## Supplementary material for "Comprehensive profiling of the human intestinal DNA virome and prediction of disease-associated bacterial hosts in severe Myalgic Encephalomyelitis/Chronic Fatigue Syndrome (ME/CFS)": S Table 5 WMS OTUs

| contig_id | contig_length | provirus | proviral_length | gene_count | viral_genes | host_genes | checkv_quality | miuvig_quality | completeness | completeness_method | contamination |
|---|---|---|---|---|---|---|---|---|---|---|---|
| S9-final-viral-combined_k141_18\|\|full | 2748 | No | NA | 2 | 0 | 0 | Not-determined | Genome-fragment | NA | NA | 0 |
| S9-final-viral-combined_k141_27311\|\|full | 10561 | No | NA | 12 | 0 | 2 | Medium-quality | Genome-fragment | 80.96 | AAI-based (high-confidence) | 0 |
| S9-final-viral-combined_k141_61921\|\|full | 3541 | No | NA | 3 | 0 | 1 | Not-determined | Genome-fragment | NA | NA | 0 |
| S9-final-viral-combined_k141_70652\|\|lt2gene | 1000 | No | NA | 2 | 1 | 0 | Low-quality | Genome-fragment | 1.86 | HMM-based (lower-bound) | 0 |
| S9-final-viral-combined_k141_128240\|\|full | 2114 | No | NA | 3 | 0 | 0 | Not-determined | Genome-fragment | NA | NA | 0 |
| S9-final-viral-combined_k141_80221\|\|full | 1203 | No | NA | 5 | 0 | 0 | Not-determined | Genome-fragment | NA | NA | 0 |
| S11-final-viral-combined_k141_221480\|\|full | 1000 | No | NA | 2 | 0 | 1 | Low-quality | Genome-fragment | 2.34 | AAI-based (medium-confidence) | 0 |
| S1-final-viral-combined_k141_132027\|\|lt2gene | 1000 | No | NA | 1 | 1 | 0 | Low-quality | Genome-fragment | 2.77 | AAI-based (high-confidence) | 0 |
| S9-final-viral-combined_k141_101618\|\|full | 2377 | No | NA | 2 | 0 | 2 | Not-determined | Genome-fragment | NA | NA | 0 |
| S1_k141_254098 | 1000 | No | NA | 1 | 0 | 1 | Low-quality | Genome-fragment | 0.55 | AAI-based (high-confidence) | 0 |
| S9-final-viral-combined_k141_21414\|\|full | 1549 | No | NA | 2 | 0 | 1 | Not-determined | Genome-fragment | NA | NA | 0 |
| S3-final-viral-combined_k141_78230\|\|full | 1000 | No | NA | 4 | 0 | 0 | Low-quality | Genome-fragment | 4.52 | AAI-based (high-confidence) | 0 |
| S5_k141_49235 | 1000 | No | NA | 2 | 0 | 1 | Low-quality | Genome-fragment | 1.35 | AAI-based (medium-confidence) | 0 |
| S9-final-viral-combined_k141_101645\|\|full | 1618 | No | NA | 2 | 0 | 1 | Not-determined | Genome-fragment | NA | NA | 0 |
| S7-final-viral-combined_k141_173981\|\|full | 1000 | No | NA | 4 | 0 | 0 | Low-quality | Genome-fragment | 3.17 | AAI-based (high-confidence) | 0 |
| S9-final-viral-combined_k141_16080\|\|full | 3423 | No | NA | 3 | 0 | 2 | Not-determined | Genome-fragment | NA | NA | 0 |
| S9-final-viral-combined_k141_96317\|\|full | 2087 | No | NA | 3 | 0 | 0 | Not-determined | Genome-fragment | NA | NA | 0 |
| S7-final-viral-combined_k141_163338\|\|full | 1000 | No | NA | 3 | 0 | 1 | Low-quality | Genome-fragment | 1.44 | AAI-based (medium-confidence) | 0 |
| S9-final-viral-combined_k141_32130\|\|full | 2477 | No | NA | 4 | 0 | 3 | Not-determined | Genome-fragment | NA | NA | 0 |
| S7-final-viral-combined_k141_147972\|\|lt2gene | 1000 | No | NA | 2 | 1 | 0 | Low-quality | Genome-fragment | 2.69 | AAI-based (high-confidence) | 0 |
| S9-final-viral-combined_k141_42858\|\|full | 3399 | No | NA | 3 | 0 | 2 | Not-determined | Genome-fragment | NA | NA | 0 |
| S9-final-viral-combined_k141_16107\|\|full | 4057 | No | NA | 7 | 0 | 0 | Not-determined | Genome-fragment | NA | NA | 0 |
| S9-final-viral-combined_k141_26779\|\|full | 2234 | No | NA | 3 | 0 | 0 | Not-determined | Genome-fragment | NA | NA | 0 |
| S9-final-viral-combined_k141_74918\|\|full | 2022 | No | NA | 2 | 0 | 2 | Not-determined | Genome-fragment | NA | NA | 0 |
| S12-final-viral-combined_k141_312026\|\|full | 1000 | No | NA | 4 | 0 | 0 | Low-quality | Genome-fragment | 2.58 | AAI-based (medium-confidence) | 0 |
| S2-final-viral-combined_k141_9078\|\|full | 1000 | No | NA | 2 | 1 | 0 | Low-quality | Genome-fragment | 2.25 | AAI-based (high-confidence) | 0 |
| S4-final-viral-combined_k141_24368\|\|full | 1000 | No | NA | 2 | 0 | 0 | Low-quality | Genome-fragment | 2.12 | AAI-based (medium-confidence) | 0 |
| S9-final-viral-combined_k141_86\|\|full | 3102 | No | NA | 3 | 0 | 2 | Not-determined | Genome-fragment | NA | NA | 0 |
| S9-final-viral-combined_k141_101673\|\|full | 6045 | No | NA | 4 | 0 | 2 | Not-determined | Genome-fragment | NA | NA | 0 |
| S4-final-viral-combined_k141_71788\|\|lt2gene | 1000 | No | NA | 2 | 2 | 0 | Low-quality | Genome-fragment | 2.1 | AAI-based (medium-confidence) | 0 |
| S4_k141_145029 | 1000 | No | NA | 1 | 0 | 0 | Low-quality | Genome-fragment | 1.87 | AAI-based (high-confidence) | 0 |
| S8-final-viral-combined_k141_82360\|\|lt2gene | 1000 | No | NA | 1 | 1 | 0 | Low-quality | Genome-fragment | 2.12 | AAI-based (high-confidence) | 0 |
| S9-final-viral-combined_k141_16134\|\|full | 1525 | No | NA | 2 | 0 | 2 | Not-determined | Genome-fragment | NA | NA | 0 |
| S9-final-viral-combined_k141_9435\|\|full | 1001 | No | NA | 2 | 0 | 0 | Low-quality | Genome-fragment | 1.02 | AAI-based (medium-confidence) | 0 |
| S9-final-viral-combined_k141_26789\|\|full | 1108 | No | NA | 2 | 0 | 1 | Not-determined | Genome-fragment | NA | NA | 0 |
| S9-final-viral-combined_k141_117781\|\|full | 1622 | No | NA | 2 | 0 | 0 | Not-determined | Genome-fragment | NA | NA | 0 |
| S9-final-viral-combined_k141_26797\|\|full | 1946 | No | NA | 2 | 0 | 1 | Not-determined | Genome-fragment | NA | NA | 0 |
| S9-final-viral-combined_k141_108039\|\|lt2gene | 1001 | No | NA | 1 | 1 | 0 | Low-quality | Genome-fragment | 2.34 | AAI-based (high-confidence) | 0 |
| S9-final-viral-combined_k141_168\|\|full | 4262 | No | NA | 5 | 0 | 3 | Not-determined | Genome-fragment | NA | NA | 0 |
| S11-final-viral-combined_k141_60295\|\|full | 1001 | No | NA | 2 | 0 | 0 | Low-quality | Genome-fragment | 2.78 | AAI-based (medium-confidence) | 0 |
| S11-final-viral-combined_k141_60310\|\|full | 1001 | No | NA | 2 | 0 | 0 | Low-quality | Genome-fragment | 2.8 | AAI-based (medium-confidence) | 0 |
| S1-final-viral-combined_k141_179428\|\|full | 1001 | No | NA | 3 | 0 | 0 | Low-quality | Genome-fragment | 1.64 | AAI-based (high-confidence) | 0 |
| S9-final-viral-combined_k141_112457\|\|full | 2848 | No | NA | 4 | 0 | 2 | Not-determined | Genome-fragment | NA | NA | 0 |
| S9-final-viral-combined_k141_74987\|\|full | 2344 | No | NA | 4 | 0 | 3 | Not-determined | Genome-fragment | NA | NA | 0 |
| S9-final-viral-combined_k141_37568\|\|full | 1514 | No | NA | 4 | 0 | 0 | Not-determined | Genome-fragment | NA | NA | 0 |
| S5-final-viral-combined_k141_100625\|\|full | 1001 | No | NA | 2 | 0 | 0 | Low-quality | Genome-fragment | 1.51 | AAI-based (high-confidence) | 0 |
| S7-final-viral-combined_k141_194217\|\|full | 1001 | No | NA | 2 | 0 | 0 | Low-quality | Genome-fragment | 2.42 | AAI-based (high-confidence) | 0 |
| S9-final-viral-combined_k141_75027\|\|full | 3303 | No | NA | 3 | 0 | 3 | Not-determined | Genome-fragment | NA | NA | 0 |
| S9-final-viral-combined_k141_75033\|\|full | 1004 | No | NA | 2 | 0 | 0 | Not-determined | Genome-fragment | NA | NA | 0 |
| S7-final-viral-combined_k141_14684\|\|full | 1001 | No | NA | 3 | 0 | 1 | Low-quality | Genome-fragment | 0.75 | AAI-based (medium-confidence) | 0 |
| S7_k141_210522 | 1001 | No | NA | 1 | 0 | 0 | Low-quality | Genome-fragment | 1.2 | AAI-based (high-confidence) | 0 |
| S9-final-viral-combined_k141_96403\|\|full | 6462 | No | NA | 8 | 0 | 4 | Not-determined | Genome-fragment | NA | NA | 0 |
| S9-final-viral-combined_k141_101800\|\|full | 1511 | No | NA | 2 | 0 | 0 | Not-determined | Genome-fragment | NA | NA | 0 |
| S17-final-viral-combined_k141_256713\|\|full | 1001 | No | NA | 4 | 0 | 0 | Low-quality | Genome-fragment | 2.28 | AAI-based (medium-confidence) | 0 |
| S17-final-viral-combined_k141_209668\|\|full | 1001 | No | NA | 2 | 0 | 0 | Low-quality | Genome-fragment | 2.54 | AAI-based (medium-confidence) | 0 |
| S17-k141_145617 | 1001 | No | NA | 1 | 0 | 0 | Low-quality | Genome-fragment | 1.45 | AAI-based (high-confidence) | 0 |
| S17-k141_193417 | 1001 | No | NA | 4 | 0 | 0 | Low-quality | Genome-fragment | 2.49 | AAI-based (medium-confidence) | 0 |
| S12-final-viral-combined_k141_67449\|\|lt2gene | 1001 | No | NA | 1 | 1 | 0 | Low-quality | Genome-fragment | 2.36 | AAI-based (high-confidence) | 0 |
| S9-final-viral-combined_k141_69628\|\|full | 2855 | No | NA | 3 | 0 | 2 | Not-determined | Genome-fragment | NA | NA | 0 |
| S12-final-viral-combined_k141_337949\|\|lt2gene | 1001 | No | NA | 2 | 1 | 0 | Low-quality | Genome-fragment | 2.67 | AAI-based (medium-confidence) | 0 |
| S12_k141_357790 | 1001 | No | NA | 1 | 0 | 1 | Low-quality | Genome-fragment | 0.17 | AAI-based (medium-confidence) | 0 |
| S9-final-viral-combined_k141_85755\|\|full | 4207 | No | NA | 6 | 0 | 1 | Not-determined | Genome-fragment | NA | NA | 0 |
| S9-final-viral-combined_k141_69645\|\|full | 3178 | No | NA | 5 | 0 | 0 | Not-determined | Genome-fragment | NA | NA | 0 |
| S2_k141_87457 | 1001 | No | NA | 5 | 0 | 0 | Low-quality | Genome-fragment | 1.65 | AAI-based (high-confidence) | 0 |
| S4-final-viral-combined_k141_24500\|\|full | 1001 | No | NA | 2 | 1 | 0 | Low-quality | Genome-fragment | 2.27 | AAI-based (high-confidence) | 0 |
| S9-final-viral-combined_k141_43024\|\|full | 1925 | No | NA | 3 | 0 | 1 | Not-determined | Genome-fragment | NA | NA | 0 |
| S9-final-viral-combined_k141_287\|\|full | 5659 | No | NA | 4 | 0 | 2 | Not-determined | Genome-fragment | NA | NA | 0 |
| S6-final-viral-combined_k141_10749\|\|full | 1001 | No | NA | 3 | 0 | 1 | Low-quality | Genome-fragment | 2.35 | AAI-based (high-confidence) | 0 |
| S9-final-viral-combined_k141_5476\|\|full | 2190 | No | NA | 5 | 0 | 0 | Not-determined | Genome-fragment | NA | NA | 0 |
| S9-final-viral-combined_k141_299\|\|full | 1375 | No | NA | 2 | 0 | 1 | Not-determined | Genome-fragment | NA | NA | 0 |
| S9-final-viral-combined_k141_305\|\|full | 3295 | No | NA | 4 | 0 | 2 | Not-determined | Genome-fragment | NA | NA | 0 |
| S8-final-viral-combined_k141_37709\|\|full | 1001 | No | NA | 2 | 0 | 0 | Low-quality | Genome-fragment | 0.66 | AAI-based (medium-confidence) | 0 |
| S9-final-viral-combined_k141_26934\|\|full | 2820 | No | NA | 3 | 0 | 1 | Not-determined | Genome-fragment | NA | NA | 0 |
| S9-final-viral-combined_k141_32282\|\|full | 1099 | No | NA | 3 | 0 | 1 | Not-determined | Genome-fragment | NA | NA | 0 |
| S8_k141_166590 | 1001 | No | NA | 1 | 0 | 1 | Low-quality | Genome-fragment | 0.71 | AAI-based (high-confidence) | 0 |
| S9-final-viral-combined_k141_32287\|\|full | 2131 | No | NA | 6 | 0 | 1 | Not-determined | Genome-fragment | NA | NA | 0 |
| S9-final-viral-combined_k141_32289\|\|full | 1042 | No | NA | 2 | 0 | 1 | Not-determined | Genome-fragment | NA | NA | 0 |
| S9-final-viral-combined_k141_123194\|\|full | 1827 | No | NA | 2 | 0 | 1 | Not-determined | Genome-fragment | NA | NA | 0 |
| S9-final-viral-combined_k141_48351\|\|full | 2893 | No | NA | 7 | 0 | 1 | Not-determined | Genome-fragment | NA | NA | 0 |
| S11-final-viral-combined_k141_10324\|\|full | 1002 | No | NA | 3 | 0 | 0 | Low-quality | Genome-fragment | 1.63 | AAI-based (medium-confidence) | 0 |
| S9-final-viral-combined_k141_16253\|\|full | 2326 | No | NA | 4 | 0 | 2 | Not-determined | Genome-fragment | NA | NA | 0 |
| S9-final-viral-combined_k141_117850\|\|full | 4441 | No | NA | 3 | 0 | 3 | Not-determined | Genome-fragment | NA | NA | 0 |
| S9-final-viral-combined_k141_64277\|\|full | 95138 | Yes | 90419 | 119 | 34 | 12 | High-quality | High-quality | 100 | AAI-based (high-confidence) | 4.96 |
| S11_k141_218887 | 1002 | No | NA | 3 | 0 | 0 | Low-quality | Genome-fragment | 2.05 | AAI-based (high-confidence) | 0 |
| S9-final-viral-combined_k141_80423\|\|full | 1883 | No | NA | 3 | 0 | 2 | Not-determined | Genome-fragment | NA | NA | 0 |
| S1-final-viral-combined_k141_238768\|\|full | 1002 | No | NA | 3 | 1 | 0 | Low-quality | Genome-fragment | 1.5 | AAI-based (medium-confidence) | 0 |
| S5-final-viral-combined_k141_187922\|\|full | 1002 | No | NA | 3 | 0 | 1 | Low-quality | Genome-fragment | 2.16 | AAI-based (medium-confidence) | 0 |
| S9-final-viral-combined_k141_27000\|\|full | 3168 | No | NA | 3 | 0 | 1 | Not-determined | Genome-fragment | NA | NA | 0 |
| S9-final-viral-combined_k141_101848\|\|full | 3941 | No | NA | 5 | 0 | 3 | Not-determined | Genome-fragment | NA | NA | 0 |
| S9-final-viral-combined_k141_27009\|\|full | 5257 | No | NA | 5 | 0 | 4 | Not-determined | Genome-fragment | NA | NA | 0 |
| S9-final-viral-combined_k141_96528\|\|full | 1011 | No | NA | 3 | 0 | 0 | Not-determined | Genome-fragment | NA | NA | 0 |
| S5-final-viral-combined_k141_26622\|\|full | 1002 | No | NA | 2 | 1 | 0 | Low-quality | Genome-fragment | 0.61 | AAI-based (high-confidence) | 0 |
| S5_k141_74139 | 1002 | No | NA | 1 | 0 | 1 | Low-quality | Genome-fragment | 0.42 | AAI-based (medium-confidence) | 0 |
| S9-final-viral-combined_k141_21587\|\|full | 1630 | No | NA | 2 | 0 | 1 | Not-determined | Genome-fragment | NA | NA | 0 |
| S7_k141_184241 | 1002 | No | NA | 2 | 0 | 0 | Low-quality | Genome-fragment | 1.39 | AAI-based (medium-confidence) | 0 |
| S7_k141_156568 | 1002 | No | NA | 2 | 0 | 1 | Low-quality | Genome-fragment | 0.66 | AAI-based (medium-confidence) | 0 |
| S9-final-viral-combined_k141_80509\|\|full | 2958 | No | NA | 4 | 0 | 1 | Not-determined | Genome-fragment | NA | NA | 0 |
| S17-final-viral-combined_k141_2883\|\|full | 1002 | Yes | 504 | 2 | 1 | 1 | Low-quality | Genome-fragment | 0.69 | AAI-based (high-confidence) | 49.7 |
| S9-final-viral-combined_k141_32337\|\|full | 1910 | No | NA | 3 | 0 | 0 | Not-determined | Genome-fragment | NA | NA | 0 |
| S9-final-viral-combined_k141_117949\|\|full | 1389 | No | NA | 3 | 0 | 1 | Not-determined | Genome-fragment | NA | NA | 0 |
| S9-final-viral-combined_k141_16354\|\|full | 1703 | No | NA | 2 | 0 | 1 | Not-determined | Genome-fragment | NA | NA | 0 |
| S9-final-viral-combined_k141_48425\|\|full | 5789 | No | NA | 8 | 0 | 4 | Not-determined | Genome-fragment | NA | NA | 0 |
| S12_k141_390149 | 1002 | No | NA | 2 | 2 | 0 | Low-quality | Genome-fragment | 2.2 | AAI-based (high-confidence) | 0 |
| S8-final-viral-combined_k141_195215\|\|full | 1002 | No | NA | 4 | 0 | 0 | Low-quality | Genome-fragment | 1 | AAI-based (high-confidence) | 0 |
| S9-final-viral-combined_k141_37598\|\|full | 2183 | No | NA | 2 | 0 | 0 | Not-determined | Genome-fragment | NA | NA | 0 |
| S8_k141_95325 | 1002 | No | NA | 3 | 0 | 0 | Low-quality | Genome-fragment | 1.73 | AAI-based (high-confidence) | 0 |
| S8_k141_187889 | 1002 | No | NA | 2 | 0 | 1 | Low-quality | Genome-fragment | 3.26 | AAI-based (high-confidence) | 0 |
| S1-final-viral-combined_k141_151461\|\|full | 1003 | No | NA | 3 | 0 | 1 | Low-quality | Genome-fragment | 0.69 | AAI-based (medium-confidence) | 0 |
| S9-final-viral-combined_k141_48451\|\|full | 2296 | No | NA | 3 | 0 | 2 | Not-determined | Genome-fragment | NA | NA | 0 |
| S9-final-viral-combined_k141_64400\|\|full | 3263 | No | NA | 4 | 0 | 2 | Not-determined | Genome-fragment | NA | NA | 0 |
| S9-final-viral-combined_k141_85928\|\|full | 2736 | No | NA | 2 | 0 | 1 | Not-determined | Genome-fragment | NA | NA | 0 |
| S9-final-viral-combined_k141_112628\|\|full | 1497 | No | NA | 2 | 0 | 1 | Not-determined | Genome-fragment | NA | NA | 0 |
| S1-final-viral-combined_k141_16219\|\|lt2gene | 1003 | No | NA | 1 | 1 | 0 | Low-quality | Genome-fragment | 2.63 | AAI-based (high-confidence) | 0 |
| S3-final-viral-combined_k141_136120\|\|full | 1003 | No | NA | 3 | 0 | 0 | Low-quality | Genome-fragment | 3.04 | AAI-based (medium-confidence) | 0 |
| S9-final-viral-combined_k141_479\|\|full | 2809 | No | NA | 4 | 0 | 2 | Not-determined | Genome-fragment | NA | NA | 0 |
| S9-final-viral-combined_k141_117972\|\|full | 1924 | No | NA | 2 | 0 | 2 | Not-determined | Genome-fragment | NA | NA | 0 |
| S9-final-viral-combined_k141_27116\|\|full | 6599 | No | NA | 5 | 0 | 2 | Not-determined | Genome-fragment | NA | NA | 0 |
| S9-final-viral-combined_k141_59132\|\|full | 1486 | No | NA | 2 | 0 | 1 | Not-determined | Genome-fragment | NA | NA | 0 |
| S5_k141_199576 | 1003 | No | NA | 1 | 0 | 1 | Low-quality | Genome-fragment | 1.88 | AAI-based (high-confidence) | 0 |
| S7-final-viral-combined_k141_66882\|\|lt2gene | 1003 | No | NA | 1 | 1 | 0 | Low-quality | Genome-fragment | 2.19 | AAI-based (high-confidence) | 0 |
| S17-final-viral-combined_k141_80973\|\|full | 1003 | No | NA | 4 | 1 | 0 | Low-quality | Genome-fragment | 6.66 | AAI-based (high-confidence) | 0 |
| S9-final-viral-combined_k141_112695\|\|full | 2514 | No | NA | 4 | 0 | 1 | Not-determined | Genome-fragment | NA | NA | 0 |
| S9-final-viral-combined_k141_80603\|\|full | 1005 | No | NA | 3 | 0 | 0 | Not-determined | Genome-fragment | NA | NA | 0 |
| S9-final-viral-combined_k141_59179\|\|full | 2886 | No | NA | 5 | 0 | 4 | Not-determined | Genome-fragment | NA | NA | 0 |
| S9-final-viral-combined_k141_53731\|\|full | 1687 | No | NA | 3 | 0 | 3 | Not-determined | Genome-fragment | NA | NA | 0 |
| S9-final-viral-combined_k141_96645\|\|full | 1076 | No | NA | 3 | 0 | 1 | Not-determined | Genome-fragment | NA | NA | 0 |
| S9-final-viral-combined_k141_48495\|\|full | 1411 | No | NA | 3 | 0 | 0 | Not-determined | Genome-fragment | NA | NA | 0 |
| S12-final-viral-combined_k141_135765\|\|full | 1003 | No | NA | 3 | 2 | 0 | Low-quality | Genome-fragment | 1.77 | AAI-based (high-confidence) | 0 |
| S12_k141_47120 | 1003 | No | NA | 2 | 0 | 0 | Low-quality | Genome-fragment | 0.94 | AAI-based (high-confidence) | 0 |
| S9-final-viral-combined_k141_37676\|\|full | 3769 | No | NA | 5 | 0 | 2 | Not-determined | Genome-fragment | NA | NA | 0 |
| S9-final-viral-combined_k141_16439\|\|full | 1359 | No | NA | 3 | 0 | 3 | Not-determined | Genome-fragment | NA | NA | 0 |
| S9-final-viral-combined_k141_21601\|\|full | 1568 | No | NA | 3 | 0 | 0 | Not-determined | Genome-fragment | NA | NA | 0 |
| S2-final-viral-combined_k141_191035\|\|full | 1003 | No | NA | 2 | 0 | 0 | Low-quality | Genome-fragment | 1.24 | AAI-based (medium-confidence) | 0 |
| S2-final-viral-combined_k141_208406\|\|lt2gene | 1003 | No | NA | 2 | 1 | 0 | Low-quality | Genome-fragment | 2.55 | AAI-based (medium-confidence) | 0 |
| S4-final-viral-combined_k141_127725\|\|full | 1003 | No | NA | 3 | 1 | 0 | Low-quality | Genome-fragment | 2.47 | AAI-based (high-confidence) | 0 |
| S9-final-viral-combined_k141_96686\|\|full | 1116 | No | NA | 3 | 0 | 0 | Not-determined | Genome-fragment | NA | NA | 0 |
| S4_k141_28814 | 1003 | No | NA | 1 | 0 | 0 | Low-quality | Genome-fragment | 0.75 | AAI-based (high-confidence) | 0 |
| S9-final-viral-combined_k141_5572\|\|full | 1160 | No | NA | 2 | 0 | 0 | Not-determined | Genome-fragment | NA | NA | 0 |
| S9-final-viral-combined_k141_107113\|\|full | 2137 | No | NA | 2 | 0 | 2 | Not-determined | Genome-fragment | NA | NA | 0 |
| S8-final-viral-combined_k141_244332\|\|full | 1003 | No | NA | 2 | 0 | 0 | Low-quality | Genome-fragment | 2.39 | AAI-based (high-confidence) | 0 |
| S9-final-viral-combined_k141_102119\|\|full | 1892 | No | NA | 1 | 0 | 0 | Not-determined | Genome-fragment | NA | NA | 0 |
| S8-final-viral-combined_k141_167785\|\|full | 1003 | No | NA | 2 | 0 | 0 | Low-quality | Genome-fragment | 1.79 | AAI-based (high-confidence) | 0 |
| S9-final-viral-combined_k141_59247\|\|full | 1630 | No | NA | 3 | 0 | 2 | Not-determined | Genome-fragment | NA | NA | 0 |
| S8-final-viral-combined_k141_128674\|\|lt2gene | 1003 | No | NA | 2 | 2 | 0 | Low-quality | Genome-fragment | 1.75 | HMM-based (lower-bound) | 0 |
| S9-final-viral-combined_k141_21506\|\|lt2gene | 1004 | No | NA | 1 | 1 | 0 | Low-quality | Genome-fragment | 2.61 | AAI-based (high-confidence) | 0 |
| S9-final-viral-combined_k141_386\|\|lt2gene | 1004 | No | NA | 2 | 0 | 1 | Low-quality | Genome-fragment | 1.61 | AAI-based (medium-confidence) | 0 |
| S9-final-viral-combined_k141_102132\|\|full | 1576 | No | NA | 3 | 0 | 1 | Not-determined | Genome-fragment | NA | NA | 0 |
| S9-final-viral-combined_k141_59250\|\|full | 2764 | No | NA | 3 | 0 | 2 | Not-determined | Genome-fragment | NA | NA | 0 |
| S9-final-viral-combined_k141_96745\|\|full | 1846 | No | NA | 3 | 0 | 0 | Not-determined | Genome-fragment | NA | NA | 0 |
| S11_k141_155867 | 1004 | No | NA | 2 | 0 | 0 | Low-quality | Genome-fragment | 2.51 | AAI-based (high-confidence) | 0 |
| S9-final-viral-combined_k141_48628\|\|full | 1890 | No | NA | 2 | 0 | 2 | Not-determined | Genome-fragment | NA | NA | 0 |
| S1-final-viral-combined_k141_238667\|\|full | 1004 | No | NA | 3 | 1 | 0 | Low-quality | Genome-fragment | 0.61 | HMM-based (lower-bound) | 0 |
| S1-final-viral-combined_k141_13666\|\|lt2gene | 1004 | No | NA | 1 | 1 | 0 | Low-quality | Genome-fragment | 3.21 | AAI-based (high-confidence) | 0 |
| S1_k141_242902 | 1004 | No | NA | 2 | 0 | 0 | Low-quality | Genome-fragment | 2.36 | AAI-based (high-confidence) | 0 |
| S9-final-viral-combined_k141_610\|\|full | 1479 | No | NA | 2 | 0 | 0 | Not-determined | Genome-fragment | NA | NA | 0 |
| S9-final-viral-combined_k141_5668\|\|full | 1594 | No | NA | 4 | 0 | 0 | Not-determined | Genome-fragment | NA | NA | 0 |
| S1_k141_309685 | 1004 | No | NA | 3 | 2 | 0 | Low-quality | Genome-fragment | 2.51 | AAI-based (high-confidence) | 0 |
| S9-final-viral-combined_k141_59282\|\|full | 1078 | No | NA | 3 | 0 | 0 | Not-determined | Genome-fragment | NA | NA | 0 |
| S9-final-viral-combined_k141_59296\|\|full | 1097 | No | NA | 3 | 0 | 2 | Not-determined | Genome-fragment | NA | NA | 0 |
| S9-final-viral-combined_k141_118134\|\|full | 1988 | No | NA | 3 | 0 | 2 | Not-determined | Genome-fragment | NA | NA | 0 |
| S9-final-viral-combined_k141_118149\|\|full | 2527 | No | NA | 3 | 0 | 3 | Not-determined | Genome-fragment | NA | NA | 0 |
| S9-final-viral-combined_k141_107208\|\|full | 5224 | No | NA | 8 | 0 | 0 | Not-determined | Genome-fragment | NA | NA | 0 |
| S3-final-viral-combined_k141_37307\|\|lt2gene | 1004 | No | NA | 1 | 1 | 0 | Low-quality | Genome-fragment | 2.29 | AAI-based (medium-confidence) | 0 |
| S9-final-viral-combined_k141_96864\|\|full | 2790 | No | NA | 3 | 0 | 2 | Not-determined | Genome-fragment | NA | NA | 0 |
| S9-final-viral-combined_k141_32383\|\|full | 5423 | No | NA | 4 | 0 | 3 | Not-determined | Genome-fragment | NA | NA | 0 |
| S5-final-viral-combined_k141_78615\|\|full | 1004 | No | NA | 2 | 0 | 0 | Low-quality | Genome-fragment | 1.9 | AAI-based (high-confidence) | 0 |
| S9-final-viral-combined_k141_10972\|\|full | 1531 | No | NA | 2 | 0 | 1 | Not-determined | Genome-fragment | NA | NA | 0 |
| S9-final-viral-combined_k141_59345\|\|full | 1827 | No | NA | 3 | 0 | 2 | Not-determined | Genome-fragment | NA | NA | 0 |
| S12-final-viral-combined_k141_261345\|\|full | 1004 | No | NA | 3 | 0 | 0 | Low-quality | Genome-fragment | 1.06 | AAI-based (high-confidence) | 0 |
| S12-final-viral-combined_k141_315632\|\|full | 1004 | No | NA | 2 | 1 | 0 | Low-quality | Genome-fragment | 2.19 | AAI-based (high-confidence) | 0 |
| S9-final-viral-combined_k141_32398\|\|full | 2574 | No | NA | 2 | 0 | 2 | Not-determined | Genome-fragment | NA | NA | 0 |
| S12-final-viral-combined_k141_217859\|\|lt2gene | 1004 | No | NA | 2 | 1 | 0 | Low-quality | Genome-fragment | 0.82 | HMM-based (lower-bound) | 0 |
| S9-final-viral-combined_k141_86103\|\|full | 2608 | No | NA | 3 | 0 | 2 | Not-determined | Genome-fragment | NA | NA | 0 |
| S9-final-viral-combined_k141_702\|\|full | 3727 | No | NA | 3 | 0 | 2 | Not-determined | Genome-fragment | NA | NA | 0 |
| S9-final-viral-combined_k141_86118\|\|full | 2382 | No | NA | 2 | 0 | 2 | Not-determined | Genome-fragment | NA | NA | 0 |
| S9-final-viral-combined_k141_37803\|\|full | 1048 | No | NA | 3 | 0 | 1 | Not-determined | Genome-fragment | NA | NA | 0 |
| S2-final-viral-combined_k141_42687\|\|full | 1004 | No | NA | 5 | 3 | 0 | Low-quality | Genome-fragment | 1.65 | AAI-based (high-confidence) | 0 |
| S2-final-viral-combined_k141_70451\|\|full | 1004 | No | NA | 3 | 0 | 0 | Low-quality | Genome-fragment | 2.24 | AAI-based (medium-confidence) | 0 |
| S9-final-viral-combined_k141_102219\|\|full | 1520 | No | NA | 2 | 0 | 1 | Not-determined | Genome-fragment | NA | NA | 0 |
| S4-final-viral-combined_k141_87892\|\|lt2gene | 1004 | No | NA | 2 | 1 | 0 | Low-quality | Genome-fragment | 2.64 | AAI-based (high-confidence) | 0 |
| S9-final-viral-combined_k141_48768\|\|full | 4645 | No | NA | 3 | 0 | 3 | Not-determined | Genome-fragment | NA | NA | 0 |
| S8-final-viral-combined_k141_147665\|\|full | 1004 | No | NA | 3 | 0 | 0 | Low-quality | Genome-fragment | 0.8 | AAI-based (high-confidence) | 0 |
| S9-final-viral-combined_k141_108241\|\|full | 1005 | No | NA | 3 | 0 | 2 | Low-quality | Genome-fragment | 1.84 | AAI-based (medium-confidence) | 0 |
| S11-final-viral-combined_k141_241219\|\|full | 1005 | No | NA | 3 | 0 | 0 | Low-quality | Genome-fragment | 0.89 | AAI-based (high-confidence) | 0 |
| S9-final-viral-combined_k141_53913\|\|full | 2392 | No | NA | 2 | 0 | 2 | Not-determined | Genome-fragment | NA | NA | 0 |
| S9-final-viral-combined_k141_118237\|\|full | 2111 | No | NA | 4 | 0 | 1 | Not-determined | Genome-fragment | NA | NA | 0 |
| S11_k141_257486 | 1005 | No | NA | 2 | 0 | 0 | Low-quality | Genome-fragment | 2.26 | AAI-based (high-confidence) | 0 |
| S9-final-viral-combined_k141_21772\|\|full | 2196 | No | NA | 3 | 0 | 0 | Not-determined | Genome-fragment | NA | NA | 0 |
| S9-final-viral-combined_k141_16513\|\|full | 1923 | No | NA | 5 | 0 | 2 | Not-determined | Genome-fragment | NA | NA | 0 |
| S9-final-viral-combined_k141_5767\|\|full | 4900 | No | NA | 5 | 0 | 0 | Not-determined | Genome-fragment | NA | NA | 0 |
| S9-final-viral-combined_k141_10990\|\|full | 52978 | No | NA | 50 | 1 | 18 | Medium-quality | Genome-fragment | 80.16 | HMM-based (lower-bound) | 0 |
| S5_k141_167266 | 1005 | No | NA | 3 | 0 | 0 | Low-quality | Genome-fragment | 2.63 | AAI-based (high-confidence) | 0 |
| S9-final-viral-combined_k141_27269\|\|full | 2697 | No | NA | 3 | 0 | 1 | Not-determined | Genome-fragment | NA | NA | 0 |
| S17-final-viral-combined_k141_139274\|\|full | 1005 | No | NA | 2 | 0 | 1 | Low-quality | Genome-fragment | 0.45 | AAI-based (medium-confidence) | 0 |
| S9-final-viral-combined_k141_37880\|\|full | 2114 | No | NA | 3 | 0 | 2 | Not-determined | Genome-fragment | NA | NA | 0 |
| S9-final-viral-combined_k141_27278\|\|full | 2464 | No | NA | 4 | 0 | 1 | Not-determined | Genome-fragment | NA | NA | 0 |
| S12-final-viral-combined_k141_150834\|\|full | 1005 | No | NA | 3 | 0 | 0 | Low-quality | Genome-fragment | 2.39 | AAI-based (medium-confidence) | 0 |
| S12-final-viral-combined_k141_356355\|\|lt2gene | 1005 | No | NA | 1 | 1 | 0 | Low-quality | Genome-fragment | 0.62 | HMM-based (lower-bound) | 0 |
| S9-final-viral-combined_k141_107318\|\|full | 2853 | No | NA | 4 | 0 | 0 | Not-determined | Genome-fragment | NA | NA | 0 |
| S9-final-viral-combined_k141_16580\|\|full | 2707 | No | NA | 2 | 0 | 0 | Not-determined | Genome-fragment | NA | NA | 0 |
| S12_k141_160583 | 1005 | No | NA | 1 | 0 | 1 | Low-quality | Genome-fragment | 1.04 | AAI-based (medium-confidence) | 0 |
| S9-final-viral-combined_k141_80897\|\|full | 4099 | No | NA | 5 | 0 | 0 | Not-determined | Genome-fragment | NA | NA | 0 |
| S9-final-viral-combined_k141_102297\|\|full | 2760 | No | NA | 5 | 0 | 2 | Not-determined | Genome-fragment | NA | NA | 0 |
| S9-final-viral-combined_k141_118292\|\|full | 4656 | No | NA | 4 | 0 | 3 | Not-determined | Genome-fragment | NA | NA | 0 |
| S9-final-viral-combined_k141_59492\|\|full | 1146 | No | NA | 3 | 0 | 1 | Not-determined | Genome-fragment | NA | NA | 0 |
| S9-final-viral-combined_k141_112903\|\|full | 2427 | No | NA | 2 | 0 | 2 | Not-determined | Genome-fragment | NA | NA | 0 |
| S4_k141_132142 | 1005 | No | NA | 3 | 0 | 0 | Low-quality | Genome-fragment | 2.35 | AAI-based (medium-confidence) | 0 |
| S9-final-viral-combined_k141_824\|\|full | 2295 | No | NA | 3 | 0 | 1 | Not-determined | Genome-fragment | NA | NA | 0 |
| S9-final-viral-combined_k141_112916\|\|full | 2348 | No | NA | 4 | 0 | 3 | Not-determined | Genome-fragment | NA | NA | 0 |
| S9-final-viral-combined_k141_53991\|\|full | 61938 | Yes | 38377 | 68 | 26 | 13 | High-quality | High-quality | 90.69 | AAI-based (medium-confidence) | 38.04 |
| S6-final-viral-combined_k141_130583\|\|lt2gene | 1005 | No | NA | 1 | 1 | 0 | Low-quality | Genome-fragment | 1.32 | HMM-based (lower-bound) | 0 |
| S9-final-viral-combined_k141_102344\|\|full | 3777 | No | NA | 2 | 0 | 0 | Not-determined | Genome-fragment | NA | NA | 0 |
| S9-final-viral-combined_k141_86247\|\|full | 1010 | No | NA | 3 | 0 | 0 | Not-determined | Genome-fragment | NA | NA | 0 |
| S8-final-viral-combined_k141_120656\|\|full | 1005 | No | NA | 3 | 1 | 0 | Low-quality | Genome-fragment | 0.55 | HMM-based (lower-bound) | 0 |
| S8_k141_234378 | 1005 | No | NA | 2 | 0 | 0 | Low-quality | Genome-fragment | 0.68 | AAI-based (medium-confidence) | 0 |
| S9-final-viral-combined_k141_80975\|\|full | 1121 | No | NA | 2 | 0 | 0 | Not-determined | Genome-fragment | NA | NA | 0 |
| S9-final-viral-combined_k141_898\|\|full | 2197 | No | NA | 3 | 0 | 1 | Not-determined | Genome-fragment | NA | NA | 0 |
| S9-final-viral-combined_k141_59595\|\|full | 4898 | No | NA | 4 | 0 | 2 | Not-determined | Genome-fragment | NA | NA | 0 |
| S8_k141_22586 | 1005 | No | NA | 2 | 0 | 0 | Low-quality | Genome-fragment | 0.64 | AAI-based (high-confidence) | 0 |
| S9-final-viral-combined_k141_102361\|\|full | 5078 | No | NA | 7 | 0 | 6 | Not-determined | Genome-fragment | NA | NA | 0 |
| S9-final-viral-combined_k141_92846\|\|full | 1006 | No | NA | 3 | 0 | 0 | Low-quality | Genome-fragment | 2.55 | AAI-based (high-confidence) | 0 |
| S9_k141_5414 | 1006 | No | NA | 2 | 0 | 0 | Low-quality | Genome-fragment | 0.56 | AAI-based (medium-confidence) | 0 |
| S9-final-viral-combined_k141_102368\|\|full | 2183 | No | NA | 3 | 0 | 1 | Not-determined | Genome-fragment | NA | NA | 0 |
| S9-final-viral-combined_k141_32630\|\|full | 6808 | No | NA | 7 | 0 | 4 | Not-determined | Genome-fragment | NA | NA | 0 |
| S9-final-viral-combined_k141_916\|\|full | 5764 | No | NA | 4 | 0 | 1 | Not-determined | Genome-fragment | NA | NA | 0 |
| S9-final-viral-combined_k141_48856\|\|full | 2275 | No | NA | 3 | 0 | 0 | Not-determined | Genome-fragment | NA | NA | 0 |
| S11_k141_96511 | 1006 | No | NA | 1 | 0 | 0 | Low-quality | Genome-fragment | 1.69 | AAI-based (high-confidence) | 0 |
| S9-final-viral-combined_k141_5932\|\|full | 3430 | No | NA | 5 | 0 | 0 | Not-determined | Genome-fragment | NA | NA | 0 |
| S9-final-viral-combined_k141_64659\|\|full | 1171 | No | NA | 2 | 0 | 1 | Not-determined | Genome-fragment | NA | NA | 0 |
| S9-final-viral-combined_k141_5937\|\|full | 1378 | No | NA | 3 | 0 | 0 | Not-determined | Genome-fragment | NA | NA | 0 |
| S9-final-viral-combined_k141_38008\|\|full | 2032 | No | NA | 4 | 0 | 2 | Not-determined | Genome-fragment | NA | NA | 0 |
| S9-final-viral-combined_k141_64676\|\|full | 1246 | No | NA | 2 | 0 | 1 | Not-determined | Genome-fragment | NA | NA | 0 |
| S9-final-viral-combined_k141_97142\|\|full | 7110 | No | NA | 5 | 0 | 5 | Not-determined | Genome-fragment | NA | NA | 0 |
| S9-final-viral-combined_k141_32669\|\|full | 3323 | No | NA | 3 | 0 | 1 | Not-determined | Genome-fragment | NA | NA | 0 |
| S1-final-viral-combined_k141_70920\|\|full | 1006 | No | NA | 2 | 0 | 0 | Low-quality | Genome-fragment | 1.51 | AAI-based (medium-confidence) | 0 |
| S5_k141_67176 | 1006 | No | NA | 2 | 0 | 0 | Low-quality | Genome-fragment | 2.06 | AAI-based (medium-confidence) | 0 |
| S9-final-viral-combined_k141_48891\|\|full | 2210 | No | NA | 2 | 0 | 1 | Not-determined | Genome-fragment | NA | NA | 0 |
| S7-final-viral-combined_k141_54060\|\|full | 1006 | No | NA | 2 | 0 | 0 | Low-quality | Genome-fragment | 1.59 | AAI-based (medium-confidence) | 0 |
| S12-final-viral-combined_k141_9666\|\|full | 1006 | No | NA | 3 | 0 | 0 | Low-quality | Genome-fragment | 2.07 | AAI-based (medium-confidence) | 0 |
| S9-final-viral-combined_k141_32690\|\|full | 2110 | No | NA | 3 | 0 | 1 | Not-determined | Genome-fragment | NA | NA | 0 |
| S9-final-viral-combined_k141_64721\|\|full | 2037 | No | NA | 3 | 0 | 3 | Not-determined | Genome-fragment | NA | NA | 0 |
| S12-final-viral-combined_k141_199360\|\|full | 1006 | No | NA | 2 | 2 | 0 | Low-quality | Genome-fragment | 1.78 | AAI-based (high-confidence) | 0 |
| S9-final-viral-combined_k141_11101\|\|full | 16618 | No | NA | 19 | 0 | 6 | Not-determined | Genome-fragment | NA | NA | 0 |
| S9-final-viral-combined_k141_48911\|\|full | 4270 | No | NA | 4 | 0 | 1 | Not-determined | Genome-fragment | NA | NA | 0 |
| S9-final-viral-combined_k141_81063\|\|full | 1841 | No | NA | 2 | 0 | 2 | Not-determined | Genome-fragment | NA | NA | 0 |
| S9-final-viral-combined_k141_69955\|\|full | 4458 | No | NA | 5 | 0 | 0 | Not-determined | Genome-fragment | NA | NA | 0 |
| S9-final-viral-combined_k141_11112\|\|full | 1735 | No | NA | 2 | 0 | 1 | Not-determined | Genome-fragment | NA | NA | 0 |
| S9-final-viral-combined_k141_91374\|\|full | 1755 | No | NA | 2 | 0 | 1 | Not-determined | Genome-fragment | NA | NA | 0 |
| S2-final-viral-combined_k141_13601\|\|lt2gene | 1006 | No | NA | 2 | 0 | 0 | Low-quality | Genome-fragment | 1.54 | AAI-based (high-confidence) | 0 |
| S4_k141_62838 | 1006 | No | NA | 1 | 0 | 0 | Low-quality | Genome-fragment | 2.52 | AAI-based (medium-confidence) | 0 |
| S4_k141_114413 | 1006 | No | NA | 2 | 0 | 0 | Low-quality | Genome-fragment | 1.67 | AAI-based (high-confidence) | 0 |
| S4_k141_13643 | 1006 | No | NA | 1 | 0 | 0 | Low-quality | Genome-fragment | 2.26 | AAI-based (high-confidence) | 0 |
| S6_k141_76011 | 1006 | No | NA | 3 | 1 | 0 | Low-quality | Genome-fragment | 2.51 | AAI-based (high-confidence) | 0 |
| S9-final-viral-combined_k141_82433\|\|full | 1007 | No | NA | 2 | 2 | 0 | Low-quality | Genome-fragment | 2.23 | AAI-based (high-confidence) | 0 |
| S9-final-viral-combined_k141_107416\|\|full | 2090 | No | NA | 4 | 0 | 1 | Not-determined | Genome-fragment | NA | NA | 0 |
| S9-final-viral-combined_k141_70007\|\|full | 2867 | No | NA | 3 | 0 | 2 | Not-determined | Genome-fragment | NA | NA | 0 |
| S9-final-viral-combined_k141_38048\|\|full | 1195 | No | NA | 3 | 0 | 0 | Not-determined | Genome-fragment | NA | NA | 0 |
| S9-final-viral-combined_k141_11167\|\|full | 1509 | No | NA | 2 | 0 | 2 | Not-determined | Genome-fragment | NA | NA | 0 |
| S9-final-viral-combined_k141_86397\|\|full | 4826 | No | NA | 3 | 0 | 2 | Not-determined | Genome-fragment | NA | NA | 0 |
| S9-final-viral-combined_k141_38061\|\|full | 2053 | No | NA | 3 | 0 | 2 | Not-determined | Genome-fragment | NA | NA | 0 |
| S9-final-viral-combined_k141_48980\|\|full | 4670 | No | NA | 7 | 0 | 5 | Not-determined | Genome-fragment | NA | NA | 0 |
| S9_k141_25337 | 1007 | No | NA | 1 | 0 | 1 | Low-quality | Genome-fragment | 2.25 | AAI-based (medium-confidence) | 0 |
| S9-final-viral-combined_k141_48999\|\|full | 2400 | No | NA | 3 | 0 | 3 | Not-determined | Genome-fragment | NA | NA | 0 |
| S9-final-viral-combined_k141_27431\|\|full | 1719 | No | NA | 4 | 0 | 0 | Not-determined | Genome-fragment | NA | NA | 0 |
| S11-final-viral-combined_k141_132281\|\|full | 1007 | No | NA | 2 | 0 | 1 | Low-quality | Genome-fragment | 0.84 | AAI-based (medium-confidence) | 0 |
| S9-final-viral-combined_k141_1115\|\|full | 3274 | No | NA | 6 | 0 | 0 | Not-determined | Genome-fragment | NA | NA | 0 |
| S9-final-viral-combined_k141_64752\|\|full | 4374 | No | NA | 5 | 0 | 5 | Not-determined | Genome-fragment | NA | NA | 0 |
| S9-final-viral-combined_k141_6012\|\|full | 3639 | No | NA | 4 | 0 | 2 | Not-determined | Genome-fragment | NA | NA | 0 |
| S1-final-viral-combined_k141_363837\|\|full | 1007 | No | NA | 2 | 0 | 0 | Low-quality | Genome-fragment | 0.76 | AAI-based (medium-confidence) | 0 |
| S9-final-viral-combined_k141_27448\|\|full | 3836 | No | NA | 3 | 0 | 3 | Not-determined | Genome-fragment | NA | NA | 0 |
| S1-final-viral-combined_k141_236843\|\|lt2gene | 1007 | No | NA | 1 | 1 | 0 | Low-quality | Genome-fragment | 1.79 | HMM-based (lower-bound) | 0 |
| S9-final-viral-combined_k141_27471\|\|full | 2939 | No | NA | 2 | 0 | 1 | Not-determined | Genome-fragment | NA | NA | 0 |
| S9-final-viral-combined_k141_64784\|\|full | 1769 | No | NA | 5 | 0 | 0 | Not-determined | Genome-fragment | NA | NA | 0 |
| S9-final-viral-combined_k141_64785\|\|full | 1564 | No | NA | 2 | 0 | 0 | Not-determined | Genome-fragment | NA | NA | 0 |
| S17-final-viral-combined_k141_35307\|\|full | 1007 | No | NA | 2 | 0 | 0 | Low-quality | Genome-fragment | 0.72 | AAI-based (medium-confidence) | 0 |
| S9-final-viral-combined_k141_81192\|\|full | 2057 | No | NA | 2 | 0 | 1 | Not-determined | Genome-fragment | NA | NA | 0 |
| S9-final-viral-combined_k141_54087\|\|full | 3583 | No | NA | 4 | 0 | 3 | Not-determined | Genome-fragment | NA | NA | 0 |
| S9-final-viral-combined_k141_97219\|\|full | 3835 | No | NA | 3 | 0 | 2 | Not-determined | Genome-fragment | NA | NA | 0 |
| S9-final-viral-combined_k141_54102\|\|full | 1748 | No | NA | 4 | 0 | 3 | Not-determined | Genome-fragment | NA | NA | 0 |
| S9-final-viral-combined_k141_38176\|\|full | 1562 | No | NA | 2 | 0 | 1 | Not-determined | Genome-fragment | NA | NA | 0 |
| S9-final-viral-combined_k141_38183\|\|full | 1405 | No | NA | 3 | 0 | 1 | Not-determined | Genome-fragment | NA | NA | 0 |
| S9-final-viral-combined_k141_81263\|\|full | 1016 | No | NA | 2 | 0 | 0 | Not-determined | Genome-fragment | NA | NA | 0 |
| S17-final-viral-combined_k141_5221\|\|full | 1007 | No | NA | 3 | 0 | 0 | Low-quality | Genome-fragment | 1.96 | AAI-based (high-confidence) | 0 |
| S9-final-viral-combined_k141_54136\|\|full | 2200 | No | NA | 4 | 0 | 0 | Not-determined | Genome-fragment | NA | NA | 0 |
| S9-final-viral-combined_k141_107449\|\|full | 2976 | No | NA | 5 | 0 | 0 | Not-determined | Genome-fragment | NA | NA | 0 |
| S12-final-viral-combined_k141_390112\|\|full | 1007 | No | NA | 2 | 1 | 0 | Low-quality | Genome-fragment | 2.52 | AAI-based (high-confidence) | 0 |
| S9-final-viral-combined_k141_91559\|\|full | 2029 | No | NA | 2 | 0 | 1 | Not-determined | Genome-fragment | NA | NA | 0 |
| S9-final-viral-combined_k141_27506\|\|full | 7113 | No | NA | 8 | 0 | 0 | Not-determined | Genome-fragment | NA | NA | 0 |
| S9-final-viral-combined_k141_1255\|\|full | 1565 | No | NA | 3 | 0 | 1 | Not-determined | Genome-fragment | NA | NA | 0 |
| S9-final-viral-combined_k141_123614\|\|full | 1022 | No | NA | 3 | 0 | 3 | Not-determined | Genome-fragment | NA | NA | 0 |
| S9-final-viral-combined_k141_118553\|\|full | 2676 | No | NA | 5 | 0 | 0 | Not-determined | Genome-fragment | NA | NA | 0 |
| S12_k141_293816 | 1007 | No | NA | 3 | 1 | 0 | Low-quality | Genome-fragment | 0.54 | AAI-based (high-confidence) | 0 |
| S9-final-viral-combined_k141_91580\|\|full | 5090 | No | NA | 5 | 0 | 1 | Not-determined | Genome-fragment | NA | NA | 0 |
| S9-final-viral-combined_k141_54162\|\|full | 2009 | No | NA | 3 | 0 | 0 | Not-determined | Genome-fragment | NA | NA | 0 |
| S9-final-viral-combined_k141_91604\|\|full | 6465 | No | NA | 8 | 0 | 6 | Not-determined | Genome-fragment | NA | NA | 0 |
| S12_k141_176746 | 1007 | Yes | 376 | 2 | 1 | 1 | Low-quality | Genome-fragment | 1.05 | AAI-based (high-confidence) | 62.66 |
| S2_k141_69923 | 1007 | No | NA | 3 | 0 | 2 | Low-quality | Genome-fragment | 2.36 | AAI-based (high-confidence) | 0 |
| S9-final-viral-combined_k141_59785\|\|full | 1475 | No | NA | 3 | 0 | 1 | Not-determined | Genome-fragment | NA | NA | 0 |
| S9-final-viral-combined_k141_123667\|\|full | 1457 | No | NA | 2 | 0 | 1 | Not-determined | Genome-fragment | NA | NA | 0 |
| S8-final-viral-combined_k141_94524\|\|full | 1007 | No | NA | 4 | 1 | 0 | Low-quality | Genome-fragment | 3.24 | AAI-based (high-confidence) | 0 |
| S9-final-viral-combined_k141_107528\|\|full | 1534 | No | NA | 3 | 0 | 0 | Not-determined | Genome-fragment | NA | NA | 0 |
| S9-final-viral-combined_k141_113103\|\|full | 1735 | No | NA | 3 | 0 | 2 | Not-determined | Genome-fragment | NA | NA | 0 |
| S8-final-viral-combined_k141_112398\|\|lt2gene | 1007 | No | NA | 1 | 1 | 0 | Low-quality | Genome-fragment | 0.7 | HMM-based (lower-bound) | 0 |
| S8_k141_221455 | 1007 | No | NA | 1 | 0 | 1 | Low-quality | Genome-fragment | 0.61 | AAI-based (medium-confidence) | 0 |
| S9-final-viral-combined_k141_64845\|\|full | 1340 | No | NA | 3 | 0 | 2 | Not-determined | Genome-fragment | NA | NA | 0 |
| S9-final-viral-combined_k141_90196\|\|full | 1008 | No | NA | 4 | 0 | 0 | Low-quality | Genome-fragment | 1.44 | AAI-based (medium-confidence) | 0 |
| S9-final-viral-combined_k141_86572\|\|full | 2154 | No | NA | 2 | 0 | 1 | Not-determined | Genome-fragment | NA | NA | 0 |
| S9-final-viral-combined_k141_22091\|\|full | 1425 | No | NA | 3 | 0 | 2 | Not-determined | Genome-fragment | NA | NA | 0 |
| S9-final-viral-combined_k141_38052\|\|lt2gene | 1008 | No | NA | 1 | 1 | 0 | Low-quality | Genome-fragment | 2.4 | AAI-based (high-confidence) | 0 |
| S9-final-viral-combined_k141_64859\|\|full | 9084 | No | NA | 14 | 0 | 6 | Not-determined | Genome-fragment | NA | NA | 0 |
| S9-final-viral-combined_k141_86580\|\|full | 2124 | No | NA | 3 | 0 | 3 | Not-determined | Genome-fragment | NA | NA | 0 |
| S9-final-viral-combined_k141_1334\|\|full | 1560 | No | NA | 2 | 0 | 1 | Not-determined | Genome-fragment | NA | NA | 0 |
| S11-final-viral-combined_k141_46292\|\|full | 1008 | No | NA | 3 | 0 | 0 | Low-quality | Genome-fragment | 2.2 | AAI-based (medium-confidence) | 0 |
| S11-final-viral-combined_k141_78559\|\|full | 1008 | No | NA | 2 | 0 | 1 | Low-quality | Genome-fragment | 1.88 | AAI-based (medium-confidence) | 0 |
| S9-final-viral-combined_k141_102631\|\|full | 2780 | No | NA | 3 | 0 | 3 | Not-determined | Genome-fragment | NA | NA | 0 |
| S9-final-viral-combined_k141_123754\|\|full | 1303 | No | NA | 2 | 0 | 1 | Not-determined | Genome-fragment | NA | NA | 0 |
| S9-final-viral-combined_k141_91678\|\|full | 2498 | No | NA | 3 | 0 | 0 | Not-determined | Genome-fragment | NA | NA | 0 |
| S9-final-viral-combined_k141_102639\|\|full | 3116 | No | NA | 3 | 0 | 3 | Not-determined | Genome-fragment | NA | NA | 0 |
| S9-final-viral-combined_k141_27618\|\|full | 1749 | No | NA | 2 | 0 | 2 | Not-determined | Genome-fragment | NA | NA | 0 |
| S3-final-viral-combined_k141_116063\|\|lt2gene | 1008 | No | NA | 2 | 1 | 0 | Low-quality | Genome-fragment | 2.66 | AAI-based (medium-confidence) | 0 |
| S9-final-viral-combined_k141_64935\|\|full | 1443 | No | NA | 2 | 0 | 0 | Not-determined | Genome-fragment | NA | NA | 0 |
| S9-final-viral-combined_k141_6108\|\|full | 1243 | No | NA | 4 | 0 | 0 | Not-determined | Genome-fragment | NA | NA | 0 |
| S7-final-viral-combined_k141_47370\|\|full | 1008 | No | NA | 3 | 1 | 0 | Low-quality | Genome-fragment | 2.48 | AAI-based (high-confidence) | 0 |
| S9-final-viral-combined_k141_64944\|\|full | 3466 | No | NA | 5 | 0 | 4 | Not-determined | Genome-fragment | NA | NA | 0 |
| S9-final-viral-combined_k141_118657\|\|full | 1782 | No | NA | 2 | 0 | 0 | Not-determined | Genome-fragment | NA | NA | 0 |
| S7_k141_74505 | 1008 | No | NA | 2 | 0 | 1 | Low-quality | Genome-fragment | 1.45 | AAI-based (medium-confidence) | 0 |
| S9-final-viral-combined_k141_11400\|\|full | 1535 | No | NA | 3 | 0 | 0 | Not-determined | Genome-fragment | NA | NA | 0 |
| S9-final-viral-combined_k141_27687\|\|full | 1024 | No | NA | 3 | 0 | 1 | Not-determined | Genome-fragment | NA | NA | 0 |
| S9-final-viral-combined_k141_33106\|\|full | 1673 | No | NA | 2 | 0 | 0 | Not-determined | Genome-fragment | NA | NA | 0 |
| S9-final-viral-combined_k141_59889\|\|full | 1384 | No | NA | 1 | 0 | 1 | Not-determined | Genome-fragment | NA | NA | 0 |
| S7_k141_108024 | 1008 | No | NA | 2 | 0 | 1 | Low-quality | Genome-fragment | 2.06 | AAI-based (medium-confidence) | 0 |
| S7_k141_220303 | 1008 | No | NA | 2 | 0 | 1 | Low-quality | Genome-fragment | 0.41 | AAI-based (medium-confidence) | 0 |
| S9-final-viral-combined_k141_22136\|\|full | 2657 | No | NA | 3 | 0 | 1 | Not-determined | Genome-fragment | NA | NA | 0 |
| S12-final-viral-combined_k141_366295\|\|lt2gene | 1008 | No | NA | 2 | 2 | 0 | Low-quality | Genome-fragment | 2.07 | AAI-based (medium-confidence) | 0 |
| S9-final-viral-combined_k141_54363\|\|full | 1061 | No | NA | 2 | 0 | 2 | Not-determined | Genome-fragment | NA | NA | 0 |
| S2_k141_139658 | 1008 | No | NA | 1 | 0 | 1 | Low-quality | Genome-fragment | 0.57 | AAI-based (medium-confidence) | 0 |
| S9-final-viral-combined_k141_33158\|\|full | 2590 | No | NA | 5 | 0 | 4 | Not-determined | Genome-fragment | NA | NA | 0 |
| S9-final-viral-combined_k141_11423\|\|full | 1776 | No | NA | 4 | 0 | 0 | Not-determined | Genome-fragment | NA | NA | 0 |
| S4-final-viral-combined_k141_123075\|\|full | 1008 | No | NA | 3 | 0 | 0 | Low-quality | Genome-fragment | 1.79 | AAI-based (medium-confidence) | 0 |
| S9-final-viral-combined_k141_11424\|\|full | 2810 | No | NA | 4 | 0 | 0 | Not-determined | Genome-fragment | NA | NA | 0 |
| S6-final-viral-combined_k141_67902\|\|lt2gene | 1008 | No | NA | 2 | 1 | 0 | Low-quality | Genome-fragment | 2.13 | AAI-based (high-confidence) | 0 |
| S9-final-viral-combined_k141_102708\|\|full | 1836 | No | NA | 3 | 0 | 2 | Not-determined | Genome-fragment | NA | NA | 0 |
| S11-final-viral-combined_k141_1909\|\|full | 1009 | No | NA | 2 | 0 | 0 | Low-quality | Genome-fragment | 1.52 | AAI-based (medium-confidence) | 0 |
| S9-final-viral-combined_k141_11440\|\|full | 1662 | No | NA | 2 | 0 | 0 | Not-determined | Genome-fragment | NA | NA | 0 |
| S11-final-viral-combined_k141_126314\|\|full | 1009 | No | NA | 3 | 0 | 1 | Low-quality | Genome-fragment | 2.87 | AAI-based (high-confidence) | 0 |
| S11-final-viral-combined_k141_147480\|\|full | 1009 | No | NA | 4 | 1 | 0 | Low-quality | Genome-fragment | 1.76 | AAI-based (medium-confidence) | 0 |
| S1-final-viral-combined_k141_268120\|\|full | 1009 | No | NA | 3 | 1 | 0 | Low-quality | Genome-fragment | 2.73 | AAI-based (high-confidence) | 0 |
| S5_k141_71145 | 1009 | No | NA | 3 | 0 | 0 | Low-quality | Genome-fragment | 0.4 | AAI-based (medium-confidence) | 0 |
| S12-final-viral-combined_k141_136420\|\|full | 1009 | No | NA | 2 | 0 | 0 | Low-quality | Genome-fragment | 2.57 | AAI-based (high-confidence) | 0 |
| S12-final-viral-combined_k141_89972\|\|lt2gene | 1009 | No | NA | 2 | 2 | 0 | Low-quality | Genome-fragment | 1.43 | HMM-based (lower-bound) | 0 |
| S9-final-viral-combined_k141_27794\|\|full | 4228 | No | NA | 4 | 0 | 4 | Not-determined | Genome-fragment | NA | NA | 0 |
| S9-final-viral-combined_k141_43624\|\|full | 2884 | No | NA | 2 | 0 | 2 | Not-determined | Genome-fragment | NA | NA | 0 |
| S9-final-viral-combined_k141_27824\|\|full | 3150 | No | NA | 5 | 0 | 3 | Not-determined | Genome-fragment | NA | NA | 0 |
| S9-final-viral-combined_k141_43635\|\|full | 11936 | No | NA | 5 | 0 | 1 | Not-determined | Genome-fragment | NA | NA | 0 |
| S12-final-viral-combined_k141_297935\|\|lt2gene | 1009 | No | NA | 2 | 2 | 0 | Low-quality | Genome-fragment | 2.72 | AAI-based (medium-confidence) | 0 |
| S12_k141_199802 | 1009 | No | NA | 1 | 1 | 0 | Low-quality | Genome-fragment | 0.47 | AAI-based (high-confidence) | 0 |
| S9-final-viral-combined_k141_75548\|\|full | 6295 | No | NA | 3 | 0 | 3 | Not-determined | Genome-fragment | NA | NA | 0 |
| S12_k141_317852 | 1009 | No | NA | 2 | 0 | 0 | Low-quality | Genome-fragment | 0.54 | AAI-based (high-confidence) | 0 |
| S2-final-viral-combined_k141_125920\|\|full | 1009 | No | NA | 2 | 1 | 0 | Low-quality | Genome-fragment | 1.41 | AAI-based (medium-confidence) | 0 |
| S6-final-viral-combined_k141_38689\|\|full | 1009 | No | NA | 2 | 0 | 0 | Low-quality | Genome-fragment | 1.38 | AAI-based (medium-confidence) | 0 |
| S9-final-viral-combined_k141_16775\|\|full | 2111 | No | NA | 3 | 0 | 1 | Not-determined | Genome-fragment | NA | NA | 0 |
| S9-final-viral-combined_k141_86752\|\|full | 2899 | No | NA | 4 | 0 | 0 | Not-determined | Genome-fragment | NA | NA | 0 |
| S9-final-viral-combined_k141_91837\|\|full | 2703 | No | NA | 3 | 0 | 3 | Not-determined | Genome-fragment | NA | NA | 0 |
| S8_k141_19242 | 1009 | No | NA | 1 | 0 | 1 | Low-quality | Genome-fragment | 0.59 | AAI-based (medium-confidence) | 0 |
| S1_k141_202753 | 1010 | No | NA | 2 | 0 | 0 | Low-quality | Genome-fragment | 2.41 | AAI-based (medium-confidence) | 0 |
| S1_k141_73154 | 1010 | No | NA | 1 | 0 | 1 | Low-quality | Genome-fragment | 0.67 | AAI-based (medium-confidence) | 0 |
| S1_k141_390043 | 1010 | No | NA | 2 | 0 | 1 | Low-quality | Genome-fragment | 0.44 | AAI-based (medium-confidence) | 0 |
| S9-final-viral-combined_k141_102840\|\|full | 1404 | No | NA | 3 | 0 | 0 | Not-determined | Genome-fragment | NA | NA | 0 |
| S5-final-viral-combined_k141_45398\|\|full | 1010 | No | NA | 3 | 3 | 0 | Low-quality | Genome-fragment | 2.64 | AAI-based (high-confidence) | 0 |
| S5_k141_208360 | 1010 | No | NA | 1 | 0 | 0 | Low-quality | Genome-fragment | 0.66 | AAI-based (medium-confidence) | 0 |
| S9-final-viral-combined_k141_97506\|\|full | 4155 | No | NA | 5 | 0 | 3 | Not-determined | Genome-fragment | NA | NA | 0 |
| S12_k141_133365 | 1010 | No | NA | 2 | 0 | 2 | Low-quality | Genome-fragment | 0.8 | AAI-based (medium-confidence) | 0 |
| S4-final-viral-combined_k141_22390\|\|full | 1010 | No | NA | 3 | 0 | 1 | Low-quality | Genome-fragment | 0.44 | AAI-based (medium-confidence) | 0 |
| S9-final-viral-combined_k141_43653\|\|full | 2225 | No | NA | 2 | 0 | 1 | Not-determined | Genome-fragment | NA | NA | 0 |
| S9-final-viral-combined_k141_22387\|\|full | 2989 | No | NA | 4 | 0 | 3 | Not-determined | Genome-fragment | NA | NA | 0 |
| S4_k141_102294 | 1010 | No | NA | 2 | 0 | 0 | Low-quality | Genome-fragment | 1.06 | AAI-based (high-confidence) | 0 |
| S8_k141_43513 | 1010 | No | NA | 2 | 0 | 0 | Low-quality | Genome-fragment | 1.08 | AAI-based (high-confidence) | 0 |
| S9-final-viral-combined_k141_70220\|\|full | 1928 | No | NA | 3 | 0 | 2 | Not-determined | Genome-fragment | NA | NA | 0 |
| S9-final-viral-combined_k141_127926\|\|full | 1011 | No | NA | 4 | 1 | 0 | Low-quality | Genome-fragment | 2.16 | AAI-based (medium-confidence) | 0 |
| S11-final-viral-combined_k141_266226\|\|full | 1011 | No | NA | 3 | 2 | 0 | Low-quality | Genome-fragment | 2.38 | AAI-based (high-confidence) | 0 |
| S9-final-viral-combined_k141_22427\|\|full | 1219 | No | NA | 4 | 0 | 0 | Not-determined | Genome-fragment | NA | NA | 0 |
| S9-final-viral-combined_k141_11542\|\|full | 2125 | No | NA | 4 | 0 | 0 | Not-determined | Genome-fragment | NA | NA | 0 |
| S11-final-viral-combined_k141_260242\|\|full | 1011 | No | NA | 2 | 0 | 0 | Low-quality | Genome-fragment | 1.8 | AAI-based (high-confidence) | 0 |
| S9-final-viral-combined_k141_22437\|\|full | 1655 | No | NA | 3 | 0 | 0 | Not-determined | Genome-fragment | NA | NA | 0 |
| S9-final-viral-combined_k141_6387\|\|full | 2354 | No | NA | 3 | 0 | 1 | Not-determined | Genome-fragment | NA | NA | 0 |
| S9-final-viral-combined_k141_75612\|\|full | 1665 | No | NA | 3 | 0 | 1 | Not-determined | Genome-fragment | NA | NA | 0 |
| S11-final-viral-combined_k141_312627\|\|lt2gene | 1011 | No | NA | 1 | 1 | 0 | Low-quality | Genome-fragment | 2.28 | AAI-based (high-confidence) | 0 |
| S9-final-viral-combined_k141_123995\|\|full | 1106 | No | NA | 2 | 0 | 1 | Not-determined | Genome-fragment | NA | NA | 0 |
| S11-final-viral-combined_k141_54431\|\|lt2gene | 1011 | No | NA | 1 | 1 | 0 | Low-quality | Genome-fragment | 0.65 | HMM-based (lower-bound) | 0 |
| S9-final-viral-combined_k141_65132\|\|full | 1728 | No | NA | 2 | 0 | 2 | Not-determined | Genome-fragment | NA | NA | 0 |
| S9-final-viral-combined_k141_124013\|\|full | 3148 | No | NA | 4 | 0 | 1 | Not-determined | Genome-fragment | NA | NA | 0 |
| S9-final-viral-combined_k141_118836\|\|full | 6761 | No | NA | 7 | 0 | 4 | Not-determined | Genome-fragment | NA | NA | 0 |
| S11_k141_19356 | 1011 | No | NA | 1 | 0 | 0 | Low-quality | Genome-fragment | 0.7 | AAI-based (medium-confidence) | 0 |
| S1-final-viral-combined_k141_19176\|\|full | 1011 | No | NA | 2 | 0 | 1 | Low-quality | Genome-fragment | 2.38 | AAI-based (medium-confidence) | 0 |
| S9-final-viral-combined_k141_16859\|\|full | 2757 | No | NA | 4 | 0 | 0 | Not-determined | Genome-fragment | NA | NA | 0 |
| S9-final-viral-combined_k141_102893\|\|full | 2884 | No | NA | 3 | 0 | 1 | Not-determined | Genome-fragment | NA | NA | 0 |
| S1-final-viral-combined_k141_39556\|\|full | 1011 | No | NA | 3 | 0 | 0 | Low-quality | Genome-fragment | 1.93 | AAI-based (medium-confidence) | 0 |
| S9-final-viral-combined_k141_70344\|\|full | 1724 | No | NA | 3 | 0 | 2 | Not-determined | Genome-fragment | NA | NA | 0 |
| S9-final-viral-combined_k141_124050\|\|full | 3234 | No | NA | 4 | 0 | 3 | Not-determined | Genome-fragment | NA | NA | 0 |
| S9-final-viral-combined_k141_22475\|\|full | 2857 | No | NA | 4 | 0 | 1 | Not-determined | Genome-fragment | NA | NA | 0 |
| S9-final-viral-combined_k141_22476\|\|full | 1912 | No | NA | 2 | 0 | 1 | Not-determined | Genome-fragment | NA | NA | 0 |
| S9-final-viral-combined_k141_6416\|\|full | 6509 | No | NA | 9 | 0 | 1 | Not-determined | Genome-fragment | NA | NA | 0 |
| S9-final-viral-combined_k141_6434\|\|full | 5896 | No | NA | 4 | 0 | 4 | Not-determined | Genome-fragment | NA | NA | 0 |
| S1-final-viral-combined_k141_73347\|\|full | 1011 | No | NA | 3 | 1 | 0 | Low-quality | Genome-fragment | 2.19 | AAI-based (medium-confidence) | 0 |
| S9-final-viral-combined_k141_118884\|\|full | 2150 | No | NA | 3 | 0 | 2 | Not-determined | Genome-fragment | NA | NA | 0 |
| S9-final-viral-combined_k141_6448\|\|full | 1203 | No | NA | 2 | 0 | 1 | Not-determined | Genome-fragment | NA | NA | 0 |
| S1-final-viral-combined_k141_73801\|\|full | 1011 | No | NA | 4 | 0 | 0 | Low-quality | Genome-fragment | 2.59 | AAI-based (high-confidence) | 0 |
| S9-final-viral-combined_k141_81619\|\|full | 1677 | No | NA | 3 | 0 | 0 | Not-determined | Genome-fragment | NA | NA | 0 |
| S9-final-viral-combined_k141_22494\|\|full | 2907 | No | NA | 5 | 0 | 3 | Not-determined | Genome-fragment | NA | NA | 0 |
| S9-final-viral-combined_k141_124081\|\|full | 1809 | No | NA | 2 | 0 | 1 | Not-determined | Genome-fragment | NA | NA | 0 |
| S3_k141_21907 | 1011 | No | NA | 3 | 0 | 2 | Low-quality | Genome-fragment | 1.57 | AAI-based (medium-confidence) | 0 |
| S7-final-viral-combined_k141_93462\|\|full | 1011 | No | NA | 3 | 3 | 0 | Low-quality | Genome-fragment | 2.21 | AAI-based (high-confidence) | 0 |

| | | | | | | | | | | | |
|---|---|---|---|---|---|---|---|---|---|---|---|
| S7_k141_60893 | 1011 | No | NA | 2 | 2 | 0 | Low-quality | Genome-fragment | 1.94 | AAI-based (high-confidence) | 0 |
| S12_k141_179759 | 1011 | No | NA | 2 | 2 | 0 | Low-quality | Genome-fragment | 1.54 | HMM-based (lower-bound) | 0 |
| S9-final-viral-combined_k141_113286\|\|full | 5219 | No | NA | 9 | 0 | 0 | Not-determined | Genome-fragment | NA | NA | 0 |
| S2-final-viral-combined_k141_121831\|\|full | 1011 | No | NA | 2 | 0 | 1 | Low-quality | Genome-fragment | 0.74 | AAI-based (medium-confidence) | 0 |
| S9-final-viral-combined_k141_97550\|\|full | 6576 | No | NA | 9 | 0 | 4 | Not-determined | Genome-fragment | NA | NA | 0 |
| S9-final-viral-combined_k141_49339\|\|full | 1607 | No | NA | 2 | 0 | 2 | Not-determined | Genome-fragment | NA | NA | 0 |
| S9-final-viral-combined_k141_124119\|\|full | 2659 | No | NA | 5 | 0 | 0 | Not-determined | Genome-fragment | NA | NA | 0 |
| S9-final-viral-combined_k141_1700\|\|full | 1288 | No | NA | 2 | 0 | 1 | Not-determined | Genome-fragment | NA | NA | 0 |
| S9-final-viral-combined_k141_91982\|\|full | 2373 | No | NA | 3 | 0 | 2 | Not-determined | Genome-fragment | NA | NA | 0 |
| S9-final-viral-combined_k141_113325\|\|full | 1661 | No | NA | 2 | 0 | 0 | Not-determined | Genome-fragment | NA | NA | 0 |
| S9-final-viral-combined_k141_49367\|\|full | 2232 | No | NA | 3 | 0 | 0 | Not-determined | Genome-fragment | NA | NA | 0 |
| S2-final-viral-combined_k141_48877\|\|lt2gene | 1011 | No | NA | 1 | 1 | 0 | Low-quality | Genome-fragment | 3.46 | HMM-based (lower-bound) | 0 |
| S9-final-viral-combined_k141_113334\|\|full | 2106 | No | NA | 2 | 0 | 2 | Not-determined | Genome-fragment | NA | NA | 0 |
| S9-final-viral-combined_k141_81683\|\|full | 1872 | No | NA | 4 | 0 | 2 | Not-determined | Genome-fragment | NA | NA | 0 |
| S9-final-viral-combined_k141_54741\|\|full | 3801 | No | NA | 6 | 0 | 0 | Not-determined | Genome-fragment | NA | NA | 0 |
| S9-final-viral-combined_k141_60152\|\|full | 1748 | No | NA | 3 | 0 | 2 | Not-determined | Genome-fragment | NA | NA | 0 |
| S9-final-viral-combined_k141_81767\|\|full | 1890 | No | NA | 3 | 0 | 2 | Not-determined | Genome-fragment | NA | NA | 0 |
| S9-final-viral-combined_k141_43812\|\|full | 2690 | No | NA | 4 | 0 | 3 | Not-determined | Genome-fragment | NA | NA | 0 |
| S2_k141_209746 | 1011 | No | NA | 1 | 0 | 0 | Low-quality | Genome-fragment | 2.1 | AAI-based (high-confidence) | 0 |
| S4_k141_30108 | 1011 | No | NA | 1 | 0 | 0 | Low-quality | Genome-fragment | 1.01 | AAI-based (high-confidence) | 0 |
| S4_k141_59007 | 1011 | No | NA | 1 | 0 | 0 | Low-quality | Genome-fragment | 1.08 | AAI-based (high-confidence) | 0 |
| S6_k141_38305 | 1011 | No | NA | 1 | 0 | 0 | Low-quality | Genome-fragment | 0.63 | AAI-based (high-confidence) | 0 |
| S9-final-viral-combined_k141_107986\|\|full | 1313 | No | NA | 2 | 0 | 0 | Not-determined | Genome-fragment | NA | NA | 0 |
| S9-final-viral-combined_k141_41230\|\|lt2gene | 1012 | No | NA | 2 | 2 | 0 | Low-quality | Genome-fragment | 2.2 | AAI-based (high-confidence) | 0 |
| S9-final-viral-combined_k141_119028\|\|full | 2167 | No | NA | 2 | 0 | 2 | Not-determined | Genome-fragment | NA | NA | 0 |
| S11-final-viral-combined_k141_260954\|\|full | 1012 | No | NA | 3 | 0 | 0 | Low-quality | Genome-fragment | 2.4 | AAI-based (high-confidence) | 0 |
| S1-final-viral-combined_k141_363364\|\|full | 1012 | No | NA | 3 | 0 | 1 | Low-quality | Genome-fragment | 0.83 | AAI-based (medium-confidence) | 0 |
| S9-final-viral-combined_k141_97633\|\|full | 2086 | No | NA | 3 | 0 | 2 | Not-determined | Genome-fragment | NA | NA | 0 |
| S9-final-viral-combined_k141_22701\|\|full | 4420 | No | NA | 5 | 0 | 3 | Not-determined | Genome-fragment | NA | NA | 0 |
| S9-final-viral-combined_k141_75888\|\|full | 3102 | No | NA | 3 | 0 | 2 | Not-determined | Genome-fragment | NA | NA | 0 |
| S9-final-viral-combined_k141_65378\|\|full | 4624 | No | NA | 6 | 0 | 1 | Not-determined | Genome-fragment | NA | NA | 0 |
| S1_k141_346007 | 1012 | No | NA | 1 | 0 | 1 | Low-quality | Genome-fragment | 0.38 | AAI-based (medium-confidence) | 0 |
| S9-final-viral-combined_k141_119043\|\|full | 1902 | No | NA | 4 | 0 | 2 | Not-determined | Genome-fragment | NA | NA | 0 |
| S1_k141_342765 | 1012 | No | NA | 1 | 0 | 1 | Low-quality | Genome-fragment | 0.64 | AAI-based (high-confidence) | 0 |
| S9-final-viral-combined_k141_60228\|\|full | 1964 | No | NA | 3 | 0 | 2 | Not-determined | Genome-fragment | NA | NA | 0 |
| S3-final-viral-combined_k141_98471\|\|full | 1012 | No | NA | 3 | 0 | 0 | Low-quality | Genome-fragment | 1.64 | AAI-based (medium-confidence) | 0 |
| S5-final-viral-combined_k141_163033\|\|full | 1012 | No | NA | 3 | 1 | 0 | Low-quality | Genome-fragment | 1.59 | AAI-based (high-confidence) | 0 |
| S9-final-viral-combined_k141_124178\|\|full | 1477 | No | NA | 2 | 0 | 0 | Not-determined | Genome-fragment | NA | NA | 0 |
| S9-final-viral-combined_k141_17102\|\|full | 1224 | No | NA | 3 | 0 | 0 | Not-determined | Genome-fragment | NA | NA | 0 |
| S5-final-viral-combined_k141_7633\|\|full | 1012 | No | NA | 2 | 0 | 0 | Low-quality | Genome-fragment | 0.57 | AAI-based (medium-confidence) | 0 |
| S9-final-viral-combined_k141_17104\|\|full | 2536 | No | NA | 4 | 0 | 3 | Not-determined | Genome-fragment | NA | NA | 0 |
| S9-final-viral-combined_k141_49486\|\|full | 4755 | No | NA | 6 | 0 | 0 | Not-determined | Genome-fragment | NA | NA | 0 |
| S9-final-viral-combined_k141_65428\|\|full | 1312 | No | NA | 3 | 0 | 2 | Not-determined | Genome-fragment | NA | NA | 0 |
| S5-final-viral-combined_k141_193241\|\|full | 1012 | No | NA | 3 | 1 | 0 | Low-quality | Genome-fragment | 3 | AAI-based (medium-confidence) | 0 |
| S9-final-viral-combined_k141_97696\|\|full | 1615 | No | NA | 2 | 0 | 1 | Not-determined | Genome-fragment | NA | NA | 0 |
| S9-final-viral-combined_k141_22758\|\|full | 3388 | No | NA | 3 | 0 | 3 | Not-determined | Genome-fragment | NA | NA | 0 |
| S9-final-viral-combined_k141_43917\|\|full | 1622 | No | NA | 2 | 0 | 0 | Not-determined | Genome-fragment | NA | NA | 0 |
| S9-final-viral-combined_k141_70577\|\|full | 1813 | No | NA | 3 | 0 | 3 | Not-determined | Genome-fragment | NA | NA | 0 |
| S9-final-viral-combined_k141_54870\|\|full | 6477 | No | NA | 6 | 0 | 1 | Not-determined | Genome-fragment | NA | NA | 0 |
| S5_k141_24917 | 1012 | No | NA | 1 | 0 | 0 | Low-quality | Genome-fragment | 0.57 | AAI-based (medium-confidence) | 0 |
| S7_k141_4465 | 1012 | No | NA | 3 | 0 | 0 | Low-quality | Genome-fragment | 3.05 | AAI-based (medium-confidence) | 0 |
| S9-final-viral-combined_k141_22768\|\|full | 4193 | No | NA | 6 | 0 | 3 | Not-determined | Genome-fragment | NA | NA | 0 |
| S9-final-viral-combined_k141_54874\|\|full | 4928 | No | NA | 4 | 0 | 2 | Not-determined | Genome-fragment | NA | NA | 0 |
| S2-final-viral-combined_k141_200740\|\|lt2gene | 1012 | No | NA | 1 | 1 | 0 | Low-quality | Genome-fragment | 1.74 | AAI-based (high-confidence) | 0 |
| S4-final-viral-combined_k141_132705\|\|full | 1012 | No | NA | 2 | 0 | 1 | Low-quality | Genome-fragment | 2.73 | AAI-based (medium-confidence) | 0 |
| S9-final-viral-combined_k141_124242\|\|full | 2391 | No | NA | 2 | 0 | 2 | Not-determined | Genome-fragment | NA | NA | 0 |
| S9-final-viral-combined_k141_108098\|\|full | 11780 | No | NA | 13 | 0 | 2 | Not-determined | Genome-fragment | NA | NA | 0 |
| S4-final-viral-combined_k141_155184\|\|full | 1012 | No | NA | 3 | 1 | 0 | Low-quality | Genome-fragment | 2.08 | AAI-based (high-confidence) | 0 |
| S9-final-viral-combined_k141_60336\|\|full | 1929 | No | NA | 4 | 0 | 2 | Not-determined | Genome-fragment | NA | NA | 0 |
| S9-final-viral-combined_k141_28127\|\|full | 1435 | No | NA | 3 | 0 | 0 | Not-determined | Genome-fragment | NA | NA | 0 |
| S9-final-viral-combined_k141_38556\|\|full | 1176 | No | NA | 3 | 0 | 0 | Not-determined | Genome-fragment | NA | NA | 0 |
| S4_k141_120030 | 1012 | No | NA | 1 | 0 | 1 | Low-quality | Genome-fragment | 2.26 | AAI-based (medium-confidence) | 0 |
| S4_k141_102546 | 1012 | No | NA | 2 | 0 | 0 | Low-quality | Genome-fragment | 1.62 | AAI-based (medium-confidence) | 0 |
| S9-final-viral-combined_k141_28154\|\|full | 3358 | No | NA | 4 | 0 | 2 | Not-determined | Genome-fragment | NA | NA | 0 |
| S9-final-viral-combined_k141_60361\|\|full | 1758 | No | NA | 2 | 0 | 0 | Not-determined | Genome-fragment | NA | NA | 0 |
| S9-final-viral-combined_k141_113537\|\|full | 1430 | No | NA | 2 | 0 | 1 | Not-determined | Genome-fragment | NA | NA | 0 |
| S8-final-viral-combined_k141_71875\|\|lt2gene | 1012 | No | NA | 2 | 0 | 2 | Low-quality | Genome-fragment | 0.49 | AAI-based (medium-confidence) | 0 |
| S8-final-viral-combined_k141_158363\|\|lt2gene | 1012 | No | NA | 2 | 2 | 0 | Low-quality | Genome-fragment | 1.62 | HMM-based (lower-bound) | 0 |
| S9-final-viral-combined_k141_97755\|\|full | 46121 | Yes | 27506 | 58 | 24 | 13 | Medium-quality | Genome-fragment | 60.07 | AAI-based (medium-confidence) | 40.36 |
| S9-final-viral-combined_k141_124283\|\|full | 1971 | No | NA | 3 | 0 | 2 | Not-determined | Genome-fragment | NA | NA | 0 |
| S8_k141_28646 | 1012 | No | NA | 2 | 0 | 0 | Low-quality | Genome-fragment | 2.33 | AAI-based (high-confidence) | 0 |
| S9-final-viral-combined_k141_70662\|\|full | 2885 | No | NA | 3 | 0 | 3 | Not-determined | Genome-fragment | NA | NA | 0 |
| S9-final-viral-combined_k141_28178\|\|full | 2834 | No | NA | 4 | 0 | 2 | Not-determined | Genome-fragment | NA | NA | 0 |
| S9-final-viral-combined_k141_124289\|\|full | 5708 | No | NA | 11 | 0 | 1 | Not-determined | Genome-fragment | NA | NA | 0 |
| S9-final-viral-combined_k141_65537\|\|full | 3401 | No | NA | 5 | 0 | 3 | Not-determined | Genome-fragment | NA | NA | 0 |
| S9-final-viral-combined_k141_42484\|\|full | 1013 | No | NA | 5 | 1 | 0 | Low-quality | Genome-fragment | 1.9 | AAI-based (medium-confidence) | 0 |
| S9-final-viral-combined_k141_6877\|\|full | 3760 | No | NA | 5 | 0 | 0 | Not-determined | Genome-fragment | NA | NA | 0 |
| S9-final-viral-combined_k141_14018\|\|lt2gene | 1013 | No | NA | 1 | 1 | 0 | Low-quality | Genome-fragment | 2.37 | HMM-based (lower-bound) | 0 |
| S9-final-viral-combined_k141_22906\|\|full | 1244 | No | NA | 2 | 0 | 1 | Not-determined | Genome-fragment | NA | NA | 0 |
| S1-final-viral-combined_k141_129799\|\|full | 1013 | No | NA | 3 | 0 | 2 | Low-quality | Genome-fragment | 0.45 | AAI-based (medium-confidence) | 0 |
| S9-final-viral-combined_k141_60417\|\|full | 1375 | No | NA | 3 | 0 | 3 | Not-determined | Genome-fragment | NA | NA | 0 |
| S9-final-viral-combined_k141_33230\|\|full | 41954 | Yes | 27561 | 58 | 11 | 12 | Medium-quality | Genome-fragment | 84.97 | AAI-based (high-confidence) | 34.31 |
| S3-final-viral-combined_k141_19862\|\|full | 1013 | Yes | 680 | 2 | 1 | 1 | Low-quality | Genome-fragment | 0.23 | HMM-based (lower-bound) | 32.87 |
| S9-final-viral-combined_k141_17227\|\|full | 2299 | No | NA | 2 | 0 | 1 | Not-determined | Genome-fragment | NA | NA | 0 |
| S3-final-viral-combined_k141_56360\|\|full | 1013 | No | NA | 2 | 0 | 0 | Low-quality | Genome-fragment | 0.73 | AAI-based (medium-confidence) | 0 |
| S9-final-viral-combined_k141_49568\|\|full | 4992 | No | NA | 6 | 0 | 2 | Not-determined | Genome-fragment | NA | NA | 0 |
| S9-final-viral-combined_k141_17245\|\|full | 2165 | No | NA | 3 | 0 | 0 | Not-determined | Genome-fragment | NA | NA | 0 |
| S7_k141_29961 | 1013 | No | NA | 1 | 0 | 0 | Low-quality | Genome-fragment | 0.71 | AAI-based (high-confidence) | 0 |
| S9-final-viral-combined_k141_17262\|\|full | 1438 | No | NA | 3 | 0 | 1 | Not-determined | Genome-fragment | NA | NA | 0 |
| S9-final-viral-combined_k141_17275\|\|full | 1975 | No | NA | 4 | 0 | 0 | Not-determined | Genome-fragment | NA | NA | 0 |
| S9-final-viral-combined_k141_119052\|\|full | 1806 | No | NA | 2 | 0 | 0 | Not-determined | Genome-fragment | NA | NA | 0 |
| S17-final-viral-combined_k141_69188\|\|lt2gene | 1013 | No | NA | 1 | 1 | 0 | Low-quality | Genome-fragment | 2.32 | AAI-based (high-confidence) | 0 |
| S9-final-viral-combined_k141_113626\|\|full | 1595 | No | NA | 3 | 0 | 0 | Not-determined | Genome-fragment | NA | NA | 0 |
| S12-final-viral-combined_k141_193111\|\|full | 1013 | No | NA | 2 | 0 | 0 | Low-quality | Genome-fragment | 2.8 | AAI-based (high-confidence) | 0 |
| S9-final-viral-combined_k141_124315\|\|full | 3192 | No | NA | 4 | 0 | 0 | Not-determined | Genome-fragment | NA | NA | 0 |
| S12_k141_322645 | 1013 | No | NA | 2 | 0 | 1 | Low-quality | Genome-fragment | 0.81 | AAI-based (medium-confidence) | 0 |
| S2-final-viral-combined_k141_202436\|\|full | 1013 | No | NA | 2 | 0 | 0 | Low-quality | Genome-fragment | 1.43 | AAI-based (high-confidence) | 0 |
| S9-final-viral-combined_k141_81877\|\|full | 12933 | No | NA | 18 | 0 | 5 | Not-determined | Genome-fragment | NA | NA | 0 |
| S9-final-viral-combined_k141_60466\|\|full | 14246 | No | NA | 14 | 0 | 7 | Not-determined | Genome-fragment | NA | NA | 0 |
| S9-final-viral-combined_k141_70788\|\|full | 1294 | No | NA | 3 | 0 | 0 | Not-determined | Genome-fragment | NA | NA | 0 |
| S9-final-viral-combined_k141_65634\|\|full | 1696 | No | NA | 2 | 0 | 1 | Not-determined | Genome-fragment | NA | NA | 0 |
| S9-final-viral-combined_k141_22993\|\|full | 1812 | No | NA | 2 | 0 | 0 | Not-determined | Genome-fragment | NA | NA | 0 |
| S2-final-viral-combined_k141_130641\|\|full | 1013 | No | NA | 2 | 1 | 0 | Low-quality | Genome-fragment | 2.96 | AAI-based (high-confidence) | 0 |
| S2-final-viral-combined_k141_6256\|\|full | 1013 | No | NA | 4 | 0 | 0 | Low-quality | Genome-fragment | 1.34 | AAI-based (high-confidence) | 0 |
| S9-final-viral-combined_k141_23007\|\|full | 2337 | No | NA | 3 | 0 | 2 | Not-determined | Genome-fragment | NA | NA | 0 |
| S2-final-viral-combined_k141_97382\|\|lt2gene | 1013 | No | NA | 1 | 1 | 0 | Low-quality | Genome-fragment | 0.63 | HMM-based (lower-bound) | 0 |
| S9-final-viral-combined_k141_49672\|\|full | 14140 | No | NA | 11 | 0 | 6 | Not-determined | Genome-fragment | NA | NA | 0 |
| S9-final-viral-combined_k141_103194\|\|full | 1964 | No | NA | 4 | 0 | 2 | Not-determined | Genome-fragment | NA | NA | 0 |
| S2_k141_243481 | 1013 | No | NA | 2 | 0 | 1 | Low-quality | Genome-fragment | 1.28 | AAI-based (medium-confidence) | 0 |
| S9-final-viral-combined_k141_103203\|\|full | 2290 | No | NA | 3 | 0 | 1 | Not-determined | Genome-fragment | NA | NA | 0 |
| S9-final-viral-combined_k141_87023\|\|full | 1960 | No | NA | 2 | 0 | 2 | Not-determined | Genome-fragment | NA | NA | 0 |
| S11-final-viral-combined_k141_40047\|\|full | 1014 | No | NA | 2 | 0 | 1 | Low-quality | Genome-fragment | 0.46 | AAI-based (medium-confidence) | 0 |
| S9-final-viral-combined_k141_65689\|\|full | 1506 | No | NA | 2 | 0 | 1 | Not-determined | Genome-fragment | NA | NA | 0 |
| S9-final-viral-combined_k141_55042\|\|full | 4244 | No | NA | 6 | 0 | 1 | Not-determined | Genome-fragment | NA | NA | 0 |
| S9-final-viral-combined_k141_11866\|\|full | 1988 | No | NA | 3 | 0 | 1 | Not-determined | Genome-fragment | NA | NA | 0 |
| S9-final-viral-combined_k141_87106\|\|full | 1042 | No | NA | 3 | 0 | 0 | Not-determined | Genome-fragment | NA | NA | 0 |
| S9-final-viral-combined_k141_103271\|\|full | 2102 | No | NA | 3 | 0 | 0 | Not-determined | Genome-fragment | NA | NA | 0 |
| S9-final-viral-combined_k141_49726\|\|full | 4409 | No | NA | 5 | 0 | 0 | Not-determined | Genome-fragment | NA | NA | 0 |
| S9-final-viral-combined_k141_17376\|\|full | 2169 | No | NA | 3 | 0 | 2 | Not-determined | Genome-fragment | NA | NA | 0 |
| S11-final-viral-combined_k141_242373\|\|lt2gene | 1014 | No | NA | 1 | 1 | 0 | Low-quality | Genome-fragment | 2.77 | AAI-based (medium-confidence) | 0 |
| S9-final-viral-combined_k141_28314\|\|full | 2234 | No | NA | 3 | 0 | 1 | Not-determined | Genome-fragment | NA | NA | 0 |
| S11_k141_289981 | 1014 | No | NA | 3 | 0 | 0 | Low-quality | Genome-fragment | 2.18 | AAI-based (high-confidence) | 0 |
| S9-final-viral-combined_k141_28321\|\|full | 2474 | No | NA | 4 | 0 | 2 | Not-determined | Genome-fragment | NA | NA | 0 |
| S1_k141_271796 | 1014 | No | NA | 2 | 0 | 0 | Low-quality | Genome-fragment | 0.59 | AAI-based (medium-confidence) | 0 |
| S5-final-viral-combined_k141_170026\|\|full | 1014 | No | NA | 3 | 0 | 0 | Low-quality | Genome-fragment | 2 | AAI-based (medium-confidence) | 0 |
| S5-final-viral-combined_k141_518\|\|lt2gene | 1014 | No | NA | 1 | 0 | 0 | Low-quality | Genome-fragment | 2.59 | AAI-based (high-confidence) | 0 |
| S7-final-viral-combined_k141_67860\|\|full | 1014 | No | NA | 2 | 0 | 2 | Low-quality | Genome-fragment | 0.32 | AAI-based (medium-confidence) | 0 |
| S9-final-viral-combined_k141_108227\|\|full | 1768 | No | NA | 3 | 0 | 2 | Not-determined | Genome-fragment | NA | NA | 0 |
| S9-final-viral-combined_k141_75960\|\|full | 1547 | No | NA | 3 | 0 | 3 | Not-determined | Genome-fragment | NA | NA | 0 |
| S17-k141_157531 | 1014 | No | NA | 4 | 0 | 0 | Low-quality | Genome-fragment | 2.07 | AAI-based (high-confidence) | 0 |
| S17-k141_86259 | 1014 | No | NA | 4 | 0 | 0 | Low-quality | Genome-fragment | 1.74 | AAI-based (medium-confidence) | 0 |
| S12-final-viral-combined_k141_34117\|\|full | 1014 | No | NA | 2 | 1 | 0 | Low-quality | Genome-fragment | 2.06 | AAI-based (high-confidence) | 0 |
| S12-final-viral-combined_k141_321842\|\|full | 1014 | No | NA | 4 | 0 | 0 | Low-quality | Genome-fragment | 1.71 | AAI-based (high-confidence) | 0 |
| S9-final-viral-combined_k141_81967\|\|full | 2416 | No | NA | 6 | 0 | 1 | Not-determined | Genome-fragment | NA | NA | 0 |
| S12-final-viral-combined_k141_367240\|\|lt2gene | 1014 | No | NA | 1 | 0 | 0 | Low-quality | Genome-fragment | 2.79 | AAI-based (medium-confidence) | 0 |
| S2-final-viral-combined_k141_76965\|\|lt2gene | 1014 | No | NA | 1 | 1 | 0 | Low-quality | Genome-fragment | 2.6 | AAI-based (high-confidence) | 0 |
| S9-final-viral-combined_k141_97840\|\|full | 2284 | No | NA | 3 | 0 | 2 | Not-determined | Genome-fragment | NA | NA | 0 |
| S9-final-viral-combined_k141_119138\|\|full | 19823 | No | NA | 18 | 0 | 11 | Not-determined | Genome-fragment | NA | NA | 0 |
| S9-final-viral-combined_k141_76013\|\|full | 1603 | No | NA | 3 | 0 | 1 | Not-determined | Genome-fragment | NA | NA | 0 |
| S9-final-viral-combined_k141_38821\|\|full | 2630 | No | NA | 2 | 0 | 1 | Not-determined | Genome-fragment | NA | NA | 0 |
| S2-final-viral-combined_k141_80291\|\|lt2gene | 1014 | No | NA | 2 | 1 | 0 | Low-quality | Genome-fragment | 2.74 | AAI-based (high-confidence) | 0 |
| S9-final-viral-combined_k141_108280\|\|full | 1597 | No | NA | 3 | 0 | 0 | Not-determined | Genome-fragment | NA | NA | 0 |
| S9-final-viral-combined_k141_49846\|\|full | 4402 | No | NA | 4 | 0 | 0 | Not-determined | Genome-fragment | NA | NA | 0 |
| S9-final-viral-combined_k141_65859\|\|full | 2822 | No | NA | 3 | 0 | 3 | Not-determined | Genome-fragment | NA | NA | 0 |
| S4-final-viral-combined_k141_103131\|\|full | 1014 | No | NA | 3 | 0 | 1 | Low-quality | Genome-fragment | 1.48 | AAI-based (high-confidence) | 0 |
| S4-final-viral-combined_k141_58989\|\|lt2gene | 1014 | No | NA | 2 | 1 | 0 | Low-quality | Genome-fragment | 2.28 | AAI-based (medium-confidence) | 0 |
| S9-final-viral-combined_k141_23081\|\|full | 3304 | No | NA | 3 | 0 | 3 | Not-determined | Genome-fragment | NA | NA | 0 |
| S6-final-viral-combined_k141_1339\|\|full | 1014 | No | NA | 2 | 0 | 1 | Low-quality | Genome-fragment | 0.48 | AAI-based (high-confidence) | 0 |
| S9-final-viral-combined_k141_65877\|\|full | 1974 | No | NA | 4 | 0 | 2 | Not-determined | Genome-fragment | NA | NA | 0 |
| S8-final-viral-combined_k141_83451\|\|lt2gene | 1014 | No | NA | 2 | 1 | 0 | Low-quality | Genome-fragment | 1.78 | AAI-based (high-confidence) | 0 |
| S9-final-viral-combined_k141_17525\|\|full | 2422 | No | NA | 4 | 0 | 2 | Not-determined | Genome-fragment | NA | NA | 0 |
| S8_k141_173134 | 1014 | No | NA | 1 | 0 | 0 | Low-quality | Genome-fragment | 0.75 | AAI-based (medium-confidence) | 0 |
| S9-final-viral-combined_k141_60610\|\|full | 2187 | No | NA | 3 | 0 | 1 | Not-determined | Genome-fragment | NA | NA | 0 |
| S9-final-viral-combined_k141_2042\|\|full | 38861 | Yes | 31053 | 45 | 5 | 5 | Medium-quality | Genome-fragment | 68.98 | AAI-based (high-confidence) | 20.09 |
| S9-final-viral-combined_k141_23130\|\|full | 2086 | No | NA | 2 | 0 | 0 | Not-determined | Genome-fragment | NA | NA | 0 |
| S9-final-viral-combined_k141_76068\|\|full | 2866 | No | NA | 2 | 0 | 0 | Not-determined | Genome-fragment | NA | NA | 0 |
| S9-final-viral-combined_k141_85743\|\|full | 1015 | No | NA | 4 | 4 | 0 | Low-quality | Genome-fragment | 3.19 | AAI-based (high-confidence) | 0 |
| S9-final-viral-combined_k141_115400\|\|full | 1015 | No | NA | 3 | 0 | 0 | Low-quality | Genome-fragment | 0.4 | AAI-based (medium-confidence) | 0 |
| S9-final-viral-combined_k141_23142\|\|full | 13473 | No | NA | 15 | 0 | 1 | Not-determined | Genome-fragment | NA | NA | 0 |
| S9-final-viral-combined_k141_49880\|\|full | 1553 | No | NA | 3 | 0 | 2 | Not-determined | Genome-fragment | NA | NA | 0 |
| S11-final-viral-combined_k141_59514\|\|full | 1015 | No | NA | 4 | 0 | 0 | Low-quality | Genome-fragment | 1.65 | AAI-based (high-confidence) | 0 |
| S9-final-viral-combined_k141_33405\|\|full | 1884 | No | NA | 4 | 0 | 2 | Not-determined | Genome-fragment | NA | NA | 0 |
| S11_k141_239566 | 1015 | No | NA | 1 | 0 | 0 | Low-quality | Genome-fragment | 1.67 | AAI-based (medium-confidence) | 0 |
| S1-final-viral-combined_k141_76605\|\|full | 1015 | No | NA | 2 | 0 | 0 | Low-quality | Genome-fragment | 0.76 | AAI-based (medium-confidence) | 0 |
| S1-final-viral-combined_k141_375441\|\|full | 1015 | No | NA | 2 | 0 | 1 | Low-quality | Genome-fragment | 2.17 | AAI-based (medium-confidence) | 0 |
| S9-final-viral-combined_k141_33421\|\|full | 1723 | No | NA | 3 | 0 | 3 | Not-determined | Genome-fragment | NA | NA | 0 |
| S3-final-viral-combined_k141_79746\|\|full | 1015 | No | NA | 3 | 0 | 0 | Low-quality | Genome-fragment | 2.52 | AAI-based (medium-confidence) | 0 |
| S9-final-viral-combined_k141_2130\|\|full | 2653 | No | NA | 4 | 0 | 4 | Not-determined | Genome-fragment | NA | NA | 0 |
| S9-final-viral-combined_k141_124574\|\|full | 1686 | No | NA | 3 | 0 | 3 | Not-determined | Genome-fragment | NA | NA | 0 |
| S9-final-viral-combined_k141_17589\|\|full | 2283 | No | NA | 3 | 0 | 2 | Not-determined | Genome-fragment | NA | NA | 0 |
| S9-final-viral-combined_k141_55305\|\|full | 1332 | No | NA | 5 | 0 | 1 | Not-determined | Genome-fragment | NA | NA | 0 |
| S9-final-viral-combined_k141_17591\|\|full | 4162 | No | NA | 5 | 0 | 0 | Not-determined | Genome-fragment | NA | NA | 0 |
| S3_k141_12277 | 1015 | No | NA | 1 | 0 | 1 | Low-quality | Genome-fragment | 2.44 | AAI-based (high-confidence) | 0 |
| S9-final-viral-combined_k141_103358\|\|full | 8408 | No | NA | 4 | 0 | 3 | Not-determined | Genome-fragment | NA | NA | 0 |
| S5-final-viral-combined_k141_135348\|\|full | 1015 | No | NA | 3 | 3 | 0 | Low-quality | Genome-fragment | 2.57 | AAI-based (medium-confidence) | 0 |
| S9-final-viral-combined_k141_71022\|\|full | 1018 | No | NA | 2 | 0 | 2 | Not-determined | Genome-fragment | NA | NA | 0 |
| S9-final-viral-combined_k141_55339\|\|full | 4769 | No | NA | 4 | 0 | 3 | Not-determined | Genome-fragment | NA | NA | 0 |
| S5-final-viral-combined_k141_79883\|\|lt2gene | 1015 | No | NA | 1 | 1 | 0 | Low-quality | Genome-fragment | 1.52 | AAI-based (high-confidence) | 0 |
| S9-final-viral-combined_k141_55349\|\|full | 2669 | No | NA | 4 | 0 | 1 | Not-determined | Genome-fragment | NA | NA | 0 |
| S9-final-viral-combined_k141_82135\|\|full | 2734 | No | NA | 3 | 0 | 1 | Not-determined | Genome-fragment | NA | NA | 0 |
| S9-final-viral-combined_k141_97913\|\|full | 1433 | No | NA | 3 | 0 | 2 | Not-determined | Genome-fragment | NA | NA | 0 |
| S7-final-viral-combined_k141_220984\|\|full | 1015 | Yes | 503 | 3 | 1 | 1 | Low-quality | Genome-fragment | 1.4 | AAI-based (high-confidence) | 50.44 |
| S17-final-viral-combined_k141_74417\|\|full | 1015 | No | NA | 3 | 0 | 2 | Low-quality | Genome-fragment | 0.36 | AAI-based (medium-confidence) | 0 |
| S12-final-viral-combined_k141_25826\|\|full | 1015 | No | NA | 2 | 0 | 0 | Low-quality | Genome-fragment | 1.66 | AAI-based (medium-confidence) | 0 |
| S9-final-viral-combined_k141_108379\|\|full | 2556 | No | NA | 3 | 0 | 1 | Not-determined | Genome-fragment | NA | NA | 0 |
| S9-final-viral-combined_k141_55404\|\|full | 2179 | No | NA | 2 | 0 | 1 | Not-determined | Genome-fragment | NA | NA | 0 |
| S9-final-viral-combined_k141_7225\|\|full | 1553 | No | NA | 2 | 0 | 1 | Not-determined | Genome-fragment | NA | NA | 0 |
| S12-final-viral-combined_k141_211435\|\|lt2gene | 1015 | No | NA | 2 | 1 | 0 | Low-quality | Genome-fragment | 2.56 | AAI-based (medium-confidence) | 0 |
| S9-final-viral-combined_k141_55416\|\|full | 3682 | No | NA | 4 | 0 | 4 | Not-determined | Genome-fragment | NA | NA | 0 |
| S9-final-viral-combined_k141_97945\|\|full | 1998 | No | NA | 3 | 0 | 2 | Not-determined | Genome-fragment | NA | NA | 0 |
| S12_k141_351943 | 1015 | No | NA | 2 | 0 | 0 | Low-quality | Genome-fragment | 3.86 | AAI-based (high-confidence) | 0 |
| S9-final-viral-combined_k141_97948\|\|full | 3083 | No | NA | 4 | 0 | 0 | Not-determined | Genome-fragment | NA | NA | 0 |
| S9-final-viral-combined_k141_60754\|\|full | 2282 | No | NA | 3 | 0 | 0 | Not-determined | Genome-fragment | NA | NA | 0 |
| S12_k141_81639 | 1015 | No | NA | 2 | 0 | 0 | Low-quality | Genome-fragment | 0.55 | AAI-based (high-confidence) | 0 |
| S2-final-viral-combined_k141_49613\|\|full | 1015 | No | NA | 3 | 2 | 0 | Low-quality | Genome-fragment | 1.85 | HMM-based (lower-bound) | 0 |
| S9-final-viral-combined_k141_7232\|\|full | 2610 | No | NA | 3 | 0 | 3 | Not-determined | Genome-fragment | NA | NA | 0 |
| S9-final-viral-combined_k141_82228\|\|full | 1662 | No | NA | 4 | 0 | 0 | Not-determined | Genome-fragment | NA | NA | 0 |
| S9-final-viral-combined_k141_12108\|\|full | 3730 | No | NA | 5 | 0 | 2 | Not-determined | Genome-fragment | NA | NA | 0 |
| S8-final-viral-combined_k141_107501\|\|full | 1015 | No | NA | 2 | 1 | 0 | Low-quality | Genome-fragment | 1.85 | AAI-based (medium-confidence) | 0 |
| S8-final-viral-combined_k141_271396\|\|full | 1015 | No | NA | 2 | 0 | 0 | Low-quality | Genome-fragment | 2.2 | AAI-based (medium-confidence) | 0 |
| S8-final-viral-combined_k141_10271\|\|full | 1015 | No | NA | 2 | 0 | 0 | Low-quality | Genome-fragment | 0.66 | AAI-based (high-confidence) | 0 |
| S11-final-viral-combined_k141_33192\|\|full | 1016 | No | NA | 3 | 1 | 0 | Low-quality | Genome-fragment | 2.5 | AAI-based (medium-confidence) | 0 |
| S9-final-viral-combined_k141_17744\|\|full | 2576 | No | NA | 2 | 0 | 2 | Not-determined | Genome-fragment | NA | NA | 0 |
| S9-final-viral-combined_k141_23303\|\|full | 2894 | No | NA | 4 | 0 | 3 | Not-determined | Genome-fragment | NA | NA | 0 |
| S11-final-viral-combined_k141_207281\|\|lt2gene | 1016 | No | NA | 2 | 1 | 0 | Low-quality | Genome-fragment | 1.18 | AAI-based (medium-confidence) | 0 |
| S9-final-viral-combined_k141_17759\|\|full | 4027 | No | NA | 5 | 0 | 1 | Not-determined | Genome-fragment | NA | NA | 0 |
| S9-final-viral-combined_k141_12150\|\|full | 4577 | No | NA | 4 | 0 | 4 | Not-determined | Genome-fragment | NA | NA | 0 |
| S1-final-viral-combined_k141_194280\|\|full | 1016 | No | NA | 2 | 0 | 1 | Low-quality | Genome-fragment | 15.76 | AAI-based (medium-confidence) | 0 |
| S9-final-viral-combined_k141_17788\|\|full | 11015 | No | NA | 9 | 0 | 6 | Not-determined | Genome-fragment | NA | NA | 0 |
| S9-final-viral-combined_k141_17789\|\|full | 2498 | No | NA | 2 | 0 | 1 | Not-determined | Genome-fragment | NA | NA | 0 |
| S9-final-viral-combined_k141_28427\|\|full | 1199 | No | NA | 3 | 0 | 1 | Not-determined | Genome-fragment | NA | NA | 0 |
| S9-final-viral-combined_k141_55492\|\|full | 1274 | No | NA | 3 | 0 | 2 | Not-determined | Genome-fragment | NA | NA | 0 |
| S1_k141_318985 | 1016 | No | NA | 1 | 0 | 1 | Low-quality | Genome-fragment | 0.71 | AAI-based (high-confidence) | 0 |
| S5_k141_144978 | 1016 | No | NA | 2 | 0 | 0 | Low-quality | Genome-fragment | 7.01 | AAI-based (high-confidence) | 0 |
| S9-final-viral-combined_k141_33473\|\|full | 1202 | No | NA | 2 | 0 | 1 | Not-determined | Genome-fragment | NA | NA | 0 |
| S9-final-viral-combined_k141_55507\|\|full | 1429 | No | NA | 3 | 0 | 0 | Not-determined | Genome-fragment | NA | NA | 0 |
| S9-final-viral-combined_k141_12205\|\|full | 1542 | No | NA | 3 | 0 | 2 | Not-determined | Genome-fragment | NA | NA | 0 |
| S9-final-viral-combined_k141_82293\|\|full | 19360 | No | NA | 18 | 0 | 10 | Not-determined | Genome-fragment | NA | NA | 0 |
| S7-final-viral-combined_k141_143168\|\|full | 1016 | No | NA | 3 | 1 | 0 | Low-quality | Genome-fragment | 2.6 | AAI-based (high-confidence) | 0 |
| S17-final-viral-combined_k141_110883\|\|full | 1016 | No | NA | 2 | 0 | 0 | Low-quality | Genome-fragment | 1.38 | AAI-based (medium-confidence) | 0 |
| S9-final-viral-combined_k141_119377\|\|full | 3419 | No | NA | 4 | 0 | 3 | Not-determined | Genome-fragment | NA | NA | 0 |
| S9-final-viral-combined_k141_87381\|\|full | 3519 | No | NA | 6 | 0 | 2 | Not-determined | Genome-fragment | NA | NA | 0 |
| S9-final-viral-combined_k141_2348\|\|full | 1215 | No | NA | 2 | 0 | 1 | Not-determined | Genome-fragment | NA | NA | 0 |
| S9-final-viral-combined_k141_71118\|\|full | 2771 | No | NA | 3 | 0 | 1 | Not-determined | Genome-fragment | NA | NA | 0 |
| S12-final-viral-combined_k141_240948\|\|full | 1016 | No | NA | 3 | 0 | 1 | Low-quality | Genome-fragment | 0.75 | AAI-based (medium-confidence) | 0 |
| S12-final-viral-combined_k141_309841\|\|full | 1016 | No | NA | 3 | 0 | 0 | Low-quality | Genome-fragment | 2.64 | AAI-based (high-confidence) | 0 |
| S9-final-viral-combined_k141_71124\|\|full | 7164 | No | NA | 6 | 0 | 3 | Not-determined | Genome-fragment | NA | NA | 0 |
| S9-final-viral-combined_k141_55551\|\|full | 2895 | No | NA | 3 | 0 | 2 | Not-determined | Genome-fragment | NA | NA | 0 |
| S9-final-viral-combined_k141_92671\|\|full | 1647 | No | NA | 3 | 0 | 0 | Not-determined | Genome-fragment | NA | NA | 0 |
| S12-final-viral-combined_k141_16054\|\|full | 1016 | No | NA | 2 | 0 | 0 | Low-quality | Genome-fragment | 1.73 | AAI-based (high-confidence) | 0 |
| S4_k141_114384 | 1016 | No | NA | 2 | 0 | 0 | Low-quality | Genome-fragment | 1.26 | AAI-based (high-confidence) | 0 |
| S9-final-viral-combined_k141_98074\|\|full | 1374 | No | NA | 3 | 0 | 0 | Not-determined | Genome-fragment | NA | NA | 0 |
| S9-final-viral-combined_k141_87411\|\|full | 1479 | No | NA | 3 | 0 | 0 | Not-determined | Genome-fragment | NA | NA | 0 |
| S9-final-viral-combined_k141_13693\|\|full | 1017 | No | NA | 2 | 1 | 0 | Low-quality | Genome-fragment | 2.19 | AAI-based (medium-confidence) | 0 |
| S9-final-viral-combined_k141_28569\|\|full | 2666 | No | NA | 3 | 0 | 0 | Not-determined | Genome-fragment | NA | NA | 0 |
| S9-final-viral-combined_k141_119428\|\|full | 1633 | No | NA | 4 | 0 | 1 | Not-determined | Genome-fragment | NA | NA | 0 |
| S9-final-viral-combined_k141_79301\|\|lt2gene | 1017 | No | NA | 2 | 1 | 0 | Low-quality | Genome-fragment | 2.53 | AAI-based (high-confidence) | 0 |
| S11-final-viral-combined_k141_89094\|\|full | 1017 | No | NA | 3 | 2 | 0 | Low-quality | Genome-fragment | 2.5 | AAI-based (high-confidence) | 0 |
| S1-final-viral-combined_k141_129920\|\|full | 1017 | No | NA | 3 | 0 | 0 | Low-quality | Genome-fragment | 1.55 | AAI-based (medium-confidence) | 0 |
| S9-final-viral-combined_k141_98127\|\|full | 1228 | No | NA | 3 | 0 | 0 | Not-determined | Genome-fragment | NA | NA | 0 |
| S9-final-viral-combined_k141_60894\|\|full | 3525 | No | NA | 3 | 0 | 1 | Not-determined | Genome-fragment | NA | NA | 0 |
| S9-final-viral-combined_k141_92731\|\|full | 2954 | No | NA | 5 | 0 | 0 | Not-determined | Genome-fragment | NA | NA | 0 |
| S1-final-viral-combined_k141_317633\|\|lt2gene | 1017 | No | NA | 2 | 2 | 0 | Low-quality | Genome-fragment | 1.69 | AAI-based (high-confidence) | 0 |
| S9-final-viral-combined_k141_98152\|\|full | 3335 | No | NA | 5 | 0 | 0 | Not-determined | Genome-fragment | NA | NA | 0 |
| S9-final-viral-combined_k141_71209\|\|full | 3637 | No | NA | 3 | 0 | 2 | Not-determined | Genome-fragment | NA | NA | 0 |
| S9-final-viral-combined_k141_103556\|\|full | 1043 | No | NA | 5 | 0 | 0 | Not-determined | Genome-fragment | NA | NA | 0 |
| S9-final-viral-combined_k141_33510\|\|full | 2110 | No | NA | 2 | 0 | 0 | Not-determined | Genome-fragment | NA | NA | 0 |
| S9-final-viral-combined_k141_103559\|\|full | 8547 | No | NA | 7 | 0 | 6 | Not-determined | Genome-fragment | NA | NA | 0 |
| S9-final-viral-combined_k141_39169\|\|full | 4786 | No | NA | 4 | 0 | 1 | Not-determined | Genome-fragment | NA | NA | 0 |
| S5-final-viral-combined_k141_17168\|\|full | 1017 | No | NA | 2 | 1 | 0 | Low-quality | Genome-fragment | 1.97 | AAI-based (high-confidence) | 0 |
| S7-final-viral-combined_k141_151117\|\|full | 1017 | No | NA | 4 | 1 | 0 | Low-quality | Genome-fragment | 2.58 | AAI-based (high-confidence) | 0 |
| S9-final-viral-combined_k141_39183\|\|full | 2033 | No | NA | 2 | 0 | 2 | Not-determined | Genome-fragment | NA | NA | 0 |
| S17-k141_146064 | 1017 | No | NA | 2 | 0 | 2 | Low-quality | Genome-fragment | 1.04 | AAI-based (medium-confidence) | 0 |
| S9-final-viral-combined_k141_66268\|\|full | 4333 | No | NA | 6 | 0 | 2 | Not-determined | Genome-fragment | NA | NA | 0 |
| S12-final-viral-combined_k141_331711\|\|full | 1017 | No | NA | 2 | 1 | 0 | Low-quality | Genome-fragment | 2.63 | AAI-based (medium-confidence) | 0 |
| S9-final-viral-combined_k141_7492\|\|full | 2367 | No | NA | 2 | 0 | 2 | Not-determined | Genome-fragment | NA | NA | 0 |
| S12_k141_232758 | 1017 | No | NA | 2 | 1 | 0 | Low-quality | Genome-fragment | 0.73 | AAI-based (medium-confidence) | 0 |
| S9-final-viral-combined_k141_71254\|\|full | 2761 | No | NA | 4 | 0 | 1 | Not-determined | Genome-fragment | NA | NA | 0 |
| S2-final-viral-combined_k141_222193\|\|full | 1017 | No | NA | 3 | 1 | 0 | Low-quality | Genome-fragment | 2.36 | AAI-based (medium-confidence) | 0 |
| S8-final-viral-combined_k141_91846\|\|full | 1017 | No | NA | 3 | 0 | 0 | Low-quality | Genome-fragment | 1.67 | AAI-based (medium-confidence) | 0 |
| S9-final-viral-combined_k141_28657\|\|full | 8326 | No | NA | 13 | 0 | 5 | Not-determined | Genome-fragment | NA | NA | 0 |
| S9-final-viral-combined_k141_87551\|\|full | 2492 | No | NA | 4 | 0 | 0 | Not-determined | Genome-fragment | NA | NA | 0 |
| S9-final-viral-combined_k141_28662\|\|full | 1242 | No | NA | 2 | 0 | 0 | Not-determined | Genome-fragment | NA | NA | 0 |
| S9-final-viral-combined_k141_39253\|\|full | 1416 | No | NA | 2 | 0 | 1 | Not-determined | Genome-fragment | NA | NA | 0 |
| S9-final-viral-combined_k141_17465\|\|lt2gene | 1018 | No | NA | 2 | 1 | 0 | Low-quality | Genome-fragment | 2.71 | AAI-based (high-confidence) | 0 |
| S9-final-viral-combined_k141_82397\|\|full | 1958 | No | NA | 4 | 0 | 4 | Not-determined | Genome-fragment | NA | NA | 0 |
| S9-final-viral-combined_k141_103620\|\|full | 1767 | No | NA | 3 | 0 | 1 | Not-determined | Genome-fragment | NA | NA | 0 |
| S9-final-viral-combined_k141_124733\|\|full | 3145 | No | NA | 4 | 0 | 0 | Not-determined | Genome-fragment | NA | NA | 0 |
| S9-final-viral-combined_k141_108514\|\|full | 2127 | No | NA | 4 | 0 | 1 | Not-determined | Genome-fragment | NA | NA | 0 |
| S9-final-viral-combined_k141_71300\|\|full | 1913 | No | NA | 2 | 0 | 1 | Not-determined | Genome-fragment | NA | NA | 0 |
| S9-final-viral-combined_k141_12394\|\|full | 2714 | No | NA | 3 | 0 | 0 | Not-determined | Genome-fragment | NA | NA | 0 |
| S9-final-viral-combined_k141_17959\|\|full | 2927 | No | NA | 4 | 0 | 1 | Not-determined | Genome-fragment | NA | NA | 0 |
| S11_k141_326073 | 1018 | No | NA | 2 | 0 | 1 | Low-quality | Genome-fragment | 1.28 | AAI-based (high-confidence) | 0 |
| S9-final-viral-combined_k141_50388\|\|full | 1238 | No | NA | 3 | 0 | 1 | Not-determined | Genome-fragment | NA | NA | 0 |
| S5-final-viral-combined_k141_63315\|\|full | 1018 | No | NA | 4 | 0 | 1 | Low-quality | Genome-fragment | 0.29 | AAI-based (medium-confidence) | 0 |
| S9-final-viral-combined_k141_87615\|\|full | 3540 | No | NA | 2 | 0 | 2 | Not-determined | Genome-fragment | NA | NA | 0 |
| S5-final-viral-combined_k141_22930\|\|full | 1018 | No | NA | 3 | 0 | 0 | Low-quality | Genome-fragment | 1.88 | AAI-based (medium-confidence) | 0 |
| S5-final-viral-combined_k141_133269\|\|full | 1018 | No | NA | 2 | 0 | 0 | Low-quality | Genome-fragment | 0.64 | AAI-based (medium-confidence) | 0 |
| S5-final-viral-combined_k141_26173\|\|full | 1018 | No | NA | 2 | 1 | 0 | Low-quality | Genome-fragment | 2.25 | AAI-based (high-confidence) | 0 |
| S5_k141_21929 | 1018 | No | NA | 1 | 0 | 1 | Low-quality | Genome-fragment | 2.12 | AAI-based (high-confidence) | 0 |
| S7-final-viral-combined_k141_86430\|\|full | 1018 | No | NA | 3 | 0 | 0 | Low-quality | Genome-fragment | 1.06 | AAI-based (medium-confidence) | 0 |
| S9-final-viral-combined_k141_119563\|\|full | 1441 | No | NA | 3 | 0 | 1 | Not-determined | Genome-fragment | NA | NA | 0 |
| S7-final-viral-combined_k141_244194\|\|full | 1018 | No | NA | 2 | 0 | 0 | Low-quality | Genome-fragment | 0.99 | AAI-based (high-confidence) | 0 |
| S12-final-viral-combined_k141_1130\|\|full | 1018 | No | NA | 2 | 0 | 0 | Low-quality | Genome-fragment | 1.24 | AAI-based (medium-confidence) | 0 |
| S12-final-viral-combined_k141_337148\|\|lt2gene | 1018 | No | NA | 1 | 0 | 0 | Low-quality | Genome-fragment | 6.57 | AAI-based (high-confidence) | 0 |
| S12-final-viral-combined_k141_321381\|\|lt2gene | 1018 | No | NA | 2 | 1 | 0 | Low-quality | Genome-fragment | 2.2 | AAI-based (high-confidence) | 0 |
| S12_k141_125931 | 1018 | No | NA | 1 | 1 | 0 | Low-quality | Genome-fragment | 2.42 | HMM-based (lower-bound) | 0 |
| S9-final-viral-combined_k141_108599\|\|full | 2865 | No | NA | 3 | 0 | 0 | Not-determined | Genome-fragment | NA | NA | 0 |
| S4-final-viral-combined_k141_80850\|\|full | 1018 | No | NA | 3 | 0 | 0 | Low-quality | Genome-fragment | 2.65 | AAI-based (high-confidence) | 0 |
| S9-final-viral-combined_k141_82481\|\|full | 2666 | No | NA | 3 | 0 | 2 | Not-determined | Genome-fragment | NA | NA | 0 |
| S4-final-viral-combined_k141_165078\|\|full | 1018 | No | NA | 3 | 0 | 0 | Low-quality | Genome-fragment | 2.25 | AAI-based (medium-confidence) | 0 |
| S9-final-viral-combined_k141_113920\|\|full | 1257 | No | NA | 2 | 0 | 1 | Not-determined | Genome-fragment | NA | NA | 0 |
| S9-final-viral-combined_k141_18095\|\|full | 4771 | No | NA | 4 | 0 | 1 | Not-determined | Genome-fragment | NA | NA | 0 |
| S4-final-viral-combined_k141_31960\|\|lt2gene | 1018 | No | NA | 1 | 1 | 0 | Low-quality | Genome-fragment | 3.04 | AAI-based (medium-confidence) | 0 |
| S4_k141_144510 | 1018 | No | NA | 1 | 0 | 0 | Low-quality | Genome-fragment | 2.38 | AAI-based (high-confidence) | 0 |
| S6-final-viral-combined_k141_124804\|\|full | 1018 | No | NA | 2 | 0 | 0 | Low-quality | Genome-fragment | 0.73 | AAI-based (medium-confidence) | 0 |
| S8_k141_19757 | 1018 | No | NA | 2 | 0 | 0 | Low-quality | Genome-fragment | 8.15 | AAI-based (medium-confidence) | 0 |
| S9-final-viral-combined_k141_108695\|\|full | 2308 | No | NA | 4 | 0 | 3 | Not-determined | Genome-fragment | NA | NA | 0 |
| S11-final-viral-combined_k141_22853\|\|lt2gene | 1019 | No | NA | 2 | 1 | 0 | Low-quality | Genome-fragment | 1.33 | HMM-based (lower-bound) | 0 |
| S1-final-viral-combined_k141_285168\|\|full | 1019 | No | NA | 3 | 0 | 0 | Low-quality | Genome-fragment | 2.48 | AAI-based (high-confidence) | 0 |
| S9-final-viral-combined_k141_23411\|\|full | 2327 | No | NA | 3 | 0 | 3 | Not-determined | Genome-fragment | NA | NA | 0 |
| S1_k141_17327 | 1019 | No | NA | 3 | 0 | 0 | Low-quality | Genome-fragment | 1.4 | AAI-based (medium-confidence) | 0 |
| S3-final-viral-combined_k141_51850\|\|full | 1019 | No | NA | 4 | 1 | 0 | Low-quality | Genome-fragment | 2.67 | AAI-based (medium-confidence) | 0 |
| S3_k141_47395 | 1019 | No | NA | 1 | 1 | 0 | Low-quality | Genome-fragment | 2.42 | HMM-based (lower-bound) | 0 |
| S9-final-viral-combined_k141_18274\|\|full | 7313 | No | NA | 10 | 0 | 0 | Not-determined | Genome-fragment | NA | NA | 0 |
| S9-final-viral-combined_k141_23447\|\|full | 1362 | No | NA | 2 | 0 | 0 | Not-determined | Genome-fragment | NA | NA | 0 |
| S9-final-viral-combined_k141_18287\|\|full | 1543 | No | NA | 2 | 0 | 1 | Not-determined | Genome-fragment | NA | NA | 0 |
| S7-final-viral-combined_k141_36944\|\|full | 1019 | No | NA | 3 | 0 | 0 | Low-quality | Genome-fragment | 0.72 | AAI-based (medium-confidence) | 0 |
| S9-final-viral-combined_k141_71406\|\|full | 2590 | No | NA | 3 | 0 | 1 | Not-determined | Genome-fragment | NA | NA | 0 |
| S9-final-viral-combined_k141_119617\|\|full | 1249 | No | NA | 3 | 0 | 0 | Not-determined | Genome-fragment | NA | NA | 0 |
| S17-final-viral-combined_k141_121605\|\|full | 1019 | No | NA | 2 | 1 | 0 | Low-quality | Genome-fragment | 1.63 | HMM-based (lower-bound) | 0 |
| S9-final-viral-combined_k141_23467\|\|full | 3279 | No | NA | 5 | 0 | 3 | Not-determined | Genome-fragment | NA | NA | 0 |
| S12-final-viral-combined_k141_242030\|\|full | 1019 | No | NA | 2 | 0 | 1 | Low-quality | Genome-fragment | 0.86 | AAI-based (medium-confidence) | 0 |
| S9-final-viral-combined_k141_113982\|\|full | 1078 | No | NA | 3 | 0 | 0 | Not-determined | Genome-fragment | NA | NA | 0 |
| S9-final-viral-combined_k141_113997\|\|full | 1579 | No | NA | 2 | 0 | 0 | Not-determined | Genome-fragment | NA | NA | 0 |
| S9-final-viral-combined_k141_50533\|\|full | 2206 | No | NA | 2 | 0 | 1 | Not-determined | Genome-fragment | NA | NA | 0 |
| S12-final-viral-combined_k141_362616\|\|full | 1019 | No | NA | 4 | 0 | 0 | Low-quality | Genome-fragment | 2.34 | AAI-based (medium-confidence) | 0 |
| S9-final-viral-combined_k141_12737\|\|full | 1279 | No | NA | 3 | 0 | 1 | Not-determined | Genome-fragment | NA | NA | 0 |
| S12-final-viral-combined_k141_202455\|\|lt2gene | 1019 | No | NA | 1 | 1 | 0 | Low-quality | Genome-fragment | 2.93 | AAI-based (high-confidence) | 0 |
| S9-final-viral-combined_k141_50568\|\|full | 2223 | No | NA | 3 | 0 | 2 | Not-determined | Genome-fragment | NA | NA | 0 |
| S9-final-viral-combined_k141_12746\|\|full | 5276 | No | NA | 4 | 0 | 0 | Not-determined | Genome-fragment | NA | NA | 0 |
| S12_k141_70088 | 1019 | No | NA | 2 | 1 | 0 | Low-quality | Genome-fragment | 0.55 | AAI-based (high-confidence) | 0 |
| S9-final-viral-combined_k141_44505\|\|full | 6443 | No | NA | 9 | 0 | 1 | Not-determined | Genome-fragment | NA | NA | 0 |
| S9-final-viral-combined_k141_23526\|\|full | 1166 | No | NA | 2 | 0 | 1 | Not-determined | Genome-fragment | NA | NA | 0 |
| S12_k141_63393 | 1019 | No | NA | 1 | 0 | 0 | Low-quality | Genome-fragment | 1.87 | AAI-based (medium-confidence) | 0 |
| S9-final-viral-combined_k141_124857\|\|full | 199218 | Yes | 56670 | 220 | 20 | 102 | Medium-quality | Genome-fragment | 65.17 | HMM-based (lower-bound) | 71.55 |
| S12_k141_283835 | 1019 | No | NA | 2 | 0 | 2 | Low-quality | Genome-fragment | 1.23 | AAI-based (medium-confidence) | 0 |
| S9-final-viral-combined_k141_124865\|\|full | 7139 | No | NA | 5 | 0 | 4 | Not-determined | Genome-fragment | NA | NA | 0 |
| S9-final-viral-combined_k141_55759\|\|full | 1214 | No | NA | 2 | 0 | 1 | Not-determined | Genome-fragment | NA | NA | 0 |
| S9-final-viral-combined_k141_12802\|\|full | 1448 | No | NA | 2 | 0 | 2 | Not-determined | Genome-fragment | NA | NA | 0 |
| S2-final-viral-combined_k141_182157\|\|full | 1019 | No | NA | 2 | 0 | 0 | Low-quality | Genome-fragment | 1.7 | AAI-based (medium-confidence) | 0 |
| S9-final-viral-combined_k141_124878\|\|full | 2691 | No | NA | 3 | 0 | 3 | Not-determined | Genome-fragment | NA | NA | 0 |
| S4-final-viral-combined_k141_162829\|\|full | 1019 | No | NA | 2 | 0 | 0 | Low-quality | Genome-fragment | 1.72 | AAI-based (high-confidence) | 0 |
| S9-final-viral-combined_k141_108776\|\|full | 5138 | No | NA | 4 | 0 | 3 | Not-determined | Genome-fragment | NA | NA | 0 |
| S8-final-viral-combined_k141_243842\|\|lt2gene | 1019 | No | NA | 2 | 0 | 0 | Low-quality | Genome-fragment | 2.54 | AAI-based (medium-confidence) | 0 |
| S9-final-viral-combined_k141_23618\|\|full | 2897 | No | NA | 3 | 0 | 3 | Not-determined | Genome-fragment | NA | NA | 0 |
| S9-final-viral-combined_k141_82741\|\|full | 1896 | No | NA | 2 | 0 | 1 | Not-determined | Genome-fragment | NA | NA | 0 |
| S9-final-viral-combined_k141_44547\|\|full | 1782 | No | NA | 3 | 0 | 0 | Not-determined | Genome-fragment | NA | NA | 0 |
| S8-final-viral-combined_k141_17005\|\|lt2gene | 1019 | No | NA | 1 | 1 | 0 | Low-quality | Genome-fragment | 2.37 | AAI-based (high-confidence) | 0 |
| S9-final-viral-combined_k141_87838\|\|full | 1146 | No | NA | 2 | 0 | 0 | Not-determined | Genome-fragment | NA | NA | 0 |
| S8_k141_135678 | 1019 | No | NA | 3 | 2 | 0 | Low-quality | Genome-fragment | 1.94 | AAI-based (high-confidence) | 0 |
| S9-final-viral-combined_k141_23622\|\|full | 2001 | No | NA | 3 | 0 | 1 | Not-determined | Genome-fragment | NA | NA | 0 |
| S11-final-viral-combined_k141_126019\|\|full | 1020 | No | NA | 2 | 0 | 1 | Low-quality | Genome-fragment | 1.6 | AAI-based (medium-confidence) | 0 |
| S9-final-viral-combined_k141_87857\|\|full | 5892 | No | NA | 7 | 0 | 1 | Not-determined | Genome-fragment | NA | NA | 0 |
| S9-final-viral-combined_k141_28805\|\|full | 59739 | Yes | 35476 | 65 | 10 | 11 | Medium-quality | Genome-fragment | 67.17 | HMM-based (lower-bound) | 40.62 |
| S9-final-viral-combined_k141_7880\|\|full | 4975 | No | NA | 6 | 0 | 5 | Not-determined | Genome-fragment | NA | NA | 0 |
| S11-final-viral-combined_k141_104158\|\|lt2gene | 1020 | No | NA | 1 | 1 | 0 | Low-quality | Genome-fragment | 1.49 | HMM-based (lower-bound) | 0 |
| S9-final-viral-combined_k141_33812\|\|full | 1866 | No | NA | 4 | 0 | 1 | Not-determined | Genome-fragment | NA | NA | 0 |
| S9-final-viral-combined_k141_98281\|\|full | 2342 | No | NA | 2 | 0 | 1 | Not-determined | Genome-fragment | NA | NA | 0 |
| S1-final-viral-combined_k141_109457\|\|full | 1020 | No | NA | 4 | 0 | 1 | Low-quality | Genome-fragment | 3.09 | AAI-based (high-confidence) | 0 |
| S9-final-viral-combined_k141_66432\|\|full | 2332 | No | NA | 4 | 0 | 1 | Not-determined | Genome-fragment | NA | NA | 0 |
| S5-final-viral-combined_k141_152915\|\|lt2gene | 1020 | No | NA | 1 | 1 | 0 | Low-quality | Genome-fragment | 2.71 | AAI-based (high-confidence) | 0 |
| S7_k141_95862 | 1020 | No | NA | 1 | 0 | 1 | Low-quality | Genome-fragment | 1.4 | AAI-based (medium-confidence) | 0 |
| S9-final-viral-combined_k141_82840\|\|full | 1405 | No | NA | 3 | 0 | 1 | Not-determined | Genome-fragment | NA | NA | 0 |
| S12_k141_6205 | 1020 | No | NA | 3 | 0 | 0 | Low-quality | Genome-fragment | 2.17 | AAI-based (medium-confidence) | 0 |
| S9-final-viral-combined_k141_66447\|\|full | 1112 | No | NA | 2 | 0 | 0 | Not-determined | Genome-fragment | NA | NA | 0 |
| S12_k141_211237 | 1020 | No | NA | 3 | 0 | 1 | Low-quality | Genome-fragment | 2.66 | AAI-based (high-confidence) | 0 |
| S9-final-viral-combined_k141_93068\|\|full | 1734 | No | NA | 3 | 0 | 0 | Not-determined | Genome-fragment | NA | NA | 0 |
| S9-final-viral-combined_k141_2765\|\|full | 11811 | No | NA | 8 | 0 | 6 | Not-determined | Genome-fragment | NA | NA | 0 |
| S2-final-viral-combined_k141_227005\|\|full | 1020 | No | NA | 2 | 1 | 0 | Low-quality | Genome-fragment | 0.75 | HMM-based (lower-bound) | 0 |
| S2_k141_202342 | 1020 | No | NA | 2 | 0 | 0 | Low-quality | Genome-fragment | 2.43 | AAI-based (high-confidence) | 0 |
| S4_k141_70502 | 1020 | No | NA | 2 | 0 | 0 | Low-quality | Genome-fragment | 0.51 | AAI-based (high-confidence) | 0 |
| S9-final-viral-combined_k141_28888\|\|full | 2973 | No | NA | 3 | 0 | 1 | Not-determined | Genome-fragment | NA | NA | 0 |
| S8-final-viral-combined_k141_119409\|\|lt2gene | 1020 | No | NA | 1 | 1 | 0 | Low-quality | Genome-fragment | 2.2 | AAI-based (high-confidence) | 0 |
| S9-final-viral-combined_k141_33875\|\|full | 1529 | No | NA | 2 | 0 | 1 | Not-determined | Genome-fragment | NA | NA | 0 |
| S9-final-viral-combined_k141_28895\|\|full | 1949 | No | NA | 4 | 0 | 1 | Not-determined | Genome-fragment | NA | NA | 0 |
| S9-final-viral-combined_k141_119682\|\|full | 1425 | No | NA | 3 | 0 | 0 | Not-determined | Genome-fragment | NA | NA | 0 |
| S9-final-viral-combined_k141_82431\|\|full | 1021 | No | NA | 2 | 0 | 1 | Low-quality | Genome-fragment | 1.03 | AAI-based (high-confidence) | 0 |
| S9-final-viral-combined_k141_50775\|\|full | 1502 | No | NA | 3 | 0 | 0 | Not-determined | Genome-fragment | NA | NA | 0 |
| S9-final-viral-combined_k141_125111\|\|full | 3267 | No | NA | 3 | 0 | 2 | Not-determined | Genome-fragment | NA | NA | 0 |
| S9-final-viral-combined_k141_95764\|\|full | 1021 | No | NA | 3 | 0 | 0 | Low-quality | Genome-fragment | 1.98 | AAI-based (medium-confidence) | 0 |
| S11-final-viral-combined_k141_192434\|\|lt2gene | 1021 | No | NA | 1 | 1 | 0 | Low-quality | Genome-fragment | 1.77 | HMM-based (lower-bound) | 0 |
| S1-final-viral-combined_k141_225030\|\|lt2gene | 1021 | No | NA | 1 | 1 | 0 | Low-quality | Genome-fragment | 1.72 | HMM-based (lower-bound) | 0 |
| S9-final-viral-combined_k141_44647\|\|full | 2059 | No | NA | 5 | 0 | 0 | Not-determined | Genome-fragment | NA | NA | 0 |
| S1_k141_112474 | 1021 | No | NA | 1 | 0 | 1 | Low-quality | Genome-fragment | 0.32 | AAI-based (medium-confidence) | 0 |
| S9-final-viral-combined_k141_88033\|\|full | 3145 | No | NA | 3 | 0 | 2 | Not-determined | Genome-fragment | NA | NA | 0 |
| S9-final-viral-combined_k141_50813\|\|full | 1040 | No | NA | 2 | 0 | 0 | Not-determined | Genome-fragment | NA | NA | 0 |
| S3-final-viral-combined_k141_87050\|\|full | 1021 | No | NA | 2 | 0 | 0 | Low-quality | Genome-fragment | 1.28 | AAI-based (medium-confidence) | 0 |
| S9-final-viral-combined_k141_119698\|\|full | 4567 | No | NA | 6 | 0 | 2 | Not-determined | Genome-fragment | NA | NA | 0 |
| S9-final-viral-combined_k141_39711\|\|full | 1126 | No | NA | 2 | 0 | 0 | Not-determined | Genome-fragment | NA | NA | 0 |
| S9-final-viral-combined_k141_114104\|\|full | 2300 | No | NA | 4 | 0 | 2 | Not-determined | Genome-fragment | NA | NA | 0 |
| S9-final-viral-combined_k141_119729\|\|full | 3328 | No | NA | 2 | 0 | 2 | Not-determined | Genome-fragment | NA | NA | 0 |
| S9-final-viral-combined_k141_83057\|\|full | 5541 | No | NA | 6 | 0 | 5 | Not-determined | Genome-fragment | NA | NA | 0 |
| S9-final-viral-combined_k141_71647\|\|full | 22390 | No | NA | 46 | 11 | 2 | Medium-quality | Genome-fragment | 55.46 | AAI-based (medium-confidence) | 0 |
| S9-final-viral-combined_k141_18470\|\|full | 1224 | No | NA | 3 | 0 | 1 | Not-determined | Genome-fragment | NA | NA | 0 |
| S9-final-viral-combined_k141_8026\|\|full | 2474 | No | NA | 2 | 0 | 0 | Not-determined | Genome-fragment | NA | NA | 0 |
| S9-final-viral-combined_k141_50838\|\|full | 6286 | No | NA | 8 | 0 | 0 | Not-determined | Genome-fragment | NA | NA | 0 |
| S9-final-viral-combined_k141_8034\|\|full | 1995 | No | NA | 4 | 0 | 1 | Not-determined | Genome-fragment | NA | NA | 0 |
| S9-final-viral-combined_k141_88107\|\|full | 2337 | No | NA | 3 | 0 | 2 | Not-determined | Genome-fragment | NA | NA | 0 |
| S17-final-viral-combined_k141_163289\|\|full | 1021 | No | NA | 4 | 0 | 0 | Low-quality | Genome-fragment | 2.01 | AAI-based (medium-confidence) | 0 |
| S9-final-viral-combined_k141_66624\|\|full | 1173 | No | NA | 2 | 0 | 0 | Not-determined | Genome-fragment | NA | NA | 0 |
| S9-final-viral-combined_k141_18502\|\|full | 1123 | No | NA | 2 | 0 | 1 | Not-determined | Genome-fragment | NA | NA | 0 |
| S9-final-viral-combined_k141_88130\|\|full | 1685 | No | NA | 4 | 0 | 1 | Not-determined | Genome-fragment | NA | NA | 0 |
| S9-final-viral-combined_k141_18503\|\|full | 1386 | No | NA | 3 | 0 | 0 | Not-determined | Genome-fragment | NA | NA | 0 |
| S12-final-viral-combined_k141_10219\|\|full | 1021 | No | NA | 3 | 0 | 0 | Low-quality | Genome-fragment | 2.41 | AAI-based (high-confidence) | 0 |
| S9-final-viral-combined_k141_66639\|\|full | 4820 | No | NA | 4 | 0 | 1 | Not-determined | Genome-fragment | NA | NA | 0 |
| S12_k141_10074 | 1021 | No | NA | 1 | 0 | 0 | Low-quality | Genome-fragment | 2.6 | AAI-based (high-confidence) | 0 |
| S2_k141_116060 | 1021 | No | NA | 2 | 2 | 0 | Low-quality | Genome-fragment | 2.6 | AAI-based (high-confidence) | 0 |
| S9-final-viral-combined_k141_18555\|\|full | 3614 | No | NA | 4 | 0 | 3 | Not-determined | Genome-fragment | NA | NA | 0 |
| S9-final-viral-combined_k141_29025\|\|full | 1237 | No | NA | 2 | 0 | 1 | Not-determined | Genome-fragment | NA | NA | 0 |
| S9-final-viral-combined_k141_56119\|\|full | 2288 | No | NA | 3 | 0 | 2 | Not-determined | Genome-fragment | NA | NA | 0 |
| S4-final-viral-combined_k141_45340\|\|full | 1021 | No | NA | 2 | 0 | 0 | Low-quality | Genome-fragment | 0.7 | AAI-based (high-confidence) | 0 |
| S8-final-viral-combined_k141_133333\|\|full | 1021 | No | NA | 4 | 1 | 0 | Low-quality | Genome-fragment | 0.66 | HMM-based (lower-bound) | 0 |
| S9-final-viral-combined_k141_83158\|\|full | 2228 | No | NA | 2 | 0 | 0 | Not-determined | Genome-fragment | NA | NA | 0 |
| S9-final-viral-combined_k141_18579\|\|full | 1519 | No | NA | 4 | 0 | 1 | Not-determined | Genome-fragment | NA | NA | 0 |
| S8_k141_213008 | 1021 | No | NA | 3 | 0 | 0 | Low-quality | Genome-fragment | 4.89 | AAI-based (high-confidence) | 0 |
| S9-final-viral-combined_k141_18601\|\|full | 1654 | No | NA | 2 | 0 | 1 | Not-determined | Genome-fragment | NA | NA | 0 |
| S9-final-viral-combined_k141_33862\|\|lt2gene | 1022 | No | NA | 2 | 1 | 0 | Low-quality | Genome-fragment | 2.24 | AAI-based (high-confidence) | 0 |
| S9-final-viral-combined_k141_76914\|\|full | 2127 | No | NA | 3 | 0 | 0 | Not-determined | Genome-fragment | NA | NA | 0 |
| S9-final-viral-combined_k141_23827\|\|full | 2849 | No | NA | 3 | 0 | 1 | Not-determined | Genome-fragment | NA | NA | 0 |
| S9-final-viral-combined_k141_2960\|\|full | 1872 | No | NA | 2 | 0 | 1 | Not-determined | Genome-fragment | NA | NA | 0 |
| S9-final-viral-combined_k141_8095\|\|full | 5415 | No | NA | 4 | 0 | 1 | Not-determined | Genome-fragment | NA | NA | 0 |
| S9-final-viral-combined_k141_119905\|\|full | 2428 | No | NA | 3 | 0 | 3 | Not-determined | Genome-fragment | NA | NA | 0 |

| | | | | | | | | | | | |
|---|---|---|---|---|---|---|---|---|---|---|---|
| S9-final-viral-combined_k141_83225\|\|full | 2244 | No | NA | 3 | 0 | 1 | Not-determined | Genome-fragment | NA | NA | 0 |
| S9-final-viral-combined_k141_83231\|\|full | 5975 | No | NA | 10 | 0 | 6 | Not-determined | Genome-fragment | NA | NA | 0 |
| S1-final-viral-combined_k141_149430\|\|full | 1022 | No | NA | 3 | 0 | 1 | Low-quality | Genome-fragment | 2.43 | AAI-based (high-confidence) | 0 |
| S9-final-viral-combined_k141_44815\|\|full | 1896 | No | NA | 2 | 0 | 2 | Not-determined | Genome-fragment | NA | NA | 0 |
| S9-final-viral-combined_k141_119941\|\|full | 1163 | No | NA | 2 | 0 | 0 | Not-determined | Genome-fragment | NA | NA | 0 |
| S9-final-viral-combined_k141_76946\|\|full | 2742 | No | NA | 3 | 0 | 2 | Not-determined | Genome-fragment | NA | NA | 0 |
| S9-final-viral-combined_k141_93232\|\|full | 50669 | Yes | 44439 | 62 | 33 | 6 | High-quality | High-quality | 100 | AAI-based (high-confidence) | 12.3 |
| S9-final-viral-combined_k141_29073\|\|full | 3187 | No | NA | 3 | 0 | 3 | Not-determined | Genome-fragment | NA | NA | 0 |
| S1-final-viral-combined_k141_241242\|\|full | 1022 | No | NA | 2 | 0 | 0 | Low-quality | Genome-fragment | 1.02 | AAI-based (high-confidence) | 0 |
| S1-final-viral-combined_k141_175645\|\|lt2gene | 1022 | No | NA | 1 | 1 | 0 | Low-quality | Genome-fragment | 2.78 | AAI-based (high-confidence) | 0 |
| S9-final-viral-combined_k141_29084\|\|full | 1338 | No | NA | 2 | 0 | 1 | Not-determined | Genome-fragment | NA | NA | 0 |
| S9-final-viral-combined_k141_83270\|\|full | 1178 | No | NA | 1 | 0 | 0 | Not-determined | Genome-fragment | NA | NA | 0 |
| S9-final-viral-combined_k141_119966\|\|full | 1066 | No | NA | 2 | 0 | 2 | Not-determined | Genome-fragment | NA | NA | 0 |
| S9-final-viral-combined_k141_12969\|\|full | 6441 | No | NA | 5 | 0 | 0 | Not-determined | Genome-fragment | NA | NA | 0 |
| S1_k141_160136 | 1022 | No | NA | 2 | 0 | 0 | Low-quality | Genome-fragment | 1.71 | AAI-based (medium-confidence) | 0 |
| S9-final-viral-combined_k141_18698\|\|full | 1753 | No | NA | 2 | 0 | 0 | Not-determined | Genome-fragment | NA | NA | 0 |
| S9-final-viral-combined_k141_98450\|\|full | 1192 | No | NA | 3 | 0 | 2 | Not-determined | Genome-fragment | NA | NA | 0 |
| S1_k141_391403 | 1022 | No | NA | 1 | 0 | 0 | Low-quality | Genome-fragment | 1.25 | AAI-based (high-confidence) | 0 |
| S5-final-viral-combined_k141_56158\|\|full | 1022 | No | NA | 3 | 3 | 0 | Low-quality | Genome-fragment | 3.2 | AAI-based (high-confidence) | 0 |
| S5-final-viral-combined_k141_5641\|\|full | 1022 | No | NA | 3 | 0 | 0 | Low-quality | Genome-fragment | 3 | AAI-based (high-confidence) | 0 |
| S9-final-viral-combined_k141_18726\|\|full | 2122 | No | NA | 4 | 0 | 1 | Not-determined | Genome-fragment | NA | NA | 0 |
| S5-final-viral-combined_k141_67030\|\|full | 1022 | No | NA | 3 | 2 | 0 | Low-quality | Genome-fragment | 3.03 | AAI-based (high-confidence) | 0 |
| S7-final-viral-combined_k141_151242\|\|full | 1022 | No | NA | 2 | 0 | 1 | Low-quality | Genome-fragment | 2.23 | AAI-based (medium-confidence) | 0 |
| S17-k141_11498 | 1022 | No | NA | 2 | 0 | 0 | Low-quality | Genome-fragment | 2.29 | AAI-based (high-confidence) | 0 |
| S9-final-viral-combined_k141_18730\|\|full | 1529 | No | NA | 2 | 0 | 1 | Not-determined | Genome-fragment | NA | NA | 0 |
| S17-k141_196673 | 1022 | No | NA | 2 | 1 | 0 | Low-quality | Genome-fragment | 16.44 | AAI-based (high-confidence) | 0 |
| S12-final-viral-combined_k141_89262\|\|full | 1022 | No | NA | 1 | 1 | 0 | Low-quality | Genome-fragment | 2.04 | AAI-based (medium-confidence) | 0 |
| S9-final-viral-combined_k141_18749\|\|full | 3719 | No | NA | 3 | 0 | 0 | Not-determined | Genome-fragment | NA | NA | 0 |
| S9-final-viral-combined_k141_50940\|\|full | 3017 | No | NA | 4 | 0 | 0 | Not-determined | Genome-fragment | NA | NA | 0 |
| S12-final-viral-combined_k141_279240\|\|full | 1022 | No | NA | 2 | 0 | 0 | Low-quality | Genome-fragment | 0.74 | AAI-based (medium-confidence) | 0 |
| S9-final-viral-combined_k141_114343\|\|full | 2566 | No | NA | 3 | 0 | 2 | Not-determined | Genome-fragment | NA | NA | 0 |
| S12-final-viral-combined_k141_164717\|\|full | 1022 | No | NA | 2 | 1 | 0 | Low-quality | Genome-fragment | 1.41 | AAI-based (high-confidence) | 0 |
| S9-final-viral-combined_k141_114356\|\|full | 1453 | No | NA | 2 | 0 | 2 | Not-determined | Genome-fragment | NA | NA | 0 |
| S12_k141_387881 | 1022 | No | NA | 1 | 1 | 0 | Low-quality | Genome-fragment | 2.79 | AAI-based (high-confidence) | 0 |
| S9-final-viral-combined_k141_108985\|\|full | 1433 | No | NA | 3 | 0 | 2 | Not-determined | Genome-fragment | NA | NA | 0 |
| S9-final-viral-combined_k141_61619\|\|full | 2655 | No | NA | 4 | 0 | 0 | Not-determined | Genome-fragment | NA | NA | 0 |
| S9-final-viral-combined_k141_93347\|\|full | 1293 | No | NA | 3 | 0 | 0 | Not-determined | Genome-fragment | NA | NA | 0 |
| S9-final-viral-combined_k141_83368\|\|full | 2461 | No | NA | 3 | 0 | 3 | Not-determined | Genome-fragment | NA | NA | 0 |
| S2-final-viral-combined_k141_72501\|\|lt2gene | 1022 | No | NA | 1 | 1 | 0 | Low-quality | Genome-fragment | 2.24 | AAI-based (high-confidence) | 0 |
| S2_k141_84139 | 1022 | No | NA | 4 | 0 | 0 | Low-quality | Genome-fragment | 1.69 | AAI-based (medium-confidence) | 0 |
| S8-final-viral-combined_k141_249966\|\|full | 1022 | No | NA | 3 | 0 | 0 | Low-quality | Genome-fragment | 2.23 | AAI-based (high-confidence) | 0 |
| S11-final-viral-combined_k141_255435\|\|lt2gene | 1023 | No | NA | 1 | 1 | 0 | Low-quality | Genome-fragment | 1.44 | HMM-based (lower-bound) | 0 |
| S1_k141_237366 | 1023 | No | NA | 3 | 1 | 0 | Low-quality | Genome-fragment | 0.94 | AAI-based (high-confidence) | 0 |
| S9-final-viral-combined_k141_120040\|\|full | 1487 | No | NA | 2 | 0 | 1 | Not-determined | Genome-fragment | NA | NA | 0 |
| S1_k141_129549 | 1023 | No | NA | 1 | 0 | 0 | Low-quality | Genome-fragment | 0.66 | AAI-based (high-confidence) | 0 |
| S1_k141_195477 | 1023 | No | NA | 1 | 0 | 1 | Low-quality | Genome-fragment | 0.31 | AAI-based (medium-confidence) | 0 |
| S5-final-viral-combined_k141_39408\|\|full | 1023 | No | NA | 3 | 1 | 0 | Low-quality | Genome-fragment | 2.14 | AAI-based (high-confidence) | 0 |
| S9-final-viral-combined_k141_98520\|\|full | 1402 | No | NA | 3 | 0 | 0 | Not-determined | Genome-fragment | NA | NA | 0 |
| S5_k141_238 | 1023 | No | NA | 1 | 0 | 1 | Low-quality | Genome-fragment | 0.53 | AAI-based (medium-confidence) | 0 |
| S7-final-viral-combined_k141_88405\|\|full | 1023 | No | NA | 2 | 1 | 0 | Low-quality | Genome-fragment | 2.21 | AAI-based (high-confidence) | 0 |
| S17-final-viral-combined_k141_188638\|\|full | 1023 | No | NA | 4 | 0 | 0 | Low-quality | Genome-fragment | 2.41 | AAI-based (high-confidence) | 0 |
| S9-final-viral-combined_k141_24056\|\|full | 3037 | No | NA | 3 | 0 | 1 | Not-determined | Genome-fragment | NA | NA | 0 |
| S9-final-viral-combined_k141_13151\|\|full | 1249 | No | NA | 3 | 0 | 1 | Not-determined | Genome-fragment | NA | NA | 0 |
| S9-final-viral-combined_k141_71908\|\|full | 3486 | No | NA | 5 | 0 | 5 | Not-determined | Genome-fragment | NA | NA | 0 |
| S9-final-viral-combined_k141_8347\|\|full | 1115 | No | NA | 3 | 0 | 1 | Not-determined | Genome-fragment | NA | NA | 0 |
| S17-final-viral-combined_k141_32009\|\|full | 1023 | No | NA | 3 | 1 | 0 | Low-quality | Genome-fragment | 2.75 | AAI-based (medium-confidence) | 0 |
| S17-final-viral-combined_k141_99708\|\|full | 1023 | No | NA | 2 | 0 | 0 | Low-quality | Genome-fragment | 1.06 | AAI-based (medium-confidence) | 0 |
| S9-final-viral-combined_k141_45052\|\|full | 5219 | No | NA | 6 | 0 | 5 | Not-determined | Genome-fragment | NA | NA | 0 |
| S2_k141_43142 | 1023 | No | NA | 2 | 0 | 0 | Low-quality | Genome-fragment | 1.59 | AAI-based (medium-confidence) | 0 |
| S4-final-viral-combined_k141_134748\|\|full | 1023 | No | NA | 3 | 1 | 0 | Low-quality | Genome-fragment | 1.66 | AAI-based (high-confidence) | 0 |
| S9-final-viral-combined_k141_13250\|\|full | 2950 | No | NA | 3 | 0 | 1 | Not-determined | Genome-fragment | NA | NA | 0 |
| S9-final-viral-combined_k141_18893\|\|full | 2593 | No | NA | 3 | 0 | 1 | Not-determined | Genome-fragment | NA | NA | 0 |
| S9-final-viral-combined_k141_39921\|\|full | 1353 | No | NA | 2 | 0 | 1 | Not-determined | Genome-fragment | NA | NA | 0 |
| S9-final-viral-combined_k141_61670\|\|full | 1314 | No | NA | 3 | 0 | 2 | Not-determined | Genome-fragment | NA | NA | 0 |
| S9-final-viral-combined_k141_13280\|\|full | 1198 | No | NA | 2 | 0 | 0 | Not-determined | Genome-fragment | NA | NA | 0 |
| S9-final-viral-combined_k141_77206\|\|full | 2522 | No | NA | 2 | 0 | 0 | Not-determined | Genome-fragment | NA | NA | 0 |
| S9-final-viral-combined_k141_45088\|\|full | 1498 | No | NA | 2 | 0 | 1 | Not-determined | Genome-fragment | NA | NA | 0 |
| S9-final-viral-combined_k141_98595\|\|full | 2015 | No | NA | 2 | 0 | 2 | Not-determined | Genome-fragment | NA | NA | 0 |
| S4-final-viral-combined_k141_44350\|\|full | 1023 | No | NA | 3 | 0 | 0 | Low-quality | Genome-fragment | 2.03 | AAI-based (high-confidence) | 0 |
| S9-final-viral-combined_k141_18947\|\|full | 1757 | No | NA | 2 | 0 | 0 | Not-determined | Genome-fragment | NA | NA | 0 |
| S9-final-viral-combined_k141_109090\|\|full | 3918 | No | NA | 4 | 0 | 4 | Not-determined | Genome-fragment | NA | NA | 0 |
| S9-final-viral-combined_k141_13308\|\|full | 1729 | No | NA | 3 | 0 | 0 | Not-determined | Genome-fragment | NA | NA | 0 |
| S9-final-viral-combined_k141_45113\|\|full | 1362 | No | NA | 2 | 0 | 0 | Not-determined | Genome-fragment | NA | NA | 0 |
| S9-final-viral-combined_k141_8440\|\|full | 7592 | No | NA | 6 | 0 | 1 | Not-determined | Genome-fragment | NA | NA | 0 |
| S9-final-viral-combined_k141_13325\|\|full | 3334 | No | NA | 4 | 0 | 3 | Not-determined | Genome-fragment | NA | NA | 0 |
| S4-final-viral-combined_k141_156839\|\|full | 1023 | No | NA | 2 | 0 | 2 | Low-quality | Genome-fragment | 0.56 | AAI-based (medium-confidence) | 0 |
| S4_k141_114656 | 1023 | No | NA | 3 | 2 | 0 | Low-quality | Genome-fragment | 1.46 | HMM-based (lower-bound) | 0 |
| S6-final-viral-combined_k141_82446\|\|full | 1023 | No | NA | 3 | 0 | 0 | Low-quality | Genome-fragment | 0.95 | AAI-based (high-confidence) | 0 |
| S8-final-viral-combined_k141_20976\|\|full | 1023 | No | NA | 3 | 0 | 0 | Low-quality | Genome-fragment | 2.3 | AAI-based (medium-confidence) | 0 |
| S8_k141_173869 | 1023 | No | NA | 1 | 0 | 0 | Low-quality | Genome-fragment | 2.01 | AAI-based (high-confidence) | 0 |
| S9-final-viral-combined_k141_88795\|\|full | 1024 | No | NA | 4 | 1 | 0 | Low-quality | Genome-fragment | 1.27 | HMM-based (lower-bound) | 0 |
| S9_k141_53384 | 1024 | No | NA | 3 | 2 | 0 | Low-quality | Genome-fragment | 2.66 | AAI-based (high-confidence) | 0 |
| S11-final-viral-combined_k141_294408\|\|full | 1024 | No | NA | 3 | 1 | 0 | Low-quality | Genome-fragment | 1.04 | AAI-based (medium-confidence) | 0 |
| S11_k141_225185 | 1024 | No | NA | 1 | 0 | 1 | Low-quality | Genome-fragment | 0.44 | AAI-based (medium-confidence) | 0 |
| S1-final-viral-combined_k141_247126\|\|full | 1024 | No | NA | 3 | 1 | 0 | Low-quality | Genome-fragment | 1.73 | AAI-based (medium-confidence) | 0 |
| S9-final-viral-combined_k141_51131\|\|full | 1876 | No | NA | 3 | 0 | 1 | Not-determined | Genome-fragment | NA | NA | 0 |
| S3-final-viral-combined_k141_112598\|\|lt2gene | 1024 | No | NA | 1 | 1 | 0 | Low-quality | Genome-fragment | 2.45 | AAI-based (high-confidence) | 0 |
| S9-final-viral-combined_k141_125354\|\|full | 1849 | No | NA | 3 | 0 | 1 | Not-determined | Genome-fragment | NA | NA | 0 |
| S3-final-viral-combined_k141_136988\|\|lt2gene | 1024 | No | NA | 2 | 1 | 0 | Low-quality | Genome-fragment | 2.6 | AAI-based (medium-confidence) | 0 |
| S5-final-viral-combined_k141_4356\|\|full | 1024 | No | NA | 3 | 0 | 0 | Low-quality | Genome-fragment | 1.76 | AAI-based (medium-confidence) | 0 |
| S9-final-viral-combined_k141_77343\|\|full | 1293 | No | NA | 4 | 0 | 2 | Not-determined | Genome-fragment | NA | NA | 0 |
| S9-final-viral-combined_k141_71973\|\|full | 2954 | No | NA | 4 | 0 | 2 | Not-determined | Genome-fragment | NA | NA | 0 |
| S5-final-viral-combined_k141_92790\|\|full | 1024 | No | NA | 2 | 1 | 0 | Low-quality | Genome-fragment | 2.2 | AAI-based (high-confidence) | 0 |
| S9-final-viral-combined_k141_45238\|\|full | 2265 | No | NA | 3 | 0 | 2 | Not-determined | Genome-fragment | NA | NA | 0 |
| S5-final-viral-combined_k141_40719\|\|full | 1024 | No | NA | 2 | 0 | 0 | Low-quality | Genome-fragment | 0.99 | AAI-based (high-confidence) | 0 |
| S9-final-viral-combined_k141_77355\|\|full | 5721 | No | NA | 10 | 0 | 0 | Not-determined | Genome-fragment | NA | NA | 0 |
| S9-final-viral-combined_k141_8506\|\|full | 2385 | No | NA | 2 | 0 | 1 | Not-determined | Genome-fragment | NA | NA | 0 |
| S9-final-viral-combined_k141_66732\|\|full | 1491 | No | NA | 3 | 0 | 2 | Not-determined | Genome-fragment | NA | NA | 0 |
| S5-final-viral-combined_k141_69336\|\|lt2gene | 1024 | No | NA | 1 | 1 | 0 | Low-quality | Genome-fragment | 0.63 | HMM-based (lower-bound) | 0 |
| S17-final-viral-combined_k141_207471\|\|full | 1024 | No | NA | 2 | 2 | 0 | Low-quality | Genome-fragment | 1.81 | HMM-based (lower-bound) | 0 |
| S9-final-viral-combined_k141_8514\|\|full | 2468 | No | NA | 4 | 0 | 3 | Not-determined | Genome-fragment | NA | NA | 0 |
| S17-final-viral-combined_k141_125690\|\|lt2gene | 1024 | No | NA | 2 | 1 | 0 | Low-quality | Genome-fragment | 2.33 | AAI-based (high-confidence) | 0 |
| S9-final-viral-combined_k141_56467\|\|full | 2131 | No | NA | 5 | 0 | 0 | Not-determined | Genome-fragment | NA | NA | 0 |
| S17-k141_111422 | 1024 | No | NA | 1 | 0 | 0 | Low-quality | Genome-fragment | 3.52 | AAI-based (high-confidence) | 0 |
| S12-final-viral-combined_k141_149637\|\|lt2gene | 1024 | No | NA | 2 | 0 | 2 | Low-quality | Genome-fragment | 1.1 | AAI-based (medium-confidence) | 0 |
| S9-final-viral-combined_k141_45266\|\|full | 2688 | No | NA | 2 | 0 | 2 | Not-determined | Genome-fragment | NA | NA | 0 |
| S9-final-viral-combined_k141_13390\|\|full | 5225 | No | NA | 4 | 0 | 4 | Not-determined | Genome-fragment | NA | NA | 0 |
| S9-final-viral-combined_k141_66758\|\|full | 2204 | No | NA | 3 | 0 | 2 | Not-determined | Genome-fragment | NA | NA | 0 |
| S12_k141_374763 | 1024 | No | NA | 2 | 1 | 0 | Low-quality | Genome-fragment | 3.03 | AAI-based (high-confidence) | 0 |
| S9-final-viral-combined_k141_61819\|\|full | 2252 | No | NA | 3 | 0 | 0 | Not-determined | Genome-fragment | NA | NA | 0 |
| S4-final-viral-combined_k141_152675\|\|full | 1024 | No | NA | 3 | 0 | 1 | Low-quality | Genome-fragment | 2.06 | AAI-based (high-confidence) | 0 |
| S4_k141_50719 | 1024 | No | NA | 3 | 0 | 0 | Low-quality | Genome-fragment | 8.27 | AAI-based (medium-confidence) | 0 |
| S8-final-viral-combined_k141_53162\|\|full | 1024 | No | NA | 2 | 0 | 1 | Low-quality | Genome-fragment | 2.18 | AAI-based (medium-confidence) | 0 |
| S8_k141_82387 | 1024 | No | NA | 1 | 0 | 1 | Low-quality | Genome-fragment | 0.77 | AAI-based (high-confidence) | 0 |
| S9-final-viral-combined_k141_13432\|\|full | 3168 | No | NA | 6 | 0 | 0 | Not-determined | Genome-fragment | NA | NA | 0 |
| S9-final-viral-combined_k141_93538\|\|full | 1091 | No | NA | 3 | 0 | 1 | Not-determined | Genome-fragment | NA | NA | 0 |
| S9-final-viral-combined_k141_77412\|\|full | 1525 | No | NA | 3 | 0 | 0 | Not-determined | Genome-fragment | NA | NA | 0 |
| S9-final-viral-combined_k141_99856\|\|full | 1025 | No | NA | 4 | 3 | 0 | Low-quality | Genome-fragment | 2.22 | AAI-based (medium-confidence) | 0 |
| S9-final-viral-combined_k141_47584\|\|full | 1025 | No | NA | 2 | 0 | 1 | Low-quality | Genome-fragment | 0.79 | AAI-based (medium-confidence) | 0 |
| S9-final-viral-combined_k141_19081\|\|full | 2596 | No | NA | 3 | 0 | 1 | Not-determined | Genome-fragment | NA | NA | 0 |
| S9-final-viral-combined_k141_66822\|\|full | 2123 | No | NA | 3 | 0 | 1 | Not-determined | Genome-fragment | NA | NA | 0 |
| S11-final-viral-combined_k141_235062\|\|full | 1025 | No | NA | 3 | 0 | 0 | Low-quality | Genome-fragment | 1.79 | AAI-based (medium-confidence) | 0 |
| S9-final-viral-combined_k141_29350\|\|full | 1174 | No | NA | 2 | 0 | 2 | Not-determined | Genome-fragment | NA | NA | 0 |
| S9-final-viral-combined_k141_93553\|\|full | 3376 | No | NA | 3 | 0 | 3 | Not-determined | Genome-fragment | NA | NA | 0 |
| S11-final-viral-combined_k141_180500\|\|full | 1025 | No | NA | 2 | 0 | 0 | Low-quality | Genome-fragment | 2.41 | AAI-based (high-confidence) | 0 |
| S9-final-viral-combined_k141_93556\|\|full | 1847 | No | NA | 2 | 0 | 2 | Not-determined | Genome-fragment | NA | NA | 0 |
| S9-final-viral-combined_k141_24316\|\|full | 1096 | No | NA | 3 | 0 | 0 | Not-determined | Genome-fragment | NA | NA | 0 |
| S9-final-viral-combined_k141_3210\|\|full | 2124 | No | NA | 3 | 0 | 2 | Not-determined | Genome-fragment | NA | NA | 0 |
| S1-final-viral-combined_k141_162810\|\|full | 1025 | No | NA | 2 | 1 | 0 | Low-quality | Genome-fragment | 1.9 | AAI-based (high-confidence) | 0 |
| S3-final-viral-combined_k141_35604\|\|full | 1025 | No | NA | 2 | 0 | 0 | Low-quality | Genome-fragment | 1.64 | AAI-based (high-confidence) | 0 |
| S9-final-viral-combined_k141_29363\|\|full | 1617 | No | NA | 4 | 0 | 1 | Not-determined | Genome-fragment | NA | NA | 0 |
| S9-final-viral-combined_k141_56567\|\|full | 2455 | No | NA | 3 | 0 | 2 | Not-determined | Genome-fragment | NA | NA | 0 |
| S9-final-viral-combined_k141_3232\|\|full | 1003 | No | NA | 3 | 0 | 0 | Not-determined | Genome-fragment | NA | NA | 0 |
| S9-final-viral-combined_k141_77472\|\|full | 2144 | No | NA | 4 | 0 | 3 | Not-determined | Genome-fragment | NA | NA | 0 |
| S5-final-viral-combined_k141_78822\|\|lt2gene | 1025 | No | NA | 1 | 1 | 0 | Low-quality | Genome-fragment | 2.16 | AAI-based (high-confidence) | 0 |
| S9-final-viral-combined_k141_3241\|\|full | 1726 | No | NA | 3 | 0 | 0 | Not-determined | Genome-fragment | NA | NA | 0 |
| S9-final-viral-combined_k141_109158\|\|full | 1410 | No | NA | 3 | 0 | 2 | Not-determined | Genome-fragment | NA | NA | 0 |
| S9-final-viral-combined_k141_98830\|\|full | 8992 | No | NA | 5 | 0 | 0 | Not-determined | Genome-fragment | NA | NA | 0 |
| S7-final-viral-combined_k141_196100\|\|full | 1025 | No | NA | 3 | 1 | 0 | Low-quality | Genome-fragment | 1.98 | AAI-based (high-confidence) | 0 |
| S9-final-viral-combined_k141_83693\|\|full | 1497 | No | NA | 3 | 0 | 0 | Not-determined | Genome-fragment | NA | NA | 0 |
| S9-final-viral-combined_k141_66896\|\|full | 1651 | No | NA | 3 | 0 | 0 | Not-determined | Genome-fragment | NA | NA | 0 |
| S9-final-viral-combined_k141_77512\|\|full | 2054 | No | NA | 3 | 0 | 2 | Not-determined | Genome-fragment | NA | NA | 0 |
| S17-final-viral-combined_k141_228466\|\|full | 1025 | No | NA | 3 | 0 | 0 | Low-quality | Genome-fragment | 1.41 | AAI-based (medium-confidence) | 0 |
| S9-final-viral-combined_k141_34618\|\|full | 1260 | No | NA | 2 | 0 | 1 | Not-determined | Genome-fragment | NA | NA | 0 |
| S9-final-viral-combined_k141_40227\|\|full | 14586 | No | NA | 18 | 0 | 2 | Not-determined | Genome-fragment | NA | NA | 0 |
| S9-final-viral-combined_k141_114849\|\|full | 1294 | No | NA | 2 | 0 | 0 | Not-determined | Genome-fragment | NA | NA | 0 |
| S9-final-viral-combined_k141_45423\|\|full | 2337 | No | NA | 2 | 0 | 2 | Not-determined | Genome-fragment | NA | NA | 0 |
| S12-final-viral-combined_k141_93996\|\|full | 1025 | No | NA | 2 | 0 | 1 | Low-quality | Genome-fragment | 0.66 | AAI-based (medium-confidence) | 0 |
| S12-final-viral-combined_k141_65288\|\|full | 1025 | No | NA | 3 | 1 | 0 | Low-quality | Genome-fragment | 2.79 | AAI-based (medium-confidence) | 0 |
| S12_k141_344875 | 1025 | No | NA | 2 | 0 | 0 | Low-quality | Genome-fragment | 0.2 | AAI-based (medium-confidence) | 0 |
| S2-final-viral-combined_k141_127476\|\|full | 1025 | No | NA | 3 | 0 | 2 | Low-quality | Genome-fragment | 0.48 | AAI-based (medium-confidence) | 0 |
| S9-final-viral-combined_k141_40235\|\|full | 2632 | No | NA | 6 | 0 | 2 | Not-determined | Genome-fragment | NA | NA | 0 |
| S9-final-viral-combined_k141_40244\|\|full | 1779 | No | NA | 2 | 0 | 0 | Not-determined | Genome-fragment | NA | NA | 0 |
| S4_k141_24006 | 1025 | No | NA | 2 | 1 | 0 | Low-quality | Genome-fragment | 0.51 | HMM-based (lower-bound) | 0 |
| S9-final-viral-combined_k141_114890\|\|full | 1523 | No | NA | 2 | 0 | 1 | Not-determined | Genome-fragment | NA | NA | 0 |
| S9-final-viral-combined_k141_56665\|\|full | 1414 | No | NA | 3 | 0 | 2 | Not-determined | Genome-fragment | NA | NA | 0 |
| S4_k141_143268 | 1025 | No | NA | 2 | 0 | 0 | Low-quality | Genome-fragment | 1.85 | AAI-based (high-confidence) | 0 |
| S9-final-viral-combined_k141_72175\|\|full | 3340 | No | NA | 3 | 0 | 2 | Not-determined | Genome-fragment | NA | NA | 0 |
| S4_k141_53134 | 1025 | No | NA | 2 | 0 | 1 | Low-quality | Genome-fragment | 2.34 | AAI-based (high-confidence) | 0 |
| S8-final-viral-combined_k141_26172\|\|full | 1025 | No | NA | 3 | 2 | 0 | Low-quality | Genome-fragment | 2.73 | AAI-based (medium-confidence) | 0 |
| S8-final-viral-combined_k141_189837\|\|full | 1025 | No | NA | 3 | 0 | 0 | Low-quality | Genome-fragment | 0.67 | AAI-based (high-confidence) | 0 |
| S9-final-viral-combined_k141_66940\|\|full | 4728 | No | NA | 5 | 0 | 4 | Not-determined | Genome-fragment | NA | NA | 0 |
| S9-final-viral-combined_k141_56714\|\|full | 1329 | No | NA | 3 | 0 | 1 | Not-determined | Genome-fragment | NA | NA | 0 |
| S8_k141_204889 | 1025 | No | NA | 2 | 1 | 0 | Low-quality | Genome-fragment | 1.82 | AAI-based (high-confidence) | 0 |
| S9-final-viral-combined_k141_8611\|\|full | 5725 | No | NA | 10 | 0 | 5 | Not-determined | Genome-fragment | NA | NA | 0 |
| S9-final-viral-combined_k141_56720\|\|full | 2613 | No | NA | 5 | 0 | 3 | Not-determined | Genome-fragment | NA | NA | 0 |
| S11_k141_112816 | 1026 | No | NA | 3 | 0 | 0 | Low-quality | Genome-fragment | 2.54 | AAI-based (medium-confidence) | 0 |
| S9-final-viral-combined_k141_51266\|\|full | 2680 | No | NA | 4 | 0 | 1 | Not-determined | Genome-fragment | NA | NA | 0 |
| S1-final-viral-combined_k141_22904\|\|full | 1026 | No | NA | 2 | 2 | 0 | Low-quality | Genome-fragment | 1.75 | AAI-based (high-confidence) | 0 |
| S1-final-viral-combined_k141_72382\|\|full | 1026 | No | NA | 3 | 0 | 0 | Low-quality | Genome-fragment | 0.8 | AAI-based (medium-confidence) | 0 |
| S9-final-viral-combined_k141_8653\|\|full | 1076 | No | NA | 3 | 0 | 1 | Not-determined | Genome-fragment | NA | NA | 0 |
| S1-final-viral-combined_k141_393408\|\|full | 1026 | No | NA | 3 | 1 | 0 | Low-quality | Genome-fragment | 2.31 | AAI-based (high-confidence) | 0 |
| S9-final-viral-combined_k141_98965\|\|full | 3473 | No | NA | 5 | 0 | 1 | Not-determined | Genome-fragment | NA | NA | 0 |
| S7_k141_178289 | 1026 | No | NA | 3 | 0 | 0 | Low-quality | Genome-fragment | 1.25 | AAI-based (medium-confidence) | 0 |
| S17-final-viral-combined_k141_203145\|\|full | 1026 | No | NA | 2 | 0 | 0 | Low-quality | Genome-fragment | 2.11 | AAI-based (medium-confidence) | 0 |
| S17-final-viral-combined_k141_13519\|\|full | 1026 | No | NA | 3 | 1 | 0 | Low-quality | Genome-fragment | 2.43 | AAI-based (high-confidence) | 0 |
| S17-final-viral-combined_k141_161615\|\|full | 1026 | No | NA | 2 | 1 | 0 | Low-quality | Genome-fragment | 0.79 | AAI-based (high-confidence) | 0 |
| S9-final-viral-combined_k141_120604\|\|full | 1771 | No | NA | 2 | 0 | 1 | Not-determined | Genome-fragment | NA | NA | 0 |
| S12-final-viral-combined_k141_376490\|\|full | 1026 | Yes | 336 | 2 | 1 | 1 | Low-quality | Genome-fragment | 0.56 | AAI-based (high-confidence) | 67.25 |
| S9-final-viral-combined_k141_114979\|\|full | 8497 | No | NA | 8 | 0 | 6 | Not-determined | Genome-fragment | NA | NA | 0 |
| S12-final-viral-combined_k141_308974\|\|lt2gene | 1026 | No | NA | 1 | 1 | 0 | Low-quality | Genome-fragment | 1.97 | HMM-based (lower-bound) | 0 |
| S12_k141_307864 | 1026 | No | NA | 1 | 0 | 1 | Low-quality | Genome-fragment | 2.08 | AAI-based (high-confidence) | 0 |
| S12_k141_50028 | 1026 | No | NA | 3 | 1 | 0 | Low-quality | Genome-fragment | 0.76 | AAI-based (high-confidence) | 0 |
| S9-final-viral-combined_k141_83831\|\|full | 2398 | No | NA | 4 | 0 | 2 | Not-determined | Genome-fragment | NA | NA | 0 |
| S9-final-viral-combined_k141_62019\|\|full | 1471 | No | NA | 3 | 0 | 1 | Not-determined | Genome-fragment | NA | NA | 0 |
| S9-final-viral-combined_k141_34793\|\|full | 1044 | No | NA | 2 | 0 | 0 | Not-determined | Genome-fragment | NA | NA | 0 |
| S4-final-viral-combined_k141_14051\|\|full | 1026 | No | NA | 3 | 0 | 0 | Low-quality | Genome-fragment | 0.99 | AAI-based (high-confidence) | 0 |
| S9-final-viral-combined_k141_125608\|\|full | 2340 | No | NA | 2 | 0 | 1 | Not-determined | Genome-fragment | NA | NA | 0 |
| S9-final-viral-combined_k141_34810\|\|full | 5601 | No | NA | 6 | 0 | 2 | Not-determined | Genome-fragment | NA | NA | 0 |
| S9-final-viral-combined_k141_88450\|\|full | 4599 | No | NA | 6 | 0 | 3 | Not-determined | Genome-fragment | NA | NA | 0 |
| S9-final-viral-combined_k141_34821\|\|full | 1281 | No | NA | 3 | 0 | 2 | Not-determined | Genome-fragment | NA | NA | 0 |
| S9-final-viral-combined_k141_120630\|\|full | 1928 | No | NA | 2 | 0 | 1 | Not-determined | Genome-fragment | NA | NA | 0 |
| S9-final-viral-combined_k141_13607\|\|full | 13483 | No | NA | 15 | 0 | 5 | Not-determined | Genome-fragment | NA | NA | 0 |
| S4-final-viral-combined_k141_114151\|\|full | 1026 | No | NA | 3 | 0 | 0 | Low-quality | Genome-fragment | 1.29 | AAI-based (high-confidence) | 0 |
| S4_k141_53745 | 1026 | No | NA | 2 | 0 | 1 | Low-quality | Genome-fragment | 1.94 | AAI-based (medium-confidence) | 0 |
| S9-final-viral-combined_k141_83909\|\|full | 1732 | No | NA | 4 | 0 | 2 | Not-determined | Genome-fragment | NA | NA | 0 |
| S8-final-viral-combined_k141_70284\|\|lt2gene | 1026 | No | NA | 2 | 1 | 0 | Low-quality | Genome-fragment | 2.72 | AAI-based (high-confidence) | 0 |
| S9-final-viral-combined_k141_88497\|\|full | 3192 | No | NA | 4 | 0 | 2 | Not-determined | Genome-fragment | NA | NA | 0 |
| S9-final-viral-combined_k141_120676\|\|full | 3921 | No | NA | 5 | 0 | 3 | Not-determined | Genome-fragment | NA | NA | 0 |
| S9_k141_32515 | 1027 | No | NA | 1 | 0 | 0 | Low-quality | Genome-fragment | 8.41 | AAI-based (high-confidence) | 0 |
| S11-final-viral-combined_k141_126059\|\|full | 1027 | No | NA | 3 | 1 | 0 | Low-quality | Genome-fragment | 2.35 | AAI-based (high-confidence) | 0 |
| S9-final-viral-combined_k141_104589\|\|full | 3022 | No | NA | 2 | 0 | 1 | Not-determined | Genome-fragment | NA | NA | 0 |
| S9-final-viral-combined_k141_125651\|\|full | 2483 | No | NA | 3 | 0 | 3 | Not-determined | Genome-fragment | NA | NA | 0 |
| S9-final-viral-combined_k141_109300\|\|full | 1489 | No | NA | 2 | 0 | 1 | Not-determined | Genome-fragment | NA | NA | 0 |
| S9-final-viral-combined_k141_93772\|\|full | 1090 | No | NA | 2 | 0 | 0 | Not-determined | Genome-fragment | NA | NA | 0 |
| S11-final-viral-combined_k141_78255\|\|lt2gene | 1027 | No | NA | 1 | 1 | 0 | Low-quality | Genome-fragment | 2.48 | AAI-based (high-confidence) | 0 |
| S1-final-viral-combined_k141_148858\|\|full | 1027 | No | NA | 3 | 1 | 0 | Low-quality | Genome-fragment | 2.45 | AAI-based (high-confidence) | 0 |
| S9-final-viral-combined_k141_99059\|\|full | 2339 | No | NA | 3 | 0 | 1 | Not-determined | Genome-fragment | NA | NA | 0 |
| S5-final-viral-combined_k141_111468\|\|full | 1027 | No | NA | 3 | 0 | 0 | Low-quality | Genome-fragment | 0.7 | AAI-based (high-confidence) | 0 |
| S9-final-viral-combined_k141_99107\|\|full | 2957 | No | NA | 3 | 0 | 3 | Not-determined | Genome-fragment | NA | NA | 0 |
| S9-final-viral-combined_k141_88518\|\|full | 3216 | No | NA | 4 | 0 | 0 | Not-determined | Genome-fragment | NA | NA | 0 |
| S9-final-viral-combined_k141_19273\|\|full | 1469 | No | NA | 2 | 0 | 1 | Not-determined | Genome-fragment | NA | NA | 0 |
| S5-final-viral-combined_k141_52499\|\|full | 1027 | No | NA | 3 | 0 | 0 | Low-quality | Genome-fragment | 1.95 | AAI-based (high-confidence) | 0 |
| S9-final-viral-combined_k141_24557\|\|full | 3704 | No | NA | 3 | 0 | 3 | Not-determined | Genome-fragment | NA | NA | 0 |
| S7-final-viral-combined_k141_226465\|\|full | 1027 | No | NA | 3 | 0 | 0 | Low-quality | Genome-fragment | 2.32 | AAI-based (high-confidence) | 0 |
| S9-final-viral-combined_k141_115022\|\|full | 3414 | No | NA | 4 | 0 | 2 | Not-determined | Genome-fragment | NA | NA | 0 |
| S7-final-viral-combined_k141_203658\|\|full | 1027 | No | NA | 3 | 0 | 0 | Low-quality | Genome-fragment | 2.56 | AAI-based (high-confidence) | 0 |
| S9-final-viral-combined_k141_120727\|\|full | 2149 | No | NA | 3 | 0 | 3 | Not-determined | Genome-fragment | NA | NA | 0 |
| S7-final-viral-combined_k141_67453\|\|lt2gene | 1027 | No | NA | 1 | 1 | 0 | Low-quality | Genome-fragment | 19.49 | AAI-based (high-confidence) | 0 |
| S9-final-viral-combined_k141_40359\|\|full | 1336 | No | NA | 3 | 0 | 0 | Not-determined | Genome-fragment | NA | NA | 0 |
| S9-final-viral-combined_k141_34945\|\|full | 1067 | No | NA | 2 | 0 | 1 | Not-determined | Genome-fragment | NA | NA | 0 |
| S9-final-viral-combined_k141_56822\|\|full | 1481 | No | NA | 4 | 0 | 3 | Not-determined | Genome-fragment | NA | NA | 0 |
| S17-final-viral-combined_k141_93719\|\|full | 1027 | No | NA | 3 | 0 | 0 | Low-quality | Genome-fragment | 1.18 | AAI-based (medium-confidence) | 0 |
| S17-k141_184406 | 1027 | No | NA | 2 | 0 | 1 | Low-quality | Genome-fragment | 0.49 | AAI-based (medium-confidence) | 0 |
| S9-final-viral-combined_k141_88583\|\|full | 2498 | No | NA | 3 | 0 | 0 | Not-determined | Genome-fragment | NA | NA | 0 |
| S9-final-viral-combined_k141_51484\|\|full | 1418 | No | NA | 2 | 0 | 0 | Not-determined | Genome-fragment | NA | NA | 0 |
| S9-final-viral-combined_k141_88598\|\|full | 2114 | No | NA | 4 | 0 | 0 | Not-determined | Genome-fragment | NA | NA | 0 |
| S9-final-viral-combined_k141_56855\|\|full | 2339 | No | NA | 2 | 0 | 1 | Not-determined | Genome-fragment | NA | NA | 0 |
| S9-final-viral-combined_k141_77796\|\|full | 1479 | No | NA | 3 | 0 | 1 | Not-determined | Genome-fragment | NA | NA | 0 |
| S17-k141_208747 | 1027 | No | NA | 1 | 0 | 1 | Low-quality | Genome-fragment | 1.27 | AAI-based (medium-confidence) | 0 |
| S9-final-viral-combined_k141_88632\|\|full | 1130 | No | NA | 2 | 0 | 1 | Not-determined | Genome-fragment | NA | NA | 0 |
| S9-final-viral-combined_k141_109322\|\|full | 103451 | Yes | 43439 | 113 | 27 | 51 | High-quality | High-quality | 100 | AAI-based (high-confidence) | 58.01 |
| S9-final-viral-combined_k141_51498\|\|full | 2292 | No | NA | 3 | 0 | 2 | Not-determined | Genome-fragment | NA | NA | 0 |
| S9-final-viral-combined_k141_125712\|\|full | 5977 | No | NA | 8 | 0 | 1 | Not-determined | Genome-fragment | NA | NA | 0 |
| S12-final-viral-combined_k141_71489\|\|full | 1027 | No | NA | 4 | 2 | 0 | Low-quality | Genome-fragment | 3.06 | AAI-based (high-confidence) | 0 |
| S12-final-viral-combined_k141_310156\|\|full | 1027 | No | NA | 3 | 0 | 0 | Low-quality | Genome-fragment | 2.88 | AAI-based (medium-confidence) | 0 |
| S12-final-viral-combined_k141_363056\|\|full | 1027 | No | NA | 3 | 0 | 2 | Low-quality | Genome-fragment | 1.15 | AAI-based (medium-confidence) | 0 |
| S9-final-viral-combined_k141_40392\|\|full | 2220 | No | NA | 2 | 0 | 1 | Not-determined | Genome-fragment | NA | NA | 0 |
| S9-final-viral-combined_k141_120791\|\|full | 1723 | No | NA | 3 | 0 | 0 | Not-determined | Genome-fragment | NA | NA | 0 |
| S12_k141_339725 | 1027 | No | NA | 4 | 0 | 0 | Low-quality | Genome-fragment | 2.42 | AAI-based (high-confidence) | 0 |
| S2-final-viral-combined_k141_29416\|\|full | 1027 | No | NA | 2 | 0 | 2 | Low-quality | Genome-fragment | 2.11 | AAI-based (medium-confidence) | 0 |
| S2_k141_10566 | 1027 | No | NA | 1 | 0 | 0 | Low-quality | Genome-fragment | 0.75 | AAI-based (medium-confidence) | 0 |
| S2_k141_57931 | 1027 | No | NA | 2 | 0 | 0 | Low-quality | Genome-fragment | 2.36 | AAI-based (medium-confidence) | 0 |
| S2_k141_78766 | 1027 | No | NA | 2 | 1 | 0 | Low-quality | Genome-fragment | 1.33 | AAI-based (high-confidence) | 0 |
| S9-final-viral-combined_k141_120803\|\|full | 1225 | No | NA | 2 | 0 | 1 | Not-determined | Genome-fragment | NA | NA | 0 |
| S8-final-viral-combined_k141_137477\|\|lt2gene | 1027 | No | NA | 1 | 1 | 0 | Low-quality | Genome-fragment | 2.43 | AAI-based (high-confidence) | 0 |
| S9-final-viral-combined_k141_27853\|\|full | 1028 | No | NA | 3 | 0 | 0 | Low-quality | Genome-fragment | 1.64 | AAI-based (medium-confidence) | 0 |
| S9-final-viral-combined_k141_113720\|\|full | 1028 | No | NA | 3 | 0 | 0 | Low-quality | Genome-fragment | 2.02 | AAI-based (medium-confidence) | 0 |
| S9-final-viral-combined_k141_115088\|\|full | 2297 | No | NA | 3 | 0 | 2 | Not-determined | Genome-fragment | NA | NA | 0 |
| S9-final-viral-combined_k141_115949\|\|full | 1028 | No | NA | 2 | 1 | 0 | Low-quality | Genome-fragment | 0.51 | HMM-based (lower-bound) | 0 |
| S9-final-viral-combined_k141_104675\|\|full | 3636 | No | NA | 3 | 0 | 2 | Not-determined | Genome-fragment | NA | NA | 0 |
| S9-final-viral-combined_k141_35045\|\|full | 2116 | No | NA | 3 | 0 | 2 | Not-determined | Genome-fragment | NA | NA | 0 |
| S1-final-viral-combined_k141_124731\|\|full | 1028 | No | NA | 4 | 0 | 0 | Low-quality | Genome-fragment | 1.44 | AAI-based (high-confidence) | 0 |
| S3-final-viral-combined_k141_102562\|\|full | 1028 | No | NA | 3 | 1 | 0 | Low-quality | Genome-fragment | 2.25 | AAI-based (medium-confidence) | 0 |
| S5-final-viral-combined_k141_161234\|\|full | 1028 | No | NA | 2 | 1 | 0 | Low-quality | Genome-fragment | 0.8 | AAI-based (high-confidence) | 0 |
| S5-final-viral-combined_k141_109667\|\|full | 1028 | No | NA | 3 | 0 | 0 | Low-quality | Genome-fragment | 3.01 | AAI-based (high-confidence) | 0 |
| S9-final-viral-combined_k141_120824\|\|full | 1374 | No | NA | 2 | 0 | 2 | Not-determined | Genome-fragment | NA | NA | 0 |
| S5-final-viral-combined_k141_82989\|\|lt2gene | 1028 | No | NA | 2 | 1 | 0 | Low-quality | Genome-fragment | 25.27 | AAI-based (medium-confidence) | 0 |
| S9-final-viral-combined_k141_51604\|\|full | 2810 | No | NA | 3 | 0 | 1 | Not-determined | Genome-fragment | NA | NA | 0 |
| S9-final-viral-combined_k141_77844\|\|full | 2883 | No | NA | 4 | 0 | 1 | Not-determined | Genome-fragment | NA | NA | 0 |
| S9-final-viral-combined_k141_57010\|\|full | 2456 | No | NA | 4 | 0 | 1 | Not-determined | Genome-fragment | NA | NA | 0 |
| S9-final-viral-combined_k141_19464\|\|full | 3623 | No | NA | 4 | 0 | 2 | Not-determined | Genome-fragment | NA | NA | 0 |
| S7-final-viral-combined_k141_242528\|\|full | 1028 | No | NA | 2 | 0 | 0 | Low-quality | Genome-fragment | 2.03 | AAI-based (high-confidence) | 0 |
| S9-final-viral-combined_k141_24651\|\|full | 2147 | No | NA | 3 | 0 | 2 | Not-determined | Genome-fragment | NA | NA | 0 |
| S9-final-viral-combined_k141_99255\|\|full | 1762 | No | NA | 3 | 0 | 2 | Not-determined | Genome-fragment | NA | NA | 0 |
| S9-final-viral-combined_k141_24657\|\|full | 1612 | No | NA | 3 | 0 | 1 | Not-determined | Genome-fragment | NA | NA | 0 |
| S9-final-viral-combined_k141_57026\|\|full | 3537 | No | NA | 5 | 0 | 0 | Not-determined | Genome-fragment | NA | NA | 0 |
| S9-final-viral-combined_k141_77868\|\|full | 4251 | No | NA | 7 | 0 | 1 | Not-determined | Genome-fragment | NA | NA | 0 |
| S9-final-viral-combined_k141_24661\|\|full | 2066 | No | NA | 3 | 0 | 0 | Not-determined | Genome-fragment | NA | NA | 0 |
| S7-final-viral-combined_k141_31118\|\|lt2gene | 1028 | No | NA | 1 | 1 | 0 | Low-quality | Genome-fragment | 2.31 | AAI-based (high-confidence) | 0 |
| S17-k141_139020 | 1028 | No | NA | 2 | 0 | 0 | Low-quality | Genome-fragment | 0.67 | AAI-based (medium-confidence) | 0 |
| S9-final-viral-combined_k141_35116\|\|full | 1990 | No | NA | 2 | 0 | 2 | Not-determined | Genome-fragment | NA | NA | 0 |
| S12-final-viral-combined_k141_372788\|\|full | 1028 | No | NA | 2 | 0 | 0 | Low-quality | Genome-fragment | 1.22 | AAI-based (medium-confidence) | 0 |
| S9-final-viral-combined_k141_45764\|\|full | 2367 | No | NA | 3 | 0 | 2 | Not-determined | Genome-fragment | NA | NA | 0 |
| S9-final-viral-combined_k141_62260\|\|full | 1635 | No | NA | 2 | 0 | 0 | Not-determined | Genome-fragment | NA | NA | 0 |
| S9-final-viral-combined_k141_3461\|\|full | 41772 | No | NA | 59 | 10 | 0 | High-quality | High-quality | 100 | AAI-based (high-confidence) | 0 |
| S9-final-viral-combined_k141_8891\|\|full | 1815 | No | NA | 3 | 0 | 2 | Not-determined | Genome-fragment | NA | NA | 0 |
| S12-final-viral-combined_k141_211794\|\|full | 1028 | No | NA | 2 | 0 | 0 | Low-quality | Genome-fragment | 2.03 | AAI-based (medium-confidence) | 0 |
| S12_k141_258913 | 1028 | No | NA | 2 | 0 | 0 | Low-quality | Genome-fragment | 2.26 | AAI-based (high-confidence) | 0 |
| S2-final-viral-combined_k141_196869\|\|full | 1028 | No | NA | 3 | 0 | 2 | Low-quality | Genome-fragment | 0.34 | AAI-based (medium-confidence) | 0 |
| S9-final-viral-combined_k141_45785\|\|full | 1210 | No | NA | 2 | 0 | 1 | Not-determined | Genome-fragment | NA | NA | 0 |
| S9-final-viral-combined_k141_40517\|\|full | 1605 | No | NA | 4 | 0 | 0 | Not-determined | Genome-fragment | NA | NA | 0 |
| S2-final-viral-combined_k141_86909\|\|full | 1028 | No | NA | 3 | 2 | 0 | Low-quality | Genome-fragment | 2.38 | AAI-based (high-confidence) | 0 |
| S2-final-viral-combined_k141_221808\|\|lt2gene | 1028 | No | NA | 1 | 1 | 0 | Low-quality | Genome-fragment | 0.71 | HMM-based (lower-bound) | 0 |
| S4-final-viral-combined_k141_82060\|\|full | 1028 | No | NA | 2 | 0 | 0 | Low-quality | Genome-fragment | 0.84 | AAI-based (medium-confidence) | 0 |
| S9-final-viral-combined_k141_3539\|\|full | 1141 | No | NA | 3 | 0 | 0 | Not-determined | Genome-fragment | NA | NA | 0 |
| S8-final-viral-combined_k141_232220\|\|full | 1028 | No | NA | 3 | 1 | 0 | Low-quality | Genome-fragment | 1.12 | AAI-based (medium-confidence) | 0 |
| S9-final-viral-combined_k141_120976\|\|full | 2890 | No | NA | 2 | 0 | 2 | Not-determined | Genome-fragment | NA | NA | 0 |
| S8_k141_253371 | 1028 | No | NA | 2 | 0 | 1 | Low-quality | Genome-fragment | 1.06 | AAI-based (medium-confidence) | 0 |
| S9-final-viral-combined_k141_57208\|\|full | 2046 | No | NA | 2 | 0 | 0 | Not-determined | Genome-fragment | NA | NA | 0 |
| S11-final-viral-combined_k141_10237\|\|full | 1029 | No | NA | 3 | 0 | 0 | Low-quality | Genome-fragment | 2.94 | AAI-based (medium-confidence) | 0 |
| S9-final-viral-combined_k141_35248\|\|full | 11465 | No | NA | 15 | 0 | 4 | Not-determined | Genome-fragment | NA | NA | 0 |
| S11-final-viral-combined_k141_298946\|\|full | 1029 | No | NA | 3 | 1 | 0 | Low-quality | Genome-fragment | 1.67 | AAI-based (medium-confidence) | 0 |
| S9-final-viral-combined_k141_115223\|\|full | 1858 | No | NA | 3 | 0 | 1 | Not-determined | Genome-fragment | NA | NA | 0 |
| S9-final-viral-combined_k141_78006\|\|full | 7183 | No | NA | 5 | 0 | 1 | Not-determined | Genome-fragment | NA | NA | 0 |
| S11_k141_154141 | 1029 | No | NA | 1 | 0 | 1 | Low-quality | Genome-fragment | 0.26 | AAI-based (high-confidence) | 0 |
| S9-final-viral-combined_k141_78021\|\|full | 7287 | No | NA | 7 | 0 | 7 | Not-determined | Genome-fragment | NA | NA | 0 |
| S9-final-viral-combined_k141_121049\|\|full | 1004 | No | NA | 3 | 0 | 1 | Not-determined | Genome-fragment | NA | NA | 0 |
| S9-final-viral-combined_k141_67276\|\|full | 6839 | No | NA | 9 | 0 | 4 | Not-determined | Genome-fragment | NA | NA | 0 |
| S11_k141_216465 | 1029 | No | NA | 1 | 0 | 1 | Low-quality | Genome-fragment | 0.36 | AAI-based (medium-confidence) | 0 |
| S1-final-viral-combined_k141_116216\|\|full | 1029 | No | NA | 3 | 0 | 0 | Low-quality | Genome-fragment | 2.6 | AAI-based (high-confidence) | 0 |
| S9-final-viral-combined_k141_125989\|\|full | 3298 | No | NA | 2 | 0 | 2 | Not-determined | Genome-fragment | NA | NA | 0 |
| S1-final-viral-combined_k141_302896\|\|full | 1029 | No | NA | 3 | 0 | 0 | Low-quality | Genome-fragment | 2.24 | AAI-based (high-confidence) | 0 |
| S9-final-viral-combined_k141_40544\|\|full | 5484 | No | NA | 6 | 0 | 6 | Not-determined | Genome-fragment | NA | NA | 0 |
| S1-final-viral-combined_k141_188384\|\|full | 1029 | No | NA | 3 | 1 | 0 | Low-quality | Genome-fragment | 2.8 | AAI-based (high-confidence) | 0 |
| S5-final-viral-combined_k141_109819\|\|full | 1029 | No | NA | 4 | 1 | 0 | Low-quality | Genome-fragment | 2.05 | AAI-based (medium-confidence) | 0 |
| S9-final-viral-combined_k141_29611\|\|full | 413955 | Yes | 96620 | 499 | 35 | 266 | Complete | High-quality | 100 | Provirus (high-confidence) | 76.66 |
| S9-final-viral-combined_k141_62401\|\|full | 2470 | No | NA | 3 | 0 | 2 | Not-determined | Genome-fragment | NA | NA | 0 |
| S9-final-viral-combined_k141_3609\|\|full | 1186 | No | NA | 2 | 0 | 1 | Not-determined | Genome-fragment | NA | NA | 0 |
| S9-final-viral-combined_k141_62410\|\|full | 2661 | No | NA | 3 | 0 | 0 | Not-determined | Genome-fragment | NA | NA | 0 |
| S9-final-viral-combined_k141_29645\|\|full | 3055 | No | NA | 3 | 0 | 3 | Not-determined | Genome-fragment | NA | NA | 0 |
| S5-final-viral-combined_k141_124995\|\|lt2gene | 1029 | No | NA | 2 | 1 | 0 | Low-quality | Genome-fragment | 2.65 | AAI-based (high-confidence) | 0 |
| S5-final-viral-combined_k141_198522\|\|lt2gene | 1029 | No | NA | 2 | 2 | 0 | Low-quality | Genome-fragment | 2.25 | AAI-based (high-confidence) | 0 |
| S9-final-viral-combined_k141_29664\|\|full | 1679 | No | NA | 3 | 0 | 0 | Not-determined | Genome-fragment | NA | NA | 0 |
| S7-final-viral-combined_k141_130863\|\|full | 1029 | No | NA | 3 | 1 | 0 | Low-quality | Genome-fragment | 2.34 | AAI-based (high-confidence) | 0 |
| S9-final-viral-combined_k141_51949\|\|full | 1262 | No | NA | 3 | 0 | 1 | Not-determined | Genome-fragment | NA | NA | 0 |
| S9-final-viral-combined_k141_99483\|\|full | 1502 | No | NA | 3 | 0 | 2 | Not-determined | Genome-fragment | NA | NA | 0 |
| S17-k141_254135 | 1029 | No | NA | 2 | 0 | 0 | Low-quality | Genome-fragment | 1.72 | AAI-based (high-confidence) | 0 |
| S9-final-viral-combined_k141_51956\|\|full | 2782 | No | NA | 6 | 0 | 2 | Not-determined | Genome-fragment | NA | NA | 0 |
| S12-final-viral-combined_k141_2349\|\|full | 1029 | No | NA | 3 | 0 | 1 | Low-quality | Genome-fragment | 1.62 | AAI-based (medium-confidence) | 0 |
| S9-final-viral-combined_k141_67383\|\|full | 3089 | No | NA | 4 | 0 | 4 | Not-determined | Genome-fragment | NA | NA | 0 |
| S12-final-viral-combined_k141_388047\|\|lt2gene | 1029 | No | NA | 1 | 1 | 0 | Low-quality | Genome-fragment | 0.92 | HMM-based (lower-bound) | 0 |
| S9-final-viral-combined_k141_40587\|\|full | 1997 | No | NA | 3 | 0 | 0 | Not-determined | Genome-fragment | NA | NA | 0 |
| S12-final-viral-combined_k141_92609\|\|lt2gene | 1029 | No | NA | 1 | 1 | 0 | Low-quality | Genome-fragment | 2.58 | AAI-based (high-confidence) | 0 |
| S9-final-viral-combined_k141_57377\|\|full | 2098 | No | NA | 4 | 0 | 2 | Not-determined | Genome-fragment | NA | NA | 0 |
| S9-final-viral-combined_k141_109588\|\|full | 1246 | No | NA | 2 | 0 | 1 | Not-determined | Genome-fragment | NA | NA | 0 |
| S9-final-viral-combined_k141_19728\|\|full | 4612 | No | NA | 3 | 0 | 2 | Not-determined | Genome-fragment | NA | NA | 0 |
| S12_k141_208204 | 1029 | No | NA | 2 | 0 | 1 | Low-quality | Genome-fragment | 1.44 | AAI-based (medium-confidence) | 0 |
| S2-final-viral-combined_k141_145164\|\|full | 1029 | No | NA | 3 | 0 | 0 | Low-quality | Genome-fragment | 1.79 | AAI-based (high-confidence) | 0 |
| S9-final-viral-combined_k141_62477\|\|full | 2264 | No | NA | 2 | 0 | 1 | Not-determined | Genome-fragment | NA | NA | 0 |
| S2-final-viral-combined_k141_188393\|\|full | 1029 | No | NA | 2 | 0 | 0 | Low-quality | Genome-fragment | 1.88 | AAI-based (medium-confidence) | 0 |
| S9-final-viral-combined_k141_78167\|\|full | 1313 | No | NA | 2 | 0 | 1 | Not-determined | Genome-fragment | NA | NA | 0 |
| S2_k141_70349 | 1029 | No | NA | 1 | 0 | 0 | Low-quality | Genome-fragment | 0.59 | AAI-based (high-confidence) | 0 |
| S9-final-viral-combined_k141_57395\|\|full | 1114 | No | NA | 3 | 0 | 0 | Not-determined | Genome-fragment | NA | NA | 0 |
| S2_k141_9348 | 1029 | No | NA | 1 | 0 | 1 | Low-quality | Genome-fragment | 0.25 | AAI-based (medium-confidence) | 0 |
| S9-final-viral-combined_k141_3745\|\|full | 1584 | No | NA | 3 | 0 | 0 | Not-determined | Genome-fragment | NA | NA | 0 |
| S4-final-viral-combined_k141_31600\|\|full | 1029 | No | NA | 3 | 0 | 1 | Low-quality | Genome-fragment | 0.98 | AAI-based (medium-confidence) | 0 |
| S6-final-viral-combined_k141_77448\|\|full | 1029 | No | NA | 3 | 0 | 3 | Low-quality | Genome-fragment | 0.45 | AAI-based (medium-confidence) | 0 |
| S9-final-viral-combined_k141_115419\|\|full | 2231 | No | NA | 3 | 0 | 2 | Not-determined | Genome-fragment | NA | NA | 0 |
| S9-final-viral-combined_k141_78204\|\|full | 2396 | No | NA | 4 | 0 | 1 | Not-determined | Genome-fragment | NA | NA | 0 |
| S9-final-viral-combined_k141_104856\|\|full | 2394 | No | NA | 2 | 0 | 1 | Not-determined | Genome-fragment | NA | NA | 0 |
| S9-final-viral-combined_k141_29708\|\|full | 1616 | No | NA | 2 | 0 | 1 | Not-determined | Genome-fragment | NA | NA | 0 |
| S6-final-viral-combined_k141_109873\|\|full | 1029 | No | NA | 3 | 0 | 0 | Low-quality | Genome-fragment | 1.35 | AAI-based (high-confidence) | 0 |
| S11-final-viral-combined_k141_163736\|\|lt2gene | 1030 | No | NA | 1 | 0 | 0 | Low-quality | Genome-fragment | 1.68 | AAI-based (high-confidence) | 0 |
| S1_k141_94801 | 1030 | No | NA | 2 | 0 | 1 | Low-quality | Genome-fragment | 2.18 | AAI-based (high-confidence) | 0 |
| S9-final-viral-combined_k141_24997\|\|full | 1094 | No | NA | 3 | 0 | 2 | Not-determined | Genome-fragment | NA | NA | 0 |
| S9-final-viral-combined_k141_121242\|\|full | 1408 | No | NA | 3 | 0 | 2 | Not-determined | Genome-fragment | NA | NA | 0 |
| S9-final-viral-combined_k141_57429\|\|full | 3154 | No | NA | 5 | 0 | 1 | Not-determined | Genome-fragment | NA | NA | 0 |
| S3-final-viral-combined_k141_81623\|\|full | 1030 | No | NA | 3 | 1 | 0 | Low-quality | Genome-fragment | 1.66 | AAI-based (medium-confidence) | 0 |
| S3-final-viral-combined_k141_40795\|\|lt2gene | 1030 | No | NA | 1 | 1 | 0 | Low-quality | Genome-fragment | 2.74 | AAI-based (high-confidence) | 0 |
| S9-final-viral-combined_k141_72491\|\|full | 1943 | No | NA | 3 | 0 | 3 | Not-determined | Genome-fragment | NA | NA | 0 |
| S9-final-viral-combined_k141_109643\|\|full | 3475 | No | NA | 5 | 0 | 1 | Not-determined | Genome-fragment | NA | NA | 0 |
| S9-final-viral-combined_k141_25007\|\|full | 30091 | Yes | 27739 | 36 | 20 | 5 | Medium-quality | Genome-fragment | 59.16 | AAI-based (medium-confidence) | 7.82 |
| S7-final-viral-combined_k141_159266\|\|full | 1030 | No | NA | 3 | 0 | 1 | Low-quality | Genome-fragment | 0.96 | AAI-based (medium-confidence) | 0 |
| S9-final-viral-combined_k141_89121\|\|full | 2096 | No | NA | 4 | 0 | 0 | Not-determined | Genome-fragment | NA | NA | 0 |
| S7-final-viral-combined_k141_222402\|\|full | 1030 | No | NA | 3 | 1 | 0 | Low-quality | Genome-fragment | 1.72 | HMM-based (lower-bound) | 0 |
| S9-final-viral-combined_k141_84194\|\|full | 2347 | No | NA | 2 | 0 | 0 | Not-determined | Genome-fragment | NA | NA | 0 |
| S17-final-viral-combined_k141_236903\|\|full | 1030 | No | NA | 2 | 1 | 0 | Low-quality | Genome-fragment | 3.21 | AAI-based (high-confidence) | 0 |
| S9-final-viral-combined_k141_67478\|\|full | 4359 | No | NA | 7 | 0 | 2 | Not-determined | Genome-fragment | NA | NA | 0 |
| S9-final-viral-combined_k141_67485\|\|full | 1831 | No | NA | 3 | 0 | 0 | Not-determined | Genome-fragment | NA | NA | 0 |
| S17-final-viral-combined_k141_152713\|\|full | 1030 | No | NA | 2 | 0 | 1 | Low-quality | Genome-fragment | 0.59 | AAI-based (medium-confidence) | 0 |
| S9-final-viral-combined_k141_67492\|\|full | 1694 | No | NA | 2 | 0 | 2 | Not-determined | Genome-fragment | NA | NA | 0 |
| S9-final-viral-combined_k141_9179\|\|full | 1681 | No | NA | 3 | 0 | 0 | Not-determined | Genome-fragment | NA | NA | 0 |
| S17-final-viral-combined_k141_9808\|\|full | 1030 | No | NA | 2 | 0 | 0 | Low-quality | Genome-fragment | 2.08 | AAI-based (medium-confidence) | 0 |
| S9-final-viral-combined_k141_67507\|\|full | 1496 | No | NA | 2 | 0 | 2 | Not-determined | Genome-fragment | NA | NA | 0 |
| S9-final-viral-combined_k141_115543\|\|full | 2792 | No | NA | 3 | 0 | 1 | Not-determined | Genome-fragment | NA | NA | 0 |
| S17-final-viral-combined_k141_82361\|\|lt2gene | 1030 | No | NA | 2 | 0 | 0 | Low-quality | Genome-fragment | 2.46 | AAI-based (medium-confidence) | 0 |
| S9-final-viral-combined_k141_72581\|\|full | 1259 | No | NA | 2 | 0 | 1 | Not-determined | Genome-fragment | NA | NA | 0 |
| S17-k141_79643 | 1030 | No | NA | 2 | 2 | 0 | Low-quality | Genome-fragment | 3.4 | AAI-based (high-confidence) | 0 |
| S9-final-viral-combined_k141_35415\|\|full | 3936 | No | NA | 5 | 0 | 3 | Not-determined | Genome-fragment | NA | NA | 0 |
| S9-final-viral-combined_k141_29789\|\|full | 1917 | No | NA | 3 | 0 | 3 | Not-determined | Genome-fragment | NA | NA | 0 |
| S9-final-viral-combined_k141_29806\|\|full | 1885 | No | NA | 4 | 0 | 2 | Not-determined | Genome-fragment | NA | NA | 0 |
| S9-final-viral-combined_k141_115564\|\|full | 1022 | No | NA | 2 | 0 | 1 | Not-determined | Genome-fragment | NA | NA | 0 |
| S12-final-viral-combined_k141_309457\|\|full | 1030 | No | NA | 4 | 2 | 0 | Low-quality | Genome-fragment | 1.79 | AAI-based (high-confidence) | 0 |
| S9-final-viral-combined_k141_99643\|\|full | 5489 | No | NA | 6 | 0 | 1 | Not-determined | Genome-fragment | NA | NA | 0 |
| S9-final-viral-combined_k141_25065\|\|full | 14748 | No | NA | 6 | 0 | 2 | Not-determined | Genome-fragment | NA | NA | 0 |
| S9-final-viral-combined_k141_72601\|\|full | 1878 | No | NA | 4 | 0 | 2 | Not-determined | Genome-fragment | NA | NA | 0 |
| S9-final-viral-combined_k141_45959\|\|full | 1449 | No | NA | 2 | 0 | 0 | Not-determined | Genome-fragment | NA | NA | 0 |
| S2_k141_245569 | 1030 | No | NA | 1 | 0 | 0 | Low-quality | Genome-fragment | 1.24 | AAI-based (medium-confidence) | 0 |
| S6_k141_35861 | 1030 | No | NA | 1 | 0 | 1 | Low-quality | Genome-fragment | 0.18 | AAI-based (high-confidence) | 0 |
| S9-final-viral-combined_k141_84249\|\|full | 2785 | No | NA | 3 | 0 | 1 | Not-determined | Genome-fragment | NA | NA | 0 |
| S9-final-viral-combined_k141_84250\|\|full | 1440 | No | NA | 3 | 0 | 1 | Not-determined | Genome-fragment | NA | NA | 0 |
| S9-final-viral-combined_k141_109714\|\|full | 8846 | No | NA | 8 | 0 | 7 | Not-determined | Genome-fragment | NA | NA | 0 |
| S9-final-viral-combined_k141_3878\|\|full | 5555 | No | NA | 6 | 1 | 0 | High-quality | High-quality | 100 | AAI-based (high-confidence) | 0 |
| S8-final-viral-combined_k141_186169\|\|full | 1030 | No | NA | 4 | 0 | 0 | Low-quality | Genome-fragment | 2.47 | AAI-based (high-confidence) | 0 |
| S9_k141_75911 | 1031 | No | NA | 2 | 0 | 0 | Low-quality | Genome-fragment | 1.37 | AAI-based (medium-confidence) | 0 |
| S11-final-viral-combined_k141_223346\|\|full | 1031 | No | NA | 2 | 0 | 0 | Low-quality | Genome-fragment | 2.51 | AAI-based (medium-confidence) | 0 |
| S9-final-viral-combined_k141_45998\|\|full | 1751 | No | NA | 4 | 0 | 0 | Not-determined | Genome-fragment | NA | NA | 0 |
| S9-final-viral-combined_k141_89249\|\|full | 25111 | No | NA | 42 | 12 | 0 | Medium-quality | Genome-fragment | 56.42 | AAI-based (high-confidence) | 0 |
| S9-final-viral-combined_k141_46011\|\|full | 2259 | No | NA | 3 | 0 | 1 | Not-determined | Genome-fragment | NA | NA | 0 |
| S9-final-viral-combined_k141_67544\|\|full | 1319 | No | NA | 2 | 0 | 1 | Not-determined | Genome-fragment | NA | NA | 0 |
| S9-final-viral-combined_k141_35522\|\|full | 1924 | No | NA | 2 | 0 | 0 | Not-determined | Genome-fragment | NA | NA | 0 |
| S9-final-viral-combined_k141_9230\|\|full | 2260 | No | NA | 4 | 0 | 1 | Not-determined | Genome-fragment | NA | NA | 0 |
| S9-final-viral-combined_k141_105058\|\|full | 1640 | No | NA | 3 | 0 | 3 | Not-determined | Genome-fragment | NA | NA | 0 |
| S11_k141_66707 | 1031 | No | NA | 2 | 0 | 0 | Low-quality | Genome-fragment | 1.98 | AAI-based (high-confidence) | 0 |
| S9-final-viral-combined_k141_105067\|\|full | 2911 | No | NA | 5 | 0 | 0 | Not-determined | Genome-fragment | NA | NA | 0 |
| S1_k141_222392 | 1031 | No | NA | 1 | 0 | 1 | Low-quality | Genome-fragment | 0.36 | AAI-based (medium-confidence) | 0 |
| S9-final-viral-combined_k141_121464\|\|full | 1912 | No | NA | 3 | 0 | 2 | Not-determined | Genome-fragment | NA | NA | 0 |
| S9-final-viral-combined_k141_67619\|\|full | 2278 | No | NA | 3 | 0 | 3 | Not-determined | Genome-fragment | NA | NA | 0 |
| S5_k141_205857 | 1031 | No | NA | 3 | 0 | 1 | Low-quality | Genome-fragment | 2.21 | AAI-based (medium-confidence) | 0 |
| S9-final-viral-combined_k141_89354\|\|full | 1979 | No | NA | 3 | 0 | 2 | Not-determined | Genome-fragment | NA | NA | 0 |
| S9-final-viral-combined_k141_14185\|\|full | 2561 | No | NA | 3 | 0 | 2 | Not-determined | Genome-fragment | NA | NA | 0 |
| S17-final-viral-combined_k141_39295\|\|full | 1031 | No | NA | 2 | 0 | 2 | Low-quality | Genome-fragment | 0.42 | AAI-based (medium-confidence) | 0 |
| S9-final-viral-combined_k141_78481\|\|full | 1574 | No | NA | 3 | 0 | 1 | Not-determined | Genome-fragment | NA | NA | 0 |
| S9-final-viral-combined_k141_3982\|\|full | 1683 | No | NA | 3 | 0 | 0 | Not-determined | Genome-fragment | NA | NA | 0 |
| S9-final-viral-combined_k141_115654\|\|full | 1728 | No | NA | 3 | 0 | 2 | Not-determined | Genome-fragment | NA | NA | 0 |
| S9-final-viral-combined_k141_14226\|\|full | 1199 | No | NA | 2 | 0 | 2 | Not-determined | Genome-fragment | NA | NA | 0 |
| S9-final-viral-combined_k141_25157\|\|full | 1432 | No | NA | 3 | 0 | 1 | Not-determined | Genome-fragment | NA | NA | 0 |
| S9-final-viral-combined_k141_25165\|\|full | 1363 | No | NA | 3 | 0 | 2 | Not-determined | Genome-fragment | NA | NA | 0 |
| S9-final-viral-combined_k141_126256\|\|full | 1599 | No | NA | 4 | 0 | 0 | Not-determined | Genome-fragment | NA | NA | 0 |
| S9-final-viral-combined_k141_89420\|\|full | 1437 | No | NA | 3 | 0 | 2 | Not-determined | Genome-fragment | NA | NA | 0 |
| S9-final-viral-combined_k141_72636\|\|full | 2204 | No | NA | 5 | 0 | 1 | Not-determined | Genome-fragment | NA | NA | 0 |
| S9-final-viral-combined_k141_52208\|\|full | 1745 | No | NA | 2 | 0 | 1 | Not-determined | Genome-fragment | NA | NA | 0 |

| | | | | | | | | | | | |
|---|---|---|---|---|---|---|---|---|---|---|---|
| S17-k141_207077 | 1031 | No | NA | 1 | 1 | 0 | Low-quality | Genome-fragment | 1.08 | HMM-based (lower-bound) | 0 |
| S12-final-viral-combined_k141_373554\|\|full | 1031 | No | NA | 1 | 0 | 1 | Low-quality | Genome-fragment | 0.29 | AAI-based (high-confidence) | 0 |
| S9-final-viral-combined_k141_115753\|\|full | 1316 | No | NA | 3 | 0 | 1 | Not-determined | Genome-fragment | NA | NA | 0 |
| S9-final-viral-combined_k141_67691\|\|full | 2826 | No | NA | 4 | 0 | 1 | Not-determined | Genome-fragment | NA | NA | 0 |
| S9-final-viral-combined_k141_89444\|\|full | 1618 | No | NA | 3 | 0 | 0 | Not-determined | Genome-fragment | NA | NA | 0 |
| S9-final-viral-combined_k141_89445\|\|full | 2342 | No | NA | 3 | 0 | 3 | Not-determined | Genome-fragment | NA | NA | 0 |
| S12-final-viral-combined_k141_5016\|\|full | 1031 | No | NA | 2 | 0 | 1 | Low-quality | Genome-fragment | 1.86 | AAI-based (high-confidence) | 0 |
| S9-final-viral-combined_k141_19994\|\|full | 1814 | No | NA | 3 | 0 | 1 | Not-determined | Genome-fragment | NA | NA | 0 |
| S9-final-viral-combined_k141_57504\|\|full | 1713 | No | NA | 3 | 0 | 0 | Not-determined | Genome-fragment | NA | NA | 0 |
| S9-final-viral-combined_k141_25250\|\|full | 1310 | No | NA | 2 | 0 | 1 | Not-determined | Genome-fragment | NA | NA | 0 |
| S9-final-viral-combined_k141_115776\|\|full | 2027 | No | NA | 3 | 0 | 0 | Not-determined | Genome-fragment | NA | NA | 0 |
| S9-final-viral-combined_k141_67719\|\|full | 3214 | No | NA | 5 | 0 | 5 | Not-determined | Genome-fragment | NA | NA | 0 |
| S9-final-viral-combined_k141_72667\|\|full | 2520 | No | NA | 4 | 0 | 1 | Not-determined | Genome-fragment | NA | NA | 0 |
| S12-final-viral-combined_k141_243330\|\|lt2gene | 1031 | No | NA | 1 | 1 | 0 | Low-quality | Genome-fragment | 1.79 | AAI-based (high-confidence) | 0 |
| S9-final-viral-combined_k141_29914\|\|full | 3512 | No | NA | 5 | 0 | 1 | Not-determined | Genome-fragment | NA | NA | 0 |
| S2-final-viral-combined_k141_234290\|\|lt2gene | 1031 | No | NA | 1 | 1 | 0 | Low-quality | Genome-fragment | 2.73 | AAI-based (high-confidence) | 0 |
| S4-final-viral-combined_k141_114850\|\|full | 1031 | No | NA | 4 | 1 | 0 | Low-quality | Genome-fragment | 2.38 | AAI-based (medium-confidence) | 0 |
| S9-final-viral-combined_k141_29936\|\|full | 1542 | No | NA | 3 | 0 | 1 | Not-determined | Genome-fragment | NA | NA | 0 |
| S4_k141_69250 | 1031 | No | NA | 3 | 2 | 0 | Low-quality | Genome-fragment | 0.58 | AAI-based (high-confidence) | 0 |
| S9-final-viral-combined_k141_9402\|\|full | 2727 | No | NA | 3 | 0 | 3 | Not-determined | Genome-fragment | NA | NA | 0 |
| S8_k141_216795 | 1031 | No | NA | 1 | 0 | 1 | Low-quality | Genome-fragment | 1.76 | AAI-based (high-confidence) | 0 |
| S11_k141_56944 | 1032 | No | NA | 4 | 1 | 0 | Low-quality | Genome-fragment | 1.19 | HMM-based (lower-bound) | 0 |
| S9-final-viral-combined_k141_126321\|\|full | 1888 | No | NA | 3 | 0 | 1 | Not-determined | Genome-fragment | NA | NA | 0 |
| S9-final-viral-combined_k141_35663\|\|full | 1629 | No | NA | 2 | 0 | 1 | Not-determined | Genome-fragment | NA | NA | 0 |
| S1-final-viral-combined_k141_318328\|\|full | 1032 | No | NA | 2 | 0 | 0 | Low-quality | Genome-fragment | 2.63 | AAI-based (medium-confidence) | 0 |
| S9-final-viral-combined_k141_25284\|\|full | 9080 | No | NA | 7 | 0 | 6 | Not-determined | Genome-fragment | NA | NA | 0 |
| S9-final-viral-combined_k141_121625\|\|full | 2849 | No | NA | 4 | 0 | 2 | Not-determined | Genome-fragment | NA | NA | 0 |
| S9-final-viral-combined_k141_9423\|\|full | 4377 | No | NA | 6 | 0 | 5 | Not-determined | Genome-fragment | NA | NA | 0 |
| S3-final-viral-combined_k141_7655\|\|lt2gene | 1032 | No | NA | 2 | 2 | 0 | Low-quality | Genome-fragment | 2.29 | AAI-based (high-confidence) | 0 |
| S9-final-viral-combined_k141_67797\|\|full | 3177 | No | NA | 3 | 0 | 2 | Not-determined | Genome-fragment | NA | NA | 0 |
| S9-final-viral-combined_k141_67799\|\|full | 1023 | No | NA | 2 | 0 | 0 | Not-determined | Genome-fragment | NA | NA | 0 |
| S9-final-viral-combined_k141_14418\|\|full | 3132 | No | NA | 5 | 0 | 0 | Not-determined | Genome-fragment | NA | NA | 0 |
| S9-final-viral-combined_k141_109869\|\|full | 1319 | No | NA | 3 | 0 | 1 | Not-determined | Genome-fragment | NA | NA | 0 |
| S5_k141_167550 | 1032 | No | NA | 3 | 0 | 0 | Low-quality | Genome-fragment | 0.59 | AAI-based (high-confidence) | 0 |
| S9-final-viral-combined_k141_99745\|\|full | 2363 | No | NA | 4 | 0 | 1 | Not-determined | Genome-fragment | NA | NA | 0 |
| S12-final-viral-combined_k141_211088\|\|full | 1032 | No | NA | 3 | 0 | 0 | Low-quality | Genome-fragment | 2.93 | AAI-based (high-confidence) | 0 |
| S9-final-viral-combined_k141_72726\|\|full | 2961 | No | NA | 4 | 0 | 0 | Not-determined | Genome-fragment | NA | NA | 0 |
| S9-final-viral-combined_k141_29980\|\|full | 1968 | No | NA | 3 | 0 | 0 | Not-determined | Genome-fragment | NA | NA | 0 |
| S12_k141_81584 | 1032 | No | NA | 2 | 0 | 0 | Low-quality | Genome-fragment | 0.57 | AAI-based (high-confidence) | 0 |
| S9-final-viral-combined_k141_78706\|\|full | 1963 | No | NA | 3 | 0 | 1 | Not-determined | Genome-fragment | NA | NA | 0 |
| S2-final-viral-combined_k141_209338\|\|full | 1032 | No | NA | 3 | 0 | 1 | Low-quality | Genome-fragment | 2.66 | AAI-based (medium-confidence) | 0 |
| S2_k141_223394 | 1032 | No | NA | 1 | 0 | 0 | Low-quality | Genome-fragment | 0.42 | AAI-based (medium-confidence) | 0 |
| S4-final-viral-combined_k141_78140\|\|full | 1032 | No | NA | 3 | 0 | 1 | Low-quality | Genome-fragment | 1.79 | AAI-based (medium-confidence) | 0 |
| S4_k141_153945 | 1032 | No | NA | 3 | 0 | 0 | Low-quality | Genome-fragment | 1.72 | AAI-based (high-confidence) | 0 |
| S8-final-viral-combined_k141_62520\|\|lt2gene | 1032 | No | NA | 2 | 1 | 0 | Low-quality | Genome-fragment | 0.66 | HMM-based (lower-bound) | 0 |
| S9-final-viral-combined_k141_111231\|\|full | 1033 | No | NA | 3 | 0 | 0 | Low-quality | Genome-fragment | 1.86 | AAI-based (medium-confidence) | 0 |
| S9-final-viral-combined_k141_72770\|\|full | 2037 | No | NA | 3 | 0 | 2 | Not-determined | Genome-fragment | NA | NA | 0 |
| S9-final-viral-combined_k141_78736\|\|full | 1795 | No | NA | 2 | 0 | 0 | Not-determined | Genome-fragment | NA | NA | 0 |
| S9-final-viral-combined_k141_109925\|\|full | 3081 | No | NA | 3 | 0 | 2 | Not-determined | Genome-fragment | NA | NA | 0 |
| S9-final-viral-combined_k141_84378\|\|full | 3000 | No | NA | 4 | 0 | 3 | Not-determined | Genome-fragment | NA | NA | 0 |
| S11-final-viral-combined_k141_248292\|\|full | 1033 | No | NA | 2 | 0 | 0 | Low-quality | Genome-fragment | 1.46 | AAI-based (medium-confidence) | 0 |
| S9-final-viral-combined_k141_89517\|\|full | 3137 | No | NA | 3 | 0 | 1 | Not-determined | Genome-fragment | NA | NA | 0 |
| S9-final-viral-combined_k141_121710\|\|full | 2094 | No | NA | 3 | 0 | 2 | Not-determined | Genome-fragment | NA | NA | 0 |
| S9-final-viral-combined_k141_105128\|\|full | 3298 | No | NA | 5 | 0 | 1 | Not-determined | Genome-fragment | NA | NA | 0 |
| S11-final-viral-combined_k141_187345\|\|full | 1033 | No | NA | 3 | 0 | 0 | Low-quality | Genome-fragment | 1.04 | AAI-based (medium-confidence) | 0 |
| S9-final-viral-combined_k141_4194\|\|full | 1180 | No | NA | 3 | 0 | 1 | Not-determined | Genome-fragment | NA | NA | 0 |
| S9-final-viral-combined_k141_99845\|\|full | 1943 | No | NA | 3 | 0 | 3 | Not-determined | Genome-fragment | NA | NA | 0 |
| S9-final-viral-combined_k141_40869\|\|full | 2998 | No | NA | 3 | 0 | 2 | Not-determined | Genome-fragment | NA | NA | 0 |
| S11-final-viral-combined_k141_222996\|\|lt2gene | 1033 | No | NA | 1 | 1 | 0 | Low-quality | Genome-fragment | 0.97 | HMM-based (lower-bound) | 0 |
| S9-final-viral-combined_k141_72801\|\|full | 5048 | No | NA | 4 | 0 | 1 | Not-determined | Genome-fragment | NA | NA | 0 |
| S9-final-viral-combined_k141_121754\|\|full | 1325 | No | NA | 3 | 0 | 1 | Not-determined | Genome-fragment | NA | NA | 0 |
| S9-final-viral-combined_k141_30029\|\|full | 6016 | No | NA | 11 | 0 | 1 | Not-determined | Genome-fragment | NA | NA | 0 |
| S1-final-viral-combined_k141_251192\|\|full | 1033 | No | NA | 3 | 0 | 0 | Low-quality | Genome-fragment | 2.41 | AAI-based (high-confidence) | 0 |
| S9-final-viral-combined_k141_9500\|\|full | 2304 | No | NA | 3 | 0 | 0 | Not-determined | Genome-fragment | NA | NA | 0 |
| S9-final-viral-combined_k141_67912\|\|full | 1805 | No | NA | 4 | 0 | 3 | Not-determined | Genome-fragment | NA | NA | 0 |
| S9-final-viral-combined_k141_126341\|\|full | 9842 | No | NA | 12 | 0 | 1 | Not-determined | Genome-fragment | NA | NA | 0 |
| S5_k141_74002 | 1033 | No | NA | 3 | 0 | 0 | Low-quality | Genome-fragment | 2.55 | AAI-based (medium-confidence) | 0 |
| S5_k141_30005 | 1033 | No | NA | 3 | 0 | 0 | Low-quality | Genome-fragment | 1.4 | AAI-based (medium-confidence) | 0 |
| S9-final-viral-combined_k141_30032\|\|full | 6936 | No | NA | 5 | 0 | 5 | Not-determined | Genome-fragment | NA | NA | 0 |
| S7-final-viral-combined_k141_113529\|\|full | 1033 | No | NA | 2 | 0 | 0 | Low-quality | Genome-fragment | 2.04 | AAI-based (high-confidence) | 0 |
| S7-final-viral-combined_k141_159253\|\|lt2gene | 1033 | No | NA | 2 | 2 | 0 | Low-quality | Genome-fragment | 2.79 | AAI-based (medium-confidence) | 0 |
| S7-final-viral-combined_k141_38611\|\|lt2gene | 1033 | No | NA | 1 | 1 | 0 | Low-quality | Genome-fragment | 2.15 | AAI-based (high-confidence) | 0 |
| S17-final-viral-combined_k141_160006\|\|full | 1033 | No | NA | 3 | 1 | 0 | Low-quality | Genome-fragment | 2.98 | AAI-based (high-confidence) | 0 |
| S9-final-viral-combined_k141_30041\|\|full | 2292 | No | NA | 4 | 0 | 0 | Not-determined | Genome-fragment | NA | NA | 0 |
| S9-final-viral-combined_k141_126365\|\|full | 2768 | No | NA | 4 | 0 | 3 | Not-determined | Genome-fragment | NA | NA | 0 |
| S17-final-viral-combined_k141_64213\|\|full | 1033 | No | NA | 3 | 3 | 0 | Low-quality | Genome-fragment | 2.85 | AAI-based (high-confidence) | 0 |
| S9-final-viral-combined_k141_35736\|\|full | 2578 | No | NA | 6 | 0 | 1 | Not-determined | Genome-fragment | NA | NA | 0 |
| S9-final-viral-combined_k141_109995\|\|full | 1552 | No | NA | 4 | 0 | 0 | Not-determined | Genome-fragment | NA | NA | 0 |
| S9-final-viral-combined_k141_72832\|\|full | 1078 | No | NA | 2 | 0 | 1 | Not-determined | Genome-fragment | NA | NA | 0 |
| S9-final-viral-combined_k141_84493\|\|full | 1027 | No | NA | 2 | 0 | 0 | Not-determined | Genome-fragment | NA | NA | 0 |
| S17-final-viral-combined_k141_155344\|\|lt2gene | 1033 | No | NA | 2 | 1 | 0 | Low-quality | Genome-fragment | 2.54 | AAI-based (medium-confidence) | 0 |
| S12-final-viral-combined_k141_308608\|\|full | 1033 | Yes | 423 | 2 | 1 | 1 | Low-quality | Genome-fragment | 0.55 | HMM-based (lower-bound) | 59.05 |
| S9-final-viral-combined_k141_89587\|\|full | 1160 | No | NA | 4 | 0 | 0 | Not-determined | Genome-fragment | NA | NA | 0 |
| S9-final-viral-combined_k141_126389\|\|full | 3647 | No | NA | 7 | 0 | 3 | Not-determined | Genome-fragment | NA | NA | 0 |
| S12-final-viral-combined_k141_396656\|\|full | 1033 | No | NA | 2 | 2 | 0 | Low-quality | Genome-fragment | 2.26 | AAI-based (high-confidence) | 0 |
| S9-final-viral-combined_k141_46188\|\|full | 1074 | No | NA | 3 | 0 | 0 | Not-determined | Genome-fragment | NA | NA | 0 |
| S9-final-viral-combined_k141_46203\|\|full | 1787 | No | NA | 2 | 0 | 2 | Not-determined | Genome-fragment | NA | NA | 0 |
| S9-final-viral-combined_k141_25440\|\|full | 1477 | No | NA | 2 | 0 | 1 | Not-determined | Genome-fragment | NA | NA | 0 |
| S12-final-viral-combined_k141_314659\|\|full | 1033 | No | NA | 2 | 2 | 0 | Low-quality | Genome-fragment | 2.63 | AAI-based (high-confidence) | 0 |
| S9-final-viral-combined_k141_46214\|\|full | 3186 | No | NA | 3 | 0 | 0 | Not-determined | Genome-fragment | NA | NA | 0 |
| S9-final-viral-combined_k141_30124\|\|full | 2057 | No | NA | 2 | 0 | 2 | Not-determined | Genome-fragment | NA | NA | 0 |
| S9-final-viral-combined_k141_105178\|\|full | 1494 | No | NA | 2 | 0 | 1 | Not-determined | Genome-fragment | NA | NA | 0 |
| S12-final-viral-combined_k141_81324\|\|full | 1033 | No | NA | 2 | 0 | 0 | Low-quality | Genome-fragment | 1.76 | AAI-based (high-confidence) | 0 |
| S9-final-viral-combined_k141_126467\|\|full | 1942 | No | NA | 2 | 0 | 1 | Not-determined | Genome-fragment | NA | NA | 0 |
| S9-final-viral-combined_k141_72891\|\|full | 1643 | No | NA | 2 | 0 | 2 | Not-determined | Genome-fragment | NA | NA | 0 |
| S9-final-viral-combined_k141_40988\|\|full | 3104 | No | NA | 5 | 0 | 4 | Not-determined | Genome-fragment | NA | NA | 0 |
| S9-final-viral-combined_k141_46260\|\|full | 3289 | No | NA | 5 | 0 | 2 | Not-determined | Genome-fragment | NA | NA | 0 |
| S9-final-viral-combined_k141_126480\|\|full | 3020 | No | NA | 5 | 0 | 1 | Not-determined | Genome-fragment | NA | NA | 0 |
| S9-final-viral-combined_k141_72902\|\|full | 1451 | No | NA | 2 | 0 | 1 | Not-determined | Genome-fragment | NA | NA | 0 |
| S9-final-viral-combined_k141_99943\|\|full | 3100 | No | NA | 3 | 0 | 3 | Not-determined | Genome-fragment | NA | NA | 0 |
| S12-final-viral-combined_k141_220662\|\|lt2gene | 1033 | No | NA | 2 | 2 | 0 | Low-quality | Genome-fragment | 2.37 | AAI-based (medium-confidence) | 0 |
| S12_k141_229923 | 1033 | No | NA | 2 | 1 | 0 | Low-quality | Genome-fragment | 3.5 | HMM-based (lower-bound) | 0 |
| S9-final-viral-combined_k141_30212\|\|full | 2148 | No | NA | 3 | 0 | 1 | Not-determined | Genome-fragment | NA | NA | 0 |
| S9-final-viral-combined_k141_78905\|\|full | 1111 | No | NA | 3 | 0 | 0 | Not-determined | Genome-fragment | NA | NA | 0 |
| S2-final-viral-combined_k141_117916\|\|full | 1033 | No | NA | 3 | 0 | 1 | Low-quality | Genome-fragment | 0.62 | AAI-based (medium-confidence) | 0 |
| S9-final-viral-combined_k141_41058\|\|full | 1598 | No | NA | 2 | 0 | 2 | Not-determined | Genome-fragment | NA | NA | 0 |
| S9-final-viral-combined_k141_99990\|\|full | 2222 | No | NA | 2 | 0 | 0 | Not-determined | Genome-fragment | NA | NA | 0 |
| S4-final-viral-combined_k141_12414\|\|full | 1033 | No | NA | 3 | 3 | 0 | Low-quality | Genome-fragment | 1.69 | AAI-based (high-confidence) | 0 |
| S11-final-viral-combined_k141_92607\|\|full | 1034 | No | NA | 3 | 0 | 0 | Low-quality | Genome-fragment | 0.96 | AAI-based (medium-confidence) | 0 |
| S11-final-viral-combined_k141_147842\|\|full | 1034 | No | NA | 2 | 0 | 0 | Low-quality | Genome-fragment | 1.8 | AAI-based (medium-confidence) | 0 |
| S9-final-viral-combined_k141_100007\|\|full | 3186 | No | NA | 3 | 0 | 2 | Not-determined | Genome-fragment | NA | NA | 0 |
| S9-final-viral-combined_k141_72925\|\|full | 1137 | No | NA | 3 | 0 | 0 | Not-determined | Genome-fragment | NA | NA | 0 |
| S9-final-viral-combined_k141_121920\|\|full | 1376 | No | NA | 3 | 0 | 0 | Not-determined | Genome-fragment | NA | NA | 0 |
| S1-final-viral-combined_k141_217583\|\|lt2gene | 1034 | No | NA | 2 | 1 | 0 | Low-quality | Genome-fragment | 2.88 | AAI-based (medium-confidence) | 0 |
| S9-final-viral-combined_k141_30245\|\|full | 1641 | No | NA | 3 | 0 | 2 | Not-determined | Genome-fragment | NA | NA | 0 |
| S9-final-viral-combined_k141_121927\|\|full | 9005 | No | NA | 13 | 0 | 2 | Not-determined | Genome-fragment | NA | NA | 0 |
| S1_k141_187950 | 1034 | No | NA | 3 | 1 | 0 | Low-quality | Genome-fragment | 2.14 | AAI-based (medium-confidence) | 0 |
| S3-final-viral-combined_k141_2508\|\|full | 1034 | No | NA | 5 | 0 | 0 | Low-quality | Genome-fragment | 2.71 | AAI-based (high-confidence) | 0 |
| S9-final-viral-combined_k141_100035\|\|full | 1781 | No | NA | 2 | 0 | 2 | Not-determined | Genome-fragment | NA | NA | 0 |
| S9-final-viral-combined_k141_121938\|\|full | 1486 | No | NA | 3 | 0 | 0 | Not-determined | Genome-fragment | NA | NA | 0 |
| S9-final-viral-combined_k141_84547\|\|full | 1752 | No | NA | 2 | 0 | 1 | Not-determined | Genome-fragment | NA | NA | 0 |
| S9-final-viral-combined_k141_52546\|\|full | 7164 | No | NA | 5 | 0 | 3 | Not-determined | Genome-fragment | NA | NA | 0 |
| S9-final-viral-combined_k141_52550\|\|full | 2115 | No | NA | 3 | 0 | 0 | Not-determined | Genome-fragment | NA | NA | 0 |
| S9-final-viral-combined_k141_116003\|\|full | 2045 | No | NA | 2 | 0 | 1 | Not-determined | Genome-fragment | NA | NA | 0 |
| S9-final-viral-combined_k141_30279\|\|full | 4737 | No | NA | 5 | 0 | 3 | Not-determined | Genome-fragment | NA | NA | 0 |
| S9-final-viral-combined_k141_89799\|\|full | 1985 | No | NA | 3 | 0 | 2 | Not-determined | Genome-fragment | NA | NA | 0 |
| S9-final-viral-combined_k141_73006\|\|full | 1042 | No | NA | 2 | 0 | 1 | Not-determined | Genome-fragment | NA | NA | 0 |
| S9-final-viral-combined_k141_94328\|\|full | 2610 | No | NA | 3 | 0 | 0 | Not-determined | Genome-fragment | NA | NA | 0 |
| S9-final-viral-combined_k141_14703\|\|full | 1068 | No | NA | 3 | 0 | 1 | Not-determined | Genome-fragment | NA | NA | 0 |
| S9-final-viral-combined_k141_122003\|\|full | 1051 | No | NA | 2 | 0 | 0 | Not-determined | Genome-fragment | NA | NA | 0 |
| S9-final-viral-combined_k141_105393\|\|full | 2727 | No | NA | 3 | 0 | 0 | Not-determined | Genome-fragment | NA | NA | 0 |
| S9-final-viral-combined_k141_116047\|\|full | 2108 | No | NA | 3 | 0 | 0 | Not-determined | Genome-fragment | NA | NA | 0 |
| S3-final-viral-combined_k141_49481\|\|full | 1034 | No | NA | 2 | 0 | 0 | Low-quality | Genome-fragment | 0.58 | AAI-based (medium-confidence) | 0 |
| S9-final-viral-combined_k141_94369\|\|full | 1910 | No | NA | 3 | 0 | 1 | Not-determined | Genome-fragment | NA | NA | 0 |
| S9-final-viral-combined_k141_78952\|\|full | 1942 | No | NA | 4 | 0 | 0 | Not-determined | Genome-fragment | NA | NA | 0 |
| S3_k141_85285 | 1034 | No | NA | 2 | 0 | 0 | Low-quality | Genome-fragment | 1.31 | AAI-based (high-confidence) | 0 |
| S5-final-viral-combined_k141_83545\|\|full | 1034 | No | NA | 3 | 1 | 0 | Low-quality | Genome-fragment | 1.9 | AAI-based (high-confidence) | 0 |
| S5-final-viral-combined_k141_78485\|\|full | 1034 | No | NA | 2 | 1 | 0 | Low-quality | Genome-fragment | 2.2 | AAI-based (high-confidence) | 0 |
| S9-final-viral-combined_k141_52613\|\|full | 3908 | No | NA | 5 | 0 | 0 | Not-determined | Genome-fragment | NA | NA | 0 |
| S9-final-viral-combined_k141_89907\|\|full | 2032 | No | NA | 3 | 0 | 1 | Not-determined | Genome-fragment | NA | NA | 0 |
| S9-final-viral-combined_k141_89920\|\|full | 1129 | No | NA | 2 | 0 | 2 | Not-determined | Genome-fragment | NA | NA | 0 |
| S5-final-viral-combined_k141_204500\|\|full | 1034 | No | NA | 4 | 1 | 0 | Low-quality | Genome-fragment | 2.35 | AAI-based (medium-confidence) | 0 |
| S9-final-viral-combined_k141_63174\|\|full | 3950 | No | NA | 4 | 0 | 3 | Not-determined | Genome-fragment | NA | NA | 0 |
| S9-final-viral-combined_k141_20274\|\|full | 5711 | No | NA | 8 | 0 | 2 | Not-determined | Genome-fragment | NA | NA | 0 |
| S9-final-viral-combined_k141_14752\|\|full | 4713 | No | NA | 6 | 0 | 6 | Not-determined | Genome-fragment | NA | NA | 0 |
| S9-final-viral-combined_k141_35929\|\|full | 1493 | No | NA | 3 | 0 | 2 | Not-determined | Genome-fragment | NA | NA | 0 |
| S9-final-viral-combined_k141_35930\|\|full | 1745 | No | NA | 3 | 0 | 2 | Not-determined | Genome-fragment | NA | NA | 0 |
| S9-final-viral-combined_k141_20287\|\|full | 4234 | No | NA | 7 | 0 | 1 | Not-determined | Genome-fragment | NA | NA | 0 |
| S7-final-viral-combined_k141_183295\|\|full | 1034 | No | NA | 2 | 0 | 0 | Low-quality | Genome-fragment | 2.25 | AAI-based (medium-confidence) | 0 |
| S7-final-viral-combined_k141_152322\|\|full | 1034 | No | NA | 3 | 0 | 0 | Low-quality | Genome-fragment | 1.15 | AAI-based (medium-confidence) | 0 |
| S17-final-viral-combined_k141_117954\|\|full | 1034 | No | NA | 3 | 0 | 0 | Low-quality | Genome-fragment | 2.59 | AAI-based (high-confidence) | 0 |
| S9-final-viral-combined_k141_41167\|\|full | 8576 | No | NA | 7 | 0 | 3 | Not-determined | Genome-fragment | NA | NA | 0 |
| S9-final-viral-combined_k141_116120\|\|full | 2234 | No | NA | 3 | 0 | 3 | Not-determined | Genome-fragment | NA | NA | 0 |
| S17-final-viral-combined_k141_199782\|\|full | 1034 | No | NA | 2 | 0 | 0 | Low-quality | Genome-fragment | 0.74 | AAI-based (medium-confidence) | 0 |
| S9-final-viral-combined_k141_46394\|\|full | 1393 | No | NA | 2 | 0 | 0 | Not-determined | Genome-fragment | NA | NA | 0 |
| S12-final-viral-combined_k141_235769\|\|full | 1034 | No | NA | 3 | 1 | 0 | Low-quality | Genome-fragment | 2.99 | AAI-based (medium-confidence) | 0 |
| S9-final-viral-combined_k141_41180\|\|full | 1607 | No | NA | 3 | 0 | 2 | Not-determined | Genome-fragment | NA | NA | 0 |
| S9-final-viral-combined_k141_14809\|\|full | 2058 | No | NA | 3 | 0 | 1 | Not-determined | Genome-fragment | NA | NA | 0 |
| S9-final-viral-combined_k141_57820\|\|full | 3230 | No | NA | 5 | 0 | 0 | Not-determined | Genome-fragment | NA | NA | 0 |
| S9-final-viral-combined_k141_122067\|\|full | 10762 | No | NA | 12 | 0 | 7 | Not-determined | Genome-fragment | NA | NA | 0 |
| S12-final-viral-combined_k141_21270\|\|full | 1034 | No | NA | 2 | 0 | 1 | Low-quality | Genome-fragment | 1.27 | AAI-based (medium-confidence) | 0 |
| S9-final-viral-combined_k141_52671\|\|full | 5771 | No | NA | 7 | 0 | 3 | Not-determined | Genome-fragment | NA | NA | 0 |
| S9-final-viral-combined_k141_46439\|\|full | 1298 | No | NA | 2 | 0 | 0 | Not-determined | Genome-fragment | NA | NA | 0 |
| S9-final-viral-combined_k141_35990\|\|full | 2108 | No | NA | 2 | 0 | 1 | Not-determined | Genome-fragment | NA | NA | 0 |
| S9-final-viral-combined_k141_105462\|\|full | 30714 | No | NA | 43 | 19 | 3 | Medium-quality | Genome-fragment | 68.9 | AAI-based (high-confidence) | 0 |
| S12-final-viral-combined_k141_122318\|\|full | 1034 | No | NA | 3 | 0 | 0 | Low-quality | Genome-fragment | 0.3 | AAI-based (medium-confidence) | 0 |
| S2-final-viral-combined_k141_219847\|\|full | 1034 | No | NA | 4 | 0 | 0 | Low-quality | Genome-fragment | 2.07 | AAI-based (medium-confidence) | 0 |
| S2-final-viral-combined_k141_107981\|\|full | 1034 | No | NA | 2 | 0 | 2 | Low-quality | Genome-fragment | 0.31 | AAI-based (medium-confidence) | 0 |
| S9-final-viral-combined_k141_122084\|\|full | 2786 | No | NA | 3 | 0 | 1 | Not-determined | Genome-fragment | NA | NA | 0 |
| S9-final-viral-combined_k141_84735\|\|full | 3232 | No | NA | 5 | 0 | 1 | Not-determined | Genome-fragment | NA | NA | 0 |
| S2-final-viral-combined_k141_202578\|\|full | 1034 | No | NA | 2 | 0 | 0 | Low-quality | Genome-fragment | 0.74 | AAI-based (medium-confidence) | 0 |
| S9-final-viral-combined_k141_57844\|\|full | 2487 | No | NA | 4 | 0 | 1 | Not-determined | Genome-fragment | NA | NA | 0 |
| S8-final-viral-combined_k141_164368\|\|lt2gene | 1034 | No | NA | 2 | 1 | 0 | Low-quality | Genome-fragment | 0.96 | AAI-based (medium-confidence) | 0 |
| S9-final-viral-combined_k141_36010\|\|full | 2108 | No | NA | 6 | 0 | 0 | Not-determined | Genome-fragment | NA | NA | 0 |
| S9-final-viral-combined_k141_57855\|\|full | 2717 | No | NA | 3 | 0 | 1 | Not-determined | Genome-fragment | NA | NA | 0 |
| S8_k141_114240 | 1034 | No | NA | 1 | 0 | 0 | Low-quality | Genome-fragment | 1.6 | AAI-based (medium-confidence) | 0 |
| S11-final-viral-combined_k141_136302\|\|full | 1035 | No | NA | 3 | 1 | 0 | Low-quality | Genome-fragment | 0.67 | HMM-based (lower-bound) | 0 |
| S11_k141_199534 | 1035 | No | NA | 2 | 0 | 1 | Low-quality | Genome-fragment | 2.12 | AAI-based (medium-confidence) | 0 |
| S1-final-viral-combined_k141_334103\|\|full | 1035 | No | NA | 3 | 1 | 0 | Low-quality | Genome-fragment | 1.85 | AAI-based (high-confidence) | 0 |
| S9-final-viral-combined_k141_36053\|\|full | 1242 | No | NA | 4 | 0 | 1 | Not-determined | Genome-fragment | NA | NA | 0 |
| S9-final-viral-combined_k141_110263\|\|full | 2737 | No | NA | 3 | 0 | 0 | Not-determined | Genome-fragment | NA | NA | 0 |
| S1_k141_268356 | 1035 | No | NA | 3 | 0 | 2 | Low-quality | Genome-fragment | 0.5 | AAI-based (medium-confidence) | 0 |
| S9-final-viral-combined_k141_100210\|\|full | 4272 | No | NA | 3 | 0 | 3 | Not-determined | Genome-fragment | NA | NA | 0 |
| S9-final-viral-combined_k141_94577\|\|full | 2178 | No | NA | 3 | 0 | 2 | Not-determined | Genome-fragment | NA | NA | 0 |
| S3_k141_31389 | 1035 | No | NA | 1 | 0 | 1 | Low-quality | Genome-fragment | 0.44 | AAI-based (medium-confidence) | 0 |
| S9-final-viral-combined_k141_90016\|\|full | 3753 | No | NA | 5 | 0 | 3 | Not-determined | Genome-fragment | NA | NA | 0 |
| S9-final-viral-combined_k141_46507\|\|full | 2612 | No | NA | 5 | 0 | 2 | Not-determined | Genome-fragment | NA | NA | 0 |
| S7-final-viral-combined_k141_226848\|\|full | 1035 | No | NA | 3 | 0 | 0 | Low-quality | Genome-fragment | 1.67 | AAI-based (high-confidence) | 0 |
| S9-final-viral-combined_k141_84849\|\|full | 1468 | No | NA | 2 | 0 | 1 | Not-determined | Genome-fragment | NA | NA | 0 |
| S9-final-viral-combined_k141_41220\|\|full | 1201 | No | NA | 1 | 0 | 1 | Not-determined | Genome-fragment | NA | NA | 0 |
| S9-final-viral-combined_k141_63412\|\|full | 1018 | No | NA | 2 | 0 | 2 | Not-determined | Genome-fragment | NA | NA | 0 |
| S12-final-viral-combined_k141_386017\|\|full | 1035 | No | NA | 1 | 0 | 0 | Low-quality | Genome-fragment | 1.72 | AAI-based (medium-confidence) | 0 |
| S12-final-viral-combined_k141_95332\|\|full | 1035 | No | NA | 4 | 0 | 0 | Low-quality | Genome-fragment | 2.21 | AAI-based (medium-confidence) | 0 |
| S12-final-viral-combined_k141_265465\|\|full | 1035 | No | NA | 3 | 0 | 0 | Low-quality | Genome-fragment | 2.44 | AAI-based (medium-confidence) | 0 |
| S9-final-viral-combined_k141_25704\|\|full | 1967 | No | NA | 2 | 0 | 1 | Not-determined | Genome-fragment | NA | NA | 0 |
| S9-final-viral-combined_k141_25706\|\|full | 1685 | No | NA | 2 | 0 | 1 | Not-determined | Genome-fragment | NA | NA | 0 |
| S12-final-viral-combined_k141_345613\|\|lt2gene | 1035 | No | NA | 1 | 1 | 0 | Low-quality | Genome-fragment | 1.88 | AAI-based (high-confidence) | 0 |
| S9-final-viral-combined_k141_126620\|\|full | 3278 | No | NA | 5 | 0 | 3 | Not-determined | Genome-fragment | NA | NA | 0 |
| S2_k141_158887 | 1035 | No | NA | 3 | 1 | 0 | Low-quality | Genome-fragment | 2.41 | AAI-based (high-confidence) | 0 |
| S4_k141_75982 | 1035 | No | NA | 3 | 0 | 0 | Low-quality | Genome-fragment | 1.01 | AAI-based (medium-confidence) | 0 |
| S9-final-viral-combined_k141_84913\|\|full | 4199 | No | NA | 4 | 0 | 2 | Not-determined | Genome-fragment | NA | NA | 0 |
| S4_k141_118283 | 1035 | No | NA | 2 | 0 | 0 | Low-quality | Genome-fragment | 1.5 | AAI-based (high-confidence) | 0 |
| S9-final-viral-combined_k141_57937\|\|full | 2896 | No | NA | 3 | 0 | 2 | Not-determined | Genome-fragment | NA | NA | 0 |
| S9-final-viral-combined_k141_22965\|\|full | 1036 | No | NA | 3 | 0 | 0 | Low-quality | Genome-fragment | 0.85 | AAI-based (medium-confidence) | 0 |
| S9-final-viral-combined_k141_63464\|\|full | 1581 | No | NA | 2 | 0 | 1 | Not-determined | Genome-fragment | NA | NA | 0 |
| S9-final-viral-combined_k141_9884\|\|full | 1862 | No | NA | 2 | 0 | 2 | Not-determined | Genome-fragment | NA | NA | 0 |
| S9-final-viral-combined_k141_30435\|\|full | 2066 | No | NA | 2 | 0 | 1 | Not-determined | Genome-fragment | NA | NA | 0 |
| S9-final-viral-combined_k141_23738\|\|lt2gene | 1036 | No | NA | 1 | 1 | 0 | Low-quality | Genome-fragment | 0.77 | HMM-based (lower-bound) | 0 |
| S11-final-viral-combined_k141_127675\|\|full | 1036 | No | NA | 2 | 0 | 0 | Low-quality | Genome-fragment | 2.15 | AAI-based (medium-confidence) | 0 |
| S9-final-viral-combined_k141_79115\|\|full | 2460 | No | NA | 3 | 0 | 0 | Not-determined | Genome-fragment | NA | NA | 0 |
| S9-final-viral-combined_k141_110324\|\|full | 2398 | No | NA | 4 | 0 | 4 | Not-determined | Genome-fragment | NA | NA | 0 |
| S9-final-viral-combined_k141_36196\|\|full | 1886 | No | NA | 3 | 0 | 0 | Not-determined | Genome-fragment | NA | NA | 0 |
| S11-final-viral-combined_k141_247174\|\|full | 1036 | No | NA | 3 | 0 | 0 | Low-quality | Genome-fragment | 3.14 | AAI-based (medium-confidence) | 0 |
| S7-final-viral-combined_k141_95159\|\|full | 1036 | No | NA | 3 | 0 | 0 | Low-quality | Genome-fragment | 1.78 | AAI-based (medium-confidence) | 0 |
| S7-final-viral-combined_k141_35944\|\|lt2gene | 1036 | No | NA | 2 | 0 | 0 | Low-quality | Genome-fragment | 1.92 | AAI-based (medium-confidence) | 0 |
| S9-final-viral-combined_k141_79133\|\|full | 1024 | No | NA | 2 | 0 | 1 | Not-determined | Genome-fragment | NA | NA | 0 |
| S9-final-viral-combined_k141_30472\|\|full | 4844 | No | NA | 7 | 0 | 3 | Not-determined | Genome-fragment | NA | NA | 0 |
| S17-final-viral-combined_k141_129024\|\|full | 1036 | No | NA | 3 | 1 | 0 | Low-quality | Genome-fragment | 2.37 | AAI-based (medium-confidence) | 0 |
| S12-final-viral-combined_k141_272478\|\|full | 1036 | No | NA | 3 | 0 | 0 | Low-quality | Genome-fragment | 1.2 | AAI-based (medium-confidence) | 0 |
| S9-final-viral-combined_k141_41287\|\|full | 2633 | No | NA | 5 | 0 | 3 | Not-determined | Genome-fragment | NA | NA | 0 |
| S9-final-viral-combined_k141_30483\|\|full | 2109 | No | NA | 3 | 0 | 1 | Not-determined | Genome-fragment | NA | NA | 0 |
| S9-final-viral-combined_k141_41289\|\|full | 1291 | No | NA | 1 | 0 | 0 | Not-determined | Genome-fragment | NA | NA | 0 |
| S9-final-viral-combined_k141_9918\|\|full | 1438 | No | NA | 3 | 0 | 2 | Not-determined | Genome-fragment | NA | NA | 0 |
| S9-final-viral-combined_k141_9929\|\|full | 1874 | No | NA | 3 | 0 | 3 | Not-determined | Genome-fragment | NA | NA | 0 |
| S9-final-viral-combined_k141_126656\|\|full | 7921 | No | NA | 3 | 0 | 2 | Not-determined | Genome-fragment | NA | NA | 0 |
| S12-final-viral-combined_k141_215420\|\|full | 1036 | No | NA | 3 | 0 | 0 | Low-quality | Genome-fragment | 2.56 | AAI-based (medium-confidence) | 0 |
| S2-final-viral-combined_k141_40943\|\|full | 1036 | No | NA | 2 | 0 | 0 | Low-quality | Genome-fragment | 0.69 | AAI-based (high-confidence) | 0 |
| S2-final-viral-combined_k141_195006\|\|lt2gene | 1036 | No | NA | 1 | 1 | 0 | Low-quality | Genome-fragment | 2.62 | AAI-based (high-confidence) | 0 |
| S9_k141_39619 | 1037 | No | NA | 2 | 0 | 0 | Low-quality | Genome-fragment | 1.73 | AAI-based (high-confidence) | 0 |
| S11-final-viral-combined_k141_77953\|\|full | 1037 | No | NA | 2 | 0 | 1 | Low-quality | Genome-fragment | 0.45 | AAI-based (medium-confidence) | 0 |
| S9-final-viral-combined_k141_58051\|\|full | 2115 | No | NA | 3 | 0 | 1 | Not-determined | Genome-fragment | NA | NA | 0 |
| S11-final-viral-combined_k141_305212\|\|lt2gene | 1037 | No | NA | 1 | 1 | 0 | Low-quality | Genome-fragment | 2.27 | AAI-based (high-confidence) | 0 |
| S9-final-viral-combined_k141_90167\|\|full | 1756 | No | NA | 3 | 0 | 2 | Not-determined | Genome-fragment | NA | NA | 0 |
| S9-final-viral-combined_k141_94690\|\|full | 7777 | No | NA | 14 | 0 | 6 | Not-determined | Genome-fragment | NA | NA | 0 |
| S9-final-viral-combined_k141_68596\|\|full | 2570 | No | NA | 3 | 0 | 2 | Not-determined | Genome-fragment | NA | NA | 0 |
| S9-final-viral-combined_k141_63519\|\|full | 1020 | No | NA | 2 | 0 | 1 | Not-determined | Genome-fragment | NA | NA | 0 |
| S1-final-viral-combined_k141_347725\|\|full | 1037 | No | NA | 2 | 0 | 0 | Low-quality | Genome-fragment | 2.14 | AAI-based (medium-confidence) | 0 |
| S5-final-viral-combined_k141_57499\|\|full | 1037 | No | NA | 4 | 0 | 0 | Low-quality | Genome-fragment | 1.66 | AAI-based (high-confidence) | 0 |
| S7-final-viral-combined_k141_134520\|\|full | 1037 | No | NA | 2 | 0 | 1 | Low-quality | Genome-fragment | 0.5 | AAI-based (high-confidence) | 0 |
| S9-final-viral-combined_k141_126713\|\|full | 1343 | No | NA | 3 | 0 | 1 | Not-determined | Genome-fragment | NA | NA | 0 |
| S9-final-viral-combined_k141_90185\|\|full | 4470 | No | NA | 2 | 0 | 2 | Not-determined | Genome-fragment | NA | NA | 0 |
| S7-final-viral-combined_k141_128899\|\|full | 1037 | No | NA | 3 | 1 | 0 | Low-quality | Genome-fragment | 1.85 | HMM-based (lower-bound) | 0 |
| S7_k141_131325 | 1037 | No | NA | 3 | 1 | 0 | Low-quality | Genome-fragment | 2.36 | AAI-based (high-confidence) | 0 |
| S9-final-viral-combined_k141_20485\|\|full | 90295 | Yes | 38188 | 110 | 36 | 45 | Complete | High-quality | 100 | Provirus (high-confidence) | 57.71 |
| S9-final-viral-combined_k141_122305\|\|full | 1101 | No | NA | 2 | 0 | 1 | Not-determined | Genome-fragment | NA | NA | 0 |
| S9-final-viral-combined_k141_9931\|\|full | 90630 | Yes | 43033 | 87 | 25 | 16 | Medium-quality | Genome-fragment | 63.56 | AAI-based (high-confidence) | 52.52 |
| S17-final-viral-combined_k141_260489\|\|full | 1037 | No | NA | 2 | 1 | 0 | Low-quality | Genome-fragment | 0.56 | HMM-based (lower-bound) | 0 |
| S17-final-viral-combined_k141_250368\|\|full | 1037 | No | NA | 3 | 1 | 0 | Low-quality | Genome-fragment | 1.83 | HMM-based (lower-bound) | 0 |
| S17-k141_129807 | 1037 | No | NA | 3 | 0 | 0 | Low-quality | Genome-fragment | 8.1 | AAI-based (high-confidence) | 0 |
| S9-final-viral-combined_k141_52901\|\|full | 2274 | No | NA | 3 | 0 | 3 | Not-determined | Genome-fragment | NA | NA | 0 |
| S17-k141_143946 | 1037 | No | NA | 2 | 0 | 1 | Low-quality | Genome-fragment | 1.49 | AAI-based (high-confidence) | 0 |
| S9-final-viral-combined_k141_4592\|\|full | 5015 | No | NA | 7 | 0 | 5 | Not-determined | Genome-fragment | NA | NA | 0 |
| S12-final-viral-combined_k141_59656\|\|full | 1037 | No | NA | 3 | 1 | 0 | Low-quality | Genome-fragment | 2.36 | AAI-based (medium-confidence) | 0 |
| S2-final-viral-combined_k141_187210\|\|full | 1037 | No | NA | 2 | 0 | 1 | Low-quality | Genome-fragment | 1.52 | AAI-based (medium-confidence) | 0 |
| S9-final-viral-combined_k141_52911\|\|full | 1469 | No | NA | 2 | 0 | 0 | Not-determined | Genome-fragment | NA | NA | 0 |
| S9-final-viral-combined_k141_68636\|\|full | 1051 | No | NA | 3 | 0 | 1 | Not-determined | Genome-fragment | NA | NA | 0 |
| S9-final-viral-combined_k141_94795\|\|full | 2286 | No | NA | 3 | 0 | 1 | Not-determined | Genome-fragment | NA | NA | 0 |
| S2-final-viral-combined_k141_222648\|\|full | 1037 | No | NA | 2 | 2 | 0 | Low-quality | Genome-fragment | 2.57 | AAI-based (high-confidence) | 0 |
| S9-final-viral-combined_k141_94800\|\|full | 5033 | No | NA | 6 | 0 | 5 | Not-determined | Genome-fragment | NA | NA | 0 |
| S9-final-viral-combined_k141_105561\|\|full | 3635 | No | NA | 6 | 0 | 1 | Not-determined | Genome-fragment | NA | NA | 0 |
| S9-final-viral-combined_k141_41506\|\|full | 2332 | No | NA | 3 | 0 | 0 | Not-determined | Genome-fragment | NA | NA | 0 |
| S9-final-viral-combined_k141_41507\|\|full | 1394 | No | NA | 2 | 0 | 2 | Not-determined | Genome-fragment | NA | NA | 0 |
| S9-final-viral-combined_k141_30636\|\|full | 2752 | No | NA | 4 | 0 | 0 | Not-determined | Genome-fragment | NA | NA | 0 |
| S9-final-viral-combined_k141_116398\|\|full | 5525 | No | NA | 7 | 0 | 2 | Not-determined | Genome-fragment | NA | NA | 0 |
| S9-final-viral-combined_k141_36299\|\|full | 2607 | No | NA | 5 | 0 | 2 | Not-determined | Genome-fragment | NA | NA | 0 |
| S9-final-viral-combined_k141_63666\|\|full | 1547 | No | NA | 2 | 0 | 1 | Not-determined | Genome-fragment | NA | NA | 0 |
| S9-final-viral-combined_k141_58080\|\|full | 1961 | No | NA | 2 | 0 | 2 | Not-determined | Genome-fragment | NA | NA | 0 |
| S2-final-viral-combined_k141_43047\|\|lt2gene | 1037 | No | NA | 1 | 1 | 0 | Low-quality | Genome-fragment | 2.63 | AAI-based (high-confidence) | 0 |
| S9-final-viral-combined_k141_30672\|\|full | 1974 | No | NA | 3 | 0 | 1 | Not-determined | Genome-fragment | NA | NA | 0 |
| S8_k141_22263 | 1037 | No | NA | 2 | 0 | 0 | Low-quality | Genome-fragment | 2.46 | AAI-based (medium-confidence) | 0 |
| S9-final-viral-combined_k141_20602\|\|full | 1974 | No | NA | 2 | 0 | 2 | Not-determined | Genome-fragment | NA | NA | 0 |
| S9-final-viral-combined_k141_49194\|\|full | 1038 | No | NA | 3 | 0 | 2 | Low-quality | Genome-fragment | 0.34 | AAI-based (high-confidence) | 0 |
| S9_k141_7196 | 1038 | No | NA | 5 | 0 | 0 | Low-quality | Genome-fragment | 0.7 | AAI-based (high-confidence) | 0 |
| S11-final-viral-combined_k141_21024\|\|full | 1038 | No | NA | 2 | 0 | 0 | Low-quality | Genome-fragment | 2.1 | AAI-based (high-confidence) | 0 |
| S9-final-viral-combined_k141_58120\|\|full | 1547 | No | NA | 1 | 0 | 1 | Not-determined | Genome-fragment | NA | NA | 0 |
| S11-final-viral-combined_k141_47382\|\|lt2gene | 1038 | No | NA | 1 | 0 | 0 | Low-quality | Genome-fragment | 1.82 | AAI-based (medium-confidence) | 0 |
| S9-final-viral-combined_k141_58125\|\|full | 4444 | No | NA | 5 | 0 | 0 | Not-determined | Genome-fragment | NA | NA | 0 |
| S9-final-viral-combined_k141_52999\|\|full | 2000 | No | NA | 2 | 0 | 2 | Not-determined | Genome-fragment | NA | NA | 0 |
| S9-final-viral-combined_k141_126758\|\|full | 1646 | No | NA | 2 | 0 | 0 | Not-determined | Genome-fragment | NA | NA | 0 |
| S11_k141_156595 | 1038 | No | NA | 1 | 1 | 0 | Low-quality | Genome-fragment | 2.16 | AAI-based (high-confidence) | 0 |
| S1-final-viral-combined_k141_84663\|\|full | 1038 | No | NA | 2 | 2 | 0 | Low-quality | Genome-fragment | 2.62 | AAI-based (high-confidence) | 0 |
| S1-final-viral-combined_k141_324147\|\|full | 1038 | No | NA | 2 | 0 | 1 | Low-quality | Genome-fragment | 0.78 | AAI-based (medium-confidence) | 0 |
| S9-final-viral-combined_k141_15065\|\|full | 4187 | No | NA | 6 | 0 | 2 | Not-determined | Genome-fragment | NA | NA | 0 |
| S9-final-viral-combined_k141_126769\|\|full | 4039 | No | NA | 4 | 0 | 0 | Not-determined | Genome-fragment | NA | NA | 0 |
| S9-final-viral-combined_k141_90372\|\|full | 11598 | No | NA | 11 | 0 | 8 | Not-determined | Genome-fragment | NA | NA | 0 |
| S9-final-viral-combined_k141_84979\|\|full | 1519 | No | NA | 3 | 0 | 1 | Not-determined | Genome-fragment | NA | NA | 0 |
| S9-final-viral-combined_k141_100514\|\|full | 1942 | No | NA | 3 | 0 | 0 | Not-determined | Genome-fragment | NA | NA | 0 |
| S1_k141_319682 | 1038 | No | NA | 1 | 0 | 0 | Low-quality | Genome-fragment | 0.91 | AAI-based (high-confidence) | 0 |
| S9-final-viral-combined_k141_122484\|\|full | 1385 | No | NA | 2 | 0 | 1 | Not-determined | Genome-fragment | NA | NA | 0 |
| S9-final-viral-combined_k141_4687\|\|full | 2693 | No | NA | 3 | 0 | 1 | Not-determined | Genome-fragment | NA | NA | 0 |
| S9-final-viral-combined_k141_30743\|\|full | 3177 | No | NA | 4 | 0 | 0 | Not-determined | Genome-fragment | NA | NA | 0 |
| S9-final-viral-combined_k141_110439\|\|full | 7250 | No | NA | 9 | 0 | 4 | Not-determined | Genome-fragment | NA | NA | 0 |
| S9-final-viral-combined_k141_73514\|\|full | 2315 | No | NA | 3 | 0 | 2 | Not-determined | Genome-fragment | NA | NA | 0 |
| S9-final-viral-combined_k141_79515\|\|full | 2070 | No | NA | 2 | 0 | 2 | Not-determined | Genome-fragment | NA | NA | 0 |
| S9-final-viral-combined_k141_122543\|\|full | 1388 | No | NA | 4 | 0 | 0 | Not-determined | Genome-fragment | NA | NA | 0 |
| S9-final-viral-combined_k141_10070\|\|full | 7254 | No | NA | 5 | 0 | 2 | Not-determined | Genome-fragment | NA | NA | 0 |
| S5-final-viral-combined_k141_27739\|\|full | 1038 | No | NA | 3 | 1 | 0 | Low-quality | Genome-fragment | 2.67 | AAI-based (high-confidence) | 0 |
| S9-final-viral-combined_k141_68881\|\|full | 1520 | No | NA | 2 | 0 | 2 | Not-determined | Genome-fragment | NA | NA | 0 |
| S9-final-viral-combined_k141_63829\|\|full | 12632 | No | NA | 17 | 0 | 7 | Not-determined | Genome-fragment | NA | NA | 0 |
| S5-final-viral-combined_k141_197102\|\|full | 1038 | No | NA | 3 | 1 | 0 | Low-quality | Genome-fragment | 3.21 | AAI-based (high-confidence) | 0 |
| S9-final-viral-combined_k141_68888\|\|full | 1174 | No | NA | 3 | 0 | 0 | Not-determined | Genome-fragment | NA | NA | 0 |
| S5-final-viral-combined_k141_188500\|\|full | 1038 | No | NA | 3 | 1 | 0 | Low-quality | Genome-fragment | 2.58 | HMM-based (lower-bound) | 0 |
| S5-final-viral-combined_k141_91704\|\|lt2gene | 1038 | No | NA | 2 | 0 | 0 | Low-quality | Genome-fragment | 0.64 | AAI-based (high-confidence) | 0 |
| S9-final-viral-combined_k141_85059\|\|full | 4571 | No | NA | 5 | 0 | 3 | Not-determined | Genome-fragment | NA | NA | 0 |
| S5_k141_46987 | 1038 | No | NA | 1 | 0 | 0 | Low-quality | Genome-fragment | 1.44 | AAI-based (high-confidence) | 0 |
| S9-final-viral-combined_k141_79532\|\|full | 6693 | No | NA | 5 | 0 | 1 | Not-determined | Genome-fragment | NA | NA | 0 |
| S5_k141_4139 | 1038 | No | NA | 3 | 0 | 0 | Low-quality | Genome-fragment | 7.12 | AAI-based (high-confidence) | 0 |
| S9-final-viral-combined_k141_110458\|\|full | 1458 | No | NA | 3 | 0 | 2 | Not-determined | Genome-fragment | NA | NA | 0 |
| S7_k141_226762 | 1038 | No | NA | 2 | 0 | 1 | Low-quality | Genome-fragment | 0.9 | AAI-based (high-confidence) | 0 |
| S17-final-viral-combined_k141_138139\|\|full | 1038 | No | NA | 4 | 2 | 0 | Low-quality | Genome-fragment | 2.39 | AAI-based (high-confidence) | 0 |
| S9-final-viral-combined_k141_90479\|\|full | 4547 | No | NA | 4 | 0 | 4 | Not-determined | Genome-fragment | NA | NA | 0 |
| S9-final-viral-combined_k141_36459\|\|full | 29514 | No | NA | 51 | 19 | 0 | Medium-quality | Genome-fragment | 89.69 | AAI-based (high-confidence) | 0 |
| S17-final-viral-combined_k141_152578\|\|full | 1038 | No | NA | 4 | 0 | 0 | Low-quality | Genome-fragment | 2.45 | AAI-based (high-confidence) | 0 |
| S9-final-viral-combined_k141_53137\|\|full | 1298 | No | NA | 2 | 0 | 1 | Not-determined | Genome-fragment | NA | NA | 0 |
| S9-final-viral-combined_k141_95014\|\|full | 2998 | No | NA | 3 | 0 | 3 | Not-determined | Genome-fragment | NA | NA | 0 |
| S12-final-viral-combined_k141_159271\|\|full | 1038 | No | NA | 3 | 1 | 0 | Low-quality | Genome-fragment | 0.63 | HMM-based (lower-bound) | 0 |
| S9-final-viral-combined_k141_10141\|\|full | 6594 | No | NA | 6 | 0 | 3 | Not-determined | Genome-fragment | NA | NA | 0 |
| S9-final-viral-combined_k141_46771\|\|full | 2292 | No | NA | 2 | 0 | 2 | Not-determined | Genome-fragment | NA | NA | 0 |
| S12-final-viral-combined_k141_32091\|\|lt2gene | 1038 | No | NA | 1 | 1 | 0 | Low-quality | Genome-fragment | 0.72 | HMM-based (lower-bound) | 0 |
| S12_k141_122990 | 1038 | No | NA | 1 | 0 | 1 | Low-quality | Genome-fragment | 2.5 | AAI-based (high-confidence) | 0 |
| S12_k141_224244 | 1038 | No | NA | 2 | 0 | 1 | Low-quality | Genome-fragment | 0.64 | AAI-based (medium-confidence) | 0 |
| S9-final-viral-combined_k141_63904\|\|full | 1130 | No | NA | 2 | 0 | 2 | Not-determined | Genome-fragment | NA | NA | 0 |
| S12_k141_395526 | 1038 | No | NA | 1 | 0 | 0 | Low-quality | Genome-fragment | 1.83 | AAI-based (high-confidence) | 0 |
| S9-final-viral-combined_k141_79607\|\|full | 1252 | No | NA | 3 | 0 | 0 | Not-determined | Genome-fragment | NA | NA | 0 |
| S9-final-viral-combined_k141_46804\|\|full | 8520 | No | NA | 10 | 0 | 8 | Not-determined | Genome-fragment | NA | NA | 0 |
| S2-final-viral-combined_k141_220097\|\|full | 1038 | No | NA | 3 | 1 | 0 | Low-quality | Genome-fragment | 2.64 | AAI-based (medium-confidence) | 0 |
| S9-final-viral-combined_k141_36526\|\|full | 1124 | No | NA | 2 | 0 | 1 | Not-determined | Genome-fragment | NA | NA | 0 |
| S2_k141_243759 | 1038 | No | NA | 3 | 1 | 0 | Low-quality | Genome-fragment | 1.14 | AAI-based (high-confidence) | 0 |
| S4-final-viral-combined_k141_106357\|\|full | 1038 | No | NA | 2 | 0 | 1 | Low-quality | Genome-fragment | 0.37 | AAI-based (medium-confidence) | 0 |
| S9-final-viral-combined_k141_100710\|\|full | 1092 | No | NA | 1 | 0 | 1 | Not-determined | Genome-fragment | NA | NA | 0 |
| S4_k141_49856 | 1038 | No | NA | 4 | 1 | 0 | Low-quality | Genome-fragment | 1.56 | AAI-based (medium-confidence) | 0 |
| S9-final-viral-combined_k141_41615\|\|full | 2346 | No | NA | 2 | 0 | 2 | Not-determined | Genome-fragment | NA | NA | 0 |
| S9-final-viral-combined_k141_116492\|\|full | 1714 | No | NA | 3 | 0 | 2 | Not-determined | Genome-fragment | NA | NA | 0 |
| S6-final-viral-combined_k141_81723\|\|full | 1038 | No | NA | 3 | 1 | 0 | Low-quality | Genome-fragment | 1.54 | HMM-based (lower-bound) | 0 |
| S9-final-viral-combined_k141_41644\|\|full | 1790 | No | NA | 4 | 0 | 0 | Not-determined | Genome-fragment | NA | NA | 0 |
| S9-final-viral-combined_k141_110571\|\|full | 3823 | No | NA | 5 | 0 | 2 | Not-determined | Genome-fragment | NA | NA | 0 |
| S8-final-viral-combined_k141_121582\|\|full | 1038 | No | NA | 3 | 0 | 0 | Low-quality | Genome-fragment | 1.39 | AAI-based (medium-confidence) | 0 |
| S9-final-viral-combined_k141_26173\|\|full | 1965 | No | NA | 3 | 0 | 1 | Not-determined | Genome-fragment | NA | NA | 0 |
| S9-final-viral-combined_k141_10213\|\|full | 1186 | No | NA | 2 | 0 | 1 | Not-determined | Genome-fragment | NA | NA | 0 |
| S8_k141_24031 | 1038 | No | NA | 2 | 0 | 1 | Low-quality | Genome-fragment | 2.17 | AAI-based (high-confidence) | 0 |
| S9-final-viral-combined_k141_53178\|\|full | 1387 | No | NA | 2 | 0 | 1 | Not-determined | Genome-fragment | NA | NA | 0 |
| S9-final-viral-combined_k141_95072\|\|full | 2479 | No | NA | 2 | 0 | 2 | Not-determined | Genome-fragment | NA | NA | 0 |
| S9-final-viral-combined_k141_97083\|\|full | 1039 | No | NA | 3 | 2 | 0 | Low-quality | Genome-fragment | 2.88 | AAI-based (high-confidence) | 0 |
| S9-final-viral-combined_k141_60207\|\|full | 1039 | No | NA | 3 | 0 | 0 | Low-quality | Genome-fragment | 2.28 | AAI-based (high-confidence) | 0 |
| S9-final-viral-combined_k141_88708\|\|full | 1039 | No | NA | 3 | 0 | 0 | Low-quality | Genome-fragment | 2.23 | AAI-based (medium-confidence) | 0 |
| S9_k141_30942 | 1039 | No | NA | 1 | 0 | 0 | Low-quality | Genome-fragment | 0.66 | AAI-based (high-confidence) | 0 |
| S9-final-viral-combined_k141_100783\|\|full | 1594 | No | NA | 2 | 0 | 1 | Not-determined | Genome-fragment | NA | NA | 0 |
| S9-final-viral-combined_k141_69102\|\|full | 1342 | No | NA | 2 | 0 | 2 | Not-determined | Genome-fragment | NA | NA | 0 |
| S11-final-viral-combined_k141_60999\|\|full | 1039 | No | NA | 3 | 0 | 0 | Low-quality | Genome-fragment | 0.79 | AAI-based (high-confidence) | 0 |
| S9-final-viral-combined_k141_69103\|\|full | 5438 | No | NA | 7 | 0 | 3 | Not-determined | Genome-fragment | NA | NA | 0 |
| S1-final-viral-combined_k141_150039\|\|full | 1039 | No | NA | 4 | 0 | 0 | Low-quality | Genome-fragment | 2.42 | AAI-based (high-confidence) | 0 |
| S9-final-viral-combined_k141_36694\|\|full | 1421 | No | NA | 3 | 0 | 1 | Not-determined | Genome-fragment | NA | NA | 0 |
| S1_k141_109164 | 1039 | No | NA | 2 | 0 | 0 | Low-quality | Genome-fragment | 1.92 | AAI-based (medium-confidence) | 0 |
| S9-final-viral-combined_k141_126981\|\|full | 4142 | No | NA | 4 | 0 | 2 | Not-determined | Genome-fragment | NA | NA | 0 |
| S3_k141_81461 | 1039 | No | NA | 1 | 1 | 0 | Low-quality | Genome-fragment | 1.1 | AAI-based (high-confidence) | 0 |
| S7-final-viral-combined_k141_226730\|\|full | 1039 | No | NA | 2 | 0 | 0 | Low-quality | Genome-fragment | 1.55 | AAI-based (medium-confidence) | 0 |
| S17-final-viral-combined_k141_210775\|\|full | 1039 | No | NA | 3 | 0 | 0 | Low-quality | Genome-fragment | 2.38 | AAI-based (medium-confidence) | 0 |
| S2-final-viral-combined_k141_172383\|\|lt2gene | 1039 | No | NA | 1 | 1 | 0 | Low-quality | Genome-fragment | 2.25 | AAI-based (high-confidence) | 0 |
| S2_k141_241185 | 1039 | No | NA | 2 | 0 | 2 | Low-quality | Genome-fragment | 0.94 | AAI-based (medium-confidence) | 0 |
| S8_k141_204886 | 1039 | No | NA | 2 | 0 | 2 | Low-quality | Genome-fragment | 0.8 | AAI-based (medium-confidence) | 0 |
| S9-final-viral-combined_k141_19063\|\|lt2gene | 1040 | No | NA | 2 | 1 | 0 | Low-quality | Genome-fragment | 2.24 | AAI-based (high-confidence) | 0 |
| S9-final-viral-combined_k141_4936\|\|full | 2115 | No | NA | 2 | 0 | 0 | Not-determined | Genome-fragment | NA | NA | 0 |
| S9-final-viral-combined_k141_36747\|\|full | 5815 | No | NA | 6 | 0 | 1 | Not-determined | Genome-fragment | NA | NA | 0 |
| S1-final-viral-combined_k141_217186\|\|full | 1040 | No | NA | 3 | 0 | 0 | Low-quality | Genome-fragment | 2.47 | AAI-based (high-confidence) | 0 |
| S9-final-viral-combined_k141_53295\|\|full | 1578 | No | NA | 3 | 0 | 1 | Not-determined | Genome-fragment | NA | NA | 0 |
| S9-final-viral-combined_k141_85328\|\|full | 4066 | No | NA | 4 | 0 | 3 | Not-determined | Genome-fragment | NA | NA | 0 |
| S9-final-viral-combined_k141_116694\|\|full | 2187 | No | NA | 3 | 0 | 2 | Not-determined | Genome-fragment | NA | NA | 0 |
| S1_k141_379001 | 1040 | No | NA | 1 | 0 | 1 | Low-quality | Genome-fragment | 0.5 | AAI-based (medium-confidence) | 0 |
| S9-final-viral-combined_k141_41802\|\|full | 4608 | No | NA | 9 | 0 | 1 | Not-determined | Genome-fragment | NA | NA | 0 |
| S9-final-viral-combined_k141_26287\|\|full | 1879 | No | NA | 3 | 0 | 2 | Not-determined | Genome-fragment | NA | NA | 0 |
| S7_k141_99963 | 1040 | No | NA | 2 | 0 | 1 | Low-quality | Genome-fragment | 0.16 | AAI-based (medium-confidence) | 0 |
| S9-final-viral-combined_k141_41818\|\|full | 2621 | No | NA | 3 | 0 | 2 | Not-determined | Genome-fragment | NA | NA | 0 |
| S17-final-viral-combined_k141_11390\|\|full | 1040 | No | NA | 3 | 2 | 0 | Low-quality | Genome-fragment | 2.23 | AAI-based (high-confidence) | 0 |
| S9-final-viral-combined_k141_90808\|\|full | 1222 | No | NA | 2 | 0 | 1 | Not-determined | Genome-fragment | NA | NA | 0 |
| S9-final-viral-combined_k141_30979\|\|full | 3417 | No | NA | 4 | 0 | 3 | Not-determined | Genome-fragment | NA | NA | 0 |
| S9-final-viral-combined_k141_73784\|\|full | 1465 | No | NA | 3 | 0 | 1 | Not-determined | Genome-fragment | NA | NA | 0 |
| S9-final-viral-combined_k141_90818\|\|full | 1486 | No | NA | 3 | 0 | 0 | Not-determined | Genome-fragment | NA | NA | 0 |
| S9-final-viral-combined_k141_69175\|\|full | 4213 | No | NA | 3 | 0 | 1 | Not-determined | Genome-fragment | NA | NA | 0 |
| S9-final-viral-combined_k141_73804\|\|full | 17806 | No | NA | 18 | 0 | 9 | Not-determined | Genome-fragment | NA | NA | 0 |
| S17-final-viral-combined_k141_105895\|\|full | 1040 | No | NA | 2 | 0 | 0 | Low-quality | Genome-fragment | 2.04 | AAI-based (high-confidence) | 0 |
| S9-final-viral-combined_k141_58366\|\|full | 1277 | No | NA | 3 | 0 | 1 | Not-determined | Genome-fragment | NA | NA | 0 |
| S9-final-viral-combined_k141_36873\|\|full | 1689 | No | NA | 3 | 0 | 0 | Not-determined | Genome-fragment | NA | NA | 0 |
| S17-final-viral-combined_k141_150482\|\|lt2gene | 1040 | No | NA | 2 | 0 | 0 | Low-quality | Genome-fragment | 1.92 | AAI-based (medium-confidence) | 0 |
| S9-final-viral-combined_k141_85431\|\|full | 3215 | No | NA | 6 | 0 | 1 | Not-determined | Genome-fragment | NA | NA | 0 |
| S9-final-viral-combined_k141_31057\|\|full | 3222 | No | NA | 3 | 0 | 2 | Not-determined | Genome-fragment | NA | NA | 0 |
| S9-final-viral-combined_k141_116758\|\|full | 2439 | No | NA | 3 | 0 | 2 | Not-determined | Genome-fragment | NA | NA | 0 |
| S9-final-viral-combined_k141_85438\|\|full | 2835 | No | NA | 3 | 0 | 2 | Not-determined | Genome-fragment | NA | NA | 0 |
| S9-final-viral-combined_k141_116765\|\|full | 5298 | No | NA | 9 | 0 | 4 | Not-determined | Genome-fragment | NA | NA | 0 |
| S9-final-viral-combined_k141_41907\|\|full | 1695 | No | NA | 3 | 0 | 0 | Not-determined | Genome-fragment | NA | NA | 0 |
| S17-k141_196905 | 1040 | No | NA | 2 | 0 | 0 | Low-quality | Genome-fragment | 2.29 | AAI-based (high-confidence) | 0 |
| S9-final-viral-combined_k141_105822\|\|full | 43006 | No | NA | 59 | 32 | 0 | High-quality | High-quality | 97.86 | AAI-based (high-confidence) | 0 |
| S17-k141_266339 | 1040 | No | NA | 2 | 0 | 1 | Low-quality | Genome-fragment | 0.33 | AAI-based (medium-confidence) | 0 |
| S9-final-viral-combined_k141_116787\|\|full | 1193 | No | NA | 2 | 0 | 1 | Not-determined | Genome-fragment | NA | NA | 0 |
| S9-final-viral-combined_k141_90897\|\|full | 1854 | No | NA | 4 | 0 | 0 | Not-determined | Genome-fragment | NA | NA | 0 |
| S12-final-viral-combined_k141_220734\|\|full | 1040 | No | NA | 3 | 1 | 0 | Low-quality | Genome-fragment | 0.66 | HMM-based (lower-bound) | 0 |
| S9-final-viral-combined_k141_15463\|\|full | 1189 | No | NA | 3 | 0 | 1 | Not-determined | Genome-fragment | NA | NA | 0 |
| S12-final-viral-combined_k141_75489\|\|full | 1040 | No | NA | 3 | 3 | 0 | Low-quality | Genome-fragment | 1.82 | HMM-based (lower-bound) | 0 |
| S9-final-viral-combined_k141_69210\|\|full | 2624 | No | NA | 2 | 0 | 2 | Not-determined | Genome-fragment | NA | NA | 0 |
| S9-final-viral-combined_k141_105854\|\|full | 1308 | No | NA | 2 | 0 | 1 | Not-determined | Genome-fragment | NA | NA | 0 |
| S12-final-viral-combined_k141_29418\|\|full | 1040 | No | NA | 3 | 0 | 1 | Low-quality | Genome-fragment | 0.27 | AAI-based (medium-confidence) | 0 |
| S9-final-viral-combined_k141_53426\|\|full | 1787 | No | NA | 3 | 0 | 2 | Not-determined | Genome-fragment | NA | NA | 0 |
| S12-final-viral-combined_k141_367961\|\|lt2gene | 1040 | No | NA | 2 | 1 | 0 | Low-quality | Genome-fragment | 2.52 | AAI-based (high-confidence) | 0 |
| S2-final-viral-combined_k141_56058\|\|lt2gene | 1040 | No | NA | 1 | 1 | 0 | Low-quality | Genome-fragment | 2.21 | AAI-based (high-confidence) | 0 |
| S9-final-viral-combined_k141_79889\|\|full | 2416 | No | NA | 6 | 0 | 1 | Not-determined | Genome-fragment | NA | NA | 0 |
| S9-final-viral-combined_k141_31066\|\|full | 3896 | No | NA | 3 | 0 | 1 | Not-determined | Genome-fragment | NA | NA | 0 |
| S9-final-viral-combined_k141_93448\|\|full | 1041 | No | NA | 3 | 0 | 0 | Low-quality | Genome-fragment | 2.63 | AAI-based (high-confidence) | 0 |
| S9-final-viral-combined_k141_58458\|\|full | 1893 | No | NA | 4 | 0 | 1 | Not-determined | Genome-fragment | NA | NA | 0 |
| S9-final-viral-combined_k141_26410\|\|full | 63551 | No | NA | 95 | 33 | 2 | High-quality | High-quality | 100 | AAI-based (high-confidence) | 0 |
| S1-final-viral-combined_k141_3224\|\|lt2gene | 1041 | No | NA | 2 | 1 | 0 | Low-quality | Genome-fragment | 1.9 | AAI-based (medium-confidence) | 0 |
| S5-final-viral-combined_k141_155346\|\|full | 1041 | No | NA | 3 | 2 | 0 | Low-quality | Genome-fragment | 3.07 | AAI-based (high-confidence) | 0 |
| S5-final-viral-combined_k141_16432\|\|lt2gene | 1041 | No | NA | 2 | 2 | 0 | Low-quality | Genome-fragment | 1.5 | AAI-based (high-confidence) | 0 |
| S9-final-viral-combined_k141_122746\|\|full | 1987 | No | NA | 3 | 0 | 1 | Not-determined | Genome-fragment | NA | NA | 0 |
| S9-final-viral-combined_k141_122747\|\|full | 2196 | No | NA | 4 | 0 | 1 | Not-determined | Genome-fragment | NA | NA | 0 |

| | | | | | | | | | | | |
|---|---|---|---|---|---|---|---|---|---|---|---|
| S9-final-viral-combined_k141_101111\|\|full | 1634 | No | NA | 3 | 0 | 1 | Not-determined | Genome-fragment | NA | NA | 0 |
| S9-final-viral-combined_k141_10362\|\|full | 1328 | No | NA | 3 | 0 | 1 | Not-determined | Genome-fragment | NA | NA | 0 |
| S9-final-viral-combined_k141_26458\|\|full | 1282 | No | NA | 3 | 0 | 0 | Not-determined | Genome-fragment | NA | NA | 0 |
| S9-final-viral-combined_k141_15629\|\|full | 2450 | No | NA | 2 | 0 | 2 | Not-determined | Genome-fragment | NA | NA | 0 |
| S7-final-viral-combined_k141_121472\|\|lt2gene | 1041 | No | NA | 2 | 1 | 0 | Low-quality | Genome-fragment | 2.19 | AAI-based (high-confidence) | 0 |
| S9-final-viral-combined_k141_58515\|\|full | 1459 | No | NA | 3 | 0 | 2 | Not-determined | Genome-fragment | NA | NA | 0 |
| S12-final-viral-combined_k141_352541\|\|full | 1041 | No | NA | 3 | 0 | 0 | Low-quality | Genome-fragment | 1.52 | AAI-based (high-confidence) | 0 |
| S9-final-viral-combined_k141_37121\|\|full | 1587 | No | NA | 3 | 0 | 0 | Not-determined | Genome-fragment | NA | NA | 0 |
| S9-final-viral-combined_k141_37126\|\|full | 1601 | No | NA | 3 | 0 | 0 | Not-determined | Genome-fragment | NA | NA | 0 |
| S9-final-viral-combined_k141_26533\|\|full | 1823 | No | NA | 2 | 0 | 1 | Not-determined | Genome-fragment | NA | NA | 0 |
| S9-final-viral-combined_k141_101207\|\|full | 1217 | No | NA | 2 | 0 | 1 | Not-determined | Genome-fragment | NA | NA | 0 |
| S9-final-viral-combined_k141_26559\|\|full | 1948 | No | NA | 3 | 0 | 2 | Not-determined | Genome-fragment | NA | NA | 0 |
| S9-final-viral-combined_k141_47018\|\|full | 1183 | No | NA | 4 | 0 | 0 | Not-determined | Genome-fragment | NA | NA | 0 |
| S12_k141_256968 | 1041 | No | NA | 3 | 0 | 2 | Low-quality | Genome-fragment | 2.12 | AAI-based (medium-confidence) | 0 |
| S2-final-viral-combined_k141_52837\|\|full | 1041 | No | NA | 3 | 0 | 0 | Low-quality | Genome-fragment | 1.28 | AAI-based (medium-confidence) | 0 |
| S2-final-viral-combined_k141_240189\|\|full | 1041 | No | NA | 2 | 0 | 0 | Low-quality | Genome-fragment | 1.83 | AAI-based (high-confidence) | 0 |
| S9-final-viral-combined_k141_127086\|\|full | 1014 | No | NA | 2 | 0 | 1 | Not-determined | Genome-fragment | NA | NA | 0 |
| S9-final-viral-combined_k141_5130\|\|full | 1336 | No | NA | 2 | 0 | 1 | Not-determined | Genome-fragment | NA | NA | 0 |
| S9-final-viral-combined_k141_26615\|\|full | 3058 | No | NA | 5 | 0 | 0 | Not-determined | Genome-fragment | NA | NA | 0 |
| S4_k141_76287 | 1041 | No | NA | 2 | 0 | 1 | Low-quality | Genome-fragment | 0.14 | AAI-based (medium-confidence) | 0 |
| S9-final-viral-combined_k141_15729\|\|full | 3159 | No | NA | 4 | 0 | 2 | Not-determined | Genome-fragment | NA | NA | 0 |
| S11-final-viral-combined_k141_245289\|\|full | 1042 | No | NA | 2 | 0 | 0 | Low-quality | Genome-fragment | 2.06 | AAI-based (medium-confidence) | 0 |
| S9-final-viral-combined_k141_74040\|\|full | 5135 | No | NA | 4 | 0 | 2 | Not-determined | Genome-fragment | NA | NA | 0 |
| S1-final-viral-combined_k141_342141\|\|full | 1042 | No | NA | 3 | 0 | 0 | Low-quality | Genome-fragment | 2.15 | AAI-based (medium-confidence) | 0 |
| S9-final-viral-combined_k141_10460\|\|full | 3549 | No | NA | 3 | 0 | 1 | Not-determined | Genome-fragment | NA | NA | 0 |
| S1-final-viral-combined_k141_146027\|\|full | 1042 | No | NA | 4 | 1 | 0 | Low-quality | Genome-fragment | 2.36 | AAI-based (high-confidence) | 0 |
| S9-final-viral-combined_k141_74063\|\|full | 4092 | No | NA | 6 | 0 | 1 | Not-determined | Genome-fragment | NA | NA | 0 |
| S9-final-viral-combined_k141_21198\|\|full | 1041 | No | NA | 2 | 0 | 1 | Not-determined | Genome-fragment | NA | NA | 0 |
| S9-final-viral-combined_k141_15776\|\|full | 2609 | No | NA | 3 | 0 | 3 | Not-determined | Genome-fragment | NA | NA | 0 |
| S9-final-viral-combined_k141_26652\|\|full | 13994 | No | NA | 11 | 0 | 4 | Not-determined | Genome-fragment | NA | NA | 0 |
| S9-final-viral-combined_k141_26654\|\|full | 3200 | No | NA | 4 | 0 | 3 | Not-determined | Genome-fragment | NA | NA | 0 |
| S9-final-viral-combined_k141_26656\|\|full | 1343 | No | NA | 3 | 0 | 0 | Not-determined | Genome-fragment | NA | NA | 0 |
| S9-final-viral-combined_k141_101270\|\|full | 1105 | No | NA | 2 | 0 | 2 | Not-determined | Genome-fragment | NA | NA | 0 |
| S9-final-viral-combined_k141_26666\|\|full | 2981 | No | NA | 4 | 0 | 3 | Not-determined | Genome-fragment | NA | NA | 0 |
| S9-final-viral-combined_k141_58608\|\|full | 4746 | No | NA | 9 | 0 | 1 | Not-determined | Genome-fragment | NA | NA | 0 |
| S9-final-viral-combined_k141_47096\|\|full | 1379 | No | NA | 3 | 0 | 1 | Not-determined | Genome-fragment | NA | NA | 0 |
| S9-final-viral-combined_k141_80036\|\|full | 2664 | No | NA | 5 | 0 | 0 | Not-determined | Genome-fragment | NA | NA | 0 |
| S9-final-viral-combined_k141_95399\|\|full | 1338 | No | NA | 2 | 0 | 0 | Not-determined | Genome-fragment | NA | NA | 0 |
| S1_k141_38 | 1042 | No | NA | 3 | 0 | 1 | Low-quality | Genome-fragment | 0.76 | AAI-based (medium-confidence) | 0 |
| S9-final-viral-combined_k141_74121\|\|full | 2460 | No | NA | 5 | 0 | 1 | Not-determined | Genome-fragment | NA | NA | 0 |
| S5-final-viral-combined_k141_67484\|\|full | 1042 | No | NA | 4 | 1 | 0 | Low-quality | Genome-fragment | 0.63 | AAI-based (medium-confidence) | 0 |
| S9-final-viral-combined_k141_10518\|\|full | 1133 | No | NA | 3 | 0 | 0 | Not-determined | Genome-fragment | NA | NA | 0 |
| S5-final-viral-combined_k141_64131\|\|lt2gene | 1042 | No | NA | 1 | 1 | 0 | Low-quality | Genome-fragment | 1.86 | HMM-based (lower-bound) | 0 |
| S9-final-viral-combined_k141_37257\|\|full | 2774 | No | NA | 4 | 0 | 4 | Not-determined | Genome-fragment | NA | NA | 0 |
| S7_k141_80582 | 1042 | No | NA | 2 | 0 | 2 | Low-quality | Genome-fragment | 0.78 | AAI-based (medium-confidence) | 0 |
| S9-final-viral-combined_k141_37289\|\|full | 2196 | No | NA | 3 | 0 | 1 | Not-determined | Genome-fragment | NA | NA | 0 |
| S9-final-viral-combined_k141_31296\|\|full | 1591 | No | NA | 3 | 0 | 2 | Not-determined | Genome-fragment | NA | NA | 0 |
| S9-final-viral-combined_k141_21301\|\|full | 1444 | No | NA | 1 | 0 | 0 | Not-determined | Genome-fragment | NA | NA | 0 |
| S17-final-viral-combined_k141_103019\|\|full | 1042 | No | NA | 2 | 0 | 1 | Low-quality | Genome-fragment | 1.66 | AAI-based (medium-confidence) | 0 |
| S9-final-viral-combined_k141_21310\|\|full | 6308 | No | NA | 6 | 0 | 3 | Not-determined | Genome-fragment | NA | NA | 0 |
| S17-final-viral-combined_k141_140102\|\|full | 1042 | No | NA | 3 | 0 | 0 | Low-quality | Genome-fragment | 2.61 | AAI-based (medium-confidence) | 0 |
| S9-final-viral-combined_k141_42180\|\|full | 7539 | No | NA | 4 | 0 | 4 | Not-determined | Genome-fragment | NA | NA | 0 |
| S17-k141_44388 | 1042 | No | NA | 2 | 0 | 0 | Low-quality | Genome-fragment | 0.77 | AAI-based (medium-confidence) | 0 |
| S9-final-viral-combined_k141_80110\|\|full | 3636 | No | NA | 2 | 0 | 1 | Not-determined | Genome-fragment | NA | NA | 0 |
| S9-final-viral-combined_k141_116905\|\|full | 3364 | No | NA | 3 | 0 | 2 | Not-determined | Genome-fragment | NA | NA | 0 |
| S12-final-viral-combined_k141_3132\|\|full | 1042 | No | NA | 3 | 1 | 0 | Low-quality | Genome-fragment | 1.73 | AAI-based (high-confidence) | 0 |
| S12-final-viral-combined_k141_324970\|\|full | 1042 | No | NA | 2 | 1 | 0 | Low-quality | Genome-fragment | 1.71 | AAI-based (high-confidence) | 0 |
| S9-final-viral-combined_k141_116938\|\|full | 1094 | No | NA | 2 | 0 | 0 | Not-determined | Genome-fragment | NA | NA | 0 |
| S9-final-viral-combined_k141_42214\|\|full | 5813 | No | NA | 2 | 0 | 0 | Not-determined | Genome-fragment | NA | NA | 0 |
| S12_k141_343032 | 1042 | No | NA | 1 | 0 | 1 | Low-quality | Genome-fragment | 0.17 | AAI-based (high-confidence) | 0 |
| S12_k141_260513 | 1042 | No | NA | 2 | 0 | 0 | Low-quality | Genome-fragment | 2.13 | AAI-based (high-confidence) | 0 |
| S9-final-viral-combined_k141_47111\|\|full | 1510 | No | NA | 2 | 0 | 1 | Not-determined | Genome-fragment | NA | NA | 0 |
| S9-final-viral-combined_k141_47115\|\|full | 2687 | No | NA | 4 | 0 | 1 | Not-determined | Genome-fragment | NA | NA | 0 |
| S9-final-viral-combined_k141_47119\|\|full | 2589 | No | NA | 2 | 0 | 2 | Not-determined | Genome-fragment | NA | NA | 0 |
| S9-final-viral-combined_k141_127159\|\|full | 1755 | No | NA | 3 | 0 | 1 | Not-determined | Genome-fragment | NA | NA | 0 |
| S2-final-viral-combined_k141_54858\|\|full | 1042 | No | NA | 3 | 0 | 0 | Low-quality | Genome-fragment | 2.14 | AAI-based (high-confidence) | 0 |
| S2-final-viral-combined_k141_154491\|\|lt2gene | 1042 | No | NA | 1 | 1 | 0 | Low-quality | Genome-fragment | 2.2 | AAI-based (high-confidence) | 0 |
| S9-final-viral-combined_k141_42267\|\|full | 2251 | No | NA | 4 | 0 | 0 | Not-determined | Genome-fragment | NA | NA | 0 |
| S2_k141_119555 | 1042 | No | NA | 2 | 0 | 0 | Low-quality | Genome-fragment | 1.09 | AAI-based (high-confidence) | 0 |
| S9-final-viral-combined_k141_31425\|\|full | 2543 | No | NA | 2 | 0 | 1 | Not-determined | Genome-fragment | NA | NA | 0 |
| S4-final-viral-combined_k141_113983\|\|full | 1042 | No | NA | 3 | 0 | 1 | Low-quality | Genome-fragment | 1.01 | AAI-based (medium-confidence) | 0 |
| S9-final-viral-combined_k141_31440\|\|full | 4603 | No | NA | 5 | 0 | 1 | Not-determined | Genome-fragment | NA | NA | 0 |
| S4_k141_147591 | 1042 | No | NA | 2 | 0 | 0 | Low-quality | Genome-fragment | 1.09 | AAI-based (high-confidence) | 0 |
| S4_k141_24386 | 1042 | No | NA | 1 | 0 | 0 | Low-quality | Genome-fragment | 0.74 | AAI-based (high-confidence) | 0 |
| S9-final-viral-combined_k141_117052\|\|full | 1328 | No | NA | 5 | 0 | 0 | Not-determined | Genome-fragment | NA | NA | 0 |
| S9-final-viral-combined_k141_95550\|\|full | 1423 | No | NA | 2 | 0 | 2 | Not-determined | Genome-fragment | NA | NA | 0 |
| S9-final-viral-combined_k141_16017\|\|full | 2960 | No | NA | 3 | 0 | 1 | Not-determined | Genome-fragment | NA | NA | 0 |
| S9-final-viral-combined_k141_117093\|\|full | 3091 | No | NA | 3 | 0 | 0 | Not-determined | Genome-fragment | NA | NA | 0 |
| S6_k141_65318 | 1042 | No | NA | 3 | 0 | 0 | Low-quality | Genome-fragment | 2.77 | AAI-based (medium-confidence) | 0 |
| S9-final-viral-combined_k141_117094\|\|full | 2018 | No | NA | 2 | 0 | 1 | Not-determined | Genome-fragment | NA | NA | 0 |
| S8_k141_255585 | 1042 | No | NA | 1 | 0 | 0 | Low-quality | Genome-fragment | 0.68 | AAI-based (high-confidence) | 0 |
| S9-final-viral-combined_k141_117124\|\|full | 2120 | No | NA | 2 | 0 | 2 | Not-determined | Genome-fragment | NA | NA | 0 |
| S9-final-viral-combined_k141_31456\|\|full | 48386 | Yes | 34943 | 54 | 9 | 6 | Medium-quality | Genome-fragment | 72.49 | AAI-based (medium-confidence) | 27.78 |
| S9-final-viral-combined_k141_74315\|\|full | 4687 | No | NA | 5 | 0 | 4 | Not-determined | Genome-fragment | NA | NA | 0 |
| S9-final-viral-combined_k141_101527\|\|full | 2567 | No | NA | 2 | 0 | 2 | Not-determined | Genome-fragment | NA | NA | 0 |
| S9-final-viral-combined_k141_64950\|\|lt2gene | 1043 | No | NA | 2 | 1 | 0 | Low-quality | Genome-fragment | 2.23 | AAI-based (high-confidence) | 0 |
| S9-final-viral-combined_k141_106161\|\|full | 2404 | No | NA | 4 | 0 | 1 | Not-determined | Genome-fragment | NA | NA | 0 |
| S9-final-viral-combined_k141_127273\|\|full | 1035 | No | NA | 2 | 0 | 2 | Not-determined | Genome-fragment | NA | NA | 0 |
| S9-final-viral-combined_k141_106166\|\|full | 1679 | No | NA | 2 | 0 | 0 | Not-determined | Genome-fragment | NA | NA | 0 |
| S9-final-viral-combined_k141_117152\|\|full | 1720 | No | NA | 5 | 0 | 2 | Not-determined | Genome-fragment | NA | NA | 0 |
| S9-final-viral-combined_k141_127281\|\|full | 2522 | No | NA | 3 | 0 | 3 | Not-determined | Genome-fragment | NA | NA | 0 |
| S9-final-viral-combined_k141_101551\|\|full | 4535 | No | NA | 6 | 0 | 0 | Not-determined | Genome-fragment | NA | NA | 0 |
| S11-final-viral-combined_k141_42262\|\|full | 1043 | No | NA | 3 | 0 | 0 | Low-quality | Genome-fragment | 2.29 | AAI-based (high-confidence) | 0 |
| S11-final-viral-combined_k141_329087\|\|full | 1043 | No | NA | 3 | 0 | 0 | Low-quality | Genome-fragment | 2.11 | AAI-based (medium-confidence) | 0 |
| S11-final-viral-combined_k141_135144\|\|lt2gene | 1043 | No | NA | 2 | 2 | 0 | Low-quality | Genome-fragment | 1.68 | HMM-based (lower-bound) | 0 |
| S1-final-viral-combined_k141_245604\|\|lt2gene | 1043 | No | NA | 1 | 1 | 0 | Low-quality | Genome-fragment | 20.33 | AAI-based (medium-confidence) | 0 |
| S5-final-viral-combined_k141_209279\|\|full | 1043 | No | NA | 2 | 0 | 0 | Low-quality | Genome-fragment | 1.91 | AAI-based (medium-confidence) | 0 |
| S9-final-viral-combined_k141_95624\|\|full | 3522 | No | NA | 3 | 0 | 1 | Not-determined | Genome-fragment | NA | NA | 0 |
| S17-final-viral-combined_k141_90077\|\|full | 1043 | No | NA | 2 | 1 | 0 | Low-quality | Genome-fragment | 2.64 | AAI-based (high-confidence) | 0 |
| S17-final-viral-combined_k141_98610\|\|lt2gene | 1043 | No | NA | 2 | 1 | 0 | Low-quality | Genome-fragment | 0.63 | HMM-based (lower-bound) | 0 |
| S9-final-viral-combined_k141_106258\|\|full | 1149 | No | NA | 2 | 0 | 0 | Not-determined | Genome-fragment | NA | NA | 0 |
| S9-final-viral-combined_k141_101602\|\|full | 2492 | No | NA | 3 | 0 | 1 | Not-determined | Genome-fragment | NA | NA | 0 |
| S12-final-viral-combined_k141_54795\|\|full | 1043 | No | NA | 2 | 0 | 0 | Low-quality | Genome-fragment | 2.44 | AAI-based (medium-confidence) | 0 |
| S9-final-viral-combined_k141_127319\|\|full | 7358 | No | NA | 7 | 0 | 6 | Not-determined | Genome-fragment | NA | NA | 0 |
| S12-final-viral-combined_k141_229714\|\|full | 1043 | No | NA | 3 | 0 | 1 | Low-quality | Genome-fragment | 0.65 | AAI-based (medium-confidence) | 0 |
| S2-final-viral-combined_k141_126249\|\|full | 1043 | No | NA | 3 | 0 | 0 | Low-quality | Genome-fragment | 0.59 | AAI-based (medium-confidence) | 0 |
| S9-final-viral-combined_k141_31614\|\|full | 2041 | No | NA | 2 | 0 | 1 | Not-determined | Genome-fragment | NA | NA | 0 |
| S9-final-viral-combined_k141_42439\|\|full | 1387 | No | NA | 2 | 0 | 1 | Not-determined | Genome-fragment | NA | NA | 0 |
| S9-final-viral-combined_k141_106308\|\|full | 3315 | No | NA | 4 | 0 | 1 | Not-determined | Genome-fragment | NA | NA | 0 |
| S9-final-viral-combined_k141_31682\|\|full | 2326 | No | NA | 2 | 0 | 2 | Not-determined | Genome-fragment | NA | NA | 0 |
| S9-final-viral-combined_k141_110827\|\|full | 3816 | No | NA | 5 | 0 | 2 | Not-determined | Genome-fragment | NA | NA | 0 |
| S4-final-viral-combined_k141_30678\|\|full | 1043 | No | NA | 3 | 0 | 1 | Low-quality | Genome-fragment | 0.38 | AAI-based (medium-confidence) | 0 |
| S9-final-viral-combined_k141_74484\|\|full | 2643 | No | NA | 4 | 0 | 2 | Not-determined | Genome-fragment | NA | NA | 0 |
| S6-final-viral-combined_k141_125401\|\|full | 1043 | No | NA | 4 | 3 | 0 | Low-quality | Genome-fragment | 1.67 | HMM-based (lower-bound) | 0 |
| S9-final-viral-combined_k141_110846\|\|full | 5478 | No | NA | 5 | 0 | 4 | Not-determined | Genome-fragment | NA | NA | 0 |
| S9-final-viral-combined_k141_106379\|\|full | 2048 | No | NA | 4 | 0 | 0 | Not-determined | Genome-fragment | NA | NA | 0 |
| S8-final-viral-combined_k141_237713\|\|lt2gene | 1043 | No | NA | 1 | 1 | 0 | Low-quality | Genome-fragment | 2.48 | AAI-based (high-confidence) | 0 |
| S8_k141_267644 | 1043 | No | NA | 1 | 1 | 0 | Low-quality | Genome-fragment | 0.75 | AAI-based (high-confidence) | 0 |
| S9-final-viral-combined_k141_48028\|\|lt2gene | 1044 | No | NA | 2 | 1 | 0 | Low-quality | Genome-fragment | 2.27 | AAI-based (high-confidence) | 0 |
| S9-final-viral-combined_k141_110875\|\|full | 1505 | No | NA | 2 | 0 | 1 | Not-determined | Genome-fragment | NA | NA | 0 |
| S9-final-viral-combined_k141_47311\|\|full | 1851 | No | NA | 3 | 0 | 1 | Not-determined | Genome-fragment | NA | NA | 0 |
| S9-final-viral-combined_k141_74507\|\|full | 3556 | No | NA | 4 | 0 | 1 | Not-determined | Genome-fragment | NA | NA | 0 |
| S11-final-viral-combined_k141_95743\|\|lt2gene | 1044 | No | NA | 2 | 1 | 0 | Low-quality | Genome-fragment | 2.67 | AAI-based (high-confidence) | 0 |
| S11_k141_310817 | 1044 | No | NA | 4 | 0 | 0 | Low-quality | Genome-fragment | 1.69 | AAI-based (high-confidence) | 0 |
| S9-final-viral-combined_k141_110908\|\|full | 5545 | No | NA | 5 | 0 | 3 | Not-determined | Genome-fragment | NA | NA | 0 |
| S9-final-viral-combined_k141_47363\|\|full | 1151 | No | NA | 4 | 0 | 1 | Not-determined | Genome-fragment | NA | NA | 0 |
| S1-final-viral-combined_k141_159859\|\|full | 1044 | No | NA | 2 | 1 | 0 | Low-quality | Genome-fragment | 2.11 | AAI-based (medium-confidence) | 0 |
| S5-final-viral-combined_k141_2706\|\|full | 1044 | No | NA | 3 | 1 | 0 | Low-quality | Genome-fragment | 2.16 | AAI-based (medium-confidence) | 0 |
| S9-final-viral-combined_k141_117317\|\|full | 1167 | No | NA | 1 | 0 | 0 | Not-determined | Genome-fragment | NA | NA | 0 |
| S9-final-viral-combined_k141_95825\|\|full | 1052 | No | NA | 2 | 0 | 1 | Not-determined | Genome-fragment | NA | NA | 0 |
| S5-final-viral-combined_k141_203745\|\|full | 1044 | No | NA | 3 | 0 | 1 | Low-quality | Genome-fragment | 0.66 | AAI-based (medium-confidence) | 0 |
| S17-k141_125977 | 1044 | No | NA | 2 | 1 | 0 | Low-quality | Genome-fragment | 2.1 | AAI-based (high-confidence) | 0 |
| S9-final-viral-combined_k141_106504\|\|full | 4219 | No | NA | 3 | 0 | 1 | Not-determined | Genome-fragment | NA | NA | 0 |
| S12-final-viral-combined_k141_359752\|\|lt2gene | 1044 | No | NA | 1 | 0 | 0 | Low-quality | Genome-fragment | 2.13 | AAI-based (high-confidence) | 0 |
| S2_k141_94686 | 1044 | No | NA | 1 | 0 | 1 | Low-quality | Genome-fragment | 0.69 | AAI-based (high-confidence) | 0 |
| S9-final-viral-combined_k141_117424\|\|full | 2978 | No | NA | 4 | 0 | 1 | Not-determined | Genome-fragment | NA | NA | 0 |
| S4-final-viral-combined_k141_105304\|\|full | 1044 | No | NA | 3 | 1 | 0 | Low-quality | Genome-fragment | 0.62 | AAI-based (high-confidence) | 0 |
| S9-final-viral-combined_k141_110937\|\|full | 1160 | No | NA | 3 | 0 | 2 | Not-determined | Genome-fragment | NA | NA | 0 |
| S9-final-viral-combined_k141_95959\|\|full | 2967 | No | NA | 1 | 0 | 1 | Not-determined | Genome-fragment | NA | NA | 0 |
| S9-final-viral-combined_k141_106570\|\|full | 1029 | No | NA | 2 | 0 | 1 | Not-determined | Genome-fragment | NA | NA | 0 |
| S9-final-viral-combined_k141_96018\|\|full | 4555 | No | NA | 3 | 0 | 3 | Not-determined | Genome-fragment | NA | NA | 0 |
| S9-final-viral-combined_k141_106607\|\|full | 3665 | No | NA | 4 | 0 | 3 | Not-determined | Genome-fragment | NA | NA | 0 |
| S9-final-viral-combined_k141_106617\|\|full | 3274 | No | NA | 3 | 0 | 2 | Not-determined | Genome-fragment | NA | NA | 0 |
| S9-final-viral-combined_k141_111003\|\|full | 1225 | No | NA | 3 | 0 | 1 | Not-determined | Genome-fragment | NA | NA | 0 |
| S9-final-viral-combined_k141_127537\|\|full | 3128 | No | NA | 3 | 0 | 2 | Not-determined | Genome-fragment | NA | NA | 0 |
| S9-final-viral-combined_k141_127540\|\|full | 1113 | No | NA | 2 | 0 | 1 | Not-determined | Genome-fragment | NA | NA | 0 |
| S9-final-viral-combined_k141_127545\|\|full | 2989 | No | NA | 3 | 0 | 3 | Not-determined | Genome-fragment | NA | NA | 0 |
| S9-final-viral-combined_k141_106662\|\|full | 2471 | No | NA | 4 | 0 | 2 | Not-determined | Genome-fragment | NA | NA | 0 |
| S4_k141_73093 | 1044 | No | NA | 1 | 0 | 0 | Low-quality | Genome-fragment | 1.2 | AAI-based (high-confidence) | 0 |
| S9-final-viral-combined_k141_42719\|\|full | 4225 | No | NA | 9 | 0 | 0 | Not-determined | Genome-fragment | NA | NA | 0 |
| S6-final-viral-combined_k141_104815\|\|full | 1044 | No | NA | 2 | 0 | 1 | Low-quality | Genome-fragment | 1.7 | AAI-based (high-confidence) | 0 |
| S9-final-viral-combined_k141_31834\|\|full | 3549 | No | NA | 6 | 0 | 4 | Not-determined | Genome-fragment | NA | NA | 0 |
| S9_k141_62479 | 1045 | No | NA | 2 | 0 | 0 | Low-quality | Genome-fragment | 2.8 | AAI-based (high-confidence) | 0 |
| S11-final-viral-combined_k141_62145\|\|lt2gene | 1045 | No | NA | 1 | 1 | 0 | Low-quality | Genome-fragment | 2.45 | AAI-based (high-confidence) | 0 |
| S11-final-viral-combined_k141_301018\|\|lt2gene | 1045 | No | NA | 1 | 1 | 0 | Low-quality | Genome-fragment | 19.47 | AAI-based (high-confidence) | 0 |
| S9-final-viral-combined_k141_31854\|\|full | 2735 | No | NA | 3 | 0 | 3 | Not-determined | Genome-fragment | NA | NA | 0 |
| S11_k141_255351 | 1045 | No | NA | 2 | 0 | 0 | Low-quality | Genome-fragment | 0.68 | AAI-based (high-confidence) | 0 |
| S1-final-viral-combined_k141_297508\|\|full | 1045 | No | NA | 4 | 1 | 0 | Low-quality | Genome-fragment | 1.96 | AAI-based (high-confidence) | 0 |
| S1-final-viral-combined_k141_335887\|\|full | 1045 | No | NA | 3 | 1 | 0 | Low-quality | Genome-fragment | 1.8 | AAI-based (medium-confidence) | 0 |
| S1-final-viral-combined_k141_243642\|\|full | 1045 | No | NA | 3 | 0 | 0 | Low-quality | Genome-fragment | 2.57 | AAI-based (high-confidence) | 0 |
| S9-final-viral-combined_k141_106733\|\|full | 1880 | No | NA | 2 | 0 | 2 | Not-determined | Genome-fragment | NA | NA | 0 |
| S1-final-viral-combined_k141_111128\|\|full | 1045 | No | NA | 3 | 0 | 0 | Low-quality | Genome-fragment | 1.76 | AAI-based (high-confidence) | 0 |
| S9-final-viral-combined_k141_106751\|\|full | 4826 | No | NA | 7 | 0 | 2 | Not-determined | Genome-fragment | NA | NA | 0 |
| S5_k141_21553 | 1045 | No | NA | 2 | 0 | 2 | Low-quality | Genome-fragment | 3.44 | AAI-based (medium-confidence) | 0 |
| S17-final-viral-combined_k141_214599\|\|full | 1045 | No | NA | 3 | 0 | 0 | Low-quality | Genome-fragment | 2.93 | AAI-based (high-confidence) | 0 |
| S9-final-viral-combined_k141_127771\|\|full | 2179 | No | NA | 3 | 0 | 1 | Not-determined | Genome-fragment | NA | NA | 0 |
| S9-final-viral-combined_k141_111151\|\|full | 1410 | No | NA | 3 | 0 | 0 | Not-determined | Genome-fragment | NA | NA | 0 |
| S17-final-viral-combined_k141_195949\|\|full | 1045 | No | NA | 2 | 0 | 0 | Low-quality | Genome-fragment | 0.78 | AAI-based (medium-confidence) | 0 |
| S12-final-viral-combined_k141_51000\|\|full | 1045 | No | NA | 2 | 0 | 1 | Low-quality | Genome-fragment | 1.91 | AAI-based (medium-confidence) | 0 |
| S12-final-viral-combined_k141_133897\|\|full | 1045 | No | NA | 3 | 1 | 0 | Low-quality | Genome-fragment | 1.67 | HMM-based (lower-bound) | 0 |
| S9-final-viral-combined_k141_111171\|\|full | 1729 | No | NA | 2 | 0 | 2 | Not-determined | Genome-fragment | NA | NA | 0 |
| S9-final-viral-combined_k141_127812\|\|full | 1005 | No | NA | 2 | 0 | 1 | Not-determined | Genome-fragment | NA | NA | 0 |
| S12-final-viral-combined_k141_93914\|\|lt2gene | 1045 | No | NA | 1 | 1 | 0 | Low-quality | Genome-fragment | 2.91 | AAI-based (high-confidence) | 0 |
| S9-final-viral-combined_k141_32027\|\|full | 1929 | No | NA | 5 | 0 | 0 | Not-determined | Genome-fragment | NA | NA | 0 |
| S12_k141_127957 | 1045 | No | NA | 1 | 0 | 0 | Low-quality | Genome-fragment | 0.57 | AAI-based (high-confidence) | 0 |
| S9-final-viral-combined_k141_47774\|\|full | 3253 | No | NA | 4 | 0 | 3 | Not-determined | Genome-fragment | NA | NA | 0 |
| S9-final-viral-combined_k141_111201\|\|full | 1334 | No | NA | 2 | 0 | 0 | Not-determined | Genome-fragment | NA | NA | 0 |
| S9-final-viral-combined_k141_32086\|\|full | 1575 | No | NA | 2 | 0 | 1 | Not-determined | Genome-fragment | NA | NA | 0 |
| S12_k141_96585 | 1045 | No | NA | 3 | 0 | 0 | Low-quality | Genome-fragment | 2.04 | AAI-based (medium-confidence) | 0 |
| S2-final-viral-combined_k141_66574\|\|lt2gene | 1045 | No | NA | 2 | 2 | 0 | Low-quality | Genome-fragment | 2 | AAI-based (high-confidence) | 0 |
| S9-final-viral-combined_k141_111243\|\|full | 1673 | No | NA | 2 | 0 | 2 | Not-determined | Genome-fragment | NA | NA | 0 |
| S2_k141_135819 | 1045 | No | NA | 1 | 0 | 0 | Low-quality | Genome-fragment | 1.33 | AAI-based (high-confidence) | 0 |
| S2_k141_32438 | 1045 | No | NA | 1 | 0 | 0 | Low-quality | Genome-fragment | 1.75 | AAI-based (high-confidence) | 0 |
| S9-final-viral-combined_k141_111336\|\|full | 1995 | No | NA | 2 | 0 | 1 | Not-determined | Genome-fragment | NA | NA | 0 |
| S4-final-viral-combined_k141_102810\|\|lt2gene | 1045 | No | NA | 2 | 0 | 0 | Low-quality | Genome-fragment | 2.53 | AAI-based (high-confidence) | 0 |
| S9-final-viral-combined_k141_48030\|\|full | 3739 | No | NA | 3 | 0 | 2 | Not-determined | Genome-fragment | NA | NA | 0 |
| S9-final-viral-combined_k141_111515\|\|full | 2295 | No | NA | 4 | 0 | 0 | Not-determined | Genome-fragment | NA | NA | 0 |
| S8_k141_214262 | 1045 | No | NA | 2 | 0 | 1 | Low-quality | Genome-fragment | 2.13 | AAI-based (medium-confidence) | 0 |
| S9-final-viral-combined_k141_94387\|\|full | 1046 | No | NA | 2 | 0 | 0 | Low-quality | Genome-fragment | 1.63 | AAI-based (medium-confidence) | 0 |
| S9_k141_78394 | 1046 | No | NA | 2 | 0 | 0 | Low-quality | Genome-fragment | 1.75 | AAI-based (medium-confidence) | 0 |
| S9-final-viral-combined_k141_111791\|\|full | 2896 | No | NA | 4 | 0 | 2 | Not-determined | Genome-fragment | NA | NA | 0 |
| S9-final-viral-combined_k141_111884\|\|full | 1984 | No | NA | 2 | 0 | 1 | Not-determined | Genome-fragment | NA | NA | 0 |
| S9-final-viral-combined_k141_111935\|\|full | 3079 | No | NA | 3 | 0 | 2 | Not-determined | Genome-fragment | NA | NA | 0 |
| S11-final-viral-combined_k141_127570\|\|full | 1046 | No | NA | 3 | 0 | 1 | Low-quality | Genome-fragment | 0.44 | AAI-based (medium-confidence) | 0 |
| S9-final-viral-combined_k141_112067\|\|full | 1840 | No | NA | 2 | 0 | 1 | Not-determined | Genome-fragment | NA | NA | 0 |
| S11-final-viral-combined_k141_259849\|\|full | 1046 | No | NA | 2 | 0 | 1 | Low-quality | Genome-fragment | 0.76 | AAI-based (medium-confidence) | 0 |
| S11-final-viral-combined_k141_82237\|\|full | 1046 | No | NA | 2 | 0 | 0 | Low-quality | Genome-fragment | 2.78 | AAI-based (medium-confidence) | 0 |
| S9-final-viral-combined_k141_37587\|\|0_partial | 82228 | Yes | 46493 | 73 | 10 | 17 | Medium-quality | Genome-fragment | 85.36 | AAI-based (high-confidence) | 43.46 |
| S9-final-viral-combined_k141_43110\|\|0_partial | 15458 | No | NA | 18 | 0 | 7 | Not-determined | Genome-fragment | NA | NA | 0 |
| S9-final-viral-combined_k141_5526\|\|0_partial | 51870 | Yes | 36902 | 63 | 7 | 16 | Medium-quality | Genome-fragment | 66.25 | AAI-based (high-confidence) | 28.86 |
| S1-final-viral-combined_k141_94227\|\|full | 1046 | No | NA | 3 | 0 | 0 | Low-quality | Genome-fragment | 2.06 | AAI-based (medium-confidence) | 0 |
| S9-final-viral-combined_k141_5696\|\|0_partial | 14816 | No | NA | 17 | 0 | 13 | Not-determined | Genome-fragment | NA | NA | 0 |
| S1-final-viral-combined_k141_164467\|\|full | 1046 | No | NA | 4 | 1 | 0 | Low-quality | Genome-fragment | 2.57 | AAI-based (medium-confidence) | 0 |
| S1_k141_159538 | 1046 | No | NA | 3 | 0 | 0 | Low-quality | Genome-fragment | 2.54 | AAI-based (high-confidence) | 0 |
| S5-final-viral-combined_k141_25072\|\|lt2gene | 1046 | No | NA | 1 | 1 | 0 | Low-quality | Genome-fragment | 0.56 | HMM-based (lower-bound) | 0 |
| S9-final-viral-combined_k141_33229\|\|0_partial | 23889 | No | NA | 36 | 10 | 0 | Medium-quality | Genome-fragment | 62.78 | AAI-based (medium-confidence) | 0 |
| S9-final-viral-combined_k141_54941\|\|0_partial | 19411 | No | NA | 11 | 0 | 6 | Not-determined | Genome-fragment | NA | NA | 0 |
| S9-final-viral-combined_k141_44292\|\|0_partial | 19773 | No | NA | 16 | 0 | 10 | Not-determined | Genome-fragment | NA | NA | 0 |
| S17-final-viral-combined_k141_200117\|\|full | 1046 | No | NA | 2 | 0 | 1 | Low-quality | Genome-fragment | 0.54 | AAI-based (medium-confidence) | 0 |
| S9-final-viral-combined_k141_76274\|\|0_partial | 68824 | Yes | 31753 | 78 | 23 | 20 | Complete | High-quality | 100 | Provirus (medium-confidence) | 53.86 |
| S9-final-viral-combined_k141_23386\|\|0_partial | 21837 | No | NA | 23 | 0 | 9 | Not-determined | Genome-fragment | NA | NA | 0 |
| S9-final-viral-combined_k141_44402\|\|0_partial | 32331 | No | NA | 46 | 9 | 1 | Medium-quality | Genome-fragment | 76.65 | AAI-based (medium-confidence) | 0 |
| S12-final-viral-combined_k141_209813\|\|full | 1046 | No | NA | 3 | 1 | 0 | Low-quality | Genome-fragment | 1.64 | AAI-based (high-confidence) | 0 |
| S9-final-viral-combined_k141_93117\|\|0_partial | 19133 | No | NA | 13 | 0 | 7 | Not-determined | Genome-fragment | NA | NA | 0 |
| S12-final-viral-combined_k141_378296\|\|full | 1046 | No | NA | 3 | 0 | 0 | Low-quality | Genome-fragment | 2.12 | AAI-based (medium-confidence) | 0 |
| S12-final-viral-combined_k141_114797\|\|full | 1046 | No | NA | 3 | 1 | 0 | Low-quality | Genome-fragment | 3.01 | AAI-based (high-confidence) | 0 |
| S12_k141_94395 | 1046 | No | NA | 2 | 0 | 1 | Low-quality | Genome-fragment | 1.85 | AAI-based (medium-confidence) | 0 |
| S2-final-viral-combined_k141_198209\|\|full | 1046 | No | NA | 4 | 1 | 0 | Low-quality | Genome-fragment | 3.01 | AAI-based (high-confidence) | 0 |
| S9-final-viral-combined_k141_61898\|\|0_partial | 22191 | No | NA | 17 | 0 | 8 | Not-determined | Genome-fragment | NA | NA | 0 |
| S9-final-viral-combined_k141_56773\|\|0_partial | 5561 | Yes | 2977 | 7 | 1 | 1 | Medium-quality | Genome-fragment | 71.54 | HMM-based (lower-bound) | 46.47 |
| S2_k141_35532 | 1046 | No | NA | 3 | 0 | 0 | Low-quality | Genome-fragment | 1.08 | AAI-based (medium-confidence) | 0 |
| S8_k141_40800 | 1046 | No | NA | 1 | 0 | 0 | Low-quality | Genome-fragment | 2.84 | AAI-based (high-confidence) | 0 |
| S9-final-viral-combined_k141_4767\|\|full | 1047 | No | NA | 2 | 0 | 0 | Low-quality | Genome-fragment | 8.14 | AAI-based (medium-confidence) | 0 |
| S11-final-viral-combined_k141_20828\|\|full | 1047 | No | NA | 2 | 1 | 0 | Low-quality | Genome-fragment | 1.74 | HMM-based (lower-bound) | 0 |
| S9-final-viral-combined_k141_19806\|\|0_partial | 11167 | No | NA | 11 | 0 | 6 | Not-determined | Genome-fragment | NA | NA | 0 |
| S3-final-viral-combined_k141_37833\|\|full | 1047 | No | NA | 3 | 0 | 0 | Low-quality | Genome-fragment | 2.48 | AAI-based (high-confidence) | 0 |
| S3_k141_108175 | 1047 | No | NA | 3 | 0 | 0 | Low-quality | Genome-fragment | 2.32 | AAI-based (high-confidence) | 0 |
| S7-final-viral-combined_k141_44401\|\|full | 1047 | No | NA | 3 | 0 | 0 | Low-quality | Genome-fragment | 1.79 | AAI-based (medium-confidence) | 0 |
| S17-final-viral-combined_k141_77806\|\|full | 1047 | No | NA | 3 | 1 | 0 | Low-quality | Genome-fragment | 2.72 | AAI-based (high-confidence) | 0 |
| S9-final-viral-combined_k141_14742\|\|0_partial | 11051 | No | NA | 9 | 0 | 7 | Not-determined | Genome-fragment | NA | NA | 0 |
| S9-final-viral-combined_k141_9647\|\|0_partial | 12997 | No | NA | 11 | 0 | 5 | Not-determined | Genome-fragment | NA | NA | 0 |
| S9-final-viral-combined_k141_4518\|\|0_partial | 33577 | No | NA | 17 | 0 | 11 | Not-determined | Genome-fragment | NA | NA | 0 |
| S9-final-viral-combined_k141_58078\|\|0_partial | 31656 | No | NA | 20 | 0 | 9 | Not-determined | Genome-fragment | NA | NA | 0 |
| S9-final-viral-combined_k141_63934\|\|0_partial | 19182 | No | NA | 11 | 0 | 9 | Not-determined | Genome-fragment | NA | NA | 0 |
| S17-final-viral-combined_k141_230779\|\|full | 1047 | No | NA | 3 | 0 | 0 | Low-quality | Genome-fragment | 1.69 | AAI-based (high-confidence) | 0 |
| S9-final-viral-combined_k141_10274\|\|0_partial | 16360 | No | NA | 14 | 0 | 11 | Not-determined | Genome-fragment | NA | NA | 0 |
| S9-final-viral-combined_k141_122742\|\|0_partial | 24352 | No | NA | 16 | 0 | 4 | Medium-quality | Genome-fragment | 73.74 | AAI-based (medium-confidence) | 0 |
| S17-k141_4339 | 1047 | No | NA | 2 | 0 | 0 | Low-quality | Genome-fragment | 15.95 | AAI-based (medium-confidence) | 0 |
| S9-final-viral-combined_k141_10665\|\|0_partial | 10566 | No | NA | 11 | 0 | 7 | Not-determined | Genome-fragment | NA | NA | 0 |
| S9-final-viral-combined_k141_96070\|\|0_partial | 11396 | No | NA | 11 | 0 | 9 | Not-determined | Genome-fragment | NA | NA | 0 |
| S9-final-viral-combined_k141_106708\|\|0_partial | 13339 | No | NA | 10 | 0 | 8 | Not-determined | Genome-fragment | NA | NA | 0 |
| S12-final-viral-combined_k141_359392\|\|full | 1047 | No | NA | 3 | 0 | 0 | Low-quality | Genome-fragment | 2.06 | AAI-based (medium-confidence) | 0 |
| S2_k141_18565 | 1047 | No | NA | 1 | 0 | 1 | Low-quality | Genome-fragment | 0.3 | AAI-based (medium-confidence) | 0 |
| S11-final-viral-combined_k141_248108\|\|lt2gene | 1048 | No | NA | 1 | 1 | 0 | Low-quality | Genome-fragment | 2.34 | AAI-based (high-confidence) | 0 |
| S11-final-viral-combined_k141_182330\|\|lt2gene | 1048 | No | NA | 2 | 1 | 0 | Low-quality | Genome-fragment | 1.97 | AAI-based (high-confidence) | 0 |
| S5_k141_135256 | 1048 | No | NA | 1 | 0 | 0 | Low-quality | Genome-fragment | 0.81 | AAI-based (high-confidence) | 0 |
| S9-final-viral-combined_k141_75345\|\|lt2gene | 2631 | No | NA | 1 | 0 | 0 | Not-determined | Genome-fragment | NA | NA | 0 |
| S17-final-viral-combined_k141_253535\|\|full | 1048 | No | NA | 4 | 0 | 0 | Low-quality | Genome-fragment | 1.3 | AAI-based (high-confidence) | 0 |
| S8-final-viral-combined_k141_158152\|\|full | 1048 | No | NA | 2 | 2 | 0 | Low-quality | Genome-fragment | 1.75 | AAI-based (high-confidence) | 0 |
| S11_k141_38222 | 1049 | No | NA | 4 | 0 | 0 | Low-quality | Genome-fragment | 1.42 | AAI-based (high-confidence) | 0 |
| S1-final-viral-combined_k141_16500\|\|full | 1049 | No | NA | 2 | 0 | 1 | Low-quality | Genome-fragment | 2.69 | AAI-based (high-confidence) | 0 |
| S9-final-viral-combined_k141_10986\|\|lt2gene | 1368 | No | NA | 2 | 0 | 1 | Not-determined | Genome-fragment | NA | NA | 0 |
| S1-final-viral-combined_k141_297613\|\|full | 1049 | No | NA | 2 | 0 | 0 | Low-quality | Genome-fragment | 1.22 | AAI-based (medium-confidence) | 0 |
| S1-final-viral-combined_k141_111833\|\|full | 1049 | No | NA | 2 | 1 | 0 | Low-quality | Genome-fragment | 2.03 | AAI-based (high-confidence) | 0 |
| S3-final-viral-combined_k141_1865\|\|full | 1049 | No | NA | 2 | 1 | 0 | Low-quality | Genome-fragment | 1.84 | AAI-based (medium-confidence) | 0 |
| S9-final-viral-combined_k141_123509\|\|lt2gene | 1585 | No | NA | 1 | 0 | 0 | Not-determined | Genome-fragment | NA | NA | 0 |
| S5-final-viral-combined_k141_112418\|\|full | 1049 | No | NA | 2 | 0 | 2 | Low-quality | Genome-fragment | 0.45 | AAI-based (medium-confidence) | 0 |
| S5-final-viral-combined_k141_147508\|\|lt2gene | 1049 | No | NA | 2 | 2 | 0 | Low-quality | Genome-fragment | 2.95 | AAI-based (high-confidence) | 0 |
| S7-final-viral-combined_k141_159815\|\|full | 1049 | No | NA | 4 | 1 | 0 | Low-quality | Genome-fragment | 2.02 | AAI-based (medium-confidence) | 0 |
| S9-final-viral-combined_k141_16684\|\|lt2gene | 3277 | No | NA | 1 | 0 | 1 | Not-determined | Genome-fragment | NA | NA | 0 |
| S7_k141_33911 | 1049 | No | NA | 1 | 0 | 0 | Low-quality | Genome-fragment | 1.88 | AAI-based (high-confidence) | 0 |
| S17-k141_80826 | 1049 | No | NA | 2 | 0 | 0 | Low-quality | Genome-fragment | 3.23 | AAI-based (medium-confidence) | 0 |
| S17-k141_59189 | 1049 | No | NA | 1 | 0 | 1 | Low-quality | Genome-fragment | 1.86 | AAI-based (high-confidence) | 0 |
| S17-k141_17225 | 1049 | No | NA | 2 | 1 | 0 | Low-quality | Genome-fragment | 2.01 | AAI-based (high-confidence) | 0 |
| S12_k141_192265 | 1049 | No | NA | 1 | 0 | 0 | Low-quality | Genome-fragment | 1.43 | AAI-based (medium-confidence) | 0 |
| S2-final-viral-combined_k141_210509\|\|full | 1049 | No | NA | 4 | 0 | 0 | Low-quality | Genome-fragment | 2.7 | AAI-based (high-confidence) | 0 |
| S2-final-viral-combined_k141_108558\|\|full | 1049 | No | NA | 3 | 0 | 0 | Low-quality | Genome-fragment | 2.07 | AAI-based (medium-confidence) | 0 |
| S2-final-viral-combined_k141_108573\|\|full | 1049 | No | NA | 3 | 0 | 0 | Low-quality | Genome-fragment | 2.08 | AAI-based (medium-confidence) | 0 |
| S2_k141_163296 | 1049 | No | NA | 3 | 1 | 0 | Low-quality | Genome-fragment | 15.7 | AAI-based (high-confidence) | 0 |
| S4-final-viral-combined_k141_76065\|\|full | 1049 | No | NA | 2 | 0 | 0 | Low-quality | Genome-fragment | 1.44 | AAI-based (high-confidence) | 0 |
| S11-final-viral-combined_k141_213662\|\|lt2gene | 1050 | No | NA | 2 | 0 | 0 | Low-quality | Genome-fragment | 1.95 | AAI-based (high-confidence) | 0 |
| S11_k141_241660 | 1050 | No | NA | 2 | 0 | 0 | Low-quality | Genome-fragment | 2.62 | AAI-based (high-confidence) | 0 |
| S11_k141_160266 | 1050 | No | NA | 1 | 0 | 1 | Low-quality | Genome-fragment | 2.07 | AAI-based (high-confidence) | 0 |
| S1-final-viral-combined_k141_178240\|\|full | 1050 | No | NA | 3 | 1 | 0 | Low-quality | Genome-fragment | 2.48 | AAI-based (high-confidence) | 0 |
| S1-final-viral-combined_k141_243219\|\|lt2gene | 1050 | No | NA | 1 | 1 | 0 | Low-quality | Genome-fragment | 1.74 | AAI-based (high-confidence) | 0 |
| S3-final-viral-combined_k141_58118\|\|lt2gene | 1050 | No | NA | 1 | 1 | 0 | Low-quality | Genome-fragment | 0.96 | HMM-based (lower-bound) | 0 |
| S7_k141_185264 | 1050 | No | NA | 3 | 1 | 0 | Low-quality | Genome-fragment | 2.03 | AAI-based (high-confidence) | 0 |
| S9-final-viral-combined_k141_23006\|\|lt2gene | 1314 | No | NA | 1 | 0 | 0 | Not-determined | Genome-fragment | NA | NA | 0 |
| S17-k141_28237 | 1050 | No | NA | 2 | 0 | 1 | Low-quality | Genome-fragment | 2.75 | AAI-based (medium-confidence) | 0 |
| S12-final-viral-combined_k141_46232\|\|full | 1050 | No | NA | 3 | 1 | 0 | Low-quality | Genome-fragment | 2.37 | AAI-based (high-confidence) | 0 |
| S12-final-viral-combined_k141_200616\|\|full | 1050 | No | NA | 3 | 2 | 0 | Low-quality | Genome-fragment | 2.32 | HMM-based (lower-bound) | 0 |
| S2-final-viral-combined_k141_207420\|\|full | 1050 | No | NA | 3 | 0 | 0 | Low-quality | Genome-fragment | 1.89 | AAI-based (medium-confidence) | 0 |
| S6-final-viral-combined_k141_44630\|\|full | 1050 | No | NA | 3 | 0 | 0 | Low-quality | Genome-fragment | 4.12 | AAI-based (medium-confidence) | 0 |
| S8_k141_69815 | 1050 | No | NA | 2 | 1 | 0 | Low-quality | Genome-fragment | 2.41 | AAI-based (medium-confidence) | 0 |
| S1-final-viral-combined_k141_174246\|\|lt2gene | 1051 | No | NA | 1 | 1 | 0 | Low-quality | Genome-fragment | 2.68 | AAI-based (high-confidence) | 0 |
| S5-final-viral-combined_k141_37104\|\|full | 1051 | No | NA | 3 | 1 | 0 | Low-quality | Genome-fragment | 2.07 | AAI-based (high-confidence) | 0 |
| S5-final-viral-combined_k141_190157\|\|full | 1051 | No | NA | 2 | 0 | 0 | Low-quality | Genome-fragment | 1.76 | AAI-based (medium-confidence) | 0 |
| S5-final-viral-combined_k141_35794\|\|lt2gene | 1051 | No | NA | 2 | 0 | 1 | Low-quality | Genome-fragment | 1.43 | AAI-based (medium-confidence) | 0 |
| S17-k141_262360 | 1051 | No | NA | 2 | 0 | 0 | Low-quality | Genome-fragment | 1.97 | AAI-based (medium-confidence) | 0 |
| S12-final-viral-combined_k141_21768\|\|full | 1051 | No | NA | 2 | 0 | 0 | Low-quality | Genome-fragment | 1.75 | AAI-based (high-confidence) | 0 |
| S4-final-viral-combined_k141_25324\|\|full | 1051 | No | NA | 4 | 0 | 0 | Low-quality | Genome-fragment | 1.62 | AAI-based (high-confidence) | 0 |
| S8-final-viral-combined_k141_244643\|\|full | 1051 | No | NA | 3 | 0 | 0 | Low-quality | Genome-fragment | 0.77 | AAI-based (medium-confidence) | 0 |
| S9-final-viral-combined_k141_103726\|\|lt2gene | 1158 | No | NA | 2 | 0 | 1 | Not-determined | Genome-fragment | NA | NA | 0 |
| S8-final-viral-combined_k141_54304\|\|full | 1051 | No | NA | 2 | 0 | 0 | Low-quality | Genome-fragment | 1.44 | AAI-based (medium-confidence) | 0 |
| S8_k141_126400 | 1051 | No | NA | 1 | 0 | 0 | Low-quality | Genome-fragment | 1.74 | AAI-based (high-confidence) | 0 |
| S9-final-viral-combined_k141_66679\|\|full | 1052 | No | NA | 3 | 0 | 0 | Low-quality | Genome-fragment | 2.53 | AAI-based (medium-confidence) | 0 |
| S9_k141_31178 | 1052 | No | NA | 1 | 1 | 0 | Low-quality | Genome-fragment | 1.48 | HMM-based (lower-bound) | 0 |
| S11_k141_196535 | 1052 | No | NA | 4 | 0 | 0 | Low-quality | Genome-fragment | 1.98 | AAI-based (medium-confidence) | 0 |
| S3-final-viral-combined_k141_3448\|\|full | 1052 | No | NA | 2 | 0 | 0 | Low-quality | Genome-fragment | 1.68 | AAI-based (high-confidence) | 0 |
| S3-final-viral-combined_k141_4994\|\|full | 1052 | No | NA | 2 | 0 | 0 | Low-quality | Genome-fragment | 1.3 | AAI-based (high-confidence) | 0 |
| S5-final-viral-combined_k141_200618\|\|full | 1052 | No | NA | 2 | 1 | 0 | Low-quality | Genome-fragment | 2.55 | AAI-based (high-confidence) | 0 |
| S7-final-viral-combined_k141_93489\|\|full | 1052 | No | NA | 3 | 0 | 0 | Low-quality | Genome-fragment | 1.33 | AAI-based (high-confidence) | 0 |
| S9-final-viral-combined_k141_93198\|\|lt2gene | 2167 | No | NA | 2 | 0 | 0 | Not-determined | Genome-fragment | NA | NA | 0 |
| S7-final-viral-combined_k141_8725\|\|full | 1052 | No | NA | 2 | 0 | 0 | Low-quality | Genome-fragment | 2.05 | AAI-based (medium-confidence) | 0 |
| S17-final-viral-combined_k141_22787\|\|full | 1052 | No | NA | 3 | 1 | 0 | Low-quality | Genome-fragment | 3.04 | AAI-based (medium-confidence) | 0 |
| S17-final-viral-combined_k141_58063\|\|full | 1052 | No | NA | 2 | 2 | 0 | Low-quality | Genome-fragment | 1.9 | HMM-based (lower-bound) | 0 |
| S17-final-viral-combined_k141_137374\|\|full | 1052 | No | NA | 3 | 0 | 0 | Low-quality | Genome-fragment | 2.43 | AAI-based (high-confidence) | 0 |
| S17-final-viral-combined_k141_49364\|\|full | 1052 | No | NA | 2 | 0 | 0 | Low-quality | Genome-fragment | 2.06 | AAI-based (medium-confidence) | 0 |
| S17-k141_169645 | 1052 | No | NA | 1 | 0 | 0 | Low-quality | Genome-fragment | 1.25 | AAI-based (high-confidence) | 0 |
| S12_k141_72011 | 1052 | No | NA | 3 | 0 | 0 | Low-quality | Genome-fragment | 2.17 | AAI-based (medium-confidence) | 0 |
| S2_k141_144452 | 1052 | No | NA | 2 | 0 | 0 | Low-quality | Genome-fragment | 0.48 | AAI-based (medium-confidence) | 0 |
| S2_k141_133307 | 1052 | No | NA | 3 | 0 | 0 | Low-quality | Genome-fragment | 2.92 | AAI-based (medium-confidence) | 0 |
| S6-final-viral-combined_k141_30548\|\|full | 1052 | No | NA | 4 | 0 | 0 | Low-quality | Genome-fragment | 2.94 | AAI-based (medium-confidence) | 0 |
| S8-final-viral-combined_k141_104886\|\|lt2gene | 1052 | No | NA | 2 | 1 | 0 | Low-quality | Genome-fragment | 2.69 | AAI-based (high-confidence) | 0 |
| S11-final-viral-combined_k141_308804\|\|full | 1053 | No | NA | 2 | 0 | 0 | Low-quality | Genome-fragment | 1.99 | AAI-based (medium-confidence) | 0 |
| S11-final-viral-combined_k141_325920\|\|full | 1053 | No | NA | 3 | 0 | 0 | Low-quality | Genome-fragment | 2.49 | AAI-based (medium-confidence) | 0 |
| S11-final-viral-combined_k141_301515\|\|full | 1053 | No | NA | 3 | 0 | 0 | Low-quality | Genome-fragment | 2.48 | AAI-based (medium-confidence) | 0 |
| S11_k141_278563 | 1053 | No | NA | 2 | 0 | 0 | Low-quality | Genome-fragment | 1.96 | AAI-based (high-confidence) | 0 |
| S11_k141_226113 | 1053 | No | NA | 3 | 0 | 0 | Low-quality | Genome-fragment | 2.9 | AAI-based (high-confidence) | 0 |
| S1-final-viral-combined_k141_151614\|\|full | 1053 | No | NA | 3 | 0 | 2 | Low-quality | Genome-fragment | 0.78 | AAI-based (medium-confidence) | 0 |
| S1-final-viral-combined_k141_188168\|\|full | 1053 | No | NA | 2 | 0 | 0 | Low-quality | Genome-fragment | 2.92 | AAI-based (high-confidence) | 0 |
| S9-final-viral-combined_k141_9291\|\|lt2gene | 1495 | No | NA | 1 | 0 | 0 | Not-determined | Genome-fragment | NA | NA | 0 |
| S9-final-viral-combined_k141_46147\|\|lt2gene | 1402 | No | NA | 1 | 0 | 1 | Not-determined | Genome-fragment | NA | NA | 0 |
| S1_k141_188431 | 1053 | No | NA | 1 | 0 | 1 | Low-quality | Genome-fragment | 0.49 | AAI-based (medium-confidence) | 0 |
| S5_k141_148552 | 1053 | No | NA | 1 | 1 | 0 | Low-quality | Genome-fragment | 2.26 | HMM-based (lower-bound) | 0 |
| S7-final-viral-combined_k141_137255\|\|lt2gene | 1053 | No | NA | 1 | 1 | 0 | Low-quality | Genome-fragment | 1.95 | HMM-based (lower-bound) | 0 |
| S7-final-viral-combined_k141_130441\|\|lt2gene | 1053 | No | NA | 2 | 2 | 0 | Low-quality | Genome-fragment | 0.79 | AAI-based (medium-confidence) | 0 |
| S9-final-viral-combined_k141_72790\|\|lt2gene | 1816 | No | NA | 1 | 0 | 0 | Not-determined | Genome-fragment | NA | NA | 0 |
| S17-final-viral-combined_k141_240538\|\|full | 1053 | No | NA | 3 | 0 | 0 | Low-quality | Genome-fragment | 1.82 | AAI-based (high-confidence) | 0 |
| S12-final-viral-combined_k141_18163\|\|full | 1053 | No | NA | 2 | 0 | 1 | Low-quality | Genome-fragment | 0.6 | AAI-based (medium-confidence) | 0 |
| S12-final-viral-combined_k141_234346\|\|lt2gene | 1053 | No | NA | 2 | 1 | 0 | Low-quality | Genome-fragment | 2.15 | AAI-based (high-confidence) | 0 |
| S2-final-viral-combined_k141_171111\|\|full | 1053 | No | NA | 4 | 1 | 0 | Low-quality | Genome-fragment | 2.61 | AAI-based (high-confidence) | 0 |
| S2-final-viral-combined_k141_214785\|\|full | 1053 | No | NA | 3 | 0 | 1 | Low-quality | Genome-fragment | 0.48 | AAI-based (medium-confidence) | 0 |
| S2-final-viral-combined_k141_170854\|\|lt2gene | 1053 | No | NA | 2 | 1 | 0 | Low-quality | Genome-fragment | 2.98 | AAI-based (high-confidence) | 0 |
| S4_k141_115323 | 1053 | No | NA | 1 | 0 | 0 | Low-quality | Genome-fragment | 0.93 | AAI-based (high-confidence) | 0 |
| S8-final-viral-combined_k141_23687\|\|full | 1053 | No | NA | 4 | 0 | 0 | Low-quality | Genome-fragment | 0.93 | AAI-based (medium-confidence) | 0 |
| S9-final-viral-combined_k141_52810\|\|lt2gene | 2091 | No | NA | 1 | 0 | 0 | Not-determined | Genome-fragment | NA | NA | 0 |
| S8-final-viral-combined_k141_191056\|\|full | 1053 | No | NA | 3 | 0 | 0 | Low-quality | Genome-fragment | 2.48 | AAI-based (high-confidence) | 0 |
| S8-final-viral-combined_k141_215251\|\|full | 1053 | No | NA | 2 | 0 | 0 | Low-quality | Genome-fragment | 2.08 | AAI-based (medium-confidence) | 0 |
| S1-final-viral-combined_k141_387282\|\|full | 1054 | No | NA | 3 | 1 | 0 | Low-quality | Genome-fragment | 2.48 | AAI-based (high-confidence) | 0 |
| S1-final-viral-combined_k141_377851\|\|full | 1054 | No | NA | 2 | 0 | 0 | Low-quality | Genome-fragment | 2.05 | AAI-based (medium-confidence) | 0 |
| S5-final-viral-combined_k141_130839\|\|lt2gene | 1054 | No | NA | 1 | 1 | 0 | Low-quality | Genome-fragment | 2.44 | AAI-based (high-confidence) | 0 |
| S17-final-viral-combined_k141_63866\|\|lt2gene | 1054 | No | NA | 2 | 1 | 0 | Low-quality | Genome-fragment | 1.92 | HMM-based (lower-bound) | 0 |
| S17-final-viral-combined_k141_241837\|\|lt2gene | 1054 | No | NA | 1 | 1 | 0 | Low-quality | Genome-fragment | 2.82 | AAI-based (high-confidence) | 0 |
| S12-final-viral-combined_k141_13200\|\|full | 1054 | No | NA | 4 | 1 | 0 | Low-quality | Genome-fragment | 2.54 | AAI-based (high-confidence) | 0 |
| S12_k141_385619 | 1054 | No | NA | 3 | 0 | 2 | Low-quality | Genome-fragment | 2.23 | AAI-based (high-confidence) | 0 |
| S9-final-viral-combined_k141_58343\|\|lt2gene | 1520 | No | NA | 1 | 0 | 0 | Not-determined | Genome-fragment | NA | NA | 0 |
| S4-final-viral-combined_k141_121588\|\|lt2gene | 1054 | No | NA | 2 | 2 | 0 | Low-quality | Genome-fragment | 0.39 | HMM-based (lower-bound) | 0 |
| S4_k141_76767 | 1054 | No | NA | 3 | 1 | 0 | Low-quality | Genome-fragment | 1.06 | AAI-based (high-confidence) | 0 |
| S6-final-viral-combined_k141_99443\|\|full | 1054 | No | NA | 2 | 0 | 1 | Low-quality | Genome-fragment | 0.47 | AAI-based (medium-confidence) | 0 |
| S6-final-viral-combined_k141_74481\|\|lt2gene | 1054 | No | NA | 2 | 1 | 0 | Low-quality | Genome-fragment | 3.88 | HMM-based (lower-bound) | 0 |
| S8-final-viral-combined_k141_202649\|\|full | 1054 | No | NA | 3 | 0 | 0 | Low-quality | Genome-fragment | 2.29 | AAI-based (medium-confidence) | 0 |
| S8-final-viral-combined_k141_135778\|\|full | 1054 | No | NA | 2 | 0 | 1 | Low-quality | Genome-fragment | 1.43 | AAI-based (medium-confidence) | 0 |
| S8_k141_152419 | 1054 | No | NA | 1 | 1 | 0 | Low-quality | Genome-fragment | 0.93 | HMM-based (lower-bound) | 0 |
| S9_k141_112305 | 3113 | No | NA | 2 | 0 | 0 | Not-determined | Genome-fragment | NA | NA | 0 |
| S9_k141_122993 | 8129 | No | NA | 18 | 0 | 18 | Not-determined | Genome-fragment | NA | NA | 0 |
| S9_k141_53485 | 1440 | No | NA | 1 | 0 | 0 | Not-determined | Genome-fragment | NA | NA | 0 |
| S9_k141_106986 | 1614 | No | NA | 3 | 0 | 0 | Not-determined | Genome-fragment | NA | NA | 0 |
| S9_k141_101639 | 1372 | No | NA | 1 | 0 | 1 | Not-determined | Genome-fragment | NA | NA | 0 |
| S9_k141_112338 | 1072 | No | NA | 2 | 0 | 2 | Not-determined | Genome-fragment | NA | NA | 0 |
| S9_k141_10739 | 1112 | No | NA | 1 | 0 | 0 | Not-determined | Genome-fragment | NA | NA | 0 |
| S11-final-viral-combined_k141_58489\|\|full | 1055 | No | NA | 2 | 0 | 0 | Low-quality | Genome-fragment | 0.68 | AAI-based (medium-confidence) | 0 |
| S9_k141_64239 | 1127 | No | NA | 3 | 0 | 0 | Not-determined | Genome-fragment | NA | NA | 0 |
| S9_k141_10765 | 2555 | No | NA | 4 | 0 | 1 | Not-determined | Genome-fragment | NA | NA | 0 |
| S11-final-viral-combined_k141_91999\|\|full | 1055 | No | NA | 3 | 0 | 0 | Low-quality | Genome-fragment | 1.5 | AAI-based (medium-confidence) | 0 |
| S9_k141_97 | 1094 | No | NA | 1 | 0 | 1 | Not-determined | Genome-fragment | NA | NA | 0 |
| S11-final-viral-combined_k141_301696\|\|lt2gene | 1055 | No | NA | 1 | 1 | 0 | Low-quality | Genome-fragment | 3.01 | AAI-based (high-confidence) | 0 |
| S1_k141_285035 | 1055 | No | NA | 1 | 0 | 1 | Low-quality | Genome-fragment | 0.1 | AAI-based (medium-confidence) | 0 |
| S9_k141_48218 | 3813 | No | NA | 3 | 0 | 3 | Not-determined | Genome-fragment | NA | NA | 0 |
| S3-final-viral-combined_k141_62294\|\|full | 1055 | No | NA | 3 | 0 | 0 | Low-quality | Genome-fragment | 2.39 | AAI-based (medium-confidence) | 0 |
| S5-final-viral-combined_k141_145529\|\|lt2gene | 1055 | No | NA | 2 | 1 | 0 | Low-quality | Genome-fragment | 3.06 | AAI-based (high-confidence) | 0 |
| S9_k141_21476 | 9252 | No | NA | 10 | 0 | 1 | Not-determined | Genome-fragment | NA | NA | 0 |
| S9_k141_37559 | 1304 | No | NA | 1 | 0 | 0 | Not-determined | Genome-fragment | NA | NA | 0 |
| S9_k141_183 | 4319 | No | NA | 3 | 0 | 1 | Not-determined | Genome-fragment | NA | NA | 0 |
| S9_k141_74993 | 2033 | No | NA | 2 | 0 | 1 | Not-determined | Genome-fragment | NA | NA | 0 |
| S9_k141_197 | 2199 | No | NA | 2 | 0 | 1 | Not-determined | Genome-fragment | NA | NA | 0 |
| S9_k141_112490 | 2523 | No | NA | 1 | 0 | 1 | Not-determined | Genome-fragment | NA | NA | 0 |
| S9_k141_117800 | 1795 | No | NA | 3 | 0 | 2 | Not-determined | Genome-fragment | NA | NA | 0 |
| S9_k141_69615 | 2008 | No | NA | 1 | 0 | 1 | Not-determined | Genome-fragment | NA | NA | 0 |
| S9_k141_32208 | 1323 | No | NA | 1 | 0 | 0 | Not-determined | Genome-fragment | NA | NA | 0 |
| S5_k141_152706 | 1055 | No | NA | 1 | 0 | 0 | Low-quality | Genome-fragment | 1.38 | AAI-based (high-confidence) | 0 |
| S7-final-viral-combined_k141_169125\|\|full | 1055 | No | NA | 2 | 0 | 1 | Low-quality | Genome-fragment | 1.64 | AAI-based (medium-confidence) | 0 |
| S9_k141_267 | 3385 | No | NA | 3 | 0 | 2 | Not-determined | Genome-fragment | NA | NA | 0 |
| S9_k141_112569 | 2011 | No | NA | 3 | 0 | 2 | Not-determined | Genome-fragment | NA | NA | 0 |
| S7-final-viral-combined_k141_190750\|\|full | 1055 | No | NA | 4 | 0 | 0 | Low-quality | Genome-fragment | 2.5 | AAI-based (medium-confidence) | 0 |
| S9_k141_48326 | 1248 | No | NA | 1 | 0 | 0 | Not-determined | Genome-fragment | NA | NA | 0 |
| S7_k141_5065 | 1055 | No | NA | 1 | 0 | 1 | Low-quality | Genome-fragment | 0.67 | AAI-based (high-confidence) | 0 |
| S9_k141_117823 | 1373 | No | NA | 1 | 0 | 1 | Not-determined | Genome-fragment | NA | NA | 0 |
| S17-k141_45507 | 1055 | No | NA | 2 | 0 | 0 | Low-quality | Genome-fragment | 1.64 | AAI-based (high-confidence) | 0 |
| S9_k141_59026 | 1149 | No | NA | 1 | 0 | 0 | Not-determined | Genome-fragment | NA | NA | 0 |
| S9_k141_101841 | 3731 | No | NA | 3 | 0 | 0 | Not-determined | Genome-fragment | NA | NA | 0 |
| S12_k141_225280 | 1055 | No | NA | 2 | 1 | 0 | Low-quality | Genome-fragment | 0.57 | AAI-based (high-confidence) | 0 |
| S2-final-viral-combined_k141_1668\|\|full | 1055 | No | NA | 3 | 1 | 0 | Low-quality | Genome-fragment | 1.96 | AAI-based (medium-confidence) | 0 |
| S9_k141_75180 | 1973 | No | NA | 2 | 0 | 1 | Not-determined | Genome-fragment | NA | NA | 0 |
| S9_k141_101845 | 3595 | No | NA | 2 | 0 | 2 | Not-determined | Genome-fragment | NA | NA | 0 |
| S9_k141_96501 | 1266 | No | NA | 1 | 0 | 1 | Not-determined | Genome-fragment | NA | NA | 0 |
| S4-final-viral-combined_k141_75388\|\|full | 1055 | No | NA | 3 | 0 | 0 | Low-quality | Genome-fragment | 2.52 | AAI-based (high-confidence) | 0 |
| S8-final-viral-combined_k141_60335\|\|full | 1055 | No | NA | 3 | 1 | 0 | Low-quality | Genome-fragment | 2.27 | AAI-based (medium-confidence) | 0 |
| S9_k141_27022 | 1219 | No | NA | 1 | 0 | 0 | Not-determined | Genome-fragment | NA | NA | 0 |
| S9_k141_117902 | 2257 | No | NA | 3 | 0 | 2 | Not-determined | Genome-fragment | NA | NA | 0 |
| S8-final-viral-combined_k141_182756\|\|lt2gene | 1055 | No | NA | 2 | 1 | 0 | Low-quality | Genome-fragment | 2.09 | AAI-based (medium-confidence) | 0 |
| S9_k141_101876 | 6057 | No | NA | 12 | 0 | 10 | Not-determined | Genome-fragment | NA | NA | 0 |
| S9_k141_64387 | 1092 | No | NA | 1 | 0 | 1 | Not-determined | Genome-fragment | NA | NA | 0 |
| S9_k141_117947 | 3656 | No | NA | 3 | 0 | 3 | Not-determined | Genome-fragment | NA | NA | 0 |
| S9_k141_85874 | 1733 | No | NA | 3 | 0 | 2 | Not-determined | Genome-fragment | NA | NA | 0 |
| S9_k141_101966 | 3218 | No | NA | 5 | 0 | 5 | Not-determined | Genome-fragment | NA | NA | 0 |
| S9_k141_96593 | 10757 | No | NA | 3 | 0 | 2 | Not-determined | Genome-fragment | NA | NA | 0 |
| S8_k141_14240 | 1055 | No | NA | 1 | 0 | 0 | Low-quality | Genome-fragment | 0.63 | AAI-based (high-confidence) | 0 |
| S1-final-viral-combined_k141_65890\|\|full | 1056 | No | NA | 2 | 1 | 0 | Low-quality | Genome-fragment | 0.8 | HMM-based (lower-bound) | 0 |
| S9_k141_69773 | 2475 | No | NA | 1 | 0 | 1 | Not-determined | Genome-fragment | NA | NA | 0 |
| S3-final-viral-combined_k141_51566\|\|full | 1056 | No | NA | 2 | 0 | 1 | Low-quality | Genome-fragment | 0.48 | AAI-based (medium-confidence) | 0 |
| S9_k141_59140 | 1614 | No | NA | 2 | 0 | 1 | Not-determined | Genome-fragment | NA | NA | 0 |
| S9_k141_102037 | 3441 | No | NA | 2 | 0 | 2 | Not-determined | Genome-fragment | NA | NA | 0 |

| | | | | | | | | | | | |
|---|---|---|---|---|---|---|---|---|---|---|---|
| S9_k141_53692 | 9365 | No | NA | 1 | 0 | 1 | Not-determined | Genome-fragment | NA | NA | 0 |
| S7-final-viral-combined_k141_227167\|\|full | 1056 | No | NA | 2 | 0 | 0 | Low-quality | Genome-fragment | 1.9 | AAI-based (medium-confidence) | 0 |
| S9_k141_75318 | 3810 | No | NA | 3 | 0 | 2 | Not-determined | Genome-fragment | NA | NA | 0 |
| S9_k141_59158 | 1200 | No | NA | 1 | 0 | 1 | Not-determined | Genome-fragment | NA | NA | 0 |
| S7-final-viral-combined_k141_175244\|\|lt2gene | 1056 | No | NA | 2 | 1 | 0 | Low-quality | Genome-fragment | 2.29 | AAI-based (high-confidence) | 0 |
| S9_k141_59165 | 1155 | No | NA | 1 | 0 | 1 | Not-determined | Genome-fragment | NA | NA | 0 |
| S9_k141_16429 | 3574 | No | NA | 4 | 0 | 4 | Not-determined | Genome-fragment | NA | NA | 0 |
| S7_k141_36178 | 1056 | No | NA | 2 | 0 | 0 | Low-quality | Genome-fragment | 0.86 | AAI-based (medium-confidence) | 0 |
| S17-final-viral-combined_k141_137215\|\|full | 1056 | No | NA | 4 | 0 | 0 | Low-quality | Genome-fragment | 2.65 | AAI-based (medium-confidence) | 0 |
| S17-k141_28011 | 1056 | No | NA | 2 | 1 | 0 | Low-quality | Genome-fragment | 2.4 | AAI-based (high-confidence) | 0 |
| S9_k141_80637 | 3724 | No | NA | 4 | 0 | 3 | Not-determined | Genome-fragment | NA | NA | 0 |
| S9_k141_37691 | 1187 | No | NA | 2 | 0 | 0 | Not-determined | Genome-fragment | NA | NA | 0 |
| S9_k141_75361 | 6033 | No | NA | 2 | 0 | 2 | Not-determined | Genome-fragment | NA | NA | 0 |
| S9_k141_102082 | 3832 | No | NA | 5 | 0 | 0 | Not-determined | Genome-fragment | NA | NA | 0 |
| S12-final-viral-combined_k141_267922\|\|lt2gene | 1056 | No | NA | 2 | 1 | 0 | Low-quality | Genome-fragment | 2.75 | AAI-based (high-confidence) | 0 |
| S4-final-viral-combined_k141_34284\|\|full | 1056 | No | NA | 2 | 0 | 0 | Low-quality | Genome-fragment | 1.56 | AAI-based (medium-confidence) | 0 |
| S9_k141_64472 | 5450 | No | NA | 7 | 1 | 0 | High-quality | High-quality | 90.91 | AAI-based (medium-confidence) | 0 |
| S4_k141_154182 | 1056 | No | NA | 2 | 0 | 0 | Low-quality | Genome-fragment | 2.36 | AAI-based (medium-confidence) | 0 |
| S9_k141_598 | 1054 | No | NA | 1 | 0 | 1 | Not-determined | Genome-fragment | NA | NA | 0 |
| S9_k141_96751 | 6695 | No | NA | 6 | 0 | 3 | Not-determined | Genome-fragment | NA | NA | 0 |
| S9_k141_53853 | 2062 | No | NA | 1 | 0 | 1 | Not-determined | Genome-fragment | NA | NA | 0 |
| S9_k141_86071 | 1022 | No | NA | 1 | 0 | 1 | Not-determined | Genome-fragment | NA | NA | 0 |
| S9_k141_59317 | 1501 | No | NA | 1 | 0 | 1 | Not-determined | Genome-fragment | NA | NA | 0 |
| S9_k141_48692 | 2306 | No | NA | 2 | 0 | 1 | Not-determined | Genome-fragment | NA | NA | 0 |
| S9_k141_107209 | 4298 | No | NA | 1 | 0 | 1 | Not-determined | Genome-fragment | NA | NA | 0 |
| S4_k141_2243 | 1056 | No | NA | 2 | 0 | 1 | Low-quality | Genome-fragment | 2.37 | AAI-based (medium-confidence) | 0 |
| S9_k141_48699 | 5833 | No | NA | 3 | 0 | 3 | Not-determined | Genome-fragment | NA | NA | 0 |
| S9_k141_21434 | 1057 | No | NA | 2 | 0 | 0 | Low-quality | Genome-fragment | 0.97 | AAI-based (high-confidence) | 0 |
| S11-final-viral-combined_k141_138489\|\|full | 1057 | No | NA | 3 | 1 | 0 | Low-quality | Genome-fragment | 1.3 | AAI-based (medium-confidence) | 0 |
| S9_k141_86102 | 1496 | No | NA | 2 | 0 | 2 | Not-determined | Genome-fragment | NA | NA | 0 |
| S9_k141_43271 | 2194 | No | NA | 1 | 0 | 1 | Not-determined | Genome-fragment | NA | NA | 0 |
| S9_k141_37805 | 1106 | No | NA | 1 | 0 | 1 | Not-determined | Genome-fragment | NA | NA | 0 |
| S9_k141_123378 | 1295 | No | NA | 1 | 0 | 1 | Not-determined | Genome-fragment | NA | NA | 0 |
| S9_k141_43281 | 1445 | No | NA | 1 | 0 | 0 | Not-determined | Genome-fragment | NA | NA | 0 |
| S9_k141_32452 | 2186 | No | NA | 1 | 0 | 0 | Not-determined | Genome-fragment | NA | NA | 0 |
| S11-final-viral-combined_k141_183264\|\|full | 1057 | No | NA | 2 | 0 | 2 | Low-quality | Genome-fragment | 0.51 | AAI-based (medium-confidence) | 0 |
| S1-final-viral-combined_k141_134958\|\|full | 1057 | No | NA | 3 | 0 | 0 | Low-quality | Genome-fragment | 3.17 | AAI-based (high-confidence) | 0 |
| S9_k141_37826 | 4171 | No | NA | 1 | 1 | 0 | Not-determined | Genome-fragment | NA | NA | 0 |
| S9_k141_80781 | 1126 | No | NA | 1 | 0 | 1 | Not-determined | Genome-fragment | NA | NA | 0 |
| S1_k141_318752 | 1057 | No | NA | 2 | 0 | 1 | Low-quality | Genome-fragment | 2.77 | AAI-based (medium-confidence) | 0 |
| S9_k141_86196 | 1498 | No | NA | 2 | 0 | 1 | Not-determined | Genome-fragment | NA | NA | 0 |
| S9_k141_118238 | 4356 | No | NA | 3 | 0 | 2 | Not-determined | Genome-fragment | NA | NA | 0 |
| S9_k141_80803 | 2123 | No | NA | 1 | 0 | 0 | Not-determined | Genome-fragment | NA | NA | 0 |
| S9_k141_107272 | 1775 | No | NA | 2 | 0 | 2 | Not-determined | Genome-fragment | NA | NA | 0 |
| S9_k141_37869 | 3947 | No | NA | 6 | 0 | 1 | Not-determined | Genome-fragment | NA | NA | 0 |
| S9_k141_5775 | 1879 | No | NA | 2 | 0 | 1 | Not-determined | Genome-fragment | NA | NA | 0 |
| S9_k141_80830 | 7294 | No | NA | 11 | 0 | 3 | Not-determined | Genome-fragment | NA | NA | 0 |
| S9_k141_123395 | 5834 | No | NA | 8 | 0 | 4 | Not-determined | Genome-fragment | NA | NA | 0 |
| S9_k141_80854 | 4924 | No | NA | 6 | 0 | 2 | Not-determined | Genome-fragment | NA | NA | 0 |
| S9_k141_789 | 1830 | No | NA | 1 | 0 | 0 | Not-determined | Genome-fragment | NA | NA | 0 |
| S1_k141_108947 | 1057 | No | NA | 2 | 0 | 0 | Low-quality | Genome-fragment | 3.45 | AAI-based (high-confidence) | 0 |
| S7_k141_209108 | 1057 | No | NA | 3 | 0 | 0 | Low-quality | Genome-fragment | 5.04 | AAI-based (high-confidence) | 0 |
| S7_k141_15667 | 1057 | No | NA | 2 | 0 | 0 | Low-quality | Genome-fragment | 2.33 | AAI-based (high-confidence) | 0 |
| S9_k141_59493 | 1454 | No | NA | 1 | 0 | 0 | Not-determined | Genome-fragment | NA | NA | 0 |
| S7_k141_224227 | 1057 | No | NA | 2 | 0 | 0 | Low-quality | Genome-fragment | 0.76 | AAI-based (medium-confidence) | 0 |
| S9_k141_97062 | 2878 | No | NA | 3 | 0 | 2 | Not-determined | Genome-fragment | NA | NA | 0 |
| S12-final-viral-combined_k141_35943\|\|lt2gene | 1057 | No | NA | 2 | 1 | 0 | Low-quality | Genome-fragment | 0.94 | HMM-based (lower-bound) | 0 |
| S9_k141_43428 | 1381 | No | NA | 2 | 0 | 0 | Not-determined | Genome-fragment | NA | NA | 0 |
| S9_k141_37964 | 1108 | No | NA | 2 | 0 | 0 | Not-determined | Genome-fragment | NA | NA | 0 |
| S9_k141_59591 | 1319 | No | NA | 4 | 0 | 2 | Not-determined | Genome-fragment | NA | NA | 0 |
| S9_k141_54007 | 2761 | No | NA | 2 | 0 | 1 | Not-determined | Genome-fragment | NA | NA | 0 |
| S9_k141_86296 | 1078 | No | NA | 1 | 0 | 1 | Not-determined | Genome-fragment | NA | NA | 0 |
| S12-final-viral-combined_k141_311249\|\|lt2gene | 1057 | No | NA | 2 | 1 | 0 | Low-quality | Genome-fragment | 2.39 | AAI-based (medium-confidence) | 0 |
| S2-final-viral-combined_k141_20708\|\|lt2gene | 1057 | No | NA | 1 | 1 | 0 | Low-quality | Genome-fragment | 1.54 | HMM-based (lower-bound) | 0 |
| S9_k141_11080 | 1000 | No | NA | 1 | 0 | 0 | Not-determined | Genome-fragment | NA | NA | 0 |
| S9_k141_11081 | 1029 | No | NA | 1 | 0 | 1 | Not-determined | Genome-fragment | NA | NA | 0 |
| S4-final-viral-combined_k141_68479\|\|full | 1057 | No | NA | 2 | 0 | 0 | Low-quality | Genome-fragment | 2.77 | AAI-based (medium-confidence) | 0 |
| S9_k141_80996 | 4337 | No | NA | 6 | 0 | 6 | Not-determined | Genome-fragment | NA | NA | 0 |
| S6_k141_58266 | 1057 | No | NA | 2 | 0 | 2 | Low-quality | Genome-fragment | 0.37 | AAI-based (medium-confidence) | 0 |
| S9_k141_123466 | 2023 | No | NA | 1 | 0 | 1 | Not-determined | Genome-fragment | NA | NA | 0 |
| S9_k141_118421 | 2030 | No | NA | 2 | 0 | 0 | Not-determined | Genome-fragment | NA | NA | 0 |
| S9_k141_5951 | 5033 | No | NA | 4 | 0 | 0 | Not-determined | Genome-fragment | NA | NA | 0 |
| S9_k141_59618 | 3377 | No | NA | 1 | 0 | 1 | Not-determined | Genome-fragment | NA | NA | 0 |
| S9_k141_5959 | 1090 | No | NA | 1 | 0 | 1 | Not-determined | Genome-fragment | NA | NA | 0 |
| S9_k141_951 | 1641 | No | NA | 2 | 0 | 1 | Not-determined | Genome-fragment | NA | NA | 0 |
| S9_k141_953 | 2140 | No | NA | 1 | 0 | 0 | Not-determined | Genome-fragment | NA | NA | 0 |
| S9_k141_958 | 1299 | No | NA | 1 | 0 | 1 | Not-determined | Genome-fragment | NA | NA | 0 |
| S9_k141_971 | 3933 | No | NA | 4 | 0 | 1 | Not-determined | Genome-fragment | NA | NA | 0 |
| S9_k141_123492 | 11321 | No | NA | 14 | 0 | 14 | Not-determined | Genome-fragment | NA | NA | 0 |
| S9_k141_64706 | 4164 | No | NA | 7 | 0 | 0 | Not-determined | Genome-fragment | NA | NA | 0 |
| S9_k141_11095 | 6723 | No | NA | 12 | 0 | 1 | Not-determined | Genome-fragment | NA | NA | 0 |
| S8_k141_181469 | 1057 | No | NA | 2 | 0 | 0 | Low-quality | Genome-fragment | 0.69 | AAI-based (high-confidence) | 0 |
| S8_k141_8344 | 1057 | No | NA | 1 | 0 | 0 | Low-quality | Genome-fragment | 1.06 | AAI-based (high-confidence) | 0 |
| S9-final-viral-combined_k141_6290\|\|full | 1058 | No | NA | 3 | 1 | 0 | Low-quality | Genome-fragment | 3.03 | AAI-based (high-confidence) | 0 |
| S9_k141_102486 | 1073 | No | NA | 1 | 0 | 0 | Not-determined | Genome-fragment | NA | NA | 0 |
| S11-final-viral-combined_k141_65965\|\|full | 1058 | No | NA | 3 | 0 | 1 | Low-quality | Genome-fragment | 0.55 | AAI-based (medium-confidence) | 0 |
| S9_k141_91379 | 4998 | No | NA | 1 | 0 | 0 | Not-determined | Genome-fragment | NA | NA | 0 |
| S9_k141_69979 | 1195 | No | NA | 3 | 0 | 2 | Not-determined | Genome-fragment | NA | NA | 0 |
| S1-final-viral-combined_k141_142819\|\|full | 1058 | No | NA | 2 | 0 | 2 | Low-quality | Genome-fragment | 0.38 | AAI-based (medium-confidence) | 0 |
| S9_k141_118481 | 1331 | No | NA | 1 | 0 | 0 | Not-determined | Genome-fragment | NA | NA | 0 |
| S1-final-viral-combined_k141_96163\|\|full | 1058 | No | NA | 2 | 2 | 0 | Low-quality | Genome-fragment | 2.92 | AAI-based (high-confidence) | 0 |
| S9_k141_102542 | 3076 | No | NA | 2 | 0 | 2 | Not-determined | Genome-fragment | NA | NA | 0 |
| S9_k141_123545 | 1038 | No | NA | 1 | 0 | 1 | Not-determined | Genome-fragment | NA | NA | 0 |
| S1_k141_68868 | 1058 | No | NA | 1 | 0 | 0 | Low-quality | Genome-fragment | 0.86 | AAI-based (high-confidence) | 0 |
| S9_k141_48989 | 1571 | No | NA | 2 | 0 | 1 | Not-determined | Genome-fragment | NA | NA | 0 |
| S3-final-viral-combined_k141_89943\|\|full | 1058 | No | NA | 2 | 0 | 0 | Low-quality | Genome-fragment | 2.09 | AAI-based (medium-confidence) | 0 |
| S3-final-viral-combined_k141_33719\|\|full | 1058 | No | NA | 3 | 0 | 0 | Low-quality | Genome-fragment | 0.73 | AAI-based (medium-confidence) | 0 |
| S5-final-viral-combined_k141_201112\|\|full | 1058 | No | NA | 3 | 0 | 1 | Low-quality | Genome-fragment | 0.58 | AAI-based (high-confidence) | 0 |
| S9_k141_27452 | 3548 | No | NA | 1 | 0 | 1 | Not-determined | Genome-fragment | NA | NA | 0 |
| S9_k141_91499 | 1578 | No | NA | 1 | 0 | 0 | Not-determined | Genome-fragment | NA | NA | 0 |
| S9_k141_32850 | 1404 | No | NA | 2 | 0 | 0 | Not-determined | Genome-fragment | NA | NA | 0 |
| S9_k141_38162 | 1172 | No | NA | 1 | 0 | 1 | Not-determined | Genome-fragment | NA | NA | 0 |
| S9_k141_97227 | 1359 | No | NA | 2 | 0 | 1 | Not-determined | Genome-fragment | NA | NA | 0 |
| S9_k141_1205 | 1415 | No | NA | 1 | 0 | 1 | Not-determined | Genome-fragment | NA | NA | 0 |
| S9_k141_38166 | 1652 | No | NA | 2 | 0 | 1 | Not-determined | Genome-fragment | NA | NA | 0 |
| S9_k141_38172 | 1033 | No | NA | 1 | 0 | 0 | Not-determined | Genome-fragment | NA | NA | 0 |
| S5-final-viral-combined_k141_76987\|\|full | 1058 | No | NA | 3 | 1 | 0 | Low-quality | Genome-fragment | 7.84 | AAI-based (high-confidence) | 0 |
| S9_k141_22024 | 1575 | No | NA | 2 | 0 | 1 | Not-determined | Genome-fragment | NA | NA | 0 |
| S9_k141_97251 | 1475 | No | NA | 2 | 0 | 0 | Not-determined | Genome-fragment | NA | NA | 0 |
| S9_k141_22027 | 4155 | No | NA | 5 | 0 | 1 | Not-determined | Genome-fragment | NA | NA | 0 |
| S9_k141_22030 | 5466 | No | NA | 6 | 0 | 4 | Not-determined | Genome-fragment | NA | NA | 0 |
| S9_k141_86492 | 1424 | No | NA | 2 | 0 | 0 | Not-determined | Genome-fragment | NA | NA | 0 |
| S5-final-viral-combined_k141_68012\|\|full | 1058 | No | NA | 4 | 2 | 0 | Low-quality | Genome-fragment | 1.79 | AAI-based (medium-confidence) | 0 |
| S9_k141_81301 | 4909 | No | NA | 8 | 0 | 6 | Not-determined | Genome-fragment | NA | NA | 0 |
| S9_k141_59749 | 2284 | No | NA | 4 | 0 | 2 | Not-determined | Genome-fragment | NA | NA | 0 |
| S9_k141_118569 | 5511 | No | NA | 3 | 0 | 3 | Not-determined | Genome-fragment | NA | NA | 0 |
| S7-final-viral-combined_k141_205129\|\|full | 1058 | No | NA | 3 | 0 | 0 | Low-quality | Genome-fragment | 2.63 | AAI-based (high-confidence) | 0 |
| S9_k141_123637 | 1066 | No | NA | 2 | 0 | 1 | Not-determined | Genome-fragment | NA | NA | 0 |
| S9_k141_59772 | 1037 | No | NA | 1 | 0 | 0 | Not-determined | Genome-fragment | NA | NA | 0 |
| S9_k141_123663 | 1024 | No | NA | 1 | 0 | 1 | Not-determined | Genome-fragment | NA | NA | 0 |
| S9_k141_54171 | 1175 | No | NA | 2 | 0 | 2 | Not-determined | Genome-fragment | NA | NA | 0 |
| S7-final-viral-combined_k141_238295\|\|full | 1058 | No | NA | 1 | 0 | 0 | Low-quality | Genome-fragment | 0.58 | AAI-based (high-confidence) | 0 |
| S9_k141_123693 | 1430 | No | NA | 2 | 0 | 1 | Not-determined | Genome-fragment | NA | NA | 0 |
| S9_k141_86539 | 1139 | No | NA | 1 | 0 | 1 | Not-determined | Genome-fragment | NA | NA | 0 |
| S7-final-viral-combined_k141_118290\|\|full | 1058 | Yes | 794 | 2 | 1 | 1 | Low-quality | Genome-fragment | 1.11 | HMM-based (lower-bound) | 24.95 |
| S7_k141_14923 | 1058 | No | NA | 2 | 0 | 1 | Low-quality | Genome-fragment | 1.65 | AAI-based (medium-confidence) | 0 |
| S9_k141_6049 | 1095 | No | NA | 1 | 0 | 1 | Not-determined | Genome-fragment | NA | NA | 0 |
| S9_k141_1336 | 1152 | No | NA | 1 | 0 | 1 | Not-determined | Genome-fragment | NA | NA | 0 |
| S9_k141_1339 | 1223 | No | NA | 1 | 0 | 1 | Not-determined | Genome-fragment | NA | NA | 0 |
| S9_k141_64874 | 1571 | No | NA | 2 | 0 | 1 | Not-determined | Genome-fragment | NA | NA | 0 |
| S12-final-viral-combined_k141_234699\|\|full | 1058 | No | NA | 3 | 0 | 0 | Low-quality | Genome-fragment | 2.37 | AAI-based (high-confidence) | 0 |
| S12-final-viral-combined_k141_335766\|\|full | 1058 | No | NA | 2 | 0 | 1 | Low-quality | Genome-fragment | 1.23 | AAI-based (medium-confidence) | 0 |
| S9_k141_86600 | 1035 | No | NA | 4 | 0 | 4 | Not-determined | Genome-fragment | NA | NA | 0 |
| S9_k141_54302 | 1355 | No | NA | 1 | 0 | 1 | Not-determined | Genome-fragment | NA | NA | 0 |
| S12-final-viral-combined_k141_52191\|\|full | 1058 | No | NA | 2 | 0 | 0 | Low-quality | Genome-fragment | 1.65 | AAI-based (medium-confidence) | 0 |
| S9_k141_102650 | 3487 | No | NA | 5 | 0 | 1 | Not-determined | Genome-fragment | NA | NA | 0 |
| S9_k141_33050 | 4061 | No | NA | 6 | 0 | 1 | Not-determined | Genome-fragment | NA | NA | 0 |
| S9_k141_113139 | 5222 | No | NA | 5 | 0 | 4 | Not-determined | Genome-fragment | NA | NA | 0 |
| S9_k141_27633 | 1469 | No | NA | 2 | 0 | 1 | Not-determined | Genome-fragment | NA | NA | 0 |
| S12-final-viral-combined_k141_337974\|\|full | 1058 | No | NA | 3 | 0 | 0 | Low-quality | Genome-fragment | 2.72 | AAI-based (medium-confidence) | 0 |
| S9_k141_123793 | 1026 | No | NA | 1 | 0 | 1 | Not-determined | Genome-fragment | NA | NA | 0 |
| S9_k141_59838 | 1160 | No | NA | 1 | 0 | 1 | Not-determined | Genome-fragment | NA | NA | 0 |
| S9_k141_118687 | 1578 | No | NA | 2 | 0 | 1 | Not-determined | Genome-fragment | NA | NA | 0 |
| S9_k141_33128 | 1205 | No | NA | 1 | 0 | 1 | Not-determined | Genome-fragment | NA | NA | 0 |
| S9_k141_43541 | 1018 | No | NA | 2 | 0 | 1 | Not-determined | Genome-fragment | NA | NA | 0 |
| S9_k141_59921 | 3731 | No | NA | 5 | 0 | 3 | Not-determined | Genome-fragment | NA | NA | 0 |
| S9_k141_102682 | 1105 | No | NA | 2 | 0 | 1 | Not-determined | Genome-fragment | NA | NA | 0 |
| S9_k141_33164 | 1509 | No | NA | 2 | 0 | 1 | Not-determined | Genome-fragment | NA | NA | 0 |
| S12-final-viral-combined_k141_240745\|\|full | 1058 | No | NA | 1 | 0 | 0 | Low-quality | Genome-fragment | 2.52 | AAI-based (medium-confidence) | 0 |
| S6-final-viral-combined_k141_117069\|\|full | 1058 | No | NA | 4 | 0 | 0 | Low-quality | Genome-fragment | 1.98 | AAI-based (medium-confidence) | 0 |
| S9_k141_6194 | 5172 | No | NA | 5 | 0 | 2 | Not-determined | Genome-fragment | NA | NA | 0 |
| S9_k141_11430 | 1026 | No | NA | 2 | 0 | 1 | Not-determined | Genome-fragment | NA | NA | 0 |
| S8-final-viral-combined_k141_237272\|\|lt2gene | 1058 | No | NA | 2 | 0 | 0 | Low-quality | Genome-fragment | 39.56 | AAI-based (medium-confidence) | 0 |
| S9_k141_107701 | 3063 | No | NA | 3 | 0 | 3 | Not-determined | Genome-fragment | NA | NA | 0 |
| S9-final-viral-combined_k141_127677\|\|full | 1059 | No | NA | 3 | 0 | 0 | Low-quality | Genome-fragment | 2.36 | AAI-based (high-confidence) | 0 |
| S9_k141_16756 | 1246 | No | NA | 1 | 0 | 0 | Not-determined | Genome-fragment | NA | NA | 0 |
| S9_k141_38310 | 2758 | No | NA | 2 | 0 | 0 | Not-determined | Genome-fragment | NA | NA | 0 |
| S9_k141_38323 | 1320 | No | NA | 2 | 0 | 1 | Not-determined | Genome-fragment | NA | NA | 0 |
| S9_k141_43638 | 1385 | No | NA | 1 | 0 | 0 | Not-determined | Genome-fragment | NA | NA | 0 |
| S9-final-viral-combined_k141_67851\|\|lt2gene | 1059 | No | NA | 2 | 2 | 0 | Low-quality | Genome-fragment | 1.34 | HMM-based (lower-bound) | 0 |
| S9_k141_97479 | 1845 | No | NA | 2 | 0 | 1 | Not-determined | Genome-fragment | NA | NA | 0 |
| S9_k141_70197 | 1063 | No | NA | 1 | 0 | 1 | Not-determined | Genome-fragment | NA | NA | 0 |
| S9_k141_54511 | 1728 | No | NA | 2 | 0 | 0 | Not-determined | Genome-fragment | NA | NA | 0 |
| S11-final-viral-combined_k141_240025\|\|full | 1059 | No | NA | 3 | 1 | 0 | Low-quality | Genome-fragment | 3.31 | AAI-based (high-confidence) | 0 |
| S9_k141_1524 | 1193 | No | NA | 4 | 0 | 2 | Not-determined | Genome-fragment | NA | NA | 0 |
| S9_k141_123938 | 1264 | No | NA | 1 | 0 | 1 | Not-determined | Genome-fragment | NA | NA | 0 |
| S9_k141_70231 | 5111 | No | NA | 6 | 0 | 0 | Not-determined | Genome-fragment | NA | NA | 0 |
| S9_k141_16829 | 2054 | No | NA | 3 | 0 | 2 | Not-determined | Genome-fragment | NA | NA | 0 |
| S9_k141_123953 | 1610 | No | NA | 2 | 0 | 1 | Not-determined | Genome-fragment | NA | NA | 0 |
| S9_k141_11551 | 1131 | No | NA | 2 | 0 | 0 | Not-determined | Genome-fragment | NA | NA | 0 |
| S9_k141_27947 | 1331 | No | NA | 1 | 0 | 0 | Not-determined | Genome-fragment | NA | NA | 0 |
| S9_k141_65129 | 1092 | No | NA | 1 | 0 | 1 | Not-determined | Genome-fragment | NA | NA | 0 |
| S9_k141_16843 | 1274 | No | NA | 1 | 0 | 1 | Not-determined | Genome-fragment | NA | NA | 0 |
| S9_k141_81552 | 1227 | No | NA | 1 | 0 | 1 | Not-determined | Genome-fragment | NA | NA | 0 |
| S11-final-viral-combined_k141_321788\|\|full | 1059 | No | NA | 2 | 0 | 0 | Low-quality | Genome-fragment | 2.05 | AAI-based (medium-confidence) | 0 |
| S9_k141_86814 | 3669 | No | NA | 5 | 0 | 5 | Not-determined | Genome-fragment | NA | NA | 0 |
| S9_k141_118854 | 1047 | No | NA | 1 | 0 | 1 | Not-determined | Genome-fragment | NA | NA | 0 |
| S9_k141_113242 | 3263 | No | NA | 3 | 0 | 2 | Not-determined | Genome-fragment | NA | NA | 0 |
| S11_k141_181159 | 1059 | No | NA | 2 | 0 | 0 | Low-quality | Genome-fragment | 0.68 | AAI-based (medium-confidence) | 0 |
| S11_k141_321180 | 1059 | No | NA | 2 | 0 | 0 | Low-quality | Genome-fragment | 2.75 | AAI-based (high-confidence) | 0 |
| S1-final-viral-combined_k141_150710\|\|full | 1059 | No | NA | 2 | 0 | 0 | Low-quality | Genome-fragment | 0.86 | AAI-based (high-confidence) | 0 |
| S9_k141_113259 | 3241 | No | NA | 2 | 0 | 2 | Not-determined | Genome-fragment | NA | NA | 0 |
| S9_k141_81627 | 2208 | No | NA | 3 | 0 | 1 | Not-determined | Genome-fragment | NA | NA | 0 |
| S9_k141_54668 | 1029 | No | NA | 2 | 0 | 1 | Not-determined | Genome-fragment | NA | NA | 0 |
| S9_k141_43722 | 1060 | No | NA | 1 | 0 | 1 | Not-determined | Genome-fragment | NA | NA | 0 |
| S1-final-viral-combined_k141_276406\|\|full | 1059 | No | NA | 2 | 0 | 1 | Low-quality | Genome-fragment | 0.52 | AAI-based (medium-confidence) | 0 |
| S9_k141_113309 | 2001 | No | NA | 1 | 0 | 1 | Not-determined | Genome-fragment | NA | NA | 0 |
| S5-final-viral-combined_k141_29060\|\|full | 1059 | No | NA | 3 | 1 | 0 | Low-quality | Genome-fragment | 2.22 | AAI-based (medium-confidence) | 0 |
| S9_k141_75751 | 1078 | No | NA | 1 | 0 | 1 | Not-determined | Genome-fragment | NA | NA | 0 |
| S9_k141_75769 | 1066 | No | NA | 4 | 0 | 0 | Not-determined | Genome-fragment | NA | NA | 0 |
| S9_k141_75781 | 3822 | No | NA | 4 | 0 | 4 | Not-determined | Genome-fragment | NA | NA | 0 |
| S9_k141_92051 | 3213 | No | NA | 3 | 0 | 3 | Not-determined | Genome-fragment | NA | NA | 0 |
| S9_k141_75803 | 6719 | No | NA | 9 | 0 | 2 | Not-determined | Genome-fragment | NA | NA | 0 |
| S9_k141_22608 | 3349 | No | NA | 3 | 0 | 1 | Not-determined | Genome-fragment | NA | NA | 0 |
| S9_k141_38425 | 3815 | No | NA | 1 | 0 | 1 | Not-determined | Genome-fragment | NA | NA | 0 |
| S9_k141_60164 | 1056 | No | NA | 2 | 0 | 2 | Not-determined | Genome-fragment | NA | NA | 0 |
| S9_k141_1736 | 1085 | No | NA | 1 | 0 | 1 | Not-determined | Genome-fragment | NA | NA | 0 |
| S9_k141_1739 | 1336 | No | NA | 1 | 0 | 0 | Not-determined | Genome-fragment | NA | NA | 0 |
| S9_k141_70472 | 3335 | No | NA | 3 | 0 | 1 | Not-determined | Genome-fragment | NA | NA | 0 |
| S9_k141_22640 | 1045 | No | NA | 3 | 0 | 0 | Not-determined | Genome-fragment | NA | NA | 0 |
| S5-final-viral-combined_k141_31776\|\|full | 1059 | No | NA | 2 | 1 | 0 | Low-quality | Genome-fragment | 2.2 | AAI-based (high-confidence) | 0 |
| S9_k141_92124 | 1041 | No | NA | 1 | 0 | 1 | Not-determined | Genome-fragment | NA | NA | 0 |
| S9_k141_86857 | 1307 | No | NA | 2 | 0 | 0 | Not-determined | Genome-fragment | NA | NA | 0 |
| S7_k141_218025 | 1059 | No | NA | 2 | 0 | 0 | Low-quality | Genome-fragment | 1 | AAI-based (high-confidence) | 0 |
| S17-final-viral-combined_k141_196696\|\|full | 1059 | No | NA | 5 | 1 | 0 | Low-quality | Genome-fragment | 2.8 | AAI-based (high-confidence) | 0 |
| S9_k141_28051 | 1605 | No | NA | 4 | 0 | 1 | Not-determined | Genome-fragment | NA | NA | 0 |
| S12-final-viral-combined_k141_43924\|\|full | 1059 | No | NA | 3 | 1 | 0 | Low-quality | Genome-fragment | 0.64 | HMM-based (lower-bound) | 0 |
| S12_k141_125950 | 1059 | No | NA | 1 | 0 | 0 | Low-quality | Genome-fragment | 0.47 | AAI-based (high-confidence) | 0 |
| S4_k141_150536 | 1059 | No | NA | 2 | 0 | 2 | Low-quality | Genome-fragment | 0.55 | AAI-based (medium-confidence) | 0 |
| S9_k141_65420 | 1384 | No | NA | 2 | 0 | 0 | Not-determined | Genome-fragment | NA | NA | 0 |
| S9_k141_86878 | 28068 | No | NA | 1 | 0 | 0 | Not-determined | Genome-fragment | NA | NA | 0 |
| S9_k141_113498 | 1854 | No | NA | 3 | 0 | 1 | Not-determined | Genome-fragment | NA | NA | 0 |
| S11-final-viral-combined_k141_99537\|\|full | 1060 | No | NA | 3 | 1 | 0 | Low-quality | Genome-fragment | 1.4 | HMM-based (lower-bound) | 0 |
| S9_k141_70589 | 1121 | No | NA | 1 | 0 | 1 | Not-determined | Genome-fragment | NA | NA | 0 |
| S3_k141_16513 | 1060 | No | NA | 2 | 0 | 1 | Low-quality | Genome-fragment | 0.26 | AAI-based (medium-confidence) | 0 |
| S9_k141_70594 | 1177 | No | NA | 1 | 0 | 1 | Not-determined | Genome-fragment | NA | NA | 0 |
| S9_k141_81797 | 1003 | No | NA | 1 | 0 | 1 | Not-determined | Genome-fragment | NA | NA | 0 |
| S9_k141_54891 | 2082 | No | NA | 4 | 0 | 0 | Not-determined | Genome-fragment | NA | NA | 0 |
| S9_k141_103009 | 1031 | No | NA | 1 | 0 | 0 | Not-determined | Genome-fragment | NA | NA | 0 |
| S9_k141_38535 | 3310 | No | NA | 2 | 0 | 1 | Not-determined | Genome-fragment | NA | NA | 0 |
| S5-final-viral-combined_k141_171858\|\|full | 1060 | No | NA | 2 | 0 | 0 | Low-quality | Genome-fragment | 2.09 | AAI-based (medium-confidence) | 0 |
| S9_k141_54925 | 1365 | No | NA | 2 | 0 | 1 | Not-determined | Genome-fragment | NA | NA | 0 |
| S5-final-viral-combined_k141_162102\|\|lt2gene | 1060 | No | NA | 2 | 1 | 0 | Low-quality | Genome-fragment | 2.28 | AAI-based (high-confidence) | 0 |
| S9_k141_113532 | 3423 | No | NA | 3 | 0 | 2 | Not-determined | Genome-fragment | NA | NA | 0 |
| S9_k141_6819 | 1252 | No | NA | 1 | 0 | 1 | Not-determined | Genome-fragment | NA | NA | 0 |
| S9_k141_22868 | 3916 | No | NA | 2 | 0 | 1 | Not-determined | Genome-fragment | NA | NA | 0 |
| S9_k141_70683 | 5586 | No | NA | 2 | 0 | 0 | Not-determined | Genome-fragment | NA | NA | 0 |
| S9_k141_81841 | 4518 | No | NA | 2 | 0 | 1 | Not-determined | Genome-fragment | NA | NA | 0 |
| S9_k141_70750 | 2664 | No | NA | 2 | 0 | 2 | Not-determined | Genome-fragment | NA | NA | 0 |
| S9_k141_28235 | 2403 | No | NA | 2 | 0 | 0 | Not-determined | Genome-fragment | NA | NA | 0 |
| S9_k141_28236 | 1096 | No | NA | 1 | 0 | 1 | Not-determined | Genome-fragment | NA | NA | 0 |
| S9_k141_38578 | 1213 | No | NA | 1 | 0 | 0 | Not-determined | Genome-fragment | NA | NA | 0 |
| S9_k141_55012 | 1336 | No | NA | 1 | 0 | 0 | Not-determined | Genome-fragment | NA | NA | 0 |
| S7-final-viral-combined_k141_146205\|\|full | 1060 | No | NA | 3 | 1 | 0 | Low-quality | Genome-fragment | 2.29 | AAI-based (medium-confidence) | 0 |
| S17-final-viral-combined_k141_21063\|\|full | 1060 | No | NA | 3 | 0 | 1 | Low-quality | Genome-fragment | 0.54 | AAI-based (medium-confidence) | 0 |
| S9_k141_6983 | 1004 | No | NA | 2 | 0 | 0 | Not-determined | Genome-fragment | NA | NA | 0 |
| S9_k141_28275 | 1368 | No | NA | 2 | 0 | 0 | Not-determined | Genome-fragment | NA | NA | 0 |
| S9_k141_38657 | 1312 | No | NA | 1 | 0 | 1 | Not-determined | Genome-fragment | NA | NA | 0 |
| S9_k141_81948 | 1184 | No | NA | 2 | 0 | 1 | Not-determined | Genome-fragment | NA | NA | 0 |
| S9_k141_124410 | 3270 | No | NA | 3 | 0 | 3 | Not-determined | Genome-fragment | NA | NA | 0 |
| S9_k141_70808 | 6028 | No | NA | 5 | 0 | 5 | Not-determined | Genome-fragment | NA | NA | 0 |
| S9_k141_17342 | 1023 | No | NA | 1 | 0 | 1 | Not-determined | Genome-fragment | NA | NA | 0 |
| S9_k141_49731 | 3475 | No | NA | 3 | 0 | 2 | Not-determined | Genome-fragment | NA | NA | 0 |
| S9_k141_108215 | 1262 | No | NA | 1 | 0 | 1 | Not-determined | Genome-fragment | NA | NA | 0 |
| S9_k141_103303 | 1014 | No | NA | 2 | 0 | 1 | Not-determined | Genome-fragment | NA | NA | 0 |
| S9_k141_44188 | 2099 | No | NA | 2 | 0 | 1 | Not-determined | Genome-fragment | NA | NA | 0 |
| S9_k141_44189 | 1012 | No | NA | 2 | 0 | 1 | Not-determined | Genome-fragment | NA | NA | 0 |
| S9_k141_97816 | 33242 | No | NA | 17 | 0 | 5 | Not-determined | Genome-fragment | NA | NA | 0 |
| S9_k141_33372 | 1507 | No | NA | 1 | 0 | 1 | Not-determined | Genome-fragment | NA | NA | 0 |
| S9_k141_119113 | 1244 | No | NA | 3 | 0 | 1 | Not-determined | Genome-fragment | NA | NA | 0 |
| S9_k141_75989 | 1117 | No | NA | 1 | 0 | 0 | Not-determined | Genome-fragment | NA | NA | 0 |
| S9_k141_65768 | 1590 | No | NA | 1 | 0 | 0 | Not-determined | Genome-fragment | NA | NA | 0 |
| S9_k141_75990 | 6391 | No | NA | 4 | 0 | 3 | Not-determined | Genome-fragment | NA | NA | 0 |
| S9_k141_87187 | 25238 | No | NA | 39 | 0 | 31 | Not-determined | Genome-fragment | NA | NA | 0 |
| S12_k141_36777 | 1060 | No | NA | 1 | 0 | 1 | Low-quality | Genome-fragment | 2.99 | AAI-based (medium-confidence) | 0 |
| S9_k141_33392 | 8414 | No | NA | 8 | 0 | 8 | Not-determined | Genome-fragment | NA | NA | 0 |
| S9_k141_70929 | 1877 | No | NA | 3 | 0 | 2 | Not-determined | Genome-fragment | NA | NA | 0 |
| S9_k141_17501 | 1763 | No | NA | 2 | 0 | 0 | Not-determined | Genome-fragment | NA | NA | 0 |
| S9_k141_60586 | 1308 | No | NA | 2 | 0 | 1 | Not-determined | Genome-fragment | NA | NA | 0 |
| S9_k141_55198 | 6796 | No | NA | 5 | 0 | 1 | Not-determined | Genome-fragment | NA | NA | 0 |
| S9_k141_124517 | 2220 | No | NA | 1 | 0 | 0 | Not-determined | Genome-fragment | NA | NA | 0 |
| S9_k141_33397 | 1458 | No | NA | 1 | 0 | 1 | Not-determined | Genome-fragment | NA | NA | 0 |
| S12_k141_93685 | 1060 | No | NA | 2 | 0 | 0 | Low-quality | Genome-fragment | 0.57 | AAI-based (high-confidence) | 0 |
| S2-final-viral-combined_k141_85104\|\|full | 1060 | No | NA | 2 | 0 | 0 | Low-quality | Genome-fragment | 1.52 | AAI-based (medium-confidence) | 0 |
| S9_k141_2097 | 1565 | No | NA | 2 | 0 | 2 | Not-determined | Genome-fragment | NA | NA | 0 |
| S2_k141_11508 | 1060 | No | NA | 3 | 0 | 0 | Low-quality | Genome-fragment | 2.73 | AAI-based (high-confidence) | 0 |
| S9_k141_49893 | 9651 | No | NA | 2 | 0 | 0 | Not-determined | Genome-fragment | NA | NA | 0 |
| S9_k141_2112 | 2358 | No | NA | 3 | 0 | 2 | Not-determined | Genome-fragment | NA | NA | 0 |
| S2_k141_16178 | 1060 | No | NA | 3 | 0 | 0 | Low-quality | Genome-fragment | 2.2 | AAI-based (medium-confidence) | 0 |
| S9_k141_33426 | 9829 | No | NA | 7 | 0 | 2 | Not-determined | Genome-fragment | NA | NA | 0 |
| S9_k141_44300 | 3468 | No | NA | 2 | 0 | 1 | Not-determined | Genome-fragment | NA | NA | 0 |
| S9_k141_33437 | 5306 | No | NA | 3 | 0 | 2 | Not-determined | Genome-fragment | NA | NA | 0 |
| S9_k141_92533 | 1627 | No | NA | 2 | 0 | 1 | Not-determined | Genome-fragment | NA | NA | 0 |
| S2_k141_6263 | 1060 | No | NA | 1 | 0 | 1 | Low-quality | Genome-fragment | 0.37 | AAI-based (medium-confidence) | 0 |
| S4-final-viral-combined_k141_22818\|\|full | 1060 | No | NA | 3 | 0 | 2 | Low-quality | Genome-fragment | 1.55 | AAI-based (high-confidence) | 0 |
| S9_k141_23223 | 3014 | No | NA | 1 | 0 | 1 | Not-determined | Genome-fragment | NA | NA | 0 |
| S9_k141_55340 | 1519 | No | NA | 3 | 0 | 0 | Not-determined | Genome-fragment | NA | NA | 0 |
| S9_k141_92567 | 1123 | No | NA | 2 | 0 | 0 | Not-determined | Genome-fragment | NA | NA | 0 |
| S9_k141_66026 | 4639 | No | NA | 5 | 0 | 3 | Not-determined | Genome-fragment | NA | NA | 0 |
| S9_k141_108358 | 1454 | No | NA | 2 | 0 | 0 | Not-determined | Genome-fragment | NA | NA | 0 |
| S9_k141_76180 | 1109 | No | NA | 1 | 0 | 0 | Not-determined | Genome-fragment | NA | NA | 0 |
| S4-final-viral-combined_k141_90288\|\|lt2gene | 1060 | No | NA | 1 | 1 | 0 | Low-quality | Genome-fragment | 1.72 | HMM-based (lower-bound) | 0 |
| S4_k141_156157 | 1060 | No | NA | 2 | 0 | 0 | Low-quality | Genome-fragment | 1.05 | AAI-based (high-confidence) | 0 |
| S9_k141_12052 | 1238 | No | NA | 2 | 0 | 1 | Not-determined | Genome-fragment | NA | NA | 0 |
| S9_k141_60730 | 1210 | No | NA | 1 | 0 | 0 | Not-determined | Genome-fragment | NA | NA | 0 |
| S9-final-viral-combined_k141_67516\|\|lt2gene | 1061 | No | NA | 2 | 1 | 0 | Low-quality | Genome-fragment | 1.57 | HMM-based (lower-bound) | 0 |
| S9_k141_87287 | 1097 | No | NA | 1 | 0 | 1 | Not-determined | Genome-fragment | NA | NA | 0 |
| S9_k141_55446 | 2161 | No | NA | 1 | 0 | 1 | Not-determined | Genome-fragment | NA | NA | 0 |
| S11-final-viral-combined_k141_170906\|\|full | 1061 | No | NA | 3 | 0 | 0 | Low-quality | Genome-fragment | 2.05 | AAI-based (high-confidence) | 0 |
| S9_k141_23306 | 2070 | No | NA | 2 | 0 | 1 | Not-determined | Genome-fragment | NA | NA | 0 |
| S9_k141_119339 | 6441 | No | NA | 1 | 0 | 1 | Not-determined | Genome-fragment | NA | NA | 0 |
| S11_k141_138123 | 1061 | No | NA | 3 | 0 | 0 | Low-quality | Genome-fragment | 0.8 | AAI-based (high-confidence) | 0 |
| S9_k141_50122 | 1281 | No | NA | 2 | 0 | 1 | Not-determined | Genome-fragment | NA | NA | 0 |
| S11_k141_304463 | 1061 | No | NA | 2 | 0 | 1 | Low-quality | Genome-fragment | 0.37 | AAI-based (medium-confidence) | 0 |
| S1-final-viral-combined_k141_229995\|\|full | 1061 | No | NA | 3 | 0 | 0 | Low-quality | Genome-fragment | 1.6 | AAI-based (high-confidence) | 0 |
| S5_k141_42642 | 1061 | No | NA | 1 | 0 | 1 | Low-quality | Genome-fragment | 0.57 | AAI-based (high-confidence) | 0 |
| S9_k141_28449 | 1722 | No | NA | 2 | 0 | 1 | Not-determined | Genome-fragment | NA | NA | 0 |
| S9_k141_23356 | 1218 | No | NA | 1 | 0 | 1 | Not-determined | Genome-fragment | NA | NA | 0 |
| S9_k141_28454 | 1642 | No | NA | 2 | 0 | 2 | Not-determined | Genome-fragment | NA | NA | 0 |
| S17-final-viral-combined_k141_182615\|\|full | 1061 | No | NA | 3 | 1 | 0 | Low-quality | Genome-fragment | 2.69 | AAI-based (medium-confidence) | 0 |
| S9_k141_12231 | 1252 | No | NA | 2 | 0 | 2 | Not-determined | Genome-fragment | NA | NA | 0 |
| S17-final-viral-combined_k141_141281\|\|full | 1061 | No | NA | 2 | 0 | 0 | Low-quality | Genome-fragment | 0.58 | AAI-based (medium-confidence) | 0 |
| S12-final-viral-combined_k141_173536\|\|lt2gene | 1061 | No | NA | 2 | 2 | 0 | Low-quality | Genome-fragment | 2.63 | AAI-based (high-confidence) | 0 |
| S2-final-viral-combined_k141_152804\|\|full | 1061 | No | NA | 2 | 0 | 0 | Low-quality | Genome-fragment | 2.54 | AAI-based (medium-confidence) | 0 |
| S2-final-viral-combined_k141_71170\|\|full | 1061 | No | NA | 2 | 0 | 0 | Low-quality | Genome-fragment | 2.28 | AAI-based (high-confidence) | 0 |
| S9_k141_28581 | 1403 | No | NA | 1 | 0 | 0 | Not-determined | Genome-fragment | NA | NA | 0 |
| S9_k141_33527 | 1190 | No | NA | 1 | 0 | 1 | Not-determined | Genome-fragment | NA | NA | 0 |
| S2_k141_151235 | 1061 | No | NA | 2 | 0 | 0 | Low-quality | Genome-fragment | 1.96 | AAI-based (medium-confidence) | 0 |
| S9_k141_71225 | 5795 | No | NA | 10 | 0 | 1 | Not-determined | Genome-fragment | NA | NA | 0 |
| S4-final-viral-combined_k141_165568\|\|full | 1061 | No | NA | 3 | 0 | 0 | Low-quality | Genome-fragment | 1.73 | AAI-based (high-confidence) | 0 |
| S9_k141_7489 | 1343 | No | NA | 1 | 0 | 0 | Not-determined | Genome-fragment | NA | NA | 0 |
| S4-final-viral-combined_k141_153421\|\|lt2gene | 1061 | No | NA | 1 | 1 | 0 | Low-quality | Genome-fragment | 2.54 | AAI-based (high-confidence) | 0 |
| S9_k141_60965 | 1507 | No | NA | 4 | 0 | 0 | Not-determined | Genome-fragment | NA | NA | 0 |
| S9_k141_66281 | 1274 | No | NA | 2 | 0 | 0 | Not-determined | Genome-fragment | NA | NA | 0 |
| S9_k141_12372 | 1234 | No | NA | 2 | 0 | 0 | Not-determined | Genome-fragment | NA | NA | 0 |
| S9_k141_2448 | 1359 | No | NA | 1 | 0 | 0 | Not-determined | Genome-fragment | NA | NA | 0 |
| S9_k141_17925 | 1426 | No | NA | 3 | 0 | 1 | Not-determined | Genome-fragment | NA | NA | 0 |
| S6-final-viral-combined_k141_49802\|\|full | 1061 | No | NA | 3 | 0 | 0 | Low-quality | Genome-fragment | 2.29 | AAI-based (high-confidence) | 0 |
| S9_k141_39264 | 1526 | No | NA | 2 | 0 | 2 | Not-determined | Genome-fragment | NA | NA | 0 |
| S9_k141_119531 | 6014 | No | NA | 4 | 0 | 1 | Not-determined | Genome-fragment | NA | NA | 0 |
| S9_k141_87590 | 1378 | No | NA | 2 | 0 | 1 | Not-determined | Genome-fragment | NA | NA | 0 |
| S9_k141_12403 | 4026 | No | NA | 5 | 0 | 2 | Not-determined | Genome-fragment | NA | NA | 0 |
| S8_k141_110912 | 1061 | No | NA | 4 | 0 | 0 | Low-quality | Genome-fragment | 3.21 | AAI-based (medium-confidence) | 0 |
| S9_k141_92913 | 4464 | No | NA | 5 | 0 | 4 | Not-determined | Genome-fragment | NA | NA | 0 |
| S9_k141_12445 | 1652 | No | NA | 1 | 0 | 1 | Not-determined | Genome-fragment | NA | NA | 0 |
| S9_k141_66317 | 1309 | No | NA | 1 | 0 | 1 | Not-determined | Genome-fragment | NA | NA | 0 |
| S9_k141_61053 | 1475 | No | NA | 1 | 0 | 1 | Not-determined | Genome-fragment | NA | NA | 0 |
| S9-final-viral-combined_k141_90970\|\|full | 1062 | No | NA | 2 | 0 | 0 | Low-quality | Genome-fragment | 2.64 | AAI-based (medium-confidence) | 0 |
| S9_k141_12462 | 4158 | No | NA | 3 | 0 | 3 | Not-determined | Genome-fragment | NA | NA | 0 |
| S9_k141_119581 | 1793 | No | NA | 2 | 0 | 0 | Not-determined | Genome-fragment | NA | NA | 0 |
| S1-final-viral-combined_k141_120326\|\|full | 1062 | No | NA | 3 | 0 | 0 | Low-quality | Genome-fragment | 0.81 | AAI-based (high-confidence) | 0 |
| S9_k141_98240 | 3086 | No | NA | 5 | 0 | 0 | Not-determined | Genome-fragment | NA | NA | 0 |
| S1-final-viral-combined_k141_96959\|\|full | 1062 | No | NA | 4 | 0 | 0 | Low-quality | Genome-fragment | 1.82 | AAI-based (high-confidence) | 0 |
| S9_k141_76397 | 1702 | No | NA | 2 | 0 | 2 | Not-determined | Genome-fragment | NA | NA | 0 |
| S1-final-viral-combined_k141_179759\|\|full | 1062 | No | NA | 3 | 0 | 0 | Low-quality | Genome-fragment | 3.44 | AAI-based (high-confidence) | 0 |
| S9_k141_76419 | 6893 | No | NA | 7 | 0 | 2 | Not-determined | Genome-fragment | NA | NA | 0 |
| S9_k141_50434 | 1127 | No | NA | 1 | 0 | 1 | Not-determined | Genome-fragment | NA | NA | 0 |
| S3-final-viral-combined_k141_127141\|\|full | 1062 | No | NA | 2 | 0 | 1 | Low-quality | Genome-fragment | 0.63 | AAI-based (medium-confidence) | 0 |
| S9_k141_18090 | 4913 | No | NA | 3 | 0 | 1 | Not-determined | Genome-fragment | NA | NA | 0 |
| S9_k141_82487 | 1602 | No | NA | 2 | 0 | 0 | Not-determined | Genome-fragment | NA | NA | 0 |
| S3-final-viral-combined_k141_4667\|\|full | 1062 | No | NA | 3 | 0 | 0 | Low-quality | Genome-fragment | 1.92 | AAI-based (high-confidence) | 0 |
| S7-final-viral-combined_k141_211396\|\|lt2gene | 1062 | No | NA | 2 | 2 | 0 | Low-quality | Genome-fragment | 1.51 | HMM-based (lower-bound) | 0 |
| S9_k141_61124 | 1359 | No | NA | 2 | 0 | 2 | Not-determined | Genome-fragment | NA | NA | 0 |
| S7-final-viral-combined_k141_7465\|\|lt2gene | 1062 | No | NA | 2 | 2 | 0 | Low-quality | Genome-fragment | 2.36 | AAI-based (high-confidence) | 0 |
| S9_k141_113957 | 4144 | No | NA | 5 | 0 | 0 | Not-determined | Genome-fragment | NA | NA | 0 |
| S9_k141_33732 | 3512 | No | NA | 3 | 0 | 3 | Not-determined | Genome-fragment | NA | NA | 0 |
| S9_k141_71360 | 2194 | No | NA | 3 | 0 | 1 | Not-determined | Genome-fragment | NA | NA | 0 |
| S17-final-viral-combined_k141_170653\|\|full | 1062 | No | NA | 5 | 0 | 0 | Low-quality | Genome-fragment | 2.26 | AAI-based (high-confidence) | 0 |
| S9_k141_71370 | 5727 | No | NA | 2 | 0 | 1 | Not-determined | Genome-fragment | NA | NA | 0 |
| S9_k141_12629 | 1779 | No | NA | 3 | 0 | 3 | Not-determined | Genome-fragment | NA | NA | 0 |
| S9_k141_87685 | 1031 | No | NA | 2 | 0 | 0 | Not-determined | Genome-fragment | NA | NA | 0 |
| S9_k141_39420 | 4716 | No | NA | 5 | 0 | 1 | Not-determined | Genome-fragment | NA | NA | 0 |
| S17-final-viral-combined_k141_84422\|\|full | 1062 | No | NA | 2 | 1 | 0 | Low-quality | Genome-fragment | 2.52 | AAI-based (high-confidence) | 0 |
| S9_k141_71400 | 4373 | No | NA | 4 | 0 | 4 | Not-determined | Genome-fragment | NA | NA | 0 |
| S9_k141_55641 | 6750 | No | NA | 4 | 0 | 3 | Not-determined | Genome-fragment | NA | NA | 0 |
| S9_k141_55661 | 1038 | No | NA | 2 | 0 | 2 | Not-determined | Genome-fragment | NA | NA | 0 |
| S9_k141_7677 | 1785 | No | NA | 2 | 0 | 1 | Not-determined | Genome-fragment | NA | NA | 0 |
| S17-k141_193759 | 1062 | No | NA | 1 | 0 | 1 | Low-quality | Genome-fragment | 0.7 | AAI-based (high-confidence) | 0 |
| S9_k141_50511 | 1282 | No | NA | 2 | 0 | 2 | Not-determined | Genome-fragment | NA | NA | 0 |
| S9_k141_114004 | 1656 | No | NA | 1 | 0 | 0 | Not-determined | Genome-fragment | NA | NA | 0 |
| S9_k141_55681 | 2667 | No | NA | 3 | 0 | 0 | Not-determined | Genome-fragment | NA | NA | 0 |
| S9_k141_44490 | 2524 | No | NA | 1 | 0 | 0 | Not-determined | Genome-fragment | NA | NA | 0 |
| S12-final-viral-combined_k141_208372\|\|full | 1062 | No | NA | 2 | 0 | 1 | Low-quality | Genome-fragment | 1.28 | AAI-based (high-confidence) | 0 |
| S9_k141_50588 | 4444 | No | NA | 6 | 0 | 5 | Not-determined | Genome-fragment | NA | NA | 0 |
| S9_k141_7733 | 2998 | No | NA | 2 | 0 | 0 | Not-determined | Genome-fragment | NA | NA | 0 |
| S9_k141_12789 | 1016 | No | NA | 1 | 0 | 0 | Not-determined | Genome-fragment | NA | NA | 0 |
| S9_k141_61267 | 2568 | No | NA | 1 | 0 | 1 | Not-determined | Genome-fragment | NA | NA | 0 |
| S12_k141_98457 | 1062 | No | NA | 1 | 0 | 0 | Low-quality | Genome-fragment | 1.81 | AAI-based (high-confidence) | 0 |
| S9_k141_66373 | 1293 | No | NA | 2 | 0 | 2 | Not-determined | Genome-fragment | NA | NA | 0 |
| S9_k141_87792 | 9049 | No | NA | 8 | 0 | 8 | Not-determined | Genome-fragment | NA | NA | 0 |
| S9_k141_108773 | 1038 | No | NA | 1 | 0 | 1 | Not-determined | Genome-fragment | NA | NA | 0 |
| S9_k141_44541 | 1094 | No | NA | 1 | 0 | 0 | Not-determined | Genome-fragment | NA | NA | 0 |
| S9_k141_108787 | 1114 | No | NA | 1 | 0 | 0 | Not-determined | Genome-fragment | NA | NA | 0 |
| S9_k141_61303 | 1064 | No | NA | 1 | 0 | 1 | Not-determined | Genome-fragment | NA | NA | 0 |
| S9_k141_12839 | 1446 | No | NA | 1 | 0 | 1 | Not-determined | Genome-fragment | NA | NA | 0 |
| S9_k141_108798 | 5124 | No | NA | 7 | 0 | 0 | Not-determined | Genome-fragment | NA | NA | 0 |
| S9_k141_124914 | 1182 | No | NA | 2 | 0 | 1 | Not-determined | Genome-fragment | NA | NA | 0 |
| S9_k141_98264 | 9643 | No | NA | 2 | 0 | 1 | Not-determined | Genome-fragment | NA | NA | 0 |
| S9_k141_82766 | 3429 | No | NA | 4 | 0 | 3 | Not-determined | Genome-fragment | NA | NA | 0 |
| S12_k141_33022 | 1062 | No | NA | 2 | 1 | 0 | Low-quality | Genome-fragment | 2.93 | AAI-based (high-confidence) | 0 |
| S9_k141_98279 | 1997 | No | NA | 2 | 0 | 1 | Not-determined | Genome-fragment | NA | NA | 0 |
| S4-final-viral-combined_k141_128781\|\|lt2gene | 1062 | No | NA | 2 | 1 | 0 | Low-quality | Genome-fragment | 0.69 | HMM-based (lower-bound) | 0 |
| S9_k141_2748 | 3030 | No | NA | 5 | 0 | 0 | Not-determined | Genome-fragment | NA | NA | 0 |
| S9_k141_82844 | 1005 | No | NA | 1 | 0 | 0 | Not-determined | Genome-fragment | NA | NA | 0 |
| S9_k141_2754 | 2974 | No | NA | 3 | 0 | 2 | Not-determined | Genome-fragment | NA | NA | 0 |
| S8-final-viral-combined_k141_32038\|\|full | 1062 | No | NA | 3 | 0 | 0 | Low-quality | Genome-fragment | 1.76 | AAI-based (high-confidence) | 0 |
| S9_k141_93069 | 3343 | No | NA | 4 | 0 | 2 | Not-determined | Genome-fragment | NA | NA | 0 |
| S9_k141_7913 | 1643 | No | NA | 1 | 0 | 1 | Not-determined | Genome-fragment | NA | NA | 0 |
| S9_k141_103983 | 1214 | No | NA | 3 | 0 | 0 | Not-determined | Genome-fragment | NA | NA | 0 |
| S9_k141_2790 | 1210 | No | NA | 1 | 0 | 1 | Not-determined | Genome-fragment | NA | NA | 0 |
| S9-final-viral-combined_k141_48284\|\|full | 1063 | No | NA | 3 | 1 | 0 | Low-quality | Genome-fragment | 1.91 | AAI-based (high-confidence) | 0 |
| S11-final-viral-combined_k141_162437\|\|full | 1063 | No | NA | 4 | 1 | 0 | Low-quality | Genome-fragment | 2.18 | AAI-based (medium-confidence) | 0 |
| S9_k141_82893 | 3598 | No | NA | 5 | 0 | 4 | Not-determined | Genome-fragment | NA | NA | 0 |
| S11-final-viral-combined_k141_314483\|\|full | 1063 | No | NA | 2 | 0 | 1 | Low-quality | Genome-fragment | 0.73 | AAI-based (medium-confidence) | 0 |
| S17-final-viral-combined_k141_256032\|\|full | 1063 | No | NA | 3 | 2 | 0 | Low-quality | Genome-fragment | 2.33 | AAI-based (high-confidence) | 0 |
| S9_k141_93095 | 1130 | No | NA | 3 | 0 | 3 | Not-determined | Genome-fragment | NA | NA | 0 |
| S9_k141_87967 | 1378 | No | NA | 2 | 0 | 1 | Not-determined | Genome-fragment | NA | NA | 0 |
| S9_k141_114036 | 2305 | No | NA | 1 | 0 | 0 | Not-determined | Genome-fragment | NA | NA | 0 |
| S17-final-viral-combined_k141_181839\|\|full | 1063 | No | NA | 3 | 2 | 1 | Low-quality | Genome-fragment | 2.46 | AAI-based (high-confidence) | 0 |
| S9_k141_23694 | 1062 | No | NA | 3 | 0 | 2 | Not-determined | Genome-fragment | NA | NA | 0 |
| S9_k141_83003 | 1130 | No | NA | 1 | 0 | 0 | Not-determined | Genome-fragment | NA | NA | 0 |
| S9_k141_50811 | 1471 | No | NA | 1 | 0 | 1 | Not-determined | Genome-fragment | NA | NA | 0 |
| S9_k141_55972 | 1862 | No | NA | 4 | 0 | 1 | Not-determined | Genome-fragment | NA | NA | 0 |
| S9_k141_66557 | 4224 | No | NA | 2 | 0 | 0 | Not-determined | Genome-fragment | NA | NA | 0 |
| S9_k141_50826 | 4490 | No | NA | 5 | 0 | 3 | Not-determined | Genome-fragment | NA | NA | 0 |
| S17-k141_186583 | 1063 | No | NA | 3 | 0 | 0 | Low-quality | Genome-fragment | 2.12 | AAI-based (medium-confidence) | 0 |
| S9_k141_33954 | 2554 | No | NA | 1 | 0 | 1 | Not-determined | Genome-fragment | NA | NA | 0 |
| S9_k141_83062 | 2004 | No | NA | 3 | 0 | 1 | Not-determined | Genome-fragment | NA | NA | 0 |
| S9_k141_61332 | 3472 | No | NA | 3 | 0 | 2 | Not-determined | Genome-fragment | NA | NA | 0 |
| S9_k141_50839 | 1036 | No | NA | 2 | 0 | 0 | Not-determined | Genome-fragment | NA | NA | 0 |
| S9_k141_104090 | 1106 | No | NA | 1 | 0 | 1 | Not-determined | Genome-fragment | NA | NA | 0 |
| S9_k141_2862 | 3386 | No | NA | 3 | 0 | 3 | Not-determined | Genome-fragment | NA | NA | 0 |
| S9_k141_88115 | 1049 | No | NA | 1 | 0 | 0 | Not-determined | Genome-fragment | NA | NA | 0 |

| | | | | | | | | | | | |
|---|---|---|---|---|---|---|---|---|---|---|---|
| S9_k141_76822 | 1405 | No | NA | 1 | 0 | 0 | Not-determined | Genome-fragment | NA | NA | 0 |
| S9_k141_98391 | 3054 | No | NA | 4 | 0 | 3 | Not-determined | Genome-fragment | NA | NA | 0 |
| S12-final-viral-combined_k141_24813\|\|full | 1063 | No | NA | 2 | 0 | 1 | Low-quality | Genome-fragment | 1.82 | AAI-based (medium-confidence) | 0 |
| S9_k141_93181 | 1145 | No | NA | 2 | 0 | 0 | Not-determined | Genome-fragment | NA | NA | 0 |
| S9_k141_104169 | 4074 | No | NA | 8 | 0 | 0 | Medium-quality | Genome-fragment | 69.89 | AAI-based (medium-confidence) | 0 |
| S9_k141_104188 | 3112 | No | NA | 4 | 0 | 0 | Not-determined | Genome-fragment | NA | NA | 0 |
| S12_k141_211540 | 1063 | No | NA | 2 | 0 | 0 | Low-quality | Genome-fragment | 0.71 | AAI-based (medium-confidence) | 0 |
| S9_k141_39771 | 1355 | No | NA | 1 | 0 | 1 | Not-determined | Genome-fragment | NA | NA | 0 |
| S9_k141_29045 | 9750 | No | NA | 18 | 0 | 17 | Not-determined | Genome-fragment | NA | NA | 0 |
| S9_k141_12895 | 18327 | No | NA | 18 | 0 | 4 | Not-determined | Genome-fragment | NA | NA | 0 |
| S12_k141_348550 | 1063 | No | NA | 2 | 0 | 0 | Low-quality | Genome-fragment | 1.88 | AAI-based (high-confidence) | 0 |
| S9_k141_39785 | 1318 | No | NA | 1 | 0 | 0 | Not-determined | Genome-fragment | NA | NA | 0 |
| S9_k141_56161 | 4176 | No | NA | 5 | 0 | 2 | Not-determined | Genome-fragment | NA | NA | 0 |
| S9_k141_39790 | 1207 | No | NA | 3 | 0 | 1 | Not-determined | Genome-fragment | NA | NA | 0 |
| S9_k141_56162 | 2275 | No | NA | 1 | 0 | 1 | Not-determined | Genome-fragment | NA | NA | 0 |
| S9_k141_12927 | 1263 | No | NA | 2 | 0 | 0 | Not-determined | Genome-fragment | NA | NA | 0 |
| S2-final-viral-combined_k141_180558\|\|full | 1063 | No | NA | 3 | 0 | 1 | Low-quality | Genome-fragment | 1.56 | AAI-based (high-confidence) | 0 |
| S9_k141_119951 | 1000 | No | NA | 1 | 0 | 0 | Not-determined | Genome-fragment | NA | NA | 0 |
| S2-final-viral-combined_k141_131353\|\|full | 1063 | No | NA | 2 | 0 | 1 | Low-quality | Genome-fragment | 2.54 | AAI-based (medium-confidence) | 0 |
| S2_k141_126931 | 1063 | No | NA | 3 | 0 | 0 | Low-quality | Genome-fragment | 1.11 | AAI-based (high-confidence) | 0 |
| S9_k141_98412 | 1396 | No | NA | 2 | 0 | 1 | Not-determined | Genome-fragment | NA | NA | 0 |
| S4-final-viral-combined_k141_40094\|\|full | 1063 | No | NA | 2 | 1 | 0 | Low-quality | Genome-fragment | 2.71 | AAI-based (high-confidence) | 0 |
| S9_k141_8199 | 3319 | No | NA | 3 | 0 | 1 | Not-determined | Genome-fragment | NA | NA | 0 |
| S8_k141_6037 | 1063 | No | NA | 3 | 1 | 0 | Low-quality | Genome-fragment | 2.42 | AAI-based (high-confidence) | 0 |
| S9_k141_71790 | 3250 | No | NA | 3 | 0 | 1 | Not-determined | Genome-fragment | NA | NA | 0 |
| S11-final-viral-combined_k141_264272\|\|lt2gene | 1064 | No | NA | 1 | 1 | 0 | Low-quality | Genome-fragment | 2.62 | AAI-based (high-confidence) | 0 |
| S9_k141_3045 | 2063 | No | NA | 2 | 0 | 2 | Not-determined | Genome-fragment | NA | NA | 0 |
| S9_k141_13047 | 6352 | No | NA | 1 | 0 | 1 | Not-determined | Genome-fragment | NA | NA | 0 |
| S9_k141_76995 | 2378 | No | NA | 1 | 0 | 1 | Not-determined | Genome-fragment | NA | NA | 0 |
| S1-final-viral-combined_k141_313304\|\|full | 1064 | No | NA | 3 | 1 | 0 | Low-quality | Genome-fragment | 1.9 | AAI-based (high-confidence) | 0 |
| S9_k141_66684 | 1375 | No | NA | 1 | 0 | 0 | Not-determined | Genome-fragment | NA | NA | 0 |
| S9_k141_61598 | 1141 | No | NA | 1 | 0 | 0 | Not-determined | Genome-fragment | NA | NA | 0 |
| S9_k141_8258 | 1117 | No | NA | 1 | 0 | 1 | Not-determined | Genome-fragment | NA | NA | 0 |
| S9_k141_50936 | 1089 | No | NA | 1 | 0 | 1 | Not-determined | Genome-fragment | NA | NA | 0 |
| S9_k141_114339 | 1574 | No | NA | 2 | 0 | 1 | Not-determined | Genome-fragment | NA | NA | 0 |
| S9_k141_50939 | 1108 | No | NA | 3 | 0 | 1 | Not-determined | Genome-fragment | NA | NA | 0 |
| S9_k141_39845 | 1048 | No | NA | 2 | 0 | 0 | Not-determined | Genome-fragment | NA | NA | 0 |
| S1-final-viral-combined_k141_210059\|\|full | 1064 | No | NA | 3 | 1 | 0 | Low-quality | Genome-fragment | 2.32 | AAI-based (high-confidence) | 0 |
| S9_k141_23972 | 1432 | No | NA | 1 | 0 | 1 | Not-determined | Genome-fragment | NA | NA | 0 |
| S9_k141_93348 | 1409 | No | NA | 3 | 0 | 0 | Not-determined | Genome-fragment | NA | NA | 0 |
| S9_k141_3089 | 1134 | No | NA | 1 | 0 | 1 | Not-determined | Genome-fragment | NA | NA | 0 |
| S9_k141_34191 | 1154 | No | NA | 1 | 0 | 0 | Not-determined | Genome-fragment | NA | NA | 0 |
| S9_k141_56250 | 3112 | No | NA | 5 | 0 | 2 | Not-determined | Genome-fragment | NA | NA | 0 |
| S9_k141_18785 | 1057 | No | NA | 2 | 0 | 1 | Not-determined | Genome-fragment | NA | NA | 0 |
| S1-final-viral-combined_k141_300325\|\|lt2gene | 1064 | No | NA | 2 | 1 | 0 | Low-quality | Genome-fragment | 2.05 | HMM-based (lower-bound) | 0 |
| S9_k141_83401 | 1265 | No | NA | 1 | 0 | 1 | Not-determined | Genome-fragment | NA | NA | 0 |
| S9_k141_8287 | 1213 | No | NA | 1 | 0 | 0 | Not-determined | Genome-fragment | NA | NA | 0 |
| S9_k141_120036 | 1345 | No | NA | 2 | 0 | 2 | Not-determined | Genome-fragment | NA | NA | 0 |
| S9_k141_98502 | 1687 | No | NA | 5 | 0 | 4 | Not-determined | Genome-fragment | NA | NA | 0 |
| S9_k141_77109 | 1283 | No | NA | 1 | 0 | 0 | Not-determined | Genome-fragment | NA | NA | 0 |
| S9_k141_114421 | 1045 | No | NA | 1 | 0 | 1 | Not-determined | Genome-fragment | NA | NA | 0 |
| S9_k141_8313 | 1179 | No | NA | 1 | 0 | 0 | Not-determined | Genome-fragment | NA | NA | 0 |
| S9_k141_18802 | 1867 | No | NA | 3 | 0 | 0 | Not-determined | Genome-fragment | NA | NA | 0 |
| S9_k141_125240 | 1094 | No | NA | 2 | 0 | 0 | Not-determined | Genome-fragment | NA | NA | 0 |
| S9_k141_120085 | 1118 | No | NA | 1 | 0 | 1 | Not-determined | Genome-fragment | NA | NA | 0 |
| S9_k141_29161 | 4733 | No | NA | 2 | 0 | 2 | Not-determined | Genome-fragment | NA | NA | 0 |
| S9_k141_8337 | 2978 | No | NA | 3 | 0 | 3 | Not-determined | Genome-fragment | NA | NA | 0 |
| S9_k141_120117 | 1382 | No | NA | 2 | 0 | 0 | Not-determined | Genome-fragment | NA | NA | 0 |
| S9_k141_120142 | 1322 | No | NA | 1 | 0 | 1 | Not-determined | Genome-fragment | NA | NA | 0 |
| S9_k141_18844 | 5638 | No | NA | 3 | 0 | 2 | Not-determined | Genome-fragment | NA | NA | 0 |
| S9_k141_24126 | 6203 | No | NA | 5 | 0 | 1 | Not-determined | Genome-fragment | NA | NA | 0 |
| S9_k141_13215 | 5801 | No | NA | 4 | 0 | 2 | Not-determined | Genome-fragment | NA | NA | 0 |
| S1_k141_376641 | 1064 | No | NA | 1 | 0 | 0 | Low-quality | Genome-fragment | 2.23 | AAI-based (high-confidence) | 0 |
| S9_k141_77185 | 1007 | No | NA | 1 | 0 | 1 | Not-determined | Genome-fragment | NA | NA | 0 |
| S9_k141_13266 | 2229 | No | NA | 3 | 0 | 1 | Not-determined | Genome-fragment | NA | NA | 0 |
| S9_k141_18916 | 1636 | No | NA | 2 | 0 | 2 | Not-determined | Genome-fragment | NA | NA | 0 |
| S9_k141_45093 | 1068 | No | NA | 1 | 0 | 0 | Not-determined | Genome-fragment | NA | NA | 0 |
| S9_k141_29245 | 2208 | No | NA | 2 | 0 | 1 | Not-determined | Genome-fragment | NA | NA | 0 |
| S9_k141_13297 | 1264 | No | NA | 2 | 0 | 0 | Not-determined | Genome-fragment | NA | NA | 0 |
| S9_k141_39962 | 1976 | No | NA | 1 | 0 | 1 | Not-determined | Genome-fragment | NA | NA | 0 |
| S5-final-viral-combined_k141_116021\|\|full | 1064 | No | NA | 3 | 1 | 0 | Low-quality | Genome-fragment | 1.97 | AAI-based (high-confidence) | 0 |
| S9_k141_51090 | 2970 | No | NA | 3 | 0 | 1 | Not-determined | Genome-fragment | NA | NA | 0 |
| S9_k141_61735 | 1532 | No | NA | 2 | 0 | 0 | Not-determined | Genome-fragment | NA | NA | 0 |
| S5_k141_1696 | 1064 | No | NA | 1 | 0 | 1 | Low-quality | Genome-fragment | 0.4 | AAI-based (high-confidence) | 0 |
| S9_k141_120262 | 1217 | No | NA | 1 | 0 | 0 | Not-determined | Genome-fragment | NA | NA | 0 |
| S9_k141_125363 | 2091 | No | NA | 4 | 0 | 4 | Not-determined | Genome-fragment | NA | NA | 0 |
| S9_k141_24232 | 3251 | No | NA | 9 | 0 | 7 | Not-determined | Genome-fragment | NA | NA | 0 |
| S9_k141_120272 | 11828 | No | NA | 18 | 0 | 9 | Not-determined | Genome-fragment | NA | NA | 0 |
| S9_k141_120278 | 1802 | No | NA | 3 | 0 | 3 | Not-determined | Genome-fragment | NA | NA | 0 |
| S9_k141_40039 | 2964 | No | NA | 2 | 0 | 0 | Not-determined | Genome-fragment | NA | NA | 0 |
| S9_k141_45253 | 1497 | No | NA | 1 | 0 | 0 | Not-determined | Genome-fragment | NA | NA | 0 |
| S9_k141_13379 | 12264 | No | NA | 1 | 0 | 1 | Not-determined | Genome-fragment | NA | NA | 0 |
| S9_k141_19025 | 55093 | No | NA | 70 | 9 | 0 | High-quality | High-quality | 90.22 | AAI-based (high-confidence) | 0 |
| S9_k141_24282 | 6325 | No | NA | 3 | 0 | 0 | Not-determined | Genome-fragment | NA | NA | 0 |
| S7-final-viral-combined_k141_53652\|\|full | 1064 | No | NA | 4 | 1 | 0 | Low-quality | Genome-fragment | 2.09 | AAI-based (high-confidence) | 0 |
| S9_k141_120313 | 1056 | No | NA | 1 | 0 | 0 | Not-determined | Genome-fragment | NA | NA | 0 |
| S12-final-viral-combined_k141_17272\|\|full | 1064 | No | NA | 3 | 1 | 0 | Low-quality | Genome-fragment | 1.41 | HMM-based (lower-bound) | 0 |
| S9_k141_120333 | 1124 | No | NA | 2 | 0 | 2 | Not-determined | Genome-fragment | NA | NA | 0 |
| S12-final-viral-combined_k141_1769\|\|full | 1064 | No | NA | 4 | 0 | 2 | Low-quality | Genome-fragment | 0.31 | AAI-based (medium-confidence) | 0 |
| S9_k141_125422 | 31732 | No | NA | 27 | 0 | 18 | Not-determined | Genome-fragment | NA | NA | 0 |
| S9_k141_72086 | 1219 | No | NA | 2 | 0 | 1 | Not-determined | Genome-fragment | NA | NA | 0 |
| S12_k141_144277 | 1064 | No | NA | 3 | 0 | 0 | Low-quality | Genome-fragment | 0.56 | AAI-based (high-confidence) | 0 |
| S9_k141_45348 | 3273 | No | NA | 2 | 0 | 1 | Not-determined | Genome-fragment | NA | NA | 0 |
| S2-final-viral-combined_k141_129330\|\|full | 1064 | No | NA | 3 | 0 | 0 | Low-quality | Genome-fragment | 2.43 | AAI-based (medium-confidence) | 0 |
| S9_k141_120427 | 1373 | No | NA | 1 | 0 | 1 | Not-determined | Genome-fragment | NA | NA | 0 |
| S9_k141_93563 | 3155 | No | NA | 4 | 0 | 1 | Not-determined | Genome-fragment | NA | NA | 0 |
| S4-final-viral-combined_k141_117952\|\|full | 1064 | No | NA | 3 | 1 | 0 | Low-quality | Genome-fragment | 1.61 | HMM-based (lower-bound) | 0 |
| S9_k141_24348 | 6316 | No | NA | 7 | 0 | 2 | Not-determined | Genome-fragment | NA | NA | 0 |
| S6_k141_52463 | 1064 | No | NA | 3 | 0 | 0 | Low-quality | Genome-fragment | 1.26 | AAI-based (high-confidence) | 0 |
| S9-final-viral-combined_k141_122966\|\|full | 1065 | No | NA | 3 | 0 | 0 | Low-quality | Genome-fragment | 2.9 | AAI-based (medium-confidence) | 0 |
| S9_k141_83667 | 1007 | No | NA | 1 | 0 | 1 | Not-determined | Genome-fragment | NA | NA | 0 |
| S9_k141_40215 | 4880 | No | NA | 4 | 0 | 2 | Not-determined | Genome-fragment | NA | NA | 0 |
| S9_k141_34586 | 2323 | No | NA | 2 | 0 | 2 | Not-determined | Genome-fragment | NA | NA | 0 |
| S9_k141_120458 | 1197 | No | NA | 1 | 0 | 1 | Not-determined | Genome-fragment | NA | NA | 0 |
| S9_k141_34630 | 1159 | No | NA | 2 | 0 | 1 | Not-determined | Genome-fragment | NA | NA | 0 |
| S11-final-viral-combined_k141_87737\|\|full | 1065 | No | NA | 5 | 1 | 0 | Low-quality | Genome-fragment | 2.19 | AAI-based (high-confidence) | 0 |
| S9_k141_45466 | 1246 | No | NA | 1 | 0 | 0 | Not-determined | Genome-fragment | NA | NA | 0 |
| S9_k141_120509 | 1258 | No | NA | 1 | 0 | 0 | Not-determined | Genome-fragment | NA | NA | 0 |
| S9_k141_83744 | 1913 | No | NA | 4 | 0 | 1 | Not-determined | Genome-fragment | NA | NA | 0 |
| S9_k141_13537 | 3594 | No | NA | 3 | 0 | 0 | Not-determined | Genome-fragment | NA | NA | 0 |
| S11_k141_317021 | 1065 | No | NA | 2 | 1 | 0 | Low-quality | Genome-fragment | 1.26 | AAI-based (medium-confidence) | 0 |
| S9_k141_61970 | 1038 | No | NA | 1 | 0 | 1 | Not-determined | Genome-fragment | NA | NA | 0 |
| S9_k141_77570 | 1225 | No | NA | 1 | 0 | 0 | Not-determined | Genome-fragment | NA | NA | 0 |
| S9_k141_93669 | 1688 | No | NA | 1 | 0 | 0 | Not-determined | Genome-fragment | NA | NA | 0 |
| S9_k141_34729 | 1097 | No | NA | 3 | 0 | 0 | Not-determined | Genome-fragment | NA | NA | 0 |
| S9_k141_114967 | 2505 | No | NA | 3 | 0 | 2 | Not-determined | Genome-fragment | NA | NA | 0 |
| S9_k141_61987 | 1941 | No | NA | 1 | 0 | 1 | Not-determined | Genome-fragment | NA | NA | 0 |
| S11_k141_91541 | 1065 | No | NA | 2 | 0 | 0 | Low-quality | Genome-fragment | 1.27 | AAI-based (medium-confidence) | 0 |
| S9_k141_114971 | 26413 | No | NA | 37 | 22 | 2 | Medium-quality | Genome-fragment | 61.92 | AAI-based (high-confidence) | 0 |
| S9_k141_24388 | 5019 | No | NA | 8 | 0 | 3 | Not-determined | Genome-fragment | NA | NA | 0 |
| S9_k141_88387 | 1860 | No | NA | 2 | 0 | 1 | Not-determined | Genome-fragment | NA | NA | 0 |
| S11_k141_259055 | 1065 | No | NA | 2 | 0 | 0 | Low-quality | Genome-fragment | 2.44 | AAI-based (high-confidence) | 0 |
| S9_k141_62015 | 1328 | No | NA | 1 | 0 | 0 | Not-determined | Genome-fragment | NA | NA | 0 |
| S9_k141_109222 | 2150 | No | NA | 1 | 0 | 1 | Not-determined | Genome-fragment | NA | NA | 0 |
| S1-final-viral-combined_k141_362148\|\|lt2gene | 1065 | No | NA | 2 | 1 | 0 | Low-quality | Genome-fragment | 2.78 | AAI-based (high-confidence) | 0 |
| S1_k141_183006 | 1065 | No | NA | 1 | 0 | 1 | Low-quality | Genome-fragment | 1.78 | AAI-based (medium-confidence) | 0 |
| S9_k141_125607 | 1485 | No | NA | 1 | 0 | 1 | Not-determined | Genome-fragment | NA | NA | 0 |
| S9_k141_109234 | 3380 | No | NA | 2 | 0 | 0 | Not-determined | Genome-fragment | NA | NA | 0 |
| S9_k141_125616 | 2038 | No | NA | 2 | 0 | 2 | Not-determined | Genome-fragment | NA | NA | 0 |
| S3-final-viral-combined_k141_99924\|\|full | 1065 | No | NA | 2 | 0 | 1 | Low-quality | Genome-fragment | 0.63 | AAI-based (medium-confidence) | 0 |
| S9_k141_34843 | 4927 | No | NA | 2 | 0 | 1 | Not-determined | Genome-fragment | NA | NA | 0 |
| S17-final-viral-combined_k141_156754\|\|full | 1065 | No | NA | 3 | 1 | 0 | Low-quality | Genome-fragment | 2.01 | AAI-based (medium-confidence) | 0 |
| S9_k141_93758 | 9212 | No | NA | 8 | 0 | 8 | Not-determined | Genome-fragment | NA | NA | 0 |
| S9_k141_34892 | 1872 | No | NA | 2 | 0 | 2 | Not-determined | Genome-fragment | NA | NA | 0 |
| S9_k141_104620 | 2587 | No | NA | 1 | 0 | 0 | Not-determined | Genome-fragment | NA | NA | 0 |
| S9_k141_88517 | 1389 | No | NA | 1 | 0 | 1 | Not-determined | Genome-fragment | NA | NA | 0 |
| S9_k141_77712 | 1172 | No | NA | 1 | 0 | 0 | Not-determined | Genome-fragment | NA | NA | 0 |
| S9_k141_120695 | 3989 | No | NA | 4 | 0 | 4 | Not-determined | Genome-fragment | NA | NA | 0 |
| S9_k141_34898 | 1384 | No | NA | 1 | 0 | 0 | Not-determined | Genome-fragment | NA | NA | 0 |
| S9_k141_88526 | 9856 | No | NA | 9 | 0 | 9 | Not-determined | Genome-fragment | NA | NA | 0 |
| S9_k141_72217 | 4695 | No | NA | 1 | 0 | 0 | Not-determined | Genome-fragment | NA | NA | 0 |
| S9_k141_8789 | 2848 | No | NA | 1 | 0 | 1 | Not-determined | Genome-fragment | NA | NA | 0 |
| S17_k141_74923 | 1065 | No | NA | 2 | 1 | 0 | Low-quality | Genome-fragment | 0.67 | HMM-based (lower-bound) | 0 |
| S12-final-viral-combined_k141_332320\|\|full | 1065 | No | NA | 2 | 0 | 0 | Low-quality | Genome-fragment | 2.84 | AAI-based (medium-confidence) | 0 |
| S9_k141_83927 | 1626 | No | NA | 1 | 0 | 0 | Not-determined | Genome-fragment | NA | NA | 0 |
| S9_k141_72260 | 1319 | No | NA | 1 | 0 | 1 | Not-determined | Genome-fragment | NA | NA | 0 |
| S9_k141_88605 | 1093 | No | NA | 1 | 0 | 1 | Not-determined | Genome-fragment | NA | NA | 0 |
| S12_k141_314102 | 1065 | No | NA | 3 | 0 | 0 | Low-quality | Genome-fragment | 0.64 | AAI-based (medium-confidence) | 0 |
| S2_k141_14561 | 1065 | No | NA | 2 | 0 | 1 | Low-quality | Genome-fragment | 0.35 | AAI-based (high-confidence) | 0 |
| S4_k141_29454 | 1065 | No | NA | 2 | 0 | 0 | Low-quality | Genome-fragment | 2.11 | AAI-based (high-confidence) | 0 |
| S9_k141_115050 | 1488 | No | NA | 1 | 0 | 1 | Not-determined | Genome-fragment | NA | NA | 0 |
| S9_k141_51500 | 1355 | No | NA | 2 | 0 | 0 | Not-determined | Genome-fragment | NA | NA | 0 |
| S9_k141_45635 | 1393 | No | NA | 2 | 0 | 0 | Not-determined | Genome-fragment | NA | NA | 0 |
| S9_k141_77823 | 1385 | No | NA | 1 | 0 | 0 | Not-determined | Genome-fragment | NA | NA | 0 |
| S9_k141_125773 | 1268 | No | NA | 1 | 0 | 1 | Not-determined | Genome-fragment | NA | NA | 0 |
| S9_k141_24647 | 1005 | No | NA | 2 | 0 | 2 | Not-determined | Genome-fragment | NA | NA | 0 |
| S9_k141_40477 | 1083 | No | NA | 2 | 0 | 1 | Not-determined | Genome-fragment | NA | NA | 0 |
| S9_k141_77857 | 3712 | No | NA | 5 | 0 | 0 | Not-determined | Genome-fragment | NA | NA | 0 |
| S8_k141_1732 | 1065 | No | NA | 1 | 1 | 0 | Low-quality | Genome-fragment | 0.69 | AAI-based (high-confidence) | 0 |
| S9-final-viral-combined_k141_41787\|\|full | 1066 | No | NA | 2 | 0 | 0 | Low-quality | Genome-fragment | 2.83 | AAI-based (high-confidence) | 0 |
| S9-final-viral-combined_k141_95869\|\|full | 1066 | No | NA | 2 | 0 | 0 | Low-quality | Genome-fragment | 2.03 | AAI-based (high-confidence) | 0 |
| S9_k141_104703 | 1473 | No | NA | 1 | 0 | 1 | Not-determined | Genome-fragment | NA | NA | 0 |
| S9_k141_88725 | 1538 | No | NA | 1 | 0 | 1 | Not-determined | Genome-fragment | NA | NA | 0 |
| S9_k141_13878 | 1293 | No | NA | 1 | 0 | 1 | Not-determined | Genome-fragment | NA | NA | 0 |
| S9_k141_83978 | 1564 | No | NA | 2 | 0 | 2 | Not-determined | Genome-fragment | NA | NA | 0 |
| S9_k141_120958 | 1668 | No | NA | 1 | 0 | 1 | Not-determined | Genome-fragment | NA | NA | 0 |
| S11-final-viral-combined_k141_322719\|\|full | 1066 | No | NA | 2 | 0 | 0 | Low-quality | Genome-fragment | 0.75 | AAI-based (medium-confidence) | 0 |
| S9_k141_62279 | 15206 | No | NA | 17 | 0 | 7 | Not-determined | Genome-fragment | NA | NA | 0 |
| S9_k141_35211 | 1114 | No | NA | 2 | 0 | 0 | Not-determined | Genome-fragment | NA | NA | 0 |
| S9_k141_24745 | 3919 | No | NA | 3 | 0 | 1 | Not-determined | Genome-fragment | NA | NA | 0 |
| S9_k141_8971 | 3000 | No | NA | 2 | 0 | 0 | Not-determined | Genome-fragment | NA | NA | 0 |
| S9_k141_9031 | 2229 | No | NA | 1 | 0 | 1 | Not-determined | Genome-fragment | NA | NA | 0 |
| S1-final-viral-combined_k141_346882\|\|full | 1066 | No | NA | 3 | 1 | 0 | Low-quality | Genome-fragment | 1.91 | AAI-based (medium-confidence) | 0 |
| S9_k141_35280 | 11818 | No | NA | 10 | 0 | 9 | Not-determined | Genome-fragment | NA | NA | 0 |
| S9_k141_57251 | 15440 | No | NA | 31 | 0 | 30 | Not-determined | Genome-fragment | NA | NA | 0 |
| S9_k141_57274 | 1952 | No | NA | 4 | 0 | 2 | Not-determined | Genome-fragment | NA | NA | 0 |
| S9_k141_78031 | 2178 | No | NA | 1 | 0 | 1 | Not-determined | Genome-fragment | NA | NA | 0 |
| S9_k141_121078 | 1353 | No | NA | 1 | 0 | 0 | Not-determined | Genome-fragment | NA | NA | 0 |
| S9_k141_3581 | 1680 | No | NA | 1 | 0 | 0 | Not-determined | Genome-fragment | NA | NA | 0 |
| S9_k141_67314 | 1559 | No | NA | 3 | 0 | 1 | Not-determined | Genome-fragment | NA | NA | 0 |
| S1-final-viral-combined_k141_19422\|\|full | 1066 | No | NA | 2 | 0 | 1 | Low-quality | Genome-fragment | 1.67 | AAI-based (medium-confidence) | 0 |
| S9_k141_88971 | 6836 | No | NA | 1 | 0 | 1 | Not-determined | Genome-fragment | NA | NA | 0 |
| S9_k141_3629 | 1051 | No | NA | 1 | 0 | 1 | Not-determined | Genome-fragment | NA | NA | 0 |
| S9_k141_45866 | 4630 | No | NA | 3 | 0 | 3 | Not-determined | Genome-fragment | NA | NA | 0 |
| S1-final-viral-combined_k141_173169\|\|full | 1066 | No | NA | 3 | 0 | 0 | Low-quality | Genome-fragment | 2.66 | AAI-based (medium-confidence) | 0 |
| S9_k141_72404 | 4699 | No | NA | 4 | 0 | 4 | Not-determined | Genome-fragment | NA | NA | 0 |
| S3-final-viral-combined_k141_69815\|\|full | 1066 | No | NA | 4 | 0 | 0 | Low-quality | Genome-fragment | 1.6 | AAI-based (high-confidence) | 0 |
| S9_k141_45892 | 1492 | No | NA | 1 | 0 | 1 | Not-determined | Genome-fragment | NA | NA | 0 |
| S9_k141_109580 | 1349 | No | NA | 1 | 0 | 0 | Not-determined | Genome-fragment | NA | NA | 0 |
| S3_k141_80874 | 1066 | No | NA | 3 | 0 | 0 | Low-quality | Genome-fragment | 1.35 | AAI-based (high-confidence) | 0 |
| S5_k141_64072 | 1066 | No | NA | 1 | 0 | 1 | Low-quality | Genome-fragment | 3.54 | AAI-based (high-confidence) | 0 |
| S9_k141_3725 | 3217 | No | NA | 2 | 0 | 2 | Not-determined | Genome-fragment | NA | NA | 0 |
| S7_k141_78129 | 1066 | No | NA | 3 | 0 | 0 | Low-quality | Genome-fragment | 2.4 | AAI-based (high-confidence) | 0 |
| S9_k141_84138 | 7744 | No | NA | 5 | 0 | 1 | Not-determined | Genome-fragment | NA | NA | 0 |
| S17-final-viral-combined_k141_38771\|\|full | 1066 | No | NA | 3 | 1 | 0 | Low-quality | Genome-fragment | 3.72 | AAI-based (high-confidence) | 0 |
| S9_k141_89053 | 1365 | No | NA | 2 | 0 | 0 | Not-determined | Genome-fragment | NA | NA | 0 |
| S12-final-viral-combined_k141_339232\|\|full | 1066 | No | NA | 4 | 0 | 0 | Low-quality | Genome-fragment | 2.94 | AAI-based (high-confidence) | 0 |
| S12-final-viral-combined_k141_23361\|\|full | 1066 | No | NA | 3 | 0 | 1 | Low-quality | Genome-fragment | 1.37 | AAI-based (medium-confidence) | 0 |
| S9_k141_45915 | 2674 | No | NA | 2 | 0 | 0 | Not-determined | Genome-fragment | NA | NA | 0 |
| S9_k141_93843 | 2424 | No | NA | 2 | 0 | 2 | Not-determined | Genome-fragment | NA | NA | 0 |
| S9_k141_121222 | 1697 | No | NA | 1 | 0 | 1 | Not-determined | Genome-fragment | NA | NA | 0 |
| S9_k141_52017 | 2018 | No | NA | 1 | 0 | 1 | Not-determined | Genome-fragment | NA | NA | 0 |
| S9_k141_62516 | 2163 | No | NA | 2 | 0 | 1 | Not-determined | Genome-fragment | NA | NA | 0 |
| S12-final-viral-combined_k141_248808\|\|full | 1066 | No | NA | 2 | 0 | 0 | Low-quality | Genome-fragment | 1.29 | AAI-based (high-confidence) | 0 |
| S9_k141_78223 | 1093 | No | NA | 1 | 0 | 1 | Not-determined | Genome-fragment | NA | NA | 0 |
| S9_k141_67426 | 1683 | No | NA | 1 | 0 | 0 | Not-determined | Genome-fragment | NA | NA | 0 |
| S9_k141_72487 | 1203 | No | NA | 2 | 0 | 0 | Not-determined | Genome-fragment | NA | NA | 0 |
| S9_k141_45935 | 2254 | No | NA | 2 | 0 | 0 | Not-determined | Genome-fragment | NA | NA | 0 |
| S9_k141_3814 | 3043 | No | NA | 3 | 0 | 2 | Not-determined | Genome-fragment | NA | NA | 0 |
| S9_k141_45936 | 1197 | No | NA | 1 | 0 | 0 | Not-determined | Genome-fragment | NA | NA | 0 |
| S9_k141_104932 | 2823 | No | NA | 3 | 0 | 2 | Not-determined | Genome-fragment | NA | NA | 0 |
| S12-final-viral-combined_k141_400887\|\|full | 1066 | No | NA | 3 | 0 | 0 | Low-quality | Genome-fragment | 1.2 | AAI-based (high-confidence) | 0 |
| S9_k141_40601 | 1429 | No | NA | 2 | 0 | 2 | Not-determined | Genome-fragment | NA | NA | 0 |
| S9_k141_14004 | 6947 | No | NA | 6 | 0 | 3 | Not-determined | Genome-fragment | NA | NA | 0 |
| S12-final-viral-combined_k141_3458\|\|lt2gene | 1066 | No | NA | 2 | 0 | 1 | Low-quality | Genome-fragment | 1.43 | AAI-based (medium-confidence) | 0 |
| S9_k141_3842 | 22440 | No | NA | 22 | 0 | 8 | Not-determined | Genome-fragment | NA | NA | 0 |
| S9_k141_109676 | 2208 | No | NA | 3 | 0 | 2 | Not-determined | Genome-fragment | NA | NA | 0 |
| S12_k141_122436 | 1066 | No | NA | 3 | 0 | 0 | Low-quality | Genome-fragment | 2.28 | AAI-based (high-confidence) | 0 |
| S9_k141_93943 | 4238 | No | NA | 5 | 0 | 2 | Not-determined | Genome-fragment | NA | NA | 0 |
| S9_k141_29785 | 4514 | No | NA | 10 | 0 | 9 | Not-determined | Genome-fragment | NA | NA | 0 |
| S9_k141_115550 | 1162 | No | NA | 1 | 0 | 1 | Not-determined | Genome-fragment | NA | NA | 0 |
| S2-final-viral-combined_k141_48890\|\|full | 1066 | No | NA | 2 | 0 | 1 | Low-quality | Genome-fragment | 0.87 | AAI-based (medium-confidence) | 0 |
| S9_k141_126104 | 1147 | No | NA | 1 | 0 | 1 | Not-determined | Genome-fragment | NA | NA | 0 |
| S2_k141_83974 | 1066 | No | NA | 2 | 1 | 0 | Low-quality | Genome-fragment | 3.8 | HMM-based (lower-bound) | 0 |
| S9_k141_84236 | 22854 | No | NA | 18 | 0 | 1 | Not-determined | Genome-fragment | NA | NA | 0 |
| S9_k141_45964 | 2472 | No | NA | 2 | 0 | 1 | Not-determined | Genome-fragment | NA | NA | 0 |
| S9_k141_3864 | 1129 | No | NA | 1 | 0 | 0 | Not-determined | Genome-fragment | NA | NA | 0 |
| S9_k141_72611 | 6580 | No | NA | 5 | 0 | 2 | Not-determined | Genome-fragment | NA | NA | 0 |
| S9_k141_9222 | 1358 | No | NA | 3 | 0 | 2 | Not-determined | Genome-fragment | NA | NA | 0 |
| S6-final-viral-combined_k141_51172\|\|full | 1066 | No | NA | 2 | 0 | 0 | Low-quality | Genome-fragment | 0.91 | AAI-based (high-confidence) | 0 |
| S9_k141_67536 | 6390 | No | NA | 5 | 0 | 1 | Not-determined | Genome-fragment | NA | NA | 0 |
| S9_k141_67537 | 1365 | No | NA | 2 | 0 | 1 | Not-determined | Genome-fragment | NA | NA | 0 |
| S8-final-viral-combined_k141_5303\|\|full | 1066 | No | NA | 3 | 2 | 0 | Low-quality | Genome-fragment | 1.3 | HMM-based (lower-bound) | 0 |
| S9-final-viral-combined_k141_87001\|\|full | 1067 | No | NA | 3 | 0 | 0 | Low-quality | Genome-fragment | 2.4 | AAI-based (high-confidence) | 0 |
| S9_k141_78416 | 6381 | No | NA | 9 | 0 | 9 | Not-determined | Genome-fragment | NA | NA | 0 |
| S9_k141_67584 | 2197 | No | NA | 1 | 0 | 0 | Not-determined | Genome-fragment | NA | NA | 0 |
| S9_k141_9237 | 1101 | No | NA | 1 | 0 | 1 | Not-determined | Genome-fragment | NA | NA | 0 |
| S9_k141_14170 | 1316 | No | NA | 2 | 0 | 1 | Not-determined | Genome-fragment | NA | NA | 0 |
| S9_k141_115677 | 3425 | No | NA | 4 | 0 | 1 | Not-determined | Genome-fragment | NA | NA | 0 |
| S9_k141_78516 | 1373 | No | NA | 1 | 0 | 1 | Not-determined | Genome-fragment | NA | NA | 0 |
| S9_k141_14246 | 1753 | No | NA | 2 | 0 | 1 | Not-determined | Genome-fragment | NA | NA | 0 |
| S7-final-viral-combined_k141_219231\|\|lt2gene | 1067 | No | NA | 2 | 1 | 0 | Low-quality | Genome-fragment | 2.88 | AAI-based (high-confidence) | 0 |
| S9_k141_84314 | 1286 | No | NA | 1 | 0 | 0 | Not-determined | Genome-fragment | NA | NA | 0 |
| S17-final-viral-combined_k141_53502\|\|lt2gene | 1067 | No | NA | 2 | 1 | 0 | Low-quality | Genome-fragment | 2.47 | AAI-based (medium-confidence) | 0 |
| S9_k141_52237 | 1678 | No | NA | 1 | 0 | 1 | Not-determined | Genome-fragment | NA | NA | 0 |
| S9_k141_57500 | 1967 | No | NA | 5 | 0 | 5 | Not-determined | Genome-fragment | NA | NA | 0 |
| S12-final-viral-combined_k141_362570\|\|full | 1067 | No | NA | 2 | 0 | 0 | Low-quality | Genome-fragment | 1.6 | AAI-based (medium-confidence) | 0 |
| S9_k141_40736 | 1367 | No | NA | 1 | 0 | 1 | Not-determined | Genome-fragment | NA | NA | 0 |
| S2-final-viral-combined_k141_186346\|\|full | 1067 | No | NA | 3 | 0 | 2 | Low-quality | Genome-fragment | 0.29 | AAI-based (medium-confidence) | 0 |
| S9_k141_121605 | 1132 | No | NA | 1 | 0 | 1 | Not-determined | Genome-fragment | NA | NA | 0 |
| S9_k141_78631 | 1003 | No | NA | 1 | 0 | 0 | Not-determined | Genome-fragment | NA | NA | 0 |
| S9_k141_9398 | 1248 | No | NA | 1 | 0 | 0 | Not-determined | Genome-fragment | NA | NA | 0 |
| S9_k141_20045 | 1015 | No | NA | 2 | 0 | 1 | Not-determined | Genome-fragment | NA | NA | 0 |
| S9_k141_4059 | 1387 | No | NA | 1 | 0 | 0 | Not-determined | Genome-fragment | NA | NA | 0 |
| S9_k141_14434 | 1523 | No | NA | 2 | 0 | 1 | Not-determined | Genome-fragment | NA | NA | 0 |
| S4-final-viral-combined_k141_139665\|\|full | 1067 | No | NA | 3 | 2 | 0 | Low-quality | Genome-fragment | 2.52 | AAI-based (high-confidence) | 0 |
| S9_k141_72715 | 3091 | No | NA | 5 | 0 | 0 | Not-determined | Genome-fragment | NA | NA | 0 |
| S9_k141_14445 | 2176 | No | NA | 1 | 0 | 0 | Not-determined | Genome-fragment | NA | NA | 0 |
| S9_k141_40764 | 1327 | No | NA | 1 | 0 | 1 | Not-determined | Genome-fragment | NA | NA | 0 |
| S9_k141_109894 | 1153 | No | NA | 1 | 0 | 1 | Not-determined | Genome-fragment | NA | NA | 0 |
| S9_k141_14447 | 3643 | No | NA | 3 | 0 | 2 | Not-determined | Genome-fragment | NA | NA | 0 |
| S9_k141_72722 | 1027 | No | NA | 3 | 0 | 1 | Not-determined | Genome-fragment | NA | NA | 0 |
| S4-final-viral-combined_k141_81329\|\|full | 1067 | No | NA | 2 | 0 | 0 | Low-quality | Genome-fragment | 1.65 | AAI-based (medium-confidence) | 0 |
| S9_k141_14455 | 2037 | No | NA | 4 | 0 | 0 | Not-determined | Genome-fragment | NA | NA | 0 |
| S4-final-viral-combined_k141_34298\|\|full | 1067 | No | NA | 2 | 1 | 0 | Low-quality | Genome-fragment | 3.13 | AAI-based (high-confidence) | 0 |
| S9_k141_78718 | 1518 | No | NA | 2 | 0 | 2 | Not-determined | Genome-fragment | NA | NA | 0 |
| S9_k141_52329 | 1100 | No | NA | 2 | 0 | 1 | Not-determined | Genome-fragment | NA | NA | 0 |
| S9_k141_14481 | 1637 | No | NA | 2 | 0 | 2 | Not-determined | Genome-fragment | NA | NA | 0 |
| S9_k141_52338 | 1029 | No | NA | 1 | 0 | 0 | Not-determined | Genome-fragment | NA | NA | 0 |
| S9_k141_78754 | 1391 | No | NA | 1 | 0 | 0 | Not-determined | Genome-fragment | NA | NA | 0 |
| S9_k141_9484 | 1489 | No | NA | 1 | 0 | 0 | Not-determined | Genome-fragment | NA | NA | 0 |
| S9_k141_72798 | 1476 | No | NA | 1 | 0 | 1 | Not-determined | Genome-fragment | NA | NA | 0 |
| S9_k141_99830 | 1745 | No | NA | 1 | 0 | 0 | Not-determined | Genome-fragment | NA | NA | 0 |
| S9-final-viral-combined_k141_101566\|\|lt2gene | 1068 | No | NA | 2 | 1 | 0 | Low-quality | Genome-fragment | 2.49 | AAI-based (medium-confidence) | 0 |
| S9_k141_20112 | 1103 | No | NA | 2 | 0 | 2 | Not-determined | Genome-fragment | NA | NA | 0 |
| S9_k141_40874 | 4042 | No | NA | 5 | 0 | 3 | Not-determined | Genome-fragment | NA | NA | 0 |
| S9_k141_7495 | 1068 | No | NA | 2 | 0 | 0 | Low-quality | Genome-fragment | 0.88 | AAI-based (medium-confidence) | 0 |
| S9_k141_25359 | 3472 | No | NA | 3 | 0 | 3 | Not-determined | Genome-fragment | NA | NA | 0 |
| S9_k141_126358 | 2741 | No | NA | 6 | 0 | 6 | Not-determined | Genome-fragment | NA | NA | 0 |
| S9_k141_9516 | 3953 | No | NA | 7 | 0 | 7 | Not-determined | Genome-fragment | NA | NA | 0 |
| S11-final-viral-combined_k141_183284\|\|full | 1068 | No | NA | 3 | 0 | 0 | Low-quality | Genome-fragment | 2.07 | AAI-based (medium-confidence) | 0 |
| S9_k141_89584 | 3146 | No | NA | 4 | 0 | 3 | Not-determined | Genome-fragment | NA | NA | 0 |
| S9_k141_89597 | 1592 | No | NA | 1 | 0 | 0 | Not-determined | Genome-fragment | NA | NA | 0 |
| S9_k141_94189 | 2523 | No | NA | 1 | 0 | 1 | Not-determined | Genome-fragment | NA | NA | 0 |
| S9_k141_9558 | 2989 | No | NA | 3 | 0 | 0 | Not-determined | Genome-fragment | NA | NA | 0 |
| S9_k141_94199 | 1164 | No | NA | 2 | 0 | 1 | Not-determined | Genome-fragment | NA | NA | 0 |
| S9_k141_25475 | 1461 | No | NA | 3 | 0 | 2 | Not-determined | Genome-fragment | NA | NA | 0 |
| S9_k141_9603 | 1696 | No | NA | 2 | 0 | 0 | Not-determined | Genome-fragment | NA | NA | 0 |
| S9_k141_30129 | 1176 | No | NA | 1 | 0 | 1 | Not-determined | Genome-fragment | NA | NA | 0 |
| S9_k141_78889 | 1605 | No | NA | 2 | 0 | 1 | Not-determined | Genome-fragment | NA | NA | 0 |
| S11-final-viral-combined_k141_61543\|\|full | 1068 | No | NA | 3 | 0 | 0 | Low-quality | Genome-fragment | 2.52 | AAI-based (high-confidence) | 0 |
| S9_k141_46255 | 1287 | No | NA | 1 | 0 | 1 | Not-determined | Genome-fragment | NA | NA | 0 |
| S11-final-viral-combined_k141_241328\|\|full | 1068 | No | NA | 3 | 1 | 0 | Low-quality | Genome-fragment | 1.4 | HMM-based (lower-bound) | 0 |
| S9_k141_68032 | 1214 | No | NA | 3 | 0 | 1 | Not-determined | Genome-fragment | NA | NA | 0 |
| S11-final-viral-combined_k141_186929\|\|full | 1068 | No | NA | 2 | 1 | 0 | Low-quality | Genome-fragment | 1.63 | HMM-based (lower-bound) | 0 |
| S11-final-viral-combined_k141_215863\|\|full | 1068 | No | NA | 3 | 0 | 0 | Low-quality | Genome-fragment | 2.36 | AAI-based (medium-confidence) | 0 |
| S9_k141_121879 | 7736 | No | NA | 5 | 0 | 3 | Not-determined | Genome-fragment | NA | NA | 0 |
| S9_k141_78899 | 5779 | No | NA | 8 | 0 | 6 | Not-determined | Genome-fragment | NA | NA | 0 |
| S9_k141_121900 | 1019 | No | NA | 1 | 0 | 1 | Not-determined | Genome-fragment | NA | NA | 0 |
| S9_k141_89699 | 2119 | No | NA | 2 | 0 | 2 | Not-determined | Genome-fragment | NA | NA | 0 |
| S9_k141_4292 | 3669 | No | NA | 3 | 0 | 1 | Not-determined | Genome-fragment | NA | NA | 0 |
| S9_k141_20227 | 2265 | No | NA | 5 | 0 | 1 | Not-determined | Genome-fragment | NA | NA | 0 |
| S11-final-viral-combined_k141_269272\|\|lt2gene | 1068 | No | NA | 2 | 1 | 0 | Low-quality | Genome-fragment | 3.19 | AAI-based (medium-confidence) | 0 |
| S9_k141_25606 | 1002 | No | NA | 1 | 0 | 0 | Not-determined | Genome-fragment | NA | NA | 0 |
| S9_k141_25609 | 3250 | No | NA | 3 | 0 | 3 | Not-determined | Genome-fragment | NA | NA | 0 |
| S9_k141_100042 | 4220 | No | NA | 10 | 0 | 9 | Not-determined | Genome-fragment | NA | NA | 0 |
| S9_k141_41073 | 8200 | No | NA | 6 | 0 | 5 | Not-determined | Genome-fragment | NA | NA | 0 |
| S9_k141_35853 | 1792 | No | NA | 3 | 0 | 1 | Not-determined | Genome-fragment | NA | NA | 0 |
| S9_k141_73005 | 1012 | No | NA | 1 | 0 | 1 | Not-determined | Genome-fragment | NA | NA | 0 |
| S9_k141_57764 | 1055 | No | NA | 1 | 0 | 1 | Not-determined | Genome-fragment | NA | NA | 0 |
| S9_k141_110142 | 1294 | No | NA | 1 | 0 | 1 | Not-determined | Genome-fragment | NA | NA | 0 |
| S9_k141_126533 | 1083 | No | NA | 2 | 0 | 1 | Not-determined | Genome-fragment | NA | NA | 0 |
| S11_k141_19439 | 1068 | No | NA | 1 | 0 | 0 | Low-quality | Genome-fragment | 0.51 | AAI-based (medium-confidence) | 0 |
| S1-final-viral-combined_k141_118705\|\|full | 1068 | No | NA | 4 | 0 | 0 | Low-quality | Genome-fragment | 2.47 | AAI-based (high-confidence) | 0 |
| S9_k141_94378 | 4509 | No | NA | 5 | 0 | 5 | Not-determined | Genome-fragment | NA | NA | 0 |
| S9_k141_73103 | 1329 | No | NA | 2 | 0 | 1 | Not-determined | Genome-fragment | NA | NA | 0 |
| S7-final-viral-combined_k141_114641\|\|full | 1068 | No | NA | 2 | 0 | 1 | Low-quality | Genome-fragment | 1.9 | AAI-based (medium-confidence) | 0 |
| S7-final-viral-combined_k141_16034\|\|full | 1068 | No | NA | 3 | 0 | 0 | Low-quality | Genome-fragment | 4.06 | AAI-based (high-confidence) | 0 |
| S9_k141_116116 | 1007 | No | NA | 1 | 0 | 1 | Not-determined | Genome-fragment | NA | NA | 0 |
| S7_k141_154351 | 1068 | No | NA | 2 | 0 | 0 | Low-quality | Genome-fragment | 2.29 | AAI-based (medium-confidence) | 0 |
| S9_k141_73144 | 1018 | No | NA | 1 | 0 | 0 | Not-determined | Genome-fragment | NA | NA | 0 |
| S9_k141_35952 | 11525 | No | NA | 25 | 0 | 24 | Not-determined | Genome-fragment | NA | NA | 0 |
| S9_k141_94481 | 1797 | No | NA | 2 | 0 | 1 | Not-determined | Genome-fragment | NA | NA | 0 |
| S9_k141_9685 | 1081 | No | NA | 1 | 0 | 0 | Not-determined | Genome-fragment | NA | NA | 0 |
| S9_k141_126566 | 2636 | No | NA | 2 | 0 | 1 | Not-determined | Genome-fragment | NA | NA | 0 |
| S9_k141_84727 | 1071 | No | NA | 2 | 0 | 2 | Not-determined | Genome-fragment | NA | NA | 0 |
| S9_k141_9717 | 2121 | No | NA | 1 | 0 | 1 | Not-determined | Genome-fragment | NA | NA | 0 |
| S9_k141_30378 | 1142 | No | NA | 3 | 0 | 0 | Not-determined | Genome-fragment | NA | NA | 0 |
| S17-final-viral-combined_k141_81874\|\|full | 1068 | No | NA | 4 | 1 | 0 | Low-quality | Genome-fragment | 2.52 | AAI-based (high-confidence) | 0 |
| S9_k141_57848 | 1073 | No | NA | 1 | 0 | 1 | Not-determined | Genome-fragment | NA | NA | 0 |
| S9_k141_4442 | 2893 | No | NA | 1 | 0 | 0 | Not-determined | Genome-fragment | NA | NA | 0 |
| S9_k141_122107 | 1244 | No | NA | 2 | 0 | 0 | Not-determined | Genome-fragment | NA | NA | 0 |
| S9_k141_94521 | 3163 | No | NA | 4 | 0 | 3 | Not-determined | Genome-fragment | NA | NA | 0 |
| S9_k141_73186 | 1367 | No | NA | 2 | 0 | 1 | Not-determined | Genome-fragment | NA | NA | 0 |
| S9_k141_122119 | 1472 | No | NA | 2 | 0 | 2 | Not-determined | Genome-fragment | NA | NA | 0 |
| S9_k141_63353 | 1017 | No | NA | 1 | 0 | 0 | Not-determined | Genome-fragment | NA | NA | 0 |
| S9_k141_90013 | 1302 | No | NA | 1 | 0 | 1 | Not-determined | Genome-fragment | NA | NA | 0 |
| S9_k141_20372 | 1123 | No | NA | 1 | 0 | 0 | Not-determined | Genome-fragment | NA | NA | 0 |
| S17-final-viral-combined_k141_74577\|\|full | 1068 | No | NA | 3 | 0 | 1 | Low-quality | Genome-fragment | 0.75 | AAI-based (high-confidence) | 0 |
| S9_k141_116247 | 3713 | No | NA | 3 | 0 | 3 | Not-determined | Genome-fragment | NA | NA | 0 |
| S9_k141_20377 | 3184 | No | NA | 1 | 0 | 0 | Not-determined | Genome-fragment | NA | NA | 0 |
| S12_k141_228689 | 1068 | No | NA | 2 | 0 | 2 | Low-quality | Genome-fragment | 0.32 | AAI-based (medium-confidence) | 0 |
| S9_k141_52748 | 1213 | No | NA | 5 | 0 | 3 | Not-determined | Genome-fragment | NA | NA | 0 |
| S9_k141_52759 | 1161 | No | NA | 2 | 0 | 1 | Not-determined | Genome-fragment | NA | NA | 0 |
| S8-final-viral-combined_k141_264432\|\|full | 1068 | No | NA | 3 | 0 | 1 | Low-quality | Genome-fragment | 0.91 | AAI-based (medium-confidence) | 0 |
| S9_k141_14831 | 1010 | No | NA | 1 | 0 | 0 | Not-determined | Genome-fragment | NA | NA | 0 |
| S9_k141_25687 | 1094 | No | NA | 2 | 0 | 0 | Not-determined | Genome-fragment | NA | NA | 0 |
| S9_k141_90075 | 4065 | No | NA | 6 | 0 | 2 | Not-determined | Genome-fragment | NA | NA | 0 |
| S9-final-viral-combined_k141_36387\|\|full | 1069 | No | NA | 2 | 0 | 1 | Low-quality | Genome-fragment | 8.32 | AAI-based (high-confidence) | 0 |
| S9_k141_122207 | 1515 | No | NA | 2 | 0 | 2 | Not-determined | Genome-fragment | NA | NA | 0 |
| S11-final-viral-combined_k141_33341\|\|full | 1069 | No | NA | 2 | 1 | 0 | Low-quality | Genome-fragment | 1.78 | AAI-based (high-confidence) | 0 |
| S11-final-viral-combined_k141_219851\|\|full | 1069 | No | NA | 3 | 0 | 1 | Low-quality | Genome-fragment | 0.44 | AAI-based (medium-confidence) | 0 |
| S9_k141_68476 | 1399 | No | NA | 1 | 0 | 0 | Not-determined | Genome-fragment | NA | NA | 0 |
| S9_k141_84937 | 1140 | No | NA | 1 | 0 | 1 | Not-determined | Genome-fragment | NA | NA | 0 |
| S9_k141_52780 | 2869 | No | NA | 3 | 0 | 1 | Not-determined | Genome-fragment | NA | NA | 0 |
| S1_k141_337552 | 1069 | No | NA | 1 | 0 | 1 | Low-quality | Genome-fragment | 0.18 | AAI-based (high-confidence) | 0 |
| S9_k141_110319 | 1003 | No | NA | 1 | 0 | 0 | Not-determined | Genome-fragment | NA | NA | 0 |
| S9_k141_41272 | 1066 | No | NA | 1 | 0 | 1 | Not-determined | Genome-fragment | NA | NA | 0 |
| S9_k141_57964 | 1182 | No | NA | 2 | 0 | 0 | Not-determined | Genome-fragment | NA | NA | 0 |
| S9_k141_79122 | 1195 | No | NA | 2 | 0 | 0 | Not-determined | Genome-fragment | NA | NA | 0 |
| S5-final-viral-combined_k141_211127\|\|full | 1069 | No | NA | 2 | 0 | 0 | Low-quality | Genome-fragment | 1.99 | AAI-based (medium-confidence) | 0 |
| S5-final-viral-combined_k141_33780\|\|full | 1069 | No | NA | 3 | 0 | 1 | Low-quality | Genome-fragment | 0.91 | AAI-based (medium-confidence) | 0 |
| S9_k141_25724 | 1058 | No | NA | 3 | 0 | 0 | Not-determined | Genome-fragment | NA | NA | 0 |
| S9_k141_68526 | 3507 | No | NA | 2 | 0 | 2 | Not-determined | Genome-fragment | NA | NA | 0 |
| S5-final-viral-combined_k141_25411\|\|full | 1069 | No | NA | 2 | 0 | 0 | Low-quality | Genome-fragment | 1.87 | AAI-based (high-confidence) | 0 |
| S9_k141_68542 | 1072 | No | NA | 2 | 0 | 2 | Not-determined | Genome-fragment | NA | NA | 0 |
| S9_k141_68549 | 1275 | No | NA | 2 | 0 | 1 | Not-determined | Genome-fragment | NA | NA | 0 |
| S9_k141_46627 | 1174 | No | NA | 1 | 0 | 1 | Not-determined | Genome-fragment | NA | NA | 0 |
| S9_k141_73409 | 6462 | No | NA | 3 | 0 | 3 | Not-determined | Genome-fragment | NA | NA | 0 |
| S9_k141_126662 | 1273 | No | NA | 3 | 0 | 0 | Not-determined | Genome-fragment | NA | NA | 0 |
| S5_k141_207017 | 1069 | No | NA | 1 | 0 | 0 | Low-quality | Genome-fragment | 1.79 | AAI-based (medium-confidence) | 0 |
| S9_k141_122290 | 1118 | No | NA | 2 | 0 | 0 | Not-determined | Genome-fragment | NA | NA | 0 |
| S9_k141_36230 | 1176 | No | NA | 1 | 0 | 1 | Not-determined | Genome-fragment | NA | NA | 0 |
| S9_k141_36238 | 1256 | No | NA | 1 | 0 | 1 | Not-determined | Genome-fragment | NA | NA | 0 |
| S9_k141_116337 | 1603 | No | NA | 1 | 0 | 0 | Not-determined | Genome-fragment | NA | NA | 0 |
| S9_k141_90190 | 5336 | No | NA | 1 | 0 | 0 | Not-determined | Genome-fragment | NA | NA | 0 |
| S5_k141_202930 | 1069 | No | NA | 1 | 0 | 1 | Low-quality | Genome-fragment | 0.5 | AAI-based (medium-confidence) | 0 |
| S17-final-viral-combined_k141_61342\|\|lt2gene | 1069 | No | NA | 2 | 1 | 0 | Low-quality | Genome-fragment | 2.34 | AAI-based (medium-confidence) | 0 |
| S12-final-viral-combined_k141_110038\|\|full | 1069 | No | NA | 2 | 0 | 0 | Low-quality | Genome-fragment | 1.69 | AAI-based (high-confidence) | 0 |
| S9_k141_4577 | 1421 | No | NA | 1 | 0 | 0 | Not-determined | Genome-fragment | NA | NA | 0 |
| S6-final-viral-combined_k141_21155\|\|full | 1069 | No | NA | 3 | 1 | 0 | Low-quality | Genome-fragment | 1.76 | AAI-based (high-confidence) | 0 |
| S9_k141_100349 | 3838 | No | NA | 4 | 0 | 3 | Not-determined | Genome-fragment | NA | NA | 0 |
| S9_k141_52872 | 1218 | No | NA | 1 | 0 | 1 | Not-determined | Genome-fragment | NA | NA | 0 |
| S8_k141_243856 | 1069 | No | NA | 1 | 0 | 1 | Low-quality | Genome-fragment | 0.73 | AAI-based (high-confidence) | 0 |
| S9_k141_94761 | 5607 | No | NA | 6 | 0 | 5 | Not-determined | Genome-fragment | NA | NA | 0 |
| S9_k141_4584 | 1161 | No | NA | 1 | 0 | 0 | Not-determined | Genome-fragment | NA | NA | 0 |
| S9_k141_90214 | 3450 | No | NA | 5 | 0 | 5 | Not-determined | Genome-fragment | NA | NA | 0 |
| S9_k141_110408 | 1152 | No | NA | 1 | 0 | 0 | Not-determined | Genome-fragment | NA | NA | 0 |
| S9_k141_122314 | 1169 | No | NA | 2 | 0 | 1 | Not-determined | Genome-fragment | NA | NA | 0 |
| S9_k141_46708 | 4237 | No | NA | 2 | 0 | 0 | Not-determined | Genome-fragment | NA | NA | 0 |
| S9_k141_79332 | 1449 | No | NA | 3 | 0 | 0 | Not-determined | Genome-fragment | NA | NA | 0 |
| S9_k141_36269 | 1069 | No | NA | 4 | 0 | 0 | Not-determined | Genome-fragment | NA | NA | 0 |
| S9_k141_49616 | 1070 | No | NA | 1 | 0 | 1 | Low-quality | Genome-fragment | 2.55 | AAI-based (medium-confidence) | 0 |
| S11_k141_271466 | 1070 | No | NA | 2 | 1 | 0 | Low-quality | Genome-fragment | 0.62 | HMM-based (lower-bound) | 0 |
| S9_k141_79373 | 7864 | No | NA | 5 | 0 | 5 | Not-determined | Genome-fragment | NA | NA | 0 |
| S9_k141_73468 | 2553 | No | NA | 2 | 0 | 2 | Not-determined | Genome-fragment | NA | NA | 0 |
| S9_k141_15040 | 1072 | No | NA | 1 | 0 | 0 | Not-determined | Genome-fragment | NA | NA | 0 |
| S9_k141_20599 | 1502 | No | NA | 2 | 0 | 1 | Not-determined | Genome-fragment | NA | NA | 0 |
| S9_k141_25887 | 6057 | No | NA | 7 | 0 | 6 | Not-determined | Genome-fragment | NA | NA | 0 |
| S9_k141_30689 | 1077 | No | NA | 1 | 0 | 1 | Not-determined | Genome-fragment | NA | NA | 0 |
| S9_k141_20618 | 4564 | No | NA | 5 | 0 | 0 | Not-determined | Genome-fragment | NA | NA | 0 |
| S9_k141_25911 | 1039 | No | NA | 2 | 0 | 1 | Not-determined | Genome-fragment | NA | NA | 0 |
| S9_k141_63720 | 6385 | No | NA | 5 | 0 | 4 | Not-determined | Genome-fragment | NA | NA | 0 |
| S9_k141_126748 | 1460 | No | NA | 2 | 0 | 1 | Not-determined | Genome-fragment | NA | NA | 0 |
| S1-final-viral-combined_k141_104636\|\|full | 1070 | No | NA | 3 | 1 | 0 | Low-quality | Genome-fragment | 1.68 | HMM-based (lower-bound) | 0 |
| S9_k141_20655 | 4108 | No | NA | 1 | 0 | 0 | Not-determined | Genome-fragment | NA | NA | 0 |
| S9_k141_126750 | 3047 | No | NA | 4 | 0 | 4 | Not-determined | Genome-fragment | NA | NA | 0 |
| S1-final-viral-combined_k141_125132\|\|full | 1070 | No | NA | 3 | 0 | 0 | Low-quality | Genome-fragment | 0.81 | AAI-based (high-confidence) | 0 |
| S9_k141_79460 | 1496 | No | NA | 3 | 0 | 0 | Not-determined | Genome-fragment | NA | NA | 0 |
| S9_k141_84972 | 1733 | No | NA | 2 | 0 | 1 | Not-determined | Genome-fragment | NA | NA | 0 |
| S9_k141_36371 | 1947 | No | NA | 3 | 0 | 0 | Not-determined | Genome-fragment | NA | NA | 0 |
| S9_k141_36377 | 1253 | No | NA | 2 | 0 | 0 | Not-determined | Genome-fragment | NA | NA | 0 |
| S9_k141_58187 | 1576 | No | NA | 2 | 0 | 1 | Not-determined | Genome-fragment | NA | NA | 0 |

| | | | | | | | | | | | |
|---|---|---|---|---|---|---|---|---|---|---|---|
| S9_k141_122479 | 5221 | No | NA | 6 | 0 | 0 | Not-determined | Genome-fragment | NA | NA | 0 |
| S1-final-viral-combined_k141_266863\|\|lt2gene | 1070 | No | NA | 2 | 1 | 0 | Low-quality | Genome-fragment | 3.17 | AAI-based (high-confidence) | 0 |
| S9_k141_68832 | 6574 | No | NA | 1 | 0 | 1 | Not-determined | Genome-fragment | NA | NA | 0 |
| S9_k141_73523 | 4115 | No | NA | 5 | 0 | 0 | Not-determined | Genome-fragment | NA | NA | 0 |
| S9_k141_94935 | 1209 | No | NA | 1 | 0 | 1 | Not-determined | Genome-fragment | NA | NA | 0 |
| S9_k141_36423 | 3183 | No | NA | 4 | 0 | 4 | Not-determined | Genome-fragment | NA | NA | 0 |
| S9_k141_25999 | 1171 | No | NA | 1 | 0 | 0 | Not-determined | Genome-fragment | NA | NA | 0 |
| S9_k141_53058 | 3310 | No | NA | 2 | 0 | 2 | Not-determined | Genome-fragment | NA | NA | 0 |
| S9_k141_20698 | 1106 | No | NA | 2 | 0 | 0 | Not-determined | Genome-fragment | NA | NA | 0 |
| S9_k141_30756 | 3188 | No | NA | 2 | 0 | 2 | Not-determined | Genome-fragment | NA | NA | 0 |
| S9_k141_4718 | 1392 | No | NA | 2 | 0 | 0 | Not-determined | Genome-fragment | NA | NA | 0 |
| S9_k141_85049 | 1654 | No | NA | 3 | 0 | 2 | Not-determined | Genome-fragment | NA | NA | 0 |
| S9_k141_94970 | 1834 | No | NA | 2 | 0 | 2 | Not-determined | Genome-fragment | NA | NA | 0 |
| S9_k141_94986 | 3938 | No | NA | 2 | 0 | 0 | Not-determined | Genome-fragment | NA | NA | 0 |
| S9_k141_68908 | 1212 | No | NA | 3 | 0 | 0 | Not-determined | Genome-fragment | NA | NA | 0 |
| S1-final-viral-combined_k141_160650\|\|lt2gene | 1070 | No | NA | 1 | 1 | 0 | Low-quality | Genome-fragment | 2.18 | AAI-based (medium-confidence) | 0 |
| S9_k141_10126 | 2121 | No | NA | 1 | 0 | 1 | Not-determined | Genome-fragment | NA | NA | 0 |
| S9_k141_53114 | 2871 | No | NA | 2 | 0 | 1 | Not-determined | Genome-fragment | NA | NA | 0 |
| S9_k141_94997 | 6999 | No | NA | 13 | 0 | 2 | Not-determined | Genome-fragment | NA | NA | 0 |
| S7-final-viral-combined_k141_136498\|\|full | 1070 | No | NA | 1 | 0 | 1 | Low-quality | Genome-fragment | 2.26 | AAI-based (high-confidence) | 0 |
| S9_k141_20764 | 1879 | No | NA | 2 | 0 | 1 | Not-determined | Genome-fragment | NA | NA | 0 |
| S7-final-viral-combined_k141_45468\|\|full | 1070 | No | NA | 2 | 0 | 0 | Low-quality | Genome-fragment | 1.96 | AAI-based (high-confidence) | 0 |
| S9_k141_90492 | 3505 | No | NA | 1 | 0 | 1 | Not-determined | Genome-fragment | NA | NA | 0 |
| S9_k141_20777 | 4115 | No | NA | 5 | 0 | 2 | Not-determined | Genome-fragment | NA | NA | 0 |
| S17-final-viral-combined_k141_59998\|\|lt2gene | 1070 | No | NA | 1 | 1 | 0 | Low-quality | Genome-fragment | 0.74 | HMM-based (lower-bound) | 0 |
| S17-k141_158541 | 1070 | No | NA | 1 | 0 | 1 | Low-quality | Genome-fragment | 0.75 | AAI-based (high-confidence) | 0 |
| S9_k141_41592 | 1001 | No | NA | 1 | 0 | 0 | Not-determined | Genome-fragment | NA | NA | 0 |
| S12-final-viral-combined_k141_153084\|\|lt2gene | 1070 | No | NA | 2 | 1 | 0 | Low-quality | Genome-fragment | 0.74 | HMM-based (lower-bound) | 0 |
| S9_k141_15199 | 1036 | No | NA | 1 | 0 | 1 | Not-determined | Genome-fragment | NA | NA | 0 |
| S9_k141_15230 | 9630 | No | NA | 7 | 0 | 3 | Not-determined | Genome-fragment | NA | NA | 0 |
| S9_k141_85198 | 1310 | No | NA | 1 | 0 | 1 | Not-determined | Genome-fragment | NA | NA | 0 |
| S9_k141_110589 | 3615 | No | NA | 1 | 0 | 1 | Not-determined | Genome-fragment | NA | NA | 0 |
| S9_k141_95068 | 1427 | No | NA | 2 | 0 | 0 | Not-determined | Genome-fragment | NA | NA | 0 |
| S9_k141_15250 | 1512 | No | NA | 1 | 0 | 1 | Not-determined | Genome-fragment | NA | NA | 0 |
| S9_k141_73608 | 15847 | No | NA | 16 | 0 | 10 | Not-determined | Genome-fragment | NA | NA | 0 |
| S12-final-viral-combined_k141_375264\|\|lt2gene | 1070 | No | NA | 2 | 1 | 0 | Low-quality | Genome-fragment | 1.79 | AAI-based (high-confidence) | 0 |
| S9_k141_20862 | 3613 | No | NA | 6 | 0 | 3 | Not-determined | Genome-fragment | NA | NA | 0 |
| S12_k141_311026 | 1070 | No | NA | 2 | 1 | 0 | Low-quality | Genome-fragment | 1.91 | HMM-based (lower-bound) | 0 |
| S9_k141_116571 | 5288 | No | NA | 6 | 0 | 3 | Not-determined | Genome-fragment | NA | NA | 0 |
| S9_k141_95089 | 1355 | No | NA | 1 | 0 | 0 | Not-determined | Genome-fragment | NA | NA | 0 |
| S9_k141_90693 | 1088 | No | NA | 1 | 0 | 1 | Not-determined | Genome-fragment | NA | NA | 0 |
| S9_k141_20883 | 7317 | No | NA | 6 | 0 | 5 | Not-determined | Genome-fragment | NA | NA | 0 |
| S9_k141_126967 | 1338 | No | NA | 1 | 0 | 0 | Not-determined | Genome-fragment | NA | NA | 0 |
| S4-final-viral-combined_k141_160099\|\|full | 1070 | No | NA | 3 | 1 | 0 | Low-quality | Genome-fragment | 1.71 | AAI-based (medium-confidence) | 0 |
| S4_k141_49365 | 1070 | No | NA | 2 | 0 | 0 | Low-quality | Genome-fragment | 0.99 | AAI-based (medium-confidence) | 0 |
| S6-final-viral-combined_k141_15389\|\|full | 1070 | No | NA | 2 | 2 | 0 | Low-quality | Genome-fragment | 1.31 | HMM-based (lower-bound) | 0 |
| S9_k141_85311 | 3213 | No | NA | 3 | 0 | 0 | Not-determined | Genome-fragment | NA | NA | 0 |
| S9_k141_15358 | 2692 | No | NA | 1 | 1 | 0 | Not-determined | Genome-fragment | NA | NA | 0 |
| S9_k141_73744 | 1578 | No | NA | 1 | 0 | 1 | Not-determined | Genome-fragment | NA | NA | 0 |
| S9_k141_116666 | 3076 | No | NA | 3 | 0 | 2 | Not-determined | Genome-fragment | NA | NA | 0 |
| S9_k141_73750 | 3316 | No | NA | 2 | 0 | 2 | Not-determined | Genome-fragment | NA | NA | 0 |
| S6-final-viral-combined_k141_25954\|\|lt2gene | 1070 | No | NA | 1 | 1 | 0 | Low-quality | Genome-fragment | 2.56 | AAI-based (high-confidence) | 0 |
| S9_k141_100905 | 3204 | No | NA | 4 | 0 | 2 | Not-determined | Genome-fragment | NA | NA | 0 |
| S9_k141_101281 | 1071 | No | NA | 1 | 0 | 1 | Low-quality | Genome-fragment | 3.6 | AAI-based (medium-confidence) | 0 |
| S9_k141_41847 | 1272 | No | NA | 2 | 0 | 0 | Not-determined | Genome-fragment | NA | NA | 0 |
| S9_k141_69184 | 1772 | No | NA | 1 | 0 | 1 | Not-determined | Genome-fragment | NA | NA | 0 |
| S9_k141_36851 | 1183 | No | NA | 1 | 0 | 1 | Not-determined | Genome-fragment | NA | NA | 0 |
| S9_k141_5020 | 1114 | No | NA | 1 | 0 | 0 | Not-determined | Genome-fragment | NA | NA | 0 |
| S9_k141_73819 | 1498 | No | NA | 1 | 0 | 1 | Not-determined | Genome-fragment | NA | NA | 0 |
| S9_k141_116742 | 1540 | No | NA | 2 | 0 | 1 | Not-determined | Genome-fragment | NA | NA | 0 |
| S9_k141_73837 | 2323 | No | NA | 1 | 0 | 1 | Not-determined | Genome-fragment | NA | NA | 0 |
| S9_k141_110702 | 2084 | No | NA | 3 | 0 | 2 | Not-determined | Genome-fragment | NA | NA | 0 |
| S9_k141_41947 | 1269 | No | NA | 1 | 0 | 0 | Not-determined | Genome-fragment | NA | NA | 0 |
| S9_k141_20945 | 10667 | No | NA | 10 | 0 | 9 | Not-determined | Genome-fragment | NA | NA | 0 |
| S9_k141_85471 | 1071 | No | NA | 1 | 0 | 0 | Not-determined | Genome-fragment | NA | NA | 0 |
| S1-final-viral-combined_k141_129997\|\|full | 1071 | No | NA | 2 | 0 | 1 | Low-quality | Genome-fragment | 0.68 | AAI-based (high-confidence) | 0 |
| S9_k141_58405 | 1866 | No | NA | 2 | 0 | 0 | Not-determined | Genome-fragment | NA | NA | 0 |
| S9_k141_53398 | 4849 | No | NA | 2 | 0 | 2 | Not-determined | Genome-fragment | NA | NA | 0 |
| S9_k141_5083 | 1712 | No | NA | 1 | 0 | 1 | Not-determined | Genome-fragment | NA | NA | 0 |
| S9_k141_105843 | 1204 | No | NA | 2 | 0 | 2 | Not-determined | Genome-fragment | NA | NA | 0 |
| S9_k141_10313 | 3213 | No | NA | 3 | 0 | 0 | Not-determined | Genome-fragment | NA | NA | 0 |
| S9_k141_15499 | 5249 | No | NA | 4 | 0 | 3 | Not-determined | Genome-fragment | NA | NA | 0 |
| S9_k141_41969 | 2462 | No | NA | 1 | 0 | 0 | Not-determined | Genome-fragment | NA | NA | 0 |
| S9_k141_10326 | 1100 | No | NA | 1 | 0 | 0 | Not-determined | Genome-fragment | NA | NA | 0 |
| S9_k141_15514 | 1009 | No | NA | 1 | 0 | 0 | Not-determined | Genome-fragment | NA | NA | 0 |
| S9_k141_122739 | 1684 | No | NA | 2 | 0 | 1 | Not-determined | Genome-fragment | NA | NA | 0 |
| S9_k141_31062 | 1170 | No | NA | 1 | 0 | 0 | Not-determined | Genome-fragment | NA | NA | 0 |
| S9_k141_101055 | 2937 | No | NA | 3 | 0 | 3 | Not-determined | Genome-fragment | NA | NA | 0 |
| S9_k141_79893 | 1903 | No | NA | 3 | 0 | 2 | Not-determined | Genome-fragment | NA | NA | 0 |
| S9_k141_95175 | 1471 | No | NA | 1 | 0 | 1 | Not-determined | Genome-fragment | NA | NA | 0 |
| S9_k141_101079 | 3259 | No | NA | 4 | 0 | 4 | Not-determined | Genome-fragment | NA | NA | 0 |
| S9_k141_53450 | 4512 | No | NA | 4 | 0 | 2 | Not-determined | Genome-fragment | NA | NA | 0 |
| S9_k141_21049 | 7788 | No | NA | 1 | 0 | 1 | Not-determined | Genome-fragment | NA | NA | 0 |
| S1-final-viral-combined_k141_392331\|\|full | 1071 | No | NA | 4 | 1 | 0 | Low-quality | Genome-fragment | 1.71 | HMM-based (lower-bound) | 0 |
| S3-final-viral-combined_k141_88484\|\|full | 1071 | No | NA | 2 | 0 | 1 | Low-quality | Genome-fragment | 0.9 | AAI-based (medium-confidence) | 0 |
| S9_k141_69296 | 1166 | No | NA | 2 | 0 | 2 | Not-determined | Genome-fragment | NA | NA | 0 |
| S9_k141_53470 | 1592 | No | NA | 2 | 0 | 1 | Not-determined | Genome-fragment | NA | NA | 0 |
| S9_k141_37086 | 1104 | No | NA | 1 | 0 | 0 | Not-determined | Genome-fragment | NA | NA | 0 |
| S9_k141_79953 | 1583 | No | NA | 1 | 0 | 1 | Not-determined | Genome-fragment | NA | NA | 0 |
| S9_k141_10407 | 2275 | No | NA | 2 | 0 | 2 | Not-determined | Genome-fragment | NA | NA | 0 |
| S9_k141_95241 | 1572 | No | NA | 2 | 0 | 1 | Not-determined | Genome-fragment | NA | NA | 0 |
| S9_k141_79980 | 1233 | No | NA | 1 | 0 | 1 | Not-determined | Genome-fragment | NA | NA | 0 |
| S9_k141_42021 | 3782 | No | NA | 2 | 0 | 2 | Not-determined | Genome-fragment | NA | NA | 0 |
| S9_k141_122809 | 3496 | No | NA | 6 | 0 | 4 | Not-determined | Genome-fragment | NA | NA | 0 |
| S9_k141_37153 | 1175 | No | NA | 1 | 0 | 1 | Not-determined | Genome-fragment | NA | NA | 0 |
| S9_k141_106039 | 4247 | No | NA | 6 | 0 | 3 | Not-determined | Genome-fragment | NA | NA | 0 |
| S3-final-viral-combined_k141_107381\|\|full | 1071 | No | NA | 3 | 0 | 1 | Low-quality | Genome-fragment | 0.87 | AAI-based (medium-confidence) | 0 |
| S9_k141_26614 | 1174 | No | NA | 1 | 0 | 0 | Not-determined | Genome-fragment | NA | NA | 0 |
| S7_k141_239658 | 1071 | No | NA | 3 | 0 | 0 | Low-quality | Genome-fragment | 1.94 | AAI-based (high-confidence) | 0 |
| S9_k141_21161 | 2323 | No | NA | 1 | 0 | 1 | Not-determined | Genome-fragment | NA | NA | 0 |
| S7_k141_182147 | 1071 | No | NA | 2 | 1 | 0 | Low-quality | Genome-fragment | 2.74 | AAI-based (high-confidence) | 0 |
| S9_k141_106067 | 3084 | No | NA | 2 | 0 | 2 | Not-determined | Genome-fragment | NA | NA | 0 |
| S12-final-viral-combined_k141_207024\|\|full | 1071 | No | NA | 4 | 0 | 0 | Low-quality | Genome-fragment | 2.45 | AAI-based (high-confidence) | 0 |
| S12-final-viral-combined_k141_347575\|\|full | 1071 | No | NA | 3 | 1 | 0 | Low-quality | Genome-fragment | 2.65 | AAI-based (high-confidence) | 0 |
| S9_k141_10480 | 3938 | No | NA | 7 | 1 | 0 | High-quality | High-quality | 94.63 | HMM-based (lower-bound) | 0 |
| S9_k141_58594 | 1420 | No | NA | 3 | 0 | 3 | Not-determined | Genome-fragment | NA | NA | 0 |
| S9_k141_80008 | 2006 | No | NA | 4 | 0 | 2 | Not-determined | Genome-fragment | NA | NA | 0 |
| S9_k141_69458 | 1210 | No | NA | 1 | 0 | 0 | Not-determined | Genome-fragment | NA | NA | 0 |
| S12-final-viral-combined_k141_203417\|\|lt2gene | 1071 | No | NA | 2 | 2 | 0 | Low-quality | Genome-fragment | 1.77 | AAI-based (high-confidence) | 0 |
| S9_k141_69465 | 1512 | No | NA | 1 | 0 | 1 | Not-determined | Genome-fragment | NA | NA | 0 |
| S9_k141_74088 | 1335 | No | NA | 1 | 0 | 0 | Not-determined | Genome-fragment | NA | NA | 0 |
| S12-final-viral-combined_k141_277230\|\|lt2gene | 1071 | No | NA | 1 | 1 | 0 | Low-quality | Genome-fragment | 0.88 | HMM-based (lower-bound) | 0 |
| S9_k141_31251 | 1104 | No | NA | 2 | 0 | 0 | Not-determined | Genome-fragment | NA | NA | 0 |
| S2-final-viral-combined_k141_194534\|\|full | 1071 | No | NA | 2 | 0 | 0 | Low-quality | Genome-fragment | 2.87 | AAI-based (high-confidence) | 0 |
| S9_k141_15849 | 5805 | No | NA | 10 | 0 | 1 | Not-determined | Genome-fragment | NA | NA | 0 |
| S9_k141_101377 | 6564 | No | NA | 8 | 0 | 3 | Not-determined | Genome-fragment | NA | NA | 0 |
| S9_k141_42187 | 1127 | No | NA | 1 | 0 | 1 | Not-determined | Genome-fragment | NA | NA | 0 |
| S9_k141_80116 | 3701 | No | NA | 2 | 0 | 2 | Not-determined | Genome-fragment | NA | NA | 0 |
| S8_k141_11460 | 1071 | No | NA | 1 | 0 | 0 | Low-quality | Genome-fragment | 0.75 | AAI-based (medium-confidence) | 0 |
| S9_k141_37339 | 1076 | No | NA | 4 | 0 | 0 | Not-determined | Genome-fragment | NA | NA | 0 |
| S11-final-viral-combined_k141_10708\|\|full | 1072 | No | NA | 3 | 0 | 0 | Low-quality | Genome-fragment | 1.1 | AAI-based (medium-confidence) | 0 |
| S9_k141_127153 | 1426 | No | NA | 1 | 0 | 1 | Not-determined | Genome-fragment | NA | NA | 0 |
| S9_k141_31404 | 1197 | No | NA | 3 | 0 | 3 | Not-determined | Genome-fragment | NA | NA | 0 |
| S9_k141_10635 | 1015 | No | NA | 1 | 0 | 0 | Not-determined | Genome-fragment | NA | NA | 0 |
| S1-final-viral-combined_k141_7169\|\|full | 1072 | No | NA | 3 | 0 | 0 | Low-quality | Genome-fragment | 2.05 | AAI-based (medium-confidence) | 0 |
| S9_k141_31434 | 4399 | No | NA | 2 | 0 | 2 | Not-determined | Genome-fragment | NA | NA | 0 |
| S9_k141_95543 | 3372 | No | NA | 4 | 0 | 4 | Not-determined | Genome-fragment | NA | NA | 0 |
| S9_k141_101466 | 1176 | No | NA | 3 | 0 | 1 | Not-determined | Genome-fragment | NA | NA | 0 |
| S3-final-viral-combined_k141_73907\|\|full | 1072 | No | NA | 4 | 0 | 0 | Low-quality | Genome-fragment | 1.01 | AAI-based (medium-confidence) | 0 |
| S9_k141_127258 | 2589 | No | NA | 3 | 0 | 1 | Not-determined | Genome-fragment | NA | NA | 0 |
| S9_k141_16032 | 2373 | No | NA | 5 | 0 | 0 | Not-determined | Genome-fragment | NA | NA | 0 |
| S9_k141_47196 | 1044 | No | NA | 2 | 0 | 0 | Not-determined | Genome-fragment | NA | NA | 0 |
| S9_k141_101559 | 1337 | No | NA | 1 | 0 | 1 | Not-determined | Genome-fragment | NA | NA | 0 |
| S9_k141_31504 | 1697 | No | NA | 2 | 0 | 2 | Not-determined | Genome-fragment | NA | NA | 0 |
| S9_k141_106243 | 7672 | No | NA | 9 | 0 | 6 | Not-determined | Genome-fragment | NA | NA | 0 |
| S7-final-viral-combined_k141_180497\|\|lt2gene | 1072 | No | NA | 1 | 1 | 0 | Low-quality | Genome-fragment | 2.03 | HMM-based (lower-bound) | 0 |
| S9_k141_42452 | 4460 | No | NA | 2 | 0 | 1 | Not-determined | Genome-fragment | NA | NA | 0 |
| S9_k141_31703 | 3531 | No | NA | 1 | 0 | 0 | Not-determined | Genome-fragment | NA | NA | 0 |
| S17-final-viral-combined_k141_187613\|\|full | 1072 | No | NA | 3 | 0 | 1 | Low-quality | Genome-fragment | 0.56 | AAI-based (medium-confidence) | 0 |
| S17-k141_187897 | 1072 | No | NA | 1 | 0 | 1 | Low-quality | Genome-fragment | 0.6 | AAI-based (medium-confidence) | 0 |
| S9_k141_106461 | 1175 | No | NA | 1 | 0 | 1 | Not-determined | Genome-fragment | NA | NA | 0 |
| S9_k141_31739 | 3184 | No | NA | 4 | 0 | 4 | Not-determined | Genome-fragment | NA | NA | 0 |
| S9_k141_117390 | 1045 | No | NA | 1 | 0 | 1 | Not-determined | Genome-fragment | NA | NA | 0 |
| S12-final-viral-combined_k141_360696\|\|lt2gene | 1072 | No | NA | 1 | 1 | 0 | Low-quality | Genome-fragment | 0.59 | HMM-based (lower-bound) | 0 |
| S9_k141_106520 | 1093 | No | NA | 1 | 0 | 0 | Not-determined | Genome-fragment | NA | NA | 0 |
| S9_k141_106559 | 7978 | No | NA | 6 | 0 | 5 | Not-determined | Genome-fragment | NA | NA | 0 |
| S9_k141_117520 | 1082 | No | NA | 4 | 0 | 0 | Not-determined | Genome-fragment | NA | NA | 0 |
| S9_k141_42648 | 2201 | No | NA | 1 | 0 | 1 | Not-determined | Genome-fragment | NA | NA | 0 |
| S9_k141_110997 | 3617 | No | NA | 2 | 0 | 0 | Not-determined | Genome-fragment | NA | NA | 0 |
| S9_k141_74703 | 4620 | No | NA | 5 | 0 | 2 | Not-determined | Genome-fragment | NA | NA | 0 |
| S9_k141_127536 | 1313 | No | NA | 1 | 0 | 1 | Not-determined | Genome-fragment | NA | NA | 0 |
| S12_k141_155025 | 1072 | No | NA | 2 | 0 | 0 | Low-quality | Genome-fragment | 0.78 | AAI-based (medium-confidence) | 0 |
| S9_k141_42663 | 2212 | No | NA | 2 | 0 | 2 | Not-determined | Genome-fragment | NA | NA | 0 |
| S9_k141_47550 | 1760 | No | NA | 3 | 0 | 0 | Not-determined | Genome-fragment | NA | NA | 0 |
| S9_k141_127559 | 1324 | No | NA | 2 | 0 | 2 | Not-determined | Genome-fragment | NA | NA | 0 |
| S9_k141_127565 | 1200 | No | NA | 2 | 0 | 0 | Not-determined | Genome-fragment | NA | NA | 0 |
| S2-final-viral-combined_k141_105778\|\|lt2gene | 1072 | No | NA | 2 | 1 | 0 | Low-quality | Genome-fragment | 1.79 | HMM-based (lower-bound) | 0 |
| S2_k141_135760 | 1072 | No | NA | 3 | 0 | 0 | Low-quality | Genome-fragment | 1.05 | AAI-based (high-confidence) | 0 |
| S9_k141_42707 | 6015 | No | NA | 6 | 0 | 1 | Not-determined | Genome-fragment | NA | NA | 0 |
| S9_k141_47603 | 1343 | No | NA | 2 | 0 | 0 | Not-determined | Genome-fragment | NA | NA | 0 |
| S9_k141_127625 | 1425 | No | NA | 2 | 0 | 0 | Not-determined | Genome-fragment | NA | NA | 0 |
| S4-final-viral-combined_k141_83897\|\|full | 1072 | No | NA | 2 | 2 | 0 | Low-quality | Genome-fragment | 2.97 | AAI-based (high-confidence) | 0 |
| S9_k141_31844 | 1111 | No | NA | 2 | 0 | 1 | Not-determined | Genome-fragment | NA | NA | 0 |
| S9_k141_96224 | 1264 | No | NA | 1 | 0 | 1 | Not-determined | Genome-fragment | NA | NA | 0 |
| S4_k141_137455 | 1072 | No | NA | 1 | 0 | 1 | Low-quality | Genome-fragment | 0.65 | AAI-based (medium-confidence) | 0 |
| S9_k141_32004 | 2893 | No | NA | 4 | 0 | 2 | Not-determined | Genome-fragment | NA | NA | 0 |
| S9_k141_32026 | 5039 | No | NA | 10 | 0 | 9 | Not-determined | Genome-fragment | NA | NA | 0 |
| S9_k141_111192 | 1061 | No | NA | 1 | 0 | 1 | Not-determined | Genome-fragment | NA | NA | 0 |
| S6_k141_28189 | 1072 | No | NA | 2 | 0 | 0 | Low-quality | Genome-fragment | 2.13 | AAI-based (high-confidence) | 0 |
| S9_k141_127892 | 1043 | No | NA | 2 | 0 | 1 | Not-determined | Genome-fragment | NA | NA | 0 |
| S9_k141_111207 | 1211 | No | NA | 1 | 0 | 0 | Not-determined | Genome-fragment | NA | NA | 0 |
| S8-final-viral-combined_k141_54946\|\|full | 1072 | No | NA | 3 | 1 | 0 | Low-quality | Genome-fragment | 2.68 | AAI-based (high-confidence) | 0 |
| S9_k141_111237 | 3424 | No | NA | 2 | 0 | 1 | Not-determined | Genome-fragment | NA | NA | 0 |
| S9_k141_47867 | 11339 | No | NA | 2 | 0 | 0 | Not-determined | Genome-fragment | NA | NA | 0 |
| S9_k141_47937 | 1795 | No | NA | 2 | 0 | 2 | Not-determined | Genome-fragment | NA | NA | 0 |
| S9_k141_111454 | 1096 | No | NA | 2 | 0 | 1 | Not-determined | Genome-fragment | NA | NA | 0 |
| S9_k141_111488 | 2167 | No | NA | 4 | 0 | 1 | Not-determined | Genome-fragment | NA | NA | 0 |
| S9_k141_111537 | 1522 | No | NA | 1 | 0 | 0 | Not-determined | Genome-fragment | NA | NA | 0 |
| S9_k141_111575 | 1254 | No | NA | 1 | 0 | 1 | Not-determined | Genome-fragment | NA | NA | 0 |
| S11-final-viral-combined_k141_318520\|\|full | 1073 | No | NA | 2 | 0 | 1 | Low-quality | Genome-fragment | 0.33 | AAI-based (medium-confidence) | 0 |
| S9_k141_48112 | 2968 | No | NA | 3 | 0 | 0 | Not-determined | Genome-fragment | NA | NA | 0 |
| S9_k141_128151 | 1116 | No | NA | 2 | 0 | 1 | Not-determined | Genome-fragment | NA | NA | 0 |
| S9_k141_128198 | 1195 | No | NA | 3 | 0 | 0 | Not-determined | Genome-fragment | NA | NA | 0 |
| S9_k141_128211 | 1203 | No | NA | 2 | 0 | 2 | Not-determined | Genome-fragment | NA | NA | 0 |
| S9_k141_128232 | 5983 | No | NA | 10 | 3 | 0 | Complete | High-quality | 100 | DTR (high-confidence) | 0 |
| S9_k141_128248 | 3515 | No | NA | 6 | 0 | 2 | Not-determined | Genome-fragment | NA | NA | 0 |
| S11-final-viral-combined_k141_130664\|\|full | 1073 | No | NA | 2 | 0 | 0 | Low-quality | Genome-fragment | 1.09 | AAI-based (medium-confidence) | 0 |
| S11-final-viral-combined_k141_204529\|\|lt2gene | 1073 | No | NA | 2 | 2 | 0 | Low-quality | Genome-fragment | 2.35 | AAI-based (high-confidence) | 0 |
| S1-final-viral-combined_k141_233149\|\|full | 1073 | No | NA | 3 | 0 | 2 | Low-quality | Genome-fragment | 0.95 | AAI-based (medium-confidence) | 0 |
| S9_k141_111769 | 1014 | No | NA | 3 | 0 | 1 | Not-determined | Genome-fragment | NA | NA | 0 |
| S9_k141_111947 | 1718 | No | NA | 2 | 0 | 2 | Not-determined | Genome-fragment | NA | NA | 0 |
| S1-final-viral-combined_k141_13762\|\|lt2gene | 1073 | No | NA | 1 | 1 | 0 | Low-quality | Genome-fragment | 1.51 | HMM-based (lower-bound) | 0 |
| S9_k141_112192 | 1043 | No | NA | 1 | 0 | 0 | Not-determined | Genome-fragment | NA | NA | 0 |
| S9_k141_112196 | 1631 | No | NA | 1 | 0 | 1 | Not-determined | Genome-fragment | NA | NA | 0 |
| S9_k141_112201 | 4184 | No | NA | 5 | 0 | 4 | Not-determined | Genome-fragment | NA | NA | 0 |
| S1_k141_133010 | 1073 | No | NA | 3 | 2 | 0 | Low-quality | Genome-fragment | 9.98 | AAI-based (high-confidence) | 0 |
| S11-final-viral-combined_k141_190241\|\|full | 1349 | No | NA | 5 | 0 | 0 | Not-determined | Genome-fragment | NA | NA | 0 |
| S1_k141_122264 | 1073 | No | NA | 2 | 0 | 0 | Low-quality | Genome-fragment | 0.61 | AAI-based (medium-confidence) | 0 |
| S17-final-viral-combined_k141_193187\|\|full | 1073 | No | NA | 3 | 0 | 1 | Low-quality | Genome-fragment | 0.64 | AAI-based (medium-confidence) | 0 |
| S11-final-viral-combined_k141_179156\|\|full | 1688 | No | NA | 2 | 0 | 2 | Not-determined | Genome-fragment | NA | NA | 0 |
| S11-final-viral-combined_k141_55135\|\|full | 1888 | No | NA | 3 | 0 | 0 | Not-determined | Genome-fragment | NA | NA | 0 |
| S11-final-viral-combined_k141_96474\|\|full | 2250 | No | NA | 4 | 0 | 0 | Not-determined | Genome-fragment | NA | NA | 0 |
| S11-final-viral-combined_k141_137838\|\|full | 1703 | No | NA | 3 | 0 | 0 | Not-determined | Genome-fragment | NA | NA | 0 |
| S17-final-viral-combined_k141_71414\|\|full | 1073 | No | NA | 2 | 0 | 1 | Low-quality | Genome-fragment | 1.46 | AAI-based (medium-confidence) | 0 |
| S11-final-viral-combined_k141_110257\|\|full | 1844 | No | NA | 3 | 0 | 2 | Not-determined | Genome-fragment | NA | NA | 0 |
| S11-final-viral-combined_k141_27578\|\|full | 1291 | No | NA | 2 | 0 | 0 | Not-determined | Genome-fragment | NA | NA | 0 |
| S17-k141_88347 | 1073 | No | NA | 1 | 0 | 0 | Low-quality | Genome-fragment | 1.27 | AAI-based (high-confidence) | 0 |
| S11-final-viral-combined_k141_206749\|\|full | 1071 | No | NA | 2 | 0 | 0 | Not-determined | Genome-fragment | NA | NA | 0 |
| S4-final-viral-combined_k141_117215\|\|full | 1073 | No | NA | 2 | 0 | 0 | Low-quality | Genome-fragment | 2.47 | AAI-based (medium-confidence) | 0 |
| S11-final-viral-combined_k141_137877\|\|full | 1139 | No | NA | 2 | 0 | 0 | Not-determined | Genome-fragment | NA | NA | 0 |
| S6-final-viral-combined_k141_19288\|\|full | 1073 | No | NA | 2 | 0 | 0 | Low-quality | Genome-fragment | 1.79 | AAI-based (high-confidence) | 0 |
| S11-final-viral-combined_k141_13874\|\|full | 2756 | No | NA | 3 | 0 | 2 | Not-determined | Genome-fragment | NA | NA | 0 |
| S11-final-viral-combined_k141_317024\|\|full | 2208 | No | NA | 3 | 0 | 2 | Not-determined | Genome-fragment | NA | NA | 0 |
| S11-final-viral-combined_k141_137899\|\|full | 4922 | No | NA | 7 | 0 | 5 | Not-determined | Genome-fragment | NA | NA | 0 |
| S11-final-viral-combined_k141_206772\|\|full | 4054 | No | NA | 4 | 0 | 3 | Not-determined | Genome-fragment | NA | NA | 0 |
| S11-final-viral-combined_k141_303240\|\|full | 7401 | No | NA | 7 | 0 | 1 | Not-determined | Genome-fragment | NA | NA | 0 |
| S11-final-viral-combined_k141_151648\|\|full | 2560 | No | NA | 3 | 0 | 0 | Not-determined | Genome-fragment | NA | NA | 0 |
| S11-final-viral-combined_k141_248132\|\|full | 2705 | No | NA | 4 | 0 | 1 | Not-determined | Genome-fragment | NA | NA | 0 |
| S11-final-viral-combined_k141_317033\|\|full | 9395 | No | NA | 8 | 0 | 7 | Not-determined | Genome-fragment | NA | NA | 0 |
| S6_k141_110998 | 1073 | No | NA | 2 | 0 | 1 | Low-quality | Genome-fragment | 0.84 | AAI-based (high-confidence) | 0 |
| S1-final-viral-combined_k141_14767\|\|full | 1074 | No | NA | 3 | 0 | 0 | Low-quality | Genome-fragment | 1.69 | AAI-based (high-confidence) | 0 |
| S1-final-viral-combined_k141_277753\|\|lt2gene | 1074 | No | NA | 2 | 2 | 0 | Low-quality | Genome-fragment | 2.3 | AAI-based (high-confidence) | 0 |
| S11-final-viral-combined_k141_41367\|\|full | 39181 | No | NA | 57 | 12 | 2 | Medium-quality | Genome-fragment | 60.35 | AAI-based (high-confidence) | 0 |
| S11-final-viral-combined_k141_317054\|\|full | 2873 | No | NA | 4 | 0 | 1 | Not-determined | Genome-fragment | NA | NA | 0 |
| S1_k141_209565 | 1074 | No | NA | 1 | 0 | 1 | Low-quality | Genome-fragment | 1.8 | AAI-based (high-confidence) | 0 |
| S7_k141_228500 | 1074 | No | NA | 2 | 0 | 0 | Low-quality | Genome-fragment | 2.17 | AAI-based (high-confidence) | 0 |
| S12-final-viral-combined_k141_389889\|\|full | 1074 | No | NA | 3 | 0 | 0 | Low-quality | Genome-fragment | 2.06 | AAI-based (medium-confidence) | 0 |
| S11-final-viral-combined_k141_192971\|\|full | 2196 | No | NA | 4 | 0 | 4 | Not-determined | Genome-fragment | NA | NA | 0 |
| S11-final-viral-combined_k141_82799\|\|full | 1401 | No | NA | 3 | 0 | 0 | Not-determined | Genome-fragment | NA | NA | 0 |
| S12-final-viral-combined_k141_205760\|\|full | 1074 | No | NA | 3 | 0 | 2 | Low-quality | Genome-fragment | 0.53 | AAI-based (medium-confidence) | 0 |
| S11-final-viral-combined_k141_110327\|\|full | 5631 | No | NA | 8 | 0 | 7 | Not-determined | Genome-fragment | NA | NA | 0 |
| S11-final-viral-combined_k141_192994\|\|full | 1494 | No | NA | 2 | 0 | 1 | Not-determined | Genome-fragment | NA | NA | 0 |
| S11-final-viral-combined_k141_206830\|\|full | 1223 | No | NA | 3 | 0 | 0 | Not-determined | Genome-fragment | NA | NA | 0 |
| S12-final-viral-combined_k141_104751\|\|lt2gene | 1074 | No | NA | 1 | 1 | 0 | Low-quality | Genome-fragment | 1.69 | HMM-based (lower-bound) | 0 |
| S11-final-viral-combined_k141_41400\|\|full | 2528 | No | NA | 4 | 0 | 2 | Not-determined | Genome-fragment | NA | NA | 0 |
| S12_k141_289597 | 1074 | No | NA | 2 | 0 | 1 | Low-quality | Genome-fragment | 0.94 | AAI-based (medium-confidence) | 0 |
| S12_k141_145908 | 1074 | No | NA | 3 | 0 | 0 | Low-quality | Genome-fragment | 3.18 | AAI-based (medium-confidence) | 0 |
| S11-final-viral-combined_k141_124198\|\|full | 2965 | No | NA | 3 | 0 | 0 | Not-determined | Genome-fragment | NA | NA | 0 |
| S9-final-viral-combined_k141_112634\|\|full | 1075 | No | NA | 3 | 0 | 0 | Low-quality | Genome-fragment | 2.13 | AAI-based (high-confidence) | 0 |
| S11-final-viral-combined_k141_289520\|\|full | 4615 | No | NA | 4 | 0 | 3 | Not-determined | Genome-fragment | NA | NA | 0 |
| S11-final-viral-combined_k141_289537\|\|full | 1272 | No | NA | 3 | 0 | 1 | Not-determined | Genome-fragment | NA | NA | 0 |
| S11-final-viral-combined_k141_99776\|\|full | 1075 | No | NA | 2 | 0 | 0 | Low-quality | Genome-fragment | 1.31 | AAI-based (high-confidence) | 0 |
| S11-final-viral-combined_k141_206880\|\|full | 3453 | No | NA | 4 | 0 | 2 | Not-determined | Genome-fragment | NA | NA | 0 |
| S1-final-viral-combined_k141_357319\|\|full | 1075 | No | NA | 3 | 0 | 0 | Low-quality | Genome-fragment | 2.22 | AAI-based (high-confidence) | 0 |
| S1-final-viral-combined_k141_63314\|\|lt2gene | 1075 | No | NA | 1 | 1 | 0 | Low-quality | Genome-fragment | 2.27 | AAI-based (high-confidence) | 0 |
| S1_k141_290381 | 1075 | No | NA | 1 | 0 | 0 | Low-quality | Genome-fragment | 0.42 | AAI-based (high-confidence) | 0 |
| S11-final-viral-combined_k141_69075\|\|full | 1749 | No | NA | 2 | 0 | 0 | Not-determined | Genome-fragment | NA | NA | 0 |
| S3_k141_100007 | 1075 | No | NA | 3 | 1 | 0 | Low-quality | Genome-fragment | 0.49 | AAI-based (high-confidence) | 0 |
| S5-final-viral-combined_k141_46073\|\|full | 1075 | No | NA | 3 | 1 | 0 | Low-quality | Genome-fragment | 2.49 | AAI-based (high-confidence) | 0 |
| S17-final-viral-combined_k141_55739\|\|full | 1075 | No | NA | 2 | 0 | 0 | Low-quality | Genome-fragment | 0.64 | AAI-based (high-confidence) | 0 |
| S17-k141_13701 | 1075 | No | NA | 2 | 0 | 0 | Low-quality | Genome-fragment | 0.81 | AAI-based (high-confidence) | 0 |
| S17-k141_58578 | 1075 | No | NA | 2 | 0 | 2 | Low-quality | Genome-fragment | 0.74 | AAI-based (medium-confidence) | 0 |
| S11-final-viral-combined_k141_234385\|\|full | 3633 | No | NA | 6 | 0 | 0 | Not-determined | Genome-fragment | NA | NA | 0 |
| S11-final-viral-combined_k141_165609\|\|full | 2474 | No | NA | 4 | 0 | 0 | Not-determined | Genome-fragment | NA | NA | 0 |
| S12-final-viral-combined_k141_126693\|\|lt2gene | 1075 | No | NA | 2 | 2 | 0 | Low-quality | Genome-fragment | 3.11 | AAI-based (high-confidence) | 0 |
| S12_k141_381496 | 1075 | No | NA | 2 | 0 | 0 | Low-quality | Genome-fragment | 2.08 | AAI-based (medium-confidence) | 0 |
| S2-final-viral-combined_k141_84787\|\|full | 1075 | No | NA | 3 | 0 | 0 | Low-quality | Genome-fragment | 1.83 | AAI-based (medium-confidence) | 0 |
| S2-final-viral-combined_k141_107010\|\|full | 1075 | No | NA | 3 | 0 | 0 | Low-quality | Genome-fragment | 1.58 | AAI-based (high-confidence) | 0 |
| S11-final-viral-combined_k141_151854\|\|full | 2466 | No | NA | 3 | 0 | 3 | Not-determined | Genome-fragment | NA | NA | 0 |
| S11-final-viral-combined_k141_165646\|\|full | 1294 | No | NA | 2 | 0 | 1 | Not-determined | Genome-fragment | NA | NA | 0 |
| S2-final-viral-combined_k141_89812\|\|lt2gene | 1075 | No | NA | 1 | 1 | 0 | Low-quality | Genome-fragment | 2.55 | AAI-based (high-confidence) | 0 |
| S11-final-viral-combined_k141_193148\|\|full | 39231 | Yes | 30175 | 35 | 5 | 4 | Medium-quality | Genome-fragment | 67.75 | AAI-based (high-confidence) | 23.08 |
| S2_k141_42050 | 1075 | No | NA | 1 | 1 | 0 | Low-quality | Genome-fragment | 1.61 | HMM-based (lower-bound) | 0 |
| S11-final-viral-combined_k141_124324\|\|full | 3304 | No | NA | 4 | 0 | 4 | Not-determined | Genome-fragment | NA | NA | 0 |
| S8-final-viral-combined_k141_96731\|\|full | 1075 | No | NA | 4 | 0 | 0 | Low-quality | Genome-fragment | 1.59 | AAI-based (medium-confidence) | 0 |
| S9_k141_907 | 1076 | No | NA | 4 | 1 | 0 | Low-quality | Genome-fragment | 11.43 | AAI-based (high-confidence) | 0 |
| S11-final-viral-combined_k141_324496\|\|full | 1076 | No | NA | 3 | 0 | 0 | Low-quality | Genome-fragment | 2.69 | AAI-based (high-confidence) | 0 |
| S11-final-viral-combined_k141_234426\|\|full | 13340 | No | NA | 11 | 0 | 9 | Not-determined | Genome-fragment | NA | NA | 0 |
| S1-final-viral-combined_k141_365998\|\|full | 1076 | No | NA | 2 | 0 | 0 | Low-quality | Genome-fragment | 0.69 | AAI-based (medium-confidence) | 0 |
| S11-final-viral-combined_k141_69182\|\|full | 1057 | No | NA | 3 | 0 | 1 | Not-determined | Genome-fragment | NA | NA | 0 |
| S11-final-viral-combined_k141_124393\|\|full | 2429 | No | NA | 6 | 0 | 0 | Not-determined | Genome-fragment | NA | NA | 0 |
| S1-final-viral-combined_k141_285671\|\|full | 1076 | No | NA | 3 | 0 | 0 | Low-quality | Genome-fragment | 0.85 | AAI-based (medium-confidence) | 0 |
| S11-final-viral-combined_k141_220808\|\|full | 7884 | No | NA | 6 | 0 | 0 | Not-determined | Genome-fragment | NA | NA | 0 |
| S3-final-viral-combined_k141_116328\|\|full | 1076 | No | NA | 2 | 1 | 0 | Low-quality | Genome-fragment | 1.78 | AAI-based (high-confidence) | 0 |
| S11-final-viral-combined_k141_138099\|\|full | 1288 | No | NA | 3 | 0 | 1 | Not-determined | Genome-fragment | NA | NA | 0 |
| S5-final-viral-combined_k141_72904\|\|full | 1076 | No | NA | 3 | 3 | 0 | Low-quality | Genome-fragment | 2.36 | AAI-based (high-confidence) | 0 |
| S11-final-viral-combined_k141_14092\|\|full | 2575 | No | NA | 3 | 0 | 0 | Not-determined | Genome-fragment | NA | NA | 0 |
| S11-final-viral-combined_k141_165709\|\|full | 2871 | No | NA | 3 | 0 | 0 | Not-determined | Genome-fragment | NA | NA | 0 |
| S7-final-viral-combined_k141_40078\|\|full | 1076 | No | NA | 3 | 1 | 0 | Low-quality | Genome-fragment | 0.88 | HMM-based (lower-bound) | 0 |
| S11-final-viral-combined_k141_207007\|\|full | 52392 | No | NA | 64 | 0 | 23 | Not-determined | Genome-fragment | NA | NA | 0 |
| S7_k141_143163 | 1076 | No | NA | 2 | 2 | 0 | Low-quality | Genome-fragment | 2.23 | AAI-based (high-confidence) | 0 |
| S11-final-viral-combined_k141_96692\|\|full | 2102 | No | NA | 2 | 0 | 2 | Not-determined | Genome-fragment | NA | NA | 0 |
| S11-final-viral-combined_k141_262077\|\|full | 1984 | No | NA | 5 | 0 | 0 | Not-determined | Genome-fragment | NA | NA | 0 |
| S11-final-viral-combined_k141_110485\|\|full | 2196 | No | NA | 3 | 0 | 2 | Not-determined | Genome-fragment | NA | NA | 0 |
| S11-final-viral-combined_k141_55409\|\|full | 1926 | No | NA | 2 | 0 | 2 | Not-determined | Genome-fragment | NA | NA | 0 |
| S17-final-viral-combined_k141_104028\|\|full | 1076 | No | NA | 3 | 0 | 2 | Low-quality | Genome-fragment | 1.42 | AAI-based (medium-confidence) | 0 |
| S17-final-viral-combined_k141_104099\|\|full | 1076 | No | NA | 2 | 1 | 0 | Low-quality | Genome-fragment | 1.94 | AAI-based (medium-confidence) | 0 |
| S11-final-viral-combined_k141_289605\|\|full | 1941 | No | NA | 4 | 0 | 3 | Not-determined | Genome-fragment | NA | NA | 0 |
| S11-final-viral-combined_k141_41510\|\|full | 6143 | No | NA | 8 | 0 | 6 | Not-determined | Genome-fragment | NA | NA | 0 |
| S12-final-viral-combined_k141_66048\|\|full | 1076 | No | NA | 4 | 2 | 0 | Low-quality | Genome-fragment | 1.72 | AAI-based (medium-confidence) | 0 |
| S12-final-viral-combined_k141_248145\|\|lt2gene | 1076 | No | NA | 2 | 1 | 0 | Low-quality | Genome-fragment | 1.84 | AAI-based (medium-confidence) | 0 |
| S11-final-viral-combined_k141_220854\|\|full | 1989 | No | NA | 2 | 0 | 1 | Not-determined | Genome-fragment | NA | NA | 0 |
| S11-final-viral-combined_k141_69207\|\|full | 8496 | No | NA | 9 | 0 | 6 | Not-determined | Genome-fragment | NA | NA | 0 |
| S2-final-viral-combined_k141_211951\|\|lt2gene | 1076 | No | NA | 1 | 1 | 0 | Low-quality | Genome-fragment | 0.58 | HMM-based (lower-bound) | 0 |
| S2_k141_64120 | 1076 | No | NA | 1 | 0 | 0 | Low-quality | Genome-fragment | 1.48 | AAI-based (high-confidence) | 0 |
| S11-final-viral-combined_k141_165785\|\|full | 1869 | No | NA | 4 | 0 | 0 | Not-determined | Genome-fragment | NA | NA | 0 |
| S4-final-viral-combined_k141_111719\|\|full | 1076 | No | NA | 3 | 0 | 0 | Low-quality | Genome-fragment | 2.05 | AAI-based (medium-confidence) | 0 |
| S11-final-viral-combined_k141_96737\|\|full | 2752 | No | NA | 2 | 0 | 1 | Not-determined | Genome-fragment | NA | NA | 0 |
| S11-final-viral-combined_k141_124446\|\|full | 4103 | No | NA | 5 | 0 | 3 | Not-determined | Genome-fragment | NA | NA | 0 |
| S8-final-viral-combined_k141_258279\|\|full | 1076 | No | NA | 2 | 1 | 0 | Low-quality | Genome-fragment | 1.65 | AAI-based (medium-confidence) | 0 |
| S11-final-viral-combined_k141_193271\|\|full | 2104 | No | NA | 4 | 0 | 2 | Not-determined | Genome-fragment | NA | NA | 0 |
| S11_k141_319123 | 1077 | No | NA | 3 | 0 | 0 | Low-quality | Genome-fragment | 2.37 | AAI-based (high-confidence) | 0 |
| S11_k141_119984 | 1077 | No | NA | 4 | 0 | 1 | Low-quality | Genome-fragment | 2.13 | AAI-based (medium-confidence) | 0 |
| S1_k141_64782 | 1077 | No | NA | 3 | 1 | 0 | Low-quality | Genome-fragment | 0.28 | HMM-based (lower-bound) | 0 |
| S17-final-viral-combined_k141_201062\|\|full | 1077 | No | NA | 2 | 0 | 0 | Low-quality | Genome-fragment | 3.05 | AAI-based (medium-confidence) | 0 |
| S11-final-viral-combined_k141_303534\|\|full | 2821 | No | NA | 4 | 0 | 3 | Not-determined | Genome-fragment | NA | NA | 0 |
| S17-k141_200767 | 1077 | No | NA | 2 | 0 | 0 | Low-quality | Genome-fragment | 1.67 | AAI-based (medium-confidence) | 0 |
| S11-final-viral-combined_k141_151974\|\|full | 6276 | No | NA | 11 | 0 | 1 | Not-determined | Genome-fragment | NA | NA | 0 |
| S12_k141_361606 | 1077 | No | NA | 4 | 0 | 1 | Low-quality | Genome-fragment | 2.47 | AAI-based (medium-confidence) | 0 |
| S11-final-viral-combined_k141_248401\|\|full | 1839 | No | NA | 2 | 0 | 1 | Not-determined | Genome-fragment | NA | NA | 0 |
| S2-final-viral-combined_k141_127089\|\|full | 1077 | No | NA | 3 | 0 | 0 | Low-quality | Genome-fragment | 0.66 | AAI-based (medium-confidence) | 0 |
| S2-final-viral-combined_k141_99321\|\|full | 1077 | No | NA | 3 | 0 | 2 | Low-quality | Genome-fragment | 1.44 | AAI-based (high-confidence) | 0 |
| S2-final-viral-combined_k141_193804\|\|full | 1077 | No | NA | 3 | 0 | 1 | Low-quality | Genome-fragment | 2.47 | AAI-based (medium-confidence) | 0 |
| S11-final-viral-combined_k141_110566\|\|full | 1929 | No | NA | 4 | 0 | 1 | Not-determined | Genome-fragment | NA | NA | 0 |
| S11-final-viral-combined_k141_138247\|\|full | 2105 | No | NA | 4 | 0 | 2 | Not-determined | Genome-fragment | NA | NA | 0 |
| S2-final-viral-combined_k141_112723\|\|full | 1077 | No | NA | 3 | 3 | 0 | Low-quality | Genome-fragment | 1.97 | AAI-based (high-confidence) | 0 |
| S11-final-viral-combined_k141_152012\|\|full | 1485 | No | NA | 3 | 0 | 0 | Not-determined | Genome-fragment | NA | NA | 0 |
| S11-final-viral-combined_k141_83070\|\|full | 3223 | No | NA | 2 | 0 | 1 | Not-determined | Genome-fragment | NA | NA | 0 |
| S6-final-viral-combined_k141_119045\|\|full | 1077 | No | NA | 2 | 0 | 1 | Low-quality | Genome-fragment | 0.51 | AAI-based (medium-confidence) | 0 |
| S6-final-viral-combined_k141_3121\|\|full | 1077 | No | NA | 1 | 0 | 0 | Low-quality | Genome-fragment | 2.14 | AAI-based (high-confidence) | 0 |
| S11-final-viral-combined_k141_124507\|\|full | 5044 | No | NA | 10 | 0 | 1 | Not-determined | Genome-fragment | NA | NA | 0 |
| S11-final-viral-combined_k141_152036\|\|full | 3174 | No | NA | 4 | 0 | 0 | Not-determined | Genome-fragment | NA | NA | 0 |
| S11-final-viral-combined_k141_289712\|\|full | 18580 | No | NA | 18 | 0 | 11 | Not-determined | Genome-fragment | NA | NA | 0 |
| S8-final-viral-combined_k141_187774\|\|full | 1077 | No | NA | 2 | 0 | 0 | Low-quality | Genome-fragment | 0.66 | AAI-based (high-confidence) | 0 |
| S11-final-viral-combined_k141_165943\|\|full | 1150 | No | NA | 2 | 0 | 0 | Not-determined | Genome-fragment | NA | NA | 0 |
| S8-final-viral-combined_k141_155984\|\|full | 1077 | No | NA | 3 | 0 | 0 | Low-quality | Genome-fragment | 0.7 | AAI-based (high-confidence) | 0 |
| S8_k141_79854 | 1077 | No | NA | 2 | 0 | 0 | Low-quality | Genome-fragment | 0.86 | AAI-based (high-confidence) | 0 |
| S11_k141_319543 | 1078 | No | NA | 3 | 2 | 0 | Low-quality | Genome-fragment | 2.94 | AAI-based (high-confidence) | 0 |
| S11_k141_6006 | 1078 | No | NA | 1 | 1 | 0 | Low-quality | Genome-fragment | 0.57 | HMM-based (lower-bound) | 0 |
| S1-final-viral-combined_k141_142958\|\|full | 1078 | No | NA | 2 | 0 | 0 | Low-quality | Genome-fragment | 2.35 | AAI-based (medium-confidence) | 0 |
| S11-final-viral-combined_k141_28013\|\|full | 1332 | No | NA | 3 | 0 | 0 | Not-determined | Genome-fragment | NA | NA | 0 |
| S11-final-viral-combined_k141_14266\|\|full | 1946 | No | NA | 3 | 0 | 1 | Not-determined | Genome-fragment | NA | NA | 0 |
| S11-final-viral-combined_k141_289730\|\|full | 1482 | No | NA | 2 | 0 | 0 | Not-determined | Genome-fragment | NA | NA | 0 |
| S11-final-viral-combined_k141_138299\|\|full | 2429 | No | NA | 3 | 0 | 3 | Not-determined | Genome-fragment | NA | NA | 0 |
| S11-final-viral-combined_k141_41659\|\|full | 1428 | No | NA | 2 | 0 | 1 | Not-determined | Genome-fragment | NA | NA | 0 |
| S11-final-viral-combined_k141_289732\|\|full | 1308 | No | NA | 2 | 0 | 1 | Not-determined | Genome-fragment | NA | NA | 0 |
| S5-final-viral-combined_k141_89346\|\|full | 1078 | No | NA | 3 | 2 | 0 | Low-quality | Genome-fragment | 2.31 | AAI-based (high-confidence) | 0 |
| S5_k141_183108 | 1078 | No | NA | 3 | 0 | 0 | Low-quality | Genome-fragment | 2.66 | AAI-based (high-confidence) | 0 |
| S7-final-viral-combined_k141_15790\|\|full | 1078 | No | NA | 2 | 1 | 0 | Low-quality | Genome-fragment | 1.83 | AAI-based (medium-confidence) | 0 |
| S11-final-viral-combined_k141_110636\|\|full | 2419 | No | NA | 4 | 0 | 1 | Not-determined | Genome-fragment | NA | NA | 0 |
| S11-final-viral-combined_k141_69335\|\|full | 2329 | No | NA | 2 | 0 | 1 | Not-determined | Genome-fragment | NA | NA | 0 |
| S11-final-viral-combined_k141_83107\|\|full | 1314 | No | NA | 2 | 0 | 0 | Not-determined | Genome-fragment | NA | NA | 0 |
| S11-final-viral-combined_k141_69337\|\|full | 1968 | No | NA | 2 | 0 | 2 | Not-determined | Genome-fragment | NA | NA | 0 |
| S7-final-viral-combined_k141_27956\|\|full | 1078 | No | NA | 3 | 0 | 0 | Low-quality | Genome-fragment | 0.72 | AAI-based (medium-confidence) | 0 |
| S11-final-viral-combined_k141_14318\|\|full | 1999 | No | NA | 2 | 0 | 1 | Not-determined | Genome-fragment | NA | NA | 0 |
| S11-final-viral-combined_k141_303682\|\|full | 5729 | No | NA | 5 | 0 | 3 | Not-determined | Genome-fragment | NA | NA | 0 |
| S11-final-viral-combined_k141_69357\|\|full | 5297 | No | NA | 4 | 0 | 1 | Not-determined | Genome-fragment | NA | NA | 0 |
| S11-final-viral-combined_k141_220937\|\|full | 2503 | No | NA | 5 | 0 | 1 | Not-determined | Genome-fragment | NA | NA | 0 |
| S11-final-viral-combined_k141_83135\|\|full | 2540 | No | NA | 2 | 0 | 1 | Not-determined | Genome-fragment | NA | NA | 0 |
| S11-final-viral-combined_k141_69376\|\|full | 1833 | No | NA | 2 | 0 | 1 | Not-determined | Genome-fragment | NA | NA | 0 |
| S11-final-viral-combined_k141_248565\|\|full | 5089 | No | NA | 5 | 0 | 3 | Not-determined | Genome-fragment | NA | NA | 0 |
| S11-final-viral-combined_k141_276066\|\|full | 2238 | No | NA | 2 | 0 | 1 | Not-determined | Genome-fragment | NA | NA | 0 |
| S11-final-viral-combined_k141_96991\|\|full | 1429 | No | NA | 2 | 0 | 2 | Not-determined | Genome-fragment | NA | NA | 0 |
| S11-final-viral-combined_k141_179605\|\|full | 43615 | Yes | 22617 | 47 | 12 | 17 | Medium-quality | Genome-fragment | 54 | AAI-based (high-confidence) | 48.14 |
| S7_k141_20563 | 1078 | No | NA | 1 | 0 | 0 | Low-quality | Genome-fragment | 1.8 | AAI-based (medium-confidence) | 0 |
| S11-final-viral-combined_k141_14365\|\|full | 2206 | No | NA | 3 | 0 | 2 | Not-determined | Genome-fragment | NA | NA | 0 |
| S11-final-viral-combined_k141_234757\|\|full | 3517 | No | NA | 3 | 0 | 1 | Not-determined | Genome-fragment | NA | NA | 0 |
| S17-final-viral-combined_k141_138877\|\|full | 1078 | No | NA | 2 | 2 | 0 | Low-quality | Genome-fragment | 3.16 | AAI-based (high-confidence) | 0 |
| S11-final-viral-combined_k141_234790\|\|full | 1037 | No | NA | 3 | 0 | 1 | Not-determined | Genome-fragment | NA | NA | 0 |
| S17-final-viral-combined_k141_141231\|\|full | 1078 | No | NA | 3 | 0 | 0 | Low-quality | Genome-fragment | 2.88 | AAI-based (medium-confidence) | 0 |
| S11-final-viral-combined_k141_152167\|\|full | 1048 | No | NA | 3 | 0 | 2 | Not-determined | Genome-fragment | NA | NA | 0 |
| S11-final-viral-combined_k141_28104\|\|full | 1684 | No | NA | 3 | 0 | 2 | Not-determined | Genome-fragment | NA | NA | 0 |
| S11-final-viral-combined_k141_193408\|\|full | 27118 | No | NA | 23 | 0 | 21 | Not-determined | Genome-fragment | NA | NA | 0 |
| S11-final-viral-combined_k141_193418\|\|full | 1502 | No | NA | 2 | 0 | 0 | Not-determined | Genome-fragment | NA | NA | 0 |
| S17-k141_263484 | 1078 | No | NA | 2 | 0 | 0 | Low-quality | Genome-fragment | 0.86 | AAI-based (high-confidence) | 0 |
| S11-final-viral-combined_k141_152188\|\|full | 1580 | No | NA | 3 | 0 | 2 | Not-determined | Genome-fragment | NA | NA | 0 |
| S11-final-viral-combined_k141_193419\|\|full | 1960 | No | NA | 2 | 0 | 1 | Not-determined | Genome-fragment | NA | NA | 0 |
| S12-final-viral-combined_k141_219655\|\|full | 1078 | No | NA | 3 | 0 | 2 | Low-quality | Genome-fragment | 0.34 | AAI-based (medium-confidence) | 0 |
| S11-final-viral-combined_k141_138346\|\|full | 1540 | No | NA | 3 | 0 | 1 | Not-determined | Genome-fragment | NA | NA | 0 |
| S12-final-viral-combined_k141_195978\|\|lt2gene | 1078 | No | NA | 1 | 1 | 0 | Low-quality | Genome-fragment | 2.38 | AAI-based (high-confidence) | 0 |
| S4-final-viral-combined_k141_151424\|\|full | 1078 | No | NA | 3 | 2 | 0 | Low-quality | Genome-fragment | 1.59 | AAI-based (medium-confidence) | 0 |
| S11-final-viral-combined_k141_97034\|\|full | 2698 | No | NA | 3 | 0 | 2 | Not-determined | Genome-fragment | NA | NA | 0 |
| S6-final-viral-combined_k141_124395\|\|full | 1078 | No | NA | 2 | 0 | 2 | Low-quality | Genome-fragment | 0.78 | AAI-based (medium-confidence) | 0 |
| S11-final-viral-combined_k141_41853\|\|full | 1711 | No | NA | 3 | 0 | 3 | Not-determined | Genome-fragment | NA | NA | 0 |
| S11-final-viral-combined_k141_248582\|\|full | 14636 | No | NA | 7 | 0 | 3 | Not-determined | Genome-fragment | NA | NA | 0 |
| S11-final-viral-combined_k141_110756\|\|full | 1467 | No | NA | 2 | 0 | 0 | Not-determined | Genome-fragment | NA | NA | 0 |
| S11-final-viral-combined_k141_221028\|\|full | 1021 | No | NA | 3 | 0 | 0 | Not-determined | Genome-fragment | NA | NA | 0 |
| S11-final-viral-combined_k141_289855\|\|full | 4843 | No | NA | 6 | 0 | 6 | Not-determined | Genome-fragment | NA | NA | 0 |
| S11-final-viral-combined_k141_28164\|\|full | 1133 | No | NA | 2 | 0 | 1 | Not-determined | Genome-fragment | NA | NA | 0 |
| S11-final-viral-combined_k141_97068\|\|full | 2884 | No | NA | 3 | 0 | 1 | Not-determined | Genome-fragment | NA | NA | 0 |
| S8_k141_59308 | 1078 | No | NA | 2 | 0 | 0 | Low-quality | Genome-fragment | 2.77 | AAI-based (medium-confidence) | 0 |
| S11-final-viral-combined_k141_97086\|\|full | 3110 | No | NA | 2 | 0 | 1 | Not-determined | Genome-fragment | NA | NA | 0 |
| S11-final-viral-combined_k141_96787\|\|lt2gene | 1079 | No | NA | 2 | 2 | 0 | Low-quality | Genome-fragment | 2.48 | HMM-based (lower-bound) | 0 |
| S11_k141_119503 | 1079 | No | NA | 1 | 1 | 0 | Low-quality | Genome-fragment | 2.76 | AAI-based (high-confidence) | 0 |
| S11-final-viral-combined_k141_166152\|\|full | 1902 | No | NA | 4 | 0 | 1 | Not-determined | Genome-fragment | NA | NA | 0 |
| S11-final-viral-combined_k141_138475\|\|full | 2004 | No | NA | 3 | 0 | 0 | Not-determined | Genome-fragment | NA | NA | 0 |
| S11-final-viral-combined_k141_138485\|\|full | 1253 | No | NA | 3 | 0 | 2 | Not-determined | Genome-fragment | NA | NA | 0 |
| S3_k141_128208 | 1079 | No | NA | 1 | 0 | 1 | Low-quality | Genome-fragment | 3.24 | AAI-based (medium-confidence) | 0 |
| S11-final-viral-combined_k141_248664\|\|full | 1516 | No | NA | 3 | 0 | 0 | Not-determined | Genome-fragment | NA | NA | 0 |
| S11-final-viral-combined_k141_14466\|\|full | 4424 | No | NA | 5 | 0 | 2 | Not-determined | Genome-fragment | NA | NA | 0 |
| S11-final-viral-combined_k141_289941\|\|full | 2447 | No | NA | 2 | 0 | 1 | Not-determined | Genome-fragment | NA | NA | 0 |
| S11-final-viral-combined_k141_207291\|\|full | 3391 | No | NA | 6 | 0 | 2 | Not-determined | Genome-fragment | NA | NA | 0 |
| S5-final-viral-combined_k141_136162\|\|lt2gene | 1079 | No | NA | 2 | 2 | 0 | Low-quality | Genome-fragment | 1.66 | AAI-based (medium-confidence) | 0 |
| S11-final-viral-combined_k141_97132\|\|full | 2253 | No | NA | 3 | 0 | 1 | Not-determined | Genome-fragment | NA | NA | 0 |
| S11-final-viral-combined_k141_289967\|\|full | 2622 | No | NA | 3 | 0 | 2 | Not-determined | Genome-fragment | NA | NA | 0 |
| S7-final-viral-combined_k141_62448\|\|full | 1079 | No | NA | 2 | 1 | 0 | Low-quality | Genome-fragment | 1.55 | HMM-based (lower-bound) | 0 |
| S11-final-viral-combined_k141_124799\|\|full | 1338 | No | NA | 3 | 0 | 0 | Not-determined | Genome-fragment | NA | NA | 0 |
| S11-final-viral-combined_k141_83281\|\|full | 4130 | No | NA | 4 | 0 | 3 | Not-determined | Genome-fragment | NA | NA | 0 |
| S7-final-viral-combined_k141_44478\|\|full | 1079 | No | NA | 2 | 0 | 0 | Low-quality | Genome-fragment | 3.06 | AAI-based (medium-confidence) | 0 |
| S11-final-viral-combined_k141_110814\|\|full | 1123 | No | NA | 3 | 0 | 1 | Not-determined | Genome-fragment | NA | NA | 0 |
| S17-final-viral-combined_k141_111172\|\|full | 1079 | No | NA | 3 | 0 | 0 | Low-quality | Genome-fragment | 2.13 | AAI-based (medium-confidence) | 0 |
| S12-final-viral-combined_k141_39316\|\|full | 1079 | No | NA | 3 | 0 | 0 | Low-quality | Genome-fragment | 3.39 | AAI-based (high-confidence) | 0 |
| S12-final-viral-combined_k141_247451\|\|lt2gene | 1079 | No | NA | 2 | 1 | 0 | Low-quality | Genome-fragment | 2.12 | AAI-based (high-confidence) | 0 |
| S11-final-viral-combined_k141_235008\|\|full | 1079 | No | NA | 2 | 0 | 1 | Not-determined | Genome-fragment | NA | NA | 0 |
| S2_k141_177254 | 1079 | No | NA | 2 | 0 | 0 | Low-quality | Genome-fragment | 3.7 | AAI-based (high-confidence) | 0 |
| S11-final-viral-combined_k141_303892\|\|full | 2867 | No | NA | 5 | 0 | 1 | Not-determined | Genome-fragment | NA | NA | 0 |
| S11-final-viral-combined_k141_290010\|\|full | 1002 | No | NA | 3 | 0 | 0 | Not-determined | Genome-fragment | NA | NA | 0 |
| S11-final-viral-combined_k141_221167\|\|full | 1555 | No | NA | 3 | 0 | 0 | Not-determined | Genome-fragment | NA | NA | 0 |
| S1-final-viral-combined_k141_122213\|\|full | 1080 | No | NA | 3 | 0 | 2 | Low-quality | Genome-fragment | 0.37 | AAI-based (medium-confidence) | 0 |
| S11-final-viral-combined_k141_138530\|\|full | 1407 | No | NA | 3 | 0 | 2 | Not-determined | Genome-fragment | NA | NA | 0 |
| S3-final-viral-combined_k141_10580\|\|full | 1080 | No | NA | 4 | 2 | 0 | Low-quality | Genome-fragment | 2.6 | AAI-based (high-confidence) | 0 |
| S11-final-viral-combined_k141_14547\|\|full | 7471 | No | NA | 7 | 0 | 1 | Not-determined | Genome-fragment | NA | NA | 0 |
| S11-final-viral-combined_k141_124887\|\|full | 1112 | No | NA | 2 | 0 | 1 | Not-determined | Genome-fragment | NA | NA | 0 |

| | | | | | | | | | | | |
|---|---|---|---|---|---|---|---|---|---|---|---|
| S11-final-viral-combined_k141_290030\|\|full | 1365 | No | NA | 3 | 0 | 1 | Not-determined | Genome-fragment | NA | NA | 0 |
| S5-final-viral-combined_k141_3580\|\|full | 1080 | No | NA | 3 | 0 | 0 | Low-quality | Genome-fragment | 2.12 | AAI-based (medium-confidence) | 0 |
| S5-final-viral-combined_k141_85088\|\|full | 1080 | No | NA | 3 | 2 | 0 | Low-quality | Genome-fragment | 2.38 | AAI-based (medium-confidence) | 0 |
| S11-final-viral-combined_k141_152339\|\|full | 7421 | No | NA | 11 | 0 | 2 | Not-determined | Genome-fragment | NA | NA | 0 |
| S5-final-viral-combined_k141_21801\|\|full | 1080 | No | NA | 2 | 0 | 0 | Low-quality | Genome-fragment | 1.86 | AAI-based (medium-confidence) | 0 |
| S17-final-viral-combined_k141_47928\|\|full | 1080 | No | NA | 2 | 0 | 0 | Low-quality | Genome-fragment | 2.8 | AAI-based (medium-confidence) | 0 |
| S11-final-viral-combined_k141_124914\|\|full | 2484 | No | NA | 2 | 0 | 2 | Not-determined | Genome-fragment | NA | NA | 0 |
| S11-final-viral-combined_k141_28316\|\|full | 2324 | No | NA | 3 | 0 | 0 | Not-determined | Genome-fragment | NA | NA | 0 |
| S17-final-viral-combined_k141_265642\|\|lt2gene | 1080 | No | NA | 2 | 1 | 0 | Low-quality | Genome-fragment | 1.66 | HMM-based (lower-bound) | 0 |
| S11-final-viral-combined_k141_152356\|\|full | 3180 | No | NA | 5 | 0 | 0 | Not-determined | Genome-fragment | NA | NA | 0 |
| S2-final-viral-combined_k141_40394\|\|full | 1080 | No | NA | 3 | 0 | 0 | Low-quality | Genome-fragment | 2.05 | AAI-based (medium-confidence) | 0 |
| S11-final-viral-combined_k141_276341\|\|full | 2914 | No | NA | 5 | 0 | 0 | Not-determined | Genome-fragment | NA | NA | 0 |
| S11-final-viral-combined_k141_28317\|\|full | 6207 | No | NA | 5 | 0 | 4 | Not-determined | Genome-fragment | NA | NA | 0 |
| S11-final-viral-combined_k141_221231\|\|full | 2137 | No | NA | 4 | 0 | 0 | Not-determined | Genome-fragment | NA | NA | 0 |
| S11-final-viral-combined_k141_276342\|\|full | 1349 | No | NA | 2 | 0 | 1 | Not-determined | Genome-fragment | NA | NA | 0 |
| S11-final-viral-combined_k141_678\|\|full | 7158 | No | NA | 9 | 0 | 2 | Not-determined | Genome-fragment | NA | NA | 0 |
| S11-final-viral-combined_k141_193702\|\|full | 3304 | No | NA | 3 | 0 | 2 | Not-determined | Genome-fragment | NA | NA | 0 |
| S4-final-viral-combined_k141_10744\|\|full | 1080 | No | NA | 3 | 1 | 0 | Low-quality | Genome-fragment | 1.31 | AAI-based (high-confidence) | 0 |
| S11-final-viral-combined_k141_303944\|\|full | 1673 | No | NA | 3 | 0 | 1 | Not-determined | Genome-fragment | NA | NA | 0 |
| S8-final-viral-combined_k141_266458\|\|full | 1080 | No | NA | 3 | 1 | 0 | Low-quality | Genome-fragment | 0.82 | AAI-based (high-confidence) | 0 |
| S11-final-viral-combined_k141_97292\|\|full | 2922 | No | NA | 3 | 0 | 1 | Not-determined | Genome-fragment | NA | NA | 0 |
| S8-final-viral-combined_k141_245828\|\|full | 1080 | No | NA | 4 | 1 | 0 | Low-quality | Genome-fragment | 1.95 | AAI-based (high-confidence) | 0 |
| S8_k141_35867 | 1080 | No | NA | 2 | 0 | 0 | Low-quality | Genome-fragment | 2.45 | AAI-based (high-confidence) | 0 |
| S9-final-viral-combined_k141_8864\|\|lt2gene | 1081 | No | NA | 1 | 1 | 0 | Low-quality | Genome-fragment | 2.81 | AAI-based (high-confidence) | 0 |
| S11-final-viral-combined_k141_235142\|\|full | 2992 | No | NA | 2 | 0 | 2 | Not-determined | Genome-fragment | NA | NA | 0 |
| S11-final-viral-combined_k141_297626\|\|lt2gene | 1081 | No | NA | 2 | 1 | 0 | Low-quality | Genome-fragment | 2.47 | AAI-based (high-confidence) | 0 |
| S11-final-viral-combined_k141_55982\|\|full | 1896 | No | NA | 3 | 0 | 0 | Not-determined | Genome-fragment | NA | NA | 0 |
| S1_k141_169781 | 1081 | No | NA | 2 | 0 | 1 | Low-quality | Genome-fragment | 2.77 | AAI-based (medium-confidence) | 0 |
| S5-final-viral-combined_k141_122425\|\|lt2gene | 1081 | No | NA | 2 | 2 | 0 | Low-quality | Genome-fragment | 2.41 | AAI-based (high-confidence) | 0 |
| S11-final-viral-combined_k141_235186\|\|full | 1539 | No | NA | 2 | 0 | 1 | Not-determined | Genome-fragment | NA | NA | 0 |
| S11-final-viral-combined_k141_166423\|\|full | 1429 | No | NA | 2 | 0 | 1 | Not-determined | Genome-fragment | NA | NA | 0 |
| S11-final-viral-combined_k141_110981\|\|full | 1763 | No | NA | 2 | 0 | 1 | Not-determined | Genome-fragment | NA | NA | 0 |
| S17-final-viral-combined_k141_206409\|\|full | 1081 | No | NA | 3 | 2 | 0 | Low-quality | Genome-fragment | 2.71 | AAI-based (high-confidence) | 0 |
| S17-k141_121614 | 1081 | No | NA | 2 | 0 | 0 | Low-quality | Genome-fragment | 0.93 | AAI-based (high-confidence) | 0 |
| S11-final-viral-combined_k141_207506\|\|full | 1835 | No | NA | 2 | 0 | 2 | Not-determined | Genome-fragment | NA | NA | 0 |
| S12-final-viral-combined_k141_152557\|\|full | 1081 | No | NA | 3 | 0 | 0 | Low-quality | Genome-fragment | 0.86 | AAI-based (medium-confidence) | 0 |
| S11-final-viral-combined_k141_28397\|\|full | 1134 | No | NA | 3 | 0 | 0 | Not-determined | Genome-fragment | NA | NA | 0 |
| S11-final-viral-combined_k141_221310\|\|full | 32507 | No | NA | 40 | 11 | 1 | High-quality | High-quality | 91.28 | AAI-based (high-confidence) | 0 |
| S11-final-viral-combined_k141_317802\|\|full | 1124 | No | NA | 2 | 0 | 1 | Not-determined | Genome-fragment | NA | NA | 0 |
| S12-final-viral-combined_k141_144712\|\|full | 1081 | No | NA | 3 | 0 | 0 | Low-quality | Genome-fragment | 2.27 | AAI-based (high-confidence) | 0 |
| S11-final-viral-combined_k141_235212\|\|full | 2812 | No | NA | 3 | 0 | 2 | Not-determined | Genome-fragment | NA | NA | 0 |
| S11-final-viral-combined_k141_317806\|\|full | 3456 | No | NA | 2 | 0 | 2 | Not-determined | Genome-fragment | NA | NA | 0 |
| S11-final-viral-combined_k141_276481\|\|full | 1948 | No | NA | 3 | 0 | 2 | Not-determined | Genome-fragment | NA | NA | 0 |
| S2-final-viral-combined_k141_85560\|\|full | 1081 | No | NA | 3 | 0 | 0 | Low-quality | Genome-fragment | 2.6 | AAI-based (medium-confidence) | 0 |
| S11-final-viral-combined_k141_304041\|\|full | 2567 | No | NA | 4 | 0 | 0 | Not-determined | Genome-fragment | NA | NA | 0 |
| S11-final-viral-combined_k141_69727\|\|full | 1233 | No | NA | 3 | 0 | 0 | Not-determined | Genome-fragment | NA | NA | 0 |
| S11-final-viral-combined_k141_193830\|\|full | 2039 | No | NA | 3 | 0 | 3 | Not-determined | Genome-fragment | NA | NA | 0 |
| S11-final-viral-combined_k141_83551\|\|full | 1533 | No | NA | 3 | 0 | 1 | Not-determined | Genome-fragment | NA | NA | 0 |
| S11-final-viral-combined_k141_221357\|\|full | 1644 | No | NA | 3 | 0 | 1 | Not-determined | Genome-fragment | NA | NA | 0 |
| S11-final-viral-combined_k141_166477\|\|full | 5569 | No | NA | 6 | 0 | 3 | Not-determined | Genome-fragment | NA | NA | 0 |
| S11-final-viral-combined_k141_14636\|\|full | 35938 | No | NA | 38 | 20 | 0 | High-quality | High-quality | 92.99 | AAI-based (high-confidence) | 0 |
| S11-final-viral-combined_k141_304103\|\|full | 2158 | No | NA | 3 | 0 | 1 | Not-determined | Genome-fragment | NA | NA | 0 |
| S2-final-viral-combined_k141_214961\|\|full | 1081 | No | NA | 3 | 1 | 0 | Low-quality | Genome-fragment | 2.54 | AAI-based (medium-confidence) | 0 |
| S4-final-viral-combined_k141_96024\|\|full | 1081 | No | NA | 3 | 0 | 0 | Low-quality | Genome-fragment | 0.79 | AAI-based (high-confidence) | 0 |
| S6-final-viral-combined_k141_44715\|\|full | 1081 | No | NA | 2 | 1 | 0 | Low-quality | Genome-fragment | 2.25 | AAI-based (high-confidence) | 0 |
| S11-final-viral-combined_k141_290135\|\|full | 2427 | No | NA | 3 | 0 | 1 | Not-determined | Genome-fragment | NA | NA | 0 |
| S6-final-viral-combined_k141_69940\|\|full | 1081 | No | NA | 2 | 1 | 0 | Low-quality | Genome-fragment | 0.59 | HMM-based (lower-bound) | 0 |
| S6-final-viral-combined_k141_137290\|\|lt2gene | 1081 | No | NA | 1 | 1 | 0 | Low-quality | Genome-fragment | 2.62 | AAI-based (high-confidence) | 0 |
| S9-final-viral-combined_k141_121678\|\|full | 1082 | No | NA | 3 | 0 | 0 | Low-quality | Genome-fragment | 2.82 | AAI-based (high-confidence) | 0 |
| S11-final-viral-combined_k141_83571\|\|full | 40621 | Yes | 32207 | 51 | 12 | 6 | Medium-quality | Genome-fragment | 82.38 | AAI-based (medium-confidence) | 20.71 |
| S11-final-viral-combined_k141_14663\|\|full | 1393 | No | NA | 3 | 0 | 0 | Not-determined | Genome-fragment | NA | NA | 0 |
| S11-final-viral-combined_k141_166490\|\|full | 1913 | No | NA | 3 | 0 | 3 | Not-determined | Genome-fragment | NA | NA | 0 |
| S9-final-viral-combined_k141_89878\|\|full | 1082 | No | NA | 4 | 0 | 0 | Low-quality | Genome-fragment | 1.03 | AAI-based (medium-confidence) | 0 |
| S11-final-viral-combined_k141_125034\|\|full | 2448 | No | NA | 4 | 0 | 1 | Not-determined | Genome-fragment | NA | NA | 0 |
| S9-final-viral-combined_k141_64016\|\|full | 1082 | No | NA | 2 | 0 | 0 | Low-quality | Genome-fragment | 2.05 | AAI-based (high-confidence) | 0 |
| S9_k141_28071 | 1082 | No | NA | 1 | 0 | 1 | Low-quality | Genome-fragment | 0.31 | AAI-based (high-confidence) | 0 |
| S11-final-viral-combined_k141_83624\|\|full | 4525 | No | NA | 4 | 0 | 4 | Not-determined | Genome-fragment | NA | NA | 0 |
| S11-final-viral-combined_k141_189933\|\|lt2gene | 1082 | No | NA | 2 | 1 | 0 | Low-quality | Genome-fragment | 0.7 | HMM-based (lower-bound) | 0 |
| S11-final-viral-combined_k141_304139\|\|full | 1834 | No | NA | 3 | 0 | 2 | Not-determined | Genome-fragment | NA | NA | 0 |
| S1-final-viral-combined_k141_137762\|\|full | 1082 | No | NA | 2 | 2 | 0 | Low-quality | Genome-fragment | 2.35 | AAI-based (high-confidence) | 0 |
| S1-final-viral-combined_k141_221507\|\|full | 1082 | No | NA | 3 | 0 | 0 | Low-quality | Genome-fragment | 0.81 | AAI-based (high-confidence) | 0 |
| S11-final-viral-combined_k141_125119\|\|full | 7617 | No | NA | 6 | 0 | 5 | Not-determined | Genome-fragment | NA | NA | 0 |
| S11-final-viral-combined_k141_248996\|\|full | 4568 | No | NA | 4 | 0 | 3 | Not-determined | Genome-fragment | NA | NA | 0 |
| S11-final-viral-combined_k141_152628\|\|full | 2165 | No | NA | 3 | 0 | 0 | Not-determined | Genome-fragment | NA | NA | 0 |
| S11-final-viral-combined_k141_977\|\|full | 3504 | No | NA | 2 | 0 | 1 | Not-determined | Genome-fragment | NA | NA | 0 |
| S11-final-viral-combined_k141_276570\|\|full | 17411 | No | NA | 15 | 0 | 6 | Not-determined | Genome-fragment | NA | NA | 0 |
| S1-final-viral-combined_k141_106745\|\|full | 1082 | No | NA | 3 | 0 | 1 | Low-quality | Genome-fragment | 1.09 | AAI-based (high-confidence) | 0 |
| S1_k141_112514 | 1082 | No | NA | 5 | 0 | 0 | Low-quality | Genome-fragment | 2.11 | AAI-based (high-confidence) | 0 |
| S11-final-viral-combined_k141_83703\|\|full | 1215 | No | NA | 3 | 0 | 0 | Not-determined | Genome-fragment | NA | NA | 0 |
| S11-final-viral-combined_k141_28483\|\|full | 6699 | No | NA | 7 | 0 | 5 | Not-determined | Genome-fragment | NA | NA | 0 |
| S7_k141_236602 | 1082 | No | NA | 3 | 1 | 0 | Low-quality | Genome-fragment | 1.86 | AAI-based (medium-confidence) | 0 |
| S11-final-viral-combined_k141_166608\|\|full | 1533 | No | NA | 3 | 0 | 2 | Not-determined | Genome-fragment | NA | NA | 0 |
| S11-final-viral-combined_k141_207691\|\|full | 2158 | No | NA | 2 | 0 | 2 | Not-determined | Genome-fragment | NA | NA | 0 |
| S11-final-viral-combined_k141_180040\|\|full | 1278 | No | NA | 3 | 0 | 0 | Not-determined | Genome-fragment | NA | NA | 0 |
| S11-final-viral-combined_k141_304194\|\|full | 3101 | No | NA | 3 | 0 | 0 | Not-determined | Genome-fragment | NA | NA | 0 |
| S11-final-viral-combined_k141_152665\|\|full | 2272 | No | NA | 3 | 0 | 1 | Not-determined | Genome-fragment | NA | NA | 0 |
| S11-final-viral-combined_k141_152666\|\|full | 4114 | No | NA | 4 | 0 | 3 | Not-determined | Genome-fragment | NA | NA | 0 |
| S11-final-viral-combined_k141_14777\|\|full | 3326 | No | NA | 4 | 0 | 4 | Not-determined | Genome-fragment | NA | NA | 0 |
| S11-final-viral-combined_k141_83742\|\|full | 1727 | No | NA | 2 | 0 | 2 | Not-determined | Genome-fragment | NA | NA | 0 |
| S11-final-viral-combined_k141_97609\|\|full | 2449 | No | NA | 2 | 0 | 2 | Not-determined | Genome-fragment | NA | NA | 0 |
| S11-final-viral-combined_k141_180077\|\|full | 2558 | No | NA | 2 | 0 | 1 | Not-determined | Genome-fragment | NA | NA | 0 |
| S17-final-viral-combined_k141_178340\|\|full | 1082 | No | NA | 3 | 2 | 0 | Low-quality | Genome-fragment | 2.84 | AAI-based (high-confidence) | 0 |
| S17-final-viral-combined_k141_179667\|\|full | 1082 | No | NA | 3 | 1 | 0 | Low-quality | Genome-fragment | 2.54 | AAI-based (high-confidence) | 0 |
| S12-final-viral-combined_k141_116436\|\|full | 1082 | No | NA | 3 | 0 | 0 | Low-quality | Genome-fragment | 2.07 | AAI-based (high-confidence) | 0 |
| S11-final-viral-combined_k141_221462\|\|full | 3657 | No | NA | 6 | 0 | 0 | Not-determined | Genome-fragment | NA | NA | 0 |
| S11-final-viral-combined_k141_290235\|\|full | 3874 | No | NA | 4 | 0 | 3 | Not-determined | Genome-fragment | NA | NA | 0 |
| S12-final-viral-combined_k141_285014\|\|lt2gene | 1082 | No | NA | 1 | 1 | 0 | Low-quality | Genome-fragment | 3.04 | AAI-based (high-confidence) | 0 |
| S2-final-viral-combined_k141_168749\|\|full | 1082 | No | NA | 2 | 0 | 0 | Low-quality | Genome-fragment | 2.39 | AAI-based (medium-confidence) | 0 |
| S8-final-viral-combined_k141_485\|\|full | 1082 | No | NA | 3 | 0 | 0 | Low-quality | Genome-fragment | 1.75 | AAI-based (medium-confidence) | 0 |
| S8-final-viral-combined_k141_228090\|\|full | 1082 | No | NA | 3 | 1 | 0 | Low-quality | Genome-fragment | 2.73 | AAI-based (medium-confidence) | 0 |
| S11-final-viral-combined_k141_180807\|\|full | 1083 | No | NA | 2 | 0 | 2 | Low-quality | Genome-fragment | 0.54 | AAI-based (medium-confidence) | 0 |
| S11-final-viral-combined_k141_42215\|\|full | 2113 | No | NA | 3 | 0 | 3 | Not-determined | Genome-fragment | NA | NA | 0 |
| S11-final-viral-combined_k141_276638\|\|full | 1891 | No | NA | 2 | 0 | 2 | Not-determined | Genome-fragment | NA | NA | 0 |
| S11-final-viral-combined_k141_167741\|\|full | 1083 | No | NA | 2 | 0 | 2 | Low-quality | Genome-fragment | 0.4 | AAI-based (medium-confidence) | 0 |
| S3-final-viral-combined_k141_63671\|\|full | 1083 | No | NA | 3 | 1 | 0 | Low-quality | Genome-fragment | 1.34 | HMM-based (lower-bound) | 0 |
| S11-final-viral-combined_k141_125176\|\|full | 67341 | Yes | 22887 | 66 | 3 | 25 | Medium-quality | Genome-fragment | 56.45 | AAI-based (high-confidence) | 66.01 |
| S11-final-viral-combined_k141_304239\|\|full | 44706 | No | NA | 66 | 17 | 0 | High-quality | High-quality | 99.61 | AAI-based (high-confidence) | 0 |
| S11-final-viral-combined_k141_83805\|\|full | 1108 | No | NA | 2 | 0 | 2 | Not-determined | Genome-fragment | NA | NA | 0 |
| S11-final-viral-combined_k141_56123\|\|full | 1493 | No | NA | 3 | 0 | 1 | Not-determined | Genome-fragment | NA | NA | 0 |
| S11-final-viral-combined_k141_152773\|\|full | 4344 | No | NA | 3 | 0 | 2 | Not-determined | Genome-fragment | NA | NA | 0 |
| S11-final-viral-combined_k141_290278\|\|full | 1406 | No | NA | 2 | 0 | 1 | Not-determined | Genome-fragment | NA | NA | 0 |
| S3_k141_34196 | 1083 | No | NA | 2 | 0 | 2 | Low-quality | Genome-fragment | 0.67 | AAI-based (medium-confidence) | 0 |
| S5-final-viral-combined_k141_153436\|\|full | 1083 | No | NA | 3 | 1 | 0 | Low-quality | Genome-fragment | 2.46 | AAI-based (high-confidence) | 0 |
| S11-final-viral-combined_k141_304274\|\|full | 4945 | No | NA | 5 | 0 | 1 | Not-determined | Genome-fragment | NA | NA | 0 |
| S5_k141_198233 | 1083 | No | NA | 2 | 0 | 0 | Low-quality | Genome-fragment | 3.02 | AAI-based (high-confidence) | 0 |
| S11-final-viral-combined_k141_207831\|\|full | 2553 | No | NA | 3 | 0 | 1 | Not-determined | Genome-fragment | NA | NA | 0 |
| S11-final-viral-combined_k141_249131\|\|full | 1946 | No | NA | 3 | 0 | 1 | Not-determined | Genome-fragment | NA | NA | 0 |
| S11-final-viral-combined_k141_193956\|\|full | 3898 | No | NA | 5 | 0 | 1 | Not-determined | Genome-fragment | NA | NA | 0 |
| S11-final-viral-combined_k141_235594\|\|full | 4229 | No | NA | 3 | 0 | 2 | Not-determined | Genome-fragment | NA | NA | 0 |
| S11-final-viral-combined_k141_152802\|\|full | 1326 | No | NA | 2 | 0 | 1 | Not-determined | Genome-fragment | NA | NA | 0 |
| S11-final-viral-combined_k141_14892\|\|full | 1933 | No | NA | 2 | 0 | 2 | Not-determined | Genome-fragment | NA | NA | 0 |
| S11-final-viral-combined_k141_221578\|\|full | 1748 | No | NA | 3 | 0 | 1 | Not-determined | Genome-fragment | NA | NA | 0 |
| S7-final-viral-combined_k141_34868\|\|full | 1083 | No | NA | 3 | 3 | 0 | Low-quality | Genome-fragment | 2.31 | AAI-based (high-confidence) | 0 |
| S11-final-viral-combined_k141_28535\|\|full | 1398 | No | NA | 3 | 0 | 0 | Not-determined | Genome-fragment | NA | NA | 0 |
| S11-final-viral-combined_k141_193989\|\|full | 10709 | No | NA | 12 | 0 | 7 | Not-determined | Genome-fragment | NA | NA | 0 |
| S11-final-viral-combined_k141_14951\|\|full | 1625 | No | NA | 2 | 0 | 0 | Not-determined | Genome-fragment | NA | NA | 0 |
| S7_k141_44748 | 1083 | No | NA | 2 | 0 | 0 | Low-quality | Genome-fragment | 1.34 | AAI-based (high-confidence) | 0 |
| S7_k141_147372 | 1083 | No | NA | 1 | 0 | 0 | Low-quality | Genome-fragment | 2.53 | AAI-based (high-confidence) | 0 |
| S7_k141_40399 | 1083 | No | NA | 4 | 1 | 0 | Low-quality | Genome-fragment | 2.14 | AAI-based (high-confidence) | 0 |
| S11-final-viral-combined_k141_28579\|\|full | 1687 | No | NA | 3 | 0 | 1 | Not-determined | Genome-fragment | NA | NA | 0 |
| S17-k141_89315 | 1083 | No | NA | 3 | 0 | 0 | Low-quality | Genome-fragment | 1.21 | AAI-based (high-confidence) | 0 |
| S17-k141_86918 | 1083 | No | NA | 1 | 0 | 1 | Low-quality | Genome-fragment | 0.58 | AAI-based (medium-confidence) | 0 |
| S11-final-viral-combined_k141_262998\|\|full | 1937 | No | NA | 3 | 0 | 1 | Not-determined | Genome-fragment | NA | NA | 0 |
| S17-k141_21943 | 1083 | No | NA | 1 | 0 | 1 | Low-quality | Genome-fragment | 0.6 | AAI-based (medium-confidence) | 0 |
| S12-final-viral-combined_k141_268303\|\|full | 1083 | No | NA | 2 | 0 | 2 | Low-quality | Genome-fragment | 1 | AAI-based (medium-confidence) | 0 |
| S2-final-viral-combined_k141_198037\|\|full | 1083 | No | NA | 2 | 0 | 1 | Low-quality | Genome-fragment | 0.51 | AAI-based (medium-confidence) | 0 |
| S11-final-viral-combined_k141_235715\|\|full | 3285 | No | NA | 4 | 0 | 4 | Not-determined | Genome-fragment | NA | NA | 0 |
| S11-final-viral-combined_k141_138897\|\|full | 1390 | No | NA | 2 | 0 | 1 | Not-determined | Genome-fragment | NA | NA | 0 |
| S11-final-viral-combined_k141_56201\|\|full | 1662 | No | NA | 3 | 0 | 0 | Not-determined | Genome-fragment | NA | NA | 0 |
| S6-final-viral-combined_k141_64187\|\|lt2gene | 1083 | No | NA | 2 | 2 | 0 | Low-quality | Genome-fragment | 2.45 | AAI-based (medium-confidence) | 0 |
| S9_k141_69999 | 1084 | No | NA | 2 | 0 | 0 | Low-quality | Genome-fragment | 2.08 | AAI-based (high-confidence) | 0 |
| S11-final-viral-combined_k141_42398\|\|full | 2063 | No | NA | 3 | 0 | 0 | Not-determined | Genome-fragment | NA | NA | 0 |
| S11-final-viral-combined_k141_284655\|\|full | 1084 | No | NA | 4 | 0 | 0 | Low-quality | Genome-fragment | 1.3 | AAI-based (high-confidence) | 0 |
| S11-final-viral-combined_k141_249267\|\|full | 4115 | No | NA | 8 | 0 | 3 | Not-determined | Genome-fragment | NA | NA | 0 |
| S11-final-viral-combined_k141_304378\|\|full | 4477 | No | NA | 4 | 0 | 3 | Not-determined | Genome-fragment | NA | NA | 0 |
| S11-final-viral-combined_k141_44971\|\|lt2gene | 1084 | No | NA | 2 | 2 | 0 | Low-quality | Genome-fragment | 1.89 | HMM-based (lower-bound) | 0 |
| S11-final-viral-combined_k141_125333\|\|full | 1107 | No | NA | 3 | 0 | 2 | Not-determined | Genome-fragment | NA | NA | 0 |
| S11-final-viral-combined_k141_208008\|\|full | 2441 | No | NA | 3 | 0 | 3 | Not-determined | Genome-fragment | NA | NA | 0 |
| S11-final-viral-combined_k141_276868\|\|full | 1085 | No | NA | 2 | 0 | 2 | Not-determined | Genome-fragment | NA | NA | 0 |
| S11-final-viral-combined_k141_180286\|\|full | 1851 | No | NA | 3 | 0 | 1 | Not-determined | Genome-fragment | NA | NA | 0 |
| S11-final-viral-combined_k141_208028\|\|full | 2676 | No | NA | 5 | 0 | 3 | Not-determined | Genome-fragment | NA | NA | 0 |
| S1-final-viral-combined_k141_180381\|\|full | 1084 | No | NA | 2 | 0 | 2 | Low-quality | Genome-fragment | 0.36 | AAI-based (medium-confidence) | 0 |
| S5-final-viral-combined_k141_130777\|\|lt2gene | 1084 | No | NA | 2 | 2 | 0 | Low-quality | Genome-fragment | 1.51 | HMM-based (lower-bound) | 0 |
| S11-final-viral-combined_k141_304450\|\|full | 4346 | No | NA | 5 | 0 | 2 | Not-determined | Genome-fragment | NA | NA | 0 |
| S5-final-viral-combined_k141_177341\|\|lt2gene | 1084 | No | NA | 2 | 2 | 0 | Low-quality | Genome-fragment | 1.34 | HMM-based (lower-bound) | 0 |
| S11-final-viral-combined_k141_290474\|\|full | 2726 | No | NA | 5 | 0 | 2 | Not-determined | Genome-fragment | NA | NA | 0 |
| S7-final-viral-combined_k141_131833\|\|full | 1084 | No | NA | 3 | 0 | 0 | Low-quality | Genome-fragment | 2.16 | AAI-based (medium-confidence) | 0 |
| S12_k141_271346 | 1084 | No | NA | 1 | 0 | 1 | Low-quality | Genome-fragment | 1.08 | AAI-based (high-confidence) | 0 |
| S12_k141_325839 | 1084 | No | NA | 2 | 0 | 0 | Low-quality | Genome-fragment | 1.65 | AAI-based (medium-confidence) | 0 |
| S2-final-viral-combined_k141_197679\|\|full | 1084 | No | NA | 2 | 1 | 0 | Low-quality | Genome-fragment | 24.52 | AAI-based (medium-confidence) | 0 |
| S6-final-viral-combined_k141_120952\|\|full | 1084 | No | NA | 2 | 2 | 0 | Low-quality | Genome-fragment | 1.01 | AAI-based (high-confidence) | 0 |
| S8_k141_262734 | 1084 | No | NA | 2 | 0 | 0 | Low-quality | Genome-fragment | 0.81 | AAI-based (high-confidence) | 0 |
| S9-final-viral-combined_k141_18395\|\|full | 1085 | No | NA | 4 | 3 | 0 | Low-quality | Genome-fragment | 2.44 | AAI-based (medium-confidence) | 0 |
| S11-final-viral-combined_k141_111363\|\|full | 4188 | No | NA | 6 | 0 | 1 | Not-determined | Genome-fragment | NA | NA | 0 |
| S11-final-viral-combined_k141_290506\|\|full | 1120 | No | NA | 2 | 0 | 0 | Not-determined | Genome-fragment | NA | NA | 0 |
| S9_k141_49467 | 1085 | No | NA | 2 | 0 | 0 | Low-quality | Genome-fragment | 2.18 | AAI-based (high-confidence) | 0 |
| S1-final-viral-combined_k141_136303\|\|full | 1085 | No | NA | 3 | 0 | 1 | Low-quality | Genome-fragment | 1.74 | AAI-based (medium-confidence) | 0 |
| S11-final-viral-combined_k141_28648\|\|full | 2878 | No | NA | 4 | 0 | 1 | Not-determined | Genome-fragment | NA | NA | 0 |
| S1_k141_286825 | 1085 | No | NA | 3 | 0 | 1 | Low-quality | Genome-fragment | 0.68 | AAI-based (high-confidence) | 0 |
| S1_k141_207196 | 1085 | No | NA | 1 | 0 | 0 | Low-quality | Genome-fragment | 1.01 | AAI-based (high-confidence) | 0 |
| S11-final-viral-combined_k141_125424\|\|full | 2776 | No | NA | 5 | 0 | 1 | Not-determined | Genome-fragment | NA | NA | 0 |
| S11-final-viral-combined_k141_70034\|\|full | 1178 | No | NA | 2 | 0 | 1 | Not-determined | Genome-fragment | NA | NA | 0 |
| S11-final-viral-combined_k141_42544\|\|full | 1389 | No | NA | 3 | 0 | 1 | Not-determined | Genome-fragment | NA | NA | 0 |
| S11-final-viral-combined_k141_221651\|\|full | 1919 | No | NA | 2 | 0 | 2 | Not-determined | Genome-fragment | NA | NA | 0 |
| S11-final-viral-combined_k141_125431\|\|full | 1526 | No | NA | 3 | 0 | 2 | Not-determined | Genome-fragment | NA | NA | 0 |
| S3-final-viral-combined_k141_3009\|\|full | 1085 | No | NA | 3 | 0 | 0 | Low-quality | Genome-fragment | 0.49 | AAI-based (medium-confidence) | 0 |
| S11-final-viral-combined_k141_304521\|\|full | 1473 | No | NA | 3 | 0 | 1 | Not-determined | Genome-fragment | NA | NA | 0 |
| S17-final-viral-combined_k141_248136\|\|full | 1085 | No | NA | 2 | 1 | 0 | Low-quality | Genome-fragment | 2.78 | AAI-based (high-confidence) | 0 |
| S12-final-viral-combined_k141_311471\|\|full | 1085 | No | NA | 3 | 2 | 0 | Low-quality | Genome-fragment | 2.95 | AAI-based (high-confidence) | 0 |
| S12-final-viral-combined_k141_95213\|\|full | 1085 | No | NA | 3 | 2 | 0 | Low-quality | Genome-fragment | 2.3 | AAI-based (medium-confidence) | 0 |
| S11-final-viral-combined_k141_98028\|\|full | 1612 | No | NA | 2 | 0 | 0 | Not-determined | Genome-fragment | NA | NA | 0 |
| S11-final-viral-combined_k141_98034\|\|full | 2475 | No | NA | 2 | 0 | 2 | Not-determined | Genome-fragment | NA | NA | 0 |
| S11-final-viral-combined_k141_208056\|\|full | 3203 | No | NA | 5 | 0 | 0 | Not-determined | Genome-fragment | NA | NA | 0 |
| S11-final-viral-combined_k141_98039\|\|full | 2192 | No | NA | 2 | 0 | 0 | Not-determined | Genome-fragment | NA | NA | 0 |
| S12-final-viral-combined_k141_113943\|\|full | 1085 | No | NA | 3 | 0 | 0 | Low-quality | Genome-fragment | 1.89 | AAI-based (high-confidence) | 0 |
| S2-final-viral-combined_k141_233930\|\|full | 1085 | No | NA | 2 | 0 | 0 | Low-quality | Genome-fragment | 2.27 | AAI-based (high-confidence) | 0 |
| S11-final-viral-combined_k141_15104\|\|full | 1995 | No | NA | 3 | 0 | 0 | Not-determined | Genome-fragment | NA | NA | 0 |
| S11-final-viral-combined_k141_180367\|\|full | 2786 | No | NA | 5 | 0 | 0 | Not-determined | Genome-fragment | NA | NA | 0 |
| S11-final-viral-combined_k141_263208\|\|full | 3620 | No | NA | 4 | 0 | 2 | Not-determined | Genome-fragment | NA | NA | 0 |
| S2-final-viral-combined_k141_202799\|\|full | 1085 | No | NA | 3 | 1 | 0 | Low-quality | Genome-fragment | 3.04 | AAI-based (high-confidence) | 0 |
| S2_k141_31574 | 1085 | No | NA | 1 | 0 | 0 | Low-quality | Genome-fragment | 0.1 | AAI-based (medium-confidence) | 0 |
| S11-final-viral-combined_k141_318116\|\|full | 2577 | No | NA | 2 | 0 | 0 | Not-determined | Genome-fragment | NA | NA | 0 |
| S8-final-viral-combined_k141_147097\|\|full | 1085 | No | NA | 3 | 0 | 1 | Low-quality | Genome-fragment | 1.61 | AAI-based (medium-confidence) | 0 |
| S9-final-viral-combined_k141_114357\|\|full | 1086 | No | NA | 3 | 0 | 1 | Low-quality | Genome-fragment | 0.86 | AAI-based (medium-confidence) | 0 |
| S1-final-viral-combined_k141_283497\|\|full | 1086 | No | NA | 2 | 2 | 0 | Low-quality | Genome-fragment | 3.28 | AAI-based (medium-confidence) | 0 |
| S1_k141_68546 | 1086 | No | NA | 3 | 0 | 1 | Low-quality | Genome-fragment | 2.16 | AAI-based (high-confidence) | 0 |
| S1_k141_344699 | 1086 | No | NA | 1 | 0 | 1 | Low-quality | Genome-fragment | 0.73 | AAI-based (high-confidence) | 0 |
| S7-final-viral-combined_k141_216341\|\|full | 1086 | No | NA | 3 | 0 | 1 | Low-quality | Genome-fragment | 1.82 | AAI-based (medium-confidence) | 0 |
| S2-final-viral-combined_k141_175796\|\|full | 1086 | No | NA | 3 | 2 | 0 | Low-quality | Genome-fragment | 1.55 | HMM-based (lower-bound) | 0 |
| S11-final-viral-combined_k141_194178\|\|full | 2474 | No | NA | 5 | 0 | 1 | Not-determined | Genome-fragment | NA | NA | 0 |
| S11-final-viral-combined_k141_194179\|\|full | 1721 | No | NA | 2 | 0 | 0 | Not-determined | Genome-fragment | NA | NA | 0 |
| S2_k141_189078 | 1086 | No | NA | 1 | 0 | 0 | Low-quality | Genome-fragment | 0.7 | AAI-based (high-confidence) | 0 |
| S11-final-viral-combined_k141_70147\|\|full | 1798 | No | NA | 3 | 0 | 0 | Not-determined | Genome-fragment | NA | NA | 0 |
| S4-final-viral-combined_k141_149035\|\|lt2gene | 1086 | No | NA | 1 | 1 | 0 | Low-quality | Genome-fragment | 2.19 | AAI-based (high-confidence) | 0 |
| S8_k141_221822 | 1086 | No | NA | 1 | 0 | 1 | Low-quality | Genome-fragment | 0.18 | AAI-based (high-confidence) | 0 |
| S11-final-viral-combined_k141_1327\|\|full | 11792 | No | NA | 8 | 0 | 6 | Not-determined | Genome-fragment | NA | NA | 0 |
| S11-final-viral-combined_k141_180409\|\|full | 3918 | No | NA | 4 | 0 | 3 | Not-determined | Genome-fragment | NA | NA | 0 |
| S8_k141_264579 | 1086 | No | NA | 1 | 0 | 0 | Low-quality | Genome-fragment | 2.4 | AAI-based (medium-confidence) | 0 |
| S9-final-viral-combined_k141_32282\|\|full | 1087 | No | NA | 2 | 0 | 1 | Low-quality | Genome-fragment | 2.3 | AAI-based (high-confidence) | 0 |
| S11-final-viral-combined_k141_84094\|\|full | 1791 | No | NA | 2 | 0 | 1 | Not-determined | Genome-fragment | NA | NA | 0 |
| S9-final-viral-combined_k141_102275\|\|full | 1087 | No | NA | 2 | 0 | 0 | Low-quality | Genome-fragment | 1.42 | AAI-based (medium-confidence) | 0 |
| S11-final-viral-combined_k141_135080\|\|full | 1087 | No | NA | 2 | 0 | 1 | Low-quality | Genome-fragment | 0.85 | AAI-based (medium-confidence) | 0 |
| S11-final-viral-combined_k141_98124\|\|full | 1543 | No | NA | 2 | 0 | 2 | Not-determined | Genome-fragment | NA | NA | 0 |
| S11-final-viral-combined_k141_235970\|\|full | 1630 | No | NA | 4 | 0 | 0 | Not-determined | Genome-fragment | NA | NA | 0 |
| S11-final-viral-combined_k141_138983\|\|full | 9845 | No | NA | 10 | 0 | 5 | Not-determined | Genome-fragment | NA | NA | 0 |
| S11-final-viral-combined_k141_191990\|\|full | 1087 | Yes | 883 | 4 | 1 | 1 | Low-quality | Genome-fragment | 1.34 | AAI-based (high-confidence) | 18.77 |
| S11-final-viral-combined_k141_138987\|\|full | 7709 | No | NA | 9 | 0 | 9 | Not-determined | Genome-fragment | NA | NA | 0 |
| S1-final-viral-combined_k141_105608\|\|full | 1087 | No | NA | 3 | 0 | 0 | Low-quality | Genome-fragment | 1.85 | AAI-based (high-confidence) | 0 |
| S11-final-viral-combined_k141_194205\|\|full | 35517 | No | NA | 43 | 20 | 1 | Medium-quality | Genome-fragment | 57.83 | AAI-based (high-confidence) | 0 |
| S11-final-viral-combined_k141_318179\|\|full | 1099 | No | NA | 2 | 0 | 1 | Not-determined | Genome-fragment | NA | NA | 0 |
| S3-final-viral-combined_k141_51927\|\|full | 1087 | No | NA | 3 | 2 | 0 | Low-quality | Genome-fragment | 2.18 | AAI-based (medium-confidence) | 0 |
| S11-final-viral-combined_k141_180451\|\|full | 2010 | No | NA | 2 | 0 | 2 | Not-determined | Genome-fragment | NA | NA | 0 |
| S11-final-viral-combined_k141_318209\|\|full | 3508 | No | NA | 4 | 0 | 2 | Not-determined | Genome-fragment | NA | NA | 0 |
| S11-final-viral-combined_k141_180462\|\|full | 1056 | No | NA | 2 | 0 | 0 | Not-determined | Genome-fragment | NA | NA | 0 |
| S11-final-viral-combined_k141_304728\|\|full | 1610 | No | NA | 2 | 0 | 1 | Not-determined | Genome-fragment | NA | NA | 0 |
| S11-final-viral-combined_k141_290674\|\|full | 4647 | No | NA | 5 | 0 | 3 | Not-determined | Genome-fragment | NA | NA | 0 |
| S7-final-viral-combined_k141_47377\|\|lt2gene | 1087 | No | NA | 1 | 1 | 0 | Low-quality | Genome-fragment | 1.02 | HMM-based (lower-bound) | 0 |
| S11-final-viral-combined_k141_277120\|\|full | 1646 | No | NA | 2 | 0 | 1 | Not-determined | Genome-fragment | NA | NA | 0 |
| S17-final-viral-combined_k141_126659\|\|full | 1087 | No | NA | 3 | 1 | 0 | Low-quality | Genome-fragment | 2.53 | AAI-based (medium-confidence) | 0 |
| S11-final-viral-combined_k141_111441\|\|full | 2336 | No | NA | 3 | 0 | 2 | Not-determined | Genome-fragment | NA | NA | 0 |
| S12-final-viral-combined_k141_225744\|\|full | 1087 | No | NA | 3 | 0 | 0 | Low-quality | Genome-fragment | 2.32 | AAI-based (high-confidence) | 0 |
| S11-final-viral-combined_k141_304737\|\|full | 13474 | No | NA | 7 | 0 | 6 | Not-determined | Genome-fragment | NA | NA | 0 |
| S12_k141_188795 | 1087 | No | NA | 1 | 0 | 0 | Low-quality | Genome-fragment | 0.69 | AAI-based (high-confidence) | 0 |
| S11-final-viral-combined_k141_318242\|\|full | 5155 | No | NA | 5 | 0 | 3 | Not-determined | Genome-fragment | NA | NA | 0 |
| S11-final-viral-combined_k141_167054\|\|full | 2831 | No | NA | 3 | 0 | 1 | Not-determined | Genome-fragment | NA | NA | 0 |
| S11-final-viral-combined_k141_277171\|\|full | 3497 | No | NA | 4 | 0 | 3 | Not-determined | Genome-fragment | NA | NA | 0 |
| S2-final-viral-combined_k141_114733\|\|full | 1087 | No | NA | 3 | 0 | 0 | Low-quality | Genome-fragment | 2.55 | AAI-based (medium-confidence) | 0 |
| S11-final-viral-combined_k141_318255\|\|full | 6149 | No | NA | 5 | 0 | 2 | Not-determined | Genome-fragment | NA | NA | 0 |
| S2_k141_176191 | 1087 | No | NA | 4 | 0 | 0 | Low-quality | Genome-fragment | 1.68 | AAI-based (high-confidence) | 0 |
| S6-final-viral-combined_k141_31686\|\|full | 1087 | No | NA | 2 | 0 | 1 | Low-quality | Genome-fragment | 0.77 | AAI-based (medium-confidence) | 0 |
| S8-final-viral-combined_k141_108470\|\|full | 1087 | No | NA | 3 | 0 | 0 | Low-quality | Genome-fragment | 3.26 | AAI-based (high-confidence) | 0 |
| S8_k141_217131 | 1087 | No | NA | 2 | 0 | 0 | Low-quality | Genome-fragment | 2.28 | AAI-based (high-confidence) | 0 |
| S11-final-viral-combined_k141_111189\|\|full | 1088 | No | NA | 3 | 0 | 0 | Low-quality | Genome-fragment | 2.5 | AAI-based (medium-confidence) | 0 |
| S11-final-viral-combined_k141_290741\|\|full | 3672 | No | NA | 6 | 0 | 4 | Not-determined | Genome-fragment | NA | NA | 0 |
| S11-final-viral-combined_k141_249558\|\|full | 2159 | No | NA | 2 | 0 | 2 | Not-determined | Genome-fragment | NA | NA | 0 |
| S11-final-viral-combined_k141_159708\|\|lt2gene | 1088 | No | NA | 2 | 1 | 0 | Low-quality | Genome-fragment | 2.87 | AAI-based (medium-confidence) | 0 |
| S11_k141_148037 | 1088 | No | NA | 2 | 0 | 1 | Low-quality | Genome-fragment | 1.03 | AAI-based (medium-confidence) | 0 |
| S11-final-viral-combined_k141_221958\|\|full | 5904 | No | NA | 5 | 0 | 4 | Not-determined | Genome-fragment | NA | NA | 0 |
| S1-final-viral-combined_k141_47855\|\|full | 1088 | No | NA | 3 | 1 | 0 | Low-quality | Genome-fragment | 2.01 | HMM-based (lower-bound) | 0 |
| S11-final-viral-combined_k141_208250\|\|full | 7473 | No | NA | 7 | 0 | 1 | Not-determined | Genome-fragment | NA | NA | 0 |
| S1-final-viral-combined_k141_182328\|\|lt2gene | 1088 | No | NA | 2 | 1 | 0 | Low-quality | Genome-fragment | 1.76 | HMM-based (lower-bound) | 0 |
| S1_k141_332685 | 1088 | No | NA | 4 | 0 | 0 | Low-quality | Genome-fragment | 1.79 | AAI-based (high-confidence) | 0 |
| S11-final-viral-combined_k141_249597\|\|full | 1223 | No | NA | 2 | 0 | 0 | Not-determined | Genome-fragment | NA | NA | 0 |
| S11-final-viral-combined_k141_304791\|\|full | 1029 | No | NA | 2 | 0 | 1 | Not-determined | Genome-fragment | NA | NA | 0 |
| S3_k141_58021 | 1088 | No | NA | 2 | 0 | 0 | Low-quality | Genome-fragment | 3.14 | AAI-based (high-confidence) | 0 |
| S11-final-viral-combined_k141_42825\|\|full | 1129 | No | NA | 2 | 0 | 0 | Not-determined | Genome-fragment | NA | NA | 0 |
| S11-final-viral-combined_k141_249624\|\|full | 1192 | No | NA | 3 | 0 | 1 | Not-determined | Genome-fragment | NA | NA | 0 |
| S11-final-viral-combined_k141_1461\|\|full | 2336 | No | NA | 2 | 0 | 0 | Not-determined | Genome-fragment | NA | NA | 0 |
| S11-final-viral-combined_k141_167194\|\|full | 1582 | No | NA | 5 | 0 | 0 | Not-determined | Genome-fragment | NA | NA | 0 |
| S11-final-viral-combined_k141_111503\|\|full | 5123 | No | NA | 5 | 0 | 5 | Not-determined | Genome-fragment | NA | NA | 0 |
| S11-final-viral-combined_k141_290825\|\|full | 2823 | No | NA | 3 | 0 | 2 | Not-determined | Genome-fragment | NA | NA | 0 |
| S11-final-viral-combined_k141_318352\|\|full | 1465 | No | NA | 2 | 0 | 1 | Not-determined | Genome-fragment | NA | NA | 0 |
| S11-final-viral-combined_k141_236146\|\|full | 2472 | No | NA | 4 | 0 | 0 | Not-determined | Genome-fragment | NA | NA | 0 |
| S11-final-viral-combined_k141_236165\|\|full | 2736 | No | NA | 3 | 0 | 2 | Not-determined | Genome-fragment | NA | NA | 0 |
| S11-final-viral-combined_k141_125755\|\|full | 5174 | No | NA | 5 | 0 | 4 | Not-determined | Genome-fragment | NA | NA | 0 |
| S11-final-viral-combined_k141_180631\|\|full | 1580 | No | NA | 4 | 0 | 0 | Not-determined | Genome-fragment | NA | NA | 0 |
| S11-final-viral-combined_k141_318374\|\|full | 1204 | No | NA | 2 | 0 | 0 | Not-determined | Genome-fragment | NA | NA | 0 |
| S11-final-viral-combined_k141_208315\|\|full | 8699 | No | NA | 8 | 0 | 3 | Not-determined | Genome-fragment | NA | NA | 0 |
| S11-final-viral-combined_k141_290886\|\|full | 1082 | No | NA | 3 | 0 | 1 | Not-determined | Genome-fragment | NA | NA | 0 |
| S11-final-viral-combined_k141_29048\|\|full | 1418 | No | NA | 4 | 0 | 1 | Not-determined | Genome-fragment | NA | NA | 0 |
| S11-final-viral-combined_k141_180646\|\|full | 4824 | No | NA | 3 | 0 | 2 | Not-determined | Genome-fragment | NA | NA | 0 |
| S3_k141_54716 | 1088 | No | NA | 2 | 0 | 0 | Low-quality | Genome-fragment | 2.65 | AAI-based (high-confidence) | 0 |
| S11-final-viral-combined_k141_222067\|\|full | 7653 | No | NA | 8 | 0 | 4 | Not-determined | Genome-fragment | NA | NA | 0 |
| S11-final-viral-combined_k141_98321\|\|full | 1071 | No | NA | 3 | 0 | 3 | Not-determined | Genome-fragment | NA | NA | 0 |
| S7-final-viral-combined_k141_113827\|\|full | 1088 | No | NA | 2 | 0 | 0 | Low-quality | Genome-fragment | 1.48 | AAI-based (medium-confidence) | 0 |
| S11-final-viral-combined_k141_194482\|\|full | 2147 | No | NA | 3 | 0 | 1 | Not-determined | Genome-fragment | NA | NA | 0 |
| S11-final-viral-combined_k141_277321\|\|full | 7007 | No | NA | 12 | 0 | 0 | Not-determined | Genome-fragment | NA | NA | 0 |
| S11-final-viral-combined_k141_277325\|\|full | 2426 | No | NA | 3 | 0 | 1 | Not-determined | Genome-fragment | NA | NA | 0 |
| S11-final-viral-combined_k141_1594\|\|full | 7166 | No | NA | 7 | 0 | 3 | Not-determined | Genome-fragment | NA | NA | 0 |
| S11-final-viral-combined_k141_84326\|\|full | 23591 | No | NA | 38 | 16 | 0 | Medium-quality | Genome-fragment | 53.16 | AAI-based (medium-confidence) | 0 |
| S11-final-viral-combined_k141_263485\|\|full | 3947 | No | NA | 2 | 0 | 1 | Not-determined | Genome-fragment | NA | NA | 0 |
| S11-final-viral-combined_k141_139196\|\|full | 2052 | No | NA | 3 | 0 | 2 | Not-determined | Genome-fragment | NA | NA | 0 |
| S7-final-viral-combined_k141_218744\|\|full | 1088 | No | NA | 3 | 0 | 0 | Low-quality | Genome-fragment | 2.55 | AAI-based (high-confidence) | 0 |
| S11-final-viral-combined_k141_290937\|\|full | 1379 | No | NA | 3 | 0 | 0 | Not-determined | Genome-fragment | NA | NA | 0 |
| S11-final-viral-combined_k141_304863\|\|full | 1960 | No | NA | 3 | 0 | 0 | Not-determined | Genome-fragment | NA | NA | 0 |
| S11-final-viral-combined_k141_180685\|\|full | 9701 | No | NA | 14 | 0 | 6 | Not-determined | Genome-fragment | NA | NA | 0 |
| S11-final-viral-combined_k141_15480\|\|full | 1784 | No | NA | 2 | 0 | 1 | Not-determined | Genome-fragment | NA | NA | 0 |
| S11-final-viral-combined_k141_194525\|\|full | 2249 | No | NA | 3 | 0 | 0 | Not-determined | Genome-fragment | NA | NA | 0 |
| S11-final-viral-combined_k141_84363\|\|full | 1336 | No | NA | 2 | 0 | 2 | Not-determined | Genome-fragment | NA | NA | 0 |
| S11-final-viral-combined_k141_208394\|\|full | 2383 | No | NA | 3 | 0 | 0 | Not-determined | Genome-fragment | NA | NA | 0 |
| S11-final-viral-combined_k141_263516\|\|full | 1363 | No | NA | 4 | 0 | 0 | Not-determined | Genome-fragment | NA | NA | 0 |
| S11-final-viral-combined_k141_84378\|\|full | 5350 | No | NA | 5 | 0 | 3 | Not-determined | Genome-fragment | NA | NA | 0 |
| S11-final-viral-combined_k141_290980\|\|full | 3348 | No | NA | 4 | 0 | 1 | Not-determined | Genome-fragment | NA | NA | 0 |
| S7-final-viral-combined_k141_118922\|\|full | 1088 | No | NA | 3 | 1 | 0 | Low-quality | Genome-fragment | 2.46 | AAI-based (medium-confidence) | 0 |
| S11-final-viral-combined_k141_222198\|\|full | 4449 | No | NA | 3 | 0 | 0 | Not-determined | Genome-fragment | NA | NA | 0 |
| S7-final-viral-combined_k141_149122\|\|lt2gene | 1088 | No | NA | 1 | 1 | 0 | Low-quality | Genome-fragment | 0.97 | HMM-based (lower-bound) | 0 |
| S17-final-viral-combined_k141_27848\|\|full | 1088 | No | NA | 2 | 0 | 0 | Low-quality | Genome-fragment | 2.68 | AAI-based (high-confidence) | 0 |
| S12-final-viral-combined_k141_252526\|\|full | 1088 | No | NA | 2 | 0 | 0 | Low-quality | Genome-fragment | 1.63 | AAI-based (medium-confidence) | 0 |
| S12-final-viral-combined_k141_262263\|\|full | 1088 | No | NA | 3 | 0 | 0 | Low-quality | Genome-fragment | 1.99 | AAI-based (high-confidence) | 0 |
| S11-final-viral-combined_k141_236214\|\|full | 93358 | Yes | 32851 | 103 | 22 | 50 | Medium-quality | Genome-fragment | 54.08 | AAI-based (high-confidence) | 64.81 |
| S11-final-viral-combined_k141_291042\|\|full | 4908 | No | NA | 3 | 0 | 2 | Not-determined | Genome-fragment | NA | NA | 0 |
| S12-final-viral-combined_k141_63862\|\|full | 1088 | No | NA | 3 | 1 | 0 | Low-quality | Genome-fragment | 1.77 | AAI-based (medium-confidence) | 0 |
| S12-final-viral-combined_k141_204828\|\|lt2gene | 1088 | No | NA | 2 | 1 | 0 | Low-quality | Genome-fragment | 1.99 | HMM-based (lower-bound) | 0 |
| S2-final-viral-combined_k141_16642\|\|full | 1088 | No | NA | 2 | 0 | 1 | Low-quality | Genome-fragment | 1.18 | AAI-based (high-confidence) | 0 |
| S11-final-viral-combined_k141_15586\|\|full | 3064 | No | NA | 4 | 0 | 2 | Not-determined | Genome-fragment | NA | NA | 0 |
| S11-final-viral-combined_k141_84433\|\|full | 1136 | No | NA | 2 | 0 | 1 | Not-determined | Genome-fragment | NA | NA | 0 |
| S2-final-viral-combined_k141_62857\|\|lt2gene | 1088 | No | NA | 1 | 1 | 0 | Low-quality | Genome-fragment | 1.42 | HMM-based (lower-bound) | 0 |
| S11-final-viral-combined_k141_42928\|\|full | 2884 | No | NA | 5 | 0 | 0 | Not-determined | Genome-fragment | NA | NA | 0 |
| S4-final-viral-combined_k141_107661\|\|full | 1088 | No | NA | 3 | 0 | 1 | Low-quality | Genome-fragment | 0.93 | AAI-based (medium-confidence) | 0 |
| S8-final-viral-combined_k141_83045\|\|full | 1088 | No | NA | 2 | 2 | 0 | Low-quality | Genome-fragment | 2.66 | AAI-based (medium-confidence) | 0 |
| S9-final-viral-combined_k141_96349\|\|full | 1089 | No | NA | 2 | 2 | 0 | Low-quality | Genome-fragment | 3.02 | AAI-based (high-confidence) | 0 |
| S9-final-viral-combined_k141_118173\|\|lt2gene | 1089 | No | NA | 2 | 1 | 0 | Low-quality | Genome-fragment | 2.21 | AAI-based (medium-confidence) | 0 |
| S11-final-viral-combined_k141_227860\|\|full | 1089 | No | NA | 2 | 0 | 0 | Low-quality | Genome-fragment | 1.27 | AAI-based (high-confidence) | 0 |
| S11_k141_166508 | 1089 | No | NA | 2 | 0 | 2 | Low-quality | Genome-fragment | 2.84 | AAI-based (high-confidence) | 0 |
| S1-final-viral-combined_k141_57600\|\|full | 1089 | No | NA | 3 | 0 | 1 | Low-quality | Genome-fragment | 0.84 | AAI-based (medium-confidence) | 0 |
| S11-final-viral-combined_k141_236254\|\|full | 5795 | No | NA | 10 | 0 | 0 | Not-determined | Genome-fragment | NA | NA | 0 |
| S11-final-viral-combined_k141_70349\|\|full | 1364 | No | NA | 3 | 0 | 2 | Not-determined | Genome-fragment | NA | NA | 0 |
| S11-final-viral-combined_k141_139335\|\|full | 3106 | No | NA | 3 | 0 | 3 | Not-determined | Genome-fragment | NA | NA | 0 |
| S11-final-viral-combined_k141_1702\|\|full | 1755 | No | NA | 3 | 0 | 0 | Not-determined | Genome-fragment | NA | NA | 0 |
| S11-final-viral-combined_k141_167421\|\|full | 1227 | No | NA | 2 | 0 | 0 | Not-determined | Genome-fragment | NA | NA | 0 |
| S7-final-viral-combined_k141_127004\|\|full | 1089 | No | NA | 3 | 0 | 0 | Low-quality | Genome-fragment | 1.87 | AAI-based (high-confidence) | 0 |
| S7_k141_153329 | 1089 | No | NA | 1 | 0 | 1 | Low-quality | Genome-fragment | 2.78 | AAI-based (medium-confidence) | 0 |
| S17-final-viral-combined_k141_190984\|\|full | 1089 | No | NA | 3 | 0 | 0 | Low-quality | Genome-fragment | 1.09 | AAI-based (medium-confidence) | 0 |
| S17-final-viral-combined_k141_224844\|\|full | 1089 | No | NA | 2 | 0 | 1 | Low-quality | Genome-fragment | 0.95 | AAI-based (medium-confidence) | 0 |
| S11-final-viral-combined_k141_304985\|\|full | 20916 | No | NA | 25 | 0 | 12 | Not-determined | Genome-fragment | NA | NA | 0 |
| S17-k141_102882 | 1089 | No | NA | 3 | 1 | 0 | Low-quality | Genome-fragment | 1.47 | AAI-based (high-confidence) | 0 |
| S11-final-viral-combined_k141_222289\|\|full | 3755 | No | NA | 4 | 0 | 4 | Not-determined | Genome-fragment | NA | NA | 0 |
| S4-final-viral-combined_k141_66946\|\|lt2gene | 1089 | No | NA | 2 | 0 | 0 | Low-quality | Genome-fragment | 2.44 | AAI-based (medium-confidence) | 0 |
| S11-final-viral-combined_k141_167448\|\|full | 2472 | No | NA | 2 | 0 | 2 | Not-determined | Genome-fragment | NA | NA | 0 |
| S4_k141_140390 | 1089 | No | NA | 1 | 0 | 0 | Low-quality | Genome-fragment | 2.76 | AAI-based (high-confidence) | 0 |
| S11-final-viral-combined_k141_98435\|\|full | 1658 | No | NA | 3 | 0 | 2 | Not-determined | Genome-fragment | NA | NA | 0 |
| S11-final-viral-combined_k141_43002\|\|full | 1570 | No | NA | 3 | 0 | 0 | Not-determined | Genome-fragment | NA | NA | 0 |
| S4_k141_127999 | 1089 | No | NA | 2 | 0 | 0 | Low-quality | Genome-fragment | 2.29 | AAI-based (medium-confidence) | 0 |
| S11-final-viral-combined_k141_43020\|\|full | 1574 | No | NA | 3 | 0 | 0 | Not-determined | Genome-fragment | NA | NA | 0 |
| S8-final-viral-combined_k141_142404\|\|full | 1089 | No | NA | 2 | 0 | 1 | Low-quality | Genome-fragment | 1.64 | AAI-based (medium-confidence) | 0 |
| S11-final-viral-combined_k141_1715\|\|full | 2274 | No | NA | 4 | 0 | 2 | Not-determined | Genome-fragment | NA | NA | 0 |
| S11-final-viral-combined_k141_98447\|\|full | 8569 | No | NA | 10 | 0 | 4 | Not-determined | Genome-fragment | NA | NA | 0 |
| S11-final-viral-combined_k141_56859\|\|full | 3570 | No | NA | 5 | 0 | 1 | Not-determined | Genome-fragment | NA | NA | 0 |
| S11-final-viral-combined_k141_111715\|\|full | 2297 | No | NA | 3 | 0 | 1 | Not-determined | Genome-fragment | NA | NA | 0 |
| S9_k141_47737 | 1090 | No | NA | 1 | 0 | 0 | Low-quality | Genome-fragment | 0.85 | AAI-based (high-confidence) | 0 |
| S11-final-viral-combined_k141_180865\|\|full | 3308 | No | NA | 3 | 0 | 1 | Not-determined | Genome-fragment | NA | NA | 0 |
| S11-final-viral-combined_k141_98465\|\|full | 5033 | No | NA | 6 | 0 | 6 | Not-determined | Genome-fragment | NA | NA | 0 |
| S11-final-viral-combined_k141_246626\|\|full | 1090 | No | NA | 2 | 0 | 0 | Low-quality | Genome-fragment | 0.98 | AAI-based (medium-confidence) | 0 |
| S1-final-viral-combined_k141_355791\|\|full | 1090 | No | NA | 3 | 1 | 0 | Low-quality | Genome-fragment | 2.02 | AAI-based (high-confidence) | 0 |
| S1-final-viral-combined_k141_224840\|\|full | 1090 | No | NA | 2 | 1 | 0 | Low-quality | Genome-fragment | 1.25 | HMM-based (lower-bound) | 0 |
| S1_k141_247084 | 1090 | No | NA | 3 | 0 | 0 | Low-quality | Genome-fragment | 2.33 | AAI-based (high-confidence) | 0 |
| S1_k141_361946 | 1090 | No | NA | 1 | 0 | 0 | Low-quality | Genome-fragment | 0.85 | AAI-based (medium-confidence) | 0 |
| S5-final-viral-combined_k141_67957\|\|full | 1090 | No | NA | 2 | 0 | 1 | Low-quality | Genome-fragment | 1.33 | AAI-based (medium-confidence) | 0 |
| S11-final-viral-combined_k141_125887\|\|full | 2383 | No | NA | 4 | 0 | 4 | Not-determined | Genome-fragment | NA | NA | 0 |
| S7-final-viral-combined_k141_58301\|\|full | 1090 | No | NA | 2 | 0 | 0 | Low-quality | Genome-fragment | 0.42 | AAI-based (high-confidence) | 0 |
| S11-final-viral-combined_k141_84587\|\|full | 3623 | No | NA | 5 | 0 | 3 | Not-determined | Genome-fragment | NA | NA | 0 |
| S7-final-viral-combined_k141_220265\|\|lt2gene | 1090 | No | NA | 2 | 1 | 0 | Low-quality | Genome-fragment | 3.05 | AAI-based (high-confidence) | 0 |
| S11-final-viral-combined_k141_98540\|\|full | 3233 | No | NA | 2 | 0 | 0 | Not-determined | Genome-fragment | NA | NA | 0 |
| S11-final-viral-combined_k141_305018\|\|full | 3869 | No | NA | 7 | 0 | 4 | Not-determined | Genome-fragment | NA | NA | 0 |
| S7-final-viral-combined_k141_60396\|\|lt2gene | 1090 | No | NA | 1 | 1 | 0 | Low-quality | Genome-fragment | 1.43 | HMM-based (lower-bound) | 0 |
| S17-final-viral-combined_k141_214800\|\|lt2gene | 1090 | No | NA | 2 | 1 | 0 | Low-quality | Genome-fragment | 2.44 | AAI-based (medium-confidence) | 0 |
| S11-final-viral-combined_k141_222372\|\|full | 1061 | No | NA | 2 | 0 | 0 | Not-determined | Genome-fragment | NA | NA | 0 |
| S11-final-viral-combined_k141_222378\|\|full | 1909 | No | NA | 2 | 0 | 2 | Not-determined | Genome-fragment | NA | NA | 0 |
| S11-final-viral-combined_k141_29236\|\|full | 3373 | No | NA | 3 | 0 | 2 | Not-determined | Genome-fragment | NA | NA | 0 |
| S11-final-viral-combined_k141_236390\|\|full | 1922 | No | NA | 3 | 0 | 3 | Not-determined | Genome-fragment | NA | NA | 0 |
| S12-final-viral-combined_k141_361210\|\|full | 1090 | No | NA | 2 | 0 | 0 | Low-quality | Genome-fragment | 1.99 | AAI-based (high-confidence) | 0 |
| S11-final-viral-combined_k141_153428\|\|full | 2225 | No | NA | 2 | 0 | 1 | Not-determined | Genome-fragment | NA | NA | 0 |
| S11-final-viral-combined_k141_277598\|\|full | 1242 | No | NA | 3 | 0 | 1 | Not-determined | Genome-fragment | NA | NA | 0 |
| S11-final-viral-combined_k141_208690\|\|full | 3963 | No | NA | 4 | 0 | 3 | Not-determined | Genome-fragment | NA | NA | 0 |
| S11-final-viral-combined_k141_291200\|\|full | 6314 | No | NA | 3 | 0 | 0 | Not-determined | Genome-fragment | NA | NA | 0 |
| S11-final-viral-combined_k141_153451\|\|full | 4906 | No | NA | 3 | 0 | 1 | Not-determined | Genome-fragment | NA | NA | 0 |
| S11-final-viral-combined_k141_139530\|\|full | 3908 | No | NA | 6 | 0 | 2 | Not-determined | Genome-fragment | NA | NA | 0 |
| S12_k141_369377 | 1090 | No | NA | 3 | 0 | 1 | Low-quality | Genome-fragment | 2.08 | AAI-based (medium-confidence) | 0 |
| S11-final-viral-combined_k141_194736\|\|full | 2452 | No | NA | 4 | 0 | 2 | Not-determined | Genome-fragment | NA | NA | 0 |
| S11-final-viral-combined_k141_98600\|\|full | 1525 | No | NA | 4 | 0 | 0 | Not-determined | Genome-fragment | NA | NA | 0 |
| S2-final-viral-combined_k141_21370\|\|full | 1090 | No | NA | 2 | 0 | 1 | Low-quality | Genome-fragment | 2.82 | AAI-based (medium-confidence) | 0 |
| S11-final-viral-combined_k141_15754\|\|full | 1853 | No | NA | 3 | 0 | 3 | Not-determined | Genome-fragment | NA | NA | 0 |
| S6-final-viral-combined_k141_17019\|\|full | 1090 | No | NA | 2 | 0 | 1 | Low-quality | Genome-fragment | 1.52 | AAI-based (medium-confidence) | 0 |
| S9-final-viral-combined_k141_84388\|\|full | 1091 | No | NA | 3 | 1 | 0 | Low-quality | Genome-fragment | 2.64 | AAI-based (high-confidence) | 0 |
| S11-final-viral-combined_k141_249860\|\|full | 1702 | No | NA | 1 | 0 | 1 | Not-determined | Genome-fragment | NA | NA | 0 |
| S11-final-viral-combined_k141_111844\|\|full | 5929 | No | NA | 5 | 0 | 4 | Not-determined | Genome-fragment | NA | NA | 0 |
| S11-final-viral-combined_k141_62717\|\|full | 1091 | No | NA | 3 | 0 | 0 | Low-quality | Genome-fragment | 2.46 | AAI-based (high-confidence) | 0 |
| S11-final-viral-combined_k141_43189\|\|full | 1365 | No | NA | 3 | 0 | 1 | Not-determined | Genome-fragment | NA | NA | 0 |
| S11-final-viral-combined_k141_291272\|\|full | 2536 | No | NA | 2 | 0 | 2 | Not-determined | Genome-fragment | NA | NA | 0 |
| S11-final-viral-combined_k141_57082\|\|full | 1033 | No | NA | 2 | 0 | 2 | Not-determined | Genome-fragment | NA | NA | 0 |
| S1-final-viral-combined_k141_329671\|\|full | 1091 | No | NA | 3 | 1 | 0 | Low-quality | Genome-fragment | 0.67 | HMM-based (lower-bound) | 0 |
| S11-final-viral-combined_k141_167608\|\|full | 2858 | No | NA | 6 | 0 | 5 | Not-determined | Genome-fragment | NA | NA | 0 |
| S1-final-viral-combined_k141_53686\|\|full | 1091 | No | NA | 2 | 2 | 0 | Low-quality | Genome-fragment | 2.25 | AAI-based (high-confidence) | 0 |
| S11-final-viral-combined_k141_15775\|\|full | 17898 | No | NA | 15 | 0 | 6 | Not-determined | Genome-fragment | NA | NA | 0 |
| S11-final-viral-combined_k141_277658\|\|full | 2099 | No | NA | 2 | 0 | 2 | Not-determined | Genome-fragment | NA | NA | 0 |
| S5-final-viral-combined_k141_187736\|\|lt2gene | 1091 | No | NA | 2 | 1 | 0 | Low-quality | Genome-fragment | 2.67 | AAI-based (high-confidence) | 0 |
| S17-final-viral-combined_k141_213443\|\|full | 1091 | No | NA | 2 | 1 | 0 | Low-quality | Genome-fragment | 2.67 | AAI-based (high-confidence) | 0 |
| S11-final-viral-combined_k141_111868\|\|full | 2379 | No | NA | 3 | 0 | 0 | Not-determined | Genome-fragment | NA | NA | 0 |
| S17-final-viral-combined_k141_118195\|\|full | 1091 | No | NA | 3 | 0 | 0 | Low-quality | Genome-fragment | 1.33 | AAI-based (high-confidence) | 0 |
| S11-final-viral-combined_k141_181029\|\|full | 2467 | No | NA | 3 | 0 | 1 | Not-determined | Genome-fragment | NA | NA | 0 |
| S11-final-viral-combined_k141_249929\|\|full | 3063 | No | NA | 3 | 0 | 1 | Not-determined | Genome-fragment | NA | NA | 0 |
| S12-final-viral-combined_k141_117327\|\|lt2gene | 1091 | No | NA | 2 | 1 | 0 | Low-quality | Genome-fragment | 1.79 | AAI-based (high-confidence) | 0 |
| S11-final-viral-combined_k141_305209\|\|full | 1496 | No | NA | 3 | 0 | 3 | Not-determined | Genome-fragment | NA | NA | 0 |
| S11-final-viral-combined_k141_291313\|\|full | 10306 | No | NA | 4 | 0 | 1 | Not-determined | Genome-fragment | NA | NA | 0 |
| S11-final-viral-combined_k141_98673\|\|full | 2957 | No | NA | 3 | 0 | 3 | Not-determined | Genome-fragment | NA | NA | 0 |
| S12-final-viral-combined_k141_16617\|\|lt2gene | 1091 | No | NA | 2 | 2 | 0 | Low-quality | Genome-fragment | 1.96 | AAI-based (high-confidence) | 0 |
| S11-final-viral-combined_k141_1906\|\|full | 1647 | No | NA | 3 | 0 | 0 | Not-determined | Genome-fragment | NA | NA | 0 |
| S2-final-viral-combined_k141_244126\|\|full | 1091 | No | NA | 2 | 0 | 0 | Low-quality | Genome-fragment | 0.74 | AAI-based (medium-confidence) | 0 |
| S2_k141_129732 | 1091 | No | NA | 3 | 0 | 0 | Low-quality | Genome-fragment | 1.82 | AAI-based (high-confidence) | 0 |
| S11-final-viral-combined_k141_318741\|\|full | 1926 | No | NA | 4 | 0 | 0 | Not-determined | Genome-fragment | NA | NA | 0 |
| S4-final-viral-combined_k141_152272\|\|full | 1091 | No | NA | 2 | 0 | 0 | Low-quality | Genome-fragment | 2.51 | AAI-based (medium-confidence) | 0 |
| S11-final-viral-combined_k141_236538\|\|full | 1224 | No | NA | 2 | 0 | 0 | Not-determined | Genome-fragment | NA | NA | 0 |
| S11-final-viral-combined_k141_318748\|\|full | 2564 | No | NA | 4 | 0 | 1 | Not-determined | Genome-fragment | NA | NA | 0 |
| S11-final-viral-combined_k141_249989\|\|full | 1055 | No | NA | 2 | 0 | 0 | Not-determined | Genome-fragment | NA | NA | 0 |
| S11-final-viral-combined_k141_318761\|\|full | 2304 | No | NA | 2 | 0 | 1 | Not-determined | Genome-fragment | NA | NA | 0 |
| S11-final-viral-combined_k141_167712\|\|full | 2996 | No | NA | 3 | 0 | 1 | Not-determined | Genome-fragment | NA | NA | 0 |
| S4_k141_49353 | 1091 | No | NA | 1 | 0 | 0 | Low-quality | Genome-fragment | 0.44 | AAI-based (medium-confidence) | 0 |
| S6_k141_100262 | 1091 | No | NA | 3 | 1 | 0 | Low-quality | Genome-fragment | 2.25 | AAI-based (medium-confidence) | 0 |
| S11-final-viral-combined_k141_98730\|\|full | 5675 | No | NA | 6 | 0 | 3 | Not-determined | Genome-fragment | NA | NA | 0 |
| S8-final-viral-combined_k141_174057\|\|full | 1091 | No | NA | 3 | 0 | 0 | Low-quality | Genome-fragment | 2.06 | AAI-based (medium-confidence) | 0 |
| S11-final-viral-combined_k141_84828\|\|full | 1111 | No | NA | 3 | 0 | 2 | Not-determined | Genome-fragment | NA | NA | 0 |
| S9-final-viral-combined_k141_64693\|\|lt2gene | 1092 | No | NA | 2 | 1 | 0 | Low-quality | Genome-fragment | 2.96 | AAI-based (high-confidence) | 0 |
| S11-final-viral-combined_k141_263865\|\|full | 1537 | No | NA | 2 | 0 | 2 | Not-determined | Genome-fragment | NA | NA | 0 |

| | | | | | | | | | | | |
|---|---|---|---|---|---|---|---|---|---|---|---|
| S9_k141_59339 | 1092 | No | NA | 2 | 0 | 0 | Low-quality | Genome-fragment | 1.82 | AAI-based (high-confidence) | 0 |
| S11-final-viral-combined_k141_208956\|\|full | 2340 | No | NA | 4 | 0 | 0 | Not-determined | Genome-fragment | NA | NA | 0 |
| S11-final-viral-combined_k141_291410\|\|full | 1489 | No | NA | 2 | 0 | 1 | Not-determined | Genome-fragment | NA | NA | 0 |
| S9_k141_12863 | 1092 | No | NA | 3 | 2 | 0 | Low-quality | Genome-fragment | 2.66 | AAI-based (high-confidence) | 0 |
| S11-final-viral-combined_k141_43337\|\|full | 4912 | No | NA | 5 | 0 | 2 | Not-determined | Genome-fragment | NA | NA | 0 |
| S11-final-viral-combined_k141_263889\|\|full | 4397 | No | NA | 6 | 0 | 2 | Not-determined | Genome-fragment | NA | NA | 0 |
| S11-final-viral-combined_k141_160037\|\|lt2gene | 1092 | No | NA | 2 | 1 | 0 | Low-quality | Genome-fragment | 2.74 | AAI-based (high-confidence) | 0 |
| S11-final-viral-combined_k141_98795\|\|full | 2152 | No | NA | 3 | 0 | 0 | Not-determined | Genome-fragment | NA | NA | 0 |
| S11_k141_55103 | 1092 | No | NA | 2 | 0 | 0 | Low-quality | Genome-fragment | 1.64 | AAI-based (medium-confidence) | 0 |
| S11-final-viral-combined_k141_236617\|\|full | 1236 | No | NA | 2 | 0 | 0 | Not-determined | Genome-fragment | NA | NA | 0 |
| S1-final-viral-combined_k141_154027\|\|lt2gene | 1092 | No | NA | 1 | 1 | 0 | Low-quality | Genome-fragment | 2.06 | AAI-based (high-confidence) | 0 |
| S11-final-viral-combined_k141_305298\|\|full | 1117 | No | NA | 2 | 0 | 0 | Not-determined | Genome-fragment | NA | NA | 0 |
| S11-final-viral-combined_k141_305321\|\|full | 1003 | No | NA | 3 | 0 | 1 | Not-determined | Genome-fragment | NA | NA | 0 |
| S11-final-viral-combined_k141_43400\|\|full | 3299 | No | NA | 5 | 0 | 2 | Not-determined | Genome-fragment | NA | NA | 0 |
| S11-final-viral-combined_k141_70453\|\|full | 1029 | No | NA | 4 | 0 | 0 | Not-determined | Genome-fragment | NA | NA | 0 |
| S11-final-viral-combined_k141_277905\|\|full | 2919 | No | NA | 3 | 0 | 2 | Not-determined | Genome-fragment | NA | NA | 0 |
| S11-final-viral-combined_k141_167763\|\|full | 1135 | No | NA | 3 | 0 | 0 | Not-determined | Genome-fragment | NA | NA | 0 |
| S11-final-viral-combined_k141_291465\|\|full | 1188 | No | NA | 3 | 0 | 1 | Not-determined | Genome-fragment | NA | NA | 0 |
| S1_k141_159061 | 1092 | No | NA | 2 | 0 | 0 | Low-quality | Genome-fragment | 0.26 | AAI-based (medium-confidence) | 0 |
| S11-final-viral-combined_k141_209025\|\|full | 1289 | No | NA | 3 | 0 | 2 | Not-determined | Genome-fragment | NA | NA | 0 |
| S11-final-viral-combined_k141_43402\|\|full | 12229 | No | NA | 11 | 0 | 4 | Not-determined | Genome-fragment | NA | NA | 0 |
| S11-final-viral-combined_k141_236712\|\|full | 1082 | No | NA | 2 | 0 | 1 | Not-determined | Genome-fragment | NA | NA | 0 |
| S3_k141_45297 | 1092 | No | NA | 2 | 0 | 0 | Low-quality | Genome-fragment | 0.5 | AAI-based (high-confidence) | 0 |
| S11-final-viral-combined_k141_236721\|\|full | 2957 | No | NA | 2 | 0 | 1 | Not-determined | Genome-fragment | NA | NA | 0 |
| S11-final-viral-combined_k141_305367\|\|full | 9795 | No | NA | 9 | 0 | 6 | Not-determined | Genome-fragment | NA | NA | 0 |
| S5-final-viral-combined_k141_17875\|\|lt2gene | 1092 | No | NA | 1 | 1 | 0 | Low-quality | Genome-fragment | 1.81 | HMM-based (lower-bound) | 0 |
| S11-final-viral-combined_k141_195011\|\|full | 1524 | No | NA | 3 | 0 | 1 | Not-determined | Genome-fragment | NA | NA | 0 |
| S5-final-viral-combined_k141_189541\|\|lt2gene | 1092 | No | NA | 1 | 1 | 0 | Low-quality | Genome-fragment | 0.65 | AAI-based (high-confidence) | 0 |
| S17-k141_129812 | 1092 | No | NA | 3 | 3 | 0 | Low-quality | Genome-fragment | 2.55 | AAI-based (high-confidence) | 0 |
| S11-final-viral-combined_k141_70468\|\|full | 2208 | No | NA | 2 | 0 | 1 | Not-determined | Genome-fragment | NA | NA | 0 |
| S11-final-viral-combined_k141_291493\|\|full | 11081 | No | NA | 10 | 0 | 8 | Not-determined | Genome-fragment | NA | NA | 0 |
| S11-final-viral-combined_k141_139879\|\|full | 6832 | No | NA | 5 | 0 | 3 | Not-determined | Genome-fragment | NA | NA | 0 |
| S11-final-viral-combined_k141_57355\|\|full | 4273 | No | NA | 8 | 0 | 1 | Not-determined | Genome-fragment | NA | NA | 0 |
| S11-final-viral-combined_k141_195033\|\|full | 1242 | No | NA | 2 | 0 | 0 | Not-determined | Genome-fragment | NA | NA | 0 |
| S11-final-viral-combined_k141_84925\|\|full | 2743 | No | NA | 3 | 0 | 0 | Not-determined | Genome-fragment | NA | NA | 0 |
| S11-final-viral-combined_k141_181217\|\|full | 2259 | No | NA | 3 | 0 | 3 | Not-determined | Genome-fragment | NA | NA | 0 |
| S11-final-viral-combined_k141_153678\|\|full | 4348 | No | NA | 3 | 0 | 2 | Not-determined | Genome-fragment | NA | NA | 0 |
| S11-final-viral-combined_k141_16103\|\|full | 2391 | No | NA | 2 | 0 | 1 | Not-determined | Genome-fragment | NA | NA | 0 |
| S2-final-viral-combined_k141_101970\|\|full | 1092 | No | NA | 2 | 0 | 0 | Low-quality | Genome-fragment | 0.94 | AAI-based (high-confidence) | 0 |
| S11-final-viral-combined_k141_84956\|\|full | 1375 | No | NA | 2 | 0 | 2 | Not-determined | Genome-fragment | NA | NA | 0 |
| S2-final-viral-combined_k141_194839\|\|full | 1092 | No | NA | 3 | 0 | 1 | Low-quality | Genome-fragment | 1.36 | AAI-based (medium-confidence) | 0 |
| S11-final-viral-combined_k141_112100\|\|full | 1866 | No | NA | 3 | 0 | 2 | Not-determined | Genome-fragment | NA | NA | 0 |
| S11-final-viral-combined_k141_222729\|\|full | 1271 | No | NA | 2 | 0 | 1 | Not-determined | Genome-fragment | NA | NA | 0 |
| S11-final-viral-combined_k141_167828\|\|full | 1709 | No | NA | 2 | 0 | 1 | Not-determined | Genome-fragment | NA | NA | 0 |
| S2-final-viral-combined_k141_111400\|\|full | 1092 | No | NA | 3 | 0 | 0 | Low-quality | Genome-fragment | 2.67 | AAI-based (medium-confidence) | 0 |
| S11-final-viral-combined_k141_278092\|\|full | 1577 | No | NA | 2 | 0 | 0 | Not-determined | Genome-fragment | NA | NA | 0 |
| S2_k141_238357 | 1092 | No | NA | 3 | 1 | 0 | Low-quality | Genome-fragment | 2.81 | AAI-based (high-confidence) | 0 |
| S11-final-viral-combined_k141_57407\|\|full | 7371 | No | NA | 8 | 0 | 5 | Not-determined | Genome-fragment | NA | NA | 0 |
| S11-final-viral-combined_k141_139912\|\|full | 1417 | No | NA | 3 | 0 | 0 | Not-determined | Genome-fragment | NA | NA | 0 |
| S11-final-viral-combined_k141_319018\|\|full | 3487 | No | NA | 3 | 0 | 0 | Not-determined | Genome-fragment | NA | NA | 0 |
| S11-final-viral-combined_k141_57412\|\|full | 1996 | No | NA | 3 | 0 | 2 | Not-determined | Genome-fragment | NA | NA | 0 |
| S11-final-viral-combined_k141_70570\|\|full | 1184 | No | NA | 3 | 0 | 0 | Not-determined | Genome-fragment | NA | NA | 0 |
| S11-final-viral-combined_k141_236884\|\|full | 1821 | No | NA | 3 | 0 | 2 | Not-determined | Genome-fragment | NA | NA | 0 |
| S11-final-viral-combined_k141_236894\|\|full | 2215 | No | NA | 3 | 0 | 1 | Not-determined | Genome-fragment | NA | NA | 0 |
| S11-final-viral-combined_k141_264050\|\|full | 97251 | Yes | 49308 | 116 | 22 | 32 | Complete | High-quality | 100 | Provirus (high-confidence) | 49.3 |
| S4-final-viral-combined_k141_120179\|\|full | 1092 | No | NA | 3 | 0 | 2 | Low-quality | Genome-fragment | 1.48 | AAI-based (medium-confidence) | 0 |
| S11-final-viral-combined_k141_139935\|\|full | 10462 | No | NA | 8 | 0 | 3 | Not-determined | Genome-fragment | NA | NA | 0 |
| S11-final-viral-combined_k141_98951\|\|full | 1074 | No | NA | 3 | 0 | 1 | Not-determined | Genome-fragment | NA | NA | 0 |
| S8_k141_255645 | 1092 | No | NA | 1 | 0 | 1 | Low-quality | Genome-fragment | 0.39 | AAI-based (medium-confidence) | 0 |
| S9-final-viral-combined_k141_98497\|\|full | 1093 | No | NA | 3 | 1 | 0 | Low-quality | Genome-fragment | 2.9 | AAI-based (high-confidence) | 0 |
| S11-final-viral-combined_k141_156475\|\|full | 1093 | No | NA | 3 | 1 | 0 | Low-quality | Genome-fragment | 3.04 | AAI-based (medium-confidence) | 0 |
| S11-final-viral-combined_k141_133892\|\|full | 1093 | No | NA | 3 | 0 | 0 | Low-quality | Genome-fragment | 2.94 | AAI-based (medium-confidence) | 0 |
| S11-final-viral-combined_k141_2197\|\|full | 1493 | No | NA | 2 | 0 | 0 | Not-determined | Genome-fragment | NA | NA | 0 |
| S1-final-viral-combined_k141_257785\|\|full | 1093 | Yes | 880 | 4 | 1 | 1 | Low-quality | Genome-fragment | 2.66 | AAI-based (medium-confidence) | 19.49 |
| S11-final-viral-combined_k141_139941\|\|full | 20245 | No | NA | 17 | 0 | 11 | Not-determined | Genome-fragment | NA | NA | 0 |
| S1-final-viral-combined_k141_94176\|\|full | 1093 | No | NA | 2 | 0 | 1 | Low-quality | Genome-fragment | 1.37 | AAI-based (medium-confidence) | 0 |
| S3_k141_117633 | 1093 | No | NA | 4 | 0 | 0 | Low-quality | Genome-fragment | 2.22 | AAI-based (high-confidence) | 0 |
| S11-final-viral-combined_k141_209287\|\|full | 1199 | No | NA | 2 | 0 | 1 | Not-determined | Genome-fragment | NA | NA | 0 |
| S3_k141_78672 | 1093 | No | NA | 3 | 0 | 0 | Low-quality | Genome-fragment | 1.83 | AAI-based (medium-confidence) | 0 |
| S11-final-viral-combined_k141_209295\|\|full | 3044 | No | NA | 3 | 0 | 2 | Not-determined | Genome-fragment | NA | NA | 0 |
| S11-final-viral-combined_k141_126308\|\|full | 3276 | No | NA | 2 | 0 | 2 | Not-determined | Genome-fragment | NA | NA | 0 |
| S12-final-viral-combined_k141_151925\|\|full | 1093 | No | NA | 3 | 0 | 0 | Low-quality | Genome-fragment | 2.39 | AAI-based (high-confidence) | 0 |
| S12-final-viral-combined_k141_221613\|\|full | 1093 | No | NA | 3 | 0 | 0 | Low-quality | Genome-fragment | 2.2 | AAI-based (medium-confidence) | 0 |
| S11-final-viral-combined_k141_112147\|\|full | 19697 | No | NA | 16 | 3 | 0 | Medium-quality | Genome-fragment | 50.22 | AAI-based (high-confidence) | 0 |
| S2-final-viral-combined_k141_198978\|\|full | 1093 | No | NA | 2 | 0 | 1 | Low-quality | Genome-fragment | 0.36 | AAI-based (medium-confidence) | 0 |
| S11-final-viral-combined_k141_70697\|\|full | 2112 | No | NA | 2 | 0 | 2 | Not-determined | Genome-fragment | NA | NA | 0 |
| S11-final-viral-combined_k141_16155\|\|full | 3812 | No | NA | 5 | 0 | 3 | Not-determined | Genome-fragment | NA | NA | 0 |
| S2-final-viral-combined_k141_56830\|\|full | 1093 | No | NA | 2 | 0 | 0 | Low-quality | Genome-fragment | 1.93 | AAI-based (high-confidence) | 0 |
| S8-final-viral-combined_k141_243307\|\|full | 1093 | No | NA | 2 | 2 | 0 | Low-quality | Genome-fragment | 2.12 | AAI-based (medium-confidence) | 0 |
| S11-final-viral-combined_k141_126341\|\|full | 2990 | No | NA | 4 | 0 | 2 | Not-determined | Genome-fragment | NA | NA | 0 |
| S11-final-viral-combined_k141_126352\|\|full | 1957 | No | NA | 2 | 0 | 1 | Not-determined | Genome-fragment | NA | NA | 0 |
| S11-final-viral-combined_k141_319045\|\|full | 16235 | No | NA | 14 | 0 | 10 | Not-determined | Genome-fragment | NA | NA | 0 |
| S11-final-viral-combined_k141_2297\|\|full | 2622 | No | NA | 3 | 0 | 2 | Not-determined | Genome-fragment | NA | NA | 0 |
| S11-final-viral-combined_k141_264168\|\|full | 5957 | No | NA | 6 | 0 | 4 | Not-determined | Genome-fragment | NA | NA | 0 |
| S11-final-viral-combined_k141_305611\|\|full | 1626 | No | NA | 3 | 0 | 0 | Not-determined | Genome-fragment | NA | NA | 0 |
| S11-final-viral-combined_k141_29788\|\|full | 3329 | No | NA | 2 | 0 | 2 | Not-determined | Genome-fragment | NA | NA | 0 |
| S8-final-viral-combined_k141_67710\|\|lt2gene | 1093 | No | NA | 1 | 1 | 0 | Low-quality | Genome-fragment | 2.92 | AAI-based (high-confidence) | 0 |
| S11-final-viral-combined_k141_140009\|\|full | 54887 | Yes | 33397 | 59 | 22 | 14 | High-quality | High-quality | 96.44 | AAI-based (high-confidence) | 39.15 |
| S11-final-viral-combined_k141_2336\|\|full | 2731 | No | NA | 3 | 0 | 2 | Not-determined | Genome-fragment | NA | NA | 0 |
| S11-final-viral-combined_k141_4744\|\|full | 1094 | No | NA | 3 | 0 | 0 | Low-quality | Genome-fragment | 0.7 | AAI-based (medium-confidence) | 0 |
| S11-final-viral-combined_k141_29819\|\|full | 3968 | No | NA | 4 | 0 | 0 | Not-determined | Genome-fragment | NA | NA | 0 |
| S11-final-viral-combined_k141_209357\|\|full | 1440 | No | NA | 2 | 0 | 2 | Not-determined | Genome-fragment | NA | NA | 0 |
| S11-final-viral-combined_k141_140040\|\|full | 6389 | No | NA | 4 | 0 | 3 | Not-determined | Genome-fragment | NA | NA | 0 |
| S11-final-viral-combined_k141_222766\|\|full | 1186 | No | NA | 3 | 0 | 1 | Not-determined | Genome-fragment | NA | NA | 0 |
| S11_k141_282279 | 1094 | No | NA | 4 | 0 | 0 | Low-quality | Genome-fragment | 2.71 | AAI-based (medium-confidence) | 0 |
| S1-final-viral-combined_k141_268607\|\|full | 1094 | No | NA | 2 | 0 | 0 | Low-quality | Genome-fragment | 2.13 | AAI-based (high-confidence) | 0 |
| S1_k141_166642 | 1094 | No | NA | 2 | 0 | 0 | Low-quality | Genome-fragment | 1.16 | AAI-based (medium-confidence) | 0 |
| S11-final-viral-combined_k141_209410\|\|full | 2029 | No | NA | 2 | 0 | 1 | Not-determined | Genome-fragment | NA | NA | 0 |
| S3-final-viral-combined_k141_72673\|\|full | 1094 | No | NA | 3 | 0 | 0 | Low-quality | Genome-fragment | 1.87 | AAI-based (high-confidence) | 0 |
| S3-final-viral-combined_k141_67912\|\|full | 1094 | No | NA | 3 | 0 | 0 | Low-quality | Genome-fragment | 1.88 | AAI-based (high-confidence) | 0 |
| S5-final-viral-combined_k141_11939\|\|lt2gene | 1094 | No | NA | 1 | 1 | 0 | Low-quality | Genome-fragment | 1.62 | HMM-based (lower-bound) | 0 |
| S7_k141_78925 | 1094 | No | NA | 4 | 0 | 0 | Low-quality | Genome-fragment | 1.84 | AAI-based (medium-confidence) | 0 |
| S12-final-viral-combined_k141_120319\|\|full | 1094 | No | NA | 3 | 1 | 0 | Low-quality | Genome-fragment | 0.77 | AAI-based (medium-confidence) | 0 |
| S12_k141_380517 | 1094 | No | NA | 2 | 0 | 0 | Low-quality | Genome-fragment | 3.14 | AAI-based (medium-confidence) | 0 |
| S12_k141_28743 | 1094 | No | NA | 3 | 0 | 0 | Low-quality | Genome-fragment | 2 | AAI-based (high-confidence) | 0 |
| S11-final-viral-combined_k141_2432\|\|full | 2553 | No | NA | 2 | 0 | 2 | Not-determined | Genome-fragment | NA | NA | 0 |
| S11-final-viral-combined_k141_29856\|\|full | 1175 | No | NA | 2 | 0 | 2 | Not-determined | Genome-fragment | NA | NA | 0 |
| S11-final-viral-combined_k141_43692\|\|full | 1612 | No | NA | 3 | 0 | 2 | Not-determined | Genome-fragment | NA | NA | 0 |
| S4_k141_161777 | 1094 | No | NA | 3 | 0 | 0 | Low-quality | Genome-fragment | 0.88 | AAI-based (medium-confidence) | 0 |
| S8-final-viral-combined_k141_108354\|\|full | 1094 | No | NA | 3 | 1 | 0 | Low-quality | Genome-fragment | 0.47 | HMM-based (lower-bound) | 0 |
| S11-final-viral-combined_k141_70845\|\|full | 2785 | No | NA | 3 | 0 | 0 | Not-determined | Genome-fragment | NA | NA | 0 |
| S8-final-viral-combined_k141_49344\|\|lt2gene | 1094 | No | NA | 2 | 0 | 1 | Low-quality | Genome-fragment | 1.74 | AAI-based (medium-confidence) | 0 |
| S11-final-viral-combined_k141_305809\|\|full | 2121 | No | NA | 3 | 0 | 0 | Not-determined | Genome-fragment | NA | NA | 0 |
| S11-final-viral-combined_k141_85099\|\|full | 1802 | No | NA | 4 | 0 | 0 | Not-determined | Genome-fragment | NA | NA | 0 |
| S11-final-viral-combined_k141_278316\|\|full | 1475 | No | NA | 2 | 0 | 1 | Not-determined | Genome-fragment | NA | NA | 0 |
| S11-final-viral-combined_k141_250516\|\|full | 1573 | No | NA | 3 | 0 | 1 | Not-determined | Genome-fragment | NA | NA | 0 |
| S11-final-viral-combined_k141_209521\|\|full | 1256 | No | NA | 2 | 0 | 1 | Not-determined | Genome-fragment | NA | NA | 0 |
| S9-final-viral-combined_k141_22349\|\|full | 1095 | No | NA | 3 | 1 | 0 | Low-quality | Genome-fragment | 2.37 | AAI-based (medium-confidence) | 0 |
| S11-final-viral-combined_k141_209524\|\|full | 4955 | No | NA | 4 | 0 | 2 | Not-determined | Genome-fragment | NA | NA | 0 |
| S11-final-viral-combined_k141_126580\|\|full | 1776 | No | NA | 2 | 0 | 2 | Not-determined | Genome-fragment | NA | NA | 0 |
| S11-final-viral-combined_k141_168048\|\|full | 52585 | Yes | 29594 | 59 | 20 | 10 | Complete | High-quality | 100 | Provirus (medium-confidence) | 43.72 |
| S11-final-viral-combined_k141_278337\|\|full | 3373 | No | NA | 5 | 0 | 0 | Not-determined | Genome-fragment | NA | NA | 0 |
| S11-final-viral-combined_k141_321266\|\|full | 1095 | No | NA | 2 | 0 | 0 | Low-quality | Genome-fragment | 1.32 | AAI-based (high-confidence) | 0 |
| S1-final-viral-combined_k141_234097\|\|full | 1095 | No | NA | 3 | 0 | 3 | Low-quality | Genome-fragment | 0.32 | AAI-based (medium-confidence) | 0 |
| S1-final-viral-combined_k141_123886\|\|full | 1095 | No | NA | 4 | 0 | 0 | Low-quality | Genome-fragment | 2.59 | AAI-based (high-confidence) | 0 |
| S11-final-viral-combined_k141_222957\|\|full | 4457 | No | NA | 6 | 0 | 6 | Not-determined | Genome-fragment | NA | NA | 0 |
| S5-final-viral-combined_k141_167942\|\|full | 1095 | No | NA | 3 | 0 | 0 | Low-quality | Genome-fragment | 2.25 | AAI-based (medium-confidence) | 0 |
| S11-final-viral-combined_k141_168091\|\|full | 2826 | No | NA | 6 | 0 | 2 | Not-determined | Genome-fragment | NA | NA | 0 |
| S11-final-viral-combined_k141_140237\|\|full | 1643 | No | NA | 3 | 0 | 2 | Not-determined | Genome-fragment | NA | NA | 0 |
| S7-final-viral-combined_k141_239555\|\|full | 1095 | No | NA | 3 | 1 | 0 | Low-quality | Genome-fragment | 0.77 | HMM-based (lower-bound) | 0 |
| S11-final-viral-combined_k141_57802\|\|full | 1624 | No | NA | 2 | 0 | 1 | Not-determined | Genome-fragment | NA | NA | 0 |
| S11-final-viral-combined_k141_140250\|\|full | 6783 | No | NA | 6 | 0 | 3 | Not-determined | Genome-fragment | NA | NA | 0 |
| S7_k141_28532 | 1095 | No | NA | 3 | 0 | 0 | Low-quality | Genome-fragment | 2.63 | AAI-based (high-confidence) | 0 |
| S12-final-viral-combined_k141_188308\|\|lt2gene | 1095 | No | NA | 1 | 1 | 0 | Low-quality | Genome-fragment | 1.2 | HMM-based (lower-bound) | 0 |
| S11-final-viral-combined_k141_2587\|\|full | 1570 | No | NA | 3 | 0 | 2 | Not-determined | Genome-fragment | NA | NA | 0 |
| S11-final-viral-combined_k141_181414\|\|full | 18242 | No | NA | 19 | 0 | 14 | Not-determined | Genome-fragment | NA | NA | 0 |
| S11-final-viral-combined_k141_140272\|\|full | 1717 | No | NA | 2 | 0 | 2 | Not-determined | Genome-fragment | NA | NA | 0 |
| S11-final-viral-combined_k141_209597\|\|full | 1279 | No | NA | 2 | 0 | 1 | Not-determined | Genome-fragment | NA | NA | 0 |
| S12-final-viral-combined_k141_71082\|\|lt2gene | 1095 | No | NA | 2 | 1 | 0 | Low-quality | Genome-fragment | 1.97 | AAI-based (medium-confidence) | 0 |
| S11-final-viral-combined_k141_43818\|\|full | 3280 | No | NA | 2 | 0 | 2 | Not-determined | Genome-fragment | NA | NA | 0 |
| S11-final-viral-combined_k141_154076\|\|full | 1004 | No | NA | 2 | 0 | 0 | Not-determined | Genome-fragment | NA | NA | 0 |
| S11-final-viral-combined_k141_30034\|\|full | 1935 | No | NA | 4 | 0 | 2 | Not-determined | Genome-fragment | NA | NA | 0 |
| S11-final-viral-combined_k141_99216\|\|full | 1503 | No | NA | 4 | 0 | 1 | Not-determined | Genome-fragment | NA | NA | 0 |
| S11-final-viral-combined_k141_237088\|\|full | 1058 | No | NA | 3 | 0 | 0 | Not-determined | Genome-fragment | NA | NA | 0 |
| S11-final-viral-combined_k141_57868\|\|full | 6395 | No | NA | 7 | 0 | 4 | Not-determined | Genome-fragment | NA | NA | 0 |
| S11-final-viral-combined_k141_2619\|\|full | 1832 | No | NA | 4 | 0 | 0 | Not-determined | Genome-fragment | NA | NA | 0 |
| S12-final-viral-combined_k141_356955\|\|lt2gene | 1095 | No | NA | 2 | 1 | 0 | Low-quality | Genome-fragment | 1.48 | HMM-based (lower-bound) | 0 |
| S11-final-viral-combined_k141_168179\|\|full | 1797 | No | NA | 2 | 0 | 1 | Not-determined | Genome-fragment | NA | NA | 0 |
| S11-final-viral-combined_k141_71008\|\|full | 4359 | No | NA | 4 | 0 | 4 | Not-determined | Genome-fragment | NA | NA | 0 |
| S12_k141_137048 | 1095 | No | NA | 2 | 1 | 0 | Low-quality | Genome-fragment | 2.13 | AAI-based (high-confidence) | 0 |
| S12_k141_304895 | 1095 | No | NA | 3 | 0 | 0 | Low-quality | Genome-fragment | 2.3 | AAI-based (medium-confidence) | 0 |
| S2-final-viral-combined_k141_130159\|\|full | 1095 | No | NA | 4 | 2 | 0 | Low-quality | Genome-fragment | 1.47 | AAI-based (medium-confidence) | 0 |
| S11-final-viral-combined_k141_195395\|\|full | 1126 | No | NA | 3 | 0 | 1 | Not-determined | Genome-fragment | NA | NA | 0 |
| S11-final-viral-combined_k141_112367\|\|full | 1498 | No | NA | 2 | 0 | 0 | Not-determined | Genome-fragment | NA | NA | 0 |
| S11-final-viral-combined_k141_16242\|\|full | 4543 | No | NA | 3 | 0 | 3 | Not-determined | Genome-fragment | NA | NA | 0 |
| S2-final-viral-combined_k141_61690\|\|full | 1095 | No | NA | 3 | 0 | 0 | Low-quality | Genome-fragment | 2.11 | AAI-based (medium-confidence) | 0 |
| S2-final-viral-combined_k141_174231\|\|lt2gene | 1095 | No | NA | 1 | 1 | 0 | Low-quality | Genome-fragment | 1.23 | HMM-based (lower-bound) | 0 |
| S11-final-viral-combined_k141_305945\|\|full | 16105 | No | NA | 13 | 0 | 4 | Not-determined | Genome-fragment | NA | NA | 0 |
| S11-final-viral-combined_k141_264325\|\|full | 1355 | No | NA | 3 | 0 | 2 | Not-determined | Genome-fragment | NA | NA | 0 |
| S11-final-viral-combined_k141_278498\|\|full | 1730 | No | NA | 2 | 0 | 0 | Not-determined | Genome-fragment | NA | NA | 0 |
| S11-final-viral-combined_k141_278504\|\|full | 1838 | No | NA | 2 | 0 | 0 | Not-determined | Genome-fragment | NA | NA | 0 |
| S11-final-viral-combined_k141_43904\|\|full | 1967 | No | NA | 4 | 0 | 1 | Not-determined | Genome-fragment | NA | NA | 0 |
| S11-final-viral-combined_k141_126619\|\|full | 2005 | No | NA | 3 | 0 | 0 | Not-determined | Genome-fragment | NA | NA | 0 |
| S11-final-viral-combined_k141_126622\|\|full | 1951 | No | NA | 2 | 0 | 2 | Not-determined | Genome-fragment | NA | NA | 0 |
| S2_k141_232206 | 1095 | No | NA | 1 | 0 | 1 | Low-quality | Genome-fragment | 0.29 | AAI-based (medium-confidence) | 0 |
| S4-final-viral-combined_k141_85774\|\|full | 1095 | No | NA | 4 | 0 | 0 | Low-quality | Genome-fragment | 2.15 | AAI-based (medium-confidence) | 0 |
| S4-final-viral-combined_k141_60163\|\|full | 1095 | No | NA | 5 | 0 | 0 | Low-quality | Genome-fragment | 2.35 | AAI-based (high-confidence) | 0 |
| S11-final-viral-combined_k141_305980\|\|full | 2416 | No | NA | 3 | 0 | 3 | Not-determined | Genome-fragment | NA | NA | 0 |
| S4_k141_126461 | 1095 | No | NA | 1 | 0 | 0 | Low-quality | Genome-fragment | 1.15 | AAI-based (high-confidence) | 0 |
| S11-final-viral-combined_k141_264382\|\|full | 1876 | No | NA | 3 | 0 | 1 | Not-determined | Genome-fragment | NA | NA | 0 |
| S9-final-viral-combined_k141_88662\|\|full | 1096 | No | NA | 3 | 0 | 0 | Low-quality | Genome-fragment | 2.7 | AAI-based (high-confidence) | 0 |
| S11-final-viral-combined_k141_81233\|\|full | 1096 | No | NA | 2 | 1 | 0 | Low-quality | Genome-fragment | 2.03 | HMM-based (lower-bound) | 0 |
| S1-final-viral-combined_k141_280478\|\|full | 1096 | No | NA | 3 | 3 | 0 | Low-quality | Genome-fragment | 3.13 | AAI-based (high-confidence) | 0 |
| S1_k141_393818 | 1096 | No | NA | 2 | 0 | 0 | Low-quality | Genome-fragment | 0.69 | AAI-based (high-confidence) | 0 |
| S11-final-viral-combined_k141_140338\|\|full | 2324 | No | NA | 3 | 0 | 3 | Not-determined | Genome-fragment | NA | NA | 0 |
| S11-final-viral-combined_k141_112434\|\|full | 2033 | No | NA | 4 | 0 | 2 | Not-determined | Genome-fragment | NA | NA | 0 |
| S3-final-viral-combined_k141_6930\|\|full | 1096 | No | NA | 2 | 0 | 0 | Low-quality | Genome-fragment | 1.76 | AAI-based (medium-confidence) | 0 |
| S11-final-viral-combined_k141_181538\|\|full | 4750 | No | NA | 3 | 0 | 2 | Not-determined | Genome-fragment | NA | NA | 0 |
| S11-final-viral-combined_k141_2743\|\|full | 4672 | No | NA | 8 | 0 | 0 | Not-determined | Genome-fragment | NA | NA | 0 |
| S5-final-viral-combined_k141_135712\|\|full | 1096 | No | NA | 3 | 0 | 0 | Low-quality | Genome-fragment | 1.33 | AAI-based (medium-confidence) | 0 |
| S11-final-viral-combined_k141_305996\|\|full | 1018 | No | NA | 3 | 0 | 1 | Not-determined | Genome-fragment | NA | NA | 0 |
| S11-final-viral-combined_k141_112475\|\|full | 1090 | No | NA | 2 | 0 | 1 | Not-determined | Genome-fragment | NA | NA | 0 |
| S5-final-viral-combined_k141_64376\|\|full | 1096 | No | NA | 3 | 1 | 0 | Low-quality | Genome-fragment | 2.38 | AAI-based (high-confidence) | 0 |
| S11-final-viral-combined_k141_195425\|\|full | 29097 | No | NA | 25 | 6 | 2 | Medium-quality | Genome-fragment | 67.93 | AAI-based (high-confidence) | 0 |
| S5-final-viral-combined_k141_209623\|\|full | 1096 | No | NA | 3 | 1 | 0 | Low-quality | Genome-fragment | 1.88 | AAI-based (medium-confidence) | 0 |
| S11-final-viral-combined_k141_209769\|\|full | 2583 | No | NA | 2 | 0 | 0 | Not-determined | Genome-fragment | NA | NA | 0 |
| S5-final-viral-combined_k141_58369\|\|full | 1096 | No | NA | 2 | 0 | 0 | Low-quality | Genome-fragment | 2.58 | AAI-based (medium-confidence) | 0 |
| S11-final-viral-combined_k141_223216\|\|full | 1246 | No | NA | 2 | 0 | 0 | Not-determined | Genome-fragment | NA | NA | 0 |
| S5-final-viral-combined_k141_181491\|\|lt2gene | 1096 | No | NA | 2 | 2 | 0 | Low-quality | Genome-fragment | 1.35 | HMM-based (lower-bound) | 0 |
| S7-final-viral-combined_k141_170368\|\|full | 1096 | No | NA | 2 | 0 | 0 | Low-quality | Genome-fragment | 0.57 | AAI-based (medium-confidence) | 0 |
| S7-final-viral-combined_k141_51532\|\|lt2gene | 1096 | No | NA | 1 | 1 | 0 | Low-quality | Genome-fragment | 0.76 | HMM-based (lower-bound) | 0 |
| S11-final-viral-combined_k141_30178\|\|full | 1697 | No | NA | 3 | 0 | 0 | Not-determined | Genome-fragment | NA | NA | 0 |
| S17-final-viral-combined_k141_85241\|\|full | 1096 | No | NA | 2 | 2 | 0 | Low-quality | Genome-fragment | 2.92 | AAI-based (high-confidence) | 0 |
| S11-final-viral-combined_k141_319432\|\|full | 1999 | No | NA | 5 | 0 | 1 | Not-determined | Genome-fragment | NA | NA | 0 |
| S17-final-viral-combined_k141_263280\|\|lt2gene | 1096 | No | NA | 2 | 1 | 0 | Low-quality | Genome-fragment | 2.33 | AAI-based (high-confidence) | 0 |
| S12-final-viral-combined_k141_35770\|\|full | 1096 | No | NA | 3 | 2 | 0 | Low-quality | Genome-fragment | 2.17 | AAI-based (high-confidence) | 0 |
| S11-final-viral-combined_k141_112535\|\|full | 12484 | No | NA | 10 | 0 | 9 | Not-determined | Genome-fragment | NA | NA | 0 |
| S11-final-viral-combined_k141_58063\|\|full | 5498 | No | NA | 7 | 0 | 6 | Not-determined | Genome-fragment | NA | NA | 0 |
| S11-final-viral-combined_k141_209823\|\|full | 2018 | No | NA | 2 | 0 | 2 | Not-determined | Genome-fragment | NA | NA | 0 |
| S12-final-viral-combined_k141_216548\|\|full | 1096 | No | NA | 2 | 0 | 1 | Low-quality | Genome-fragment | 0.82 | AAI-based (medium-confidence) | 0 |
| S11-final-viral-combined_k141_278616\|\|full | 6260 | No | NA | 8 | 0 | 3 | Not-determined | Genome-fragment | NA | NA | 0 |
| S11-final-viral-combined_k141_278622\|\|full | 2558 | No | NA | 4 | 0 | 3 | Not-determined | Genome-fragment | NA | NA | 0 |
| S12-final-viral-combined_k141_310809\|\|lt2gene | 1096 | No | NA | 2 | 2 | 0 | Low-quality | Genome-fragment | 1.35 | HMM-based (lower-bound) | 0 |
| S11-final-viral-combined_k141_85317\|\|full | 4878 | No | NA | 4 | 0 | 3 | Not-determined | Genome-fragment | NA | NA | 0 |
| S11-final-viral-combined_k141_168339\|\|full | 5631 | No | NA | 4 | 0 | 2 | Not-determined | Genome-fragment | NA | NA | 0 |
| S12_k141_59398 | 1096 | No | NA | 2 | 0 | 1 | Low-quality | Genome-fragment | 1.21 | AAI-based (medium-confidence) | 0 |
| S11-final-viral-combined_k141_112546\|\|full | 3378 | No | NA | 5 | 0 | 3 | Not-determined | Genome-fragment | NA | NA | 0 |
| S11-final-viral-combined_k141_154333\|\|full | 3327 | No | NA | 3 | 0 | 2 | Not-determined | Genome-fragment | NA | NA | 0 |
| S4-final-viral-combined_k141_27992\|\|full | 1096 | No | NA | 4 | 0 | 0 | Low-quality | Genome-fragment | 3.17 | AAI-based (high-confidence) | 0 |
| S11-final-viral-combined_k141_250800\|\|full | 4425 | No | NA | 5 | 0 | 3 | Not-determined | Genome-fragment | NA | NA | 0 |
| S4-final-viral-combined_k141_144693\|\|full | 1096 | No | NA | 2 | 2 | 0 | Low-quality | Genome-fragment | 2.77 | AAI-based (high-confidence) | 0 |
| S6_k141_66036 | 1096 | No | NA | 3 | 0 | 2 | Low-quality | Genome-fragment | 1.39 | AAI-based (medium-confidence) | 0 |
| S8-final-viral-combined_k141_232863\|\|full | 1096 | No | NA | 2 | 0 | 1 | Low-quality | Genome-fragment | 1.43 | AAI-based (medium-confidence) | 0 |
| S8-final-viral-combined_k141_168306\|\|full | 1096 | No | NA | 3 | 0 | 3 | Low-quality | Genome-fragment | 1.34 | AAI-based (medium-confidence) | 0 |
| S1-final-viral-combined_k141_8013\|\|full | 1097 | No | NA | 2 | 0 | 1 | Low-quality | Genome-fragment | 0.8 | AAI-based (medium-confidence) | 0 |
| S11-final-viral-combined_k141_126689\|\|full | 1068 | No | NA | 3 | 0 | 0 | Not-determined | Genome-fragment | NA | NA | 0 |
| S11-final-viral-combined_k141_44034\|\|full | 1979 | No | NA | 4 | 0 | 0 | Not-determined | Genome-fragment | NA | NA | 0 |
| S3_k141_43350 | 1097 | No | NA | 1 | 0 | 1 | Low-quality | Genome-fragment | 0.98 | AAI-based (medium-confidence) | 0 |
| S11-final-viral-combined_k141_168389\|\|full | 1864 | No | NA | 2 | 0 | 1 | Not-determined | Genome-fragment | NA | NA | 0 |
| S11-final-viral-combined_k141_99466\|\|full | 1287 | No | NA | 3 | 0 | 1 | Not-determined | Genome-fragment | NA | NA | 0 |
| S11-final-viral-combined_k141_264551\|\|full | 2864 | No | NA | 3 | 0 | 0 | Not-determined | Genome-fragment | NA | NA | 0 |
| S5_k141_149634 | 1097 | No | NA | 4 | 2 | 0 | Low-quality | Genome-fragment | 2.04 | AAI-based (medium-confidence) | 0 |
| S11-final-viral-combined_k141_44071\|\|full | 1884 | No | NA | 3 | 0 | 2 | Not-determined | Genome-fragment | NA | NA | 0 |
| S7-final-viral-combined_k141_163862\|\|lt2gene | 1097 | No | NA | 2 | 0 | 0 | Low-quality | Genome-fragment | 2.65 | AAI-based (high-confidence) | 0 |
| S17-final-viral-combined_k141_58415\|\|lt2gene | 1097 | No | NA | 1 | 1 | 0 | Low-quality | Genome-fragment | 2.82 | AAI-based (high-confidence) | 0 |
| S12_k141_236282 | 1097 | No | NA | 2 | 0 | 1 | Low-quality | Genome-fragment | 1.89 | AAI-based (medium-confidence) | 0 |
| S11-final-viral-combined_k141_44125\|\|full | 3365 | No | NA | 4 | 0 | 2 | Not-determined | Genome-fragment | NA | NA | 0 |
| S11-final-viral-combined_k141_126781\|\|full | 2606 | No | NA | 4 | 0 | 2 | Not-determined | Genome-fragment | NA | NA | 0 |
| S2-final-viral-combined_k141_46532\|\|lt2gene | 1097 | No | NA | 2 | 1 | 0 | Low-quality | Genome-fragment | 2.23 | AAI-based (high-confidence) | 0 |
| S2-final-viral-combined_k141_216099\|\|lt2gene | 1097 | No | NA | 1 | 1 | 0 | Low-quality | Genome-fragment | 2.73 | AAI-based (high-confidence) | 0 |
| S11-final-viral-combined_k141_154394\|\|full | 1445 | No | NA | 2 | 0 | 2 | Not-determined | Genome-fragment | NA | NA | 0 |
| S8-final-viral-combined_k141_200502\|\|lt2gene | 1097 | No | NA | 2 | 1 | 0 | Low-quality | Genome-fragment | 2.67 | AAI-based (high-confidence) | 0 |
| S11-final-viral-combined_k141_223391\|\|full | 1480 | No | NA | 2 | 0 | 0 | Not-determined | Genome-fragment | NA | NA | 0 |
| S11-final-viral-combined_k141_168515\|\|full | 1873 | No | NA | 4 | 0 | 0 | Not-determined | Genome-fragment | NA | NA | 0 |
| S11-final-viral-combined_k141_16494\|\|full | 6970 | No | NA | 8 | 0 | 6 | Not-determined | Genome-fragment | NA | NA | 0 |
| S11-final-viral-combined_k141_292043\|\|full | 2728 | No | NA | 3 | 0 | 3 | Not-determined | Genome-fragment | NA | NA | 0 |
| S11-final-viral-combined_k141_58228\|\|full | 1536 | No | NA | 3 | 0 | 1 | Not-determined | Genome-fragment | NA | NA | 0 |
| S8_k141_201130 | 1097 | No | NA | 4 | 0 | 0 | Low-quality | Genome-fragment | 0.78 | AAI-based (high-confidence) | 0 |
| S11-final-viral-combined_k141_98309\|\|full | 1098 | No | NA | 5 | 0 | 0 | Low-quality | Genome-fragment | 2.48 | AAI-based (high-confidence) | 0 |
| S11-final-viral-combined_k141_122375\|\|full | 1098 | No | NA | 2 | 0 | 0 | Low-quality | Genome-fragment | 2.3 | AAI-based (high-confidence) | 0 |
| S11-final-viral-combined_k141_85484\|\|full | 4792 | No | NA | 5 | 0 | 4 | Not-determined | Genome-fragment | NA | NA | 0 |
| S11_k141_138549 | 1098 | No | NA | 3 | 1 | 0 | Low-quality | Genome-fragment | 1.75 | HMM-based (lower-bound) | 0 |
| S11-final-viral-combined_k141_223435\|\|full | 3352 | No | NA | 6 | 0 | 4 | Not-determined | Genome-fragment | NA | NA | 0 |
| S11-final-viral-combined_k141_306288\|\|full | 4708 | No | NA | 3 | 0 | 3 | Not-determined | Genome-fragment | NA | NA | 0 |
| S11-final-viral-combined_k141_126840\|\|full | 4587 | No | NA | 6 | 0 | 4 | Not-determined | Genome-fragment | NA | NA | 0 |
| S11-final-viral-combined_k141_181717\|\|full | 1917 | No | NA | 3 | 0 | 1 | Not-determined | Genome-fragment | NA | NA | 0 |
| S1-final-viral-combined_k141_11249\|\|full | 1098 | No | NA | 3 | 1 | 0 | Low-quality | Genome-fragment | 2.61 | AAI-based (medium-confidence) | 0 |
| S5-final-viral-combined_k141_11926\|\|full | 1098 | No | NA | 3 | 0 | 0 | Low-quality | Genome-fragment | 2.84 | AAI-based (high-confidence) | 0 |
| S11-final-viral-combined_k141_237557\|\|full | 8994 | No | NA | 9 | 0 | 6 | Not-determined | Genome-fragment | NA | NA | 0 |
| S11-final-viral-combined_k141_181745\|\|full | 1799 | No | NA | 5 | 0 | 0 | Not-determined | Genome-fragment | NA | NA | 0 |
| S11-final-viral-combined_k141_126850\|\|full | 2725 | No | NA | 3 | 0 | 1 | Not-determined | Genome-fragment | NA | NA | 0 |
| S7_k141_219939 | 1098 | No | NA | 1 | 0 | 0 | Low-quality | Genome-fragment | 0.82 | AAI-based (medium-confidence) | 0 |
| S11-final-viral-combined_k141_181770\|\|full | 1638 | No | NA | 3 | 0 | 0 | Not-determined | Genome-fragment | NA | NA | 0 |
| S17-k141_74001 | 1098 | No | NA | 3 | 0 | 3 | Low-quality | Genome-fragment | 3.1 | AAI-based (medium-confidence) | 0 |
| S11-final-viral-combined_k141_58323\|\|full | 2916 | No | NA | 4 | 0 | 3 | Not-determined | Genome-fragment | NA | NA | 0 |
| S11-final-viral-combined_k141_126864\|\|full | 2595 | No | NA | 3 | 0 | 2 | Not-determined | Genome-fragment | NA | NA | 0 |
| S11-final-viral-combined_k141_30396\|\|full | 2842 | No | NA | 6 | 0 | 1 | Not-determined | Genome-fragment | NA | NA | 0 |
| S11-final-viral-combined_k141_112733\|\|full | 34643 | No | NA | 49 | 22 | 1 | High-quality | High-quality | 95.1 | AAI-based (medium-confidence) | 0 |
| S12-final-viral-combined_k141_383132\|\|lt2gene | 1098 | No | NA | 2 | 1 | 0 | Low-quality | Genome-fragment | 2.55 | AAI-based (high-confidence) | 0 |
| S12_k141_59332 | 1098 | No | NA | 1 | 0 | 1 | Low-quality | Genome-fragment | 0.37 | AAI-based (medium-confidence) | 0 |
| S11-final-viral-combined_k141_210043\|\|full | 1013 | No | NA | 2 | 0 | 2 | Not-determined | Genome-fragment | NA | NA | 0 |
| S4-final-viral-combined_k141_107094\|\|lt2gene | 1098 | No | NA | 1 | 1 | 0 | Low-quality | Genome-fragment | 6.43 | AAI-based (high-confidence) | 0 |
| S11-final-viral-combined_k141_168654\|\|full | 2833 | No | NA | 3 | 0 | 2 | Not-determined | Genome-fragment | NA | NA | 0 |
| S9_k141_105499 | 1099 | No | NA | 2 | 0 | 0 | Low-quality | Genome-fragment | 2.56 | AAI-based (high-confidence) | 0 |
| S11-final-viral-combined_k141_3041\|\|full | 1292 | No | NA | 2 | 0 | 1 | Not-determined | Genome-fragment | NA | NA | 0 |
| S11-final-viral-combined_k141_238404\|\|full | 1099 | No | NA | 3 | 0 | 0 | Low-quality | Genome-fragment | 2.73 | AAI-based (medium-confidence) | 0 |
| S11-final-viral-combined_k141_238790\|\|full | 1099 | No | NA | 2 | 0 | 1 | Low-quality | Genome-fragment | 0.82 | AAI-based (high-confidence) | 0 |
| S11-final-viral-combined_k141_140706\|\|full | 3472 | No | NA | 4 | 0 | 3 | Not-determined | Genome-fragment | NA | NA | 0 |
| S11_k141_265804 | 1099 | No | NA | 2 | 1 | 0 | Low-quality | Genome-fragment | 2.99 | AAI-based (high-confidence) | 0 |
| S11-final-viral-combined_k141_58388\|\|full | 6454 | No | NA | 4 | 0 | 4 | Not-determined | Genome-fragment | NA | NA | 0 |
| S11_k141_173161 | 1099 | No | NA | 2 | 0 | 0 | Low-quality | Genome-fragment | 0.62 | AAI-based (medium-confidence) | 0 |
| S11_k141_173719 | 1099 | No | NA | 1 | 1 | 0 | Low-quality | Genome-fragment | 1.83 | HMM-based (lower-bound) | 0 |
| S1-final-viral-combined_k141_363124\|\|full | 1099 | No | NA | 2 | 1 | 0 | Low-quality | Genome-fragment | 2.43 | AAI-based (high-confidence) | 0 |
| S11-final-viral-combined_k141_250998\|\|full | 1961 | No | NA | 2 | 0 | 0 | Not-determined | Genome-fragment | NA | NA | 0 |
| S11-final-viral-combined_k141_210116\|\|full | 2869 | No | NA | 3 | 0 | 3 | Not-determined | Genome-fragment | NA | NA | 0 |
| S11-final-viral-combined_k141_223530\|\|full | 5243 | No | NA | 5 | 0 | 4 | Not-determined | Genome-fragment | NA | NA | 0 |
| S1-final-viral-combined_k141_275741\|\|full | 1099 | No | NA | 2 | 2 | 0 | Low-quality | Genome-fragment | 2.51 | AAI-based (high-confidence) | 0 |
| S11-final-viral-combined_k141_126984\|\|full | 5222 | No | NA | 6 | 0 | 4 | Not-determined | Genome-fragment | NA | NA | 0 |
| S3-final-viral-combined_k141_43716\|\|full | 1099 | No | NA | 2 | 0 | 0 | Low-quality | Genome-fragment | 1.95 | AAI-based (high-confidence) | 0 |
| S5_k141_125357 | 1099 | No | NA | 1 | 1 | 0 | Low-quality | Genome-fragment | 2.57 | AAI-based (high-confidence) | 0 |
| S11-final-viral-combined_k141_140743\|\|full | 2718 | No | NA | 4 | 0 | 0 | Not-determined | Genome-fragment | NA | NA | 0 |
| S11-final-viral-combined_k141_112847\|\|full | 3397 | No | NA | 4 | 0 | 3 | Not-determined | Genome-fragment | NA | NA | 0 |
| S11-final-viral-combined_k141_168697\|\|full | 3683 | No | NA | 4 | 0 | 2 | Not-determined | Genome-fragment | NA | NA | 0 |
| S7_k141_96765 | 1099 | No | NA | 3 | 0 | 0 | Low-quality | Genome-fragment | 1.13 | AAI-based (medium-confidence) | 0 |
| S12-final-viral-combined_k141_323559\|\|lt2gene | 1099 | No | NA | 1 | 1 | 0 | Low-quality | Genome-fragment | 2.41 | AAI-based (high-confidence) | 0 |
| S2-final-viral-combined_k141_4517\|\|full | 1099 | No | NA | 4 | 1 | 0 | Low-quality | Genome-fragment | 2.74 | AAI-based (high-confidence) | 0 |
| S11-final-viral-combined_k141_306508\|\|full | 3742 | No | NA | 5 | 0 | 2 | Not-determined | Genome-fragment | NA | NA | 0 |
| S11-final-viral-combined_k141_30534\|\|full | 1796 | No | NA | 3 | 0 | 0 | Not-determined | Genome-fragment | NA | NA | 0 |
| S11-final-viral-combined_k141_30538\|\|full | 1441 | No | NA | 3 | 0 | 0 | Not-determined | Genome-fragment | NA | NA | 0 |
| S2_k141_230241 | 1099 | No | NA | 1 | 0 | 0 | Low-quality | Genome-fragment | 1.57 | AAI-based (high-confidence) | 0 |
| S2_k141_79670 | 1099 | No | NA | 2 | 0 | 0 | Low-quality | Genome-fragment | 0.52 | AAI-based (medium-confidence) | 0 |
| S11-final-viral-combined_k141_237709\|\|full | 1256 | No | NA | 2 | 0 | 0 | Not-determined | Genome-fragment | NA | NA | 0 |
| S4-final-viral-combined_k141_21354\|\|lt2gene | 1099 | No | NA | 2 | 1 | 0 | Low-quality | Genome-fragment | 1.48 | AAI-based (high-confidence) | 0 |
| S11-final-viral-combined_k141_99757\|\|full | 15586 | No | NA | 16 | 0 | 13 | Not-determined | Genome-fragment | NA | NA | 0 |
| S11-final-viral-combined_k141_99758\|\|full | 1109 | No | NA | 3 | 0 | 1 | Not-determined | Genome-fragment | NA | NA | 0 |
| S6-final-viral-combined_k141_66711\|\|full | 1099 | No | NA | 3 | 1 | 0 | Low-quality | Genome-fragment | 1.98 | HMM-based (lower-bound) | 0 |
| S8-final-viral-combined_k141_25523\|\|lt2gene | 1099 | No | NA | 2 | 0 | 0 | Low-quality | Genome-fragment | 1.55 | AAI-based (medium-confidence) | 0 |
| S11-final-viral-combined_k141_30567\|\|full | 1946 | No | NA | 2 | 0 | 2 | Not-determined | Genome-fragment | NA | NA | 0 |
| S8-final-viral-combined_k141_132765\|\|lt2gene | 1099 | No | NA | 1 | 1 | 0 | Low-quality | Genome-fragment | 1.23 | HMM-based (lower-bound) | 0 |
| S11-final-viral-combined_k141_140835\|\|full | 4220 | No | NA | 5 | 0 | 4 | Not-determined | Genome-fragment | NA | NA | 0 |
| S11-final-viral-combined_k141_99763\|\|full | 3416 | No | NA | 3 | 0 | 0 | Not-determined | Genome-fragment | NA | NA | 0 |
| S3_k141_65342 | 1100 | No | NA | 2 | 0 | 0 | Low-quality | Genome-fragment | 2.37 | AAI-based (medium-confidence) | 0 |
| S11-final-viral-combined_k141_210173\|\|full | 2708 | No | NA | 3 | 0 | 2 | Not-determined | Genome-fragment | NA | NA | 0 |
| S11-final-viral-combined_k141_210183\|\|full | 1568 | No | NA | 3 | 0 | 1 | Not-determined | Genome-fragment | NA | NA | 0 |
| S5-final-viral-combined_k141_206150\|\|full | 1100 | No | NA | 4 | 0 | 1 | Low-quality | Genome-fragment | 1.57 | AAI-based (medium-confidence) | 0 |
| S5-final-viral-combined_k141_38691\|\|full | 1100 | No | NA | 2 | 0 | 0 | Low-quality | Genome-fragment | 2.44 | AAI-based (high-confidence) | 0 |
| S5_k141_106412 | 1100 | No | NA | 1 | 1 | 0 | Low-quality | Genome-fragment | 0.74 | HMM-based (lower-bound) | 0 |
| S11-final-viral-combined_k141_237759\|\|full | 3709 | No | NA | 3 | 0 | 3 | Not-determined | Genome-fragment | NA | NA | 0 |
| S7-final-viral-combined_k141_120762\|\|full | 1100 | No | NA | 2 | 0 | 0 | Low-quality | Genome-fragment | 2.35 | AAI-based (medium-confidence) | 0 |
| S17-final-viral-combined_k141_81699\|\|full | 1100 | No | NA | 2 | 0 | 1 | Low-quality | Genome-fragment | 1.83 | AAI-based (medium-confidence) | 0 |
| S11-final-viral-combined_k141_292215\|\|full | 1036 | No | NA | 2 | 0 | 1 | Not-determined | Genome-fragment | NA | NA | 0 |
| S11-final-viral-combined_k141_168794\|\|full | 2853 | No | NA | 4 | 0 | 2 | Not-determined | Genome-fragment | NA | NA | 0 |
| S11-final-viral-combined_k141_154801\|\|full | 2012 | No | NA | 2 | 0 | 0 | Not-determined | Genome-fragment | NA | NA | 0 |
| S11-final-viral-combined_k141_58538\|\|full | 35663 | No | NA | 52 | 19 | 1 | Medium-quality | Genome-fragment | 68.08 | AAI-based (medium-confidence) | 0 |
| S17-final-viral-combined_k141_140852\|\|full | 1100 | No | NA | 2 | 0 | 2 | Low-quality | Genome-fragment | 1.4 | AAI-based (medium-confidence) | 0 |
| S17-final-viral-combined_k141_211305\|\|lt2gene | 1100 | No | NA | 1 | 1 | 0 | Low-quality | Genome-fragment | 1.77 | HMM-based (lower-bound) | 0 |
| S11-final-viral-combined_k141_251133\|\|full | 1404 | No | NA | 2 | 0 | 2 | Not-determined | Genome-fragment | NA | NA | 0 |
| S11-final-viral-combined_k141_279015\|\|full | 1557 | No | NA | 3 | 0 | 2 | Not-determined | Genome-fragment | NA | NA | 0 |
| S17-k141_141849 | 1100 | No | NA | 2 | 0 | 0 | Low-quality | Genome-fragment | 2.17 | AAI-based (medium-confidence) | 0 |
| S11-final-viral-combined_k141_264960\|\|full | 1323 | No | NA | 3 | 0 | 2 | Not-determined | Genome-fragment | NA | NA | 0 |
| S12-final-viral-combined_k141_18825\|\|full | 1100 | No | NA | 3 | 1 | 0 | Low-quality | Genome-fragment | 2.58 | AAI-based (medium-confidence) | 0 |
| S11-final-viral-combined_k141_3171\|\|full | 1639 | No | NA | 4 | 0 | 2 | Not-determined | Genome-fragment | NA | NA | 0 |
| S12-final-viral-combined_k141_270434\|\|full | 1100 | No | NA | 4 | 0 | 1 | Low-quality | Genome-fragment | 1.31 | AAI-based (high-confidence) | 0 |
| S12-final-viral-combined_k141_213960\|\|full | 1100 | No | NA | 2 | 0 | 0 | Low-quality | Genome-fragment | 0.64 | AAI-based (high-confidence) | 0 |
| S11-final-viral-combined_k141_71617\|\|full | 1489 | No | NA | 2 | 0 | 1 | Not-determined | Genome-fragment | NA | NA | 0 |
| S11-final-viral-combined_k141_16852\|\|full | 2856 | No | NA | 3 | 0 | 0 | Not-determined | Genome-fragment | NA | NA | 0 |
| S11-final-viral-combined_k141_195895\|\|full | 3898 | No | NA | 3 | 0 | 0 | Not-determined | Genome-fragment | NA | NA | 0 |
| S11-final-viral-combined_k141_127152\|\|full | 1173 | No | NA | 3 | 0 | 0 | Not-determined | Genome-fragment | NA | NA | 0 |
| S4-final-viral-combined_k141_118932\|\|full | 1100 | No | NA | 3 | 0 | 1 | Low-quality | Genome-fragment | 0.66 | AAI-based (medium-confidence) | 0 |
| S4-final-viral-combined_k141_59354\|\|lt2gene | 1100 | No | NA | 2 | 2 | 0 | Low-quality | Genome-fragment | 3.68 | AAI-based (high-confidence) | 0 |
| S11-final-viral-combined_k141_319717\|\|full | 1902 | No | NA | 3 | 0 | 1 | Not-determined | Genome-fragment | NA | NA | 0 |
| S11-final-viral-combined_k141_168886\|\|full | 3414 | No | NA | 4 | 0 | 2 | Not-determined | Genome-fragment | NA | NA | 0 |
| S4_k141_137683 | 1100 | No | NA | 1 | 1 | 0 | Low-quality | Genome-fragment | 0.42 | HMM-based (lower-bound) | 0 |
| S6-final-viral-combined_k141_123476\|\|full | 1100 | No | NA | 2 | 0 | 0 | Low-quality | Genome-fragment | 1.28 | AAI-based (medium-confidence) | 0 |
| S8_k141_273351 | 1100 | No | NA | 2 | 0 | 0 | Low-quality | Genome-fragment | 1.17 | AAI-based (high-confidence) | 0 |
| S11-final-viral-combined_k141_168894\|\|full | 1469 | No | NA | 2 | 0 | 1 | Not-determined | Genome-fragment | NA | NA | 0 |
| S1-final-viral-combined_k141_191475\|\|full | 1101 | No | NA | 2 | 1 | 0 | Low-quality | Genome-fragment | 2.24 | AAI-based (medium-confidence) | 0 |
| S11-final-viral-combined_k141_195935\|\|full | 2459 | No | NA | 3 | 0 | 0 | Not-determined | Genome-fragment | NA | NA | 0 |
| S1-final-viral-combined_k141_198022\|\|lt2gene | 1101 | No | NA | 1 | 1 | 0 | Low-quality | Genome-fragment | 2.52 | AAI-based (high-confidence) | 0 |
| S1_k141_275097 | 1101 | No | NA | 2 | 0 | 0 | Low-quality | Genome-fragment | 2.8 | AAI-based (medium-confidence) | 0 |
| S7-final-viral-combined_k141_185762\|\|lt2gene | 1101 | No | NA | 2 | 1 | 0 | Low-quality | Genome-fragment | 2.72 | AAI-based (high-confidence) | 0 |
| S11-final-viral-combined_k141_319774\|\|full | 1705 | No | NA | 4 | 0 | 2 | Not-determined | Genome-fragment | NA | NA | 0 |
| S17-final-viral-combined_k141_151021\|\|full | 1101 | No | NA | 2 | 0 | 2 | Low-quality | Genome-fragment | 2.51 | AAI-based (medium-confidence) | 0 |
| S11-final-viral-combined_k141_99916\|\|full | 3504 | No | NA | 5 | 0 | 0 | Not-determined | Genome-fragment | NA | NA | 0 |
| S12-final-viral-combined_k141_116057\|\|full | 1101 | No | NA | 4 | 0 | 2 | Low-quality | Genome-fragment | 2.31 | AAI-based (medium-confidence) | 0 |
| S2-final-viral-combined_k141_101010\|\|full | 1101 | No | NA | 2 | 0 | 0 | Low-quality | Genome-fragment | 2.24 | AAI-based (high-confidence) | 0 |
| S6-final-viral-combined_k141_84259\|\|full | 1101 | No | NA | 2 | 0 | 1 | Low-quality | Genome-fragment | 1.59 | AAI-based (high-confidence) | 0 |
| S6-final-viral-combined_k141_92039\|\|full | 1101 | No | NA | 3 | 0 | 1 | Low-quality | Genome-fragment | 1.27 | AAI-based (medium-confidence) | 0 |
| S11-final-viral-combined_k141_58607\|\|full | 1644 | No | NA | 2 | 0 | 1 | Not-determined | Genome-fragment | NA | NA | 0 |
| S11-final-viral-combined_k141_279160\|\|full | 3143 | No | NA | 3 | 0 | 1 | Not-determined | Genome-fragment | NA | NA | 0 |
| S6-final-viral-combined_k141_121166\|\|full | 1101 | No | NA | 3 | 0 | 0 | Low-quality | Genome-fragment | 2.68 | AAI-based (high-confidence) | 0 |
| S11-final-viral-combined_k141_127241\|\|full | 3269 | No | NA | 2 | 0 | 1 | Not-determined | Genome-fragment | NA | NA | 0 |
| S8-final-viral-combined_k141_224221\|\|lt2gene | 1101 | No | NA | 1 | 1 | 0 | Low-quality | Genome-fragment | 1.03 | AAI-based (high-confidence) | 0 |
| S11-final-viral-combined_k141_155656\|\|lt2gene | 1102 | No | NA | 2 | 0 | 1 | Low-quality | Genome-fragment | 1.2 | AAI-based (medium-confidence) | 0 |
| S11-final-viral-combined_k141_210192\|\|full | 1368 | No | NA | 2 | 0 | 1 | Not-determined | Genome-fragment | NA | NA | 0 |
| S11-final-viral-combined_k141_237896\|\|full | 7225 | No | NA | 8 | 0 | 1 | Not-determined | Genome-fragment | NA | NA | 0 |
| S11-final-viral-combined_k141_306746\|\|full | 1397 | No | NA | 3 | 0 | 1 | Not-determined | Genome-fragment | NA | NA | 0 |
| S11-final-viral-combined_k141_196003\|\|full | 25024 | No | NA | 22 | 0 | 16 | Not-determined | Genome-fragment | NA | NA | 0 |
| S11-final-viral-combined_k141_237902\|\|full | 2731 | No | NA | 2 | 0 | 2 | Not-determined | Genome-fragment | NA | NA | 0 |
| S11-final-viral-combined_k141_30808\|\|full | 2259 | No | NA | 3 | 0 | 0 | Not-determined | Genome-fragment | NA | NA | 0 |
| S11-final-viral-combined_k141_265060\|\|full | 1768 | No | NA | 4 | 0 | 2 | Not-determined | Genome-fragment | NA | NA | 0 |
| S11-final-viral-combined_k141_58622\|\|full | 2061 | No | NA | 2 | 0 | 1 | Not-determined | Genome-fragment | NA | NA | 0 |
| S1-final-viral-combined_k141_168174\|\|full | 1102 | No | NA | 2 | 0 | 2 | Low-quality | Genome-fragment | 0.53 | AAI-based (medium-confidence) | 0 |
| S11-final-viral-combined_k141_154845\|\|full | 25037 | No | NA | 34 | 22 | 0 | Medium-quality | Genome-fragment | 84.36 | AAI-based (high-confidence) | 0 |
| S3_k141_89452 | 1102 | No | NA | 2 | 0 | 0 | Low-quality | Genome-fragment | 0.93 | AAI-based (high-confidence) | 0 |
| S7-final-viral-combined_k141_138383\|\|full | 1102 | No | NA | 4 | 1 | 0 | Low-quality | Genome-fragment | 2.9 | AAI-based (high-confidence) | 0 |
| S11-final-viral-combined_k141_279215\|\|full | 7111 | No | NA | 7 | 0 | 5 | Not-determined | Genome-fragment | NA | NA | 0 |
| S11-final-viral-combined_k141_237940\|\|full | 3000 | No | NA | 3 | 0 | 0 | Not-determined | Genome-fragment | NA | NA | 0 |
| S11-final-viral-combined_k141_154875\|\|full | 2638 | No | NA | 4 | 0 | 0 | Not-determined | Genome-fragment | NA | NA | 0 |
| S7-final-viral-combined_k141_109828\|\|full | 1102 | No | NA | 3 | 0 | 0 | Low-quality | Genome-fragment | 2.46 | AAI-based (medium-confidence) | 0 |
| S11-final-viral-combined_k141_306804\|\|full | 3154 | No | NA | 6 | 0 | 0 | Not-determined | Genome-fragment | NA | NA | 0 |
| S11-final-viral-combined_k141_100032\|\|full | 1272 | No | NA | 2 | 0 | 0 | Not-determined | Genome-fragment | NA | NA | 0 |
| S11-final-viral-combined_k141_85931\|\|full | 2164 | No | NA | 2 | 0 | 2 | Not-determined | Genome-fragment | NA | NA | 0 |
| S11-final-viral-combined_k141_100035\|\|full | 1369 | No | NA | 2 | 0 | 1 | Not-determined | Genome-fragment | NA | NA | 0 |
| S7_k141_213637 | 1102 | No | NA | 2 | 0 | 1 | Low-quality | Genome-fragment | 2.21 | AAI-based (medium-confidence) | 0 |
| S17-final-viral-combined_k141_5177\|\|full | 1102 | No | NA | 2 | 0 | 0 | Low-quality | Genome-fragment | 2.18 | AAI-based (medium-confidence) | 0 |
| S17-final-viral-combined_k141_252406\|\|full | 1102 | No | NA | 3 | 0 | 1 | Low-quality | Genome-fragment | 1.55 | AAI-based (medium-confidence) | 0 |
| S11-final-viral-combined_k141_279251\|\|full | 2228 | No | NA | 2 | 0 | 2 | Not-determined | Genome-fragment | NA | NA | 0 |
| S11-final-viral-combined_k141_154907\|\|full | 1422 | No | NA | 2 | 0 | 0 | Not-determined | Genome-fragment | NA | NA | 0 |
| S12_k141_189259 | 1102 | No | NA | 1 | 0 | 0 | Low-quality | Genome-fragment | 2.11 | AAI-based (high-confidence) | 0 |
| S11-final-viral-combined_k141_279272\|\|full | 8112 | No | NA | 7 | 0 | 4 | Not-determined | Genome-fragment | NA | NA | 0 |
| S11-final-viral-combined_k141_319895\|\|full | 2995 | No | NA | 2 | 0 | 1 | Not-determined | Genome-fragment | NA | NA | 0 |
| S12_k141_284395 | 1102 | No | NA | 3 | 0 | 0 | Low-quality | Genome-fragment | 16 | AAI-based (medium-confidence) | 0 |
| S11-final-viral-combined_k141_100079\|\|full | 7099 | No | NA | 6 | 0 | 5 | Not-determined | Genome-fragment | NA | NA | 0 |
| S2_k141_115476 | 1102 | No | NA | 3 | 0 | 0 | Low-quality | Genome-fragment | 3.34 | AAI-based (medium-confidence) | 0 |
| S11-final-viral-combined_k141_265141\|\|full | 2626 | No | NA | 3 | 0 | 1 | Not-determined | Genome-fragment | NA | NA | 0 |
| S6-final-viral-combined_k141_91275\|\|full | 1102 | No | NA | 2 | 0 | 0 | Low-quality | Genome-fragment | 1.74 | AAI-based (high-confidence) | 0 |
| S11-final-viral-combined_k141_100091\|\|full | 5189 | No | NA | 5 | 0 | 4 | Not-determined | Genome-fragment | NA | NA | 0 |
| S8-final-viral-combined_k141_219217\|\|full | 1102 | No | NA | 2 | 0 | 1 | Low-quality | Genome-fragment | 1.96 | AAI-based (medium-confidence) | 0 |
| S8-final-viral-combined_k141_225844\|\|lt2gene | 1102 | No | NA | 1 | 1 | 0 | Low-quality | Genome-fragment | 2.53 | AAI-based (high-confidence) | 0 |
| S9-final-viral-combined_k141_51973\|\|full | 1103 | No | NA | 3 | 1 | 0 | Low-quality | Genome-fragment | 1.36 | AAI-based (high-confidence) | 0 |
| S11-final-viral-combined_k141_198523\|\|full | 1103 | No | NA | 2 | 0 | 0 | Low-quality | Genome-fragment | 0.73 | AAI-based (high-confidence) | 0 |
| S11-final-viral-combined_k141_141082\|\|full | 1695 | No | NA | 4 | 0 | 1 | Not-determined | Genome-fragment | NA | NA | 0 |
| S11-final-viral-combined_k141_30888\|\|full | 2978 | No | NA | 3 | 0 | 3 | Not-determined | Genome-fragment | NA | NA | 0 |
| S11-final-viral-combined_k141_154976\|\|full | 1699 | No | NA | 4 | 0 | 2 | Not-determined | Genome-fragment | NA | NA | 0 |
| S11-final-viral-combined_k141_100124\|\|full | 2476 | No | NA | 3 | 0 | 0 | Not-determined | Genome-fragment | NA | NA | 0 |
| S1-final-viral-combined_k141_7191\|\|full | 1103 | No | NA | 3 | 0 | 3 | Low-quality | Genome-fragment | 0.84 | AAI-based (medium-confidence) | 0 |
| S11-final-viral-combined_k141_3381\|\|full | 6826 | No | NA | 9 | 0 | 3 | Not-determined | Genome-fragment | NA | NA | 0 |
| S11-final-viral-combined_k141_17029\|\|full | 30219 | No | NA | 51 | 5 | 1 | Medium-quality | Genome-fragment | 54.28 | AAI-based (high-confidence) | 0 |
| S1_k141_234796 | 1103 | No | NA | 1 | 0 | 0 | Low-quality | Genome-fragment | 2.58 | AAI-based (medium-confidence) | 0 |
| S5-final-viral-combined_k141_169196\|\|full | 1103 | No | NA | 2 | 1 | 0 | Low-quality | Genome-fragment | 0.52 | AAI-based (medium-confidence) | 0 |
| S5-final-viral-combined_k141_20205\|\|lt2gene | 1103 | No | NA | 2 | 2 | 0 | Low-quality | Genome-fragment | 2.17 | AAI-based (medium-confidence) | 0 |
| S11-final-viral-combined_k141_127327\|\|full | 2000 | No | NA | 3 | 0 | 2 | Not-determined | Genome-fragment | NA | NA | 0 |
| S7-final-viral-combined_k141_80103\|\|lt2gene | 1103 | No | NA | 2 | 1 | 0 | Low-quality | Genome-fragment | 1.86 | AAI-based (high-confidence) | 0 |
| S11-final-viral-combined_k141_292437\|\|full | 1860 | No | NA | 4 | 0 | 1 | Not-determined | Genome-fragment | NA | NA | 0 |
| S11-final-viral-combined_k141_320010\|\|full | 1810 | No | NA | 2 | 0 | 0 | Not-determined | Genome-fragment | NA | NA | 0 |
| S11-final-viral-combined_k141_44721\|\|full | 2058 | No | NA | 2 | 0 | 2 | Not-determined | Genome-fragment | NA | NA | 0 |
| S11-final-viral-combined_k141_210369\|\|full | 6782 | No | NA | 4 | 0 | 3 | Not-determined | Genome-fragment | NA | NA | 0 |
| S11-final-viral-combined_k141_100171\|\|full | 5209 | No | NA | 7 | 0 | 5 | Not-determined | Genome-fragment | NA | NA | 0 |
| S7_k141_20925 | 1103 | Yes | 617 | 2 | 1 | 1 | Low-quality | Genome-fragment | 1.52 | AAI-based (high-confidence) | 44.06 |
| S11-final-viral-combined_k141_306971\|\|full | 2249 | No | NA | 3 | 0 | 1 | Not-determined | Genome-fragment | NA | NA | 0 |
| S12_k141_167148 | 1103 | No | NA | 1 | 1 | 0 | Low-quality | Genome-fragment | 1.9 | AAI-based (high-confidence) | 0 |
| S3-final-viral-combined_k141_23875\|\|full | 1104 | No | NA | 3 | 1 | 0 | Low-quality | Genome-fragment | 1.82 | AAI-based (high-confidence) | 0 |

| | | | | | | | | | | | |
|---|---|---|---|---|---|---|---|---|---|---|---|
| S11-final-viral-combined_k141_210383\|\|full | 3325 | No | NA | 4 | 0 | 2 | Not-determined | Genome-fragment | NA | NA | 0 |
| S3-final-viral-combined_k141_56749\|\|full | 1104 | No | NA | 2 | 0 | 1 | Low-quality | Genome-fragment | 1.25 | AAI-based (medium-confidence) | 0 |
| S11-final-viral-combined_k141_44751\|\|full | 19980 | No | NA | 19 | 0 | 16 | Not-determined | Genome-fragment | NA | NA | 0 |
| S11-final-viral-combined_k141_169244\|\|full | 1716 | No | NA | 3 | 0 | 0 | Not-determined | Genome-fragment | NA | NA | 0 |
| S11-final-viral-combined_k141_169247\|\|full | 2445 | No | NA | 3 | 0 | 3 | Not-determined | Genome-fragment | NA | NA | 0 |
| S5-final-viral-combined_k141_125167\|\|full | 1104 | No | NA | 3 | 1 | 0 | Low-quality | Genome-fragment | 1.22 | HMM-based (lower-bound) | 0 |
| S11-final-viral-combined_k141_292514\|\|full | 3758 | No | NA | 4 | 0 | 0 | Not-determined | Genome-fragment | NA | NA | 0 |
| S11-final-viral-combined_k141_113239\|\|full | 2196 | No | NA | 3 | 0 | 1 | Not-determined | Genome-fragment | NA | NA | 0 |
| S5-final-viral-combined_k141_135395\|\|full | 1104 | No | NA | 3 | 0 | 2 | Low-quality | Genome-fragment | 0.32 | AAI-based (medium-confidence) | 0 |
| S7-final-viral-combined_k141_60979\|\|lt2gene | 1104 | No | NA | 1 | 1 | 0 | Low-quality | Genome-fragment | 4.41 | AAI-based (high-confidence) | 0 |
| S17-final-viral-combined_k141_72412\|\|full | 1104 | No | NA | 2 | 0 | 0 | Low-quality | Genome-fragment | 2.43 | AAI-based (medium-confidence) | 0 |
| S17-k141_164075 | 1104 | No | NA | 1 | 0 | 1 | Low-quality | Genome-fragment | 2.26 | AAI-based (high-confidence) | 0 |
| S11-final-viral-combined_k141_141197\|\|full | 1868 | No | NA | 3 | 0 | 2 | Not-determined | Genome-fragment | NA | NA | 0 |
| S11-final-viral-combined_k141_238131\|\|full | 1505 | No | NA | 3 | 0 | 2 | Not-determined | Genome-fragment | NA | NA | 0 |
| S12-final-viral-combined_k141_50847\|\|full | 1104 | No | NA | 3 | 1 | 0 | Low-quality | Genome-fragment | 1.01 | HMM-based (lower-bound) | 0 |
| S12-final-viral-combined_k141_230375\|\|lt2gene | 1104 | No | NA | 1 | 1 | 0 | Low-quality | Genome-fragment | 0.76 | HMM-based (lower-bound) | 0 |
| S12_k141_322180 | 1104 | No | NA | 2 | 0 | 0 | Low-quality | Genome-fragment | 0.62 | AAI-based (medium-confidence) | 0 |
| S11-final-viral-combined_k141_169317\|\|full | 5954 | No | NA | 4 | 0 | 3 | Not-determined | Genome-fragment | NA | NA | 0 |
| S11-final-viral-combined_k141_31018\|\|full | 2045 | No | NA | 3 | 0 | 1 | Not-determined | Genome-fragment | NA | NA | 0 |
| S11-final-viral-combined_k141_71915\|\|full | 9424 | No | NA | 5 | 0 | 1 | Not-determined | Genome-fragment | NA | NA | 0 |
| S2-final-viral-combined_k141_20766\|\|full | 1104 | No | NA | 3 | 1 | 0 | Low-quality | Genome-fragment | 3.76 | AAI-based (medium-confidence) | 0 |
| S2-final-viral-combined_k141_114710\|\|full | 1104 | No | NA | 3 | 0 | 0 | Low-quality | Genome-fragment | 0.81 | AAI-based (high-confidence) | 0 |
| S11-final-viral-combined_k141_292605\|\|full | 1474 | No | NA | 3 | 0 | 0 | Not-determined | Genome-fragment | NA | NA | 0 |
| S11-final-viral-combined_k141_182617\|\|full | 4298 | No | NA | 9 | 0 | 0 | Not-determined | Genome-fragment | NA | NA | 0 |
| S2_k141_212493 | 1104 | No | NA | 2 | 0 | 0 | Low-quality | Genome-fragment | 1.75 | AAI-based (medium-confidence) | 0 |
| S11-final-viral-combined_k141_100323\|\|full | 1922 | No | NA | 3 | 0 | 1 | Not-determined | Genome-fragment | NA | NA | 0 |
| S11-final-viral-combined_k141_155163\|\|full | 1164 | No | NA | 3 | 0 | 1 | Not-determined | Genome-fragment | NA | NA | 0 |
| S11-final-viral-combined_k141_279469\|\|full | 3712 | No | NA | 7 | 0 | 0 | Not-determined | Genome-fragment | NA | NA | 0 |
| S11-final-viral-combined_k141_113315\|\|full | 5021 | No | NA | 4 | 0 | 3 | Not-determined | Genome-fragment | NA | NA | 0 |
| S11-final-viral-combined_k141_3507\|\|full | 82880 | Yes | 41645 | 117 | 18 | 35 | Complete | High-quality | 100 | Provirus (high-confidence) | 49.75 |
| S11-final-viral-combined_k141_71977\|\|full | 3178 | No | NA | 6 | 0 | 2 | Not-determined | Genome-fragment | NA | NA | 0 |
| S11-final-viral-combined_k141_224183\|\|full | 1045 | No | NA | 2 | 0 | 0 | Not-determined | Genome-fragment | NA | NA | 0 |
| S2_k141_26004 | 1104 | No | NA | 1 | 0 | 1 | Low-quality | Genome-fragment | 0.26 | AAI-based (medium-confidence) | 0 |
| S6-final-viral-combined_k141_41071\|\|full | 1104 | No | NA | 3 | 2 | 0 | Low-quality | Genome-fragment | 2 | AAI-based (high-confidence) | 0 |
| S11-final-viral-combined_k141_3516\|\|full | 1204 | No | NA | 2 | 0 | 1 | Not-determined | Genome-fragment | NA | NA | 0 |
| S11-final-viral-combined_k141_251437\|\|full | 10600 | No | NA | 12 | 0 | 9 | Not-determined | Genome-fragment | NA | NA | 0 |
| S11-final-viral-combined_k141_44934\|\|full | 2353 | No | NA | 2 | 0 | 0 | Not-determined | Genome-fragment | NA | NA | 0 |
| S11-final-viral-combined_k141_320191\|\|full | 4334 | No | NA | 5 | 0 | 2 | Not-determined | Genome-fragment | NA | NA | 0 |
| S6_k141_60055 | 1104 | Yes | 527 | 2 | 1 | 1 | Low-quality | Genome-fragment | 0.15 | HMM-based (lower-bound) | 52.26 |
| S11-final-viral-combined_k141_44944\|\|full | 2892 | No | NA | 3 | 0 | 2 | Not-determined | Genome-fragment | NA | NA | 0 |
| S8_k141_156309 | 1104 | No | NA | 2 | 1 | 0 | Low-quality | Genome-fragment | 1.88 | HMM-based (lower-bound) | 0 |
| S1-final-viral-combined_k141_102578\|\|full | 1105 | No | NA | 2 | 0 | 0 | Low-quality | Genome-fragment | 1.32 | AAI-based (medium-confidence) | 0 |
| S1-final-viral-combined_k141_131229\|\|lt2gene | 1105 | No | NA | 1 | 1 | 0 | Low-quality | Genome-fragment | 1.82 | HMM-based (lower-bound) | 0 |
| S11-final-viral-combined_k141_210623\|\|full | 1395 | No | NA | 3 | 0 | 0 | Not-determined | Genome-fragment | NA | NA | 0 |
| S11-final-viral-combined_k141_31075\|\|full | 1227 | No | NA | 3 | 0 | 1 | Not-determined | Genome-fragment | NA | NA | 0 |
| S1-final-viral-combined_k141_147009\|\|lt2gene | 1105 | No | NA | 2 | 1 | 0 | Low-quality | Genome-fragment | 3.02 | AAI-based (high-confidence) | 0 |
| S11-final-viral-combined_k141_224240\|\|full | 1252 | No | NA | 3 | 0 | 0 | Not-determined | Genome-fragment | NA | NA | 0 |
| S11-final-viral-combined_k141_196187\|\|full | 2285 | No | NA | 3 | 0 | 2 | Not-determined | Genome-fragment | NA | NA | 0 |
| S1_k141_314062 | 1105 | No | NA | 2 | 0 | 2 | Low-quality | Genome-fragment | 0.35 | AAI-based (medium-confidence) | 0 |
| S1_k141_134301 | 1105 | No | NA | 2 | 0 | 1 | Low-quality | Genome-fragment | 2.47 | AAI-based (medium-confidence) | 0 |
| S11-final-viral-combined_k141_44977\|\|full | 14535 | No | NA | 18 | 0 | 6 | Not-determined | Genome-fragment | NA | NA | 0 |
| S11-final-viral-combined_k141_86440\|\|full | 1117 | No | NA | 3 | 0 | 1 | Not-determined | Genome-fragment | NA | NA | 0 |
| S11-final-viral-combined_k141_279539\|\|full | 4149 | No | NA | 3 | 0 | 3 | Not-determined | Genome-fragment | NA | NA | 0 |
| S11-final-viral-combined_k141_210655\|\|full | 1658 | No | NA | 2 | 0 | 2 | Not-determined | Genome-fragment | NA | NA | 0 |
| S11-final-viral-combined_k141_141351\|\|full | 4306 | No | NA | 6 | 0 | 6 | Not-determined | Genome-fragment | NA | NA | 0 |
| S3-final-viral-combined_k141_55832\|\|full | 1105 | No | NA | 4 | 0 | 0 | Low-quality | Genome-fragment | 2.03 | AAI-based (high-confidence) | 0 |
| S11-final-viral-combined_k141_320266\|\|full | 1154 | No | NA | 3 | 0 | 0 | Not-determined | Genome-fragment | NA | NA | 0 |
| S5-final-viral-combined_k141_205106\|\|lt2gene | 1105 | No | NA | 2 | 2 | 0 | Low-quality | Genome-fragment | 2.38 | AAI-based (high-confidence) | 0 |
| S5-final-viral-combined_k141_173360\|\|lt2gene | 1105 | No | NA | 2 | 2 | 0 | Low-quality | Genome-fragment | 1.34 | HMM-based (lower-bound) | 0 |
| S12-final-viral-combined_k141_80405\|\|full | 1105 | No | NA | 2 | 1 | 0 | Low-quality | Genome-fragment | 1.68 | HMM-based (lower-bound) | 0 |
| S11-final-viral-combined_k141_210668\|\|full | 2311 | No | NA | 4 | 0 | 3 | Not-determined | Genome-fragment | NA | NA | 0 |
| S12-final-viral-combined_k141_50052\|\|full | 1105 | No | NA | 2 | 0 | 0 | Low-quality | Genome-fragment | 1.49 | AAI-based (medium-confidence) | 0 |
| S12-final-viral-combined_k141_158420\|\|lt2gene | 1105 | No | NA | 1 | 1 | 0 | Low-quality | Genome-fragment | 1.97 | HMM-based (lower-bound) | 0 |
| S11-final-viral-combined_k141_100495\|\|full | 1573 | No | NA | 3 | 0 | 0 | Not-determined | Genome-fragment | NA | NA | 0 |
| S2-final-viral-combined_k141_235562\|\|full | 1105 | No | NA | 2 | 1 | 0 | Low-quality | Genome-fragment | 1.71 | AAI-based (high-confidence) | 0 |
| S6-final-viral-combined_k141_111708\|\|full | 1105 | No | NA | 4 | 0 | 0 | Low-quality | Genome-fragment | 0.64 | AAI-based (medium-confidence) | 0 |
| S11-final-viral-combined_k141_196225\|\|full | 3954 | No | NA | 7 | 0 | 1 | Not-determined | Genome-fragment | NA | NA | 0 |
| S8-final-viral-combined_k141_118713\|\|full | 1105 | No | NA | 3 | 0 | 0 | Low-quality | Genome-fragment | 0.99 | AAI-based (high-confidence) | 0 |
| S11-final-viral-combined_k141_182785\|\|full | 4590 | No | NA | 7 | 0 | 1 | Not-determined | Genome-fragment | NA | NA | 0 |
| S8-final-viral-combined_k141_82407\|\|lt2gene | 1105 | No | NA | 2 | 1 | 0 | Low-quality | Genome-fragment | 1.27 | HMM-based (lower-bound) | 0 |
| S11-final-viral-combined_k141_101819\|\|lt2gene | 1106 | No | NA | 1 | 1 | 0 | Low-quality | Genome-fragment | 1.02 | HMM-based (lower-bound) | 0 |
| S11-final-viral-combined_k141_279617\|\|full | 2506 | No | NA | 4 | 0 | 2 | Not-determined | Genome-fragment | NA | NA | 0 |
| S11-final-viral-combined_k141_141366\|\|full | 1487 | No | NA | 2 | 0 | 0 | Not-determined | Genome-fragment | NA | NA | 0 |
| S11-final-viral-combined_k141_141367\|\|full | 1212 | No | NA | 3 | 0 | 0 | Not-determined | Genome-fragment | NA | NA | 0 |
| S11-final-viral-combined_k141_127635\|\|full | 2045 | No | NA | 4 | 0 | 2 | Not-determined | Genome-fragment | NA | NA | 0 |
| S11-final-viral-combined_k141_3589\|\|full | 25278 | No | NA | 27 | 0 | 15 | Not-determined | Genome-fragment | NA | NA | 0 |
| S11-final-viral-combined_k141_123914\|\|lt2gene | 1106 | No | NA | 2 | 1 | 0 | Low-quality | Genome-fragment | 3.11 | AAI-based (medium-confidence) | 0 |
| S11-final-viral-combined_k141_210725\|\|full | 4802 | No | NA | 3 | 0 | 2 | Not-determined | Genome-fragment | NA | NA | 0 |
| S1_k141_37854 | 1106 | No | NA | 1 | 0 | 0 | Low-quality | Genome-fragment | 0.63 | AAI-based (medium-confidence) | 0 |
| S11-final-viral-combined_k141_141402\|\|full | 2618 | No | NA | 7 | 0 | 1 | Not-determined | Genome-fragment | NA | NA | 0 |
| S11-final-viral-combined_k141_292874\|\|full | 10593 | No | NA | 12 | 0 | 5 | Not-determined | Genome-fragment | NA | NA | 0 |
| S11-final-viral-combined_k141_141417\|\|full | 1948 | No | NA | 3 | 0 | 2 | Not-determined | Genome-fragment | NA | NA | 0 |
| S11-final-viral-combined_k141_279651\|\|full | 2028 | No | NA | 3 | 0 | 1 | Not-determined | Genome-fragment | NA | NA | 0 |
| S1_k141_338709 | 1106 | No | NA | 2 | 0 | 1 | Low-quality | Genome-fragment | 2.42 | AAI-based (high-confidence) | 0 |
| S3-final-viral-combined_k141_134633\|\|lt2gene | 1106 | No | NA | 2 | 1 | 0 | Low-quality | Genome-fragment | 1.78 | HMM-based (lower-bound) | 0 |
| S7_k141_189616 | 1106 | No | NA | 3 | 0 | 0 | Low-quality | Genome-fragment | 1.05 | AAI-based (high-confidence) | 0 |
| S11-final-viral-combined_k141_113444\|\|full | 4636 | No | NA | 5 | 0 | 3 | Not-determined | Genome-fragment | NA | NA | 0 |
| S12-final-viral-combined_k141_65313\|\|full | 1106 | No | NA | 4 | 2 | 0 | Low-quality | Genome-fragment | 2.73 | AAI-based (medium-confidence) | 0 |
| S11-final-viral-combined_k141_113448\|\|full | 6419 | No | NA | 9 | 0 | 1 | Not-determined | Genome-fragment | NA | NA | 0 |
| S11-final-viral-combined_k141_86587\|\|full | 5982 | No | NA | 5 | 0 | 3 | Not-determined | Genome-fragment | NA | NA | 0 |
| S12-final-viral-combined_k141_378751\|\|lt2gene | 1106 | No | NA | 1 | 1 | 0 | Low-quality | Genome-fragment | 2.45 | AAI-based (high-confidence) | 0 |
| S11-final-viral-combined_k141_155350\|\|full | 1383 | No | NA | 3 | 0 | 0 | Not-determined | Genome-fragment | NA | NA | 0 |
| S12_k141_271250 | 1106 | No | NA | 3 | 0 | 0 | Low-quality | Genome-fragment | 1.05 | AAI-based (high-confidence) | 0 |
| S11-final-viral-combined_k141_127706\|\|full | 1152 | No | NA | 2 | 0 | 1 | Not-determined | Genome-fragment | NA | NA | 0 |
| S11-final-viral-combined_k141_265453\|\|full | 4483 | No | NA | 8 | 1 | 0 | High-quality | High-quality | 100 | HMM-based (lower-bound) | 0 |
| S11-final-viral-combined_k141_320409\|\|full | 2418 | No | NA | 2 | 0 | 1 | Not-determined | Genome-fragment | NA | NA | 0 |
| S11-final-viral-combined_k141_141474\|\|full | 3338 | No | NA | 5 | 0 | 0 | Not-determined | Genome-fragment | NA | NA | 0 |
| S12_k141_125715 | 1106 | No | NA | 2 | 0 | 0 | Low-quality | Genome-fragment | 1.8 | AAI-based (medium-confidence) | 0 |
| S11-final-viral-combined_k141_155379\|\|full | 1244 | No | NA | 3 | 0 | 1 | Not-determined | Genome-fragment | NA | NA | 0 |
| S2-final-viral-combined_k141_171994\|\|full | 1106 | No | NA | 2 | 0 | 1 | Low-quality | Genome-fragment | 1.47 | AAI-based (medium-confidence) | 0 |
| S6_k141_124363 | 1106 | No | NA | 1 | 0 | 1 | Low-quality | Genome-fragment | 0.41 | AAI-based (medium-confidence) | 0 |
| S8-final-viral-combined_k141_235206\|\|full | 1106 | No | NA | 3 | 1 | 0 | Low-quality | Genome-fragment | 2.8 | AAI-based (medium-confidence) | 0 |
| S8-final-viral-combined_k141_205250\|\|full | 1106 | No | NA | 2 | 0 | 1 | Low-quality | Genome-fragment | 0.69 | AAI-based (high-confidence) | 0 |
| S11-final-viral-combined_k141_265511\|\|full | 1870 | No | NA | 3 | 0 | 1 | Not-determined | Genome-fragment | NA | NA | 0 |
| S11-final-viral-combined_k141_155429\|\|full | 2755 | No | NA | 4 | 0 | 4 | Not-determined | Genome-fragment | NA | NA | 0 |
| S11-final-viral-combined_k141_3692\|\|full | 7314 | No | NA | 6 | 0 | 4 | Not-determined | Genome-fragment | NA | NA | 0 |
| S9-final-viral-combined_k141_103096\|\|full | 1107 | No | NA | 5 | 0 | 0 | Low-quality | Genome-fragment | 3.02 | AAI-based (medium-confidence) | 0 |
| S11-final-viral-combined_k141_31984\|\|full | 1107 | No | NA | 3 | 2 | 0 | Low-quality | Genome-fragment | 1.97 | AAI-based (high-confidence) | 0 |
| S1_k141_269605 | 1107 | No | NA | 2 | 0 | 1 | Low-quality | Genome-fragment | 0.31 | AAI-based (high-confidence) | 0 |
| S11-final-viral-combined_k141_86699\|\|full | 1544 | No | NA | 3 | 0 | 1 | Not-determined | Genome-fragment | NA | NA | 0 |
| S11-final-viral-combined_k141_141550\|\|full | 1770 | No | NA | 4 | 0 | 1 | Not-determined | Genome-fragment | NA | NA | 0 |
| S3_k141_100439 | 1107 | Yes | 517 | 2 | 1 | 1 | Low-quality | Genome-fragment | 0.24 | HMM-based (lower-bound) | 53.3 |
| S7-final-viral-combined_k141_127462\|\|lt2gene | 1107 | No | NA | 2 | 2 | 0 | Low-quality | Genome-fragment | 2.33 | AAI-based (high-confidence) | 0 |
| S7_k141_102034 | 1107 | No | NA | 1 | 0 | 0 | Low-quality | Genome-fragment | 1.04 | AAI-based (high-confidence) | 0 |
| S11-final-viral-combined_k141_141573\|\|full | 1900 | No | NA | 5 | 0 | 3 | Not-determined | Genome-fragment | NA | NA | 0 |
| S17-final-viral-combined_k141_160582\|\|full | 1107 | No | NA | 6 | 0 | 0 | Low-quality | Genome-fragment | 2.04 | AAI-based (medium-confidence) | 0 |
| S12-final-viral-combined_k141_122837\|\|full | 1107 | No | NA | 3 | 0 | 0 | Low-quality | Genome-fragment | 3.91 | AAI-based (high-confidence) | 0 |
| S12-final-viral-combined_k141_224893\|\|full | 1107 | No | NA | 3 | 0 | 0 | Low-quality | Genome-fragment | 0.49 | AAI-based (medium-confidence) | 0 |
| S12-final-viral-combined_k141_208239\|\|lt2gene | 1107 | No | NA | 1 | 1 | 0 | Low-quality | Genome-fragment | 3.23 | AAI-based (high-confidence) | 0 |
| S12_k141_371686 | 1107 | No | NA | 1 | 0 | 0 | Low-quality | Genome-fragment | 0.61 | AAI-based (high-confidence) | 0 |
| S2-final-viral-combined_k141_244626\|\|full | 1107 | No | NA | 4 | 1 | 0 | Low-quality | Genome-fragment | 2.38 | AAI-based (high-confidence) | 0 |
| S11-final-viral-combined_k141_86725\|\|full | 3391 | No | NA | 4 | 0 | 1 | Not-determined | Genome-fragment | NA | NA | 0 |
| S4-final-viral-combined_k141_121387\|\|full | 1107 | No | NA | 1 | 0 | 1 | Low-quality | Genome-fragment | 0.6 | AAI-based (medium-confidence) | 0 |
| S9-final-viral-combined_k141_71606\|\|full | 1108 | No | NA | 3 | 0 | 0 | Low-quality | Genome-fragment | 1.36 | AAI-based (medium-confidence) | 0 |
| S11-final-viral-combined_k141_293050\|\|full | 1160 | No | NA | 3 | 0 | 3 | Not-determined | Genome-fragment | NA | NA | 0 |
| S11-final-viral-combined_k141_279848\|\|full | 1012 | No | NA | 2 | 0 | 1 | Not-determined | Genome-fragment | NA | NA | 0 |
| S11-final-viral-combined_k141_155521\|\|full | 3228 | No | NA | 7 | 0 | 0 | Not-determined | Genome-fragment | NA | NA | 0 |
| S11-final-viral-combined_k141_128336\|\|full | 1108 | No | NA | 3 | 0 | 0 | Low-quality | Genome-fragment | 2.22 | AAI-based (high-confidence) | 0 |
| S11-final-viral-combined_k141_31377\|\|full | 2221 | No | NA | 4 | 0 | 3 | Not-determined | Genome-fragment | NA | NA | 0 |
| S11-final-viral-combined_k141_279870\|\|full | 1116 | No | NA | 3 | 0 | 1 | Not-determined | Genome-fragment | NA | NA | 0 |
| S11-final-viral-combined_k141_155555\|\|full | 1946 | No | NA | 2 | 0 | 1 | Not-determined | Genome-fragment | NA | NA | 0 |
| S11-final-viral-combined_k141_224464\|\|full | 2882 | No | NA | 4 | 0 | 3 | Not-determined | Genome-fragment | NA | NA | 0 |
| S11-final-viral-combined_k141_59289\|\|full | 2632 | No | NA | 3 | 0 | 1 | Not-determined | Genome-fragment | NA | NA | 0 |
| S11-final-viral-combined_k141_261821\|\|full | 1108 | No | NA | 2 | 0 | 1 | Low-quality | Genome-fragment | 1.65 | AAI-based (medium-confidence) | 0 |
| S11-final-viral-combined_k141_17428\|\|full | 1254 | No | NA | 3 | 0 | 0 | Not-determined | Genome-fragment | NA | NA | 0 |
| S11-final-viral-combined_k141_196476\|\|full | 1552 | No | NA | 4 | 0 | 2 | Not-determined | Genome-fragment | NA | NA | 0 |
| S11-final-viral-combined_k141_293133\|\|full | 3673 | No | NA | 6 | 0 | 2 | Not-determined | Genome-fragment | NA | NA | 0 |
| S11-final-viral-combined_k141_59315\|\|full | 1446 | No | NA | 2 | 0 | 1 | Not-determined | Genome-fragment | NA | NA | 0 |
| S1-final-viral-combined_k141_159363\|\|full | 1108 | No | NA | 3 | 2 | 0 | Low-quality | Genome-fragment | 2.44 | AAI-based (high-confidence) | 0 |
| S11-final-viral-combined_k141_100734\|\|full | 2012 | No | NA | 3 | 0 | 1 | Not-determined | Genome-fragment | NA | NA | 0 |
| S11-final-viral-combined_k141_238424\|\|full | 1120 | No | NA | 2 | 0 | 2 | Not-determined | Genome-fragment | NA | NA | 0 |
| S11-final-viral-combined_k141_320569\|\|full | 4590 | No | NA | 3 | 0 | 1 | Not-determined | Genome-fragment | NA | NA | 0 |
| S11-final-viral-combined_k141_72318\|\|full | 1930 | No | NA | 3 | 0 | 2 | Not-determined | Genome-fragment | NA | NA | 0 |
| S11-final-viral-combined_k141_17449\|\|full | 1319 | No | NA | 2 | 0 | 0 | Not-determined | Genome-fragment | NA | NA | 0 |
| S5-final-viral-combined_k141_94235\|\|full | 1108 | No | NA | 4 | 1 | 0 | Low-quality | Genome-fragment | 2.83 | AAI-based (high-confidence) | 0 |
| S5-final-viral-combined_k141_39700\|\|lt2gene | 1108 | No | NA | 2 | 1 | 0 | Low-quality | Genome-fragment | 2.55 | AAI-based (high-confidence) | 0 |
| S11-final-viral-combined_k141_113569\|\|full | 1309 | No | NA | 3 | 0 | 0 | Not-determined | Genome-fragment | NA | NA | 0 |
| S11-final-viral-combined_k141_31399\|\|full | 1307 | No | NA | 2 | 0 | 2 | Not-determined | Genome-fragment | NA | NA | 0 |
| S11-final-viral-combined_k141_86834\|\|full | 3422 | No | NA | 3 | 0 | 2 | Not-determined | Genome-fragment | NA | NA | 0 |
| S11-final-viral-combined_k141_293157\|\|full | 8579 | No | NA | 13 | 0 | 1 | Not-determined | Genome-fragment | NA | NA | 0 |
| S5_k141_113537 | 1108 | No | NA | 2 | 0 | 0 | Low-quality | Genome-fragment | 2.11 | AAI-based (high-confidence) | 0 |
| S11-final-viral-combined_k141_127870\|\|full | 3563 | No | NA | 5 | 0 | 5 | Not-determined | Genome-fragment | NA | NA | 0 |
| S11-final-viral-combined_k141_100749\|\|full | 2523 | No | NA | 2 | 0 | 2 | Not-determined | Genome-fragment | NA | NA | 0 |
| S11-final-viral-combined_k141_45309\|\|full | 1551 | No | NA | 3 | 0 | 0 | Not-determined | Genome-fragment | NA | NA | 0 |
| S11-final-viral-combined_k141_251870\|\|full | 5498 | No | NA | 4 | 0 | 0 | Not-determined | Genome-fragment | NA | NA | 0 |
| S11-final-viral-combined_k141_320601\|\|full | 5306 | No | NA | 9 | 0 | 1 | Not-determined | Genome-fragment | NA | NA | 0 |
| S7-final-viral-combined_k141_63944\|\|full | 1108 | No | NA | 2 | 0 | 0 | Low-quality | Genome-fragment | 2.82 | AAI-based (medium-confidence) | 0 |
| S11-final-viral-combined_k141_251886\|\|full | 4078 | No | NA | 4 | 0 | 3 | Not-determined | Genome-fragment | NA | NA | 0 |
| S7-final-viral-combined_k141_55363\|\|full | 1108 | No | NA | 2 | 1 | 0 | Low-quality | Genome-fragment | 0.69 | AAI-based (medium-confidence) | 0 |
| S11-final-viral-combined_k141_224496\|\|full | 2155 | No | NA | 4 | 0 | 1 | Not-determined | Genome-fragment | NA | NA | 0 |
| S17-final-viral-combined_k141_247066\|\|full | 1108 | No | NA | 3 | 0 | 0 | Low-quality | Genome-fragment | 5.23 | AAI-based (high-confidence) | 0 |
| S17-final-viral-combined_k141_237991\|\|full | 1108 | No | NA | 2 | 0 | 1 | Low-quality | Genome-fragment | 0.85 | AAI-based (medium-confidence) | 0 |
| S11-final-viral-combined_k141_3889\|\|full | 1371 | No | NA | 3 | 0 | 1 | Not-determined | Genome-fragment | NA | NA | 0 |
| S11-final-viral-combined_k141_113605\|\|full | 1691 | No | NA | 3 | 0 | 0 | Not-determined | Genome-fragment | NA | NA | 0 |
| S12_k141_302691 | 1108 | No | NA | 2 | 0 | 1 | Low-quality | Genome-fragment | 0.63 | AAI-based (medium-confidence) | 0 |
| S11-final-viral-combined_k141_279980\|\|full | 7494 | No | NA | 8 | 0 | 6 | Not-determined | Genome-fragment | NA | NA | 0 |
| S11-final-viral-combined_k141_72366\|\|full | 8725 | No | NA | 10 | 0 | 8 | Not-determined | Genome-fragment | NA | NA | 0 |
| S4-final-viral-combined_k141_88055\|\|lt2gene | 1108 | No | NA | 2 | 0 | 0 | Low-quality | Genome-fragment | 2.56 | AAI-based (high-confidence) | 0 |
| S11-final-viral-combined_k141_113627\|\|full | 5037 | No | NA | 5 | 0 | 2 | Not-determined | Genome-fragment | NA | NA | 0 |
| S11-final-viral-combined_k141_224538\|\|full | 5670 | No | NA | 8 | 0 | 1 | Not-determined | Genome-fragment | NA | NA | 0 |
| S6-final-viral-combined_k141_140197\|\|full | 1108 | No | NA | 2 | 0 | 0 | Low-quality | Genome-fragment | 0.71 | AAI-based (high-confidence) | 0 |
| S11-final-viral-combined_k141_100782\|\|full | 3276 | No | NA | 4 | 0 | 2 | Not-determined | Genome-fragment | NA | NA | 0 |
| S11-final-viral-combined_k141_280054\|\|full | 4466 | No | NA | 4 | 0 | 3 | Not-determined | Genome-fragment | NA | NA | 0 |
| S11-final-viral-combined_k141_300273\|\|full | 1109 | No | NA | 3 | 1 | 0 | Low-quality | Genome-fragment | 2.94 | AAI-based (medium-confidence) | 0 |
| S11-final-viral-combined_k141_265795\|\|full | 1457 | No | NA | 2 | 0 | 0 | Not-determined | Genome-fragment | NA | NA | 0 |
| S11-final-viral-combined_k141_72418\|\|full | 6660 | No | NA | 8 | 0 | 7 | Not-determined | Genome-fragment | NA | NA | 0 |
| S11-final-viral-combined_k141_84132\|\|lt2gene | 1109 | No | NA | 2 | 2 | 0 | Low-quality | Genome-fragment | 3.27 | AAI-based (medium-confidence) | 0 |
| S11-final-viral-combined_k141_169829\|\|full | 6475 | No | NA | 6 | 0 | 4 | Not-determined | Genome-fragment | NA | NA | 0 |
| S1-final-viral-combined_k141_362249\|\|full | 1109 | No | NA | 2 | 0 | 0 | Low-quality | Genome-fragment | 0.83 | AAI-based (high-confidence) | 0 |
| S11-final-viral-combined_k141_45457\|\|full | 1515 | No | NA | 3 | 0 | 1 | Not-determined | Genome-fragment | NA | NA | 0 |
| S11-final-viral-combined_k141_141868\|\|full | 2893 | No | NA | 2 | 0 | 2 | Not-determined | Genome-fragment | NA | NA | 0 |
| S11-final-viral-combined_k141_211311\|\|full | 2345 | No | NA | 3 | 0 | 3 | Not-determined | Genome-fragment | NA | NA | 0 |
| S11-final-viral-combined_k141_141880\|\|full | 4066 | No | NA | 6 | 0 | 5 | Not-determined | Genome-fragment | NA | NA | 0 |
| S11-final-viral-combined_k141_196625\|\|full | 2544 | No | NA | 5 | 0 | 3 | Not-determined | Genome-fragment | NA | NA | 0 |
| S11-final-viral-combined_k141_211342\|\|full | 1681 | No | NA | 2 | 0 | 0 | Not-determined | Genome-fragment | NA | NA | 0 |
| S1-final-viral-combined_k141_15988\|\|full | 1109 | No | NA | 4 | 0 | 0 | Low-quality | Genome-fragment | 2.46 | AAI-based (high-confidence) | 0 |
| S11-final-viral-combined_k141_141901\|\|full | 2500 | No | NA | 2 | 0 | 2 | Not-determined | Genome-fragment | NA | NA | 0 |
| S11-final-viral-combined_k141_238736\|\|full | 3794 | No | NA | 4 | 0 | 4 | Not-determined | Genome-fragment | NA | NA | 0 |
| S11-final-viral-combined_k141_252044\|\|full | 1278 | No | NA | 3 | 0 | 0 | Not-determined | Genome-fragment | NA | NA | 0 |
| S11-final-viral-combined_k141_307549\|\|full | 1042 | No | NA | 3 | 0 | 1 | Not-determined | Genome-fragment | NA | NA | 0 |
| S11-final-viral-combined_k141_265892\|\|full | 3137 | No | NA | 3 | 0 | 3 | Not-determined | Genome-fragment | NA | NA | 0 |
| S11-final-viral-combined_k141_293302\|\|full | 3103 | No | NA | 3 | 0 | 1 | Not-determined | Genome-fragment | NA | NA | 0 |
| S11-final-viral-combined_k141_59424\|\|full | 3312 | No | NA | 3 | 0 | 2 | Not-determined | Genome-fragment | NA | NA | 0 |
| S11-final-viral-combined_k141_72444\|\|full | 2445 | No | NA | 3 | 0 | 0 | Not-determined | Genome-fragment | NA | NA | 0 |
| S11-final-viral-combined_k141_293323\|\|full | 1882 | No | NA | 4 | 0 | 1 | Not-determined | Genome-fragment | NA | NA | 0 |
| S3-final-viral-combined_k141_104631\|\|lt2gene | 1109 | No | NA | 2 | 1 | 0 | Low-quality | Genome-fragment | 4.08 | HMM-based (lower-bound) | 0 |
| S11-final-viral-combined_k141_224695\|\|full | 1757 | No | NA | 3 | 0 | 0 | Not-determined | Genome-fragment | NA | NA | 0 |
| S11-final-viral-combined_k141_224703\|\|full | 1602 | No | NA | 3 | 0 | 1 | Not-determined | Genome-fragment | NA | NA | 0 |
| S5-final-viral-combined_k141_125717\|\|full | 1109 | No | NA | 2 | 0 | 1 | Low-quality | Genome-fragment | 0.86 | AAI-based (medium-confidence) | 0 |
| S5-final-viral-combined_k141_136352\|\|full | 1109 | No | NA | 2 | 1 | 0 | Low-quality | Genome-fragment | 3.19 | AAI-based (high-confidence) | 0 |
| S17-final-viral-combined_k141_249961\|\|full | 1109 | No | NA | 2 | 0 | 0 | Low-quality | Genome-fragment | 1.31 | AAI-based (high-confidence) | 0 |
| S17-final-viral-combined_k141_55532\|\|full | 1109 | No | NA | 2 | 0 | 2 | Low-quality | Genome-fragment | 0.42 | AAI-based (medium-confidence) | 0 |
| S17-final-viral-combined_k141_69863\|\|lt2gene | 1109 | No | NA | 1 | 1 | 0 | Low-quality | Genome-fragment | 3.24 | AAI-based (high-confidence) | 0 |
| S11-final-viral-combined_k141_252077\|\|full | 2818 | No | NA | 3 | 0 | 1 | Not-determined | Genome-fragment | NA | NA | 0 |
| S17-k141_247035 | 1109 | No | NA | 3 | 0 | 0 | Low-quality | Genome-fragment | 2.35 | AAI-based (medium-confidence) | 0 |
| S8-final-viral-combined_k141_243785\|\|full | 1109 | No | NA | 2 | 0 | 1 | Low-quality | Genome-fragment | 0.95 | AAI-based (medium-confidence) | 0 |
| S11-final-viral-combined_k141_4024\|\|full | 3783 | No | NA | 3 | 0 | 3 | Not-determined | Genome-fragment | NA | NA | 0 |
| S11-final-viral-combined_k141_211449\|\|full | 1155 | No | NA | 2 | 0 | 1 | Not-determined | Genome-fragment | NA | NA | 0 |
| S11-final-viral-combined_k141_141980\|\|full | 5595 | No | NA | 3 | 0 | 2 | Not-determined | Genome-fragment | NA | NA | 0 |
| S8-final-viral-combined_k141_180108\|\|lt2gene | 1109 | No | NA | 1 | 1 | 0 | Low-quality | Genome-fragment | 2.17 | AAI-based (high-confidence) | 0 |
| S11-final-viral-combined_k141_238840\|\|full | 1170 | No | NA | 3 | 0 | 1 | Not-determined | Genome-fragment | NA | NA | 0 |
| S9-final-viral-combined_k141_22147\|\|full | 1110 | No | NA | 3 | 2 | 0 | Low-quality | Genome-fragment | 1.82 | HMM-based (lower-bound) | 0 |
| S11-final-viral-combined_k141_196723\|\|full | 4995 | No | NA | 3 | 0 | 2 | Not-determined | Genome-fragment | NA | NA | 0 |
| S11_k141_3965 | 1110 | No | NA | 4 | 1 | 0 | Low-quality | Genome-fragment | 1.37 | HMM-based (lower-bound) | 0 |
| S11-final-viral-combined_k141_155744\|\|full | 4127 | No | NA | 4 | 0 | 3 | Not-determined | Genome-fragment | NA | NA | 0 |
| S11-final-viral-combined_k141_142011\|\|full | 3762 | No | NA | 3 | 0 | 2 | Not-determined | Genome-fragment | NA | NA | 0 |
| S5-final-viral-combined_k141_136852\|\|full | 1110 | No | NA | 3 | 0 | 3 | Low-quality | Genome-fragment | 1.51 | AAI-based (medium-confidence) | 0 |
| S11-final-viral-combined_k141_142024\|\|full | 1402 | No | NA | 3 | 0 | 0 | Not-determined | Genome-fragment | NA | NA | 0 |
| S11-final-viral-combined_k141_128089\|\|full | 4590 | No | NA | 2 | 0 | 2 | Not-determined | Genome-fragment | NA | NA | 0 |
| S11-final-viral-combined_k141_128090\|\|full | 5380 | No | NA | 6 | 0 | 1 | Not-determined | Genome-fragment | NA | NA | 0 |
| S11-final-viral-combined_k141_320710\|\|full | 44314 | No | NA | 64 | 7 | 0 | High-quality | High-quality | 99.26 | AAI-based (high-confidence) | 0 |
| S7-final-viral-combined_k141_142477\|\|full | 1110 | No | NA | 3 | 0 | 0 | Low-quality | Genome-fragment | 2.4 | AAI-based (high-confidence) | 0 |
| S17-final-viral-combined_k141_240018\|\|full | 1110 | No | NA | 4 | 0 | 0 | Low-quality | Genome-fragment | 2.06 | AAI-based (medium-confidence) | 0 |
| S11-final-viral-combined_k141_59465\|\|full | 1441 | No | NA | 5 | 0 | 0 | Not-determined | Genome-fragment | NA | NA | 0 |
| S11-final-viral-combined_k141_87174\|\|full | 1997 | No | NA | 3 | 0 | 1 | Not-determined | Genome-fragment | NA | NA | 0 |
| S17-final-viral-combined_k141_41741\|\|lt2gene | 1110 | No | NA | 2 | 0 | 0 | Low-quality | Genome-fragment | 2.05 | AAI-based (high-confidence) | 0 |
| S11-final-viral-combined_k141_59468\|\|full | 4626 | No | NA | 4 | 0 | 4 | Not-determined | Genome-fragment | NA | NA | 0 |
| S11-final-viral-combined_k141_293463\|\|full | 3246 | No | NA | 5 | 0 | 3 | Not-determined | Genome-fragment | NA | NA | 0 |
| S11-final-viral-combined_k141_224781\|\|full | 1613 | No | NA | 2 | 0 | 1 | Not-determined | Genome-fragment | NA | NA | 0 |
| S11-final-viral-combined_k141_196765\|\|full | 2632 | No | NA | 3 | 0 | 1 | Not-determined | Genome-fragment | NA | NA | 0 |
| S11-final-viral-combined_k141_4071\|\|full | 26339 | No | NA | 25 | 0 | 18 | Not-determined | Genome-fragment | NA | NA | 0 |
| S8-final-viral-combined_k141_166079\|\|full | 1110 | No | NA | 3 | 1 | 0 | Low-quality | Genome-fragment | 2.98 | AAI-based (medium-confidence) | 0 |
| S8-final-viral-combined_k141_75485\|\|full | 1110 | No | NA | 3 | 0 | 0 | Low-quality | Genome-fragment | 0.58 | AAI-based (medium-confidence) | 0 |
| S11-final-viral-combined_k141_87209\|\|full | 1872 | No | NA | 5 | 0 | 0 | Not-determined | Genome-fragment | NA | NA | 0 |
| S11-final-viral-combined_k141_183386\|\|full | 2873 | No | NA | 6 | 0 | 2 | Not-determined | Genome-fragment | NA | NA | 0 |
| S8-final-viral-combined_k141_200966\|\|full | 1110 | No | NA | 2 | 0 | 0 | Low-quality | Genome-fragment | 1.78 | AAI-based (medium-confidence) | 0 |
| S9-final-viral-combined_k141_70257\|\|full | 1111 | No | NA | 3 | 0 | 2 | Low-quality | Genome-fragment | 0.34 | AAI-based (medium-confidence) | 0 |
| S9-final-viral-combined_k141_91066\|\|lt2gene | 1111 | No | NA | 2 | 1 | 0 | Low-quality | Genome-fragment | 1.65 | HMM-based (lower-bound) | 0 |
| S11-final-viral-combined_k141_59520\|\|full | 1269 | No | NA | 3 | 0 | 3 | Not-determined | Genome-fragment | NA | NA | 0 |
| S9_k141_7934 | 1111 | No | NA | 3 | 0 | 0 | Low-quality | Genome-fragment | 2.73 | AAI-based (high-confidence) | 0 |
| S17-final-viral-combined_k141_196092\|\|lt2gene | 1111 | No | NA | 2 | 0 | 0 | Low-quality | Genome-fragment | 2.64 | AAI-based (high-confidence) | 0 |
| S11-final-viral-combined_k141_224831\|\|full | 1431 | No | NA | 2 | 0 | 1 | Not-determined | Genome-fragment | NA | NA | 0 |
| S12-final-viral-combined_k141_84515\|\|full | 1111 | No | NA | 2 | 2 | 0 | Low-quality | Genome-fragment | 1.18 | HMM-based (lower-bound) | 0 |
| S11-final-viral-combined_k141_211569\|\|full | 2831 | No | NA | 3 | 0 | 0 | Not-determined | Genome-fragment | NA | NA | 0 |
| S11-final-viral-combined_k141_211570\|\|full | 1226 | No | NA | 2 | 0 | 1 | Not-determined | Genome-fragment | NA | NA | 0 |
| S11-final-viral-combined_k141_238964\|\|full | 3568 | No | NA | 6 | 0 | 0 | Not-determined | Genome-fragment | NA | NA | 0 |
| S12-final-viral-combined_k141_213761\|\|full | 1111 | No | NA | 3 | 2 | 0 | Low-quality | Genome-fragment | 1.38 | AAI-based (high-confidence) | 0 |
| S11-final-viral-combined_k141_87242\|\|full | 1783 | No | NA | 5 | 0 | 0 | Not-determined | Genome-fragment | NA | NA | 0 |
| S4_k141_118996 | 1111 | No | NA | 2 | 0 | 1 | Low-quality | Genome-fragment | 2.5 | AAI-based (medium-confidence) | 0 |
| S11-final-viral-combined_k141_45766\|\|full | 1375 | No | NA | 3 | 0 | 1 | Not-determined | Genome-fragment | NA | NA | 0 |
| S11-final-viral-combined_k141_31735\|\|full | 1804 | No | NA | 3 | 0 | 1 | Not-determined | Genome-fragment | NA | NA | 0 |
| S8_k141_220634 | 1111 | No | NA | 4 | 1 | 0 | Low-quality | Genome-fragment | 1.12 | AAI-based (high-confidence) | 0 |
| S11-final-viral-combined_k141_19002\|\|full | 1112 | No | NA | 1 | 0 | 1 | Low-quality | Genome-fragment | 1.71 | AAI-based (high-confidence) | 0 |
| S11-final-viral-combined_k141_196849\|\|full | 1255 | No | NA | 2 | 0 | 1 | Not-determined | Genome-fragment | NA | NA | 0 |
| S11_k141_9081 | 1112 | No | NA | 2 | 0 | 0 | Low-quality | Genome-fragment | 1.73 | AAI-based (high-confidence) | 0 |
| S1-final-viral-combined_k141_1801\|\|full | 1112 | No | NA | 2 | 0 | 0 | Low-quality | Genome-fragment | 1.56 | AAI-based (high-confidence) | 0 |
| S5-final-viral-combined_k141_157644\|\|full | 1112 | No | NA | 3 | 2 | 0 | Low-quality | Genome-fragment | 2.3 | AAI-based (high-confidence) | 0 |
| S5_k141_24110 | 1112 | No | NA | 2 | 2 | 0 | Low-quality | Genome-fragment | 3.41 | AAI-based (high-confidence) | 0 |
| S17-final-viral-combined_k141_138583\|\|full | 1112 | No | NA | 3 | 0 | 0 | Low-quality | Genome-fragment | 1.62 | AAI-based (medium-confidence) | 0 |
| S17-k141_72106 | 1112 | No | NA | 2 | 0 | 0 | Low-quality | Genome-fragment | 0.94 | AAI-based (medium-confidence) | 0 |
| S11-final-viral-combined_k141_128238\|\|full | 1578 | No | NA | 2 | 0 | 0 | Not-determined | Genome-fragment | NA | NA | 0 |
| S11-final-viral-combined_k141_224892\|\|full | 3009 | No | NA | 6 | 0 | 2 | Not-determined | Genome-fragment | NA | NA | 0 |
| S11-final-viral-combined_k141_101124\|\|full | 2664 | No | NA | 2 | 0 | 2 | Not-determined | Genome-fragment | NA | NA | 0 |
| S11-final-viral-combined_k141_280409\|\|full | 5485 | No | NA | 6 | 0 | 5 | Not-determined | Genome-fragment | NA | NA | 0 |
| S11-final-viral-combined_k141_252337\|\|full | 1729 | No | NA | 2 | 0 | 1 | Not-determined | Genome-fragment | NA | NA | 0 |
| S11-final-viral-combined_k141_4199\|\|full | 2023 | No | NA | 4 | 0 | 0 | Not-determined | Genome-fragment | NA | NA | 0 |
| S12-final-viral-combined_k141_357122\|\|full | 1112 | No | NA | 4 | 0 | 0 | Low-quality | Genome-fragment | 2.23 | AAI-based (high-confidence) | 0 |
| S11-final-viral-combined_k141_293637\|\|full | 1292 | No | NA | 2 | 0 | 0 | Not-determined | Genome-fragment | NA | NA | 0 |
| S11-final-viral-combined_k141_183471\|\|full | 3734 | No | NA | 3 | 0 | 2 | Not-determined | Genome-fragment | NA | NA | 0 |
| S12-final-viral-combined_k141_66758\|\|full | 1112 | No | NA | 2 | 0 | 0 | Low-quality | Genome-fragment | 0.85 | AAI-based (medium-confidence) | 0 |
| S11-final-viral-combined_k141_239067\|\|full | 1660 | No | NA | 4 | 0 | 2 | Not-determined | Genome-fragment | NA | NA | 0 |
| S11-final-viral-combined_k141_59649\|\|full | 3961 | No | NA | 4 | 0 | 2 | Not-determined | Genome-fragment | NA | NA | 0 |
| S12_k141_320047 | 1112 | No | NA | 1 | 0 | 0 | Low-quality | Genome-fragment | 0.98 | AAI-based (high-confidence) | 0 |
| S4-final-viral-combined_k141_108662\|\|full | 1112 | No | NA | 2 | 0 | 1 | Low-quality | Genome-fragment | 0.83 | AAI-based (medium-confidence) | 0 |
| S4-final-viral-combined_k141_18445\|\|full | 1112 | No | NA | 3 | 0 | 0 | Low-quality | Genome-fragment | 1.09 | AAI-based (medium-confidence) | 0 |
| S6-final-viral-combined_k141_74577\|\|full | 1112 | No | NA | 2 | 1 | 0 | Low-quality | Genome-fragment | 1.03 | AAI-based (high-confidence) | 0 |
| S11-final-viral-combined_k141_224945\|\|full | 3849 | No | NA | 6 | 0 | 4 | Not-determined | Genome-fragment | NA | NA | 0 |
| S11-final-viral-combined_k141_266239\|\|full | 1191 | No | NA | 4 | 0 | 0 | Not-determined | Genome-fragment | NA | NA | 0 |
| S8-final-viral-combined_k141_41875\|\|full | 1112 | No | NA | 3 | 0 | 0 | Low-quality | Genome-fragment | 1.36 | AAI-based (medium-confidence) | 0 |
| S11-final-viral-combined_k141_239117\|\|full | 3082 | No | NA | 5 | 0 | 3 | Not-determined | Genome-fragment | NA | NA | 0 |
| S8-final-viral-combined_k141_275718\|\|lt2gene | 1112 | No | NA | 1 | 1 | 0 | Low-quality | Genome-fragment | 0.47 | HMM-based (lower-bound) | 0 |
| S11-final-viral-combined_k141_266244\|\|full | 1275 | No | NA | 2 | 0 | 1 | Not-determined | Genome-fragment | NA | NA | 0 |
| S11-final-viral-combined_k141_183534\|\|full | 2312 | No | NA | 6 | 0 | 0 | Not-determined | Genome-fragment | NA | NA | 0 |
| S11-final-viral-combined_k141_155959\|\|full | 2869 | No | NA | 2 | 0 | 2 | Not-determined | Genome-fragment | NA | NA | 0 |
| S11-final-viral-combined_k141_4269\|\|full | 1274 | No | NA | 2 | 0 | 1 | Not-determined | Genome-fragment | NA | NA | 0 |
| S9_k141_30345 | 1113 | No | NA | 3 | 1 | 0 | Low-quality | Genome-fragment | 3.54 | AAI-based (high-confidence) | 0 |
| S11-final-viral-combined_k141_72737\|\|full | 5612 | No | NA | 6 | 0 | 5 | Not-determined | Genome-fragment | NA | NA | 0 |
| S11-final-viral-combined_k141_4274\|\|full | 3639 | No | NA | 4 | 0 | 1 | Not-determined | Genome-fragment | NA | NA | 0 |
| S11-final-viral-combined_k141_17703\|\|full | 1967 | No | NA | 2 | 0 | 2 | Not-determined | Genome-fragment | NA | NA | 0 |
| S11-final-viral-combined_k141_307896\|\|full | 1123 | No | NA | 3 | 0 | 0 | Not-determined | Genome-fragment | NA | NA | 0 |
| S11_k141_31639 | 1113 | No | NA | 2 | 1 | 0 | Low-quality | Genome-fragment | 2.48 | AAI-based (high-confidence) | 0 |
| S1-final-viral-combined_k141_320066\|\|full | 1113 | No | NA | 2 | 1 | 0 | Low-quality | Genome-fragment | 2.36 | AAI-based (high-confidence) | 0 |
| S5_k141_169765 | 1113 | No | NA | 2 | 2 | 0 | Low-quality | Genome-fragment | 2.72 | AAI-based (medium-confidence) | 0 |
| S7-final-viral-combined_k141_222901\|\|full | 1113 | No | NA | 3 | 2 | 0 | Low-quality | Genome-fragment | 2.65 | AAI-based (high-confidence) | 0 |
| S11-final-viral-combined_k141_196991\|\|full | 11885 | No | NA | 8 | 0 | 6 | Not-determined | Genome-fragment | NA | NA | 0 |
| S11-final-viral-combined_k141_128342\|\|full | 8797 | No | NA | 6 | 0 | 5 | Not-determined | Genome-fragment | NA | NA | 0 |
| S17-final-viral-combined_k141_95250\|\|full | 1113 | No | NA | 2 | 0 | 0 | Low-quality | Genome-fragment | 1.36 | AAI-based (medium-confidence) | 0 |
| S17-final-viral-combined_k141_30386\|\|full | 1113 | No | NA | 3 | 0 | 1 | Low-quality | Genome-fragment | 2.18 | AAI-based (medium-confidence) | 0 |
| S11-final-viral-combined_k141_101322\|\|full | 1262 | No | NA | 2 | 0 | 1 | Not-determined | Genome-fragment | NA | NA | 0 |
| S11-final-viral-combined_k141_320937\|\|full | 3301 | No | NA | 4 | 0 | 0 | Not-determined | Genome-fragment | NA | NA | 0 |
| S17-final-viral-combined_k141_210921\|\|full | 1113 | No | NA | 3 | 1 | 0 | Low-quality | Genome-fragment | 3.27 | AAI-based (high-confidence) | 0 |
| S17-k141_171356 | 1113 | No | NA | 2 | 2 | 0 | Low-quality | Genome-fragment | 2.75 | AAI-based (high-confidence) | 0 |
| S11-final-viral-combined_k141_128374\|\|full | 1526 | No | NA | 3 | 0 | 1 | Not-determined | Genome-fragment | NA | NA | 0 |
| S11-final-viral-combined_k141_239191\|\|full | 1094 | No | NA | 3 | 0 | 1 | Not-determined | Genome-fragment | NA | NA | 0 |
| S11-final-viral-combined_k141_239194\|\|full | 3607 | No | NA | 5 | 0 | 0 | Not-determined | Genome-fragment | NA | NA | 0 |
| S11-final-viral-combined_k141_45984\|\|full | 1285 | No | NA | 2 | 0 | 1 | Not-determined | Genome-fragment | NA | NA | 0 |
| S11-final-viral-combined_k141_156020\|\|full | 6290 | No | NA | 4 | 0 | 3 | Not-determined | Genome-fragment | NA | NA | 0 |
| S11-final-viral-combined_k141_31957\|\|full | 1173 | No | NA | 2 | 0 | 1 | Not-determined | Genome-fragment | NA | NA | 0 |
| S17-k141_243312 | 1113 | No | NA | 2 | 0 | 0 | Low-quality | Genome-fragment | 1.71 | AAI-based (medium-confidence) | 0 |
| S11-final-viral-combined_k141_128396\|\|full | 2171 | No | NA | 2 | 0 | 0 | Not-determined | Genome-fragment | NA | NA | 0 |
| S11-final-viral-combined_k141_72833\|\|full | 2651 | No | NA | 2 | 0 | 1 | Not-determined | Genome-fragment | NA | NA | 0 |
| S11-final-viral-combined_k141_72837\|\|full | 5810 | No | NA | 3 | 0 | 3 | Not-determined | Genome-fragment | NA | NA | 0 |
| S12-final-viral-combined_k141_23492\|\|full | 1113 | No | NA | 3 | 0 | 0 | Low-quality | Genome-fragment | 1.54 | AAI-based (medium-confidence) | 0 |
| S12-final-viral-combined_k141_11517\|\|lt2gene | 1113 | No | NA | 1 | 1 | 0 | Low-quality | Genome-fragment | 2.44 | HMM-based (lower-bound) | 0 |
| S12_k141_365228 | 1113 | No | NA | 2 | 0 | 2 | Low-quality | Genome-fragment | 0.41 | AAI-based (medium-confidence) | 0 |
| S11-final-viral-combined_k141_280643\|\|full | 4488 | No | NA | 7 | 0 | 7 | Not-determined | Genome-fragment | NA | NA | 0 |
| S2-final-viral-combined_k141_214238\|\|lt2gene | 1113 | No | NA | 1 | 1 | 0 | Low-quality | Genome-fragment | 3.18 | AAI-based (high-confidence) | 0 |
| S4-final-viral-combined_k141_98497\|\|full | 1113 | No | NA | 2 | 0 | 0 | Low-quality | Genome-fragment | 5.13 | AAI-based (high-confidence) | 0 |
| S6_k141_100515 | 1113 | No | NA | 4 | 0 | 0 | Low-quality | Genome-fragment | 3.29 | AAI-based (high-confidence) | 0 |
| S11-final-viral-combined_k141_17823\|\|full | 1120 | No | NA | 2 | 0 | 1 | Not-determined | Genome-fragment | NA | NA | 0 |
| S11-final-viral-combined_k141_72881\|\|full | 3767 | No | NA | 8 | 0 | 1 | Not-determined | Genome-fragment | NA | NA | 0 |
| S9_k141_34500 | 1114 | No | NA | 1 | 0 | 1 | Low-quality | Genome-fragment | 0.79 | AAI-based (high-confidence) | 0 |
| S11-final-viral-combined_k141_101337\|\|full | 78390 | Yes | 25287 | 84 | 13 | 28 | Medium-quality | Genome-fragment | 66.28 | AAI-based (high-confidence) | 67.74 |
| S5-final-viral-combined_k141_197499\|\|full | 1114 | No | NA | 3 | 0 | 1 | Low-quality | Genome-fragment | 2.35 | AAI-based (high-confidence) | 0 |
| S5-final-viral-combined_k141_140133\|\|full | 1114 | No | NA | 3 | 0 | 1 | Low-quality | Genome-fragment | 3.56 | AAI-based (medium-confidence) | 0 |
| S11-final-viral-combined_k141_59771\|\|full | 2047 | No | NA | 2 | 0 | 0 | Not-determined | Genome-fragment | NA | NA | 0 |
| S17-k141_61519 | 1114 | No | NA | 2 | 0 | 0 | Low-quality | Genome-fragment | 2.23 | AAI-based (medium-confidence) | 0 |
| S11-final-viral-combined_k141_211748\|\|full | 1271 | No | NA | 3 | 0 | 0 | Not-determined | Genome-fragment | NA | NA | 0 |
| S12-final-viral-combined_k141_120861\|\|full | 1114 | No | NA | 3 | 0 | 0 | Low-quality | Genome-fragment | 2.7 | AAI-based (medium-confidence) | 0 |
| S11-final-viral-combined_k141_239251\|\|full | 2923 | No | NA | 3 | 0 | 2 | Not-determined | Genome-fragment | NA | NA | 0 |
| S11-final-viral-combined_k141_225160\|\|full | 2855 | No | NA | 4 | 0 | 2 | Not-determined | Genome-fragment | NA | NA | 0 |
| S11-final-viral-combined_k141_142382\|\|full | 2907 | No | NA | 3 | 0 | 3 | Not-determined | Genome-fragment | NA | NA | 0 |
| S11-final-viral-combined_k141_17859\|\|full | 1461 | No | NA | 3 | 0 | 1 | Not-determined | Genome-fragment | NA | NA | 0 |
| S11-final-viral-combined_k141_321043\|\|full | 3471 | No | NA | 3 | 0 | 2 | Not-determined | Genome-fragment | NA | NA | 0 |
| S11-final-viral-combined_k141_72904\|\|full | 3507 | No | NA | 5 | 0 | 0 | Not-determined | Genome-fragment | NA | NA | 0 |
| S12-final-viral-combined_k141_372428\|\|lt2gene | 1114 | No | NA | 1 | 1 | 0 | Low-quality | Genome-fragment | 26.77 | HMM-based (lower-bound) | 0 |
| S11-final-viral-combined_k141_170361\|\|full | 20669 | No | NA | 19 | 0 | 13 | Not-determined | Genome-fragment | NA | NA | 0 |
| S12-final-viral-combined_k141_5533\|\|lt2gene | 1114 | No | NA | 1 | 1 | 0 | Low-quality | Genome-fragment | 2.62 | AAI-based (medium-confidence) | 0 |
| S11-final-viral-combined_k141_114169\|\|full | 3584 | No | NA | 3 | 0 | 0 | Not-determined | Genome-fragment | NA | NA | 0 |
| S11-final-viral-combined_k141_252598\|\|full | 1660 | No | NA | 2 | 0 | 0 | Not-determined | Genome-fragment | NA | NA | 0 |
| S11-final-viral-combined_k141_101386\|\|full | 5797 | No | NA | 5 | 0 | 4 | Not-determined | Genome-fragment | NA | NA | 0 |
| S12_k141_332850 | 1114 | No | NA | 1 | 0 | 0 | Low-quality | Genome-fragment | 2.71 | AAI-based (high-confidence) | 0 |
| S11-final-viral-combined_k141_114181\|\|full | 2508 | No | NA | 4 | 0 | 3 | Not-determined | Genome-fragment | NA | NA | 0 |
| S11-final-viral-combined_k141_142436\|\|full | 1232 | No | NA | 3 | 0 | 0 | Not-determined | Genome-fragment | NA | NA | 0 |
| S11-final-viral-combined_k141_87425\|\|full | 1359 | No | NA | 2 | 0 | 1 | Not-determined | Genome-fragment | NA | NA | 0 |
| S11-final-viral-combined_k141_128480\|\|full | 1608 | No | NA | 4 | 0 | 0 | Not-determined | Genome-fragment | NA | NA | 0 |
| S2-final-viral-combined_k141_204760\|\|lt2gene | 1114 | No | NA | 2 | 1 | 0 | Low-quality | Genome-fragment | 2.35 | AAI-based (medium-confidence) | 0 |
| S11-final-viral-combined_k141_183782\|\|full | 5951 | No | NA | 5 | 0 | 3 | Not-determined | Genome-fragment | NA | NA | 0 |
| S11-final-viral-combined_k141_197126\|\|full | 1541 | No | NA | 3 | 0 | 1 | Not-determined | Genome-fragment | NA | NA | 0 |
| S2_k141_35286 | 1114 | No | NA | 1 | 1 | 0 | Low-quality | Genome-fragment | 2.45 | AAI-based (high-confidence) | 0 |
| S11-final-viral-combined_k141_321069\|\|full | 1554 | No | NA | 3 | 0 | 0 | Not-determined | Genome-fragment | NA | NA | 0 |
| S11-final-viral-combined_k141_32064\|\|full | 9622 | No | NA | 10 | 0 | 5 | Not-determined | Genome-fragment | NA | NA | 0 |
| S2_k141_57237 | 1114 | No | NA | 2 | 0 | 0 | Low-quality | Genome-fragment | 2.04 | AAI-based (high-confidence) | 0 |
| S11-final-viral-combined_k141_239278\|\|full | 1369 | No | NA | 3 | 0 | 1 | Not-determined | Genome-fragment | NA | NA | 0 |
| S11-final-viral-combined_k141_59819\|\|full | 2970 | No | NA | 3 | 0 | 3 | Not-determined | Genome-fragment | NA | NA | 0 |
| S11-final-viral-combined_k141_225258\|\|full | 1838 | No | NA | 2 | 0 | 1 | Not-determined | Genome-fragment | NA | NA | 0 |
| S11-final-viral-combined_k141_239286\|\|full | 8211 | No | NA | 6 | 0 | 5 | Not-determined | Genome-fragment | NA | NA | 0 |
| S11-final-viral-combined_k141_225273\|\|full | 3414 | No | NA | 4 | 0 | 0 | Not-determined | Genome-fragment | NA | NA | 0 |
| S8-final-viral-combined_k141_190417\|\|full | 1114 | No | NA | 3 | 0 | 2 | Low-quality | Genome-fragment | 0.92 | AAI-based (medium-confidence) | 0 |
| S1-final-viral-combined_k141_334434\|\|full | 1115 | No | NA | 2 | 1 | 0 | Low-quality | Genome-fragment | 2.56 | AAI-based (medium-confidence) | 0 |
| S1-final-viral-combined_k141_306287\|\|full | 1115 | No | NA | 2 | 2 | 0 | Low-quality | Genome-fragment | 8.96 | AAI-based (high-confidence) | 0 |
| S1-final-viral-combined_k141_372179\|\|full | 1115 | No | NA | 3 | 0 | 0 | Low-quality | Genome-fragment | 2.14 | AAI-based (high-confidence) | 0 |
| S1-final-viral-combined_k141_123028\|\|lt2gene | 1115 | No | NA | 1 | 1 | 0 | Low-quality | Genome-fragment | 1.63 | HMM-based (lower-bound) | 0 |
| S11-final-viral-combined_k141_128542\|\|full | 1966 | No | NA | 2 | 0 | 0 | Not-determined | Genome-fragment | NA | NA | 0 |
| S11-final-viral-combined_k141_59850\|\|full | 1708 | No | NA | 3 | 0 | 3 | Not-determined | Genome-fragment | NA | NA | 0 |
| S1_k141_67728 | 1115 | No | NA | 1 | 0 | 1 | Low-quality | Genome-fragment | 0.63 | AAI-based (high-confidence) | 0 |
| S11-final-viral-combined_k141_308080\|\|full | 4058 | No | NA | 4 | 0 | 3 | Not-determined | Genome-fragment | NA | NA | 0 |
| S3-final-viral-combined_k141_29177\|\|lt2gene | 1115 | No | NA | 2 | 0 | 0 | Low-quality | Genome-fragment | 0.73 | AAI-based (high-confidence) | 0 |
| S11-final-viral-combined_k141_32126\|\|full | 3174 | No | NA | 3 | 0 | 2 | Not-determined | Genome-fragment | NA | NA | 0 |
| S11-final-viral-combined_k141_17913\|\|full | 1858 | No | NA | 2 | 0 | 2 | Not-determined | Genome-fragment | NA | NA | 0 |
| S11-final-viral-combined_k141_197192\|\|full | 2678 | No | NA | 3 | 0 | 2 | Not-determined | Genome-fragment | NA | NA | 0 |
| S11-final-viral-combined_k141_239323\|\|full | 1434 | No | NA | 3 | 0 | 2 | Not-determined | Genome-fragment | NA | NA | 0 |
| S11-final-viral-combined_k141_128548\|\|full | 7927 | No | NA | 5 | 0 | 4 | Not-determined | Genome-fragment | NA | NA | 0 |
| S11-final-viral-combined_k141_170434\|\|full | 1582 | No | NA | 3 | 0 | 3 | Not-determined | Genome-fragment | NA | NA | 0 |
| S3_k141_79065 | 1115 | No | NA | 2 | 0 | 0 | Low-quality | Genome-fragment | 1.65 | AAI-based (medium-confidence) | 0 |
| S3_k141_16261 | 1115 | No | NA | 1 | 0 | 0 | Low-quality | Genome-fragment | 1.88 | AAI-based (high-confidence) | 0 |
| S11-final-viral-combined_k141_321139\|\|full | 1354 | No | NA | 3 | 0 | 3 | Not-determined | Genome-fragment | NA | NA | 0 |
| S11-final-viral-combined_k141_142548\|\|full | 2196 | No | NA | 2 | 0 | 2 | Not-determined | Genome-fragment | NA | NA | 0 |
| S5-final-viral-combined_k141_109930\|\|full | 1115 | No | NA | 3 | 0 | 0 | Low-quality | Genome-fragment | 1.6 | AAI-based (medium-confidence) | 0 |
| S7-final-viral-combined_k141_70850\|\|full | 1115 | No | NA | 2 | 0 | 0 | Low-quality | Genome-fragment | 2.92 | AAI-based (medium-confidence) | 0 |
| S11-final-viral-combined_k141_225377\|\|full | 2149 | No | NA | 4 | 0 | 2 | Not-determined | Genome-fragment | NA | NA | 0 |
| S7_k141_201950 | 1115 | No | NA | 1 | 0 | 0 | Low-quality | Genome-fragment | 0.84 | AAI-based (high-confidence) | 0 |
| S11-final-viral-combined_k141_17943\|\|full | 12845 | No | NA | 12 | 0 | 2 | Not-determined | Genome-fragment | NA | NA | 0 |
| S17-final-viral-combined_k141_215753\|\|full | 1115 | No | NA | 3 | 0 | 1 | Low-quality | Genome-fragment | 3.16 | AAI-based (medium-confidence) | 0 |
| S17-final-viral-combined_k141_92726\|\|full | 1115 | No | NA | 2 | 0 | 0 | Low-quality | Genome-fragment | 3.08 | AAI-based (medium-confidence) | 0 |
| S11-final-viral-combined_k141_266567\|\|full | 2760 | No | NA | 3 | 0 | 3 | Not-determined | Genome-fragment | NA | NA | 0 |
| S17-k141_187981 | 1115 | No | NA | 3 | 0 | 1 | Low-quality | Genome-fragment | 2.15 | AAI-based (medium-confidence) | 0 |
| S12-final-viral-combined_k141_156144\|\|lt2gene | 1115 | No | NA | 1 | 1 | 0 | Low-quality | Genome-fragment | 2.08 | HMM-based (lower-bound) | 0 |
| S11-final-viral-combined_k141_225408\|\|full | 1306 | No | NA | 2 | 0 | 2 | Not-determined | Genome-fragment | NA | NA | 0 |
| S11-final-viral-combined_k141_266569\|\|full | 2128 | No | NA | 3 | 0 | 0 | Not-determined | Genome-fragment | NA | NA | 0 |
| S11-final-viral-combined_k141_252701\|\|full | 3423 | No | NA | 4 | 0 | 1 | Not-determined | Genome-fragment | NA | NA | 0 |
| S4-final-viral-combined_k141_20921\|\|full | 1115 | No | NA | 3 | 0 | 0 | Low-quality | Genome-fragment | 2.38 | AAI-based (medium-confidence) | 0 |
| S4-final-viral-combined_k141_152897\|\|full | 1115 | No | NA | 3 | 0 | 0 | Low-quality | Genome-fragment | 0.66 | AAI-based (high-confidence) | 0 |
| S11-final-viral-combined_k141_321203\|\|full | 11275 | No | NA | 12 | 0 | 7 | Not-determined | Genome-fragment | NA | NA | 0 |
| S8-final-viral-combined_k141_76406\|\|lt2gene | 1115 | No | NA | 2 | 1 | 0 | Low-quality | Genome-fragment | 2.26 | AAI-based (medium-confidence) | 0 |
| S11-final-viral-combined_k141_293937\|\|full | 1658 | No | NA | 3 | 0 | 1 | Not-determined | Genome-fragment | NA | NA | 0 |
| S11-final-viral-combined_k141_32211\|\|full | 1310 | No | NA | 3 | 0 | 0 | Not-determined | Genome-fragment | NA | NA | 0 |
| S11-final-viral-combined_k141_280838\|\|full | 3539 | No | NA | 3 | 0 | 2 | Not-determined | Genome-fragment | NA | NA | 0 |
| S9-final-viral-combined_k141_86507\|\|lt2gene | 1116 | No | NA | 2 | 1 | 0 | Low-quality | Genome-fragment | 2.44 | AAI-based (medium-confidence) | 0 |
| S11-final-viral-combined_k141_117161\|\|full | 1116 | No | NA | 2 | 0 | 0 | Low-quality | Genome-fragment | 3.03 | AAI-based (medium-confidence) | 0 |
| S11-final-viral-combined_k141_293955\|\|full | 1458 | No | NA | 2 | 0 | 1 | Not-determined | Genome-fragment | NA | NA | 0 |
| S11-final-viral-combined_k141_280851\|\|full | 2343 | No | NA | 3 | 0 | 2 | Not-determined | Genome-fragment | NA | NA | 0 |
| S1-final-viral-combined_k141_388509\|\|full | 1116 | No | NA | 3 | 0 | 0 | Low-quality | Genome-fragment | 2.87 | AAI-based (high-confidence) | 0 |
| S11-final-viral-combined_k141_59983\|\|full | 8832 | No | NA | 12 | 0 | 2 | Not-determined | Genome-fragment | NA | NA | 0 |
| S1_k141_121468 | 1116 | No | NA | 1 | 0 | 1 | Low-quality | Genome-fragment | 2.18 | AAI-based (high-confidence) | 0 |
| S5_k141_165069 | 1116 | No | NA | 3 | 0 | 3 | Low-quality | Genome-fragment | 3.27 | AAI-based (medium-confidence) | 0 |
| S11-final-viral-combined_k141_18011\|\|full | 2225 | No | NA | 2 | 0 | 1 | Not-determined | Genome-fragment | NA | NA | 0 |
| S7-final-viral-combined_k141_72885\|\|full | 1116 | No | NA | 3 | 1 | 0 | Low-quality | Genome-fragment | 2.87 | AAI-based (medium-confidence) | 0 |
| S17-final-viral-combined_k141_254372\|\|full | 1116 | No | NA | 2 | 1 | 0 | Low-quality | Genome-fragment | 3.43 | AAI-based (high-confidence) | 0 |
| S11-final-viral-combined_k141_46317\|\|full | 1098 | No | NA | 2 | 0 | 1 | Not-determined | Genome-fragment | NA | NA | 0 |
| S11-final-viral-combined_k141_73113\|\|full | 2972 | No | NA | 3 | 0 | 2 | Not-determined | Genome-fragment | NA | NA | 0 |
| S11-final-viral-combined_k141_252803\|\|full | 1158 | No | NA | 3 | 0 | 1 | Not-determined | Genome-fragment | NA | NA | 0 |
| S11-final-viral-combined_k141_18042\|\|full | 3092 | No | NA | 4 | 0 | 1 | Not-determined | Genome-fragment | NA | NA | 0 |

| | | | | | | | | | | | |
|---|---|---|---|---|---|---|---|---|---|---|---|
| S11-final-viral-combined_k141_46321\|\|full | 1617 | No | NA | 3 | 0 | 1 | Not-determined | Genome-fragment | NA | NA | 0 |
| S12-final-viral-combined_k141_18685\|\|full | 1116 | No | NA | 3 | 0 | 2 | Low-quality | Genome-fragment | 0.35 | AAI-based (medium-confidence) | 0 |
| S12_k141_260654 | 1116 | No | NA | 1 | 0 | 1 | Low-quality | Genome-fragment | 0.54 | AAI-based (high-confidence) | 0 |
| S11-final-viral-combined_k141_197365\|\|full | 1134 | No | NA | 3 | 0 | 1 | Not-determined | Genome-fragment | NA | NA | 0 |
| S11-final-viral-combined_k141_156384\|\|full | 2394 | No | NA | 3 | 0 | 0 | Not-determined | Genome-fragment | NA | NA | 0 |
| S11-final-viral-combined_k141_321320\|\|full | 8480 | No | NA | 6 | 0 | 2 | Not-determined | Genome-fragment | NA | NA | 0 |
| S11-final-viral-combined_k141_18057\|\|full | 3008 | No | NA | 2 | 0 | 2 | Not-determined | Genome-fragment | NA | NA | 0 |
| S2-final-viral-combined_k141_113103\|\|full | 1116 | No | NA | 3 | 0 | 0 | Low-quality | Genome-fragment | 2.63 | AAI-based (medium-confidence) | 0 |
| S11-final-viral-combined_k141_114373\|\|full | 1697 | No | NA | 4 | 0 | 0 | Not-determined | Genome-fragment | NA | NA | 0 |
| S2-final-viral-combined_k141_136713\|\|lt2gene | 1116 | No | NA | 2 | 1 | 0 | Low-quality | Genome-fragment | 3.6 | HMM-based (lower-bound) | 0 |
| S2-final-viral-combined_k141_91452\|\|lt2gene | 1116 | No | NA | 1 | 1 | 0 | Low-quality | Genome-fragment | 2.41 | AAI-based (high-confidence) | 0 |
| S2_k141_105983 | 1116 | No | NA | 2 | 0 | 1 | Low-quality | Genome-fragment | 0.73 | AAI-based (high-confidence) | 0 |
| S11-final-viral-combined_k141_73170\|\|full | 2674 | No | NA | 4 | 0 | 1 | Not-determined | Genome-fragment | NA | NA | 0 |
| S11-final-viral-combined_k141_32294\|\|full | 1992 | No | NA | 2 | 0 | 0 | Not-determined | Genome-fragment | NA | NA | 0 |
| S11-final-viral-combined_k141_170679\|\|full | 1548 | No | NA | 2 | 0 | 1 | Not-determined | Genome-fragment | NA | NA | 0 |
| S9-final-viral-combined_k141_121607\|\|full | 1117 | No | NA | 2 | 1 | 0 | Low-quality | Genome-fragment | 2.4 | AAI-based (high-confidence) | 0 |
| S9-final-viral-combined_k141_120499\|\|lt2gene | 1117 | No | NA | 1 | 1 | 0 | Low-quality | Genome-fragment | 3.18 | AAI-based (high-confidence) | 0 |
| S11-final-viral-combined_k141_225613\|\|full | 2008 | No | NA | 3 | 0 | 2 | Not-determined | Genome-fragment | NA | NA | 0 |
| S11-final-viral-combined_k141_101738\|\|full | 2146 | No | NA | 2 | 0 | 2 | Not-determined | Genome-fragment | NA | NA | 0 |
| S9-final-viral-combined_k141_14919\|\|lt2gene | 1117 | No | NA | 2 | 2 | 0 | Low-quality | Genome-fragment | 1.62 | HMM-based (lower-bound) | 0 |
| S11-final-viral-combined_k141_219267\|\|full | 1117 | No | NA | 2 | 0 | 0 | Low-quality | Genome-fragment | 0.8 | AAI-based (medium-confidence) | 0 |
| S11-final-viral-combined_k141_4781\|\|full | 7037 | No | NA | 7 | 0 | 6 | Not-determined | Genome-fragment | NA | NA | 0 |
| S11-final-viral-combined_k141_290097\|\|lt2gene | 1117 | No | NA | 1 | 1 | 0 | Low-quality | Genome-fragment | 3.18 | AAI-based (medium-confidence) | 0 |
| S11-final-viral-combined_k141_184590\|\|lt2gene | 1117 | No | NA | 2 | 2 | 0 | Low-quality | Genome-fragment | 1.36 | HMM-based (lower-bound) | 0 |
| S11-final-viral-combined_k141_118549\|\|lt2gene | 1117 | No | NA | 2 | 1 | 0 | Low-quality | Genome-fragment | 0.43 | HMM-based (lower-bound) | 0 |
| S11-final-viral-combined_k141_73204\|\|full | 68219 | No | NA | 63 | 3 | 13 | Medium-quality | Genome-fragment | 61.42 | AAI-based (medium-confidence) | 0 |
| S1-final-viral-combined_k141_229217\|\|full | 1117 | No | NA | 2 | 0 | 1 | Low-quality | Genome-fragment | 0.66 | AAI-based (medium-confidence) | 0 |
| S1-final-viral-combined_k141_109906\|\|lt2gene | 1117 | No | NA | 2 | 2 | 0 | Low-quality | Genome-fragment | 3.41 | AAI-based (high-confidence) | 0 |
| S11-final-viral-combined_k141_128850\|\|full | 1278 | No | NA | 3 | 0 | 1 | Not-determined | Genome-fragment | NA | NA | 0 |
| S11-final-viral-combined_k141_87734\|\|full | 1461 | No | NA | 2 | 0 | 0 | Not-determined | Genome-fragment | NA | NA | 0 |
| S1_k141_177163 | 1117 | No | NA | 1 | 0 | 1 | Low-quality | Genome-fragment | 0.83 | AAI-based (high-confidence) | 0 |
| S3-final-viral-combined_k141_12098\|\|full | 1117 | No | NA | 3 | 0 | 0 | Low-quality | Genome-fragment | 0.84 | AAI-based (high-confidence) | 0 |
| S3_k141_92974 | 1117 | No | NA | 3 | 1 | 0 | Low-quality | Genome-fragment | 1.86 | AAI-based (high-confidence) | 0 |
| S11-final-viral-combined_k141_281156\|\|full | 2820 | No | NA | 3 | 0 | 2 | Not-determined | Genome-fragment | NA | NA | 0 |
| S11-final-viral-combined_k141_60101\|\|full | 1853 | No | NA | 5 | 0 | 1 | Not-determined | Genome-fragment | NA | NA | 0 |
| S5-final-viral-combined_k141_71676\|\|full | 1117 | No | NA | 3 | 0 | 0 | Low-quality | Genome-fragment | 0.58 | AAI-based (medium-confidence) | 0 |
| S11-final-viral-combined_k141_308351\|\|full | 7416 | No | NA | 7 | 0 | 0 | Not-determined | Genome-fragment | NA | NA | 0 |
| S7-final-viral-combined_k141_197113\|\|full | 1117 | No | NA | 3 | 0 | 0 | Low-quality | Genome-fragment | 1.37 | AAI-based (medium-confidence) | 0 |
| S7_k141_220450 | 1117 | No | NA | 4 | 0 | 0 | Low-quality | Genome-fragment | 2.43 | AAI-based (high-confidence) | 0 |
| S17-final-viral-combined_k141_189489\|\|full | 1117 | No | NA | 4 | 0 | 0 | Low-quality | Genome-fragment | 2.58 | AAI-based (high-confidence) | 0 |
| S11-final-viral-combined_k141_142849\|\|full | 1321 | No | NA | 3 | 0 | 1 | Not-determined | Genome-fragment | NA | NA | 0 |
| S11-final-viral-combined_k141_114605\|\|full | 2373 | No | NA | 3 | 0 | 2 | Not-determined | Genome-fragment | NA | NA | 0 |
| S11-final-viral-combined_k141_60166\|\|full | 1586 | No | NA | 3 | 0 | 1 | Not-determined | Genome-fragment | NA | NA | 0 |
| S11-final-viral-combined_k141_239689\|\|full | 1577 | No | NA | 3 | 0 | 1 | Not-determined | Genome-fragment | NA | NA | 0 |
| S17-final-viral-combined_k141_34137\|\|full | 1117 | No | NA | 2 | 0 | 0 | Low-quality | Genome-fragment | 1.48 | AAI-based (medium-confidence) | 0 |
| S12-final-viral-combined_k141_206145\|\|lt2gene | 1117 | No | NA | 1 | 1 | 0 | Low-quality | Genome-fragment | 2.04 | HMM-based (lower-bound) | 0 |
| S11-final-viral-combined_k141_184106\|\|full | 2715 | No | NA | 5 | 0 | 0 | Not-determined | Genome-fragment | NA | NA | 0 |
| S2-final-viral-combined_k141_99034\|\|full | 1117 | No | NA | 4 | 1 | 0 | Low-quality | Genome-fragment | 3.2 | AAI-based (high-confidence) | 0 |
| S11-final-viral-combined_k141_60207\|\|full | 1289 | No | NA | 3 | 0 | 1 | Not-determined | Genome-fragment | NA | NA | 0 |
| S11-final-viral-combined_k141_308412\|\|full | 1855 | No | NA | 2 | 0 | 1 | Not-determined | Genome-fragment | NA | NA | 0 |
| S11-final-viral-combined_k141_73299\|\|full | 3029 | No | NA | 4 | 0 | 2 | Not-determined | Genome-fragment | NA | NA | 0 |
| S11-final-viral-combined_k141_239757\|\|full | 1506 | No | NA | 2 | 0 | 0 | Not-determined | Genome-fragment | NA | NA | 0 |
| S11-final-viral-combined_k141_101935\|\|full | 1397 | No | NA | 2 | 0 | 0 | Not-determined | Genome-fragment | NA | NA | 0 |
| S11-final-viral-combined_k141_128971\|\|full | 6508 | No | NA | 4 | 0 | 2 | Not-determined | Genome-fragment | NA | NA | 0 |
| S4-final-viral-combined_k141_149870\|\|full | 1117 | No | NA | 3 | 0 | 0 | Low-quality | Genome-fragment | 2.88 | AAI-based (high-confidence) | 0 |
| S11-final-viral-combined_k141_212302\|\|full | 4453 | No | NA | 4 | 0 | 3 | Not-determined | Genome-fragment | NA | NA | 0 |
| S11-final-viral-combined_k141_18264\|\|full | 1985 | No | NA | 4 | 0 | 2 | Not-determined | Genome-fragment | NA | NA | 0 |
| S11-final-viral-combined_k141_294209\|\|full | 1300 | No | NA | 3 | 0 | 1 | Not-determined | Genome-fragment | NA | NA | 0 |
| S11-final-viral-combined_k141_128982\|\|full | 1716 | No | NA | 3 | 0 | 1 | Not-determined | Genome-fragment | NA | NA | 0 |
| S11-final-viral-combined_k141_101945\|\|full | 1171 | No | NA | 2 | 0 | 1 | Not-determined | Genome-fragment | NA | NA | 0 |
| S4-final-viral-combined_k141_73918\|\|full | 1117 | No | NA | 2 | 0 | 1 | Low-quality | Genome-fragment | 1.63 | AAI-based (high-confidence) | 0 |
| S4_k141_75828 | 1117 | No | NA | 2 | 0 | 0 | Low-quality | Genome-fragment | 2.32 | AAI-based (high-confidence) | 0 |
| S11-final-viral-combined_k141_270124\|\|lt2gene | 1118 | No | NA | 2 | 1 | 0 | Low-quality | Genome-fragment | 2.44 | AAI-based (high-confidence) | 0 |
| S11-final-viral-combined_k141_170863\|\|full | 2819 | No | NA | 4 | 0 | 3 | Not-determined | Genome-fragment | NA | NA | 0 |
| S5_k141_152380 | 1118 | No | NA | 1 | 0 | 0 | Low-quality | Genome-fragment | 1.49 | AAI-based (high-confidence) | 0 |
| S11-final-viral-combined_k141_46519\|\|full | 4119 | No | NA | 4 | 0 | 1 | Not-determined | Genome-fragment | NA | NA | 0 |
| S11-final-viral-combined_k141_308486\|\|full | 2291 | No | NA | 3 | 0 | 0 | Not-determined | Genome-fragment | NA | NA | 0 |
| S11-final-viral-combined_k141_101985\|\|full | 1526 | No | NA | 2 | 0 | 1 | Not-determined | Genome-fragment | NA | NA | 0 |
| S5_k141_140802 | 1118 | No | NA | 1 | 0 | 0 | Low-quality | Genome-fragment | 1.88 | AAI-based (medium-confidence) | 0 |
| S11-final-viral-combined_k141_321507\|\|full | 5593 | No | NA | 5 | 0 | 4 | Not-determined | Genome-fragment | NA | NA | 0 |
| S7-final-viral-combined_k141_235223\|\|full | 1118 | No | NA | 2 | 0 | 1 | Low-quality | Genome-fragment | 3.57 | AAI-based (medium-confidence) | 0 |
| S7-final-viral-combined_k141_108001\|\|full | 1118 | No | NA | 1 | 0 | 0 | Low-quality | Genome-fragment | 2.75 | AAI-based (medium-confidence) | 0 |
| S11-final-viral-combined_k141_32620\|\|full | 1682 | No | NA | 6 | 0 | 0 | Not-determined | Genome-fragment | NA | NA | 0 |
| S7-final-viral-combined_k141_99458\|\|full | 1118 | No | NA | 3 | 1 | 0 | Low-quality | Genome-fragment | 2.51 | AAI-based (high-confidence) | 0 |
| S7_k141_162243 | 1118 | No | NA | 2 | 0 | 0 | Low-quality | Genome-fragment | 0.33 | AAI-based (medium-confidence) | 0 |
| S17-final-viral-combined_k141_152278\|\|full | 1118 | No | NA | 3 | 1 | 0 | Low-quality | Genome-fragment | 1.96 | AAI-based (medium-confidence) | 0 |
| S11-final-viral-combined_k141_321533\|\|full | 1280 | No | NA | 2 | 0 | 2 | Not-determined | Genome-fragment | NA | NA | 0 |
| S12-final-viral-combined_k141_235099\|\|full | 1118 | No | NA | 3 | 0 | 0 | Low-quality | Genome-fragment | 2.15 | AAI-based (medium-confidence) | 0 |
| S6-final-viral-combined_k141_34400\|\|full | 1118 | No | NA | 3 | 0 | 0 | Low-quality | Genome-fragment | 1.33 | AAI-based (high-confidence) | 0 |
| S6-final-viral-combined_k141_102260\|\|lt2gene | 1118 | No | NA | 2 | 1 | 0 | Low-quality | Genome-fragment | 3.47 | AAI-based (high-confidence) | 0 |
| S8-final-viral-combined_k141_260314\|\|full | 1118 | No | NA | 2 | 2 | 0 | Low-quality | Genome-fragment | 2.72 | AAI-based (high-confidence) | 0 |
| S11-final-viral-combined_k141_143006\|\|full | 1860 | No | NA | 2 | 0 | 1 | Not-determined | Genome-fragment | NA | NA | 0 |
| S8_k141_90445 | 1118 | No | NA | 2 | 0 | 0 | Low-quality | Genome-fragment | 2.58 | AAI-based (high-confidence) | 0 |
| S11_k141_18771 | 1119 | No | NA | 1 | 0 | 1 | Low-quality | Genome-fragment | 0.52 | AAI-based (medium-confidence) | 0 |
| S11-final-viral-combined_k141_225789\|\|full | 3044 | No | NA | 5 | 0 | 2 | Not-determined | Genome-fragment | NA | NA | 0 |
| S11_k141_40108 | 1119 | No | NA | 1 | 0 | 1 | Low-quality | Genome-fragment | 1.34 | AAI-based (medium-confidence) | 0 |
| S11-final-viral-combined_k141_281391\|\|full | 2907 | No | NA | 5 | 0 | 2 | Not-determined | Genome-fragment | NA | NA | 0 |
| S3_k141_91325 | 1119 | No | NA | 2 | 0 | 0 | Low-quality | Genome-fragment | 1.07 | AAI-based (high-confidence) | 0 |
| S5-final-viral-combined_k141_184785\|\|lt2gene | 1119 | No | NA | 2 | 2 | 0 | Low-quality | Genome-fragment | 2.54 | AAI-based (high-confidence) | 0 |
| S11-final-viral-combined_k141_239905\|\|full | 3719 | No | NA | 4 | 0 | 1 | Not-determined | Genome-fragment | NA | NA | 0 |
| S11-final-viral-combined_k141_281396\|\|full | 2329 | No | NA | 3 | 0 | 0 | Not-determined | Genome-fragment | NA | NA | 0 |
| S11-final-viral-combined_k141_281397\|\|full | 2075 | No | NA | 2 | 0 | 2 | Not-determined | Genome-fragment | NA | NA | 0 |
| S11-final-viral-combined_k141_266936\|\|full | 3232 | No | NA | 4 | 0 | 0 | Not-determined | Genome-fragment | NA | NA | 0 |
| S5_k141_113752 | 1119 | Yes | 503 | 2 | 1 | 1 | Low-quality | Genome-fragment | 1.12 | AAI-based (high-confidence) | 55.05 |
| S11-final-viral-combined_k141_253181\|\|full | 1885 | No | NA | 2 | 0 | 0 | Not-determined | Genome-fragment | NA | NA | 0 |
| S11-final-viral-combined_k141_87960\|\|full | 2559 | No | NA | 3 | 0 | 2 | Not-determined | Genome-fragment | NA | NA | 0 |
| S11-final-viral-combined_k141_4989\|\|full | 1456 | No | NA | 2 | 0 | 1 | Not-determined | Genome-fragment | NA | NA | 0 |
| S7-final-viral-combined_k141_37013\|\|full | 1119 | No | NA | 3 | 0 | 0 | Low-quality | Genome-fragment | 0.77 | AAI-based (medium-confidence) | 0 |
| S11-final-viral-combined_k141_170949\|\|full | 3632 | No | NA | 3 | 0 | 3 | Not-determined | Genome-fragment | NA | NA | 0 |
| S12-final-viral-combined_k141_129767\|\|lt2gene | 1119 | No | NA | 1 | 1 | 0 | Low-quality | Genome-fragment | 1 | HMM-based (lower-bound) | 0 |
| S2_k141_97706 | 1119 | No | NA | 1 | 0 | 0 | Low-quality | Genome-fragment | 2.47 | AAI-based (medium-confidence) | 0 |
| S4-final-viral-combined_k141_49501\|\|full | 1119 | No | NA | 3 | 0 | 0 | Low-quality | Genome-fragment | 2.61 | AAI-based (medium-confidence) | 0 |
| S11-final-viral-combined_k141_212400\|\|full | 2151 | No | NA | 4 | 0 | 0 | Not-determined | Genome-fragment | NA | NA | 0 |
| S11-final-viral-combined_k141_102128\|\|full | 1781 | No | NA | 2 | 0 | 1 | Not-determined | Genome-fragment | NA | NA | 0 |
| S11-final-viral-combined_k141_88009\|\|full | 1624 | No | NA | 2 | 0 | 0 | Not-determined | Genome-fragment | NA | NA | 0 |
| S11-final-viral-combined_k141_143062\|\|full | 1460 | No | NA | 3 | 0 | 2 | Not-determined | Genome-fragment | NA | NA | 0 |
| S8_k141_113601 | 1119 | No | NA | 2 | 0 | 1 | Low-quality | Genome-fragment | 1.72 | AAI-based (medium-confidence) | 0 |
| S9-final-viral-combined_k141_90448\|\|lt2gene | 1120 | No | NA | 2 | 1 | 0 | Low-quality | Genome-fragment | 2.43 | AAI-based (medium-confidence) | 0 |
| S9_k141_72399 | 1120 | No | NA | 2 | 0 | 0 | Low-quality | Genome-fragment | 1.87 | AAI-based (medium-confidence) | 0 |
| S11-final-viral-combined_k141_184209\|\|full | 1461 | No | NA | 3 | 0 | 1 | Not-determined | Genome-fragment | NA | NA | 0 |
| S11-final-viral-combined_k141_5089\|\|full | 2305 | No | NA | 3 | 0 | 3 | Not-determined | Genome-fragment | NA | NA | 0 |
| S11-final-viral-combined_k141_267016\|\|full | 8364 | No | NA | 4 | 0 | 0 | Not-determined | Genome-fragment | NA | NA | 0 |
| S11-final-viral-combined_k141_51087\|\|full | 1120 | No | NA | 3 | 1 | 0 | Low-quality | Genome-fragment | 0.68 | HMM-based (lower-bound) | 0 |
| S11_k141_86251 | 1120 | No | NA | 3 | 1 | 0 | Low-quality | Genome-fragment | 2.61 | AAI-based (high-confidence) | 0 |
| S1_k141_141084 | 1120 | No | NA | 1 | 0 | 1 | Low-quality | Genome-fragment | 0.52 | AAI-based (high-confidence) | 0 |
| S11-final-viral-combined_k141_212474\|\|full | 2949 | No | NA | 4 | 0 | 2 | Not-determined | Genome-fragment | NA | NA | 0 |
| S11-final-viral-combined_k141_156734\|\|full | 1820 | No | NA | 2 | 0 | 1 | Not-determined | Genome-fragment | NA | NA | 0 |
| S1_k141_79330 | 1120 | No | NA | 3 | 0 | 0 | Low-quality | Genome-fragment | 2.05 | AAI-based (medium-confidence) | 0 |
| S3-final-viral-combined_k141_33599\|\|full | 1120 | No | NA | 2 | 1 | 0 | Low-quality | Genome-fragment | 2.91 | AAI-based (high-confidence) | 0 |
| S11-final-viral-combined_k141_156746\|\|full | 2407 | No | NA | 3 | 0 | 3 | Not-determined | Genome-fragment | NA | NA | 0 |
| S3_k141_98656 | 1120 | No | NA | 1 | 0 | 0 | Low-quality | Genome-fragment | 2.88 | AAI-based (high-confidence) | 0 |
| S5-final-viral-combined_k141_71291\|\|full | 1120 | No | NA | 2 | 0 | 0 | Low-quality | Genome-fragment | 2.71 | AAI-based (high-confidence) | 0 |
| S17-final-viral-combined_k141_173708\|\|full | 1120 | No | NA | 2 | 0 | 0 | Low-quality | Genome-fragment | 2.48 | AAI-based (high-confidence) | 0 |
| S17-final-viral-combined_k141_232695\|\|full | 1120 | No | NA | 3 | 0 | 0 | Low-quality | Genome-fragment | 2.08 | AAI-based (high-confidence) | 0 |
| S11-final-viral-combined_k141_114879\|\|full | 1871 | No | NA | 2 | 0 | 2 | Not-determined | Genome-fragment | NA | NA | 0 |
| S11-final-viral-combined_k141_267118\|\|full | 3038 | No | NA | 4 | 0 | 2 | Not-determined | Genome-fragment | NA | NA | 0 |
| S11-final-viral-combined_k141_294458\|\|full | 1620 | No | NA | 4 | 0 | 0 | Not-determined | Genome-fragment | NA | NA | 0 |
| S11-final-viral-combined_k141_60490\|\|full | 6613 | No | NA | 5 | 0 | 3 | Not-determined | Genome-fragment | NA | NA | 0 |
| S11-final-viral-combined_k141_267120\|\|full | 1600 | No | NA | 3 | 0 | 1 | Not-determined | Genome-fragment | NA | NA | 0 |
| S11-final-viral-combined_k141_143164\|\|full | 1040 | No | NA | 2 | 0 | 0 | Not-determined | Genome-fragment | NA | NA | 0 |
| S11-final-viral-combined_k141_156752\|\|full | 2539 | No | NA | 3 | 0 | 0 | Not-determined | Genome-fragment | NA | NA | 0 |
| S11-final-viral-combined_k141_294469\|\|full | 2621 | No | NA | 3 | 0 | 1 | Not-determined | Genome-fragment | NA | NA | 0 |
| S11-final-viral-combined_k141_281599\|\|full | 1175 | No | NA | 2 | 0 | 0 | Not-determined | Genome-fragment | NA | NA | 0 |
| S11-final-viral-combined_k141_184317\|\|full | 1218 | No | NA | 3 | 0 | 2 | Not-determined | Genome-fragment | NA | NA | 0 |
| S12_k141_280272 | 1120 | No | NA | 3 | 0 | 0 | Low-quality | Genome-fragment | 3.04 | AAI-based (high-confidence) | 0 |
| S11-final-viral-combined_k141_32725\|\|full | 2628 | No | NA | 5 | 0 | 2 | Not-determined | Genome-fragment | NA | NA | 0 |
| S11-final-viral-combined_k141_308741\|\|full | 1062 | No | NA | 3 | 0 | 0 | Not-determined | Genome-fragment | NA | NA | 0 |
| S11-final-viral-combined_k141_143193\|\|full | 3726 | No | NA | 3 | 0 | 1 | Not-determined | Genome-fragment | NA | NA | 0 |
| S2-final-viral-combined_k141_170669\|\|full | 1120 | No | NA | 2 | 0 | 0 | Low-quality | Genome-fragment | 2.17 | AAI-based (medium-confidence) | 0 |
| S11-final-viral-combined_k141_102257\|\|full | 1366 | No | NA | 4 | 0 | 1 | Not-determined | Genome-fragment | NA | NA | 0 |
| S2-final-viral-combined_k141_245134\|\|full | 1120 | No | NA | 2 | 2 | 0 | Low-quality | Genome-fragment | 2.42 | AAI-based (high-confidence) | 0 |
| S11-final-viral-combined_k141_18533\|\|full | 2368 | No | NA | 3 | 0 | 2 | Not-determined | Genome-fragment | NA | NA | 0 |
| S11-final-viral-combined_k141_281637\|\|full | 3665 | No | NA | 5 | 0 | 4 | Not-determined | Genome-fragment | NA | NA | 0 |
| S11-final-viral-combined_k141_143204\|\|full | 6095 | No | NA | 3 | 0 | 2 | Not-determined | Genome-fragment | NA | NA | 0 |
| S2_k141_170009 | 1120 | No | NA | 2 | 0 | 1 | Low-quality | Genome-fragment | 2.5 | AAI-based (high-confidence) | 0 |
| S11-final-viral-combined_k141_102266\|\|full | 9586 | No | NA | 12 | 0 | 6 | Not-determined | Genome-fragment | NA | NA | 0 |
| S11-final-viral-combined_k141_114954\|\|full | 1192 | No | NA | 3 | 0 | 3 | Not-determined | Genome-fragment | NA | NA | 0 |
| S11-final-viral-combined_k141_253255\|\|full | 1743 | No | NA | 3 | 0 | 3 | Not-determined | Genome-fragment | NA | NA | 0 |
| S11-final-viral-combined_k141_281649\|\|full | 2587 | No | NA | 2 | 0 | 1 | Not-determined | Genome-fragment | NA | NA | 0 |
| S11-final-viral-combined_k141_197938\|\|full | 10593 | No | NA | 7 | 0 | 2 | Not-determined | Genome-fragment | NA | NA | 0 |
| S4_k141_161422 | 1120 | No | NA | 2 | 0 | 0 | Low-quality | Genome-fragment | 1.87 | AAI-based (medium-confidence) | 0 |
| S8-final-viral-combined_k141_272835\|\|lt2gene | 1120 | No | NA | 2 | 1 | 0 | Low-quality | Genome-fragment | 1.64 | HMM-based (lower-bound) | 0 |
| S3_k141_1207 | 1121 | No | NA | 2 | 0 | 0 | Low-quality | Genome-fragment | 0.52 | AAI-based (high-confidence) | 0 |
| S5-final-viral-combined_k141_50475\|\|full | 1121 | No | NA | 2 | 1 | 0 | Low-quality | Genome-fragment | 0.77 | HMM-based (lower-bound) | 0 |
| S11-final-viral-combined_k141_18562\|\|full | 2755 | No | NA | 2 | 0 | 2 | Not-determined | Genome-fragment | NA | NA | 0 |
| S11-final-viral-combined_k141_5246\|\|full | 1235 | No | NA | 3 | 0 | 1 | Not-determined | Genome-fragment | NA | NA | 0 |
| S5_k141_83478 | 1121 | No | NA | 1 | 0 | 1 | Low-quality | Genome-fragment | 3.58 | AAI-based (medium-confidence) | 0 |
| S7-final-viral-combined_k141_12371\|\|full | 1121 | No | NA | 3 | 1 | 0 | Low-quality | Genome-fragment | 1.61 | HMM-based (lower-bound) | 0 |
| S11-final-viral-combined_k141_184382\|\|full | 1577 | No | NA | 2 | 0 | 1 | Not-determined | Genome-fragment | NA | NA | 0 |
| S11-final-viral-combined_k141_240139\|\|full | 7259 | No | NA | 2 | 0 | 2 | Not-determined | Genome-fragment | NA | NA | 0 |
| S7-final-viral-combined_k141_198265\|\|full | 1121 | No | NA | 2 | 1 | 0 | Low-quality | Genome-fragment | 2.75 | AAI-based (medium-confidence) | 0 |
| S7_k141_126320 | 1121 | No | NA | 1 | 0 | 1 | Low-quality | Genome-fragment | 0.37 | AAI-based (medium-confidence) | 0 |
| S11-final-viral-combined_k141_32810\|\|full | 5526 | No | NA | 6 | 0 | 5 | Not-determined | Genome-fragment | NA | NA | 0 |
| S11-final-viral-combined_k141_156843\|\|full | 2314 | No | NA | 4 | 0 | 1 | Not-determined | Genome-fragment | NA | NA | 0 |
| S11-final-viral-combined_k141_321706\|\|full | 2631 | No | NA | 3 | 0 | 2 | Not-determined | Genome-fragment | NA | NA | 0 |
| S17-final-viral-combined_k141_171932\|\|full | 1121 | No | NA | 3 | 0 | 0 | Low-quality | Genome-fragment | 3.04 | AAI-based (medium-confidence) | 0 |
| S12-final-viral-combined_k141_43283\|\|full | 1121 | No | NA | 4 | 1 | 0 | Low-quality | Genome-fragment | 2.23 | AAI-based (medium-confidence) | 0 |
| S11-final-viral-combined_k141_212683\|\|full | 2859 | No | NA | 4 | 0 | 0 | Not-determined | Genome-fragment | NA | NA | 0 |
| S11-final-viral-combined_k141_129337\|\|full | 1715 | No | NA | 2 | 0 | 1 | Not-determined | Genome-fragment | NA | NA | 0 |
| S11-final-viral-combined_k141_267154\|\|full | 1019 | No | NA | 2 | 0 | 2 | Not-determined | Genome-fragment | NA | NA | 0 |
| S11-final-viral-combined_k141_184420\|\|full | 2304 | No | NA | 3 | 0 | 1 | Not-determined | Genome-fragment | NA | NA | 0 |
| S12-final-viral-combined_k141_9685\|\|full | 1121 | No | NA | 3 | 0 | 0 | Low-quality | Genome-fragment | 2.14 | AAI-based (medium-confidence) | 0 |
| S12-final-viral-combined_k141_49973\|\|full | 1121 | No | NA | 2 | 0 | 1 | Low-quality | Genome-fragment | 0.58 | AAI-based (medium-confidence) | 0 |
| S11-final-viral-combined_k141_73611\|\|full | 2491 | No | NA | 3 | 0 | 0 | Not-determined | Genome-fragment | NA | NA | 0 |
| S12-final-viral-combined_k141_290242\|\|lt2gene | 1121 | No | NA | 1 | 1 | 0 | Low-quality | Genome-fragment | 1.47 | HMM-based (lower-bound) | 0 |
| S12_k141_284464 | 1121 | No | NA | 2 | 0 | 0 | Low-quality | Genome-fragment | 0.52 | AAI-based (high-confidence) | 0 |
| S11-final-viral-combined_k141_294568\|\|full | 1201 | No | NA | 2 | 0 | 1 | Not-determined | Genome-fragment | NA | NA | 0 |
| S2-final-viral-combined_k141_104771\|\|full | 1121 | No | NA | 4 | 2 | 0 | Low-quality | Genome-fragment | 2.69 | AAI-based (high-confidence) | 0 |
| S2-final-viral-combined_k141_171337\|\|full | 1121 | No | NA | 3 | 0 | 0 | Low-quality | Genome-fragment | 0.62 | AAI-based (high-confidence) | 0 |
| S11-final-viral-combined_k141_46816\|\|full | 29694 | No | NA | 41 | 8 | 0 | Medium-quality | Genome-fragment | 62.99 | AAI-based (high-confidence) | 0 |
| S11-final-viral-combined_k141_171066\|\|full | 3031 | No | NA | 3 | 0 | 1 | Not-determined | Genome-fragment | NA | NA | 0 |
| S11-final-viral-combined_k141_32902\|\|full | 5899 | No | NA | 7 | 0 | 3 | Not-determined | Genome-fragment | NA | NA | 0 |
| S2-final-viral-combined_k141_90845\|\|full | 1121 | No | NA | 2 | 0 | 0 | Low-quality | Genome-fragment | 3.6 | AAI-based (high-confidence) | 0 |
| S4_k141_163236 | 1121 | No | NA | 2 | 0 | 0 | Low-quality | Genome-fragment | 0.93 | AAI-based (high-confidence) | 0 |
| S11-final-viral-combined_k141_73663\|\|full | 1350 | No | NA | 3 | 0 | 0 | Not-determined | Genome-fragment | NA | NA | 0 |
| S11-final-viral-combined_k141_46838\|\|full | 3952 | No | NA | 4 | 0 | 2 | Not-determined | Genome-fragment | NA | NA | 0 |
| S11-final-viral-combined_k141_115152\|\|full | 1453 | No | NA | 3 | 0 | 0 | Not-determined | Genome-fragment | NA | NA | 0 |
| S11-final-viral-combined_k141_171084\|\|full | 3657 | No | NA | 3 | 0 | 3 | Not-determined | Genome-fragment | NA | NA | 0 |
| S11-final-viral-combined_k141_156954\|\|full | 5358 | No | NA | 3 | 0 | 2 | Not-determined | Genome-fragment | NA | NA | 0 |
| S11-final-viral-combined_k141_294612\|\|full | 1328 | No | NA | 3 | 0 | 0 | Not-determined | Genome-fragment | NA | NA | 0 |
| S6_k141_139142 | 1121 | No | NA | 3 | 0 | 0 | Low-quality | Genome-fragment | 2.23 | AAI-based (high-confidence) | 0 |
| S8-final-viral-combined_k141_176008\|\|full | 1121 | No | NA | 4 | 0 | 1 | Low-quality | Genome-fragment | 2.92 | AAI-based (medium-confidence) | 0 |
| S11-final-viral-combined_k141_46844\|\|full | 7709 | No | NA | 7 | 0 | 7 | Not-determined | Genome-fragment | NA | NA | 0 |
| S11-final-viral-combined_k141_143285\|\|full | 1249 | No | NA | 2 | 0 | 1 | Not-determined | Genome-fragment | NA | NA | 0 |
| S11-final-viral-combined_k141_156974\|\|full | 1185 | No | NA | 2 | 0 | 0 | Not-determined | Genome-fragment | NA | NA | 0 |
| S11-final-viral-combined_k141_115172\|\|full | 3313 | No | NA | 3 | 0 | 3 | Not-determined | Genome-fragment | NA | NA | 0 |
| S11-final-viral-combined_k141_294627\|\|full | 3425 | No | NA | 4 | 0 | 3 | Not-determined | Genome-fragment | NA | NA | 0 |
| S11-final-viral-combined_k141_5402\|\|full | 1572 | No | NA | 2 | 0 | 1 | Not-determined | Genome-fragment | NA | NA | 0 |
| S8-final-viral-combined_k141_4556\|\|lt2gene | 1121 | No | NA | 2 | 1 | 0 | Low-quality | Genome-fragment | 2.31 | AAI-based (high-confidence) | 0 |
| S8_k141_35475 | 1121 | No | NA | 2 | 0 | 1 | Low-quality | Genome-fragment | 0.4 | AAI-based (medium-confidence) | 0 |
| S11-final-viral-combined_k141_198080\|\|full | 21682 | No | NA | 18 | 0 | 13 | Not-determined | Genome-fragment | NA | NA | 0 |
| S11-final-viral-combined_k141_294666\|\|full | 1076 | No | NA | 2 | 0 | 0 | Not-determined | Genome-fragment | NA | NA | 0 |
| S11-final-viral-combined_k141_156986\|\|full | 2721 | No | NA | 4 | 0 | 3 | Not-determined | Genome-fragment | NA | NA | 0 |
| S11-final-viral-combined_k141_294673\|\|full | 1440 | No | NA | 2 | 0 | 1 | Not-determined | Genome-fragment | NA | NA | 0 |
| S11-final-viral-combined_k141_245733\|\|full | 1122 | No | NA | 5 | 2 | 0 | Low-quality | Genome-fragment | 3.02 | AAI-based (high-confidence) | 0 |
| S11_k141_223054 | 1122 | No | NA | 2 | 1 | 0 | Low-quality | Genome-fragment | 0.4 | HMM-based (lower-bound) | 0 |
| S11_k141_50654 | 1122 | No | NA | 2 | 0 | 1 | Low-quality | Genome-fragment | 1.88 | AAI-based (medium-confidence) | 0 |
| S5-final-viral-combined_k141_105411\|\|full | 1122 | No | NA | 3 | 1 | 0 | Low-quality | Genome-fragment | 2.49 | AAI-based (medium-confidence) | 0 |
| S7-final-viral-combined_k141_49363\|\|full | 1122 | No | NA | 2 | 1 | 0 | Low-quality | Genome-fragment | 1.68 | HMM-based (lower-bound) | 0 |
| S11-final-viral-combined_k141_294692\|\|full | 1634 | No | NA | 3 | 0 | 0 | Not-determined | Genome-fragment | NA | NA | 0 |
| S7-final-viral-combined_k141_146858\|\|lt2gene | 1122 | No | NA | 2 | 0 | 0 | Low-quality | Genome-fragment | 2.67 | AAI-based (medium-confidence) | 0 |
| S11-final-viral-combined_k141_294704\|\|full | 2735 | No | NA | 2 | 0 | 2 | Not-determined | Genome-fragment | NA | NA | 0 |
| S11-final-viral-combined_k141_88295\|\|full | 1400 | No | NA | 2 | 0 | 2 | Not-determined | Genome-fragment | NA | NA | 0 |
| S17-final-viral-combined_k141_188794\|\|lt2gene | 1122 | No | NA | 1 | 1 | 0 | Low-quality | Genome-fragment | 3.62 | HMM-based (lower-bound) | 0 |
| S17-k141_192648 | 1122 | No | NA | 2 | 0 | 1 | Low-quality | Genome-fragment | 0.49 | AAI-based (medium-confidence) | 0 |
| S17-k141_260762 | 1122 | No | NA | 3 | 0 | 0 | Low-quality | Genome-fragment | 3.11 | AAI-based (high-confidence) | 0 |
| S17-k141_52707 | 1122 | No | NA | 2 | 0 | 1 | Low-quality | Genome-fragment | 0.79 | AAI-based (high-confidence) | 0 |
| S11-final-viral-combined_k141_240405\|\|full | 4886 | No | NA | 4 | 0 | 1 | Not-determined | Genome-fragment | NA | NA | 0 |
| S11-final-viral-combined_k141_171175\|\|full | 2002 | No | NA | 3 | 0 | 0 | Not-determined | Genome-fragment | NA | NA | 0 |
| S11-final-viral-combined_k141_321834\|\|full | 2398 | No | NA | 3 | 0 | 2 | Not-determined | Genome-fragment | NA | NA | 0 |
| S12_k141_132836 | 1122 | No | NA | 2 | 0 | 1 | Low-quality | Genome-fragment | 0.38 | AAI-based (medium-confidence) | 0 |
| S11-final-viral-combined_k141_88332\|\|full | 6184 | No | NA | 9 | 0 | 1 | Not-determined | Genome-fragment | NA | NA | 0 |
| S4-final-viral-combined_k141_158481\|\|lt2gene | 1122 | No | NA | 1 | 1 | 0 | Low-quality | Genome-fragment | 3.7 | AAI-based (high-confidence) | 0 |
| S6_k141_79827 | 1122 | No | NA | 1 | 0 | 1 | Low-quality | Genome-fragment | 0.73 | AAI-based (high-confidence) | 0 |
| S11-final-viral-combined_k141_71774\|\|lt2gene | 1123 | No | NA | 2 | 1 | 0 | Low-quality | Genome-fragment | 2.85 | AAI-based (high-confidence) | 0 |
| S11_k141_63829 | 1123 | No | NA | 2 | 0 | 1 | Low-quality | Genome-fragment | 1.42 | AAI-based (high-confidence) | 0 |
| S1-final-viral-combined_k141_40606\|\|full | 1123 | No | NA | 4 | 1 | 0 | Low-quality | Genome-fragment | 2.78 | AAI-based (high-confidence) | 0 |
| S11-final-viral-combined_k141_253582\|\|full | 1998 | No | NA | 2 | 0 | 1 | Not-determined | Genome-fragment | NA | NA | 0 |
| S1-final-viral-combined_k141_10739\|\|lt2gene | 1123 | No | NA | 2 | 1 | 0 | Low-quality | Genome-fragment | 2.62 | AAI-based (high-confidence) | 0 |
| S1-final-viral-combined_k141_262888\|\|lt2gene | 1123 | No | NA | 1 | 1 | 0 | Low-quality | Genome-fragment | 1.83 | HMM-based (lower-bound) | 0 |
| S11-final-viral-combined_k141_226135\|\|full | 3347 | No | NA | 3 | 0 | 2 | Not-determined | Genome-fragment | NA | NA | 0 |
| S11-final-viral-combined_k141_184668\|\|full | 4157 | No | NA | 5 | 0 | 1 | Not-determined | Genome-fragment | NA | NA | 0 |
| S11-final-viral-combined_k141_294784\|\|full | 1267 | No | NA | 3 | 0 | 1 | Not-determined | Genome-fragment | NA | NA | 0 |
| S11-final-viral-combined_k141_267318\|\|full | 1848 | No | NA | 5 | 0 | 2 | Not-determined | Genome-fragment | NA | NA | 0 |
| S11-final-viral-combined_k141_321860\|\|full | 2792 | No | NA | 3 | 0 | 1 | Not-determined | Genome-fragment | NA | NA | 0 |
| S5-final-viral-combined_k141_7351\|\|full | 1123 | No | NA | 2 | 1 | 0 | Low-quality | Genome-fragment | 2.49 | HMM-based (lower-bound) | 0 |
| S11-final-viral-combined_k141_115309\|\|full | 1433 | No | NA | 3 | 0 | 1 | Not-determined | Genome-fragment | NA | NA | 0 |
| S11-final-viral-combined_k141_129506\|\|full | 1692 | No | NA | 2 | 0 | 1 | Not-determined | Genome-fragment | NA | NA | 0 |
| S5-final-viral-combined_k141_16226\|\|full | 1123 | No | NA | 4 | 1 | 0 | Low-quality | Genome-fragment | 0.77 | HMM-based (lower-bound) | 0 |
| S5-final-viral-combined_k141_170243\|\|lt2gene | 1123 | No | NA | 1 | 1 | 0 | Low-quality | Genome-fragment | 0.61 | HMM-based (lower-bound) | 0 |
| S11-final-viral-combined_k141_129543\|\|full | 2241 | No | NA | 2 | 0 | 2 | Not-determined | Genome-fragment | NA | NA | 0 |
| S5-final-viral-combined_k141_91722\|\|lt2gene | 1123 | No | NA | 1 | 1 | 0 | Low-quality | Genome-fragment | 2.84 | AAI-based (high-confidence) | 0 |
| S11-final-viral-combined_k141_33110\|\|full | 4127 | No | NA | 3 | 0 | 1 | Not-determined | Genome-fragment | NA | NA | 0 |
| S17-final-viral-combined_k141_185565\|\|full | 1123 | No | NA | 3 | 1 | 0 | Low-quality | Genome-fragment | 2.06 | HMM-based (lower-bound) | 0 |
| S11-final-viral-combined_k141_47099\|\|full | 2004 | No | NA | 3 | 0 | 3 | Not-determined | Genome-fragment | NA | NA | 0 |
| S11-final-viral-combined_k141_226165\|\|full | 33207 | No | NA | 40 | 8 | 3 | Medium-quality | Genome-fragment | 78.45 | AAI-based (medium-confidence) | 0 |
| S11-final-viral-combined_k141_33113\|\|full | 6894 | No | NA | 11 | 0 | 7 | Not-determined | Genome-fragment | NA | NA | 0 |
| S12-final-viral-combined_k141_67946\|\|full | 1123 | No | NA | 3 | 1 | 0 | Low-quality | Genome-fragment | 0.71 | HMM-based (lower-bound) | 0 |
| S11-final-viral-combined_k141_5563\|\|full | 4271 | No | NA | 4 | 0 | 3 | Not-determined | Genome-fragment | NA | NA | 0 |
| S12-final-viral-combined_k141_371844\|\|full | 1123 | No | NA | 2 | 0 | 0 | Low-quality | Genome-fragment | 1.93 | AAI-based (medium-confidence) | 0 |
| S11-final-viral-combined_k141_47141\|\|full | 2093 | No | NA | 2 | 0 | 1 | Not-determined | Genome-fragment | NA | NA | 0 |
| S12-final-viral-combined_k141_379252\|\|full | 1123 | No | NA | 2 | 0 | 1 | Low-quality | Genome-fragment | 1.84 | AAI-based (medium-confidence) | 0 |
| S12-final-viral-combined_k141_300508\|\|full | 1123 | No | NA | 2 | 0 | 0 | Low-quality | Genome-fragment | 0.76 | AAI-based (high-confidence) | 0 |
| S11-final-viral-combined_k141_267363\|\|full | 2317 | No | NA | 5 | 0 | 0 | Not-determined | Genome-fragment | NA | NA | 0 |
| S12-final-viral-combined_k141_387549\|\|lt2gene | 1123 | No | NA | 2 | 1 | 0 | Low-quality | Genome-fragment | 2.75 | AAI-based (medium-confidence) | 0 |
| S11-final-viral-combined_k141_129593\|\|full | 1972 | No | NA | 2 | 0 | 1 | Not-determined | Genome-fragment | NA | NA | 0 |
| S11-final-viral-combined_k141_171340\|\|full | 1388 | No | NA | 3 | 0 | 0 | Not-determined | Genome-fragment | NA | NA | 0 |
| S12_k141_180810 | 1123 | No | NA | 3 | 0 | 0 | Low-quality | Genome-fragment | 0.6 | AAI-based (high-confidence) | 0 |
| S11-final-viral-combined_k141_184770\|\|full | 3195 | No | NA | 3 | 0 | 1 | Not-determined | Genome-fragment | NA | NA | 0 |
| S2-final-viral-combined_k141_152660\|\|full | 1123 | No | NA | 2 | 0 | 0 | Low-quality | Genome-fragment | 2.23 | AAI-based (medium-confidence) | 0 |
| S2_k141_4420 | 1123 | No | NA | 1 | 0 | 0 | Low-quality | Genome-fragment | 2.71 | AAI-based (high-confidence) | 0 |
| S11-final-viral-combined_k141_240616\|\|full | 1668 | No | NA | 2 | 0 | 1 | Not-determined | Genome-fragment | NA | NA | 0 |
| S11-final-viral-combined_k141_184791\|\|full | 8204 | No | NA | 6 | 0 | 6 | Not-determined | Genome-fragment | NA | NA | 0 |
| S8_k141_97849 | 1123 | No | NA | 2 | 0 | 0 | Low-quality | Genome-fragment | 1.12 | AAI-based (high-confidence) | 0 |
| S9-final-viral-combined_k141_123687\|\|lt2gene | 1124 | No | NA | 1 | 1 | 0 | Low-quality | Genome-fragment | 2.83 | AAI-based (high-confidence) | 0 |
| S11-final-viral-combined_k141_130410\|\|full | 1124 | No | NA | 3 | 0 | 0 | Low-quality | Genome-fragment | 2.32 | AAI-based (medium-confidence) | 0 |
| S11-final-viral-combined_k141_309270\|\|full | 4510 | No | NA | 4 | 0 | 3 | Not-determined | Genome-fragment | NA | NA | 0 |
| S11-final-viral-combined_k141_321938\|\|full | 42506 | Yes | 39787 | 45 | 7 | 7 | High-quality | High-quality | 92.89 | AAI-based (medium-confidence) | 6.4 |
| S11-final-viral-combined_k141_242641\|\|full | 1124 | No | NA | 3 | 0 | 0 | Low-quality | Genome-fragment | 2.73 | AAI-based (high-confidence) | 0 |
| S11-final-viral-combined_k141_226236\|\|full | 3272 | No | NA | 5 | 0 | 4 | Not-determined | Genome-fragment | NA | NA | 0 |
| S11-final-viral-combined_k141_283895\|\|full | 1124 | No | NA | 3 | 0 | 0 | Low-quality | Genome-fragment | 1.46 | AAI-based (medium-confidence) | 0 |
| S11_k141_119680 | 1124 | No | NA | 2 | 0 | 0 | Low-quality | Genome-fragment | 2.98 | AAI-based (high-confidence) | 0 |
| S11-final-viral-combined_k141_33212\|\|full | 1762 | No | NA | 2 | 0 | 1 | Not-determined | Genome-fragment | NA | NA | 0 |
| S1-final-viral-combined_k141_152916\|\|full | 1124 | No | NA | 2 | 0 | 0 | Low-quality | Genome-fragment | 1.31 | AAI-based (medium-confidence) | 0 |
| S11-final-viral-combined_k141_129642\|\|full | 3035 | No | NA | 2 | 0 | 2 | Not-determined | Genome-fragment | NA | NA | 0 |
| S11-final-viral-combined_k141_267475\|\|full | 5970 | No | NA | 5 | 0 | 2 | Not-determined | Genome-fragment | NA | NA | 0 |
| S11-final-viral-combined_k141_226248\|\|full | 2138 | No | NA | 1 | 0 | 0 | Not-determined | Genome-fragment | NA | NA | 0 |
| S11-final-viral-combined_k141_33239\|\|full | 3753 | No | NA | 6 | 0 | 0 | Not-determined | Genome-fragment | NA | NA | 0 |
| S1-final-viral-combined_k141_5534\|\|full | 1124 | No | NA | 3 | 0 | 0 | Low-quality | Genome-fragment | 1.89 | AAI-based (high-confidence) | 0 |
| S11-final-viral-combined_k141_226260\|\|full | 1096 | No | NA | 3 | 0 | 3 | Not-determined | Genome-fragment | NA | NA | 0 |
| S11-final-viral-combined_k141_253685\|\|full | 1124 | No | NA | 3 | 1 | 0 | Low-quality | Genome-fragment | 1.47 | HMM-based (lower-bound) | 0 |
| S1-final-viral-combined_k141_347276\|\|lt2gene | 1124 | No | NA | 1 | 1 | 0 | Low-quality | Genome-fragment | 3.15 | AAI-based (high-confidence) | 0 |
| S1_k141_165134 | 1124 | No | NA | 2 | 0 | 0 | Low-quality | Genome-fragment | 1.87 | AAI-based (medium-confidence) | 0 |
| S3-final-viral-combined_k141_38501\|\|full | 1124 | No | NA | 2 | 0 | 1 | Low-quality | Genome-fragment | 0.69 | AAI-based (high-confidence) | 0 |
| S11-final-viral-combined_k141_102820\|\|full | 3130 | No | NA | 3 | 0 | 3 | Not-determined | Genome-fragment | NA | NA | 0 |
| S11-final-viral-combined_k141_143622\|\|full | 4110 | No | NA | 3 | 0 | 2 | Not-determined | Genome-fragment | NA | NA | 0 |
| S5-final-viral-combined_k141_108527\|\|lt2gene | 1124 | No | NA | 2 | 0 | 0 | Low-quality | Genome-fragment | 2.81 | AAI-based (high-confidence) | 0 |
| S17-final-viral-combined_k141_99861\|\|full | 1124 | No | NA | 3 | 0 | 1 | Low-quality | Genome-fragment | 0.55 | AAI-based (medium-confidence) | 0 |
| S12-final-viral-combined_k141_222173\|\|lt2gene | 1124 | No | NA | 2 | 1 | 0 | Low-quality | Genome-fragment | 1.42 | AAI-based (high-confidence) | 0 |
| S12-final-viral-combined_k141_191767\|\|lt2gene | 1124 | No | NA | 2 | 0 | 0 | Low-quality | Genome-fragment | 2 | AAI-based (high-confidence) | 0 |
| S11-final-viral-combined_k141_253869\|\|full | 1539 | No | NA | 2 | 0 | 0 | Not-determined | Genome-fragment | NA | NA | 0 |
| S11-final-viral-combined_k141_295020\|\|full | 1278 | No | NA | 2 | 0 | 2 | Not-determined | Genome-fragment | NA | NA | 0 |
| S11-final-viral-combined_k141_213079\|\|full | 1387 | No | NA | 2 | 0 | 2 | Not-determined | Genome-fragment | NA | NA | 0 |
| S12-final-viral-combined_k141_145140\|\|lt2gene | 1124 | No | NA | 2 | 1 | 0 | Low-quality | Genome-fragment | 2.91 | AAI-based (high-confidence) | 0 |
| S12-final-viral-combined_k141_216401\|\|lt2gene | 1124 | No | NA | 2 | 1 | 0 | Low-quality | Genome-fragment | 0.73 | HMM-based (lower-bound) | 0 |
| S11-final-viral-combined_k141_240699\|\|full | 2872 | No | NA | 4 | 0 | 2 | Not-determined | Genome-fragment | NA | NA | 0 |
| S11-final-viral-combined_k141_213099\|\|full | 1737 | No | NA | 2 | 0 | 2 | Not-determined | Genome-fragment | NA | NA | 0 |
| S11-final-viral-combined_k141_309356\|\|full | 3029 | No | NA | 3 | 0 | 1 | Not-determined | Genome-fragment | NA | NA | 0 |
| S11-final-viral-combined_k141_5677\|\|full | 27110 | No | NA | 20 | 0 | 13 | Not-determined | Genome-fragment | NA | NA | 0 |
| S12_k141_306609 | 1124 | No | NA | 2 | 2 | 0 | Low-quality | Genome-fragment | 2.45 | AAI-based (high-confidence) | 0 |
| S2-final-viral-combined_k141_140001\|\|full | 1124 | No | NA | 3 | 0 | 1 | Low-quality | Genome-fragment | 0.98 | AAI-based (medium-confidence) | 0 |
| S4-final-viral-combined_k141_123459\|\|full | 1124 | No | NA | 3 | 0 | 0 | Low-quality | Genome-fragment | 2.29 | AAI-based (medium-confidence) | 0 |
| S11-final-viral-combined_k141_5688\|\|full | 5923 | No | NA | 6 | 0 | 2 | Not-determined | Genome-fragment | NA | NA | 0 |
| S4-final-viral-combined_k141_124102\|\|full | 1124 | No | NA | 2 | 1 | 0 | Low-quality | Genome-fragment | 0.99 | AAI-based (high-confidence) | 0 |
| S11-final-viral-combined_k141_88560\|\|full | 1958 | No | NA | 3 | 0 | 2 | Not-determined | Genome-fragment | NA | NA | 0 |
| S11-final-viral-combined_k141_309384\|\|full | 2203 | No | NA | 2 | 0 | 0 | Not-determined | Genome-fragment | NA | NA | 0 |
| S11-final-viral-combined_k141_5699\|\|full | 1302 | No | NA | 3 | 0 | 2 | Not-determined | Genome-fragment | NA | NA | 0 |
| S11-final-viral-combined_k141_282052\|\|full | 3434 | No | NA | 3 | 0 | 2 | Not-determined | Genome-fragment | NA | NA | 0 |
| S11-final-viral-combined_k141_226380\|\|full | 2118 | No | NA | 3 | 0 | 3 | Not-determined | Genome-fragment | NA | NA | 0 |
| S8-final-viral-combined_k141_197780\|\|full | 1124 | No | NA | 3 | 0 | 0 | Low-quality | Genome-fragment | 1.19 | AAI-based (high-confidence) | 0 |
| S11-final-viral-combined_k141_253934\|\|full | 6029 | No | NA | 3 | 0 | 1 | Not-determined | Genome-fragment | NA | NA | 0 |
| S11-final-viral-combined_k141_322062\|\|full | 1482 | No | NA | 3 | 0 | 2 | Not-determined | Genome-fragment | NA | NA | 0 |
| S11-final-viral-combined_k141_253948\|\|full | 1824 | No | NA | 2 | 0 | 1 | Not-determined | Genome-fragment | NA | NA | 0 |
| S11-final-viral-combined_k141_61033\|\|full | 2335 | No | NA | 3 | 0 | 2 | Not-determined | Genome-fragment | NA | NA | 0 |
| S11-final-viral-combined_k141_213161\|\|full | 3307 | No | NA | 3 | 0 | 0 | Not-determined | Genome-fragment | NA | NA | 0 |
| S11-final-viral-combined_k141_253144\|\|full | 1125 | No | NA | 4 | 2 | 0 | Low-quality | Genome-fragment | 1.73 | AAI-based (medium-confidence) | 0 |
| S11-final-viral-combined_k141_129751\|\|full | 1367 | No | NA | 3 | 0 | 1 | Not-determined | Genome-fragment | NA | NA | 0 |
| S11-final-viral-combined_k141_240783\|\|full | 1770 | No | NA | 3 | 0 | 0 | Not-determined | Genome-fragment | NA | NA | 0 |
| S11-final-viral-combined_k141_47377\|\|full | 15742 | No | NA | 12 | 0 | 11 | Not-determined | Genome-fragment | NA | NA | 0 |
| S11-final-viral-combined_k141_7949\|\|full | 1125 | No | NA | 3 | 0 | 0 | Low-quality | Genome-fragment | 2.89 | AAI-based (high-confidence) | 0 |
| S11-final-viral-combined_k141_18915\|\|full | 3029 | No | NA | 2 | 0 | 1 | Not-determined | Genome-fragment | NA | NA | 0 |
| S11-final-viral-combined_k141_187736\|\|full | 1125 | No | NA | 3 | 0 | 0 | Low-quality | Genome-fragment | 2.45 | AAI-based (medium-confidence) | 0 |
| S11-final-viral-combined_k141_240798\|\|full | 3716 | No | NA | 4 | 0 | 1 | Not-determined | Genome-fragment | NA | NA | 0 |
| S11-final-viral-combined_k141_115425\|\|full | 1700 | No | NA | 2 | 0 | 1 | Not-determined | Genome-fragment | NA | NA | 0 |
| S1-final-viral-combined_k141_46294\|\|full | 1125 | No | NA | 2 | 0 | 0 | Low-quality | Genome-fragment | 1.24 | AAI-based (medium-confidence) | 0 |
| S1-final-viral-combined_k141_328239\|\|full | 1125 | No | NA | 4 | 0 | 0 | Low-quality | Genome-fragment | 2.17 | AAI-based (high-confidence) | 0 |
| S11-final-viral-combined_k141_171499\|\|full | 2649 | No | NA | 3 | 0 | 3 | Not-determined | Genome-fragment | NA | NA | 0 |
| S1_k141_300389 | 1125 | No | NA | 2 | 0 | 0 | Low-quality | Genome-fragment | 2.06 | AAI-based (medium-confidence) | 0 |
| S11-final-viral-combined_k141_143768\|\|full | 2984 | No | NA | 3 | 0 | 2 | Not-determined | Genome-fragment | NA | NA | 0 |
| S11-final-viral-combined_k141_198512\|\|full | 41546 | Yes | 36129 | 60 | 9 | 3 | Medium-quality | Genome-fragment | 66.2 | AAI-based (high-confidence) | 13.04 |
| S11-final-viral-combined_k141_171514\|\|full | 3260 | No | NA | 5 | 0 | 2 | Not-determined | Genome-fragment | NA | NA | 0 |
| S1_k141_45831 | 1125 | No | NA | 2 | 0 | 1 | Low-quality | Genome-fragment | 3.72 | AAI-based (medium-confidence) | 0 |
| S5-final-viral-combined_k141_108199\|\|lt2gene | 1125 | No | NA | 1 | 1 | 0 | Low-quality | Genome-fragment | 3.33 | AAI-based (high-confidence) | 0 |
| S11-final-viral-combined_k141_115461\|\|full | 1087 | No | NA | 2 | 0 | 0 | Not-determined | Genome-fragment | NA | NA | 0 |
| S7-final-viral-combined_k141_26649\|\|full | 1125 | No | NA | 3 | 0 | 2 | Low-quality | Genome-fragment | 2.59 | AAI-based (high-confidence) | 0 |
| S11-final-viral-combined_k141_226492\|\|full | 1329 | No | NA | 3 | 0 | 0 | Not-determined | Genome-fragment | NA | NA | 0 |
| S11-final-viral-combined_k141_282155\|\|full | 2555 | No | NA | 2 | 0 | 1 | Not-determined | Genome-fragment | NA | NA | 0 |
| S11-final-viral-combined_k141_171529\|\|full | 2066 | No | NA | 3 | 0 | 3 | Not-determined | Genome-fragment | NA | NA | 0 |
| S11-final-viral-combined_k141_184917\|\|full | 1710 | No | NA | 3 | 0 | 1 | Not-determined | Genome-fragment | NA | NA | 0 |
| S11-final-viral-combined_k141_322136\|\|full | 1337 | No | NA | 2 | 0 | 0 | Not-determined | Genome-fragment | NA | NA | 0 |
| S11-final-viral-combined_k141_322149\|\|full | 2473 | No | NA | 3 | 0 | 2 | Not-determined | Genome-fragment | NA | NA | 0 |
| S11-final-viral-combined_k141_157537\|\|full | 1219 | No | NA | 2 | 0 | 1 | Not-determined | Genome-fragment | NA | NA | 0 |
| S17-final-viral-combined_k141_232319\|\|lt2gene | 1125 | No | NA | 1 | 1 | 0 | Low-quality | Genome-fragment | 2.54 | AAI-based (high-confidence) | 0 |
| S11-final-viral-combined_k141_198559\|\|full | 8454 | No | NA | 8 | 0 | 7 | Not-determined | Genome-fragment | NA | NA | 0 |
| S11-final-viral-combined_k141_226519\|\|full | 1349 | No | NA | 2 | 0 | 0 | Not-determined | Genome-fragment | NA | NA | 0 |
| S12-final-viral-combined_k141_184339\|\|full | 1125 | No | NA | 2 | 0 | 0 | Low-quality | Genome-fragment | 0.82 | AAI-based (medium-confidence) | 0 |
| S11-final-viral-combined_k141_267814\|\|full | 2597 | No | NA | 3 | 0 | 2 | Not-determined | Genome-fragment | NA | NA | 0 |
| S12-final-viral-combined_k141_259427\|\|full | 1125 | No | NA | 2 | 0 | 0 | Low-quality | Genome-fragment | 0.87 | AAI-based (high-confidence) | 0 |
| S11-final-viral-combined_k141_157590\|\|full | 4156 | No | NA | 6 | 0 | 1 | Not-determined | Genome-fragment | NA | NA | 0 |
| S12-final-viral-combined_k141_210363\|\|full | 1125 | No | NA | 4 | 1 | 0 | Low-quality | Genome-fragment | 1.81 | AAI-based (high-confidence) | 0 |
| S11-final-viral-combined_k141_322218\|\|full | 1688 | No | NA | 3 | 0 | 1 | Not-determined | Genome-fragment | NA | NA | 0 |
| S11-final-viral-combined_k141_19057\|\|full | 1050 | No | NA | 3 | 0 | 1 | Not-determined | Genome-fragment | NA | NA | 0 |
| S4-final-viral-combined_k141_122922\|\|lt2gene | 1125 | No | NA | 1 | 1 | 0 | Low-quality | Genome-fragment | 2.89 | AAI-based (medium-confidence) | 0 |
| S11-final-viral-combined_k141_184984\|\|full | 2428 | No | NA | 2 | 0 | 2 | Not-determined | Genome-fragment | NA | NA | 0 |
| S11-final-viral-combined_k141_240890\|\|full | 24596 | No | NA | 30 | 0 | 18 | Not-determined | Genome-fragment | NA | NA | 0 |
| S11-final-viral-combined_k141_282227\|\|full | 27969 | No | NA | 36 | 16 | 3 | Medium-quality | Genome-fragment | 59.35 | AAI-based (medium-confidence) | 0 |
| S11-final-viral-combined_k141_103072\|\|full | 2025 | No | NA | 3 | 0 | 1 | Not-determined | Genome-fragment | NA | NA | 0 |
| S9-final-viral-combined_k141_67816\|\|full | 1126 | No | NA | 3 | 0 | 0 | Low-quality | Genome-fragment | 0.63 | AAI-based (medium-confidence) | 0 |
| S11-final-viral-combined_k141_213368\|\|full | 4328 | No | NA | 3 | 0 | 3 | Not-determined | Genome-fragment | NA | NA | 0 |
| S11-final-viral-combined_k141_322286\|\|full | 6907 | No | NA | 3 | 0 | 3 | Not-determined | Genome-fragment | NA | NA | 0 |
| S11-final-viral-combined_k141_306546\|\|full | 1126 | No | NA | 3 | 0 | 0 | Low-quality | Genome-fragment | 2.63 | AAI-based (high-confidence) | 0 |
| S11-final-viral-combined_k141_157704\|\|full | 1671 | No | NA | 4 | 0 | 1 | Not-determined | Genome-fragment | NA | NA | 0 |
| S1-final-viral-combined_k141_369485\|\|lt2gene | 1126 | No | NA | 1 | 1 | 0 | Low-quality | Genome-fragment | 18.95 | AAI-based (high-confidence) | 0 |
| S1-final-viral-combined_k141_206538\|\|lt2gene | 1126 | No | NA | 2 | 2 | 0 | Low-quality | Genome-fragment | 2.44 | AAI-based (high-confidence) | 0 |
| S1_k141_270609 | 1126 | No | NA | 1 | 0 | 0 | Low-quality | Genome-fragment | 0.91 | AAI-based (high-confidence) | 0 |
| S11-final-viral-combined_k141_61241\|\|full | 6106 | No | NA | 6 | 0 | 4 | Not-determined | Genome-fragment | NA | NA | 0 |
| S11-final-viral-combined_k141_295252\|\|full | 1729 | No | NA | 2 | 0 | 2 | Not-determined | Genome-fragment | NA | NA | 0 |
| S11-final-viral-combined_k141_19156\|\|full | 1372 | No | NA | 6 | 0 | 0 | Not-determined | Genome-fragment | NA | NA | 0 |
| S11-final-viral-combined_k141_282320\|\|full | 1145 | No | NA | 2 | 0 | 0 | Not-determined | Genome-fragment | NA | NA | 0 |
| S11-final-viral-combined_k141_226624\|\|full | 1965 | No | NA | 2 | 0 | 2 | Not-determined | Genome-fragment | NA | NA | 0 |
| S11-final-viral-combined_k141_198700\|\|full | 1241 | No | NA | 2 | 0 | 1 | Not-determined | Genome-fragment | NA | NA | 0 |
| S3-final-viral-combined_k141_43122\|\|full | 1126 | No | NA | 2 | 0 | 0 | Low-quality | Genome-fragment | 2.25 | AAI-based (high-confidence) | 0 |
| S11-final-viral-combined_k141_322338\|\|full | 3842 | No | NA | 3 | 0 | 2 | Not-determined | Genome-fragment | NA | NA | 0 |
| S11-final-viral-combined_k141_254282\|\|full | 4445 | No | NA | 3 | 0 | 2 | Not-determined | Genome-fragment | NA | NA | 0 |
| S11-final-viral-combined_k141_322354\|\|full | 1799 | No | NA | 2 | 0 | 0 | Not-determined | Genome-fragment | NA | NA | 0 |
| S5-final-viral-combined_k141_681\|\|full | 1126 | Yes | 723 | 3 | 1 | 1 | Low-quality | Genome-fragment | 1.8 | HMM-based (lower-bound) | 35.79 |
| S5-final-viral-combined_k141_58271\|\|full | 1126 | No | NA | 2 | 0 | 1 | Low-quality | Genome-fragment | 0.47 | AAI-based (medium-confidence) | 0 |
| S11-final-viral-combined_k141_33645\|\|full | 1113 | No | NA | 3 | 0 | 1 | Not-determined | Genome-fragment | NA | NA | 0 |
| S7-final-viral-combined_k141_122794\|\|full | 1126 | No | NA | 3 | 1 | 0 | Low-quality | Genome-fragment | 3.06 | AAI-based (high-confidence) | 0 |
| S7-final-viral-combined_k141_53857\|\|full | 1126 | No | NA | 2 | 0 | 1 | Low-quality | Genome-fragment | 5.91 | AAI-based (medium-confidence) | 0 |
| S11-final-viral-combined_k141_254333\|\|full | 1996 | No | NA | 2 | 0 | 1 | Not-determined | Genome-fragment | NA | NA | 0 |
| S11-final-viral-combined_k141_61339\|\|full | 2617 | No | NA | 2 | 0 | 1 | Not-determined | Genome-fragment | NA | NA | 0 |
| S7-final-viral-combined_k141_9110\|\|full | 1126 | No | NA | 2 | 1 | 0 | Low-quality | Genome-fragment | 0.69 | HMM-based (lower-bound) | 0 |
| S11-final-viral-combined_k141_47668\|\|full | 1338 | No | NA | 2 | 0 | 1 | Not-determined | Genome-fragment | NA | NA | 0 |
| S7_k141_44491 | 1126 | No | NA | 2 | 1 | 0 | Low-quality | Genome-fragment | 1.4 | AAI-based (high-confidence) | 0 |
| S17-final-viral-combined_k141_109193\|\|full | 1126 | No | NA | 2 | 1 | 0 | Low-quality | Genome-fragment | 2.06 | AAI-based (medium-confidence) | 0 |
| S11-final-viral-combined_k141_295357\|\|full | 3206 | No | NA | 4 | 0 | 2 | Not-determined | Genome-fragment | NA | NA | 0 |
| S17-k141_127114 | 1126 | No | NA | 5 | 5 | 0 | Low-quality | Genome-fragment | 3.71 | AAI-based (high-confidence) | 0 |
| S11-final-viral-combined_k141_322466\|\|full | 1661 | No | NA | 3 | 0 | 0 | Not-determined | Genome-fragment | NA | NA | 0 |
| S11-final-viral-combined_k141_144014\|\|full | 4971 | No | NA | 4 | 0 | 2 | Not-determined | Genome-fragment | NA | NA | 0 |
| S17-k141_128814 | 1126 | No | NA | 1 | 0 | 1 | Low-quality | Genome-fragment | 0.69 | AAI-based (medium-confidence) | 0 |
| S11-final-viral-combined_k141_213459\|\|full | 2362 | No | NA | 3 | 0 | 2 | Not-determined | Genome-fragment | NA | NA | 0 |
| S11-final-viral-combined_k141_213474\|\|full | 2424 | No | NA | 3 | 0 | 1 | Not-determined | Genome-fragment | NA | NA | 0 |
| S11-final-viral-combined_k141_185140\|\|full | 3038 | No | NA | 3 | 0 | 2 | Not-determined | Genome-fragment | NA | NA | 0 |
| S11-final-viral-combined_k141_33758\|\|full | 1141 | No | NA | 2 | 0 | 0 | Not-determined | Genome-fragment | NA | NA | 0 |
| S12-final-viral-combined_k141_132699\|\|full | 1126 | No | NA | 2 | 1 | 0 | Low-quality | Genome-fragment | 2.21 | AAI-based (medium-confidence) | 0 |
| S2-final-viral-combined_k141_168373\|\|full | 1126 | No | NA | 3 | 1 | 0 | Low-quality | Genome-fragment | 2.73 | AAI-based (high-confidence) | 0 |
| S2-final-viral-combined_k141_244329\|\|full | 1126 | No | NA | 2 | 0 | 1 | Low-quality | Genome-fragment | 1.57 | AAI-based (medium-confidence) | 0 |
| S11-final-viral-combined_k141_144079\|\|full | 1180 | No | NA | 2 | 0 | 0 | Not-determined | Genome-fragment | NA | NA | 0 |
| S4-final-viral-combined_k141_57995\|\|lt2gene | 1126 | No | NA | 2 | 1 | 0 | Low-quality | Genome-fragment | 2.93 | AAI-based (medium-confidence) | 0 |
| S6-final-viral-combined_k141_53903\|\|full | 1126 | No | NA | 4 | 0 | 0 | Low-quality | Genome-fragment | 1.34 | AAI-based (high-confidence) | 0 |
| S8-final-viral-combined_k141_250707\|\|lt2gene | 1126 | No | NA | 2 | 0 | 0 | Low-quality | Genome-fragment | 2.44 | AAI-based (medium-confidence) | 0 |
| S11-final-viral-combined_k141_267993\|\|full | 74748 | Yes | 64139 | 97 | 26 | 5 | High-quality | High-quality | 100 | AAI-based (medium-confidence) | 14.19 |
| S9-final-viral-combined_k141_33673\|\|full | 1127 | No | NA | 3 | 2 | 0 | Low-quality | Genome-fragment | 2.5 | AAI-based (high-confidence) | 0 |
| S11_k141_297500 | 1127 | No | NA | 2 | 1 | 0 | Low-quality | Genome-fragment | 1.78 | AAI-based (medium-confidence) | 0 |
| S11-final-viral-combined_k141_198863\|\|full | 2613 | No | NA | 3 | 0 | 2 | Not-determined | Genome-fragment | NA | NA | 0 |
| S11-final-viral-combined_k141_241192\|\|full | 1646 | No | NA | 3 | 0 | 1 | Not-determined | Genome-fragment | NA | NA | 0 |
| S11-final-viral-combined_k141_157977\|\|full | 5705 | No | NA | 6 | 0 | 4 | Not-determined | Genome-fragment | NA | NA | 0 |
| S11-final-viral-combined_k141_88966\|\|full | 1506 | No | NA | 3 | 0 | 1 | Not-determined | Genome-fragment | NA | NA | 0 |
| S1-final-viral-combined_k141_362117\|\|full | 1127 | No | NA | 3 | 1 | 0 | Low-quality | Genome-fragment | 2.2 | AAI-based (medium-confidence) | 0 |
| S11-final-viral-combined_k141_295490\|\|full | 1685 | No | NA | 3 | 0 | 0 | Not-determined | Genome-fragment | NA | NA | 0 |
| S1-final-viral-combined_k141_350441\|\|full | 1127 | No | NA | 2 | 0 | 0 | Low-quality | Genome-fragment | 0.9 | AAI-based (medium-confidence) | 0 |
| S5-final-viral-combined_k141_112842\|\|full | 1127 | No | NA | 4 | 0 | 2 | Low-quality | Genome-fragment | 0.78 | AAI-based (high-confidence) | 0 |

| | | | | | | | | | | | |
|---|---|---|---|---|---|---|---|---|---|---|---|
| S11-final-viral-combined_k141_144137\|\|full | 3229 | No | NA | 3 | 0 | 1 | Not-determined | Genome-fragment | NA | NA | 0 |
| S7-final-viral-combined_k141_187752\|\|full | 1127 | No | NA | 2 | 0 | 0 | Low-quality | Genome-fragment | 1.91 | AAI-based (medium-confidence) | 0 |
| S7-final-viral-combined_k141_171838\|\|full | 1127 | No | NA | 2 | 0 | 0 | Low-quality | Genome-fragment | 2.75 | AAI-based (medium-confidence) | 0 |
| S11-final-viral-combined_k141_103309\|\|full | 1662 | No | NA | 3 | 0 | 1 | Not-determined | Genome-fragment | NA | NA | 0 |
| S11-final-viral-combined_k141_254464\|\|full | 5746 | No | NA | 3 | 0 | 1 | Not-determined | Genome-fragment | NA | NA | 0 |
| S12_k141_191382 | 1127 | No | NA | 2 | 0 | 1 | Low-quality | Genome-fragment | 2.46 | AAI-based (medium-confidence) | 0 |
| S11-final-viral-combined_k141_171863\|\|full | 6406 | No | NA | 11 | 0 | 2 | Not-determined | Genome-fragment | NA | NA | 0 |
| S11-final-viral-combined_k141_158015\|\|full | 1731 | No | NA | 3 | 0 | 2 | Not-determined | Genome-fragment | NA | NA | 0 |
| S11-final-viral-combined_k141_309920\|\|full | 2898 | No | NA | 2 | 0 | 0 | Not-determined | Genome-fragment | NA | NA | 0 |
| S11-final-viral-combined_k141_171865\|\|full | 2401 | No | NA | 3 | 0 | 3 | Not-determined | Genome-fragment | NA | NA | 0 |
| S2-final-viral-combined_k141_173907\|\|full | 1127 | No | NA | 2 | 0 | 1 | Low-quality | Genome-fragment | 0.52 | AAI-based (medium-confidence) | 0 |
| S11-final-viral-combined_k141_268061\|\|full | 2823 | No | NA | 3 | 0 | 2 | Not-determined | Genome-fragment | NA | NA | 0 |
| S11-final-viral-combined_k141_282615\|\|full | 1661 | No | NA | 3 | 0 | 0 | Not-determined | Genome-fragment | NA | NA | 0 |
| S2-final-viral-combined_k141_196382\|\|full | 1127 | No | NA | 2 | 0 | 0 | Low-quality | Genome-fragment | 1.95 | AAI-based (medium-confidence) | 0 |
| S2-final-viral-combined_k141_82409\|\|full | 1127 | No | NA | 1 | 0 | 0 | Low-quality | Genome-fragment | 1.76 | AAI-based (high-confidence) | 0 |
| S11-final-viral-combined_k141_116018\|\|full | 1531 | No | NA | 2 | 0 | 1 | Not-determined | Genome-fragment | NA | NA | 0 |
| S11-final-viral-combined_k141_268086\|\|full | 1783 | No | NA | 4 | 0 | 0 | Not-determined | Genome-fragment | NA | NA | 0 |
| S11-final-viral-combined_k141_282630\|\|full | 6359 | No | NA | 5 | 0 | 3 | Not-determined | Genome-fragment | NA | NA | 0 |
| S11-final-viral-combined_k141_19371\|\|full | 4078 | No | NA | 4 | 0 | 2 | Not-determined | Genome-fragment | NA | NA | 0 |
| S2-final-viral-combined_k141_180386\|\|lt2gene | 1127 | No | NA | 1 | 1 | 0 | Low-quality | Genome-fragment | 2.58 | AAI-based (high-confidence) | 0 |
| S6-final-viral-combined_k141_64605\|\|full | 1127 | No | NA | 3 | 1 | 0 | Low-quality | Genome-fragment | 2.23 | AAI-based (medium-confidence) | 0 |
| S8-final-viral-combined_k141_146599\|\|full | 1127 | No | NA | 4 | 0 | 0 | Low-quality | Genome-fragment | 3.54 | AAI-based (medium-confidence) | 0 |
| S8_k141_17927 | 1127 | No | NA | 2 | 0 | 0 | Low-quality | Genome-fragment | 1.97 | AAI-based (medium-confidence) | 0 |
| S11-final-viral-combined_k141_47739\|\|full | 1820 | No | NA | 3 | 0 | 1 | Not-determined | Genome-fragment | NA | NA | 0 |
| S11-final-viral-combined_k141_6271\|\|full | 1154 | No | NA | 2 | 0 | 1 | Not-determined | Genome-fragment | NA | NA | 0 |
| S11-final-viral-combined_k141_74670\|\|full | 1141 | No | NA | 2 | 0 | 2 | Not-determined | Genome-fragment | NA | NA | 0 |
| S9-final-viral-combined_k141_78073\|\|lt2gene | 1128 | No | NA | 1 | 1 | 0 | Low-quality | Genome-fragment | 1.04 | HMM-based (lower-bound) | 0 |
| S1-final-viral-combined_k141_214642\|\|lt2gene | 1128 | No | NA | 2 | 1 | 0 | Low-quality | Genome-fragment | 2.32 | AAI-based (high-confidence) | 0 |
| S11-final-viral-combined_k141_309974\|\|full | 7093 | No | NA | 9 | 0 | 9 | Not-determined | Genome-fragment | NA | NA | 0 |
| S1-final-viral-combined_k141_2632\|\|lt2gene | 1128 | No | NA | 1 | 1 | 0 | Low-quality | Genome-fragment | 0.4 | HMM-based (lower-bound) | 0 |
| S11-final-viral-combined_k141_171951\|\|full | 1896 | No | NA | 3 | 0 | 3 | Not-determined | Genome-fragment | NA | NA | 0 |
| S11-final-viral-combined_k141_74678\|\|full | 2952 | No | NA | 4 | 0 | 2 | Not-determined | Genome-fragment | NA | NA | 0 |
| S11-final-viral-combined_k141_61692\|\|full | 2279 | No | NA | 3 | 0 | 1 | Not-determined | Genome-fragment | NA | NA | 0 |
| S1_k141_222745 | 1128 | No | NA | 3 | 1 | 0 | Low-quality | Genome-fragment | 2.41 | AAI-based (medium-confidence) | 0 |
| S11-final-viral-combined_k141_310011\|\|full | 1398 | No | NA | 3 | 0 | 0 | Not-determined | Genome-fragment | NA | NA | 0 |
| S11-final-viral-combined_k141_61709\|\|full | 1025 | No | NA | 3 | 0 | 0 | Not-determined | Genome-fragment | NA | NA | 0 |
| S11-final-viral-combined_k141_241434\|\|full | 1269 | No | NA | 2 | 0 | 1 | Not-determined | Genome-fragment | NA | NA | 0 |
| S3-final-viral-combined_k141_70592\|\|full | 1128 | No | NA | 3 | 0 | 0 | Low-quality | Genome-fragment | 1.37 | AAI-based (medium-confidence) | 0 |
| S11-final-viral-combined_k141_213710\|\|full | 5127 | No | NA | 6 | 0 | 1 | Not-determined | Genome-fragment | NA | NA | 0 |
| S11-final-viral-combined_k141_310045\|\|full | 2762 | No | NA | 3 | 0 | 1 | Not-determined | Genome-fragment | NA | NA | 0 |
| S11-final-viral-combined_k141_227013\|\|full | 10273 | No | NA | 6 | 0 | 4 | Not-determined | Genome-fragment | NA | NA | 0 |
| S11-final-viral-combined_k141_222787\|\|full | 1470 | No | NA | 3 | 0 | 2 | Not-determined | Genome-fragment | NA | NA | 0 |
| S11-final-viral-combined_k141_89079\|\|full | 1945 | No | NA | 3 | 0 | 2 | Not-determined | Genome-fragment | NA | NA | 0 |
| S11-final-viral-combined_k141_198995\|\|full | 6653 | No | NA | 3 | 0 | 1 | Not-determined | Genome-fragment | NA | NA | 0 |
| S11-final-viral-combined_k141_19470\|\|full | 2769 | No | NA | 4 | 0 | 1 | Not-determined | Genome-fragment | NA | NA | 0 |
| S5-final-viral-combined_k141_101636\|\|lt2gene | 1128 | No | NA | 2 | 2 | 0 | Low-quality | Genome-fragment | 2.52 | AAI-based (high-confidence) | 0 |
| S11-final-viral-combined_k141_241481\|\|full | 1992 | No | NA | 1 | 0 | 1 | Not-determined | Genome-fragment | NA | NA | 0 |
| S7-final-viral-combined_k141_138248\|\|full | 1128 | No | NA | 2 | 0 | 1 | Low-quality | Genome-fragment | 0.82 | AAI-based (medium-confidence) | 0 |
| S11-final-viral-combined_k141_6394\|\|full | 1941 | No | NA | 2 | 0 | 1 | Not-determined | Genome-fragment | NA | NA | 0 |
| S2-final-viral-combined_k141_172619\|\|lt2gene | 1128 | No | NA | 2 | 1 | 0 | Low-quality | Genome-fragment | 1.41 | HMM-based (lower-bound) | 0 |
| S11-final-viral-combined_k141_213736\|\|full | 1360 | No | NA | 3 | 0 | 2 | Not-determined | Genome-fragment | NA | NA | 0 |
| S4_k141_121786 | 1128 | No | NA | 2 | 0 | 2 | Low-quality | Genome-fragment | 0.65 | AAI-based (medium-confidence) | 0 |
| S11-final-viral-combined_k141_322809\|\|full | 1438 | No | NA | 3 | 0 | 0 | Not-determined | Genome-fragment | NA | NA | 0 |
| S11-final-viral-combined_k141_130247\|\|full | 2196 | No | NA | 2 | 0 | 0 | Not-determined | Genome-fragment | NA | NA | 0 |
| S9-final-viral-combined_k141_67856\|\|full | 1129 | No | NA | 3 | 0 | 0 | Low-quality | Genome-fragment | 2.15 | AAI-based (medium-confidence) | 0 |
| S11-final-viral-combined_k141_268286\|\|full | 2820 | No | NA | 4 | 0 | 0 | Not-determined | Genome-fragment | NA | NA | 0 |
| S11-final-viral-combined_k141_227046\|\|full | 3420 | No | NA | 4 | 0 | 0 | Not-determined | Genome-fragment | NA | NA | 0 |
| S11-final-viral-combined_k141_137956\|\|full | 1129 | No | NA | 3 | 0 | 0 | Low-quality | Genome-fragment | 2.81 | AAI-based (medium-confidence) | 0 |
| S11-final-viral-combined_k141_154096\|\|lt2gene | 1129 | No | NA | 2 | 1 | 0 | Low-quality | Genome-fragment | 2.48 | AAI-based (high-confidence) | 0 |
| S11-final-viral-combined_k141_103483\|\|full | 1169 | No | NA | 2 | 0 | 1 | Not-determined | Genome-fragment | NA | NA | 0 |
| S11_k141_260207 | 1129 | No | NA | 2 | 0 | 1 | Low-quality | Genome-fragment | 1.53 | AAI-based (medium-confidence) | 0 |
| S1-final-viral-combined_k141_273282\|\|full | 1129 | No | NA | 3 | 0 | 1 | Low-quality | Genome-fragment | 2.37 | AAI-based (medium-confidence) | 0 |
| S1-final-viral-combined_k141_12549\|\|full | 1129 | No | NA | 2 | 0 | 1 | Low-quality | Genome-fragment | 0.88 | AAI-based (medium-confidence) | 0 |
| S11-final-viral-combined_k141_34002\|\|full | 3042 | No | NA | 3 | 0 | 2 | Not-determined | Genome-fragment | NA | NA | 0 |
| S1_k141_264555 | 1129 | No | NA | 1 | 0 | 0 | Low-quality | Genome-fragment | 1.94 | AAI-based (high-confidence) | 0 |
| S3-final-viral-combined_k141_65566\|\|full | 1129 | No | NA | 2 | 0 | 0 | Low-quality | Genome-fragment | 1.88 | AAI-based (high-confidence) | 0 |
| S11-final-viral-combined_k141_322857\|\|full | 1115 | No | NA | 2 | 0 | 0 | Not-determined | Genome-fragment | NA | NA | 0 |
| S3-final-viral-combined_k141_101540\|\|lt2gene | 1129 | No | NA | 2 | 1 | 0 | Low-quality | Genome-fragment | 2.47 | AAI-based (high-confidence) | 0 |
| S7-final-viral-combined_k141_51463\|\|full | 1129 | No | NA | 2 | 0 | 1 | Low-quality | Genome-fragment | 2.11 | AAI-based (high-confidence) | 0 |
| S11-final-viral-combined_k141_103559\|\|full | 2430 | No | NA | 3 | 0 | 2 | Not-determined | Genome-fragment | NA | NA | 0 |
| S7_k141_188268 | 1129 | No | NA | 2 | 1 | 0 | Low-quality | Genome-fragment | 1.72 | AAI-based (high-confidence) | 0 |
| S11-final-viral-combined_k141_158306\|\|full | 3402 | No | NA | 3 | 0 | 3 | Not-determined | Genome-fragment | NA | NA | 0 |
| S11-final-viral-combined_k141_61885\|\|full | 2853 | No | NA | 4 | 0 | 1 | Not-determined | Genome-fragment | NA | NA | 0 |
| S17-k141_179063 | 1129 | No | NA | 2 | 0 | 0 | Low-quality | Genome-fragment | 0.58 | AAI-based (medium-confidence) | 0 |
| S12-final-viral-combined_k141_190870\|\|full | 1129 | No | NA | 3 | 0 | 0 | Low-quality | Genome-fragment | 0.54 | AAI-based (medium-confidence) | 0 |
| S11-final-viral-combined_k141_254765\|\|full | 4084 | No | NA | 3 | 0 | 3 | Not-determined | Genome-fragment | NA | NA | 0 |
| S12-final-viral-combined_k141_371092\|\|lt2gene | 1129 | No | NA | 2 | 1 | 0 | Low-quality | Genome-fragment | 2.54 | AAI-based (high-confidence) | 0 |
| S11-final-viral-combined_k141_74851\|\|full | 2387 | No | NA | 2 | 0 | 2 | Not-determined | Genome-fragment | NA | NA | 0 |
| S12-final-viral-combined_k141_140149\|\|lt2gene | 1129 | No | NA | 2 | 1 | 0 | Low-quality | Genome-fragment | 1.85 | AAI-based (high-confidence) | 0 |
| S8-final-viral-combined_k141_34114\|\|full | 1129 | No | NA | 2 | 0 | 0 | Low-quality | Genome-fragment | 1.16 | AAI-based (high-confidence) | 0 |
| S11-final-viral-combined_k141_61897\|\|full | 4819 | No | NA | 4 | 0 | 3 | Not-determined | Genome-fragment | NA | NA | 0 |
| S11_k141_127515 | 1130 | No | NA | 1 | 0 | 1 | Low-quality | Genome-fragment | 0.42 | AAI-based (medium-confidence) | 0 |
| S11-final-viral-combined_k141_254779\|\|full | 4367 | No | NA | 4 | 0 | 3 | Not-determined | Genome-fragment | NA | NA | 0 |
| S11-final-viral-combined_k141_213863\|\|full | 2154 | No | NA | 2 | 0 | 1 | Not-determined | Genome-fragment | NA | NA | 0 |
| S11-final-viral-combined_k141_172205\|\|full | 4086 | No | NA | 3 | 0 | 3 | Not-determined | Genome-fragment | NA | NA | 0 |
| S11-final-viral-combined_k141_310229\|\|full | 4643 | No | NA | 4 | 0 | 3 | Not-determined | Genome-fragment | NA | NA | 0 |
| S1-final-viral-combined_k141_202534\|\|full | 1130 | No | NA | 2 | 1 | 0 | Low-quality | Genome-fragment | 2.24 | AAI-based (high-confidence) | 0 |
| S11-final-viral-combined_k141_103605\|\|full | 1753 | No | NA | 2 | 0 | 2 | Not-determined | Genome-fragment | NA | NA | 0 |
| S11-final-viral-combined_k141_158379\|\|full | 1782 | No | NA | 3 | 0 | 3 | Not-determined | Genome-fragment | NA | NA | 0 |
| S1-final-viral-combined_k141_30840\|\|full | 1130 | No | NA | 3 | 2 | 0 | Low-quality | Genome-fragment | 2.5 | AAI-based (high-confidence) | 0 |
| S7-final-viral-combined_k141_57550\|\|full | 1130 | No | NA | 2 | 0 | 0 | Low-quality | Genome-fragment | 3.25 | AAI-based (high-confidence) | 0 |
| S17-final-viral-combined_k141_11208\|\|full | 1130 | No | NA | 4 | 0 | 0 | Low-quality | Genome-fragment | 2.61 | AAI-based (high-confidence) | 0 |
| S11-final-viral-combined_k141_268379\|\|full | 1564 | No | NA | 2 | 0 | 1 | Not-determined | Genome-fragment | NA | NA | 0 |
| S2-final-viral-combined_k141_110588\|\|full | 1130 | No | NA | 2 | 0 | 0 | Low-quality | Genome-fragment | 2.14 | AAI-based (medium-confidence) | 0 |
| S6-final-viral-combined_k141_58691\|\|full | 1130 | No | NA | 3 | 0 | 0 | Low-quality | Genome-fragment | 2.74 | AAI-based (medium-confidence) | 0 |
| S8-final-viral-combined_k141_140201\|\|full | 1130 | No | NA | 4 | 0 | 1 | Low-quality | Genome-fragment | 2.19 | AAI-based (high-confidence) | 0 |
| S11-final-viral-combined_k141_310317\|\|full | 1729 | No | NA | 3 | 0 | 2 | Not-determined | Genome-fragment | NA | NA | 0 |
| S8-final-viral-combined_k141_33846\|\|lt2gene | 1130 | No | NA | 1 | 1 | 0 | Low-quality | Genome-fragment | 2.4 | AAI-based (high-confidence) | 0 |
| S9-final-viral-combined_k141_67159\|\|full | 1131 | No | NA | 2 | 0 | 0 | Low-quality | Genome-fragment | 2.25 | AAI-based (medium-confidence) | 0 |
| S11-final-viral-combined_k141_74947\|\|full | 1510 | No | NA | 3 | 0 | 0 | Not-determined | Genome-fragment | NA | NA | 0 |
| S11-final-viral-combined_k141_310323\|\|full | 3595 | No | NA | 4 | 0 | 4 | Not-determined | Genome-fragment | NA | NA | 0 |
| S11-final-viral-combined_k141_32838\|\|full | 1131 | No | NA | 2 | 0 | 1 | Low-quality | Genome-fragment | 1.16 | AAI-based (medium-confidence) | 0 |
| S11-final-viral-combined_k141_20009\|\|full | 1131 | No | NA | 2 | 0 | 1 | Low-quality | Genome-fragment | 0.76 | AAI-based (medium-confidence) | 0 |
| S11-final-viral-combined_k141_89287\|\|full | 1039 | No | NA | 3 | 0 | 1 | Not-determined | Genome-fragment | NA | NA | 0 |
| S1-final-viral-combined_k141_188668\|\|full | 1131 | No | NA | 4 | 1 | 0 | Low-quality | Genome-fragment | 2.07 | HMM-based (lower-bound) | 0 |
| S11-final-viral-combined_k141_241674\|\|full | 1996 | No | NA | 2 | 0 | 1 | Not-determined | Genome-fragment | NA | NA | 0 |
| S11-final-viral-combined_k141_19660\|\|full | 2202 | No | NA | 2 | 0 | 2 | Not-determined | Genome-fragment | NA | NA | 0 |
| S11-final-viral-combined_k141_61989\|\|full | 2112 | No | NA | 2 | 0 | 2 | Not-determined | Genome-fragment | NA | NA | 0 |
| S11-final-viral-combined_k141_116251\|\|full | 13998 | No | NA | 13 | 0 | 12 | Not-determined | Genome-fragment | NA | NA | 0 |
| S1-final-viral-combined_k141_28615\|\|full | 1131 | No | NA | 4 | 0 | 0 | Low-quality | Genome-fragment | 2.74 | AAI-based (high-confidence) | 0 |
| S1-final-viral-combined_k141_72490\|\|lt2gene | 1131 | No | NA | 2 | 2 | 0 | Low-quality | Genome-fragment | 1.79 | AAI-based (high-confidence) | 0 |
| S11-final-viral-combined_k141_172346\|\|full | 1674 | No | NA | 2 | 0 | 0 | Not-determined | Genome-fragment | NA | NA | 0 |
| S5_k141_21009 | 1131 | No | NA | 1 | 0 | 0 | Low-quality | Genome-fragment | 2.01 | AAI-based (high-confidence) | 0 |
| S5_k141_104942 | 1131 | No | NA | 1 | 0 | 1 | Low-quality | Genome-fragment | 2.37 | AAI-based (high-confidence) | 0 |
| S11-final-viral-combined_k141_282865\|\|full | 2412 | No | NA | 3 | 0 | 2 | Not-determined | Genome-fragment | NA | NA | 0 |
| S11-final-viral-combined_k141_47974\|\|full | 2931 | No | NA | 7 | 0 | 3 | Not-determined | Genome-fragment | NA | NA | 0 |
| S11-final-viral-combined_k141_6644\|\|full | 8228 | No | NA | 4 | 0 | 2 | Not-determined | Genome-fragment | NA | NA | 0 |
| S7-final-viral-combined_k141_227077\|\|full | 1131 | No | NA | 3 | 0 | 0 | Low-quality | Genome-fragment | 0.9 | AAI-based (medium-confidence) | 0 |
| S11-final-viral-combined_k141_47980\|\|full | 2776 | No | NA | 3 | 0 | 2 | Not-determined | Genome-fragment | NA | NA | 0 |
| S11-final-viral-combined_k141_254913\|\|full | 1401 | No | NA | 3 | 0 | 1 | Not-determined | Genome-fragment | NA | NA | 0 |
| S11-final-viral-combined_k141_62015\|\|full | 1794 | No | NA | 3 | 0 | 0 | Not-determined | Genome-fragment | NA | NA | 0 |
| S11-final-viral-combined_k141_74983\|\|full | 49793 | No | NA | 72 | 20 | 0 | High-quality | High-quality | 99.25 | AAI-based (high-confidence) | 0 |
| S11-final-viral-combined_k141_34202\|\|full | 11240 | No | NA | 12 | 0 | 0 | Not-determined | Genome-fragment | NA | NA | 0 |
| S12-final-viral-combined_k141_382454\|\|full | 1131 | No | NA | 3 | 0 | 1 | Low-quality | Genome-fragment | 1.6 | AAI-based (high-confidence) | 0 |
| S11-final-viral-combined_k141_103695\|\|full | 6662 | No | NA | 4 | 0 | 0 | Not-determined | Genome-fragment | NA | NA | 0 |
| S11-final-viral-combined_k141_254940\|\|full | 4438 | No | NA | 5 | 0 | 5 | Not-determined | Genome-fragment | NA | NA | 0 |
| S11-final-viral-combined_k141_144563\|\|full | 3065 | No | NA | 5 | 0 | 3 | Not-determined | Genome-fragment | NA | NA | 0 |
| S12-final-viral-combined_k141_267302\|\|full | 1131 | No | NA | 2 | 0 | 0 | Low-quality | Genome-fragment | 2.69 | AAI-based (high-confidence) | 0 |
| S11-final-viral-combined_k141_282903\|\|full | 9631 | No | NA | 7 | 0 | 3 | Not-determined | Genome-fragment | NA | NA | 0 |
| S11-final-viral-combined_k141_116333\|\|full | 2336 | No | NA | 2 | 0 | 2 | Not-determined | Genome-fragment | NA | NA | 0 |
| S11-final-viral-combined_k141_310363\|\|full | 9043 | No | NA | 7 | 0 | 2 | Not-determined | Genome-fragment | NA | NA | 0 |
| S11-final-viral-combined_k141_241759\|\|full | 2958 | No | NA | 2 | 0 | 2 | Not-determined | Genome-fragment | NA | NA | 0 |
| S12_k141_260067 | 1131 | No | NA | 2 | 0 | 0 | Low-quality | Genome-fragment | 2.22 | AAI-based (high-confidence) | 0 |
| S2-final-viral-combined_k141_101145\|\|lt2gene | 1131 | No | NA | 2 | 1 | 0 | Low-quality | Genome-fragment | 1.64 | AAI-based (high-confidence) | 0 |
| S4-final-viral-combined_k141_150867\|\|full | 1131 | No | NA | 3 | 0 | 0 | Low-quality | Genome-fragment | 0.99 | AAI-based (medium-confidence) | 0 |
| S8-final-viral-combined_k141_66369\|\|full | 1131 | No | NA | 3 | 1 | 0 | Low-quality | Genome-fragment | 3.43 | AAI-based (high-confidence) | 0 |
| S11-final-viral-combined_k141_227368\|\|full | 1659 | No | NA | 3 | 0 | 1 | Not-determined | Genome-fragment | NA | NA | 0 |
| S8_k141_116247 | 1131 | No | NA | 2 | 0 | 0 | Low-quality | Genome-fragment | 1.69 | AAI-based (medium-confidence) | 0 |
| S11-final-viral-combined_k141_89382\|\|full | 1922 | No | NA | 3 | 0 | 3 | Not-determined | Genome-fragment | NA | NA | 0 |
| S11-final-viral-combined_k141_212064\|\|full | 1132 | No | NA | 3 | 0 | 0 | Low-quality | Genome-fragment | 1.99 | AAI-based (high-confidence) | 0 |
| S11-final-viral-combined_k141_6725\|\|full | 2162 | No | NA | 3 | 0 | 1 | Not-determined | Genome-fragment | NA | NA | 0 |
| S11-final-viral-combined_k141_213977\|\|full | 5453 | No | NA | 2 | 0 | 0 | Not-determined | Genome-fragment | NA | NA | 0 |
| S1-final-viral-combined_k141_134118\|\|full | 1132 | No | NA | 2 | 1 | 0 | Low-quality | Genome-fragment | 2.86 | AAI-based (medium-confidence) | 0 |
| S1-final-viral-combined_k141_315802\|\|full | 1132 | No | NA | 3 | 0 | 0 | Low-quality | Genome-fragment | 2.64 | AAI-based (high-confidence) | 0 |
| S11-final-viral-combined_k141_199259\|\|full | 4112 | No | NA | 6 | 0 | 5 | Not-determined | Genome-fragment | NA | NA | 0 |
| S5-final-viral-combined_k141_99962\|\|full | 1132 | No | NA | 2 | 0 | 0 | Low-quality | Genome-fragment | 3.25 | AAI-based (high-confidence) | 0 |
| S5-final-viral-combined_k141_194184\|\|full | 1132 | No | NA | 2 | 0 | 0 | Low-quality | Genome-fragment | 1.72 | AAI-based (high-confidence) | 0 |
| S11-final-viral-combined_k141_34236\|\|full | 1809 | No | NA | 3 | 0 | 1 | Not-determined | Genome-fragment | NA | NA | 0 |
| S7-final-viral-combined_k141_135524\|\|full | 1132 | No | NA | 2 | 0 | 1 | Low-quality | Genome-fragment | 2.14 | AAI-based (medium-confidence) | 0 |
| S11-final-viral-combined_k141_144596\|\|full | 2080 | No | NA | 3 | 0 | 1 | Not-determined | Genome-fragment | NA | NA | 0 |
| S11-final-viral-combined_k141_227391\|\|full | 4881 | No | NA | 8 | 0 | 2 | Not-determined | Genome-fragment | NA | NA | 0 |
| S7-final-viral-combined_k141_145732\|\|full | 1132 | No | NA | 2 | 0 | 0 | Low-quality | Genome-fragment | 2.38 | AAI-based (medium-confidence) | 0 |
| S7_k141_158372 | 1132 | No | NA | 4 | 0 | 0 | Low-quality | Genome-fragment | 0.74 | AAI-based (high-confidence) | 0 |
| S12_k141_138998 | 1132 | No | NA | 2 | 2 | 0 | Low-quality | Genome-fragment | 2.51 | AAI-based (high-confidence) | 0 |
| S4-final-viral-combined_k141_123827\|\|lt2gene | 1132 | No | NA | 1 | 0 | 0 | Low-quality | Genome-fragment | 2.23 | AAI-based (high-confidence) | 0 |
| S11-final-viral-combined_k141_322973\|\|full | 2030 | No | NA | 2 | 0 | 1 | Not-determined | Genome-fragment | NA | NA | 0 |
| S11-final-viral-combined_k141_62134\|\|full | 2457 | No | NA | 1 | 0 | 1 | Not-determined | Genome-fragment | NA | NA | 0 |
| S6-final-viral-combined_k141_38998\|\|full | 1132 | No | NA | 3 | 0 | 0 | Low-quality | Genome-fragment | 3.43 | AAI-based (medium-confidence) | 0 |
| S8-final-viral-combined_k141_142329\|\|full | 1132 | No | NA | 2 | 0 | 0 | Low-quality | Genome-fragment | 2.51 | AAI-based (medium-confidence) | 0 |
| S11-final-viral-combined_k141_62139\|\|full | 1475 | No | NA | 2 | 0 | 0 | Not-determined | Genome-fragment | NA | NA | 0 |
| S11-final-viral-combined_k141_322981\|\|full | 3603 | No | NA | 5 | 0 | 1 | Not-determined | Genome-fragment | NA | NA | 0 |
| S11-final-viral-combined_k141_99648\|\|full | 1133 | No | NA | 2 | 1 | 0 | Low-quality | Genome-fragment | 1.62 | AAI-based (high-confidence) | 0 |
| S11-final-viral-combined_k141_307155\|\|full | 1133 | No | NA | 2 | 0 | 2 | Low-quality | Genome-fragment | 0.82 | AAI-based (medium-confidence) | 0 |
| S11-final-viral-combined_k141_295868\|\|full | 1924 | No | NA | 2 | 0 | 2 | Not-determined | Genome-fragment | NA | NA | 0 |
| S11-final-viral-combined_k141_33655\|\|lt2gene | 1133 | No | NA | 2 | 1 | 0 | Low-quality | Genome-fragment | 0.69 | HMM-based (lower-bound) | 0 |
| S11-final-viral-combined_k141_34302\|\|full | 1129 | No | NA | 2 | 0 | 2 | Not-determined | Genome-fragment | NA | NA | 0 |
| S1-final-viral-combined_k141_314547\|\|full | 1133 | No | NA | 4 | 1 | 0 | Low-quality | Genome-fragment | 2.02 | AAI-based (medium-confidence) | 0 |
| S11-final-viral-combined_k141_34306\|\|full | 2000 | No | NA | 3 | 0 | 1 | Not-determined | Genome-fragment | NA | NA | 0 |
| S11-final-viral-combined_k141_255061\|\|full | 3198 | No | NA | 2 | 0 | 0 | Not-determined | Genome-fragment | NA | NA | 0 |
| S11-final-viral-combined_k141_268581\|\|full | 1291 | No | NA | 2 | 0 | 1 | Not-determined | Genome-fragment | NA | NA | 0 |
| S1-final-viral-combined_k141_373640\|\|full | 1133 | No | NA | 3 | 0 | 0 | Low-quality | Genome-fragment | 1 | AAI-based (medium-confidence) | 0 |
| S11-final-viral-combined_k141_6801\|\|full | 2415 | No | NA | 2 | 0 | 1 | Not-determined | Genome-fragment | NA | NA | 0 |
| S1-final-viral-combined_k141_277046\|\|full | 1133 | No | NA | 3 | 0 | 0 | Low-quality | Genome-fragment | 2.35 | AAI-based (high-confidence) | 0 |
| S11-final-viral-combined_k141_89478\|\|full | 2183 | No | NA | 3 | 0 | 0 | Not-determined | Genome-fragment | NA | NA | 0 |
| S11-final-viral-combined_k141_89482\|\|full | 3493 | No | NA | 3 | 0 | 2 | Not-determined | Genome-fragment | NA | NA | 0 |
| S1_k141_199478 | 1133 | No | NA | 3 | 0 | 0 | Low-quality | Genome-fragment | 2.07 | AAI-based (high-confidence) | 0 |
| S17-final-viral-combined_k141_78042\|\|full | 1133 | No | NA | 2 | 0 | 2 | Low-quality | Genome-fragment | 2.53 | AAI-based (medium-confidence) | 0 |
| S11-final-viral-combined_k141_323003\|\|full | 6416 | No | NA | 7 | 0 | 1 | Not-determined | Genome-fragment | NA | NA | 0 |
| S11-final-viral-combined_k141_310499\|\|full | 2198 | No | NA | 3 | 0 | 1 | Not-determined | Genome-fragment | NA | NA | 0 |
| S17-final-viral-combined_k141_171657\|\|full | 1133 | No | NA | 2 | 0 | 0 | Low-quality | Genome-fragment | 2.11 | AAI-based (high-confidence) | 0 |
| S11-final-viral-combined_k141_310510\|\|full | 1018 | No | NA | 3 | 0 | 1 | Not-determined | Genome-fragment | NA | NA | 0 |
| S17-final-viral-combined_k141_205645\|\|full | 1133 | No | NA | 2 | 0 | 0 | Low-quality | Genome-fragment | 0.66 | AAI-based (medium-confidence) | 0 |
| S11-final-viral-combined_k141_310515\|\|full | 1088 | No | NA | 1 | 0 | 0 | Not-determined | Genome-fragment | NA | NA | 0 |
| S17-k141_53661 | 1133 | No | NA | 1 | 0 | 0 | Low-quality | Genome-fragment | 0.72 | AAI-based (high-confidence) | 0 |
| S11-final-viral-combined_k141_158709\|\|full | 2265 | No | NA | 3 | 0 | 2 | Not-determined | Genome-fragment | NA | NA | 0 |
| S11-final-viral-combined_k141_172444\|\|full | 1961 | No | NA | 3 | 0 | 2 | Not-determined | Genome-fragment | NA | NA | 0 |
| S11-final-viral-combined_k141_158713\|\|full | 1627 | No | NA | 3 | 0 | 2 | Not-determined | Genome-fragment | NA | NA | 0 |
| S11-final-viral-combined_k141_116484\|\|full | 1937 | No | NA | 2 | 0 | 1 | Not-determined | Genome-fragment | NA | NA | 0 |
| S11-final-viral-combined_k141_75185\|\|full | 1237 | No | NA | 3 | 0 | 1 | Not-determined | Genome-fragment | NA | NA | 0 |
| S12-final-viral-combined_k141_174841\|\|lt2gene | 1133 | No | NA | 1 | 1 | 0 | Low-quality | Genome-fragment | 1.59 | HMM-based (lower-bound) | 0 |
| S11-final-viral-combined_k141_116489\|\|full | 1715 | No | NA | 3 | 0 | 1 | Not-determined | Genome-fragment | NA | NA | 0 |
| S12-final-viral-combined_k141_116452\|\|lt2gene | 1133 | No | NA | 2 | 1 | 0 | Low-quality | Genome-fragment | 2.43 | AAI-based (high-confidence) | 0 |
| S11-final-viral-combined_k141_268653\|\|full | 1356 | No | NA | 2 | 0 | 0 | Not-determined | Genome-fragment | NA | NA | 0 |
| S11-final-viral-combined_k141_48135\|\|full | 2578 | No | NA | 2 | 0 | 2 | Not-determined | Genome-fragment | NA | NA | 0 |
| S11-final-viral-combined_k141_323046\|\|full | 2840 | No | NA | 5 | 0 | 2 | Not-determined | Genome-fragment | NA | NA | 0 |
| S11-final-viral-combined_k141_144734\|\|full | 2510 | No | NA | 4 | 0 | 2 | Not-determined | Genome-fragment | NA | NA | 0 |
| S12_k141_160604 | 1133 | No | NA | 2 | 2 | 0 | Low-quality | Genome-fragment | 2.6 | HMM-based (lower-bound) | 0 |
| S11-final-viral-combined_k141_323051\|\|full | 4670 | No | NA | 4 | 0 | 3 | Not-determined | Genome-fragment | NA | NA | 0 |
| S11-final-viral-combined_k141_310594\|\|full | 2109 | No | NA | 3 | 0 | 3 | Not-determined | Genome-fragment | NA | NA | 0 |
| S2-final-viral-combined_k141_14270\|\|full | 1133 | No | NA | 3 | 0 | 1 | Low-quality | Genome-fragment | 2.3 | AAI-based (medium-confidence) | 0 |
| S11-final-viral-combined_k141_214134\|\|full | 6240 | No | NA | 6 | 0 | 3 | Not-determined | Genome-fragment | NA | NA | 0 |
| S9-final-viral-combined_k141_15894\|\|full | 1134 | No | NA | 2 | 0 | 1 | Low-quality | Genome-fragment | 0.68 | AAI-based (high-confidence) | 0 |
| S11-final-viral-combined_k141_19943\|\|full | 8416 | No | NA | 4 | 0 | 2 | Not-determined | Genome-fragment | NA | NA | 0 |
| S11-final-viral-combined_k141_130522\|\|full | 5428 | No | NA | 5 | 0 | 3 | Not-determined | Genome-fragment | NA | NA | 0 |
| S9-final-viral-combined_k141_76899\|\|lt2gene | 1134 | No | NA | 2 | 2 | 0 | Low-quality | Genome-fragment | 0.67 | HMM-based (lower-bound) | 0 |
| S11-final-viral-combined_k141_199387\|\|full | 1709 | No | NA | 3 | 0 | 2 | Not-determined | Genome-fragment | NA | NA | 0 |
| S11-final-viral-combined_k141_6909\|\|full | 4035 | No | NA | 4 | 0 | 3 | Not-determined | Genome-fragment | NA | NA | 0 |
| S11-final-viral-combined_k141_185731\|\|full | 2307 | No | NA | 2 | 0 | 1 | Not-determined | Genome-fragment | NA | NA | 0 |
| S11-final-viral-combined_k141_130546\|\|full | 1132 | No | NA | 3 | 0 | 0 | Not-determined | Genome-fragment | NA | NA | 0 |
| S11-final-viral-combined_k141_135167\|\|lt2gene | 1134 | No | NA | 2 | 1 | 0 | Low-quality | Genome-fragment | 2.63 | AAI-based (medium-confidence) | 0 |
| S11-final-viral-combined_k141_130565\|\|full | 1658 | No | NA | 2 | 0 | 0 | Not-determined | Genome-fragment | NA | NA | 0 |
| S1-final-viral-combined_k141_330154\|\|lt2gene | 1134 | No | NA | 2 | 1 | 0 | Low-quality | Genome-fragment | 2.84 | AAI-based (high-confidence) | 0 |
| S11-final-viral-combined_k141_323081\|\|full | 1340 | No | NA | 3 | 0 | 0 | Not-determined | Genome-fragment | NA | NA | 0 |
| S12-final-viral-combined_k141_393326\|\|full | 1134 | No | NA | 3 | 0 | 0 | Low-quality | Genome-fragment | 1.35 | AAI-based (high-confidence) | 0 |
| S11-final-viral-combined_k141_89595\|\|full | 1540 | No | NA | 2 | 0 | 0 | Not-determined | Genome-fragment | NA | NA | 0 |
| S11-final-viral-combined_k141_62349\|\|full | 2023 | No | NA | 2 | 0 | 2 | Not-determined | Genome-fragment | NA | NA | 0 |
| S11-final-viral-combined_k141_241917\|\|full | 6489 | No | NA | 4 | 0 | 3 | Not-determined | Genome-fragment | NA | NA | 0 |
| S4-final-viral-combined_k141_154972\|\|full | 1134 | No | NA | 3 | 0 | 1 | Low-quality | Genome-fragment | 0.56 | AAI-based (medium-confidence) | 0 |
| S8-final-viral-combined_k141_149766\|\|full | 1134 | No | NA | 2 | 0 | 0 | Low-quality | Genome-fragment | 2.62 | AAI-based (medium-confidence) | 0 |
| S11-final-viral-combined_k141_6937\|\|full | 4593 | No | NA | 5 | 0 | 0 | Not-determined | Genome-fragment | NA | NA | 0 |
| S11-final-viral-combined_k141_20013\|\|full | 2995 | No | NA | 5 | 0 | 2 | Not-determined | Genome-fragment | NA | NA | 0 |
| S9-final-viral-combined_k141_122830\|\|lt2gene | 1135 | No | NA | 2 | 1 | 0 | Low-quality | Genome-fragment | 2.9 | AAI-based (high-confidence) | 0 |
| S11-final-viral-combined_k141_89616\|\|full | 2105 | No | NA | 2 | 0 | 2 | Not-determined | Genome-fragment | NA | NA | 0 |
| S11-final-viral-combined_k141_310684\|\|full | 4421 | No | NA | 3 | 0 | 2 | Not-determined | Genome-fragment | NA | NA | 0 |
| S11-final-viral-combined_k141_227630\|\|full | 1118 | No | NA | 3 | 0 | 0 | Not-determined | Genome-fragment | NA | NA | 0 |
| S9_k141_28102 | 1135 | No | NA | 2 | 0 | 0 | Low-quality | Genome-fragment | 2.89 | AAI-based (high-confidence) | 0 |
| S11-final-viral-combined_k141_241944\|\|full | 3301 | No | NA | 2 | 0 | 2 | Not-determined | Genome-fragment | NA | NA | 0 |
| S11-final-viral-combined_k141_20038\|\|full | 1622 | No | NA | 3 | 0 | 0 | Not-determined | Genome-fragment | NA | NA | 0 |
| S11-final-viral-combined_k141_172559\|\|full | 1362 | No | NA | 1 | 0 | 1 | Not-determined | Genome-fragment | NA | NA | 0 |
| S11-final-viral-combined_k141_20045\|\|full | 2709 | No | NA | 3 | 0 | 0 | Not-determined | Genome-fragment | NA | NA | 0 |
| S11-final-viral-combined_k141_310738\|\|full | 3034 | No | NA | 6 | 0 | 0 | Not-determined | Genome-fragment | NA | NA | 0 |
| S11-final-viral-combined_k141_116589\|\|full | 25615 | No | NA | 38 | 1 | 11 | Medium-quality | Genome-fragment | 70.25 | AAI-based (medium-confidence) | 0 |
| S11-final-viral-combined_k141_289690\|\|full | 1135 | No | NA | 3 | 2 | 0 | Low-quality | Genome-fragment | 1.76 | HMM-based (lower-bound) | 0 |
| S11-final-viral-combined_k141_20063\|\|full | 2862 | No | NA | 3 | 0 | 3 | Not-determined | Genome-fragment | NA | NA | 0 |
| S1-final-viral-combined_k141_91357\|\|lt2gene | 1135 | No | NA | 2 | 2 | 0 | Low-quality | Genome-fragment | 2.73 | AAI-based (high-confidence) | 0 |
| S11-final-viral-combined_k141_268767\|\|full | 2437 | No | NA | 2 | 0 | 2 | Not-determined | Genome-fragment | NA | NA | 0 |
| S5-final-viral-combined_k141_84835\|\|full | 1135 | No | NA | 3 | 1 | 0 | Low-quality | Genome-fragment | 2.83 | AAI-based (high-confidence) | 0 |
| S5-final-viral-combined_k141_165525\|\|full | 1135 | No | NA | 4 | 2 | 0 | Low-quality | Genome-fragment | 2.75 | AAI-based (high-confidence) | 0 |
| S7-final-viral-combined_k141_77260\|\|full | 1135 | No | NA | 4 | 1 | 0 | Low-quality | Genome-fragment | 2.29 | AAI-based (high-confidence) | 0 |
| S2-final-viral-combined_k141_211975\|\|full | 1135 | No | NA | 3 | 0 | 1 | Low-quality | Genome-fragment | 1.65 | AAI-based (medium-confidence) | 0 |
| S11-final-viral-combined_k141_255255\|\|full | 1784 | No | NA | 3 | 0 | 1 | Not-determined | Genome-fragment | NA | NA | 0 |
| S2-final-viral-combined_k141_194282\|\|lt2gene | 1135 | No | NA | 2 | 2 | 0 | Low-quality | Genome-fragment | 1.8 | HMM-based (lower-bound) | 0 |
| S11-final-viral-combined_k141_296048\|\|full | 2381 | No | NA | 3 | 0 | 1 | Not-determined | Genome-fragment | NA | NA | 0 |
| S11-final-viral-combined_k141_214256\|\|full | 2163 | No | NA | 3 | 0 | 1 | Not-determined | Genome-fragment | NA | NA | 0 |
| S11-final-viral-combined_k141_130609\|\|full | 2879 | No | NA | 4 | 0 | 0 | Not-determined | Genome-fragment | NA | NA | 0 |
| S11-final-viral-combined_k141_172648\|\|full | 1087 | No | NA | 2 | 0 | 1 | Not-determined | Genome-fragment | NA | NA | 0 |
| S4-final-viral-combined_k141_75661\|\|full | 1135 | No | NA | 3 | 1 | 0 | Low-quality | Genome-fragment | 2.42 | AAI-based (high-confidence) | 0 |
| S8-final-viral-combined_k141_254566\|\|full | 1135 | No | NA | 4 | 0 | 0 | Low-quality | Genome-fragment | 1.23 | AAI-based (medium-confidence) | 0 |
| S11-final-viral-combined_k141_34581\|\|full | 2812 | No | NA | 2 | 0 | 2 | Not-determined | Genome-fragment | NA | NA | 0 |
| S11-final-viral-combined_k141_283148\|\|full | 2380 | No | NA | 3 | 0 | 0 | Not-determined | Genome-fragment | NA | NA | 0 |
| S8-final-viral-combined_k141_32326\|\|full | 1135 | No | NA | 3 | 2 | 0 | Low-quality | Genome-fragment | 2.82 | AAI-based (high-confidence) | 0 |
| S9-final-viral-combined_k141_37250\|\|full | 1136 | No | NA | 4 | 0 | 0 | Low-quality | Genome-fragment | 2.03 | AAI-based (medium-confidence) | 0 |
| S11-final-viral-combined_k141_62468\|\|full | 2054 | No | NA | 3 | 0 | 2 | Not-determined | Genome-fragment | NA | NA | 0 |
| S11-final-viral-combined_k141_145751\|\|full | 1136 | No | NA | 3 | 1 | 0 | Low-quality | Genome-fragment | 1.38 | AAI-based (high-confidence) | 0 |
| S11-final-viral-combined_k141_54951\|\|full | 1136 | No | NA | 2 | 1 | 0 | Low-quality | Genome-fragment | 2.5 | AAI-based (high-confidence) | 0 |
| S1-final-viral-combined_k141_39174\|\|full | 1136 | No | NA | 5 | 0 | 0 | Low-quality | Genome-fragment | 2.92 | AAI-based (high-confidence) | 0 |
| S11-final-viral-combined_k141_7054\|\|full | 4526 | No | NA | 4 | 0 | 2 | Not-determined | Genome-fragment | NA | NA | 0 |
| S11-final-viral-combined_k141_283178\|\|full | 1004 | No | NA | 2 | 0 | 0 | Not-determined | Genome-fragment | NA | NA | 0 |
| S11-final-viral-combined_k141_62481\|\|full | 2617 | No | NA | 5 | 0 | 1 | Not-determined | Genome-fragment | NA | NA | 0 |
| S11-final-viral-combined_k141_75358\|\|full | 1428 | No | NA | 4 | 0 | 0 | Not-determined | Genome-fragment | NA | NA | 0 |
| S1-final-viral-combined_k141_262209\|\|lt2gene | 1136 | No | NA | 2 | 1 | 0 | Low-quality | Genome-fragment | 2.9 | AAI-based (high-confidence) | 0 |
| S11-final-viral-combined_k141_310841\|\|full | 3506 | No | NA | 5 | 0 | 0 | Not-determined | Genome-fragment | NA | NA | 0 |
| S11-final-viral-combined_k141_48246\|\|full | 48975 | Yes | 41339 | 68 | 9 | 5 | Medium-quality | Genome-fragment | 71 | AAI-based (high-confidence) | 15.59 |
| S3_k141_126467 | 1136 | No | NA | 2 | 1 | 0 | Low-quality | Genome-fragment | 0.88 | AAI-based (high-confidence) | 0 |
| S5-final-viral-combined_k141_207864\|\|full | 1136 | No | NA | 3 | 0 | 0 | Low-quality | Genome-fragment | 2.61 | AAI-based (high-confidence) | 0 |
| S11-final-viral-combined_k141_283206\|\|full | 4754 | No | NA | 4 | 0 | 2 | Not-determined | Genome-fragment | NA | NA | 0 |
| S11-final-viral-combined_k141_296138\|\|full | 2463 | No | NA | 4 | 0 | 0 | Not-determined | Genome-fragment | NA | NA | 0 |
| S11-final-viral-combined_k141_255314\|\|full | 2981 | No | NA | 2 | 0 | 2 | Not-determined | Genome-fragment | NA | NA | 0 |
| S11-final-viral-combined_k141_75384\|\|full | 1197 | No | NA | 2 | 0 | 1 | Not-determined | Genome-fragment | NA | NA | 0 |
| S11-final-viral-combined_k141_48303\|\|full | 1118 | No | NA | 2 | 0 | 1 | Not-determined | Genome-fragment | NA | NA | 0 |
| S11-final-viral-combined_k141_199584\|\|full | 2017 | No | NA | 2 | 0 | 1 | Not-determined | Genome-fragment | NA | NA | 0 |
| S11-final-viral-combined_k141_89746\|\|full | 2645 | No | NA | 2 | 0 | 2 | Not-determined | Genome-fragment | NA | NA | 0 |
| S5-final-viral-combined_k141_146271\|\|full | 1136 | No | NA | 2 | 2 | 0 | Low-quality | Genome-fragment | 2.96 | AAI-based (high-confidence) | 0 |
| S17-final-viral-combined_k141_193410\|\|full | 1136 | No | NA | 2 | 1 | 0 | Low-quality | Genome-fragment | 2.03 | AAI-based (high-confidence) | 0 |
| S17-final-viral-combined_k141_94826\|\|full | 1136 | No | NA | 2 | 0 | 0 | Low-quality | Genome-fragment | 2.24 | AAI-based (high-confidence) | 0 |
| S17-final-viral-combined_k141_117455\|\|lt2gene | 1136 | No | NA | 1 | 1 | 0 | Low-quality | Genome-fragment | 1.68 | HMM-based (lower-bound) | 0 |
| S11-final-viral-combined_k141_283220\|\|full | 34186 | No | NA | 50 | 16 | 1 | High-quality | High-quality | 95.51 | AAI-based (medium-confidence) | 0 |
| S17-k141_194942 | 1136 | No | NA | 2 | 0 | 1 | Low-quality | Genome-fragment | 0.89 | AAI-based (high-confidence) | 0 |
| S11-final-viral-combined_k141_7143\|\|full | 2117 | No | NA | 3 | 0 | 1 | Not-determined | Genome-fragment | NA | NA | 0 |
| S12-final-viral-combined_k141_353053\|\|full | 1136 | No | NA | 4 | 0 | 0 | Low-quality | Genome-fragment | 1.67 | AAI-based (high-confidence) | 0 |
| S4-final-viral-combined_k141_159776\|\|full | 1136 | No | NA | 4 | 0 | 0 | Low-quality | Genome-fragment | 2.94 | AAI-based (medium-confidence) | 0 |
| S11-final-viral-combined_k141_214365\|\|full | 1426 | No | NA | 3 | 0 | 0 | Not-determined | Genome-fragment | NA | NA | 0 |
| S11-final-viral-combined_k141_62588\|\|full | 1396 | No | NA | 2 | 0 | 1 | Not-determined | Genome-fragment | NA | NA | 0 |
| S6-final-viral-combined_k141_82804\|\|lt2gene | 1136 | No | NA | 2 | 1 | 0 | Low-quality | Genome-fragment | 1.31 | HMM-based (lower-bound) | 0 |
| S11-final-viral-combined_k141_34753\|\|full | 1418 | No | NA | 3 | 0 | 0 | Not-determined | Genome-fragment | NA | NA | 0 |
| S11-final-viral-combined_k141_214378\|\|full | 1514 | No | NA | 2 | 0 | 2 | Not-determined | Genome-fragment | NA | NA | 0 |
| S11-final-viral-combined_k141_104191\|\|full | 11212 | No | NA | 12 | 0 | 5 | Not-determined | Genome-fragment | NA | NA | 0 |
| S11-final-viral-combined_k141_75425\|\|full | 2411 | No | NA | 5 | 0 | 0 | Not-determined | Genome-fragment | NA | NA | 0 |
| S11-final-viral-combined_k141_242159\|\|full | 1392 | No | NA | 2 | 0 | 1 | Not-determined | Genome-fragment | NA | NA | 0 |
| S11-final-viral-combined_k141_212074\|\|full | 1137 | No | NA | 3 | 0 | 0 | Low-quality | Genome-fragment | 2.47 | AAI-based (high-confidence) | 0 |
| S1-final-viral-combined_k141_88030\|\|full | 1137 | No | NA | 3 | 0 | 0 | Low-quality | Genome-fragment | 2.76 | AAI-based (high-confidence) | 0 |
| S11-final-viral-combined_k141_7201\|\|full | 1488 | No | NA | 5 | 0 | 1 | Not-determined | Genome-fragment | NA | NA | 0 |
| S1-final-viral-combined_k141_192250\|\|full | 1137 | No | NA | 3 | 0 | 1 | Low-quality | Genome-fragment | 0.95 | AAI-based (medium-confidence) | 0 |
| S11-final-viral-combined_k141_296193\|\|full | 1027 | No | NA | 3 | 0 | 0 | Not-determined | Genome-fragment | NA | NA | 0 |
| S1_k141_158589 | 1137 | No | NA | 3 | 2 | 0 | Low-quality | Genome-fragment | 2.09 | AAI-based (high-confidence) | 0 |
| S11-final-viral-combined_k141_62642\|\|full | 1283 | No | NA | 3 | 0 | 0 | Not-determined | Genome-fragment | NA | NA | 0 |
| S5_k141_150561 | 1137 | No | NA | 1 | 0 | 1 | Low-quality | Genome-fragment | 0.76 | AAI-based (high-confidence) | 0 |
| S11-final-viral-combined_k141_242200\|\|full | 7074 | No | NA | 7 | 0 | 7 | Not-determined | Genome-fragment | NA | NA | 0 |
| S11-final-viral-combined_k141_116729\|\|full | 1169 | No | NA | 2 | 0 | 2 | Not-determined | Genome-fragment | NA | NA | 0 |
| S11-final-viral-combined_k141_34814\|\|full | 1159 | No | NA | 2 | 0 | 1 | Not-determined | Genome-fragment | NA | NA | 0 |
| S11-final-viral-combined_k141_145114\|\|full | 2950 | No | NA | 4 | 0 | 3 | Not-determined | Genome-fragment | NA | NA | 0 |
| S11-final-viral-combined_k141_283286\|\|full | 1313 | No | NA | 3 | 0 | 2 | Not-determined | Genome-fragment | NA | NA | 0 |
| S7-final-viral-combined_k141_100936\|\|lt2gene | 1137 | No | NA | 2 | 2 | 0 | Low-quality | Genome-fragment | 2.56 | AAI-based (high-confidence) | 0 |
| S11-final-viral-combined_k141_172889\|\|full | 29228 | No | NA | 37 | 9 | 0 | Medium-quality | Genome-fragment | 55.57 | HMM-based (lower-bound) | 0 |
| S17-final-viral-combined_k141_243866\|\|full | 1137 | No | NA | 3 | 0 | 0 | Low-quality | Genome-fragment | 0.63 | AAI-based (medium-confidence) | 0 |
| S11-final-viral-combined_k141_214450\|\|full | 1334 | No | NA | 3 | 0 | 1 | Not-determined | Genome-fragment | NA | NA | 0 |
| S17-final-viral-combined_k141_99992\|\|full | 1137 | No | NA | 4 | 0 | 3 | Low-quality | Genome-fragment | 0.33 | AAI-based (medium-confidence) | 0 |
| S17-final-viral-combined_k141_197082\|\|lt2gene | 1137 | No | NA | 2 | 1 | 0 | Low-quality | Genome-fragment | 3.12 | AAI-based (high-confidence) | 0 |
| S11-final-viral-combined_k141_130829\|\|full | 3411 | No | NA | 5 | 0 | 0 | Not-determined | Genome-fragment | NA | NA | 0 |
| S12-final-viral-combined_k141_18143\|\|full | 1137 | No | NA | 3 | 0 | 1 | Low-quality | Genome-fragment | 2.04 | AAI-based (high-confidence) | 0 |
| S11-final-viral-combined_k141_62713\|\|full | 2241 | No | NA | 3 | 0 | 3 | Not-determined | Genome-fragment | NA | NA | 0 |
| S2-final-viral-combined_k141_137135\|\|full | 1137 | No | NA | 1 | 0 | 0 | Low-quality | Genome-fragment | 2.81 | AAI-based (high-confidence) | 0 |
| S6-final-viral-combined_k141_109618\|\|full | 1137 | No | NA | 3 | 1 | 0 | Low-quality | Genome-fragment | 4 | AAI-based (medium-confidence) | 0 |
| S11-final-viral-combined_k141_20306\|\|full | 2660 | No | NA | 3 | 0 | 1 | Not-determined | Genome-fragment | NA | NA | 0 |
| S11-final-viral-combined_k141_255547\|\|full | 1815 | No | NA | 3 | 0 | 2 | Not-determined | Genome-fragment | NA | NA | 0 |
| S11-final-viral-combined_k141_130848\|\|full | 6171 | No | NA | 5 | 0 | 5 | Not-determined | Genome-fragment | NA | NA | 0 |
| S11-final-viral-combined_k141_158946\|\|full | 3921 | No | NA | 3 | 0 | 2 | Not-determined | Genome-fragment | NA | NA | 0 |
| S11-final-viral-combined_k141_89888\|\|full | 1621 | No | NA | 2 | 0 | 2 | Not-determined | Genome-fragment | NA | NA | 0 |
| S11-final-viral-combined_k141_296287\|\|full | 1707 | No | NA | 3 | 0 | 1 | Not-determined | Genome-fragment | NA | NA | 0 |
| S11-final-viral-combined_k141_145188\|\|full | 1882 | No | NA | 3 | 0 | 2 | Not-determined | Genome-fragment | NA | NA | 0 |
| S11-final-viral-combined_k141_145195\|\|full | 1171 | No | NA | 2 | 0 | 2 | Not-determined | Genome-fragment | NA | NA | 0 |
| S11-final-viral-combined_k141_62740\|\|full | 3443 | No | NA | 4 | 0 | 2 | Not-determined | Genome-fragment | NA | NA | 0 |
| S11-final-viral-combined_k141_172953\|\|full | 3342 | No | NA | 3 | 0 | 2 | Not-determined | Genome-fragment | NA | NA | 0 |
| S11-final-viral-combined_k141_268967\|\|full | 17804 | No | NA | 21 | 11 | 0 | Medium-quality | Genome-fragment | 50.48 | AAI-based (medium-confidence) | 0 |
| S11-final-viral-combined_k141_114309\|\|full | 1138 | No | NA | 3 | 0 | 1 | Low-quality | Genome-fragment | 2.37 | AAI-based (medium-confidence) | 0 |
| S1-final-viral-combined_k141_95384\|\|lt2gene | 1138 | No | NA | 1 | 1 | 0 | Low-quality | Genome-fragment | 2.62 | AAI-based (high-confidence) | 0 |
| S11-final-viral-combined_k141_199715\|\|full | 1469 | No | NA | 4 | 0 | 1 | Not-determined | Genome-fragment | NA | NA | 0 |
| S11-final-viral-combined_k141_130858\|\|full | 4004 | No | NA | 3 | 0 | 1 | Not-determined | Genome-fragment | NA | NA | 0 |
| S11-final-viral-combined_k141_34911\|\|full | 2471 | No | NA | 2 | 0 | 1 | Not-determined | Genome-fragment | NA | NA | 0 |
| S11-final-viral-combined_k141_172968\|\|full | 4215 | No | NA | 2 | 0 | 2 | Not-determined | Genome-fragment | NA | NA | 0 |
| S11-final-viral-combined_k141_75613\|\|full | 7842 | No | NA | 6 | 0 | 5 | Not-determined | Genome-fragment | NA | NA | 0 |
| S11-final-viral-combined_k141_62752\|\|full | 1426 | No | NA | 1 | 0 | 0 | Not-determined | Genome-fragment | NA | NA | 0 |
| S7_k141_129436 | 1138 | No | NA | 1 | 0 | 0 | Low-quality | Genome-fragment | 8.43 | AAI-based (high-confidence) | 0 |
| S11-final-viral-combined_k141_116860\|\|full | 4752 | No | NA | 5 | 0 | 2 | Not-determined | Genome-fragment | NA | NA | 0 |
| S11-final-viral-combined_k141_255630\|\|full | 1254 | No | NA | 2 | 0 | 0 | Not-determined | Genome-fragment | NA | NA | 0 |
| S17-final-viral-combined_k141_143518\|\|full | 1138 | No | NA | 3 | 1 | 0 | Low-quality | Genome-fragment | 3.18 | AAI-based (high-confidence) | 0 |
| S12-final-viral-combined_k141_224009\|\|full | 1138 | No | NA | 3 | 0 | 0 | Low-quality | Genome-fragment | 2.08 | AAI-based (high-confidence) | 0 |
| S12-final-viral-combined_k141_192011\|\|lt2gene | 1138 | No | NA | 1 | 1 | 0 | Low-quality | Genome-fragment | 2.39 | AAI-based (high-confidence) | 0 |
| S11-final-viral-combined_k141_173006\|\|full | 1185 | No | NA | 2 | 0 | 1 | Not-determined | Genome-fragment | NA | NA | 0 |
| S11-final-viral-combined_k141_255672\|\|full | 1737 | No | NA | 3 | 0 | 1 | Not-determined | Genome-fragment | NA | NA | 0 |
| S11-final-viral-combined_k141_199781\|\|full | 3534 | No | NA | 5 | 0 | 2 | Not-determined | Genome-fragment | NA | NA | 0 |
| S11-final-viral-combined_k141_296376\|\|full | 2750 | No | NA | 3 | 0 | 2 | Not-determined | Genome-fragment | NA | NA | 0 |
| S11-final-viral-combined_k141_48581\|\|full | 2864 | No | NA | 7 | 0 | 0 | Not-determined | Genome-fragment | NA | NA | 0 |
| S8_k141_243122 | 1138 | No | NA | 2 | 0 | 0 | Low-quality | Genome-fragment | 2.38 | AAI-based (medium-confidence) | 0 |
| S11-final-viral-combined_k141_130913\|\|full | 1689 | No | NA | 3 | 0 | 0 | Not-determined | Genome-fragment | NA | NA | 0 |
| S11-final-viral-combined_k141_186079\|\|full | 1778 | No | NA | 3 | 0 | 2 | Not-determined | Genome-fragment | NA | NA | 0 |
| S11-final-viral-combined_k141_104323\|\|full | 10685 | No | NA | 9 | 0 | 6 | Not-determined | Genome-fragment | NA | NA | 0 |
| S9-final-viral-combined_k141_125289\|\|full | 1139 | No | NA | 3 | 1 | 0 | Low-quality | Genome-fragment | 0.78 | HMM-based (lower-bound) | 0 |
| S11-final-viral-combined_k141_35001\|\|full | 5316 | No | NA | 7 | 0 | 0 | Not-determined | Genome-fragment | NA | NA | 0 |
| S11-final-viral-combined_k141_255685\|\|full | 22563 | No | NA | 19 | 0 | 18 | Not-determined | Genome-fragment | NA | NA | 0 |
| S11-final-viral-combined_k141_104339\|\|full | 3068 | No | NA | 5 | 0 | 2 | Not-determined | Genome-fragment | NA | NA | 0 |
| S11-final-viral-combined_k141_283452\|\|full | 1630 | No | NA | 3 | 0 | 0 | Not-determined | Genome-fragment | NA | NA | 0 |
| S11-final-viral-combined_k141_201510\|\|full | 1139 | No | NA | 3 | 0 | 0 | Low-quality | Genome-fragment | 1.31 | AAI-based (medium-confidence) | 0 |
| S11-final-viral-combined_k141_242389\|\|full | 3488 | No | NA | 4 | 0 | 2 | Not-determined | Genome-fragment | NA | NA | 0 |
| S11-final-viral-combined_k141_246599\|\|full | 1139 | No | NA | 4 | 0 | 0 | Low-quality | Genome-fragment | 2.47 | AAI-based (medium-confidence) | 0 |
| S11-final-viral-combined_k141_75697\|\|full | 1284 | No | NA | 3 | 0 | 1 | Not-determined | Genome-fragment | NA | NA | 0 |
| S11-final-viral-combined_k141_116954\|\|full | 1970 | No | NA | 4 | 0 | 0 | Not-determined | Genome-fragment | NA | NA | 0 |
| S11-final-viral-combined_k141_199834\|\|full | 1121 | No | NA | 3 | 0 | 0 | Not-determined | Genome-fragment | NA | NA | 0 |
| S11-final-viral-combined_k141_186127\|\|full | 1205 | No | NA | 2 | 0 | 1 | Not-determined | Genome-fragment | NA | NA | 0 |
| S11-final-viral-combined_k141_255749\|\|full | 2763 | No | NA | 4 | 0 | 1 | Not-determined | Genome-fragment | NA | NA | 0 |
| S11-final-viral-combined_k141_104371\|\|full | 2909 | No | NA | 3 | 0 | 0 | Not-determined | Genome-fragment | NA | NA | 0 |
| S11_k141_244342 | 1139 | No | NA | 1 | 0 | 0 | Low-quality | Genome-fragment | 3.8 | AAI-based (high-confidence) | 0 |
| S11-final-viral-combined_k141_131016\|\|full | 2256 | No | NA | 2 | 0 | 1 | Not-determined | Genome-fragment | NA | NA | 0 |
| S5_k141_186479 | 1139 | No | NA | 2 | 2 | 0 | Low-quality | Genome-fragment | 1.89 | AAI-based (medium-confidence) | 0 |
| S11-final-viral-combined_k141_323536\|\|full | 5205 | No | NA | 4 | 0 | 3 | Not-determined | Genome-fragment | NA | NA | 0 |
| S17-final-viral-combined_k141_201413\|\|full | 1139 | No | NA | 3 | 2 | 0 | Low-quality | Genome-fragment | 2.6 | AAI-based (high-confidence) | 0 |
| S17-k141_158073 | 1139 | No | NA | 3 | 2 | 0 | Low-quality | Genome-fragment | 2.23 | AAI-based (high-confidence) | 0 |
| S11-final-viral-combined_k141_7443\|\|full | 1460 | No | NA | 3 | 0 | 2 | Not-determined | Genome-fragment | NA | NA | 0 |
| S11-final-viral-combined_k141_311182\|\|full | 2012 | No | NA | 2 | 0 | 0 | Not-determined | Genome-fragment | NA | NA | 0 |
| S12-final-viral-combined_k141_21269\|\|lt2gene | 1139 | No | NA | 2 | 1 | 0 | Low-quality | Genome-fragment | 1.98 | AAI-based (medium-confidence) | 0 |
| S11-final-viral-combined_k141_35124\|\|full | 3379 | No | NA | 4 | 0 | 1 | Not-determined | Genome-fragment | NA | NA | 0 |
| S12_k141_352897 | 1139 | No | NA | 2 | 0 | 0 | Low-quality | Genome-fragment | 4.1 | AAI-based (high-confidence) | 0 |
| S12_k141_36578 | 1139 | No | NA | 1 | 1 | 0 | Low-quality | Genome-fragment | 15.26 | AAI-based (medium-confidence) | 0 |
| S11-final-viral-combined_k141_35138\|\|full | 1928 | No | NA | 3 | 0 | 1 | Not-determined | Genome-fragment | NA | NA | 0 |
| S11-final-viral-combined_k141_296536\|\|full | 2632 | No | NA | 5 | 0 | 1 | Not-determined | Genome-fragment | NA | NA | 0 |
| S11-final-viral-combined_k141_75797\|\|full | 2701 | No | NA | 3 | 0 | 2 | Not-determined | Genome-fragment | NA | NA | 0 |
| S2-final-viral-combined_k141_189136\|\|lt2gene | 1139 | No | NA | 1 | 1 | 0 | Low-quality | Genome-fragment | 1.66 | HMM-based (lower-bound) | 0 |
| S2_k141_218123 | 1139 | No | NA | 1 | 0 | 0 | Low-quality | Genome-fragment | 2.22 | AAI-based (high-confidence) | 0 |
| S11-final-viral-combined_k141_214759\|\|full | 1140 | No | NA | 4 | 0 | 0 | Low-quality | Genome-fragment | 2.52 | AAI-based (medium-confidence) | 0 |
| S11_k141_313893 | 1140 | No | NA | 1 | 0 | 1 | Low-quality | Genome-fragment | 1.1 | AAI-based (medium-confidence) | 0 |
| S1_k141_143386 | 1140 | No | NA | 1 | 0 | 0 | Low-quality | Genome-fragment | 3.01 | AAI-based (high-confidence) | 0 |
| S11-final-viral-combined_k141_35175\|\|full | 1963 | No | NA | 3 | 0 | 1 | Not-determined | Genome-fragment | NA | NA | 0 |
| S5-final-viral-combined_k141_138505\|\|full | 1140 | No | NA | 2 | 0 | 0 | Low-quality | Genome-fragment | 0.72 | AAI-based (medium-confidence) | 0 |
| S7-final-viral-combined_k141_194127\|\|full | 1140 | No | NA | 3 | 1 | 0 | Low-quality | Genome-fragment | 1.72 | HMM-based (lower-bound) | 0 |
| S11-final-viral-combined_k141_173219\|\|full | 2964 | No | NA | 3 | 0 | 3 | Not-determined | Genome-fragment | NA | NA | 0 |
| S11-final-viral-combined_k141_323612\|\|full | 5764 | No | NA | 7 | 0 | 3 | Not-determined | Genome-fragment | NA | NA | 0 |
| S11-final-viral-combined_k141_173230\|\|full | 1664 | No | NA | 2 | 0 | 2 | Not-determined | Genome-fragment | NA | NA | 0 |
| S7-final-viral-combined_k141_76010\|\|full | 1140 | No | NA | 3 | 0 | 0 | Low-quality | Genome-fragment | 1.75 | AAI-based (high-confidence) | 0 |
| S7-final-viral-combined_k141_234223\|\|lt2gene | 1140 | No | NA | 2 | 1 | 0 | Low-quality | Genome-fragment | 3.34 | AAI-based (high-confidence) | 0 |
| S11-final-viral-combined_k141_117121\|\|full | 4319 | No | NA | 6 | 0 | 2 | Not-determined | Genome-fragment | NA | NA | 0 |
| S17-final-viral-combined_k141_152468\|\|full | 1140 | No | NA | 3 | 0 | 0 | Low-quality | Genome-fragment | 1.43 | AAI-based (high-confidence) | 0 |
| S11-final-viral-combined_k141_159198\|\|full | 11160 | No | NA | 12 | 0 | 9 | Not-determined | Genome-fragment | NA | NA | 0 |
| S11-final-viral-combined_k141_214692\|\|full | 5415 | No | NA | 8 | 0 | 2 | Not-determined | Genome-fragment | NA | NA | 0 |
| S17-final-viral-combined_k141_188853\|\|full | 1140 | No | NA | 3 | 0 | 1 | Low-quality | Genome-fragment | 1.46 | AAI-based (medium-confidence) | 0 |
| S17-k141_133285 | 1140 | No | NA | 1 | 0 | 1 | Low-quality | Genome-fragment | 0.37 | AAI-based (medium-confidence) | 0 |
| S11-final-viral-combined_k141_35275\|\|full | 1198 | No | NA | 3 | 0 | 2 | Not-determined | Genome-fragment | NA | NA | 0 |
| S11-final-viral-combined_k141_242624\|\|full | 6531 | No | NA | 7 | 0 | 4 | Not-determined | Genome-fragment | NA | NA | 0 |
| S12-final-viral-combined_k141_324669\|\|full | 1140 | No | NA | 3 | 0 | 0 | Low-quality | Genome-fragment | 1.94 | AAI-based (high-confidence) | 0 |
| S12-final-viral-combined_k141_299995\|\|full | 1140 | No | NA | 2 | 0 | 1 | Low-quality | Genome-fragment | 0.34 | AAI-based (medium-confidence) | 0 |
| S11-final-viral-combined_k141_228262\|\|full | 1537 | No | NA | 2 | 0 | 1 | Not-determined | Genome-fragment | NA | NA | 0 |
| S11-final-viral-combined_k141_311294\|\|full | 1112 | No | NA | 2 | 0 | 0 | Not-determined | Genome-fragment | NA | NA | 0 |
| S2-final-viral-combined_k141_234088\|\|full | 1140 | No | NA | 3 | 0 | 2 | Low-quality | Genome-fragment | 0.64 | AAI-based (medium-confidence) | 0 |
| S4-final-viral-combined_k141_62016\|\|full | 1140 | No | NA | 3 | 0 | 0 | Low-quality | Genome-fragment | 3.04 | AAI-based (high-confidence) | 0 |
| S11-final-viral-combined_k141_173341\|\|full | 5297 | No | NA | 4 | 0 | 3 | Not-determined | Genome-fragment | NA | NA | 0 |
| S4_k141_155938 | 1140 | No | NA | 3 | 1 | 0 | Low-quality | Genome-fragment | 1.38 | AAI-based (high-confidence) | 0 |
| S8-final-viral-combined_k141_184476\|\|full | 1140 | No | NA | 2 | 0 | 1 | Low-quality | Genome-fragment | 1.67 | AAI-based (medium-confidence) | 0 |
| S8_k141_40114 | 1140 | No | NA | 1 | 0 | 1 | Low-quality | Genome-fragment | 0.7 | AAI-based (medium-confidence) | 0 |
| S11-final-viral-combined_k141_159271\|\|full | 2979 | No | NA | 4 | 0 | 4 | Not-determined | Genome-fragment | NA | NA | 0 |
| S11-final-viral-combined_k141_200084\|\|full | 38510 | No | NA | 52 | 15 | 1 | Medium-quality | Genome-fragment | 63.01 | AAI-based (high-confidence) | 0 |
| S8_k141_110884 | 1140 | No | NA | 3 | 0 | 1 | Low-quality | Genome-fragment | 3.64 | AAI-based (medium-confidence) | 0 |
| S9-final-viral-combined_k141_37848\|\|full | 1141 | No | NA | 3 | 0 | 2 | Low-quality | Genome-fragment | 0.84 | AAI-based (medium-confidence) | 0 |
| S9-final-viral-combined_k141_87806\|\|full | 1141 | No | NA | 2 | 0 | 1 | Low-quality | Genome-fragment | 1.91 | AAI-based (high-confidence) | 0 |
| S11_k141_92491 | 1141 | No | NA | 3 | 0 | 0 | Low-quality | Genome-fragment | 1.91 | AAI-based (high-confidence) | 0 |

| | | | | | | | | | | | | | |
|---|---|---|---|---|---|---|---|---|---|---|---|---|---|
| S1-final-viral-combined_k141_86126\|\|full | 1141 | No | NA | | 3 | 0 | 1 | Low-quality | Genome-fragment | | 2.37 | AAI-based (medium-confidence) | 0 |
| S11-final-viral-combined_k141_311331\|\|full | 1470 | No | NA | | 3 | 0 | 0 | Not-determined | Genome-fragment | NA | | NA | 0 |
| S7-final-viral-combined_k141_179007\|\|lt2gene | 1141 | No | NA | | 2 | 0 | 0 | Low-quality | Genome-fragment | | 2.68 | AAI-based (high-confidence) | 0 |
| S17-k141_46904 | 1141 | No | NA | | 2 | 0 | 2 | Low-quality | Genome-fragment | | 0.22 | AAI-based (medium-confidence) | 0 |
| S12-final-viral-combined_k141_14199\|\|full | 1141 | No | NA | | 3 | 0 | 0 | Low-quality | Genome-fragment | | 2.37 | AAI-based (medium-confidence) | 0 |
| S11-final-viral-combined_k141_90352\|\|full | 6629 | No | NA | | 6 | 0 | 2 | Not-determined | Genome-fragment | NA | | NA | 0 |
| S11-final-viral-combined_k141_159321\|\|full | 9146 | No | NA | | 11 | 0 | 3 | Not-determined | Genome-fragment | NA | | NA | 0 |
| S12-final-viral-combined_k141_122243\|\|lt2gene | 1141 | No | NA | | 2 | 2 | 0 | Low-quality | Genome-fragment | | 3.02 | AAI-based (high-confidence) | 0 |
| S11-final-viral-combined_k141_186413\|\|full | 3351 | No | NA | | 4 | 0 | 3 | Not-determined | Genome-fragment | NA | | NA | 0 |
| S2-final-viral-combined_k141_215443\|\|full | 1141 | No | NA | | 2 | 0 | 0 | Low-quality | Genome-fragment | | 2.58 | AAI-based (medium-confidence) | 0 |
| S11-final-viral-combined_k141_63110\|\|full | 2038 | No | NA | | 2 | 0 | 1 | Not-determined | Genome-fragment | NA | | NA | 0 |
| S11-final-viral-combined_k141_173471\|\|full | 3445 | No | NA | | 4 | 0 | 2 | Not-determined | Genome-fragment | NA | | NA | 0 |
| S6_k141_126190 | 1141 | No | NA | | 1 | 0 | 1 | Low-quality | Genome-fragment | | 0.39 | AAI-based (medium-confidence) | 0 |
| S8-final-viral-combined_k141_119403\|\|full | 1141 | No | NA | | 2 | 0 | 0 | Low-quality | Genome-fragment | | 2.01 | AAI-based (medium-confidence) | 0 |
| S8-final-viral-combined_k141_247991\|\|full | 1141 | No | NA | | 3 | 0 | 0 | Low-quality | Genome-fragment | | 1.79 | AAI-based (medium-confidence) | 0 |
| S11-final-viral-combined_k141_296760\|\|full | 1091 | No | NA | | 2 | 0 | 0 | Not-determined | Genome-fragment | NA | | NA | 0 |
| S11-final-viral-combined_k141_7641\|\|full | 4477 | No | NA | | 4 | 0 | 2 | Not-determined | Genome-fragment | NA | | NA | 0 |
| S11-final-viral-combined_k141_256055\|\|full | 1980 | No | NA | | 2 | 0 | 1 | Not-determined | Genome-fragment | NA | | NA | 0 |
| S11-final-viral-combined_k141_269433\|\|full | 2748 | No | NA | | 3 | 0 | 1 | Not-determined | Genome-fragment | NA | | NA | 0 |
| S8_k141_90868 | 1141 | No | NA | | 2 | 0 | 0 | Low-quality | Genome-fragment | | 1.27 | AAI-based (high-confidence) | 0 |
| S11-final-viral-combined_k141_214847\|\|full | 1586 | No | NA | | 2 | 0 | 1 | Not-determined | Genome-fragment | NA | | NA | 0 |
| S11-final-viral-combined_k141_145490\|\|full | 1342 | No | NA | | 2 | 0 | 1 | Not-determined | Genome-fragment | NA | | NA | 0 |
| S11-final-viral-combined_k141_228403\|\|full | 5657 | No | NA | | 6 | 0 | 3 | Not-determined | Genome-fragment | NA | | NA | 0 |
| S11-final-viral-combined_k141_35492\|\|full | 2701 | No | NA | | 4 | 0 | 2 | Not-determined | Genome-fragment | NA | | NA | 0 |
| S11-final-viral-combined_k141_173511\|\|full | 1852 | No | NA | | 3 | 0 | 2 | Not-determined | Genome-fragment | NA | | NA | 0 |
| S11-final-viral-combined_k141_7686\|\|full | 1358 | No | NA | | 4 | 0 | 1 | Not-determined | Genome-fragment | NA | | NA | 0 |
| S11-final-viral-combined_k141_214891\|\|full | 1593 | No | NA | | 1 | 0 | 1 | Not-determined | Genome-fragment | NA | | NA | 0 |
| S9-final-viral-combined_k141_91439\|\|full | 1142 | Yes | | 553 | 3 | 2 | 1 | Low-quality | Genome-fragment | | 1.46 | AAI-based (medium-confidence) | 51.58 |
| S11-final-viral-combined_k141_63199\|\|full | 1240 | No | NA | | 3 | 0 | 0 | Not-determined | Genome-fragment | NA | | NA | 0 |
| S11-final-viral-combined_k141_231969\|\|full | 1142 | No | NA | | 2 | 0 | 1 | Low-quality | Genome-fragment | | 1.95 | AAI-based (medium-confidence) | 0 |
| S11-final-viral-combined_k141_104672\|\|full | 1662 | No | NA | | 1 | 0 | 1 | Not-determined | Genome-fragment | NA | | NA | 0 |
| S11-final-viral-combined_k141_228417\|\|full | 3415 | No | NA | | 6 | 0 | 0 | Not-determined | Genome-fragment | NA | | NA | 0 |
| S11-final-viral-combined_k141_283660\|\|full | 1688 | No | NA | | 2 | 0 | 1 | Not-determined | Genome-fragment | NA | | NA | 0 |
| S1_k141_335417 | 1142 | No | NA | | 1 | 0 | 1 | Low-quality | Genome-fragment | | 0.45 | AAI-based (medium-confidence) | 0 |
| S11-final-viral-combined_k141_35521\|\|full | 1395 | No | NA | | 2 | 0 | 1 | Not-determined | Genome-fragment | NA | | NA | 0 |
| S11-final-viral-combined_k141_104730\|\|full | 2241 | No | NA | | 5 | 0 | 3 | Not-determined | Genome-fragment | NA | | NA | 0 |
| S11-final-viral-combined_k141_296844\|\|full | 2650 | No | NA | | 3 | 0 | 1 | Not-determined | Genome-fragment | NA | | NA | 0 |
| S11-final-viral-combined_k141_35532\|\|full | 2050 | No | NA | | 2 | 0 | 1 | Not-determined | Genome-fragment | NA | | NA | 0 |
| S11-final-viral-combined_k141_117370\|\|full | 2550 | No | NA | | 4 | 0 | 1 | Not-determined | Genome-fragment | NA | | NA | 0 |
| S11-final-viral-combined_k141_159502\|\|full | 1705 | No | NA | | 3 | 0 | 2 | Not-determined | Genome-fragment | NA | | NA | 0 |
| S5-final-viral-combined_k141_142196\|\|full | 1142 | No | NA | | 2 | 0 | 1 | Low-quality | Genome-fragment | | 0.75 | AAI-based (medium-confidence) | 0 |
| S11-final-viral-combined_k141_173590\|\|full | 17118 | No | NA | | 6 | 0 | 5 | Not-determined | Genome-fragment | NA | | NA | 0 |
| S11-final-viral-combined_k141_117394\|\|full | 2322 | No | NA | | 2 | 0 | 2 | Not-determined | Genome-fragment | NA | | NA | 0 |
| S11-final-viral-combined_k141_228485\|\|full | 2400 | No | NA | | 3 | 0 | 2 | Not-determined | Genome-fragment | NA | | NA | 0 |
| S11-final-viral-combined_k141_145644\|\|full | 3018 | No | NA | | 3 | 0 | 2 | Not-determined | Genome-fragment | NA | | NA | 0 |
| S7-final-viral-combined_k141_47799\|\|full | 1142 | No | NA | | 3 | 0 | 0 | Low-quality | Genome-fragment | | 2.24 | AAI-based (medium-confidence) | 0 |
| [illegible] | [illegible] | [illegible] | [illegible] | [illegible] | [illegible] | [illegible] | [illegible] | [illegible] | [illegible] | [illegible] | [illegible] | [illegible] | [illegible] |

| | | | | | | | | | | | |
|---|---|---|---|---|---|---|---|---|---|---|---|
| S11-final-viral-combined_k141_216014\|\|full | 3309 | No | NA | 3 | 0 | 2 | Not-determined | Genome-fragment | NA | NA | 0 |
| S11-final-viral-combined_k141_132236\|\|full | 4274 | No | NA | 4 | 0 | 4 | Not-determined | Genome-fragment | NA | NA | 0 |
| S12-final-viral-combined_k141_324172\|\|full | 1155 | No | NA | 3 | 2 | 0 | Low-quality | Genome-fragment | 1.97 | AAI-based (high-confidence) | 0 |
| S11-final-viral-combined_k141_285099\|\|full | 5957 | No | NA | 7 | 0 | 1 | Not-determined | Genome-fragment | NA | NA | 0 |
| S12_k141_220389 | 1155 | No | NA | 1 | 1 | 0 | Low-quality | Genome-fragment | 0.73 | HMM-based (lower-bound) | 0 |
| S12_k141_45481 | 1155 | No | NA | 2 | 1 | 0 | Low-quality | Genome-fragment | 0.81 | HMM-based (lower-bound) | 0 |
| S11-final-viral-combined_k141_325095\|\|full | 3168 | No | NA | 5 | 0 | 1 | Not-determined | Genome-fragment | NA | NA | 0 |
| S2-final-viral-combined_k141_12618\|\|lt2gene | 1155 | No | NA | 1 | 1 | 0 | Low-quality | Genome-fragment | 2.39 | AAI-based (high-confidence) | 0 |
| S11-final-viral-combined_k141_187998\|\|full | 1639 | No | NA | 2 | 0 | 0 | Not-determined | Genome-fragment | NA | NA | 0 |
| S11-final-viral-combined_k141_325131\|\|full | 1030 | No | NA | 2 | 0 | 0 | Not-determined | Genome-fragment | NA | NA | 0 |
| S11-final-viral-combined_k141_174806\|\|full | 1428 | No | NA | 2 | 0 | 1 | Not-determined | Genome-fragment | NA | NA | 0 |
| S11-final-viral-combined_k141_216094\|\|full | 4020 | No | NA | 5 | 0 | 5 | Not-determined | Genome-fragment | NA | NA | 0 |
| S11-final-viral-combined_k141_312775\|\|full | 2477 | No | NA | 5 | 0 | 1 | Not-determined | Genome-fragment | NA | NA | 0 |
| S6-final-viral-combined_k141_106465\|\|lt2gene | 1155 | No | NA | 2 | 1 | 0 | Low-quality | Genome-fragment | 1.89 | HMM-based (lower-bound) | 0 |
| S11-final-viral-combined_k141_321695\|\|full | 1156 | No | NA | 3 | 0 | 0 | Low-quality | Genome-fragment | 1.96 | AAI-based (high-confidence) | 0 |
| S11-final-viral-combined_k141_257621\|\|full | 1611 | No | NA | 2 | 0 | 1 | Not-determined | Genome-fragment | NA | NA | 0 |
| S5-final-viral-combined_k141_156005\|\|full | 1156 | No | NA | 3 | 3 | 0 | Low-quality | Genome-fragment | 3.54 | AAI-based (medium-confidence) | 0 |
| S11-final-viral-combined_k141_188033\|\|full | 1140 | No | NA | 3 | 0 | 1 | Not-determined | Genome-fragment | NA | NA | 0 |
| S11-final-viral-combined_k141_147034\|\|full | 1570 | No | NA | 3 | 0 | 2 | Not-determined | Genome-fragment | NA | NA | 0 |
| S7-final-viral-combined_k141_14247\|\|full | 1156 | No | NA | 3 | 0 | 0 | Low-quality | Genome-fragment | 2.49 | AAI-based (medium-confidence) | 0 |
| S7-final-viral-combined_k141_163345\|\|full | 1156 | No | NA | 3 | 0 | 0 | Low-quality | Genome-fragment | 2.81 | AAI-based (high-confidence) | 0 |
| S11-final-viral-combined_k141_285215\|\|full | 4845 | No | NA | 5 | 0 | 2 | Not-determined | Genome-fragment | NA | NA | 0 |
| S11-final-viral-combined_k141_64392\|\|full | 42456 | No | NA | 43 | 9 | 3 | High-quality | High-quality | 99.12 | AAI-based (medium-confidence) | 0 |
| S11-final-viral-combined_k141_161005\|\|full | 4739 | No | NA | 5 | 0 | 1 | Not-determined | Genome-fragment | NA | NA | 0 |
| S7-final-viral-combined_k141_129787\|\|lt2gene | 1156 | No | NA | 2 | 1 | 0 | Low-quality | Genome-fragment | 2.73 | HMM-based (lower-bound) | 0 |
| S17-final-viral-combined_k141_23886\|\|full | 1156 | No | NA | 2 | 0 | 1 | Low-quality | Genome-fragment | 1.75 | AAI-based (medium-confidence) | 0 |
| S11-final-viral-combined_k141_77553\|\|full | 2160 | No | NA | 4 | 0 | 1 | Not-determined | Genome-fragment | NA | NA | 0 |
| S8-final-viral-combined_k141_66537\|\|full | 1156 | No | NA | 3 | 1 | 0 | Low-quality | Genome-fragment | 2.62 | AAI-based (high-confidence) | 0 |
| S8_k141_23147 | 1156 | No | NA | 1 | 0 | 0 | Low-quality | Genome-fragment | 1.41 | AAI-based (high-confidence) | 0 |
| S11-final-viral-combined_k141_312811\|\|full | 1626 | No | NA | 3 | 0 | 0 | Not-determined | Genome-fragment | NA | NA | 0 |
| S9_k141_118315 | 1157 | No | NA | 2 | 0 | 2 | Low-quality | Genome-fragment | 0.64 | AAI-based (medium-confidence) | 0 |
| S11-final-viral-combined_k141_244239\|\|full | 1662 | No | NA | 3 | 0 | 1 | Not-determined | Genome-fragment | NA | NA | 0 |
| S11-final-viral-combined_k141_125399\|\|lt2gene | 1157 | No | NA | 1 | 0 | 0 | Low-quality | Genome-fragment | 2.15 | AAI-based (high-confidence) | 0 |
| S11-final-viral-combined_k141_9732\|\|full | 1217 | No | NA | 2 | 0 | 2 | Not-determined | Genome-fragment | NA | NA | 0 |
| S1-final-viral-combined_k141_348384\|\|full | 1157 | No | NA | 3 | 0 | 0 | Low-quality | Genome-fragment | 2.42 | AAI-based (medium-confidence) | 0 |
| S11-final-viral-combined_k141_50768\|\|full | 1810 | No | NA | 2 | 0 | 0 | Not-determined | Genome-fragment | NA | NA | 0 |
| S11-final-viral-combined_k141_64448\|\|full | 4342 | No | NA | 5 | 0 | 3 | Not-determined | Genome-fragment | NA | NA | 0 |
| S11-final-viral-combined_k141_270847\|\|full | 1979 | No | NA | 4 | 0 | 0 | Not-determined | Genome-fragment | NA | NA | 0 |
| S11-final-viral-combined_k141_132290\|\|full | 1537 | No | NA | 3 | 0 | 0 | Not-determined | Genome-fragment | NA | NA | 0 |
| S1-final-viral-combined_k141_184627\|\|full | 1157 | No | NA | 3 | 0 | 1 | Low-quality | Genome-fragment | 0.68 | AAI-based (medium-confidence) | 0 |
| S11-final-viral-combined_k141_91959\|\|full | 1493 | No | NA | 3 | 0 | 2 | Not-determined | Genome-fragment | NA | NA | 0 |
| S11-final-viral-combined_k141_244273\|\|full | 2926 | No | NA | 4 | 0 | 2 | Not-determined | Genome-fragment | NA | NA | 0 |
| S11-final-viral-combined_k141_216193\|\|full | 1197 | No | NA | 2 | 0 | 2 | Not-determined | Genome-fragment | NA | NA | 0 |
| S11-final-viral-combined_k141_50800\|\|full | 9429 | No | NA | 8 | 0 | 6 | Not-determined | Genome-fragment | NA | NA | 0 |
| S1-final-viral-combined_k141_111446\|\|full | 1157 | No | NA | 2 | 1 | 0 | Low-quality | Genome-fragment | 0.73 | HMM-based (lower-bound) | 0 |
| S5-final-viral-combined_k141_6831\|\|full | 1157 | No | NA | 4 | 1 | 0 | Low-quality | Genome-fragment | 2.47 | HMM-based (lower-bound) | 0 |
| S5-final-viral-combined_k141_109528\|\|lt2gene | 1157 | No | NA | 2 | 2 | 0 | Low-quality | Genome-fragment | 3.11 | AAI-based (high-confidence) | 0 |
| S5_k141_109261 | 1157 | No | NA | 3 | 0 | 0 | Low-quality | Genome-fragment | 9.48 | AAI-based (medium-confidence) | 0 |
| S11-final-viral-combined_k141_106428\|\|full | 8622 | No | NA | 9 | 0 | 5 | Not-determined | Genome-fragment | NA | NA | 0 |
| S11-final-viral-combined_k141_9778\|\|full | 1495 | No | NA | 3 | 0 | 3 | Not-determined | Genome-fragment | NA | NA | 0 |
| S11-final-viral-combined_k141_229981\|\|full | 1457 | No | NA | 3 | 0 | 0 | Not-determined | Genome-fragment | NA | NA | 0 |
| S12-final-viral-combined_k141_77285\|\|full | 1157 | No | NA | 2 | 0 | 0 | Low-quality | Genome-fragment | 0.75 | AAI-based (medium-confidence) | 0 |
| S11-final-viral-combined_k141_298241\|\|full | 1233 | No | NA | 2 | 0 | 1 | Not-determined | Genome-fragment | NA | NA | 0 |
| S11-final-viral-combined_k141_325227\|\|full | 1584 | No | NA | 3 | 0 | 0 | Not-determined | Genome-fragment | NA | NA | 0 |
| S11-final-viral-combined_k141_92019\|\|full | 4681 | No | NA | 7 | 0 | 3 | Not-determined | Genome-fragment | NA | NA | 0 |
| S11-final-viral-combined_k141_174976\|\|full | 1004 | No | NA | 2 | 0 | 1 | Not-determined | Genome-fragment | NA | NA | 0 |
| S11-final-viral-combined_k141_50841\|\|full | 3452 | No | NA | 3 | 0 | 3 | Not-determined | Genome-fragment | NA | NA | 0 |
| S11-final-viral-combined_k141_161111\|\|full | 4925 | No | NA | 5 | 0 | 2 | Not-determined | Genome-fragment | NA | NA | 0 |
| S11-final-viral-combined_k141_77639\|\|full | 1039 | No | NA | 2 | 0 | 1 | Not-determined | Genome-fragment | NA | NA | 0 |
| S11-final-viral-combined_k141_106448\|\|full | 1676 | No | NA | 2 | 0 | 2 | Not-determined | Genome-fragment | NA | NA | 0 |
| S11-final-viral-combined_k141_132383\|\|full | 2414 | No | NA | 2 | 0 | 2 | Not-determined | Genome-fragment | NA | NA | 0 |
| S11-final-viral-combined_k141_77680\|\|full | 1123 | No | NA | 2 | 0 | 2 | Not-determined | Genome-fragment | NA | NA | 0 |
| S11-final-viral-combined_k141_64569\|\|full | 3171 | No | NA | 4 | 0 | 0 | Not-determined | Genome-fragment | NA | NA | 0 |
| S2_k141_180733 | 1157 | No | NA | 3 | 2 | 0 | Low-quality | Genome-fragment | 3.03 | AAI-based (medium-confidence) | 0 |
| S8-final-viral-combined_k141_185608\|\|full | 1157 | No | NA | 4 | 0 | 0 | Low-quality | Genome-fragment | 3.49 | AAI-based (high-confidence) | 0 |
| S11-final-viral-combined_k141_77717\|\|full | 3687 | No | NA | 4 | 0 | 2 | Not-determined | Genome-fragment | NA | NA | 0 |
| S11-final-viral-combined_k141_77720\|\|full | 1903 | No | NA | 2 | 0 | 2 | Not-determined | Genome-fragment | NA | NA | 0 |
| S11-final-viral-combined_k141_257839\|\|full | 1393 | No | NA | 2 | 0 | 1 | Not-determined | Genome-fragment | NA | NA | 0 |
| S11-final-viral-combined_k141_244352\|\|full | 2472 | No | NA | 3 | 0 | 1 | Not-determined | Genome-fragment | NA | NA | 0 |
| S5_k141_74 | 1158 | No | NA | 1 | 0 | 1 | Low-quality | Genome-fragment | 0.75 | AAI-based (high-confidence) | 0 |
| S11-final-viral-combined_k141_92058\|\|full | 1062 | No | NA | 2 | 0 | 1 | Not-determined | Genome-fragment | NA | NA | 0 |
| S7-final-viral-combined_k141_202583\|\|lt2gene | 1158 | No | NA | 1 | 1 | 0 | Low-quality | Genome-fragment | 2.64 | AAI-based (medium-confidence) | 0 |
| S11-final-viral-combined_k141_37075\|\|full | 1347 | No | NA | 3 | 0 | 1 | Not-determined | Genome-fragment | NA | NA | 0 |
| S11-final-viral-combined_k141_21950\|\|full | 4432 | No | NA | 6 | 0 | 6 | Not-determined | Genome-fragment | NA | NA | 0 |
| S17-final-viral-combined_k141_22030\|\|full | 1158 | No | NA | 4 | 0 | 0 | Low-quality | Genome-fragment | 2.58 | AAI-based (high-confidence) | 0 |
| S11-final-viral-combined_k141_325295\|\|full | 2238 | No | NA | 3 | 0 | 2 | Not-determined | Genome-fragment | NA | NA | 0 |
| S11-final-viral-combined_k141_298381\|\|full | 1573 | No | NA | 3 | 0 | 0 | Not-determined | Genome-fragment | NA | NA | 0 |
| S12-final-viral-combined_k141_298635\|\|full | 1158 | No | NA | 4 | 4 | 0 | Low-quality | Genome-fragment | 3.35 | AAI-based (high-confidence) | 0 |
| S11-final-viral-combined_k141_147287\|\|full | 1219 | No | NA | 1 | 0 | 0 | Not-determined | Genome-fragment | NA | NA | 0 |
| S11-final-viral-combined_k141_202201\|\|full | 1257 | No | NA | 3 | 0 | 2 | Not-determined | Genome-fragment | NA | NA | 0 |
| S12_k141_154407 | 1158 | No | NA | 3 | 1 | 0 | Low-quality | Genome-fragment | 1.89 | AAI-based (medium-confidence) | 0 |
| S11-final-viral-combined_k141_202207\|\|full | 7291 | No | NA | 7 | 0 | 4 | Not-determined | Genome-fragment | NA | NA | 0 |
| S11-final-viral-combined_k141_9897\|\|full | 1536 | No | NA | 3 | 0 | 2 | Not-determined | Genome-fragment | NA | NA | 0 |
| S11-final-viral-combined_k141_257911\|\|full | 3067 | No | NA | 4 | 0 | 4 | Not-determined | Genome-fragment | NA | NA | 0 |
| S2-final-viral-combined_k141_3888\|\|full | 1158 | No | NA | 2 | 1 | 0 | Low-quality | Genome-fragment | 2.49 | AAI-based (high-confidence) | 0 |
| S11-final-viral-combined_k141_270995\|\|full | 2308 | No | NA | 4 | 0 | 0 | Not-determined | Genome-fragment | NA | NA | 0 |
| S11-final-viral-combined_k141_257931\|\|full | 2201 | No | NA | 3 | 0 | 2 | Not-determined | Genome-fragment | NA | NA | 0 |
| S2-final-viral-combined_k141_26346\|\|full | 1158 | No | NA | 3 | 1 | 0 | Low-quality | Genome-fragment | 1.72 | AAI-based (high-confidence) | 0 |
| S2-final-viral-combined_k141_184830\|\|lt2gene | 1158 | No | NA | 2 | 1 | 0 | Low-quality | Genome-fragment | 2.91 | AAI-based (medium-confidence) | 0 |
| S4_k141_550 | 1158 | No | NA | 3 | 0 | 0 | Low-quality | Genome-fragment | 2.97 | AAI-based (high-confidence) | 0 |
| S11-final-viral-combined_k141_119150\|\|full | 3434 | No | NA | 3 | 0 | 1 | Not-determined | Genome-fragment | NA | NA | 0 |
| S4_k141_101770 | 1158 | No | NA | 2 | 0 | 1 | Low-quality | Genome-fragment | 3.14 | AAI-based (medium-confidence) | 0 |
| S11-final-viral-combined_k141_51036\|\|full | 1884 | No | NA | 2 | 0 | 2 | Not-determined | Genome-fragment | NA | NA | 0 |
| S11-final-viral-combined_k141_51040\|\|full | 2345 | No | NA | 3 | 0 | 3 | Not-determined | Genome-fragment | NA | NA | 0 |
| S11-final-viral-combined_k141_92114\|\|full | 48959 | Yes | 26924 | 55 | 7 | 13 | Medium-quality | Genome-fragment | 58.85 | AAI-based (high-confidence) | 45.01 |
| S11-final-viral-combined_k141_313604\|\|full | 1159 | No | NA | 3 | 0 | 0 | Low-quality | Genome-fragment | 1.98 | AAI-based (medium-confidence) | 0 |
| S11-final-viral-combined_k141_202257\|\|full | 4943 | No | NA | 4 | 0 | 4 | Not-determined | Genome-fragment | NA | NA | 0 |
| S11-final-viral-combined_k141_191509\|\|lt2gene | 1159 | No | NA | 1 | 1 | 0 | Low-quality | Genome-fragment | 1.93 | HMM-based (lower-bound) | 0 |
| S1-final-viral-combined_k141_151228\|\|full | 1159 | No | NA | 2 | 0 | 0 | Low-quality | Genome-fragment | 2.18 | AAI-based (medium-confidence) | 0 |
| S1-final-viral-combined_k141_88160\|\|full | 1159 | No | NA | 3 | 0 | 0 | Low-quality | Genome-fragment | 2.49 | AAI-based (medium-confidence) | 0 |
| S1-final-viral-combined_k141_241218\|\|full | 1159 | No | NA | 3 | 1 | 0 | Low-quality | Genome-fragment | 2.98 | AAI-based (high-confidence) | 0 |
| S1-final-viral-combined_k141_161477\|\|full | 1159 | No | NA | 3 | 0 | 1 | Low-quality | Genome-fragment | 3.19 | AAI-based (medium-confidence) | 0 |
| S1-final-viral-combined_k141_223079\|\|lt2gene | 1159 | No | NA | 1 | 1 | 0 | Low-quality | Genome-fragment | 1.06 | HMM-based (lower-bound) | 0 |
| S3_k141_16872 | 1159 | No | NA | 3 | 0 | 1 | Low-quality | Genome-fragment | 1.57 | AAI-based (medium-confidence) | 0 |
| S5-final-viral-combined_k141_96000\|\|full | 1159 | No | NA | 3 | 1 | 0 | Low-quality | Genome-fragment | 0.75 | HMM-based (lower-bound) | 0 |
| S11-final-viral-combined_k141_77799\|\|full | 2061 | No | NA | 3 | 0 | 2 | Not-determined | Genome-fragment | NA | NA | 0 |
| S11-final-viral-combined_k141_92172\|\|full | 2155 | No | NA | 2 | 0 | 2 | Not-determined | Genome-fragment | NA | NA | 0 |
| S11-final-viral-combined_k141_258048\|\|full | 1724 | No | NA | 2 | 0 | 0 | Not-determined | Genome-fragment | NA | NA | 0 |
| S11-final-viral-combined_k141_119215\|\|full | 27769 | No | NA | 41 | 14 | 2 | Medium-quality | Genome-fragment | 73.83 | AAI-based (medium-confidence) | 0 |
| S11-final-viral-combined_k141_106590\|\|full | 1708 | No | NA | 3 | 0 | 2 | Not-determined | Genome-fragment | NA | NA | 0 |
| S11-final-viral-combined_k141_119233\|\|full | 2005 | No | NA | 4 | 0 | 0 | Not-determined | Genome-fragment | NA | NA | 0 |
| S11-final-viral-combined_k141_188358\|\|full | 4547 | No | NA | 7 | 0 | 3 | Not-determined | Genome-fragment | NA | NA | 0 |
| S11-final-viral-combined_k141_216453\|\|full | 5389 | No | NA | 6 | 0 | 4 | Not-determined | Genome-fragment | NA | NA | 0 |
| S7-final-viral-combined_k141_119201\|\|full | 1159 | No | NA | 3 | 0 | 0 | Low-quality | Genome-fragment | 1.22 | AAI-based (medium-confidence) | 0 |
| S11-final-viral-combined_k141_106610\|\|full | 1242 | No | NA | 3 | 0 | 0 | Not-determined | Genome-fragment | NA | NA | 0 |
| S11-final-viral-combined_k141_230211\|\|full | 1068 | No | NA | 2 | 0 | 1 | Not-determined | Genome-fragment | NA | NA | 0 |
| S11-final-viral-combined_k141_147471\|\|full | 1109 | No | NA | 2 | 0 | 1 | Not-determined | Genome-fragment | NA | NA | 0 |
| S7-final-viral-combined_k141_9628\|\|lt2gene | 1159 | No | NA | 1 | 1 | 0 | Low-quality | Genome-fragment | 0.18 | HMM-based (lower-bound) | 0 |
| S11-final-viral-combined_k141_298516\|\|full | 1726 | No | NA | 3 | 0 | 3 | Not-determined | Genome-fragment | NA | NA | 0 |
| S11-final-viral-combined_k141_175235\|\|full | 3864 | No | NA | 4 | 0 | 1 | Not-determined | Genome-fragment | NA | NA | 0 |
| S11-final-viral-combined_k141_92234\|\|full | 2810 | No | NA | 4 | 0 | 2 | Not-determined | Genome-fragment | NA | NA | 0 |
| S7_k141_136557 | 1159 | No | NA | 1 | 0 | 1 | Low-quality | Genome-fragment | 0.59 | AAI-based (medium-confidence) | 0 |
| S7_k141_159273 | 1159 | No | NA | 2 | 0 | 0 | Low-quality | Genome-fragment | 0.48 | AAI-based (medium-confidence) | 0 |
| S11-final-viral-combined_k141_119250\|\|full | 3278 | No | NA | 4 | 0 | 2 | Not-determined | Genome-fragment | NA | NA | 0 |
| S17-final-viral-combined_k141_90127\|\|full | 1159 | No | NA | 4 | 2 | 0 | Low-quality | Genome-fragment | 2.76 | AAI-based (medium-confidence) | 0 |
| S11-final-viral-combined_k141_202349\|\|full | 2955 | No | NA | 3 | 0 | 2 | Not-determined | Genome-fragment | NA | NA | 0 |
| S11-final-viral-combined_k141_132591\|\|full | 5418 | No | NA | 3 | 0 | 1 | Not-determined | Genome-fragment | NA | NA | 0 |
| S11-final-viral-combined_k141_230239\|\|full | 3186 | No | NA | 2 | 0 | 2 | Not-determined | Genome-fragment | NA | NA | 0 |
| S17-final-viral-combined_k141_42955\|\|full | 1159 | No | NA | 4 | 0 | 0 | Low-quality | Genome-fragment | 2.31 | AAI-based (high-confidence) | 0 |
| S11-final-viral-combined_k141_132604\|\|full | 3501 | No | NA | 6 | 0 | 0 | Not-determined | Genome-fragment | NA | NA | 0 |
| S11-final-viral-combined_k141_325514\|\|full | 3886 | No | NA | 5 | 0 | 0 | Not-determined | Genome-fragment | NA | NA | 0 |
| S17-final-viral-combined_k141_159988\|\|lt2gene | 1159 | No | NA | 2 | 2 | 0 | Low-quality | Genome-fragment | 3.83 | AAI-based (high-confidence) | 0 |
| S11-final-viral-combined_k141_202383\|\|full | 5218 | No | NA | 5 | 0 | 4 | Not-determined | Genome-fragment | NA | NA | 0 |
| S12-final-viral-combined_k141_239852\|\|lt2gene | 1159 | No | NA | 2 | 1 | 0 | Low-quality | Genome-fragment | 2.39 | AAI-based (high-confidence) | 0 |
| S11-final-viral-combined_k141_271085\|\|full | 2928 | No | NA | 2 | 0 | 1 | Not-determined | Genome-fragment | NA | NA | 0 |
| S11-final-viral-combined_k141_51234\|\|full | 1728 | No | NA | 2 | 0 | 2 | Not-determined | Genome-fragment | NA | NA | 0 |
| S11-final-viral-combined_k141_271088\|\|full | 1958 | No | NA | 4 | 0 | 0 | Not-determined | Genome-fragment | NA | NA | 0 |
| S11-final-viral-combined_k141_92282\|\|full | 2399 | No | NA | 4 | 0 | 1 | Not-determined | Genome-fragment | NA | NA | 0 |
| S11-final-viral-combined_k141_202396\|\|full | 1371 | No | NA | 2 | 0 | 1 | Not-determined | Genome-fragment | NA | NA | 0 |
| S2_k141_47323 | 1159 | No | NA | 2 | 0 | 1 | Low-quality | Genome-fragment | 0.55 | AAI-based (medium-confidence) | 0 |
| S8-final-viral-combined_k141_15694\|\|full | 1159 | No | NA | 3 | 0 | 0 | Low-quality | Genome-fragment | 1.94 | AAI-based (medium-confidence) | 0 |
| S11-final-viral-combined_k141_202401\|\|full | 2760 | No | NA | 3 | 0 | 2 | Not-determined | Genome-fragment | NA | NA | 0 |
| S9_k141_63565 | 1160 | No | NA | 3 | 0 | 0 | Low-quality | Genome-fragment | 2.87 | AAI-based (high-confidence) | 0 |
| S11-final-viral-combined_k141_271097\|\|full | 1154 | No | NA | 2 | 0 | 1 | Not-determined | Genome-fragment | NA | NA | 0 |
| S11-final-viral-combined_k141_92308\|\|full | 1669 | No | NA | 2 | 0 | 2 | Not-determined | Genome-fragment | NA | NA | 0 |
| S11-final-viral-combined_k141_204555\|\|full | 1160 | No | NA | 3 | 0 | 0 | Low-quality | Genome-fragment | 2.78 | AAI-based (medium-confidence) | 0 |
| S11-final-viral-combined_k141_64820\|\|full | 1679 | No | NA | 3 | 0 | 3 | Not-determined | Genome-fragment | NA | NA | 0 |
| S11-final-viral-combined_k141_313267\|\|full | 1000 | No | NA | 3 | 0 | 0 | Not-determined | Genome-fragment | NA | NA | 0 |
| S11-final-viral-combined_k141_291012\|\|lt2gene | 1160 | No | NA | 2 | 1 | 0 | Low-quality | Genome-fragment | 1.52 | HMM-based (lower-bound) | 0 |
| S11-final-viral-combined_k141_51270\|\|full | 3471 | No | NA | 4 | 0 | 1 | Not-determined | Genome-fragment | NA | NA | 0 |
| S11-final-viral-combined_k141_64832\|\|full | 1886 | No | NA | 4 | 0 | 2 | Not-determined | Genome-fragment | NA | NA | 0 |
| S11-final-viral-combined_k141_285651\|\|full | 3006 | No | NA | 3 | 0 | 1 | Not-determined | Genome-fragment | NA | NA | 0 |
| S11-final-viral-combined_k141_77906\|\|full | 2251 | No | NA | 3 | 0 | 2 | Not-determined | Genome-fragment | NA | NA | 0 |
| S11-final-viral-combined_k141_64878\|\|full | 4049 | No | NA | 6 | 0 | 5 | Not-determined | Genome-fragment | NA | NA | 0 |
| S1-final-viral-combined_k141_32380\|\|lt2gene | 1160 | No | NA | 1 | 1 | 0 | Low-quality | Genome-fragment | 3.07 | AAI-based (medium-confidence) | 0 |
| S1_k141_213359 | 1160 | No | NA | 3 | 0 | 2 | Low-quality | Genome-fragment | 0.88 | AAI-based (medium-confidence) | 0 |
| S3-final-viral-combined_k141_30952\|\|lt2gene | 1160 | No | NA | 1 | 1 | 0 | Low-quality | Genome-fragment | 3 | AAI-based (high-confidence) | 0 |
| S11-final-viral-combined_k141_161396\|\|full | 2317 | No | NA | 2 | 0 | 1 | Not-determined | Genome-fragment | NA | NA | 0 |
| S11-final-viral-combined_k141_258190\|\|full | 7507 | No | NA | 6 | 0 | 6 | Not-determined | Genome-fragment | NA | NA | 0 |
| S5_k141_190372 | 1160 | No | NA | 2 | 0 | 1 | Low-quality | Genome-fragment | 1.03 | AAI-based (medium-confidence) | 0 |
| S11-final-viral-combined_k141_147637\|\|full | 4936 | No | NA | 7 | 0 | 2 | Not-determined | Genome-fragment | NA | NA | 0 |
| S11-final-viral-combined_k141_132714\|\|full | 3398 | No | NA | 4 | 0 | 1 | Not-determined | Genome-fragment | NA | NA | 0 |
| S17-final-viral-combined_k141_233749\|\|lt2gene | 1160 | No | NA | 2 | 1 | 0 | Low-quality | Genome-fragment | 2.86 | AAI-based (medium-confidence) | 0 |
| S11-final-viral-combined_k141_147642\|\|full | 1426 | No | NA | 3 | 0 | 0 | Not-determined | Genome-fragment | NA | NA | 0 |
| S12-final-viral-combined_k141_380417\|\|full | 1160 | No | NA | 2 | 0 | 0 | Low-quality | Genome-fragment | 1.75 | AAI-based (high-confidence) | 0 |
| S8-final-viral-combined_k141_52757\|\|full | 1160 | No | NA | 2 | 0 | 0 | Low-quality | Genome-fragment | 5.46 | AAI-based (high-confidence) | 0 |
| S11-final-viral-combined_k141_161427\|\|full | 8073 | No | NA | 12 | 0 | 3 | Not-determined | Genome-fragment | NA | NA | 0 |
| S8-final-viral-combined_k141_78336\|\|full | 1160 | No | NA | 4 | 1 | 0 | Low-quality | Genome-fragment | 2.14 | HMM-based (lower-bound) | 0 |
| S9_k141_9380 | 1161 | No | NA | 2 | 0 | 2 | Low-quality | Genome-fragment | 2.43 | AAI-based (medium-confidence) | 0 |
| S11_k141_311053 | 1161 | No | NA | 2 | 2 | 0 | Low-quality | Genome-fragment | 2.47 | AAI-based (high-confidence) | 0 |
| S11_k141_133361 | 1161 | No | NA | 3 | 0 | 0 | Low-quality | Genome-fragment | 2.95 | AAI-based (high-confidence) | 0 |
| S11-final-viral-combined_k141_132765\|\|full | 1155 | No | NA | 3 | 0 | 1 | Not-determined | Genome-fragment | NA | NA | 0 |
| S11-final-viral-combined_k141_64962\|\|full | 2617 | No | NA | 3 | 0 | 3 | Not-determined | Genome-fragment | NA | NA | 0 |
| S1-final-viral-combined_k141_24380\|\|full | 1161 | No | NA | 3 | 0 | 1 | Low-quality | Genome-fragment | 2.73 | AAI-based (medium-confidence) | 0 |
| S11-final-viral-combined_k141_298764\|\|full | 7041 | No | NA | 5 | 0 | 4 | Not-determined | Genome-fragment | NA | NA | 0 |
| S11-final-viral-combined_k141_175474\|\|full | 2364 | No | NA | 5 | 0 | 2 | Not-determined | Genome-fragment | NA | NA | 0 |
| S11-final-viral-combined_k141_258291\|\|full | 1361 | No | NA | 3 | 0 | 0 | Not-determined | Genome-fragment | NA | NA | 0 |
| S11-final-viral-combined_k141_106846\|\|full | 2976 | No | NA | 4 | 0 | 2 | Not-determined | Genome-fragment | NA | NA | 0 |
| S1_k141_291706 | 1161 | No | NA | 2 | 0 | 0 | Low-quality | Genome-fragment | 1.39 | AAI-based (medium-confidence) | 0 |
| S5-final-viral-combined_k141_2297\|\|full | 1161 | No | NA | 2 | 0 | 2 | Low-quality | Genome-fragment | 0.65 | AAI-based (medium-confidence) | 0 |
| S7-final-viral-combined_k141_50240\|\|full | 1161 | No | NA | 2 | 0 | 0 | Low-quality | Genome-fragment | 1.79 | AAI-based (medium-confidence) | 0 |
| S11-final-viral-combined_k141_78056\|\|full | 4060 | No | NA | 3 | 0 | 1 | Not-determined | Genome-fragment | NA | NA | 0 |
| S11-final-viral-combined_k141_64994\|\|full | 2752 | No | NA | 3 | 0 | 2 | Not-determined | Genome-fragment | NA | NA | 0 |
| S11-final-viral-combined_k141_161520\|\|full | 4379 | No | NA | 3 | 0 | 3 | Not-determined | Genome-fragment | NA | NA | 0 |
| S11-final-viral-combined_k141_202544\|\|full | 2510 | No | NA | 2 | 0 | 2 | Not-determined | Genome-fragment | NA | NA | 0 |
| S11-final-viral-combined_k141_65004\|\|full | 6394 | No | NA | 8 | 0 | 4 | Not-determined | Genome-fragment | NA | NA | 0 |
| S7-final-viral-combined_k141_160921\|\|lt2gene | 1161 | No | NA | 2 | 1 | 0 | Low-quality | Genome-fragment | 1.7 | HMM-based (lower-bound) | 0 |
| S11-final-viral-combined_k141_132826\|\|full | 1711 | No | NA | 3 | 0 | 2 | Not-determined | Genome-fragment | NA | NA | 0 |
| S17-final-viral-combined_k141_70231\|\|full | 1161 | No | NA | 2 | 0 | 1 | Low-quality | Genome-fragment | 2.52 | AAI-based (medium-confidence) | 0 |
| S11-final-viral-combined_k141_78101\|\|full | 1491 | No | NA | 2 | 0 | 2 | Not-determined | Genome-fragment | NA | NA | 0 |
| S11-final-viral-combined_k141_22350\|\|full | 1184 | No | NA | 2 | 0 | 0 | Not-determined | Genome-fragment | NA | NA | 0 |
| S11-final-viral-combined_k141_119495\|\|full | 1748 | No | NA | 3 | 0 | 1 | Not-determined | Genome-fragment | NA | NA | 0 |
| S11-final-viral-combined_k141_298795\|\|full | 1504 | No | NA | 3 | 0 | 2 | Not-determined | Genome-fragment | NA | NA | 0 |
| S17-final-viral-combined_k141_73393\|\|full | 1161 | No | NA | 3 | 1 | 0 | Low-quality | Genome-fragment | 2.42 | AAI-based (high-confidence) | 0 |
| S11-final-viral-combined_k141_132854\|\|full | 1045 | No | NA | 2 | 0 | 1 | Not-determined | Genome-fragment | NA | NA | 0 |
| S11-final-viral-combined_k141_216811\|\|full | 2117 | No | NA | 4 | 0 | 3 | Not-determined | Genome-fragment | NA | NA | 0 |
| S11-final-viral-combined_k141_37534\|\|full | 5111 | No | NA | 7 | 0 | 1 | Not-determined | Genome-fragment | NA | NA | 0 |
| S12-final-viral-combined_k141_265119\|\|full | 1161 | No | NA | 2 | 0 | 0 | Low-quality | Genome-fragment | 2.49 | AAI-based (high-confidence) | 0 |
| S11-final-viral-combined_k141_37546\|\|full | 1492 | No | NA | 3 | 0 | 1 | Not-determined | Genome-fragment | NA | NA | 0 |
| S11-final-viral-combined_k141_313437\|\|full | 1116 | No | NA | 3 | 0 | 2 | Not-determined | Genome-fragment | NA | NA | 0 |
| S12-final-viral-combined_k141_13613\|\|lt2gene | 1161 | No | NA | 1 | 1 | 0 | Low-quality | Genome-fragment | 2.61 | AAI-based (high-confidence) | 0 |
| S2-final-viral-combined_k141_229361\|\|full | 1161 | No | NA | 2 | 0 | 0 | Low-quality | Genome-fragment | 0.64 | AAI-based (high-confidence) | 0 |
| S2-final-viral-combined_k141_189009\|\|full | 1161 | No | NA | 2 | 0 | 1 | Low-quality | Genome-fragment | 0.71 | AAI-based (medium-confidence) | 0 |
| S11-final-viral-combined_k141_132895\|\|full | 10610 | No | NA | 8 | 0 | 7 | Not-determined | Genome-fragment | NA | NA | 0 |
| S8-final-viral-combined_k141_109685\|\|full | 1161 | No | NA | 2 | 0 | 0 | Low-quality | Genome-fragment | 2.8 | AAI-based (high-confidence) | 0 |
| S8_k141_258310 | 1161 | No | NA | 2 | 0 | 0 | Low-quality | Genome-fragment | 2.63 | AAI-based (medium-confidence) | 0 |
| S9-final-viral-combined_k141_56932\|\|full | 1162 | No | NA | 3 | 0 | 0 | Low-quality | Genome-fragment | 0.81 | AAI-based (medium-confidence) | 0 |
| S3-final-viral-combined_k141_37267\|\|full | 1162 | No | NA | 3 | 0 | 1 | Low-quality | Genome-fragment | 0.42 | AAI-based (medium-confidence) | 0 |
| S5-final-viral-combined_k141_65518\|\|full | 1162 | No | NA | 2 | 0 | 0 | Low-quality | Genome-fragment | 1.93 | AAI-based (medium-confidence) | 0 |
| S11-final-viral-combined_k141_78177\|\|full | 10409 | No | NA | 6 | 0 | 5 | Not-determined | Genome-fragment | NA | NA | 0 |
| S11-final-viral-combined_k141_92581\|\|full | 1269 | No | NA | 3 | 0 | 1 | Not-determined | Genome-fragment | NA | NA | 0 |
| S11-final-viral-combined_k141_22448\|\|full | 1406 | No | NA | 2 | 0 | 2 | Not-determined | Genome-fragment | NA | NA | 0 |
| S11-final-viral-combined_k141_244878\|\|full | 1148 | No | NA | 3 | 0 | 1 | Not-determined | Genome-fragment | NA | NA | 0 |
| S11-final-viral-combined_k141_325689\|\|full | 1791 | No | NA | 3 | 0 | 1 | Not-determined | Genome-fragment | NA | NA | 0 |
| S17-final-viral-combined_k141_6437\|\|full | 1162 | No | NA | 2 | 0 | 1 | Low-quality | Genome-fragment | 0.93 | AAI-based (high-confidence) | 0 |
| S11-final-viral-combined_k141_244885\|\|full | 2818 | No | NA | 3 | 0 | 2 | Not-determined | Genome-fragment | NA | NA | 0 |
| S11-final-viral-combined_k141_78203\|\|full | 12085 | No | NA | 14 | 0 | 10 | Not-determined | Genome-fragment | NA | NA | 0 |
| S12-final-viral-combined_k141_82658\|\|full | 1162 | No | NA | 3 | 0 | 1 | Low-quality | Genome-fragment | 0.54 | AAI-based (high-confidence) | 0 |
| S11-final-viral-combined_k141_92612\|\|full | 2229 | No | NA | 3 | 0 | 0 | Not-determined | Genome-fragment | NA | NA | 0 |
| S11-final-viral-combined_k141_106993\|\|full | 3344 | No | NA | 4 | 0 | 0 | Not-determined | Genome-fragment | NA | NA | 0 |
| S2_k141_192361 | 1162 | No | NA | 3 | 0 | 0 | Low-quality | Genome-fragment | 2.12 | AAI-based (medium-confidence) | 0 |
| S11-final-viral-combined_k141_271415\|\|full | 1428 | No | NA | 3 | 0 | 2 | Not-determined | Genome-fragment | NA | NA | 0 |
| S4_k141_107021 | 1162 | No | NA | 3 | 0 | 1 | Low-quality | Genome-fragment | 1.71 | AAI-based (medium-confidence) | 0 |
| S11-final-viral-combined_k141_298862\|\|full | 2146 | No | NA | 3 | 0 | 1 | Not-determined | Genome-fragment | NA | NA | 0 |
| S11-final-viral-combined_k141_313520\|\|full | 3493 | No | NA | 3 | 0 | 2 | Not-determined | Genome-fragment | NA | NA | 0 |
| S11-final-viral-combined_k141_188652\|\|full | 1744 | No | NA | 3 | 0 | 1 | Not-determined | Genome-fragment | NA | NA | 0 |
| S11-final-viral-combined_k141_10225\|\|full | 2503 | No | NA | 3 | 0 | 0 | Not-determined | Genome-fragment | NA | NA | 0 |
| S8-final-viral-combined_k141_55024\|\|full | 1162 | No | NA | 4 | 0 | 0 | Low-quality | Genome-fragment | 0.74 | AAI-based (high-confidence) | 0 |
| S8_k141_85761 | 1162 | No | NA | 2 | 0 | 0 | Low-quality | Genome-fragment | 1.56 | AAI-based (high-confidence) | 0 |
| S1-final-viral-combined_k141_104591\|\|lt2gene | 1163 | No | NA | 2 | 0 | 1 | Low-quality | Genome-fragment | 6.93 | AAI-based (medium-confidence) | 0 |
| S5-final-viral-combined_k141_175117\|\|full | 1163 | No | NA | 3 | 2 | 0 | Low-quality | Genome-fragment | 2.39 | AAI-based (medium-confidence) | 0 |
| S5_k141_33909 | 1163 | No | NA | 2 | 0 | 0 | Low-quality | Genome-fragment | 0.47 | AAI-based (medium-confidence) | 0 |
| S7-final-viral-combined_k141_72393\|\|full | 1163 | No | NA | 3 | 1 | 0 | Low-quality | Genome-fragment | 3.09 | AAI-based (medium-confidence) | 0 |
| S11-final-viral-combined_k141_202748\|\|full | 2379 | No | NA | 3 | 0 | 1 | Not-determined | Genome-fragment | NA | NA | 0 |
| S12-final-viral-combined_k141_309396\|\|full | 1163 | No | NA | 2 | 0 | 0 | Low-quality | Genome-fragment | 2.48 | AAI-based (high-confidence) | 0 |
| S6-final-viral-combined_k141_132470\|\|full | 1163 | No | NA | 3 | 0 | 1 | Low-quality | Genome-fragment | 0.54 | AAI-based (medium-confidence) | 0 |
| S11-final-viral-combined_k141_313553\|\|full | 4095 | No | NA | 5 | 0 | 3 | Not-determined | Genome-fragment | NA | NA | 0 |
| S11-final-viral-combined_k141_216898\|\|full | 1209 | No | NA | 3 | 0 | 0 | Not-determined | Genome-fragment | NA | NA | 0 |
| S6_k141_19090 | 1163 | No | NA | 2 | 1 | 0 | Low-quality | Genome-fragment | 3.56 | AAI-based (high-confidence) | 0 |
| S11-final-viral-combined_k141_161719\|\|full | 2818 | No | NA | 4 | 0 | 2 | Not-determined | Genome-fragment | NA | NA | 0 |
| S6_k141_21461 | 1163 | No | NA | 1 | 0 | 1 | Low-quality | Genome-fragment | 3.86 | AAI-based (medium-confidence) | 0 |
| S9-final-viral-combined_k141_57841\|\|full | 1164 | No | NA | 2 | 1 | 0 | Low-quality | Genome-fragment | 2.08 | AAI-based (medium-confidence) | 0 |
| S11-final-viral-combined_k141_170146\|\|full | 1164 | No | NA | 3 | 0 | 0 | Low-quality | Genome-fragment | 1.78 | AAI-based (high-confidence) | 0 |
| S1-final-viral-combined_k141_102501\|\|full | 1164 | No | NA | 2 | 0 | 1 | Low-quality | Genome-fragment | 1.42 | AAI-based (medium-confidence) | 0 |
| S11-final-viral-combined_k141_202796\|\|full | 2395 | No | NA | 2 | 0 | 1 | Not-determined | Genome-fragment | NA | NA | 0 |
| S1-final-viral-combined_k141_27965\|\|full | 1164 | No | NA | 3 | 0 | 0 | Low-quality | Genome-fragment | 0.8 | AAI-based (medium-confidence) | 0 |
| S11-final-viral-combined_k141_107078\|\|full | 3355 | No | NA | 3 | 0 | 1 | Not-determined | Genome-fragment | NA | NA | 0 |
| S11-final-viral-combined_k141_147914\|\|full | 4338 | No | NA | 3 | 0 | 2 | Not-determined | Genome-fragment | NA | NA | 0 |
| S11-final-viral-combined_k141_65264\|\|full | 2830 | No | NA | 3 | 0 | 1 | Not-determined | Genome-fragment | NA | NA | 0 |
| S1_k141_292778 | 1164 | No | NA | 3 | 0 | 0 | Low-quality | Genome-fragment | 2.43 | AAI-based (high-confidence) | 0 |
| S11-final-viral-combined_k141_230669\|\|full | 2712 | No | NA | 3 | 0 | 2 | Not-determined | Genome-fragment | NA | NA | 0 |
| S5-final-viral-combined_k141_35864\|\|lt2gene | 1164 | No | NA | 2 | 1 | 0 | Low-quality | Genome-fragment | 2.61 | AAI-based (high-confidence) | 0 |
| S5_k141_110042 | 1164 | No | NA | 2 | 0 | 1 | Low-quality | Genome-fragment | 0.74 | AAI-based (high-confidence) | 0 |
| S6-final-viral-combined_k141_66055\|\|full | 1164 | No | NA | 2 | 2 | 0 | Low-quality | Genome-fragment | 1.95 | AAI-based (high-confidence) | 0 |
| S11-final-viral-combined_k141_328530\|\|full | 1165 | No | NA | 2 | 2 | 0 | Low-quality | Genome-fragment | 3.39 | AAI-based (high-confidence) | 0 |
| S11-final-viral-combined_k141_78354\|\|full | 2297 | No | NA | 3 | 0 | 2 | Not-determined | Genome-fragment | NA | NA | 0 |
| S11-final-viral-combined_k141_258156\|\|lt2gene | 1165 | No | NA | 2 | 1 | 0 | Low-quality | Genome-fragment | 2.53 | AAI-based (high-confidence) | 0 |
| S11-final-viral-combined_k141_325817\|\|full | 2866 | No | NA | 4 | 0 | 0 | Not-determined | Genome-fragment | NA | NA | 0 |
| S1-final-viral-combined_k141_162643\|\|full | 1165 | No | NA | 3 | 0 | 0 | Low-quality | Genome-fragment | 2.49 | AAI-based (medium-confidence) | 0 |
| S11-final-viral-combined_k141_51768\|\|full | 7033 | No | NA | 7 | 0 | 3 | Not-determined | Genome-fragment | NA | NA | 0 |
| S11-final-viral-combined_k141_325821\|\|full | 3108 | No | NA | 2 | 0 | 2 | Not-determined | Genome-fragment | NA | NA | 0 |
| S11-final-viral-combined_k141_78380\|\|full | 1014 | No | NA | 2 | 0 | 0 | Not-determined | Genome-fragment | NA | NA | 0 |
| S11-final-viral-combined_k141_202859\|\|full | 2400 | No | NA | 2 | 0 | 0 | Not-determined | Genome-fragment | NA | NA | 0 |
| S5-final-viral-combined_k141_93876\|\|full | 1165 | No | NA | 3 | 1 | 0 | Low-quality | Genome-fragment | 2.1 | AAI-based (high-confidence) | 0 |
| S11-final-viral-combined_k141_188809\|\|full | 4400 | No | NA | 5 | 0 | 1 | Not-determined | Genome-fragment | NA | NA | 0 |
| S11-final-viral-combined_k141_147972\|\|full | 1574 | No | NA | 2 | 0 | 1 | Not-determined | Genome-fragment | NA | NA | 0 |
| S11-final-viral-combined_k141_51791\|\|full | 2345 | No | NA | 4 | 0 | 0 | Not-determined | Genome-fragment | NA | NA | 0 |
| S11-final-viral-combined_k141_299014\|\|full | 2029 | No | NA | 2 | 0 | 1 | Not-determined | Genome-fragment | NA | NA | 0 |
| S7_k141_32133 | 1165 | No | NA | 2 | 0 | 0 | Low-quality | Genome-fragment | 2.58 | AAI-based (high-confidence) | 0 |
| S17-final-viral-combined_k141_248504\|\|full | 1165 | No | NA | 2 | 0 | 2 | Low-quality | Genome-fragment | 0.48 | AAI-based (medium-confidence) | 0 |
| S11-final-viral-combined_k141_299036\|\|full | 2012 | No | NA | 3 | 0 | 3 | Not-determined | Genome-fragment | NA | NA | 0 |
| S11-final-viral-combined_k141_37809\|\|full | 1084 | No | NA | 3 | 0 | 0 | Not-determined | Genome-fragment | NA | NA | 0 |
| S17-final-viral-combined_k141_180515\|\|lt2gene | 1165 | No | NA | 2 | 1 | 0 | Low-quality | Genome-fragment | 2.45 | AAI-based (high-confidence) | 0 |
| S12_k141_5436 | 1165 | No | NA | 2 | 0 | 0 | Low-quality | Genome-fragment | 1.24 | AAI-based (high-confidence) | 0 |
| S2-final-viral-combined_k141_219676\|\|full | 1165 | No | NA | 4 | 1 | 0 | Low-quality | Genome-fragment | 2.81 | AAI-based (high-confidence) | 0 |
| S11-final-viral-combined_k141_78453\|\|full | 2591 | No | NA | 3 | 0 | 0 | Not-determined | Genome-fragment | NA | NA | 0 |
| S11-final-viral-combined_k141_325889\|\|full | 6911 | No | NA | 8 | 0 | 3 | Not-determined | Genome-fragment | NA | NA | 0 |
| S11-final-viral-combined_k141_202958\|\|full | 2142 | No | NA | 2 | 0 | 2 | Not-determined | Genome-fragment | NA | NA | 0 |
| S2-final-viral-combined_k141_107128\|\|full | 1165 | No | NA | 2 | 0 | 0 | Low-quality | Genome-fragment | 0.69 | AAI-based (medium-confidence) | 0 |
| S2_k141_122135 | 1165 | No | NA | 1 | 0 | 1 | Low-quality | Genome-fragment | 2.41 | AAI-based (medium-confidence) | 0 |
| S11-final-viral-combined_k141_161836\|\|full | 2527 | No | NA | 4 | 0 | 2 | Not-determined | Genome-fragment | NA | NA | 0 |
| S6-final-viral-combined_k141_92592\|\|full | 1165 | No | NA | 3 | 0 | 0 | Low-quality | Genome-fragment | 1.47 | AAI-based (high-confidence) | 0 |
| S8-final-viral-combined_k141_222829\|\|full | 1165 | No | NA | 3 | 2 | 0 | Low-quality | Genome-fragment | 1.25 | AAI-based (high-confidence) | 0 |
| S11-final-viral-combined_k141_107223\|\|full | 2402 | No | NA | 3 | 0 | 1 | Not-determined | Genome-fragment | NA | NA | 0 |
| S9-final-viral-combined_k141_63461\|\|full | 1166 | No | NA | 3 | 1 | 0 | Low-quality | Genome-fragment | 2.13 | HMM-based (lower-bound) | 0 |
| S11-final-viral-combined_k141_313750\|\|full | 2903 | No | NA | 6 | 0 | 2 | Not-determined | Genome-fragment | NA | NA | 0 |
| S9-final-viral-combined_k141_55045\|\|lt2gene | 1166 | No | NA | 1 | 1 | 0 | Low-quality | Genome-fragment | 2.87 | AAI-based (high-confidence) | 0 |
| S11-final-viral-combined_k141_51877\|\|full | 5668 | No | NA | 4 | 0 | 4 | Not-determined | Genome-fragment | NA | NA | 0 |
| S11-final-viral-combined_k141_114811\|\|full | 1166 | No | NA | 3 | 0 | 1 | Low-quality | Genome-fragment | 3.3 | AAI-based (medium-confidence) | 0 |
| S11-final-viral-combined_k141_51893\|\|full | 1132 | No | NA | 4 | 0 | 0 | Not-determined | Genome-fragment | NA | NA | 0 |
| S1-final-viral-combined_k141_132131\|\|full | 1166 | No | NA | 4 | 2 | 0 | Low-quality | Genome-fragment | 2.77 | AAI-based (high-confidence) | 0 |
| S11-final-viral-combined_k141_107248\|\|full | 2085 | No | NA | 3 | 0 | 1 | Not-determined | Genome-fragment | NA | NA | 0 |
| S11-final-viral-combined_k141_230836\|\|full | 1440 | No | NA | 5 | 0 | 0 | Not-determined | Genome-fragment | NA | NA | 0 |
| S1-final-viral-combined_k141_227209\|\|full | 1166 | No | NA | 2 | 0 | 2 | Low-quality | Genome-fragment | 0.37 | AAI-based (medium-confidence) | 0 |
| S1-final-viral-combined_k141_98472\|\|full | 1166 | No | NA | 2 | 0 | 0 | Low-quality | Genome-fragment | 0.69 | AAI-based (high-confidence) | 0 |
| S5-final-viral-combined_k141_8651\|\|full | 1166 | No | NA | 4 | 1 | 0 | Low-quality | Genome-fragment | 1.86 | AAI-based (high-confidence) | 0 |
| S5_k141_83186 | 1166 | No | NA | 2 | 0 | 0 | Low-quality | Genome-fragment | 2.37 | AAI-based (high-confidence) | 0 |
| S9-final-viral-combined_k141_82483\|\|full | 1167 | No | NA | 2 | 0 | 1 | Low-quality | Genome-fragment | 0.65 | AAI-based (high-confidence) | 0 |
| S11_k141_151010 | 1167 | No | NA | 2 | 0 | 0 | Low-quality | Genome-fragment | 2.15 | AAI-based (high-confidence) | 0 |
| S1-final-viral-combined_k141_61896\|\|full | 1167 | No | NA | 3 | 0 | 0 | Low-quality | Genome-fragment | 1.9 | AAI-based (high-confidence) | 0 |
| S1_k141_70609 | 1167 | No | NA | 3 | 1 | 0 | Low-quality | Genome-fragment | 1.92 | AAI-based (high-confidence) | 0 |
| S11-final-viral-combined_k141_107280\|\|full | 1312 | No | NA | 2 | 0 | 0 | Not-determined | Genome-fragment | NA | NA | 0 |
| S5-final-viral-combined_k141_80306\|\|full | 1167 | No | NA | 3 | 1 | 0 | Low-quality | Genome-fragment | 1.67 | AAI-based (high-confidence) | 0 |
| S11-final-viral-combined_k141_133201\|\|full | 3030 | No | NA | 3 | 0 | 2 | Not-determined | Genome-fragment | NA | NA | 0 |
| S11-final-viral-combined_k141_161935\|\|full | 1045 | No | NA | 3 | 0 | 0 | Not-determined | Genome-fragment | NA | NA | 0 |
| S17-final-viral-combined_k141_206937\|\|lt2gene | 1167 | No | NA | 1 | 1 | 0 | Low-quality | Genome-fragment | 3.48 | AAI-based (medium-confidence) | 0 |
| S11-final-viral-combined_k141_51946\|\|full | 9480 | No | NA | 9 | 0 | 4 | Not-determined | Genome-fragment | NA | NA | 0 |
| S11-final-viral-combined_k141_188983\|\|full | 1518 | No | NA | 2 | 0 | 0 | Not-determined | Genome-fragment | NA | NA | 0 |
| S12-final-viral-combined_k141_88246\|\|full | 1167 | No | NA | 2 | 0 | 2 | Low-quality | Genome-fragment | 0.68 | AAI-based (medium-confidence) | 0 |
| S11-final-viral-combined_k141_313814\|\|full | 1343 | No | NA | 2 | 0 | 0 | Not-determined | Genome-fragment | NA | NA | 0 |
| S12-final-viral-combined_k141_343602\|\|full | 1167 | No | NA | 3 | 1 | 0 | Low-quality | Genome-fragment | 1.53 | HMM-based (lower-bound) | 0 |
| S11-final-viral-combined_k141_92934\|\|full | 1815 | No | NA | 4 | 0 | 0 | Not-determined | Genome-fragment | NA | NA | 0 |
| S11-final-viral-combined_k141_299255\|\|full | 1297 | No | NA | 3 | 0 | 2 | Not-determined | Genome-fragment | NA | NA | 0 |
| S12-final-viral-combined_k141_345117\|\|full | 1167 | No | NA | 3 | 0 | 2 | Low-quality | Genome-fragment | 0.06 | AAI-based (medium-confidence) | 0 |
| S8-final-viral-combined_k141_210\|\|full | 1167 | No | NA | 3 | 0 | 0 | Low-quality | Genome-fragment | 2.45 | AAI-based (medium-confidence) | 0 |
| S9-final-viral-combined_k141_33556\|\|lt2gene | 1168 | No | NA | 2 | 1 | 0 | Low-quality | Genome-fragment | 0.75 | HMM-based (lower-bound) | 0 |
| S11-final-viral-combined_k141_319394\|\|full | 1168 | No | NA | 3 | 1 | 0 | Low-quality | Genome-fragment | 2.57 | AAI-based (high-confidence) | 0 |
| S11-final-viral-combined_k141_299292\|\|full | 1978 | No | NA | 3 | 0 | 1 | Not-determined | Genome-fragment | NA | NA | 0 |
| S11-final-viral-combined_k141_119944\|\|full | 3663 | No | NA | 4 | 0 | 2 | Not-determined | Genome-fragment | NA | NA | 0 |
| S11-final-viral-combined_k141_258785\|\|full | 2268 | No | NA | 3 | 0 | 1 | Not-determined | Genome-fragment | NA | NA | 0 |
| S11-final-viral-combined_k141_90103\|\|full | 1168 | No | NA | 3 | 0 | 0 | Low-quality | Genome-fragment | 2.01 | AAI-based (medium-confidence) | 0 |
| S11-final-viral-combined_k141_286138\|\|full | 2337 | No | NA | 3 | 0 | 1 | Not-determined | Genome-fragment | NA | NA | 0 |
| S11-final-viral-combined_k141_299304\|\|full | 2123 | No | NA | 3 | 0 | 0 | Not-determined | Genome-fragment | NA | NA | 0 |
| S1-final-viral-combined_k141_111992\|\|full | 1168 | No | NA | 2 | 0 | 1 | Low-quality | Genome-fragment | 0.71 | AAI-based (medium-confidence) | 0 |
| S1_k141_376664 | 1168 | No | NA | 3 | 1 | 0 | Low-quality | Genome-fragment | 0.59 | HMM-based (lower-bound) | 0 |
| S5-final-viral-combined_k141_206022\|\|full | 1168 | No | NA | 3 | 1 | 0 | Low-quality | Genome-fragment | 2 | AAI-based (high-confidence) | 0 |
| S5-final-viral-combined_k141_7867\|\|full | 1168 | No | NA | 3 | 1 | 0 | Low-quality | Genome-fragment | 2.45 | AAI-based (medium-confidence) | 0 |
| S11-final-viral-combined_k141_22874\|\|full | 4029 | No | NA | 3 | 0 | 2 | Not-determined | Genome-fragment | NA | NA | 0 |
| S5-final-viral-combined_k141_94860\|\|full | 1168 | No | NA | 2 | 0 | 1 | Low-quality | Genome-fragment | 1.59 | AAI-based (high-confidence) | 0 |
| S5-final-viral-combined_k141_137060\|\|lt2gene | 1168 | No | NA | 2 | 2 | 0 | Low-quality | Genome-fragment | 2.77 | AAI-based (medium-confidence) | 0 |
| S17-final-viral-combined_k141_78491\|\|full | 1168 | Yes | 769 | 3 | 2 | 1 | Low-quality | Genome-fragment | 1.59 | AAI-based (medium-confidence) | 34.16 |
| S17-final-viral-combined_k141_51250\|\|full | 1168 | No | NA | 4 | 1 | 0 | Low-quality | Genome-fragment | 2.83 | AAI-based (medium-confidence) | 0 |
| S11-final-viral-combined_k141_271772\|\|full | 4643 | No | NA | 4 | 0 | 4 | Not-determined | Genome-fragment | NA | NA | 0 |
| S11-final-viral-combined_k141_22908\|\|full | 1571 | No | NA | 2 | 0 | 2 | Not-determined | Genome-fragment | NA | NA | 0 |
| S11-final-viral-combined_k141_78633\|\|full | 2775 | No | NA | 3 | 0 | 3 | Not-determined | Genome-fragment | NA | NA | 0 |
| S11-final-viral-combined_k141_258844\|\|full | 5543 | No | NA | 5 | 0 | 3 | Not-determined | Genome-fragment | NA | NA | 0 |
| S11-final-viral-combined_k141_107400\|\|full | 3653 | No | NA | 3 | 0 | 2 | Not-determined | Genome-fragment | NA | NA | 0 |
| S11-final-viral-combined_k141_245309\|\|full | 2514 | No | NA | 3 | 0 | 2 | Not-determined | Genome-fragment | NA | NA | 0 |
| S11-final-viral-combined_k141_148191\|\|full | 6996 | No | NA | 5 | 0 | 3 | Not-determined | Genome-fragment | NA | NA | 0 |
| S11-final-viral-combined_k141_120030\|\|full | 2932 | No | NA | 3 | 0 | 0 | Not-determined | Genome-fragment | NA | NA | 0 |
| S11-final-viral-combined_k141_38161\|\|full | 3140 | No | NA | 2 | 0 | 2 | Not-determined | Genome-fragment | NA | NA | 0 |
| S11-final-viral-combined_k141_286234\|\|full | 5886 | No | NA | 6 | 0 | 4 | Not-determined | Genome-fragment | NA | NA | 0 |
| S11-final-viral-combined_k141_107461\|\|full | 2948 | No | NA | 4 | 0 | 4 | Not-determined | Genome-fragment | NA | NA | 0 |
| S11-final-viral-combined_k141_52134\|\|full | 2643 | No | NA | 4 | 0 | 0 | Not-determined | Genome-fragment | NA | NA | 0 |
| S11-final-viral-combined_k141_10605\|\|full | 1344 | No | NA | 3 | 0 | 1 | Not-determined | Genome-fragment | NA | NA | 0 |
| S17-k141_222528 | 1168 | No | NA | 2 | 0 | 0 | Low-quality | Genome-fragment | 0.72 | AAI-based (medium-confidence) | 0 |
| S12-final-viral-combined_k141_331766\|\|full | 1168 | No | NA | 2 | 0 | 1 | Low-quality | Genome-fragment | 0.37 | AAI-based (medium-confidence) | 0 |
| S11-final-viral-combined_k141_22952\|\|full | 1505 | No | NA | 3 | 0 | 1 | Not-determined | Genome-fragment | NA | NA | 0 |
| S2-final-viral-combined_k141_40290\|\|full | 1168 | No | NA | 2 | 0 | 0 | Low-quality | Genome-fragment | 2.54 | AAI-based (medium-confidence) | 0 |
| S11-final-viral-combined_k141_133385\|\|full | 1385 | No | NA | 2 | 0 | 1 | Not-determined | Genome-fragment | NA | NA | 0 |
| S11-final-viral-combined_k141_231071\|\|full | 1529 | No | NA | 3 | 0 | 0 | Not-determined | Genome-fragment | NA | NA | 0 |
| S11-final-viral-combined_k141_52162\|\|full | 1155 | No | NA | 2 | 0 | 1 | Not-determined | Genome-fragment | NA | NA | 0 |
| S11-final-viral-combined_k141_133410\|\|full | 3031 | No | NA | 2 | 0 | 1 | Not-determined | Genome-fragment | NA | NA | 0 |
| S11-final-viral-combined_k141_38224\|\|full | 2374 | No | NA | 5 | 0 | 3 | Not-determined | Genome-fragment | NA | NA | 0 |
| S2_k141_148825 | 1168 | No | NA | 1 | 0 | 0 | Low-quality | Genome-fragment | 2.73 | AAI-based (high-confidence) | 0 |
| S11-final-viral-combined_k141_217300\|\|full | 2111 | No | NA | 4 | 0 | 2 | Not-determined | Genome-fragment | NA | NA | 0 |
| S11-final-viral-combined_k141_52209\|\|full | 2261 | No | NA | 3 | 0 | 2 | Not-determined | Genome-fragment | NA | NA | 0 |
| S4_k141_166750 | 1168 | No | NA | 3 | 2 | 0 | Low-quality | Genome-fragment | 0.67 | AAI-based (high-confidence) | 0 |
| S11-final-viral-combined_k141_258961\|\|full | 1665 | No | NA | 2 | 0 | 1 | Not-determined | Genome-fragment | NA | NA | 0 |
| S11-final-viral-combined_k141_23003\|\|full | 2179 | No | NA | 3 | 0 | 1 | Not-determined | Genome-fragment | NA | NA | 0 |
| S8-final-viral-combined_k141_184719\|\|lt2gene | 1168 | No | NA | 2 | 1 | 0 | Low-quality | Genome-fragment | 2.14 | HMM-based (lower-bound) | 0 |
| S11-final-viral-combined_k141_299519\|\|full | 1423 | No | NA | 3 | 0 | 2 | Not-determined | Genome-fragment | NA | NA | 0 |
| S11-final-viral-combined_k141_258998\|\|full | 2696 | No | NA | 2 | 0 | 2 | Not-determined | Genome-fragment | NA | NA | 0 |
| S11-final-viral-combined_k141_52256\|\|full | 1178 | No | NA | 3 | 0 | 0 | Not-determined | Genome-fragment | NA | NA | 0 |
| S11-final-viral-combined_k141_52257\|\|full | 4068 | No | NA | 6 | 0 | 2 | Not-determined | Genome-fragment | NA | NA | 0 |
| S11-final-viral-combined_k141_65801\|\|full | 1230 | No | NA | 3 | 0 | 0 | Not-determined | Genome-fragment | NA | NA | 0 |
| S8-final-viral-combined_k141_241257\|\|lt2gene | 1168 | No | NA | 1 | 1 | 0 | Low-quality | Genome-fragment | 1.1 | HMM-based (lower-bound) | 0 |
| S11-final-viral-combined_k141_23019\|\|full | 1345 | No | NA | 3 | 0 | 0 | Not-determined | Genome-fragment | NA | NA | 0 |
| S11-final-viral-combined_k141_93196\|\|full | 1640 | No | NA | 2 | 0 | 1 | Not-determined | Genome-fragment | NA | NA | 0 |
| S9-final-viral-combined_k141_104791\|\|full | 1169 | No | NA | 3 | 1 | 0 | Low-quality | Genome-fragment | 2.79 | AAI-based (high-confidence) | 0 |
| S11-final-viral-combined_k141_65829\|\|full | 10638 | No | NA | 10 | 0 | 8 | Not-determined | Genome-fragment | NA | NA | 0 |
| S11-final-viral-combined_k141_326164\|\|full | 2250 | No | NA | 3 | 0 | 2 | Not-determined | Genome-fragment | NA | NA | 0 |
| S9-final-viral-combined_k141_21289\|\|full | 1169 | No | NA | 3 | 1 | 0 | Low-quality | Genome-fragment | 2.54 | AAI-based (high-confidence) | 0 |
| S11-final-viral-combined_k141_38308\|\|full | 1488 | No | NA | 2 | 0 | 0 | Not-determined | Genome-fragment | NA | NA | 0 |
| S11-final-viral-combined_k141_133539\|\|full | 1058 | No | NA | 3 | 0 | 1 | Not-determined | Genome-fragment | NA | NA | 0 |
| S11_k141_24683 | 1169 | No | NA | 3 | 2 | 0 | Low-quality | Genome-fragment | 2.06 | AAI-based (high-confidence) | 0 |
| S1-final-viral-combined_k141_205908\|\|full | 1169 | No | NA | 3 | 3 | 0 | Low-quality | Genome-fragment | 2.49 | AAI-based (high-confidence) | 0 |
| S1-final-viral-combined_k141_371924\|\|full | 1169 | No | NA | 3 | 0 | 0 | Low-quality | Genome-fragment | 1.58 | AAI-based (medium-confidence) | 0 |
| S11-final-viral-combined_k141_286378\|\|full | 2079 | No | NA | 3 | 0 | 2 | Not-determined | Genome-fragment | NA | NA | 0 |
| S11-final-viral-combined_k141_148345\|\|full | 3428 | No | NA | 5 | 0 | 2 | Not-determined | Genome-fragment | NA | NA | 0 |
| S11-final-viral-combined_k141_326184\|\|full | 1763 | No | NA | 5 | 0 | 1 | Not-determined | Genome-fragment | NA | NA | 0 |
| S11-final-viral-combined_k141_78790\|\|full | 1032 | No | NA | 3 | 0 | 1 | Not-determined | Genome-fragment | NA | NA | 0 |
| S11-final-viral-combined_k141_93262\|\|full | 2691 | No | NA | 3 | 0 | 2 | Not-determined | Genome-fragment | NA | NA | 0 |
| S11-final-viral-combined_k141_148351\|\|full | 1415 | No | NA | 2 | 0 | 1 | Not-determined | Genome-fragment | NA | NA | 0 |
| S1-final-viral-combined_k141_78730\|\|lt2gene | 1169 | No | NA | 2 | 1 | 0 | Low-quality | Genome-fragment | 2.05 | HMM-based (lower-bound) | 0 |
| S11-final-viral-combined_k141_176210\|\|full | 12254 | No | NA | 11 | 0 | 10 | Not-determined | Genome-fragment | NA | NA | 0 |
| S11-final-viral-combined_k141_162328\|\|full | 2803 | No | NA | 2 | 0 | 2 | Not-determined | Genome-fragment | NA | NA | 0 |
| S5-final-viral-combined_k141_189616\|\|full | 1169 | No | NA | 2 | 0 | 2 | Low-quality | Genome-fragment | 0.53 | AAI-based (high-confidence) | 0 |
| S7-final-viral-combined_k141_226015\|\|full | 1169 | No | NA | 3 | 0 | 0 | Low-quality | Genome-fragment | 2.97 | AAI-based (high-confidence) | 0 |
| S11-final-viral-combined_k141_93325\|\|full | 3002 | No | NA | 5 | 0 | 4 | Not-determined | Genome-fragment | NA | NA | 0 |
| S11-final-viral-combined_k141_148412\|\|full | 3186 | No | NA | 5 | 0 | 2 | Not-determined | Genome-fragment | NA | NA | 0 |
| S11-final-viral-combined_k141_245608\|\|full | 3718 | No | NA | 4 | 0 | 3 | Not-determined | Genome-fragment | NA | NA | 0 |
| S11-final-viral-combined_k141_326255\|\|full | 2117 | No | NA | 3 | 0 | 0 | Not-determined | Genome-fragment | NA | NA | 0 |
| S17-final-viral-combined_k141_185368\|\|full | 1169 | No | NA | 3 | 0 | 1 | Low-quality | Genome-fragment | 2.55 | AAI-based (medium-confidence) | 0 |
| S11-final-viral-combined_k141_299603\|\|full | 1820 | No | NA | 4 | 0 | 0 | Not-determined | Genome-fragment | NA | NA | 0 |
| S2-final-viral-combined_k141_88013\|\|full | 1169 | No | NA | 2 | 0 | 1 | Low-quality | Genome-fragment | 0.6 | AAI-based (medium-confidence) | 0 |
| S2-final-viral-combined_k141_195413\|\|lt2gene | 1169 | No | NA | 1 | 1 | 0 | Low-quality | Genome-fragment | 2.58 | AAI-based (high-confidence) | 0 |
| S2_k141_125372 | 1169 | No | NA | 3 | 0 | 3 | Low-quality | Genome-fragment | 3.05 | AAI-based (high-confidence) | 0 |
| S2_k141_234573 | 1169 | No | NA | 3 | 0 | 3 | Low-quality | Genome-fragment | 1 | AAI-based (medium-confidence) | 0 |
| S11-final-viral-combined_k141_314054\|\|full | 1078 | No | NA | 2 | 0 | 1 | Not-determined | Genome-fragment | NA | NA | 0 |
| S4_k141_70074 | 1169 | No | NA | 2 | 0 | 0 | Low-quality | Genome-fragment | 1.64 | AAI-based (medium-confidence) | 0 |
| S11-final-viral-combined_k141_65983\|\|full | 1258 | No | NA | 3 | 0 | 1 | Not-determined | Genome-fragment | NA | NA | 0 |
| S11-final-viral-combined_k141_299635\|\|full | 9977 | No | NA | 12 | 0 | 2 | Not-determined | Genome-fragment | NA | NA | 0 |
| S11-final-viral-combined_k141_314066\|\|full | 3355 | No | NA | 3 | 0 | 1 | Not-determined | Genome-fragment | NA | NA | 0 |
| S8_k141_147792 | 1169 | No | NA | 1 | 0 | 0 | Low-quality | Genome-fragment | 1.88 | AAI-based (medium-confidence) | 0 |
| S11-final-viral-combined_k141_93388\|\|full | 4997 | No | NA | 4 | 0 | 3 | Not-determined | Genome-fragment | NA | NA | 0 |
| S8_k141_57259 | 1169 | No | NA | 2 | 0 | 0 | Low-quality | Genome-fragment | 2.25 | AAI-based (high-confidence) | 0 |
| S9-final-viral-combined_k141_25771\|\|full | 1170 | No | NA | 2 | 0 | 2 | Low-quality | Genome-fragment | 1.96 | AAI-based (medium-confidence) | 0 |
| S9-final-viral-combined_k141_46702\|\|full | 1170 | No | NA | 2 | 0 | 0 | Low-quality | Genome-fragment | 0.8 | AAI-based (high-confidence) | 0 |
| S9_k141_12982 | 1170 | No | NA | 1 | 1 | 0 | Low-quality | Genome-fragment | 2.56 | AAI-based (high-confidence) | 0 |
| S11-final-viral-combined_k141_217499\|\|full | 5368 | No | NA | 7 | 0 | 2 | Not-determined | Genome-fragment | NA | NA | 0 |
| S11-final-viral-combined_k141_176324\|\|full | 5945 | No | NA | 8 | 0 | 5 | Not-determined | Genome-fragment | NA | NA | 0 |
| S11-final-viral-combined_k141_184028\|\|lt2gene | 1170 | No | NA | 1 | 1 | 0 | Low-quality | Genome-fragment | 2.54 | AAI-based (high-confidence) | 0 |

| | | | | | | | | | | | |
|---|---|---|---|---|---|---|---|---|---|---|---|
| S11-final-viral-combined_k141_10780\|\|full | 2101 | No | NA | 2 | 0 | 2 | Not-determined | Genome-fragment | NA | NA | 0 |
| S11-final-viral-combined_k141_25850\|\|lt2gene | 1170 | No | NA | 2 | 0 | 0 | Low-quality | Genome-fragment | 3.07 | AAI-based (medium-confidence) | 0 |
| S11_k141_245645 | 1170 | No | NA | 2 | 0 | 0 | Low-quality | Genome-fragment | 1.75 | AAI-based (medium-confidence) | 0 |
| S11-final-viral-combined_k141_286527\|\|full | 3901 | No | NA | 6 | 0 | 2 | Not-determined | Genome-fragment | NA | NA | 0 |
| S1-final-viral-combined_k141_159835\|\|full | 1170 | No | NA | 4 | 0 | 0 | Low-quality | Genome-fragment | 2.75 | AAI-based (medium-confidence) | 0 |
| S11-final-viral-combined_k141_162474\|\|full | 2666 | No | NA | 4 | 0 | 0 | Not-determined | Genome-fragment | NA | NA | 0 |
| S11-final-viral-combined_k141_66052\|\|full | 7901 | No | NA | 7 | 0 | 6 | Not-determined | Genome-fragment | NA | NA | 0 |
| S1_k141_152318 | 1170 | No | NA | 1 | 0 | 0 | Low-quality | Genome-fragment | 2.09 | AAI-based (medium-confidence) | 0 |
| S1_k141_236392 | 1170 | No | NA | 2 | 0 | 0 | Low-quality | Genome-fragment | 2.23 | AAI-based (medium-confidence) | 0 |
| S11-final-viral-combined_k141_217555\|\|full | 1013 | No | NA | 2 | 0 | 0 | Not-determined | Genome-fragment | NA | NA | 0 |
| S11-final-viral-combined_k141_217557\|\|full | 2425 | No | NA | 4 | 0 | 0 | Not-determined | Genome-fragment | NA | NA | 0 |
| S5-final-viral-combined_k141_166393\|\|full | 1170 | No | NA | 3 | 2 | 0 | Low-quality | Genome-fragment | 2.69 | HMM-based (lower-bound) | 0 |
| S11-final-viral-combined_k141_217568\|\|full | 1066 | No | NA | 3 | 0 | 0 | Not-determined | Genome-fragment | NA | NA | 0 |
| S11-final-viral-combined_k141_259209\|\|full | 1274 | No | NA | 3 | 0 | 0 | Not-determined | Genome-fragment | NA | NA | 0 |
| S11-final-viral-combined_k141_286549\|\|full | 3433 | No | NA | 3 | 0 | 3 | Not-determined | Genome-fragment | NA | NA | 0 |
| S11-final-viral-combined_k141_176392\|\|full | 3156 | No | NA | 4 | 0 | 2 | Not-determined | Genome-fragment | NA | NA | 0 |
| S5-final-viral-combined_k141_789\|\|lt2gene | 1170 | No | NA | 1 | 1 | 0 | Low-quality | Genome-fragment | 0.84 | HMM-based (lower-bound) | 0 |
| S7-final-viral-combined_k141_130309\|\|full | 1170 | No | NA | 2 | 0 | 0 | Low-quality | Genome-fragment | 2.9 | AAI-based (medium-confidence) | 0 |
| S11-final-viral-combined_k141_314161\|\|full | 1610 | No | NA | 2 | 0 | 1 | Not-determined | Genome-fragment | NA | NA | 0 |
| S11-final-viral-combined_k141_245701\|\|full | 1938 | No | NA | 3 | 0 | 2 | Not-determined | Genome-fragment | NA | NA | 0 |
| S17-final-viral-combined_k141_20297\|\|full | 1170 | No | NA | 3 | 1 | 0 | Low-quality | Genome-fragment | 2.31 | AAI-based (high-confidence) | 0 |
| S17-final-viral-combined_k141_197638\|\|lt2gene | 1170 | No | NA | 2 | 1 | 0 | Low-quality | Genome-fragment | 4.01 | HMM-based (lower-bound) | 0 |
| S11-final-viral-combined_k141_148553\|\|full | 3622 | No | NA | 5 | 0 | 3 | Not-determined | Genome-fragment | NA | NA | 0 |
| S2_k141_228768 | 1170 | No | NA | 3 | 1 | 0 | Low-quality | Genome-fragment | 1.15 | AAI-based (high-confidence) | 0 |
| S11-final-viral-combined_k141_52476\|\|full | 2382 | No | NA | 2 | 0 | 1 | Not-determined | Genome-fragment | NA | NA | 0 |
| S2_k141_90985 | 1170 | No | NA | 1 | 0 | 0 | Low-quality | Genome-fragment | 1 | AAI-based (medium-confidence) | 0 |
| S4-final-viral-combined_k141_16991\|\|full | 1170 | No | NA | 2 | 0 | 0 | Low-quality | Genome-fragment | 3.53 | AAI-based (medium-confidence) | 0 |
| S8-final-viral-combined_k141_156366\|\|full | 1170 | No | NA | 3 | 0 | 0 | Low-quality | Genome-fragment | 1.7 | AAI-based (high-confidence) | 0 |
| S11-final-viral-combined_k141_326367\|\|full | 1495 | No | NA | 2 | 0 | 2 | Not-determined | Genome-fragment | NA | NA | 0 |
| S9-final-viral-combined_k141_120263\|\|lt2gene | 1171 | No | NA | 1 | 1 | 0 | Low-quality | Genome-fragment | 1.53 | HMM-based (lower-bound) | 0 |
| S11-final-viral-combined_k141_185615\|\|lt2gene | 1171 | No | NA | 1 | 1 | 0 | Low-quality | Genome-fragment | 2.7 | AAI-based (high-confidence) | 0 |
| S11-final-viral-combined_k141_10923\|\|full | 1563 | No | NA | 2 | 0 | 0 | Not-determined | Genome-fragment | NA | NA | 0 |
| S11-final-viral-combined_k141_217663\|\|lt2gene | 1171 | No | NA | 2 | 2 | 0 | Low-quality | Genome-fragment | 2.63 | AAI-based (high-confidence) | 0 |
| S11-final-viral-combined_k141_107873\|\|full | 4457 | No | NA | 4 | 0 | 2 | Not-determined | Genome-fragment | NA | NA | 0 |
| S3-final-viral-combined_k141_106458\|\|full | 1171 | No | NA | 3 | 1 | 0 | Low-quality | Genome-fragment | 2.64 | AAI-based (medium-confidence) | 0 |
| S5-final-viral-combined_k141_202083\|\|full | 1171 | No | NA | 2 | 0 | 0 | Low-quality | Genome-fragment | 2.89 | AAI-based (high-confidence) | 0 |
| S17-final-viral-combined_k141_185396\|\|full | 1171 | No | NA | 3 | 1 | 0 | Low-quality | Genome-fragment | 1.66 | HMM-based (lower-bound) | 0 |
| S12-final-viral-combined_k141_187898\|\|full | 1171 | No | NA | 2 | 0 | 1 | Low-quality | Genome-fragment | 2.51 | AAI-based (high-confidence) | 0 |
| S2-final-viral-combined_k141_12552\|\|full | 1171 | No | NA | 5 | 1 | 0 | Low-quality | Genome-fragment | 2.66 | AAI-based (high-confidence) | 0 |
| S11-final-viral-combined_k141_299828\|\|full | 2445 | No | NA | 3 | 0 | 1 | Not-determined | Genome-fragment | NA | NA | 0 |
| S11-final-viral-combined_k141_259323\|\|full | 1130 | No | NA | 3 | 0 | 2 | Not-determined | Genome-fragment | NA | NA | 0 |
| S11-final-viral-combined_k141_148618\|\|full | 2983 | No | NA | 3 | 0 | 1 | Not-determined | Genome-fragment | NA | NA | 0 |
| S11-final-viral-combined_k141_38556\|\|full | 5955 | No | NA | 7 | 0 | 4 | Not-determined | Genome-fragment | NA | NA | 0 |
| S2-final-viral-combined_k141_241261\|\|lt2gene | 1171 | No | NA | 1 | 1 | 0 | Low-quality | Genome-fragment | 2.47 | AAI-based (high-confidence) | 0 |
| S11-final-viral-combined_k141_79002\|\|full | 4180 | No | NA | 6 | 0 | 1 | Not-determined | Genome-fragment | NA | NA | 0 |
| S6-final-viral-combined_k141_126628\|\|lt2gene | 1171 | No | NA | 2 | 0 | 0 | Low-quality | Genome-fragment | 2.91 | AAI-based (medium-confidence) | 0 |
| S11-final-viral-combined_k141_120242\|\|full | 1342 | No | NA | 3 | 0 | 1 | Not-determined | Genome-fragment | NA | NA | 0 |
| S11-final-viral-combined_k141_52586\|\|full | 1432 | No | NA | 2 | 0 | 1 | Not-determined | Genome-fragment | NA | NA | 0 |
| S6-final-viral-combined_k141_121394\|\|lt2gene | 1171 | No | NA | 1 | 1 | 0 | Low-quality | Genome-fragment | 3.14 | AAI-based (high-confidence) | 0 |
| S11-final-viral-combined_k141_286692\|\|full | 1772 | No | NA | 2 | 0 | 1 | Not-determined | Genome-fragment | NA | NA | 0 |
| S11-final-viral-combined_k141_299858\|\|full | 2725 | No | NA | 4 | 0 | 2 | Not-determined | Genome-fragment | NA | NA | 0 |
| S11-final-viral-combined_k141_231430\|\|full | 4783 | No | NA | 4 | 0 | 1 | Not-determined | Genome-fragment | NA | NA | 0 |
| S11-final-viral-combined_k141_326483\|\|full | 2401 | No | NA | 3 | 0 | 2 | Not-determined | Genome-fragment | NA | NA | 0 |
| S11-final-viral-combined_k141_314220\|\|full | 1026 | No | NA | 2 | 0 | 2 | Not-determined | Genome-fragment | NA | NA | 0 |
| S8-final-viral-combined_k141_263253\|\|lt2gene | 1171 | No | NA | 2 | 1 | 0 | Low-quality | Genome-fragment | 0.76 | AAI-based (high-confidence) | 0 |
| S11-final-viral-combined_k141_87047\|\|full | 1172 | No | NA | 3 | 3 | 0 | Low-quality | Genome-fragment | 2.47 | AAI-based (high-confidence) | 0 |
| S1-final-viral-combined_k141_2758\|\|full | 1172 | No | NA | 3 | 0 | 0 | Low-quality | Genome-fragment | 0.65 | AAI-based (high-confidence) | 0 |
| S11-final-viral-combined_k141_38625\|\|full | 4074 | No | NA | 8 | 0 | 0 | Not-determined | Genome-fragment | NA | NA | 0 |
| S11-final-viral-combined_k141_217681\|\|full | 1108 | No | NA | 2 | 0 | 2 | Not-determined | Genome-fragment | NA | NA | 0 |
| S11-final-viral-combined_k141_120257\|\|full | 35507 | Yes | 27209 | 53 | 14 | 12 | Medium-quality | Genome-fragment | 57.27 | AAI-based (high-confidence) | 23.37 |
| S11-final-viral-combined_k141_272238\|\|full | 5660 | No | NA | 7 | 0 | 1 | Not-determined | Genome-fragment | NA | NA | 0 |
| S1-final-viral-combined_k141_261273\|\|full | 1172 | No | NA | 3 | 2 | 0 | Low-quality | Genome-fragment | 1.6 | AAI-based (high-confidence) | 0 |
| S1-final-viral-combined_k141_97527\|\|full | 1172 | No | NA | 3 | 0 | 0 | Low-quality | Genome-fragment | 2.83 | AAI-based (high-confidence) | 0 |
| S11-final-viral-combined_k141_79075\|\|full | 2988 | No | NA | 4 | 0 | 3 | Not-determined | Genome-fragment | NA | NA | 0 |
| S17-final-viral-combined_k141_89137\|\|full | 1172 | No | NA | 3 | 0 | 0 | Low-quality | Genome-fragment | 3.14 | AAI-based (medium-confidence) | 0 |
| S11-final-viral-combined_k141_314244\|\|full | 1838 | No | NA | 3 | 0 | 2 | Not-determined | Genome-fragment | NA | NA | 0 |
| S11-final-viral-combined_k141_52713\|\|full | 1895 | No | NA | 2 | 0 | 2 | Not-determined | Genome-fragment | NA | NA | 0 |
| S2-final-viral-combined_k141_79365\|\|full | 1172 | No | NA | 5 | 1 | 0 | Low-quality | Genome-fragment | 3.32 | AAI-based (high-confidence) | 0 |
| S2-final-viral-combined_k141_175769\|\|full | 1172 | No | NA | 2 | 0 | 1 | Low-quality | Genome-fragment | 0.89 | AAI-based (medium-confidence) | 0 |
| S11-final-viral-combined_k141_52722\|\|full | 1438 | No | NA | 3 | 0 | 1 | Not-determined | Genome-fragment | NA | NA | 0 |
| S4-final-viral-combined_k141_32145\|\|lt2gene | 1172 | No | NA | 2 | 0 | 1 | Low-quality | Genome-fragment | 1.27 | AAI-based (medium-confidence) | 0 |
| S8-final-viral-combined_k141_234012\|\|full | 1172 | No | NA | 4 | 0 | 0 | Low-quality | Genome-fragment | 2.52 | AAI-based (high-confidence) | 0 |
| S11-final-viral-combined_k141_11116\|\|full | 2205 | No | NA | 3 | 0 | 2 | Not-determined | Genome-fragment | NA | NA | 0 |
| S11-final-viral-combined_k141_299954\|\|full | 26312 | No | NA | 41 | 7 | 0 | Medium-quality | Genome-fragment | 70.68 | AAI-based (high-confidence) | 0 |
| S11-final-viral-combined_k141_314271\|\|full | 1493 | No | NA | 2 | 0 | 0 | Not-determined | Genome-fragment | NA | NA | 0 |
| S11-final-viral-combined_k141_286765\|\|full | 2758 | No | NA | 3 | 0 | 2 | Not-determined | Genome-fragment | NA | NA | 0 |
| S11-final-viral-combined_k141_176572\|\|full | 4084 | No | NA | 6 | 0 | 2 | Not-determined | Genome-fragment | NA | NA | 0 |
| S11-final-viral-combined_k141_176586\|\|full | 2459 | No | NA | 4 | 0 | 0 | Not-determined | Genome-fragment | NA | NA | 0 |
| S11-final-viral-combined_k141_162800\|\|full | 1576 | No | NA | 3 | 0 | 0 | Not-determined | Genome-fragment | NA | NA | 0 |
| S11-final-viral-combined_k141_314292\|\|full | 2791 | No | NA | 3 | 0 | 1 | Not-determined | Genome-fragment | NA | NA | 0 |
| S8-final-viral-combined_k141_216775\|\|lt2gene | 1172 | No | NA | 1 | 1 | 0 | Low-quality | Genome-fragment | 2.54 | AAI-based (high-confidence) | 0 |
| S11-final-viral-combined_k141_271676\|\|full | 1173 | No | NA | 3 | 3 | 0 | Low-quality | Genome-fragment | 2.53 | AAI-based (medium-confidence) | 0 |
| S1_k141_268405 | 1173 | No | NA | 2 | 0 | 1 | Low-quality | Genome-fragment | 0.47 | AAI-based (medium-confidence) | 0 |
| S1_k141_141330 | 1173 | No | NA | 1 | 0 | 1 | Low-quality | Genome-fragment | 3.31 | AAI-based (medium-confidence) | 0 |
| S11-final-viral-combined_k141_162850\|\|full | 2655 | No | NA | 4 | 0 | 4 | Not-determined | Genome-fragment | NA | NA | 0 |
| S3_k141_58161 | 1173 | No | NA | 4 | 0 | 0 | Low-quality | Genome-fragment | 1.9 | AAI-based (high-confidence) | 0 |
| S11-final-viral-combined_k141_176636\|\|full | 4554 | No | NA | 6 | 0 | 1 | Not-determined | Genome-fragment | NA | NA | 0 |
| S5-final-viral-combined_k141_61727\|\|full | 1173 | No | NA | 3 | 0 | 0 | Low-quality | Genome-fragment | 2.99 | AAI-based (high-confidence) | 0 |
| S5_k141_141932 | 1173 | No | NA | 1 | 0 | 1 | Low-quality | Genome-fragment | 0.32 | AAI-based (medium-confidence) | 0 |
| S11-final-viral-combined_k141_23496\|\|full | 1005 | No | NA | 3 | 0 | 2 | Not-determined | Genome-fragment | NA | NA | 0 |
| S12-final-viral-combined_k141_251464\|\|full | 1173 | No | NA | 3 | 1 | 0 | Low-quality | Genome-fragment | 2.54 | AAI-based (medium-confidence) | 0 |
| S11-final-viral-combined_k141_120395\|\|full | 3673 | No | NA | 5 | 0 | 3 | Not-determined | Genome-fragment | NA | NA | 0 |
| S11-final-viral-combined_k141_108088\|\|full | 3610 | No | NA | 5 | 0 | 3 | Not-determined | Genome-fragment | NA | NA | 0 |
| S12-final-viral-combined_k141_3454\|\|full | 1173 | No | NA | 2 | 0 | 0 | Low-quality | Genome-fragment | 2.59 | AAI-based (high-confidence) | 0 |
| S11-final-viral-combined_k141_52861\|\|full | 2944 | No | NA | 3 | 0 | 1 | Not-determined | Genome-fragment | NA | NA | 0 |
| S12-final-viral-combined_k141_206205\|\|full | 1173 | No | NA | 3 | 0 | 1 | Low-quality | Genome-fragment | 1.14 | AAI-based (medium-confidence) | 0 |
| S12_k141_145657 | 1173 | No | NA | 1 | 0 | 1 | Low-quality | Genome-fragment | 0.77 | AAI-based (high-confidence) | 0 |
| S11-final-viral-combined_k141_203779\|\|full | 1368 | No | NA | 3 | 0 | 0 | Not-determined | Genome-fragment | NA | NA | 0 |
| S2_k141_151046 | 1173 | No | NA | 4 | 0 | 0 | Low-quality | Genome-fragment | 3.55 | AAI-based (medium-confidence) | 0 |
| S11-final-viral-combined_k141_259526\|\|full | 1555 | No | NA | 3 | 0 | 2 | Not-determined | Genome-fragment | NA | NA | 0 |
| S9-final-viral-combined_k141_11877\|\|full | 1174 | No | NA | 2 | 0 | 1 | Low-quality | Genome-fragment | 0.92 | AAI-based (medium-confidence) | 0 |
| S9_k141_15183 | 1174 | No | NA | 3 | 0 | 2 | Low-quality | Genome-fragment | 0.63 | AAI-based (medium-confidence) | 0 |
| S5-final-viral-combined_k141_169526\|\|full | 1174 | No | NA | 3 | 0 | 1 | Low-quality | Genome-fragment | 3.1 | AAI-based (medium-confidence) | 0 |
| S11-final-viral-combined_k141_326648\|\|full | 1569 | No | NA | 2 | 0 | 2 | Not-determined | Genome-fragment | NA | NA | 0 |
| S5_k141_53117 | 1174 | No | NA | 1 | 0 | 1 | Low-quality | Genome-fragment | 3 | AAI-based (medium-confidence) | 0 |
| S7-final-viral-combined_k141_39488\|\|lt2gene | 1174 | No | NA | 1 | 1 | 0 | Low-quality | Genome-fragment | 0.37 | HMM-based (lower-bound) | 0 |
| S12-final-viral-combined_k141_2844\|\|full | 1174 | No | NA | 3 | 0 | 2 | Low-quality | Genome-fragment | 1.81 | AAI-based (medium-confidence) | 0 |
| S11-final-viral-combined_k141_108155\|\|full | 2332 | No | NA | 2 | 0 | 1 | Not-determined | Genome-fragment | NA | NA | 0 |
| S12-final-viral-combined_k141_365411\|\|lt2gene | 1174 | No | NA | 2 | 1 | 0 | Low-quality | Genome-fragment | 1.51 | HMM-based (lower-bound) | 0 |
| S2-final-viral-combined_k141_147301\|\|full | 1174 | No | NA | 4 | 1 | 0 | Low-quality | Genome-fragment | 2.56 | AAI-based (high-confidence) | 0 |
| S11-final-viral-combined_k141_79302\|\|full | 2189 | No | NA | 3 | 0 | 1 | Not-determined | Genome-fragment | NA | NA | 0 |
| S11-final-viral-combined_k141_108180\|\|full | 1168 | No | NA | 2 | 0 | 0 | Not-determined | Genome-fragment | NA | NA | 0 |
| S11-final-viral-combined_k141_217790\|\|full | 54204 | Yes | 40062 | 39 | 5 | 6 | Medium-quality | Genome-fragment | 66.2 | AAI-based (high-confidence) | 26.09 |
| S2-final-viral-combined_k141_128817\|\|full | 1174 | No | NA | 2 | 2 | 0 | Low-quality | Genome-fragment | 1.94 | AAI-based (high-confidence) | 0 |
| S2-final-viral-combined_k141_245271\|\|full | 1174 | No | NA | 2 | 1 | 0 | Low-quality | Genome-fragment | 2.75 | AAI-based (high-confidence) | 0 |
| S4-final-viral-combined_k141_122412\|\|full | 1174 | No | NA | 3 | 1 | 0 | Low-quality | Genome-fragment | 2.31 | AAI-based (high-confidence) | 0 |
| S4_k141_107795 | 1174 | No | NA | 2 | 0 | 1 | Low-quality | Genome-fragment | 0.9 | AAI-based (high-confidence) | 0 |
| S4_k141_117836 | 1174 | No | NA | 2 | 0 | 1 | Low-quality | Genome-fragment | 0.71 | AAI-based (high-confidence) | 0 |
| S11-final-viral-combined_k141_148887\|\|full | 13818 | No | NA | 6 | 0 | 6 | Not-determined | Genome-fragment | NA | NA | 0 |
| S11-final-viral-combined_k141_300173\|\|full | 3838 | No | NA | 4 | 0 | 2 | Not-determined | Genome-fragment | NA | NA | 0 |
| S6-final-viral-combined_k141_3811\|\|full | 1174 | No | NA | 4 | 1 | 0 | Low-quality | Genome-fragment | 1.68 | AAI-based (medium-confidence) | 0 |
| S8-final-viral-combined_k141_163927\|\|full | 1174 | No | NA | 3 | 0 | 0 | Low-quality | Genome-fragment | 2.44 | AAI-based (high-confidence) | 0 |
| S11-final-viral-combined_k141_38926\|\|full | 2361 | No | NA | 3 | 0 | 0 | Not-determined | Genome-fragment | NA | NA | 0 |
| S8-final-viral-combined_k141_192886\|\|full | 1174 | No | NA | 2 | 0 | 0 | Low-quality | Genome-fragment | 2.05 | AAI-based (medium-confidence) | 0 |
| S11-final-viral-combined_k141_66410\|\|full | 2340 | No | NA | 3 | 0 | 2 | Not-determined | Genome-fragment | NA | NA | 0 |
| S11-final-viral-combined_k141_300220\|\|full | 5841 | No | NA | 7 | 0 | 1 | Not-determined | Genome-fragment | NA | NA | 0 |
| S11-final-viral-combined_k141_93844\|\|full | 2080 | No | NA | 3 | 0 | 1 | Not-determined | Genome-fragment | NA | NA | 0 |
| S8_k141_84575 | 1174 | No | NA | 2 | 1 | 0 | Low-quality | Genome-fragment | 2.17 | AAI-based (medium-confidence) | 0 |
| S11-final-viral-combined_k141_231847\|\|full | 3277 | No | NA | 5 | 0 | 3 | Not-determined | Genome-fragment | NA | NA | 0 |
| S11-final-viral-combined_k141_52991\|\|full | 2605 | No | NA | 4 | 0 | 2 | Not-determined | Genome-fragment | NA | NA | 0 |
| S9-final-viral-combined_k141_43179\|\|full | 1175 | No | NA | 2 | 0 | 0 | Low-quality | Genome-fragment | 2.02 | AAI-based (high-confidence) | 0 |
| S11-final-viral-combined_k141_246160\|\|full | 3172 | No | NA | 3 | 0 | 3 | Not-determined | Genome-fragment | NA | NA | 0 |
| S11-final-viral-combined_k141_234411\|\|full | 1175 | No | NA | 3 | 1 | 0 | Low-quality | Genome-fragment | 2.1 | AAI-based (medium-confidence) | 0 |
| S11-final-viral-combined_k141_66488\|\|full | 1922 | No | NA | 6 | 0 | 0 | Not-determined | Genome-fragment | NA | NA | 0 |
| S11-final-viral-combined_k141_163036\|\|full | 1810 | No | NA | 3 | 0 | 1 | Not-determined | Genome-fragment | NA | NA | 0 |
| S11-final-viral-combined_k141_93898\|\|full | 2959 | No | NA | 3 | 0 | 2 | Not-determined | Genome-fragment | NA | NA | 0 |
| S11-final-viral-combined_k141_189723\|\|full | 7594 | No | NA | 8 | 0 | 7 | Not-determined | Genome-fragment | NA | NA | 0 |
| S11-final-viral-combined_k141_66558\|\|full | 2522 | No | NA | 5 | 0 | 3 | Not-determined | Genome-fragment | NA | NA | 0 |
| S11-final-viral-combined_k141_23676\|\|full | 2425 | No | NA | 3 | 0 | 3 | Not-determined | Genome-fragment | NA | NA | 0 |
| S11-final-viral-combined_k141_300270\|\|full | 1877 | No | NA | 3 | 0 | 2 | Not-determined | Genome-fragment | NA | NA | 0 |
| S11-final-viral-combined_k141_120632\|\|full | 1499 | No | NA | 3 | 0 | 0 | Not-determined | Genome-fragment | NA | NA | 0 |
| S11-final-viral-combined_k141_302505\|\|full | 1175 | No | NA | 4 | 0 | 0 | Low-quality | Genome-fragment | 0.92 | AAI-based (high-confidence) | 0 |
| S1-final-viral-combined_k141_38241\|\|full | 1175 | No | NA | 2 | 1 | 0 | Low-quality | Genome-fragment | 0.47 | HMM-based (lower-bound) | 0 |
| S1_k141_366987 | 1175 | No | NA | 1 | 0 | 0 | Low-quality | Genome-fragment | 1.76 | AAI-based (high-confidence) | 0 |
| S11-final-viral-combined_k141_259765\|\|full | 1340 | No | NA | 3 | 0 | 1 | Not-determined | Genome-fragment | NA | NA | 0 |
| S11-final-viral-combined_k141_38992\|\|full | 1122 | No | NA | 3 | 0 | 1 | Not-determined | Genome-fragment | NA | NA | 0 |
| S1_k141_246642 | 1175 | No | NA | 2 | 0 | 1 | Low-quality | Genome-fragment | 2.68 | AAI-based (medium-confidence) | 0 |
| S11-final-viral-combined_k141_39009\|\|full | 2521 | No | NA | 3 | 0 | 2 | Not-determined | Genome-fragment | NA | NA | 0 |
| S11-final-viral-combined_k141_93928\|\|full | 5712 | No | NA | 8 | 0 | 6 | Not-determined | Genome-fragment | NA | NA | 0 |
| S3-final-viral-combined_k141_51419\|\|full | 1175 | No | NA | 4 | 0 | 0 | Low-quality | Genome-fragment | 2.7 | AAI-based (medium-confidence) | 0 |
| S3-final-viral-combined_k141_17879\|\|lt2gene | 1175 | No | NA | 1 | 0 | 0 | Low-quality | Genome-fragment | 3.22 | AAI-based (high-confidence) | 0 |
| S11-final-viral-combined_k141_189779\|\|full | 20977 | No | NA | 20 | 0 | 13 | Not-determined | Genome-fragment | NA | NA | 0 |
| S5-final-viral-combined_k141_50421\|\|lt2gene | 1175 | No | NA | 2 | 1 | 0 | Low-quality | Genome-fragment | 2.84 | AAI-based (medium-confidence) | 0 |
| S11-final-viral-combined_k141_272702\|\|full | 1010 | No | NA | 2 | 0 | 1 | Not-determined | Genome-fragment | NA | NA | 0 |
| S11-final-viral-combined_k141_53200\|\|full | 7248 | No | NA | 11 | 0 | 1 | Not-determined | Genome-fragment | NA | NA | 0 |
| S11-final-viral-combined_k141_120734\|\|full | 1859 | No | NA | 3 | 0 | 1 | Not-determined | Genome-fragment | NA | NA | 0 |
| S11-final-viral-combined_k141_149038\|\|full | 1432 | No | NA | 3 | 0 | 1 | Not-determined | Genome-fragment | NA | NA | 0 |
| S7-final-viral-combined_k141_128569\|\|full | 1175 | No | NA | 3 | 0 | 0 | Low-quality | Genome-fragment | 1.31 | AAI-based (medium-confidence) | 0 |
| S11-final-viral-combined_k141_39068\|\|full | 5045 | No | NA | 4 | 0 | 2 | Not-determined | Genome-fragment | NA | NA | 0 |
| S7-final-viral-combined_k141_222100\|\|full | 1175 | No | NA | 2 | 1 | 0 | Low-quality | Genome-fragment | 2.76 | AAI-based (high-confidence) | 0 |
| S7_k141_219413 | 1175 | No | NA | 3 | 1 | 0 | Low-quality | Genome-fragment | 0.74 | HMM-based (lower-bound) | 0 |
| S11-final-viral-combined_k141_11421\|\|full | 1180 | No | NA | 2 | 0 | 0 | Not-determined | Genome-fragment | NA | NA | 0 |
| S11-final-viral-combined_k141_11424\|\|full | 5971 | No | NA | 5 | 0 | 5 | Not-determined | Genome-fragment | NA | NA | 0 |
| S11-final-viral-combined_k141_189844\|\|full | 10912 | No | NA | 11 | 0 | 6 | Not-determined | Genome-fragment | NA | NA | 0 |
| S11-final-viral-combined_k141_326909\|\|full | 5257 | No | NA | 5 | 0 | 3 | Not-determined | Genome-fragment | NA | NA | 0 |
| S17-final-viral-combined_k141_24759\|\|lt2gene | 1175 | No | NA | 2 | 2 | 0 | Low-quality | Genome-fragment | 3.13 | AAI-based (medium-confidence) | 0 |
| S17-k141_115246 | 1175 | No | NA | 1 | 0 | 0 | Low-quality | Genome-fragment | 0.19 | AAI-based (high-confidence) | 0 |
| S11-final-viral-combined_k141_287220\|\|full | 1270 | No | NA | 2 | 0 | 0 | Not-determined | Genome-fragment | NA | NA | 0 |
| S11-final-viral-combined_k141_134462\|\|full | 1721 | No | NA | 5 | 0 | 2 | Not-determined | Genome-fragment | NA | NA | 0 |
| S11-final-viral-combined_k141_300466\|\|full | 3280 | No | NA | 4 | 0 | 3 | Not-determined | Genome-fragment | NA | NA | 0 |
| S12-final-viral-combined_k141_376910\|\|full | 1175 | No | NA | 3 | 0 | 0 | Low-quality | Genome-fragment | 2.15 | AAI-based (medium-confidence) | 0 |
| S11-final-viral-combined_k141_176995\|\|full | 2799 | No | NA | 3 | 0 | 1 | Not-determined | Genome-fragment | NA | NA | 0 |
| S11-final-viral-combined_k141_66735\|\|full | 3356 | No | NA | 4 | 0 | 3 | Not-determined | Genome-fragment | NA | NA | 0 |
| S11-final-viral-combined_k141_189869\|\|full | 1058 | No | NA | 2 | 0 | 1 | Not-determined | Genome-fragment | NA | NA | 0 |
| S11-final-viral-combined_k141_232051\|\|full | 3535 | No | NA | 4 | 0 | 2 | Not-determined | Genome-fragment | NA | NA | 0 |
| S11-final-viral-combined_k141_108513\|\|full | 2955 | No | NA | 3 | 0 | 2 | Not-determined | Genome-fragment | NA | NA | 0 |
| S11-final-viral-combined_k141_120785\|\|full | 1363 | No | NA | 2 | 0 | 0 | Not-determined | Genome-fragment | NA | NA | 0 |
| S11-final-viral-combined_k141_108514\|\|full | 4966 | No | NA | 4 | 0 | 2 | Not-determined | Genome-fragment | NA | NA | 0 |
| S11-final-viral-combined_k141_177022\|\|full | 1248 | No | NA | 3 | 0 | 1 | Not-determined | Genome-fragment | NA | NA | 0 |
| S12-final-viral-combined_k141_399617\|\|full | 1175 | No | NA | 2 | 0 | 0 | Low-quality | Genome-fragment | 2.68 | AAI-based (high-confidence) | 0 |
| S2_k141_104226 | 1175 | No | NA | 1 | 0 | 0 | Low-quality | Genome-fragment | 3.26 | AAI-based (high-confidence) | 0 |
| S4_k141_75956 | 1175 | No | NA | 2 | 1 | 0 | Low-quality | Genome-fragment | 1.19 | AAI-based (high-confidence) | 0 |
| S8-final-viral-combined_k141_186354\|\|full | 1175 | No | NA | 2 | 2 | 0 | Low-quality | Genome-fragment | 0.86 | HMM-based (lower-bound) | 0 |
| S11-final-viral-combined_k141_149186\|\|full | 1141 | No | NA | 1 | 0 | 1 | Not-determined | Genome-fragment | NA | NA | 0 |
| S9-final-viral-combined_k141_98560\|\|full | 1176 | No | NA | 2 | 0 | 0 | Low-quality | Genome-fragment | 0.44 | AAI-based (medium-confidence) | 0 |
| S9-final-viral-combined_k141_40981\|\|full | 1176 | No | NA | 3 | 2 | 0 | Low-quality | Genome-fragment | 3.85 | AAI-based (high-confidence) | 0 |
| S9-final-viral-combined_k141_113251\|\|lt2gene | 1176 | No | NA | 1 | 1 | 0 | Low-quality | Genome-fragment | 2.49 | AAI-based (high-confidence) | 0 |
| S11-final-viral-combined_k141_301502\|\|full | 1176 | No | NA | 4 | 3 | 0 | Low-quality | Genome-fragment | 1.87 | AAI-based (high-confidence) | 0 |
| S1-final-viral-combined_k141_11153\|\|full | 1176 | No | NA | 2 | 0 | 0 | Low-quality | Genome-fragment | 1.54 | AAI-based (medium-confidence) | 0 |
| S3-final-viral-combined_k141_126242\|\|lt2gene | 1176 | No | NA | 1 | 1 | 0 | Low-quality | Genome-fragment | 1.65 | HMM-based (lower-bound) | 0 |
| S7-final-viral-combined_k141_237370\|\|full | 1176 | No | NA | 2 | 0 | 0 | Low-quality | Genome-fragment | 1 | AAI-based (medium-confidence) | 0 |
| S7-final-viral-combined_k141_187254\|\|full | 1176 | No | NA | 3 | 0 | 0 | Low-quality | Genome-fragment | 1.14 | AAI-based (high-confidence) | 0 |
| S12-final-viral-combined_k141_18122\|\|lt2gene | 1176 | No | NA | 2 | 2 | 0 | Low-quality | Genome-fragment | 3.14 | AAI-based (medium-confidence) | 0 |
| S11-final-viral-combined_k141_11572\|\|full | 2683 | No | NA | 3 | 0 | 3 | Not-determined | Genome-fragment | NA | NA | 0 |
| S11-final-viral-combined_k141_149267\|\|full | 2734 | No | NA | 4 | 0 | 3 | Not-determined | Genome-fragment | NA | NA | 0 |
| S12_k141_78446 | 1176 | No | NA | 1 | 0 | 1 | Low-quality | Genome-fragment | 0.77 | AAI-based (high-confidence) | 0 |
| S11-final-viral-combined_k141_163336\|\|full | 1607 | No | NA | 3 | 0 | 1 | Not-determined | Genome-fragment | NA | NA | 0 |
| S2-final-viral-combined_k141_177677\|\|lt2gene | 1176 | No | NA | 1 | 1 | 0 | Low-quality | Genome-fragment | 1.32 | HMM-based (lower-bound) | 0 |
| S11-final-viral-combined_k141_23942\|\|full | 2155 | No | NA | 3 | 0 | 2 | Not-determined | Genome-fragment | NA | NA | 0 |
| S11-final-viral-combined_k141_177123\|\|full | 1972 | No | NA | 3 | 0 | 0 | Not-determined | Genome-fragment | NA | NA | 0 |
| S8-final-viral-combined_k141_90048\|\|full | 1176 | No | NA | 3 | 0 | 1 | Low-quality | Genome-fragment | 2.03 | AAI-based (medium-confidence) | 0 |
| S11-final-viral-combined_k141_23949\|\|full | 2660 | No | NA | 4 | 0 | 2 | Not-determined | Genome-fragment | NA | NA | 0 |
| S11-final-viral-combined_k141_39279\|\|full | 2899 | No | NA | 3 | 0 | 1 | Not-determined | Genome-fragment | NA | NA | 0 |
| S8-final-viral-combined_k141_119208\|\|lt2gene | 1176 | No | NA | 1 | 1 | 0 | Low-quality | Genome-fragment | 2.13 | AAI-based (high-confidence) | 0 |
| S11-final-viral-combined_k141_53292\|\|full | 3475 | No | NA | 3 | 0 | 3 | Not-determined | Genome-fragment | NA | NA | 0 |
| S11-final-viral-combined_k141_79765\|\|full | 1297 | No | NA | 3 | 0 | 0 | Not-determined | Genome-fragment | NA | NA | 0 |
| S11-final-viral-combined_k141_120946\|\|full | 1240 | No | NA | 3 | 0 | 0 | Not-determined | Genome-fragment | NA | NA | 0 |
| S11-final-viral-combined_k141_246608\|\|full | 1079 | No | NA | 3 | 0 | 1 | Not-determined | Genome-fragment | NA | NA | 0 |
| S11-final-viral-combined_k141_39310\|\|full | 2144 | No | NA | 3 | 0 | 2 | Not-determined | Genome-fragment | NA | NA | 0 |
| S8_k141_269704 | 1176 | No | NA | 1 | 0 | 0 | Low-quality | Genome-fragment | 0.69 | AAI-based (medium-confidence) | 0 |
| S11-final-viral-combined_k141_79794\|\|full | 1192 | No | NA | 2 | 0 | 1 | Not-determined | Genome-fragment | NA | NA | 0 |
| S9-final-viral-combined_k141_74455\|\|full | 1177 | No | NA | 3 | 0 | 0 | Low-quality | Genome-fragment | 1.8 | AAI-based (medium-confidence) | 0 |
| S11-final-viral-combined_k141_200093\|\|full | 1177 | No | NA | 3 | 0 | 0 | Low-quality | Genome-fragment | 2.65 | AAI-based (medium-confidence) | 0 |
| S1-final-viral-combined_k141_49114\|\|full | 1177 | No | NA | 3 | 3 | 0 | Low-quality | Genome-fragment | 3.05 | AAI-based (high-confidence) | 0 |
| S11-final-viral-combined_k141_189966\|\|full | 1248 | No | NA | 2 | 0 | 1 | Not-determined | Genome-fragment | NA | NA | 0 |
| S1-final-viral-combined_k141_336052\|\|lt2gene | 1177 | No | NA | 1 | 1 | 0 | Low-quality | Genome-fragment | 2.61 | AAI-based (medium-confidence) | 0 |
| S11-final-viral-combined_k141_218090\|\|full | 1508 | No | NA | 3 | 0 | 0 | Not-determined | Genome-fragment | NA | NA | 0 |
| S11-final-viral-combined_k141_314613\|\|full | 1103 | No | NA | 1 | 0 | 0 | Not-determined | Genome-fragment | NA | NA | 0 |
| S11-final-viral-combined_k141_190000\|\|full | 3884 | No | NA | 5 | 0 | 0 | Not-determined | Genome-fragment | NA | NA | 0 |
| S11-final-viral-combined_k141_314635\|\|full | 5451 | No | NA | 5 | 0 | 1 | Not-determined | Genome-fragment | NA | NA | 0 |
| S11-final-viral-combined_k141_108744\|\|full | 7120 | No | NA | 6 | 0 | 1 | Not-determined | Genome-fragment | NA | NA | 0 |
| S11-final-viral-combined_k141_287393\|\|full | 1190 | No | NA | 2 | 0 | 2 | Not-determined | Genome-fragment | NA | NA | 0 |
| S1_k141_346805 | 1177 | No | NA | 3 | 0 | 0 | Low-quality | Genome-fragment | 2.29 | AAI-based (medium-confidence) | 0 |
| S11-final-viral-combined_k141_24088\|\|full | 2916 | No | NA | 5 | 0 | 2 | Not-determined | Genome-fragment | NA | NA | 0 |
| S3_k141_80913 | 1177 | No | NA | 1 | 0 | 1 | Low-quality | Genome-fragment | 0.76 | AAI-based (medium-confidence) | 0 |
| S11-final-viral-combined_k141_177232\|\|full | 2724 | No | NA | 3 | 0 | 3 | Not-determined | Genome-fragment | NA | NA | 0 |
| S7_k141_32103 | 1177 | No | NA | 2 | 0 | 0 | Low-quality | Genome-fragment | 1.92 | AAI-based (medium-confidence) | 0 |
| S11-final-viral-combined_k141_163470\|\|full | 5623 | No | NA | 4 | 0 | 4 | Not-determined | Genome-fragment | NA | NA | 0 |
| S7_k141_197928 | 1177 | No | NA | 1 | 0 | 0 | Low-quality | Genome-fragment | 0.96 | AAI-based (high-confidence) | 0 |
| S11-final-viral-combined_k141_190052\|\|full | 1597 | No | NA | 3 | 0 | 0 | Not-determined | Genome-fragment | NA | NA | 0 |
| S17-final-viral-combined_k141_56950\|\|full | 1177 | No | NA | 2 | 0 | 0 | Low-quality | Genome-fragment | 2.79 | AAI-based (high-confidence) | 0 |
| S17-final-viral-combined_k141_115570\|\|full | 1177 | No | NA | 3 | 0 | 1 | Low-quality | Genome-fragment | 0.98 | AAI-based (medium-confidence) | 0 |
| S12-final-viral-combined_k141_322764\|\|full | 1177 | No | NA | 2 | 0 | 2 | Low-quality | Genome-fragment | 2.22 | AAI-based (medium-confidence) | 0 |
| S2_k141_219310 | 1177 | No | NA | 3 | 0 | 0 | Low-quality | Genome-fragment | 3.55 | AAI-based (medium-confidence) | 0 |
| S11-final-viral-combined_k141_149463\|\|full | 3628 | No | NA | 4 | 0 | 3 | Not-determined | Genome-fragment | NA | NA | 0 |
| S11-final-viral-combined_k141_300632\|\|full | 1971 | No | NA | 4 | 0 | 2 | Not-determined | Genome-fragment | NA | NA | 0 |
| S11-final-viral-combined_k141_163520\|\|full | 2002 | No | NA | 3 | 0 | 0 | Not-determined | Genome-fragment | NA | NA | 0 |
| S11-final-viral-combined_k141_134772\|\|full | 1489 | No | NA | 2 | 0 | 1 | Not-determined | Genome-fragment | NA | NA | 0 |
| S11-final-viral-combined_k141_24156\|\|full | 1089 | No | NA | 2 | 0 | 0 | Not-determined | Genome-fragment | NA | NA | 0 |
| S8-final-viral-combined_k141_97281\|\|full | 1177 | No | NA | 3 | 1 | 0 | Low-quality | Genome-fragment | 2.02 | AAI-based (medium-confidence) | 0 |
| S8_k141_37330 | 1177 | No | NA | 4 | 1 | 0 | Low-quality | Genome-fragment | 1.91 | AAI-based (medium-confidence) | 0 |
| S9-final-viral-combined_k141_33112\|\|full | 1178 | No | NA | 2 | 1 | 0 | Low-quality | Genome-fragment | 2.62 | AAI-based (medium-confidence) | 0 |
| S11-final-viral-combined_k141_94135\|\|full | 1889 | No | NA | 4 | 0 | 1 | Not-determined | Genome-fragment | NA | NA | 0 |
| S9-final-viral-combined_k141_33313\|\|full | 1178 | No | NA | 2 | 0 | 0 | Low-quality | Genome-fragment | 2.85 | AAI-based (high-confidence) | 0 |
| S11-final-viral-combined_k141_287511\|\|full | 1662 | No | NA | 4 | 0 | 1 | Not-determined | Genome-fragment | NA | NA | 0 |
| S11-final-viral-combined_k141_207475\|\|full | 1178 | No | NA | 4 | 0 | 1 | Low-quality | Genome-fragment | 0.73 | AAI-based (medium-confidence) | 0 |
| S11-final-viral-combined_k141_10419\|\|full | 1178 | No | NA | 2 | 0 | 0 | Low-quality | Genome-fragment | 2.12 | AAI-based (high-confidence) | 0 |
| S1-final-viral-combined_k141_327445\|\|full | 1178 | No | NA | 3 | 0 | 2 | Low-quality | Genome-fragment | 2.16 | AAI-based (medium-confidence) | 0 |
| S11-final-viral-combined_k141_149549\|\|full | 1604 | No | NA | 2 | 0 | 1 | Not-determined | Genome-fragment | NA | NA | 0 |
| S5-final-viral-combined_k141_9588\|\|full | 1178 | No | NA | 3 | 1 | 0 | Low-quality | Genome-fragment | 2.15 | AAI-based (high-confidence) | 0 |
| S11-final-viral-combined_k141_327235\|\|full | 1106 | No | NA | 3 | 0 | 1 | Not-determined | Genome-fragment | NA | NA | 0 |
| S11-final-viral-combined_k141_67020\|\|full | 1961 | No | NA | 2 | 0 | 1 | Not-determined | Genome-fragment | NA | NA | 0 |
| S11-final-viral-combined_k141_218247\|\|full | 1220 | No | NA | 3 | 0 | 0 | Not-determined | Genome-fragment | NA | NA | 0 |
| S11-final-viral-combined_k141_287555\|\|full | 4921 | No | NA | 3 | 0 | 0 | Not-determined | Genome-fragment | NA | NA | 0 |
| S5-final-viral-combined_k141_110414\|\|full | 1178 | No | NA | 5 | 1 | 0 | Low-quality | Genome-fragment | 1.85 | HMM-based (lower-bound) | 0 |
| S11-final-viral-combined_k141_149563\|\|full | 9238 | No | NA | 6 | 0 | 5 | Not-determined | Genome-fragment | NA | NA | 0 |
| S5-final-viral-combined_k141_117603\|\|lt2gene | 1178 | No | NA | 1 | 1 | 0 | Low-quality | Genome-fragment | 2.23 | HMM-based (lower-bound) | 0 |
| S11-final-viral-combined_k141_11859\|\|full | 1098 | No | NA | 2 | 0 | 0 | Not-determined | Genome-fragment | NA | NA | 0 |
| S11-final-viral-combined_k141_218283\|\|full | 1329 | No | NA | 3 | 0 | 0 | Not-determined | Genome-fragment | NA | NA | 0 |
| S11-final-viral-combined_k141_149569\|\|full | 5724 | No | NA | 5 | 0 | 3 | Not-determined | Genome-fragment | NA | NA | 0 |
| S11-final-viral-combined_k141_300761\|\|full | 3392 | No | NA | 3 | 0 | 3 | Not-determined | Genome-fragment | NA | NA | 0 |
| S11-final-viral-combined_k141_232358\|\|full | 1286 | No | NA | 3 | 0 | 1 | Not-determined | Genome-fragment | NA | NA | 0 |
| S11-final-viral-combined_k141_300762\|\|full | 4623 | No | NA | 5 | 0 | 5 | Not-determined | Genome-fragment | NA | NA | 0 |
| S5-final-viral-combined_k141_129991\|\|lt2gene | 1178 | No | NA | 2 | 2 | 0 | Low-quality | Genome-fragment | 2.01 | AAI-based (medium-confidence) | 0 |
| S11-final-viral-combined_k141_149573\|\|full | 1785 | No | NA | 3 | 0 | 1 | Not-determined | Genome-fragment | NA | NA | 0 |
| S11-final-viral-combined_k141_94206\|\|full | 1225 | No | NA | 3 | 0 | 0 | Not-determined | Genome-fragment | NA | NA | 0 |
| S11-final-viral-combined_k141_218294\|\|full | 1241 | No | NA | 2 | 0 | 0 | Not-determined | Genome-fragment | NA | NA | 0 |
| S11-final-viral-combined_k141_300777\|\|full | 2895 | No | NA | 3 | 0 | 3 | Not-determined | Genome-fragment | NA | NA | 0 |
| S7-final-viral-combined_k141_221855\|\|full | 1178 | No | NA | 3 | 0 | 0 | Low-quality | Genome-fragment | 1.24 | AAI-based (high-confidence) | 0 |
| S7-final-viral-combined_k141_243737\|\|full | 1178 | No | NA | 4 | 0 | 1 | Low-quality | Genome-fragment | 2.52 | AAI-based (medium-confidence) | 0 |
| S7_k141_5955 | 1178 | No | NA | 3 | 0 | 0 | Low-quality | Genome-fragment | 2.87 | AAI-based (high-confidence) | 0 |
| S11-final-viral-combined_k141_232381\|\|full | 2659 | No | NA | 3 | 0 | 1 | Not-determined | Genome-fragment | NA | NA | 0 |
| S17-final-viral-combined_k141_225934\|\|lt2gene | 1178 | No | NA | 2 | 1 | 0 | Low-quality | Genome-fragment | 3.88 | AAI-based (high-confidence) | 0 |
| S11-final-viral-combined_k141_39616\|\|full | 3186 | No | NA | 3 | 0 | 2 | Not-determined | Genome-fragment | NA | NA | 0 |
| S11-final-viral-combined_k141_300795\|\|full | 1967 | No | NA | 3 | 0 | 0 | Not-determined | Genome-fragment | NA | NA | 0 |
| S11-final-viral-combined_k141_218314\|\|full | 3111 | No | NA | 3 | 0 | 2 | Not-determined | Genome-fragment | NA | NA | 0 |
| S11-final-viral-combined_k141_67111\|\|full | 3318 | No | NA | 4 | 0 | 3 | Not-determined | Genome-fragment | NA | NA | 0 |
| S11-final-viral-combined_k141_246860\|\|full | 2451 | No | NA | 3 | 0 | 1 | Not-determined | Genome-fragment | NA | NA | 0 |
| S11-final-viral-combined_k141_11918\|\|full | 2298 | No | NA | 3 | 0 | 3 | Not-determined | Genome-fragment | NA | NA | 0 |
| S2_k141_48539 | 1178 | No | NA | 1 | 0 | 0 | Low-quality | Genome-fragment | 3.22 | AAI-based (high-confidence) | 0 |
| S2_k141_89081 | 1178 | No | NA | 1 | 0 | 1 | Low-quality | Genome-fragment | 3.96 | AAI-based (medium-confidence) | 0 |
| S4_k141_36994 | 1178 | No | NA | 5 | 0 | 0 | Low-quality | Genome-fragment | 3.2 | AAI-based (high-confidence) | 0 |
| S6-final-viral-combined_k141_113300\|\|lt2gene | 1178 | No | NA | 2 | 1 | 0 | Low-quality | Genome-fragment | 2.69 | AAI-based (medium-confidence) | 0 |
| S11-final-viral-combined_k141_149626\|\|full | 1818 | No | NA | 3 | 0 | 2 | Not-determined | Genome-fragment | NA | NA | 0 |
| S11-final-viral-combined_k141_273141\|\|full | 1454 | No | NA | 2 | 0 | 2 | Not-determined | Genome-fragment | NA | NA | 0 |
| S11-final-viral-combined_k141_314841\|\|full | 3042 | No | NA | 4 | 0 | 3 | Not-determined | Genome-fragment | NA | NA | 0 |
| S8-final-viral-combined_k141_37209\|\|full | 1178 | No | NA | 3 | 0 | 0 | Low-quality | Genome-fragment | 2.89 | AAI-based (medium-confidence) | 0 |
| S9-final-viral-combined_k141_14380\|\|full | 1179 | No | NA | 2 | 0 | 0 | Low-quality | Genome-fragment | 2.52 | AAI-based (medium-confidence) | 0 |
| S9_k141_102230 | 1179 | No | NA | 1 | 0 | 0 | Low-quality | Genome-fragment | 1.97 | AAI-based (high-confidence) | 0 |
| S11-final-viral-combined_k141_53659\|\|full | 2212 | No | NA | 4 | 0 | 2 | Not-determined | Genome-fragment | NA | NA | 0 |
| S1-final-viral-combined_k141_18177\|\|full | 1179 | No | NA | 3 | 0 | 1 | Low-quality | Genome-fragment | 0.59 | AAI-based (medium-confidence) | 0 |
| S3-final-viral-combined_k141_32553\|\|full | 1179 | No | NA | 3 | 0 | 0 | Low-quality | Genome-fragment | 2.49 | AAI-based (medium-confidence) | 0 |
| S11-final-viral-combined_k141_163688\|\|full | 1239 | No | NA | 3 | 0 | 1 | Not-determined | Genome-fragment | NA | NA | 0 |
| S11-final-viral-combined_k141_53678\|\|full | 1079 | No | NA | 2 | 0 | 1 | Not-determined | Genome-fragment | NA | NA | 0 |
| S11-final-viral-combined_k141_327401\|\|full | 2976 | No | NA | 3 | 0 | 3 | Not-determined | Genome-fragment | NA | NA | 0 |
| S11-final-viral-combined_k141_177536\|\|full | 1356 | No | NA | 3 | 0 | 0 | Not-determined | Genome-fragment | NA | NA | 0 |
| S3-final-viral-combined_k141_45698\|\|full | 1179 | No | NA | 2 | 0 | 0 | Low-quality | Genome-fragment | 0.84 | AAI-based (medium-confidence) | 0 |
| S7-final-viral-combined_k141_3097\|\|full | 1179 | No | NA | 3 | 1 | 0 | Low-quality | Genome-fragment | 2.92 | AAI-based (high-confidence) | 0 |
| S7-final-viral-combined_k141_209842\|\|full | 1179 | No | NA | 3 | 0 | 0 | Low-quality | Genome-fragment | 2.22 | AAI-based (medium-confidence) | 0 |
| S2-final-viral-combined_k141_142120\|\|full | 1179 | No | NA | 3 | 0 | 0 | Low-quality | Genome-fragment | 2.08 | AAI-based (medium-confidence) | 0 |
| S11-final-viral-combined_k141_67193\|\|full | 1489 | No | NA | 2 | 0 | 2 | Not-determined | Genome-fragment | NA | NA | 0 |
| S11-final-viral-combined_k141_67204\|\|full | 1531 | No | NA | 3 | 0 | 3 | Not-determined | Genome-fragment | NA | NA | 0 |
| S4_k141_116818 | 1179 | No | NA | 3 | 0 | 0 | Low-quality | Genome-fragment | 2.43 | AAI-based (medium-confidence) | 0 |
| S8-final-viral-combined_k141_62412\|\|full | 1179 | No | NA | 2 | 0 | 0 | Low-quality | Genome-fragment | 1.38 | AAI-based (medium-confidence) | 0 |
| S1-final-viral-combined_k141_239630\|\|full | 1180 | No | NA | 3 | 0 | 1 | Low-quality | Genome-fragment | 0.54 | AAI-based (medium-confidence) | 0 |
| S11-final-viral-combined_k141_135052\|\|full | 1804 | No | NA | 2 | 0 | 2 | Not-determined | Genome-fragment | NA | NA | 0 |
| S1-final-viral-combined_k141_98282\|\|full | 1180 | No | NA | 4 | 0 | 0 | Low-quality | Genome-fragment | 0.79 | AAI-based (medium-confidence) | 0 |
| S3-final-viral-combined_k141_86000\|\|lt2gene | 1180 | No | NA | 2 | 2 | 0 | Low-quality | Genome-fragment | 2.54 | AAI-based (high-confidence) | 0 |
| S11-final-viral-combined_k141_135076\|\|full | 2620 | No | NA | 3 | 0 | 1 | Not-determined | Genome-fragment | NA | NA | 0 |
| S17-final-viral-combined_k141_71068\|\|lt2gene | 1180 | No | NA | 2 | 2 | 0 | Low-quality | Genome-fragment | 3.05 | AAI-based (high-confidence) | 0 |
| S17-k141_204970 | 1180 | No | NA | 2 | 0 | 1 | Low-quality | Genome-fragment | 1.11 | AAI-based (high-confidence) | 0 |
| S11-final-viral-combined_k141_287787\|\|full | 3604 | No | NA | 6 | 0 | 0 | Not-determined | Genome-fragment | NA | NA | 0 |
| S12-final-viral-combined_k141_189237\|\|full | 1180 | No | NA | 2 | 0 | 0 | Low-quality | Genome-fragment | 2.03 | AAI-based (high-confidence) | 0 |
| S11-final-viral-combined_k141_327506\|\|full | 1833 | No | NA | 4 | 0 | 0 | Not-determined | Genome-fragment | NA | NA | 0 |
| S12-final-viral-combined_k141_379109\|\|full | 1180 | No | NA | 5 | 1 | 0 | Low-quality | Genome-fragment | 2.67 | AAI-based (high-confidence) | 0 |
| S11-final-viral-combined_k141_327515\|\|full | 5378 | No | NA | 6 | 0 | 2 | Not-determined | Genome-fragment | NA | NA | 0 |
| S11-final-viral-combined_k141_135121\|\|full | 1836 | No | NA | 3 | 0 | 1 | Not-determined | Genome-fragment | NA | NA | 0 |
| S11-final-viral-combined_k141_300884\|\|full | 1226 | No | NA | 3 | 0 | 1 | Not-determined | Genome-fragment | NA | NA | 0 |
| S2-final-viral-combined_k141_55686\|\|full | 1180 | No | NA | 3 | 0 | 0 | Low-quality | Genome-fragment | 2.31 | AAI-based (medium-confidence) | 0 |
| S11-final-viral-combined_k141_177619\|\|full | 5825 | No | NA | 5 | 0 | 1 | Not-determined | Genome-fragment | NA | NA | 0 |
| S11-final-viral-combined_k141_247052\|\|full | 1889 | No | NA | 2 | 0 | 1 | Not-determined | Genome-fragment | NA | NA | 0 |
| S4-final-viral-combined_k141_71645\|\|full | 1180 | No | NA | 2 | 0 | 0 | Low-quality | Genome-fragment | 2.76 | AAI-based (high-confidence) | 0 |
| S11_k141_145671 | 1181 | No | NA | 5 | 0 | 0 | Low-quality | Genome-fragment | 2.5 | AAI-based (high-confidence) | 0 |
| S11-final-viral-combined_k141_109071\|\|full | 3147 | No | NA | 4 | 0 | 1 | Not-determined | Genome-fragment | NA | NA | 0 |
| S1-final-viral-combined_k141_360582\|\|full | 1181 | No | NA | 2 | 0 | 2 | Low-quality | Genome-fragment | 0.75 | AAI-based (medium-confidence) | 0 |
| S11-final-viral-combined_k141_53770\|\|full | 1760 | No | NA | 2 | 0 | 1 | Not-determined | Genome-fragment | NA | NA | 0 |
| S11-final-viral-combined_k141_190420\|\|full | 1243 | No | NA | 2 | 0 | 0 | Not-determined | Genome-fragment | NA | NA | 0 |
| S11-final-viral-combined_k141_327565\|\|full | 1983 | No | NA | 3 | 0 | 0 | Not-determined | Genome-fragment | NA | NA | 0 |
| S1_k141_51933 | 1181 | No | NA | 2 | 0 | 0 | Low-quality | Genome-fragment | 1.75 | AAI-based (high-confidence) | 0 |
| S11-final-viral-combined_k141_190424\|\|full | 5442 | No | NA | 4 | 0 | 1 | Not-determined | Genome-fragment | NA | NA | 0 |
| S11-final-viral-combined_k141_163916\|\|full | 4063 | No | NA | 4 | 0 | 3 | Not-determined | Genome-fragment | NA | NA | 0 |
| S1_k141_193370 | 1181 | No | NA | 3 | 1 | 0 | Low-quality | Genome-fragment | 1.43 | AAI-based (high-confidence) | 0 |
| S3-final-viral-combined_k141_105525\|\|full | 1181 | No | NA | 4 | 0 | 0 | Low-quality | Genome-fragment | 3.21 | AAI-based (high-confidence) | 0 |
| S5-final-viral-combined_k141_77674\|\|full | 1181 | No | NA | 3 | 3 | 0 | Low-quality | Genome-fragment | 3.21 | AAI-based (medium-confidence) | 0 |
| S11-final-viral-combined_k141_39912\|\|full | 1553 | No | NA | 4 | 0 | 0 | Not-determined | Genome-fragment | NA | NA | 0 |
| S5-final-viral-combined_k141_160659\|\|lt2gene | 1181 | No | NA | 2 | 1 | 0 | Low-quality | Genome-fragment | 1.77 | AAI-based (medium-confidence) | 0 |
| S11-final-viral-combined_k141_39918\|\|full | 2254 | No | NA | 2 | 0 | 1 | Not-determined | Genome-fragment | NA | NA | 0 |
| S7-final-viral-combined_k141_234938\|\|full | 1181 | No | NA | 2 | 0 | 0 | Low-quality | Genome-fragment | 2.46 | AAI-based (medium-confidence) | 0 |
| S11-final-viral-combined_k141_315046\|\|full | 1471 | No | NA | 3 | 0 | 0 | Not-determined | Genome-fragment | NA | NA | 0 |
| S17-final-viral-combined_k141_33740\|\|full | 1181 | No | NA | 3 | 0 | 0 | Low-quality | Genome-fragment | 1.89 | AAI-based (medium-confidence) | 0 |
| S11-final-viral-combined_k141_39930\|\|full | 1404 | No | NA | 2 | 0 | 0 | Not-determined | Genome-fragment | NA | NA | 0 |
| S11-final-viral-combined_k141_94498\|\|full | 3503 | No | NA | 4 | 0 | 4 | Not-determined | Genome-fragment | NA | NA | 0 |
| S11-final-viral-combined_k141_204522\|\|full | 30685 | No | NA | 35 | 12 | 0 | Medium-quality | Genome-fragment | 66.54 | AAI-based (high-confidence) | 0 |
| S11-final-viral-combined_k141_67279\|\|full | 3404 | No | NA | 3 | 0 | 1 | Not-determined | Genome-fragment | NA | NA | 0 |
| S12-final-viral-combined_k141_121545\|\|full | 1181 | No | NA | 4 | 1 | 0 | Low-quality | Genome-fragment | 1.91 | AAI-based (high-confidence) | 0 |
| S12-final-viral-combined_k141_361985\|\|full | 1181 | No | NA | 5 | 0 | 0 | Low-quality | Genome-fragment | 1.88 | AAI-based (high-confidence) | 0 |
| S12-final-viral-combined_k141_249522\|\|full | 1181 | No | NA | 2 | 0 | 1 | Low-quality | Genome-fragment | 0.36 | AAI-based (medium-confidence) | 0 |
| S12_k141_232515 | 1181 | No | NA | 1 | 0 | 0 | Low-quality | Genome-fragment | 3.02 | AAI-based (high-confidence) | 0 |
| S12_k141_114490 | 1181 | No | NA | 2 | 0 | 2 | Low-quality | Genome-fragment | 0.49 | AAI-based (medium-confidence) | 0 |
| S11-final-viral-combined_k141_164006\|\|full | 1523 | No | NA | 3 | 0 | 0 | Not-determined | Genome-fragment | NA | NA | 0 |
| S4-final-viral-combined_k141_134635\|\|full | 1181 | No | NA | 2 | 0 | 1 | Low-quality | Genome-fragment | 1.63 | AAI-based (medium-confidence) | 0 |
| S11-final-viral-combined_k141_232649\|\|full | 1890 | No | NA | 2 | 0 | 0 | Not-determined | Genome-fragment | NA | NA | 0 |
| S11-final-viral-combined_k141_190522\|\|full | 1894 | No | NA | 3 | 0 | 1 | Not-determined | Genome-fragment | NA | NA | 0 |
| S11-final-viral-combined_k141_247182\|\|full | 2407 | No | NA | 2 | 0 | 1 | Not-determined | Genome-fragment | NA | NA | 0 |
| S11-final-viral-combined_k141_190526\|\|full | 1183 | No | NA | 2 | 0 | 1 | Not-determined | Genome-fragment | NA | NA | 0 |
| S11-final-viral-combined_k141_288028\|\|full | 1221 | No | NA | 2 | 0 | 0 | Not-determined | Genome-fragment | NA | NA | 0 |
| S4-final-viral-combined_k141_13785\|\|full | 1181 | No | NA | 2 | 0 | 1 | Low-quality | Genome-fragment | 1.28 | AAI-based (high-confidence) | 0 |
| S11-final-viral-combined_k141_80324\|\|full | 1372 | No | NA | 4 | 0 | 0 | Not-determined | Genome-fragment | NA | NA | 0 |
| S6-final-viral-combined_k141_76479\|\|full | 1181 | No | NA | 3 | 0 | 0 | Low-quality | Genome-fragment | 0.78 | AAI-based (medium-confidence) | 0 |
| S11-final-viral-combined_k141_288046\|\|full | 1549 | No | NA | 2 | 0 | 2 | Not-determined | Genome-fragment | NA | NA | 0 |
| S11-final-viral-combined_k141_135223\|\|full | 1464 | No | NA | 2 | 0 | 0 | Not-determined | Genome-fragment | NA | NA | 0 |
| S11-final-viral-combined_k141_53880\|\|full | 1701 | No | NA | 3 | 0 | 2 | Not-determined | Genome-fragment | NA | NA | 0 |
| S11-final-viral-combined_k141_94598\|\|full | 2479 | No | NA | 4 | 0 | 3 | Not-determined | Genome-fragment | NA | NA | 0 |
| S11-final-viral-combined_k141_164043\|\|full | 2099 | No | NA | 1 | 0 | 0 | Not-determined | Genome-fragment | NA | NA | 0 |
| S11-final-viral-combined_k141_247220\|\|full | 1640 | No | NA | 2 | 0 | 1 | Not-determined | Genome-fragment | NA | NA | 0 |
| S11-final-viral-combined_k141_149928\|\|full | 1639 | No | NA | 2 | 0 | 0 | Not-determined | Genome-fragment | NA | NA | 0 |
| S11-final-viral-combined_k141_121616\|\|full | 1471 | No | NA | 4 | 0 | 1 | Not-determined | Genome-fragment | NA | NA | 0 |
| S8-final-viral-combined_k141_176625\|\|full | 1181 | No | NA | 2 | 1 | 0 | Low-quality | Genome-fragment | 0.71 | HMM-based (lower-bound) | 0 |
| S8-final-viral-combined_k141_96488\|\|full | 1181 | No | NA | 2 | 0 | 0 | Low-quality | Genome-fragment | 2.63 | AAI-based (high-confidence) | 0 |
| S8-final-viral-combined_k141_146960\|\|full | 1181 | No | NA | 2 | 1 | 0 | Low-quality | Genome-fragment | 2.6 | AAI-based (high-confidence) | 0 |
| S11-final-viral-combined_k141_190594\|\|full | 2720 | No | NA | 4 | 0 | 0 | Not-determined | Genome-fragment | NA | NA | 0 |
| S11-final-viral-combined_k141_149961\|\|full | 1388 | No | NA | 2 | 0 | 1 | Not-determined | Genome-fragment | NA | NA | 0 |
| S8_k141_12422 | 1181 | No | NA | 2 | 0 | 0 | Low-quality | Genome-fragment | 0.44 | AAI-based (medium-confidence) | 0 |
| S11-final-viral-combined_k141_173658\|\|full | 1182 | No | NA | 2 | 0 | 1 | Low-quality | Genome-fragment | 0.54 | AAI-based (medium-confidence) | 0 |
| S11-final-viral-combined_k141_176502\|\|full | 1182 | No | NA | 2 | 1 | 0 | Low-quality | Genome-fragment | 2.01 | AAI-based (medium-confidence) | 0 |
| S1-final-viral-combined_k141_35082\|\|full | 1182 | No | NA | 2 | 0 | 0 | Low-quality | Genome-fragment | 1.44 | AAI-based (high-confidence) | 0 |
| S1_k141_100511 | 1182 | No | NA | 1 | 0 | 1 | Low-quality | Genome-fragment | 0.79 | AAI-based (high-confidence) | 0 |
| S11-final-viral-combined_k141_190624\|\|full | 2130 | No | NA | 5 | 0 | 1 | Not-determined | Genome-fragment | NA | NA | 0 |
| S11-final-viral-combined_k141_80385\|\|full | 1952 | No | NA | 2 | 0 | 2 | Not-determined | Genome-fragment | NA | NA | 0 |
| S5-final-viral-combined_k141_23788\|\|full | 1182 | No | NA | 4 | 2 | 0 | Low-quality | Genome-fragment | 2.21 | AAI-based (high-confidence) | 0 |
| S11-final-viral-combined_k141_232774\|\|full | 2128 | No | NA | 3 | 0 | 1 | Not-determined | Genome-fragment | NA | NA | 0 |
| S5-final-viral-combined_k141_77934\|\|lt2gene | 1182 | No | NA | 2 | 2 | 0 | Low-quality | Genome-fragment | 2.63 | AAI-based (medium-confidence) | 0 |
| S11-final-viral-combined_k141_12240\|\|full | 1372 | No | NA | 2 | 0 | 0 | Not-determined | Genome-fragment | NA | NA | 0 |
| S11-final-viral-combined_k141_177894\|\|full | 2476 | No | NA | 3 | 0 | 1 | Not-determined | Genome-fragment | NA | NA | 0 |
| S11-final-viral-combined_k141_315198\|\|full | 1285 | No | NA | 5 | 0 | 0 | Not-determined | Genome-fragment | NA | NA | 0 |
| S5_k141_20540 | 1182 | No | NA | 2 | 0 | 0 | Low-quality | Genome-fragment | 2.39 | AAI-based (medium-confidence) | 0 |
| S7-final-viral-combined_k141_209631\|\|full | 1182 | No | NA | 3 | 2 | 0 | Low-quality | Genome-fragment | 1.91 | AAI-based (medium-confidence) | 0 |
| S11-final-viral-combined_k141_109231\|\|full | 4063 | No | NA | 4 | 0 | 2 | Not-determined | Genome-fragment | NA | NA | 0 |
| S17-k141_258606 | 1182 | No | NA | 4 | 0 | 0 | Low-quality | Genome-fragment | 1.66 | AAI-based (high-confidence) | 0 |
| S11-final-viral-combined_k141_260603\|\|full | 2783 | No | NA | 4 | 0 | 1 | Not-determined | Genome-fragment | NA | NA | 0 |
| S2-final-viral-combined_k141_133608\|\|full | 1182 | No | NA | 3 | 0 | 2 | Low-quality | Genome-fragment | 1.65 | AAI-based (medium-confidence) | 0 |
| S11-final-viral-combined_k141_247323\|\|full | 5327 | No | NA | 6 | 0 | 4 | Not-determined | Genome-fragment | NA | NA | 0 |
| S2_k141_128440 | 1182 | No | NA | 2 | 1 | 0 | Low-quality | Genome-fragment | 2.4 | AAI-based (high-confidence) | 0 |
| S11-final-viral-combined_k141_327765\|\|full | 1003 | No | NA | 3 | 0 | 0 | Not-determined | Genome-fragment | NA | NA | 0 |
| S11-final-viral-combined_k141_247333\|\|full | 2049 | No | NA | 3 | 0 | 1 | Not-determined | Genome-fragment | NA | NA | 0 |
| S4_k141_99229 | 1182 | No | NA | 2 | 0 | 1 | Low-quality | Genome-fragment | 2.57 | AAI-based (medium-confidence) | 0 |
| S11-final-viral-combined_k141_273513\|\|full | 1644 | No | NA | 2 | 0 | 2 | Not-determined | Genome-fragment | NA | NA | 0 |
| S6_k141_124448 | 1182 | No | NA | 2 | 1 | 0 | Low-quality | Genome-fragment | 0.75 | HMM-based (lower-bound) | 0 |
| S11-final-viral-combined_k141_273529\|\|full | 3632 | No | NA | 4 | 0 | 2 | Not-determined | Genome-fragment | NA | NA | 0 |
| S11-final-viral-combined_k141_218752\|\|full | 2069 | No | NA | 4 | 0 | 0 | Not-determined | Genome-fragment | NA | NA | 0 |
| S11-final-viral-combined_k141_218769\|\|full | 1351 | No | NA | 4 | 0 | 0 | Not-determined | Genome-fragment | NA | NA | 0 |
| S11-final-viral-combined_k141_12389\|\|full | 4400 | No | NA | 5 | 0 | 2 | Not-determined | Genome-fragment | NA | NA | 0 |
| S11-final-viral-combined_k141_24779\|\|full | 2881 | No | NA | 3 | 0 | 1 | Not-determined | Genome-fragment | NA | NA | 0 |
| S11-final-viral-combined_k141_301122\|\|full | 1125 | No | NA | 2 | 0 | 1 | Not-determined | Genome-fragment | NA | NA | 0 |
| S9-final-viral-combined_k141_43712\|\|full | 1183 | No | NA | 3 | 0 | 0 | Low-quality | Genome-fragment | 2.9 | AAI-based (medium-confidence) | 0 |
| S11-final-viral-combined_k141_178041\|\|full | 1342 | No | NA | 2 | 0 | 1 | Not-determined | Genome-fragment | NA | NA | 0 |
| S9_k141_111764 | 1183 | No | NA | 4 | 0 | 0 | Low-quality | Genome-fragment | 1.59 | AAI-based (medium-confidence) | 0 |
| S11-final-viral-combined_k141_190789\|\|full | 2101 | No | NA | 2 | 0 | 1 | Not-determined | Genome-fragment | NA | NA | 0 |
| S11-final-viral-combined_k141_40252\|\|full | 1105 | No | NA | 2 | 0 | 1 | Not-determined | Genome-fragment | NA | NA | 0 |
| S11-final-viral-combined_k141_199640\|\|full | 1183 | No | NA | 3 | 1 | 0 | Low-quality | Genome-fragment | 1.89 | AAI-based (high-confidence) | 0 |
| S11-final-viral-combined_k141_40256\|\|full | 2396 | No | NA | 2 | 0 | 1 | Not-determined | Genome-fragment | NA | NA | 0 |
| S11-final-viral-combined_k141_269969\|\|full | 1183 | No | NA | 2 | 1 | 0 | Low-quality | Genome-fragment | 0.75 | HMM-based (lower-bound) | 0 |
| S11-final-viral-combined_k141_40274\|\|full | 1992 | No | NA | 3 | 0 | 1 | Not-determined | Genome-fragment | NA | NA | 0 |

| | | | | | | | | | | | |
|---|---|---|---|---|---|---|---|---|---|---|---|
| S11-final-viral-combined_k141_54027\|\|full | 6540 | No | NA | 7 | 0 | 6 | Not-determined | Genome-fragment | NA | NA | 0 |
| S11-final-viral-combined_k141_259984\|\|lt2gene | 1183 | No | NA | 2 | 2 | 0 | Low-quality | Genome-fragment | 3.19 | AAI-based (high-confidence) | 0 |
| S11-final-viral-combined_k141_273619\|\|full | 17297 | No | NA | 13 | 0 | 9 | Not-determined | Genome-fragment | NA | NA | 0 |
| S11-final-viral-combined_k141_40306\|\|full | 1937 | No | NA | 2 | 0 | 2 | Not-determined | Genome-fragment | NA | NA | 0 |
| S1_k141_193675 | 1183 | No | NA | 2 | 0 | 0 | Low-quality | Genome-fragment | 0.77 | AAI-based (medium-confidence) | 0 |
| S11-final-viral-combined_k141_247376\|\|full | 1327 | No | NA | 3 | 0 | 0 | Not-determined | Genome-fragment | NA | NA | 0 |
| S11-final-viral-combined_k141_150192\|\|full | 4416 | No | NA | 7 | 0 | 0 | Not-determined | Genome-fragment | NA | NA | 0 |
| S11-final-viral-combined_k141_273646\|\|full | 3588 | No | NA | 3 | 0 | 1 | Not-determined | Genome-fragment | NA | NA | 0 |
| S17-final-viral-combined_k141_258258\|\|full | 1183 | No | NA | 4 | 1 | 0 | Low-quality | Genome-fragment | 2.87 | AAI-based (medium-confidence) | 0 |
| S11-final-viral-combined_k141_150210\|\|full | 1758 | No | NA | 4 | 0 | 2 | Not-determined | Genome-fragment | NA | NA | 0 |
| S11-final-viral-combined_k141_67643\|\|full | 7699 | No | NA | 3 | 0 | 2 | Not-determined | Genome-fragment | NA | NA | 0 |
| S17-k141_115366 | 1183 | No | NA | 1 | 0 | 0 | Low-quality | Genome-fragment | 0.82 | AAI-based (high-confidence) | 0 |
| S12_k141_70007 | 1183 | No | NA | 2 | 0 | 0 | Low-quality | Genome-fragment | 0.67 | AAI-based (medium-confidence) | 0 |
| S11-final-viral-combined_k141_260837\|\|full | 9683 | No | NA | 8 | 0 | 4 | Not-determined | Genome-fragment | NA | NA | 0 |
| S2-final-viral-combined_k141_214549\|\|full | 1183 | No | NA | 2 | 0 | 0 | Low-quality | Genome-fragment | 2.19 | AAI-based (high-confidence) | 0 |
| S2-final-viral-combined_k141_185872\|\|lt2gene | 1183 | No | NA | 2 | 0 | 0 | Low-quality | Genome-fragment | 2.76 | AAI-based (medium-confidence) | 0 |
| S2_k141_63579 | 1183 | No | NA | 1 | 0 | 1 | Low-quality | Genome-fragment | 0.31 | AAI-based (high-confidence) | 0 |
| S11-final-viral-combined_k141_301161\|\|full | 3371 | No | NA | 3 | 0 | 2 | Not-determined | Genome-fragment | NA | NA | 0 |
| S11-final-viral-combined_k141_121821\|\|full | 1965 | No | NA | 2 | 0 | 0 | Not-determined | Genome-fragment | NA | NA | 0 |
| S2_k141_131801 | 1183 | No | NA | 2 | 0 | 0 | Low-quality | Genome-fragment | 1.23 | AAI-based (high-confidence) | 0 |
| S6-final-viral-combined_k141_103191\|\|full | 1183 | No | NA | 4 | 0 | 0 | Low-quality | Genome-fragment | 0.91 | AAI-based (high-confidence) | 0 |
| S8_k141_102680 | 1183 | No | NA | 2 | 0 | 0 | Low-quality | Genome-fragment | 7.61 | AAI-based (medium-confidence) | 0 |
| S11-final-viral-combined_k141_178107\|\|full | 1125 | No | NA | 3 | 0 | 1 | Not-determined | Genome-fragment | NA | NA | 0 |
| S11-final-viral-combined_k141_40383\|\|full | 1214 | No | NA | 4 | 0 | 0 | Not-determined | Genome-fragment | NA | NA | 0 |
| S11-final-viral-combined_k141_200302\|\|full | 1184 | No | NA | 3 | 0 | 0 | Low-quality | Genome-fragment | 2.07 | AAI-based (high-confidence) | 0 |
| S11-final-viral-combined_k141_12564\|\|full | 2530 | No | NA | 2 | 0 | 2 | Not-determined | Genome-fragment | NA | NA | 0 |
| S11-final-viral-combined_k141_135631\|\|full | 2304 | No | NA | 2 | 0 | 2 | Not-determined | Genome-fragment | NA | NA | 0 |
| S11-final-viral-combined_k141_327900\|\|full | 3173 | No | NA | 2 | 0 | 2 | Not-determined | Genome-fragment | NA | NA | 0 |
| S11-final-viral-combined_k141_327903\|\|full | 1782 | No | NA | 3 | 0 | 2 | Not-determined | Genome-fragment | NA | NA | 0 |
| S1-final-viral-combined_k141_250855\|\|full | 1184 | No | NA | 3 | 3 | 0 | Low-quality | Genome-fragment | 2.95 | AAI-based (high-confidence) | 0 |
| S11-final-viral-combined_k141_288277\|\|full | 5935 | No | NA | 4 | 0 | 4 | Not-determined | Genome-fragment | NA | NA | 0 |
| S1-final-viral-combined_k141_303952\|\|full | 1184 | No | NA | 2 | 0 | 0 | Low-quality | Genome-fragment | 2.89 | AAI-based (medium-confidence) | 0 |
| S11-final-viral-combined_k141_301257\|\|full | 2657 | No | NA | 2 | 0 | 2 | Not-determined | Genome-fragment | NA | NA | 0 |
| S1-final-viral-combined_k141_109605\|\|full | 1184 | No | NA | 3 | 0 | 0 | Low-quality | Genome-fragment | 1.81 | AAI-based (high-confidence) | 0 |
| S11-final-viral-combined_k141_135685\|\|full | 1881 | No | NA | 3 | 0 | 3 | Not-determined | Genome-fragment | NA | NA | 0 |
| S11-final-viral-combined_k141_190972\|\|full | 1391 | No | NA | 2 | 0 | 2 | Not-determined | Genome-fragment | NA | NA | 0 |
| S11-final-viral-combined_k141_327931\|\|full | 1145 | No | NA | 2 | 0 | 2 | Not-determined | Genome-fragment | NA | NA | 0 |
| S11-final-viral-combined_k141_190975\|\|full | 1973 | No | NA | 2 | 0 | 1 | Not-determined | Genome-fragment | NA | NA | 0 |
| S1-final-viral-combined_k141_91380\|\|lt2gene | 1184 | No | NA | 1 | 0 | 0 | Low-quality | Genome-fragment | 2.46 | AAI-based (medium-confidence) | 0 |
| S1-final-viral-combined_k141_390483\|\|lt2gene | 1184 | No | NA | 2 | 1 | 0 | Low-quality | Genome-fragment | 2.79 | AAI-based (high-confidence) | 0 |
| S1_k141_170410 | 1184 | No | NA | 1 | 0 | 1 | Low-quality | Genome-fragment | 0.44 | AAI-based (high-confidence) | 0 |
| S11-final-viral-combined_k141_109637\|\|full | 1292 | No | NA | 2 | 0 | 2 | Not-determined | Genome-fragment | NA | NA | 0 |
| S11-final-viral-combined_k141_150384\|\|full | 1727 | No | NA | 2 | 0 | 0 | Not-determined | Genome-fragment | NA | NA | 0 |
| S11-final-viral-combined_k141_150388\|\|full | 1844 | No | NA | 2 | 0 | 1 | Not-determined | Genome-fragment | NA | NA | 0 |
| S7_k141_75336 | 1184 | No | NA | 2 | 0 | 2 | Low-quality | Genome-fragment | 0.77 | AAI-based (medium-confidence) | 0 |
| S17-k141_193920 | 1184 | No | NA | 3 | 0 | 0 | Low-quality | Genome-fragment | 1.61 | AAI-based (high-confidence) | 0 |
| S11-final-viral-combined_k141_164284\|\|full | 1228 | No | NA | 3 | 0 | 3 | Not-determined | Genome-fragment | NA | NA | 0 |
| S12-final-viral-combined_k141_81888\|\|full | 1184 | No | NA | 3 | 3 | 0 | Low-quality | Genome-fragment | 3.17 | AAI-based (medium-confidence) | 0 |
| S11-final-viral-combined_k141_12637\|\|full | 1567 | No | NA | 3 | 0 | 3 | Not-determined | Genome-fragment | NA | NA | 0 |
| S11-final-viral-combined_k141_67845\|\|full | 1609 | No | NA | 3 | 0 | 1 | Not-determined | Genome-fragment | NA | NA | 0 |
| S12-final-viral-combined_k141_350420\|\|full | 1184 | No | NA | 3 | 1 | 0 | Low-quality | Genome-fragment | 2.52 | AAI-based (medium-confidence) | 0 |
| S11-final-viral-combined_k141_80600\|\|full | 1465 | No | NA | 4 | 0 | 1 | Not-determined | Genome-fragment | NA | NA | 0 |
| S11-final-viral-combined_k141_191059\|\|full | 1791 | No | NA | 3 | 0 | 1 | Not-determined | Genome-fragment | NA | NA | 0 |
| S11-final-viral-combined_k141_273868\|\|full | 1985 | No | NA | 3 | 0 | 2 | Not-determined | Genome-fragment | NA | NA | 0 |
| S12_k141_342329 | 1184 | No | NA | 2 | 0 | 2 | Low-quality | Genome-fragment | 1.96 | AAI-based (high-confidence) | 0 |
| S11-final-viral-combined_k141_204970\|\|full | 2173 | No | NA | 3 | 0 | 1 | Not-determined | Genome-fragment | NA | NA | 0 |
| S2-final-viral-combined_k141_219177\|\|full | 1184 | No | NA | 2 | 1 | 0 | Low-quality | Genome-fragment | 1.73 | HMM-based (lower-bound) | 0 |
| S11-final-viral-combined_k141_247594\|\|full | 6155 | No | NA | 6 | 0 | 3 | Not-determined | Genome-fragment | NA | NA | 0 |
| S8-final-viral-combined_k141_108233\|\|full | 1184 | No | NA | 3 | 0 | 0 | Low-quality | Genome-fragment | 0.68 | AAI-based (high-confidence) | 0 |
| S11-final-viral-combined_k141_109719\|\|full | 1728 | No | NA | 2 | 0 | 1 | Not-determined | Genome-fragment | NA | NA | 0 |
| S11-final-viral-combined_k141_40545\|\|full | 1185 | No | NA | 2 | 0 | 1 | Low-quality | Genome-fragment | 1.97 | AAI-based (medium-confidence) | 0 |
| S11-final-viral-combined_k141_122010\|\|full | 7761 | No | NA | 8 | 0 | 8 | Not-determined | Genome-fragment | NA | NA | 0 |
| S11-final-viral-combined_k141_54280\|\|full | 3638 | No | NA | 6 | 0 | 0 | Not-determined | Genome-fragment | NA | NA | 0 |
| S11-final-viral-combined_k141_328025\|\|full | 5836 | No | NA | 5 | 0 | 4 | Not-determined | Genome-fragment | NA | NA | 0 |
| S11-final-viral-combined_k141_25022\|\|full | 2420 | No | NA | 4 | 0 | 0 | Not-determined | Genome-fragment | NA | NA | 0 |
| S11_k141_30556 | 1185 | No | NA | 3 | 1 | 0 | Low-quality | Genome-fragment | 2.04 | AAI-based (high-confidence) | 0 |
| S11-final-viral-combined_k141_288422\|\|full | 2361 | No | NA | 3 | 0 | 1 | Not-determined | Genome-fragment | NA | NA | 0 |
| S11-final-viral-combined_k141_67915\|\|full | 1158 | No | NA | 3 | 0 | 0 | Not-determined | Genome-fragment | NA | NA | 0 |
| S11-final-viral-combined_k141_80708\|\|full | 1024 | No | NA | 2 | 0 | 2 | Not-determined | Genome-fragment | NA | NA | 0 |
| S11-final-viral-combined_k141_315601\|\|full | 2054 | No | NA | 4 | 0 | 1 | Not-determined | Genome-fragment | NA | NA | 0 |
| S1-final-viral-combined_k141_348217\|\|full | 1185 | No | NA | 3 | 0 | 0 | Low-quality | Genome-fragment | 1.02 | AAI-based (medium-confidence) | 0 |
| S11-final-viral-combined_k141_219078\|\|full | 2330 | No | NA | 2 | 0 | 2 | Not-determined | Genome-fragment | NA | NA | 0 |
| S11-final-viral-combined_k141_135832\|\|full | 1541 | No | NA | 3 | 0 | 1 | Not-determined | Genome-fragment | NA | NA | 0 |
| S11-final-viral-combined_k141_301389\|\|full | 3150 | No | NA | 4 | 0 | 2 | Not-determined | Genome-fragment | NA | NA | 0 |
| S11-final-viral-combined_k141_274039\|\|full | 3273 | No | NA | 4 | 0 | 3 | Not-determined | Genome-fragment | NA | NA | 0 |
| S1-final-viral-combined_k141_372440\|\|full | 1185 | No | NA | 3 | 0 | 1 | Low-quality | Genome-fragment | 9.49 | AAI-based (medium-confidence) | 0 |
| S11-final-viral-combined_k141_122075\|\|full | 1078 | No | NA | 2 | 0 | 0 | Not-determined | Genome-fragment | NA | NA | 0 |
| S11-final-viral-combined_k141_315651\|\|full | 2619 | No | NA | 4 | 0 | 3 | Not-determined | Genome-fragment | NA | NA | 0 |
| S5-final-viral-combined_k141_82132\|\|full | 1185 | No | NA | 4 | 1 | 0 | Low-quality | Genome-fragment | 0.78 | HMM-based (lower-bound) | 0 |
| S11-final-viral-combined_k141_12804\|\|full | 3022 | No | NA | 3 | 0 | 1 | Not-determined | Genome-fragment | NA | NA | 0 |
| S11-final-viral-combined_k141_233322\|\|full | 3682 | No | NA | 6 | 0 | 6 | Not-determined | Genome-fragment | NA | NA | 0 |
| S5-final-viral-combined_k141_158247\|\|full | 1185 | No | NA | 3 | 0 | 0 | Low-quality | Genome-fragment | 0.68 | AAI-based (high-confidence) | 0 |
| S11-final-viral-combined_k141_315674\|\|full | 1778 | No | NA | 4 | 0 | 1 | Not-determined | Genome-fragment | NA | NA | 0 |
| S11-final-viral-combined_k141_301444\|\|full | 2584 | No | NA | 3 | 0 | 3 | Not-determined | Genome-fragment | NA | NA | 0 |
| S11-final-viral-combined_k141_54454\|\|full | 1702 | No | NA | 2 | 0 | 2 | Not-determined | Genome-fragment | NA | NA | 0 |
| S5_k141_126809 | 1185 | No | NA | 1 | 0 | 0 | Low-quality | Genome-fragment | 2.86 | AAI-based (high-confidence) | 0 |
| S11-final-viral-combined_k141_135934\|\|full | 5264 | No | NA | 6 | 0 | 4 | Not-determined | Genome-fragment | NA | NA | 0 |
| S11-final-viral-combined_k141_288610\|\|full | 2083 | No | NA | 3 | 0 | 1 | Not-determined | Genome-fragment | NA | NA | 0 |
| S7_k141_150040 | 1185 | No | NA | 3 | 0 | 0 | Low-quality | Genome-fragment | 2.17 | AAI-based (medium-confidence) | 0 |
| S11-final-viral-combined_k141_233370\|\|full | 2004 | No | NA | 3 | 0 | 2 | Not-determined | Genome-fragment | NA | NA | 0 |
| S11-final-viral-combined_k141_247835\|\|full | 2222 | No | NA | 2 | 0 | 2 | Not-determined | Genome-fragment | NA | NA | 0 |
| S11-final-viral-combined_k141_135946\|\|full | 2674 | No | NA | 2 | 0 | 2 | Not-determined | Genome-fragment | NA | NA | 0 |
| S11-final-viral-combined_k141_301482\|\|full | 3005 | No | NA | 3 | 0 | 1 | Not-determined | Genome-fragment | NA | NA | 0 |
| S11-final-viral-combined_k141_219172\|\|full | 3655 | No | NA | 4 | 0 | 3 | Not-determined | Genome-fragment | NA | NA | 0 |
| S11-final-viral-combined_k141_315728\|\|full | 3274 | No | NA | 6 | 0 | 3 | Not-determined | Genome-fragment | NA | NA | 0 |
| S11-final-viral-combined_k141_261182\|\|full | 7049 | No | NA | 6 | 0 | 4 | Not-determined | Genome-fragment | NA | NA | 0 |
| S11-final-viral-combined_k141_219179\|\|full | 2822 | No | NA | 2 | 0 | 1 | Not-determined | Genome-fragment | NA | NA | 0 |
| S12-final-viral-combined_k141_69674\|\|full | 1185 | No | NA | 3 | 1 | 0 | Low-quality | Genome-fragment | 2.17 | AAI-based (medium-confidence) | 0 |
| S11-final-viral-combined_k141_205155\|\|full | 1801 | No | NA | 3 | 0 | 0 | Not-determined | Genome-fragment | NA | NA | 0 |
| S12-final-viral-combined_k141_311441\|\|full | 1185 | No | NA | 3 | 0 | 0 | Low-quality | Genome-fragment | 1.1 | AAI-based (medium-confidence) | 0 |
| S8-final-viral-combined_k141_137774\|\|full | 1185 | No | NA | 4 | 0 | 0 | Low-quality | Genome-fragment | 0.95 | AAI-based (high-confidence) | 0 |
| S11-final-viral-combined_k141_40799\|\|full | 1318 | No | NA | 3 | 0 | 1 | Not-determined | Genome-fragment | NA | NA | 0 |
| S11-final-viral-combined_k141_25222\|\|full | 1720 | No | NA | 2 | 0 | 2 | Not-determined | Genome-fragment | NA | NA | 0 |
| S11-final-viral-combined_k141_219200\|\|full | 1149 | No | NA | 2 | 0 | 1 | Not-determined | Genome-fragment | NA | NA | 0 |
| S11-final-viral-combined_k141_274122\|\|full | 1238 | No | NA | 3 | 0 | 1 | Not-determined | Genome-fragment | NA | NA | 0 |
| S8-final-viral-combined_k141_186398\|\|lt2gene | 1185 | No | NA | 2 | 2 | 0 | Low-quality | Genome-fragment | 2.19 | HMM-based (lower-bound) | 0 |
| S11-final-viral-combined_k141_68126\|\|full | 1152 | No | NA | 3 | 0 | 1 | Not-determined | Genome-fragment | NA | NA | 0 |
| S11-final-viral-combined_k141_219208\|\|full | 1622 | No | NA | 3 | 0 | 1 | Not-determined | Genome-fragment | NA | NA | 0 |
| S9-final-viral-combined_k141_94703\|\|full | 1186 | No | NA | 3 | 0 | 0 | Low-quality | Genome-fragment | 2.99 | AAI-based (medium-confidence) | 0 |
| S7-final-viral-combined_k141_125265\|\|lt2gene | 1186 | No | NA | 2 | 1 | 0 | Low-quality | Genome-fragment | 0.68 | HMM-based (lower-bound) | 0 |
| S17-k141_171229 | 1186 | No | NA | 4 | 2 | 0 | Low-quality | Genome-fragment | 3.16 | AAI-based (high-confidence) | 0 |
| S11-final-viral-combined_k141_95220\|\|full | 1475 | No | NA | 3 | 0 | 0 | Not-determined | Genome-fragment | NA | NA | 0 |
| S12-final-viral-combined_k141_163170\|\|full | 1186 | No | NA | 3 | 2 | 0 | Low-quality | Genome-fragment | 2.92 | AAI-based (medium-confidence) | 0 |
| S4-final-viral-combined_k141_136223\|\|full | 1186 | No | NA | 2 | 0 | 1 | Low-quality | Genome-fragment | 0.69 | AAI-based (medium-confidence) | 0 |
| S8-final-viral-combined_k141_77581\|\|full | 1186 | No | NA | 2 | 0 | 1 | Low-quality | Genome-fragment | 7.56 | AAI-based (medium-confidence) | 0 |
| S11-final-viral-combined_k141_95225\|\|full | 8766 | No | NA | 14 | 0 | 2 | Not-determined | Genome-fragment | NA | NA | 0 |
| S11-final-viral-combined_k141_25252\|\|full | 2644 | No | NA | 2 | 0 | 2 | Not-determined | Genome-fragment | NA | NA | 0 |
| S11-final-viral-combined_k141_68174\|\|full | 3936 | No | NA | 3 | 0 | 2 | Not-determined | Genome-fragment | NA | NA | 0 |
| S11-final-viral-combined_k141_191337\|\|full | 3240 | No | NA | 6 | 0 | 0 | Not-determined | Genome-fragment | NA | NA | 0 |
| S11-final-viral-combined_k141_150758\|\|full | 1550 | No | NA | 2 | 0 | 2 | Not-determined | Genome-fragment | NA | NA | 0 |
| S8-final-viral-combined_k141_204506\|\|full | 1186 | No | NA | 3 | 0 | 0 | Low-quality | Genome-fragment | 2.34 | AAI-based (high-confidence) | 0 |
| S11-final-viral-combined_k141_294583\|\|full | 1187 | No | NA | 2 | 1 | 0 | Low-quality | Genome-fragment | 2.99 | AAI-based (medium-confidence) | 0 |
| S11-final-viral-combined_k141_219258\|\|full | 1344 | No | NA | 3 | 0 | 1 | Not-determined | Genome-fragment | NA | NA | 0 |
| S11-final-viral-combined_k141_315820\|\|full | 1640 | No | NA | 2 | 0 | 2 | Not-determined | Genome-fragment | NA | NA | 0 |
| S11-final-viral-combined_k141_193435\|\|lt2gene | 1187 | No | NA | 1 | 1 | 0 | Low-quality | Genome-fragment | 1.07 | HMM-based (lower-bound) | 0 |
| S11-final-viral-combined_k141_136036\|\|full | 2158 | No | NA | 3 | 0 | 0 | Not-determined | Genome-fragment | NA | NA | 0 |
| S11-final-viral-combined_k141_95263\|\|full | 1209 | No | NA | 3 | 0 | 1 | Not-determined | Genome-fragment | NA | NA | 0 |
| S11_k141_276873 | 1187 | No | NA | 1 | 0 | 1 | Low-quality | Genome-fragment | 0.52 | AAI-based (high-confidence) | 0 |
| S11-final-viral-combined_k141_219271\|\|full | 1316 | No | NA | 3 | 0 | 0 | Not-determined | Genome-fragment | NA | NA | 0 |
| S11-final-viral-combined_k141_191365\|\|full | 1753 | No | NA | 2 | 0 | 0 | Not-determined | Genome-fragment | NA | NA | 0 |
| S11-final-viral-combined_k141_164615\|\|full | 1202 | No | NA | 2 | 0 | 0 | Not-determined | Genome-fragment | NA | NA | 0 |
| S11-final-viral-combined_k141_261270\|\|full | 2406 | No | NA | 2 | 0 | 1 | Not-determined | Genome-fragment | NA | NA | 0 |
| S11-final-viral-combined_k141_150816\|\|full | 1012 | No | NA | 2 | 0 | 1 | Not-determined | Genome-fragment | NA | NA | 0 |
| S11-final-viral-combined_k141_288693\|\|full | 2536 | No | NA | 3 | 0 | 1 | Not-determined | Genome-fragment | NA | NA | 0 |
| S11-final-viral-combined_k141_328271\|\|full | 3002 | No | NA | 2 | 0 | 2 | Not-determined | Genome-fragment | NA | NA | 0 |
| S1-final-viral-combined_k141_199166\|\|lt2gene | 1187 | No | NA | 1 | 1 | 0 | Low-quality | Genome-fragment | 2.56 | AAI-based (medium-confidence) | 0 |
| S11-final-viral-combined_k141_261302\|\|full | 4488 | No | NA | 4 | 0 | 2 | Not-determined | Genome-fragment | NA | NA | 0 |
| S11-final-viral-combined_k141_13067\|\|full | 2890 | No | NA | 2 | 0 | 0 | Not-determined | Genome-fragment | NA | NA | 0 |
| S1-final-viral-combined_k141_51000\|\|lt2gene | 1187 | No | NA | 1 | 1 | 0 | Low-quality | Genome-fragment | 2.7 | AAI-based (high-confidence) | 0 |
| S11-final-viral-combined_k141_95372\|\|full | 1515 | No | NA | 3 | 0 | 1 | Not-determined | Genome-fragment | NA | NA | 0 |
| S1_k141_296370 | 1187 | No | NA | 1 | 0 | 1 | Low-quality | Genome-fragment | 0.66 | AAI-based (medium-confidence) | 0 |
| S5_k141_47642 | 1187 | No | NA | 4 | 0 | 0 | Low-quality | Genome-fragment | 0.65 | AAI-based (medium-confidence) | 0 |
| S7-final-viral-combined_k141_14181\|\|full | 1187 | No | NA | 2 | 0 | 1 | Low-quality | Genome-fragment | 0.54 | AAI-based (medium-confidence) | 0 |
| S7-final-viral-combined_k141_58578\|\|full | 1187 | No | NA | 3 | 0 | 0 | Low-quality | Genome-fragment | 2.72 | AAI-based (high-confidence) | 0 |
| S11-final-viral-combined_k141_248034\|\|full | 5985 | No | NA | 6 | 0 | 3 | Not-determined | Genome-fragment | NA | NA | 0 |
| S12_k141_191473 | 1187 | No | NA | 2 | 0 | 1 | Low-quality | Genome-fragment | 1.33 | AAI-based (high-confidence) | 0 |
| S11-final-viral-combined_k141_110185\|\|full | 1587 | No | NA | 3 | 0 | 0 | Not-determined | Genome-fragment | NA | NA | 0 |
| S11-final-viral-combined_k141_301630\|\|full | 5034 | No | NA | 4 | 0 | 4 | Not-determined | Genome-fragment | NA | NA | 0 |
| S11-final-viral-combined_k141_261397\|\|full | 1434 | No | NA | 2 | 0 | 0 | Not-determined | Genome-fragment | NA | NA | 0 |
| S2-final-viral-combined_k141_482\|\|full | 1187 | No | NA | 2 | 1 | 0 | Low-quality | Genome-fragment | 1.9 | AAI-based (high-confidence) | 0 |
| S2-final-viral-combined_k141_137315\|\|full | 1187 | No | NA | 2 | 0 | 0 | Low-quality | Genome-fragment | 1.75 | AAI-based (high-confidence) | 0 |
| S11-final-viral-combined_k141_191463\|\|full | 1691 | No | NA | 4 | 0 | 1 | Not-determined | Genome-fragment | NA | NA | 0 |
| S2-final-viral-combined_k141_142357\|\|full | 1187 | No | NA | 5 | 0 | 0 | Low-quality | Genome-fragment | 2.75 | AAI-based (medium-confidence) | 0 |
| S4-final-viral-combined_k141_162395\|\|full | 1187 | No | NA | 2 | 0 | 1 | Low-quality | Genome-fragment | 0.57 | AAI-based (high-confidence) | 0 |
| S9-final-viral-combined_k141_82726\|\|full | 1188 | No | NA | 3 | 0 | 0 | Low-quality | Genome-fragment | 1.8 | AAI-based (medium-confidence) | 0 |
| S11-final-viral-combined_k141_191472\|\|full | 1153 | No | NA | 2 | 0 | 1 | Not-determined | Genome-fragment | NA | NA | 0 |
| S11-final-viral-combined_k141_25450\|\|full | 1119 | No | NA | 3 | 0 | 1 | Not-determined | Genome-fragment | NA | NA | 0 |
| S11-final-viral-combined_k141_95439\|\|full | 4285 | No | NA | 4 | 0 | 3 | Not-determined | Genome-fragment | NA | NA | 0 |
| S9_k141_73694 | 1188 | No | NA | 2 | 0 | 0 | Low-quality | Genome-fragment | 0.46 | AAI-based (medium-confidence) | 0 |
| S1-final-viral-combined_k141_160413\|\|lt2gene | 1188 | No | NA | 2 | 1 | 0 | Low-quality | Genome-fragment | 2.87 | AAI-based (medium-confidence) | 0 |
| S5_k141_47667 | 1188 | No | NA | 3 | 0 | 1 | Low-quality | Genome-fragment | 1.27 | AAI-based (medium-confidence) | 0 |
| S17-final-viral-combined_k141_2901\|\|full | 1188 | Yes | 492 | 2 | 1 | 1 | Low-quality | Genome-fragment | 0.6 | AAI-based (high-confidence) | 58.59 |
| S17-k141_236914 | 1188 | No | NA | 1 | 0 | 1 | Low-quality | Genome-fragment | 0.31 | AAI-based (high-confidence) | 0 |
| S11-final-viral-combined_k141_274349\|\|full | 4427 | No | NA | 4 | 0 | 3 | Not-determined | Genome-fragment | NA | NA | 0 |
| S12-final-viral-combined_k141_231325\|\|full | 1188 | No | NA | 2 | 1 | 0 | Low-quality | Genome-fragment | 2.27 | AAI-based (high-confidence) | 0 |
| S12_k141_321567 | 1188 | No | NA | 4 | 0 | 0 | Low-quality | Genome-fragment | 2.54 | AAI-based (high-confidence) | 0 |
| S11-final-viral-combined_k141_136252\|\|full | 1241 | No | NA | 2 | 0 | 0 | Not-determined | Genome-fragment | NA | NA | 0 |
| S4_k141_112641 | 1188 | No | NA | 2 | 1 | 0 | Low-quality | Genome-fragment | 1.17 | AAI-based (high-confidence) | 0 |
| S8-final-viral-combined_k141_77506\|\|full | 1188 | No | NA | 4 | 2 | 0 | Low-quality | Genome-fragment | 3.28 | AAI-based (high-confidence) | 0 |
| S11-final-viral-combined_k141_41093\|\|full | 2508 | No | NA | 3 | 0 | 2 | Not-determined | Genome-fragment | NA | NA | 0 |
| S9-final-viral-combined_k141_98596\|\|full | 1189 | No | NA | 3 | 0 | 1 | Low-quality | Genome-fragment | 0.99 | AAI-based (medium-confidence) | 0 |
| S11-final-viral-combined_k141_117912\|\|full | 1189 | No | NA | 2 | 0 | 2 | Low-quality | Genome-fragment | 0.9 | AAI-based (medium-confidence) | 0 |
| S1-final-viral-combined_k141_297322\|\|lt2gene | 1189 | No | NA | 2 | 1 | 0 | Low-quality | Genome-fragment | 2.81 | HMM-based (lower-bound) | 0 |
| S11-final-viral-combined_k141_41114\|\|full | 4969 | No | NA | 7 | 0 | 2 | Not-determined | Genome-fragment | NA | NA | 0 |
| S11-final-viral-combined_k141_178794\|\|full | 1489 | No | NA | 2 | 0 | 1 | Not-determined | Genome-fragment | NA | NA | 0 |
| S7-final-viral-combined_k141_45526\|\|full | 1189 | No | NA | 2 | 0 | 0 | Low-quality | Genome-fragment | 0.75 | AAI-based (medium-confidence) | 0 |
| S11-final-viral-combined_k141_219584\|\|full | 1755 | No | NA | 3 | 0 | 1 | Not-determined | Genome-fragment | NA | NA | 0 |
| S4-final-viral-combined_k141_95728\|\|lt2gene | 1189 | No | NA | 1 | 1 | 0 | Low-quality | Genome-fragment | 1.64 | AAI-based (high-confidence) | 0 |
| S4-final-viral-combined_k141_62003\|\|lt2gene | 1189 | No | NA | 1 | 1 | 0 | Low-quality | Genome-fragment | 2.5 | HMM-based (lower-bound) | 0 |
| S11-final-viral-combined_k141_233842\|\|full | 2020 | No | NA | 3 | 0 | 2 | Not-determined | Genome-fragment | NA | NA | 0 |
| S6_k141_110392 | 1189 | No | NA | 2 | 0 | 0 | Low-quality | Genome-fragment | 1.36 | AAI-based (high-confidence) | 0 |
| S11-final-viral-combined_k141_301929\|\|full | 1271 | No | NA | 1 | 0 | 0 | Not-determined | Genome-fragment | NA | NA | 0 |
| S11-final-viral-combined_k141_32453\|\|full | 1190 | No | NA | 2 | 0 | 0 | Low-quality | Genome-fragment | 2.33 | AAI-based (high-confidence) | 0 |
| S11-final-viral-combined_k141_301938\|\|full | 1386 | No | NA | 3 | 0 | 0 | Not-determined | Genome-fragment | NA | NA | 0 |
| S11-final-viral-combined_k141_316033\|\|full | 2732 | No | NA | 3 | 0 | 0 | Not-determined | Genome-fragment | NA | NA | 0 |
| S11_k141_142811 | 1190 | No | NA | 2 | 0 | 1 | Low-quality | Genome-fragment | 1.14 | AAI-based (high-confidence) | 0 |
| S11-final-viral-combined_k141_191723\|\|full | 3240 | No | NA | 4 | 0 | 3 | Not-determined | Genome-fragment | NA | NA | 0 |
| S1-final-viral-combined_k141_287671\|\|full | 1190 | No | NA | 2 | 0 | 2 | Low-quality | Genome-fragment | 0.64 | AAI-based (medium-confidence) | 0 |
| S1-final-viral-combined_k141_276653\|\|full | 1190 | No | NA | 2 | 0 | 1 | Low-quality | Genome-fragment | 0.67 | AAI-based (high-confidence) | 0 |
| S11-final-viral-combined_k141_328443\|\|full | 1925 | No | NA | 3 | 0 | 1 | Not-determined | Genome-fragment | NA | NA | 0 |
| S11-final-viral-combined_k141_233923\|\|full | 1017 | No | NA | 3 | 0 | 1 | Not-determined | Genome-fragment | NA | NA | 0 |
| S5-final-viral-combined_k141_78977\|\|lt2gene | 1190 | No | NA | 1 | 1 | 0 | Low-quality | Genome-fragment | 2.85 | AAI-based (medium-confidence) | 0 |
| S7-final-viral-combined_k141_220653\|\|full | 1190 | No | NA | 2 | 0 | 1 | Low-quality | Genome-fragment | 0.7 | AAI-based (medium-confidence) | 0 |
| S11-final-viral-combined_k141_233930\|\|full | 1526 | No | NA | 2 | 0 | 0 | Not-determined | Genome-fragment | NA | NA | 0 |
| S12-final-viral-combined_k141_374465\|\|lt2gene | 1190 | No | NA | 1 | 1 | 0 | Low-quality | Genome-fragment | 0.46 | HMM-based (lower-bound) | 0 |
| S2-final-viral-combined_k141_228742\|\|full | 1190 | No | NA | 2 | 1 | 0 | Low-quality | Genome-fragment | 2.96 | AAI-based (medium-confidence) | 0 |
| S11-final-viral-combined_k141_68502\|\|full | 2346 | No | NA | 2 | 0 | 0 | Not-determined | Genome-fragment | NA | NA | 0 |
| S11-final-viral-combined_k141_219686\|\|full | 3860 | No | NA | 4 | 0 | 3 | Not-determined | Genome-fragment | NA | NA | 0 |
| S11-final-viral-combined_k141_289006\|\|full | 1139 | No | NA | 2 | 0 | 0 | Not-determined | Genome-fragment | NA | NA | 0 |
| S11-final-viral-combined_k141_191787\|\|full | 4495 | No | NA | 4 | 0 | 1 | Not-determined | Genome-fragment | NA | NA | 0 |
| S11-final-viral-combined_k141_81271\|\|full | 2545 | No | NA | 3 | 0 | 2 | Not-determined | Genome-fragment | NA | NA | 0 |
| S11-final-viral-combined_k141_289023\|\|full | 1281 | No | NA | 3 | 0 | 1 | Not-determined | Genome-fragment | NA | NA | 0 |
| S11-final-viral-combined_k141_122693\|\|full | 2094 | No | NA | 2 | 0 | 2 | Not-determined | Genome-fragment | NA | NA | 0 |
| S11-final-viral-combined_k141_136444\|\|full | 16632 | No | NA | 15 | 0 | 3 | Not-determined | Genome-fragment | NA | NA | 0 |
| S2-final-viral-combined_k141_182956\|\|full | 1190 | No | NA | 2 | 0 | 0 | Low-quality | Genome-fragment | 2.22 | AAI-based (high-confidence) | 0 |
| S11-final-viral-combined_k141_328498\|\|full | 3237 | No | NA | 3 | 0 | 1 | Not-determined | Genome-fragment | NA | NA | 0 |
| S11-final-viral-combined_k141_302027\|\|full | 2079 | No | NA | 3 | 0 | 3 | Not-determined | Genome-fragment | NA | NA | 0 |
| S11-final-viral-combined_k141_219733\|\|full | 1323 | No | NA | 2 | 0 | 2 | Not-determined | Genome-fragment | NA | NA | 0 |
| S11-final-viral-combined_k141_55003\|\|full | 1996 | No | NA | 4 | 0 | 3 | Not-determined | Genome-fragment | NA | NA | 0 |
| S2-final-viral-combined_k141_123516\|\|full | 1190 | No | NA | 3 | 1 | 0 | Low-quality | Genome-fragment | 2.09 | AAI-based (high-confidence) | 0 |
| S2-final-viral-combined_k141_162342\|\|lt2gene | 1190 | No | NA | 2 | 2 | 0 | Low-quality | Genome-fragment | 2.47 | AAI-based (high-confidence) | 0 |
| S2_k141_208006 | 1190 | No | NA | 3 | 0 | 2 | Low-quality | Genome-fragment | 2.36 | AAI-based (high-confidence) | 0 |
| S11-final-viral-combined_k141_68527\|\|full | 2390 | No | NA | 4 | 0 | 3 | Not-determined | Genome-fragment | NA | NA | 0 |
| S8_k141_186645 | 1190 | No | NA | 3 | 0 | 0 | Low-quality | Genome-fragment | 0.95 | AAI-based (high-confidence) | 0 |
| S8_k141_211098 | 1190 | No | NA | 1 | 0 | 0 | Low-quality | Genome-fragment | 1.72 | AAI-based (medium-confidence) | 0 |
| S9_k141_106062 | 1191 | No | NA | 3 | 0 | 1 | Low-quality | Genome-fragment | 0.8 | AAI-based (medium-confidence) | 0 |
| S11-final-viral-combined_k141_194006\|\|full | 1191 | No | NA | 3 | 1 | 0 | Low-quality | Genome-fragment | 1.33 | HMM-based (lower-bound) | 0 |
| S11-final-viral-combined_k141_20181\|\|full | 1191 | No | NA | 3 | 0 | 1 | Low-quality | Genome-fragment | 0.71 | AAI-based (medium-confidence) | 0 |
| S11-final-viral-combined_k141_261762\|\|full | 1472 | No | NA | 2 | 0 | 2 | Not-determined | Genome-fragment | NA | NA | 0 |
| S11-final-viral-combined_k141_151165\|\|full | 9167 | No | NA | 7 | 0 | 6 | Not-determined | Genome-fragment | NA | NA | 0 |
| S11-final-viral-combined_k141_274574\|\|full | 1220 | No | NA | 3 | 0 | 0 | Not-determined | Genome-fragment | NA | NA | 0 |
| S1-final-viral-combined_k141_200911\|\|lt2gene | 1191 | No | NA | 2 | 1 | 0 | Low-quality | Genome-fragment | 2.54 | AAI-based (high-confidence) | 0 |
| S11-final-viral-combined_k141_328565\|\|full | 4168 | No | NA | 3 | 0 | 1 | Not-determined | Genome-fragment | NA | NA | 0 |
| S11-final-viral-combined_k141_55077\|\|full | 1706 | No | NA | 2 | 0 | 0 | Not-determined | Genome-fragment | NA | NA | 0 |
| S11-final-viral-combined_k141_274602\|\|full | 1562 | No | NA | 3 | 0 | 1 | Not-determined | Genome-fragment | NA | NA | 0 |
| S11-final-viral-combined_k141_274604\|\|full | 1978 | No | NA | 3 | 0 | 0 | Not-determined | Genome-fragment | NA | NA | 0 |
| S1_k141_301193 | 1191 | No | NA | 1 | 0 | 1 | Low-quality | Genome-fragment | 0.29 | AAI-based (high-confidence) | 0 |
| S11-final-viral-combined_k141_178953\|\|full | 6371 | No | NA | 6 | 0 | 2 | Not-determined | Genome-fragment | NA | NA | 0 |
| S11-final-viral-combined_k141_219811\|\|full | 2467 | No | NA | 7 | 0 | 0 | Not-determined | Genome-fragment | NA | NA | 0 |
| S11-final-viral-combined_k141_136561\|\|full | 3583 | No | NA | 4 | 0 | 2 | Not-determined | Genome-fragment | NA | NA | 0 |
| S11-final-viral-combined_k141_205702\|\|full | 1178 | No | NA | 3 | 0 | 0 | Not-determined | Genome-fragment | NA | NA | 0 |
| S3_k141_124910 | 1191 | No | NA | 2 | 0 | 0 | Low-quality | Genome-fragment | 2.22 | AAI-based (high-confidence) | 0 |
| S5-final-viral-combined_k141_146839\|\|full | 1191 | No | NA | 4 | 0 | 0 | Low-quality | Genome-fragment | 3.47 | AAI-based (high-confidence) | 0 |
| S5_k141_88083 | 1191 | No | NA | 2 | 0 | 0 | Low-quality | Genome-fragment | 2.48 | AAI-based (high-confidence) | 0 |
| S11-final-viral-combined_k141_25886\|\|full | 2033 | No | NA | 4 | 0 | 0 | Not-determined | Genome-fragment | NA | NA | 0 |
| S11-final-viral-combined_k141_151242\|\|full | 3679 | No | NA | 5 | 0 | 4 | Not-determined | Genome-fragment | NA | NA | 0 |
| S11-final-viral-combined_k141_95855\|\|full | 2712 | No | NA | 4 | 0 | 0 | Not-determined | Genome-fragment | NA | NA | 0 |
| S12_k141_4486 | 1191 | No | NA | 3 | 1 | 0 | Low-quality | Genome-fragment | 3.29 | AAI-based (high-confidence) | 0 |
| S4-final-viral-combined_k141_78182\|\|full | 1191 | No | NA | 4 | 0 | 0 | Low-quality | Genome-fragment | 1.45 | AAI-based (high-confidence) | 0 |
| S4_k141_36250 | 1191 | No | NA | 2 | 0 | 0 | Low-quality | Genome-fragment | 1.19 | AAI-based (medium-confidence) | 0 |
| S6-final-viral-combined_k141_139316\|\|lt2gene | 1191 | No | NA | 1 | 1 | 0 | Low-quality | Genome-fragment | 2.16 | AAI-based (high-confidence) | 0 |
| S11-final-viral-combined_k141_136596\|\|full | 5533 | No | NA | 3 | 0 | 1 | Not-determined | Genome-fragment | NA | NA | 0 |
| S8-final-viral-combined_k141_131304\|\|full | 1191 | No | NA | 2 | 0 | 1 | Low-quality | Genome-fragment | 0.64 | AAI-based (medium-confidence) | 0 |
| S11-final-viral-combined_k141_289191\|\|full | 5275 | No | NA | 6 | 0 | 3 | Not-determined | Genome-fragment | NA | NA | 0 |
| S8-final-viral-combined_k141_145326\|\|full | 1191 | No | NA | 4 | 0 | 0 | Low-quality | Genome-fragment | 1.6 | AAI-based (medium-confidence) | 0 |
| S9-final-viral-combined_k141_74074\|\|lt2gene | 1192 | No | NA | 2 | 1 | 0 | Low-quality | Genome-fragment | 3.3 | AAI-based (high-confidence) | 0 |
| S11-final-viral-combined_k141_165226\|\|full | 2058 | No | NA | 3 | 0 | 2 | Not-determined | Genome-fragment | NA | NA | 0 |
| S9_k141_58903 | 1192 | No | NA | 1 | 0 | 0 | Low-quality | Genome-fragment | 1.22 | AAI-based (high-confidence) | 0 |
| S11-final-viral-combined_k141_25941\|\|full | 3404 | No | NA | 4 | 0 | 1 | Not-determined | Genome-fragment | NA | NA | 0 |
| S11-final-viral-combined_k141_98316\|\|lt2gene | 1192 | No | NA | 1 | 1 | 0 | Low-quality | Genome-fragment | 3.1 | AAI-based (high-confidence) | 0 |
| S11-final-viral-combined_k141_136615\|\|full | 5494 | No | NA | 6 | 0 | 4 | Not-determined | Genome-fragment | NA | NA | 0 |
| S11-final-viral-combined_k141_136618\|\|full | 1900 | No | NA | 3 | 0 | 3 | Not-determined | Genome-fragment | NA | NA | 0 |
| S11-final-viral-combined_k141_328682\|\|full | 6458 | No | NA | 6 | 0 | 5 | Not-determined | Genome-fragment | NA | NA | 0 |
| S1-final-viral-combined_k141_141716\|\|full | 1192 | Yes | 384 | 2 | 1 | 1 | Low-quality | Genome-fragment | 0.71 | AAI-based (high-confidence) | 67.79 |
| S11-final-viral-combined_k141_328693\|\|full | 1128 | No | NA | 4 | 0 | 1 | Not-determined | Genome-fragment | NA | NA | 0 |
| S5-final-viral-combined_k141_125162\|\|full | 1192 | No | NA | 4 | 3 | 0 | Low-quality | Genome-fragment | 2.56 | AAI-based (medium-confidence) | 0 |
| S11-final-viral-combined_k141_179090\|\|full | 2107 | No | NA | 2 | 0 | 2 | Not-determined | Genome-fragment | NA | NA | 0 |
| S12-final-viral-combined_k141_103187\|\|lt2gene | 1192 | No | NA | 2 | 1 | 0 | Low-quality | Genome-fragment | 0.77 | HMM-based (lower-bound) | 0 |
| S6-final-viral-combined_k141_47267\|\|full | 1192 | No | NA | 2 | 2 | 0 | Low-quality | Genome-fragment | 2.47 | HMM-based (lower-bound) | 0 |
| S9-final-viral-combined_k141_102627\|\|full | 1193 | No | NA | 2 | 2 | 0 | Low-quality | Genome-fragment | 1.93 | AAI-based (high-confidence) | 0 |
| S11-final-viral-combined_k141_294678\|\|lt2gene | 1193 | No | NA | 2 | 1 | 0 | Low-quality | Genome-fragment | 3.34 | AAI-based (high-confidence) | 0 |
| S1-final-viral-combined_k141_65482\|\|full | 1193 | No | NA | 2 | 0 | 0 | Low-quality | Genome-fragment | 1.8 | AAI-based (high-confidence) | 0 |
| S7-final-viral-combined_k141_175465\|\|full | 1193 | No | NA | 2 | 0 | 0 | Low-quality | Genome-fragment | 2.35 | AAI-based (high-confidence) | 0 |
| S11-final-viral-combined_k141_26014\|\|full | 1423 | No | NA | 2 | 0 | 0 | Not-determined | Genome-fragment | NA | NA | 0 |
| S11-final-viral-combined_k141_191955\|\|full | 2430 | No | NA | 3 | 0 | 2 | Not-determined | Genome-fragment | NA | NA | 0 |
| S11-final-viral-combined_k141_81598\|\|full | 1993 | No | NA | 3 | 0 | 2 | Not-determined | Genome-fragment | NA | NA | 0 |
| S7-final-viral-combined_k141_198945\|\|full | 1193 | No | NA | 2 | 0 | 0 | Low-quality | Genome-fragment | 2.06 | AAI-based (high-confidence) | 0 |
| S7_k141_51282 | 1193 | No | NA | 2 | 0 | 1 | Low-quality | Genome-fragment | 0.52 | AAI-based (medium-confidence) | 0 |
| S12-final-viral-combined_k141_258059\|\|full | 1193 | No | NA | 3 | 1 | 0 | Low-quality | Genome-fragment | 2.81 | AAI-based (high-confidence) | 0 |
| S12-final-viral-combined_k141_275929\|\|full | 1193 | No | NA | 2 | 0 | 0 | Low-quality | Genome-fragment | 0.86 | AAI-based (medium-confidence) | 0 |
| S12-final-viral-combined_k141_336782\|\|lt2gene | 1193 | No | NA | 1 | 1 | 0 | Low-quality | Genome-fragment | 1.19 | HMM-based (lower-bound) | 0 |
| S11-final-viral-combined_k141_136744\|\|full | 3205 | No | NA | 4 | 0 | 2 | Not-determined | Genome-fragment | NA | NA | 0 |
| S8-final-viral-combined_k141_217599\|\|full | 1193 | No | NA | 3 | 0 | 1 | Low-quality | Genome-fragment | 2.02 | AAI-based (medium-confidence) | 0 |
| S9-final-viral-combined_k141_101861\|\|full | 1194 | No | NA | 2 | 0 | 0 | Low-quality | Genome-fragment | 2.86 | AAI-based (high-confidence) | 0 |
| S11-final-viral-combined_k141_26097\|\|full | 1608 | No | NA | 2 | 0 | 2 | Not-determined | Genome-fragment | NA | NA | 0 |
| S11-final-viral-combined_k141_13761\|\|full | 2478 | No | NA | 2 | 0 | 1 | Not-determined | Genome-fragment | NA | NA | 0 |
| S9_k141_82087 | 1194 | No | NA | 3 | 0 | 0 | Low-quality | Genome-fragment | 2.88 | AAI-based (high-confidence) | 0 |
| S11-final-viral-combined_k141_124687\|\|full | 1194 | No | NA | 2 | 0 | 0 | Low-quality | Genome-fragment | 0.32 | AAI-based (medium-confidence) | 0 |
| S11-final-viral-combined_k141_81646\|\|full | 2258 | No | NA | 4 | 0 | 2 | Not-determined | Genome-fragment | NA | NA | 0 |
| S1-final-viral-combined_k141_136345\|\|full | 1194 | No | NA | 3 | 0 | 1 | Low-quality | Genome-fragment | 0.85 | AAI-based (medium-confidence) | 0 |
| S11-final-viral-combined_k141_136796\|\|full | 2167 | No | NA | 3 | 0 | 0 | Not-determined | Genome-fragment | NA | NA | 0 |
| S1-final-viral-combined_k141_276092\|\|full | 1194 | No | NA | 3 | 1 | 0 | Low-quality | Genome-fragment | 0.77 | HMM-based (lower-bound) | 0 |
| S1-final-viral-combined_k141_20116\|\|lt2gene | 1194 | No | NA | 2 | 2 | 0 | Low-quality | Genome-fragment | 2.57 | AAI-based (medium-confidence) | 0 |
| S11-final-viral-combined_k141_302484\|\|full | 1306 | No | NA | 3 | 0 | 0 | Not-determined | Genome-fragment | NA | NA | 0 |
| S1_k141_58778 | 1194 | No | NA | 1 | 0 | 1 | Low-quality | Genome-fragment | 2.44 | AAI-based (medium-confidence) | 0 |
| S5-final-viral-combined_k141_106260\|\|full | 1194 | No | NA | 3 | 1 | 0 | Low-quality | Genome-fragment | 1.89 | AAI-based (high-confidence) | 0 |
| S11-final-viral-combined_k141_328798\|\|full | 2610 | No | NA | 5 | 0 | 0 | Not-determined | Genome-fragment | NA | NA | 0 |
| S17-final-viral-combined_k141_78993\|\|lt2gene | 1194 | No | NA | 2 | 1 | 0 | Low-quality | Genome-fragment | 0.74 | HMM-based (lower-bound) | 0 |
| S12_k141_300859 | 1194 | No | NA | 1 | 0 | 1 | Low-quality | Genome-fragment | 3.37 | AAI-based (medium-confidence) | 0 |
| S2-final-viral-combined_k141_244388\|\|full | 1194 | No | NA | 3 | 0 | 1 | Low-quality | Genome-fragment | 2.85 | AAI-based (high-confidence) | 0 |
| S2-final-viral-combined_k141_7392\|\|full | 1194 | No | NA | 2 | 1 | 0 | Low-quality | Genome-fragment | 2.57 | AAI-based (high-confidence) | 0 |
| S11-final-viral-combined_k141_328840\|\|full | 4511 | No | NA | 4 | 0 | 3 | Not-determined | Genome-fragment | NA | NA | 0 |
| S4_k141_13000 | 1194 | No | NA | 2 | 2 | 0 | Low-quality | Genome-fragment | 1.91 | AAI-based (high-confidence) | 0 |
| S11-final-viral-combined_k141_151446\|\|full | 2083 | No | NA | 3 | 0 | 3 | Not-determined | Genome-fragment | NA | NA | 0 |
| S11-final-viral-combined_k141_302584\|\|full | 1106 | No | NA | 2 | 0 | 0 | Not-determined | Genome-fragment | NA | NA | 0 |
| S8_k141_50788 | 1194 | No | NA | 1 | 0 | 0 | Low-quality | Genome-fragment | 1.55 | AAI-based (high-confidence) | 0 |
| S11-final-viral-combined_k141_151488\|\|full | 1018 | No | NA | 2 | 0 | 1 | Not-determined | Genome-fragment | NA | NA | 0 |
| S11-final-viral-combined_k141_136951\|\|full | 1692 | No | NA | 2 | 0 | 2 | Not-determined | Genome-fragment | NA | NA | 0 |
| S9_k141_123755 | 1195 | No | NA | 2 | 0 | 2 | Low-quality | Genome-fragment | 0.37 | AAI-based (medium-confidence) | 0 |
| S11-final-viral-combined_k141_275068\|\|full | 2749 | No | NA | 3 | 0 | 3 | Not-determined | Genome-fragment | NA | NA | 0 |
| S9_k141_119413 | 1195 | No | NA | 4 | 0 | 0 | Low-quality | Genome-fragment | 2.4 | AAI-based (high-confidence) | 0 |
| S11-final-viral-combined_k141_207825\|\|full | 1195 | No | NA | 3 | 1 | 0 | Low-quality | Genome-fragment | 2.26 | AAI-based (high-confidence) | 0 |
| S11_k141_253537 | 1195 | No | NA | 2 | 0 | 0 | Low-quality | Genome-fragment | 1.81 | AAI-based (medium-confidence) | 0 |
| S11-final-viral-combined_k141_81877\|\|full | 4097 | No | NA | 3 | 0 | 3 | Not-determined | Genome-fragment | NA | NA | 0 |
| S11-final-viral-combined_k141_68874\|\|full | 2190 | No | NA | 3 | 0 | 1 | Not-determined | Genome-fragment | NA | NA | 0 |
| S11-final-viral-combined_k141_123287\|\|full | 2795 | No | NA | 4 | 0 | 0 | Not-determined | Genome-fragment | NA | NA | 0 |
| S11-final-viral-combined_k141_137033\|\|full | 6213 | No | NA | 8 | 0 | 7 | Not-determined | Genome-fragment | NA | NA | 0 |
| S1_k141_364305 | 1195 | No | NA | 1 | 0 | 1 | Low-quality | Genome-fragment | 0.95 | AAI-based (high-confidence) | 0 |
| S11-final-viral-combined_k141_137044\|\|full | 1053 | No | NA | 3 | 0 | 0 | Not-determined | Genome-fragment | NA | NA | 0 |
| S11-final-viral-combined_k141_81916\|\|full | 2654 | No | NA | 4 | 0 | 2 | Not-determined | Genome-fragment | NA | NA | 0 |
| S5-final-viral-combined_k141_5385\|\|lt2gene | 1195 | No | NA | 1 | 0 | 0 | Low-quality | Genome-fragment | 2.47 | AAI-based (high-confidence) | 0 |
| S7-final-viral-combined_k141_13842\|\|full | 1195 | No | NA | 3 | 0 | 1 | Low-quality | Genome-fragment | 1.27 | AAI-based (high-confidence) | 0 |
| S11-final-viral-combined_k141_137073\|\|full | 2055 | No | NA | 3 | 0 | 0 | Not-determined | Genome-fragment | NA | NA | 0 |
| S11-final-viral-combined_k141_26402\|\|full | 2251 | No | NA | 4 | 0 | 1 | Not-determined | Genome-fragment | NA | NA | 0 |
| S7_k141_107846 | 1195 | No | NA | 2 | 0 | 0 | Low-quality | Genome-fragment | 2.2 | AAI-based (high-confidence) | 0 |
| S11-final-viral-combined_k141_96357\|\|full | 2237 | No | NA | 3 | 0 | 1 | Not-determined | Genome-fragment | NA | NA | 0 |
| S17-final-viral-combined_k141_257058\|\|full | 1195 | No | NA | 3 | 1 | 0 | Low-quality | Genome-fragment | 2.43 | AAI-based (high-confidence) | 0 |
| S17-k141_8364 | 1195 | No | NA | 2 | 0 | 0 | Low-quality | Genome-fragment | 2.6 | AAI-based (high-confidence) | 0 |
| S11-final-viral-combined_k141_192361\|\|full | 2335 | No | NA | 3 | 0 | 1 | Not-determined | Genome-fragment | NA | NA | 0 |
| S11-final-viral-combined_k141_192363\|\|full | 1790 | No | NA | 3 | 0 | 2 | Not-determined | Genome-fragment | NA | NA | 0 |
| S11-final-viral-combined_k141_123409\|\|full | 2848 | No | NA | 3 | 0 | 1 | Not-determined | Genome-fragment | NA | NA | 0 |
| S12-final-viral-combined_k141_104024\|\|lt2gene | 1195 | No | NA | 1 | 1 | 0 | Low-quality | Genome-fragment | 2.56 | AAI-based (high-confidence) | 0 |
| S11-final-viral-combined_k141_316608\|\|full | 1399 | No | NA | 2 | 0 | 2 | Not-determined | Genome-fragment | NA | NA | 0 |
| S12_k141_246229 | 1195 | No | NA | 5 | 0 | 0 | Low-quality | Genome-fragment | 1.97 | AAI-based (high-confidence) | 0 |
| S2-final-viral-combined_k141_65502\|\|full | 1195 | Yes | 335 | 3 | 1 | 1 | Low-quality | Genome-fragment | 0.17 | AAI-based (medium-confidence) | 71.97 |
| S2-final-viral-combined_k141_3663\|\|lt2gene | 1195 | No | NA | 2 | 1 | 0 | Low-quality | Genome-fragment | 2.41 | AAI-based (high-confidence) | 0 |
| S11-final-viral-combined_k141_26585\|\|full | 10287 | No | NA | 10 | 0 | 8 | Not-determined | Genome-fragment | NA | NA | 0 |
| S2-final-viral-combined_k141_27746\|\|lt2gene | 1195 | No | NA | 2 | 1 | 0 | Low-quality | Genome-fragment | 2.97 | AAI-based (high-confidence) | 0 |
| S11-final-viral-combined_k141_329005\|\|full | 1086 | No | NA | 2 | 0 | 2 | Not-determined | Genome-fragment | NA | NA | 0 |
| S11-final-viral-combined_k141_329008\|\|full | 1770 | No | NA | 2 | 0 | 1 | Not-determined | Genome-fragment | NA | NA | 0 |
| S11-final-viral-combined_k141_192453\|\|full | 2940 | No | NA | 3 | 0 | 1 | Not-determined | Genome-fragment | NA | NA | 0 |
| S4_k141_21422 | 1195 | No | NA | 1 | 0 | 0 | Low-quality | Genome-fragment | 1.99 | AAI-based (medium-confidence) | 0 |
| S11-final-viral-combined_k141_275323\|\|full | 11311 | No | NA | 9 | 0 | 9 | Not-determined | Genome-fragment | NA | NA | 0 |
| S11-final-viral-combined_k141_26628\|\|full | 2788 | No | NA | 3 | 0 | 1 | Not-determined | Genome-fragment | NA | NA | 0 |
| S8_k141_125646 | 1195 | No | NA | 1 | 1 | 0 | Low-quality | Genome-fragment | 1.19 | AAI-based (medium-confidence) | 0 |
| S8_k141_92110 | 1195 | No | NA | 3 | 2 | 0 | Low-quality | Genome-fragment | 3.71 | AAI-based (high-confidence) | 0 |
| S1-final-viral-combined_k141_151648\|\|full | 1196 | No | NA | 3 | 0 | 0 | Low-quality | Genome-fragment | 3.39 | AAI-based (high-confidence) | 0 |
| S11-final-viral-combined_k141_26670\|\|full | 1103 | No | NA | 2 | 0 | 1 | Not-determined | Genome-fragment | NA | NA | 0 |
| S11-final-viral-combined_k141_275376\|\|full | 1837 | No | NA | 3 | 0 | 1 | Not-determined | Genome-fragment | NA | NA | 0 |
| S1-final-viral-combined_k141_218381\|\|full | 1196 | No | NA | 2 | 0 | 2 | Low-quality | Genome-fragment | 1.75 | AAI-based (high-confidence) | 0 |
| S11-final-viral-combined_k141_82137\|\|full | 2940 | No | NA | 3 | 0 | 3 | Not-determined | Genome-fragment | NA | NA | 0 |
| S1-final-viral-combined_k141_329009\|\|full | 1196 | No | NA | 2 | 0 | 0 | Low-quality | Genome-fragment | 2.51 | AAI-based (medium-confidence) | 0 |
| S11-final-viral-combined_k141_123643\|\|full | 2085 | No | NA | 3 | 0 | 2 | Not-determined | Genome-fragment | NA | NA | 0 |
| S11-final-viral-combined_k141_220472\|\|full | 1306 | No | NA | 3 | 0 | 1 | Not-determined | Genome-fragment | NA | NA | 0 |
| S11-final-viral-combined_k141_82177\|\|full | 1015 | No | NA | 2 | 0 | 2 | Not-determined | Genome-fragment | NA | NA | 0 |
| S11-final-viral-combined_k141_26706\|\|full | 1622 | No | NA | 2 | 0 | 1 | Not-determined | Genome-fragment | NA | NA | 0 |
| S11-final-viral-combined_k141_123658\|\|full | 1860 | No | NA | 2 | 0 | 1 | Not-determined | Genome-fragment | NA | NA | 0 |
| S5_k141_125564 | 1196 | No | NA | 1 | 0 | 1 | Low-quality | Genome-fragment | 0.79 | AAI-based (high-confidence) | 0 |
| S17-final-viral-combined_k141_51629\|\|full | 1196 | No | NA | 3 | 0 | 0 | Low-quality | Genome-fragment | 0.79 | AAI-based (medium-confidence) | 0 |
| S11-final-viral-combined_k141_137427\|\|full | 3130 | No | NA | 3 | 0 | 3 | Not-determined | Genome-fragment | NA | NA | 0 |
| S12_k141_5446 | 1196 | No | NA | 2 | 1 | 0 | Low-quality | Genome-fragment | 1.37 | AAI-based (medium-confidence) | 0 |
| S11-final-viral-combined_k141_303066\|\|full | 2255 | No | NA | 3 | 0 | 3 | Not-determined | Genome-fragment | NA | NA | 0 |
| S12_k141_362016 | 1196 | No | NA | 4 | 1 | 0 | Low-quality | Genome-fragment | 2.97 | AAI-based (medium-confidence) | 0 |
| S2-final-viral-combined_k141_57049\|\|full | 1196 | No | NA | 3 | 0 | 1 | Low-quality | Genome-fragment | 2.45 | AAI-based (high-confidence) | 0 |
| S11-final-viral-combined_k141_137498\|\|full | 4177 | No | NA | 3 | 0 | 1 | Not-determined | Genome-fragment | NA | NA | 0 |
| S2-final-viral-combined_k141_196488\|\|full | 1196 | No | NA | 2 | 0 | 0 | Low-quality | Genome-fragment | 1.46 | AAI-based (high-confidence) | 0 |
| S11-final-viral-combined_k141_137509\|\|full | 2420 | No | NA | 3 | 0 | 1 | Not-determined | Genome-fragment | NA | NA | 0 |
| S11-final-viral-combined_k141_192708\|\|full | 1096 | No | NA | 2 | 0 | 1 | Not-determined | Genome-fragment | NA | NA | 0 |
| S11-final-viral-combined_k141_137547\|\|full | 1126 | No | NA | 2 | 0 | 1 | Not-determined | Genome-fragment | NA | NA | 0 |
| S2-final-viral-combined_k141_127322\|\|lt2gene | 1196 | No | NA | 1 | 1 | 0 | Low-quality | Genome-fragment | 2.83 | AAI-based (high-confidence) | 0 |
| S11-final-viral-combined_k141_329193\|\|full | 2169 | No | NA | 3 | 0 | 1 | Not-determined | Genome-fragment | NA | NA | 0 |
| S11-final-viral-combined_k141_82361\|\|full | 14273 | No | NA | 13 | 0 | 12 | Not-determined | Genome-fragment | NA | NA | 0 |
| S11-final-viral-combined_k141_316880\|\|full | 2304 | No | NA | 2 | 0 | 2 | Not-determined | Genome-fragment | NA | NA | 0 |
| S11-final-viral-combined_k141_192773\|\|full | 5206 | No | NA | 8 | 0 | 2 | Not-determined | Genome-fragment | NA | NA | 0 |
| S8-final-viral-combined_k141_115793\|\|full | 1196 | No | NA | 3 | 0 | 0 | Low-quality | Genome-fragment | 1 | AAI-based (high-confidence) | 0 |
| S11-final-viral-combined_k141_206350\|\|full | 12767 | No | NA | 12 | 0 | 8 | Not-determined | Genome-fragment | NA | NA | 0 |
| S11-final-viral-combined_k141_275525\|\|full | 11803 | No | NA | 12 | 0 | 9 | Not-determined | Genome-fragment | NA | NA | 0 |
| S11-final-viral-combined_k141_329260\|\|full | 9328 | No | NA | 8 | 0 | 3 | Not-determined | Genome-fragment | NA | NA | 0 |
| S8-final-viral-combined_k141_87995\|\|full | 1196 | No | NA | 2 | 0 | 1 | Low-quality | Genome-fragment | 2.27 | AAI-based (medium-confidence) | 0 |
| S11-final-viral-combined_k141_26933\|\|full | 1043 | No | NA | 3 | 0 | 0 | Not-determined | Genome-fragment | NA | NA | 0 |
| S11-final-viral-combined_k141_26956\|\|full | 1366 | No | NA | 3 | 0 | 0 | Not-determined | Genome-fragment | NA | NA | 0 |
| S8-final-viral-combined_k141_67175\|\|full | 1196 | No | NA | 3 | 0 | 2 | Low-quality | Genome-fragment | 2.75 | AAI-based (medium-confidence) | 0 |
| S11-final-viral-combined_k141_206462\|\|full | 2926 | No | NA | 2 | 0 | 2 | Not-determined | Genome-fragment | NA | NA | 0 |
| S11-final-viral-combined_k141_329343\|\|full | 1303 | No | NA | 2 | 0 | 1 | Not-determined | Genome-fragment | NA | NA | 0 |
| S9-final-viral-combined_k141_100194\|\|full | 1197 | No | NA | 3 | 1 | 0 | Low-quality | Genome-fragment | 2.69 | AAI-based (medium-confidence) | 0 |
| S11-final-viral-combined_k141_329421\|\|full | 2051 | No | NA | 3 | 0 | 3 | Not-determined | Genome-fragment | NA | NA | 0 |
| S9-final-viral-combined_k141_28565\|\|lt2gene | 1197 | No | NA | 1 | 1 | 0 | Low-quality | Genome-fragment | 1.79 | HMM-based (lower-bound) | 0 |
| S11_k141_295013 | 1197 | No | NA | 3 | 0 | 2 | Low-quality | Genome-fragment | 0.71 | AAI-based (medium-confidence) | 0 |
| S11-final-viral-combined_k141_27115\|\|full | 1681 | No | NA | 2 | 0 | 1 | Not-determined | Genome-fragment | NA | NA | 0 |
| S11-final-viral-combined_k141_206655\|\|full | 2551 | No | NA | 2 | 0 | 0 | Not-determined | Genome-fragment | NA | NA | 0 |
| S11-final-viral-combined_k141_27141\|\|full | 3494 | No | NA | 4 | 0 | 4 | Not-determined | Genome-fragment | NA | NA | 0 |
| S11-final-viral-combined_k141_329493\|\|full | 51811 | No | NA | 41 | 1 | 29 | High-quality | High-quality | 100 | HMM-based (lower-bound) | 0 |
| S1_k141_107624 | 1197 | No | NA | 1 | 0 | 0 | Low-quality | Genome-fragment | 3.07 | AAI-based (medium-confidence) | 0 |
| S11-final-viral-combined_k141_329522\|\|full | 7380 | No | NA | 6 | 0 | 6 | Not-determined | Genome-fragment | NA | NA | 0 |
| S11-final-viral-combined_k141_27202\|\|full | 1973 | No | NA | 4 | 0 | 2 | Not-determined | Genome-fragment | NA | NA | 0 |
| S11-final-viral-combined_k141_27219\|\|full | 3045 | No | NA | 7 | 0 | 1 | Not-determined | Genome-fragment | NA | NA | 0 |
| S11-final-viral-combined_k141_27282\|\|full | 2612 | No | NA | 3 | 0 | 0 | Not-determined | Genome-fragment | NA | NA | 0 |
| S1_k141_193419 | 1197 | No | NA | 1 | 0 | 1 | Low-quality | Genome-fragment | 0.67 | AAI-based (high-confidence) | 0 |
| S11-final-viral-combined_k141_329809\|\|full | 1281 | No | NA | 3 | 0 | 0 | Not-determined | Genome-fragment | NA | NA | 0 |
| S11-final-viral-combined_k141_329864\|\|full | 3938 | No | NA | 5 | 0 | 2 | Not-determined | Genome-fragment | NA | NA | 0 |
| S3-final-viral-combined_k141_22812\|\|lt2gene | 1197 | No | NA | 2 | 0 | 0 | Low-quality | Genome-fragment | 2.11 | AAI-based (high-confidence) | 0 |
| S11-final-viral-combined_k141_330012\|\|full | 1157 | No | NA | 3 | 0 | 1 | Not-determined | Genome-fragment | NA | NA | 0 |
| S11-final-viral-combined_k141_330119\|\|full | 1613 | No | NA | 3 | 0 | 1 | Not-determined | Genome-fragment | NA | NA | 0 |
| S11-final-viral-combined_k141_330252\|\|full | 6534 | No | NA | 10 | 0 | 0 | Not-determined | Genome-fragment | NA | NA | 0 |
| S11-final-viral-combined_k141_330345\|\|full | 1780 | No | NA | 2 | 0 | 2 | Not-determined | Genome-fragment | NA | NA | 0 |
| S11-final-viral-combined_k141_330368\|\|full | 4243 | No | NA | 3 | 0 | 1 | Not-determined | Genome-fragment | NA | NA | 0 |
| S3_k141_130305 | 1197 | No | NA | 2 | 0 | 2 | Low-quality | Genome-fragment | 1.57 | AAI-based (medium-confidence) | 0 |
| S11-final-viral-combined_k141_124528\|\|0_partial | 12304 | No | NA | 11 | 0 | 4 | Not-determined | Genome-fragment | NA | NA | 0 |
| S11-final-viral-combined_k141_96978\|\|0_partial | 16088 | No | NA | 14 | 0 | 12 | Not-determined | Genome-fragment | NA | NA | 0 |
| S3_k141_124419 | 1197 | No | NA | 2 | 0 | 0 | Low-quality | Genome-fragment | 0.55 | AAI-based (high-confidence) | 0 |
| S11-final-viral-combined_k141_248668\|\|0_partial | 10925 | No | NA | 9 | 0 | 6 | Not-determined | Genome-fragment | NA | NA | 0 |
| S5-final-viral-combined_k141_21394\|\|full | 1197 | No | NA | 2 | 0 | 1 | Low-quality | Genome-fragment | 2.11 | AAI-based (high-confidence) | 0 |
| S11-final-viral-combined_k141_236046\|\|0_partial | 22654 | No | NA | 20 | 0 | 16 | Not-determined | Genome-fragment | NA | NA | 0 |
| S17-final-viral-combined_k141_167568\|\|full | 1197 | No | NA | 2 | 0 | 2 | Low-quality | Genome-fragment | 0.46 | AAI-based (medium-confidence) | 0 |
| S11-final-viral-combined_k141_153467\|\|0_partial | 25494 | No | NA | 19 | 0 | 15 | Not-determined | Genome-fragment | NA | NA | 0 |
| S17-final-viral-combined_k141_108127\|\|full | 1197 | No | NA | 2 | 0 | 0 | Low-quality | Genome-fragment | 0.78 | AAI-based (high-confidence) | 0 |
| S12-final-viral-combined_k141_271871\|\|full | 1197 | No | NA | 2 | 0 | 0 | Low-quality | Genome-fragment | 2.72 | AAI-based (medium-confidence) | 0 |
| S12_k141_87408 | 1197 | No | NA | 2 | 0 | 0 | Low-quality | Genome-fragment | 0.65 | AAI-based (high-confidence) | 0 |
| S12_k141_144612 | 1197 | No | NA | 3 | 0 | 0 | Low-quality | Genome-fragment | 2.05 | AAI-based (high-confidence) | 0 |
| S2-final-viral-combined_k141_104938\|\|lt2gene | 1197 | No | NA | 1 | 1 | 0 | Low-quality | Genome-fragment | 1.57 | HMM-based (lower-bound) | 0 |
| S6-final-viral-combined_k141_78058\|\|full | 1197 | No | NA | 2 | 0 | 0 | Low-quality | Genome-fragment | 2.77 | AAI-based (high-confidence) | 0 |
| S8-final-viral-combined_k141_231507\|\|full | 1197 | No | NA | 3 | 0 | 1 | Low-quality | Genome-fragment | 0.78 | AAI-based (medium-confidence) | 0 |
| S8-final-viral-combined_k141_212425\|\|full | 1197 | No | NA | 4 | 0 | 0 | Low-quality | Genome-fragment | 0.78 | AAI-based (high-confidence) | 0 |

| | | | | | | | | | | | |
|---|---|---|---|---|---|---|---|---|---|---|---|
| S8_k141_87954 | 1197 | No | NA | 2 | 1 | 0 | Low-quality | Genome-fragment | 1.91 | AAI-based (high-confidence) | 0 |
| S9-final-viral-combined_k141_76842||full | 1198 | No | NA | 3 | 0 | 0 | Low-quality | Genome-fragment | 2.8 | AAI-based (medium-confidence) | 0 |
| S11-final-viral-combined_k141_18338||0_partial | 13664 | No | NA | 11 | 0 | 8 | Not-determined | Genome-fragment | NA | NA | 0 |
| S9-final-viral-combined_k141_89724||full | 1198 | No | NA | 3 | 0 | 0 | Low-quality | Genome-fragment | 0.35 | AAI-based (medium-confidence) | 0 |
| S11-final-viral-combined_k141_36901||full | 1198 | No | NA | 3 | 0 | 0 | Low-quality | Genome-fragment | 1.8 | AAI-based (medium-confidence) | 0 |
| S11_k141_191183 | 1198 | No | NA | 2 | 0 | 0 | Low-quality | Genome-fragment | 2.12 | AAI-based (high-confidence) | 0 |
| S11-final-viral-combined_k141_129668||0_partial | 15245 | No | NA | 11 | 0 | 11 | Not-determined | Genome-fragment | NA | NA | 0 |
| S1-final-viral-combined_k141_197635||full | 1198 | No | NA | 3 | 0 | 1 | Low-quality | Genome-fragment | 1.64 | AAI-based (medium-confidence) | 0 |
| S1-final-viral-combined_k141_369279||full | 1198 | No | NA | 5 | 0 | 0 | Low-quality | Genome-fragment | 2.05 | AAI-based (high-confidence) | 0 |
| S11-final-viral-combined_k141_47427||0_partial | 30930 | No | NA | 30 | 0 | 16 | Not-determined | Genome-fragment | NA | NA | 0 |
| S11-final-viral-combined_k141_199102||0_partial | 21178 | No | NA | 21 | 0 | 17 | Not-determined | Genome-fragment | NA | NA | 0 |
| S1-final-viral-combined_k141_289473||full | 1198 | No | NA | 3 | 0 | 1 | Low-quality | Genome-fragment | 0.7 | AAI-based (medium-confidence) | 0 |
| S11-final-viral-combined_k141_283224||0_partial | 11396 | No | NA | 9 | 0 | 7 | Not-determined | Genome-fragment | NA | NA | 0 |
| S12-final-viral-combined_k141_325590||full | 1198 | No | NA | 2 | 0 | 1 | Low-quality | Genome-fragment | 0.47 | AAI-based (medium-confidence) | 0 |
| S11-final-viral-combined_k141_200131||0_partial | 13297 | No | NA | 11 | 0 | 8 | Not-determined | Genome-fragment | NA | NA | 0 |
| S11-final-viral-combined_k141_242954||0_partial | 13557 | No | NA | 11 | 0 | 9 | Not-determined | Genome-fragment | NA | NA | 0 |
| S12_k141_137877 | 1198 | No | NA | 3 | 0 | 0 | Low-quality | Genome-fragment | 2.76 | AAI-based (medium-confidence) | 0 |
| S11-final-viral-combined_k141_146499||0_partial | 10736 | No | NA | 11 | 0 | 3 | Not-determined | Genome-fragment | NA | NA | 0 |
| S2-final-viral-combined_k141_151538||full | 1198 | No | NA | 2 | 0 | 1 | Low-quality | Genome-fragment | 1.63 | AAI-based (medium-confidence) | 0 |
| S6_k141_118051 | 1198 | No | NA | 2 | 0 | 1 | Low-quality | Genome-fragment | 4 | AAI-based (medium-confidence) | 0 |
| S8-final-viral-combined_k141_179335||full | 1198 | No | NA | 3 | 1 | 0 | Low-quality | Genome-fragment | 2.71 | AAI-based (medium-confidence) | 0 |
| S11-final-viral-combined_k141_4559||full | 1199 | No | NA | 2 | 0 | 0 | Low-quality | Genome-fragment | 2.36 | AAI-based (high-confidence) | 0 |
| S11-final-viral-combined_k141_313109||full | 1199 | No | NA | 3 | 0 | 1 | Low-quality | Genome-fragment | 2.2 | AAI-based (medium-confidence) | 0 |
| S11-final-viral-combined_k141_161582||full | 1199 | No | NA | 2 | 0 | 0 | Low-quality | Genome-fragment | 2.12 | AAI-based (high-confidence) | 0 |
| S11_k141_22994 | 1199 | No | NA | 1 | 0 | 0 | Low-quality | Genome-fragment | 0.9 | AAI-based (high-confidence) | 0 |
| S1-final-viral-combined_k141_134486||full | 1199 | No | NA | 3 | 0 | 0 | Low-quality | Genome-fragment | 2.66 | AAI-based (medium-confidence) | 0 |
| S11-final-viral-combined_k141_275441||0_partial | 11597 | No | NA | 8 | 0 | 6 | Not-determined | Genome-fragment | NA | NA | 0 |
| S1-final-viral-combined_k141_173800||full | 1199 | No | NA | 4 | 3 | 0 | Low-quality | Genome-fragment | 3.2 | AAI-based (medium-confidence) | 0 |
| S1_k141_15332 | 1199 | No | NA | 3 | 2 | 0 | Low-quality | Genome-fragment | 1.44 | AAI-based (high-confidence) | 0 |
| S11-final-viral-combined_k141_234401||lt2gene | 1243 | No | NA | 2 | 0 | 0 | Not-determined | Genome-fragment | NA | NA | 0 |
| S11-final-viral-combined_k141_55359||lt2gene | 1106 | No | NA | 1 | 0 | 0 | Not-determined | Genome-fragment | NA | NA | 0 |
| S17-final-viral-combined_k141_206889||lt2gene | 1199 | No | NA | 2 | 1 | 0 | Low-quality | Genome-fragment | 31.1 | AAI-based (medium-confidence) | 0 |
| S17-k141_11545 | 1199 | No | NA | 1 | 0 | 1 | Low-quality | Genome-fragment | 0.45 | AAI-based (high-confidence) | 0 |
| S2-final-viral-combined_k141_91251||full | 1199 | No | NA | 3 | 0 | 0 | Low-quality | Genome-fragment | 2.59 | AAI-based (medium-confidence) | 0 |
| S11-final-viral-combined_k141_317354||lt2gene | 1102 | No | NA | 1 | 0 | 0 | Not-determined | Genome-fragment | NA | NA | 0 |
| S11-final-viral-combined_k141_152129||lt2gene | 1111 | No | NA | 2 | 0 | 0 | Not-determined | Genome-fragment | NA | NA | 0 |
| S4-final-viral-combined_k141_72548||full | 1199 | No | NA | 3 | 0 | 0 | Low-quality | Genome-fragment | 2.79 | AAI-based (high-confidence) | 0 |
| S8_k141_210104 | 1199 | No | NA | 2 | 0 | 2 | Low-quality | Genome-fragment | 0.6 | AAI-based (medium-confidence) | 0 |
| S9-final-viral-combined_k141_32219||full | 1200 | No | NA | 4 | 3 | 0 | Low-quality | Genome-fragment | 2.6 | AAI-based (medium-confidence) | 0 |
| S11-final-viral-combined_k141_78924||full | 1200 | No | NA | 2 | 0 | 2 | Low-quality | Genome-fragment | 0.75 | AAI-based (medium-confidence) | 0 |
| S1-final-viral-combined_k141_162482||full | 1200 | No | NA | 3 | 1 | 0 | Low-quality | Genome-fragment | 2.2 | AAI-based (medium-confidence) | 0 |
| S11-final-viral-combined_k141_110829||lt2gene | 1730 | No | NA | 1 | 0 | 0 | Not-determined | Genome-fragment | NA | NA | 0 |
| S11-final-viral-combined_k141_166256||lt2gene | 1091 | No | NA | 2 | 0 | 0 | Not-determined | Genome-fragment | NA | NA | 0 |
| S5-final-viral-combined_k141_49840||full | 1200 | No | NA | 3 | 1 | 0 | Low-quality | Genome-fragment | 2.44 | AAI-based (high-confidence) | 0 |
| S11-final-viral-combined_k141_166357||lt2gene | 1285 | No | NA | 1 | 0 | 0 | Not-determined | Genome-fragment | NA | NA | 0 |
| S11-final-viral-combined_k141_179840||lt2gene | 1264 | No | NA | 1 | 0 | 0 | Not-determined | Genome-fragment | NA | NA | 0 |
| S7_k141_121767 | 1200 | No | NA | 2 | 0 | 0 | Low-quality | Genome-fragment | 0.97 | AAI-based (medium-confidence) | 0 |
| S11-final-viral-combined_k141_262751||lt2gene | 1425 | No | NA | 2 | 0 | 0 | Not-determined | Genome-fragment | NA | NA | 0 |
| S11-final-viral-combined_k141_42167||lt2gene | 1013 | No | NA | 1 | 0 | 1 | Not-determined | Genome-fragment | NA | NA | 0 |
| S11-final-viral-combined_k141_262805||lt2gene | 1482 | No | NA | 1 | 0 | 0 | Not-determined | Genome-fragment | NA | NA | 0 |
| S17-final-viral-combined_k141_121124||full | 1200 | No | NA | 3 | 1 | 0 | Low-quality | Genome-fragment | 2.78 | AAI-based (medium-confidence) | 0 |
| S17-k141_255435 | 1200 | No | NA | 2 | 1 | 0 | Low-quality | Genome-fragment | 2.23 | AAI-based (high-confidence) | 0 |
| S8-final-viral-combined_k141_252531||full | 1200 | No | NA | 3 | 0 | 0 | Low-quality | Genome-fragment | 2.34 | AAI-based (high-confidence) | 0 |
| S9-final-viral-combined_k141_104994||full | 1201 | No | NA | 2 | 0 | 0 | Low-quality | Genome-fragment | 3.02 | AAI-based (medium-confidence) | 0 |
| S11-final-viral-combined_k141_30592||full | 1201 | No | NA | 3 | 0 | 1 | Low-quality | Genome-fragment | 2.24 | AAI-based (medium-confidence) | 0 |
| S11-final-viral-combined_k141_30348||lt2gene | 1201 | No | NA | 2 | 1 | 0 | Low-quality | Genome-fragment | 0.69 | HMM-based (lower-bound) | 0 |
| S11-final-viral-combined_k141_70145||lt2gene | 1122 | No | NA | 1 | 0 | 0 | Not-determined | Genome-fragment | NA | NA | 0 |
| S1-final-viral-combined_k141_323914||lt2gene | 1201 | No | NA | 2 | 2 | 0 | Low-quality | Genome-fragment | 2.14 | AAI-based (high-confidence) | 0 |
| S1-final-viral-combined_k141_30235||lt2gene | 1201 | No | NA | 1 | 1 | 0 | Low-quality | Genome-fragment | 3.74 | AAI-based (high-confidence) | 0 |
| S3-final-viral-combined_k141_51708||full | 1201 | No | NA | 2 | 0 | 0 | Low-quality | Genome-fragment | 1.08 | AAI-based (medium-confidence) | 0 |
| S11-final-viral-combined_k141_263698||lt2gene | 1086 | No | NA | 1 | 0 | 0 | Not-determined | Genome-fragment | NA | NA | 0 |
| S5-final-viral-combined_k141_17230||full | 1201 | No | NA | 3 | 0 | 0 | Low-quality | Genome-fragment | 2.14 | AAI-based (medium-confidence) | 0 |
| S11-final-viral-combined_k141_208973||lt2gene | 2022 | No | NA | 2 | 0 | 0 | Not-determined | Genome-fragment | NA | NA | 0 |
| S11-final-viral-combined_k141_209406||lt2gene | 1223 | No | NA | 1 | 0 | 0 | Not-determined | Genome-fragment | NA | NA | 0 |
| S7_k141_76279 | 1201 | No | NA | 1 | 0 | 0 | Low-quality | Genome-fragment | 2.79 | AAI-based (high-confidence) | 0 |
| S11-final-viral-combined_k141_264263||lt2gene | 3639 | No | NA | 2 | 0 | 0 | Not-determined | Genome-fragment | NA | NA | 0 |
| S12-final-viral-combined_k141_175188||full | 1201 | No | NA | 2 | 0 | 0 | Low-quality | Genome-fragment | 1.28 | AAI-based (high-confidence) | 0 |
| S4-final-viral-combined_k141_4296||full | 1201 | No | NA | 3 | 0 | 1 | Low-quality | Genome-fragment | 0.8 | AAI-based (medium-confidence) | 0 |
| S4-final-viral-combined_k141_27134||full | 1201 | No | NA | 3 | 0 | 2 | Low-quality | Genome-fragment | 0.75 | AAI-based (medium-confidence) | 0 |
| S8-final-viral-combined_k141_172713||lt2gene | 1201 | No | NA | 1 | 1 | 0 | Low-quality | Genome-fragment | 1.87 | HMM-based (lower-bound) | 0 |
| S8_k141_184903 | 1201 | No | NA | 1 | 0 | 1 | Low-quality | Genome-fragment | 0.52 | AAI-based (high-confidence) | 0 |
| S9-final-viral-combined_k141_28563||full | 1202 | No | NA | 2 | 0 | 0 | Low-quality | Genome-fragment | 2.44 | AAI-based (medium-confidence) | 0 |
| S11-final-viral-combined_k141_154342||lt2gene | 1435 | No | NA | 1 | 0 | 1 | Not-determined | Genome-fragment | NA | NA | 0 |
| S11-final-viral-combined_k141_223315||lt2gene | 1141 | No | NA | 1 | 0 | 1 | Not-determined | Genome-fragment | NA | NA | 0 |
| S9-final-viral-combined_k141_124894||full | 1202 | No | NA | 2 | 0 | 1 | Low-quality | Genome-fragment | 0.95 | AAI-based (medium-confidence) | 0 |
| S11-final-viral-combined_k141_182608||lt2gene | 1202 | No | NA | 1 | 1 | 0 | Low-quality | Genome-fragment | 2.63 | AAI-based (medium-confidence) | 0 |
| S11_k141_180688 | 1202 | No | NA | 2 | 0 | 1 | Low-quality | Genome-fragment | 0.38 | AAI-based (medium-confidence) | 0 |
| S1_k141_3150 | 1202 | No | NA | 4 | 0 | 2 | Low-quality | Genome-fragment | 1.83 | AAI-based (high-confidence) | 0 |
| S3-final-viral-combined_k141_130553||full | 1202 | No | NA | 3 | 0 | 0 | Low-quality | Genome-fragment | 2.62 | AAI-based (high-confidence) | 0 |
| S11-final-viral-combined_k141_168705||lt2gene | 1904 | No | NA | 1 | 0 | 1 | Not-determined | Genome-fragment | NA | NA | 0 |
| S5-final-viral-combined_k141_134950||full | 1202 | No | NA | 2 | 1 | 0 | Low-quality | Genome-fragment | 2.71 | AAI-based (medium-confidence) | 0 |
| S5-final-viral-combined_k141_79767||lt2gene | 1202 | No | NA | 2 | 2 | 0 | Low-quality | Genome-fragment | 2.57 | AAI-based (high-confidence) | 0 |
| S5_k141_110240 | 1202 | No | NA | 3 | 3 | 0 | Low-quality | Genome-fragment | 3.4 | AAI-based (high-confidence) | 0 |
| S7-final-viral-combined_k141_104701||full | 1202 | No | NA | 3 | 0 | 0 | Low-quality | Genome-fragment | 1.66 | AAI-based (medium-confidence) | 0 |
| S17-final-viral-combined_k141_181424||full | 1202 | No | NA | 2 | 1 | 0 | Low-quality | Genome-fragment | 3.03 | AAI-based (medium-confidence) | 0 |
| S17-final-viral-combined_k141_188106||full | 1202 | No | NA | 3 | 0 | 0 | Low-quality | Genome-fragment | 2.06 | AAI-based (medium-confidence) | 0 |
| S17-final-viral-combined_k141_19169||full | 1202 | No | NA | 2 | 0 | 0 | Low-quality | Genome-fragment | 0.97 | AAI-based (medium-confidence) | 0 |
| S12-final-viral-combined_k141_131712||full | 1202 | No | NA | 5 | 0 | 0 | Low-quality | Genome-fragment | 3.52 | AAI-based (high-confidence) | 0 |
| S2_k141_132694 | 1202 | No | NA | 4 | 0 | 0 | Low-quality | Genome-fragment | 2.08 | AAI-based (high-confidence) | 0 |
| S6-final-viral-combined_k141_99955||full | 1202 | No | NA | 3 | 0 | 0 | Low-quality | Genome-fragment | 1.56 | AAI-based (high-confidence) | 0 |
| S6-final-viral-combined_k141_39108||full | 1202 | No | NA | 3 | 0 | 0 | Low-quality | Genome-fragment | 1.66 | AAI-based (high-confidence) | 0 |
| S8-final-viral-combined_k141_74933||full | 1202 | No | NA | 2 | 1 | 0 | Low-quality | Genome-fragment | 2.41 | AAI-based (high-confidence) | 0 |
| S9-final-viral-combined_k141_49619||lt2gene | 1203 | No | NA | 2 | 1 | 0 | Low-quality | Genome-fragment | 2.64 | AAI-based (medium-confidence) | 0 |
| S11-final-viral-combined_k141_72347||lt2gene | 1576 | No | NA | 2 | 0 | 2 | Not-determined | Genome-fragment | NA | NA | 0 |
| S11-final-viral-combined_k141_228326||full | 1203 | No | NA | 3 | 0 | 0 | Low-quality | Genome-fragment | 0.49 | AAI-based (medium-confidence) | 0 |
| S3-final-viral-combined_k141_98328||full | 1203 | No | NA | 3 | 0 | 0 | Low-quality | Genome-fragment | 2.67 | AAI-based (high-confidence) | 0 |
| S3-final-viral-combined_k141_51666||lt2gene | 1203 | No | NA | 1 | 1 | 0 | Low-quality | Genome-fragment | 1.93 | HMM-based (lower-bound) | 0 |
| S5-final-viral-combined_k141_197232||full | 1203 | No | NA | 4 | 0 | 0 | Low-quality | Genome-fragment | 2.67 | AAI-based (high-confidence) | 0 |
| S7-final-viral-combined_k141_88778||full | 1203 | No | NA | 2 | 0 | 2 | Low-quality | Genome-fragment | 0.41 | AAI-based (high-confidence) | 0 |
| S12-final-viral-combined_k141_173332||full | 1203 | No | NA | 3 | 0 | 0 | Low-quality | Genome-fragment | 3.41 | AAI-based (high-confidence) | 0 |
| S11-final-viral-combined_k141_252296||lt2gene | 1107 | No | NA | 2 | 0 | 0 | Not-determined | Genome-fragment | NA | NA | 0 |
| S2-final-viral-combined_k141_24291||lt2gene | 1203 | No | NA | 1 | 1 | 0 | Low-quality | Genome-fragment | 2.7 | AAI-based (medium-confidence) | 0 |
| S2_k141_128097 | 1203 | No | NA | 1 | 0 | 1 | Low-quality | Genome-fragment | 4.03 | AAI-based (high-confidence) | 0 |
| S11-final-viral-combined_k141_183775||lt2gene | 1116 | No | NA | 1 | 0 | 1 | Not-determined | Genome-fragment | NA | NA | 0 |
| S8_k141_248379 | 1203 | No | NA | 2 | 0 | 0 | Low-quality | Genome-fragment | 1.72 | AAI-based (medium-confidence) | 0 |
| S11-final-viral-combined_k141_114239||lt2gene | 1021 | No | NA | 1 | 0 | 0 | Not-determined | Genome-fragment | NA | NA | 0 |
| S9-final-viral-combined_k141_4206||full | 1204 | No | NA | 2 | 0 | 0 | Low-quality | Genome-fragment | 1.77 | AAI-based (medium-confidence) | 0 |
| S11-final-viral-combined_k141_146157||full | 1204 | No | NA | 4 | 1 | 0 | Low-quality | Genome-fragment | 1.97 | AAI-based (high-confidence) | 0 |
| S11-final-viral-combined_k141_212001||lt2gene | 1415 | No | NA | 1 | 0 | 0 | Not-determined | Genome-fragment | NA | NA | 0 |
| S11-final-viral-combined_k141_294012||lt2gene | 1252 | No | NA | 1 | 0 | 0 | Not-determined | Genome-fragment | NA | NA | 0 |
| S11-final-viral-combined_k141_174757||full | 1204 | No | NA | 2 | 0 | 0 | Low-quality | Genome-fragment | 1.71 | AAI-based (high-confidence) | 0 |
| S11-final-viral-combined_k141_212232||lt2gene | 1077 | No | NA | 1 | 0 | 0 | Not-determined | Genome-fragment | NA | NA | 0 |
| S1-final-viral-combined_k141_343573||full | 1204 | No | NA | 3 | 0 | 1 | Low-quality | Genome-fragment | 1.64 | AAI-based (medium-confidence) | 0 |
| S1_k141_153504 | 1204 | No | NA | 3 | 0 | 2 | Low-quality | Genome-fragment | 3.01 | AAI-based (high-confidence) | 0 |
| S5-final-viral-combined_k141_60746||lt2gene | 1204 | No | NA | 2 | 1 | 0 | Low-quality | Genome-fragment | 1.96 | HMM-based (lower-bound) | 0 |
| S11-final-viral-combined_k141_267134||lt2gene | 2447 | No | NA | 2 | 0 | 1 | Not-determined | Genome-fragment | NA | NA | 0 |
| S11-final-viral-combined_k141_253375||lt2gene | 2268 | No | NA | 2 | 0 | 0 | Not-determined | Genome-fragment | NA | NA | 0 |
| S5_k141_134041 | 1204 | No | NA | 1 | 0 | 0 | Low-quality | Genome-fragment | 2.8 | AAI-based (medium-confidence) | 0 |
| S5_k141_32244 | 1204 | No | NA | 1 | 0 | 1 | Low-quality | Genome-fragment | 2.02 | AAI-based (high-confidence) | 0 |
| S5_k141_42187 | 1204 | No | NA | 2 | 0 | 2 | Low-quality | Genome-fragment | 2.16 | AAI-based (medium-confidence) | 0 |
| S7-final-viral-combined_k141_170268||full | 1204 | No | NA | 3 | 1 | 0 | Low-quality | Genome-fragment | 3.6 | AAI-based (high-confidence) | 0 |
| S7_k141_211687 | 1204 | No | NA | 1 | 0 | 1 | Low-quality | Genome-fragment | 0.32 | AAI-based (high-confidence) | 0 |
| S11-final-viral-combined_k141_102591||lt2gene | 1241 | No | NA | 1 | 0 | 0 | Not-determined | Genome-fragment | NA | NA | 0 |
| S17-final-viral-combined_k141_245967||lt2gene | 1204 | No | NA | 2 | 1 | 0 | Low-quality | Genome-fragment | 2.21 | HMM-based (lower-bound) | 0 |
| S17-k141_10510 | 1204 | No | NA | 1 | 0 | 0 | Low-quality | Genome-fragment | 1.16 | AAI-based (high-confidence) | 0 |
| S11-final-viral-combined_k141_74132||lt2gene | 1171 | No | NA | 2 | 0 | 1 | Not-determined | Genome-fragment | NA | NA | 0 |
| S11-final-viral-combined_k141_157344||lt2gene | 1022 | No | NA | 1 | 0 | 0 | Not-determined | Genome-fragment | NA | NA | 0 |
| S12-final-viral-combined_k141_50442||full | 1204 | No | NA | 4 | 0 | 0 | Low-quality | Genome-fragment | 2.06 | AAI-based (high-confidence) | 0 |
| S12-final-viral-combined_k141_314578||full | 1204 | No | NA | 2 | 0 | 0 | Low-quality | Genome-fragment | 2.76 | AAI-based (high-confidence) | 0 |
| S12_k141_238449 | 1204 | No | NA | 1 | 0 | 0 | Low-quality | Genome-fragment | 1.69 | AAI-based (high-confidence) | 0 |
| S11-final-viral-combined_k141_309559||lt2gene | 2216 | No | NA | 1 | 0 | 0 | Not-determined | Genome-fragment | NA | NA | 0 |
| S2_k141_40416 | 1204 | No | NA | 1 | 0 | 1 | Low-quality | Genome-fragment | 0.65 | AAI-based (medium-confidence) | 0 |
| S11-final-viral-combined_k141_74444||lt2gene | 1628 | No | NA | 1 | 0 | 0 | Not-determined | Genome-fragment | NA | NA | 0 |
| S8-final-viral-combined_k141_70213||full | 1204 | No | NA | 2 | 0 | 0 | Low-quality | Genome-fragment | 0.9 | AAI-based (medium-confidence) | 0 |
| S11-final-viral-combined_k141_295345||lt2gene | 1385 | No | NA | 2 | 0 | 1 | Not-determined | Genome-fragment | NA | NA | 0 |
| S11-final-viral-combined_k141_309868||lt2gene | 1267 | No | NA | 2 | 0 | 1 | Not-determined | Genome-fragment | NA | NA | 0 |
| S11-final-viral-combined_k141_236973||full | 1205 | No | NA | 2 | 1 | 0 | Low-quality | Genome-fragment | 3.2 | AAI-based (medium-confidence) | 0 |
| S11-final-viral-combined_k141_100255||full | 1205 | No | NA | 2 | 1 | 0 | Low-quality | Genome-fragment | 1.86 | HMM-based (lower-bound) | 0 |
| S11-final-viral-combined_k141_198511||full | 1205 | No | NA | 2 | 0 | 2 | Low-quality | Genome-fragment | 0.36 | AAI-based (medium-confidence) | 0 |
| S11-final-viral-combined_k141_326167||full | 1205 | No | NA | 2 | 1 | 0 | Low-quality | Genome-fragment | 2.55 | AAI-based (high-confidence) | 0 |
| S11_k141_321186 | 1205 | No | NA | 1 | 0 | 1 | Low-quality | Genome-fragment | 1.46 | AAI-based (high-confidence) | 0 |
| S1-final-viral-combined_k141_120667||full | 1205 | No | NA | 4 | 1 | 0 | Low-quality | Genome-fragment | 0.48 | HMM-based (lower-bound) | 0 |
| S11-final-viral-combined_k141_172134||lt2gene | 1981 | No | NA | 2 | 0 | 0 | Not-determined | Genome-fragment | NA | NA | 0 |
| S11-final-viral-combined_k141_172285||lt2gene | 1141 | No | NA | 2 | 0 | 1 | Not-determined | Genome-fragment | NA | NA | 0 |
| S11-final-viral-combined_k141_48007||lt2gene | 1708 | No | NA | 2 | 0 | 0 | Not-determined | Genome-fragment | NA | NA | 0 |
| S1-final-viral-combined_k141_230061||full | 1205 | No | NA | 3 | 0 | 2 | Low-quality | Genome-fragment | 2.1 | AAI-based (high-confidence) | 0 |
| S1-final-viral-combined_k141_32504||full | 1205 | No | NA | 4 | 0 | 0 | Low-quality | Genome-fragment | 2.78 | AAI-based (high-confidence) | 0 |
| S5-final-viral-combined_k141_142545||full | 1205 | No | NA | 2 | 0 | 0 | Low-quality | Genome-fragment | 0.75 | AAI-based (high-confidence) | 0 |
| S11-final-viral-combined_k141_199423||lt2gene | 2659 | No | NA | 1 | 0 | 1 | Not-determined | Genome-fragment | NA | NA | 0 |
| S7-final-viral-combined_k141_48740||full | 1205 | No | NA | 3 | 2 | 0 | Low-quality | Genome-fragment | 3.35 | AAI-based (high-confidence) | 0 |
| S7-final-viral-combined_k141_185308||lt2gene | 1205 | No | NA | 2 | 2 | 0 | Low-quality | Genome-fragment | 3.49 | AAI-based (high-confidence) | 0 |
| S7-final-viral-combined_k141_49041||lt2gene | 1205 | No | NA | 2 | 2 | 0 | Low-quality | Genome-fragment | 2.43 | AAI-based (high-confidence) | 0 |
| S17-final-viral-combined_k141_219670||full | 1205 | No | NA | 3 | 0 | 0 | Low-quality | Genome-fragment | 1.87 | AAI-based (high-confidence) | 0 |
| S12_k141_151420 | 1205 | No | NA | 3 | 1 | 0 | Low-quality | Genome-fragment | 3.14 | AAI-based (high-confidence) | 0 |
| S11-final-viral-combined_k141_283307||lt2gene | 1079 | No | NA | 1 | 0 | 1 | Not-determined | Genome-fragment | NA | NA | 0 |
| S12_k141_97556 | 1205 | No | NA | 2 | 0 | 0 | Low-quality | Genome-fragment | 1.22 | AAI-based (high-confidence) | 0 |
| S11-final-viral-combined_k141_214660||lt2gene | 1179 | No | NA | 2 | 0 | 1 | Not-determined | Genome-fragment | NA | NA | 0 |
| S8-final-viral-combined_k141_195815||full | 1205 | No | NA | 2 | 0 | 0 | Low-quality | Genome-fragment | 2.9 | AAI-based (medium-confidence) | 0 |
| S9-final-viral-combined_k141_17156||full | 1206 | No | NA | 4 | 0 | 0 | Low-quality | Genome-fragment | 2.78 | AAI-based (high-confidence) | 0 |
| S9-final-viral-combined_k141_89485||lt2gene | 1206 | No | NA | 2 | 0 | 1 | Low-quality | Genome-fragment | 2.16 | AAI-based (medium-confidence) | 0 |
| S9-final-viral-combined_k141_9730||lt2gene | 1206 | No | NA | 2 | 1 | 0 | Low-quality | Genome-fragment | 2.93 | AAI-based (high-confidence) | 0 |
| S9_k141_99754 | 1206 | No | NA | 2 | 0 | 0 | Low-quality | Genome-fragment | 0.13 | AAI-based (medium-confidence) | 0 |
| S11_k141_221243 | 1206 | No | NA | 3 | 2 | 0 | Low-quality | Genome-fragment | 2.47 | AAI-based (medium-confidence) | 0 |
| S5-final-viral-combined_k141_46648||full | 1206 | No | NA | 2 | 0 | 0 | Low-quality | Genome-fragment | 0.61 | AAI-based (medium-confidence) | 0 |
| S5-final-viral-combined_k141_56475||full | 1206 | No | NA | 4 | 0 | 4 | Low-quality | Genome-fragment | 0.35 | AAI-based (medium-confidence) | 0 |
| S5_k141_154735 | 1206 | No | NA | 3 | 0 | 0 | Low-quality | Genome-fragment | 2.01 | AAI-based (high-confidence) | 0 |
| S5_k141_164351 | 1206 | No | NA | 1 | 0 | 1 | Low-quality | Genome-fragment | 1.83 | AAI-based (medium-confidence) | 0 |
| S17-final-viral-combined_k141_119274||full | 1206 | No | NA | 2 | 1 | 0 | Low-quality | Genome-fragment | 2.39 | AAI-based (high-confidence) | 0 |
| S2-final-viral-combined_k141_236495||full | 1206 | No | NA | 3 | 0 | 1 | Low-quality | Genome-fragment | 1.97 | AAI-based (medium-confidence) | 0 |
| S2-final-viral-combined_k141_61183||full | 1206 | No | NA | 2 | 0 | 2 | Low-quality | Genome-fragment | 3.07 | AAI-based (medium-confidence) | 0 |
| S2-final-viral-combined_k141_52815||lt2gene | 1206 | No | NA | 2 | 1 | 0 | Low-quality | Genome-fragment | 2.23 | HMM-based (lower-bound) | 0 |
| S2_k141_67303 | 1206 | No | NA | 2 | 0 | 2 | Low-quality | Genome-fragment | 1.15 | AAI-based (medium-confidence) | 0 |
| S9-final-viral-combined_k141_104303||full | 1207 | No | NA | 2 | 0 | 1 | Low-quality | Genome-fragment | 1 | AAI-based (medium-confidence) | 0 |
| S11-final-viral-combined_k141_49820||lt2gene | 3119 | No | NA | 1 | 0 | 0 | Not-determined | Genome-fragment | NA | NA | 0 |
| S11-final-viral-combined_k141_246168||full | 1207 | No | NA | 3 | 1 | 0 | Low-quality | Genome-fragment | 0.63 | HMM-based (lower-bound) | 0 |
| S11-final-viral-combined_k141_3364||lt2gene | 1207 | No | NA | 1 | 1 | 0 | Low-quality | Genome-fragment | 3.27 | AAI-based (high-confidence) | 0 |
| S11-final-viral-combined_k141_215778||lt2gene | 1695 | No | NA | 2 | 0 | 0 | Not-determined | Genome-fragment | NA | NA | 0 |
| S11-final-viral-combined_k141_297882||lt2gene | 1474 | No | NA | 1 | 0 | 0 | Not-determined | Genome-fragment | NA | NA | 0 |
| S1-final-viral-combined_k141_379162||lt2gene | 1207 | No | NA | 1 | 1 | 0 | Low-quality | Genome-fragment | 3.11 | AAI-based (high-confidence) | 0 |
| S1-final-viral-combined_k141_28659||lt2gene | 1207 | No | NA | 2 | 2 | 0 | Low-quality | Genome-fragment | 2.7 | AAI-based (high-confidence) | 0 |
| S5-final-viral-combined_k141_60396||full | 1207 | No | NA | 3 | 0 | 0 | Low-quality | Genome-fragment | 0.86 | AAI-based (high-confidence) | 0 |
| S11-final-viral-combined_k141_187742||lt2gene | 1650 | No | NA | 2 | 0 | 1 | Not-determined | Genome-fragment | NA | NA | 0 |
| S17-final-viral-combined_k141_55864||full | 1207 | No | NA | 2 | 0 | 0 | Low-quality | Genome-fragment | 2.84 | AAI-based (high-confidence) | 0 |
| S17-final-viral-combined_k141_117005||lt2gene | 1207 | No | NA | 2 | 1 | 0 | Low-quality | Genome-fragment | 2.99 | AAI-based (medium-confidence) | 0 |
| S12-final-viral-combined_k141_137414||full | 1207 | No | NA | 3 | 2 | 0 | Low-quality | Genome-fragment | 1.21 | HMM-based (lower-bound) | 0 |
| S2_k141_54522 | 1207 | No | NA | 1 | 0 | 1 | Low-quality | Genome-fragment | 0.53 | AAI-based (medium-confidence) | 0 |
| S11-final-viral-combined_k141_92255||lt2gene | 1012 | No | NA | 2 | 0 | 0 | Not-determined | Genome-fragment | NA | NA | 0 |
| S6_k141_81841 | 1207 | No | NA | 2 | 1 | 0 | Low-quality | Genome-fragment | 2.24 | AAI-based (high-confidence) | 0 |
| S8-final-viral-combined_k141_238854||full | 1207 | No | NA | 3 | 0 | 0 | Low-quality | Genome-fragment | 1.77 | AAI-based (medium-confidence) | 0 |
| S11-final-viral-combined_k141_92505||lt2gene | 1884 | No | NA | 1 | 0 | 1 | Not-determined | Genome-fragment | NA | NA | 0 |
| S8-final-viral-combined_k141_68664||full | 1207 | No | NA | 2 | 0 | 0 | Low-quality | Genome-fragment | 2.64 | AAI-based (high-confidence) | 0 |
| S9-final-viral-combined_k141_106750||full | 1208 | No | NA | 2 | 0 | 1 | Low-quality | Genome-fragment | 0.59 | AAI-based (high-confidence) | 0 |
| S11_k141_95751 | 1208 | No | NA | 3 | 0 | 0 | Low-quality | Genome-fragment | 2.41 | AAI-based (medium-confidence) | 0 |
| S1-final-viral-combined_k141_319764||full | 1208 | No | NA | 3 | 0 | 0 | Low-quality | Genome-fragment | 3.12 | AAI-based (medium-confidence) | 0 |
| S1_k141_221333 | 1208 | No | NA | 1 | 0 | 0 | Low-quality | Genome-fragment | 6.92 | AAI-based (high-confidence) | 0 |
| S5-final-viral-combined_k141_58107||full | 1208 | No | NA | 2 | 1 | 0 | Low-quality | Genome-fragment | 0.83 | HMM-based (lower-bound) | 0 |
| S5_k141_36810 | 1208 | No | NA | 2 | 0 | 0 | Low-quality | Genome-fragment | 1.85 | AAI-based (medium-confidence) | 0 |
| S5_k141_5150 | 1208 | No | NA | 1 | 0 | 1 | Low-quality | Genome-fragment | 2.59 | AAI-based (high-confidence) | 0 |
| S17-k141_223433 | 1208 | No | NA | 3 | 1 | 0 | Low-quality | Genome-fragment | 1.19 | AAI-based (medium-confidence) | 0 |
| S12-final-viral-combined_k141_52293||lt2gene | 1208 | No | NA | 1 | 1 | 0 | Low-quality | Genome-fragment | 2.43 | AAI-based (medium-confidence) | 0 |
| S11-final-viral-combined_k141_133362||lt2gene | 1466 | No | NA | 1 | 0 | 1 | Not-determined | Genome-fragment | NA | NA | 0 |
| S11-final-viral-combined_k141_133368||lt2gene | 1322 | No | NA | 2 | 0 | 0 | Not-determined | Genome-fragment | NA | NA | 0 |
| S11-final-viral-combined_k141_299486||lt2gene | 1534 | No | NA | 2 | 0 | 0 | Not-determined | Genome-fragment | NA | NA | 0 |
| S2-final-viral-combined_k141_238444||full | 1208 | No | NA | 3 | 0 | 1 | Low-quality | Genome-fragment | 8.93 | AAI-based (medium-confidence) | 0 |
| S11-final-viral-combined_k141_245491||lt2gene | 1159 | No | NA | 1 | 0 | 1 | Not-determined | Genome-fragment | NA | NA | 0 |
| S2_k141_3467 | 1208 | No | NA | 1 | 0 | 0 | Low-quality | Genome-fragment | 1.57 | AAI-based (high-confidence) | 0 |
| S4-final-viral-combined_k141_92344||lt2gene | 1208 | No | NA | 2 | 1 | 0 | Low-quality | Genome-fragment | 1.95 | HMM-based (lower-bound) | 0 |
| S6-final-viral-combined_k141_87499||full | 1208 | No | NA | 3 | 0 | 1 | Low-quality | Genome-fragment | 0.77 | AAI-based (medium-confidence) | 0 |
| S11-final-viral-combined_k141_52520||lt2gene | 1289 | No | NA | 1 | 0 | 0 | Not-determined | Genome-fragment | NA | NA | 0 |
| S9-final-viral-combined_k141_105401||lt2gene | 1209 | No | NA | 1 | 1 | 0 | Low-quality | Genome-fragment | 1.41 | AAI-based (high-confidence) | 0 |
| S11-final-viral-combined_k141_323955||full | 1209 | No | NA | 3 | 1 | 0 | Low-quality | Genome-fragment | 2.73 | AAI-based (medium-confidence) | 0 |
| S1-final-viral-combined_k141_151522||full | 1209 | No | NA | 2 | 0 | 1 | Low-quality | Genome-fragment | 0.7 | AAI-based (medium-confidence) | 0 |
| S5-final-viral-combined_k141_31755||full | 1209 | No | NA | 4 | 0 | 0 | Low-quality | Genome-fragment | 2.53 | AAI-based (medium-confidence) | 0 |
| S11-final-viral-combined_k141_203764||lt2gene | 3361 | No | NA | 1 | 0 | 1 | Not-determined | Genome-fragment | NA | NA | 0 |
| S5-final-viral-combined_k141_127001||lt2gene | 1209 | No | NA | 2 | 1 | 0 | Low-quality | Genome-fragment | 2.74 | AAI-based (medium-confidence) | 0 |
| S7-final-viral-combined_k141_105038||full | 1209 | No | NA | 2 | 0 | 0 | Low-quality | Genome-fragment | 2.54 | AAI-based (high-confidence) | 0 |
| S7_k141_188626 | 1209 | No | NA | 2 | 0 | 0 | Low-quality | Genome-fragment | 2.75 | AAI-based (high-confidence) | 0 |
| S12-final-viral-combined_k141_61143||full | 1209 | No | NA | 4 | 0 | 0 | Low-quality | Genome-fragment | 1.57 | AAI-based (medium-confidence) | 0 |
| S12-final-viral-combined_k141_94434||full | 1209 | No | NA | 2 | 0 | 0 | Low-quality | Genome-fragment | 3.56 | AAI-based (high-confidence) | 0 |
| S12-final-viral-combined_k141_356060||lt2gene | 1209 | No | NA | 1 | 1 | 0 | Low-quality | Genome-fragment | 1.32 | HMM-based (lower-bound) | 0 |
| S11-final-viral-combined_k141_259887||lt2gene | 1084 | No | NA | 2 | 0 | 0 | Not-determined | Genome-fragment | NA | NA | 0 |
| S11-final-viral-combined_k141_326970||lt2gene | 2305 | No | NA | 2 | 0 | 1 | Not-determined | Genome-fragment | NA | NA | 0 |
| S12-final-viral-combined_k141_14071||lt2gene | 1209 | No | NA | 1 | 1 | 0 | Low-quality | Genome-fragment | 3.71 | AAI-based (medium-confidence) | 0 |
| S8-final-viral-combined_k141_174862||full | 1209 | No | NA | 3 | 0 | 0 | Low-quality | Genome-fragment | 8.37 | AAI-based (medium-confidence) | 0 |
| S11-final-viral-combined_k141_314649||lt2gene | 1725 | No | NA | 2 | 0 | 1 | Not-determined | Genome-fragment | NA | NA | 0 |
| S11-final-viral-combined_k141_94169||lt2gene | 1261 | No | NA | 1 | 0 | 0 | Not-determined | Genome-fragment | NA | NA | 0 |
| S8-final-viral-combined_k141_245753||full | 1209 | No | NA | 4 | 1 | 0 | Low-quality | Genome-fragment | 2.48 | AAI-based (medium-confidence) | 0 |
| S11-final-viral-combined_k141_287719||lt2gene | 1740 | No | NA | 2 | 0 | 0 | Not-determined | Genome-fragment | NA | NA | 0 |
| S8-final-viral-combined_k141_120292||lt2gene | 1209 | No | NA | 1 | 1 | 0 | Low-quality | Genome-fragment | 1.29 | AAI-based (high-confidence) | 0 |
| S11-final-viral-combined_k141_224003||full | 1210 | No | NA | 3 | 1 | 0 | Low-quality | Genome-fragment | 2.75 | AAI-based (high-confidence) | 0 |
| S11-final-viral-combined_k141_174266||full | 1210 | No | NA | 4 | 0 | 0 | Low-quality | Genome-fragment | 2.64 | AAI-based (high-confidence) | 0 |
| S5-final-viral-combined_k141_109141||full | 1210 | No | NA | 3 | 0 | 0 | Low-quality | Genome-fragment | 1.9 | AAI-based (medium-confidence) | 0 |
| S7-final-viral-combined_k141_147086||full | 1210 | No | NA | 2 | 0 | 0 | Low-quality | Genome-fragment | 2.17 | AAI-based (medium-confidence) | 0 |
| S12-final-viral-combined_k141_342425||lt2gene | 1210 | No | NA | 2 | 1 | 0 | Low-quality | Genome-fragment | 2.04 | HMM-based (lower-bound) | 0 |
| S12_k141_372566 | 1210 | No | NA | 1 | 1 | 0 | Low-quality | Genome-fragment | 0.65 | AAI-based (high-confidence) | 0 |
| S2-final-viral-combined_k141_209302||lt2gene | 1210 | No | NA | 2 | 1 | 0 | Low-quality | Genome-fragment | 0.78 | HMM-based (lower-bound) | 0 |
| S2_k141_115807 | 1210 | No | NA | 2 | 0 | 0 | Low-quality | Genome-fragment | 1.72 | AAI-based (high-confidence) | 0 |
| S4_k141_40472 | 1210 | No | NA | 3 | 0 | 0 | Low-quality | Genome-fragment | 1.96 | AAI-based (medium-confidence) | 0 |
| S11-final-viral-combined_k141_315339||lt2gene | 1276 | No | NA | 2 | 0 | 0 | Not-determined | Genome-fragment | NA | NA | 0 |
| S11-final-viral-combined_k141_109764||lt2gene | 1329 | No | NA | 1 | 0 | 0 | Not-determined | Genome-fragment | NA | NA | 0 |
| S11-final-viral-combined_k141_247683||lt2gene | 1036 | No | NA | 2 | 0 | 0 | Not-determined | Genome-fragment | NA | NA | 0 |
| S11-final-viral-combined_k141_54378||lt2gene | 1003 | No | NA | 1 | 0 | 0 | Not-determined | Genome-fragment | NA | NA | 0 |
| S11-final-viral-combined_k141_247740||lt2gene | 1382 | No | NA | 1 | 0 | 0 | Not-determined | Genome-fragment | NA | NA | 0 |
| S11-final-viral-combined_k141_301420||lt2gene | 2040 | No | NA | 1 | 0 | 1 | Not-determined | Genome-fragment | NA | NA | 0 |
| S11-final-viral-combined_k141_71632||full | 1211 | No | NA | 2 | 1 | 0 | Low-quality | Genome-fragment | 3.04 | AAI-based (high-confidence) | 0 |
| S3-final-viral-combined_k141_35845||full | 1211 | No | NA | 3 | 0 | 1 | Low-quality | Genome-fragment | 1.65 | AAI-based (medium-confidence) | 0 |
| S11-final-viral-combined_k141_219235||lt2gene | 1365 | No | NA | 1 | 0 | 0 | Not-determined | Genome-fragment | NA | NA | 0 |
| S11-final-viral-combined_k141_95305||lt2gene | 1332 | No | NA | 2 | 0 | 0 | Not-determined | Genome-fragment | NA | NA | 0 |
| S3_k141_125143 | 1211 | No | NA | 3 | 0 | 0 | Low-quality | Genome-fragment | 2.43 | AAI-based (medium-confidence) | 0 |
| S5-final-viral-combined_k141_119200||full | 1211 | No | NA | 2 | 1 | 0 | Low-quality | Genome-fragment | 1.74 | HMM-based (lower-bound) | 0 |
| S11-final-viral-combined_k141_178905||lt2gene | 3008 | No | NA | 1 | 0 | 1 | Not-determined | Genome-fragment | NA | NA | 0 |
| S7-final-viral-combined_k141_39329||lt2gene | 1211 | No | NA | 2 | 1 | 0 | Low-quality | Genome-fragment | 3.01 | AAI-based (high-confidence) | 0 |
| S17-final-viral-combined_k141_217916||full | 1211 | No | NA | 4 | 1 | 0 | Low-quality | Genome-fragment | 2.72 | AAI-based (high-confidence) | 0 |
| S17-final-viral-combined_k141_30210||full | 1211 | No | NA | 2 | 0 | 0 | Low-quality | Genome-fragment | 2.17 | AAI-based (high-confidence) | 0 |
| S12-final-viral-combined_k141_139857||full | 1211 | No | NA | 3 | 0 | 0 | Low-quality | Genome-fragment | 2.02 | AAI-based (high-confidence) | 0 |
| S2-final-viral-combined_k141_130015||full | 1211 | No | NA | 2 | 0 | 0 | Low-quality | Genome-fragment | 0.89 | AAI-based (high-confidence) | 0 |
| S4-final-viral-combined_k141_42772||full | 1211 | Yes | 497 | 5 | 1 | 1 | Low-quality | Genome-fragment | 1.25 | AAI-based (medium-confidence) | 58.96 |
| S4-final-viral-combined_k141_156790||lt2gene | 1211 | No | NA | 2 | 1 | 0 | Low-quality | Genome-fragment | 3.22 | AAI-based (high-confidence) | 0 |
| S6-final-viral-combined_k141_57653||full | 1211 | No | NA | 1 | 0 | 0 | Low-quality | Genome-fragment | 3.01 | AAI-based (high-confidence) | 0 |
| S6_k141_47697 | 1211 | No | NA | 1 | 0 | 1 | Low-quality | Genome-fragment | 0.52 | AAI-based (medium-confidence) | 0 |
| S8-final-viral-combined_k141_190664||full | 1211 | No | NA | 3 | 0 | 0 | Low-quality | Genome-fragment | 1.16 | AAI-based (medium-confidence) | 0 |
| S8-final-viral-combined_k141_195067||full | 1211 | No | NA | 2 | 0 | 0 | Low-quality | Genome-fragment | 3.04 | AAI-based (medium-confidence) | 0 |
| S11-final-viral-combined_k141_27367||lt2gene | 1325 | No | NA | 1 | 0 | 1 | Not-determined | Genome-fragment | NA | NA | 0 |
| S11_k141_13825 | 1783 | No | NA | 2 | 0 | 0 | Not-determined | Genome-fragment | NA | NA | 0 |
| S11_k141_275662 | 3899 | No | NA | 3 | 0 | 3 | Not-determined | Genome-fragment | NA | NA | 0 |
| S8_k141_223889 | 1211 | No | NA | 2 | 0 | 0 | Low-quality | Genome-fragment | 1.88 | AAI-based (high-confidence) | 0 |
| S11_k141_96490 | 1396 | No | NA | 2 | 0 | 2 | Not-determined | Genome-fragment | NA | NA | 0 |
| S11_k141_55146 | 1286 | No | NA | 1 | 0 | 0 | Not-determined | Genome-fragment | NA | NA | 0 |
| S8_k141_112434 | 1211 | No | NA | 2 | 1 | 0 | Low-quality | Genome-fragment | 3.75 | AAI-based (high-confidence) | 0 |
| S11_k141_289478 | 1132 | No | NA | 1 | 0 | 1 | Not-determined | Genome-fragment | NA | NA | 0 |
| S11_k141_248134 | 1243 | No | NA | 1 | 0 | 1 | Not-determined | Genome-fragment | NA | NA | 0 |
| S9_k141_110372 | 1212 | No | NA | 2 | 0 | 0 | Low-quality | Genome-fragment | 2.06 | AAI-based (high-confidence) | 0 |
| S11_k141_137920 | 1224 | No | NA | 3 | 0 | 3 | Not-determined | Genome-fragment | NA | NA | 0 |
| S11_k141_151683 | 1402 | No | NA | 1 | 0 | 1 | Not-determined | Genome-fragment | NA | NA | 0 |
| S11_k141_110332 | 1156 | No | NA | 1 | 0 | 0 | Not-determined | Genome-fragment | NA | NA | 0 |
| S11_k141_192995 | 1334 | No | NA | 1 | 0 | 0 | Not-determined | Genome-fragment | NA | NA | 0 |
| S11_k141_220598 | 4952 | No | NA | 5 | 0 | 5 | Not-determined | Genome-fragment | NA | NA | 0 |
| S11-final-viral-combined_k141_209317||full | 1212 | No | NA | 2 | 0 | 0 | Low-quality | Genome-fragment | 3.16 | AAI-based (high-confidence) | 0 |
| S11_k141_289518 | 1491 | No | NA | 1 | 0 | 1 | Not-determined | Genome-fragment | NA | NA | 0 |
| S11_k141_137951 | 1147 | No | NA | 1 | 0 | 0 | Not-determined | Genome-fragment | NA | NA | 0 |
| S11_k141_220623 | 4368 | No | NA | 4 | 0 | 3 | Not-determined | Genome-fragment | NA | NA | 0 |
| S1-final-viral-combined_k141_109767||full | 1212 | No | NA | 2 | 0 | 1 | Low-quality | Genome-fragment | 0.82 | AAI-based (medium-confidence) | 0 |
| S1_k141_100081 | 1212 | No | NA | 1 | 0 | 1 | Low-quality | Genome-fragment | 2.08 | AAI-based (high-confidence) | 0 |
| S11_k141_27600 | 1926 | No | NA | 1 | 0 | 1 | Not-determined | Genome-fragment | NA | NA | 0 |
| S11_k141_220641 | 1986 | No | NA | 2 | 0 | 1 | Not-determined | Genome-fragment | NA | NA | 0 |
| S5-final-viral-combined_k141_95856||full | 1212 | No | NA | 2 | 0 | 0 | Low-quality | Genome-fragment | 2.02 | AAI-based (medium-confidence) | 0 |
| S12-final-viral-combined_k141_74596||full | 1212 | No | NA | 3 | 1 | 0 | Low-quality | Genome-fragment | 2.45 | AAI-based (medium-confidence) | 0 |
| S11_k141_69070 | 3140 | No | NA | 2 | 0 | 2 | Not-determined | Genome-fragment | NA | NA | 0 |
| S11_k141_317161 | 1391 | No | NA | 2 | 0 | 1 | Not-determined | Genome-fragment | NA | NA | 0 |
| S2-final-viral-combined_k141_233876||full | 1212 | No | NA | 3 | 1 | 0 | Low-quality | Genome-fragment | 2.73 | AAI-based (high-confidence) | 0 |
| S11_k141_234402 | 1150 | No | NA | 2 | 0 | 1 | Not-determined | Genome-fragment | NA | NA | 0 |
| S11_k141_124294 | 1285 | No | NA | 1 | 0 | 0 | Not-determined | Genome-fragment | NA | NA | 0 |
| S11_k141_69147 | 2861 | No | NA | 1 | 0 | 1 | Not-determined | Genome-fragment | NA | NA | 0 |
| S11_k141_55340 | 1176 | No | NA | 1 | 0 | 1 | Not-determined | Genome-fragment | NA | NA | 0 |
| S11_k141_303398 | 25741 | No | NA | 27 | 4 | 0 | Medium-quality | Genome-fragment | 59.61 | AAI-based (high-confidence) | 0 |
| S11_k141_317187 | 6935 | No | NA | 6 | 0 | 5 | Not-determined | Genome-fragment | NA | NA | 0 |
| S11_k141_317193 | 1152 | No | NA | 2 | 0 | 0 | Not-determined | Genome-fragment | NA | NA | 0 |
| S11_k141_193176 | 5286 | No | NA | 5 | 0 | 4 | Not-determined | Genome-fragment | NA | NA | 0 |
| S11_k141_289586 | 1460 | No | NA | 1 | 0 | 1 | Not-determined | Genome-fragment | NA | NA | 0 |
| S11_k141_69159 | 2835 | No | NA | 4 | 0 | 0 | Not-determined | Genome-fragment | NA | NA | 0 |
| S2_k141_41821 | 1212 | No | NA | 1 | 0 | 1 | Low-quality | Genome-fragment | 0.53 | AAI-based (medium-confidence) | 0 |
| S4_k141_68919 | 1212 | No | NA | 3 | 0 | 0 | Low-quality | Genome-fragment | 1.39 | AAI-based (medium-confidence) | 0 |
| S8-final-viral-combined_k141_95571||full | 1212 | No | NA | 3 | 0 | 2 | Low-quality | Genome-fragment | 1.22 | AAI-based (medium-confidence) | 0 |
| S11_k141_151897 | 1009 | No | NA | 2 | 0 | 1 | Not-determined | Genome-fragment | NA | NA | 0 |
| S11_k141_234460 | 1275 | No | NA | 3 | 0 | 0 | Not-determined | Genome-fragment | NA | NA | 0 |
| S9-final-viral-combined_k141_79144||full | 1213 | No | NA | 3 | 0 | 0 | Low-quality | Genome-fragment | 3.12 | AAI-based (high-confidence) | 0 |
| S11_k141_179422 | 1007 | No | NA | 3 | 0 | 0 | Not-determined | Genome-fragment | NA | NA | 0 |
| S11_k141_110470 | 1558 | No | NA | 1 | 0 | 0 | Not-determined | Genome-fragment | NA | NA | 0 |
| S1_k141_18532 | 1213 | No | NA | 4 | 1 | 0 | Low-quality | Genome-fragment | 3.6 | AAI-based (high-confidence) | 0 |
| S11_k141_165734 | 1329 | No | NA | 1 | 0 | 1 | Not-determined | Genome-fragment | NA | NA | 0 |
| S17-final-viral-combined_k141_22629||lt2gene | 1213 | No | NA | 2 | 1 | 0 | Low-quality | Genome-fragment | 2.95 | AAI-based (high-confidence) | 0 |
| S11_k141_193248 | 1488 | No | NA | 2 | 0 | 2 | Not-determined | Genome-fragment | NA | NA | 0 |
| S11_k141_165752 | 1250 | No | NA | 3 | 0 | 3 | Not-determined | Genome-fragment | NA | NA | 0 |
| S11_k141_220873 | 1120 | No | NA | 3 | 0 | 1 | Not-determined | Genome-fragment | NA | NA | 0 |
| S11_k141_27835 | 1258 | No | NA | 1 | 0 | 1 | Not-determined | Genome-fragment | NA | NA | 0 |
| S11_k141_124458 | 4269 | No | NA | 5 | 0 | 3 | Not-determined | Genome-fragment | NA | NA | 0 |
| S17-k141_34166 | 1213 | No | NA | 1 | 0 | 0 | Low-quality | Genome-fragment | 2.84 | AAI-based (medium-confidence) | 0 |
| S11_k141_165812 | 5881 | No | NA | 5 | 0 | 5 | Not-determined | Genome-fragment | NA | NA | 0 |
| S17-k141_239172 | 1213 | No | NA | 3 | 0 | 1 | Low-quality | Genome-fragment | 0.83 | AAI-based (medium-confidence) | 0 |
| S11_k141_207078 | 1128 | No | NA | 1 | 0 | 1 | Not-determined | Genome-fragment | NA | NA | 0 |
| S11_k141_165831 | 1826 | No | NA | 1 | 0 | 1 | Not-determined | Genome-fragment | NA | NA | 0 |
| S11_k141_193295 | 1164 | No | NA | 1 | 0 | 1 | Not-determined | Genome-fragment | NA | NA | 0 |
| S11_k141_248378 | 1318 | No | NA | 1 | 0 | 0 | Not-determined | Genome-fragment | NA | NA | 0 |
| S17-k141_175284 | 1213 | No | NA | 3 | 0 | 0 | Low-quality | Genome-fragment | 2.79 | AAI-based (high-confidence) | 0 |
| S11_k141_110548 | 1585 | No | NA | 2 | 0 | 0 | Not-determined | Genome-fragment | NA | NA | 0 |
| S12-final-viral-combined_k141_228179||full | 1213 | No | NA | 3 | 1 | 0 | Low-quality | Genome-fragment | 3.04 | AAI-based (high-confidence) | 0 |
| S11_k141_96843 | 1112 | No | NA | 2 | 0 | 1 | Not-determined | Genome-fragment | NA | NA | 0 |
| S11_k141_152004 | 1191 | No | NA | 3 | 0 | 0 | Not-determined | Genome-fragment | NA | NA | 0 |
| S11_k141_165906 | 9212 | No | NA | 18 | 0 | 3 | Not-determined | Genome-fragment | NA | NA | 0 |
| S11_k141_193357 | 4053 | No | NA | 5 | 0 | 1 | Not-determined | Genome-fragment | NA | NA | 0 |
| S11_k141_41607 | 9671 | No | NA | 10 | 0 | 6 | Not-determined | Genome-fragment | NA | NA | 0 |
| S2-final-viral-combined_k141_210184||full | 1213 | No | NA | 3 | 2 | 0 | Low-quality | Genome-fragment | 2.59 | AAI-based (medium-confidence) | 0 |
| S11_k141_152031 | 1170 | No | NA | 1 | 0 | 0 | Not-determined | Genome-fragment | NA | NA | 0 |
| S11_k141_124508 | 1684 | No | NA | 1 | 0 | 0 | Not-determined | Genome-fragment | NA | NA | 0 |
| S11_k141_28007 | 1114 | No | NA | 1 | 0 | 0 | Not-determined | Genome-fragment | NA | NA | 0 |
| S11_k141_110599 | 2991 | No | NA | 2 | 0 | 2 | Not-determined | Genome-fragment | NA | NA | 0 |
| S2-final-viral-combined_k141_78542||full | 1213 | No | NA | 3 | 1 | 0 | Low-quality | Genome-fragment | 2.68 | AAI-based (medium-confidence) | 0 |
| S11_k141_124518 | 6073 | No | NA | 9 | 0 | 2 | Not-determined | Genome-fragment | NA | NA | 0 |
| S11_k141_275977 | 1071 | No | NA | 2 | 0 | 0 | Not-determined | Genome-fragment | NA | NA | 0 |
| S9-final-viral-combined_k141_35052||full | 1214 | No | NA | 3 | 1 | 0 | Low-quality | Genome-fragment | 2.7 | AAI-based (medium-confidence) | 0 |
| S11_k141_165970 | 1327 | No | NA | 2 | 0 | 0 | Not-determined | Genome-fragment | NA | NA | 0 |
| S11_k141_152076 | 8724 | No | NA | 6 | 0 | 5 | Not-determined | Genome-fragment | NA | NA | 0 |
| S11_k141_276013 | 1247 | No | NA | 1 | 0 | 0 | Not-determined | Genome-fragment | NA | NA | 0 |
| S11_k141_220939 | 1624 | No | NA | 2 | 0 | 1 | Not-determined | Genome-fragment | NA | NA | 0 |
| S11_k141_276045 | 2511 | No | NA | 3 | 0 | 1 | Not-determined | Genome-fragment | NA | NA | 0 |
| S11_k141_41710 | 5452 | No | NA | 7 | 0 | 0 | Not-determined | Genome-fragment | NA | NA | 0 |
| S11_k141_124601 | 6510 | No | NA | 7 | 0 | 2 | Not-determined | Genome-fragment | NA | NA | 0 |
| S9_k141_4083 | 1214 | No | NA | 2 | 0 | 0 | Low-quality | Genome-fragment | 2.41 | AAI-based (high-confidence) | 0 |
| S11_k141_193395 | 1600 | No | NA | 4 | 0 | 0 | Not-determined | Genome-fragment | NA | NA | 0 |
| S11_k141_474 | 6135 | No | NA | 2 | 0 | 0 | Not-determined | Genome-fragment | NA | NA | 0 |
| S11_k141_152143 | 1816 | No | NA | 1 | 0 | 1 | Not-determined | Genome-fragment | NA | NA | 0 |
| S11_k141_317508 | 2225 | No | NA | 3 | 0 | 1 | Not-determined | Genome-fragment | NA | NA | 0 |
| S11-final-viral-combined_k141_323594||full | 1214 | No | NA | 3 | 0 | 0 | Low-quality | Genome-fragment | 2.06 | AAI-based (high-confidence) | 0 |
| S11_k141_152145 | 5554 | No | NA | 4 | 0 | 2 | Not-determined | Genome-fragment | NA | NA | 0 |
| S11_k141_55610 | 58459 | No | NA | 102 | 27 | 2 | Medium-quality | Genome-fragment | 78.18 | AAI-based (high-confidence) | 0 |
| S11_k141_28096 | 5032 | No | NA | 5 | 0 | 4 | Not-determined | Genome-fragment | NA | NA | 0 |
| S11-final-viral-combined_k141_132632||full | 1214 | No | NA | 3 | 3 | 0 | Low-quality | Genome-fragment | 2.68 | AAI-based (high-confidence) | 0 |
| S11-final-viral-combined_k141_303499||lt2gene | 1214 | No | NA | 2 | 2 | 0 | Low-quality | Genome-fragment | 1.73 | HMM-based (lower-bound) | 0 |
| S11_k141_289837 | 2224 | No | NA | 4 | 0 | 0 | Not-determined | Genome-fragment | NA | NA | 0 |
| S11_k141_234833 | 7240 | No | NA | 7 | 0 | 4 | Not-determined | Genome-fragment | NA | NA | 0 |
| S11_k141_14390 | 52675 | No | NA | 77 | 16 | 2 | Complete | High-quality | 100 | DTR (high-confidence) | 0 |
| S11_k141_289849 | 4677 | No | NA | 7 | 0 | 1 | Not-determined | Genome-fragment | NA | NA | 0 |
| S11_k141_193489 | 5847 | No | NA | 3 | 0 | 3 | Not-determined | Genome-fragment | NA | NA | 0 |
| S1-final-viral-combined_k141_377767||full | 1214 | No | NA | 3 | 1 | 0 | Low-quality | Genome-fragment | 2.86 | AAI-based (high-confidence) | 0 |
| S11_k141_234871 | 1358 | No | NA | 2 | 0 | 0 | Not-determined | Genome-fragment | NA | NA | 0 |
| S1-final-viral-combined_k141_161820||lt2gene | 1214 | No | NA | 2 | 2 | 0 | Low-quality | Genome-fragment | 2.99 | AAI-based (high-confidence) | 0 |
| S3_k141_78616 | 1214 | No | NA | 3 | 1 | 0 | Low-quality | Genome-fragment | 3.15 | AAI-based (high-confidence) | 0 |
| S7-final-viral-combined_k141_93029||full | 1214 | No | NA | 2 | 0 | 0 | Low-quality | Genome-fragment | 1.87 | AAI-based (high-confidence) | 0 |
| S11_k141_193551 | 1387 | No | NA | 1 | 0 | 0 | Not-determined | Genome-fragment | NA | NA | 0 |
| S11_k141_276200 | 1533 | No | NA | 1 | 0 | 1 | Not-determined | Genome-fragment | NA | NA | 0 |
| S11_k141_193566 | 3730 | No | NA | 2 | 0 | 1 | Not-determined | Genome-fragment | NA | NA | 0 |
| S7-final-viral-combined_k141_141821||full | 1214 | No | NA | 2 | 0 | 2 | Low-quality | Genome-fragment | 0.39 | AAI-based (medium-confidence) | 0 |
| S11_k141_152284 | 1756 | No | NA | 1 | 0 | 0 | Not-determined | Genome-fragment | NA | NA | 0 |
| S11_k141_124809 | 1603 | No | NA | 2 | 0 | 2 | Not-determined | Genome-fragment | NA | NA | 0 |
| S11_k141_625 | 1073 | No | NA | 2 | 0 | 1 | Not-determined | Genome-fragment | NA | NA | 0 |
| S17-final-viral-combined_k141_191529||full | 1214 | No | NA | 2 | 1 | 0 | Low-quality | Genome-fragment | 1.7 | HMM-based (lower-bound) | 0 |
| S11_k141_289996 | 1033 | No | NA | 1 | 0 | 1 | Not-determined | Genome-fragment | NA | NA | 0 |
| S11_k141_97176 | 1176 | No | NA | 1 | 0 | 1 | Not-determined | Genome-fragment | NA | NA | 0 |
| S17-final-viral-combined_k141_98303||full | 1214 | No | NA | 4 | 1 | 0 | Low-quality | Genome-fragment | 2.47 | AAI-based (high-confidence) | 0 |
| S11_k141_83308 | 4753 | No | NA | 5 | 0 | 1 | Not-determined | Genome-fragment | NA | NA | 0 |
| S11_k141_69569 | 2665 | No | NA | 2 | 0 | 2 | Not-determined | Genome-fragment | NA | NA | 0 |
| S11_k141_193635 | 1856 | No | NA | 2 | 0 | 2 | Not-determined | Genome-fragment | NA | NA | 0 |
| S11_k141_193655 | 1045 | No | NA | 1 | 0 | 1 | Not-determined | Genome-fragment | NA | NA | 0 |
| S11_k141_97223 | 1191 | No | NA | 1 | 0 | 1 | Not-determined | Genome-fragment | NA | NA | 0 |
| S11_k141_276339 | 2177 | No | NA | 2 | 0 | 1 | Not-determined | Genome-fragment | NA | NA | 0 |
| S11_k141_110886 | 1342 | No | NA | 1 | 0 | 1 | Not-determined | Genome-fragment | NA | NA | 0 |
| S12_k141_98019 | 1214 | No | NA | 2 | 0 | 0 | Low-quality | Genome-fragment | 2.61 | AAI-based (high-confidence) | 0 |
| S11_k141_207380 | 1321 | No | NA | 1 | 0 | 1 | Not-determined | Genome-fragment | NA | NA | 0 |
| S2-final-viral-combined_k141_85209||full | 1214 | No | NA | 3 | 0 | 0 | Low-quality | Genome-fragment | 2.93 | AAI-based (high-confidence) | 0 |
| S11_k141_290078 | 1232 | No | NA | 1 | 0 | 1 | Not-determined | Genome-fragment | NA | NA | 0 |
| S11_k141_69651 | 7232 | No | NA | 6 | 0 | 6 | Not-determined | Genome-fragment | NA | NA | 0 |
| S11_k141_193716 | 1942 | No | NA | 2 | 0 | 2 | Not-determined | Genome-fragment | NA | NA | 0 |
| S11_k141_138544 | 1039 | No | NA | 2 | 0 | 2 | Not-determined | Genome-fragment | NA | NA | 0 |
| S2-final-viral-combined_k141_8661||full | 1214 | No | NA | 5 | 0 | 0 | Low-quality | Genome-fragment | 2.15 | AAI-based (medium-confidence) | 0 |
| S11_k141_152423 | 1036 | No | NA | 1 | 0 | 1 | Not-determined | Genome-fragment | NA | NA | 0 |
| S11_k141_303977 | 4802 | No | NA | 5 | 0 | 2 | Not-determined | Genome-fragment | NA | NA | 0 |
| S11_k141_28333 | 4189 | No | NA | 5 | 0 | 4 | Not-determined | Genome-fragment | NA | NA | 0 |
| S11_k141_758 | 4660 | No | NA | 3 | 0 | 1 | Not-determined | Genome-fragment | NA | NA | 0 |
| S11_k141_97342 | 1323 | No | NA | 1 | 0 | 1 | Not-determined | Genome-fragment | NA | NA | 0 |
| S11_k141_790 | 1099 | No | NA | 1 | 0 | 0 | Not-determined | Genome-fragment | NA | NA | 0 |
| S11_k141_262664 | 1305 | No | NA | 1 | 0 | 1 | Not-determined | Genome-fragment | NA | NA | 0 |
| S11_k141_820 | 3390 | No | NA | 3 | 0 | 2 | Not-determined | Genome-fragment | NA | NA | 0 |
| S11_k141_276471 | 4483 | No | NA | 3 | 0 | 0 | Not-determined | Genome-fragment | NA | NA | 0 |
| S11_k141_138641 | 1219 | No | NA | 2 | 0 | 1 | Not-determined | Genome-fragment | NA | NA | 0 |
| S4-final-viral-combined_k141_23531||full | 1214 | No | NA | 2 | 0 | 0 | Low-quality | Genome-fragment | 1.57 | AAI-based (medium-confidence) | 0 |
| S11_k141_28409 | 4185 | No | NA | 3 | 0 | 3 | Not-determined | Genome-fragment | NA | NA | 0 |
| S4_k141_128382 | 1214 | No | NA | 1 | 0 | 1 | Low-quality | Genome-fragment | 0.43 | AAI-based (medium-confidence) | 0 |
| S11_k141_179873 | 2831 | No | NA | 2 | 0 | 2 | Not-determined | Genome-fragment | NA | NA | 0 |
| S11_k141_28421 | 1068 | No | NA | 1 | 0 | 1 | Not-determined | Genome-fragment | NA | NA | 0 |
| S6-final-viral-combined_k141_44845||full | 1214 | No | NA | 3 | 0 | 0 | Low-quality | Genome-fragment | 3.05 | AAI-based (medium-confidence) | 0 |
| S11_k141_235285 | 1394 | No | NA | 3 | 0 | 1 | Not-determined | Genome-fragment | NA | NA | 0 |
| S6-final-viral-combined_k141_134369||full | 1214 | No | NA | 2 | 0 | 1 | Low-quality | Genome-fragment | 0.44 | AAI-based (medium-confidence) | 0 |
| S11_k141_235313 | 2204 | No | NA | 2 | 0 | 2 | Not-determined | Genome-fragment | NA | NA | 0 |

| | | | | | | | | | | | | |
|---|---|---|---|---|---|---|---|---|---|---|---|---|
| S11_k141_207625 | 4441 | No | NA | 6 | 0 | 3 | Not-determined | Genome-fragment | NA | | NA | 0 |
| S11_k141_111136 | 2575 | No | NA | 2 | 0 | 0 | Not-determined | Genome-fragment | NA | | NA | 0 |
| S11_k141_179994 | 2102 | No | NA | 2 | 0 | 1 | Not-determined | Genome-fragment | NA | | NA | 0 |
| S11_k141_249009 | 1842 | No | NA | 1 | 0 | 1 | Not-determined | Genome-fragment | NA | | NA | 0 |
| S11_k141_97574 | 1111 | No | NA | 1 | 0 | 0 | Not-determined | Genome-fragment | NA | | NA | 0 |
| S11_k141_166613 | 2755 | No | NA | 3 | 0 | 3 | Not-determined | Genome-fragment | NA | | NA | 0 |
| S11_k141_304203 | 3181 | No | NA | 4 | 0 | 2 | Not-determined | Genome-fragment | NA | | NA | 0 |
| S11_k141_42194 | 4668 | No | NA | 2 | 0 | 2 | Not-determined | Genome-fragment | NA | | NA | 0 |
| S11_k141_249062 | 1408 | No | NA | 1 | 0 | 1 | Not-determined | Genome-fragment | NA | | NA | 0 |
| S11_k141_166679 | 1160 | No | NA | 1 | 0 | 1 | Not-determined | Genome-fragment | NA | | NA | 0 |
| S11_k141_1030 | 2785 | No | NA | 2 | 0 | 0 | Not-determined | Genome-fragment | NA | | NA | 0 |
| S11_k141_152723 | 1070 | No | NA | 1 | 0 | 0 | Not-determined | Genome-fragment | NA | | NA | 0 |
| S11_k141_152748 | 3936 | No | NA | 4 | 0 | 2 | Not-determined | Genome-fragment | NA | | NA | 0 |
| S11_k141_14851 | 1738 | No | NA | 1 | 0 | 1 | Not-determined | Genome-fragment | NA | | NA | 0 |
| S11_k141_262907 | 1019 | No | NA | 3 | 0 | 0 | Not-determined | Genome-fragment | NA | | NA | 0 |
| S11_k141_221549 | 1436 | No | NA | 2 | 0 | 1 | Not-determined | Genome-fragment | NA | | NA | 0 |
| S11_k141_249127 | 1611 | No | NA | 1 | 0 | 1 | Not-determined | Genome-fragment | NA | | NA | 0 |
| S6-final-viral-combined_k141_134838\|\|full | 1214 | No | NA | 3 | 0 | 0 | Low-quality | Genome-fragment | | 1.45 | AAI-based (high-confidence) | 0 |
| S11_k141_180181 | 14561 | No | NA | 18 | 0 | 3 | Not-determined | Genome-fragment | NA | | NA | 0 |
| S11_k141_28549 | 1207 | No | NA | 1 | 0 | 0 | Not-determined | Genome-fragment | NA | | NA | 0 |
| S8-final-viral-combined_k141_115210\|\|full | 1214 | No | NA | 2 | 0 | 0 | Low-quality | Genome-fragment | | 2.02 | AAI-based (medium-confidence) | 0 |
| S11_k141_262965 | 1248 | No | NA | 2 | 0 | 0 | Not-determined | Genome-fragment | NA | | NA | 0 |
| S11_k141_42346 | 1255 | No | NA | 2 | 0 | 2 | Not-determined | Genome-fragment | NA | | NA | 0 |
| S8-final-viral-combined_k141_78306\|\|full | 1214 | No | NA | 3 | 1 | 0 | Low-quality | Genome-fragment | | 2.5 | AAI-based (medium-confidence) | 0 |
| S11_k141_111273 | 1804 | No | NA | 2 | 0 | 2 | Not-determined | Genome-fragment | NA | | NA | 0 |
| S11_k141_1167 | 3161 | No | NA | 5 | 0 | 0 | Not-determined | Genome-fragment | NA | | NA | 0 |
| S11_k141_194015 | 1776 | No | NA | 2 | 0 | 0 | Not-determined | Genome-fragment | NA | | NA | 0 |
| S11_k141_207945 | 1782 | No | NA | 2 | 0 | 1 | Not-determined | Genome-fragment | NA | | NA | 0 |
| S11-final-viral-combined_k141_180782\|\|full | 1215 | No | NA | 3 | 0 | 0 | Low-quality | Genome-fragment | | 2.99 | AAI-based (high-confidence) | 0 |
| S11_k141_249236 | 1047 | No | NA | 2 | 0 | 1 | Not-determined | Genome-fragment | NA | | NA | 0 |
| S11_k141_56214 | 2090 | No | NA | 1 | 0 | 0 | Not-determined | Genome-fragment | NA | | NA | 0 |
| S11_k141_207992 | 1144 | No | NA | 1 | 0 | 1 | Not-determined | Genome-fragment | NA | | NA | 0 |
| S11_k141_180264 | 1799 | No | NA | 2 | 0 | 2 | Not-determined | Genome-fragment | NA | | NA | 0 |
| S11_k141_263076 | 3498 | No | NA | 5 | 0 | 5 | Not-determined | Genome-fragment | NA | | NA | 0 |
| S11_k141_235809 | 1325 | No | NA | 2 | 0 | 1 | Not-determined | Genome-fragment | NA | | NA | 0 |
| S11-final-viral-combined_k141_24371\|\|full | 1215 | No | NA | 5 | 0 | 0 | Low-quality | Genome-fragment | | 3.53 | AAI-based (high-confidence) | 0 |
| S11_k141_166890 | 1119 | No | NA | 1 | 0 | 1 | Not-determined | Genome-fragment | NA | | NA | 0 |
| S11_k141_125365 | 1798 | No | NA | 2 | 0 | 1 | Not-determined | Genome-fragment | NA | | NA | 0 |
| S1_k141_265845 | 1215 | No | NA | 2 | 1 | 0 | Low-quality | Genome-fragment | | 0.52 | HMM-based (lower-bound) | 0 |
| S11_k141_15040 | 2347 | No | NA | 2 | 0 | 1 | Not-determined | Genome-fragment | NA | | NA | 0 |
| S11_k141_249332 | 1170 | No | NA | 1 | 0 | 1 | Not-determined | Genome-fragment | NA | | NA | 0 |
| S3_k141_104129 | 1215 | No | NA | 3 | 3 | 0 | Low-quality | Genome-fragment | | 3.21 | AAI-based (high-confidence) | 0 |
| S5-final-viral-combined_k141_206875\|\|full | 1215 | Yes | 807 | 3 | 1 | 1 | Low-quality | Genome-fragment | | 2.06 | AAI-based (medium-confidence) | 33.58 |
| S5-final-viral-combined_k141_94215\|\|lt2gene | 1215 | No | NA | 1 | 1 | 0 | Low-quality | Genome-fragment | | 2.95 | AAI-based (high-confidence) | 0 |
| S11_k141_70036 | 3240 | No | NA | 3 | 0 | 0 | Not-determined | Genome-fragment | NA | | NA | 0 |
| S11_k141_221652 | 3998 | No | NA | 3 | 0 | 3 | Not-determined | Genome-fragment | NA | | NA | 0 |
| S7-final-viral-combined_k141_99942\|\|full | 1215 | No | NA | 4 | 1 | 0 | Low-quality | Genome-fragment | | 2.62 | AAI-based (medium-confidence) | 0 |
| S11_k141_70078 | 2142 | No | NA | 1 | 0 | 1 | Not-determined | Genome-fragment | NA | | NA | 0 |
| S11_k141_28735 | 1724 | No | NA | 1 | 0 | 1 | Not-determined | Genome-fragment | NA | | NA | 0 |
| S7-final-viral-combined_k141_4571\|\|lt2gene | 1215 | No | NA | 1 | 1 | 0 | Low-quality | Genome-fragment | | 2.59 | AAI-based (high-confidence) | 0 |
| S11_k141_318103 | 2153 | No | NA | 3 | 0 | 2 | Not-determined | Genome-fragment | NA | | NA | 0 |
| S11_k141_208089 | 1048 | No | NA | 1 | 0 | 1 | Not-determined | Genome-fragment | NA | | NA | 0 |
| S11_k141_263223 | 2307 | No | NA | 1 | 0 | 1 | Not-determined | Genome-fragment | NA | | NA | 0 |
| S11_k141_152933 | 2432 | No | NA | 2 | 0 | 2 | Not-determined | Genome-fragment | NA | | NA | 0 |
| S11_k141_290586 | 1327 | No | NA | 3 | 0 | 2 | Not-determined | Genome-fragment | NA | | NA | 0 |
| S17-final-viral-combined_k141_15164\|\|full | 1215 | No | NA | 3 | 1 | 0 | Low-quality | Genome-fragment | | 0.43 | HMM-based (lower-bound) | 0 |
| S11_k141_180387 | 1046 | No | NA | 1 | 0 | 0 | Not-determined | Genome-fragment | NA | | NA | 0 |
| S11_k141_70149 | 1258 | No | NA | 1 | 0 | 0 | Not-determined | Genome-fragment | NA | | NA | 0 |
| S11_k141_290598 | 1650 | No | NA | 2 | 0 | 0 | Not-determined | Genome-fragment | NA | | NA | 0 |
| S11_k141_180423 | 4813 | No | NA | 4 | 0 | 4 | Not-determined | Genome-fragment | NA | | NA | 0 |
| S11_k141_15162 | 3300 | No | NA | 5 | 0 | 2 | Not-determined | Genome-fragment | NA | | NA | 0 |
| S11_k141_166981 | 1136 | No | NA | 1 | 0 | 1 | Not-determined | Genome-fragment | NA | | NA | 0 |
| S11_k141_249504 | 4957 | No | NA | 5 | 0 | 2 | Not-determined | Genome-fragment | NA | | NA | 0 |
| S11_k141_15196 | 1887 | No | NA | 3 | 0 | 2 | Not-determined | Genome-fragment | NA | | NA | 0 |
| S11_k141_249511 | 1029 | No | NA | 2 | 0 | 2 | Not-determined | Genome-fragment | NA | | NA | 0 |
| S11_k141_318192 | 1129 | No | NA | 2 | 0 | 1 | Not-determined | Genome-fragment | NA | | NA | 0 |
| S17-final-viral-combined_k141_163542\|\|full | 1215 | Yes | 471 | 2 | 1 | 1 | Low-quality | Genome-fragment | | 0.98 | AAI-based (medium-confidence) | 61.23 |
| S12-final-viral-combined_k141_124016\|\|full | 1215 | No | NA | 3 | 1 | 0 | Low-quality | Genome-fragment | | 2.23 | AAI-based (high-confidence) | 0 |
| S12-final-viral-combined_k141_95069\|\|lt2gene | 1215 | No | NA | 2 | 1 | 0 | Low-quality | Genome-fragment | | 3.45 | AAI-based (high-confidence) | 0 |
| S11_k141_15269 | 1035 | No | NA | 1 | 0 | 1 | Not-determined | Genome-fragment | NA | | NA | 0 |
| S11_k141_139058 | 1122 | No | NA | 2 | 0 | 2 | Not-determined | Genome-fragment | NA | | NA | 0 |
| S11_k141_194326 | 15603 | No | NA | 13 | 0 | 7 | Not-determined | Genome-fragment | NA | | NA | 0 |
| S11_k141_42731 | 5776 | No | NA | 5 | 0 | 3 | Not-determined | Genome-fragment | NA | | NA | 0 |
| S11_k141_249555 | 1852 | No | NA | 1 | 0 | 0 | Not-determined | Genome-fragment | NA | | NA | 0 |
| S11_k141_1421 | 1335 | No | NA | 2 | 0 | 0 | Not-determined | Genome-fragment | NA | | NA | 0 |
| S11_k141_180532 | 1028 | No | NA | 1 | 0 | 1 | Not-determined | Genome-fragment | NA | | NA | 0 |
| S2-final-viral-combined_k141_75390\|\|full | 1215 | No | NA | 3 | 1 | 0 | Low-quality | Genome-fragment | | 2.48 | AAI-based (medium-confidence) | 0 |
| S11_k141_1432 | 12745 | No | NA | 10 | 0 | 5 | Not-determined | Genome-fragment | NA | | NA | 0 |
| S11_k141_221981 | 1096 | No | NA | 1 | 0 | 0 | Not-determined | Genome-fragment | NA | | NA | 0 |
| S11_k141_263351 | 1698 | No | NA | 4 | 0 | 3 | Not-determined | Genome-fragment | NA | | NA | 0 |
| S11_k141_139113 | 3497 | No | NA | 3 | 0 | 3 | Not-determined | Genome-fragment | NA | | NA | 0 |
| S11_k141_167160 | 2555 | No | NA | 1 | 0 | 0 | Not-determined | Genome-fragment | NA | | NA | 0 |
| S11_k141_153073 | 3250 | No | NA | 2 | 0 | 0 | Not-determined | Genome-fragment | NA | | NA | 0 |
| S11_k141_111498 | 1369 | No | NA | 1 | 0 | 0 | Not-determined | Genome-fragment | NA | | NA | 0 |
| S11_k141_208306 | 4111 | No | NA | 1 | 0 | 1 | Not-determined | Genome-fragment | NA | | NA | 0 |
| S11_k141_153097 | 2029 | No | NA | 3 | 0 | 2 | Not-determined | Genome-fragment | NA | | NA | 0 |
| S2_k141_23464 | 1215 | No | NA | 2 | 1 | 0 | Low-quality | Genome-fragment | | 1.37 | AAI-based (medium-confidence) | 0 |
| S11_k141_84301 | 3462 | No | NA | 5 | 0 | 1 | Not-determined | Genome-fragment | NA | | NA | 0 |
| S8-final-viral-combined_k141_274295\|\|full | 1215 | No | NA | 3 | 0 | 0 | Low-quality | Genome-fragment | | 2.83 | AAI-based (medium-confidence) | 0 |
| S11_k141_111551 | 3702 | No | NA | 2 | 0 | 1 | Not-determined | Genome-fragment | NA | | NA | 0 |
| S11_k141_56661 | 3355 | No | NA | 2 | 0 | 2 | Not-determined | Genome-fragment | NA | | NA | 0 |
| S8-final-viral-combined_k141_218676\|\|full | 1215 | No | NA | 3 | 0 | 0 | Low-quality | Genome-fragment | | 1.93 | AAI-based (medium-confidence) | 0 |
| S9-final-viral-combined_k141_85819\|\|full | 1216 | No | NA | 4 | 0 | 0 | Low-quality | Genome-fragment | | 2.48 | AAI-based (medium-confidence) | 0 |
| S11_k141_139232 | 1195 | No | NA | 1 | 0 | 1 | Not-determined | Genome-fragment | NA | | NA | 0 |
| S11-final-viral-combined_k141_43924\|\|full | 1216 | No | NA | 3 | 0 | 2 | Low-quality | Genome-fragment | | 4.01 | AAI-based (high-confidence) | 0 |
| S11_k141_318440 | 1621 | No | NA | 1 | 0 | 0 | Not-determined | Genome-fragment | NA | | NA | 0 |
| S11-final-viral-combined_k141_256929\|\|full | 1216 | No | NA | 4 | 1 | 0 | Low-quality | Genome-fragment | | 0.79 | HMM-based (lower-bound) | 0 |
| S11_k141_153247 | 3195 | No | NA | 5 | 0 | 1 | Not-determined | Genome-fragment | NA | | NA | 0 |
| S11_k141_84404 | 1152 | No | NA | 2 | 0 | 1 | Not-determined | Genome-fragment | NA | | NA | 0 |
| S11-final-viral-combined_k141_149692\|\|full | 1216 | No | NA | 4 | 0 | 0 | Low-quality | Genome-fragment | | 0.79 | AAI-based (high-confidence) | 0 |
| S11_k141_208431 | 1138 | No | NA | 1 | 0 | 1 | Not-determined | Genome-fragment | NA | | NA | 0 |
| S11_k141_111626 | 1262 | No | NA | 1 | 0 | 1 | Not-determined | Genome-fragment | NA | | NA | 0 |
| S11_k141_236242 | 2678 | No | NA | 5 | 0 | 2 | Not-determined | Genome-fragment | NA | | NA | 0 |
| S11_k141_180781 | 1460 | No | NA | 2 | 0 | 0 | Not-determined | Genome-fragment | NA | | NA | 0 |
| S11_k141_56762 | 7865 | No | NA | 13 | 0 | 3 | Not-determined | Genome-fragment | NA | | NA | 0 |
| S11_k141_222262 | 1124 | No | NA | 2 | 0 | 0 | Not-determined | Genome-fragment | NA | | NA | 0 |
| S11_k141_194597 | 1175 | No | NA | 3 | 0 | 0 | Not-determined | Genome-fragment | NA | | NA | 0 |
| S11_k141_56786 | 1630 | No | NA | 1 | 0 | 0 | Not-determined | Genome-fragment | NA | | NA | 0 |
| S11_k141_277464 | 1101 | No | NA | 1 | 0 | 0 | Not-determined | Genome-fragment | NA | | NA | 0 |
| S11_k141_208515 | 5999 | No | NA | 2 | 0 | 2 | Not-determined | Genome-fragment | NA | | NA | 0 |
| S1-final-viral-combined_k141_202036\|\|full | 1216 | No | NA | 2 | 0 | 0 | Low-quality | Genome-fragment | | 2.61 | AAI-based (medium-confidence) | 0 |
| S11_k141_249794 | 3833 | No | NA | 2 | 0 | 1 | Not-determined | Genome-fragment | NA | | NA | 0 |
| S1-final-viral-combined_k141_344835\|\|full | 1216 | No | NA | 4 | 0 | 1 | Low-quality | Genome-fragment | | 2.87 | AAI-based (medium-confidence) | 0 |
| S1-final-viral-combined_k141_122176\|\|lt2gene | 1216 | No | NA | 2 | 1 | 0 | Low-quality | Genome-fragment | | 2.26 | HMM-based (lower-bound) | 0 |
| S11_k141_98487 | 1102 | No | NA | 1 | 0 | 1 | Not-determined | Genome-fragment | NA | | NA | 0 |
| S11_k141_111752 | 2358 | No | NA | 2 | 0 | 1 | Not-determined | Genome-fragment | NA | | NA | 0 |
| S11_k141_125896 | 1377 | No | NA | 1 | 0 | 0 | Not-determined | Genome-fragment | NA | | NA | 0 |
| S11_k141_153395 | 1021 | No | NA | 1 | 0 | 1 | Not-determined | Genome-fragment | NA | | NA | 0 |
| S1_k141_174599 | 1216 | No | NA | 1 | 0 | 0 | Low-quality | Genome-fragment | | 2.03 | AAI-based (medium-confidence) | 0 |
| S11_k141_222396 | 1108 | No | NA | 2 | 0 | 0 | Not-determined | Genome-fragment | NA | | NA | 0 |
| S3_k141_41591 | 1216 | No | NA | 2 | 0 | 0 | Low-quality | Genome-fragment | | 7.33 | AAI-based (high-confidence) | 0 |
| S11_k141_84637 | 1546 | No | NA | 3 | 0 | 3 | Not-determined | Genome-fragment | NA | | NA | 0 |
| S7-final-viral-combined_k141_71384\|\|full | 1216 | No | NA | 3 | 1 | 0 | Low-quality | Genome-fragment | | 3.22 | AAI-based (high-confidence) | 0 |
| S11_k141_194754 | 1109 | No | NA | 2 | 0 | 0 | Not-determined | Genome-fragment | NA | | NA | 0 |
| S11_k141_249838 | 1064 | No | NA | 1 | 0 | 0 | Not-determined | Genome-fragment | NA | | NA | 0 |
| S17-final-viral-combined_k141_142573\|\|full | 1216 | No | NA | 4 | 1 | 0 | Low-quality | Genome-fragment | | 2.08 | AAI-based (high-confidence) | 0 |
| S11_k141_167569 | 5025 | No | NA | 4 | 0 | 3 | Not-determined | Genome-fragment | NA | | NA | 0 |
| S11_k141_125994 | 1989 | No | NA | 1 | 0 | 0 | Not-determined | Genome-fragment | NA | | NA | 0 |
| S11_k141_249883 | 1147 | No | NA | 3 | 0 | 0 | Not-determined | Genome-fragment | NA | | NA | 0 |
| S11_k141_236471 | 2019 | No | NA | 2 | 0 | 0 | Not-determined | Genome-fragment | NA | | NA | 0 |
| S11_k141_98648 | 1078 | No | NA | 3 | 0 | 2 | Not-determined | Genome-fragment | NA | | NA | 0 |
| S11_k141_208787 | 1058 | No | NA | 2 | 0 | 1 | Not-determined | Genome-fragment | NA | | NA | 0 |
| S11_k141_98658 | 1159 | No | NA | 1 | 0 | 1 | Not-determined | Genome-fragment | NA | | NA | 0 |
| S6-final-viral-combined_k141_110879\|\|full | 1216 | No | NA | 3 | 0 | 2 | Low-quality | Genome-fragment | | 0.54 | AAI-based (medium-confidence) | 0 |
| S11_k141_84775 | 1568 | No | NA | 2 | 0 | 0 | Not-determined | Genome-fragment | NA | | NA | 0 |
| S11_k141_277740 | 1610 | No | NA | 2 | 0 | 0 | Not-determined | Genome-fragment | NA | | NA | 0 |
| S11_k141_1921 | 1631 | No | NA | 2 | 0 | 1 | Not-determined | Genome-fragment | NA | | NA | 0 |
| S8-final-viral-combined_k141_215\|\|full | 1216 | No | NA | 3 | 2 | 0 | Low-quality | Genome-fragment | | 2.2 | HMM-based (lower-bound) | 0 |
| S11_k141_15863 | 1657 | No | NA | 2 | 0 | 2 | Not-determined | Genome-fragment | NA | | NA | 0 |
| S11_k141_318755 | 1234 | No | NA | 1 | 0 | 1 | Not-determined | Genome-fragment | NA | | NA | 0 |
| S11-final-viral-combined_k141_46076\|\|full | 1217 | No | NA | 5 | 0 | 0 | Low-quality | Genome-fragment | | 2.96 | AAI-based (high-confidence) | 0 |
| S11_k141_181062 | 1027 | No | NA | 2 | 0 | 0 | Not-determined | Genome-fragment | NA | | NA | 0 |
| S11_k141_98728 | 1152 | No | NA | 1 | 0 | 1 | Not-determined | Genome-fragment | NA | | NA | 0 |
| S11_k141_263848 | 2028 | No | NA | 1 | 0 | 1 | Not-determined | Genome-fragment | NA | | NA | 0 |
| S11_k141_263867 | 1551 | No | NA | 2 | 0 | 1 | Not-determined | Genome-fragment | NA | | NA | 0 |
| S11_k141_117873 | 1217 | No | NA | 3 | 0 | 0 | Low-quality | Genome-fragment | | 3.05 | AAI-based (high-confidence) | 0 |
| S11_k141_277834 | 2450 | No | NA | 1 | 0 | 0 | Not-determined | Genome-fragment | NA | | NA | 0 |
| S11_k141_318838 | 1024 | No | NA | 1 | 0 | 1 | Not-determined | Genome-fragment | NA | | NA | 0 |
| S11_k141_181129 | 2531 | No | NA | 2 | 0 | 2 | Not-determined | Genome-fragment | NA | | NA | 0 |
| S1-final-viral-combined_k141_92728\|\|full | 1217 | No | NA | 3 | 2 | 0 | Low-quality | Genome-fragment | | 2.21 | AAI-based (medium-confidence) | 0 |
| S11_k141_208972 | 1128 | No | NA | 2 | 0 | 2 | Not-determined | Genome-fragment | NA | | NA | 0 |
| S11_k141_139747 | 1202 | No | NA | 2 | 0 | 2 | Not-determined | Genome-fragment | NA | | NA | 0 |
| S11_k141_277865 | 1136 | No | NA | 2 | 0 | 1 | Not-determined | Genome-fragment | NA | | NA | 0 |
| S11_k141_126172 | 1030 | No | NA | 2 | 0 | 2 | Not-determined | Genome-fragment | NA | | NA | 0 |
| S11_k141_2055 | 1538 | No | NA | 2 | 0 | 1 | Not-determined | Genome-fragment | NA | | NA | 0 |
| S11_k141_153579 | 6515 | No | NA | 7 | 0 | 0 | Not-determined | Genome-fragment | NA | | NA | 0 |
| S11_k141_318865 | 1134 | No | NA | 1 | 0 | 1 | Not-determined | Genome-fragment | NA | | NA | 0 |
| S11_k141_209016 | 3466 | No | NA | 3 | 0 | 2 | Not-determined | Genome-fragment | NA | | NA | 0 |
| S11_k141_305338 | 1326 | No | NA | 1 | 0 | 1 | Not-determined | Genome-fragment | NA | | NA | 0 |
| S1-final-viral-combined_k141_301569\|\|lt2gene | 1217 | No | NA | 1 | 1 | 0 | Low-quality | Genome-fragment | | 2.03 | HMM-based (lower-bound) | 0 |
| S11_k141_236685 | 1542 | No | NA | 2 | 0 | 2 | Not-determined | Genome-fragment | NA | | NA | 0 |
| S11_k141_139811 | 1232 | No | NA | 1 | 0 | 0 | Not-determined | Genome-fragment | NA | | NA | 0 |
| S11_k141_250126 | 1331 | No | NA | 3 | 0 | 1 | Not-determined | Genome-fragment | NA | | NA | 0 |
| S3-final-viral-combined_k141_54798\|\|full | 1217 | No | NA | 4 | 0 | 0 | Low-quality | Genome-fragment | | 0.79 | AAI-based (high-confidence) | 0 |
| S11_k141_277988 | 2095 | No | NA | 2 | 0 | 2 | Not-determined | Genome-fragment | NA | | NA | 0 |
| S11_k141_57339 | 1130 | No | NA | 1 | 0 | 1 | Not-determined | Genome-fragment | NA | | NA | 0 |
| S11_k141_318965 | 3063 | No | NA | 7 | 0 | 0 | Not-determined | Genome-fragment | NA | | NA | 0 |
| S11_k141_16088 | 1157 | No | NA | 2 | 0 | 2 | Not-determined | Genome-fragment | NA | | NA | 0 |
| S11_k141_98841 | 1075 | No | NA | 1 | 0 | 1 | Not-determined | Genome-fragment | NA | | NA | 0 |
| S11_k141_222692 | 3870 | No | NA | 3 | 0 | 3 | Not-determined | Genome-fragment | NA | | NA | 0 |
| S11_k141_70520 | 1679 | No | NA | 3 | 0 | 1 | Not-determined | Genome-fragment | NA | | NA | 0 |
| S11_k141_209154 | 1679 | No | NA | 2 | 0 | 1 | Not-determined | Genome-fragment | NA | | NA | 0 |
| S11_k141_70565 | 1331 | No | NA | 1 | 0 | 1 | Not-determined | Genome-fragment | NA | | NA | 0 |
| S3_k141_61697 | 1217 | No | NA | 2 | 0 | 0 | Low-quality | Genome-fragment | | 0.94 | AAI-based (high-confidence) | 0 |
| S11_k141_209181 | 4757 | No | NA | 10 | 0 | 1 | Not-determined | Genome-fragment | NA | | NA | 0 |
| S11_k141_43504 | 1737 | No | NA | 1 | 0 | 0 | Not-determined | Genome-fragment | NA | | NA | 0 |
| S5-final-viral-combined_k141_154951\|\|lt2gene | 1217 | No | NA | 2 | 2 | 0 | Low-quality | Genome-fragment | | 2.97 | AAI-based (medium-confidence) | 0 |
| S11_k141_43509 | 1699 | No | NA | 3 | 0 | 1 | Not-determined | Genome-fragment | NA | | NA | 0 |
| S11_k141_57493 | 1800 | No | NA | 2 | 0 | 0 | Not-determined | Genome-fragment | NA | | NA | 0 |
| S11_k141_305507 | 3402 | No | NA | 4 | 0 | 2 | Not-determined | Genome-fragment | NA | | NA | 0 |
| S11_k141_85034 | 1039 | No | NA | 2 | 0 | 2 | Not-determined | Genome-fragment | NA | | NA | 0 |
| S11_k141_209262 | 3321 | No | NA | 5 | 0 | 1 | Not-determined | Genome-fragment | NA | | NA | 0 |
| S7-final-viral-combined_k141_228197\|\|full | 1217 | No | NA | 2 | 0 | 1 | Low-quality | Genome-fragment | | 0.72 | AAI-based (medium-confidence) | 0 |
| S11_k141_278183 | 3744 | No | NA | 4 | 0 | 4 | Not-determined | Genome-fragment | NA | | NA | 0 |
| S11_k141_126326 | 1038 | No | NA | 3 | 0 | 1 | Not-determined | Genome-fragment | NA | | NA | 0 |
| S11_k141_195214 | 1499 | No | NA | 2 | 0 | 1 | Not-determined | Genome-fragment | NA | | NA | 0 |
| S11_k141_278198 | 1449 | No | NA | 2 | 0 | 0 | Not-determined | Genome-fragment | NA | | NA | 0 |
| S11_k141_264171 | 1365 | No | NA | 1 | 0 | 0 | Not-determined | Genome-fragment | NA | | NA | 0 |
| S11_k141_57568 | 1008 | No | NA | 1 | 0 | 1 | Not-determined | Genome-fragment | NA | | NA | 0 |
| S7-final-viral-combined_k141_50814\|\|full | 1217 | No | NA | 3 | 1 | 0 | Low-quality | Genome-fragment | | 3.19 | AAI-based (high-confidence) | 0 |
| S11_k141_153822 | 3123 | No | NA | 1 | 0 | 0 | Not-determined | Genome-fragment | NA | | NA | 0 |
| S11_k141_29815 | 4198 | No | NA | 3 | 0 | 3 | Not-determined | Genome-fragment | NA | | NA | 0 |
| S11_k141_250423 | 1169 | No | NA | 1 | 0 | 0 | Not-determined | Genome-fragment | NA | | NA | 0 |
| S11_k141_112187 | 1205 | No | NA | 1 | 0 | 1 | Not-determined | Genome-fragment | NA | | NA | 0 |
| S7_k141_191781 | 1217 | No | NA | 3 | 2 | 0 | Low-quality | Genome-fragment | | 3.35 | AAI-based (high-confidence) | 0 |
| S11_k141_70787 | 1309 | No | NA | 1 | 0 | 0 | Not-determined | Genome-fragment | NA | | NA | 0 |
| S12-final-viral-combined_k141_236477\|\|full | 1217 | No | NA | 3 | 0 | 0 | Low-quality | Genome-fragment | | 1.87 | AAI-based (high-confidence) | 0 |
| S11_k141_236984 | 2322 | No | NA | 2 | 0 | 1 | Not-determined | Genome-fragment | NA | | NA | 0 |
| S12-final-viral-combined_k141_80236\|\|full | 1217 | No | NA | 3 | 2 | 0 | Low-quality | Genome-fragment | | 3.08 | AAI-based (medium-confidence) | 0 |
| S12-final-viral-combined_k141_367305\|\|full | 1217 | No | NA | 3 | 1 | 0 | Low-quality | Genome-fragment | | 1.49 | AAI-based (medium-confidence) | 0 |
| S12_k141_380729 | 1217 | No | NA | 1 | 0 | 1 | Low-quality | Genome-fragment | | 0.74 | AAI-based (medium-confidence) | 0 |
| S11_k141_2406 | 1163 | No | NA | 1 | 0 | 1 | Not-determined | Genome-fragment | NA | | NA | 0 |
| S11_k141_305745 | 1357 | No | NA | 2 | 0 | 1 | Not-determined | Genome-fragment | NA | | NA | 0 |
| S11_k141_126480 | 3315 | No | NA | 1 | 0 | 1 | Not-determined | Genome-fragment | NA | | NA | 0 |
| S11_k141_43679 | 1075 | No | NA | 2 | 0 | 0 | Not-determined | Genome-fragment | NA | | NA | 0 |
| S11_k141_305773 | 3434 | No | NA | 2 | 0 | 1 | Not-determined | Genome-fragment | NA | | NA | 0 |
| S11_k141_112253 | 1109 | No | NA | 2 | 0 | 2 | Not-determined | Genome-fragment | NA | | NA | 0 |
| S11_k141_126517 | 1084 | No | NA | 1 | 0 | 1 | Not-determined | Genome-fragment | NA | | NA | 0 |
| S2-final-viral-combined_k141_199793\|\|full | 1217 | No | NA | 2 | 0 | 1 | Low-quality | Genome-fragment | | 1.56 | AAI-based (medium-confidence) | 0 |
| S11_k141_2515 | 1425 | No | NA | 1 | 0 | 1 | Not-determined | Genome-fragment | NA | | NA | 0 |
| S4-final-viral-combined_k141_102863\|\|full | 1217 | No | NA | 2 | 0 | 1 | Low-quality | Genome-fragment | | 1.99 | AAI-based (medium-confidence) | 0 |
| S4-final-viral-combined_k141_161639\|\|lt2gene | 1217 | No | NA | 2 | 1 | 0 | Low-quality | Genome-fragment | | 0.75 | HMM-based (lower-bound) | 0 |
| S11_k141_70903 | 1426 | No | NA | 1 | 0 | 1 | Not-determined | Genome-fragment | NA | | NA | 0 |
| S11_k141_319255 | 1683 | No | NA | 2 | 0 | 2 | Not-determined | Genome-fragment | NA | | NA | 0 |
| S11_k141_99161 | 1534 | No | NA | 1 | 0 | 0 | Not-determined | Genome-fragment | NA | | NA | 0 |
| S11_k141_209564 | 1061 | No | NA | 4 | 0 | 0 | Not-determined | Genome-fragment | NA | | NA | 0 |
| S11_k141_29974 | 1833 | No | NA | 1 | 0 | 1 | Not-determined | Genome-fragment | NA | | NA | 0 |
| S11_k141_209579 | 1917 | No | NA | 2 | 0 | 1 | Not-determined | Genome-fragment | NA | | NA | 0 |
| S11_k141_250572 | 9488 | No | NA | 5 | 0 | 5 | Not-determined | Genome-fragment | NA | | NA | 0 |
| S11_k141_57846 | 1905 | No | NA | 1 | 0 | 1 | Not-determined | Genome-fragment | NA | | NA | 0 |
| S11_k141_278441 | 1905 | No | NA | 2 | 0 | 0 | Not-determined | Genome-fragment | NA | | NA | 0 |
| S11_k141_319308 | 3376 | No | NA | 1 | 0 | 1 | Not-determined | Genome-fragment | NA | | NA | 0 |
| S11_k141_154099 | 1173 | No | NA | 2 | 0 | 1 | Not-determined | Genome-fragment | NA | | NA | 0 |
| S8-final-viral-combined_k141_19941\|\|full | 1217 | No | NA | 4 | 2 | 0 | Low-quality | Genome-fragment | | 3.53 | AAI-based (high-confidence) | 0 |
| S8_k141_195681 | 1217 | No | NA | 2 | 0 | 0 | Low-quality | Genome-fragment | | 2.1 | AAI-based (high-confidence) | 0 |
| S11_k141_30058 | 1017 | No | NA | 1 | 0 | 0 | Not-determined | Genome-fragment | NA | | NA | 0 |
| S11_k141_16241 | 1120 | No | NA | 1 | 0 | 1 | Not-determined | Genome-fragment | NA | | NA | 0 |
| S11-final-viral-combined_k141_300241\|\|full | 1218 | No | NA | 3 | 0 | 0 | Low-quality | Genome-fragment | | 3.92 | AAI-based (high-confidence) | 0 |
| S11_k141_209642 | 1295 | No | NA | 1 | 0 | 1 | Not-determined | Genome-fragment | NA | | NA | 0 |
| S11_k141_223124 | 4336 | No | NA | 4 | 0 | 1 | Not-determined | Genome-fragment | NA | | NA | 0 |
| S11_k141_237173 | 1059 | No | NA | 2 | 0 | 0 | Not-determined | Genome-fragment | NA | | NA | 0 |
| S11_k141_43929 | 1632 | No | NA | 1 | 0 | 1 | Not-determined | Genome-fragment | NA | | NA | 0 |
| S11_k141_223143 | 1132 | No | NA | 1 | 0 | 1 | Not-determined | Genome-fragment | NA | | NA | 0 |
| S11_k141_57964 | 1503 | No | NA | 2 | 0 | 1 | Not-determined | Genome-fragment | NA | | NA | 0 |
| S11_k141_264378 | 1757 | No | NA | 3 | 0 | 0 | Not-determined | Genome-fragment | NA | | NA | 0 |
| S11_k141_278558 | 3324 | No | NA | 2 | 0 | 0 | Not-determined | Genome-fragment | NA | | NA | 0 |
| S11-final-viral-combined_k141_41053\|\|full | 1218 | No | NA | 2 | 0 | 1 | Low-quality | Genome-fragment | | 1.6 | AAI-based (medium-confidence) | 0 |
| S11_k141_319376 | 1456 | No | NA | 2 | 0 | 2 | Not-determined | Genome-fragment | NA | | NA | 0 |
| S1-final-viral-combined_k141_193472\|\|full | 1218 | No | NA | 2 | 1 | 0 | Low-quality | Genome-fragment | | 3.7 | AAI-based (high-confidence) | 0 |
| S11_k141_16327 | 1769 | No | NA | 2 | 0 | 0 | Not-determined | Genome-fragment | NA | | NA | 0 |
| S11_k141_99393 | 5795 | No | NA | 5 | 0 | 4 | Not-determined | Genome-fragment | NA | | NA | 0 |
| S11_k141_2787 | 1274 | No | NA | 2 | 0 | 1 | Not-determined | Genome-fragment | NA | | NA | 0 |
| S11_k141_181594 | 2639 | No | NA | 3 | 0 | 2 | Not-determined | Genome-fragment | NA | | NA | 0 |
| S11_k141_154329 | 1922 | No | NA | 2 | 0 | 2 | Not-determined | Genome-fragment | NA | | NA | 0 |
| S11_k141_154330 | 1094 | No | NA | 1 | 0 | 1 | Not-determined | Genome-fragment | NA | | NA | 0 |
| S11_k141_85351 | 1962 | No | NA | 1 | 0 | 1 | Not-determined | Genome-fragment | NA | | NA | 0 |
| S11_k141_140485 | 1457 | No | NA | 1 | 0 | 0 | Not-determined | Genome-fragment | NA | | NA | 0 |
| S11_k141_223290 | 1377 | No | NA | 1 | 0 | 1 | Not-determined | Genome-fragment | NA | | NA | 0 |
| S1_k141_346203 | 1218 | No | NA | 3 | 0 | 0 | Low-quality | Genome-fragment | | 2.9 | AAI-based (medium-confidence) | 0 |
| S11_k141_264539 | 3271 | No | NA | 2 | 0 | 1 | Not-determined | Genome-fragment | NA | | NA | 0 |
| S11_k141_58172 | 1304 | No | NA | 1 | 0 | 1 | Not-determined | Genome-fragment | NA | | NA | 0 |
| S11_k141_44081 | 1038 | No | NA | 1 | 0 | 1 | Not-determined | Genome-fragment | NA | | NA | 0 |
| S11_k141_44095 | 2471 | No | NA | 3 | 0 | 1 | Not-determined | Genome-fragment | NA | | NA | 0 |
| S11_k141_264583 | 1023 | No | NA | 2 | 0 | 2 | Not-determined | Genome-fragment | NA | | NA | 0 |
| S7-final-viral-combined_k141_2982\|\|lt2gene | 1218 | No | NA | 2 | 0 | 1 | Low-quality | Genome-fragment | | 0.29 | AAI-based (medium-confidence) | 0 |
| S17-final-viral-combined_k141_179024\|\|full | 1218 | No | NA | 3 | 0 | 0 | Low-quality | Genome-fragment | | 2.23 | AAI-based (high-confidence) | 0 |
| S11_k141_58212 | 3061 | No | NA | 1 | 0 | 1 | Not-determined | Genome-fragment | NA | | NA | 0 |
| S11_k141_44116 | 1354 | No | NA | 1 | 0 | 1 | Not-determined | Genome-fragment | NA | | NA | 0 |
| S11_k141_168454 | 2691 | No | NA | 5 | 0 | 0 | Not-determined | Genome-fragment | NA | | NA | 0 |
| S11_k141_291961 | 6857 | No | NA | 6 | 0 | 6 | Not-determined | Genome-fragment | NA | | NA | 0 |
| S11_k141_223347 | 5407 | No | NA | 7 | 0 | 3 | Not-determined | Genome-fragment | NA | | NA | 0 |
| S17-final-viral-combined_k141_227022\|\|full | 1218 | No | NA | 3 | 3 | 0 | Low-quality | Genome-fragment | | 1.71 | AAI-based (high-confidence) | 0 |
| S17-final-viral-combined_k141_231452\|\|full | 1218 | No | NA | 3 | 0 | 0 | Low-quality | Genome-fragment | | 15.2 | AAI-based (medium-confidence) | 0 |
| S11_k141_71311 | 1317 | No | NA | 2 | 0 | 0 | Not-determined | Genome-fragment | NA | | NA | 0 |
| S11_k141_154373 | 2068 | No | NA | 3 | 0 | 1 | Not-determined | Genome-fragment | NA | | NA | 0 |
| S11_k141_2947 | 1071 | No | NA | 1 | 0 | 0 | Not-determined | Genome-fragment | NA | | NA | 0 |
| S11_k141_306219 | 2009 | No | NA | 2 | 0 | 2 | Not-determined | Genome-fragment | NA | | NA | 0 |
| S11_k141_292003 | 2158 | No | NA | 1 | 0 | 1 | Not-determined | Genome-fragment | NA | | NA | 0 |
| S17-final-viral-combined_k141_254342\|\|full | 1218 | No | NA | 3 | 0 | 1 | Low-quality | Genome-fragment | | 2.66 | AAI-based (medium-confidence) | 0 |
| S11_k141_237505 | 1582 | No | NA | 2 | 0 | 0 | Not-determined | Genome-fragment | NA | | NA | 0 |
| S11_k141_16505 | 1233 | No | NA | 2 | 0 | 0 | Not-determined | Genome-fragment | NA | | NA | 0 |
| S2-final-viral-combined_k141_62558\|\|lt2gene | 1218 | No | NA | 1 | 1 | 0 | Low-quality | Genome-fragment | | 3.93 | HMM-based (lower-bound) | 0 |
| S11_k141_181710 | 1646 | No | NA | 1 | 0 | 0 | Not-determined | Genome-fragment | NA | | NA | 0 |
| S11_k141_112724 | 3824 | No | NA | 4 | 0 | 2 | Not-determined | Genome-fragment | NA | | NA | 0 |
| S11_k141_44247 | 1681 | No | NA | 2 | 0 | 1 | Not-determined | Genome-fragment | NA | | NA | 0 |
| S11_k141_85518 | 1329 | No | NA | 2 | 0 | 1 | Not-determined | Genome-fragment | NA | | NA | 0 |
| S11_k141_30366 | 1567 | No | NA | 3 | 0 | 1 | Not-determined | Genome-fragment | NA | | NA | 0 |
| S11_k141_223450 | 2771 | No | NA | 2 | 0 | 1 | Not-determined | Genome-fragment | NA | | NA | 0 |
| S11_k141_168612 | 1111 | No | NA | 1 | 0 | 1 | Not-determined | Genome-fragment | NA | | NA | 0 |
| S11_k141_140666 | 1089 | No | NA | 1 | 0 | 1 | Not-determined | Genome-fragment | NA | | NA | 0 |
| S11_k141_3034 | 3115 | No | NA | 5 | 0 | 0 | Not-determined | Genome-fragment | NA | | NA | 0 |
| S11_k141_319535 | 3696 | No | NA | 2 | 0 | 1 | Not-determined | Genome-fragment | NA | | NA | 0 |
| S4-final-viral-combined_k141_160754\|\|full | 1218 | No | NA | 3 | 0 | 1 | Low-quality | Genome-fragment | | 1 | AAI-based (medium-confidence) | 0 |
| S11_k141_168651 | 1122 | No | NA | 2 | 0 | 2 | Not-determined | Genome-fragment | NA | | NA | 0 |
| S11_k141_210060 | 3461 | No | NA | 2 | 0 | 1 | Not-determined | Genome-fragment | NA | | NA | 0 |
| S8-final-viral-combined_k141_198167\|\|full | 1218 | No | NA | 3 | 0 | 1 | Low-quality | Genome-fragment | | 1.19 | AAI-based (medium-confidence) | 0 |
| S11_k141_195720 | 1373 | No | NA | 3 | 0 | 0 | Not-determined | Genome-fragment | NA | | NA | 0 |
| S11_k141_154513 | 1003 | No | NA | 1 | 0 | 0 | Not-determined | Genome-fragment | NA | | NA | 0 |
| S8-final-viral-combined_k141_233020\|\|full | 1218 | No | NA | 2 | 0 | 0 | Low-quality | Genome-fragment | | 2.77 | AAI-based (medium-confidence) | 0 |
| S8-final-viral-combined_k141_201046\|\|full | 1218 | No | NA | 2 | 0 | 2 | Low-quality | Genome-fragment | | 1.06 | AAI-based (medium-confidence) | 0 |
| S11_k141_112808 | 15664 | No | NA | 14 | 0 | 10 | Not-determined | Genome-fragment | NA | | NA | 0 |
| S8-final-viral-combined_k141_223139\|\|lt2gene | 1218 | Yes | 617 | 2 | 1 | 1 | Low-quality | Genome-fragment | | 0.97 | HMM-based (lower-bound) | 49.34 |
| S11_k141_306479 | 1338 | No | NA | 1 | 0 | 0 | Not-determined | Genome-fragment | NA | | NA | 0 |
| S11_k141_168694 | 1533 | No | NA | 1 | 0 | 0 | Not-determined | Genome-fragment | NA | | NA | 0 |
| S11_k141_278950 | 5061 | No | NA | 3 | 0 | 1 | Not-determined | Genome-fragment | NA | | NA | 0 |
| S11_k141_58482 | 1290 | No | NA | 2 | 0 | 2 | Not-determined | Genome-fragment | NA | | NA | 0 |
| S9-final-viral-combined_k141_11266\|\|full | 1219 | No | NA | 3 | 1 | 0 | Low-quality | Genome-fragment | | 3.09 | AAI-based (high-confidence) | 0 |
| S11_k141_112976 | 1013 | No | NA | 3 | 0 | 2 | Not-determined | Genome-fragment | NA | | NA | 0 |
| S11_k141_154748 | 6306 | No | NA | 12 | 0 | 12 | Not-determined | Genome-fragment | NA | | NA | 0 |
| S11_k141_195808 | 4951 | No | NA | 2 | 0 | 2 | Not-determined | Genome-fragment | NA | | NA | 0 |
| S11_k141_210187 | 3650 | No | NA | 4 | 0 | 1 | Not-determined | Genome-fragment | NA | | NA | 0 |
| S11_k141_3116 | 38537 | Yes | 31746 | 49 | 12 | 4 | Medium-quality | Genome-fragment | | 88.7 | AAI-based (medium-confidence) | 17.62 |
| S11_k141_30609 | 1078 | No | NA | 1 | 0 | 0 | Not-determined | Genome-fragment | NA | | NA | 0 |
| S9_k141_91796 | 1219 | No | NA | 2 | 0 | 0 | Low-quality | Genome-fragment | | 1.63 | AAI-based (high-confidence) | 0 |
| S11_k141_251106 | 1058 | No | NA | 2 | 0 | 1 | Not-determined | Genome-fragment | NA | | NA | 0 |
| S11_k141_182093 | 8205 | No | NA | 5 | 0 | 0 | Not-determined | Genome-fragment | NA | | NA | 0 |
| S11_k141_44351 | 2276 | No | NA | 2 | 0 | 2 | Not-determined | Genome-fragment | NA | | NA | 0 |
| S11_k141_251140 | 1083 | No | NA | 1 | 0 | 1 | Not-determined | Genome-fragment | NA | | NA | 0 |
| S11_k141_30653 | 3047 | No | NA | 4 | 0 | 2 | Not-determined | Genome-fragment | NA | | NA | 0 |
| S11_k141_195898 | 1057 | No | NA | 1 | 0 | 1 | Not-determined | Genome-fragment | NA | | NA | 0 |
| S11_k141_71644 | 1257 | No | NA | 1 | 0 | 0 | Not-determined | Genome-fragment | NA | | NA | 0 |
| S11_k141_127188 | 1269 | No | NA | 2 | 0 | 1 | Not-determined | Genome-fragment | NA | | NA | 0 |
| S11_k141_279117 | 5563 | No | NA | 3 | 0 | 1 | Not-determined | Genome-fragment | NA | | NA | 0 |
| S11_k141_3241 | 1081 | No | NA | 1 | 0 | 0 | Not-determined | Genome-fragment | NA | | NA | 0 |
| S11_k141_58605 | 1034 | No | NA | 1 | 0 | 0 | Not-determined | Genome-fragment | NA | | NA | 0 |
| S11_k141_279202 | 1741 | No | NA | 3 | 0 | 0 | Not-determined | Genome-fragment | NA | | NA | 0 |
| S11-final-viral-combined_k141_80519\|\|full | 1219 | No | NA | 4 | 0 | 0 | Low-quality | Genome-fragment | | 0.92 | AAI-based (high-confidence) | 0 |
| S11_k141_210218 | 3456 | No | NA | 2 | 0 | 1 | Not-determined | Genome-fragment | NA | | NA | 0 |
| S11_k141_251212 | 1117 | No | NA | 1 | 0 | 1 | Not-determined | Genome-fragment | NA | | NA | 0 |
| S11_k141_58639 | 1731 | No | NA | 2 | 0 | 1 | Not-determined | Genome-fragment | NA | | NA | 0 |
| S11_k141_85898 | 4738 | No | NA | 7 | 0 | 0 | Not-determined | Genome-fragment | NA | | NA | 0 |
| S11_k141_265092 | 1390 | No | NA | 3 | 0 | 2 | Not-determined | Genome-fragment | NA | | NA | 0 |
| S11_k141_210222 | 3775 | No | NA | 4 | 0 | 2 | Not-determined | Genome-fragment | NA | | NA | 0 |
| S11_k141_265104 | 6150 | No | NA | 4 | 0 | 0 | Not-determined | Genome-fragment | NA | | NA | 0 |
| S11_k141_182252 | 1829 | No | NA | 3 | 0 | 2 | Not-determined | Genome-fragment | NA | | NA | 0 |
| S11_k141_85929 | 1347 | No | NA | 2 | 0 | 1 | Not-determined | Genome-fragment | NA | | NA | 0 |
| S11_k141_154887 | 1187 | No | NA | 3 | 0 | 0 | Not-determined | Genome-fragment | NA | | NA | 0 |
| S11_k141_223884 | 3636 | No | NA | 3 | 0 | 3 | Not-determined | Genome-fragment | NA | | NA | 0 |
| S11_k141_16953 | 2819 | No | NA | 2 | 0 | 0 | Not-determined | Genome-fragment | NA | | NA | 0 |
| S11-final-viral-combined_k141_173797\|\|lt2gene | 1219 | No | NA | 2 | 1 | 0 | Low-quality | Genome-fragment | | 3.3 | AAI-based (high-confidence) | 0 |
| S11_k141_154938 | 2514 | No | NA | 2 | 0 | 0 | Not-determined | Genome-fragment | NA | | NA | 0 |
| S11_k141_265167 | 3571 | No | NA | 2 | 0 | 2 | Not-determined | Genome-fragment | NA | | NA | 0 |
| S11_k141_319945 | 1791 | No | NA | 2 | 0 | 0 | Not-determined | Genome-fragment | NA | | NA | 0 |
| S11_k141_141104 | 1304 | No | NA | 2 | 0 | 1 | Not-determined | Genome-fragment | NA | | NA | 0 |
| S11_k141_231620 | 1219 | No | NA | 2 | 0 | 0 | Low-quality | Genome-fragment | | 2.04 | AAI-based (medium-confidence) | 0 |
| S11_k141_279354 | 1093 | No | NA | 2 | 0 | 1 | Not-determined | Genome-fragment | NA | | NA | 0 |
| S11_k141_113146 | 5316 | No | NA | 3 | 0 | 0 | Not-determined | Genome-fragment | NA | | NA | 0 |
| S1_k141_186486 | 1219 | No | NA | 2 | 0 | 2 | Low-quality | Genome-fragment | | 0.36 | AAI-based (medium-confidence) | 0 |
| S11_k141_319985 | 1267 | No | NA | 1 | 0 | 1 | Not-determined | Genome-fragment | NA | | NA | 0 |
| S11_k141_319990 | 2451 | No | NA | 1 | 0 | 1 | Not-determined | Genome-fragment | NA | | NA | 0 |
| S11_k141_292421 | 6384 | No | NA | 8 | 0 | 1 | Not-determined | Genome-fragment | NA | | NA | 0 |
| S5-final-viral-combined_k141_202004\|\|full | 1219 | No | NA | 2 | 0 | 2 | Low-quality | Genome-fragment | | 0.81 | AAI-based (high-confidence) | 0 |
| S11_k141_210374 | 1060 | No | NA | 1 | 0 | 1 | Not-determined | Genome-fragment | NA | | NA | 0 |
| S5-final-viral-combined_k141_102114\|\|lt2gene | 1219 | No | NA | 2 | 1 | 0 | Low-quality | Genome-fragment | | 3.06 | AAI-based (high-confidence) | 0 |
| S11_k141_58743 | 1063 | No | NA | 3 | 0 | 0 | Not-determined | Genome-fragment | NA | | NA | 0 |
| S11_k141_86123 | 1750 | No | NA | 1 | 0 | 1 | Not-determined | Genome-fragment | NA | | NA | 0 |
| S5-final-viral-combined_k141_129123\|\|lt2gene | 1219 | No | NA | 2 | 1 | 0 | Low-quality | Genome-fragment | | 2.78 | AAI-based (medium-confidence) | 0 |
| S12-final-viral-combined_k141_352593\|\|full | 1219 | No | NA | 2 | 0 | 1 | Low-quality | Genome-fragment | | 1.42 | AAI-based (high-confidence) | 0 |
| S11_k141_292520 | 3131 | No | NA | 4 | 0 | 0 | Not-determined | Genome-fragment | NA | | NA | 0 |
| S11_k141_30972 | 1261 | No | NA | 1 | 0 | 1 | Not-determined | Genome-fragment | NA | | NA | 0 |
| S12-final-viral-combined_k141_173829\|\|lt2gene | 1219 | No | NA | 2 | 1 | 0 | Low-quality | Genome-fragment | | 2.9 | AAI-based (high-confidence) | 0 |
| S11_k141_86198 | 1026 | No | NA | 1 | 0 | 0 | Not-determined | Genome-fragment | NA | | NA | 0 |
| S2_k141_247298 | 1219 | No | NA | 1 | 0 | 0 | Low-quality | Genome-fragment | | 2.94 | AAI-based (medium-confidence) | 0 |
| S11_k141_155154 | 1071 | No | NA | 2 | 0 | 0 | Not-determined | Genome-fragment | NA | | NA | 0 |
| S11_k141_279460 | 1069 | No | NA | 3 | 0 | 3 | Not-determined | Genome-fragment | NA | | NA | 0 |
| S11_k141_86310 | 2719 | No | NA | 3 | 0 | 2 | Not-determined | Genome-fragment | NA | | NA | 0 |
| S6_k141_23701 | 1219 | No | NA | 1 | 0 | 0 | Low-quality | Genome-fragment | | 3.6 | AAI-based (high-confidence) | 0 |
| S11_k141_113322 | 4061 | No | NA | 1 | 0 | 0 | Not-determined | Genome-fragment | NA | | NA | 0 |
| S11_k141_196153 | 1310 | No | NA | 1 | 0 | 0 | Not-determined | Genome-fragment | NA | | NA | 0 |
| S11_k141_141303 | 1307 | No | NA | 1 | 0 | 1 | Not-determined | Genome-fragment | NA | | NA | 0 |
| S11_k141_251462 | 7019 | No | NA | 11 | 0 | 7 | Not-determined | Genome-fragment | NA | | NA | 0 |
| S8-final-viral-combined_k141_201964\|\|full | 1219 | No | NA | 2 | 0 | 1 | Low-quality | Genome-fragment | | 0.83 | AAI-based (medium-confidence) | 0 |
| S11_k141_31076 | 1162 | No | NA | 2 | 0 | 2 | Not-determined | Genome-fragment | NA | | NA | 0 |
| S11_k141_58900 | 3738 | No | NA | 2 | 0 | 1 | Not-determined | Genome-fragment | NA | | NA | 0 |
| S11_k141_320258 | 3012 | No | NA | 2 | 0 | 1 | Not-determined | Genome-fragment | NA | | NA | 0 |
| S11_k141_292829 | 2807 | No | NA | 2 | 0 | 1 | Not-determined | Genome-fragment | NA | | NA | 0 |
| S11_k141_265304 | 2729 | No | NA | 1 | 0 | 0 | Not-determined | Genome-fragment | NA | | NA | 0 |
| S11_k141_100489 | 1449 | No | NA | 2 | 0 | 0 | Not-determined | Genome-fragment | NA | | NA | 0 |
| S9_k141_92492 | 1220 | No | NA | 1 | 0 | 1 | Low-quality | Genome-fragment | | 0.81 | AAI-based (high-confidence) | 0 |
| S11-final-viral-combined_k141_217260\|\|lt2gene | 1220 | No | NA | 2 | 0 | 0 | Low-quality | Genome-fragment | | 2.61 | AAI-based (medium-confidence) | 0 |
| S11_k141_196245 | 8940 | No | NA | 6 | 0 | 4 | Not-determined | Genome-fragment | NA | | NA | 0 |
| S11_k141_86545 | 1444 | No | NA | 1 | 0 | 1 | Not-determined | Genome-fragment | NA | | NA | 0 |
| S1_k141_113604 | 1220 | No | NA | 2 | 1 | 0 | Low-quality | Genome-fragment | | 2.56 | AAI-based (high-confidence) | 0 |
| S11_k141_251681 | 1225 | No | NA | 1 | 0 | 1 | Not-determined | Genome-fragment | NA | | NA | 0 |
| S11_k141_72149 | 1562 | No | NA | 1 | 0 | 1 | Not-determined | Genome-fragment | NA | | NA | 0 |
| S11_k141_224395 | 3132 | No | NA | 3 | 0 | 1 | Not-determined | Genome-fragment | NA | | NA | 0 |
| S11_k141_31250 | 1488 | No | NA | 2 | 0 | 1 | Not-determined | Genome-fragment | NA | | NA | 0 |
| S3-final-viral-combined_k141_49729\|\|full | 1220 | No | NA | 4 | 1 | 0 | Low-quality | Genome-fragment | | 3.2 | AAI-based (high-confidence) | 0 |
| S11_k141_279802 | 1083 | No | NA | 2 | 0 | 2 | Not-determined | Genome-fragment | NA | | NA | 0 |
| S11_k141_113483 | 3983 | No | NA | 5 | 0 | 3 | Not-determined | Genome-fragment | NA | | NA | 0 |
| S11_k141_141553 | 4341 | No | NA | 6 | 0 | 0 | Not-determined | Genome-fragment | NA | | NA | 0 |
| S11_k141_210963 | 1071 | No | NA | 2 | 0 | 1 | Not-determined | Genome-fragment | NA | | NA | 0 |
| S3-final-viral-combined_k141_44024\|\|lt2gene | 1220 | No | NA | 1 | 1 | 0 | Low-quality | Genome-fragment | | 3.17 | AAI-based (high-confidence) | 0 |
| S11_k141_3723 | 1039 | No | NA | 3 | 0 | 1 | Not-determined | Genome-fragment | NA | | NA | 0 |
| S11_k141_265545 | 8265 | No | NA | 8 | 0 | 3 | Not-determined | Genome-fragment | NA | | NA | 0 |
| S3_k141_54194 | 1220 | No | NA | 5 | 0 | 0 | Low-quality | Genome-fragment | | 2.29 | AAI-based (high-confidence) | 0 |
| S7-final-viral-combined_k141_132857\|\|full | 1220 | No | NA | 3 | 0 | 0 | Low-quality | Genome-fragment | | 3.21 | AAI-based (high-confidence) | 0 |
| S11_k141_224458 | 1187 | No | NA | 2 | 0 | 2 | Not-determined | Genome-fragment | NA | | NA | 0 |
| S11_k141_224460 | 1244 | No | NA | 1 | 0 | 1 | Not-determined | Genome-fragment | NA | | NA | 0 |
| S7-final-viral-combined_k141_70210\|\|full | 1220 | No | NA | 3 | 1 | 0 | Low-quality | Genome-fragment | | 19.08 | AAI-based (high-confidence) | 0 |
| S11_k141_320521 | 4453 | No | NA | 7 | 0 | 1 | Not-determined | Genome-fragment | NA | | NA | 0 |
| S7-final-viral-combined_k141_41988\|\|lt2gene | 1220 | No | NA | 1 | 0 | 0 | Low-quality | Genome-fragment | | 2.61 | AAI-based (high-confidence) | 0 |
| S17-k141_82930 | 1220 | No | NA | 2 | 0 | 0 | Low-quality | Genome-fragment | | 0.97 | AAI-based (high-confidence) | 0 |
| S11_k141_86807 | 1144 | No | NA | 2 | 0 | 2 | Not-determined | Genome-fragment | NA | | NA | 0 |
| S4_k141_43994 | 1220 | No | NA | 2 | 0 | 0 | Low-quality | Genome-fragment | | 1.23 | AAI-based (high-confidence) | 0 |
| S11_k141_320555 | 31066 | No | NA | 38 | 11 | 1 | Medium-quality | Genome-fragment | | 60.9 | AAI-based (high-confidence) | 0 |
| S11_k141_211099 | 4665 | No | NA | 5 | 0 | 5 | Not-determined | Genome-fragment | NA | | NA | 0 |
| S11_k141_265652 | 3153 | No | NA | 2 | 0 | 2 | Not-determined | Genome-fragment | NA | | NA | 0 |
| S11_k141_31428 | 1027 | No | NA | 1 | 0 | 0 | Not-determined | Genome-fragment | NA | | NA | 0 |
| S8-final-viral-combined_k141_93421\|\|full | 1220 | No | NA | 3 | 0 | 0 | Low-quality | Genome-fragment | | 2.34 | AAI-based (medium-confidence) | 0 |
| S11_k141_155622 | 1835 | No | NA | 5 | 0 | 4 | Not-determined | Genome-fragment | NA | | NA | 0 |
| S9-final-viral-combined_k141_83714\|\|full | 1221 | No | NA | 3 | 0 | 0 | Low-quality | Genome-fragment | | 3.07 | AAI-based (medium-confidence) | 0 |
| S11_k141_127920 | 3011 | No | NA | 1 | 0 | 1 | Not-determined | Genome-fragment | NA | | NA | 0 |
| S11-final-viral-combined_k141_21893\|\|full | 1221 | No | NA | 3 | 0 | 0 | Low-quality | Genome-fragment | | 3.15 | AAI-based (medium-confidence) | 0 |
| S11_k141_3906 | 1309 | No | NA | 2 | 0 | 1 | Not-determined | Genome-fragment | NA | | NA | 0 |
| S11_k141_141799 | 2136 | No | NA | 1 | 0 | 1 | Not-determined | Genome-fragment | NA | | NA | 0 |
| S11_k141_155670 | 1614 | No | NA | 1 | 0 | 1 | Not-determined | Genome-fragment | NA | | NA | 0 |
| S11_k141_72396 | 3201 | No | NA | 2 | 0 | 2 | Not-determined | Genome-fragment | NA | | NA | 0 |
| S11-final-viral-combined_k141_313619\|\|lt2gene | 1221 | No | NA | 1 | 1 | 0 | Low-quality | Genome-fragment | | 0.84 | HMM-based (lower-bound) | 0 |
| S11_k141_265796 | 4641 | No | NA | 6 | 0 | 6 | Not-determined | Genome-fragment | NA | | NA | 0 |
| S11_k141_178787 | 1221 | No | NA | 3 | 1 | 0 | Low-quality | Genome-fragment | | 3.22 | AAI-based (high-confidence) | 0 |
| S1-final-viral-combined_k141_39209\|\|full | 1221 | No | NA | 2 | 1 | 0 | Low-quality | Genome-fragment | | 2.87 | AAI-based (high-confidence) | 0 |
| S1-final-viral-combined_k141_174326\|\|full | 1221 | No | NA | 4 | 0 | 0 | Low-quality | Genome-fragment | | 4.86 | AAI-based (high-confidence) | 0 |

| | | | | | | | | | | | |
|---|---|---|---|---|---|---|---|---|---|---|---|
| S1-final-viral-combined_k141_201392\|\|lt2gene | 1221 | No | NA | 2 | 0 | 1 | Low-quality | Genome-fragment | 1.67 | AAI-based (medium-confidence) | 0 |
| S11_k141_87006 | 2723 | No | NA | 2 | 0 | 2 | Not-determined | Genome-fragment | NA | NA | 0 |
| S1_k141_300402 | 1221 | No | NA | 2 | 2 | 0 | Low-quality | Genome-fragment | 0.92 | AAI-based (high-confidence) | 0 |
| S11_k141_141895 | 1029 | No | NA | 1 | 0 | 1 | Not-determined | Genome-fragment | NA | NA | 0 |
| S11_k141_252028 | 1718 | No | NA | 1 | 0 | 1 | Not-determined | Genome-fragment | NA | NA | 0 |
| S11_k141_265877 | 1001 | No | NA | 1 | 0 | 1 | Not-determined | Genome-fragment | NA | NA | 0 |
| S11_k141_211356 | 2508 | No | NA | 1 | 0 | 0 | Not-determined | Genome-fragment | NA | NA | 0 |
| S11_k141_224654 | 1991 | No | NA | 1 | 0 | 1 | Not-determined | Genome-fragment | NA | NA | 0 |
| S11_k141_211370 | 1906 | No | NA | 1 | 0 | 1 | Not-determined | Genome-fragment | NA | NA | 0 |
| S11_k141_169920 | 2932 | No | NA | 2 | 0 | 2 | Not-determined | Genome-fragment | NA | NA | 0 |
| S11_k141_238785 | 1590 | No | NA | 1 | 0 | 1 | Not-determined | Genome-fragment | NA | NA | 0 |
| S11_k141_183224 | 1460 | No | NA | 3 | 0 | 1 | Not-determined | Genome-fragment | NA | NA | 0 |
| S7-final-viral-combined_k141_2625\|\|full | 1221 | No | NA | 3 | 1 | 0 | Low-quality | Genome-fragment | 3 | AAI-based (high-confidence) | 0 |
| S11_k141_4026 | 1113 | No | NA | 1 | 0 | 0 | Not-determined | Genome-fragment | NA | NA | 0 |
| S7-final-viral-combined_k141_157950\|\|full | 1221 | No | NA | 2 | 0 | 0 | Low-quality | Genome-fragment | 0.98 | AAI-based (high-confidence) | 0 |
| S11_k141_100981 | 1049 | No | NA | 1 | 0 | 1 | Not-determined | Genome-fragment | NA | NA | 0 |
| S7_k141_188805 | 1221 | No | NA | 3 | 0 | 0 | Low-quality | Genome-fragment | 2.5 | AAI-based (medium-confidence) | 0 |
| S11_k141_155745 | 1688 | No | NA | 2 | 0 | 2 | Not-determined | Genome-fragment | NA | NA | 0 |
| S12-final-viral-combined_k141_200672\|\|lt2gene | 1221 | No | NA | 2 | 1 | 0 | Low-quality | Genome-fragment | 2.62 | AAI-based (high-confidence) | 0 |
| S11_k141_307602 | 3067 | No | NA | 2 | 0 | 2 | Not-determined | Genome-fragment | NA | NA | 0 |
| S11_k141_169996 | 2388 | No | NA | 1 | 0 | 1 | Not-determined | Genome-fragment | NA | NA | 0 |
| S11_k141_252146 | 3010 | No | NA | 3 | 0 | 0 | Not-determined | Genome-fragment | NA | NA | 0 |
| S11_k141_252148 | 5297 | No | NA | 7 | 0 | 0 | Not-determined | Genome-fragment | NA | NA | 0 |
| S12_k141_253233 | 1221 | No | NA | 1 | 0 | 0 | Low-quality | Genome-fragment | 1.24 | AAI-based (high-confidence) | 0 |
| S11_k141_31644 | 1019 | No | NA | 1 | 0 | 0 | Not-determined | Genome-fragment | NA | NA | 0 |
| S11_k141_320738 | 1430 | No | NA | 2 | 0 | 1 | Not-determined | Genome-fragment | NA | NA | 0 |
| S11_k141_155790 | 1856 | No | NA | 2 | 0 | 2 | Not-determined | Genome-fragment | NA | NA | 0 |
| S4_k141_135533 | 1221 | No | NA | 2 | 0 | 0 | Low-quality | Genome-fragment | 1.87 | AAI-based (medium-confidence) | 0 |
| S11_k141_238874 | 1573 | No | NA | 2 | 0 | 2 | Not-determined | Genome-fragment | NA | NA | 0 |
| S9-final-viral-combined_k141_32868\|\|lt2gene | 1222 | No | NA | 2 | 1 | 0 | Low-quality | Genome-fragment | 2.7 | AAI-based (high-confidence) | 0 |
| S11_k141_252222 | 4432 | No | NA | 5 | 0 | 2 | Not-determined | Genome-fragment | NA | NA | 0 |
| S11_k141_142080 | 1832 | No | NA | 2 | 0 | 0 | Not-determined | Genome-fragment | NA | NA | 0 |
| S11-final-viral-combined_k141_302966\|\|full | 1222 | Yes | 727 | 3 | 1 | 1 | Low-quality | Genome-fragment | 1.15 | AAI-based (medium-confidence) | 40.51 |
| S11-final-viral-combined_k141_46804\|\|lt2gene | 1222 | No | NA | 2 | 1 | 0 | Low-quality | Genome-fragment | 0.74 | HMM-based (lower-bound) | 0 |
| S11_k141_252272 | 4954 | No | NA | 6 | 0 | 5 | Not-determined | Genome-fragment | NA | NA | 0 |
| S11_k141_101057 | 1713 | No | NA | 1 | 0 | 1 | Not-determined | Genome-fragment | NA | NA | 0 |
| S11_k141_87265 | 1142 | No | NA | 3 | 0 | 1 | Not-determined | Genome-fragment | NA | NA | 0 |
| S11_k141_266138 | 1289 | No | NA | 1 | 0 | 0 | Not-determined | Genome-fragment | NA | NA | 0 |
| S11-final-viral-combined_k141_200031\|\|lt2gene | 1222 | No | NA | 2 | 1 | 0 | Low-quality | Genome-fragment | 2.57 | AAI-based (medium-confidence) | 0 |
| S1-final-viral-combined_k141_246721\|\|full | 1222 | No | NA | 3 | 0 | 1 | Low-quality | Genome-fragment | 1.12 | AAI-based (high-confidence) | 0 |
| S11_k141_101116 | 2250 | No | NA | 2 | 0 | 1 | Not-determined | Genome-fragment | NA | NA | 0 |
| S11_k141_72605 | 1190 | No | NA | 3 | 0 | 0 | Not-determined | Genome-fragment | NA | NA | 0 |
| S11_k141_87293 | 37508 | No | NA | 46 | 10 | 0 | Complete | High-quality | 100 | DTR (high-confidence) | 0 |
| S11_k141_59660 | 3319 | No | NA | 5 | 0 | 3 | Not-determined | Genome-fragment | NA | NA | 0 |
| S11_k141_280452 | 1134 | No | NA | 1 | 0 | 1 | Not-determined | Genome-fragment | NA | NA | 0 |
| S11_k141_87300 | 1186 | No | NA | 2 | 0 | 0 | Not-determined | Genome-fragment | NA | NA | 0 |
| S11_k141_266247 | 1079 | No | NA | 2 | 0 | 2 | Not-determined | Genome-fragment | NA | NA | 0 |
| S3_k141_9753 | 1222 | No | NA | 5 | 0 | 0 | Low-quality | Genome-fragment | 1.96 | AAI-based (high-confidence) | 0 |
| S11_k141_266315 | 2193 | No | NA | 3 | 0 | 1 | Not-determined | Genome-fragment | NA | NA | 0 |
| S11_k141_211662 | 1182 | No | NA | 1 | 0 | 0 | Not-determined | Genome-fragment | NA | NA | 0 |
| S11_k141_31865 | 3709 | No | NA | 3 | 0 | 0 | Not-determined | Genome-fragment | NA | NA | 0 |
| S11_k141_266343 | 1497 | No | NA | 2 | 0 | 2 | Not-determined | Genome-fragment | NA | NA | 0 |
| S12-final-viral-combined_k141_326325\|\|full | 1222 | No | NA | 5 | 0 | 0 | Low-quality | Genome-fragment | 3.8 | AAI-based (high-confidence) | 0 |
| S11_k141_266396 | 3009 | No | NA | 1 | 0 | 0 | Not-determined | Genome-fragment | NA | NA | 0 |
| S11_k141_45976 | 1036 | No | NA | 1 | 0 | 1 | Not-determined | Genome-fragment | NA | NA | 0 |
| S11_k141_320969 | 1452 | No | NA | 1 | 0 | 1 | Not-determined | Genome-fragment | NA | NA | 0 |
| S11_k141_72821 | 1086 | No | NA | 1 | 0 | 1 | Not-determined | Genome-fragment | NA | NA | 0 |
| S12-final-viral-combined_k141_82962\|\|full | 1222 | No | NA | 3 | 0 | 0 | Low-quality | Genome-fragment | 3.73 | AAI-based (high-confidence) | 0 |
| S11_k141_183640 | 1671 | No | NA | 2 | 0 | 2 | Not-determined | Genome-fragment | NA | NA | 0 |
| S12_k141_56236 | 1222 | No | NA | 4 | 0 | 0 | Low-quality | Genome-fragment | 1.88 | AAI-based (high-confidence) | 0 |
| S11_k141_183699 | 2495 | No | NA | 2 | 0 | 0 | Not-determined | Genome-fragment | NA | NA | 0 |
| S11_k141_87374 | 1206 | No | NA | 2 | 0 | 1 | Not-determined | Genome-fragment | NA | NA | 0 |
| S11_k141_183717 | 1219 | No | NA | 3 | 0 | 2 | Not-determined | Genome-fragment | NA | NA | 0 |
| S4-final-viral-combined_k141_133668\|\|lt2gene | 1222 | No | NA | 1 | 1 | 0 | Low-quality | Genome-fragment | 1.64 | AAI-based (high-confidence) | 0 |
| S11_k141_46106 | 2136 | No | NA | 5 | 0 | 4 | Not-determined | Genome-fragment | NA | NA | 0 |
| S11_k141_4475 | 1968 | No | NA | 2 | 0 | 2 | Not-determined | Genome-fragment | NA | NA | 0 |
| S11_k141_128477 | 3377 | No | NA | 4 | 0 | 3 | Not-determined | Genome-fragment | NA | NA | 0 |
| S11_k141_225246 | 1037 | No | NA | 1 | 0 | 0 | Not-determined | Genome-fragment | NA | NA | 0 |
| S11_k141_4510 | 1051 | No | NA | 1 | 0 | 1 | Not-determined | Genome-fragment | NA | NA | 0 |
| S11_k141_4545 | 1280 | No | NA | 1 | 0 | 1 | Not-determined | Genome-fragment | NA | NA | 0 |
| S4_k141_37182 | 1222 | No | NA | 2 | 1 | 0 | Low-quality | Genome-fragment | 2.14 | AAI-based (high-confidence) | 0 |
| S11_k141_156190 | 1146 | No | NA | 1 | 0 | 0 | Not-determined | Genome-fragment | NA | NA | 0 |
| S11_k141_183803 | 3487 | No | NA | 3 | 0 | 3 | Not-determined | Genome-fragment | NA | NA | 0 |
| S11_k141_32131 | 1085 | No | NA | 1 | 0 | 0 | Not-determined | Genome-fragment | NA | NA | 0 |
| S11_k141_170443 | 4149 | No | NA | 3 | 0 | 2 | Not-determined | Genome-fragment | NA | NA | 0 |
| S8-final-viral-combined_k141_54260\|\|full | 1222 | No | NA | 2 | 0 | 0 | Low-quality | Genome-fragment | 0.72 | AAI-based (medium-confidence) | 0 |
| S11_k141_321171 | 1031 | No | NA | 3 | 0 | 2 | Not-determined | Genome-fragment | NA | NA | 0 |
| S11_k141_46222 | 1051 | No | NA | 1 | 0 | 0 | Not-determined | Genome-fragment | NA | NA | 0 |
| S11_k141_225393 | 1104 | No | NA | 1 | 0 | 1 | Not-determined | Genome-fragment | NA | NA | 0 |
| S11_k141_183845 | 4229 | No | NA | 4 | 0 | 0 | Not-determined | Genome-fragment | NA | NA | 0 |
| S8_k141_166767 | 1222 | No | NA | 2 | 0 | 1 | Low-quality | Genome-fragment | 2.2 | AAI-based (high-confidence) | 0 |
| S11_k141_239363 | 3153 | No | NA | 3 | 0 | 3 | Not-determined | Genome-fragment | NA | NA | 0 |
| S11-final-viral-combined_k141_305022\|\|full | 1223 | No | NA | 2 | 0 | 0 | Low-quality | Genome-fragment | 2.09 | AAI-based (high-confidence) | 0 |
| S11-final-viral-combined_k141_23352\|\|lt2gene | 1223 | No | NA | 2 | 0 | 0 | Low-quality | Genome-fragment | 0.8 | AAI-based (high-confidence) | 0 |
| S11_k141_170502 | 1590 | No | NA | 2 | 0 | 1 | Not-determined | Genome-fragment | NA | NA | 0 |
| S11_k141_128609 | 1260 | No | NA | 2 | 0 | 1 | Not-determined | Genome-fragment | NA | NA | 0 |
| S11_k141_168491 | 1223 | No | NA | 2 | 0 | 0 | Low-quality | Genome-fragment | 2.66 | AAI-based (medium-confidence) | 0 |
| S11_k141_114260 | 3455 | No | NA | 2 | 0 | 2 | Not-determined | Genome-fragment | NA | NA | 0 |
| S11_k141_308167 | 3271 | No | NA | 4 | 0 | 3 | Not-determined | Genome-fragment | NA | NA | 0 |
| S11_k141_321198 | 1508 | No | NA | 1 | 0 | 1 | Not-determined | Genome-fragment | NA | NA | 0 |
| S11_k141_114275 | 6262 | No | NA | 10 | 0 | 1 | Not-determined | Genome-fragment | NA | NA | 0 |
| S5-final-viral-combined_k141_207620\|\|full | 1223 | No | NA | 2 | 0 | 0 | Low-quality | Genome-fragment | 1.42 | AAI-based (medium-confidence) | 0 |
| S11_k141_225481 | 3302 | No | NA | 5 | 0 | 3 | Not-determined | Genome-fragment | NA | NA | 0 |
| S11_k141_183913 | 1565 | No | NA | 1 | 0 | 1 | Not-determined | Genome-fragment | NA | NA | 0 |
| S11_k141_142640 | 3076 | No | NA | 4 | 0 | 2 | Not-determined | Genome-fragment | NA | NA | 0 |
| S11_k141_4688 | 1683 | No | NA | 1 | 0 | 0 | Not-determined | Genome-fragment | NA | NA | 0 |
| S11_k141_252777 | 1200 | No | NA | 2 | 0 | 2 | Not-determined | Genome-fragment | NA | NA | 0 |
| S11_k141_142680 | 1844 | No | NA | 2 | 0 | 0 | Not-determined | Genome-fragment | NA | NA | 0 |
| S12-final-viral-combined_k141_184160\|\|full | 1223 | No | NA | 3 | 0 | 0 | Low-quality | Genome-fragment | 2.34 | AAI-based (medium-confidence) | 0 |
| S11_k141_142681 | 3539 | No | NA | 6 | 0 | 0 | Not-determined | Genome-fragment | NA | NA | 0 |
| S12-final-viral-combined_k141_294318\|\|full | 1223 | No | NA | 3 | 1 | 0 | Low-quality | Genome-fragment | 2.52 | AAI-based (high-confidence) | 0 |
| S11_k141_239475 | 1145 | No | NA | 1 | 0 | 0 | Not-determined | Genome-fragment | NA | NA | 0 |
| S11_k141_18045 | 13513 | No | NA | 16 | 0 | 0 | Not-determined | Genome-fragment | NA | NA | 0 |
| S11_k141_142705 | 1219 | No | NA | 2 | 0 | 2 | Not-determined | Genome-fragment | NA | NA | 0 |
| S11_k141_321349 | 1090 | No | NA | 2 | 0 | 1 | Not-determined | Genome-fragment | NA | NA | 0 |
| S11_k141_294099 | 1453 | No | NA | 1 | 0 | 1 | Not-determined | Genome-fragment | NA | NA | 0 |
| S12-final-viral-combined_k141_77365\|\|full | 1223 | No | NA | 2 | 1 | 0 | Low-quality | Genome-fragment | 1.88 | AAI-based (high-confidence) | 0 |
| S2-final-viral-combined_k141_211108\|\|full | 1223 | No | NA | 3 | 0 | 0 | Low-quality | Genome-fragment | 2.81 | AAI-based (medium-confidence) | 0 |
| S11_k141_197417 | 1107 | No | NA | 1 | 0 | 0 | Not-determined | Genome-fragment | NA | NA | 0 |
| S11_k141_252896 | 1101 | No | NA | 1 | 0 | 0 | Not-determined | Genome-fragment | NA | NA | 0 |
| S11_k141_225596 | 1673 | No | NA | 2 | 0 | 0 | Not-determined | Genome-fragment | NA | NA | 0 |
| S11_k141_321378 | 1395 | No | NA | 3 | 0 | 1 | Not-determined | Genome-fragment | NA | NA | 0 |
| S11_k141_294125 | 1420 | No | NA | 3 | 0 | 1 | Not-determined | Genome-fragment | NA | NA | 0 |
| S2-final-viral-combined_k141_179439\|\|lt2gene | 1223 | No | NA | 2 | 2 | 0 | Low-quality | Genome-fragment | 3.59 | AAI-based (high-confidence) | 0 |
| S11_k141_156492 | 1498 | No | NA | 1 | 0 | 1 | Not-determined | Genome-fragment | NA | NA | 0 |
| S11_k141_183997 | 1568 | No | NA | 2 | 0 | 1 | Not-determined | Genome-fragment | NA | NA | 0 |
| S4_k141_153718 | 1223 | No | NA | 1 | 1 | 0 | Low-quality | Genome-fragment | 0.7 | HMM-based (lower-bound) | 0 |
| S11_k141_114533 | 2678 | No | NA | 2 | 0 | 0 | Not-determined | Genome-fragment | NA | NA | 0 |
| S11_k141_18130 | 1055 | No | NA | 1 | 0 | 1 | Not-determined | Genome-fragment | NA | NA | 0 |
| S11_k141_32418 | 2610 | No | NA | 3 | 0 | 0 | Not-determined | Genome-fragment | NA | NA | 0 |
| S11_k141_197501 | 1104 | No | NA | 2 | 0 | 1 | Not-determined | Genome-fragment | NA | NA | 0 |
| S11_k141_142842 | 4890 | No | NA | 3 | 0 | 2 | Not-determined | Genome-fragment | NA | NA | 0 |
| S11_k141_73232 | 5174 | No | NA | 4 | 0 | 2 | Not-determined | Genome-fragment | NA | NA | 0 |
| S11_k141_32479 | 10399 | No | NA | 11 | 0 | 4 | Not-determined | Genome-fragment | NA | NA | 0 |
| S11_k141_142867 | 1304 | No | NA | 2 | 0 | 1 | Not-determined | Genome-fragment | NA | NA | 0 |
| S11_k141_101907 | 3524 | No | NA | 3 | 0 | 3 | Not-determined | Genome-fragment | NA | NA | 0 |
| S6_k141_190 | 1223 | No | NA | 2 | 0 | 0 | Low-quality | Genome-fragment | 2.98 | AAI-based (medium-confidence) | 0 |
| S11-final-viral-combined_k141_156536\|\|full | 1224 | No | NA | 3 | 1 | 0 | Low-quality | Genome-fragment | 3.38 | AAI-based (high-confidence) | 0 |
| S11-final-viral-combined_k141_324622\|\|full | 1224 | No | NA | 3 | 1 | 0 | Low-quality | Genome-fragment | 3.43 | AAI-based (high-confidence) | 0 |
| S1-final-viral-combined_k141_329369\|\|full | 1224 | No | NA | 2 | 0 | 0 | Low-quality | Genome-fragment | 1.91 | AAI-based (medium-confidence) | 0 |
| S11_k141_156566 | 1236 | No | NA | 1 | 0 | 1 | Not-determined | Genome-fragment | NA | NA | 0 |
| S11_k141_142919 | 1248 | No | NA | 1 | 0 | 1 | Not-determined | Genome-fragment | NA | NA | 0 |
| S1-final-viral-combined_k141_124331\|\|full | 1224 | No | NA | 2 | 0 | 1 | Low-quality | Genome-fragment | 1.88 | AAI-based (medium-confidence) | 0 |
| S11_k141_266915 | 2339 | No | NA | 1 | 0 | 0 | Not-determined | Genome-fragment | NA | NA | 0 |
| S11_k141_321484 | 1295 | No | NA | 2 | 0 | 0 | Not-determined | Genome-fragment | NA | NA | 0 |
| S11_k141_142963 | 1321 | No | NA | 1 | 0 | 1 | Not-determined | Genome-fragment | NA | NA | 0 |
| S3-final-viral-combined_k141_96267\|\|full | 1224 | No | NA | 3 | 1 | 0 | Low-quality | Genome-fragment | 2.62 | AAI-based (high-confidence) | 0 |
| S7-final-viral-combined_k141_188352\|\|full | 1224 | No | NA | 3 | 0 | 0 | Low-quality | Genome-fragment | 9.54 | AAI-based (medium-confidence) | 0 |
| S11_k141_281348 | 3528 | No | NA | 4 | 0 | 2 | Not-determined | Genome-fragment | NA | NA | 0 |
| S11_k141_143005 | 1216 | No | NA | 2 | 0 | 0 | Not-determined | Genome-fragment | NA | NA | 0 |
| S17-k141_173260 | 1224 | No | NA | 2 | 0 | 0 | Low-quality | Genome-fragment | 4.23 | AAI-based (medium-confidence) | 0 |
| S11_k141_4976 | 3308 | No | NA | 5 | 0 | 3 | Not-determined | Genome-fragment | NA | NA | 0 |
| S11_k141_87956 | 1195 | No | NA | 3 | 0 | 0 | Not-determined | Genome-fragment | NA | NA | 0 |
| S11_k141_266944 | 1653 | No | NA | 1 | 0 | 0 | Not-determined | Genome-fragment | NA | NA | 0 |
| S11_k141_321582 | 3158 | No | NA | 2 | 0 | 2 | Not-determined | Genome-fragment | NA | NA | 0 |
| S11_k141_294337 | 2458 | No | NA | 4 | 0 | 0 | Not-determined | Genome-fragment | NA | NA | 0 |
| S12-final-viral-combined_k141_51258\|\|full | 1224 | No | NA | 3 | 1 | 0 | Low-quality | Genome-fragment | 2.67 | AAI-based (high-confidence) | 0 |
| S11_k141_281447 | 1492 | No | NA | 2 | 0 | 1 | Not-determined | Genome-fragment | NA | NA | 0 |
| S11_k141_294349 | 1651 | No | NA | 2 | 0 | 0 | Not-determined | Genome-fragment | NA | NA | 0 |
| S11_k141_294353 | 1954 | No | NA | 1 | 0 | 0 | Not-determined | Genome-fragment | NA | NA | 0 |
| S11_k141_239961 | 2373 | No | NA | 2 | 0 | 1 | Not-determined | Genome-fragment | NA | NA | 0 |
| S11_k141_102155 | 1310 | No | NA | 1 | 0 | 1 | Not-determined | Genome-fragment | NA | NA | 0 |
| S12-final-viral-combined_k141_219819\|\|full | 1224 | No | NA | 4 | 2 | 0 | Low-quality | Genome-fragment | 2.01 | HMM-based (lower-bound) | 0 |
| S12_k141_378331 | 1224 | No | NA | 2 | 0 | 2 | Low-quality | Genome-fragment | 1.58 | AAI-based (medium-confidence) | 0 |
| S12_k141_244439 | 1224 | No | NA | 3 | 0 | 0 | Low-quality | Genome-fragment | 2.7 | AAI-based (medium-confidence) | 0 |
| S11_k141_88048 | 3569 | No | NA | 4 | 0 | 2 | Not-determined | Genome-fragment | NA | NA | 0 |
| S11_k141_294430 | 1953 | No | NA | 1 | 0 | 1 | Not-determined | Genome-fragment | NA | NA | 0 |
| S11_k141_308659 | 3793 | No | NA | 4 | 0 | 1 | Not-determined | Genome-fragment | NA | NA | 0 |
| S11_k141_156721 | 1036 | No | NA | 1 | 0 | 1 | Not-determined | Genome-fragment | NA | NA | 0 |
| S11_k141_197822 | 1035 | No | NA | 2 | 0 | 0 | Not-determined | Genome-fragment | NA | NA | 0 |
| S11_k141_308703 | 2105 | No | NA | 5 | 0 | 0 | Not-determined | Genome-fragment | NA | NA | 0 |
| S11_k141_32691 | 155831 | No | NA | 188 | 1 | 11 | Medium-quality | Genome-fragment | 70.25 | HMM-based (lower-bound) | 0 |
| S11_k141_308724 | 1366 | No | NA | 1 | 0 | 1 | Not-determined | Genome-fragment | NA | NA | 0 |
| S2-final-viral-combined_k141_140603\|\|full | 1224 | No | NA | 4 | 4 | 0 | Low-quality | Genome-fragment | 2.28 | AAI-based (high-confidence) | 0 |
| S11_k141_32727 | 1431 | No | NA | 2 | 0 | 1 | Not-determined | Genome-fragment | NA | NA | 0 |
| S11_k141_281617 | 1419 | No | NA | 2 | 0 | 1 | Not-determined | Genome-fragment | NA | NA | 0 |
| S11_k141_129201 | 1070 | No | NA | 1 | 0 | 1 | Not-determined | Genome-fragment | NA | NA | 0 |
| S6_k141_57252 | 1224 | No | NA | 2 | 0 | 0 | Low-quality | Genome-fragment | 1.45 | AAI-based (high-confidence) | 0 |
| S11_k141_212577 | 3105 | No | NA | 2 | 0 | 2 | Not-determined | Genome-fragment | NA | NA | 0 |
| S11_k141_18554 | 1010 | No | NA | 2 | 0 | 2 | Not-determined | Genome-fragment | NA | NA | 0 |
| S11_k141_88119 | 1496 | No | NA | 1 | 0 | 1 | Not-determined | Genome-fragment | NA | NA | 0 |
| S11_k141_5222 | 5412 | No | NA | 4 | 0 | 4 | Not-determined | Genome-fragment | NA | NA | 0 |
| S11_k141_32788 | 1488 | No | NA | 2 | 0 | 2 | Not-determined | Genome-fragment | NA | NA | 0 |
| S11_k141_184366 | 1541 | No | NA | 1 | 0 | 1 | Not-determined | Genome-fragment | NA | NA | 0 |
| S11_k141_240138 | 8458 | No | NA | 4 | 0 | 2 | Not-determined | Genome-fragment | NA | NA | 0 |
| S11_k141_60538 | 1124 | No | NA | 1 | 0 | 0 | Not-determined | Genome-fragment | NA | NA | 0 |
| S8_k141_83765 | 1224 | No | NA | 4 | 0 | 0 | Low-quality | Genome-fragment | 0.87 | AAI-based (high-confidence) | 0 |
| S11_k141_156845 | 1662 | No | NA | 1 | 0 | 0 | Not-determined | Genome-fragment | NA | NA | 0 |
| S11_k141_143274 | 1338 | No | NA | 1 | 0 | 1 | Not-determined | Genome-fragment | NA | NA | 0 |
| S11-final-viral-combined_k141_176248\|\|full | 1225 | No | NA | 4 | 0 | 0 | Low-quality | Genome-fragment | 0.95 | AAI-based (high-confidence) | 0 |
| S11_k141_102357 | 7962 | No | NA | 2 | 0 | 1 | Not-determined | Genome-fragment | NA | NA | 0 |
| S3_k141_97369 | 1225 | No | NA | 3 | 0 | 0 | Low-quality | Genome-fragment | 2.13 | AAI-based (medium-confidence) | 0 |
| S11_k141_226077 | 2750 | No | NA | 2 | 0 | 0 | Not-determined | Genome-fragment | NA | NA | 0 |
| S11_k141_198020 | 1369 | No | NA | 1 | 0 | 1 | Not-determined | Genome-fragment | NA | NA | 0 |
| S12_k141_246534 | 1225 | No | NA | 4 | 0 | 0 | Low-quality | Genome-fragment | 3.04 | AAI-based (high-confidence) | 0 |
| S4_k141_29360 | 1225 | No | NA | 1 | 0 | 1 | Low-quality | Genome-fragment | 0.66 | AAI-based (medium-confidence) | 0 |
| S11_k141_102416 | 2974 | No | NA | 3 | 0 | 0 | Not-determined | Genome-fragment | NA | NA | 0 |
| S11_k141_321756 | 1083 | No | NA | 1 | 0 | 1 | Not-determined | Genome-fragment | NA | NA | 0 |
| S6-final-viral-combined_k141_32751\|\|full | 1225 | No | NA | 4 | 4 | 0 | Low-quality | Genome-fragment | 2.67 | AAI-based (high-confidence) | 0 |
| S11_k141_102437 | 3217 | No | NA | 5 | 0 | 3 | Not-determined | Genome-fragment | NA | NA | 0 |
| S11_k141_253455 | 1372 | No | NA | 2 | 0 | 2 | Not-determined | Genome-fragment | NA | NA | 0 |
| S6-final-viral-combined_k141_128555\|\|full | 1225 | No | NA | 4 | 0 | 0 | Low-quality | Genome-fragment | 2.59 | AAI-based (medium-confidence) | 0 |
| S11_k141_32975 | 1165 | No | NA | 1 | 0 | 1 | Not-determined | Genome-fragment | NA | NA | 0 |
| S11_k141_171112 | 4208 | No | NA | 3 | 0 | 2 | Not-determined | Genome-fragment | NA | NA | 0 |
| S11_k141_32994 | 1305 | No | NA | 3 | 0 | 0 | Not-determined | Genome-fragment | NA | NA | 0 |
| S11_k141_171119 | 1642 | No | NA | 1 | 0 | 1 | Not-determined | Genome-fragment | NA | NA | 0 |
| S11_k141_171121 | 2648 | No | NA | 3 | 0 | 1 | Not-determined | Genome-fragment | NA | NA | 0 |
| S11_k141_308958 | 3355 | No | NA | 2 | 0 | 2 | Not-determined | Genome-fragment | NA | NA | 0 |
| S11_k141_294719 | 1092 | No | NA | 2 | 0 | 0 | Not-determined | Genome-fragment | NA | NA | 0 |
| S1-final-viral-combined_k141_149079\|\|full | 1226 | No | NA | 3 | 2 | 0 | Low-quality | Genome-fragment | 2.75 | AAI-based (medium-confidence) | 0 |
| S11_k141_157024 | 1568 | No | NA | 1 | 0 | 1 | Not-determined | Genome-fragment | NA | NA | 0 |
| S11_k141_309009 | 1066 | No | NA | 1 | 0 | 1 | Not-determined | Genome-fragment | NA | NA | 0 |
| S1-final-viral-combined_k141_142473\|\|full | 1226 | No | NA | 3 | 0 | 0 | Low-quality | Genome-fragment | 1.69 | AAI-based (high-confidence) | 0 |
| S11_k141_129476 | 2116 | No | NA | 1 | 0 | 0 | Not-determined | Genome-fragment | NA | NA | 0 |
| S5-final-viral-combined_k141_213093\|\|full | 1226 | No | NA | 3 | 1 | 0 | Low-quality | Genome-fragment | 2.54 | AAI-based (high-confidence) | 0 |
| S11_k141_157076 | 5853 | No | NA | 8 | 0 | 1 | Not-determined | Genome-fragment | NA | NA | 0 |
| S5_k141_65740 | 1226 | No | NA | 3 | 0 | 0 | Low-quality | Genome-fragment | 19.96 | AAI-based (high-confidence) | 0 |
| S7-final-viral-combined_k141_244769\|\|lt2gene | 1226 | No | NA | 1 | 1 | 0 | Low-quality | Genome-fragment | 1.79 | HMM-based (lower-bound) | 0 |
| S11_k141_226153 | 1271 | No | NA | 1 | 0 | 1 | Not-determined | Genome-fragment | NA | NA | 0 |
| S11_k141_143496 | 4427 | No | NA | 5 | 0 | 3 | Not-determined | Genome-fragment | NA | NA | 0 |
| S17-final-viral-combined_k141_13609\|\|full | 1226 | No | NA | 2 | 1 | 0 | Low-quality | Genome-fragment | 0.74 | HMM-based (lower-bound) | 0 |
| S17-final-viral-combined_k141_105096\|\|full | 1226 | No | NA | 2 | 0 | 1 | Low-quality | Genome-fragment | 2.15 | AAI-based (medium-confidence) | 0 |
| S11_k141_171300 | 9777 | No | NA | 6 | 0 | 4 | Not-determined | Genome-fragment | NA | NA | 0 |
| S11_k141_226170 | 3873 | No | NA | 3 | 0 | 3 | Not-determined | Genome-fragment | NA | NA | 0 |
| S11_k141_33141 | 1292 | No | NA | 2 | 0 | 2 | Not-determined | Genome-fragment | NA | NA | 0 |
| S11_k141_171317 | 3793 | No | NA | 2 | 0 | 2 | Not-determined | Genome-fragment | NA | NA | 0 |
| S11_k141_294900 | 3329 | No | NA | 2 | 0 | 1 | Not-determined | Genome-fragment | NA | NA | 0 |
| S11_k141_88493 | 1353 | No | NA | 1 | 0 | 1 | Not-determined | Genome-fragment | NA | NA | 0 |
| S17-final-viral-combined_k141_186042\|\|full | 1226 | No | NA | 3 | 1 | 0 | Low-quality | Genome-fragment | 2.51 | HMM-based (lower-bound) | 0 |
| S17-final-viral-combined_k141_232422\|\|full | 1226 | No | NA | 3 | 1 | 0 | Low-quality | Genome-fragment | 3.35 | AAI-based (high-confidence) | 0 |
| S11_k141_74031 | 3426 | No | NA | 5 | 0 | 3 | Not-determined | Genome-fragment | NA | NA | 0 |
| S11_k141_253793 | 1077 | No | NA | 3 | 0 | 1 | Not-determined | Genome-fragment | NA | NA | 0 |
| S11_k141_88512 | 1633 | No | NA | 2 | 0 | 1 | Not-determined | Genome-fragment | NA | NA | 0 |
| S11_k141_281933 | 1815 | No | NA | 4 | 0 | 1 | Not-determined | Genome-fragment | NA | NA | 0 |
| S11_k141_33254 | 1392 | No | NA | 1 | 0 | 0 | Not-determined | Genome-fragment | NA | NA | 0 |
| S12-final-viral-combined_k141_248892\|\|full | 1226 | No | NA | 2 | 0 | 0 | Low-quality | Genome-fragment | 2.39 | AAI-based (medium-confidence) | 0 |
| S11_k141_213055 | 2472 | No | NA | 2 | 0 | 2 | Not-determined | Genome-fragment | NA | NA | 0 |
| S6-final-viral-combined_k141_76992\|\|full | 1226 | No | NA | 2 | 0 | 1 | Low-quality | Genome-fragment | 2.09 | AAI-based (medium-confidence) | 0 |
| S11_k141_157290 | 1284 | No | NA | 1 | 0 | 1 | Not-determined | Genome-fragment | NA | NA | 0 |
| S11_k141_240687 | 1091 | No | NA | 3 | 0 | 0 | Not-determined | Genome-fragment | NA | NA | 0 |
| S11_k141_213082 | 1201 | No | NA | 1 | 0 | 1 | Not-determined | Genome-fragment | NA | NA | 0 |
| S11_k141_198458 | 1077 | No | NA | 1 | 0 | 1 | Not-determined | Genome-fragment | NA | NA | 0 |
| S11_k141_47298 | 17525 | No | NA | 16 | 0 | 7 | Not-determined | Genome-fragment | NA | NA | 0 |
| S11_k141_157328 | 3563 | No | NA | 3 | 0 | 3 | Not-determined | Genome-fragment | NA | NA | 0 |
| S11_k141_102849 | 3427 | No | NA | 1 | 0 | 1 | Not-determined | Genome-fragment | NA | NA | 0 |
| S11_k141_129680 | 1620 | No | NA | 2 | 0 | 1 | Not-determined | Genome-fragment | NA | NA | 0 |
| S11_k141_47307 | 1283 | No | NA | 2 | 0 | 1 | Not-determined | Genome-fragment | NA | NA | 0 |
| S5-final-viral-combined_k141_194785\|\|lt2gene | 1227 | No | NA | 1 | 1 | 0 | Low-quality | Genome-fragment | 0.94 | HMM-based (lower-bound) | 0 |
| S7-final-viral-combined_k141_193534\|\|full | 1227 | No | NA | 2 | 0 | 1 | Low-quality | Genome-fragment | 1.63 | AAI-based (high-confidence) | 0 |
| S11_k141_5726 | 2291 | No | NA | 1 | 0 | 1 | Not-determined | Genome-fragment | NA | NA | 0 |
| S11_k141_184834 | 1088 | No | NA | 1 | 0 | 0 | Not-determined | Genome-fragment | NA | NA | 0 |
| S17-k141_243019 | 1227 | No | NA | 2 | 0 | 0 | Low-quality | Genome-fragment | 2.33 | AAI-based (medium-confidence) | 0 |
| S11_k141_171487 | 6578 | No | NA | 6 | 0 | 5 | Not-determined | Genome-fragment | NA | NA | 0 |
| S11_k141_129813 | 1093 | No | NA | 1 | 0 | 0 | Not-determined | Genome-fragment | NA | NA | 0 |
| S11_k141_129827 | 1694 | No | NA | 1 | 0 | 0 | Not-determined | Genome-fragment | NA | NA | 0 |
| S11_k141_171503 | 2184 | No | NA | 2 | 0 | 0 | Not-determined | Genome-fragment | NA | NA | 0 |
| S11_k141_33446 | 1072 | No | NA | 1 | 0 | 1 | Not-determined | Genome-fragment | NA | NA | 0 |
| S11_k141_47398 | 3222 | No | NA | 3 | 0 | 2 | Not-determined | Genome-fragment | NA | NA | 0 |
| S11_k141_254067 | 1672 | No | NA | 1 | 0 | 1 | Not-determined | Genome-fragment | NA | NA | 0 |
| S11_k141_184893 | 1886 | No | NA | 1 | 0 | 1 | Not-determined | Genome-fragment | NA | NA | 0 |
| S12-final-viral-combined_k141_126541\|\|full | 1227 | No | NA | 3 | 2 | 0 | Low-quality | Genome-fragment | 2.92 | AAI-based (medium-confidence) | 0 |
| S11_k141_184924 | 2102 | No | NA | 6 | 0 | 0 | Not-determined | Genome-fragment | NA | NA | 0 |
| S11_k141_74301 | 3031 | No | NA | 2 | 0 | 1 | Not-determined | Genome-fragment | NA | NA | 0 |
| S11_k141_115492 | 2418 | No | NA | 2 | 0 | 2 | Not-determined | Genome-fragment | NA | NA | 0 |
| S11_k141_254133 | 3537 | No | NA | 3 | 0 | 3 | Not-determined | Genome-fragment | NA | NA | 0 |
| S11_k141_5827 | 1001 | No | NA | 2 | 0 | 1 | Not-determined | Genome-fragment | NA | NA | 0 |
| S11_k141_322217 | 2517 | No | NA | 2 | 0 | 2 | Not-determined | Genome-fragment | NA | NA | 0 |
| S11_k141_322234 | 1020 | No | NA | 2 | 0 | 0 | Not-determined | Genome-fragment | NA | NA | 0 |
| S11_k141_143870 | 1004 | No | NA | 1 | 0 | 1 | Not-determined | Genome-fragment | NA | NA | 0 |
| S12-final-viral-combined_k141_250065\|\|full | 1227 | No | NA | 2 | 0 | 0 | Low-quality | Genome-fragment | 2.3 | AAI-based (medium-confidence) | 0 |
| S12-final-viral-combined_k141_73995\|\|lt2gene | 1227 | No | NA | 2 | 1 | 0 | Low-quality | Genome-fragment | 0.79 | HMM-based (lower-bound) | 0 |
| S11_k141_226583 | 1276 | No | NA | 1 | 0 | 1 | Not-determined | Genome-fragment | NA | NA | 0 |
| S2-final-viral-combined_k141_107872\|\|full | 1227 | No | NA | 1 | 0 | 1 | Low-quality | Genome-fragment | 2.68 | AAI-based (high-confidence) | 0 |
| S11_k141_267863 | 1329 | No | NA | 1 | 0 | 1 | Not-determined | Genome-fragment | NA | NA | 0 |
| S11_k141_309645 | 3952 | No | NA | 2 | 0 | 2 | Not-determined | Genome-fragment | NA | NA | 0 |
| S11_k141_47569 | 5513 | No | NA | 6 | 0 | 4 | Not-determined | Genome-fragment | NA | NA | 0 |
| S11_k141_198693 | 1056 | No | NA | 3 | 0 | 0 | Not-determined | Genome-fragment | NA | NA | 0 |
| S11_k141_5945 | 1774 | No | NA | 1 | 0 | 1 | Not-determined | Genome-fragment | NA | NA | 0 |
| S11_k141_171685 | 3820 | No | NA | 3 | 0 | 3 | Not-determined | Genome-fragment | NA | NA | 0 |
| S11_k141_185049 | 3118 | No | NA | 4 | 0 | 3 | Not-determined | Genome-fragment | NA | NA | 0 |
| S11_k141_295305 | 12142 | No | NA | 12 | 0 | 7 | Not-determined | Genome-fragment | NA | NA | 0 |
| S11_k141_33647 | 1992 | No | NA | 3 | 0 | 2 | Not-determined | Genome-fragment | NA | NA | 0 |
| S11_k141_88781 | 3666 | No | NA | 2 | 0 | 2 | Not-determined | Genome-fragment | NA | NA | 0 |
| S4_k141_16729 | 1227 | No | NA | 4 | 0 | 0 | Low-quality | Genome-fragment | 2.15 | AAI-based (medium-confidence) | 0 |
| S8-final-viral-combined_k141_135273\|\|full | 1227 | No | NA | 2 | 0 | 0 | Low-quality | Genome-fragment | 2.28 | AAI-based (high-confidence) | 0 |
| S11_k141_226672 | 2197 | No | NA | 3 | 0 | 2 | Not-determined | Genome-fragment | NA | NA | 0 |
| S11_k141_6021 | 1027 | No | NA | 1 | 0 | 1 | Not-determined | Genome-fragment | NA | NA | 0 |
| S11_k141_33670 | 1537 | No | NA | 2 | 0 | 2 | Not-determined | Genome-fragment | NA | NA | 0 |
| S8-final-viral-combined_k141_148082\|\|full | 1227 | No | NA | 2 | 0 | 0 | Low-quality | Genome-fragment | 1.62 | AAI-based (medium-confidence) | 0 |
| S11_k141_19215 | 8203 | No | NA | 5 | 0 | 3 | Not-determined | Genome-fragment | NA | NA | 0 |
| S11_k141_267972 | 3581 | No | NA | 3 | 0 | 3 | Not-determined | Genome-fragment | NA | NA | 0 |
| S11_k141_309756 | 1094 | No | NA | 1 | 0 | 0 | Not-determined | Genome-fragment | NA | NA | 0 |
| S11_k141_267973 | 1517 | No | NA | 1 | 0 | 1 | Not-determined | Genome-fragment | NA | NA | 0 |
| S11_k141_115749 | 1228 | No | NA | 3 | 0 | 1 | Not-determined | Genome-fragment | NA | NA | 0 |
| S11_k141_33688 | 6066 | No | NA | 5 | 0 | 2 | Not-determined | Genome-fragment | NA | NA | 0 |
| S11_k141_267987 | 2295 | No | NA | 3 | 0 | 0 | Not-determined | Genome-fragment | NA | NA | 0 |
| S11_k141_295349 | 6316 | No | NA | 6 | 0 | 2 | Not-determined | Genome-fragment | NA | NA | 0 |
| S8-final-viral-combined_k141_183418\|\|lt2gene | 1227 | No | NA | 1 | 0 | 0 | Low-quality | Genome-fragment | 2.47 | AAI-based (high-confidence) | 0 |
| S11_k141_115796 | 1477 | No | NA | 2 | 0 | 1 | Not-determined | Genome-fragment | NA | NA | 0 |
| S11_k141_144040 | 1234 | No | NA | 2 | 0 | 1 | Not-determined | Genome-fragment | NA | NA | 0 |
| S11_k141_144059 | 1198 | No | NA | 2 | 0 | 2 | Not-determined | Genome-fragment | NA | NA | 0 |
| S11_k141_198852 | 5054 | No | NA | 3 | 0 | 2 | Not-determined | Genome-fragment | NA | NA | 0 |
| S11_k141_115886 | 9283 | No | NA | 7 | 0 | 4 | Not-determined | Genome-fragment | NA | NA | 0 |
| S11_k141_322586 | 1154 | No | NA | 2 | 0 | 1 | Not-determined | Genome-fragment | NA | NA | 0 |
| S11_k141_171838 | 1158 | No | NA | 1 | 0 | 0 | Not-determined | Genome-fragment | NA | NA | 0 |
| S11_k141_241187 | 1379 | No | NA | 2 | 0 | 1 | Not-determined | Genome-fragment | NA | NA | 0 |
| S11_k141_144145 | 1250 | No | NA | 1 | 0 | 1 | Not-determined | Genome-fragment | NA | NA | 0 |
| S11_k141_144163 | 1126 | No | NA | 3 | 0 | 0 | Not-determined | Genome-fragment | NA | NA | 0 |
| S11_k141_61574 | 1296 | No | NA | 1 | 0 | 1 | Not-determined | Genome-fragment | NA | NA | 0 |
| S11_k141_6203 | 45102 | No | NA | 41 | 1 | 9 | Medium-quality | Genome-fragment | 50.63 | HMM-based (lower-bound) | 0 |
| S11_k141_158032 | 2830 | No | NA | 2 | 0 | 1 | Not-determined | Genome-fragment | NA | NA | 0 |
| S9-final-viral-combined_k141_106820\|\|full | 1228 | No | NA | 3 | 0 | 0 | Low-quality | Genome-fragment | 0.69 | AAI-based (medium-confidence) | 0 |
| S11_k141_74646 | 2112 | No | NA | 2 | 0 | 1 | Not-determined | Genome-fragment | NA | NA | 0 |
| S11_k141_309932 | 3747 | No | NA | 5 | 0 | 2 | Not-determined | Genome-fragment | NA | NA | 0 |
| S11_k141_61636 | 1114 | No | NA | 1 | 0 | 1 | Not-determined | Genome-fragment | NA | NA | 0 |
| S11_k141_19395 | 2167 | No | NA | 4 | 0 | 2 | Not-determined | Genome-fragment | NA | NA | 0 |
| S11_k141_47732 | 3945 | No | NA | 4 | 0 | 0 | Not-determined | Genome-fragment | NA | NA | 0 |
| S11_k141_171937 | 1192 | No | NA | 3 | 0 | 1 | Not-determined | Genome-fragment | NA | NA | 0 |
| S11_k141_130151 | 1414 | No | NA | 2 | 0 | 2 | Not-determined | Genome-fragment | NA | NA | 0 |
| S11-final-viral-combined_k141_187000\|\|full | 1228 | No | NA | 3 | 0 | 0 | Low-quality | Genome-fragment | 1.27 | AAI-based (medium-confidence) | 0 |
| S11_k141_185280 | 3636 | No | NA | 5 | 0 | 2 | Not-determined | Genome-fragment | NA | NA | 0 |
| S11_k141_198945 | 3317 | No | NA | 3 | 0 | 2 | Not-determined | Genome-fragment | NA | NA | 0 |
| S11-final-viral-combined_k141_312468\|\|full | 1228 | No | NA | 2 | 0 | 1 | Low-quality | Genome-fragment | 2.96 | AAI-based (high-confidence) | 0 |
| S11_k141_268174 | 1195 | No | NA | 3 | 0 | 0 | Not-determined | Genome-fragment | NA | NA | 0 |
| S11_k141_144291 | 2479 | No | NA | 5 | 0 | 0 | Not-determined | Genome-fragment | NA | NA | 0 |
| S11_k141_185318 | 1741 | No | NA | 3 | 0 | 1 | Not-determined | Genome-fragment | NA | NA | 0 |
| S11_k141_310018 | 1254 | No | NA | 1 | 0 | 1 | Not-determined | Genome-fragment | NA | NA | 0 |
| S11_k141_6375 | 1093 | No | NA | 2 | 0 | 1 | Not-determined | Genome-fragment | NA | NA | 0 |
| S11_k141_241483 | 1285 | No | NA | 2 | 0 | 2 | Not-determined | Genome-fragment | NA | NA | 0 |
| S11_k141_268261 | 1051 | No | NA | 1 | 0 | 1 | Not-determined | Genome-fragment | NA | NA | 0 |
| S11_k141_310089 | 1408 | No | NA | 1 | 0 | 1 | Not-determined | Genome-fragment | NA | NA | 0 |
| S11_k141_322807 | 1090 | No | NA | 1 | 0 | 1 | Not-determined | Genome-fragment | NA | NA | 0 |
| S11-final-viral-combined_k141_137806\|\|full | 1228 | No | NA | 3 | 0 | 0 | Low-quality | Genome-fragment | 2.48 | AAI-based (medium-confidence) | 0 |
| S11_k141_19499 | 6399 | No | NA | 4 | 0 | 3 | Not-determined | Genome-fragment | NA | NA | 0 |
| S11_k141_310100 | 1113 | No | NA | 1 | 0 | 1 | Not-determined | Genome-fragment | NA | NA | 0 |
| S11_k141_172103 | 1235 | No | NA | 2 | 0 | 2 | Not-determined | Genome-fragment | NA | NA | 0 |
| S11_k141_89113 | 1322 | No | NA | 1 | 0 | 0 | Not-determined | Genome-fragment | NA | NA | 0 |
| S11_k141_310110 | 1484 | No | NA | 1 | 0 | 1 | Not-determined | Genome-fragment | NA | NA | 0 |
| S1-final-viral-combined_k141_285189\|\|full | 1228 | No | NA | 3 | 0 | 0 | Low-quality | Genome-fragment | 0.83 | AAI-based (medium-confidence) | 0 |
| S11_k141_213759 | 2562 | No | NA | 1 | 0 | 1 | Not-determined | Genome-fragment | NA | NA | 0 |
| S11_k141_61811 | 4298 | No | NA | 5 | 0 | 3 | Not-determined | Genome-fragment | NA | NA | 0 |
| S1-final-viral-combined_k141_194928\|\|full | 1228 | No | NA | 3 | 0 | 0 | Low-quality | Genome-fragment | 3.28 | AAI-based (high-confidence) | 0 |
| S11_k141_295614 | 4760 | No | NA | 2 | 0 | 0 | Not-determined | Genome-fragment | NA | NA | 0 |
| S11_k141_295622 | 1096 | No | NA | 1 | 0 | 1 | Not-determined | Genome-fragment | NA | NA | 0 |
| S11_k141_227122 | 1106 | No | NA | 1 | 0 | 1 | Not-determined | Genome-fragment | NA | NA | 0 |
| S11_k141_74823 | 1186 | No | NA | 4 | 0 | 3 | Not-determined | Genome-fragment | NA | NA | 0 |
| S11_k141_213817 | 1116 | No | NA | 2 | 0 | 0 | Not-determined | Genome-fragment | NA | NA | 0 |
| S11_k141_295627 | 3074 | No | NA | 2 | 0 | 2 | Not-determined | Genome-fragment | NA | NA | 0 |
| S1_k141_346514 | 1228 | No | NA | 4 | 0 | 0 | Low-quality | Genome-fragment | 22.21 | AAI-based (high-confidence) | 0 |
| S11_k141_116154 | 2118 | No | NA | 3 | 0 | 1 | Not-determined | Genome-fragment | NA | NA | 0 |
| S11_k141_241558 | 1612 | No | NA | 2 | 0 | 2 | Not-determined | Genome-fragment | NA | NA | 0 |
| S7-final-viral-combined_k141_49830\|\|full | 1228 | No | NA | 5 | 0 | 0 | Low-quality | Genome-fragment | 5.83 | AAI-based (high-confidence) | 0 |
| S17-final-viral-combined_k141_8181\|\|full | 1228 | No | NA | 2 | 0 | 0 | Low-quality | Genome-fragment | 2.76 | AAI-based (high-confidence) | 0 |
| S11_k141_310268 | 1331 | No | NA | 2 | 0 | 1 | Not-determined | Genome-fragment | NA | NA | 0 |
| S11_k141_268382 | 1524 | No | NA | 2 | 0 | 1 | Not-determined | Genome-fragment | NA | NA | 0 |
| S11_k141_213877 | 1050 | No | NA | 2 | 0 | 2 | Not-determined | Genome-fragment | NA | NA | 0 |
| S11_k141_34127 | 1028 | No | NA | 2 | 0 | 2 | Not-determined | Genome-fragment | NA | NA | 0 |
| S11_k141_158427 | 3104 | No | NA | 2 | 0 | 0 | Not-determined | Genome-fragment | NA | NA | 0 |
| S2-final-viral-combined_k141_28343\|\|full | 1228 | No | NA | 3 | 0 | 0 | Low-quality | Genome-fragment | 2.13 | AAI-based (high-confidence) | 0 |
| S4-final-viral-combined_k141_40149\|\|full | 1228 | No | NA | 2 | 0 | 1 | Low-quality | Genome-fragment | 0.71 | AAI-based (medium-confidence) | 0 |
| S4_k141_146941 | 1228 | No | NA | 1 | 0 | 0 | Low-quality | Genome-fragment | 0.58 | AAI-based (high-confidence) | 0 |
| S11_k141_172281 | 1007 | No | NA | 1 | 0 | 1 | Not-determined | Genome-fragment | NA | NA | 0 |
| S11_k141_89268 | 1684 | No | NA | 4 | 0 | 0 | Not-determined | Genome-fragment | NA | NA | 0 |
| S11_k141_268393 | 1846 | No | NA | 4 | 0 | 0 | Not-determined | Genome-fragment | NA | NA | 0 |
| S11_k141_74957 | 1393 | No | NA | 1 | 0 | 0 | Not-determined | Genome-fragment | NA | NA | 0 |
| S4_k141_18733 | 1228 | No | NA | 1 | 0 | 1 | Low-quality | Genome-fragment | 0.86 | AAI-based (high-confidence) | 0 |
| S11_k141_322932 | 1304 | No | NA | 1 | 0 | 1 | Not-determined | Genome-fragment | NA | NA | 0 |
| S11_k141_144480 | 2122 | No | NA | 4 | 0 | 3 | Not-determined | Genome-fragment | NA | NA | 0 |
| S11_k141_103668 | 1533 | No | NA | 2 | 0 | 0 | Not-determined | Genome-fragment | NA | NA | 0 |
| S6_k141_66074 | 1228 | No | NA | 2 | 0 | 1 | Low-quality | Genome-fragment | 2.38 | AAI-based (medium-confidence) | 0 |
| S11_k141_213922 | 1078 | No | NA | 2 | 0 | 1 | Not-determined | Genome-fragment | NA | NA | 0 |
| S8-final-viral-combined_k141_31823\|\|full | 1228 | No | NA | 3 | 2 | 0 | Low-quality | Genome-fragment | 1.86 | HMM-based (lower-bound) | 0 |
| S11_k141_241744 | 1013 | No | NA | 1 | 0 | 0 | Not-determined | Genome-fragment | NA | NA | 0 |
| S9-final-viral-combined_k141_35731\|\|full | 1229 | No | NA | 2 | 0 | 0 | Low-quality | Genome-fragment | 1.1 | AAI-based (medium-confidence) | 0 |
| S11_k141_75002 | 2947 | No | NA | 1 | 0 | 1 | Not-determined | Genome-fragment | NA | NA | 0 |
| S9-final-viral-combined_k141_97417\|\|lt2gene | 1229 | No | NA | 1 | 1 | 0 | Low-quality | Genome-fragment | 1.61 | HMM-based (lower-bound) | 0 |
| S11_k141_116361 | 1005 | No | NA | 2 | 0 | 1 | Not-determined | Genome-fragment | NA | NA | 0 |
| S11_k141_116381 | 3407 | No | NA | 1 | 0 | 0 | Not-determined | Genome-fragment | NA | NA | 0 |
| S11_k141_199273 | 1006 | No | NA | 1 | 0 | 1 | Not-determined | Genome-fragment | NA | NA | 0 |
| S11_k141_19801 | 1479 | No | NA | 2 | 0 | 2 | Not-determined | Genome-fragment | NA | NA | 0 |
| S11_k141_268564 | 3320 | No | NA | 6 | 0 | 0 | Not-determined | Genome-fragment | NA | NA | 0 |
| S11_k141_268578 | 9152 | No | NA | 5 | 0 | 3 | Not-determined | Genome-fragment | NA | NA | 0 |
| S11_k141_185659 | 1157 | No | NA | 1 | 0 | 1 | Not-determined | Genome-fragment | NA | NA | 0 |
| S11_k141_75075 | 1615 | No | NA | 1 | 0 | 0 | Not-determined | Genome-fragment | NA | NA | 0 |
| S1-final-viral-combined_k141_200958\|\|full | 1229 | No | NA | 2 | 1 | 0 | Low-quality | Genome-fragment | 4.28 | AAI-based (high-confidence) | 0 |
| S11_k141_227508 | 3797 | No | NA | 1 | 0 | 1 | Not-determined | Genome-fragment | NA | NA | 0 |
| S11_k141_214083 | 6181 | No | NA | 4 | 0 | 4 | Not-determined | Genome-fragment | NA | NA | 0 |
| S11_k141_34410 | 1059 | No | NA | 3 | 0 | 0 | Not-determined | Genome-fragment | NA | NA | 0 |
| S1-final-viral-combined_k141_270021\|\|lt2gene | 1229 | No | NA | 2 | 1 | 0 | Low-quality | Genome-fragment | 3.52 | AAI-based (high-confidence) | 0 |
| S11_k141_48130 | 1605 | No | NA | 3 | 0 | 2 | Not-determined | Genome-fragment | NA | NA | 0 |
| S5_k141_64176 | 1229 | No | NA | 2 | 1 | 0 | Low-quality | Genome-fragment | 2.45 | AAI-based (medium-confidence) | 0 |
| S11_k141_19927 | 1620 | No | NA | 2 | 0 | 1 | Not-determined | Genome-fragment | NA | NA | 0 |
| S11_k141_116530 | 1983 | No | NA | 1 | 0 | 1 | Not-determined | Genome-fragment | NA | NA | 0 |
| S11_k141_130559 | 1620 | No | NA | 3 | 0 | 2 | Not-determined | Genome-fragment | NA | NA | 0 |
| S11_k141_172510 | 1397 | No | NA | 2 | 0 | 1 | Not-determined | Genome-fragment | NA | NA | 0 |
| S11_k141_310651 | 2998 | No | NA | 1 | 0 | 1 | Not-determined | Genome-fragment | NA | NA | 0 |
| S11_k141_214185 | 2246 | No | NA | 1 | 0 | 0 | Not-determined | Genome-fragment | NA | NA | 0 |
| S11_k141_295963 | 1103 | No | NA | 1 | 0 | 0 | Not-determined | Genome-fragment | NA | NA | 0 |
| S11_k141_144801 | 1123 | No | NA | 2 | 0 | 0 | Not-determined | Genome-fragment | NA | NA | 0 |
| S11_k141_62383 | 1062 | No | NA | 1 | 0 | 1 | Not-determined | Genome-fragment | NA | NA | 0 |
| S11_k141_323109 | 1261 | No | NA | 1 | 0 | 1 | Not-determined | Genome-fragment | NA | NA | 0 |
| S17-k141_106903 | 1229 | No | NA | 1 | 0 | 1 | Low-quality | Genome-fragment | 0.33 | AAI-based (high-confidence) | 0 |
| S4_k141_31795 | 1229 | No | NA | 2 | 0 | 0 | Low-quality | Genome-fragment | 1.3 | AAI-based (high-confidence) | 0 |
| S11_k141_283072 | 1101 | No | NA | 2 | 0 | 1 | Not-determined | Genome-fragment | NA | NA | 0 |
| S8-final-viral-combined_k141_268542\|\|full | 1229 | No | NA | 2 | 0 | 1 | Low-quality | Genome-fragment | 0.86 | AAI-based (medium-confidence) | 0 |
| S11_k141_323163 | 1514 | No | NA | 1 | 0 | 1 | Not-determined | Genome-fragment | NA | NA | 0 |
| S8-final-viral-combined_k141_273811\|\|full | 1229 | No | NA | 3 | 0 | 0 | Low-quality | Genome-fragment | 0.8 | AAI-based (high-confidence) | 0 |
| S11_k141_283116 | 1342 | No | NA | 2 | 0 | 0 | Not-determined | Genome-fragment | NA | NA | 0 |
| S11_k141_89678 | 1087 | No | NA | 1 | 0 | 0 | Not-determined | Genome-fragment | NA | NA | 0 |
| S11_k141_34576 | 1117 | No | NA | 2 | 0 | 0 | Not-determined | Genome-fragment | NA | NA | 0 |
| S8-final-viral-combined_k141_265227\|\|lt2gene | 1229 | No | NA | 2 | 1 | 0 | Low-quality | Genome-fragment | 0.69 | HMM-based (lower-bound) | 0 |
| S9-final-viral-combined_k141_40092\|\|full | 1230 | No | NA | 2 | 0 | 1 | Low-quality | Genome-fragment | 1.52 | AAI-based (medium-confidence) | 0 |
| S11_k141_20129 | 3620 | No | NA | 4 | 0 | 3 | Not-determined | Genome-fragment | NA | NA | 0 |
| S11_k141_62504 | 1660 | No | NA | 2 | 0 | 0 | Not-determined | Genome-fragment | NA | NA | 0 |
| S11-final-viral-combined_k141_983\|\|full | 1230 | No | NA | 3 | 0 | 0 | Low-quality | Genome-fragment | 2.42 | AAI-based (high-confidence) | 0 |
| S11_k141_255327 | 1003 | No | NA | 1 | 0 | 0 | Not-determined | Genome-fragment | NA | NA | 0 |
| S11_k141_214335 | 4386 | No | NA | 5 | 0 | 4 | Not-determined | Genome-fragment | NA | NA | 0 |
| S11_k141_227747 | 3210 | No | NA | 2 | 0 | 1 | Not-determined | Genome-fragment | NA | NA | 0 |
| S11-final-viral-combined_k141_211973\|\|lt2gene | 1230 | No | NA | 1 | 1 | 0 | Low-quality | Genome-fragment | 0.48 | HMM-based (lower-bound) | 0 |
| S11_k141_145003 | 1991 | No | NA | 1 | 0 | 0 | Not-determined | Genome-fragment | NA | NA | 0 |
| S11_k141_268890 | 1413 | No | NA | 2 | 0 | 1 | Not-determined | Genome-fragment | NA | NA | 0 |
| S11_k141_62576 | 4067 | No | NA | 2 | 0 | 0 | Not-determined | Genome-fragment | NA | NA | 0 |
| S11_k141_48366 | 3355 | No | NA | 2 | 0 | 2 | Not-determined | Genome-fragment | NA | NA | 0 |

| | | | | | | | | | | | | |
|---|---|---|---|---|---|---|---|---|---|---|---|---|
| S11_k141_20234 | 2744 | No | NA | 3 | 0 | 2 | Not-determined | Genome-fragment | NA | | NA | 0 |
| S11_k141_242148 | 3701 | No | NA | 2 | 0 | 1 | Not-determined | Genome-fragment | NA | | NA | 0 |
| S11_k141_62593 | 1264 | No | NA | 3 | 0 | 3 | Not-determined | Genome-fragment | NA | | NA | 0 |
| S11_k141_80131 | 1230 | No | NA | 3 | 1 | 0 | Low-quality | Genome-fragment | | 2.66 | AAI-based (medium-confidence) | 0 |
| S5-final-viral-combined_k141_37717\|\|full | 1230 | No | NA | 3 | 1 | 0 | Low-quality | Genome-fragment | | 2.76 | AAI-based (high-confidence) | 0 |
| S11_k141_20253 | 1247 | No | NA | 3 | 0 | 1 | Not-determined | Genome-fragment | NA | | NA | 0 |
| S11_k141_268920 | 1702 | No | NA | 1 | 0 | 1 | Not-determined | Genome-fragment | NA | | NA | 0 |
| S11_k141_75440 | 3710 | No | NA | 3 | 0 | 3 | Not-determined | Genome-fragment | NA | | NA | 0 |
| S11_k141_130764 | 1683 | No | NA | 2 | 0 | 2 | Not-determined | Genome-fragment | NA | | NA | 0 |
| S11_k141_75470 | 1179 | No | NA | 1 | 0 | 1 | Not-determined | Genome-fragment | NA | | NA | 0 |
| S5-final-viral-combined_k141_22282\|\|full | 1230 | No | NA | 4 | 0 | 0 | Low-quality | Genome-fragment | | 1.67 | AAI-based (high-confidence) | 0 |
| S11_k141_323330 | 3156 | No | NA | 5 | 0 | 2 | Not-determined | Genome-fragment | NA | | NA | 0 |
| S11_k141_130808 | 1808 | No | NA | 3 | 0 | 0 | Not-determined | Genome-fragment | NA | | NA | 0 |
| S11_k141_323355 | 1833 | No | NA | 2 | 0 | 2 | Not-determined | Genome-fragment | NA | | NA | 0 |
| S11_k141_255519 | 1452 | No | NA | 1 | 0 | 1 | Not-determined | Genome-fragment | NA | | NA | 0 |
| S11_k141_242259 | 1192 | No | NA | 2 | 0 | 1 | Not-determined | Genome-fragment | NA | | NA | 0 |
| S11_k141_172904 | 1178 | No | NA | 1 | 0 | 0 | Not-determined | Genome-fragment | NA | | NA | 0 |
| S5_k141_17740 | 1230 | No | NA | 3 | 1 | 0 | Low-quality | Genome-fragment | | 3.6 | AAI-based (high-confidence) | 0 |
| S11_k141_130846 | 3405 | No | NA | 3 | 0 | 0 | Not-determined | Genome-fragment | NA | | NA | 0 |
| S7-final-viral-combined_k141_127117\|\|full | 1230 | No | NA | 3 | 0 | 0 | Low-quality | Genome-fragment | | 20.86 | AAI-based (medium-confidence) | 0 |
| S11_k141_89875 | 6523 | No | NA | 2 | 0 | 2 | Not-determined | Genome-fragment | NA | | NA | 0 |
| S11_k141_104253 | 1145 | No | NA | 1 | 0 | 0 | Not-determined | Genome-fragment | NA | | NA | 0 |
| S12_k141_318936 | 1230 | No | NA | 2 | 0 | 1 | Low-quality | Genome-fragment | | 0.8 | AAI-based (high-confidence) | 0 |
| S11_k141_34921 | 3306 | No | NA | 3 | 0 | 1 | Not-determined | Genome-fragment | NA | | NA | 0 |
| S11_k141_62773 | 1091 | No | NA | 2 | 0 | 0 | Not-determined | Genome-fragment | NA | | NA | 0 |
| S11_k141_228018 | 1115 | No | NA | 1 | 0 | 0 | Not-determined | Genome-fragment | NA | | NA | 0 |
| S11_k141_228021 | 1191 | No | NA | 3 | 0 | 0 | Not-determined | Genome-fragment | NA | | NA | 0 |
| S11_k141_62802 | 1105 | No | NA | 2 | 0 | 1 | Not-determined | Genome-fragment | NA | | NA | 0 |
| S8_k141_218140 | 1230 | No | NA | 2 | 0 | 0 | Low-quality | Genome-fragment | | 0.75 | AAI-based (medium-confidence) | 0 |
| S11_k141_116893 | 1458 | No | NA | 1 | 0 | 0 | Not-determined | Genome-fragment | NA | | NA | 0 |
| S11_k141_228072 | 1606 | No | NA | 1 | 0 | 1 | Not-determined | Genome-fragment | NA | | NA | 0 |
| S9-final-viral-combined_k141_86625\|\|full | 1231 | No | NA | 3 | 0 | 0 | Low-quality | Genome-fragment | | 3.15 | AAI-based (high-confidence) | 0 |
| S11_k141_130959 | 4611 | No | NA | 3 | 0 | 2 | Not-determined | Genome-fragment | NA | | NA | 0 |
| S11_k141_323507 | 3255 | No | NA | 2 | 0 | 1 | Not-determined | Genome-fragment | NA | | NA | 0 |
| S11_k141_255720 | 4258 | No | NA | 3 | 0 | 3 | Not-determined | Genome-fragment | NA | | NA | 0 |
| S11_k141_311103 | 1207 | No | NA | 2 | 0 | 1 | Not-determined | Genome-fragment | NA | | NA | 0 |
| S11_k141_323522 | 1631 | No | NA | 1 | 0 | 1 | Not-determined | Genome-fragment | NA | | NA | 0 |
| S11-final-viral-combined_k141_259525\|\|full | 1231 | No | NA | 2 | 0 | 1 | Low-quality | Genome-fragment | | 2.19 | AAI-based (medium-confidence) | 0 |
| S11-final-viral-combined_k141_126464\|\|lt2gene | 1231 | No | NA | 1 | 1 | 0 | Low-quality | Genome-fragment | | 2.77 | AAI-based (medium-confidence) | 0 |
| S11_k141_48693 | 1304 | No | NA | 2 | 0 | 1 | Not-determined | Genome-fragment | NA | | NA | 0 |
| S3-final-viral-combined_k141_61607\|\|full | 1231 | No | NA | 2 | 0 | 0 | Low-quality | Genome-fragment | | 1.28 | AAI-based (medium-confidence) | 0 |
| S11_k141_90106 | 1449 | No | NA | 1 | 0 | 1 | Not-determined | Genome-fragment | NA | | NA | 0 |
| S11_k141_104393 | 2290 | No | NA | 1 | 0 | 1 | Not-determined | Genome-fragment | NA | | NA | 0 |
| S5-final-viral-combined_k141_90994\|\|full | 1231 | No | NA | 2 | 1 | 0 | Low-quality | Genome-fragment | | 2.94 | AAI-based (medium-confidence) | 0 |
| S11_k141_48732 | 1525 | No | NA | 3 | 0 | 2 | Not-determined | Genome-fragment | NA | | NA | 0 |
| S5-final-viral-combined_k141_42096\|\|full | 1231 | No | NA | 3 | 0 | 0 | Low-quality | Genome-fragment | | 1.19 | AAI-based (medium-confidence) | 0 |
| S11_k141_62950 | 3859 | No | NA | 5 | 0 | 2 | Not-determined | Genome-fragment | NA | | NA | 0 |
| S5-final-viral-combined_k141_87428\|\|full | 1231 | No | NA | 3 | 0 | 0 | Low-quality | Genome-fragment | | 2.68 | AAI-based (medium-confidence) | 0 |
| S11_k141_242547 | 1964 | No | NA | 1 | 0 | 1 | Not-determined | Genome-fragment | NA | | NA | 0 |
| S11_k141_35219 | 1206 | No | NA | 2 | 0 | 0 | Not-determined | Genome-fragment | NA | | NA | 0 |
| S11_k141_186225 | 3100 | No | NA | 2 | 0 | 2 | Not-determined | Genome-fragment | NA | | NA | 0 |
| S7-final-viral-combined_k141_227967\|\|full | 1231 | No | NA | 3 | 0 | 0 | Low-quality | Genome-fragment | | 2.33 | AAI-based (high-confidence) | 0 |
| S11_k141_228205 | 1058 | No | NA | 1 | 0 | 1 | Not-determined | Genome-fragment | NA | | NA | 0 |
| S11_k141_200073 | 1173 | No | NA | 2 | 0 | 1 | Not-determined | Genome-fragment | NA | | NA | 0 |
| S11_k141_311320 | 1820 | No | NA | 2 | 0 | 0 | Not-determined | Genome-fragment | NA | | NA | 0 |
| S11_k141_159258 | 2012 | No | NA | 5 | 0 | 0 | Not-determined | Genome-fragment | NA | | NA | 0 |
| S11_k141_104432 | 3538 | No | NA | 2 | 0 | 2 | Not-determined | Genome-fragment | NA | | NA | 0 |
| S11_k141_145452 | 1035 | No | NA | 1 | 0 | 0 | Not-determined | Genome-fragment | NA | | NA | 0 |
| S11_k141_131220 | 1287 | No | NA | 2 | 0 | 2 | Not-determined | Genome-fragment | NA | | NA | 0 |
| S12-final-viral-combined_k141_146895\|\|full | 1231 | No | NA | 2 | 0 | 0 | Low-quality | Genome-fragment | | 2.64 | AAI-based (medium-confidence) | 0 |
| S11_k141_283597 | 1811 | No | NA | 1 | 0 | 0 | Not-determined | Genome-fragment | NA | | NA | 0 |
| S12-final-viral-combined_k141_309233\|\|lt2gene | 1231 | No | NA | 2 | 2 | 0 | Low-quality | Genome-fragment | | 2.5 | AAI-based (medium-confidence) | 0 |
| S11_k141_214826 | 1014 | No | NA | 1 | 0 | 1 | Not-determined | Genome-fragment | NA | | NA | 0 |
| S11_k141_269403 | 1395 | No | NA | 1 | 0 | 1 | Not-determined | Genome-fragment | NA | | NA | 0 |
| S11_k141_145471 | 1393 | No | NA | 2 | 0 | 2 | Not-determined | Genome-fragment | NA | | NA | 0 |
| S11_k141_186473 | 3066 | No | NA | 2 | 0 | 1 | Not-determined | Genome-fragment | NA | | NA | 0 |
| S11_k141_159380 | 1196 | No | NA | 2 | 0 | 2 | Not-determined | Genome-fragment | NA | | NA | 0 |
| S2-final-viral-combined_k141_75160\|\|full | 1231 | No | NA | 3 | 0 | 0 | Low-quality | Genome-fragment | | 3.39 | AAI-based (medium-confidence) | 0 |
| S6-final-viral-combined_k141_80841\|\|full | 1231 | No | NA | 2 | 0 | 0 | Low-quality | Genome-fragment | | 3.93 | AAI-based (medium-confidence) | 0 |
| S11_k141_173510 | 1720 | No | NA | 2 | 0 | 2 | Not-determined | Genome-fragment | NA | | NA | 0 |
| S11_k141_159404 | 1255 | No | NA | 2 | 0 | 0 | Not-determined | Genome-fragment | NA | | NA | 0 |
| S11-final-viral-combined_k141_86451\|\|full | 1232 | No | NA | 2 | 0 | 0 | Low-quality | Genome-fragment | | 2.62 | AAI-based (medium-confidence) | 0 |
| S11_k141_296800 | 1296 | No | NA | 1 | 0 | 1 | Not-determined | Genome-fragment | NA | | NA | 0 |
| S11_k141_214893 | 1021 | No | NA | 1 | 0 | 1 | Not-determined | Genome-fragment | NA | | NA | 0 |
| S11_k141_20701 | 1138 | No | NA | 2 | 0 | 1 | Not-determined | Genome-fragment | NA | | NA | 0 |
| S11_k141_200254 | 1164 | No | NA | 2 | 0 | 1 | Not-determined | Genome-fragment | NA | | NA | 0 |
| S11_k141_173540 | 1347 | No | NA | 1 | 0 | 1 | Not-determined | Genome-fragment | NA | | NA | 0 |
| S11-final-viral-combined_k141_214944\|\|full | 1232 | No | NA | 3 | 1 | 0 | Low-quality | Genome-fragment | | 2.93 | AAI-based (medium-confidence) | 0 |
| S11_k141_269510 | 1018 | No | NA | 1 | 0 | 1 | Not-determined | Genome-fragment | NA | | NA | 0 |
| S11_k141_35522 | 1412 | No | NA | 1 | 0 | 0 | Not-determined | Genome-fragment | NA | | NA | 0 |
| S11_k141_269523 | 1214 | No | NA | 1 | 0 | 1 | Not-determined | Genome-fragment | NA | | NA | 0 |
| S11_k141_242882 | 2548 | No | NA | 2 | 0 | 2 | Not-determined | Genome-fragment | NA | | NA | 0 |
| S11_k141_7859 | 1043 | No | NA | 1 | 0 | 0 | Not-determined | Genome-fragment | NA | | NA | 0 |
| S1-final-viral-combined_k141_352017\|\|full | 1232 | No | NA | 4 | 0 | 0 | Low-quality | Genome-fragment | | 3.05 | AAI-based (high-confidence) | 0 |
| S11_k141_269614 | 1018 | No | NA | 1 | 0 | 1 | Not-determined | Genome-fragment | NA | | NA | 0 |
| S11_k141_104841 | 4064 | No | NA | 3 | 0 | 3 | Not-determined | Genome-fragment | NA | | NA | 0 |
| S11_k141_117411 | 1039 | No | NA | 1 | 0 | 0 | Not-determined | Genome-fragment | NA | | NA | 0 |
| S11_k141_311538 | 4706 | No | NA | 3 | 0 | 3 | Not-determined | Genome-fragment | NA | | NA | 0 |
| S11_k141_76117 | 4320 | No | NA | 4 | 0 | 3 | Not-determined | Genome-fragment | NA | | NA | 0 |
| S11_k141_7952 | 2240 | No | NA | 2 | 0 | 0 | Not-determined | Genome-fragment | NA | | NA | 0 |
| S11_k141_256225 | 1127 | No | NA | 1 | 0 | 1 | Not-determined | Genome-fragment | NA | | NA | 0 |
| S11_k141_200413 | 1606 | No | NA | 3 | 0 | 0 | Not-determined | Genome-fragment | NA | | NA | 0 |
| S11_k141_49195 | 1120 | No | NA | 1 | 0 | 0 | Not-determined | Genome-fragment | NA | | NA | 0 |
| S11_k141_228642 | 1021 | No | NA | 2 | 0 | 0 | Not-determined | Genome-fragment | NA | | NA | 0 |
| S11_k141_323959 | 1157 | No | NA | 1 | 0 | 1 | Not-determined | Genome-fragment | NA | | NA | 0 |
| S7_k141_184430 | 1232 | No | NA | 1 | 0 | 0 | Low-quality | Genome-fragment | | 1.2 | AAI-based (high-confidence) | 0 |
| S11_k141_49202 | 1449 | No | NA | 2 | 0 | 2 | Not-determined | Genome-fragment | NA | | NA | 0 |
| S17-final-viral-combined_k141_109987\|\|full | 1232 | No | NA | 3 | 0 | 1 | Low-quality | Genome-fragment | | 1.67 | AAI-based (high-confidence) | 0 |
| S12-final-viral-combined_k141_305606\|\|full | 1232 | No | NA | 2 | 0 | 0 | Low-quality | Genome-fragment | | 1.97 | AAI-based (medium-confidence) | 0 |
| S11_k141_117607 | 1281 | No | NA | 1 | 0 | 0 | Not-determined | Genome-fragment | NA | | NA | 0 |
| S11_k141_63390 | 2892 | No | NA | 1 | 0 | 0 | Not-determined | Genome-fragment | NA | | NA | 0 |
| S12_k141_101223 | 1232 | No | NA | 3 | 2 | 0 | Low-quality | Genome-fragment | | 2.93 | AAI-based (high-confidence) | 0 |
| S11_k141_269752 | 1811 | No | NA | 1 | 0 | 1 | Not-determined | Genome-fragment | NA | | NA | 0 |
| S4-final-viral-combined_k141_166272\|\|full | 1232 | No | NA | 4 | 0 | 0 | Low-quality | Genome-fragment | | 3.31 | AAI-based (medium-confidence) | 0 |
| S11_k141_131501 | 1074 | No | NA | 2 | 0 | 0 | Not-determined | Genome-fragment | NA | | NA | 0 |
| S11_k141_311662 | 2152 | No | NA | 3 | 0 | 2 | Not-determined | Genome-fragment | NA | | NA | 0 |
| S4-final-viral-combined_k141_12297\|\|lt2gene | 1232 | No | NA | 1 | 1 | 0 | Low-quality | Genome-fragment | | 1.62 | AAI-based (high-confidence) | 0 |
| S11_k141_104981 | 2075 | No | NA | 2 | 0 | 1 | Not-determined | Genome-fragment | NA | | NA | 0 |
| S4_k141_158992 | 1232 | No | NA | 2 | 1 | 0 | Low-quality | Genome-fragment | | 0.71 | AAI-based (high-confidence) | 0 |
| S4_k141_27810 | 1232 | No | NA | 1 | 1 | 0 | Low-quality | Genome-fragment | | 0.4 | HMM-based (lower-bound) | 0 |
| S11_k141_173730 | 1104 | No | NA | 2 | 0 | 1 | Not-determined | Genome-fragment | NA | | NA | 0 |
| S11_k141_311717 | 5215 | No | NA | 1 | 0 | 1 | Not-determined | Genome-fragment | NA | | NA | 0 |
| S11_k141_311737 | 3593 | No | NA | 2 | 0 | 2 | Not-determined | Genome-fragment | NA | | NA | 0 |
| S11-final-viral-combined_k141_279270\|\|full | 1233 | No | NA | 2 | 0 | 0 | Low-quality | Genome-fragment | | 3.85 | AAI-based (high-confidence) | 0 |
| S11_k141_131558 | 1191 | No | NA | 1 | 0 | 1 | Not-determined | Genome-fragment | NA | | NA | 0 |
| S11_k141_269860 | 1127 | No | NA | 1 | 0 | 0 | Not-determined | Genome-fragment | NA | | NA | 0 |
| S11_k141_8187 | 1026 | No | NA | 2 | 0 | 0 | Not-determined | Genome-fragment | NA | | NA | 0 |
| S11_k141_49279 | 3264 | No | NA | 2 | 0 | 1 | Not-determined | Genome-fragment | NA | | NA | 0 |
| S11_k141_311789 | 1247 | No | NA | 2 | 0 | 1 | Not-determined | Genome-fragment | NA | | NA | 0 |
| S11_k141_269885 | 1289 | No | NA | 1 | 0 | 0 | Not-determined | Genome-fragment | NA | | NA | 0 |
| S11_k141_49322 | 4555 | No | NA | 3 | 0 | 1 | Not-determined | Genome-fragment | NA | | NA | 0 |
| S11_k141_311801 | 1723 | No | NA | 2 | 0 | 2 | Not-determined | Genome-fragment | NA | | NA | 0 |
| S11-final-viral-combined_k141_197395\|\|lt2gene | 1233 | No | NA | 1 | 1 | 0 | Low-quality | Genome-fragment | | 1.13 | HMM-based (lower-bound) | 0 |
| S11_k141_284004 | 1349 | No | NA | 2 | 0 | 1 | Not-determined | Genome-fragment | NA | | NA | 0 |
| S11_k141_76406 | 1123 | No | NA | 1 | 0 | 1 | Not-determined | Genome-fragment | NA | | NA | 0 |
| S1-final-viral-combined_k141_192956\|\|full | 1233 | No | NA | 3 | 0 | 0 | Low-quality | Genome-fragment | | 0.76 | AAI-based (medium-confidence) | 0 |
| S11_k141_269922 | 1066 | No | NA | 1 | 0 | 0 | Not-determined | Genome-fragment | NA | | NA | 0 |
| S11_k141_215201 | 2009 | No | NA | 2 | 0 | 2 | Not-determined | Genome-fragment | NA | | NA | 0 |
| S11_k141_105099 | 5115 | No | NA | 2 | 0 | 2 | Not-determined | Genome-fragment | NA | | NA | 0 |
| S11_k141_243219 | 4802 | No | NA | 3 | 0 | 2 | Not-determined | Genome-fragment | NA | | NA | 0 |
| S5-final-viral-combined_k141_198660\|\|lt2gene | 1233 | No | NA | 1 | 1 | 0 | Low-quality | Genome-fragment | | 0.76 | HMM-based (lower-bound) | 0 |
| S11_k141_173885 | 4421 | No | NA | 3 | 0 | 3 | Not-determined | Genome-fragment | NA | | NA | 0 |
| S11_k141_284136 | 1600 | No | NA | 1 | 0 | 0 | Not-determined | Genome-fragment | NA | | NA | 0 |
| S7-final-viral-combined_k141_11020\|\|full | 1233 | No | NA | 2 | 0 | 2 | Low-quality | Genome-fragment | | 2.72 | AAI-based (medium-confidence) | 0 |
| S11_k141_243270 | 2788 | No | NA | 4 | 0 | 0 | Not-determined | Genome-fragment | NA | | NA | 0 |
| S11_k141_90940 | 4050 | No | NA | 2 | 0 | 2 | Not-determined | Genome-fragment | NA | | NA | 0 |
| S11_k141_49483 | 2669 | No | NA | 4 | 0 | 4 | Not-determined | Genome-fragment | NA | | NA | 0 |
| S11_k141_187007 | 1190 | No | NA | 1 | 0 | 0 | Not-determined | Genome-fragment | NA | | NA | 0 |
| S11_k141_105237 | 3589 | No | NA | 1 | 0 | 1 | Not-determined | Genome-fragment | NA | | NA | 0 |
| S17-final-viral-combined_k141_167710\|\|full | 1233 | No | NA | 2 | 0 | 0 | Low-quality | Genome-fragment | | 1.93 | AAI-based (medium-confidence) | 0 |
| S17-final-viral-combined_k141_17736\|\|full | 1233 | No | NA | 3 | 0 | 0 | Low-quality | Genome-fragment | | 3.08 | AAI-based (high-confidence) | 0 |
| S11_k141_215367 | 4872 | No | NA | 4 | 0 | 3 | Not-determined | Genome-fragment | NA | | NA | 0 |
| S11_k141_187075 | 1151 | No | NA | 2 | 0 | 0 | Not-determined | Genome-fragment | NA | | NA | 0 |
| S11_k141_297373 | 1431 | No | NA | 1 | 0 | 0 | Not-determined | Genome-fragment | NA | | NA | 0 |
| S11_k141_36067 | 5147 | No | NA | 5 | 0 | 4 | Not-determined | Genome-fragment | NA | | NA | 0 |
| S17-final-viral-combined_k141_182284\|\|lt2gene | 1233 | No | NA | 2 | 1 | 0 | Low-quality | Genome-fragment | | 3.08 | AAI-based (medium-confidence) | 0 |
| S11_k141_117946 | 1182 | No | NA | 1 | 0 | 1 | Not-determined | Genome-fragment | NA | | NA | 0 |
| S11_k141_284305 | 4821 | No | NA | 4 | 0 | 3 | Not-determined | Genome-fragment | NA | | NA | 0 |
| S11_k141_228983 | 1778 | No | NA | 1 | 0 | 1 | Not-determined | Genome-fragment | NA | | NA | 0 |
| S17_k141_152454 | 1233 | No | NA | 2 | 0 | 0 | Low-quality | Genome-fragment | | 2.06 | AAI-based (medium-confidence) | 0 |
| S11_k141_63750 | 1094 | No | NA | 2 | 0 | 1 | Not-determined | Genome-fragment | NA | | NA | 0 |
| S11_k141_131817 | 2154 | No | NA | 2 | 0 | 0 | Not-determined | Genome-fragment | NA | | NA | 0 |
| S11_k141_187160 | 1357 | No | NA | 2 | 0 | 1 | Not-determined | Genome-fragment | NA | | NA | 0 |
| S11_k141_297407 | 8265 | No | NA | 5 | 0 | 4 | Not-determined | Genome-fragment | NA | | NA | 0 |
| S11_k141_256724 | 1692 | No | NA | 2 | 0 | 1 | Not-determined | Genome-fragment | NA | | NA | 0 |
| S11_k141_131829 | 2112 | No | NA | 2 | 0 | 2 | Not-determined | Genome-fragment | NA | | NA | 0 |
| S11_k141_284360 | 1068 | No | NA | 3 | 0 | 0 | Not-determined | Genome-fragment | NA | | NA | 0 |
| S11_k141_146200 | 1380 | No | NA | 2 | 0 | 1 | Not-determined | Genome-fragment | NA | | NA | 0 |
| S12-final-viral-combined_k141_218539\|\|full | 1233 | No | NA | 3 | 0 | 0 | Low-quality | Genome-fragment | | 1.7 | AAI-based (medium-confidence) | 0 |
| S11_k141_311999 | 3547 | No | NA | 3 | 0 | 3 | Not-determined | Genome-fragment | NA | | NA | 0 |
| S11_k141_297437 | 1958 | No | NA | 2 | 0 | 1 | Not-determined | Genome-fragment | NA | | NA | 0 |
| S11_k141_200935 | 6390 | No | NA | 3 | 0 | 2 | Not-determined | Genome-fragment | NA | | NA | 0 |
| S11_k141_324354 | 1024 | No | NA | 4 | 0 | 4 | Not-determined | Genome-fragment | NA | | NA | 0 |
| S11_k141_63767 | 4145 | No | NA | 3 | 0 | 2 | Not-determined | Genome-fragment | NA | | NA | 0 |
| S11_k141_118037 | 1133 | No | NA | 3 | 0 | 2 | Not-determined | Genome-fragment | NA | | NA | 0 |
| S11_k141_200951 | 1149 | No | NA | 2 | 0 | 1 | Not-determined | Genome-fragment | NA | | NA | 0 |
| S11_k141_200999 | 4704 | No | NA | 2 | 0 | 2 | Not-determined | Genome-fragment | NA | | NA | 0 |
| S11_k141_201004 | 1767 | No | NA | 2 | 0 | 2 | Not-determined | Genome-fragment | NA | | NA | 0 |
| S11_k141_76788 | 1046 | No | NA | 1 | 0 | 1 | Not-determined | Genome-fragment | NA | | NA | 0 |
| S11_k141_229105 | 1193 | No | NA | 1 | 0 | 1 | Not-determined | Genome-fragment | NA | | NA | 0 |
| S11_k141_256886 | 1529 | No | NA | 3 | 0 | 1 | Not-determined | Genome-fragment | NA | | NA | 0 |
| S11_k141_229129 | 1190 | No | NA | 1 | 0 | 1 | Not-determined | Genome-fragment | NA | | NA | 0 |
| S12-final-viral-combined_k141_231558\|\|full | 1233 | No | NA | 4 | 0 | 0 | Low-quality | Genome-fragment | | 1.02 | AAI-based (medium-confidence) | 0 |
| S11_k141_324401 | 4511 | No | NA | 2 | 0 | 1 | Not-determined | Genome-fragment | NA | | NA | 0 |
| S11_k141_76846 | 1173 | No | NA | 1 | 0 | 0 | Not-determined | Genome-fragment | NA | | NA | 0 |
| S11_k141_21408 | 1377 | No | NA | 2 | 0 | 1 | Not-determined | Genome-fragment | NA | | NA | 0 |
| S11_k141_146378 | 1090 | No | NA | 1 | 0 | 0 | Not-determined | Genome-fragment | NA | | NA | 0 |
| S11_k141_91215 | 1489 | No | NA | 2 | 0 | 1 | Not-determined | Genome-fragment | NA | | NA | 0 |
| S12-final-viral-combined_k141_231530\|\|lt2gene | 1233 | No | NA | 1 | 1 | 0 | Low-quality | Genome-fragment | | 0.47 | HMM-based (lower-bound) | 0 |
| S4-final-viral-combined_k141_148690\|\|full | 1233 | No | NA | 2 | 0 | 0 | Low-quality | Genome-fragment | | 2.13 | AAI-based (high-confidence) | 0 |
| S4_k141_117760 | 1233 | No | NA | 2 | 0 | 0 | Low-quality | Genome-fragment | | 0.53 | AAI-based (medium-confidence) | 0 |
| S11_k141_324474 | 3179 | No | NA | 4 | 0 | 4 | Not-determined | Genome-fragment | NA | | NA | 0 |
| S11_k141_49875 | 1025 | No | NA | 1 | 0 | 1 | Not-determined | Genome-fragment | NA | | NA | 0 |
| S11_k141_8849 | 3857 | No | NA | 2 | 0 | 1 | Not-determined | Genome-fragment | NA | | NA | 0 |
| S8-final-viral-combined_k141_271227\|\|full | 1233 | No | NA | 2 | 0 | 0 | Low-quality | Genome-fragment | | 1.79 | AAI-based (medium-confidence) | 0 |
| S9-final-viral-combined_k141_82235\|\|full | 1234 | No | NA | 3 | 1 | 0 | Low-quality | Genome-fragment | | 1.99 | AAI-based (high-confidence) | 0 |
| S17-final-viral-combined_k141_116998\|\|full | 1234 | No | NA | 3 | 0 | 0 | Low-quality | Genome-fragment | | 2 | AAI-based (medium-confidence) | 0 |
| S4-final-viral-combined_k141_160636\|\|full | 1234 | No | NA | 3 | 0 | 2 | Low-quality | Genome-fragment | | 0.87 | AAI-based (medium-confidence) | 0 |
| S11_k141_63849 | 1999 | No | NA | 2 | 0 | 1 | Not-determined | Genome-fragment | NA | | NA | 0 |
| S11_k141_8895 | 3624 | No | NA | 3 | 0 | 3 | Not-determined | Genome-fragment | NA | | NA | 0 |
| S11_k141_118189 | 1502 | No | NA | 4 | 0 | 4 | Not-determined | Genome-fragment | NA | | NA | 0 |
| S11_k141_160324 | 1117 | No | NA | 2 | 0 | 1 | Not-determined | Genome-fragment | NA | | NA | 0 |
| S11_k141_91268 | 7536 | No | NA | 3 | 0 | 2 | Not-determined | Genome-fragment | NA | | NA | 0 |
| S8-final-viral-combined_k141_172064\|\|full | 1234 | No | NA | 1 | 0 | 1 | Low-quality | Genome-fragment | | 2.32 | AAI-based (high-confidence) | 0 |
| S11_k141_243628 | 1214 | No | NA | 2 | 0 | 2 | Not-determined | Genome-fragment | NA | | NA | 0 |
| S11_k141_243649 | 3228 | No | NA | 2 | 0 | 1 | Not-determined | Genome-fragment | NA | | NA | 0 |
| S11_k141_105653 | 1479 | No | NA | 3 | 0 | 1 | Not-determined | Genome-fragment | NA | | NA | 0 |
| S9-final-viral-combined_k141_79303\|\|full | 1235 | No | NA | 2 | 0 | 0 | Low-quality | Genome-fragment | | 2.79 | AAI-based (medium-confidence) | 0 |
| S11_k141_257056 | 1053 | No | NA | 2 | 0 | 1 | Not-determined | Genome-fragment | NA | | NA | 0 |
| S11-final-viral-combined_k141_399\|\|full | 1235 | No | NA | 3 | 1 | 0 | Low-quality | Genome-fragment | | 3.39 | AAI-based (medium-confidence) | 0 |
| S11_k141_284533 | 1000 | No | NA | 1 | 0 | 0 | Not-determined | Genome-fragment | NA | | NA | 0 |
| S11_k141_270247 | 1198 | No | NA | 2 | 0 | 1 | Not-determined | Genome-fragment | NA | | NA | 0 |
| S11_k141_49994 | 3046 | No | NA | 4 | 0 | 1 | Not-determined | Genome-fragment | NA | | NA | 0 |
| S11_k141_312264 | 1312 | No | NA | 1 | 0 | 0 | Not-determined | Genome-fragment | NA | | NA | 0 |
| S11_k141_324527 | 1002 | No | NA | 1 | 0 | 1 | Not-determined | Genome-fragment | NA | | NA | 0 |
| S11_k141_201265 | 1629 | No | NA | 3 | 0 | 0 | Not-determined | Genome-fragment | NA | | NA | 0 |
| S1-final-viral-combined_k141_54451\|\|full | 1235 | No | NA | 2 | 0 | 2 | Low-quality | Genome-fragment | | 0.39 | AAI-based (medium-confidence) | 0 |
| S11_k141_187442 | 1720 | No | NA | 3 | 0 | 2 | Not-determined | Genome-fragment | NA | | NA | 0 |
| S1-final-viral-combined_k141_386392\|\|full | 1235 | No | NA | 4 | 1 | 0 | Low-quality | Genome-fragment | | 1.86 | AAI-based (high-confidence) | 0 |
| S11_k141_312324 | 1047 | No | NA | 1 | 0 | 0 | Not-determined | Genome-fragment | NA | | NA | 0 |
| S11_k141_105758 | 4371 | No | NA | 10 | 0 | 0 | Not-determined | Genome-fragment | NA | | NA | 0 |
| S11_k141_105766 | 1141 | No | NA | 1 | 0 | 1 | Not-determined | Genome-fragment | NA | | NA | 0 |
| S11_k141_21535 | 1015 | No | NA | 2 | 0 | 1 | Not-determined | Genome-fragment | NA | | NA | 0 |
| S1_k141_177159 | 1235 | No | NA | 3 | 2 | 0 | Low-quality | Genome-fragment | | 1.99 | AAI-based (high-confidence) | 0 |
| S11_k141_201344 | 6828 | No | NA | 8 | 0 | 3 | Not-determined | Genome-fragment | NA | | NA | 0 |
| S11_k141_21572 | 3393 | No | NA | 4 | 0 | 4 | Not-determined | Genome-fragment | NA | | NA | 0 |
| S5-final-viral-combined_k141_22059\|\|full | 1235 | No | NA | 3 | 1 | 0 | Low-quality | Genome-fragment | | 3.52 | AAI-based (high-confidence) | 0 |
| S11_k141_21622 | 5846 | No | NA | 7 | 0 | 1 | Not-determined | Genome-fragment | NA | | NA | 0 |
| S11_k141_50239 | 1176 | No | NA | 1 | 0 | 0 | Not-determined | Genome-fragment | NA | | NA | 0 |
| S11_k141_297793 | 1658 | No | NA | 1 | 0 | 1 | Not-determined | Genome-fragment | NA | | NA | 0 |
| S11_k141_201429 | 1129 | No | NA | 1 | 0 | 1 | Not-determined | Genome-fragment | NA | | NA | 0 |
| S5-final-viral-combined_k141_131787\|\|full | 1235 | No | NA | 4 | 3 | 0 | Low-quality | Genome-fragment | | 2.89 | AAI-based (high-confidence) | 0 |
| S11_k141_297808 | 3713 | No | NA | 3 | 0 | 3 | Not-determined | Genome-fragment | NA | | NA | 0 |
| S11_k141_270427 | 1605 | No | NA | 2 | 0 | 0 | Not-determined | Genome-fragment | NA | | NA | 0 |
| S11_k141_297837 | 2502 | No | NA | 2 | 0 | 1 | Not-determined | Genome-fragment | NA | | NA | 0 |
| S11_k141_36536 | 1022 | No | NA | 2 | 0 | 1 | Not-determined | Genome-fragment | NA | | NA | 0 |
| S11_k141_284762 | 2754 | No | NA | 1 | 0 | 0 | Not-determined | Genome-fragment | NA | | NA | 0 |
| S11_k141_270475 | 1157 | No | NA | 2 | 0 | 0 | Not-determined | Genome-fragment | NA | | NA | 0 |
| S17-final-viral-combined_k141_112910\|\|full | 1235 | No | NA | 3 | 0 | 0 | Low-quality | Genome-fragment | | 2.22 | AAI-based (high-confidence) | 0 |
| S11_k141_229574 | 1466 | No | NA | 3 | 0 | 1 | Not-determined | Genome-fragment | NA | | NA | 0 |
| S11_k141_91546 | 1237 | No | NA | 1 | 0 | 1 | Not-determined | Genome-fragment | NA | | NA | 0 |
| S17-final-viral-combined_k141_195822\|\|full | 1235 | No | NA | 4 | 0 | 0 | Low-quality | Genome-fragment | | 3.16 | AAI-based (medium-confidence) | 0 |
| S11_k141_324811 | 1613 | No | NA | 1 | 0 | 1 | Not-determined | Genome-fragment | NA | | NA | 0 |
| S2-final-viral-combined_k141_101394\|\|full | 1235 | No | NA | 2 | 0 | 0 | Low-quality | Genome-fragment | | 2.29 | AAI-based (high-confidence) | 0 |
| S11_k141_284840 | 1018 | No | NA | 1 | 0 | 0 | Not-determined | Genome-fragment | NA | | NA | 0 |
| S11_k141_229649 | 1359 | No | NA | 1 | 0 | 1 | Not-determined | Genome-fragment | NA | | NA | 0 |
| S11_k141_118583 | 3069 | No | NA | 3 | 0 | 2 | Not-determined | Genome-fragment | NA | | NA | 0 |
| S11_k141_229654 | 1511 | No | NA | 1 | 0 | 1 | Not-determined | Genome-fragment | NA | | NA | 0 |
| S6_k141_127704 | 1235 | No | NA | 3 | 0 | 1 | Low-quality | Genome-fragment | | 1.56 | AAI-based (medium-confidence) | 0 |
| S11_k141_64247 | 1008 | No | NA | 2 | 0 | 1 | Not-determined | Genome-fragment | NA | | NA | 0 |
| S11_k141_270561 | 1015 | No | NA | 2 | 0 | 1 | Not-determined | Genome-fragment | NA | | NA | 0 |
| S11_k141_160723 | 3362 | No | NA | 3 | 0 | 3 | Not-determined | Genome-fragment | NA | | NA | 0 |
| S11_k141_77228 | 3519 | No | NA | 3 | 0 | 3 | Not-determined | Genome-fragment | NA | | NA | 0 |
| S8-final-viral-combined_k141_172723\|\|full | 1235 | No | NA | 3 | 0 | 0 | Low-quality | Genome-fragment | | 3.15 | AAI-based (medium-confidence) | 0 |
| S11_k141_174658 | 1257 | No | NA | 2 | 0 | 1 | Not-determined | Genome-fragment | NA | | NA | 0 |
| S11_k141_324915 | 6418 | No | NA | 2 | 0 | 1 | Not-determined | Genome-fragment | NA | | NA | 0 |
| S8-final-viral-combined_k141_11041\|\|full | 1235 | No | NA | 3 | 0 | 1 | Low-quality | Genome-fragment | | 0.86 | AAI-based (medium-confidence) | 0 |
| S11_k141_243985 | 4033 | No | NA | 1 | 0 | 0 | Not-determined | Genome-fragment | NA | | NA | 0 |
| S11_k141_257478 | 3505 | No | NA | 3 | 0 | 0 | Not-determined | Genome-fragment | NA | | NA | 0 |
| S11_k141_201669 | 1005 | No | NA | 1 | 0 | 0 | Not-determined | Genome-fragment | NA | | NA | 0 |
| S11_k141_118665 | 1907 | No | NA | 2 | 0 | 2 | Not-determined | Genome-fragment | NA | | NA | 0 |
| S9-final-viral-combined_k141_79124\|\|lt2gene | 1236 | No | NA | 1 | 1 | 0 | Low-quality | Genome-fragment | | 3.07 | AAI-based (high-confidence) | 0 |
| S5-final-viral-combined_k141_16579\|\|full | 1236 | No | NA | 2 | 1 | 0 | Low-quality | Genome-fragment | | 2.34 | AAI-based (medium-confidence) | 0 |
| S11_k141_257502 | 3124 | No | NA | 4 | 0 | 1 | Not-determined | Genome-fragment | NA | | NA | 0 |
| S11_k141_257508 | 6092 | No | NA | 7 | 0 | 3 | Not-determined | Genome-fragment | NA | | NA | 0 |
| S7-final-viral-combined_k141_27129\|\|full | 1236 | No | NA | 4 | 1 | 0 | Low-quality | Genome-fragment | | 2.83 | AAI-based (high-confidence) | 0 |
| S11_k141_201716 | 1211 | No | NA | 1 | 0 | 1 | Not-determined | Genome-fragment | NA | | NA | 0 |
| S7_k141_54765 | 1236 | No | NA | 2 | 1 | 0 | Low-quality | Genome-fragment | | 2.24 | HMM-based (lower-bound) | 0 |
| S11_k141_174743 | 1008 | No | NA | 1 | 0 | 1 | Not-determined | Genome-fragment | NA | | NA | 0 |
| S11_k141_312656 | 1020 | No | NA | 1 | 0 | 1 | Not-determined | Genome-fragment | NA | | NA | 0 |
| S11_k141_36785 | 1176 | No | NA | 2 | 0 | 1 | Not-determined | Genome-fragment | NA | | NA | 0 |
| S2-final-viral-combined_k141_67502\|\|full | 1236 | No | NA | 3 | 0 | 2 | Low-quality | Genome-fragment | | 0.5 | AAI-based (medium-confidence) | 0 |
| S11_k141_216016 | 1025 | No | NA | 1 | 0 | 0 | Not-determined | Genome-fragment | NA | | NA | 0 |
| S11_k141_146961 | 1042 | No | NA | 2 | 0 | 2 | Not-determined | Genome-fragment | NA | | NA | 0 |
| S11_k141_21847 | 1521 | No | NA | 1 | 0 | 1 | Not-determined | Genome-fragment | NA | | NA | 0 |
| S11_k141_270774 | 3079 | No | NA | 3 | 0 | 0 | Not-determined | Genome-fragment | NA | | NA | 0 |
| S11_k141_36840 | 1042 | No | NA | 1 | 0 | 0 | Not-determined | Genome-fragment | NA | | NA | 0 |
| S11_k141_118792 | 6573 | No | NA | 7 | 0 | 5 | Not-determined | Genome-fragment | NA | | NA | 0 |
| S8_k141_27172 | 1236 | No | NA | 3 | 0 | 0 | Low-quality | Genome-fragment | | 2 | AAI-based (medium-confidence) | 0 |
| S11_k141_147008 | 1005 | No | NA | 1 | 0 | 1 | Not-determined | Genome-fragment | NA | | NA | 0 |
| S11-final-viral-combined_k141_231733\|\|full | 1237 | No | NA | 3 | 1 | 0 | Low-quality | Genome-fragment | | 0.75 | HMM-based (lower-bound) | 0 |
| S11_k141_312781 | 1087 | No | NA | 1 | 0 | 1 | Not-determined | Genome-fragment | NA | | NA | 0 |
| S1-final-viral-combined_k141_29523\|\|full | 1237 | No | NA | 3 | 0 | 0 | Low-quality | Genome-fragment | | 2.49 | AAI-based (high-confidence) | 0 |
| S11_k141_201880 | 13760 | No | NA | 13 | 0 | 12 | Not-determined | Genome-fragment | NA | | NA | 0 |
| S11_k141_201884 | 1002 | No | NA | 1 | 0 | 1 | Not-determined | Genome-fragment | NA | | NA | 0 |
| S5-final-viral-combined_k141_121263\|\|full | 1237 | No | NA | 3 | 1 | 0 | Low-quality | Genome-fragment | | 2.24 | AAI-based (medium-confidence) | 0 |
| S11_k141_64400 | 1114 | No | NA | 3 | 0 | 0 | Not-determined | Genome-fragment | NA | | NA | 0 |
| S11_k141_64410 | 1660 | No | NA | 1 | 0 | 1 | Not-determined | Genome-fragment | NA | | NA | 0 |
| S11_k141_161025 | 1355 | No | NA | 2 | 0 | 1 | Not-determined | Genome-fragment | NA | | NA | 0 |
| S11_k141_312812 | 3782 | No | NA | 2 | 0 | 0 | Not-determined | Genome-fragment | NA | | NA | 0 |
| S11_k141_201984 | 3899 | No | NA | 4 | 0 | 2 | Not-determined | Genome-fragment | NA | | NA | 0 |
| S11_k141_106378 | 1378 | No | NA | 2 | 0 | 1 | Not-determined | Genome-fragment | NA | | NA | 0 |
| S11_k141_201993 | 1111 | No | NA | 1 | 0 | 1 | Not-determined | Genome-fragment | NA | | NA | 0 |
| S11_k141_132332 | 1319 | No | NA | 3 | 0 | 0 | Not-determined | Genome-fragment | NA | | NA | 0 |
| S11_k141_325187 | 6715 | No | NA | 9 | 0 | 3 | Not-determined | Genome-fragment | NA | | NA | 0 |
| S5-final-viral-combined_k141_68743\|\|lt2gene | 1237 | No | NA | 1 | 1 | 0 | Low-quality | Genome-fragment | | 2.15 | HMM-based (lower-bound) | 0 |
| S11_k141_270881 | 1093 | No | NA | 1 | 0 | 1 | Not-determined | Genome-fragment | NA | | NA | 0 |
| S11_k141_312906 | 1055 | No | NA | 2 | 0 | 1 | Not-determined | Genome-fragment | NA | | NA | 0 |
| S11_k141_64506 | 1322 | No | NA | 1 | 0 | 0 | Not-determined | Genome-fragment | NA | | NA | 0 |
| S5_k141_98771 | 1237 | No | NA | 1 | 0 | 0 | Low-quality | Genome-fragment | | 2.53 | AAI-based (medium-confidence) | 0 |
| S11_k141_188172 | 1027 | No | NA | 2 | 0 | 2 | Not-determined | Genome-fragment | NA | | NA | 0 |
| S11_k141_230016 | 4361 | No | NA | 3 | 0 | 2 | Not-determined | Genome-fragment | NA | | NA | 0 |
| S11_k141_106474 | 8121 | No | NA | 8 | 0 | 5 | Not-determined | Genome-fragment | NA | | NA | 0 |
| S11_k141_106491 | 1286 | No | NA | 1 | 0 | 0 | Not-determined | Genome-fragment | NA | | NA | 0 |
| S11_k141_257827 | 1074 | No | NA | 2 | 0 | 1 | Not-determined | Genome-fragment | NA | | NA | 0 |
| S2-final-viral-combined_k141_33422\|\|full | 1237 | No | NA | 3 | 0 | 1 | Low-quality | Genome-fragment | | 1.69 | AAI-based (medium-confidence) | 0 |
| S11_k141_270907 | 1173 | No | NA | 1 | 0 | 1 | Not-determined | Genome-fragment | NA | | NA | 0 |
| S11_k141_147244 | 1608 | No | NA | 2 | 0 | 0 | Not-determined | Genome-fragment | NA | | NA | 0 |
| S11_k141_202183 | 1826 | No | NA | 1 | 0 | 1 | Not-determined | Genome-fragment | NA | | NA | 0 |
| S11_k141_325317 | 1668 | No | NA | 2 | 0 | 2 | Not-determined | Genome-fragment | NA | | NA | 0 |
| S9_k141_9494 | 1238 | No | NA | 4 | 0 | 0 | Low-quality | Genome-fragment | | 3.78 | AAI-based (medium-confidence) | 0 |
| S11_k141_285473 | 3504 | No | NA | 3 | 0 | 1 | Not-determined | Genome-fragment | NA | | NA | 0 |
| S11-final-viral-combined_k141_39438\|\|full | 1238 | No | NA | 3 | 0 | 2 | Low-quality | Genome-fragment | | 1.95 | AAI-based (medium-confidence) | 0 |
| S11_k141_257935 | 1556 | No | NA | 2 | 0 | 1 | Not-determined | Genome-fragment | NA | | NA | 0 |
| S11_k141_244498 | 1498 | No | NA | 2 | 0 | 0 | Not-determined | Genome-fragment | NA | | NA | 0 |
| S11_k141_175166 | 1098 | No | NA | 1 | 0 | 1 | Not-determined | Genome-fragment | NA | | NA | 0 |
| S11_k141_302054 | 1238 | No | NA | 3 | 0 | 0 | Low-quality | Genome-fragment | | 2.27 | AAI-based (medium-confidence) | 0 |
| S11_k141_161264 | 1090 | No | NA | 1 | 0 | 1 | Not-determined | Genome-fragment | NA | | NA | 0 |
| S11_k141_325406 | 1087 | No | NA | 1 | 0 | 1 | Not-determined | Genome-fragment | NA | | NA | 0 |
| S11_k141_92143 | 1224 | No | NA | 2 | 0 | 0 | Not-determined | Genome-fragment | NA | | NA | 0 |
| S1-final-viral-combined_k141_329408\|\|full | 1238 | No | NA | 3 | 0 | 0 | Low-quality | Genome-fragment | | 2.85 | AAI-based (medium-confidence) | 0 |
| S11_k141_132554 | 3759 | No | NA | 4 | 0 | 3 | Not-determined | Genome-fragment | NA | | NA | 0 |
| S11_k141_77822 | 1217 | No | NA | 1 | 0 | 1 | Not-determined | Genome-fragment | NA | | NA | 0 |
| S3_k141_93549 | 1238 | No | NA | 1 | 0 | 1 | Low-quality | Genome-fragment | | 0.33 | AAI-based (medium-confidence) | 0 |
| S5-final-viral-combined_k141_187618\|\|full | 1238 | No | NA | 4 | 1 | 0 | Low-quality | Genome-fragment | | 2.85 | AAI-based (medium-confidence) | 0 |
| S7-final-viral-combined_k141_46216\|\|full | 1238 | No | NA | 4 | 0 | 1 | Low-quality | Genome-fragment | | 1.11 | AAI-based (medium-confidence) | 0 |
| S11_k141_175239 | 1008 | No | NA | 1 | 0 | 1 | Not-determined | Genome-fragment | NA | | NA | 0 |
| S11_k141_64765 | 2631 | No | NA | 3 | 0 | 0 | Not-determined | Genome-fragment | NA | | NA | 0 |
| S11_k141_51214 | 7051 | No | NA | 9 | 0 | 6 | Not-determined | Genome-fragment | NA | | NA | 0 |
| S11_k141_51219 | 5074 | No | NA | 3 | 0 | 0 | Not-determined | Genome-fragment | NA | | NA | 0 |
| S11_k141_244593 | 1142 | No | NA | 4 | 0 | 0 | Not-determined | Genome-fragment | NA | | NA | 0 |
| S11_k141_64777 | 1798 | No | NA | 1 | 0 | 0 | Not-determined | Genome-fragment | NA | | NA | 0 |
| S11_k141_298585 | 1125 | No | NA | 1 | 0 | 0 | Not-determined | Genome-fragment | NA | | NA | 0 |
| S11_k141_175294 | 1003 | No | NA | 1 | 0 | 1 | Not-determined | Genome-fragment | NA | | NA | 0 |
| S11_k141_106732 | 3295 | No | NA | 2 | 0 | 2 | Not-determined | Genome-fragment | NA | | NA | 0 |
| S11_k141_10024 | 2283 | No | NA | 2 | 0 | 2 | Not-determined | Genome-fragment | NA | | NA | 0 |
| S11_k141_92388 | 1471 | No | NA | 1 | 0 | 1 | Not-determined | Genome-fragment | NA | | NA | 0 |
| S11_k141_119381 | 1046 | No | NA | 2 | 0 | 1 | Not-determined | Genome-fragment | NA | | NA | 0 |
| S11_k141_188491 | 1075 | No | NA | 2 | 0 | 1 | Not-determined | Genome-fragment | NA | | NA | 0 |
| S11_k141_325535 | 2195 | No | NA | 2 | 0 | 1 | Not-determined | Genome-fragment | NA | | NA | 0 |
| S11_k141_77928 | 1448 | No | NA | 3 | 0 | 1 | Not-determined | Genome-fragment | NA | | NA | 0 |
| S11_k141_106772 | 7204 | No | NA | 5 | 0 | 0 | Not-determined | Genome-fragment | NA | | NA | 0 |
| S11_k141_37405 | 1282 | No | NA | 1 | 0 | 0 | Not-determined | Genome-fragment | NA | | NA | 0 |
| S11_k141_22205 | 3540 | No | NA | 5 | 0 | 4 | Not-determined | Genome-fragment | NA | | NA | 0 |
| S7_k141_243336 | 1238 | No | NA | 3 | 0 | 3 | Low-quality | Genome-fragment | | 3.23 | AAI-based (high-confidence) | 0 |
| S11_k141_106812 | 2519 | No | NA | 1 | 0 | 1 | Not-determined | Genome-fragment | NA | | NA | 0 |
| S11_k141_175439 | 1012 | No | NA | 3 | 0 | 1 | Not-determined | Genome-fragment | NA | | NA | 0 |
| S17-final-viral-combined_k141_125357\|\|full | 1238 | No | NA | 3 | 0 | 2 | Low-quality | Genome-fragment | | 0.9 | AAI-based (medium-confidence) | 0 |
| S11_k141_106832 | 3963 | No | NA | 3 | 0 | 1 | Not-determined | Genome-fragment | NA | | NA | 0 |
| S17-final-viral-combined_k141_83549\|\|full | 1238 | No | NA | 2 | 0 | 0 | Low-quality | Genome-fragment | | 1.35 | AAI-based (medium-confidence) | 0 |
| S11_k141_51400 | 5370 | No | NA | 3 | 0 | 3 | Not-determined | Genome-fragment | NA | | NA | 0 |
| S17-final-viral-combined_k141_60519\|\|full | 1238 | No | NA | 2 | 1 | 0 | Low-quality | Genome-fragment | | 2.55 | AAI-based (high-confidence) | 0 |
| S11_k141_313382 | 2284 | No | NA | 2 | 0 | 2 | Not-determined | Genome-fragment | NA | | NA | 0 |
| S12-final-viral-combined_k141_331875\|\|lt2gene | 1238 | No | NA | 2 | 2 | 0 | Low-quality | Genome-fragment | | 9.62 | AAI-based (medium-confidence) | 0 |
| S11_k141_298787 | 1911 | No | NA | 2 | 0 | 2 | Not-determined | Genome-fragment | NA | | NA | 0 |
| S11_k141_22345 | 1135 | No | NA | 2 | 0 | 2 | Not-determined | Genome-fragment | NA | | NA | 0 |
| S4-final-viral-combined_k141_34844\|\|full | 1238 | No | NA | 3 | 2 | 0 | Low-quality | Genome-fragment | | 3.47 | AAI-based (high-confidence) | 0 |
| S11_k141_313402 | 2209 | No | NA | 4 | 0 | 2 | Not-determined | Genome-fragment | NA | | NA | 0 |
| S11_k141_10123 | 1229 | No | NA | 3 | 0 | 1 | Not-determined | Genome-fragment | NA | | NA | 0 |
| S11_k141_230488 | 1161 | No | NA | 1 | 0 | 0 | Not-determined | Genome-fragment | NA | | NA | 0 |
| S11_k141_10144 | 1180 | No | NA | 1 | 0 | 1 | Not-determined | Genome-fragment | NA | | NA | 0 |
| S11_k141_119535 | 4371 | No | NA | 4 | 0 | 2 | Not-determined | Genome-fragment | NA | | NA | 0 |
| S11_k141_313467 | 1041 | No | NA | 1 | 0 | 0 | Not-determined | Genome-fragment | NA | | NA | 0 |
| S11_k141_258391 | 1027 | No | NA | 2 | 0 | 1 | Not-determined | Genome-fragment | NA | | NA | 0 |
| S11_k141_271385 | 1935 | No | NA | 1 | 0 | 0 | Not-determined | Genome-fragment | NA | | NA | 0 |
| S11_k141_22475 | 1231 | No | NA | 2 | 0 | 2 | Not-determined | Genome-fragment | NA | | NA | 0 |
| S11_k141_78266 | 1817 | No | NA | 1 | 0 | 0 | Not-determined | Genome-fragment | NA | | NA | 0 |
| S11_k141_271461 | 1320 | No | NA | 1 | 0 | 1 | Not-determined | Genome-fragment | NA | | NA | 0 |
| S4-final-viral-combined_k141_147053\|\|full | 1238 | No | NA | 3 | 0 | 2 | Low-quality | Genome-fragment | | 1.16 | AAI-based (medium-confidence) | 0 |
| S8-final-viral-combined_k141_98942\|\|full | 1238 | No | NA | 4 | 1 | 0 | Low-quality | Genome-fragment | | 2.97 | AAI-based (high-confidence) | 0 |
| S11_k141_188691 | 1448 | No | NA | 2 | 0 | 1 | Not-determined | Genome-fragment | NA | | NA | 0 |
| S11_k141_188697 | 1069 | No | NA | 2 | 0 | 1 | Not-determined | Genome-fragment | NA | | NA | 0 |
| S11_k141_325756 | 1462 | No | NA | 1 | 0 | 1 | Not-determined | Genome-fragment | NA | | NA | 0 |
| S11_k141_175671 | 1024 | No | NA | 1 | 0 | 0 | Not-determined | Genome-fragment | NA | | NA | 0 |
| S11_k141_258513 | 1008 | No | NA | 2 | 0 | 0 | Not-determined | Genome-fragment | NA | | NA | 0 |
| S8-final-viral-combined_k141_113379\|\|full | 1238 | No | NA | 3 | 0 | 2 | Low-quality | Genome-fragment | | 0.66 | AAI-based (medium-confidence) | 0 |
| S11_k141_51724 | 1547 | No | NA | 3 | 0 | 1 | Not-determined | Genome-fragment | NA | | NA | 0 |
| S11_k141_78315 | 6288 | No | NA | 7 | 0 | 3 | Not-determined | Genome-fragment | NA | | NA | 0 |
| S11_k141_188763 | 1305 | No | NA | 1 | 0 | 1 | Not-determined | Genome-fragment | NA | | NA | 0 |
| S11_k141_271551 | 1033 | No | NA | 2 | 0 | 1 | Not-determined | Genome-fragment | NA | | NA | 0 |
| S9-final-viral-combined_k141_55092\|\|full | 1239 | No | NA | 3 | 0 | 0 | Low-quality | Genome-fragment | | 1.91 | AAI-based (medium-confidence) | 0 |
| S11_k141_230715 | 1042 | No | NA | 1 | 0 | 0 | Not-determined | Genome-fragment | NA | | NA | 0 |
| S11_k141_133089 | 2354 | No | NA | 2 | 0 | 0 | Not-determined | Genome-fragment | NA | | NA | 0 |
| S11_k141_299016 | 2991 | No | NA | 1 | 0 | 1 | Not-determined | Genome-fragment | NA | | NA | 0 |
| S11_k141_217004 | 5728 | No | NA | 4 | 0 | 3 | Not-determined | Genome-fragment | NA | | NA | 0 |
| S11-final-viral-combined_k141_95848\|\|full | 1239 | No | NA | 3 | 0 | 0 | Low-quality | Genome-fragment | | 2.31 | AAI-based (medium-confidence) | 0 |
| S11_k141_107189 | 3327 | No | NA | 5 | 0 | 1 | Not-determined | Genome-fragment | NA | | NA | 0 |
| S11_k141_175792 | 2178 | No | NA | 2 | 0 | 1 | Not-determined | Genome-fragment | NA | | NA | 0 |
| S17-final-viral-combined_k141_257896\|\|full | 1239 | No | NA | 3 | 0 | 1 | Low-quality | Genome-fragment | | 0.59 | AAI-based (medium-confidence) | 0 |
| S11_k141_37858 | 1140 | No | NA | 1 | 0 | 1 | Not-determined | Genome-fragment | NA | | NA | 0 |
| S11_k141_202984 | 1905 | No | NA | 4 | 0 | 1 | Not-determined | Genome-fragment | NA | | NA | 0 |
| S17-final-viral-combined_k141_163119\|\|full | 1239 | No | NA | 3 | 0 | 0 | Low-quality | Genome-fragment | | 0.89 | AAI-based (medium-confidence) | 0 |
| S11_k141_286045 | 1177 | No | NA | 3 | 0 | 0 | Not-determined | Genome-fragment | NA | | NA | 0 |
| S11_k141_22797 | 1122 | No | NA | 2 | 0 | 1 | Not-determined | Genome-fragment | NA | | NA | 0 |
| S11_k141_286065 | 1066 | No | NA | 1 | 0 | 0 | Not-determined | Genome-fragment | NA | | NA | 0 |
| S11_k141_188942 | 2904 | No | NA | 4 | 0 | 0 | Not-determined | Genome-fragment | NA | | NA | 0 |
| S11_k141_203053 | 1141 | No | NA | 1 | 0 | 1 | Not-determined | Genome-fragment | NA | | NA | 0 |
| S11_k141_188974 | 1130 | No | NA | 2 | 0 | 1 | Not-determined | Genome-fragment | NA | | NA | 0 |
| S11_k141_325956 | 2321 | No | NA | 1 | 0 | 0 | Not-determined | Genome-fragment | NA | | NA | 0 |
| S12-final-viral-combined_k141_20393\|\|full | 1239 | No | NA | 2 | 0 | 1 | Low-quality | Genome-fragment | | 0.87 | AAI-based (medium-confidence) | 0 |
| S11_k141_325968 | 13756 | No | NA | 14 | 0 | 4 | Not-determined | Genome-fragment | NA | | NA | 0 |
| S11_k141_217188 | 1483 | No | NA | 1 | 0 | 1 | Not-determined | Genome-fragment | NA | | NA | 0 |
| S11_k141_258765 | 1086 | No | NA | 1 | 0 | 0 | Not-determined | Genome-fragment | NA | | NA | 0 |
| S11_k141_299273 | 1141 | No | NA | 1 | 0 | 1 | Not-determined | Genome-fragment | NA | | NA | 0 |
| S11_k141_245235 | 1687 | No | NA | 4 | 0 | 3 | Not-determined | Genome-fragment | NA | | NA | 0 |
| S11_k141_325998 | 1041 | No | NA | 2 | 0 | 2 | Not-determined | Genome-fragment | NA | | NA | 0 |
| S11_k141_119974 | 1245 | No | NA | 2 | 0 | 1 | Not-determined | Genome-fragment | NA | | NA | 0 |
| S12_k141_287867 | 1239 | No | NA | 2 | 0 | 0 | Low-quality | Genome-fragment | | 2.2 | AAI-based (high-confidence) | 0 |
| S2-final-viral-combined_k141_17602\|\|lt2gene | 1239 | No | NA | 2 | 2 | 0 | Low-quality | Genome-fragment | | 4.51 | HMM-based (lower-bound) | 0 |
| S11_k141_189035 | 1041 | No | NA | 2 | 0 | 0 | Not-determined | Genome-fragment | NA | | NA | 0 |
| S11_k141_22882 | 3084 | No | NA | 3 | 0 | 3 | Not-determined | Genome-fragment | NA | | NA | 0 |
| S11_k141_10588 | 1409 | No | NA | 1 | 0 | 1 | Not-determined | Genome-fragment | NA | | NA | 0 |
| S11_k141_78616 | 1378 | No | NA | 2 | 0 | 0 | Not-determined | Genome-fragment | NA | | NA | 0 |
| S2_k141_157116 | 1239 | No | NA | 2 | 1 | 0 | Low-quality | Genome-fragment | | 4.2 | HMM-based (lower-bound) | 0 |
| S11_k141_93056 | 1790 | No | NA | 2 | 0 | 0 | Not-determined | Genome-fragment | NA | | NA | 0 |
| S4-final-viral-combined_k141_29409\|\|full | 1239 | No | NA | 4 | 1 | 0 | Low-quality | Genome-fragment | | 3.56 | AAI-based (high-confidence) | 0 |
| S11_k141_52143 | 1047 | No | NA | 2 | 0 | 0 | Not-determined | Genome-fragment | NA | | NA | 0 |
| S4_k141_149820 | 1239 | No | NA | 2 | 0 | 2 | Low-quality | Genome-fragment | | 2.08 | AAI-based (medium-confidence) | 0 |
| S8_k141_26796 | 1239 | No | NA | 2 | 0 | 0 | Low-quality | Genome-fragment | | 1.16 | AAI-based (high-confidence) | 0 |
| S8_k141_10245 | 1239 | No | NA | 1 | 0 | 0 | Low-quality | Genome-fragment | | 1.26 | AAI-based (high-confidence) | 0 |
| S11_k141_176121 | 1238 | No | NA | 2 | 0 | 2 | Not-determined | Genome-fragment | NA | | NA | 0 |
| S11-final-viral-combined_k141_153026\|\|full | 1240 | No | NA | 5 | 0 | 0 | Low-quality | Genome-fragment | | 2.71 | AAI-based (high-confidence) | 0 |
| S11_k141_162197 | 1150 | No | NA | 1 | 0 | 1 | Not-determined | Genome-fragment | NA | | NA | 0 |

| | | | | | | | | | | | |
|---|---|---|---|---|---|---|---|---|---|---|---|
| S11_k141_286285 | 1267 | No | NA | 3 | 0 | 0 | Not-determined | Genome-fragment | NA | NA | 0 |
| S11-final-viral-combined_k141_268505\|\|full | 1240 | No | NA | 2 | 0 | 1 | Low-quality | Genome-fragment | 0.73 | AAI-based (medium-confidence) | 0 |
| S11_k141_271886 | 1529 | No | NA | 1 | 0 | 1 | Not-determined | Genome-fragment | NA | NA | 0 |
| S11-final-viral-combined_k141_160226\|\|full | 1240 | No | NA | 4 | 1 | 0 | Low-quality | Genome-fragment | 1.61 | AAI-based (high-confidence) | 0 |
| S11_k141_299501 | 3074 | No | NA | 4 | 0 | 3 | Not-determined | Genome-fragment | NA | NA | 0 |
| S1-final-viral-combined_k141_33912\|\|full | 1240 | No | NA | 3 | 0 | 0 | Low-quality | Genome-fragment | 0.98 | AAI-based (medium-confidence) | 0 |
| S3_k141_99966 | 1240 | No | NA | 2 | 0 | 0 | Low-quality | Genome-fragment | 0.81 | AAI-based (high-confidence) | 0 |
| S3_k141_8814 | 1240 | No | NA | 1 | 0 | 1 | Low-quality | Genome-fragment | 0.87 | AAI-based (high-confidence) | 0 |
| S5-final-viral-combined_k141_168657\|\|full | 1240 | No | NA | 5 | 4 | 0 | Low-quality | Genome-fragment | 3.07 | AAI-based (medium-confidence) | 0 |
| S11_k141_107618 | 11448 | No | NA | 11 | 0 | 10 | Not-determined | Genome-fragment | NA | NA | 0 |
| S5-final-viral-combined_k141_204359\|\|full | 1240 | No | NA | 6 | 1 | 0 | Low-quality | Genome-fragment | 2.87 | AAI-based (medium-confidence) | 0 |
| S11_k141_133535 | 3719 | No | NA | 2 | 0 | 1 | Not-determined | Genome-fragment | NA | NA | 0 |
| S11_k141_107672 | 1446 | No | NA | 1 | 0 | 1 | Not-determined | Genome-fragment | NA | NA | 0 |
| S11_k141_245533 | 1072 | No | NA | 1 | 0 | 1 | Not-determined | Genome-fragment | NA | NA | 0 |
| S11_k141_231187 | 1353 | No | NA | 2 | 0 | 2 | Not-determined | Genome-fragment | NA | NA | 0 |
| S5-final-viral-combined_k141_136011\|\|lt2gene | 1240 | No | NA | 2 | 2 | 0 | Low-quality | Genome-fragment | 2.96 | AAI-based (medium-confidence) | 0 |
| S12-final-viral-combined_k141_153804\|\|full | 1240 | No | NA | 3 | 0 | 1 | Low-quality | Genome-fragment | 2.84 | AAI-based (high-confidence) | 0 |
| S11_k141_245620 | 2657 | No | NA | 4 | 0 | 2 | Not-determined | Genome-fragment | NA | NA | 0 |
| S11_k141_162416 | 1047 | No | NA | 1 | 0 | 0 | Not-determined | Genome-fragment | NA | NA | 0 |
| S11_k141_286497 | 5594 | No | NA | 5 | 0 | 3 | Not-determined | Genome-fragment | NA | NA | 0 |
| S12-final-viral-combined_k141_276585\|\|full | 1240 | No | NA | 2 | 0 | 2 | Low-quality | Genome-fragment | 1.65 | AAI-based (medium-confidence) | 0 |
| S2-final-viral-combined_k141_148107\|\|full | 1240 | No | NA | 2 | 2 | 0 | Low-quality | Genome-fragment | 3.07 | AAI-based (high-confidence) | 0 |
| S11_k141_65988 | 1066 | No | NA | 1 | 0 | 0 | Not-determined | Genome-fragment | NA | NA | 0 |
| S11_k141_259105 | 1206 | No | NA | 3 | 0 | 2 | Not-determined | Genome-fragment | NA | NA | 0 |
| S2-final-viral-combined_k141_81580\|\|full | 1240 | No | NA | 3 | 0 | 1 | Low-quality | Genome-fragment | 1.57 | AAI-based (medium-confidence) | 0 |
| S4-final-viral-combined_k141_159098\|\|full | 1240 | No | NA | 3 | 1 | 0 | Low-quality | Genome-fragment | 2.96 | AAI-based (high-confidence) | 0 |
| S11_k141_176327 | 1857 | No | NA | 2 | 0 | 1 | Not-determined | Genome-fragment | NA | NA | 0 |
| S11_k141_272042 | 7974 | No | NA | 12 | 0 | 3 | Not-determined | Genome-fragment | NA | NA | 0 |
| S4_k141_35287 | 1240 | No | NA | 4 | 0 | 0 | Low-quality | Genome-fragment | 1.24 | AAI-based (high-confidence) | 0 |
| S8-final-viral-combined_k141_245197\|\|full | 1240 | No | NA | 5 | 0 | 0 | Low-quality | Genome-fragment | 3.41 | AAI-based (high-confidence) | 0 |
| S11_k141_38455 | 1202 | No | NA | 1 | 0 | 0 | Not-determined | Genome-fragment | NA | NA | 0 |
| S11_k141_217562 | 1315 | No | NA | 3 | 0 | 2 | Not-determined | Genome-fragment | NA | NA | 0 |
| S11_k141_78886 | 1145 | No | NA | 1 | 0 | 1 | Not-determined | Genome-fragment | NA | NA | 0 |
| S1-final-viral-combined_k141_321653\|\|full | 1241 | No | NA | 3 | 0 | 0 | Low-quality | Genome-fragment | 3.35 | AAI-based (high-confidence) | 0 |
| S11_k141_217596 | 1542 | No | NA | 1 | 0 | 0 | Not-determined | Genome-fragment | NA | NA | 0 |
| S1-final-viral-combined_k141_8498\|\|full | 1241 | No | NA | 3 | 0 | 0 | Low-quality | Genome-fragment | 2.2 | AAI-based (high-confidence) | 0 |
| S11_k141_272105 | 16657 | No | NA | 14 | 0 | 12 | Not-determined | Genome-fragment | NA | NA | 0 |
| S11_k141_93539 | 1030 | No | NA | 1 | 0 | 1 | Not-determined | Genome-fragment | NA | NA | 0 |
| S11_k141_10911 | 1074 | No | NA | 1 | 0 | 1 | Not-determined | Genome-fragment | NA | NA | 0 |
| S11_k141_189371 | 3670 | No | NA | 4 | 0 | 1 | Not-determined | Genome-fragment | NA | NA | 0 |
| S11_k141_38531 | 1353 | No | NA | 2 | 0 | 2 | Not-determined | Genome-fragment | NA | NA | 0 |
| S11_k141_299810 | 1469 | No | NA | 2 | 0 | 0 | Not-determined | Genome-fragment | NA | NA | 0 |
| S11_k141_38554 | 1443 | No | NA | 5 | 0 | 4 | Not-determined | Genome-fragment | NA | NA | 0 |
| S11_k141_162643 | 1000 | No | NA | 2 | 0 | 2 | Not-determined | Genome-fragment | NA | NA | 0 |
| S11_k141_79032 | 1006 | No | NA | 1 | 0 | 1 | Not-determined | Genome-fragment | NA | NA | 0 |
| S11_k141_299864 | 2172 | No | NA | 2 | 0 | 1 | Not-determined | Genome-fragment | NA | NA | 0 |
| S11_k141_259360 | 1401 | No | NA | 4 | 0 | 4 | Not-determined | Genome-fragment | NA | NA | 0 |
| S11_k141_176490 | 1571 | No | NA | 3 | 0 | 1 | Not-determined | Genome-fragment | NA | NA | 0 |
| S11_k141_120259 | 2207 | No | NA | 3 | 0 | 0 | Not-determined | Genome-fragment | NA | NA | 0 |
| S11_k141_133991 | 1098 | No | NA | 1 | 0 | 1 | Not-determined | Genome-fragment | NA | NA | 0 |
| S17-final-viral-combined_k141_119017\|\|full | 1241 | No | NA | 3 | 0 | 0 | Low-quality | Genome-fragment | 0.82 | AAI-based (medium-confidence) | 0 |
| S11_k141_134004 | 1710 | No | NA | 1 | 0 | 1 | Not-determined | Genome-fragment | NA | NA | 0 |
| S17-final-viral-combined_k141_143004\|\|full | 1241 | No | NA | 3 | 1 | 0 | Low-quality | Genome-fragment | 3.49 | AAI-based (medium-confidence) | 0 |
| S11_k141_286748 | 2022 | No | NA | 2 | 0 | 2 | Not-determined | Genome-fragment | NA | NA | 0 |
| S17-final-viral-combined_k141_109414\|\|full | 1241 | No | NA | 4 | 1 | 0 | Low-quality | Genome-fragment | 2.63 | AAI-based (medium-confidence) | 0 |
| S11_k141_217700 | 1149 | No | NA | 1 | 0 | 0 | Not-determined | Genome-fragment | NA | NA | 0 |
| S11_k141_52760 | 1009 | No | NA | 2 | 0 | 1 | Not-determined | Genome-fragment | NA | NA | 0 |
| S11_k141_23477 | 2050 | No | NA | 1 | 0 | 1 | Not-determined | Genome-fragment | NA | NA | 0 |
| S12-final-viral-combined_k141_61405\|\|full | 1241 | No | NA | 3 | 2 | 0 | Low-quality | Genome-fragment | 2.66 | AAI-based (high-confidence) | 0 |
| S11_k141_79182 | 1244 | No | NA | 1 | 0 | 0 | Not-determined | Genome-fragment | NA | NA | 0 |
| S11_k141_176640 | 1149 | No | NA | 2 | 0 | 2 | Not-determined | Genome-fragment | NA | NA | 0 |
| S12-final-viral-combined_k141_212896\|\|full | 1241 | No | NA | 2 | 0 | 0 | Low-quality | Genome-fragment | 3.07 | AAI-based (medium-confidence) | 0 |
| S11_k141_108102 | 3027 | No | NA | 1 | 0 | 1 | Not-determined | Genome-fragment | NA | NA | 0 |
| S12_k141_323563 | 1241 | No | NA | 1 | 0 | 1 | Low-quality | Genome-fragment | 0.8 | AAI-based (high-confidence) | 0 |
| S11_k141_11210 | 1254 | No | NA | 2 | 0 | 0 | Not-determined | Genome-fragment | NA | NA | 0 |
| S11_k141_134180 | 1829 | No | NA | 2 | 0 | 0 | Not-determined | Genome-fragment | NA | NA | 0 |
| S11_k141_203817 | 1266 | No | NA | 1 | 0 | 1 | Not-determined | Genome-fragment | NA | NA | 0 |
| S11_k141_326644 | 1001 | No | NA | 1 | 0 | 0 | Not-determined | Genome-fragment | NA | NA | 0 |
| S11_k141_326661 | 3827 | No | NA | 4 | 0 | 4 | Not-determined | Genome-fragment | NA | NA | 0 |
| S11_k141_120505 | 4952 | No | NA | 3 | 0 | 2 | Not-determined | Genome-fragment | NA | NA | 0 |
| S2_k141_99960 | 1241 | No | NA | 4 | 0 | 0 | Low-quality | Genome-fragment | 2.99 | AAI-based (medium-confidence) | 0 |
| S11_k141_79316 | 1050 | No | NA | 1 | 0 | 0 | Not-determined | Genome-fragment | NA | NA | 0 |
| S11_k141_300179 | 1398 | No | NA | 1 | 0 | 1 | Not-determined | Genome-fragment | NA | NA | 0 |
| S11_k141_286979 | 5397 | No | NA | 3 | 0 | 2 | Not-determined | Genome-fragment | NA | NA | 0 |
| S11_k141_286984 | 1043 | No | NA | 2 | 0 | 0 | Not-determined | Genome-fragment | NA | NA | 0 |
| S11_k141_203924 | 1313 | No | NA | 1 | 0 | 0 | Not-determined | Genome-fragment | NA | NA | 0 |
| S11_k141_38940 | 1192 | No | NA | 1 | 0 | 0 | Not-determined | Genome-fragment | NA | NA | 0 |
| S11_k141_148902 | 1362 | No | NA | 1 | 0 | 1 | Not-determined | Genome-fragment | NA | NA | 0 |
| S11_k141_108236 | 2090 | No | NA | 5 | 0 | 0 | Not-determined | Genome-fragment | NA | NA | 0 |
| S11_k141_23607 | 1377 | No | NA | 1 | 0 | 1 | Not-determined | Genome-fragment | NA | NA | 0 |
| S11_k141_53001 | 1750 | No | NA | 2 | 0 | 1 | Not-determined | Genome-fragment | NA | NA | 0 |
| S11_k141_246159 | 1143 | No | NA | 1 | 0 | 1 | Not-determined | Genome-fragment | NA | NA | 0 |
| S11_k141_66478 | 1039 | No | NA | 1 | 0 | 0 | Not-determined | Genome-fragment | NA | NA | 0 |
| S11_k141_148956 | 1051 | No | NA | 1 | 0 | 0 | Not-determined | Genome-fragment | NA | NA | 0 |
| S2_k141_196633 | 1241 | No | NA | 1 | 0 | 0 | Low-quality | Genome-fragment | 0.8 | AAI-based (high-confidence) | 0 |
| S4-final-viral-combined_k141_143599\|\|full | 1241 | No | NA | 2 | 1 | 0 | Low-quality | Genome-fragment | 2.94 | AAI-based (high-confidence) | 0 |
| S11_k141_246210 | 1212 | No | NA | 2 | 0 | 0 | Not-determined | Genome-fragment | NA | NA | 0 |
| S11_k141_326784 | 4573 | No | NA | 3 | 0 | 2 | Not-determined | Genome-fragment | NA | NA | 0 |
| S8-final-viral-combined_k141_78024\|\|lt2gene | 1241 | No | NA | 2 | 2 | 0 | Low-quality | Genome-fragment | 2.05 | AAI-based (high-confidence) | 0 |
| S11_k141_319127 | 1242 | No | NA | 3 | 0 | 0 | Low-quality | Genome-fragment | 2.13 | AAI-based (high-confidence) | 0 |
| S1-final-viral-combined_k141_100712\|\|full | 1242 | No | NA | 4 | 0 | 0 | Low-quality | Genome-fragment | 2.51 | AAI-based (medium-confidence) | 0 |
| S11_k141_53137 | 3481 | No | NA | 2 | 0 | 2 | Not-determined | Genome-fragment | NA | NA | 0 |
| S1-final-viral-combined_k141_102843\|\|full | 1242 | No | NA | 3 | 0 | 0 | Low-quality | Genome-fragment | 5 | AAI-based (medium-confidence) | 0 |
| S3-final-viral-combined_k141_85250\|\|full | 1242 | No | NA | 2 | 0 | 0 | Low-quality | Genome-fragment | 0.99 | AAI-based (medium-confidence) | 0 |
| S7-final-viral-combined_k141_120748\|\|full | 1242 | No | NA | 3 | 0 | 3 | Low-quality | Genome-fragment | 0.98 | AAI-based (medium-confidence) | 0 |
| S11_k141_287146 | 2322 | No | NA | 2 | 0 | 2 | Not-determined | Genome-fragment | NA | NA | 0 |
| S7_k141_99173 | 1242 | No | NA | 1 | 0 | 1 | Low-quality | Genome-fragment | 0.33 | AAI-based (high-confidence) | 0 |
| S11_k141_108393 | 1420 | No | NA | 3 | 0 | 3 | Not-determined | Genome-fragment | NA | NA | 0 |
| S11_k141_120710 | 3682 | No | NA | 2 | 0 | 0 | Not-determined | Genome-fragment | NA | NA | 0 |
| S11_k141_23784 | 4481 | No | NA | 4 | 0 | 3 | Not-determined | Genome-fragment | NA | NA | 0 |
| S17-final-viral-combined_k141_202776\|\|full | 1242 | No | NA | 2 | 0 | 2 | Low-quality | Genome-fragment | 0.86 | AAI-based (medium-confidence) | 0 |
| S11_k141_300384 | 2412 | No | NA | 4 | 0 | 1 | Not-determined | Genome-fragment | NA | NA | 0 |
| S11_k141_272738 | 1302 | No | NA | 2 | 0 | 1 | Not-determined | Genome-fragment | NA | NA | 0 |
| S11_k141_246377 | 1124 | No | NA | 2 | 0 | 0 | Not-determined | Genome-fragment | NA | NA | 0 |
| S11_k141_176949 | 5860 | No | NA | 4 | 0 | 3 | Not-determined | Genome-fragment | NA | NA | 0 |
| S17-final-viral-combined_k141_5417\|\|full | 1242 | No | NA | 4 | 0 | 0 | Low-quality | Genome-fragment | 1.89 | AAI-based (high-confidence) | 0 |
| S17-final-viral-combined_k141_66351\|\|lt2gene | 1242 | No | NA | 2 | 1 | 0 | Low-quality | Genome-fragment | 2 | HMM-based (lower-bound) | 0 |
| S11_k141_300440 | 1305 | No | NA | 1 | 0 | 1 | Not-determined | Genome-fragment | NA | NA | 0 |
| S11_k141_314514 | 1025 | No | NA | 2 | 0 | 0 | Not-determined | Genome-fragment | NA | NA | 0 |
| S11_k141_108469 | 1040 | No | NA | 1 | 0 | 1 | Not-determined | Genome-fragment | NA | NA | 0 |
| S11_k141_39149 | 2533 | No | NA | 3 | 0 | 0 | Not-determined | Genome-fragment | NA | NA | 0 |
| S11_k141_246471 | 1127 | No | NA | 1 | 0 | 1 | Not-determined | Genome-fragment | NA | NA | 0 |
| S11_k141_287287 | 1266 | No | NA | 2 | 0 | 1 | Not-determined | Genome-fragment | NA | NA | 0 |
| S11_k141_79665 | 1159 | No | NA | 1 | 0 | 1 | Not-determined | Genome-fragment | NA | NA | 0 |
| S11_k141_66793 | 1224 | No | NA | 2 | 0 | 1 | Not-determined | Genome-fragment | NA | NA | 0 |
| S11_k141_11514 | 2155 | No | NA | 1 | 0 | 1 | Not-determined | Genome-fragment | NA | NA | 0 |
| S17_k141_247546 | 1242 | No | NA | 1 | 0 | 0 | Low-quality | Genome-fragment | 2.92 | AAI-based (medium-confidence) | 0 |
| S11_k141_79679 | 4049 | No | NA | 6 | 0 | 0 | Not-determined | Genome-fragment | NA | NA | 0 |
| S11_k141_11529 | 1012 | No | NA | 2 | 0 | 2 | Not-determined | Genome-fragment | NA | NA | 0 |
| S11_k141_177089 | 3450 | No | NA | 3 | 0 | 3 | Not-determined | Genome-fragment | NA | NA | 0 |
| S11_k141_246544 | 1906 | No | NA | 3 | 0 | 2 | Not-determined | Genome-fragment | NA | NA | 0 |
| S11_k141_79732 | 1279 | No | NA | 1 | 0 | 0 | Not-determined | Genome-fragment | NA | NA | 0 |
| S11_k141_108644 | 1397 | No | NA | 1 | 0 | 1 | Not-determined | Genome-fragment | NA | NA | 0 |
| S11_k141_53283 | 4331 | No | NA | 6 | 0 | 3 | Not-determined | Genome-fragment | NA | NA | 0 |
| S11_k141_120934 | 1735 | No | NA | 3 | 0 | 0 | Not-determined | Genome-fragment | NA | NA | 0 |
| S11_k141_246602 | 1147 | No | NA | 1 | 0 | 1 | Not-determined | Genome-fragment | NA | NA | 0 |
| S2-final-viral-combined_k141_192277\|\|full | 1242 | No | NA | 3 | 1 | 0 | Low-quality | Genome-fragment | 2.53 | AAI-based (high-confidence) | 0 |
| S4-final-viral-combined_k141_164207\|\|full | 1242 | No | NA | 4 | 0 | 0 | Low-quality | Genome-fragment | 3.13 | AAI-based (medium-confidence) | 0 |
| S11_k141_218069 | 1480 | No | NA | 1 | 0 | 1 | Not-determined | Genome-fragment | NA | NA | 0 |
| S6-final-viral-combined_k141_129259\|\|full | 1242 | No | NA | 1 | 0 | 0 | Low-quality | Genome-fragment | 0.53 | AAI-based (medium-confidence) | 0 |
| S11_k141_177145 | 1188 | No | NA | 2 | 0 | 2 | Not-determined | Genome-fragment | NA | NA | 0 |
| S11_k141_23973 | 1101 | No | NA | 1 | 0 | 1 | Not-determined | Genome-fragment | NA | NA | 0 |
| S11_k141_121000 | 2073 | No | NA | 3 | 0 | 3 | Not-determined | Genome-fragment | NA | NA | 0 |
| S8_k141_54615 | 1242 | No | NA | 2 | 1 | 0 | Low-quality | Genome-fragment | 2.22 | AAI-based (high-confidence) | 0 |
| S11_k141_246639 | 3850 | No | NA | 2 | 0 | 0 | Not-determined | Genome-fragment | NA | NA | 0 |
| S11_k141_24017 | 3174 | No | NA | 1 | 0 | 1 | Not-determined | Genome-fragment | NA | NA | 0 |
| S11_k141_177192 | 1158 | No | NA | 1 | 0 | 1 | Not-determined | Genome-fragment | NA | NA | 0 |
| S11_k141_260030 | 1184 | No | NA | 1 | 0 | 1 | Not-determined | Genome-fragment | NA | NA | 0 |
| S11_k141_246666 | 1248 | No | NA | 2 | 0 | 0 | Not-determined | Genome-fragment | NA | NA | 0 |
| S11_k141_121021 | 1060 | No | NA | 1 | 0 | 1 | Not-determined | Genome-fragment | NA | NA | 0 |
| S11_k141_121026 | 1159 | No | NA | 1 | 0 | 0 | Not-determined | Genome-fragment | NA | NA | 0 |
| S11_k141_177243 | 1947 | No | NA | 1 | 0 | 1 | Not-determined | Genome-fragment | NA | NA | 0 |
| S11_k141_108786 | 3065 | No | NA | 3 | 0 | 3 | Not-determined | Genome-fragment | NA | NA | 0 |
| S11_k141_190050 | 13936 | No | NA | 11 | 0 | 4 | Not-determined | Genome-fragment | NA | NA | 0 |
| S11_k141_314693 | 1348 | No | NA | 1 | 0 | 0 | Not-determined | Genome-fragment | NA | NA | 0 |
| S11_k141_11780 | 1830 | No | NA | 3 | 0 | 0 | Not-determined | Genome-fragment | NA | NA | 0 |
| S11_k141_11789 | 1537 | No | NA | 2 | 0 | 0 | Not-determined | Genome-fragment | NA | NA | 0 |
| S9-final-viral-combined_k141_26848\|\|full | 1243 | No | NA | 3 | 0 | 0 | Low-quality | Genome-fragment | 1.75 | AAI-based (medium-confidence) | 0 |
| S11_k141_108832 | 2630 | No | NA | 3 | 0 | 3 | Not-determined | Genome-fragment | NA | NA | 0 |
| S9_k141_12534 | 1243 | No | NA | 1 | 0 | 1 | Low-quality | Genome-fragment | 0.3 | AAI-based (medium-confidence) | 0 |
| S11-final-viral-combined_k141_84223\|\|full | 1243 | No | NA | 3 | 0 | 0 | Low-quality | Genome-fragment | 2.59 | AAI-based (high-confidence) | 0 |
| S11_k141_94148 | 1394 | No | NA | 1 | 0 | 1 | Not-determined | Genome-fragment | NA | NA | 0 |
| S11_k141_134846 | 1074 | No | NA | 1 | 0 | 1 | Not-determined | Genome-fragment | NA | NA | 0 |
| S11_k141_163577 | 1006 | No | NA | 3 | 0 | 1 | Not-determined | Genome-fragment | NA | NA | 0 |
| S11_k141_79996 | 2385 | No | NA | 2 | 0 | 1 | Not-determined | Genome-fragment | NA | NA | 0 |
| S11_k141_24255 | 4156 | No | NA | 6 | 0 | 1 | Not-determined | Genome-fragment | NA | NA | 0 |
| S11_k141_134883 | 3690 | No | NA | 6 | 0 | 1 | Not-determined | Genome-fragment | NA | NA | 0 |
| S11-final-viral-combined_k141_274560\|\|full | 1243 | No | NA | 3 | 2 | 0 | Low-quality | Genome-fragment | 2.81 | AAI-based (medium-confidence) | 0 |
| S11_k141_306871 | 1243 | No | NA | 2 | 0 | 1 | Low-quality | Genome-fragment | 0.59 | AAI-based (medium-confidence) | 0 |
| S11_k141_260222 | 1331 | No | NA | 1 | 0 | 1 | Not-determined | Genome-fragment | NA | NA | 0 |
| S11_k141_80060 | 1126 | No | NA | 1 | 0 | 0 | Not-determined | Genome-fragment | NA | NA | 0 |
| S11_k141_287630 | 11510 | No | NA | 15 | 0 | 2 | Not-determined | Genome-fragment | NA | NA | 0 |
| S11_k141_39648 | 1580 | No | NA | 2 | 0 | 1 | Not-determined | Genome-fragment | NA | NA | 0 |
| S1_k141_313080 | 1243 | No | NA | 2 | 0 | 0 | Low-quality | Genome-fragment | 1.19 | AAI-based (medium-confidence) | 0 |
| S11_k141_53654 | 2340 | No | NA | 5 | 0 | 0 | Not-determined | Genome-fragment | NA | NA | 0 |
| S5-final-viral-combined_k141_56285\|\|full | 1243 | No | NA | 3 | 0 | 0 | Low-quality | Genome-fragment | 1.86 | AAI-based (medium-confidence) | 0 |
| S11_k141_134977 | 5032 | No | NA | 9 | 0 | 1 | Not-determined | Genome-fragment | NA | NA | 0 |
| S11_k141_163684 | 9784 | No | NA | 5 | 0 | 3 | Not-determined | Genome-fragment | NA | NA | 0 |
| S7-final-viral-combined_k141_243892\|\|full | 1243 | No | NA | 3 | 0 | 0 | Low-quality | Genome-fragment | 1.69 | AAI-based (high-confidence) | 0 |
| S11_k141_135007 | 4722 | No | NA | 3 | 0 | 3 | Not-determined | Genome-fragment | NA | NA | 0 |
| S11_k141_135014 | 1216 | No | NA | 2 | 0 | 0 | Not-determined | Genome-fragment | NA | NA | 0 |
| S11_k141_327436 | 1029 | No | NA | 1 | 0 | 0 | Not-determined | Genome-fragment | NA | NA | 0 |
| S11_k141_260322 | 1221 | No | NA | 1 | 0 | 1 | Not-determined | Genome-fragment | NA | NA | 0 |
| S11_k141_260356 | 1331 | No | NA | 2 | 0 | 2 | Not-determined | Genome-fragment | NA | NA | 0 |
| S11_k141_24458 | 3620 | No | NA | 4 | 0 | 2 | Not-determined | Genome-fragment | NA | NA | 0 |
| S11_k141_39777 | 1056 | No | NA | 2 | 0 | 1 | Not-determined | Genome-fragment | NA | NA | 0 |
| S11_k141_314959 | 1097 | No | NA | 1 | 0 | 0 | Not-determined | Genome-fragment | NA | NA | 0 |
| S11_k141_39828 | 1090 | No | NA | 2 | 0 | 0 | Not-determined | Genome-fragment | NA | NA | 0 |
| S11_k141_327547 | 1045 | No | NA | 1 | 0 | 1 | Not-determined | Genome-fragment | NA | NA | 0 |
| S11_k141_327564 | 1469 | No | NA | 2 | 0 | 0 | Not-determined | Genome-fragment | NA | NA | 0 |
| S7_k141_57380 | 1243 | No | NA | 1 | 0 | 1 | Low-quality | Genome-fragment | 2.28 | AAI-based (medium-confidence) | 0 |
| S17-final-viral-combined_k141_248207\|\|full | 1243 | No | NA | 3 | 0 | 1 | Low-quality | Genome-fragment | 1.83 | AAI-based (medium-confidence) | 0 |
| S11_k141_190467 | 1585 | No | NA | 1 | 0 | 1 | Not-determined | Genome-fragment | NA | NA | 0 |
| S17-final-viral-combined_k141_195811\|\|full | 1243 | No | NA | 2 | 0 | 0 | Low-quality | Genome-fragment | 0.74 | AAI-based (high-confidence) | 0 |
| S11_k141_232619 | 1689 | No | NA | 2 | 0 | 2 | Not-determined | Genome-fragment | NA | NA | 0 |
| S11_k141_301005 | 1225 | No | NA | 3 | 0 | 0 | Not-determined | Genome-fragment | NA | NA | 0 |
| S17-final-viral-combined_k141_85517\|\|full | 1243 | No | NA | 3 | 0 | 0 | Low-quality | Genome-fragment | 1.74 | AAI-based (high-confidence) | 0 |
| S11_k141_121560 | 1625 | No | NA | 2 | 0 | 2 | Not-determined | Genome-fragment | NA | NA | 0 |
| S11_k141_24622 | 1336 | No | NA | 2 | 0 | 2 | Not-determined | Genome-fragment | NA | NA | 0 |
| S11_k141_53862 | 1191 | No | NA | 1 | 0 | 0 | Not-determined | Genome-fragment | NA | NA | 0 |
| S12-final-viral-combined_k141_126075\|\|lt2gene | 1243 | No | NA | 2 | 1 | 0 | Low-quality | Genome-fragment | 2.45 | AAI-based (high-confidence) | 0 |
| S11_k141_149896 | 7229 | No | NA | 10 | 0 | 2 | Not-determined | Genome-fragment | NA | NA | 0 |
| S11_k141_204594 | 4009 | No | NA | 3 | 0 | 2 | Not-determined | Genome-fragment | NA | NA | 0 |
| S11_k141_164040 | 4136 | No | NA | 7 | 0 | 7 | Not-determined | Genome-fragment | NA | NA | 0 |
| S2_k141_146210 | 1243 | No | NA | 1 | 0 | 0 | Low-quality | Genome-fragment | 2.11 | AAI-based (high-confidence) | 0 |
| S8-final-viral-combined_k141_173973\|\|full | 1243 | No | NA | 3 | 0 | 0 | Low-quality | Genome-fragment | 0.81 | AAI-based (high-confidence) | 0 |
| S9-final-viral-combined_k141_36470\|\|full | 1244 | No | NA | 3 | 0 | 0 | Low-quality | Genome-fragment | 0.8 | AAI-based (medium-confidence) | 0 |
| S11_k141_190581 | 2082 | No | NA | 3 | 0 | 1 | Not-determined | Genome-fragment | NA | NA | 0 |
| S11_k141_260516 | 1175 | No | NA | 1 | 0 | 0 | Not-determined | Genome-fragment | NA | NA | 0 |
| S11-final-viral-combined_k141_136643\|\|full | 1244 | No | NA | 3 | 0 | 2 | Low-quality | Genome-fragment | 0.91 | AAI-based (medium-confidence) | 0 |
| S1-final-viral-combined_k141_304828\|\|full | 1244 | No | NA | 5 | 0 | 0 | Low-quality | Genome-fragment | 2.15 | AAI-based (high-confidence) | 0 |
| S11_k141_327708 | 3343 | No | NA | 3 | 0 | 2 | Not-determined | Genome-fragment | NA | NA | 0 |
| S1-final-viral-combined_k141_178208\|\|full | 1244 | No | NA | 3 | 0 | 0 | Low-quality | Genome-fragment | 2.98 | AAI-based (medium-confidence) | 0 |
| S11_k141_12231 | 1540 | No | NA | 1 | 0 | 1 | Not-determined | Genome-fragment | NA | NA | 0 |
| S11_k141_121674 | 1372 | No | NA | 1 | 0 | 1 | Not-determined | Genome-fragment | NA | NA | 0 |
| S1_k141_363294 | 1244 | No | NA | 2 | 0 | 0 | Low-quality | Genome-fragment | 1.42 | AAI-based (medium-confidence) | 0 |
| S1_k141_209013 | 1244 | No | NA | 1 | 0 | 0 | Low-quality | Genome-fragment | 3.37 | AAI-based (medium-confidence) | 0 |
| S11_k141_40114 | 3697 | No | NA | 3 | 0 | 2 | Not-determined | Genome-fragment | NA | NA | 0 |
| S11_k141_327748 | 1510 | No | NA | 4 | 0 | 0 | Not-determined | Genome-fragment | NA | NA | 0 |
| S11_k141_260610 | 1142 | No | NA | 1 | 0 | 0 | Not-determined | Genome-fragment | NA | NA | 0 |
| S11_k141_24709 | 2224 | No | NA | 1 | 0 | 1 | Not-determined | Genome-fragment | NA | NA | 0 |
| S11_k141_24733 | 1574 | No | NA | 2 | 0 | 2 | Not-determined | Genome-fragment | NA | NA | 0 |
| S11_k141_67487 | 2666 | No | NA | 5 | 0 | 2 | Not-determined | Genome-fragment | NA | NA | 0 |
| S11_k141_327800 | 2100 | No | NA | 2 | 0 | 1 | Not-determined | Genome-fragment | NA | NA | 0 |
| S11_k141_218783 | 4240 | No | NA | 3 | 0 | 3 | Not-determined | Genome-fragment | NA | NA | 0 |
| S11_k141_260706 | 1194 | No | NA | 1 | 0 | 0 | Not-determined | Genome-fragment | NA | NA | 0 |
| S11_k141_24801 | 1167 | No | NA | 2 | 0 | 2 | Not-determined | Genome-fragment | NA | NA | 0 |
| S11_k141_260725 | 1408 | No | NA | 1 | 0 | 1 | Not-determined | Genome-fragment | NA | NA | 0 |
| S11_k141_67558 | 1462 | No | NA | 2 | 0 | 1 | Not-determined | Genome-fragment | NA | NA | 0 |
| S11_k141_12432 | 1530 | No | NA | 2 | 0 | 0 | Not-determined | Genome-fragment | NA | NA | 0 |
| S11_k141_53993 | 1065 | No | NA | 2 | 0 | 2 | Not-determined | Genome-fragment | NA | NA | 0 |
| S11_k141_260756 | 1044 | No | NA | 1 | 0 | 0 | Not-determined | Genome-fragment | NA | NA | 0 |
| S11_k141_247363 | 1125 | No | NA | 1 | 0 | 1 | Not-determined | Genome-fragment | NA | NA | 0 |
| S5-final-viral-combined_k141_94576\|\|full | 1244 | No | NA | 3 | 1 | 0 | Low-quality | Genome-fragment | 0.85 | HMM-based (lower-bound) | 0 |
| S11_k141_54105 | 1209 | No | NA | 2 | 0 | 0 | Not-determined | Genome-fragment | NA | NA | 0 |
| S7-final-viral-combined_k141_189787\|\|full | 1244 | No | NA | 3 | 0 | 0 | Low-quality | Genome-fragment | 2.44 | AAI-based (medium-confidence) | 0 |
| S11_k141_109518 | 2607 | No | NA | 1 | 0 | 1 | Not-determined | Genome-fragment | NA | NA | 0 |
| S11_k141_54123 | 1616 | No | NA | 2 | 0 | 0 | Not-determined | Genome-fragment | NA | NA | 0 |
| S11_k141_327862 | 1456 | No | NA | 2 | 0 | 1 | Not-determined | Genome-fragment | NA | NA | 0 |
| S11_k141_301185 | 1244 | No | NA | 2 | 0 | 2 | Not-determined | Genome-fragment | NA | NA | 0 |
| S11_k141_178105 | 1791 | No | NA | 3 | 0 | 2 | Not-determined | Genome-fragment | NA | NA | 0 |
| S11_k141_150292 | 1531 | No | NA | 2 | 0 | 0 | Not-determined | Genome-fragment | NA | NA | 0 |
| S11_k141_273733 | 1161 | No | NA | 2 | 0 | 0 | Not-determined | Genome-fragment | NA | NA | 0 |
| S7_k141_166795 | 1244 | No | NA | 3 | 0 | 0 | Low-quality | Genome-fragment | 1.55 | AAI-based (high-confidence) | 0 |
| S17-final-viral-combined_k141_253647\|\|full | 1244 | No | NA | 2 | 1 | 0 | Low-quality | Genome-fragment | 2.45 | HMM-based (lower-bound) | 0 |
| S11_k141_24911 | 1272 | No | NA | 1 | 0 | 0 | Not-determined | Genome-fragment | NA | NA | 0 |
| S11_k141_12603 | 1988 | No | NA | 3 | 0 | 2 | Not-determined | Genome-fragment | NA | NA | 0 |
| S11_k141_233179 | 12033 | No | NA | 9 | 0 | 4 | Not-determined | Genome-fragment | NA | NA | 0 |
| S17_k141_254861 | 1244 | No | NA | 3 | 0 | 0 | Low-quality | Genome-fragment | 1.74 | AAI-based (high-confidence) | 0 |
| S12-final-viral-combined_k141_387081\|\|full | 1244 | No | NA | 2 | 0 | 0 | Low-quality | Genome-fragment | 2.34 | AAI-based (high-confidence) | 0 |
| S11_k141_288352 | 1180 | No | NA | 2 | 0 | 2 | Not-determined | Genome-fragment | NA | NA | 0 |
| S11_k141_191030 | 1830 | No | NA | 2 | 0 | 2 | Not-determined | Genome-fragment | NA | NA | 0 |
| S11_k141_288369 | 1710 | No | NA | 1 | 0 | 1 | Not-determined | Genome-fragment | NA | NA | 0 |
| S11_k141_12639 | 1211 | No | NA | 1 | 0 | 1 | Not-determined | Genome-fragment | NA | NA | 0 |
| S12-final-viral-combined_k141_243985\|\|full | 1244 | No | NA | 3 | 0 | 0 | Low-quality | Genome-fragment | 0.89 | AAI-based (medium-confidence) | 0 |
| S11_k141_109691 | 1486 | No | NA | 1 | 0 | 1 | Not-determined | Genome-fragment | NA | NA | 0 |
| S11_k141_164316 | 1740 | No | NA | 1 | 0 | 0 | Not-determined | Genome-fragment | NA | NA | 0 |
| S11_k141_261021 | 2870 | No | NA | 3 | 0 | 1 | Not-determined | Genome-fragment | NA | NA | 0 |
| S11_k141_12683 | 3017 | No | NA | 2 | 0 | 2 | Not-determined | Genome-fragment | NA | NA | 0 |
| S11_k141_67891 | 3072 | No | NA | 5 | 0 | 3 | Not-determined | Genome-fragment | NA | NA | 0 |
| S11_k141_219045 | 4213 | No | NA | 4 | 0 | 2 | Not-determined | Genome-fragment | NA | NA | 0 |
| S11_k141_67909 | 1198 | No | NA | 1 | 0 | 1 | Not-determined | Genome-fragment | NA | NA | 0 |
| S11_k141_233239 | 1442 | No | NA | 1 | 0 | 0 | Not-determined | Genome-fragment | NA | NA | 0 |
| S11_k141_328028 | 3809 | No | NA | 1 | 0 | 0 | Not-determined | Genome-fragment | NA | NA | 0 |
| S12-final-viral-combined_k141_69093\|\|lt2gene | 1244 | No | NA | 2 | 1 | 0 | Low-quality | Genome-fragment | 2.68 | AAI-based (high-confidence) | 0 |
| S11_k141_164413 | 1272 | No | NA | 3 | 0 | 3 | Not-determined | Genome-fragment | NA | NA | 0 |
| S11_k141_40617 | 1109 | No | NA | 1 | 0 | 1 | Not-determined | Genome-fragment | NA | NA | 0 |
| S11_k141_301394 | 2043 | No | NA | 2 | 0 | 2 | Not-determined | Genome-fragment | NA | NA | 0 |
| S11_k141_205109 | 1162 | No | NA | 2 | 0 | 2 | Not-determined | Genome-fragment | NA | NA | 0 |
| S11_k141_191150 | 1255 | No | NA | 1 | 0 | 0 | Not-determined | Genome-fragment | NA | NA | 0 |
| S12_k141_90111 | 1244 | No | NA | 4 | 1 | 0 | Low-quality | Genome-fragment | 3.22 | AAI-based (high-confidence) | 0 |
| S11_k141_150562 | 2144 | No | NA | 2 | 0 | 1 | Not-determined | Genome-fragment | NA | NA | 0 |
| S11_k141_178374 | 1812 | No | NA | 2 | 0 | 0 | Not-determined | Genome-fragment | NA | NA | 0 |
| S11_k141_233331 | 9311 | No | NA | 19 | 0 | 18 | Not-determined | Genome-fragment | NA | NA | 0 |
| S2-final-viral-combined_k141_7737\|\|full | 1244 | No | NA | 3 | 2 | 0 | Low-quality | Genome-fragment | 2.8 | AAI-based (high-confidence) | 0 |
| S11_k141_164441 | 1094 | No | NA | 1 | 0 | 1 | Not-determined | Genome-fragment | NA | NA | 0 |
| S4_k141_29431 | 1244 | No | NA | 4 | 1 | 0 | Low-quality | Genome-fragment | 3.57 | AAI-based (high-confidence) | 0 |
| S11_k141_135922 | 1593 | No | NA | 2 | 0 | 0 | Not-determined | Genome-fragment | NA | NA | 0 |
| S6-final-viral-combined_k141_85567\|\|full | 1244 | No | NA | 3 | 0 | 1 | Low-quality | Genome-fragment | 2.49 | AAI-based (high-confidence) | 0 |
| S11_k141_95138 | 3834 | No | NA | 2 | 0 | 2 | Not-determined | Genome-fragment | NA | NA | 0 |
| S11_k141_178425 | 1872 | No | NA | 1 | 0 | 0 | Not-determined | Genome-fragment | NA | NA | 0 |
| S8-final-viral-combined_k141_13154\|\|full | 1244 | No | NA | 3 | 0 | 0 | Low-quality | Genome-fragment | 1.76 | AAI-based (medium-confidence) | 0 |
| S11_k141_68095 | 1512 | No | NA | 1 | 0 | 0 | Not-determined | Genome-fragment | NA | NA | 0 |
| S8-final-viral-combined_k141_108645\|\|full | 1244 | No | NA | 2 | 0 | 0 | Low-quality | Genome-fragment | 0.78 | AAI-based (medium-confidence) | 0 |
| S9_k141_23515 | 1245 | No | NA | 2 | 0 | 0 | Low-quality | Genome-fragment | 2.98 | AAI-based (medium-confidence) | 0 |
| S11-final-viral-combined_k141_278457\|\|full | 1245 | No | NA | 3 | 0 | 1 | Low-quality | Genome-fragment | 1.54 | AAI-based (high-confidence) | 0 |
| S1-final-viral-combined_k141_194042\|\|full | 1245 | No | NA | 2 | 0 | 1 | Low-quality | Genome-fragment | 7.48 | AAI-based (medium-confidence) | 0 |
| S11_k141_288655 | 1776 | No | NA | 1 | 0 | 1 | Not-determined | Genome-fragment | NA | NA | 0 |
| S11_k141_68181 | 1268 | No | NA | 2 | 0 | 1 | Not-determined | Genome-fragment | NA | NA | 0 |
| S11_k141_150759 | 2653 | No | NA | 2 | 0 | 1 | Not-determined | Genome-fragment | NA | NA | 0 |
| S11_k141_219251 | 1074 | No | NA | 1 | 0 | 1 | Not-determined | Genome-fragment | NA | NA | 0 |
| S11_k141_261243 | 3031 | No | NA | 1 | 0 | 0 | Not-determined | Genome-fragment | NA | NA | 0 |
| S11_k141_164606 | 1393 | No | NA | 1 | 0 | 1 | Not-determined | Genome-fragment | NA | NA | 0 |
| S11_k141_40904 | 1913 | No | NA | 2 | 0 | 1 | Not-determined | Genome-fragment | NA | NA | 0 |
| S1-final-viral-combined_k141_376156\|\|full | 1245 | No | NA | 3 | 0 | 0 | Low-quality | Genome-fragment | 0.86 | AAI-based (medium-confidence) | 0 |
| S11_k141_274188 | 1892 | No | NA | 1 | 0 | 1 | Not-determined | Genome-fragment | NA | NA | 0 |
| S1-final-viral-combined_k141_15883\|\|lt2gene | 1245 | No | NA | 2 | 2 | 0 | Low-quality | Genome-fragment | 3.16 | AAI-based (high-confidence) | 0 |
| S11_k141_315827 | 1368 | No | NA | 1 | 0 | 1 | Not-determined | Genome-fragment | NA | NA | 0 |
| S1_k141_306582 | 1245 | No | NA | 2 | 0 | 2 | Low-quality | Genome-fragment | 0.58 | AAI-based (medium-confidence) | 0 |
| S11_k141_233587 | 1308 | No | NA | 1 | 0 | 0 | Not-determined | Genome-fragment | NA | NA | 0 |
| S17-final-viral-combined_k141_203812\|\|full | 1245 | No | NA | 3 | 0 | 0 | Low-quality | Genome-fragment | 2.08 | AAI-based (medium-confidence) | 0 |
| S17-final-viral-combined_k141_38866\|\|lt2gene | 1245 | No | NA | 2 | 1 | 0 | Low-quality | Genome-fragment | 3.66 | AAI-based (high-confidence) | 0 |
| S17_k141_237720 | 1245 | No | NA | 4 | 0 | 0 | Low-quality | Genome-fragment | 1.59 | AAI-based (high-confidence) | 0 |
| S11_k141_80997 | 15746 | No | NA | 10 | 0 | 6 | Not-determined | Genome-fragment | NA | NA | 0 |
| S11_k141_54715 | 1880 | No | NA | 1 | 0 | 1 | Not-determined | Genome-fragment | NA | NA | 0 |
| S17_k141_108394 | 1245 | No | NA | 1 | 0 | 0 | Low-quality | Genome-fragment | 1.87 | AAI-based (medium-confidence) | 0 |
| S11_k141_54756 | 1589 | No | NA | 1 | 0 | 1 | Not-determined | Genome-fragment | NA | NA | 0 |
| S11_k141_288772 | 1642 | No | NA | 2 | 0 | 0 | Not-determined | Genome-fragment | NA | NA | 0 |
| S11_k141_122402 | 7036 | No | NA | 9 | 0 | 2 | Not-determined | Genome-fragment | NA | NA | 0 |
| S12-final-viral-combined_k141_335813\|\|full | 1245 | No | NA | 3 | 0 | 0 | Low-quality | Genome-fragment | 1.71 | AAI-based (medium-confidence) | 0 |
| S11_k141_150932 | 2684 | No | NA | 3 | 0 | 1 | Not-determined | Genome-fragment | NA | NA | 0 |
| S12-final-viral-combined_k141_379982\|\|full | 1245 | No | NA | 2 | 0 | 0 | Low-quality | Genome-fragment | 2.48 | AAI-based (high-confidence) | 0 |
| S11_k141_191526 | 1052 | No | NA | 2 | 0 | 1 | Not-determined | Genome-fragment | NA | NA | 0 |
| S11_k141_95486 | 1338 | No | NA | 1 | 0 | 1 | Not-determined | Genome-fragment | NA | NA | 0 |
| S12_k141_316090 | 1245 | No | NA | 1 | 1 | 0 | Low-quality | Genome-fragment | 2.96 | HMM-based (lower-bound) | 0 |
| S11_k141_13259 | 1908 | No | NA | 3 | 0 | 1 | Not-determined | Genome-fragment | NA | NA | 0 |
| S11_k141_95499 | 1502 | No | NA | 1 | 0 | 0 | Not-determined | Genome-fragment | NA | NA | 0 |
| S11_k141_164833 | 4605 | No | NA | 4 | 0 | 4 | Not-determined | Genome-fragment | NA | NA | 0 |
| S11_k141_191560 | 3972 | No | NA | 2 | 0 | 2 | Not-determined | Genome-fragment | NA | NA | 0 |
| S11_k141_191569 | 1207 | No | NA | 1 | 0 | 0 | Not-determined | Genome-fragment | NA | NA | 0 |
| S2_k141_240119 | 1245 | No | NA | 2 | 0 | 0 | Low-quality | Genome-fragment | 2.44 | AAI-based (high-confidence) | 0 |
| S6-final-viral-combined_k141_3558\|\|full | 1245 | No | NA | 2 | 0 | 2 | Low-quality | Genome-fragment | 1.52 | AAI-based (medium-confidence) | 0 |
| S11_k141_136316 | 1132 | No | NA | 2 | 0 | 2 | Not-determined | Genome-fragment | NA | NA | 0 |
| S11_k141_122517 | 1872 | No | NA | 2 | 0 | 2 | Not-determined | Genome-fragment | NA | NA | 0 |
| S9-final-viral-combined_k141_70474\|\|full | 1246 | No | NA | 2 | 0 | 0 | Low-quality | Genome-fragment | 2.08 | AAI-based (high-confidence) | 0 |
| S11_k141_54883 | 1018 | No | NA | 2 | 0 | 0 | Not-determined | Genome-fragment | NA | NA | 0 |
| S11_k141_205476 | 1159 | No | NA | 1 | 0 | 0 | Not-determined | Genome-fragment | NA | NA | 0 |
| S11-final-viral-combined_k141_97491\|\|full | 1246 | No | NA | 3 | 0 | 0 | Low-quality | Genome-fragment | 2.57 | AAI-based (high-confidence) | 0 |
| S11_k141_54900 | 3683 | No | NA | 3 | 0 | 3 | Not-determined | Genome-fragment | NA | NA | 0 |
| S11_k141_81187 | 1003 | No | NA | 1 | 0 | 0 | Not-determined | Genome-fragment | NA | NA | 0 |
| S11_k141_191712 | 1724 | No | NA | 1 | 0 | 0 | Not-determined | Genome-fragment | NA | NA | 0 |
| S11_k141_288977 | 22884 | No | NA | 9 | 0 | 4 | Not-determined | Genome-fragment | NA | NA | 0 |
| S1-final-viral-combined_k141_193460\|\|full | 1246 | No | NA | 2 | 2 | 0 | Low-quality | Genome-fragment | 2.92 | AAI-based (high-confidence) | 0 |
| S11_k141_41200 | 1700 | No | NA | 1 | 0 | 0 | Not-determined | Genome-fragment | NA | NA | 0 |
| S11_k141_191816 | 1261 | No | NA | 2 | 0 | 1 | Not-determined | Genome-fragment | NA | NA | 0 |
| S1-final-viral-combined_k141_150924\|\|lt2gene | 1246 | No | NA | 2 | 1 | 0 | Low-quality | Genome-fragment | 2.86 | AAI-based (high-confidence) | 0 |
| S1_k141_131115 | 1246 | No | NA | 2 | 0 | 0 | Low-quality | Genome-fragment | 1.01 | AAI-based (high-confidence) | 0 |
| S11_k141_178902 | 3006 | No | NA | 5 | 0 | 2 | Not-determined | Genome-fragment | NA | NA | 0 |
| S3-final-viral-combined_k141_39415\|\|lt2gene | 1246 | No | NA | 2 | 1 | 0 | Low-quality | Genome-fragment | 2.52 | AAI-based (high-confidence) | 0 |
| S11_k141_178908 | 1006 | No | NA | 1 | 0 | 0 | Not-determined | Genome-fragment | NA | NA | 0 |
| S3_k141_127769 | 1246 | No | NA | 1 | 0 | 1 | Low-quality | Genome-fragment | 0.62 | AAI-based (medium-confidence) | 0 |
| S11_k141_234077 | 2675 | No | NA | 1 | 0 | 1 | Not-determined | Genome-fragment | NA | NA | 0 |
| S11_k141_41317 | 1232 | No | NA | 1 | 0 | 1 | Not-determined | Genome-fragment | NA | NA | 0 |
| S11_k141_136549 | 1032 | No | NA | 2 | 0 | 1 | Not-determined | Genome-fragment | NA | NA | 0 |
| S11_k141_328624 | 3795 | No | NA | 5 | 0 | 5 | Not-determined | Genome-fragment | NA | NA | 0 |
| S11_k141_316247 | 1046 | No | NA | 1 | 0 | 1 | Not-determined | Genome-fragment | NA | NA | 0 |
| S5-final-viral-combined_k141_4633\|\|full | 1246 | No | NA | 3 | 3 | 0 | Low-quality | Genome-fragment | 3.11 | AAI-based (high-confidence) | 0 |
| S12-final-viral-combined_k141_351680\|\|full | 1246 | No | NA | 2 | 0 | 1 | Low-quality | Genome-fragment | 0.57 | AAI-based (medium-confidence) | 0 |
| S11_k141_13630 | 1089 | No | NA | 1 | 0 | 0 | Not-determined | Genome-fragment | NA | NA | 0 |
| S12-final-viral-combined_k141_303671\|\|full | 1246 | No | NA | 2 | 0 | 0 | Low-quality | Genome-fragment | 2.72 | AAI-based (medium-confidence) | 0 |
| S8-final-viral-combined_k141_68815\|\|full | 1246 | No | NA | 3 | 1 | 0 | Low-quality | Genome-fragment | 2.18 | AAI-based (high-confidence) | 0 |
| S11_k141_165277 | 1492 | No | NA | 3 | 0 | 2 | Not-determined | Genome-fragment | NA | NA | 0 |
| S9-final-viral-combined_k141_35256\|\|full | 1247 | No | NA | 3 | 0 | 1 | Low-quality | Genome-fragment | 1.69 | AAI-based (medium-confidence) | 0 |
| S11-final-viral-combined_k141_250804\|\|full | 1247 | No | NA | 2 | 0 | 0 | Low-quality | Genome-fragment | 1.92 | AAI-based (high-confidence) | 0 |
| S11_k141_316317 | 1122 | No | NA | 2 | 0 | 1 | Not-determined | Genome-fragment | NA | NA | 0 |
| S11_k141_165319 | 1119 | No | NA | 2 | 0 | 2 | Not-determined | Genome-fragment | NA | NA | 0 |
| S11_k141_151398 | 1119 | No | NA | 2 | 0 | 0 | Not-determined | Genome-fragment | NA | NA | 0 |
| S11_k141_26046 | 1232 | No | NA | 1 | 0 | 1 | Not-determined | Genome-fragment | NA | NA | 0 |
| S11_k141_316359 | 7547 | No | NA | 10 | 0 | 4 | Not-determined | Genome-fragment | NA | NA | 0 |
| S11_k141_289343 | 3212 | No | NA | 7 | 0 | 1 | Not-determined | Genome-fragment | NA | NA | 0 |
| S11_k141_289388 | 1515 | No | NA | 2 | 0 | 0 | Not-determined | Genome-fragment | NA | NA | 0 |
| S11_k141_274853 | 2575 | No | NA | 2 | 0 | 0 | Not-determined | Genome-fragment | NA | NA | 0 |
| S11_k141_96102 | 5004 | No | NA | 7 | 0 | 0 | Not-determined | Genome-fragment | NA | NA | 0 |
| S11-final-viral-combined_k141_38922\|\|full | 1247 | No | NA | 3 | 0 | 0 | Low-quality | Genome-fragment | 3.02 | AAI-based (high-confidence) | 0 |
| S11_k141_68780 | 5674 | No | NA | 3 | 0 | 0 | Not-determined | Genome-fragment | NA | NA | 0 |
| S11_k141_26243 | 1265 | No | NA | 3 | 0 | 0 | Not-determined | Genome-fragment | NA | NA | 0 |
| S11_k141_302544 | 3691 | No | NA | 4 | 0 | 2 | Not-determined | Genome-fragment | NA | NA | 0 |
| S11_k141_81762 | 1104 | No | NA | 1 | 0 | 0 | Not-determined | Genome-fragment | NA | NA | 0 |
| S11_k141_192129 | 2969 | No | NA | 5 | 0 | 1 | Not-determined | Genome-fragment | NA | NA | 0 |
| S11_k141_136900 | 1266 | No | NA | 2 | 0 | 1 | Not-determined | Genome-fragment | NA | NA | 0 |
| S11_k141_81783 | 1101 | No | NA | 1 | 0 | 0 | Not-determined | Genome-fragment | NA | NA | 0 |
| S11_k141_220170 | 1674 | No | NA | 3 | 0 | 2 | Not-determined | Genome-fragment | NA | NA | 0 |
| S11_k141_206083 | 1216 | No | NA | 1 | 0 | 1 | Not-determined | Genome-fragment | NA | NA | 0 |
| S1_k141_309870 | 1247 | No | NA | 3 | 0 | 1 | Low-quality | Genome-fragment | 0.7 | AAI-based (medium-confidence) | 0 |
| S11_k141_26312 | 1051 | No | NA | 4 | 0 | 3 | Not-determined | Genome-fragment | NA | NA | 0 |
| S11_k141_26327 | 1035 | No | NA | 2 | 0 | 0 | Not-determined | Genome-fragment | NA | NA | 0 |
| S11_k141_328875 | 3328 | No | NA | 5 | 0 | 1 | Not-determined | Genome-fragment | NA | NA | 0 |
| S11_k141_151552 | 1034 | No | NA | 1 | 0 | 1 | Not-determined | Genome-fragment | NA | NA | 0 |
| S11_k141_192272 | 3356 | No | NA | 3 | 0 | 1 | Not-determined | Genome-fragment | NA | NA | 0 |
| S11_k141_328921 | 1236 | No | NA | 1 | 0 | 0 | Not-determined | Genome-fragment | NA | NA | 0 |
| S3_k141_66996 | 1247 | No | NA | 1 | 1 | 0 | Low-quality | Genome-fragment | 3.43 | HMM-based (lower-bound) | 0 |
| S11_k141_137054 | 1409 | No | NA | 2 | 0 | 1 | Not-determined | Genome-fragment | NA | NA | 0 |
| S11_k141_206131 | 1349 | No | NA | 1 | 0 | 0 | Not-determined | Genome-fragment | NA | NA | 0 |
| S11_k141_302808 | 6197 | No | NA | 5 | 0 | 4 | Not-determined | Genome-fragment | NA | NA | 0 |
| S11_k141_137136 | 8446 | No | NA | 5 | 0 | 4 | Not-determined | Genome-fragment | NA | NA | 0 |
| S11_k141_123428 | 1129 | No | NA | 1 | 0 | 0 | Not-determined | Genome-fragment | NA | NA | 0 |
| S5-final-viral-combined_k141_136927\|\|full | 1247 | No | NA | 2 | 0 | 1 | Low-quality | Genome-fragment | 1.5 | AAI-based (high-confidence) | 0 |
| S5-final-viral-combined_k141_67965\|\|full | 1247 | No | NA | 4 | 1 | 0 | Low-quality | Genome-fragment | 2.33 | AAI-based (high-confidence) | 0 |
| S11_k141_316632 | 3745 | No | NA | 4 | 0 | 3 | Not-determined | Genome-fragment | NA | NA | 0 |
| S11_k141_328990 | 1019 | No | NA | 1 | 0 | 0 | Not-determined | Genome-fragment | NA | NA | 0 |
| S11_k141_192429 | 1355 | No | NA | 2 | 0 | 2 | Not-determined | Genome-fragment | NA | NA | 0 |
| S11_k141_68900 | 1105 | No | NA | 1 | 0 | 0 | Not-determined | Genome-fragment | NA | NA | 0 |
| S11_k141_26594 | 4478 | No | NA | 2 | 0 | 2 | Not-determined | Genome-fragment | NA | NA | 0 |
| S11_k141_137281 | 1273 | No | NA | 1 | 0 | 0 | Not-determined | Genome-fragment | NA | NA | 0 |
| S11_k141_302950 | 1000 | No | NA | 1 | 0 | 0 | Not-determined | Genome-fragment | NA | NA | 0 |
| S11_k141_220436 | 2012 | No | NA | 3 | 0 | 1 | Not-determined | Genome-fragment | NA | NA | 0 |
| S5-final-viral-combined_k141_185175\|\|full | 1247 | No | NA | 2 | 0 | 2 | Low-quality | Genome-fragment | 2.79 | AAI-based (medium-confidence) | 0 |
| S11_k141_137441 | 1570 | No | NA | 1 | 0 | 0 | Not-determined | Genome-fragment | NA | NA | 0 |
| S11_k141_275463 | 1521 | No | NA | 2 | 0 | 2 | Not-determined | Genome-fragment | NA | NA | 0 |
| S11_k141_316875 | 1199 | No | NA | 1 | 0 | 1 | Not-determined | Genome-fragment | NA | NA | 0 |
| S7-final-viral-combined_k141_236659\|\|lt2gene | 1247 | No | NA | 2 | 2 | 0 | Low-quality | Genome-fragment | 2.51 | AAI-based (high-confidence) | 0 |
| S7-final-viral-combined_k141_55397\|\|lt2gene | 1247 | No | NA | 2 | 0 | 0 | Low-quality | Genome-fragment | 3.64 | AAI-based (medium-confidence) | 0 |
| S11_k141_26900 | 5247 | No | NA | 4 | 0 | 2 | Not-determined | Genome-fragment | NA | NA | 0 |
| S7-final-viral-combined_k141_120003\|\|lt2gene | 1247 | No | NA | 2 | 2 | 0 | Low-quality | Genome-fragment | 2.01 | HMM-based (lower-bound) | 0 |
| S11_k141_275614 | 2814 | No | NA | 7 | 0 | 0 | Not-determined | Genome-fragment | NA | NA | 0 |
| S17-final-viral-combined_k141_6034\|\|full | 1247 | No | NA | 2 | 0 | 0 | Low-quality | Genome-fragment | 0.79 | AAI-based (medium-confidence) | 0 |
| S11_k141_124028 | 1592 | No | NA | 3 | 0 | 2 | Not-determined | Genome-fragment | NA | NA | 0 |
| S11_k141_137801 | 1195 | No | NA | 1 | 0 | 0 | Not-determined | Genome-fragment | NA | NA | 0 |
| S11_k141_27139 | 1111 | No | NA | 2 | 0 | 1 | Not-determined | Genome-fragment | NA | NA | 0 |
| S11_k141_330074 | 1008 | No | NA | 1 | 0 | 1 | Not-determined | Genome-fragment | NA | NA | 0 |
| S4-final-viral-combined_k141_121544\|\|full | 1247 | No | NA | 5 | 2 | 0 | Low-quality | Genome-fragment | 1.9 | AAI-based (medium-confidence) | 0 |
| S11_k141_330477 | 5123 | No | NA | 9 | 0 | 0 | Not-determined | Genome-fragment | NA | NA | 0 |
| S11_k141_330479 | 36558 | No | NA | 42 | 8 | 0 | Complete | High-quality | 100 | DTR (medium-confidence) | 0 |
| S11_k141_330484 | 4823 | No | NA | 7 | 0 | 0 | Not-determined | Genome-fragment | NA | NA | 0 |
| S8-final-viral-combined_k141_79910\|\|full | 1247 | No | NA | 2 | 0 | 0 | Low-quality | Genome-fragment | 1.91 | AAI-based (medium-confidence) | 0 |
| S11_k141_330585 | 1029 | No | NA | 2 | 0 | 0 | Not-determined | Genome-fragment | NA | NA | 0 |
| S8-final-viral-combined_k141_241728\|\|full | 1247 | No | NA | 2 | 0 | 0 | Low-quality | Genome-fragment | 0.81 | AAI-based (high-confidence) | 0 |
| S1-final-viral-combined_k141_70298\|\|full | 2245 | No | NA | 5 | 0 | 0 | Not-determined | Genome-fragment | NA | NA | 0 |
| S1-final-viral-combined_k141_324053\|\|full | 41421 | No | NA | 60 | 23 | 0 | High-quality | High-quality | 96.35 | AAI-based (high-confidence) | 0 |
| S1-final-viral-combined_k141_359778\|\|full | 6875 | No | NA | 11 | 0 | 0 | Not-determined | Genome-fragment | NA | NA | 0 |
| S8_k141_113905 | 1247 | No | NA | 1 | 0 | 0 | Low-quality | Genome-fragment | 2.79 | AAI-based (high-confidence) | 0 |
| S1-final-viral-combined_k141_394572\|\|full | 4760 | No | NA | 10 | 0 | 0 | Not-determined | Genome-fragment | NA | NA | 0 |
| S1-final-viral-combined_k141_394574\|\|full | 46833 | No | NA | 61 | 15 | 1 | High-quality | High-quality | 100 | AAI-based (high-confidence) | 0 |

| | | | | | | | | | | | |
|---|---|---|---|---|---|---|---|---|---|---|---|
| S11-final-viral-combined_k141_79427\|\|lt2gene | 1248 | No | NA | 2 | 2 | 0 | Low-quality | Genome-fragment | 3.15 | AAI-based (medium-confidence) | 0 |
| S1-final-viral-combined_k141_394628\|\|full | 10807 | No | NA | 13 | 0 | 1 | Medium-quality | Genome-fragment | 73.96 | AAI-based (high-confidence) | 0 |
| S11-final-viral-combined_k141_150592\|\|lt2gene | 1248 | No | NA | 1 | 1 | 0 | Low-quality | Genome-fragment | 3.07 | AAI-based (high-confidence) | 0 |
| S1-final-viral-combined_k141_180985\|\|full | 9945 | No | NA | 11 | 0 | 6 | Not-determined | Genome-fragment | NA | NA | 0 |
| S1-final-viral-combined_k141_206076\|\|full | 1248 | No | NA | 3 | 0 | 0 | Low-quality | Genome-fragment | 2.57 | AAI-based (medium-confidence) | 0 |
| S1-final-viral-combined_k141_106935\|\|full | 1248 | No | NA | 3 | 0 | 0 | Low-quality | Genome-fragment | 2.69 | AAI-based (high-confidence) | 0 |
| S1-final-viral-combined_k141_164556\|\|full | 4037 | No | NA | 3 | 0 | 1 | Not-determined | Genome-fragment | NA | NA | 0 |
| S1-final-viral-combined_k141_279715\|\|full | 9462 | No | NA | 8 | 0 | 7 | Not-determined | Genome-fragment | NA | NA | 0 |
| S1-final-viral-combined_k141_131658\|\|full | 1645 | No | NA | 2 | 0 | 0 | Not-determined | Genome-fragment | NA | NA | 0 |
| S1-final-viral-combined_k141_49387\|\|full | 3092 | No | NA | 3 | 0 | 1 | Not-determined | Genome-fragment | NA | NA | 0 |
| S1-final-viral-combined_k141_93906\|\|full | 1248 | No | NA | 3 | 0 | 0 | Low-quality | Genome-fragment | 2.6 | AAI-based (medium-confidence) | 0 |
| S1-final-viral-combined_k141_345549\|\|full | 1238 | No | NA | 2 | 0 | 1 | Not-determined | Genome-fragment | NA | NA | 0 |
| S1-final-viral-combined_k141_361999\|\|full | 1012 | No | NA | 2 | 0 | 1 | Not-determined | Genome-fragment | NA | NA | 0 |
| S1-final-viral-combined_k141_179474\|\|full | 1248 | No | NA | 2 | 0 | 0 | Low-quality | Genome-fragment | 0.92 | AAI-based (medium-confidence) | 0 |
| S1-final-viral-combined_k141_164566\|\|full | 2328 | No | NA | 3 | 0 | 1 | Not-determined | Genome-fragment | NA | NA | 0 |
| S1-final-viral-combined_k141_263274\|\|full | 1900 | No | NA | 3 | 0 | 0 | Not-determined | Genome-fragment | NA | NA | 0 |
| S1-final-viral-combined_k141_16472\|\|full | 3693 | No | NA | 4 | 0 | 2 | Not-determined | Genome-fragment | NA | NA | 0 |
| S1-final-viral-combined_k141_31527\|\|full | 1248 | No | NA | 4 | 0 | 0 | Low-quality | Genome-fragment | 1.31 | AAI-based (high-confidence) | 0 |
| S1-final-viral-combined_k141_49429\|\|full | 6245 | No | NA | 7 | 0 | 0 | Not-determined | Genome-fragment | NA | NA | 0 |
| S1-final-viral-combined_k141_32969\|\|full | 2815 | No | NA | 4 | 0 | 1 | Not-determined | Genome-fragment | NA | NA | 0 |
| S1-final-viral-combined_k141_148133\|\|full | 4223 | No | NA | 5 | 0 | 5 | Not-determined | Genome-fragment | NA | NA | 0 |
| S1-final-viral-combined_k141_82325\|\|full | 1223 | No | NA | 3 | 0 | 2 | Not-determined | Genome-fragment | NA | NA | 0 |
| S7-final-viral-combined_k141_220032\|\|full | 1248 | No | NA | 4 | 1 | 0 | Low-quality | Genome-fragment | 3.08 | AAI-based (high-confidence) | 0 |
| S7-final-viral-combined_k141_141731\|\|full | 1248 | No | NA | 3 | 1 | 0 | Low-quality | Genome-fragment | 3.16 | AAI-based (high-confidence) | 0 |
| S7-final-viral-combined_k141_204679\|\|lt2gene | 1248 | No | NA | 1 | 1 | 0 | Low-quality | Genome-fragment | 2.08 | HMM-based (lower-bound) | 0 |
| S7_k141_60452 | 1248 | No | NA | 3 | 1 | 0 | Low-quality | Genome-fragment | 3.44 | AAI-based (medium-confidence) | 0 |
| S2-final-viral-combined_k141_157508\|\|full | 1248 | No | NA | 2 | 0 | 0 | Low-quality | Genome-fragment | 3.05 | AAI-based (medium-confidence) | 0 |
| S1-final-viral-combined_k141_345591\|\|full | 6096 | No | NA | 6 | 0 | 4 | Not-determined | Genome-fragment | NA | NA | 0 |
| S1-final-viral-combined_k141_100\|\|full | 1612 | No | NA | 3 | 0 | 3 | Not-determined | Genome-fragment | NA | NA | 0 |
| S1-final-viral-combined_k141_329133\|\|full | 1097 | No | NA | 2 | 0 | 0 | Not-determined | Genome-fragment | NA | NA | 0 |
| S1-final-viral-combined_k141_263299\|\|full | 1643 | No | NA | 3 | 0 | 2 | Not-determined | Genome-fragment | NA | NA | 0 |
| S1-final-viral-combined_k141_131740\|\|full | 1336 | No | NA | 2 | 0 | 1 | Not-determined | Genome-fragment | NA | NA | 0 |
| S2-final-viral-combined_k141_86410\|\|full | 1248 | No | NA | 2 | 0 | 1 | Low-quality | Genome-fragment | 0.95 | AAI-based (medium-confidence) | 0 |
| S2_k141_137370 | 1248 | No | NA | 2 | 1 | 0 | Low-quality | Genome-fragment | 2.46 | AAI-based (high-confidence) | 0 |
| S1-final-viral-combined_k141_123\|\|full | 4501 | No | NA | 6 | 0 | 0 | Not-determined | Genome-fragment | NA | NA | 0 |
| S4_k141_81945 | 1248 | No | NA | 1 | 0 | 1 | Low-quality | Genome-fragment | 0.42 | AAI-based (medium-confidence) | 0 |
| S4_k141_54047 | 1248 | No | NA | 3 | 1 | 0 | Low-quality | Genome-fragment | 0.7 | HMM-based (lower-bound) | 0 |
| S1-final-viral-combined_k141_362058\|\|full | 2733 | No | NA | 4 | 0 | 1 | Not-determined | Genome-fragment | NA | NA | 0 |
| S1-final-viral-combined_k141_115273\|\|full | 1534 | No | NA | 2 | 0 | 1 | Not-determined | Genome-fragment | NA | NA | 0 |
| S1-final-viral-combined_k141_246845\|\|full | 1418 | No | NA | 3 | 0 | 2 | Not-determined | Genome-fragment | NA | NA | 0 |
| S1-final-viral-combined_k141_33039\|\|full | 1619 | No | NA | 2 | 0 | 1 | Not-determined | Genome-fragment | NA | NA | 0 |
| S11-final-viral-combined_k141_125856\|\|full | 1249 | No | NA | 3 | 2 | 0 | Low-quality | Genome-fragment | 2.48 | AAI-based (high-confidence) | 0 |
| S11-final-viral-combined_k141_142090\|\|full | 1249 | No | NA | 3 | 0 | 0 | Low-quality | Genome-fragment | 2.49 | AAI-based (high-confidence) | 0 |
| S1-final-viral-combined_k141_230440\|\|full | 4314 | No | NA | 6 | 0 | 2 | Not-determined | Genome-fragment | NA | NA | 0 |
| S1-final-viral-combined_k141_246847\|\|full | 8531 | No | NA | 12 | 0 | 0 | Not-determined | Genome-fragment | NA | NA | 0 |
| S1-final-viral-combined_k141_39539\|\|lt2gene | 1249 | No | NA | 2 | 0 | 0 | Low-quality | Genome-fragment | 2.29 | AAI-based (high-confidence) | 0 |
| S1_k141_178479 | 1249 | No | NA | 1 | 0 | 0 | Low-quality | Genome-fragment | 0.21 | AAI-based (high-confidence) | 0 |
| S1-final-viral-combined_k141_115301\|\|full | 1168 | No | NA | 3 | 0 | 0 | Not-determined | Genome-fragment | NA | NA | 0 |
| S1-final-viral-combined_k141_115308\|\|full | 1495 | No | NA | 1 | 0 | 1 | Not-determined | Genome-fragment | NA | NA | 0 |
| S1-final-viral-combined_k141_213971\|\|full | 1146 | No | NA | 2 | 0 | 0 | Not-determined | Genome-fragment | NA | NA | 0 |
| S1-final-viral-combined_k141_164618\|\|full | 2006 | No | NA | 3 | 0 | 0 | Not-determined | Genome-fragment | NA | NA | 0 |
| S7-final-viral-combined_k141_236250\|\|full | 1249 | No | NA | 3 | 0 | 0 | Low-quality | Genome-fragment | 1.56 | AAI-based (high-confidence) | 0 |
| S12-final-viral-combined_k141_325856\|\|full | 1249 | No | NA | 2 | 0 | 0 | Low-quality | Genome-fragment | 2.78 | AAI-based (medium-confidence) | 0 |
| S1-final-viral-combined_k141_345645\|\|full | 7264 | No | NA | 6 | 0 | 4 | Not-determined | Genome-fragment | NA | NA | 0 |
| S1-final-viral-combined_k141_345650\|\|full | 4889 | No | NA | 7 | 0 | 3 | Not-determined | Genome-fragment | NA | NA | 0 |
| S12_k141_331028 | 1249 | No | NA | 3 | 1 | 0 | Low-quality | Genome-fragment | 2.28 | AAI-based (medium-confidence) | 0 |
| S1-final-viral-combined_k141_164641\|\|full | 1522 | No | NA | 3 | 0 | 2 | Not-determined | Genome-fragment | NA | NA | 0 |
| S1-final-viral-combined_k141_49525\|\|full | 6238 | No | NA | 6 | 0 | 3 | Not-determined | Genome-fragment | NA | NA | 0 |
| S2-final-viral-combined_k141_115957\|\|lt2gene | 1249 | No | NA | 2 | 2 | 0 | Low-quality | Genome-fragment | 2.73 | AAI-based (high-confidence) | 0 |
| S8-final-viral-combined_k141_232367\|\|full | 1249 | No | NA | 2 | 0 | 1 | Low-quality | Genome-fragment | 1.98 | AAI-based (medium-confidence) | 0 |
| S8-final-viral-combined_k141_186988\|\|lt2gene | 1249 | No | NA | 2 | 2 | 0 | Low-quality | Genome-fragment | 24.05 | AAI-based (high-confidence) | 0 |
| S1-final-viral-combined_k141_279838\|\|full | 1865 | No | NA | 2 | 0 | 0 | Not-determined | Genome-fragment | NA | NA | 0 |
| S1-final-viral-combined_k141_296311\|\|full | 2788 | No | NA | 4 | 0 | 1 | Not-determined | Genome-fragment | NA | NA | 0 |
| S1-final-viral-combined_k141_312828\|\|full | 4702 | No | NA | 8 | 0 | 1 | Not-determined | Genome-fragment | NA | NA | 0 |
| S8_k141_63002 | 1249 | No | NA | 2 | 1 | 0 | Low-quality | Genome-fragment | 3.65 | AAI-based (medium-confidence) | 0 |
| S1-final-viral-combined_k141_98846\|\|full | 2623 | No | NA | 5 | 0 | 2 | Not-determined | Genome-fragment | NA | NA | 0 |
| S1-final-viral-combined_k141_312847\|\|full | 4048 | No | NA | 4 | 0 | 2 | Not-determined | Genome-fragment | NA | NA | 0 |
| S9_k141_127222 | 1250 | No | NA | 3 | 1 | 0 | Low-quality | Genome-fragment | 3.02 | AAI-based (high-confidence) | 0 |
| S1-final-viral-combined_k141_329213\|\|full | 1017 | No | NA | 1 | 0 | 0 | Not-determined | Genome-fragment | NA | NA | 0 |
| S1-final-viral-combined_k141_131842\|\|full | 1897 | No | NA | 2 | 0 | 2 | Not-determined | Genome-fragment | NA | NA | 0 |
| S1-final-viral-combined_k141_197551\|\|full | 1738 | No | NA | 4 | 0 | 1 | Not-determined | Genome-fragment | NA | NA | 0 |
| S1-final-viral-combined_k141_246918\|\|full | 1139 | No | NA | 2 | 0 | 0 | Not-determined | Genome-fragment | NA | NA | 0 |
| S1-final-viral-combined_k141_16599\|\|full | 6002 | No | NA | 9 | 0 | 2 | Not-determined | Genome-fragment | NA | NA | 0 |
| S1-final-viral-combined_k141_230510\|\|full | 1180 | No | NA | 3 | 0 | 1 | Not-determined | Genome-fragment | NA | NA | 0 |
| S1-final-viral-combined_k141_131856\|\|full | 1634 | No | NA | 2 | 0 | 2 | Not-determined | Genome-fragment | NA | NA | 0 |
| S11-final-viral-combined_k141_299497\|\|full | 1250 | No | NA | 3 | 0 | 1 | Low-quality | Genome-fragment | 1.85 | AAI-based (medium-confidence) | 0 |
| S11-final-viral-combined_k141_163914\|\|full | 1250 | No | NA | 2 | 0 | 2 | Low-quality | Genome-fragment | 2.35 | AAI-based (medium-confidence) | 0 |
| S1-final-viral-combined_k141_197561\|\|full | 2858 | No | NA | 4 | 0 | 0 | Not-determined | Genome-fragment | NA | NA | 0 |
| S1-final-viral-combined_k141_65855\|\|full | 5397 | No | NA | 6 | 0 | 2 | Not-determined | Genome-fragment | NA | NA | 0 |
| S11-final-viral-combined_k141_32982\|\|lt2gene | 1250 | No | NA | 2 | 2 | 0 | Low-quality | Genome-fragment | 2.92 | AAI-based (medium-confidence) | 0 |
| S1-final-viral-combined_k141_312862\|\|full | 1523 | No | NA | 3 | 0 | 2 | Not-determined | Genome-fragment | NA | NA | 0 |
| S1-final-viral-combined_k141_362174\|\|full | 4671 | No | NA | 2 | 0 | 2 | Not-determined | Genome-fragment | NA | NA | 0 |
| S1-final-viral-combined_k141_65862\|\|full | 2109 | No | NA | 3 | 0 | 2 | Not-determined | Genome-fragment | NA | NA | 0 |
| S1-final-viral-combined_k141_16621\|\|full | 2421 | No | NA | 3 | 0 | 2 | Not-determined | Genome-fragment | NA | NA | 0 |
| S1-final-viral-combined_k141_246923\|\|full | 30540 | No | NA | 42 | 9 | 0 | Medium-quality | Genome-fragment | 58.68 | HMM-based (lower-bound) | 0 |
| S1-final-viral-combined_k141_164693\|\|full | 31466 | No | NA | 23 | 0 | 17 | Not-determined | Genome-fragment | NA | NA | 0 |
| S1-final-viral-combined_k141_362181\|\|full | 1359 | No | NA | 1 | 0 | 1 | Not-determined | Genome-fragment | NA | NA | 0 |
| S1-final-viral-combined_k141_296349\|\|full | 1975 | No | NA | 3 | 0 | 2 | Not-determined | Genome-fragment | NA | NA | 0 |
| S1-final-viral-combined_k141_104145\|\|full | 1250 | No | NA | 2 | 0 | 1 | Low-quality | Genome-fragment | 3.76 | AAI-based (medium-confidence) | 0 |
| S1-final-viral-combined_k141_6228\|\|full | 1250 | No | NA | 4 | 4 | 0 | Low-quality | Genome-fragment | 2.84 | HMM-based (lower-bound) | 0 |
| S1-final-viral-combined_k141_131904\|\|full | 1036 | No | NA | 3 | 0 | 0 | Not-determined | Genome-fragment | NA | NA | 0 |
| S1-final-viral-combined_k141_205779\|\|full | 1250 | No | NA | 3 | 2 | 0 | Low-quality | Genome-fragment | 0.92 | AAI-based (high-confidence) | 0 |
| S1_k141_182070 | 1250 | No | NA | 2 | 0 | 0 | Low-quality | Genome-fragment | 2.99 | AAI-based (high-confidence) | 0 |
| S3-final-viral-combined_k141_58480\|\|full | 1250 | No | NA | 3 | 0 | 0 | Low-quality | Genome-fragment | 2.46 | AAI-based (high-confidence) | 0 |
| S1-final-viral-combined_k141_197622\|\|full | 3109 | No | NA | 2 | 0 | 2 | Not-determined | Genome-fragment | NA | NA | 0 |
| S3-final-viral-combined_k141_36652\|\|lt2gene | 1250 | No | NA | 2 | 2 | 0 | Low-quality | Genome-fragment | 2.76 | AAI-based (high-confidence) | 0 |
| S5-final-viral-combined_k141_157670\|\|lt2gene | 1250 | No | NA | 1 | 1 | 0 | Low-quality | Genome-fragment | 1.97 | HMM-based (lower-bound) | 0 |
| S1-final-viral-combined_k141_296379\|\|full | 3834 | No | NA | 4 | 0 | 2 | Not-determined | Genome-fragment | NA | NA | 0 |
| S1-final-viral-combined_k141_263400\|\|full | 1731 | No | NA | 3 | 0 | 0 | Not-determined | Genome-fragment | NA | NA | 0 |
| S7-final-viral-combined_k141_185228\|\|full | 1250 | No | NA | 2 | 0 | 0 | Low-quality | Genome-fragment | 3 | AAI-based (high-confidence) | 0 |
| S1-final-viral-combined_k141_279969\|\|full | 1850 | No | NA | 3 | 0 | 0 | Not-determined | Genome-fragment | NA | NA | 0 |
| S1-final-viral-combined_k141_296413\|\|full | 2249 | No | NA | 4 | 0 | 0 | Not-determined | Genome-fragment | NA | NA | 0 |
| S1-final-viral-combined_k141_345719\|\|full | 1304 | No | NA | 3 | 0 | 0 | Not-determined | Genome-fragment | NA | NA | 0 |
| S7-final-viral-combined_k141_219151\|\|full | 1250 | No | NA | 3 | 1 | 0 | Low-quality | Genome-fragment | 2.92 | AAI-based (high-confidence) | 0 |
| S1-final-viral-combined_k141_115431\|\|full | 5502 | No | NA | 6 | 0 | 3 | Not-determined | Genome-fragment | NA | NA | 0 |
| S1-final-viral-combined_k141_345728\|\|full | 1759 | No | NA | 3 | 0 | 3 | Not-determined | Genome-fragment | NA | NA | 0 |
| S17-final-viral-combined_k141_120805\|\|full | 1250 | No | NA | 2 | 0 | 0 | Low-quality | Genome-fragment | 2.11 | AAI-based (medium-confidence) | 0 |
| S12_k141_39131 | 1250 | No | NA | 3 | 0 | 0 | Low-quality | Genome-fragment | 2.77 | AAI-based (medium-confidence) | 0 |
| S8-final-viral-combined_k141_180127\|\|lt2gene | 1250 | No | NA | 1 | 1 | 0 | Low-quality | Genome-fragment | 2.84 | AAI-based (high-confidence) | 0 |
| S1-final-viral-combined_k141_164775\|\|full | 2449 | No | NA | 4 | 0 | 0 | Not-determined | Genome-fragment | NA | NA | 0 |
| S1-final-viral-combined_k141_131979\|\|full | 1309 | No | NA | 3 | 0 | 0 | Not-determined | Genome-fragment | NA | NA | 0 |
| S1-final-viral-combined_k141_98968\|\|full | 2423 | No | NA | 3 | 0 | 1 | Not-determined | Genome-fragment | NA | NA | 0 |
| S1-final-viral-combined_k141_131989\|\|full | 2979 | No | NA | 2 | 0 | 2 | Not-determined | Genome-fragment | NA | NA | 0 |
| S11-final-viral-combined_k141_321053\|\|lt2gene | 1251 | No | NA | 2 | 1 | 0 | Low-quality | Genome-fragment | 0.81 | AAI-based (high-confidence) | 0 |
| S1-final-viral-combined_k141_59467\|\|full | 1251 | No | NA | 4 | 1 | 0 | Low-quality | Genome-fragment | 2.58 | AAI-based (medium-confidence) | 0 |
| S1-final-viral-combined_k141_258321\|\|full | 1251 | No | NA | 2 | 0 | 0 | Low-quality | Genome-fragment | 2.29 | AAI-based (medium-confidence) | 0 |
| S1-final-viral-combined_k141_49597\|\|full | 2649 | No | NA | 3 | 0 | 3 | Not-determined | Genome-fragment | NA | NA | 0 |
| S1-final-viral-combined_k141_230689\|\|full | 1097 | No | NA | 3 | 0 | 0 | Not-determined | Genome-fragment | NA | NA | 0 |
| S7-final-viral-combined_k141_16549\|\|full | 1251 | No | NA | 2 | 0 | 0 | Low-quality | Genome-fragment | 2.93 | AAI-based (medium-confidence) | 0 |
| S1-final-viral-combined_k141_16783\|\|full | 3449 | No | NA | 3 | 0 | 1 | Not-determined | Genome-fragment | NA | NA | 0 |
| S1-final-viral-combined_k141_164818\|\|full | 2083 | No | NA | 3 | 0 | 2 | Not-determined | Genome-fragment | NA | NA | 0 |
| S17-k141_76634 | 1251 | No | NA | 1 | 0 | 1 | Low-quality | Genome-fragment | 0.33 | AAI-based (medium-confidence) | 0 |
| S12-final-viral-combined_k141_98847\|\|full | 1251 | No | NA | 2 | 0 | 1 | Low-quality | Genome-fragment | 0.51 | AAI-based (medium-confidence) | 0 |
| S1-final-viral-combined_k141_181277\|\|full | 6877 | No | NA | 8 | 0 | 1 | Not-determined | Genome-fragment | NA | NA | 0 |
| S1-final-viral-combined_k141_115473\|\|full | 1841 | No | NA | 3 | 0 | 2 | Not-determined | Genome-fragment | NA | NA | 0 |
| S1-final-viral-combined_k141_329318\|\|full | 2509 | No | NA | 4 | 0 | 1 | Not-determined | Genome-fragment | NA | NA | 0 |
| S1-final-viral-combined_k141_520\|\|full | 5155 | No | NA | 3 | 0 | 3 | Not-determined | Genome-fragment | NA | NA | 0 |
| S1-final-viral-combined_k141_49641\|\|full | 5667 | No | NA | 6 | 0 | 2 | Not-determined | Genome-fragment | NA | NA | 0 |
| S1-final-viral-combined_k141_329334\|\|full | 1549 | No | NA | 2 | 0 | 1 | Not-determined | Genome-fragment | NA | NA | 0 |
| S1-final-viral-combined_k141_148362\|\|full | 4458 | No | NA | 5 | 0 | 5 | Not-determined | Genome-fragment | NA | NA | 0 |
| S12-final-viral-combined_k141_384594\|\|full | 1251 | No | NA | 3 | 1 | 0 | Low-quality | Genome-fragment | 1.96 | AAI-based (medium-confidence) | 0 |
| S1-final-viral-combined_k141_378728\|\|full | 3309 | No | NA | 4 | 0 | 1 | Not-determined | Genome-fragment | NA | NA | 0 |
| S1-final-viral-combined_k141_132065\|\|full | 1111 | No | NA | 3 | 0 | 1 | Not-determined | Genome-fragment | NA | NA | 0 |
| S1-final-viral-combined_k141_230768\|\|full | 7742 | No | NA | 10 | 0 | 1 | Not-determined | Genome-fragment | NA | NA | 0 |
| S1-final-viral-combined_k141_345795\|\|full | 1614 | No | NA | 2 | 0 | 2 | Not-determined | Genome-fragment | NA | NA | 0 |
| S1-final-viral-combined_k141_181306\|\|full | 1755 | No | NA | 2 | 0 | 2 | Not-determined | Genome-fragment | NA | NA | 0 |
| S2-final-viral-combined_k141_172083\|\|full | 1251 | No | NA | 3 | 0 | 0 | Low-quality | Genome-fragment | 2.58 | AAI-based (medium-confidence) | 0 |
| S4-final-viral-combined_k141_9637\|\|full | 1251 | No | NA | 3 | 0 | 0 | Low-quality | Genome-fragment | 3.26 | AAI-based (medium-confidence) | 0 |
| S1-final-viral-combined_k141_296563\|\|full | 1392 | No | NA | 3 | 0 | 0 | Not-determined | Genome-fragment | NA | NA | 0 |
| S8-final-viral-combined_k141_40783\|\|full | 1251 | No | NA | 3 | 0 | 0 | Low-quality | Genome-fragment | 2.38 | AAI-based (medium-confidence) | 0 |
| S8_k141_1032 | 1251 | No | NA | 1 | 0 | 0 | Low-quality | Genome-fragment | 0.9 | AAI-based (high-confidence) | 0 |
| S1-final-viral-combined_k141_569\|\|full | 3426 | No | NA | 2 | 0 | 2 | Not-determined | Genome-fragment | NA | NA | 0 |
| S1-final-viral-combined_k141_362379\|\|full | 5941 | No | NA | 5 | 0 | 4 | Not-determined | Genome-fragment | NA | NA | 0 |
| S9-final-viral-combined_k141_14472\|\|full | 1252 | No | NA | 3 | 0 | 1 | Low-quality | Genome-fragment | 0.55 | AAI-based (medium-confidence) | 0 |
| S9_k141_8396 | 1252 | No | NA | 1 | 0 | 0 | Low-quality | Genome-fragment | 1.51 | AAI-based (high-confidence) | 0 |
| S1-final-viral-combined_k141_313044\|\|full | 2141 | No | NA | 4 | 0 | 3 | Not-determined | Genome-fragment | NA | NA | 0 |
| S1-final-viral-combined_k141_362385\|\|full | 4873 | No | NA | 5 | 0 | 3 | Not-determined | Genome-fragment | NA | NA | 0 |
| S11-final-viral-combined_k141_3767\|\|lt2gene | 1252 | No | NA | 2 | 0 | 0 | Low-quality | Genome-fragment | 2.83 | AAI-based (medium-confidence) | 0 |
| S1_k141_341663 | 1252 | No | NA | 5 | 0 | 0 | Low-quality | Genome-fragment | 2.02 | AAI-based (high-confidence) | 0 |
| S1-final-viral-combined_k141_263562\|\|full | 29679 | No | NA | 44 | 4 | 0 | Medium-quality | Genome-fragment | 64.61 | AAI-based (high-confidence) | 0 |
| S1-final-viral-combined_k141_115571\|\|full | 1874 | No | NA | 4 | 0 | 3 | Not-determined | Genome-fragment | NA | NA | 0 |
| S1-final-viral-combined_k141_33145\|\|full | 4351 | No | NA | 4 | 0 | 2 | Not-determined | Genome-fragment | NA | NA | 0 |
| S1-final-viral-combined_k141_49700\|\|full | 2906 | No | NA | 5 | 0 | 0 | Not-determined | Genome-fragment | NA | NA | 0 |
| S1-final-viral-combined_k141_362395\|\|full | 5235 | No | NA | 4 | 0 | 3 | Not-determined | Genome-fragment | NA | NA | 0 |
| S1-final-viral-combined_k141_230822\|\|full | 3064 | No | NA | 2 | 0 | 1 | Not-determined | Genome-fragment | NA | NA | 0 |
| S1-final-viral-combined_k141_132120\|\|full | 1370 | No | NA | 2 | 0 | 2 | Not-determined | Genome-fragment | NA | NA | 0 |
| S1-final-viral-combined_k141_197846\|\|full | 1026 | No | NA | 2 | 0 | 2 | Not-determined | Genome-fragment | NA | NA | 0 |
| S1-final-viral-combined_k141_99095\|\|full | 1314 | No | NA | 3 | 0 | 2 | Not-determined | Genome-fragment | NA | NA | 0 |
| S1-final-viral-combined_k141_164894\|\|full | 29496 | No | NA | 46 | 21 | 0 | Medium-quality | Genome-fragment | 85.65 | AAI-based (high-confidence) | 0 |
| S1-final-viral-combined_k141_280157\|\|full | 2889 | No | NA | 2 | 0 | 1 | Not-determined | Genome-fragment | NA | NA | 0 |
| S1-final-viral-combined_k141_280172\|\|full | 3616 | No | NA | 6 | 0 | 1 | Not-determined | Genome-fragment | NA | NA | 0 |
| S7-final-viral-combined_k141_22946\|\|full | 1252 | No | NA | 3 | 1 | 0 | Low-quality | Genome-fragment | 5.94 | AAI-based (high-confidence) | 0 |
| S17-final-viral-combined_k141_55103\|\|full | 1252 | No | NA | 2 | 0 | 1 | Low-quality | Genome-fragment | 2.17 | AAI-based (medium-confidence) | 0 |
| S1-final-viral-combined_k141_33184\|\|full | 1076 | No | NA | 2 | 0 | 1 | Not-determined | Genome-fragment | NA | NA | 0 |
| S12-final-viral-combined_k141_156170\|\|full | 1252 | No | NA | 2 | 1 | 0 | Low-quality | Genome-fragment | 1.72 | AAI-based (medium-confidence) | 0 |
| S12-final-viral-combined_k141_332767\|\|lt2gene | 1252 | No | NA | 1 | 1 | 0 | Low-quality | Genome-fragment | 2.64 | AAI-based (high-confidence) | 0 |
| S12_k141_76718 | 1252 | No | NA | 2 | 0 | 0 | Low-quality | Genome-fragment | 2.41 | AAI-based (high-confidence) | 0 |
| S12_k141_365527 | 1252 | No | NA | 3 | 0 | 0 | Low-quality | Genome-fragment | 2.31 | AAI-based (medium-confidence) | 0 |
| S1-final-viral-combined_k141_263600\|\|full | 1973 | No | NA | 3 | 0 | 1 | Not-determined | Genome-fragment | NA | NA | 0 |
| S1-final-viral-combined_k141_66039\|\|full | 1937 | No | NA | 3 | 0 | 1 | Not-determined | Genome-fragment | NA | NA | 0 |
| S2-final-viral-combined_k141_29076\|\|full | 1252 | No | NA | 4 | 2 | 0 | Low-quality | Genome-fragment | 2 | AAI-based (high-confidence) | 0 |
| S1-final-viral-combined_k141_132134\|\|full | 9922 | No | NA | 10 | 0 | 9 | Not-determined | Genome-fragment | NA | NA | 0 |
| S1-final-viral-combined_k141_33199\|\|full | 1519 | No | NA | 3 | 0 | 1 | Not-determined | Genome-fragment | NA | NA | 0 |
| S1-final-viral-combined_k141_181436\|\|full | 1352 | No | NA | 3 | 0 | 1 | Not-determined | Genome-fragment | NA | NA | 0 |
| S1-final-viral-combined_k141_49766\|\|full | 1294 | No | NA | 2 | 0 | 0 | Not-determined | Genome-fragment | NA | NA | 0 |
| S8-final-viral-combined_k141_241935\|\|full | 1252 | No | NA | 3 | 0 | 0 | Low-quality | Genome-fragment | 2.75 | AAI-based (high-confidence) | 0 |
| S1-final-viral-combined_k141_329445\|\|full | 1792 | No | NA | 4 | 0 | 0 | Not-determined | Genome-fragment | NA | NA | 0 |
| S1-final-viral-combined_k141_280210\|\|full | 1105 | No | NA | 2 | 0 | 1 | Not-determined | Genome-fragment | NA | NA | 0 |
| S1-final-viral-combined_k141_181457\|\|full | 1798 | No | NA | 3 | 0 | 1 | Not-determined | Genome-fragment | NA | NA | 0 |
| S1-final-viral-combined_k141_280217\|\|full | 1290 | No | NA | 3 | 0 | 0 | Not-determined | Genome-fragment | NA | NA | 0 |
| S1-final-viral-combined_k141_230849\|\|full | 1348 | No | NA | 3 | 0 | 0 | Not-determined | Genome-fragment | NA | NA | 0 |
| S1-final-viral-combined_k141_230856\|\|full | 1528 | No | NA | 2 | 0 | 1 | Not-determined | Genome-fragment | NA | NA | 0 |
| S1-final-viral-combined_k141_115656\|\|full | 2196 | No | NA | 3 | 0 | 1 | Not-determined | Genome-fragment | NA | NA | 0 |
| S11_k141_135261 | 1253 | No | NA | 2 | 0 | 0 | Low-quality | Genome-fragment | 2.08 | AAI-based (medium-confidence) | 0 |
| S1-final-viral-combined_k141_296624\|\|full | 1938 | No | NA | 2 | 0 | 1 | Not-determined | Genome-fragment | NA | NA | 0 |
| S1_k141_83431 | 1253 | No | NA | 1 | 0 | 1 | Low-quality | Genome-fragment | 0.22 | AAI-based (high-confidence) | 0 |
| S1-final-viral-combined_k141_313129\|\|full | 2009 | No | NA | 2 | 0 | 1 | Not-determined | Genome-fragment | NA | NA | 0 |
| S1-final-viral-combined_k141_378876\|\|full | 2433 | No | NA | 4 | 0 | 2 | Not-determined | Genome-fragment | NA | NA | 0 |
| S1-final-viral-combined_k141_378882\|\|full | 1069 | No | NA | 3 | 0 | 0 | Not-determined | Genome-fragment | NA | NA | 0 |
| S1-final-viral-combined_k141_148488\|\|full | 14737 | No | NA | 15 | 0 | 11 | Not-determined | Genome-fragment | NA | NA | 0 |
| S1-final-viral-combined_k141_230872\|\|full | 1919 | No | NA | 3 | 0 | 2 | Not-determined | Genome-fragment | NA | NA | 0 |
| S2_k141_185164 | 1253 | No | NA | 1 | 0 | 1 | Low-quality | Genome-fragment | 1.08 | AAI-based (high-confidence) | 0 |
| S1-final-viral-combined_k141_99204\|\|full | 8186 | No | NA | 8 | 0 | 5 | Not-determined | Genome-fragment | NA | NA | 0 |
| S1-final-viral-combined_k141_115697\|\|full | 4304 | No | NA | 5 | 0 | 1 | Not-determined | Genome-fragment | NA | NA | 0 |
| S1-final-viral-combined_k141_16922\|\|full | 1604 | No | NA | 3 | 0 | 0 | Not-determined | Genome-fragment | NA | NA | 0 |
| S1-final-viral-combined_k141_132190\|\|full | 2921 | No | NA | 4 | 0 | 0 | Not-determined | Genome-fragment | NA | NA | 0 |
| S4-final-viral-combined_k141_40084\|\|full | 1253 | No | NA | 3 | 0 | 0 | Low-quality | Genome-fragment | 0.78 | AAI-based (medium-confidence) | 0 |
| S1-final-viral-combined_k141_115702\|\|full | 8565 | No | NA | 10 | 0 | 6 | Not-determined | Genome-fragment | NA | NA | 0 |
| S1-final-viral-combined_k141_329493\|\|full | 3415 | No | NA | 5 | 0 | 1 | Not-determined | Genome-fragment | NA | NA | 0 |
| S1-final-viral-combined_k141_82836\|\|full | 3954 | No | NA | 5 | 0 | 4 | Not-determined | Genome-fragment | NA | NA | 0 |
| S1-final-viral-combined_k141_99231\|\|full | 1200 | No | NA | 2 | 0 | 1 | Not-determined | Genome-fragment | NA | NA | 0 |
| S1-final-viral-combined_k141_132226\|\|full | 1685 | No | NA | 2 | 0 | 2 | Not-determined | Genome-fragment | NA | NA | 0 |
| S1-final-viral-combined_k141_66078\|\|full | 7206 | No | NA | 7 | 0 | 4 | Not-determined | Genome-fragment | NA | NA | 0 |
| S1-final-viral-combined_k141_296707\|\|full | 1491 | No | NA | 3 | 0 | 0 | Not-determined | Genome-fragment | NA | NA | 0 |
| S8-final-viral-combined_k141_94913\|\|full | 1253 | No | NA | 2 | 0 | 0 | Low-quality | Genome-fragment | 3 | AAI-based (high-confidence) | 0 |
| S1-final-viral-combined_k141_181565\|\|full | 1189 | No | NA | 2 | 0 | 1 | Not-determined | Genome-fragment | NA | NA | 0 |
| S11-final-viral-combined_k141_267883\|\|full | 1254 | No | NA | 3 | 0 | 1 | Low-quality | Genome-fragment | 2.76 | AAI-based (medium-confidence) | 0 |
| S1_k141_213612 | 1254 | No | NA | 3 | 0 | 0 | Low-quality | Genome-fragment | 3.8 | AAI-based (medium-confidence) | 0 |
| S1_k141_148001 | 1254 | No | NA | 3 | 0 | 1 | Low-quality | Genome-fragment | 4 | AAI-based (medium-confidence) | 0 |
| S17-final-viral-combined_k141_37822\|\|lt2gene | 1254 | No | NA | 2 | 0 | 2 | Low-quality | Genome-fragment | 0.95 | AAI-based (medium-confidence) | 0 |
| S12-final-viral-combined_k141_3232\|\|full | 1254 | No | NA | 3 | 0 | 2 | Low-quality | Genome-fragment | 1.72 | AAI-based (medium-confidence) | 0 |
| S1-final-viral-combined_k141_66136\|\|full | 1579 | No | NA | 4 | 0 | 0 | Not-determined | Genome-fragment | NA | NA | 0 |
| S2-final-viral-combined_k141_99010\|\|full | 1254 | No | NA | 3 | 0 | 0 | Low-quality | Genome-fragment | 2.69 | AAI-based (medium-confidence) | 0 |
| S1-final-viral-combined_k141_296744\|\|full | 1077 | No | NA | 2 | 0 | 0 | Not-determined | Genome-fragment | NA | NA | 0 |
| S1-final-viral-combined_k141_66159\|\|full | 1712 | No | NA | 1 | 0 | 0 | Not-determined | Genome-fragment | NA | NA | 0 |
| S1-final-viral-combined_k141_181626\|\|full | 4731 | No | NA | 5 | 0 | 3 | Not-determined | Genome-fragment | NA | NA | 0 |
| S1-final-viral-combined_k141_346049\|\|full | 7612 | No | NA | 7 | 0 | 7 | Not-determined | Genome-fragment | NA | NA | 0 |
| S1-final-viral-combined_k141_115820\|\|full | 1216 | No | NA | 2 | 0 | 0 | Not-determined | Genome-fragment | NA | NA | 0 |
| S1-final-viral-combined_k141_66176\|\|full | 8491 | No | NA | 6 | 0 | 2 | Not-determined | Genome-fragment | NA | NA | 0 |
| S1-final-viral-combined_k141_296777\|\|full | 1489 | No | NA | 2 | 0 | 0 | Not-determined | Genome-fragment | NA | NA | 0 |
| S1-final-viral-combined_k141_132334\|\|full | 3908 | No | NA | 5 | 0 | 1 | Not-determined | Genome-fragment | NA | NA | 0 |
| S2-final-viral-combined_k141_184686\|\|full | 1254 | No | NA | 3 | 1 | 0 | Low-quality | Genome-fragment | 0.83 | AAI-based (high-confidence) | 0 |
| S1-final-viral-combined_k141_148617\|\|full | 2261 | No | NA | 3 | 0 | 2 | Not-determined | Genome-fragment | NA | NA | 0 |
| S6_k141_66521 | 1254 | No | NA | 1 | 0 | 0 | Low-quality | Genome-fragment | 2.45 | AAI-based (high-confidence) | 0 |
| S1-final-viral-combined_k141_198030\|\|full | 1150 | No | NA | 2 | 0 | 2 | Not-determined | Genome-fragment | NA | NA | 0 |
| S8-final-viral-combined_k141_8549\|\|lt2gene | 1254 | No | NA | 2 | 2 | 0 | Low-quality | Genome-fragment | 1.96 | HMM-based (lower-bound) | 0 |
| S9_k141_125496 | 1255 | No | NA | 1 | 0 | 1 | Low-quality | Genome-fragment | 3.54 | AAI-based (medium-confidence) | 0 |
| S11-final-viral-combined_k141_111032\|\|full | 1255 | No | NA | 2 | 1 | 0 | Low-quality | Genome-fragment | 2.63 | AAI-based (high-confidence) | 0 |
| S1-final-viral-combined_k141_115877\|\|full | 1096 | No | NA | 2 | 0 | 2 | Not-determined | Genome-fragment | NA | NA | 0 |
| S1-final-viral-combined_k141_82984\|\|full | 1845 | No | NA | 2 | 0 | 1 | Not-determined | Genome-fragment | NA | NA | 0 |
| S1-final-viral-combined_k141_17057\|\|full | 2661 | No | NA | 5 | 0 | 0 | Not-determined | Genome-fragment | NA | NA | 0 |
| S11_k141_208527 | 1255 | No | NA | 3 | 0 | 0 | Low-quality | Genome-fragment | 2.76 | AAI-based (high-confidence) | 0 |
| S1-final-viral-combined_k141_198055\|\|full | 2480 | No | NA | 3 | 0 | 3 | Not-determined | Genome-fragment | NA | NA | 0 |
| S1-final-viral-combined_k141_17065\|\|full | 1807 | No | NA | 4 | 0 | 0 | Not-determined | Genome-fragment | NA | NA | 0 |
| S1_k141_271227 | 1255 | No | NA | 4 | 1 | 0 | Low-quality | Genome-fragment | 2.67 | AAI-based (medium-confidence) | 0 |
| S3-final-viral-combined_k141_60057\|\|full | 1255 | No | NA | 3 | 1 | 0 | Low-quality | Genome-fragment | 2.28 | AAI-based (high-confidence) | 0 |
| S1-final-viral-combined_k141_263835\|\|full | 6120 | No | NA | 6 | 0 | 3 | Not-determined | Genome-fragment | NA | NA | 0 |
| S5-final-viral-combined_k141_194600\|\|lt2gene | 1255 | No | NA | 1 | 1 | 0 | Low-quality | Genome-fragment | 0.73 | AAI-based (high-confidence) | 0 |
| S1-final-viral-combined_k141_362573\|\|full | 1045 | No | NA | 2 | 0 | 2 | Not-determined | Genome-fragment | NA | NA | 0 |
| S7-final-viral-combined_k141_150578\|\|full | 1255 | No | NA | 4 | 0 | 0 | Low-quality | Genome-fragment | 2.82 | AAI-based (medium-confidence) | 0 |
| S1-final-viral-combined_k141_247416\|\|full | 3573 | No | NA | 3 | 0 | 1 | Not-determined | Genome-fragment | NA | NA | 0 |
| S2-final-viral-combined_k141_82342\|\|lt2gene | 1255 | No | NA | 2 | 1 | 0 | Low-quality | Genome-fragment | 2.42 | AAI-based (medium-confidence) | 0 |
| S4-final-viral-combined_k141_84318\|\|full | 1255 | No | NA | 3 | 1 | 0 | Low-quality | Genome-fragment | 2.95 | AAI-based (medium-confidence) | 0 |
| S6-final-viral-combined_k141_132593\|\|full | 1255 | No | NA | 2 | 0 | 1 | Low-quality | Genome-fragment | 2.1 | AAI-based (medium-confidence) | 0 |
| S8-final-viral-combined_k141_75795\|\|full | 1255 | No | NA | 3 | 1 | 0 | Low-quality | Genome-fragment | 2.08 | AAI-based (high-confidence) | 0 |
| S1-final-viral-combined_k141_115950\|\|full | 3853 | No | NA | 6 | 0 | 0 | Not-determined | Genome-fragment | NA | NA | 0 |
| S1-final-viral-combined_k141_165159\|\|full | 4317 | No | NA | 6 | 0 | 1 | Not-determined | Genome-fragment | NA | NA | 0 |
| S1-final-viral-combined_k141_346214\|\|full | 1734 | No | NA | 2 | 0 | 2 | Not-determined | Genome-fragment | NA | NA | 0 |
| S1-final-viral-combined_k141_137654\|\|full | 1256 | No | NA | 3 | 0 | 0 | Low-quality | Genome-fragment | 1.5 | AAI-based (medium-confidence) | 0 |
| S1_k141_212297 | 1256 | No | NA | 1 | 0 | 1 | Low-quality | Genome-fragment | 0.47 | AAI-based (medium-confidence) | 0 |
| S17-k141_24160 | 1256 | No | NA | 2 | 0 | 2 | Low-quality | Genome-fragment | 0.36 | AAI-based (medium-confidence) | 0 |
| S12-final-viral-combined_k141_393318\|\|full | 1256 | No | NA | 3 | 1 | 0 | Low-quality | Genome-fragment | 2.11 | AAI-based (medium-confidence) | 0 |
| S1-final-viral-combined_k141_346236\|\|full | 7326 | No | NA | 8 | 0 | 5 | Not-determined | Genome-fragment | NA | NA | 0 |
| S1-final-viral-combined_k141_329694\|\|full | 1051 | No | NA | 3 | 0 | 0 | Not-determined | Genome-fragment | NA | NA | 0 |
| S2-final-viral-combined_k141_92149\|\|full | 1256 | No | NA | 3 | 0 | 0 | Low-quality | Genome-fragment | 3.38 | AAI-based (medium-confidence) | 0 |
| S1-final-viral-combined_k141_346243\|\|full | 1951 | No | NA | 3 | 0 | 0 | Not-determined | Genome-fragment | NA | NA | 0 |
| S4-final-viral-combined_k141_72799\|\|full | 1256 | No | NA | 2 | 0 | 0 | Low-quality | Genome-fragment | 1.94 | AAI-based (high-confidence) | 0 |
| S6-final-viral-combined_k141_133667\|\|full | 1256 | No | NA | 4 | 1 | 0 | Low-quality | Genome-fragment | 3.45 | HMM-based (lower-bound) | 0 |
| S9_k141_117264 | 1257 | No | NA | 2 | 0 | 0 | Low-quality | Genome-fragment | 2.03 | AAI-based (medium-confidence) | 0 |
| S11_k141_162534 | 1257 | No | NA | 2 | 0 | 0 | Low-quality | Genome-fragment | 1.95 | AAI-based (high-confidence) | 0 |
| S1-final-viral-combined_k141_165204\|\|full | 19280 | No | NA | 19 | 0 | 13 | Not-determined | Genome-fragment | NA | NA | 0 |
| S1-final-viral-combined_k141_296801\|\|full | 1488 | No | NA | 2 | 0 | 0 | Not-determined | Genome-fragment | NA | NA | 0 |
| S1-final-viral-combined_k141_292365\|\|full | 1257 | No | NA | 5 | 1 | 0 | Low-quality | Genome-fragment | 3.35 | AAI-based (medium-confidence) | 0 |
| S1_k141_92191 | 1257 | No | NA | 3 | 2 | 0 | Low-quality | Genome-fragment | 3.5 | AAI-based (high-confidence) | 0 |
| S1-final-viral-combined_k141_148726\|\|full | 2511 | No | NA | 3 | 0 | 2 | Not-determined | Genome-fragment | NA | NA | 0 |
| S1-final-viral-combined_k141_66238\|\|full | 4280 | No | NA | 5 | 0 | 4 | Not-determined | Genome-fragment | NA | NA | 0 |
| S1-final-viral-combined_k141_379037\|\|full | 3595 | No | NA | 3 | 0 | 1 | Not-determined | Genome-fragment | NA | NA | 0 |
| S1_k141_78736 | 1257 | No | NA | 2 | 0 | 1 | Low-quality | Genome-fragment | 0.42 | AAI-based (medium-confidence) | 0 |
| S1-final-viral-combined_k141_362683\|\|full | 1893 | No | NA | 3 | 0 | 0 | Not-determined | Genome-fragment | NA | NA | 0 |
| S5-final-viral-combined_k141_6870\|\|full | 1257 | No | NA | 3 | 0 | 0 | Low-quality | Genome-fragment | 3.16 | AAI-based (medium-confidence) | 0 |
| S5-final-viral-combined_k141_158416\|\|lt2gene | 1257 | No | NA | 2 | 1 | 0 | Low-quality | Genome-fragment | 0.69 | HMM-based (lower-bound) | 0 |
| S1-final-viral-combined_k141_231001\|\|full | 5638 | No | NA | 5 | 0 | 3 | Not-determined | Genome-fragment | NA | NA | 0 |
| S1-final-viral-combined_k141_927\|\|full | 1954 | No | NA | 4 | 0 | 2 | Not-determined | Genome-fragment | NA | NA | 0 |
| S7-final-viral-combined_k141_203239\|\|full | 1257 | No | NA | 3 | 2 | 0 | Low-quality | Genome-fragment | 3.39 | AAI-based (high-confidence) | 0 |
| S1-final-viral-combined_k141_17181\|\|full | 1423 | No | NA | 4 | 0 | 0 | Not-determined | Genome-fragment | NA | NA | 0 |
| S1-final-viral-combined_k141_83124\|\|full | 1993 | No | NA | 4 | 0 | 0 | Not-determined | Genome-fragment | NA | NA | 0 |
| S12-final-viral-combined_k141_22510\|\|full | 1257 | No | NA | 4 | 1 | 0 | Low-quality | Genome-fragment | 3.67 | AAI-based (high-confidence) | 0 |
| S12-final-viral-combined_k141_162794\|\|full | 1257 | No | NA | 3 | 0 | 0 | Low-quality | Genome-fragment | 1.84 | AAI-based (medium-confidence) | 0 |
| S12-final-viral-combined_k141_197981\|\|full | 1257 | No | NA | 3 | 2 | 0 | Low-quality | Genome-fragment | 2.49 | AAI-based (medium-confidence) | 0 |
| S12_k141_77351 | 1257 | No | NA | 2 | 0 | 1 | Low-quality | Genome-fragment | 0.43 | AAI-based (high-confidence) | 0 |
| S1-final-viral-combined_k141_116093\|\|full | 2600 | No | NA | 2 | 0 | 2 | Not-determined | Genome-fragment | NA | NA | 0 |
| S1-final-viral-combined_k141_198227\|\|full | 2396 | No | NA | 4 | 0 | 2 | Not-determined | Genome-fragment | NA | NA | 0 |
| S1-final-viral-combined_k141_247557\|\|full | 3920 | No | NA | 5 | 0 | 1 | Not-determined | Genome-fragment | NA | NA | 0 |
| S1-final-viral-combined_k141_313279\|\|full | 2341 | No | NA | 5 | 0 | 1 | Not-determined | Genome-fragment | NA | NA | 0 |
| S1-final-viral-combined_k141_280547\|\|full | 3342 | No | NA | 4 | 0 | 2 | Not-determined | Genome-fragment | NA | NA | 0 |
| S1-final-viral-combined_k141_264022\|\|full | 1672 | No | NA | 4 | 0 | 3 | Not-determined | Genome-fragment | NA | NA | 0 |
| S2-final-viral-combined_k141_54558\|\|lt2gene | 1257 | No | NA | 2 | 2 | 0 | Low-quality | Genome-fragment | 3.29 | AAI-based (high-confidence) | 0 |
| S1-final-viral-combined_k141_247577\|\|full | 3867 | No | NA | 3 | 0 | 3 | Not-determined | Genome-fragment | NA | NA | 0 |
| S1-final-viral-combined_k141_247578\|\|full | 4106 | No | NA | 7 | 0 | 3 | Not-determined | Genome-fragment | NA | NA | 0 |
| S1-final-viral-combined_k141_346382\|\|full | 9721 | No | NA | 8 | 0 | 5 | Not-determined | Genome-fragment | NA | NA | 0 |
| S2_k141_71516 | 1257 | No | NA | 3 | 0 | 1 | Low-quality | Genome-fragment | 0.78 | AAI-based (medium-confidence) | 0 |
| S1-final-viral-combined_k141_346392\|\|full | 1568 | No | NA | 2 | 0 | 1 | Not-determined | Genome-fragment | NA | NA | 0 |
| S1-final-viral-combined_k141_1014\|\|full | 2051 | No | NA | 2 | 0 | 2 | Not-determined | Genome-fragment | NA | NA | 0 |
| S1-final-viral-combined_k141_313366\|\|full | 1089 | No | NA | 3 | 0 | 0 | Not-determined | Genome-fragment | NA | NA | 0 |
| S1-final-viral-combined_k141_313369\|\|full | 2881 | No | NA | 3 | 0 | 3 | Not-determined | Genome-fragment | NA | NA | 0 |
| S1-final-viral-combined_k141_296921\|\|full | 3878 | No | NA | 3 | 0 | 3 | Not-determined | Genome-fragment | NA | NA | 0 |
| S1-final-viral-combined_k141_231084\|\|full | 2193 | No | NA | 2 | 0 | 2 | Not-determined | Genome-fragment | NA | NA | 0 |
| S1-final-viral-combined_k141_148824\|\|full | 3095 | No | NA | 4 | 0 | 2 | Not-determined | Genome-fragment | NA | NA | 0 |
| S4-final-viral-combined_k141_74858\|\|full | 1257 | No | NA | 2 | 0 | 2 | Low-quality | Genome-fragment | 2.09 | AAI-based (medium-confidence) | 0 |
| S9-final-viral-combined_k141_27549\|\|full | 1258 | Yes | 773 | 4 | 2 | 1 | Low-quality | Genome-fragment | 1.75 | AAI-based (medium-confidence) | 38.55 |
| S9-final-viral-combined_k141_92090\|\|full | 1258 | No | NA | 3 | 0 | 0 | Low-quality | Genome-fragment | 2.19 | AAI-based (high-confidence) | 0 |
| S1-final-viral-combined_k141_214898\|\|full | 2240 | No | NA | 2 | 0 | 0 | Not-determined | Genome-fragment | NA | NA | 0 |
| S1-final-viral-combined_k141_329783\|\|full | 2111 | No | NA | 2 | 0 | 1 | Not-determined | Genome-fragment | NA | NA | 0 |
| S1-final-viral-combined_k141_64309\|\|full | 1258 | No | NA | 4 | 1 | 0 | Low-quality | Genome-fragment | 3.57 | AAI-based (high-confidence) | 0 |
| S1-final-viral-combined_k141_1080\|\|full | 4002 | No | NA | 6 | 0 | 1 | Not-determined | Genome-fragment | NA | NA | 0 |
| S3-final-viral-combined_k141_1896\|\|full | 1258 | No | NA | 4 | 1 | 0 | Low-quality | Genome-fragment | 2.2 | AAI-based (high-confidence) | 0 |
| S5-final-viral-combined_k141_170758\|\|full | 1258 | No | NA | 2 | 1 | 0 | Low-quality | Genome-fragment | 2.12 | AAI-based (medium-confidence) | 0 |
| S1-final-viral-combined_k141_231131\|\|full | 4792 | No | NA | 6 | 0 | 1 | Not-determined | Genome-fragment | NA | NA | 0 |
| S1-final-viral-combined_k141_296985\|\|full | 3839 | No | NA | 3 | 0 | 0 | Not-determined | Genome-fragment | NA | NA | 0 |
| S5-final-viral-combined_k141_129244\|\|full | 1258 | No | NA | 4 | 1 | 0 | Low-quality | Genome-fragment | 8.68 | AAI-based (high-confidence) | 0 |
| S1-final-viral-combined_k141_1114\|\|full | 5544 | No | NA | 7 | 0 | 3 | Not-determined | Genome-fragment | NA | NA | 0 |
| S12-final-viral-combined_k141_169520\|\|full | 1258 | No | NA | 4 | 0 | 0 | Low-quality | Genome-fragment | 3.44 | AAI-based (medium-confidence) | 0 |
| S1-final-viral-combined_k141_346507\|\|full | 1520 | No | NA | 4 | 0 | 0 | Not-determined | Genome-fragment | NA | NA | 0 |
| S1-final-viral-combined_k141_379264\|\|full | 1178 | No | NA | 2 | 0 | 2 | Not-determined | Genome-fragment | NA | NA | 0 |
| S12-final-viral-combined_k141_182199\|\|full | 1258 | No | NA | 3 | 2 | 0 | Low-quality | Genome-fragment | 2.79 | AAI-based (high-confidence) | 0 |
| S2-final-viral-combined_k141_57188\|\|full | 1258 | No | NA | 4 | 2 | 0 | Low-quality | Genome-fragment | 2.92 | AAI-based (high-confidence) | 0 |
| S2-final-viral-combined_k141_238112\|\|full | 1258 | No | NA | 2 | 0 | 2 | Low-quality | Genome-fragment | 2.85 | AAI-based (medium-confidence) | 0 |
| S4-final-viral-combined_k141_21756\|\|full | 1258 | No | NA | 3 | 0 | 0 | Low-quality | Genome-fragment | 0.79 | AAI-based (medium-confidence) | 0 |
| S1-final-viral-combined_k141_132662\|\|full | 8323 | No | NA | 10 | 0 | 10 | Not-determined | Genome-fragment | NA | NA | 0 |
| S6-final-viral-combined_k141_136267\|\|full | 1258 | No | NA | 4 | 1 | 0 | Low-quality | Genome-fragment | 1.46 | AAI-based (high-confidence) | 0 |
| S6-final-viral-combined_k141_97993\|\|full | 1258 | No | NA | 3 | 1 | 0 | Low-quality | Genome-fragment | 2.76 | AAI-based (high-confidence) | 0 |
| S8-final-viral-combined_k141_218896\|\|full | 1258 | No | NA | 3 | 0 | 1 | Low-quality | Genome-fragment | 1.62 | AAI-based (medium-confidence) | 0 |
| S1-final-viral-combined_k141_231210\|\|full | 1596 | No | NA | 3 | 0 | 0 | Not-determined | Genome-fragment | NA | NA | 0 |
| S1-final-viral-combined_k141_379289\|\|full | 6739 | No | NA | 7 | 0 | 6 | Not-determined | Genome-fragment | NA | NA | 0 |
| S8-final-viral-combined_k141_128089\|\|full | 1258 | No | NA | 2 | 0 | 1 | Low-quality | Genome-fragment | 2.97 | AAI-based (medium-confidence) | 0 |
| S11-final-viral-combined_k141_314082\|\|full | 1259 | No | NA | 4 | 0 | 0 | Low-quality | Genome-fragment | 2.03 | AAI-based (high-confidence) | 0 |
| S1-final-viral-combined_k141_147340\|\|full | 1259 | No | NA | 4 | 0 | 0 | Low-quality | Genome-fragment | 2.93 | AAI-based (high-confidence) | 0 |
| S1-final-viral-combined_k141_362955\|\|full | 1349 | No | NA | 2 | 0 | 0 | Not-determined | Genome-fragment | NA | NA | 0 |
| S1-final-viral-combined_k141_1173\|\|full | 1774 | No | NA | 3 | 0 | 2 | Not-determined | Genome-fragment | NA | NA | 0 |
| S1-final-viral-combined_k141_329842\|\|full | 2244 | No | NA | 2 | 0 | 1 | Not-determined | Genome-fragment | NA | NA | 0 |
| S1-final-viral-combined_k141_215037\|\|full | 2623 | No | NA | 3 | 0 | 2 | Not-determined | Genome-fragment | NA | NA | 0 |
| S1-final-viral-combined_k141_171238\|\|lt2gene | 1259 | No | NA | 2 | 2 | 0 | Low-quality | Genome-fragment | 2.25 | AAI-based (high-confidence) | 0 |
| S1-final-viral-combined_k141_66406\|\|full | 7675 | No | NA | 11 | 0 | 8 | Not-determined | Genome-fragment | NA | NA | 0 |
| S1_k141_174759 | 1259 | No | NA | 3 | 1 | 0 | Low-quality | Genome-fragment | 3.57 | AAI-based (high-confidence) | 0 |
| S3_k141_111915 | 1259 | No | NA | 1 | 0 | 1 | Low-quality | Genome-fragment | 0.47 | AAI-based (medium-confidence) | 0 |
| S5-final-viral-combined_k141_24518\|\|full | 1259 | No | NA | 3 | 0 | 0 | Low-quality | Genome-fragment | 0.78 | AAI-based (medium-confidence) | 0 |
| S1-final-viral-combined_k141_231234\|\|full | 26780 | No | NA | 26 | 0 | 0 | Medium-quality | Genome-fragment | 65.27 | AAI-based (high-confidence) | 0 |
| S17-k141_238843 | 1259 | No | NA | 3 | 1 | 0 | Low-quality | Genome-fragment | 2.67 | AAI-based (high-confidence) | 0 |
| S1-final-viral-combined_k141_66458\|\|full | 4385 | No | NA | 5 | 0 | 4 | Not-determined | Genome-fragment | NA | NA | 0 |
| S1-final-viral-combined_k141_66459\|\|full | 3202 | No | NA | 3 | 0 | 1 | Not-determined | Genome-fragment | NA | NA | 0 |
| S1-final-viral-combined_k141_363002\|\|full | 3731 | No | NA | 5 | 0 | 2 | Not-determined | Genome-fragment | NA | NA | 0 |
| S12-final-viral-combined_k141_318265\|\|full | 1259 | No | NA | 2 | 1 | 0 | Low-quality | Genome-fragment | 1.43 | HMM-based (lower-bound) | 0 |
| S12_k141_186392 | 1259 | No | NA | 3 | 0 | 1 | Low-quality | Genome-fragment | 1.74 | AAI-based (medium-confidence) | 0 |
| S1-final-viral-combined_k141_198425\|\|full | 1762 | No | NA | 3 | 0 | 1 | Not-determined | Genome-fragment | NA | NA | 0 |
| S1-final-viral-combined_k141_99599\|\|full | 1235 | No | NA | 3 | 0 | 1 | Not-determined | Genome-fragment | NA | NA | 0 |
| S12_k141_198242 | 1259 | No | NA | 1 | 0 | 0 | Low-quality | Genome-fragment | 1.92 | AAI-based (medium-confidence) | 0 |
| S2-final-viral-combined_k141_53599\|\|full | 1259 | No | NA | 3 | 0 | 0 | Low-quality | Genome-fragment | 2.93 | AAI-based (medium-confidence) | 0 |
| S11-final-viral-combined_k141_209477\|\|full | 1260 | No | NA | 3 | 2 | 0 | Low-quality | Genome-fragment | 2.61 | AAI-based (medium-confidence) | 0 |
| S1-final-viral-combined_k141_66476\|\|full | 2057 | No | NA | 4 | 0 | 0 | Not-determined | Genome-fragment | NA | NA | 0 |
| S1-final-viral-combined_k141_66480\|\|full | 1278 | No | NA | 3 | 0 | 1 | Not-determined | Genome-fragment | NA | NA | 0 |
| S11-final-viral-combined_k141_255447\|\|full | 1260 | No | NA | 3 | 0 | 0 | Low-quality | Genome-fragment | 1.7 | AAI-based (high-confidence) | 0 |
| S1-final-viral-combined_k141_33611\|\|full | 1255 | No | NA | 3 | 0 | 0 | Not-determined | Genome-fragment | NA | NA | 0 |
| S1-final-viral-combined_k141_17366\|\|full | 1716 | No | NA | 2 | 0 | 0 | Not-determined | Genome-fragment | NA | NA | 0 |
| S1-final-viral-combined_k141_346613\|\|full | 2854 | No | NA | 2 | 0 | 2 | Not-determined | Genome-fragment | NA | NA | 0 |
| S1-final-viral-combined_k141_17380\|\|full | 2636 | No | NA | 3 | 0 | 1 | Not-determined | Genome-fragment | NA | NA | 0 |
| S3-final-viral-combined_k141_8866\|\|lt2gene | 1260 | No | NA | 2 | 2 | 0 | Low-quality | Genome-fragment | 2.36 | AAI-based (high-confidence) | 0 |
| S1-final-viral-combined_k141_33639\|\|full | 1070 | No | NA | 3 | 0 | 0 | Not-determined | Genome-fragment | NA | NA | 0 |
| S1-final-viral-combined_k141_17397\|\|full | 1952 | No | NA | 3 | 0 | 2 | Not-determined | Genome-fragment | NA | NA | 0 |
| S5-final-viral-combined_k141_54800\|\|full | 1260 | No | NA | 3 | 0 | 0 | Low-quality | Genome-fragment | 1.96 | AAI-based (medium-confidence) | 0 |
| S1-final-viral-combined_k141_1271\|\|full | 2498 | No | NA | 3 | 0 | 0 | Not-determined | Genome-fragment | NA | NA | 0 |
| S5_k141_158512 | 1260 | No | NA | 3 | 0 | 0 | Low-quality | Genome-fragment | 2.43 | AAI-based (medium-confidence) | 0 |
| S7-final-viral-combined_k141_129716\|\|full | 1260 | No | NA | 4 | 0 | 0 | Low-quality | Genome-fragment | 1.68 | AAI-based (high-confidence) | 0 |
| S1-final-viral-combined_k141_83362\|\|full | 4290 | No | NA | 3 | 0 | 2 | Not-determined | Genome-fragment | NA | NA | 0 |
| S1-final-viral-combined_k141_165469\|\|full | 2502 | No | NA | 4 | 0 | 0 | Not-determined | Genome-fragment | NA | NA | 0 |
| S1-final-viral-combined_k141_264264\|\|full | 4517 | No | NA | 6 | 0 | 0 | Not-determined | Genome-fragment | NA | NA | 0 |
| S1-final-viral-combined_k141_49936\|\|full | 3706 | No | NA | 3 | 0 | 3 | Not-determined | Genome-fragment | NA | NA | 0 |
| S1-final-viral-combined_k141_329891\|\|full | 2841 | No | NA | 3 | 0 | 1 | Not-determined | Genome-fragment | NA | NA | 0 |
| S1-final-viral-combined_k141_280807\|\|full | 9154 | No | NA | 9 | 0 | 0 | Not-determined | Genome-fragment | NA | NA | 0 |
| S17-final-viral-combined_k141_51532\|\|full | 1260 | No | NA | 4 | 1 | 0 | Low-quality | Genome-fragment | 2.59 | AAI-based (high-confidence) | 0 |
| S1-final-viral-combined_k141_280811\|\|full | 1108 | No | NA | 2 | 0 | 0 | Not-determined | Genome-fragment | NA | NA | 0 |
| S1-final-viral-combined_k141_346665\|\|full | 6147 | No | NA | 9 | 0 | 1 | Not-determined | Genome-fragment | NA | NA | 0 |
| S2_k141_229975 | 1260 | No | NA | 2 | 0 | 1 | Low-quality | Genome-fragment | 2.1 | AAI-based (high-confidence) | 0 |
| S1-final-viral-combined_k141_1318\|\|full | 2439 | No | NA | 4 | 0 | 1 | Not-determined | Genome-fragment | NA | NA | 0 |
| S8-final-viral-combined_k141_92304\|\|lt2gene | 1260 | No | NA | 2 | 1 | 0 | Low-quality | Genome-fragment | 2.89 | AAI-based (high-confidence) | 0 |
| S1-final-viral-combined_k141_231354\|\|full | 3544 | No | NA | 5 | 0 | 0 | Not-determined | Genome-fragment | NA | NA | 0 |
| S1_k141_69059 | 1261 | No | NA | 2 | 0 | 0 | Low-quality | Genome-fragment | 0.12 | AAI-based (medium-confidence) | 0 |
| S3_k141_47123 | 1261 | No | NA | 3 | 0 | 0 | Low-quality | Genome-fragment | 3.34 | AAI-based (high-confidence) | 0 |
| S1-final-viral-combined_k141_231366\|\|full | 1547 | No | NA | 2 | 0 | 1 | Not-determined | Genome-fragment | NA | NA | 0 |
| S1-final-viral-combined_k141_1349\|\|full | 2020 | No | NA | 3 | 0 | 1 | Not-determined | Genome-fragment | NA | NA | 0 |
| S1-final-viral-combined_k141_83405\|\|full | 1705 | No | NA | 3 | 0 | 1 | Not-determined | Genome-fragment | NA | NA | 0 |
| S1-final-viral-combined_k141_264291\|\|full | 22782 | No | NA | 27 | 0 | 3 | Not-determined | Genome-fragment | NA | NA | 0 |
| S5-final-viral-combined_k141_151518\|\|full | 1261 | No | NA | 3 | 2 | 0 | Low-quality | Genome-fragment | 3.11 | AAI-based (high-confidence) | 0 |
| S1-final-viral-combined_k141_297288\|\|full | 1838 | No | NA | 3 | 0 | 1 | Not-determined | Genome-fragment | NA | NA | 0 |
| S1-final-viral-combined_k141_198541\|\|full | 6626 | No | NA | 6 | 0 | 3 | Not-determined | Genome-fragment | NA | NA | 0 |
| S1-final-viral-combined_k141_17449\|\|full | 4689 | No | NA | 5 | 0 | 3 | Not-determined | Genome-fragment | NA | NA | 0 |
| S17-final-viral-combined_k141_38687\|\|full | 1261 | No | NA | 2 | 0 | 2 | Low-quality | Genome-fragment | 0.6 | AAI-based (medium-confidence) | 0 |
| S1-final-viral-combined_k141_280825\|\|full | 4670 | No | NA | 6 | 0 | 4 | Not-determined | Genome-fragment | NA | NA | 0 |
| S17-final-viral-combined_k141_263398\|\|full | 1261 | No | NA | 3 | 0 | 2 | Low-quality | Genome-fragment | 0.62 | AAI-based (medium-confidence) | 0 |
| S17-k141_75494 | 1261 | No | NA | 1 | 0 | 1 | Low-quality | Genome-fragment | 1.12 | AAI-based (medium-confidence) | 0 |
| S1-final-viral-combined_k141_247921\|\|full | 2429 | No | NA | 4 | 0 | 1 | Not-determined | Genome-fragment | NA | NA | 0 |
| S12-final-viral-combined_k141_89539\|\|full | 1261 | No | NA | 4 | 0 | 0 | Low-quality | Genome-fragment | 0.75 | AAI-based (medium-confidence) | 0 |
| S12-final-viral-combined_k141_142756\|\|full | 1261 | No | NA | 3 | 0 | 0 | Low-quality | Genome-fragment | 0.63 | AAI-based (medium-confidence) | 0 |
| S12_k141_75666 | 1261 | No | NA | 2 | 0 | 1 | Low-quality | Genome-fragment | 0.65 | AAI-based (medium-confidence) | 0 |
| S12_k141_11294 | 1261 | No | NA | 1 | 0 | 1 | Low-quality | Genome-fragment | 0.7 | AAI-based (medium-confidence) | 0 |
| S1-final-viral-combined_k141_1415\|\|full | 2973 | No | NA | 3 | 0 | 1 | Not-determined | Genome-fragment | NA | NA | 0 |
| S1-final-viral-combined_k141_280879\|\|full | 4978 | No | NA | 2 | 0 | 2 | Not-determined | Genome-fragment | NA | NA | 0 |
| S1-final-viral-combined_k141_231411\|\|full | 12380 | No | NA | 12 | 1 | 0 | Medium-quality | Genome-fragment | 83.34 | AAI-based (high-confidence) | 0 |
| S4-final-viral-combined_k141_8479\|\|full | 1261 | No | NA | 2 | 0 | 0 | Low-quality | Genome-fragment | 1.34 | AAI-based (medium-confidence) | 0 |
| S4-final-viral-combined_k141_94351\|\|full | 1261 | No | NA | 4 | 0 | 0 | Low-quality | Genome-fragment | 2.08 | AAI-based (high-confidence) | 0 |
| S1-final-viral-combined_k141_198600\|\|full | 6249 | No | NA | 2 | 0 | 1 | Not-determined | Genome-fragment | NA | NA | 0 |
| S1-final-viral-combined_k141_280887\|\|full | 1568 | No | NA | 3 | 0 | 0 | Not-determined | Genome-fragment | NA | NA | 0 |
| S1-final-viral-combined_k141_99749\|\|full | 1461 | No | NA | 3 | 0 | 2 | Not-determined | Genome-fragment | NA | NA | 0 |
| S1-final-viral-combined_k141_132934\|\|full | 51171 | Yes | 43190 | 72 | 16 | 7 | Complete | High-quality | 100 | Provirus (high-confidence) | 15.6 |
| S1-final-viral-combined_k141_99759\|\|full | 2867 | No | NA | 4 | 0 | 0 | Not-determined | Genome-fragment | NA | NA | 0 |
| S1-final-viral-combined_k141_66554\|\|full | 2727 | No | NA | 3 | 0 | 2 | Not-determined | Genome-fragment | NA | NA | 0 |
| S1-final-viral-combined_k141_198626\|\|full | 1157 | No | NA | 3 | 0 | 1 | Not-determined | Genome-fragment | NA | NA | 0 |
| S4-final-viral-combined_k141_74761\|\|full | 1261 | No | NA | 3 | 0 | 1 | Low-quality | Genome-fragment | 1.2 | AAI-based (high-confidence) | 0 |
| S1-final-viral-combined_k141_248007\|\|full | 3599 | No | NA | 3 | 0 | 2 | Not-determined | Genome-fragment | NA | NA | 0 |
| S1-final-viral-combined_k141_33813\|\|full | 16823 | No | NA | 18 | 0 | 12 | Not-determined | Genome-fragment | NA | NA | 0 |
| S9-final-viral-combined_k141_105521\|\|full | 1262 | No | NA | 4 | 0 | 0 | Low-quality | Genome-fragment | 2.51 | AAI-based (medium-confidence) | 0 |
| S9-final-viral-combined_k141_92151\|\|lt2gene | 1262 | No | NA | 1 | 1 | 0 | Low-quality | Genome-fragment | 2.12 | HMM-based (lower-bound) | 0 |
| S1-final-viral-combined_k141_132983\|\|full | 4415 | No | NA | 4 | 0 | 4 | Not-determined | Genome-fragment | NA | NA | 0 |
| S7-final-viral-combined_k141_9721\|\|full | 1262 | No | NA | 3 | 0 | 0 | Low-quality | Genome-fragment | 2.13 | AAI-based (high-confidence) | 0 |

| | | | | | | | | | | | | | |
|---|---|---|---|---|---|---|---|---|---|---|---|---|---|
| S1-final-viral-combined_k141_215337\|\|full | 6484 | No | NA | | 5 | 0 | 5 | Not-determined | Genome-fragment | NA | | NA | 0 |
| S17-final-viral-combined_k141_102433\|\|full | 1262 | No | NA | | 5 | 1 | 0 | Low-quality | Genome-fragment | | 1.2 | AAI-based (high-confidence) | 0 |
| S1-final-viral-combined_k141_182154\|\|full | 1442 | No | NA | | 2 | 0 | 1 | Not-determined | Genome-fragment | NA | | NA | 0 |
| S1-final-viral-combined_k141_50047\|\|full | 4594 | No | NA | | 4 | 0 | 4 | Not-determined | Genome-fragment | NA | | NA | 0 |
| S17-final-viral-combined_k141_248665\|\|full | 1262 | No | NA | | 3 | 1 | 0 | Low-quality | Genome-fragment | | 0.81 | HMM-based (lower-bound) | 0 |
| S1-final-viral-combined_k141_215349\|\|full | 8512 | No | NA | | 6 | 0 | 5 | Not-determined | Genome-fragment | NA | | NA | 0 |
| S12-final-viral-combined_k141_138509\|\|full | 1262 | No | NA | | 2 | 0 | 0 | Low-quality | Genome-fragment | | 2.36 | AAI-based (high-confidence) | 0 |
| S12-final-viral-combined_k141_86548\|\|lt2gene | 1262 | No | NA | | 1 | 1 | 0 | Low-quality | Genome-fragment | | 3.02 | AAI-based (high-confidence) | 0 |
| S2-final-viral-combined_k141_87233\|\|full | 1262 | No | NA | | 2 | 0 | 0 | Low-quality | Genome-fragment | | 2.44 | AAI-based (medium-confidence) | 0 |
| S2-final-viral-combined_k141_110072\|\|full | 1262 | No | NA | | 3 | 1 | 0 | Low-quality | Genome-fragment | | 2.85 | AAI-based (high-confidence) | 0 |
| S1-final-viral-combined_k141_248087\|\|full | 2419 | No | NA | | 2 | 0 | 1 | Not-determined | Genome-fragment | NA | | NA | 0 |
| S4_k141_142401 | 1262 | No | NA | | 3 | 2 | 0 | Low-quality | Genome-fragment | | 0.73 | AAI-based (high-confidence) | 0 |
| S1-final-viral-combined_k141_182241\|\|full | 1177 | No | NA | | 3 | 0 | 1 | Not-determined | Genome-fragment | NA | | NA | 0 |
| S1-final-viral-combined_k141_33908\|\|full | 1454 | No | NA | | 3 | 0 | 1 | Not-determined | Genome-fragment | NA | | NA | 0 |
| S1-final-viral-combined_k141_313824\|\|full | 4085 | No | NA | | 6 | 0 | 4 | Not-determined | Genome-fragment | NA | | NA | 0 |
| S6_k141_111742 | 1262 | No | NA | | 5 | 0 | 0 | Low-quality | Genome-fragment | | 1.32 | AAI-based (medium-confidence) | 0 |
| S1-final-viral-combined_k141_281012\|\|full | 3269 | No | NA | | 3 | 0 | 1 | Not-determined | Genome-fragment | NA | | NA | 0 |
| S1-final-viral-combined_k141_165561\|\|full | 2062 | No | NA | | 2 | 0 | 0 | Not-determined | Genome-fragment | NA | | NA | 0 |
| S1-final-viral-combined_k141_281018\|\|full | 5269 | No | NA | | 5 | 0 | 4 | Not-determined | Genome-fragment | NA | | NA | 0 |
| S1-final-viral-combined_k141_346881\|\|full | 2845 | No | NA | | 4 | 0 | 0 | Not-determined | Genome-fragment | NA | | NA | 0 |
| S9-final-viral-combined_k141_16039\|\|full | 1263 | No | NA | | 2 | 1 | 0 | Low-quality | Genome-fragment | | 1.77 | HMM-based (lower-bound) | 0 |
| S1-final-viral-combined_k141_281033\|\|full | 1082 | No | NA | | 2 | 0 | 1 | Not-determined | Genome-fragment | NA | | NA | 0 |
| S1-final-viral-combined_k141_165607\|\|full | 2641 | No | NA | | 3 | 0 | 0 | Not-determined | Genome-fragment | NA | | NA | 0 |
| S1-final-viral-combined_k141_165608\|\|full | 1401 | No | NA | | 3 | 0 | 0 | Not-determined | Genome-fragment | NA | | NA | 0 |
| S11-final-viral-combined_k141_187844\|\|full | 1263 | No | NA | | 3 | 1 | 0 | Low-quality | Genome-fragment | | 2.8 | AAI-based (high-confidence) | 0 |
| S1-final-viral-combined_k141_116554\|\|full | 1799 | No | NA | | 2 | 0 | 0 | Not-determined | Genome-fragment | NA | | NA | 0 |
| S1-final-viral-combined_k141_116556\|\|full | 1108 | No | NA | | 2 | 0 | 0 | Not-determined | Genome-fragment | NA | | NA | 0 |
| S1-final-viral-combined_k141_72777\|\|full | 1263 | No | NA | | 2 | 0 | 0 | Low-quality | Genome-fragment | | 1.91 | AAI-based (high-confidence) | 0 |
| S1-final-viral-combined_k141_328207\|\|lt2gene | 1263 | No | NA | | 1 | 1 | 0 | Low-quality | Genome-fragment | | 1.18 | HMM-based (lower-bound) | 0 |
| S1_k141_218752 | 1263 | No | NA | | 2 | 0 | 0 | Low-quality | Genome-fragment | | 2.72 | AAI-based (high-confidence) | 0 |
| S1-final-viral-combined_k141_231492\|\|full | 24462 | No | NA | | 23 | 0 | 13 | Not-determined | Genome-fragment | NA | | NA | 0 |
| S1_k141_73160 | 1263 | No | NA | | 2 | 0 | 0 | Low-quality | Genome-fragment | | 3.28 | AAI-based (high-confidence) | 0 |
| S1-final-viral-combined_k141_281062\|\|full | 4248 | No | NA | | 3 | 0 | 1 | Not-determined | Genome-fragment | NA | | NA | 0 |
| S1-final-viral-combined_k141_17651\|\|full | 7507 | No | NA | | 11 | 1 | 0 | High-quality | High-quality | | 100 | HMM-based (lower-bound) | 0 |
| S1-final-viral-combined_k141_165641\|\|full | 3142 | No | NA | | 7 | 0 | 2 | Not-determined | Genome-fragment | NA | | NA | 0 |
| S1-final-viral-combined_k141_297485\|\|full | 9631 | No | NA | | 8 | 0 | 6 | Not-determined | Genome-fragment | NA | | NA | 0 |
| S3_k141_118011 | 1263 | No | NA | | 1 | 0 | 0 | Low-quality | Genome-fragment | | 0.95 | AAI-based (high-confidence) | 0 |
| S1-final-viral-combined_k141_313851\|\|full | 13334 | No | NA | | 20 | 5 | 0 | High-quality | High-quality | | 96 | AAI-based (medium-confidence) | 0 |
| S7-final-viral-combined_k141_70245\|\|lt2gene | 1263 | No | NA | | 1 | 1 | 0 | Low-quality | Genome-fragment | | 0.87 | HMM-based (lower-bound) | 0 |
| S17-final-viral-combined_k141_228696\|\|lt2gene | 1263 | No | NA | | 2 | 2 | 0 | Low-quality | Genome-fragment | | 2.65 | AAI-based (medium-confidence) | 0 |
| S17-k141_228362 | 1263 | No | NA | | 2 | 0 | 0 | Low-quality | Genome-fragment | | 3.1 | AAI-based (medium-confidence) | 0 |
| S1-final-viral-combined_k141_313865\|\|full | 1924 | No | NA | | 3 | 0 | 2 | Not-determined | Genome-fragment | NA | | NA | 0 |
| S1-final-viral-combined_k141_1583\|\|full | 2544 | No | NA | | 2 | 0 | 1 | Not-determined | Genome-fragment | NA | | NA | 0 |
| S1-final-viral-combined_k141_17683\|\|full | 3754 | No | NA | | 5 | 0 | 4 | Not-determined | Genome-fragment | NA | | NA | 0 |
| S1-final-viral-combined_k141_99918\|\|full | 1945 | No | NA | | 5 | 0 | 0 | Not-determined | Genome-fragment | NA | | NA | 0 |
| S1-final-viral-combined_k141_17689\|\|full | 1087 | No | NA | | 2 | 0 | 1 | Not-determined | Genome-fragment | NA | | NA | 0 |
| S1-final-viral-combined_k141_50160\|\|full | 3125 | No | NA | | 3 | 0 | 0 | Not-determined | Genome-fragment | NA | | NA | 0 |
| S12-final-viral-combined_k141_88161\|\|full | 1263 | No | NA | | 4 | 1 | 0 | Low-quality | Genome-fragment | | 3.08 | AAI-based (medium-confidence) | 0 |
| S12-final-viral-combined_k141_360711\|\|lt2gene | 1263 | No | NA | | 2 | 2 | 0 | Low-quality | Genome-fragment | | 2.06 | HMM-based (lower-bound) | 0 |
| S1-final-viral-combined_k141_231562\|\|full | 1779 | No | NA | | 7 | 0 | 0 | Not-determined | Genome-fragment | NA | | NA | 0 |
| S1-final-viral-combined_k141_116633\|\|full | 1623 | No | NA | | 3 | 0 | 2 | Not-determined | Genome-fragment | NA | | NA | 0 |
| S1-final-viral-combined_k141_297511\|\|full | 1326 | No | NA | | 2 | 0 | 2 | Not-determined | Genome-fragment | NA | | NA | 0 |
| S1-final-viral-combined_k141_1602\|\|full | 2568 | No | NA | | 6 | 0 | 1 | Not-determined | Genome-fragment | NA | | NA | 0 |
| S1-final-viral-combined_k141_133144\|\|full | 7309 | No | NA | | 8 | 0 | 1 | Not-determined | Genome-fragment | NA | | NA | 0 |
| S1-final-viral-combined_k141_182370\|\|full | 40907 | No | NA | | 29 | 0 | 18 | Not-determined | Genome-fragment | NA | | NA | 0 |
| S1-final-viral-combined_k141_182372\|\|full | 1768 | No | NA | | 4 | 0 | 1 | Not-determined | Genome-fragment | NA | | NA | 0 |
| S8-final-viral-combined_k141_151700\|\|full | 1263 | No | NA | | 3 | 0 | 0 | Low-quality | Genome-fragment | | 2.09 | AAI-based (high-confidence) | 0 |
| S8-final-viral-combined_k141_77966\|\|full | 1263 | No | NA | | 3 | 0 | 1 | Low-quality | Genome-fragment | | 1.36 | AAI-based (medium-confidence) | 0 |
| S8_k141_126740 | 1263 | No | NA | | 2 | 0 | 2 | Low-quality | Genome-fragment | | 2.61 | AAI-based (high-confidence) | 0 |
| S1-final-viral-combined_k141_149272\|\|full | 7892 | No | NA | | 7 | 0 | 3 | Not-determined | Genome-fragment | NA | | NA | 0 |
| S11-final-viral-combined_k141_329672\|\|full | 1264 | No | NA | | 3 | 0 | 0 | Low-quality | Genome-fragment | | 2.89 | AAI-based (high-confidence) | 0 |
| S1-final-viral-combined_k141_50193\|\|full | 14977 | No | NA | | 12 | 0 | 10 | Not-determined | Genome-fragment | NA | | NA | 0 |
| S1-final-viral-combined_k141_133193\|\|full | 2543 | No | NA | | 2 | 0 | 2 | Not-determined | Genome-fragment | NA | | NA | 0 |
| S1-final-viral-combined_k141_379564\|\|full | 1660 | No | NA | | 3 | 0 | 2 | Not-determined | Genome-fragment | NA | | NA | 0 |
| S11_k141_213778 | 1264 | No | NA | | 2 | 1 | 0 | Low-quality | Genome-fragment | | 2.93 | AAI-based (medium-confidence) | 0 |
| S1-final-viral-combined_k141_83571\|\|full | 1142 | No | NA | | 2 | 0 | 0 | Not-determined | Genome-fragment | NA | | NA | 0 |
| S1-final-viral-combined_k141_242032\|\|lt2gene | 1264 | No | NA | | 2 | 1 | 0 | Low-quality | Genome-fragment | | 2.21 | AAI-based (high-confidence) | 0 |
| S5-final-viral-combined_k141_66500\|\|full | 1264 | No | NA | | 3 | 1 | 0 | Low-quality | Genome-fragment | | 2.41 | HMM-based (lower-bound) | 0 |
| S1-final-viral-combined_k141_83585\|\|full | 1408 | No | NA | | 1 | 0 | 1 | Not-determined | Genome-fragment | NA | | NA | 0 |
| S1-final-viral-combined_k141_198931\|\|full | 1623 | No | NA | | 2 | 0 | 0 | Not-determined | Genome-fragment | NA | | NA | 0 |
| S1-final-viral-combined_k141_379598\|\|full | 3489 | No | NA | | 5 | 0 | 3 | Not-determined | Genome-fragment | NA | | NA | 0 |
| S1-final-viral-combined_k141_116691\|\|full | 2948 | No | NA | | 2 | 0 | 2 | Not-determined | Genome-fragment | NA | | NA | 0 |
| S5_k141_64805 | 1264 | No | NA | | 4 | 1 | 0 | Low-quality | Genome-fragment | | 1.85 | AAI-based (high-confidence) | 0 |
| S2-final-viral-combined_k141_6993\|\|full | 1264 | No | NA | | 3 | 0 | 0 | Low-quality | Genome-fragment | | 2.2 | AAI-based (medium-confidence) | 0 |
| S1-final-viral-combined_k141_231572\|\|full | 4555 | No | NA | | 7 | 0 | 1 | Not-determined | Genome-fragment | NA | | NA | 0 |
| S2_k141_183774 | 1264 | No | NA | | 5 | 0 | 0 | Low-quality | Genome-fragment | | 0.78 | AAI-based (high-confidence) | 0 |
| S1-final-viral-combined_k141_314006\|\|full | 1934 | No | NA | | 3 | 0 | 2 | Not-determined | Genome-fragment | NA | | NA | 0 |
| S1-final-viral-combined_k141_17781\|\|full | 2233 | No | NA | | 2 | 0 | 1 | Not-determined | Genome-fragment | NA | | NA | 0 |
| S1-final-viral-combined_k141_149340\|\|full | 5690 | No | NA | | 6 | 0 | 4 | Not-determined | Genome-fragment | NA | | NA | 0 |
| S8-final-viral-combined_k141_49187\|\|full | 1264 | No | NA | | 3 | 0 | 0 | Low-quality | Genome-fragment | | 2.71 | AAI-based (medium-confidence) | 0 |
| S9_k141_90026 | 1265 | No | NA | | 2 | 2 | 0 | Low-quality | Genome-fragment | | 3.15 | AAI-based (medium-confidence) | 0 |
| S11-final-viral-combined_k141_175283\|\|full | 1265 | No | NA | | 4 | 2 | 0 | Low-quality | Genome-fragment | | 1.1 | HMM-based (lower-bound) | 0 |
| S1-final-viral-combined_k141_34176\|\|full | 1578 | No | NA | | 3 | 0 | 2 | Not-determined | Genome-fragment | NA | | NA | 0 |
| S1-final-viral-combined_k141_347139\|\|full | 5145 | No | NA | | 4 | 0 | 4 | Not-determined | Genome-fragment | NA | | NA | 0 |
| S1-final-viral-combined_k141_21789\|\|full | 1265 | No | NA | | 4 | 1 | 0 | Low-quality | Genome-fragment | | 3.57 | AAI-based (medium-confidence) | 0 |
| S1-final-viral-combined_k141_50280\|\|full | 2116 | No | NA | | 3 | 0 | 3 | Not-determined | Genome-fragment | NA | | NA | 0 |
| S1-final-viral-combined_k141_330329\|\|full | 33155 | No | NA | | 36 | 7 | 4 | Medium-quality | Genome-fragment | | 77.41 | AAI-based (medium-confidence) | 0 |
| S1-final-viral-combined_k141_141459\|\|full | 1265 | No | NA | | 3 | 0 | 1 | Low-quality | Genome-fragment | | 1.64 | AAI-based (high-confidence) | 0 |
| S5-final-viral-combined_k141_178108\|\|full | 1265 | No | NA | | 4 | 1 | 0 | Low-quality | Genome-fragment | | 1.42 | HMM-based (lower-bound) | 0 |
| S1-final-viral-combined_k141_182446\|\|full | 1108 | No | NA | | 3 | 0 | 0 | Not-determined | Genome-fragment | NA | | NA | 0 |
| S1-final-viral-combined_k141_50297\|\|full | 1910 | No | NA | | 4 | 0 | 0 | Not-determined | Genome-fragment | NA | | NA | 0 |
| S1-final-viral-combined_k141_50304\|\|full | 1883 | No | NA | | 2 | 0 | 2 | Not-determined | Genome-fragment | NA | | NA | 0 |
| S7-final-viral-combined_k141_218510\|\|full | 1265 | No | NA | | 4 | 0 | 1 | Low-quality | Genome-fragment | | 0.99 | AAI-based (medium-confidence) | 0 |
| S1-final-viral-combined_k141_50306\|\|full | 2559 | No | NA | | 3 | 0 | 0 | Not-determined | Genome-fragment | NA | | NA | 0 |
| S1-final-viral-combined_k141_215482\|\|full | 3289 | No | NA | | 2 | 0 | 2 | Not-determined | Genome-fragment | NA | | NA | 0 |
| S1-final-viral-combined_k141_133285\|\|full | 1286 | No | NA | | 2 | 0 | 1 | Not-determined | Genome-fragment | NA | | NA | 0 |
| S1-final-viral-combined_k141_149378\|\|full | 2141 | No | NA | | 2 | 0 | 2 | Not-determined | Genome-fragment | NA | | NA | 0 |
| S7-final-viral-combined_k141_180068\|\|full | 1265 | No | NA | | 3 | 0 | 1 | Low-quality | Genome-fragment | | 2.75 | AAI-based (medium-confidence) | 0 |
| S7-final-viral-combined_k141_243108\|\|full | 1265 | No | NA | | 2 | 0 | 1 | Low-quality | Genome-fragment | | 1.95 | AAI-based (medium-confidence) | 0 |
| S1-final-viral-combined_k141_297659\|\|full | 13147 | No | NA | | 14 | 0 | 7 | Not-determined | Genome-fragment | NA | | NA | 0 |
| S1-final-viral-combined_k141_215511\|\|full | 3293 | No | NA | | 3 | 0 | 2 | Not-determined | Genome-fragment | NA | | NA | 0 |
| S1-final-viral-combined_k141_34278\|\|full | 1536 | No | NA | | 3 | 0 | 2 | Not-determined | Genome-fragment | NA | | NA | 0 |
| S17-final-viral-combined_k141_189598\|\|lt2gene | 1265 | No | NA | | 1 | 1 | 0 | Low-quality | Genome-fragment | | 1.19 | HMM-based (lower-bound) | 0 |
| S1-final-viral-combined_k141_83726\|\|full | 3759 | No | NA | | 3 | 0 | 3 | Not-determined | Genome-fragment | NA | | NA | 0 |
| S12-final-viral-combined_k141_179970\|\|full | 1265 | No | NA | | 2 | 0 | 1 | Low-quality | Genome-fragment | | 1.5 | AAI-based (high-confidence) | 0 |
| S1-final-viral-combined_k141_149420\|\|full | 3087 | No | NA | | 2 | 0 | 2 | Not-determined | Genome-fragment | NA | | NA | 0 |
| S12-final-viral-combined_k141_32847\|\|full | 1265 | No | NA | | 3 | 1 | 0 | Low-quality | Genome-fragment | | 2.04 | AAI-based (high-confidence) | 0 |
| S2-final-viral-combined_k141_227638\|\|lt2gene | 1265 | No | NA | | 2 | 2 | 0 | Low-quality | Genome-fragment | | 4.44 | HMM-based (lower-bound) | 0 |
| S1-final-viral-combined_k141_215575\|\|full | 2424 | No | NA | | 2 | 0 | 1 | Not-determined | Genome-fragment | NA | | NA | 0 |
| S6-final-viral-combined_k141_35870\|\|full | 1265 | No | NA | | 3 | 0 | 0 | Low-quality | Genome-fragment | | 0.9 | AAI-based (medium-confidence) | 0 |
| S9-final-viral-combined_k141_63495\|\|full | 1266 | No | NA | | 3 | 0 | 1 | Low-quality | Genome-fragment | | 0.91 | AAI-based (medium-confidence) | 0 |
| S1-final-viral-combined_k141_248221\|\|full | 3655 | No | NA | | 6 | 0 | 2 | Not-determined | Genome-fragment | NA | | NA | 0 |
| S1-final-viral-combined_k141_83783\|\|full | 4480 | No | NA | | 4 | 0 | 0 | Not-determined | Genome-fragment | NA | | NA | 0 |
| S1-final-viral-combined_k141_165771\|\|full | 2697 | No | NA | | 3 | 0 | 0 | Not-determined | Genome-fragment | NA | | NA | 0 |
| S1-final-viral-combined_k141_50386\|\|full | 1062 | No | NA | | 2 | 0 | 0 | Not-determined | Genome-fragment | NA | | NA | 0 |
| S1-final-viral-combined_k141_231792\|\|full | 3453 | No | NA | | 4 | 0 | 2 | Not-determined | Genome-fragment | NA | | NA | 0 |
| S1-final-viral-combined_k141_297754\|\|full | 2068 | No | NA | | 3 | 0 | 0 | Not-determined | Genome-fragment | NA | | NA | 0 |
| S1-final-viral-combined_k141_231798\|\|full | 6162 | No | NA | | 5 | 0 | 5 | Not-determined | Genome-fragment | NA | | NA | 0 |
| S11-final-viral-combined_k141_96324\|\|full | 1266 | No | NA | | 4 | 0 | 1 | Low-quality | Genome-fragment | | 1.08 | AAI-based (medium-confidence) | 0 |
| S1-final-viral-combined_k141_50405\|\|full | 4526 | No | NA | | 6 | 0 | 4 | Not-determined | Genome-fragment | NA | | NA | 0 |
| S1-final-viral-combined_k141_215650\|\|full | 3054 | No | NA | | 4 | 0 | 3 | Not-determined | Genome-fragment | NA | | NA | 0 |
| S7_k141_224000 | 1266 | No | NA | | 2 | 0 | 0 | Low-quality | Genome-fragment | | 2.17 | AAI-based (medium-confidence) | 0 |
| S1-final-viral-combined_k141_248293\|\|full | 1693 | No | NA | | 3 | 0 | 2 | Not-determined | Genome-fragment | NA | | NA | 0 |
| S1-final-viral-combined_k141_149472\|\|full | 23957 | No | NA | | 24 | 0 | 12 | Not-determined | Genome-fragment | NA | | NA | 0 |
| S1-final-viral-combined_k141_281370\|\|full | 3032 | No | NA | | 4 | 0 | 3 | Not-determined | Genome-fragment | NA | | NA | 0 |
| S1-final-viral-combined_k141_314247\|\|full | 1717 | No | NA | | 3 | 0 | 0 | Not-determined | Genome-fragment | NA | | NA | 0 |
| S1-final-viral-combined_k141_248315\|\|full | 3898 | No | NA | | 4 | 0 | 2 | Not-determined | Genome-fragment | NA | | NA | 0 |
| S1-final-viral-combined_k141_330509\|\|full | 3074 | No | NA | | 3 | 0 | 1 | Not-determined | Genome-fragment | NA | | NA | 0 |
| S1-final-viral-combined_k141_149498\|\|full | 1593 | No | NA | | 4 | 0 | 0 | Not-determined | Genome-fragment | NA | | NA | 0 |
| S1-final-viral-combined_k141_330512\|\|full | 1601 | No | NA | | 2 | 0 | 1 | Not-determined | Genome-fragment | NA | | NA | 0 |
| S17-final-viral-combined_k141_258108\|\|full | 1266 | No | NA | | 2 | 0 | 0 | Low-quality | Genome-fragment | | 0.79 | AAI-based (medium-confidence) | 0 |
| S1-final-viral-combined_k141_379919\|\|full | 5614 | No | NA | | 6 | 0 | 1 | Not-determined | Genome-fragment | NA | | NA | 0 |
| S17-final-viral-combined_k141_94140\|\|full | 1266 | No | NA | | 3 | 0 | 1 | Low-quality | Genome-fragment | | 1.8 | AAI-based (high-confidence) | 0 |
| S1-final-viral-combined_k141_149506\|\|full | 4629 | No | NA | | 5 | 0 | 3 | Not-determined | Genome-fragment | NA | | NA | 0 |
| S17-final-viral-combined_k141_63298\|\|lt2gene | 1266 | No | NA | | 2 | 2 | 0 | Low-quality | Genome-fragment | | 2.74 | AAI-based (high-confidence) | 0 |
| S1-final-viral-combined_k141_149508\|\|full | 1199 | No | NA | | 2 | 0 | 1 | Not-determined | Genome-fragment | NA | | NA | 0 |
| S1-final-viral-combined_k141_379959\|\|full | 2161 | No | NA | | 3 | 0 | 2 | Not-determined | Genome-fragment | NA | | NA | 0 |
| S1-final-viral-combined_k141_50436\|\|full | 5139 | No | NA | | 7 | 0 | 1 | Not-determined | Genome-fragment | NA | | NA | 0 |
| S1-final-viral-combined_k141_34429\|\|full | 2830 | No | NA | | 4 | 0 | 3 | Not-determined | Genome-fragment | NA | | NA | 0 |
| S1-final-viral-combined_k141_281425\|\|full | 1255 | No | NA | | 2 | 0 | 0 | Not-determined | Genome-fragment | NA | | NA | 0 |
| S1-final-viral-combined_k141_182634\|\|full | 1867 | No | NA | | 2 | 0 | 1 | Not-determined | Genome-fragment | NA | | NA | 0 |
| S1-final-viral-combined_k141_248392\|\|full | 10209 | No | NA | | 7 | 0 | 3 | Not-determined | Genome-fragment | NA | | NA | 0 |
| S1-final-viral-combined_k141_248394\|\|full | 2444 | No | NA | | 6 | 0 | 0 | Not-determined | Genome-fragment | NA | | NA | 0 |
| S12-final-viral-combined_k141_340915\|\|full | 1266 | No | NA | | 2 | 1 | 0 | Low-quality | Genome-fragment | | 1.3 | AAI-based (high-confidence) | 0 |
| S1-final-viral-combined_k141_2019\|\|full | 2850 | No | NA | | 4 | 0 | 0 | Not-determined | Genome-fragment | NA | | NA | 0 |
| S12-final-viral-combined_k141_359373\|\|full | 1266 | No | NA | | 4 | 3 | 0 | Low-quality | Genome-fragment | | 2.72 | AAI-based (high-confidence) | 0 |
| S1-final-viral-combined_k141_133538\|\|full | 2158 | No | NA | | 2 | 0 | 1 | Not-determined | Genome-fragment | NA | | NA | 0 |
| S1-final-viral-combined_k141_2030\|\|full | 7214 | No | NA | | 6 | 0 | 3 | Not-determined | Genome-fragment | NA | | NA | 0 |
| S1-final-viral-combined_k141_182676\|\|full | 2046 | No | NA | | 3 | 0 | 1 | Not-determined | Genome-fragment | NA | | NA | 0 |
| S1-final-viral-combined_k141_199239\|\|full | 1356 | No | NA | | 3 | 0 | 2 | Not-determined | Genome-fragment | NA | | NA | 0 |
| S1-final-viral-combined_k141_18089\|\|full | 3427 | No | NA | | 2 | 0 | 2 | Not-determined | Genome-fragment | NA | | NA | 0 |
| S1-final-viral-combined_k141_133562\|\|full | 1398 | No | NA | | 3 | 0 | 0 | Not-determined | Genome-fragment | NA | | NA | 0 |
| S1-final-viral-combined_k141_330606\|\|full | 2498 | No | NA | | 2 | 0 | 2 | Not-determined | Genome-fragment | NA | | NA | 0 |
| S1-final-viral-combined_k141_50483\|\|full | 1952 | No | NA | | 2 | 0 | 1 | Not-determined | Genome-fragment | NA | | NA | 0 |
| S1-final-viral-combined_k141_83918\|\|full | 2606 | No | NA | | 3 | 0 | 3 | Not-determined | Genome-fragment | NA | | NA | 0 |
| S1-final-viral-combined_k141_116855\|\|full | 1907 | No | NA | | 4 | 0 | 1 | Not-determined | Genome-fragment | NA | | NA | 0 |
| S1-final-viral-combined_k141_231924\|\|full | 7654 | No | NA | | 4 | 0 | 4 | Not-determined | Genome-fragment | NA | | NA | 0 |
| S1-final-viral-combined_k141_231927\|\|full | 1181 | No | NA | | 3 | 0 | 1 | Not-determined | Genome-fragment | NA | | NA | 0 |
| S2-final-viral-combined_k141_206725\|\|full | 1266 | No | NA | | 3 | 0 | 0 | Low-quality | Genome-fragment | | 1.87 | AAI-based (medium-confidence) | 0 |
| S4-final-viral-combined_k141_119859\|\|full | 1266 | No | NA | | 3 | 2 | 0 | Low-quality | Genome-fragment | | 2.86 | AAI-based (medium-confidence) | 0 |
| S1-final-viral-combined_k141_231935\|\|full | 1137 | No | NA | | 3 | 0 | 0 | Not-determined | Genome-fragment | NA | | NA | 0 |
| S1-final-viral-combined_k141_116870\|\|full | 1422 | No | NA | | 4 | 0 | 1 | Not-determined | Genome-fragment | NA | | NA | 0 |
| S1-final-viral-combined_k141_18104\|\|full | 4658 | No | NA | | 6 | 0 | 0 | Not-determined | Genome-fragment | NA | | NA | 0 |
| S1-final-viral-combined_k141_199268\|\|full | 1580 | No | NA | | 2 | 0 | 1 | Not-determined | Genome-fragment | NA | | NA | 0 |
| S1-final-viral-combined_k141_50508\|\|full | 4230 | No | NA | | 3 | 0 | 0 | Not-determined | Genome-fragment | NA | | NA | 0 |
| S1-final-viral-combined_k141_100196\|\|full | 1288 | No | NA | | 2 | 0 | 1 | Not-determined | Genome-fragment | NA | | NA | 0 |
| S1-final-viral-combined_k141_133606\|\|full | 2147 | No | NA | | 3 | 0 | 2 | Not-determined | Genome-fragment | NA | | NA | 0 |
| S1-final-viral-combined_k141_149656\|\|full | 1023 | No | NA | | 3 | 0 | 1 | Not-determined | Genome-fragment | NA | | NA | 0 |
| S4-final-viral-combined_k141_59766\|\|full | 1266 | No | NA | | 2 | 0 | 1 | Low-quality | Genome-fragment | | 1.97 | AAI-based (medium-confidence) | 0 |
| S11-final-viral-combined_k141_295353\|\|full | 1267 | No | NA | | 4 | 1 | 0 | Low-quality | Genome-fragment | | 2.61 | AAI-based (high-confidence) | 0 |
| S1-final-viral-combined_k141_363666\|\|full | 3581 | No | NA | | 2 | 0 | 2 | Not-determined | Genome-fragment | NA | | NA | 0 |
| S1-final-viral-combined_k141_49448\|\|full | 1267 | No | NA | | 2 | 0 | 0 | Low-quality | Genome-fragment | | 1.91 | AAI-based (medium-confidence) | 0 |
| S1-final-viral-combined_k141_1861\|\|full | 1267 | No | NA | | 2 | 2 | 0 | Low-quality | Genome-fragment | | 3.37 | AAI-based (high-confidence) | 0 |
| S1-final-viral-combined_k141_167084\|\|full | 1267 | No | NA | | 4 | 2 | 0 | Low-quality | Genome-fragment | | 2.59 | AAI-based (medium-confidence) | 0 |
| S1-final-viral-combined_k141_373692\|\|full | 1267 | No | NA | | 4 | 1 | 0 | Low-quality | Genome-fragment | | 2.68 | AAI-based (high-confidence) | 0 |
| S1-final-viral-combined_k141_165809\|\|full | 1287 | No | NA | | 3 | 0 | 1 | Not-determined | Genome-fragment | NA | | NA | 0 |
| S1-final-viral-combined_k141_116927\|\|full | 2484 | No | NA | | 4 | 0 | 3 | Not-determined | Genome-fragment | NA | | NA | 0 |
| S1-final-viral-combined_k141_215796\|\|full | 2222 | No | NA | | 3 | 0 | 2 | Not-determined | Genome-fragment | NA | | NA | 0 |
| S1-final-viral-combined_k141_363693\|\|full | 1100 | No | NA | | 2 | 0 | 1 | Not-determined | Genome-fragment | NA | | NA | 0 |
| S1-final-viral-combined_k141_100258\|\|full | 1129 | No | NA | | 3 | 0 | 1 | Not-determined | Genome-fragment | NA | | NA | 0 |
| S5-final-viral-combined_k141_209650\|\|full | 1267 | No | NA | | 3 | 1 | 0 | Low-quality | Genome-fragment | | 0.92 | AAI-based (medium-confidence) | 0 |
| S1-final-viral-combined_k141_231995\|\|full | 1557 | No | NA | | 3 | 0 | 2 | Not-determined | Genome-fragment | NA | | NA | 0 |
| S17-final-viral-combined_k141_123339\|\|full | 1267 | No | NA | | 4 | 2 | 0 | Low-quality | Genome-fragment | | 3.48 | AAI-based (high-confidence) | 0 |
| S1-final-viral-combined_k141_215806\|\|full | 12770 | No | NA | | 16 | 0 | 10 | Not-determined | Genome-fragment | NA | | NA | 0 |
| S1-final-viral-combined_k141_330694\|\|full | 4167 | No | NA | | 4 | 0 | 2 | Not-determined | Genome-fragment | NA | | NA | 0 |
| S12_k141_116012 | 1267 | No | NA | | 2 | 0 | 0 | Low-quality | Genome-fragment | | 0.76 | AAI-based (medium-confidence) | 0 |
| S1-final-viral-combined_k141_165834\|\|full | 3191 | No | NA | | 4 | 0 | 2 | Not-determined | Genome-fragment | NA | | NA | 0 |
| S1-final-viral-combined_k141_100287\|\|full | 1261 | No | NA | | 3 | 0 | 1 | Not-determined | Genome-fragment | NA | | NA | 0 |
| S1-final-viral-combined_k141_215846\|\|full | 3825 | No | NA | | 7 | 0 | 1 | Not-determined | Genome-fragment | NA | | NA | 0 |
| S8-final-viral-combined_k141_50868\|\|lt2gene | 1267 | No | NA | | 2 | 2 | 0 | Low-quality | Genome-fragment | | 1.98 | HMM-based (lower-bound) | 0 |
| S11-final-viral-combined_k141_293636\|\|full | 1268 | No | NA | | 3 | 0 | 0 | Low-quality | Genome-fragment | | 2.12 | AAI-based (high-confidence) | 0 |
| S11-final-viral-combined_k141_189106\|\|lt2gene | 1268 | No | NA | | 2 | 2 | 0 | Low-quality | Genome-fragment | | 3.71 | AAI-based (high-confidence) | 0 |
| S1-final-viral-combined_k141_284382\|\|full | 1268 | No | NA | | 2 | 0 | 1 | Low-quality | Genome-fragment | | 1.5 | AAI-based (medium-confidence) | 0 |
| S1-final-viral-combined_k141_116938\|\|full | 24124 | No | NA | | 33 | 18 | 1 | Medium-quality | Genome-fragment | | 68.29 | AAI-based (high-confidence) | 0 |
| S1-final-viral-combined_k141_363720\|\|full | 3342 | No | NA | | 4 | 0 | 0 | Not-determined | Genome-fragment | NA | | NA | 0 |
| S1-final-viral-combined_k141_100316\|\|full | 1494 | No | NA | | 4 | 0 | 1 | Not-determined | Genome-fragment | NA | | NA | 0 |
| S1-final-viral-combined_k141_281674\|\|full | 1491 | No | NA | | 3 | 0 | 1 | Not-determined | Genome-fragment | NA | | NA | 0 |
| S1-final-viral-combined_k141_2186\|\|full | 1001 | No | NA | | 2 | 0 | 0 | Not-determined | Genome-fragment | NA | | NA | 0 |
| S1-final-viral-combined_k141_281685\|\|full | 2332 | No | NA | | 4 | 0 | 0 | Not-determined | Genome-fragment | NA | | NA | 0 |
| S1-final-viral-combined_k141_215880\|\|full | 5433 | No | NA | | 6 | 0 | 5 | Not-determined | Genome-fragment | NA | | NA | 0 |
| S1-final-viral-combined_k141_182835\|\|full | 1166 | No | NA | | 1 | 0 | 0 | Not-determined | Genome-fragment | NA | | NA | 0 |
| S1-final-viral-combined_k141_330737\|\|full | 4652 | No | NA | | 4 | 0 | 3 | Not-determined | Genome-fragment | NA | | NA | 0 |
| S1-final-viral-combined_k141_2222\|\|full | 1255 | No | NA | | 2 | 0 | 1 | Not-determined | Genome-fragment | NA | | NA | 0 |
| S1-final-viral-combined_k141_330745\|\|full | 4079 | No | NA | | 3 | 0 | 0 | Not-determined | Genome-fragment | NA | | NA | 0 |
| S1-final-viral-combined_k141_232732\|\|lt2gene | 1268 | No | NA | | 2 | 0 | 0 | Low-quality | Genome-fragment | | 9.37 | AAI-based (high-confidence) | 0 |
| S1-final-viral-combined_k141_264709\|\|full | 1460 | No | NA | | 2 | 0 | 1 | Not-determined | Genome-fragment | NA | | NA | 0 |
| S1-final-viral-combined_k141_248622\|\|full | 22409 | No | NA | | 11 | 0 | 3 | Medium-quality | Genome-fragment | | 64.62 | AAI-based (high-confidence) | 0 |
| S1-final-viral-combined_k141_100347\|\|full | 1286 | No | NA | | 3 | 0 | 1 | Not-determined | Genome-fragment | NA | | NA | 0 |
| S1-final-viral-combined_k141_199398\|\|full | 2876 | No | NA | | 7 | 0 | 0 | Not-determined | Genome-fragment | NA | | NA | 0 |
| S1_k141_345241 | 1268 | No | NA | | 1 | 0 | 1 | Low-quality | Genome-fragment | | 1.99 | AAI-based (medium-confidence) | 0 |
| S1-final-viral-combined_k141_66977\|\|full | 2000 | No | NA | | 4 | 0 | 2 | Not-determined | Genome-fragment | NA | | NA | 0 |
| S5_k141_86862 | 1268 | No | NA | | 3 | 0 | 0 | Low-quality | Genome-fragment | | 0.75 | AAI-based (high-confidence) | 0 |
| S1-final-viral-combined_k141_116993\|\|full | 2178 | No | NA | | 3 | 0 | 2 | Not-determined | Genome-fragment | NA | | NA | 0 |
| S1-final-viral-combined_k141_182900\|\|full | 1624 | No | NA | | 3 | 0 | 2 | Not-determined | Genome-fragment | NA | | NA | 0 |
| S7-final-viral-combined_k141_143503\|\|full | 1268 | Yes | | 902 | 2 | 1 | 1 | Low-quality | Genome-fragment | | 0.07 | HMM-based (lower-bound) | 28.86 |
| S1-final-viral-combined_k141_165889\|\|full | 1234 | No | NA | | 2 | 0 | 0 | Not-determined | Genome-fragment | NA | | NA | 0 |
| S1-final-viral-combined_k141_50640\|\|full | 2374 | No | NA | | 2 | 0 | 2 | Not-determined | Genome-fragment | NA | | NA | 0 |
| S1-final-viral-combined_k141_347563\|\|full | 2261 | No | NA | | 3 | 0 | 1 | Not-determined | Genome-fragment | NA | | NA | 0 |
| S1-final-viral-combined_k141_314438\|\|full | 12494 | No | NA | | 13 | 0 | 7 | Not-determined | Genome-fragment | NA | | NA | 0 |
| S7-final-viral-combined_k141_5570\|\|full | 1268 | No | NA | | 3 | 0 | 0 | Low-quality | Genome-fragment | | 2.37 | AAI-based (high-confidence) | 0 |
| S1-final-viral-combined_k141_18312\|\|full | 1560 | No | NA | | 5 | 0 | 1 | Not-determined | Genome-fragment | NA | | NA | 0 |
| S12-final-viral-combined_k141_112953\|\|lt2gene | 1268 | No | NA | | 1 | 1 | 0 | Low-quality | Genome-fragment | | 0.38 | HMM-based (lower-bound) | 0 |
| S1-final-viral-combined_k141_67035\|\|full | 2328 | No | NA | | 3 | 0 | 0 | Not-determined | Genome-fragment | NA | | NA | 0 |
| S1-final-viral-combined_k141_133794\|\|full | 1244 | No | NA | | 2 | 0 | 0 | Not-determined | Genome-fragment | NA | | NA | 0 |
| S1-final-viral-combined_k141_133795\|\|full | 1381 | No | NA | | 3 | 0 | 2 | Not-determined | Genome-fragment | NA | | NA | 0 |
| S1-final-viral-combined_k141_380184\|\|full | 6487 | No | NA | | 6 | 0 | 3 | Not-determined | Genome-fragment | NA | | NA | 0 |
| S1-final-viral-combined_k141_67049\|\|full | 4664 | No | NA | | 5 | 0 | 3 | Not-determined | Genome-fragment | NA | | NA | 0 |
| S1-final-viral-combined_k141_50688\|\|full | 6550 | No | NA | | 5 | 0 | 5 | Not-determined | Genome-fragment | NA | | NA | 0 |
| S1-final-viral-combined_k141_133811\|\|full | 2234 | No | NA | | 4 | 0 | 1 | Not-determined | Genome-fragment | NA | | NA | 0 |
| S1-final-viral-combined_k141_380204\|\|full | 1244 | No | NA | | 3 | 0 | 0 | Not-determined | Genome-fragment | NA | | NA | 0 |
| S6-final-viral-combined_k141_6951\|\|full | 1268 | No | NA | | 2 | 0 | 0 | Low-quality | Genome-fragment | | 2.67 | AAI-based (medium-confidence) | 0 |
| S8-final-viral-combined_k141_107694\|\|full | 1268 | No | NA | | 4 | 1 | 0 | Low-quality | Genome-fragment | | 3.19 | AAI-based (high-confidence) | 0 |
| S11_k141_156038 | 1269 | No | NA | | 3 | 0 | 0 | Low-quality | Genome-fragment | | 0.89 | AAI-based (medium-confidence) | 0 |
| S1-final-viral-combined_k141_304890\|\|full | 1269 | No | NA | | 4 | 0 | 0 | Low-quality | Genome-fragment | | 2.28 | AAI-based (high-confidence) | 0 |
| S1-final-viral-combined_k141_387981\|\|lt2gene | 1269 | No | NA | | 1 | 1 | 0 | Low-quality | Genome-fragment | | 3.37 | AAI-based (high-confidence) | 0 |
| S1-final-viral-combined_k141_100430\|\|full | 33842 | No | NA | | 46 | 19 | 1 | Medium-quality | Genome-fragment | | 71.89 | AAI-based (high-confidence) | 0 |
| S1-final-viral-combined_k141_84201\|\|full | 2307 | No | NA | | 3 | 0 | 2 | Not-determined | Genome-fragment | NA | | NA | 0 |
| S5-final-viral-combined_k141_144533\|\|lt2gene | 1269 | No | NA | | 2 | 1 | 0 | Low-quality | Genome-fragment | | 3.64 | AAI-based (medium-confidence) | 0 |
| S7-final-viral-combined_k141_5746\|\|full | 1269 | No | NA | | 3 | 0 | 1 | Low-quality | Genome-fragment | | 3.18 | AAI-based (high-confidence) | 0 |
| S1-final-viral-combined_k141_182958\|\|full | 2801 | No | NA | | 3 | 0 | 2 | Not-determined | Genome-fragment | NA | | NA | 0 |
| S17-final-viral-combined_k141_115186\|\|full | 1269 | No | NA | | 4 | 0 | 0 | Low-quality | Genome-fragment | | 2.54 | AAI-based (medium-confidence) | 0 |
| S1-final-viral-combined_k141_363851\|\|full | 4337 | No | NA | | 4 | 0 | 3 | Not-determined | Genome-fragment | NA | | NA | 0 |
| S17-k141_17326 | 1269 | No | NA | | 2 | 0 | 1 | Low-quality | Genome-fragment | | 3.82 | AAI-based (medium-confidence) | 0 |
| S1-final-viral-combined_k141_18367\|\|full | 6352 | No | NA | | 6 | 0 | 2 | Not-determined | Genome-fragment | NA | | NA | 0 |
| S1-final-viral-combined_k141_298237\|\|full | 2076 | No | NA | | 2 | 0 | 2 | Not-determined | Genome-fragment | NA | | NA | 0 |
| S1-final-viral-combined_k141_232159\|\|full | 6812 | No | NA | | 9 | 0 | 6 | Not-determined | Genome-fragment | NA | | NA | 0 |
| S1-final-viral-combined_k141_347641\|\|full | 2238 | No | NA | | 3 | 0 | 1 | Not-determined | Genome-fragment | NA | | NA | 0 |
| S12-final-viral-combined_k141_363815\|\|lt2gene | 1269 | No | NA | | 2 | 1 | 0 | Low-quality | Genome-fragment | | 3.15 | AAI-based (medium-confidence) | 0 |
| S1-final-viral-combined_k141_149844\|\|full | 1817 | No | NA | | 3 | 0 | 1 | Not-determined | Genome-fragment | NA | | NA | 0 |
| S1-final-viral-combined_k141_84250\|\|full | 1770 | No | NA | | 3 | 0 | 0 | Not-determined | Genome-fragment | NA | | NA | 0 |
| S2_k141_23623 | 1269 | No | NA | | 2 | 0 | 0 | Low-quality | Genome-fragment | | 1.3 | AAI-based (medium-confidence) | 0 |
| S1-final-viral-combined_k141_363895\|\|full | 1868 | No | NA | | 3 | 0 | 2 | Not-determined | Genome-fragment | NA | | NA | 0 |
| S1-final-viral-combined_k141_50804\|\|full | 1474 | No | NA | | 2 | 0 | 1 | Not-determined | Genome-fragment | NA | | NA | 0 |
| S1-final-viral-combined_k141_84278\|\|full | 2303 | No | NA | | 3 | 0 | 1 | Not-determined | Genome-fragment | NA | | NA | 0 |
| S4-final-viral-combined_k141_112866\|\|full | 1269 | No | NA | | 2 | 1 | 0 | Low-quality | Genome-fragment | | 9.92 | AAI-based (high-confidence) | 0 |
| S6-final-viral-combined_k141_2095\|\|full | 1269 | No | NA | | 2 | 0 | 0 | Low-quality | Genome-fragment | | 0.81 | AAI-based (medium-confidence) | 0 |
| S6_k141_33901 | 1269 | No | NA | | 2 | 0 | 1 | Low-quality | Genome-fragment | | 0.61 | AAI-based (medium-confidence) | 0 |
| S1-final-viral-combined_k141_166012\|\|full | 1602 | No | NA | | 2 | 0 | 1 | Not-determined | Genome-fragment | NA | | NA | 0 |
| S9-final-viral-combined_k141_47314\|\|full | 1270 | No | NA | | 3 | 0 | 0 | Low-quality | Genome-fragment | | 3.2 | AAI-based (medium-confidence) | 0 |
| S1-final-viral-combined_k141_330799\|\|full | 1587 | No | NA | | 3 | 0 | 1 | Not-determined | Genome-fragment | NA | | NA | 0 |
| S11-final-viral-combined_k141_72132\|\|full | 1270 | No | NA | | 3 | 1 | 0 | Low-quality | Genome-fragment | | 2.67 | AAI-based (high-confidence) | 0 |
| S11-final-viral-combined_k141_241991\|\|full | 1270 | No | NA | | 3 | 0 | 0 | Low-quality | Genome-fragment | | 2.58 | AAI-based (medium-confidence) | 0 |
| S11_k141_196667 | 1270 | No | NA | | 3 | 3 | 0 | Low-quality | Genome-fragment | | 3.05 | AAI-based (high-confidence) | 0 |
| S1-final-viral-combined_k141_84323\|\|full | 2179 | No | NA | | 3 | 0 | 1 | Not-determined | Genome-fragment | NA | | NA | 0 |
| S1-final-viral-combined_k141_133924\|\|full | 2043 | No | NA | | 2 | 0 | 2 | Not-determined | Genome-fragment | NA | | NA | 0 |
| S7-final-viral-combined_k141_143953\|\|full | 1270 | No | NA | | 3 | 0 | 0 | Low-quality | Genome-fragment | | 2 | AAI-based (medium-confidence) | 0 |
| S1-final-viral-combined_k141_183059\|\|full | 1145 | No | NA | | 3 | 0 | 0 | Not-determined | Genome-fragment | NA | | NA | 0 |
| S7_k141_237871 | 1270 | No | NA | | 2 | 0 | 1 | Low-quality | Genome-fragment | | 0.3 | AAI-based (medium-confidence) | 0 |
| S12-final-viral-combined_k141_68224\|\|full | 1270 | No | NA | | 3 | 1 | 0 | Low-quality | Genome-fragment | | 2.03 | AAI-based (medium-confidence) | 0 |
| S12-final-viral-combined_k141_1557\|\|full | 1270 | No | NA | | 2 | 0 | 2 | Low-quality | Genome-fragment | | 0.57 | AAI-based (medium-confidence) | 0 |
| S1-final-viral-combined_k141_84331\|\|full | 1456 | No | NA | | 4 | 0 | 1 | Not-determined | Genome-fragment | NA | | NA | 0 |
| S1-final-viral-combined_k141_248802\|\|full | 2049 | No | NA | | 3 | 0 | 2 | Not-determined | Genome-fragment | NA | | NA | 0 |
| S1-final-viral-combined_k141_347705\|\|full | 1947 | No | NA | | 2 | 0 | 2 | Not-determined | Genome-fragment | NA | | NA | 0 |
| S1-final-viral-combined_k141_298364\|\|full | 8315 | No | NA | | 10 | 0 | 6 | Not-determined | Genome-fragment | NA | | NA | 0 |
| S2-final-viral-combined_k141_149185\|\|full | 1270 | No | NA | | 3 | 0 | 2 | Low-quality | Genome-fragment | | 3.04 | AAI-based (medium-confidence) | 0 |
| S2-final-viral-combined_k141_77122\|\|full | 1270 | No | NA | | 2 | 0 | 0 | Low-quality | Genome-fragment | | 0.66 | AAI-based (medium-confidence) | 0 |
| S2-final-viral-combined_k141_144077\|\|full | 1270 | No | NA | | 3 | 0 | 0 | Low-quality | Genome-fragment | | 2.36 | AAI-based (high-confidence) | 0 |
| S1-final-viral-combined_k141_363959\|\|full | 1139 | No | NA | | 2 | 0 | 1 | Not-determined | Genome-fragment | NA | | NA | 0 |
| S1-final-viral-combined_k141_298369\|\|full | 4282 | No | NA | | 5 | 0 | 4 | Not-determined | Genome-fragment | NA | | NA | 0 |
| S2_k141_217002 | 1270 | No | NA | | 1 | 0 | 1 | Low-quality | Genome-fragment | | 0.45 | AAI-based (medium-confidence) | 0 |
| S1-final-viral-combined_k141_298371\|\|full | 1669 | No | NA | | 3 | 0 | 1 | Not-determined | Genome-fragment | NA | | NA | 0 |
| S6-final-viral-combined_k141_74650\|\|full | 1270 | No | NA | | 2 | 0 | 0 | Low-quality | Genome-fragment | | 0.28 | AAI-based (medium-confidence) | 0 |
| S11-final-viral-combined_k141_146178\|\|full | 1271 | No | NA | | 2 | 0 | 0 | Low-quality | Genome-fragment | | 0.84 | AAI-based (high-confidence) | 0 |
| S1-final-viral-combined_k141_50899\|\|full | 2221 | No | NA | | 2 | 0 | 1 | Not-determined | Genome-fragment | NA | | NA | 0 |
| S1-final-viral-combined_k141_264932\|\|full | 1841 | No | NA | | 2 | 0 | 1 | Not-determined | Genome-fragment | NA | | NA | 0 |
| S1-final-viral-combined_k141_149946\|\|full | 1764 | No | NA | | 3 | 0 | 1 | Not-determined | Genome-fragment | NA | | NA | 0 |
| S1-final-viral-combined_k141_134002\|\|full | 2384 | No | NA | | 2 | 0 | 2 | Not-determined | Genome-fragment | NA | | NA | 0 |
| S11_k141_54609 | 1271 | No | NA | | 1 | 0 | 0 | Low-quality | Genome-fragment | | 1.1 | AAI-based (medium-confidence) | 0 |
| S1-final-viral-combined_k141_100575\|\|full | 2589 | No | NA | | 3 | 0 | 0 | Not-determined | Genome-fragment | NA | | NA | 0 |
| S1-final-viral-combined_k141_216142\|\|full | 1587 | No | NA | | 3 | 0 | 1 | Not-determined | Genome-fragment | NA | | NA | 0 |
| S1-final-viral-combined_k141_218209\|\|lt2gene | 1271 | No | NA | | 1 | 0 | 0 | Low-quality | Genome-fragment | | 2.74 | AAI-based (high-confidence) | 0 |
| S1_k141_383660 | 1271 | No | NA | | 1 | 0 | 0 | Low-quality | Genome-fragment | | 5.56 | AAI-based (high-confidence) | 0 |
| S1-final-viral-combined_k141_248922\|\|full | 1475 | No | NA | | 4 | 0 | 0 | Not-determined | Genome-fragment | NA | | NA | 0 |
| S1-final-viral-combined_k141_232342\|\|full | 1878 | No | NA | | 2 | 0 | 1 | Not-determined | Genome-fragment | NA | | NA | 0 |
| S1-final-viral-combined_k141_380353\|\|full | 3713 | No | NA | | 5 | 0 | 5 | Not-determined | Genome-fragment | NA | | NA | 0 |
| S1-final-viral-combined_k141_150026\|\|full | 2384 | No | NA | | 4 | 0 | 1 | Not-determined | Genome-fragment | NA | | NA | 0 |
| S17-final-viral-combined_k141_136747\|\|full | 1271 | No | NA | | 3 | 1 | 0 | Low-quality | Genome-fragment | | 0.82 | HMM-based (lower-bound) | 0 |
| S17-final-viral-combined_k141_84807\|\|lt2gene | 1271 | No | NA | | 1 | 1 | 0 | Low-quality | Genome-fragment | | 2.86 | AAI-based (high-confidence) | 0 |
| S1-final-viral-combined_k141_232363\|\|full | 1320 | No | NA | | 4 | 0 | 0 | Not-determined | Genome-fragment | NA | | NA | 0 |
| S17-final-viral-combined_k141_230229\|\|lt2gene | 1271 | No | NA | | 2 | 2 | 0 | Low-quality | Genome-fragment | | 2.29 | AAI-based (high-confidence) | 0 |
| S12-final-viral-combined_k141_202568\|\|full | 1271 | No | NA | | 4 | 1 | 0 | Low-quality | Genome-fragment | | 2.27 | AAI-based (high-confidence) | 0 |
| S1-final-viral-combined_k141_248961\|\|full | 1870 | No | NA | | 2 | 0 | 0 | Not-determined | Genome-fragment | NA | | NA | 0 |
| S4_k141_52887 | 1271 | No | NA | | 3 | 0 | 0 | Low-quality | Genome-fragment | | 2.34 | AAI-based (medium-confidence) | 0 |
| S1-final-viral-combined_k141_248980\|\|full | 3016 | No | NA | | 4 | 0 | 0 | Not-determined | Genome-fragment | NA | | NA | 0 |
| S8-final-viral-combined_k141_202732\|\|full | 1271 | No | NA | | 3 | 0 | 0 | Low-quality | Genome-fragment | | 3.09 | AAI-based (high-confidence) | 0 |
| S9-final-viral-combined_k141_117119\|\|full | 1272 | No | NA | | 3 | 1 | 0 | Low-quality | Genome-fragment | | 2.6 | AAI-based (medium-confidence) | 0 |
| S1-final-viral-combined_k141_364030\|\|full | 1950 | No | NA | | 2 | 0 | 1 | Not-determined | Genome-fragment | NA | | NA | 0 |
| S1-final-viral-combined_k141_183180\|\|full | 2674 | No | NA | | 3 | 0 | 1 | Not-determined | Genome-fragment | NA | | NA | 0 |
| S11-final-viral-combined_k141_276370\|\|full | 1272 | No | NA | | 4 | 2 | 0 | Low-quality | Genome-fragment | | 3.54 | AAI-based (high-confidence) | 0 |
| S1-final-viral-combined_k141_314820\|\|full | 7045 | No | NA | | 7 | 0 | 3 | Not-determined | Genome-fragment | NA | | NA | 0 |
| S1-final-viral-combined_k141_216260\|\|full | 10510 | No | NA | | 14 | 0 | 2 | Not-determined | Genome-fragment | NA | | NA | 0 |
| S11_k141_310567 | 1272 | No | NA | | 1 | 0 | 0 | Low-quality | Genome-fragment | | 1.82 | AAI-based (high-confidence) | 0 |
| S1-final-viral-combined_k141_6053\|\|full | 1272 | No | NA | | 3 | 0 | 0 | Low-quality | Genome-fragment | | 3.41 | AAI-based (high-confidence) | 0 |
| S1-final-viral-combined_k141_134149\|\|full | 1014 | No | NA | | 2 | 0 | 1 | Not-determined | Genome-fragment | NA | | NA | 0 |
| S1-final-viral-combined_k141_51109\|\|full | 13768 | No | NA | | 12 | 0 | 11 | Not-determined | Genome-fragment | NA | | NA | 0 |
| S1-final-viral-combined_k141_330923\|\|full | 1360 | No | NA | | 2 | 0 | 1 | Not-determined | Genome-fragment | NA | | NA | 0 |
| S1-final-viral-combined_k141_78447\|\|full | 1272 | No | NA | | 3 | 0 | 1 | Low-quality | Genome-fragment | | 1.08 | AAI-based (medium-confidence) | 0 |
| S1-final-viral-combined_k141_2529\|\|full | 3354 | No | NA | | 5 | 0 | 1 | Not-determined | Genome-fragment | NA | | NA | 0 |
| S1-final-viral-combined_k141_298421\|\|full | 1191 | No | NA | | 3 | 0 | 2 | Not-determined | Genome-fragment | NA | | NA | 0 |
| S1-final-viral-combined_k141_282012\|\|full | 2092 | No | NA | | 3 | 0 | 1 | Not-determined | Genome-fragment | NA | | NA | 0 |
| S1-final-viral-combined_k141_84504\|\|full | 2605 | No | NA | | 3 | 0 | 1 | Not-determined | Genome-fragment | NA | | NA | 0 |
| S1-final-viral-combined_k141_249068\|\|full | 2202 | No | NA | | 3 | 0 | 1 | Not-determined | Genome-fragment | NA | | NA | 0 |
| S1-final-viral-combined_k141_216301\|\|full | 3221 | No | NA | | 4 | 0 | 1 | Not-determined | Genome-fragment | NA | | NA | 0 |
| S1-final-viral-combined_k141_183239\|\|full | 2621 | No | NA | | 4 | 0 | 3 | Not-determined | Genome-fragment | NA | | NA | 0 |
| S1-final-viral-combined_k141_364086\|\|full | 2317 | No | NA | | 5 | 0 | 0 | Not-determined | Genome-fragment | NA | | NA | 0 |
| S1-final-viral-combined_k141_199714\|\|full | 20970 | No | NA | | 16 | 0 | 10 | Not-determined | Genome-fragment | NA | | NA | 0 |
| S1-final-viral-combined_k141_364091\|\|full | 1930 | No | NA | | 2 | 0 | 0 | Not-determined | Genome-fragment | NA | | NA | 0 |
| S3-final-viral-combined_k141_47567\|\|full | 1272 | No | NA | | 2 | 0 | 0 | Low-quality | Genome-fragment | | 1.14 | AAI-based (medium-confidence) | 0 |
| S1-final-viral-combined_k141_265058\|\|full | 1656 | No | NA | | 2 | 0 | 2 | Not-determined | Genome-fragment | NA | | NA | 0 |
| S3-final-viral-combined_k141_100208\|\|full | 1272 | No | NA | | 2 | 0 | 0 | Low-quality | Genome-fragment | | 0.82 | AAI-based (high-confidence) | 0 |
| S12-final-viral-combined_k141_303071\|\|full | 1272 | No | NA | | 4 | 1 | 0 | Low-quality | Genome-fragment | | 2.98 | AAI-based (high-confidence) | 0 |
| S1-final-viral-combined_k141_216341\|\|full | 1234 | No | NA | | 2 | 0 | 2 | Not-determined | Genome-fragment | NA | | NA | 0 |
| S1-final-viral-combined_k141_298484\|\|full | 6273 | No | NA | | 8 | 0 | 4 | Not-determined | Genome-fragment | NA | | NA | 0 |
| S12-final-viral-combined_k141_230220\|\|full | 1272 | No | NA | | 2 | 1 | 0 | Low-quality | Genome-fragment | | 0.78 | AAI-based (medium-confidence) | 0 |
| S1-final-viral-combined_k141_298499\|\|full | 1444 | No | NA | | 2 | 0 | 0 | Not-determined | Genome-fragment | NA | | NA | 0 |
| S1-final-viral-combined_k141_100703\|\|full | 1037 | No | NA | | 2 | 0 | 2 | Not-determined | Genome-fragment | NA | | NA | 0 |
| S1-final-viral-combined_k141_100709\|\|full | 1086 | No | NA | | 3 | 0 | 1 | Not-determined | Genome-fragment | NA | | NA | 0 |
| S2-final-viral-combined_k141_101933\|\|lt2gene | 1272 | No | NA | | 2 | 1 | 0 | Low-quality | Genome-fragment | | 0.82 | HMM-based (lower-bound) | 0 |
| S8-final-viral-combined_k141_197749\|\|full | 1272 | No | NA | | 4 | 0 | 0 | Low-quality | Genome-fragment | | 0.97 | AAI-based (medium-confidence) | 0 |
| S9-final-viral-combined_k141_24029\|\|full | 1273 | No | NA | | 2 | 0 | 2 | Low-quality | Genome-fragment | | 0.4 | AAI-based (medium-confidence) | 0 |
| S1_k141_331007 | 1273 | No | NA | | 3 | 0 | 0 | Low-quality | Genome-fragment | | 1.94 | AAI-based (medium-confidence) | 0 |
| S3-final-viral-combined_k141_117359\|\|full | 1273 | No | NA | | 3 | 0 | 0 | Low-quality | Genome-fragment | | 1.29 | AAI-based (medium-confidence) | 0 |
| S3_k141_33476 | 1273 | No | NA | | 1 | 0 | 0 | Low-quality | Genome-fragment | | 2.74 | AAI-based (high-confidence) | 0 |
| S1-final-viral-combined_k141_298524\|\|full | 1858 | No | NA | | 2 | 0 | 2 | Not-determined | Genome-fragment | NA | | NA | 0 |
| S7-final-viral-combined_k141_4826\|\|lt2gene | 1273 | No | NA | | 2 | 1 | 0 | Low-quality | Genome-fragment | | 3.06 | AAI-based (high-confidence) | 0 |
| S12_k141_340996 | 1273 | No | NA | | 3 | 3 | 0 | Low-quality | Genome-fragment | | 2.72 | AAI-based (medium-confidence) | 0 |
| S1-final-viral-combined_k141_18536\|\|full | 2350 | No | NA | | 2 | 0 | 1 | Not-determined | Genome-fragment | NA | | NA | 0 |
| S1-final-viral-combined_k141_314909\|\|full | 2072 | No | NA | | 2 | 0 | 2 | Not-determined | Genome-fragment | NA | | NA | 0 |
| S1-final-viral-combined_k141_232545\|\|full | 2315 | No | NA | | 3 | 0 | 3 | Not-determined | Genome-fragment | NA | | NA | 0 |
| S1-final-viral-combined_k141_166301\|\|full | 4165 | No | NA | | 4 | 0 | 4 | Not-determined | Genome-fragment | NA | | NA | 0 |
| S2-final-viral-combined_k141_218351\|\|full | 1273 | No | NA | | 3 | 1 | 0 | Low-quality | Genome-fragment | | 2.19 | AAI-based (medium-confidence) | 0 |
| S2-final-viral-combined_k141_75321\|\|full | 1273 | No | NA | | 5 | 1 | 0 | Low-quality | Genome-fragment | | 3.05 | AAI-based (high-confidence) | 0 |
| S1-final-viral-combined_k141_51246\|\|full | 8429 | No | NA | | 2 | 0 | 2 | Not-determined | Genome-fragment | NA | | NA | 0 |
| S1-final-viral-combined_k141_51259\|\|full | 2502 | No | NA | | 2 | 0 | 1 | Not-determined | Genome-fragment | NA | | NA | 0 |
| S2-final-viral-combined_k141_159714\|\|full | 1273 | No | NA | | 3 | 1 | 0 | Low-quality | Genome-fragment | | 3.64 | AAI-based (medium-confidence) | 0 |
| S1-final-viral-combined_k141_314950\|\|full | 7784 | No | NA | | 8 | 0 | 7 | Not-determined | Genome-fragment | NA | | NA | 0 |
| S1-final-viral-combined_k141_93536\|\|full | 1274 | No | NA | | 1 | 0 | 0 | Low-quality | Genome-fragment | | 1.8 | AAI-based (medium-confidence) | 0 |
| S1-final-viral-combined_k141_314958\|\|full | 1718 | No | NA | | 4 | 0 | 0 | Not-determined | Genome-fragment | NA | | NA | 0 |
| S5_k141_3420 | 1274 | No | NA | | 3 | 0 | 0 | Low-quality | Genome-fragment | | 2.01 | AAI-based (medium-confidence) | 0 |
| S1-final-viral-combined_k141_380551\|\|full | 8050 | No | NA | | 7 | 0 | 5 | Not-determined | Genome-fragment | NA | | NA | 0 |
| S7-final-viral-combined_k141_34292\|\|full | 1274 | No | NA | | 3 | 0 | 0 | Low-quality | Genome-fragment | | 3.21 | AAI-based (high-confidence) | 0 |
| S1-final-viral-combined_k141_18586\|\|full | 7915 | No | NA | | 9 | 0 | 8 | Not-determined | Genome-fragment | NA | | NA | 0 |
| S1-final-viral-combined_k141_216457\|\|full | 3126 | No | NA | | 5 | 0 | 3 | Not-determined | Genome-fragment | NA | | NA | 0 |
| S7_k141_191709 | 1274 | No | NA | | 2 | 0 | 1 | Low-quality | Genome-fragment | | 2.88 | AAI-based (medium-confidence) | 0 |
| S17-final-viral-combined_k141_166828\|\|lt2gene | 1274 | No | NA | | 2 | 1 | 0 | Low-quality | Genome-fragment | | 1.86 | HMM-based (lower-bound) | 0 |
| S1-final-viral-combined_k141_117457\|\|full | 5007 | No | NA | | 4 | 0 | 3 | Not-determined | Genome-fragment | NA | | NA | 0 |
| S1-final-viral-combined_k141_18591\|\|full | 3694 | No | NA | | 2 | 0 | 1 | Not-determined | Genome-fragment | NA | | NA | 0 |
| S1-final-viral-combined_k141_380562\|\|full | 2154 | No | NA | | 3 | 0 | 3 | Not-determined | Genome-fragment | NA | | NA | 0 |
| S1-final-viral-combined_k141_183359\|\|full | 1487 | No | NA | | 3 | 0 | 0 | Not-determined | Genome-fragment | NA | | NA | 0 |
| S1-final-viral-combined_k141_117464\|\|full | 3600 | No | NA | | 3 | 0 | 3 | Not-determined | Genome-fragment | NA | | NA | 0 |
| S17-final-viral-combined_k141_227178\|\|lt2gene | 1274 | No | NA | | 1 | 1 | 0 | Low-quality | Genome-fragment | | 3.12 | AAI-based (high-confidence) | 0 |
| S1-final-viral-combined_k141_35165\|\|full | 1984 | No | NA | | 3 | 0 | 0 | Not-determined | Genome-fragment | NA | | NA | 0 |
| S1-final-viral-combined_k141_67547\|\|full | 8953 | No | NA | | 8 | 0 | 5 | Not-determined | Genome-fragment | NA | | NA | 0 |
| S12-final-viral-combined_k141_93953\|\|full | 1274 | No | NA | | 3 | 0 | 0 | Low-quality | Genome-fragment | | 3.37 | AAI-based (medium-confidence) | 0 |
| S12-final-viral-combined_k141_143423\|\|lt2gene | 1274 | No | NA | | 2 | 1 | 0 | Low-quality | Genome-fragment | | 2.57 | AAI-based (high-confidence) | 0 |
| S12_k141_391441 | 1274 | No | NA | | 2 | 0 | 0 | Low-quality | Genome-fragment | | 1.92 | AAI-based (medium-confidence) | 0 |
| S1-final-viral-combined_k141_265210\|\|full | 5624 | No | NA | | 5 | 0 | 3 | Not-determined | Genome-fragment | NA | | NA | 0 |
| S1-final-viral-combined_k141_51345\|\|full | 3565 | No | NA | | 5 | 0 | 3 | Not-determined | Genome-fragment | NA | | NA | 0 |
| S1-final-viral-combined_k141_18646\|\|full | 1326 | No | NA | | 3 | 0 | 2 | Not-determined | Genome-fragment | NA | | NA | 0 |
| S2-final-viral-combined_k141_100377\|\|lt2gene | 1274 | No | NA | | 2 | 1 | 0 | Low-quality | Genome-fragment | | 3.59 | AAI-based (high-confidence) | 0 |
| S1-final-viral-combined_k141_216511\|\|full | 1899 | No | NA | | 3 | 0 | 1 | Not-determined | Genome-fragment | NA | | NA | 0 |
| S1-final-viral-combined_k141_380613\|\|full | 1442 | No | NA | | 2 | 0 | 1 | Not-determined | Genome-fragment | NA | | NA | 0 |
| S1-final-viral-combined_k141_364260\|\|full | 2646 | No | NA | | 4 | 0 | 3 | Not-determined | Genome-fragment | NA | | NA | 0 |
| S1-final-viral-combined_k141_166375\|\|full | 13085 | No | NA | | 9 | 0 | 5 | Not-determined | Genome-fragment | NA | | NA | 0 |
| S1-final-viral-combined_k141_232661\|\|full | 1185 | No | NA | | 3 | 0 | 2 | Not-determined | Genome-fragment | NA | | NA | 0 |
| S2_k141_53791 | 1274 | No | NA | | 3 | 1 | 0 | Low-quality | Genome-fragment | | 2.74 | AAI-based (high-confidence) | 0 |
| S4-final-viral-combined_k141_57793\|\|full | 1274 | No | NA | | 3 | 0 | 1 | Low-quality | Genome-fragment | | 0.74 | AAI-based (medium-confidence) | 0 |
| S1-final-viral-combined_k141_18667\|\|full | 3195 | No | NA | | 5 | 0 | 3 | Not-determined | Genome-fragment | NA | | NA | 0 |
| S4_k141_65180 | 1274 | No | NA | | 2 | 0 | 0 | Low-quality | Genome-fragment | | 1.39 | AAI-based (medium-confidence) | 0 |
| S1-final-viral-combined_k141_35203\|\|full | 1571 | No | NA | | 2 | 0 | 1 | Not-determined | Genome-fragment | NA | | NA | 0 |
| S8-final-viral-combined_k141_219661\|\|full | 1274 | No | NA | | 2 | 0 | 1 | Low-quality | Genome-fragment | | 2.25 | AAI-based (high-confidence) | 0 |
| S1-final-viral-combined_k141_265224\|\|full | 13415 | No | NA | | 12 | 0 | 4 | Not-determined | Genome-fragment | NA | | NA | 0 |
| S1-final-viral-combined_k141_249288\|\|full | 1911 | No | NA | | 3 | 0 | 1 | Not-determined | Genome-fragment | NA | | NA | 0 |
| S1-final-viral-combined_k141_67591\|\|full | 1622 | No | NA | | 4 | 0 | 0 | Not-determined | Genome-fragment | NA | | NA | 0 |
| S1-final-viral-combined_k141_298661\|\|full | 1890 | No | NA | | 3 | 0 | 0 | Not-determined | Genome-fragment | NA | | NA | 0 |
| S1-final-viral-combined_k141_249299\|\|full | 7775 | No | NA | | 10 | 0 | 0 | Not-determined | Genome-fragment | NA | | NA | 0 |
| S1-final-viral-combined_k141_166392\|\|full | 19346 | No | NA | | 31 | 10 | 0 | Medium-quality | Genome-fragment | | 50.16 | AAI-based (high-confidence) | 0 |
| S8-final-viral-combined_k141_30683\|\|full | 1274 | No | NA | | 2 | 0 | 0 | Low-quality | Genome-fragment | | 2.27 | AAI-based (high-confidence) | 0 |
| S1-final-viral-combined_k141_67598\|\|full | 1226 | No | NA | | 3 | 0 | 0 | Not-determined | Genome-fragment | NA | | NA | 0 |
| S1-final-viral-combined_k141_2710\|\|full | 2471 | No | NA | | 2 | 0 | 0 | Not-determined | Genome-fragment | NA | | NA | 0 |
| S8-final-viral-combined_k141_68778\|\|full | 1274 | No | NA | | 4 | 0 | 0 | Low-quality | Genome-fragment | | 3.42 | AAI-based (medium-confidence) | 0 |
| S1-final-viral-combined_k141_51405\|\|full | 2195 | No | NA | | 2 | 0 | 1 | Not-determined | Genome-fragment | NA | | NA | 0 |
| S1-final-viral-combined_k141_249364\|\|full | 1332 | No | NA | | 2 | 0 | 0 | Not-determined | Genome-fragment | NA | | NA | 0 |
| S11-final-viral-combined_k141_58996\|\|full | 1275 | No | NA | | 3 | 0 | 0 | Low-quality | Genome-fragment | | 1.41 | AAI-based (medium-confidence) | 0 |
| S17-final-viral-combined_k141_173968\|\|full | 1275 | No | NA | | 3 | 0 | 1 | Low-quality | Genome-fragment | | 2.23 | AAI-based (medium-confidence) | 0 |
| S17-k141_215519 | 1275 | No | NA | | 2 | 1 | 0 | Low-quality | Genome-fragment | | 2.36 | HMM-based (lower-bound) | 0 |
| S12-final-viral-combined_k141_84953\|\|full | 1275 | No | NA | | 2 | 0 | 0 | Low-quality | Genome-fragment | | 2.25 | AAI-based (high-confidence) | 0 |
| S1-final-viral-combined_k141_315090\|\|full | 3547 | No | NA | | 6 | 0 | 0 | Not-determined | Genome-fragment | NA | | NA | 0 |
| S12-final-viral-combined_k141_44620\|\|full | 1275 | No | NA | | 4 | 3 | 0 | Low-quality | Genome-fragment | | 3.73 | AAI-based (high-confidence) | 0 |
| S1-final-viral-combined_k141_232722\|\|full | 1797 | No | NA | | 2 | 0 | 1 | Not-determined | Genome-fragment | NA | | NA | 0 |
| S2_k141_122549 | 1275 | No | NA | | 2 | 0 | 0 | Low-quality | Genome-fragment | | 1.65 | AAI-based (high-confidence) | 0 |
| S4_k141_147136 | 1275 | No | NA | | 1 | 0 | 0 | Low-quality | Genome-fragment | | 0.84 | AAI-based (high-confidence) | 0 |
| S1-final-viral-combined_k141_84824\|\|full | 7870 | No | NA | | 7 | 0 | 5 | Not-determined | Genome-fragment | NA | | NA | 0 |

| | | | | | | | | | | | |
|---|---|---|---|---|---|---|---|---|---|---|---|
| S8_k141_214746 | 1275 | No | NA | 1 | 0 | 1 | Low-quality | Genome-fragment | 4.07 | AAI-based (medium-confidence) | 0 |
| S1-final-viral-combined_k141_2792\|\|full | 3573 | No | NA | 3 | 0 | 1 | Not-determined | Genome-fragment | NA | NA | 0 |
| S9-final-viral-combined_k141_89872\|\|full | 1276 | No | NA | 3 | 0 | 0 | Low-quality | Genome-fragment | 1.55 | AAI-based (medium-confidence) | 0 |
| S1-final-viral-combined_k141_331161\|\|full | 7521 | No | NA | 7 | 0 | 1 | Not-determined | Genome-fragment | NA | NA | 0 |
| S1-final-viral-combined_k141_364387\|\|full | 2514 | No | NA | 4 | 0 | 0 | Not-determined | Genome-fragment | NA | NA | 0 |
| S9-final-viral-combined_k141_13135\|\|lt2gene | 1276 | No | NA | 1 | 1 | 0 | Low-quality | Genome-fragment | 4.04 | AAI-based (high-confidence) | 0 |
| S1-final-viral-combined_k141_2801\|\|full | 2086 | No | NA | 4 | 0 | 3 | Not-determined | Genome-fragment | NA | NA | 0 |
| S1-final-viral-combined_k141_380734\|\|full | 1827 | No | NA | 3 | 0 | 0 | Not-determined | Genome-fragment | NA | NA | 0 |
| S9_k141_97622 | 1276 | No | NA | 3 | 0 | 1 | Low-quality | Genome-fragment | 3.14 | AAI-based (medium-confidence) | 0 |
| S1-final-viral-combined_k141_298784\|\|full | 1141 | No | NA | 2 | 0 | 0 | Not-determined | Genome-fragment | NA | NA | 0 |
| S1-final-viral-combined_k141_232747\|\|full | 3481 | No | NA | 5 | 0 | 3 | Not-determined | Genome-fragment | NA | NA | 0 |
| S1-final-viral-combined_k141_265302\|\|full | 4589 | No | NA | 4 | 0 | 2 | Not-determined | Genome-fragment | NA | NA | 0 |
| S1-final-viral-combined_k141_150348\|\|full | 2404 | No | NA | 4 | 0 | 1 | Not-determined | Genome-fragment | NA | NA | 0 |
| S11_k141_144273 | 1276 | No | NA | 4 | 1 | 0 | Low-quality | Genome-fragment | 2.28 | AAI-based (high-confidence) | 0 |
| S1-final-viral-combined_k141_298822\|\|full | 4036 | No | NA | 3 | 0 | 3 | Not-determined | Genome-fragment | NA | NA | 0 |
| S7-final-viral-combined_k141_73133\|\|full | 1276 | No | NA | 4 | 1 | 0 | Low-quality | Genome-fragment | 2.69 | AAI-based (high-confidence) | 0 |
| S12-final-viral-combined_k141_228962\|\|full | 1276 | No | NA | 2 | 0 | 0 | Low-quality | Genome-fragment | 1.13 | AAI-based (medium-confidence) | 0 |
| S4_k141_161294 | 1276 | No | NA | 1 | 0 | 1 | Low-quality | Genome-fragment | 0.21 | AAI-based (high-confidence) | 0 |
| S1-final-viral-combined_k141_166494\|\|full | 2066 | No | NA | 3 | 0 | 1 | Not-determined | Genome-fragment | NA | NA | 0 |
| S1-final-viral-combined_k141_331225\|\|full | 2170 | No | NA | 2 | 0 | 1 | Not-determined | Genome-fragment | NA | NA | 0 |
| S6-final-viral-combined_k141_132581\|\|full | 1276 | No | NA | 3 | 0 | 0 | Low-quality | Genome-fragment | 0.81 | AAI-based (high-confidence) | 0 |
| S1-final-viral-combined_k141_100856\|\|full | 3924 | No | NA | 5 | 0 | 4 | Not-determined | Genome-fragment | NA | NA | 0 |
| S8-final-viral-combined_k141_199135\|\|full | 1276 | No | NA | 3 | 3 | 0 | Low-quality | Genome-fragment | 2.61 | HMM-based (lower-bound) | 0 |
| S1-final-viral-combined_k141_15910\|\|full | 1277 | No | NA | 3 | 0 | 0 | Low-quality | Genome-fragment | 2.43 | AAI-based (medium-confidence) | 0 |
| S1-final-viral-combined_k141_282300\|\|full | 2805 | No | NA | 4 | 0 | 3 | Not-determined | Genome-fragment | NA | NA | 0 |
| S5-final-viral-combined_k141_155443\|\|full | 1277 | No | NA | 3 | 0 | 0 | Low-quality | Genome-fragment | 2 | AAI-based (medium-confidence) | 0 |
| S7-final-viral-combined_k141_207165\|\|full | 1277 | No | NA | 2 | 1 | 0 | Low-quality | Genome-fragment | 3.42 | AAI-based (high-confidence) | 0 |
| S1-final-viral-combined_k141_298862\|\|full | 1153 | No | NA | 3 | 0 | 1 | Not-determined | Genome-fragment | NA | NA | 0 |
| S7-final-viral-combined_k141_40177\|\|full | 1277 | No | NA | 3 | 1 | 0 | Low-quality | Genome-fragment | 3.87 | AAI-based (medium-confidence) | 0 |
| S1-final-viral-combined_k141_282312\|\|full | 1880 | No | NA | 2 | 0 | 0 | Not-determined | Genome-fragment | NA | NA | 0 |
| S1-final-viral-combined_k141_166512\|\|full | 2342 | No | NA | 3 | 0 | 3 | Not-determined | Genome-fragment | NA | NA | 0 |
| S7-final-viral-combined_k141_79832\|\|full | 1277 | No | NA | 4 | 0 | 1 | Low-quality | Genome-fragment | 1.85 | AAI-based (medium-confidence) | 0 |
| S1-final-viral-combined_k141_232865\|\|full | 1514 | No | NA | 3 | 0 | 2 | Not-determined | Genome-fragment | NA | NA | 0 |
| S1-final-viral-combined_k141_134565\|\|full | 2563 | No | NA | 5 | 0 | 2 | Not-determined | Genome-fragment | NA | NA | 0 |
| S12-final-viral-combined_k141_282931\|\|full | 1277 | No | NA | 3 | 0 | 1 | Low-quality | Genome-fragment | 0.75 | AAI-based (medium-confidence) | 0 |
| S8-final-viral-combined_k141_164626\|\|full | 1277 | No | NA | 4 | 0 | 0 | Low-quality | Genome-fragment | 2.78 | AAI-based (high-confidence) | 0 |
| S1-final-viral-combined_k141_282352\|\|full | 2157 | No | NA | 2 | 0 | 0 | Not-determined | Genome-fragment | NA | NA | 0 |
| S9-final-viral-combined_k141_30932\|\|full | 1278 | No | NA | 3 | 0 | 1 | Low-quality | Genome-fragment | 2.56 | AAI-based (medium-confidence) | 0 |
| S9_k141_97946 | 1278 | No | NA | 2 | 2 | 0 | Low-quality | Genome-fragment | 3.75 | AAI-based (high-confidence) | 0 |
| S9_k141_28440 | 1278 | No | NA | 3 | 0 | 0 | Low-quality | Genome-fragment | 1.37 | AAI-based (high-confidence) | 0 |
| S1-final-viral-combined_k141_315214\|\|full | 1904 | No | NA | 3 | 0 | 2 | Not-determined | Genome-fragment | NA | NA | 0 |
| S1-final-viral-combined_k141_134610\|\|full | 2055 | No | NA | 2 | 0 | 1 | Not-determined | Genome-fragment | NA | NA | 0 |
| S1-final-viral-combined_k141_100929\|\|full | 8310 | No | NA | 6 | 0 | 5 | Not-determined | Genome-fragment | NA | NA | 0 |
| S1-final-viral-combined_k141_200146\|\|full | 1553 | No | NA | 2 | 0 | 0 | Not-determined | Genome-fragment | NA | NA | 0 |
| S1-final-viral-combined_k141_100934\|\|full | 1982 | No | NA | 4 | 0 | 0 | Not-determined | Genome-fragment | NA | NA | 0 |
| S1-final-viral-combined_k141_35429\|\|full | 10905 | No | NA | 11 | 0 | 8 | Not-determined | Genome-fragment | NA | NA | 0 |
| S1-final-viral-combined_k141_35438\|\|full | 1072 | No | NA | 3 | 0 | 0 | Not-determined | Genome-fragment | NA | NA | 0 |
| S1-final-viral-combined_k141_315245\|\|full | 1040 | No | NA | 2 | 0 | 1 | Not-determined | Genome-fragment | NA | NA | 0 |
| S11-final-viral-combined_k141_277392\|\|full | 1278 | No | NA | 3 | 2 | 0 | Low-quality | Genome-fragment | 2.52 | HMM-based (lower-bound) | 0 |
| S1-final-viral-combined_k141_18900\|\|full | 2155 | No | NA | 2 | 0 | 1 | Not-determined | Genome-fragment | NA | NA | 0 |
| S11-final-viral-combined_k141_215820\|\|full | 1278 | No | NA | 4 | 1 | 0 | Low-quality | Genome-fragment | 2.47 | AAI-based (medium-confidence) | 0 |
| S1-final-viral-combined_k141_85004\|\|full | 9532 | No | NA | 11 | 0 | 8 | Not-determined | Genome-fragment | NA | NA | 0 |
| S11_k141_34522 | 1278 | No | NA | 3 | 2 | 0 | Low-quality | Genome-fragment | 1.55 | AAI-based (high-confidence) | 0 |
| S1-final-viral-combined_k141_117799\|\|full | 1339 | No | NA | 4 | 0 | 0 | Not-determined | Genome-fragment | NA | NA | 0 |
| S1-final-viral-combined_k141_163768\|\|full | 1278 | No | NA | 2 | 0 | 1 | Low-quality | Genome-fragment | 1.74 | AAI-based (high-confidence) | 0 |
| S1-final-viral-combined_k141_117811\|\|full | 2672 | No | NA | 3 | 0 | 2 | Not-determined | Genome-fragment | NA | NA | 0 |
| S1-final-viral-combined_k141_18934\|\|full | 3151 | No | NA | 4 | 0 | 2 | Not-determined | Genome-fragment | NA | NA | 0 |
| S1-final-viral-combined_k141_323733\|\|lt2gene | 1278 | No | NA | 1 | 1 | 0 | Low-quality | Genome-fragment | 2.01 | AAI-based (high-confidence) | 0 |
| S1-final-viral-combined_k141_265385\|\|full | 7166 | No | NA | 5 | 0 | 2 | Not-determined | Genome-fragment | NA | NA | 0 |
| S1-final-viral-combined_k141_348182\|\|full | 1133 | No | NA | 2 | 0 | 2 | Not-determined | Genome-fragment | NA | NA | 0 |
| S3-final-viral-combined_k141_40112\|\|full | 1278 | No | NA | 3 | 0 | 0 | Low-quality | Genome-fragment | 1.52 | AAI-based (medium-confidence) | 0 |
| S3-final-viral-combined_k141_130695\|\|lt2gene | 1278 | No | NA | 1 | 1 | 0 | Low-quality | Genome-fragment | 2.89 | AAI-based (high-confidence) | 0 |
| S1-final-viral-combined_k141_282406\|\|full | 1849 | No | NA | 3 | 0 | 2 | Not-determined | Genome-fragment | NA | NA | 0 |
| S5-final-viral-combined_k141_154669\|\|full | 1278 | No | NA | 4 | 1 | 0 | Low-quality | Genome-fragment | 0.73 | AAI-based (high-confidence) | 0 |
| S1-final-viral-combined_k141_331403\|\|full | 1776 | No | NA | 2 | 0 | 2 | Not-determined | Genome-fragment | NA | NA | 0 |
| S5_k141_148029 | 1278 | No | NA | 1 | 0 | 1 | Low-quality | Genome-fragment | 3.08 | AAI-based (high-confidence) | 0 |
| S1-final-viral-combined_k141_67893\|\|full | 1732 | No | NA | 2 | 0 | 0 | Not-determined | Genome-fragment | NA | NA | 0 |
| S17-k141_126021 | 1278 | No | NA | 3 | 0 | 0 | Low-quality | Genome-fragment | 2.24 | AAI-based (medium-confidence) | 0 |
| S12-final-viral-combined_k141_236201\|\|full | 1278 | No | NA | 3 | 0 | 0 | Low-quality | Genome-fragment | 2.87 | AAI-based (medium-confidence) | 0 |
| S12_k141_116336 | 1278 | No | NA | 3 | 0 | 0 | Low-quality | Genome-fragment | 2.36 | AAI-based (medium-confidence) | 0 |
| S1-final-viral-combined_k141_364641\|\|full | 12703 | No | NA | 11 | 0 | 5 | Not-determined | Genome-fragment | NA | NA | 0 |
| S1-final-viral-combined_k141_298963\|\|full | 3330 | No | NA | 3 | 0 | 3 | Not-determined | Genome-fragment | NA | NA | 0 |
| S2-final-viral-combined_k141_94967\|\|full | 1278 | No | NA | 3 | 0 | 0 | Low-quality | Genome-fragment | 1.1 | AAI-based (high-confidence) | 0 |
| S1-final-viral-combined_k141_216905\|\|full | 2094 | No | NA | 4 | 0 | 0 | Not-determined | Genome-fragment | NA | NA | 0 |
| S2-final-viral-combined_k141_7633\|\|full | 1278 | No | NA | 3 | 0 | 0 | Low-quality | Genome-fragment | 1.89 | AAI-based (medium-confidence) | 0 |
| S1-final-viral-combined_k141_249603\|\|full | 2015 | No | NA | 2 | 0 | 0 | Not-determined | Genome-fragment | NA | NA | 0 |
| S4-final-viral-combined_k141_61823\|\|full | 1278 | No | NA | 4 | 0 | 2 | Low-quality | Genome-fragment | 0.93 | AAI-based (medium-confidence) | 0 |
| S4_k141_69956 | 1278 | No | NA | 1 | 0 | 0 | Low-quality | Genome-fragment | 2.31 | AAI-based (medium-confidence) | 0 |
| S9-final-viral-combined_k141_110446\|\|full | 1279 | No | NA | 2 | 0 | 0 | Low-quality | Genome-fragment | 2.56 | AAI-based (medium-confidence) | 0 |
| S1-final-viral-combined_k141_7635\|\|full | 1279 | No | NA | 2 | 1 | 0 | Low-quality | Genome-fragment | 0.83 | HMM-based (lower-bound) | 0 |
| S1-final-viral-combined_k141_19017\|\|full | 9596 | No | NA | 8 | 0 | 6 | Not-determined | Genome-fragment | NA | NA | 0 |
| S1-final-viral-combined_k141_35544\|\|full | 2790 | No | NA | 2 | 0 | 1 | Not-determined | Genome-fragment | NA | NA | 0 |
| S1-final-viral-combined_k141_282465\|\|full | 16050 | No | NA | 18 | 0 | 13 | Not-determined | Genome-fragment | NA | NA | 0 |
| S1-final-viral-combined_k141_364710\|\|full | 5270 | No | NA | 7 | 0 | 4 | Not-determined | Genome-fragment | NA | NA | 0 |
| S1-final-viral-combined_k141_19031\|\|full | 32923 | No | NA | 52 | 24 | 0 | High-quality | High-quality | 90.89 | AAI-based (medium-confidence) | 0 |
| S1-final-viral-combined_k141_35585\|\|full | 1758 | No | NA | 3 | 0 | 0 | Not-determined | Genome-fragment | NA | NA | 0 |
| S1-final-viral-combined_k141_134769\|\|full | 2160 | No | NA | 2 | 0 | 2 | Not-determined | Genome-fragment | NA | NA | 0 |
| S1-final-viral-combined_k141_361745\|\|full | 1279 | No | NA | 2 | 0 | 1 | Low-quality | Genome-fragment | 2.27 | AAI-based (medium-confidence) | 0 |
| S1-final-viral-combined_k141_101131\|\|full | 2438 | No | NA | 4 | 0 | 0 | Not-determined | Genome-fragment | NA | NA | 0 |
| S1_k141_197480 | 1279 | No | NA | 2 | 0 | 1 | Low-quality | Genome-fragment | 4.46 | AAI-based (high-confidence) | 0 |
| S3-final-viral-combined_k141_118590\|\|lt2gene | 1279 | No | NA | 2 | 1 | 0 | Low-quality | Genome-fragment | 3.87 | AAI-based (high-confidence) | 0 |
| S1-final-viral-combined_k141_117936\|\|full | 3822 | No | NA | 3 | 0 | 3 | Not-determined | Genome-fragment | NA | NA | 0 |
| S1-final-viral-combined_k141_150685\|\|full | 1465 | No | NA | 2 | 0 | 2 | Not-determined | Genome-fragment | NA | NA | 0 |
| S5-final-viral-combined_k141_193833\|\|full | 1279 | No | NA | 2 | 0 | 0 | Low-quality | Genome-fragment | 1.13 | AAI-based (high-confidence) | 0 |
| S1-final-viral-combined_k141_282546\|\|full | 1444 | No | NA | 3 | 0 | 1 | Not-determined | Genome-fragment | NA | NA | 0 |
| S5-final-viral-combined_k141_105197\|\|lt2gene | 1279 | No | NA | 1 | 1 | 0 | Low-quality | Genome-fragment | 1.8 | HMM-based (lower-bound) | 0 |
| S1-final-viral-combined_k141_117951\|\|full | 1724 | No | NA | 6 | 0 | 0 | Not-determined | Genome-fragment | NA | NA | 0 |
| S1-final-viral-combined_k141_85124\|\|full | 1783 | No | NA | 3 | 0 | 1 | Not-determined | Genome-fragment | NA | NA | 0 |
| S17-final-viral-combined_k141_249103\|\|full | 1279 | No | NA | 3 | 0 | 1 | Low-quality | Genome-fragment | 0.78 | AAI-based (medium-confidence) | 0 |
| S1-final-viral-combined_k141_67972\|\|full | 1606 | No | NA | 3 | 0 | 1 | Not-determined | Genome-fragment | NA | NA | 0 |
| S1-final-viral-combined_k141_233047\|\|full | 7150 | No | NA | 9 | 0 | 0 | Not-determined | Genome-fragment | NA | NA | 0 |
| S1-final-viral-combined_k141_348379\|\|full | 3411 | No | NA | 5 | 0 | 1 | Not-determined | Genome-fragment | NA | NA | 0 |
| S17-final-viral-combined_k141_114672\|\|lt2gene | 1279 | No | NA | 1 | 1 | 0 | Low-quality | Genome-fragment | 2.13 | HMM-based (lower-bound) | 0 |
| S2-final-viral-combined_k141_38518\|\|lt2gene | 1279 | No | NA | 2 | 1 | 0 | Low-quality | Genome-fragment | 3.78 | AAI-based (high-confidence) | 0 |
| S11-final-viral-combined_k141_200768\|\|full | 1280 | No | NA | 3 | 0 | 1 | Low-quality | Genome-fragment | 1.04 | AAI-based (medium-confidence) | 0 |
| S11-final-viral-combined_k141_165338\|\|lt2gene | 1280 | No | NA | 2 | 1 | 0 | Low-quality | Genome-fragment | 2.33 | AAI-based (medium-confidence) | 0 |
| S1-final-viral-combined_k141_299172\|\|full | 1093 | No | NA | 3 | 0 | 2 | Not-determined | Genome-fragment | NA | NA | 0 |
| S1-final-viral-combined_k141_192703\|\|full | 1280 | No | NA | 3 | 0 | 1 | Low-quality | Genome-fragment | 2.28 | AAI-based (medium-confidence) | 0 |
| S1-final-viral-combined_k141_166678\|\|full | 1838 | No | NA | 2 | 0 | 0 | Not-determined | Genome-fragment | NA | NA | 0 |
| S1-final-viral-combined_k141_51737\|\|full | 1002 | No | NA | 2 | 0 | 1 | Not-determined | Genome-fragment | NA | NA | 0 |
| S1-final-viral-combined_k141_249800\|\|full | 1242 | No | NA | 3 | 0 | 1 | Not-determined | Genome-fragment | NA | NA | 0 |
| S5-final-viral-combined_k141_117598\|\|full | 1280 | No | NA | 2 | 0 | 0 | Low-quality | Genome-fragment | 2.24 | AAI-based (medium-confidence) | 0 |
| S1-final-viral-combined_k141_35647\|\|full | 2276 | No | NA | 3 | 0 | 3 | Not-determined | Genome-fragment | NA | NA | 0 |
| S1-final-viral-combined_k141_364786\|\|full | 1786 | No | NA | 4 | 0 | 0 | Not-determined | Genome-fragment | NA | NA | 0 |
| S1-final-viral-combined_k141_348455\|\|full | 2650 | No | NA | 2 | 0 | 1 | Not-determined | Genome-fragment | NA | NA | 0 |
| S1-final-viral-combined_k141_35652\|\|full | 1658 | No | NA | 2 | 0 | 2 | Not-determined | Genome-fragment | NA | NA | 0 |
| S1-final-viral-combined_k141_315474\|\|full | 3917 | No | NA | 3 | 0 | 2 | Not-determined | Genome-fragment | NA | NA | 0 |
| S17-final-viral-combined_k141_5963\|\|full | 1280 | No | NA | 2 | 1 | 0 | Low-quality | Genome-fragment | 1.83 | AAI-based (high-confidence) | 0 |
| S1-final-viral-combined_k141_200283\|\|full | 2335 | No | NA | 6 | 0 | 3 | Not-determined | Genome-fragment | NA | NA | 0 |
| S1-final-viral-combined_k141_166713\|\|full | 7384 | No | NA | 11 | 0 | 1 | Not-determined | Genome-fragment | NA | NA | 0 |
| S1-final-viral-combined_k141_183965\|\|full | 1463 | No | NA | 3 | 0 | 1 | Not-determined | Genome-fragment | NA | NA | 0 |
| S1-final-viral-combined_k141_200289\|\|full | 5690 | No | NA | 7 | 0 | 1 | Not-determined | Genome-fragment | NA | NA | 0 |
| S12-final-viral-combined_k141_237871\|\|full | 1280 | No | NA | 2 | 0 | 1 | Low-quality | Genome-fragment | 2.41 | AAI-based (high-confidence) | 0 |
| S12-final-viral-combined_k141_129958\|\|full | 1280 | No | NA | 2 | 0 | 0 | Low-quality | Genome-fragment | 1.82 | AAI-based (medium-confidence) | 0 |
| S1-final-viral-combined_k141_315507\|\|full | 1661 | No | NA | 1 | 0 | 1 | Not-determined | Genome-fragment | NA | NA | 0 |
| S1-final-viral-combined_k141_183975\|\|full | 1105 | No | NA | 3 | 0 | 1 | Not-determined | Genome-fragment | NA | NA | 0 |
| S1-final-viral-combined_k141_364829\|\|full | 2216 | No | NA | 3 | 0 | 2 | Not-determined | Genome-fragment | NA | NA | 0 |
| S1-final-viral-combined_k141_35686\|\|full | 4294 | No | NA | 4 | 0 | 1 | Not-determined | Genome-fragment | NA | NA | 0 |
| S1-final-viral-combined_k141_315517\|\|full | 2443 | No | NA | 2 | 0 | 1 | Not-determined | Genome-fragment | NA | NA | 0 |
| S1-final-viral-combined_k141_282686\|\|full | 1498 | No | NA | 3 | 0 | 3 | Not-determined | Genome-fragment | NA | NA | 0 |
| S1-final-viral-combined_k141_35710\|\|full | 1666 | No | NA | 3 | 0 | 0 | Not-determined | Genome-fragment | NA | NA | 0 |
| S1-final-viral-combined_k141_348530\|\|full | 1998 | No | NA | 4 | 0 | 2 | Not-determined | Genome-fragment | NA | NA | 0 |
| S1-final-viral-combined_k141_134917\|\|full | 18137 | No | NA | 11 | 0 | 8 | Not-determined | Genome-fragment | NA | NA | 0 |
| S2-final-viral-combined_k141_222099\|\|lt2gene | 1280 | No | NA | 2 | 2 | 0 | Low-quality | Genome-fragment | 4.66 | HMM-based (lower-bound) | 0 |
| S2_k141_111917 | 1280 | No | NA | 3 | 0 | 0 | Low-quality | Genome-fragment | 1.75 | AAI-based (high-confidence) | 0 |
| S4-final-viral-combined_k141_76405\|\|full | 1280 | No | NA | 4 | 3 | 0 | Low-quality | Genome-fragment | 2.99 | AAI-based (medium-confidence) | 0 |
| S1-final-viral-combined_k141_150879\|\|full | 1424 | No | NA | 2 | 0 | 0 | Not-determined | Genome-fragment | NA | NA | 0 |
| S1-final-viral-combined_k141_118064\|\|full | 5054 | No | NA | 5 | 0 | 1 | Not-determined | Genome-fragment | NA | NA | 0 |
| S1-final-viral-combined_k141_315593\|\|full | 1263 | No | NA | 3 | 0 | 3 | Not-determined | Genome-fragment | NA | NA | 0 |
| S6_k141_7765 | 1280 | No | NA | 4 | 0 | 3 | Low-quality | Genome-fragment | 0.71 | AAI-based (medium-confidence) | 0 |
| S8-final-viral-combined_k141_221263\|\|full | 1280 | No | NA | 3 | 0 | 0 | Low-quality | Genome-fragment | 2.81 | AAI-based (medium-confidence) | 0 |
| S9-final-viral-combined_k141_70282\|\|full | 1281 | No | NA | 5 | 0 | 0 | Low-quality | Genome-fragment | 3.63 | AAI-based (high-confidence) | 0 |
| S1-final-viral-combined_k141_200349\|\|full | 3169 | No | NA | 4 | 0 | 3 | Not-determined | Genome-fragment | NA | NA | 0 |
| S11_k141_24207 | 1281 | No | NA | 2 | 0 | 0 | Low-quality | Genome-fragment | 2.82 | AAI-based (high-confidence) | 0 |
| S11-final-viral-combined_k141_239601\|\|lt2gene | 1281 | No | NA | 1 | 0 | 0 | Low-quality | Genome-fragment | 8.28 | AAI-based (high-confidence) | 0 |
| S7-final-viral-combined_k141_238797\|\|full | 1281 | No | NA | 2 | 0 | 0 | Low-quality | Genome-fragment | 2.01 | AAI-based (high-confidence) | 0 |
| S7-final-viral-combined_k141_232581\|\|full | 1281 | No | NA | 3 | 0 | 0 | Low-quality | Genome-fragment | 3.28 | AAI-based (high-confidence) | 0 |
| S1-final-viral-combined_k141_249928\|\|full | 2678 | No | NA | 3 | 0 | 1 | Not-determined | Genome-fragment | NA | NA | 0 |
| S1-final-viral-combined_k141_51810\|\|full | 32421 | No | NA | 27 | 0 | 14 | Not-determined | Genome-fragment | NA | NA | 0 |
| S1-final-viral-combined_k141_166759\|\|full | 7186 | No | NA | 9 | 0 | 2 | Not-determined | Genome-fragment | NA | NA | 0 |
| S1-final-viral-combined_k141_299404\|\|full | 6504 | No | NA | 7 | 0 | 5 | Not-determined | Genome-fragment | NA | NA | 0 |
| S1-final-viral-combined_k141_299412\|\|full | 2256 | No | NA | 3 | 0 | 0 | Not-determined | Genome-fragment | NA | NA | 0 |
| S1-final-viral-combined_k141_150933\|\|full | 2101 | No | NA | 2 | 0 | 1 | Not-determined | Genome-fragment | NA | NA | 0 |
| S12_k141_369283 | 1281 | No | NA | 4 | 0 | 0 | Low-quality | Genome-fragment | 1.33 | AAI-based (high-confidence) | 0 |
| S12_k141_392111 | 1281 | No | NA | 2 | 2 | 0 | Low-quality | Genome-fragment | 1.98 | HMM-based (lower-bound) | 0 |
| S1-final-viral-combined_k141_35798\|\|full | 1779 | No | NA | 3 | 0 | 0 | Not-determined | Genome-fragment | NA | NA | 0 |
| S2_k141_43093 | 1281 | No | NA | 3 | 0 | 0 | Low-quality | Genome-fragment | 2.14 | AAI-based (high-confidence) | 0 |
| S9-final-viral-combined_k141_38572\|\|full | 1282 | No | NA | 3 | 1 | 0 | Low-quality | Genome-fragment | 2.27 | AAI-based (medium-confidence) | 0 |
| S9-final-viral-combined_k141_6880\|\|full | 1282 | No | NA | 3 | 0 | 0 | Low-quality | Genome-fragment | 3.58 | AAI-based (medium-confidence) | 0 |
| S11-final-viral-combined_k141_329364\|\|lt2gene | 1282 | No | NA | 2 | 0 | 0 | Low-quality | Genome-fragment | 3.51 | AAI-based (medium-confidence) | 0 |
| S1-final-viral-combined_k141_51856\|\|full | 4104 | No | NA | 3 | 0 | 3 | Not-determined | Genome-fragment | NA | NA | 0 |
| S11_k141_268446 | 1282 | No | NA | 1 | 0 | 1 | Low-quality | Genome-fragment | 2.86 | AAI-based (medium-confidence) | 0 |
| S1-final-viral-combined_k141_202535\|\|full | 1282 | No | NA | 3 | 0 | 0 | Low-quality | Genome-fragment | 2.29 | AAI-based (high-confidence) | 0 |
| S1-final-viral-combined_k141_101293\|\|full | 4123 | No | NA | 3 | 0 | 0 | Not-determined | Genome-fragment | NA | NA | 0 |
| S1-final-viral-combined_k141_217105\|\|full | 1748 | No | NA | 2 | 0 | 0 | Not-determined | Genome-fragment | NA | NA | 0 |
| S1-final-viral-combined_k141_315670\|\|full | 1413 | No | NA | 2 | 0 | 0 | Not-determined | Genome-fragment | NA | NA | 0 |
| S1-final-viral-combined_k141_3084\|\|full | 2491 | No | NA | 4 | 0 | 3 | Not-determined | Genome-fragment | NA | NA | 0 |
| S1-final-viral-combined_k141_101299\|\|full | 3051 | No | NA | 4 | 0 | 2 | Not-determined | Genome-fragment | NA | NA | 0 |
| S1-final-viral-combined_k141_188400\|\|full | 1282 | No | NA | 2 | 0 | 0 | Low-quality | Genome-fragment | 3.12 | AAI-based (medium-confidence) | 0 |
| S2-final-viral-combined_k141_52921\|\|full | 1282 | No | NA | 3 | 0 | 1 | Low-quality | Genome-fragment | 2.21 | AAI-based (medium-confidence) | 0 |
| S2-final-viral-combined_k141_51420\|\|full | 1282 | No | NA | 3 | 3 | 0 | Low-quality | Genome-fragment | 2.67 | AAI-based (medium-confidence) | 0 |
| S1-final-viral-combined_k141_265741\|\|full | 4039 | No | NA | 3 | 0 | 0 | Not-determined | Genome-fragment | NA | NA | 0 |
| S2-final-viral-combined_k141_245322\|\|full | 1282 | No | NA | 5 | 0 | 0 | Low-quality | Genome-fragment | 3.74 | AAI-based (high-confidence) | 0 |
| S1-final-viral-combined_k141_19316\|\|full | 2977 | No | NA | 2 | 0 | 2 | Not-determined | Genome-fragment | NA | NA | 0 |
| S4-final-viral-combined_k141_17605\|\|full | 1282 | No | NA | 4 | 0 | 4 | Low-quality | Genome-fragment | 0.37 | AAI-based (medium-confidence) | 0 |
| S4-final-viral-combined_k141_97251\|\|full | 1282 | No | NA | 2 | 0 | 2 | Low-quality | Genome-fragment | 2.26 | AAI-based (medium-confidence) | 0 |
| S6-final-viral-combined_k141_71264\|\|full | 1282 | No | NA | 3 | 0 | 1 | Low-quality | Genome-fragment | 0.94 | AAI-based (medium-confidence) | 0 |
| S1-final-viral-combined_k141_348641\|\|full | 3050 | No | NA | 4 | 0 | 1 | Not-determined | Genome-fragment | NA | NA | 0 |
| S1-final-viral-combined_k141_51908\|\|full | 1454 | No | NA | 3 | 0 | 1 | Not-determined | Genome-fragment | NA | NA | 0 |
| S1-final-viral-combined_k141_118160\|\|full | 2847 | No | NA | 4 | 0 | 4 | Not-determined | Genome-fragment | NA | NA | 0 |
| S9-final-viral-combined_k141_94893\|\|full | 1283 | No | NA | 5 | 1 | 0 | Low-quality | Genome-fragment | 3.33 | AAI-based (high-confidence) | 0 |
| S1-final-viral-combined_k141_184217\|\|full | 2419 | No | NA | 3 | 0 | 1 | Not-determined | Genome-fragment | NA | NA | 0 |
| S9-final-viral-combined_k141_419\|\|lt2gene | 1283 | No | NA | 1 | 1 | 0 | Low-quality | Genome-fragment | 2.91 | AAI-based (high-confidence) | 0 |
| S1-final-viral-combined_k141_258647\|\|full | 1283 | No | NA | 3 | 2 | 0 | Low-quality | Genome-fragment | 2.15 | AAI-based (high-confidence) | 0 |
| S1-final-viral-combined_k141_98280\|\|full | 1283 | No | NA | 3 | 0 | 0 | Low-quality | Genome-fragment | 3 | AAI-based (high-confidence) | 0 |
| S1-final-viral-combined_k141_184239\|\|full | 1975 | No | NA | 3 | 0 | 0 | Not-determined | Genome-fragment | NA | NA | 0 |
| S1-final-viral-combined_k141_44716\|\|lt2gene | 1283 | No | NA | 1 | 1 | 0 | Low-quality | Genome-fragment | 2.81 | AAI-based (high-confidence) | 0 |
| S1-final-viral-combined_k141_315698\|\|full | 2708 | No | NA | 4 | 0 | 3 | Not-determined | Genome-fragment | NA | NA | 0 |
| S1_k141_101906 | 1283 | No | NA | 3 | 1 | 0 | Low-quality | Genome-fragment | 2.97 | AAI-based (high-confidence) | 0 |
| S1_k141_40501 | 1283 | No | NA | 4 | 0 | 0 | Low-quality | Genome-fragment | 2.88 | AAI-based (high-confidence) | 0 |
| S1-final-viral-combined_k141_282791\|\|full | 2242 | No | NA | 3 | 0 | 2 | Not-determined | Genome-fragment | NA | NA | 0 |
| S1-final-viral-combined_k141_3151\|\|full | 1035 | No | NA | 3 | 0 | 0 | Not-determined | Genome-fragment | NA | NA | 0 |
| S7_k141_197203 | 1283 | No | NA | 4 | 0 | 0 | Low-quality | Genome-fragment | 3.09 | AAI-based (medium-confidence) | 0 |
| S8-final-viral-combined_k141_69892\|\|full | 1283 | No | NA | 3 | 0 | 0 | Low-quality | Genome-fragment | 2.09 | AAI-based (medium-confidence) | 0 |
| S1-final-viral-combined_k141_151032\|\|full | 1537 | No | NA | 3 | 0 | 1 | Not-determined | Genome-fragment | NA | NA | 0 |
| S1-final-viral-combined_k141_381237\|\|full | 5758 | No | NA | 5 | 0 | 2 | Not-determined | Genome-fragment | NA | NA | 0 |
| S1-final-viral-combined_k141_118167\|\|full | 50249 | No | NA | 72 | 19 | 1 | Medium-quality | Genome-fragment | 82.74 | AAI-based (high-confidence) | 0 |
| S1-final-viral-combined_k141_348744\|\|full | 6467 | No | NA | 6 | 0 | 3 | Not-determined | Genome-fragment | NA | NA | 0 |
| S1-final-viral-combined_k141_118179\|\|full | 1306 | No | NA | 3 | 0 | 2 | Not-determined | Genome-fragment | NA | NA | 0 |
| S1-final-viral-combined_k141_365071\|\|full | 2683 | No | NA | 2 | 0 | 1 | Not-determined | Genome-fragment | NA | NA | 0 |
| S1-final-viral-combined_k141_299558\|\|full | 4412 | No | NA | 7 | 0 | 0 | Not-determined | Genome-fragment | NA | NA | 0 |
| S1-final-viral-combined_k141_166838\|\|full | 2515 | No | NA | 3 | 0 | 0 | Not-determined | Genome-fragment | NA | NA | 0 |
| S1-final-viral-combined_k141_331775\|\|full | 1468 | No | NA | 3 | 0 | 0 | Not-determined | Genome-fragment | NA | NA | 0 |
| S1-final-viral-combined_k141_68288\|\|full | 2902 | No | NA | 4 | 0 | 1 | Not-determined | Genome-fragment | NA | NA | 0 |
| S1-final-viral-combined_k141_166846\|\|full | 4543 | No | NA | 9 | 0 | 7 | Not-determined | Genome-fragment | NA | NA | 0 |
| S8-final-viral-combined_k141_188585\|\|full | 1283 | No | NA | 2 | 1 | 0 | Low-quality | Genome-fragment | 2.19 | AAI-based (medium-confidence) | 0 |
| S1-final-viral-combined_k141_19376\|\|full | 2693 | No | NA | 3 | 0 | 1 | Not-determined | Genome-fragment | NA | NA | 0 |
| S1-final-viral-combined_k141_299574\|\|full | 7245 | No | NA | 7 | 0 | 7 | Not-determined | Genome-fragment | NA | NA | 0 |
| S9-final-viral-combined_k141_16105\|\|full | 1284 | No | NA | 2 | 0 | 0 | Low-quality | Genome-fragment | 0.91 | AAI-based (medium-confidence) | 0 |
| S11-final-viral-combined_k141_153401\|\|full | 1284 | No | NA | 3 | 2 | 0 | Low-quality | Genome-fragment | 2.12 | HMM-based (lower-bound) | 0 |
| S1-final-viral-combined_k141_382148\|\|full | 1284 | No | NA | 5 | 1 | 0 | Low-quality | Genome-fragment | 2.36 | AAI-based (high-confidence) | 0 |
| S1-final-viral-combined_k141_101441\|\|full | 7714 | No | NA | 9 | 0 | 7 | Not-determined | Genome-fragment | NA | NA | 0 |
| S1-final-viral-combined_k141_184317\|\|full | 2425 | No | NA | 3 | 0 | 1 | Not-determined | Genome-fragment | NA | NA | 0 |
| S1-final-viral-combined_k141_166892\|\|full | 1159 | No | NA | 3 | 0 | 1 | Not-determined | Genome-fragment | NA | NA | 0 |
| S1-final-viral-combined_k141_52059\|\|full | 1919 | No | NA | 4 | 0 | 3 | Not-determined | Genome-fragment | NA | NA | 0 |
| S1-final-viral-combined_k141_183843\|\|lt2gene | 1284 | No | NA | 2 | 1 | 0 | Low-quality | Genome-fragment | 0.69 | HMM-based (lower-bound) | 0 |
| S5_k141_27573 | 1284 | No | NA | 3 | 0 | 0 | Low-quality | Genome-fragment | 3.25 | AAI-based (medium-confidence) | 0 |
| S1-final-viral-combined_k141_151062\|\|full | 2658 | No | NA | 4 | 0 | 2 | Not-determined | Genome-fragment | NA | NA | 0 |
| S1-final-viral-combined_k141_135108\|\|full | 3830 | No | NA | 5 | 0 | 4 | Not-determined | Genome-fragment | NA | NA | 0 |
| S1-final-viral-combined_k141_135117\|\|full | 2467 | No | NA | 2 | 0 | 2 | Not-determined | Genome-fragment | NA | NA | 0 |
| S7-final-viral-combined_k141_57547\|\|full | 1284 | No | NA | 2 | 0 | 0 | Low-quality | Genome-fragment | 1.17 | AAI-based (medium-confidence) | 0 |
| S1-final-viral-combined_k141_331801\|\|full | 1086 | No | NA | 3 | 0 | 1 | Not-determined | Genome-fragment | NA | NA | 0 |
| S17-final-viral-combined_k141_117860\|\|full | 1284 | No | NA | 4 | 0 | 0 | Low-quality | Genome-fragment | 10.2 | AAI-based (high-confidence) | 0 |
| S17-k141_256949 | 1284 | No | NA | 3 | 0 | 0 | Low-quality | Genome-fragment | 2.71 | AAI-based (high-confidence) | 0 |
| S1-final-viral-combined_k141_331826\|\|full | 1787 | No | NA | 2 | 0 | 1 | Not-determined | Genome-fragment | NA | NA | 0 |
| S17-k141_1719 | 1284 | No | NA | 3 | 3 | 0 | Low-quality | Genome-fragment | 2.88 | AAI-based (high-confidence) | 0 |
| S1-final-viral-combined_k141_250104\|\|full | 3834 | No | NA | 7 | 0 | 0 | Not-determined | Genome-fragment | NA | NA | 0 |
| S1-final-viral-combined_k141_166940\|\|full | 2541 | No | NA | 4 | 0 | 2 | Not-determined | Genome-fragment | NA | NA | 0 |
| S1-final-viral-combined_k141_315820\|\|full | 3107 | No | NA | 4 | 0 | 1 | Not-determined | Genome-fragment | NA | NA | 0 |
| S1-final-viral-combined_k141_348843\|\|full | 1999 | No | NA | 2 | 0 | 2 | Not-determined | Genome-fragment | NA | NA | 0 |
| S12-final-viral-combined_k141_304077\|\|full | 1284 | No | NA | 2 | 0 | 1 | Low-quality | Genome-fragment | 0.78 | AAI-based (medium-confidence) | 0 |
| S1-final-viral-combined_k141_166959\|\|full | 6692 | No | NA | 7 | 0 | 5 | Not-determined | Genome-fragment | NA | NA | 0 |
| S12-final-viral-combined_k141_80969\|\|full | 1284 | No | NA | 3 | 1 | 0 | Low-quality | Genome-fragment | 0.59 | AAI-based (high-confidence) | 0 |
| S12_k141_362757 | 1284 | No | NA | 2 | 0 | 0 | Low-quality | Genome-fragment | 0.7 | AAI-based (high-confidence) | 0 |
| S1-final-viral-combined_k141_118306\|\|full | 1556 | No | NA | 3 | 0 | 1 | Not-determined | Genome-fragment | NA | NA | 0 |
| S1-final-viral-combined_k141_118308\|\|full | 4195 | No | NA | 6 | 0 | 5 | Not-determined | Genome-fragment | NA | NA | 0 |
| S2_k141_80903 | 1284 | No | NA | 4 | 1 | 0 | Low-quality | Genome-fragment | 3.04 | AAI-based (high-confidence) | 0 |
| S1-final-viral-combined_k141_283014\|\|full | 2199 | No | NA | 4 | 0 | 2 | Not-determined | Genome-fragment | NA | NA | 0 |
| S1-final-viral-combined_k141_381368\|\|full | 3482 | No | NA | 3 | 0 | 2 | Not-determined | Genome-fragment | NA | NA | 0 |
| S1-final-viral-combined_k141_283024\|\|full | 2296 | No | NA | 3 | 0 | 2 | Not-determined | Genome-fragment | NA | NA | 0 |
| S1-final-viral-combined_k141_118317\|\|full | 2583 | No | NA | 4 | 0 | 0 | Not-determined | Genome-fragment | NA | NA | 0 |
| S1-final-viral-combined_k141_331896\|\|full | 2459 | No | NA | 4 | 1 | 0 | Medium-quality | Genome-fragment | 82.78 | AAI-based (medium-confidence) | 0 |
| S1-final-viral-combined_k141_118338\|\|full | 3406 | No | NA | 2 | 0 | 1 | Not-determined | Genome-fragment | NA | NA | 0 |
| S6-final-viral-combined_k141_133644\|\|lt2gene | 1284 | No | NA | 2 | 1 | 0 | Low-quality | Genome-fragment | 2.4 | AAI-based (medium-confidence) | 0 |
| S1-final-viral-combined_k141_184468\|\|full | 1440 | No | NA | 2 | 0 | 1 | Not-determined | Genome-fragment | NA | NA | 0 |
| S1-final-viral-combined_k141_52159\|\|full | 1873 | No | NA | 2 | 0 | 1 | Not-determined | Genome-fragment | NA | NA | 0 |
| S1-final-viral-combined_k141_151126\|\|full | 9387 | No | NA | 10 | 0 | 9 | Not-determined | Genome-fragment | NA | NA | 0 |
| S1-final-viral-combined_k141_19500\|\|full | 8178 | No | NA | 11 | 0 | 10 | Not-determined | Genome-fragment | NA | NA | 0 |
| S9-final-viral-combined_k141_10250\|\|full | 1285 | No | NA | 3 | 1 | 0 | Low-quality | Genome-fragment | 2.72 | AAI-based (medium-confidence) | 0 |
| S9-final-viral-combined_k141_108846\|\|lt2gene | 1285 | No | NA | 2 | 0 | 0 | Low-quality | Genome-fragment | 3.13 | AAI-based (medium-confidence) | 0 |
| S9-final-viral-combined_k141_24094\|\|lt2gene | 1285 | No | NA | 2 | 1 | 0 | Low-quality | Genome-fragment | 3.46 | AAI-based (high-confidence) | 0 |
| S7-final-viral-combined_k141_151312\|\|full | 1285 | No | NA | 3 | 0 | 0 | Low-quality | Genome-fragment | 0.88 | AAI-based (medium-confidence) | 0 |
| S1-final-viral-combined_k141_250168\|\|full | 3547 | No | NA | 4 | 0 | 1 | Not-determined | Genome-fragment | NA | NA | 0 |
| S12-final-viral-combined_k141_95238\|\|full | 1285 | No | NA | 4 | 0 | 4 | Low-quality | Genome-fragment | 0.42 | AAI-based (medium-confidence) | 0 |
| S1-final-viral-combined_k141_184510\|\|full | 1893 | No | NA | 5 | 0 | 2 | Not-determined | Genome-fragment | NA | NA | 0 |
| S2-final-viral-combined_k141_75818\|\|lt2gene | 1285 | No | NA | 1 | 1 | 0 | Low-quality | Genome-fragment | 1.88 | HMM-based (lower-bound) | 0 |
| S1-final-viral-combined_k141_299723\|\|full | 2167 | No | NA | 3 | 0 | 0 | Not-determined | Genome-fragment | NA | NA | 0 |
| S1-final-viral-combined_k141_167055\|\|full | 5315 | No | NA | 5 | 0 | 3 | Not-determined | Genome-fragment | NA | NA | 0 |
| S1-final-viral-combined_k141_3323\|\|full | 2239 | No | NA | 3 | 0 | 1 | Not-determined | Genome-fragment | NA | NA | 0 |
| S1-final-viral-combined_k141_200684\|\|full | 3186 | No | NA | 3 | 0 | 1 | Not-determined | Genome-fragment | NA | NA | 0 |
| S1-final-viral-combined_k141_118476\|\|full | 1283 | No | NA | 3 | 0 | 1 | Not-determined | Genome-fragment | NA | NA | 0 |
| S1-final-viral-combined_k141_381464\|\|full | 6346 | No | NA | 5 | 0 | 2 | Not-determined | Genome-fragment | NA | NA | 0 |
| S1-final-viral-combined_k141_200690\|\|full | 5018 | No | NA | 4 | 0 | 2 | Not-determined | Genome-fragment | NA | NA | 0 |
| S9_k141_97343 | 1286 | No | NA | 1 | 0 | 0 | Low-quality | Genome-fragment | 2.74 | AAI-based (high-confidence) | 0 |
| S1-final-viral-combined_k141_19565\|\|full | 2051 | No | NA | 2 | 0 | 0 | Not-determined | Genome-fragment | NA | NA | 0 |
| S1-final-viral-combined_k141_36139\|\|full | 3978 | No | NA | 3 | 0 | 3 | Not-determined | Genome-fragment | NA | NA | 0 |
| S1-final-viral-combined_k141_283072\|\|full | 1266 | No | NA | 3 | 0 | 0 | Not-determined | Genome-fragment | NA | NA | 0 |
| S11-final-viral-combined_k141_201636\|\|full | 1286 | No | NA | 2 | 0 | 0 | Low-quality | Genome-fragment | 2.47 | AAI-based (medium-confidence) | 0 |
| S1-final-viral-combined_k141_19573\|\|full | 3658 | No | NA | 6 | 0 | 0 | Not-determined | Genome-fragment | NA | NA | 0 |
| S1-final-viral-combined_k141_331972\|\|full | 3953 | No | NA | 4 | 0 | 1 | Not-determined | Genome-fragment | NA | NA | 0 |
| S11-final-viral-combined_k141_135581\|\|full | 1286 | No | NA | 3 | 0 | 1 | Low-quality | Genome-fragment | 3.01 | AAI-based (medium-confidence) | 0 |
| S1-final-viral-combined_k141_318944\|\|full | 1286 | No | NA | 3 | 0 | 1 | Low-quality | Genome-fragment | 1.86 | AAI-based (medium-confidence) | 0 |
| S5-final-viral-combined_k141_71287\|\|full | 1286 | No | NA | 2 | 0 | 1 | Low-quality | Genome-fragment | 0.57 | AAI-based (medium-confidence) | 0 |
| S1-final-viral-combined_k141_217500\|\|full | 2633 | No | NA | 3 | 0 | 2 | Not-determined | Genome-fragment | NA | NA | 0 |
| S1-final-viral-combined_k141_331983\|\|full | 4009 | No | NA | 5 | 0 | 4 | Not-determined | Genome-fragment | NA | NA | 0 |
| S5-final-viral-combined_k141_87759\|\|full | 1286 | No | NA | 4 | 3 | 0 | Low-quality | Genome-fragment | 2.89 | AAI-based (high-confidence) | 0 |
| S1-final-viral-combined_k141_184559\|\|full | 6417 | No | NA | 8 | 0 | 5 | Not-determined | Genome-fragment | NA | NA | 0 |
| S5-final-viral-combined_k141_34548\|\|full | 1286 | No | NA | 2 | 0 | 2 | Low-quality | Genome-fragment | 0.88 | AAI-based (medium-confidence) | 0 |
| S1-final-viral-combined_k141_167127\|\|full | 1317 | No | NA | 2 | 0 | 2 | Not-determined | Genome-fragment | NA | NA | 0 |
| S1-final-viral-combined_k141_200726\|\|full | 7071 | No | NA | 5 | 0 | 5 | Not-determined | Genome-fragment | NA | NA | 0 |
| S7-final-viral-combined_k141_1275\|\|full | 1286 | No | NA | 3 | 1 | 0 | Low-quality | Genome-fragment | 0.84 | AAI-based (high-confidence) | 0 |
| S7-final-viral-combined_k141_29172\|\|full | 1286 | No | NA | 3 | 0 | 2 | Low-quality | Genome-fragment | 1.75 | AAI-based (medium-confidence) | 0 |
| S17-final-viral-combined_k141_217566\|\|full | 1286 | No | NA | 3 | 0 | 2 | Low-quality | Genome-fragment | 0.41 | AAI-based (medium-confidence) | 0 |
| S1-final-viral-combined_k141_184595\|\|full | 1751 | No | NA | 3 | 0 | 0 | Not-determined | Genome-fragment | NA | NA | 0 |
| S1-final-viral-combined_k141_349004\|\|full | 9315 | No | NA | 10 | 0 | 3 | Not-determined | Genome-fragment | NA | NA | 0 |
| S1-final-viral-combined_k141_3375\|\|full | 2695 | No | NA | 3 | 0 | 1 | Not-determined | Genome-fragment | NA | NA | 0 |
| S1-final-viral-combined_k141_365357\|\|full | 1089 | No | NA | 3 | 0 | 0 | Not-determined | Genome-fragment | NA | NA | 0 |
| S1-final-viral-combined_k141_250283\|\|full | 7171 | No | NA | 5 | 0 | 4 | Not-determined | Genome-fragment | NA | NA | 0 |
| S12-final-viral-combined_k141_274610\|\|full | 1286 | No | NA | 4 | 0 | 1 | Low-quality | Genome-fragment | 2.13 | AAI-based (medium-confidence) | 0 |
| S1-final-viral-combined_k141_381525\|\|full | 1029 | No | NA | 3 | 0 | 0 | Not-determined | Genome-fragment | NA | NA | 0 |
| S12-final-viral-combined_k141_293840\|\|full | 1286 | No | NA | 3 | 1 | 0 | Low-quality | Genome-fragment | 3.07 | AAI-based (medium-confidence) | 0 |
| S12-final-viral-combined_k141_110611\|\|lt2gene | 1286 | No | NA | 1 | 1 | 0 | Low-quality | Genome-fragment | 2.83 | AAI-based (medium-confidence) | 0 |
| S1-final-viral-combined_k141_101528\|\|full | 5060 | No | NA | 6 | 0 | 0 | Not-determined | Genome-fragment | NA | NA | 0 |
| S1-final-viral-combined_k141_184606\|\|full | 4984 | No | NA | 7 | 0 | 2 | Not-determined | Genome-fragment | NA | NA | 0 |
| S2-final-viral-combined_k141_230575\|\|full | 1286 | No | NA | 2 | 1 | 0 | Low-quality | Genome-fragment | 2.88 | AAI-based (high-confidence) | 0 |
| S1-final-viral-combined_k141_332037\|\|full | 1828 | No | NA | 4 | 0 | 0 | Not-determined | Genome-fragment | NA | NA | 0 |
| S1-final-viral-combined_k141_316024\|\|full | 2394 | No | NA | 3 | 0 | 1 | Not-determined | Genome-fragment | NA | NA | 0 |
| S1-final-viral-combined_k141_167167\|\|full | 4995 | No | NA | 6 | 0 | 5 | Not-determined | Genome-fragment | NA | NA | 0 |
| S1-final-viral-combined_k141_233529\|\|full | 1082 | No | NA | 3 | 0 | 0 | Not-determined | Genome-fragment | NA | NA | 0 |
| S2-final-viral-combined_k141_90248\|\|full | 1286 | No | NA | 4 | 1 | 0 | Low-quality | Genome-fragment | 2.84 | AAI-based (medium-confidence) | 0 |
| S4-final-viral-combined_k141_56574\|\|full | 1286 | No | NA | 4 | 1 | 0 | Low-quality | Genome-fragment | 2.14 | AAI-based (medium-confidence) | 0 |
| S1-final-viral-combined_k141_184637\|\|full | 1533 | No | NA | 3 | 0 | 0 | Not-determined | Genome-fragment | NA | NA | 0 |
| S1-final-viral-combined_k141_381578\|\|full | 5871 | No | NA | 5 | 0 | 3 | Not-determined | Genome-fragment | NA | NA | 0 |
| S1-final-viral-combined_k141_316076\|\|full | 2664 | No | NA | 4 | 0 | 1 | Not-determined | Genome-fragment | NA | NA | 0 |
| S1-final-viral-combined_k141_101601\|\|full | 3689 | No | NA | 4 | 0 | 3 | Not-determined | Genome-fragment | NA | NA | 0 |
| S1-final-viral-combined_k141_135415\|\|full | 3368 | No | NA | 5 | 0 | 1 | Not-determined | Genome-fragment | NA | NA | 0 |
| S1-final-viral-combined_k141_184685\|\|full | 1359 | No | NA | 2 | 0 | 0 | Not-determined | Genome-fragment | NA | NA | 0 |
| S1-final-viral-combined_k141_68539\|\|full | 2912 | No | NA | 2 | 0 | 1 | Not-determined | Genome-fragment | NA | NA | 0 |
| S1-final-viral-combined_k141_266136\|\|full | 1057 | No | NA | 2 | 0 | 1 | Not-determined | Genome-fragment | NA | NA | 0 |
| S1-final-viral-combined_k141_365418\|\|full | 1497 | No | NA | 2 | 0 | 0 | Not-determined | Genome-fragment | NA | NA | 0 |
| S1-final-viral-combined_k141_184708\|\|full | 2059 | No | NA | 4 | 0 | 2 | Not-determined | Genome-fragment | NA | NA | 0 |
| S4-final-viral-combined_k141_4559\|\|full | 1286 | No | NA | 2 | 0 | 0 | Low-quality | Genome-fragment | 2.69 | AAI-based (medium-confidence) | 0 |
| S1-final-viral-combined_k141_135468\|\|full | 1078 | No | NA | 2 | 0 | 2 | Not-determined | Genome-fragment | NA | NA | 0 |
| S1-final-viral-combined_k141_118613\|\|full | 3683 | No | NA | 5 | 0 | 5 | Not-determined | Genome-fragment | NA | NA | 0 |
| S4_k141_36257 | 1286 | No | NA | 3 | 0 | 0 | Low-quality | Genome-fragment | 1.56 | AAI-based (high-confidence) | 0 |
| S1-final-viral-combined_k141_36332\|\|full | 1255 | No | NA | 3 | 0 | 3 | Not-determined | Genome-fragment | NA | NA | 0 |
| S6-final-viral-combined_k141_104109\|\|lt2gene | 1286 | No | NA | 2 | 2 | 0 | Low-quality | Genome-fragment | 1.56 | HMM-based (lower-bound) | 0 |
| S1-final-viral-combined_k141_135497\|\|full | 2828 | No | NA | 5 | 0 | 0 | Not-determined | Genome-fragment | NA | NA | 0 |
| S1-final-viral-combined_k141_283237\|\|full | 7063 | No | NA | 8 | 0 | 6 | Not-determined | Genome-fragment | NA | NA | 0 |
| S8-final-viral-combined_k141_163460\|\|full | 1286 | No | NA | 3 | 1 | 0 | Low-quality | Genome-fragment | 2.46 | AAI-based (high-confidence) | 0 |
| S8-final-viral-combined_k141_94779\|\|lt2gene | 1286 | No | NA | 1 | 1 | 0 | Low-quality | Genome-fragment | 3.6 | AAI-based (high-confidence) | 0 |
| S1-final-viral-combined_k141_85495\|\|full | 1138 | No | NA | 2 | 0 | 2 | Not-determined | Genome-fragment | NA | NA | 0 |
| S11-final-viral-combined_k141_185467\|\|full | 1287 | No | NA | 3 | 0 | 1 | Low-quality | Genome-fragment | 0.78 | AAI-based (high-confidence) | 0 |
| S1-final-viral-combined_k141_316184\|\|full | 1202 | No | NA | 3 | 0 | 0 | Not-determined | Genome-fragment | NA | NA | 0 |
| S1-final-viral-combined_k141_388769\|\|full | 1287 | No | NA | 3 | 0 | 0 | Low-quality | Genome-fragment | 2.68 | AAI-based (high-confidence) | 0 |
| S1-final-viral-combined_k141_151426\|\|full | 1720 | No | NA | 4 | 0 | 2 | Not-determined | Genome-fragment | NA | NA | 0 |
| S3-final-viral-combined_k141_54011\|\|full | 1287 | No | NA | 2 | 0 | 0 | Low-quality | Genome-fragment | 2.53 | AAI-based (high-confidence) | 0 |
| S1-final-viral-combined_k141_85518\|\|full | 5965 | No | NA | 5 | 0 | 1 | Not-determined | Genome-fragment | NA | NA | 0 |
| S7-final-viral-combined_k141_160286\|\|full | 1287 | No | NA | 3 | 0 | 2 | Low-quality | Genome-fragment | 2.16 | AAI-based (medium-confidence) | 0 |
| S7-final-viral-combined_k141_143757\|\|lt2gene | 1287 | No | NA | 2 | 1 | 0 | Low-quality | Genome-fragment | 2.29 | AAI-based (medium-confidence) | 0 |
| S2-final-viral-combined_k141_151263\|\|full | 1287 | No | NA | 2 | 1 | 0 | Low-quality | Genome-fragment | 2.07 | HMM-based (lower-bound) | 0 |
| S1-final-viral-combined_k141_233690\|\|full | 1161 | No | NA | 3 | 0 | 0 | Not-determined | Genome-fragment | NA | NA | 0 |
| S1-final-viral-combined_k141_217596\|\|full | 4235 | No | NA | 6 | 0 | 2 | Not-determined | Genome-fragment | NA | NA | 0 |
| S2-final-viral-combined_k141_235916\|\|full | 1287 | No | NA | 3 | 0 | 0 | Low-quality | Genome-fragment | 0.78 | AAI-based (medium-confidence) | 0 |
| S6-final-viral-combined_k141_51920\|\|full | 1287 | No | NA | 4 | 0 | 0 | Low-quality | Genome-fragment | 2.54 | AAI-based (high-confidence) | 0 |
| S1-final-viral-combined_k141_381703\|\|full | 1847 | No | NA | 4 | 0 | 2 | Not-determined | Genome-fragment | NA | NA | 0 |
| S1-final-viral-combined_k141_349100\|\|full | 1832 | No | NA | 3 | 0 | 0 | Not-determined | Genome-fragment | NA | NA | 0 |
| S8_k141_8339 | 1287 | No | NA | 3 | 0 | 3 | Low-quality | Genome-fragment | 0.34 | AAI-based (medium-confidence) | 0 |
| S9-final-viral-combined_k141_119546\|\|full | 1288 | No | NA | 4 | 0 | 0 | Low-quality | Genome-fragment | 3.26 | AAI-based (medium-confidence) | 0 |
| S1-final-viral-combined_k141_200927\|\|full | 2511 | No | NA | 3 | 0 | 0 | Not-determined | Genome-fragment | NA | NA | 0 |
| S9-final-viral-combined_k141_19160\|\|full | 1288 | No | NA | 2 | 0 | 1 | Low-quality | Genome-fragment | 2.68 | AAI-based (medium-confidence) | 0 |
| S9_k141_35112 | 1288 | No | NA | 4 | 2 | 0 | Low-quality | Genome-fragment | 3.19 | AAI-based (high-confidence) | 0 |
| S11-final-viral-combined_k141_62334\|\|full | 1288 | No | NA | 4 | 0 | 0 | Low-quality | Genome-fragment | 1.03 | AAI-based (medium-confidence) | 0 |
| S1-final-viral-combined_k141_151508\|\|full | 1303 | No | NA | 3 | 0 | 1 | Not-determined | Genome-fragment | NA | NA | 0 |
| S1-final-viral-combined_k141_249955\|\|full | 1288 | No | NA | 2 | 0 | 1 | Low-quality | Genome-fragment | 0.65 | AAI-based (medium-confidence) | 0 |
| S1-final-viral-combined_k141_277548\|\|full | 1288 | No | NA | 3 | 0 | 2 | Low-quality | Genome-fragment | 0.41 | AAI-based (medium-confidence) | 0 |
| S5-final-viral-combined_k141_192569\|\|full | 1288 | No | NA | 3 | 1 | 0 | Low-quality | Genome-fragment | 2.6 | AAI-based (high-confidence) | 0 |
| S7-final-viral-combined_k141_62068\|\|full | 1288 | No | NA | 3 | 0 | 0 | Low-quality | Genome-fragment | 2.9 | AAI-based (high-confidence) | 0 |
| S1-final-viral-combined_k141_19809\|\|full | 1036 | No | NA | 3 | 0 | 2 | Not-determined | Genome-fragment | NA | NA | 0 |
| S1-final-viral-combined_k141_151536\|\|full | 1055 | No | NA | 2 | 0 | 1 | Not-determined | Genome-fragment | NA | NA | 0 |
| S1-final-viral-combined_k141_250503\|\|full | 1618 | No | NA | 2 | 0 | 1 | Not-determined | Genome-fragment | NA | NA | 0 |
| S1-final-viral-combined_k141_19821\|\|full | 1381 | No | NA | 2 | 0 | 1 | Not-determined | Genome-fragment | NA | NA | 0 |
| S1-final-viral-combined_k141_266361\|\|full | 1647 | No | NA | 2 | 0 | 2 | Not-determined | Genome-fragment | NA | NA | 0 |
| S1-final-viral-combined_k141_19846\|\|full | 1540 | No | NA | 3 | 0 | 0 | Not-determined | Genome-fragment | NA | NA | 0 |
| S1-final-viral-combined_k141_19850\|\|full | 1845 | No | NA | 2 | 0 | 1 | Not-determined | Genome-fragment | NA | NA | 0 |
| S1-final-viral-combined_k141_250537\|\|full | 1509 | No | NA | 2 | 0 | 0 | Not-determined | Genome-fragment | NA | NA | 0 |
| S12-final-viral-combined_k141_360730\|\|full | 1288 | No | NA | 3 | 0 | 0 | Low-quality | Genome-fragment | 2 | AAI-based (medium-confidence) | 0 |
| S12_k141_292167 | 1288 | No | NA | 2 | 0 | 0 | Low-quality | Genome-fragment | 2.79 | AAI-based (high-confidence) | 0 |
| S4-final-viral-combined_k141_40748\|\|lt2gene | 1288 | No | NA | 1 | 0 | 0 | Low-quality | Genome-fragment | 3.59 | AAI-based (medium-confidence) | 0 |
| S1-final-viral-combined_k141_151589\|\|full | 2845 | No | NA | 5 | 0 | 2 | Not-determined | Genome-fragment | NA | NA | 0 |
| S1-final-viral-combined_k141_118763\|\|full | 1787 | No | NA | 3 | 0 | 1 | Not-determined | Genome-fragment | NA | NA | 0 |
| S4_k141_43729 | 1288 | No | NA | 2 | 1 | 0 | Low-quality | Genome-fragment | 2.33 | AAI-based (medium-confidence) | 0 |
| S8-final-viral-combined_k141_228522\|\|full | 1288 | No | NA | 2 | 0 | 0 | Low-quality | Genome-fragment | 1.03 | AAI-based (medium-confidence) | 0 |
| S1-final-viral-combined_k141_118766\|\|full | 1036 | No | NA | 2 | 0 | 0 | Not-determined | Genome-fragment | NA | NA | 0 |
| S1-final-viral-combined_k141_332236\|\|full | 30000 | No | NA | 33 | 18 | 0 | Medium-quality | Genome-fragment | 55.15 | AAI-based (high-confidence) | 0 |
| S9-final-viral-combined_k141_66392\|\|full | 1289 | No | NA | 4 | 0 | 0 | Low-quality | Genome-fragment | 3.29 | AAI-based (high-confidence) | 0 |
| S1-final-viral-combined_k141_118771\|\|full | 1154 | No | NA | 2 | 0 | 2 | Not-determined | Genome-fragment | NA | NA | 0 |
| S1-final-viral-combined_k141_151600\|\|full | 1631 | No | NA | 2 | 0 | 0 | Not-determined | Genome-fragment | NA | NA | 0 |
| S1-final-viral-combined_k141_332245\|\|full | 1598 | No | NA | 2 | 0 | 1 | Not-determined | Genome-fragment | NA | NA | 0 |
| S11-final-viral-combined_k141_78557\|\|full | 1289 | No | NA | 4 | 0 | 0 | Low-quality | Genome-fragment | 1.27 | AAI-based (medium-confidence) | 0 |
| S11_k141_68054 | 1289 | No | NA | 1 | 0 | 0 | Low-quality | Genome-fragment | 1.32 | AAI-based (high-confidence) | 0 |
| S1-final-viral-combined_k141_151634\|\|full | 1626 | No | NA | 4 | 0 | 4 | Not-determined | Genome-fragment | NA | NA | 0 |
| S1-final-viral-combined_k141_52510\|\|full | 1697 | No | NA | 4 | 0 | 1 | Not-determined | Genome-fragment | NA | NA | 0 |
| S5-final-viral-combined_k141_190993\|\|full | 1289 | No | NA | 4 | 0 | 0 | Low-quality | Genome-fragment | 3.08 | AAI-based (high-confidence) | 0 |
| S1-final-viral-combined_k141_283401\|\|full | 1409 | No | NA | 2 | 0 | 0 | Not-determined | Genome-fragment | NA | NA | 0 |
| S5-final-viral-combined_k141_23745\|\|full | 1289 | No | NA | 2 | 1 | 0 | Low-quality | Genome-fragment | 1.42 | AAI-based (medium-confidence) | 0 |
| S1-final-viral-combined_k141_381793\|\|full | 1424 | No | NA | 3 | 0 | 0 | Not-determined | Genome-fragment | NA | NA | 0 |
| S1-final-viral-combined_k141_52552\|\|full | 1194 | No | NA | 3 | 0 | 0 | Not-determined | Genome-fragment | NA | NA | 0 |
| S5_k141_74201 | 1289 | No | NA | 2 | 0 | 0 | Low-quality | Genome-fragment | 2.58 | AAI-based (high-confidence) | 0 |
| S1-final-viral-combined_k141_250666\|\|full | 1888 | No | NA | 3 | 0 | 0 | Not-determined | Genome-fragment | NA | NA | 0 |
| S7_k141_124388 | 1289 | No | NA | 1 | 0 | 0 | Low-quality | Genome-fragment | 2.61 | AAI-based (medium-confidence) | 0 |
| S1-final-viral-combined_k141_19954\|\|full | 2345 | No | NA | 4 | 0 | 2 | Not-determined | Genome-fragment | NA | NA | 0 |
| S1-final-viral-combined_k141_365658\|\|full | 1132 | No | NA | 2 | 0 | 2 | Not-determined | Genome-fragment | NA | NA | 0 |
| S1-final-viral-combined_k141_365659\|\|full | 1509 | No | NA | 3 | 0 | 2 | Not-determined | Genome-fragment | NA | NA | 0 |
| S1-final-viral-combined_k141_381811\|\|full | 2194 | No | NA | 3 | 0 | 2 | Not-determined | Genome-fragment | NA | NA | 0 |
| S1-final-viral-combined_k141_151691\|\|full | 1324 | No | NA | 2 | 0 | 0 | Not-determined | Genome-fragment | NA | NA | 0 |
| S17-final-viral-combined_k141_64203\|\|full | 1289 | No | NA | 3 | 2 | 0 | Low-quality | Genome-fragment | 3.3 | AAI-based (medium-confidence) | 0 |
| S12_k141_313414 | 1289 | No | NA | 1 | 0 | 0 | Low-quality | Genome-fragment | 0.77 | AAI-based (medium-confidence) | 0 |
| S1-final-viral-combined_k141_52592\|\|full | 2694 | No | NA | 3 | 0 | 2 | Not-determined | Genome-fragment | NA | NA | 0 |
| S2-final-viral-combined_k141_53326\|\|lt2gene | 1289 | No | NA | 1 | 0 | 0 | Low-quality | Genome-fragment | 3.26 | AAI-based (high-confidence) | 0 |
| S4-final-viral-combined_k141_97248\|\|full | 1289 | No | NA | 3 | 0 | 0 | Low-quality | Genome-fragment | 0.79 | AAI-based (medium-confidence) | 0 |
| S1-final-viral-combined_k141_184924\|\|full | 2907 | No | NA | 5 | 0 | 0 | Not-determined | Genome-fragment | NA | NA | 0 |
| S1-final-viral-combined_k141_3702\|\|full | 2646 | No | NA | 3 | 0 | 1 | Not-determined | Genome-fragment | NA | NA | 0 |
| S1-final-viral-combined_k141_250698\|\|full | 1687 | No | NA | 2 | 0 | 2 | Not-determined | Genome-fragment | NA | NA | 0 |
| S1-final-viral-combined_k141_250700\|\|full | 1273 | No | NA | 2 | 0 | 2 | Not-determined | Genome-fragment | NA | NA | 0 |
| S1-final-viral-combined_k141_68649\|\|full | 7904 | No | NA | 8 | 0 | 3 | Not-determined | Genome-fragment | NA | NA | 0 |
| S8_k141_255566 | 1289 | No | NA | 2 | 0 | 1 | Low-quality | Genome-fragment | 2.03 | AAI-based (medium-confidence) | 0 |
| S1-final-viral-combined_k141_316446\|\|full | 6007 | No | NA | 7 | 0 | 4 | Not-determined | Genome-fragment | NA | NA | 0 |
| S8_k141_101791 | 1289 | No | NA | 3 | 1 | 0 | Low-quality | Genome-fragment | 1.72 | AAI-based (high-confidence) | 0 |
| S9-final-viral-combined_k141_85456\|\|full | 1290 | No | NA | 3 | 1 | 0 | Low-quality | Genome-fragment | 2.44 | AAI-based (medium-confidence) | 0 |
| S1-final-viral-combined_k141_283463\|\|full | 1831 | No | NA | 3 | 0 | 1 | Not-determined | Genome-fragment | NA | NA | 0 |
| S11-final-viral-combined_k141_76403\|\|full | 1290 | No | NA | 3 | 1 | 0 | Low-quality | Genome-fragment | 1.44 | HMM-based (lower-bound) | 0 |
| S1-final-viral-combined_k141_135673\|\|full | 4556 | No | NA | 4 | 0 | 3 | Not-determined | Genome-fragment | NA | NA | 0 |
| S1-final-viral-combined_k141_300212\|\|full | 5209 | No | NA | 2 | 0 | 0 | Not-determined | Genome-fragment | NA | NA | 0 |
| S1-final-viral-combined_k141_233933\|\|full | 1148 | No | NA | 3 | 0 | 1 | Not-determined | Genome-fragment | NA | NA | 0 |
| S11-final-viral-combined_k141_189068\|\|lt2gene | 1290 | No | NA | 2 | 1 | 0 | Low-quality | Genome-fragment | 2.95 | AAI-based (medium-confidence) | 0 |
| S11_k141_174589 | 1290 | No | NA | 2 | 0 | 2 | Low-quality | Genome-fragment | 0.21 | AAI-based (medium-confidence) | 0 |
| S1-final-viral-combined_k141_233954\|\|full | 2405 | No | NA | 3 | 0 | 1 | Not-determined | Genome-fragment | NA | NA | 0 |
| S1-final-viral-combined_k141_184976\|\|full | 3945 | No | NA | 9 | 0 | 2 | Not-determined | Genome-fragment | NA | NA | 0 |
| S1-final-viral-combined_k141_151708\|\|full | 13557 | No | NA | 13 | 2 | 1 | Medium-quality | Genome-fragment | 87.49 | AAI-based (high-confidence) | 0 |
| S1-final-viral-combined_k141_33837\|\|full | 1290 | No | NA | 3 | 0 | 0 | Low-quality | Genome-fragment | 0.82 | AAI-based (medium-confidence) | 0 |
| S1-final-viral-combined_k141_168672\|\|full | 1290 | No | NA | 3 | 0 | 1 | Low-quality | Genome-fragment | 3.33 | AAI-based (medium-confidence) | 0 |
| S7-final-viral-combined_k141_78892\|\|full | 1290 | No | NA | 2 | 0 | 0 | Low-quality | Genome-fragment | 2.14 | AAI-based (medium-confidence) | 0 |

| | | | | | | | | | | | |
|---|---|---|---|---|---|---|---|---|---|---|---|
| S1-final-viral-combined_k141_233991\|\|full | 5685 | No | NA | 6 | 0 | 4 | Not-determined | Genome-fragment | NA | NA | 0 |
| S12-final-viral-combined_k141_117211\|\|full | 1290 | No | NA | 3 | 0 | 0 | Low-quality | Genome-fragment | 2.59 | AAI-based (medium-confidence) | 0 |
| S1-final-viral-combined_k141_283520\|\|full | 2752 | No | NA | 3 | 0 | 1 | Not-determined | Genome-fragment | NA | NA | 0 |
| S1-final-viral-combined_k141_52643\|\|full | 23536 | No | NA | 30 | 8 | 0 | Medium-quality | Genome-fragment | 56.07 | AAI-based (medium-confidence) | 0 |
| S4_k141_19346 | 1290 | No | NA | 3 | 0 | 1 | Low-quality | Genome-fragment | 1.42 | AAI-based (medium-confidence) | 0 |
| S6-final-viral-combined_k141_48187\|\|full | 1290 | No | NA | 3 | 0 | 0 | Low-quality | Genome-fragment | 1.8 | AAI-based (medium-confidence) | 0 |
| S1-final-viral-combined_k141_349308\|\|full | 2168 | No | NA | 3 | 0 | 1 | Not-determined | Genome-fragment | NA | NA | 0 |
| S1-final-viral-combined_k141_217640\|\|full | 2069 | No | NA | 3 | 0 | 1 | Not-determined | Genome-fragment | NA | NA | 0 |
| S1-final-viral-combined_k141_101951\|\|full | 3580 | No | NA | 5 | 0 | 2 | Not-determined | Genome-fragment | NA | NA | 0 |
| S8-final-viral-combined_k141_12110\|\|lt2gene | 1290 | No | NA | 2 | 1 | 0 | Low-quality | Genome-fragment | 3.42 | AAI-based (high-confidence) | 0 |
| S9-final-viral-combined_k141_62825\|\|lt2gene | 1291 | No | NA | 2 | 2 | 0 | Low-quality | Genome-fragment | 3.14 | AAI-based (high-confidence) | 0 |
| S1-final-viral-combined_k141_101958\|\|full | 2110 | No | NA | 2 | 0 | 2 | Not-determined | Genome-fragment | NA | NA | 0 |
| S1-final-viral-combined_k141_118921\|\|full | 1021 | No | NA | 4 | 0 | 0 | Not-determined | Genome-fragment | NA | NA | 0 |
| S1-final-viral-combined_k141_118928\|\|full | 3159 | No | NA | 4 | 0 | 0 | Not-determined | Genome-fragment | NA | NA | 0 |
| S1-final-viral-combined_k141_283600\|\|full | 14499 | No | NA | 14 | 0 | 12 | Not-determined | Genome-fragment | NA | NA | 0 |
| S1-final-viral-combined_k141_332485\|\|full | 2344 | No | NA | 5 | 0 | 0 | Not-determined | Genome-fragment | NA | NA | 0 |
| S1-final-viral-combined_k141_266623\|\|full | 1848 | No | NA | 2 | 0 | 2 | Not-determined | Genome-fragment | NA | NA | 0 |
| S1-final-viral-combined_k141_201170\|\|full | 3655 | No | NA | 4 | 0 | 4 | Not-determined | Genome-fragment | NA | NA | 0 |
| S1-final-viral-combined_k141_52701\|\|full | 3376 | No | NA | 3 | 0 | 3 | Not-determined | Genome-fragment | NA | NA | 0 |
| S1-final-viral-combined_k141_363910\|\|full | 1291 | No | NA | 2 | 0 | 0 | Low-quality | Genome-fragment | 0.51 | AAI-based (medium-confidence) | 0 |
| S1-final-viral-combined_k141_152645\|\|full | 1291 | No | NA | 2 | 0 | 0 | Low-quality | Genome-fragment | 0.89 | AAI-based (medium-confidence) | 0 |
| S3-final-viral-combined_k141_127278\|\|full | 1291 | No | NA | 5 | 0 | 0 | Low-quality | Genome-fragment | 2.14 | AAI-based (high-confidence) | 0 |
| S1-final-viral-combined_k141_266673\|\|full | 12403 | No | NA | 15 | 0 | 3 | Not-determined | Genome-fragment | NA | NA | 0 |
| S1-final-viral-combined_k141_167554\|\|full | 7832 | No | NA | 9 | 0 | 6 | Not-determined | Genome-fragment | NA | NA | 0 |
| S1-final-viral-combined_k141_201201\|\|full | 4084 | No | NA | 3 | 0 | 3 | Not-determined | Genome-fragment | NA | NA | 0 |
| S1-final-viral-combined_k141_118971\|\|full | 1803 | No | NA | 2 | 0 | 2 | Not-determined | Genome-fragment | NA | NA | 0 |
| S1-final-viral-combined_k141_52708\|\|full | 51903 | No | NA | 48 | 3 | 34 | High-quality | High-quality | 100 | AAI-based (high-confidence) | 0 |
| S1-final-viral-combined_k141_217721\|\|full | 3624 | No | NA | 4 | 0 | 2 | Not-determined | Genome-fragment | NA | NA | 0 |
| S1-final-viral-combined_k141_185075\|\|full | 3523 | No | NA | 4 | 0 | 0 | Not-determined | Genome-fragment | NA | NA | 0 |
| S1-final-viral-combined_k141_52728\|\|full | 2957 | No | NA | 4 | 0 | 1 | Not-determined | Genome-fragment | NA | NA | 0 |
| S3-final-viral-combined_k141_60798\|\|full | 1291 | No | NA | 3 | 0 | 0 | Low-quality | Genome-fragment | 0.97 | AAI-based (medium-confidence) | 0 |
| S1-final-viral-combined_k141_250889\|\|full | 1037 | No | NA | 2 | 0 | 0 | Not-determined | Genome-fragment | NA | NA | 0 |
| S1-final-viral-combined_k141_167587\|\|full | 3081 | No | NA | 4 | 0 | 1 | Not-determined | Genome-fragment | NA | NA | 0 |
| S1-final-viral-combined_k141_234102\|\|full | 1077 | No | NA | 3 | 0 | 1 | Not-determined | Genome-fragment | NA | NA | 0 |
| S7-final-viral-combined_k141_188081\|\|lt2gene | 1291 | No | NA | 2 | 1 | 0 | Low-quality | Genome-fragment | 3.36 | AAI-based (medium-confidence) | 0 |
| S1-final-viral-combined_k141_201261\|\|full | 2229 | No | NA | 4 | 0 | 3 | Not-determined | Genome-fragment | NA | NA | 0 |
| S1-final-viral-combined_k141_266706\|\|full | 1698 | No | NA | 2 | 0 | 2 | Not-determined | Genome-fragment | NA | NA | 0 |
| S17-final-viral-combined_k141_169200\|\|full | 1291 | No | NA | 4 | 0 | 0 | Low-quality | Genome-fragment | 2.08 | AAI-based (high-confidence) | 0 |
| S1-final-viral-combined_k141_266731\|\|full | 3347 | No | NA | 4 | 0 | 1 | Not-determined | Genome-fragment | NA | NA | 0 |
| S17-k141_177889 | 1291 | No | NA | 2 | 0 | 0 | Low-quality | Genome-fragment | 2.5 | AAI-based (medium-confidence) | 0 |
| S1-final-viral-combined_k141_382081\|\|full | 3345 | No | NA | 3 | 0 | 3 | Not-determined | Genome-fragment | NA | NA | 0 |
| S17-k141_253373 | 1291 | No | NA | 3 | 0 | 3 | Low-quality | Genome-fragment | 0.12 | AAI-based (medium-confidence) | 0 |
| S4-final-viral-combined_k141_21661\|\|full | 1291 | No | NA | 2 | 0 | 0 | Low-quality | Genome-fragment | 2.61 | AAI-based (high-confidence) | 0 |
| S9-final-viral-combined_k141_34456\|\|full | 1292 | No | NA | 3 | 1 | 0 | Low-quality | Genome-fragment | 3.08 | AAI-based (medium-confidence) | 0 |
| S11-final-viral-combined_k141_130518\|\|full | 1292 | No | NA | 4 | 1 | 0 | Low-quality | Genome-fragment | 3.55 | AAI-based (high-confidence) | 0 |
| S1-final-viral-combined_k141_266749\|\|full | 2576 | No | NA | 5 | 0 | 2 | Not-determined | Genome-fragment | NA | NA | 0 |
| S11-final-viral-combined_k141_75755\|\|full | 1292 | No | NA | 2 | 0 | 0 | Low-quality | Genome-fragment | 2.82 | AAI-based (high-confidence) | 0 |
| S1-final-viral-combined_k141_300462\|\|full | 1427 | No | NA | 2 | 0 | 1 | Not-determined | Genome-fragment | NA | NA | 0 |
| S1-final-viral-combined_k141_52817\|\|full | 1182 | No | NA | 3 | 0 | 1 | Not-determined | Genome-fragment | NA | NA | 0 |
| S1-final-viral-combined_k141_332633\|\|full | 2498 | No | NA | 3 | 0 | 1 | Not-determined | Genome-fragment | NA | NA | 0 |
| S11-final-viral-combined_k141_261318\|\|full | 1292 | No | NA | 4 | 1 | 0 | Low-quality | Genome-fragment | 2.88 | AAI-based (high-confidence) | 0 |
| S1-final-viral-combined_k141_250985\|\|full | 2030 | No | NA | 2 | 0 | 1 | Not-determined | Genome-fragment | NA | NA | 0 |
| S11_k141_199458 | 1292 | No | NA | 1 | 0 | 1 | Low-quality | Genome-fragment | 0.29 | AAI-based (medium-confidence) | 0 |
| S1-final-viral-combined_k141_316835\|\|full | 2680 | No | NA | 4 | 0 | 2 | Not-determined | Genome-fragment | NA | NA | 0 |
| S1-final-viral-combined_k141_316836\|\|full | 2063 | No | NA | 3 | 0 | 1 | Not-determined | Genome-fragment | NA | NA | 0 |
| S1-final-viral-combined_k141_382136\|\|full | 4480 | No | NA | 5 | 0 | 2 | Not-determined | Genome-fragment | NA | NA | 0 |
| S1-final-viral-combined_k141_102063\|\|full | 2332 | No | NA | 3 | 0 | 2 | Not-determined | Genome-fragment | NA | NA | 0 |
| S1-final-viral-combined_k141_297503\|\|full | 1292 | No | NA | 3 | 0 | 0 | Low-quality | Genome-fragment | 1.88 | AAI-based (medium-confidence) | 0 |
| S1-final-viral-combined_k141_167618\|\|full | 1077 | No | NA | 3 | 0 | 0 | Not-determined | Genome-fragment | NA | NA | 0 |
| S1_k141_115751 | 1292 | No | NA | 2 | 0 | 1 | Low-quality | Genome-fragment | 2.72 | AAI-based (medium-confidence) | 0 |
| S1-final-viral-combined_k141_349500\|\|full | 11922 | No | NA | 18 | 0 | 0 | Not-determined | Genome-fragment | NA | NA | 0 |
| S5_k141_81734 | 1292 | No | NA | 5 | 0 | 0 | Low-quality | Genome-fragment | 3.19 | AAI-based (medium-confidence) | 0 |
| S1-final-viral-combined_k141_283694\|\|full | 1836 | No | NA | 3 | 0 | 1 | Not-determined | Genome-fragment | NA | NA | 0 |
| S7-final-viral-combined_k141_130918\|\|full | 1292 | No | NA | 3 | 0 | 1 | Low-quality | Genome-fragment | 2.89 | AAI-based (high-confidence) | 0 |
| S7_k141_115274 | 1292 | No | NA | 1 | 0 | 0 | Low-quality | Genome-fragment | 1.57 | AAI-based (high-confidence) | 0 |
| S1-final-viral-combined_k141_300514\|\|full | 1042 | No | NA | 2 | 0 | 0 | Not-determined | Genome-fragment | NA | NA | 0 |
| S1-final-viral-combined_k141_85909\|\|full | 5249 | No | NA | 3 | 0 | 3 | Not-determined | Genome-fragment | NA | NA | 0 |
| S1-final-viral-combined_k141_85916\|\|full | 1918 | No | NA | 3 | 0 | 1 | Not-determined | Genome-fragment | NA | NA | 0 |
| S12-final-viral-combined_k141_91899\|\|full | 1292 | No | NA | 3 | 0 | 1 | Low-quality | Genome-fragment | 1.52 | AAI-based (medium-confidence) | 0 |
| S1-final-viral-combined_k141_20231\|\|full | 1920 | No | NA | 5 | 0 | 0 | Not-determined | Genome-fragment | NA | NA | 0 |
| S1-final-viral-combined_k141_167648\|\|full | 1583 | No | NA | 2 | 0 | 2 | Not-determined | Genome-fragment | NA | NA | 0 |
| S2_k141_238330 | 1292 | No | NA | 1 | 0 | 1 | Low-quality | Genome-fragment | 0.84 | AAI-based (high-confidence) | 0 |
| S1-final-viral-combined_k141_217776\|\|full | 1079 | No | NA | 2 | 0 | 1 | Not-determined | Genome-fragment | NA | NA | 0 |
| S4-final-viral-combined_k141_36688\|\|full | 1292 | No | NA | 3 | 0 | 0 | Low-quality | Genome-fragment | 2.79 | AAI-based (medium-confidence) | 0 |
| S4-final-viral-combined_k141_33604\|\|full | 1292 | No | NA | 2 | 0 | 1 | Low-quality | Genome-fragment | 2.01 | AAI-based (medium-confidence) | 0 |
| S1-final-viral-combined_k141_217787\|\|full | 1681 | No | NA | 2 | 0 | 0 | Not-determined | Genome-fragment | NA | NA | 0 |
| S1-final-viral-combined_k141_167672\|\|full | 2954 | No | NA | 4 | 0 | 2 | Not-determined | Genome-fragment | NA | NA | 0 |
| S4-final-viral-combined_k141_90355\|\|full | 1292 | No | NA | 5 | 0 | 0 | Low-quality | Genome-fragment | 2.09 | AAI-based (high-confidence) | 0 |
| S4-final-viral-combined_k141_48978\|\|lt2gene | 1292 | No | NA | 2 | 1 | 0 | Low-quality | Genome-fragment | 2.07 | AAI-based (medium-confidence) | 0 |
| S4_k141_121282 | 1292 | No | NA | 1 | 0 | 0 | Low-quality | Genome-fragment | 1.21 | AAI-based (high-confidence) | 0 |
| S6-final-viral-combined_k141_21890\|\|full | 1292 | No | NA | 4 | 2 | 0 | Low-quality | Genome-fragment | 3.76 | AAI-based (high-confidence) | 0 |
| S1-final-viral-combined_k141_185282\|\|full | 1501 | No | NA | 4 | 0 | 0 | Not-determined | Genome-fragment | NA | NA | 0 |
| S1-final-viral-combined_k141_251126\|\|full | 1753 | No | NA | 3 | 0 | 2 | Not-determined | Genome-fragment | NA | NA | 0 |
| S1-final-viral-combined_k141_251139\|\|full | 2517 | No | NA | 3 | 0 | 3 | Not-determined | Genome-fragment | NA | NA | 0 |
| S1-final-viral-combined_k141_167694\|\|full | 1355 | No | NA | 4 | 0 | 0 | Not-determined | Genome-fragment | NA | NA | 0 |
| S1-final-viral-combined_k141_167711\|\|full | 3534 | No | NA | 3 | 0 | 2 | Not-determined | Genome-fragment | NA | NA | 0 |
| S1-final-viral-combined_k141_36590\|\|full | 4185 | No | NA | 6 | 0 | 1 | Not-determined | Genome-fragment | NA | NA | 0 |
| S1-final-viral-combined_k141_52983\|\|full | 1319 | No | NA | 2 | 0 | 1 | Not-determined | Genome-fragment | NA | NA | 0 |
| S1-final-viral-combined_k141_266948\|\|full | 1055 | No | NA | 2 | 0 | 1 | Not-determined | Genome-fragment | NA | NA | 0 |
| S1-final-viral-combined_k141_3928\|\|full | 2805 | No | NA | 3 | 0 | 1 | Not-determined | Genome-fragment | NA | NA | 0 |
| S1-final-viral-combined_k141_185336\|\|full | 2431 | No | NA | 4 | 1 | 0 | Medium-quality | Genome-fragment | 56.15 | AAI-based (high-confidence) | 0 |
| S11-final-viral-combined_k141_96823\|\|full | 1293 | No | NA | 3 | 1 | 0 | Low-quality | Genome-fragment | 3.31 | AAI-based (high-confidence) | 0 |
| S11-final-viral-combined_k141_177925\|\|full | 1293 | No | NA | 2 | 1 | 0 | Low-quality | Genome-fragment | 1.44 | AAI-based (medium-confidence) | 0 |
| S1-final-viral-combined_k141_281660\|\|full | 1293 | No | NA | 3 | 2 | 0 | Low-quality | Genome-fragment | 2.95 | AAI-based (medium-confidence) | 0 |
| S1-final-viral-combined_k141_5850\|\|full | 1293 | No | NA | 2 | 1 | 0 | Low-quality | Genome-fragment | 0.49 | HMM-based (lower-bound) | 0 |
| S1-final-viral-combined_k141_102192\|\|full | 4919 | No | NA | 5 | 0 | 2 | Not-determined | Genome-fragment | NA | NA | 0 |
| S1-final-viral-combined_k141_86042\|\|full | 14024 | No | NA | 17 | 0 | 2 | Not-determined | Genome-fragment | NA | NA | 0 |
| S1_k141_284281 | 1293 | No | NA | 3 | 0 | 2 | Low-quality | Genome-fragment | 2.16 | AAI-based (medium-confidence) | 0 |
| S1_k141_322530 | 1293 | No | NA | 3 | 0 | 2 | Low-quality | Genome-fragment | 2.49 | AAI-based (medium-confidence) | 0 |
| S3-final-viral-combined_k141_125594\|\|lt2gene | 1293 | No | NA | 2 | 1 | 0 | Low-quality | Genome-fragment | 3.13 | AAI-based (high-confidence) | 0 |
| S1-final-viral-combined_k141_167726\|\|full | 4106 | No | NA | 3 | 0 | 1 | Not-determined | Genome-fragment | NA | NA | 0 |
| S1-final-viral-combined_k141_300668\|\|full | 1232 | No | NA | 2 | 0 | 0 | Not-determined | Genome-fragment | NA | NA | 0 |
| S1-final-viral-combined_k141_102222\|\|full | 1021 | No | NA | 2 | 0 | 0 | Not-determined | Genome-fragment | NA | NA | 0 |
| S7-final-viral-combined_k141_159424\|\|full | 1293 | No | NA | 4 | 0 | 0 | Low-quality | Genome-fragment | 0.8 | AAI-based (medium-confidence) | 0 |
| S7_k141_30825 | 1293 | No | NA | 1 | 0 | 0 | Low-quality | Genome-fragment | 0.21 | AAI-based (high-confidence) | 0 |
| S1-final-viral-combined_k141_300679\|\|full | 1068 | No | NA | 3 | 0 | 1 | Not-determined | Genome-fragment | NA | NA | 0 |
| S17-final-viral-combined_k141_137284\|\|full | 1293 | No | NA | 3 | 1 | 0 | Low-quality | Genome-fragment | 2.3 | AAI-based (high-confidence) | 0 |
| S17-k141_258331 | 1293 | No | NA | 1 | 0 | 1 | Low-quality | Genome-fragment | 1.09 | AAI-based (high-confidence) | 0 |
| S1-final-viral-combined_k141_349643\|\|full | 2253 | No | NA | 4 | 0 | 1 | Not-determined | Genome-fragment | NA | NA | 0 |
| S4_k141_37345 | 1293 | No | NA | 2 | 0 | 0 | Low-quality | Genome-fragment | 2.74 | AAI-based (high-confidence) | 0 |
| S8-final-viral-combined_k141_210025\|\|full | 1293 | No | NA | 4 | 3 | 0 | Low-quality | Genome-fragment | 2.11 | AAI-based (medium-confidence) | 0 |
| S1-final-viral-combined_k141_152130\|\|full | 1340 | No | NA | 3 | 0 | 1 | Not-determined | Genome-fragment | NA | NA | 0 |
| S1-final-viral-combined_k141_86098\|\|full | 1049 | No | NA | 3 | 0 | 0 | Not-determined | Genome-fragment | NA | NA | 0 |
| S8_k141_50437 | 1293 | No | NA | 3 | 0 | 0 | Low-quality | Genome-fragment | 1.98 | AAI-based (medium-confidence) | 0 |
| S11-final-viral-combined_k141_138238\|\|full | 1294 | No | NA | 3 | 0 | 1 | Low-quality | Genome-fragment | 1.94 | AAI-based (medium-confidence) | 0 |
| S1-final-viral-combined_k141_119322\|\|full | 13601 | No | NA | 11 | 0 | 8 | Not-determined | Genome-fragment | NA | NA | 0 |
| S1-final-viral-combined_k141_36707\|\|full | 6204 | No | NA | 5 | 0 | 3 | Not-determined | Genome-fragment | NA | NA | 0 |
| S1-final-viral-combined_k141_362288\|\|full | 1294 | No | NA | 2 | 2 | 0 | Low-quality | Genome-fragment | 3.55 | AAI-based (high-confidence) | 0 |
| S1-final-viral-combined_k141_349701\|\|full | 1041 | No | NA | 2 | 0 | 0 | Not-determined | Genome-fragment | NA | NA | 0 |
| S1-final-viral-combined_k141_217981\|\|full | 2649 | No | NA | 3 | 0 | 0 | Not-determined | Genome-fragment | NA | NA | 0 |
| S1-final-viral-combined_k141_332863\|\|full | 3847 | No | NA | 5 | 0 | 0 | Not-determined | Genome-fragment | NA | NA | 0 |
| S1-final-viral-combined_k141_294232\|\|full | 1294 | No | NA | 5 | 0 | 0 | Low-quality | Genome-fragment | 3.18 | AAI-based (high-confidence) | 0 |
| S1-final-viral-combined_k141_185476\|\|full | 2040 | No | NA | 2 | 0 | 1 | Not-determined | Genome-fragment | NA | NA | 0 |
| S1_k141_394414 | 1294 | No | NA | 3 | 0 | 2 | Low-quality | Genome-fragment | 1.91 | AAI-based (medium-confidence) | 0 |
| S1-final-viral-combined_k141_365922\|\|full | 4433 | No | NA | 10 | 0 | 3 | Not-determined | Genome-fragment | NA | NA | 0 |
| S1-final-viral-combined_k141_349753\|\|full | 4712 | No | NA | 3 | 0 | 2 | Not-determined | Genome-fragment | NA | NA | 0 |
| S1-final-viral-combined_k141_234464\|\|full | 3147 | No | NA | 4 | 0 | 3 | Not-determined | Genome-fragment | NA | NA | 0 |
| S1-final-viral-combined_k141_68998\|\|full | 56973 | Yes | 55700 | 92 | 22 | 2 | High-quality | High-quality | 100 | AAI-based (high-confidence) | 2.23 |
| S7-final-viral-combined_k141_73265\|\|lt2gene | 1294 | No | NA | 2 | 1 | 0 | Low-quality | Genome-fragment | 3.17 | AAI-based (high-confidence) | 0 |
| S1-final-viral-combined_k141_217999\|\|full | 3033 | No | NA | 3 | 0 | 2 | Not-determined | Genome-fragment | NA | NA | 0 |
| S2-final-viral-combined_k141_177997\|\|full | 1294 | No | NA | 4 | 0 | 0 | Low-quality | Genome-fragment | 3.13 | AAI-based (high-confidence) | 0 |
| S11-final-viral-combined_k141_96575\|\|full | 1295 | No | NA | 2 | 0 | 0 | Low-quality | Genome-fragment | 1.53 | AAI-based (medium-confidence) | 0 |
| S11_k141_58805 | 1295 | No | NA | 2 | 0 | 1 | Low-quality | Genome-fragment | 0.95 | AAI-based (medium-confidence) | 0 |
| S1_k141_353480 | 1295 | No | NA | 3 | 0 | 0 | Low-quality | Genome-fragment | 0.99 | AAI-based (high-confidence) | 0 |
| S1-final-viral-combined_k141_251366\|\|full | 6845 | No | NA | 7 | 0 | 4 | Not-determined | Genome-fragment | NA | NA | 0 |
| S1-final-viral-combined_k141_36762\|\|full | 1412 | No | NA | 2 | 0 | 0 | Not-determined | Genome-fragment | NA | NA | 0 |
| S1-final-viral-combined_k141_317138\|\|full | 19449 | No | NA | 19 | 0 | 12 | Not-determined | Genome-fragment | NA | NA | 0 |
| S1-final-viral-combined_k141_167848\|\|full | 2286 | No | NA | 3 | 0 | 2 | Not-determined | Genome-fragment | NA | NA | 0 |
| S1-final-viral-combined_k141_152133\|\|full | 1595 | No | NA | 3 | 0 | 2 | Not-determined | Genome-fragment | NA | NA | 0 |
| S1-final-viral-combined_k141_349820\|\|full | 3028 | No | NA | 4 | 0 | 3 | Not-determined | Genome-fragment | NA | NA | 0 |
| S1-final-viral-combined_k141_201696\|\|full | 2966 | No | NA | 4 | 0 | 0 | Not-determined | Genome-fragment | NA | NA | 0 |
| S1-final-viral-combined_k141_69079\|\|full | 1407 | No | NA | 2 | 0 | 1 | Not-determined | Genome-fragment | NA | NA | 0 |
| S1-final-viral-combined_k141_251378\|\|full | 1333 | No | NA | 2 | 0 | 1 | Not-determined | Genome-fragment | NA | NA | 0 |
| S1_k141_338639 | 1295 | No | NA | 1 | 0 | 1 | Low-quality | Genome-fragment | 0.72 | AAI-based (medium-confidence) | 0 |
| S3-final-viral-combined_k141_133260\|\|full | 1295 | No | NA | 4 | 1 | 0 | Low-quality | Genome-fragment | 2.21 | AAI-based (high-confidence) | 0 |
| S1-final-viral-combined_k141_251387\|\|full | 3571 | No | NA | 3 | 0 | 3 | Not-determined | Genome-fragment | NA | NA | 0 |
| S1-final-viral-combined_k141_283999\|\|full | 1838 | No | NA | 3 | 0 | 1 | Not-determined | Genome-fragment | NA | NA | 0 |
| S1-final-viral-combined_k141_349869\|\|full | 2049 | No | NA | 3 | 0 | 1 | Not-determined | Genome-fragment | NA | NA | 0 |
| S1-final-viral-combined_k141_300776\|\|full | 2146 | No | NA | 4 | 0 | 0 | Not-determined | Genome-fragment | NA | NA | 0 |
| S5-final-viral-combined_k141_110719\|\|full | 1295 | No | NA | 2 | 0 | 1 | Low-quality | Genome-fragment | 1.69 | AAI-based (medium-confidence) | 0 |
| S1-final-viral-combined_k141_267198\|\|full | 4386 | No | NA | 5 | 0 | 2 | Not-determined | Genome-fragment | NA | NA | 0 |
| S7-final-viral-combined_k141_51555\|\|full | 1295 | No | NA | 2 | 0 | 2 | Low-quality | Genome-fragment | 0.82 | AAI-based (medium-confidence) | 0 |
| S7-final-viral-combined_k141_120959\|\|lt2gene | 1295 | No | NA | 2 | 2 | 0 | Low-quality | Genome-fragment | 2.85 | AAI-based (medium-confidence) | 0 |
| S12-final-viral-combined_k141_372250\|\|full | 1295 | Yes | 504 | 2 | 1 | 1 | Low-quality | Genome-fragment | 0.62 | HMM-based (lower-bound) | 61.08 |
| S1-final-viral-combined_k141_4054\|\|full | 6441 | No | NA | 7 | 0 | 1 | Not-determined | Genome-fragment | NA | NA | 0 |
| S1-final-viral-combined_k141_349888\|\|full | 2505 | No | NA | 4 | 0 | 2 | Not-determined | Genome-fragment | NA | NA | 0 |
| S12-final-viral-combined_k141_92224\|\|full | 1295 | No | NA | 2 | 0 | 0 | Low-quality | Genome-fragment | 3.9 | AAI-based (high-confidence) | 0 |
| S1-final-viral-combined_k141_167919\|\|full | 1848 | No | NA | 3 | 0 | 1 | Not-determined | Genome-fragment | NA | NA | 0 |
| S1-final-viral-combined_k141_36852\|\|full | 3191 | No | NA | 2 | 0 | 2 | Not-determined | Genome-fragment | NA | NA | 0 |
| S1-final-viral-combined_k141_382380\|\|full | 1780 | No | NA | 4 | 0 | 0 | Not-determined | Genome-fragment | NA | NA | 0 |
| S1-final-viral-combined_k141_201769\|\|full | 2819 | No | NA | 2 | 0 | 2 | Not-determined | Genome-fragment | NA | NA | 0 |
| S12-final-viral-combined_k141_367716\|\|full | 1295 | No | NA | 2 | 0 | 2 | Low-quality | Genome-fragment | 0.64 | AAI-based (medium-confidence) | 0 |
| S1-final-viral-combined_k141_185613\|\|full | 1174 | No | NA | 4 | 0 | 1 | Not-determined | Genome-fragment | NA | NA | 0 |
| S2-final-viral-combined_k141_16746\|\|full | 1295 | No | NA | 4 | 1 | 0 | Low-quality | Genome-fragment | 2.21 | AAI-based (high-confidence) | 0 |
| S1-final-viral-combined_k141_300893\|\|full | 5554 | No | NA | 8 | 0 | 0 | Not-determined | Genome-fragment | NA | NA | 0 |
| S1-final-viral-combined_k141_284081\|\|full | 6143 | No | NA | 5 | 0 | 4 | Not-determined | Genome-fragment | NA | NA | 0 |
| S1-final-viral-combined_k141_152265\|\|full | 1631 | No | NA | 2 | 0 | 0 | Not-determined | Genome-fragment | NA | NA | 0 |
| S1-final-viral-combined_k141_251587\|\|full | 1388 | No | NA | 4 | 0 | 1 | Not-determined | Genome-fragment | NA | NA | 0 |
| S6-final-viral-combined_k141_59075\|\|full | 1295 | No | NA | 4 | 1 | 0 | Low-quality | Genome-fragment | 2.54 | AAI-based (medium-confidence) | 0 |
| S1-final-viral-combined_k141_366151\|\|full | 1085 | No | NA | 2 | 0 | 1 | Not-determined | Genome-fragment | NA | NA | 0 |
| S6-final-viral-combined_k141_50310\|\|full | 1295 | No | NA | 3 | 0 | 2 | Low-quality | Genome-fragment | 0.65 | AAI-based (medium-confidence) | 0 |
| S11-final-viral-combined_k141_91228\|\|full | 1296 | No | NA | 4 | 1 | 0 | Low-quality | Genome-fragment | 3.7 | AAI-based (medium-confidence) | 0 |
| S1-final-viral-combined_k141_251599\|\|full | 7549 | No | NA | 7 | 0 | 4 | Not-determined | Genome-fragment | NA | NA | 0 |
| S11-final-viral-combined_k141_68813\|\|lt2gene | 1296 | No | NA | 2 | 1 | 0 | Low-quality | Genome-fragment | 2.91 | AAI-based (high-confidence) | 0 |
| S1-final-viral-combined_k141_317268\|\|full | 2956 | No | NA | 3 | 0 | 2 | Not-determined | Genome-fragment | NA | NA | 0 |
| S1-final-viral-combined_k141_382442\|\|full | 2054 | No | NA | 4 | 0 | 1 | Not-determined | Genome-fragment | NA | NA | 0 |
| S11_k141_218652 | 1296 | No | NA | 3 | 0 | 0 | Low-quality | Genome-fragment | 2.2 | AAI-based (high-confidence) | 0 |
| S1-final-viral-combined_k141_333127\|\|full | 1808 | No | NA | 3 | 0 | 2 | Not-determined | Genome-fragment | NA | NA | 0 |
| S3_k141_51247 | 1296 | No | NA | 2 | 0 | 0 | Low-quality | Genome-fragment | 2.99 | AAI-based (medium-confidence) | 0 |
| S1-final-viral-combined_k141_284134\|\|full | 1722 | No | NA | 4 | 0 | 1 | Not-determined | Genome-fragment | NA | NA | 0 |
| S1-final-viral-combined_k141_350110\|\|full | 1209 | No | NA | 3 | 0 | 0 | Not-determined | Genome-fragment | NA | NA | 0 |
| S1-final-viral-combined_k141_333140\|\|full | 4169 | No | NA | 4 | 0 | 2 | Not-determined | Genome-fragment | NA | NA | 0 |
| S1-final-viral-combined_k141_119692\|\|full | 4446 | No | NA | 3 | 0 | 0 | Not-determined | Genome-fragment | NA | NA | 0 |
| S17-final-viral-combined_k141_100062\|\|full | 1296 | No | NA | 3 | 0 | 0 | Low-quality | Genome-fragment | 0.63 | AAI-based (medium-confidence) | 0 |
| S1-final-viral-combined_k141_350141\|\|full | 2972 | No | NA | 3 | 0 | 1 | Not-determined | Genome-fragment | NA | NA | 0 |
| S17-final-viral-combined_k141_102652\|\|full | 1296 | No | NA | 3 | 0 | 0 | Low-quality | Genome-fragment | 1.85 | AAI-based (medium-confidence) | 0 |
| S17-final-viral-combined_k141_172669\|\|full | 1296 | No | NA | 2 | 0 | 2 | Low-quality | Genome-fragment | 0.61 | AAI-based (medium-confidence) | 0 |
| S1-final-viral-combined_k141_102476\|\|full | 1613 | No | NA | 5 | 0 | 0 | Not-determined | Genome-fragment | NA | NA | 0 |
| S1-final-viral-combined_k141_382517\|\|full | 1815 | No | NA | 3 | 0 | 3 | Not-determined | Genome-fragment | NA | NA | 0 |
| S1-final-viral-combined_k141_185766\|\|full | 1701 | No | NA | 2 | 0 | 1 | Not-determined | Genome-fragment | NA | NA | 0 |
| S12-final-viral-combined_k141_366846\|\|full | 1296 | No | NA | 2 | 0 | 1 | Low-quality | Genome-fragment | 0.66 | AAI-based (medium-confidence) | 0 |
| S2-final-viral-combined_k141_241957\|\|full | 1296 | No | NA | 3 | 0 | 0 | Low-quality | Genome-fragment | 1.63 | AAI-based (medium-confidence) | 0 |
| S1-final-viral-combined_k141_251694\|\|full | 2833 | No | NA | 5 | 0 | 0 | Not-determined | Genome-fragment | NA | NA | 0 |
| S1-final-viral-combined_k141_152349\|\|full | 1809 | No | NA | 2 | 0 | 1 | Not-determined | Genome-fragment | NA | NA | 0 |
| S1-final-viral-combined_k141_317361\|\|full | 1597 | No | NA | 3 | 0 | 1 | Not-determined | Genome-fragment | NA | NA | 0 |
| S4_k141_23528 | 1296 | No | NA | 1 | 0 | 0 | Low-quality | Genome-fragment | 2.59 | AAI-based (high-confidence) | 0 |
| S1-final-viral-combined_k141_284189\|\|full | 4651 | No | NA | 6 | 0 | 0 | Not-determined | Genome-fragment | NA | NA | 0 |
| S1-final-viral-combined_k141_37083\|\|full | 2826 | No | NA | 3 | 0 | 0 | Not-determined | Genome-fragment | NA | NA | 0 |
| S1-final-viral-combined_k141_152380\|\|full | 6320 | No | NA | 8 | 0 | 1 | Not-determined | Genome-fragment | NA | NA | 0 |
| S8-final-viral-combined_k141_74832\|\|full | 1296 | No | NA | 2 | 0 | 1 | Low-quality | Genome-fragment | 0.85 | AAI-based (medium-confidence) | 0 |
| S1-final-viral-combined_k141_20801\|\|full | 1892 | No | NA | 3 | 0 | 1 | Not-determined | Genome-fragment | NA | NA | 0 |
| S1-final-viral-combined_k141_323250\|\|full | 1297 | No | NA | 3 | 0 | 0 | Low-quality | Genome-fragment | 2.92 | AAI-based (high-confidence) | 0 |
| S1-final-viral-combined_k141_382567\|\|full | 1862 | No | NA | 3 | 0 | 1 | Not-determined | Genome-fragment | NA | NA | 0 |
| S1-final-viral-combined_k141_317391\|\|full | 7534 | No | NA | 11 | 0 | 0 | Not-determined | Genome-fragment | NA | NA | 0 |
| S1_k141_233371 | 1297 | No | NA | 3 | 0 | 0 | Low-quality | Genome-fragment | 3.41 | AAI-based (high-confidence) | 0 |
| S1-final-viral-combined_k141_102499\|\|full | 2667 | No | NA | 3 | 0 | 3 | Not-determined | Genome-fragment | NA | NA | 0 |
| S3_k141_109237 | 1297 | No | NA | 2 | 0 | 0 | Low-quality | Genome-fragment | 3.09 | AAI-based (high-confidence) | 0 |
| S7-final-viral-combined_k141_506\|\|full | 1297 | No | NA | 4 | 0 | 0 | Low-quality | Genome-fragment | 1.64 | AAI-based (high-confidence) | 0 |
| S1-final-viral-combined_k141_284219\|\|full | 3774 | No | NA | 4 | 0 | 4 | Not-determined | Genome-fragment | NA | NA | 0 |
| S1-final-viral-combined_k141_382586\|\|full | 2080 | No | NA | 3 | 0 | 2 | Not-determined | Genome-fragment | NA | NA | 0 |
| S7-final-viral-combined_k141_64158\|\|full | 1297 | No | NA | 3 | 0 | 0 | Low-quality | Genome-fragment | 2.2 | AAI-based (medium-confidence) | 0 |
| S7_k141_204446 | 1297 | No | NA | 3 | 0 | 0 | Low-quality | Genome-fragment | 1.22 | AAI-based (high-confidence) | 0 |
| S1-final-viral-combined_k141_152405\|\|full | 1345 | No | NA | 2 | 0 | 2 | Not-determined | Genome-fragment | NA | NA | 0 |
| S1-final-viral-combined_k141_37145\|\|full | 1967 | No | NA | 3 | 0 | 0 | Not-determined | Genome-fragment | NA | NA | 0 |
| S1-final-viral-combined_k141_119827\|\|full | 6494 | No | NA | 7 | 0 | 4 | Not-determined | Genome-fragment | NA | NA | 0 |
| S17-final-viral-combined_k141_59498\|\|full | 1297 | No | NA | 3 | 0 | 2 | Low-quality | Genome-fragment | 1 | AAI-based (medium-confidence) | 0 |
| S17-final-viral-combined_k141_199368\|\|full | 1297 | No | NA | 2 | 0 | 0 | Low-quality | Genome-fragment | 3.24 | AAI-based (high-confidence) | 0 |
| S4-final-viral-combined_k141_166683\|\|full | 1297 | No | NA | 3 | 0 | 2 | Low-quality | Genome-fragment | 2.39 | AAI-based (medium-confidence) | 0 |
| S1-final-viral-combined_k141_382618\|\|full | 1159 | No | NA | 2 | 0 | 2 | Not-determined | Genome-fragment | NA | NA | 0 |
| S1-final-viral-combined_k141_53438\|\|full | 1362 | No | NA | 4 | 0 | 3 | Not-determined | Genome-fragment | NA | NA | 0 |
| S1-final-viral-combined_k141_69226\|\|full | 4163 | No | NA | 5 | 0 | 3 | Not-determined | Genome-fragment | NA | NA | 0 |
| S6-final-viral-combined_k141_100526\|\|lt2gene | 1297 | No | NA | 2 | 2 | 0 | Low-quality | Genome-fragment | 4.03 | AAI-based (high-confidence) | 0 |
| S1-final-viral-combined_k141_69229\|\|full | 1348 | No | NA | 2 | 0 | 0 | Not-determined | Genome-fragment | NA | NA | 0 |
| S1-final-viral-combined_k141_37155\|\|full | 2040 | No | NA | 2 | 0 | 1 | Not-determined | Genome-fragment | NA | NA | 0 |
| S1-final-viral-combined_k141_202031\|\|full | 2428 | No | NA | 3 | 0 | 3 | Not-determined | Genome-fragment | NA | NA | 0 |
| S6_k141_130921 | 1297 | No | NA | 2 | 0 | 0 | Low-quality | Genome-fragment | 0.77 | AAI-based (high-confidence) | 0 |
| S1-final-viral-combined_k141_267514\|\|full | 1609 | No | NA | 4 | 0 | 2 | Not-determined | Genome-fragment | NA | NA | 0 |
| S1-final-viral-combined_k141_53455\|\|full | 2567 | No | NA | 4 | 0 | 0 | Not-determined | Genome-fragment | NA | NA | 0 |
| S1-final-viral-combined_k141_136479\|\|full | 2497 | No | NA | 2 | 0 | 2 | Not-determined | Genome-fragment | NA | NA | 0 |
| S11-final-viral-combined_k141_108122\|\|full | 1298 | No | NA | 3 | 0 | 0 | Low-quality | Genome-fragment | 2.67 | AAI-based (high-confidence) | 0 |
| S11_k141_240901 | 1298 | No | NA | 3 | 0 | 0 | Low-quality | Genome-fragment | 1.39 | AAI-based (high-confidence) | 0 |
| S1-final-viral-combined_k141_202043\|\|full | 6612 | No | NA | 9 | 0 | 2 | Not-determined | Genome-fragment | NA | NA | 0 |
| S1-final-viral-combined_k141_168253\|\|full | 1466 | No | NA | 2 | 0 | 1 | Not-determined | Genome-fragment | NA | NA | 0 |
| S1-final-viral-combined_k141_318313\|\|full | 1298 | No | NA | 3 | 1 | 0 | Low-quality | Genome-fragment | 0.79 | HMM-based (lower-bound) | 0 |
| S1-final-viral-combined_k141_69258\|\|full | 2359 | No | NA | 3 | 0 | 2 | Not-determined | Genome-fragment | NA | NA | 0 |
| S1-final-viral-combined_k141_333282\|\|full | 3042 | No | NA | 4 | 0 | 2 | Not-determined | Genome-fragment | NA | NA | 0 |
| S1-final-viral-combined_k141_69264\|\|full | 2270 | No | NA | 2 | 0 | 1 | Not-determined | Genome-fragment | NA | NA | 0 |
| S1-final-viral-combined_k141_284284\|\|full | 6439 | No | NA | 5 | 0 | 2 | Not-determined | Genome-fragment | NA | NA | 0 |
| S1-final-viral-combined_k141_119899\|\|full | 1647 | No | NA | 3 | 0 | 1 | Not-determined | Genome-fragment | NA | NA | 0 |
| S5_k141_71222 | 1298 | No | NA | 3 | 0 | 1 | Low-quality | Genome-fragment | 2.84 | AAI-based (medium-confidence) | 0 |
| S7-final-viral-combined_k141_118310\|\|full | 1298 | No | NA | 3 | 1 | 0 | Low-quality | Genome-fragment | 3.32 | AAI-based (high-confidence) | 0 |
| S1-final-viral-combined_k141_53493\|\|full | 1764 | No | NA | 3 | 0 | 2 | Not-determined | Genome-fragment | NA | NA | 0 |
| S7-final-viral-combined_k141_97333\|\|full | 1298 | No | NA | 2 | 1 | 0 | Low-quality | Genome-fragment | 1.69 | AAI-based (medium-confidence) | 0 |
| S1-final-viral-combined_k141_86424\|\|full | 3212 | No | NA | 6 | 0 | 2 | Not-determined | Genome-fragment | NA | NA | 0 |
| S1-final-viral-combined_k141_168280\|\|full | 2887 | No | NA | 2 | 0 | 2 | Not-determined | Genome-fragment | NA | NA | 0 |
| S7-final-viral-combined_k141_100964\|\|full | 1298 | No | NA | 2 | 0 | 2 | Low-quality | Genome-fragment | 0.78 | AAI-based (medium-confidence) | 0 |
| S17-k141_103544 | 1298 | No | NA | 2 | 0 | 0 | Low-quality | Genome-fragment | 5.16 | AAI-based (medium-confidence) | 0 |
| S1-final-viral-combined_k141_53509\|\|full | 1206 | No | NA | 3 | 0 | 3 | Not-determined | Genome-fragment | NA | NA | 0 |
| S1-final-viral-combined_k141_69280\|\|full | 7371 | No | NA | 6 | 0 | 5 | Not-determined | Genome-fragment | NA | NA | 0 |
| S1-final-viral-combined_k141_4229\|\|full | 1382 | No | NA | 2 | 0 | 0 | Not-determined | Genome-fragment | NA | NA | 0 |
| S1-final-viral-combined_k141_53519\|\|full | 2423 | No | NA | 3 | 0 | 0 | Not-determined | Genome-fragment | NA | NA | 0 |
| S1-final-viral-combined_k141_333331\|\|full | 1443 | No | NA | 3 | 0 | 1 | Not-determined | Genome-fragment | NA | NA | 0 |
| S2-final-viral-combined_k141_117106\|\|lt2gene | 1298 | No | NA | 2 | 2 | 0 | Low-quality | Genome-fragment | 2.16 | AAI-based (high-confidence) | 0 |
| S1-final-viral-combined_k141_168302\|\|full | 5547 | No | NA | 4 | 0 | 0 | Not-determined | Genome-fragment | NA | NA | 0 |
| S6_k141_130749 | 1298 | No | NA | 2 | 1 | 0 | Low-quality | Genome-fragment | 1.62 | HMM-based (lower-bound) | 0 |
| S8-final-viral-combined_k141_197826\|\|full | 1298 | No | NA | 2 | 0 | 0 | Low-quality | Genome-fragment | 2.04 | AAI-based (high-confidence) | 0 |
| S1-final-viral-combined_k141_317531\|\|full | 4932 | No | NA | 4 | 0 | 3 | Not-determined | Genome-fragment | NA | NA | 0 |
| S8-final-viral-combined_k141_152396\|\|full | 1298 | No | NA | 2 | 0 | 1 | Low-quality | Genome-fragment | 0.82 | AAI-based (medium-confidence) | 0 |
| S1-final-viral-combined_k141_53562\|\|full | 2021 | No | NA | 3 | 0 | 0 | Not-determined | Genome-fragment | NA | NA | 0 |
| S9-final-viral-combined_k141_14862\|\|lt2gene | 1299 | No | NA | 1 | 1 | 0 | Low-quality | Genome-fragment | 2.99 | AAI-based (medium-confidence) | 0 |
| S11-final-viral-combined_k141_162382\|\|full | 1299 | No | NA | 3 | 1 | 0 | Low-quality | Genome-fragment | 2.14 | AAI-based (medium-confidence) | 0 |
| S1-final-viral-combined_k141_152507\|\|full | 1688 | No | NA | 3 | 0 | 1 | Not-determined | Genome-fragment | NA | NA | 0 |
| S1-final-viral-combined_k141_53574\|\|full | 2147 | No | NA | 2 | 0 | 2 | Not-determined | Genome-fragment | NA | NA | 0 |
| S11-final-viral-combined_k141_93487\|\|lt2gene | 1299 | No | NA | 2 | 1 | 0 | Low-quality | Genome-fragment | 0.87 | AAI-based (medium-confidence) | 0 |
| S1-final-viral-combined_k141_382736\|\|full | 1024 | No | NA | 2 | 0 | 0 | Not-determined | Genome-fragment | NA | NA | 0 |
| S1-final-viral-combined_k141_86472\|\|full | 3924 | No | NA | 5 | 0 | 4 | Not-determined | Genome-fragment | NA | NA | 0 |
| S1-final-viral-combined_k141_374301\|\|full | 1299 | No | NA | 4 | 2 | 0 | Low-quality | Genome-fragment | 2.88 | AAI-based (high-confidence) | 0 |
| S1-final-viral-combined_k141_301348\|\|full | 2190 | No | NA | 4 | 0 | 1 | Not-determined | Genome-fragment | NA | NA | 0 |
| S1-final-viral-combined_k141_69297\|\|full | 3193 | No | NA | 3 | 0 | 1 | Not-determined | Genome-fragment | NA | NA | 0 |
| S1-final-viral-combined_k141_317584\|\|full | 3768 | No | NA | 4 | 0 | 0 | Not-determined | Genome-fragment | NA | NA | 0 |
| S5-final-viral-combined_k141_90123\|\|full | 1299 | No | NA | 3 | 2 | 0 | Low-quality | Genome-fragment | 2.99 | AAI-based (high-confidence) | 0 |
| S1-final-viral-combined_k141_317585\|\|full | 10279 | No | NA | 6 | 0 | 3 | Not-determined | Genome-fragment | NA | NA | 0 |
| S1-final-viral-combined_k141_37288\|\|full | 3487 | No | NA | 3 | 0 | 2 | Not-determined | Genome-fragment | NA | NA | 0 |
| S1-final-viral-combined_k141_136614\|\|full | 23699 | No | NA | 22 | 0 | 17 | Not-determined | Genome-fragment | NA | NA | 0 |
| S1-final-viral-combined_k141_284391\|\|full | 1829 | No | NA | 3 | 0 | 0 | Not-determined | Genome-fragment | NA | NA | 0 |
| S1-final-viral-combined_k141_37297\|\|full | 1592 | No | NA | 3 | 0 | 1 | Not-determined | Genome-fragment | NA | NA | 0 |
| S7-final-viral-combined_k141_42926\|\|lt2gene | 1299 | No | NA | 1 | 1 | 0 | Low-quality | Genome-fragment | 2.7 | AAI-based (high-confidence) | 0 |
| S17-k141_95955 | 1299 | No | NA | 2 | 0 | 0 | Low-quality | Genome-fragment | 2.17 | AAI-based (medium-confidence) | 0 |
| S12_k141_208274 | 1299 | No | NA | 3 | 0 | 0 | Low-quality | Genome-fragment | 1.76 | AAI-based (high-confidence) | 0 |
| S12_k141_361979 | 1299 | No | NA | 2 | 1 | 0 | Low-quality | Genome-fragment | 0.1 | HMM-based (lower-bound) | 0 |
| S8-final-viral-combined_k141_240124\|\|full | 1299 | No | NA | 3 | 1 | 0 | Low-quality | Genome-fragment | 2.14 | AAI-based (high-confidence) | 0 |
| S1-final-viral-combined_k141_382784\|\|full | 3661 | No | NA | 4 | 0 | 1 | Not-determined | Genome-fragment | NA | NA | 0 |
| S1-final-viral-combined_k141_69323\|\|full | 2788 | No | NA | 4 | 0 | 2 | Not-determined | Genome-fragment | NA | NA | 0 |
| S1-final-viral-combined_k141_218372\|\|full | 1901 | No | NA | 3 | 0 | 1 | Not-determined | Genome-fragment | NA | NA | 0 |
| S1-final-viral-combined_k141_382799\|\|full | 1658 | No | NA | 2 | 0 | 0 | Not-determined | Genome-fragment | NA | NA | 0 |
| S1-final-viral-combined_k141_86509\|\|full | 3386 | No | NA | 3 | 0 | 2 | Not-determined | Genome-fragment | NA | NA | 0 |
| S8-final-viral-combined_k141_57428\|\|full | 1299 | No | NA | 3 | 0 | 1 | Low-quality | Genome-fragment | 0.6 | AAI-based (medium-confidence) | 0 |
| S1-final-viral-combined_k141_53652\|\|full | 6252 | No | NA | 6 | 0 | 4 | Not-determined | Genome-fragment | NA | NA | 0 |
| S8-final-viral-combined_k141_138770\|\|lt2gene | 1299 | No | NA | 2 | 0 | 2 | Low-quality | Genome-fragment | 1.85 | AAI-based (medium-confidence) | 0 |
| S9-final-viral-combined_k141_97179\|\|full | 1300 | No | NA | 2 | 0 | 0 | Low-quality | Genome-fragment | 3.49 | AAI-based (medium-confidence) | 0 |
| S11-final-viral-combined_k141_153243\|\|full | 1300 | No | NA | 4 | 2 | 0 | Low-quality | Genome-fragment | 1.03 | AAI-based (high-confidence) | 0 |
| S11-final-viral-combined_k141_95483\|\|full | 1300 | No | NA | 3 | 1 | 0 | Low-quality | Genome-fragment | 1.54 | HMM-based (lower-bound) | 0 |
| S11_k141_251661 | 1300 | No | NA | 3 | 2 | 0 | Low-quality | Genome-fragment | 2.67 | AAI-based (high-confidence) | 0 |
| S1-final-viral-combined_k141_238583\|\|full | 1300 | No | NA | 3 | 0 | 0 | Low-quality | Genome-fragment | 4.26 | AAI-based (high-confidence) | 0 |
| S1-final-viral-combined_k141_382828\|\|full | 3208 | No | NA | 3 | 0 | 2 | Not-determined | Genome-fragment | NA | NA | 0 |
| S1-final-viral-combined_k141_53661\|\|full | 1245 | No | NA | 2 | 0 | 0 | Not-determined | Genome-fragment | NA | NA | 0 |
| S1-final-viral-combined_k141_366471\|\|full | 3089 | No | NA | 2 | 0 | 2 | Not-determined | Genome-fragment | NA | NA | 0 |
| S1-final-viral-combined_k141_382860\|\|full | 5283 | No | NA | 6 | 0 | 1 | Not-determined | Genome-fragment | NA | NA | 0 |
| S3-final-viral-combined_k141_10332\|\|full | 1300 | No | NA | 3 | 0 | 1 | Low-quality | Genome-fragment | 1.23 | AAI-based (high-confidence) | 0 |
| S1-final-viral-combined_k141_366492\|\|full | 1134 | No | NA | 2 | 0 | 0 | Not-determined | Genome-fragment | NA | NA | 0 |
| S7_k141_8047 | 1300 | No | NA | 3 | 2 | 0 | Low-quality | Genome-fragment | 2.94 | AAI-based (high-confidence) | 0 |
| S1-final-viral-combined_k141_366494\|\|full | 3124 | No | NA | 3 | 0 | 3 | Not-determined | Genome-fragment | NA | NA | 0 |
| S12-final-viral-combined_k141_154155\|\|lt2gene | 1300 | No | NA | 2 | 2 | 0 | Low-quality | Genome-fragment | 2.82 | AAI-based (medium-confidence) | 0 |
| S1-final-viral-combined_k141_120067\|\|full | 2792 | No | NA | 3 | 0 | 2 | Not-determined | Genome-fragment | NA | NA | 0 |
| S2-final-viral-combined_k141_184482\|\|full | 1300 | No | NA | 3 | 1 | 0 | Low-quality | Genome-fragment | 2.83 | HMM-based (lower-bound) | 0 |
| S2-final-viral-combined_k141_132115\|\|full | 1300 | No | NA | 4 | 1 | 0 | Low-quality | Genome-fragment | 3.48 | AAI-based (medium-confidence) | 0 |
| S4-final-viral-combined_k141_18294\|\|full | 1300 | No | NA | 4 | 0 | 0 | Low-quality | Genome-fragment | 0.88 | AAI-based (medium-confidence) | 0 |
| S6-final-viral-combined_k141_77130\|\|full | 1300 | No | NA | 2 | 0 | 0 | Low-quality | Genome-fragment | 1.02 | AAI-based (medium-confidence) | 0 |
| S1-final-viral-combined_k141_382889\|\|full | 1417 | No | NA | 2 | 0 | 0 | Not-determined | Genome-fragment | NA | NA | 0 |
| S1-final-viral-combined_k141_350434\|\|full | 3818 | No | NA | 3 | 0 | 2 | Not-determined | Genome-fragment | NA | NA | 0 |
| S1-final-viral-combined_k141_185969\|\|full | 1079 | No | NA | 3 | 0 | 0 | Not-determined | Genome-fragment | NA | NA | 0 |
| S6-final-viral-combined_k141_75414\|\|full | 1300 | No | NA | 2 | 0 | 0 | Low-quality | Genome-fragment | 1.89 | AAI-based (medium-confidence) | 0 |
| S6-final-viral-combined_k141_58560\|\|full | 1300 | No | NA | 3 | 0 | 0 | Low-quality | Genome-fragment | 0.74 | AAI-based (medium-confidence) | 0 |
| S1-final-viral-combined_k141_350443\|\|full | 1810 | No | NA | 3 | 0 | 2 | Not-determined | Genome-fragment | NA | NA | 0 |
| S1-final-viral-combined_k141_317684\|\|full | 3134 | No | NA | 3 | 0 | 3 | Not-determined | Genome-fragment | NA | NA | 0 |
| S8-final-viral-combined_k141_69362\|\|full | 1300 | No | NA | 3 | 0 | 1 | Low-quality | Genome-fragment | 0.73 | AAI-based (medium-confidence) | 0 |
| S8-final-viral-combined_k141_120468\|\|full | 1300 | No | NA | 2 | 1 | 0 | Low-quality | Genome-fragment | 2.48 | AAI-based (medium-confidence) | 0 |
| S1-final-viral-combined_k141_37364\|\|full | 15419 | No | NA | 13 | 0 | 9 | Not-determined | Genome-fragment | NA | NA | 0 |
| S1-final-viral-combined_k141_102779\|\|full | 5413 | No | NA | 4 | 0 | 4 | Not-determined | Genome-fragment | NA | NA | 0 |
| S1-final-viral-combined_k141_102791\|\|full | 1640 | No | NA | 6 | 0 | 2 | Not-determined | Genome-fragment | NA | NA | 0 |
| S1-final-viral-combined_k141_102797\|\|full | 2497 | No | NA | 7 | 0 | 5 | Not-determined | Genome-fragment | NA | NA | 0 |
| S1-final-viral-combined_k141_53711\|\|full | 1716 | No | NA | 3 | 0 | 2 | Not-determined | Genome-fragment | NA | NA | 0 |
| S8-final-viral-combined_k141_228279\|\|full | 1300 | No | NA | 2 | 0 | 0 | Low-quality | Genome-fragment | 1.32 | AAI-based (high-confidence) | 0 |
| S11-final-viral-combined_k141_98398\|\|full | 1301 | No | NA | 3 | 1 | 0 | Low-quality | Genome-fragment | 2.99 | AAI-based (medium-confidence) | 0 |
| S1-final-viral-combined_k141_6403\|\|full | 16152 | No | NA | 15 | 0 | 10 | Not-determined | Genome-fragment | NA | NA | 0 |
| S11_k141_299058 | 1301 | No | NA | 2 | 0 | 2 | Low-quality | Genome-fragment | 1.74 | AAI-based (medium-confidence) | 0 |
| S1-final-viral-combined_k141_44703\|\|lt2gene | 1301 | No | NA | 1 | 1 | 0 | Low-quality | Genome-fragment | 2.88 | AAI-based (high-confidence) | 0 |
| S1-final-viral-combined_k141_218466\|\|full | 1369 | No | NA | 2 | 0 | 1 | Not-determined | Genome-fragment | NA | NA | 0 |
| S1-final-viral-combined_k141_69467\|\|full | 2541 | No | NA | 2 | 0 | 2 | Not-determined | Genome-fragment | NA | NA | 0 |
| S3-final-viral-combined_k141_34956\|\|full | 1301 | No | NA | 3 | 1 | 0 | Low-quality | Genome-fragment | 0.96 | HMM-based (lower-bound) | 0 |
| S1-final-viral-combined_k141_102801\|\|full | 28850 | No | NA | 39 | 0 | 3 | Medium-quality | Genome-fragment | 55.22 | AAI-based (medium-confidence) | 0 |
| S5-final-viral-combined_k141_8900\|\|full | 1301 | No | NA | 3 | 1 | 0 | Low-quality | Genome-fragment | 2.66 | AAI-based (high-confidence) | 0 |
| S1-final-viral-combined_k141_234992\|\|full | 1248 | No | NA | 2 | 0 | 1 | Not-determined | Genome-fragment | NA | NA | 0 |
| S12-final-viral-combined_k141_217780\|\|full | 1301 | No | NA | 2 | 0 | 1 | Low-quality | Genome-fragment | 0.49 | AAI-based (medium-confidence) | 0 |
| S12-final-viral-combined_k141_235250\|\|full | 1301 | No | NA | 3 | 1 | 0 | Low-quality | Genome-fragment | 0.84 | HMM-based (lower-bound) | 0 |
| S1-final-viral-combined_k141_136798\|\|full | 1573 | No | NA | 2 | 0 | 1 | Not-determined | Genome-fragment | NA | NA | 0 |
| S1-final-viral-combined_k141_120181\|\|full | 1313 | No | NA | 3 | 0 | 1 | Not-determined | Genome-fragment | NA | NA | 0 |
| S12-final-viral-combined_k141_288074\|\|full | 1301 | No | NA | 3 | 0 | 0 | Low-quality | Genome-fragment | 2.5 | AAI-based (high-confidence) | 0 |
| S1-final-viral-combined_k141_350557\|\|full | 1872 | No | NA | 2 | 0 | 1 | Not-determined | Genome-fragment | NA | NA | 0 |
| S12-final-viral-combined_k141_262697\|\|full | 1301 | No | NA | 3 | 1 | 0 | Low-quality | Genome-fragment | 2.94 | AAI-based (medium-confidence) | 0 |
| S1-final-viral-combined_k141_218529\|\|full | 2312 | No | NA | 3 | 0 | 2 | Not-determined | Genome-fragment | NA | NA | 0 |
| S1-final-viral-combined_k141_69541\|\|full | 1501 | No | NA | 2 | 0 | 1 | Not-determined | Genome-fragment | NA | NA | 0 |
| S1-final-viral-combined_k141_37450\|\|full | 2048 | No | NA | 4 | 0 | 1 | Not-determined | Genome-fragment | NA | NA | 0 |
| S4-final-viral-combined_k141_73128\|\|full | 1301 | No | NA | 1 | 0 | 1 | Low-quality | Genome-fragment | 3.4 | AAI-based (medium-confidence) | 0 |
| S1-final-viral-combined_k141_37455\|\|full | 1585 | No | NA | 3 | 0 | 2 | Not-determined | Genome-fragment | NA | NA | 0 |
| S6-final-viral-combined_k141_42525\|\|lt2gene | 1301 | No | NA | 2 | 1 | 0 | Low-quality | Genome-fragment | 4.2 | HMM-based (lower-bound) | 0 |
| S1-final-viral-combined_k141_168526\|\|full | 4162 | No | NA | 4 | 0 | 0 | Not-determined | Genome-fragment | NA | NA | 0 |
| S8-final-viral-combined_k141_198255\|\|full | 1301 | No | NA | 4 | 0 | 1 | Low-quality | Genome-fragment | 2.97 | AAI-based (medium-confidence) | 0 |
| S1-final-viral-combined_k141_37463\|\|full | 6272 | No | NA | 7 | 0 | 4 | Not-determined | Genome-fragment | NA | NA | 0 |
| S1-final-viral-combined_k141_301545\|\|full | 3874 | No | NA | 6 | 0 | 2 | Not-determined | Genome-fragment | NA | NA | 0 |
| S1-final-viral-combined_k141_235063\|\|full | 1857 | No | NA | 3 | 0 | 1 | Not-determined | Genome-fragment | NA | NA | 0 |
| S1-final-viral-combined_k141_267757\|\|full | 2010 | No | NA | 4 | 0 | 0 | Not-determined | Genome-fragment | NA | NA | 0 |
| S1-final-viral-combined_k141_86625\|\|full | 1930 | No | NA | 4 | 0 | 0 | Not-determined | Genome-fragment | NA | NA | 0 |
| S1-final-viral-combined_k141_69579\|\|full | 3336 | No | NA | 3 | 0 | 3 | Not-determined | Genome-fragment | NA | NA | 0 |
| S1-final-viral-combined_k141_53859\|\|full | 2583 | No | NA | 4 | 0 | 2 | Not-determined | Genome-fragment | NA | NA | 0 |
| S1-final-viral-combined_k141_252016\|\|full | 1028 | No | NA | 2 | 0 | 1 | Not-determined | Genome-fragment | NA | NA | 0 |
| S1-final-viral-combined_k141_168587\|\|full | 3460 | No | NA | 3 | 0 | 1 | Not-determined | Genome-fragment | NA | NA | 0 |
| S1-final-viral-combined_k141_218581\|\|full | 2391 | No | NA | 3 | 0 | 0 | Not-determined | Genome-fragment | NA | NA | 0 |
| S9-final-viral-combined_k141_97200\|\|lt2gene | 1302 | No | NA | 2 | 1 | 0 | Low-quality | Genome-fragment | 3.3 | AAI-based (high-confidence) | 0 |
| S1-final-viral-combined_k141_4564\|\|full | 1165 | No | NA | 2 | 0 | 1 | Not-determined | Genome-fragment | NA | NA | 0 |
| S1-final-viral-combined_k141_389538\|\|full | 1302 | No | NA | 3 | 0 | 1 | Low-quality | Genome-fragment | 0.63 | AAI-based (medium-confidence) | 0 |
| S1-final-viral-combined_k141_64679\|\|full | 1302 | No | NA | 3 | 0 | 1 | Low-quality | Genome-fragment | 0.89 | AAI-based (medium-confidence) | 0 |
| S3-final-viral-combined_k141_37123\|\|full | 1302 | No | NA | 2 | 0 | 0 | Low-quality | Genome-fragment | 3.61 | AAI-based (medium-confidence) | 0 |
| S1-final-viral-combined_k141_152756\|\|full | 27818 | No | NA | 20 | 0 | 16 | Not-determined | Genome-fragment | NA | NA | 0 |
| S1-final-viral-combined_k141_120296\|\|full | 3574 | No | NA | 4 | 0 | 0 | Not-determined | Genome-fragment | NA | NA | 0 |
| S1-final-viral-combined_k141_366632\|\|full | 5534 | No | NA | 4 | 0 | 3 | Not-determined | Genome-fragment | NA | NA | 0 |
| S1-final-viral-combined_k141_120300\|\|full | 2008 | No | NA | 3 | 0 | 1 | Not-determined | Genome-fragment | NA | NA | 0 |
| S1-final-viral-combined_k141_102920\|\|full | 1296 | No | NA | 2 | 0 | 0 | Not-determined | Genome-fragment | NA | NA | 0 |
| S1-final-viral-combined_k141_120311\|\|full | 1672 | No | NA | 3 | 0 | 3 | Not-determined | Genome-fragment | NA | NA | 0 |
| S1-final-viral-combined_k141_21333\|\|full | 2458 | No | NA | 3 | 0 | 2 | Not-determined | Genome-fragment | NA | NA | 0 |
| S3-final-viral-combined_k141_106407\|\|full | 1302 | No | NA | 2 | 0 | 2 | Low-quality | Genome-fragment | 0.96 | AAI-based (high-confidence) | 0 |
| S5-final-viral-combined_k141_169014\|\|full | 1302 | No | NA | 3 | 1 | 2 | Low-quality | Genome-fragment | 1.39 | AAI-based (medium-confidence) | 0 |
| S7-final-viral-combined_k141_116722\|\|lt2gene | 1302 | No | NA | 2 | 1 | 0 | Low-quality | Genome-fragment | 2.62 | AAI-based (high-confidence) | 0 |
| S7_k141_226280 | 1302 | No | NA | 3 | 0 | 0 | Low-quality | Genome-fragment | 2.73 | AAI-based (high-confidence) | 0 |
| S1-final-viral-combined_k141_53869\|\|full | 1742 | No | NA | 2 | 0 | 2 | Not-determined | Genome-fragment | NA | NA | 0 |
| S1-final-viral-combined_k141_317890\|\|full | 11345 | No | NA | 9 | 0 | 8 | Not-determined | Genome-fragment | NA | NA | 0 |
| S17-k141_95930 | 1302 | No | NA | 2 | 1 | 0 | Low-quality | Genome-fragment | 3.39 | AAI-based (medium-confidence) | 0 |
| S4-final-viral-combined_k141_138100\|\|full | 1302 | No | NA | 1 | 1 | 0 | Low-quality | Genome-fragment | 3.11 | AAI-based (medium-confidence) | 0 |
| S1-final-viral-combined_k141_235083\|\|full | 3280 | No | NA | 6 | 0 | 1 | Not-determined | Genome-fragment | NA | NA | 0 |
| S1-final-viral-combined_k141_120348\|\|full | 2511 | No | NA | 5 | 0 | 0 | Not-determined | Genome-fragment | NA | NA | 0 |
| S1-final-viral-combined_k141_383133\|\|full | 1121 | No | NA | 2 | 0 | 1 | Not-determined | Genome-fragment | NA | NA | 0 |
| S1-final-viral-combined_k141_152828\|\|full | 5176 | No | NA | 5 | 0 | 5 | Not-determined | Genome-fragment | NA | NA | 0 |
| S8-final-viral-combined_k141_266813\|\|full | 1302 | No | NA | 3 | 3 | 0 | Low-quality | Genome-fragment | 2.85 | AAI-based (high-confidence) | 0 |
| S1-final-viral-combined_k141_267840\|\|full | 1685 | No | NA | 3 | 0 | 2 | Not-determined | Genome-fragment | NA | NA | 0 |
| S1-final-viral-combined_k141_301628\|\|full | 8878 | No | NA | 6 | 0 | 4 | Not-determined | Genome-fragment | NA | NA | 0 |
| S8_k141_105122 | 1302 | No | NA | 1 | 0 | 1 | Low-quality | Genome-fragment | 0.48 | AAI-based (medium-confidence) | 0 |
| S1-final-viral-combined_k141_202311\|\|full | 2215 | No | NA | 3 | 0 | 1 | Not-determined | Genome-fragment | NA | NA | 0 |
| S1-final-viral-combined_k141_284761\|\|full | 1127 | No | NA | 3 | 0 | 1 | Not-determined | Genome-fragment | NA | NA | 0 |
| S8_k141_151317 | 1302 | No | NA | 1 | 0 | 0 | Low-quality | Genome-fragment | 0.91 | AAI-based (medium-confidence) | 0 |
| S9-final-viral-combined_k141_93907\|\|full | 1303 | No | NA | 6 | 0 | 0 | Low-quality | Genome-fragment | 2.12 | AAI-based (medium-confidence) | 0 |
| S1-final-viral-combined_k141_350691\|\|full | 1753 | No | NA | 2 | 0 | 2 | Not-determined | Genome-fragment | NA | NA | 0 |
| S1-final-viral-combined_k141_383172\|\|full | 1799 | No | NA | 3 | 0 | 0 | Not-determined | Genome-fragment | NA | NA | 0 |
| S11-final-viral-combined_k141_122369\|\|full | 1303 | No | NA | 3 | 1 | 0 | Low-quality | Genome-fragment | 2.61 | AAI-based (high-confidence) | 0 |
| S1-final-viral-combined_k141_366710\|\|full | 13558 | No | NA | 13 | 0 | 6 | Not-determined | Genome-fragment | NA | NA | 0 |
| S1-final-viral-combined_k141_218650\|\|full | 15962 | No | NA | 13 | 0 | 9 | Not-determined | Genome-fragment | NA | NA | 0 |
| S1-final-viral-combined_k141_202359\|\|full | 2107 | No | NA | 3 | 0 | 1 | Not-determined | Genome-fragment | NA | NA | 0 |
| S1-final-viral-combined_k141_37418\|\|full | 1303 | No | NA | 2 | 0 | 1 | Low-quality | Genome-fragment | 1.77 | AAI-based (medium-confidence) | 0 |
| S1-final-viral-combined_k141_225546\|\|full | 1303 | No | NA | 3 | 0 | 0 | Low-quality | Genome-fragment | 2.25 | AAI-based (medium-confidence) | 0 |
| S5-final-viral-combined_k141_34793\|\|full | 1303 | No | NA | 4 | 3 | 0 | Low-quality | Genome-fragment | 3.34 | AAI-based (high-confidence) | 0 |
| S7-final-viral-combined_k141_95365\|\|full | 1303 | No | NA | 2 | 0 | 2 | Low-quality | Genome-fragment | 2.18 | AAI-based (medium-confidence) | 0 |
| S1-final-viral-combined_k141_366730\|\|full | 2575 | No | NA | 4 | 0 | 2 | Not-determined | Genome-fragment | NA | NA | 0 |
| S1-final-viral-combined_k141_86725\|\|full | 2748 | No | NA | 3 | 0 | 0 | Not-determined | Genome-fragment | NA | NA | 0 |
| S7-final-viral-combined_k141_224150\|\|full | 1303 | No | NA | 4 | 0 | 0 | Low-quality | Genome-fragment | 3.59 | AAI-based (medium-confidence) | 0 |
| S1-final-viral-combined_k141_4679\|\|full | 8732 | No | NA | 12 | 0 | 9 | Not-determined | Genome-fragment | NA | NA | 0 |
| S1-final-viral-combined_k141_120456\|\|full | 5308 | No | NA | 5 | 0 | 4 | Not-determined | Genome-fragment | NA | NA | 0 |
| S17-final-viral-combined_k141_256960\|\|lt2gene | 1303 | No | NA | 1 | 1 | 0 | Low-quality | Genome-fragment | 2.85 | AAI-based (high-confidence) | 0 |
| S12-final-viral-combined_k141_358321\|\|full | 1303 | No | NA | 3 | 0 | 1 | Low-quality | Genome-fragment | 3.15 | AAI-based (medium-confidence) | 0 |
| S12-final-viral-combined_k141_77027\|\|full | 1303 | No | NA | 5 | 0 | 0 | Low-quality | Genome-fragment | 2.72 | AAI-based (high-confidence) | 0 |
| S1-final-viral-combined_k141_202425\|\|full | 2201 | No | NA | 4 | 0 | 1 | Not-determined | Genome-fragment | NA | NA | 0 |
| S1-final-viral-combined_k141_235209\|\|full | 2188 | No | NA | 3 | 0 | 1 | Not-determined | Genome-fragment | NA | NA | 0 |
| S12-final-viral-combined_k141_331371\|\|full | 1303 | No | NA | 2 | 1 | 0 | Low-quality | Genome-fragment | 0.5 | AAI-based (medium-confidence) | 0 |
| S1-final-viral-combined_k141_218702\|\|full | 4002 | No | NA | 5 | 0 | 2 | Not-determined | Genome-fragment | NA | NA | 0 |
| S12-final-viral-combined_k141_103156\|\|lt2gene | 1303 | No | NA | 2 | 0 | 0 | Low-quality | Genome-fragment | 2.56 | AAI-based (high-confidence) | 0 |

| | | | | | | | | | | | |
|---|---|---|---|---|---|---|---|---|---|---|---|
| S12_k141_323514 | 1303 | No | NA | 2 | 0 | 0 | Low-quality | Genome-fragment | 3.07 | AAI-based (medium-confidence) | 0 |
| S1-final-viral-combined_k141_202439\|\|full | 6438 | No | NA | 7 | 0 | 2 | Not-determined | Genome-fragment | NA | NA | 0 |
| S1-final-viral-combined_k141_186188\|\|full | 4922 | No | NA | 2 | 0 | 2 | Not-determined | Genome-fragment | NA | NA | 0 |
| S2-final-viral-combined_k141_180810\|\|lt2gene | 1303 | No | NA | 1 | 1 | 0 | Low-quality | Genome-fragment | 0.85 | AAI-based (high-confidence) | 0 |
| S8-final-viral-combined_k141_219820\|\|full | 1303 | No | NA | 5 | 0 | 0 | Low-quality | Genome-fragment | 3.78 | AAI-based (high-confidence) | 0 |
| S8-final-viral-combined_k141_11332\|\|lt2gene | 1303 | No | NA | 2 | 1 | 0 | Low-quality | Genome-fragment | 2.24 | AAI-based (high-confidence) | 0 |
| S1-final-viral-combined_k141_333564\|\|full | 7240 | No | NA | 6 | 0 | 5 | Not-determined | Genome-fragment | NA | NA | 0 |
| S1-final-viral-combined_k141_120533\|\|full | 8687 | Yes | 5570 | 7 | 1 | 1 | High-quality | High-quality | 100 | HMM-based (lower-bound) | 35.88 |
| S1-final-viral-combined_k141_120545\|\|full | 1123 | No | NA | 2 | 0 | 0 | Not-determined | Genome-fragment | NA | NA | 0 |
| S1-final-viral-combined_k141_267892\|\|full | 4423 | No | NA | 7 | 0 | 6 | Not-determined | Genome-fragment | NA | NA | 0 |
| S11-final-viral-combined_k141_110579\|\|full | 1304 | No | NA | 2 | 0 | 0 | Low-quality | Genome-fragment | 2.52 | AAI-based (medium-confidence) | 0 |
| S1-final-viral-combined_k141_333589\|\|full | 1690 | No | NA | 3 | 0 | 2 | Not-determined | Genome-fragment | NA | NA | 0 |
| S1-final-viral-combined_k141_230977\|\|full | 1304 | No | NA | 3 | 2 | 0 | Low-quality | Genome-fragment | 1.82 | HMM-based (lower-bound) | 0 |
| S1-final-viral-combined_k141_149040\|\|full | 1304 | No | NA | 2 | 0 | 0 | Low-quality | Genome-fragment | 1.66 | AAI-based (high-confidence) | 0 |
| S1-final-viral-combined_k141_19076\|\|full | 1304 | No | NA | 2 | 0 | 0 | Low-quality | Genome-fragment | 3.22 | AAI-based (medium-confidence) | 0 |
| S1_k141_231223 | 1304 | No | NA | 2 | 0 | 1 | Low-quality | Genome-fragment | 0.7 | AAI-based (medium-confidence) | 0 |
| S1-final-viral-combined_k141_267920\|\|full | 2449 | No | NA | 4 | 0 | 2 | Not-determined | Genome-fragment | NA | NA | 0 |
| S7-final-viral-combined_k141_234962\|\|full | 1304 | No | NA | 2 | 0 | 0 | Low-quality | Genome-fragment | 3.66 | AAI-based (high-confidence) | 0 |
| S1-final-viral-combined_k141_202576\|\|full | 2500 | No | NA | 2 | 0 | 2 | Not-determined | Genome-fragment | NA | NA | 0 |
| S1-final-viral-combined_k141_4847\|\|full | 11103 | No | NA | 11 | 0 | 10 | Not-determined | Genome-fragment | NA | NA | 0 |
| S1-final-viral-combined_k141_69837\|\|full | 4079 | No | NA | 6 | 0 | 0 | Not-determined | Genome-fragment | NA | NA | 0 |
| S12-final-viral-combined_k141_150338\|\|full | 1304 | No | NA | 4 | 0 | 2 | Low-quality | Genome-fragment | 2.92 | AAI-based (medium-confidence) | 0 |
| S1-final-viral-combined_k141_4867\|\|full | 2952 | No | NA | 4 | 0 | 1 | Not-determined | Genome-fragment | NA | NA | 0 |
| S1-final-viral-combined_k141_86880\|\|full | 3353 | No | NA | 4 | 0 | 2 | Not-determined | Genome-fragment | NA | NA | 0 |
| S4_k141_49987 | 1304 | No | NA | 2 | 1 | 0 | Low-quality | Genome-fragment | 0.75 | AAI-based (high-confidence) | 0 |
| S11-final-viral-combined_k141_98023\|\|full | 1305 | No | NA | 3 | 0 | 0 | Low-quality | Genome-fragment | 3.51 | AAI-based (medium-confidence) | 0 |
| S1-final-viral-combined_k141_169012\|\|full | 2570 | No | NA | 3 | 0 | 0 | Not-determined | Genome-fragment | NA | NA | 0 |
| S1-final-viral-combined_k141_239263\|\|full | 1305 | No | NA | 3 | 0 | 0 | Low-quality | Genome-fragment | 1.77 | AAI-based (medium-confidence) | 0 |
| S1-final-viral-combined_k141_302001\|\|full | 7775 | No | NA | 3 | 0 | 3 | Not-determined | Genome-fragment | NA | NA | 0 |
| S1-final-viral-combined_k141_218938\|\|full | 1239 | No | NA | 3 | 0 | 0 | Not-determined | Genome-fragment | NA | NA | 0 |
| S1-final-viral-combined_k141_103146\|\|full | 3003 | No | NA | 4 | 0 | 3 | Not-determined | Genome-fragment | NA | NA | 0 |
| S1-final-viral-combined_k141_302003\|\|full | 3242 | No | NA | 5 | 0 | 0 | Not-determined | Genome-fragment | NA | NA | 0 |
| S1-final-viral-combined_k141_292363\|\|lt2gene | 1305 | No | NA | 2 | 1 | 0 | Low-quality | Genome-fragment | 25.65 | AAI-based (high-confidence) | 0 |
| S12-final-viral-combined_k141_9231\|\|full | 1305 | No | NA | 3 | 3 | 0 | Low-quality | Genome-fragment | 2.78 | AAI-based (high-confidence) | 0 |
| S1-final-viral-combined_k141_284782\|\|full | 1165 | No | NA | 4 | 0 | 0 | Not-determined | Genome-fragment | NA | NA | 0 |
| S1-final-viral-combined_k141_103157\|\|full | 2664 | No | NA | 5 | 0 | 2 | Not-determined | Genome-fragment | NA | NA | 0 |
| S12_k141_358732 | 1305 | No | NA | 1 | 1 | 0 | Low-quality | Genome-fragment | 2.93 | AAI-based (high-confidence) | 0 |
| S6-final-viral-combined_k141_71736\|\|full | 1305 | No | NA | 3 | 0 | 0 | Low-quality | Genome-fragment | 3.69 | AAI-based (high-confidence) | 0 |
| S1-final-viral-combined_k141_17010\|\|full | 1306 | No | NA | 4 | 0 | 0 | Low-quality | Genome-fragment | 1.01 | AAI-based (medium-confidence) | 0 |
| S1-final-viral-combined_k141_318268\|\|full | 3755 | No | NA | 6 | 0 | 6 | Not-determined | Genome-fragment | NA | NA | 0 |
| S5-final-viral-combined_k141_145209\|\|full | 1306 | No | NA | 2 | 2 | 0 | Low-quality | Genome-fragment | 0.69 | HMM-based (lower-bound) | 0 |
| S1-final-viral-combined_k141_202678\|\|full | 2525 | No | NA | 3 | 0 | 1 | Not-determined | Genome-fragment | NA | NA | 0 |
| S1-final-viral-combined_k141_318278\|\|full | 1072 | No | NA | 2 | 0 | 1 | Not-determined | Genome-fragment | NA | NA | 0 |
| S1-final-viral-combined_k141_4934\|\|full | 1453 | No | NA | 2 | 0 | 0 | Not-determined | Genome-fragment | NA | NA | 0 |
| S7_k141_124620 | 1306 | No | NA | 1 | 0 | 0 | Low-quality | Genome-fragment | 2.85 | AAI-based (high-confidence) | 0 |
| S1-final-viral-combined_k141_284811\|\|full | 3450 | No | NA | 3 | 0 | 3 | Not-determined | Genome-fragment | NA | NA | 0 |
| [illegible] | | | | | | | | | | | |

| | | | | | | | | | | | |
|---|---|---|---|---|---|---|---|---|---|---|---|
| S1-final-viral-combined_k141_253642\|\|full | 1489 | No | NA | 3 | 0 | 2 | Not-determined | Genome-fragment | NA | NA | 0 |
| S1_k141_291681 | 1320 | No | NA | 3 | 1 | 0 | Low-quality | Genome-fragment | 2.26 | AAI-based (high-confidence) | 0 |
| S3-final-viral-combined_k141_59265\|\|lt2gene | 1320 | No | NA | 1 | 1 | 0 | Low-quality | Genome-fragment | 2.6 | AAI-based (medium-confidence) | 0 |
| S1-final-viral-combined_k141_352329\|\|full | 1448 | No | NA | 2 | 0 | 1 | Not-determined | Genome-fragment | NA | NA | 0 |
| S7-final-viral-combined_k141_134296\|\|full | 1320 | No | NA | 3 | 0 | 0 | Low-quality | Genome-fragment | 3.15 | AAI-based (medium-confidence) | 0 |
| S1-final-viral-combined_k141_55415\|\|full | 2894 | No | NA | 2 | 0 | 2 | Not-determined | Genome-fragment | NA | NA | 0 |
| S7-final-viral-combined_k141_155553\|\|lt2gene | 1320 | No | NA | 2 | 1 | 0 | Low-quality | Genome-fragment | 2.78 | AAI-based (high-confidence) | 0 |
| S17-final-viral-combined_k141_222413\|\|lt2gene | 1320 | No | NA | 2 | 1 | 0 | Low-quality | Genome-fragment | 2.92 | AAI-based (high-confidence) | 0 |
| S12-final-viral-combined_k141_138323\|\|full | 1320 | No | NA | 3 | 0 | 0 | Low-quality | Genome-fragment | 2.03 | AAI-based (medium-confidence) | 0 |
| S1-final-viral-combined_k141_6158\|\|full | 1751 | No | NA | 4 | 0 | 2 | Not-determined | Genome-fragment | NA | NA | 0 |
| S1-final-viral-combined_k141_154109\|\|full | 2476 | No | NA | 4 | 0 | 0 | Not-determined | Genome-fragment | NA | NA | 0 |
| S1-final-viral-combined_k141_352371\|\|full | 2414 | No | NA | 3 | 0 | 2 | Not-determined | Genome-fragment | NA | NA | 0 |
| S8-final-viral-combined_k141_105590\|\|full | 1320 | No | NA | 2 | 0 | 0 | Low-quality | Genome-fragment | 0.45 | AAI-based (medium-confidence) | 0 |
| S1-final-viral-combined_k141_204097\|\|full | 1027 | No | NA | 3 | 0 | 0 | Not-determined | Genome-fragment | NA | NA | 0 |
| S1-final-viral-combined_k141_154138\|\|full | 6770 | No | NA | 7 | 0 | 4 | Not-determined | Genome-fragment | NA | NA | 0 |
| S1-final-viral-combined_k141_88224\|\|full | 2599 | No | NA | 4 | 0 | 0 | Not-determined | Genome-fragment | NA | NA | 0 |
| S11-final-viral-combined_k141_171872\|\|full | 1321 | No | NA | 4 | 0 | 1 | Low-quality | Genome-fragment | 1.55 | AAI-based (medium-confidence) | 0 |
| S1-final-viral-combined_k141_170125\|\|full | 2362 | No | NA | 3 | 0 | 3 | Not-determined | Genome-fragment | NA | NA | 0 |
| S11_k141_153219 | 1321 | No | NA | 4 | 0 | 1 | Low-quality | Genome-fragment | 0.36 | AAI-based (medium-confidence) | 0 |
| S1-final-viral-combined_k141_347072\|\|full | 1321 | No | NA | 3 | 0 | 0 | Low-quality | Genome-fragment | 3.06 | AAI-based (medium-confidence) | 0 |
| S1-final-viral-combined_k141_187867\|\|full | 1373 | No | NA | 2 | 0 | 2 | Not-determined | Genome-fragment | NA | NA | 0 |
| S1-final-viral-combined_k141_334900\|\|full | 1360 | No | NA | 3 | 0 | 1 | Not-determined | Genome-fragment | NA | NA | 0 |
| S1-final-viral-combined_k141_71304\|\|full | 3044 | No | NA | 3 | 0 | 1 | Not-determined | Genome-fragment | NA | NA | 0 |
| S5-final-viral-combined_k141_56100\|\|full | 1321 | No | NA | 3 | 1 | 0 | Low-quality | Genome-fragment | 3.42 | AAI-based (medium-confidence) | 0 |
| S12-final-viral-combined_k141_293130\|\|full | 1321 | No | NA | 1 | 0 | 1 | Low-quality | Genome-fragment | 2.2 | AAI-based (high-confidence) | 0 |
| S12_k141_366528 | 1321 | No | NA | 2 | 0 | 1 | Low-quality | Genome-fragment | 0.25 | AAI-based (high-confidence) | 0 |
| S1-final-viral-combined_k141_236738\|\|full | 1035 | No | NA | 2 | 0 | 0 | Not-determined | Genome-fragment | NA | NA | 0 |
| S2_k141_1430 | 1321 | No | NA | 3 | 1 | 0 | Low-quality | Genome-fragment | 3.19 | AAI-based (high-confidence) | 0 |
| S1-final-viral-combined_k141_71342\|\|full | 1080 | No | NA | 3 | 0 | 0 | Not-determined | Genome-fragment | NA | NA | 0 |
| S4_k141_62876 | 1321 | No | NA | 2 | 0 | 0 | Low-quality | Genome-fragment | 2.58 | AAI-based (medium-confidence) | 0 |
| S1-final-viral-combined_k141_269442\|\|full | 2346 | No | NA | 3 | 0 | 2 | Not-determined | Genome-fragment | NA | NA | 0 |
| S1-final-viral-combined_k141_6241\|\|full | 1168 | No | NA | 2 | 0 | 1 | Not-determined | Genome-fragment | NA | NA | 0 |
| S1-final-viral-combined_k141_71349\|\|full | 6290 | No | NA | 5 | 0 | 5 | Not-determined | Genome-fragment | NA | NA | 0 |
| S6-final-viral-combined_k141_37217\|\|full | 1321 | No | NA | 4 | 1 | 0 | Low-quality | Genome-fragment | 4.44 | AAI-based (high-confidence) | 0 |
| S11-final-viral-combined_k141_71780\|\|full | 1322 | No | NA | 3 | 0 | 1 | Low-quality | Genome-fragment | 1.96 | AAI-based (medium-confidence) | 0 |
| S1-final-viral-combined_k141_334958\|\|full | 3247 | No | NA | 3 | 0 | 2 | Not-determined | Genome-fragment | NA | NA | 0 |
| S1-final-viral-combined_k141_6287\|\|full | 2265 | No | NA | 2 | 0 | 2 | Not-determined | Genome-fragment | NA | NA | 0 |
| S11-final-viral-combined_k141_161250\|\|full | 1322 | No | NA | 4 | 0 | 0 | Low-quality | Genome-fragment | 1.92 | AAI-based (high-confidence) | 0 |
| S1-final-viral-combined_k141_368164\|\|full | 5465 | No | NA | 5 | 0 | 1 | Not-determined | Genome-fragment | NA | NA | 0 |
| S11-final-viral-combined_k141_320817\|\|lt2gene | 1322 | No | NA | 2 | 2 | 0 | Low-quality | Genome-fragment | 1.88 | HMM-based (lower-bound) | 0 |
| S1-final-viral-combined_k141_138381\|\|full | 1960 | No | NA | 2 | 0 | 2 | Not-determined | Genome-fragment | NA | NA | 0 |
| S11_k141_249518 | 1322 | No | NA | 3 | 0 | 0 | Low-quality | Genome-fragment | 2.64 | AAI-based (medium-confidence) | 0 |
| S1-final-viral-combined_k141_241671\|\|lt2gene | 1322 | No | NA | 1 | 1 | 0 | Low-quality | Genome-fragment | 2.16 | HMM-based (lower-bound) | 0 |
| S1-final-viral-combined_k141_88337\|\|full | 2282 | No | NA | 4 | 0 | 0 | Not-determined | Genome-fragment | NA | NA | 0 |
| S1-final-viral-combined_k141_138393\|\|full | 3534 | No | NA | 5 | 0 | 0 | Not-determined | Genome-fragment | NA | NA | 0 |
| S7-final-viral-combined_k141_205879\|\|full | 1322 | No | NA | 2 | 0 | 2 | Low-quality | Genome-fragment | 0.78 | AAI-based (medium-confidence) | 0 |
| S1-final-viral-combined_k141_88380\|\|full | 1579 | No | NA | 3 | 0 | 1 | Not-determined | Genome-fragment | NA | NA | 0 |
| S17-final-viral-combined_k141_239966\|\|full | 1322 | No | NA | 2 | 0 | 0 | Low-quality | Genome-fragment | 2.37 | AAI-based (medium-confidence) | 0 |
| S1-final-viral-combined_k141_204228\|\|full | 3133 | No | NA | 4 | 0 | 2 | Not-determined | Genome-fragment | NA | NA | 0 |
| S4-final-viral-combined_k141_80025\|\|full | 1322 | No | NA | 3 | 2 | 0 | Low-quality | Genome-fragment | 2.41 | AAI-based (high-confidence) | 0 |
| S1-final-viral-combined_k141_71457\|\|full | 2191 | No | NA | 3 | 0 | 2 | Not-determined | Genome-fragment | NA | NA | 0 |
| S1-final-viral-combined_k141_71467\|\|full | 1335 | No | NA | 2 | 0 | 1 | Not-determined | Genome-fragment | NA | NA | 0 |
| S1-final-viral-combined_k141_335033\|\|full | 3609 | No | NA | 3 | 0 | 3 | Not-determined | Genome-fragment | NA | NA | 0 |
| S6-final-viral-combined_k141_27244\|\|full | 1322 | No | NA | 3 | 0 | 0 | Low-quality | Genome-fragment | 2.81 | AAI-based (medium-confidence) | 0 |
| S9_k141_77050 | 1323 | No | NA | 1 | 0 | 0 | Low-quality | Genome-fragment | 2.21 | AAI-based (high-confidence) | 0 |
| S1-final-viral-combined_k141_188004\|\|full | 2333 | No | NA | 3 | 0 | 1 | Not-determined | Genome-fragment | NA | NA | 0 |
| S1-final-viral-combined_k141_269559\|\|full | 3015 | No | NA | 3 | 0 | 1 | Not-determined | Genome-fragment | NA | NA | 0 |
| S11-final-viral-combined_k141_195284\|\|full | 1323 | No | NA | 3 | 1 | 0 | Low-quality | Genome-fragment | 2.09 | AAI-based (high-confidence) | 0 |
| S5-final-viral-combined_k141_173411\|\|full | 1323 | No | NA | 3 | 0 | 0 | Low-quality | Genome-fragment | 1.18 | AAI-based (high-confidence) | 0 |
| S1-final-viral-combined_k141_319772\|\|full | 2371 | No | NA | 3 | 0 | 3 | Not-determined | Genome-fragment | NA | NA | 0 |
| S1-final-viral-combined_k141_385001\|\|full | 3902 | No | NA | 7 | 0 | 0 | Not-determined | Genome-fragment | NA | NA | 0 |
| S1-final-viral-combined_k141_253948\|\|full | 1905 | No | NA | 2 | 0 | 1 | Not-determined | Genome-fragment | NA | NA | 0 |
| S5-final-viral-combined_k141_200211\|\|full | 1323 | No | NA | 2 | 0 | 0 | Low-quality | Genome-fragment | 2.97 | AAI-based (medium-confidence) | 0 |
| S1-final-viral-combined_k141_236949\|\|full | 2730 | No | NA | 4 | 0 | 0 | Not-determined | Genome-fragment | NA | NA | 0 |
| S7-final-viral-combined_k141_164830\|\|full | 1323 | No | NA | 3 | 0 | 1 | Low-quality | Genome-fragment | 2.27 | AAI-based (medium-confidence) | 0 |
| S12-final-viral-combined_k141_314404\|\|lt2gene | 1323 | No | NA | 1 | 1 | 0 | Low-quality | Genome-fragment | 0.72 | AAI-based (high-confidence) | 0 |
| S1-final-viral-combined_k141_236957\|\|full | 3898 | No | NA | 3 | 0 | 3 | Not-determined | Genome-fragment | NA | NA | 0 |
| S12-final-viral-combined_k141_33182\|\|lt2gene | 1323 | No | NA | 1 | 1 | 0 | Low-quality | Genome-fragment | 0.91 | HMM-based (lower-bound) | 0 |
| S1-final-viral-combined_k141_204356\|\|full | 1105 | No | NA | 4 | 0 | 2 | Not-determined | Genome-fragment | NA | NA | 0 |
| S1-final-viral-combined_k141_236972\|\|full | 1777 | No | NA | 3 | 0 | 0 | Not-determined | Genome-fragment | NA | NA | 0 |
| S17-final-viral-combined_k141_30320\|\|full | 1324 | No | NA | 2 | 0 | 0 | Low-quality | Genome-fragment | 2.73 | AAI-based (medium-confidence) | 0 |
| S1-final-viral-combined_k141_236997\|\|full | 3461 | No | NA | 4 | 0 | 1 | Not-determined | Genome-fragment | NA | NA | 0 |
| S1-final-viral-combined_k141_269581\|\|full | 3561 | No | NA | 3 | 0 | 2 | Not-determined | Genome-fragment | NA | NA | 0 |
| S12-final-viral-combined_k141_203407\|\|full | 1324 | No | NA | 3 | 0 | 0 | Low-quality | Genome-fragment | 1.25 | AAI-based (medium-confidence) | 0 |
| S1-final-viral-combined_k141_39344\|\|full | 1041 | No | NA | 3 | 0 | 0 | Not-determined | Genome-fragment | NA | NA | 0 |
| S1-final-viral-combined_k141_368329\|\|full | 1188 | No | NA | 2 | 0 | 1 | Not-determined | Genome-fragment | NA | NA | 0 |
| S1-final-viral-combined_k141_286280\|\|full | 2694 | No | NA | 6 | 0 | 0 | Not-determined | Genome-fragment | NA | NA | 0 |
| S1-final-viral-combined_k141_22802\|\|full | 1482 | No | NA | 3 | 0 | 1 | Not-determined | Genome-fragment | NA | NA | 0 |
| S1-final-viral-combined_k141_122415\|\|full | 3573 | No | NA | 4 | 0 | 2 | Not-determined | Genome-fragment | NA | NA | 0 |
| S12_k141_398855 | 1324 | No | NA | 2 | 0 | 2 | Low-quality | Genome-fragment | 0.9 | AAI-based (medium-confidence) | 0 |
| S1-final-viral-combined_k141_55716\|\|full | 2494 | No | NA | 6 | 0 | 0 | Not-determined | Genome-fragment | NA | NA | 0 |
| S1-final-viral-combined_k141_269609\|\|full | 1403 | No | NA | 3 | 0 | 2 | Not-determined | Genome-fragment | NA | NA | 0 |
| S1-final-viral-combined_k141_220465\|\|full | 1161 | No | NA | 3 | 0 | 0 | Not-determined | Genome-fragment | NA | NA | 0 |
| S2-final-viral-combined_k141_42847\|\|full | 1324 | No | NA | 4 | 0 | 0 | Low-quality | Genome-fragment | 3.7 | AAI-based (high-confidence) | 0 |
| S1-final-viral-combined_k141_220467\|\|full | 1491 | No | NA | 3 | 0 | 0 | Not-determined | Genome-fragment | NA | NA | 0 |
| S1-final-viral-combined_k141_269614\|\|full | 3384 | No | NA | 5 | 0 | 0 | Not-determined | Genome-fragment | NA | NA | 0 |
| S2_k141_55909 | 1324 | No | NA | 2 | 0 | 0 | Low-quality | Genome-fragment | 2.13 | AAI-based (medium-confidence) | 0 |
| S1-final-viral-combined_k141_204444\|\|full | 3789 | No | NA | 7 | 0 | 3 | Not-determined | Genome-fragment | NA | NA | 0 |
| S1-final-viral-combined_k141_6516\|\|full | 3905 | No | NA | 5 | 0 | 1 | Not-determined | Genome-fragment | NA | NA | 0 |
| S1-final-viral-combined_k141_237077\|\|full | 3517 | No | NA | 3 | 0 | 2 | Not-determined | Genome-fragment | NA | NA | 0 |
| S4-final-viral-combined_k141_93550\|\|full | 1324 | No | NA | 2 | 0 | 1 | Low-quality | Genome-fragment | 2.5 | AAI-based (medium-confidence) | 0 |
| S11-final-viral-combined_k141_216036\|\|full | 1325 | No | NA | 4 | 3 | 0 | Low-quality | Genome-fragment | 6.25 | AAI-based (medium-confidence) | 0 |
| S1-final-viral-combined_k141_55767\|\|full | 1984 | No | NA | 3 | 0 | 0 | Not-determined | Genome-fragment | NA | NA | 0 |
| S1-final-viral-combined_k141_4446\|\|full | 1325 | No | NA | 3 | 0 | 0 | Low-quality | Genome-fragment | 0.62 | AAI-based (medium-confidence) | 0 |
| S1-final-viral-combined_k141_154436\|\|full | 1325 | No | NA | 3 | 1 | 0 | Low-quality | Genome-fragment | 2.37 | AAI-based (high-confidence) | 0 |
| S1_k141_321268 | 1325 | No | NA | 4 | 0 | 0 | Low-quality | Genome-fragment | 2.92 | AAI-based (high-confidence) | 0 |
| S1-final-viral-combined_k141_6542\|\|full | 1314 | No | NA | 3 | 0 | 0 | Not-determined | Genome-fragment | NA | NA | 0 |
| S1-final-viral-combined_k141_319959\|\|full | 1412 | No | NA | 3 | 0 | 1 | Not-determined | Genome-fragment | NA | NA | 0 |
| S3_k141_92733 | 1325 | No | NA | 2 | 1 | 0 | Low-quality | Genome-fragment | 0.61 | AAI-based (high-confidence) | 0 |
| S1-final-viral-combined_k141_55824\|\|full | 1234 | No | NA | 2 | 0 | 1 | Not-determined | Genome-fragment | NA | NA | 0 |
| S1-final-viral-combined_k141_71645\|\|full | 1476 | No | NA | 3 | 0 | 0 | Not-determined | Genome-fragment | NA | NA | 0 |
| S5-final-viral-combined_k141_129521\|\|full | 1325 | No | NA | 3 | 0 | 0 | Low-quality | Genome-fragment | 1.73 | AAI-based (high-confidence) | 0 |
| S17-k141_164207 | 1325 | No | NA | 2 | 1 | 0 | Low-quality | Genome-fragment | 1.95 | AAI-based (high-confidence) | 0 |
| S4-final-viral-combined_k141_69730\|\|full | 1325 | No | NA | 2 | 0 | 0 | Low-quality | Genome-fragment | 0.77 | AAI-based (medium-confidence) | 0 |
| S1-final-viral-combined_k141_319988\|\|full | 2929 | No | NA | 4 | 0 | 2 | Not-determined | Genome-fragment | NA | NA | 0 |
| S1-final-viral-combined_k141_39461\|\|full | 2406 | No | NA | 3 | 0 | 2 | Not-determined | Genome-fragment | NA | NA | 0 |
| S1-final-viral-combined_k141_237175\|\|full | 3330 | No | NA | 4 | 0 | 4 | Not-determined | Genome-fragment | NA | NA | 0 |
| S4-final-viral-combined_k141_55369\|\|full | 1325 | Yes | 1053 | 2 | 1 | 1 | Low-quality | Genome-fragment | 0.42 | HMM-based (lower-bound) | 20.53 |
| S1-final-viral-combined_k141_279200\|\|lt2gene | 1326 | No | NA | 2 | 0 | 0 | Low-quality | Genome-fragment | 2.29 | AAI-based (medium-confidence) | 0 |
| S1-final-viral-combined_k141_154423\|\|full | 3502 | No | NA | 7 | 0 | 0 | Not-determined | Genome-fragment | NA | NA | 0 |
| S12-final-viral-combined_k141_173211\|\|full | 1326 | No | NA | 3 | 0 | 0 | Low-quality | Genome-fragment | 2.81 | AAI-based (high-confidence) | 0 |
| S1-final-viral-combined_k141_385327\|\|full | 1122 | No | NA | 3 | 0 | 0 | Not-determined | Genome-fragment | NA | NA | 0 |
| S12-final-viral-combined_k141_6284\|\|full | 1326 | No | NA | 3 | 0 | 0 | Low-quality | Genome-fragment | 1.61 | AAI-based (high-confidence) | 0 |
| S1-final-viral-combined_k141_335205\|\|full | 1369 | No | NA | 2 | 0 | 0 | Not-determined | Genome-fragment | NA | NA | 0 |
| S1-final-viral-combined_k141_71697\|\|full | 2815 | No | NA | 5 | 0 | 1 | Not-determined | Genome-fragment | NA | NA | 0 |
| S12-final-viral-combined_k141_195791\|\|lt2gene | 1326 | No | NA | 2 | 1 | 0 | Low-quality | Genome-fragment | 2.85 | AAI-based (medium-confidence) | 0 |
| S6-final-viral-combined_k141_119352\|\|full | 1326 | No | NA | 3 | 0 | 0 | Low-quality | Genome-fragment | 1.39 | AAI-based (medium-confidence) | 0 |
| S8-final-viral-combined_k141_125689\|\|full | 1326 | No | NA | 3 | 0 | 0 | Low-quality | Genome-fragment | 3.08 | AAI-based (high-confidence) | 0 |
| S1-final-viral-combined_k141_204572\|\|full | 2335 | No | NA | 3 | 0 | 1 | Not-determined | Genome-fragment | NA | NA | 0 |
| S1-final-viral-combined_k141_71711\|\|full | 2555 | No | NA | 3 | 0 | 1 | Not-determined | Genome-fragment | NA | NA | 0 |
| S1-final-viral-combined_k141_385364\|\|full | 3816 | No | NA | 6 | 0 | 1 | Not-determined | Genome-fragment | NA | NA | 0 |
| S1-final-viral-combined_k141_154463\|\|full | 1601 | No | NA | 3 | 0 | 0 | Not-determined | Genome-fragment | NA | NA | 0 |
| S1-final-viral-combined_k141_335225\|\|full | 2051 | No | NA | 3 | 0 | 3 | Not-determined | Genome-fragment | NA | NA | 0 |
| S1-final-viral-combined_k141_352880\|\|full | 3316 | No | NA | 4 | 0 | 3 | Not-determined | Genome-fragment | NA | NA | 0 |
| S1-final-viral-combined_k141_320039\|\|full | 11688 | No | NA | 7 | 0 | 1 | Not-determined | Genome-fragment | NA | NA | 0 |
| S9-final-viral-combined_k141_83619\|\|full | 1327 | No | NA | 3 | 0 | 0 | Low-quality | Genome-fragment | 3.3 | AAI-based (high-confidence) | 0 |
| S1-final-viral-combined_k141_303562\|\|full | 1469 | No | NA | 2 | 0 | 0 | Not-determined | Genome-fragment | NA | NA | 0 |
| S1-final-viral-combined_k141_138779\|\|full | 2099 | No | NA | 3 | 0 | 1 | Not-determined | Genome-fragment | NA | NA | 0 |
| S1-final-viral-combined_k141_55885\|\|full | 1331 | No | NA | 2 | 0 | 2 | Not-determined | Genome-fragment | NA | NA | 0 |
| S1-final-viral-combined_k141_237252\|\|full | 2423 | No | NA | 2 | 0 | 2 | Not-determined | Genome-fragment | NA | NA | 0 |
| S9_k141_109611 | 1327 | No | NA | 2 | 0 | 1 | Low-quality | Genome-fragment | 2.22 | AAI-based (medium-confidence) | 0 |
| S1-final-viral-combined_k141_320070\|\|full | 2528 | No | NA | 4 | 0 | 3 | Not-determined | Genome-fragment | NA | NA | 0 |
| S1-final-viral-combined_k141_88754\|\|full | 1340 | No | NA | 2 | 0 | 1 | Not-determined | Genome-fragment | NA | NA | 0 |
| S11-final-viral-combined_k141_292739\|\|full | 1327 | No | NA | 2 | 0 | 0 | Low-quality | Genome-fragment | 3.27 | AAI-based (high-confidence) | 0 |
| S11-final-viral-combined_k141_41873\|\|lt2gene | 1327 | No | NA | 2 | 1 | 0 | Low-quality | Genome-fragment | 2.75 | AAI-based (high-confidence) | 0 |
| S3-final-viral-combined_k141_28006\|\|full | 1327 | No | NA | 3 | 0 | 1 | Low-quality | Genome-fragment | 1.22 | AAI-based (medium-confidence) | 0 |
| S1-final-viral-combined_k141_71754\|\|full | 8817 | No | NA | 3 | 0 | 0 | Not-determined | Genome-fragment | NA | NA | 0 |
| S1-final-viral-combined_k141_368528\|\|full | 3359 | No | NA | 4 | 0 | 0 | Not-determined | Genome-fragment | NA | NA | 0 |
| S5-final-viral-combined_k141_100193\|\|full | 1327 | No | NA | 2 | 0 | 0 | Low-quality | Genome-fragment | 2.19 | AAI-based (high-confidence) | 0 |
| S1-final-viral-combined_k141_104641\|\|full | 5201 | No | NA | 4 | 0 | 4 | Not-determined | Genome-fragment | NA | NA | 0 |
| S17-final-viral-combined_k141_113062\|\|full | 1327 | No | NA | 4 | 0 | 0 | Low-quality | Genome-fragment | 3.89 | AAI-based (high-confidence) | 0 |
| S1-final-viral-combined_k141_204651\|\|full | 1418 | No | NA | 2 | 0 | 0 | Not-determined | Genome-fragment | NA | NA | 0 |
| S1-final-viral-combined_k141_254239\|\|full | 1270 | No | NA | 3 | 0 | 1 | Not-determined | Genome-fragment | NA | NA | 0 |
| S1-final-viral-combined_k141_55914\|\|full | 1016 | No | NA | 2 | 0 | 1 | Not-determined | Genome-fragment | NA | NA | 0 |
| S12-final-viral-combined_k141_254145\|\|full | 1327 | No | NA | 3 | 0 | 0 | Low-quality | Genome-fragment | 1.89 | AAI-based (medium-confidence) | 0 |
| S12_k141_316447 | 1327 | No | NA | 2 | 0 | 0 | Low-quality | Genome-fragment | 0.71 | AAI-based (high-confidence) | 0 |
| S1-final-viral-combined_k141_88783\|\|full | 4582 | No | NA | 4 | 0 | 3 | Not-determined | Genome-fragment | NA | NA | 0 |
| S1-final-viral-combined_k141_138826\|\|full | 12830 | No | NA | 14 | 0 | 11 | Not-determined | Genome-fragment | NA | NA | 0 |
| S1-final-viral-combined_k141_6714\|\|full | 1729 | No | NA | 2 | 0 | 1 | Not-determined | Genome-fragment | NA | NA | 0 |
| S1-final-viral-combined_k141_269868\|\|full | 1295 | No | NA | 2 | 0 | 1 | Not-determined | Genome-fragment | NA | NA | 0 |
| S1-final-viral-combined_k141_122680\|\|full | 1316 | No | NA | 3 | 0 | 0 | Not-determined | Genome-fragment | NA | NA | 0 |
| S1-final-viral-combined_k141_23049\|\|full | 2383 | No | NA | 2 | 0 | 2 | Not-determined | Genome-fragment | NA | NA | 0 |
| S1-final-viral-combined_k141_23065\|\|full | 1347 | No | NA | 4 | 0 | 0 | Not-determined | Genome-fragment | NA | NA | 0 |
| S4_k141_128961 | 1327 | No | NA | 2 | 1 | 0 | Low-quality | Genome-fragment | 1.11 | AAI-based (medium-confidence) | 0 |
| S1-final-viral-combined_k141_104689\|\|full | 2863 | No | NA | 3 | 0 | 0 | Not-determined | Genome-fragment | NA | NA | 0 |
| S1-final-viral-combined_k141_320142\|\|full | 1054 | No | NA | 2 | 0 | 1 | Not-determined | Genome-fragment | NA | NA | 0 |
| S1-final-viral-combined_k141_320146\|\|full | 2087 | No | NA | 2 | 0 | 1 | Not-determined | Genome-fragment | NA | NA | 0 |
| S1-final-viral-combined_k141_286411\|\|full | 1822 | No | NA | 3 | 0 | 3 | Not-determined | Genome-fragment | NA | NA | 0 |
| S8_k141_271 | 1327 | No | NA | 4 | 0 | 0 | Low-quality | Genome-fragment | 2.15 | AAI-based (high-confidence) | 0 |
| S1-final-viral-combined_k141_170521\|\|full | 2425 | No | NA | 3 | 0 | 3 | Not-determined | Genome-fragment | NA | NA | 0 |
| S9-final-viral-combined_k141_18028\|\|full | 1328 | No | NA | 4 | 1 | 0 | Low-quality | Genome-fragment | 2.89 | AAI-based (medium-confidence) | 0 |
| S1-final-viral-combined_k141_170529\|\|full | 6376 | No | NA | 7 | 0 | 4 | Not-determined | Genome-fragment | NA | NA | 0 |
| S1-final-viral-combined_k141_220777\|\|full | 1798 | No | NA | 3 | 0 | 1 | Not-determined | Genome-fragment | NA | NA | 0 |
| S1-final-viral-combined_k141_104716\|\|full | 2363 | No | NA | 4 | 0 | 2 | Not-determined | Genome-fragment | NA | NA | 0 |
| S11-final-viral-combined_k141_172357\|\|full | 1328 | No | NA | 3 | 0 | 0 | Low-quality | Genome-fragment | 1.77 | AAI-based (medium-confidence) | 0 |
| S1-final-viral-combined_k141_170552\|\|full | 7229 | No | NA | 12 | 0 | 0 | Not-determined | Genome-fragment | NA | NA | 0 |
| S11-final-viral-combined_k141_254981\|\|full | 1328 | No | NA | 6 | 0 | 0 | Low-quality | Genome-fragment | 1.17 | AAI-based (medium-confidence) | 0 |
| S1-final-viral-combined_k141_368653\|\|full | 1596 | No | NA | 3 | 0 | 0 | Not-determined | Genome-fragment | NA | NA | 0 |
| S11_k141_256167 | 1328 | No | NA | 3 | 1 | 0 | Low-quality | Genome-fragment | 1.53 | AAI-based (high-confidence) | 0 |
| S1-final-viral-combined_k141_56006\|\|full | 1810 | No | NA | 2 | 0 | 1 | Not-determined | Genome-fragment | NA | NA | 0 |
| S1-final-viral-combined_k141_56014\|\|full | 1565 | No | NA | 2 | 0 | 2 | Not-determined | Genome-fragment | NA | NA | 0 |
| S3-final-viral-combined_k141_134054\|\|full | 1328 | No | NA | 2 | 0 | 0 | Low-quality | Genome-fragment | 3.43 | AAI-based (medium-confidence) | 0 |
| S1-final-viral-combined_k141_122761\|\|full | 9263 | No | NA | 9 | 0 | 5 | Not-determined | Genome-fragment | NA | NA | 0 |
| S1-final-viral-combined_k141_254315\|\|full | 1057 | No | NA | 2 | 0 | 1 | Not-determined | Genome-fragment | NA | NA | 0 |
| S7-final-viral-combined_k141_77223\|\|full | 1328 | No | NA | 3 | 0 | 1 | Low-quality | Genome-fragment | 0.73 | AAI-based (medium-confidence) | 0 |
| S12-final-viral-combined_k141_40353\|\|full | 1328 | No | NA | 3 | 1 | 0 | Low-quality | Genome-fragment | 3.84 | AAI-based (medium-confidence) | 0 |
| S1-final-viral-combined_k141_56069\|\|full | 2100 | No | NA | 2 | 0 | 1 | Not-determined | Genome-fragment | NA | NA | 0 |
| S12-final-viral-combined_k141_394747\|\|full | 1328 | No | NA | 3 | 0 | 0 | Low-quality | Genome-fragment | 2.57 | AAI-based (high-confidence) | 0 |
| S1-final-viral-combined_k141_88935\|\|full | 1795 | No | NA | 3 | 0 | 0 | Not-determined | Genome-fragment | NA | NA | 0 |
| S2-final-viral-combined_k141_132507\|\|full | 1328 | No | NA | 3 | 0 | 1 | Low-quality | Genome-fragment | 1.09 | AAI-based (high-confidence) | 0 |
| S11-final-viral-combined_k141_269216\|\|full | 1329 | No | NA | 3 | 0 | 1 | Low-quality | Genome-fragment | 2.62 | AAI-based (medium-confidence) | 0 |
| S1-final-viral-combined_k141_220828\|\|full | 1551 | No | NA | 3 | 0 | 1 | Not-determined | Genome-fragment | NA | NA | 0 |
| S11_k141_225105 | 1329 | No | NA | 2 | 0 | 1 | Low-quality | Genome-fragment | 2.17 | AAI-based (medium-confidence) | 0 |
| S1-final-viral-combined_k141_39616\|\|full | 1892 | No | NA | 2 | 0 | 1 | Not-determined | Genome-fragment | NA | NA | 0 |
| S1-final-viral-combined_k141_320324\|\|full | 2336 | No | NA | 4 | 0 | 3 | Not-determined | Genome-fragment | NA | NA | 0 |
| S1-final-viral-combined_k141_181133\|\|full | 1329 | No | NA | 3 | 0 | 2 | Low-quality | Genome-fragment | 1 | AAI-based (medium-confidence) | 0 |
| S1-final-viral-combined_k141_353113\|\|full | 2736 | No | NA | 6 | 0 | 1 | Not-determined | Genome-fragment | NA | NA | 0 |
| S1-final-viral-combined_k141_320339\|\|full | 1584 | No | NA | 3 | 0 | 1 | Not-determined | Genome-fragment | NA | NA | 0 |
| S1-final-viral-combined_k141_39641\|\|full | 4304 | No | NA | 6 | 0 | 3 | Not-determined | Genome-fragment | NA | NA | 0 |
| S1-final-viral-combined_k141_353591\|\|lt2gene | 1329 | No | NA | 2 | 1 | 0 | Low-quality | Genome-fragment | 3.08 | AAI-based (high-confidence) | 0 |
| S1-final-viral-combined_k141_220843\|\|full | 2294 | No | NA | 4 | 0 | 0 | Not-determined | Genome-fragment | NA | NA | 0 |
| S3_k141_3140 | 1329 | No | NA | 1 | 0 | 0 | Low-quality | Genome-fragment | 3.28 | AAI-based (medium-confidence) | 0 |
| S5-final-viral-combined_k141_85091\|\|full | 1329 | No | NA | 3 | 0 | 0 | Low-quality | Genome-fragment | 1.13 | AAI-based (medium-confidence) | 0 |
| S5-final-viral-combined_k141_43874\|\|full | 1329 | No | NA | 3 | 0 | 2 | Low-quality | Genome-fragment | 3.95 | AAI-based (medium-confidence) | 0 |
| S1-final-viral-combined_k141_56158\|\|full | 3130 | No | NA | 5 | 0 | 1 | Not-determined | Genome-fragment | NA | NA | 0 |
| S1-final-viral-combined_k141_138938\|\|full | 3370 | No | NA | 6 | 0 | 1 | Not-determined | Genome-fragment | NA | NA | 0 |
| S1-final-viral-combined_k141_6880\|\|full | 2517 | No | NA | 2 | 0 | 2 | Not-determined | Genome-fragment | NA | NA | 0 |
| S1-final-viral-combined_k141_368751\|\|full | 4795 | No | NA | 5 | 0 | 1 | Not-determined | Genome-fragment | NA | NA | 0 |
| S2_k141_134693 | 1329 | No | NA | 3 | 0 | 1 | Low-quality | Genome-fragment | 3.52 | AAI-based (high-confidence) | 0 |
| S1-final-viral-combined_k141_335455\|\|full | 4119 | No | NA | 4 | 0 | 3 | Not-determined | Genome-fragment | NA | NA | 0 |
| S8_k141_127274 | 1329 | No | NA | 2 | 0 | 1 | Low-quality | Genome-fragment | 2.6 | AAI-based (medium-confidence) | 0 |
| S7-final-viral-combined_k141_140089\|\|full | 1330 | No | NA | 3 | 1 | 0 | Low-quality | Genome-fragment | 2.58 | AAI-based (medium-confidence) | 0 |
| S7_k141_197583 | 1330 | No | NA | 2 | 0 | 2 | Low-quality | Genome-fragment | 0.3 | AAI-based (medium-confidence) | 0 |
| S1-final-viral-combined_k141_353148\|\|full | 8373 | No | NA | 5 | 0 | 4 | Not-determined | Genome-fragment | NA | NA | 0 |
| S1-final-viral-combined_k141_188439\|\|full | 1975 | No | NA | 3 | 0 | 1 | Not-determined | Genome-fragment | NA | NA | 0 |
| S1-final-viral-combined_k141_303886\|\|full | 1647 | No | NA | 3 | 0 | 2 | Not-determined | Genome-fragment | NA | NA | 0 |
| S1-final-viral-combined_k141_154710\|\|full | 1716 | No | NA | 3 | 0 | 2 | Not-determined | Genome-fragment | NA | NA | 0 |
| S1-final-viral-combined_k141_88984\|\|full | 2221 | No | NA | 3 | 0 | 2 | Not-determined | Genome-fragment | NA | NA | 0 |
| S7_k141_122529 | 1330 | No | NA | 2 | 0 | 1 | Low-quality | Genome-fragment | 2.64 | AAI-based (high-confidence) | 0 |
| S1-final-viral-combined_k141_188475\|\|full | 2154 | No | NA | 2 | 0 | 1 | Not-determined | Genome-fragment | NA | NA | 0 |
| S1-final-viral-combined_k141_286614\|\|full | 3265 | No | NA | 6 | 0 | 0 | Not-determined | Genome-fragment | NA | NA | 0 |
| S17-final-viral-combined_k141_114252\|\|lt2gene | 1330 | No | NA | 1 | 1 | 0 | Low-quality | Genome-fragment | 3.88 | AAI-based (high-confidence) | 0 |
| S12-final-viral-combined_k141_51789\|\|full | 1330 | No | NA | 3 | 0 | 0 | Low-quality | Genome-fragment | 2.56 | AAI-based (high-confidence) | 0 |
| S2-final-viral-combined_k141_110530\|\|full | 1330 | No | NA | 4 | 0 | 0 | Low-quality | Genome-fragment | 3.63 | AAI-based (medium-confidence) | 0 |
| S6_k141_52926 | 1330 | No | NA | 5 | 1 | 0 | Low-quality | Genome-fragment | 1.06 | HMM-based (lower-bound) | 0 |
| S1-final-viral-combined_k141_23248\|\|full | 83257 | Yes | 29089 | 90 | 23 | 35 | Complete | High-quality | 100 | Provirus (medium-confidence) | 65.06 |
| S1-final-viral-combined_k141_139011\|\|full | 1462 | No | NA | 3 | 0 | 1 | Not-determined | Genome-fragment | NA | NA | 0 |
| S1-final-viral-combined_k141_204943\|\|full | 1817 | No | NA | 3 | 0 | 1 | Not-determined | Genome-fragment | NA | NA | 0 |
| S1-final-viral-combined_k141_188509\|\|full | 3297 | No | NA | 5 | 0 | 2 | Not-determined | Genome-fragment | NA | NA | 0 |
| S6_k141_20299 | 1330 | No | NA | 2 | 0 | 0 | Low-quality | Genome-fragment | 1.5 | AAI-based (high-confidence) | 0 |
| S1-final-viral-combined_k141_102768\|\|full | 1331 | No | NA | 4 | 0 | 0 | Low-quality | Genome-fragment | 2.18 | AAI-based (high-confidence) | 0 |
| S1-final-viral-combined_k141_335515\|\|full | 1905 | No | NA | 3 | 0 | 0 | Not-determined | Genome-fragment | NA | NA | 0 |
| S1-final-viral-combined_k141_303928\|\|full | 3706 | No | NA | 7 | 0 | 1 | Not-determined | Genome-fragment | NA | NA | 0 |
| S1-final-viral-combined_k141_324904\|\|full | 1331 | No | NA | 3 | 0 | 0 | Low-quality | Genome-fragment | 4.04 | AAI-based (high-confidence) | 0 |
| S1-final-viral-combined_k141_62540\|\|full | 1331 | No | NA | 3 | 1 | 0 | Low-quality | Genome-fragment | 3.04 | AAI-based (high-confidence) | 0 |
| S1-final-viral-combined_k141_286676\|\|full | 1902 | No | NA | 4 | 0 | 0 | Not-determined | Genome-fragment | NA | NA | 0 |
| S1-final-viral-combined_k141_154779\|\|full | 2656 | No | NA | 3 | 0 | 1 | Not-determined | Genome-fragment | NA | NA | 0 |
| S3-final-viral-combined_k141_121393\|\|full | 1331 | No | NA | 3 | 0 | 0 | Low-quality | Genome-fragment | 1.87 | AAI-based (medium-confidence) | 0 |
| S7-final-viral-combined_k141_196516\|\|full | 1331 | No | NA | 2 | 0 | 0 | Low-quality | Genome-fragment | 0.82 | AAI-based (high-confidence) | 0 |
| S17-final-viral-combined_k141_111542\|\|full | 1331 | No | NA | 2 | 1 | 0 | Low-quality | Genome-fragment | 2.7 | AAI-based (high-confidence) | 0 |
| S1-final-viral-combined_k141_188545\|\|full | 10480 | No | NA | 13 | 0 | 7 | Not-determined | Genome-fragment | NA | NA | 0 |
| S1-final-viral-combined_k141_353246\|\|full | 1788 | No | NA | 2 | 0 | 1 | Not-determined | Genome-fragment | NA | NA | 0 |
| S1-final-viral-combined_k141_7081\|\|full | 3028 | No | NA | 4 | 0 | 2 | Not-determined | Genome-fragment | NA | NA | 0 |
| S1-final-viral-combined_k141_286747\|\|full | 1768 | No | NA | 3 | 0 | 1 | Not-determined | Genome-fragment | NA | NA | 0 |
| S1-final-viral-combined_k141_237642\|\|full | 15115 | No | NA | 16 | 0 | 11 | Not-determined | Genome-fragment | NA | NA | 0 |
| S1-final-viral-combined_k141_335579\|\|full | 2820 | No | NA | 4 | 0 | 2 | Not-determined | Genome-fragment | NA | NA | 0 |
| S1-final-viral-combined_k141_188615\|\|full | 1636 | No | NA | 3 | 0 | 0 | Not-determined | Genome-fragment | NA | NA | 0 |
| S1-final-viral-combined_k141_286810\|\|full | 1208 | No | NA | 2 | 0 | 0 | Not-determined | Genome-fragment | NA | NA | 0 |
| S1-final-viral-combined_k141_353292\|\|full | 1044 | No | NA | 4 | 0 | 0 | Not-determined | Genome-fragment | NA | NA | 0 |
| S1-final-viral-combined_k141_237668\|\|full | 1040 | No | NA | 2 | 0 | 0 | Not-determined | Genome-fragment | NA | NA | 0 |
| S1-final-viral-combined_k141_385639\|\|full | 4015 | No | NA | 5 | 0 | 3 | Not-determined | Genome-fragment | NA | NA | 0 |
| S17-final-viral-combined_k141_249778\|\|full | 1331 | No | NA | 4 | 0 | 0 | Low-quality | Genome-fragment | 2.27 | AAI-based (medium-confidence) | 0 |
| S1-final-viral-combined_k141_72128\|\|full | 1570 | No | NA | 3 | 0 | 1 | Not-determined | Genome-fragment | NA | NA | 0 |
| S1-final-viral-combined_k141_254645\|\|full | 2462 | No | NA | 4 | 0 | 3 | Not-determined | Genome-fragment | NA | NA | 0 |
| S8_k141_249435 | 1331 | No | NA | 2 | 1 | 0 | Low-quality | Genome-fragment | 1.59 | HMM-based (lower-bound) | 0 |
| S1-final-viral-combined_k141_254654\|\|full | 3063 | No | NA | 6 | 0 | 0 | Not-determined | Genome-fragment | NA | NA | 0 |
| S1-final-viral-combined_k141_188651\|\|full | 3225 | No | NA | 6 | 0 | 0 | Not-determined | Genome-fragment | NA | NA | 0 |
| S1-final-viral-combined_k141_254665\|\|full | 8648 | No | NA | 11 | 0 | 7 | Not-determined | Genome-fragment | NA | NA | 0 |
| S1-final-viral-combined_k141_72143\|\|full | 2209 | No | NA | 3 | 0 | 2 | Not-determined | Genome-fragment | NA | NA | 0 |
| S1-final-viral-combined_k141_72148\|\|full | 1751 | No | NA | 2 | 0 | 1 | Not-determined | Genome-fragment | NA | NA | 0 |
| S1-final-viral-combined_k141_237340\|\|full | 1332 | Yes | 675 | 2 | 1 | 1 | Low-quality | Genome-fragment | 1.12 | AAI-based (high-confidence) | 49.32 |
| S1-final-viral-combined_k141_353336\|\|full | 1297 | No | NA | 2 | 0 | 0 | Not-determined | Genome-fragment | NA | NA | 0 |
| S1-final-viral-combined_k141_205068\|\|full | 1739 | No | NA | 2 | 0 | 0 | Not-determined | Genome-fragment | NA | NA | 0 |
| S1-final-viral-combined_k141_312413\|\|full | 1332 | No | NA | 4 | 0 | 0 | Low-quality | Genome-fragment | 2.49 | AAI-based (high-confidence) | 0 |
| S1-final-viral-combined_k141_188697\|\|full | 14428 | No | NA | 11 | 0 | 9 | Not-determined | Genome-fragment | NA | NA | 0 |
| S1-final-viral-combined_k141_72167\|\|full | 34592 | No | NA | 52 | 13 | 1 | High-quality | High-quality | 100 | AAI-based (high-confidence) | 0 |
| S1_k141_188975 | 1332 | No | NA | 3 | 0 | 1 | Low-quality | Genome-fragment | 0.96 | AAI-based (medium-confidence) | 0 |
| S1_k141_241860 | 1332 | No | NA | 3 | 0 | 0 | Low-quality | Genome-fragment | 0.94 | AAI-based (high-confidence) | 0 |
| S1-final-viral-combined_k141_104942\|\|full | 7518 | No | NA | 8 | 0 | 4 | Not-determined | Genome-fragment | NA | NA | 0 |
| S1-final-viral-combined_k141_39805\|\|full | 1251 | No | NA | 2 | 0 | 2 | Not-determined | Genome-fragment | NA | NA | 0 |
| S1-final-viral-combined_k141_7160\|\|full | 2078 | No | NA | 3 | 0 | 2 | Not-determined | Genome-fragment | NA | NA | 0 |
| S5-final-viral-combined_k141_91744\|\|full | 1332 | No | NA | 3 | 0 | 0 | Low-quality | Genome-fragment | 1.42 | AAI-based (medium-confidence) | 0 |
| S5-final-viral-combined_k141_75981\|\|full | 1332 | No | NA | 5 | 0 | 0 | Low-quality | Genome-fragment | 2.68 | AAI-based (medium-confidence) | 0 |
| S1-final-viral-combined_k141_56490\|\|full | 1911 | No | NA | 2 | 0 | 1 | Not-determined | Genome-fragment | NA | NA | 0 |
| S1-final-viral-combined_k141_385740\|\|full | 1176 | No | NA | 2 | 0 | 0 | Not-determined | Genome-fragment | NA | NA | 0 |
| S5-final-viral-combined_k141_131244\|\|full | 1332 | No | NA | 3 | 1 | 0 | Low-quality | Genome-fragment | 3.19 | AAI-based (medium-confidence) | 0 |
| S1-final-viral-combined_k141_23477\|\|full | 1462 | No | NA | 2 | 0 | 2 | Not-determined | Genome-fragment | NA | NA | 0 |
| S1-final-viral-combined_k141_23489\|\|full | 1008 | No | NA | 2 | 0 | 1 | Not-determined | Genome-fragment | NA | NA | 0 |
| S5-final-viral-combined_k141_60096\|\|lt2gene | 1332 | No | NA | 2 | 1 | 0 | Low-quality | Genome-fragment | 1.73 | AAI-based (medium-confidence) | 0 |
| S17-final-viral-combined_k141_16443\|\|full | 1332 | No | NA | 3 | 0 | 0 | Low-quality | Genome-fragment | 0.83 | AAI-based (medium-confidence) | 0 |
| S12-final-viral-combined_k141_66062\|\|full | 1332 | No | NA | 4 | 0 | 1 | Low-quality | Genome-fragment | 3.34 | AAI-based (medium-confidence) | 0 |
| S1-final-viral-combined_k141_39814\|\|full | 4005 | No | NA | 6 | 0 | 4 | Not-determined | Genome-fragment | NA | NA | 0 |
| S1-final-viral-combined_k141_23497\|\|full | 1630 | No | NA | 3 | 0 | 0 | Not-determined | Genome-fragment | NA | NA | 0 |
| S1-final-viral-combined_k141_139377\|\|full | 11313 | No | NA | 11 | 0 | 9 | Not-determined | Genome-fragment | NA | NA | 0 |
| S2-final-viral-combined_k141_74119\|\|full | 1332 | No | NA | 2 | 0 | 0 | Low-quality | Genome-fragment | 1.31 | AAI-based (high-confidence) | 0 |
| S2_k141_129198 | 1332 | No | NA | 3 | 0 | 0 | Low-quality | Genome-fragment | 1.4 | AAI-based (high-confidence) | 0 |
| S4-final-viral-combined_k141_10555\|\|full | 1332 | No | NA | 4 | 2 | 0 | Low-quality | Genome-fragment | 2.91 | AAI-based (high-confidence) | 0 |
| S4_k141_127944 | 1332 | No | NA | 2 | 0 | 1 | Low-quality | Genome-fragment | 0.76 | AAI-based (medium-confidence) | 0 |
| S1-final-viral-combined_k141_39849\|\|full | 1612 | No | NA | 3 | 0 | 2 | Not-determined | Genome-fragment | NA | NA | 0 |
| S1-final-viral-combined_k141_205225\|\|full | 5252 | No | NA | 4 | 0 | 2 | Not-determined | Genome-fragment | NA | NA | 0 |
| S8-final-viral-combined_k141_269324\|\|full | 1332 | No | NA | 3 | 1 | 0 | Low-quality | Genome-fragment | 2.38 | AAI-based (high-confidence) | 0 |
| S1-final-viral-combined_k141_72339\|\|full | 2731 | No | NA | 5 | 0 | 0 | Not-determined | Genome-fragment | NA | NA | 0 |
| S9-final-viral-combined_k141_17103\|\|full | 1333 | No | NA | 3 | 0 | 1 | Low-quality | Genome-fragment | 2.28 | AAI-based (medium-confidence) | 0 |
| S1-final-viral-combined_k141_155036\|\|full | 7408 | No | NA | 7 | 0 | 5 | Not-determined | Genome-fragment | NA | NA | 0 |
| S1-final-viral-combined_k141_23553\|\|full | 1214 | No | NA | 3 | 0 | 2 | Not-determined | Genome-fragment | NA | NA | 0 |
| S1-final-viral-combined_k141_171103\|\|full | 4032 | No | NA | 6 | 0 | 1 | Not-determined | Genome-fragment | NA | NA | 0 |
| S1-final-viral-combined_k141_64499\|\|full | 1333 | No | NA | 3 | 0 | 0 | Low-quality | Genome-fragment | 3.14 | AAI-based (high-confidence) | 0 |
| S7-final-viral-combined_k141_83292\|\|full | 1333 | No | NA | 2 | 0 | 0 | Low-quality | Genome-fragment | 1.07 | AAI-based (medium-confidence) | 0 |
| S17-final-viral-combined_k141_47009\|\|full | 1333 | Yes | 757 | 2 | 1 | 1 | Low-quality | Genome-fragment | 0.52 | AAI-based (high-confidence) | 43.21 |
| S1-final-viral-combined_k141_320968\|\|full | 2408 | No | NA | 3 | 0 | 0 | Not-determined | Genome-fragment | NA | NA | 0 |
| S12-final-viral-combined_k141_14311\|\|full | 1333 | No | NA | 2 | 0 | 1 | Low-quality | Genome-fragment | 2.45 | AAI-based (medium-confidence) | 0 |
| S2-final-viral-combined_k141_231917\|\|lt2gene | 1333 | No | NA | 1 | 1 | 0 | Low-quality | Genome-fragment | 1.75 | HMM-based (lower-bound) | 0 |
| S1-final-viral-combined_k141_39945\|\|full | 1218 | No | NA | 4 | 0 | 0 | Not-determined | Genome-fragment | NA | NA | 0 |
| S1-final-viral-combined_k141_335855\|\|full | 1963 | No | NA | 3 | 0 | 0 | Not-determined | Genome-fragment | NA | NA | 0 |
| S1-final-viral-combined_k141_304191\|\|full | 1531 | No | NA | 3 | 0 | 1 | Not-determined | Genome-fragment | NA | NA | 0 |
| S2_k141_101837 | 1333 | No | NA | 2 | 0 | 0 | Low-quality | Genome-fragment | 2.13 | AAI-based (medium-confidence) | 0 |
| S8-final-viral-combined_k141_204738\|\|full | 1333 | No | NA | 2 | 0 | 0 | Low-quality | Genome-fragment | 2.97 | AAI-based (medium-confidence) | 0 |
| S1-final-viral-combined_k141_254840\|\|full | 1907 | No | NA | 3 | 0 | 1 | Not-determined | Genome-fragment | NA | NA | 0 |
| S1-final-viral-combined_k141_104412\|\|full | 1334 | No | NA | 2 | 0 | 0 | Low-quality | Genome-fragment | 3.79 | AAI-based (medium-confidence) | 0 |
| S1-final-viral-combined_k141_353510\|\|full | 1792 | No | NA | 2 | 0 | 2 | Not-determined | Genome-fragment | NA | NA | 0 |
| S1-final-viral-combined_k141_178184\|\|full | 1334 | No | NA | 3 | 1 | 0 | Low-quality | Genome-fragment | 2.86 | AAI-based (high-confidence) | 0 |
| S1-final-viral-combined_k141_47860\|\|full | 1334 | No | NA | 4 | 3 | 0 | Low-quality | Genome-fragment | 2.71 | AAI-based (medium-confidence) | 0 |
| S1-final-viral-combined_k141_23590\|\|full | 2577 | No | NA | 2 | 0 | 1 | Not-determined | Genome-fragment | NA | NA | 0 |
| S1-final-viral-combined_k141_304236\|\|full | 3128 | No | NA | 4 | 0 | 4 | Not-determined | Genome-fragment | NA | NA | 0 |
| S1-final-viral-combined_k141_123387\|\|full | 2810 | No | NA | 3 | 0 | 2 | Not-determined | Genome-fragment | NA | NA | 0 |
| S1-final-viral-combined_k141_254858\|\|full | 1904 | No | NA | 2 | 0 | 0 | Not-determined | Genome-fragment | NA | NA | 0 |
| S1-final-viral-combined_k141_221262\|\|full | 1237 | No | NA | 2 | 0 | 1 | Not-determined | Genome-fragment | NA | NA | 0 |
| S5_k141_101927 | 1334 | No | NA | 1 | 0 | 1 | Low-quality | Genome-fragment | 0.39 | AAI-based (medium-confidence) | 0 |
| S7-final-viral-combined_k141_9901\|\|full | 1334 | No | NA | 3 | 1 | 0 | Low-quality | Genome-fragment | 2.47 | AAI-based (medium-confidence) | 0 |
| S1-final-viral-combined_k141_155113\|\|full | 2028 | No | NA | 2 | 0 | 0 | Not-determined | Genome-fragment | NA | NA | 0 |
| S17-k141_158645 | 1334 | No | NA | 1 | 0 | 1 | Low-quality | Genome-fragment | 3.18 | AAI-based (high-confidence) | 0 |
| S1-final-viral-combined_k141_89478\|\|full | 2734 | No | NA | 3 | 0 | 2 | Not-determined | Genome-fragment | NA | NA | 0 |
| S12-final-viral-combined_k141_301791\|\|full | 1334 | No | NA | 2 | 1 | 0 | Low-quality | Genome-fragment | 2.18 | AAI-based (high-confidence) | 0 |
| S12-final-viral-combined_k141_229955\|\|lt2gene | 1334 | No | NA | 1 | 1 | 0 | Low-quality | Genome-fragment | 0.99 | HMM-based (lower-bound) | 0 |
| S1-final-viral-combined_k141_105166\|\|full | 1366 | No | NA | 3 | 0 | 0 | Not-determined | Genome-fragment | NA | NA | 0 |
| S1-final-viral-combined_k141_40023\|\|full | 4256 | No | NA | 4 | 0 | 4 | Not-determined | Genome-fragment | NA | NA | 0 |
| S1-final-viral-combined_k141_40025\|\|full | 1292 | No | NA | 4 | 0 | 1 | Not-determined | Genome-fragment | NA | NA | 0 |
| S12_k141_227250 | 1334 | No | NA | 1 | 0 | 1 | Low-quality | Genome-fragment | 0.88 | AAI-based (high-confidence) | 0 |
| S1-final-viral-combined_k141_7397\|\|full | 4165 | No | NA | 4 | 0 | 3 | Not-determined | Genome-fragment | NA | NA | 0 |
| S1-final-viral-combined_k141_254935\|\|full | 1911 | No | NA | 4 | 0 | 1 | Not-determined | Genome-fragment | NA | NA | 0 |
| S1-final-viral-combined_k141_238091\|\|full | 3362 | No | NA | 4 | 0 | 2 | Not-determined | Genome-fragment | NA | NA | 0 |
| S1-final-viral-combined_k141_7400\|\|full | 1186 | No | NA | 2 | 0 | 0 | Not-determined | Genome-fragment | NA | NA | 0 |
| S8-final-viral-combined_k141_149016\|\|full | 1334 | No | NA | 3 | 1 | 0 | Low-quality | Genome-fragment | 0.95 | HMM-based (lower-bound) | 0 |
| S1-final-viral-combined_k141_171227\|\|full | 1061 | No | NA | 2 | 0 | 1 | Not-determined | Genome-fragment | NA | NA | 0 |
| S9-final-viral-combined_k141_95758\|\|full | 1335 | No | NA | 3 | 0 | 0 | Low-quality | Genome-fragment | 3.29 | AAI-based (medium-confidence) | 0 |
| S1-final-viral-combined_k141_238125\|\|full | 3581 | No | NA | 6 | 0 | 2 | Not-determined | Genome-fragment | NA | NA | 0 |
| S1-final-viral-combined_k141_386046\|\|full | 1003 | No | NA | 2 | 0 | 0 | Not-determined | Genome-fragment | NA | NA | 0 |
| S11-final-viral-combined_k141_170065\|\|full | 1335 | No | NA | 4 | 0 | 1 | Low-quality | Genome-fragment | 1.11 | AAI-based (medium-confidence) | 0 |
| S1-final-viral-combined_k141_335918\|\|full | 1180 | No | NA | 3 | 0 | 1 | Not-determined | Genome-fragment | NA | NA | 0 |
| S1-final-viral-combined_k141_386079\|\|full | 1614 | No | NA | 3 | 0 | 1 | Not-determined | Genome-fragment | NA | NA | 0 |
| S1-final-viral-combined_k141_56729\|\|full | 1277 | No | NA | 4 | 0 | 1 | Not-determined | Genome-fragment | NA | NA | 0 |
| S11-final-viral-combined_k141_32505\|\|full | 1335 | No | NA | 3 | 0 | 0 | Low-quality | Genome-fragment | 2.96 | AAI-based (medium-confidence) | 0 |
| S1-final-viral-combined_k141_40115\|\|full | 2384 | No | NA | 4 | 1 | 0 | Medium-quality | Genome-fragment | 66.38 | AAI-based (high-confidence) | 0 |
| S11-final-viral-combined_k141_325950\|\|full | 1335 | No | NA | 4 | 0 | 0 | Low-quality | Genome-fragment | 2.81 | AAI-based (high-confidence) | 0 |
| S1-final-viral-combined_k141_315753\|\|lt2gene | 1335 | No | NA | 2 | 2 | 0 | Low-quality | Genome-fragment | 3.16 | AAI-based (high-confidence) | 0 |
| S1-final-viral-combined_k141_221401\|\|full | 1458 | No | NA | 3 | 0 | 2 | Not-determined | Genome-fragment | NA | NA | 0 |
| S1-final-viral-combined_k141_304343\|\|full | 1178 | No | NA | 2 | 0 | 0 | Not-determined | Genome-fragment | NA | NA | 0 |
| S1-final-viral-combined_k141_287147\|\|full | 2003 | No | NA | 3 | 0 | 3 | Not-determined | Genome-fragment | NA | NA | 0 |
| S1-final-viral-combined_k141_321236\|\|full | 1456 | No | NA | 3 | 0 | 0 | Not-determined | Genome-fragment | NA | NA | 0 |
| S1-final-viral-combined_k141_23256\|\|lt2gene | 1335 | No | NA | 1 | 0 | 0 | Low-quality | Genome-fragment | 3 | AAI-based (high-confidence) | 0 |
| S5-final-viral-combined_k141_24621\|\|full | 1335 | Yes | 873 | 2 | 1 | 1 | Low-quality | Genome-fragment | 0.99 | HMM-based (lower-bound) | 34.61 |
| S12-final-viral-combined_k141_37631\|\|full | 1335 | No | NA | 3 | 0 | 0 | Low-quality | Genome-fragment | 3.09 | AAI-based (medium-confidence) | 0 |
| S12-final-viral-combined_k141_397033\|\|full | 1335 | No | NA | 4 | 0 | 0 | Low-quality | Genome-fragment | 2.1 | AAI-based (medium-confidence) | 0 |
| S2-final-viral-combined_k141_121658\|\|lt2gene | 1335 | No | NA | 2 | 1 | 0 | Low-quality | Genome-fragment | 3.27 | AAI-based (high-confidence) | 0 |
| S2_k141_170380 | 1335 | No | NA | 2 | 0 | 1 | Low-quality | Genome-fragment | 0.46 | AAI-based (medium-confidence) | 0 |
| S1-final-viral-combined_k141_270684\|\|full | 1747 | No | NA | 5 | 0 | 2 | Not-determined | Genome-fragment | NA | NA | 0 |
| S1-final-viral-combined_k141_72579\|\|full | 1745 | No | NA | 3 | 0 | 3 | Not-determined | Genome-fragment | NA | NA | 0 |
| S1-final-viral-combined_k141_205530\|\|full | 32603 | No | NA | 38 | 0 | 8 | Not-determined | Genome-fragment | NA | NA | 0 |
| S1-final-viral-combined_k141_238241\|\|full | 2301 | No | NA | 3 | 0 | 1 | Not-determined | Genome-fragment | NA | NA | 0 |
| S1-final-viral-combined_k141_139670\|\|full | 1532 | No | NA | 3 | 0 | 1 | Not-determined | Genome-fragment | NA | NA | 0 |
| S1-final-viral-combined_k141_105385\|\|full | 1454 | No | NA | 2 | 0 | 1 | Not-determined | Genome-fragment | NA | NA | 0 |
| S1-final-viral-combined_k141_270711\|\|full | 1321 | No | NA | 3 | 0 | 0 | Not-determined | Genome-fragment | NA | NA | 0 |
| S4-final-viral-combined_k141_50123\|\|full | 1335 | No | NA | 2 | 0 | 0 | Low-quality | Genome-fragment | 0.56 | AAI-based (medium-confidence) | 0 |
| S8_k141_245075 | 1335 | No | NA | 2 | 0 | 2 | Low-quality | Genome-fragment | 0.43 | AAI-based (medium-confidence) | 0 |
| S1-final-viral-combined_k141_40166\|\|full | 2131 | No | NA | 4 | 0 | 2 | Not-determined | Genome-fragment | NA | NA | 0 |
| S1-final-viral-combined_k141_72605\|\|full | 2239 | No | NA | 3 | 0 | 0 | Not-determined | Genome-fragment | NA | NA | 0 |
| S1-final-viral-combined_k141_227156\|\|full | 1336 | No | NA | 3 | 0 | 0 | Low-quality | Genome-fragment | 2.41 | AAI-based (high-confidence) | 0 |
| S1-final-viral-combined_k141_23694\|\|full | 1730 | No | NA | 2 | 0 | 2 | Not-determined | Genome-fragment | NA | NA | 0 |
| S1_k141_307378 | 1336 | No | NA | 3 | 1 | 2 | Low-quality | Genome-fragment | 3.31 | AAI-based (high-confidence) | 0 |
| S1-final-viral-combined_k141_155280\|\|full | 1423 | No | NA | 3 | 0 | 1 | Not-determined | Genome-fragment | NA | NA | 0 |
| S1_k141_174636 | 1336 | No | NA | 2 | 0 | 0 | Low-quality | Genome-fragment | 2.92 | AAI-based (high-confidence) | 0 |
| S1-final-viral-combined_k141_369324\|\|full | 2347 | No | NA | 3 | 0 | 1 | Not-determined | Genome-fragment | NA | NA | 0 |
| S3_k141_69671 | 1336 | No | NA | 3 | 0 | 1 | Low-quality | Genome-fragment | 2.23 | AAI-based (medium-confidence) | 0 |
| S5-final-viral-combined_k141_102134\|\|full | 1336 | No | NA | 4 | 0 | 0 | Low-quality | Genome-fragment | 2.79 | AAI-based (medium-confidence) | 0 |
| S7-final-viral-combined_k141_74921\|\|full | 1336 | No | NA | 3 | 0 | 0 | Low-quality | Genome-fragment | 2.47 | AAI-based (medium-confidence) | 0 |
| S1-final-viral-combined_k141_7602\|\|full | 3549 | No | NA | 6 | 0 | 0 | Not-determined | Genome-fragment | NA | NA | 0 |
| S1-final-viral-combined_k141_40201\|\|full | 1757 | No | NA | 2 | 0 | 2 | Not-determined | Genome-fragment | NA | NA | 0 |
| S1-final-viral-combined_k141_304377\|\|full | 95231 | Yes | 58512 | 129 | 31 | 29 | Complete | High-quality | 100 | Provirus (high-confidence) | 38.56 |
| S1-final-viral-combined_k141_89651\|\|full | 2227 | No | NA | 3 | 0 | 1 | Not-determined | Genome-fragment | NA | NA | 0 |
| S1-final-viral-combined_k141_386146\|\|full | 2689 | No | NA | 2 | 0 | 2 | Not-determined | Genome-fragment | NA | NA | 0 |
| S12-final-viral-combined_k141_329447\|\|full | 1336 | No | NA | 5 | 0 | 0 | Low-quality | Genome-fragment | 3.04 | AAI-based (medium-confidence) | 0 |
| S1-final-viral-combined_k141_321302\|\|full | 1017 | No | NA | 3 | 0 | 0 | Not-determined | Genome-fragment | NA | NA | 0 |
| S1-final-viral-combined_k141_155374\|\|full | 1284 | No | NA | 3 | 0 | 1 | Not-determined | Genome-fragment | NA | NA | 0 |
| S1-final-viral-combined_k141_353757\|\|full | 35912 | No | NA | 57 | 11 | 2 | High-quality | High-quality | 98.88 | AAI-based (high-confidence) | 0 |
| S12-final-viral-combined_k141_47760\|\|lt2gene | 1336 | No | NA | 2 | 1 | 0 | Low-quality | Genome-fragment | 3.76 | AAI-based (medium-confidence) | 0 |
| S1-final-viral-combined_k141_171395\|\|full | 2456 | No | NA | 3 | 0 | 3 | Not-determined | Genome-fragment | NA | NA | 0 |
| S1-final-viral-combined_k141_221553\|\|full | 3298 | No | NA | 6 | 0 | 1 | Not-determined | Genome-fragment | NA | NA | 0 |
| S1-final-viral-combined_k141_56913\|\|full | 3428 | No | NA | 3 | 0 | 2 | Not-determined | Genome-fragment | NA | NA | 0 |
| S2-final-viral-combined_k141_4300\|\|lt2gene | 1336 | No | NA | 2 | 1 | 0 | Low-quality | Genome-fragment | 2.19 | AAI-based (medium-confidence) | 0 |
| S1-final-viral-combined_k141_72675\|\|full | 9589 | No | NA | 15 | 0 | 2 | Not-determined | Genome-fragment | NA | NA | 0 |
| S4-final-viral-combined_k141_14756\|\|full | 1336 | No | NA | 1 | 0 | 0 | Low-quality | Genome-fragment | 1.07 | AAI-based (high-confidence) | 0 |
| S1-final-viral-combined_k141_7669\|\|full | 3185 | No | NA | 4 | 0 | 1 | Not-determined | Genome-fragment | NA | NA | 0 |
| S1-final-viral-combined_k141_386207\|\|full | 1965 | No | NA | 3 | 0 | 0 | Not-determined | Genome-fragment | NA | NA | 0 |
| S8-final-viral-combined_k141_231568\|\|full | 1336 | No | NA | 2 | 0 | 2 | Low-quality | Genome-fragment | 1 | AAI-based (medium-confidence) | 0 |
| S1-final-viral-combined_k141_321361\|\|full | 3361 | No | NA | 3 | 0 | 2 | Not-determined | Genome-fragment | NA | NA | 0 |
| S11-final-viral-combined_k141_45197\|\|full | 1337 | No | NA | 3 | 1 | 0 | Low-quality | Genome-fragment | 2.64 | AAI-based (medium-confidence) | 0 |
| S1-final-viral-combined_k141_188182\|\|full | 1337 | No | NA | 4 | 1 | 0 | Low-quality | Genome-fragment | 3 | AAI-based (medium-confidence) | 0 |
| S1-final-viral-combined_k141_353801\|\|full | 2219 | No | NA | 3 | 0 | 3 | Not-determined | Genome-fragment | NA | NA | 0 |
| S1-final-viral-combined_k141_189195\|\|full | 2729 | No | NA | 2 | 0 | 2 | Not-determined | Genome-fragment | NA | NA | 0 |
| S1_k141_271463 | 1337 | No | NA | 3 | 0 | 1 | Low-quality | Genome-fragment | 1.3 | AAI-based (high-confidence) | 0 |
| S1-final-viral-combined_k141_23795\|\|full | 1091 | No | NA | 1 | 0 | 1 | Not-determined | Genome-fragment | NA | NA | 0 |
| S1_k141_57521 | 1337 | No | NA | 2 | 0 | 0 | Low-quality | Genome-fragment | 3.83 | AAI-based (medium-confidence) | 0 |
| S5-final-viral-combined_k141_18360\|\|full | 1337 | No | NA | 3 | 2 | 0 | Low-quality | Genome-fragment | 2.86 | AAI-based (high-confidence) | 0 |
| S1-final-viral-combined_k141_369407\|\|full | 3325 | No | NA | 3 | 0 | 2 | Not-determined | Genome-fragment | NA | NA | 0 |
| S1-final-viral-combined_k141_287343\|\|full | 1721 | No | NA | 3 | 0 | 0 | Not-determined | Genome-fragment | NA | NA | 0 |
| S17-final-viral-combined_k141_84512\|\|full | 1337 | No | NA | 3 | 0 | 1 | Low-quality | Genome-fragment | 3.59 | AAI-based (medium-confidence) | 0 |
| S12-final-viral-combined_k141_13815\|\|full | 1337 | No | NA | 2 | 0 | 1 | Low-quality | Genome-fragment | 3.04 | AAI-based (high-confidence) | 0 |
| S12-final-viral-combined_k141_317442\|\|full | 1337 | No | NA | 3 | 0 | 1 | Low-quality | Genome-fragment | 0.69 | AAI-based (medium-confidence) | 0 |
| S1-final-viral-combined_k141_171472\|\|full | 4811 | No | NA | 2 | 0 | 1 | Not-determined | Genome-fragment | NA | NA | 0 |
| S1-final-viral-combined_k141_23829\|\|full | 4865 | No | NA | 5 | 0 | 3 | Not-determined | Genome-fragment | NA | NA | 0 |
| S1-final-viral-combined_k141_89728\|\|full | 1564 | No | NA | 3 | 0 | 2 | Not-determined | Genome-fragment | NA | NA | 0 |
| S6-final-viral-combined_k141_17839\|\|full | 1337 | No | NA | 2 | 0 | 0 | Low-quality | Genome-fragment | 2.82 | AAI-based (medium-confidence) | 0 |
| S8_k141_3746 | 1337 | No | NA | 3 | 2 | 0 | Low-quality | Genome-fragment | 4.16 | AAI-based (high-confidence) | 0 |
| S1-final-viral-combined_k141_336087\|\|full | 2192 | No | NA | 4 | 0 | 2 | Not-determined | Genome-fragment | NA | NA | 0 |
| S1-final-viral-combined_k141_221653\|\|full | 2819 | No | NA | 3 | 0 | 1 | Not-determined | Genome-fragment | NA | NA | 0 |
| S9-final-viral-combined_k141_19115\|\|lt2gene | 1338 | No | NA | 2 | 1 | 0 | Low-quality | Genome-fragment | 3.28 | AAI-based (high-confidence) | 0 |
| S11-final-viral-combined_k141_58070\|\|full | 1338 | No | NA | 2 | 0 | 1 | Low-quality | Genome-fragment | 1.4 | AAI-based (medium-confidence) | 0 |
| S1-final-viral-combined_k141_123715\|\|full | 1571 | No | NA | 2 | 0 | 1 | Not-determined | Genome-fragment | NA | NA | 0 |
| S1-final-viral-combined_k141_221664\|\|full | 2598 | No | NA | 3 | 0 | 0 | Not-determined | Genome-fragment | NA | NA | 0 |
| S1-final-viral-combined_k141_369483\|\|full | 1669 | No | NA | 3 | 0 | 0 | Not-determined | Genome-fragment | NA | NA | 0 |
| S1-final-viral-combined_k141_114690\|\|full | 1338 | No | NA | 3 | 1 | 0 | Low-quality | Genome-fragment | 1.98 | HMM-based (lower-bound) | 0 |
| S1-final-viral-combined_k141_287402\|\|full | 1489 | No | NA | 3 | 0 | 1 | Not-determined | Genome-fragment | NA | NA | 0 |
| S7-final-viral-combined_k141_14010\|\|full | 1338 | No | NA | 2 | 0 | 0 | Low-quality | Genome-fragment | 0.84 | AAI-based (medium-confidence) | 0 |

| | | | | | | | | | | | |
|---|---|---|---|---|---|---|---|---|---|---|---|
| S17-final-viral-combined_k141_184913\|\|full | 1338 | No | NA | 3 | 0 | 1 | Low-quality | Genome-fragment | 0.84 | AAI-based (medium-confidence) | 0 |
| S17-final-viral-combined_k141_173328\|\|full | 1338 | No | NA | 3 | 1 | 0 | Low-quality | Genome-fragment | 3.07 | AAI-based (high-confidence) | 0 |
| S1-final-viral-combined_k141_386329\|\|full | 1899 | No | NA | 3 | 0 | 0 | Not-determined | Genome-fragment | NA | NA | 0 |
| S1-final-viral-combined_k141_105556\|\|full | 3361 | No | NA | 4 | 0 | 3 | Not-determined | Genome-fragment | NA | NA | 0 |
| S12-final-viral-combined_k141_177253\|\|full | 1338 | No | NA | 3 | 0 | 0 | Low-quality | Genome-fragment | 4.2 | AAI-based (high-confidence) | 0 |
| S2-final-viral-combined_k141_1715\|\|full | 1338 | No | NA | 4 | 1 | 0 | Low-quality | Genome-fragment | 3.18 | AAI-based (medium-confidence) | 0 |
| S1-final-viral-combined_k141_139808\|\|full | 1041 | No | NA | 3 | 0 | 2 | Not-determined | Genome-fragment | NA | NA | 0 |
| S1-final-viral-combined_k141_139812\|\|full | 2300 | No | NA | 5 | 0 | 2 | Not-determined | Genome-fragment | NA | NA | 0 |
| S2-final-viral-combined_k141_65541\|\|full | 1338 | No | NA | 3 | 0 | 0 | Low-quality | Genome-fragment | 1.74 | AAI-based (high-confidence) | 0 |
| S1-final-viral-combined_k141_23926\|\|full | 1824 | No | NA | 3 | 0 | 3 | Not-determined | Genome-fragment | NA | NA | 0 |
| S1-final-viral-combined_k141_369564\|\|full | 2759 | No | NA | 3 | 0 | 0 | Not-determined | Genome-fragment | NA | NA | 0 |
| S1-final-viral-combined_k141_221729\|\|full | 2369 | No | NA | 2 | 0 | 1 | Not-determined | Genome-fragment | NA | NA | 0 |
| S11-final-viral-combined_k141_13220\|\|full | 1339 | No | NA | 2 | 0 | 0 | Low-quality | Genome-fragment | 1.99 | AAI-based (medium-confidence) | 0 |
| S11-final-viral-combined_k141_120576\|\|lt2gene | 1339 | No | NA | 2 | 1 | 0 | Low-quality | Genome-fragment | 2.96 | AAI-based (high-confidence) | 0 |
| S1-final-viral-combined_k141_23944\|\|full | 3188 | No | NA | 5 | 0 | 1 | Not-determined | Genome-fragment | NA | NA | 0 |
| S1-final-viral-combined_k141_336180\|\|full | 2886 | No | NA | 3 | 0 | 2 | Not-determined | Genome-fragment | NA | NA | 0 |
| S1-final-viral-combined_k141_353978\|\|full | 7324 | No | NA | 5 | 0 | 1 | Not-determined | Genome-fragment | NA | NA | 0 |
| S1-final-viral-combined_k141_309564\|\|lt2gene | 1339 | No | NA | 1 | 1 | 0 | Low-quality | Genome-fragment | 3.08 | AAI-based (high-confidence) | 0 |
| S5_k141_200554 | 1339 | No | NA | 1 | 0 | 1 | Low-quality | Genome-fragment | 4.45 | AAI-based (high-confidence) | 0 |
| S17-final-viral-combined_k141_129508\|\|full | 1339 | No | NA | 1 | 0 | 1 | Low-quality | Genome-fragment | 2.32 | AAI-based (high-confidence) | 0 |
| S1-final-viral-combined_k141_40359\|\|full | 2256 | No | NA | 3 | 0 | 1 | Not-determined | Genome-fragment | NA | NA | 0 |
| S17-k141_87581 | 1339 | No | NA | 2 | 0 | 1 | Low-quality | Genome-fragment | 1.71 | AAI-based (medium-confidence) | 0 |
| S12-final-viral-combined_k141_34491\|\|full | 1339 | No | NA | 3 | 0 | 0 | Low-quality | Genome-fragment | 2.26 | AAI-based (medium-confidence) | 0 |
| S12-final-viral-combined_k141_365480\|\|full | 1339 | No | NA | 2 | 0 | 0 | Low-quality | Genome-fragment | 1.41 | AAI-based (medium-confidence) | 0 |
| S2-final-viral-combined_k141_160671\|\|full | 1339 | No | NA | 3 | 0 | 0 | Low-quality | Genome-fragment | 3.52 | AAI-based (high-confidence) | 0 |
| S1-final-viral-combined_k141_270983\|\|full | 5619 | No | NA | 5 | 0 | 4 | Not-determined | Genome-fragment | NA | NA | 0 |
| S1-final-viral-combined_k141_72817\|\|full | 1386 | No | NA | 4 | 0 | 1 | Not-determined | Genome-fragment | NA | NA | 0 |
| S1-final-viral-combined_k141_171534\|\|full | 1652 | No | NA | 3 | 0 | 1 | Not-determined | Genome-fragment | NA | NA | 0 |
| S1-final-viral-combined_k141_238300\|\|full | 1059 | No | NA | 2 | 0 | 1 | Not-determined | Genome-fragment | NA | NA | 0 |
| S8-final-viral-combined_k141_59194\|\|full | 1339 | No | NA | 4 | 0 | 0 | Low-quality | Genome-fragment | 0.73 | AAI-based (high-confidence) | 0 |
| S9-final-viral-combined_k141_111265\|\|full | 1340 | No | NA | 3 | 0 | 2 | Low-quality | Genome-fragment | 3.25 | AAI-based (medium-confidence) | 0 |
| S1-final-viral-combined_k141_155544\|\|full | 2159 | No | NA | 3 | 0 | 1 | Not-determined | Genome-fragment | NA | NA | 0 |
| S1-final-viral-combined_k141_386428\|\|full | 3816 | No | NA | 4 | 0 | 4 | Not-determined | Genome-fragment | NA | NA | 0 |
| S1-final-viral-combined_k141_155567\|\|full | 2203 | No | NA | 3 | 0 | 1 | Not-determined | Genome-fragment | NA | NA | 0 |
| S9_k141_81047 | 1340 | No | NA | 3 | 1 | 0 | Low-quality | Genome-fragment | 2.85 | AAI-based (high-confidence) | 0 |
| S11-final-viral-combined_k141_70008\|\|full | 1340 | No | NA | 3 | 0 | 0 | Low-quality | Genome-fragment | 1.64 | AAI-based (high-confidence) | 0 |
| S1-final-viral-combined_k141_354024\|\|full | 1931 | No | NA | 3 | 0 | 1 | Not-determined | Genome-fragment | NA | NA | 0 |
| S11-final-viral-combined_k141_130239\|\|full | 1340 | No | NA | 3 | 0 | 0 | Low-quality | Genome-fragment | 2.39 | AAI-based (high-confidence) | 0 |
| S1-final-viral-combined_k141_155602\|\|full | 1800 | No | NA | 3 | 0 | 1 | Not-determined | Genome-fragment | NA | NA | 0 |
| S1-final-viral-combined_k141_238357\|\|full | 4145 | No | NA | 5 | 0 | 2 | Not-determined | Genome-fragment | NA | NA | 0 |
| S11-final-viral-combined_k141_185532\|\|full | 1340 | No | NA | 3 | 0 | 2 | Low-quality | Genome-fragment | 0.84 | AAI-based (medium-confidence) | 0 |
| S1-final-viral-combined_k141_354048\|\|full | 2865 | No | NA | 3 | 0 | 3 | Not-determined | Genome-fragment | NA | NA | 0 |
| S1-final-viral-combined_k141_189459\|\|full | 4613 | No | NA | 4 | 0 | 1 | Not-determined | Genome-fragment | NA | NA | 0 |
| S11-final-viral-combined_k141_204859\|\|full | 1340 | No | NA | 3 | 1 | 0 | Low-quality | Genome-fragment | 1.6 | AAI-based (medium-confidence) | 0 |
| S1-final-viral-combined_k141_57147\|\|full | 10578 | No | NA | 14 | 0 | 1 | Not-determined | Genome-fragment | NA | NA | 0 |
| S1-final-viral-combined_k141_306778\|\|full | 1340 | No | NA | 3 | 1 | 0 | Low-quality | Genome-fragment | 1.22 | HMM-based (lower-bound) | 0 |
| S1-final-viral-combined_k141_143289\|\|full | 1340 | No | NA | 3 | 0 | 0 | Low-quality | Genome-fragment | 1.17 | AAI-based (medium-confidence) | 0 |
| S1-final-viral-combined_k141_155639\|\|full | 1043 | No | NA | 1 | 0 | 0 | Not-determined | Genome-fragment | NA | NA | 0 |
| S1-final-viral-combined_k141_62576\|\|full | 1340 | No | NA | 3 | 0 | 0 | Low-quality | Genome-fragment | 3.68 | AAI-based (medium-confidence) | 0 |
| S1-final-viral-combined_k141_89958\|\|full | 6602 | No | NA | 2 | 0 | 1 | Not-determined | Genome-fragment | NA | NA | 0 |
| S3-final-viral-combined_k141_100425\|\|full | 1340 | No | NA | 2 | 0 | 0 | Low-quality | Genome-fragment | 3.09 | AAI-based (medium-confidence) | 0 |
| S1-final-viral-combined_k141_238388\|\|full | 1217 | No | NA | 2 | 0 | 1 | Not-determined | Genome-fragment | NA | NA | 0 |
| S1-final-viral-combined_k141_255314\|\|full | 2027 | No | NA | 2 | 0 | 1 | Not-determined | Genome-fragment | NA | NA | 0 |
| S1-final-viral-combined_k141_271072\|\|full | 1469 | No | NA | 3 | 0 | 0 | Not-determined | Genome-fragment | NA | NA | 0 |
| S1-final-viral-combined_k141_386520\|\|full | 1654 | No | NA | 2 | 0 | 2 | Not-determined | Genome-fragment | NA | NA | 0 |
| S5-final-viral-combined_k141_169476\|\|full | 1340 | No | NA | 3 | 0 | 0 | Low-quality | Genome-fragment | 0.69 | AAI-based (high-confidence) | 0 |
| S1-final-viral-combined_k141_189507\|\|full | 1073 | No | NA | 2 | 0 | 1 | Not-determined | Genome-fragment | NA | NA | 0 |
| S1-final-viral-combined_k141_89986\|\|full | 1717 | No | NA | 2 | 0 | 0 | Not-determined | Genome-fragment | NA | NA | 0 |
| S5-final-viral-combined_k141_192161\|\|full | 1340 | No | NA | 3 | 1 | 0 | Low-quality | Genome-fragment | 2.19 | AAI-based (high-confidence) | 0 |
| S17-k141_45085 | 1340 | No | NA | 2 | 0 | 2 | Low-quality | Genome-fragment | 3.42 | AAI-based (medium-confidence) | 0 |
| S12-final-viral-combined_k141_60801\|\|full | 1340 | No | NA | 2 | 0 | 0 | Low-quality | Genome-fragment | 2.74 | AAI-based (medium-confidence) | 0 |
| S1-final-viral-combined_k141_155694\|\|full | 1999 | No | NA | 2 | 0 | 1 | Not-determined | Genome-fragment | NA | NA | 0 |
| S2-final-viral-combined_k141_35519\|\|full | 1340 | No | NA | 4 | 1 | 0 | Low-quality | Genome-fragment | 2.99 | AAI-based (high-confidence) | 0 |
| S2-final-viral-combined_k141_224440\|\|full | 1340 | No | NA | 3 | 0 | 0 | Low-quality | Genome-fragment | 0.93 | AAI-based (high-confidence) | 0 |
| S1-final-viral-combined_k141_238443\|\|full | 1149 | No | NA | 2 | 0 | 1 | Not-determined | Genome-fragment | NA | NA | 0 |
| S1-final-viral-combined_k141_123964\|\|full | 9518 | No | NA | 9 | 0 | 6 | Not-determined | Genome-fragment | NA | NA | 0 |
| S1-final-viral-combined_k141_238452\|\|full | 2185 | No | NA | 3 | 0 | 2 | Not-determined | Genome-fragment | NA | NA | 0 |
| S1-final-viral-combined_k141_386580\|\|full | 3061 | No | NA | 4 | 0 | 2 | Not-determined | Genome-fragment | NA | NA | 0 |
| S2-final-viral-combined_k141_206395\|\|full | 1340 | No | NA | 2 | 0 | 2 | Low-quality | Genome-fragment | 3.39 | AAI-based (high-confidence) | 0 |
| S6-final-viral-combined_k141_50563\|\|lt2gene | 1340 | No | NA | 1 | 1 | 0 | Low-quality | Genome-fragment | 0.56 | HMM-based (lower-bound) | 0 |
| S1-final-viral-combined_k141_123973\|\|full | 2010 | No | NA | 3 | 0 | 1 | Not-determined | Genome-fragment | NA | NA | 0 |
| S9-final-viral-combined_k141_34957\|\|full | 1341 | No | NA | 4 | 0 | 0 | Low-quality | Genome-fragment | 2.29 | AAI-based (medium-confidence) | 0 |
| S1-final-viral-combined_k141_221900\|\|full | 1132 | No | NA | 3 | 0 | 0 | Not-determined | Genome-fragment | NA | NA | 0 |
| S1-final-viral-combined_k141_255378\|\|full | 1626 | No | NA | 2 | 0 | 0 | Not-determined | Genome-fragment | NA | NA | 0 |
| S1-final-viral-combined_k141_336359\|\|full | 1603 | No | NA | 3 | 0 | 2 | Not-determined | Genome-fragment | NA | NA | 0 |
| S1-final-viral-combined_k141_369702\|\|full | 5629 | No | NA | 5 | 0 | 2 | Not-determined | Genome-fragment | NA | NA | 0 |
| S11-final-viral-combined_k141_117647\|\|full | 1341 | No | NA | 3 | 0 | 0 | Low-quality | Genome-fragment | 1.22 | AAI-based (medium-confidence) | 0 |
| S1-final-viral-combined_k141_238493\|\|full | 3455 | No | NA | 3 | 0 | 3 | Not-determined | Genome-fragment | NA | NA | 0 |
| S1-final-viral-combined_k141_321697\|\|full | 2032 | No | NA | 3 | 0 | 0 | Not-determined | Genome-fragment | NA | NA | 0 |
| S11-final-viral-combined_k141_173766\|\|full | 1341 | No | NA | 3 | 1 | 0 | Low-quality | Genome-fragment | 2.84 | AAI-based (high-confidence) | 0 |
| S11-final-viral-combined_k141_270678\|\|full | 1341 | No | NA | 3 | 0 | 1 | Low-quality | Genome-fragment | 1.97 | AAI-based (medium-confidence) | 0 |
| S11_k141_69213 | 1341 | No | NA | 1 | 0 | 0 | Low-quality | Genome-fragment | 2.32 | AAI-based (high-confidence) | 0 |
| S1-final-viral-combined_k141_287643\|\|full | 2964 | No | NA | 2 | 0 | 1 | Not-determined | Genome-fragment | NA | NA | 0 |
| S1-final-viral-combined_k141_189589\|\|full | 1445 | No | NA | 3 | 0 | 1 | Not-determined | Genome-fragment | NA | NA | 0 |
| S1-final-viral-combined_k141_8058\|\|full | 1483 | No | NA | 3 | 0 | 1 | Not-determined | Genome-fragment | NA | NA | 0 |
| S1-final-viral-combined_k141_327960\|\|full | 1341 | No | NA | 4 | 0 | 0 | Low-quality | Genome-fragment | 22.83 | AAI-based (high-confidence) | 0 |
| S1-final-viral-combined_k141_237010\|\|lt2gene | 1341 | No | NA | 1 | 1 | 0 | Low-quality | Genome-fragment | 3.28 | AAI-based (high-confidence) | 0 |
| S3-final-viral-combined_k141_7521\|\|full | 1341 | No | NA | 2 | 0 | 1 | Low-quality | Genome-fragment | 1.21 | AAI-based (medium-confidence) | 0 |
| S3-final-viral-combined_k141_2234\|\|full | 1341 | No | NA | 3 | 0 | 1 | Low-quality | Genome-fragment | 3.09 | AAI-based (high-confidence) | 0 |
| S3-final-viral-combined_k141_44100\|\|lt2gene | 1341 | No | NA | 2 | 1 | 0 | Low-quality | Genome-fragment | 3.32 | AAI-based (medium-confidence) | 0 |
| S1-final-viral-combined_k141_206015\|\|full | 1856 | No | NA | 4 | 0 | 0 | Not-determined | Genome-fragment | NA | NA | 0 |
| S5-final-viral-combined_k141_111783\|\|full | 1341 | No | NA | 2 | 0 | 0 | Low-quality | Genome-fragment | 2.77 | AAI-based (high-confidence) | 0 |
| S1-final-viral-combined_k141_189619\|\|full | 1050 | No | NA | 3 | 0 | 0 | Not-determined | Genome-fragment | NA | NA | 0 |
| S5-final-viral-combined_k141_199570\|\|lt2gene | 1341 | No | NA | 1 | 1 | 0 | Low-quality | Genome-fragment | 2.45 | AAI-based (medium-confidence) | 0 |
| S1-final-viral-combined_k141_369761\|\|full | 1546 | No | NA | 2 | 0 | 0 | Not-determined | Genome-fragment | NA | NA | 0 |
| S1-final-viral-combined_k141_304839\|\|full | 1623 | No | NA | 2 | 0 | 0 | Not-determined | Genome-fragment | NA | NA | 0 |
| S1-final-viral-combined_k141_24198\|\|full | 2006 | No | NA | 2 | 0 | 1 | Not-determined | Genome-fragment | NA | NA | 0 |
| S1-final-viral-combined_k141_73082\|\|full | 1539 | No | NA | 3 | 0 | 0 | Not-determined | Genome-fragment | NA | NA | 0 |
| S7-final-viral-combined_k141_136575\|\|full | 1341 | No | NA | 4 | 2 | 0 | Low-quality | Genome-fragment | 1.61 | AAI-based (high-confidence) | 0 |
| S1-final-viral-combined_k141_386667\|\|full | 3839 | No | NA | 5 | 0 | 3 | Not-determined | Genome-fragment | NA | NA | 0 |
| S1-final-viral-combined_k141_336466\|\|full | 4891 | No | NA | 4 | 0 | 3 | Not-determined | Genome-fragment | NA | NA | 0 |
| S7-final-viral-combined_k141_202503\|\|full | 1341 | No | NA | 5 | 0 | 0 | Low-quality | Genome-fragment | 1.46 | AAI-based (medium-confidence) | 0 |
| S7-final-viral-combined_k141_193415\|\|full | 1341 | No | NA | 2 | 1 | 0 | Low-quality | Genome-fragment | 3.08 | AAI-based (medium-confidence) | 0 |
| S1-final-viral-combined_k141_369787\|\|full | 2325 | No | NA | 2 | 0 | 2 | Not-determined | Genome-fragment | NA | NA | 0 |
| S12-final-viral-combined_k141_26968\|\|full | 1341 | No | NA | 4 | 0 | 0 | Low-quality | Genome-fragment | 3.25 | AAI-based (high-confidence) | 0 |
| S12-final-viral-combined_k141_44914\|\|full | 1341 | No | NA | 2 | 0 | 2 | Low-quality | Genome-fragment | 2.32 | AAI-based (medium-confidence) | 0 |
| S1-final-viral-combined_k141_189692\|\|full | 12207 | No | NA | 14 | 0 | 6 | Not-determined | Genome-fragment | NA | NA | 0 |
| S12-final-viral-combined_k141_100021\|\|full | 1341 | No | NA | 4 | 0 | 0 | Low-quality | Genome-fragment | 1.8 | AAI-based (medium-confidence) | 0 |
| S12-final-viral-combined_k141_286806\|\|lt2gene | 1341 | No | NA | 2 | 2 | 0 | Low-quality | Genome-fragment | 1.16 | HMM-based (lower-bound) | 0 |
| S1-final-viral-combined_k141_304895\|\|full | 1663 | No | NA | 3 | 0 | 2 | Not-determined | Genome-fragment | NA | NA | 0 |
| S12_k141_291068 | 1341 | No | NA | 2 | 0 | 1 | Low-quality | Genome-fragment | 3.8 | AAI-based (medium-confidence) | 0 |
| S1-final-viral-combined_k141_321851\|\|full | 5585 | No | NA | 3 | 0 | 2 | Not-determined | Genome-fragment | NA | NA | 0 |
| S2-final-viral-combined_k141_99389\|\|full | 1341 | No | NA | 1 | 0 | 1 | Low-quality | Genome-fragment | 1.05 | AAI-based (high-confidence) | 0 |
| S2-final-viral-combined_k141_20537\|\|full | 1341 | No | NA | 3 | 0 | 0 | Low-quality | Genome-fragment | 0.92 | AAI-based (medium-confidence) | 0 |
| S1-final-viral-combined_k141_336541\|\|full | 2578 | No | NA | 4 | 0 | 1 | Not-determined | Genome-fragment | NA | NA | 0 |
| S8-final-viral-combined_k141_70024\|\|full | 1341 | No | NA | 2 | 0 | 0 | Low-quality | Genome-fragment | 2.1 | AAI-based (high-confidence) | 0 |
| S1-final-viral-combined_k141_336555\|\|full | 1012 | No | NA | 2 | 0 | 0 | Not-determined | Genome-fragment | NA | NA | 0 |
| S9-final-viral-combined_k141_6327\|\|full | 1342 | No | NA | 4 | 2 | 0 | Low-quality | Genome-fragment | 2.27 | AAI-based (high-confidence) | 0 |
| S1-final-viral-combined_k141_296812\|\|full | 1342 | No | NA | 4 | 0 | 0 | Low-quality | Genome-fragment | 1.92 | AAI-based (high-confidence) | 0 |
| S1-final-viral-combined_k141_73131\|\|full | 4383 | No | NA | 4 | 0 | 3 | Not-determined | Genome-fragment | NA | NA | 0 |
| S1-final-viral-combined_k141_8205\|\|full | 1576 | No | NA | 3 | 0 | 0 | Not-determined | Genome-fragment | NA | NA | 0 |
| S1-final-viral-combined_k141_153857\|\|lt2gene | 1342 | No | NA | 2 | 1 | 0 | Low-quality | Genome-fragment | 3.65 | AAI-based (medium-confidence) | 0 |
| S1-final-viral-combined_k141_238680\|\|full | 1760 | No | NA | 2 | 0 | 1 | Not-determined | Genome-fragment | NA | NA | 0 |
| S5-final-viral-combined_k141_78157\|\|full | 1342 | No | NA | 2 | 1 | 0 | Low-quality | Genome-fragment | 3.03 | AAI-based (medium-confidence) | 0 |
| S5_k141_68431 | 1342 | No | NA | 1 | 0 | 1 | Low-quality | Genome-fragment | 3.78 | AAI-based (medium-confidence) | 0 |
| S1-final-viral-combined_k141_354135\|\|full | 3752 | No | NA | 7 | 0 | 1 | Not-determined | Genome-fragment | NA | NA | 0 |
| S1-final-viral-combined_k141_255501\|\|full | 1802 | No | NA | 4 | 0 | 0 | Not-determined | Genome-fragment | NA | NA | 0 |
| S1-final-viral-combined_k141_140027\|\|full | 4093 | No | NA | 3 | 0 | 3 | Not-determined | Genome-fragment | NA | NA | 0 |
| S7-final-viral-combined_k141_215465\|\|full | 1342 | No | NA | 4 | 2 | 0 | Low-quality | Genome-fragment | 3.61 | AAI-based (high-confidence) | 0 |
| S1-final-viral-combined_k141_8264\|\|full | 1300 | No | NA | 2 | 0 | 0 | Not-determined | Genome-fragment | NA | NA | 0 |
| S1-final-viral-combined_k141_336640\|\|full | 2548 | No | NA | 4 | 0 | 3 | Not-determined | Genome-fragment | NA | NA | 0 |
| S7_k141_47522 | 1342 | No | NA | 2 | 0 | 1 | Low-quality | Genome-fragment | 0.75 | AAI-based (high-confidence) | 0 |
| S12-final-viral-combined_k141_262075\|\|full | 1342 | No | NA | 3 | 0 | 2 | Low-quality | Genome-fragment | 0.4 | AAI-based (high-confidence) | 0 |
| S1-final-viral-combined_k141_189784\|\|full | 1827 | No | NA | 2 | 0 | 1 | Not-determined | Genome-fragment | NA | NA | 0 |
| S1-final-viral-combined_k141_255537\|\|full | 2488 | No | NA | 3 | 0 | 1 | Not-determined | Genome-fragment | NA | NA | 0 |
| S1-final-viral-combined_k141_271270\|\|full | 1430 | No | NA | 3 | 0 | 1 | Not-determined | Genome-fragment | NA | NA | 0 |
| S1-final-viral-combined_k141_189791\|\|full | 1663 | No | NA | 3 | 0 | 2 | Not-determined | Genome-fragment | NA | NA | 0 |
| S1-final-viral-combined_k141_287883\|\|full | 2156 | No | NA | 2 | 0 | 2 | Not-determined | Genome-fragment | NA | NA | 0 |
| S1-final-viral-combined_k141_305028\|\|full | 2397 | No | NA | 3 | 0 | 1 | Not-determined | Genome-fragment | NA | NA | 0 |
| S1-final-viral-combined_k141_124184\|\|full | 4174 | No | NA | 4 | 0 | 3 | Not-determined | Genome-fragment | NA | NA | 0 |
| S1-final-viral-combined_k141_386841\|\|full | 1742 | No | NA | 2 | 0 | 2 | Not-determined | Genome-fragment | NA | NA | 0 |
| S1-final-viral-combined_k141_271282\|\|full | 1970 | No | NA | 3 | 0 | 2 | Not-determined | Genome-fragment | NA | NA | 0 |
| S12-final-viral-combined_k141_199921\|\|full | 1342 | No | NA | 3 | 0 | 0 | Low-quality | Genome-fragment | 1.16 | AAI-based (medium-confidence) | 0 |
| S1-final-viral-combined_k141_287907\|\|full | 1965 | No | NA | 4 | 0 | 0 | Not-determined | Genome-fragment | NA | NA | 0 |
| S1-final-viral-combined_k141_8317\|\|full | 4345 | No | NA | 6 | 0 | 4 | Not-determined | Genome-fragment | NA | NA | 0 |
| S1-final-viral-combined_k141_140052\|\|full | 1950 | No | NA | 4 | 0 | 1 | Not-determined | Genome-fragment | NA | NA | 0 |
| S12-final-viral-combined_k141_231534\|\|lt2gene | 1342 | No | NA | 2 | 1 | 0 | Low-quality | Genome-fragment | 3.17 | HMM-based (lower-bound) | 0 |
| S1-final-viral-combined_k141_369976\|\|full | 1070 | No | NA | 2 | 0 | 1 | Not-determined | Genome-fragment | NA | NA | 0 |
| S1-final-viral-combined_k141_171841\|\|full | 2828 | No | NA | 3 | 0 | 0 | Not-determined | Genome-fragment | NA | NA | 0 |
| S12_k141_306697 | 1342 | No | NA | 2 | 0 | 0 | Low-quality | Genome-fragment | 2.38 | AAI-based (high-confidence) | 0 |
| S1-final-viral-combined_k141_255611\|\|full | 1549 | No | NA | 2 | 0 | 0 | Not-determined | Genome-fragment | NA | NA | 0 |
| S1-final-viral-combined_k141_189869\|\|full | 3664 | No | NA | 3 | 0 | 3 | Not-determined | Genome-fragment | NA | NA | 0 |
| S12_k141_343857 | 1342 | No | NA | 1 | 0 | 0 | Low-quality | Genome-fragment | 0.74 | AAI-based (high-confidence) | 0 |
| S2-final-viral-combined_k141_163580\|\|full | 1342 | No | NA | 2 | 0 | 0 | Low-quality | Genome-fragment | 3.2 | AAI-based (medium-confidence) | 0 |
| S1-final-viral-combined_k141_354276\|\|full | 1516 | No | NA | 3 | 0 | 1 | Not-determined | Genome-fragment | NA | NA | 0 |
| S2-final-viral-combined_k141_205721\|\|full | 1342 | No | NA | 3 | 2 | 0 | Low-quality | Genome-fragment | 2.36 | AAI-based (high-confidence) | 0 |
| S1-final-viral-combined_k141_171862\|\|full | 3750 | No | NA | 3 | 0 | 3 | Not-determined | Genome-fragment | NA | NA | 0 |
| S9-final-viral-combined_k141_19706\|\|full | 1343 | No | NA | 3 | 0 | 2 | Low-quality | Genome-fragment | 1.83 | AAI-based (medium-confidence) | 0 |
| S1-final-viral-combined_k141_222105\|\|full | 1213 | No | NA | 2 | 0 | 2 | Not-determined | Genome-fragment | NA | NA | 0 |
| S1-final-viral-combined_k141_386903\|\|full | 1297 | No | NA | 3 | 0 | 1 | Not-determined | Genome-fragment | NA | NA | 0 |
| S11-final-viral-combined_k141_229280\|\|full | 1343 | No | NA | 3 | 0 | 1 | Low-quality | Genome-fragment | 2.73 | AAI-based (medium-confidence) | 0 |
| S11_k141_264131 | 1343 | No | NA | 4 | 2 | 0 | Low-quality | Genome-fragment | 1.01 | AAI-based (high-confidence) | 0 |
| S1-final-viral-combined_k141_57370\|\|full | 24795 | Yes | 20871 | 39 | 5 | 5 | Medium-quality | Genome-fragment | 59.24 | AAI-based (high-confidence) | 15.83 |
| S1-final-viral-combined_k141_262004\|\|full | 1343 | No | NA | 4 | 0 | 0 | Low-quality | Genome-fragment | 3.76 | AAI-based (medium-confidence) | 0 |
| S1-final-viral-combined_k141_171909\|\|full | 6593 | No | NA | 10 | 0 | 3 | Not-determined | Genome-fragment | NA | NA | 0 |
| S5-final-viral-combined_k141_171092\|\|full | 1343 | No | NA | 2 | 0 | 1 | Low-quality | Genome-fragment | 1.78 | AAI-based (medium-confidence) | 0 |
| S7-final-viral-combined_k141_169322\|\|full | 1343 | No | NA | 5 | 0 | 0 | Low-quality | Genome-fragment | 1.67 | AAI-based (high-confidence) | 0 |
| S7-final-viral-combined_k141_37947\|\|full | 1343 | No | NA | 2 | 1 | 0 | Low-quality | Genome-fragment | 1.1 | HMM-based (lower-bound) | 0 |
| S1-final-viral-combined_k141_370021\|\|full | 2783 | No | NA | 3 | 0 | 2 | Not-determined | Genome-fragment | NA | NA | 0 |
| S1-final-viral-combined_k141_271402\|\|full | 1085 | No | NA | 2 | 0 | 2 | Not-determined | Genome-fragment | NA | NA | 0 |
| S1-final-viral-combined_k141_24416\|\|full | 2159 | No | NA | 4 | 0 | 1 | Not-determined | Genome-fragment | NA | NA | 0 |
| S7_k141_188001 | 1343 | No | NA | 2 | 0 | 1 | Low-quality | Genome-fragment | 2.69 | AAI-based (medium-confidence) | 0 |
| S17-final-viral-combined_k141_186608\|\|full | 1343 | No | NA | 3 | 2 | 0 | Low-quality | Genome-fragment | 3.58 | AAI-based (high-confidence) | 0 |
| S12-final-viral-combined_k141_228650\|\|full | 1343 | No | NA | 3 | 0 | 0 | Low-quality | Genome-fragment | 3.51 | AAI-based (high-confidence) | 0 |
| S1-final-viral-combined_k141_354386\|\|full | 2092 | No | NA | 2 | 0 | 1 | Not-determined | Genome-fragment | NA | NA | 0 |
| S1-final-viral-combined_k141_255737\|\|full | 6452 | No | NA | 6 | 0 | 2 | Not-determined | Genome-fragment | NA | NA | 0 |
| S1-final-viral-combined_k141_386975\|\|full | 3555 | No | NA | 4 | 0 | 3 | Not-determined | Genome-fragment | NA | NA | 0 |
| S1-final-viral-combined_k141_206381\|\|full | 2467 | No | NA | 4 | 0 | 1 | Not-determined | Genome-fragment | NA | NA | 0 |
| S2-final-viral-combined_k141_104611\|\|full | 1343 | No | NA | 4 | 0 | 0 | Low-quality | Genome-fragment | 3.7 | AAI-based (medium-confidence) | 0 |
| S1-final-viral-combined_k141_90307\|\|full | 1128 | No | NA | 2 | 0 | 0 | Not-determined | Genome-fragment | NA | NA | 0 |
| S1-final-viral-combined_k141_90311\|\|full | 1050 | No | NA | 2 | 0 | 0 | Not-determined | Genome-fragment | NA | NA | 0 |
| S1-final-viral-combined_k141_387021\|\|full | 1597 | No | NA | 2 | 0 | 1 | Not-determined | Genome-fragment | NA | NA | 0 |
| S1-final-viral-combined_k141_387026\|\|full | 1245 | No | NA | 3 | 0 | 0 | Not-determined | Genome-fragment | NA | NA | 0 |
| S1-final-viral-combined_k141_90332\|\|full | 2889 | No | NA | 5 | 0 | 1 | Not-determined | Genome-fragment | NA | NA | 0 |
| S2-final-viral-combined_k141_170777\|\|full | 1343 | No | NA | 3 | 0 | 1 | Low-quality | Genome-fragment | 1.52 | AAI-based (medium-confidence) | 0 |
| S4-final-viral-combined_k141_25684\|\|full | 1343 | No | NA | 3 | 0 | 0 | Low-quality | Genome-fragment | 2.17 | AAI-based (medium-confidence) | 0 |
| S1-final-viral-combined_k141_24494\|\|full | 4577 | No | NA | 4 | 0 | 0 | Not-determined | Genome-fragment | NA | NA | 0 |
| S9-final-viral-combined_k141_53788\|\|full | 1344 | No | NA | 3 | 0 | 0 | Low-quality | Genome-fragment | 1.5 | AAI-based (medium-confidence) | 0 |
| S1-final-viral-combined_k141_255823\|\|full | 1713 | No | NA | 3 | 0 | 1 | Not-determined | Genome-fragment | NA | NA | 0 |
| S1-final-viral-combined_k141_238910\|\|full | 1672 | No | NA | 3 | 0 | 1 | Not-determined | Genome-fragment | NA | NA | 0 |
| S1-final-viral-combined_k141_322161\|\|full | 2824 | No | NA | 3 | 0 | 2 | Not-determined | Genome-fragment | NA | NA | 0 |
| S1-final-viral-combined_k141_124452\|\|full | 1636 | No | NA | 2 | 0 | 0 | Not-determined | Genome-fragment | NA | NA | 0 |
| S11-final-viral-combined_k141_242236\|\|full | 1344 | No | NA | 2 | 0 | 1 | Low-quality | Genome-fragment | 0.31 | AAI-based (medium-confidence) | 0 |
| S1-final-viral-combined_k141_370128\|\|full | 1664 | No | NA | 2 | 0 | 2 | Not-determined | Genome-fragment | NA | NA | 0 |
| S1-final-viral-combined_k141_301690\|\|full | 1344 | No | NA | 2 | 0 | 0 | Low-quality | Genome-fragment | 7.24 | AAI-based (high-confidence) | 0 |
| S5_k141_171525 | 1344 | No | NA | 1 | 0 | 0 | Low-quality | Genome-fragment | 1.02 | AAI-based (high-confidence) | 0 |
| S7-final-viral-combined_k141_189310\|\|full | 1344 | No | NA | 3 | 0 | 3 | Low-quality | Genome-fragment | 1.19 | AAI-based (medium-confidence) | 0 |
| S1-final-viral-combined_k141_24515\|\|full | 1255 | No | NA | 2 | 0 | 1 | Not-determined | Genome-fragment | NA | NA | 0 |
| S7-final-viral-combined_k141_179549\|\|full | 1344 | No | NA | 3 | 2 | 0 | Low-quality | Genome-fragment | 1.91 | AAI-based (high-confidence) | 0 |
| S1-final-viral-combined_k141_255854\|\|full | 2165 | No | NA | 2 | 0 | 1 | Not-determined | Genome-fragment | NA | NA | 0 |
| S1-final-viral-combined_k141_288059\|\|full | 2307 | No | NA | 2 | 0 | 2 | Not-determined | Genome-fragment | NA | NA | 0 |
| S17-final-viral-combined_k141_56186\|\|full | 1344 | No | NA | 2 | 0 | 0 | Low-quality | Genome-fragment | 2.51 | AAI-based (medium-confidence) | 0 |
| S17-final-viral-combined_k141_69471\|\|full | 1344 | No | NA | 3 | 1 | 0 | Low-quality | Genome-fragment | 3.79 | AAI-based (high-confidence) | 0 |
| S1-final-viral-combined_k141_190157\|\|full | 1215 | No | NA | 2 | 0 | 1 | Not-determined | Genome-fragment | NA | NA | 0 |
| S17-final-viral-combined_k141_265542\|\|full | 1344 | No | NA | 4 | 1 | 0 | Low-quality | Genome-fragment | 3.58 | AAI-based (high-confidence) | 0 |
| S12-final-viral-combined_k141_172359\|\|full | 1344 | No | NA | 3 | 0 | 0 | Low-quality | Genome-fragment | 3.28 | AAI-based (medium-confidence) | 0 |
| S1-final-viral-combined_k141_40822\|\|full | 2397 | No | NA | 2 | 0 | 1 | Not-determined | Genome-fragment | NA | NA | 0 |
| S12-final-viral-combined_k141_383082\|\|lt2gene | 1344 | No | NA | 2 | 2 | 0 | Low-quality | Genome-fragment | 2.92 | AAI-based (high-confidence) | 0 |
| S1-final-viral-combined_k141_73338\|\|full | 2748 | No | NA | 3 | 0 | 2 | Not-determined | Genome-fragment | NA | NA | 0 |
| S2-final-viral-combined_k141_173375\|\|full | 1344 | No | NA | 4 | 1 | 0 | Low-quality | Genome-fragment | 3.21 | AAI-based (medium-confidence) | 0 |
| S4_k141_49111 | 1344 | No | NA | 4 | 0 | 0 | Low-quality | Genome-fragment | 2.33 | AAI-based (medium-confidence) | 0 |
| S6-final-viral-combined_k141_34480\|\|full | 1344 | No | NA | 3 | 0 | 0 | Low-quality | Genome-fragment | 3.86 | AAI-based (medium-confidence) | 0 |
| S1-final-viral-combined_k141_40863\|\|full | 1408 | No | NA | 3 | 0 | 2 | Not-determined | Genome-fragment | NA | NA | 0 |
| S8-final-viral-combined_k141_265370\|\|full | 1344 | No | NA | 3 | 0 | 0 | Low-quality | Genome-fragment | 2.27 | AAI-based (high-confidence) | 0 |
| S11-final-viral-combined_k141_239790\|\|full | 1345 | No | NA | 4 | 0 | 0 | Low-quality | Genome-fragment | 3.14 | AAI-based (high-confidence) | 0 |
| S11-final-viral-combined_k141_62023\|\|full | 1345 | No | NA | 4 | 1 | 0 | Low-quality | Genome-fragment | 3.22 | AAI-based (high-confidence) | 0 |
| S1-final-viral-combined_k141_24572\|\|full | 3269 | No | NA | 3 | 0 | 1 | Not-determined | Genome-fragment | NA | NA | 0 |
| S1-final-viral-combined_k141_8502\|\|full | 1679 | No | NA | 4 | 0 | 0 | Not-determined | Genome-fragment | NA | NA | 0 |
| S11-final-viral-combined_k141_227700\|\|full | 1345 | No | NA | 4 | 0 | 0 | Low-quality | Genome-fragment | 2.3 | AAI-based (high-confidence) | 0 |
| S5-final-viral-combined_k141_127424\|\|full | 1345 | No | NA | 4 | 0 | 1 | Low-quality | Genome-fragment | 1.34 | AAI-based (medium-confidence) | 0 |
| S5_k141_111090 | 1345 | No | NA | 4 | 0 | 0 | Low-quality | Genome-fragment | 2.92 | AAI-based (high-confidence) | 0 |
| S1-final-viral-combined_k141_124528\|\|full | 4577 | No | NA | 6 | 0 | 2 | Not-determined | Genome-fragment | NA | NA | 0 |
| S7-final-viral-combined_k141_151641\|\|full | 1345 | No | NA | 3 | 1 | 0 | Low-quality | Genome-fragment | 0.86 | HMM-based (lower-bound) | 0 |
| S12-final-viral-combined_k141_209557\|\|full | 1345 | No | NA | 4 | 0 | 1 | Low-quality | Genome-fragment | 0.41 | AAI-based (medium-confidence) | 0 |
| S12-final-viral-combined_k141_65170\|\|full | 1345 | No | NA | 2 | 0 | 0 | Low-quality | Genome-fragment | 3.05 | AAI-based (medium-confidence) | 0 |
| S1-final-viral-combined_k141_90416\|\|full | 9681 | No | NA | 9 | 0 | 7 | Not-determined | Genome-fragment | NA | NA | 0 |
| S12_k141_47518 | 1345 | No | NA | 1 | 0 | 0 | Low-quality | Genome-fragment | 1.37 | AAI-based (high-confidence) | 0 |
| S1-final-viral-combined_k141_155883\|\|full | 2050 | No | NA | 3 | 0 | 2 | Not-determined | Genome-fragment | NA | NA | 0 |
| S2-final-viral-combined_k141_171352\|\|full | 1345 | No | NA | 2 | 0 | 1 | Low-quality | Genome-fragment | 2.27 | AAI-based (high-confidence) | 0 |
| S4-final-viral-combined_k141_114901\|\|full | 1345 | No | NA | 2 | 1 | 0 | Low-quality | Genome-fragment | 1.93 | AAI-based (high-confidence) | 0 |
| S1-final-viral-combined_k141_305287\|\|full | 2148 | No | NA | 4 | 0 | 2 | Not-determined | Genome-fragment | NA | NA | 0 |
| S1-final-viral-combined_k141_238991\|\|full | 1823 | No | NA | 3 | 0 | 1 | Not-determined | Genome-fragment | NA | NA | 0 |
| S1-final-viral-combined_k141_206505\|\|full | 1442 | No | NA | 2 | 0 | 1 | Not-determined | Genome-fragment | NA | NA | 0 |
| S1-final-viral-combined_k141_190258\|\|full | 4449 | No | NA | 3 | 0 | 3 | Not-determined | Genome-fragment | NA | NA | 0 |
| S1-final-viral-combined_k141_73444\|\|full | 3161 | No | NA | 4 | 0 | 2 | Not-determined | Genome-fragment | NA | NA | 0 |
| S11-final-viral-combined_k141_20498\|\|full | 1346 | No | NA | 3 | 0 | 0 | Low-quality | Genome-fragment | 1.8 | AAI-based (medium-confidence) | 0 |
| S1-final-viral-combined_k141_255975\|\|full | 3193 | No | NA | 5 | 0 | 3 | Not-determined | Genome-fragment | NA | NA | 0 |
| S11-final-viral-combined_k141_65374\|\|full | 1346 | No | NA | 2 | 0 | 0 | Low-quality | Genome-fragment | 3.35 | AAI-based (high-confidence) | 0 |
| S1-final-viral-combined_k141_271693\|\|full | 1234 | No | NA | 3 | 0 | 1 | Not-determined | Genome-fragment | NA | NA | 0 |
| S1-final-viral-combined_k141_336818\|\|full | 3934 | No | NA | 5 | 0 | 3 | Not-determined | Genome-fragment | NA | NA | 0 |
| S1-final-viral-combined_k141_8576\|\|full | 2240 | No | NA | 3 | 0 | 0 | Not-determined | Genome-fragment | NA | NA | 0 |
| S3-final-viral-combined_k141_98273\|\|full | 1346 | No | NA | 4 | 0 | 0 | Low-quality | Genome-fragment | 0.85 | AAI-based (medium-confidence) | 0 |
| S3-final-viral-combined_k141_6479\|\|full | 1346 | No | NA | 3 | 0 | 0 | Low-quality | Genome-fragment | 0.95 | AAI-based (medium-confidence) | 0 |
| S3_k141_101694 | 1346 | No | NA | 3 | 0 | 0 | Low-quality | Genome-fragment | 2.25 | AAI-based (high-confidence) | 0 |
| S1-final-viral-combined_k141_336847\|\|full | 2490 | No | NA | 3 | 0 | 1 | Not-determined | Genome-fragment | NA | NA | 0 |
| S1-final-viral-combined_k141_90511\|\|full | 1748 | No | NA | 2 | 0 | 1 | Not-determined | Genome-fragment | NA | NA | 0 |
| S7-final-viral-combined_k141_215380\|\|full | 1346 | No | NA | 2 | 0 | 2 | Low-quality | Genome-fragment | 2.53 | AAI-based (medium-confidence) | 0 |
| S1-final-viral-combined_k141_40968\|\|full | 2165 | No | NA | 4 | 0 | 0 | Not-determined | Genome-fragment | NA | NA | 0 |
| S1-final-viral-combined_k141_124601\|\|full | 1852 | No | NA | 2 | 0 | 0 | Not-determined | Genome-fragment | NA | NA | 0 |
| S7_k141_118027 | 1346 | No | NA | 3 | 1 | 0 | Low-quality | Genome-fragment | 2.5 | AAI-based (high-confidence) | 0 |
| S17-final-viral-combined_k141_67500\|\|full | 1346 | No | NA | 2 | 0 | 1 | Low-quality | Genome-fragment | 0.45 | AAI-based (medium-confidence) | 0 |
| S1-final-viral-combined_k141_73519\|\|full | 2586 | No | NA | 3 | 0 | 2 | Not-determined | Genome-fragment | NA | NA | 0 |
| S12-final-viral-combined_k141_314422\|\|full | 1346 | No | NA | 4 | 0 | 0 | Low-quality | Genome-fragment | 3.82 | AAI-based (high-confidence) | 0 |
| S1-final-viral-combined_k141_322336\|\|full | 2309 | No | NA | 2 | 0 | 2 | Not-determined | Genome-fragment | NA | NA | 0 |
| S12-final-viral-combined_k141_182113\|\|full | 1346 | No | NA | 2 | 0 | 0 | Low-quality | Genome-fragment | 1.05 | AAI-based (medium-confidence) | 0 |
| S12_k141_106959 | 1346 | No | NA | 1 | 0 | 1 | Low-quality | Genome-fragment | 0.51 | AAI-based (high-confidence) | 0 |
| S1-final-viral-combined_k141_305400\|\|full | 1369 | No | NA | 2 | 0 | 1 | Not-determined | Genome-fragment | NA | NA | 0 |
| S1-final-viral-combined_k141_57613\|\|full | 6822 | No | NA | 4 | 0 | 3 | Not-determined | Genome-fragment | NA | NA | 0 |
| S1-final-viral-combined_k141_40977\|\|full | 1926 | No | NA | 2 | 0 | 2 | Not-determined | Genome-fragment | NA | NA | 0 |
| S2-final-viral-combined_k141_140664\|\|full | 1346 | No | NA | 3 | 0 | 0 | Low-quality | Genome-fragment | 3.02 | AAI-based (medium-confidence) | 0 |
| S1-final-viral-combined_k141_40979\|\|full | 3406 | No | NA | 6 | 0 | 3 | Not-determined | Genome-fragment | NA | NA | 0 |
| S6-final-viral-combined_k141_49304\|\|full | 1346 | No | NA | 5 | 0 | 0 | Low-quality | Genome-fragment | 3.28 | AAI-based (medium-confidence) | 0 |
| S1-final-viral-combined_k141_387285\|\|full | 1396 | No | NA | 4 | 0 | 1 | Not-determined | Genome-fragment | NA | NA | 0 |
| S1-final-viral-combined_k141_8717\|\|full | 1850 | No | NA | 2 | 0 | 2 | Not-determined | Genome-fragment | NA | NA | 0 |
| S1-final-viral-combined_k141_24776\|\|full | 3157 | No | NA | 3 | 0 | 2 | Not-determined | Genome-fragment | NA | NA | 0 |
| S8_k141_148762 | 1346 | No | NA | 3 | 0 | 0 | Low-quality | Genome-fragment | 1.44 | AAI-based (high-confidence) | 0 |
| S1-final-viral-combined_k141_155968\|\|full | 2204 | No | NA | 4 | 0 | 3 | Not-determined | Genome-fragment | NA | NA | 0 |
| S11-final-viral-combined_k141_213524\|\|full | 1347 | No | NA | 3 | 1 | 0 | Low-quality | Genome-fragment | 2.27 | AAI-based (high-confidence) | 0 |
| S1_k141_140091 | 1347 | No | NA | 3 | 0 | 1 | Low-quality | Genome-fragment | 0.41 | AAI-based (medium-confidence) | 0 |
| S1-final-viral-combined_k141_8748\|\|full | 2713 | No | NA | 5 | 0 | 1 | Not-determined | Genome-fragment | NA | NA | 0 |
| S3-final-viral-combined_k141_51948\|\|full | 1347 | No | NA | 2 | 0 | 0 | Low-quality | Genome-fragment | 3.03 | AAI-based (medium-confidence) | 0 |
| S1-final-viral-combined_k141_222507\|\|full | 1544 | No | NA | 3 | 0 | 0 | Not-determined | Genome-fragment | NA | NA | 0 |
| S1-final-viral-combined_k141_387310\|\|full | 3924 | No | NA | 3 | 0 | 2 | Not-determined | Genome-fragment | NA | NA | 0 |
| S1-final-viral-combined_k141_190441\|\|full | 1510 | No | NA | 3 | 0 | 0 | Not-determined | Genome-fragment | NA | NA | 0 |
| S5-final-viral-combined_k141_31297\|\|lt2gene | 1347 | No | NA | 1 | 1 | 0 | Low-quality | Genome-fragment | 2.9 | AAI-based (high-confidence) | 0 |
| S1-final-viral-combined_k141_387324\|\|full | 1596 | No | NA | 3 | 0 | 2 | Not-determined | Genome-fragment | NA | NA | 0 |
| S7-final-viral-combined_k141_30197\|\|full | 1347 | No | NA | 3 | 0 | 1 | Low-quality | Genome-fragment | 2.79 | AAI-based (high-confidence) | 0 |
| S1-final-viral-combined_k141_190451\|\|full | 1131 | No | NA | 3 | 0 | 1 | Not-determined | Genome-fragment | NA | NA | 0 |
| S17-k141_29765 | 1347 | No | NA | 2 | 0 | 0 | Low-quality | Genome-fragment | 2.51 | AAI-based (medium-confidence) | 0 |
| S1-final-viral-combined_k141_288255\|\|full | 13560 | No | NA | 14 | 0 | 12 | Not-determined | Genome-fragment | NA | NA | 0 |
| S1-final-viral-combined_k141_73562\|\|full | 2836 | No | NA | 5 | 0 | 1 | Not-determined | Genome-fragment | NA | NA | 0 |
| S1-final-viral-combined_k141_370410\|\|full | 1298 | No | NA | 3 | 0 | 0 | Not-determined | Genome-fragment | NA | NA | 0 |
| S12-final-viral-combined_k141_72442\|\|full | 1347 | No | NA | 2 | 1 | 0 | Low-quality | Genome-fragment | 0.87 | HMM-based (lower-bound) | 0 |
| S12-final-viral-combined_k141_91123\|\|full | 1347 | No | NA | 2 | 2 | 0 | Low-quality | Genome-fragment | 2.79 | HMM-based (lower-bound) | 0 |
| S1-final-viral-combined_k141_322413\|\|full | 2955 | No | NA | 2 | 0 | 1 | Not-determined | Genome-fragment | NA | NA | 0 |
| S2-final-viral-combined_k141_145007\|\|full | 1347 | No | NA | 4 | 1 | 0 | Low-quality | Genome-fragment | 1.64 | AAI-based (medium-confidence) | 0 |
| S1-final-viral-combined_k141_206672\|\|full | 2420 | No | NA | 4 | 0 | 0 | Not-determined | Genome-fragment | NA | NA | 0 |
| S1-final-viral-combined_k141_322416\|\|full | 2455 | No | NA | 2 | 0 | 1 | Not-determined | Genome-fragment | NA | NA | 0 |
| S1-final-viral-combined_k141_256029\|\|full | 4316 | No | NA | 2 | 0 | 2 | Not-determined | Genome-fragment | NA | NA | 0 |
| S1-final-viral-combined_k141_370443\|\|full | 1950 | No | NA | 3 | 0 | 1 | Not-determined | Genome-fragment | NA | NA | 0 |
| S2_k141_116220 | 1347 | No | NA | 3 | 0 | 0 | Low-quality | Genome-fragment | 11.04 | AAI-based (high-confidence) | 0 |
| S4-final-viral-combined_k141_121300\|\|full | 1347 | No | NA | 3 | 0 | 1 | Low-quality | Genome-fragment | 2.26 | AAI-based (medium-confidence) | 0 |
| S1-final-viral-combined_k141_41079\|\|full | 1293 | No | NA | 3 | 0 | 1 | Not-determined | Genome-fragment | NA | NA | 0 |
| S1-final-viral-combined_k141_24864\|\|full | 1915 | No | NA | 3 | 0 | 2 | Not-determined | Genome-fragment | NA | NA | 0 |
| S1-final-viral-combined_k141_8806\|\|full | 1105 | No | NA | 2 | 0 | 1 | Not-determined | Genome-fragment | NA | NA | 0 |
| S8-final-viral-combined_k141_6669\|\|full | 1347 | No | NA | 3 | 0 | 0 | Low-quality | Genome-fragment | 3.43 | AAI-based (medium-confidence) | 0 |
| S1-final-viral-combined_k141_106396\|\|full | 2043 | No | NA | 4 | 0 | 1 | Not-determined | Genome-fragment | NA | NA | 0 |
| S8_k141_18908 | 1347 | No | NA | 3 | 1 | 0 | Low-quality | Genome-fragment | 0.9 | AAI-based (high-confidence) | 0 |
| S1-final-viral-combined_k141_222635\|\|full | 1372 | No | NA | 3 | 0 | 0 | Not-determined | Genome-fragment | NA | NA | 0 |
| S1-final-viral-combined_k141_190531\|\|full | 2639 | No | NA | 3 | 0 | 1 | Not-determined | Genome-fragment | NA | NA | 0 |
| S11-final-viral-combined_k141_207502\|\|full | 1348 | No | NA | 3 | 1 | 0 | Low-quality | Genome-fragment | 3.02 | AAI-based (medium-confidence) | 0 |
| S11-final-viral-combined_k141_15629\|\|full | 1348 | No | NA | 3 | 0 | 0 | Low-quality | Genome-fragment | 3.03 | AAI-based (medium-confidence) | 0 |
| S1-final-viral-combined_k141_288322\|\|full | 1119 | No | NA | 3 | 0 | 0 | Not-determined | Genome-fragment | NA | NA | 0 |
| S1-final-viral-combined_k141_90688\|\|full | 1467 | No | NA | 2 | 0 | 1 | Not-determined | Genome-fragment | NA | NA | 0 |
| S11-final-viral-combined_k141_227330\|\|full | 1348 | No | NA | 4 | 1 | 0 | Low-quality | Genome-fragment | 2.47 | HMM-based (lower-bound) | 0 |
| S11_k141_230400 | 1348 | No | NA | 3 | 0 | 0 | Low-quality | Genome-fragment | 1.45 | AAI-based (high-confidence) | 0 |
| S1-final-viral-combined_k141_354724\|\|full | 2308 | No | NA | 3 | 0 | 2 | Not-determined | Genome-fragment | NA | NA | 0 |
| S1-final-viral-combined_k141_288357\|\|full | 1066 | No | NA | 2 | 0 | 2 | Not-determined | Genome-fragment | NA | NA | 0 |
| S1-final-viral-combined_k141_370543\|\|full | 2383 | No | NA | 2 | 0 | 0 | Not-determined | Genome-fragment | NA | NA | 0 |
| S1-final-viral-combined_k141_124910\|\|full | 1348 | No | NA | 2 | 0 | 0 | Low-quality | Genome-fragment | 0.43 | AAI-based (medium-confidence) | 0 |
| S1-final-viral-combined_k141_370551\|\|full | 2730 | No | NA | 4 | 0 | 1 | Not-determined | Genome-fragment | NA | NA | 0 |
| S1-final-viral-combined_k141_140372\|\|full | 2207 | No | NA | 5 | 0 | 1 | Not-determined | Genome-fragment | NA | NA | 0 |
| S1-final-viral-combined_k141_258316\|\|full | 1348 | No | NA | 2 | 0 | 0 | Low-quality | Genome-fragment | 2.12 | AAI-based (high-confidence) | 0 |
| S1-final-viral-combined_k141_87278\|\|lt2gene | 1348 | No | NA | 2 | 2 | 0 | Low-quality | Genome-fragment | 3.96 | AAI-based (high-confidence) | 0 |
| S1-final-viral-combined_k141_222683\|\|full | 1474 | No | NA | 2 | 0 | 0 | Not-determined | Genome-fragment | NA | NA | 0 |
| S1_k141_110103 | 1348 | No | NA | 2 | 0 | 2 | Low-quality | Genome-fragment | 0.35 | AAI-based (medium-confidence) | 0 |
| S1-final-viral-combined_k141_354739\|\|full | 2778 | No | NA | 4 | 0 | 3 | Not-determined | Genome-fragment | NA | NA | 0 |
| S7-final-viral-combined_k141_41486\|\|full | 1348 | No | NA | 3 | 1 | 0 | Low-quality | Genome-fragment | 1.71 | AAI-based (high-confidence) | 0 |
| S7_k141_184709 | 1348 | No | NA | 3 | 0 | 0 | Low-quality | Genome-fragment | 1.42 | AAI-based (medium-confidence) | 0 |
| S17-k141_6522 | 1348 | No | NA | 1 | 0 | 1 | Low-quality | Genome-fragment | 0.73 | AAI-based (medium-confidence) | 0 |
| S12-final-viral-combined_k141_23607\|\|full | 1348 | No | NA | 2 | 0 | 1 | Low-quality | Genome-fragment | 0.92 | AAI-based (medium-confidence) | 0 |
| S2-final-viral-combined_k141_51952\|\|full | 1348 | No | NA | 2 | 0 | 0 | Low-quality | Genome-fragment | 2.27 | AAI-based (high-confidence) | 0 |
| S1-final-viral-combined_k141_140412\|\|full | 1274 | No | NA | 3 | 0 | 3 | Not-determined | Genome-fragment | NA | NA | 0 |
| S1-final-viral-combined_k141_288436\|\|full | 1813 | No | NA | 2 | 0 | 1 | Not-determined | Genome-fragment | NA | NA | 0 |
| S2-final-viral-combined_k141_163550\|\|full | 1348 | No | NA | 3 | 0 | 0 | Low-quality | Genome-fragment | 2.67 | AAI-based (medium-confidence) | 0 |
| S1-final-viral-combined_k141_190626\|\|full | 3578 | No | NA | 6 | 0 | 0 | Not-determined | Genome-fragment | NA | NA | 0 |
| S1-final-viral-combined_k141_190636\|\|full | 2958 | No | NA | 3 | 0 | 1 | Not-determined | Genome-fragment | NA | NA | 0 |
| S1-final-viral-combined_k141_239362\|\|full | 2485 | No | NA | 4 | 0 | 3 | Not-determined | Genome-fragment | NA | NA | 0 |
| S1-final-viral-combined_k141_354782\|\|full | 1387 | No | NA | 3 | 0 | 2 | Not-determined | Genome-fragment | NA | NA | 0 |
| S4_k141_53873 | 1348 | No | NA | 2 | 0 | 0 | Low-quality | Genome-fragment | 1.63 | AAI-based (medium-confidence) | 0 |
| S9-final-viral-combined_k141_24458\|\|full | 1349 | No | NA | 3 | 0 | 0 | Low-quality | Genome-fragment | 2.25 | AAI-based (high-confidence) | 0 |
| S9-final-viral-combined_k141_76805\|\|lt2gene | 1349 | No | NA | 1 | 1 | 0 | Low-quality | Genome-fragment | 3.8 | AAI-based (high-confidence) | 0 |
| S1-final-viral-combined_k141_172284\|\|full | 3383 | No | NA | 3 | 0 | 1 | Not-determined | Genome-fragment | NA | NA | 0 |
| S11_k141_242987 | 1349 | No | NA | 1 | 1 | 0 | Low-quality | Genome-fragment | 0.92 | HMM-based (lower-bound) | 0 |
| S1-final-viral-combined_k141_41288\|\|full | 3092 | No | NA | 3 | 0 | 2 | Not-determined | Genome-fragment | NA | NA | 0 |
| S1-final-viral-combined_k141_370684\|\|full | 1207 | No | NA | 3 | 0 | 1 | Not-determined | Genome-fragment | NA | NA | 0 |
| S1-final-viral-combined_k141_271958\|\|full | 1890 | No | NA | 2 | 0 | 0 | Not-determined | Genome-fragment | NA | NA | 0 |
| S1-final-viral-combined_k141_352733\|\|full | 1349 | No | NA | 3 | 0 | 0 | Low-quality | Genome-fragment | 2.74 | AAI-based (medium-confidence) | 0 |
| S1-final-viral-combined_k141_270381\|\|full | 1349 | No | NA | 6 | 0 | 0 | Low-quality | Genome-fragment | 3.52 | AAI-based (high-confidence) | 0 |
| S3-final-viral-combined_k141_66341\|\|full | 1349 | No | NA | 2 | 0 | 0 | Low-quality | Genome-fragment | 2.69 | AAI-based (medium-confidence) | 0 |
| S7-final-viral-combined_k141_8593\|\|lt2gene | 1349 | No | NA | 1 | 1 | 0 | Low-quality | Genome-fragment | 0.93 | HMM-based (lower-bound) | 0 |
| S7_k141_49775 | 1349 | No | NA | 3 | 0 | 0 | Low-quality | Genome-fragment | 2.79 | AAI-based (medium-confidence) | 0 |
| S17-final-viral-combined_k141_44881\|\|full | 1349 | No | NA | 3 | 0 | 1 | Low-quality | Genome-fragment | 0.79 | AAI-based (medium-confidence) | 0 |
| S12-final-viral-combined_k141_62323\|\|full | 1349 | No | NA | 3 | 2 | 0 | Low-quality | Genome-fragment | 1.36 | AAI-based (high-confidence) | 0 |
| S1-final-viral-combined_k141_57839\|\|full | 2059 | No | NA | 3 | 0 | 2 | Not-determined | Genome-fragment | NA | NA | 0 |
| S12-final-viral-combined_k141_216329\|\|full | 1349 | No | NA | 4 | 0 | 1 | Low-quality | Genome-fragment | 1.88 | AAI-based (medium-confidence) | 0 |
| S1-final-viral-combined_k141_140577\|\|full | 1142 | No | NA | 2 | 0 | 0 | Not-determined | Genome-fragment | NA | NA | 0 |
| S1-final-viral-combined_k141_354899\|\|full | 18395 | No | NA | 26 | 16 | 1 | Medium-quality | Genome-fragment | 72.57 | AAI-based (high-confidence) | 0 |
| S1-final-viral-combined_k141_322501\|\|full | 3431 | No | NA | 4 | 0 | 3 | Not-determined | Genome-fragment | NA | NA | 0 |
| S1-final-viral-combined_k141_190702\|\|full | 3774 | No | NA | 5 | 0 | 4 | Not-determined | Genome-fragment | NA | NA | 0 |
| S1-final-viral-combined_k141_25092\|\|full | 2101 | No | NA | 3 | 0 | 2 | Not-determined | Genome-fragment | NA | NA | 0 |
| S1-final-viral-combined_k141_370755\|\|full | 3345 | No | NA | 3 | 0 | 2 | Not-determined | Genome-fragment | NA | NA | 0 |
| S1-final-viral-combined_k141_156205\|\|full | 1018 | No | NA | 3 | 0 | 0 | Not-determined | Genome-fragment | NA | NA | 0 |
| S1-final-viral-combined_k141_354936\|\|full | 2127 | No | NA | 3 | 0 | 1 | Not-determined | Genome-fragment | NA | NA | 0 |
| S1-final-viral-combined_k141_73770\|\|full | 3573 | No | NA | 4 | 0 | 2 | Not-determined | Genome-fragment | NA | NA | 0 |
| S1-final-viral-combined_k141_106563\|\|full | 3362 | No | NA | 3 | 0 | 3 | Not-determined | Genome-fragment | NA | NA | 0 |
| S1-final-viral-combined_k141_156234\|\|full | 6549 | No | NA | 6 | 0 | 0 | Not-determined | Genome-fragment | NA | NA | 0 |
| S1-final-viral-combined_k141_41369\|\|full | 36028 | No | NA | 60 | 17 | 1 | Medium-quality | Genome-fragment | 83.64 | AAI-based (high-confidence) | 0 |
| S2-final-viral-combined_k141_15836\|\|full | 1349 | No | NA | 4 | 3 | 0 | Low-quality | Genome-fragment | 2.04 | HMM-based (lower-bound) | 0 |
| S2-final-viral-combined_k141_130272\|\|full | 1349 | No | NA | 4 | 1 | 0 | Low-quality | Genome-fragment | 3.79 | AAI-based (high-confidence) | 0 |
| S9-final-viral-combined_k141_124536\|\|full | 1350 | No | NA | 3 | 1 | 0 | Low-quality | Genome-fragment | 2.73 | AAI-based (medium-confidence) | 0 |
| S11-final-viral-combined_k141_18742\|\|lt2gene | 1350 | No | NA | 2 | 2 | 0 | Low-quality | Genome-fragment | 5.94 | AAI-based (high-confidence) | 0 |
| S7-final-viral-combined_k141_133769\|\|full | 1350 | No | NA | 3 | 0 | 1 | Low-quality | Genome-fragment | 0.46 | AAI-based (medium-confidence) | 0 |
| S1-final-viral-combined_k141_370831\|\|full | 1410 | No | NA | 2 | 0 | 0 | Not-determined | Genome-fragment | NA | NA | 0 |
| S17-final-viral-combined_k141_135993\|\|full | 1350 | No | NA | 3 | 0 | 0 | Low-quality | Genome-fragment | 3.68 | AAI-based (medium-confidence) | 0 |
| S1-final-viral-combined_k141_256361\|\|full | 5195 | No | NA | 6 | 0 | 2 | Not-determined | Genome-fragment | NA | NA | 0 |
| S1-final-viral-combined_k141_125057\|\|full | 1650 | No | NA | 3 | 0 | 0 | Not-determined | Genome-fragment | NA | NA | 0 |
| S2_k141_237059 | 1350 | No | NA | 2 | 0 | 0 | Low-quality | Genome-fragment | 2.66 | AAI-based (medium-confidence) | 0 |
| S1-final-viral-combined_k141_91001\|\|full | 3962 | No | NA | 4 | 0 | 4 | Not-determined | Genome-fragment | NA | NA | 0 |
| S1-final-viral-combined_k141_140700\|\|full | 4058 | No | NA | 3 | 0 | 2 | Not-determined | Genome-fragment | NA | NA | 0 |
| S8-final-viral-combined_k141_133682\|\|full | 1350 | No | NA | 3 | 0 | 0 | Low-quality | Genome-fragment | 1.44 | AAI-based (medium-confidence) | 0 |
| S1-final-viral-combined_k141_125075\|\|full | 5175 | No | NA | 9 | 0 | 3 | Not-determined | Genome-fragment | NA | NA | 0 |
| S1-final-viral-combined_k141_41379\|\|full | 10985 | No | NA | 10 | 0 | 8 | Not-determined | Genome-fragment | NA | NA | 0 |
| S1-final-viral-combined_k141_288677\|\|full | 1883 | No | NA | 3 | 0 | 2 | Not-determined | Genome-fragment | NA | NA | 0 |
| S1-final-viral-combined_k141_41387\|\|full | 3852 | No | NA | 6 | 0 | 0 | Not-determined | Genome-fragment | NA | NA | 0 |
| S1-final-viral-combined_k141_156272\|\|full | 1191 | No | NA | 2 | 0 | 1 | Not-determined | Genome-fragment | NA | NA | 0 |
| S11-final-viral-combined_k141_67256\|\|full | 1351 | No | NA | 2 | 0 | 1 | Low-quality | Genome-fragment | 0.45 | AAI-based (medium-confidence) | 0 |
| S1-final-viral-combined_k141_322615\|\|full | 2495 | No | NA | 3 | 0 | 2 | Not-determined | Genome-fragment | NA | NA | 0 |
| S1-final-viral-combined_k141_9055\|\|full | 6453 | No | NA | 5 | 0 | 2 | Not-determined | Genome-fragment | NA | NA | 0 |
| S1-final-viral-combined_k141_120927\|\|full | 1351 | No | NA | 2 | 0 | 0 | Low-quality | Genome-fragment | 2.35 | AAI-based (medium-confidence) | 0 |
| S1-final-viral-combined_k141_387706\|\|full | 2088 | No | NA | 4 | 0 | 1 | Not-determined | Genome-fragment | NA | NA | 0 |
| S1-final-viral-combined_k141_73863\|\|full | 1208 | No | NA | 3 | 0 | 0 | Not-determined | Genome-fragment | NA | NA | 0 |
| S1-final-viral-combined_k141_207052\|\|full | 2177 | No | NA | 3 | 0 | 3 | Not-determined | Genome-fragment | NA | NA | 0 |
| S1-final-viral-combined_k141_73865\|\|full | 1058 | No | NA | 2 | 0 | 1 | Not-determined | Genome-fragment | NA | NA | 0 |
| S5-final-viral-combined_k141_180510\|\|full | 1351 | No | NA | 1 | 0 | 1 | Low-quality | Genome-fragment | 1.75 | AAI-based (medium-confidence) | 0 |
| S1-final-viral-combined_k141_239603\|\|full | 1244 | No | NA | 2 | 0 | 1 | Not-determined | Genome-fragment | NA | NA | 0 |
| S1-final-viral-combined_k141_256430\|\|full | 2323 | No | NA | 2 | 0 | 2 | Not-determined | Genome-fragment | NA | NA | 0 |
| S1-final-viral-combined_k141_9085\|\|full | 1355 | No | NA | 2 | 0 | 1 | Not-determined | Genome-fragment | NA | NA | 0 |
| S1-final-viral-combined_k141_125146\|\|full | 3434 | No | NA | 2 | 0 | 2 | Not-determined | Genome-fragment | NA | NA | 0 |
| S1-final-viral-combined_k141_272193\|\|full | 7269 | No | NA | 8 | 0 | 6 | Not-determined | Genome-fragment | NA | NA | 0 |
| S5-final-viral-combined_k141_19329\|\|lt2gene | 1351 | No | NA | 2 | 1 | 0 | Low-quality | Genome-fragment | 3.02 | AAI-based (high-confidence) | 0 |
| S1-final-viral-combined_k141_387752\|\|full | 1907 | No | NA | 3 | 0 | 3 | Not-determined | Genome-fragment | NA | NA | 0 |
| S1-final-viral-combined_k141_305953\|\|full | 1861 | No | NA | 2 | 0 | 0 | Not-determined | Genome-fragment | NA | NA | 0 |
| S1-final-viral-combined_k141_156356\|\|full | 2977 | No | NA | 3 | 0 | 1 | Not-determined | Genome-fragment | NA | NA | 0 |
| S7_k141_125183 | 1351 | No | NA | 2 | 0 | 0 | Low-quality | Genome-fragment | 2.26 | AAI-based (high-confidence) | 0 |
| S17-final-viral-combined_k141_184683\|\|full | 1351 | No | NA | 2 | 1 | 0 | Low-quality | Genome-fragment | 26.52 | AAI-based (medium-confidence) | 0 |
| S17-final-viral-combined_k141_140012\|\|full | 1351 | No | NA | 4 | 0 | 0 | Low-quality | Genome-fragment | 2.1 | AAI-based (high-confidence) | 0 |
| S1-final-viral-combined_k141_370973\|\|full | 5800 | No | NA | 6 | 0 | 4 | Not-determined | Genome-fragment | NA | NA | 0 |
| S1-final-viral-combined_k141_25263\|\|full | 3557 | No | NA | 6 | 0 | 3 | Not-determined | Genome-fragment | NA | NA | 0 |
| S1-final-viral-combined_k141_25270\|\|full | 2212 | No | NA | 3 | 0 | 0 | Not-determined | Genome-fragment | NA | NA | 0 |
| S1-final-viral-combined_k141_106706\|\|full | 2003 | No | NA | 3 | 0 | 2 | Not-determined | Genome-fragment | NA | NA | 0 |
| S17-final-viral-combined_k141_86136\|\|full | 1351 | No | NA | 3 | 0 | 0 | Low-quality | Genome-fragment | 3.11 | AAI-based (high-confidence) | 0 |
| S1-final-viral-combined_k141_322691\|\|full | 1730 | No | NA | 3 | 0 | 0 | Not-determined | Genome-fragment | NA | NA | 0 |
| S1-final-viral-combined_k141_337370\|\|full | 1371 | No | NA | 3 | 0 | 1 | Not-determined | Genome-fragment | NA | NA | 0 |
| S1-final-viral-combined_k141_322699\|\|full | 1207 | No | NA | 2 | 0 | 0 | Not-determined | Genome-fragment | NA | NA | 0 |
| S1-final-viral-combined_k141_322706\|\|full | 5676 | No | NA | 9 | 0 | 3 | Not-determined | Genome-fragment | NA | NA | 0 |
| S1-final-viral-combined_k141_106730\|\|full | 6847 | No | NA | 6 | 0 | 1 | Not-determined | Genome-fragment | NA | NA | 0 |
| S1-final-viral-combined_k141_9182\|\|full | 1683 | No | NA | 3 | 0 | 2 | Not-determined | Genome-fragment | NA | NA | 0 |
| S1-final-viral-combined_k141_106732\|\|full | 4934 | No | NA | 3 | 0 | 3 | Not-determined | Genome-fragment | NA | NA | 0 |
| S17-final-viral-combined_k141_264920\|\|full | 1351 | No | NA | 3 | 0 | 0 | Low-quality | Genome-fragment | 1.75 | AAI-based (medium-confidence) | 0 |
| S12-final-viral-combined_k141_68396\|\|full | 1351 | No | NA | 2 | 0 | 2 | Low-quality | Genome-fragment | 0.43 | AAI-based (medium-confidence) | 0 |
| S1-final-viral-combined_k141_125205\|\|full | 1862 | No | NA | 2 | 0 | 1 | Not-determined | Genome-fragment | NA | NA | 0 |
| S1-final-viral-combined_k141_305987\|\|full | 2649 | No | NA | 3 | 0 | 3 | Not-determined | Genome-fragment | NA | NA | 0 |

| | | | | | | | | | | | | |
|---|---|---|---|---|---|---|---|---|---|---|---|---|
| S12_k141_266953 | 1351 | No | NA | | 3 | 1 | 0 | Low-quality | Genome-fragment | 2.73 | AAI-based (medium-confidence) | 0 |
| S1-final-viral-combined_k141_140868\|\|full | 1163 | No | NA | | 2 | 0 | 1 | Not-determined | Genome-fragment | NA | NA | 0 |
| S1-final-viral-combined_k141_222988\|\|full | 1539 | No | NA | | 3 | 0 | 2 | Not-determined | Genome-fragment | NA | NA | 0 |
| S2-final-viral-combined_k141_78900\|\|full | 1351 | No | NA | | 3 | 1 | 0 | Low-quality | Genome-fragment | 2.17 | AAI-based (high-confidence) | 0 |
| S4_k141_147421 | 1351 | No | NA | | 1 | 0 | 1 | Low-quality | Genome-fragment | 0.75 | AAI-based (medium-confidence) | 0 |
| S1-final-viral-combined_k141_25376\|\|full | 3237 | No | NA | | 6 | 0 | 2 | Not-determined | Genome-fragment | NA | NA | 0 |
| S8-final-viral-combined_k141_85286\|\|full | 1351 | No | NA | | 2 | 1 | 0 | Low-quality | Genome-fragment | 2.04 | HMM-based (lower-bound) | 0 |
| S1-final-viral-combined_k141_222997\|\|full | 6376 | No | NA | | 6 | 0 | 4 | Not-determined | Genome-fragment | NA | NA | 0 |
| S1-final-viral-combined_k141_74001\|\|full | 1009 | No | NA | | 2 | 0 | 1 | Not-determined | Genome-fragment | NA | NA | 0 |
| S9_k141_127364 | 1352 | No | NA | | 2 | 2 | 0 | Low-quality | Genome-fragment | 3.33 | AAI-based (medium-confidence) | 0 |
| S11-final-viral-combined_k141_194298\|\|full | 1352 | No | NA | | 3 | 0 | 0 | Low-quality | Genome-fragment | 3.09 | AAI-based (medium-confidence) | 0 |
| S1-final-viral-combined_k141_223024\|\|full | 1994 | No | NA | | 2 | 0 | 2 | Not-determined | Genome-fragment | NA | NA | 0 |
| S1-final-viral-combined_k141_288806\|\|full | 4902 | No | NA | | 4 | 0 | 2 | Not-determined | Genome-fragment | NA | NA | 0 |
| S1-final-viral-combined_k141_387896\|\|full | 1626 | No | NA | | 2 | 0 | 1 | Not-determined | Genome-fragment | NA | NA | 0 |
| S1-final-viral-combined_k141_223052\|\|full | 2683 | No | NA | | 5 | 0 | 1 | Not-determined | Genome-fragment | NA | NA | 0 |
| S11-final-viral-combined_k141_174057\|\|full | 1352 | No | NA | | 3 | 0 | 0 | Low-quality | Genome-fragment | 2.59 | AAI-based (medium-confidence) | 0 |
| S1-final-viral-combined_k141_352357\|\|full | 1352 | No | NA | | 3 | 0 | 2 | Low-quality | Genome-fragment | 0.46 | AAI-based (medium-confidence) | 0 |
| S1-final-viral-combined_k141_288835\|\|full | 1899 | No | NA | | 2 | 0 | 2 | Not-determined | Genome-fragment | NA | NA | 0 |
| S1-final-viral-combined_k141_89876\|\|full | 1352 | No | NA | | 2 | 0 | 2 | Low-quality | Genome-fragment | 2.26 | AAI-based (medium-confidence) | 0 |
| S1-final-viral-combined_k141_58197\|\|full | 11554 | No | NA | | 11 | 0 | 7 | Not-determined | Genome-fragment | NA | NA | 0 |
| S1-final-viral-combined_k141_337458\|\|full | 3360 | No | NA | | 3 | 0 | 2 | Not-determined | Genome-fragment | NA | NA | 0 |
| S1-final-viral-combined_k141_106797\|\|full | 1357 | No | NA | | 3 | 0 | 3 | Not-determined | Genome-fragment | NA | NA | 0 |
| S1-final-viral-combined_k141_156506\|\|full | 1441 | No | NA | | 3 | 0 | 2 | Not-determined | Genome-fragment | NA | NA | 0 |
| S1-final-viral-combined_k141_207263\|\|full | 1834 | No | NA | | 2 | 0 | 2 | Not-determined | Genome-fragment | NA | NA | 0 |
| S1-final-viral-combined_k141_106810\|\|full | 1165 | No | NA | | 3 | 0 | 1 | Not-determined | Genome-fragment | NA | NA | 0 |
| S1-final-viral-combined_k141_156514\|\|full | 1614 | No | NA | | 4 | 0 | 0 | Not-determined | Genome-fragment | NA | NA | 0 |
| S1-final-viral-combined_k141_156523\|\|full | 4747 | No | NA | | 13 | 0 | 0 | Not-determined | Genome-fragment | NA | NA | 0 |
| S3-final-viral-combined_k141_56462\|\|full | 1352 | No | NA | | 3 | 0 | 1 | Low-quality | Genome-fragment | 0.75 | AAI-based (high-confidence) | 0 |
| S1-final-viral-combined_k141_172632\|\|full | 3069 | No | NA | | 3 | 0 | 1 | Not-determined | Genome-fragment | NA | NA | 0 |
| S1-final-viral-combined_k141_239790\|\|full | 4990 | No | NA | | 7 | 1 | 0 | Not-determined | Genome-fragment | NA | NA | 0 |
| S5_k141_63420 | 1352 | No | NA | | 1 | 0 | 0 | Low-quality | Genome-fragment | 2.81 | AAI-based (medium-confidence) | 0 |
| S1-final-viral-combined_k141_172636\|\|full | 4551 | No | NA | | 3 | 0 | 3 | Not-determined | Genome-fragment | NA | NA | 0 |
| S7-final-viral-combined_k141_23194\|\|full | 1352 | No | NA | | 3 | 1 | 0 | Low-quality | Genome-fragment | 1.57 | AAI-based (medium-confidence) | 0 |
| S1-final-viral-combined_k141_306142\|\|full | 1797 | No | NA | | 3 | 0 | 2 | Not-determined | Genome-fragment | NA | NA | 0 |
| S1-final-viral-combined_k141_106867\|\|full | 2344 | No | NA | | 3 | 0 | 1 | Not-determined | Genome-fragment | NA | NA | 0 |
| S1-final-viral-combined_k141_9350\|\|full | 13160 | No | NA | | 11 | 0 | 5 | Not-determined | Genome-fragment | NA | NA | 0 |
| S1-final-viral-combined_k141_223081\|\|full | 1704 | No | NA | | 3 | 0 | 0 | Not-determined | Genome-fragment | NA | NA | 0 |
| S1-final-viral-combined_k141_207327\|\|full | 2608 | No | NA | | 2 | 0 | 1 | Not-determined | Genome-fragment | NA | NA | 0 |
| S1-final-viral-combined_k141_91342\|\|full | 7354 | No | NA | | 6 | 0 | 1 | Not-determined | Genome-fragment | NA | NA | 0 |
| S1-final-viral-combined_k141_337549\|\|full | 1411 | No | NA | | 3 | 0 | 2 | Not-determined | Genome-fragment | NA | NA | 0 |
| S1-final-viral-combined_k141_337559\|\|full | 2228 | No | NA | | 3 | 0 | 2 | Not-determined | Genome-fragment | NA | NA | 0 |
| S17-final-viral-combined_k141_68903\|\|full | 1352 | No | NA | | 3 | 2 | 0 | Low-quality | Genome-fragment | 3.94 | AAI-based (medium-confidence) | 0 |
| S17-k141_90264 | 1352 | No | NA | | 1 | 0 | 0 | Low-quality | Genome-fragment | 0.88 | AAI-based (high-confidence) | 0 |
| S1-final-viral-combined_k141_156571\|\|full | 2115 | No | NA | | 4 | 0 | 2 | Not-determined | Genome-fragment | NA | NA | 0 |
| S12_k141_364594 | 1352 | No | NA | | 2 | 0 | 0 | Low-quality | Genome-fragment | 2.44 | AAI-based (high-confidence) | 0 |
| S2-final-viral-combined_k141_69242\|\|full | 1352 | No | NA | | 3 | 0 | 0 | Low-quality | Genome-fragment | 1.93 | AAI-based (high-confidence) | 0 |
| S1-final-viral-combined_k141_322919\|\|full | 3817 | No | NA | | 6 | 0 | 3 | Not-determined | Genome-fragment | NA | NA | 0 |
| S11_k141_206084 | 1353 | No | NA | | 4 | 0 | 0 | Low-quality | Genome-fragment | 2.21 | AAI-based (high-confidence) | 0 |
| S1-final-viral-combined_k141_355243\|\|full | 4048 | No | NA | | 5 | 0 | 4 | Not-determined | Genome-fragment | NA | NA | 0 |
| S1-final-viral-combined_k141_371190\|\|full | 3691 | No | NA | | 2 | 0 | 2 | Not-determined | Genome-fragment | NA | NA | 0 |
| S1-final-viral-combined_k141_41664\|\|full | 1064 | No | NA | | 3 | 0 | 0 | Not-determined | Genome-fragment | NA | NA | 0 |
| S1-final-viral-combined_k141_66473\|\|full | 1353 | No | NA | | 4 | 0 | 0 | Low-quality | Genome-fragment | 2.63 | AAI-based (high-confidence) | 0 |
| S1-final-viral-combined_k141_384458\|\|full | 1353 | No | NA | | 2 | 0 | 1 | Low-quality | Genome-fragment | 2.24 | AAI-based (medium-confidence) | 0 |
| S1-final-viral-combined_k141_306247\|\|full | 7697 | No | NA | | 7 | 0 | 7 | Not-determined | Genome-fragment | NA | NA | 0 |
| S3-final-viral-combined_k141_76608\|\|full | 1353 | No | NA | | 2 | 0 | 1 | Low-quality | Genome-fragment | 1.41 | AAI-based (medium-confidence) | 0 |
| S5-final-viral-combined_k141_163063\|\|full | 1353 | No | NA | | 3 | 0 | 1 | Low-quality | Genome-fragment | 3.1 | AAI-based (medium-confidence) | 0 |
| S1-final-viral-combined_k141_223210\|\|full | 4751 | No | NA | | 4 | 0 | 2 | Not-determined | Genome-fragment | NA | NA | 0 |
| S1-final-viral-combined_k141_156637\|\|full | 11186 | No | NA | | 20 | 5 | 1 | Medium-quality | Genome-fragment | 87.8 | AAI-based (high-confidence) | 0 |
| S1-final-viral-combined_k141_9480\|\|full | 3989 | No | NA | | 3 | 0 | 2 | Not-determined | Genome-fragment | NA | NA | 0 |
| S1-final-viral-combined_k141_74233\|\|full | 1521 | No | NA | | 2 | 0 | 0 | Not-determined | Genome-fragment | NA | NA | 0 |
| S1-final-viral-combined_k141_41739\|\|full | 3473 | No | NA | | 4 | 0 | 1 | Not-determined | Genome-fragment | NA | NA | 0 |
| S1-final-viral-combined_k141_141152\|\|full | 1503 | No | NA | | 3 | 0 | 2 | Not-determined | Genome-fragment | NA | NA | 0 |
| S5-final-viral-combined_k141_115663\|\|full | 1353 | No | NA | | 2 | 0 | 0 | Low-quality | Genome-fragment | 3.41 | AAI-based (high-confidence) | 0 |
| S1-final-viral-combined_k141_223253\|\|full | 1149 | No | NA | | 2 | 0 | 0 | Not-determined | Genome-fragment | NA | NA | 0 |
| S1-final-viral-combined_k141_387998\|\|full | 1351 | No | NA | | 2 | 0 | 1 | Not-determined | Genome-fragment | NA | NA | 0 |
| S1-final-viral-combined_k141_74265\|\|full | 3061 | No | NA | | 2 | 0 | 2 | Not-determined | Genome-fragment | NA | NA | 0 |
| S12-final-viral-combined_k141_93654\|\|full | 1353 | No | NA | | 3 | 1 | 0 | Low-quality | Genome-fragment | 2.78 | AAI-based (high-confidence) | 0 |
| S1-final-viral-combined_k141_322982\|\|full | 1745 | No | NA | | 3 | 0 | 2 | Not-determined | Genome-fragment | NA | NA | 0 |
| S12-final-viral-combined_k141_92833\|\|lt2gene | 1353 | No | NA | | 2 | 2 | 0 | Low-quality | Genome-fragment | 3.14 | AAI-based (high-confidence) | 0 |
| S1-final-viral-combined_k141_223296\|\|full | 1717 | No | NA | | 3 | 0 | 0 | Not-determined | Genome-fragment | NA | NA | 0 |
| S1-final-viral-combined_k141_388049\|\|full | 1066 | No | NA | | 2 | 0 | 1 | Not-determined | Genome-fragment | NA | NA | 0 |
| S1-final-viral-combined_k141_125392\|\|full | 2693 | No | NA | | 3 | 0 | 1 | Not-determined | Genome-fragment | NA | NA | 0 |
| S1-final-viral-combined_k141_58377\|\|full | 3266 | No | NA | | 3 | 0 | 2 | Not-determined | Genome-fragment | NA | NA | 0 |
| S1-final-viral-combined_k141_107040\|\|full | 2210 | No | NA | | 3 | 0 | 2 | Not-determined | Genome-fragment | NA | NA | 0 |
| S4-final-viral-combined_k141_147200\|\|full | 1353 | No | NA | | 4 | 0 | 0 | Low-quality | Genome-fragment | 3.69 | AAI-based (medium-confidence) | 0 |
| S4-final-viral-combined_k141_94077\|\|full | 1353 | No | NA | | 3 | 0 | 0 | Low-quality | Genome-fragment | 2.72 | AAI-based (medium-confidence) | 0 |
| S1-final-viral-combined_k141_9511\|\|full | 4921 | No | NA | | 3 | 0 | 1 | Not-determined | Genome-fragment | NA | NA | 0 |
| S4-final-viral-combined_k141_71212\|\|lt2gene | 1353 | No | NA | | 2 | 2 | 0 | Low-quality | Genome-fragment | 2.82 | AAI-based (high-confidence) | 0 |
| S9_k141_2222 | 1354 | No | NA | | 3 | 0 | 0 | Low-quality | Genome-fragment | 4.1 | AAI-based (medium-confidence) | 0 |
| S11-final-viral-combined_k141_210789\|\|full | 1354 | No | NA | | 4 | 0 | 0 | Low-quality | Genome-fragment | 3.84 | AAI-based (medium-confidence) | 0 |
| S1-final-viral-combined_k141_58411\|\|full | 4030 | No | NA | | 3 | 0 | 3 | Not-determined | Genome-fragment | NA | NA | 0 |
| S1-final-viral-combined_k141_9534\|\|full | 4899 | No | NA | | 5 | 0 | 3 | Not-determined | Genome-fragment | NA | NA | 0 |
| S1-final-viral-combined_k141_249563\|\|full | 1354 | No | NA | | 3 | 0 | 0 | Low-quality | Genome-fragment | 2.44 | AAI-based (medium-confidence) | 0 |
| S5-final-viral-combined_k141_145589\|\|lt2gene | 1354 | No | NA | | 2 | 2 | 0 | Low-quality | Genome-fragment | 2.66 | AAI-based (high-confidence) | 0 |
| S17-final-viral-combined_k141_83516\|\|full | 1354 | No | NA | | 3 | 0 | 0 | Low-quality | Genome-fragment | 3.12 | AAI-based (medium-confidence) | 0 |
| S17-k141_129806 | 1354 | No | NA | | 2 | 0 | 0 | Low-quality | Genome-fragment | 3.29 | AAI-based (high-confidence) | 0 |
| S1-final-viral-combined_k141_107073\|\|full | 21709 | No | NA | | 31 | 13 | 0 | Medium-quality | Genome-fragment | 56.42 | AAI-based (high-confidence) | 0 |
| S6-final-viral-combined_k141_89390\|\|full | 1354 | No | NA | | 2 | 0 | 0 | Low-quality | Genome-fragment | 2.88 | AAI-based (high-confidence) | 0 |
| S1-final-viral-combined_k141_107102\|\|full | 2689 | No | NA | | 4 | 0 | 0 | Not-determined | Genome-fragment | NA | NA | 0 |
| S1-final-viral-combined_k141_9568\|\|full | 1437 | No | NA | | 2 | 0 | 0 | Not-determined | Genome-fragment | NA | NA | 0 |
| S8-final-viral-combined_k141_155484\|\|full | 1354 | No | NA | | 4 | 0 | 1 | Low-quality | Genome-fragment | 1.32 | AAI-based (medium-confidence) | 0 |
| S1-final-viral-combined_k141_74361\|\|full | 4014 | No | NA | | 5 | 0 | 3 | Not-determined | Genome-fragment | NA | NA | 0 |
| S1-final-viral-combined_k141_388132\|\|full | 5532 | No | NA | | 4 | 0 | 4 | Not-determined | Genome-fragment | NA | NA | 0 |
| S8_k141_239573 | 1354 | No | NA | | 2 | 0 | 0 | Low-quality | Genome-fragment | 0.88 | AAI-based (medium-confidence) | 0 |
| S1-final-viral-combined_k141_191183\|\|full | 1771 | No | NA | | 2 | 0 | 1 | Not-determined | Genome-fragment | NA | NA | 0 |
| S1-final-viral-combined_k141_236127\|\|full | 1355 | No | NA | | 3 | 0 | 1 | Low-quality | Genome-fragment | 0.72 | AAI-based (medium-confidence) | 0 |
| S5-final-viral-combined_k141_4241\|\|full | 1355 | No | NA | | 4 | 0 | 0 | Low-quality | Genome-fragment | 0.85 | AAI-based (medium-confidence) | 0 |
| S5-final-viral-combined_k141_143873\|\|lt2gene | 1355 | No | NA | | 2 | 1 | 0 | Low-quality | Genome-fragment | 1.52 | HMM-based (lower-bound) | 0 |
| S1-final-viral-combined_k141_74412\|\|full | 1165 | No | NA | | 2 | 0 | 1 | Not-determined | Genome-fragment | NA | NA | 0 |
| S7-final-viral-combined_k141_55930\|\|full | 1355 | No | NA | | 4 | 1 | 0 | Low-quality | Genome-fragment | 3.82 | AAI-based (high-confidence) | 0 |
| S1-final-viral-combined_k141_156796\|\|full | 4204 | No | NA | | 5 | 0 | 2 | Not-determined | Genome-fragment | NA | NA | 0 |
| S17-k141_155801 | 1355 | No | NA | | 4 | 0 | 2 | Low-quality | Genome-fragment | 2.79 | AAI-based (high-confidence) | 0 |
| S1-final-viral-combined_k141_141285\|\|full | 3915 | No | NA | | 4 | 0 | 2 | Not-determined | Genome-fragment | NA | NA | 0 |
| S1-final-viral-combined_k141_306336\|\|full | 1582 | No | NA | | 3 | 0 | 2 | Not-determined | Genome-fragment | NA | NA | 0 |
| S12-final-viral-combined_k141_374418\|\|full | 1355 | No | NA | | 4 | 0 | 0 | Low-quality | Genome-fragment | 2.66 | AAI-based (high-confidence) | 0 |
| S12-final-viral-combined_k141_160769\|\|full | 1355 | No | NA | | 2 | 0 | 1 | Low-quality | Genome-fragment | 2.13 | AAI-based (medium-confidence) | 0 |
| S1-final-viral-combined_k141_125544\|\|full | 5186 | No | NA | | 5 | 0 | 4 | Not-determined | Genome-fragment | NA | NA | 0 |
| S1-final-viral-combined_k141_371448\|\|full | 5455 | No | NA | | 6 | 0 | 3 | Not-determined | Genome-fragment | NA | NA | 0 |
| S1-final-viral-combined_k141_191206\|\|full | 1634 | No | NA | | 2 | 0 | 2 | Not-determined | Genome-fragment | NA | NA | 0 |
| S1-final-viral-combined_k141_355665\|\|full | 2602 | No | NA | | 3 | 0 | 3 | Not-determined | Genome-fragment | NA | NA | 0 |
| S1-final-viral-combined_k141_388241\|\|full | 2572 | No | NA | | 3 | 0 | 3 | Not-determined | Genome-fragment | NA | NA | 0 |
| S12-final-viral-combined_k141_237135\|\|lt2gene | 1355 | No | NA | | 2 | 1 | 0 | Low-quality | Genome-fragment | 0.75 | HMM-based (lower-bound) | 0 |
| S2-final-viral-combined_k141_35834\|\|full | 1355 | No | NA | | 3 | 0 | 1 | Low-quality | Genome-fragment | 3.17 | AAI-based (medium-confidence) | 0 |
| S1-final-viral-combined_k141_371493\|\|full | 5659 | No | NA | | 9 | 0 | 7 | Not-determined | Genome-fragment | NA | NA | 0 |
| S8-final-viral-combined_k141_265009\|\|full | 1355 | No | NA | | 3 | 0 | 0 | Low-quality | Genome-fragment | 5.39 | AAI-based (high-confidence) | 0 |
| S1-final-viral-combined_k141_41971\|\|full | 1643 | No | NA | | 3 | 0 | 3 | Not-determined | Genome-fragment | NA | NA | 0 |
| S1-final-viral-combined_k141_323197\|\|full | 4581 | No | NA | | 4 | 0 | 4 | Not-determined | Genome-fragment | NA | NA | 0 |
| S1-final-viral-combined_k141_9760\|\|full | 1316 | No | NA | | 2 | 0 | 1 | Not-determined | Genome-fragment | NA | NA | 0 |
| S1-final-viral-combined_k141_91800\|\|full | 1833 | No | NA | | 3 | 0 | 0 | Not-determined | Genome-fragment | NA | NA | 0 |
| S1-final-viral-combined_k141_173115\|\|full | 1511 | No | NA | | 3 | 0 | 1 | Not-determined | Genome-fragment | NA | NA | 0 |
| S1-final-viral-combined_k141_91809\|\|full | 4005 | No | NA | | 3 | 0 | 3 | Not-determined | Genome-fragment | NA | NA | 0 |
| S1-final-viral-combined_k141_289223\|\|full | 1207 | No | NA | | 4 | 0 | 2 | Not-determined | Genome-fragment | NA | NA | 0 |
| S1-final-viral-combined_k141_74545\|\|full | 10676 | No | NA | | 6 | 0 | 6 | Not-determined | Genome-fragment | NA | NA | 0 |
| S1-final-viral-combined_k141_323240\|\|full | 1798 | No | NA | | 3 | 0 | 2 | Not-determined | Genome-fragment | NA | NA | 0 |
| S8-final-viral-combined_k141_56067\|\|full | 1355 | No | NA | | 3 | 2 | 0 | Low-quality | Genome-fragment | 2.4 | AAI-based (high-confidence) | 0 |
| S11-final-viral-combined_k141_20096\|\|full | 1356 | No | NA | | 2 | 0 | 0 | Low-quality | Genome-fragment | 2.41 | AAI-based (medium-confidence) | 0 |
| S1-final-viral-combined_k141_191296\|\|full | 2033 | No | NA | | 2 | 0 | 2 | Not-determined | Genome-fragment | NA | NA | 0 |
| S1-final-viral-combined_k141_156897\|\|full | 1184 | No | NA | | 2 | 0 | 2 | Not-determined | Genome-fragment | NA | NA | 0 |
| S1-final-viral-combined_k141_42023\|\|full | 1313 | No | NA | | 1 | 0 | 1 | Not-determined | Genome-fragment | NA | NA | 0 |
| S1-final-viral-combined_k141_289252\|\|full | 1023 | No | NA | | 2 | 0 | 1 | Not-determined | Genome-fragment | NA | NA | 0 |
| S1-final-viral-combined_k141_173137\|\|full | 1157 | No | NA | | 2 | 0 | 0 | Not-determined | Genome-fragment | NA | NA | 0 |
| S1-final-viral-combined_k141_55688\|\|full | 1356 | No | NA | | 4 | 0 | 0 | Low-quality | Genome-fragment | 2.79 | AAI-based (medium-confidence) | 0 |
| S1-final-viral-combined_k141_257156\|\|full | 1449 | No | NA | | 3 | 0 | 3 | Not-determined | Genome-fragment | NA | NA | 0 |
| S1-final-viral-combined_k141_223295\|\|full | 1010 | No | NA | | 2 | 0 | 2 | Not-determined | Genome-fragment | NA | NA | 0 |
| S1-final-viral-combined_k141_141431\|\|full | 3026 | No | NA | | 3 | 0 | 2 | Not-determined | Genome-fragment | NA | NA | 0 |
| S1-final-viral-combined_k141_289291\|\|full | 1221 | No | NA | | 2 | 0 | 1 | Not-determined | Genome-fragment | NA | NA | 0 |
| S1-final-viral-combined_k141_146617\|\|full | 1356 | No | NA | | 3 | 1 | 0 | Low-quality | Genome-fragment | 2.27 | AAI-based (high-confidence) | 0 |
| S1-final-viral-combined_k141_306481\|\|full | 1438 | No | NA | | 3 | 0 | 2 | Not-determined | Genome-fragment | NA | NA | 0 |
| S1-final-viral-combined_k141_25859\|\|full | 5118 | No | NA | | 6 | 0 | 6 | Not-determined | Genome-fragment | NA | NA | 0 |
| S1-final-viral-combined_k141_289301\|\|full | 1635 | No | NA | | 3 | 0 | 1 | Not-determined | Genome-fragment | NA | NA | 0 |
| S1-final-viral-combined_k141_16170\|\|lt2gene | 1356 | No | NA | | 1 | 1 | 0 | Low-quality | Genome-fragment | 1.24 | HMM-based (lower-bound) | 0 |
| S7_k141_90886 | 1356 | No | NA | | 4 | 0 | 0 | Low-quality | Genome-fragment | 1.72 | AAI-based (high-confidence) | 0 |
| S17-final-viral-combined_k141_91780\|\|full | 1356 | No | NA | | 3 | 0 | 0 | Low-quality | Genome-fragment | 2.57 | AAI-based (medium-confidence) | 0 |
| S1-final-viral-combined_k141_257165\|\|full | 1088 | No | NA | | 2 | 0 | 2 | Not-determined | Genome-fragment | NA | NA | 0 |
| S1-final-viral-combined_k141_141449\|\|full | 6302 | No | NA | | 10 | 0 | 0 | Not-determined | Genome-fragment | NA | NA | 0 |
| S1-final-viral-combined_k141_306499\|\|full | 2406 | No | NA | | 3 | 0 | 2 | Not-determined | Genome-fragment | NA | NA | 0 |
| S17-k141_33681 | 1356 | No | NA | | 1 | 0 | 1 | Low-quality | Genome-fragment | 0.47 | AAI-based (medium-confidence) | 0 |
| S12-final-viral-combined_k141_211436\|\|lt2gene | 1356 | No | NA | | 2 | 1 | 0 | Low-quality | Genome-fragment | 2.26 | AAI-based (high-confidence) | 0 |
| S1-final-viral-combined_k141_257176\|\|full | 1276 | No | NA | | 3 | 0 | 0 | Not-determined | Genome-fragment | NA | NA | 0 |
| S1-final-viral-combined_k141_323361\|\|full | 2268 | No | NA | | 3 | 0 | 2 | Not-determined | Genome-fragment | NA | NA | 0 |
| S12_k141_127844 | 1356 | No | NA | | 2 | 0 | 0 | Low-quality | Genome-fragment | 1.14 | AAI-based (high-confidence) | 0 |
| S1-final-viral-combined_k141_289369\|\|full | 4427 | No | NA | | 3 | 0 | 2 | Not-determined | Genome-fragment | NA | NA | 0 |
| S2-final-viral-combined_k141_142957\|\|full | 1356 | No | NA | | 3 | 0 | 0 | Low-quality | Genome-fragment | 2.8 | AAI-based (medium-confidence) | 0 |
| S2_k141_217159 | 1356 | No | NA | | 4 | 0 | 0 | Low-quality | Genome-fragment | 1.93 | AAI-based (high-confidence) | 0 |
| S1-final-viral-combined_k141_306555\|\|full | 5943 | No | NA | | 5 | 0 | 4 | Not-determined | Genome-fragment | NA | NA | 0 |
| S1-final-viral-combined_k141_74656\|\|full | 1282 | No | NA | | 2 | 0 | 2 | Not-determined | Genome-fragment | NA | NA | 0 |
| S1-final-viral-combined_k141_240282\|\|full | 2968 | No | NA | | 3 | 0 | 1 | Not-determined | Genome-fragment | NA | NA | 0 |
| S1-final-viral-combined_k141_337961\|\|full | 1168 | No | NA | | 2 | 0 | 2 | Not-determined | Genome-fragment | NA | NA | 0 |
| S1-final-viral-combined_k141_337972\|\|full | 2006 | No | NA | | 2 | 0 | 0 | Not-determined | Genome-fragment | NA | NA | 0 |
| S1-final-viral-combined_k141_173207\|\|full | 1270 | No | NA | | 3 | 0 | 0 | Not-determined | Genome-fragment | NA | NA | 0 |
| S1-final-viral-combined_k141_191447\|\|full | 1539 | No | NA | | 2 | 0 | 1 | Not-determined | Genome-fragment | NA | NA | 0 |
| S11-final-viral-combined_k141_170025\|\|lt2gene | 1357 | No | NA | | 2 | 0 | 0 | Low-quality | Genome-fragment | 3.63 | AAI-based (medium-confidence) | 0 |
| S1-final-viral-combined_k141_223426\|\|full | 8237 | No | NA | | 8 | 0 | 6 | Not-determined | Genome-fragment | NA | NA | 0 |
| S1-final-viral-combined_k141_191473\|\|full | 1902 | No | NA | | 3 | 0 | 1 | Not-determined | Genome-fragment | NA | NA | 0 |
| S1-final-viral-combined_k141_381385\|\|full | 1357 | No | NA | | 2 | 0 | 1 | Low-quality | Genome-fragment | 1.15 | AAI-based (medium-confidence) | 0 |
| S1-final-viral-combined_k141_223439\|\|full | 1898 | No | NA | | 4 | 0 | 2 | Not-determined | Genome-fragment | NA | NA | 0 |
| S1-final-viral-combined_k141_23246\|\|full | 1357 | No | NA | | 3 | 0 | 0 | Low-quality | Genome-fragment | 2.28 | AAI-based (high-confidence) | 0 |
| S1-final-viral-combined_k141_63497\|\|full | 1357 | No | NA | | 3 | 0 | 0 | Low-quality | Genome-fragment | 0.92 | AAI-based (high-confidence) | 0 |
| S1-final-viral-combined_k141_289478\|\|full | 2737 | No | NA | | 4 | 0 | 1 | Not-determined | Genome-fragment | NA | NA | 0 |
| S1-final-viral-combined_k141_125700\|\|full | 3378 | No | NA | | 4 | 0 | 3 | Not-determined | Genome-fragment | NA | NA | 0 |
| S1_k141_305954 | 1357 | No | NA | | 3 | 3 | 0 | Low-quality | Genome-fragment | 1.89 | AAI-based (high-confidence) | 0 |
| S3-final-viral-combined_k141_66423\|\|lt2gene | 1357 | No | NA | | 1 | 1 | 0 | Low-quality | Genome-fragment | 3.68 | AAI-based (high-confidence) | 0 |
| S5-final-viral-combined_k141_99641\|\|full | 1357 | No | NA | | 2 | 0 | 2 | Low-quality | Genome-fragment | 0.84 | AAI-based (high-confidence) | 0 |
| S7_k141_113061 | 1357 | No | NA | | 2 | 0 | 0 | Low-quality | Genome-fragment | 0.52 | AAI-based (medium-confidence) | 0 |
| S1-final-viral-combined_k141_371637\|\|full | 1592 | No | NA | | 2 | 0 | 1 | Not-determined | Genome-fragment | NA | NA | 0 |
| S1-final-viral-combined_k141_371653\|\|full | 1413 | No | NA | | 2 | 0 | 0 | Not-determined | Genome-fragment | NA | NA | 0 |
| S1-final-viral-combined_k141_257326\|\|full | 5277 | No | NA | | 5 | 0 | 4 | Not-determined | Genome-fragment | NA | NA | 0 |
| S12-final-viral-combined_k141_82537\|\|full | 1357 | No | NA | | 4 | 0 | 0 | Low-quality | Genome-fragment | 2.92 | AAI-based (high-confidence) | 0 |
| S12-final-viral-combined_k141_334592\|\|full | 1357 | No | NA | | 3 | 1 | 0 | Low-quality | Genome-fragment | 2.44 | AAI-based (medium-confidence) | 0 |
| S1-final-viral-combined_k141_191590\|\|full | 1673 | No | NA | | 3 | 0 | 0 | Not-determined | Genome-fragment | NA | NA | 0 |
| S1-final-viral-combined_k141_371685\|\|full | 1294 | No | NA | | 3 | 0 | 0 | Not-determined | Genome-fragment | NA | NA | 0 |
| S12_k141_302033 | 1357 | No | NA | | 2 | 1 | 0 | Low-quality | Genome-fragment | 2 | HMM-based (lower-bound) | 0 |
| S1-final-viral-combined_k141_92003\|\|full | 2440 | No | NA | | 3 | 0 | 3 | Not-determined | Genome-fragment | NA | NA | 0 |
| S1-final-viral-combined_k141_306743\|\|full | 5882 | No | NA | | 4 | 0 | 4 | Not-determined | Genome-fragment | NA | NA | 0 |
| S1-final-viral-combined_k141_42225\|\|full | 6330 | No | NA | | 5 | 0 | 4 | Not-determined | Genome-fragment | NA | NA | 0 |
| S1-final-viral-combined_k141_92013\|\|full | 1035 | No | NA | | 3 | 0 | 0 | Not-determined | Genome-fragment | NA | NA | 0 |
| S1-final-viral-combined_k141_26125\|\|full | 17878 | No | NA | | 14 | 0 | 13 | Not-determined | Genome-fragment | NA | NA | 0 |
| S1-final-viral-combined_k141_92035\|\|full | 5893 | No | NA | | 7 | 0 | 3 | Not-determined | Genome-fragment | NA | NA | 0 |
| S1-final-viral-combined_k141_10093\|\|full | 1982 | No | NA | | 2 | 0 | 2 | Not-determined | Genome-fragment | NA | NA | 0 |
| S1-final-viral-combined_k141_338175\|\|full | 1683 | No | NA | | 3 | 0 | 2 | Not-determined | Genome-fragment | NA | NA | 0 |
| S2-final-viral-combined_k141_38194\|\|full | 1357 | No | NA | | 4 | 0 | 0 | Low-quality | Genome-fragment | 2.72 | AAI-based (medium-confidence) | 0 |
| S2_k141_49806 | 1357 | No | NA | | 3 | 0 | 0 | Low-quality | Genome-fragment | 1.06 | AAI-based (medium-confidence) | 0 |
| S6_k141_26029 | 1357 | No | NA | | 2 | 0 | 2 | Low-quality | Genome-fragment | 1.06 | AAI-based (medium-confidence) | 0 |
| S1-final-viral-combined_k141_107632\|\|full | 1398 | No | NA | | 3 | 0 | 1 | Not-determined | Genome-fragment | NA | NA | 0 |
| S8-final-viral-combined_k141_65422\|\|full | 1357 | No | NA | | 3 | 0 | 0 | Low-quality | Genome-fragment | 3.45 | AAI-based (medium-confidence) | 0 |
| S1-final-viral-combined_k141_323584\|\|full | 4093 | No | NA | | 5 | 0 | 3 | Not-determined | Genome-fragment | NA | NA | 0 |
| S1-final-viral-combined_k141_173326\|\|full | 7068 | No | NA | | 6 | 0 | 5 | Not-determined | Genome-fragment | NA | NA | 0 |
| S1-final-viral-combined_k141_173328\|\|full | 1519 | No | NA | | 2 | 0 | 0 | Not-determined | Genome-fragment | NA | NA | 0 |
| S1-final-viral-combined_k141_10116\|\|full | 1127 | No | NA | | 3 | 0 | 0 | Not-determined | Genome-fragment | NA | NA | 0 |
| S1-final-viral-combined_k141_289664\|\|full | 1391 | No | NA | | 2 | 0 | 2 | Not-determined | Genome-fragment | NA | NA | 0 |
| S11-final-viral-combined_k141_227054\|\|full | 1358 | No | NA | | 3 | 0 | 0 | Low-quality | Genome-fragment | 2.8 | AAI-based (high-confidence) | 0 |
| S11_k141_113780 | 1358 | No | NA | | 1 | 0 | 1 | Low-quality | Genome-fragment | 0.82 | AAI-based (medium-confidence) | 0 |
| S1-final-viral-combined_k141_125837\|\|full | 1053 | No | NA | | 2 | 0 | 0 | Not-determined | Genome-fragment | NA | NA | 0 |
| S1-final-viral-combined_k141_173350\|\|full | 1381 | No | NA | | 3 | 0 | 0 | Not-determined | Genome-fragment | NA | NA | 0 |
| S1-final-viral-combined_k141_125841\|\|full | 4904 | No | NA | | 7 | 0 | 2 | Not-determined | Genome-fragment | NA | NA | 0 |
| S1-final-viral-combined_k141_150632\|\|lt2gene | 1358 | No | NA | | 1 | 1 | 0 | Low-quality | Genome-fragment | 3.57 | AAI-based (high-confidence) | 0 |
| S1-final-viral-combined_k141_338224\|\|full | 1431 | No | NA | | 3 | 0 | 1 | Not-determined | Genome-fragment | NA | NA | 0 |
| S1-final-viral-combined_k141_371753\|\|full | 2158 | No | NA | | 4 | 0 | 3 | Not-determined | Genome-fragment | NA | NA | 0 |
| S1-final-viral-combined_k141_173376\|\|full | 6546 | No | NA | | 7 | 0 | 7 | Not-determined | Genome-fragment | NA | NA | 0 |
| S1-final-viral-combined_k141_191640\|\|full | 9295 | No | NA | | 5 | 0 | 4 | Not-determined | Genome-fragment | NA | NA | 0 |
| S3-final-viral-combined_k141_89933\|\|full | 1358 | No | NA | | 2 | 0 | 2 | Low-quality | Genome-fragment | 0.94 | AAI-based (medium-confidence) | 0 |
| S1-final-viral-combined_k141_157270\|\|full | 3973 | No | NA | | 3 | 0 | 2 | Not-determined | Genome-fragment | NA | NA | 0 |
| S1-final-viral-combined_k141_356053\|\|full | 1357 | No | NA | | 2 | 0 | 1 | Not-determined | Genome-fragment | NA | NA | 0 |
| S1-final-viral-combined_k141_289704\|\|full | 2015 | No | NA | | 3 | 0 | 2 | Not-determined | Genome-fragment | NA | NA | 0 |
| S5-final-viral-combined_k141_181804\|\|lt2gene | 1358 | No | NA | | 1 | 1 | 0 | Low-quality | Genome-fragment | 1.01 | HMM-based (lower-bound) | 0 |
| S7-final-viral-combined_k141_174592\|\|full | 1358 | No | NA | | 2 | 0 | 0 | Low-quality | Genome-fragment | 2.62 | AAI-based (medium-confidence) | 0 |
| S1-final-viral-combined_k141_107666\|\|full | 1513 | No | NA | | 3 | 0 | 1 | Not-determined | Genome-fragment | NA | NA | 0 |
| S17-final-viral-combined_k141_54603\|\|full | 1358 | No | NA | | 3 | 0 | 1 | Low-quality | Genome-fragment | 0.83 | AAI-based (medium-confidence) | 0 |
| S2-final-viral-combined_k141_74650\|\|full | 1358 | No | NA | | 3 | 1 | 0 | Low-quality | Genome-fragment | 3.01 | AAI-based (high-confidence) | 0 |
| S9-final-viral-combined_k141_71895\|\|full | 1359 | No | NA | | 4 | 1 | 0 | Low-quality | Genome-fragment | 4.04 | AAI-based (high-confidence) | 0 |
| S1-final-viral-combined_k141_257496\|\|full | 1589 | No | NA | | 4 | 0 | 1 | Not-determined | Genome-fragment | NA | NA | 0 |
| S1-final-viral-combined_k141_306863\|\|full | 6578 | No | NA | | 3 | 0 | 3 | Not-determined | Genome-fragment | NA | NA | 0 |
| S11-final-viral-combined_k141_323947\|\|lt2gene | 1359 | No | NA | | 2 | 1 | 0 | Low-quality | Genome-fragment | 2.19 | HMM-based (lower-bound) | 0 |
| S1-final-viral-combined_k141_289714\|\|full | 2537 | No | NA | | 2 | 0 | 2 | Not-determined | Genome-fragment | NA | NA | 0 |
| S11-final-viral-combined_k141_11921\|\|lt2gene | 1359 | No | NA | | 2 | 1 | 0 | Low-quality | Genome-fragment | 3.01 | AAI-based (high-confidence) | 0 |
| S1-final-viral-combined_k141_356106\|\|full | 1123 | No | NA | | 3 | 0 | 0 | Not-determined | Genome-fragment | NA | NA | 0 |
| S1-final-viral-combined_k141_240544\|\|full | 2192 | No | NA | | 4 | 0 | 3 | Not-determined | Genome-fragment | NA | NA | 0 |
| S1-final-viral-combined_k141_371791\|\|full | 9824 | No | NA | | 9 | 0 | 1 | Not-determined | Genome-fragment | NA | NA | 0 |
| S1-final-viral-combined_k141_257516\|\|full | 3121 | No | NA | | 3 | 0 | 0 | Not-determined | Genome-fragment | NA | NA | 0 |
| S1_k141_387052 | 1359 | No | NA | | 2 | 0 | 0 | Low-quality | Genome-fragment | 2.43 | AAI-based (high-confidence) | 0 |
| S1-final-viral-combined_k141_208276\|\|full | 3147 | No | NA | | 6 | 0 | 0 | Not-determined | Genome-fragment | NA | NA | 0 |
| S5-final-viral-combined_k141_34106\|\|full | 1359 | No | NA | | 3 | 0 | 0 | Low-quality | Genome-fragment | 3.1 | AAI-based (medium-confidence) | 0 |
| S5-final-viral-combined_k141_97665\|\|full | 1359 | No | NA | | 3 | 3 | 0 | Low-quality | Genome-fragment | 2.03 | AAI-based (high-confidence) | 0 |
| S7-final-viral-combined_k141_89791\|\|full | 1359 | No | NA | | 3 | 0 | 0 | Low-quality | Genome-fragment | 3.2 | AAI-based (medium-confidence) | 0 |
| S1-final-viral-combined_k141_42397\|\|full | 2257 | No | NA | | 2 | 0 | 0 | Not-determined | Genome-fragment | NA | NA | 0 |
| S1-final-viral-combined_k141_74781\|\|full | 1052 | No | NA | | 2 | 0 | 2 | Not-determined | Genome-fragment | NA | NA | 0 |
| S1-final-viral-combined_k141_74782\|\|full | 1311 | No | NA | | 3 | 0 | 0 | Not-determined | Genome-fragment | NA | NA | 0 |
| S1-final-viral-combined_k141_42403\|\|full | 1920 | No | NA | | 2 | 0 | 1 | Not-determined | Genome-fragment | NA | NA | 0 |
| S7_k141_200361 | 1359 | No | NA | | 2 | 0 | 1 | Low-quality | Genome-fragment | 0.65 | AAI-based (medium-confidence) | 0 |
| S1-final-viral-combined_k141_141627\|\|full | 2149 | No | NA | | 4 | 0 | 0 | Not-determined | Genome-fragment | NA | NA | 0 |
| S1-final-viral-combined_k141_191743\|\|full | 1109 | No | NA | | 2 | 0 | 0 | Not-determined | Genome-fragment | NA | NA | 0 |
| S17-final-viral-combined_k141_113710\|\|full | 1359 | No | NA | | 3 | 0 | 2 | Low-quality | Genome-fragment | 4.12 | AAI-based (medium-confidence) | 0 |
| S17-final-viral-combined_k141_96062\|\|full | 1359 | No | NA | | 2 | 0 | 0 | Low-quality | Genome-fragment | 1.74 | AAI-based (high-confidence) | 0 |
| S1-final-viral-combined_k141_289787\|\|full | 2484 | No | NA | | 5 | 0 | 1 | Not-determined | Genome-fragment | NA | NA | 0 |
| S12-final-viral-combined_k141_72428\|\|full | 1359 | No | NA | | 3 | 2 | 0 | Low-quality | Genome-fragment | 3.18 | AAI-based (medium-confidence) | 0 |
| S1-final-viral-combined_k141_273054\|\|full | 1917 | No | NA | | 2 | 0 | 2 | Not-determined | Genome-fragment | NA | NA | 0 |
| S12-final-viral-combined_k141_275023\|\|full | 1359 | No | NA | | 3 | 2 | 0 | Low-quality | Genome-fragment | 3.19 | AAI-based (medium-confidence) | 0 |
| S1-final-viral-combined_k141_74851\|\|full | 3100 | No | NA | | 4 | 0 | 3 | Not-determined | Genome-fragment | NA | NA | 0 |
| S1-final-viral-combined_k141_356210\|\|full | 4888 | No | NA | | 2 | 0 | 2 | Not-determined | Genome-fragment | NA | NA | 0 |
| S1-final-viral-combined_k141_257617\|\|full | 9271 | No | NA | | 9 | 0 | 1 | Not-determined | Genome-fragment | NA | NA | 0 |
| S1-final-viral-combined_k141_173467\|\|full | 3287 | No | NA | | 3 | 0 | 1 | Not-determined | Genome-fragment | NA | NA | 0 |
| S1-final-viral-combined_k141_338393\|\|full | 3233 | No | NA | | 4 | 0 | 2 | Not-determined | Genome-fragment | NA | NA | 0 |
| S12-final-viral-combined_k141_232241\|\|lt2gene | 1359 | No | NA | | 2 | 2 | 0 | Low-quality | Genome-fragment | 3.64 | AAI-based (medium-confidence) | 0 |
| S4-final-viral-combined_k141_58563\|\|full | 1359 | No | NA | | 3 | 0 | 1 | Low-quality | Genome-fragment | 1.49 | AAI-based (medium-confidence) | 0 |
| S1-final-viral-combined_k141_107766\|\|full | 53116 | Yes | | 36184 | 61 | 10 | 13 | Medium-quality | Genome-fragment | 86.89 | AAI-based (high-confidence) | 31.88 |
| S1-final-viral-combined_k141_240655\|\|full | 11704 | No | NA | | 10 | 0 | 4 | Not-determined | Genome-fragment | NA | NA | 0 |
| S1-final-viral-combined_k141_356255\|\|full | 2072 | No | NA | | 4 | 0 | 0 | Not-determined | Genome-fragment | NA | NA | 0 |
| S1-final-viral-combined_k141_92329\|\|full | 8493 | No | NA | | 8 | 0 | 7 | Not-determined | Genome-fragment | NA | NA | 0 |
| S4-final-viral-combined_k141_163862\|\|full | 1359 | No | NA | | 2 | 1 | 0 | Low-quality | Genome-fragment | 0.86 | HMM-based (lower-bound) | 0 |
| S1-final-viral-combined_k141_240679\|\|full | 2128 | No | NA | | 2 | 0 | 1 | Not-determined | Genome-fragment | NA | NA | 0 |
| S4-final-viral-combined_k141_95825\|\|full | 1359 | No | NA | | 2 | 0 | 2 | Low-quality | Genome-fragment | 2.25 | AAI-based (medium-confidence) | 0 |
| S8_k141_10999 | 1359 | No | NA | | 1 | 0 | 0 | Low-quality | Genome-fragment | 1.72 | AAI-based (medium-confidence) | 0 |
| S1-final-viral-combined_k141_257700\|\|full | 2642 | No | NA | | 4 | 0 | 2 | Not-determined | Genome-fragment | NA | NA | 0 |
| S9-final-viral-combined_k141_52301\|\|full | 1360 | No | NA | | 3 | 1 | 0 | Low-quality | Genome-fragment | 2.98 | AAI-based (high-confidence) | 0 |
| S1-final-viral-combined_k141_141883\|\|full | 1360 | No | NA | | 4 | 2 | 0 | Low-quality | Genome-fragment | 3.25 | AAI-based (high-confidence) | 0 |
| S1-final-viral-combined_k141_59018\|\|full | 7793 | No | NA | | 6 | 0 | 5 | Not-determined | Genome-fragment | NA | NA | 0 |
| S1-final-viral-combined_k141_376631\|\|full | 1360 | No | NA | | 2 | 0 | 0 | Low-quality | Genome-fragment | 2.74 | AAI-based (high-confidence) | 0 |
| S1-final-viral-combined_k141_59021\|\|full | 1806 | No | NA | | 3 | 0 | 2 | Not-determined | Genome-fragment | NA | NA | 0 |
| S1-final-viral-combined_k141_10264\|\|full | 8107 | No | NA | | 7 | 0 | 1 | Not-determined | Genome-fragment | NA | NA | 0 |
| S1-final-viral-combined_k141_356345\|\|full | 1377 | No | NA | | 2 | 0 | 1 | Not-determined | Genome-fragment | NA | NA | 0 |
| S1-final-viral-combined_k141_191846\|\|full | 12676 | No | NA | | 11 | 0 | 9 | Not-determined | Genome-fragment | NA | NA | 0 |
| S7-final-viral-combined_k141_148260\|\|full | 1360 | No | NA | | 2 | 0 | 0 | Low-quality | Genome-fragment | 2.36 | AAI-based (high-confidence) | 0 |
| S7_k141_130141 | 1360 | No | NA | | 2 | 0 | 0 | Low-quality | Genome-fragment | 1.08 | AAI-based (high-confidence) | 0 |
| S1-final-viral-combined_k141_356352\|\|full | 4104 | No | NA | | 5 | 0 | 3 | Not-determined | Genome-fragment | NA | NA | 0 |
| S1-final-viral-combined_k141_289954\|\|full | 1967 | No | NA | | 2 | 0 | 2 | Not-determined | Genome-fragment | NA | NA | 0 |
| S17-final-viral-combined_k141_250269\|\|full | 1360 | No | NA | | 3 | 1 | 0 | Low-quality | Genome-fragment | 2.42 | AAI-based (medium-confidence) | 0 |
| S17-k141_256459 | 1360 | No | NA | | 1 | 0 | 0 | Low-quality | Genome-fragment | 1.89 | AAI-based (medium-confidence) | 0 |
| S12-final-viral-combined_k141_186985\|\|full | 1360 | No | NA | | 2 | 1 | 0 | Low-quality | Genome-fragment | 3 | AAI-based (medium-confidence) | 0 |
| S2-final-viral-combined_k141_231133\|\|full | 1360 | No | NA | | 3 | 0 | 1 | Low-quality | Genome-fragment | 3.35 | AAI-based (medium-confidence) | 0 |
| S1-final-viral-combined_k141_141832\|\|full | 2495 | No | NA | | 4 | 0 | 1 | Not-determined | Genome-fragment | NA | NA | 0 |
| S1-final-viral-combined_k141_289965\|\|full | 2219 | No | NA | | 3 | 0 | 2 | Not-determined | Genome-fragment | NA | NA | 0 |
| S1-final-viral-combined_k141_173568\|\|full | 1414 | No | NA | | 2 | 0 | 0 | Not-determined | Genome-fragment | NA | NA | 0 |
| S1-final-viral-combined_k141_26417\|\|full | 14494 | No | NA | | 14 | 0 | 10 | Not-determined | Genome-fragment | NA | NA | 0 |
| S1-final-viral-combined_k141_126047\|\|full | 3054 | No | NA | | 5 | 0 | 0 | Not-determined | Genome-fragment | NA | NA | 0 |
| S2-final-viral-combined_k141_60621\|\|full | 1360 | No | NA | | 2 | 0 | 0 | Low-quality | Genome-fragment | 2.92 | AAI-based (medium-confidence) | 0 |
| S1-final-viral-combined_k141_126049\|\|full | 2171 | No | NA | | 3 | 0 | 1 | Not-determined | Genome-fragment | NA | NA | 0 |
| S8-final-viral-combined_k141_124314\|\|lt2gene | 1360 | No | NA | | 2 | 1 | 0 | Low-quality | Genome-fragment | 2.92 | AAI-based (high-confidence) | 0 |
| S1-final-viral-combined_k141_59084\|\|full | 4879 | No | NA | | 7 | 0 | 0 | Not-determined | Genome-fragment | NA | NA | 0 |
| S1_k141_127924 | 1361 | No | NA | | 2 | 0 | 1 | Low-quality | Genome-fragment | 2.26 | AAI-based (high-confidence) | 0 |
| S1-final-viral-combined_k141_59085\|\|full | 4630 | No | NA | | 5 | 0 | 1 | Not-determined | Genome-fragment | NA | NA | 0 |
| S7-final-viral-combined_k141_26078\|\|full | 1361 | No | NA | | 5 | 1 | 0 | Low-quality | Genome-fragment | 1.22 | HMM-based (lower-bound) | 0 |
| S17-final-viral-combined_k141_152452\|\|full | 1361 | No | NA | | 5 | 2 | 0 | Low-quality | Genome-fragment | 2.62 | AAI-based (medium-confidence) | 0 |
| S1-final-viral-combined_k141_10286\|\|full | 36514 | No | NA | | 31 | 0 | 20 | Not-determined | Genome-fragment | NA | NA | 0 |
| S17-k141_222839 | 1361 | No | NA | | 1 | 0 | 1 | Low-quality | Genome-fragment | 4.57 | AAI-based (medium-confidence) | 0 |
| S1-final-viral-combined_k141_372074\|\|full | 1598 | No | NA | | 2 | 0 | 0 | Not-determined | Genome-fragment | NA | NA | 0 |
| S17-k141_57873 | 1361 | No | NA | | 2 | 0 | 0 | Low-quality | Genome-fragment | 2.29 | AAI-based (high-confidence) | 0 |
| S1-final-viral-combined_k141_372078\|\|full | 1065 | No | NA | | 4 | 0 | 0 | Not-determined | Genome-fragment | NA | NA | 0 |
| S12-final-viral-combined_k141_58370\|\|full | 1361 | No | NA | | 2 | 0 | 0 | Low-quality | Genome-fragment | 2.19 | AAI-based (medium-confidence) | 0 |
| S1-final-viral-combined_k141_307184\|\|full | 2618 | No | NA | | 2 | 0 | 2 | Not-determined | Genome-fragment | NA | NA | 0 |
| S1-final-viral-combined_k141_126103\|\|full | 5176 | No | NA | | 4 | 0 | 3 | Not-determined | Genome-fragment | NA | NA | 0 |
| S1-final-viral-combined_k141_42645\|\|full | 3411 | No | NA | | 4 | 0 | 3 | Not-determined | Genome-fragment | NA | NA | 0 |
| S1-final-viral-combined_k141_26465\|\|full | 1153 | No | NA | | 2 | 0 | 0 | Not-determined | Genome-fragment | NA | NA | 0 |
| S1-final-viral-combined_k141_257851\|\|full | 2363 | No | NA | | 3 | 0 | 1 | Not-determined | Genome-fragment | NA | NA | 0 |
| S12-final-viral-combined_k141_283639\|\|full | 1361 | No | NA | | 3 | 0 | 0 | Low-quality | Genome-fragment | 2.39 | AAI-based (medium-confidence) | 0 |
| S6-final-viral-combined_k141_36703\|\|full | 1361 | No | NA | | 2 | 0 | 0 | Low-quality | Genome-fragment | 1.28 | AAI-based (high-confidence) | 0 |
| S1-final-viral-combined_k141_257856\|\|full | 6862 | No | NA | | 6 | 0 | 4 | Not-determined | Genome-fragment | NA | NA | 0 |
| S1-final-viral-combined_k141_372134\|\|full | 2541 | No | NA | | 4 | 0 | 0 | Not-determined | Genome-fragment | NA | NA | 0 |
| S6-final-viral-combined_k141_14070\|\|full | 1361 | No | NA | | 2 | 0 | 0 | Low-quality | Genome-fragment | 3.29 | AAI-based (medium-confidence) | 0 |
| S1-final-viral-combined_k141_338549\|\|full | 5377 | No | NA | | 6 | 0 | 5 | Not-determined | Genome-fragment | NA | NA | 0 |
| S1-final-viral-combined_k141_257885\|\|full | 6549 | No | NA | | 7 | 0 | 4 | Not-determined | Genome-fragment | NA | NA | 0 |
| S1-final-viral-combined_k141_307207\|\|full | 3451 | No | NA | | 3 | 0 | 2 | Not-determined | Genome-fragment | NA | NA | 0 |
| S8-final-viral-combined_k141_77169\|\|full | 1361 | No | NA | | 3 | 0 | 0 | Low-quality | Genome-fragment | 0.98 | AAI-based (medium-confidence) | 0 |
| S1-final-viral-combined_k141_338569\|\|full | 1554 | No | NA | | 1 | 0 | 0 | Not-determined | Genome-fragment | NA | NA | 0 |
| S11-final-viral-combined_k141_278870\|\|full | 1362 | No | NA | | 3 | 0 | 0 | Low-quality | Genome-fragment | 0.48 | AAI-based (medium-confidence) | 0 |
| S11-final-viral-combined_k141_200627\|\|full | 1362 | No | NA | | 3 | 3 | 0 | Low-quality | Genome-fragment | 4.57 | AAI-based (high-confidence) | 0 |
| S1-final-viral-combined_k141_26544\|\|full | 3373 | No | NA | | 4 | 0 | 2 | Not-determined | Genome-fragment | NA | NA | 0 |
| S5-final-viral-combined_k141_189045\|\|full | 1362 | No | NA | | 3 | 3 | 0 | Low-quality | Genome-fragment | 1.97 | AAI-based (high-confidence) | 0 |
| S1-final-viral-combined_k141_338582\|\|full | 3030 | No | NA | | 5 | 0 | 0 | Not-determined | Genome-fragment | NA | NA | 0 |
| S1-final-viral-combined_k141_42694\|\|full | 1185 | No | NA | | 3 | 0 | 2 | Not-determined | Genome-fragment | NA | NA | 0 |
| S7-final-viral-combined_k141_205419\|\|full | 1362 | No | NA | | 2 | 0 | 1 | Low-quality | Genome-fragment | 1.65 | AAI-based (medium-confidence) | 0 |
| S1-final-viral-combined_k141_208550\|\|full | 2897 | No | NA | | 4 | 0 | 2 | Not-determined | Genome-fragment | NA | NA | 0 |
| S1-final-viral-combined_k141_141993\|\|full | 1324 | No | NA | | 3 | 0 | 0 | Not-determined | Genome-fragment | NA | NA | 0 |
| S1-final-viral-combined_k141_92523\|\|full | 31793 | Yes | | 21321 | 39 | 10 | 8 | Medium-quality | Genome-fragment | 61.28 | AAI-based (high-confidence) | 32.94 |
| S1-final-viral-combined_k141_224136\|\|full | 1047 | No | NA | | 3 | 0 | 0 | Not-determined | Genome-fragment | NA | NA | 0 |
| S1-final-viral-combined_k141_75145\|\|full | 1030 | No | NA | | 2 | 0 | 1 | Not-determined | Genome-fragment | NA | NA | 0 |
| S1-final-viral-combined_k141_257962\|\|full | 9387 | No | NA | | 11 | 0 | 7 | Not-determined | Genome-fragment | NA | NA | 0 |
| S1-final-viral-combined_k141_224160\|\|full | 1863 | No | NA | | 3 | 0 | 0 | Not-determined | Genome-fragment | NA | NA | 0 |
| S1-final-viral-combined_k141_208588\|\|full | 6162 | No | NA | | 6 | 0 | 3 | Not-determined | Genome-fragment | NA | NA | 0 |
| S1-final-viral-combined_k141_173782\|\|full | 2331 | No | NA | | 4 | 0 | 1 | Not-determined | Genome-fragment | NA | NA | 0 |
| S1-final-viral-combined_k141_126302\|\|full | 1213 | No | NA | | 2 | 0 | 0 | Not-determined | Genome-fragment | NA | NA | 0 |
| S12-final-viral-combined_k141_338824\|\|full | 1362 | No | NA | | 3 | 0 | 0 | Low-quality | Genome-fragment | 2.67 | AAI-based (high-confidence) | 0 |
| S1-final-viral-combined_k141_372268\|\|full | 2254 | No | NA | | 5 | 0 | 1 | Not-determined | Genome-fragment | NA | NA | 0 |
| S1-final-viral-combined_k141_59189\|\|full | 1792 | No | NA | | 3 | 0 | 0 | Not-determined | Genome-fragment | NA | NA | 0 |
| S8_k141_201770 | 1362 | No | NA | | 2 | 0 | 0 | Low-quality | Genome-fragment | 1.75 | AAI-based (high-confidence) | 0 |
| S9_k141_125582 | 1363 | No | NA | | 1 | 1 | 0 | Low-quality | Genome-fragment | 2.5 | AAI-based (high-confidence) | 0 |
| S1-final-viral-combined_k141_59204\|\|full | 2603 | No | NA | | 2 | 0 | 2 | Not-determined | Genome-fragment | NA | NA | 0 |
| S1-final-viral-combined_k141_324115\|\|full | 1649 | No | NA | | 3 | 0 | 3 | Not-determined | Genome-fragment | NA | NA | 0 |
| S1-final-viral-combined_k141_388583\|\|full | 6837 | No | NA | | 4 | 0 | 2 | Not-determined | Genome-fragment | NA | NA | 0 |
| S1-final-viral-combined_k141_307299\|\|full | 3198 | No | NA | | 3 | 0 | 1 | Not-determined | Genome-fragment | NA | NA | 0 |
| S1-final-viral-combined_k141_388584\|\|full | 5613 | No | NA | | 7 | 0 | 5 | Not-determined | Genome-fragment | NA | NA | 0 |
| S1-final-viral-combined_k141_290203\|\|full | 1976 | No | NA | | 3 | 0 | 1 | Not-determined | Genome-fragment | NA | NA | 0 |
| S1-final-viral-combined_k141_224189\|\|full | 3708 | No | NA | | 5 | 0 | 0 | Not-determined | Genome-fragment | NA | NA | 0 |
| S1-final-viral-combined_k141_26615\|\|full | 1487 | No | NA | | 3 | 0 | 2 | Not-determined | Genome-fragment | NA | NA | 0 |
| S1-final-viral-combined_k141_324141\|\|full | 2377 | No | NA | | 3 | 0 | 3 | Not-determined | Genome-fragment | NA | NA | 0 |
| S1-final-viral-combined_k141_208661\|\|full | 6595 | No | NA | | 9 | 0 | 1 | Not-determined | Genome-fragment | NA | NA | 0 |
| S1-final-viral-combined_k141_92636\|\|full | 1470 | No | NA | | 3 | 0 | 0 | Not-determined | Genome-fragment | NA | NA | 0 |
| S1-final-viral-combined_k141_142067\|\|full | 1200 | No | NA | | 2 | 0 | 1 | Not-determined | Genome-fragment | NA | NA | 0 |
| S1-final-viral-combined_k141_388606\|\|full | 2761 | No | NA | | 3 | 0 | 0 | Not-determined | Genome-fragment | NA | NA | 0 |
| S11-final-viral-combined_k141_63822\|\|full | 1363 | No | NA | | 2 | 0 | 1 | Low-quality | Genome-fragment | 0.3 | AAI-based (medium-confidence) | 0 |
| S1-final-viral-combined_k141_356572\|\|full | 1989 | No | NA | | 3 | 0 | 2 | Not-determined | Genome-fragment | NA | NA | 0 |
| S1-final-viral-combined_k141_240987\|\|full | 1298 | No | NA | | 2 | 0 | 1 | Not-determined | Genome-fragment | NA | NA | 0 |
| S1-final-viral-combined_k141_240990\|\|full | 1070 | No | NA | | 3 | 0 | 1 | Not-determined | Genome-fragment | NA | NA | 0 |
| S1-final-viral-combined_k141_192004\|\|full | 1033 | No | NA | | 2 | 0 | 2 | Not-determined | Genome-fragment | NA | NA | 0 |
| S1-final-viral-combined_k141_59261\|\|full | 3074 | No | NA | | 4 | 0 | 2 | Not-determined | Genome-fragment | NA | NA | 0 |
| S1-final-viral-combined_k141_42351\|\|lt2gene | 1363 | No | NA | | 1 | 1 | 0 | Low-quality | Genome-fragment | 2.23 | HMM-based (lower-bound) | 0 |
| S1-final-viral-combined_k141_208687\|\|full | 5658 | No | NA | | 5 | 0 | 3 | Not-determined | Genome-fragment | NA | NA | 0 |
| S1-final-viral-combined_k141_338697\|\|full | 1991 | No | NA | | 4 | 0 | 1 | Not-determined | Genome-fragment | NA | NA | 0 |
| S1-final-viral-combined_k141_208701\|\|full | 1833 | No | NA | | 3 | 0 | 0 | Not-determined | Genome-fragment | NA | NA | 0 |
| S1-final-viral-combined_k141_208706\|\|full | 1015 | No | NA | | 3 | 0 | 3 | Not-determined | Genome-fragment | NA | NA | 0 |
| S5-final-viral-combined_k141_110799\|\|full | 1363 | No | NA | | 2 | 1 | 0 | Low-quality | Genome-fragment | 3.01 | AAI-based (medium-confidence) | 0 |
| S1-final-viral-combined_k141_258096\|\|full | 2122 | No | NA | | 2 | 0 | 0 | Not-determined | Genome-fragment | NA | NA | 0 |
| S17-final-viral-combined_k141_31591\|\|full | 1363 | Yes | | 378 | 3 | 1 | 1 | Low-quality | Genome-fragment | 0.79 | AAI-based (medium-confidence) | 72.27 |
| S12-final-viral-combined_k141_84908\|\|lt2gene | 1363 | No | NA | | 1 | 1 | 0 | Low-quality | Genome-fragment | 0.92 | HMM-based (lower-bound) | 0 |
| S1-final-viral-combined_k141_208724\|\|full | 1617 | No | NA | | 3 | 0 | 1 | Not-determined | Genome-fragment | NA | NA | 0 |
| S1-final-viral-combined_k141_192089\|\|full | 1952 | No | NA | | 2 | 0 | 2 | Not-determined | Genome-fragment | NA | NA | 0 |
| S12_k141_9733 | 1363 | No | NA | | 2 | 0 | 0 | Low-quality | Genome-fragment | 2.38 | AAI-based (high-confidence) | 0 |
| S6-final-viral-combined_k141_85634\|\|full | 1363 | No | NA | | 3 | 0 | 0 | Low-quality | Genome-fragment | 2.05 | AAI-based (medium-confidence) | 0 |
| S1-final-viral-combined_k141_59315\|\|full | 3298 | No | NA | | 3 | 0 | 1 | Not-determined | Genome-fragment | NA | NA | 0 |
| S6-final-viral-combined_k141_40550\|\|lt2gene | 1363 | No | NA | | 1 | 1 | 0 | Low-quality | Genome-fragment | 4.59 | AAI-based (high-confidence) | 0 |
| S8-final-viral-combined_k141_102766\|\|full | 1363 | No | NA | | 2 | 0 | 0 | Low-quality | Genome-fragment | 2.5 | AAI-based (high-confidence) | 0 |
| S1-final-viral-combined_k141_338748\|\|full | 3819 | No | NA | | 4 | 0 | 1 | Not-determined | Genome-fragment | NA | NA | 0 |
| S1-final-viral-combined_k141_157822\|\|full | 4452 | No | NA | | 4 | 0 | 2 | Not-determined | Genome-fragment | NA | NA | 0 |
| S9-final-viral-combined_k141_108283\|\|full | 1364 | No | NA | | 3 | 0 | 1 | Low-quality | Genome-fragment | 1.26 | AAI-based (medium-confidence) | 0 |
| S1-final-viral-combined_k141_157834\|\|full | 1451 | No | NA | | 3 | 0 | 0 | Not-determined | Genome-fragment | NA | NA | 0 |
| S1-final-viral-combined_k141_126470\|\|full | 3043 | No | NA | | 3 | 0 | 3 | Not-determined | Genome-fragment | NA | NA | 0 |
| S1-final-viral-combined_k141_208764\|\|full | 2000 | No | NA | | 2 | 0 | 1 | Not-determined | Genome-fragment | NA | NA | 0 |
| S9_k141_84895 | 1364 | No | NA | | 1 | 0 | 1 | Low-quality | Genome-fragment | 0.48 | AAI-based (medium-confidence) | 0 |
| S1-final-viral-combined_k141_208775\|\|full | 4059 | No | NA | | 8 | 0 | 0 | Not-determined | Genome-fragment | NA | NA | 0 |
| S1-final-viral-combined_k141_241111\|\|full | 1438 | No | NA | | 4 | 0 | 1 | Not-determined | Genome-fragment | NA | NA | 0 |
| S11-final-viral-combined_k141_55419\|\|full | 1364 | No | NA | | 3 | 1 | 0 | Low-quality | Genome-fragment | 0.83 | HMM-based (lower-bound) | 0 |
| S1-final-viral-combined_k141_338785\|\|full | 1235 | No | NA | | 2 | 0 | 0 | Not-determined | Genome-fragment | NA | NA | 0 |
| S1-final-viral-combined_k141_185200\|\|full | 1364 | No | NA | | 3 | 0 | 0 | Low-quality | Genome-fragment | 3.74 | AAI-based (medium-confidence) | 0 |
| S1-final-viral-combined_k141_388731\|\|full | 5268 | No | NA | | 9 | 0 | 5 | Not-determined | Genome-fragment | NA | NA | 0 |
| S1-final-viral-combined_k141_258197\|\|full | 2226 | No | NA | | 2 | 0 | 2 | Not-determined | Genome-fragment | NA | NA | 0 |
| S7-final-viral-combined_k141_239352\|\|full | 1364 | No | NA | | 5 | 0 | 0 | Low-quality | Genome-fragment | 3.04 | AAI-based (medium-confidence) | 0 |
| S17-k141_149286 | 1364 | No | NA | | 5 | 2 | 0 | Low-quality | Genome-fragment | 1.72 | AAI-based (medium-confidence) | 0 |
| S12-final-viral-combined_k141_376748\|\|full | 1364 | No | NA | | 4 | 0 | 0 | Low-quality | Genome-fragment | 2.24 | AAI-based (high-confidence) | 0 |
| S12-final-viral-combined_k141_265283\|\|full | 1364 | No | NA | | 4 | 1 | 0 | Low-quality | Genome-fragment | 2.84 | AAI-based (high-confidence) | 0 |
| S12_k141_124575 | 1364 | No | NA | | 3 | 0 | 0 | Low-quality | Genome-fragment | 3.46 | AAI-based (medium-confidence) | 0 |
| S1-final-viral-combined_k141_338837\|\|full | 6024 | No | NA | | 9 | 0 | 6 | Not-determined | Genome-fragment | NA | NA | 0 |
| S2-final-viral-combined_k141_55720\|\|full | 1364 | No | NA | | 2 | 0 | 0 | Low-quality | Genome-fragment | 2.4 | AAI-based (medium-confidence) | 0 |
| S1-final-viral-combined_k141_208837\|\|full | 1137 | No | NA | | 2 | 0 | 0 | Not-determined | Genome-fragment | NA | NA | 0 |
| S1-final-viral-combined_k141_290347\|\|full | 2652 | No | NA | | 4 | 0 | 0 | Not-determined | Genome-fragment | NA | NA | 0 |
| S1-final-viral-combined_k141_290350\|\|full | 2575 | No | NA | | 4 | 0 | 0 | Not-determined | Genome-fragment | NA | NA | 0 |
| S11-final-viral-combined_k141_310043\|\|full | 1365 | No | NA | | 2 | 0 | 0 | Low-quality | Genome-fragment | 1.51 | AAI-based (medium-confidence) | 0 |
| S1-final-viral-combined_k141_165878\|\|full | 1365 | No | NA | | 3 | 0 | 1 | Low-quality | Genome-fragment | 1.12 | AAI-based (medium-confidence) | 0 |
| S1-final-viral-combined_k141_324293\|\|full | 1480 | No | NA | | 3 | 0 | 1 | Not-determined | Genome-fragment | NA | NA | 0 |
| S1-final-viral-combined_k141_87489\|\|full | 1365 | No | NA | | 2 | 0 | 2 | Low-quality | Genome-fragment | 0.71 | AAI-based (medium-confidence) | 0 |
| S17-k141_220745 | 1365 | No | NA | | 2 | 0 | 2 | Low-quality | Genome-fragment | 0.15 | AAI-based (medium-confidence) | 0 |
| S1-final-viral-combined_k141_307324\|\|full | 5594 | No | NA | | 6 | 0 | 1 | Not-determined | Genome-fragment | NA | NA | 0 |
| S1-final-viral-combined_k141_258249\|\|full | 1265 | No | NA | | 2 | 0 | 0 | Not-determined | Genome-fragment | NA | NA | 0 |
| S12-final-viral-combined_k141_86776\|\|full | 1365 | No | NA | | 4 | 0 | 0 | Low-quality | Genome-fragment | 3.59 | AAI-based (high-confidence) | 0 |
| S12-final-viral-combined_k141_157135\|\|full | 1365 | No | NA | | 3 | 2 | 0 | Low-quality | Genome-fragment | 3.44 | AAI-based (medium-confidence) | 0 |
| S1-final-viral-combined_k141_126617\|\|full | 1255 | No | NA | | 2 | 0 | 1 | Not-determined | Genome-fragment | NA | NA | 0 |
| S1-final-viral-combined_k141_126618\|\|full | 1177 | No | NA | | 3 | 0 | 2 | Not-determined | Genome-fragment | NA | NA | 0 |
| S1-final-viral-combined_k141_258282\|\|full | 3193 | No | NA | | 2 | 0 | 0 | Not-determined | Genome-fragment | NA | NA | 0 |
| S1-final-viral-combined_k141_142368\|\|full | 1347 | No | NA | | 2 | 0 | 1 | Not-determined | Genome-fragment | NA | NA | 0 |
| S1-final-viral-combined_k141_142374\|\|full | 4294 | No | NA | | 6 | 0 | 3 | Not-determined | Genome-fragment | NA | NA | 0 |
| S12-final-viral-combined_k141_80641\|\|full | 1365 | No | NA | | 5 | 0 | 0 | Low-quality | Genome-fragment | 3.1 | AAI-based (medium-confidence) | 0 |
| S1-final-viral-combined_k141_59551\|\|full | 1063 | No | NA | | 3 | 0 | 1 | Not-determined | Genome-fragment | NA | NA | 0 |
| S1-final-viral-combined_k141_10502\|\|full | 7776 | No | NA | | 7 | 0 | 4 | Not-determined | Genome-fragment | NA | NA | 0 |
| S2-final-viral-combined_k141_40814\|\|full | 1365 | No | NA | | 2 | 2 | 0 | Low-quality | Genome-fragment | 2.97 | AAI-based (high-confidence) | 0 |
| S1-final-viral-combined_k141_192321\|\|full | 5826 | No | NA | | 6 | 0 | 1 | Not-determined | Genome-fragment | NA | NA | 0 |

| | | | | | | | | | | | |
|---|---|---|---|---|---|---|---|---|---|---|---|
| S1-final-viral-combined_k141_290423\|\|full | 3760 | No | NA | 3 | 0 | 3 | Not-determined | Genome-fragment | NA | NA | 0 |
| S1-final-viral-combined_k141_372596\|\|full | 1144 | No | NA | 3 | 0 | 1 | Not-determined | Genome-fragment | NA | NA | 0 |
| S1-final-viral-combined_k141_307388\|\|full | 3367 | No | NA | 2 | 0 | 1 | Not-determined | Genome-fragment | NA | NA | 0 |
| S6-final-viral-combined_k141_22231\|\|full | 1365 | No | NA | 4 | 1 | 0 | Low-quality | Genome-fragment | 1.05 | AAI-based (medium-confidence) | 0 |
| S1-final-viral-combined_k141_273637\|\|full | 1063 | No | NA | 2 | 0 | 0 | Not-determined | Genome-fragment | NA | NA | 0 |
| S1-final-viral-combined_k141_356823\|\|full | 2818 | No | NA | 3 | 0 | 1 | Not-determined | Genome-fragment | NA | NA | 0 |
| S6_k141_30238 | 1365 | No | NA | 5 | 0 | 0 | Low-quality | Genome-fragment | 3.26 | AAI-based (medium-confidence) | 0 |
| S8-final-viral-combined_k141_245992\|\|full | 1365 | No | NA | 5 | 0 | 0 | Low-quality | Genome-fragment | 3.52 | AAI-based (high-confidence) | 0 |
| S1-final-viral-combined_k141_258317\|\|full | 10332 | No | NA | 10 | 0 | 7 | Not-determined | Genome-fragment | NA | NA | 0 |
| S1-final-viral-combined_k141_75464\|\|full | 3366 | No | NA | 3 | 0 | 2 | Not-determined | Genome-fragment | NA | NA | 0 |
| S8-final-viral-combined_k141_248152\|\|full | 1365 | No | NA | 2 | 0 | 0 | Low-quality | Genome-fragment | 3.43 | AAI-based (medium-confidence) | 0 |
| S9-final-viral-combined_k141_117660\|\|full | 1366 | No | NA | 3 | 0 | 0 | Low-quality | Genome-fragment | 4.32 | AAI-based (high-confidence) | 0 |
| S1-final-viral-combined_k141_26768\|\|full | 4603 | No | NA | 3 | 0 | 2 | Not-determined | Genome-fragment | NA | NA | 0 |
| S1-final-viral-combined_k141_93038\|\|full | 2996 | No | NA | 4 | 0 | 0 | Not-determined | Genome-fragment | NA | NA | 0 |
| S11-final-viral-combined_k141_142\|\|full | 1366 | No | NA | 3 | 0 | 0 | Low-quality | Genome-fragment | 3.09 | AAI-based (medium-confidence) | 0 |
| S1-final-viral-combined_k141_43167\|\|full | 4573 | No | NA | 4 | 0 | 3 | Not-determined | Genome-fragment | NA | NA | 0 |
| S1_k141_223601 | 1366 | No | NA | 3 | 2 | 0 | Low-quality | Genome-fragment | 2.98 | AAI-based (high-confidence) | 0 |
| S1-final-viral-combined_k141_93047\|\|full | 1103 | No | NA | 3 | 0 | 0 | Not-determined | Genome-fragment | NA | NA | 0 |
| S1-final-viral-combined_k141_307405\|\|full | 6449 | No | NA | 4 | 0 | 2 | Not-determined | Genome-fragment | NA | NA | 0 |
| S1-final-viral-combined_k141_142454\|\|full | 2101 | No | NA | 4 | 0 | 2 | Not-determined | Genome-fragment | NA | NA | 0 |
| S1-final-viral-combined_k141_43178\|\|full | 1469 | No | NA | 2 | 0 | 1 | Not-determined | Genome-fragment | NA | NA | 0 |
| S7-final-viral-combined_k141_113697\|\|full | 1366 | No | NA | 5 | 2 | 0 | Low-quality | Genome-fragment | 2.93 | AAI-based (high-confidence) | 0 |
| S1-final-viral-combined_k141_174173\|\|full | 1844 | No | NA | 3 | 0 | 0 | Not-determined | Genome-fragment | NA | NA | 0 |
| S1-final-viral-combined_k141_290494\|\|full | 1908 | No | NA | 2 | 0 | 1 | Not-determined | Genome-fragment | NA | NA | 0 |
| S1-final-viral-combined_k141_324368\|\|full | 2937 | No | NA | 3 | 0 | 2 | Not-determined | Genome-fragment | NA | NA | 0 |
| S1-final-viral-combined_k141_174209\|\|full | 1162 | No | NA | 3 | 0 | 0 | Not-determined | Genome-fragment | NA | NA | 0 |
| S1-final-viral-combined_k141_26840\|\|full | 1184 | No | NA | 3 | 0 | 0 | Not-determined | Genome-fragment | NA | NA | 0 |
| S1-final-viral-combined_k141_142461\|\|full | 3111 | No | NA | 4 | 0 | 0 | Not-determined | Genome-fragment | NA | NA | 0 |
| S7-final-viral-combined_k141_4090\|\|full | 1366 | No | NA | 3 | 0 | 1 | Low-quality | Genome-fragment | 2.35 | AAI-based (high-confidence) | 0 |
| S1-final-viral-combined_k141_126778\|\|full | 2211 | No | NA | 5 | 0 | 0 | Not-determined | Genome-fragment | NA | NA | 0 |
| S1-final-viral-combined_k141_10609\|\|full | 1942 | No | NA | 5 | 0 | 0 | Not-determined | Genome-fragment | NA | NA | 0 |
| S1-final-viral-combined_k141_388920\|\|full | 2501 | No | NA | 6 | 0 | 0 | Not-determined | Genome-fragment | NA | NA | 0 |
| S7-final-viral-combined_k141_17553\|\|full | 1366 | No | NA | 4 | 1 | 0 | Low-quality | Genome-fragment | 3.68 | AAI-based (medium-confidence) | 0 |
| S1-final-viral-combined_k141_372636\|\|full | 2537 | No | NA | 4 | 0 | 2 | Not-determined | Genome-fragment | NA | NA | 0 |
| S7_k141_223601 | 1366 | No | NA | 1 | 0 | 0 | Low-quality | Genome-fragment | 0.89 | AAI-based (high-confidence) | 0 |
| S1-final-viral-combined_k141_126796\|\|full | 7642 | No | NA | 10 | 0 | 4 | Not-determined | Genome-fragment | NA | NA | 0 |
| S17-final-viral-combined_k141_174723\|\|full | 1366 | No | NA | 2 | 0 | 2 | Low-quality | Genome-fragment | 0.94 | AAI-based (medium-confidence) | 0 |
| S1-final-viral-combined_k141_356908\|\|full | 2769 | No | NA | 4 | 0 | 2 | Not-determined | Genome-fragment | NA | NA | 0 |
| S1-final-viral-combined_k141_273732\|\|full | 2507 | No | NA | 2 | 0 | 0 | Not-determined | Genome-fragment | NA | NA | 0 |
| S1-final-viral-combined_k141_10650\|\|full | 2964 | No | NA | 3 | 0 | 3 | Not-determined | Genome-fragment | NA | NA | 0 |
| S12-final-viral-combined_k141_385633\|\|full | 1366 | No | NA | 2 | 0 | 1 | Low-quality | Genome-fragment | 1.49 | AAI-based (high-confidence) | 0 |
| S12_k141_173641 | 1366 | No | NA | 1 | 0 | 0 | Low-quality | Genome-fragment | 0.63 | AAI-based (high-confidence) | 0 |
| S1-final-viral-combined_k141_224578\|\|full | 25656 | No | NA | 36 | 10 | 0 | Medium-quality | Genome-fragment | 59.81 | AAI-based (high-confidence) | 0 |
| S1-final-viral-combined_k141_338951\|\|full | 1137 | No | NA | 2 | 0 | 0 | Not-determined | Genome-fragment | NA | NA | 0 |
| S12_k141_31593 | 1366 | No | NA | 3 | 0 | 0 | Low-quality | Genome-fragment | 1.36 | AAI-based (high-confidence) | 0 |
| S1-final-viral-combined_k141_307508\|\|full | 1131 | No | NA | 3 | 0 | 0 | Not-determined | Genome-fragment | NA | NA | 0 |
| S1-final-viral-combined_k141_192472\|\|full | 2734 | No | NA | 4 | 0 | 3 | Not-determined | Genome-fragment | NA | NA | 0 |
| S2-final-viral-combined_k141_46120\|\|full | 1366 | No | NA | 3 | 0 | 0 | Low-quality | Genome-fragment | 2.69 | AAI-based (medium-confidence) | 0 |
| S2-final-viral-combined_k141_80025\|\|full | 1366 | No | NA | 3 | 0 | 1 | Low-quality | Genome-fragment | 0.72 | AAI-based (high-confidence) | 0 |
| S1-final-viral-combined_k141_258450\|\|full | 14490 | No | NA | 17 | 0 | 11 | Not-determined | Genome-fragment | NA | NA | 0 |
| S1-final-viral-combined_k141_126855\|\|full | 3339 | No | NA | 4 | 0 | 1 | Not-determined | Genome-fragment | NA | NA | 0 |
| S1-final-viral-combined_k141_200032\|\|full | 1367 | No | NA | 2 | 0 | 0 | Low-quality | Genome-fragment | 0.91 | AAI-based (medium-confidence) | 0 |
| S1-final-viral-combined_k141_10701\|\|full | 1774 | No | NA | 3 | 0 | 1 | Not-determined | Genome-fragment | NA | NA | 0 |
| S1-final-viral-combined_k141_356923\|\|full | 2853 | No | NA | 3 | 0 | 3 | Not-determined | Genome-fragment | NA | NA | 0 |
| S1-final-viral-combined_k141_319785\|\|full | 1367 | No | NA | 3 | 0 | 2 | Low-quality | Genome-fragment | 2.47 | AAI-based (medium-confidence) | 0 |
| S1-final-viral-combined_k141_344624\|\|full | 1367 | No | NA | 2 | 0 | 1 | Low-quality | Genome-fragment | 2.55 | AAI-based (medium-confidence) | 0 |
| S1-final-viral-combined_k141_126927\|\|full | 2900 | No | NA | 2 | 0 | 1 | Not-determined | Genome-fragment | NA | NA | 0 |
| S1-final-viral-combined_k141_307608\|\|full | 2871 | No | NA | 3 | 0 | 1 | Not-determined | Genome-fragment | NA | NA | 0 |
| S1-final-viral-combined_k141_372747\|\|full | 1108 | No | NA | 3 | 0 | 2 | Not-determined | Genome-fragment | NA | NA | 0 |
| S1-final-viral-combined_k141_10770\|\|full | 3126 | No | NA | 5 | 0 | 2 | Not-determined | Genome-fragment | NA | NA | 0 |
| S1-final-viral-combined_k141_75632\|\|full | 2241 | No | NA | 4 | 0 | 2 | Not-determined | Genome-fragment | NA | NA | 0 |
| S3-final-viral-combined_k141_101559\|\|full | 1367 | No | NA | 2 | 0 | 1 | Low-quality | Genome-fragment | 0.72 | AAI-based (medium-confidence) | 0 |
| S5-final-viral-combined_k141_71535\|\|full | 1367 | No | NA | 4 | 0 | 1 | Low-quality | Genome-fragment | 1.78 | AAI-based (high-confidence) | 0 |
| S5-final-viral-combined_k141_80262\|\|lt2gene | 1367 | No | NA | 1 | 1 | 0 | Low-quality | Genome-fragment | 3.49 | AAI-based (medium-confidence) | 0 |
| S7-final-viral-combined_k141_74437\|\|full | 1367 | No | NA | 4 | 1 | 0 | Low-quality | Genome-fragment | 0.76 | HMM-based (lower-bound) | 0 |
| S1-final-viral-combined_k141_290621\|\|full | 2136 | No | NA | 2 | 0 | 1 | Not-determined | Genome-fragment | NA | NA | 0 |
| S1-final-viral-combined_k141_93226\|\|full | 1250 | No | NA | 3 | 0 | 2 | Not-determined | Genome-fragment | NA | NA | 0 |
| S1-final-viral-combined_k141_10801\|\|full | 1596 | No | NA | 2 | 0 | 1 | Not-determined | Genome-fragment | NA | NA | 0 |
| S1-final-viral-combined_k141_126948\|\|full | 2468 | No | NA | 3 | 0 | 1 | Not-determined | Genome-fragment | NA | NA | 0 |
| S17-k141_228107 | 1367 | No | NA | 2 | 0 | 0 | Low-quality | Genome-fragment | 0.78 | AAI-based (medium-confidence) | 0 |
| S1-final-viral-combined_k141_209205\|\|full | 8161 | No | NA | 5 | 0 | 4 | Not-determined | Genome-fragment | NA | NA | 0 |
| S1-final-viral-combined_k141_290643\|\|full | 2193 | No | NA | 3 | 0 | 0 | Not-determined | Genome-fragment | NA | NA | 0 |
| S12-final-viral-combined_k141_300415\|\|full | 1367 | No | NA | 4 | 0 | 0 | Low-quality | Genome-fragment | 0.86 | AAI-based (medium-confidence) | 0 |
| S1-final-viral-combined_k141_224718\|\|full | 2108 | No | NA | 2 | 0 | 1 | Not-determined | Genome-fragment | NA | NA | 0 |
| S1-final-viral-combined_k141_158309\|\|full | 4305 | No | NA | 6 | 0 | 4 | Not-determined | Genome-fragment | NA | NA | 0 |
| S12_k141_274374 | 1367 | No | NA | 3 | 3 | 0 | Low-quality | Genome-fragment | 2.64 | AAI-based (medium-confidence) | 0 |
| S1-final-viral-combined_k141_241508\|\|full | 3962 | No | NA | 3 | 0 | 2 | Not-determined | Genome-fragment | NA | NA | 0 |
| S1-final-viral-combined_k141_43378\|\|full | 1444 | No | NA | 3 | 0 | 0 | Not-determined | Genome-fragment | NA | NA | 0 |
| S1-final-viral-combined_k141_126965\|\|full | 3275 | No | NA | 3 | 0 | 3 | Not-determined | Genome-fragment | NA | NA | 0 |
| S12_k141_96259 | 1367 | No | NA | 1 | 0 | 0 | Low-quality | Genome-fragment | 0.91 | AAI-based (high-confidence) | 0 |
| S2-final-viral-combined_k141_44276\|\|full | 1367 | No | NA | 2 | 0 | 0 | Low-quality | Genome-fragment | 1.64 | AAI-based (high-confidence) | 0 |
| S2-final-viral-combined_k141_168180\|\|full | 1367 | No | NA | 3 | 0 | 1 | Low-quality | Genome-fragment | 2.31 | AAI-based (medium-confidence) | 0 |
| S1-final-viral-combined_k141_372867\|\|full | 3087 | No | NA | 3 | 0 | 1 | Not-determined | Genome-fragment | NA | NA | 0 |
| S1-final-viral-combined_k141_27092\|\|full | 1930 | No | NA | 3 | 0 | 1 | Not-determined | Genome-fragment | NA | NA | 0 |
| S2-final-viral-combined_k141_226294\|\|lt2gene | 1367 | No | NA | 2 | 1 | 0 | Low-quality | Genome-fragment | 4.63 | HMM-based (lower-bound) | 0 |
| S1-final-viral-combined_k141_59847\|\|full | 2058 | No | NA | 2 | 0 | 1 | Not-determined | Genome-fragment | NA | NA | 0 |
| S1-final-viral-combined_k141_10888\|\|full | 4172 | No | NA | 3 | 0 | 2 | Not-determined | Genome-fragment | NA | NA | 0 |
| S2_k141_142111 | 1367 | No | NA | 4 | 0 | 2 | Low-quality | Genome-fragment | 0.68 | AAI-based (medium-confidence) | 0 |
| S1-final-viral-combined_k141_324647\|\|full | 2415 | No | NA | 3 | 0 | 1 | Not-determined | Genome-fragment | NA | NA | 0 |
| S1-final-viral-combined_k141_357034\|\|full | 1499 | No | NA | 2 | 0 | 0 | Not-determined | Genome-fragment | NA | NA | 0 |
| S4-final-viral-combined_k141_22531\|\|full | 1367 | No | NA | 2 | 0 | 0 | Low-quality | Genome-fragment | 0.86 | AAI-based (high-confidence) | 0 |
| S1-final-viral-combined_k141_93331\|\|full | 1920 | No | NA | 3 | 0 | 0 | Not-determined | Genome-fragment | NA | NA | 0 |
| S1-final-viral-combined_k141_59870\|\|full | 8442 | No | NA | 8 | 0 | 7 | Not-determined | Genome-fragment | NA | NA | 0 |
| S1-final-viral-combined_k141_158350\|\|full | 2154 | No | NA | 2 | 0 | 0 | Not-determined | Genome-fragment | NA | NA | 0 |
| S1-final-viral-combined_k141_389161\|\|full | 3173 | No | NA | 3 | 0 | 0 | Not-determined | Genome-fragment | NA | NA | 0 |
| S8-final-viral-combined_k141_214967\|\|full | 1367 | No | NA | 2 | 0 | 0 | Low-quality | Genome-fragment | 2.04 | AAI-based (medium-confidence) | 0 |
| S11_k141_311240 | 1368 | No | NA | 1 | 0 | 0 | Low-quality | Genome-fragment | 0.79 | AAI-based (medium-confidence) | 0 |
| S1-final-viral-combined_k141_372880\|\|full | 2053 | No | NA | 2 | 0 | 2 | Not-determined | Genome-fragment | NA | NA | 0 |
| S1-final-viral-combined_k141_241575\|\|full | 3262 | No | NA | 5 | 0 | 2 | Not-determined | Genome-fragment | NA | NA | 0 |
| S1-final-viral-combined_k141_258671\|\|full | 2040 | No | NA | 4 | 0 | 2 | Not-determined | Genome-fragment | NA | NA | 0 |
| S1-final-viral-combined_k141_357066\|\|full | 1502 | No | NA | 3 | 0 | 0 | Not-determined | Genome-fragment | NA | NA | 0 |
| S1-final-viral-combined_k141_339243\|\|full | 10674 | No | NA | 6 | 0 | 5 | Not-determined | Genome-fragment | NA | NA | 0 |
| S3-final-viral-combined_k141_32075\|\|full | 1368 | No | NA | 3 | 2 | 0 | Low-quality | Genome-fragment | 4.66 | HMM-based (lower-bound) | 0 |
| S1-final-viral-combined_k141_273920\|\|full | 2281 | No | NA | 3 | 0 | 3 | Not-determined | Genome-fragment | NA | NA | 0 |
| S1-final-viral-combined_k141_258713\|\|full | 2322 | No | NA | 3 | 0 | 1 | Not-determined | Genome-fragment | NA | NA | 0 |
| S3_k141_132228 | 1368 | No | NA | 3 | 0 | 0 | Low-quality | Genome-fragment | 0.89 | AAI-based (high-confidence) | 0 |
| S5_k141_93291 | 1368 | No | NA | 2 | 0 | 1 | Low-quality | Genome-fragment | 0.44 | AAI-based (medium-confidence) | 0 |
| S1-final-viral-combined_k141_127056\|\|full | 1213 | No | NA | 2 | 0 | 1 | Not-determined | Genome-fragment | NA | NA | 0 |
| S12-final-viral-combined_k141_315485\|\|full | 1368 | No | NA | 2 | 0 | 0 | Low-quality | Genome-fragment | 1.04 | AAI-based (high-confidence) | 0 |
| S1-final-viral-combined_k141_241618\|\|full | 1226 | No | NA | 3 | 0 | 0 | Not-determined | Genome-fragment | NA | NA | 0 |
| S1-final-viral-combined_k141_158428\|\|full | 3046 | No | NA | 3 | 0 | 3 | Not-determined | Genome-fragment | NA | NA | 0 |
| S1-final-viral-combined_k141_258750\|\|full | 2296 | No | NA | 3 | 0 | 1 | Not-determined | Genome-fragment | NA | NA | 0 |
| S1-final-viral-combined_k141_11011\|\|full | 2573 | No | NA | 5 | 0 | 1 | Not-determined | Genome-fragment | NA | NA | 0 |
| S1-final-viral-combined_k141_142721\|\|full | 14643 | No | NA | 11 | 0 | 6 | Not-determined | Genome-fragment | NA | NA | 0 |
| S1-final-viral-combined_k141_339289\|\|full | 2443 | No | NA | 2 | 0 | 2 | Not-determined | Genome-fragment | NA | NA | 0 |
| S12-final-viral-combined_k141_156627\|\|lt2gene | 1368 | No | NA | 2 | 1 | 0 | Low-quality | Genome-fragment | 2.15 | AAI-based (high-confidence) | 0 |
| S1-final-viral-combined_k141_258764\|\|full | 3699 | No | NA | 5 | 0 | 4 | Not-determined | Genome-fragment | NA | NA | 0 |
| S4-final-viral-combined_k141_49493\|\|full | 1368 | No | NA | 3 | 0 | 1 | Low-quality | Genome-fragment | 1.11 | AAI-based (medium-confidence) | 0 |
| S1-final-viral-combined_k141_324699\|\|full | 6150 | No | NA | 6 | 0 | 0 | Not-determined | Genome-fragment | NA | NA | 0 |
| S6-final-viral-combined_k141_107922\|\|lt2gene | 1368 | No | NA | 2 | 2 | 0 | Low-quality | Genome-fragment | 4.08 | AAI-based (high-confidence) | 0 |
| S8-final-viral-combined_k141_274472\|\|full | 1368 | No | NA | 4 | 0 | 0 | Low-quality | Genome-fragment | 3.01 | AAI-based (medium-confidence) | 0 |
| S1-final-viral-combined_k141_224870\|\|full | 2416 | No | NA | 3 | 0 | 3 | Not-determined | Genome-fragment | NA | NA | 0 |
| S8_k141_162993 | 1368 | No | NA | 1 | 0 | 1 | Low-quality | Genome-fragment | 0.76 | AAI-based (high-confidence) | 0 |
| S11-final-viral-combined_k141_242248\|\|full | 1369 | No | NA | 2 | 0 | 0 | Low-quality | Genome-fragment | 3.07 | AAI-based (high-confidence) | 0 |
| S1-final-viral-combined_k141_59925\|\|full | 52971 | No | NA | 46 | 0 | 39 | Not-determined | Genome-fragment | NA | NA | 0 |
| S11-final-viral-combined_k141_11434\|\|full | 1369 | No | NA | 2 | 0 | 2 | Low-quality | Genome-fragment | 0.43 | AAI-based (medium-confidence) | 0 |
| S1-final-viral-combined_k141_324717\|\|full | 1048 | No | NA | 2 | 0 | 2 | Not-determined | Genome-fragment | NA | NA | 0 |
| S1_k141_295172 | 1369 | No | NA | 3 | 0 | 2 | Low-quality | Genome-fragment | 0.88 | AAI-based (medium-confidence) | 0 |
| S1-final-viral-combined_k141_209411\|\|full | 1230 | No | NA | 3 | 0 | 2 | Not-determined | Genome-fragment | NA | NA | 0 |
| S1-final-viral-combined_k141_224901\|\|full | 1283 | No | NA | 2 | 0 | 1 | Not-determined | Genome-fragment | NA | NA | 0 |
| S5-final-viral-combined_k141_102894\|\|full | 1369 | No | NA | 2 | 0 | 0 | Low-quality | Genome-fragment | 2.09 | AAI-based (medium-confidence) | 0 |
| S1-final-viral-combined_k141_75831\|\|full | 1886 | No | NA | 4 | 0 | 0 | Not-determined | Genome-fragment | NA | NA | 0 |
| S1-final-viral-combined_k141_108788\|\|full | 1310 | No | NA | 3 | 0 | 1 | Not-determined | Genome-fragment | NA | NA | 0 |
| S5-final-viral-combined_k141_156960\|\|lt2gene | 1369 | No | NA | 2 | 1 | 0 | Low-quality | Genome-fragment | 5.61 | AAI-based (high-confidence) | 0 |
| S1-final-viral-combined_k141_273985\|\|full | 1495 | No | NA | 2 | 0 | 2 | Not-determined | Genome-fragment | NA | NA | 0 |
| S1-final-viral-combined_k141_75865\|\|full | 1677 | No | NA | 2 | 0 | 1 | Not-determined | Genome-fragment | NA | NA | 0 |
| S1-final-viral-combined_k141_93481\|\|full | 2468 | No | NA | 3 | 0 | 2 | Not-determined | Genome-fragment | NA | NA | 0 |
| S1-final-viral-combined_k141_389319\|\|full | 1129 | No | NA | 2 | 0 | 0 | Not-determined | Genome-fragment | NA | NA | 0 |
| S5_k141_14870 | 1369 | No | NA | 1 | 0 | 1 | Low-quality | Genome-fragment | 1.36 | AAI-based (high-confidence) | 0 |
| S7-final-viral-combined_k141_228383\|\|full | 1369 | No | NA | 3 | 1 | 0 | Low-quality | Genome-fragment | 3.56 | AAI-based (medium-confidence) | 0 |
| S1-final-viral-combined_k141_224943\|\|full | 1659 | No | NA | 3 | 0 | 0 | Not-determined | Genome-fragment | NA | NA | 0 |
| S12-final-viral-combined_k141_339533\|\|full | 1369 | No | NA | 2 | 1 | 0 | Low-quality | Genome-fragment | 3.05 | HMM-based (lower-bound) | 0 |
| S12-final-viral-combined_k141_194155\|\|full | 1369 | No | NA | 3 | 0 | 0 | Low-quality | Genome-fragment | 3.56 | AAI-based (medium-confidence) | 0 |
| S1-final-viral-combined_k141_224958\|\|full | 1971 | No | NA | 3 | 0 | 3 | Not-determined | Genome-fragment | NA | NA | 0 |
| S1-final-viral-combined_k141_27227\|\|full | 2195 | No | NA | 4 | 0 | 3 | Not-determined | Genome-fragment | NA | NA | 0 |
| S12-final-viral-combined_k141_311372\|\|full | 1369 | No | NA | 2 | 0 | 0 | Low-quality | Genome-fragment | 3.22 | AAI-based (high-confidence) | 0 |
| S1-final-viral-combined_k141_290967\|\|full | 2186 | No | NA | 4 | 0 | 2 | Not-determined | Genome-fragment | NA | NA | 0 |
| S1-final-viral-combined_k141_59989\|\|full | 4408 | No | NA | 6 | 0 | 2 | Not-determined | Genome-fragment | NA | NA | 0 |
| S12_k141_307545 | 1369 | No | NA | 2 | 0 | 0 | Low-quality | Genome-fragment | 1.39 | AAI-based (high-confidence) | 0 |
| S2-final-viral-combined_k141_60484\|\|full | 1369 | No | NA | 2 | 0 | 0 | Low-quality | Genome-fragment | 1.28 | AAI-based (medium-confidence) | 0 |
| S2-final-viral-combined_k141_132864\|\|full | 1369 | No | NA | 2 | 1 | 0 | Low-quality | Genome-fragment | 1.02 | HMM-based (lower-bound) | 0 |
| S6-final-viral-combined_k141_109738\|\|full | 1369 | No | NA | 3 | 0 | 1 | Low-quality | Genome-fragment | 2.75 | AAI-based (medium-confidence) | 0 |
| S11-final-viral-combined_k141_104404\|\|full | 1370 | No | NA | 2 | 0 | 1 | Low-quality | Genome-fragment | 0.98 | AAI-based (high-confidence) | 0 |
| S1-final-viral-combined_k141_43587\|\|full | 1040 | No | NA | 2 | 0 | 2 | Not-determined | Genome-fragment | NA | NA | 0 |
| S1-final-viral-combined_k141_357178\|\|full | 3218 | No | NA | 6 | 0 | 1 | Not-determined | Genome-fragment | NA | NA | 0 |
| S1-final-viral-combined_k141_339423\|\|full | 1635 | No | NA | 3 | 0 | 1 | Not-determined | Genome-fragment | NA | NA | 0 |
| S11_k141_310315 | 1370 | No | NA | 2 | 0 | 0 | Low-quality | Genome-fragment | 1.12 | AAI-based (medium-confidence) | 0 |
| S1-final-viral-combined_k141_364587\|\|full | 1370 | No | NA | 2 | 0 | 1 | Low-quality | Genome-fragment | 0.98 | AAI-based (high-confidence) | 0 |
| S1-final-viral-combined_k141_93574\|\|full | 1175 | No | NA | 2 | 0 | 1 | Not-determined | Genome-fragment | NA | NA | 0 |
| S1-final-viral-combined_k141_366313\|\|full | 1370 | No | NA | 4 | 1 | 0 | Low-quality | Genome-fragment | 2.63 | AAI-based (high-confidence) | 0 |
| S1-final-viral-combined_k141_43610\|\|full | 1916 | No | NA | 2 | 0 | 0 | Not-determined | Genome-fragment | NA | NA | 0 |
| S1-final-viral-combined_k141_284338\|\|full | 1370 | No | NA | 3 | 0 | 0 | Low-quality | Genome-fragment | 3.19 | AAI-based (medium-confidence) | 0 |
| S1-final-viral-combined_k141_80577\|\|full | 1370 | No | NA | 4 | 2 | 0 | Low-quality | Genome-fragment | 3.1 | AAI-based (high-confidence) | 0 |
| S1-final-viral-combined_k141_43614\|\|full | 8848 | No | NA | 8 | 0 | 7 | Not-determined | Genome-fragment | NA | NA | 0 |
| S1_k141_142451 | 1370 | No | NA | 2 | 2 | 0 | Low-quality | Genome-fragment | 3.41 | AAI-based (high-confidence) | 0 |
| S5-final-viral-combined_k141_152610\|\|full | 1370 | No | NA | 2 | 1 | 0 | Low-quality | Genome-fragment | 3.23 | AAI-based (high-confidence) | 0 |
| S1-final-viral-combined_k141_225049\|\|full | 2716 | No | NA | 3 | 0 | 2 | Not-determined | Genome-fragment | NA | NA | 0 |
| S7-final-viral-combined_k141_10772\|\|full | 1370 | No | NA | 4 | 1 | 0 | Low-quality | Genome-fragment | 3.3 | AAI-based (high-confidence) | 0 |
| S7-final-viral-combined_k141_50217\|\|full | 1370 | No | NA | 3 | 0 | 0 | Low-quality | Genome-fragment | 2.11 | AAI-based (medium-confidence) | 0 |
| S7-final-viral-combined_k141_244328\|\|full | 1370 | No | NA | 4 | 0 | 0 | Low-quality | Genome-fragment | 1.3 | AAI-based (high-confidence) | 0 |
| S6-final-viral-combined_k141_33383\|\|full | 1370 | No | NA | 2 | 0 | 0 | Low-quality | Genome-fragment | 3.99 | AAI-based (high-confidence) | 0 |
| S8-final-viral-combined_k141_106898\|\|full | 1370 | No | NA | 3 | 0 | 0 | Low-quality | Genome-fragment | 3.17 | AAI-based (high-confidence) | 0 |
| S1-final-viral-combined_k141_43654\|\|full | 4173 | No | NA | 3 | 0 | 0 | Not-determined | Genome-fragment | NA | NA | 0 |
| S8-final-viral-combined_k141_6781\|\|full | 1370 | No | NA | 2 | 0 | 1 | Low-quality | Genome-fragment | 2.62 | AAI-based (medium-confidence) | 0 |
| S1-final-viral-combined_k141_339468\|\|full | 9421 | No | NA | 8 | 0 | 7 | Not-determined | Genome-fragment | NA | NA | 0 |
| S1-final-viral-combined_k141_11155\|\|full | 1816 | No | NA | 2 | 0 | 1 | Not-determined | Genome-fragment | NA | NA | 0 |
| S1-final-viral-combined_k141_259053\|\|full | 2814 | No | NA | 3 | 0 | 1 | Not-determined | Genome-fragment | NA | NA | 0 |
| S3-final-viral-combined_k141_62370\|\|lt2gene | 1371 | No | NA | 1 | 0 | 0 | Low-quality | Genome-fragment | 3.19 | AAI-based (medium-confidence) | 0 |
| S1-final-viral-combined_k141_76038\|\|full | 2678 | No | NA | 2 | 0 | 2 | Not-determined | Genome-fragment | NA | NA | 0 |
| S1-final-viral-combined_k141_291106\|\|full | 1048 | No | NA | 2 | 0 | 2 | Not-determined | Genome-fragment | NA | NA | 0 |
| S1-final-viral-combined_k141_339483\|\|full | 1437 | No | NA | 3 | 0 | 0 | Not-determined | Genome-fragment | NA | NA | 0 |
| S5-final-viral-combined_k141_140824\|\|full | 1371 | No | NA | 2 | 0 | 0 | Low-quality | Genome-fragment | 3.41 | AAI-based (medium-confidence) | 0 |
| S12-final-viral-combined_k141_106240\|\|full | 1371 | No | NA | 3 | 0 | 0 | Low-quality | Genome-fragment | 0.7 | AAI-based (high-confidence) | 0 |
| S1-final-viral-combined_k141_192842\|\|full | 1530 | No | NA | 3 | 0 | 1 | Not-determined | Genome-fragment | NA | NA | 0 |
| S8-final-viral-combined_k141_246208\|\|full | 1371 | No | NA | 3 | 0 | 0 | Low-quality | Genome-fragment | 0.74 | AAI-based (high-confidence) | 0 |
| S1-final-viral-combined_k141_291127\|\|full | 1345 | No | NA | 3 | 0 | 2 | Not-determined | Genome-fragment | NA | NA | 0 |
| S1-final-viral-combined_k141_241935\|\|full | 2116 | No | NA | 3 | 0 | 2 | Not-determined | Genome-fragment | NA | NA | 0 |
| S8-final-viral-combined_k141_272042\|\|lt2gene | 1371 | No | NA | 1 | 1 | 0 | Low-quality | Genome-fragment | 2.86 | AAI-based (medium-confidence) | 0 |
| S1-final-viral-combined_k141_142869\|\|full | 3199 | No | NA | 3 | 0 | 2 | Not-determined | Genome-fragment | NA | NA | 0 |
| S1-final-viral-combined_k141_291153\|\|full | 3375 | No | NA | 3 | 0 | 2 | Not-determined | Genome-fragment | NA | NA | 0 |
| S11-final-viral-combined_k141_197562\|\|full | 1372 | No | NA | 4 | 0 | 0 | Low-quality | Genome-fragment | 1.09 | AAI-based (medium-confidence) | 0 |
| S11-final-viral-combined_k141_33155\|\|full | 1372 | No | NA | 4 | 0 | 0 | Low-quality | Genome-fragment | 2.44 | AAI-based (medium-confidence) | 0 |
| S1-final-viral-combined_k141_308135\|\|full | 7330 | No | NA | 6 | 0 | 5 | Not-determined | Genome-fragment | NA | NA | 0 |
| S11-final-viral-combined_k141_50081\|\|full | 1372 | No | NA | 3 | 2 | 0 | Low-quality | Genome-fragment | 0.86 | AAI-based (medium-confidence) | 0 |
| S1-final-viral-combined_k141_324961\|\|full | 1745 | No | NA | 2 | 0 | 1 | Not-determined | Genome-fragment | NA | NA | 0 |
| S11-final-viral-combined_k141_116114\|\|lt2gene | 1372 | No | NA | 1 | 1 | 0 | Low-quality | Genome-fragment | 2.56 | HMM-based (lower-bound) | 0 |
| S5-final-viral-combined_k141_206459\|\|full | 1372 | No | NA | 2 | 1 | 0 | Low-quality | Genome-fragment | 2.55 | AAI-based (medium-confidence) | 0 |
| S1-final-viral-combined_k141_209536\|\|full | 1246 | No | NA | 3 | 0 | 2 | Not-determined | Genome-fragment | NA | NA | 0 |
| S1-final-viral-combined_k141_27327\|\|full | 1287 | No | NA | 3 | 0 | 0 | Not-determined | Genome-fragment | NA | NA | 0 |
| S1-final-viral-combined_k141_225210\|\|full | 2134 | No | NA | 3 | 0 | 3 | Not-determined | Genome-fragment | NA | NA | 0 |
| S17-final-viral-combined_k141_172215\|\|lt2gene | 1372 | No | NA | 1 | 1 | 0 | Low-quality | Genome-fragment | 1.22 | HMM-based (lower-bound) | 0 |
| S17-final-viral-combined_k141_73341\|\|lt2gene | 1372 | No | NA | 1 | 1 | 0 | Low-quality | Genome-fragment | 1.56 | HMM-based (lower-bound) | 0 |
| S1-final-viral-combined_k141_308160\|\|full | 7772 | No | NA | 7 | 0 | 4 | Not-determined | Genome-fragment | NA | NA | 0 |
| S17-k141_137054 | 1372 | No | NA | 1 | 0 | 0 | Low-quality | Genome-fragment | 3.23 | AAI-based (medium-confidence) | 0 |
| S1-final-viral-combined_k141_43787\|\|full | 1058 | No | NA | 2 | 0 | 2 | Not-determined | Genome-fragment | NA | NA | 0 |
| S1-final-viral-combined_k141_209557\|\|full | 1260 | No | NA | 3 | 0 | 0 | Not-determined | Genome-fragment | NA | NA | 0 |
| S12-final-viral-combined_k141_47685\|\|full | 1372 | Yes | 406 | 3 | 1 | 1 | Low-quality | Genome-fragment | 1.07 | AAI-based (high-confidence) | 70.41 |
| S1-final-viral-combined_k141_158676\|\|full | 3323 | No | NA | 3 | 0 | 1 | Not-determined | Genome-fragment | NA | NA | 0 |
| S1-final-viral-combined_k141_357373\|\|full | 2966 | No | NA | 5 | 0 | 0 | Medium-quality | Genome-fragment | 53.65 | AAI-based (high-confidence) | 0 |
| S6_k141_67651 | 1372 | No | NA | 4 | 0 | 0 | Low-quality | Genome-fragment | 1.08 | AAI-based (medium-confidence) | 0 |
| S1-final-viral-combined_k141_357408\|\|full | 2120 | No | NA | 3 | 0 | 1 | Not-determined | Genome-fragment | NA | NA | 0 |
| S8-final-viral-combined_k141_245718\|\|full | 1372 | No | NA | 2 | 0 | 0 | Low-quality | Genome-fragment | 3.12 | AAI-based (medium-confidence) | 0 |
| S1-final-viral-combined_k141_339560\|\|full | 2117 | No | NA | 4 | 0 | 0 | Not-determined | Genome-fragment | NA | NA | 0 |
| S1-final-viral-combined_k141_225273\|\|full | 1510 | No | NA | 4 | 0 | 1 | Not-determined | Genome-fragment | NA | NA | 0 |
| S1-final-viral-combined_k141_373226\|\|full | 1167 | No | NA | 2 | 0 | 0 | Not-determined | Genome-fragment | NA | NA | 0 |
| S8-final-viral-combined_k141_52337\|\|full | 1372 | No | NA | 2 | 1 | 0 | Low-quality | Genome-fragment | 3.08 | AAI-based (medium-confidence) | 0 |
| S1-final-viral-combined_k141_158702\|\|full | 3677 | No | NA | 3 | 0 | 0 | Not-determined | Genome-fragment | NA | NA | 0 |
| S1-final-viral-combined_k141_357424\|\|full | 2032 | No | NA | 4 | 0 | 0 | Not-determined | Genome-fragment | NA | NA | 0 |
| S8_k141_63009 | 1372 | No | NA | 2 | 0 | 1 | Low-quality | Genome-fragment | 2.27 | AAI-based (high-confidence) | 0 |
| S1-final-viral-combined_k141_49799\|\|full | 1373 | No | NA | 5 | 0 | 0 | Low-quality | Genome-fragment | 4.02 | AAI-based (high-confidence) | 0 |
| S1-final-viral-combined_k141_124868\|\|full | 1373 | No | NA | 3 | 0 | 2 | Low-quality | Genome-fragment | 1.87 | AAI-based (medium-confidence) | 0 |
| S3_k141_119212 | 1373 | No | NA | 1 | 0 | 1 | Low-quality | Genome-fragment | 2.96 | AAI-based (high-confidence) | 0 |
| S1-final-viral-combined_k141_109055\|\|full | 2786 | No | NA | 3 | 0 | 3 | Not-determined | Genome-fragment | NA | NA | 0 |
| S1-final-viral-combined_k141_373236\|\|full | 1699 | No | NA | 2 | 0 | 1 | Not-determined | Genome-fragment | NA | NA | 0 |
| S5-final-viral-combined_k141_137903\|\|lt2gene | 1373 | No | NA | 1 | 1 | 0 | Low-quality | Genome-fragment | 3.04 | AAI-based (medium-confidence) | 0 |
| S7-final-viral-combined_k141_219758\|\|full | 1373 | No | NA | 4 | 0 | 0 | Low-quality | Genome-fragment | 2.44 | AAI-based (medium-confidence) | 0 |
| S17-k141_8488 | 1373 | No | NA | 1 | 1 | 0 | Low-quality | Genome-fragment | 0.79 | HMM-based (lower-bound) | 0 |
| S1-final-viral-combined_k141_11293\|\|full | 1122 | No | NA | 3 | 0 | 0 | Not-determined | Genome-fragment | NA | NA | 0 |
| S1-final-viral-combined_k141_142988\|\|full | 1586 | No | NA | 2 | 0 | 0 | Not-determined | Genome-fragment | NA | NA | 0 |
| S1-final-viral-combined_k141_142995\|\|full | 1088 | No | NA | 3 | 0 | 0 | Not-determined | Genome-fragment | NA | NA | 0 |
| S12-final-viral-combined_k141_398105\|\|lt2gene | 1373 | No | NA | 2 | 1 | 0 | Low-quality | Genome-fragment | 2.24 | AAI-based (high-confidence) | 0 |
| S2-final-viral-combined_k141_15122\|\|full | 1373 | No | NA | 5 | 1 | 0 | Low-quality | Genome-fragment | 0.89 | HMM-based (lower-bound) | 0 |
| S1-final-viral-combined_k141_27433\|\|full | 1540 | No | NA | 3 | 0 | 0 | Not-determined | Genome-fragment | NA | NA | 0 |
| S8-final-viral-combined_k141_201866\|\|full | 1373 | No | NA | 3 | 0 | 0 | Low-quality | Genome-fragment | 2.14 | AAI-based (high-confidence) | 0 |
| S8_k141_40984 | 1373 | No | NA | 2 | 0 | 1 | Low-quality | Genome-fragment | 1.3 | AAI-based (medium-confidence) | 0 |
| S9-final-viral-combined_k141_12024\|\|full | 1374 | No | NA | 2 | 1 | 0 | Low-quality | Genome-fragment | 1.58 | HMM-based (lower-bound) | 0 |
| S1-final-viral-combined_k141_127534\|\|full | 1327 | No | NA | 4 | 0 | 0 | Not-determined | Genome-fragment | NA | NA | 0 |
| S1-final-viral-combined_k141_389654\|\|full | 1464 | No | NA | 2 | 0 | 1 | Not-determined | Genome-fragment | NA | NA | 0 |
| S11_k141_17713 | 1374 | No | NA | 3 | 0 | 1 | Low-quality | Genome-fragment | 1.42 | AAI-based (medium-confidence) | 0 |
| S1-final-viral-combined_k141_127181\|\|full | 1374 | No | NA | 3 | 0 | 1 | Low-quality | Genome-fragment | 0.35 | AAI-based (medium-confidence) | 0 |
| S1-final-viral-combined_k141_96885\|\|full | 1374 | No | NA | 5 | 1 | 0 | Low-quality | Genome-fragment | 4.24 | AAI-based (high-confidence) | 0 |
| S1-final-viral-combined_k141_174918\|\|full | 1044 | No | NA | 2 | 0 | 1 | Not-determined | Genome-fragment | NA | NA | 0 |
| S1-final-viral-combined_k141_76289\|\|full | 1406 | No | NA | 2 | 0 | 0 | Not-determined | Genome-fragment | NA | NA | 0 |
| S1-final-viral-combined_k141_389674\|\|full | 1513 | No | NA | 4 | 0 | 0 | Not-determined | Genome-fragment | NA | NA | 0 |
| S1-final-viral-combined_k141_291405\|\|full | 1808 | No | NA | 2 | 0 | 2 | Not-determined | Genome-fragment | NA | NA | 0 |
| S1_k141_251887 | 1374 | No | NA | 7 | 0 | 0 | Low-quality | Genome-fragment | 1.3 | AAI-based (medium-confidence) | 0 |
| S1-final-viral-combined_k141_373308\|\|full | 14841 | No | NA | 17 | 1 | 6 | High-quality | High-quality | 100 | HMM-based (lower-bound) | 0 |
| S7-final-viral-combined_k141_97086\|\|full | 1374 | No | NA | 2 | 0 | 0 | Low-quality | Genome-fragment | 3.19 | AAI-based (medium-confidence) | 0 |
| S1-final-viral-combined_k141_93949\|\|full | 2152 | No | NA | 2 | 0 | 1 | Not-determined | Genome-fragment | NA | NA | 0 |
| S1-final-viral-combined_k141_93952\|\|full | 1245 | No | NA | 3 | 0 | 3 | Not-determined | Genome-fragment | NA | NA | 0 |
| S1-final-viral-combined_k141_325093\|\|full | 3682 | No | NA | 3 | 0 | 2 | Not-determined | Genome-fragment | NA | NA | 0 |
| S17-final-viral-combined_k141_186018\|\|full | 1374 | No | NA | 3 | 0 | 0 | Low-quality | Genome-fragment | 1.63 | AAI-based (high-confidence) | 0 |
| S1-final-viral-combined_k141_373316\|\|full | 1320 | No | NA | 3 | 0 | 0 | Not-determined | Genome-fragment | NA | NA | 0 |
| S12-final-viral-combined_k141_157741\|\|full | 1374 | No | NA | 3 | 0 | 0 | Low-quality | Genome-fragment | 0.4 | AAI-based (medium-confidence) | 0 |
| S1-final-viral-combined_k141_225467\|\|full | 1651 | No | NA | 3 | 0 | 1 | Not-determined | Genome-fragment | NA | NA | 0 |
| S12-final-viral-combined_k141_313514\|\|lt2gene | 1374 | No | NA | 2 | 1 | 0 | Low-quality | Genome-fragment | 3.42 | AAI-based (medium-confidence) | 0 |
| S1-final-viral-combined_k141_357597\|\|full | 2856 | No | NA | 4 | 0 | 0 | Not-determined | Genome-fragment | NA | NA | 0 |
| S12_k141_251889 | 1374 | No | NA | 1 | 0 | 1 | Low-quality | Genome-fragment | 1.09 | AAI-based (high-confidence) | 0 |
| S1-final-viral-combined_k141_109189\|\|full | 1068 | No | NA | 3 | 0 | 0 | Not-determined | Genome-fragment | NA | NA | 0 |
| S1-final-viral-combined_k141_93978\|\|full | 2082 | No | NA | 6 | 0 | 1 | Not-determined | Genome-fragment | NA | NA | 0 |
| S1-final-viral-combined_k141_308339\|\|full | 1051 | No | NA | 2 | 0 | 1 | Not-determined | Genome-fragment | NA | NA | 0 |
| S1-final-viral-combined_k141_357608\|\|full | 1136 | No | NA | 3 | 0 | 1 | Not-determined | Genome-fragment | NA | NA | 0 |
| S1-final-viral-combined_k141_325139\|\|full | 1318 | No | NA | 2 | 0 | 0 | Not-determined | Genome-fragment | NA | NA | 0 |
| S12_k141_290378 | 1374 | No | NA | 2 | 0 | 0 | Low-quality | Genome-fragment | 0.83 | AAI-based (medium-confidence) | 0 |
| S12_k141_263654 | 1374 | No | NA | 2 | 0 | 0 | Low-quality | Genome-fragment | 3.09 | AAI-based (high-confidence) | 0 |
| S1-final-viral-combined_k141_109209\|\|full | 1184 | No | NA | 2 | 0 | 0 | Not-determined | Genome-fragment | NA | NA | 0 |
| S1-final-viral-combined_k141_242176\|\|full | 1837 | No | NA | 3 | 0 | 0 | Not-determined | Genome-fragment | NA | NA | 0 |
| S1-final-viral-combined_k141_242177\|\|full | 2512 | No | NA | 3 | 0 | 2 | Not-determined | Genome-fragment | NA | NA | 0 |
| S2-final-viral-combined_k141_134788\|\|full | 1374 | No | NA | 3 | 0 | 0 | Low-quality | Genome-fragment | 3.3 | AAI-based (medium-confidence) | 0 |
| S1-final-viral-combined_k141_259295\|\|full | 1521 | No | NA | 3 | 0 | 1 | Not-determined | Genome-fragment | NA | NA | 0 |
| S1-final-viral-combined_k141_389753\|\|full | 6271 | No | NA | 7 | 0 | 5 | Not-determined | Genome-fragment | NA | NA | 0 |
| S1-final-viral-combined_k141_143184\|\|full | 1064 | No | NA | 2 | 0 | 1 | Not-determined | Genome-fragment | NA | NA | 0 |
| S1-final-viral-combined_k141_225513\|\|full | 1708 | No | NA | 2 | 0 | 0 | Not-determined | Genome-fragment | NA | NA | 0 |
| S1-final-viral-combined_k141_94055\|\|full | 3142 | No | NA | 3 | 0 | 3 | Not-determined | Genome-fragment | NA | NA | 0 |
| S2-final-viral-combined_k141_36066\|\|full | 1374 | No | NA | 3 | 0 | 1 | Low-quality | Genome-fragment | 1.35 | AAI-based (high-confidence) | 0 |
| S2-final-viral-combined_k141_71630\|\|lt2gene | 1374 | No | NA | 2 | 2 | 0 | Low-quality | Genome-fragment | 3.21 | AAI-based (medium-confidence) | 0 |
| S2_k141_148713 | 1374 | No | NA | 1 | 0 | 1 | Low-quality | Genome-fragment | 0.45 | AAI-based (medium-confidence) | 0 |
| S1-final-viral-combined_k141_373408\|\|full | 1444 | No | NA | 2 | 0 | 0 | Not-determined | Genome-fragment | NA | NA | 0 |
| S1-final-viral-combined_k141_94073\|\|full | 1416 | No | NA | 2 | 0 | 0 | Not-determined | Genome-fragment | NA | NA | 0 |
| S2_k141_124059 | 1374 | No | NA | 5 | 1 | 0 | Low-quality | Genome-fragment | 3.76 | AAI-based (high-confidence) | 0 |
| S4-final-viral-combined_k141_20824\|\|full | 1374 | No | NA | 2 | 0 | 1 | Low-quality | Genome-fragment | 0.77 | AAI-based (medium-confidence) | 0 |
| S1-final-viral-combined_k141_43992\|\|full | 16717 | No | NA | 21 | 0 | 13 | Not-determined | Genome-fragment | NA | NA | 0 |
| S1-final-viral-combined_k141_158942\|\|full | 2370 | No | NA | 2 | 0 | 0 | Not-determined | Genome-fragment | NA | NA | 0 |
| S4-final-viral-combined_k141_22017\|\|lt2gene | 1374 | No | NA | 1 | 1 | 0 | Low-quality | Genome-fragment | 1.29 | AAI-based (high-confidence) | 0 |
| S1-final-viral-combined_k141_94103\|\|full | 1658 | No | NA | 2 | 0 | 1 | Not-determined | Genome-fragment | NA | NA | 0 |
| S1-final-viral-combined_k141_158950\|\|full | 3983 | No | NA | 3 | 0 | 3 | Not-determined | Genome-fragment | NA | NA | 0 |
| S6-final-viral-combined_k141_27266\|\|lt2gene | 1374 | No | NA | 2 | 1 | 0 | Low-quality | Genome-fragment | 2.92 | AAI-based (high-confidence) | 0 |
| S8_k141_127143 | 1374 | No | NA | 1 | 0 | 1 | Low-quality | Genome-fragment | 1.34 | AAI-based (medium-confidence) | 0 |
| S1-final-viral-combined_k141_239248\|\|full | 1375 | No | NA | 4 | 2 | 0 | Low-quality | Genome-fragment | 3.59 | AAI-based (high-confidence) | 0 |
| S1-final-viral-combined_k141_325276\|\|full | 2940 | No | NA | 4 | 0 | 3 | Not-determined | Genome-fragment | NA | NA | 0 |
| S1-final-viral-combined_k141_193296\|\|full | 1601 | No | NA | 2 | 0 | 2 | Not-determined | Genome-fragment | NA | NA | 0 |
| S1-final-viral-combined_k141_78618\|\|full | 1375 | No | NA | 3 | 0 | 1 | Low-quality | Genome-fragment | 3.58 | AAI-based (medium-confidence) | 0 |
| S1-final-viral-combined_k141_37171\|\|lt2gene | 1375 | No | NA | 1 | 1 | 0 | Low-quality | Genome-fragment | 2.6 | HMM-based (lower-bound) | 0 |
| S1-final-viral-combined_k141_209886\|\|full | 1925 | No | NA | 3 | 0 | 1 | Not-determined | Genome-fragment | NA | NA | 0 |
| S1-final-viral-combined_k141_193300\|\|full | 3063 | No | NA | 3 | 0 | 3 | Not-determined | Genome-fragment | NA | NA | 0 |
| S1-final-viral-combined_k141_158978\|\|full | 2188 | No | NA | 3 | 0 | 2 | Not-determined | Genome-fragment | NA | NA | 0 |
| S1-final-viral-combined_k141_274508\|\|full | 3144 | No | NA | 4 | 0 | 2 | Not-determined | Genome-fragment | NA | NA | 0 |
| S3_k141_121074 | 1375 | No | NA | 2 | 0 | 0 | Low-quality | Genome-fragment | 1.74 | AAI-based (high-confidence) | 0 |
| S17-final-viral-combined_k141_77050\|\|full | 1375 | No | NA | 3 | 1 | 0 | Low-quality | Genome-fragment | 3.27 | AAI-based (medium-confidence) | 0 |
| S12-final-viral-combined_k141_302956\|\|lt2gene | 1375 | No | NA | 2 | 2 | 0 | Low-quality | Genome-fragment | 4.29 | AAI-based (medium-confidence) | 0 |
| S1-final-viral-combined_k141_60615\|\|full | 1445 | No | NA | 3 | 0 | 0 | Not-determined | Genome-fragment | NA | NA | 0 |
| S1-final-viral-combined_k141_193328\|\|full | 2848 | No | NA | 3 | 0 | 1 | Not-determined | Genome-fragment | NA | NA | 0 |
| S12-final-viral-combined_k141_233648\|\|lt2gene | 1375 | No | NA | 2 | 1 | 0 | Low-quality | Genome-fragment | 3.34 | AAI-based (high-confidence) | 0 |
| S2-final-viral-combined_k141_53778\|\|full | 1375 | No | NA | 4 | 0 | 0 | Low-quality | Genome-fragment | 3.03 | AAI-based (high-confidence) | 0 |
| S1-final-viral-combined_k141_143322\|\|full | 3643 | No | NA | 6 | 0 | 4 | Not-determined | Genome-fragment | NA | NA | 0 |
| S1-final-viral-combined_k141_373481\|\|full | 1268 | No | NA | 2 | 0 | 1 | Not-determined | Genome-fragment | NA | NA | 0 |
| S1-final-viral-combined_k141_357765\|\|full | 2439 | No | NA | 3 | 0 | 1 | Not-determined | Genome-fragment | NA | NA | 0 |
| S2_k141_101096 | 1375 | No | NA | 3 | 1 | 0 | Low-quality | Genome-fragment | 2.34 | AAI-based (medium-confidence) | 0 |
| S2_k141_165353 | 1375 | No | NA | 4 | 0 | 0 | Low-quality | Genome-fragment | 1.43 | AAI-based (high-confidence) | 0 |
| S1-final-viral-combined_k141_159030\|\|full | 3408 | No | NA | 4 | 0 | 1 | Not-determined | Genome-fragment | NA | NA | 0 |
| S11-final-viral-combined_k141_265617\|\|full | 1376 | No | NA | 2 | 0 | 1 | Low-quality | Genome-fragment | 2.61 | AAI-based (medium-confidence) | 0 |
| S1-final-viral-combined_k141_291710\|\|full | 3103 | No | NA | 4 | 0 | 1 | Not-determined | Genome-fragment | NA | NA | 0 |
| S1-final-viral-combined_k141_259475\|\|full | 1284 | No | NA | 4 | 0 | 0 | Not-determined | Genome-fragment | NA | NA | 0 |
| S1-final-viral-combined_k141_175193\|\|full | 1291 | No | NA | 2 | 0 | 1 | Not-determined | Genome-fragment | NA | NA | 0 |
| S11-final-viral-combined_k141_322564\|\|lt2gene | 1376 | No | NA | 1 | 1 | 0 | Low-quality | Genome-fragment | 0.95 | HMM-based (lower-bound) | 0 |
| S1-final-viral-combined_k141_325353\|\|full | 1454 | No | NA | 3 | 0 | 1 | Not-determined | Genome-fragment | NA | NA | 0 |
| S3_k141_17429 | 1376 | No | NA | 4 | 0 | 2 | Low-quality | Genome-fragment | 4.14 | AAI-based (medium-confidence) | 0 |
| S1-final-viral-combined_k141_193427\|\|full | 1260 | No | NA | 3 | 0 | 0 | Not-determined | Genome-fragment | NA | NA | 0 |
| S1-final-viral-combined_k141_259508\|\|full | 1693 | No | NA | 4 | 0 | 2 | Not-determined | Genome-fragment | NA | NA | 0 |
| S5-final-viral-combined_k141_4460\|\|full | 1376 | No | NA | 3 | 0 | 0 | Low-quality | Genome-fragment | 3.76 | AAI-based (high-confidence) | 0 |
| S1-final-viral-combined_k141_94257\|\|full | 6577 | No | NA | 5 | 0 | 5 | Not-determined | Genome-fragment | NA | NA | 0 |
| S7-final-viral-combined_k141_70393\|\|full | 1376 | Yes | 1127 | 3 | 1 | 1 | Low-quality | Genome-fragment | 2.26 | HMM-based (lower-bound) | 18.1 |
| S1-final-viral-combined_k141_109429\|\|full | 2798 | No | NA | 2 | 0 | 1 | Not-determined | Genome-fragment | NA | NA | 0 |
| S2-final-viral-combined_k141_134805\|\|full | 1376 | No | NA | 3 | 0 | 0 | Low-quality | Genome-fragment | 3.26 | AAI-based (medium-confidence) | 0 |
| S1-final-viral-combined_k141_143382\|\|full | 5793 | No | NA | 9 | 0 | 1 | Not-determined | Genome-fragment | NA | NA | 0 |
| S1-final-viral-combined_k141_94270\|\|full | 1817 | No | NA | 3 | 0 | 3 | Not-determined | Genome-fragment | NA | NA | 0 |
| S1-final-viral-combined_k141_308595\|\|full | 2257 | No | NA | 3 | 0 | 3 | Not-determined | Genome-fragment | NA | NA | 0 |
| S1-final-viral-combined_k141_291803\|\|full | 1933 | No | NA | 4 | 0 | 0 | Not-determined | Genome-fragment | NA | NA | 0 |
| S1-final-viral-combined_k141_143404\|\|full | 1635 | No | NA | 2 | 0 | 1 | Not-determined | Genome-fragment | NA | NA | 0 |
| S2-final-viral-combined_k141_93690\|\|full | 1376 | No | NA | 3 | 1 | 0 | Low-quality | Genome-fragment | 3.03 | AAI-based (medium-confidence) | 0 |
| S2_k141_134971 | 1376 | No | NA | 3 | 0 | 0 | Low-quality | Genome-fragment | 2.24 | AAI-based (high-confidence) | 0 |
| S1-final-viral-combined_k141_11616\|\|full | 8251 | No | NA | 7 | 0 | 2 | Not-determined | Genome-fragment | NA | NA | 0 |
| S4-final-viral-combined_k141_70266\|\|full | 1376 | No | NA | 4 | 0 | 0 | Low-quality | Genome-fragment | 2.02 | AAI-based (medium-confidence) | 0 |
| S1-final-viral-combined_k141_259602\|\|full | 3074 | No | NA | 8 | 0 | 0 | Not-determined | Genome-fragment | NA | NA | 0 |
| S8-final-viral-combined_k141_242716\|\|full | 1376 | No | NA | 2 | 0 | 1 | Low-quality | Genome-fragment | 0.44 | AAI-based (medium-confidence) | 0 |
| S11-final-viral-combined_k141_91314\|\|full | 1377 | No | NA | 3 | 0 | 0 | Low-quality | Genome-fragment | 3.22 | AAI-based (high-confidence) | 0 |
| S11-final-viral-combined_k141_268295\|\|lt2gene | 1377 | No | NA | 2 | 1 | 0 | Low-quality | Genome-fragment | 3.83 | AAI-based (high-confidence) | 0 |
| S1-final-viral-combined_k141_76546\|\|full | 17519 | No | NA | 14 | 0 | 10 | Not-determined | Genome-fragment | NA | NA | 0 |
| S1-final-viral-combined_k141_76548\|\|full | 1438 | No | NA | 3 | 0 | 0 | Not-determined | Genome-fragment | NA | NA | 0 |
| S1-final-viral-combined_k141_133439\|\|lt2gene | 1377 | No | NA | 1 | 1 | 0 | Low-quality | Genome-fragment | 3.05 | AAI-based (high-confidence) | 0 |
| S1-final-viral-combined_k141_255271\|\|lt2gene | 1377 | No | NA | 1 | 0 | 0 | Low-quality | Genome-fragment | 3.9 | AAI-based (high-confidence) | 0 |
| S5-final-viral-combined_k141_90268\|\|full | 1377 | No | NA | 3 | 0 | 0 | Low-quality | Genome-fragment | 3.13 | AAI-based (medium-confidence) | 0 |
| S1-final-viral-combined_k141_143459\|\|full | 35960 | No | NA | 50 | 24 | 1 | Medium-quality | Genome-fragment | 77.24 | HMM-based (lower-bound) | 0 |
| S5-final-viral-combined_k141_161464\|\|full | 1377 | No | NA | 3 | 1 | 0 | Low-quality | Genome-fragment | 1.26 | HMM-based (lower-bound) | 0 |
| S7_k141_193569 | 1377 | No | NA | 1 | 0 | 1 | Low-quality | Genome-fragment | 2.09 | AAI-based (high-confidence) | 0 |
| S1-final-viral-combined_k141_193534\|\|full | 2395 | No | NA | 2 | 0 | 2 | Not-determined | Genome-fragment | NA | NA | 0 |
| S1-final-viral-combined_k141_390031\|\|full | 1120 | No | NA | 2 | 0 | 0 | Not-determined | Genome-fragment | NA | NA | 0 |
| S17-final-viral-combined_k141_148219\|\|full | 1377 | No | NA | 3 | 0 | 0 | Low-quality | Genome-fragment | 3.54 | AAI-based (high-confidence) | 0 |
| S17-final-viral-combined_k141_194216\|\|full | 1377 | No | NA | 2 | 0 | 1 | Low-quality | Genome-fragment | 0.86 | AAI-based (medium-confidence) | 0 |
| S12-final-viral-combined_k141_396379\|\|full | 1377 | No | NA | 4 | 0 | 0 | Low-quality | Genome-fragment | 2.1 | AAI-based (medium-confidence) | 0 |
| S1-final-viral-combined_k141_175346\|\|full | 1046 | No | NA | 2 | 0 | 0 | Not-determined | Genome-fragment | NA | NA | 0 |
| S1-final-viral-combined_k141_11704\|\|full | 1795 | No | NA | 3 | 0 | 0 | Not-determined | Genome-fragment | NA | NA | 0 |
| S1-final-viral-combined_k141_325510\|\|full | 1748 | No | NA | 3 | 0 | 2 | Not-determined | Genome-fragment | NA | NA | 0 |
| S1-final-viral-combined_k141_109586\|\|full | 5043 | No | NA | 4 | 0 | 4 | Not-determined | Genome-fragment | NA | NA | 0 |
| S1-final-viral-combined_k141_259664\|\|full | 5143 | No | NA | 10 | 0 | 0 | Not-determined | Genome-fragment | NA | NA | 0 |
| S1-final-viral-combined_k141_259667\|\|full | 2235 | No | NA | 3 | 0 | 0 | Not-determined | Genome-fragment | NA | NA | 0 |
| S1-final-viral-combined_k141_274732\|\|full | 3235 | No | NA | 3 | 0 | 1 | Not-determined | Genome-fragment | NA | NA | 0 |
| S2-final-viral-combined_k141_137449\|\|full | 1377 | No | NA | 2 | 0 | 0 | Low-quality | Genome-fragment | 1.24 | AAI-based (medium-confidence) | 0 |
| S1-final-viral-combined_k141_94446\|\|full | 1551 | No | NA | 3 | 0 | 0 | Not-determined | Genome-fragment | NA | NA | 0 |
| S1-final-viral-combined_k141_339904\|\|full | 2733 | No | NA | 3 | 0 | 2 | Not-determined | Genome-fragment | NA | NA | 0 |
| S2-final-viral-combined_k141_75075\|\|full | 1377 | No | NA | 4 | 0 | 1 | Low-quality | Genome-fragment | 3.35 | AAI-based (medium-confidence) | 0 |
| S11-final-viral-combined_k141_22425\|\|full | 1378 | No | NA | 2 | 0 | 2 | Low-quality | Genome-fragment | 0.97 | AAI-based (medium-confidence) | 0 |
| S1-final-viral-combined_k141_370906\|\|full | 1378 | No | NA | 3 | 0 | 2 | Low-quality | Genome-fragment | 0.8 | AAI-based (medium-confidence) | 0 |
| S5-final-viral-combined_k141_129310\|\|full | 1378 | No | NA | 2 | 0 | 1 | Low-quality | Genome-fragment | 0.71 | AAI-based (medium-confidence) | 0 |
| S1-final-viral-combined_k141_308752\|\|full | 1352 | No | NA | 2 | 0 | 2 | Not-determined | Genome-fragment | NA | NA | 0 |
| S1-final-viral-combined_k141_11754\|\|full | 1912 | No | NA | 4 | 0 | 1 | Not-determined | Genome-fragment | NA | NA | 0 |
| S1-final-viral-combined_k141_94487\|\|full | 1246 | No | NA | 3 | 0 | 1 | Not-determined | Genome-fragment | NA | NA | 0 |
| S1-final-viral-combined_k141_339949\|\|full | 2358 | No | NA | 3 | 0 | 1 | Not-determined | Genome-fragment | NA | NA | 0 |
| S7_k141_243494 | 1378 | No | NA | 2 | 0 | 0 | Low-quality | Genome-fragment | 0.52 | AAI-based (medium-confidence) | 0 |
| S1-final-viral-combined_k141_76632\|\|full | 1084 | No | NA | 2 | 0 | 1 | Not-determined | Genome-fragment | NA | NA | 0 |
| S1-final-viral-combined_k141_11768\|\|full | 2216 | No | NA | 3 | 0 | 1 | Not-determined | Genome-fragment | NA | NA | 0 |
| S1-final-viral-combined_k141_259730\|\|full | 1325 | No | NA | 2 | 0 | 0 | Not-determined | Genome-fragment | NA | NA | 0 |
| S1-final-viral-combined_k141_325590\|\|full | 2718 | No | NA | 5 | 0 | 0 | Not-determined | Genome-fragment | NA | NA | 0 |
| S12-final-viral-combined_k141_225055\|\|full | 1378 | No | NA | 2 | 0 | 0 | Low-quality | Genome-fragment | 1.97 | AAI-based (medium-confidence) | 0 |
| S1-final-viral-combined_k141_292019\|\|full | 1599 | No | NA | 2 | 0 | 1 | Not-determined | Genome-fragment | NA | NA | 0 |
| S1-final-viral-combined_k141_339976\|\|full | 4809 | No | NA | 5 | 0 | 4 | Not-determined | Genome-fragment | NA | NA | 0 |
| S1-final-viral-combined_k141_159242\|\|full | 1339 | No | NA | 2 | 0 | 2 | Not-determined | Genome-fragment | NA | NA | 0 |
| S1-final-viral-combined_k141_225915\|\|full | 1333 | No | NA | 2 | 0 | 1 | Not-determined | Genome-fragment | NA | NA | 0 |
| S1-final-viral-combined_k141_325624\|\|full | 2429 | No | NA | 2 | 0 | 1 | Not-determined | Genome-fragment | NA | NA | 0 |
| S1-final-viral-combined_k141_242525\|\|full | 1657 | No | NA | 3 | 0 | 2 | Not-determined | Genome-fragment | NA | NA | 0 |
| S1-final-viral-combined_k141_340006\|\|full | 1004 | No | NA | 3 | 0 | 1 | Not-determined | Genome-fragment | NA | NA | 0 |
| S1-final-viral-combined_k141_109670\|\|full | 1883 | No | NA | 2 | 0 | 0 | Not-determined | Genome-fragment | NA | NA | 0 |
| S1-final-viral-combined_k141_175395\|\|full | 4979 | No | NA | 3 | 0 | 2 | Not-determined | Genome-fragment | NA | NA | 0 |
| S1-final-viral-combined_k141_44463\|\|full | 1910 | No | NA | 3 | 0 | 3 | Not-determined | Genome-fragment | NA | NA | 0 |
| S2-final-viral-combined_k141_10340\|\|lt2gene | 1378 | No | NA | 1 | 1 | 0 | Low-quality | Genome-fragment | 2.77 | AAI-based (high-confidence) | 0 |
| S1-final-viral-combined_k141_28078\|\|full | 1615 | No | NA | 2 | 0 | 0 | Not-determined | Genome-fragment | NA | NA | 0 |
| S1-final-viral-combined_k141_193687\|\|full | 5738 | No | NA | 8 | 0 | 5 | Not-determined | Genome-fragment | NA | NA | 0 |
| S6-final-viral-combined_k141_30098\|\|full | 1378 | No | NA | 3 | 1 | 0 | Low-quality | Genome-fragment | 2.03 | AAI-based (high-confidence) | 0 |
| S8_k141_117153 | 1378 | No | NA | 3 | 1 | 0 | Low-quality | Genome-fragment | 1.89 | AAI-based (high-confidence) | 0 |
| S1-final-viral-combined_k141_94540\|\|full | 3204 | No | NA | 4 | 0 | 1 | Not-determined | Genome-fragment | NA | NA | 0 |
| S1-final-viral-combined_k141_390159\|\|full | 8356 | No | NA | 10 | 0 | 7 | Not-determined | Genome-fragment | NA | NA | 0 |
| S8_k141_129494 | 1378 | No | NA | 6 | 0 | 0 | Low-quality | Genome-fragment | 0.73 | AAI-based (medium-confidence) | 0 |
| S1-final-viral-combined_k141_159314\|\|full | 8804 | No | NA | 8 | 0 | 3 | Not-determined | Genome-fragment | NA | NA | 0 |
| S1-final-viral-combined_k141_175425\|\|full | 3175 | No | NA | 3 | 0 | 3 | Not-determined | Genome-fragment | NA | NA | 0 |
| S9-final-viral-combined_k141_74571\|\|full | 1379 | No | NA | 4 | 3 | 0 | Low-quality | Genome-fragment | 4.33 | AAI-based (high-confidence) | 0 |
| S1-final-viral-combined_k141_308848\|\|full | 21928 | No | NA | 24 | 2 | 0 | Medium-quality | Genome-fragment | 66.34 | AAI-based (high-confidence) | 0 |
| S1-final-viral-combined_k141_175439\|\|full | 1526 | No | NA | 2 | 0 | 0 | Not-determined | Genome-fragment | NA | NA | 0 |
| S11-final-viral-combined_k141_23772\|\|full | 1379 | No | NA | 2 | 0 | 0 | Low-quality | Genome-fragment | 2.18 | AAI-based (high-confidence) | 0 |
| S1-final-viral-combined_k141_188216\|\|full | 1379 | No | NA | 3 | 0 | 0 | Low-quality | Genome-fragment | 3.27 | AAI-based (medium-confidence) | 0 |
| S1-final-viral-combined_k141_373837\|\|full | 2562 | No | NA | 4 | 0 | 2 | Not-determined | Genome-fragment | NA | NA | 0 |
| S7-final-viral-combined_k141_172094\|\|full | 1379 | No | NA | 3 | 0 | 0 | Low-quality | Genome-fragment | 2.1 | AAI-based (medium-confidence) | 0 |
| S7_k141_155732 | 1379 | No | NA | 3 | 0 | 0 | Low-quality | Genome-fragment | 1.3 | AAI-based (high-confidence) | 0 |
| S2_k141_98521 | 1379 | No | NA | 1 | 0 | 1 | Low-quality | Genome-fragment | 1.37 | AAI-based (medium-confidence) | 0 |
| S6-final-viral-combined_k141_77524\|\|full | 1379 | No | NA | 4 | 0 | 0 | Low-quality | Genome-fragment | 1.64 | AAI-based (high-confidence) | 0 |
| S1-final-viral-combined_k141_308897\|\|full | 1427 | No | NA | 2 | 0 | 2 | Not-determined | Genome-fragment | NA | NA | 0 |
| S1-final-viral-combined_k141_94600\|\|full | 4172 | No | NA | 5 | 0 | 0 | Not-determined | Genome-fragment | NA | NA | 0 |
| S9-final-viral-combined_k141_54509\|\|full | 1380 | No | NA | 3 | 0 | 0 | Low-quality | Genome-fragment | 2.94 | AAI-based (high-confidence) | 0 |
| S1-final-viral-combined_k141_292137\|\|full | 2464 | No | NA | 2 | 0 | 1 | Not-determined | Genome-fragment | NA | NA | 0 |
| S1-final-viral-combined_k141_259876\|\|full | 2698 | No | NA | 3 | 0 | 2 | Not-determined | Genome-fragment | NA | NA | 0 |
| S1-final-viral-combined_k141_143736\|\|full | 1458 | No | NA | 2 | 0 | 0 | Not-determined | Genome-fragment | NA | NA | 0 |
| S3-final-viral-combined_k141_39223\|\|lt2gene | 1380 | No | NA | 2 | 1 | 0 | Low-quality | Genome-fragment | 1.29 | HMM-based (lower-bound) | 0 |
| S1-final-viral-combined_k141_76819\|\|full | 3399 | No | NA | 3 | 0 | 1 | Not-determined | Genome-fragment | NA | NA | 0 |
| S7_k141_134603 | 1380 | No | NA | 4 | 0 | 3 | Low-quality | Genome-fragment | 3.07 | AAI-based (high-confidence) | 0 |

| | | | | | | | | | | | |
|---|---|---|---|---|---|---|---|---|---|---|---|
| S1-final-viral-combined_k141_340119\|\|full | 2185 | No | NA | 4 | 0 | 0 | Not-determined | Genome-fragment | NA | NA | 0 |
| S1-final-viral-combined_k141_358290\|\|full | 7925 | No | NA | 12 | 0 | 2 | Not-determined | Genome-fragment | NA | NA | 0 |
| S17-final-viral-combined_k141_124786\|\|full | 1380 | No | NA | 4 | 1 | 0 | Low-quality | Genome-fragment | 1.8 | AAI-based (high-confidence) | 0 |
| S1-final-viral-combined_k141_325744\|\|full | 2600 | No | NA | 4 | 0 | 0 | Not-determined | Genome-fragment | NA | NA | 0 |
| S1-final-viral-combined_k141_292172\|\|full | 3553 | No | NA | 5 | 0 | 0 | Not-determined | Genome-fragment | NA | NA | 0 |
| S1-final-viral-combined_k141_109782\|\|full | 1092 | No | NA | 3 | 0 | 1 | Not-determined | Genome-fragment | NA | NA | 0 |
| S17-final-viral-combined_k141_117919\|\|full | 1380 | No | NA | 3 | 2 | 0 | Low-quality | Genome-fragment | 1.47 | HMM-based (lower-bound) | 0 |
| S1-final-viral-combined_k141_94640\|\|full | 1858 | No | NA | 3 | 0 | 2 | Not-determined | Genome-fragment | NA | NA | 0 |
| S1-final-viral-combined_k141_325770\|\|full | 1456 | No | NA | 4 | 0 | 0 | Not-determined | Genome-fragment | NA | NA | 0 |
| S1-final-viral-combined_k141_11974\|\|full | 2796 | No | NA | 3 | 0 | 2 | Not-determined | Genome-fragment | NA | NA | 0 |
| S17-final-viral-combined_k141_132250\|\|full | 1380 | No | NA | 2 | 0 | 0 | Low-quality | Genome-fragment | 3.38 | AAI-based (medium-confidence) | 0 |
| S1-final-viral-combined_k141_226060\|\|full | 2384 | No | NA | 5 | 0 | 0 | Not-determined | Genome-fragment | NA | NA | 0 |
| S1-final-viral-combined_k141_274988\|\|full | 3811 | No | NA | 3 | 0 | 2 | Not-determined | Genome-fragment | NA | NA | 0 |
| S1-final-viral-combined_k141_76919\|\|full | 1837 | No | NA | 2 | 0 | 0 | Not-determined | Genome-fragment | NA | NA | 0 |
| S17-k141_144452 | 1380 | No | NA | 4 | 0 | 0 | Low-quality | Genome-fragment | 3 | AAI-based (high-confidence) | 0 |
| S1-final-viral-combined_k141_275003\|\|full | 2284 | No | NA | 3 | 0 | 2 | Not-determined | Genome-fragment | NA | NA | 0 |
| S1-final-viral-combined_k141_292254\|\|full | 2197 | No | NA | 3 | 0 | 2 | Not-determined | Genome-fragment | NA | NA | 0 |
| S1-final-viral-combined_k141_109895\|\|full | 2438 | No | NA | 2 | 0 | 2 | Not-determined | Genome-fragment | NA | NA | 0 |
| S12-final-viral-combined_k141_143272\|\|full | 1380 | No | NA | 2 | 0 | 1 | Low-quality | Genome-fragment | 1.24 | AAI-based (medium-confidence) | 0 |
| S1-final-viral-combined_k141_128107\|\|full | 1594 | No | NA | 3 | 0 | 0 | Not-determined | Genome-fragment | NA | NA | 0 |
| S1-final-viral-combined_k141_340220\|\|full | 1364 | No | NA | 3 | 0 | 1 | Not-determined | Genome-fragment | NA | NA | 0 |
| S2-final-viral-combined_k141_74355\|\|full | 1380 | No | NA | 3 | 0 | 0 | Low-quality | Genome-fragment | 1.29 | AAI-based (medium-confidence) | 0 |
| S1-final-viral-combined_k141_325836\|\|full | 1102 | No | NA | 2 | 0 | 1 | Not-determined | Genome-fragment | NA | NA | 0 |
| S1-final-viral-combined_k141_374057\|\|full | 4007 | No | NA | 4 | 0 | 2 | Not-determined | Genome-fragment | NA | NA | 0 |
| S1-final-viral-combined_k141_259967\|\|full | 1129 | No | NA | 2 | 0 | 2 | Not-determined | Genome-fragment | NA | NA | 0 |
| S1-final-viral-combined_k141_76963\|\|full | 6215 | No | NA | 8 | 0 | 0 | Not-determined | Genome-fragment | NA | NA | 0 |
| S2-final-viral-combined_k141_55168\|\|full | 1380 | No | NA | 3 | 0 | 2 | Low-quality | Genome-fragment | 1.88 | AAI-based (high-confidence) | 0 |
| S1-final-viral-combined_k141_358456\|\|full | 3028 | No | NA | 3 | 0 | 2 | Not-determined | Genome-fragment | NA | NA | 0 |
| S1-final-viral-combined_k141_143889\|\|full | 2155 | No | NA | 2 | 0 | 1 | Not-determined | Genome-fragment | NA | NA | 0 |
| S1-final-viral-combined_k141_175599\|\|full | 3562 | No | NA | 3 | 0 | 3 | Not-determined | Genome-fragment | NA | NA | 0 |
| S8-final-viral-combined_k141_26546\|\|full | 1380 | No | NA | 3 | 1 | 0 | Low-quality | Genome-fragment | 3.41 | AAI-based (high-confidence) | 0 |
| S1-final-viral-combined_k141_159466\|\|full | 1360 | No | NA | 2 | 0 | 2 | Not-determined | Genome-fragment | NA | NA | 0 |
| S1-final-viral-combined_k141_44761\|\|full | 3695 | No | NA | 4 | 0 | 0 | Not-determined | Genome-fragment | NA | NA | 0 |
| S1-final-viral-combined_k141_390370\|\|full | 1380 | No | NA | 3 | 0 | 0 | Not-determined | Genome-fragment | NA | NA | 0 |
| S1-final-viral-combined_k141_159490\|\|full | 1027 | No | NA | 2 | 0 | 1 | Not-determined | Genome-fragment | NA | NA | 0 |
| S8_k141_148396 | 1380 | No | NA | 1 | 0 | 0 | Low-quality | Genome-fragment | 2.18 | AAI-based (high-confidence) | 0 |
| S1-final-viral-combined_k141_226179\|\|full | 5210 | No | NA | 5 | 0 | 5 | Not-determined | Genome-fragment | NA | NA | 0 |
| S11_k141_125990 | 1381 | No | NA | 2 | 0 | 0 | Low-quality | Genome-fragment | 2.3 | AAI-based (high-confidence) | 0 |
| S1-final-viral-combined_k141_340271\|\|full | 2316 | No | NA | 3 | 0 | 1 | Not-determined | Genome-fragment | NA | NA | 0 |
| S1-final-viral-combined_k141_366264\|\|full | 1381 | No | NA | 2 | 0 | 0 | Low-quality | Genome-fragment | 3.71 | AAI-based (medium-confidence) | 0 |
| S1-final-viral-combined_k141_94699\|\|full | 1305 | No | NA | 3 | 0 | 0 | Not-determined | Genome-fragment | NA | NA | 0 |
| S1-final-viral-combined_k141_374121\|\|full | 2485 | No | NA | 3 | 0 | 2 | Not-determined | Genome-fragment | NA | NA | 0 |
| S1-final-viral-combined_k141_210573\|\|full | 2846 | No | NA | 4 | 0 | 3 | Not-determined | Genome-fragment | NA | NA | 0 |
| S1-final-viral-combined_k141_242800\|\|full | 1477 | No | NA | 2 | 0 | 1 | Not-determined | Genome-fragment | NA | NA | 0 |
| S1-final-viral-combined_k141_16926\|\|lt2gene | 1381 | No | NA | 1 | 1 | 0 | Low-quality | Genome-fragment | 1.81 | HMM-based (lower-bound) | 0 |
| S1-final-viral-combined_k141_12222\|\|full | 1097 | No | NA | 2 | 0 | 0 | Not-determined | Genome-fragment | NA | NA | 0 |
| S1_k141_149379 | 1381 | No | NA | 3 | 0 | 1 | Low-quality | Genome-fragment | 0.41 | AAI-based (medium-confidence) | 0 |
| S1-final-viral-combined_k141_226252\|\|full | 1960 | No | NA | 4 | 0 | 1 | Not-determined | Genome-fragment | NA | NA | 0 |
| S1-final-viral-combined_k141_159520\|\|full | 1025 | No | NA | 1 | 0 | 0 | Not-determined | Genome-fragment | NA | NA | 0 |
| S1-final-viral-combined_k141_390446\|\|full | 1343 | No | NA | 2 | 0 | 1 | Not-determined | Genome-fragment | NA | NA | 0 |
| S1_k141_344850 | 1381 | No | NA | 2 | 0 | 1 | Low-quality | Genome-fragment | 0.46 | AAI-based (medium-confidence) | 0 |
| S1-final-viral-combined_k141_77053\|\|full | 5133 | No | NA | 3 | 0 | 2 | Not-determined | Genome-fragment | NA | NA | 0 |
| S1-final-viral-combined_k141_159544\|\|full | 21136 | No | NA | 31 | 15 | 0 | Medium-quality | Genome-fragment | 58.23 | AAI-based (high-confidence) | 0 |
| S1-final-viral-combined_k141_242858\|\|full | 2447 | No | NA | 3 | 0 | 0 | Not-determined | Genome-fragment | NA | NA | 0 |
| S5-final-viral-combined_k141_11066\|\|full | 1381 | No | NA | 2 | 0 | 1 | Low-quality | Genome-fragment | 0.72 | AAI-based (medium-confidence) | 0 |
| S7-final-viral-combined_k141_164457\|\|full | 1381 | No | NA | 3 | 0 | 0 | Low-quality | Genome-fragment | 2.46 | AAI-based (high-confidence) | 0 |
| S7-final-viral-combined_k141_196228\|\|full | 1381 | No | NA | 2 | 0 | 0 | Low-quality | Genome-fragment | 0.86 | AAI-based (medium-confidence) | 0 |
| S1-final-viral-combined_k141_94756\|\|full | 3255 | No | NA | 3 | 0 | 1 | Not-determined | Genome-fragment | NA | NA | 0 |
| S1-final-viral-combined_k141_309197\|\|full | 1447 | No | NA | 4 | 0 | 0 | Not-determined | Genome-fragment | NA | NA | 0 |
| S1-final-viral-combined_k141_61490\|\|full | 2132 | No | NA | 3 | 0 | 3 | Not-determined | Genome-fragment | NA | NA | 0 |
| S1-final-viral-combined_k141_260188\|\|full | 1262 | No | NA | 2 | 0 | 1 | Not-determined | Genome-fragment | NA | NA | 0 |
| S12-final-viral-combined_k141_115055\|\|full | 1381 | No | NA | 4 | 1 | 0 | Low-quality | Genome-fragment | 2.29 | AAI-based (medium-confidence) | 0 |
| S1-final-viral-combined_k141_226314\|\|full | 1444 | No | NA | 2 | 0 | 2 | Not-determined | Genome-fragment | NA | NA | 0 |
| S1-final-viral-combined_k141_28458\|\|full | 1182 | No | NA | 2 | 0 | 0 | Not-determined | Genome-fragment | NA | NA | 0 |
| S1-final-viral-combined_k141_77099\|\|full | 8957 | No | NA | 8 | 0 | 6 | Not-determined | Genome-fragment | NA | NA | 0 |
| S1-final-viral-combined_k141_226324\|\|full | 2543 | No | NA | 3 | 0 | 0 | Not-determined | Genome-fragment | NA | NA | 0 |
| S1-final-viral-combined_k141_77105\|\|full | 2083 | No | NA | 2 | 0 | 2 | Not-determined | Genome-fragment | NA | NA | 0 |
| S1-final-viral-combined_k141_326008\|\|full | 1115 | No | NA | 3 | 0 | 0 | Not-determined | Genome-fragment | NA | NA | 0 |
| S1-final-viral-combined_k141_374219\|\|full | 19640 | No | NA | 32 | 12 | 0 | Medium-quality | Genome-fragment | 50.09 | AAI-based (high-confidence) | 0 |
| S1-final-viral-combined_k141_226358\|\|full | 6207 | No | NA | 6 | 0 | 4 | Not-determined | Genome-fragment | NA | NA | 0 |
| S8-final-viral-combined_k141_156843\|\|full | 1381 | No | NA | 3 | 1 | 0 | Low-quality | Genome-fragment | 4.41 | AAI-based (medium-confidence) | 0 |
| S11-final-viral-combined_k141_267283\|\|full | 1382 | No | NA | 2 | 0 | 1 | Low-quality | Genome-fragment | 1.3 | AAI-based (medium-confidence) | 0 |
| S1-final-viral-combined_k141_292516\|\|full | 4391 | No | NA | 4 | 0 | 3 | Not-determined | Genome-fragment | NA | NA | 0 |
| S1-final-viral-combined_k141_358683\|\|full | 3643 | No | NA | 4 | 0 | 0 | Not-determined | Genome-fragment | NA | NA | 0 |
| S1-final-viral-combined_k141_144089\|\|full | 1089 | No | NA | 2 | 0 | 1 | Not-determined | Genome-fragment | NA | NA | 0 |
| S1-final-viral-combined_k141_242932\|\|full | 1500 | No | NA | 4 | 0 | 0 | Not-determined | Genome-fragment | NA | NA | 0 |
| S1-final-viral-combined_k141_144092\|\|full | 4427 | No | NA | 4 | 0 | 4 | Not-determined | Genome-fragment | NA | NA | 0 |
| S1-final-viral-combined_k141_390566\|\|full | 1063 | No | NA | 3 | 0 | 0 | Not-determined | Genome-fragment | NA | NA | 0 |
| S11-final-viral-combined_k141_288604\|\|full | 1382 | No | NA | 2 | 0 | 1 | Low-quality | Genome-fragment | 0.99 | AAI-based (medium-confidence) | 0 |
| S1-final-viral-combined_k141_193884\|\|full | 3087 | No | NA | 2 | 0 | 1 | Not-determined | Genome-fragment | NA | NA | 0 |
| S1-final-viral-combined_k141_36657\|\|full | 1382 | No | NA | 4 | 2 | 0 | Low-quality | Genome-fragment | 2.36 | AAI-based (medium-confidence) | 0 |
| S1-final-viral-combined_k141_340436\|\|full | 1295 | No | NA | 3 | 0 | 0 | Not-determined | Genome-fragment | NA | NA | 0 |
| S1-final-viral-combined_k141_77188\|\|full | 11572 | No | NA | 7 | 0 | 0 | Not-determined | Genome-fragment | NA | NA | 0 |
| S1-final-viral-combined_k141_90233\|\|full | 1382 | No | NA | 4 | 0 | 0 | Low-quality | Genome-fragment | 2.47 | AAI-based (high-confidence) | 0 |
| S3_k141_53944 | 1382 | No | NA | 3 | 1 | 0 | Low-quality | Genome-fragment | 0.8 | AAI-based (high-confidence) | 0 |
| S3_k141_96758 | 1382 | No | NA | 2 | 0 | 1 | Low-quality | Genome-fragment | 0.87 | AAI-based (medium-confidence) | 0 |
| S3_k141_120095 | 1382 | No | NA | 4 | 0 | 0 | Low-quality | Genome-fragment | 0.64 | AAI-based (medium-confidence) | 0 |
| S1-final-viral-combined_k141_61626\|\|full | 7167 | No | NA | 7 | 0 | 7 | Not-determined | Genome-fragment | NA | NA | 0 |
| S7-final-viral-combined_k141_179588\|\|full | 1382 | No | NA | 3 | 1 | 0 | Low-quality | Genome-fragment | 1.73 | AAI-based (high-confidence) | 0 |
| S1-final-viral-combined_k141_12453\|\|full | 2574 | No | NA | 3 | 0 | 1 | Not-determined | Genome-fragment | NA | NA | 0 |
| S1-final-viral-combined_k141_128366\|\|full | 1936 | No | NA | 2 | 0 | 0 | Not-determined | Genome-fragment | NA | NA | 0 |
| S1-final-viral-combined_k141_159670\|\|full | 6533 | No | NA | 5 | 0 | 3 | Not-determined | Genome-fragment | NA | NA | 0 |
| S7-final-viral-combined_k141_9328\|\|full | 1382 | No | NA | 2 | 0 | 2 | Low-quality | Genome-fragment | 0.44 | AAI-based (medium-confidence) | 0 |
| S17-final-viral-combined_k141_29847\|\|full | 1382 | No | NA | 3 | 0 | 2 | Low-quality | Genome-fragment | 0.49 | AAI-based (medium-confidence) | 0 |
| S1-final-viral-combined_k141_260367\|\|full | 1150 | No | NA | 3 | 0 | 0 | Not-determined | Genome-fragment | NA | NA | 0 |
| S1-final-viral-combined_k141_358777\|\|full | 1773 | No | NA | 2 | 0 | 1 | Not-determined | Genome-fragment | NA | NA | 0 |
| S2-final-viral-combined_k141_128912\|\|lt2gene | 1382 | No | NA | 2 | 0 | 1 | Low-quality | Genome-fragment | 1.68 | AAI-based (medium-confidence) | 0 |
| S8-final-viral-combined_k141_201279\|\|full | 1382 | No | NA | 2 | 0 | 0 | Low-quality | Genome-fragment | 2.69 | AAI-based (medium-confidence) | 0 |
| S1-final-viral-combined_k141_61657\|\|full | 4586 | No | NA | 4 | 0 | 2 | Not-determined | Genome-fragment | NA | NA | 0 |
| S1-final-viral-combined_k141_390677\|\|full | 2078 | No | NA | 4 | 0 | 1 | Not-determined | Genome-fragment | NA | NA | 0 |
| S1-final-viral-combined_k141_144171\|\|full | 3511 | No | NA | 4 | 0 | 4 | Not-determined | Genome-fragment | NA | NA | 0 |
| S1-final-viral-combined_k141_94851\|\|full | 1587 | No | NA | 3 | 0 | 0 | Not-determined | Genome-fragment | NA | NA | 0 |
| S11-final-viral-combined_k141_201164\|\|lt2gene | 1383 | No | NA | 2 | 2 | 0 | Low-quality | Genome-fragment | 4.31 | AAI-based (medium-confidence) | 0 |
| S1-final-viral-combined_k141_326103\|\|full | 9513 | No | NA | 10 | 0 | 7 | Not-determined | Genome-fragment | NA | NA | 0 |
| S1-final-viral-combined_k141_358813\|\|full | 1412 | No | NA | 2 | 0 | 0 | Not-determined | Genome-fragment | NA | NA | 0 |
| S7-final-viral-combined_k141_56638\|\|full | 1383 | No | NA | 1 | 0 | 0 | Low-quality | Genome-fragment | 2.16 | AAI-based (medium-confidence) | 0 |
| S1-final-viral-combined_k141_159711\|\|full | 1615 | No | NA | 3 | 0 | 1 | Not-determined | Genome-fragment | NA | NA | 0 |
| S1-final-viral-combined_k141_45039\|\|full | 1776 | No | NA | 3 | 0 | 1 | Not-determined | Genome-fragment | NA | NA | 0 |
| S17-final-viral-combined_k141_180519\|\|full | 1383 | No | NA | 3 | 0 | 0 | Low-quality | Genome-fragment | 3.27 | AAI-based (medium-confidence) | 0 |
| S1-final-viral-combined_k141_226517\|\|full | 1232 | No | NA | 2 | 0 | 1 | Not-determined | Genome-fragment | NA | NA | 0 |
| S1-final-viral-combined_k141_110004\|\|full | 2056 | No | NA | 3 | 0 | 2 | Not-determined | Genome-fragment | NA | NA | 0 |
| S17-final-viral-combined_k141_39093\|\|full | 1383 | No | NA | 3 | 2 | 0 | Low-quality | Genome-fragment | 3.18 | AAI-based (high-confidence) | 0 |
| S17-k141_30728 | 1383 | No | NA | 3 | 1 | 0 | Low-quality | Genome-fragment | 3.89 | AAI-based (high-confidence) | 0 |
| S1-final-viral-combined_k141_374392\|\|full | 1200 | No | NA | 3 | 0 | 1 | Not-determined | Genome-fragment | NA | NA | 0 |
| S1-final-viral-combined_k141_159733\|\|full | 1456 | No | NA | 3 | 0 | 1 | Not-determined | Genome-fragment | NA | NA | 0 |
| S2-final-viral-combined_k141_134900\|\|full | 1383 | No | NA | 2 | 0 | 1 | Low-quality | Genome-fragment | 1.04 | AAI-based (medium-confidence) | 0 |
| S2-final-viral-combined_k141_231194\|\|full | 1383 | No | NA | 2 | 0 | 0 | Low-quality | Genome-fragment | 3.54 | AAI-based (high-confidence) | 0 |
| S1-final-viral-combined_k141_374410\|\|full | 1754 | No | NA | 2 | 0 | 2 | Not-determined | Genome-fragment | NA | NA | 0 |
| S1-final-viral-combined_k141_110023\|\|full | 3147 | No | NA | 2 | 0 | 1 | Not-determined | Genome-fragment | NA | NA | 0 |
| S2_k141_88626 | 1383 | No | NA | 2 | 0 | 1 | Low-quality | Genome-fragment | 0.82 | AAI-based (medium-confidence) | 0 |
| S8_k141_201051 | 1383 | No | NA | 2 | 1 | 0 | Low-quality | Genome-fragment | 0.88 | HMM-based (lower-bound) | 0 |
| S1-final-viral-combined_k141_159746\|\|full | 1733 | No | NA | 3 | 0 | 1 | Not-determined | Genome-fragment | NA | NA | 0 |
| S1-final-viral-combined_k141_45089\|\|full | 2294 | No | NA | 3 | 0 | 0 | Not-determined | Genome-fragment | NA | NA | 0 |
| S11-final-viral-combined_k141_165966\|\|full | 1384 | No | NA | 3 | 0 | 2 | Low-quality | Genome-fragment | 0.83 | AAI-based (high-confidence) | 0 |
| S1-final-viral-combined_k141_374442\|\|full | 1794 | No | NA | 3 | 0 | 2 | Not-determined | Genome-fragment | NA | NA | 0 |
| S1-final-viral-combined_k141_110042\|\|full | 3863 | No | NA | 4 | 0 | 2 | Not-determined | Genome-fragment | NA | NA | 0 |
| S7-final-viral-combined_k141_198085\|\|full | 1384 | No | NA | 3 | 0 | 1 | Low-quality | Genome-fragment | 1.64 | AAI-based (medium-confidence) | 0 |
| S1-final-viral-combined_k141_94880\|\|full | 4909 | No | NA | 5 | 0 | 0 | Not-determined | Genome-fragment | NA | NA | 0 |
| S17-k141_76197 | 1384 | No | NA | 1 | 1 | 0 | Low-quality | Genome-fragment | 1.26 | HMM-based (lower-bound) | 0 |
| S1-final-viral-combined_k141_340546\|\|full | 14534 | No | NA | 15 | 0 | 11 | Not-determined | Genome-fragment | NA | NA | 0 |
| S1-final-viral-combined_k141_275374\|\|full | 7885 | No | NA | 10 | 0 | 7 | Not-determined | Genome-fragment | NA | NA | 0 |
| S1-final-viral-combined_k141_326225\|\|full | 2199 | No | NA | 4 | 0 | 2 | Not-determined | Genome-fragment | NA | NA | 0 |
| S1-final-viral-combined_k141_243144\|\|full | 8426 | No | NA | 6 | 0 | 3 | Not-determined | Genome-fragment | NA | NA | 0 |
| S1-final-viral-combined_k141_77320\|\|full | 4371 | No | NA | 5 | 0 | 1 | Not-determined | Genome-fragment | NA | NA | 0 |
| S1-final-viral-combined_k141_326250\|\|full | 1072 | No | NA | 3 | 0 | 1 | Not-determined | Genome-fragment | NA | NA | 0 |
| S1-final-viral-combined_k141_340558\|\|full | 27649 | No | NA | 35 | 8 | 0 | Medium-quality | Genome-fragment | 52.94 | HMM-based (lower-bound) | 0 |
| S1-final-viral-combined_k141_194124\|\|full | 14062 | No | NA | 17 | 0 | 3 | Not-determined | Genome-fragment | NA | NA | 0 |
| S1-final-viral-combined_k141_77327\|\|full | 2946 | No | NA | 5 | 0 | 1 | Not-determined | Genome-fragment | NA | NA | 0 |
| S1-final-viral-combined_k141_28676\|\|full | 4247 | No | NA | 8 | 0 | 3 | Not-determined | Genome-fragment | NA | NA | 0 |
| S1-final-viral-combined_k141_358934\|\|full | 1964 | No | NA | 3 | 0 | 0 | Not-determined | Genome-fragment | NA | NA | 0 |
| S1-final-viral-combined_k141_275444\|\|full | 1161 | No | NA | 2 | 0 | 1 | Not-determined | Genome-fragment | NA | NA | 0 |
| S1-final-viral-combined_k141_77329\|\|full | 7173 | No | NA | 11 | 0 | 0 | Not-determined | Genome-fragment | NA | NA | 0 |
| S1-final-viral-combined_k141_292811\|\|full | 3284 | No | NA | 6 | 0 | 3 | Not-determined | Genome-fragment | NA | NA | 0 |
| S8-final-viral-combined_k141_76422\|\|full | 1384 | No | NA | 3 | 0 | 0 | Low-quality | Genome-fragment | 3.28 | AAI-based (medium-confidence) | 0 |
| S1-final-viral-combined_k141_340600\|\|full | 4058 | No | NA | 5 | 0 | 5 | Not-determined | Genome-fragment | NA | NA | 0 |
| S9_k141_76135 | 1385 | No | NA | 2 | 0 | 2 | Low-quality | Genome-fragment | 2.17 | AAI-based (medium-confidence) | 0 |
| S1-final-viral-combined_k141_332161\|\|full | 1385 | No | NA | 3 | 1 | 0 | Low-quality | Genome-fragment | 1.02 | HMM-based (lower-bound) | 0 |
| S1-final-viral-combined_k141_222761\|\|full | 1385 | No | NA | 3 | 0 | 0 | Low-quality | Genome-fragment | 2.65 | AAI-based (high-confidence) | 0 |
| S1-final-viral-combined_k141_77380\|\|full | 3789 | No | NA | 5 | 0 | 2 | Not-determined | Genome-fragment | NA | NA | 0 |
| S5_k141_192284 | 1385 | No | NA | 1 | 0 | 1 | Low-quality | Genome-fragment | 0.59 | AAI-based (medium-confidence) | 0 |
| S1-final-viral-combined_k141_340616\|\|full | 1638 | No | NA | 3 | 0 | 0 | Not-determined | Genome-fragment | NA | NA | 0 |
| S1-final-viral-combined_k141_77389\|\|full | 1645 | No | NA | 3 | 0 | 0 | Not-determined | Genome-fragment | NA | NA | 0 |
| S17-final-viral-combined_k141_135615\|\|full | 1385 | No | NA | 3 | 1 | 0 | Low-quality | Genome-fragment | 3.2 | AAI-based (high-confidence) | 0 |
| S12_k141_385376 | 1385 | No | NA | 2 | 1 | 0 | Low-quality | Genome-fragment | 2.75 | AAI-based (high-confidence) | 0 |
| S1-final-viral-combined_k141_12688\|\|full | 2585 | No | NA | 4 | 0 | 3 | Not-determined | Genome-fragment | NA | NA | 0 |
| S1-final-viral-combined_k141_340631\|\|full | 2990 | No | NA | 4 | 0 | 1 | Not-determined | Genome-fragment | NA | NA | 0 |
| S12_k141_213592 | 1385 | No | NA | 6 | 0 | 0 | Low-quality | Genome-fragment | 2.35 | AAI-based (high-confidence) | 0 |
| S1-final-viral-combined_k141_194239\|\|full | 3406 | No | NA | 5 | 0 | 0 | Not-determined | Genome-fragment | NA | NA | 0 |
| S2-final-viral-combined_k141_211250\|\|full | 1385 | No | NA | 4 | 1 | 0 | Low-quality | Genome-fragment | 2.9 | AAI-based (high-confidence) | 0 |
| S2-final-viral-combined_k141_246421\|\|full | 1385 | No | NA | 2 | 0 | 2 | Low-quality | Genome-fragment | 0.44 | AAI-based (medium-confidence) | 0 |
| S2_k141_114514 | 1385 | No | NA | 2 | 0 | 0 | Low-quality | Genome-fragment | 3.28 | AAI-based (medium-confidence) | 0 |
| S4-final-viral-combined_k141_42788\|\|full | 1385 | No | NA | 2 | 0 | 0 | Low-quality | Genome-fragment | 2.71 | AAI-based (medium-confidence) | 0 |
| S4-final-viral-combined_k141_94099\|\|full | 1385 | No | NA | 4 | 1 | 0 | Low-quality | Genome-fragment | 2.61 | AAI-based (medium-confidence) | 0 |
| S1-final-viral-combined_k141_374616\|\|full | 1030 | No | NA | 2 | 0 | 1 | Not-determined | Genome-fragment | NA | NA | 0 |
| S1-final-viral-combined_k141_340654\|\|full | 3865 | No | NA | 5 | 0 | 3 | Not-determined | Genome-fragment | NA | NA | 0 |
| S1-final-viral-combined_k141_374624\|\|full | 1262 | No | NA | 2 | 0 | 0 | Not-determined | Genome-fragment | NA | NA | 0 |
| S4-final-viral-combined_k141_161890\|\|lt2gene | 1385 | No | NA | 1 | 1 | 0 | Low-quality | Genome-fragment | 2.02 | HMM-based (lower-bound) | 0 |
| S8-final-viral-combined_k141_257513\|\|full | 1385 | No | NA | 3 | 0 | 0 | Low-quality | Genome-fragment | 0.56 | AAI-based (medium-confidence) | 0 |
| S8-final-viral-combined_k141_11209\|\|full | 1385 | No | NA | 3 | 1 | 0 | Low-quality | Genome-fragment | 3.61 | AAI-based (medium-confidence) | 0 |
| S1-final-viral-combined_k141_61974\|\|full | 4390 | No | NA | 4 | 0 | 3 | Not-determined | Genome-fragment | NA | NA | 0 |
| S1-final-viral-combined_k141_77544\|\|full | 1008 | No | NA | 2 | 0 | 0 | Not-determined | Genome-fragment | NA | NA | 0 |
| S1-final-viral-combined_k141_28797\|\|full | 2571 | No | NA | 7 | 0 | 0 | Not-determined | Genome-fragment | NA | NA | 0 |
| S1-final-viral-combined_k141_236027\|\|full | 1386 | No | NA | 4 | 1 | 0 | Low-quality | Genome-fragment | 3.69 | AAI-based (medium-confidence) | 0 |
| S1-final-viral-combined_k141_77560\|\|full | 2921 | No | NA | 3 | 0 | 2 | Not-determined | Genome-fragment | NA | NA | 0 |
| S1-final-viral-combined_k141_292934\|\|full | 2316 | No | NA | 2 | 0 | 2 | Not-determined | Genome-fragment | NA | NA | 0 |
| S1-final-viral-combined_k141_243343\|\|full | 2084 | No | NA | 2 | 0 | 0 | Not-determined | Genome-fragment | NA | NA | 0 |
| S1-final-viral-combined_k141_28829\|\|full | 2137 | No | NA | 2 | 0 | 0 | Not-determined | Genome-fragment | NA | NA | 0 |
| S1-final-viral-combined_k141_41148\|\|full | 1386 | No | NA | 4 | 0 | 1 | Low-quality | Genome-fragment | 4.14 | AAI-based (medium-confidence) | 0 |
| S1-final-viral-combined_k141_77591\|\|full | 1496 | No | NA | 2 | 0 | 1 | Not-determined | Genome-fragment | NA | NA | 0 |
| S1_k141_189145 | 1386 | No | NA | 2 | 0 | 0 | Low-quality | Genome-fragment | 1.31 | AAI-based (high-confidence) | 0 |
| S1-final-viral-combined_k141_77598\|\|full | 1349 | No | NA | 2 | 0 | 1 | Not-determined | Genome-fragment | NA | NA | 0 |
| S5_k141_96058 | 1386 | No | NA | 3 | 0 | 3 | Low-quality | Genome-fragment | 1.4 | AAI-based (medium-confidence) | 0 |
| S7-final-viral-combined_k141_95151\|\|full | 1386 | No | NA | 3 | 0 | 0 | Low-quality | Genome-fragment | 2.9 | AAI-based (medium-confidence) | 0 |
| S1-final-viral-combined_k141_292971\|\|full | 4485 | No | NA | 4 | 0 | 4 | Not-determined | Genome-fragment | NA | NA | 0 |
| S12-final-viral-combined_k141_32216\|\|full | 1386 | No | NA | 3 | 0 | 0 | Low-quality | Genome-fragment | 1.08 | AAI-based (medium-confidence) | 0 |
| S12_k141_214225 | 1386 | No | NA | 2 | 0 | 2 | Low-quality | Genome-fragment | 0.71 | AAI-based (medium-confidence) | 0 |
| S2-final-viral-combined_k141_157549\|\|full | 1386 | No | NA | 3 | 2 | 0 | Low-quality | Genome-fragment | 0.66 | HMM-based (lower-bound) | 0 |
| S8-final-viral-combined_k141_96491\|\|full | 1386 | Yes | 339 | 3 | 1 | 2 | Low-quality | Genome-fragment | 0.68 | HMM-based (lower-bound) | 75.54 |
| S8_k141_68222 | 1386 | No | NA | 1 | 1 | 0 | Low-quality | Genome-fragment | 1.58 | HMM-based (lower-bound) | 0 |
| S9-final-viral-combined_k141_127871\|\|full | 1387 | No | NA | 3 | 2 | 0 | Low-quality | Genome-fragment | 2.15 | HMM-based (lower-bound) | 0 |
| S1-final-viral-combined_k141_391029\|\|full | 1199 | No | NA | 2 | 0 | 2 | Not-determined | Genome-fragment | NA | NA | 0 |
| S1-final-viral-combined_k141_293030\|\|full | 1006 | No | NA | 3 | 0 | 1 | Not-determined | Genome-fragment | NA | NA | 0 |
| S11-final-viral-combined_k141_321873\|\|full | 1387 | No | NA | 2 | 0 | 1 | Low-quality | Genome-fragment | 0.15 | AAI-based (medium-confidence) | 0 |
| S5-final-viral-combined_k141_126023\|\|lt2gene | 1387 | No | NA | 2 | 2 | 0 | Low-quality | Genome-fragment | 3.16 | AAI-based (high-confidence) | 0 |
| S1-final-viral-combined_k141_293050\|\|full | 2844 | No | NA | 5 | 0 | 3 | Not-determined | Genome-fragment | NA | NA | 0 |
| S1-final-viral-combined_k141_95099\|\|full | 2114 | No | NA | 2 | 0 | 2 | Not-determined | Genome-fragment | NA | NA | 0 |
| S1-final-viral-combined_k141_194461\|\|full | 5898 | No | NA | 5 | 0 | 2 | Not-determined | Genome-fragment | NA | NA | 0 |
| S1-final-viral-combined_k141_243443\|\|full | 1814 | No | NA | 3 | 0 | 0 | Not-determined | Genome-fragment | NA | NA | 0 |
| S1-final-viral-combined_k141_159970\|\|full | 1530 | No | NA | 3 | 0 | 2 | Not-determined | Genome-fragment | NA | NA | 0 |
| S1-final-viral-combined_k141_128847\|\|full | 3204 | No | NA | 5 | 0 | 4 | Not-determined | Genome-fragment | NA | NA | 0 |
| S17-final-viral-combined_k141_199451\|\|full | 1387 | No | NA | 4 | 1 | 0 | Low-quality | Genome-fragment | 3.06 | AAI-based (high-confidence) | 0 |
| S1-final-viral-combined_k141_62134\|\|full | 1775 | No | NA | 3 | 0 | 1 | Not-determined | Genome-fragment | NA | NA | 0 |
| S1-final-viral-combined_k141_95140\|\|full | 2151 | No | NA | 2 | 0 | 1 | Not-determined | Genome-fragment | NA | NA | 0 |
| S1-final-viral-combined_k141_211062\|\|full | 1340 | No | NA | 2 | 0 | 2 | Not-determined | Genome-fragment | NA | NA | 0 |
| S1-final-viral-combined_k141_326505\|\|full | 1334 | No | NA | 2 | 0 | 1 | Not-determined | Genome-fragment | NA | NA | 0 |
| S12-final-viral-combined_k141_319292\|\|full | 1387 | No | NA | 4 | 0 | 0 | Low-quality | Genome-fragment | 2.31 | AAI-based (medium-confidence) | 0 |
| S1-final-viral-combined_k141_128895\|\|full | 1241 | No | NA | 2 | 0 | 1 | Not-determined | Genome-fragment | NA | NA | 0 |
| S1-final-viral-combined_k141_374730\|\|full | 2357 | No | NA | 2 | 0 | 1 | Not-determined | Genome-fragment | NA | NA | 0 |
| S1-final-viral-combined_k141_340882\|\|full | 5939 | No | NA | 5 | 0 | 2 | Not-determined | Genome-fragment | NA | NA | 0 |
| S1-final-viral-combined_k141_194503\|\|full | 2758 | No | NA | 3 | 0 | 2 | Not-determined | Genome-fragment | NA | NA | 0 |
| S1-final-viral-combined_k141_211079\|\|full | 1567 | No | NA | 3 | 0 | 1 | Not-determined | Genome-fragment | NA | NA | 0 |
| S1-final-viral-combined_k141_144429\|\|full | 3019 | No | NA | 3 | 0 | 3 | Not-determined | Genome-fragment | NA | NA | 0 |
| S12-final-viral-combined_k141_227239\|\|full | 1387 | No | NA | 2 | 0 | 1 | Low-quality | Genome-fragment | 2.14 | AAI-based (high-confidence) | 0 |
| S1-final-viral-combined_k141_144432\|\|full | 1108 | No | NA | 3 | 0 | 1 | Not-determined | Genome-fragment | NA | NA | 0 |
| S1-final-viral-combined_k141_293113\|\|full | 2934 | No | NA | 3 | 0 | 3 | Not-determined | Genome-fragment | NA | NA | 0 |
| S12-final-viral-combined_k141_50002\|\|full | 1387 | No | NA | 4 | 2 | 0 | Low-quality | Genome-fragment | 2.92 | AAI-based (medium-confidence) | 0 |
| S12_k141_47899 | 1387 | No | NA | 3 | 1 | 0 | Low-quality | Genome-fragment | 2.69 | AAI-based (high-confidence) | 0 |
| S2-final-viral-combined_k141_239246\|\|full | 1387 | No | NA | 3 | 0 | 1 | Low-quality | Genome-fragment | 3.29 | AAI-based (medium-confidence) | 0 |
| S4-final-viral-combined_k141_54466\|\|full | 1387 | No | NA | 4 | 0 | 0 | Low-quality | Genome-fragment | 3.71 | AAI-based (high-confidence) | 0 |
| S8-final-viral-combined_k141_247928\|\|full | 1387 | No | NA | 3 | 0 | 0 | Low-quality | Genome-fragment | 2.61 | AAI-based (medium-confidence) | 0 |
| S1-final-viral-combined_k141_391075\|\|full | 2676 | No | NA | 3 | 0 | 1 | Not-determined | Genome-fragment | NA | NA | 0 |
| S1-final-viral-combined_k141_226939\|\|full | 2300 | No | NA | 4 | 0 | 1 | Not-determined | Genome-fragment | NA | NA | 0 |
| S1-final-viral-combined_k141_194589\|\|full | 6538 | No | NA | 3 | 0 | 3 | Not-determined | Genome-fragment | NA | NA | 0 |
| S9_k141_36117 | 1388 | No | NA | 3 | 0 | 0 | Low-quality | Genome-fragment | 2.36 | AAI-based (medium-confidence) | 0 |
| S1-final-viral-combined_k141_95226\|\|full | 1003 | No | NA | 3 | 0 | 1 | Not-determined | Genome-fragment | NA | NA | 0 |
| S1-final-viral-combined_k141_194599\|\|full | 2503 | No | NA | 2 | 0 | 1 | Not-determined | Genome-fragment | NA | NA | 0 |
| S1-final-viral-combined_k141_275787\|\|full | 3339 | No | NA | 6 | 0 | 4 | Not-determined | Genome-fragment | NA | NA | 0 |
| S1-final-viral-combined_k141_77779\|\|full | 8290 | No | NA | 4 | 0 | 1 | Not-determined | Genome-fragment | NA | NA | 0 |
| S1-final-viral-combined_k141_211188\|\|full | 5708 | No | NA | 7 | 0 | 6 | Not-determined | Genome-fragment | NA | NA | 0 |
| S11-final-viral-combined_k141_65472\|\|full | 1388 | No | NA | 2 | 0 | 0 | Low-quality | Genome-fragment | 0.55 | AAI-based (medium-confidence) | 0 |
| S1-final-viral-combined_k141_243589\|\|full | 2888 | No | NA | 3 | 0 | 2 | Not-determined | Genome-fragment | NA | NA | 0 |
| S1-final-viral-combined_k141_144502\|\|full | 6069 | No | NA | 7 | 0 | 2 | Not-determined | Genome-fragment | NA | NA | 0 |
| S1-final-viral-combined_k141_144535\|\|full | 3158 | No | NA | 4 | 0 | 3 | Not-determined | Genome-fragment | NA | NA | 0 |
| S1-final-viral-combined_k141_160108\|\|full | 3566 | No | NA | 4 | 0 | 2 | Not-determined | Genome-fragment | NA | NA | 0 |
| S1-final-viral-combined_k141_110220\|\|full | 7891 | No | NA | 5 | 0 | 4 | Not-determined | Genome-fragment | NA | NA | 0 |
| S1-final-viral-combined_k141_227003\|\|full | 25087 | No | NA | 34 | 9 | 1 | Medium-quality | Genome-fragment | 56.45 | AAI-based (high-confidence) | 0 |
| S1-final-viral-combined_k141_341003\|\|full | 2453 | No | NA | 2 | 0 | 2 | Not-determined | Genome-fragment | NA | NA | 0 |
| S1-final-viral-combined_k141_374867\|\|full | 1196 | No | NA | 2 | 0 | 1 | Not-determined | Genome-fragment | NA | NA | 0 |
| S1-final-viral-combined_k141_243633\|\|full | 1652 | No | NA | 2 | 0 | 1 | Not-determined | Genome-fragment | NA | NA | 0 |
| S11_k141_255229 | 1388 | No | NA | 3 | 0 | 3 | Low-quality | Genome-fragment | 1.75 | AAI-based (medium-confidence) | 0 |
| S1-final-viral-combined_k141_260913\|\|full | 2089 | No | NA | 3 | 0 | 2 | Not-determined | Genome-fragment | NA | NA | 0 |
| S1-final-viral-combined_k141_260914\|\|full | 3318 | No | NA | 3 | 0 | 1 | Not-determined | Genome-fragment | NA | NA | 0 |
| S1-final-viral-combined_k141_236835\|\|full | 1388 | No | NA | 3 | 0 | 0 | Low-quality | Genome-fragment | 1.64 | AAI-based (medium-confidence) | 0 |
| S17-final-viral-combined_k141_11109\|\|full | 1388 | No | NA | 3 | 0 | 0 | Low-quality | Genome-fragment | 1.87 | AAI-based (high-confidence) | 0 |
| S1-final-viral-combined_k141_243666\|\|full | 2772 | No | NA | 3 | 0 | 2 | Not-determined | Genome-fragment | NA | NA | 0 |
| S1-final-viral-combined_k141_160176\|\|full | 6577 | No | NA | 5 | 0 | 4 | Not-determined | Genome-fragment | NA | NA | 0 |
| S1-final-viral-combined_k141_211260\|\|full | 1853 | No | NA | 2 | 0 | 2 | Not-determined | Genome-fragment | NA | NA | 0 |
| S1-final-viral-combined_k141_211270\|\|full | 1163 | No | NA | 2 | 0 | 2 | Not-determined | Genome-fragment | NA | NA | 0 |
| S1-final-viral-combined_k141_12952\|\|full | 5661 | No | NA | 5 | 0 | 4 | Not-determined | Genome-fragment | NA | NA | 0 |
| S1-final-viral-combined_k141_326687\|\|full | 4194 | No | NA | 6 | 0 | 0 | Not-determined | Genome-fragment | NA | NA | 0 |
| S12-final-viral-combined_k141_241872\|\|full | 1388 | No | NA | 3 | 0 | 0 | Low-quality | Genome-fragment | 1.05 | AAI-based (medium-confidence) | 0 |
| S1-final-viral-combined_k141_12958\|\|full | 1640 | No | NA | 3 | 0 | 2 | Not-determined | Genome-fragment | NA | NA | 0 |
| S12-final-viral-combined_k141_165702\|\|full | 1388 | No | NA | 3 | 0 | 1 | Low-quality | Genome-fragment | 1.33 | AAI-based (high-confidence) | 0 |
| S1-final-viral-combined_k141_45410\|\|full | 3754 | No | NA | 8 | 0 | 5 | Not-determined | Genome-fragment | NA | NA | 0 |
| S12-final-viral-combined_k141_219422\|\|lt2gene | 1388 | No | NA | 1 | 1 | 0 | Low-quality | Genome-fragment | 4.51 | AAI-based (high-confidence) | 0 |
| S6-final-viral-combined_k141_12825\|\|full | 1388 | No | NA | 4 | 0 | 0 | Low-quality | Genome-fragment | 4.01 | AAI-based (high-confidence) | 0 |
| S1-final-viral-combined_k141_176160\|\|full | 5334 | No | NA | 4 | 0 | 3 | Not-determined | Genome-fragment | NA | NA | 0 |
| S6_k141_18691 | 1388 | No | NA | 4 | 0 | 0 | Low-quality | Genome-fragment | 0.74 | AAI-based (high-confidence) | 0 |
| S1-final-viral-combined_k141_309869\|\|full | 3454 | No | NA | 2 | 0 | 1 | Not-determined | Genome-fragment | NA | NA | 0 |
| S1-final-viral-combined_k141_359429\|\|full | 2840 | No | NA | 2 | 0 | 2 | Not-determined | Genome-fragment | NA | NA | 0 |
| S3-final-viral-combined_k141_11545\|\|full | 1389 | No | NA | 3 | 0 | 0 | Low-quality | Genome-fragment | 2.93 | AAI-based (medium-confidence) | 0 |
| S17-final-viral-combined_k141_6027\|\|full | 1389 | No | NA | 2 | 0 | 2 | Low-quality | Genome-fragment | 1.13 | AAI-based (medium-confidence) | 0 |
| S12-final-viral-combined_k141_103812\|\|full | 1389 | No | NA | 4 | 1 | 0 | Low-quality | Genome-fragment | 3.02 | AAI-based (high-confidence) | 0 |
| S2-final-viral-combined_k141_145329\|\|full | 1389 | No | NA | 3 | 0 | 0 | Low-quality | Genome-fragment | 2.82 | AAI-based (high-confidence) | 0 |
| S1-final-viral-combined_k141_77881\|\|full | 1455 | No | NA | 3 | 0 | 0 | Not-determined | Genome-fragment | NA | NA | 0 |
| S1-final-viral-combined_k141_293333\|\|full | 1512 | No | NA | 3 | 0 | 1 | Not-determined | Genome-fragment | NA | NA | 0 |
| S4-final-viral-combined_k141_154077\|\|full | 1389 | No | NA | 4 | 0 | 1 | Low-quality | Genome-fragment | 6.61 | AAI-based (medium-confidence) | 0 |
| S4_k141_102034 | 1389 | No | NA | 3 | 1 | 0 | Low-quality | Genome-fragment | 3.48 | AAI-based (high-confidence) | 0 |
| S6_k141_139679 | 1389 | No | NA | 4 | 2 | 0 | Low-quality | Genome-fragment | 2.16 | HMM-based (lower-bound) | 0 |
| S1-final-viral-combined_k141_45489\|\|full | 5207 | No | NA | 4 | 0 | 2 | Not-determined | Genome-fragment | NA | NA | 0 |
| S1-final-viral-combined_k141_326800\|\|full | 2165 | No | NA | 2 | 0 | 2 | Not-determined | Genome-fragment | NA | NA | 0 |
| S1-final-viral-combined_k141_144713\|\|full | 1031 | No | NA | 3 | 0 | 1 | Not-determined | Genome-fragment | NA | NA | 0 |
| S1-final-viral-combined_k141_211406\|\|full | 6409 | No | NA | 11 | 0 | 1 | Not-determined | Genome-fragment | NA | NA | 0 |
| S8_k141_142840 | 1389 | No | NA | 1 | 0 | 0 | Low-quality | Genome-fragment | 0.98 | AAI-based (medium-confidence) | 0 |
| S1-final-viral-combined_k141_309943\|\|full | 11578 | No | NA | 9 | 0 | 8 | Not-determined | Genome-fragment | NA | NA | 0 |
| S8_k141_271865 | 1389 | No | NA | 1 | 0 | 0 | Low-quality | Genome-fragment | 1.68 | AAI-based (high-confidence) | 0 |
| S11-final-viral-combined_k141_111656\|\|full | 1390 | No | NA | 2 | 0 | 0 | Low-quality | Genome-fragment | 3.2 | AAI-based (medium-confidence) | 0 |
| S1-final-viral-combined_k141_360449\|\|full | 1390 | No | NA | 2 | 0 | 0 | Low-quality | Genome-fragment | 1.91 | AAI-based (medium-confidence) | 0 |
| S3-final-viral-combined_k141_19147\|\|full | 1390 | No | NA | 2 | 0 | 2 | Low-quality | Genome-fragment | 0.5 | AAI-based (medium-confidence) | 0 |
| S17-final-viral-combined_k141_254900\|\|full | 1390 | No | NA | 2 | 0 | 2 | Low-quality | Genome-fragment | 0.87 | AAI-based (medium-confidence) | 0 |
| S12-final-viral-combined_k141_313859\|\|full | 1390 | No | NA | 3 | 0 | 0 | Low-quality | Genome-fragment | 4.22 | AAI-based (medium-confidence) | 0 |
| S2-final-viral-combined_k141_8038\|\|full | 1390 | No | NA | 5 | 0 | 0 | Low-quality | Genome-fragment | 3.89 | AAI-based (high-confidence) | 0 |
| S9-final-viral-combined_k141_12352\|\|full | 1391 | No | NA | 3 | 1 | 0 | Low-quality | Genome-fragment | 0.96 | HMM-based (lower-bound) | 0 |
| S1-final-viral-combined_k141_227203\|\|full | 4267 | No | NA | 7 | 0 | 1 | Not-determined | Genome-fragment | NA | NA | 0 |
| S1-final-viral-combined_k141_275961\|\|full | 2403 | No | NA | 3 | 0 | 2 | Not-determined | Genome-fragment | NA | NA | 0 |
| S1-final-viral-combined_k141_227206\|\|full | 1566 | No | NA | 1 | 0 | 0 | Not-determined | Genome-fragment | NA | NA | 0 |
| S11-final-viral-combined_k141_29300\|\|full | 1391 | No | NA | 3 | 1 | 0 | Low-quality | Genome-fragment | 3.05 | AAI-based (high-confidence) | 0 |
| S1-final-viral-combined_k141_275978\|\|full | 1039 | No | NA | 3 | 0 | 1 | Not-determined | Genome-fragment | NA | NA | 0 |
| S1-final-viral-combined_k141_194848\|\|full | 3888 | No | NA | 4 | 0 | 3 | Not-determined | Genome-fragment | NA | NA | 0 |
| S1-final-viral-combined_k141_173515\|\|full | 1391 | No | NA | 4 | 2 | 0 | Low-quality | Genome-fragment | 3.33 | AAI-based (medium-confidence) | 0 |
| S3-final-viral-combined_k141_17845\|\|full | 1391 | No | NA | 4 | 0 | 0 | Low-quality | Genome-fragment | 9.49 | AAI-based (high-confidence) | 0 |
| S1-final-viral-combined_k141_95486\|\|full | 2101 | No | NA | 2 | 0 | 2 | Not-determined | Genome-fragment | NA | NA | 0 |
| S1-final-viral-combined_k141_176320\|\|full | 3528 | No | NA | 3 | 0 | 2 | Not-determined | Genome-fragment | NA | NA | 0 |
| S1-final-viral-combined_k141_243919\|\|full | 7021 | No | NA | 6 | 0 | 0 | Not-determined | Genome-fragment | NA | NA | 0 |
| S1-final-viral-combined_k141_144772\|\|full | 3714 | No | NA | 2 | 0 | 1 | Not-determined | Genome-fragment | NA | NA | 0 |
| S1-final-viral-combined_k141_129288\|\|full | 1697 | No | NA | 2 | 0 | 1 | Not-determined | Genome-fragment | NA | NA | 0 |
| S1-final-viral-combined_k141_77993\|\|full | 1768 | No | NA | 3 | 0 | 1 | Not-determined | Genome-fragment | NA | NA | 0 |
| S1-final-viral-combined_k141_194879\|\|full | 1682 | No | NA | 2 | 0 | 0 | Not-determined | Genome-fragment | NA | NA | 0 |
| S1-final-viral-combined_k141_194880\|\|full | 1197 | No | NA | 2 | 0 | 2 | Not-determined | Genome-fragment | NA | NA | 0 |
| S1-final-viral-combined_k141_77995\|\|full | 1663 | No | NA | 2 | 0 | 1 | Not-determined | Genome-fragment | NA | NA | 0 |
| S1-final-viral-combined_k141_176340\|\|full | 1658 | No | NA | 2 | 0 | 2 | Not-determined | Genome-fragment | NA | NA | 0 |
| S5-final-viral-combined_k141_178336\|\|full | 1391 | No | NA | 2 | 0 | 0 | Low-quality | Genome-fragment | 0.69 | AAI-based (medium-confidence) | 0 |
| S5-final-viral-combined_k141_18560\|\|full | 1391 | No | NA | 3 | 0 | 0 | Low-quality | Genome-fragment | 0.84 | AAI-based (medium-confidence) | 0 |
| S1-final-viral-combined_k141_211506\|\|full | 7339 | No | NA | 10 | 0 | 1 | Not-determined | Genome-fragment | NA | NA | 0 |
| S1-final-viral-combined_k141_310044\|\|full | 3514 | No | NA | 4 | 0 | 1 | Not-determined | Genome-fragment | NA | NA | 0 |
| S1-final-viral-combined_k141_227265\|\|full | 2707 | No | NA | 3 | 0 | 2 | Not-determined | Genome-fragment | NA | NA | 0 |
| S2-final-viral-combined_k141_233342\|\|full | 1391 | No | NA | 3 | 1 | 0 | Low-quality | Genome-fragment | 20.82 | AAI-based (high-confidence) | 0 |
| S1-final-viral-combined_k141_129338\|\|full | 4388 | No | NA | 8 | 0 | 0 | Not-determined | Genome-fragment | NA | NA | 0 |
| S1-final-viral-combined_k141_341227\|\|full | 1670 | No | NA | 2 | 0 | 2 | Not-determined | Genome-fragment | NA | NA | 0 |
| S1-final-viral-combined_k141_45594\|\|full | 2018 | No | NA | 3 | 0 | 0 | Not-determined | Genome-fragment | NA | NA | 0 |
| S6-final-viral-combined_k141_132374\|\|full | 1391 | No | NA | 3 | 0 | 0 | Low-quality | Genome-fragment | 2.78 | AAI-based (high-confidence) | 0 |
| S8-final-viral-combined_k141_236227\|\|full | 1391 | No | NA | 2 | 1 | 0 | Low-quality | Genome-fragment | 3.18 | AAI-based (high-confidence) | 0 |
| S8-final-viral-combined_k141_86842\|\|full | 1391 | No | NA | 3 | 0 | 1 | Low-quality | Genome-fragment | 2.39 | AAI-based (medium-confidence) | 0 |
| S1-final-viral-combined_k141_211536\|\|full | 1260 | No | NA | 2 | 0 | 1 | Not-determined | Genome-fragment | NA | NA | 0 |
| S9-final-viral-combined_k141_46556\|\|full | 1392 | No | NA | 2 | 0 | 0 | Low-quality | Genome-fragment | 3.35 | AAI-based (medium-confidence) | 0 |
| S11-final-viral-combined_k141_187869\|\|full | 1392 | No | NA | 3 | 0 | 0 | Low-quality | Genome-fragment | 3.49 | AAI-based (medium-confidence) | 0 |
| S1-final-viral-combined_k141_129384\|\|full | 1395 | No | NA | 3 | 0 | 1 | Not-determined | Genome-fragment | NA | NA | 0 |
| S1-final-viral-combined_k141_364145\|\|full | 1392 | No | NA | 4 | 1 | 0 | Low-quality | Genome-fragment | 1.91 | AAI-based (high-confidence) | 0 |
| S1-final-viral-combined_k141_326953\|\|full | 4893 | No | NA | 5 | 0 | 3 | Not-determined | Genome-fragment | NA | NA | 0 |
| S1-final-viral-combined_k141_13200\|\|full | 2387 | No | NA | 2 | 0 | 2 | Not-determined | Genome-fragment | NA | NA | 0 |
| S1-final-viral-combined_k141_261287\|\|full | 6326 | No | NA | 6 | 0 | 3 | Not-determined | Genome-fragment | NA | NA | 0 |
| S1-final-viral-combined_k141_13210\|\|full | 2614 | No | NA | 4 | 0 | 1 | Not-determined | Genome-fragment | NA | NA | 0 |
| S1-final-viral-combined_k141_326978\|\|full | 1762 | No | NA | 2 | 0 | 1 | Not-determined | Genome-fragment | NA | NA | 0 |
| S1-final-viral-combined_k141_375225\|\|full | 2161 | No | NA | 4 | 0 | 3 | Not-determined | Genome-fragment | NA | NA | 0 |
| S1-final-viral-combined_k141_95553\|\|full | 2461 | No | NA | 2 | 0 | 0 | Not-determined | Genome-fragment | NA | NA | 0 |
| S1-final-viral-combined_k141_45647\|\|full | 2024 | No | NA | 3 | 0 | 1 | Not-determined | Genome-fragment | NA | NA | 0 |
| S1-final-viral-combined_k141_375241\|\|full | 2027 | No | NA | 2 | 0 | 2 | Not-determined | Genome-fragment | NA | NA | 0 |
| S1-final-viral-combined_k141_78024\|\|full | 1440 | No | NA | 1 | 0 | 0 | Not-determined | Genome-fragment | NA | NA | 0 |
| S3-final-viral-combined_k141_112493\|\|full | 1392 | No | NA | 3 | 0 | 0 | Low-quality | Genome-fragment | 2.86 | AAI-based (medium-confidence) | 0 |
| S1-final-viral-combined_k141_13242\|\|full | 3388 | No | NA | 3 | 0 | 1 | Not-determined | Genome-fragment | NA | NA | 0 |
| S1-final-viral-combined_k141_310152\|\|full | 1567 | No | NA | 3 | 0 | 1 | Not-determined | Genome-fragment | NA | NA | 0 |
| S1-final-viral-combined_k141_95582\|\|full | 1138 | No | NA | 2 | 0 | 0 | Not-determined | Genome-fragment | NA | NA | 0 |
| S1-final-viral-combined_k141_144933\|\|full | 2901 | No | NA | 4 | 0 | 4 | Not-determined | Genome-fragment | NA | NA | 0 |
| S7-final-viral-combined_k141_211524\|\|full | 1392 | No | NA | 2 | 0 | 0 | Low-quality | Genome-fragment | 2.73 | AAI-based (medium-confidence) | 0 |
| S1-final-viral-combined_k141_195010\|\|full | 1349 | No | NA | 3 | 0 | 1 | Not-determined | Genome-fragment | NA | NA | 0 |
| S1-final-viral-combined_k141_176469\|\|full | 7374 | No | NA | 8 | 0 | 5 | Not-determined | Genome-fragment | NA | NA | 0 |
| S1-final-viral-combined_k141_211726\|\|full | 1197 | No | NA | 2 | 0 | 2 | Not-determined | Genome-fragment | NA | NA | 0 |
| S1-final-viral-combined_k141_29201\|\|full | 3024 | No | NA | 2 | 0 | 0 | Not-determined | Genome-fragment | NA | NA | 0 |
| S7-final-viral-combined_k141_113437\|\|lt2gene | 1392 | No | NA | 1 | 1 | 0 | Low-quality | Genome-fragment | 0.96 | HMM-based (lower-bound) | 0 |
| S1-final-viral-combined_k141_211738\|\|full | 1098 | No | NA | 3 | 0 | 1 | Not-determined | Genome-fragment | NA | NA | 0 |
| S17-final-viral-combined_k141_57682\|\|full | 1392 | No | NA | 5 | 1 | 0 | Low-quality | Genome-fragment | 2.52 | HMM-based (lower-bound) | 0 |
| S17-final-viral-combined_k141_120268\|\|full | 1392 | No | NA | 3 | 0 | 0 | Low-quality | Genome-fragment | 2.65 | AAI-based (medium-confidence) | 0 |
| S17-final-viral-combined_k141_152774\|\|full | 1392 | No | NA | 3 | 0 | 1 | Low-quality | Genome-fragment | 0.94 | AAI-based (medium-confidence) | 0 |
| S12_k141_35626 | 1392 | No | NA | 3 | 0 | 1 | Low-quality | Genome-fragment | 0.68 | AAI-based (medium-confidence) | 0 |
| S1-final-viral-combined_k141_276250\|\|full | 1538 | No | NA | 2 | 0 | 0 | Not-determined | Genome-fragment | NA | NA | 0 |
| S1-final-viral-combined_k141_160638\|\|full | 4579 | No | NA | 7 | 0 | 1 | Not-determined | Genome-fragment | NA | NA | 0 |
| S1-final-viral-combined_k141_145002\|\|full | 3910 | No | NA | 5 | 0 | 0 | Not-determined | Genome-fragment | NA | NA | 0 |
| S2_k141_25223 | 1392 | No | NA | 2 | 1 | 0 | Low-quality | Genome-fragment | 3.05 | AAI-based (medium-confidence) | 0 |
| S1-final-viral-combined_k141_359658\|\|full | 1125 | No | NA | 2 | 0 | 1 | Not-determined | Genome-fragment | NA | NA | 0 |
| S4-final-viral-combined_k141_124979\|\|full | 1392 | No | NA | 2 | 1 | 0 | Low-quality | Genome-fragment | 2.75 | AAI-based (medium-confidence) | 0 |
| S4-final-viral-combined_k141_62557\|\|full | 1392 | No | NA | 3 | 0 | 0 | Low-quality | Genome-fragment | 3.75 | AAI-based (high-confidence) | 0 |
| S1-final-viral-combined_k141_375331\|\|full | 2857 | No | NA | 3 | 0 | 0 | Not-determined | Genome-fragment | NA | NA | 0 |
| S9-final-viral-combined_k141_10887\|\|full | 1393 | No | NA | 3 | 1 | 0 | Low-quality | Genome-fragment | 1.72 | HMM-based (lower-bound) | 0 |
| S9-final-viral-combined_k141_108965\|\|full | 1393 | No | NA | 4 | 0 | 3 | Low-quality | Genome-fragment | 0.35 | AAI-based (medium-confidence) | 0 |
| S11-final-viral-combined_k141_110701\|\|full | 1393 | No | NA | 4 | 0 | 0 | Low-quality | Genome-fragment | 3.65 | AAI-based (medium-confidence) | 0 |
| S1-final-viral-combined_k141_244188\|\|full | 2690 | No | NA | 2 | 0 | 2 | Not-determined | Genome-fragment | NA | NA | 0 |
| S1-final-viral-combined_k141_341429\|\|full | 1603 | No | NA | 3 | 0 | 1 | Not-determined | Genome-fragment | NA | NA | 0 |
| S1-final-viral-combined_k141_391471\|\|full | 1393 | No | NA | 3 | 0 | 1 | Low-quality | Genome-fragment | 2.42 | AAI-based (medium-confidence) | 0 |
| S1-final-viral-combined_k141_238969\|\|lt2gene | 1393 | No | NA | 1 | 1 | 0 | Low-quality | Genome-fragment | 3.31 | AAI-based (medium-confidence) | 0 |
| S5-final-viral-combined_k141_64358\|\|full | 1393 | No | NA | 3 | 3 | 0 | Low-quality | Genome-fragment | 2.26 | AAI-based (high-confidence) | 0 |
| S1-final-viral-combined_k141_176552\|\|full | 1403 | No | NA | 2 | 0 | 0 | Not-determined | Genome-fragment | NA | NA | 0 |
| S1-final-viral-combined_k141_160702\|\|full | 1434 | No | NA | 3 | 0 | 2 | Not-determined | Genome-fragment | NA | NA | 0 |
| S1-final-viral-combined_k141_227480\|\|full | 1630 | No | NA | 2 | 0 | 2 | Not-determined | Genome-fragment | NA | NA | 0 |
| S7-final-viral-combined_k141_190937\|\|full | 1393 | No | NA | 6 | 0 | 0 | Low-quality | Genome-fragment | 3.06 | AAI-based (medium-confidence) | 0 |
| S7-final-viral-combined_k141_24719\|\|lt2gene | 1393 | No | NA | 2 | 1 | 0 | Low-quality | Genome-fragment | 3.62 | AAI-based (medium-confidence) | 0 |
| S1-final-viral-combined_k141_244241\|\|full | 6631 | No | NA | 6 | 0 | 5 | Not-determined | Genome-fragment | NA | NA | 0 |
| S12-final-viral-combined_k141_26460\|\|full | 1393 | No | NA | 3 | 1 | 0 | Low-quality | Genome-fragment | 1.94 | AAI-based (medium-confidence) | 0 |
| S1-final-viral-combined_k141_45768\|\|full | 1400 | No | NA | 3 | 0 | 1 | Not-determined | Genome-fragment | NA | NA | 0 |
| S1-final-viral-combined_k141_261473\|\|full | 3267 | No | NA | 6 | 0 | 0 | Not-determined | Genome-fragment | NA | NA | 0 |
| S1-final-viral-combined_k141_78110\|\|full | 2783 | No | NA | 4 | 0 | 1 | Not-determined | Genome-fragment | NA | NA | 0 |
| S1-final-viral-combined_k141_276375\|\|full | 4618 | No | NA | 4 | 0 | 3 | Not-determined | Genome-fragment | NA | NA | 0 |
| S2-final-viral-combined_k141_228507\|\|full | 1393 | No | NA | 3 | 0 | 1 | Low-quality | Genome-fragment | 2.33 | AAI-based (high-confidence) | 0 |
| S1-final-viral-combined_k141_195174\|\|full | 2046 | No | NA | 3 | 0 | 1 | Not-determined | Genome-fragment | NA | NA | 0 |
| S1-final-viral-combined_k141_341508\|\|full | 5068 | No | NA | 7 | 0 | 0 | Not-determined | Genome-fragment | NA | NA | 0 |
| S2-final-viral-combined_k141_22016\|\|full | 1393 | No | NA | 3 | 0 | 0 | Low-quality | Genome-fragment | 1.79 | AAI-based (high-confidence) | 0 |
| S1-final-viral-combined_k141_211897\|\|full | 1033 | No | NA | 2 | 0 | 1 | Not-determined | Genome-fragment | NA | NA | 0 |
| S2-final-viral-combined_k141_57801\|\|lt2gene | 1393 | No | NA | 2 | 1 | 0 | Low-quality | Genome-fragment | 23.47 | AAI-based (high-confidence) | 0 |
| S8-final-viral-combined_k141_71481\|\|full | 1393 | No | NA | 3 | 1 | 0 | Low-quality | Genome-fragment | 3.26 | AAI-based (high-confidence) | 0 |
| S11-final-viral-combined_k141_251369\|\|full | 1394 | No | NA | 3 | 0 | 0 | Low-quality | Genome-fragment | 2.41 | AAI-based (medium-confidence) | 0 |
| S1-final-viral-combined_k141_129690\|\|full | 9792 | No | NA | 9 | 0 | 6 | Not-determined | Genome-fragment | NA | NA | 0 |
| S11-final-viral-combined_k141_68864\|\|full | 1394 | No | NA | 3 | 0 | 2 | Low-quality | Genome-fragment | 0.51 | AAI-based (medium-confidence) | 0 |
| S1-final-viral-combined_k141_145154\|\|full | 3138 | No | NA | 2 | 0 | 2 | Not-determined | Genome-fragment | NA | NA | 0 |
| S1-final-viral-combined_k141_110752\|\|full | 2287 | No | NA | 3 | 0 | 1 | Not-determined | Genome-fragment | NA | NA | 0 |
| S1-final-viral-combined_k141_276419\|\|full | 7039 | No | NA | 7 | 0 | 3 | Not-determined | Genome-fragment | NA | NA | 0 |
| S1-final-viral-combined_k141_145157\|\|full | 1885 | No | NA | 3 | 0 | 1 | Not-determined | Genome-fragment | NA | NA | 0 |
| S11-final-viral-combined_k141_73465\|\|lt2gene | 1394 | No | NA | 2 | 1 | 0 | Low-quality | Genome-fragment | 0.53 | HMM-based (lower-bound) | 0 |
| S1-final-viral-combined_k141_327085\|\|full | 2197 | No | NA | 3 | 0 | 0 | Not-determined | Genome-fragment | NA | NA | 0 |
| S1-final-viral-combined_k141_176656\|\|full | 1495 | No | NA | 3 | 0 | 2 | Not-determined | Genome-fragment | NA | NA | 0 |
| S1-final-viral-combined_k141_375389\|\|full | 1066 | No | NA | 3 | 0 | 0 | Not-determined | Genome-fragment | NA | NA | 0 |
| S1-final-viral-combined_k141_176664\|\|full | 1637 | No | NA | 2 | 0 | 0 | Not-determined | Genome-fragment | NA | NA | 0 |
| S1-final-viral-combined_k141_293800\|\|full | 1516 | No | NA | 2 | 0 | 1 | Not-determined | Genome-fragment | NA | NA | 0 |
| S1-final-viral-combined_k141_45826\|\|full | 1894 | No | NA | 2 | 0 | 1 | Not-determined | Genome-fragment | NA | NA | 0 |
| S1-final-viral-combined_k141_375397\|\|full | 3445 | No | NA | 3 | 0 | 3 | Not-determined | Genome-fragment | NA | NA | 0 |
| S1-final-viral-combined_k141_280149\|\|full | 1394 | No | NA | 2 | 1 | 0 | Low-quality | Genome-fragment | 3.31 | AAI-based (medium-confidence) | 0 |
| S1-final-viral-combined_k141_17061\|\|full | 1394 | No | NA | 3 | 0 | 1 | Low-quality | Genome-fragment | 3.09 | AAI-based (high-confidence) | 0 |
| S1-final-viral-combined_k141_62680\|\|full | 8004 | No | NA | 4 | 0 | 1 | Not-determined | Genome-fragment | NA | NA | 0 |
| S1-final-viral-combined_k141_62703\|\|full | 2980 | No | NA | 3 | 0 | 2 | Not-determined | Genome-fragment | NA | NA | 0 |
| S1-final-viral-combined_k141_244380\|\|full | 2220 | No | NA | 2 | 0 | 0 | Not-determined | Genome-fragment | NA | NA | 0 |
| S1-final-viral-combined_k141_195247\|\|full | 1993 | No | NA | 3 | 0 | 3 | Not-determined | Genome-fragment | NA | NA | 0 |
| S1_k141_18432 | 1394 | No | NA | 5 | 1 | 0 | Low-quality | Genome-fragment | 3.09 | AAI-based (high-confidence) | 0 |
| S1-final-viral-combined_k141_293843\|\|full | 1283 | No | NA | 2 | 0 | 1 | Not-determined | Genome-fragment | NA | NA | 0 |
| S1_k141_108487 | 1394 | No | NA | 2 | 1 | 0 | Low-quality | Genome-fragment | 2.27 | AAI-based (high-confidence) | 0 |
| S5-final-viral-combined_k141_58671\|\|full | 1394 | No | NA | 2 | 0 | 0 | Low-quality | Genome-fragment | 3.09 | AAI-based (medium-confidence) | 0 |
| S1-final-viral-combined_k141_293849\|\|full | 2971 | No | NA | 5 | 0 | 3 | Not-determined | Genome-fragment | NA | NA | 0 |
| S1-final-viral-combined_k141_95908\|\|full | 1328 | No | NA | 2 | 0 | 1 | Not-determined | Genome-fragment | NA | NA | 0 |
| S1-final-viral-combined_k141_391684\|\|full | 1456 | No | NA | 5 | 0 | 0 | Not-determined | Genome-fragment | NA | NA | 0 |
| S1-final-viral-combined_k141_293865\|\|full | 1028 | No | NA | 2 | 0 | 1 | Not-determined | Genome-fragment | NA | NA | 0 |
| S17-final-viral-combined_k141_174083\|\|full | 1394 | No | NA | 2 | 0 | 2 | Low-quality | Genome-fragment | 2.62 | AAI-based (high-confidence) | 0 |
| S8-final-viral-combined_k141_13623\|\|full | 1394 | Yes | 1139 | 3 | 1 | 1 | Low-quality | Genome-fragment | 2.37 | AAI-based (high-confidence) | 18.29 |
| S1-final-viral-combined_k141_62759\|\|full | 1023 | No | NA | 2 | 0 | 1 | Not-determined | Genome-fragment | NA | NA | 0 |
| S1-final-viral-combined_k141_244453\|\|full | 1230 | No | NA | 3 | 0 | 0 | Not-determined | Genome-fragment | NA | NA | 0 |
| S1-final-viral-combined_k141_129804\|\|full | 3564 | No | NA | 4 | 0 | 3 | Not-determined | Genome-fragment | NA | NA | 0 |
| S1-final-viral-combined_k141_176753\|\|full | 2490 | No | NA | 2 | 0 | 1 | Not-determined | Genome-fragment | NA | NA | 0 |

| | | | | | | | | | | | |
|---|---|---|---|---|---|---|---|---|---|---|---|
| S8-final-viral-combined_k141_268108\|\|lt2gene | 1394 | No | NA | 2 | 1 | 0 | Low-quality | Genome-fragment | 3.47 | AAI-based (high-confidence) | 0 |
| S5-final-viral-combined_k141_32112\|\|full | 1395 | No | NA | 2 | 0 | 0 | Low-quality | Genome-fragment | 0.82 | AAI-based (high-confidence) | 0 |
| S1-final-viral-combined_k141_95965\|\|full | 1171 | No | NA | 2 | 0 | 0 | Not-determined | Genome-fragment | NA | NA | 0 |
| S1-final-viral-combined_k141_227663\|\|full | 1474 | No | NA | 4 | 0 | 0 | Not-determined | Genome-fragment | NA | NA | 0 |
| S1-final-viral-combined_k141_29364\|\|full | 1510 | No | NA | 3 | 0 | 1 | Not-determined | Genome-fragment | NA | NA | 0 |
| S1-final-viral-combined_k141_13617\|\|full | 1620 | No | NA | 3 | 0 | 1 | Not-determined | Genome-fragment | NA | NA | 0 |
| S1-final-viral-combined_k141_244499\|\|full | 2123 | No | NA | 4 | 0 | 1 | Not-determined | Genome-fragment | NA | NA | 0 |
| S1-final-viral-combined_k141_78357\|\|full | 2025 | No | NA | 2 | 0 | 1 | Not-determined | Genome-fragment | NA | NA | 0 |
| S1-final-viral-combined_k141_110909\|\|full | 1558 | No | NA | 2 | 0 | 0 | Not-determined | Genome-fragment | NA | NA | 0 |
| S1-final-viral-combined_k141_78362\|\|full | 3283 | No | NA | 5 | 0 | 0 | Not-determined | Genome-fragment | NA | NA | 0 |
| S1-final-viral-combined_k141_96007\|\|full | 1398 | No | NA | 3 | 0 | 0 | Not-determined | Genome-fragment | NA | NA | 0 |
| S1-final-viral-combined_k141_244507\|\|full | 5859 | No | NA | 8 | 0 | 4 | Not-determined | Genome-fragment | NA | NA | 0 |
| S17-final-viral-combined_k141_73315\|\|full | 1395 | No | NA | 2 | 0 | 0 | Low-quality | Genome-fragment | 2.21 | AAI-based (high-confidence) | 0 |
| S1-final-viral-combined_k141_310323\|\|full | 1010 | No | NA | 3 | 0 | 1 | Not-determined | Genome-fragment | NA | NA | 0 |
| S1-final-viral-combined_k141_13643\|\|full | 1999 | No | NA | 3 | 0 | 2 | Not-determined | Genome-fragment | NA | NA | 0 |
| S1-final-viral-combined_k141_212110\|\|full | 3464 | No | NA | 5 | 0 | 3 | Not-determined | Genome-fragment | NA | NA | 0 |
| S1-final-viral-combined_k141_160964\|\|full | 16754 | No | NA | 17 | 0 | 9 | Not-determined | Genome-fragment | NA | NA | 0 |
| S12-final-viral-combined_k141_148867\|\|full | 1395 | Yes | 996 | 2 | 1 | 1 | Low-quality | Genome-fragment | 1.78 | AAI-based (medium-confidence) | 28.6 |
| S1-final-viral-combined_k141_359901\|\|full | 2255 | No | NA | 2 | 0 | 2 | Not-determined | Genome-fragment | NA | NA | 0 |
| S1-final-viral-combined_k141_78379\|\|full | 3396 | No | NA | 4 | 0 | 0 | Not-determined | Genome-fragment | NA | NA | 0 |
| S1-final-viral-combined_k141_78382\|\|full | 1435 | No | NA | 2 | 0 | 2 | Not-determined | Genome-fragment | NA | NA | 0 |
| S1-final-viral-combined_k141_129871\|\|full | 1876 | No | NA | 5 | 0 | 0 | Not-determined | Genome-fragment | NA | NA | 0 |
| S2-final-viral-combined_k141_113474\|\|full | 1395 | No | NA | 3 | 0 | 1 | Low-quality | Genome-fragment | 0.72 | AAI-based (medium-confidence) | 0 |
| S1-final-viral-combined_k141_46048\|\|full | 1880 | No | NA | 3 | 0 | 2 | Not-determined | Genome-fragment | NA | NA | 0 |
| S4-final-viral-combined_k141_30438\|\|lt2gene | 1395 | No | NA | 2 | 2 | 0 | Low-quality | Genome-fragment | 2.09 | HMM-based (lower-bound) | 0 |
| S1-final-viral-combined_k141_46059\|\|full | 2879 | No | NA | 2 | 0 | 2 | Not-determined | Genome-fragment | NA | NA | 0 |
| S1-final-viral-combined_k141_227745\|\|full | 3662 | No | NA | 3 | 0 | 2 | Not-determined | Genome-fragment | NA | NA | 0 |
| S8-final-viral-combined_k141_207258\|\|full | 1395 | No | NA | 3 | 0 | 2 | Low-quality | Genome-fragment | 0.75 | AAI-based (medium-confidence) | 0 |
| S1-final-viral-combined_k141_261620\|\|full | 2151 | No | NA | 2 | 0 | 2 | Not-determined | Genome-fragment | NA | NA | 0 |
| S11-final-viral-combined_k141_124189\|\|full | 1396 | No | NA | 2 | 0 | 1 | Low-quality | Genome-fragment | 0.79 | AAI-based (medium-confidence) | 0 |
| S1-final-viral-combined_k141_195464\|\|full | 7560 | No | NA | 7 | 0 | 6 | Not-determined | Genome-fragment | NA | NA | 0 |
| S1-final-viral-combined_k141_110985\|\|full | 20869 | No | NA | 19 | 0 | 15 | Not-determined | Genome-fragment | NA | NA | 0 |
| S1-final-viral-combined_k141_227795\|\|full | 5097 | No | NA | 6 | 0 | 2 | Not-determined | Genome-fragment | NA | NA | 0 |
| S11-final-viral-combined_k141_96640\|\|full | 1396 | No | NA | 3 | 0 | 0 | Low-quality | Genome-fragment | 3.29 | AAI-based (high-confidence) | 0 |
| S11-final-viral-combined_k141_58764\|\|full | 1396 | No | NA | 3 | 1 | 0 | Low-quality | Genome-fragment | 2.97 | AAI-based (high-confidence) | 0 |
| S1-final-viral-combined_k141_244629\|\|full | 1202 | No | NA | 3 | 0 | 0 | Not-determined | Genome-fragment | NA | NA | 0 |
| S11_k141_227042 | 1396 | No | NA | 1 | 0 | 0 | Low-quality | Genome-fragment | 2.41 | AAI-based (high-confidence) | 0 |
| S1-final-viral-combined_k141_244654\|\|full | 2958 | No | NA | 3 | 0 | 3 | Not-determined | Genome-fragment | NA | NA | 0 |
| S1-final-viral-combined_k141_293998\|\|full | 1456 | No | NA | 2 | 0 | 0 | Not-determined | Genome-fragment | NA | NA | 0 |
| S3_k141_36125 | 1396 | No | NA | 2 | 0 | 0 | Low-quality | Genome-fragment | 2.33 | AAI-based (medium-confidence) | 0 |
| S1-final-viral-combined_k141_261681\|\|full | 1244 | No | NA | 3 | 0 | 2 | Not-determined | Genome-fragment | NA | NA | 0 |
| S1-final-viral-combined_k141_62943\|\|full | 1380 | No | NA | 2 | 0 | 0 | Not-determined | Genome-fragment | NA | NA | 0 |
| S1-final-viral-combined_k141_111063\|\|full | 3028 | No | NA | 4 | 0 | 0 | Not-determined | Genome-fragment | NA | NA | 0 |
| S5-final-viral-combined_k141_194871\|\|full | 1396 | No | NA | 3 | 0 | 2 | Low-quality | Genome-fragment | 0.85 | AAI-based (medium-confidence) | 0 |
| S5_k141_11531 | 1396 | No | NA | 4 | 1 | 0 | Low-quality | Genome-fragment | 0.9 | HMM-based (lower-bound) | 0 |
| S17-final-viral-combined_k141_226139\|\|full | 1396 | No | NA | 4 | 0 | 1 | Low-quality | Genome-fragment | 0.44 | AAI-based (medium-confidence) | 0 |
| S1-final-viral-combined_k141_62976\|\|full | 1676 | No | NA | 4 | 0 | 1 | Not-determined | Genome-fragment | NA | NA | 0 |
| S1-final-viral-combined_k141_195557\|\|full | 1598 | No | NA | 3 | 0 | 1 | Not-determined | Genome-fragment | NA | NA | 0 |
| S1-final-viral-combined_k141_391816\|\|full | 2178 | No | NA | 3 | 0 | 1 | Not-determined | Genome-fragment | NA | NA | 0 |
| S1-final-viral-combined_k141_391822\|\|full | 1460 | No | NA | 3 | 0 | 2 | Not-determined | Genome-fragment | NA | NA | 0 |
| S1-final-viral-combined_k141_195570\|\|full | 3113 | No | NA | 2 | 0 | 2 | Not-determined | Genome-fragment | NA | NA | 0 |
| S1-final-viral-combined_k141_63027\|\|full | 1695 | No | NA | 3 | 0 | 2 | Not-determined | Genome-fragment | NA | NA | 0 |
| S1-final-viral-combined_k141_130071\|\|full | 6617 | No | NA | 5 | 0 | 3 | Not-determined | Genome-fragment | NA | NA | 0 |
| S1-final-viral-combined_k141_244754\|\|full | 2296 | No | NA | 4 | 0 | 0 | Not-determined | Genome-fragment | NA | NA | 0 |
| S17-final-viral-combined_k141_216990\|\|full | 1396 | No | NA | 3 | 1 | 0 | Low-quality | Genome-fragment | 3.49 | AAI-based (medium-confidence) | 0 |
| S1-final-viral-combined_k141_195608\|\|full | 2408 | No | NA | 5 | 0 | 3 | Not-determined | Genome-fragment | NA | NA | 0 |
| S1-final-viral-combined_k141_375745\|\|full | 2017 | No | NA | 2 | 0 | 2 | Not-determined | Genome-fragment | NA | NA | 0 |
| S12-final-viral-combined_k141_385080\|\|full | 1396 | No | NA | 3 | 0 | 2 | Low-quality | Genome-fragment | 1.86 | AAI-based (medium-confidence) | 0 |
| S12_k141_391482 | 1396 | No | NA | 4 | 0 | 0 | Low-quality | Genome-fragment | 3.57 | AAI-based (medium-confidence) | 0 |
| S1-final-viral-combined_k141_375782\|\|full | 1375 | No | NA | 3 | 0 | 1 | Not-determined | Genome-fragment | NA | NA | 0 |
| S1-final-viral-combined_k141_111169\|\|full | 2231 | No | NA | 4 | 0 | 0 | Not-determined | Genome-fragment | NA | NA | 0 |
| S1-final-viral-combined_k141_130175\|\|full | 1428 | No | NA | 3 | 0 | 1 | Not-determined | Genome-fragment | NA | NA | 0 |
| S1-final-viral-combined_k141_341944\|\|full | 2525 | No | NA | 4 | 0 | 2 | Not-determined | Genome-fragment | NA | NA | 0 |
| S2-final-viral-combined_k141_52343\|\|full | 1396 | No | NA | 4 | 1 | 0 | Low-quality | Genome-fragment | 3.32 | AAI-based (high-confidence) | 0 |
| S1-final-viral-combined_k141_111214\|\|full | 1093 | No | NA | 3 | 0 | 0 | Not-determined | Genome-fragment | NA | NA | 0 |
| S2-final-viral-combined_k141_146419\|\|full | 1396 | No | NA | 1 | 0 | 0 | Low-quality | Genome-fragment | 3.04 | AAI-based (high-confidence) | 0 |
| S1-final-viral-combined_k141_391910\|\|full | 7107 | No | NA | 8 | 0 | 5 | Not-determined | Genome-fragment | NA | NA | 0 |
| S1-final-viral-combined_k141_63151\|\|full | 2518 | No | NA | 3 | 0 | 0 | Not-determined | Genome-fragment | NA | NA | 0 |
| S2_k141_63752 | 1396 | No | NA | 4 | 0 | 0 | Low-quality | Genome-fragment | 2.33 | AAI-based (high-confidence) | 0 |
| S1-final-viral-combined_k141_391913\|\|full | 1051 | No | NA | 2 | 0 | 2 | Not-determined | Genome-fragment | NA | NA | 0 |
| S4-final-viral-combined_k141_114998\|\|full | 1396 | No | NA | 3 | 0 | 0 | Low-quality | Genome-fragment | 2.69 | AAI-based (high-confidence) | 0 |
| S1-final-viral-combined_k141_212378\|\|full | 1772 | No | NA | 2 | 0 | 1 | Not-determined | Genome-fragment | NA | NA | 0 |
| S1-final-viral-combined_k141_276816\|\|full | 1225 | No | NA | 2 | 0 | 0 | Not-determined | Genome-fragment | NA | NA | 0 |
| S1-final-viral-combined_k141_276817\|\|full | 1535 | No | NA | 3 | 0 | 0 | Not-determined | Genome-fragment | NA | NA | 0 |
| S1-final-viral-combined_k141_96143\|\|full | 1130 | No | NA | 2 | 0 | 0 | Not-determined | Genome-fragment | NA | NA | 0 |
| S1-final-viral-combined_k141_227917\|\|full | 18499 | No | NA | 26 | 7 | 1 | Medium-quality | Genome-fragment | 50.48 | AAI-based (high-confidence) | 0 |
| S1-final-viral-combined_k141_227929\|\|full | 2381 | No | NA | 2 | 0 | 1 | Not-determined | Genome-fragment | NA | NA | 0 |
| S1-final-viral-combined_k141_244857\|\|full | 6672 | No | NA | 5 | 0 | 5 | Not-determined | Genome-fragment | NA | NA | 0 |
| S4-final-viral-combined_k141_160390\|\|lt2gene | 1396 | No | NA | 1 | 1 | 0 | Low-quality | Genome-fragment | 2.25 | AAI-based (high-confidence) | 0 |
| S8-final-viral-combined_k141_184391\|\|full | 1396 | No | NA | 3 | 1 | 0 | Low-quality | Genome-fragment | 3.4 | AAI-based (medium-confidence) | 0 |
| S1-final-viral-combined_k141_360189\|\|full | 2983 | No | NA | 4 | 0 | 3 | Not-determined | Genome-fragment | NA | NA | 0 |
| S1-final-viral-combined_k141_244871\|\|full | 1869 | No | NA | 3 | 0 | 0 | Not-determined | Genome-fragment | NA | NA | 0 |
| S8-final-viral-combined_k141_257723\|\|full | 1396 | No | NA | 3 | 0 | 0 | Low-quality | Genome-fragment | 1.25 | AAI-based (high-confidence) | 0 |
| S1-final-viral-combined_k141_347093\|\|full | 1397 | No | NA | 3 | 0 | 0 | Low-quality | Genome-fragment | 2.96 | AAI-based (medium-confidence) | 0 |
| S5_k141_54507 | 1397 | No | NA | 2 | 1 | 0 | Low-quality | Genome-fragment | 3.25 | AAI-based (medium-confidence) | 0 |
| S1-final-viral-combined_k141_161386\|\|full | 3092 | No | NA | 3 | 0 | 2 | Not-determined | Genome-fragment | NA | NA | 0 |
| S17-final-viral-combined_k141_52862\|\|lt2gene | 1397 | No | NA | 2 | 1 | 0 | Low-quality | Genome-fragment | 3.46 | AAI-based (high-confidence) | 0 |
| S1-final-viral-combined_k141_29625\|\|full | 4934 | No | NA | 4 | 0 | 0 | Not-determined | Genome-fragment | NA | NA | 0 |
| S1-final-viral-combined_k141_391970\|\|full | 1650 | No | NA | 2 | 0 | 0 | Not-determined | Genome-fragment | NA | NA | 0 |
| S1-final-viral-combined_k141_342044\|\|full | 1582 | No | NA | 3 | 0 | 0 | Not-determined | Genome-fragment | NA | NA | 0 |
| S1-final-viral-combined_k141_111297\|\|full | 1918 | No | NA | 4 | 0 | 2 | Not-determined | Genome-fragment | NA | NA | 0 |
| S12-final-viral-combined_k141_205905\|\|full | 1397 | No | NA | 2 | 1 | 0 | Low-quality | Genome-fragment | 3.3 | AAI-based (medium-confidence) | 0 |
| S1-final-viral-combined_k141_391974\|\|full | 4486 | No | NA | 6 | 0 | 0 | Not-determined | Genome-fragment | NA | NA | 0 |
| S1-final-viral-combined_k141_212493\|\|full | 1886 | No | NA | 3 | 0 | 0 | Not-determined | Genome-fragment | NA | NA | 0 |
| S12-final-viral-combined_k141_56856\|\|full | 1397 | No | NA | 2 | 1 | 0 | Low-quality | Genome-fragment | 2.6 | AAI-based (medium-confidence) | 0 |
| S6-final-viral-combined_k141_36578\|\|full | 1397 | No | NA | 3 | 0 | 2 | Low-quality | Genome-fragment | 1.79 | AAI-based (medium-confidence) | 0 |
| S1-final-viral-combined_k141_262040\|\|full | 2262 | No | NA | 5 | 0 | 1 | Not-determined | Genome-fragment | NA | NA | 0 |
| S1-final-viral-combined_k141_276911\|\|full | 7487 | No | NA | 8 | 0 | 1 | Not-determined | Genome-fragment | NA | NA | 0 |
| S1-final-viral-combined_k141_212516\|\|full | 1184 | No | NA | 2 | 0 | 1 | Not-determined | Genome-fragment | NA | NA | 0 |
| S8-final-viral-combined_k141_71446\|\|full | 1397 | No | NA | 3 | 0 | 0 | Low-quality | Genome-fragment | 2.88 | AAI-based (high-confidence) | 0 |
| S1-final-viral-combined_k141_63216\|\|full | 5240 | No | NA | 5 | 0 | 4 | Not-determined | Genome-fragment | NA | NA | 0 |
| S1-final-viral-combined_k141_14015\|\|full | 5176 | No | NA | 3 | 0 | 1 | Not-determined | Genome-fragment | NA | NA | 0 |
| S1-final-viral-combined_k141_195867\|\|full | 1579 | No | NA | 3 | 0 | 0 | Not-determined | Genome-fragment | NA | NA | 0 |
| S9-final-viral-combined_k141_85306\|\|full | 1398 | No | NA | 4 | 0 | 0 | Low-quality | Genome-fragment | 3.32 | AAI-based (medium-confidence) | 0 |
| S11-final-viral-combined_k141_11005\|\|full | 1398 | No | NA | 5 | 1 | 0 | Low-quality | Genome-fragment | 3.15 | AAI-based (high-confidence) | 0 |
| S1-final-viral-combined_k141_111366\|\|full | 1731 | No | NA | 3 | 0 | 0 | Not-determined | Genome-fragment | NA | NA | 0 |
| S11-final-viral-combined_k141_54592\|\|full | 1398 | No | NA | 3 | 0 | 0 | Low-quality | Genome-fragment | 2.94 | AAI-based (medium-confidence) | 0 |
| S1-final-viral-combined_k141_161472\|\|full | 1471 | No | NA | 1 | 0 | 0 | Not-determined | Genome-fragment | NA | NA | 0 |
| S1-final-viral-combined_k141_244986\|\|full | 1754 | No | NA | 4 | 0 | 0 | Not-determined | Genome-fragment | NA | NA | 0 |
| S3-final-viral-combined_k141_55328\|\|full | 1398 | No | NA | 4 | 0 | 1 | Low-quality | Genome-fragment | 0.66 | AAI-based (medium-confidence) | 0 |
| S1-final-viral-combined_k141_195909\|\|full | 1267 | No | NA | 2 | 0 | 2 | Not-determined | Genome-fragment | NA | NA | 0 |
| S5-final-viral-combined_k141_47566\|\|full | 1398 | No | NA | 3 | 0 | 0 | Low-quality | Genome-fragment | 1.99 | AAI-based (medium-confidence) | 0 |
| S1-final-viral-combined_k141_327240\|\|full | 3339 | No | NA | 3 | 0 | 0 | Not-determined | Genome-fragment | NA | NA | 0 |
| S1-final-viral-combined_k141_360285\|\|full | 40364 | No | NA | 53 | 17 | 4 | Medium-quality | Genome-fragment | 81.89 | AAI-based (medium-confidence) | 0 |
| S1-final-viral-combined_k141_111382\|\|full | 2261 | No | NA | 3 | 0 | 2 | Not-determined | Genome-fragment | NA | NA | 0 |
| S1-final-viral-combined_k141_262091\|\|full | 12847 | No | NA | 9 | 0 | 7 | Not-determined | Genome-fragment | NA | NA | 0 |
| S5-final-viral-combined_k141_32405\|\|full | 1398 | No | NA | 3 | 2 | 0 | Low-quality | Genome-fragment | 2.25 | AAI-based (high-confidence) | 0 |
| S1-final-viral-combined_k141_130297\|\|full | 1936 | No | NA | 2 | 0 | 0 | Not-determined | Genome-fragment | NA | NA | 0 |
| S5-final-viral-combined_k141_194497\|\|full | 1398 | No | NA | 4 | 4 | 0 | Low-quality | Genome-fragment | 3.49 | AAI-based (medium-confidence) | 0 |
| S1-final-viral-combined_k141_29703\|\|full | 1147 | No | NA | 2 | 0 | 0 | Not-determined | Genome-fragment | NA | NA | 0 |
| S1-final-viral-combined_k141_78621\|\|full | 1694 | No | NA | 3 | 0 | 2 | Not-determined | Genome-fragment | NA | NA | 0 |
| S17-final-viral-combined_k141_179235\|\|full | 1398 | No | NA | 3 | 0 | 0 | Low-quality | Genome-fragment | 3.48 | AAI-based (high-confidence) | 0 |
| S12-final-viral-combined_k141_262316\|\|full | 1398 | No | NA | 3 | 0 | 1 | Low-quality | Genome-fragment | 2.03 | AAI-based (medium-confidence) | 0 |
| S12-final-viral-combined_k141_66411\|\|lt2gene | 1398 | No | NA | 2 | 1 | 0 | Low-quality | Genome-fragment | 2.56 | AAI-based (medium-confidence) | 0 |
| S2-final-viral-combined_k141_138318\|\|full | 1398 | No | NA | 3 | 0 | 0 | Low-quality | Genome-fragment | 2.4 | AAI-based (medium-confidence) | 0 |
| S6-final-viral-combined_k141_70158\|\|full | 1398 | No | NA | 2 | 0 | 1 | Low-quality | Genome-fragment | 1.66 | AAI-based (high-confidence) | 0 |
| S11-final-viral-combined_k141_304497\|\|full | 1399 | No | NA | 3 | 0 | 0 | Low-quality | Genome-fragment | 1.64 | AAI-based (medium-confidence) | 0 |
| S11-final-viral-combined_k141_239327\|\|full | 1399 | No | NA | 3 | 1 | 0 | Low-quality | Genome-fragment | 2.53 | AAI-based (high-confidence) | 0 |
| S1-final-viral-combined_k141_145813\|\|full | 2205 | No | NA | 2 | 0 | 1 | Not-determined | Genome-fragment | NA | NA | 0 |
| S1-final-viral-combined_k141_104718\|\|full | 1399 | No | NA | 2 | 2 | 0 | Low-quality | Genome-fragment | 2.29 | AAI-based (high-confidence) | 0 |
| S1-final-viral-combined_k141_14061\|\|full | 2093 | No | NA | 3 | 0 | 2 | Not-determined | Genome-fragment | NA | NA | 0 |
| S1-final-viral-combined_k141_376024\|\|full | 4398 | No | NA | 5 | 0 | 5 | Not-determined | Genome-fragment | NA | NA | 0 |
| S3_k141_71076 | 1399 | No | NA | 1 | 0 | 0 | Low-quality | Genome-fragment | 2.84 | AAI-based (high-confidence) | 0 |
| S1-final-viral-combined_k141_310716\|\|full | 2560 | No | NA | 5 | 0 | 1 | Not-determined | Genome-fragment | NA | NA | 0 |
| S1-final-viral-combined_k141_294277\|\|full | 1729 | No | NA | 2 | 0 | 1 | Not-determined | Genome-fragment | NA | NA | 0 |
| S1-final-viral-combined_k141_29754\|\|full | 2482 | No | NA | 4 | 0 | 3 | Not-determined | Genome-fragment | NA | NA | 0 |
| S1-final-viral-combined_k141_212638\|\|full | 2887 | No | NA | 3 | 0 | 3 | Not-determined | Genome-fragment | NA | NA | 0 |
| S1-final-viral-combined_k141_310739\|\|full | 1218 | No | NA | 3 | 0 | 1 | Not-determined | Genome-fragment | NA | NA | 0 |
| S5-final-viral-combined_k141_81182\|\|full | 1399 | No | NA | 2 | 1 | 0 | Low-quality | Genome-fragment | 2.85 | AAI-based (medium-confidence) | 0 |
| S1-final-viral-combined_k141_195996\|\|full | 1841 | No | NA | 3 | 0 | 1 | Not-determined | Genome-fragment | NA | NA | 0 |
| S1-final-viral-combined_k141_262173\|\|full | 2923 | No | NA | 5 | 0 | 3 | Not-determined | Genome-fragment | NA | NA | 0 |
| S1-final-viral-combined_k141_294318\|\|full | 4880 | No | NA | 4 | 0 | 3 | Not-determined | Genome-fragment | NA | NA | 0 |
| S1-final-viral-combined_k141_310784\|\|full | 2162 | No | NA | 4 | 0 | 0 | Not-determined | Genome-fragment | NA | NA | 0 |
| S7-final-viral-combined_k141_55126\|\|full | 1399 | No | NA | 2 | 0 | 2 | Low-quality | Genome-fragment | 3.59 | AAI-based (medium-confidence) | 0 |
| S1-final-viral-combined_k141_228215\|\|full | 1126 | No | NA | 2 | 0 | 2 | Not-determined | Genome-fragment | NA | NA | 0 |
| S17-final-viral-combined_k141_156640\|\|full | 1399 | No | NA | 1 | 0 | 0 | Low-quality | Genome-fragment | 2.43 | AAI-based (medium-confidence) | 0 |
| S1-final-viral-combined_k141_96322\|\|full | 7654 | No | NA | 8 | 0 | 5 | Not-determined | Genome-fragment | NA | NA | 0 |
| S17-final-viral-combined_k141_115031\|\|full | 1399 | Yes | 633 | 2 | 1 | 1 | Low-quality | Genome-fragment | 1.77 | AAI-based (high-confidence) | 54.75 |
| S1-final-viral-combined_k141_130382\|\|full | 1803 | No | NA | 3 | 0 | 0 | Not-determined | Genome-fragment | NA | NA | 0 |
| S1-final-viral-combined_k141_14169\|\|full | 3598 | No | NA | 2 | 0 | 2 | Not-determined | Genome-fragment | NA | NA | 0 |
| S1-final-viral-combined_k141_46389\|\|full | 5208 | No | NA | 7 | 0 | 7 | Not-determined | Genome-fragment | NA | NA | 0 |
| S1-final-viral-combined_k141_14173\|\|full | 1397 | No | NA | 4 | 0 | 1 | Not-determined | Genome-fragment | NA | NA | 0 |
| S17-final-viral-combined_k141_148647\|\|full | 1399 | No | NA | 3 | 0 | 1 | Low-quality | Genome-fragment | 0.47 | AAI-based (medium-confidence) | 0 |
| S1-final-viral-combined_k141_111476\|\|full | 2056 | No | NA | 3 | 0 | 1 | Not-determined | Genome-fragment | NA | NA | 0 |
| S1-final-viral-combined_k141_294390\|\|full | 2646 | No | NA | 3 | 0 | 2 | Not-determined | Genome-fragment | NA | NA | 0 |
| S17-final-viral-combined_k141_197240\|\|full | 1399 | No | NA | 5 | 0 | 0 | Low-quality | Genome-fragment | 3.06 | AAI-based (medium-confidence) | 0 |
| S12-final-viral-combined_k141_38355\|\|full | 1399 | No | NA | 6 | 0 | 0 | Low-quality | Genome-fragment | 1.74 | AAI-based (high-confidence) | 0 |
| S1-final-viral-combined_k141_310870\|\|full | 2227 | No | NA | 4 | 0 | 0 | Not-determined | Genome-fragment | NA | NA | 0 |
| S2-final-viral-combined_k141_117584\|\|full | 1399 | No | NA | 2 | 0 | 0 | Low-quality | Genome-fragment | 2.56 | AAI-based (medium-confidence) | 0 |
| S2-final-viral-combined_k141_30471\|\|lt2gene | 1399 | No | NA | 1 | 1 | 0 | Low-quality | Genome-fragment | 4.51 | HMM-based (lower-bound) | 0 |
| S4-final-viral-combined_k141_121007\|\|full | 1399 | No | NA | 2 | 0 | 0 | Low-quality | Genome-fragment | 1.9 | AAI-based (medium-confidence) | 0 |
| S9-final-viral-combined_k141_81461\|\|full | 1400 | No | NA | 2 | 1 | 0 | Low-quality | Genome-fragment | 1.76 | AAI-based (high-confidence) | 0 |
| S1-final-viral-combined_k141_78848\|\|full | 1825 | No | NA | 2 | 0 | 2 | Not-determined | Genome-fragment | NA | NA | 0 |
| S9-final-viral-combined_k141_58435\|\|full | 1400 | No | NA | 3 | 0 | 0 | Low-quality | Genome-fragment | 3.13 | AAI-based (high-confidence) | 0 |
| S5-final-viral-combined_k141_194557\|\|full | 1400 | No | NA | 2 | 1 | 0 | Low-quality | Genome-fragment | 2.35 | HMM-based (lower-bound) | 0 |
| S1-final-viral-combined_k141_196149\|\|full | 1759 | No | NA | 4 | 0 | 2 | Not-determined | Genome-fragment | NA | NA | 0 |
| S5-final-viral-combined_k141_123129\|\|full | 1400 | No | NA | 4 | 0 | 0 | Low-quality | Genome-fragment | 1.75 | AAI-based (medium-confidence) | 0 |
| S1-final-viral-combined_k141_327482\|\|full | 3683 | No | NA | 6 | 0 | 0 | Not-determined | Genome-fragment | NA | NA | 0 |
| S1-final-viral-combined_k141_63515\|\|full | 4684 | No | NA | 4 | 0 | 2 | Not-determined | Genome-fragment | NA | NA | 0 |
| S17-final-viral-combined_k141_222894\|\|full | 1400 | No | NA | 6 | 0 | 0 | Low-quality | Genome-fragment | 3.89 | AAI-based (medium-confidence) | 0 |
| S17-final-viral-combined_k141_71269\|\|full | 1400 | No | NA | 2 | 1 | 0 | Low-quality | Genome-fragment | 3.22 | AAI-based (high-confidence) | 0 |
| S1-final-viral-combined_k141_360494\|\|full | 5356 | No | NA | 6 | 0 | 6 | Not-determined | Genome-fragment | NA | NA | 0 |
| S1-final-viral-combined_k141_46509\|\|full | 1197 | No | NA | 3 | 0 | 1 | Not-determined | Genome-fragment | NA | NA | 0 |
| S12-final-viral-combined_k141_264015\|\|full | 1400 | No | NA | 3 | 2 | 0 | Low-quality | Genome-fragment | 3.12 | AAI-based (medium-confidence) | 0 |
| S1-final-viral-combined_k141_63549\|\|full | 2599 | No | NA | 2 | 0 | 2 | Not-determined | Genome-fragment | NA | NA | 0 |
| S12-final-viral-combined_k141_84111\|\|lt2gene | 1400 | No | NA | 2 | 1 | 0 | Low-quality | Genome-fragment | 2.74 | AAI-based (high-confidence) | 0 |
| S12-final-viral-combined_k141_93445\|\|lt2gene | 1400 | No | NA | 2 | 1 | 0 | Low-quality | Genome-fragment | 3.59 | AAI-based (high-confidence) | 0 |
| S1-final-viral-combined_k141_294545\|\|full | 2722 | No | NA | 4 | 0 | 1 | Not-determined | Genome-fragment | NA | NA | 0 |
| S1-final-viral-combined_k141_245194\|\|full | 2435 | No | NA | 4 | 0 | 1 | Not-determined | Genome-fragment | NA | NA | 0 |
| S2-final-viral-combined_k141_236801\|\|full | 1400 | No | NA | 2 | 0 | 0 | Low-quality | Genome-fragment | 2.33 | AAI-based (medium-confidence) | 0 |
| S6-final-viral-combined_k141_44423\|\|full | 1400 | No | NA | 4 | 0 | 3 | Low-quality | Genome-fragment | 0.41 | AAI-based (medium-confidence) | 0 |
| S9-final-viral-combined_k141_54866\|\|full | 1401 | No | NA | 3 | 1 | 0 | Low-quality | Genome-fragment | 3.52 | AAI-based (high-confidence) | 0 |
| S1-final-viral-combined_k141_342349\|\|full | 9185 | No | NA | 8 | 0 | 5 | Not-determined | Genome-fragment | NA | NA | 0 |
| S11-final-viral-combined_k141_181809\|\|full | 1401 | No | NA | 3 | 0 | 0 | Low-quality | Genome-fragment | 2.22 | AAI-based (high-confidence) | 0 |
| S1-final-viral-combined_k141_262365\|\|full | 1712 | No | NA | 2 | 0 | 2 | Not-determined | Genome-fragment | NA | NA | 0 |
| S1-final-viral-combined_k141_31434\|\|full | 1401 | No | NA | 2 | 0 | 1 | Low-quality | Genome-fragment | 1.08 | AAI-based (high-confidence) | 0 |
| S1-final-viral-combined_k141_392237\|\|full | 2003 | No | NA | 2 | 0 | 1 | Not-determined | Genome-fragment | NA | NA | 0 |
| S1-final-viral-combined_k141_196245\|\|full | 1752 | No | NA | 3 | 0 | 2 | Not-determined | Genome-fragment | NA | NA | 0 |
| S3-final-viral-combined_k141_79537\|\|lt2gene | 1401 | No | NA | 2 | 1 | 0 | Low-quality | Genome-fragment | 3.25 | AAI-based (high-confidence) | 0 |
| S1-final-viral-combined_k141_177679\|\|full | 1057 | No | NA | 2 | 0 | 2 | Not-determined | Genome-fragment | NA | NA | 0 |
| S1-final-viral-combined_k141_130517\|\|full | 1803 | No | NA | 3 | 0 | 2 | Not-determined | Genome-fragment | NA | NA | 0 |
| S7-final-viral-combined_k141_228835\|\|full | 1401 | No | NA | 3 | 2 | 0 | Low-quality | Genome-fragment | 2.5 | HMM-based (lower-bound) | 0 |
| S1-final-viral-combined_k141_161787\|\|full | 1046 | No | NA | 2 | 0 | 1 | Not-determined | Genome-fragment | NA | NA | 0 |
| S1-final-viral-combined_k141_294609\|\|full | 1940 | No | NA | 2 | 0 | 0 | Not-determined | Genome-fragment | NA | NA | 0 |
| S1-final-viral-combined_k141_342407\|\|full | 6208 | No | NA | 6 | 0 | 2 | Not-determined | Genome-fragment | NA | NA | 0 |
| S1-final-viral-combined_k141_161810\|\|full | 2700 | No | NA | 5 | 0 | 1 | Not-determined | Genome-fragment | NA | NA | 0 |
| S17-k141_143001 | 1401 | No | NA | 2 | 0 | 2 | Low-quality | Genome-fragment | 2.77 | AAI-based (medium-confidence) | 0 |
| S1-final-viral-combined_k141_130541\|\|full | 2838 | No | NA | 7 | 0 | 1 | Not-determined | Genome-fragment | NA | NA | 0 |
| S1-final-viral-combined_k141_342429\|\|full | 5111 | No | NA | 6 | 0 | 3 | Not-determined | Genome-fragment | NA | NA | 0 |
| S1-final-viral-combined_k141_63621\|\|full | 9431 | No | NA | 9 | 0 | 4 | Not-determined | Genome-fragment | NA | NA | 0 |
| S1-final-viral-combined_k141_96569\|\|full | 2603 | No | NA | 3 | 0 | 0 | Not-determined | Genome-fragment | NA | NA | 0 |
| S12-final-viral-combined_k141_335881\|\|full | 1401 | No | NA | 3 | 1 | 0 | Low-quality | Genome-fragment | 2.37 | AAI-based (high-confidence) | 0 |
| S1-final-viral-combined_k141_392304\|\|full | 3564 | No | NA | 5 | 0 | 3 | Not-determined | Genome-fragment | NA | NA | 0 |
| S1-final-viral-combined_k141_30095\|\|full | 1682 | No | NA | 3 | 0 | 0 | Not-determined | Genome-fragment | NA | NA | 0 |
| S1-final-viral-combined_k141_212892\|\|full | 22088 | No | NA | 28 | 9 | 0 | Medium-quality | Genome-fragment | 56.33 | AAI-based (high-confidence) | 0 |
| S2-final-viral-combined_k141_20217\|\|full | 1401 | No | NA | 3 | 1 | 0 | Low-quality | Genome-fragment | 2.24 | AAI-based (medium-confidence) | 0 |
| S1-final-viral-combined_k141_342497\|\|full | 1736 | No | NA | 4 | 0 | 2 | Not-determined | Genome-fragment | NA | NA | 0 |
| S1-final-viral-combined_k141_294694\|\|full | 2242 | No | NA | 3 | 0 | 1 | Not-determined | Genome-fragment | NA | NA | 0 |
| S11-final-viral-combined_k141_235220\|\|full | 1402 | No | NA | 2 | 1 | 0 | Low-quality | Genome-fragment | 2.17 | AAI-based (medium-confidence) | 0 |
| S11-final-viral-combined_k141_214665\|\|full | 1402 | No | NA | 4 | 0 | 0 | Low-quality | Genome-fragment | 3.81 | AAI-based (medium-confidence) | 0 |
| S1-final-viral-combined_k141_196382\|\|full | 19410 | No | NA | 21 | 0 | 5 | Not-determined | Genome-fragment | NA | NA | 0 |
| S11_k141_253637 | 1402 | No | NA | 1 | 0 | 1 | Low-quality | Genome-fragment | 3.95 | AAI-based (medium-confidence) | 0 |
| S1-final-viral-combined_k141_342553\|\|full | 2033 | No | NA | 2 | 0 | 2 | Not-determined | Genome-fragment | NA | NA | 0 |
| S1-final-viral-combined_k141_294729\|\|full | 6787 | No | NA | 7 | 0 | 6 | Not-determined | Genome-fragment | NA | NA | 0 |
| S1-final-viral-combined_k141_30137\|\|full | 1901 | No | NA | 3 | 0 | 2 | Not-determined | Genome-fragment | NA | NA | 0 |
| S1-final-viral-combined_k141_262512\|\|full | 8540 | No | NA | 9 | 0 | 5 | Not-determined | Genome-fragment | NA | NA | 0 |
| S1-final-viral-combined_k141_96704\|\|full | 1770 | No | NA | 2 | 0 | 1 | Not-determined | Genome-fragment | NA | NA | 0 |
| S5-final-viral-combined_k141_163326\|\|full | 1402 | No | NA | 3 | 1 | 0 | Low-quality | Genome-fragment | 1.57 | AAI-based (high-confidence) | 0 |
| S1-final-viral-combined_k141_96708\|\|full | 4235 | No | NA | 5 | 0 | 3 | Not-determined | Genome-fragment | NA | NA | 0 |
| S5-final-viral-combined_k141_120206\|\|full | 1402 | No | NA | 4 | 0 | 1 | Low-quality | Genome-fragment | 2.87 | AAI-based (medium-confidence) | 0 |
| S7-final-viral-combined_k141_154160\|\|full | 1402 | No | NA | 2 | 0 | 0 | Low-quality | Genome-fragment | 2.73 | AAI-based (high-confidence) | 0 |
| S17-final-viral-combined_k141_146853\|\|full | 1402 | No | NA | 2 | 0 | 2 | Low-quality | Genome-fragment | 3.27 | AAI-based (medium-confidence) | 0 |
| S12-final-viral-combined_k141_262365\|\|full | 1402 | No | NA | 3 | 1 | 0 | Low-quality | Genome-fragment | 4.38 | AAI-based (medium-confidence) | 0 |
| S1-final-viral-combined_k141_30190\|\|full | 2281 | No | NA | 2 | 0 | 1 | Not-determined | Genome-fragment | NA | NA | 0 |
| S4_k141_25661 | 1402 | No | NA | 1 | 0 | 1 | Low-quality | Genome-fragment | 2.39 | AAI-based (high-confidence) | 0 |
| S1-final-viral-combined_k141_96744\|\|full | 1305 | No | NA | 3 | 0 | 3 | Not-determined | Genome-fragment | NA | NA | 0 |
| S1-final-viral-combined_k141_14442\|\|full | 2168 | No | NA | 3 | 0 | 2 | Not-determined | Genome-fragment | NA | NA | 0 |
| S8-final-viral-combined_k141_93827\|\|full | 1402 | No | NA | 4 | 0 | 2 | Low-quality | Genome-fragment | 0.48 | AAI-based (medium-confidence) | 0 |
| S1-final-viral-combined_k141_177882\|\|full | 2511 | No | NA | 3 | 0 | 0 | Not-determined | Genome-fragment | NA | NA | 0 |
| S1-final-viral-combined_k141_130673\|\|full | 1372 | No | NA | 3 | 0 | 1 | Not-determined | Genome-fragment | NA | NA | 0 |
| S1-final-viral-combined_k141_327763\|\|full | 1665 | No | NA | 2 | 0 | 1 | Not-determined | Genome-fragment | NA | NA | 0 |
| S9_k141_71579 | 1403 | No | NA | 3 | 1 | 0 | Low-quality | Genome-fragment | 3.9 | AAI-based (medium-confidence) | 0 |
| S1-final-viral-combined_k141_30268\|\|full | 1399 | No | NA | 2 | 0 | 1 | Not-determined | Genome-fragment | NA | NA | 0 |
| S11_k141_256064 | 1403 | No | NA | 2 | 1 | 0 | Low-quality | Genome-fragment | 1.49 | AAI-based (high-confidence) | 0 |
| S1-final-viral-combined_k141_85652\|\|full | 1403 | No | NA | 3 | 0 | 2 | Low-quality | Genome-fragment | 0.9 | AAI-based (medium-confidence) | 0 |
| S5-final-viral-combined_k141_123314\|\|full | 1403 | No | NA | 4 | 1 | 0 | Low-quality | Genome-fragment | 0.87 | AAI-based (medium-confidence) | 0 |
| S1-final-viral-combined_k141_96848\|\|full | 1534 | No | NA | 2 | 0 | 1 | Not-determined | Genome-fragment | NA | NA | 0 |
| S1-final-viral-combined_k141_14519\|\|full | 3505 | No | NA | 4 | 0 | 0 | Not-determined | Genome-fragment | NA | NA | 0 |
| S1-final-viral-combined_k141_63789\|\|full | 7809 | No | NA | 3 | 0 | 2 | Not-determined | Genome-fragment | NA | NA | 0 |
| S5-final-viral-combined_k141_9241\|\|lt2gene | 1403 | No | NA | 2 | 1 | 0 | Low-quality | Genome-fragment | 0.91 | HMM-based (lower-bound) | 0 |
| S1-final-viral-combined_k141_277430\|\|full | 9829 | No | NA | 9 | 0 | 5 | Not-determined | Genome-fragment | NA | NA | 0 |
| S7_k141_235448 | 1403 | No | NA | 1 | 0 | 1 | Low-quality | Genome-fragment | 0.37 | AAI-based (high-confidence) | 0 |
| S1-final-viral-combined_k141_14545\|\|full | 1915 | No | NA | 2 | 0 | 1 | Not-determined | Genome-fragment | NA | NA | 0 |
| S17-final-viral-combined_k141_175998\|\|full | 1403 | No | NA | 3 | 1 | 0 | Low-quality | Genome-fragment | 1.03 | HMM-based (lower-bound) | 0 |
| S1-final-viral-combined_k141_162071\|\|full | 7200 | No | NA | 8 | 0 | 4 | Not-determined | Genome-fragment | NA | NA | 0 |
| S17-k141_187443 | 1403 | No | NA | 2 | 0 | 0 | Low-quality | Genome-fragment | 3.14 | AAI-based (medium-confidence) | 0 |
| S1-final-viral-combined_k141_245656\|\|full | 2212 | No | NA | 3 | 0 | 1 | Not-determined | Genome-fragment | NA | NA | 0 |
| S1-final-viral-combined_k141_111757\|\|full | 2907 | No | NA | 3 | 0 | 2 | Not-determined | Genome-fragment | NA | NA | 0 |
| S1-final-viral-combined_k141_245665\|\|full | 2715 | No | NA | 3 | 0 | 2 | Not-determined | Genome-fragment | NA | NA | 0 |
| S1-final-viral-combined_k141_14568\|\|full | 2291 | No | NA | 3 | 0 | 0 | Not-determined | Genome-fragment | NA | NA | 0 |
| S12_k141_167245 | 1403 | No | NA | 3 | 0 | 1 | Low-quality | Genome-fragment | 0.87 | AAI-based (medium-confidence) | 0 |
| S1-final-viral-combined_k141_360771\|\|full | 15514 | No | NA | 15 | 0 | 12 | Not-determined | Genome-fragment | NA | NA | 0 |
| S1-final-viral-combined_k141_262668\|\|full | 7816 | No | NA | 5 | 0 | 3 | Not-determined | Genome-fragment | NA | NA | 0 |
| S2-final-viral-combined_k141_159159\|\|full | 1403 | No | NA | 3 | 0 | 0 | Low-quality | Genome-fragment | 3.27 | AAI-based (high-confidence) | 0 |
| S2-final-viral-combined_k141_68212\|\|full | 1403 | No | NA | 2 | 0 | 1 | Low-quality | Genome-fragment | 2.52 | AAI-based (high-confidence) | 0 |
| S1-final-viral-combined_k141_360796\|\|full | 1400 | No | NA | 3 | 0 | 0 | Not-determined | Genome-fragment | NA | NA | 0 |
| S1-final-viral-combined_k141_277494\|\|full | 2251 | No | NA | 3 | 0 | 0 | Not-determined | Genome-fragment | NA | NA | 0 |
| S4-final-viral-combined_k141_109724\|\|full | 1403 | No | NA | 2 | 0 | 0 | Low-quality | Genome-fragment | 2.83 | AAI-based (medium-confidence) | 0 |
| S1-final-viral-combined_k141_162158\|\|full | 4450 | No | NA | 4 | 0 | 3 | Not-determined | Genome-fragment | NA | NA | 0 |
| S1-final-viral-combined_k141_311423\|\|full | 10389 | No | NA | 12 | 0 | 11 | Not-determined | Genome-fragment | NA | NA | 0 |
| S6-final-viral-combined_k141_92261\|\|lt2gene | 1403 | No | NA | 1 | 1 | 0 | Low-quality | Genome-fragment | 2.82 | HMM-based (lower-bound) | 0 |
| S1-final-viral-combined_k141_178042\|\|full | 2484 | No | NA | 3 | 0 | 1 | Not-determined | Genome-fragment | NA | NA | 0 |
| S8-final-viral-combined_k141_162590\|\|full | 1403 | No | NA | 5 | 0 | 0 | Low-quality | Genome-fragment | 1.9 | AAI-based (high-confidence) | 0 |
| S8-final-viral-combined_k141_197065\|\|full | 1403 | No | NA | 3 | 0 | 0 | Low-quality | Genome-fragment | 2.8 | AAI-based (medium-confidence) | 0 |
| S1-final-viral-combined_k141_392563\|\|full | 1935 | No | NA | 2 | 0 | 2 | Not-determined | Genome-fragment | NA | NA | 0 |
| S8-final-viral-combined_k141_250362\|\|full | 1403 | No | NA | 2 | 0 | 1 | Low-quality | Genome-fragment | 2.56 | AAI-based (medium-confidence) | 0 |
| S1-final-viral-combined_k141_96933\|\|full | 2481 | No | NA | 4 | 0 | 2 | Not-determined | Genome-fragment | NA | NA | 0 |
| S11-final-viral-combined_k141_138104\|\|full | 1404 | No | NA | 4 | 0 | 0 | Low-quality | Genome-fragment | 1.08 | AAI-based (high-confidence) | 0 |
| S1-final-viral-combined_k141_14620\|\|full | 2688 | No | NA | 4 | 0 | 3 | Not-determined | Genome-fragment | NA | NA | 0 |
| S1-final-viral-combined_k141_277574\|\|full | 1274 | No | NA | 2 | 0 | 1 | Not-determined | Genome-fragment | NA | NA | 0 |
| S1-final-viral-combined_k141_342839\|\|full | 6548 | No | NA | 6 | 0 | 3 | Not-determined | Genome-fragment | NA | NA | 0 |
| S1-final-viral-combined_k141_241449\|\|full | 1404 | No | NA | 3 | 0 | 1 | Low-quality | Genome-fragment | 2.57 | AAI-based (medium-confidence) | 0 |
| S17-final-viral-combined_k141_32825\|\|full | 1404 | No | NA | 3 | 0 | 1 | Low-quality | Genome-fragment | 0.68 | AAI-based (medium-confidence) | 0 |
| S12-final-viral-combined_k141_256488\|\|full | 1404 | No | NA | 4 | 2 | 0 | Low-quality | Genome-fragment | 2.78 | AAI-based (medium-confidence) | 0 |
| S12_k141_286254 | 1404 | No | NA | 2 | 0 | 0 | Low-quality | Genome-fragment | 1.39 | AAI-based (high-confidence) | 0 |
| S4_k141_153089 | 1404 | No | NA | 2 | 0 | 0 | Low-quality | Genome-fragment | 1.41 | AAI-based (medium-confidence) | 0 |
| S9_k141_64718 | 1405 | No | NA | 3 | 1 | 0 | Low-quality | Genome-fragment | 2.79 | AAI-based (high-confidence) | 0 |
| S1-final-viral-combined_k141_299371\|\|full | 1405 | No | NA | 3 | 0 | 0 | Low-quality | Genome-fragment | 3.65 | AAI-based (high-confidence) | 0 |
| S5-final-viral-combined_k141_52073\|\|full | 1405 | No | NA | 4 | 0 | 0 | Low-quality | Genome-fragment | 2.45 | AAI-based (high-confidence) | 0 |
| S1-final-viral-combined_k141_30471\|\|full | 3012 | No | NA | 3 | 0 | 2 | Not-determined | Genome-fragment | NA | NA | 0 |
| S1-final-viral-combined_k141_96973\|\|full | 2784 | No | NA | 4 | 0 | 2 | Not-determined | Genome-fragment | NA | NA | 0 |
| S1-final-viral-combined_k141_360908\|\|full | 2331 | No | NA | 2 | 0 | 1 | Not-determined | Genome-fragment | NA | NA | 0 |
| S1-final-viral-combined_k141_130899\|\|full | 1532 | No | NA | 3 | 0 | 0 | Not-determined | Genome-fragment | NA | NA | 0 |
| S7_k141_32386 | 1405 | No | NA | 2 | 0 | 0 | Low-quality | Genome-fragment | 1.33 | AAI-based (high-confidence) | 0 |
| S1-final-viral-combined_k141_96982\|\|full | 1183 | No | NA | 3 | 0 | 1 | Not-determined | Genome-fragment | NA | NA | 0 |
| S1-final-viral-combined_k141_262781\|\|full | 2045 | No | NA | 3 | 0 | 2 | Not-determined | Genome-fragment | NA | NA | 0 |
| S17-final-viral-combined_k141_67205\|\|lt2gene | 1405 | No | NA | 1 | 1 | 0 | Low-quality | Genome-fragment | 3.83 | AAI-based (medium-confidence) | 0 |
| S17-final-viral-combined_k141_204368\|\|lt2gene | 1405 | No | NA | 1 | 1 | 0 | Low-quality | Genome-fragment | 2.19 | AAI-based (high-confidence) | 0 |
| S12_k141_198456 | 1405 | No | NA | 1 | 0 | 0 | Low-quality | Genome-fragment | 0.77 | AAI-based (high-confidence) | 0 |
| S2-final-viral-combined_k141_176384\|\|full | 1405 | No | NA | 1 | 0 | 0 | Low-quality | Genome-fragment | 2.87 | AAI-based (medium-confidence) | 0 |
| S1-final-viral-combined_k141_111905\|\|full | 8392 | No | NA | 10 | 0 | 6 | Not-determined | Genome-fragment | NA | NA | 0 |
| S2-final-viral-combined_k141_75930\|\|full | 1405 | Yes | 346 | 2 | 1 | 1 | Low-quality | Genome-fragment | 0.14 | HMM-based (lower-bound) | 75.37 |
| S2-final-viral-combined_k141_246569\|\|full | 1405 | No | NA | 4 | 0 | 2 | Low-quality | Genome-fragment | 0.45 | AAI-based (medium-confidence) | 0 |
| S1-final-viral-combined_k141_342937\|\|full | 1493 | No | NA | 2 | 0 | 0 | Not-determined | Genome-fragment | NA | NA | 0 |
| S4-final-viral-combined_k141_130968\|\|full | 1405 | No | NA | 2 | 0 | 0 | Low-quality | Genome-fragment | 3.67 | AAI-based (high-confidence) | 0 |
| S1-final-viral-combined_k141_392674\|\|full | 8872 | No | NA | 10 | 0 | 6 | Not-determined | Genome-fragment | NA | NA | 0 |
| S4-final-viral-combined_k141_34100\|\|full | 1405 | No | NA | 3 | 3 | 0 | Low-quality | Genome-fragment | 2.33 | AAI-based (high-confidence) | 0 |
| S8-final-viral-combined_k141_196854\|\|full | 1405 | No | NA | 2 | 0 | 2 | Low-quality | Genome-fragment | 1.01 | AAI-based (high-confidence) | 0 |
| S8_k141_63621 | 1405 | No | NA | 3 | 0 | 0 | Low-quality | Genome-fragment | 0.91 | AAI-based (high-confidence) | 0 |
| S1-final-viral-combined_k141_111943\|\|full | 3930 | No | NA | 4 | 0 | 4 | Not-determined | Genome-fragment | NA | NA | 0 |
| S9-final-viral-combined_k141_73568\|\|full | 1406 | No | NA | 3 | 0 | 2 | Low-quality | Genome-fragment | 0.36 | AAI-based (medium-confidence) | 0 |
| S3-final-viral-combined_k141_70155\|\|full | 1406 | No | NA | 3 | 0 | 2 | Low-quality | Genome-fragment | 1.23 | AAI-based (medium-confidence) | 0 |
| S7-final-viral-combined_k141_233893\|\|full | 1406 | No | NA | 2 | 0 | 0 | Low-quality | Genome-fragment | 3.25 | AAI-based (medium-confidence) | 0 |
| S12-final-viral-combined_k141_152710\|\|full | 1406 | No | NA | 4 | 2 | 0 | Low-quality | Genome-fragment | 2.51 | AAI-based (high-confidence) | 0 |
| S12-final-viral-combined_k141_106609\|\|full | 1406 | No | NA | 2 | 0 | 1 | Low-quality | Genome-fragment | 1.03 | AAI-based (medium-confidence) | 0 |
| S1-final-viral-combined_k141_196594\|\|full | 18806 | No | NA | 14 | 0 | 10 | Not-determined | Genome-fragment | NA | NA | 0 |
| S1-final-viral-combined_k141_14799\|\|full | 1184 | No | NA | 2 | 0 | 2 | Not-determined | Genome-fragment | NA | NA | 0 |
| S12-final-viral-combined_k141_326650\|\|full | 1406 | No | NA | 3 | 2 | 0 | Low-quality | Genome-fragment | 2.96 | AAI-based (high-confidence) | 0 |
| S12_k141_249948 | 1406 | No | NA | 2 | 0 | 1 | Low-quality | Genome-fragment | 0.42 | AAI-based (medium-confidence) | 0 |
| S1-final-viral-combined_k141_196600\|\|full | 2384 | No | NA | 3 | 0 | 1 | Not-determined | Genome-fragment | NA | NA | 0 |
| S2-final-viral-combined_k141_124498\|\|full | 1406 | No | NA | 5 | 0 | 0 | Low-quality | Genome-fragment | 1.33 | AAI-based (medium-confidence) | 0 |
| S1-final-viral-combined_k141_213047\|\|full | 5460 | No | NA | 8 | 0 | 1 | Not-determined | Genome-fragment | NA | NA | 0 |
| S4-final-viral-combined_k141_146310\|\|full | 1406 | No | NA | 4 | 0 | 0 | Low-quality | Genome-fragment | 2.41 | AAI-based (medium-confidence) | 0 |
| S4-final-viral-combined_k141_7350\|\|full | 1406 | No | NA | 3 | 0 | 0 | Low-quality | Genome-fragment | 0.9 | AAI-based (high-confidence) | 0 |
| S4-final-viral-combined_k141_58287\|\|full | 1406 | No | NA | 4 | 1 | 0 | Low-quality | Genome-fragment | 1.81 | AAI-based (medium-confidence) | 0 |
| S1-final-viral-combined_k141_376712\|\|full | 3755 | No | NA | 5 | 0 | 0 | Not-determined | Genome-fragment | NA | NA | 0 |
| S1-final-viral-combined_k141_146421\|\|full | 3926 | No | NA | 5 | 0 | 3 | Not-determined | Genome-fragment | NA | NA | 0 |
| S4-final-viral-combined_k141_149485\|\|lt2gene | 1406 | No | NA | 2 | 2 | 0 | Low-quality | Genome-fragment | 2.85 | AAI-based (medium-confidence) | 0 |
| S1-final-viral-combined_k141_178244\|\|full | 13070 | No | NA | 12 | 0 | 9 | Not-determined | Genome-fragment | NA | NA | 0 |
| S1-final-viral-combined_k141_162364\|\|full | 1666 | No | NA | 3 | 0 | 0 | Not-determined | Genome-fragment | NA | NA | 0 |
| S1-final-viral-combined_k141_282570\|\|full | 1407 | No | NA | 5 | 1 | 0 | Low-quality | Genome-fragment | 2.54 | AAI-based (high-confidence) | 0 |
| S1-final-viral-combined_k141_208248\|\|full | 1407 | No | NA | 3 | 1 | 0 | Low-quality | Genome-fragment | 1.48 | HMM-based (lower-bound) | 0 |
| S1-final-viral-combined_k141_228729\|\|full | 3059 | No | NA | 6 | 0 | 1 | Not-determined | Genome-fragment | NA | NA | 0 |
| S1-final-viral-combined_k141_97138\|\|full | 1304 | No | NA | 2 | 0 | 1 | Not-determined | Genome-fragment | NA | NA | 0 |
| S1-final-viral-combined_k141_361004\|\|full | 2927 | No | NA | 3 | 0 | 0 | Not-determined | Genome-fragment | NA | NA | 0 |
| S1-final-viral-combined_k141_162420\|\|full | 1768 | No | NA | 2 | 0 | 1 | Not-determined | Genome-fragment | NA | NA | 0 |
| S1-final-viral-combined_k141_311738\|\|full | 5704 | No | NA | 5 | 0 | 3 | Not-determined | Genome-fragment | NA | NA | 0 |
| S1-final-viral-combined_k141_392782\|\|full | 1378 | No | NA | 2 | 0 | 1 | Not-determined | Genome-fragment | NA | NA | 0 |
| S7-final-viral-combined_k141_43126\|\|lt2gene | 1407 | No | NA | 1 | 1 | 0 | Low-quality | Genome-fragment | 3.59 | AAI-based (high-confidence) | 0 |
| S1-final-viral-combined_k141_262826\|\|full | 4131 | No | NA | 3 | 0 | 2 | Not-determined | Genome-fragment | NA | NA | 0 |
| S1-final-viral-combined_k141_262833\|\|full | 2349 | No | NA | 5 | 0 | 3 | Not-determined | Genome-fragment | NA | NA | 0 |
| S1-final-viral-combined_k141_262837\|\|full | 2044 | No | NA | 3 | 0 | 2 | Not-determined | Genome-fragment | NA | NA | 0 |
| S1-final-viral-combined_k141_361045\|\|full | 1143 | No | NA | 3 | 0 | 0 | Not-determined | Genome-fragment | NA | NA | 0 |
| S6-final-viral-combined_k141_8089\|\|full | 1407 | No | NA | 2 | 0 | 0 | Low-quality | Genome-fragment | 0.54 | AAI-based (high-confidence) | 0 |
| S6_k141_71719 | 1407 | No | NA | 2 | 0 | 0 | Low-quality | Genome-fragment | 4.53 | AAI-based (medium-confidence) | 0 |
| S8_k141_236701 | 1407 | No | NA | 2 | 1 | 0 | Low-quality | Genome-fragment | 1.1 | AAI-based (high-confidence) | 0 |
| S1-final-viral-combined_k141_131088\|\|full | 2101 | No | NA | 2 | 0 | 1 | Not-determined | Genome-fragment | NA | NA | 0 |
| S1-final-viral-combined_k141_293308\|\|full | 1408 | No | NA | 3 | 0 | 2 | Low-quality | Genome-fragment | 0.79 | AAI-based (medium-confidence) | 0 |
| S1-final-viral-combined_k141_229196\|\|full | 1408 | No | NA | 2 | 1 | 0 | Low-quality | Genome-fragment | 3.32 | AAI-based (high-confidence) | 0 |
| S1-final-viral-combined_k141_213093\|\|full | 2618 | No | NA | 3 | 0 | 2 | Not-determined | Genome-fragment | NA | NA | 0 |
| S1-final-viral-combined_k141_14924\|\|full | 1701 | No | NA | 3 | 0 | 2 | Not-determined | Genome-fragment | NA | NA | 0 |
| S7-final-viral-combined_k141_153946\|\|full | 1408 | No | NA | 3 | 0 | 0 | Low-quality | Genome-fragment | 1.32 | AAI-based (high-confidence) | 0 |
| S1-final-viral-combined_k141_311832\|\|full | 1962 | No | NA | 2 | 0 | 0 | Not-determined | Genome-fragment | NA | NA | 0 |
| S1-final-viral-combined_k141_131134\|\|full | 2251 | No | NA | 2 | 0 | 1 | Not-determined | Genome-fragment | NA | NA | 0 |
| S1-final-viral-combined_k141_146542\|\|full | 1289 | No | NA | 2 | 0 | 1 | Not-determined | Genome-fragment | NA | NA | 0 |
| S7-final-viral-combined_k141_189269\|\|full | 1408 | No | NA | 4 | 1 | 0 | Low-quality | Genome-fragment | 3.85 | AAI-based (medium-confidence) | 0 |
| S17-final-viral-combined_k141_39513\|\|full | 1408 | No | NA | 4 | 0 | 0 | Low-quality | Genome-fragment | 2.82 | AAI-based (high-confidence) | 0 |
| S17-final-viral-combined_k141_63315\|\|lt2gene | 1408 | No | NA | 1 | 1 | 0 | Low-quality | Genome-fragment | 2.06 | HMM-based (lower-bound) | 0 |
| S1-final-viral-combined_k141_277867\|\|full | 5089 | No | NA | 4 | 0 | 1 | Not-determined | Genome-fragment | NA | NA | 0 |
| S1-final-viral-combined_k141_277869\|\|full | 1217 | No | NA | 3 | 0 | 0 | Not-determined | Genome-fragment | NA | NA | 0 |
| S17-k141_98910 | 1408 | No | NA | 2 | 0 | 1 | Low-quality | Genome-fragment | 1.8 | AAI-based (high-confidence) | 0 |
| S2-final-viral-combined_k141_5247\|\|full | 1408 | No | NA | 2 | 0 | 2 | Low-quality | Genome-fragment | 2.16 | AAI-based (high-confidence) | 0 |
| S1-final-viral-combined_k141_178504\|\|full | 1005 | No | NA | 2 | 0 | 0 | Not-determined | Genome-fragment | NA | NA | 0 |
| S2_k141_230770 | 1408 | No | NA | 1 | 0 | 0 | Low-quality | Genome-fragment | 3.02 | AAI-based (high-confidence) | 0 |
| S9-final-viral-combined_k141_125114\|\|full | 1409 | No | NA | 3 | 2 | 0 | Low-quality | Genome-fragment | 2.77 | AAI-based (medium-confidence) | 0 |
| S1-final-viral-combined_k141_198889\|\|full | 1409 | No | NA | 3 | 1 | 0 | Low-quality | Genome-fragment | 3.73 | AAI-based (high-confidence) | 0 |
| S1-final-viral-combined_k141_95686\|\|full | 1409 | No | NA | 4 | 1 | 0 | Low-quality | Genome-fragment | 0.91 | HMM-based (lower-bound) | 0 |
| S3-final-viral-combined_k141_117599\|\|full | 1409 | No | NA | 3 | 0 | 0 | Low-quality | Genome-fragment | 30.22 | AAI-based (medium-confidence) | 0 |
| S1-final-viral-combined_k141_15006\|\|full | 2288 | No | NA | 2 | 0 | 1 | Not-determined | Genome-fragment | NA | NA | 0 |
| S1-final-viral-combined_k141_311924\|\|full | 1070 | No | NA | 2 | 0 | 2 | Not-determined | Genome-fragment | NA | NA | 0 |
| S1-final-viral-combined_k141_277956\|\|full | 1334 | No | NA | 3 | 0 | 1 | Not-determined | Genome-fragment | NA | NA | 0 |
| S1-final-viral-combined_k141_30797\|\|full | 2608 | No | NA | 2 | 0 | 2 | Not-determined | Genome-fragment | NA | NA | 0 |
| S5-final-viral-combined_k141_41053\|\|full | 1409 | No | NA | 2 | 0 | 1 | Low-quality | Genome-fragment | 3.41 | AAI-based (medium-confidence) | 0 |
| S1-final-viral-combined_k141_246071\|\|full | 1277 | No | NA | 2 | 0 | 2 | Not-determined | Genome-fragment | NA | NA | 0 |
| S1-final-viral-combined_k141_377005\|\|full | 5271 | No | NA | 9 | 0 | 5 | Not-determined | Genome-fragment | NA | NA | 0 |
| S1-final-viral-combined_k141_162599\|\|full | 1421 | No | NA | 2 | 0 | 2 | Not-determined | Genome-fragment | NA | NA | 0 |
| S7-final-viral-combined_k141_234312\|\|full | 1409 | No | NA | 2 | 0 | 0 | Low-quality | Genome-fragment | 2.92 | AAI-based (medium-confidence) | 0 |
| S17-final-viral-combined_k141_48340\|\|full | 1409 | No | NA | 4 | 1 | 0 | Low-quality | Genome-fragment | 3.07 | HMM-based (lower-bound) | 0 |
| S12-final-viral-combined_k141_323151\|\|full | 1409 | No | NA | 3 | 1 | 0 | Low-quality | Genome-fragment | 1.8 | AAI-based (high-confidence) | 0 |
| S12-final-viral-combined_k141_246838\|\|full | 1409 | No | NA | 2 | 0 | 1 | Low-quality | Genome-fragment | 0.89 | AAI-based (high-confidence) | 0 |
| S1-final-viral-combined_k141_196823\|\|full | 2800 | No | NA | 3 | 0 | 3 | Not-determined | Genome-fragment | NA | NA | 0 |
| S1-final-viral-combined_k141_246087\|\|full | 1274 | No | NA | 2 | 0 | 2 | Not-determined | Genome-fragment | NA | NA | 0 |
| S1-final-viral-combined_k141_196828\|\|full | 7360 | No | NA | 5 | 0 | 4 | Not-determined | Genome-fragment | NA | NA | 0 |
| S1-final-viral-combined_k141_343254\|\|full | 5265 | No | NA | 7 | 0 | 3 | Not-determined | Genome-fragment | NA | NA | 0 |
| S1-final-viral-combined_k141_263032\|\|full | 4804 | No | NA | 5 | 0 | 0 | Not-determined | Genome-fragment | NA | NA | 0 |
| S12-final-viral-combined_k141_50133\|\|full | 1409 | No | NA | 2 | 0 | 2 | Low-quality | Genome-fragment | 0.45 | AAI-based (medium-confidence) | 0 |
| S1-final-viral-combined_k141_46780\|\|full | 1133 | No | NA | 3 | 0 | 1 | Not-determined | Genome-fragment | NA | NA | 0 |
| S1-final-viral-combined_k141_377044\|\|full | 2306 | No | NA | 3 | 0 | 2 | Not-determined | Genome-fragment | NA | NA | 0 |
| S4-final-viral-combined_k141_17882\|\|lt2gene | 1409 | No | NA | 2 | 1 | 0 | Low-quality | Genome-fragment | 3.28 | AAI-based (medium-confidence) | 0 |
| S1-final-viral-combined_k141_343285\|\|full | 1188 | No | NA | 3 | 0 | 0 | Not-determined | Genome-fragment | NA | NA | 0 |
| S1-final-viral-combined_k141_79745\|\|full | 3058 | No | NA | 4 | 0 | 0 | Not-determined | Genome-fragment | NA | NA | 0 |
| S6-final-viral-combined_k141_21410\|\|full | 1409 | No | NA | 3 | 1 | 0 | Low-quality | Genome-fragment | 2.37 | HMM-based (lower-bound) | 0 |
| S8-final-viral-combined_k141_206698\|\|full | 1409 | No | NA | 4 | 0 | 0 | Low-quality | Genome-fragment | 2.93 | AAI-based (medium-confidence) | 0 |
| S8-final-viral-combined_k141_184955\|\|lt2gene | 1409 | No | NA | 2 | 2 | 0 | Low-quality | Genome-fragment | 2.36 | HMM-based (lower-bound) | 0 |
| S8_k141_169428 | 1409 | No | NA | 3 | 0 | 0 | Low-quality | Genome-fragment | 2.27 | AAI-based (high-confidence) | 0 |
| S1-final-viral-combined_k141_295384\|\|full | 3321 | No | NA | 3 | 0 | 3 | Not-determined | Genome-fragment | NA | NA | 0 |
| S1-final-viral-combined_k141_392866\|\|full | 2404 | No | NA | 2 | 0 | 1 | Not-determined | Genome-fragment | NA | NA | 0 |
| S9_k141_30140 | 1410 | No | NA | 3 | 1 | 0 | Low-quality | Genome-fragment | 2.87 | AAI-based (medium-confidence) | 0 |
| S1-final-viral-combined_k141_97358\|\|full | 4144 | No | NA | 3 | 0 | 1 | Not-determined | Genome-fragment | NA | NA | 0 |
| S5-final-viral-combined_k141_205934\|\|full | 1410 | No | NA | 6 | 3 | 0 | Low-quality | Genome-fragment | 4.4 | AAI-based (high-confidence) | 0 |
| S1-final-viral-combined_k141_343320\|\|full | 1332 | No | NA | 3 | 0 | 1 | Not-determined | Genome-fragment | NA | NA | 0 |
| S1-final-viral-combined_k141_131270\|\|full | 38955 | No | NA | 66 | 18 | 0 | High-quality | High-quality | 92.32 | AAI-based (high-confidence) | 0 |
| S1-final-viral-combined_k141_278071\|\|full | 2025 | No | NA | 2 | 0 | 1 | Not-determined | Genome-fragment | NA | NA | 0 |
| S1-final-viral-combined_k141_15111\|\|full | 11982 | No | NA | 11 | 0 | 9 | Not-determined | Genome-fragment | NA | NA | 0 |
| S17-final-viral-combined_k141_39129\|\|full | 1410 | No | NA | 3 | 0 | 0 | Low-quality | Genome-fragment | 4.46 | AAI-based (medium-confidence) | 0 |
| S17-k141_188948 | 1410 | No | NA | 4 | 1 | 0 | Low-quality | Genome-fragment | 4.34 | AAI-based (medium-confidence) | 0 |
| S1-final-viral-combined_k141_328479\|\|full | 6665 | No | NA | 4 | 0 | 2 | Not-determined | Genome-fragment | NA | NA | 0 |
| S1-final-viral-combined_k141_263102\|\|full | 1171 | No | NA | 2 | 0 | 2 | Not-determined | Genome-fragment | NA | NA | 0 |
| S1-final-viral-combined_k141_30961\|\|full | 8961 | No | NA | 10 | 0 | 5 | Not-determined | Genome-fragment | NA | NA | 0 |
| S1-final-viral-combined_k141_97406\|\|full | 6420 | No | NA | 4 | 0 | 3 | Not-determined | Genome-fragment | NA | NA | 0 |
| S17-k141_41652 | 1410 | No | NA | 2 | 0 | 2 | Low-quality | Genome-fragment | 0.79 | AAI-based (medium-confidence) | 0 |
| S12-final-viral-combined_k141_138893\|\|full | 1410 | No | NA | 3 | 0 | 0 | Low-quality | Genome-fragment | 3.59 | AAI-based (medium-confidence) | 0 |
| S1-final-viral-combined_k141_361307\|\|full | 2051 | No | NA | 4 | 0 | 0 | Not-determined | Genome-fragment | NA | NA | 0 |
| S4-final-viral-combined_k141_119987\|\|full | 1410 | No | NA | 4 | 0 | 0 | Low-quality | Genome-fragment | 2.04 | AAI-based (medium-confidence) | 0 |
| S1-final-viral-combined_k141_328503\|\|full | 7085 | No | NA | 4 | 0 | 3 | Not-determined | Genome-fragment | NA | NA | 0 |
| S4-final-viral-combined_k141_30509\|\|full | 1410 | No | NA | 4 | 0 | 0 | Low-quality | Genome-fragment | 2.72 | AAI-based (high-confidence) | 0 |
| S1-final-viral-combined_k141_97446\|\|full | 1581 | No | NA | 1 | 0 | 1 | Not-determined | Genome-fragment | NA | NA | 0 |
| S1-final-viral-combined_k141_162832\|\|full | 3707 | No | NA | 4 | 0 | 2 | Not-determined | Genome-fragment | NA | NA | 0 |
| S4_k141_58524 | 1410 | No | NA | 5 | 0 | 0 | Low-quality | Genome-fragment | 1.37 | AAI-based (medium-confidence) | 0 |
| S8-final-viral-combined_k141_247623\|\|full | 1410 | No | NA | 2 | 0 | 2 | Low-quality | Genome-fragment | 2.36 | AAI-based (medium-confidence) | 0 |
| S1-final-viral-combined_k141_295527\|\|full | 1996 | No | NA | 3 | 0 | 0 | Not-determined | Genome-fragment | NA | NA | 0 |
| S1-final-viral-combined_k141_79885\|\|full | 2466 | No | NA | 3 | 0 | 1 | Not-determined | Genome-fragment | NA | NA | 0 |
| S11-final-viral-combined_k141_2552\|\|full | 1411 | No | NA | 2 | 0 | 0 | Low-quality | Genome-fragment | 1.75 | AAI-based (high-confidence) | 0 |

| | | | | | | | | | | | |
|---|---|---|---|---|---|---|---|---|---|---|---|
| S1-final-viral-combined_k141_278178\|\|full | 2993 | No | NA | 2 | 0 | 2 | Not-determined | Genome-fragment | NA | NA | 0 |
| S1-final-viral-combined_k141_213333\|\|full | 21039 | No | NA | 13 | 0 | 7 | Not-determined | Genome-fragment | NA | NA | 0 |
| S11-final-viral-combined_k141_99632\|\|full | 1411 | No | NA | 2 | 0 | 0 | Low-quality | Genome-fragment | 1.86 | AAI-based (medium-confidence) | 0 |
| S1-final-viral-combined_k141_146820\|\|full | 1665 | No | NA | 3 | 0 | 1 | Not-determined | Genome-fragment | NA | NA | 0 |
| S1-final-viral-combined_k141_219971\|\|full | 1411 | No | NA | 3 | 0 | 1 | Low-quality | Genome-fragment | 2.55 | AAI-based (medium-confidence) | 0 |
| S17-final-viral-combined_k141_11887\|\|full | 1411 | No | NA | 2 | 0 | 0 | Low-quality | Genome-fragment | 2 | AAI-based (medium-confidence) | 0 |
| S1-final-viral-combined_k141_343426\|\|full | 2842 | No | NA | 3 | 0 | 3 | Not-determined | Genome-fragment | NA | NA | 0 |
| S1-final-viral-combined_k141_295578\|\|full | 1682 | No | NA | 2 | 0 | 1 | Not-determined | Genome-fragment | NA | NA | 0 |
| S12-final-viral-combined_k141_151482\|\|full | 1411 | No | NA | 5 | 0 | 0 | Low-quality | Genome-fragment | 3.9 | AAI-based (high-confidence) | 0 |
| S12-final-viral-combined_k141_220976\|\|full | 1411 | No | NA | 2 | 0 | 0 | Low-quality | Genome-fragment | 2.9 | AAI-based (medium-confidence) | 0 |
| S12-final-viral-combined_k141_21678\|\|full | 1411 | No | NA | 2 | 0 | 2 | Low-quality | Genome-fragment | 0.83 | AAI-based (medium-confidence) | 0 |
| S1-final-viral-combined_k141_131364\|\|full | 3594 | No | NA | 4 | 0 | 2 | Not-determined | Genome-fragment | NA | NA | 0 |
| S12-final-viral-combined_k141_276744\|\|full | 1411 | No | NA | 2 | 0 | 0 | Low-quality | Genome-fragment | 2.59 | AAI-based (high-confidence) | 0 |
| S1-final-viral-combined_k141_229228\|\|full | 1698 | No | NA | 2 | 0 | 1 | Not-determined | Genome-fragment | NA | NA | 0 |
| S1-final-viral-combined_k141_328545\|\|full | 1454 | No | NA | 2 | 0 | 1 | Not-determined | Genome-fragment | NA | NA | 0 |
| S1-final-viral-combined_k141_64440\|\|full | 2187 | No | NA | 5 | 0 | 3 | Not-determined | Genome-fragment | NA | NA | 0 |
| S1-final-viral-combined_k141_178932\|\|full | 4991 | No | NA | 6 | 0 | 4 | Not-determined | Genome-fragment | NA | NA | 0 |
| S12-final-viral-combined_k141_46389\|\|full | 1411 | No | NA | 1 | 0 | 0 | Low-quality | Genome-fragment | 1.71 | AAI-based (high-confidence) | 0 |
| S1-final-viral-combined_k141_213445\|\|full | 1736 | No | NA | 3 | 0 | 1 | Not-determined | Genome-fragment | NA | NA | 0 |
| S1-final-viral-combined_k141_15256\|\|full | 1758 | No | NA | 3 | 0 | 2 | Not-determined | Genome-fragment | NA | NA | 0 |
| S1-final-viral-combined_k141_377333\|\|full | 2265 | No | NA | 2 | 0 | 2 | Not-determined | Genome-fragment | NA | NA | 0 |
| S12_k141_396602 | 1411 | No | NA | 1 | 0 | 1 | Low-quality | Genome-fragment | 2.4 | AAI-based (high-confidence) | 0 |
| S1-final-viral-combined_k141_112235\|\|full | 1577 | No | NA | 2 | 0 | 1 | Not-determined | Genome-fragment | NA | NA | 0 |
| S2-final-viral-combined_k141_121736\|\|full | 1411 | No | NA | 3 | 0 | 0 | Low-quality | Genome-fragment | 1.65 | AAI-based (medium-confidence) | 0 |
| S1-final-viral-combined_k141_361494\|\|full | 3565 | No | NA | 3 | 0 | 3 | Not-determined | Genome-fragment | NA | NA | 0 |
| S2-final-viral-combined_k141_9760\|\|full | 1411 | No | NA | 2 | 0 | 1 | Low-quality | Genome-fragment | 1.84 | AAI-based (medium-confidence) | 0 |
| S1-final-viral-combined_k141_229306\|\|full | 2127 | No | NA | 3 | 0 | 0 | Not-determined | Genome-fragment | NA | NA | 0 |
| S1-final-viral-combined_k141_328594\|\|full | 1584 | No | NA | 2 | 0 | 1 | Not-determined | Genome-fragment | NA | NA | 0 |
| S1-final-viral-combined_k141_246266\|\|full | 1756 | No | NA | 3 | 0 | 2 | Not-determined | Genome-fragment | NA | NA | 0 |
| S1-final-viral-combined_k141_178979\|\|full | 3064 | No | NA | 3 | 0 | 2 | Not-determined | Genome-fragment | NA | NA | 0 |
| S1-final-viral-combined_k141_263231\|\|full | 9264 | No | NA | 11 | 0 | 3 | Not-determined | Genome-fragment | NA | NA | 0 |
| S1-final-viral-combined_k141_97684\|\|full | 1982 | No | NA | 3 | 0 | 1 | Not-determined | Genome-fragment | NA | NA | 0 |
| S1-final-viral-combined_k141_393095\|\|full | 2552 | No | NA | 3 | 0 | 1 | Not-determined | Genome-fragment | NA | NA | 0 |
| S1-final-viral-combined_k141_213472\|\|full | 1728 | No | NA | 3 | 0 | 1 | Not-determined | Genome-fragment | NA | NA | 0 |
| S1-final-viral-combined_k141_31166\|\|full | 1633 | No | NA | 3 | 0 | 0 | Not-determined | Genome-fragment | NA | NA | 0 |
| S2-final-viral-combined_k141_33223\|\|lt2gene | 1411 | No | NA | 1 | 1 | 0 | Low-quality | Genome-fragment | 4.78 | HMM-based (lower-bound) | 0 |
| S4_k141_157726 | 1411 | No | NA | 2 | 0 | 0 | Low-quality | Genome-fragment | 2.17 | AAI-based (medium-confidence) | 0 |
| S1-final-viral-combined_k141_31177\|\|full | 1327 | No | NA | 2 | 0 | 1 | Not-determined | Genome-fragment | NA | NA | 0 |
| S1-final-viral-combined_k141_229344\|\|full | 5530 | No | NA | 5 | 0 | 2 | Not-determined | Genome-fragment | NA | NA | 0 |
| S9-final-viral-combined_k141_45440\|\|full | 1412 | No | NA | 2 | 0 | 1 | Low-quality | Genome-fragment | 2.93 | AAI-based (medium-confidence) | 0 |
| S11-final-viral-combined_k141_263877\|\|full | 1412 | No | NA | 1 | 0 | 1 | Low-quality | Genome-fragment | 2.37 | AAI-based (high-confidence) | 0 |
| S11-final-viral-combined_k141_313222\|\|full | 1412 | No | NA | 2 | 0 | 1 | Low-quality | Genome-fragment | 1.92 | AAI-based (medium-confidence) | 0 |
| S11-final-viral-combined_k141_247574\|\|full | 1412 | No | NA | 4 | 0 | 0 | Low-quality | Genome-fragment | 2.19 | AAI-based (medium-confidence) | 0 |
| S1-final-viral-combined_k141_163070\|\|full | 6143 | No | NA | 10 | 0 | 0 | Not-determined | Genome-fragment | NA | NA | 0 |
| S11-final-viral-combined_k141_48295\|\|lt2gene | 1412 | No | NA | 1 | 1 | 0 | Low-quality | Genome-fragment | 2.47 | HMM-based (lower-bound) | 0 |
| S1-final-viral-combined_k141_112310\|\|full | 1788 | No | NA | 2 | 0 | 2 | Not-determined | Genome-fragment | NA | NA | 0 |
| S1-final-viral-combined_k141_64598\|\|full | 4833 | No | NA | 4 | 0 | 1 | Not-determined | Genome-fragment | NA | NA | 0 |
| S5-final-viral-combined_k141_10916\|\|full | 1412 | No | NA | 3 | 2 | 0 | Low-quality | Genome-fragment | 3.86 | AAI-based (high-confidence) | 0 |
| S7-final-viral-combined_k141_5962\|\|lt2gene | 1412 | No | NA | 2 | 1 | 0 | Low-quality | Genome-fragment | 0.81 | HMM-based (lower-bound) | 0 |
| S17-final-viral-combined_k141_263632\|\|lt2gene | 1412 | No | NA | 2 | 1 | 0 | Low-quality | Genome-fragment | 0.91 | HMM-based (lower-bound) | 0 |
| S1-final-viral-combined_k141_64632\|\|full | 1708 | No | NA | 3 | 0 | 0 | Not-determined | Genome-fragment | NA | NA | 0 |
| S12-final-viral-combined_k141_228435\|\|full | 1412 | No | NA | 3 | 2 | 0 | Low-quality | Genome-fragment | 2.26 | HMM-based (lower-bound) | 0 |
| S12-final-viral-combined_k141_283526\|\|full | 1412 | No | NA | 4 | 4 | 0 | Low-quality | Genome-fragment | 3.97 | AAI-based (medium-confidence) | 0 |
| S1-final-viral-combined_k141_64646\|\|full | 2442 | No | NA | 4 | 0 | 2 | Not-determined | Genome-fragment | NA | NA | 0 |
| S4-final-viral-combined_k141_121890\|\|lt2gene | 1412 | No | NA | 2 | 1 | 0 | Low-quality | Genome-fragment | 3.11 | AAI-based (high-confidence) | 0 |
| S1-final-viral-combined_k141_312273\|\|full | 4589 | No | NA | 5 | 0 | 3 | Not-determined | Genome-fragment | NA | NA | 0 |
| S1-final-viral-combined_k141_47220\|\|full | 3629 | No | NA | 3 | 0 | 2 | Not-determined | Genome-fragment | NA | NA | 0 |
| S1-final-viral-combined_k141_179105\|\|full | 2751 | No | NA | 4 | 0 | 1 | Not-determined | Genome-fragment | NA | NA | 0 |
| S1-final-viral-combined_k141_377516\|\|full | 4950 | No | NA | 4 | 0 | 4 | Not-determined | Genome-fragment | NA | NA | 0 |
| S4_k141_116765 | 1412 | No | NA | 3 | 0 | 0 | Low-quality | Genome-fragment | 1.93 | AAI-based (medium-confidence) | 0 |
| S9-final-viral-combined_k141_607\|\|full | 1413 | No | NA | 2 | 1 | 0 | Low-quality | Genome-fragment | 0.84 | AAI-based (medium-confidence) | 0 |
| S11-final-viral-combined_k141_126391\|\|full | 1413 | No | NA | 4 | 1 | 0 | Low-quality | Genome-fragment | 1.93 | AAI-based (high-confidence) | 0 |
| S11-final-viral-combined_k141_45066\|\|full | 1413 | No | NA | 3 | 0 | 0 | Low-quality | Genome-fragment | 2.99 | AAI-based (high-confidence) | 0 |
| S11-final-viral-combined_k141_87218\|\|full | 1413 | No | NA | 2 | 0 | 0 | Low-quality | Genome-fragment | 1.13 | AAI-based (high-confidence) | 0 |
| S1-final-viral-combined_k141_393186\|\|full | 1084 | No | NA | 2 | 0 | 0 | Not-determined | Genome-fragment | NA | NA | 0 |
| S11-final-viral-combined_k141_10579\|\|full | 1413 | No | NA | 2 | 0 | 0 | Low-quality | Genome-fragment | 0.99 | AAI-based (medium-confidence) | 0 |
| S11_k141_217532 | 1413 | No | NA | 3 | 1 | 0 | Low-quality | Genome-fragment | 33.95 | HMM-based (lower-bound) | 0 |
| S1-final-viral-combined_k141_47266\|\|full | 1802 | No | NA | 3 | 0 | 0 | Not-determined | Genome-fragment | NA | NA | 0 |
| S1-final-viral-combined_k141_264703\|\|full | 1413 | No | NA | 3 | 0 | 0 | Low-quality | Genome-fragment | 3.93 | AAI-based (high-confidence) | 0 |
| S1-final-viral-combined_k141_213652\|\|full | 1610 | No | NA | 4 | 0 | 2 | Not-determined | Genome-fragment | NA | NA | 0 |
| S1-final-viral-combined_k141_156495\|\|full | 1413 | No | NA | 4 | 0 | 0 | Low-quality | Genome-fragment | 1.04 | AAI-based (medium-confidence) | 0 |
| S1-final-viral-combined_k141_97901\|\|full | 3768 | No | NA | 4 | 0 | 2 | Not-determined | Genome-fragment | NA | NA | 0 |
| S5-final-viral-combined_k141_43177\|\|full | 1413 | No | NA | 4 | 1 | 0 | Low-quality | Genome-fragment | 3.2 | AAI-based (high-confidence) | 0 |
| S1-final-viral-combined_k141_328849\|\|full | 5550 | No | NA | 5 | 0 | 3 | Not-determined | Genome-fragment | NA | NA | 0 |
| S7_k141_33379 | 1413 | No | NA | 3 | 1 | 0 | Low-quality | Genome-fragment | 3.36 | AAI-based (high-confidence) | 0 |
| S1-final-viral-combined_k141_163172\|\|full | 3059 | No | NA | 3 | 0 | 1 | Not-determined | Genome-fragment | NA | NA | 0 |
| S17-final-viral-combined_k141_165682\|\|full | 1413 | No | NA | 3 | 0 | 2 | Low-quality | Genome-fragment | 0.93 | AAI-based (medium-confidence) | 0 |
| S12_k141_159583 | 1413 | No | NA | 2 | 0 | 0 | Low-quality | Genome-fragment | 0.65 | AAI-based (high-confidence) | 0 |
| S1-final-viral-combined_k141_295836\|\|full | 9231 | No | NA | 9 | 0 | 7 | Not-determined | Genome-fragment | NA | NA | 0 |
| S12_k141_384040 | 1413 | No | NA | 3 | 0 | 1 | Low-quality | Genome-fragment | 2.49 | AAI-based (high-confidence) | 0 |
| S1-final-viral-combined_k141_15568\|\|full | 5924 | No | NA | 4 | 0 | 4 | Not-determined | Genome-fragment | NA | NA | 0 |
| S1-final-viral-combined_k141_147093\|\|full | 5411 | No | NA | 8 | 0 | 1 | Not-determined | Genome-fragment | NA | NA | 0 |
| S2-final-viral-combined_k141_190714\|\|full | 1413 | Yes | 861 | 3 | 1 | 1 | Low-quality | Genome-fragment | 0.35 | HMM-based (lower-bound) | 39.07 |
| S1-final-viral-combined_k141_343723\|\|full | 3737 | No | NA | 4 | 0 | 3 | Not-determined | Genome-fragment | NA | NA | 0 |
| S1-final-viral-combined_k141_179302\|\|full | 1280 | No | NA | 2 | 0 | 1 | Not-determined | Genome-fragment | NA | NA | 0 |
| S1-final-viral-combined_k141_131545\|\|full | 3674 | No | NA | 4 | 0 | 4 | Not-determined | Genome-fragment | NA | NA | 0 |
| S1-final-viral-combined_k141_15606\|\|full | 3910 | No | NA | 5 | 0 | 2 | Not-determined | Genome-fragment | NA | NA | 0 |
| S4-final-viral-combined_k141_45389\|\|full | 1413 | No | NA | 3 | 2 | 0 | Low-quality | Genome-fragment | 2.64 | HMM-based (lower-bound) | 0 |
| S1-final-viral-combined_k141_278337\|\|full | 1813 | No | NA | 2 | 0 | 1 | Not-determined | Genome-fragment | NA | NA | 0 |
| S4-final-viral-combined_k141_40460\|\|full | 1413 | No | NA | 2 | 0 | 0 | Low-quality | Genome-fragment | 3.46 | AAI-based (high-confidence) | 0 |
| S11-final-viral-combined_k141_223232\|\|full | 1414 | No | NA | 2 | 1 | 0 | Low-quality | Genome-fragment | 2.24 | AAI-based (medium-confidence) | 0 |
| S1-final-viral-combined_k141_163265\|\|full | 3138 | No | NA | 3 | 0 | 3 | Not-determined | Genome-fragment | NA | NA | 0 |
| S1-final-viral-combined_k141_98058\|\|full | 4343 | No | NA | 7 | 0 | 5 | Not-determined | Genome-fragment | NA | NA | 0 |
| S11-final-viral-combined_k141_294429\|\|full | 1414 | No | NA | 2 | 0 | 1 | Low-quality | Genome-fragment | 2.44 | AAI-based (medium-confidence) | 0 |
| S1-final-viral-combined_k141_64844\|\|full | 1754 | No | NA | 3 | 0 | 2 | Not-determined | Genome-fragment | NA | NA | 0 |
| S11_k141_263031 | 1414 | No | NA | 3 | 1 | 0 | Low-quality | Genome-fragment | 2.52 | AAI-based (high-confidence) | 0 |
| S1-final-viral-combined_k141_39170\|\|full | 1414 | No | NA | 2 | 0 | 1 | Low-quality | Genome-fragment | 2.57 | AAI-based (medium-confidence) | 0 |
| S1-final-viral-combined_k141_80424\|\|full | 1240 | No | NA | 2 | 0 | 1 | Not-determined | Genome-fragment | NA | NA | 0 |
| S1-final-viral-combined_k141_278405\|\|full | 14224 | No | NA | 14 | 0 | 11 | Not-determined | Genome-fragment | NA | NA | 0 |
| S1-final-viral-combined_k141_312504\|\|full | 7191 | No | NA | 3 | 0 | 0 | Not-determined | Genome-fragment | NA | NA | 0 |
| S5-final-viral-combined_k141_113617\|\|full | 1414 | No | NA | 4 | 2 | 0 | Low-quality | Genome-fragment | 3.55 | AAI-based (high-confidence) | 0 |
| S17-final-viral-combined_k141_141200\|\|full | 1414 | No | NA | 3 | 0 | 0 | Low-quality | Genome-fragment | 0.78 | AAI-based (high-confidence) | 0 |
| S17-final-viral-combined_k141_84342\|\|lt2gene | 1414 | No | NA | 2 | 1 | 0 | Low-quality | Genome-fragment | 0.87 | HMM-based (lower-bound) | 0 |
| S1-final-viral-combined_k141_312515\|\|full | 1665 | No | NA | 2 | 0 | 2 | Not-determined | Genome-fragment | NA | NA | 0 |
| S1-final-viral-combined_k141_197351\|\|full | 1492 | No | NA | 2 | 0 | 1 | Not-determined | Genome-fragment | NA | NA | 0 |
| S1-final-viral-combined_k141_361735\|\|full | 2477 | No | NA | 2 | 0 | 0 | Not-determined | Genome-fragment | NA | NA | 0 |
| S17-k141_222205 | 1414 | No | NA | 2 | 0 | 1 | Low-quality | Genome-fragment | 2.47 | AAI-based (medium-confidence) | 0 |
| S2_k141_64847 | 1414 | No | NA | 1 | 0 | 0 | Low-quality | Genome-fragment | 0.76 | AAI-based (medium-confidence) | 0 |
| S4-final-viral-combined_k141_151552\|\|full | 1414 | No | NA | 5 | 1 | 0 | Low-quality | Genome-fragment | 2.13 | HMM-based (lower-bound) | 0 |
| S1-final-viral-combined_k141_31355\|\|full | 2282 | No | NA | 2 | 0 | 1 | Not-determined | Genome-fragment | NA | NA | 0 |
| S1-final-viral-combined_k141_361752\|\|full | 6111 | No | NA | 14 | 0 | 0 | Not-determined | Genome-fragment | NA | NA | 0 |
| S8-final-viral-combined_k141_211075\|\|full | 1414 | No | NA | 3 | 0 | 2 | Low-quality | Genome-fragment | 2.17 | AAI-based (high-confidence) | 0 |
| S11_k141_124461 | 1415 | No | NA | 4 | 0 | 0 | Low-quality | Genome-fragment | 1.5 | AAI-based (high-confidence) | 0 |
| S1-final-viral-combined_k141_64942\|\|full | 2878 | No | NA | 4 | 0 | 3 | Not-determined | Genome-fragment | NA | NA | 0 |
| S1-final-viral-combined_k141_229531\|\|full | 1339 | No | NA | 3 | 0 | 1 | Not-determined | Genome-fragment | NA | NA | 0 |
| S1-final-viral-combined_k141_197382\|\|full | 2266 | No | NA | 5 | 0 | 0 | Not-determined | Genome-fragment | NA | NA | 0 |
| S1-final-viral-combined_k141_31407\|\|full | 1428 | No | NA | 2 | 0 | 1 | Not-determined | Genome-fragment | NA | NA | 0 |
| S1-final-viral-combined_k141_15714\|\|full | 1355 | No | NA | 3 | 0 | 3 | Not-determined | Genome-fragment | NA | NA | 0 |
| S1-final-viral-combined_k141_181844\|\|lt2gene | 1415 | No | NA | 2 | 0 | 0 | Low-quality | Genome-fragment | 2.08 | AAI-based (medium-confidence) | 0 |
| S1-final-viral-combined_k141_64969\|\|full | 6637 | No | NA | 5 | 0 | 3 | Not-determined | Genome-fragment | NA | NA | 0 |
| S5_k141_79694 | 1415 | No | NA | 3 | 0 | 0 | Low-quality | Genome-fragment | 5.63 | AAI-based (high-confidence) | 0 |
| S1-final-viral-combined_k141_343831\|\|full | 10144 | No | NA | 7 | 0 | 6 | Not-determined | Genome-fragment | NA | NA | 0 |
| S7-final-viral-combined_k141_155719\|\|full | 1415 | No | NA | 3 | 1 | 0 | Low-quality | Genome-fragment | 3.43 | AAI-based (high-confidence) | 0 |
| S17-final-viral-combined_k141_188874\|\|lt2gene | 1415 | No | NA | 2 | 0 | 0 | Low-quality | Genome-fragment | 3.45 | AAI-based (medium-confidence) | 0 |
| S1-final-viral-combined_k141_377814\|\|full | 1699 | No | NA | 3 | 0 | 0 | Not-determined | Genome-fragment | NA | NA | 0 |
| S1-final-viral-combined_k141_163363\|\|full | 1845 | No | NA | 2 | 0 | 0 | Not-determined | Genome-fragment | NA | NA | 0 |
| S12-final-viral-combined_k141_51066\|\|full | 1415 | No | NA | 3 | 1 | 0 | Low-quality | Genome-fragment | 4.16 | AAI-based (high-confidence) | 0 |
| S1-final-viral-combined_k141_197401\|\|full | 53457 | No | NA | 66 | 4 | 4 | Medium-quality | Genome-fragment | 76.98 | HMM-based (lower-bound) | 0 |
| S12-final-viral-combined_k141_34755\|\|full | 1415 | No | NA | 3 | 0 | 2 | Low-quality | Genome-fragment | 1.33 | AAI-based (medium-confidence) | 0 |
| S2-final-viral-combined_k141_197665\|\|full | 1415 | No | NA | 2 | 0 | 0 | Low-quality | Genome-fragment | 2.44 | AAI-based (high-confidence) | 0 |
| S1-final-viral-combined_k141_296047\|\|full | 2583 | No | NA | 3 | 0 | 0 | Not-determined | Genome-fragment | NA | NA | 0 |
| S2-final-viral-combined_k141_127633\|\|full | 1415 | No | NA | 4 | 2 | 0 | Low-quality | Genome-fragment | 3.23 | AAI-based (high-confidence) | 0 |
| S1-final-viral-combined_k141_179395\|\|full | 1544 | No | NA | 2 | 0 | 1 | Not-determined | Genome-fragment | NA | NA | 0 |
| S4_k141_130909 | 1415 | No | NA | 4 | 0 | 0 | Low-quality | Genome-fragment | 3.59 | AAI-based (high-confidence) | 0 |
| S1-final-viral-combined_k141_163388\|\|full | 2196 | No | NA | 3 | 0 | 2 | Not-determined | Genome-fragment | NA | NA | 0 |
| S1-final-viral-combined_k141_361829\|\|full | 6103 | No | NA | 5 | 0 | 0 | Not-determined | Genome-fragment | NA | NA | 0 |
| S9-final-viral-combined_k141_18144\|\|full | 1416 | No | NA | 2 | 0 | 0 | Low-quality | Genome-fragment | 2.77 | AAI-based (medium-confidence) | 0 |
| S11-final-viral-combined_k141_321652\|\|full | 1416 | No | NA | 1 | 0 | 1 | Low-quality | Genome-fragment | 1.65 | AAI-based (high-confidence) | 0 |
| S11-final-viral-combined_k141_227464\|\|full | 1416 | No | NA | 2 | 1 | 0 | Low-quality | Genome-fragment | 2.38 | HMM-based (lower-bound) | 0 |
| S1-final-viral-combined_k141_163434\|\|full | 1415 | No | NA | 3 | 0 | 1 | Not-determined | Genome-fragment | NA | NA | 0 |
| S1-final-viral-combined_k141_147332\|\|full | 1690 | No | NA | 3 | 0 | 1 | Not-determined | Genome-fragment | NA | NA | 0 |
| S1_k141_197509 | 1416 | No | NA | 1 | 0 | 1 | Low-quality | Genome-fragment | 2.39 | AAI-based (high-confidence) | 0 |
| S3-final-viral-combined_k141_59119\|\|full | 1416 | No | NA | 3 | 0 | 0 | Low-quality | Genome-fragment | 1.13 | AAI-based (medium-confidence) | 0 |
| S1-final-viral-combined_k141_15827\|\|full | 2315 | No | NA | 3 | 0 | 1 | Not-determined | Genome-fragment | NA | NA | 0 |
| S1-final-viral-combined_k141_377904\|\|full | 1493 | No | NA | 2 | 0 | 1 | Not-determined | Genome-fragment | NA | NA | 0 |
| S1-final-viral-combined_k141_47549\|\|full | 1457 | No | NA | 3 | 0 | 0 | Not-determined | Genome-fragment | NA | NA | 0 |
| S12-final-viral-combined_k141_136425\|\|full | 1416 | No | NA | 2 | 0 | 1 | Low-quality | Genome-fragment | 1.22 | AAI-based (medium-confidence) | 0 |
| S4_k141_99721 | 1416 | No | NA | 2 | 0 | 2 | Low-quality | Genome-fragment | 0.34 | AAI-based (medium-confidence) | 0 |
| S6-final-viral-combined_k141_136388\|\|full | 1416 | No | NA | 2 | 0 | 1 | Low-quality | Genome-fragment | 3.02 | AAI-based (medium-confidence) | 0 |
| S1-final-viral-combined_k141_278676\|\|full | 4967 | No | NA | 7 | 0 | 0 | Not-determined | Genome-fragment | NA | NA | 0 |
| S8_k141_228984 | 1416 | No | NA | 2 | 0 | 0 | Low-quality | Genome-fragment | 2.69 | AAI-based (high-confidence) | 0 |
| S1-final-viral-combined_k141_361884\|\|full | 5842 | No | NA | 5 | 0 | 2 | Not-determined | Genome-fragment | NA | NA | 0 |
| S1-final-viral-combined_k141_31461\|\|full | 6984 | No | NA | 7 | 0 | 5 | Not-determined | Genome-fragment | NA | NA | 0 |
| S11-final-viral-combined_k141_240644\|\|lt2gene | 1417 | No | NA | 2 | 2 | 0 | Low-quality | Genome-fragment | 2.2 | AAI-based (high-confidence) | 0 |
| S11_k141_273849 | 1417 | No | NA | 2 | 0 | 0 | Low-quality | Genome-fragment | 0.92 | AAI-based (high-confidence) | 0 |
| S1-final-viral-combined_k141_19104\|\|full | 1417 | No | NA | 2 | 0 | 0 | Low-quality | Genome-fragment | 2.98 | AAI-based (medium-confidence) | 0 |
| S1-final-viral-combined_k141_19507\|\|full | 1417 | No | NA | 3 | 0 | 1 | Low-quality | Genome-fragment | 4.04 | AAI-based (medium-confidence) | 0 |
| S1-final-viral-combined_k141_112660\|\|full | 2902 | No | NA | 4 | 0 | 3 | Not-determined | Genome-fragment | NA | NA | 0 |
| S1-final-viral-combined_k141_163526\|\|full | 2018 | No | NA | 3 | 0 | 0 | Not-determined | Genome-fragment | NA | NA | 0 |
| S5-final-viral-combined_k141_155366\|\|full | 1417 | No | NA | 3 | 0 | 2 | Low-quality | Genome-fragment | 2.87 | AAI-based (medium-confidence) | 0 |
| S5-final-viral-combined_k141_24199\|\|full | 1417 | No | NA | 3 | 0 | 2 | Low-quality | Genome-fragment | 1.16 | AAI-based (medium-confidence) | 0 |
| S1-final-viral-combined_k141_31487\|\|full | 2413 | No | NA | 3 | 0 | 2 | Not-determined | Genome-fragment | NA | NA | 0 |
| S17-final-viral-combined_k141_230847\|\|full | 1417 | No | NA | 3 | 1 | 0 | Low-quality | Genome-fragment | 2.1 | AAI-based (high-confidence) | 0 |
| S2-final-viral-combined_k141_115757\|\|full | 1417 | No | NA | 3 | 0 | 1 | Low-quality | Genome-fragment | 3.09 | AAI-based (medium-confidence) | 0 |
| S11-final-viral-combined_k141_138732\|\|lt2gene | 1418 | No | NA | 2 | 0 | 0 | Low-quality | Genome-fragment | 3 | AAI-based (high-confidence) | 0 |
| S1-final-viral-combined_k141_15947\|\|full | 1432 | No | NA | 4 | 0 | 0 | Not-determined | Genome-fragment | NA | NA | 0 |
| S11_k141_257500 | 1418 | No | NA | 1 | 0 | 1 | Low-quality | Genome-fragment | 0.4 | AAI-based (high-confidence) | 0 |
| S1-final-viral-combined_k141_62595\|\|full | 1418 | No | NA | 3 | 0 | 0 | Low-quality | Genome-fragment | 3.26 | AAI-based (medium-confidence) | 0 |
| S1-final-viral-combined_k141_278812\|\|full | 5625 | No | NA | 6 | 0 | 2 | Not-determined | Genome-fragment | NA | NA | 0 |
| S1-final-viral-combined_k141_278018\|\|full | 2935 | No | NA | 3 | 0 | 3 | Not-determined | Genome-fragment | NA | NA | 0 |
| S1-final-viral-combined_k141_246683\|\|full | 3237 | No | NA | 4 | 0 | 0 | Not-determined | Genome-fragment | NA | NA | 0 |
| S3-final-viral-combined_k141_100413\|\|lt2gene | 1418 | No | NA | 2 | 1 | 0 | Low-quality | Genome-fragment | 3.21 | AAI-based (medium-confidence) | 0 |
| S1-final-viral-combined_k141_278831\|\|full | 2926 | No | NA | 3 | 0 | 1 | Not-determined | Genome-fragment | NA | NA | 0 |
| S17-final-viral-combined_k141_201383\|\|full | 1418 | No | NA | 2 | 0 | 1 | Low-quality | Genome-fragment | 0.54 | AAI-based (medium-confidence) | 0 |
| S1-final-viral-combined_k141_246694\|\|full | 10495 | No | NA | 6 | 0 | 5 | Not-determined | Genome-fragment | NA | NA | 0 |
| S1-final-viral-combined_k141_163625\|\|full | 1814 | No | NA | 3 | 0 | 1 | Not-determined | Genome-fragment | NA | NA | 0 |
| S1-final-viral-combined_k141_393651\|\|full | 3295 | No | NA | 2 | 0 | 2 | Not-determined | Genome-fragment | NA | NA | 0 |
| S1-final-viral-combined_k141_47745\|\|full | 1241 | No | NA | 2 | 0 | 2 | Not-determined | Genome-fragment | NA | NA | 0 |
| S1-final-viral-combined_k141_112803\|\|full | 2216 | No | NA | 3 | 0 | 0 | Not-determined | Genome-fragment | NA | NA | 0 |
| S1-final-viral-combined_k141_147553\|\|full | 2070 | No | NA | 3 | 0 | 2 | Not-determined | Genome-fragment | NA | NA | 0 |
| S17-final-viral-combined_k141_179834\|\|full | 1418 | No | NA | 3 | 0 | 0 | Low-quality | Genome-fragment | 1.4 | AAI-based (medium-confidence) | 0 |
| S1-final-viral-combined_k141_147557\|\|full | 1515 | No | NA | 2 | 0 | 1 | Not-determined | Genome-fragment | NA | NA | 0 |
| S1-final-viral-combined_k141_344130\|\|full | 4503 | No | NA | 3 | 0 | 2 | Not-determined | Genome-fragment | NA | NA | 0 |
| S17-k141_21015 | 1418 | No | NA | 2 | 0 | 0 | Low-quality | Genome-fragment | 2.54 | AAI-based (medium-confidence) | 0 |
| S1-final-viral-combined_k141_112833\|\|full | 3460 | No | NA | 5 | 0 | 5 | Not-determined | Genome-fragment | NA | NA | 0 |
| S1-final-viral-combined_k141_278877\|\|full | 1488 | No | NA | 3 | 0 | 0 | Not-determined | Genome-fragment | NA | NA | 0 |
| S1-final-viral-combined_k141_163677\|\|full | 1944 | No | NA | 3 | 0 | 1 | Not-determined | Genome-fragment | NA | NA | 0 |
| S1-final-viral-combined_k141_229765\|\|full | 1745 | No | NA | 2 | 0 | 2 | Not-determined | Genome-fragment | NA | NA | 0 |
| S12-final-viral-combined_k141_355952\|\|full | 1418 | No | NA | 2 | 0 | 1 | Low-quality | Genome-fragment | 0.72 | AAI-based (medium-confidence) | 0 |
| S1-final-viral-combined_k141_393680\|\|full | 2520 | No | NA | 3 | 0 | 3 | Not-determined | Genome-fragment | NA | NA | 0 |
| S12-final-viral-combined_k141_164783\|\|full | 1418 | No | NA | 3 | 0 | 2 | Low-quality | Genome-fragment | 0.77 | AAI-based (medium-confidence) | 0 |
| S1-final-viral-combined_k141_393733\|\|full | 1566 | No | NA | 2 | 0 | 1 | Not-determined | Genome-fragment | NA | NA | 0 |
| S12_k141_153770 | 1418 | No | NA | 2 | 0 | 1 | Low-quality | Genome-fragment | 1.01 | AAI-based (high-confidence) | 0 |
| S2_k141_150337 | 1418 | No | NA | 3 | 0 | 2 | Low-quality | Genome-fragment | 2.8 | AAI-based (medium-confidence) | 0 |
| S1-final-viral-combined_k141_112909\|\|full | 8288 | No | NA | 9 | 0 | 1 | Not-determined | Genome-fragment | NA | NA | 0 |
| S4_k141_134795 | 1418 | No | NA | 4 | 0 | 0 | Low-quality | Genome-fragment | 3.9 | AAI-based (high-confidence) | 0 |
| S4_k141_3573 | 1418 | No | NA | 3 | 0 | 0 | Low-quality | Genome-fragment | 3.66 | AAI-based (medium-confidence) | 0 |
| S4_k141_66216 | 1418 | No | NA | 3 | 0 | 1 | Low-quality | Genome-fragment | 1.72 | AAI-based (high-confidence) | 0 |
| S1-final-viral-combined_k141_65340\|\|full | 2116 | No | NA | 5 | 0 | 2 | Not-determined | Genome-fragment | NA | NA | 0 |
| S1-final-viral-combined_k141_112929\|\|full | 1558 | No | NA | 2 | 0 | 1 | Not-determined | Genome-fragment | NA | NA | 0 |
| S1-final-viral-combined_k141_229859\|\|full | 2572 | No | NA | 3 | 0 | 2 | Not-determined | Genome-fragment | NA | NA | 0 |
| S9-final-viral-combined_k141_116329\|\|full | 1419 | No | NA | 3 | 0 | 1 | Low-quality | Genome-fragment | 2.26 | AAI-based (medium-confidence) | 0 |
| S9_k141_106044 | 1419 | No | NA | 1 | 0 | 1 | Low-quality | Genome-fragment | 3.28 | AAI-based (medium-confidence) | 0 |
| S11-final-viral-combined_k141_65485\|\|full | 1419 | No | NA | 3 | 0 | 1 | Low-quality | Genome-fragment | 1.08 | AAI-based (medium-confidence) | 0 |
| S1-final-viral-combined_k141_16114\|\|full | 1347 | No | NA | 2 | 0 | 1 | Not-determined | Genome-fragment | NA | NA | 0 |
| S1-final-viral-combined_k141_344339\|\|full | 1223 | No | NA | 3 | 0 | 0 | Not-determined | Genome-fragment | NA | NA | 0 |
| S1-final-viral-combined_k141_112967\|\|full | 2238 | No | NA | 4 | 0 | 4 | Not-determined | Genome-fragment | NA | NA | 0 |
| S5-final-viral-combined_k141_29255\|\|full | 1419 | No | NA | 3 | 2 | 0 | Low-quality | Genome-fragment | 2.34 | AAI-based (high-confidence) | 0 |
| S1-final-viral-combined_k141_47967\|\|full | 1727 | No | NA | 3 | 0 | 0 | Not-determined | Genome-fragment | NA | NA | 0 |
| S1-final-viral-combined_k141_16175\|\|full | 3217 | No | NA | 3 | 0 | 2 | Not-determined | Genome-fragment | NA | NA | 0 |
| S17-final-viral-combined_k141_27309\|\|full | 1419 | No | NA | 3 | 0 | 0 | Low-quality | Genome-fragment | 0.69 | AAI-based (medium-confidence) | 0 |
| S17-k141_258737 | 1419 | No | NA | 4 | 1 | 0 | Low-quality | Genome-fragment | 1.87 | AAI-based (high-confidence) | 0 |
| S1-final-viral-combined_k141_393855\|\|full | 5875 | No | NA | 7 | 0 | 2 | Not-determined | Genome-fragment | NA | NA | 0 |
| S6-final-viral-combined_k141_65946\|\|full | 1419 | No | NA | 3 | 0 | 0 | Low-quality | Genome-fragment | 2.79 | AAI-based (medium-confidence) | 0 |
| S6-final-viral-combined_k141_104907\|\|full | 1419 | No | NA | 3 | 2 | 0 | Low-quality | Genome-fragment | 3.11 | AAI-based (medium-confidence) | 0 |
| S1-final-viral-combined_k141_81266\|\|full | 3600 | No | NA | 3 | 0 | 2 | Not-determined | Genome-fragment | NA | NA | 0 |
| S1-final-viral-combined_k141_393889\|\|full | 3128 | No | NA | 5 | 0 | 2 | Not-determined | Genome-fragment | NA | NA | 0 |
| S9-final-viral-combined_k141_121686\|\|full | 1420 | No | NA | 3 | 2 | 0 | Low-quality | Genome-fragment | 3.02 | AAI-based (medium-confidence) | 0 |
| S1-final-viral-combined_k141_230006\|\|full | 2257 | No | NA | 2 | 0 | 1 | Not-determined | Genome-fragment | NA | NA | 0 |
| S11-final-viral-combined_k141_83043\|\|full | 1420 | No | NA | 3 | 2 | 0 | Low-quality | Genome-fragment | 3.06 | AAI-based (medium-confidence) | 0 |
| S1-final-viral-combined_k141_393911\|\|full | 3184 | No | NA | 3 | 0 | 2 | Not-determined | Genome-fragment | NA | NA | 0 |
| S11-final-viral-combined_k141_79313\|\|full | 1420 | No | NA | 2 | 0 | 0 | Low-quality | Genome-fragment | 1.26 | AAI-based (medium-confidence) | 0 |
| S1-final-viral-combined_k141_300391\|\|full | 1420 | No | NA | 3 | 1 | 0 | Low-quality | Genome-fragment | 2.93 | AAI-based (medium-confidence) | 0 |
| S1-final-viral-combined_k141_98643\|\|full | 5861 | No | NA | 7 | 0 | 1 | Not-determined | Genome-fragment | NA | NA | 0 |
| S1-final-viral-combined_k141_356848\|\|full | 1420 | No | NA | 2 | 0 | 1 | Low-quality | Genome-fragment | 2.4 | AAI-based (medium-confidence) | 0 |
| S1-final-viral-combined_k141_53379\|\|lt2gene | 1420 | No | NA | 2 | 2 | 0 | Low-quality | Genome-fragment | 3.76 | AAI-based (high-confidence) | 0 |
| S7-final-viral-combined_k141_177496\|\|full | 1420 | No | NA | 4 | 1 | 0 | Low-quality | Genome-fragment | 2.19 | HMM-based (lower-bound) | 0 |
| S1-final-viral-combined_k141_344487\|\|full | 2801 | No | NA | 3 | 0 | 1 | Not-determined | Genome-fragment | NA | NA | 0 |
| S1-final-viral-combined_k141_98669\|\|full | 1048 | No | NA | 1 | 0 | 0 | Not-determined | Genome-fragment | NA | NA | 0 |
| S7_k141_59063 | 1420 | No | NA | 5 | 0 | 0 | Low-quality | Genome-fragment | 3.45 | AAI-based (medium-confidence) | 0 |
| S17-final-viral-combined_k141_38196\|\|full | 1420 | No | NA | 2 | 0 | 0 | Low-quality | Genome-fragment | 4.01 | AAI-based (medium-confidence) | 0 |
| S1-final-viral-combined_k141_344538\|\|full | 5812 | No | NA | 3 | 0 | 2 | Not-determined | Genome-fragment | NA | NA | 0 |
| S1-final-viral-combined_k141_65577\|\|full | 2304 | No | NA | 2 | 0 | 2 | Not-determined | Genome-fragment | NA | NA | 0 |
| S1-final-viral-combined_k141_230112\|\|full | 2050 | No | NA | 1 | 0 | 1 | Not-determined | Genome-fragment | NA | NA | 0 |
| S1-final-viral-combined_k141_98682\|\|full | 1334 | No | NA | 2 | 0 | 0 | Not-determined | Genome-fragment | NA | NA | 0 |
| S1-final-viral-combined_k141_344559\|\|full | 10060 | No | NA | 5 | 0 | 1 | Not-determined | Genome-fragment | NA | NA | 0 |
| S1-final-viral-combined_k141_394044\|\|full | 1624 | No | NA | 2 | 0 | 1 | Not-determined | Genome-fragment | NA | NA | 0 |
| S2_k141_89273 | 1420 | No | NA | 3 | 0 | 0 | Low-quality | Genome-fragment | 2.83 | AAI-based (medium-confidence) | 0 |
| S1-final-viral-combined_k141_147807\|\|full | 6130 | No | NA | 7 | 0 | 5 | Not-determined | Genome-fragment | NA | NA | 0 |
| S1-final-viral-combined_k141_16338\|\|full | 5343 | No | NA | 5 | 0 | 3 | Not-determined | Genome-fragment | NA | NA | 0 |
| S1-final-viral-combined_k141_16340\|\|full | 1439 | No | NA | 2 | 0 | 0 | Not-determined | Genome-fragment | NA | NA | 0 |
| S1-final-viral-combined_k141_65587\|\|full | 1537 | No | NA | 2 | 0 | 1 | Not-determined | Genome-fragment | NA | NA | 0 |
| S8-final-viral-combined_k141_250821\|\|full | 1420 | No | NA | 3 | 0 | 1 | Low-quality | Genome-fragment | 3.98 | AAI-based (medium-confidence) | 0 |
| S1-final-viral-combined_k141_31984\|\|full | 1592 | No | NA | 2 | 0 | 1 | Not-determined | Genome-fragment | NA | NA | 0 |
| S1-final-viral-combined_k141_16347\|\|full | 2347 | No | NA | 4 | 0 | 0 | Not-determined | Genome-fragment | NA | NA | 0 |
| S11-final-viral-combined_k141_117611\|\|full | 1421 | No | NA | 4 | 1 | 0 | Low-quality | Genome-fragment | 3.1 | HMM-based (lower-bound) | 0 |
| S1-final-viral-combined_k141_230199\|\|full | 4661 | No | NA | 5 | 0 | 4 | Not-determined | Genome-fragment | NA | NA | 0 |
| S1-final-viral-combined_k141_279260\|\|full | 1230 | No | NA | 3 | 0 | 0 | Not-determined | Genome-fragment | NA | NA | 0 |
| S1-final-viral-combined_k141_16388\|\|full | 9905 | No | NA | 9 | 0 | 4 | Not-determined | Genome-fragment | NA | NA | 0 |
| S1-final-viral-combined_k141_180198\|\|full | 1850 | No | NA | 2 | 0 | 1 | Not-determined | Genome-fragment | NA | NA | 0 |
| S1-final-viral-combined_k141_279287\|\|full | 1062 | No | NA | 3 | 0 | 0 | Not-determined | Genome-fragment | NA | NA | 0 |
| S11-final-viral-combined_k141_160367\|\|full | 1421 | No | NA | 3 | 1 | 0 | Low-quality | Genome-fragment | 3.97 | AAI-based (medium-confidence) | 0 |
| S1-final-viral-combined_k141_164133\|\|full | 1232 | No | NA | 2 | 0 | 0 | Not-determined | Genome-fragment | NA | NA | 0 |
| S1-final-viral-combined_k141_81501\|\|full | 1355 | No | NA | 2 | 0 | 0 | Not-determined | Genome-fragment | NA | NA | 0 |
| S1-final-viral-combined_k141_180265\|\|full | 6112 | No | NA | 8 | 0 | 5 | Not-determined | Genome-fragment | NA | NA | 0 |
| S1-final-viral-combined_k141_81544\|\|full | 3598 | No | NA | 3 | 0 | 1 | Not-determined | Genome-fragment | NA | NA | 0 |
| S11-final-viral-combined_k141_151217\|\|full | 1421 | No | NA | 4 | 0 | 2 | Low-quality | Genome-fragment | 4.43 | AAI-based (medium-confidence) | 0 |
| S1-final-viral-combined_k141_150725\|\|full | 1421 | No | NA | 4 | 2 | 0 | Low-quality | Genome-fragment | 2.53 | AAI-based (high-confidence) | 0 |
| S1-final-viral-combined_k141_32084\|\|full | 1598 | No | NA | 3 | 0 | 2 | Not-determined | Genome-fragment | NA | NA | 0 |
| S1-final-viral-combined_k141_232700\|\|lt2gene | 1421 | No | NA | 2 | 1 | 0 | Low-quality | Genome-fragment | 3.38 | AAI-based (high-confidence) | 0 |
| S1-final-viral-combined_k141_164218\|\|full | 4350 | No | NA | 5 | 0 | 5 | Not-determined | Genome-fragment | NA | NA | 0 |
| S5-final-viral-combined_k141_32249\|\|lt2gene | 1421 | No | NA | 1 | 1 | 0 | Low-quality | Genome-fragment | 0.76 | HMM-based (lower-bound) | 0 |
| S5-final-viral-combined_k141_211676\|\|lt2gene | 1421 | Yes | 767 | 2 | 1 | 1 | Low-quality | Genome-fragment | 1.97 | AAI-based (high-confidence) | 46.02 |
| S1-final-viral-combined_k141_32158\|\|full | 2006 | No | NA | 2 | 0 | 0 | Not-determined | Genome-fragment | NA | NA | 0 |
| S7-final-viral-combined_k141_239085\|\|full | 1421 | No | NA | 3 | 2 | 0 | Low-quality | Genome-fragment | 3.32 | AAI-based (medium-confidence) | 0 |
| S17-final-viral-combined_k141_214846\|\|full | 1421 | No | NA | 3 | 0 | 0 | Low-quality | Genome-fragment | 0.9 | AAI-based (medium-confidence) | 0 |
| S1-final-viral-combined_k141_32191\|\|full | 4281 | No | NA | 5 | 0 | 3 | Not-determined | Genome-fragment | NA | NA | 0 |
| S17-final-viral-combined_k141_228804\|\|full | 1421 | No | NA | 2 | 0 | 2 | Low-quality | Genome-fragment | 1.14 | AAI-based (medium-confidence) | 0 |
| S2-final-viral-combined_k141_83679\|\|full | 1421 | No | NA | 2 | 0 | 1 | Low-quality | Genome-fragment | 1.93 | AAI-based (high-confidence) | 0 |
| S1-final-viral-combined_k141_180399\|\|full | 2259 | No | NA | 3 | 0 | 0 | Not-determined | Genome-fragment | NA | NA | 0 |
| S6-final-viral-combined_k141_17667\|\|full | 1421 | No | NA | 3 | 2 | 0 | Low-quality | Genome-fragment | 2.88 | AAI-based (medium-confidence) | 0 |
| S1-final-viral-combined_k141_180418\|\|full | 3326 | No | NA | 7 | 0 | 1 | Not-determined | Genome-fragment | NA | NA | 0 |
| S8_k141_15634 | 1421 | No | NA | 4 | 0 | 0 | Low-quality | Genome-fragment | 1.41 | AAI-based (high-confidence) | 0 |
| S1-final-viral-combined_k141_394327\|\|full | 2698 | No | NA | 3 | 0 | 1 | Not-determined | Genome-fragment | NA | NA | 0 |
| S1-final-viral-combined_k141_344889\|\|full | 1003 | No | NA | 2 | 0 | 0 | Not-determined | Genome-fragment | NA | NA | 0 |
| S1-final-viral-combined_k141_394332\|\|full | 3514 | No | NA | 4 | 0 | 2 | Not-determined | Genome-fragment | NA | NA | 0 |
| S1-final-viral-combined_k141_81739\|\|full | 2093 | No | NA | 3 | 0 | 2 | Not-determined | Genome-fragment | NA | NA | 0 |
| S1-final-viral-combined_k141_164401\|\|full | 3099 | No | NA | 4 | 0 | 3 | Not-determined | Genome-fragment | NA | NA | 0 |
| S1-final-viral-combined_k141_180468\|\|full | 2632 | No | NA | 2 | 0 | 1 | Not-determined | Genome-fragment | NA | NA | 0 |
| S9-final-viral-combined_k141_16364\|\|full | 1422 | No | NA | 4 | 0 | 0 | Low-quality | Genome-fragment | 1.38 | AAI-based (medium-confidence) | 0 |
| S1-final-viral-combined_k141_113633\|\|full | 1532 | No | NA | 3 | 0 | 0 | Not-determined | Genome-fragment | NA | NA | 0 |
| S1-final-viral-combined_k141_48389\|\|full | 3839 | No | NA | 4 | 0 | 3 | Not-determined | Genome-fragment | NA | NA | 0 |
| S1-final-viral-combined_k141_388921\|\|full | 1422 | No | NA | 3 | 1 | 0 | Low-quality | Genome-fragment | 0.96 | HMM-based (lower-bound) | 0 |
| S3-final-viral-combined_k141_95249\|\|full | 1422 | No | NA | 3 | 1 | 0 | Low-quality | Genome-fragment | 3.5 | AAI-based (high-confidence) | 0 |
| S1-final-viral-combined_k141_394391\|\|full | 2595 | No | NA | 3 | 0 | 2 | Not-determined | Genome-fragment | NA | NA | 0 |
| S1-final-viral-combined_k141_394396\|\|full | 2809 | No | NA | 3 | 0 | 3 | Not-determined | Genome-fragment | NA | NA | 0 |
| S1-final-viral-combined_k141_279492\|\|full | 29085 | No | NA | 48 | 15 | 1 | Medium-quality | Genome-fragment | 81.84 | AAI-based (high-confidence) | 0 |
| S1-final-viral-combined_k141_81854\|\|full | 1747 | No | NA | 3 | 0 | 1 | Not-determined | Genome-fragment | NA | NA | 0 |
| S2-final-viral-combined_k141_57371\|\|full | 1422 | No | NA | 3 | 1 | 0 | Low-quality | Genome-fragment | 0.23 | HMM-based (lower-bound) | 0 |
| S1-final-viral-combined_k141_345059\|\|full | 8482 | No | NA | 3 | 0 | 1 | Not-determined | Genome-fragment | NA | NA | 0 |
| S1-final-viral-combined_k141_180603\|\|full | 2608 | No | NA | 3 | 0 | 0 | Not-determined | Genome-fragment | NA | NA | 0 |
| S2-final-viral-combined_k141_71970\|\|full | 1422 | No | NA | 3 | 0 | 1 | Low-quality | Genome-fragment | 1.89 | AAI-based (medium-confidence) | 0 |
| S1-final-viral-combined_k141_32497\|\|full | 2082 | No | NA | 2 | 0 | 1 | Not-determined | Genome-fragment | NA | NA | 0 |
| S1-final-viral-combined_k141_48530\|\|full | 1964 | No | NA | 3 | 0 | 3 | Not-determined | Genome-fragment | NA | NA | 0 |
| S4-final-viral-combined_k141_15020\|\|full | 1422 | Yes | 750 | 2 | 1 | 1 | Low-quality | Genome-fragment | 1.06 | AAI-based (high-confidence) | 47.26 |
| S9_k141_32953 | 1423 | No | NA | 5 | 0 | 0 | Low-quality | Genome-fragment | 2.38 | AAI-based (medium-confidence) | 0 |
| S5-final-viral-combined_k141_129375\|\|full | 1423 | No | NA | 3 | 0 | 2 | Low-quality | Genome-fragment | 0.4 | AAI-based (medium-confidence) | 0 |
| S5-final-viral-combined_k141_104587\|\|full | 1423 | No | NA | 2 | 0 | 0 | Low-quality | Genome-fragment | 2.05 | AAI-based (medium-confidence) | 0 |
| S7_k141_145339 | 1423 | No | NA | 1 | 0 | 0 | Low-quality | Genome-fragment | 3.1 | AAI-based (high-confidence) | 0 |
| S1-final-viral-combined_k141_32547\|\|full | 1576 | No | NA | 3 | 0 | 0 | Not-determined | Genome-fragment | NA | NA | 0 |
| S1-final-viral-combined_k141_345112\|\|full | 1311 | No | NA | 3 | 0 | 0 | Not-determined | Genome-fragment | NA | NA | 0 |
| S1-final-viral-combined_k141_164436\|\|full | 5584 | No | NA | 3 | 0 | 3 | Not-determined | Genome-fragment | NA | NA | 0 |
| S1-final-viral-combined_k141_164439\|\|full | 2106 | No | NA | 3 | 0 | 1 | Not-determined | Genome-fragment | NA | NA | 0 |
| S17-final-viral-combined_k141_84431\|\|full | 1423 | No | NA | 3 | 2 | 0 | Low-quality | Genome-fragment | 3.52 | AAI-based (medium-confidence) | 0 |
| S1-final-viral-combined_k141_164485\|\|full | 13714 | No | NA | 18 | 1 | 2 | Medium-quality | Genome-fragment | 89.89 | AAI-based (high-confidence) | 0 |
| S1-final-viral-combined_k141_48676\|\|full | 1195 | No | NA | 2 | 0 | 1 | Not-determined | Genome-fragment | NA | NA | 0 |
| S17-final-viral-combined_k141_230826\|\|full | 1423 | No | NA | 3 | 1 | 0 | Low-quality | Genome-fragment | 2.18 | AAI-based (high-confidence) | 0 |
| S12-final-viral-combined_k141_140064\|\|full | 1423 | No | NA | 2 | 0 | 0 | Low-quality | Genome-fragment | 2.17 | AAI-based (high-confidence) | 0 |
| S1-final-viral-combined_k141_279611\|\|full | 1716 | No | NA | 3 | 0 | 1 | Not-determined | Genome-fragment | NA | NA | 0 |
| S1-final-viral-combined_k141_180779\|\|full | 1388 | No | NA | 3 | 0 | 0 | Not-determined | Genome-fragment | NA | NA | 0 |
| S1-final-viral-combined_k141_180781\|\|full | 1442 | No | NA | 2 | 0 | 0 | Not-determined | Genome-fragment | NA | NA | 0 |
| S2-final-viral-combined_k141_230261\|\|full | 1423 | No | NA | 3 | 0 | 0 | Low-quality | Genome-fragment | 3.71 | AAI-based (medium-confidence) | 0 |
| S1-final-viral-combined_k141_345168\|\|full | 1677 | No | NA | 4 | 0 | 1 | Not-determined | Genome-fragment | NA | NA | 0 |
| S1-final-viral-combined_k141_32738\|\|full | 3179 | No | NA | 4 | 0 | 3 | Not-determined | Genome-fragment | NA | NA | 0 |
| S1-final-viral-combined_k141_180806\|\|full | 2018 | No | NA | 2 | 0 | 1 | Not-determined | Genome-fragment | NA | NA | 0 |
| S1-final-viral-combined_k141_180807\|\|full | 2337 | No | NA | 5 | 0 | 0 | Not-determined | Genome-fragment | NA | NA | 0 |
| S1-final-viral-combined_k141_345238\|\|full | 7040 | No | NA | 6 | 0 | 2 | Not-determined | Genome-fragment | NA | NA | 0 |
| S2-final-viral-combined_k141_122264\|\|full | 1423 | No | NA | 2 | 0 | 0 | Low-quality | Genome-fragment | 0.94 | AAI-based (medium-confidence) | 0 |
| S1-final-viral-combined_k141_345249\|\|full | 2666 | No | NA | 4 | 0 | 1 | Not-determined | Genome-fragment | NA | NA | 0 |
| S1-final-viral-combined_k141_82151\|\|full | 2117 | No | NA | 2 | 0 | 2 | Not-determined | Genome-fragment | NA | NA | 0 |
| S1-final-viral-combined_k141_48809\|\|full | 2078 | No | NA | 3 | 0 | 3 | Not-determined | Genome-fragment | NA | NA | 0 |
| S2_k141_23862 | 1423 | No | NA | 5 | 0 | 0 | Low-quality | Genome-fragment | 3.67 | AAI-based (medium-confidence) | 0 |
| S1-final-viral-combined_k141_114145\|\|full | 2105 | No | NA | 2 | 0 | 2 | Not-determined | Genome-fragment | NA | NA | 0 |
| S1-final-viral-combined_k141_82187\|\|full | 2508 | No | NA | 3 | 0 | 0 | Not-determined | Genome-fragment | NA | NA | 0 |
| S2_k141_143844 | 1423 | No | NA | 1 | 0 | 1 | Low-quality | Genome-fragment | 0.86 | AAI-based (high-confidence) | 0 |
| S1-final-viral-combined_k141_345325\|\|full | 2041 | No | NA | 4 | 0 | 0 | Not-determined | Genome-fragment | NA | NA | 0 |
| S1-final-viral-combined_k141_82205\|\|full | 1118 | No | NA | 2 | 0 | 1 | Not-determined | Genome-fragment | NA | NA | 0 |
| S1-final-viral-combined_k141_48883\|\|full | 9453 | No | NA | 9 | 0 | 7 | Not-determined | Genome-fragment | NA | NA | 0 |
| S1-final-viral-combined_k141_345351\|\|full | 3352 | No | NA | 2 | 0 | 2 | Not-determined | Genome-fragment | NA | NA | 0 |
| S1-final-viral-combined_k141_345383\|\|full | 1542 | No | NA | 2 | 0 | 1 | Not-determined | Genome-fragment | NA | NA | 0 |
| S4-final-viral-combined_k141_10245\|\|full | 1423 | No | NA | 3 | 0 | 0 | Low-quality | Genome-fragment | 1.87 | AAI-based (high-confidence) | 0 |
| S1-final-viral-combined_k141_32863\|\|full | 15902 | No | NA | 20 | 0 | 10 | Medium-quality | Genome-fragment | 50.78 | AAI-based (medium-confidence) | 0 |
| S1-final-viral-combined_k141_48966\|\|full | 3073 | No | NA | 3 | 0 | 1 | Not-determined | Genome-fragment | NA | NA | 0 |
| S1-final-viral-combined_k141_32868\|\|full | 6744 | No | NA | 8 | 0 | 5 | Not-determined | Genome-fragment | NA | NA | 0 |
| S1-final-viral-combined_k141_114282\|\|full | 2133 | No | NA | 3 | 0 | 2 | Not-determined | Genome-fragment | NA | NA | 0 |
| S1-final-viral-combined_k141_345404\|\|full | 1314 | No | NA | 2 | 0 | 0 | Not-determined | Genome-fragment | NA | NA | 0 |
| S1-final-viral-combined_k141_345429\|\|full | 1091 | No | NA | 2 | 0 | 0 | Not-determined | Genome-fragment | NA | NA | 0 |
| S8-final-viral-combined_k141_246658\|\|full | 1423 | No | NA | 2 | 0 | 1 | Low-quality | Genome-fragment | 2.18 | AAI-based (medium-confidence) | 0 |
| S1-final-viral-combined_k141_32883\|\|full | 3720 | No | NA | 4 | 0 | 0 | Not-determined | Genome-fragment | NA | NA | 0 |
| S1-final-viral-combined_k141_114382\|\|full | 1539 | No | NA | 3 | 0 | 1 | Not-determined | Genome-fragment | NA | NA | 0 |
| S1-final-viral-combined_k141_114474\|\|full | 2678 | No | NA | 3 | 0 | 2 | Not-determined | Genome-fragment | NA | NA | 0 |
| S9-final-viral-combined_k141_75458\|\|full | 1424 | No | NA | 3 | 1 | 0 | Low-quality | Genome-fragment | 0.54 | HMM-based (lower-bound) | 0 |
| S9-final-viral-combined_k141_56527\|\|full | 1424 | No | NA | 3 | 1 | 0 | Low-quality | Genome-fragment | 3.16 | AAI-based (high-confidence) | 0 |
| S9_k141_32613 | 1424 | No | NA | 1 | 0 | 0 | Low-quality | Genome-fragment | 2.38 | AAI-based (high-confidence) | 0 |
| S9_k141_21954 | 1424 | No | NA | 4 | 0 | 1 | Low-quality | Genome-fragment | 3.35 | AAI-based (high-confidence) | 0 |
| S1-final-viral-combined_k141_49249\|\|full | 2946 | No | NA | 6 | 0 | 3 | Not-determined | Genome-fragment | NA | NA | 0 |
| S11-final-viral-combined_k141_195610\|\|full | 1424 | No | NA | 6 | 1 | 0 | Low-quality | Genome-fragment | 1.03 | AAI-based (medium-confidence) | 0 |
| S1-final-viral-combined_k141_49284\|\|full | 2005 | No | NA | 3 | 0 | 1 | Not-determined | Genome-fragment | NA | NA | 0 |
| S1-final-viral-combined_k141_284667\|\|full | 1424 | No | NA | 2 | 0 | 1 | Low-quality | Genome-fragment | 2.21 | AAI-based (medium-confidence) | 0 |
| S5_k141_85022 | 1424 | No | NA | 2 | 1 | 0 | Low-quality | Genome-fragment | 3.26 | HMM-based (lower-bound) | 0 |
| S17-final-viral-combined_k141_47580\|\|full | 1424 | No | NA | 2 | 0 | 1 | Low-quality | Genome-fragment | 0.71 | AAI-based (high-confidence) | 0 |
| S12-final-viral-combined_k141_188638\|\|full | 1424 | No | NA | 4 | 1 | 0 | Low-quality | Genome-fragment | 3.19 | AAI-based (medium-confidence) | 0 |
| S1-final-viral-combined_k141_114978\|\|full | 2619 | No | NA | 3 | 0 | 3 | Not-determined | Genome-fragment | NA | NA | 0 |
| S1-final-viral-combined_k141_115088\|\|full | 3609 | No | NA | 4 | 0 | 1 | Not-determined | Genome-fragment | NA | NA | 0 |
| S1-final-viral-combined_k141_115104\|\|full | 1284 | No | NA | 3 | 0 | 2 | Not-determined | Genome-fragment | NA | NA | 0 |
| S2-final-viral-combined_k141_238421\|\|full | 1424 | No | NA | 3 | 0 | 2 | Low-quality | Genome-fragment | 0.75 | AAI-based (medium-confidence) | 0 |
| S9-final-viral-combined_k141_89585\|\|full | 1425 | No | NA | 2 | 0 | 0 | Low-quality | Genome-fragment | 0.8 | AAI-based (medium-confidence) | 0 |
| S1-final-viral-combined_k141_34689\|\|full | 1425 | No | NA | 2 | 0 | 0 | Low-quality | Genome-fragment | 3.78 | AAI-based (high-confidence) | 0 |
| S1-final-viral-combined_k141_125241\|\|full | 1425 | No | NA | 3 | 0 | 0 | Low-quality | Genome-fragment | 2.22 | AAI-based (medium-confidence) | 0 |
| S1-final-viral-combined_k141_2551\|\|0_partial | 17087 | No | NA | 15 | 0 | 10 | Not-determined | Genome-fragment | NA | NA | 0 |
| S3-final-viral-combined_k141_85139\|\|lt2gene | 1425 | No | NA | 1 | 1 | 0 | Low-quality | Genome-fragment | 2.08 | HMM-based (lower-bound) | 0 |
| S7-final-viral-combined_k141_128011\|\|lt2gene | 1425 | No | NA | 1 | 1 | 0 | Low-quality | Genome-fragment | 1.34 | HMM-based (lower-bound) | 0 |
| S7_k141_17556 | 1425 | No | NA | 1 | 0 | 0 | Low-quality | Genome-fragment | 1.76 | AAI-based (medium-confidence) | 0 |
| S1-final-viral-combined_k141_331544\|\|0_partial | 26230 | Yes | 14540 | 33 | 7 | 8 | Complete | High-quality | 100 | Provirus (medium-confidence) | 44.57 |
| S7_k141_60291 | 1425 | No | NA | 2 | 0 | 0 | Low-quality | Genome-fragment | 3.23 | AAI-based (medium-confidence) | 0 |
| S1-final-viral-combined_k141_68332\|\|0_partial | 12047 | No | NA | 10 | 0 | 7 | Not-determined | Genome-fragment | NA | NA | 0 |
| S1-final-viral-combined_k141_265860\|\|0_partial | 23566 | No | NA | 22 | 0 | 13 | Not-determined | Genome-fragment | NA | NA | 0 |
| S1-final-viral-combined_k141_85225\|\|0_partial | 67554 | Yes | 41415 | 86 | 21 | 20 | High-quality | High-quality | 100 | AAI-based (high-confidence) | 38.69 |
| S17-final-viral-combined_k141_218694\|\|full | 1425 | No | NA | 4 | 2 | 0 | Low-quality | Genome-fragment | 2.54 | AAI-based (high-confidence) | 0 |
| S1-final-viral-combined_k141_36114\|\|0_partial | 10668 | No | NA | 10 | 0 | 9 | Not-determined | Genome-fragment | NA | NA | 0 |
| S12-final-viral-combined_k141_243470\|\|full | 1425 | No | NA | 5 | 0 | 0 | Low-quality | Genome-fragment | 3.44 | AAI-based (medium-confidence) | 0 |
| S2-final-viral-combined_k141_47288\|\|full | 1425 | No | NA | 3 | 0 | 0 | Low-quality | Genome-fragment | 23.23 | AAI-based (medium-confidence) | 0 |
| S1-final-viral-combined_k141_136097\|\|0_partial | 19804 | No | NA | 23 | 1 | 6 | Medium-quality | Genome-fragment | 50.57 | AAI-based (high-confidence) | 0 |
| S1-final-viral-combined_k141_168463\|\|0_partial | 23512 | No | NA | 28 | 0 | 7 | Not-determined | Genome-fragment | NA | NA | 0 |
| S1-final-viral-combined_k141_284778\|\|0_partial | 14860 | No | NA | 11 | 0 | 8 | Not-determined | Genome-fragment | NA | NA | 0 |
| S2-final-viral-combined_k141_144466\|\|lt2gene | 1425 | No | NA | 2 | 0 | 0 | Low-quality | Genome-fragment | 2.98 | AAI-based (medium-confidence) | 0 |
| S1-final-viral-combined_k141_169272\|\|0_partial | 12125 | No | NA | 14 | 0 | 12 | Not-determined | Genome-fragment | NA | NA | 0 |
| S1-final-viral-combined_k141_121278\|\|0_partial | 11574 | No | NA | 10 | 0 | 9 | Not-determined | Genome-fragment | NA | NA | 0 |
| S4-final-viral-combined_k141_104762\|\|full | 1425 | No | NA | 3 | 0 | 0 | Low-quality | Genome-fragment | 1.13 | AAI-based (medium-confidence) | 0 |
| S8-final-viral-combined_k141_109996\|\|full | 1425 | No | NA | 3 | 0 | 0 | Low-quality | Genome-fragment | 2.6 | AAI-based (medium-confidence) | 0 |
| S9-final-viral-combined_k141_91942\|\|lt2gene | 1426 | No | NA | 1 | 1 | 0 | Low-quality | Genome-fragment | 3.34 | AAI-based (high-confidence) | 0 |
| S11-final-viral-combined_k141_255992\|\|full | 1426 | No | NA | 3 | 2 | 0 | Low-quality | Genome-fragment | 3.34 | AAI-based (medium-confidence) | 0 |
| S1-final-viral-combined_k141_138958\|\|0_partial | 13426 | No | NA | 16 | 0 | 10 | Not-determined | Genome-fragment | NA | NA | 0 |
| S1-final-viral-combined_k141_255289\|\|1_partial | 11584 | No | NA | 7 | 0 | 5 | Not-determined | Genome-fragment | NA | NA | 0 |
| S11_k141_165217 | 1426 | No | NA | 3 | 0 | 1 | Low-quality | Genome-fragment | 2.86 | AAI-based (medium-confidence) | 0 |
| S1-final-viral-combined_k141_155743\|\|0_partial | 15180 | No | NA | 15 | 0 | 11 | Not-determined | Genome-fragment | NA | NA | 0 |
| S7-final-viral-combined_k141_86098\|\|full | 1426 | No | NA | 3 | 0 | 0 | Low-quality | Genome-fragment | 2.45 | AAI-based (medium-confidence) | 0 |
| S1-final-viral-combined_k141_273012\|\|0_partial | 28053 | No | NA | 26 | 0 | 18 | Not-determined | Genome-fragment | NA | NA | 0 |
| S1-final-viral-combined_k141_307323\|\|0_partial | 70097 | Yes | 33763 | 80 | 16 | 19 | Medium-quality | Genome-fragment | 88.54 | AAI-based (high-confidence) | 51.83 |
| S17-final-viral-combined_k141_70831\|\|full | 1426 | No | NA | 2 | 0 | 0 | Low-quality | Genome-fragment | 3.05 | AAI-based (medium-confidence) | 0 |
| S17-k141_49920 | 1426 | No | NA | 1 | 0 | 1 | Low-quality | Genome-fragment | 4.02 | AAI-based (medium-confidence) | 0 |
| S1-final-viral-combined_k141_326034\|\|0_partial | 18298 | No | NA | 23 | 0 | 9 | Not-determined | Genome-fragment | NA | NA | 0 |
| S1-final-viral-combined_k141_94988\|\|0_partial | 39556 | Yes | 28256 | 44 | 7 | 19 | Medium-quality | Genome-fragment | 68.23 | AAI-based (medium-confidence) | 28.57 |
| S12-final-viral-combined_k141_201923\|\|full | 1426 | No | NA | 2 | 0 | 0 | Low-quality | Genome-fragment | 2.83 | AAI-based (medium-confidence) | 0 |
| S1-final-viral-combined_k141_375477\|\|0_partial | 12579 | No | NA | 6 | 0 | 5 | Not-determined | Genome-fragment | NA | NA | 0 |
| S12_k141_122617 | 1426 | No | NA | 2 | 1 | 0 | Low-quality | Genome-fragment | 0.78 | AAI-based (high-confidence) | 0 |
| S1-final-viral-combined_k141_294159\|\|0_partial | 18405 | No | NA | 14 | 0 | 9 | Not-determined | Genome-fragment | NA | NA | 0 |
| S1-final-viral-combined_k141_294159\|\|1_partial | 13349 | No | NA | 8 | 0 | 6 | Not-determined | Genome-fragment | NA | NA | 0 |
| S2-final-viral-combined_k141_114024\|\|full | 1426 | No | NA | 3 | 1 | 0 | Low-quality | Genome-fragment | 1.67 | AAI-based (high-confidence) | 0 |
| S2-final-viral-combined_k141_126674\|\|full | 1426 | No | NA | 3 | 0 | 0 | Low-quality | Genome-fragment | 2.6 | AAI-based (high-confidence) | 0 |
| S1-final-viral-combined_k141_115303\|\|lt2gene | 1159 | No | NA | 1 | 0 | 0 | Not-determined | Genome-fragment | NA | NA | 0 |
| S11-final-viral-combined_k141_281579\|\|full | 1427 | No | NA | 2 | 0 | 0 | Low-quality | Genome-fragment | 3.42 | AAI-based (high-confidence) | 0 |
| S1-final-viral-combined_k141_164803\|\|full | 1427 | No | NA | 3 | 0 | 0 | Low-quality | Genome-fragment | 1.46 | AAI-based (high-confidence) | 0 |
| S1-final-viral-combined_k141_296380\|\|lt2gene | 1223 | No | NA | 2 | 0 | 2 | Not-determined | Genome-fragment | NA | NA | 0 |
| S5-final-viral-combined_k141_70563\|\|lt2gene | 1427 | No | NA | 1 | 1 | 0 | Low-quality | Genome-fragment | 3.13 | AAI-based (medium-confidence) | 0 |
| S7_k141_244184 | 1427 | No | NA | 3 | 0 | 0 | Low-quality | Genome-fragment | 2.79 | AAI-based (medium-confidence) | 0 |
| S17-k141_143253 | 1427 | No | NA | 1 | 0 | 1 | Low-quality | Genome-fragment | 0.53 | AAI-based (medium-confidence) | 0 |
| S4-final-viral-combined_k141_29280\|\|full | 1427 | No | NA | 2 | 0 | 0 | Low-quality | Genome-fragment | 2.17 | AAI-based (medium-confidence) | 0 |
| S4-final-viral-combined_k141_166564\|\|full | 1427 | No | NA | 3 | 1 | 0 | Low-quality | Genome-fragment | 2.85 | AAI-based (high-confidence) | 0 |
| S8-final-viral-combined_k141_187829\|\|full | 1427 | No | NA | 2 | 0 | 0 | Low-quality | Genome-fragment | 0.92 | AAI-based (high-confidence) | 0 |
| S8_k141_51616 | 1427 | No | NA | 4 | 1 | 0 | Low-quality | Genome-fragment | 4.04 | AAI-based (high-confidence) | 0 |
| S11-final-viral-combined_k141_80281\|\|full | 1428 | No | NA | 3 | 1 | 0 | Low-quality | Genome-fragment | 3.7 | AAI-based (high-confidence) | 0 |
| S3-final-viral-combined_k141_15094\|\|lt2gene | 1428 | No | NA | 2 | 2 | 0 | Low-quality | Genome-fragment | 4.08 | AAI-based (high-confidence) | 0 |

| | | | | | | | | | | | |
|---|---|---|---|---|---|---|---|---|---|---|---|
| S3_k141_25468 | 1428 | No | NA | 2 | 0 | 1 | Low-quality | Genome-fragment | 3.02 | AAI-based (medium-confidence) | 0 |
| S1-final-viral-combined_k141_296926\|\|lt2gene | 1405 | No | NA | 2 | 0 | 2 | Not-determined | Genome-fragment | NA | NA | 0 |
| S1-final-viral-combined_k141_264200\|\|lt2gene | 1778 | No | NA | 2 | 0 | 0 | Not-determined | Genome-fragment | NA | NA | 0 |
| S1-final-viral-combined_k141_329884\|\|lt2gene | 1175 | No | NA | 1 | 0 | 0 | Not-determined | Genome-fragment | NA | NA | 0 |
| S5-final-viral-combined_k141_184449\|\|full | 1428 | No | NA | 3 | 3 | 0 | Low-quality | Genome-fragment | 3.56 | AAI-based (high-confidence) | 0 |
| S5-final-viral-combined_k141_20609\|\|lt2gene | 1428 | No | NA | 1 | 1 | 0 | Low-quality | Genome-fragment | 2.48 | HMM-based (lower-bound) | 0 |
| S7-final-viral-combined_k141_84874\|\|full | 1428 | No | NA | 3 | 0 | 0 | Low-quality | Genome-fragment | 3.28 | AAI-based (medium-confidence) | 0 |
| S17-final-viral-combined_k141_28222\|\|full | 1428 | No | NA | 3 | 3 | 0 | Low-quality | Genome-fragment | 3.06 | AAI-based (medium-confidence) | 0 |
| S1-final-viral-combined_k141_165741\|\|lt2gene | 1364 | No | NA | 1 | 0 | 0 | Not-determined | Genome-fragment | NA | NA | 0 |
| S1-final-viral-combined_k141_17936\|\|lt2gene | 1082 | No | NA | 1 | 0 | 0 | Not-determined | Genome-fragment | NA | NA | 0 |
| S12-final-viral-combined_k141_268597\|\|full | 1428 | No | NA | 7 | 0 | 0 | Low-quality | Genome-fragment | 3.48 | AAI-based (medium-confidence) | 0 |
| S12-final-viral-combined_k141_228184\|\|full | 1428 | No | NA | 4 | 0 | 0 | Low-quality | Genome-fragment | 0.96 | AAI-based (medium-confidence) | 0 |
| S6-final-viral-combined_k141_113492\|\|lt2gene | 1428 | No | NA | 1 | 1 | 0 | Low-quality | Genome-fragment | 4.33 | AAI-based (high-confidence) | 0 |
| S1-final-viral-combined_k141_264563\|\|lt2gene | 1433 | No | NA | 1 | 0 | 0 | Not-determined | Genome-fragment | NA | NA | 0 |
| S1-final-viral-combined_k141_314281\|\|lt2gene | 1802 | No | NA | 2 | 0 | 0 | Not-determined | Genome-fragment | NA | NA | 0 |
| S1-final-viral-combined_k141_100187\|\|lt2gene | 1441 | No | NA | 2 | 0 | 2 | Not-determined | Genome-fragment | NA | NA | 0 |
| S6_k141_33179 | 1428 | No | NA | 1 | 0 | 1 | Low-quality | Genome-fragment | 4.03 | AAI-based (medium-confidence) | 0 |
| S1-final-viral-combined_k141_2463\|\|lt2gene | 1055 | No | NA | 1 | 0 | 0 | Not-determined | Genome-fragment | NA | NA | 0 |
| S8-final-viral-combined_k141_17333\|\|full | 1428 | No | NA | 4 | 0 | 0 | Low-quality | Genome-fragment | 3.83 | AAI-based (medium-confidence) | 0 |
| S9_k141_62998 | 1429 | No | NA | 2 | 1 | 0 | Low-quality | Genome-fragment | 2.47 | HMM-based (lower-bound) | 0 |
| S11-final-viral-combined_k141_17850\|\|full | 1429 | No | NA | 3 | 0 | 3 | Low-quality | Genome-fragment | 0.84 | AAI-based (medium-confidence) | 0 |
| S1-final-viral-combined_k141_315185\|\|lt2gene | 1705 | No | NA | 2 | 0 | 1 | Not-determined | Genome-fragment | NA | NA | 0 |
| S1-final-viral-combined_k141_364578\|\|lt2gene | 1903 | No | NA | 2 | 0 | 0 | Not-determined | Genome-fragment | NA | NA | 0 |
| S1-final-viral-combined_k141_331427\|\|lt2gene | 1451 | No | NA | 2 | 0 | 1 | Not-determined | Genome-fragment | NA | NA | 0 |
| S1-final-viral-combined_k141_119081\|\|full | 1429 | No | NA | 3 | 0 | 2 | Low-quality | Genome-fragment | 1.94 | AAI-based (medium-confidence) | 0 |
| S3-final-viral-combined_k141_85640\|\|full | 1429 | No | NA | 2 | 0 | 0 | Low-quality | Genome-fragment | 3.08 | AAI-based (medium-confidence) | 0 |
| S5-final-viral-combined_k141_141693\|\|full | 1429 | No | NA | 2 | 0 | 0 | Low-quality | Genome-fragment | 0.8 | AAI-based (medium-confidence) | 0 |
| S7-final-viral-combined_k141_223893\|\|full | 1429 | No | NA | 2 | 0 | 0 | Low-quality | Genome-fragment | 4.17 | AAI-based (medium-confidence) | 0 |
| S12-final-viral-combined_k141_88600\|\|full | 1429 | No | NA | 2 | 0 | 0 | Low-quality | Genome-fragment | 1.05 | AAI-based (medium-confidence) | 0 |
| S12_k141_141966 | 1429 | No | NA | 2 | 0 | 0 | Low-quality | Genome-fragment | 2.35 | AAI-based (high-confidence) | 0 |
| S4_k141_103831 | 1429 | No | NA | 4 | 0 | 0 | Low-quality | Genome-fragment | 1.08 | AAI-based (medium-confidence) | 0 |
| S1-final-viral-combined_k141_200516\|\|lt2gene | 1187 | No | NA | 2 | 0 | 0 | Not-determined | Genome-fragment | NA | NA | 0 |
| S6-final-viral-combined_k141_107985\|\|full | 1429 | No | NA | 3 | 0 | 0 | Low-quality | Genome-fragment | 2.67 | AAI-based (medium-confidence) | 0 |
| S9-final-viral-combined_k141_2500\|\|full | 1430 | No | NA | 2 | 0 | 0 | Low-quality | Genome-fragment | 1.2 | AAI-based (high-confidence) | 0 |
| S9-final-viral-combined_k141_12642\|\|full | 1430 | No | NA | 3 | 2 | 0 | Low-quality | Genome-fragment | 3.72 | AAI-based (high-confidence) | 0 |
| S11-final-viral-combined_k141_114741\|\|full | 1430 | No | NA | 2 | 0 | 0 | Low-quality | Genome-fragment | 0.16 | AAI-based (medium-confidence) | 0 |
| S1-final-viral-combined_k141_151873\|\|full | 1430 | No | NA | 3 | 0 | 2 | Low-quality | Genome-fragment | 1.74 | AAI-based (medium-confidence) | 0 |
| S1-final-viral-combined_k141_167241\|\|lt2gene | 1092 | No | NA | 2 | 0 | 0 | Not-determined | Genome-fragment | NA | NA | 0 |
| S1-final-viral-combined_k141_381630\|\|lt2gene | 1422 | No | NA | 2 | 0 | 0 | Not-determined | Genome-fragment | NA | NA | 0 |
| S1-final-viral-combined_k141_250422\|\|lt2gene | 1072 | No | NA | 2 | 0 | 0 | Not-determined | Genome-fragment | NA | NA | 0 |
| S1-final-viral-combined_k141_193282\|\|full | 1430 | No | NA | 3 | 0 | 2 | Low-quality | Genome-fragment | 0.8 | AAI-based (medium-confidence) | 0 |
| S1-final-viral-combined_k141_167259\|\|lt2gene | 1084 | No | NA | 2 | 0 | 0 | Not-determined | Genome-fragment | NA | NA | 0 |
| S1_k141_374469 | 1430 | No | NA | 1 | 0 | 1 | Low-quality | Genome-fragment | 4.64 | AAI-based (high-confidence) | 0 |
| S1-final-viral-combined_k141_349201\|\|lt2gene | 1349 | No | NA | 2 | 0 | 0 | Not-determined | Genome-fragment | NA | NA | 0 |
| S7-final-viral-combined_k141_113747\|\|full | 1430 | No | NA | 3 | 0 | 1 | Low-quality | Genome-fragment | 0.14 | AAI-based (medium-confidence) | 0 |
| S12_k141_158026 | 1430 | No | NA | 1 | 0 | 1 | Low-quality | Genome-fragment | 3.14 | AAI-based (medium-confidence) | 0 |
| S12_k141_213044 | 1430 | No | NA | 1 | 0 | 1 | Low-quality | Genome-fragment | 1.62 | AAI-based (medium-confidence) | 0 |
| S4-final-viral-combined_k141_50697\|\|full | 1430 | No | NA | 2 | 0 | 0 | Low-quality | Genome-fragment | 3.56 | AAI-based (medium-confidence) | 0 |
| S4-final-viral-combined_k141_73867\|\|full | 1430 | No | NA | 3 | 1 | 0 | Low-quality | Genome-fragment | 2.46 | HMM-based (lower-bound) | 0 |
| S4_k141_92764 | 1430 | No | NA | 1 | 0 | 0 | Low-quality | Genome-fragment | 0.24 | AAI-based (high-confidence) | 0 |
| S6-final-viral-combined_k141_9720\|\|lt2gene | 1430 | No | NA | 2 | 0 | 0 | Low-quality | Genome-fragment | 2.61 | AAI-based (high-confidence) | 0 |
| S8-final-viral-combined_k141_9687\|\|full | 1430 | No | NA | 2 | 0 | 0 | Low-quality | Genome-fragment | 3.75 | AAI-based (medium-confidence) | 0 |
| S11-final-viral-combined_k141_140585\|\|full | 1431 | No | NA | 2 | 0 | 0 | Low-quality | Genome-fragment | 2.86 | AAI-based (high-confidence) | 0 |
| S1-final-viral-combined_k141_3923\|\|lt2gene | 1091 | No | NA | 1 | 0 | 0 | Not-determined | Genome-fragment | NA | NA | 0 |
| S1-final-viral-combined_k141_168077\|\|lt2gene | 1585 | No | NA | 2 | 0 | 1 | Not-determined | Genome-fragment | NA | NA | 0 |
| S1-final-viral-combined_k141_185804\|\|lt2gene | 2118 | No | NA | 2 | 0 | 0 | Not-determined | Genome-fragment | NA | NA | 0 |
| S1-final-viral-combined_k141_284006\|\|full | 1431 | No | NA | 2 | 0 | 1 | Low-quality | Genome-fragment | 0.45 | AAI-based (medium-confidence) | 0 |
| S5-final-viral-combined_k141_45645\|\|full | 1431 | No | NA | 3 | 0 | 1 | Low-quality | Genome-fragment | 2.6 | AAI-based (medium-confidence) | 0 |
| S12-final-viral-combined_k141_232380\|\|full | 1431 | No | NA | 4 | 0 | 1 | Low-quality | Genome-fragment | 1.2 | AAI-based (medium-confidence) | 0 |
| S12-final-viral-combined_k141_364929\|\|full | 1431 | No | NA | 3 | 1 | 0 | Low-quality | Genome-fragment | 3.19 | AAI-based (medium-confidence) | 0 |
| S6-final-viral-combined_k141_140622\|\|full | 1431 | No | NA | 3 | 3 | 0 | Low-quality | Genome-fragment | 2.37 | AAI-based (high-confidence) | 0 |
| S9-final-viral-combined_k141_104199\|\|full | 1432 | No | NA | 6 | 1 | 0 | Low-quality | Genome-fragment | 3.51 | AAI-based (high-confidence) | 0 |
| S9-final-viral-combined_k141_127682\|\|full | 1432 | No | NA | 4 | 1 | 0 | Low-quality | Genome-fragment | 2.41 | HMM-based (lower-bound) | 0 |
| S11-final-viral-combined_k141_146760\|\|lt2gene | 1432 | No | NA | 2 | 1 | 0 | Low-quality | Genome-fragment | 3.26 | AAI-based (medium-confidence) | 0 |
| S1-final-viral-combined_k141_21210\|\|lt2gene | 1432 | No | NA | 1 | 1 | 0 | Low-quality | Genome-fragment | 2.24 | AAI-based (medium-confidence) | 0 |
| S1-final-viral-combined_k141_168797\|\|lt2gene | 1142 | No | NA | 1 | 0 | 0 | Not-determined | Genome-fragment | NA | NA | 0 |
| S7-final-viral-combined_k141_126180\|\|full | 1432 | No | NA | 3 | 0 | 0 | Low-quality | Genome-fragment | 0.83 | AAI-based (medium-confidence) | 0 |
| S1-final-viral-combined_k141_4853\|\|lt2gene | 7370 | No | NA | 1 | 0 | 1 | Not-determined | Genome-fragment | NA | NA | 0 |
| S12-final-viral-combined_k141_251840\|\|full | 1432 | No | NA | 3 | 0 | 1 | Low-quality | Genome-fragment | 2.5 | AAI-based (medium-confidence) | 0 |
| S1-final-viral-combined_k141_5016\|\|lt2gene | 1588 | No | NA | 1 | 0 | 1 | Not-determined | Genome-fragment | NA | NA | 0 |
| S12-final-viral-combined_k141_331259\|\|full | 1432 | No | NA | 3 | 0 | 2 | Low-quality | Genome-fragment | 2.37 | AAI-based (medium-confidence) | 0 |
| S1-final-viral-combined_k141_302302\|\|lt2gene | 1208 | No | NA | 1 | 0 | 0 | Not-determined | Genome-fragment | NA | NA | 0 |
| S2-final-viral-combined_k141_174596\|\|full | 1432 | No | NA | 3 | 0 | 0 | Low-quality | Genome-fragment | 1.08 | AAI-based (medium-confidence) | 0 |
| S9_k141_69702 | 1433 | No | NA | 4 | 0 | 0 | Low-quality | Genome-fragment | 2.39 | AAI-based (medium-confidence) | 0 |
| S1-final-viral-combined_k141_169564\|\|lt2gene | 1161 | No | NA | 2 | 0 | 1 | Not-determined | Genome-fragment | NA | NA | 0 |
| S1-final-viral-combined_k141_152652\|\|full | 1433 | No | NA | 5 | 1 | 0 | Low-quality | Genome-fragment | 2.96 | AAI-based (high-confidence) | 0 |
| S1-final-viral-combined_k141_219842\|\|lt2gene | 1013 | No | NA | 1 | 0 | 0 | Not-determined | Genome-fragment | NA | NA | 0 |
| S1-final-viral-combined_k141_137939\|\|lt2gene | 1401 | No | NA | 1 | 0 | 0 | Not-determined | Genome-fragment | NA | NA | 0 |
| S7-final-viral-combined_k141_119825\|\|full | 1433 | No | NA | 2 | 0 | 0 | Low-quality | Genome-fragment | 2.35 | AAI-based (medium-confidence) | 0 |
| S1-final-viral-combined_k141_319129\|\|lt2gene | 3578 | No | NA | 1 | 0 | 0 | Not-determined | Genome-fragment | NA | NA | 0 |
| S12-final-viral-combined_k141_178275\|\|lt2gene | 1433 | No | NA | 2 | 1 | 0 | Low-quality | Genome-fragment | 3.63 | AAI-based (high-confidence) | 0 |
| S1-final-viral-combined_k141_121748\|\|lt2gene | 1116 | No | NA | 1 | 0 | 0 | Not-determined | Genome-fragment | NA | NA | 0 |
| S1-final-viral-combined_k141_384707\|\|lt2gene | 1387 | No | NA | 2 | 0 | 2 | Not-determined | Genome-fragment | NA | NA | 0 |
| S2_k141_159607 | 1433 | No | NA | 1 | 0 | 1 | Low-quality | Genome-fragment | 0.45 | AAI-based (medium-confidence) | 0 |
| S4_k141_59304 | 1433 | No | NA | 4 | 0 | 0 | Low-quality | Genome-fragment | 2.08 | AAI-based (medium-confidence) | 0 |
| S8-final-viral-combined_k141_245074\|\|full | 1433 | No | NA | 3 | 1 | 0 | Low-quality | Genome-fragment | 4.38 | AAI-based (high-confidence) | 0 |
| S9-final-viral-combined_k141_7398\|\|full | 1434 | No | NA | 3 | 0 | 0 | Low-quality | Genome-fragment | 3.12 | AAI-based (high-confidence) | 0 |
| S1-final-viral-combined_k141_301253\|\|full | 1434 | No | NA | 2 | 0 | 0 | Low-quality | Genome-fragment | 3.93 | AAI-based (high-confidence) | 0 |
| S1-final-viral-combined_k141_55595\|\|full | 1434 | No | NA | 3 | 2 | 0 | Low-quality | Genome-fragment | 2.83 | AAI-based (medium-confidence) | 0 |
| S5-final-viral-combined_k141_84977\|\|full | 1434 | No | NA | 2 | 0 | 1 | Low-quality | Genome-fragment | 0.64 | AAI-based (medium-confidence) | 0 |
| S17-final-viral-combined_k141_190807\|\|full | 1434 | No | NA | 3 | 2 | 0 | Low-quality | Genome-fragment | 3.66 | AAI-based (high-confidence) | 0 |
| S17-final-viral-combined_k141_59779\|\|full | 1434 | No | NA | 4 | 0 | 0 | Low-quality | Genome-fragment | 3.66 | AAI-based (medium-confidence) | 0 |
| S1-final-viral-combined_k141_254354\|\|lt2gene | 1030 | No | NA | 2 | 0 | 0 | Not-determined | Genome-fragment | NA | NA | 0 |
| S1-final-viral-combined_k141_71935\|\|lt2gene | 1665 | No | NA | 2 | 0 | 0 | Not-determined | Genome-fragment | NA | NA | 0 |
| S2-final-viral-combined_k141_150199\|\|full | 1434 | No | NA | 2 | 0 | 2 | Low-quality | Genome-fragment | 0.86 | AAI-based (medium-confidence) | 0 |
| S8-final-viral-combined_k141_247523\|\|full | 1434 | No | NA | 2 | 0 | 0 | Low-quality | Genome-fragment | 1.22 | AAI-based (high-confidence) | 0 |
| S11-final-viral-combined_k141_54116\|\|full | 1435 | No | NA | 2 | 0 | 0 | Low-quality | Genome-fragment | 3.49 | AAI-based (high-confidence) | 0 |
| S1-final-viral-combined_k141_67577\|\|full | 1435 | No | NA | 2 | 0 | 1 | Low-quality | Genome-fragment | 0.52 | AAI-based (medium-confidence) | 0 |
| S1-final-viral-combined_k141_70953\|\|full | 1435 | No | NA | 4 | 0 | 0 | Low-quality | Genome-fragment | 3.36 | AAI-based (high-confidence) | 0 |
| S1-final-viral-combined_k141_222638\|\|full | 1435 | No | NA | 3 | 1 | 0 | Low-quality | Genome-fragment | 2.38 | AAI-based (high-confidence) | 0 |
| S17-final-viral-combined_k141_43956\|\|full | 1435 | No | NA | 3 | 0 | 0 | Low-quality | Genome-fragment | 2.65 | AAI-based (medium-confidence) | 0 |
| S1-final-viral-combined_k141_89267\|\|lt2gene | 3957 | No | NA | 2 | 0 | 1 | Not-determined | Genome-fragment | NA | NA | 0 |
| S1-final-viral-combined_k141_221209\|\|lt2gene | 1753 | No | NA | 1 | 0 | 0 | Not-determined | Genome-fragment | NA | NA | 0 |
| S1-final-viral-combined_k141_188898\|\|lt2gene | 3077 | No | NA | 1 | 0 | 0 | Not-determined | Genome-fragment | NA | NA | 0 |
| S12-final-viral-combined_k141_175032\|\|full | 1435 | No | NA | 3 | 3 | 0 | Low-quality | Genome-fragment | 2.81 | AAI-based (high-confidence) | 0 |
| S2-final-viral-combined_k141_125071\|\|full | 1435 | No | NA | 4 | 3 | 0 | Low-quality | Genome-fragment | 4.01 | AAI-based (high-confidence) | 0 |
| S4_k141_164089 | 1435 | No | NA | 1 | 0 | 0 | Low-quality | Genome-fragment | 2.4 | AAI-based (high-confidence) | 0 |
| S6_k141_36596 | 1435 | No | NA | 3 | 0 | 0 | Low-quality | Genome-fragment | 1.7 | AAI-based (high-confidence) | 0 |
| S8-final-viral-combined_k141_178292\|\|full | 1435 | No | NA | 4 | 0 | 0 | Low-quality | Genome-fragment | 2.96 | AAI-based (medium-confidence) | 0 |
| S11_k141_260199 | 1436 | No | NA | 1 | 0 | 0 | Low-quality | Genome-fragment | 1.58 | AAI-based (high-confidence) | 0 |
| S1_k141_96876 | 1436 | No | NA | 1 | 0 | 0 | Low-quality | Genome-fragment | 2.87 | AAI-based (high-confidence) | 0 |
| S5-final-viral-combined_k141_188391\|\|full | 1436 | No | NA | 2 | 0 | 0 | Low-quality | Genome-fragment | 1.2 | AAI-based (medium-confidence) | 0 |
| S1-final-viral-combined_k141_369669\|\|lt2gene | 2403 | No | NA | 1 | 0 | 0 | Not-determined | Genome-fragment | NA | NA | 0 |
| S1-final-viral-combined_k141_369676\|\|lt2gene | 2031 | No | NA | 2 | 0 | 0 | Not-determined | Genome-fragment | NA | NA | 0 |
| S7-final-viral-combined_k141_200930\|\|full | 1436 | No | NA | 5 | 1 | 0 | Low-quality | Genome-fragment | 2.29 | AAI-based (high-confidence) | 0 |
| S1-final-viral-combined_k141_336489\|\|lt2gene | 1108 | No | NA | 2 | 0 | 2 | Not-determined | Genome-fragment | NA | NA | 0 |
| S1-final-viral-combined_k141_386715\|\|lt2gene | 2317 | No | NA | 1 | 0 | 1 | Not-determined | Genome-fragment | NA | NA | 0 |
| S12_k141_93753 | 1436 | No | NA | 3 | 0 | 0 | Low-quality | Genome-fragment | 1.43 | AAI-based (high-confidence) | 0 |
| S2_k141_79681 | 1436 | No | NA | 7 | 1 | 0 | Low-quality | Genome-fragment | 2.68 | AAI-based (medium-confidence) | 0 |
| S8-final-viral-combined_k141_208289\|\|full | 1436 | No | NA | 3 | 0 | 0 | Low-quality | Genome-fragment | 3.39 | AAI-based (medium-confidence) | 0 |
| S9-final-viral-combined_k141_85194\|\|full | 1437 | No | NA | 2 | 0 | 0 | Low-quality | Genome-fragment | 1.74 | AAI-based (medium-confidence) | 0 |
| S1-final-viral-combined_k141_354598\|\|lt2gene | 1508 | No | NA | 1 | 0 | 0 | Not-determined | Genome-fragment | NA | NA | 0 |
| S11-final-viral-combined_k141_196784\|\|full | 1437 | No | NA | 3 | 0 | 2 | Low-quality | Genome-fragment | 0.96 | AAI-based (medium-confidence) | 0 |
| S11-final-viral-combined_k141_59852\|\|full | 1437 | No | NA | 3 | 0 | 0 | Low-quality | Genome-fragment | 2.63 | AAI-based (medium-confidence) | 0 |
| S11-final-viral-combined_k141_326912\|\|lt2gene | 1437 | No | NA | 2 | 1 | 0 | Low-quality | Genome-fragment | 3.68 | AAI-based (medium-confidence) | 0 |
| S1_k141_377661 | 1437 | No | NA | 3 | 0 | 0 | Low-quality | Genome-fragment | 1.92 | AAI-based (high-confidence) | 0 |
| S5-final-viral-combined_k141_190554\|\|full | 1437 | No | NA | 4 | 1 | 0 | Low-quality | Genome-fragment | 3.11 | AAI-based (high-confidence) | 0 |
| S1-final-viral-combined_k141_337301\|\|lt2gene | 5748 | No | NA | 1 | 0 | 1 | Not-determined | Genome-fragment | NA | NA | 0 |
| S5-final-viral-combined_k141_14430\|\|full | 1437 | No | NA | 3 | 2 | 0 | Low-quality | Genome-fragment | 2.88 | AAI-based (high-confidence) | 0 |
| S1-final-viral-combined_k141_322652\|\|lt2gene | 1580 | No | NA | 1 | 0 | 0 | Not-determined | Genome-fragment | NA | NA | 0 |
| S17-final-viral-combined_k141_166893\|\|full | 1437 | No | NA | 4 | 1 | 0 | Low-quality | Genome-fragment | 3.36 | AAI-based (high-confidence) | 0 |
| S1-final-viral-combined_k141_355110\|\|lt2gene | 1189 | No | NA | 1 | 0 | 0 | Not-determined | Genome-fragment | NA | NA | 0 |
| S17-final-viral-combined_k141_81451\|\|full | 1437 | No | NA | 2 | 0 | 0 | Low-quality | Genome-fragment | 1 | AAI-based (high-confidence) | 0 |
| S6-final-viral-combined_k141_138698\|\|full | 1437 | No | NA | 3 | 0 | 1 | Low-quality | Genome-fragment | 2.11 | AAI-based (medium-confidence) | 0 |
| S8-final-viral-combined_k141_74322\|\|full | 1437 | No | NA | 4 | 0 | 0 | Low-quality | Genome-fragment | 3.07 | AAI-based (medium-confidence) | 0 |
| S9-final-viral-combined_k141_56382\|\|full | 1438 | No | NA | 3 | 0 | 0 | Low-quality | Genome-fragment | 5.05 | AAI-based (high-confidence) | 0 |
| S9-final-viral-combined_k141_4589\|\|full | 1438 | No | NA | 3 | 1 | 0 | Low-quality | Genome-fragment | 2.04 | AAI-based (high-confidence) | 0 |
| S9_k141_76277 | 1438 | No | NA | 2 | 0 | 0 | Low-quality | Genome-fragment | 0.58 | AAI-based (medium-confidence) | 0 |
| S1-final-viral-combined_k141_289087\|\|lt2gene | 1056 | No | NA | 1 | 0 | 0 | Not-determined | Genome-fragment | NA | NA | 0 |
| S11_k141_142257 | 1438 | No | NA | 2 | 0 | 1 | Low-quality | Genome-fragment | 2.81 | AAI-based (medium-confidence) | 0 |
| S1-final-viral-combined_k141_377976\|\|full | 1438 | No | NA | 4 | 0 | 0 | Low-quality | Genome-fragment | 3.04 | AAI-based (high-confidence) | 0 |
| S1-final-viral-combined_k141_49157\|\|full | 1438 | No | NA | 4 | 0 | 1 | Low-quality | Genome-fragment | 0.45 | AAI-based (medium-confidence) | 0 |
| S3-final-viral-combined_k141_6823\|\|full | 1438 | No | NA | 5 | 0 | 0 | Low-quality | Genome-fragment | 2.24 | AAI-based (medium-confidence) | 0 |
| S5-final-viral-combined_k141_71431\|\|full | 1438 | No | NA | 4 | 1 | 0 | Low-quality | Genome-fragment | 2.15 | AAI-based (medium-confidence) | 0 |
| S1-final-viral-combined_k141_58869\|\|lt2gene | 1039 | No | NA | 1 | 0 | 0 | Not-determined | Genome-fragment | NA | NA | 0 |
| S1-final-viral-combined_k141_157389\|\|lt2gene | 2225 | No | NA | 2 | 0 | 0 | Not-determined | Genome-fragment | NA | NA | 0 |
| S5_k141_202695 | 1438 | No | NA | 1 | 0 | 1 | Low-quality | Genome-fragment | 4.68 | AAI-based (high-confidence) | 0 |
| S7-final-viral-combined_k141_167275\|\|full | 1438 | No | NA | 2 | 0 | 0 | Low-quality | Genome-fragment | 2.07 | AAI-based (medium-confidence) | 0 |
| S1-final-viral-combined_k141_191955\|\|lt2gene | 1952 | No | NA | 2 | 0 | 2 | Not-determined | Genome-fragment | NA | NA | 0 |
| S17-final-viral-combined_k141_67371\|\|full | 1438 | No | NA | 3 | 1 | 0 | Low-quality | Genome-fragment | 0.93 | HMM-based (lower-bound) | 0 |
| S1-final-viral-combined_k141_173913\|\|lt2gene | 2100 | No | NA | 1 | 0 | 0 | Not-determined | Genome-fragment | NA | NA | 0 |
| S1-final-viral-combined_k141_142217\|\|lt2gene | 1077 | No | NA | 2 | 0 | 1 | Not-determined | Genome-fragment | NA | NA | 0 |
| S1-final-viral-combined_k141_224355\|\|lt2gene | 1142 | No | NA | 1 | 0 | 1 | Not-determined | Genome-fragment | NA | NA | 0 |
| S17-final-viral-combined_k141_163596\|\|full | 1438 | No | NA | 1 | 0 | 0 | Low-quality | Genome-fragment | 3.09 | AAI-based (high-confidence) | 0 |
| S12-final-viral-combined_k141_224226\|\|full | 1438 | No | NA | 2 | 0 | 1 | Low-quality | Genome-fragment | 0.41 | AAI-based (medium-confidence) | 0 |
| S4_k141_115441 | 1438 | No | NA | 2 | 0 | 0 | Low-quality | Genome-fragment | 3.35 | AAI-based (medium-confidence) | 0 |
| S6_k141_98240 | 1438 | No | NA | 2 | 0 | 2 | Low-quality | Genome-fragment | 0.36 | AAI-based (medium-confidence) | 0 |
| S1-final-viral-combined_k141_273937\|\|lt2gene | 1060 | No | NA | 1 | 0 | 1 | Not-determined | Genome-fragment | NA | NA | 0 |
| S1-final-viral-combined_k141_192678\|\|lt2gene | 1017 | No | NA | 1 | 0 | 1 | Not-determined | Genome-fragment | NA | NA | 0 |
| S9-final-viral-combined_k141_12886\|\|full | 1439 | No | NA | 3 | 0 | 0 | Low-quality | Genome-fragment | 1.77 | AAI-based (medium-confidence) | 0 |
| S11-final-viral-combined_k141_223596\|\|full | 1439 | No | NA | 5 | 1 | 0 | Low-quality | Genome-fragment | 2.94 | AAI-based (high-confidence) | 0 |
| S11-final-viral-combined_k141_319754\|\|full | 1439 | No | NA | 2 | 0 | 0 | Low-quality | Genome-fragment | 2.06 | AAI-based (medium-confidence) | 0 |
| S7-final-viral-combined_k141_224211\|\|full | 1439 | No | NA | 3 | 0 | 2 | Low-quality | Genome-fragment | 0.45 | AAI-based (medium-confidence) | 0 |
| S17-final-viral-combined_k141_189176\|\|lt2gene | 1439 | No | NA | 2 | 0 | 0 | Low-quality | Genome-fragment | 2.01 | AAI-based (medium-confidence) | 0 |
| S1-final-viral-combined_k141_325182\|\|lt2gene | 2435 | No | NA | 2 | 0 | 2 | Not-determined | Genome-fragment | NA | NA | 0 |
| S12-final-viral-combined_k141_19548\|\|full | 1439 | No | NA | 5 | 0 | 2 | Low-quality | Genome-fragment | 1 | AAI-based (medium-confidence) | 0 |
| S2-final-viral-combined_k141_137432\|\|full | 1439 | No | NA | 3 | 0 | 1 | Low-quality | Genome-fragment | 0.77 | AAI-based (medium-confidence) | 0 |
| S8-final-viral-combined_k141_157432\|\|lt2gene | 1439 | No | NA | 2 | 1 | 0 | Low-quality | Genome-fragment | 3.14 | AAI-based (high-confidence) | 0 |
| S1-final-viral-combined_k141_340043\|\|lt2gene | 1042 | No | NA | 1 | 0 | 1 | Not-determined | Genome-fragment | NA | NA | 0 |
| S1-final-viral-combined_k141_283217\|\|lt2gene | 1440 | No | NA | 1 | 1 | 0 | Low-quality | Genome-fragment | 3.17 | AAI-based (high-confidence) | 0 |
| S7_k141_242461 | 1440 | No | NA | 3 | 0 | 0 | Low-quality | Genome-fragment | 4.06 | AAI-based (medium-confidence) | 0 |
| S17-final-viral-combined_k141_216559\|\|lt2gene | 1440 | No | NA | 1 | 1 | 0 | Low-quality | Genome-fragment | 3.62 | AAI-based (medium-confidence) | 0 |
| S12-final-viral-combined_k141_76724\|\|lt2gene | 1440 | No | NA | 2 | 1 | 0 | Low-quality | Genome-fragment | 1.28 | HMM-based (lower-bound) | 0 |
| S12-final-viral-combined_k141_109395\|\|lt2gene | 1440 | No | NA | 2 | 1 | 0 | Low-quality | Genome-fragment | 3.6 | AAI-based (high-confidence) | 0 |
| S12_k141_266257 | 1440 | No | NA | 3 | 1 | 0 | Low-quality | Genome-fragment | 1.33 | AAI-based (high-confidence) | 0 |
| S2-final-viral-combined_k141_137082\|\|full | 1440 | No | NA | 2 | 0 | 1 | Low-quality | Genome-fragment | 1.34 | AAI-based (medium-confidence) | 0 |
| S2_k141_1397 | 1440 | No | NA | 2 | 0 | 0 | Low-quality | Genome-fragment | 2.87 | AAI-based (high-confidence) | 0 |
| S1-final-viral-combined_k141_94751\|\|lt2gene | 2734 | No | NA | 1 | 0 | 0 | Not-determined | Genome-fragment | NA | NA | 0 |
| S2_k141_155308 | 1440 | No | NA | 2 | 0 | 0 | Low-quality | Genome-fragment | 3.28 | AAI-based (medium-confidence) | 0 |
| S9-final-viral-combined_k141_81970\|\|full | 1441 | No | NA | 5 | 3 | 0 | Low-quality | Genome-fragment | 3.09 | AAI-based (high-confidence) | 0 |
| S1-final-viral-combined_k141_374489\|\|lt2gene | 1432 | No | NA | 1 | 0 | 1 | Not-determined | Genome-fragment | NA | NA | 0 |
| S1-final-viral-combined_k141_210819\|\|lt2gene | 1117 | No | NA | 1 | 0 | 0 | Not-determined | Genome-fragment | NA | NA | 0 |
| S9-final-viral-combined_k141_60427\|\|lt2gene | 1441 | No | NA | 2 | 2 | 0 | Low-quality | Genome-fragment | 3.36 | AAI-based (high-confidence) | 0 |
| S11_k141_81558 | 1441 | No | NA | 2 | 1 | 0 | Low-quality | Genome-fragment | 3.38 | AAI-based (high-confidence) | 0 |
| S1-final-viral-combined_k141_217253\|\|full | 1441 | No | NA | 3 | 0 | 0 | Low-quality | Genome-fragment | 1.75 | AAI-based (high-confidence) | 0 |
| S1-final-viral-combined_k141_392369\|\|full | 1441 | No | NA | 3 | 0 | 0 | Low-quality | Genome-fragment | 1.29 | AAI-based (medium-confidence) | 0 |
| S1-final-viral-combined_k141_164517\|\|full | 1441 | No | NA | 3 | 0 | 0 | Low-quality | Genome-fragment | 1.45 | AAI-based (high-confidence) | 0 |
| S1-final-viral-combined_k141_283192\|\|lt2gene | 1441 | No | NA | 1 | 1 | 0 | Low-quality | Genome-fragment | 3.16 | AAI-based (high-confidence) | 0 |
| S1-final-viral-combined_k141_160504\|\|lt2gene | 2007 | No | NA | 2 | 0 | 2 | Not-determined | Genome-fragment | NA | NA | 0 |
| S3-final-viral-combined_k141_120133\|\|full | 1441 | No | NA | 5 | 0 | 0 | Low-quality | Genome-fragment | 3.7 | AAI-based (high-confidence) | 0 |
| S3-final-viral-combined_k141_30993\|\|lt2gene | 1441 | No | NA | 2 | 0 | 0 | Low-quality | Genome-fragment | 9.31 | AAI-based (high-confidence) | 0 |
| S5_k141_18681 | 1441 | No | NA | 1 | 1 | 0 | Low-quality | Genome-fragment | 1.63 | HMM-based (lower-bound) | 0 |
| S7-final-viral-combined_k141_197354\|\|full | 1441 | No | NA | 2 | 0 | 2 | Low-quality | Genome-fragment | 1.04 | AAI-based (high-confidence) | 0 |
| S7-final-viral-combined_k141_169261\|\|full | 1441 | No | NA | 3 | 0 | 0 | Low-quality | Genome-fragment | 2.54 | AAI-based (high-confidence) | 0 |
| S17-final-viral-combined_k141_37689\|\|full | 1441 | No | NA | 2 | 1 | 0 | Low-quality | Genome-fragment | 11.27 | AAI-based (high-confidence) | 0 |
| S12-final-viral-combined_k141_84541\|\|full | 1441 | No | NA | 3 | 0 | 0 | Low-quality | Genome-fragment | 5.06 | AAI-based (high-confidence) | 0 |
| S12-final-viral-combined_k141_386907\|\|full | 1441 | No | NA | 4 | 2 | 0 | Low-quality | Genome-fragment | 4.16 | AAI-based (medium-confidence) | 0 |
| S1-final-viral-combined_k141_46442\|\|lt2gene | 1282 | No | NA | 1 | 0 | 0 | Not-determined | Genome-fragment | NA | NA | 0 |
| S12-final-viral-combined_k141_342475\|\|lt2gene | 1441 | No | NA | 1 | 1 | 0 | Low-quality | Genome-fragment | 2.98 | AAI-based (high-confidence) | 0 |
| S2-final-viral-combined_k141_40053\|\|full | 1441 | No | NA | 4 | 1 | 0 | Low-quality | Genome-fragment | 3.16 | AAI-based (medium-confidence) | 0 |
| S1-final-viral-combined_k141_212895\|\|lt2gene | 1124 | No | NA | 1 | 0 | 0 | Not-determined | Genome-fragment | NA | NA | 0 |
| S2-final-viral-combined_k141_56373\|\|lt2gene | 1441 | No | NA | 2 | 0 | 0 | Low-quality | Genome-fragment | 3.29 | AAI-based (medium-confidence) | 0 |
| S4_k141_108597 | 1441 | No | NA | 6 | 0 | 0 | Low-quality | Genome-fragment | 3.24 | AAI-based (high-confidence) | 0 |
| S1-final-viral-combined_k141_30341\|\|lt2gene | 3423 | No | NA | 1 | 0 | 0 | Not-determined | Genome-fragment | NA | NA | 0 |
| S6-final-viral-combined_k141_65897\|\|full | 1441 | No | NA | 4 | 0 | 0 | Low-quality | Genome-fragment | 2.19 | AAI-based (medium-confidence) | 0 |
| S1-final-viral-combined_k141_160972\|\|full | 1442 | No | NA | 2 | 0 | 0 | Low-quality | Genome-fragment | 2.11 | AAI-based (medium-confidence) | 0 |
| S1-final-viral-combined_k141_296762\|\|lt2gene | 1442 | No | NA | 2 | 1 | 0 | Low-quality | Genome-fragment | 3.09 | AAI-based (high-confidence) | 0 |
| S1-final-viral-combined_k141_146608\|\|lt2gene | 1697 | No | NA | 1 | 0 | 0 | Not-determined | Genome-fragment | NA | NA | 0 |
| S1_k141_304077 | 1442 | No | NA | 1 | 0 | 0 | Low-quality | Genome-fragment | 2.76 | AAI-based (medium-confidence) | 0 |
| S1-final-viral-combined_k141_178609\|\|lt2gene | 1602 | No | NA | 2 | 0 | 0 | Not-determined | Genome-fragment | NA | NA | 0 |
| S1-final-viral-combined_k141_278030\|\|lt2gene | 1042 | No | NA | 2 | 0 | 0 | Not-determined | Genome-fragment | NA | NA | 0 |
| S1-final-viral-combined_k141_97776\|\|lt2gene | 1148 | No | NA | 1 | 0 | 0 | Not-determined | Genome-fragment | NA | NA | 0 |
| S3-final-viral-combined_k141_43729\|\|full | 1442 | No | NA | 2 | 0 | 0 | Low-quality | Genome-fragment | 2.71 | AAI-based (medium-confidence) | 0 |
| S3-final-viral-combined_k141_21607\|\|full | 1442 | No | NA | 3 | 0 | 0 | Low-quality | Genome-fragment | 3.18 | AAI-based (medium-confidence) | 0 |
| S1-final-viral-combined_k141_278470\|\|lt2gene | 3557 | No | NA | 2 | 0 | 0 | Not-determined | Genome-fragment | NA | NA | 0 |
| S5-final-viral-combined_k141_132615\|\|full | 1442 | No | NA | 3 | 0 | 1 | Low-quality | Genome-fragment | 0.31 | AAI-based (medium-confidence) | 0 |
| S5-final-viral-combined_k141_150349\|\|full | 1442 | No | NA | 3 | 0 | 0 | Low-quality | Genome-fragment | 2.65 | AAI-based (high-confidence) | 0 |
| S7-final-viral-combined_k141_139907\|\|full | 1442 | No | NA | 3 | 1 | 0 | Low-quality | Genome-fragment | 3.43 | AAI-based (high-confidence) | 0 |
| S17-final-viral-combined_k141_229164\|\|full | 1442 | No | NA | 2 | 0 | 0 | Low-quality | Genome-fragment | 2.85 | AAI-based (medium-confidence) | 0 |
| S12_k141_346626 | 1442 | No | NA | 4 | 0 | 0 | Low-quality | Genome-fragment | 3.91 | AAI-based (high-confidence) | 0 |
| S1-final-viral-combined_k141_31879\|\|lt2gene | 1084 | No | NA | 2 | 0 | 0 | Not-determined | Genome-fragment | NA | NA | 0 |
| S9-final-viral-combined_k141_26784\|\|full | 1443 | No | NA | 2 | 0 | 2 | Low-quality | Genome-fragment | 0.86 | AAI-based (medium-confidence) | 0 |
| S11-final-viral-combined_k141_37556\|\|full | 1443 | No | NA | 2 | 0 | 0 | Low-quality | Genome-fragment | 1.75 | AAI-based (high-confidence) | 0 |
| S1-final-viral-combined_k141_33113\|\|full | 1443 | No | NA | 2 | 0 | 2 | Low-quality | Genome-fragment | 0.66 | AAI-based (medium-confidence) | 0 |
| S1-final-viral-combined_k141_83735\|\|full | 1443 | No | NA | 2 | 0 | 0 | Low-quality | Genome-fragment | 1.05 | AAI-based (high-confidence) | 0 |
| S1-final-viral-combined_k141_114999\|\|lt2gene | 2803 | No | NA | 1 | 0 | 0 | Not-determined | Genome-fragment | NA | NA | 0 |
| S1_k141_49376 | 3307 | No | NA | 5 | 0 | 0 | Not-determined | Genome-fragment | NA | NA | 0 |
| S1_k141_378447 | 1365 | No | NA | 2 | 0 | 2 | Not-determined | Genome-fragment | NA | NA | 0 |
| S1_k141_329087 | 1020 | No | NA | 1 | 0 | 1 | Not-determined | Genome-fragment | NA | NA | 0 |
| S1_k141_312637 | 1403 | No | NA | 2 | 0 | 0 | Not-determined | Genome-fragment | NA | NA | 0 |
| S1_k141_345535 | 1064 | No | NA | 1 | 0 | 1 | Not-determined | Genome-fragment | NA | NA | 0 |
| S1_k141_16464 | 3900 | No | NA | 2 | 0 | 2 | Not-determined | Genome-fragment | NA | NA | 0 |
| S5-final-viral-combined_k141_67929\|\|full | 1443 | No | NA | 3 | 0 | 2 | Low-quality | Genome-fragment | 3.41 | AAI-based (medium-confidence) | 0 |
| S1_k141_32948 | 1152 | No | NA | 2 | 0 | 1 | Not-determined | Genome-fragment | NA | NA | 0 |
| S1_k141_312671 | 1025 | No | NA | 2 | 0 | 1 | Not-determined | Genome-fragment | NA | NA | 0 |
| S1_k141_197462 | 3644 | No | NA | 8 | 0 | 8 | Not-determined | Genome-fragment | NA | NA | 0 |
| S17-final-viral-combined_k141_201716\|\|full | 1443 | No | NA | 2 | 1 | 0 | Low-quality | Genome-fragment | 2.13 | HMM-based (lower-bound) | 0 |
| S1_k141_246805 | 1324 | No | NA | 3 | 0 | 1 | Not-determined | Genome-fragment | NA | NA | 0 |
| S1_k141_378494 | 2025 | No | NA | 2 | 0 | 0 | Not-determined | Genome-fragment | NA | NA | 0 |
| S1_k141_329114 | 2306 | No | NA | 2 | 0 | 2 | Not-determined | Genome-fragment | NA | NA | 0 |
| S1_k141_33005 | 1003 | No | NA | 2 | 0 | 2 | Not-determined | Genome-fragment | NA | NA | 0 |
| S12-final-viral-combined_k141_329084\|\|full | 1443 | No | NA | 3 | 2 | 0 | Low-quality | Genome-fragment | 2.65 | HMM-based (lower-bound) | 0 |
| S1_k141_263297 | 1310 | No | NA | 1 | 0 | 1 | Not-determined | Genome-fragment | NA | NA | 0 |
| S1_k141_148157 | 2635 | No | NA | 1 | 0 | 0 | Not-determined | Genome-fragment | NA | NA | 0 |
| S12-final-viral-combined_k141_45789\|\|full | 1443 | No | NA | 1 | 0 | 0 | Low-quality | Genome-fragment | 2.18 | AAI-based (high-confidence) | 0 |
| S1_k141_181066 | 1039 | No | NA | 1 | 0 | 0 | Not-determined | Genome-fragment | NA | NA | 0 |
| S1_k141_378514 | 1203 | No | NA | 2 | 0 | 2 | Not-determined | Genome-fragment | NA | NA | 0 |
| S1_k141_329152 | 1221 | No | NA | 1 | 0 | 1 | Not-determined | Genome-fragment | NA | NA | 0 |
| S1_k141_329161 | 1527 | No | NA | 1 | 0 | 0 | Not-determined | Genome-fragment | NA | NA | 0 |
| S1_k141_152 | 1493 | No | NA | 1 | 0 | 0 | Not-determined | Genome-fragment | NA | NA | 0 |
| S1_k141_296285 | 3781 | No | NA | 3 | 0 | 1 | Not-determined | Genome-fragment | NA | NA | 0 |
| S1_k141_345622 | 1420 | No | NA | 1 | 0 | 0 | Not-determined | Genome-fragment | NA | NA | 0 |
| S1_k141_296300 | 1409 | No | NA | 1 | 0 | 1 | Not-determined | Genome-fragment | NA | NA | 0 |
| S1_k141_115317 | 1086 | No | NA | 1 | 0 | 1 | Not-determined | Genome-fragment | NA | NA | 0 |
| S1_k141_164646 | 1545 | No | NA | 1 | 0 | 0 | Not-determined | Genome-fragment | NA | NA | 0 |
| S12_k141_316218 | 1443 | No | NA | 2 | 0 | 2 | Low-quality | Genome-fragment | 0.72 | AAI-based (medium-confidence) | 0 |
| S1_k141_98823 | 1712 | No | NA | 1 | 0 | 0 | Not-determined | Genome-fragment | NA | NA | 0 |
| S1_k141_164656 | 1051 | No | NA | 1 | 0 | 0 | Not-determined | Genome-fragment | NA | NA | 0 |
| S1_k141_16574 | 1056 | No | NA | 1 | 0 | 1 | Not-determined | Genome-fragment | NA | NA | 0 |
| S1_k141_263323 | 2916 | No | NA | 3 | 0 | 0 | Not-determined | Genome-fragment | NA | NA | 0 |
| S1_k141_148259 | 1581 | No | NA | 1 | 0 | 0 | Not-determined | Genome-fragment | NA | NA | 0 |
| S1_k141_197558 | 2032 | No | NA | 2 | 0 | 0 | Not-determined | Genome-fragment | NA | NA | 0 |
| S4-final-viral-combined_k141_16737\|\|full | 1443 | No | NA | 4 | 1 | 0 | Low-quality | Genome-fragment | 2.78 | AAI-based (medium-confidence) | 0 |
| S1_k141_263333 | 1395 | No | NA | 1 | 0 | 1 | Not-determined | Genome-fragment | NA | NA | 0 |
| S1_k141_362171 | 1136 | No | NA | 1 | 0 | 0 | Not-determined | Genome-fragment | NA | NA | 0 |
| S1_k141_181157 | 7315 | No | NA | 7 | 0 | 0 | Not-determined | Genome-fragment | NA | NA | 0 |
| S1_k141_181161 | 1161 | No | NA | 2 | 0 | 1 | Not-determined | Genome-fragment | NA | NA | 0 |
| S1_k141_148277 | 1204 | No | NA | 2 | 0 | 2 | Not-determined | Genome-fragment | NA | NA | 0 |
| S1_k141_131885 | 2046 | No | NA | 4 | 0 | 1 | Not-determined | Genome-fragment | NA | NA | 0 |
| S6-final-viral-combined_k141_44970\|\|full | 1443 | No | NA | 3 | 0 | 2 | Low-quality | Genome-fragment | 0.6 | AAI-based (medium-confidence) | 0 |
| S1_k141_148294 | 1041 | No | NA | 2 | 0 | 2 | Not-determined | Genome-fragment | NA | NA | 0 |
| S1_k141_263360 | 1054 | No | NA | 1 | 0 | 0 | Not-determined | Genome-fragment | NA | NA | 0 |
| S1_k141_197596 | 1059 | No | NA | 2 | 0 | 0 | Not-determined | Genome-fragment | NA | NA | 0 |
| S1_k141_279900 | 4048 | No | NA | 2 | 0 | 0 | Not-determined | Genome-fragment | NA | NA | 0 |
| S1_k141_181180 | 1088 | No | NA | 1 | 0 | 0 | Not-determined | Genome-fragment | NA | NA | 0 |
| S1_k141_230587 | 1208 | No | NA | 1 | 0 | 1 | Not-determined | Genome-fragment | NA | NA | 0 |
| S8-final-viral-combined_k141_271824\|\|full | 1443 | No | NA | 4 | 0 | 0 | Low-quality | Genome-fragment | 1.02 | AAI-based (medium-confidence) | 0 |
| S8_k141_267963 | 1443 | No | NA | 1 | 0 | 1 | Low-quality | Genome-fragment | 4.07 | AAI-based (medium-confidence) | 0 |
| S1_k141_181198 | 3673 | No | NA | 8 | 0 | 8 | Not-determined | Genome-fragment | NA | NA | 0 |
| S1_k141_329277 | 1169 | No | NA | 2 | 0 | 1 | Not-determined | Genome-fragment | NA | NA | 0 |
| S1_k141_353 | 1393 | No | NA | 2 | 0 | 2 | Not-determined | Genome-fragment | NA | NA | 0 |
| S1_k141_230595 | 1170 | No | NA | 1 | 0 | 0 | Not-determined | Genome-fragment | NA | NA | 0 |
| S1_k141_16701 | 4312 | No | NA | 5 | 0 | 3 | Not-determined | Genome-fragment | NA | NA | 0 |
| S11-final-viral-combined_k141_27757\|\|full | 1444 | No | NA | 1 | 0 | 1 | Low-quality | Genome-fragment | 2.04 | AAI-based (high-confidence) | 0 |
| S1_k141_362207 | 2010 | No | NA | 4 | 0 | 3 | Not-determined | Genome-fragment | NA | NA | 0 |
| S11-final-viral-combined_k141_149277\|\|full | 1444 | No | NA | 3 | 0 | 0 | Low-quality | Genome-fragment | 0.57 | AAI-based (high-confidence) | 0 |
| S1_k141_230630 | 3470 | No | NA | 3 | 0 | 1 | Not-determined | Genome-fragment | NA | NA | 0 |
| S1_k141_410 | 2008 | No | NA | 3 | 0 | 2 | Not-determined | Genome-fragment | NA | NA | 0 |
| S1_k141_280017 | 1118 | No | NA | 1 | 0 | 1 | Not-determined | Genome-fragment | NA | NA | 0 |
| S1_k141_65939 | 1198 | No | NA | 1 | 0 | 1 | Not-determined | Genome-fragment | NA | NA | 0 |
| S1_k141_65940 | 1273 | No | NA | 1 | 0 | 1 | Not-determined | Genome-fragment | NA | NA | 0 |
| S1_k141_230702 | 4605 | No | NA | 1 | 0 | 1 | Not-determined | Genome-fragment | NA | NA | 0 |
| S1_k141_312980 | 1292 | No | NA | 1 | 0 | 0 | Not-determined | Genome-fragment | NA | NA | 0 |
| S1-final-viral-combined_k141_386694\|\|full | 1444 | No | NA | 3 | 0 | 0 | Low-quality | Genome-fragment | 1.26 | AAI-based (high-confidence) | 0 |
| S1_k141_115468 | 1089 | No | NA | 1 | 0 | 0 | Not-determined | Genome-fragment | NA | NA | 0 |
| S1-final-viral-combined_k141_236275\|\|lt2gene | 1444 | No | NA | 2 | 2 | 0 | Low-quality | Genome-fragment | 3.1 | AAI-based (high-confidence) | 0 |
| S1_k141_362303 | 1103 | No | NA | 1 | 0 | 1 | Not-determined | Genome-fragment | NA | NA | 0 |
| S1_k141_230767 | 2393 | No | NA | 1 | 0 | 1 | Not-determined | Genome-fragment | NA | NA | 0 |
| S1_k141_132060 | 1269 | No | NA | 1 | 0 | 1 | Not-determined | Genome-fragment | NA | NA | 0 |
| S3-final-viral-combined_k141_2170\|\|full | 1444 | No | NA | 3 | 1 | 0 | Low-quality | Genome-fragment | 2.69 | AAI-based (high-confidence) | 0 |
| S1_k141_148367 | 2326 | No | NA | 3 | 0 | 3 | Not-determined | Genome-fragment | NA | NA | 0 |
| S1_k141_296543 | 1032 | No | NA | 1 | 0 | 1 | Not-determined | Genome-fragment | NA | NA | 0 |
| S1_k141_296545 | 2004 | No | NA | 1 | 0 | 1 | Not-determined | Genome-fragment | NA | NA | 0 |
| S7-final-viral-combined_k141_18334\|\|full | 1444 | No | NA | 5 | 0 | 0 | Low-quality | Genome-fragment | 1.81 | AAI-based (high-confidence) | 0 |
| S1_k141_362352 | 1801 | No | NA | 2 | 0 | 1 | Not-determined | Genome-fragment | NA | NA | 0 |
| S1_k141_148425 | 1073 | No | NA | 2 | 0 | 1 | Not-determined | Genome-fragment | NA | NA | 0 |
| S1_k141_82755 | 3302 | No | NA | 3 | 0 | 3 | Not-determined | Genome-fragment | NA | NA | 0 |
| S7-final-viral-combined_k141_193996\|\|full | 1444 | No | NA | 2 | 0 | 1 | Low-quality | Genome-fragment | 1.4 | AAI-based (medium-confidence) | 0 |
| S1_k141_247158 | 1102 | No | NA | 1 | 0 | 1 | Not-determined | Genome-fragment | NA | NA | 0 |
| S1_k141_49689 | 1359 | No | NA | 2 | 0 | 1 | Not-determined | Genome-fragment | NA | NA | 0 |
| S1_k141_33138 | 4323 | No | NA | 6 | 0 | 6 | Not-determined | Genome-fragment | NA | NA | 0 |
| S1_k141_33140 | 3356 | No | NA | 1 | 0 | 0 | Not-determined | Genome-fragment | NA | NA | 0 |
| S17-k141_165843 | 1444 | No | NA | 3 | 0 | 1 | Low-quality | Genome-fragment | 3.27 | AAI-based (medium-confidence) | 0 |
| S1_k141_247167 | 2280 | No | NA | 2 | 0 | 1 | Not-determined | Genome-fragment | NA | NA | 0 |
| S1_k141_345875 | 3968 | No | NA | 4 | 0 | 3 | Not-determined | Genome-fragment | NA | NA | 0 |
| S1_k141_263569 | 4738 | No | NA | 7 | 0 | 0 | Not-determined | Genome-fragment | NA | NA | 0 |
| S1_k141_263570 | 2641 | No | NA | 6 | 0 | 6 | Not-determined | Genome-fragment | NA | NA | 0 |
| S1_k141_230826 | 1106 | No | NA | 1 | 0 | 1 | Not-determined | Genome-fragment | NA | NA | 0 |
| S1_k141_99097 | 2270 | No | NA | 1 | 0 | 1 | Not-determined | Genome-fragment | NA | NA | 0 |
| S2-final-viral-combined_k141_140621\|\|full | 1444 | No | NA | 2 | 0 | 1 | Low-quality | Genome-fragment | 4.34 | AAI-based (medium-confidence) | 0 |
| S1_k141_115609 | 2774 | No | NA | 1 | 0 | 1 | Not-determined | Genome-fragment | NA | NA | 0 |
| S1_k141_49730 | 3742 | No | NA | 1 | 0 | 0 | Not-determined | Genome-fragment | NA | NA | 0 |
| S4-final-viral-combined_k141_46786\|\|full | 1444 | No | NA | 2 | 0 | 0 | Low-quality | Genome-fragment | 0.95 | AAI-based (medium-confidence) | 0 |
| S1_k141_378796 | 1733 | No | NA | 1 | 0 | 0 | Not-determined | Genome-fragment | NA | NA | 0 |
| S1_k141_16854 | 1404 | No | NA | 1 | 0 | 1 | Not-determined | Genome-fragment | NA | NA | 0 |
| S1_k141_197883 | 1816 | No | NA | 3 | 0 | 0 | Not-determined | Genome-fragment | NA | NA | 0 |
| S1_k141_99134 | 1481 | No | NA | 2 | 0 | 2 | Not-determined | Genome-fragment | NA | NA | 0 |
| S1_k141_313098 | 1046 | No | NA | 1 | 0 | 0 | Not-determined | Genome-fragment | NA | NA | 0 |
| S1_k141_230843 | 1922 | No | NA | 3 | 0 | 1 | Not-determined | Genome-fragment | NA | NA | 0 |
| S1_k141_214325 | 2067 | No | NA | 4 | 0 | 4 | Not-determined | Genome-fragment | NA | NA | 0 |
| S1_k141_329453 | 7011 | No | NA | 8 | 0 | 4 | Not-determined | Genome-fragment | NA | NA | 0 |
| S11-final-viral-combined_k141_42755\|\|full | 1445 | No | NA | 3 | 1 | 0 | Low-quality | Genome-fragment | 3.61 | AAI-based (medium-confidence) | 0 |
| S1_k141_378877 | 1294 | No | NA | 3 | 0 | 2 | Not-determined | Genome-fragment | NA | NA | 0 |
| S1_k141_230864 | 5119 | No | NA | 2 | 0 | 2 | Not-determined | Genome-fragment | NA | NA | 0 |
| S1_k141_164963 | 1307 | No | NA | 1 | 0 | 1 | Not-determined | Genome-fragment | NA | NA | 0 |
| S1_k141_378891 | 3188 | No | NA | 7 | 0 | 1 | Not-determined | Genome-fragment | NA | NA | 0 |
| S1_k141_197951 | 1278 | No | NA | 2 | 0 | 1 | Not-determined | Genome-fragment | NA | NA | 0 |
| S1_k141_296663 | 1428 | No | NA | 2 | 0 | 0 | Not-determined | Genome-fragment | NA | NA | 0 |
| S11-final-viral-combined_k141_50886\|\|full | 1445 | No | NA | 2 | 0 | 1 | Low-quality | Genome-fragment | 1.37 | AAI-based (medium-confidence) | 0 |
| S1-final-viral-combined_k141_24921\|\|full | 1445 | No | NA | 2 | 0 | 0 | Low-quality | Genome-fragment | 2.83 | AAI-based (high-confidence) | 0 |
| S1_k141_345988 | 1304 | No | NA | 1 | 0 | 0 | Not-determined | Genome-fragment | NA | NA | 0 |
| S1_k141_378918 | 3846 | No | NA | 3 | 0 | 2 | Not-determined | Genome-fragment | NA | NA | 0 |
| S1_k141_378927 | 1126 | No | NA | 1 | 0 | 0 | Not-determined | Genome-fragment | NA | NA | 0 |
| S1_k141_115723 | 3860 | No | NA | 5 | 0 | 0 | Not-determined | Genome-fragment | NA | NA | 0 |
| S1_k141_214385 | 1203 | No | NA | 1 | 0 | 0 | Not-determined | Genome-fragment | NA | NA | 0 |
| S1_k141_66082 | 1347 | No | NA | 2 | 0 | 1 | Not-determined | Genome-fragment | NA | NA | 0 |
| S1-final-viral-combined_k141_156970\|\|lt2gene | 1445 | No | NA | 2 | 1 | 0 | Low-quality | Genome-fragment | 0.78 | HMM-based (lower-bound) | 0 |
| S3_k141_83786 | 1445 | No | NA | 3 | 1 | 0 | Low-quality | Genome-fragment | 2.03 | AAI-based (high-confidence) | 0 |
| S1_k141_263698 | 1284 | No | NA | 2 | 0 | 0 | Not-determined | Genome-fragment | NA | NA | 0 |
| S5-final-viral-combined_k141_1305\|\|lt2gene | 1445 | No | NA | 2 | 1 | 0 | Low-quality | Genome-fragment | 3.73 | AAI-based (high-confidence) | 0 |
| S1_k141_230917 | 5964 | No | NA | 4 | 0 | 1 | Not-determined | Genome-fragment | NA | NA | 0 |
| S5_k141_16453 | 1445 | No | NA | 1 | 0 | 0 | Low-quality | Genome-fragment | 2.35 | AAI-based (high-confidence) | 0 |
| S1_k141_247360 | 4461 | No | NA | 4 | 0 | 0 | Not-determined | Genome-fragment | NA | NA | 0 |
| S1_k141_263779 | 3050 | No | NA | 3 | 0 | 2 | Not-determined | Genome-fragment | NA | NA | 0 |
| S1_k141_115810 | 1082 | No | NA | 1 | 0 | 1 | Not-determined | Genome-fragment | NA | NA | 0 |
| S12-final-viral-combined_k141_74326\|\|full | 1445 | No | NA | 2 | 0 | 0 | Low-quality | Genome-fragment | 2.49 | AAI-based (medium-confidence) | 0 |
| S1_k141_214521 | 3187 | No | NA | 3 | 0 | 2 | Not-determined | Genome-fragment | NA | NA | 0 |
| S1_k141_115861 | 1105 | No | NA | 1 | 0 | 1 | Not-determined | Genome-fragment | NA | NA | 0 |
| S4-final-viral-combined_k141_98590\|\|full | 1445 | Yes | 731 | 2 | 1 | 1 | Low-quality | Genome-fragment | 1.67 | AAI-based (medium-confidence) | 49.41 |
| S1_k141_176510 | 1446 | No | NA | 1 | 0 | 1 | Low-quality | Genome-fragment | 2.47 | AAI-based (high-confidence) | 0 |
| S1_k141_66204 | 4540 | No | NA | 3 | 0 | 1 | Not-determined | Genome-fragment | NA | NA | 0 |
| S1_k141_247400 | 1008 | No | NA | 1 | 0 | 1 | Not-determined | Genome-fragment | NA | NA | 0 |
| S5_k141_63606 | 1446 | No | NA | 3 | 0 | 2 | Low-quality | Genome-fragment | 2.86 | AAI-based (medium-confidence) | 0 |
| S1_k141_148650 | 1375 | No | NA | 3 | 0 | 2 | Not-determined | Genome-fragment | NA | NA | 0 |
| S1_k141_198088 | 7010 | No | NA | 4 | 0 | 2 | Not-determined | Genome-fragment | NA | NA | 0 |
| S2-final-viral-combined_k141_243742\|\|full | 1446 | Yes | 410 | 3 | 1 | 1 | Low-quality | Genome-fragment | 1.02 | AAI-based (medium-confidence) | 71.65 |
| S1_k141_214691 | 1015 | No | NA | 2 | 0 | 1 | Not-determined | Genome-fragment | NA | NA | 0 |
| S1_k141_296810 | 1210 | No | NA | 2 | 0 | 2 | Not-determined | Genome-fragment | NA | NA | 0 |
| S1_k141_165231 | 1007 | No | NA | 1 | 0 | 1 | Not-determined | Genome-fragment | NA | NA | 0 |
| S1_k141_132457 | 1007 | No | NA | 2 | 0 | 0 | Not-determined | Genome-fragment | NA | NA | 0 |
| S1_k141_247527 | 1374 | No | NA | 3 | 0 | 1 | Not-determined | Genome-fragment | NA | NA | 0 |
| S1_k141_280492 | 1064 | No | NA | 1 | 0 | 0 | Not-determined | Genome-fragment | NA | NA | 0 |
| S1_k141_296850 | 49921 | No | NA | 68 | 19 | 1 | High-quality | High-quality | 94.65 | AAI-based (high-confidence) | 0 |
| S1_k141_132530 | 1342 | No | NA | 1 | 0 | 1 | Not-determined | Genome-fragment | NA | NA | 0 |
| S1_k141_148775 | 6970 | No | NA | 10 | 0 | 1 | Not-determined | Genome-fragment | NA | NA | 0 |
| S1_k141_181903 | 1102 | No | NA | 1 | 0 | 1 | Not-determined | Genome-fragment | NA | NA | 0 |
| S2-final-viral-combined_k141_224531\|\|full | 1446 | No | NA | 3 | 0 | 1 | Low-quality | Genome-fragment | 1.39 | AAI-based (medium-confidence) | 0 |
| S1_k141_313388 | 11884 | No | NA | 12 | 0 | 6 | Not-determined | Genome-fragment | NA | NA | 0 |
| S1_k141_198298 | 4491 | No | NA | 6 | 0 | 1 | Not-determined | Genome-fragment | NA | NA | 0 |
| S8-final-viral-combined_k141_90330\|\|full | 1446 | No | NA | 4 | 2 | 0 | Low-quality | Genome-fragment | 4.35 | AAI-based (high-confidence) | 0 |
| S1_k141_1081 | 1191 | No | NA | 2 | 0 | 0 | Not-determined | Genome-fragment | NA | NA | 0 |
| S8-final-viral-combined_k141_275979\|\|full | 1446 | No | NA | 2 | 0 | 1 | Low-quality | Genome-fragment | 0.61 | AAI-based (medium-confidence) | 0 |
| S1_k141_214936 | 1425 | No | NA | 1 | 0 | 0 | Not-determined | Genome-fragment | NA | NA | 0 |
| S1_k141_198315 | 7127 | No | NA | 5 | 0 | 5 | Not-determined | Genome-fragment | NA | NA | 0 |
| S1_k141_198318 | 1562 | No | NA | 2 | 0 | 0 | Not-determined | Genome-fragment | NA | NA | 0 |
| S1_k141_280653 | 1651 | No | NA | 3 | 0 | 0 | Not-determined | Genome-fragment | NA | NA | 0 |
| S8-final-viral-combined_k141_130050\|\|lt2gene | 1446 | No | NA | 2 | 1 | 0 | Low-quality | Genome-fragment | 1.36 | HMM-based (lower-bound) | 0 |
| S1_k141_379272 | 11296 | No | NA | 8 | 0 | 6 | Not-determined | Genome-fragment | NA | NA | 0 |
| S3-final-viral-combined_k141_77674\|\|full | 1447 | No | NA | 2 | 1 | 0 | Low-quality | Genome-fragment | 3.12 | AAI-based (high-confidence) | 0 |
| S1_k141_280703 | 1322 | No | NA | 2 | 0 | 1 | Not-determined | Genome-fragment | NA | NA | 0 |
| S3_k141_66527 | 1447 | No | NA | 3 | 0 | 0 | Low-quality | Genome-fragment | 1.32 | AAI-based (medium-confidence) | 0 |
| S1_k141_116337 | 3592 | No | NA | 3 | 0 | 3 | Not-determined | Genome-fragment | NA | NA | 0 |
| S1_k141_17283 | 1087 | No | NA | 2 | 0 | 2 | Not-determined | Genome-fragment | NA | NA | 0 |
| S5-final-viral-combined_k141_56605\|\|full | 1447 | No | NA | 4 | 0 | 1 | Low-quality | Genome-fragment | 0.85 | AAI-based (high-confidence) | 0 |
| S1_k141_198387 | 1039 | No | NA | 1 | 0 | 0 | Not-determined | Genome-fragment | NA | NA | 0 |
| S17-final-viral-combined_k141_34419\|\|full | 1447 | No | NA | 3 | 0 | 0 | Low-quality | Genome-fragment | 2.2 | AAI-based (medium-confidence) | 0 |
| S1_k141_1192 | 1094 | No | NA | 1 | 0 | 1 | Not-determined | Genome-fragment | NA | NA | 0 |
| S4-final-viral-combined_k141_112823\|\|full | 1447 | No | NA | 3 | 0 | 1 | Low-quality | Genome-fragment | 11.25 | AAI-based (high-confidence) | 0 |
| S8_k141_86261 | 1447 | No | NA | 2 | 0 | 0 | Low-quality | Genome-fragment | 3.65 | AAI-based (high-confidence) | 0 |
| S11-final-viral-combined_k141_212727\|\|full | 1448 | No | NA | 2 | 0 | 1 | Low-quality | Genome-fragment | 0.75 | AAI-based (medium-confidence) | 0 |
| S1_k141_362985 | 1255 | No | NA | 3 | 0 | 0 | Not-determined | Genome-fragment | NA | NA | 0 |
| S11-final-viral-combined_k141_77871\|\|full | 1448 | No | NA | 4 | 0 | 2 | Low-quality | Genome-fragment | 3.68 | AAI-based (high-confidence) | 0 |
| S1_k141_297144 | 1091 | No | NA | 3 | 0 | 2 | Not-determined | Genome-fragment | NA | NA | 0 |
| S11-final-viral-combined_k141_115652\|\|lt2gene | 1448 | No | NA | 2 | 2 | 0 | Low-quality | Genome-fragment | 3.8 | AAI-based (high-confidence) | 0 |
| S1_k141_247808 | 1842 | No | NA | 3 | 0 | 1 | Not-determined | Genome-fragment | NA | NA | 0 |
| S1_k141_264193 | 1124 | No | NA | 2 | 0 | 2 | Not-determined | Genome-fragment | NA | NA | 0 |
| S1_k141_379343 | 4168 | No | NA | 2 | 0 | 0 | Not-determined | Genome-fragment | NA | NA | 0 |
| S1_k141_264205 | 1160 | No | NA | 1 | 0 | 1 | Not-determined | Genome-fragment | NA | NA | 0 |
| S1_k141_280758 | 1839 | No | NA | 1 | 0 | 1 | Not-determined | Genome-fragment | NA | NA | 0 |
| S1_k141_1244 | 1507 | No | NA | 1 | 0 | 1 | Not-determined | Genome-fragment | NA | NA | 0 |
| S1_k141_215101 | 1069 | No | NA | 1 | 0 | 1 | Not-determined | Genome-fragment | NA | NA | 0 |
| S1_k141_379361 | 1117 | No | NA | 1 | 0 | 0 | Not-determined | Genome-fragment | NA | NA | 0 |
| S1-final-viral-combined_k141_353806\|\|full | 1448 | No | NA | 4 | 0 | 0 | Low-quality | Genome-fragment | 1.35 | AAI-based (high-confidence) | 0 |

| | | | | | | | | | | | |
|---|---|---|---|---|---|---|---|---|---|---|---|
| S1_k141_280780 | 1268 | No | NA | 2 | 0 | 1 | Not-determined | Genome-fragment | NA | NA | 0 |
| S1_k141_313558 | 7695 | No | NA | 5 | 0 | 4 | Not-determined | Genome-fragment | NA | NA | 0 |
| S1_k141_280793 | 1330 | No | NA | 2 | 0 | 0 | Not-determined | Genome-fragment | NA | NA | 0 |
| S1_k141_1282 | 1346 | No | NA | 2 | 0 | 1 | Not-determined | Genome-fragment | NA | NA | 0 |
| S1_k141_379373 | 2984 | No | NA | 2 | 0 | 2 | Not-determined | Genome-fragment | NA | NA | 0 |
| S1-final-viral-combined_k141_324199\|\|full | 1448 | No | NA | 4 | 0 | 0 | Low-quality | Genome-fragment | 2.38 | AAI-based (medium-confidence) | 0 |
| S1-final-viral-combined_k141_393222\|\|full | 1448 | No | NA | 3 | 0 | 0 | Low-quality | Genome-fragment | 2.37 | AAI-based (medium-confidence) | 0 |
| S1_k141_17423 | 1478 | No | NA | 1 | 0 | 0 | Not-determined | Genome-fragment | NA | NA | 0 |
| S1_k141_215157 | 1302 | No | NA | 2 | 0 | 1 | Not-determined | Genome-fragment | NA | NA | 0 |
| S1_k141_379405 | 8039 | No | NA | 5 | 0 | 3 | Not-determined | Genome-fragment | NA | NA | 0 |
| S1_k141_136803 | 1448 | No | NA | 1 | 1 | 0 | Low-quality | Genome-fragment | 1.02 | HMM-based (lower-bound) | 0 |
| S1_k141_33707 | 1571 | No | NA | 3 | 0 | 2 | Not-determined | Genome-fragment | NA | NA | 0 |
| S1_k141_1353 | 1104 | No | NA | 1 | 0 | 1 | Not-determined | Genome-fragment | NA | NA | 0 |
| S1_k141_264294 | 1280 | No | NA | 1 | 0 | 0 | Not-determined | Genome-fragment | NA | NA | 0 |
| S7-final-viral-combined_k141_127306\|\|full | 1448 | No | NA | 3 | 0 | 1 | Low-quality | Genome-fragment | 3.95 | AAI-based (high-confidence) | 0 |
| S1_k141_329973 | 1314 | No | NA | 1 | 0 | 0 | Not-determined | Genome-fragment | NA | NA | 0 |
| S7_k141_207728 | 1448 | No | NA | 2 | 1 | 0 | Low-quality | Genome-fragment | 1.13 | AAI-based (medium-confidence) | 0 |
| S1_k141_247964 | 1056 | No | NA | 1 | 0 | 1 | Not-determined | Genome-fragment | NA | NA | 0 |
| S17-final-viral-combined_k141_39597\|\|full | 1448 | No | NA | 3 | 2 | 0 | Low-quality | Genome-fragment | 2.94 | AAI-based (high-confidence) | 0 |
| S1_k141_363087 | 2027 | No | NA | 3 | 0 | 2 | Not-determined | Genome-fragment | NA | NA | 0 |
| S17-k141_125546 | 1448 | No | NA | 3 | 1 | 0 | Low-quality | Genome-fragment | 1.5 | AAI-based (high-confidence) | 0 |
| S1_k141_149099 | 1287 | No | NA | 3 | 0 | 0 | Not-determined | Genome-fragment | NA | NA | 0 |
| S12-final-viral-combined_k141_242906\|\|full | 1448 | No | NA | 3 | 1 | 0 | Low-quality | Genome-fragment | 0.79 | AAI-based (medium-confidence) | 0 |
| S12_k141_375002 | 1448 | No | NA | 2 | 1 | 0 | Low-quality | Genome-fragment | 0.78 | AAI-based (high-confidence) | 0 |
| S1_k141_33806 | 3408 | No | NA | 6 | 0 | 1 | Not-determined | Genome-fragment | NA | NA | 0 |
| S1_k141_363106 | 7523 | No | NA | 8 | 0 | 1 | Not-determined | Genome-fragment | NA | NA | 0 |
| S2-final-viral-combined_k141_196986\|\|full | 1448 | No | NA | 4 | 0 | 0 | Low-quality | Genome-fragment | 2.96 | AAI-based (medium-confidence) | 0 |
| S4_k141_111661 | 1448 | No | NA | 3 | 1 | 0 | Low-quality | Genome-fragment | 1.95 | HMM-based (lower-bound) | 0 |
| S1_k141_379465 | 1196 | No | NA | 1 | 0 | 1 | Not-determined | Genome-fragment | NA | NA | 0 |
| S1_k141_313751 | 1654 | No | NA | 1 | 0 | 0 | Not-determined | Genome-fragment | NA | NA | 0 |
| S1_k141_198678 | 3495 | No | NA | 4 | 0 | 0 | Not-determined | Genome-fragment | NA | NA | 0 |
| S1_k141_66572 | 3948 | No | NA | 6 | 0 | 2 | Not-determined | Genome-fragment | NA | NA | 0 |
| S1_k141_248067 | 1604 | No | NA | 3 | 0 | 1 | Not-determined | Genome-fragment | NA | NA | 0 |
| S1_k141_379498 | 5113 | No | NA | 2 | 0 | 2 | Not-determined | Genome-fragment | NA | NA | 0 |
| S8-final-viral-combined_k141_10336\|\|full | 1448 | No | NA | 3 | 1 | 0 | Low-quality | Genome-fragment | 2.87 | AAI-based (high-confidence) | 0 |
| S11-final-viral-combined_k141_319881\|\|full | 1449 | No | NA | 3 | 0 | 0 | Low-quality | Genome-fragment | 1.44 | AAI-based (medium-confidence) | 0 |
| S1_k141_346852 | 1005 | No | NA | 3 | 0 | 0 | Not-determined | Genome-fragment | NA | NA | 0 |
| S1_k141_17614 | 1633 | No | NA | 2 | 0 | 1 | Not-determined | Genome-fragment | NA | NA | 0 |
| S1_k141_363193 | 5948 | No | NA | 3 | 0 | 2 | Not-determined | Genome-fragment | NA | NA | 0 |
| S11-final-viral-combined_k141_160341\|\|full | 1449 | Yes | 1249 | 3 | 1 | 1 | Low-quality | Genome-fragment | 1.92 | HMM-based (lower-bound) | 13.8 |
| S1-final-viral-combined_k141_162165\|\|full | 1449 | No | NA | 3 | 0 | 2 | Low-quality | Genome-fragment | 1.03 | AAI-based (medium-confidence) | 0 |
| S1_k141_17628 | 1589 | No | NA | 4 | 0 | 0 | Not-determined | Genome-fragment | NA | NA | 0 |
| S1_k141_116555 | 2359 | No | NA | 4 | 0 | 0 | Not-determined | Genome-fragment | NA | NA | 0 |
| S1_k141_248108 | 1682 | No | NA | 2 | 0 | 1 | Not-determined | Genome-fragment | NA | NA | 0 |
| S1_k141_50131 | 3336 | No | NA | 2 | 0 | 2 | Not-determined | Genome-fragment | NA | NA | 0 |
| S1_k141_363254 | 1098 | No | NA | 2 | 0 | 1 | Not-determined | Genome-fragment | NA | NA | 0 |
| S1_k141_50140 | 1057 | No | NA | 2 | 0 | 0 | Not-determined | Genome-fragment | NA | NA | 0 |
| S1_k141_281104 | 1023 | No | NA | 1 | 0 | 1 | Not-determined | Genome-fragment | NA | NA | 0 |
| S1_k141_363266 | 3186 | No | NA | 6 | 0 | 1 | Not-determined | Genome-fragment | NA | NA | 0 |
| S1_k141_165673 | 1031 | No | NA | 1 | 0 | 1 | Not-determined | Genome-fragment | NA | NA | 0 |
| S1_k141_17719 | 3571 | No | NA | 1 | 0 | 1 | Not-determined | Genome-fragment | NA | NA | 0 |
| S1_k141_281128 | 1510 | No | NA | 2 | 0 | 0 | Not-determined | Genome-fragment | NA | NA | 0 |
| S1_k141_34021 | 1456 | No | NA | 2 | 0 | 2 | Not-determined | Genome-fragment | NA | NA | 0 |
| S1-final-viral-combined_k141_187745\|\|lt2gene | 1449 | No | NA | 1 | 1 | 0 | Low-quality | Genome-fragment | 2.12 | HMM-based (lower-bound) | 0 |
| S1_k141_354483 | 1449 | No | NA | 1 | 0 | 1 | Low-quality | Genome-fragment | 1.29 | AAI-based (medium-confidence) | 0 |
| S1_k141_379562 | 4587 | No | NA | 3 | 0 | 2 | Not-determined | Genome-fragment | NA | NA | 0 |
| S1_k141_264373 | 1102 | No | NA | 1 | 0 | 0 | Not-determined | Genome-fragment | NA | NA | 0 |
| S1_k141_330273 | 1585 | No | NA | 2 | 0 | 1 | Not-determined | Genome-fragment | NA | NA | 0 |
| S7_k141_141066 | 1449 | No | NA | 2 | 0 | 0 | Low-quality | Genome-fragment | 1.65 | AAI-based (medium-confidence) | 0 |
| S1_k141_83582 | 1238 | No | NA | 1 | 0 | 1 | Not-determined | Genome-fragment | NA | NA | 0 |
| S1_k141_198939 | 1399 | No | NA | 3 | 0 | 2 | Not-determined | Genome-fragment | NA | NA | 0 |
| S1_k141_165709 | 1337 | No | NA | 1 | 0 | 0 | Not-determined | Genome-fragment | NA | NA | 0 |
| S1_k141_264419 | 11105 | No | NA | 8 | 0 | 7 | Not-determined | Genome-fragment | NA | NA | 0 |
| S1_k141_313991 | 11007 | No | NA | 15 | 4 | 0 | Medium-quality | Genome-fragment | 80.73 | AAI-based (medium-confidence) | 0 |
| S17-final-viral-combined_k141_112600\|\|full | 1449 | No | NA | 3 | 0 | 0 | Low-quality | Genome-fragment | 3.81 | AAI-based (high-confidence) | 0 |
| S1_k141_264426 | 1298 | No | NA | 3 | 0 | 2 | Not-determined | Genome-fragment | NA | NA | 0 |
| S17-final-viral-combined_k141_114548\|\|full | 1449 | No | NA | 4 | 1 | 0 | Low-quality | Genome-fragment | 3.37 | AAI-based (medium-confidence) | 0 |
| S1_k141_34159 | 1654 | No | NA | 1 | 0 | 1 | Not-determined | Genome-fragment | NA | NA | 0 |
| S1_k141_379651 | 1044 | No | NA | 2 | 0 | 0 | Not-determined | Genome-fragment | NA | NA | 0 |
| S12-final-viral-combined_k141_308675\|\|full | 1449 | No | NA | 2 | 2 | 0 | Low-quality | Genome-fragment | 3.11 | AAI-based (high-confidence) | 0 |
| S12_k141_140012 | 1449 | No | NA | 1 | 0 | 1 | Low-quality | Genome-fragment | 0.88 | AAI-based (high-confidence) | 0 |
| S1_k141_330348 | 1189 | No | NA | 1 | 0 | 0 | Not-determined | Genome-fragment | NA | NA | 0 |
| S1_k141_379734 | 1233 | No | NA | 1 | 0 | 1 | Not-determined | Genome-fragment | NA | NA | 0 |
| S2_k141_133130 | 1449 | No | NA | 3 | 2 | 0 | Low-quality | Genome-fragment | 2.45 | AAI-based (high-confidence) | 0 |
| S6-final-viral-combined_k141_71302\|\|lt2gene | 1449 | No | NA | 1 | 1 | 0 | Low-quality | Genome-fragment | 1.9 | HMM-based (lower-bound) | 0 |
| S1_k141_1775 | 4505 | No | NA | 10 | 0 | 7 | Not-determined | Genome-fragment | NA | NA | 0 |
| S1_k141_34276 | 1056 | No | NA | 1 | 0 | 1 | Not-determined | Genome-fragment | NA | NA | 0 |
| S1_k141_297677 | 1359 | No | NA | 2 | 0 | 2 | Not-determined | Genome-fragment | NA | NA | 0 |
| S1_k141_182490 | 2817 | No | NA | 3 | 0 | 1 | Not-determined | Genome-fragment | NA | NA | 0 |
| S1_k141_363350 | 1471 | No | NA | 4 | 0 | 0 | Not-determined | Genome-fragment | NA | NA | 0 |
| S1_k141_363359 | 1604 | No | NA | 1 | 0 | 0 | Not-determined | Genome-fragment | NA | NA | 0 |
| S1_k141_66750 | 1139 | No | NA | 3 | 0 | 0 | Not-determined | Genome-fragment | NA | NA | 0 |
| S1_k141_83750 | 1180 | No | NA | 1 | 0 | 0 | Not-determined | Genome-fragment | NA | NA | 0 |
| S1_k141_182522 | 1819 | No | NA | 1 | 0 | 0 | Not-determined | Genome-fragment | NA | NA | 0 |
| S1_k141_116736 | 2340 | No | NA | 1 | 0 | 1 | Not-determined | Genome-fragment | NA | NA | 0 |
| S1_k141_314147 | 4185 | No | NA | 5 | 0 | 1 | Not-determined | Genome-fragment | NA | NA | 0 |
| S1_k141_116753 | 1052 | No | NA | 3 | 0 | 2 | Not-determined | Genome-fragment | NA | NA | 0 |
| S1_k141_231737 | 2532 | No | NA | 3 | 0 | 0 | Not-determined | Genome-fragment | NA | NA | 0 |
| S1_k141_165760 | 1475 | No | NA | 1 | 0 | 1 | Not-determined | Genome-fragment | NA | NA | 0 |
| S1_k141_363413 | 1105 | No | NA | 1 | 0 | 1 | Not-determined | Genome-fragment | NA | NA | 0 |
| S1_k141_248227 | 6517 | No | NA | 5 | 0 | 1 | Not-determined | Genome-fragment | NA | NA | 0 |
| S1_k141_314183 | 1977 | No | NA | 2 | 0 | 1 | Not-determined | Genome-fragment | NA | NA | 0 |
| S11-final-viral-combined_k141_321116\|\|full | 1450 | No | NA | 2 | 0 | 1 | Low-quality | Genome-fragment | 1.84 | AAI-based (medium-confidence) | 0 |
| S1_k141_314187 | 5510 | No | NA | 2 | 0 | 0 | Not-determined | Genome-fragment | NA | NA | 0 |
| S1-final-viral-combined_k141_223663\|\|full | 1450 | No | NA | 2 | 0 | 1 | Low-quality | Genome-fragment | 1.76 | AAI-based (medium-confidence) | 0 |
| S1_k141_379883 | 1020 | No | NA | 2 | 0 | 1 | Not-determined | Genome-fragment | NA | NA | 0 |
| S1_k141_149474 | 1246 | No | NA | 1 | 0 | 1 | Not-determined | Genome-fragment | NA | NA | 0 |
| S1_k141_215661 | 1388 | No | NA | 2 | 0 | 1 | Not-determined | Genome-fragment | NA | NA | 0 |
| S1_k141_17967 | 2605 | No | NA | 3 | 0 | 0 | Not-determined | Genome-fragment | NA | NA | 0 |
| S17-final-viral-combined_k141_247219\|\|full | 1450 | No | NA | 3 | 0 | 0 | Low-quality | Genome-fragment | 4.35 | AAI-based (medium-confidence) | 0 |
| S1_k141_100010 | 45981 | No | NA | 66 | 10 | 0 | High-quality | High-quality | 100 | AAI-based (high-confidence) | 0 |
| S1_k141_379917 | 1231 | No | NA | 2 | 0 | 0 | Not-determined | Genome-fragment | NA | NA | 0 |
| S2-final-viral-combined_k141_13826\|\|full | 1450 | No | NA | 3 | 0 | 1 | Low-quality | Genome-fragment | 3.49 | AAI-based (medium-confidence) | 0 |
| S1_k141_116805 | 3376 | No | NA | 1 | 0 | 1 | Not-determined | Genome-fragment | NA | NA | 0 |
| S1_k141_116808 | 2143 | No | NA | 2 | 0 | 2 | Not-determined | Genome-fragment | NA | NA | 0 |
| S1_k141_379925 | 1724 | No | NA | 1 | 0 | 1 | Not-determined | Genome-fragment | NA | NA | 0 |
| S1_k141_281408 | 1181 | No | NA | 1 | 0 | 1 | Not-determined | Genome-fragment | NA | NA | 0 |
| S1_k141_100052 | 1039 | No | NA | 2 | 0 | 2 | Not-determined | Genome-fragment | NA | NA | 0 |
| S1_k141_264541 | 2294 | No | NA | 1 | 0 | 0 | Not-determined | Genome-fragment | NA | NA | 0 |
| S1_k141_133501 | 3270 | No | NA | 2 | 0 | 1 | Not-determined | Genome-fragment | NA | NA | 0 |
| S2_k141_247491 | 1450 | No | NA | 3 | 0 | 0 | Low-quality | Genome-fragment | 8.03 | AAI-based (high-confidence) | 0 |
| S4-final-viral-combined_k141_86285\|\|full | 1450 | No | NA | 2 | 0 | 0 | Low-quality | Genome-fragment | 3.07 | AAI-based (high-confidence) | 0 |
| S1_k141_1974 | 4146 | No | NA | 5 | 0 | 2 | Not-determined | Genome-fragment | NA | NA | 0 |
| S1_k141_281443 | 3039 | No | NA | 2 | 0 | 1 | Not-determined | Genome-fragment | NA | NA | 0 |
| S1_k141_281452 | 1093 | No | NA | 1 | 0 | 0 | Not-determined | Genome-fragment | NA | NA | 0 |
| S4-final-viral-combined_k141_72035\|\|lt2gene | 1450 | No | NA | 2 | 0 | 1 | Low-quality | Genome-fragment | 1.68 | AAI-based (medium-confidence) | 0 |
| S4_k141_36792 | 1450 | No | NA | 3 | 0 | 0 | Low-quality | Genome-fragment | 1.46 | AAI-based (high-confidence) | 0 |
| S1_k141_314308 | 1681 | No | NA | 1 | 0 | 0 | Not-determined | Genome-fragment | NA | NA | 0 |
| S1_k141_18075 | 1751 | No | NA | 1 | 0 | 1 | Not-determined | Genome-fragment | NA | NA | 0 |
| S1_k141_330591 | 1236 | No | NA | 1 | 0 | 0 | Not-determined | Genome-fragment | NA | NA | 0 |
| S1_k141_330599 | 1520 | No | NA | 2 | 0 | 1 | Not-determined | Genome-fragment | NA | NA | 0 |
| S1_k141_231917 | 2123 | No | NA | 2 | 0 | 0 | Not-determined | Genome-fragment | NA | NA | 0 |
| S1_k141_100137 | 1501 | No | NA | 2 | 0 | 2 | Not-determined | Genome-fragment | NA | NA | 0 |
| S1_k141_149619 | 1075 | No | NA | 1 | 0 | 0 | Not-determined | Genome-fragment | NA | NA | 0 |
| S1_k141_363612 | 1526 | No | NA | 2 | 0 | 2 | Not-determined | Genome-fragment | NA | NA | 0 |
| S1_k141_330613 | 5616 | No | NA | 4 | 0 | 1 | Not-determined | Genome-fragment | NA | NA | 0 |
| S1_k141_248457 | 1683 | No | NA | 2 | 0 | 1 | Not-determined | Genome-fragment | NA | NA | 0 |
| S1_k141_2081 | 11897 | No | NA | 8 | 0 | 8 | Not-determined | Genome-fragment | NA | NA | 0 |
| S1_k141_297985 | 1036 | No | NA | 1 | 0 | 1 | Not-determined | Genome-fragment | NA | NA | 0 |
| S1_k141_330669 | 5062 | No | NA | 3 | 0 | 2 | Not-determined | Genome-fragment | NA | NA | 0 |
| S1_k141_100205 | 3115 | No | NA | 2 | 0 | 1 | Not-determined | Genome-fragment | NA | NA | 0 |
| S1_k141_380049 | 1070 | No | NA | 2 | 0 | 2 | Not-determined | Genome-fragment | NA | NA | 0 |
| S1_k141_133629 | 1691 | No | NA | 1 | 0 | 1 | Not-determined | Genome-fragment | NA | NA | 0 |
| S1_k141_330680 | 3474 | No | NA | 7 | 0 | 4 | Not-determined | Genome-fragment | NA | NA | 0 |
| S1_k141_215765 | 1055 | No | NA | 1 | 0 | 0 | Not-determined | Genome-fragment | NA | NA | 0 |
| S1_k141_380063 | 5052 | No | NA | 4 | 0 | 3 | Not-determined | Genome-fragment | NA | NA | 0 |
| S1_k141_2118 | 1417 | No | NA | 1 | 0 | 1 | Not-determined | Genome-fragment | NA | NA | 0 |
| S1_k141_199294 | 3462 | No | NA | 4 | 0 | 1 | Not-determined | Genome-fragment | NA | NA | 0 |
| S1_k141_50538 | 1205 | No | NA | 1 | 0 | 1 | Not-determined | Genome-fragment | NA | NA | 0 |
| S1_k141_149674 | 1630 | No | NA | 1 | 0 | 0 | Not-determined | Genome-fragment | NA | NA | 0 |
| S1_k141_149683 | 1668 | No | NA | 1 | 0 | 0 | Not-determined | Genome-fragment | NA | NA | 0 |
| S1_k141_199324 | 1226 | No | NA | 1 | 0 | 1 | Not-determined | Genome-fragment | NA | NA | 0 |
| S1_k141_264627 | 1460 | No | NA | 3 | 0 | 0 | Not-determined | Genome-fragment | NA | NA | 0 |
| S1_k141_182788 | 2143 | No | NA | 1 | 0 | 1 | Not-determined | Genome-fragment | NA | NA | 0 |
| S6-final-viral-combined_k141_95829\|\|full | 1450 | No | NA | 3 | 0 | 2 | Low-quality | Genome-fragment | 0.64 | AAI-based (medium-confidence) | 0 |
| S1_k141_66889 | 1139 | No | NA | 2 | 0 | 0 | Not-determined | Genome-fragment | NA | NA | 0 |
| S1_k141_380136 | 1056 | No | NA | 2 | 0 | 2 | Not-determined | Genome-fragment | NA | NA | 0 |
| S1_k141_165873 | 3046 | No | NA | 2 | 0 | 2 | Not-determined | Genome-fragment | NA | NA | 0 |
| S1_k141_264655 | 1154 | No | NA | 2 | 0 | 0 | Not-determined | Genome-fragment | NA | NA | 0 |
| S1_k141_100319 | 1465 | No | NA | 1 | 0 | 1 | Not-determined | Genome-fragment | NA | NA | 0 |
| S1_k141_2194 | 1120 | No | NA | 1 | 0 | 0 | Not-determined | Genome-fragment | NA | NA | 0 |
| S6-final-viral-combined_k141_62300\|\|full | 1450 | No | NA | 3 | 0 | 0 | Low-quality | Genome-fragment | 2.28 | AAI-based (medium-confidence) | 0 |
| S1_k141_182830 | 1456 | No | NA | 2 | 0 | 2 | Not-determined | Genome-fragment | NA | NA | 0 |
| S8-final-viral-combined_k141_224303\|\|lt2gene | 1450 | No | NA | 1 | 1 | 0 | Low-quality | Genome-fragment | 2.53 | AAI-based (medium-confidence) | 0 |
| S8-final-viral-combined_k141_181655\|\|lt2gene | 1450 | No | NA | 2 | 1 | 0 | Low-quality | Genome-fragment | 2.45 | AAI-based (high-confidence) | 0 |
| S1_k141_34669 | 1371 | No | NA | 1 | 0 | 1 | Not-determined | Genome-fragment | NA | NA | 0 |
| S1_k141_182855 | 4604 | No | NA | 5 | 0 | 1 | Not-determined | Genome-fragment | NA | NA | 0 |
| S9-final-viral-combined_k141_57824\|\|full | 1451 | No | NA | 2 | 0 | 1 | Low-quality | Genome-fragment | 1.71 | AAI-based (medium-confidence) | 0 |
| S1_k141_149761 | 1864 | No | NA | 2 | 0 | 1 | Not-determined | Genome-fragment | NA | NA | 0 |
| S1_k141_149782 | 2412 | No | NA | 2 | 0 | 2 | Not-determined | Genome-fragment | NA | NA | 0 |
| S1_k141_314455 | 1823 | No | NA | 1 | 0 | 0 | Not-determined | Genome-fragment | NA | NA | 0 |
| S9-final-viral-combined_k141_68467\|\|full | 1451 | No | NA | 3 | 1 | 0 | Low-quality | Genome-fragment | 2.39 | AAI-based (medium-confidence) | 0 |
| S1_k141_100412 | 3309 | No | NA | 2 | 0 | 2 | Not-determined | Genome-fragment | NA | NA | 0 |
| S1_k141_67027 | 2823 | No | NA | 4 | 0 | 2 | Not-determined | Genome-fragment | NA | NA | 0 |
| S1_k141_314467 | 1283 | No | NA | 1 | 0 | 1 | Not-determined | Genome-fragment | NA | NA | 0 |
| S1_k141_264767 | 1195 | No | NA | 2 | 0 | 1 | Not-determined | Genome-fragment | NA | NA | 0 |
| S1_k141_363799 | 8029 | No | NA | 10 | 0 | 3 | Not-determined | Genome-fragment | NA | NA | 0 |
| S1_k141_18342 | 1545 | No | NA | 2 | 0 | 1 | Not-determined | Genome-fragment | NA | NA | 0 |
| S1_k141_165946 | 3053 | No | NA | 2 | 0 | 1 | Not-determined | Genome-fragment | NA | NA | 0 |
| S1_k141_149824 | 1801 | No | NA | 6 | 0 | 1 | Not-determined | Genome-fragment | NA | NA | 0 |
| S11-final-viral-combined_k141_318046\|\|full | 1451 | No | NA | 3 | 0 | 1 | Low-quality | Genome-fragment | 3.09 | AAI-based (medium-confidence) | 0 |
| S1_k141_50723 | 1426 | No | NA | 3 | 0 | 0 | Not-determined | Genome-fragment | NA | NA | 0 |
| S1_k141_18358 | 1973 | No | NA | 1 | 0 | 0 | Not-determined | Genome-fragment | NA | NA | 0 |
| S11-final-viral-combined_k141_162604\|\|full | 1451 | No | NA | 4 | 1 | 0 | Low-quality | Genome-fragment | 4.05 | AAI-based (high-confidence) | 0 |
| S1_k141_84213 | 3808 | No | NA | 1 | 0 | 0 | Not-determined | Genome-fragment | NA | NA | 0 |
| S1_k141_18362 | 3772 | No | NA | 5 | 0 | 4 | Not-determined | Genome-fragment | NA | NA | 0 |
| S1_k141_182975 | 2938 | No | NA | 1 | 0 | 1 | Not-determined | Genome-fragment | NA | NA | 0 |
| S1_k141_84236 | 1069 | No | NA | 1 | 0 | 1 | Not-determined | Genome-fragment | NA | NA | 0 |
| S1_k141_307242 | 1451 | No | NA | 3 | 0 | 0 | Low-quality | Genome-fragment | 1.54 | AAI-based (medium-confidence) | 0 |
| S1_k141_182981 | 1496 | No | NA | 1 | 0 | 1 | Not-determined | Genome-fragment | NA | NA | 0 |
| S1_k141_363880 | 1131 | No | NA | 1 | 0 | 1 | Not-determined | Genome-fragment | NA | NA | 0 |
| S1_k141_281789 | 3119 | No | NA | 2 | 0 | 1 | Not-determined | Genome-fragment | NA | NA | 0 |
| S3-final-viral-combined_k141_46198\|\|full | 1451 | No | NA | 3 | 0 | 1 | Low-quality | Genome-fragment | 0.84 | AAI-based (medium-confidence) | 0 |
| S1_k141_216057 | 1034 | No | NA | 2 | 0 | 0 | Not-determined | Genome-fragment | NA | NA | 0 |
| S1_k141_347671 | 1609 | No | NA | 2 | 0 | 0 | Not-determined | Genome-fragment | NA | NA | 0 |
| S5-final-viral-combined_k141_112040\|\|full | 1451 | No | NA | 5 | 3 | 0 | Low-quality | Genome-fragment | 3.79 | AAI-based (high-confidence) | 0 |
| S1_k141_50867 | 1028 | No | NA | 2 | 0 | 1 | Not-determined | Genome-fragment | NA | NA | 0 |
| S5_k141_102696 | 1451 | No | NA | 4 | 1 | 0 | Low-quality | Genome-fragment | 2.66 | AAI-based (medium-confidence) | 0 |
| S1_k141_347723 | 3193 | No | NA | 3 | 0 | 3 | Not-determined | Genome-fragment | NA | NA | 0 |
| S1_k141_100554 | 2956 | No | NA | 1 | 0 | 0 | Not-determined | Genome-fragment | NA | NA | 0 |
| S1_k141_264900 | 2156 | No | NA | 3 | 0 | 2 | Not-determined | Genome-fragment | NA | NA | 0 |
| S1_k141_232280 | 1220 | No | NA | 1 | 0 | 1 | Not-determined | Genome-fragment | NA | NA | 0 |
| S1_k141_232309 | 1601 | No | NA | 1 | 0 | 1 | Not-determined | Genome-fragment | NA | NA | 0 |
| S17-final-viral-combined_k141_176353\|\|full | 1451 | No | NA | 3 | 0 | 0 | Low-quality | Genome-fragment | 1.14 | AAI-based (high-confidence) | 0 |
| S12_k141_17531 | 1451 | Yes | 1014 | 2 | 1 | 1 | Low-quality | Genome-fragment | 0.61 | HMM-based (lower-bound) | 30.12 |
| S2-final-viral-combined_k141_219962\|\|lt2gene | 1451 | No | NA | 1 | 1 | 0 | Low-quality | Genome-fragment | 2.42 | HMM-based (lower-bound) | 0 |
| S1_k141_34881 | 3329 | No | NA | 7 | 0 | 0 | Not-determined | Genome-fragment | NA | NA | 0 |
| S1_k141_380339 | 1409 | No | NA | 2 | 0 | 2 | Not-determined | Genome-fragment | NA | NA | 0 |
| S4-final-viral-combined_k141_154216\|\|full | 1451 | No | NA | 4 | 0 | 0 | Low-quality | Genome-fragment | 1.95 | AAI-based (medium-confidence) | 0 |
| S1_k141_232385 | 1126 | No | NA | 1 | 0 | 0 | Not-determined | Genome-fragment | NA | NA | 0 |
| S1_k141_134063 | 1977 | No | NA | 2 | 0 | 1 | Not-determined | Genome-fragment | NA | NA | 0 |
| S1_k141_51057 | 1541 | No | NA | 1 | 0 | 0 | Not-determined | Genome-fragment | NA | NA | 0 |
| S1_k141_380378 | 3583 | No | NA | 3 | 0 | 3 | Not-determined | Genome-fragment | NA | NA | 0 |
| S1_k141_67296 | 1154 | No | NA | 1 | 0 | 0 | Not-determined | Genome-fragment | NA | NA | 0 |
| S1_k141_183171 | 1052 | No | NA | 2 | 0 | 0 | Not-determined | Genome-fragment | NA | NA | 0 |
| S1_k141_281927 | 4114 | No | NA | 6 | 0 | 6 | Not-determined | Genome-fragment | NA | NA | 0 |
| S4-final-viral-combined_k141_87860\|\|full | 1451 | No | NA | 2 | 0 | 1 | Low-quality | Genome-fragment | 0.9 | AAI-based (medium-confidence) | 0 |
| S4-final-viral-combined_k141_23385\|\|lt2gene | 1451 | No | NA | 2 | 0 | 0 | Low-quality | Genome-fragment | 1.44 | AAI-based (high-confidence) | 0 |
| S1_k141_298418 | 4244 | No | NA | 3 | 0 | 3 | Not-determined | Genome-fragment | NA | NA | 0 |
| S17-final-viral-combined_k141_226377\|\|full | 1452 | No | NA | 3 | 1 | 0 | Low-quality | Genome-fragment | 3.31 | AAI-based (high-confidence) | 0 |
| S1_k141_34981 | 1307 | No | NA | 4 | 0 | 0 | Not-determined | Genome-fragment | NA | NA | 0 |
| S1_k141_265012 | 2578 | No | NA | 2 | 0 | 1 | Not-determined | Genome-fragment | NA | NA | 0 |
| S1_k141_298457 | 1694 | No | NA | 3 | 0 | 1 | Not-determined | Genome-fragment | NA | NA | 0 |
| S1_k141_282043 | 4788 | No | NA | 6 | 0 | 2 | Not-determined | Genome-fragment | NA | NA | 0 |
| S1_k141_199721 | 1655 | No | NA | 2 | 0 | 0 | Not-determined | Genome-fragment | NA | NA | 0 |
| S1_k141_249091 | 1176 | No | NA | 2 | 0 | 0 | Not-determined | Genome-fragment | NA | NA | 0 |
| S1_k141_265070 | 1264 | No | NA | 1 | 0 | 1 | Not-determined | Genome-fragment | NA | NA | 0 |
| S17-k141_88476 | 1452 | No | NA | 4 | 1 | 0 | Low-quality | Genome-fragment | 3.47 | AAI-based (medium-confidence) | 0 |
| S1_k141_84536 | 1293 | No | NA | 2 | 0 | 1 | Not-determined | Genome-fragment | NA | NA | 0 |
| S1_k141_282073 | 1003 | No | NA | 2 | 0 | 1 | Not-determined | Genome-fragment | NA | NA | 0 |
| S1_k141_347874 | 1830 | No | NA | 3 | 0 | 3 | Not-determined | Genome-fragment | NA | NA | 0 |
| S1_k141_298525 | 1033 | No | NA | 2 | 0 | 0 | Not-determined | Genome-fragment | NA | NA | 0 |
| S12-final-viral-combined_k141_137896\|\|full | 1452 | No | NA | 3 | 0 | 0 | Low-quality | Genome-fragment | 3.39 | AAI-based (high-confidence) | 0 |
| S1_k141_166290 | 1550 | No | NA | 1 | 0 | 0 | Not-determined | Genome-fragment | NA | NA | 0 |
| S2-final-viral-combined_k141_30449\|\|full | 1452 | No | NA | 3 | 1 | 0 | Low-quality | Genome-fragment | 3.85 | AAI-based (high-confidence) | 0 |
| S1_k141_347910 | 2793 | No | NA | 2 | 0 | 0 | Not-determined | Genome-fragment | NA | NA | 0 |
| S1_k141_380529 | 1130 | No | NA | 1 | 0 | 0 | Not-determined | Genome-fragment | NA | NA | 0 |
| S1_k141_364203 | 2099 | No | NA | 3 | 0 | 0 | Not-determined | Genome-fragment | NA | NA | 0 |
| S1_k141_84655 | 1220 | No | NA | 2 | 0 | 2 | Not-determined | Genome-fragment | NA | NA | 0 |
| S1_k141_183343 | 1102 | No | NA | 1 | 0 | 0 | Not-determined | Genome-fragment | NA | NA | 0 |
| S1_k141_331074 | 1144 | No | NA | 2 | 0 | 0 | Not-determined | Genome-fragment | NA | NA | 0 |
| S1_k141_282083 | 2576 | No | NA | 4 | 0 | 0 | Not-determined | Genome-fragment | NA | NA | 0 |
| S1_k141_232626 | 1424 | No | NA | 1 | 0 | 1 | Not-determined | Genome-fragment | NA | NA | 0 |
| S4-final-viral-combined_k141_113593\|\|full | 1452 | No | NA | 3 | 0 | 0 | Low-quality | Genome-fragment | 0.82 | AAI-based (medium-confidence) | 0 |
| S1_k141_117474 | 7870 | No | NA | 9 | 0 | 1 | Not-determined | Genome-fragment | NA | NA | 0 |
| S1_k141_249224 | 1893 | No | NA | 2 | 0 | 2 | Not-determined | Genome-fragment | NA | NA | 0 |
| S1_k141_331086 | 1517 | No | NA | 1 | 0 | 0 | Not-determined | Genome-fragment | NA | NA | 0 |
| S1_k141_298619 | 4821 | No | NA | 5 | 0 | 4 | Not-determined | Genome-fragment | NA | NA | 0 |
| S1_k141_51322 | 1056 | No | NA | 2 | 0 | 0 | Not-determined | Genome-fragment | NA | NA | 0 |
| S1_k141_282133 | 2254 | No | NA | 2 | 0 | 2 | Not-determined | Genome-fragment | NA | NA | 0 |
| S1_k141_216487 | 4121 | No | NA | 6 | 0 | 1 | Not-determined | Genome-fragment | NA | NA | 0 |
| S4-final-viral-combined_k141_166529\|\|full | 1452 | No | NA | 2 | 0 | 2 | Low-quality | Genome-fragment | 0.86 | AAI-based (medium-confidence) | 0 |
| S1_k141_150207 | 3054 | No | NA | 6 | 0 | 3 | Not-determined | Genome-fragment | NA | NA | 0 |
| S1_k141_2669 | 2193 | No | NA | 2 | 0 | 2 | Not-determined | Genome-fragment | NA | NA | 0 |
| S1_k141_18704 | 1024 | No | NA | 2 | 0 | 2 | Not-determined | Genome-fragment | NA | NA | 0 |
| S6_k141_66944 | 1452 | No | NA | 5 | 1 | 0 | Low-quality | Genome-fragment | 0.78 | AAI-based (high-confidence) | 0 |
| S1_k141_2699 | 4456 | No | NA | 3 | 0 | 0 | Not-determined | Genome-fragment | NA | NA | 0 |
| S1_k141_364312 | 1342 | No | NA | 2 | 0 | 2 | Not-determined | Genome-fragment | NA | NA | 0 |
| S6_k141_19955 | 1452 | No | NA | 3 | 0 | 1 | Low-quality | Genome-fragment | 0.37 | AAI-based (high-confidence) | 0 |
| S1_k141_18714 | 1477 | No | NA | 1 | 0 | 1 | Not-determined | Genome-fragment | NA | NA | 0 |
| S1_k141_265274 | 1099 | No | NA | 1 | 0 | 1 | Not-determined | Genome-fragment | NA | NA | 0 |
| S1_k141_84776 | 1413 | No | NA | 3 | 0 | 3 | Not-determined | Genome-fragment | NA | NA | 0 |
| S8-final-viral-combined_k141_209529\|\|full | 1452 | No | NA | 2 | 0 | 0 | Low-quality | Genome-fragment | 3.23 | AAI-based (high-confidence) | 0 |
| S8_k141_103828 | 1452 | No | NA | 3 | 0 | 0 | Low-quality | Genome-fragment | 0.68 | AAI-based (medium-confidence) | 0 |
| S1_k141_100773 | 12895 | No | NA | 10 | 0 | 6 | Not-determined | Genome-fragment | NA | NA | 0 |
| S1_k141_216642 | 7322 | No | NA | 5 | 0 | 2 | Not-determined | Genome-fragment | NA | NA | 0 |
| S9-final-viral-combined_k141_79931\|\|full | 1453 | No | NA | 4 | 0 | 0 | Low-quality | Genome-fragment | 2.27 | AAI-based (medium-confidence) | 0 |
| S1_k141_134452 | 1759 | No | NA | 1 | 0 | 1 | Not-determined | Genome-fragment | NA | NA | 0 |
| S1_k141_265285 | 39134 | No | NA | 65 | 24 | 1 | High-quality | High-quality | 100 | AAI-based (high-confidence) | 0 |
| S1_k141_380711 | 3410 | No | NA | 4 | 0 | 4 | Not-determined | Genome-fragment | NA | NA | 0 |
| S1_k141_84826 | 3675 | No | NA | 1 | 0 | 1 | Not-determined | Genome-fragment | NA | NA | 0 |
| S1_k141_331162 | 1379 | No | NA | 2 | 0 | 1 | Not-determined | Genome-fragment | NA | NA | 0 |
| S1_k141_282238 | 1263 | No | NA | 2 | 0 | 0 | Not-determined | Genome-fragment | NA | NA | 0 |
| S1_k141_166470 | 3136 | No | NA | 4 | 0 | 3 | Not-determined | Genome-fragment | NA | NA | 0 |
| S1_k141_298799 | 1111 | No | NA | 1 | 0 | 0 | Not-determined | Genome-fragment | NA | NA | 0 |
| S1_k141_331173 | 3080 | No | NA | 4 | 0 | 1 | Not-determined | Genome-fragment | NA | NA | 0 |
| S1_k141_265308 | 1106 | No | NA | 1 | 0 | 0 | Not-determined | Genome-fragment | NA | NA | 0 |
| S1_k141_166490 | 1680 | No | NA | 1 | 0 | 1 | Not-determined | Genome-fragment | NA | NA | 0 |
| S1_k141_150361 | 1375 | No | NA | 1 | 0 | 0 | Not-determined | Genome-fragment | NA | NA | 0 |
| S1_k141_84890 | 2597 | No | NA | 2 | 0 | 2 | Not-determined | Genome-fragment | NA | NA | 0 |
| S1_k141_2845 | 1130 | No | NA | 1 | 0 | 1 | Not-determined | Genome-fragment | NA | NA | 0 |
| S9_k141_21474 | 1453 | No | NA | 4 | 0 | 0 | Low-quality | Genome-fragment | 2.43 | AAI-based (high-confidence) | 0 |
| S1_k141_200044 | 1216 | No | NA | 2 | 0 | 0 | Not-determined | Genome-fragment | NA | NA | 0 |
| S1_k141_84931 | 1377 | No | NA | 2 | 0 | 2 | Not-determined | Genome-fragment | NA | NA | 0 |
| S11-final-viral-combined_k141_35338\|\|full | 1453 | No | NA | 3 | 0 | 1 | Low-quality | Genome-fragment | 1.9 | AAI-based (medium-confidence) | 0 |
| S11_k141_2200 | 1453 | No | NA | 1 | 0 | 1 | Low-quality | Genome-fragment | 2.74 | AAI-based (medium-confidence) | 0 |
| S1_k141_18826 | 3555 | No | NA | 4 | 0 | 3 | Not-determined | Genome-fragment | NA | NA | 0 |
| S1-final-viral-combined_k141_244617\|\|full | 1453 | No | NA | 4 | 0 | 0 | Low-quality | Genome-fragment | 3.48 | AAI-based (high-confidence) | 0 |
| S1_k141_216774 | 1944 | No | NA | 1 | 0 | 1 | Not-determined | Genome-fragment | NA | NA | 0 |
| S1_k141_380817 | 1484 | No | NA | 2 | 0 | 1 | Not-determined | Genome-fragment | NA | NA | 0 |
| S1-final-viral-combined_k141_377526\|\|full | 1453 | No | NA | 5 | 0 | 0 | Low-quality | Genome-fragment | 4.57 | AAI-based (high-confidence) | 0 |
| S1-final-viral-combined_k141_387434\|\|lt2gene | 1453 | No | NA | 2 | 2 | 0 | Low-quality | Genome-fragment | 3.2 | AAI-based (high-confidence) | 0 |
| S1_k141_100919 | 1292 | No | NA | 3 | 0 | 2 | Not-determined | Genome-fragment | NA | NA | 0 |
| S1_k141_51467 | 1735 | No | NA | 1 | 0 | 0 | Not-determined | Genome-fragment | NA | NA | 0 |
| S1_k141_150484 | 1261 | No | NA | 3 | 0 | 0 | Not-determined | Genome-fragment | NA | NA | 0 |
| S1_k141_364542 | 1934 | No | NA | 2 | 0 | 1 | Not-determined | Genome-fragment | NA | NA | 0 |
| S1_k141_18884 | 1272 | No | NA | 1 | 0 | 0 | Not-determined | Genome-fragment | NA | NA | 0 |
| S1_k141_183626 | 1044 | No | NA | 4 | 0 | 1 | Not-determined | Genome-fragment | NA | NA | 0 |
| S1_k141_216827 | 3124 | No | NA | 4 | 0 | 3 | Not-determined | Genome-fragment | NA | NA | 0 |
| S17-final-viral-combined_k141_213547\|\|full | 1453 | No | NA | 5 | 0 | 0 | Low-quality | Genome-fragment | 11.46 | AAI-based (medium-confidence) | 0 |
| S1_k141_51531 | 3667 | No | NA | 3 | 0 | 1 | Not-determined | Genome-fragment | NA | NA | 0 |
| S1_k141_265362 | 1283 | No | NA | 1 | 0 | 0 | Not-determined | Genome-fragment | NA | NA | 0 |
| S1_k141_134643 | 1059 | No | NA | 1 | 0 | 1 | Not-determined | Genome-fragment | NA | NA | 0 |
| S1_k141_67874 | 1450 | No | NA | 2 | 0 | 2 | Not-determined | Genome-fragment | NA | NA | 0 |
| S1_k141_51565 | 5463 | No | NA | 3 | 0 | 3 | Not-determined | Genome-fragment | NA | NA | 0 |
| S17-final-viral-combined_k141_216708\|\|lt2gene | 1453 | No | NA | 1 | 1 | 0 | Low-quality | Genome-fragment | 3.86 | AAI-based (high-confidence) | 0 |
| S1_k141_183694 | 1187 | No | NA | 4 | 0 | 4 | Not-determined | Genome-fragment | NA | NA | 0 |
| S1_k141_380881 | 3026 | No | NA | 5 | 0 | 2 | Not-determined | Genome-fragment | NA | NA | 0 |
| S12-final-viral-combined_k141_133547\|\|full | 1453 | No | NA | 2 | 0 | 1 | Low-quality | Genome-fragment | 2.68 | AAI-based (medium-confidence) | 0 |
| S1_k141_232991 | 3397 | No | NA | 2 | 0 | 1 | Not-determined | Genome-fragment | NA | NA | 0 |
| S1_k141_2995 | 1685 | No | NA | 3 | 0 | 3 | Not-determined | Genome-fragment | NA | NA | 0 |
| S1_k141_249638 | 1983 | No | NA | 3 | 0 | 0 | Not-determined | Genome-fragment | NA | NA | 0 |
| S1_k141_331483 | 7120 | No | NA | 16 | 0 | 16 | Not-determined | Genome-fragment | NA | NA | 0 |
| S1_k141_249663 | 1888 | No | NA | 2 | 0 | 1 | Not-determined | Genome-fragment | NA | NA | 0 |
| S1_k141_265501 | 1296 | No | NA | 4 | 0 | 3 | Not-determined | Genome-fragment | NA | NA | 0 |
| S1_k141_101108 | 4047 | No | NA | 6 | 0 | 1 | Not-determined | Genome-fragment | NA | NA | 0 |
| S12_k141_80099 | 1453 | No | NA | 3 | 1 | 0 | Low-quality | Genome-fragment | 2.62 | AAI-based (high-confidence) | 0 |
| S1_k141_134772 | 1062 | No | NA | 1 | 0 | 0 | Not-determined | Genome-fragment | NA | NA | 0 |
| S4-final-viral-combined_k141_56055\|\|full | 1453 | No | NA | 3 | 0 | 0 | Low-quality | Genome-fragment | 0.87 | AAI-based (medium-confidence) | 0 |
| S1_k141_19046 | 3956 | No | NA | 6 | 0 | 1 | Not-determined | Genome-fragment | NA | NA | 0 |
| S1_k141_249732 | 2104 | No | NA | 2 | 0 | 0 | Not-determined | Genome-fragment | NA | NA | 0 |
| S1_k141_85108 | 1484 | No | NA | 2 | 0 | 2 | Not-determined | Genome-fragment | NA | NA | 0 |
| S1_k141_101164 | 1673 | No | NA | 2 | 0 | 1 | Not-determined | Genome-fragment | NA | NA | 0 |
| S1_k141_85132 | 1070 | No | NA | 1 | 0 | 1 | Not-determined | Genome-fragment | NA | NA | 0 |
| S8-final-viral-combined_k141_127658\|\|lt2gene | 1453 | No | NA | 1 | 1 | 0 | Low-quality | Genome-fragment | 1 | HMM-based (lower-bound) | 0 |
| S1_k141_67976 | 2578 | No | NA | 3 | 0 | 1 | Not-determined | Genome-fragment | NA | NA | 0 |
| S8_k141_276011 | 1453 | No | NA | 3 | 0 | 3 | Low-quality | Genome-fragment | 0.49 | AAI-based (medium-confidence) | 0 |
| S1_k141_249771 | 1022 | No | NA | 2 | 0 | 0 | Not-determined | Genome-fragment | NA | NA | 0 |
| S1_k141_117975 | 11716 | No | NA | 9 | 0 | 4 | Not-determined | Genome-fragment | NA | NA | 0 |
| S1_k141_249825 | 1220 | No | NA | 2 | 0 | 0 | Not-determined | Genome-fragment | NA | NA | 0 |
| S1_k141_150790 | 4814 | No | NA | 4 | 0 | 3 | Not-determined | Genome-fragment | NA | NA | 0 |
| S1_k141_381023 | 1777 | No | NA | 1 | 0 | 1 | Not-determined | Genome-fragment | NA | NA | 0 |
| S11_k141_125010 | 1454 | No | NA | 3 | 0 | 0 | Low-quality | Genome-fragment | 3.78 | AAI-based (medium-confidence) | 0 |
| S1-final-viral-combined_k141_306652\|\|full | 1454 | No | NA | 3 | 0 | 0 | Low-quality | Genome-fragment | 3.34 | AAI-based (medium-confidence) | 0 |
| S1_k141_299239 | 5622 | No | NA | 10 | 0 | 3 | Not-determined | Genome-fragment | NA | NA | 0 |
| S1_k141_249863 | 1134 | No | NA | 1 | 0 | 1 | Not-determined | Genome-fragment | NA | NA | 0 |
| S1_k141_348487 | 5167 | No | NA | 7 | 0 | 5 | Not-determined | Genome-fragment | NA | NA | 0 |
| S1_k141_249880 | 1425 | No | NA | 2 | 0 | 2 | Not-determined | Genome-fragment | NA | NA | 0 |
| S1_k141_118005 | 3785 | No | NA | 4 | 0 | 1 | Not-determined | Genome-fragment | NA | NA | 0 |
| S12-final-viral-combined_k141_366114\|\|full | 1454 | No | NA | 2 | 1 | 0 | Low-quality | Genome-fragment | 3.27 | AAI-based (high-confidence) | 0 |
| S1_k141_200310 | 2167 | No | NA | 4 | 0 | 1 | Not-determined | Genome-fragment | NA | NA | 0 |
| S1_k141_265588 | 1441 | No | NA | 1 | 0 | 1 | Not-determined | Genome-fragment | NA | NA | 0 |
| S1_k141_233106 | 3485 | No | NA | 1 | 0 | 0 | Not-determined | Genome-fragment | NA | NA | 0 |
| S2_k141_216316 | 1454 | No | NA | 3 | 0 | 0 | Low-quality | Genome-fragment | 2.83 | AAI-based (medium-confidence) | 0 |
| S1_k141_315574 | 3599 | No | NA | 3 | 0 | 2 | Not-determined | Genome-fragment | NA | NA | 0 |
| S1_k141_315578 | 1104 | No | NA | 2 | 0 | 1 | Not-determined | Genome-fragment | NA | NA | 0 |
| S4-final-viral-combined_k141_138662\|\|full | 1454 | No | NA | 2 | 0 | 0 | Low-quality | Genome-fragment | 0.9 | AAI-based (medium-confidence) | 0 |
| S1_k141_282743 | 1737 | No | NA | 2 | 0 | 1 | Not-determined | Genome-fragment | NA | NA | 0 |
| S6-final-viral-combined_k141_41527\|\|full | 1454 | No | NA | 3 | 0 | 1 | Low-quality | Genome-fragment | 0.94 | AAI-based (high-confidence) | 0 |
| S1_k141_35744 | 1602 | No | NA | 2 | 0 | 2 | Not-determined | Genome-fragment | NA | NA | 0 |
| S1_k141_331580 | 4461 | No | NA | 6 | 0 | 4 | Not-determined | Genome-fragment | NA | NA | 0 |
| S8-final-viral-combined_k141_135637\|\|lt2gene | 1454 | Yes | 1260 | 2 | 1 | 1 | Low-quality | Genome-fragment | 1.22 | AAI-based (high-confidence) | 13.34 |
| S1_k141_299419 | 1497 | No | NA | 1 | 0 | 1 | Not-determined | Genome-fragment | NA | NA | 0 |
| S1_k141_200403 | 1175 | No | NA | 1 | 0 | 1 | Not-determined | Genome-fragment | NA | NA | 0 |
| S1_k141_101280 | 11549 | No | NA | 13 | 0 | 1 | Not-determined | Genome-fragment | NA | NA | 0 |
| S1_k141_381142 | 3254 | No | NA | 3 | 0 | 2 | Not-determined | Genome-fragment | NA | NA | 0 |
| S1_k141_134954 | 1763 | No | NA | 2 | 0 | 1 | Not-determined | Genome-fragment | NA | NA | 0 |
| S1_k141_184173 | 1403 | No | NA | 1 | 0 | 1 | Not-determined | Genome-fragment | NA | NA | 0 |
| S9-final-viral-combined_k141_55427\|\|full | 1455 | No | NA | 3 | 0 | 1 | Low-quality | Genome-fragment | 3.76 | AAI-based (medium-confidence) | 0 |
| S1_k141_299455 | 1358 | No | NA | 1 | 0 | 1 | Not-determined | Genome-fragment | NA | NA | 0 |
| S9-final-viral-combined_k141_50996\|\|full | 1455 | No | NA | 3 | 0 | 0 | Low-quality | Genome-fragment | 24.07 | AAI-based (medium-confidence) | 0 |
| S3-final-viral-combined_k141_72261\|\|full | 1455 | No | NA | 2 | 0 | 1 | Low-quality | Genome-fragment | 1.02 | AAI-based (medium-confidence) | 0 |
| S1_k141_233209 | 2430 | No | NA | 2 | 0 | 0 | Not-determined | Genome-fragment | NA | NA | 0 |
| S1_k141_184212 | 1590 | No | NA | 2 | 0 | 0 | Not-determined | Genome-fragment | NA | NA | 0 |
| S1_k141_3120 | 2323 | No | NA | 4 | 0 | 2 | Not-determined | Genome-fragment | NA | NA | 0 |
| S1_k141_118162 | 1399 | No | NA | 1 | 0 | 1 | Not-determined | Genome-fragment | NA | NA | 0 |
| S1_k141_299482 | 6573 | No | NA | 4 | 0 | 4 | Not-determined | Genome-fragment | NA | NA | 0 |
| S1_k141_365034 | 1025 | No | NA | 2 | 0 | 0 | Not-determined | Genome-fragment | NA | NA | 0 |
| S1_k141_35852 | 4374 | No | NA | 2 | 0 | 1 | Not-determined | Genome-fragment | NA | NA | 0 |
| S7-final-viral-combined_k141_146633\|\|full | 1455 | No | NA | 4 | 0 | 0 | Low-quality | Genome-fragment | 4.16 | AAI-based (medium-confidence) | 0 |
| S1_k141_184235 | 1102 | No | NA | 1 | 0 | 0 | Not-determined | Genome-fragment | NA | NA | 0 |
| S1_k141_184236 | 1315 | No | NA | 1 | 0 | 0 | Not-determined | Genome-fragment | NA | NA | 0 |
| S1_k141_365063 | 2173 | No | NA | 2 | 0 | 2 | Not-determined | Genome-fragment | NA | NA | 0 |
| S7-final-viral-combined_k141_105982\|\|full | 1455 | No | NA | 3 | 0 | 2 | Low-quality | Genome-fragment | 0.76 | AAI-based (medium-confidence) | 0 |
| S1_k141_315699 | 1212 | No | NA | 1 | 0 | 0 | Not-determined | Genome-fragment | NA | NA | 0 |
| S7_k141_25010 | 1455 | No | NA | 1 | 0 | 1 | Low-quality | Genome-fragment | 0.4 | AAI-based (high-confidence) | 0 |
| S17-k141_189390 | 1455 | No | NA | 1 | 0 | 0 | Low-quality | Genome-fragment | 2.86 | AAI-based (high-confidence) | 0 |
| S1_k141_184249 | 1600 | No | NA | 1 | 0 | 1 | Not-determined | Genome-fragment | NA | NA | 0 |
| S17-k141_216899 | 1455 | No | NA | 1 | 0 | 0 | Low-quality | Genome-fragment | 1.78 | AAI-based (high-confidence) | 0 |
| S1_k141_200489 | 7746 | No | NA | 10 | 0 | 0 | Not-determined | Genome-fragment | NA | NA | 0 |
| S2-final-viral-combined_k141_219098\|\|full | 1455 | No | NA | 5 | 0 | 0 | Low-quality | Genome-fragment | 2.7 | AAI-based (medium-confidence) | 0 |
| S1_k141_282869 | 1406 | No | NA | 2 | 0 | 0 | Not-determined | Genome-fragment | NA | NA | 0 |
| S1_k141_348764 | 4484 | No | NA | 4 | 0 | 4 | Not-determined | Genome-fragment | NA | NA | 0 |
| S1_k141_166857 | 2060 | No | NA | 2 | 0 | 1 | Not-determined | Genome-fragment | NA | NA | 0 |
| S1_k141_217265 | 6420 | No | NA | 4 | 0 | 4 | Not-determined | Genome-fragment | NA | NA | 0 |
| S1_k141_52061 | 3024 | No | NA | 2 | 0 | 2 | Not-determined | Genome-fragment | NA | NA | 0 |
| S2-final-viral-combined_k141_163891\|\|full | 1455 | No | NA | 2 | 0 | 0 | Low-quality | Genome-fragment | 3.01 | AAI-based (high-confidence) | 0 |
| S8-final-viral-combined_k141_245448\|\|full | 1455 | No | NA | 4 | 0 | 0 | Low-quality | Genome-fragment | 3.34 | AAI-based (high-confidence) | 0 |
| S1_k141_265870 | 4535 | No | NA | 6 | 0 | 4 | Not-determined | Genome-fragment | NA | NA | 0 |
| S11-final-viral-combined_k141_69208\|\|full | 1456 | No | NA | 2 | 0 | 1 | Low-quality | Genome-fragment | 4.65 | AAI-based (high-confidence) | 0 |
| S11-final-viral-combined_k141_108853\|\|full | 1456 | No | NA | 2 | 1 | 0 | Low-quality | Genome-fragment | 0.9 | AAI-based (high-confidence) | 0 |
| S1_k141_233385 | 3681 | No | NA | 3 | 0 | 2 | Not-determined | Genome-fragment | NA | NA | 0 |
| S1_k141_118297 | 1107 | No | NA | 2 | 0 | 2 | Not-determined | Genome-fragment | NA | NA | 0 |
| S1_k141_331873 | 3627 | No | NA | 3 | 0 | 2 | Not-determined | Genome-fragment | NA | NA | 0 |
| S1_k141_265905 | 1094 | No | NA | 1 | 0 | 1 | Not-determined | Genome-fragment | NA | NA | 0 |
| S1_k141_200564 | 1440 | No | NA | 2 | 0 | 2 | Not-determined | Genome-fragment | NA | NA | 0 |
| S1-final-viral-combined_k141_121334\|\|full | 1456 | No | NA | 5 | 0 | 0 | Low-quality | Genome-fragment | 3.02 | AAI-based (high-confidence) | 0 |
| S1_k141_135202 | 1905 | No | NA | 2 | 0 | 1 | Not-determined | Genome-fragment | NA | NA | 0 |
| S3-final-viral-combined_k141_52734\|\|full | 1456 | No | NA | 3 | 1 | 0 | Low-quality | Genome-fragment | 3.09 | AAI-based (medium-confidence) | 0 |
| S1_k141_118344 | 1346 | No | NA | 1 | 0 | 0 | Not-determined | Genome-fragment | NA | NA | 0 |
| S1_k141_151125 | 1530 | No | NA | 2 | 0 | 2 | Not-determined | Genome-fragment | NA | NA | 0 |
| S1_k141_52154 | 1298 | No | NA | 1 | 0 | 0 | Not-determined | Genome-fragment | NA | NA | 0 |
| S1_k141_315901 | 2168 | No | NA | 3 | 0 | 1 | Not-determined | Genome-fragment | NA | NA | 0 |
| S1_k141_118391 | 1275 | No | NA | 1 | 0 | 1 | Not-determined | Genome-fragment | NA | NA | 0 |
| S1_k141_381436 | 1173 | No | NA | 1 | 0 | 0 | Not-determined | Genome-fragment | NA | NA | 0 |
| S1_k141_331935 | 3644 | No | NA | 5 | 0 | 0 | Not-determined | Genome-fragment | NA | NA | 0 |
| S12_k141_184196 | 1456 | No | NA | 2 | 0 | 1 | Low-quality | Genome-fragment | 0.74 | AAI-based (medium-confidence) | 0 |
| S1_k141_217470 | 7997 | No | NA | 15 | 0 | 1 | Not-determined | Genome-fragment | NA | NA | 0 |
| S1_k141_85381 | 1267 | No | NA | 1 | 0 | 1 | Not-determined | Genome-fragment | NA | NA | 0 |
| S1_k141_52234 | 1618 | No | NA | 1 | 0 | 1 | Not-determined | Genome-fragment | NA | NA | 0 |
| S1_k141_283070 | 2049 | No | NA | 1 | 0 | 0 | Not-determined | Genome-fragment | NA | NA | 0 |
| S1_k141_250266 | 1047 | No | NA | 1 | 0 | 1 | Not-determined | Genome-fragment | NA | NA | 0 |
| S1_k141_52265 | 1314 | No | NA | 2 | 0 | 2 | Not-determined | Genome-fragment | NA | NA | 0 |
| S2-final-viral-combined_k141_242108\|\|full | 1456 | No | NA | 2 | 0 | 0 | Low-quality | Genome-fragment | 1.14 | AAI-based (high-confidence) | 0 |
| S1_k141_52314 | 2197 | No | NA | 3 | 0 | 3 | Not-determined | Genome-fragment | NA | NA | 0 |
| S1_k141_266028 | 3587 | No | NA | 5 | 0 | 2 | Not-determined | Genome-fragment | NA | NA | 0 |
| S1_k141_381523 | 1531 | No | NA | 1 | 0 | 1 | Not-determined | Genome-fragment | NA | NA | 0 |
| S1_k141_118552 | 2074 | No | NA | 5 | 0 | 5 | Not-determined | Genome-fragment | NA | NA | 0 |
| S1_k141_167168 | 1159 | No | NA | 2 | 0 | 1 | Not-determined | Genome-fragment | NA | NA | 0 |
| S1_k141_283139 | 4712 | No | NA | 6 | 0 | 0 | Not-determined | Genome-fragment | NA | NA | 0 |
| S1_k141_151295 | 1012 | No | NA | 2 | 0 | 1 | Not-determined | Genome-fragment | NA | NA | 0 |
| S1_k141_299787 | 1337 | No | NA | 1 | 0 | 1 | Not-determined | Genome-fragment | NA | NA | 0 |
| S1_k141_184636 | 1432 | No | NA | 2 | 0 | 1 | Not-determined | Genome-fragment | NA | NA | 0 |

| | | | | | | | | | | | |
|---|---|---|---|---|---|---|---|---|---|---|---|
| S1_k141_36257 | 1275 | No | NA | 1 | 0 | 0 | Not-determined | Genome-fragment | NA | NA | 0 |
| S1_k141_266051 | 3974 | No | NA | 5 | 0 | 0 | Not-determined | Genome-fragment | NA | NA | 0 |
| S1_k141_250318 | 1594 | No | NA | 3 | 0 | 0 | Not-determined | Genome-fragment | NA | NA | 0 |
| S1_k141_283179 | 1175 | No | NA | 2 | 0 | 1 | Not-determined | Genome-fragment | NA | NA | 0 |
| S6-final-viral-combined_k141_57036\|\|full | 1456 | No | NA | 4 | 0 | 0 | Low-quality | Genome-fragment | 3.41 | AAI-based (high-confidence) | 0 |
| S1_k141_68505 | 6935 | No | NA | 10 | 0 | 5 | Not-determined | Genome-fragment | NA | NA | 0 |
| S11-final-viral-combined_k141_29467\|\|full | 1457 | No | NA | 5 | 0 | 0 | Low-quality | Genome-fragment | 3.72 | AAI-based (high-confidence) | 0 |
| S1_k141_68526 | 1251 | No | NA | 2 | 0 | 2 | Not-determined | Genome-fragment | NA | NA | 0 |
| S1_k141_101632 | 1330 | No | NA | 2 | 0 | 1 | Not-determined | Genome-fragment | NA | NA | 0 |
| S1-final-viral-combined_k141_99541\|\|full | 1457 | No | NA | 3 | 0 | 0 | Low-quality | Genome-fragment | 1.05 | AAI-based (high-confidence) | 0 |
| S1_k141_101635 | 1033 | No | NA | 3 | 0 | 0 | Not-determined | Genome-fragment | NA | NA | 0 |
| S1_k141_151389 | 1789 | No | NA | 4 | 0 | 4 | Not-determined | Genome-fragment | NA | NA | 0 |
| S1-final-viral-combined_k141_323644\|\|full | 1457 | No | NA | 4 | 4 | 0 | Low-quality | Genome-fragment | 3.43 | AAI-based (high-confidence) | 0 |
| S1_k141_266226 | 1489 | No | NA | 2 | 0 | 0 | Not-determined | Genome-fragment | NA | NA | 0 |
| S1_k141_200857 | 5149 | No | NA | 5 | 0 | 5 | Not-determined | Genome-fragment | NA | NA | 0 |
| S1_k141_200890 | 1329 | No | NA | 1 | 0 | 0 | Not-determined | Genome-fragment | NA | NA | 0 |
| S7-final-viral-combined_k141_71380\|\|full | 1457 | No | NA | 3 | 0 | 2 | Low-quality | Genome-fragment | 1.57 | AAI-based (high-confidence) | 0 |
| S12-final-viral-combined_k141_102677\|\|full | 1457 | No | NA | 4 | 0 | 0 | Low-quality | Genome-fragment | 1.16 | AAI-based (medium-confidence) | 0 |
| S1_k141_266278 | 1268 | No | NA | 2 | 0 | 2 | Not-determined | Genome-fragment | NA | NA | 0 |
| S1_k141_200923 | 1910 | No | NA | 2 | 0 | 1 | Not-determined | Genome-fragment | NA | NA | 0 |
| S1_k141_266289 | 1192 | No | NA | 1 | 0 | 0 | Not-determined | Genome-fragment | NA | NA | 0 |
| S1_k141_381734 | 1338 | No | NA | 2 | 0 | 1 | Not-determined | Genome-fragment | NA | NA | 0 |
| S1_k141_68607 | 1697 | No | NA | 1 | 0 | 0 | Not-determined | Genome-fragment | NA | NA | 0 |
| S12-final-viral-combined_k141_220303\|\|full | 1457 | No | NA | 2 | 0 | 1 | Low-quality | Genome-fragment | 0.63 | AAI-based (medium-confidence) | 0 |
| S1_k141_85577 | 1591 | No | NA | 2 | 0 | 1 | Not-determined | Genome-fragment | NA | NA | 0 |
| S1_k141_349160 | 1225 | No | NA | 2 | 0 | 2 | Not-determined | Genome-fragment | NA | NA | 0 |
| S12_k141_192967 | 1457 | No | NA | 4 | 1 | 0 | Low-quality | Genome-fragment | 0.79 | AAI-based (high-confidence) | 0 |
| S9-final-viral-combined_k141_26573\|\|full | 1458 | No | NA | 4 | 4 | 0 | Low-quality | Genome-fragment | 3.5 | AAI-based (medium-confidence) | 0 |
| S1_k141_167311 | 9702 | No | NA | 7 | 0 | 6 | Not-determined | Genome-fragment | NA | NA | 0 |
| S1_k141_365621 | 1092 | No | NA | 1 | 0 | 1 | Not-determined | Genome-fragment | NA | NA | 0 |
| S1_k141_184868 | 2362 | No | NA | 2 | 0 | 0 | Not-determined | Genome-fragment | NA | NA | 0 |
| S9_k141_34362 | 1458 | No | NA | 3 | 2 | 0 | Low-quality | Genome-fragment | 2.61 | AAI-based (high-confidence) | 0 |
| S11-final-viral-combined_k141_227607\|\|full | 1458 | No | NA | 3 | 1 | 0 | Low-quality | Genome-fragment | 2.63 | AAI-based (medium-confidence) | 0 |
| S1_k141_135652 | 1160 | No | NA | 1 | 0 | 1 | Not-determined | Genome-fragment | NA | NA | 0 |
| S1_k141_151630 | 2805 | No | NA | 5 | 0 | 0 | Not-determined | Genome-fragment | NA | NA | 0 |
| S1-final-viral-combined_k141_313743\|\|full | 1458 | No | NA | 3 | 1 | 0 | Low-quality | Genome-fragment | 3.6 | AAI-based (high-confidence) | 0 |
| S1_k141_52521 | 1338 | No | NA | 3 | 0 | 0 | Not-determined | Genome-fragment | NA | NA | 0 |
| S1_k141_167371 | 1025 | No | NA | 2 | 0 | 1 | Not-determined | Genome-fragment | NA | NA | 0 |
| S1_k141_365640 | 1620 | No | NA | 3 | 0 | 3 | Not-determined | Genome-fragment | NA | NA | 0 |
| S1_k141_52538 | 1012 | No | NA | 1 | 0 | 0 | Not-determined | Genome-fragment | NA | NA | 0 |
| S1-final-viral-combined_k141_188532\|\|full | 1458 | No | NA | 2 | 0 | 1 | Low-quality | Genome-fragment | 3.52 | AAI-based (high-confidence) | 0 |
| S1_k141_283403 | 4574 | No | NA | 3 | 0 | 1 | Not-determined | Genome-fragment | NA | NA | 0 |
| S1_k141_201009 | 1109 | No | NA | 1 | 0 | 1 | Not-determined | Genome-fragment | NA | NA | 0 |
| S1_k141_19946 | 1206 | No | NA | 1 | 0 | 1 | Not-determined | Genome-fragment | NA | NA | 0 |
| S1_k141_184905 | 3195 | No | NA | 4 | 0 | 2 | Not-determined | Genome-fragment | NA | NA | 0 |
| S1_k141_316417 | 1145 | No | NA | 2 | 0 | 2 | Not-determined | Genome-fragment | NA | NA | 0 |
| S1-final-viral-combined_k141_44035\|\|lt2gene | 1458 | No | NA | 2 | 1 | 0 | Low-quality | Genome-fragment | 3.82 | AAI-based (high-confidence) | 0 |
| S1_k141_36486 | 1103 | No | NA | 1 | 0 | 0 | Not-determined | Genome-fragment | NA | NA | 0 |
| S17-final-viral-combined_k141_188786\|\|full | 1458 | No | NA | 2 | 0 | 1 | Low-quality | Genome-fragment | 1.17 | AAI-based (medium-confidence) | 0 |
| S1_k141_266500 | 4818 | No | NA | 5 | 0 | 5 | Not-determined | Genome-fragment | NA | NA | 0 |
| S1_k141_266510 | 1026 | No | NA | 1 | 0 | 1 | Not-determined | Genome-fragment | NA | NA | 0 |
| S1_k141_85698 | 7226 | No | NA | 4 | 0 | 4 | Not-determined | Genome-fragment | NA | NA | 0 |
| S1_k141_283468 | 1266 | No | NA | 3 | 0 | 0 | Not-determined | Genome-fragment | NA | NA | 0 |
| S1_k141_52608 | 3408 | No | NA | 3 | 0 | 3 | Not-determined | Genome-fragment | NA | NA | 0 |
| S1_k141_85717 | 1653 | No | NA | 2 | 0 | 0 | Not-determined | Genome-fragment | NA | NA | 0 |
| S1_k141_135692 | 6092 | No | NA | 7 | 0 | 7 | Not-determined | Genome-fragment | NA | NA | 0 |
| S1_k141_85727 | 1106 | No | NA | 2 | 0 | 2 | Not-determined | Genome-fragment | NA | NA | 0 |
| S1_k141_381907 | 1011 | No | NA | 1 | 0 | 0 | Not-determined | Genome-fragment | NA | NA | 0 |
| S1_k141_349276 | 3120 | No | NA | 4 | 0 | 1 | Not-determined | Genome-fragment | NA | NA | 0 |
| S1_k141_185014 | 1915 | No | NA | 1 | 0 | 0 | Not-determined | Genome-fragment | NA | NA | 0 |
| S12_k141_189780 | 1458 | No | NA | 1 | 1 | 0 | Low-quality | Genome-fragment | 1.54 | AAI-based (high-confidence) | 0 |
| S2-final-viral-combined_k141_178421\|\|full | 1458 | No | NA | 4 | 0 | 0 | Low-quality | Genome-fragment | 2.7 | AAI-based (medium-confidence) | 0 |
| S1_k141_233994 | 4499 | No | NA | 5 | 0 | 2 | Not-determined | Genome-fragment | NA | NA | 0 |
| S1_k141_332409 | 3011 | No | NA | 3 | 0 | 1 | Not-determined | Genome-fragment | NA | NA | 0 |
| S1_k141_101929 | 1176 | No | NA | 1 | 0 | 1 | Not-determined | Genome-fragment | NA | NA | 0 |
| S1_k141_234010 | 1587 | No | NA | 2 | 0 | 2 | Not-determined | Genome-fragment | NA | NA | 0 |
| S1_k141_332422 | 2836 | No | NA | 3 | 0 | 3 | Not-determined | Genome-fragment | NA | NA | 0 |
| S1_k141_151740 | 5898 | No | NA | 4 | 0 | 3 | Not-determined | Genome-fragment | NA | NA | 0 |
| S1_k141_332451 | 3877 | No | NA | 1 | 0 | 0 | Not-determined | Genome-fragment | NA | NA | 0 |
| S1_k141_316607 | 3410 | No | NA | 5 | 0 | 4 | Not-determined | Genome-fragment | NA | NA | 0 |
| S1_k141_85804 | 1081 | No | NA | 2 | 0 | 1 | Not-determined | Genome-fragment | NA | NA | 0 |
| S1_k141_381997 | 2151 | No | NA | 2 | 0 | 2 | Not-determined | Genome-fragment | NA | NA | 0 |
| S1_k141_135733 | 1274 | No | NA | 2 | 0 | 2 | Not-determined | Genome-fragment | NA | NA | 0 |
| S1_k141_266619 | 1677 | No | NA | 3 | 0 | 0 | Not-determined | Genome-fragment | NA | NA | 0 |
| S1_k141_185046 | 29667 | No | NA | 26 | 6 | 1 | Medium-quality | Genome-fragment | 70.38 | AAI-based (high-confidence) | 0 |
| S2-final-viral-combined_k141_232280\|\|full | 1458 | No | NA | 2 | 1 | 0 | Low-quality | Genome-fragment | 3.46 | AAI-based (medium-confidence) | 0 |
| S9-final-viral-combined_k141_31489\|\|full | 1459 | No | NA | 3 | 0 | 1 | Low-quality | Genome-fragment | 0.72 | AAI-based (medium-confidence) | 0 |
| S11-final-viral-combined_k141_153743\|\|full | 1459 | No | NA | 2 | 0 | 1 | Low-quality | Genome-fragment | 1.87 | AAI-based (high-confidence) | 0 |
| S1-final-viral-combined_k141_335374\|\|full | 1459 | No | NA | 1 | 0 | 0 | Low-quality | Genome-fragment | 3.38 | AAI-based (medium-confidence) | 0 |
| S1_k141_300411 | 1206 | No | NA | 1 | 0 | 0 | Not-determined | Genome-fragment | NA | NA | 0 |
| S1_k141_332502 | 1561 | No | NA | 1 | 0 | 1 | Not-determined | Genome-fragment | NA | NA | 0 |
| S5_k141_51165 | 1459 | No | NA | 3 | 0 | 0 | Low-quality | Genome-fragment | 3.07 | AAI-based (high-confidence) | 0 |
| S4-final-viral-combined_k141_127807\|\|full | 1459 | No | NA | 5 | 0 | 0 | Low-quality | Genome-fragment | 3.52 | AAI-based (high-confidence) | 0 |
| S1_k141_349415 | 2564 | No | NA | 4 | 0 | 0 | Not-determined | Genome-fragment | NA | NA | 0 |
| S1_k141_185095 | 3431 | No | NA | 4 | 0 | 3 | Not-determined | Genome-fragment | NA | NA | 0 |
| S1_k141_119026 | 1844 | No | NA | 2 | 0 | 2 | Not-determined | Genome-fragment | NA | NA | 0 |
| S1_k141_167612 | 1012 | No | NA | 1 | 0 | 0 | Not-determined | Genome-fragment | NA | NA | 0 |
| S1_k141_119047 | 1643 | No | NA | 2 | 0 | 1 | Not-determined | Genome-fragment | NA | NA | 0 |
| S1_k141_300450 | 1202 | No | NA | 2 | 0 | 1 | Not-determined | Genome-fragment | NA | NA | 0 |
| S1_k141_316800 | 1003 | No | NA | 1 | 0 | 1 | Not-determined | Genome-fragment | NA | NA | 0 |
| S1_k141_151897 | 1222 | No | NA | 2 | 0 | 0 | Not-determined | Genome-fragment | NA | NA | 0 |
| S1_k141_234158 | 1411 | No | NA | 1 | 0 | 1 | Not-determined | Genome-fragment | NA | NA | 0 |
| S1_k141_52845 | 1217 | No | NA | 1 | 0 | 1 | Not-determined | Genome-fragment | NA | NA | 0 |
| S1_k141_300485 | 3963 | No | NA | 2 | 0 | 1 | Not-determined | Genome-fragment | NA | NA | 0 |
| S1_k141_36560 | 9361 | No | NA | 9 | 0 | 3 | Not-determined | Genome-fragment | NA | NA | 0 |
| S1_k141_85890 | 1117 | No | NA | 1 | 0 | 1 | Not-determined | Genome-fragment | NA | NA | 0 |
| S1_k141_300504 | 1187 | No | NA | 2 | 0 | 2 | Not-determined | Genome-fragment | NA | NA | 0 |
| S6_k141_104818 | 1459 | No | NA | 1 | 0 | 0 | Low-quality | Genome-fragment | 2.13 | AAI-based (high-confidence) | 0 |
| S1_k141_310340 | 1460 | No | NA | 2 | 0 | 1 | Low-quality | Genome-fragment | 2.87 | AAI-based (medium-confidence) | 0 |
| S5-final-viral-combined_k141_19352\|\|full | 1460 | No | NA | 3 | 0 | 0 | Low-quality | Genome-fragment | 2.93 | AAI-based (medium-confidence) | 0 |
| S1_k141_217774 | 2073 | No | NA | 4 | 0 | 0 | Not-determined | Genome-fragment | NA | NA | 0 |
| S1_k141_52922 | 1223 | No | NA | 1 | 0 | 1 | Not-determined | Genome-fragment | NA | NA | 0 |
| S1_k141_135974 | 1048 | No | NA | 1 | 0 | 1 | Not-determined | Genome-fragment | NA | NA | 0 |
| S5-final-viral-combined_k141_63937\|\|full | 1460 | No | NA | 4 | 0 | 0 | Low-quality | Genome-fragment | 2.64 | AAI-based (high-confidence) | 0 |
| S1_k141_217819 | 1389 | No | NA | 2 | 0 | 0 | Not-determined | Genome-fragment | NA | NA | 0 |
| S1_k141_185265 | 11712 | No | NA | 10 | 0 | 4 | Not-determined | Genome-fragment | NA | NA | 0 |
| S1_k141_102124 | 1261 | No | NA | 1 | 0 | 1 | Not-determined | Genome-fragment | NA | NA | 0 |
| S1_k141_85998 | 3338 | No | NA | 3 | 0 | 2 | Not-determined | Genome-fragment | NA | NA | 0 |
| S1_k141_349540 | 1080 | No | NA | 1 | 0 | 1 | Not-determined | Genome-fragment | NA | NA | 0 |
| S1_k141_152030 | 1380 | No | NA | 1 | 0 | 1 | Not-determined | Genome-fragment | NA | NA | 0 |
| S5-final-viral-combined_k141_111255\|\|full | 1460 | No | NA | 4 | 0 | 0 | Low-quality | Genome-fragment | 1.64 | AAI-based (high-confidence) | 0 |
| S1_k141_349544 | 1126 | No | NA | 1 | 0 | 0 | Not-determined | Genome-fragment | NA | NA | 0 |
| S1_k141_365799 | 1590 | No | NA | 1 | 0 | 1 | Not-determined | Genome-fragment | NA | NA | 0 |
| S7-final-viral-combined_k141_180484\|\|full | 1460 | No | NA | 3 | 0 | 1 | Low-quality | Genome-fragment | 1.56 | AAI-based (medium-confidence) | 0 |
| S1_k141_217850 | 1054 | No | NA | 3 | 0 | 1 | Not-determined | Genome-fragment | NA | NA | 0 |
| S1_k141_36582 | 1525 | No | NA | 1 | 0 | 0 | Not-determined | Genome-fragment | NA | NA | 0 |
| S1_k141_365806 | 1487 | No | NA | 2 | 0 | 1 | Not-determined | Genome-fragment | NA | NA | 0 |
| S12-final-viral-combined_k141_382642\|\|full | 1460 | No | NA | 2 | 0 | 0 | Low-quality | Genome-fragment | 3.43 | AAI-based (medium-confidence) | 0 |
| S12-final-viral-combined_k141_148869\|\|full | 1460 | No | NA | 2 | 0 | 1 | Low-quality | Genome-fragment | 0.74 | AAI-based (high-confidence) | 0 |
| S1_k141_53006 | 4438 | No | NA | 11 | 0 | 11 | Not-determined | Genome-fragment | NA | NA | 0 |
| S1_k141_36621 | 3463 | No | NA | 4 | 0 | 0 | Not-determined | Genome-fragment | NA | NA | 0 |
| S12-final-viral-combined_k141_34333\|\|lt2gene | 1460 | No | NA | 1 | 1 | 0 | Low-quality | Genome-fragment | 2.46 | HMM-based (lower-bound) | 0 |
| S2-final-viral-combined_k141_157787\|\|full | 1460 | No | NA | 3 | 3 | 0 | Low-quality | Genome-fragment | 4.18 | AAI-based (medium-confidence) | 0 |
| S1_k141_68926 | 1168 | No | NA | 1 | 0 | 1 | Not-determined | Genome-fragment | NA | NA | 0 |
| S1_k141_201544 | 2005 | No | NA | 1 | 0 | 0 | Not-determined | Genome-fragment | NA | NA | 0 |
| S1_k141_185395 | 3594 | No | NA | 4 | 0 | 4 | Not-determined | Genome-fragment | NA | NA | 0 |
| S1_k141_251246 | 1024 | No | NA | 2 | 0 | 2 | Not-determined | Genome-fragment | NA | NA | 0 |
| S1_k141_86082 | 1161 | No | NA | 1 | 0 | 1 | Not-determined | Genome-fragment | NA | NA | 0 |
| S1_k141_68980 | 2234 | No | NA | 2 | 0 | 1 | Not-determined | Genome-fragment | NA | NA | 0 |
| S2-final-viral-combined_k141_216555\|\|full | 1460 | No | NA | 3 | 0 | 0 | Low-quality | Genome-fragment | 0.99 | AAI-based (medium-confidence) | 0 |
| S2-final-viral-combined_k141_59696\|\|lt2gene | 1460 | No | NA | 2 | 1 | 0 | Low-quality | Genome-fragment | 3.02 | AAI-based (high-confidence) | 0 |
| S8-final-viral-combined_k141_82017\|\|full | 1460 | No | NA | 3 | 0 | 0 | Low-quality | Genome-fragment | 2.57 | AAI-based (medium-confidence) | 0 |
| S11-final-viral-combined_k141_15646\|\|full | 1461 | No | NA | 5 | 1 | 0 | Low-quality | Genome-fragment | 2.64 | AAI-based (high-confidence) | 0 |
| S1_k141_349728 | 1024 | No | NA | 1 | 0 | 0 | Not-determined | Genome-fragment | NA | NA | 0 |
| S1_k141_20467 | 3988 | No | NA | 5 | 0 | 0 | Not-determined | Genome-fragment | NA | NA | 0 |
| S1_k141_283882 | 1223 | No | NA | 4 | 0 | 0 | Not-determined | Genome-fragment | NA | NA | 0 |
| S1_k141_185489 | 4074 | No | NA | 3 | 0 | 1 | Not-determined | Genome-fragment | NA | NA | 0 |
| S1_k141_201641 | 3597 | No | NA | 4 | 0 | 1 | Not-determined | Genome-fragment | NA | NA | 0 |
| S1_k141_317129 | 3268 | No | NA | 4 | 0 | 3 | Not-determined | Genome-fragment | NA | NA | 0 |
| S1_k141_234465 | 1037 | No | NA | 1 | 0 | 1 | Not-determined | Genome-fragment | NA | NA | 0 |
| S1_k141_317134 | 1507 | No | NA | 1 | 0 | 1 | Not-determined | Genome-fragment | NA | NA | 0 |
| S1_k141_283925 | 3005 | No | NA | 2 | 0 | 2 | Not-determined | Genome-fragment | NA | NA | 0 |
| S1-final-viral-combined_k141_159306\|\|full | 1461 | No | NA | 3 | 0 | 1 | Low-quality | Genome-fragment | 2.53 | AAI-based (medium-confidence) | 0 |
| S1_k141_300716 | 1379 | No | NA | 1 | 0 | 1 | Not-determined | Genome-fragment | NA | NA | 0 |
| S5_k141_165774 | 1461 | No | NA | 1 | 0 | 1 | Low-quality | Genome-fragment | 0.84 | AAI-based (medium-confidence) | 0 |
| S7-final-viral-combined_k141_86304\|\|full | 1461 | No | NA | 2 | 0 | 1 | Low-quality | Genome-fragment | 1.21 | AAI-based (medium-confidence) | 0 |
| S1_k141_36778 | 2536 | No | NA | 3 | 0 | 1 | Not-determined | Genome-fragment | NA | NA | 0 |
| S1_k141_365974 | 1639 | No | NA | 1 | 0 | 1 | Not-determined | Genome-fragment | NA | NA | 0 |
| S1_k141_382330 | 1081 | No | NA | 1 | 0 | 1 | Not-determined | Genome-fragment | NA | NA | 0 |
| S1_k141_20539 | 1028 | No | NA | 2 | 0 | 2 | Not-determined | Genome-fragment | NA | NA | 0 |
| S1_k141_267188 | 1466 | No | NA | 1 | 0 | 1 | Not-determined | Genome-fragment | NA | NA | 0 |
| S17-final-viral-combined_k141_161285\|\|full | 1461 | No | NA | 4 | 1 | 0 | Low-quality | Genome-fragment | 2.52 | AAI-based (medium-confidence) | 0 |
| S1_k141_332917 | 1129 | No | NA | 3 | 0 | 0 | Not-determined | Genome-fragment | NA | NA | 0 |
| S1_k141_185562 | 1016 | No | NA | 1 | 0 | 1 | Not-determined | Genome-fragment | NA | NA | 0 |
| S1_k141_152143 | 1153 | No | NA | 1 | 0 | 0 | Not-determined | Genome-fragment | NA | NA | 0 |
| S1_k141_152153 | 1375 | No | NA | 2 | 0 | 2 | Not-determined | Genome-fragment | NA | NA | 0 |
| S1_k141_349910 | 1144 | No | NA | 2 | 0 | 0 | Not-determined | Genome-fragment | NA | NA | 0 |
| S1_k141_152170 | 3313 | No | NA | 5 | 0 | 1 | Not-determined | Genome-fragment | NA | NA | 0 |
| S2_k141_115581 | 1461 | No | NA | 1 | 0 | 0 | Low-quality | Genome-fragment | 0.25 | AAI-based (high-confidence) | 0 |
| S1_k141_152179 | 1498 | No | NA | 1 | 0 | 1 | Not-determined | Genome-fragment | NA | NA | 0 |
| S1_k141_300845 | 1138 | No | NA | 1 | 0 | 1 | Not-determined | Genome-fragment | NA | NA | 0 |
| S9-final-viral-combined_k141_36940\|\|full | 1462 | No | NA | 2 | 0 | 0 | Low-quality | Genome-fragment | 1.77 | AAI-based (high-confidence) | 0 |
| S1_k141_251477 | 1520 | No | NA | 2 | 0 | 0 | Not-determined | Genome-fragment | NA | NA | 0 |
| S1_k141_332973 | 1406 | No | NA | 2 | 0 | 0 | Not-determined | Genome-fragment | NA | NA | 0 |
| S1_k141_332982 | 3498 | No | NA | 3 | 0 | 1 | Not-determined | Genome-fragment | NA | NA | 0 |
| S1_k141_349956 | 1180 | No | NA | 2 | 0 | 1 | Not-determined | Genome-fragment | NA | NA | 0 |
| S1_k141_218112 | 2429 | No | NA | 4 | 0 | 2 | Not-determined | Genome-fragment | NA | NA | 0 |
| S1_k141_119545 | 2926 | No | NA | 2 | 0 | 2 | Not-determined | Genome-fragment | NA | NA | 0 |
| S1_k141_167983 | 4055 | No | NA | 5 | 0 | 3 | Not-determined | Genome-fragment | NA | NA | 0 |
| S1_k141_102342 | 79954 | No | NA | 125 | 20 | 4 | Medium-quality | Genome-fragment | 58.02 | AAI-based (high-confidence) | 0 |
| S1_k141_218148 | 1738 | No | NA | 1 | 0 | 0 | Not-determined | Genome-fragment | NA | NA | 0 |
| S1_k141_36928 | 1067 | No | NA | 2 | 0 | 2 | Not-determined | Genome-fragment | NA | NA | 0 |
| S1_k141_366116 | 4705 | No | NA | 2 | 0 | 2 | Not-determined | Genome-fragment | NA | NA | 0 |
| S1_k141_36932 | 1006 | No | NA | 1 | 0 | 0 | Not-determined | Genome-fragment | NA | NA | 0 |
| S1_k141_86210 | 1807 | No | NA | 2 | 0 | 0 | Not-determined | Genome-fragment | NA | NA | 0 |
| S1_k141_350011 | 1045 | No | NA | 1 | 0 | 0 | Not-determined | Genome-fragment | NA | NA | 0 |
| S11-final-viral-combined_k141_229040\|\|full | 1462 | No | NA | 2 | 0 | 1 | Low-quality | Genome-fragment | 1.9 | AAI-based (medium-confidence) | 0 |
| S11-final-viral-combined_k141_215961\|\|full | 1462 | No | NA | 3 | 0 | 0 | Low-quality | Genome-fragment | 3.5 | AAI-based (medium-confidence) | 0 |
| S1_k141_119607 | 1160 | No | NA | 2 | 0 | 2 | Not-determined | Genome-fragment | NA | NA | 0 |
| S11-final-viral-combined_k141_325553\|\|lt2gene | 1462 | No | NA | 2 | 0 | 0 | Low-quality | Genome-fragment | 0.95 | AAI-based (high-confidence) | 0 |
| S1_k141_36959 | 1002 | No | NA | 2 | 0 | 2 | Not-determined | Genome-fragment | NA | NA | 0 |
| S1_k141_152303 | 1761 | No | NA | 2 | 0 | 0 | Not-determined | Genome-fragment | NA | NA | 0 |
| S1_k141_185686 | 1007 | No | NA | 1 | 0 | 0 | Not-determined | Genome-fragment | NA | NA | 0 |
| S1_k141_301017 | 1060 | No | NA | 1 | 0 | 1 | Not-determined | Genome-fragment | NA | NA | 0 |
| S1-final-viral-combined_k141_89367\|\|full | 1462 | No | NA | 3 | 0 | 0 | Low-quality | Genome-fragment | 0.5 | AAI-based (medium-confidence) | 0 |
| S1_k141_267383 | 1115 | No | NA | 2 | 0 | 2 | Not-determined | Genome-fragment | NA | NA | 0 |
| S7-final-viral-combined_k141_30799\|\|full | 1462 | No | NA | 3 | 2 | 0 | Low-quality | Genome-fragment | 3.33 | AAI-based (high-confidence) | 0 |
| S1_k141_119658 | 5080 | No | NA | 4 | 0 | 3 | Not-determined | Genome-fragment | NA | NA | 0 |
| S1_k141_20724 | 1470 | No | NA | 1 | 0 | 1 | Not-determined | Genome-fragment | NA | NA | 0 |
| S1_k141_102432 | 1731 | No | NA | 3 | 0 | 2 | Not-determined | Genome-fragment | NA | NA | 0 |
| S1_k141_136283 | 1663 | No | NA | 2 | 0 | 2 | Not-determined | Genome-fragment | NA | NA | 0 |
| S1_k141_317295 | 1849 | No | NA | 2 | 0 | 2 | Not-determined | Genome-fragment | NA | NA | 0 |
| S1_k141_366216 | 1352 | No | NA | 1 | 0 | 1 | Not-determined | Genome-fragment | NA | NA | 0 |
| S1_k141_317309 | 6320 | No | NA | 5 | 0 | 5 | Not-determined | Genome-fragment | NA | NA | 0 |
| S1_k141_234637 | 1797 | No | NA | 3 | 0 | 3 | Not-determined | Genome-fragment | NA | NA | 0 |
| S1_k141_333166 | 1168 | No | NA | 1 | 0 | 0 | Not-determined | Genome-fragment | NA | NA | 0 |
| S1_k141_37050 | 1337 | No | NA | 2 | 0 | 1 | Not-determined | Genome-fragment | NA | NA | 0 |
| S1_k141_152347 | 2026 | No | NA | 1 | 0 | 0 | Not-determined | Genome-fragment | NA | NA | 0 |
| S1_k141_102483 | 9764 | No | NA | 3 | 0 | 2 | Not-determined | Genome-fragment | NA | NA | 0 |
| S1_k141_69195 | 4476 | No | NA | 3 | 0 | 0 | Not-determined | Genome-fragment | NA | NA | 0 |
| S7_k141_209053 | 1462 | No | NA | 5 | 0 | 0 | Low-quality | Genome-fragment | 2.41 | AAI-based (high-confidence) | 0 |
| S1_k141_382563 | 1750 | No | NA | 1 | 0 | 1 | Not-determined | Genome-fragment | NA | NA | 0 |
| S1_k141_69203 | 7419 | No | NA | 4 | 0 | 3 | Not-determined | Genome-fragment | NA | NA | 0 |
| S1_k141_4131 | 1686 | No | NA | 1 | 0 | 1 | Not-determined | Genome-fragment | NA | NA | 0 |
| S1_k141_136383 | 4193 | No | NA | 2 | 0 | 0 | Not-determined | Genome-fragment | NA | NA | 0 |
| S1_k141_382582 | 1948 | No | NA | 2 | 0 | 2 | Not-determined | Genome-fragment | NA | NA | 0 |
| S12-final-viral-combined_k141_8598\|\|full | 1462 | No | NA | 2 | 0 | 1 | Low-quality | Genome-fragment | 1.9 | AAI-based (medium-confidence) | 0 |
| S4_k141_14074 | 1462 | No | NA | 3 | 0 | 0 | Low-quality | Genome-fragment | 0.99 | AAI-based (medium-confidence) | 0 |
| S1_k141_37125 | 1303 | No | NA | 2 | 0 | 1 | Not-determined | Genome-fragment | NA | NA | 0 |
| S6-final-viral-combined_k141_113942\|\|full | 1462 | No | NA | 1 | 0 | 1 | Low-quality | Genome-fragment | 1.04 | AAI-based (high-confidence) | 0 |
| S1_k141_350231 | 1921 | No | NA | 2 | 0 | 2 | Not-determined | Genome-fragment | NA | NA | 0 |
| S1_k141_168183 | 1025 | No | NA | 1 | 0 | 1 | Not-determined | Genome-fragment | NA | NA | 0 |
| S1_k141_201990 | 3869 | No | NA | 6 | 0 | 2 | Not-determined | Genome-fragment | NA | NA | 0 |
| S1_k141_382610 | 1299 | No | NA | 1 | 0 | 1 | Not-determined | Genome-fragment | NA | NA | 0 |
| S6_k141_75354 | 1462 | No | NA | 1 | 0 | 0 | Low-quality | Genome-fragment | 0.96 | AAI-based (high-confidence) | 0 |
| S1_k141_53441 | 3677 | No | NA | 8 | 0 | 2 | Not-determined | Genome-fragment | NA | NA | 0 |
| S1_k141_317471 | 1112 | No | NA | 3 | 0 | 1 | Not-determined | Genome-fragment | NA | NA | 0 |
| S1_k141_382624 | 1036 | No | NA | 2 | 0 | 1 | Not-determined | Genome-fragment | NA | NA | 0 |
| S1_k141_251749 | 4007 | No | NA | 7 | 0 | 0 | Not-determined | Genome-fragment | NA | NA | 0 |
| S1_k141_317475 | 1669 | No | NA | 1 | 0 | 1 | Not-determined | Genome-fragment | NA | NA | 0 |
| S1_k141_37184 | 1326 | No | NA | 2 | 0 | 0 | Not-determined | Genome-fragment | NA | NA | 0 |
| S1_k141_53474 | 1580 | No | NA | 1 | 0 | 1 | Not-determined | Genome-fragment | NA | NA | 0 |
| S1_k141_119887 | 1085 | No | NA | 1 | 0 | 0 | Not-determined | Genome-fragment | NA | NA | 0 |
| S5-final-viral-combined_k141_143301\|\|full | 1463 | No | NA | 2 | 0 | 0 | Low-quality | Genome-fragment | 2.28 | AAI-based (medium-confidence) | 0 |
| S1_k141_69266 | 3067 | No | NA | 4 | 0 | 0 | Not-determined | Genome-fragment | NA | NA | 0 |
| S1_k141_168270 | 1355 | No | NA | 1 | 0 | 1 | Not-determined | Genome-fragment | NA | NA | 0 |
| S1_k141_301289 | 5492 | No | NA | 3 | 0 | 1 | Not-determined | Genome-fragment | NA | NA | 0 |
| S7-final-viral-combined_k141_218195\|\|full | 1463 | No | NA | 3 | 0 | 1 | Low-quality | Genome-fragment | 2.45 | AAI-based (medium-confidence) | 0 |
| S1_k141_366348 | 1502 | No | NA | 1 | 0 | 0 | Not-determined | Genome-fragment | NA | NA | 0 |
| S1_k141_333340 | 1086 | No | NA | 1 | 0 | 1 | Not-determined | Genome-fragment | NA | NA | 0 |
| S1_k141_136572 | 1262 | No | NA | 1 | 0 | 1 | Not-determined | Genome-fragment | NA | NA | 0 |
| S1_k141_333353 | 3304 | No | NA | 6 | 0 | 1 | Not-determined | Genome-fragment | NA | NA | 0 |
| S1_k141_218289 | 1692 | No | NA | 2 | 0 | 1 | Not-determined | Genome-fragment | NA | NA | 0 |
| S1_k141_4253 | 4416 | No | NA | 3 | 0 | 0 | Not-determined | Genome-fragment | NA | NA | 0 |
| S1_k141_53581 | 1055 | No | NA | 2 | 0 | 1 | Not-determined | Genome-fragment | NA | NA | 0 |
| S1_k141_317580 | 1210 | No | NA | 2 | 0 | 1 | Not-determined | Genome-fragment | NA | NA | 0 |
| S1_k141_267614 | 1414 | No | NA | 1 | 0 | 1 | Not-determined | Genome-fragment | NA | NA | 0 |
| S1_k141_152540 | 1101 | No | NA | 3 | 0 | 3 | Not-determined | Genome-fragment | NA | NA | 0 |
| S1_k141_350367 | 1264 | No | NA | 1 | 0 | 1 | Not-determined | Genome-fragment | NA | NA | 0 |
| S1_k141_317590 | 1144 | No | NA | 1 | 0 | 0 | Not-determined | Genome-fragment | NA | NA | 0 |
| S1_k141_152559 | 1189 | No | NA | 1 | 0 | 1 | Not-determined | Genome-fragment | NA | NA | 0 |
| S7-final-viral-combined_k141_71005\|\|full | 1463 | No | NA | 2 | 0 | 2 | Low-quality | Genome-fragment | 1.48 | AAI-based (high-confidence) | 0 |
| S12-final-viral-combined_k141_183815\|\|full | 1463 | No | NA | 2 | 0 | 0 | Low-quality | Genome-fragment | 1.09 | AAI-based (medium-confidence) | 0 |
| S1_k141_4277 | 31752 | Yes | 29534 | 46 | 14 | 3 | Medium-quality | Genome-fragment | 82.52 | AAI-based (medium-confidence) | 6.99 |
| S1_k141_317638 | 1310 | No | NA | 2 | 0 | 1 | Not-determined | Genome-fragment | NA | NA | 0 |
| S1_k141_120017 | 1056 | No | NA | 2 | 0 | 0 | Not-determined | Genome-fragment | NA | NA | 0 |
| S12-final-viral-combined_k141_149701\|\|full | 1463 | No | NA | 3 | 1 | 0 | Low-quality | Genome-fragment | 2.51 | HMM-based (lower-bound) | 0 |
| S1_k141_350405 | 3848 | No | NA | 5 | 0 | 4 | Not-determined | Genome-fragment | NA | NA | 0 |
| S1_k141_53670 | 1358 | No | NA | 2 | 0 | 2 | Not-determined | Genome-fragment | NA | NA | 0 |
| S1_k141_37344 | 1372 | No | NA | 1 | 0 | 0 | Not-determined | Genome-fragment | NA | NA | 0 |
| S1_k141_120062 | 1396 | No | NA | 3 | 0 | 1 | Not-determined | Genome-fragment | NA | NA | 0 |
| S1_k141_102729 | 1011 | No | NA | 1 | 0 | 1 | Not-determined | Genome-fragment | NA | NA | 0 |
| S1_k141_234869 | 5796 | No | NA | 5 | 0 | 1 | Not-determined | Genome-fragment | NA | NA | 0 |
| S1_k141_185953 | 2473 | No | NA | 4 | 0 | 3 | Not-determined | Genome-fragment | NA | NA | 0 |
| S1_k141_382901 | 4073 | No | NA | 6 | 0 | 1 | Not-determined | Genome-fragment | NA | NA | 0 |
| S1_k141_234894 | 1498 | No | NA | 3 | 0 | 2 | Not-determined | Genome-fragment | NA | NA | 0 |
| S1_k141_234896 | 1376 | No | NA | 3 | 0 | 0 | Not-determined | Genome-fragment | NA | NA | 0 |
| S2-final-viral-combined_k141_95065\|\|full | 1463 | No | NA | 2 | 1 | 0 | Low-quality | Genome-fragment | 2.17 | HMM-based (lower-bound) | 0 |
| S2-final-viral-combined_k141_214462\|\|full | 1463 | No | NA | 3 | 0 | 2 | Low-quality | Genome-fragment | 3.35 | AAI-based (medium-confidence) | 0 |
| S2_k141_166881 | 1463 | No | NA | 2 | 1 | 0 | Low-quality | Genome-fragment | 5.22 | HMM-based (lower-bound) | 0 |
| S1_k141_350466 | 1028 | No | NA | 1 | 0 | 0 | Not-determined | Genome-fragment | NA | NA | 0 |
| S1_k141_234917 | 1249 | No | NA | 1 | 0 | 0 | Not-determined | Genome-fragment | NA | NA | 0 |
| S8-final-viral-combined_k141_263569\|\|full | 1463 | No | NA | 3 | 1 | 0 | Low-quality | Genome-fragment | 1.31 | HMM-based (lower-bound) | 0 |
| S1_k141_202160 | 1208 | No | NA | 1 | 0 | 0 | Not-determined | Genome-fragment | NA | NA | 0 |
| S1_k141_37385 | 1092 | No | NA | 1 | 0 | 0 | Not-determined | Genome-fragment | NA | NA | 0 |
| S1_k141_234979 | 1303 | No | NA | 2 | 0 | 1 | Not-determined | Genome-fragment | NA | NA | 0 |
| S1_k141_234983 | 1133 | No | NA | 1 | 0 | 0 | Not-determined | Genome-fragment | NA | NA | 0 |
| S8-final-viral-combined_k141_121668\|\|lt2gene | 1463 | No | NA | 2 | 1 | 0 | Low-quality | Genome-fragment | 4.55 | AAI-based (high-confidence) | 0 |
| S1_k141_350520 | 3045 | No | NA | 4 | 0 | 0 | Not-determined | Genome-fragment | NA | NA | 0 |
| S8_k141_106522 | 1463 | No | NA | 6 | 0 | 0 | Low-quality | Genome-fragment | 1.11 | AAI-based (high-confidence) | 0 |
| S1_k141_317798 | 1127 | No | NA | 1 | 0 | 0 | Not-determined | Genome-fragment | NA | NA | 0 |
| S1_k141_4458 | 4426 | No | NA | 2 | 0 | 0 | Not-determined | Genome-fragment | NA | NA | 0 |
| S1_k141_366607 | 6406 | No | NA | 2 | 0 | 1 | Not-determined | Genome-fragment | NA | NA | 0 |
| S9-final-viral-combined_k141_53253\|\|full | 1464 | No | NA | 3 | 1 | 0 | Low-quality | Genome-fragment | 3.53 | AAI-based (high-confidence) | 0 |
| S1_k141_37442 | 1050 | No | NA | 3 | 0 | 2 | Not-determined | Genome-fragment | NA | NA | 0 |
| S1_k141_284621 | 1137 | No | NA | 3 | 0 | 1 | Not-determined | Genome-fragment | NA | NA | 0 |
| S1-final-viral-combined_k141_216456\|\|full | 1464 | No | NA | 4 | 1 | 0 | Low-quality | Genome-fragment | 3.58 | AAI-based (medium-confidence) | 0 |
| S1_k141_136857 | 1059 | No | NA | 2 | 0 | 2 | Not-determined | Genome-fragment | NA | NA | 0 |
| S1_k141_383005 | 3024 | No | NA | 1 | 0 | 1 | Not-determined | Genome-fragment | NA | NA | 0 |
| S1_k141_251999 | 1594 | No | NA | 2 | 0 | 2 | Not-determined | Genome-fragment | NA | NA | 0 |
| S1_k141_366619 | 1515 | No | NA | 1 | 0 | 0 | Not-determined | Genome-fragment | NA | NA | 0 |
| S3-final-viral-combined_k141_127910\|\|full | 1464 | No | NA | 3 | 0 | 0 | Low-quality | Genome-fragment | 1.78 | AAI-based (high-confidence) | 0 |
| S5-final-viral-combined_k141_53073\|\|full | 1464 | No | NA | 5 | 0 | 0 | Low-quality | Genome-fragment | 1.96 | AAI-based (high-confidence) | 0 |
| S5-final-viral-combined_k141_39548\|\|lt2gene | 1464 | No | NA | 1 | 1 | 0 | Low-quality | Genome-fragment | 1.42 | HMM-based (lower-bound) | 0 |
| S1_k141_102898 | 4006 | No | NA | 3 | 0 | 0 | Not-determined | Genome-fragment | NA | NA | 0 |
| S7-final-viral-combined_k141_86208\|\|full | 1464 | No | NA | 3 | 1 | 0 | Low-quality | Genome-fragment | 0.93 | HMM-based (lower-bound) | 0 |
| S1_k141_21315 | 3898 | No | NA | 4 | 0 | 3 | Not-determined | Genome-fragment | NA | NA | 0 |
| S1_k141_152782 | 1116 | No | NA | 2 | 0 | 1 | Not-determined | Genome-fragment | NA | NA | 0 |
| S1_k141_202297 | 1071 | No | NA | 1 | 0 | 0 | Not-determined | Genome-fragment | NA | NA | 0 |
| S1_k141_267845 | 3644 | No | NA | 3 | 0 | 0 | Not-determined | Genome-fragment | NA | NA | 0 |
| S17-final-viral-combined_k141_39939\|\|full | 1464 | No | NA | 3 | 2 | 0 | Low-quality | Genome-fragment | 4.04 | AAI-based (high-confidence) | 0 |
| S1_k141_136913 | 3043 | No | NA | 3 | 0 | 1 | Not-determined | Genome-fragment | NA | NA | 0 |
| S1_k141_86692 | 3662 | No | NA | 6 | 0 | 0 | Not-determined | Genome-fragment | NA | NA | 0 |
| S1_k141_168735 | 2187 | No | NA | 1 | 0 | 0 | Not-determined | Genome-fragment | NA | NA | 0 |
| S17-final-viral-combined_k141_177319\|\|full | 1464 | No | NA | 3 | 0 | 2 | Low-quality | Genome-fragment | 0.43 | AAI-based (medium-confidence) | 0 |
| S1_k141_136963 | 4921 | No | NA | 7 | 0 | 2 | Not-determined | Genome-fragment | NA | NA | 0 |
| S17-k141_702 | 1464 | No | NA | 1 | 0 | 0 | Low-quality | Genome-fragment | 2.98 | AAI-based (high-confidence) | 0 |
| S1_k141_383227 | 1103 | No | NA | 2 | 0 | 2 | Not-determined | Genome-fragment | NA | NA | 0 |
| S1_k141_69659 | 5005 | No | NA | 5 | 0 | 4 | Not-determined | Genome-fragment | NA | NA | 0 |
| S1_k141_318017 | 9706 | No | NA | 13 | 0 | 4 | Not-determined | Genome-fragment | NA | NA | 0 |
| S1_k141_383266 | 1432 | No | NA | 2 | 0 | 1 | Not-determined | Genome-fragment | NA | NA | 0 |
| S1_k141_218736 | 1644 | No | NA | 2 | 0 | 0 | Not-determined | Genome-fragment | NA | NA | 0 |
| S1_k141_186227 | 1143 | No | NA | 1 | 0 | 1 | Not-determined | Genome-fragment | NA | NA | 0 |
| S12-final-viral-combined_k141_122696\|\|full | 1464 | No | NA | 3 | 3 | 0 | Low-quality | Genome-fragment | 3.04 | AAI-based (high-confidence) | 0 |
| S1_k141_252229 | 2720 | No | NA | 2 | 0 | 2 | Not-determined | Genome-fragment | NA | NA | 0 |
| S1_k141_235294 | 4527 | No | NA | 8 | 0 | 0 | Not-determined | Genome-fragment | NA | NA | 0 |
| S1_k141_366807 | 3664 | No | NA | 2 | 0 | 1 | Not-determined | Genome-fragment | NA | NA | 0 |
| S1_k141_69779 | 10151 | No | NA | 11 | 0 | 10 | Not-determined | Genome-fragment | NA | NA | 0 |
| S1_k141_153002 | 2227 | No | NA | 2 | 0 | 0 | Not-determined | Genome-fragment | NA | NA | 0 |
| S1_k141_318132 | 1000 | No | NA | 2 | 0 | 1 | Not-determined | Genome-fragment | NA | NA | 0 |
| S1_k141_366827 | 1661 | No | NA | 1 | 0 | 0 | Not-determined | Genome-fragment | NA | NA | 0 |
| S1_k141_69795 | 4133 | No | NA | 3 | 0 | 0 | Not-determined | Genome-fragment | NA | NA | 0 |
| S1_k141_186314 | 1174 | No | NA | 1 | 0 | 1 | Not-determined | Genome-fragment | NA | NA | 0 |
| S1_k141_252304 | 1046 | No | NA | 2 | 0 | 1 | Not-determined | Genome-fragment | NA | NA | 0 |
| S8-final-viral-combined_k141_112653\|\|full | 1464 | No | NA | 2 | 1 | 0 | Low-quality | Genome-fragment | 2.22 | HMM-based (lower-bound) | 0 |
| S1_k141_202556 | 3310 | No | NA | 2 | 0 | 0 | Not-determined | Genome-fragment | NA | NA | 0 |
| S1_k141_318181 | 1108 | No | NA | 2 | 0 | 0 | Not-determined | Genome-fragment | NA | NA | 0 |
| S1_k141_120575 | 1446 | No | NA | 2 | 0 | 2 | Not-determined | Genome-fragment | NA | NA | 0 |
| S1_k141_86836 | 1037 | No | NA | 2 | 0 | 1 | Not-determined | Genome-fragment | NA | NA | 0 |
| S1_k141_168966 | 3115 | No | NA | 3 | 0 | 3 | Not-determined | Genome-fragment | NA | NA | 0 |
| S1_k141_235363 | 1051 | No | NA | 1 | 0 | 1 | Not-determined | Genome-fragment | NA | NA | 0 |
| S1_k141_4874 | 1839 | No | NA | 1 | 0 | 1 | Not-determined | Genome-fragment | NA | NA | 0 |
| S8_k141_60784 | 1464 | No | NA | 1 | 0 | 1 | Low-quality | Genome-fragment | 1.15 | AAI-based (high-confidence) | 0 |
| S1_k141_153095 | 2947 | No | NA | 3 | 0 | 2 | Not-determined | Genome-fragment | NA | NA | 0 |
| S1_k141_37873 | 1037 | No | NA | 2 | 0 | 1 | Not-determined | Genome-fragment | NA | NA | 0 |
| S1_k141_137199 | 1014 | No | NA | 2 | 0 | 0 | Not-determined | Genome-fragment | NA | NA | 0 |
| S12-final-viral-combined_k141_399875\|\|lt2gene | 1465 | No | NA | 2 | 1 | 0 | Low-quality | Genome-fragment | 3.58 | AAI-based (medium-confidence) | 0 |
| S1_k141_235413 | 1079 | No | NA | 1 | 0 | 1 | Not-determined | Genome-fragment | NA | NA | 0 |
| S1_k141_54049 | 1627 | No | NA | 1 | 0 | 1 | Not-determined | Genome-fragment | NA | NA | 0 |
| S1_k141_302058 | 1010 | No | NA | 1 | 0 | 0 | Not-determined | Genome-fragment | NA | NA | 0 |
| S1_k141_333652 | 1769 | No | NA | 3 | 0 | 1 | Not-determined | Genome-fragment | NA | NA | 0 |
| S1_k141_367048 | 1316 | No | NA | 1 | 0 | 1 | Not-determined | Genome-fragment | NA | NA | 0 |
| S1_k141_120729 | 4968 | No | NA | 7 | 0 | 1 | Not-determined | Genome-fragment | NA | NA | 0 |
| S1_k141_268097 | 11452 | No | NA | 10 | 0 | 6 | Not-determined | Genome-fragment | NA | NA | 0 |
| S1_k141_302105 | 2033 | No | NA | 2 | 0 | 0 | Not-determined | Genome-fragment | NA | NA | 0 |
| S1_k141_284894 | 1136 | No | NA | 1 | 0 | 0 | Not-determined | Genome-fragment | NA | NA | 0 |
| S1_k141_103317 | 1087 | No | NA | 1 | 0 | 1 | Not-determined | Genome-fragment | NA | NA | 0 |
| S1_k141_383572 | 2032 | No | NA | 1 | 0 | 1 | Not-determined | Genome-fragment | NA | NA | 0 |
| S1_k141_367114 | 8354 | No | NA | 5 | 0 | 1 | Not-determined | Genome-fragment | NA | NA | 0 |
| S1_k141_120791 | 1339 | No | NA | 1 | 0 | 0 | Not-determined | Genome-fragment | NA | NA | 0 |
| S1_k141_219071 | 2798 | No | NA | 3 | 0 | 1 | Not-determined | Genome-fragment | NA | NA | 0 |
| S1_k141_252429 | 2258 | No | NA | 1 | 0 | 1 | Not-determined | Genome-fragment | NA | NA | 0 |
| S1_k141_268158 | 1080 | No | NA | 1 | 0 | 1 | Not-determined | Genome-fragment | NA | NA | 0 |
| S1_k141_235525 | 2985 | No | NA | 1 | 0 | 0 | Not-determined | Genome-fragment | NA | NA | 0 |
| S1_k141_120821 | 7741 | No | NA | 6 | 0 | 3 | Not-determined | Genome-fragment | NA | NA | 0 |
| S1_k141_268174 | 1042 | No | NA | 1 | 0 | 0 | Not-determined | Genome-fragment | NA | NA | 0 |
| S1_k141_87035 | 3281 | No | NA | 5 | 0 | 2 | Not-determined | Genome-fragment | NA | NA | 0 |
| S12_k141_368088 | 1465 | No | NA | 2 | 0 | 0 | Low-quality | Genome-fragment | 3.73 | AAI-based (high-confidence) | 0 |
| S1_k141_333767 | 4247 | No | NA | 6 | 0 | 1 | Not-determined | Genome-fragment | NA | NA | 0 |
| S1_k141_284942 | 1096 | No | NA | 1 | 0 | 1 | Not-determined | Genome-fragment | NA | NA | 0 |
| S1_k141_284952 | 1184 | No | NA | 2 | 0 | 1 | Not-determined | Genome-fragment | NA | NA | 0 |
| S6-final-viral-combined_k141_6156\|\|full | 1465 | No | NA | 3 | 0 | 1 | Low-quality | Genome-fragment | 0.76 | AAI-based (medium-confidence) | 0 |
| S6-final-viral-combined_k141_74076\|\|full | 1465 | No | NA | 3 | 0 | 0 | Low-quality | Genome-fragment | 3.71 | AAI-based (medium-confidence) | 0 |
| S1_k141_302150 | 1058 | No | NA | 2 | 0 | 0 | Not-determined | Genome-fragment | NA | NA | 0 |
| S1_k141_54188 | 3572 | No | NA | 3 | 0 | 2 | Not-determined | Genome-fragment | NA | NA | 0 |
| S1_k141_153229 | 1471 | No | NA | 2 | 0 | 1 | Not-determined | Genome-fragment | NA | NA | 0 |
| S1_k141_137383 | 1014 | No | NA | 1 | 0 | 1 | Not-determined | Genome-fragment | NA | NA | 0 |
| S9-final-viral-combined_k141_32442\|\|full | 1466 | No | NA | 4 | 0 | 0 | Low-quality | Genome-fragment | 3.36 | AAI-based (high-confidence) | 0 |
| S1_k141_137402 | 3583 | No | NA | 1 | 0 | 1 | Not-determined | Genome-fragment | NA | NA | 0 |
| S11_k141_52237 | 1466 | No | NA | 1 | 1 | 0 | Low-quality | Genome-fragment | 0.72 | HMM-based (lower-bound) | 0 |
| S1_k141_137425 | 3567 | No | NA | 3 | 0 | 0 | Not-determined | Genome-fragment | NA | NA | 0 |
| S1_k141_21516 | 1068 | No | NA | 1 | 0 | 0 | Not-determined | Genome-fragment | NA | NA | 0 |
| S1_k141_120934 | 1323 | No | NA | 3 | 0 | 0 | Not-determined | Genome-fragment | NA | NA | 0 |
| S1_k141_268283 | 3444 | No | NA | 4 | 0 | 3 | Not-determined | Genome-fragment | NA | NA | 0 |
| S1_k141_252532 | 3347 | No | NA | 1 | 0 | 1 | Not-determined | Genome-fragment | NA | NA | 0 |
| S1_k141_284997 | 1458 | No | NA | 1 | 0 | 1 | Not-determined | Genome-fragment | NA | NA | 0 |
| S1_k141_54271 | 2269 | No | NA | 2 | 0 | 2 | Not-determined | Genome-fragment | NA | NA | 0 |
| S1_k141_252540 | 7494 | No | NA | 7 | 0 | 6 | Not-determined | Genome-fragment | NA | NA | 0 |
| S1_k141_54291 | 1279 | No | NA | 1 | 0 | 1 | Not-determined | Genome-fragment | NA | NA | 0 |
| S1_k141_54303 | 5149 | No | NA | 5 | 0 | 5 | Not-determined | Genome-fragment | NA | NA | 0 |
| S3-final-viral-combined_k141_43945\|\|full | 1466 | No | NA | 4 | 1 | 0 | Low-quality | Genome-fragment | 3.51 | AAI-based (high-confidence) | 0 |
| S1_k141_351239 | 1451 | No | NA | 2 | 0 | 2 | Not-determined | Genome-fragment | NA | NA | 0 |
| S1_k141_318549 | 7120 | No | NA | 2 | 0 | 1 | Not-determined | Genome-fragment | NA | NA | 0 |
| S1_k141_351273 | 1315 | No | NA | 1 | 0 | 0 | Not-determined | Genome-fragment | NA | NA | 0 |
| S1_k141_169175 | 1321 | No | NA | 1 | 0 | 1 | Not-determined | Genome-fragment | NA | NA | 0 |
| S1_k141_202895 | 1645 | No | NA | 3 | 0 | 1 | Not-determined | Genome-fragment | NA | NA | 0 |
| S3-final-viral-combined_k141_29758\|\|lt2gene | 1466 | No | NA | 1 | 1 | 0 | Low-quality | Genome-fragment | 0.8 | HMM-based (lower-bound) | 0 |
| S5-final-viral-combined_k141_28559\|\|full | 1466 | No | NA | 3 | 1 | 0 | Low-quality | Genome-fragment | 2.36 | AAI-based (medium-confidence) | 0 |
| S1_k141_318558 | 3368 | No | NA | 1 | 0 | 0 | Not-determined | Genome-fragment | NA | NA | 0 |
| S5-final-viral-combined_k141_28805\|\|lt2gene | 1466 | No | NA | 1 | 1 | 0 | Low-quality | Genome-fragment | 2.14 | HMM-based (lower-bound) | 0 |
| S1_k141_351286 | 3791 | No | NA | 2 | 0 | 1 | Not-determined | Genome-fragment | NA | NA | 0 |
| S7-final-viral-combined_k141_92065\|\|full | 1466 | Yes | 615 | 3 | 1 | 1 | Low-quality | Genome-fragment | 1.45 | AAI-based (medium-confidence) | 58.05 |
| S7_k141_103936 | 1466 | No | NA | 1 | 0 | 0 | Low-quality | Genome-fragment | 0.25 | AAI-based (high-confidence) | 0 |
| S1_k141_252615 | 5087 | No | NA | 3 | 0 | 2 | Not-determined | Genome-fragment | NA | NA | 0 |
| S1_k141_186739 | 1635 | No | NA | 1 | 0 | 1 | Not-determined | Genome-fragment | NA | NA | 0 |
| S1_k141_351336 | 1552 | No | NA | 1 | 0 | 1 | Not-determined | Genome-fragment | NA | NA | 0 |
| S12-final-viral-combined_k141_180247\|\|lt2gene | 1466 | No | NA | 2 | 1 | 0 | Low-quality | Genome-fragment | 2.22 | AAI-based (medium-confidence) | 0 |
| S4-final-viral-combined_k141_117416\|\|full | 1466 | No | NA | 3 | 0 | 1 | Low-quality | Genome-fragment | 2.07 | AAI-based (high-confidence) | 0 |
| S1_k141_351360 | 1248 | No | NA | 2 | 0 | 0 | Not-determined | Genome-fragment | NA | NA | 0 |
| S1-final-viral-combined_k141_182581\|\|full | 1467 | No | NA | 2 | 0 | 0 | Low-quality | Genome-fragment | 2.48 | AAI-based (high-confidence) | 0 |
| S1-final-viral-combined_k141_62116\|\|lt2gene | 1467 | No | NA | 1 | 1 | 0 | Low-quality | Genome-fragment | 1.01 | HMM-based (lower-bound) | 0 |
| S1_k141_252633 | 1010 | No | NA | 1 | 0 | 1 | Not-determined | Genome-fragment | NA | NA | 0 |
| S1_k141_285081 | 2769 | No | NA | 2 | 0 | 0 | Not-determined | Genome-fragment | NA | NA | 0 |
| S1_k141_268402 | 1950 | No | NA | 1 | 0 | 0 | Not-determined | Genome-fragment | NA | NA | 0 |
| S3_k141_37487 | 1467 | No | NA | 2 | 1 | 0 | Low-quality | Genome-fragment | 0.95 | AAI-based (high-confidence) | 0 |
| S1_k141_235774 | 1109 | No | NA | 2 | 0 | 2 | Not-determined | Genome-fragment | NA | NA | 0 |
| S1_k141_351382 | 1035 | No | NA | 1 | 0 | 1 | Not-determined | Genome-fragment | NA | NA | 0 |
| S1_k141_252699 | 2352 | No | NA | 2 | 0 | 2 | Not-determined | Genome-fragment | NA | NA | 0 |
| S1_k141_87272 | 1196 | No | NA | 2 | 0 | 2 | Not-determined | Genome-fragment | NA | NA | 0 |
| S1_k141_235809 | 1084 | No | NA | 1 | 0 | 1 | Not-determined | Genome-fragment | NA | NA | 0 |
| S1_k141_121177 | 1088 | No | NA | 2 | 0 | 0 | Not-determined | Genome-fragment | NA | NA | 0 |
| S5-final-viral-combined_k141_39181\|\|full | 1467 | No | NA | 3 | 0 | 0 | Low-quality | Genome-fragment | 3.57 | AAI-based (medium-confidence) | 0 |
| S7-final-viral-combined_k141_6007\|\|full | 1467 | No | NA | 2 | 0 | 0 | Low-quality | Genome-fragment | 0.58 | AAI-based (medium-confidence) | 0 |
| S1_k141_54432 | 1065 | No | NA | 3 | 0 | 3 | Not-determined | Genome-fragment | NA | NA | 0 |
| S1_k141_367456 | 1437 | No | NA | 1 | 0 | 1 | Not-determined | Genome-fragment | NA | NA | 0 |
| S1_k141_121207 | 1678 | No | NA | 1 | 0 | 1 | Not-determined | Genome-fragment | NA | NA | 0 |
| S7-final-viral-combined_k141_7091\|\|full | 1467 | No | NA | 3 | 0 | 0 | Low-quality | Genome-fragment | 1.72 | AAI-based (medium-confidence) | 0 |
| S1_k141_5310 | 3206 | No | NA | 2 | 0 | 2 | Not-determined | Genome-fragment | NA | NA | 0 |
| S1_k141_285235 | 2423 | No | NA | 2 | 0 | 1 | Not-determined | Genome-fragment | NA | NA | 0 |
| S1_k141_169279 | 3212 | No | NA | 1 | 0 | 1 | Not-determined | Genome-fragment | NA | NA | 0 |
| S1_k141_54503 | 1142 | No | NA | 1 | 0 | 1 | Not-determined | Genome-fragment | NA | NA | 0 |
| S1_k141_268484 | 4310 | No | NA | 3 | 0 | 1 | Not-determined | Genome-fragment | NA | NA | 0 |
| S1_k141_367503 | 1069 | No | NA | 1 | 0 | 1 | Not-determined | Genome-fragment | NA | NA | 0 |

| | | | | | | | | | | | |
|---|---|---|---|---|---|---|---|---|---|---|---|
| S7-final-viral-combined_k141_109227\|\|full | 1467 | No | NA | 4 | 1 | 0 | Low-quality | Genome-fragment | 3.68 | AAI-based (high-confidence) | 0 |
| S1_k141_351537 | 1075 | No | NA | 2 | 0 | 1 | Not-determined | Genome-fragment | NA | NA | 0 |
| S1_k141_70248 | 2192 | No | NA | 3 | 0 | 3 | Not-determined | Genome-fragment | NA | NA | 0 |
| S1_k141_87368 | 1008 | No | NA | 1 | 0 | 0 | Not-determined | Genome-fragment | NA | NA | 0 |
| S1_k141_103383 | 1121 | No | NA | 1 | 0 | 1 | Not-determined | Genome-fragment | NA | NA | 0 |
| S1_k141_121267 | 1096 | No | NA | 3 | 0 | 1 | Not-determined | Genome-fragment | NA | NA | 0 |
| S1_k141_38248 | 1872 | No | NA | 2 | 0 | 2 | Not-determined | Genome-fragment | NA | NA | 0 |
| S7-final-viral-combined_k141_227626\|\|lt2gene | 1467 | No | NA | 1 | 1 | 0 | Low-quality | Genome-fragment | 3.71 | AAI-based (high-confidence) | 0 |
| S1_k141_87387 | 1056 | No | NA | 1 | 0 | 0 | Not-determined | Genome-fragment | NA | NA | 0 |
| S1_k141_137751 | 12244 | No | NA | 15 | 0 | 3 | Not-determined | Genome-fragment | NA | NA | 0 |
| S1_k141_54523 | 1393 | No | NA | 2 | 0 | 2 | Not-determined | Genome-fragment | NA | NA | 0 |
| S1_k141_103432 | 3256 | No | NA | 2 | 0 | 1 | Not-determined | Genome-fragment | NA | NA | 0 |
| S1_k141_235966 | 1610 | No | NA | 1 | 0 | 1 | Not-determined | Genome-fragment | NA | NA | 0 |
| S1_k141_268563 | 3972 | No | NA | 4 | 0 | 3 | Not-determined | Genome-fragment | NA | NA | 0 |
| S1_k141_383973 | 1955 | No | NA | 4 | 0 | 1 | Not-determined | Genome-fragment | NA | NA | 0 |
| S1_k141_153615 | 2783 | No | NA | 4 | 1 | 0 | Medium-quality | Genome-fragment | 50.34 | AAI-based (high-confidence) | 0 |
| S12-final-viral-combined_k141_280661\|\|full | 1467 | No | NA | 2 | 0 | 0 | Low-quality | Genome-fragment | 2.46 | AAI-based (high-confidence) | 0 |
| S1_k141_137796 | 1550 | No | NA | 3 | 0 | 2 | Not-determined | Genome-fragment | NA | NA | 0 |
| S1_k141_169394 | 1008 | No | NA | 1 | 0 | 1 | Not-determined | Genome-fragment | NA | NA | 0 |
| S1_k141_5403 | 1869 | No | NA | 2 | 0 | 2 | Not-determined | Genome-fragment | NA | NA | 0 |
| S1_k141_87450 | 2148 | No | NA | 6 | 0 | 3 | Not-determined | Genome-fragment | NA | NA | 0 |
| S1_k141_38364 | 4443 | No | NA | 3 | 0 | 0 | Not-determined | Genome-fragment | NA | NA | 0 |
| S6-final-viral-combined_k141_51819\|\|full | 1467 | No | NA | 4 | 0 | 0 | Low-quality | Genome-fragment | 1.32 | AAI-based (high-confidence) | 0 |
| S1_k141_5448 | 1353 | No | NA | 1 | 0 | 1 | Not-determined | Genome-fragment | NA | NA | 0 |
| S1_k141_137804 | 1096 | No | NA | 2 | 0 | 1 | Not-determined | Genome-fragment | NA | NA | 0 |
| S1_k141_187188 | 1134 | No | NA | 1 | 0 | 0 | Not-determined | Genome-fragment | NA | NA | 0 |
| S1_k141_334270 | 1116 | No | NA | 2 | 0 | 0 | Not-determined | Genome-fragment | NA | NA | 0 |
| S1_k141_203320 | 2221 | No | NA | 2 | 0 | 1 | Not-determined | Genome-fragment | NA | NA | 0 |
| S1_k141_367781 | 1720 | No | NA | 2 | 0 | 2 | Not-determined | Genome-fragment | NA | NA | 0 |
| S1_k141_103524 | 1434 | No | NA | 3 | 0 | 0 | Not-determined | Genome-fragment | NA | NA | 0 |
| S1_k141_384095 | 1004 | No | NA | 3 | 0 | 3 | Not-determined | Genome-fragment | NA | NA | 0 |
| S1_k141_121337 | 1031 | No | NA | 1 | 0 | 1 | Not-determined | Genome-fragment | NA | NA | 0 |
| S1_k141_169516 | 3753 | No | NA | 3 | 0 | 0 | Not-determined | Genome-fragment | NA | NA | 0 |
| S1_k141_236105 | 22362 | No | NA | 36 | 13 | 0 | Medium-quality | Genome-fragment | 50.89 | AAI-based (high-confidence) | 0 |
| S11-final-viral-combined_k141_168293\|\|full | 1468 | No | NA | 4 | 0 | 0 | Low-quality | Genome-fragment | 3.69 | AAI-based (medium-confidence) | 0 |
| S1_k141_367840 | 2361 | No | NA | 1 | 0 | 1 | Not-determined | Genome-fragment | NA | NA | 0 |
| S1_k141_103570 | 1097 | No | NA | 2 | 0 | 1 | Not-determined | Genome-fragment | NA | NA | 0 |
| S1_k141_187282 | 1120 | No | NA | 1 | 0 | 0 | Not-determined | Genome-fragment | NA | NA | 0 |
| S1_k141_121370 | 1038 | No | NA | 1 | 0 | 1 | Not-determined | Genome-fragment | NA | NA | 0 |
| S1_k141_103608 | 2956 | No | NA | 1 | 0 | 1 | Not-determined | Genome-fragment | NA | NA | 0 |
| S1_k141_318883 | 1191 | No | NA | 1 | 0 | 0 | Not-determined | Genome-fragment | NA | NA | 0 |
| S1_k141_351743 | 1523 | No | NA | 2 | 0 | 1 | Not-determined | Genome-fragment | NA | NA | 0 |
| S1_k141_384200 | 1364 | No | NA | 4 | 0 | 4 | Not-determined | Genome-fragment | NA | NA | 0 |
| S11_k141_75278 | 1468 | No | NA | 3 | 0 | 0 | Low-quality | Genome-fragment | 3.19 | AAI-based (medium-confidence) | 0 |
| S1_k141_302752 | 1678 | No | NA | 1 | 0 | 1 | Not-determined | Genome-fragment | NA | NA | 0 |
| S1_k141_203470 | 1217 | No | NA | 2 | 0 | 1 | Not-determined | Genome-fragment | NA | NA | 0 |
| S1_k141_38547 | 1898 | No | NA | 3 | 0 | 0 | Not-determined | Genome-fragment | NA | NA | 0 |
| S1_k141_302776 | 1115 | No | NA | 1 | 0 | 1 | Not-determined | Genome-fragment | NA | NA | 0 |
| S1_k141_367916 | 1750 | No | NA | 2 | 0 | 0 | Not-determined | Genome-fragment | NA | NA | 0 |
| S1_k141_219826 | 1172 | No | NA | 2 | 0 | 2 | Not-determined | Genome-fragment | NA | NA | 0 |
| S1_k141_103649 | 2515 | No | NA | 3 | 0 | 0 | Not-determined | Genome-fragment | NA | NA | 0 |
| S11_k141_299006 | 1468 | No | NA | 2 | 1 | 0 | Low-quality | Genome-fragment | 2.81 | AAI-based (high-confidence) | 0 |
| S1_k141_236216 | 1369 | No | NA | 1 | 0 | 1 | Not-determined | Genome-fragment | NA | NA | 0 |
| S1_k141_54804 | 1104 | No | NA | 1 | 0 | 1 | Not-determined | Genome-fragment | NA | NA | 0 |
| S1_k141_70674 | 1438 | No | NA | 1 | 0 | 1 | Not-determined | Genome-fragment | NA | NA | 0 |
| S1_k141_384300 | 1225 | No | NA | 2 | 0 | 1 | Not-determined | Genome-fragment | NA | NA | 0 |
| S5-final-viral-combined_k141_85210\|\|full | 1468 | No | NA | 3 | 0 | 0 | Low-quality | Genome-fragment | 1.19 | AAI-based (medium-confidence) | 0 |
| S5-final-viral-combined_k141_123419\|\|lt2gene | 1468 | No | NA | 1 | 1 | 0 | Low-quality | Genome-fragment | 23.76 | AAI-based (high-confidence) | 0 |
| S1_k141_54839 | 1252 | No | NA | 1 | 0 | 1 | Not-determined | Genome-fragment | NA | NA | 0 |
| S1_k141_54842 | 1793 | No | NA | 1 | 0 | 1 | Not-determined | Genome-fragment | NA | NA | 0 |
| S1_k141_54859 | 1680 | No | NA | 3 | 0 | 0 | Not-determined | Genome-fragment | NA | NA | 0 |
| S1_k141_334293 | 1073 | No | NA | 1 | 0 | 0 | Not-determined | Genome-fragment | NA | NA | 0 |
| S1_k141_268854 | 1704 | No | NA | 3 | 0 | 3 | Not-determined | Genome-fragment | NA | NA | 0 |
| S1_k141_38644 | 1710 | No | NA | 1 | 0 | 1 | Not-determined | Genome-fragment | NA | NA | 0 |
| S1_k141_351829 | 6288 | No | NA | 1 | 0 | 1 | Not-determined | Genome-fragment | NA | NA | 0 |
| S1_k141_121498 | 6289 | No | NA | 1 | 0 | 0 | Not-determined | Genome-fragment | NA | NA | 0 |
| S1_k141_351853 | 1108 | No | NA | 1 | 0 | 1 | Not-determined | Genome-fragment | NA | NA | 0 |
| S1_k141_70716 | 1422 | No | NA | 2 | 0 | 2 | Not-determined | Genome-fragment | NA | NA | 0 |
| S1_k141_137976 | 1587 | No | NA | 1 | 0 | 1 | Not-determined | Genome-fragment | NA | NA | 0 |
| S12-final-viral-combined_k141_163046\|\|full | 1468 | No | NA | 3 | 2 | 0 | Low-quality | Genome-fragment | 2.51 | AAI-based (high-confidence) | 0 |
| S1_k141_319020 | 2421 | No | NA | 3 | 0 | 2 | Not-determined | Genome-fragment | NA | NA | 0 |
| S1_k141_368049 | 1041 | No | NA | 1 | 0 | 1 | Not-determined | Genome-fragment | NA | NA | 0 |
| S1_k141_169680 | 1236 | No | NA | 1 | 0 | 1 | Not-determined | Genome-fragment | NA | NA | 0 |
| S1_k141_236280 | 2813 | No | NA | 2 | 0 | 0 | Not-determined | Genome-fragment | NA | NA | 0 |
| S1_k141_169697 | 3932 | No | NA | 4 | 0 | 3 | Not-determined | Genome-fragment | NA | NA | 0 |
| S1_k141_153725 | 3139 | No | NA | 4 | 0 | 0 | Not-determined | Genome-fragment | NA | NA | 0 |
| S1_k141_220010 | 1873 | No | NA | 2 | 0 | 1 | Not-determined | Genome-fragment | NA | NA | 0 |
| S1_k141_253171 | 1240 | No | NA | 1 | 0 | 1 | Not-determined | Genome-fragment | NA | NA | 0 |
| S1_k141_87772 | 1466 | No | NA | 2 | 0 | 2 | Not-determined | Genome-fragment | NA | NA | 0 |
| S4_k141_51607 | 1468 | No | NA | 3 | 2 | 0 | Low-quality | Genome-fragment | 0.86 | AAI-based (high-confidence) | 0 |
| S1_k141_70814 | 1328 | No | NA | 3 | 0 | 1 | Not-determined | Genome-fragment | NA | NA | 0 |
| S1_k141_268946 | 1272 | No | NA | 2 | 0 | 1 | Not-determined | Genome-fragment | NA | NA | 0 |
| S8-final-viral-combined_k141_149008\|\|full | 1468 | No | NA | 3 | 1 | 0 | Low-quality | Genome-fragment | 0.93 | HMM-based (lower-bound) | 0 |
| S1_k141_54992 | 1089 | No | NA | 2 | 0 | 2 | Not-determined | Genome-fragment | NA | NA | 0 |
| S11-final-viral-combined_k141_170229\|\|full | 1469 | No | NA | 4 | 1 | 0 | Low-quality | Genome-fragment | 2.49 | AAI-based (medium-confidence) | 0 |
| S1-final-viral-combined_k141_12158\|\|full | 1469 | No | NA | 6 | 0 | 0 | Low-quality | Genome-fragment | 4.3 | AAI-based (high-confidence) | 0 |
| S1_k141_121653 | 1392 | No | NA | 2 | 0 | 1 | Not-determined | Genome-fragment | NA | NA | 0 |
| S3_k141_99778 | 1469 | No | NA | 3 | 1 | 0 | Low-quality | Genome-fragment | 0.68 | AAI-based (high-confidence) | 0 |
| S1_k141_334426 | 7546 | No | NA | 3 | 0 | 1 | Not-determined | Genome-fragment | NA | NA | 0 |
| S5-final-viral-combined_k141_117469\|\|full | 1469 | No | NA | 2 | 2 | 0 | Low-quality | Genome-fragment | 4.18 | AAI-based (high-confidence) | 0 |
| S5-final-viral-combined_k141_4725\|\|full | 1469 | No | NA | 4 | 1 | 0 | Low-quality | Genome-fragment | 1.87 | AAI-based (high-confidence) | 0 |
| S5-final-viral-combined_k141_148888\|\|full | 1469 | No | NA | 4 | 0 | 0 | Low-quality | Genome-fragment | 2.89 | AAI-based (medium-confidence) | 0 |
| S1_k141_351966 | 1049 | No | NA | 1 | 0 | 1 | Not-determined | Genome-fragment | NA | NA | 0 |
| S1_k141_203697 | 1001 | No | NA | 3 | 0 | 0 | Not-determined | Genome-fragment | NA | NA | 0 |
| S1_k141_103925 | 1040 | No | NA | 2 | 0 | 0 | Not-determined | Genome-fragment | NA | NA | 0 |
| S1_k141_169822 | 1517 | No | NA | 3 | 0 | 2 | Not-determined | Genome-fragment | NA | NA | 0 |
| S1_k141_236431 | 1154 | No | NA | 1 | 0 | 1 | Not-determined | Genome-fragment | NA | NA | 0 |
| S1_k141_253303 | 1804 | No | NA | 2 | 0 | 1 | Not-determined | Genome-fragment | NA | NA | 0 |
| S1_k141_187510 | 4562 | No | NA | 1 | 0 | 1 | Not-determined | Genome-fragment | NA | NA | 0 |
| S1_k141_38875 | 1095 | No | NA | 2 | 0 | 1 | Not-determined | Genome-fragment | NA | NA | 0 |
| S7_k141_217874 | 1469 | No | NA | 2 | 1 | 0 | Low-quality | Genome-fragment | 2.51 | AAI-based (medium-confidence) | 0 |
| S1_k141_103998 | 1098 | No | NA | 1 | 0 | 1 | Not-determined | Genome-fragment | NA | NA | 0 |
| S1_k141_22222 | 4563 | No | NA | 6 | 0 | 0 | Not-determined | Genome-fragment | NA | NA | 0 |
| S1_k141_38913 | 1719 | No | NA | 2 | 0 | 2 | Not-determined | Genome-fragment | NA | NA | 0 |
| S1_k141_187541 | 9571 | No | NA | 6 | 0 | 4 | Not-determined | Genome-fragment | NA | NA | 0 |
| S1_k141_384637 | 6675 | No | NA | 6 | 0 | 2 | Not-determined | Genome-fragment | NA | NA | 0 |
| S1_k141_334597 | 1044 | No | NA | 1 | 0 | 0 | Not-determined | Genome-fragment | NA | NA | 0 |
| S1_k141_285846 | 3371 | No | NA | 2 | 0 | 1 | Not-determined | Genome-fragment | NA | NA | 0 |
| S1_k141_187556 | 1349 | No | NA | 1 | 0 | 1 | Not-determined | Genome-fragment | NA | NA | 0 |
| S1_k141_384647 | 1486 | No | NA | 1 | 0 | 0 | Not-determined | Genome-fragment | NA | NA | 0 |
| S1_k141_121818 | 3012 | No | NA | 4 | 0 | 2 | Not-determined | Genome-fragment | NA | NA | 0 |
| S1_k141_319350 | 1072 | No | NA | 1 | 0 | 0 | Not-determined | Genome-fragment | NA | NA | 0 |
| S1_k141_253406 | 1067 | No | NA | 2 | 0 | 1 | Not-determined | Genome-fragment | NA | NA | 0 |
| S1_k141_236542 | 2105 | No | NA | 1 | 0 | 1 | Not-determined | Genome-fragment | NA | NA | 0 |
| S1_k141_22265 | 1059 | No | NA | 1 | 0 | 1 | Not-determined | Genome-fragment | NA | NA | 0 |
| S1_k141_5961 | 3223 | No | NA | 1 | 0 | 1 | Not-determined | Genome-fragment | NA | NA | 0 |
| S1_k141_87984 | 1458 | No | NA | 1 | 0 | 1 | Not-determined | Genome-fragment | NA | NA | 0 |
| S17-final-viral-combined_k141_17213\|\|full | 1469 | No | NA | 2 | 0 | 0 | Low-quality | Genome-fragment | 1.09 | AAI-based (medium-confidence) | 0 |
| S1_k141_5984 | 1055 | No | NA | 1 | 0 | 1 | Not-determined | Genome-fragment | NA | NA | 0 |
| S1_k141_319423 | 7338 | No | NA | 5 | 0 | 5 | Not-determined | Genome-fragment | NA | NA | 0 |
| S1_k141_352192 | 1187 | No | NA | 1 | 0 | 1 | Not-determined | Genome-fragment | NA | NA | 0 |
| S1_k141_6005 | 1297 | No | NA | 2 | 0 | 0 | Not-determined | Genome-fragment | NA | NA | 0 |
| S1_k141_121849 | 2063 | No | NA | 1 | 0 | 1 | Not-determined | Genome-fragment | NA | NA | 0 |
| S1_k141_38983 | 4772 | No | NA | 6 | 0 | 5 | Not-determined | Genome-fragment | NA | NA | 0 |
| S12-final-viral-combined_k141_251216\|\|full | 1469 | No | NA | 2 | 0 | 1 | Low-quality | Genome-fragment | 1.09 | AAI-based (medium-confidence) | 0 |
| S1_k141_203893 | 2759 | No | NA | 3 | 0 | 2 | Not-determined | Genome-fragment | NA | NA | 0 |
| S1_k141_121869 | 2243 | No | NA | 3 | 0 | 2 | Not-determined | Genome-fragment | NA | NA | 0 |
| S1_k141_352211 | 2015 | No | NA | 2 | 0 | 2 | Not-determined | Genome-fragment | NA | NA | 0 |
| S1_k141_22313 | 1046 | No | NA | 1 | 0 | 1 | Not-determined | Genome-fragment | NA | NA | 0 |
| S1_k141_121909 | 1347 | No | NA | 2 | 0 | 0 | Not-determined | Genome-fragment | NA | NA | 0 |
| S2-final-viral-combined_k141_26756\|\|full | 1469 | No | NA | 3 | 1 | 0 | Low-quality | Genome-fragment | 0.96 | AAI-based (medium-confidence) | 0 |
| S2_k141_115357 | 1469 | No | NA | 1 | 0 | 0 | Low-quality | Genome-fragment | 2.45 | AAI-based (high-confidence) | 0 |
| S1_k141_121935 | 1585 | No | NA | 1 | 0 | 0 | Not-determined | Genome-fragment | NA | NA | 0 |
| S1_k141_187737 | 1290 | No | NA | 2 | 0 | 1 | Not-determined | Genome-fragment | NA | NA | 0 |
| S1_k141_253540 | 1182 | No | NA | 3 | 0 | 0 | Not-determined | Genome-fragment | NA | NA | 0 |
| S1_k141_269273 | 1259 | No | NA | 1 | 0 | 1 | Not-determined | Genome-fragment | NA | NA | 0 |
| S1_k141_71175 | 1379 | No | NA | 1 | 0 | 1 | Not-determined | Genome-fragment | NA | NA | 0 |
| S1_k141_138279 | 2062 | No | NA | 2 | 0 | 1 | Not-determined | Genome-fragment | NA | NA | 0 |
| S11-final-viral-combined_k141_37059\|\|full | 1470 | No | NA | 3 | 0 | 0 | Low-quality | Genome-fragment | 3.22 | AAI-based (high-confidence) | 0 |
| S1-final-viral-combined_k141_316121\|\|full | 1470 | No | NA | 4 | 0 | 0 | Low-quality | Genome-fragment | 3.02 | AAI-based (medium-confidence) | 0 |
| S1_k141_154035 | 1397 | No | NA | 1 | 0 | 0 | Not-determined | Genome-fragment | NA | NA | 0 |
| S1_k141_352297 | 3322 | No | NA | 1 | 0 | 1 | Not-determined | Genome-fragment | NA | NA | 0 |
| S1_k141_6096 | 1293 | No | NA | 1 | 0 | 0 | Not-determined | Genome-fragment | NA | NA | 0 |
| S1_k141_71200 | 2519 | No | NA | 2 | 0 | 0 | Not-determined | Genome-fragment | NA | NA | 0 |
| S1_k141_88155 | 1347 | No | NA | 1 | 0 | 1 | Not-determined | Genome-fragment | NA | NA | 0 |
| S1_k141_6131 | 1059 | No | NA | 2 | 0 | 2 | Not-determined | Genome-fragment | NA | NA | 0 |
| S1_k141_204065 | 3505 | No | NA | 4 | 0 | 1 | Not-determined | Genome-fragment | NA | NA | 0 |
| S17-final-viral-combined_k141_51751\|\|full | 1470 | Yes | 294 | 2 | 1 | 1 | Low-quality | Genome-fragment | 0.15 | AAI-based (medium-confidence) | 80 |
| S17-final-viral-combined_k141_194185\|\|lt2gene | 1470 | No | NA | 1 | 1 | 0 | Low-quality | Genome-fragment | 1.38 | HMM-based (lower-bound) | 0 |
| S1_k141_236658 | 1232 | No | NA | 2 | 0 | 2 | Not-determined | Genome-fragment | NA | NA | 0 |
| S2-final-viral-combined_k141_182620\|\|full | 1470 | No | NA | 3 | 0 | 1 | Low-quality | Genome-fragment | 3.1 | AAI-based (high-confidence) | 0 |
| S1_k141_269335 | 1796 | No | NA | 2 | 0 | 2 | Not-determined | Genome-fragment | NA | NA | 0 |
| S2-final-viral-combined_k141_37172\|\|lt2gene | 1470 | No | NA | 2 | 2 | 0 | Low-quality | Genome-fragment | 3.83 | AAI-based (high-confidence) | 0 |
| S1_k141_39114 | 1901 | No | NA | 2 | 0 | 0 | Not-determined | Genome-fragment | NA | NA | 0 |
| S1_k141_334791 | 3819 | No | NA | 4 | 0 | 4 | Not-determined | Genome-fragment | NA | NA | 0 |
| S1_k141_154140 | 3044 | No | NA | 5 | 0 | 3 | Not-determined | Genome-fragment | NA | NA | 0 |
| S1_k141_303172 | 1405 | No | NA | 4 | 0 | 0 | Not-determined | Genome-fragment | NA | NA | 0 |
| S1_k141_269372 | 1013 | No | NA | 1 | 0 | 1 | Not-determined | Genome-fragment | NA | NA | 0 |
| S1_k141_352408 | 1365 | No | NA | 2 | 0 | 0 | Not-determined | Genome-fragment | NA | NA | 0 |
| S1_k141_253771 | 1211 | No | NA | 3 | 0 | 0 | Not-determined | Genome-fragment | NA | NA | 0 |
| S1_k141_170115 | 40097 | No | NA | 51 | 23 | 1 | High-quality | High-quality | 100 | AAI-based (high-confidence) | 0 |
| S1_k141_204136 | 2968 | No | NA | 3 | 0 | 2 | Not-determined | Genome-fragment | NA | NA | 0 |
| S1_k141_71292 | 4202 | No | NA | 8 | 0 | 0 | Not-determined | Genome-fragment | NA | NA | 0 |
| S1_k141_204142 | 1034 | No | NA | 2 | 0 | 2 | Not-determined | Genome-fragment | NA | NA | 0 |
| S1_k141_88280 | 3161 | No | NA | 2 | 0 | 2 | Not-determined | Genome-fragment | NA | NA | 0 |
| S1_k141_170161 | 1499 | No | NA | 2 | 0 | 0 | Not-determined | Genome-fragment | NA | NA | 0 |
| S8-final-viral-combined_k141_155536\|\|full | 1470 | No | NA | 2 | 0 | 0 | Low-quality | Genome-fragment | 1.06 | AAI-based (medium-confidence) | 0 |
| S11-final-viral-combined_k141_324075\|\|full | 1471 | No | NA | 3 | 0 | 0 | Low-quality | Genome-fragment | 3.18 | AAI-based (high-confidence) | 0 |
| S1_k141_352438 | 1120 | No | NA | 3 | 0 | 0 | Not-determined | Genome-fragment | NA | NA | 0 |
| S3-final-viral-combined_k141_81836\|\|full | 1471 | No | NA | 4 | 0 | 0 | Low-quality | Genome-fragment | 1.66 | AAI-based (medium-confidence) | 0 |
| S5-final-viral-combined_k141_174766\|\|full | 1471 | No | NA | 4 | 0 | 0 | Low-quality | Genome-fragment | 2.37 | AAI-based (medium-confidence) | 0 |
| S7-final-viral-combined_k141_143996\|\|full | 1471 | No | NA | 3 | 0 | 0 | Low-quality | Genome-fragment | 2.44 | AAI-based (high-confidence) | 0 |
| S1_k141_204192 | 1754 | No | NA | 2 | 0 | 1 | Not-determined | Genome-fragment | NA | NA | 0 |
| S1_k141_6291 | 1397 | No | NA | 1 | 0 | 1 | Not-determined | Genome-fragment | NA | NA | 0 |
| S7-final-viral-combined_k141_23365\|\|full | 1471 | No | NA | 2 | 0 | 1 | Low-quality | Genome-fragment | 2.77 | AAI-based (high-confidence) | 0 |
| S1_k141_334971 | 1262 | No | NA | 2 | 0 | 2 | Not-determined | Genome-fragment | NA | NA | 0 |
| S1_k141_170214 | 1687 | No | NA | 1 | 0 | 0 | Not-determined | Genome-fragment | NA | NA | 0 |
| S1_k141_334981 | 3349 | No | NA | 2 | 0 | 2 | Not-determined | Genome-fragment | NA | NA | 0 |
| S7-final-viral-combined_k141_162765\|\|full | 1471 | No | NA | 3 | 0 | 1 | Low-quality | Genome-fragment | 0.84 | AAI-based (medium-confidence) | 0 |
| S1_k141_71398 | 1237 | No | NA | 1 | 0 | 0 | Not-determined | Genome-fragment | NA | NA | 0 |
| S1_k141_104388 | 1300 | No | NA | 2 | 0 | 2 | Not-determined | Genome-fragment | NA | NA | 0 |
| S12-final-viral-combined_k141_253887\|\|full | 1471 | No | NA | 4 | 0 | 0 | Low-quality | Genome-fragment | 3.26 | AAI-based (high-confidence) | 0 |
| S1_k141_187939 | 3278 | No | NA | 4 | 0 | 1 | Not-determined | Genome-fragment | NA | NA | 0 |
| S12-final-viral-combined_k141_240998\|\|full | 1471 | No | NA | 3 | 0 | 0 | Low-quality | Genome-fragment | 1.09 | AAI-based (medium-confidence) | 0 |
| S1_k141_352525 | 1250 | No | NA | 2 | 0 | 1 | Not-determined | Genome-fragment | NA | NA | 0 |
| S1_k141_236890 | 1145 | No | NA | 2 | 0 | 0 | Not-determined | Genome-fragment | NA | NA | 0 |
| S4-final-viral-combined_k141_85766\|\|full | 1471 | No | NA | 2 | 0 | 0 | Low-quality | Genome-fragment | 1.03 | AAI-based (high-confidence) | 0 |
| S1_k141_220391 | 1146 | No | NA | 1 | 0 | 0 | Not-determined | Genome-fragment | NA | NA | 0 |
| S8_k141_119825 | 1471 | No | NA | 4 | 1 | 0 | Low-quality | Genome-fragment | 0.95 | HMM-based (lower-bound) | 0 |
| S1_k141_55663 | 1360 | No | NA | 1 | 0 | 0 | Not-determined | Genome-fragment | NA | NA | 0 |
| S1_k141_138520 | 1632 | No | NA | 1 | 0 | 0 | Not-determined | Genome-fragment | NA | NA | 0 |
| S1_k141_204318 | 4452 | No | NA | 4 | 0 | 4 | Not-determined | Genome-fragment | NA | NA | 0 |
| S1_k141_6409 | 1036 | No | NA | 2 | 0 | 2 | Not-determined | Genome-fragment | NA | NA | 0 |
| S11-final-viral-combined_k141_264353\|\|full | 1472 | No | NA | 3 | 0 | 2 | Low-quality | Genome-fragment | 1.21 | AAI-based (medium-confidence) | 0 |
| S1_k141_236970 | 2478 | No | NA | 2 | 0 | 0 | Not-determined | Genome-fragment | NA | NA | 0 |
| S1_k141_104465 | 1574 | No | NA | 2 | 0 | 2 | Not-determined | Genome-fragment | NA | NA | 0 |
| S1_k141_39334 | 1285 | No | NA | 1 | 0 | 1 | Not-determined | Genome-fragment | NA | NA | 0 |
| S1_k141_303335 | 2425 | No | NA | 1 | 0 | 1 | Not-determined | Genome-fragment | NA | NA | 0 |
| S1_k141_286268 | 6154 | No | NA | 5 | 0 | 3 | Not-determined | Genome-fragment | NA | NA | 0 |
| S1_k141_188124 | 1645 | No | NA | 1 | 0 | 0 | Not-determined | Genome-fragment | NA | NA | 0 |
| S1_k141_6504 | 1115 | No | NA | 1 | 0 | 0 | Not-determined | Genome-fragment | NA | NA | 0 |
| S11-final-viral-combined_k141_302483\|\|full | 1472 | No | NA | 3 | 0 | 0 | Low-quality | Genome-fragment | 2.09 | AAI-based (medium-confidence) | 0 |
| S1_k141_6512 | 1592 | No | NA | 2 | 0 | 2 | Not-determined | Genome-fragment | NA | NA | 0 |
| S1_k141_55729 | 1484 | No | NA | 1 | 0 | 1 | Not-determined | Genome-fragment | NA | NA | 0 |
| S1_k141_335113 | 5677 | No | NA | 5 | 0 | 1 | Not-determined | Genome-fragment | NA | NA | 0 |
| S11-final-viral-combined_k141_42488\|\|lt2gene | 1472 | No | NA | 2 | 1 | 0 | Low-quality | Genome-fragment | 0.91 | HMM-based (lower-bound) | 0 |
| S1_k141_138643 | 1635 | No | NA | 1 | 0 | 0 | Not-determined | Genome-fragment | NA | NA | 0 |
| S1_k141_254037 | 1083 | No | NA | 1 | 0 | 0 | Not-determined | Genome-fragment | NA | NA | 0 |
| S1_k141_303394 | 1257 | No | NA | 3 | 0 | 3 | Not-determined | Genome-fragment | NA | NA | 0 |
| S1_k141_204488 | 1015 | No | NA | 1 | 0 | 1 | Not-determined | Genome-fragment | NA | NA | 0 |
| S1_k141_335170 | 1139 | No | NA | 4 | 0 | 4 | Not-determined | Genome-fragment | NA | NA | 0 |
| S1_k141_335171 | 3374 | No | NA | 1 | 0 | 1 | Not-determined | Genome-fragment | NA | NA | 0 |
| S1-final-viral-combined_k141_366109\|\|full | 1472 | No | NA | 2 | 0 | 0 | Low-quality | Genome-fragment | 1.98 | AAI-based (medium-confidence) | 0 |
| S1_k141_385234 | 1385 | No | NA | 1 | 0 | 0 | Not-determined | Genome-fragment | NA | NA | 0 |
| S1_k141_220544 | 1049 | No | NA | 1 | 0 | 1 | Not-determined | Genome-fragment | NA | NA | 0 |
| S1_k141_104516 | 1394 | No | NA | 1 | 0 | 0 | Not-determined | Genome-fragment | NA | NA | 0 |
| S1_k141_188204 | 1058 | No | NA | 2 | 0 | 0 | Not-determined | Genome-fragment | NA | NA | 0 |
| S1_k141_204519 | 1038 | No | NA | 1 | 0 | 1 | Not-determined | Genome-fragment | NA | NA | 0 |
| S1-final-viral-combined_k141_389726\|\|full | 1472 | No | NA | 2 | 1 | 0 | Low-quality | Genome-fragment | 3.22 | AAI-based (high-confidence) | 0 |
| S1_k141_220553 | 1169 | No | NA | 1 | 0 | 1 | Not-determined | Genome-fragment | NA | NA | 0 |
| S3-final-viral-combined_k141_118477\|\|full | 1472 | No | NA | 2 | 2 | 0 | Low-quality | Genome-fragment | 3.75 | AAI-based (high-confidence) | 0 |
| S1_k141_122535 | 3589 | No | NA | 3 | 0 | 2 | Not-determined | Genome-fragment | NA | NA | 0 |
| S1_k141_138742 | 1292 | No | NA | 1 | 0 | 1 | Not-determined | Genome-fragment | NA | NA | 0 |
| S1_k141_254106 | 2975 | No | NA | 2 | 0 | 2 | Not-determined | Genome-fragment | NA | NA | 0 |
| S1_k141_88645 | 2135 | No | NA | 2 | 0 | 2 | Not-determined | Genome-fragment | NA | NA | 0 |
| S5_k141_68232 | 1472 | No | NA | 1 | 0 | 0 | Low-quality | Genome-fragment | 2.53 | AAI-based (medium-confidence) | 0 |
| S1_k141_71673 | 1048 | No | NA | 2 | 0 | 1 | Not-determined | Genome-fragment | NA | NA | 0 |
| S1_k141_254138 | 1933 | No | NA | 1 | 0 | 1 | Not-determined | Genome-fragment | NA | NA | 0 |
| S1_k141_22920 | 4483 | No | NA | 11 | 0 | 10 | Not-determined | Genome-fragment | NA | NA | 0 |
| S1_k141_320024 | 3372 | No | NA | 8 | 0 | 1 | Not-determined | Genome-fragment | NA | NA | 0 |
| S1_k141_220637 | 1174 | No | NA | 1 | 0 | 1 | Not-determined | Genome-fragment | NA | NA | 0 |
| S1_k141_22951 | 4311 | No | NA | 8 | 0 | 2 | Not-determined | Genome-fragment | NA | NA | 0 |
| S1_k141_303546 | 3401 | No | NA | 5 | 0 | 2 | Not-determined | Genome-fragment | NA | NA | 0 |
| S1_k141_104617 | 7989 | No | NA | 7 | 0 | 5 | Not-determined | Genome-fragment | NA | NA | 0 |
| S1_k141_122647 | 1448 | No | NA | 2 | 0 | 0 | Not-determined | Genome-fragment | NA | NA | 0 |
| S1_k141_254254 | 1463 | No | NA | 2 | 0 | 1 | Not-determined | Genome-fragment | NA | NA | 0 |
| S7-final-viral-combined_k141_189969\|\|full | 1472 | No | NA | 3 | 1 | 0 | Low-quality | Genome-fragment | 1.7 | AAI-based (medium-confidence) | 0 |
| S17-final-viral-combined_k141_205240\|\|full | 1472 | No | NA | 2 | 0 | 0 | Low-quality | Genome-fragment | 2.63 | AAI-based (medium-confidence) | 0 |
| S1_k141_55933 | 3084 | No | NA | 1 | 0 | 1 | Not-determined | Genome-fragment | NA | NA | 0 |
| S1_k141_104665 | 1323 | No | NA | 1 | 0 | 1 | Not-determined | Genome-fragment | NA | NA | 0 |
| S1_k141_368586 | 1162 | No | NA | 1 | 0 | 0 | Not-determined | Genome-fragment | NA | NA | 0 |
| S1_k141_254300 | 1583 | No | NA | 1 | 0 | 0 | Not-determined | Genome-fragment | NA | NA | 0 |
| S1_k141_104710 | 2023 | No | NA | 1 | 0 | 1 | Not-determined | Genome-fragment | NA | NA | 0 |
| S1_k141_353013 | 1053 | No | NA | 2 | 0 | 2 | Not-determined | Genome-fragment | NA | NA | 0 |
| S1_k141_335317 | 6501 | No | NA | 8 | 0 | 6 | Not-determined | Genome-fragment | NA | NA | 0 |
| S1_k141_368616 | 1000 | No | NA | 2 | 0 | 1 | Not-determined | Genome-fragment | NA | NA | 0 |
| S1_k141_71855 | 2036 | No | NA | 1 | 0 | 1 | Not-determined | Genome-fragment | NA | NA | 0 |
| S1_k141_286456 | 1590 | No | NA | 2 | 0 | 0 | Not-determined | Genome-fragment | NA | NA | 0 |
| S1_k141_303680 | 1103 | No | NA | 1 | 0 | 1 | Not-determined | Genome-fragment | NA | NA | 0 |
| S1_k141_6742 | 1276 | No | NA | 3 | 0 | 3 | Not-determined | Genome-fragment | NA | NA | 0 |
| S1_k141_353026 | 5866 | No | NA | 6 | 0 | 0 | Not-determined | Genome-fragment | NA | NA | 0 |
| S12-final-viral-combined_k141_47381\|\|full | 1472 | No | NA | 1 | 0 | 1 | Low-quality | Genome-fragment | 1.69 | AAI-based (high-confidence) | 0 |
| S2-final-viral-combined_k141_229970\|\|full | 1472 | No | NA | 4 | 0 | 0 | Low-quality | Genome-fragment | 2.61 | AAI-based (high-confidence) | 0 |
| S4-final-viral-combined_k141_153121\|\|full | 1472 | No | NA | 3 | 1 | 0 | Low-quality | Genome-fragment | 3.25 | AAI-based (medium-confidence) | 0 |
| S1_k141_122789 | 3353 | No | NA | 4 | 0 | 2 | Not-determined | Genome-fragment | NA | NA | 0 |
| S9_k141_127904 | 1473 | No | NA | 1 | 0 | 1 | Low-quality | Genome-fragment | 0.25 | AAI-based (high-confidence) | 0 |
| S1_k141_170614 | 4193 | No | NA | 5 | 0 | 4 | Not-determined | Genome-fragment | NA | NA | 0 |
| S1_k141_23148 | 1013 | No | NA | 1 | 0 | 1 | Not-determined | Genome-fragment | NA | NA | 0 |
| S1_k141_385473 | 1072 | No | NA | 1 | 0 | 0 | Not-determined | Genome-fragment | NA | NA | 0 |
| S1_k141_254362 | 1249 | No | NA | 2 | 0 | 2 | Not-determined | Genome-fragment | NA | NA | 0 |
| S11-final-viral-combined_k141_270484\|\|full | 1473 | No | NA | 3 | 2 | 0 | Low-quality | Genome-fragment | 1.78 | HMM-based (lower-bound) | 0 |
| S1_k141_56099 | 3969 | No | NA | 4 | 0 | 1 | Not-determined | Genome-fragment | NA | NA | 0 |
| S1_k141_23185 | 1105 | No | NA | 1 | 0 | 0 | Not-determined | Genome-fragment | NA | NA | 0 |
| S1_k141_353103 | 1665 | No | NA | 3 | 0 | 0 | Not-determined | Genome-fragment | NA | NA | 0 |
| S11_k141_44174 | 1473 | No | NA | 1 | 0 | 0 | Low-quality | Genome-fragment | 1.2 | AAI-based (medium-confidence) | 0 |
| S1_k141_39650 | 1173 | No | NA | 1 | 0 | 1 | Not-determined | Genome-fragment | NA | NA | 0 |
| S1_k141_154632 | 1875 | No | NA | 1 | 0 | 1 | Not-determined | Genome-fragment | NA | NA | 0 |
| S1_k141_188426 | 1168 | No | NA | 2 | 0 | 1 | Not-determined | Genome-fragment | NA | NA | 0 |
| S1_k141_204866 | 4425 | No | NA | 5 | 0 | 5 | Not-determined | Genome-fragment | NA | NA | 0 |
| S1-final-viral-combined_k141_108867\|\|full | 1473 | No | NA | 2 | 0 | 0 | Low-quality | Genome-fragment | 2.36 | AAI-based (medium-confidence) | 0 |
| S1_k141_320365 | 4138 | No | NA | 6 | 0 | 1 | Not-determined | Genome-fragment | NA | NA | 0 |
| S17-final-viral-combined_k141_195469\|\|full | 1473 | No | NA | 4 | 1 | 0 | Low-quality | Genome-fragment | 3.93 | AAI-based (high-confidence) | 0 |
| S1_k141_170734 | 1139 | No | NA | 1 | 0 | 0 | Not-determined | Genome-fragment | NA | NA | 0 |
| S1_k141_385550 | 1196 | No | NA | 2 | 0 | 0 | Not-determined | Genome-fragment | NA | NA | 0 |
| S1_k141_122980 | 1481 | No | NA | 1 | 0 | 1 | Not-determined | Genome-fragment | NA | NA | 0 |
| S1_k141_23261 | 2711 | No | NA | 1 | 0 | 1 | Not-determined | Genome-fragment | NA | NA | 0 |
| S1_k141_237594 | 3659 | No | NA | 1 | 0 | 0 | Not-determined | Genome-fragment | NA | NA | 0 |
| S1_k141_335507 | 7287 | No | NA | 7 | 0 | 5 | Not-determined | Genome-fragment | NA | NA | 0 |
| S1_k141_170828 | 1367 | No | NA | 4 | 0 | 0 | Not-determined | Genome-fragment | NA | NA | 0 |
| S1_k141_270060 | 2270 | No | NA | 5 | 0 | 2 | Not-determined | Genome-fragment | NA | NA | 0 |
| S1_k141_7044 | 4460 | No | NA | 7 | 0 | 2 | Not-determined | Genome-fragment | NA | NA | 0 |
| S1_k141_353255 | 1275 | No | NA | 2 | 0 | 1 | Not-determined | Genome-fragment | NA | NA | 0 |
| S1_k141_188585 | 1009 | No | NA | 2 | 0 | 1 | Not-determined | Genome-fragment | NA | NA | 0 |
| S1_k141_123088 | 1726 | No | NA | 3 | 0 | 1 | Not-determined | Genome-fragment | NA | NA | 0 |
| S1_k141_188613 | 4496 | No | NA | 2 | 0 | 2 | Not-determined | Genome-fragment | NA | NA | 0 |
| S1_k141_353282 | 1246 | No | NA | 1 | 0 | 1 | Not-determined | Genome-fragment | NA | NA | 0 |
| S1_k141_170908 | 1176 | No | NA | 4 | 0 | 1 | Not-determined | Genome-fragment | NA | NA | 0 |
| S2-final-viral-combined_k141_208625\|\|full | 1473 | No | NA | 2 | 0 | 0 | Low-quality | Genome-fragment | 3.69 | AAI-based (medium-confidence) | 0 |
| S1_k141_254633 | 2237 | No | NA | 1 | 0 | 1 | Not-determined | Genome-fragment | NA | NA | 0 |
| S2-final-viral-combined_k141_95005\|\|full | 1473 | No | NA | 4 | 2 | 0 | Low-quality | Genome-fragment | 3.31 | AAI-based (high-confidence) | 0 |
| S1_k141_320615 | 1044 | No | NA | 1 | 0 | 1 | Not-determined | Genome-fragment | NA | NA | 0 |
| S1_k141_170958 | 1742 | No | NA | 1 | 0 | 1 | Not-determined | Genome-fragment | NA | NA | 0 |
| S1_k141_139213 | 2406 | No | NA | 1 | 0 | 1 | Not-determined | Genome-fragment | NA | NA | 0 |
| S1_k141_270220 | 1557 | No | NA | 1 | 0 | 1 | Not-determined | Genome-fragment | NA | NA | 0 |
| S8-final-viral-combined_k141_119505\|\|lt2gene | 1473 | No | NA | 1 | 1 | 0 | Low-quality | Genome-fragment | 2.15 | HMM-based (lower-bound) | 0 |
| S11-final-viral-combined_k141_279556\|\|full | 1474 | No | NA | 2 | 0 | 1 | Low-quality | Genome-fragment | 2.43 | AAI-based (medium-confidence) | 0 |
| S1_k141_56428 | 1034 | No | NA | 2 | 0 | 0 | Not-determined | Genome-fragment | NA | NA | 0 |
| S1_k141_89218 | 1268 | No | NA | 3 | 0 | 0 | Not-determined | Genome-fragment | NA | NA | 0 |
| S1_k141_205108 | 1152 | No | NA | 1 | 0 | 1 | Not-determined | Genome-fragment | NA | NA | 0 |
| S1_k141_56462 | 2102 | No | NA | 1 | 0 | 0 | Not-determined | Genome-fragment | NA | NA | 0 |
| S1_k141_353371 | 1974 | No | NA | 2 | 0 | 1 | Not-determined | Genome-fragment | NA | NA | 0 |
| S1_k141_89220 | 1116 | No | NA | 2 | 0 | 1 | Not-determined | Genome-fragment | NA | NA | 0 |
| S11-final-viral-combined_k141_81374\|\|lt2gene | 1474 | No | NA | 2 | 1 | 0 | Low-quality | Genome-fragment | 4 | AAI-based (high-confidence) | 0 |
| S3-final-viral-combined_k141_20806\|\|full | 1474 | No | NA | 4 | 0 | 0 | Low-quality | Genome-fragment | 4.12 | AAI-based (medium-confidence) | 0 |
| S1_k141_353413 | 2106 | No | NA | 1 | 0 | 1 | Not-determined | Genome-fragment | NA | NA | 0 |
| S1_k141_353416 | 1221 | No | NA | 2 | 0 | 0 | Not-determined | Genome-fragment | NA | NA | 0 |
| S1_k141_154959 | 1198 | No | NA | 2 | 0 | 0 | Not-determined | Genome-fragment | NA | NA | 0 |
| S1_k141_123240 | 2041 | No | NA | 5 | 0 | 0 | Not-determined | Genome-fragment | NA | NA | 0 |
| S1_k141_154968 | 1054 | No | NA | 2 | 0 | 2 | Not-determined | Genome-fragment | NA | NA | 0 |
| S1_k141_320856 | 8935 | No | NA | 11 | 0 | 8 | Not-determined | Genome-fragment | NA | NA | 0 |
| S1_k141_368991 | 4794 | No | NA | 7 | 0 | 5 | Not-determined | Genome-fragment | NA | NA | 0 |
| S1_k141_72322 | 3078 | No | NA | 3 | 0 | 1 | Not-determined | Genome-fragment | NA | NA | 0 |
| S1_k141_89329 | 1487 | No | NA | 1 | 0 | 1 | Not-determined | Genome-fragment | NA | NA | 0 |
| S1_k141_139416 | 2867 | No | NA | 1 | 0 | 0 | Not-determined | Genome-fragment | NA | NA | 0 |
| S1_k141_320873 | 7594 | No | NA | 5 | 0 | 2 | Not-determined | Genome-fragment | NA | NA | 0 |
| S3-final-viral-combined_k141_62630\|\|full | 1474 | No | NA | 3 | 0 | 0 | Low-quality | Genome-fragment | 2.92 | AAI-based (medium-confidence) | 0 |
| S1_k141_89349 | 1061 | No | NA | 2 | 0 | 1 | Not-determined | Genome-fragment | NA | NA | 0 |
| S1_k141_39883 | 1545 | No | NA | 2 | 0 | 1 | Not-determined | Genome-fragment | NA | NA | 0 |
| S1_k141_72360 | 1015 | No | NA | 2 | 0 | 0 | Not-determined | Genome-fragment | NA | NA | 0 |
| S1_k141_7235 | 1445 | No | NA | 2 | 0 | 0 | Not-determined | Genome-fragment | NA | NA | 0 |
| S7-final-viral-combined_k141_156394\|\|full | 1474 | No | NA | 3 | 0 | 1 | Low-quality | Genome-fragment | 3.02 | AAI-based (medium-confidence) | 0 |
| S1_k141_304196 | 1101 | No | NA | 1 | 0 | 0 | Not-determined | Genome-fragment | NA | NA | 0 |
| S17-final-viral-combined_k141_218427\|\|full | 1474 | No | NA | 4 | 2 | 0 | Low-quality | Genome-fragment | 2.85 | AAI-based (high-confidence) | 0 |
| S17-k141_193249 | 1474 | No | NA | 3 | 2 | 0 | Low-quality | Genome-fragment | 3.44 | AAI-based (high-confidence) | 0 |
| S1_k141_369054 | 1209 | No | NA | 1 | 0 | 1 | Not-determined | Genome-fragment | NA | NA | 0 |
| S1_k141_123357 | 1371 | No | NA | 2 | 0 | 1 | Not-determined | Genome-fragment | NA | NA | 0 |
| S12-final-viral-combined_k141_53177\|\|lt2gene | 1474 | No | NA | 2 | 0 | 0 | Low-quality | Genome-fragment | 1.82 | AAI-based (high-confidence) | 0 |
| S1_k141_188885 | 1497 | No | NA | 1 | 0 | 1 | Not-determined | Genome-fragment | NA | NA | 0 |
| S1_k141_221240 | 1014 | No | NA | 2 | 0 | 0 | Not-determined | Genome-fragment | NA | NA | 0 |
| S1_k141_123406 | 4346 | No | NA | 9 | 0 | 0 | Not-determined | Genome-fragment | NA | NA | 0 |
| S1_k141_39984 | 4909 | No | NA | 2 | 0 | 1 | Not-determined | Genome-fragment | NA | NA | 0 |
| S1_k141_139473 | 2852 | No | NA | 4 | 0 | 4 | Not-determined | Genome-fragment | NA | NA | 0 |
| S1_k141_23609 | 1076 | No | NA | 2 | 0 | 2 | Not-determined | Genome-fragment | NA | NA | 0 |
| S1_k141_56603 | 1077 | No | NA | 3 | 0 | 0 | Not-determined | Genome-fragment | NA | NA | 0 |
| S1_k141_369111 | 3402 | No | NA | 3 | 0 | 2 | Not-determined | Genome-fragment | NA | NA | 0 |
| S1_k141_254890 | 3240 | No | NA | 3 | 0 | 3 | Not-determined | Genome-fragment | NA | NA | 0 |
| S1_k141_321061 | 1013 | No | NA | 3 | 0 | 0 | Not-determined | Genome-fragment | NA | NA | 0 |
| S1_k141_353564 | 1375 | No | NA | 1 | 0 | 0 | Not-determined | Genome-fragment | NA | NA | 0 |
| S1_k141_40044 | 9105 | No | NA | 6 | 0 | 3 | Not-determined | Genome-fragment | NA | NA | 0 |
| S1_k141_321108 | 1013 | No | NA | 1 | 0 | 1 | Not-determined | Genome-fragment | NA | NA | 0 |
| S1_k141_56640 | 2535 | No | NA | 2 | 0 | 2 | Not-determined | Genome-fragment | NA | NA | 0 |
| S1_k141_221330 | 1389 | No | NA | 1 | 0 | 0 | Not-determined | Genome-fragment | NA | NA | 0 |
| S12_k141_77159 | 1474 | No | NA | 2 | 0 | 1 | Low-quality | Genome-fragment | 0.6 | AAI-based (medium-confidence) | 0 |
| S1_k141_254959 | 3919 | No | NA | 10 | 0 | 10 | Not-determined | Genome-fragment | NA | NA | 0 |
| S1_k141_238113 | 1010 | No | NA | 2 | 0 | 0 | Not-determined | Genome-fragment | NA | NA | 0 |
| S1_k141_7437 | 5808 | No | NA | 8 | 0 | 4 | Not-determined | Genome-fragment | NA | NA | 0 |
| S2-final-viral-combined_k141_204279\|\|lt2gene | 1474 | No | NA | 2 | 2 | 0 | Low-quality | Genome-fragment | 3.12 | AAI-based (high-confidence) | 0 |
| S4-final-viral-combined_k141_17812\|\|full | 1474 | No | NA | 4 | 0 | 2 | Low-quality | Genome-fragment | 0.4 | AAI-based (medium-confidence) | 0 |
| S1_k141_72528 | 1990 | No | NA | 4 | 0 | 4 | Not-determined | Genome-fragment | NA | NA | 0 |
| S8-final-viral-combined_k141_220108\|\|full | 1474 | No | NA | 4 | 1 | 0 | Low-quality | Genome-fragment | 0.99 | HMM-based (lower-bound) | 0 |
| S11-final-viral-combined_k141_72266\|\|full | 1475 | No | NA | 3 | 1 | 0 | Low-quality | Genome-fragment | 2.86 | AAI-based (high-confidence) | 0 |
| S1_k141_270631 | 2072 | No | NA | 3 | 0 | 2 | Not-determined | Genome-fragment | NA | NA | 0 |
| S11_k141_83002 | 1475 | No | NA | 4 | 0 | 2 | Low-quality | Genome-fragment | 0.48 | AAI-based (medium-confidence) | 0 |
| S1_k141_238176 | 1817 | No | NA | 2 | 0 | 1 | Not-determined | Genome-fragment | NA | NA | 0 |
| S1-final-viral-combined_k141_341574\|\|full | 1475 | No | NA | 3 | 0 | 0 | Low-quality | Genome-fragment | 2.47 | AAI-based (medium-confidence) | 0 |
| S1_k141_105330 | 1313 | No | NA | 2 | 0 | 0 | Not-determined | Genome-fragment | NA | NA | 0 |
| S1_k141_238213 | 8233 | No | NA | 8 | 0 | 8 | Not-determined | Genome-fragment | NA | NA | 0 |
| S1_k141_304374 | 1380 | No | NA | 2 | 0 | 2 | Not-determined | Genome-fragment | NA | NA | 0 |
| S1_k141_367376 | 1475 | No | NA | 1 | 0 | 0 | Low-quality | Genome-fragment | 3.96 | AAI-based (medium-confidence) | 0 |
| S1_k141_221425 | 1480 | No | NA | 1 | 0 | 1 | Not-determined | Genome-fragment | NA | NA | 0 |
| S1_k141_171323 | 1025 | No | NA | 3 | 0 | 1 | Not-determined | Genome-fragment | NA | NA | 0 |
| S7-final-viral-combined_k141_69851\|\|full | 1475 | No | NA | 5 | 0 | 3 | Low-quality | Genome-fragment | 0.45 | AAI-based (medium-confidence) | 0 |
| S1_k141_188985 | 1558 | No | NA | 1 | 0 | 1 | Not-determined | Genome-fragment | NA | NA | 0 |
| S1_k141_369263 | 1324 | No | NA | 2 | 0 | 1 | Not-determined | Genome-fragment | NA | NA | 0 |
| S1_k141_238218 | 6530 | No | NA | 6 | 0 | 1 | Not-determined | Genome-fragment | NA | NA | 0 |
| S1_k141_155219 | 1115 | No | NA | 4 | 0 | 0 | Not-determined | Genome-fragment | NA | NA | 0 |
| S17-k141_263437 | 1475 | No | NA | 2 | 0 | 0 | Low-quality | Genome-fragment | 0.35 | AAI-based (medium-confidence) | 0 |
| S1_k141_72575 | 1114 | No | NA | 3 | 0 | 1 | Not-determined | Genome-fragment | NA | NA | 0 |
| S1_k141_7546 | 1528 | No | NA | 1 | 0 | 1 | Not-determined | Genome-fragment | NA | NA | 0 |
| S1_k141_56843 | 1451 | No | NA | 1 | 0 | 1 | Not-determined | Genome-fragment | NA | NA | 0 |
| S1_k141_255099 | 2000 | No | NA | 1 | 0 | 1 | Not-determined | Genome-fragment | NA | NA | 0 |
| S1_k141_155250 | 1073 | No | NA | 2 | 0 | 2 | Not-determined | Genome-fragment | NA | NA | 0 |
| S1_k141_270713 | 1398 | No | NA | 2 | 0 | 0 | Not-determined | Genome-fragment | NA | NA | 0 |
| S2-final-viral-combined_k141_242102\|\|full | 1475 | No | NA | 3 | 0 | 1 | Low-quality | Genome-fragment | 0.84 | AAI-based (medium-confidence) | 0 |
| S8-final-viral-combined_k141_223006\|\|full | 1475 | No | NA | 2 | 0 | 0 | Low-quality | Genome-fragment | 3.42 | AAI-based (high-confidence) | 0 |
| S1_k141_287245 | 1474 | No | NA | 1 | 0 | 1 | Not-determined | Genome-fragment | NA | NA | 0 |
| S1_k141_123596 | 5818 | No | NA | 6 | 0 | 5 | Not-determined | Genome-fragment | NA | NA | 0 |
| S1_k141_221519 | 1690 | No | NA | 2 | 0 | 1 | Not-determined | Genome-fragment | NA | NA | 0 |
| S1_k141_321309 | 1261 | No | NA | 1 | 0 | 1 | Not-determined | Genome-fragment | NA | NA | 0 |
| S1_k141_123616 | 1501 | No | NA | 1 | 0 | 1 | Not-determined | Genome-fragment | NA | NA | 0 |
| S1_k141_155378 | 6534 | No | NA | 3 | 0 | 3 | Not-determined | Genome-fragment | NA | NA | 0 |
| S1_k141_89666 | 1515 | No | NA | 2 | 0 | 2 | Not-determined | Genome-fragment | NA | NA | 0 |
| S8-final-viral-combined_k141_216696\|\|full | 1475 | No | NA | 2 | 1 | 0 | Low-quality | Genome-fragment | 2.84 | AAI-based (medium-confidence) | 0 |
| S1_k141_56921 | 1050 | No | NA | 1 | 0 | 1 | Not-determined | Genome-fragment | NA | NA | 0 |
| S1_k141_221595 | 1925 | No | NA | 2 | 0 | 1 | Not-determined | Genome-fragment | NA | NA | 0 |
| S1_k141_304463 | 3168 | No | NA | 3 | 0 | 1 | Not-determined | Genome-fragment | NA | NA | 0 |
| S1_k141_56941 | 1481 | No | NA | 1 | 0 | 0 | Not-determined | Genome-fragment | NA | NA | 0 |
| S1_k141_189199 | 1222 | No | NA | 2 | 0 | 0 | Not-determined | Genome-fragment | NA | NA | 0 |
| S1_k141_386234 | 1573 | No | NA | 1 | 0 | 1 | Not-determined | Genome-fragment | NA | NA | 0 |
| S1_k141_321392 | 4265 | No | NA | 4 | 0 | 3 | Not-determined | Genome-fragment | NA | NA | 0 |
| S1_k141_304489 | 1726 | No | NA | 1 | 0 | 1 | Not-determined | Genome-fragment | NA | NA | 0 |
| S1_k141_369418 | 1538 | No | NA | 2 | 0 | 0 | Not-determined | Genome-fragment | NA | NA | 0 |
| S1_k141_270869 | 1922 | No | NA | 2 | 0 | 0 | Not-determined | Genome-fragment | NA | NA | 0 |
| S1_k141_221626 | 10940 | No | NA | 11 | 0 | 3 | Not-determined | Genome-fragment | NA | NA | 0 |
| S1_k141_72730 | 1064 | No | NA | 1 | 0 | 0 | Not-determined | Genome-fragment | NA | NA | 0 |
| S1_k141_7700 | 30471 | No | NA | 29 | 0 | 8 | Not-determined | Genome-fragment | NA | NA | 0 |
| S1_k141_189234 | 13335 | No | NA | 9 | 0 | 7 | Not-determined | Genome-fragment | NA | NA | 0 |
| S8_k141_64681 | 1475 | No | NA | 3 | 2 | 0 | Low-quality | Genome-fragment | 3.08 | AAI-based (high-confidence) | 0 |
| S1_k141_353862 | 1174 | No | NA | 2 | 0 | 1 | Not-determined | Genome-fragment | NA | NA | 0 |
| S1_k141_155467 | 1363 | No | NA | 1 | 0 | 1 | Not-determined | Genome-fragment | NA | NA | 0 |
| S1_k141_57068 | 1137 | No | NA | 1 | 0 | 0 | Not-determined | Genome-fragment | NA | NA | 0 |
| S1_k141_189258 | 2813 | No | NA | 2 | 0 | 2 | Not-determined | Genome-fragment | NA | NA | 0 |
| S1_k141_205794 | 1427 | No | NA | 1 | 0 | 1 | Not-determined | Genome-fragment | NA | NA | 0 |
| S1_k141_89770 | 1280 | No | NA | 1 | 0 | 1 | Not-determined | Genome-fragment | NA | NA | 0 |
| S1_k141_386360 | 3950 | No | NA | 5 | 0 | 0 | Not-determined | Genome-fragment | NA | NA | 0 |
| S1_k141_40330 | 1724 | No | NA | 1 | 0 | 0 | Not-determined | Genome-fragment | NA | NA | 0 |
| S1_k141_353950 | 1158 | No | NA | 2 | 0 | 2 | Not-determined | Genome-fragment | NA | NA | 0 |
| S1_k141_221731 | 1042 | No | NA | 2 | 0 | 2 | Not-determined | Genome-fragment | NA | NA | 0 |
| S1_k141_386385 | 1834 | No | NA | 4 | 0 | 0 | Not-determined | Genome-fragment | NA | NA | 0 |
| S1_k141_336177 | 1278 | No | NA | 2 | 0 | 0 | Not-determined | Genome-fragment | NA | NA | 0 |
| S1_k141_72760 | 2876 | No | NA | 2 | 0 | 1 | Not-determined | Genome-fragment | NA | NA | 0 |
| S1_k141_221757 | 2496 | No | NA | 4 | 0 | 0 | Not-determined | Genome-fragment | NA | NA | 0 |
| S1_k141_270973 | 3317 | No | NA | 4 | 0 | 2 | Not-determined | Genome-fragment | NA | NA | 0 |

| | | | | | | | | | | | |
|---|---|---|---|---|---|---|---|---|---|---|---|
| S1_k141_105675 | 1308 | No | NA | 1 | 0 | 1 | Not-determined | Genome-fragment | NA | NA | 0 |
| S1_k141_238292 | 4146 | No | NA | 3 | 0 | 0 | Not-determined | Genome-fragment | NA | NA | 0 |
| S1_k141_105702 | 3054 | No | NA | 4 | 0 | 2 | Not-determined | Genome-fragment | NA | NA | 0 |
| S9-final-viral-combined_k141_29136\|\|full | 1476 | No | NA | 3 | 0 | 0 | Low-quality | Genome-fragment | 3.57 | AAI-based (medium-confidence) | 0 |
| S1_k141_354014 | 5130 | No | NA | 6 | 0 | 3 | Not-determined | Genome-fragment | NA | NA | 0 |
| S1_k141_189419 | 1382 | No | NA | 1 | 0 | 1 | Not-determined | Genome-fragment | NA | NA | 0 |
| S1_k141_139920 | 1803 | No | NA | 2 | 0 | 1 | Not-determined | Genome-fragment | NA | NA | 0 |
| S1_k141_139931 | 1534 | No | NA | 1 | 0 | 0 | Not-determined | Genome-fragment | NA | NA | 0 |
| S1_k141_40429 | 4226 | No | NA | 3 | 0 | 2 | Not-determined | Genome-fragment | NA | NA | 0 |
| S1_k141_221834 | 1953 | No | NA | 3 | 0 | 0 | Not-determined | Genome-fragment | NA | NA | 0 |
| S9-final-viral-combined_k141_110343\|\|full | 1476 | No | NA | 4 | 2 | 0 | Low-quality | Genome-fragment | 3.21 | AAI-based (high-confidence) | 0 |
| S1-final-viral-combined_k141_294977\|\|full | 1476 | No | NA | 3 | 0 | 0 | Low-quality | Genome-fragment | 4.22 | AAI-based (high-confidence) | 0 |
| S1_k141_155604 | 2156 | No | NA | 3 | 0 | 0 | Not-determined | Genome-fragment | NA | NA | 0 |
| S1_k141_24041 | 1062 | No | NA | 1 | 0 | 0 | Not-determined | Genome-fragment | NA | NA | 0 |
| S1_k141_51945 | 1476 | No | NA | 2 | 0 | 0 | Low-quality | Genome-fragment | 3.3 | AAI-based (high-confidence) | 0 |
| S1_k141_7963 | 2989 | No | NA | 3 | 0 | 1 | Not-determined | Genome-fragment | NA | NA | 0 |
| S1_k141_369649 | 1309 | No | NA | 3 | 0 | 0 | Not-determined | Genome-fragment | NA | NA | 0 |
| S1_k141_155672 | 1050 | No | NA | 1 | 0 | 1 | Not-determined | Genome-fragment | NA | NA | 0 |
| S1_k141_7979 | 3491 | No | NA | 3 | 0 | 3 | Not-determined | Genome-fragment | NA | NA | 0 |
| S1_k141_171611 | 1586 | No | NA | 1 | 0 | 0 | Not-determined | Genome-fragment | NA | NA | 0 |
| S1_k141_221879 | 1256 | No | NA | 1 | 0 | 1 | Not-determined | Genome-fragment | NA | NA | 0 |
| S5-final-viral-combined_k141_141931\|\|full | 1476 | No | NA | 4 | 2 | 0 | Low-quality | Genome-fragment | 2.44 | AAI-based (high-confidence) | 0 |
| S1_k141_155712 | 3508 | No | NA | 4 | 0 | 1 | Not-determined | Genome-fragment | NA | NA | 0 |
| S1_k141_369684 | 4047 | No | NA | 8 | 0 | 8 | Not-determined | Genome-fragment | NA | NA | 0 |
| S1_k141_238468 | 1431 | No | NA | 2 | 0 | 1 | Not-determined | Genome-fragment | NA | NA | 0 |
| S1_k141_105829 | 1451 | No | NA | 1 | 0 | 0 | Not-determined | Genome-fragment | NA | NA | 0 |
| S1_k141_336387 | 2133 | No | NA | 4 | 0 | 2 | Not-determined | Genome-fragment | NA | NA | 0 |
| S1_k141_287648 | 1845 | No | NA | 2 | 0 | 2 | Not-determined | Genome-fragment | NA | NA | 0 |
| S7_k141_104547 | 1476 | No | NA | 5 | 0 | 0 | Low-quality | Genome-fragment | 2.14 | AAI-based (high-confidence) | 0 |
| S1_k141_271238 | 1160 | No | NA | 1 | 0 | 0 | Not-determined | Genome-fragment | NA | NA | 0 |
| S17-final-viral-combined_k141_222385\|\|full | 1476 | No | NA | 2 | 0 | 0 | Low-quality | Genome-fragment | 3.63 | AAI-based (medium-confidence) | 0 |
| S1_k141_255433 | 1262 | No | NA | 1 | 0 | 1 | Not-determined | Genome-fragment | NA | NA | 0 |
| S1_k141_171695 | 1323 | No | NA | 2 | 0 | 0 | Not-determined | Genome-fragment | NA | NA | 0 |
| S1_k141_57248 | 1259 | No | NA | 2 | 0 | 0 | Not-determined | Genome-fragment | NA | NA | 0 |
| S17-k141_31485 | 1476 | No | NA | 2 | 2 | 0 | Low-quality | Genome-fragment | 5.38 | AAI-based (high-confidence) | 0 |
| S1-final-viral-combined_k141_289920\|\|full | 1477 | No | NA | 4 | 0 | 0 | Low-quality | Genome-fragment | 3.79 | AAI-based (medium-confidence) | 0 |
| S1_k141_321801 | 1111 | No | NA | 2 | 0 | 1 | Not-determined | Genome-fragment | NA | NA | 0 |
| S1_k141_304861 | 9729 | No | NA | 2 | 0 | 1 | Not-determined | Genome-fragment | NA | NA | 0 |
| S1_k141_171726 | 1286 | No | NA | 3 | 0 | 1 | Not-determined | Genome-fragment | NA | NA | 0 |
| S1_k141_369797 | 1034 | No | NA | 1 | 0 | 0 | Not-determined | Genome-fragment | NA | NA | 0 |
| S1_k141_321835 | 2189 | No | NA | 3 | 0 | 2 | Not-determined | Genome-fragment | NA | NA | 0 |
| S5-final-viral-combined_k141_207005\|\|full | 1477 | No | NA | 3 | 0 | 0 | Low-quality | Genome-fragment | 2.82 | AAI-based (high-confidence) | 0 |
| S1_k141_386714 | 1266 | No | NA | 2 | 0 | 1 | Not-determined | Genome-fragment | NA | NA | 0 |
| S1_k141_105940 | 3570 | No | NA | 5 | 0 | 4 | Not-determined | Genome-fragment | NA | NA | 0 |
| S1_k141_8164 | 1583 | No | NA | 3 | 0 | 1 | Not-determined | Genome-fragment | NA | NA | 0 |
| S1_k141_206094 | 1523 | No | NA | 1 | 0 | 1 | Not-determined | Genome-fragment | NA | NA | 0 |
| S1_k141_304925 | 1308 | No | NA | 1 | 0 | 1 | Not-determined | Genome-fragment | NA | NA | 0 |
| S1_k141_90126 | 3432 | No | NA | 4 | 0 | 1 | Not-determined | Genome-fragment | NA | NA | 0 |
| S1_k141_90161 | 1145 | No | NA | 3 | 0 | 2 | Not-determined | Genome-fragment | NA | NA | 0 |
| S1_k141_304960 | 3045 | No | NA | 6 | 0 | 2 | Not-determined | Genome-fragment | NA | NA | 0 |
| S7-final-viral-combined_k141_243164\|\|full | 1477 | No | NA | 3 | 0 | 0 | Low-quality | Genome-fragment | 3.43 | AAI-based (high-confidence) | 0 |
| S1_k141_287830 | 1014 | No | NA | 1 | 0 | 1 | Not-determined | Genome-fragment | NA | NA | 0 |
| S1_k141_171785 | 1032 | No | NA | 2 | 0 | 1 | Not-determined | Genome-fragment | NA | NA | 0 |
| S17-final-viral-combined_k141_156193\|\|full | 1477 | No | NA | 4 | 1 | 0 | Low-quality | Genome-fragment | 2.1 | AAI-based (high-confidence) | 0 |
| S17-final-viral-combined_k141_227357\|\|full | 1477 | No | NA | 3 | 0 | 1 | Low-quality | Genome-fragment | 0.62 | AAI-based (medium-confidence) | 0 |
| S1_k141_287842 | 1428 | No | NA | 2 | 0 | 1 | Not-determined | Genome-fragment | NA | NA | 0 |
| S1_k141_369862 | 1205 | No | NA | 2 | 0 | 1 | Not-determined | Genome-fragment | NA | NA | 0 |
| S17-final-viral-combined_k141_210942\|\|full | 1477 | No | NA | 3 | 0 | 0 | Low-quality | Genome-fragment | 2.44 | AAI-based (medium-confidence) | 0 |
| S17-final-viral-combined_k141_97288\|\|full | 1477 | No | NA | 3 | 0 | 0 | Low-quality | Genome-fragment | 2.75 | AAI-based (high-confidence) | 0 |
| S1_k141_73166 | 9961 | No | NA | 7 | 0 | 7 | Not-determined | Genome-fragment | NA | NA | 0 |
| S1_k141_222008 | 1023 | No | NA | 2 | 0 | 1 | Not-determined | Genome-fragment | NA | NA | 0 |
| S1_k141_305016 | 1232 | No | NA | 2 | 0 | 0 | Not-determined | Genome-fragment | NA | NA | 0 |
| S1_k141_354176 | 1105 | No | NA | 1 | 0 | 1 | Not-determined | Genome-fragment | NA | NA | 0 |
| S1_k141_189831 | 1775 | No | NA | 2 | 0 | 0 | Not-determined | Genome-fragment | NA | NA | 0 |
| S1_k141_73204 | 1151 | No | NA | 2 | 0 | 0 | Not-determined | Genome-fragment | NA | NA | 0 |
| S1_k141_189847 | 1138 | No | NA | 1 | 0 | 1 | Not-determined | Genome-fragment | NA | NA | 0 |
| S1_k141_140080 | 1216 | No | NA | 1 | 0 | 0 | Not-determined | Genome-fragment | NA | NA | 0 |
| S12-final-viral-combined_k141_361968\|\|full | 1477 | No | NA | 4 | 0 | 0 | Low-quality | Genome-fragment | 2.24 | AAI-based (high-confidence) | 0 |
| S1_k141_171835 | 1028 | No | NA | 1 | 0 | 0 | Not-determined | Genome-fragment | NA | NA | 0 |
| S1_k141_321969 | 7531 | No | NA | 11 | 0 | 8 | Not-determined | Genome-fragment | NA | NA | 0 |
| S1_k141_124253 | 1171 | No | NA | 1 | 0 | 1 | Not-determined | Genome-fragment | NA | NA | 0 |
| S1_k141_354275 | 1128 | No | NA | 1 | 0 | 1 | Not-determined | Genome-fragment | NA | NA | 0 |
| S1_k141_222104 | 1372 | No | NA | 1 | 0 | 1 | Not-determined | Genome-fragment | NA | NA | 0 |
| S1_k141_124278 | 2302 | No | NA | 2 | 0 | 2 | Not-determined | Genome-fragment | NA | NA | 0 |
| S1_k141_386908 | 1769 | No | NA | 2 | 0 | 1 | Not-determined | Genome-fragment | NA | NA | 0 |
| S8-final-viral-combined_k141_57029\|\|full | 1477 | No | NA | 3 | 0 | 1 | Low-quality | Genome-fragment | 0.72 | AAI-based (medium-confidence) | 0 |
| S8-final-viral-combined_k141_47399\|\|lt2gene | 1477 | No | NA | 1 | 1 | 0 | Low-quality | Genome-fragment | 3.26 | AAI-based (high-confidence) | 0 |
| S11-final-viral-combined_k141_89401\|\|full | 1478 | No | NA | 3 | 0 | 2 | Low-quality | Genome-fragment | 1.39 | AAI-based (medium-confidence) | 0 |
| S1_k141_238887 | 1168 | No | NA | 3 | 0 | 1 | Not-determined | Genome-fragment | NA | NA | 0 |
| S1_k141_124340 | 1051 | No | NA | 1 | 0 | 0 | Not-determined | Genome-fragment | NA | NA | 0 |
| S1_k141_73288 | 2363 | No | NA | 1 | 0 | 1 | Not-determined | Genome-fragment | NA | NA | 0 |
| S1_k141_90282 | 3097 | No | NA | 3 | 0 | 3 | Not-determined | Genome-fragment | NA | NA | 0 |
| S1_k141_322070 | 1038 | No | NA | 1 | 0 | 1 | Not-determined | Genome-fragment | NA | NA | 0 |
| S1_k141_106115 | 26905 | No | NA | 27 | 7 | 0 | Medium-quality | Genome-fragment | 78.86 | AAI-based (high-confidence) | 0 |
| S1-final-viral-combined_k141_299305\|\|lt2gene | 1478 | No | NA | 2 | 1 | 0 | Low-quality | Genome-fragment | 1.02 | HMM-based (lower-bound) | 0 |
| S1_k141_255757 | 1163 | No | NA | 1 | 0 | 0 | Not-determined | Genome-fragment | NA | NA | 0 |
| S1_k141_57430 | 6268 | No | NA | 6 | 0 | 2 | Not-determined | Genome-fragment | NA | NA | 0 |
| S1_k141_305123 | 1108 | No | NA | 4 | 0 | 3 | Not-determined | Genome-fragment | NA | NA | 0 |
| S1_k141_222240 | 1580 | No | NA | 1 | 0 | 1 | Not-determined | Genome-fragment | NA | NA | 0 |
| S1_k141_206400 | 1266 | No | NA | 1 | 0 | 1 | Not-determined | Genome-fragment | NA | NA | 0 |
| S1_k141_171938 | 6123 | No | NA | 8 | 0 | 1 | Not-determined | Genome-fragment | NA | NA | 0 |
| S5_k141_62303 | 1478 | No | NA | 1 | 0 | 0 | Low-quality | Genome-fragment | 0.97 | AAI-based (high-confidence) | 0 |
| S1_k141_40761 | 3297 | No | NA | 1 | 0 | 1 | Not-determined | Genome-fragment | NA | NA | 0 |
| S7-final-viral-combined_k141_158064\|\|full | 1478 | No | NA | 3 | 0 | 0 | Low-quality | Genome-fragment | 2.67 | AAI-based (high-confidence) | 0 |
| S7-final-viral-combined_k141_46309\|\|full | 1478 | No | NA | 4 | 0 | 0 | Low-quality | Genome-fragment | 3.06 | AAI-based (medium-confidence) | 0 |
| S7_k141_235807 | 1478 | No | NA | 2 | 0 | 1 | Low-quality | Genome-fragment | 2.6 | AAI-based (medium-confidence) | 0 |
| S1_k141_73305 | 2220 | No | NA | 5 | 0 | 2 | Not-determined | Genome-fragment | NA | NA | 0 |
| S17-final-viral-combined_k141_111596\|\|full | 1478 | No | NA | 3 | 0 | 0 | Low-quality | Genome-fragment | 3.64 | AAI-based (medium-confidence) | 0 |
| S17-final-viral-combined_k141_135192\|\|full | 1478 | No | NA | 4 | 0 | 2 | Low-quality | Genome-fragment | 3.73 | AAI-based (high-confidence) | 0 |
| S1_k141_387063 | 3415 | No | NA | 3 | 0 | 0 | Not-determined | Genome-fragment | NA | NA | 0 |
| S1_k141_387064 | 1347 | No | NA | 2 | 0 | 0 | Not-determined | Genome-fragment | NA | NA | 0 |
| S1_k141_90345 | 1291 | No | NA | 3 | 0 | 2 | Not-determined | Genome-fragment | NA | NA | 0 |
| S17-final-viral-combined_k141_16176\|\|full | 1478 | No | NA | 3 | 0 | 0 | Low-quality | Genome-fragment | 3.62 | AAI-based (medium-confidence) | 0 |
| S1_k141_106193 | 2243 | No | NA | 1 | 0 | 0 | Not-determined | Genome-fragment | NA | NA | 0 |
| S1_k141_106201 | 1657 | No | NA | 1 | 0 | 1 | Not-determined | Genome-fragment | NA | NA | 0 |
| S1_k141_354498 | 1648 | No | NA | 2 | 0 | 2 | Not-determined | Genome-fragment | NA | NA | 0 |
| S1_k141_24530 | 4667 | No | NA | 3 | 0 | 1 | Not-determined | Genome-fragment | NA | NA | 0 |
| S1_k141_57495 | 3248 | No | NA | 3 | 0 | 1 | Not-determined | Genome-fragment | NA | NA | 0 |
| S1_k141_106233 | 1014 | No | NA | 2 | 0 | 0 | Not-determined | Genome-fragment | NA | NA | 0 |
| S1_k141_90382 | 2958 | No | NA | 7 | 0 | 2 | Not-determined | Genome-fragment | NA | NA | 0 |
| S1_k141_40850 | 1027 | No | NA | 1 | 0 | 1 | Not-determined | Genome-fragment | NA | NA | 0 |
| S1_k141_155841 | 5048 | No | NA | 5 | 0 | 1 | Not-determined | Genome-fragment | NA | NA | 0 |
| S17-final-viral-combined_k141_63314\|\|full | 1478 | No | NA | 2 | 1 | 0 | Low-quality | Genome-fragment | 3 | AAI-based (high-confidence) | 0 |
| S2_k141_220206 | 1478 | No | NA | 2 | 0 | 0 | Low-quality | Genome-fragment | 0.85 | AAI-based (high-confidence) | 0 |
| S1_k141_172005 | 3574 | No | NA | 3 | 0 | 2 | Not-determined | Genome-fragment | NA | NA | 0 |
| S1_k141_24580 | 6468 | No | NA | 1 | 0 | 1 | Not-determined | Genome-fragment | NA | NA | 0 |
| S4-final-viral-combined_k141_49781\|\|full | 1478 | No | NA | 4 | 1 | 0 | Low-quality | Genome-fragment | 0.95 | HMM-based (lower-bound) | 0 |
| S4-final-viral-combined_k141_120456\|\|full | 1478 | Yes | 1115 | 3 | 1 | 1 | Low-quality | Genome-fragment | 2.25 | HMM-based (lower-bound) | 24.56 |
| S1_k141_370220 | 1862 | No | NA | 2 | 0 | 2 | Not-determined | Genome-fragment | NA | NA | 0 |
| S1_k141_354553 | 1391 | No | NA | 1 | 0 | 0 | Not-determined | Genome-fragment | NA | NA | 0 |
| S1_k141_255961 | 1098 | No | NA | 1 | 0 | 0 | Not-determined | Genome-fragment | NA | NA | 0 |
| S1_k141_155909 | 3318 | No | NA | 3 | 0 | 1 | Not-determined | Genome-fragment | NA | NA | 0 |
| S1_k141_239043 | 1172 | No | NA | 1 | 0 | 0 | Not-determined | Genome-fragment | NA | NA | 0 |
| S1_k141_40943 | 4073 | No | NA | 2 | 0 | 2 | Not-determined | Genome-fragment | NA | NA | 0 |
| S8_k141_70646 | 1478 | No | NA | 1 | 0 | 0 | Low-quality | Genome-fragment | 2.06 | AAI-based (high-confidence) | 0 |
| S1_k141_24717 | 1071 | No | NA | 2 | 0 | 0 | Not-determined | Genome-fragment | NA | NA | 0 |
| S1_k141_82882 | 1479 | No | NA | 3 | 0 | 0 | Low-quality | Genome-fragment | 3.14 | AAI-based (high-confidence) | 0 |
| S1_k141_305376 | 3881 | No | NA | 2 | 0 | 1 | Not-determined | Genome-fragment | NA | NA | 0 |
| S17-final-viral-combined_k141_122518\|\|full | 1479 | No | NA | 3 | 0 | 0 | Low-quality | Genome-fragment | 3.32 | AAI-based (medium-confidence) | 0 |
| S17-final-viral-combined_k141_36242\|\|full | 1479 | No | NA | 3 | 2 | 0 | Low-quality | Genome-fragment | 3.91 | AAI-based (high-confidence) | 0 |
| S12-final-viral-combined_k141_109682\|\|full | 1479 | No | NA | 2 | 0 | 0 | Low-quality | Genome-fragment | 1.64 | AAI-based (medium-confidence) | 0 |
| S1_k141_90597 | 2892 | No | NA | 1 | 0 | 0 | Not-determined | Genome-fragment | NA | NA | 0 |
| S1_k141_8763 | 1173 | No | NA | 1 | 0 | 0 | Not-determined | Genome-fragment | NA | NA | 0 |
| S1_k141_24816 | 2092 | No | NA | 2 | 0 | 1 | Not-determined | Genome-fragment | NA | NA | 0 |
| S1_k141_73561 | 1903 | No | NA | 2 | 0 | 2 | Not-determined | Genome-fragment | NA | NA | 0 |
| S1_k141_41041 | 3984 | No | NA | 6 | 0 | 3 | Not-determined | Genome-fragment | NA | NA | 0 |
| S1_k141_222550 | 1228 | No | NA | 2 | 0 | 2 | Not-determined | Genome-fragment | NA | NA | 0 |
| S1_k141_41052 | 1397 | No | NA | 3 | 0 | 0 | Not-determined | Genome-fragment | NA | NA | 0 |
| S1_k141_337072 | 1202 | No | NA | 2 | 0 | 2 | Not-determined | Genome-fragment | NA | NA | 0 |
| S1_k141_24860 | 1129 | No | NA | 2 | 0 | 0 | Not-determined | Genome-fragment | NA | NA | 0 |
| S1_k141_239228 | 1082 | No | NA | 1 | 0 | 1 | Not-determined | Genome-fragment | NA | NA | 0 |
| S1_k141_206693 | 3467 | No | NA | 4 | 0 | 2 | Not-determined | Genome-fragment | NA | NA | 0 |
| S12-final-viral-combined_k141_366666\|\|full | 1479 | No | NA | 4 | 2 | 0 | Low-quality | Genome-fragment | 3.41 | AAI-based (medium-confidence) | 0 |
| S1_k141_206720 | 1809 | No | NA | 1 | 0 | 0 | Not-determined | Genome-fragment | NA | NA | 0 |
| S1_k141_370496 | 1243 | No | NA | 2 | 0 | 2 | Not-determined | Genome-fragment | NA | NA | 0 |
| S1_k141_305535 | 1399 | No | NA | 2 | 0 | 0 | Not-determined | Genome-fragment | NA | NA | 0 |
| S2-final-viral-combined_k141_171239\|\|full | 1479 | No | NA | 3 | 1 | 0 | Low-quality | Genome-fragment | 4.64 | AAI-based (medium-confidence) | 0 |
| S1_k141_24885 | 3173 | No | NA | 2 | 0 | 2 | Not-determined | Genome-fragment | NA | NA | 0 |
| S4-final-viral-combined_k141_105080\|\|full | 1479 | No | NA | 2 | 0 | 1 | Low-quality | Genome-fragment | 1.1 | AAI-based (medium-confidence) | 0 |
| S3-final-viral-combined_k141_57879\|\|full | 1480 | No | NA | 3 | 0 | 1 | Low-quality | Genome-fragment | 0.91 | AAI-based (medium-confidence) | 0 |
| S1_k141_90701 | 2632 | No | NA | 2 | 0 | 0 | Not-determined | Genome-fragment | NA | NA | 0 |
| S1_k141_156050 | 6820 | No | NA | 10 | 0 | 0 | Not-determined | Genome-fragment | NA | NA | 0 |
| S1_k141_354719 | 1305 | No | NA | 1 | 0 | 0 | Not-determined | Genome-fragment | NA | NA | 0 |
| S1_k141_305564 | 1780 | No | NA | 1 | 0 | 1 | Not-determined | Genome-fragment | NA | NA | 0 |
| S1_k141_370527 | 4004 | No | NA | 2 | 0 | 2 | Not-determined | Genome-fragment | NA | NA | 0 |
| S1_k141_24915 | 1049 | No | NA | 1 | 0 | 1 | Not-determined | Genome-fragment | NA | NA | 0 |
| S1_k141_124821 | 1716 | No | NA | 1 | 0 | 0 | Not-determined | Genome-fragment | NA | NA | 0 |
| S1_k141_206805 | 1070 | No | NA | 2 | 0 | 0 | Not-determined | Genome-fragment | NA | NA | 0 |
| S1_k141_271859 | 1267 | No | NA | 1 | 0 | 1 | Not-determined | Genome-fragment | NA | NA | 0 |
| S1_k141_57719 | 1953 | No | NA | 1 | 0 | 1 | Not-determined | Genome-fragment | NA | NA | 0 |
| S1_k141_206811 | 4634 | No | NA | 2 | 0 | 2 | Not-determined | Genome-fragment | NA | NA | 0 |
| S5-final-viral-combined_k141_135667\|\|full | 1480 | No | NA | 2 | 0 | 0 | Low-quality | Genome-fragment | 1.64 | AAI-based (high-confidence) | 0 |
| S7-final-viral-combined_k141_131406\|\|full | 1480 | No | NA | 6 | 0 | 0 | Low-quality | Genome-fragment | 3.7 | AAI-based (high-confidence) | 0 |
| S1_k141_206864 | 10080 | No | NA | 8 | 0 | 6 | Not-determined | Genome-fragment | NA | NA | 0 |
| S1_k141_57781 | 2569 | No | NA | 1 | 0 | 1 | Not-determined | Genome-fragment | NA | NA | 0 |
| S17-final-viral-combined_k141_174031\|\|full | 1480 | No | NA | 3 | 0 | 1 | Low-quality | Genome-fragment | 0.88 | AAI-based (medium-confidence) | 0 |
| S1_k141_41293 | 1145 | No | NA | 3 | 0 | 2 | Not-determined | Genome-fragment | NA | NA | 0 |
| S1_k141_239426 | 1256 | No | NA | 1 | 0 | 0 | Not-determined | Genome-fragment | NA | NA | 0 |
| S1_k141_73727 | 1608 | No | NA | 1 | 0 | 1 | Not-determined | Genome-fragment | NA | NA | 0 |
| S1_k141_222733 | 1095 | No | NA | 1 | 0 | 1 | Not-determined | Genome-fragment | NA | NA | 0 |
| S1_k141_239438 | 1152 | No | NA | 1 | 0 | 1 | Not-determined | Genome-fragment | NA | NA | 0 |
| S12-final-viral-combined_k141_175444\|\|full | 1480 | No | NA | 3 | 0 | 2 | Low-quality | Genome-fragment | 0.81 | AAI-based (medium-confidence) | 0 |
| S1_k141_288518 | 1875 | No | NA | 2 | 0 | 1 | Not-determined | Genome-fragment | NA | NA | 0 |
| S4-final-viral-combined_k141_82837\|\|full | 1480 | No | NA | 3 | 0 | 0 | Low-quality | Genome-fragment | 3.57 | AAI-based (medium-confidence) | 0 |
| S1_k141_354894 | 1252 | No | NA | 1 | 0 | 1 | Not-determined | Genome-fragment | NA | NA | 0 |
| S1_k141_140569 | 1074 | No | NA | 1 | 0 | 1 | Not-determined | Genome-fragment | NA | NA | 0 |
| S1_k141_124961 | 1158 | No | NA | 1 | 0 | 0 | Not-determined | Genome-fragment | NA | NA | 0 |
| S1_k141_124968 | 4343 | No | NA | 1 | 0 | 0 | Not-determined | Genome-fragment | NA | NA | 0 |
| S1_k141_8955 | 3790 | No | NA | 1 | 0 | 1 | Not-determined | Genome-fragment | NA | NA | 0 |
| S1_k141_370747 | 1115 | No | NA | 1 | 0 | 0 | Not-determined | Genome-fragment | NA | NA | 0 |
| S4_k141_95147 | 1480 | No | NA | 2 | 0 | 1 | Low-quality | Genome-fragment | 0.71 | AAI-based (medium-confidence) | 0 |
| S8-final-viral-combined_k141_231764\|\|full | 1480 | No | NA | 3 | 0 | 0 | Low-quality | Genome-fragment | 3.41 | AAI-based (medium-confidence) | 0 |
| S1_k141_206968 | 5324 | No | NA | 5 | 0 | 1 | Not-determined | Genome-fragment | NA | NA | 0 |
| S1_k141_73747 | 1874 | No | NA | 3 | 0 | 2 | Not-determined | Genome-fragment | NA | NA | 0 |
| S1_k141_8998 | 3058 | No | NA | 3 | 0 | 3 | Not-determined | Genome-fragment | NA | NA | 0 |
| S1_k141_305822 | 2640 | No | NA | 1 | 0 | 1 | Not-determined | Genome-fragment | NA | NA | 0 |
| S1_k141_125031 | 1116 | No | NA | 1 | 0 | 1 | Not-determined | Genome-fragment | NA | NA | 0 |
| S1_k141_57924 | 1015 | No | NA | 2 | 0 | 2 | Not-determined | Genome-fragment | NA | NA | 0 |
| S1_k141_90974 | 1031 | No | NA | 2 | 0 | 0 | Not-determined | Genome-fragment | NA | NA | 0 |
| S1_k141_106579 | 4621 | No | NA | 3 | 0 | 0 | Not-determined | Genome-fragment | NA | NA | 0 |
| S1_k141_239545 | 1200 | No | NA | 2 | 0 | 0 | Not-determined | Genome-fragment | NA | NA | 0 |
| S1_k141_288652 | 1040 | No | NA | 2 | 0 | 0 | Not-determined | Genome-fragment | NA | NA | 0 |
| S8-final-viral-combined_k141_60347\|\|full | 1480 | No | NA | 4 | 0 | 1 | Low-quality | Genome-fragment | 2.75 | AAI-based (medium-confidence) | 0 |
| S1_k141_73834 | 1219 | No | NA | 2 | 0 | 1 | Not-determined | Genome-fragment | NA | NA | 0 |
| S1_k141_25184 | 1012 | No | NA | 1 | 0 | 1 | Not-determined | Genome-fragment | NA | NA | 0 |
| S1_k141_222845 | 1820 | No | NA | 5 | 0 | 4 | Not-determined | Genome-fragment | NA | NA | 0 |
| S1_k141_125103 | 1170 | No | NA | 2 | 0 | 0 | Not-determined | Genome-fragment | NA | NA | 0 |
| S1_k141_322616 | 1395 | No | NA | 1 | 0 | 0 | Not-determined | Genome-fragment | NA | NA | 0 |
| S8-final-viral-combined_k141_264487\|\|full | 1480 | No | NA | 2 | 0 | 0 | Low-quality | Genome-fragment | 2.22 | AAI-based (medium-confidence) | 0 |
| S1_k141_305911 | 1149 | No | NA | 1 | 0 | 1 | Not-determined | Genome-fragment | NA | NA | 0 |
| S1_k141_288699 | 1488 | No | NA | 2 | 0 | 1 | Not-determined | Genome-fragment | NA | NA | 0 |
| S1_k141_207046 | 1076 | No | NA | 2 | 0 | 1 | Not-determined | Genome-fragment | NA | NA | 0 |
| S1_k141_73859 | 1341 | No | NA | 3 | 0 | 0 | Not-determined | Genome-fragment | NA | NA | 0 |
| S1_k141_370915 | 1157 | No | NA | 3 | 0 | 0 | Not-determined | Genome-fragment | NA | NA | 0 |
| S1_k141_125128 | 1464 | No | NA | 1 | 0 | 1 | Not-determined | Genome-fragment | NA | NA | 0 |
| S1_k141_222898 | 1212 | No | NA | 3 | 0 | 0 | Not-determined | Genome-fragment | NA | NA | 0 |
| S8_k141_4615 | 1480 | No | NA | 3 | 1 | 0 | Low-quality | Genome-fragment | 2.37 | AAI-based (high-confidence) | 0 |
| S1_k141_272176 | 1431 | No | NA | 2 | 0 | 1 | Not-determined | Genome-fragment | NA | NA | 0 |
| S1_k141_58013 | 1182 | No | NA | 1 | 0 | 1 | Not-determined | Genome-fragment | NA | NA | 0 |
| S1_k141_25217 | 1505 | No | NA | 2 | 0 | 1 | Not-determined | Genome-fragment | NA | NA | 0 |
| S1_k141_256451 | 1253 | No | NA | 2 | 0 | 0 | Not-determined | Genome-fragment | NA | NA | 0 |
| S7-final-viral-combined_k141_13781\|\|full | 1481 | No | NA | 3 | 1 | 0 | Low-quality | Genome-fragment | 2.76 | AAI-based (high-confidence) | 0 |
| S17-final-viral-combined_k141_113623\|\|full | 1481 | No | NA | 4 | 1 | 0 | Low-quality | Genome-fragment | 2.52 | AAI-based (high-confidence) | 0 |
| S1_k141_156369 | 1034 | No | NA | 3 | 0 | 0 | Not-determined | Genome-fragment | NA | NA | 0 |
| S17-final-viral-combined_k141_251966\|\|full | 1481 | No | NA | 2 | 1 | 0 | Low-quality | Genome-fragment | 3.22 | AAI-based (medium-confidence) | 0 |
| S1_k141_125195 | 1521 | No | NA | 2 | 0 | 1 | Not-determined | Genome-fragment | NA | NA | 0 |
| S1_k141_106726 | 1315 | No | NA | 3 | 0 | 1 | Not-determined | Genome-fragment | NA | NA | 0 |
| S1_k141_387799 | 3676 | No | NA | 4 | 0 | 1 | Not-determined | Genome-fragment | NA | NA | 0 |
| S1_k141_239674 | 1384 | No | NA | 1 | 0 | 0 | Not-determined | Genome-fragment | NA | NA | 0 |
| S1_k141_207110 | 1326 | No | NA | 1 | 0 | 0 | Not-determined | Genome-fragment | NA | NA | 0 |
| S1_k141_190812 | 1412 | No | NA | 2 | 0 | 0 | Not-determined | Genome-fragment | NA | NA | 0 |
| S1_k141_256554 | 1692 | No | NA | 2 | 0 | 2 | Not-determined | Genome-fragment | NA | NA | 0 |
| S2_k141_127978 | 1481 | No | NA | 3 | 0 | 1 | Low-quality | Genome-fragment | 0.96 | AAI-based (high-confidence) | 0 |
| S1_k141_322729 | 1140 | No | NA | 1 | 0 | 0 | Not-determined | Genome-fragment | NA | NA | 0 |
| S1_k141_73961 | 1159 | No | NA | 2 | 0 | 0 | Not-determined | Genome-fragment | NA | NA | 0 |
| S1_k141_172518 | 1077 | No | NA | 1 | 0 | 1 | Not-determined | Genome-fragment | NA | NA | 0 |
| S1_k141_172528 | 3362 | No | NA | 4 | 0 | 3 | Not-determined | Genome-fragment | NA | NA | 0 |
| S1_k141_272270 | 1858 | No | NA | 1 | 0 | 1 | Not-determined | Genome-fragment | NA | NA | 0 |
| S1_k141_272272 | 1330 | No | NA | 1 | 0 | 0 | Not-determined | Genome-fragment | NA | NA | 0 |
| S1_k141_73986 | 2810 | No | NA | 1 | 0 | 1 | Not-determined | Genome-fragment | NA | NA | 0 |
| S1_k141_305996 | 1569 | No | NA | 1 | 0 | 1 | Not-determined | Genome-fragment | NA | NA | 0 |
| S1_k141_222958 | 1222 | No | NA | 2 | 0 | 0 | Not-determined | Genome-fragment | NA | NA | 0 |
| S8-final-viral-combined_k141_153736\|\|full | 1481 | No | NA | 4 | 1 | 0 | Low-quality | Genome-fragment | 2.39 | AAI-based (medium-confidence) | 0 |
| S1_k141_106752 | 1307 | No | NA | 1 | 0 | 1 | Not-determined | Genome-fragment | NA | NA | 0 |
| S8-final-viral-combined_k141_28866\|\|full | 1481 | No | NA | 3 | 0 | 0 | Low-quality | Genome-fragment | 2.67 | AAI-based (medium-confidence) | 0 |
| S11-final-viral-combined_k141_146725\|\|lt2gene | 1482 | No | NA | 1 | 1 | 0 | Low-quality | Genome-fragment | 1.94 | HMM-based (lower-bound) | 0 |
| S1_k141_41534 | 1029 | No | NA | 1 | 0 | 1 | Not-determined | Genome-fragment | NA | NA | 0 |
| S1_k141_272325 | 1576 | No | NA | 3 | 0 | 0 | Not-determined | Genome-fragment | NA | NA | 0 |
| S1-final-viral-combined_k141_4299\|\|full | 1482 | No | NA | 3 | 0 | 0 | Low-quality | Genome-fragment | 1.21 | AAI-based (high-confidence) | 0 |
| S1_k141_172591 | 1078 | No | NA | 1 | 0 | 1 | Not-determined | Genome-fragment | NA | NA | 0 |
| S1_k141_337422 | 1094 | No | NA | 2 | 0 | 1 | Not-determined | Genome-fragment | NA | NA | 0 |
| S1_k141_91234 | 1051 | No | NA | 2 | 0 | 2 | Not-determined | Genome-fragment | NA | NA | 0 |
| S1_k141_223034 | 1063 | No | NA | 3 | 0 | 0 | Not-determined | Genome-fragment | NA | NA | 0 |
| S1_k141_91241 | 1257 | No | NA | 1 | 0 | 1 | Not-determined | Genome-fragment | NA | NA | 0 |
| S1_k141_74036 | 3340 | No | NA | 4 | 0 | 1 | Not-determined | Genome-fragment | NA | NA | 0 |
| S1_k141_306084 | 1918 | No | NA | 1 | 0 | 1 | Not-determined | Genome-fragment | NA | NA | 0 |
| S1_k141_207244 | 1136 | No | NA | 1 | 0 | 1 | Not-determined | Genome-fragment | NA | NA | 0 |
| S1_k141_387910 | 7215 | No | NA | 4 | 0 | 3 | Not-determined | Genome-fragment | NA | NA | 0 |
| S1-final-viral-combined_k141_340191\|\|full | 1482 | No | NA | 3 | 0 | 0 | Low-quality | Genome-fragment | 3.13 | AAI-based (medium-confidence) | 0 |
| S1_k141_9327 | 1239 | No | NA | 2 | 0 | 1 | Not-determined | Genome-fragment | NA | NA | 0 |
| S1-final-viral-combined_k141_122299\|\|lt2gene | 1482 | No | NA | 2 | 1 | 0 | Low-quality | Genome-fragment | 3.29 | AAI-based (medium-confidence) | 0 |
| S1_k141_371121 | 4139 | No | NA | 2 | 0 | 1 | Not-determined | Genome-fragment | NA | NA | 0 |
| S1_k141_272400 | 1401 | No | NA | 1 | 0 | 0 | Not-determined | Genome-fragment | NA | NA | 0 |
| S1_k141_280898 | 1482 | No | NA | 2 | 0 | 0 | Low-quality | Genome-fragment | 3.6 | AAI-based (high-confidence) | 0 |
| S1_k141_190948 | 1069 | No | NA | 2 | 0 | 1 | Not-determined | Genome-fragment | NA | NA | 0 |
| S1_k141_25488 | 1501 | No | NA | 2 | 0 | 0 | Not-determined | Genome-fragment | NA | NA | 0 |
| S1_k141_140975 | 1380 | No | NA | 2 | 0 | 2 | Not-determined | Genome-fragment | NA | NA | 0 |
| S3_k141_134249 | 1482 | No | NA | 1 | 0 | 0 | Low-quality | Genome-fragment | 1.88 | AAI-based (medium-confidence) | 0 |
| S5-final-viral-combined_k141_77911\|\|full | 1482 | No | NA | 4 | 0 | 0 | Low-quality | Genome-fragment | 2.87 | AAI-based (medium-confidence) | 0 |
| S1_k141_239821 | 1011 | No | NA | 2 | 0 | 1 | Not-determined | Genome-fragment | NA | NA | 0 |
| S1_k141_207358 | 3856 | No | NA | 3 | 0 | 0 | Not-determined | Genome-fragment | NA | NA | 0 |
| S1_k141_106929 | 1192 | No | NA | 2 | 0 | 0 | Not-determined | Genome-fragment | NA | NA | 0 |
| S5_k141_208592 | 1482 | No | NA | 1 | 0 | 1 | Low-quality | Genome-fragment | 0.71 | AAI-based (medium-confidence) | 0 |
| S1_k141_125355 | 1115 | No | NA | 1 | 0 | 1 | Not-determined | Genome-fragment | NA | NA | 0 |
| S7-final-viral-combined_k141_40206\|\|full | 1482 | No | NA | 2 | 0 | 0 | Low-quality | Genome-fragment | 2.08 | AAI-based (high-confidence) | 0 |
| S1_k141_355256 | 1116 | No | NA | 2 | 0 | 0 | Not-determined | Genome-fragment | NA | NA | 0 |
| S17-final-viral-combined_k141_55769\|\|full | 1482 | No | NA | 3 | 1 | 0 | Low-quality | Genome-fragment | 1.19 | HMM-based (lower-bound) | 0 |
| S1_k141_191014 | 1157 | No | NA | 2 | 0 | 1 | Not-determined | Genome-fragment | NA | NA | 0 |
| S1_k141_337616 | 2548 | No | NA | 1 | 0 | 1 | Not-determined | Genome-fragment | NA | NA | 0 |
| S1_k141_91409 | 1218 | No | NA | 1 | 0 | 1 | Not-determined | Genome-fragment | NA | NA | 0 |
| S1_k141_172792 | 2121 | No | NA | 2 | 0 | 1 | Not-determined | Genome-fragment | NA | NA | 0 |
| S1_k141_288954 | 1548 | No | NA | 3 | 0 | 0 | Not-determined | Genome-fragment | NA | NA | 0 |
| S1_k141_239871 | 1039 | No | NA | 1 | 0 | 1 | Not-determined | Genome-fragment | NA | NA | 0 |
| S1_k141_207480 | 1039 | No | NA | 2 | 0 | 0 | Not-determined | Genome-fragment | NA | NA | 0 |
| S1_k141_371256 | 1039 | No | NA | 2 | 0 | 0 | Not-determined | Genome-fragment | NA | NA | 0 |
| S1_k141_141160 | 1684 | No | NA | 1 | 0 | 1 | Not-determined | Genome-fragment | NA | NA | 0 |
| S12-final-viral-combined_k141_343635\|\|full | 1482 | No | NA | 3 | 1 | 0 | Low-quality | Genome-fragment | 3.7 | AAI-based (medium-confidence) | 0 |
| S2-final-viral-combined_k141_60674\|\|full | 1482 | No | NA | 3 | 0 | 1 | Low-quality | Genome-fragment | 3.22 | AAI-based (medium-confidence) | 0 |
| S11-final-viral-combined_k141_195273\|\|full | 1483 | No | NA | 3 | 0 | 0 | Low-quality | Genome-fragment | 2.09 | AAI-based (high-confidence) | 0 |
| S1_k141_371273 | 1051 | No | NA | 2 | 0 | 1 | Not-determined | Genome-fragment | NA | NA | 0 |
| S11-final-viral-combined_k141_155774\|\|full | 1483 | No | NA | 2 | 0 | 0 | Low-quality | Genome-fragment | 2.62 | AAI-based (medium-confidence) | 0 |
| S1_k141_58342 | 1183 | No | NA | 2 | 0 | 0 | Not-determined | Genome-fragment | NA | NA | 0 |
| S1_k141_207507 | 2216 | No | NA | 3 | 0 | 2 | Not-determined | Genome-fragment | NA | NA | 0 |
| S1_k141_74275 | 2242 | No | NA | 3 | 0 | 2 | Not-determined | Genome-fragment | NA | NA | 0 |
| S1_k141_322996 | 2117 | No | NA | 2 | 0 | 2 | Not-determined | Genome-fragment | NA | NA | 0 |
| S1_k141_156695 | 1629 | No | NA | 2 | 0 | 1 | Not-determined | Genome-fragment | NA | NA | 0 |
| S1_k141_107027 | 2324 | No | NA | 3 | 0 | 0 | Not-determined | Genome-fragment | NA | NA | 0 |
| S1_k141_355457 | 1013 | No | NA | 2 | 0 | 1 | Not-determined | Genome-fragment | NA | NA | 0 |
| S1_k141_91595 | 1214 | No | NA | 1 | 0 | 1 | Not-determined | Genome-fragment | NA | NA | 0 |
| S11-final-viral-combined_k141_231498\|\|full | 1483 | No | NA | 2 | 0 | 1 | Low-quality | Genome-fragment | 0.95 | AAI-based (medium-confidence) | 0 |
| S1_k141_74329 | 1086 | No | NA | 1 | 0 | 1 | Not-determined | Genome-fragment | NA | NA | 0 |
| S1_k141_207582 | 2502 | No | NA | 3 | 0 | 2 | Not-determined | Genome-fragment | NA | NA | 0 |
| S1_k141_355494 | 3355 | No | NA | 2 | 0 | 1 | Not-determined | Genome-fragment | NA | NA | 0 |
| S1_k141_172935 | 1499 | No | NA | 1 | 0 | 1 | Not-determined | Genome-fragment | NA | NA | 0 |
| S11-final-viral-combined_k141_232305\|\|full | 1483 | No | NA | 4 | 0 | 2 | Low-quality | Genome-fragment | 0.4 | AAI-based (medium-confidence) | 0 |
| S1_k141_337651 | 1148 | No | NA | 1 | 0 | 1 | Not-determined | Genome-fragment | NA | NA | 0 |
| S1_k141_9547 | 1559 | No | NA | 2 | 0 | 1 | Not-determined | Genome-fragment | NA | NA | 0 |
| S1_k141_9575 | 1678 | No | NA | 2 | 0 | 1 | Not-determined | Genome-fragment | NA | NA | 0 |
| S1_k141_156772 | 1450 | No | NA | 1 | 0 | 1 | Not-determined | Genome-fragment | NA | NA | 0 |
| S1_k141_107118 | 2950 | No | NA | 4 | 0 | 2 | Not-determined | Genome-fragment | NA | NA | 0 |
| S1_k141_256991 | 1428 | No | NA | 2 | 0 | 1 | Not-determined | Genome-fragment | NA | NA | 0 |
| S1_k141_371383 | 1692 | No | NA | 2 | 0 | 1 | Not-determined | Genome-fragment | NA | NA | 0 |
| S1_k141_9594 | 1573 | No | NA | 1 | 0 | 0 | Not-determined | Genome-fragment | NA | NA | 0 |
| S1_k141_388137 | 1998 | No | NA | 2 | 0 | 2 | Not-determined | Genome-fragment | NA | NA | 0 |
| S1_k141_41848 | 1003 | No | NA | 1 | 0 | 1 | Not-determined | Genome-fragment | NA | NA | 0 |
| S1_k141_91637 | 3450 | No | NA | 1 | 0 | 1 | Not-determined | Genome-fragment | NA | NA | 0 |
| S1_k141_323078 | 1383 | No | NA | 2 | 0 | 1 | Not-determined | Genome-fragment | NA | NA | 0 |
| S1-final-viral-combined_k141_5235\|\|full | 1483 | Yes | 1217 | 3 | 1 | 1 | Low-quality | Genome-fragment | 1.38 | AAI-based (medium-confidence) | 17.94 |
| S1_k141_323082 | 3149 | No | NA | 2 | 0 | 2 | Not-determined | Genome-fragment | NA | NA | 0 |
| S5-final-viral-combined_k141_7180\|\|full | 1483 | No | NA | 2 | 1 | 0 | Low-quality | Genome-fragment | 3.8 | AAI-based (medium-confidence) | 0 |
| S1_k141_25702 | 1293 | No | NA | 1 | 0 | 1 | Not-determined | Genome-fragment | NA | NA | 0 |
| S1_k141_257035 | 1320 | No | NA | 3 | 0 | 1 | Not-determined | Genome-fragment | NA | NA | 0 |
| S1_k141_9663 | 8777 | No | NA | 7 | 0 | 5 | Not-determined | Genome-fragment | NA | NA | 0 |
| S1_k141_388205 | 1048 | No | NA | 1 | 0 | 0 | Not-determined | Genome-fragment | NA | NA | 0 |
| S1_k141_289195 | 1933 | No | NA | 4 | 0 | 0 | Not-determined | Genome-fragment | NA | NA | 0 |
| S1_k141_289196 | 1142 | No | NA | 1 | 0 | 1 | Not-determined | Genome-fragment | NA | NA | 0 |
| S1_k141_41918 | 4137 | No | NA | 3 | 0 | 2 | Not-determined | Genome-fragment | NA | NA | 0 |
| S5-final-viral-combined_k141_130470\|\|full | 1483 | No | NA | 3 | 0 | 0 | Low-quality | Genome-fragment | 3.19 | AAI-based (medium-confidence) | 0 |
| S5_k141_74763 | 1483 | No | NA | 2 | 0 | 0 | Low-quality | Genome-fragment | 2 | AAI-based (medium-confidence) | 0 |
| S1_k141_355675 | 2083 | No | NA | 2 | 0 | 2 | Not-determined | Genome-fragment | NA | NA | 0 |
| S17-final-viral-combined_k141_259285\|\|full | 1483 | No | NA | 2 | 0 | 0 | Low-quality | Genome-fragment | 2.39 | AAI-based (high-confidence) | 0 |
| S1_k141_91732 | 1035 | No | NA | 2 | 0 | 0 | Not-determined | Genome-fragment | NA | NA | 0 |
| S12-final-viral-combined_k141_114366\|\|full | 1483 | No | NA | 4 | 0 | 0 | Low-quality | Genome-fragment | 1.01 | AAI-based (medium-confidence) | 0 |
| S2-final-viral-combined_k141_125971\|\|full | 1483 | No | NA | 3 | 0 | 1 | Low-quality | Genome-fragment | 1.96 | AAI-based (high-confidence) | 0 |
| S1_k141_25780 | 3684 | No | NA | 1 | 0 | 1 | Not-determined | Genome-fragment | NA | NA | 0 |
| S1_k141_207672 | 3005 | No | NA | 3 | 0 | 1 | Not-determined | Genome-fragment | NA | NA | 0 |
| S1_k141_388292 | 3129 | No | NA | 8 | 0 | 0 | Medium-quality | Genome-fragment | 51.19 | AAI-based (high-confidence) | 0 |
| S1_k141_74530 | 1051 | No | NA | 1 | 0 | 1 | Not-determined | Genome-fragment | NA | NA | 0 |
| S1_k141_289208 | 3276 | No | NA | 3 | 0 | 3 | Not-determined | Genome-fragment | NA | NA | 0 |
| S2-final-viral-combined_k141_181770\|\|full | 1483 | No | NA | 3 | 0 | 0 | Low-quality | Genome-fragment | 2.62 | AAI-based (high-confidence) | 0 |
| S1_k141_257095 | 1027 | No | NA | 1 | 0 | 0 | Not-determined | Genome-fragment | NA | NA | 0 |
| S1_k141_306402 | 1113 | No | NA | 4 | 0 | 1 | Not-determined | Genome-fragment | NA | NA | 0 |
| S1_k141_58476 | 5638 | No | NA | 6 | 0 | 4 | Not-determined | Genome-fragment | NA | NA | 0 |
| S1_k141_306403 | 1850 | No | NA | 1 | 0 | 0 | Not-determined | Genome-fragment | NA | NA | 0 |
| S1_k141_289249 | 3509 | No | NA | 4 | 0 | 4 | Not-determined | Genome-fragment | NA | NA | 0 |
| S1_k141_257134 | 3141 | No | NA | 2 | 0 | 0 | Not-determined | Genome-fragment | NA | NA | 0 |
| S1_k141_257143 | 1005 | No | NA | 1 | 0 | 1 | Not-determined | Genome-fragment | NA | NA | 0 |
| S1_k141_173160 | 1067 | No | NA | 1 | 0 | 0 | Not-determined | Genome-fragment | NA | NA | 0 |
| S1_k141_141458 | 1087 | No | NA | 2 | 0 | 0 | Not-determined | Genome-fragment | NA | NA | 0 |
| S1_k141_191366 | 3662 | No | NA | 4 | 0 | 4 | Not-determined | Genome-fragment | NA | NA | 0 |
| S1_k141_25921 | 1753 | No | NA | 1 | 0 | 1 | Not-determined | Genome-fragment | NA | NA | 0 |
| S1_k141_9923 | 3088 | No | NA | 4 | 0 | 2 | Not-determined | Genome-fragment | NA | NA | 0 |
| S1_k141_25953 | 3215 | No | NA | 3 | 0 | 2 | Not-determined | Genome-fragment | NA | NA | 0 |
| S1_k141_141492 | 3183 | No | NA | 2 | 0 | 1 | Not-determined | Genome-fragment | NA | NA | 0 |
| S1_k141_306561 | 1332 | No | NA | 3 | 0 | 0 | Not-determined | Genome-fragment | NA | NA | 0 |
| S1_k141_58596 | 1275 | No | NA | 1 | 0 | 1 | Not-determined | Genome-fragment | NA | NA | 0 |
| S1_k141_223414 | 3342 | No | NA | 5 | 0 | 0 | Not-determined | Genome-fragment | NA | NA | 0 |
| S1_k141_42112 | 1018 | No | NA | 2 | 0 | 1 | Not-determined | Genome-fragment | NA | NA | 0 |
| S1_k141_125683 | 1132 | No | NA | 2 | 0 | 1 | Not-determined | Genome-fragment | NA | NA | 0 |
| S6-final-viral-combined_k141_64198\|\|full | 1483 | No | NA | 3 | 1 | 0 | Low-quality | Genome-fragment | 1.94 | HMM-based (lower-bound) | 0 |
| S6_k141_77104 | 1483 | No | NA | 4 | 0 | 0 | Low-quality | Genome-fragment | 1.37 | AAI-based (high-confidence) | 0 |
| S1_k141_306588 | 1071 | No | NA | 1 | 0 | 1 | Not-determined | Genome-fragment | NA | NA | 0 |
| S1_k141_9977 | 1154 | No | NA | 2 | 0 | 0 | Not-determined | Genome-fragment | NA | NA | 0 |
| S1_k141_223487 | 3234 | No | NA | 3 | 0 | 2 | Not-determined | Genome-fragment | NA | NA | 0 |
| S1_k141_157028 | 1208 | No | NA | 3 | 0 | 2 | Not-determined | Genome-fragment | NA | NA | 0 |
| S1_k141_107506 | 1128 | No | NA | 2 | 0 | 0 | Not-determined | Genome-fragment | NA | NA | 0 |
| S1-final-viral-combined_k141_18251\|\|full | 1484 | No | NA | 3 | 1 | 0 | Low-quality | Genome-fragment | 42.99 | AAI-based (medium-confidence) | 0 |
| S1_k141_240361 | 1128 | No | NA | 3 | 0 | 2 | Not-determined | Genome-fragment | NA | NA | 0 |
| S1_k141_91966 | 1395 | No | NA | 1 | 0 | 1 | Not-determined | Genome-fragment | NA | NA | 0 |
| S1_k141_240383 | 1109 | No | NA | 1 | 0 | 1 | Not-determined | Genome-fragment | NA | NA | 0 |
| S1_k141_134637 | 1484 | No | NA | 4 | 0 | 0 | Low-quality | Genome-fragment | 3.22 | AAI-based (high-confidence) | 0 |
| S1_k141_323487 | 1775 | No | NA | 3 | 0 | 3 | Not-determined | Genome-fragment | NA | NA | 0 |
| S1_k141_208064 | 5094 | No | NA | 5 | 0 | 3 | Not-determined | Genome-fragment | NA | NA | 0 |
| S7-final-viral-combined_k141_167095\|\|full | 1484 | No | NA | 4 | 0 | 0 | Low-quality | Genome-fragment | 2.91 | AAI-based (medium-confidence) | 0 |
| S1_k141_157105 | 1255 | No | NA | 2 | 0 | 0 | Not-determined | Genome-fragment | NA | NA | 0 |
| S1_k141_289593 | 1319 | No | NA | 1 | 0 | 0 | Not-determined | Genome-fragment | NA | NA | 0 |
| S1_k141_191613 | 1190 | No | NA | 1 | 0 | 1 | Not-determined | Genome-fragment | NA | NA | 0 |
| S7-final-viral-combined_k141_240531\|\|lt2gene | 1484 | No | NA | 1 | 1 | 0 | Low-quality | Genome-fragment | 1.39 | HMM-based (lower-bound) | 0 |
| S1_k141_223605 | 1323 | No | NA | 1 | 0 | 1 | Not-determined | Genome-fragment | NA | NA | 0 |
| S1_k141_157173 | 1172 | No | NA | 1 | 0 | 1 | Not-determined | Genome-fragment | NA | NA | 0 |
| S12-final-viral-combined_k141_109501\|\|full | 1484 | No | NA | 5 | 0 | 0 | Low-quality | Genome-fragment | 0.86 | AAI-based (medium-confidence) | 0 |
| S1_k141_208136 | 1001 | No | NA | 1 | 0 | 1 | Not-determined | Genome-fragment | NA | NA | 0 |
| S2-final-viral-combined_k141_111915\|\|full | 1484 | No | NA | 5 | 1 | 0 | Low-quality | Genome-fragment | 2.61 | AAI-based (medium-confidence) | 0 |
| S1_k141_42248 | 1028 | No | NA | 2 | 0 | 0 | Not-determined | Genome-fragment | NA | NA | 0 |
| S1_k141_257394 | 1276 | No | NA | 1 | 0 | 1 | Not-determined | Genome-fragment | NA | NA | 0 |
| S1_k141_173357 | 1633 | No | NA | 4 | 0 | 0 | Not-determined | Genome-fragment | NA | NA | 0 |
| S1_k141_42290 | 4600 | No | NA | 1 | 0 | 1 | Not-determined | Genome-fragment | NA | NA | 0 |
| S1_k141_223620 | 31814 | No | NA | 40 | 15 | 0 | High-quality | High-quality | 96.36 | AAI-based (high-confidence) | 0 |
| S1_k141_223643 | 3175 | No | NA | 4 | 0 | 1 | Not-determined | Genome-fragment | NA | NA | 0 |
| S1_k141_92117 | 2526 | No | NA | 2 | 0 | 2 | Not-determined | Genome-fragment | NA | NA | 0 |
| S1_k141_208193 | 1187 | No | NA | 1 | 0 | 1 | Not-determined | Genome-fragment | NA | NA | 0 |
| S1_k141_58830 | 3914 | No | NA | 6 | 0 | 0 | Not-determined | Genome-fragment | NA | NA | 0 |
| S1-final-viral-combined_k141_116860\|\|full | 1485 | No | NA | 4 | 0 | 0 | Low-quality | Genome-fragment | 3.62 | AAI-based (medium-confidence) | 0 |
| S1_k141_157271 | 1818 | No | NA | 1 | 0 | 1 | Not-determined | Genome-fragment | NA | NA | 0 |
| S1_k141_26266 | 1552 | No | NA | 1 | 0 | 1 | Not-determined | Genome-fragment | NA | NA | 0 |
| S1_k141_323641 | 1534 | No | NA | 1 | 0 | 0 | Not-determined | Genome-fragment | NA | NA | 0 |
| S1_k141_240529 | 1497 | No | NA | 1 | 0 | 1 | Not-determined | Genome-fragment | NA | NA | 0 |
| S1_k141_107682 | 1457 | No | NA | 2 | 0 | 0 | Not-determined | Genome-fragment | NA | NA | 0 |
| S1-final-viral-combined_k141_333605\|\|full | 1485 | No | NA | 2 | 0 | 2 | Low-quality | Genome-fragment | 2.16 | AAI-based (high-confidence) | 0 |
| S1_k141_356094 | 1260 | No | NA | 2 | 0 | 1 | Not-determined | Genome-fragment | NA | NA | 0 |
| S1_k141_306879 | 1017 | No | NA | 2 | 0 | 1 | Not-determined | Genome-fragment | NA | NA | 0 |
| S1_k141_157322 | 1479 | No | NA | 2 | 0 | 2 | Not-determined | Genome-fragment | NA | NA | 0 |
| S1_k141_248018 | 1485 | No | NA | 3 | 1 | 0 | Low-quality | Genome-fragment | 5.09 | HMM-based (lower-bound) | 0 |
| S1_k141_223682 | 10659 | No | NA | 7 | 0 | 5 | Not-determined | Genome-fragment | NA | NA | 0 |
| S1_k141_289730 | 1715 | No | NA | 1 | 0 | 0 | Not-determined | Genome-fragment | NA | NA | 0 |
| S1_k141_107726 | 3387 | No | NA | 1 | 0 | 1 | Not-determined | Genome-fragment | NA | NA | 0 |
| S1_k141_42385 | 1483 | No | NA | 1 | 0 | 1 | Not-determined | Genome-fragment | NA | NA | 0 |
| S5-final-viral-combined_k141_108018\|\|full | 1485 | No | NA | 4 | 0 | 1 | Low-quality | Genome-fragment | 3.3 | AAI-based (medium-confidence) | 0 |
| S1_k141_26335 | 3465 | No | NA | 3 | 0 | 3 | Not-determined | Genome-fragment | NA | NA | 0 |
| S1_k141_26350 | 1108 | No | NA | 1 | 0 | 0 | Not-determined | Genome-fragment | NA | NA | 0 |
| S1_k141_208307 | 3094 | No | NA | 4 | 0 | 0 | Not-determined | Genome-fragment | NA | NA | 0 |
| S1_k141_74789 | 1266 | No | NA | 1 | 0 | 0 | Not-determined | Genome-fragment | NA | NA | 0 |
| S1_k141_289757 | 1236 | No | NA | 1 | 0 | 1 | Not-determined | Genome-fragment | NA | NA | 0 |
| S5-final-viral-combined_k141_150713\|\|full | 1485 | No | NA | 4 | 0 | 0 | Low-quality | Genome-fragment | 3.54 | AAI-based (high-confidence) | 0 |
| S1_k141_42425 | 1182 | No | NA | 2 | 0 | 1 | Not-determined | Genome-fragment | NA | NA | 0 |
| S1_k141_141629 | 1631 | No | NA | 1 | 0 | 1 | Not-determined | Genome-fragment | NA | NA | 0 |
| S1_k141_371835 | 1330 | No | NA | 2 | 0 | 1 | Not-determined | Genome-fragment | NA | NA | 0 |
| S17-final-viral-combined_k141_224926\|\|full | 1485 | No | NA | 3 | 0 | 0 | Low-quality | Genome-fragment | 3.6 | AAI-based (high-confidence) | 0 |
| S1_k141_240622 | 1683 | No | NA | 1 | 0 | 0 | Not-determined | Genome-fragment | NA | NA | 0 |
| S1_k141_273080 | 1752 | No | NA | 2 | 0 | 2 | Not-determined | Genome-fragment | NA | NA | 0 |
| S1_k141_208338 | 1023 | No | NA | 1 | 0 | 1 | Not-determined | Genome-fragment | NA | NA | 0 |
| S1_k141_157417 | 1595 | No | NA | 1 | 0 | 1 | Not-determined | Genome-fragment | NA | NA | 0 |
| S12-final-viral-combined_k141_400839\|\|full | 1485 | No | NA | 3 | 0 | 0 | Low-quality | Genome-fragment | 3.08 | AAI-based (medium-confidence) | 0 |

| | | | | | | | | | | | |
|---|---|---|---|---|---|---|---|---|---|---|---|
| S1_k141_208343 | 3119 | No | NA | 5 | 0 | 1 | Not-determined | Genome-fragment | NA | NA | 0 |
| S1_k141_306977 | 1129 | No | NA | 1 | 0 | 1 | Not-determined | Genome-fragment | NA | NA | 0 |
| S1_k141_58992 | 1046 | No | NA | 1 | 0 | 1 | Not-determined | Genome-fragment | NA | NA | 0 |
| S1_k141_42475 | 5283 | No | NA | 1 | 0 | 1 | Not-determined | Genome-fragment | NA | NA | 0 |
| S2_k141_115289 | 1485 | No | NA | 2 | 0 | 1 | Low-quality | Genome-fragment | 0.89 | AAI-based (medium-confidence) | 0 |
| S1_k141_42482 | 8193 | No | NA | 7 | 0 | 0 | Not-determined | Genome-fragment | NA | NA | 0 |
| S1_k141_157462 | 1148 | No | NA | 1 | 0 | 1 | Not-determined | Genome-fragment | NA | NA | 0 |
| S1_k141_141757 | 3622 | No | NA | 3 | 0 | 2 | Not-determined | Genome-fragment | NA | NA | 0 |
| S1_k141_223868 | 1290 | No | NA | 1 | 0 | 1 | Not-determined | Genome-fragment | NA | NA | 0 |
| S1_k141_356304 | 1154 | No | NA | 1 | 0 | 0 | Not-determined | Genome-fragment | NA | NA | 0 |
| S1_k141_371966 | 1090 | No | NA | 2 | 0 | 0 | Not-determined | Genome-fragment | NA | NA | 0 |
| S1_k141_26400 | 1066 | No | NA | 1 | 0 | 0 | Not-determined | Genome-fragment | NA | NA | 0 |
| S1_k141_157496 | 3001 | No | NA | 2 | 0 | 1 | Not-determined | Genome-fragment | NA | NA | 0 |
| S9-final-viral-combined_k141_71998\|\|full | 1486 | No | NA | 3 | 0 | 2 | Low-quality | Genome-fragment | 1.2 | AAI-based (medium-confidence) | 0 |
| S1_k141_338427 | 1135 | No | NA | 2 | 0 | 1 | Not-determined | Genome-fragment | NA | NA | 0 |
| S1_k141_323844 | 2260 | No | NA | 1 | 0 | 1 | Not-determined | Genome-fragment | NA | NA | 0 |
| S9_k141_105694 | 1486 | No | NA | 2 | 0 | 1 | Low-quality | Genome-fragment | 0.95 | AAI-based (medium-confidence) | 0 |
| S1_k141_323856 | 1321 | No | NA | 2 | 0 | 0 | Not-determined | Genome-fragment | NA | NA | 0 |
| S1_k141_257772 | 1107 | No | NA | 1 | 0 | 1 | Not-determined | Genome-fragment | NA | NA | 0 |
| S1_k141_372014 | 1848 | No | NA | 1 | 0 | 1 | Not-determined | Genome-fragment | NA | NA | 0 |
| S1_k141_173555 | 1307 | No | NA | 2 | 0 | 2 | Not-determined | Genome-fragment | NA | NA | 0 |
| S1_k141_289967 | 3525 | No | NA | 4 | 0 | 1 | Not-determined | Genome-fragment | NA | NA | 0 |
| S1_k141_240790 | 2960 | No | NA | 1 | 0 | 1 | Not-determined | Genome-fragment | NA | NA | 0 |
| S1_k141_157576 | 1046 | No | NA | 1 | 0 | 0 | Not-determined | Genome-fragment | NA | NA | 0 |
| S1_k141_356368 | 4225 | No | NA | 6 | 0 | 5 | Not-determined | Genome-fragment | NA | NA | 0 |
| S1-final-viral-combined_k141_264947\|\|full | 1486 | No | NA | 3 | 1 | 0 | Low-quality | Genome-fragment | 2.75 | AAI-based (high-confidence) | 0 |
| S1_k141_92459 | 6325 | No | NA | 1 | 0 | 1 | Not-determined | Genome-fragment | NA | NA | 0 |
| S1-final-viral-combined_k141_328153\|\|full | 1486 | No | NA | 4 | 1 | 0 | Low-quality | Genome-fragment | 11.52 | AAI-based (medium-confidence) | 0 |
| S1-final-viral-combined_k141_81669\|\|full | 1486 | No | NA | 3 | 1 | 0 | Low-quality | Genome-fragment | 3.77 | AAI-based (high-confidence) | 0 |
| S12-final-viral-combined_k141_3675\|\|full | 1486 | No | NA | 2 | 0 | 0 | Low-quality | Genome-fragment | 0.93 | AAI-based (medium-confidence) | 0 |
| S1_k141_323949 | 1176 | No | NA | 2 | 0 | 2 | Not-determined | Genome-fragment | NA | NA | 0 |
| S1_k141_26472 | 1024 | No | NA | 1 | 0 | 0 | Not-determined | Genome-fragment | NA | NA | 0 |
| S2-final-viral-combined_k141_176727\|\|full | 1486 | No | NA | 5 | 2 | 0 | Low-quality | Genome-fragment | 3.3 | AAI-based (high-confidence) | 0 |
| S1_k141_26481 | 1001 | No | NA | 3 | 0 | 3 | Not-determined | Genome-fragment | NA | NA | 0 |
| S1_k141_75051 | 1656 | No | NA | 3 | 0 | 0 | Not-determined | Genome-fragment | NA | NA | 0 |
| S1_k141_290068 | 1210 | No | NA | 2 | 0 | 0 | Not-determined | Genome-fragment | NA | NA | 0 |
| S1_k141_10353 | 1124 | No | NA | 1 | 0 | 1 | Not-determined | Genome-fragment | NA | NA | 0 |
| S1_k141_141935 | 1159 | No | NA | 1 | 0 | 1 | Not-determined | Genome-fragment | NA | NA | 0 |
| S1_k141_307219 | 1142 | No | NA | 1 | 0 | 0 | Not-determined | Genome-fragment | NA | NA | 0 |
| S1_k141_356427 | 1202 | No | NA | 2 | 0 | 0 | Not-determined | Genome-fragment | NA | NA | 0 |
| S1_k141_26531 | 1069 | No | NA | 1 | 0 | 1 | Not-determined | Genome-fragment | NA | NA | 0 |
| S1_k141_173711 | 1203 | No | NA | 2 | 0 | 1 | Not-determined | Genome-fragment | NA | NA | 0 |
| S1_k141_157678 | 1484 | No | NA | 1 | 0 | 1 | Not-determined | Genome-fragment | NA | NA | 0 |
| S1_k141_141947 | 1925 | No | NA | 1 | 0 | 1 | Not-determined | Genome-fragment | NA | NA | 0 |
| S1_k141_224118 | 1121 | No | NA | 1 | 0 | 0 | Not-determined | Genome-fragment | NA | NA | 0 |
| S1_k141_75122 | 3142 | No | NA | 2 | 0 | 2 | Not-determined | Genome-fragment | NA | NA | 0 |
| S1_k141_356460 | 1037 | No | NA | 1 | 0 | 0 | Not-determined | Genome-fragment | NA | NA | 0 |
| S2_k141_72586 | 1486 | No | NA | 2 | 1 | 0 | Low-quality | Genome-fragment | 2.13 | AAI-based (high-confidence) | 0 |
| S1_k141_324088 | 1397 | No | NA | 2 | 0 | 0 | Not-determined | Genome-fragment | NA | NA | 0 |
| S8-final-viral-combined_k141_81964\|\|full | 1486 | No | NA | 3 | 0 | 0 | Low-quality | Genome-fragment | 2.81 | AAI-based (medium-confidence) | 0 |
| S8-final-viral-combined_k141_2539\|\|full | 1486 | No | NA | 4 | 1 | 0 | Low-quality | Genome-fragment | 1.13 | AAI-based (high-confidence) | 0 |
| S8-final-viral-combined_k141_200646\|\|full | 1486 | No | NA | 2 | 0 | 1 | Low-quality | Genome-fragment | 3.63 | AAI-based (high-confidence) | 0 |
| S1_k141_307304 | 1272 | No | NA | 2 | 0 | 2 | Not-determined | Genome-fragment | NA | NA | 0 |
| S1_k141_356557 | 1299 | No | NA | 2 | 0 | 1 | Not-determined | Genome-fragment | NA | NA | 0 |
| S1_k141_75214 | 2137 | No | NA | 1 | 0 | 0 | Not-determined | Genome-fragment | NA | NA | 0 |
| S1_k141_59226 | 1260 | No | NA | 3 | 0 | 0 | Not-determined | Genome-fragment | NA | NA | 0 |
| S9-final-viral-combined_k141_43150\|\|lt2gene | 1487 | No | NA | 2 | 1 | 0 | Low-quality | Genome-fragment | 3.86 | AAI-based (high-confidence) | 0 |
| S1_k141_142071 | 1000 | No | NA | 1 | 0 | 1 | Not-determined | Genome-fragment | NA | NA | 0 |
| S1_k141_92656 | 1193 | No | NA | 1 | 0 | 1 | Not-determined | Genome-fragment | NA | NA | 0 |
| S1_k141_224204 | 1811 | No | NA | 1 | 0 | 1 | Not-determined | Genome-fragment | NA | NA | 0 |
| S1_k141_92661 | 1109 | No | NA | 1 | 0 | 0 | Not-determined | Genome-fragment | NA | NA | 0 |
| S1_k141_108097 | 3945 | No | NA | 1 | 0 | 0 | Not-determined | Genome-fragment | NA | NA | 0 |
| S5-final-viral-combined_k141_136357\|\|full | 1487 | No | NA | 3 | 0 | 0 | Low-quality | Genome-fragment | 2.06 | AAI-based (high-confidence) | 0 |
| S1_k141_75230 | 1122 | No | NA | 1 | 0 | 1 | Not-determined | Genome-fragment | NA | NA | 0 |
| S7_k141_210657 | 1487 | No | NA | 2 | 0 | 0 | Low-quality | Genome-fragment | 2.93 | AAI-based (medium-confidence) | 0 |
| S1_k141_192053 | 3034 | No | NA | 2 | 0 | 1 | Not-determined | Genome-fragment | NA | NA | 0 |
| S7_k141_190897 | 1487 | No | NA | 1 | 0 | 0 | Low-quality | Genome-fragment | 1.45 | AAI-based (high-confidence) | 0 |
| S1_k141_324203 | 1187 | No | NA | 1 | 0 | 1 | Not-determined | Genome-fragment | NA | NA | 0 |
| S1_k141_75247 | 1329 | No | NA | 2 | 0 | 2 | Not-determined | Genome-fragment | NA | NA | 0 |
| S1_k141_42837 | 1104 | No | NA | 1 | 0 | 1 | Not-determined | Genome-fragment | NA | NA | 0 |
| S1_k141_92721 | 1174 | No | NA | 2 | 0 | 1 | Not-determined | Genome-fragment | NA | NA | 0 |
| S1_k141_273434 | 1770 | No | NA | 1 | 0 | 1 | Not-determined | Genome-fragment | NA | NA | 0 |
| S1_k141_356613 | 1038 | No | NA | 1 | 0 | 0 | Not-determined | Genome-fragment | NA | NA | 0 |
| S1_k141_142140 | 1964 | No | NA | 4 | 0 | 2 | Not-determined | Genome-fragment | NA | NA | 0 |
| S1_k141_324248 | 1018 | No | NA | 1 | 0 | 1 | Not-determined | Genome-fragment | NA | NA | 0 |
| S1_k141_192117 | 4494 | No | NA | 1 | 0 | 1 | Not-determined | Genome-fragment | NA | NA | 0 |
| S1_k141_126468 | 6594 | No | NA | 5 | 0 | 5 | Not-determined | Genome-fragment | NA | NA | 0 |
| S1_k141_42911 | 1071 | No | NA | 1 | 0 | 1 | Not-determined | Genome-fragment | NA | NA | 0 |
| S17-final-viral-combined_k141_35700\|\|full | 1487 | No | NA | 2 | 0 | 2 | Low-quality | Genome-fragment | 1.7 | AAI-based (medium-confidence) | 0 |
| S1_k141_241091 | 1085 | No | NA | 2 | 0 | 2 | Not-determined | Genome-fragment | NA | NA | 0 |
| S17-final-viral-combined_k141_48656\|\|full | 1487 | No | NA | 3 | 0 | 0 | Low-quality | Genome-fragment | 3.15 | AAI-based (medium-confidence) | 0 |
| S1_k141_192164 | 1020 | No | NA | 2 | 0 | 2 | Not-determined | Genome-fragment | NA | NA | 0 |
| S17-final-viral-combined_k141_7838\|\|full | 1487 | Yes | 423 | 2 | 1 | 1 | Low-quality | Genome-fragment | 0.2 | HMM-based (lower-bound) | 71.55 |
| S1_k141_241131 | 3465 | No | NA | 3 | 0 | 3 | Not-determined | Genome-fragment | NA | NA | 0 |
| S1_k141_92809 | 10998 | No | NA | 12 | 0 | 8 | Not-determined | Genome-fragment | NA | NA | 0 |
| S1_k141_142238 | 1605 | No | NA | 1 | 0 | 1 | Not-determined | Genome-fragment | NA | NA | 0 |
| S17-final-viral-combined_k141_252727\|\|full | 1487 | No | NA | 2 | 2 | 0 | Low-quality | Genome-fragment | 3.18 | AAI-based (high-confidence) | 0 |
| S1_k141_92852 | 1155 | No | NA | 3 | 0 | 1 | Not-determined | Genome-fragment | NA | NA | 0 |
| S12-final-viral-combined_k141_49238\|\|full | 1487 | No | NA | 2 | 0 | 2 | Low-quality | Genome-fragment | 2.49 | AAI-based (medium-confidence) | 0 |
| S1_k141_388771 | 2977 | No | NA | 5 | 0 | 0 | Not-determined | Genome-fragment | NA | NA | 0 |
| S1_k141_388774 | 1210 | No | NA | 3 | 0 | 0 | Not-determined | Genome-fragment | NA | NA | 0 |
| S1_k141_356733 | 4811 | No | NA | 4 | 0 | 2 | Not-determined | Genome-fragment | NA | NA | 0 |
| S1_k141_208811 | 1114 | No | NA | 2 | 0 | 1 | Not-determined | Genome-fragment | NA | NA | 0 |
| S1_k141_208814 | 1729 | No | NA | 4 | 0 | 0 | Not-determined | Genome-fragment | NA | NA | 0 |
| S1_k141_273527 | 2210 | No | NA | 4 | 0 | 0 | Not-determined | Genome-fragment | NA | NA | 0 |
| S1_k141_92901 | 3381 | No | NA | 5 | 0 | 4 | Not-determined | Genome-fragment | NA | NA | 0 |
| S1_k141_10426 | 2939 | No | NA | 3 | 0 | 2 | Not-determined | Genome-fragment | NA | NA | 0 |
| S1_k141_208863 | 1044 | No | NA | 1 | 0 | 1 | Not-determined | Genome-fragment | NA | NA | 0 |
| S1_k141_157984 | 3441 | No | NA | 4 | 0 | 1 | Not-determined | Genome-fragment | NA | NA | 0 |
| S2_k141_213031 | 1487 | No | NA | 1 | 0 | 1 | Low-quality | Genome-fragment | 1.76 | AAI-based (high-confidence) | 0 |
| S1_k141_241248 | 2422 | No | NA | 2 | 0 | 2 | Not-determined | Genome-fragment | NA | NA | 0 |
| S1_k141_241263 | 2132 | No | NA | 1 | 0 | 1 | Not-determined | Genome-fragment | NA | NA | 0 |
| S1_k141_241267 | 1336 | No | NA | 1 | 0 | 1 | Not-determined | Genome-fragment | NA | NA | 0 |
| S1_k141_75381 | 1077 | No | NA | 1 | 0 | 1 | Not-determined | Genome-fragment | NA | NA | 0 |
| S1_k141_208875 | 18901 | No | NA | 22 | 3 | 0 | Medium-quality | Genome-fragment | 50.43 | AAI-based (high-confidence) | 0 |
| S1_k141_290405 | 1022 | No | NA | 1 | 0 | 1 | Not-determined | Genome-fragment | NA | NA | 0 |
| S1_k141_43071 | 1237 | No | NA | 1 | 0 | 1 | Not-determined | Genome-fragment | NA | NA | 0 |
| S1_k141_43079 | 3362 | No | NA | 4 | 0 | 3 | Not-determined | Genome-fragment | NA | NA | 0 |
| S6_k141_122835 | 1487 | No | NA | 1 | 0 | 0 | Low-quality | Genome-fragment | 1.05 | AAI-based (medium-confidence) | 0 |
| S8-final-viral-combined_k141_195921\|\|full | 1487 | No | NA | 3 | 0 | 1 | Low-quality | Genome-fragment | 1.67 | AAI-based (medium-confidence) | 0 |
| S1_k141_224464 | 1013 | No | NA | 3 | 0 | 2 | Not-determined | Genome-fragment | NA | NA | 0 |
| S1_k141_388864 | 1084 | No | NA | 2 | 0 | 2 | Not-determined | Genome-fragment | NA | NA | 0 |
| S1_k141_273594 | 1873 | No | NA | 2 | 0 | 0 | Not-determined | Genome-fragment | NA | NA | 0 |
| S1_k141_158041 | 1064 | No | NA | 2 | 0 | 1 | Not-determined | Genome-fragment | NA | NA | 0 |
| S1_k141_241333 | 1411 | No | NA | 1 | 0 | 1 | Not-determined | Genome-fragment | NA | NA | 0 |
| S1_k141_59568 | 4455 | No | NA | 3 | 0 | 3 | Not-determined | Genome-fragment | NA | NA | 0 |
| S1_k141_93004 | 1013 | No | NA | 2 | 0 | 1 | Not-determined | Genome-fragment | NA | NA | 0 |
| S1_k141_356819 | 1005 | No | NA | 1 | 0 | 0 | Not-determined | Genome-fragment | NA | NA | 0 |
| S1_k141_290442 | 1285 | No | NA | 4 | 0 | 0 | Not-determined | Genome-fragment | NA | NA | 0 |
| S1_k141_174152 | 4004 | No | NA | 5 | 0 | 2 | Not-determined | Genome-fragment | NA | NA | 0 |
| S1_k141_290464 | 1282 | No | NA | 4 | 0 | 0 | Not-determined | Genome-fragment | NA | NA | 0 |
| S8-final-viral-combined_k141_236609\|\|full | 1487 | No | NA | 2 | 0 | 0 | Low-quality | Genome-fragment | 0.97 | AAI-based (high-confidence) | 0 |
| S1_k141_356859 | 3153 | No | NA | 3 | 0 | 0 | Not-determined | Genome-fragment | NA | NA | 0 |
| S8_k141_253395 | 1487 | No | NA | 1 | 0 | 1 | Low-quality | Genome-fragment | 0.52 | AAI-based (medium-confidence) | 0 |
| S1_k141_126735 | 3038 | No | NA | 2 | 0 | 1 | Not-determined | Genome-fragment | NA | NA | 0 |
| S9-final-viral-combined_k141_10324\|\|full | 1488 | No | NA | 3 | 2 | 0 | Low-quality | Genome-fragment | 3.35 | AAI-based (medium-confidence) | 0 |
| S1_k141_43186 | 3920 | No | NA | 5 | 0 | 0 | Not-determined | Genome-fragment | NA | NA | 0 |
| S1_k141_75498 | 1208 | No | NA | 1 | 0 | 0 | Not-determined | Genome-fragment | NA | NA | 0 |
| S1_k141_192409 | 1024 | No | NA | 1 | 0 | 1 | Not-determined | Genome-fragment | NA | NA | 0 |
| S1_k141_43187 | 10589 | No | NA | 10 | 0 | 10 | Not-determined | Genome-fragment | NA | NA | 0 |
| S1_k141_174210 | 1444 | No | NA | 5 | 0 | 5 | Not-determined | Genome-fragment | NA | NA | 0 |
| S1-final-viral-combined_k141_87181\|\|full | 1488 | No | NA | 2 | 1 | 0 | Low-quality | Genome-fragment | 4.38 | AAI-based (high-confidence) | 0 |
| S1_k141_43205 | 1390 | No | NA | 1 | 0 | 0 | Not-determined | Genome-fragment | NA | NA | 0 |
| S1_k141_26850 | 5057 | No | NA | 1 | 0 | 1 | Not-determined | Genome-fragment | NA | NA | 0 |
| S1_k141_10635 | 1461 | No | NA | 3 | 0 | 0 | Not-determined | Genome-fragment | NA | NA | 0 |
| S1_k141_388936 | 1248 | No | NA | 1 | 0 | 0 | Not-determined | Genome-fragment | NA | NA | 0 |
| S3_k141_92635 | 1488 | No | NA | 2 | 0 | 1 | Low-quality | Genome-fragment | 0.97 | AAI-based (high-confidence) | 0 |
| S1_k141_372652 | 1626 | No | NA | 1 | 0 | 0 | Not-determined | Genome-fragment | NA | NA | 0 |
| S1_k141_126824 | 1267 | No | NA | 2 | 0 | 0 | Not-determined | Genome-fragment | NA | NA | 0 |
| S1_k141_388959 | 2065 | No | NA | 1 | 0 | 0 | Not-determined | Genome-fragment | NA | NA | 0 |
| S1_k141_224579 | 11026 | No | NA | 9 | 0 | 7 | Not-determined | Genome-fragment | NA | NA | 0 |
| S1_k141_93131 | 13737 | No | NA | 4 | 0 | 3 | Not-determined | Genome-fragment | NA | NA | 0 |
| S5_k141_39897 | 1488 | No | NA | 6 | 0 | 0 | Low-quality | Genome-fragment | 2.76 | AAI-based (medium-confidence) | 0 |
| S1_k141_209086 | 1433 | No | NA | 2 | 0 | 1 | Not-determined | Genome-fragment | NA | NA | 0 |
| S12-final-viral-combined_k141_301624\|\|full | 1488 | No | NA | 3 | 0 | 0 | Low-quality | Genome-fragment | 1.61 | AAI-based (high-confidence) | 0 |
| S1_k141_241380 | 3616 | No | NA | 3 | 0 | 2 | Not-determined | Genome-fragment | NA | NA | 0 |
| S1_k141_26965 | 4474 | No | NA | 14 | 0 | 0 | Not-determined | Genome-fragment | NA | NA | 0 |
| S1_k141_224641 | 1292 | No | NA | 3 | 0 | 2 | Not-determined | Genome-fragment | NA | NA | 0 |
| S1_k141_142569 | 3543 | No | NA | 7 | 0 | 1 | Not-determined | Genome-fragment | NA | NA | 0 |
| S1_k141_43321 | 1582 | No | NA | 2 | 0 | 1 | Not-determined | Genome-fragment | NA | NA | 0 |
| S1_k141_126934 | 1221 | No | NA | 2 | 0 | 2 | Not-determined | Genome-fragment | NA | NA | 0 |
| S1_k141_192547 | 3269 | No | NA | 2 | 0 | 2 | Not-determined | Genome-fragment | NA | NA | 0 |
| S1_k141_126943 | 10456 | No | NA | 6 | 0 | 6 | Not-determined | Genome-fragment | NA | NA | 0 |
| S1_k141_27013 | 1113 | No | NA | 1 | 0 | 0 | Not-determined | Genome-fragment | NA | NA | 0 |
| S1_k141_324588 | 1594 | No | NA | 2 | 0 | 1 | Not-determined | Genome-fragment | NA | NA | 0 |
| S1_k141_372816 | 1177 | No | NA | 2 | 0 | 2 | Not-determined | Genome-fragment | NA | NA | 0 |
| S1_k141_258556 | 1487 | No | NA | 2 | 0 | 0 | Not-determined | Genome-fragment | NA | NA | 0 |
| S1_k141_258560 | 2554 | No | NA | 2 | 0 | 2 | Not-determined | Genome-fragment | NA | NA | 0 |
| S1_k141_241501 | 4128 | No | NA | 5 | 0 | 1 | Not-determined | Genome-fragment | NA | NA | 0 |
| S12-final-viral-combined_k141_101135\|\|full | 1488 | No | NA | 2 | 1 | 0 | Low-quality | Genome-fragment | 0.68 | AAI-based (high-confidence) | 0 |
| S1_k141_241509 | 1147 | No | NA | 2 | 0 | 0 | Not-determined | Genome-fragment | NA | NA | 0 |
| S1_k141_290680 | 1789 | No | NA | 2 | 0 | 2 | Not-determined | Genome-fragment | NA | NA | 0 |
| S12-final-viral-combined_k141_106113\|\|full | 1488 | No | NA | 4 | 0 | 0 | Low-quality | Genome-fragment | 3.16 | AAI-based (high-confidence) | 0 |
| S1_k141_59810 | 1327 | No | NA | 1 | 0 | 1 | Not-determined | Genome-fragment | NA | NA | 0 |
| S1_k141_389132 | 2625 | No | NA | 2 | 0 | 1 | Not-determined | Genome-fragment | NA | NA | 0 |
| S1_k141_126977 | 1553 | No | NA | 2 | 0 | 2 | Not-determined | Genome-fragment | NA | NA | 0 |
| S1_k141_27101 | 1328 | No | NA | 1 | 0 | 0 | Not-determined | Genome-fragment | NA | NA | 0 |
| S1_k141_290704 | 1137 | No | NA | 2 | 0 | 2 | Not-determined | Genome-fragment | NA | NA | 0 |
| S1_k141_389143 | 1781 | No | NA | 1 | 0 | 1 | Not-determined | Genome-fragment | NA | NA | 0 |
| S12-final-viral-combined_k141_130126\|\|lt2gene | 1488 | No | NA | 2 | 1 | 0 | Low-quality | Genome-fragment | 3.61 | AAI-based (high-confidence) | 0 |
| S12_k141_73745 | 1488 | No | NA | 3 | 0 | 1 | Low-quality | Genome-fragment | 1.5 | AAI-based (medium-confidence) | 0 |
| S1_k141_209311 | 3120 | No | NA | 1 | 0 | 1 | Not-determined | Genome-fragment | NA | NA | 0 |
| S1_k141_192631 | 1243 | No | NA | 1 | 0 | 1 | Not-determined | Genome-fragment | NA | NA | 0 |
| S11-final-viral-combined_k141_181773\|\|lt2gene | 1489 | No | NA | 1 | 1 | 0 | Low-quality | Genome-fragment | 2.35 | HMM-based (lower-bound) | 0 |
| S1_k141_389196 | 1383 | No | NA | 2 | 0 | 0 | Not-determined | Genome-fragment | NA | NA | 0 |
| S1_k141_10975 | 1706 | No | NA | 1 | 0 | 0 | Not-determined | Genome-fragment | NA | NA | 0 |
| S1-final-viral-combined_k141_253917\|\|full | 1489 | No | NA | 2 | 0 | 0 | Low-quality | Genome-fragment | 1.58 | AAI-based (medium-confidence) | 0 |
| S1_k141_224821 | 8426 | No | NA | 9 | 0 | 8 | Not-determined | Genome-fragment | NA | NA | 0 |
| S1_k141_174403 | 1607 | No | NA | 1 | 0 | 1 | Not-determined | Genome-fragment | NA | NA | 0 |
| S1_k141_258722 | 1264 | No | NA | 2 | 0 | 1 | Not-determined | Genome-fragment | NA | NA | 0 |
| S1_k141_75727 | 1164 | No | NA | 1 | 0 | 0 | Not-determined | Genome-fragment | NA | NA | 0 |
| S1_k141_258741 | 1074 | No | NA | 1 | 0 | 0 | Not-determined | Genome-fragment | NA | NA | 0 |
| S1_k141_142731 | 1035 | No | NA | 1 | 0 | 1 | Not-determined | Genome-fragment | NA | NA | 0 |
| S1_k141_389237 | 1125 | No | NA | 1 | 0 | 1 | Not-determined | Genome-fragment | NA | NA | 0 |
| S1_k141_62638 | 1489 | No | NA | 3 | 0 | 0 | Low-quality | Genome-fragment | 3.05 | AAI-based (high-confidence) | 0 |
| S1_k141_75779 | 5450 | No | NA | 4 | 0 | 0 | Not-determined | Genome-fragment | NA | NA | 0 |
| S1_k141_258805 | 1052 | No | NA | 2 | 0 | 2 | Not-determined | Genome-fragment | NA | NA | 0 |
| S1_k141_307855 | 4218 | No | NA | 2 | 0 | 1 | Not-determined | Genome-fragment | NA | NA | 0 |
| S1_k141_142757 | 1062 | No | NA | 1 | 0 | 0 | Not-determined | Genome-fragment | NA | NA | 0 |
| S1_k141_373012 | 1485 | No | NA | 1 | 0 | 1 | Not-determined | Genome-fragment | NA | NA | 0 |
| S1_k141_389270 | 1455 | No | NA | 1 | 0 | 0 | Not-determined | Genome-fragment | NA | NA | 0 |
| S1_k141_209421 | 1782 | No | NA | 1 | 0 | 1 | Not-determined | Genome-fragment | NA | NA | 0 |
| S1_k141_373022 | 1086 | No | NA | 1 | 0 | 0 | Not-determined | Genome-fragment | NA | NA | 0 |
| S1_k141_174477 | 3046 | No | NA | 3 | 0 | 2 | Not-determined | Genome-fragment | NA | NA | 0 |
| S5-final-viral-combined_k141_185792\|\|full | 1489 | No | NA | 4 | 0 | 0 | Low-quality | Genome-fragment | 3.65 | AAI-based (high-confidence) | 0 |
| S1_k141_108805 | 1857 | No | NA | 1 | 0 | 1 | Not-determined | Genome-fragment | NA | NA | 0 |
| S1_k141_290928 | 1114 | No | NA | 1 | 0 | 1 | Not-determined | Genome-fragment | NA | NA | 0 |
| S1_k141_209461 | 1222 | No | NA | 1 | 0 | 0 | Not-determined | Genome-fragment | NA | NA | 0 |
| S7-final-viral-combined_k141_143106\|\|full | 1489 | No | NA | 3 | 0 | 0 | Low-quality | Genome-fragment | 0.94 | AAI-based (medium-confidence) | 0 |
| S1_k141_43552 | 1046 | No | NA | 1 | 0 | 0 | Not-determined | Genome-fragment | NA | NA | 0 |
| S1_k141_75911 | 3407 | No | NA | 5 | 0 | 4 | Not-determined | Genome-fragment | NA | NA | 0 |
| S1_k141_307980 | 3178 | No | NA | 2 | 0 | 1 | Not-determined | Genome-fragment | NA | NA | 0 |
| S1_k141_373087 | 2051 | No | NA | 1 | 0 | 0 | Not-determined | Genome-fragment | NA | NA | 0 |
| S1_k141_357148 | 3556 | No | NA | 4 | 0 | 1 | Not-determined | Genome-fragment | NA | NA | 0 |
| S12-final-viral-combined_k141_354878\|\|full | 1489 | No | NA | 3 | 0 | 0 | Low-quality | Genome-fragment | 2.25 | AAI-based (medium-confidence) | 0 |
| S12-final-viral-combined_k141_254701\|\|full | 1489 | No | NA | 3 | 0 | 0 | Low-quality | Genome-fragment | 2.94 | AAI-based (medium-confidence) | 0 |
| S2-final-viral-combined_k141_122368\|\|full | 1489 | No | NA | 3 | 0 | 0 | Low-quality | Genome-fragment | 2.97 | AAI-based (medium-confidence) | 0 |
| S1_k141_307994 | 1152 | No | NA | 1 | 0 | 1 | Not-determined | Genome-fragment | NA | NA | 0 |
| S1_k141_324816 | 1651 | No | NA | 1 | 0 | 1 | Not-determined | Genome-fragment | NA | NA | 0 |
| S1_k141_373101 | 1168 | No | NA | 1 | 0 | 0 | Not-determined | Genome-fragment | NA | NA | 0 |
| S1_k141_127210 | 1043 | No | NA | 2 | 0 | 0 | Not-determined | Genome-fragment | NA | NA | 0 |
| S1_k141_60032 | 3075 | No | NA | 4 | 0 | 4 | Not-determined | Genome-fragment | NA | NA | 0 |
| S1_k141_241813 | 2133 | No | NA | 3 | 0 | 1 | Not-determined | Genome-fragment | NA | NA | 0 |
| S1_k141_225015 | 1336 | No | NA | 1 | 0 | 1 | Not-determined | Genome-fragment | NA | NA | 0 |
| S1_k141_389394 | 1091 | No | NA | 1 | 0 | 1 | Not-determined | Genome-fragment | NA | NA | 0 |
| S1_k141_75950 | 7752 | No | NA | 8 | 0 | 1 | Not-determined | Genome-fragment | NA | NA | 0 |
| S4-final-viral-combined_k141_53315\|\|full | 1489 | No | NA | 2 | 0 | 0 | Low-quality | Genome-fragment | 3.03 | AAI-based (medium-confidence) | 0 |
| S8-final-viral-combined_k141_198670\|\|full | 1489 | No | NA | 2 | 0 | 0 | Low-quality | Genome-fragment | 3.1 | AAI-based (medium-confidence) | 0 |
| S1_k141_339440 | 1967 | No | NA | 2 | 0 | 0 | Not-determined | Genome-fragment | NA | NA | 0 |
| S1_k141_274045 | 5612 | No | NA | 2 | 0 | 1 | Not-determined | Genome-fragment | NA | NA | 0 |
| S11-final-viral-combined_k141_238357\|\|full | 1490 | No | NA | 4 | 1 | 0 | Low-quality | Genome-fragment | 2.65 | AAI-based (high-confidence) | 0 |
| S1_k141_192769 | 1486 | No | NA | 2 | 0 | 2 | Not-determined | Genome-fragment | NA | NA | 0 |
| S11-final-viral-combined_k141_55037\|\|full | 1490 | No | NA | 4 | 1 | 0 | Low-quality | Genome-fragment | 3.65 | AAI-based (medium-confidence) | 0 |
| S11-final-viral-combined_k141_276620\|\|lt2gene | 1490 | No | NA | 2 | 1 | 0 | Low-quality | Genome-fragment | 3.12 | AAI-based (high-confidence) | 0 |
| S1_k141_274084 | 1034 | No | NA | 1 | 0 | 0 | Not-determined | Genome-fragment | NA | NA | 0 |
| S11-final-viral-combined_k141_276648\|\|lt2gene | 1490 | No | NA | 2 | 1 | 0 | Low-quality | Genome-fragment | 3.12 | AAI-based (high-confidence) | 0 |
| S1_k141_60093 | 3884 | No | NA | 6 | 0 | 1 | Not-determined | Genome-fragment | NA | NA | 0 |
| S1-final-viral-combined_k141_44771\|\|lt2gene | 1490 | No | NA | 2 | 0 | 0 | Low-quality | Genome-fragment | 2.81 | AAI-based (high-confidence) | 0 |
| S1_k141_324892 | 1101 | No | NA | 1 | 0 | 1 | Not-determined | Genome-fragment | NA | NA | 0 |
| S1_k141_191225 | 1490 | No | NA | 2 | 0 | 0 | Low-quality | Genome-fragment | 2.55 | AAI-based (medium-confidence) | 0 |
| S17-final-viral-combined_k141_33676\|\|full | 1490 | No | NA | 3 | 0 | 0 | Low-quality | Genome-fragment | 2.62 | AAI-based (medium-confidence) | 0 |
| S1_k141_43695 | 1056 | No | NA | 1 | 0 | 1 | Not-determined | Genome-fragment | NA | NA | 0 |
| S17-final-viral-combined_k141_240555\|\|full | 1490 | No | NA | 2 | 1 | 0 | Low-quality | Genome-fragment | 1.71 | HMM-based (lower-bound) | 0 |
| S1_k141_43719 | 4945 | No | NA | 9 | 0 | 8 | Not-determined | Genome-fragment | NA | NA | 0 |
| S1_k141_373176 | 1241 | No | NA | 1 | 0 | 0 | Not-determined | Genome-fragment | NA | NA | 0 |
| S17-final-viral-combined_k141_231437\|\|full | 1490 | No | NA | 2 | 0 | 0 | Low-quality | Genome-fragment | 2.2 | AAI-based (medium-confidence) | 0 |
| S1_k141_108998 | 1316 | No | NA | 3 | 0 | 2 | Not-determined | Genome-fragment | NA | NA | 0 |
| S17-final-viral-combined_k141_77406\|\|full | 1490 | No | NA | 3 | 2 | 0 | Low-quality | Genome-fragment | 3.76 | AAI-based (medium-confidence) | 0 |
| S17-k141_92619 | 1490 | Yes | 308 | 2 | 1 | 1 | Low-quality | Genome-fragment | 0.2 | AAI-based (medium-confidence) | 79.33 |
| S12-final-viral-combined_k141_244433\|\|full | 1490 | No | NA | 3 | 0 | 0 | Low-quality | Genome-fragment | 2.73 | AAI-based (high-confidence) | 0 |
| S8-final-viral-combined_k141_206785\|\|full | 1490 | No | NA | 2 | 0 | 1 | Low-quality | Genome-fragment | 0.88 | AAI-based (medium-confidence) | 0 |
| S1_k141_27329 | 3239 | No | NA | 2 | 0 | 2 | Not-determined | Genome-fragment | NA | NA | 0 |
| S1_k141_43776 | 2402 | No | NA | 3 | 0 | 0 | Not-determined | Genome-fragment | NA | NA | 0 |
| S1_k141_27338 | 1740 | No | NA | 2 | 0 | 1 | Not-determined | Genome-fragment | NA | NA | 0 |
| S9-final-viral-combined_k141_48062\|\|full | 1491 | No | NA | 4 | 1 | 0 | Low-quality | Genome-fragment | 3.99 | AAI-based (medium-confidence) | 0 |
| S1-final-viral-combined_k141_288174\|\|full | 1491 | No | NA | 3 | 0 | 0 | Low-quality | Genome-fragment | 3.49 | AAI-based (medium-confidence) | 0 |
| S1_k141_291267 | 1076 | No | NA | 1 | 0 | 1 | Not-determined | Genome-fragment | NA | NA | 0 |
| S1_k141_274252 | 8363 | No | NA | 3 | 0 | 1 | Not-determined | Genome-fragment | NA | NA | 0 |
| S1-final-viral-combined_k141_126361\|\|lt2gene | 1491 | No | NA | 2 | 1 | 0 | Low-quality | Genome-fragment | 5.05 | HMM-based (lower-bound) | 0 |
| S1_k141_109053 | 3785 | No | NA | 4 | 0 | 1 | Not-determined | Genome-fragment | NA | NA | 0 |
| S1_k141_93802 | 2098 | No | NA | 2 | 0 | 2 | Not-determined | Genome-fragment | NA | NA | 0 |
| S1_k141_357447 | 1250 | No | NA | 1 | 0 | 0 | Not-determined | Genome-fragment | NA | NA | 0 |
| S1_k141_76213 | 1515 | No | NA | 1 | 0 | 1 | Not-determined | Genome-fragment | NA | NA | 0 |
| S1_k141_209643 | 1680 | No | NA | 3 | 0 | 1 | Not-determined | Genome-fragment | NA | NA | 0 |
| S1_k141_225303 | 2504 | No | NA | 1 | 0 | 0 | Not-determined | Genome-fragment | NA | NA | 0 |
| S1_k141_158711 | 1310 | No | NA | 1 | 0 | 1 | Not-determined | Genome-fragment | NA | NA | 0 |
| S1_k141_76234 | 1318 | No | NA | 1 | 0 | 0 | Not-determined | Genome-fragment | NA | NA | 0 |
| S1_k141_259199 | 1309 | No | NA | 1 | 0 | 1 | Not-determined | Genome-fragment | NA | NA | 0 |
| S1_k141_60337 | 1915 | No | NA | 4 | 0 | 4 | Not-determined | Genome-fragment | NA | NA | 0 |
| S1_k141_259204 | 6960 | No | NA | 1 | 0 | 1 | Not-determined | Genome-fragment | NA | NA | 0 |
| S7-final-viral-combined_k141_10277\|\|full | 1491 | Yes | 666 | 5 | 1 | 1 | Low-quality | Genome-fragment | 1.75 | AAI-based (high-confidence) | 55.33 |
| S1_k141_274333 | 1299 | No | NA | 2 | 0 | 1 | Not-determined | Genome-fragment | NA | NA | 0 |
| S1_k141_339597 | 4674 | No | NA | 3 | 0 | 3 | Not-determined | Genome-fragment | NA | NA | 0 |
| S7-final-viral-combined_k141_176302\|\|full | 1491 | No | NA | 3 | 0 | 2 | Low-quality | Genome-fragment | 2.06 | AAI-based (medium-confidence) | 0 |
| S1_k141_339606 | 6414 | No | NA | 7 | 0 | 4 | Not-determined | Genome-fragment | NA | NA | 0 |
| S1_k141_76266 | 1245 | No | NA | 1 | 0 | 1 | Not-determined | Genome-fragment | NA | NA | 0 |
| S1_k141_193029 | 1522 | No | NA | 1 | 0 | 0 | Not-determined | Genome-fragment | NA | NA | 0 |
| S1_k141_109140 | 1224 | No | NA | 1 | 0 | 0 | Not-determined | Genome-fragment | NA | NA | 0 |
| S1_k141_109149 | 1443 | No | NA | 2 | 0 | 1 | Not-determined | Genome-fragment | NA | NA | 0 |
| S17-final-viral-combined_k141_231082\|\|full | 1491 | No | NA | 3 | 2 | 0 | Low-quality | Genome-fragment | 3.49 | AAI-based (medium-confidence) | 0 |
| S1_k141_93935 | 1010 | No | NA | 1 | 0 | 1 | Not-determined | Genome-fragment | NA | NA | 0 |
| S1_k141_11367 | 1197 | No | NA | 1 | 0 | 1 | Not-determined | Genome-fragment | NA | NA | 0 |
| S2-final-viral-combined_k141_190946\|\|full | 1491 | No | NA | 3 | 0 | 0 | Low-quality | Genome-fragment | 1.01 | AAI-based (medium-confidence) | 0 |
| S1_k141_291415 | 1201 | No | NA | 1 | 0 | 1 | Not-determined | Genome-fragment | NA | NA | 0 |
| S1_k141_274374 | 1045 | No | NA | 1 | 0 | 1 | Not-determined | Genome-fragment | NA | NA | 0 |
| S8_k141_53529 | 1491 | No | NA | 3 | 1 | 0 | Low-quality | Genome-fragment | 0.97 | AAI-based (high-confidence) | 0 |
| S1_k141_174969 | 5579 | No | NA | 7 | 0 | 4 | Not-determined | Genome-fragment | NA | NA | 0 |
| S11-final-viral-combined_k141_23872\|\|full | 1492 | No | NA | 3 | 0 | 3 | Low-quality | Genome-fragment | 0.88 | AAI-based (medium-confidence) | 0 |
| S1_k141_308340 | 1015 | No | NA | 2 | 0 | 1 | Not-determined | Genome-fragment | NA | NA | 0 |
| S11_k141_230203 | 1492 | No | NA | 1 | 0 | 0 | Low-quality | Genome-fragment | 3.49 | AAI-based (medium-confidence) | 0 |
| S1_k141_175006 | 1165 | No | NA | 1 | 0 | 0 | Not-determined | Genome-fragment | NA | NA | 0 |
| S1_k141_308384 | 2552 | No | NA | 3 | 0 | 3 | Not-determined | Genome-fragment | NA | NA | 0 |
| S1-final-viral-combined_k141_114663\|\|full | 1492 | No | NA | 2 | 1 | 0 | Low-quality | Genome-fragment | 2.21 | HMM-based (lower-bound) | 0 |
| S1_k141_193195 | 2063 | No | NA | 3 | 0 | 0 | Not-determined | Genome-fragment | NA | NA | 0 |
| S3-final-viral-combined_k141_135079\|\|full | 1492 | No | NA | 6 | 1 | 0 | Low-quality | Genome-fragment | 3.13 | AAI-based (high-confidence) | 0 |
| S1_k141_259302 | 1637 | No | NA | 1 | 0 | 0 | Not-determined | Genome-fragment | NA | NA | 0 |
| S1_k141_291539 | 2220 | No | NA | 3 | 0 | 3 | Not-determined | Genome-fragment | NA | NA | 0 |
| S1_k141_143226 | 1125 | No | NA | 1 | 0 | 0 | Not-determined | Genome-fragment | NA | NA | 0 |
| S1_k141_325228 | 1177 | No | NA | 3 | 0 | 1 | Not-determined | Genome-fragment | NA | NA | 0 |
| S1_k141_158951 | 4704 | No | NA | 4 | 0 | 3 | Not-determined | Genome-fragment | NA | NA | 0 |
| S1_k141_109303 | 1555 | No | NA | 1 | 0 | 1 | Not-determined | Genome-fragment | NA | NA | 0 |
| S1_k141_143258 | 1034 | No | NA | 1 | 0 | 0 | Not-determined | Genome-fragment | NA | NA | 0 |
| S1_k141_158977 | 1846 | No | NA | 2 | 0 | 2 | Not-determined | Genome-fragment | NA | NA | 0 |
| S5_k141_163756 | 1492 | No | NA | 3 | 0 | 1 | Low-quality | Genome-fragment | 3.47 | AAI-based (medium-confidence) | 0 |
| S1_k141_76412 | 2109 | No | NA | 2 | 0 | 0 | Not-determined | Genome-fragment | NA | NA | 0 |
| S1_k141_109335 | 1050 | No | NA | 3 | 0 | 0 | Not-determined | Genome-fragment | NA | NA | 0 |
| S1_k141_143312 | 1103 | No | NA | 2 | 0 | 0 | Not-determined | Genome-fragment | NA | NA | 0 |
| S12-final-viral-combined_k141_95642\|\|full | 1492 | No | NA | 2 | 0 | 2 | Low-quality | Genome-fragment | 2.58 | AAI-based (medium-confidence) | 0 |
| S1_k141_159003 | 1183 | No | NA | 1 | 0 | 0 | Not-determined | Genome-fragment | NA | NA | 0 |
| S1_k141_159006 | 1341 | No | NA | 2 | 0 | 0 | Not-determined | Genome-fragment | NA | NA | 0 |
| S1_k141_274533 | 4039 | No | NA | 6 | 0 | 0 | Not-determined | Genome-fragment | NA | NA | 0 |
| S1_k141_27659 | 1175 | No | NA | 1 | 0 | 0 | Not-determined | Genome-fragment | NA | NA | 0 |
| S1_k141_127712 | 1722 | No | NA | 2 | 0 | 0 | Not-determined | Genome-fragment | NA | NA | 0 |
| S12_k141_299931 | 1492 | No | NA | 1 | 0 | 1 | Low-quality | Genome-fragment | 2.21 | AAI-based (medium-confidence) | 0 |
| S8_k141_59304 | 1492 | No | NA | 3 | 0 | 0 | Low-quality | Genome-fragment | 1.12 | AAI-based (high-confidence) | 0 |
| S1_k141_209913 | 1493 | No | NA | 1 | 0 | 1 | Not-determined | Genome-fragment | NA | NA | 0 |
| S1_k141_373509 | 2312 | No | NA | 3 | 0 | 1 | Not-determined | Genome-fragment | NA | NA | 0 |
| S1_k141_339808 | 6975 | No | NA | 9 | 0 | 1 | Not-determined | Genome-fragment | NA | NA | 0 |
| S1_k141_159033 | 1078 | No | NA | 1 | 0 | 1 | Not-determined | Genome-fragment | NA | NA | 0 |
| S8_k141_144085 | 1492 | No | NA | 3 | 0 | 1 | Low-quality | Genome-fragment | 2.05 | AAI-based (medium-confidence) | 0 |
| S1_k141_175176 | 1428 | No | NA | 2 | 0 | 1 | Not-determined | Genome-fragment | NA | NA | 0 |
| S1_k141_109376 | 1514 | No | NA | 3 | 0 | 0 | Not-determined | Genome-fragment | NA | NA | 0 |
| S1_k141_389841 | 1145 | No | NA | 1 | 0 | 1 | Not-determined | Genome-fragment | NA | NA | 0 |
| S9-final-viral-combined_k141_48117\|\|full | 1493 | No | NA | 4 | 1 | 0 | Low-quality | Genome-fragment | 3.61 | AAI-based (medium-confidence) | 0 |
| S9_k141_122205 | 1493 | No | NA | 5 | 2 | 0 | Low-quality | Genome-fragment | 3.57 | AAI-based (high-confidence) | 0 |
| S1_k141_357842 | 1587 | No | NA | 1 | 0 | 1 | Not-determined | Genome-fragment | NA | NA | 0 |
| S1_k141_357850 | 5169 | No | NA | 3 | 0 | 3 | Not-determined | Genome-fragment | NA | NA | 0 |
| S1_k141_159102 | 1186 | No | NA | 3 | 0 | 2 | Not-determined | Genome-fragment | NA | NA | 0 |
| S7-final-viral-combined_k141_88925\|\|full | 1493 | No | NA | 2 | 0 | 1 | Low-quality | Genome-fragment | 0.76 | AAI-based (medium-confidence) | 0 |
| S1_k141_11575 | 1669 | No | NA | 1 | 0 | 0 | Not-determined | Genome-fragment | NA | NA | 0 |
| S12-final-viral-combined_k141_36040\|\|full | 1493 | No | NA | 3 | 1 | 0 | Low-quality | Genome-fragment | 2.26 | HMM-based (lower-bound) | 0 |
| S1_k141_11592 | 1006 | No | NA | 1 | 0 | 1 | Not-determined | Genome-fragment | NA | NA | 0 |
| S1_k141_308603 | 2217 | No | NA | 4 | 0 | 2 | Not-determined | Genome-fragment | NA | NA | 0 |
| S1_k141_291815 | 1177 | No | NA | 2 | 0 | 2 | Not-determined | Genome-fragment | NA | NA | 0 |
| S12-final-viral-combined_k141_384563\|\|full | 1493 | No | NA | 3 | 0 | 1 | Low-quality | Genome-fragment | 3.69 | AAI-based (medium-confidence) | 0 |
| S1_k141_94286 | 1208 | No | NA | 1 | 0 | 1 | Not-determined | Genome-fragment | NA | NA | 0 |
| S1_k141_291847 | 1608 | No | NA | 1 | 0 | 0 | Not-determined | Genome-fragment | NA | NA | 0 |
| S1_k141_143448 | 1140 | No | NA | 1 | 0 | 1 | Not-determined | Genome-fragment | NA | NA | 0 |
| S1_k141_325457 | 1126 | No | NA | 1 | 0 | 0 | Not-determined | Genome-fragment | NA | NA | 0 |
| S1_k141_109464 | 1066 | No | NA | 1 | 0 | 1 | Not-determined | Genome-fragment | NA | NA | 0 |
| S1_k141_76561 | 1133 | No | NA | 2 | 0 | 0 | Not-determined | Genome-fragment | NA | NA | 0 |
| S1_k141_389988 | 1268 | No | NA | 2 | 0 | 2 | Not-determined | Genome-fragment | NA | NA | 0 |
| S1_k141_225750 | 6459 | No | NA | 4 | 0 | 4 | Not-determined | Genome-fragment | NA | NA | 0 |
| S1_k141_109499 | 1106 | No | NA | 1 | 0 | 1 | Not-determined | Genome-fragment | NA | NA | 0 |
| S1_k141_193504 | 1058 | No | NA | 2 | 0 | 1 | Not-determined | Genome-fragment | NA | NA | 0 |
| S1_k141_225766 | 1356 | No | NA | 2 | 0 | 1 | Not-determined | Genome-fragment | NA | NA | 0 |
| S2-final-viral-combined_k141_165526\|\|full | 1493 | No | NA | 3 | 1 | 0 | Low-quality | Genome-fragment | 1 | HMM-based (lower-bound) | 0 |
| S1_k141_210083 | 1334 | No | NA | 2 | 0 | 2 | Not-determined | Genome-fragment | NA | NA | 0 |
| S1_k141_357957 | 1389 | No | NA | 2 | 0 | 1 | Not-determined | Genome-fragment | NA | NA | 0 |
| S11-final-viral-combined_k141_86223\|\|full | 1494 | No | NA | 3 | 0 | 0 | Low-quality | Genome-fragment | 1.78 | AAI-based (medium-confidence) | 0 |
| S1-final-viral-combined_k141_130710\|\|full | 1494 | No | NA | 5 | 0 | 0 | Low-quality | Genome-fragment | 3.52 | AAI-based (medium-confidence) | 0 |
| S1_k141_291907 | 1215 | No | NA | 1 | 0 | 0 | Not-determined | Genome-fragment | NA | NA | 0 |
| S1_k141_308662 | 1031 | No | NA | 1 | 0 | 0 | Not-determined | Genome-fragment | NA | NA | 0 |
| S5-final-viral-combined_k141_155455\|\|full | 1494 | No | NA | 3 | 1 | 0 | Low-quality | Genome-fragment | 3.73 | AAI-based (high-confidence) | 0 |
| S1_k141_242477 | 1298 | No | NA | 2 | 0 | 1 | Not-determined | Genome-fragment | NA | NA | 0 |
| S1_k141_339905 | 1326 | No | NA | 1 | 0 | 1 | Not-determined | Genome-fragment | NA | NA | 0 |
| S1_k141_373728 | 1043 | No | NA | 1 | 0 | 1 | Not-determined | Genome-fragment | NA | NA | 0 |
| S1_k141_193604 | 3500 | No | NA | 4 | 0 | 2 | Not-determined | Genome-fragment | NA | NA | 0 |
| S1_k141_358048 | 2235 | No | NA | 2 | 0 | 2 | Not-determined | Genome-fragment | NA | NA | 0 |
| S1_k141_358056 | 3829 | No | NA | 3 | 0 | 3 | Not-determined | Genome-fragment | NA | NA | 0 |
| S1_k141_109628 | 3387 | No | NA | 2 | 0 | 2 | Not-determined | Genome-fragment | NA | NA | 0 |
| S1_k141_60916 | 1612 | No | NA | 1 | 0 | 1 | Not-determined | Genome-fragment | NA | NA | 0 |
| S5-final-viral-combined_k141_132286\|\|full | 1494 | No | NA | 3 | 1 | 0 | Low-quality | Genome-fragment | 2.77 | AAI-based (medium-confidence) | 0 |
| S1_k141_143576 | 1709 | No | NA | 4 | 0 | 2 | Not-determined | Genome-fragment | NA | NA | 0 |
| S1_k141_292020 | 1399 | No | NA | 1 | 0 | 0 | Not-determined | Genome-fragment | NA | NA | 0 |
| S1_k141_159243 | 1790 | No | NA | 1 | 0 | 0 | Not-determined | Genome-fragment | NA | NA | 0 |
| S1_k141_11820 | 1586 | No | NA | 2 | 0 | 1 | Not-determined | Genome-fragment | NA | NA | 0 |
| S1_k141_390116 | 1125 | No | NA | 1 | 0 | 1 | Not-determined | Genome-fragment | NA | NA | 0 |
| S1_k141_358139 | 4234 | No | NA | 6 | 0 | 1 | Not-determined | Genome-fragment | NA | NA | 0 |
| S5-final-viral-combined_k141_175812\|\|lt2gene | 1494 | No | NA | 1 | 1 | 0 | Low-quality | Genome-fragment | 1.03 | HMM-based (lower-bound) | 0 |
| S1_k141_210214 | 2023 | No | NA | 1 | 0 | 1 | Not-determined | Genome-fragment | NA | NA | 0 |
| S1_k141_143660 | 4048 | No | NA | 6 | 0 | 3 | Not-determined | Genome-fragment | NA | NA | 0 |
| S1_k141_373808 | 1022 | No | NA | 1 | 0 | 0 | Not-determined | Genome-fragment | NA | NA | 0 |
| S7-final-viral-combined_k141_175685\|\|full | 1494 | No | NA | 5 | 0 | 0 | Low-quality | Genome-fragment | 2.27 | AAI-based (medium-confidence) | 0 |
| S1_k141_109720 | 3350 | No | NA | 3 | 0 | 2 | Not-determined | Genome-fragment | NA | NA | 0 |
| S7_k141_240983 | 1494 | No | NA | 4 | 0 | 0 | Low-quality | Genome-fragment | 2.5 | AAI-based (medium-confidence) | 0 |
| S1_k141_61070 | 1752 | No | NA | 2 | 0 | 2 | Not-determined | Genome-fragment | NA | NA | 0 |
| S1_k141_44536 | 1150 | No | NA | 1 | 0 | 1 | Not-determined | Genome-fragment | NA | NA | 0 |
| S1_k141_159358 | 1871 | No | NA | 4 | 0 | 0 | Not-determined | Genome-fragment | NA | NA | 0 |
| S1_k141_274896 | 3203 | No | NA | 4 | 0 | 3 | Not-determined | Genome-fragment | NA | NA | 0 |
| S1_k141_242627 | 1405 | No | NA | 1 | 0 | 1 | Not-determined | Genome-fragment | NA | NA | 0 |
| S1_k141_358305 | 1254 | No | NA | 1 | 0 | 1 | Not-determined | Genome-fragment | NA | NA | 0 |
| S1_k141_225995 | 47567 | No | NA | 68 | 14 | 1 | High-quality | High-quality | 100 | AAI-based (high-confidence) | 0 |
| S1_k141_274929 | 3591 | No | NA | 1 | 0 | 1 | Not-determined | Genome-fragment | NA | NA | 0 |
| S1_k141_373919 | 3488 | No | NA | 2 | 0 | 2 | Not-determined | Genome-fragment | NA | NA | 0 |
| S1_k141_11964 | 2780 | No | NA | 2 | 0 | 0 | Not-determined | Genome-fragment | NA | NA | 0 |
| S1_k141_76856 | 1075 | No | NA | 1 | 0 | 1 | Not-determined | Genome-fragment | NA | NA | 0 |
| S1_k141_390214 | 1155 | No | NA | 2 | 0 | 2 | Not-determined | Genome-fragment | NA | NA | 0 |
| S1_k141_11979 | 3124 | No | NA | 2 | 0 | 1 | Not-determined | Genome-fragment | NA | NA | 0 |
| S1_k141_226027 | 1542 | No | NA | 1 | 0 | 0 | Not-determined | Genome-fragment | NA | NA | 0 |
| S1_k141_226043 | 1693 | No | NA | 2 | 0 | 1 | Not-determined | Genome-fragment | NA | NA | 0 |
| S1_k141_340164 | 1099 | No | NA | 2 | 0 | 0 | Not-determined | Genome-fragment | NA | NA | 0 |
| S1_k141_292231 | 1264 | No | NA | 2 | 0 | 1 | Not-determined | Genome-fragment | NA | NA | 0 |
| S1_k141_76896 | 1128 | No | NA | 1 | 0 | 1 | Not-determined | Genome-fragment | NA | NA | 0 |
| S1_k141_226055 | 1073 | No | NA | 1 | 0 | 0 | Not-determined | Genome-fragment | NA | NA | 0 |
| S1_k141_242656 | 1016 | No | NA | 2 | 0 | 1 | Not-determined | Genome-fragment | NA | NA | 0 |
| S1_k141_374008 | 1055 | No | NA | 1 | 0 | 1 | Not-determined | Genome-fragment | NA | NA | 0 |
| S1_k141_340185 | 1148 | No | NA | 1 | 0 | 0 | Not-determined | Genome-fragment | NA | NA | 0 |
| S1_k141_44664 | 1754 | No | NA | 3 | 0 | 1 | Not-determined | Genome-fragment | NA | NA | 0 |
| S1_k141_143848 | 1414 | No | NA | 1 | 0 | 1 | Not-determined | Genome-fragment | NA | NA | 0 |
| S1_k141_128123 | 1091 | No | NA | 1 | 0 | 0 | Not-determined | Genome-fragment | NA | NA | 0 |
| S1_k141_358418 | 4069 | No | NA | 5 | 0 | 3 | Not-determined | Genome-fragment | NA | NA | 0 |
| S1_k141_143858 | 1235 | No | NA | 1 | 0 | 0 | Not-determined | Genome-fragment | NA | NA | 0 |
| S1_k141_61287 | 1178 | No | NA | 2 | 0 | 1 | Not-determined | Genome-fragment | NA | NA | 0 |
| S1_k141_175556 | 1425 | No | NA | 1 | 0 | 0 | Not-determined | Genome-fragment | NA | NA | 0 |
| S1_k141_358450 | 1073 | No | NA | 1 | 0 | 1 | Not-determined | Genome-fragment | NA | NA | 0 |
| S1_k141_12095 | 2332 | No | NA | 3 | 0 | 1 | Not-determined | Genome-fragment | NA | NA | 0 |
| S17-final-viral-combined_k141_133974\|\|full | 1494 | No | NA | 3 | 0 | 0 | Low-quality | Genome-fragment | 2.64 | AAI-based (high-confidence) | 0 |
| S1_k141_76992 | 4750 | No | NA | 4 | 0 | 4 | Not-determined | Genome-fragment | NA | NA | 0 |
| S1_k141_325875 | 1404 | No | NA | 1 | 0 | 1 | Not-determined | Genome-fragment | NA | NA | 0 |
| S17-final-viral-combined_k141_44119\|\|full | 1494 | No | NA | 3 | 0 | 1 | Low-quality | Genome-fragment | 3.06 | AAI-based (medium-confidence) | 0 |
| S1_k141_374113 | 1124 | No | NA | 1 | 0 | 1 | Not-determined | Genome-fragment | NA | NA | 0 |
| S1_k141_275144 | 1036 | No | NA | 1 | 0 | 1 | Not-determined | Genome-fragment | NA | NA | 0 |
| S1_k141_374138 | 2357 | No | NA | 3 | 0 | 0 | Not-determined | Genome-fragment | NA | NA | 0 |
| S1_k141_143961 | 1463 | No | NA | 1 | 0 | 1 | Not-determined | Genome-fragment | NA | NA | 0 |
| S12-final-viral-combined_k141_176148\|\|full | 1494 | No | NA | 3 | 0 | 0 | Low-quality | Genome-fragment | 2.03 | AAI-based (medium-confidence) | 0 |
| S2-final-viral-combined_k141_4224\|\|full | 1494 | No | NA | 3 | 0 | 0 | Low-quality | Genome-fragment | 2.74 | AAI-based (high-confidence) | 0 |
| S1_k141_242821 | 2302 | No | NA | 1 | 0 | 1 | Not-determined | Genome-fragment | NA | NA | 0 |
| S2-final-viral-combined_k141_58925\|\|full | 1494 | No | NA | 4 | 0 | 0 | Low-quality | Genome-fragment | 3.54 | AAI-based (medium-confidence) | 0 |
| S1_k141_94753 | 1635 | No | NA | 2 | 0 | 1 | Not-determined | Genome-fragment | NA | NA | 0 |
| S1_k141_143985 | 1323 | No | NA | 1 | 0 | 1 | Not-determined | Genome-fragment | NA | NA | 0 |
| S1_k141_77081 | 1118 | No | NA | 1 | 0 | 1 | Not-determined | Genome-fragment | NA | NA | 0 |
| S1_k141_309189 | 1051 | No | NA | 2 | 0 | 0 | Not-determined | Genome-fragment | NA | NA | 0 |

| | | | | | | | | | | | |
|---|---|---|---|---|---|---|---|---|---|---|---|
| S9-final-viral-combined_k141_15970\|\|full | 1495 | No | NA | 6 | 0 | 0 | Low-quality | Genome-fragment | 3.61 | AAI-based (medium-confidence) | 0 |
| S1_k141_242903 | 1085 | No | NA | 2 | 0 | 1 | Not-determined | Genome-fragment | NA | NA | 0 |
| S1_k141_28450 | 1144 | No | NA | 4 | 0 | 3 | Not-determined | Genome-fragment | NA | NA | 0 |
| S9_k141_30596 | 1495 | No | NA | 2 | 0 | 0 | Low-quality | Genome-fragment | 5.67 | AAI-based (medium-confidence) | 0 |
| S1-final-viral-combined_k141_367267\|\|full | 1495 | No | NA | 3 | 0 | 1 | Low-quality | Genome-fragment | 1.18 | AAI-based (medium-confidence) | 0 |
| S1_k141_44878 | 1658 | No | NA | 1 | 0 | 1 | Not-determined | Genome-fragment | NA | NA | 0 |
| S1_k141_390541 | 6974 | No | NA | 6 | 0 | 5 | Not-determined | Genome-fragment | NA | NA | 0 |
| S1_k141_128272 | 5630 | No | NA | 6 | 0 | 5 | Not-determined | Genome-fragment | NA | NA | 0 |
| S1_k141_326028 | 2368 | No | NA | 3 | 0 | 1 | Not-determined | Genome-fragment | NA | NA | 0 |
| S1_k141_309236 | 2370 | No | NA | 3 | 0 | 1 | Not-determined | Genome-fragment | NA | NA | 0 |
| S1_k141_175711 | 1096 | No | NA | 1 | 0 | 1 | Not-determined | Genome-fragment | NA | NA | 0 |
| S1_k141_309255 | 1103 | No | NA | 1 | 0 | 0 | Not-determined | Genome-fragment | NA | NA | 0 |
| S1_k141_193848 | 1106 | No | NA | 1 | 0 | 1 | Not-determined | Genome-fragment | NA | NA | 0 |
| S1_k141_374250 | 1360 | No | NA | 1 | 0 | 1 | Not-determined | Genome-fragment | NA | NA | 0 |
| S1-final-viral-combined_k141_205299\|\|full | 1495 | No | NA | 3 | 1 | 0 | Low-quality | Genome-fragment | 3.52 | AAI-based (high-confidence) | 0 |
| S1_k141_358729 | 1022 | No | NA | 2 | 0 | 1 | Not-determined | Genome-fragment | NA | NA | 0 |
| S1_k141_128334 | 5542 | No | NA | 2 | 0 | 0 | Not-determined | Genome-fragment | NA | NA | 0 |
| S1_k141_390607 | 1062 | No | NA | 1 | 0 | 1 | Not-determined | Genome-fragment | NA | NA | 0 |
| S1_k141_374302 | 1075 | No | NA | 3 | 0 | 0 | Not-determined | Genome-fragment | NA | NA | 0 |
| S1_k141_374305 | 1053 | No | NA | 1 | 0 | 1 | Not-determined | Genome-fragment | NA | NA | 0 |
| S1_k141_326053 | 1717 | No | NA | 2 | 0 | 1 | Not-determined | Genome-fragment | NA | NA | 0 |
| S1_k141_275256 | 1181 | No | NA | 1 | 0 | 0 | Not-determined | Genome-fragment | NA | NA | 0 |
| S1_k141_260361 | 1822 | No | NA | 1 | 0 | 0 | Not-determined | Genome-fragment | NA | NA | 0 |
| S1_k141_109955 | 1045 | No | NA | 2 | 0 | 0 | Not-determined | Genome-fragment | NA | NA | 0 |
| S1_k141_292629 | 1277 | No | NA | 2 | 0 | 0 | Not-determined | Genome-fragment | NA | NA | 0 |
| S1-final-viral-combined_k141_242553\|\|full | 1495 | No | NA | 2 | 0 | 0 | Low-quality | Genome-fragment | 3.34 | AAI-based (high-confidence) | 0 |
| S1_k141_275306 | 1914 | No | NA | 1 | 0 | 0 | Not-determined | Genome-fragment | NA | NA | 0 |
| S1_k141_128402 | 2058 | No | NA | 2 | 0 | 1 | Not-determined | Genome-fragment | NA | NA | 0 |
| S1_k141_102204 | 1495 | No | NA | 2 | 0 | 0 | Low-quality | Genome-fragment | 3.64 | AAI-based (high-confidence) | 0 |
| S1_k141_292688 | 1043 | No | NA | 1 | 0 | 0 | Not-determined | Genome-fragment | NA | NA | 0 |
| S1_k141_109987 | 4035 | No | NA | 1 | 0 | 0 | Not-determined | Genome-fragment | NA | NA | 0 |
| S1_k141_12510 | 1665 | No | NA | 1 | 0 | 1 | Not-determined | Genome-fragment | NA | NA | 0 |
| S1_k141_292693 | 1181 | No | NA | 2 | 0 | 1 | Not-determined | Genome-fragment | NA | NA | 0 |
| S1_k141_159720 | 1073 | No | NA | 2 | 0 | 0 | Not-determined | Genome-fragment | NA | NA | 0 |
| S1_k141_226516 | 1112 | No | NA | 2 | 0 | 0 | Not-determined | Genome-fragment | NA | NA | 0 |
| S5-final-viral-combined_k141_141440\|\|full | 1495 | No | NA | 3 | 0 | 1 | Low-quality | Genome-fragment | 0.6 | AAI-based (high-confidence) | 0 |
| S1_k141_309349 | 2714 | No | NA | 3 | 0 | 3 | Not-determined | Genome-fragment | NA | NA | 0 |
| S1_k141_28651 | 1099 | No | NA | 2 | 0 | 0 | Not-determined | Genome-fragment | NA | NA | 0 |
| S1_k141_390771 | 3930 | No | NA | 4 | 0 | 2 | Not-determined | Genome-fragment | NA | NA | 0 |
| S1_k141_275338 | 1760 | No | NA | 3 | 0 | 2 | Not-determined | Genome-fragment | NA | NA | 0 |
| S1_k141_194079 | 1699 | No | NA | 2 | 0 | 1 | Not-determined | Genome-fragment | NA | NA | 0 |
| S1_k141_309387 | 1094 | No | NA | 1 | 0 | 0 | Not-determined | Genome-fragment | NA | NA | 0 |
| S7-final-viral-combined_k141_229060\|\|full | 1495 | No | NA | 2 | 0 | 2 | Low-quality | Genome-fragment | 0.69 | AAI-based (medium-confidence) | 0 |
| S7-final-viral-combined_k141_8849\|\|full | 1495 | No | NA | 3 | 1 | 0 | Low-quality | Genome-fragment | 2.52 | AAI-based (medium-confidence) | 0 |
| S7_k141_206056 | 1495 | No | NA | 6 | 0 | 0 | Low-quality | Genome-fragment | 2.46 | AAI-based (high-confidence) | 0 |
| S1_k141_243165 | 1095 | No | NA | 1 | 0 | 1 | Not-determined | Genome-fragment | NA | NA | 0 |
| S1_k141_12618 | 1095 | No | NA | 1 | 0 | 0 | Not-determined | Genome-fragment | NA | NA | 0 |
| S1_k141_358941 | 1218 | No | NA | 1 | 0 | 1 | Not-determined | Genome-fragment | NA | NA | 0 |
| S1_k141_374529 | 5331 | No | NA | 5 | 0 | 0 | Not-determined | Genome-fragment | NA | NA | 0 |
| S1_k141_292821 | 3612 | No | NA | 2 | 0 | 1 | Not-determined | Genome-fragment | NA | NA | 0 |
| S1_k141_275457 | 1833 | No | NA | 2 | 0 | 0 | Not-determined | Genome-fragment | NA | NA | 0 |
| S1_k141_61850 | 1529 | No | NA | 3 | 0 | 2 | Not-determined | Genome-fragment | NA | NA | 0 |
| S1_k141_260486 | 2048 | No | NA | 2 | 0 | 1 | Not-determined | Genome-fragment | NA | NA | 0 |
| S1_k141_309489 | 15553 | No | NA | 30 | 0 | 29 | Not-determined | Genome-fragment | NA | NA | 0 |
| S17-final-viral-combined_k141_212373\|\|full | 1495 | No | NA | 3 | 1 | 0 | Low-quality | Genome-fragment | 2.81 | AAI-based (medium-confidence) | 0 |
| S1_k141_260562 | 1821 | No | NA | 2 | 0 | 0 | Not-determined | Genome-fragment | NA | NA | 0 |
| S17-final-viral-combined_k141_42262\|\|lt2gene | 1495 | No | NA | 2 | 0 | 0 | Low-quality | Genome-fragment | 3.6 | AAI-based (medium-confidence) | 0 |
| S1_k141_275573 | 3191 | No | NA | 5 | 0 | 1 | Not-determined | Genome-fragment | NA | NA | 0 |
| S1_k141_374634 | 4127 | No | NA | 4 | 0 | 3 | Not-determined | Genome-fragment | NA | NA | 0 |
| S1_k141_374645 | 1118 | No | NA | 1 | 0 | 0 | Not-determined | Genome-fragment | NA | NA | 0 |
| S1_k141_226733 | 1245 | No | NA | 1 | 0 | 1 | Not-determined | Genome-fragment | NA | NA | 0 |
| S1_k141_390889 | 3201 | No | NA | 2 | 0 | 1 | Not-determined | Genome-fragment | NA | NA | 0 |
| S1_k141_45228 | 3358 | No | NA | 4 | 0 | 1 | Not-determined | Genome-fragment | NA | NA | 0 |
| S1_k141_243304 | 3768 | No | NA | 1 | 0 | 0 | Not-determined | Genome-fragment | NA | NA | 0 |
| S1_k141_210922 | 2912 | No | NA | 2 | 0 | 0 | Not-determined | Genome-fragment | NA | NA | 0 |
| S1_k141_260609 | 1515 | No | NA | 1 | 0 | 1 | Not-determined | Genome-fragment | NA | NA | 0 |
| S1_k141_390926 | 1012 | No | NA | 2 | 0 | 0 | Not-determined | Genome-fragment | NA | NA | 0 |
| S1_k141_62006 | 1017 | No | NA | 1 | 0 | 1 | Not-determined | Genome-fragment | NA | NA | 0 |
| S1_k141_326413 | 2039 | No | NA | 3 | 0 | 2 | Not-determined | Genome-fragment | NA | NA | 0 |
| S1_k141_359149 | 1786 | No | NA | 3 | 0 | 1 | Not-determined | Genome-fragment | NA | NA | 0 |
| S1_k141_28838 | 4081 | No | NA | 3 | 0 | 3 | Not-determined | Genome-fragment | NA | NA | 0 |
| S1_k141_77616 | 3172 | No | NA | 3 | 0 | 1 | Not-determined | Genome-fragment | NA | NA | 0 |
| S1_k141_326431 | 2272 | No | NA | 2 | 0 | 1 | Not-determined | Genome-fragment | NA | NA | 0 |
| S1_k141_175978 | 1363 | No | NA | 2 | 0 | 2 | Not-determined | Genome-fragment | NA | NA | 0 |
| S1_k141_293004 | 1210 | No | NA | 2 | 0 | 1 | Not-determined | Genome-fragment | NA | NA | 0 |
| S12-final-viral-combined_k141_112142\|\|full | 1495 | No | NA | 3 | 1 | 0 | Low-quality | Genome-fragment | 3.83 | AAI-based (medium-confidence) | 0 |
| S1_k141_293017 | 1059 | No | NA | 1 | 0 | 1 | Not-determined | Genome-fragment | NA | NA | 0 |
| S1_k141_110068 | 1237 | No | NA | 2 | 0 | 1 | Not-determined | Genome-fragment | NA | NA | 0 |
| S1_k141_391039 | 1165 | No | NA | 2 | 0 | 1 | Not-determined | Genome-fragment | NA | NA | 0 |
| S12-final-viral-combined_k141_199378\|\|lt2gene | 1495 | No | NA | 1 | 1 | 0 | Low-quality | Genome-fragment | 1.03 | HMM-based (lower-bound) | 0 |
| S1_k141_309655 | 1303 | No | NA | 1 | 0 | 1 | Not-determined | Genome-fragment | NA | NA | 0 |
| S4-final-viral-combined_k141_117761\|\|full | 1495 | No | NA | 3 | 1 | 0 | Low-quality | Genome-fragment | 3.06 | HMM-based (lower-bound) | 0 |
| S1_k141_62127 | 2399 | No | NA | 2 | 0 | 2 | Not-determined | Genome-fragment | NA | NA | 0 |
| S1_k141_293083 | 1890 | No | NA | 1 | 0 | 0 | Not-determined | Genome-fragment | NA | NA | 0 |
| S8-final-viral-combined_k141_51325\|\|full | 1495 | No | NA | 3 | 0 | 0 | Low-quality | Genome-fragment | 2.89 | AAI-based (medium-confidence) | 0 |
| S9-final-viral-combined_k141_16218\|\|full | 1496 | No | NA | 2 | 0 | 0 | Low-quality | Genome-fragment | 3.28 | AAI-based (medium-confidence) | 0 |
| S1_k141_28888 | 4476 | No | NA | 1 | 0 | 1 | Not-determined | Genome-fragment | NA | NA | 0 |
| S1_k141_128911 | 2034 | No | NA | 2 | 0 | 0 | Not-determined | Genome-fragment | NA | NA | 0 |
| S1_k141_211091 | 4322 | No | NA | 2 | 0 | 2 | Not-determined | Genome-fragment | NA | NA | 0 |
| S1_k141_62183 | 2386 | No | NA | 3 | 0 | 0 | Not-determined | Genome-fragment | NA | NA | 0 |
| S1_k141_160035 | 1102 | No | NA | 1 | 0 | 1 | Not-determined | Genome-fragment | NA | NA | 0 |
| S9_k141_54897 | 1496 | No | NA | 2 | 0 | 1 | Low-quality | Genome-fragment | 0.36 | AAI-based (medium-confidence) | 0 |
| S1_k141_95196 | 1089 | No | NA | 1 | 0 | 1 | Not-determined | Genome-fragment | NA | NA | 0 |
| S11-final-viral-combined_k141_132896\|\|full | 1496 | No | NA | 4 | 1 | 0 | Low-quality | Genome-fragment | 2.59 | AAI-based (high-confidence) | 0 |
| S1_k141_326581 | 1470 | No | NA | 2 | 0 | 0 | Not-determined | Genome-fragment | NA | NA | 0 |
| S1_k141_309751 | 1370 | No | NA | 1 | 0 | 0 | Not-determined | Genome-fragment | NA | NA | 0 |
| S1_k141_243544 | 1495 | No | NA | 1 | 0 | 0 | Not-determined | Genome-fragment | NA | NA | 0 |
| S1_k141_260827 | 6744 | No | NA | 6 | 0 | 5 | Not-determined | Genome-fragment | NA | NA | 0 |
| S1_k141_326612 | 1445 | No | NA | 2 | 0 | 1 | Not-determined | Genome-fragment | NA | NA | 0 |
| S1_k141_226951 | 1016 | No | NA | 1 | 0 | 0 | Not-determined | Genome-fragment | NA | NA | 0 |
| S1_k141_226972 | 1360 | No | NA | 1 | 0 | 0 | Not-determined | Genome-fragment | NA | NA | 0 |
| S1_k141_128959 | 1014 | No | NA | 1 | 0 | 0 | Not-determined | Genome-fragment | NA | NA | 0 |
| S1_k141_243593 | 3727 | No | NA | 3 | 0 | 2 | Not-determined | Genome-fragment | NA | NA | 0 |
| S1_k141_260869 | 1525 | No | NA | 2 | 0 | 1 | Not-determined | Genome-fragment | NA | NA | 0 |
| S1_k141_340983 | 3949 | No | NA | 7 | 0 | 2 | Not-determined | Genome-fragment | NA | NA | 0 |
| S1_k141_45377 | 3897 | No | NA | 4 | 0 | 2 | Not-determined | Genome-fragment | NA | NA | 0 |
| S1_k141_176069 | 1528 | No | NA | 1 | 0 | 1 | Not-determined | Genome-fragment | NA | NA | 0 |
| S1_k141_95310 | 1184 | No | NA | 2 | 0 | 1 | Not-determined | Genome-fragment | NA | NA | 0 |
| S1_k141_326640 | 1045 | No | NA | 1 | 0 | 1 | Not-determined | Genome-fragment | NA | NA | 0 |
| S1_k141_326641 | 2582 | No | NA | 2 | 0 | 2 | Not-determined | Genome-fragment | NA | NA | 0 |
| S1_k141_326642 | 1006 | No | NA | 2 | 0 | 1 | Not-determined | Genome-fragment | NA | NA | 0 |
| S7-final-viral-combined_k141_153928\|\|full | 1496 | No | NA | 3 | 0 | 0 | Low-quality | Genome-fragment | 3.64 | AAI-based (high-confidence) | 0 |
| S1_k141_341030 | 1113 | No | NA | 1 | 0 | 1 | Not-determined | Genome-fragment | NA | NA | 0 |
| S1_k141_275854 | 2130 | No | NA | 3 | 0 | 2 | Not-determined | Genome-fragment | NA | NA | 0 |
| S1_k141_341036 | 3201 | No | NA | 6 | 0 | 4 | Not-determined | Genome-fragment | NA | NA | 0 |
| S1_k141_129074 | 1478 | No | NA | 3 | 0 | 2 | Not-determined | Genome-fragment | NA | NA | 0 |
| S1_k141_12948 | 1213 | No | NA | 2 | 0 | 1 | Not-determined | Genome-fragment | NA | NA | 0 |
| S1_k141_341047 | 1390 | No | NA | 2 | 0 | 2 | Not-determined | Genome-fragment | NA | NA | 0 |
| S1_k141_160195 | 1298 | No | NA | 2 | 0 | 2 | Not-determined | Genome-fragment | NA | NA | 0 |
| S1_k141_374939 | 2552 | No | NA | 2 | 0 | 2 | Not-determined | Genome-fragment | NA | NA | 0 |
| S1_k141_194742 | 2163 | No | NA | 1 | 0 | 1 | Not-determined | Genome-fragment | NA | NA | 0 |
| S1_k141_110334 | 1424 | No | NA | 1 | 0 | 1 | Not-determined | Genome-fragment | NA | NA | 0 |
| S12-final-viral-combined_k141_178542\|\|full | 1496 | No | NA | 3 | 0 | 1 | Low-quality | Genome-fragment | 0.66 | AAI-based (medium-confidence) | 0 |
| S1_k141_129129 | 1026 | No | NA | 2 | 0 | 2 | Not-determined | Genome-fragment | NA | NA | 0 |
| S1_k141_62367 | 3491 | No | NA | 3 | 0 | 3 | Not-determined | Genome-fragment | NA | NA | 0 |
| S1_k141_211331 | 3503 | No | NA | 5 | 0 | 1 | Not-determined | Genome-fragment | NA | NA | 0 |
| S1_k141_391178 | 1084 | No | NA | 1 | 0 | 1 | Not-determined | Genome-fragment | NA | NA | 0 |
| S1_k141_160256 | 5257 | No | NA | 3 | 0 | 2 | Not-determined | Genome-fragment | NA | NA | 0 |
| S12-final-viral-combined_k141_367003\|\|full | 1496 | No | NA | 4 | 0 | 0 | Low-quality | Genome-fragment | 14.48 | AAI-based (high-confidence) | 0 |
| S1_k141_326769 | 1111 | No | NA | 1 | 0 | 0 | Not-determined | Genome-fragment | NA | NA | 0 |
| S2-final-viral-combined_k141_106430\|\|full | 1496 | No | NA | 3 | 0 | 0 | Low-quality | Genome-fragment | 0.94 | AAI-based (medium-confidence) | 0 |
| S1_k141_261083 | 1243 | No | NA | 2 | 0 | 0 | Not-determined | Genome-fragment | NA | NA | 0 |
| S1_k141_391222 | 1431 | No | NA | 3 | 0 | 0 | Not-determined | Genome-fragment | NA | NA | 0 |
| S1_k141_243865 | 1102 | No | NA | 2 | 0 | 0 | Not-determined | Genome-fragment | NA | NA | 0 |
| S2-final-viral-combined_k141_130373\|\|full | 1496 | No | NA | 3 | 0 | 0 | Low-quality | Genome-fragment | 1.67 | AAI-based (medium-confidence) | 0 |
| S1_k141_359534 | 1203 | No | NA | 2 | 0 | 2 | Not-determined | Genome-fragment | NA | NA | 0 |
| S4_k141_118887 | 1496 | No | NA | 2 | 0 | 0 | Low-quality | Genome-fragment | 5.13 | AAI-based (medium-confidence) | 0 |
| S9-final-viral-combined_k141_67190\|\|full | 1497 | No | NA | 2 | 2 | 0 | Low-quality | Genome-fragment | 4.36 | AAI-based (high-confidence) | 0 |
| S1_k141_391284 | 1162 | No | NA | 1 | 0 | 1 | Not-determined | Genome-fragment | NA | NA | 0 |
| S1_k141_160400 | 1039 | No | NA | 1 | 0 | 0 | Not-determined | Genome-fragment | NA | NA | 0 |
| S1-final-viral-combined_k141_164947\|\|full | 1497 | No | NA | 5 | 1 | 0 | Low-quality | Genome-fragment | 3.73 | AAI-based (medium-confidence) | 0 |
| S1_k141_160417 | 2391 | No | NA | 1 | 0 | 0 | Not-determined | Genome-fragment | NA | NA | 0 |
| S1_k141_29170 | 1027 | No | NA | 1 | 0 | 1 | Not-determined | Genome-fragment | NA | NA | 0 |
| S1-final-viral-combined_k141_225225\|\|full | 1497 | No | NA | 2 | 0 | 0 | Low-quality | Genome-fragment | 3.92 | AAI-based (high-confidence) | 0 |
| S1_k141_29173 | 1820 | No | NA | 2 | 0 | 0 | Not-determined | Genome-fragment | NA | NA | 0 |
| S1_k141_341226 | 1120 | No | NA | 1 | 0 | 1 | Not-determined | Genome-fragment | NA | NA | 0 |
| S1_k141_375151 | 5140 | No | NA | 2 | 0 | 2 | Not-determined | Genome-fragment | NA | NA | 0 |
| S1_k141_129364 | 1746 | No | NA | 2 | 0 | 1 | Not-determined | Genome-fragment | NA | NA | 0 |
| S1_k141_326930 | 1795 | No | NA | 1 | 0 | 1 | Not-determined | Genome-fragment | NA | NA | 0 |
| S5-final-viral-combined_k141_41153\|\|lt2gene | 1497 | No | NA | 2 | 1 | 0 | Low-quality | Genome-fragment | 2.82 | AAI-based (medium-confidence) | 0 |
| S1_k141_176397 | 12993 | No | NA | 13 | 0 | 9 | Not-determined | Genome-fragment | NA | NA | 0 |
| S1_k141_243978 | 1014 | No | NA | 2 | 0 | 0 | Not-determined | Genome-fragment | NA | NA | 0 |
| S1_k141_310117 | 3220 | No | NA | 3 | 0 | 2 | Not-determined | Genome-fragment | NA | NA | 0 |
| S1_k141_95542 | 1038 | No | NA | 2 | 0 | 2 | Not-determined | Genome-fragment | NA | NA | 0 |
| S1_k141_78008 | 1348 | No | NA | 2 | 0 | 0 | Not-determined | Genome-fragment | NA | NA | 0 |
| S12-final-viral-combined_k141_86122\|\|full | 1497 | No | NA | 3 | 0 | 1 | Low-quality | Genome-fragment | 1.99 | AAI-based (medium-confidence) | 0 |
| S1_k141_13227 | 1078 | No | NA | 1 | 0 | 1 | Not-determined | Genome-fragment | NA | NA | 0 |
| S1_k141_95552 | 2832 | No | NA | 1 | 0 | 1 | Not-determined | Genome-fragment | NA | NA | 0 |
| S1_k141_176444 | 1995 | No | NA | 1 | 0 | 0 | Not-determined | Genome-fragment | NA | NA | 0 |
| S1_k141_327001 | 1232 | No | NA | 2 | 0 | 0 | Not-determined | Genome-fragment | NA | NA | 0 |
| S1_k141_129464 | 1851 | No | NA | 2 | 0 | 2 | Not-determined | Genome-fragment | NA | NA | 0 |
| S12-final-viral-combined_k141_358784\|\|full | 1497 | No | NA | 2 | 0 | 2 | Low-quality | Genome-fragment | 0.45 | AAI-based (medium-confidence) | 0 |
| S12-final-viral-combined_k141_178475\|\|full | 1497 | No | NA | 2 | 0 | 1 | Low-quality | Genome-fragment | 1.14 | AAI-based (high-confidence) | 0 |
| S12-final-viral-combined_k141_360894\|\|lt2gene | 1497 | No | NA | 1 | 1 | 0 | Low-quality | Genome-fragment | 0.93 | HMM-based (lower-bound) | 0 |
| S2-final-viral-combined_k141_57555\|\|lt2gene | 1497 | Yes | 826 | 2 | 1 | 1 | Low-quality | Genome-fragment | 2.22 | AAI-based (high-confidence) | 44.82 |
| S1_k141_293585 | 1476 | No | NA | 2 | 0 | 1 | Not-determined | Genome-fragment | NA | NA | 0 |
| S1_k141_341372 | 1013 | No | NA | 2 | 0 | 2 | Not-determined | Genome-fragment | NA | NA | 0 |
| S1_k141_227380 | 1215 | No | NA | 1 | 0 | 0 | Not-determined | Genome-fragment | NA | NA | 0 |
| S1_k141_45686 | 2284 | No | NA | 2 | 0 | 0 | Not-determined | Genome-fragment | NA | NA | 0 |
| S1_k141_129513 | 1566 | No | NA | 1 | 0 | 1 | Not-determined | Genome-fragment | NA | NA | 0 |
| S1_k141_160613 | 1107 | No | NA | 3 | 0 | 0 | Not-determined | Genome-fragment | NA | NA | 0 |
| S1_k141_359628 | 1324 | No | NA | 1 | 0 | 1 | Not-determined | Genome-fragment | NA | NA | 0 |
| S1_k141_45699 | 2063 | No | NA | 6 | 0 | 5 | Not-determined | Genome-fragment | NA | NA | 0 |
| S4-final-viral-combined_k141_141426\|\|full | 1497 | No | NA | 2 | 0 | 0 | Low-quality | Genome-fragment | 2.44 | AAI-based (high-confidence) | 0 |
| S8-final-viral-combined_k141_243883\|\|full | 1497 | No | NA | 3 | 0 | 0 | Low-quality | Genome-fragment | 2.21 | AAI-based (medium-confidence) | 0 |
| S1_k141_375336 | 1872 | No | NA | 3 | 0 | 2 | Not-determined | Genome-fragment | NA | NA | 0 |
| S9-final-viral-combined_k141_28619\|\|full | 1498 | No | NA | 3 | 0 | 0 | Low-quality | Genome-fragment | 2.6 | AAI-based (high-confidence) | 0 |
| S1_k141_276289 | 1272 | No | NA | 1 | 0 | 1 | Not-determined | Genome-fragment | NA | NA | 0 |
| S1_k141_195134 | 1280 | No | NA | 1 | 0 | 1 | Not-determined | Genome-fragment | NA | NA | 0 |
| S1_k141_276317 | 1219 | No | NA | 1 | 0 | 1 | Not-determined | Genome-fragment | NA | NA | 0 |
| S1-final-viral-combined_k141_49735\|\|full | 1498 | No | NA | 3 | 0 | 2 | Low-quality | Genome-fragment | 1.2 | AAI-based (medium-confidence) | 0 |
| S1_k141_327027 | 1317 | No | NA | 2 | 0 | 0 | Not-determined | Genome-fragment | NA | NA | 0 |
| S1-final-viral-combined_k141_39761\|\|full | 1498 | No | NA | 4 | 1 | 0 | Low-quality | Genome-fragment | 3.25 | AAI-based (high-confidence) | 0 |
| S1_k141_95750 | 1603 | No | NA | 3 | 0 | 0 | Not-determined | Genome-fragment | NA | NA | 0 |
| S1-final-viral-combined_k141_156401\|\|full | 1498 | No | NA | 3 | 0 | 1 | Low-quality | Genome-fragment | 0.72 | AAI-based (medium-confidence) | 0 |
| S1_k141_293684 | 1160 | No | NA | 2 | 0 | 1 | Not-determined | Genome-fragment | NA | NA | 0 |
| S1_k141_95764 | 1266 | No | NA | 1 | 0 | 1 | Not-determined | Genome-fragment | NA | NA | 0 |
| S1_k141_13398 | 1129 | No | NA | 1 | 0 | 1 | Not-determined | Genome-fragment | NA | NA | 0 |
| S1_k141_145114 | 1036 | No | NA | 2 | 0 | 1 | Not-determined | Genome-fragment | NA | NA | 0 |
| S1_k141_126124 | 1498 | No | NA | 4 | 2 | 0 | Low-quality | Genome-fragment | 2.67 | AAI-based (high-confidence) | 0 |
| S5-final-viral-combined_k141_206570\|\|full | 1498 | No | NA | 3 | 2 | 0 | Low-quality | Genome-fragment | 2.26 | AAI-based (medium-confidence) | 0 |
| S1_k141_391556 | 4989 | No | NA | 5 | 0 | 0 | Not-determined | Genome-fragment | NA | NA | 0 |
| S1_k141_327075 | 3752 | No | NA | 3 | 0 | 3 | Not-determined | Genome-fragment | NA | NA | 0 |
| S1_k141_391561 | 1173 | No | NA | 3 | 0 | 2 | Not-determined | Genome-fragment | NA | NA | 0 |
| S17-final-viral-combined_k141_147556\|\|full | 1498 | No | NA | 2 | 0 | 0 | Low-quality | Genome-fragment | 3.03 | AAI-based (medium-confidence) | 0 |
| S17-final-viral-combined_k141_9346\|\|full | 1498 | No | NA | 3 | 1 | 0 | Low-quality | Genome-fragment | 2.79 | AAI-based (medium-confidence) | 0 |
| S1_k141_244325 | 1720 | No | NA | 2 | 0 | 0 | Not-determined | Genome-fragment | NA | NA | 0 |
| S12-final-viral-combined_k141_323134\|\|full | 1498 | No | NA | 2 | 0 | 1 | Low-quality | Genome-fragment | 1.9 | AAI-based (high-confidence) | 0 |
| S1_k141_293772 | 1081 | No | NA | 1 | 0 | 0 | Not-determined | Genome-fragment | NA | NA | 0 |
| S12-final-viral-combined_k141_139936\|\|full | 1498 | No | NA | 2 | 0 | 2 | Low-quality | Genome-fragment | 1.79 | AAI-based (medium-confidence) | 0 |
| S1_k141_391603 | 1033 | No | NA | 1 | 0 | 0 | Not-determined | Genome-fragment | NA | NA | 0 |
| S2-final-viral-combined_k141_45544\|\|full | 1498 | No | NA | 2 | 0 | 0 | Low-quality | Genome-fragment | 2.45 | AAI-based (medium-confidence) | 0 |
| S1_k141_78232 | 1459 | No | NA | 1 | 0 | 1 | Not-determined | Genome-fragment | NA | NA | 0 |
| S1_k141_176690 | 1838 | No | NA | 2 | 0 | 1 | Not-determined | Genome-fragment | NA | NA | 0 |
| S1_k141_95887 | 1241 | No | NA | 1 | 0 | 1 | Not-determined | Genome-fragment | NA | NA | 0 |
| S1_k141_375450 | 1889 | No | NA | 1 | 0 | 0 | Not-determined | Genome-fragment | NA | NA | 0 |
| S1_k141_95889 | 1060 | No | NA | 1 | 0 | 0 | Not-determined | Genome-fragment | NA | NA | 0 |
| S1_k141_62722 | 1218 | No | NA | 3 | 0 | 0 | Not-determined | Genome-fragment | NA | NA | 0 |
| S2-final-viral-combined_k141_67332\|\|lt2gene | 1498 | No | NA | 1 | 1 | 0 | Low-quality | Genome-fragment | 4.01 | AAI-based (medium-confidence) | 0 |
| S1_k141_212042 | 1251 | No | NA | 1 | 0 | 0 | Not-determined | Genome-fragment | NA | NA | 0 |
| S1_k141_29288 | 47233 | No | NA | 58 | 17 | 1 | Medium-quality | Genome-fragment | 87 | AAI-based (high-confidence) | 0 |
| S4-final-viral-combined_k141_53823\|\|full | 1498 | No | NA | 4 | 2 | 0 | Low-quality | Genome-fragment | 2.33 | AAI-based (high-confidence) | 0 |
| S8-final-viral-combined_k141_36235\|\|full | 1498 | No | NA | 2 | 0 | 0 | Low-quality | Genome-fragment | 1.74 | AAI-based (high-confidence) | 0 |
| S1_k141_276545 | 5999 | No | NA | 7 | 0 | 3 | Not-determined | Genome-fragment | NA | NA | 0 |
| S9-final-viral-combined_k141_96345\|\|full | 1499 | No | NA | 3 | 0 | 0 | Low-quality | Genome-fragment | 17.03 | AAI-based (medium-confidence) | 0 |
| S1_k141_45965 | 1039 | No | NA | 1 | 0 | 1 | Not-determined | Genome-fragment | NA | NA | 0 |
| S1_k141_227687 | 1032 | No | NA | 4 | 0 | 0 | Not-determined | Genome-fragment | NA | NA | 0 |
| S1_k141_46004 | 2690 | No | NA | 2 | 0 | 1 | Not-determined | Genome-fragment | NA | NA | 0 |
| S1_k141_78367 | 3977 | No | NA | 6 | 0 | 1 | Not-determined | Genome-fragment | NA | NA | 0 |
| S11-final-viral-combined_k141_99899\|\|full | 1499 | No | NA | 2 | 0 | 2 | Low-quality | Genome-fragment | 1.27 | AAI-based (medium-confidence) | 0 |
| S1_k141_96021 | 1215 | No | NA | 1 | 0 | 0 | Not-determined | Genome-fragment | NA | NA | 0 |
| S1_k141_244547 | 1010 | No | NA | 1 | 0 | 0 | Not-determined | Genome-fragment | NA | NA | 0 |
| S1_k141_145419 | 4246 | No | NA | 3 | 0 | 2 | Not-determined | Genome-fragment | NA | NA | 0 |
| S11-final-viral-combined_k141_257859\|\|full | 1499 | No | NA | 2 | 0 | 0 | Low-quality | Genome-fragment | 1.11 | AAI-based (medium-confidence) | 0 |
| S1_k141_195463 | 1044 | No | NA | 1 | 0 | 0 | Not-determined | Genome-fragment | NA | NA | 0 |
| S1_k141_227782 | 7723 | No | NA | 6 | 0 | 5 | Not-determined | Genome-fragment | NA | NA | 0 |
| S1_k141_227794 | 3850 | No | NA | 3 | 0 | 3 | Not-determined | Genome-fragment | NA | NA | 0 |
| S7_k141_113762 | 1499 | No | NA | 3 | 0 | 0 | Low-quality | Genome-fragment | 2.47 | AAI-based (high-confidence) | 0 |
| S1_k141_359974 | 1105 | No | NA | 3 | 0 | 0 | Not-determined | Genome-fragment | NA | NA | 0 |
| S17-k141_173680 | 1499 | No | NA | 2 | 0 | 0 | Low-quality | Genome-fragment | 2.39 | AAI-based (high-confidence) | 0 |
| S12_k141_195817 | 1499 | No | NA | 4 | 0 | 0 | Low-quality | Genome-fragment | 0.81 | AAI-based (high-confidence) | 0 |
| S1_k141_161085 | 1050 | No | NA | 1 | 0 | 1 | Not-determined | Genome-fragment | NA | NA | 0 |
| S1_k141_195503 | 1414 | No | NA | 1 | 0 | 1 | Not-determined | Genome-fragment | NA | NA | 0 |
| S1_k141_310407 | 5153 | No | NA | 4 | 0 | 1 | Not-determined | Genome-fragment | NA | NA | 0 |
| S1_k141_129998 | 2585 | No | NA | 1 | 0 | 1 | Not-determined | Genome-fragment | NA | NA | 0 |
| S12_k141_113847 | 1499 | No | NA | 2 | 0 | 0 | Low-quality | Genome-fragment | 1.06 | AAI-based (high-confidence) | 0 |
| S2-final-viral-combined_k141_90169\|\|lt2gene | 1499 | No | NA | 1 | 0 | 0 | Low-quality | Genome-fragment | 3.87 | AAI-based (high-confidence) | 0 |
| S1_k141_161120 | 1849 | No | NA | 2 | 0 | 0 | Not-determined | Genome-fragment | NA | NA | 0 |
| S1_k141_261711 | 1668 | No | NA | 1 | 0 | 0 | Not-determined | Genome-fragment | NA | NA | 0 |
| S1_k141_391799 | 1328 | No | NA | 1 | 0 | 0 | Not-determined | Genome-fragment | NA | NA | 0 |
| S1_k141_176978 | 1219 | No | NA | 1 | 0 | 0 | Not-determined | Genome-fragment | NA | NA | 0 |
| S1_k141_13787 | 2373 | No | NA | 1 | 0 | 1 | Not-determined | Genome-fragment | NA | NA | 0 |
| S4_k141_2702 | 1499 | No | NA | 1 | 0 | 1 | Low-quality | Genome-fragment | 0.42 | AAI-based (high-confidence) | 0 |
| S1_k141_176981 | 1819 | No | NA | 2 | 0 | 1 | Not-determined | Genome-fragment | NA | NA | 0 |
| S4_k141_108384 | 1499 | No | NA | 3 | 1 | 0 | Low-quality | Genome-fragment | 2.39 | AAI-based (high-confidence) | 0 |
| S1_k141_294057 | 2444 | No | NA | 3 | 0 | 1 | Not-determined | Genome-fragment | NA | NA | 0 |
| S1_k141_195565 | 2408 | No | NA | 2 | 0 | 2 | Not-determined | Genome-fragment | NA | NA | 0 |
| S1_k141_375674 | 6554 | No | NA | 6 | 0 | 5 | Not-determined | Genome-fragment | NA | NA | 0 |
| S8-final-viral-combined_k141_199834\|\|full | 1499 | No | NA | 3 | 0 | 0 | Low-quality | Genome-fragment | 1.75 | AAI-based (medium-confidence) | 0 |
| S1_k141_227834 | 1284 | No | NA | 1 | 0 | 1 | Not-determined | Genome-fragment | NA | NA | 0 |
| S1_k141_294068 | 1230 | No | NA | 1 | 0 | 1 | Not-determined | Genome-fragment | NA | NA | 0 |
| S1_k141_78484 | 1123 | No | NA | 1 | 0 | 0 | Not-determined | Genome-fragment | NA | NA | 0 |
| S1_k141_261759 | 2456 | No | NA | 4 | 0 | 0 | Not-determined | Genome-fragment | NA | NA | 0 |
| S1_k141_227847 | 1023 | No | NA | 1 | 0 | 1 | Not-determined | Genome-fragment | NA | NA | 0 |
| S9-final-viral-combined_k141_128110\|\|full | 1500 | No | NA | 3 | 1 | 0 | Low-quality | Genome-fragment | 3.78 | AAI-based (high-confidence) | 0 |
| S1_k141_176997 | 1277 | No | NA | 2 | 0 | 2 | Not-determined | Genome-fragment | NA | NA | 0 |
| S9_k141_99042 | 1500 | No | NA | 3 | 0 | 0 | Low-quality | Genome-fragment | 2.86 | AAI-based (high-confidence) | 0 |
| S1_k141_360071 | 3382 | No | NA | 1 | 0 | 1 | Not-determined | Genome-fragment | NA | NA | 0 |
| S11-final-viral-combined_k141_312160\|\|full | 1500 | No | NA | 3 | 0 | 0 | Low-quality | Genome-fragment | 4.1 | AAI-based (high-confidence) | 0 |
| S1_k141_46230 | 2004 | No | NA | 1 | 0 | 0 | Not-determined | Genome-fragment | NA | NA | 0 |
| S1_k141_145610 | 2271 | No | NA | 2 | 0 | 1 | Not-determined | Genome-fragment | NA | NA | 0 |
| S1_k141_195618 | 3248 | No | NA | 5 | 0 | 2 | Not-determined | Genome-fragment | NA | NA | 0 |
| S1_k141_111171 | 1398 | No | NA | 1 | 0 | 1 | Not-determined | Genome-fragment | NA | NA | 0 |
| S1_k141_130155 | 2432 | No | NA | 3 | 0 | 2 | Not-determined | Genome-fragment | NA | NA | 0 |
| S11-final-viral-combined_k141_232933\|\|full | 1500 | No | NA | 3 | 1 | 0 | Low-quality | Genome-fragment | 0.95 | HMM-based (lower-bound) | 0 |
| S1_k141_294133 | 3582 | No | NA | 3 | 0 | 2 | Not-determined | Genome-fragment | NA | NA | 0 |
| S1_k141_261867 | 1067 | No | NA | 1 | 0 | 1 | Not-determined | Genome-fragment | NA | NA | 0 |
| S1_k141_78535 | 1800 | No | NA | 2 | 0 | 2 | Not-determined | Genome-fragment | NA | NA | 0 |
| S1_k141_244836 | 31397 | No | NA | 35 | 9 | 0 | Medium-quality | Genome-fragment | 57.64 | AAI-based (high-confidence) | 0 |
| S1-final-viral-combined_k141_149900\|\|full | 1500 | No | NA | 3 | 1 | 0 | Low-quality | Genome-fragment | 29.92 | AAI-based (medium-confidence) | 0 |
| S1-final-viral-combined_k141_283377\|\|full | 1500 | No | NA | 3 | 0 | 0 | Low-quality | Genome-fragment | 0.84 | AAI-based (medium-confidence) | 0 |
| S3-final-viral-combined_k141_80656\|\|full | 1500 | No | NA | 5 | 1 | 0 | Low-quality | Genome-fragment | 2.48 | AAI-based (medium-confidence) | 0 |
| S1_k141_391933 | 1792 | No | NA | 2 | 0 | 1 | Not-determined | Genome-fragment | NA | NA | 0 |
| S5-final-viral-combined_k141_195413\|\|full | 1500 | No | NA | 2 | 0 | 1 | Low-quality | Genome-fragment | 3.73 | AAI-based (medium-confidence) | 0 |
| S1_k141_212405 | 1959 | No | NA | 3 | 0 | 3 | Not-determined | Genome-fragment | NA | NA | 0 |
| S7-final-viral-combined_k141_204684\|\|full | 1500 | No | NA | 5 | 0 | 1 | Low-quality | Genome-fragment | 2.07 | AAI-based (high-confidence) | 0 |
| S1_k141_276846 | 1148 | No | NA | 1 | 0 | 1 | Not-determined | Genome-fragment | NA | NA | 0 |
| S1_k141_195748 | 1074 | No | NA | 1 | 0 | 0 | Not-determined | Genome-fragment | NA | NA | 0 |
| S1_k141_212430 | 1009 | No | NA | 1 | 0 | 1 | Not-determined | Genome-fragment | NA | NA | 0 |
| S1_k141_161382 | 18199 | No | NA | 31 | 0 | 31 | Not-determined | Genome-fragment | NA | NA | 0 |
| S1_k141_391955 | 4268 | No | NA | 9 | 0 | 1 | Not-determined | Genome-fragment | NA | NA | 0 |
| S1_k141_130214 | 40887 | No | NA | 77 | 18 | 0 | Complete | High-quality | 100 | DTR (high-confidence) | 0 |
| S1_k141_261998 | 1025 | No | NA | 1 | 0 | 1 | Not-determined | Genome-fragment | NA | NA | 0 |
| S1_k141_294179 | 1674 | No | NA | 1 | 0 | 1 | Not-determined | Genome-fragment | NA | NA | 0 |
| S1_k141_227982 | 1192 | No | NA | 2 | 0 | 0 | Not-determined | Genome-fragment | NA | NA | 0 |
| S17-final-viral-combined_k141_105422\|\|full | 1500 | No | NA | 6 | 2 | 0 | Low-quality | Genome-fragment | 4.02 | AAI-based (medium-confidence) | 0 |
| S1_k141_46269 | 2494 | No | NA | 1 | 0 | 0 | Not-determined | Genome-fragment | NA | NA | 0 |
| S1_k141_145690 | 1067 | No | NA | 1 | 0 | 1 | Not-determined | Genome-fragment | NA | NA | 0 |
| S1_k141_360250 | 1158 | No | NA | 1 | 0 | 0 | Not-determined | Genome-fragment | NA | NA | 0 |
| S1_k141_111310 | 1092 | No | NA | 2 | 0 | 1 | Not-determined | Genome-fragment | NA | NA | 0 |
| S1_k141_375905 | 8836 | No | NA | 8 | 0 | 8 | Not-determined | Genome-fragment | NA | NA | 0 |
| S1_k141_276908 | 1174 | No | NA | 3 | 0 | 0 | Not-determined | Genome-fragment | NA | NA | 0 |
| S1_k141_96258 | 1165 | No | NA | 3 | 0 | 1 | Not-determined | Genome-fragment | NA | NA | 0 |
| S1_k141_310660 | 1750 | No | NA | 2 | 0 | 0 | Not-determined | Genome-fragment | NA | NA | 0 |
| S12-final-viral-combined_k141_169768\|\|full | 1500 | No | NA | 3 | 0 | 0 | Low-quality | Genome-fragment | 2.17 | AAI-based (high-confidence) | 0 |
| S1_k141_228047 | 1332 | No | NA | 2 | 0 | 0 | Not-determined | Genome-fragment | NA | NA | 0 |
| S1_k141_276965 | 1004 | No | NA | 1 | 0 | 1 | Not-determined | Genome-fragment | NA | NA | 0 |
| S12_k141_341736 | 1500 | No | NA | 2 | 0 | 0 | Low-quality | Genome-fragment | 0.79 | AAI-based (medium-confidence) | 0 |
| S1_k141_130313 | 1031 | No | NA | 4 | 0 | 3 | Not-determined | Genome-fragment | NA | NA | 0 |
| S2_k141_11138 | 1500 | No | NA | 4 | 0 | 0 | Low-quality | Genome-fragment | 3.08 | AAI-based (high-confidence) | 0 |
| S4-final-viral-combined_k141_104933\|\|full | 1500 | No | NA | 3 | 0 | 2 | Low-quality | Genome-fragment | 0.93 | AAI-based (medium-confidence) | 0 |
| S11_k141_326311 | 1501 | No | NA | 1 | 0 | 0 | Low-quality | Genome-fragment | 2.72 | AAI-based (high-confidence) | 0 |
| S1_k141_277010 | 3006 | No | NA | 2 | 0 | 2 | Not-determined | Genome-fragment | NA | NA | 0 |
| S1_k141_228123 | 1023 | No | NA | 1 | 0 | 1 | Not-determined | Genome-fragment | NA | NA | 0 |
| S1_k141_195983 | 2023 | No | NA | 4 | 0 | 0 | Not-determined | Genome-fragment | NA | NA | 0 |
| S1_k141_78677 | 6683 | No | NA | 7 | 0 | 2 | Not-determined | Genome-fragment | NA | NA | 0 |
| S1_k141_111408 | 4537 | No | NA | 4 | 0 | 4 | Not-determined | Genome-fragment | NA | NA | 0 |
| S1_k141_63344 | 1850 | No | NA | 2 | 0 | 1 | Not-determined | Genome-fragment | NA | NA | 0 |
| S3-final-viral-combined_k141_103080\|\|full | 1501 | No | NA | 2 | 0 | 0 | Low-quality | Genome-fragment | 2.29 | AAI-based (high-confidence) | 0 |
| S1_k141_245059 | 1039 | No | NA | 2 | 0 | 1 | Not-determined | Genome-fragment | NA | NA | 0 |
| S1_k141_327344 | 1012 | No | NA | 1 | 0 | 1 | Not-determined | Genome-fragment | NA | NA | 0 |
| S1_k141_46367 | 1021 | No | NA | 2 | 0 | 2 | Not-determined | Genome-fragment | NA | NA | 0 |
| S1_k141_294358 | 1383 | No | NA | 2 | 0 | 1 | Not-determined | Genome-fragment | NA | NA | 0 |
| S1_k141_376106 | 1512 | No | NA | 2 | 0 | 1 | Not-determined | Genome-fragment | NA | NA | 0 |
| S1_k141_177495 | 1135 | No | NA | 1 | 0 | 1 | Not-determined | Genome-fragment | NA | NA | 0 |
| S1_k141_145933 | 2124 | No | NA | 1 | 0 | 1 | Not-determined | Genome-fragment | NA | NA | 0 |
| S1_k141_14189 | 1082 | No | NA | 2 | 0 | 0 | Not-determined | Genome-fragment | NA | NA | 0 |
| S1_k141_392096 | 1198 | No | NA | 2 | 0 | 0 | Not-determined | Genome-fragment | NA | NA | 0 |
| S17-final-viral-combined_k141_115834\|\|full | 1501 | No | NA | 3 | 0 | 0 | Low-quality | Genome-fragment | 0.77 | AAI-based (high-confidence) | 0 |
| S1_k141_96375 | 1200 | No | NA | 3 | 0 | 2 | Not-determined | Genome-fragment | NA | NA | 0 |
| S1_k141_46476 | 1208 | No | NA | 1 | 0 | 1 | Not-determined | Genome-fragment | NA | NA | 0 |
| S1_k141_177565 | 1122 | No | NA | 1 | 0 | 0 | Not-determined | Genome-fragment | NA | NA | 0 |
| S1_k141_392141 | 1320 | No | NA | 3 | 0 | 2 | Not-determined | Genome-fragment | NA | NA | 0 |
| S1_k141_294466 | 1169 | No | NA | 1 | 0 | 0 | Not-determined | Genome-fragment | NA | NA | 0 |
| S1_k141_342311 | 1755 | No | NA | 1 | 0 | 1 | Not-determined | Genome-fragment | NA | NA | 0 |
| S1_k141_262279 | 1716 | No | NA | 1 | 0 | 1 | Not-determined | Genome-fragment | NA | NA | 0 |
| S1_k141_245143 | 1568 | No | NA | 2 | 0 | 1 | Not-determined | Genome-fragment | NA | NA | 0 |
| S1_k141_376211 | 1044 | No | NA | 1 | 0 | 1 | Not-determined | Genome-fragment | NA | NA | 0 |
| S1_k141_161698 | 4421 | No | NA | 3 | 0 | 0 | Not-determined | Genome-fragment | NA | NA | 0 |
| S17-final-viral-combined_k141_93626\|\|full | 1501 | No | NA | 3 | 0 | 0 | Low-quality | Genome-fragment | 2.65 | AAI-based (high-confidence) | 0 |
| S17-k141_170506 | 1501 | No | NA | 5 | 5 | 0 | Low-quality | Genome-fragment | 4.96 | AAI-based (high-confidence) | 0 |
| S1_k141_14279 | 1231 | No | NA | 2 | 0 | 2 | Not-determined | Genome-fragment | NA | NA | 0 |
| S12-final-viral-combined_k141_262051\|\|full | 1501 | No | NA | 3 | 0 | 1 | Low-quality | Genome-fragment | 1.27 | AAI-based (medium-confidence) | 0 |
| S1_k141_196198 | 1095 | No | NA | 2 | 0 | 0 | Not-determined | Genome-fragment | NA | NA | 0 |
| S1_k141_277239 | 3374 | No | NA | 1 | 0 | 1 | Not-determined | Genome-fragment | NA | NA | 0 |
| S1_k141_262373 | 1086 | No | NA | 1 | 0 | 1 | Not-determined | Genome-fragment | NA | NA | 0 |
| S1_k141_177647 | 17690 | No | NA | 19 | 0 | 2 | Not-determined | Genome-fragment | NA | NA | 0 |
| S12_k141_269148 | 1501 | No | NA | 1 | 0 | 1 | Low-quality | Genome-fragment | 0.39 | AAI-based (high-confidence) | 0 |
| S1_k141_196243 | 2796 | No | NA | 4 | 0 | 2 | Not-determined | Genome-fragment | NA | NA | 0 |
| S4-final-viral-combined_k141_69810\|\|full | 1501 | Yes | 1348 | 4 | 1 | 1 | Low-quality | Genome-fragment | 0.83 | HMM-based (lower-bound) | 10.19 |
| S1_k141_311020 | 1411 | No | NA | 1 | 0 | 1 | Not-determined | Genome-fragment | NA | NA | 0 |
| S1_k141_327599 | 1680 | No | NA | 1 | 0 | 0 | Not-determined | Genome-fragment | NA | NA | 0 |
| S1_k141_262465 | 1127 | No | NA | 2 | 0 | 2 | Not-determined | Genome-fragment | NA | NA | 0 |
| S1_k141_63663 | 1434 | No | NA | 1 | 0 | 1 | Not-determined | Genome-fragment | NA | NA | 0 |
| S1_k141_14348 | 1790 | No | NA | 2 | 0 | 2 | Not-determined | Genome-fragment | NA | NA | 0 |
| S1_k141_96601 | 1236 | No | NA | 1 | 0 | 0 | Not-determined | Genome-fragment | NA | NA | 0 |
| S1_k141_130561 | 1251 | No | NA | 1 | 0 | 1 | Not-determined | Genome-fragment | NA | NA | 0 |
| S8-final-viral-combined_k141_116701\|\|full | 1501 | No | NA | 3 | 0 | 0 | Low-quality | Genome-fragment | 2.52 | AAI-based (high-confidence) | 0 |
| S1_k141_245375 | 1119 | No | NA | 2 | 0 | 2 | Not-determined | Genome-fragment | NA | NA | 0 |
| S1_k141_262497 | 1172 | No | NA | 2 | 0 | 2 | Not-determined | Genome-fragment | NA | NA | 0 |
| S1_k141_342547 | 1450 | No | NA | 1 | 0 | 1 | Not-determined | Genome-fragment | NA | NA | 0 |
| S1_k141_130621 | 1353 | No | NA | 1 | 0 | 0 | Not-determined | Genome-fragment | NA | NA | 0 |
| S1_k141_262516 | 1063 | No | NA | 2 | 0 | 0 | Not-determined | Genome-fragment | NA | NA | 0 |
| S1_k141_196400 | 3263 | No | NA | 3 | 0 | 1 | Not-determined | Genome-fragment | NA | NA | 0 |
| S1_k141_14392 | 1641 | No | NA | 1 | 0 | 1 | Not-determined | Genome-fragment | NA | NA | 0 |
| S8-final-viral-combined_k141_209772\|\|full | 1501 | No | NA | 2 | 1 | 0 | Low-quality | Genome-fragment | 3.28 | AAI-based (high-confidence) | 0 |
| S1_k141_96725 | 1282 | No | NA | 1 | 0 | 0 | Not-determined | Genome-fragment | NA | NA | 0 |
| S1_k141_294751 | 1630 | No | NA | 2 | 0 | 1 | Not-determined | Genome-fragment | NA | NA | 0 |
| S1_k141_327733 | 3107 | No | NA | 2 | 0 | 0 | Not-determined | Genome-fragment | NA | NA | 0 |
| S1_k141_376420 | 2050 | No | NA | 2 | 0 | 1 | Not-determined | Genome-fragment | NA | NA | 0 |
| S1_k141_79081 | 5022 | No | NA | 5 | 0 | 2 | Not-determined | Genome-fragment | NA | NA | 0 |
| S1_k141_14441 | 1258 | No | NA | 2 | 0 | 2 | Not-determined | Genome-fragment | NA | NA | 0 |
| S9-final-viral-combined_k141_11752\|\|full | 1502 | No | NA | 1 | 0 | 0 | Low-quality | Genome-fragment | 3.88 | AAI-based (medium-confidence) | 0 |
| S1_k141_177897 | 1018 | No | NA | 1 | 0 | 1 | Not-determined | Genome-fragment | NA | NA | 0 |
| S1_k141_130686 | 1309 | No | NA | 1 | 0 | 1 | Not-determined | Genome-fragment | NA | NA | 0 |
| S1_k141_342628 | 2412 | No | NA | 1 | 0 | 1 | Not-determined | Genome-fragment | NA | NA | 0 |
| S1_k141_212940 | 59765 | No | NA | 88 | 36 | 1 | High-quality | High-quality | 99.32 | AAI-based (high-confidence) | 0 |
| S1_k141_342631 | 1772 | No | NA | 1 | 0 | 1 | Not-determined | Genome-fragment | NA | NA | 0 |
| S1_k141_63756 | 7597 | No | NA | 4 | 0 | 2 | Not-determined | Genome-fragment | NA | NA | 0 |
| S1_k141_294822 | 1089 | No | NA | 1 | 0 | 1 | Not-determined | Genome-fragment | NA | NA | 0 |
| S11-final-viral-combined_k141_190355\|\|full | 1502 | No | NA | 3 | 0 | 0 | Low-quality | Genome-fragment | 0.95 | AAI-based (medium-confidence) | 0 |
| S1_k141_360724 | 1439 | No | NA | 1 | 0 | 1 | Not-determined | Genome-fragment | NA | NA | 0 |
| S1_k141_392479 | 2312 | No | NA | 2 | 0 | 1 | Not-determined | Genome-fragment | NA | NA | 0 |
| S1_k141_360751 | 1295 | No | NA | 1 | 0 | 0 | Not-determined | Genome-fragment | NA | NA | 0 |
| S1_k141_177981 | 1124 | No | NA | 2 | 0 | 1 | Not-determined | Genome-fragment | NA | NA | 0 |
| S1-final-viral-combined_k141_271638\|\|full | 1502 | No | NA | 5 | 3 | 0 | Low-quality | Genome-fragment | 1.1 | AAI-based (medium-confidence) | 0 |
| S1_k141_311393 | 1329 | No | NA | 1 | 0 | 1 | Not-determined | Genome-fragment | NA | NA | 0 |
| S1_k141_311408 | 2492 | No | NA | 4 | 0 | 0 | Not-determined | Genome-fragment | NA | NA | 0 |
| S1_k141_327852 | 7740 | No | NA | 7 | 0 | 6 | Not-determined | Genome-fragment | NA | NA | 0 |
| S3-final-viral-combined_k141_40271\|\|full | 1502 | No | NA | 2 | 0 | 1 | Low-quality | Genome-fragment | 0.81 | AAI-based (medium-confidence) | 0 |
| S7_k141_78975 | 1502 | No | NA | 1 | 0 | 0 | Low-quality | Genome-fragment | 1.99 | AAI-based (medium-confidence) | 0 |
| S1_k141_342775 | 1033 | No | NA | 1 | 0 | 0 | Not-determined | Genome-fragment | NA | NA | 0 |
| S1_k141_277497 | 1026 | No | NA | 3 | 0 | 0 | Not-determined | Genome-fragment | NA | NA | 0 |
| S1_k141_376568 | 1194 | No | NA | 2 | 0 | 0 | Not-determined | Genome-fragment | NA | NA | 0 |
| S1_k141_327866 | 1332 | No | NA | 1 | 0 | 0 | Not-determined | Genome-fragment | NA | NA | 0 |
| S1_k141_96922 | 1200 | No | NA | 1 | 0 | 0 | Not-determined | Genome-fragment | NA | NA | 0 |
| S1_k141_327894 | 3459 | No | NA | 2 | 0 | 1 | Not-determined | Genome-fragment | NA | NA | 0 |
| S1_k141_178064 | 2554 | No | NA | 1 | 0 | 1 | Not-determined | Genome-fragment | NA | NA | 0 |
| S1_k141_360859 | 4809 | No | NA | 7 | 0 | 6 | Not-determined | Genome-fragment | NA | NA | 0 |
| S1_k141_162184 | 1000 | No | NA | 2 | 0 | 0 | Not-determined | Genome-fragment | NA | NA | 0 |
| S1_k141_360866 | 3023 | No | NA | 2 | 0 | 2 | Not-determined | Genome-fragment | NA | NA | 0 |
| S1_k141_360872 | 3325 | No | NA | 2 | 0 | 0 | Not-determined | Genome-fragment | NA | NA | 0 |
| S1_k141_162208 | 1073 | No | NA | 1 | 0 | 1 | Not-determined | Genome-fragment | NA | NA | 0 |
| S1_k141_228536 | 7871 | No | NA | 2 | 0 | 1 | Not-determined | Genome-fragment | NA | NA | 0 |
| S1_k141_162216 | 1256 | No | NA | 2 | 0 | 0 | Not-determined | Genome-fragment | NA | NA | 0 |
| S1_k141_178112 | 1000 | No | NA | 1 | 0 | 1 | Not-determined | Genome-fragment | NA | NA | 0 |
| S1_k141_327924 | 1011 | No | NA | 1 | 0 | 0 | Not-determined | Genome-fragment | NA | NA | 0 |
| S1_k141_311510 | 1184 | No | NA | 1 | 0 | 1 | Not-determined | Genome-fragment | NA | NA | 0 |
| S1_k141_392641 | 1358 | No | NA | 1 | 0 | 1 | Not-determined | Genome-fragment | NA | NA | 0 |
| S1_k141_146322 | 1602 | No | NA | 2 | 0 | 0 | Not-determined | Genome-fragment | NA | NA | 0 |
| S1_k141_327945 | 1554 | No | NA | 3 | 0 | 2 | Not-determined | Genome-fragment | NA | NA | 0 |
| S1_k141_262779 | 6432 | No | NA | 5 | 0 | 5 | Not-determined | Genome-fragment | NA | NA | 0 |
| S1_k141_376598 | 3395 | No | NA | 5 | 0 | 3 | Not-determined | Genome-fragment | NA | NA | 0 |
| S1_k141_14739 | 1084 | No | NA | 1 | 0 | 1 | Not-determined | Genome-fragment | NA | NA | 0 |
| S1_k141_311577 | 2212 | No | NA | 2 | 0 | 2 | Not-determined | Genome-fragment | NA | NA | 0 |
| S17-final-viral-combined_k141_160420\|\|full | 1502 | No | NA | 4 | 0 | 0 | Low-quality | Genome-fragment | 2.85 | AAI-based (medium-confidence) | 0 |
| S12-final-viral-combined_k141_72995\|\|full | 1502 | No | NA | 3 | 2 | 0 | Low-quality | Genome-fragment | 2.85 | AAI-based (high-confidence) | 0 |
| S1_k141_392682 | 4645 | No | NA | 5 | 0 | 3 | Not-determined | Genome-fragment | NA | NA | 0 |
| S1_k141_130956 | 1307 | No | NA | 1 | 0 | 1 | Not-determined | Genome-fragment | NA | NA | 0 |
| S1_k141_328014 | 4896 | No | NA | 1 | 0 | 0 | Not-determined | Genome-fragment | NA | NA | 0 |
| S12-final-viral-combined_k141_360622\|\|full | 1502 | No | NA | 3 | 1 | 0 | Low-quality | Genome-fragment | 1.01 | HMM-based (lower-bound) | 0 |
| S1_k141_162338 | 2051 | No | NA | 3 | 0 | 3 | Not-determined | Genome-fragment | NA | NA | 0 |
| S1_k141_14800 | 1234 | No | NA | 2 | 0 | 2 | Not-determined | Genome-fragment | NA | NA | 0 |

| | | | | | | | | | | | |
|---|---|---|---|---|---|---|---|---|---|---|---|
| S1_k141_178216 | 3099 | No | NA | 3 | 0 | 1 | Not-determined | Genome-fragment | NA | NA | 0 |
| S12_k141_251467 | 1502 | No | NA | 2 | 0 | 1 | Low-quality | Genome-fragment | 0.52 | AAI-based (high-confidence) | 0 |
| S1_k141_328051 | 1010 | No | NA | 2 | 0 | 0 | Not-determined | Genome-fragment | NA | NA | 0 |
| S1_k141_146415 | 1306 | No | NA | 1 | 0 | 0 | Not-determined | Genome-fragment | NA | NA | 0 |
| S8_k141_206093 | 1502 | No | NA | 3 | 1 | 0 | Low-quality | Genome-fragment | 1.9 | AAI-based (high-confidence) | 0 |
| S1_k141_392732 | 10728 | No | NA | 15 | 0 | 2 | Not-determined | Genome-fragment | NA | NA | 0 |
| S9-final-viral-combined_k141_7397\|\|full | 1503 | No | NA | 3 | 0 | 0 | Low-quality | Genome-fragment | 4.32 | AAI-based (medium-confidence) | 0 |
| S1_k141_328073 | 3330 | No | NA | 5 | 0 | 3 | Not-determined | Genome-fragment | NA | NA | 0 |
| S1_k141_245951 | 1837 | No | NA | 1 | 0 | 1 | Not-determined | Genome-fragment | NA | NA | 0 |
| S11_k141_265268 | 1503 | No | NA | 2 | 0 | 0 | Low-quality | Genome-fragment | 3.89 | AAI-based (high-confidence) | 0 |
| S1_k141_79418 | 1438 | No | NA | 2 | 0 | 1 | Not-determined | Genome-fragment | NA | NA | 0 |
| S1_k141_178281 | 1001 | No | NA | 2 | 0 | 0 | Not-determined | Genome-fragment | NA | NA | 0 |
| S1_k141_392753 | 1310 | No | NA | 2 | 0 | 1 | Not-determined | Genome-fragment | NA | NA | 0 |
| S1-final-viral-combined_k141_207568\|\|full | 1503 | No | NA | 4 | 0 | 2 | Low-quality | Genome-fragment | 3.41 | AAI-based (medium-confidence) | 0 |
| S1_k141_178317 | 1883 | No | NA | 1 | 0 | 1 | Not-determined | Genome-fragment | NA | NA | 0 |
| S1_k141_262816 | 1896 | No | NA | 3 | 0 | 2 | Not-determined | Genome-fragment | NA | NA | 0 |
| S1_k141_361016 | 3758 | No | NA | 1 | 0 | 1 | Not-determined | Genome-fragment | NA | NA | 0 |
| S1_k141_64130 | 1531 | No | NA | 1 | 0 | 1 | Not-determined | Genome-fragment | NA | NA | 0 |
| S1_k141_311756 | 1931 | No | NA | 2 | 0 | 0 | Not-determined | Genome-fragment | NA | NA | 0 |
| S1_k141_311758 | 1054 | No | NA | 1 | 0 | 0 | Not-determined | Genome-fragment | NA | NA | 0 |
| S3-final-viral-combined_k141_114241\|\|lt2gene | 1503 | No | NA | 1 | 1 | 0 | Low-quality | Genome-fragment | 2.84 | HMM-based (lower-bound) | 0 |
| S1_k141_178353 | 2750 | No | NA | 3 | 0 | 1 | Not-determined | Genome-fragment | NA | NA | 0 |
| S1_k141_178375 | 1341 | No | NA | 1 | 0 | 0 | Not-determined | Genome-fragment | NA | NA | 0 |
| S1_k141_262846 | 1710 | No | NA | 2 | 0 | 1 | Not-determined | Genome-fragment | NA | NA | 0 |
| S17-final-viral-combined_k141_128071\|\|full | 1503 | No | NA | 3 | 1 | 0 | Low-quality | Genome-fragment | 3.67 | AAI-based (high-confidence) | 0 |
| S4-final-viral-combined_k141_34137\|\|full | 1503 | No | NA | 2 | 0 | 0 | Low-quality | Genome-fragment | 2.45 | AAI-based (medium-confidence) | 0 |
| S1_k141_213084 | 1865 | No | NA | 1 | 0 | 1 | Not-determined | Genome-fragment | NA | NA | 0 |
| S1_k141_97218 | 1612 | No | NA | 3 | 0 | 3 | Not-determined | Genome-fragment | NA | NA | 0 |
| S8-final-viral-combined_k141_29165\|\|full | 1503 | No | NA | 4 | 1 | 0 | Low-quality | Genome-fragment | 3.7 | AAI-based (high-confidence) | 0 |
| S1_k141_112034 | 1077 | No | NA | 1 | 0 | 1 | Not-determined | Genome-fragment | NA | NA | 0 |
| S1_k141_295232 | 1568 | No | NA | 2 | 0 | 1 | Not-determined | Genome-fragment | NA | NA | 0 |
| S1_k141_46612 | 1164 | No | NA | 1 | 0 | 1 | Not-determined | Genome-fragment | NA | NA | 0 |
| S1_k141_295234 | 2726 | No | NA | 1 | 0 | 0 | Not-determined | Genome-fragment | NA | NA | 0 |
| S1_k141_79574 | 1073 | No | NA | 1 | 0 | 1 | Not-determined | Genome-fragment | NA | NA | 0 |
| S8-final-viral-combined_k141_77249\|\|full | 1503 | No | NA | 3 | 0 | 0 | Low-quality | Genome-fragment | 1.44 | AAI-based (medium-confidence) | 0 |
| S8-final-viral-combined_k141_231538\|\|lt2gene | 1503 | No | NA | 2 | 1 | 0 | Low-quality | Genome-fragment | 2.45 | HMM-based (lower-bound) | 0 |
| S1_k141_376912 | 3720 | No | NA | 2 | 0 | 2 | Not-determined | Genome-fragment | NA | NA | 0 |
| S11-final-viral-combined_k141_322119\|\|lt2gene | 1504 | No | NA | 2 | 1 | 0 | Low-quality | Genome-fragment | 1.73 | HMM-based (lower-bound) | 0 |
| S1_k141_112075 | 2854 | No | NA | 3 | 0 | 1 | Not-determined | Genome-fragment | NA | NA | 0 |
| S1_k141_14958 | 1876 | No | NA | 3 | 0 | 2 | Not-determined | Genome-fragment | NA | NA | 0 |
| S1_k141_79608 | 1258 | No | NA | 1 | 0 | 0 | Not-determined | Genome-fragment | NA | NA | 0 |
| S1_k141_97302 | 1103 | No | NA | 2 | 0 | 1 | Not-determined | Genome-fragment | NA | NA | 0 |
| S1-final-viral-combined_k141_82290\|\|full | 1504 | No | NA | 2 | 0 | 0 | Low-quality | Genome-fragment | 1.1 | AAI-based (high-confidence) | 0 |
| S1_k141_277900 | 5152 | No | NA | 7 | 0 | 1 | Not-determined | Genome-fragment | NA | NA | 0 |
| S1_k141_146593 | 1739 | No | NA | 2 | 0 | 1 | Not-determined | Genome-fragment | NA | NA | 0 |
| S1_k141_178517 | 3059 | No | NA | 4 | 0 | 3 | Not-determined | Genome-fragment | NA | NA | 0 |
| S1_k141_14995 | 1147 | No | NA | 2 | 0 | 0 | Not-determined | Genome-fragment | NA | NA | 0 |
| S1_k141_30791 | 1458 | No | NA | 1 | 0 | 0 | Not-determined | Genome-fragment | NA | NA | 0 |
| S1_k141_262957 | 1188 | No | NA | 2 | 0 | 1 | Not-determined | Genome-fragment | NA | NA | 0 |
| S1_k141_376989 | 1075 | No | NA | 1 | 0 | 0 | Not-determined | Genome-fragment | NA | NA | 0 |
| S1_k141_311941 | 6384 | Yes | 3223 | 9 | 1 | 2 | Not-determined | Genome-fragment | NA | NA | 49.51 |
| S1_k141_146653 | 1271 | No | NA | 1 | 0 | 0 | Not-determined | Genome-fragment | NA | NA | 0 |
| S1-final-viral-combined_k141_169393\|\|full | 1504 | No | NA | 3 | 0 | 0 | Low-quality | Genome-fragment | 2.8 | AAI-based (high-confidence) | 0 |
| S17-final-viral-combined_k141_81625\|\|full | 1504 | No | NA | 2 | 0 | 0 | Low-quality | Genome-fragment | 3.45 | AAI-based (medium-confidence) | 0 |
| S17-final-viral-combined_k141_91969\|\|full | 1504 | No | NA | 2 | 0 | 2 | Low-quality | Genome-fragment | 0.82 | AAI-based (high-confidence) | 0 |
| S1_k141_15092 | 1581 | No | NA | 1 | 0 | 1 | Not-determined | Genome-fragment | NA | NA | 0 |
| S1_k141_30944 | 1127 | No | NA | 1 | 0 | 1 | Not-determined | Genome-fragment | NA | NA | 0 |
| S1_k141_30954 | 1260 | No | NA | 2 | 0 | 0 | Not-determined | Genome-fragment | NA | NA | 0 |
| S1_k141_162774 | 3834 | No | NA | 4 | 0 | 1 | Not-determined | Genome-fragment | NA | NA | 0 |
| S1_k141_328489 | 1977 | No | NA | 3 | 0 | 0 | Not-determined | Genome-fragment | NA | NA | 0 |
| S17-final-viral-combined_k141_198371\|\|full | 1504 | No | NA | 3 | 0 | 0 | Low-quality | Genome-fragment | 2.07 | AAI-based (medium-confidence) | 0 |
| S1_k141_343380 | 1446 | No | NA | 1 | 0 | 1 | Not-determined | Genome-fragment | NA | NA | 0 |
| S1_k141_196965 | 1048 | No | NA | 1 | 0 | 1 | Not-determined | Genome-fragment | NA | NA | 0 |
| S1_k141_146818 | 1190 | No | NA | 1 | 0 | 0 | Not-determined | Genome-fragment | NA | NA | 0 |
| S12-final-viral-combined_k141_398640\|\|full | 1504 | No | NA | 3 | 0 | 2 | Low-quality | Genome-fragment | 2.2 | AAI-based (high-confidence) | 0 |
| S12_k141_365515 | 1504 | No | NA | 2 | 0 | 0 | Low-quality | Genome-fragment | 2.73 | AAI-based (high-confidence) | 0 |
| S1_k141_79928 | 3058 | No | NA | 2 | 0 | 1 | Not-determined | Genome-fragment | NA | NA | 0 |
| S4_k141_159594 | 1504 | No | NA | 2 | 0 | 2 | Low-quality | Genome-fragment | 0.64 | AAI-based (medium-confidence) | 0 |
| S1_k141_361406 | 3223 | No | NA | 3 | 0 | 1 | Not-determined | Genome-fragment | NA | NA | 0 |
| S1_k141_328528 | 2130 | No | NA | 4 | 0 | 2 | Not-determined | Genome-fragment | NA | NA | 0 |
| S6-final-viral-combined_k141_132416\|\|full | 1504 | No | NA | 4 | 0 | 0 | Low-quality | Genome-fragment | 1.62 | AAI-based (medium-confidence) | 0 |
| S1_k141_47001 | 1903 | No | NA | 2 | 0 | 2 | Not-determined | Genome-fragment | NA | NA | 0 |
| S1_k141_47014 | 2671 | No | NA | 4 | 0 | 0 | Not-determined | Genome-fragment | NA | NA | 0 |
| S1_k141_377299 | 2436 | No | NA | 2 | 0 | 1 | Not-determined | Genome-fragment | NA | NA | 0 |
| S9-final-viral-combined_k141_109446\|\|full | 1505 | No | NA | 3 | 1 | 0 | Low-quality | Genome-fragment | 3.9 | AAI-based (medium-confidence) | 0 |
| S1_k141_162973 | 2036 | No | NA | 3 | 0 | 1 | Not-determined | Genome-fragment | NA | NA | 0 |
| S11-final-viral-combined_k141_158405\|\|full | 1505 | No | NA | 3 | 1 | 0 | Low-quality | Genome-fragment | 4.22 | AAI-based (high-confidence) | 0 |
| S1_k141_64476 | 1560 | No | NA | 2 | 0 | 1 | Not-determined | Genome-fragment | NA | NA | 0 |
| S1_k141_361472 | 3600 | No | NA | 3 | 0 | 1 | Not-determined | Genome-fragment | NA | NA | 0 |
| S1_k141_244239 | 1505 | No | NA | 1 | 0 | 1 | Low-quality | Genome-fragment | 0.83 | AAI-based (high-confidence) | 0 |
| S1_k141_197081 | 1047 | No | NA | 1 | 0 | 0 | Not-determined | Genome-fragment | NA | NA | 0 |
| S1_k141_263215 | 7662 | No | NA | 8 | 0 | 3 | Not-determined | Genome-fragment | NA | NA | 0 |
| S1_k141_246239 | 4053 | No | NA | 4 | 0 | 3 | Not-determined | Genome-fragment | NA | NA | 0 |
| S7-final-viral-combined_k141_63313\|\|lt2gene | 1505 | No | NA | 1 | 1 | 0 | Low-quality | Genome-fragment | 3.29 | AAI-based (high-confidence) | 0 |
| S1_k141_15274 | 1039 | No | NA | 1 | 0 | 1 | Not-determined | Genome-fragment | NA | NA | 0 |
| S1_k141_64493 | 1044 | No | NA | 2 | 0 | 1 | Not-determined | Genome-fragment | NA | NA | 0 |
| S1_k141_377351 | 6497 | No | NA | 3 | 0 | 3 | Not-determined | Genome-fragment | NA | NA | 0 |
| S1_k141_31155 | 11507 | No | NA | 2 | 0 | 0 | Not-determined | Genome-fragment | NA | NA | 0 |
| S1_k141_295646 | 1204 | No | NA | 2 | 0 | 2 | Not-determined | Genome-fragment | NA | NA | 0 |
| S1_k141_328617 | 1517 | No | NA | 1 | 0 | 1 | Not-determined | Genome-fragment | NA | NA | 0 |
| S1_k141_31170 | 1047 | No | NA | 2 | 0 | 0 | Not-determined | Genome-fragment | NA | NA | 0 |
| S1_k141_328635 | 1040 | No | NA | 1 | 0 | 1 | Not-determined | Genome-fragment | NA | NA | 0 |
| S17-k141_50159 | 1505 | No | NA | 1 | 1 | 0 | Low-quality | Genome-fragment | 0.84 | HMM-based (lower-bound) | 0 |
| S12-final-viral-combined_k141_314572\|\|full | 1505 | No | NA | 2 | 0 | 1 | Low-quality | Genome-fragment | 1.92 | AAI-based (medium-confidence) | 0 |
| S1_k141_213495 | 1317 | No | NA | 2 | 0 | 0 | Not-determined | Genome-fragment | NA | NA | 0 |
| S1_k141_80104 | 3442 | No | NA | 3 | 0 | 3 | Not-determined | Genome-fragment | NA | NA | 0 |
| S1_k141_179017 | 1410 | No | NA | 1 | 0 | 1 | Not-determined | Genome-fragment | NA | NA | 0 |
| S1_k141_47121 | 1102 | No | NA | 1 | 0 | 1 | Not-determined | Genome-fragment | NA | NA | 0 |
| S4-final-viral-combined_k141_113649\|\|full | 1505 | No | NA | 2 | 0 | 0 | Low-quality | Genome-fragment | 1.08 | AAI-based (medium-confidence) | 0 |
| S1_k141_31242 | 2012 | No | NA | 3 | 0 | 1 | Not-determined | Genome-fragment | NA | NA | 0 |
| S1_k141_343594 | 1328 | No | NA | 2 | 0 | 2 | Not-determined | Genome-fragment | NA | NA | 0 |
| S1_k141_377452 | 1231 | No | NA | 2 | 0 | 1 | Not-determined | Genome-fragment | NA | NA | 0 |
| S1_k141_146985 | 3300 | No | NA | 6 | 0 | 0 | Not-determined | Genome-fragment | NA | NA | 0 |
| S1_k141_246331 | 1856 | No | NA | 2 | 0 | 1 | Not-determined | Genome-fragment | NA | NA | 0 |
| S1_k141_197176 | 1270 | No | NA | 1 | 0 | 0 | Not-determined | Genome-fragment | NA | NA | 0 |
| S1_k141_328736 | 1158 | No | NA | 2 | 0 | 1 | Not-determined | Genome-fragment | NA | NA | 0 |
| S1_k141_163103 | 1117 | No | NA | 2 | 0 | 1 | Not-determined | Genome-fragment | NA | NA | 0 |
| S8-final-viral-combined_k141_262298\|\|full | 1505 | No | NA | 4 | 2 | 0 | Low-quality | Genome-fragment | 3.91 | AAI-based (high-confidence) | 0 |
| S1_k141_213570 | 1151 | No | NA | 2 | 0 | 2 | Not-determined | Genome-fragment | NA | NA | 0 |
| S1_k141_295750 | 1269 | No | NA | 3 | 0 | 3 | Not-determined | Genome-fragment | NA | NA | 0 |
| S1_k141_229441 | 1353 | No | NA | 2 | 0 | 0 | Not-determined | Genome-fragment | NA | NA | 0 |
| S1_k141_229443 | 1047 | No | NA | 4 | 0 | 2 | Not-determined | Genome-fragment | NA | NA | 0 |
| S1_k141_328761 | 3312 | No | NA | 4 | 0 | 3 | Not-determined | Genome-fragment | NA | NA | 0 |
| S1_k141_31293 | 16897 | No | NA | 20 | 0 | 9 | Not-determined | Genome-fragment | NA | NA | 0 |
| S9-final-viral-combined_k141_57141\|\|full | 1506 | No | NA | 4 | 3 | 0 | Low-quality | Genome-fragment | 4.58 | AAI-based (high-confidence) | 0 |
| S1_k141_197257 | 3344 | No | NA | 2 | 0 | 2 | Not-determined | Genome-fragment | NA | NA | 0 |
| S9-final-viral-combined_k141_82348\|\|lt2gene | 1506 | No | NA | 2 | 2 | 0 | Low-quality | Genome-fragment | 3.71 | AAI-based (high-confidence) | 0 |
| S1_k141_246403 | 12655 | No | NA | 11 | 0 | 4 | Not-determined | Genome-fragment | NA | NA | 0 |
| S1_k141_179241 | 1949 | No | NA | 2 | 0 | 1 | Not-determined | Genome-fragment | NA | NA | 0 |
| S11-final-viral-combined_k141_323943\|\|full | 1506 | No | NA | 2 | 0 | 2 | Low-quality | Genome-fragment | 0.93 | AAI-based (high-confidence) | 0 |
| S11-final-viral-combined_k141_121633\|\|full | 1506 | No | NA | 3 | 0 | 1 | Low-quality | Genome-fragment | 2.01 | AAI-based (medium-confidence) | 0 |
| S1_k141_112452 | 1630 | No | NA | 2 | 0 | 0 | Not-determined | Genome-fragment | NA | NA | 0 |
| S1-final-viral-combined_k141_231276\|\|full | 1506 | No | NA | 3 | 0 | 1 | Low-quality | Genome-fragment | 1.02 | AAI-based (medium-confidence) | 0 |
| S1-final-viral-combined_k141_349273\|\|lt2gene | 1506 | No | NA | 1 | 0 | 0 | Low-quality | Genome-fragment | 2.71 | AAI-based (high-confidence) | 0 |
| S1_k141_80367 | 1984 | No | NA | 2 | 0 | 0 | Not-determined | Genome-fragment | NA | NA | 0 |
| S12-final-viral-combined_k141_211827\|\|full | 1506 | No | NA | 2 | 0 | 2 | Low-quality | Genome-fragment | 1.14 | AAI-based (high-confidence) | 0 |
| S1_k141_15610 | 1350 | No | NA | 2 | 0 | 0 | Not-determined | Genome-fragment | NA | NA | 0 |
| S1_k141_163280 | 2325 | No | NA | 2 | 0 | 2 | Not-determined | Genome-fragment | NA | NA | 0 |
| S2_k141_55443 | 1506 | No | NA | 3 | 0 | 1 | Low-quality | Genome-fragment | 1.11 | AAI-based (high-confidence) | 0 |
| S1_k141_361726 | 1100 | No | NA | 1 | 0 | 1 | Not-determined | Genome-fragment | NA | NA | 0 |
| S9-final-viral-combined_k141_59660\|\|full | 1507 | No | NA | 3 | 0 | 1 | Low-quality | Genome-fragment | 1.19 | AAI-based (medium-confidence) | 0 |
| S1_k141_64882 | 4207 | No | NA | 2 | 0 | 0 | Not-determined | Genome-fragment | NA | NA | 0 |
| S1_k141_47387 | 1004 | No | NA | 2 | 0 | 0 | Not-determined | Genome-fragment | NA | NA | 0 |
| S11-final-viral-combined_k141_225378\|\|full | 1507 | No | NA | 3 | 0 | 0 | Low-quality | Genome-fragment | 1.74 | AAI-based (medium-confidence) | 0 |
| S1_k141_131609 | 1128 | No | NA | 2 | 0 | 0 | Not-determined | Genome-fragment | NA | NA | 0 |
| S1_k141_278433 | 1150 | No | NA | 1 | 0 | 0 | Not-determined | Genome-fragment | NA | NA | 0 |
| S1_k141_213791 | 3333 | No | NA | 3 | 0 | 1 | Not-determined | Genome-fragment | NA | NA | 0 |
| S1_k141_179314 | 1523 | No | NA | 1 | 0 | 1 | Not-determined | Genome-fragment | NA | NA | 0 |
| S1_k141_246477 | 1043 | No | NA | 1 | 0 | 1 | Not-determined | Genome-fragment | NA | NA | 0 |
| S1_k141_98114 | 1420 | No | NA | 1 | 0 | 1 | Not-determined | Genome-fragment | NA | NA | 0 |
| S1_k141_393320 | 1464 | No | NA | 1 | 0 | 1 | Not-determined | Genome-fragment | NA | NA | 0 |
| S1_k141_179362 | 1455 | No | NA | 1 | 0 | 1 | Not-determined | Genome-fragment | NA | NA | 0 |
| S1_k141_25374 | 1507 | Yes | 237 | 3 | 1 | 1 | Low-quality | Genome-fragment | 0.13 | HMM-based (lower-bound) | 84.27 |
| S1_k141_47453 | 1335 | No | NA | 3 | 0 | 0 | Not-determined | Genome-fragment | NA | NA | 0 |
| S1_k141_393353 | 1293 | No | NA | 1 | 0 | 1 | Not-determined | Genome-fragment | NA | NA | 0 |
| S1_k141_98153 | 7386 | No | NA | 4 | 0 | 2 | Not-determined | Genome-fragment | NA | NA | 0 |
| S1_k141_377810 | 1103 | No | NA | 2 | 0 | 2 | Not-determined | Genome-fragment | NA | NA | 0 |
| S1_k141_246528 | 1121 | No | NA | 1 | 0 | 0 | Not-determined | Genome-fragment | NA | NA | 0 |
| S1_k141_278601 | 1621 | No | NA | 2 | 0 | 2 | Not-determined | Genome-fragment | NA | NA | 0 |
| S1_k141_293965 | 1507 | No | NA | 4 | 0 | 2 | Low-quality | Genome-fragment | 3.17 | AAI-based (high-confidence) | 0 |
| S1_k141_296126 | 1461 | No | NA | 3 | 0 | 0 | Not-determined | Genome-fragment | NA | NA | 0 |
| S1_k141_147362 | 1032 | No | NA | 2 | 0 | 1 | Not-determined | Genome-fragment | NA | NA | 0 |
| S1_k141_278655 | 6103 | No | NA | 8 | 0 | 7 | Not-determined | Genome-fragment | NA | NA | 0 |
| S1_k141_15829 | 1316 | No | NA | 3 | 0 | 0 | Not-determined | Genome-fragment | NA | NA | 0 |
| S1_k141_15830 | 1360 | No | NA | 1 | 0 | 1 | Not-determined | Genome-fragment | NA | NA | 0 |
| S1_k141_179582 | 1481 | No | NA | 3 | 0 | 0 | Not-determined | Genome-fragment | NA | NA | 0 |
| S1_k141_147431 | 1965 | No | NA | 2 | 0 | 1 | Not-determined | Genome-fragment | NA | NA | 0 |
| S1_k141_278720 | 1253 | No | NA | 4 | 0 | 0 | Not-determined | Genome-fragment | NA | NA | 0 |
| S3-final-viral-combined_k141_17987\|\|full | 1507 | No | NA | 2 | 2 | 0 | Low-quality | Genome-fragment | 4.05 | AAI-based (high-confidence) | 0 |
| S1_k141_246644 | 1704 | No | NA | 3 | 0 | 2 | Not-determined | Genome-fragment | NA | NA | 0 |
| S17-final-viral-combined_k141_67376\|\|full | 1507 | No | NA | 2 | 0 | 0 | Low-quality | Genome-fragment | 3.41 | AAI-based (high-confidence) | 0 |
| S12-final-viral-combined_k141_123716\|\|full | 1507 | No | NA | 4 | 1 | 0 | Low-quality | Genome-fragment | 2.29 | HMM-based (lower-bound) | 0 |
| S1_k141_361926 | 1219 | No | NA | 1 | 0 | 0 | Not-determined | Genome-fragment | NA | NA | 0 |
| S1_k141_344060 | 2604 | No | NA | 2 | 0 | 0 | Not-determined | Genome-fragment | NA | NA | 0 |
| S2-final-viral-combined_k141_33248\|\|full | 1507 | No | NA | 4 | 1 | 0 | Low-quality | Genome-fragment | 2.55 | AAI-based (high-confidence) | 0 |
| S1_k141_65210 | 5587 | No | NA | 6 | 0 | 1 | Not-determined | Genome-fragment | NA | NA | 0 |
| S2-final-viral-combined_k141_158809\|\|lt2gene | 1507 | No | NA | 2 | 2 | 0 | Low-quality | Genome-fragment | 3.18 | AAI-based (high-confidence) | 0 |
| S1_k141_112778 | 2217 | No | NA | 2 | 0 | 0 | Not-determined | Genome-fragment | NA | NA | 0 |
| S1_k141_16003 | 3289 | No | NA | 2 | 0 | 0 | Not-determined | Genome-fragment | NA | NA | 0 |
| S1_k141_163619 | 3346 | No | NA | 1 | 0 | 1 | Not-determined | Genome-fragment | NA | NA | 0 |
| S1_k141_179691 | 1329 | No | NA | 2 | 0 | 1 | Not-determined | Genome-fragment | NA | NA | 0 |
| S1_k141_163634 | 1448 | No | NA | 2 | 0 | 2 | Not-determined | Genome-fragment | NA | NA | 0 |
| S1_k141_98405 | 1488 | No | NA | 1 | 0 | 1 | Not-determined | Genome-fragment | NA | NA | 0 |
| S1_k141_378089 | 2422 | No | NA | 3 | 0 | 2 | Not-determined | Genome-fragment | NA | NA | 0 |
| S4-final-viral-combined_k141_57112\|\|full | 1507 | No | NA | 2 | 0 | 1 | Low-quality | Genome-fragment | 2.11 | AAI-based (high-confidence) | 0 |
| S9-final-viral-combined_k141_39000\|\|full | 1508 | No | NA | 2 | 0 | 0 | Low-quality | Genome-fragment | 5.9 | AAI-based (high-confidence) | 0 |
| S11-final-viral-combined_k141_127245\|\|full | 1508 | No | NA | 5 | 3 | 0 | Low-quality | Genome-fragment | 2.49 | AAI-based (high-confidence) | 0 |
| S1_k141_393696 | 4889 | No | NA | 2 | 0 | 1 | Not-determined | Genome-fragment | NA | NA | 0 |
| S1_k141_344198 | 1035 | No | NA | 1 | 0 | 1 | Not-determined | Genome-fragment | NA | NA | 0 |
| S1_k141_246760 | 1574 | No | NA | 2 | 0 | 2 | Not-determined | Genome-fragment | NA | NA | 0 |
| S1-final-viral-combined_k141_47634\|\|full | 1508 | No | NA | 3 | 0 | 1 | Low-quality | Genome-fragment | 2.84 | AAI-based (medium-confidence) | 0 |
| S17-k141_209681 | 1508 | No | NA | 4 | 4 | 0 | Low-quality | Genome-fragment | 4.96 | AAI-based (high-confidence) | 0 |
| S1_k141_31627 | 1374 | No | NA | 2 | 0 | 0 | Not-determined | Genome-fragment | NA | NA | 0 |
| S1_k141_393747 | 1982 | No | NA | 3 | 0 | 1 | Not-determined | Genome-fragment | NA | NA | 0 |
| S1_k141_393758 | 1680 | No | NA | 2 | 0 | 1 | Not-determined | Genome-fragment | NA | NA | 0 |
| S1_k141_65357 | 1278 | No | NA | 1 | 0 | 1 | Not-determined | Genome-fragment | NA | NA | 0 |
| S1_k141_229861 | 3072 | No | NA | 3 | 0 | 2 | Not-determined | Genome-fragment | NA | NA | 0 |
| S1_k141_279021 | 1730 | No | NA | 2 | 0 | 1 | Not-determined | Genome-fragment | NA | NA | 0 |
| S1_k141_31724 | 4663 | No | NA | 3 | 0 | 3 | Not-determined | Genome-fragment | NA | NA | 0 |
| S1_k141_16142 | 1325 | No | NA | 1 | 0 | 0 | Not-determined | Genome-fragment | NA | NA | 0 |
| S12-final-viral-combined_k141_257165\|\|full | 1508 | No | NA | 2 | 0 | 0 | Low-quality | Genome-fragment | 0.56 | AAI-based (medium-confidence) | 0 |
| S1_k141_31760 | 2896 | No | NA | 4 | 0 | 2 | Not-determined | Genome-fragment | NA | NA | 0 |
| S1_k141_179998 | 1037 | No | NA | 2 | 0 | 0 | Not-determined | Genome-fragment | NA | NA | 0 |
| S1_k141_344383 | 1013 | No | NA | 2 | 0 | 1 | Not-determined | Genome-fragment | NA | NA | 0 |
| S1_k141_230016 | 3470 | No | NA | 2 | 0 | 1 | Not-determined | Genome-fragment | NA | NA | 0 |
| S1_k141_344466 | 1114 | No | NA | 2 | 0 | 0 | Not-determined | Genome-fragment | NA | NA | 0 |
| S1_k141_147715 | 1203 | No | NA | 1 | 0 | 0 | Not-determined | Genome-fragment | NA | NA | 0 |
| S1_k141_147722 | 1687 | No | NA | 2 | 0 | 2 | Not-determined | Genome-fragment | NA | NA | 0 |
| S1_k141_147733 | 3026 | No | NA | 1 | 0 | 1 | Not-determined | Genome-fragment | NA | NA | 0 |
| S1_k141_163964 | 4855 | No | NA | 6 | 0 | 2 | Not-determined | Genome-fragment | NA | NA | 0 |
| S1_k141_279171 | 1072 | No | NA | 1 | 0 | 1 | Not-determined | Genome-fragment | NA | NA | 0 |
| S1_k141_344526 | 3172 | No | NA | 2 | 0 | 2 | Not-determined | Genome-fragment | NA | NA | 0 |
| S1_k141_65558 | 1000 | No | NA | 1 | 0 | 1 | Not-determined | Genome-fragment | NA | NA | 0 |
| S1_k141_230098 | 1241 | No | NA | 1 | 0 | 0 | Not-determined | Genome-fragment | NA | NA | 0 |
| S1_k141_378357 | 1165 | No | NA | 2 | 0 | 2 | Not-determined | Genome-fragment | NA | NA | 0 |
| S1_k141_394064 | 2343 | No | NA | 3 | 0 | 0 | Not-determined | Genome-fragment | NA | NA | 0 |
| S1_k141_31969 | 1328 | No | NA | 2 | 0 | 1 | Not-determined | Genome-fragment | NA | NA | 0 |
| S12_k141_17241 | 1508 | No | NA | 2 | 0 | 2 | Low-quality | Genome-fragment | 0.64 | AAI-based (medium-confidence) | 0 |
| S8_k141_267086 | 1508 | No | NA | 3 | 1 | 0 | Low-quality | Genome-fragment | 3.75 | AAI-based (high-confidence) | 0 |
| S1_k141_230253 | 1170 | No | NA | 2 | 0 | 1 | Not-determined | Genome-fragment | NA | NA | 0 |
| S1_k141_147942 | 1492 | No | NA | 2 | 0 | 1 | Not-determined | Genome-fragment | NA | NA | 0 |
| S11-final-viral-combined_k141_319373\|\|full | 1509 | No | NA | 3 | 0 | 1 | Low-quality | Genome-fragment | 0.9 | AAI-based (high-confidence) | 0 |
| S3_k141_97941 | 1509 | No | NA | 2 | 1 | 0 | Low-quality | Genome-fragment | 0.69 | AAI-based (high-confidence) | 0 |
| S1_k141_230274 | 1183 | No | NA | 3 | 0 | 3 | Not-determined | Genome-fragment | NA | NA | 0 |
| S1_k141_81554 | 1058 | No | NA | 1 | 0 | 0 | Not-determined | Genome-fragment | NA | NA | 0 |
| S7-final-viral-combined_k141_167021\|\|full | 1509 | No | NA | 3 | 1 | 0 | Low-quality | Genome-fragment | 2.5 | AAI-based (medium-confidence) | 0 |
| S1_k141_164222 | 1250 | No | NA | 1 | 0 | 0 | Not-determined | Genome-fragment | NA | NA | 0 |
| S1_k141_180302 | 2652 | No | NA | 2 | 0 | 2 | Not-determined | Genome-fragment | NA | NA | 0 |
| S12-final-viral-combined_k141_26804\|\|full | 1509 | No | NA | 4 | 0 | 1 | Low-quality | Genome-fragment | 1.09 | AAI-based (medium-confidence) | 0 |
| S1_k141_344795 | 1321 | No | NA | 1 | 0 | 0 | Not-determined | Genome-fragment | NA | NA | 0 |
| S1_k141_81665 | 1171 | No | NA | 1 | 0 | 0 | Not-determined | Genome-fragment | NA | NA | 0 |
| S1_k141_394223 | 1340 | No | NA | 2 | 0 | 1 | Not-determined | Genome-fragment | NA | NA | 0 |
| S12_k141_246857 | 1509 | No | NA | 2 | 0 | 1 | Low-quality | Genome-fragment | 0.99 | AAI-based (high-confidence) | 0 |
| S1_k141_180442 | 1191 | No | NA | 2 | 0 | 2 | Not-determined | Genome-fragment | NA | NA | 0 |
| S2_k141_224619 | 1509 | No | NA | 2 | 0 | 1 | Low-quality | Genome-fragment | 1 | AAI-based (high-confidence) | 0 |
| S1_k141_164358 | 5908 | No | NA | 5 | 0 | 3 | Not-determined | Genome-fragment | NA | NA | 0 |
| S1_k141_113579 | 1298 | No | NA | 3 | 0 | 0 | Not-determined | Genome-fragment | NA | NA | 0 |
| S1_k141_344906 | 3041 | No | NA | 5 | 0 | 3 | Not-determined | Genome-fragment | NA | NA | 0 |
| S1_k141_344921 | 3250 | No | NA | 2 | 0 | 2 | Not-determined | Genome-fragment | NA | NA | 0 |
| S1_k141_279430 | 1257 | No | NA | 1 | 0 | 0 | Not-determined | Genome-fragment | NA | NA | 0 |
| S5-final-viral-combined_k141_180244\|\|full | 1510 | No | NA | 4 | 0 | 0 | Low-quality | Genome-fragment | 2.56 | AAI-based (high-confidence) | 0 |
| S7-final-viral-combined_k141_197011\|\|full | 1510 | No | NA | 3 | 1 | 0 | Low-quality | Genome-fragment | 3.4 | AAI-based (high-confidence) | 0 |
| S1_k141_32362 | 1478 | No | NA | 2 | 0 | 1 | Not-determined | Genome-fragment | NA | NA | 0 |
| S1_k141_32371 | 3531 | No | NA | 8 | 0 | 0 | Not-determined | Genome-fragment | NA | NA | 0 |
| S1_k141_279488 | 6465 | No | NA | 4 | 0 | 1 | Not-determined | Genome-fragment | NA | NA | 0 |
| S1_k141_48434 | 3509 | No | NA | 1 | 0 | 0 | Not-determined | Genome-fragment | NA | NA | 0 |
| S7-final-viral-combined_k141_138138\|\|full | 1510 | No | NA | 4 | 1 | 0 | Low-quality | Genome-fragment | 3.25 | AAI-based (high-confidence) | 0 |
| S1_k141_48448 | 1663 | No | NA | 2 | 0 | 0 | Not-determined | Genome-fragment | NA | NA | 0 |
| S1_k141_32403 | 3012 | No | NA | 2 | 0 | 2 | Not-determined | Genome-fragment | NA | NA | 0 |
| S1_k141_48455 | 2027 | No | NA | 3 | 0 | 2 | Not-determined | Genome-fragment | NA | NA | 0 |
| S12-final-viral-combined_k141_387111\|\|full | 1510 | No | NA | 4 | 0 | 2 | Low-quality | Genome-fragment | 1.05 | AAI-based (high-confidence) | 0 |
| S1_k141_32424 | 3044 | No | NA | 2 | 0 | 1 | Not-determined | Genome-fragment | NA | NA | 0 |
| S1_k141_48488 | 1262 | No | NA | 1 | 0 | 1 | Not-determined | Genome-fragment | NA | NA | 0 |
| S1_k141_32459 | 1028 | No | NA | 2 | 0 | 1 | Not-determined | Genome-fragment | NA | NA | 0 |
| S1_k141_48513 | 1655 | No | NA | 1 | 0 | 0 | Not-determined | Genome-fragment | NA | NA | 0 |
| S1_k141_65753 | 1467 | No | NA | 1 | 0 | 0 | Not-determined | Genome-fragment | NA | NA | 0 |
| S1_k141_180663 | 1757 | No | NA | 2 | 0 | 1 | Not-determined | Genome-fragment | NA | NA | 0 |
| S1_k141_32584 | 4753 | No | NA | 5 | 0 | 3 | Not-determined | Genome-fragment | NA | NA | 0 |
| S1_k141_180699 | 12772 | No | NA | 15 | 0 | 5 | Not-determined | Genome-fragment | NA | NA | 0 |
| S1_k141_48681 | 2441 | No | NA | 5 | 0 | 5 | Not-determined | Genome-fragment | NA | NA | 0 |
| S8-final-viral-combined_k141_71798\|\|full | 1510 | No | NA | 3 | 0 | 0 | Low-quality | Genome-fragment | 2.94 | AAI-based (high-confidence) | 0 |
| S1_k141_279603 | 1225 | No | NA | 2 | 0 | 0 | Not-determined | Genome-fragment | NA | NA | 0 |
| S1_k141_48724 | 1213 | No | NA | 1 | 0 | 0 | Not-determined | Genome-fragment | NA | NA | 0 |
| S1_k141_345166 | 1025 | No | NA | 2 | 0 | 0 | Not-determined | Genome-fragment | NA | NA | 0 |
| S1_k141_32725 | 1177 | No | NA | 2 | 0 | 2 | Not-determined | Genome-fragment | NA | NA | 0 |
| S1_k141_180798 | 1077 | No | NA | 1 | 0 | 0 | Not-determined | Genome-fragment | NA | NA | 0 |
| S1_k141_32750 | 1036 | No | NA | 1 | 0 | 0 | Not-determined | Genome-fragment | NA | NA | 0 |
| S8_k141_196658 | 1510 | No | NA | 3 | 0 | 1 | Low-quality | Genome-fragment | 5.18 | AAI-based (high-confidence) | 0 |
| S1_k141_279652 | 1015 | No | NA | 1 | 0 | 1 | Not-determined | Genome-fragment | NA | NA | 0 |
| S1_k141_114025 | 3234 | No | NA | 3 | 0 | 0 | Not-determined | Genome-fragment | NA | NA | 0 |
| S1_k141_394571 | 101988 | No | NA | 124 | 17 | 3 | Complete | High-quality | 100 | DTR (high-confidence) | 0 |
| S12-final-viral-combined_k141_193312\|\|full | 1511 | No | NA | 2 | 2 | 0 | Low-quality | Genome-fragment | 2.81 | AAI-based (high-confidence) | 0 |
| S12-final-viral-combined_k141_181497\|\|full | 1511 | No | NA | 4 | 1 | 0 | Low-quality | Genome-fragment | 3.8 | AAI-based (high-confidence) | 0 |
| S1_k141_82107 | 1068 | No | NA | 1 | 0 | 1 | Not-determined | Genome-fragment | NA | NA | 0 |
| S1_k141_345285 | 1011 | No | NA | 1 | 0 | 0 | Not-determined | Genome-fragment | NA | NA | 0 |
| S1_k141_394605 | 1569 | No | NA | 1 | 0 | 0 | Not-determined | Genome-fragment | NA | NA | 0 |
| S1_k141_82162 | 7266 | No | NA | 1 | 0 | 1 | Not-determined | Genome-fragment | NA | NA | 0 |
| S1_k141_394607 | 3239 | No | NA | 5 | 0 | 1 | Not-determined | Genome-fragment | NA | NA | 0 |
| S1_k141_345311 | 12480 | No | NA | 13 | 0 | 0 | Not-determined | Genome-fragment | NA | NA | 0 |
| S1_k141_114189 | 1852 | No | NA | 1 | 0 | 1 | Not-determined | Genome-fragment | NA | NA | 0 |
| S12_k141_389763 | 1511 | No | NA | 4 | 0 | 0 | Low-quality | Genome-fragment | 2.97 | AAI-based (high-confidence) | 0 |
| S12_k141_189801 | 1511 | No | NA | 2 | 0 | 1 | Low-quality | Genome-fragment | 0.26 | AAI-based (high-confidence) | 0 |
| S1_k141_48971 | 1102 | No | NA | 2 | 0 | 2 | Not-determined | Genome-fragment | NA | NA | 0 |
| S2-final-viral-combined_k141_216748\|\|lt2gene | 1511 | No | NA | 1 | 0 | 0 | Low-quality | Genome-fragment | 2.82 | AAI-based (medium-confidence) | 0 |
| S1_k141_32878 | 18889 | No | NA | 19 | 3 | 0 | Medium-quality | Genome-fragment | 51.68 | AAI-based (high-confidence) | 0 |
| S8_k141_66168 | 1511 | No | NA | 1 | 0 | 1 | Low-quality | Genome-fragment | 1.45 | AAI-based (high-confidence) | 0 |
| S9-final-viral-combined_k141_58804\|\|full | 1512 | No | NA | 2 | 0 | 2 | Low-quality | Genome-fragment | 2.19 | AAI-based (high-confidence) | 0 |
| S1_k141_49147 | 4323 | No | NA | 4 | 0 | 0 | Not-determined | Genome-fragment | NA | NA | 0 |
| S1_k141_114574 | 1257 | No | NA | 2 | 0 | 0 | Not-determined | Genome-fragment | NA | NA | 0 |
| S1_k141_114918 | 1797 | No | NA | 2 | 0 | 0 | Not-determined | Genome-fragment | NA | NA | 0 |
| S1_k141_114986 | 1157 | No | NA | 2 | 0 | 0 | Not-determined | Genome-fragment | NA | NA | 0 |
| S3-final-viral-combined_k141_128066\|\|full | 2531 | No | NA | 2 | 0 | 0 | Not-determined | Genome-fragment | NA | NA | 0 |
| S3-final-viral-combined_k141_137874\|\|full | 8924 | No | NA | 11 | 0 | 1 | Not-determined | Genome-fragment | NA | NA | 0 |
| S9-final-viral-combined_k141_74790\|\|full | 1512 | No | NA | 2 | 0 | 2 | Low-quality | Genome-fragment | 1.06 | AAI-based (high-confidence) | 0 |
| S3-final-viral-combined_k141_132259\|\|full | 1787 | No | NA | 1 | 0 | 1 | Not-determined | Genome-fragment | NA | NA | 0 |
| S3-final-viral-combined_k141_34515\|\|full | 1965 | No | NA | 4 | 0 | 3 | Not-determined | Genome-fragment | NA | NA | 0 |
| S3-final-viral-combined_k141_97763\|\|full | 2003 | No | NA | 3 | 0 | 2 | Not-determined | Genome-fragment | NA | NA | 0 |
| S3-final-viral-combined_k141_51767\|\|full | 3838 | No | NA | 3 | 0 | 3 | Not-determined | Genome-fragment | NA | NA | 0 |
| S11_k141_196520 | 1512 | No | NA | 1 | 0 | 0 | Low-quality | Genome-fragment | 2.21 | AAI-based (high-confidence) | 0 |
| S3-final-viral-combined_k141_120780\|\|full | 6704 | No | NA | 4 | 0 | 4 | Not-determined | Genome-fragment | NA | NA | 0 |
| S3-final-viral-combined_k141_11526\|\|full | 1892 | No | NA | 3 | 0 | 3 | Not-determined | Genome-fragment | NA | NA | 0 |
| S3-final-viral-combined_k141_23051\|\|full | 2559 | No | NA | 3 | 0 | 0 | Not-determined | Genome-fragment | NA | NA | 0 |
| S12-final-viral-combined_k141_358183\|\|full | 1512 | No | NA | 3 | 1 | 0 | Low-quality | Genome-fragment | 4.15 | AAI-based (high-confidence) | 0 |
| S3-final-viral-combined_k141_51798\|\|full | 4119 | No | NA | 4 | 0 | 2 | Not-determined | Genome-fragment | NA | NA | 0 |
| S12_k141_300546 | 1512 | No | NA | 2 | 0 | 0 | Low-quality | Genome-fragment | 3.41 | AAI-based (medium-confidence) | 0 |
| S2-final-viral-combined_k141_157293\|\|full | 1512 | No | NA | 4 | 1 | 0 | Low-quality | Genome-fragment | 5.18 | HMM-based (lower-bound) | 0 |
| S3-final-viral-combined_k141_69082\|\|full | 3731 | No | NA | 3 | 0 | 2 | Not-determined | Genome-fragment | NA | NA | 0 |
| S3-final-viral-combined_k141_63347\|\|full | 1040 | No | NA | 2 | 0 | 0 | Not-determined | Genome-fragment | NA | NA | 0 |
| S2-final-viral-combined_k141_134240\|\|full | 1512 | No | NA | 2 | 0 | 1 | Low-quality | Genome-fragment | 2.01 | AAI-based (medium-confidence) | 0 |
| S3-final-viral-combined_k141_46103\|\|full | 3895 | No | NA | 5 | 0 | 3 | Not-determined | Genome-fragment | NA | NA | 0 |
| S3-final-viral-combined_k141_115104\|\|full | 6170 | No | NA | 7 | 0 | 0 | Not-determined | Genome-fragment | NA | NA | 0 |
| S3-final-viral-combined_k141_34596\|\|full | 1734 | No | NA | 2 | 0 | 2 | Not-determined | Genome-fragment | NA | NA | 0 |
| S3-final-viral-combined_k141_57565\|\|full | 1320 | No | NA | 3 | 0 | 0 | Not-determined | Genome-fragment | NA | NA | 0 |
| S6_k141_85515 | 1512 | No | NA | 1 | 0 | 0 | Low-quality | Genome-fragment | 0.66 | AAI-based (medium-confidence) | 0 |
| S8-final-viral-combined_k141_47751\|\|lt2gene | 1512 | No | NA | 1 | 1 | 0 | Low-quality | Genome-fragment | 4.44 | AAI-based (high-confidence) | 0 |
| S3-final-viral-combined_k141_115138\|\|full | 3507 | No | NA | 6 | 0 | 1 | Not-determined | Genome-fragment | NA | NA | 0 |
| S3-final-viral-combined_k141_34625\|\|full | 5349 | No | NA | 5 | 0 | 3 | Not-determined | Genome-fragment | NA | NA | 0 |
| S3-final-viral-combined_k141_86379\|\|full | 1929 | No | NA | 3 | 0 | 3 | Not-determined | Genome-fragment | NA | NA | 0 |
| S3-final-viral-combined_k141_51849\|\|full | 3389 | No | NA | 5 | 0 | 1 | Not-determined | Genome-fragment | NA | NA | 0 |
| S3-final-viral-combined_k141_5852\|\|full | 1621 | No | NA | 4 | 0 | 1 | Not-determined | Genome-fragment | NA | NA | 0 |
| S8-final-viral-combined_k141_176712\|\|lt2gene | 1512 | No | NA | 1 | 0 | 0 | Low-quality | Genome-fragment | 0.98 | AAI-based (high-confidence) | 0 |
| S3-final-viral-combined_k141_74848\|\|full | 2001 | No | NA | 4 | 0 | 0 | Not-determined | Genome-fragment | NA | NA | 0 |
| S9-final-viral-combined_k141_52797\|\|full | 1513 | No | NA | 2 | 0 | 1 | Low-quality | Genome-fragment | 2.01 | AAI-based (medium-confidence) | 0 |
| S3-final-viral-combined_k141_80585\|\|full | 1598 | No | NA | 2 | 0 | 2 | Not-determined | Genome-fragment | NA | NA | 0 |
| S3-final-viral-combined_k141_103622\|\|full | 2576 | No | NA | 4 | 0 | 1 | Not-determined | Genome-fragment | NA | NA | 0 |
| S3-final-viral-combined_k141_126694\|\|full | 1923 | No | NA | 4 | 0 | 2 | Not-determined | Genome-fragment | NA | NA | 0 |
| S5-final-viral-combined_k141_186746\|\|full | 1513 | No | NA | 2 | 0 | 2 | Low-quality | Genome-fragment | 0.5 | AAI-based (medium-confidence) | 0 |
| S3-final-viral-combined_k141_40378\|\|full | 2092 | No | NA | 4 | 0 | 0 | Not-determined | Genome-fragment | NA | NA | 0 |
| S3-final-viral-combined_k141_221\|\|full | 1790 | No | NA | 3 | 0 | 1 | Not-determined | Genome-fragment | NA | NA | 0 |
| S3-final-viral-combined_k141_103641\|\|full | 1824 | No | NA | 3 | 0 | 1 | Not-determined | Genome-fragment | NA | NA | 0 |
| S7-final-viral-combined_k141_239305\|\|full | 1513 | No | NA | 2 | 0 | 2 | Low-quality | Genome-fragment | 1.45 | AAI-based (medium-confidence) | 0 |
| S3-final-viral-combined_k141_132450\|\|full | 3199 | No | NA | 4 | 0 | 3 | Not-determined | Genome-fragment | NA | NA | 0 |
| S3-final-viral-combined_k141_40380\|\|full | 6809 | No | NA | 4 | 0 | 4 | Not-determined | Genome-fragment | NA | NA | 0 |
| S7_k141_12922 | 1513 | No | NA | 1 | 0 | 1 | Low-quality | Genome-fragment | 0.25 | AAI-based (high-confidence) | 0 |
| S3-final-viral-combined_k141_115220\|\|full | 7895 | No | NA | 11 | 0 | 1 | Not-determined | Genome-fragment | NA | NA | 0 |
| S3-final-viral-combined_k141_69190\|\|full | 2004 | No | NA | 3 | 0 | 1 | Not-determined | Genome-fragment | NA | NA | 0 |
| S3-final-viral-combined_k141_115230\|\|full | 2650 | No | NA | 6 | 0 | 0 | Not-determined | Genome-fragment | NA | NA | 0 |
| S7_k141_48136 | 1513 | No | NA | 2 | 0 | 0 | Low-quality | Genome-fragment | 2.53 | AAI-based (high-confidence) | 0 |
| S3-final-viral-combined_k141_120998\|\|full | 1317 | No | NA | 2 | 0 | 1 | Not-determined | Genome-fragment | NA | NA | 0 |
| S3-final-viral-combined_k141_40410\|\|full | 1015 | No | NA | 2 | 0 | 1 | Not-determined | Genome-fragment | NA | NA | 0 |
| S12_k141_374550 | 1513 | No | NA | 2 | 0 | 1 | Low-quality | Genome-fragment | 1.16 | AAI-based (high-confidence) | 0 |
| S3-final-viral-combined_k141_5938\|\|full | 12804 | No | NA | 11 | 0 | 9 | Not-determined | Genome-fragment | NA | NA | 0 |
| S2_k141_98024 | 1513 | No | NA | 3 | 0 | 0 | Low-quality | Genome-fragment | 3.44 | AAI-based (medium-confidence) | 0 |
| S4-final-viral-combined_k141_132515\|\|full | 1513 | No | NA | 4 | 0 | 2 | Low-quality | Genome-fragment | 0.5 | AAI-based (medium-confidence) | 0 |
| S3-final-viral-combined_k141_92204\|\|full | 2259 | No | NA | 4 | 0 | 1 | Not-determined | Genome-fragment | NA | NA | 0 |
| S3-final-viral-combined_k141_74975\|\|full | 6971 | No | NA | 9 | 0 | 7 | Not-determined | Genome-fragment | NA | NA | 0 |
| S4-final-viral-combined_k141_36331\|\|lt2gene | 1513 | No | NA | 1 | 0 | 0 | Low-quality | Genome-fragment | 2.68 | AAI-based (high-confidence) | 0 |
| S3-final-viral-combined_k141_28937\|\|full | 9541 | No | NA | 7 | 0 | 3 | Not-determined | Genome-fragment | NA | NA | 0 |
| S3-final-viral-combined_k141_57652\|\|full | 6007 | No | NA | 3 | 0 | 2 | Not-determined | Genome-fragment | NA | NA | 0 |
| S3-final-viral-combined_k141_28939\|\|full | 2734 | No | NA | 3 | 0 | 2 | Not-determined | Genome-fragment | NA | NA | 0 |
| S8_k141_110603 | 1513 | No | NA | 1 | 0 | 1 | Low-quality | Genome-fragment | 0.29 | AAI-based (medium-confidence) | 0 |
| S9-final-viral-combined_k141_108099\|\|full | 1514 | No | NA | 3 | 1 | 0 | Low-quality | Genome-fragment | 3.32 | AAI-based (high-confidence) | 0 |
| S3-final-viral-combined_k141_74992\|\|full | 1882 | No | NA | 4 | 0 | 0 | Not-determined | Genome-fragment | NA | NA | 0 |
| S9-final-viral-combined_k141_20349\|\|full | 1514 | No | NA | 2 | 0 | 1 | Low-quality | Genome-fragment | 0.73 | AAI-based (medium-confidence) | 0 |
| S1-final-viral-combined_k141_2020\|\|full | 1514 | No | NA | 6 | 1 | 0 | Low-quality | Genome-fragment | 1.99 | HMM-based (lower-bound) | 0 |
| S1_k141_155192 | 1514 | No | NA | 1 | 0 | 1 | Low-quality | Genome-fragment | 0.57 | AAI-based (high-confidence) | 0 |
| S7-final-viral-combined_k141_219822\|\|lt2gene | 1514 | No | NA | 1 | 1 | 0 | Low-quality | Genome-fragment | 2.03 | HMM-based (lower-bound) | 0 |
| S17-final-viral-combined_k141_184374\|\|full | 1514 | No | NA | 2 | 0 | 0 | Low-quality | Genome-fragment | 4.84 | AAI-based (medium-confidence) | 0 |
| S3-final-viral-combined_k141_126809\|\|full | 1897 | No | NA | 3 | 0 | 1 | Not-determined | Genome-fragment | NA | NA | 0 |
| S3-final-viral-combined_k141_126812\|\|full | 2105 | No | NA | 3 | 0 | 1 | Not-determined | Genome-fragment | NA | NA | 0 |
| S3-final-viral-combined_k141_34784\|\|full | 1094 | No | NA | 2 | 0 | 2 | Not-determined | Genome-fragment | NA | NA | 0 |
| S3-final-viral-combined_k141_17459\|\|full | 69116 | No | NA | 66 | 0 | 19 | Not-determined | Genome-fragment | NA | NA | 0 |
| S17-k141_130682 | 1514 | No | NA | 3 | 1 | 0 | Low-quality | Genome-fragment | 3.14 | AAI-based (medium-confidence) | 0 |
| S12-final-viral-combined_k141_220871\|\|lt2gene | 1514 | No | NA | 1 | 1 | 0 | Low-quality | Genome-fragment | 3.84 | AAI-based (high-confidence) | 0 |
| S3-final-viral-combined_k141_11716\|\|full | 1048 | No | NA | 3 | 0 | 0 | Not-determined | Genome-fragment | NA | NA | 0 |
| S3-final-viral-combined_k141_132594\|\|full | 6099 | No | NA | 8 | 0 | 0 | Not-determined | Genome-fragment | NA | NA | 0 |
| S3-final-viral-combined_k141_406\|\|full | 2819 | No | NA | 3 | 0 | 2 | Not-determined | Genome-fragment | NA | NA | 0 |
| S3-final-viral-combined_k141_17478\|\|full | 3953 | No | NA | 4 | 0 | 2 | Not-determined | Genome-fragment | NA | NA | 0 |
| S3-final-viral-combined_k141_46355\|\|full | 1168 | No | NA | 1 | 0 | 1 | Not-determined | Genome-fragment | NA | NA | 0 |
| S3-final-viral-combined_k141_57738\|\|full | 1378 | No | NA | 2 | 0 | 1 | Not-determined | Genome-fragment | NA | NA | 0 |
| S3-final-viral-combined_k141_52088\|\|full | 2109 | No | NA | 3 | 0 | 3 | Not-determined | Genome-fragment | NA | NA | 0 |
| S3-final-viral-combined_k141_40553\|\|full | 5218 | No | NA | 4 | 0 | 3 | Not-determined | Genome-fragment | NA | NA | 0 |
| S3-final-viral-combined_k141_63590\|\|full | 5660 | No | NA | 3 | 0 | 2 | Not-determined | Genome-fragment | NA | NA | 0 |
| S3-final-viral-combined_k141_40561\|\|full | 1591 | No | NA | 4 | 0 | 1 | Not-determined | Genome-fragment | NA | NA | 0 |
| S2_k141_131725 | 1514 | No | NA | 3 | 0 | 0 | Low-quality | Genome-fragment | 2.16 | AAI-based (high-confidence) | 0 |
| S3-final-viral-combined_k141_75035\|\|full | 2580 | No | NA | 2 | 0 | 2 | Not-determined | Genome-fragment | NA | NA | 0 |
| S3-final-viral-combined_k141_86723\|\|full | 1170 | No | NA | 3 | 0 | 1 | Not-determined | Genome-fragment | NA | NA | 0 |
| S8-final-viral-combined_k141_141912\|\|full | 1514 | No | NA | 3 | 0 | 0 | Low-quality | Genome-fragment | 1.85 | AAI-based (high-confidence) | 0 |
| S3-final-viral-combined_k141_80797\|\|full | 3146 | No | NA | 3 | 0 | 2 | Not-determined | Genome-fragment | NA | NA | 0 |
| S8-final-viral-combined_k141_259871\|\|lt2gene | 1514 | No | NA | 2 | 2 | 0 | Low-quality | Genome-fragment | 3.67 | AAI-based (high-confidence) | 0 |
| S3-final-viral-combined_k141_126929\|\|full | 1567 | No | NA | 2 | 0 | 1 | Not-determined | Genome-fragment | NA | NA | 0 |
| S3-final-viral-combined_k141_23437\|\|full | 1026 | No | NA | 2 | 0 | 1 | Not-determined | Genome-fragment | NA | NA | 0 |
| S3-final-viral-combined_k141_69383\|\|full | 1451 | No | NA | 3 | 0 | 3 | Not-determined | Genome-fragment | NA | NA | 0 |
| S11-final-viral-combined_k141_32724\|\|full | 1515 | No | NA | 2 | 0 | 0 | Low-quality | Genome-fragment | 2.45 | AAI-based (high-confidence) | 0 |
| S3-final-viral-combined_k141_17572\|\|full | 1412 | No | NA | 2 | 0 | 1 | Not-determined | Genome-fragment | NA | NA | 0 |
| S3-final-viral-combined_k141_98093\|\|full | 2209 | No | NA | 3 | 0 | 1 | Not-determined | Genome-fragment | NA | NA | 0 |
| S3-final-viral-combined_k141_132708\|\|full | 1692 | No | NA | 3 | 0 | 1 | Not-determined | Genome-fragment | NA | NA | 0 |
| S11-final-viral-combined_k141_300551\|\|full | 1515 | No | NA | 5 | 2 | 0 | Low-quality | Genome-fragment | 4.16 | AAI-based (high-confidence) | 0 |
| S3-final-viral-combined_k141_481\|\|full | 4745 | No | NA | 4 | 0 | 0 | Not-determined | Genome-fragment | NA | NA | 0 |
| S3-final-viral-combined_k141_75114\|\|full | 2171 | No | NA | 4 | 0 | 1 | Not-determined | Genome-fragment | NA | NA | 0 |
| S3-final-viral-combined_k141_490\|\|full | 1780 | No | NA | 2 | 0 | 1 | Not-determined | Genome-fragment | NA | NA | 0 |
| S3-final-viral-combined_k141_80862\|\|full | 3959 | No | NA | 3 | 0 | 2 | Not-determined | Genome-fragment | NA | NA | 0 |
| S1-final-viral-combined_k141_103787\|\|full | 1515 | No | NA | 4 | 0 | 0 | Low-quality | Genome-fragment | 3.17 | AAI-based (high-confidence) | 0 |
| S3-final-viral-combined_k141_75127\|\|full | 1080 | No | NA | 3 | 0 | 1 | Not-determined | Genome-fragment | NA | NA | 0 |
| S3-final-viral-combined_k141_36589\|\|full | 1515 | No | NA | 3 | 1 | 0 | Low-quality | Genome-fragment | 2.84 | HMM-based (lower-bound) | 0 |
| S7-final-viral-combined_k141_63872\|\|full | 1515 | No | NA | 3 | 1 | 0 | Low-quality | Genome-fragment | 2.25 | HMM-based (lower-bound) | 0 |
| S3-final-viral-combined_k141_6190\|\|full | 1421 | No | NA | 2 | 0 | 1 | Not-determined | Genome-fragment | NA | NA | 0 |
| S3-final-viral-combined_k141_34970\|\|full | 2720 | No | NA | 5 | 0 | 1 | Not-determined | Genome-fragment | NA | NA | 0 |
| S3-final-viral-combined_k141_98117\|\|full | 2071 | No | NA | 2 | 0 | 1 | Not-determined | Genome-fragment | NA | NA | 0 |
| S17-final-viral-combined_k141_93112\|\|full | 1515 | No | NA | 3 | 1 | 0 | Low-quality | Genome-fragment | 2.22 | HMM-based (lower-bound) | 0 |
| S12-final-viral-combined_k141_40393\|\|full | 1515 | No | NA | 2 | 2 | 0 | Low-quality | Genome-fragment | 3.83 | AAI-based (high-confidence) | 0 |
| S6-final-viral-combined_k141_65092\|\|full | 1515 | No | NA | 3 | 0 | 1 | Low-quality | Genome-fragment | 1.9 | AAI-based (high-confidence) | 0 |
| S3-final-viral-combined_k141_40657\|\|full | 2119 | No | NA | 6 | 0 | 2 | Not-determined | Genome-fragment | NA | NA | 0 |
| S3-final-viral-combined_k141_57849\|\|full | 1166 | No | NA | 2 | 0 | 2 | Not-determined | Genome-fragment | NA | NA | 0 |
| S8-final-viral-combined_k141_44070\|\|full | 1515 | No | NA | 3 | 1 | 0 | Low-quality | Genome-fragment | 2.77 | HMM-based (lower-bound) | 0 |
| S1-final-viral-combined_k141_350822\|\|lt2gene | 1516 | No | NA | 1 | 1 | 0 | Low-quality | Genome-fragment | 3.41 | AAI-based (high-confidence) | 0 |
| S3-final-viral-combined_k141_69470\|\|full | 1671 | No | NA | 2 | 0 | 0 | Not-determined | Genome-fragment | NA | NA | 0 |
| S1-final-viral-combined_k141_289072\|\|lt2gene | 1516 | No | NA | 1 | 1 | 0 | Low-quality | Genome-fragment | 2.22 | HMM-based (lower-bound) | 0 |
| S1_k141_381386 | 1516 | No | NA | 2 | 0 | 1 | Low-quality | Genome-fragment | 1.63 | AAI-based (high-confidence) | 0 |
| S5-final-viral-combined_k141_68040\|\|full | 1516 | No | NA | 3 | 0 | 0 | Low-quality | Genome-fragment | 2.54 | AAI-based (medium-confidence) | 0 |
| S3-final-viral-combined_k141_132783\|\|full | 1472 | No | NA | 2 | 0 | 2 | Not-determined | Genome-fragment | NA | NA | 0 |
| S3-final-viral-combined_k141_6239\|\|full | 2244 | No | NA | 3 | 0 | 2 | Not-determined | Genome-fragment | NA | NA | 0 |
| S3-final-viral-combined_k141_92473\|\|full | 2244 | No | NA | 2 | 0 | 2 | Not-determined | Genome-fragment | NA | NA | 0 |
| S3-final-viral-combined_k141_11890\|\|full | 4714 | No | NA | 7 | 0 | 2 | Not-determined | Genome-fragment | NA | NA | 0 |
| S3-final-viral-combined_k141_11897\|\|full | 2622 | No | NA | 3 | 0 | 1 | Not-determined | Genome-fragment | NA | NA | 0 |
| S3-final-viral-combined_k141_11904\|\|full | 1531 | No | NA | 3 | 0 | 1 | Not-determined | Genome-fragment | NA | NA | 0 |
| S3-final-viral-combined_k141_6271\|\|full | 3819 | No | NA | 3 | 0 | 2 | Not-determined | Genome-fragment | NA | NA | 0 |
| S3-final-viral-combined_k141_75205\|\|full | 1219 | No | NA | 3 | 0 | 0 | Not-determined | Genome-fragment | NA | NA | 0 |
| S7-final-viral-combined_k141_199330\|\|full | 1516 | No | NA | 2 | 0 | 1 | Low-quality | Genome-fragment | 0.62 | AAI-based (medium-confidence) | 0 |
| S3-final-viral-combined_k141_6278\|\|full | 2385 | No | NA | 4 | 0 | 3 | Not-determined | Genome-fragment | NA | NA | 0 |
| S3-final-viral-combined_k141_35059\|\|full | 11579 | No | NA | 11 | 0 | 7 | Not-determined | Genome-fragment | NA | NA | 0 |
| S3-final-viral-combined_k141_86910\|\|full | 3479 | No | NA | 2 | 0 | 2 | Not-determined | Genome-fragment | NA | NA | 0 |
| S7-final-viral-combined_k141_230412\|\|lt2gene | 1516 | No | NA | 2 | 0 | 0 | Low-quality | Genome-fragment | 1.67 | AAI-based (medium-confidence) | 0 |

| | | | | | | | | | | | |
|---|---|---|---|---|---|---|---|---|---|---|---|
| S3-final-viral-combined_k141_86913\|\|full | 3509 | No | NA | 5 | 0 | 2 | Not-determined | Genome-fragment | NA | NA | 0 |
| S3-final-viral-combined_k141_23593\|\|full | 2803 | No | NA | 3 | 0 | 3 | Not-determined | Genome-fragment | NA | NA | 0 |
| S3-final-viral-combined_k141_23594\|\|full | 1950 | No | NA | 3 | 0 | 1 | Not-determined | Genome-fragment | NA | NA | 0 |
| S3-final-viral-combined_k141_98216\|\|full | 7129 | No | NA | 5 | 0 | 4 | Not-determined | Genome-fragment | NA | NA | 0 |
| S3-final-viral-combined_k141_46638\|\|full | 2479 | No | NA | 3 | 0 | 1 | Not-determined | Genome-fragment | NA | NA | 0 |
| S3-final-viral-combined_k141_115597\|\|full | 5103 | No | NA | 5 | 0 | 5 | Not-determined | Genome-fragment | NA | NA | 0 |
| S3-final-viral-combined_k141_6381\|\|full | 3227 | No | NA | 3 | 0 | 1 | Not-determined | Genome-fragment | NA | NA | 0 |
| S3-final-viral-combined_k141_75264\|\|full | 8092 | No | NA | 10 | 0 | 1 | Not-determined | Genome-fragment | NA | NA | 0 |
| S17-final-viral-combined_k141_2849\|\|full | 1516 | No | NA | 2 | 1 | 0 | Low-quality | Genome-fragment | 2.4 | AAI-based (medium-confidence) | 0 |
| S12-final-viral-combined_k141_154699\|\|full | 1516 | No | NA | 5 | 0 | 0 | Low-quality | Genome-fragment | 1.96 | AAI-based (medium-confidence) | 0 |
| S3-final-viral-combined_k141_11975\|\|full | 1166 | No | NA | 2 | 0 | 2 | Not-determined | Genome-fragment | NA | NA | 0 |
| S3-final-viral-combined_k141_40838\|\|full | 2080 | No | NA | 2 | 0 | 1 | Not-determined | Genome-fragment | NA | NA | 0 |
| S3-final-viral-combined_k141_35115\|\|full | 3223 | No | NA | 2 | 0 | 1 | Not-determined | Genome-fragment | NA | NA | 0 |
| S3-final-viral-combined_k141_81080\|\|full | 1720 | No | NA | 3 | 0 | 1 | Not-determined | Genome-fragment | NA | NA | 0 |
| S3-final-viral-combined_k141_40857\|\|full | 2797 | No | NA | 3 | 0 | 1 | Not-determined | Genome-fragment | NA | NA | 0 |
| S3-final-viral-combined_k141_132922\|\|full | 2389 | No | NA | 2 | 0 | 0 | Not-determined | Genome-fragment | NA | NA | 0 |
| S3-final-viral-combined_k141_63821\|\|full | 1164 | No | NA | 3 | 0 | 0 | Not-determined | Genome-fragment | NA | NA | 0 |
| S3-final-viral-combined_k141_92586\|\|full | 2032 | No | NA | 2 | 0 | 2 | Not-determined | Genome-fragment | NA | NA | 0 |
| S3-final-viral-combined_k141_98248\|\|full | 3617 | No | NA | 4 | 0 | 1 | Not-determined | Genome-fragment | NA | NA | 0 |
| S3-final-viral-combined_k141_35146\|\|full | 1755 | No | NA | 3 | 0 | 1 | Not-determined | Genome-fragment | NA | NA | 0 |
| S3-final-viral-combined_k141_132926\|\|full | 6650 | No | NA | 6 | 0 | 4 | Not-determined | Genome-fragment | NA | NA | 0 |
| S12-final-viral-combined_k141_175404\|\|full | 1516 | No | NA | 3 | 0 | 1 | Low-quality | Genome-fragment | 5.65 | AAI-based (medium-confidence) | 0 |
| S3-final-viral-combined_k141_104145\|\|full | 2179 | No | NA | 2 | 0 | 2 | Not-determined | Genome-fragment | NA | NA | 0 |
| S12_k141_203268 | 1516 | No | NA | 1 | 0 | 0 | Low-quality | Genome-fragment | 47.39 | AAI-based (medium-confidence) | 0 |
| S2_k141_126026 | 1516 | No | NA | 4 | 3 | 0 | Low-quality | Genome-fragment | 4.21 | AAI-based (high-confidence) | 0 |
| S3-final-viral-combined_k141_109956\|\|full | 14976 | No | NA | 14 | 0 | 1 | Not-determined | Genome-fragment | NA | NA | 0 |
| S3-final-viral-combined_k141_46700\|\|full | 1731 | No | NA | 2 | 0 | 1 | Not-determined | Genome-fragment | NA | NA | 0 |
| S3-final-viral-combined_k141_40889\|\|full | 3074 | No | NA | 5 | 0 | 1 | Not-determined | Genome-fragment | NA | NA | 0 |
| S3-final-viral-combined_k141_75309\|\|full | 2606 | No | NA | 3 | 0 | 2 | Not-determined | Genome-fragment | NA | NA | 0 |
| S2_k141_193889 | 1516 | No | NA | 2 | 0 | 2 | Low-quality | Genome-fragment | 0.42 | AAI-based (high-confidence) | 0 |
| S3-final-viral-combined_k141_127304\|\|full | 1511 | No | NA | 3 | 0 | 0 | Not-determined | Genome-fragment | NA | NA | 0 |
| S6-final-viral-combined_k141_10817\|\|full | 1516 | No | NA | 3 | 1 | 0 | Low-quality | Genome-fragment | 3.91 | AAI-based (medium-confidence) | 0 |
| S8-final-viral-combined_k141_242270\|\|full | 1516 | No | NA | 2 | 0 | 1 | Low-quality | Genome-fragment | 2.64 | AAI-based (medium-confidence) | 0 |
| S9-final-viral-combined_k141_26632\|\|full | 1517 | No | NA | 4 | 0 | 0 | Low-quality | Genome-fragment | 3.04 | AAI-based (high-confidence) | 0 |
| S9-final-viral-combined_k141_98465\|\|lt2gene | 1517 | No | NA | 2 | 1 | 0 | Low-quality | Genome-fragment | 3.49 | AAI-based (high-confidence) | 0 |
| S11-final-viral-combined_k141_225585\|\|full | 1517 | No | NA | 3 | 2 | 0 | Low-quality | Genome-fragment | 3.59 | AAI-based (high-confidence) | 0 |
| S11_k141_1635 | 1517 | No | NA | 2 | 0 | 0 | Low-quality | Genome-fragment | 3.99 | AAI-based (high-confidence) | 0 |
| S1-final-viral-combined_k141_298528\|\|full | 1517 | No | NA | 4 | 2 | 0 | Low-quality | Genome-fragment | 3.76 | AAI-based (high-confidence) | 0 |
| S3-final-viral-combined_k141_12063\|\|full | 2818 | No | NA | 4 | 0 | 2 | Not-determined | Genome-fragment | NA | NA | 0 |
| S7-final-viral-combined_k141_207085\|\|full | 1517 | No | NA | 5 | 0 | 0 | Low-quality | Genome-fragment | 3.45 | AAI-based (high-confidence) | 0 |
| S7-final-viral-combined_k141_209743\|\|full | 1517 | No | NA | 4 | 1 | 0 | Low-quality | Genome-fragment | 3.43 | AAI-based (high-confidence) | 0 |
| S3-final-viral-combined_k141_6510\|\|full | 1159 | No | NA | 3 | 0 | 2 | Not-determined | Genome-fragment | NA | NA | 0 |
| S3-final-viral-combined_k141_29338\|\|full | 6194 | No | NA | 5 | 0 | 4 | Not-determined | Genome-fragment | NA | NA | 0 |
| S17-final-viral-combined_k141_262930\|\|full | 1517 | No | NA | 2 | 0 | 2 | Low-quality | Genome-fragment | 0.76 | AAI-based (medium-confidence) | 0 |
| S3-final-viral-combined_k141_121438\|\|full | 1670 | No | NA | 3 | 0 | 1 | Not-determined | Genome-fragment | NA | NA | 0 |
| S17-k141_162884 | 1517 | No | NA | 4 | 1 | 0 | Low-quality | Genome-fragment | 2.59 | AAI-based (high-confidence) | 0 |
| S12-final-viral-combined_k141_167405\|\|full | 1517 | No | NA | 4 | 0 | 0 | Low-quality | Genome-fragment | 3.16 | AAI-based (medium-confidence) | 0 |
| S3-final-viral-combined_k141_104223\|\|full | 2549 | No | NA | 3 | 0 | 0 | Not-determined | Genome-fragment | NA | NA | 0 |
| S12_k141_241359 | 1517 | No | NA | 1 | 0 | 1 | Low-quality | Genome-fragment | 0.39 | AAI-based (high-confidence) | 0 |
| S3-final-viral-combined_k141_6582\|\|full | 3084 | No | NA | 3 | 0 | 3 | Not-determined | Genome-fragment | NA | NA | 0 |
| S9-final-viral-combined_k141_32414\|\|lt2gene | 1518 | No | NA | 1 | 1 | 0 | Low-quality | Genome-fragment | 3.87 | AAI-based (high-confidence) | 0 |
| S11-final-viral-combined_k141_255129\|\|full | 1518 | No | NA | 2 | 0 | 0 | Low-quality | Genome-fragment | 2.75 | AAI-based (high-confidence) | 0 |
| S3-final-viral-combined_k141_29421\|\|full | 2192 | No | NA | 5 | 0 | 2 | Not-determined | Genome-fragment | NA | NA | 0 |
| S3-final-viral-combined_k141_104247\|\|full | 2091 | No | NA | 5 | 0 | 0 | Not-determined | Genome-fragment | NA | NA | 0 |
| S11-final-viral-combined_k141_229794\|\|full | 1518 | No | NA | 3 | 0 | 2 | Low-quality | Genome-fragment | 0.48 | AAI-based (medium-confidence) | 0 |
| S1-final-viral-combined_k141_169446\|\|lt2gene | 1518 | No | NA | 2 | 1 | 0 | Low-quality | Genome-fragment | 2.2 | AAI-based (high-confidence) | 0 |
| S5-final-viral-combined_k141_63723\|\|full | 1518 | No | NA | 4 | 0 | 1 | Low-quality | Genome-fragment | 2.77 | AAI-based (high-confidence) | 0 |
| S3-final-viral-combined_k141_829\|\|full | 4321 | No | NA | 3 | 0 | 3 | Not-determined | Genome-fragment | NA | NA | 0 |
| S3-final-viral-combined_k141_29438\|\|full | 3026 | No | NA | 2 | 0 | 1 | Not-determined | Genome-fragment | NA | NA | 0 |
| S3-final-viral-combined_k141_18003\|\|full | 1527 | No | NA | 2 | 0 | 2 | Not-determined | Genome-fragment | NA | NA | 0 |
| S7-final-viral-combined_k141_48717\|\|full | 1518 | Yes | 1287 | 3 | 1 | 1 | Low-quality | Genome-fragment | 2.58 | HMM-based (lower-bound) | 15.22 |
| S3-final-viral-combined_k141_127462\|\|full | 2131 | No | NA | 2 | 0 | 1 | Not-determined | Genome-fragment | NA | NA | 0 |
| S12-final-viral-combined_k141_128269\|\|full | 1518 | No | NA | 3 | 1 | 0 | Low-quality | Genome-fragment | 2.91 | AAI-based (medium-confidence) | 0 |
| S3-final-viral-combined_k141_46886\|\|full | 2904 | No | NA | 4 | 0 | 1 | Not-determined | Genome-fragment | NA | NA | 0 |
| S3-final-viral-combined_k141_64036\|\|full | 1864 | No | NA | 4 | 0 | 3 | Not-determined | Genome-fragment | NA | NA | 0 |
| S3-final-viral-combined_k141_110095\|\|full | 3544 | No | NA | 3 | 0 | 2 | Not-determined | Genome-fragment | NA | NA | 0 |
| S3-final-viral-combined_k141_81366\|\|full | 1844 | No | NA | 2 | 0 | 1 | Not-determined | Genome-fragment | NA | NA | 0 |
| S3-final-viral-combined_k141_898\|\|full | 3872 | No | NA | 4 | 0 | 4 | Not-determined | Genome-fragment | NA | NA | 0 |
| S3-final-viral-combined_k141_900\|\|full | 4492 | No | NA | 6 | 0 | 3 | Not-determined | Genome-fragment | NA | NA | 0 |
| S12_k141_89498 | 1518 | No | NA | 2 | 0 | 0 | Low-quality | Genome-fragment | 1.39 | AAI-based (high-confidence) | 0 |
| S3-final-viral-combined_k141_18042\|\|full | 1837 | No | NA | 2 | 0 | 0 | Not-determined | Genome-fragment | NA | NA | 0 |
| S12_k141_308865 | 1518 | No | NA | 1 | 0 | 1 | Low-quality | Genome-fragment | 0.61 | AAI-based (medium-confidence) | 0 |
| S2-final-viral-combined_k141_214434\|\|full | 1518 | No | NA | 3 | 0 | 1 | Low-quality | Genome-fragment | 2.29 | AAI-based (medium-confidence) | 0 |
| S2_k141_16560 | 1518 | No | NA | 2 | 2 | 0 | Low-quality | Genome-fragment | 3.25 | AAI-based (medium-confidence) | 0 |
| S3-final-viral-combined_k141_133131\|\|full | 2308 | No | NA | 3 | 0 | 2 | Not-determined | Genome-fragment | NA | NA | 0 |
| S3-final-viral-combined_k141_127507\|\|full | 2223 | No | NA | 2 | 0 | 2 | Not-determined | Genome-fragment | NA | NA | 0 |
| S11-final-viral-combined_k141_1294\|\|full | 1519 | No | NA | 4 | 0 | 2 | Low-quality | Genome-fragment | 0.98 | AAI-based (medium-confidence) | 0 |
| S3-final-viral-combined_k141_81435\|\|full | 1720 | No | NA | 2 | 0 | 2 | Not-determined | Genome-fragment | NA | NA | 0 |
| S3-final-viral-combined_k141_121555\|\|full | 4283 | No | NA | 7 | 0 | 3 | Not-determined | Genome-fragment | NA | NA | 0 |
| S3-final-viral-combined_k141_64120\|\|full | 2596 | No | NA | 4 | 0 | 2 | Not-determined | Genome-fragment | NA | NA | 0 |
| S11-final-viral-combined_k141_169660\|\|full | 1519 | No | NA | 3 | 1 | 0 | Low-quality | Genome-fragment | 1.71 | HMM-based (lower-bound) | 0 |
| S11-final-viral-combined_k141_40503\|\|full | 1519 | No | NA | 2 | 0 | 1 | Low-quality | Genome-fragment | 1.81 | AAI-based (medium-confidence) | 0 |
| S3-final-viral-combined_k141_81464\|\|full | 1920 | No | NA | 3 | 0 | 1 | Not-determined | Genome-fragment | NA | NA | 0 |
| S11-final-viral-combined_k141_71412\|\|lt2gene | 1519 | No | NA | 1 | 1 | 0 | Low-quality | Genome-fragment | 2.8 | AAI-based (medium-confidence) | 0 |
| S1_k141_353056 | 1519 | No | NA | 2 | 0 | 0 | Low-quality | Genome-fragment | 3.17 | AAI-based (medium-confidence) | 0 |
| S3-final-viral-combined_k141_35405\|\|full | 2080 | No | NA | 3 | 0 | 1 | Not-determined | Genome-fragment | NA | NA | 0 |
| S3-final-viral-combined_k141_5666\|\|full | 1519 | No | NA | 4 | 0 | 0 | Low-quality | Genome-fragment | 2.82 | AAI-based (high-confidence) | 0 |
| S3-final-viral-combined_k141_58437\|\|full | 5117 | No | NA | 5 | 0 | 3 | Not-determined | Genome-fragment | NA | NA | 0 |
| S5-final-viral-combined_k141_125352\|\|full | 1519 | No | NA | 3 | 0 | 0 | Low-quality | Genome-fragment | 0.87 | AAI-based (high-confidence) | 0 |
| S3-final-viral-combined_k141_75632\|\|full | 6240 | No | NA | 7 | 0 | 4 | Not-determined | Genome-fragment | NA | NA | 0 |
| S3-final-viral-combined_k141_69881\|\|full | 3591 | No | NA | 3 | 0 | 2 | Not-determined | Genome-fragment | NA | NA | 0 |
| S5-final-viral-combined_k141_156565\|\|lt2gene | 1519 | No | NA | 2 | 2 | 0 | Low-quality | Genome-fragment | 3.71 | AAI-based (high-confidence) | 0 |
| S3-final-viral-combined_k141_87399\|\|full | 1348 | No | NA | 2 | 0 | 1 | Not-determined | Genome-fragment | NA | NA | 0 |
| S17-final-viral-combined_k141_198760\|\|lt2gene | 1519 | No | NA | 2 | 1 | 0 | Low-quality | Genome-fragment | 3.81 | AAI-based (high-confidence) | 0 |
| S3-final-viral-combined_k141_127610\|\|full | 1142 | No | NA | 2 | 0 | 0 | Not-determined | Genome-fragment | NA | NA | 0 |
| S3-final-viral-combined_k141_69906\|\|full | 2877 | No | NA | 5 | 0 | 3 | Not-determined | Genome-fragment | NA | NA | 0 |
| S3-final-viral-combined_k141_58458\|\|full | 4006 | No | NA | 4 | 0 | 3 | Not-determined | Genome-fragment | NA | NA | 0 |
| S3-final-viral-combined_k141_115914\|\|full | 1187 | No | NA | 3 | 0 | 1 | Not-determined | Genome-fragment | NA | NA | 0 |
| S3-final-viral-combined_k141_18073\|\|full | 3790 | No | NA | 3 | 0 | 2 | Not-determined | Genome-fragment | NA | NA | 0 |
| S3-final-viral-combined_k141_75687\|\|full | 1165 | No | NA | 2 | 0 | 0 | Not-determined | Genome-fragment | NA | NA | 0 |
| S3-final-viral-combined_k141_6776\|\|full | 6530 | No | NA | 5 | 0 | 3 | Not-determined | Genome-fragment | NA | NA | 0 |
| S12-final-viral-combined_k141_185488\|\|full | 1519 | No | NA | 4 | 0 | 3 | Low-quality | Genome-fragment | 2.21 | AAI-based (medium-confidence) | 0 |
| S3-final-viral-combined_k141_52765\|\|full | 1601 | No | NA | 2 | 0 | 0 | Not-determined | Genome-fragment | NA | NA | 0 |
| S4-final-viral-combined_k141_22642\|\|lt2gene | 1519 | No | NA | 1 | 0 | 0 | Low-quality | Genome-fragment | 1.51 | AAI-based (high-confidence) | 0 |
| S3-final-viral-combined_k141_133261\|\|full | 2127 | No | NA | 3 | 0 | 2 | Not-determined | Genome-fragment | NA | NA | 0 |
| S3-final-viral-combined_k141_127636\|\|full | 1728 | No | NA | 4 | 0 | 0 | Not-determined | Genome-fragment | NA | NA | 0 |
| S3-final-viral-combined_k141_35483\|\|full | 2174 | No | NA | 3 | 0 | 1 | Not-determined | Genome-fragment | NA | NA | 0 |
| S3-final-viral-combined_k141_98544\|\|full | 3879 | No | NA | 4 | 0 | 3 | Not-determined | Genome-fragment | NA | NA | 0 |
| S9-final-viral-combined_k141_5895\|\|full | 1520 | No | NA | 2 | 0 | 1 | Low-quality | Genome-fragment | 0.71 | AAI-based (high-confidence) | 0 |
| S3-final-viral-combined_k141_75741\|\|full | 2810 | No | NA | 4 | 0 | 1 | Not-determined | Genome-fragment | NA | NA | 0 |
| S1-final-viral-combined_k141_213196\|\|full | 1520 | No | NA | 2 | 0 | 1 | Low-quality | Genome-fragment | 1.42 | AAI-based (medium-confidence) | 0 |
| S3-final-viral-combined_k141_64283\|\|full | 3210 | No | NA | 3 | 0 | 1 | Not-determined | Genome-fragment | NA | NA | 0 |
| S3-final-viral-combined_k141_75749\|\|full | 2690 | No | NA | 2 | 0 | 1 | Not-determined | Genome-fragment | NA | NA | 0 |
| S5-final-viral-combined_k141_205368\|\|lt2gene | 1520 | No | NA | 1 | 1 | 0 | Low-quality | Genome-fragment | 2.22 | HMM-based (lower-bound) | 0 |
| S3-final-viral-combined_k141_87519\|\|full | 1895 | No | NA | 3 | 0 | 1 | Not-determined | Genome-fragment | NA | NA | 0 |
| S3-final-viral-combined_k141_98621\|\|full | 2073 | No | NA | 3 | 0 | 1 | Not-determined | Genome-fragment | NA | NA | 0 |
| S7-final-viral-combined_k141_106347\|\|full | 1520 | No | NA | 2 | 0 | 0 | Low-quality | Genome-fragment | 1.23 | AAI-based (high-confidence) | 0 |
| S3-final-viral-combined_k141_64331\|\|full | 3862 | No | NA | 4 | 0 | 3 | Not-determined | Genome-fragment | NA | NA | 0 |
| S3-final-viral-combined_k141_6871\|\|full | 2156 | No | NA | 2 | 0 | 2 | Not-determined | Genome-fragment | NA | NA | 0 |
| S7_k141_131880 | 1520 | No | NA | 2 | 0 | 1 | Low-quality | Genome-fragment | 1.07 | AAI-based (high-confidence) | 0 |
| S3-final-viral-combined_k141_70027\|\|full | 1657 | No | NA | 3 | 0 | 1 | Not-determined | Genome-fragment | NA | NA | 0 |
| S3-final-viral-combined_k141_18197\|\|full | 1827 | No | NA | 3 | 0 | 2 | Not-determined | Genome-fragment | NA | NA | 0 |
| S3-final-viral-combined_k141_58583\|\|full | 6487 | No | NA | 5 | 0 | 5 | Not-determined | Genome-fragment | NA | NA | 0 |
| S12_k141_390299 | 1520 | No | NA | 5 | 1 | 0 | Low-quality | Genome-fragment | 2.07 | AAI-based (high-confidence) | 0 |
| S8-final-viral-combined_k141_219637\|\|full | 1520 | No | NA | 3 | 0 | 0 | Low-quality | Genome-fragment | 3.59 | AAI-based (high-confidence) | 0 |
| S1-final-viral-combined_k141_242169\|\|full | 1521 | No | NA | 3 | 0 | 0 | Low-quality | Genome-fragment | 2.87 | AAI-based (medium-confidence) | 0 |
| S3-final-viral-combined_k141_24087\|\|full | 2006 | No | NA | 4 | 0 | 2 | Not-determined | Genome-fragment | NA | NA | 0 |
| S1-final-viral-combined_k141_76974\|\|full | 1521 | No | NA | 2 | 0 | 1 | Low-quality | Genome-fragment | 1.58 | AAI-based (medium-confidence) | 0 |
| S2-final-viral-combined_k141_35871\|\|full | 1521 | No | NA | 3 | 2 | 0 | Low-quality | Genome-fragment | 4.53 | AAI-based (high-confidence) | 0 |
| S3-final-viral-combined_k141_6925\|\|full | 1014 | No | NA | 3 | 0 | 0 | Not-determined | Genome-fragment | NA | NA | 0 |
| S4-final-viral-combined_k141_142156\|\|full | 1521 | No | NA | 3 | 0 | 0 | Low-quality | Genome-fragment | 1.16 | AAI-based (high-confidence) | 0 |
| S8-final-viral-combined_k141_123556\|\|full | 1521 | No | NA | 5 | 0 | 0 | Low-quality | Genome-fragment | 3.7 | AAI-based (high-confidence) | 0 |
| S3-final-viral-combined_k141_12421\|\|full | 1284 | No | NA | 2 | 0 | 2 | Not-determined | Genome-fragment | NA | NA | 0 |
| S3-final-viral-combined_k141_133422\|\|full | 1883 | No | NA | 2 | 0 | 1 | Not-determined | Genome-fragment | NA | NA | 0 |
| S3-final-viral-combined_k141_133428\|\|full | 2244 | No | NA | 3 | 0 | 0 | Not-determined | Genome-fragment | NA | NA | 0 |
| S3-final-viral-combined_k141_70091\|\|full | 1364 | No | NA | 3 | 0 | 0 | Not-determined | Genome-fragment | NA | NA | 0 |
| S1_k141_106984 | 1522 | No | NA | 4 | 0 | 3 | Low-quality | Genome-fragment | 0.44 | AAI-based (medium-confidence) | 0 |
| S5-final-viral-combined_k141_204451\|\|full | 1522 | No | NA | 3 | 1 | 0 | Low-quality | Genome-fragment | 2.96 | AAI-based (high-confidence) | 0 |
| S3-final-viral-combined_k141_133455\|\|full | 1183 | No | NA | 3 | 0 | 1 | Not-determined | Genome-fragment | NA | NA | 0 |
| S3-final-viral-combined_k141_12479\|\|full | 2291 | No | NA | 2 | 0 | 0 | Not-determined | Genome-fragment | NA | NA | 0 |
| S3-final-viral-combined_k141_110452\|\|full | 1349 | No | NA | 2 | 0 | 2 | Not-determined | Genome-fragment | NA | NA | 0 |
| S3-final-viral-combined_k141_70129\|\|full | 1340 | No | NA | 3 | 0 | 1 | Not-determined | Genome-fragment | NA | NA | 0 |
| S7-final-viral-combined_k141_35193\|\|full | 1522 | No | NA | 3 | 0 | 0 | Low-quality | Genome-fragment | 3.97 | AAI-based (high-confidence) | 0 |
| S3-final-viral-combined_k141_24191\|\|full | 1793 | No | NA | 4 | 0 | 2 | Not-determined | Genome-fragment | NA | NA | 0 |
| S3-final-viral-combined_k141_58669\|\|full | 2809 | No | NA | 3 | 0 | 3 | Not-determined | Genome-fragment | NA | NA | 0 |
| S7_k141_157717 | 1522 | No | NA | 1 | 1 | 0 | Low-quality | Genome-fragment | 1.04 | HMM-based (lower-bound) | 0 |
| S17-final-viral-combined_k141_12785\|\|full | 1522 | No | NA | 3 | 0 | 0 | Low-quality | Genome-fragment | 3.57 | AAI-based (medium-confidence) | 0 |
| S17-final-viral-combined_k141_59855\|\|full | 1522 | No | NA | 2 | 1 | 0 | Low-quality | Genome-fragment | 3.54 | AAI-based (high-confidence) | 0 |
| S17-final-viral-combined_k141_249006\|\|full | 1522 | No | NA | 4 | 0 | 0 | Low-quality | Genome-fragment | 2.63 | AAI-based (high-confidence) | 0 |
| S3-final-viral-combined_k141_127894\|\|full | 2040 | No | NA | 4 | 0 | 1 | Not-determined | Genome-fragment | NA | NA | 0 |
| S2-final-viral-combined_k141_35397\|\|full | 1522 | No | NA | 2 | 0 | 0 | Low-quality | Genome-fragment | 1.83 | AAI-based (high-confidence) | 0 |
| S3-final-viral-combined_k141_41460\|\|full | 1068 | No | NA | 3 | 0 | 1 | Not-determined | Genome-fragment | NA | NA | 0 |
| S4-final-viral-combined_k141_110128\|\|lt2gene | 1522 | No | NA | 2 | 1 | 0 | Low-quality | Genome-fragment | 34.04 | AAI-based (high-confidence) | 0 |
| S4_k141_59811 | 1522 | No | NA | 3 | 0 | 0 | Low-quality | Genome-fragment | 2.29 | AAI-based (high-confidence) | 0 |
| S3-final-viral-combined_k141_127917\|\|full | 2825 | No | NA | 2 | 0 | 2 | Not-determined | Genome-fragment | NA | NA | 0 |
| S8_k141_235880 | 1522 | No | NA | 2 | 0 | 0 | Low-quality | Genome-fragment | 0.64 | AAI-based (medium-confidence) | 0 |
| S9-final-viral-combined_k141_112112\|\|full | 1523 | No | NA | 3 | 1 | 0 | Low-quality | Genome-fragment | 4.12 | AAI-based (high-confidence) | 0 |
| S1-final-viral-combined_k141_48813\|\|full | 1523 | No | NA | 3 | 3 | 0 | Low-quality | Genome-fragment | 2.91 | AAI-based (high-confidence) | 0 |
| S3-final-viral-combined_k141_93190\|\|full | 1774 | No | NA | 3 | 0 | 0 | Not-determined | Genome-fragment | NA | NA | 0 |
| S3_k141_116812 | 1523 | No | NA | 2 | 0 | 0 | Low-quality | Genome-fragment | 2.21 | AAI-based (high-confidence) | 0 |
| S3-final-viral-combined_k141_87700\|\|full | 1015 | No | NA | 2 | 0 | 1 | Not-determined | Genome-fragment | NA | NA | 0 |
| S5-final-viral-combined_k141_21131\|\|full | 1523 | No | NA | 3 | 0 | 3 | Low-quality | Genome-fragment | 3.07 | AAI-based (high-confidence) | 0 |
| S3-final-viral-combined_k141_133564\|\|full | 4133 | No | NA | 2 | 0 | 2 | Not-determined | Genome-fragment | NA | NA | 0 |
| S5-final-viral-combined_k141_130314\|\|full | 1523 | No | NA | 2 | 0 | 1 | Low-quality | Genome-fragment | 3.1 | AAI-based (medium-confidence) | 0 |
| S5-final-viral-combined_k141_38048\|\|lt2gene | 1523 | No | NA | 1 | 1 | 0 | Low-quality | Genome-fragment | 3.34 | AAI-based (high-confidence) | 0 |
| S12-final-viral-combined_k141_168601\|\|full | 1523 | No | NA | 5 | 0 | 0 | Low-quality | Genome-fragment | 2.45 | AAI-based (high-confidence) | 0 |
| S3-final-viral-combined_k141_76025\|\|full | 2750 | No | NA | 4 | 0 | 0 | Not-determined | Genome-fragment | NA | NA | 0 |
| S3-final-viral-combined_k141_104696\|\|full | 2206 | No | NA | 3 | 0 | 2 | Not-determined | Genome-fragment | NA | NA | 0 |
| S12-final-viral-combined_k141_315281\|\|full | 1523 | No | NA | 2 | 0 | 0 | Low-quality | Genome-fragment | 0.98 | AAI-based (high-confidence) | 0 |
| S3-final-viral-combined_k141_93232\|\|full | 2177 | No | NA | 2 | 0 | 1 | Not-determined | Genome-fragment | NA | NA | 0 |
| S12-final-viral-combined_k141_365531\|\|full | 1523 | No | NA | 2 | 0 | 0 | Low-quality | Genome-fragment | 3.94 | AAI-based (medium-confidence) | 0 |
| S3-final-viral-combined_k141_58815\|\|full | 1897 | No | NA | 2 | 0 | 0 | Not-determined | Genome-fragment | NA | NA | 0 |
| S2-final-viral-combined_k141_106609\|\|full | 1523 | No | NA | 3 | 0 | 1 | Low-quality | Genome-fragment | 2.92 | AAI-based (medium-confidence) | 0 |
| S2_k141_206297 | 1523 | No | NA | 1 | 0 | 1 | Low-quality | Genome-fragment | 1.05 | AAI-based (medium-confidence) | 0 |
| S3-final-viral-combined_k141_76067\|\|full | 2263 | No | NA | 2 | 0 | 1 | Not-determined | Genome-fragment | NA | NA | 0 |
| S8-final-viral-combined_k141_244965\|\|full | 1523 | No | NA | 3 | 0 | 2 | Low-quality | Genome-fragment | 0.97 | AAI-based (medium-confidence) | 0 |
| S3-final-viral-combined_k141_18461\|\|full | 1178 | No | NA | 2 | 0 | 0 | Not-determined | Genome-fragment | NA | NA | 0 |
| S3-final-viral-combined_k141_35783\|\|full | 4406 | No | NA | 4 | 0 | 3 | Not-determined | Genome-fragment | NA | NA | 0 |
| S3-final-viral-combined_k141_98872\|\|full | 2993 | No | NA | 3 | 0 | 2 | Not-determined | Genome-fragment | NA | NA | 0 |
| S3-final-viral-combined_k141_58871\|\|full | 7053 | No | NA | 7 | 0 | 5 | Not-determined | Genome-fragment | NA | NA | 0 |
| S3-final-viral-combined_k141_98882\|\|full | 1646 | No | NA | 3 | 0 | 1 | Not-determined | Genome-fragment | NA | NA | 0 |
| S3-final-viral-combined_k141_18468\|\|full | 8276 | No | NA | 9 | 0 | 7 | Not-determined | Genome-fragment | NA | NA | 0 |
| S3-final-viral-combined_k141_58879\|\|full | 1871 | No | NA | 2 | 0 | 2 | Not-determined | Genome-fragment | NA | NA | 0 |
| S3-final-viral-combined_k141_128107\|\|full | 1148 | No | NA | 1 | 0 | 0 | Not-determined | Genome-fragment | NA | NA | 0 |
| S11-final-viral-combined_k141_9724\|\|full | 1524 | No | NA | 4 | 0 | 0 | Low-quality | Genome-fragment | 4.12 | AAI-based (medium-confidence) | 0 |
| S12-final-viral-combined_k141_31140\|\|full | 1524 | No | NA | 3 | 1 | 0 | Low-quality | Genome-fragment | 3.1 | AAI-based (high-confidence) | 0 |
| S3-final-viral-combined_k141_64621\|\|full | 1524 | No | NA | 2 | 0 | 2 | Not-determined | Genome-fragment | NA | NA | 0 |
| S3-final-viral-combined_k141_7196\|\|full | 1512 | No | NA | 3 | 0 | 0 | Not-determined | Genome-fragment | NA | NA | 0 |
| S3-final-viral-combined_k141_87807\|\|full | 1626 | No | NA | 3 | 0 | 2 | Not-determined | Genome-fragment | NA | NA | 0 |
| S3-final-viral-combined_k141_12710\|\|full | 2022 | No | NA | 4 | 0 | 0 | Not-determined | Genome-fragment | NA | NA | 0 |
| S2-final-viral-combined_k141_105222\|\|full | 1524 | No | NA | 1 | 0 | 0 | Low-quality | Genome-fragment | 1.31 | AAI-based (high-confidence) | 0 |
| S4-final-viral-combined_k141_125902\|\|full | 1524 | No | NA | 4 | 0 | 0 | Low-quality | Genome-fragment | 3.85 | AAI-based (high-confidence) | 0 |
| S3-final-viral-combined_k141_18546\|\|full | 1081 | No | NA | 2 | 0 | 1 | Not-determined | Genome-fragment | NA | NA | 0 |
| S11-final-viral-combined_k141_41128\|\|full | 1525 | No | NA | 2 | 0 | 0 | Low-quality | Genome-fragment | 2.93 | AAI-based (high-confidence) | 0 |
| S11_k141_40807 | 1525 | No | NA | 1 | 0 | 1 | Low-quality | Genome-fragment | 2.55 | AAI-based (medium-confidence) | 0 |
| S17-final-viral-combined_k141_15517\|\|full | 1525 | No | NA | 3 | 0 | 0 | Low-quality | Genome-fragment | 3.5 | AAI-based (medium-confidence) | 0 |
| S17-k141_225370 | 1525 | No | NA | 1 | 0 | 0 | Low-quality | Genome-fragment | 0.67 | AAI-based (medium-confidence) | 0 |
| S2-final-viral-combined_k141_176263\|\|full | 1525 | No | NA | 2 | 0 | 0 | Low-quality | Genome-fragment | 2.15 | AAI-based (medium-confidence) | 0 |
| S3-final-viral-combined_k141_122123\|\|full | 1143 | No | NA | 3 | 0 | 1 | Not-determined | Genome-fragment | NA | NA | 0 |
| S3-final-viral-combined_k141_1507\|\|full | 2498 | No | NA | 3 | 0 | 2 | Not-determined | Genome-fragment | NA | NA | 0 |
| S3-final-viral-combined_k141_122145\|\|full | 1581 | No | NA | 2 | 0 | 1 | Not-determined | Genome-fragment | NA | NA | 0 |
| S3-final-viral-combined_k141_122151\|\|full | 1800 | No | NA | 2 | 0 | 2 | Not-determined | Genome-fragment | NA | NA | 0 |
| S2-final-viral-combined_k141_198429\|\|full | 1525 | No | NA | 3 | 0 | 0 | Low-quality | Genome-fragment | 2.48 | AAI-based (high-confidence) | 0 |
| S4-final-viral-combined_k141_88763\|\|full | 1525 | No | NA | 2 | 0 | 0 | Low-quality | Genome-fragment | 0.97 | AAI-based (medium-confidence) | 0 |
| S8-final-viral-combined_k141_29610\|\|lt2gene | 1525 | No | NA | 2 | 2 | 0 | Low-quality | Genome-fragment | 3.13 | AAI-based (medium-confidence) | 0 |
| S9_k141_115029 | 1526 | No | NA | 2 | 0 | 1 | Low-quality | Genome-fragment | 0.4 | AAI-based (medium-confidence) | 0 |
| S3-final-viral-combined_k141_99016\|\|full | 2108 | No | NA | 3 | 0 | 2 | Not-determined | Genome-fragment | NA | NA | 0 |
| S9_k141_127665 | 1526 | No | NA | 2 | 0 | 1 | Low-quality | Genome-fragment | 1.66 | AAI-based (medium-confidence) | 0 |
| S3-final-viral-combined_k141_12843\|\|full | 1798 | No | NA | 3 | 0 | 0 | Not-determined | Genome-fragment | NA | NA | 0 |
| S3-final-viral-combined_k141_133856\|\|full | 2189 | No | NA | 4 | 0 | 1 | Not-determined | Genome-fragment | NA | NA | 0 |
| S1-final-viral-combined_k141_84664\|\|full | 1526 | No | NA | 3 | 1 | 0 | Low-quality | Genome-fragment | 1.98 | AAI-based (medium-confidence) | 0 |
| S3-final-viral-combined_k141_99058\|\|full | 3334 | No | NA | 5 | 0 | 2 | Not-determined | Genome-fragment | NA | NA | 0 |
| S3-final-viral-combined_k141_30123\|\|full | 1352 | No | NA | 2 | 0 | 0 | Not-determined | Genome-fragment | NA | NA | 0 |
| S3-final-viral-combined_k141_110778\|\|full | 3702 | No | NA | 3 | 0 | 2 | Not-determined | Genome-fragment | NA | NA | 0 |
| S3-final-viral-combined_k141_24555\|\|full | 8023 | No | NA | 9 | 0 | 9 | Not-determined | Genome-fragment | NA | NA | 0 |
| S3-final-viral-combined_k141_24560\|\|full | 1028 | No | NA | 3 | 0 | 2 | Not-determined | Genome-fragment | NA | NA | 0 |
| S3-final-viral-combined_k141_133862\|\|full | 21160 | No | NA | 11 | 0 | 8 | Not-determined | Genome-fragment | NA | NA | 0 |
| S3-final-viral-combined_k141_128305\|\|full | 2437 | No | NA | 1 | 0 | 1 | Not-determined | Genome-fragment | NA | NA | 0 |
| S1-final-viral-combined_k141_305278\|\|full | 1526 | No | NA | 2 | 0 | 1 | Low-quality | Genome-fragment | 2.75 | AAI-based (high-confidence) | 0 |
| S1-final-viral-combined_k141_197139\|\|full | 1526 | No | NA | 2 | 0 | 1 | Low-quality | Genome-fragment | 2.01 | AAI-based (medium-confidence) | 0 |
| S3-final-viral-combined_k141_116689\|\|full | 2869 | No | NA | 2 | 0 | 1 | Not-determined | Genome-fragment | NA | NA | 0 |
| S7_k141_145470 | 1526 | No | NA | 3 | 1 | 0 | Low-quality | Genome-fragment | 3.05 | AAI-based (high-confidence) | 0 |
| S3-final-viral-combined_k141_53432\|\|full | 3454 | No | NA | 3 | 0 | 1 | Not-determined | Genome-fragment | NA | NA | 0 |
| S17-final-viral-combined_k141_17638\|\|lt2gene | 1526 | No | NA | 1 | 1 | 0 | Low-quality | Genome-fragment | 0.84 | HMM-based (lower-bound) | 0 |
| S12-final-viral-combined_k141_250760\|\|full | 1526 | No | NA | 3 | 0 | 0 | Low-quality | Genome-fragment | 3.56 | AAI-based (medium-confidence) | 0 |
| S4-final-viral-combined_k141_61032\|\|full | 1526 | No | NA | 5 | 2 | 0 | Low-quality | Genome-fragment | 4.23 | AAI-based (high-confidence) | 0 |
| S3-final-viral-combined_k141_99105\|\|full | 1347 | No | NA | 2 | 0 | 1 | Not-determined | Genome-fragment | NA | NA | 0 |
| S3-final-viral-combined_k141_70598\|\|full | 1315 | No | NA | 1 | 0 | 0 | Not-determined | Genome-fragment | NA | NA | 0 |
| S3-final-viral-combined_k141_18761\|\|full | 2163 | No | NA | 3 | 0 | 0 | Not-determined | Genome-fragment | NA | NA | 0 |
| S8-final-viral-combined_k141_164065\|\|full | 1526 | No | NA | 4 | 1 | 0 | Low-quality | Genome-fragment | 2.79 | HMM-based (lower-bound) | 0 |
| S3-final-viral-combined_k141_41827\|\|full | 1091 | No | NA | 3 | 0 | 0 | Not-determined | Genome-fragment | NA | NA | 0 |
| S3-final-viral-combined_k141_122301\|\|full | 1401 | No | NA | 2 | 0 | 1 | Not-determined | Genome-fragment | NA | NA | 0 |
| S3-final-viral-combined_k141_18781\|\|full | 2294 | No | NA | 4 | 0 | 0 | Not-determined | Genome-fragment | NA | NA | 0 |
| S1-final-viral-combined_k141_354537\|\|full | 1527 | No | NA | 3 | 2 | 0 | Low-quality | Genome-fragment | 1.03 | HMM-based (lower-bound) | 0 |
| S1-final-viral-combined_k141_305958\|\|full | 1527 | No | NA | 5 | 0 | 0 | Low-quality | Genome-fragment | 3.14 | AAI-based (medium-confidence) | 0 |
| S5-final-viral-combined_k141_132134\|\|full | 1527 | No | NA | 4 | 0 | 0 | Low-quality | Genome-fragment | 2.68 | AAI-based (medium-confidence) | 0 |
| S3-final-viral-combined_k141_59175\|\|full | 1460 | No | NA | 2 | 0 | 1 | Not-determined | Genome-fragment | NA | NA | 0 |
| S5_k141_80490 | 1527 | No | NA | 1 | 0 | 1 | Low-quality | Genome-fragment | 0.84 | AAI-based (high-confidence) | 0 |
| S3-final-viral-combined_k141_18793\|\|full | 2338 | No | NA | 2 | 0 | 1 | Not-determined | Genome-fragment | NA | NA | 0 |
| S2-final-viral-combined_k141_68175\|\|full | 1527 | No | NA | 2 | 0 | 1 | Low-quality | Genome-fragment | 5.07 | AAI-based (medium-confidence) | 0 |
| S3-final-viral-combined_k141_128406\|\|full | 1586 | No | NA | 3 | 0 | 1 | Not-determined | Genome-fragment | NA | NA | 0 |
| S2-final-viral-combined_k141_223928\|\|full | 1527 | No | NA | 5 | 0 | 0 | Low-quality | Genome-fragment | 2.1 | AAI-based (medium-confidence) | 0 |
| S3-final-viral-combined_k141_47807\|\|full | 3895 | No | NA | 5 | 0 | 0 | Not-determined | Genome-fragment | NA | NA | 0 |
| S3-final-viral-combined_k141_105176\|\|full | 3969 | No | NA | 5 | 0 | 4 | Not-determined | Genome-fragment | NA | NA | 0 |
| S3-final-viral-combined_k141_76405\|\|full | 1075 | No | NA | 2 | 0 | 0 | Not-determined | Genome-fragment | NA | NA | 0 |
| S3-final-viral-combined_k141_110979\|\|full | 1191 | No | NA | 2 | 0 | 1 | Not-determined | Genome-fragment | NA | NA | 0 |
| S8-final-viral-combined_k141_239658\|\|full | 1527 | No | NA | 4 | 0 | 0 | Low-quality | Genome-fragment | 2.83 | AAI-based (medium-confidence) | 0 |
| S3-final-viral-combined_k141_18862\|\|full | 2936 | No | NA | 3 | 0 | 2 | Not-determined | Genome-fragment | NA | NA | 0 |
| S3-final-viral-combined_k141_24705\|\|full | 1124 | No | NA | 2 | 0 | 1 | Not-determined | Genome-fragment | NA | NA | 0 |
| S11-final-viral-combined_k141_101291\|\|full | 1528 | No | NA | 3 | 1 | 0 | Low-quality | Genome-fragment | 2.73 | AAI-based (high-confidence) | 0 |
| S3-final-viral-combined_k141_122448\|\|full | 1109 | No | NA | 2 | 0 | 1 | Not-determined | Genome-fragment | NA | NA | 0 |
| S1-final-viral-combined_k141_300169\|\|full | 1528 | No | NA | 1 | 0 | 1 | Low-quality | Genome-fragment | 3.13 | AAI-based (high-confidence) | 0 |
| S3-final-viral-combined_k141_47840\|\|full | 2733 | No | NA | 3 | 0 | 0 | Not-determined | Genome-fragment | NA | NA | 0 |
| S1-final-viral-combined_k141_304778\|\|full | 1528 | No | NA | 3 | 0 | 0 | Low-quality | Genome-fragment | 1.2 | AAI-based (high-confidence) | 0 |
| S3-final-viral-combined_k141_18883\|\|full | 4133 | No | NA | 6 | 0 | 4 | Not-determined | Genome-fragment | NA | NA | 0 |
| S3-final-viral-combined_k141_93339\|\|full | 1528 | No | NA | 2 | 0 | 2 | Low-quality | Genome-fragment | 1.13 | AAI-based (medium-confidence) | 0 |
| S3_k141_64566 | 1528 | No | NA | 1 | 0 | 0 | Low-quality | Genome-fragment | 0.24 | AAI-based (high-confidence) | 0 |
| S3-final-viral-combined_k141_111048\|\|full | 3340 | No | NA | 4 | 0 | 0 | Not-determined | Genome-fragment | NA | NA | 0 |
| S3-final-viral-combined_k141_99258\|\|full | 1620 | No | NA | 3 | 0 | 1 | Not-determined | Genome-fragment | NA | NA | 0 |
| S5-final-viral-combined_k141_27827\|\|full | 1528 | No | NA | 2 | 0 | 0 | Low-quality | Genome-fragment | 2.82 | AAI-based (medium-confidence) | 0 |
| S3-final-viral-combined_k141_18936\|\|full | 6696 | No | NA | 13 | 0 | 1 | Not-determined | Genome-fragment | NA | NA | 0 |
| S7-final-viral-combined_k141_103023\|\|full | 1528 | No | NA | 3 | 2 | 0 | Low-quality | Genome-fragment | 3.53 | AAI-based (medium-confidence) | 0 |
| S3-final-viral-combined_k141_70786\|\|full | 2810 | No | NA | 3 | 0 | 2 | Not-determined | Genome-fragment | NA | NA | 0 |
| S7_k141_192064 | 1528 | Yes | 695 | 2 | 1 | 1 | Low-quality | Genome-fragment | 0.06 | HMM-based (lower-bound) | 54.52 |
| S12-final-viral-combined_k141_16923\|\|full | 1528 | No | NA | 3 | 2 | 0 | Low-quality | Genome-fragment | 3.94 | AAI-based (medium-confidence) | 0 |
| S3-final-viral-combined_k141_65118\|\|full | 1890 | No | NA | 2 | 0 | 2 | Not-determined | Genome-fragment | NA | NA | 0 |
| S9-final-viral-combined_k141_58878\|\|full | 1529 | No | NA | 4 | 2 | 0 | Low-quality | Genome-fragment | 3.17 | AAI-based (high-confidence) | 0 |
| S3-final-viral-combined_k141_1872\|\|full | 2451 | No | NA | 3 | 0 | 1 | Not-determined | Genome-fragment | NA | NA | 0 |
| S9-final-viral-combined_k141_105623\|\|full | 1529 | No | NA | 3 | 0 | 2 | Low-quality | Genome-fragment | 0.68 | AAI-based (medium-confidence) | 0 |
| S11-final-viral-combined_k141_68243\|\|full | 1529 | No | NA | 2 | 0 | 1 | Low-quality | Genome-fragment | 1.34 | AAI-based (medium-confidence) | 0 |
| S3-final-viral-combined_k141_13172\|\|full | 1261 | No | NA | 3 | 0 | 1 | Not-determined | Genome-fragment | NA | NA | 0 |
| S11-final-viral-combined_k141_137502\|\|full | 1529 | No | NA | 2 | 1 | 0 | Low-quality | Genome-fragment | 3.01 | AAI-based (medium-confidence) | 0 |
| S11-final-viral-combined_k141_66510\|\|lt2gene | 1529 | No | NA | 2 | 1 | 0 | Low-quality | Genome-fragment | 3.45 | AAI-based (medium-confidence) | 0 |
| S3-final-viral-combined_k141_124006\|\|full | 1529 | No | NA | 3 | 0 | 1 | Low-quality | Genome-fragment | 0.8 | AAI-based (medium-confidence) | 0 |
| S7-final-viral-combined_k141_115459\|\|full | 1529 | No | NA | 3 | 0 | 0 | Low-quality | Genome-fragment | 1.93 | AAI-based (high-confidence) | 0 |
| S3-final-viral-combined_k141_42068\|\|full | 2057 | No | NA | 3 | 0 | 1 | Not-determined | Genome-fragment | NA | NA | 0 |
| S17-final-viral-combined_k141_42986\|\|full | 1529 | No | NA | 2 | 0 | 0 | Low-quality | Genome-fragment | 2.45 | AAI-based (high-confidence) | 0 |
| S12-final-viral-combined_k141_127\|\|full | 1529 | No | NA | 4 | 0 | 0 | Low-quality | Genome-fragment | 2.88 | AAI-based (high-confidence) | 0 |
| S12-final-viral-combined_k141_181895\|\|full | 1529 | No | NA | 3 | 0 | 1 | Low-quality | Genome-fragment | 0.73 | AAI-based (medium-confidence) | 0 |
| S3-final-viral-combined_k141_30471\|\|full | 2255 | No | NA | 3 | 0 | 1 | Not-determined | Genome-fragment | NA | NA | 0 |
| S12_k141_194046 | 1529 | No | NA | 2 | 0 | 0 | Low-quality | Genome-fragment | 0.89 | AAI-based (high-confidence) | 0 |
| S3-final-viral-combined_k141_59390\|\|full | 1766 | No | NA | 2 | 0 | 0 | Not-determined | Genome-fragment | NA | NA | 0 |
| S3-final-viral-combined_k141_88258\|\|full | 3669 | No | NA | 3 | 0 | 2 | Not-determined | Genome-fragment | NA | NA | 0 |
| S3-final-viral-combined_k141_1943\|\|full | 1799 | No | NA | 3 | 0 | 0 | Not-determined | Genome-fragment | NA | NA | 0 |
| S11-final-viral-combined_k141_70113\|\|full | 1530 | No | NA | 3 | 1 | 0 | Low-quality | Genome-fragment | 1.15 | AAI-based (high-confidence) | 0 |
| S3-final-viral-combined_k141_70897\|\|full | 1445 | No | NA | 2 | 0 | 0 | Not-determined | Genome-fragment | NA | NA | 0 |
| S3-final-viral-combined_k141_53639\|\|full | 1517 | No | NA | 2 | 0 | 1 | Not-determined | Genome-fragment | NA | NA | 0 |
| S11_k141_124652 | 1530 | No | NA | 1 | 1 | 0 | Low-quality | Genome-fragment | 1.66 | AAI-based (medium-confidence) | 0 |
| S3-final-viral-combined_k141_59422\|\|full | 1584 | No | NA | 3 | 0 | 0 | Not-determined | Genome-fragment | NA | NA | 0 |
| S3-final-viral-combined_k141_122660\|\|full | 3130 | No | NA | 3 | 0 | 2 | Not-determined | Genome-fragment | NA | NA | 0 |
| S3-final-viral-combined_k141_13278\|\|full | 1518 | No | NA | 2 | 0 | 2 | Not-determined | Genome-fragment | NA | NA | 0 |
| S3-final-viral-combined_k141_53660\|\|full | 2379 | No | NA | 3 | 0 | 3 | Not-determined | Genome-fragment | NA | NA | 0 |
| S7-final-viral-combined_k141_153151\|\|full | 1530 | No | NA | 2 | 0 | 1 | Low-quality | Genome-fragment | 2.75 | AAI-based (medium-confidence) | 0 |
| S7_k141_61615 | 1530 | No | NA | 1 | 0 | 1 | Low-quality | Genome-fragment | 2.54 | AAI-based (medium-confidence) | 0 |
| S3-final-viral-combined_k141_105446\|\|full | 2039 | No | NA | 3 | 0 | 2 | Not-determined | Genome-fragment | NA | NA | 0 |
| S17-final-viral-combined_k141_20422\|\|lt2gene | 1530 | No | NA | 1 | 1 | 0 | Low-quality | Genome-fragment | 3.05 | AAI-based (medium-confidence) | 0 |
| S17-k141_119935 | 1530 | No | NA | 5 | 0 | 0 | Low-quality | Genome-fragment | 1.83 | AAI-based (high-confidence) | 0 |
| S3-final-viral-combined_k141_122693\|\|full | 2835 | No | NA | 2 | 0 | 1 | Not-determined | Genome-fragment | NA | NA | 0 |
| S12-final-viral-combined_k141_248918\|\|full | 1530 | No | NA | 5 | 0 | 0 | Low-quality | Genome-fragment | 3.37 | AAI-based (high-confidence) | 0 |
| S4-final-viral-combined_k141_66750\|\|full | 1530 | No | NA | 2 | 0 | 2 | Low-quality | Genome-fragment | 0.63 | AAI-based (medium-confidence) | 0 |
| S3-final-viral-combined_k141_70955\|\|full | 2555 | No | NA | 3 | 0 | 2 | Not-determined | Genome-fragment | NA | NA | 0 |
| S3-final-viral-combined_k141_13364\|\|full | 2909 | No | NA | 2 | 0 | 2 | Not-determined | Genome-fragment | NA | NA | 0 |
| S6-final-viral-combined_k141_6978\|\|full | 1530 | No | NA | 3 | 1 | 0 | Low-quality | Genome-fragment | 2.58 | AAI-based (medium-confidence) | 0 |
| S3-final-viral-combined_k141_105523\|\|full | 3335 | No | NA | 4 | 0 | 0 | Not-determined | Genome-fragment | NA | NA | 0 |
| S6_k141_59262 | 1530 | No | NA | 5 | 3 | 0 | Low-quality | Genome-fragment | 2.96 | AAI-based (high-confidence) | 0 |
| S11-final-viral-combined_k141_16866\|\|full | 1531 | No | NA | 4 | 1 | 0 | Low-quality | Genome-fragment | 1 | AAI-based (high-confidence) | 0 |
| S7-final-viral-combined_k141_211710\|\|full | 1531 | No | NA | 3 | 1 | 0 | Low-quality | Genome-fragment | 3.87 | AAI-based (medium-confidence) | 0 |
| S3-final-viral-combined_k141_117076\|\|full | 2457 | No | NA | 2 | 0 | 0 | Not-determined | Genome-fragment | NA | NA | 0 |
| S3-final-viral-combined_k141_36405\|\|full | 1477 | No | NA | 3 | 0 | 0 | Not-determined | Genome-fragment | NA | NA | 0 |
| S17-final-viral-combined_k141_58120\|\|full | 1531 | No | NA | 3 | 0 | 0 | Low-quality | Genome-fragment | 3.52 | AAI-based (medium-confidence) | 0 |
| S3-final-viral-combined_k141_111204\|\|full | 2176 | No | NA | 3 | 0 | 1 | Not-determined | Genome-fragment | NA | NA | 0 |
| S3-final-viral-combined_k141_105571\|\|full | 1471 | No | NA | 3 | 0 | 2 | Not-determined | Genome-fragment | NA | NA | 0 |
| S12-final-viral-combined_k141_300215\|\|full | 1531 | No | NA | 2 | 0 | 0 | Low-quality | Genome-fragment | 2.32 | AAI-based (high-confidence) | 0 |
| S3-final-viral-combined_k141_7700\|\|full | 1837 | No | NA | 3 | 0 | 0 | Not-determined | Genome-fragment | NA | NA | 0 |
| S2_k141_122722 | 1531 | No | NA | 3 | 0 | 0 | Low-quality | Genome-fragment | 2.61 | AAI-based (high-confidence) | 0 |
| S8-final-viral-combined_k141_221999\|\|full | 1531 | No | NA | 2 | 0 | 1 | Low-quality | Genome-fragment | 0.81 | AAI-based (medium-confidence) | 0 |
| S3-final-viral-combined_k141_134349\|\|full | 1828 | No | NA | 2 | 0 | 0 | Not-determined | Genome-fragment | NA | NA | 0 |
| S7-final-viral-combined_k141_73669\|\|full | 1532 | No | NA | 2 | 2 | 0 | Low-quality | Genome-fragment | 2.46 | HMM-based (lower-bound) | 0 |
| S3-final-viral-combined_k141_30672\|\|full | 6342 | No | NA | 4 | 0 | 4 | Not-determined | Genome-fragment | NA | NA | 0 |
| S3-final-viral-combined_k141_25069\|\|full | 22693 | No | NA | 25 | 0 | 2 | Not-determined | Genome-fragment | NA | NA | 0 |
| S17-final-viral-combined_k141_108439\|\|full | 1532 | No | NA | 1 | 0 | 1 | Low-quality | Genome-fragment | 2.01 | AAI-based (high-confidence) | 0 |
| S3-final-viral-combined_k141_134366\|\|full | 4887 | No | NA | 8 | 0 | 1 | Not-determined | Genome-fragment | NA | NA | 0 |
| S12_k141_350086 | 1532 | No | NA | 1 | 0 | 0 | Low-quality | Genome-fragment | 0.81 | AAI-based (medium-confidence) | 0 |
| S3-final-viral-combined_k141_53790\|\|full | 1290 | No | NA | 2 | 0 | 1 | Not-determined | Genome-fragment | NA | NA | 0 |
| S2-final-viral-combined_k141_53185\|\|full | 1532 | No | NA | 2 | 1 | 0 | Low-quality | Genome-fragment | 0.61 | HMM-based (lower-bound) | 0 |
| S3-final-viral-combined_k141_48185\|\|full | 1404 | No | NA | 3 | 0 | 1 | Not-determined | Genome-fragment | NA | NA | 0 |
| S3-final-viral-combined_k141_117148\|\|full | 1490 | No | NA | 3 | 0 | 2 | Not-determined | Genome-fragment | NA | NA | 0 |
| S3-final-viral-combined_k141_36493\|\|full | 2611 | No | NA | 2 | 0 | 2 | Not-determined | Genome-fragment | NA | NA | 0 |
| S2_k141_166613 | 1532 | No | NA | 3 | 0 | 0 | Low-quality | Genome-fragment | 1.38 | AAI-based (high-confidence) | 0 |
| S3-final-viral-combined_k141_71111\|\|full | 2098 | No | NA | 3 | 0 | 2 | Not-determined | Genome-fragment | NA | NA | 0 |
| S4-final-viral-combined_k141_99791\|\|full | 1532 | No | NA | 2 | 1 | 0 | Low-quality | Genome-fragment | 0.81 | HMM-based (lower-bound) | 0 |
| S4-final-viral-combined_k141_162948\|\|full | 1532 | No | NA | 2 | 0 | 1 | Low-quality | Genome-fragment | 1.74 | AAI-based (medium-confidence) | 0 |
| S6-final-viral-combined_k141_10474\|\|full | 1532 | No | NA | 3 | 0 | 0 | Low-quality | Genome-fragment | 2.76 | AAI-based (medium-confidence) | 0 |
| S3-final-viral-combined_k141_13506\|\|full | 3222 | No | NA | 4 | 0 | 2 | Not-determined | Genome-fragment | NA | NA | 0 |
| S3-final-viral-combined_k141_7765\|\|full | 1367 | No | NA | 2 | 0 | 0 | Not-determined | Genome-fragment | NA | NA | 0 |
| S3-final-viral-combined_k141_82698\|\|full | 5908 | No | NA | 5 | 0 | 5 | Not-determined | Genome-fragment | NA | NA | 0 |
| S8-final-viral-combined_k141_105698\|\|full | 1532 | No | NA | 2 | 0 | 0 | Low-quality | Genome-fragment | 1 | AAI-based (medium-confidence) | 0 |
| S8-final-viral-combined_k141_163521\|\|full | 1532 | No | NA | 3 | 0 | 1 | Low-quality | Genome-fragment | 2.28 | AAI-based (high-confidence) | 0 |
| S8-final-viral-combined_k141_146423\|\|full | 1532 | No | NA | 3 | 0 | 0 | Low-quality | Genome-fragment | 1.65 | AAI-based (high-confidence) | 0 |
| S3-final-viral-combined_k141_94100\|\|full | 2145 | No | NA | 2 | 0 | 0 | Not-determined | Genome-fragment | NA | NA | 0 |
| S1-final-viral-combined_k141_186714\|\|full | 1533 | No | NA | 3 | 1 | 0 | Low-quality | Genome-fragment | 3.56 | AAI-based (medium-confidence) | 0 |
| S3-final-viral-combined_k141_117226\|\|full | 5279 | No | NA | 4 | 0 | 1 | Not-determined | Genome-fragment | NA | NA | 0 |
| S3-final-viral-combined_k141_25185\|\|full | 1000 | No | NA | 2 | 0 | 1 | Not-determined | Genome-fragment | NA | NA | 0 |
| S5-final-viral-combined_k141_127828\|\|lt2gene | 1533 | No | NA | 1 | 1 | 0 | Low-quality | Genome-fragment | 2.24 | HMM-based (lower-bound) | 0 |
| S3-final-viral-combined_k141_76821\|\|full | 1295 | No | NA | 3 | 0 | 0 | Not-determined | Genome-fragment | NA | NA | 0 |
| S3-final-viral-combined_k141_13573\|\|full | 2060 | No | NA | 2 | 0 | 2 | Not-determined | Genome-fragment | NA | NA | 0 |
| S7-final-viral-combined_k141_144420\|\|full | 1533 | No | NA | 3 | 2 | 0 | Low-quality | Genome-fragment | 4.09 | AAI-based (high-confidence) | 0 |
| S3-final-viral-combined_k141_99600\|\|full | 1771 | No | NA | 2 | 0 | 0 | Not-determined | Genome-fragment | NA | NA | 0 |
| S3-final-viral-combined_k141_25208\|\|full | 1200 | No | NA | 3 | 0 | 2 | Not-determined | Genome-fragment | NA | NA | 0 |
| S17-final-viral-combined_k141_58156\|\|full | 1533 | No | NA | 3 | 0 | 0 | Low-quality | Genome-fragment | 1.84 | AAI-based (medium-confidence) | 0 |
| S17-final-viral-combined_k141_161521\|\|full | 1533 | No | NA | 4 | 2 | 0 | Low-quality | Genome-fragment | 3.71 | AAI-based (medium-confidence) | 0 |
| S12_k141_311753 | 1533 | No | NA | 1 | 0 | 0 | Low-quality | Genome-fragment | 2.26 | AAI-based (medium-confidence) | 0 |
| S2-final-viral-combined_k141_242420\|\|full | 1533 | No | NA | 2 | 1 | 0 | Low-quality | Genome-fragment | 5.25 | HMM-based (lower-bound) | 0 |
| S3-final-viral-combined_k141_59662\|\|full | 1634 | No | NA | 2 | 0 | 0 | Not-determined | Genome-fragment | NA | NA | 0 |
| S3-final-viral-combined_k141_94219\|\|full | 2766 | No | NA | 2 | 0 | 1 | Not-determined | Genome-fragment | NA | NA | 0 |
| S6_k141_51199 | 1533 | No | NA | 3 | 0 | 0 | Low-quality | Genome-fragment | 1.16 | AAI-based (medium-confidence) | 0 |
| S8-final-viral-combined_k141_209914\|\|full | 1533 | No | NA | 4 | 1 | 0 | Low-quality | Genome-fragment | 3.63 | AAI-based (high-confidence) | 0 |
| S8-final-viral-combined_k141_97766\|\|full | 1533 | No | NA | 5 | 0 | 0 | Low-quality | Genome-fragment | 3.72 | AAI-based (high-confidence) | 0 |
| S3-final-viral-combined_k141_36675\|\|full | 1316 | No | NA | 3 | 0 | 2 | Not-determined | Genome-fragment | NA | NA | 0 |
| S3-final-viral-combined_k141_19385\|\|full | 3629 | No | NA | 3 | 0 | 2 | Not-determined | Genome-fragment | NA | NA | 0 |
| S8_k141_126579 | 1533 | No | NA | 2 | 0 | 0 | Low-quality | Genome-fragment | 3.61 | AAI-based (high-confidence) | 0 |
| S1-final-viral-combined_k141_284569\|\|full | 1534 | No | NA | 3 | 0 | 2 | Low-quality | Genome-fragment | 0.48 | AAI-based (medium-confidence) | 0 |
| S1-final-viral-combined_k141_70016\|\|full | 1534 | No | NA | 2 | 1 | 0 | Low-quality | Genome-fragment | 3.34 | AAI-based (medium-confidence) | 0 |
| S3-final-viral-combined_k141_59703\|\|full | 1905 | No | NA | 3 | 0 | 0 | Not-determined | Genome-fragment | NA | NA | 0 |
| S3-final-viral-combined_k141_105734\|\|full | 2197 | No | NA | 3 | 0 | 1 | Not-determined | Genome-fragment | NA | NA | 0 |
| S5_k141_156133 | 1534 | No | NA | 2 | 0 | 0 | Low-quality | Genome-fragment | 10.58 | AAI-based (high-confidence) | 0 |
| S3-final-viral-combined_k141_105738\|\|full | 1686 | No | NA | 3 | 0 | 1 | Not-determined | Genome-fragment | NA | NA | 0 |
| S3-final-viral-combined_k141_53965\|\|full | 2984 | No | NA | 4 | 0 | 2 | Not-determined | Genome-fragment | NA | NA | 0 |
| S7-final-viral-combined_k141_52866\|\|full | 1534 | No | NA | 3 | 2 | 0 | Low-quality | Genome-fragment | 3.52 | AAI-based (medium-confidence) | 0 |
| S7-final-viral-combined_k141_150072\|\|full | 1534 | No | NA | 3 | 1 | 0 | Low-quality | Genome-fragment | 0.55 | HMM-based (lower-bound) | 0 |
| S3-final-viral-combined_k141_13675\|\|full | 6730 | No | NA | 6 | 0 | 4 | Not-determined | Genome-fragment | NA | NA | 0 |
| S12-final-viral-combined_k141_253154\|\|full | 1534 | Yes | 1128 | 5 | 1 | 1 | Low-quality | Genome-fragment | 0.91 | HMM-based (lower-bound) | 26.47 |
| S3-final-viral-combined_k141_105773\|\|full | 5692 | No | NA | 6 | 0 | 2 | Not-determined | Genome-fragment | NA | NA | 0 |
| S3-final-viral-combined_k141_48336\|\|full | 4054 | No | NA | 4 | 0 | 3 | Not-determined | Genome-fragment | NA | NA | 0 |
| S3-final-viral-combined_k141_71285\|\|full | 3152 | No | NA | 5 | 0 | 0 | Not-determined | Genome-fragment | NA | NA | 0 |
| S12-final-viral-combined_k141_223288\|\|full | 1534 | No | NA | 3 | 0 | 0 | Low-quality | Genome-fragment | 4.82 | AAI-based (high-confidence) | 0 |
| S12-final-viral-combined_k141_83588\|\|full | 1534 | No | NA | 3 | 0 | 1 | Low-quality | Genome-fragment | 2.08 | AAI-based (medium-confidence) | 0 |
| S12_k141_219452 | 1534 | No | NA | 1 | 0 | 0 | Low-quality | Genome-fragment | 0.84 | AAI-based (high-confidence) | 0 |
| S3-final-viral-combined_k141_76974\|\|full | 1294 | No | NA | 3 | 0 | 0 | Not-determined | Genome-fragment | NA | NA | 0 |
| S2-final-viral-combined_k141_33782\|\|full | 1534 | No | NA | 3 | 0 | 0 | Low-quality | Genome-fragment | 2.76 | AAI-based (high-confidence) | 0 |
| S3-final-viral-combined_k141_54012\|\|full | 1422 | No | NA | 2 | 0 | 1 | Not-determined | Genome-fragment | NA | NA | 0 |
| S3-final-viral-combined_k141_42533\|\|full | 1860 | No | NA | 2 | 0 | 2 | Not-determined | Genome-fragment | NA | NA | 0 |
| S8_k141_18280 | 1534 | No | NA | 2 | 0 | 0 | Low-quality | Genome-fragment | 1 | AAI-based (medium-confidence) | 0 |
| S3-final-viral-combined_k141_59773\|\|full | 1703 | No | NA | 3 | 0 | 1 | Not-determined | Genome-fragment | NA | NA | 0 |
| S3-final-viral-combined_k141_36799\|\|full | 2444 | No | NA | 4 | 0 | 2 | Not-determined | Genome-fragment | NA | NA | 0 |
| S11_k141_269759 | 1535 | No | NA | 3 | 0 | 0 | Low-quality | Genome-fragment | 2.17 | AAI-based (high-confidence) | 0 |
| S3-final-viral-combined_k141_48397\|\|full | 4145 | No | NA | 4 | 0 | 1 | Not-determined | Genome-fragment | NA | NA | 0 |
| S3-final-viral-combined_k141_88758\|\|full | 1721 | No | NA | 3 | 0 | 2 | Not-determined | Genome-fragment | NA | NA | 0 |
| S3-final-viral-combined_k141_117410\|\|full | 4974 | No | NA | 4 | 0 | 3 | Not-determined | Genome-fragment | NA | NA | 0 |
| S1-final-viral-combined_k141_141626\|\|full | 1535 | No | NA | 2 | 0 | 1 | Low-quality | Genome-fragment | 0.99 | AAI-based (high-confidence) | 0 |
| S3-final-viral-combined_k141_82975\|\|full | 1535 | No | NA | 3 | 0 | 1 | Not-determined | Genome-fragment | NA | NA | 0 |
| S1-final-viral-combined_k141_178332\|\|full | 1535 | No | NA | 4 | 0 | 1 | Low-quality | Genome-fragment | 0.72 | AAI-based (high-confidence) | 0 |
| S3-final-viral-combined_k141_2492\|\|full | 1962 | No | NA | 3 | 0 | 1 | Not-determined | Genome-fragment | NA | NA | 0 |
| S1-final-viral-combined_k141_279528\|\|full | 1535 | No | NA | 4 | 0 | 0 | Low-quality | Genome-fragment | 3.27 | AAI-based (high-confidence) | 0 |
| S17-final-viral-combined_k141_209743\|\|full | 1535 | No | NA | 4 | 1 | 0 | Low-quality | Genome-fragment | 2.91 | AAI-based (high-confidence) | 0 |
| S3-final-viral-combined_k141_111563\|\|full | 3841 | No | NA | 3 | 0 | 3 | Not-determined | Genome-fragment | NA | NA | 0 |
| S3-final-viral-combined_k141_59828\|\|full | 11258 | No | NA | 5 | 0 | 0 | Not-determined | Genome-fragment | NA | NA | 0 |
| S3-final-viral-combined_k141_19532\|\|full | 2442 | No | NA | 7 | 0 | 0 | Not-determined | Genome-fragment | NA | NA | 0 |
| S3-final-viral-combined_k141_30991\|\|full | 5822 | No | NA | 6 | 0 | 4 | Not-determined | Genome-fragment | NA | NA | 0 |
| S3-final-viral-combined_k141_77078\|\|full | 3868 | No | NA | 4 | 0 | 1 | Not-determined | Genome-fragment | NA | NA | 0 |
| S3-final-viral-combined_k141_99863\|\|full | 9447 | No | NA | 6 | 0 | 5 | Not-determined | Genome-fragment | NA | NA | 0 |
| S3-final-viral-combined_k141_65651\|\|full | 10991 | No | NA | 10 | 0 | 9 | Not-determined | Genome-fragment | NA | NA | 0 |
| S3-final-viral-combined_k141_99864\|\|full | 1316 | No | NA | 2 | 0 | 0 | Not-determined | Genome-fragment | NA | NA | 0 |
| S3-final-viral-combined_k141_134778\|\|full | 1591 | No | NA | 3 | 0 | 2 | Not-determined | Genome-fragment | NA | NA | 0 |
| S17-k141_149662 | 1535 | No | NA | 2 | 0 | 2 | Low-quality | Genome-fragment | 1.1 | AAI-based (medium-confidence) | 0 |
| S12-final-viral-combined_k141_31748\|\|full | 1535 | No | NA | 3 | 1 | 0 | Low-quality | Genome-fragment | 0.96 | HMM-based (lower-bound) | 0 |
| S3-final-viral-combined_k141_42616\|\|full | 1282 | No | NA | 2 | 0 | 1 | Not-determined | Genome-fragment | NA | NA | 0 |
| S12-final-viral-combined_k141_284524\|\|full | 1535 | No | NA | 1 | 0 | 1 | Low-quality | Genome-fragment | 2.91 | AAI-based (high-confidence) | 0 |
| S3-final-viral-combined_k141_31053\|\|full | 1287 | No | NA | 2 | 0 | 0 | Not-determined | Genome-fragment | NA | NA | 0 |
| S6-final-viral-combined_k141_64661\|\|full | 1535 | No | NA | 2 | 1 | 0 | Low-quality | Genome-fragment | 4 | AAI-based (high-confidence) | 0 |
| S3-final-viral-combined_k141_8085\|\|full | 3514 | No | NA | 4 | 0 | 2 | Not-determined | Genome-fragment | NA | NA | 0 |
| S8-final-viral-combined_k141_112912\|\|full | 1535 | No | NA | 4 | 3 | 0 | Low-quality | Genome-fragment | 1.14 | HMM-based (lower-bound) | 0 |
| S3-final-viral-combined_k141_105919\|\|full | 3046 | No | NA | 3 | 0 | 1 | Not-determined | Genome-fragment | NA | NA | 0 |

| | | | | | | | | | | | |
|---|---|---|---|---|---|---|---|---|---|---|---|
| S3-final-viral-combined_k141_134829\|\|full | 1096 | No | NA | 2 | 0 | 2 | Not-determined | Genome-fragment | NA | NA | 0 |
| S3-final-viral-combined_k141_19614\|\|full | 2259 | No | NA | 3 | 0 | 0 | Not-determined | Genome-fragment | NA | NA | 0 |
| S3-final-viral-combined_k141_8096\|\|full | 1088 | No | NA | 2 | 0 | 2 | Not-determined | Genome-fragment | NA | NA | 0 |
| S9-final-viral-combined_k141_89512\|\|full | 1536 | No | NA | 5 | 2 | 0 | Low-quality | Genome-fragment | 4.5 | AAI-based (high-confidence) | 0 |
| S11-final-viral-combined_k141_217110\|\|full | 1536 | No | NA | 3 | 2 | 0 | Low-quality | Genome-fragment | 6.29 | AAI-based (high-confidence) | 0 |
| S3-final-viral-combined_k141_83113\|\|full | 2563 | No | NA | 4 | 0 | 1 | Not-determined | Genome-fragment | NA | NA | 0 |
| S3-final-viral-combined_k141_117576\|\|full | 2452 | No | NA | 2 | 0 | 2 | Not-determined | Genome-fragment | NA | NA | 0 |
| S3-final-viral-combined_k141_37045\|\|full | 1095 | No | NA | 2 | 0 | 1 | Not-determined | Genome-fragment | NA | NA | 0 |
| S3-final-viral-combined_k141_105956\|\|full | 1362 | No | NA | 3 | 0 | 1 | Not-determined | Genome-fragment | NA | NA | 0 |
| S1-final-viral-combined_k141_106194\|\|full | 1536 | No | NA | 4 | 0 | 0 | Low-quality | Genome-fragment | 4.18 | AAI-based (high-confidence) | 0 |
| S1-final-viral-combined_k141_370308\|\|full | 1536 | No | NA | 3 | 0 | 1 | Low-quality | Genome-fragment | 1.26 | AAI-based (medium-confidence) | 0 |
| S3-final-viral-combined_k141_123287\|\|full | 1184 | No | NA | 2 | 0 | 1 | Not-determined | Genome-fragment | NA | NA | 0 |
| S3-final-viral-combined_k141_129245\|\|full | 4634 | No | NA | 5 | 0 | 4 | Not-determined | Genome-fragment | NA | NA | 0 |
| S1_k141_321779 | 1536 | No | NA | 2 | 0 | 0 | Low-quality | Genome-fragment | 3.2 | AAI-based (high-confidence) | 0 |
| S3-final-viral-combined_k141_2692\|\|full | 2226 | No | NA | 3 | 0 | 3 | Not-determined | Genome-fragment | NA | NA | 0 |
| S5-final-viral-combined_k141_105689\|\|full | 1536 | No | NA | 3 | 0 | 0 | Low-quality | Genome-fragment | 3.88 | AAI-based (medium-confidence) | 0 |
| S3-final-viral-combined_k141_111799\|\|full | 2754 | No | NA | 3 | 0 | 2 | Not-determined | Genome-fragment | NA | NA | 0 |
| S5-final-viral-combined_k141_47616\|\|lt2gene | 1536 | No | NA | 2 | 1 | 0 | Low-quality | Genome-fragment | 4.08 | AAI-based (high-confidence) | 0 |
| S7-final-viral-combined_k141_65373\|\|full | 1536 | No | NA | 3 | 0 | 1 | Low-quality | Genome-fragment | 2.05 | AAI-based (medium-confidence) | 0 |
| S7-final-viral-combined_k141_152389\|\|full | 1536 | No | NA | 2 | 1 | 0 | Low-quality | Genome-fragment | 3.55 | AAI-based (high-confidence) | 0 |
| S12-final-viral-combined_k141_100642\|\|full | 1536 | No | NA | 3 | 0 | 0 | Low-quality | Genome-fragment | 2.71 | AAI-based (medium-confidence) | 0 |
| S3-final-viral-combined_k141_123364\|\|full | 2351 | No | NA | 3 | 0 | 2 | Not-determined | Genome-fragment | NA | NA | 0 |
| S12-final-viral-combined_k141_21684\|\|full | 1536 | No | NA | 2 | 0 | 0 | Low-quality | Genome-fragment | 3.58 | AAI-based (high-confidence) | 0 |
| S12-final-viral-combined_k141_324113\|\|full | 1536 | No | NA | 2 | 0 | 0 | Low-quality | Genome-fragment | 3.55 | AAI-based (medium-confidence) | 0 |
| S4_k141_40148 | 1536 | No | NA | 2 | 1 | 0 | Low-quality | Genome-fragment | 2.65 | AAI-based (medium-confidence) | 0 |
| S3-final-viral-combined_k141_89054\|\|full | 2481 | No | NA | 3 | 0 | 0 | Not-determined | Genome-fragment | NA | NA | 0 |
| S11-final-viral-combined_k141_294648\|\|full | 1537 | No | NA | 3 | 1 | 0 | Low-quality | Genome-fragment | 0.97 | HMM-based (lower-bound) | 0 |
| S3-final-viral-combined_k141_106065\|\|full | 4333 | No | NA | 5 | 0 | 2 | Not-determined | Genome-fragment | NA | NA | 0 |
| S1-final-viral-combined_k141_168694\|\|full | 1537 | No | NA | 2 | 0 | 2 | Low-quality | Genome-fragment | 0.8 | AAI-based (medium-confidence) | 0 |
| S3-final-viral-combined_k141_134975\|\|full | 4423 | No | NA | 4 | 0 | 3 | Not-determined | Genome-fragment | NA | NA | 0 |
| S1-final-viral-combined_k141_275678\|\|full | 1537 | No | NA | 2 | 0 | 1 | Low-quality | Genome-fragment | 2 | AAI-based (medium-confidence) | 0 |
| S3-final-viral-combined_k141_13929\|\|full | 2703 | No | NA | 3 | 0 | 1 | Not-determined | Genome-fragment | NA | NA | 0 |
| S3-final-viral-combined_k141_94695\|\|full | 1877 | No | NA | 2 | 0 | 1 | Not-determined | Genome-fragment | NA | NA | 0 |
| S3-final-viral-combined_k141_42789\|\|full | 1503 | No | NA | 2 | 0 | 1 | Not-determined | Genome-fragment | NA | NA | 0 |
| S3-final-viral-combined_k141_117738\|\|full | 1032 | No | NA | 2 | 0 | 0 | Not-determined | Genome-fragment | NA | NA | 0 |
| S3-final-viral-combined_k141_129421\|\|full | 2250 | No | NA | 2 | 0 | 2 | Not-determined | Genome-fragment | NA | NA | 0 |
| S1-final-viral-combined_k141_113138\|\|full | 1537 | No | NA | 1 | 0 | 0 | Low-quality | Genome-fragment | 2.4 | AAI-based (high-confidence) | 0 |
| S1_k141_172621 | 1537 | No | NA | 2 | 0 | 0 | Low-quality | Genome-fragment | 1.04 | AAI-based (medium-confidence) | 0 |
| S3-final-viral-combined_k141_8317\|\|full | 2238 | No | NA | 3 | 0 | 1 | Not-determined | Genome-fragment | NA | NA | 0 |
| S3-final-viral-combined_k141_65873\|\|full | 2568 | No | NA | 2 | 0 | 2 | Not-determined | Genome-fragment | NA | NA | 0 |
| S3-final-viral-combined_k141_42812\|\|full | 1668 | No | NA | 2 | 0 | 1 | Not-determined | Genome-fragment | NA | NA | 0 |
| S5-final-viral-combined_k141_9832\|\|full | 1537 | No | NA | 3 | 2 | 0 | Low-quality | Genome-fragment | 2.81 | AAI-based (high-confidence) | 0 |
| S3-final-viral-combined_k141_54424\|\|full | 1849 | No | NA | 3 | 0 | 2 | Not-determined | Genome-fragment | NA | NA | 0 |
| S17-k141_7039 | 1537 | No | NA | 2 | 0 | 1 | Low-quality | Genome-fragment | 1.19 | AAI-based (high-confidence) | 0 |
| S12_k141_38187 | 1537 | No | NA | 1 | 0 | 0 | Low-quality | Genome-fragment | 2.95 | AAI-based (medium-confidence) | 0 |
| S2-final-viral-combined_k141_12191\|\|full | 1537 | No | NA | 3 | 0 | 1 | Low-quality | Genome-fragment | 4.35 | AAI-based (medium-confidence) | 0 |
| S4_k141_8407 | 1537 | No | NA | 1 | 1 | 0 | Low-quality | Genome-fragment | 1.55 | AAI-based (high-confidence) | 0 |
| S3-final-viral-combined_k141_129470\|\|full | 1895 | No | NA | 2 | 0 | 2 | Not-determined | Genome-fragment | NA | NA | 0 |
| S3-final-viral-combined_k141_8342\|\|full | 10479 | No | NA | 5 | 0 | 3 | Not-determined | Genome-fragment | NA | NA | 0 |
| S3-final-viral-combined_k141_117805\|\|full | 2968 | No | NA | 2 | 0 | 2 | Not-determined | Genome-fragment | NA | NA | 0 |
| S3-final-viral-combined_k141_135056\|\|full | 5394 | No | NA | 5 | 0 | 3 | Not-determined | Genome-fragment | NA | NA | 0 |
| S9-final-viral-combined_k141_89155\|\|full | 1538 | No | NA | 3 | 0 | 0 | Low-quality | Genome-fragment | 3.76 | AAI-based (medium-confidence) | 0 |
| S3-final-viral-combined_k141_2921\|\|full | 1541 | No | NA | 2 | 0 | 2 | Not-determined | Genome-fragment | NA | NA | 0 |
| S3-final-viral-combined_k141_42853\|\|full | 10209 | No | NA | 6 | 0 | 3 | Not-determined | Genome-fragment | NA | NA | 0 |
| S1-final-viral-combined_k141_223700\|\|full | 1538 | No | NA | 4 | 1 | 0 | Low-quality | Genome-fragment | 4.41 | AAI-based (high-confidence) | 0 |
| S3-final-viral-combined_k141_77463\|\|full | 5855 | No | NA | 6 | 0 | 4 | Not-determined | Genome-fragment | NA | NA | 0 |
| S1-final-viral-combined_k141_286817\|\|lt2gene | 1538 | No | NA | 1 | 1 | 0 | Low-quality | Genome-fragment | 3.25 | AAI-based (medium-confidence) | 0 |
| S5-final-viral-combined_k141_179389\|\|full | 1538 | No | NA | 2 | 0 | 0 | Low-quality | Genome-fragment | 2.75 | AAI-based (medium-confidence) | 0 |
| S3-final-viral-combined_k141_111996\|\|full | 2865 | No | NA | 3 | 0 | 0 | Not-determined | Genome-fragment | NA | NA | 0 |
| S7-final-viral-combined_k141_354\|\|full | 1538 | No | NA | 2 | 0 | 1 | Low-quality | Genome-fragment | 1.95 | AAI-based (high-confidence) | 0 |
| S12-final-viral-combined_k141_126192\|\|full | 1538 | No | NA | 3 | 0 | 1 | Low-quality | Genome-fragment | 3.32 | AAI-based (high-confidence) | 0 |
| S3-final-viral-combined_k141_42891\|\|full | 2603 | No | NA | 2 | 0 | 2 | Not-determined | Genome-fragment | NA | NA | 0 |
| S12-final-viral-combined_k141_325287\|\|lt2gene | 1538 | No | NA | 2 | 1 | 0 | Low-quality | Genome-fragment | 3.99 | AAI-based (high-confidence) | 0 |
| S2-final-viral-combined_k141_8304\|\|lt2gene | 1538 | No | NA | 2 | 1 | 0 | Low-quality | Genome-fragment | 4.03 | AAI-based (high-confidence) | 0 |
| S3-final-viral-combined_k141_83426\|\|full | 3099 | No | NA | 3 | 0 | 3 | Not-determined | Genome-fragment | NA | NA | 0 |
| S2_k141_237991 | 1538 | No | NA | 3 | 0 | 1 | Low-quality | Genome-fragment | 0.61 | AAI-based (medium-confidence) | 0 |
| S6_k141_37415 | 1538 | No | NA | 3 | 0 | 0 | Low-quality | Genome-fragment | 2.03 | AAI-based (medium-confidence) | 0 |
| S3-final-viral-combined_k141_123576\|\|full | 2116 | No | NA | 2 | 0 | 2 | Not-determined | Genome-fragment | NA | NA | 0 |
| S3-final-viral-combined_k141_31491\|\|full | 4838 | No | NA | 7 | 0 | 2 | Not-determined | Genome-fragment | NA | NA | 0 |
| S3-final-viral-combined_k141_65972\|\|full | 3074 | No | NA | 4 | 0 | 3 | Not-determined | Genome-fragment | NA | NA | 0 |
| S3-final-viral-combined_k141_48831\|\|full | 1250 | No | NA | 3 | 0 | 1 | Not-determined | Genome-fragment | NA | NA | 0 |
| S3-final-viral-combined_k141_89211\|\|full | 3623 | No | NA | 3 | 0 | 1 | Not-determined | Genome-fragment | NA | NA | 0 |
| S3-final-viral-combined_k141_89216\|\|full | 1000 | No | NA | 2 | 0 | 1 | Not-determined | Genome-fragment | NA | NA | 0 |
| S8_k141_180280 | 1538 | No | NA | 1 | 0 | 0 | Low-quality | Genome-fragment | 2.61 | AAI-based (medium-confidence) | 0 |
| S9-final-viral-combined_k141_64655\|\|full | 1539 | No | NA | 2 | 0 | 0 | Low-quality | Genome-fragment | 4.24 | AAI-based (medium-confidence) | 0 |
| S3-final-viral-combined_k141_48858\|\|full | 1618 | No | NA | 2 | 0 | 2 | Not-determined | Genome-fragment | NA | NA | 0 |
| S3-final-viral-combined_k141_14115\|\|full | 1154 | No | NA | 2 | 0 | 0 | Not-determined | Genome-fragment | NA | NA | 0 |
| S7-final-viral-combined_k141_137658\|\|full | 1539 | No | NA | 2 | 0 | 2 | Low-quality | Genome-fragment | 3.55 | AAI-based (medium-confidence) | 0 |
| S17-final-viral-combined_k141_174988\|\|full | 1539 | No | NA | 2 | 0 | 1 | Low-quality | Genome-fragment | 0.7 | AAI-based (medium-confidence) | 0 |
| S12-final-viral-combined_k141_351539\|\|full | 1539 | No | NA | 3 | 0 | 1 | Low-quality | Genome-fragment | 0.62 | AAI-based (medium-confidence) | 0 |
| S3-final-viral-combined_k141_129572\|\|full | 1328 | No | NA | 3 | 0 | 2 | Not-determined | Genome-fragment | NA | NA | 0 |
| S12-final-viral-combined_k141_34430\|\|full | 1539 | No | NA | 3 | 0 | 0 | Low-quality | Genome-fragment | 2.03 | AAI-based (medium-confidence) | 0 |
| S12-final-viral-combined_k141_259629\|\|full | 1539 | No | NA | 2 | 0 | 0 | Low-quality | Genome-fragment | 0.84 | AAI-based (high-confidence) | 0 |
| S2-final-viral-combined_k141_49891\|\|full | 1539 | No | NA | 4 | 0 | 0 | Low-quality | Genome-fragment | 2.7 | AAI-based (medium-confidence) | 0 |
| S3-final-viral-combined_k141_14132\|\|full | 1745 | No | NA | 3 | 0 | 0 | Not-determined | Genome-fragment | NA | NA | 0 |
| S3-final-viral-combined_k141_106306\|\|full | 2405 | No | NA | 5 | 0 | 0 | Not-determined | Genome-fragment | NA | NA | 0 |
| S2_k141_165084 | 1539 | No | NA | 1 | 0 | 1 | Low-quality | Genome-fragment | 0.41 | AAI-based (high-confidence) | 0 |
| S3-final-viral-combined_k141_106334\|\|full | 1997 | No | NA | 3 | 0 | 0 | Not-determined | Genome-fragment | NA | NA | 0 |
| S4_k141_162711 | 1539 | No | NA | 3 | 0 | 0 | Low-quality | Genome-fragment | 0.99 | AAI-based (medium-confidence) | 0 |
| S3-final-viral-combined_k141_60378\|\|full | 1291 | No | NA | 3 | 0 | 1 | Not-determined | Genome-fragment | NA | NA | 0 |
| S9_k141_117966 | 1540 | No | NA | 2 | 0 | 1 | Low-quality | Genome-fragment | 3.82 | AAI-based (high-confidence) | 0 |
| S3-final-viral-combined_k141_89305\|\|full | 1319 | No | NA | 4 | 0 | 0 | Not-determined | Genome-fragment | NA | NA | 0 |
| S3-final-viral-combined_k141_123672\|\|full | 2754 | No | NA | 4 | 0 | 0 | Not-determined | Genome-fragment | NA | NA | 0 |
| S3-final-viral-combined_k141_8524\|\|full | 1686 | No | NA | 3 | 0 | 2 | Not-determined | Genome-fragment | NA | NA | 0 |
| S11-final-viral-combined_k141_157265\|\|full | 1540 | No | NA | 3 | 0 | 0 | Low-quality | Genome-fragment | 4.22 | AAI-based (high-confidence) | 0 |
| S3-final-viral-combined_k141_89333\|\|full | 2533 | No | NA | 5 | 0 | 0 | Not-determined | Genome-fragment | NA | NA | 0 |
| S11-final-viral-combined_k141_135444\|\|lt2gene | 1540 | No | NA | 2 | 1 | 0 | Low-quality | Genome-fragment | 5.1 | AAI-based (high-confidence) | 0 |
| S3-final-viral-combined_k141_20150\|\|full | 1902 | No | NA | 4 | 0 | 0 | Not-determined | Genome-fragment | NA | NA | 0 |
| S3-final-viral-combined_k141_43064\|\|full | 2538 | No | NA | 4 | 0 | 0 | Not-determined | Genome-fragment | NA | NA | 0 |
| S11_k141_23419 | 1540 | No | NA | 4 | 1 | 0 | Low-quality | Genome-fragment | 3.24 | HMM-based (lower-bound) | 0 |
| S3-final-viral-combined_k141_135247\|\|full | 4357 | No | NA | 2 | 0 | 1 | Not-determined | Genome-fragment | NA | NA | 0 |
| S1-final-viral-combined_k141_24618\|\|lt2gene | 1540 | No | NA | 1 | 1 | 0 | Low-quality | Genome-fragment | 1.16 | AAI-based (high-confidence) | 0 |
| S3-final-viral-combined_k141_48906\|\|full | 2681 | No | NA | 4 | 0 | 1 | Not-determined | Genome-fragment | NA | NA | 0 |
| S3-final-viral-combined_k141_31597\|\|full | 14323 | No | NA | 14 | 0 | 8 | Not-determined | Genome-fragment | NA | NA | 0 |
| S1_k141_135502 | 1540 | No | NA | 2 | 0 | 1 | Low-quality | Genome-fragment | 1.92 | AAI-based (high-confidence) | 0 |
| S3-final-viral-combined_k141_48907\|\|full | 1717 | No | NA | 3 | 0 | 0 | Not-determined | Genome-fragment | NA | NA | 0 |
| S3-final-viral-combined_k141_129668\|\|full | 3002 | No | NA | 4 | 0 | 1 | Not-determined | Genome-fragment | NA | NA | 0 |
| S3-final-viral-combined_k141_8571\|\|full | 3091 | No | NA | 3 | 0 | 2 | Not-determined | Genome-fragment | NA | NA | 0 |
| S3_k141_76526 | 1540 | No | NA | 3 | 1 | 0 | Low-quality | Genome-fragment | 1.44 | AAI-based (medium-confidence) | 0 |
| S3-final-viral-combined_k141_77600\|\|full | 2241 | No | NA | 3 | 0 | 1 | Not-determined | Genome-fragment | NA | NA | 0 |
| S3-final-viral-combined_k141_123765\|\|full | 3145 | No | NA | 6 | 0 | 2 | Not-determined | Genome-fragment | NA | NA | 0 |
| S3_k141_112806 | 1540 | No | NA | 1 | 1 | 0 | Low-quality | Genome-fragment | 1.14 | HMM-based (lower-bound) | 0 |
| S3-final-viral-combined_k141_129704\|\|full | 1225 | No | NA | 3 | 0 | 1 | Not-determined | Genome-fragment | NA | NA | 0 |
| S3-final-viral-combined_k141_106453\|\|full | 2866 | No | NA | 3 | 0 | 0 | Not-determined | Genome-fragment | NA | NA | 0 |
| S5-final-viral-combined_k141_91008\|\|full | 1540 | No | NA | 4 | 0 | 0 | Low-quality | Genome-fragment | 3.62 | AAI-based (medium-confidence) | 0 |
| S3-final-viral-combined_k141_129711\|\|full | 3218 | No | NA | 4 | 0 | 1 | Not-determined | Genome-fragment | NA | NA | 0 |
| S3-final-viral-combined_k141_20213\|\|full | 2005 | No | NA | 3 | 0 | 0 | Not-determined | Genome-fragment | NA | NA | 0 |
| S3-final-viral-combined_k141_89434\|\|full | 3153 | No | NA | 6 | 0 | 0 | Not-determined | Genome-fragment | NA | NA | 0 |
| S5-final-viral-combined_k141_67775\|\|full | 1540 | No | NA | 2 | 1 | 0 | Low-quality | Genome-fragment | 2.69 | AAI-based (high-confidence) | 0 |
| S3-final-viral-combined_k141_112237\|\|full | 3139 | No | NA | 2 | 0 | 2 | Not-determined | Genome-fragment | NA | NA | 0 |
| S3-final-viral-combined_k141_89436\|\|full | 2062 | No | NA | 2 | 0 | 2 | Not-determined | Genome-fragment | NA | NA | 0 |
| S7-final-viral-combined_k141_92759\|\|full | 1540 | No | NA | 3 | 1 | 0 | Low-quality | Genome-fragment | 3.39 | AAI-based (high-confidence) | 0 |
| S3-final-viral-combined_k141_20231\|\|full | 2396 | No | NA | 3 | 0 | 1 | Not-determined | Genome-fragment | NA | NA | 0 |
| S7-final-viral-combined_k141_144877\|\|full | 1540 | No | NA | 3 | 0 | 0 | Low-quality | Genome-fragment | 2.35 | AAI-based (medium-confidence) | 0 |
| S3-final-viral-combined_k141_3273\|\|full | 3034 | No | NA | 3 | 0 | 3 | Not-determined | Genome-fragment | NA | NA | 0 |
| S3-final-viral-combined_k141_54727\|\|full | 3113 | No | NA | 3 | 0 | 1 | Not-determined | Genome-fragment | NA | NA | 0 |
| S3-final-viral-combined_k141_71896\|\|full | 2193 | No | NA | 4 | 0 | 0 | Not-determined | Genome-fragment | NA | NA | 0 |
| S17-final-viral-combined_k141_156592\|\|lt2gene | 1540 | No | NA | 1 | 1 | 0 | Low-quality | Genome-fragment | 1.97 | AAI-based (high-confidence) | 0 |
| S12-final-viral-combined_k141_109667\|\|full | 1540 | No | NA | 2 | 2 | 0 | Low-quality | Genome-fragment | 5.08 | HMM-based (lower-bound) | 0 |
| S3-final-viral-combined_k141_25991\|\|full | 2425 | No | NA | 2 | 0 | 1 | Not-determined | Genome-fragment | NA | NA | 0 |
| S3-final-viral-combined_k141_118220\|\|full | 1258 | No | NA | 3 | 0 | 1 | Not-determined | Genome-fragment | NA | NA | 0 |
| S2-final-viral-combined_k141_34047\|\|full | 1540 | No | NA | 4 | 3 | 0 | Low-quality | Genome-fragment | 2.43 | AAI-based (medium-confidence) | 0 |
| S3-final-viral-combined_k141_31684\|\|full | 5096 | No | NA | 4 | 0 | 4 | Not-determined | Genome-fragment | NA | NA | 0 |
| S2-final-viral-combined_k141_51235\|\|full | 1540 | No | NA | 3 | 0 | 1 | Low-quality | Genome-fragment | 1.09 | AAI-based (high-confidence) | 0 |
| S3-final-viral-combined_k141_14323\|\|full | 4204 | No | NA | 4 | 0 | 4 | Not-determined | Genome-fragment | NA | NA | 0 |
| S3-final-viral-combined_k141_95134\|\|full | 1940 | No | NA | 3 | 0 | 2 | Not-determined | Genome-fragment | NA | NA | 0 |
| S3-final-viral-combined_k141_83735\|\|full | 2184 | No | NA | 2 | 0 | 0 | Not-determined | Genome-fragment | NA | NA | 0 |
| S4-final-viral-combined_k141_165795\|\|full | 1540 | No | NA | 2 | 0 | 2 | Low-quality | Genome-fragment | 2.04 | AAI-based (medium-confidence) | 0 |
| S9_k141_5749 | 1541 | No | NA | 5 | 2 | 0 | Low-quality | Genome-fragment | 2.23 | AAI-based (medium-confidence) | 0 |
| S3-final-viral-combined_k141_54814\|\|full | 1937 | No | NA | 4 | 0 | 0 | Not-determined | Genome-fragment | NA | NA | 0 |
| S3-final-viral-combined_k141_49029\|\|full | 1899 | No | NA | 2 | 0 | 1 | Not-determined | Genome-fragment | NA | NA | 0 |
| S11-final-viral-combined_k141_138794\|\|full | 1541 | No | NA | 3 | 1 | 0 | Low-quality | Genome-fragment | 3.78 | AAI-based (high-confidence) | 0 |
| S3-final-viral-combined_k141_129860\|\|full | 1569 | No | NA | 3 | 0 | 1 | Not-determined | Genome-fragment | NA | NA | 0 |
| S3-final-viral-combined_k141_123911\|\|full | 1558 | No | NA | 2 | 0 | 0 | Not-determined | Genome-fragment | NA | NA | 0 |
| S3-final-viral-combined_k141_135428\|\|full | 28330 | No | NA | 47 | 19 | 0 | Medium-quality | Genome-fragment | 73.02 | AAI-based (high-confidence) | 0 |
| S3-final-viral-combined_k141_37698\|\|full | 3451 | No | NA | 5 | 0 | 1 | Not-determined | Genome-fragment | NA | NA | 0 |
| S3-final-viral-combined_k141_118338\|\|full | 3289 | No | NA | 2 | 0 | 2 | Not-determined | Genome-fragment | NA | NA | 0 |
| S3-final-viral-combined_k141_14401\|\|full | 4056 | No | NA | 5 | 0 | 0 | Not-determined | Genome-fragment | NA | NA | 0 |
| S3-final-viral-combined_k141_49076\|\|full | 1543 | No | NA | 3 | 0 | 1 | Not-determined | Genome-fragment | NA | NA | 0 |
| S11-final-viral-combined_k141_283590\|\|full | 1541 | No | NA | 3 | 0 | 1 | Low-quality | Genome-fragment | 1.2 | AAI-based (medium-confidence) | 0 |
| S3-final-viral-combined_k141_89641\|\|full | 1079 | No | NA | 3 | 0 | 1 | Not-determined | Genome-fragment | NA | NA | 0 |
| S3-final-viral-combined_k141_3421\|\|full | 2796 | No | NA | 2 | 0 | 2 | Not-determined | Genome-fragment | NA | NA | 0 |
| S3-final-viral-combined_k141_20340\|\|full | 2193 | No | NA | 5 | 0 | 0 | Not-determined | Genome-fragment | NA | NA | 0 |
| S3-final-viral-combined_k141_95241\|\|full | 2204 | No | NA | 3 | 0 | 1 | Not-determined | Genome-fragment | NA | NA | 0 |
| S11-final-viral-combined_k141_311513\|\|full | 1541 | No | NA | 2 | 1 | 0 | Low-quality | Genome-fragment | 0.13 | HMM-based (lower-bound) | 0 |
| S1-final-viral-combined_k141_272620\|\|full | 1541 | No | NA | 2 | 0 | 0 | Low-quality | Genome-fragment | 1.15 | AAI-based (medium-confidence) | 0 |
| S3-final-viral-combined_k141_123984\|\|full | 1586 | No | NA | 2 | 0 | 2 | Not-determined | Genome-fragment | NA | NA | 0 |
| S3-final-viral-combined_k141_123994\|\|full | 2789 | No | NA | 4 | 0 | 3 | Not-determined | Genome-fragment | NA | NA | 0 |
| S3-final-viral-combined_k141_72070\|\|full | 7258 | No | NA | 7 | 0 | 4 | Not-determined | Genome-fragment | NA | NA | 0 |
| S5-final-viral-combined_k141_112096\|\|full | 1541 | No | NA | 4 | 1 | 0 | Low-quality | Genome-fragment | 3.83 | AAI-based (high-confidence) | 0 |
| S7_k141_14298 | 1541 | No | NA | 2 | 0 | 1 | Low-quality | Genome-fragment | 1.01 | AAI-based (high-confidence) | 0 |
| S17-final-viral-combined_k141_7330\|\|full | 1541 | No | NA | 2 | 1 | 0 | Low-quality | Genome-fragment | 1.96 | HMM-based (lower-bound) | 0 |
| S17-final-viral-combined_k141_155440\|\|full | 1541 | No | NA | 2 | 1 | 0 | Low-quality | Genome-fragment | 0.97 | HMM-based (lower-bound) | 0 |
| S17-k141_189032 | 1541 | No | NA | 3 | 0 | 1 | Low-quality | Genome-fragment | 0.79 | AAI-based (medium-confidence) | 0 |
| S3-final-viral-combined_k141_54959\|\|full | 3458 | No | NA | 4 | 0 | 2 | Not-determined | Genome-fragment | NA | NA | 0 |
| S12-final-viral-combined_k141_387819\|\|full | 1541 | No | NA | 3 | 0 | 0 | Low-quality | Genome-fragment | 3.2 | AAI-based (medium-confidence) | 0 |
| S3-final-viral-combined_k141_106671\|\|full | 1835 | No | NA | 2 | 0 | 0 | Not-determined | Genome-fragment | NA | NA | 0 |
| S2-final-viral-combined_k141_1784\|\|full | 1541 | No | NA | 3 | 0 | 0 | Low-quality | Genome-fragment | 2.41 | AAI-based (medium-confidence) | 0 |
| S3-final-viral-combined_k141_54965\|\|full | 4932 | No | NA | 6 | 0 | 2 | Not-determined | Genome-fragment | NA | NA | 0 |
| S3-final-viral-combined_k141_66515\|\|full | 2159 | No | NA | 3 | 0 | 3 | Not-determined | Genome-fragment | NA | NA | 0 |
| S3-final-viral-combined_k141_129968\|\|full | 15044 | No | NA | 14 | 0 | 10 | Not-determined | Genome-fragment | NA | NA | 0 |
| S2-final-viral-combined_k141_211898\|\|full | 1541 | No | NA | 2 | 1 | 0 | Low-quality | Genome-fragment | 3.04 | AAI-based (high-confidence) | 0 |
| S2-final-viral-combined_k141_213400\|\|full | 1541 | No | NA | 3 | 0 | 0 | Low-quality | Genome-fragment | 2.53 | AAI-based (medium-confidence) | 0 |
| S3-final-viral-combined_k141_49197\|\|full | 1282 | No | NA | 3 | 0 | 1 | Not-determined | Genome-fragment | NA | NA | 0 |
| S6-final-viral-combined_k141_53141\|\|full | 1541 | No | NA | 4 | 0 | 2 | Low-quality | Genome-fragment | 0.76 | AAI-based (medium-confidence) | 0 |
| S8_k141_233666 | 1541 | No | NA | 2 | 0 | 0 | Low-quality | Genome-fragment | 3.79 | AAI-based (high-confidence) | 0 |
| S3-final-viral-combined_k141_129975\|\|full | 1141 | No | NA | 2 | 0 | 2 | Not-determined | Genome-fragment | NA | NA | 0 |
| S3-final-viral-combined_k141_26249\|\|full | 7527 | No | NA | 8 | 0 | 5 | Not-determined | Genome-fragment | NA | NA | 0 |
| S9_k141_34069 | 1542 | No | NA | 2 | 0 | 0 | Low-quality | Genome-fragment | 4.42 | AAI-based (high-confidence) | 0 |
| S3-final-viral-combined_k141_95336\|\|full | 1559 | No | NA | 5 | 0 | 0 | Not-determined | Genome-fragment | NA | NA | 0 |
| S1-final-viral-combined_k141_147130\|\|full | 1542 | No | NA | 1 | 0 | 1 | Low-quality | Genome-fragment | 1.84 | AAI-based (high-confidence) | 0 |
| S3-final-viral-combined_k141_3483\|\|full | 4817 | No | NA | 5 | 0 | 1 | Not-determined | Genome-fragment | NA | NA | 0 |
| S3_k141_123093 | 1542 | Yes | 566 | 2 | 1 | 1 | Low-quality | Genome-fragment | 0.7 | HMM-based (lower-bound) | 63.29 |
| S5-final-viral-combined_k141_11663\|\|full | 1542 | No | NA | 3 | 3 | 0 | Low-quality | Genome-fragment | 3.17 | AAI-based (medium-confidence) | 0 |
| S5_k141_111458 | 1542 | No | NA | 5 | 1 | 0 | Low-quality | Genome-fragment | 16.29 | AAI-based (high-confidence) | 0 |
| S3-final-viral-combined_k141_124101\|\|full | 1099 | No | NA | 3 | 0 | 0 | Not-determined | Genome-fragment | NA | NA | 0 |
| S17-final-viral-combined_k141_167062\|\|full | 1542 | No | NA | 2 | 0 | 1 | Low-quality | Genome-fragment | 1.17 | AAI-based (medium-confidence) | 0 |
| S3-final-viral-combined_k141_130018\|\|full | 2380 | No | NA | 3 | 0 | 2 | Not-determined | Genome-fragment | NA | NA | 0 |
| S3-final-viral-combined_k141_14601\|\|full | 2798 | No | NA | 3 | 0 | 2 | Not-determined | Genome-fragment | NA | NA | 0 |
| S3-final-viral-combined_k141_112613\|\|full | 3074 | No | NA | 4 | 0 | 2 | Not-determined | Genome-fragment | NA | NA | 0 |
| S3-final-viral-combined_k141_26301\|\|full | 3455 | No | NA | 3 | 0 | 3 | Not-determined | Genome-fragment | NA | NA | 0 |
| S17-final-viral-combined_k141_50590\|\|full | 1542 | No | NA | 3 | 0 | 0 | Low-quality | Genome-fragment | 3.28 | AAI-based (high-confidence) | 0 |
| S12-final-viral-combined_k141_254860\|\|full | 1542 | No | NA | 2 | 0 | 0 | Low-quality | Genome-fragment | 3.27 | AAI-based (high-confidence) | 0 |
| S3-final-viral-combined_k141_37940\|\|full | 1606 | No | NA | 3 | 0 | 0 | Not-determined | Genome-fragment | NA | NA | 0 |
| S12-final-viral-combined_k141_370474\|\|lt2gene | 1542 | No | NA | 2 | 2 | 0 | Low-quality | Genome-fragment | 3.36 | AAI-based (high-confidence) | 0 |
| S3-final-viral-combined_k141_43489\|\|full | 2577 | No | NA | 3 | 0 | 1 | Not-determined | Genome-fragment | NA | NA | 0 |
| S3-final-viral-combined_k141_72275\|\|full | 4730 | No | NA | 4 | 0 | 3 | Not-determined | Genome-fragment | NA | NA | 0 |
| S3-final-viral-combined_k141_14647\|\|full | 1041 | No | NA | 2 | 0 | 1 | Not-determined | Genome-fragment | NA | NA | 0 |
| S2-final-viral-combined_k141_243961\|\|lt2gene | 1542 | No | NA | 1 | 1 | 0 | Low-quality | Genome-fragment | 1.61 | AAI-based (high-confidence) | 0 |
| S3-final-viral-combined_k141_95465\|\|full | 4013 | No | NA | 3 | 0 | 2 | Not-determined | Genome-fragment | NA | NA | 0 |
| S8-final-viral-combined_k141_1473\|\|full | 1542 | No | NA | 3 | 2 | 0 | Low-quality | Genome-fragment | 4.28 | AAI-based (high-confidence) | 0 |
| S3-final-viral-combined_k141_95467\|\|full | 2998 | No | NA | 5 | 0 | 0 | Not-determined | Genome-fragment | NA | NA | 0 |
| S3-final-viral-combined_k141_130097\|\|full | 2831 | No | NA | 3 | 0 | 1 | Not-determined | Genome-fragment | NA | NA | 0 |
| S1_k141_73510 | 1543 | No | NA | 1 | 0 | 0 | Low-quality | Genome-fragment | 1.59 | AAI-based (high-confidence) | 0 |
| S3-final-viral-combined_k141_20592\|\|full | 2533 | No | NA | 2 | 0 | 2 | Not-determined | Genome-fragment | NA | NA | 0 |
| S3-final-viral-combined_k141_89918\|\|full | 1438 | No | NA | 5 | 0 | 0 | Not-determined | Genome-fragment | NA | NA | 0 |
| S3-final-viral-combined_k141_66690\|\|full | 1619 | No | NA | 3 | 0 | 0 | Not-determined | Genome-fragment | NA | NA | 0 |
| S3-final-viral-combined_k141_100904\|\|full | 20829 | No | NA | 19 | 0 | 16 | Not-determined | Genome-fragment | NA | NA | 0 |
| S7-final-viral-combined_k141_94661\|\|full | 1543 | No | NA | 3 | 2 | 0 | Low-quality | Genome-fragment | 3.82 | AAI-based (high-confidence) | 0 |
| S3-final-viral-combined_k141_118662\|\|full | 9295 | No | NA | 7 | 0 | 5 | Not-determined | Genome-fragment | NA | NA | 0 |
| S17-k141_206922 | 1543 | No | NA | 1 | 0 | 0 | Low-quality | Genome-fragment | 0.25 | AAI-based (high-confidence) | 0 |
| S3-final-viral-combined_k141_130163\|\|full | 1326 | No | NA | 3 | 0 | 1 | Not-determined | Genome-fragment | NA | NA | 0 |
| S3-final-viral-combined_k141_78115\|\|full | 1668 | No | NA | 3 | 0 | 1 | Not-determined | Genome-fragment | NA | NA | 0 |
| S3-final-viral-combined_k141_84052\|\|full | 1795 | No | NA | 3 | 0 | 0 | Not-determined | Genome-fragment | NA | NA | 0 |
| S3-final-viral-combined_k141_72345\|\|full | 2094 | No | NA | 3 | 0 | 3 | Not-determined | Genome-fragment | NA | NA | 0 |
| S12_k141_162170 | 1543 | No | NA | 2 | 0 | 1 | Low-quality | Genome-fragment | 1.82 | AAI-based (high-confidence) | 0 |
| S3-final-viral-combined_k141_95522\|\|full | 2085 | No | NA | 2 | 0 | 0 | Not-determined | Genome-fragment | NA | NA | 0 |
| S3-final-viral-combined_k141_130188\|\|full | 1025 | No | NA | 2 | 0 | 1 | Not-determined | Genome-fragment | NA | NA | 0 |
| S3-final-viral-combined_k141_55258\|\|full | 1844 | No | NA | 2 | 0 | 1 | Not-determined | Genome-fragment | NA | NA | 0 |
| S3-final-viral-combined_k141_43586\|\|full | 1469 | No | NA | 3 | 0 | 1 | Not-determined | Genome-fragment | NA | NA | 0 |
| S2-final-viral-combined_k141_238912\|\|full | 1543 | No | NA | 3 | 0 | 1 | Low-quality | Genome-fragment | 3 | AAI-based (high-confidence) | 0 |
| S2-final-viral-combined_k141_221676\|\|full | 1543 | No | NA | 2 | 0 | 0 | Low-quality | Genome-fragment | 1.33 | AAI-based (high-confidence) | 0 |
| S3-final-viral-combined_k141_20708\|\|full | 3231 | No | NA | 3 | 0 | 2 | Not-determined | Genome-fragment | NA | NA | 0 |
| S3-final-viral-combined_k141_32013\|\|full | 2177 | No | NA | 3 | 0 | 2 | Not-determined | Genome-fragment | NA | NA | 0 |
| S3-final-viral-combined_k141_43601\|\|full | 1433 | No | NA | 3 | 0 | 1 | Not-determined | Genome-fragment | NA | NA | 0 |
| S3-final-viral-combined_k141_32018\|\|full | 2558 | No | NA | 3 | 0 | 1 | Not-determined | Genome-fragment | NA | NA | 0 |
| S3-final-viral-combined_k141_14753\|\|full | 5484 | No | NA | 5 | 0 | 4 | Not-determined | Genome-fragment | NA | NA | 0 |
| S3-final-viral-combined_k141_14771\|\|full | 2068 | No | NA | 2 | 0 | 2 | Not-determined | Genome-fragment | NA | NA | 0 |
| S3-final-viral-combined_k141_118746\|\|full | 4678 | No | NA | 3 | 0 | 3 | Not-determined | Genome-fragment | NA | NA | 0 |
| S3-final-viral-combined_k141_38165\|\|full | 2173 | No | NA | 3 | 0 | 2 | Not-determined | Genome-fragment | NA | NA | 0 |
| S3-final-viral-combined_k141_43638\|\|full | 1401 | No | NA | 2 | 0 | 0 | Not-determined | Genome-fragment | NA | NA | 0 |
| S3-final-viral-combined_k141_112800\|\|full | 1278 | No | NA | 2 | 0 | 1 | Not-determined | Genome-fragment | NA | NA | 0 |
| S2_k141_103924 | 1543 | No | NA | 2 | 1 | 0 | Low-quality | Genome-fragment | 2.54 | AAI-based (high-confidence) | 0 |
| S3-final-viral-combined_k141_78227\|\|full | 2286 | No | NA | 4 | 0 | 0 | Not-determined | Genome-fragment | NA | NA | 0 |
| S4-final-viral-combined_k141_34168\|\|full | 1543 | No | NA | 3 | 0 | 2 | Low-quality | Genome-fragment | 1.29 | AAI-based (medium-confidence) | 0 |
| S8-final-viral-combined_k141_196277\|\|full | 1543 | No | NA | 1 | 0 | 1 | Low-quality | Genome-fragment | 0.83 | AAI-based (high-confidence) | 0 |
| S8-final-viral-combined_k141_154284\|\|full | 1543 | No | NA | 3 | 0 | 0 | Low-quality | Genome-fragment | 2.67 | AAI-based (high-confidence) | 0 |
| S9-final-viral-combined_k141_40195\|\|full | 1544 | No | NA | 3 | 1 | 0 | Low-quality | Genome-fragment | 4.03 | AAI-based (medium-confidence) | 0 |
| S11-final-viral-combined_k141_204339\|\|full | 1544 | No | NA | 3 | 1 | 0 | Low-quality | Genome-fragment | 1.08 | HMM-based (lower-bound) | 0 |
| S3-final-viral-combined_k141_9173\|\|full | 1053 | No | NA | 2 | 0 | 1 | Not-determined | Genome-fragment | NA | NA | 0 |
| S1-final-viral-combined_k141_158861\|\|lt2gene | 1544 | No | NA | 2 | 2 | 0 | Low-quality | Genome-fragment | 3.28 | AAI-based (high-confidence) | 0 |
| S3-final-viral-combined_k141_14824\|\|full | 2168 | No | NA | 3 | 0 | 1 | Not-determined | Genome-fragment | NA | NA | 0 |
| S7-final-viral-combined_k141_216666\|\|full | 1544 | No | NA | 4 | 1 | 0 | Low-quality | Genome-fragment | 0.98 | HMM-based (lower-bound) | 0 |
| S3-final-viral-combined_k141_66826\|\|full | 3194 | No | NA | 7 | 0 | 0 | Not-determined | Genome-fragment | NA | NA | 0 |
| S3-final-viral-combined_k141_130350\|\|full | 1253 | No | NA | 3 | 0 | 0 | Not-determined | Genome-fragment | NA | NA | 0 |
| S3-final-viral-combined_k141_124422\|\|full | 4625 | No | NA | 4 | 0 | 3 | Not-determined | Genome-fragment | NA | NA | 0 |
| S3-final-viral-combined_k141_3776\|\|full | 3878 | No | NA | 5 | 0 | 5 | Not-determined | Genome-fragment | NA | NA | 0 |
| S3-final-viral-combined_k141_55385\|\|full | 1497 | No | NA | 3 | 0 | 0 | Not-determined | Genome-fragment | NA | NA | 0 |
| S7-final-viral-combined_k141_150263\|\|full | 1544 | No | NA | 2 | 0 | 0 | Low-quality | Genome-fragment | 11.02 | AAI-based (medium-confidence) | 0 |
| S3-final-viral-combined_k141_112849\|\|full | 1732 | No | NA | 3 | 0 | 0 | Not-determined | Genome-fragment | NA | NA | 0 |
| S3-final-viral-combined_k141_107034\|\|full | 1700 | No | NA | 3 | 0 | 1 | Not-determined | Genome-fragment | NA | NA | 0 |
| S3-final-viral-combined_k141_14850\|\|full | 7726 | No | NA | 8 | 0 | 7 | Not-determined | Genome-fragment | NA | NA | 0 |
| S7-final-viral-combined_k141_222214\|\|full | 1544 | No | NA | 5 | 0 | 1 | Low-quality | Genome-fragment | 1.16 | AAI-based (medium-confidence) | 0 |
| S3-final-viral-combined_k141_101121\|\|full | 3195 | No | NA | 3 | 0 | 2 | Not-determined | Genome-fragment | NA | NA | 0 |
| S3-final-viral-combined_k141_20826\|\|full | 2225 | No | NA | 2 | 0 | 1 | Not-determined | Genome-fragment | NA | NA | 0 |
| S3-final-viral-combined_k141_61043\|\|full | 3516 | No | NA | 4 | 0 | 2 | Not-determined | Genome-fragment | NA | NA | 0 |
| S17-k141_4064 | 1544 | No | NA | 2 | 0 | 2 | Low-quality | Genome-fragment | 3.45 | AAI-based (high-confidence) | 0 |
| S3-final-viral-combined_k141_72541\|\|full | 1612 | No | NA | 4 | 0 | 1 | Not-determined | Genome-fragment | NA | NA | 0 |
| S12-final-viral-combined_k141_173995\|\|full | 1544 | No | NA | 4 | 0 | 0 | Low-quality | Genome-fragment | 3.06 | AAI-based (medium-confidence) | 0 |
| S12_k141_151299 | 1544 | No | NA | 1 | 0 | 0 | Low-quality | Genome-fragment | 1.91 | AAI-based (high-confidence) | 0 |
| S4_k141_34209 | 1544 | No | NA | 2 | 0 | 0 | Low-quality | Genome-fragment | 1.55 | AAI-based (high-confidence) | 0 |
| S8-final-viral-combined_k141_50713\|\|full | 1544 | No | NA | 4 | 0 | 0 | Low-quality | Genome-fragment | 3.23 | AAI-based (medium-confidence) | 0 |
| S8-final-viral-combined_k141_41413\|\|lt2gene | 1544 | No | NA | 1 | 0 | 0 | Low-quality | Genome-fragment | 3.48 | AAI-based (high-confidence) | 0 |
| S3-final-viral-combined_k141_95724\|\|full | 1329 | No | NA | 2 | 0 | 1 | Not-determined | Genome-fragment | NA | NA | 0 |
| S8-final-viral-combined_k141_271611\|\|lt2gene | 1544 | No | NA | 2 | 2 | 0 | Low-quality | Genome-fragment | 2.26 | HMM-based (lower-bound) | 0 |
| S3-final-viral-combined_k141_32194\|\|full | 2111 | No | NA | 5 | 0 | 2 | Not-determined | Genome-fragment | NA | NA | 0 |
| S9-final-viral-combined_k141_59149\|\|full | 1545 | No | NA | 2 | 0 | 1 | Low-quality | Genome-fragment | 2.13 | AAI-based (medium-confidence) | 0 |
| S11-final-viral-combined_k141_88042\|\|full | 1545 | No | NA | 3 | 2 | 0 | Low-quality | Genome-fragment | 3.86 | AAI-based (high-confidence) | 0 |
| S3-final-viral-combined_k141_135868\|\|full | 2252 | No | NA | 2 | 0 | 2 | Not-determined | Genome-fragment | NA | NA | 0 |
| S3-final-viral-combined_k141_130449\|\|full | 1276 | No | NA | 3 | 0 | 0 | Not-determined | Genome-fragment | NA | NA | 0 |
| S3-final-viral-combined_k141_3872\|\|full | 1378 | No | NA | 2 | 0 | 1 | Not-determined | Genome-fragment | NA | NA | 0 |
| S3-final-viral-combined_k141_55455\|\|full | 1087 | No | NA | 2 | 0 | 2 | Not-determined | Genome-fragment | NA | NA | 0 |
| S1-final-viral-combined_k141_318124\|\|full | 1545 | No | NA | 4 | 1 | 0 | Low-quality | Genome-fragment | 4.42 | AAI-based (high-confidence) | 0 |
| S3-final-viral-combined_k141_84344\|\|full | 1741 | No | NA | 2 | 0 | 0 | Not-determined | Genome-fragment | NA | NA | 0 |
| S3-final-viral-combined_k141_118972\|\|full | 4246 | No | NA | 3 | 0 | 1 | Not-determined | Genome-fragment | NA | NA | 0 |
| S3-final-viral-combined_k141_84355\|\|full | 1444 | No | NA | 2 | 0 | 1 | Not-determined | Genome-fragment | NA | NA | 0 |
| S5-final-viral-combined_k141_210217\|\|full | 1545 | No | NA | 3 | 2 | 0 | Low-quality | Genome-fragment | 3.14 | AAI-based (high-confidence) | 0 |
| S3-final-viral-combined_k141_20964\|\|full | 1747 | No | NA | 3 | 0 | 0 | Not-determined | Genome-fragment | NA | NA | 0 |
| S3-final-viral-combined_k141_124622\|\|full | 3951 | No | NA | 6 | 0 | 1 | Not-determined | Genome-fragment | NA | NA | 0 |
| S3-final-viral-combined_k141_67002\|\|full | 1645 | No | NA | 3 | 0 | 1 | Not-determined | Genome-fragment | NA | NA | 0 |
| S5-final-viral-combined_k141_108736\|\|lt2gene | 1545 | No | NA | 2 | 1 | 0 | Low-quality | Genome-fragment | 4.5 | AAI-based (high-confidence) | 0 |
| S3-final-viral-combined_k141_90268\|\|full | 1859 | No | NA | 2 | 0 | 2 | Not-determined | Genome-fragment | NA | NA | 0 |
| S3-final-viral-combined_k141_20990\|\|full | 2334 | No | NA | 3 | 0 | 3 | Not-determined | Genome-fragment | NA | NA | 0 |
| S3-final-viral-combined_k141_14986\|\|full | 4748 | No | NA | 6 | 0 | 1 | Not-determined | Genome-fragment | NA | NA | 0 |
| S3-final-viral-combined_k141_95827\|\|full | 1719 | No | NA | 4 | 0 | 0 | Not-determined | Genome-fragment | NA | NA | 0 |
| S3-final-viral-combined_k141_61205\|\|full | 3871 | No | NA | 5 | 0 | 2 | Not-determined | Genome-fragment | NA | NA | 0 |
| S5-final-viral-combined_k141_32420\|\|lt2gene | 1545 | No | NA | 2 | 2 | 0 | Low-quality | Genome-fragment | 0.9 | AAI-based (high-confidence) | 0 |
| S3-final-viral-combined_k141_32313\|\|full | 3977 | No | NA | 5 | 0 | 4 | Not-determined | Genome-fragment | NA | NA | 0 |
| S3-final-viral-combined_k141_26806\|\|full | 4512 | No | NA | 4 | 0 | 2 | Not-determined | Genome-fragment | NA | NA | 0 |
| S5-final-viral-combined_k141_211976\|\|lt2gene | 1545 | No | NA | 2 | 2 | 0 | Low-quality | Genome-fragment | 4.59 | AAI-based (high-confidence) | 0 |
| S3-final-viral-combined_k141_4012\|\|full | 1923 | No | NA | 2 | 0 | 0 | Not-determined | Genome-fragment | NA | NA | 0 |
| S3-final-viral-combined_k141_55604\|\|full | 2775 | No | NA | 3 | 0 | 3 | Not-determined | Genome-fragment | NA | NA | 0 |
| S3-final-viral-combined_k141_4026\|\|full | 1974 | No | NA | 2 | 0 | 1 | Not-determined | Genome-fragment | NA | NA | 0 |
| S17-final-viral-combined_k141_18932\|\|full | 1545 | No | NA | 3 | 0 | 2 | Low-quality | Genome-fragment | 0.87 | AAI-based (medium-confidence) | 0 |
| S2_k141_88690 | 1545 | No | NA | 4 | 0 | 0 | Low-quality | Genome-fragment | 4.11 | AAI-based (high-confidence) | 0 |
| S6_k141_111088 | 1545 | No | NA | 1 | 0 | 0 | Low-quality | Genome-fragment | 0.82 | AAI-based (high-confidence) | 0 |
| S3-final-viral-combined_k141_9442\|\|full | 7950 | No | NA | 6 | 0 | 5 | Not-determined | Genome-fragment | NA | NA | 0 |
| S3-final-viral-combined_k141_124751\|\|full | 7642 | No | NA | 2 | 0 | 2 | Not-determined | Genome-fragment | NA | NA | 0 |
| S3-final-viral-combined_k141_124757\|\|full | 2087 | No | NA | 2 | 0 | 0 | Not-determined | Genome-fragment | NA | NA | 0 |
| S8_k141_108514 | 1545 | No | NA | 1 | 0 | 1 | Low-quality | Genome-fragment | 0.41 | AAI-based (high-confidence) | 0 |
| S3-final-viral-combined_k141_113126\|\|full | 2724 | No | NA | 8 | 0 | 1 | Not-determined | Genome-fragment | NA | NA | 0 |
| S3-final-viral-combined_k141_15082\|\|full | 1003 | No | NA | 3 | 0 | 0 | Not-determined | Genome-fragment | NA | NA | 0 |
| S9-final-viral-combined_k141_21485\|\|full | 1546 | No | NA | 2 | 0 | 0 | Low-quality | Genome-fragment | 4.22 | AAI-based (high-confidence) | 0 |
| S9-final-viral-combined_k141_49760\|\|full | 1546 | No | NA | 3 | 0 | 1 | Low-quality | Genome-fragment | 2.25 | AAI-based (medium-confidence) | 0 |
| S12-final-viral-combined_k141_394521\|\|full | 1546 | No | NA | 1 | 0 | 0 | Low-quality | Genome-fragment | 3.38 | AAI-based (medium-confidence) | 0 |
| S12-final-viral-combined_k141_161422\|\|full | 1546 | No | NA | 5 | 0 | 0 | Low-quality | Genome-fragment | 2.78 | AAI-based (high-confidence) | 0 |
| S3-final-viral-combined_k141_90398\|\|full | 7994 | No | NA | 11 | 0 | 8 | Not-determined | Genome-fragment | NA | NA | 0 |
| S3-final-viral-combined_k141_38508\|\|full | 3622 | No | NA | 2 | 0 | 1 | Not-determined | Genome-fragment | NA | NA | 0 |
| S12-final-viral-combined_k141_175024\|\|lt2gene | 1546 | No | NA | 1 | 1 | 0 | Low-quality | Genome-fragment | 3.57 | AAI-based (high-confidence) | 0 |
| S3-final-viral-combined_k141_21163\|\|full | 1984 | No | NA | 4 | 0 | 2 | Not-determined | Genome-fragment | NA | NA | 0 |
| S3-final-viral-combined_k141_113190\|\|full | 1304 | No | NA | 2 | 0 | 1 | Not-determined | Genome-fragment | NA | NA | 0 |
| S3-final-viral-combined_k141_130681\|\|full | 1655 | No | NA | 2 | 0 | 0 | Not-determined | Genome-fragment | NA | NA | 0 |
| S2-final-viral-combined_k141_124993\|\|lt2gene | 1546 | No | NA | 1 | 1 | 0 | Low-quality | Genome-fragment | 3.8 | AAI-based (high-confidence) | 0 |
| S4_k141_120744 | 1546 | No | NA | 2 | 0 | 2 | Low-quality | Genome-fragment | 2.35 | AAI-based (high-confidence) | 0 |
| S3-final-viral-combined_k141_9561\|\|full | 1151 | No | NA | 2 | 0 | 1 | Not-determined | Genome-fragment | NA | NA | 0 |
| S3-final-viral-combined_k141_21194\|\|full | 1844 | No | NA | 3 | 0 | 1 | Not-determined | Genome-fragment | NA | NA | 0 |
| S3-final-viral-combined_k141_136168\|\|full | 1602 | No | NA | 3 | 0 | 0 | Not-determined | Genome-fragment | NA | NA | 0 |
| S3-final-viral-combined_k141_15138\|\|full | 2281 | No | NA | 4 | 0 | 3 | Not-determined | Genome-fragment | NA | NA | 0 |
| S3-final-viral-combined_k141_67242\|\|full | 1198 | No | NA | 2 | 0 | 0 | Not-determined | Genome-fragment | NA | NA | 0 |
| S9-final-viral-combined_k141_52409\|\|full | 1547 | No | NA | 4 | 0 | 1 | Low-quality | Genome-fragment | 1.82 | AAI-based (medium-confidence) | 0 |
| S3-final-viral-combined_k141_15149\|\|full | 2332 | No | NA | 3 | 0 | 1 | Not-determined | Genome-fragment | NA | NA | 0 |
| S1-final-viral-combined_k141_383571\|\|full | 1547 | No | NA | 2 | 0 | 1 | Low-quality | Genome-fragment | 0.83 | AAI-based (medium-confidence) | 0 |
| S3-final-viral-combined_k141_124878\|\|full | 1212 | No | NA | 2 | 0 | 0 | Not-determined | Genome-fragment | NA | NA | 0 |
| S3-final-viral-combined_k141_4173\|\|full | 2934 | No | NA | 3 | 0 | 1 | Not-determined | Genome-fragment | NA | NA | 0 |
| S3-final-viral-combined_k141_32500\|\|full | 3082 | No | NA | 2 | 0 | 1 | Not-determined | Genome-fragment | NA | NA | 0 |
| S3-final-viral-combined_k141_55761\|\|full | 1678 | No | NA | 3 | 0 | 0 | Not-determined | Genome-fragment | NA | NA | 0 |
| S3-final-viral-combined_k141_124907\|\|full | 1008 | No | NA | 3 | 0 | 0 | Not-determined | Genome-fragment | NA | NA | 0 |
| S3-final-viral-combined_k141_84652\|\|full | 2303 | No | NA | 3 | 0 | 3 | Not-determined | Genome-fragment | NA | NA | 0 |
| S1-final-viral-combined_k141_157544\|\|full | 1547 | No | NA | 4 | 0 | 0 | Low-quality | Genome-fragment | 1.25 | AAI-based (medium-confidence) | 0 |
| S1_k141_157453 | 1547 | No | NA | 3 | 1 | 0 | Low-quality | Genome-fragment | 32.63 | AAI-based (medium-confidence) | 0 |
| S3-final-viral-combined_k141_27092\|\|full | 2351 | No | NA | 2 | 0 | 1 | Not-determined | Genome-fragment | NA | NA | 0 |
| S3-final-viral-combined_k141_32582\|\|full | 2870 | No | NA | 3 | 0 | 0 | Not-determined | Genome-fragment | NA | NA | 0 |
| S3-final-viral-combined_k141_101434\|\|full | 2664 | No | NA | 2 | 0 | 1 | Not-determined | Genome-fragment | NA | NA | 0 |
| S3-final-viral-combined_k141_21324\|\|full | 1005 | No | NA | 1 | 0 | 1 | Not-determined | Genome-fragment | NA | NA | 0 |
| S3-final-viral-combined_k141_90520\|\|full | 2442 | No | NA | 6 | 0 | 0 | Not-determined | Genome-fragment | NA | NA | 0 |
| S3-final-viral-combined_k141_136281\|\|full | 1945 | No | NA | 4 | 0 | 1 | Not-determined | Genome-fragment | NA | NA | 0 |
| S5-final-viral-combined_k141_45275\|\|full | 1547 | No | NA | 4 | 1 | 0 | Low-quality | Genome-fragment | 4.68 | AAI-based (high-confidence) | 0 |
| S3-final-viral-combined_k141_113363\|\|full | 6572 | No | NA | 7 | 0 | 6 | Not-determined | Genome-fragment | NA | NA | 0 |
| S3-final-viral-combined_k141_90545\|\|full | 1755 | No | NA | 3 | 0 | 0 | Not-determined | Genome-fragment | NA | NA | 0 |
| S3-final-viral-combined_k141_113366\|\|full | 1125 | No | NA | 3 | 0 | 1 | Not-determined | Genome-fragment | NA | NA | 0 |
| S17-k141_246156 | 1547 | No | NA | 3 | 0 | 1 | Low-quality | Genome-fragment | 2.47 | AAI-based (medium-confidence) | 0 |
| S12-final-viral-combined_k141_1832\|\|full | 1547 | No | NA | 3 | 1 | 0 | Low-quality | Genome-fragment | 1.54 | AAI-based (medium-confidence) | 0 |
| S12-final-viral-combined_k141_199504\|\|full | 1547 | No | NA | 3 | 1 | 0 | Low-quality | Genome-fragment | 2.35 | AAI-based (high-confidence) | 0 |
| S2-final-viral-combined_k141_146212\|\|full | 1547 | No | NA | 3 | 0 | 0 | Low-quality | Genome-fragment | 2.94 | AAI-based (high-confidence) | 0 |
| S3-final-viral-combined_k141_84750\|\|full | 5924 | No | NA | 8 | 0 | 5 | Not-determined | Genome-fragment | NA | NA | 0 |
| S3-final-viral-combined_k141_72923\|\|full | 1750 | No | NA | 3 | 0 | 1 | Not-determined | Genome-fragment | NA | NA | 0 |
| S4_k141_104904 | 1547 | No | NA | 2 | 0 | 0 | Low-quality | Genome-fragment | 2.55 | AAI-based (high-confidence) | 0 |
| S3-final-viral-combined_k141_96145\|\|full | 2778 | No | NA | 4 | 0 | 1 | Not-determined | Genome-fragment | NA | NA | 0 |
| S3-final-viral-combined_k141_15254\|\|full | 1205 | No | NA | 2 | 0 | 0 | Not-determined | Genome-fragment | NA | NA | 0 |
| S3-final-viral-combined_k141_96156\|\|full | 1331 | No | NA | 3 | 0 | 1 | Not-determined | Genome-fragment | NA | NA | 0 |
| S3-final-viral-combined_k141_96174\|\|full | 3075 | No | NA | 2 | 0 | 1 | Not-determined | Genome-fragment | NA | NA | 0 |
| S3-final-viral-combined_k141_67429\|\|full | 1689 | No | NA | 3 | 0 | 1 | Not-determined | Genome-fragment | NA | NA | 0 |
| S8-final-viral-combined_k141_106540\|\|full | 1547 | No | NA | 4 | 3 | 0 | Low-quality | Genome-fragment | 2.22 | AAI-based (medium-confidence) | 0 |
| S8-final-viral-combined_k141_4111\|\|full | 1547 | No | NA | 3 | 0 | 0 | Low-quality | Genome-fragment | 1.24 | AAI-based (high-confidence) | 0 |
| S3-final-viral-combined_k141_21400\|\|full | 2637 | No | NA | 3 | 0 | 2 | Not-determined | Genome-fragment | NA | NA | 0 |
| S3-final-viral-combined_k141_61544\|\|full | 1273 | No | NA | 2 | 0 | 0 | Not-determined | Genome-fragment | NA | NA | 0 |
| S3-final-viral-combined_k141_27189\|\|full | 1636 | No | NA | 2 | 0 | 2 | Not-determined | Genome-fragment | NA | NA | 0 |
| S9_k141_28885 | 1548 | No | NA | 1 | 0 | 1 | Low-quality | Genome-fragment | 2.46 | AAI-based (medium-confidence) | 0 |
| S3-final-viral-combined_k141_113464\|\|full | 1917 | No | NA | 3 | 0 | 0 | Not-determined | Genome-fragment | NA | NA | 0 |
| S3-final-viral-combined_k141_32689\|\|full | 2593 | No | NA | 2 | 0 | 1 | Not-determined | Genome-fragment | NA | NA | 0 |
| S1-final-viral-combined_k141_228276\|\|lt2gene | 1548 | No | NA | 1 | 1 | 0 | Low-quality | Genome-fragment | 3.3 | AAI-based (high-confidence) | 0 |
| S3-final-viral-combined_k141_21421\|\|full | 1969 | No | NA | 3 | 0 | 2 | Not-determined | Genome-fragment | NA | NA | 0 |
| S3-final-viral-combined_k141_96257\|\|full | 2094 | No | NA | 3 | 0 | 1 | Not-determined | Genome-fragment | NA | NA | 0 |
| S3-final-viral-combined_k141_32712\|\|full | 2147 | No | NA | 3 | 0 | 2 | Not-determined | Genome-fragment | NA | NA | 0 |
| S17-final-viral-combined_k141_259221\|\|full | 1548 | No | NA | 4 | 0 | 0 | Low-quality | Genome-fragment | 2.11 | AAI-based (high-confidence) | 0 |
| S3-final-viral-combined_k141_15367\|\|full | 2631 | No | NA | 4 | 0 | 2 | Not-determined | Genome-fragment | NA | NA | 0 |
| S17-k141_116420 | 1548 | No | NA | 4 | 1 | 0 | Low-quality | Genome-fragment | 1.02 | HMM-based (lower-bound) | 0 |
| S3-final-viral-combined_k141_107626\|\|full | 1487 | No | NA | 2 | 0 | 2 | Not-determined | Genome-fragment | NA | NA | 0 |
| S12-final-viral-combined_k141_152122\|\|full | 1548 | No | NA | 3 | 0 | 1 | Low-quality | Genome-fragment | 3.64 | AAI-based (medium-confidence) | 0 |
| S12-final-viral-combined_k141_386882\|\|full | 1548 | No | NA | 3 | 3 | 0 | Low-quality | Genome-fragment | 2.6 | HMM-based (lower-bound) | 0 |
| S2-final-viral-combined_k141_227809\|\|full | 1548 | No | NA | 3 | 1 | 0 | Low-quality | Genome-fragment | 3.23 | AAI-based (medium-confidence) | 0 |
| S3-final-viral-combined_k141_113573\|\|full | 1690 | No | NA | 3 | 0 | 2 | Not-determined | Genome-fragment | NA | NA | 0 |
| S3-final-viral-combined_k141_9844\|\|full | 3298 | No | NA | 3 | 0 | 0 | Not-determined | Genome-fragment | NA | NA | 0 |
| S3-final-viral-combined_k141_55990\|\|full | 3644 | No | NA | 3 | 0 | 3 | Not-determined | Genome-fragment | NA | NA | 0 |
| S2_k141_226539 | 1548 | No | NA | 2 | 0 | 2 | Low-quality | Genome-fragment | 1.09 | AAI-based (high-confidence) | 0 |
| S3-final-viral-combined_k141_125226\|\|full | 1402 | No | NA | 2 | 0 | 1 | Not-determined | Genome-fragment | NA | NA | 0 |
| S4_k141_113250 | 1548 | No | NA | 3 | 0 | 0 | Low-quality | Genome-fragment | 3.28 | AAI-based (high-confidence) | 0 |
| S8-final-viral-combined_k141_239595\|\|full | 1548 | No | NA | 3 | 0 | 0 | Low-quality | Genome-fragment | 1.85 | AAI-based (medium-confidence) | 0 |
| S3-final-viral-combined_k141_119664\|\|full | 2240 | No | NA | 3 | 0 | 2 | Not-determined | Genome-fragment | NA | NA | 0 |
| S3-final-viral-combined_k141_56033\|\|full | 3155 | No | NA | 3 | 0 | 1 | Not-determined | Genome-fragment | NA | NA | 0 |
| S8_k141_137832 | 1548 | No | NA | 1 | 0 | 1 | Low-quality | Genome-fragment | 0.22 | AAI-based (medium-confidence) | 0 |
| S3-final-viral-combined_k141_125279\|\|full | 1204 | No | NA | 2 | 0 | 0 | Not-determined | Genome-fragment | NA | NA | 0 |
| S9_k141_75667 | 1549 | No | NA | 2 | 0 | 1 | Low-quality | Genome-fragment | 0.55 | AAI-based (high-confidence) | 0 |
| S3-final-viral-combined_k141_9947\|\|full | 1962 | No | NA | 2 | 0 | 1 | Not-determined | Genome-fragment | NA | NA | 0 |
| S11-final-viral-combined_k141_184557\|\|full | 1549 | No | NA | 2 | 0 | 1 | Low-quality | Genome-fragment | 3 | AAI-based (medium-confidence) | 0 |
| S11_k141_326137 | 1549 | No | NA | 1 | 0 | 0 | Low-quality | Genome-fragment | 2.8 | AAI-based (medium-confidence) | 0 |
| S3-final-viral-combined_k141_67615\|\|full | 3155 | No | NA | 3 | 0 | 1 | Not-determined | Genome-fragment | NA | NA | 0 |
| S1-final-viral-combined_k141_172038\|\|full | 1549 | No | NA | 3 | 0 | 1 | Low-quality | Genome-fragment | 0.92 | AAI-based (medium-confidence) | 0 |
| S5-final-viral-combined_k141_22775\|\|lt2gene | 1549 | No | NA | 1 | 1 | 0 | Low-quality | Genome-fragment | 3.59 | AAI-based (high-confidence) | 0 |
| S3-final-viral-combined_k141_125322\|\|full | 3176 | No | NA | 2 | 0 | 2 | Not-determined | Genome-fragment | NA | NA | 0 |
| S3-final-viral-combined_k141_85050\|\|full | 1616 | No | NA | 4 | 0 | 1 | Not-determined | Genome-fragment | NA | NA | 0 |
| S3-final-viral-combined_k141_90855\|\|full | 4412 | No | NA | 3 | 0 | 3 | Not-determined | Genome-fragment | NA | NA | 0 |
| S3-final-viral-combined_k141_107796\|\|full | 3654 | No | NA | 4 | 0 | 1 | Not-determined | Genome-fragment | NA | NA | 0 |
| S3-final-viral-combined_k141_96475\|\|full | 1015 | No | NA | 3 | 0 | 0 | Not-determined | Genome-fragment | NA | NA | 0 |
| S3-final-viral-combined_k141_131222\|\|full | 1934 | No | NA | 5 | 0 | 1 | Not-determined | Genome-fragment | NA | NA | 0 |
| S17-final-viral-combined_k141_168104\|\|full | 1549 | No | NA | 2 | 0 | 0 | Low-quality | Genome-fragment | 1.8 | AAI-based (high-confidence) | 0 |
| S3-final-viral-combined_k141_4608\|\|full | 1543 | No | NA | 2 | 0 | 0 | Not-determined | Genome-fragment | NA | NA | 0 |
| S3-final-viral-combined_k141_119792\|\|full | 2927 | No | NA | 2 | 0 | 2 | Not-determined | Genome-fragment | NA | NA | 0 |
| S3-final-viral-combined_k141_101853\|\|full | 1287 | No | NA | 2 | 0 | 0 | Not-determined | Genome-fragment | NA | NA | 0 |
| S17-final-viral-combined_k141_105720\|\|full | 1549 | No | NA | 4 | 2 | 0 | Low-quality | Genome-fragment | 3.61 | AAI-based (high-confidence) | 0 |
| S3-final-viral-combined_k141_136640\|\|full | 2836 | No | NA | 4 | 0 | 1 | Not-determined | Genome-fragment | NA | NA | 0 |
| S3-final-viral-combined_k141_96518\|\|full | 2853 | No | NA | 3 | 0 | 2 | Not-determined | Genome-fragment | NA | NA | 0 |
| S3-final-viral-combined_k141_131247\|\|full | 4089 | No | NA | 7 | 0 | 0 | Not-determined | Genome-fragment | NA | NA | 0 |
| S3-final-viral-combined_k141_96524\|\|full | 1047 | No | NA | 2 | 0 | 0 | Not-determined | Genome-fragment | NA | NA | 0 |
| S3-final-viral-combined_k141_21712\|\|full | 1328 | No | NA | 2 | 0 | 2 | Not-determined | Genome-fragment | NA | NA | 0 |
| S3-final-viral-combined_k141_44501\|\|full | 2221 | No | NA | 3 | 0 | 2 | Not-determined | Genome-fragment | NA | NA | 0 |
| S3-final-viral-combined_k141_27495\|\|full | 2279 | No | NA | 4 | 0 | 2 | Not-determined | Genome-fragment | NA | NA | 0 |
| S3-final-viral-combined_k141_50215\|\|full | 3468 | No | NA | 5 | 0 | 1 | Not-determined | Genome-fragment | NA | NA | 0 |
| S17-final-viral-combined_k141_27046\|\|lt2gene | 1549 | No | NA | 2 | 1 | 0 | Low-quality | Genome-fragment | 0.59 | HMM-based (lower-bound) | 0 |
| S3-final-viral-combined_k141_44506\|\|full | 2528 | No | NA | 4 | 0 | 2 | Not-determined | Genome-fragment | NA | NA | 0 |
| S3-final-viral-combined_k141_44508\|\|full | 2716 | No | NA | 2 | 0 | 2 | Not-determined | Genome-fragment | NA | NA | 0 |
| S3-final-viral-combined_k141_119819\|\|full | 2889 | No | NA | 3 | 0 | 2 | Not-determined | Genome-fragment | NA | NA | 0 |
| S3-final-viral-combined_k141_73262\|\|full | 33778 | No | NA | 31 | 5 | 1 | Medium-quality | Genome-fragment | 78.86 | AAI-based (high-confidence) | 0 |

| | | | | | | | | | | | |
|---|---|---|---|---|---|---|---|---|---|---|---|
| S3-final-viral-combined_k141_21739\|\|full | 1079 | No | NA | 2 | 0 | 1 | Not-determined | Genome-fragment | NA | NA | 0 |
| S17-k141_177843 | 1549 | No | NA | 3 | 0 | 0 | Low-quality | Genome-fragment | 2.37 | AAI-based (high-confidence) | 0 |
| S3-final-viral-combined_k141_32972\|\|full | 1844 | No | NA | 3 | 0 | 0 | Not-determined | Genome-fragment | NA | NA | 0 |
| S12-final-viral-combined_k141_52523\|\|full | 1549 | No | NA | 5 | 2 | 0 | Low-quality | Genome-fragment | 3.02 | AAI-based (medium-confidence) | 0 |
| S12_k141_35473 | 1549 | Yes | 1307 | 4 | 2 | 1 | Low-quality | Genome-fragment | 2.98 | AAI-based (high-confidence) | 15.62 |
| S3-final-viral-combined_k141_73308\|\|full | 2946 | No | NA | 3 | 0 | 2 | Not-determined | Genome-fragment | NA | NA | 0 |
| S3-final-viral-combined_k141_90965\|\|full | 11142 | No | NA | 11 | 0 | 7 | Not-determined | Genome-fragment | NA | NA | 0 |
| S3-final-viral-combined_k141_101946\|\|full | 2137 | No | NA | 2 | 0 | 2 | Not-determined | Genome-fragment | NA | NA | 0 |
| S3-final-viral-combined_k141_125518\|\|full | 2287 | No | NA | 4 | 0 | 0 | Not-determined | Genome-fragment | NA | NA | 0 |
| S3-final-viral-combined_k141_107958\|\|full | 3973 | No | NA | 5 | 0 | 3 | Not-determined | Genome-fragment | NA | NA | 0 |
| S3-final-viral-combined_k141_107960\|\|full | 4458 | No | NA | 5 | 0 | 0 | Not-determined | Genome-fragment | NA | NA | 0 |
| S3-final-viral-combined_k141_50344\|\|full | 1513 | No | NA | 2 | 0 | 1 | Not-determined | Genome-fragment | NA | NA | 0 |
| S3-final-viral-combined_k141_27677\|\|full | 3338 | No | NA | 4 | 0 | 2 | Not-determined | Genome-fragment | NA | NA | 0 |
| S2-final-viral-combined_k141_46557\|\|full | 1549 | No | NA | 2 | 0 | 0 | Low-quality | Genome-fragment | 3.72 | AAI-based (medium-confidence) | 0 |
| S3-final-viral-combined_k141_33060\|\|full | 2068 | No | NA | 5 | 0 | 0 | Not-determined | Genome-fragment | NA | NA | 0 |
| S8-final-viral-combined_k141_228111\|\|full | 1549 | No | NA | 3 | 2 | 0 | Low-quality | Genome-fragment | 0.86 | HMM-based (lower-bound) | 0 |
| S3-final-viral-combined_k141_50393\|\|full | 1984 | No | NA | 4 | 0 | 3 | Not-determined | Genome-fragment | NA | NA | 0 |
| S3-final-viral-combined_k141_44648\|\|full | 1633 | No | NA | 4 | 0 | 1 | Not-determined | Genome-fragment | NA | NA | 0 |
| S9_k141_108598 | 1550 | No | NA | 2 | 0 | 1 | Low-quality | Genome-fragment | 3.1 | AAI-based (medium-confidence) | 0 |
| S3-final-viral-combined_k141_15739\|\|full | 1660 | No | NA | 4 | 0 | 0 | Not-determined | Genome-fragment | NA | NA | 0 |
| S3-final-viral-combined_k141_21937\|\|full | 1665 | No | NA | 2 | 0 | 1 | Not-determined | Genome-fragment | NA | NA | 0 |
| S1-final-viral-combined_k141_258341\|\|full | 1550 | No | NA | 3 | 0 | 0 | Low-quality | Genome-fragment | 3.74 | AAI-based (medium-confidence) | 0 |
| S3-final-viral-combined_k141_62057\|\|full | 2762 | No | NA | 3 | 0 | 1 | Not-determined | Genome-fragment | NA | NA | 0 |
| S1_k141_347765 | 1550 | No | NA | 1 | 0 | 0 | Low-quality | Genome-fragment | 0.25 | AAI-based (high-confidence) | 0 |
| S3-final-viral-combined_k141_10303\|\|full | 4121 | No | NA | 4 | 0 | 1 | Not-determined | Genome-fragment | NA | NA | 0 |
| S7-final-viral-combined_k141_236578\|\|full | 1550 | No | NA | 5 | 3 | 0 | Low-quality | Genome-fragment | 3.51 | AAI-based (high-confidence) | 0 |
| S12-final-viral-combined_k141_285786\|\|full | 1550 | No | NA | 5 | 1 | 0 | Low-quality | Genome-fragment | 4.49 | AAI-based (high-confidence) | 0 |
| S3-final-viral-combined_k141_120060\|\|full | 1459 | No | NA | 2 | 0 | 1 | Not-determined | Genome-fragment | NA | NA | 0 |
| S3-final-viral-combined_k141_120071\|\|full | 1829 | No | NA | 3 | 0 | 1 | Not-determined | Genome-fragment | NA | NA | 0 |
| S12-final-viral-combined_k141_309394\|\|full | 1550 | No | NA | 3 | 0 | 0 | Low-quality | Genome-fragment | 3.34 | AAI-based (high-confidence) | 0 |
| S3-final-viral-combined_k141_85297\|\|full | 1843 | No | NA | 2 | 0 | 2 | Not-determined | Genome-fragment | NA | NA | 0 |
| S3-final-viral-combined_k141_108101\|\|full | 1930 | No | NA | 4 | 0 | 1 | Not-determined | Genome-fragment | NA | NA | 0 |
| S3-final-viral-combined_k141_131466\|\|full | 4502 | No | NA | 5 | 0 | 3 | Not-determined | Genome-fragment | NA | NA | 0 |
| S3-final-viral-combined_k141_114039\|\|full | 1615 | No | NA | 2 | 0 | 1 | Not-determined | Genome-fragment | NA | NA | 0 |
| S12-final-viral-combined_k141_175163\|\|full | 1550 | No | NA | 2 | 0 | 0 | Low-quality | Genome-fragment | 2.98 | AAI-based (medium-confidence) | 0 |
| S3-final-viral-combined_k141_125695\|\|full | 7452 | No | NA | 6 | 0 | 4 | Not-determined | Genome-fragment | NA | NA | 0 |
| S3-final-viral-combined_k141_120093\|\|full | 1010 | No | NA | 3 | 0 | 1 | Not-determined | Genome-fragment | NA | NA | 0 |
| S3-final-viral-combined_k141_85325\|\|full | 2101 | No | NA | 4 | 0 | 0 | Not-determined | Genome-fragment | NA | NA | 0 |
| S3-final-viral-combined_k141_56484\|\|full | 1411 | No | NA | 3 | 0 | 1 | Not-determined | Genome-fragment | NA | NA | 0 |
| S4-final-viral-combined_k141_98801\|\|full | 1550 | No | NA | 2 | 0 | 1 | Low-quality | Genome-fragment | 4.39 | AAI-based (medium-confidence) | 0 |
| S4_k141_158687 | 1550 | No | NA | 2 | 2 | 0 | Low-quality | Genome-fragment | 2.57 | AAI-based (high-confidence) | 0 |
| S3-final-viral-combined_k141_22066\|\|full | 1211 | No | NA | 2 | 0 | 2 | Not-determined | Genome-fragment | NA | NA | 0 |
| S11-final-viral-combined_k141_28428\|\|full | 1551 | No | NA | 4 | 3 | 0 | Low-quality | Genome-fragment | 4.76 | AAI-based (high-confidence) | 0 |
| S3-final-viral-combined_k141_27891\|\|full | 2714 | No | NA | 2 | 0 | 1 | Not-determined | Genome-fragment | NA | NA | 0 |
| S3-final-viral-combined_k141_102124\|\|full | 2103 | No | NA | 4 | 0 | 3 | Not-determined | Genome-fragment | NA | NA | 0 |
| S11-final-viral-combined_k141_181393\|\|full | 1551 | No | NA | 3 | 0 | 0 | Low-quality | Genome-fragment | 2.2 | AAI-based (high-confidence) | 0 |
| S11-final-viral-combined_k141_99386\|\|full | 1551 | No | NA | 5 | 1 | 0 | Low-quality | Genome-fragment | 4.24 | AAI-based (medium-confidence) | 0 |
| S1-final-viral-combined_k141_153559\|\|full | 1551 | No | NA | 2 | 0 | 0 | Low-quality | Genome-fragment | 1.28 | AAI-based (high-confidence) | 0 |
| S3-final-viral-combined_k141_131595\|\|full | 1121 | No | NA | 2 | 0 | 1 | Not-determined | Genome-fragment | NA | NA | 0 |
| S3-final-viral-combined_k141_27911\|\|full | 1000 | No | NA | 3 | 0 | 1 | Not-determined | Genome-fragment | NA | NA | 0 |
| S1-final-viral-combined_k141_272598\|\|full | 1551 | No | NA | 3 | 1 | 0 | Low-quality | Genome-fragment | 4.4 | AAI-based (medium-confidence) | 0 |
| S12-final-viral-combined_k141_61278\|\|full | 1551 | No | NA | 3 | 0 | 1 | Low-quality | Genome-fragment | 2.67 | AAI-based (medium-confidence) | 0 |
| S3-final-viral-combined_k141_22133\|\|full | 1812 | No | NA | 3 | 0 | 1 | Not-determined | Genome-fragment | NA | NA | 0 |
| S2-final-viral-combined_k141_103337\|\|full | 1551 | No | NA | 3 | 2 | 0 | Low-quality | Genome-fragment | 2.43 | HMM-based (lower-bound) | 0 |
| S3-final-viral-combined_k141_27928\|\|full | 2088 | No | NA | 3 | 0 | 1 | Not-determined | Genome-fragment | NA | NA | 0 |
| S3-final-viral-combined_k141_33338\|\|full | 3656 | No | NA | 2 | 0 | 2 | Not-determined | Genome-fragment | NA | NA | 0 |
| S11-final-viral-combined_k141_87287\|\|full | 1552 | No | NA | 3 | 0 | 2 | Low-quality | Genome-fragment | 1.07 | AAI-based (medium-confidence) | 0 |
| S3-final-viral-combined_k141_73613\|\|full | 1416 | No | NA | 3 | 0 | 2 | Not-determined | Genome-fragment | NA | NA | 0 |
| S3-final-viral-combined_k141_120242\|\|full | 1387 | No | NA | 2 | 0 | 2 | Not-determined | Genome-fragment | NA | NA | 0 |
| S1-final-viral-combined_k141_116651\|\|full | 1552 | No | NA | 3 | 0 | 1 | Low-quality | Genome-fragment | 2.62 | AAI-based (high-confidence) | 0 |
| S3-final-viral-combined_k141_96922\|\|full | 2452 | No | NA | 2 | 0 | 1 | Not-determined | Genome-fragment | NA | NA | 0 |
| S5_k141_114852 | 1552 | No | NA | 2 | 0 | 2 | Low-quality | Genome-fragment | 0.56 | AAI-based (high-confidence) | 0 |
| S3-final-viral-combined_k141_16010\|\|full | 1030 | No | NA | 2 | 0 | 2 | Not-determined | Genome-fragment | NA | NA | 0 |
| S7-final-viral-combined_k141_104262\|\|full | 1552 | No | NA | 4 | 1 | 0 | Low-quality | Genome-fragment | 3.56 | AAI-based (medium-confidence) | 0 |
| S3-final-viral-combined_k141_108272\|\|full | 2130 | No | NA | 3 | 0 | 1 | Not-determined | Genome-fragment | NA | NA | 0 |
| S3-final-viral-combined_k141_96945\|\|full | 2684 | No | NA | 4 | 0 | 2 | Not-determined | Genome-fragment | NA | NA | 0 |
| S3-final-viral-combined_k141_96950\|\|full | 2665 | No | NA | 4 | 0 | 2 | Not-determined | Genome-fragment | NA | NA | 0 |
| S3-final-viral-combined_k141_79630\|\|full | 2203 | No | NA | 3 | 0 | 1 | Not-determined | Genome-fragment | NA | NA | 0 |
| S3-final-viral-combined_k141_96952\|\|full | 2848 | No | NA | 2 | 0 | 2 | Not-determined | Genome-fragment | NA | NA | 0 |
| S7-final-viral-combined_k141_192016\|\|lt2gene | 1552 | No | NA | 2 | 1 | 0 | Low-quality | Genome-fragment | 2.98 | AAI-based (high-confidence) | 0 |
| S3-final-viral-combined_k141_16052\|\|full | 1346 | No | NA | 3 | 0 | 3 | Not-determined | Genome-fragment | NA | NA | 0 |
| S7-final-viral-combined_k141_18964\|\|lt2gene | 1552 | No | NA | 2 | 1 | 0 | Low-quality | Genome-fragment | 0.59 | HMM-based (lower-bound) | 0 |
| S3-final-viral-combined_k141_16053\|\|full | 1766 | No | NA | 2 | 0 | 1 | Not-determined | Genome-fragment | NA | NA | 0 |
| S12-final-viral-combined_k141_68235\|\|full | 1552 | No | NA | 2 | 1 | 0 | Low-quality | Genome-fragment | 1.74 | HMM-based (lower-bound) | 0 |
| S12-final-viral-combined_k141_13307\|\|full | 1552 | No | NA | 3 | 3 | 0 | Low-quality | Genome-fragment | 3.41 | AAI-based (high-confidence) | 0 |
| S12-final-viral-combined_k141_32826\|\|full | 1552 | No | NA | 2 | 0 | 1 | Low-quality | Genome-fragment | 2.77 | AAI-based (medium-confidence) | 0 |
| S3-final-viral-combined_k141_33438\|\|full | 1670 | No | NA | 2 | 0 | 1 | Not-determined | Genome-fragment | NA | NA | 0 |
| S3-final-viral-combined_k141_62341\|\|full | 2144 | No | NA | 2 | 0 | 1 | Not-determined | Genome-fragment | NA | NA | 0 |
| S2-final-viral-combined_k141_177659\|\|lt2gene | 1552 | No | NA | 2 | 2 | 0 | Low-quality | Genome-fragment | 3.62 | AAI-based (high-confidence) | 0 |
| S3-final-viral-combined_k141_68228\|\|full | 2844 | No | NA | 5 | 0 | 1 | Not-determined | Genome-fragment | NA | NA | 0 |
| S3-final-viral-combined_k141_39586\|\|full | 1965 | No | NA | 2 | 0 | 1 | Not-determined | Genome-fragment | NA | NA | 0 |
| S6_k141_29927 | 1552 | No | NA | 1 | 0 | 1 | Low-quality | Genome-fragment | 0.38 | AAI-based (high-confidence) | 0 |
| S3-final-viral-combined_k141_73715\|\|full | 1293 | No | NA | 1 | 0 | 1 | Not-determined | Genome-fragment | NA | NA | 0 |
| S3-final-viral-combined_k141_91336\|\|full | 4568 | No | NA | 4 | 0 | 1 | Not-determined | Genome-fragment | NA | NA | 0 |
| S3-final-viral-combined_k141_137042\|\|full | 3227 | No | NA | 4 | 0 | 3 | Not-determined | Genome-fragment | NA | NA | 0 |
| S3-final-viral-combined_k141_5137\|\|full | 6255 | No | NA | 8 | 0 | 2 | Not-determined | Genome-fragment | NA | NA | 0 |
| S3-final-viral-combined_k141_97044\|\|full | 2174 | No | NA | 4 | 0 | 2 | Not-determined | Genome-fragment | NA | NA | 0 |
| S3-final-viral-combined_k141_22301\|\|full | 2496 | No | NA | 4 | 0 | 2 | Not-determined | Genome-fragment | NA | NA | 0 |
| S3-final-viral-combined_k141_73768\|\|full | 2363 | No | NA | 3 | 0 | 1 | Not-determined | Genome-fragment | NA | NA | 0 |
| S11_k141_120422 | 1553 | No | NA | 2 | 0 | 2 | Low-quality | Genome-fragment | 3.65 | AAI-based (medium-confidence) | 0 |
| S3-final-viral-combined_k141_114351\|\|full | 2300 | No | NA | 2 | 0 | 0 | Not-determined | Genome-fragment | NA | NA | 0 |
| S1_k141_126086 | 1553 | No | NA | 4 | 0 | 0 | Low-quality | Genome-fragment | 10.43 | AAI-based (high-confidence) | 0 |
| S3-final-viral-combined_k141_45003\|\|full | 1964 | No | NA | 2 | 0 | 2 | Not-determined | Genome-fragment | NA | NA | 0 |
| S1_k141_340410 | 1553 | No | NA | 4 | 0 | 0 | Low-quality | Genome-fragment | 4.7 | AAI-based (medium-confidence) | 0 |
| S3-final-viral-combined_k141_73796\|\|full | 8556 | No | NA | 8 | 0 | 6 | Not-determined | Genome-fragment | NA | NA | 0 |
| S3-final-viral-combined_k141_45019\|\|full | 1574 | No | NA | 3 | 0 | 0 | Not-determined | Genome-fragment | NA | NA | 0 |
| S3-final-viral-combined_k141_102351\|\|full | 1177 | No | NA | 2 | 0 | 1 | Not-determined | Genome-fragment | NA | NA | 0 |
| S1_k141_31516 | 1553 | No | NA | 2 | 0 | 1 | Low-quality | Genome-fragment | 3.1 | AAI-based (medium-confidence) | 0 |
| S3-final-viral-combined_k141_85649\|\|full | 2943 | No | NA | 5 | 0 | 0 | Not-determined | Genome-fragment | NA | NA | 0 |
| S3-final-viral-combined_k141_28139\|\|full | 1126 | No | NA | 3 | 0 | 1 | Not-determined | Genome-fragment | NA | NA | 0 |
| S3-final-viral-combined_k141_91417\|\|full | 3874 | No | NA | 2 | 0 | 2 | Not-determined | Genome-fragment | NA | NA | 0 |
| S3-final-viral-combined_k141_39670\|\|full | 10064 | No | NA | 5 | 0 | 5 | Not-determined | Genome-fragment | NA | NA | 0 |
| S7-final-viral-combined_k141_97636\|\|full | 1553 | No | NA | 2 | 0 | 2 | Low-quality | Genome-fragment | 2.78 | AAI-based (medium-confidence) | 0 |
| S3-final-viral-combined_k141_10725\|\|full | 3024 | No | NA | 3 | 0 | 1 | Not-determined | Genome-fragment | NA | NA | 0 |
| S3-final-viral-combined_k141_33571\|\|full | 2956 | No | NA | 4 | 0 | 2 | Not-determined | Genome-fragment | NA | NA | 0 |
| S17-final-viral-combined_k141_203139\|\|lt2gene | 1553 | No | NA | 1 | 1 | 0 | Low-quality | Genome-fragment | 2.84 | HMM-based (lower-bound) | 0 |
| S3-final-viral-combined_k141_126118\|\|full | 1680 | No | NA | 2 | 0 | 2 | Not-determined | Genome-fragment | NA | NA | 0 |
| S3-final-viral-combined_k141_102407\|\|full | 1708 | No | NA | 3 | 0 | 0 | Not-determined | Genome-fragment | NA | NA | 0 |
| S3-final-viral-combined_k141_33577\|\|full | 1907 | No | NA | 3 | 0 | 0 | Not-determined | Genome-fragment | NA | NA | 0 |
| S3-final-viral-combined_k141_120543\|\|full | 6218 | No | NA | 5 | 0 | 4 | Not-determined | Genome-fragment | NA | NA | 0 |
| S17-final-viral-combined_k141_205939\|\|lt2gene | 1553 | No | NA | 2 | 2 | 0 | Low-quality | Genome-fragment | 4.16 | AAI-based (medium-confidence) | 0 |
| S12-final-viral-combined_k141_293415\|\|full | 1553 | No | NA | 3 | 0 | 0 | Low-quality | Genome-fragment | 4.12 | AAI-based (high-confidence) | 0 |
| S12-final-viral-combined_k141_363150\|\|full | 1553 | No | NA | 2 | 0 | 0 | Low-quality | Genome-fragment | 1.01 | AAI-based (high-confidence) | 0 |
| S3-final-viral-combined_k141_10797\|\|full | 3022 | No | NA | 3 | 0 | 1 | Not-determined | Genome-fragment | NA | NA | 0 |
| S3-final-viral-combined_k141_137146\|\|full | 1007 | No | NA | 3 | 0 | 0 | Not-determined | Genome-fragment | NA | NA | 0 |
| S3-final-viral-combined_k141_68415\|\|full | 1630 | No | NA | 3 | 0 | 0 | Not-determined | Genome-fragment | NA | NA | 0 |
| S3-final-viral-combined_k141_5362\|\|full | 1943 | No | NA | 5 | 0 | 0 | Not-determined | Genome-fragment | NA | NA | 0 |
| S6-final-viral-combined_k141_55071\|\|full | 1553 | No | NA | 4 | 1 | 0 | Low-quality | Genome-fragment | 2.57 | AAI-based (high-confidence) | 0 |
| S8-final-viral-combined_k141_142697\|\|full | 1553 | No | NA | 4 | 4 | 0 | Low-quality | Genome-fragment | 3.75 | AAI-based (high-confidence) | 0 |
| S3-final-viral-combined_k141_108433\|\|full | 1852 | No | NA | 3 | 0 | 2 | Not-determined | Genome-fragment | NA | NA | 0 |
| S8-final-viral-combined_k141_274161\|\|full | 1553 | No | NA | 4 | 1 | 0 | Low-quality | Genome-fragment | 2.69 | AAI-based (high-confidence) | 0 |
| S3-final-viral-combined_k141_5382\|\|full | 2395 | No | NA | 4 | 0 | 1 | Not-determined | Genome-fragment | NA | NA | 0 |
| S3-final-viral-combined_k141_137170\|\|full | 2139 | No | NA | 3 | 0 | 2 | Not-determined | Genome-fragment | NA | NA | 0 |
| S3-final-viral-combined_k141_10826\|\|full | 1780 | No | NA | 3 | 0 | 2 | Not-determined | Genome-fragment | NA | NA | 0 |
| S3-final-viral-combined_k141_102464\|\|full | 2218 | No | NA | 2 | 0 | 2 | Not-determined | Genome-fragment | NA | NA | 0 |
| S3-final-viral-combined_k141_131904\|\|full | 12136 | No | NA | 14 | 0 | 9 | Not-determined | Genome-fragment | NA | NA | 0 |
| S3-final-viral-combined_k141_28280\|\|full | 2057 | No | NA | 3 | 0 | 3 | Not-determined | Genome-fragment | NA | NA | 0 |
| S8_k141_157672 | 1553 | No | NA | 2 | 0 | 0 | Low-quality | Genome-fragment | 3.45 | AAI-based (high-confidence) | 0 |
| S3-final-viral-combined_k141_62594\|\|full | 1466 | No | NA | 2 | 0 | 0 | Not-determined | Genome-fragment | NA | NA | 0 |
| S3-final-viral-combined_k141_45189\|\|full | 2055 | No | NA | 2 | 0 | 1 | Not-determined | Genome-fragment | NA | NA | 0 |
| S3-final-viral-combined_k141_131927\|\|full | 1055 | No | NA | 3 | 0 | 1 | Not-determined | Genome-fragment | NA | NA | 0 |
| S3-final-viral-combined_k141_85836\|\|full | 2374 | No | NA | 5 | 0 | 2 | Not-determined | Genome-fragment | NA | NA | 0 |
| S3-final-viral-combined_k141_114533\|\|full | 1758 | No | NA | 3 | 0 | 0 | Not-determined | Genome-fragment | NA | NA | 0 |
| S9-final-viral-combined_k141_6398\|\|lt2gene | 1554 | No | NA | 2 | 2 | 0 | Low-quality | Genome-fragment | 3.94 | AAI-based (high-confidence) | 0 |
| S1-final-viral-combined_k141_247704\|\|full | 1554 | No | NA | 3 | 0 | 0 | Low-quality | Genome-fragment | 3.25 | AAI-based (medium-confidence) | 0 |
| S3-final-viral-combined_k141_99603\|\|full | 1554 | No | NA | 3 | 0 | 0 | Low-quality | Genome-fragment | 3.14 | AAI-based (medium-confidence) | 0 |
| S3-final-viral-combined_k141_45218\|\|full | 2147 | No | NA | 2 | 0 | 2 | Not-determined | Genome-fragment | NA | NA | 0 |
| S3-final-viral-combined_k141_102525\|\|full | 1761 | No | NA | 3 | 0 | 3 | Not-determined | Genome-fragment | NA | NA | 0 |
| S5-final-viral-combined_k141_26665\|\|full | 1554 | No | NA | 2 | 0 | 0 | Low-quality | Genome-fragment | 0.84 | AAI-based (high-confidence) | 0 |
| S3-final-viral-combined_k141_45242\|\|full | 1884 | No | NA | 3 | 0 | 2 | Not-determined | Genome-fragment | NA | NA | 0 |
| S7-final-viral-combined_k141_65609\|\|full | 1554 | No | NA | 3 | 0 | 0 | Low-quality | Genome-fragment | 2.97 | AAI-based (high-confidence) | 0 |
| S7_k141_185256 | 1554 | No | NA | 1 | 0 | 1 | Low-quality | Genome-fragment | 1.96 | AAI-based (high-confidence) | 0 |
| S17-final-viral-combined_k141_180449\|\|lt2gene | 1554 | No | NA | 2 | 2 | 0 | Low-quality | Genome-fragment | 5.13 | HMM-based (lower-bound) | 0 |
| S3-final-viral-combined_k141_120662\|\|full | 3475 | No | NA | 3 | 0 | 1 | Not-determined | Genome-fragment | NA | NA | 0 |
| S3-final-viral-combined_k141_137218\|\|full | 2727 | No | NA | 2 | 0 | 2 | Not-determined | Genome-fragment | NA | NA | 0 |
| S3-final-viral-combined_k141_62656\|\|full | 1233 | No | NA | 2 | 0 | 0 | Not-determined | Genome-fragment | NA | NA | 0 |
| S17-final-viral-combined_k141_228230\|\|lt2gene | 1554 | No | NA | 1 | 1 | 0 | Low-quality | Genome-fragment | 3.32 | AAI-based (high-confidence) | 0 |
| S3-final-viral-combined_k141_45292\|\|full | 5039 | No | NA | 5 | 0 | 2 | Not-determined | Genome-fragment | NA | NA | 0 |
| S12-final-viral-combined_k141_357113\|\|full | 1554 | No | NA | 3 | 0 | 0 | Low-quality | Genome-fragment | 2.59 | AAI-based (high-confidence) | 0 |
| S3-final-viral-combined_k141_102604\|\|full | 2378 | No | NA | 3 | 0 | 1 | Not-determined | Genome-fragment | NA | NA | 0 |
| S3-final-viral-combined_k141_28386\|\|full | 2583 | No | NA | 3 | 0 | 3 | Not-determined | Genome-fragment | NA | NA | 0 |
| S3-final-viral-combined_k141_108588\|\|full | 19036 | No | NA | 19 | 0 | 3 | Not-determined | Genome-fragment | NA | NA | 0 |
| S3-final-viral-combined_k141_33850\|\|full | 1019 | No | NA | 2 | 0 | 0 | Not-determined | Genome-fragment | NA | NA | 0 |
| S3-final-viral-combined_k141_62696\|\|full | 3699 | No | NA | 3 | 0 | 2 | Not-determined | Genome-fragment | NA | NA | 0 |
| S3-final-viral-combined_k141_45305\|\|full | 19171 | No | NA | 25 | 11 | 0 | Medium-quality | Genome-fragment | 51.32 | AAI-based (high-confidence) | 0 |
| S6-final-viral-combined_k141_9746\|\|full | 1554 | No | NA | 4 | 0 | 0 | Low-quality | Genome-fragment | 1.68 | AAI-based (high-confidence) | 0 |
| S3-final-viral-combined_k141_28398\|\|full | 1658 | No | NA | 2 | 0 | 1 | Not-determined | Genome-fragment | NA | NA | 0 |
| S3-final-viral-combined_k141_51133\|\|full | 3709 | No | NA | 4 | 0 | 3 | Not-determined | Genome-fragment | NA | NA | 0 |
| S3-final-viral-combined_k141_132087\|\|full | 1252 | No | NA | 3 | 0 | 1 | Not-determined | Genome-fragment | NA | NA | 0 |
| S3-final-viral-combined_k141_33903\|\|full | 1370 | No | NA | 3 | 0 | 0 | Not-determined | Genome-fragment | NA | NA | 0 |
| S3-final-viral-combined_k141_51157\|\|full | 2245 | No | NA | 2 | 0 | 1 | Not-determined | Genome-fragment | NA | NA | 0 |
| S3-final-viral-combined_k141_5572\|\|full | 2841 | No | NA | 4 | 0 | 2 | Not-determined | Genome-fragment | NA | NA | 0 |
| S3-final-viral-combined_k141_11004\|\|full | 5327 | No | NA | 7 | 0 | 2 | Not-determined | Genome-fragment | NA | NA | 0 |
| S9-final-viral-combined_k141_112863\|\|full | 1555 | No | NA | 3 | 0 | 0 | Low-quality | Genome-fragment | 2.87 | AAI-based (high-confidence) | 0 |
| S3-final-viral-combined_k141_91796\|\|full | 5281 | No | NA | 7 | 0 | 1 | Not-determined | Genome-fragment | NA | NA | 0 |
| S3-final-viral-combined_k141_86333\|\|full | 1555 | No | NA | 4 | 0 | 1 | Low-quality | Genome-fragment | 3.75 | AAI-based (high-confidence) | 0 |
| S3-final-viral-combined_k141_45373\|\|full | 2288 | No | NA | 2 | 0 | 2 | Not-determined | Genome-fragment | NA | NA | 0 |
| S3-final-viral-combined_k141_114705\|\|full | 2062 | No | NA | 3 | 0 | 1 | Not-determined | Genome-fragment | NA | NA | 0 |
| S7-final-viral-combined_k141_237921\|\|full | 1555 | No | NA | 3 | 0 | 0 | Low-quality | Genome-fragment | 1.73 | AAI-based (high-confidence) | 0 |
| S7_k141_3808 | 1555 | No | NA | 1 | 0 | 0 | Low-quality | Genome-fragment | 3.08 | AAI-based (high-confidence) | 0 |
| S3-final-viral-combined_k141_80182\|\|full | 5646 | No | NA | 5 | 0 | 4 | Not-determined | Genome-fragment | NA | NA | 0 |
| S3-final-viral-combined_k141_28454\|\|full | 1663 | No | NA | 2 | 0 | 1 | Not-determined | Genome-fragment | NA | NA | 0 |
| S3-final-viral-combined_k141_86076\|\|full | 2400 | No | NA | 2 | 0 | 1 | Not-determined | Genome-fragment | NA | NA | 0 |
| S3-final-viral-combined_k141_80194\|\|full | 2941 | No | NA | 3 | 0 | 2 | Not-determined | Genome-fragment | NA | NA | 0 |
| S3-final-viral-combined_k141_74275\|\|full | 3170 | No | NA | 3 | 0 | 2 | Not-determined | Genome-fragment | NA | NA | 0 |
| S3-final-viral-combined_k141_28477\|\|full | 1166 | No | NA | 2 | 0 | 1 | Not-determined | Genome-fragment | NA | NA | 0 |
| S2_k141_196082 | 1555 | No | NA | 3 | 0 | 1 | Low-quality | Genome-fragment | 1.11 | AAI-based (medium-confidence) | 0 |
| S3-final-viral-combined_k141_137426\|\|full | 2754 | No | NA | 3 | 0 | 1 | Not-determined | Genome-fragment | NA | NA | 0 |
| S4_k141_107346 | 1555 | No | NA | 1 | 0 | 1 | Low-quality | Genome-fragment | 2.65 | AAI-based (high-confidence) | 0 |
| S6-final-viral-combined_k141_54264\|\|full | 1555 | No | NA | 3 | 0 | 2 | Low-quality | Genome-fragment | 1.95 | AAI-based (medium-confidence) | 0 |
| S3-final-viral-combined_k141_102800\|\|full | 1313 | No | NA | 2 | 0 | 1 | Not-determined | Genome-fragment | NA | NA | 0 |
| S6-final-viral-combined_k141_37966\|\|full | 1555 | No | NA | 3 | 0 | 2 | Low-quality | Genome-fragment | 0.66 | AAI-based (medium-confidence) | 0 |
| S3-final-viral-combined_k141_57173\|\|full | 2085 | No | NA | 3 | 0 | 1 | Not-determined | Genome-fragment | NA | NA | 0 |
| S3-final-viral-combined_k141_34127\|\|full | 3008 | No | NA | 4 | 0 | 4 | Not-determined | Genome-fragment | NA | NA | 0 |
| S3-final-viral-combined_k141_51346\|\|full | 1261 | No | NA | 2 | 0 | 1 | Not-determined | Genome-fragment | NA | NA | 0 |
| S3-final-viral-combined_k141_108812\|\|full | 1722 | No | NA | 3 | 0 | 1 | Not-determined | Genome-fragment | NA | NA | 0 |
| S9-final-viral-combined_k141_54978\|\|lt2gene | 1556 | No | NA | 1 | 1 | 0 | Low-quality | Genome-fragment | 4.57 | AAI-based (medium-confidence) | 0 |
| S3-final-viral-combined_k141_28597\|\|full | 7192 | No | NA | 7 | 0 | 5 | Not-determined | Genome-fragment | NA | NA | 0 |
| S3-final-viral-combined_k141_97620\|\|full | 1210 | No | NA | 2 | 0 | 1 | Not-determined | Genome-fragment | NA | NA | 0 |
| S11-final-viral-combined_k141_135457\|\|lt2gene | 1556 | No | NA | 2 | 2 | 0 | Low-quality | Genome-fragment | 3.95 | AAI-based (high-confidence) | 0 |
| S3-final-viral-combined_k141_28618\|\|full | 4365 | No | NA | 5 | 0 | 3 | Not-determined | Genome-fragment | NA | NA | 0 |
| S3-final-viral-combined_k141_137541\|\|full | 1165 | No | NA | 3 | 0 | 0 | Not-determined | Genome-fragment | NA | NA | 0 |
| S3-final-viral-combined_k141_16731\|\|full | 5340 | No | NA | 5 | 0 | 0 | Not-determined | Genome-fragment | NA | NA | 0 |
| S3-final-viral-combined_k141_51393\|\|full | 1253 | No | NA | 2 | 0 | 0 | Not-determined | Genome-fragment | NA | NA | 0 |
| S3-final-viral-combined_k141_102922\|\|full | 1280 | No | NA | 2 | 0 | 1 | Not-determined | Genome-fragment | NA | NA | 0 |
| S3-final-viral-combined_k141_137571\|\|full | 2546 | No | NA | 5 | 0 | 0 | Not-determined | Genome-fragment | NA | NA | 0 |
| S3-final-viral-combined_k141_108852\|\|full | 2593 | No | NA | 2 | 0 | 2 | Not-determined | Genome-fragment | NA | NA | 0 |
| S3-final-viral-combined_k141_16751\|\|full | 3561 | No | NA | 4 | 0 | 2 | Not-determined | Genome-fragment | NA | NA | 0 |
| S1-final-viral-combined_k141_321354\|\|full | 1556 | No | NA | 3 | 0 | 0 | Low-quality | Genome-fragment | 2.6 | AAI-based (medium-confidence) | 0 |
| S3-final-viral-combined_k141_108891\|\|full | 3545 | No | NA | 6 | 0 | 3 | Not-determined | Genome-fragment | NA | NA | 0 |
| S3-final-viral-combined_k141_68912\|\|full | 1636 | No | NA | 2 | 0 | 1 | Not-determined | Genome-fragment | NA | NA | 0 |
| S17-final-viral-combined_k141_66866\|\|full | 1556 | No | NA | 3 | 1 | 0 | Low-quality | Genome-fragment | 3.89 | AAI-based (medium-confidence) | 0 |
| S3-final-viral-combined_k141_51441\|\|full | 7343 | No | NA | 6 | 0 | 6 | Not-determined | Genome-fragment | NA | NA | 0 |
| S3-final-viral-combined_k141_57364\|\|full | 1586 | No | NA | 3 | 0 | 1 | Not-determined | Genome-fragment | NA | NA | 0 |
| S3-final-viral-combined_k141_51465\|\|full | 1145 | No | NA | 2 | 0 | 1 | Not-determined | Genome-fragment | NA | NA | 0 |
| S3-final-viral-combined_k141_51471\|\|full | 1556 | No | NA | 3 | 0 | 0 | Not-determined | Genome-fragment | NA | NA | 0 |
| S3-final-viral-combined_k141_11357\|\|full | 1664 | No | NA | 2 | 0 | 2 | Not-determined | Genome-fragment | NA | NA | 0 |
| S3-final-viral-combined_k141_34309\|\|full | 3656 | No | NA | 4 | 0 | 2 | Not-determined | Genome-fragment | NA | NA | 0 |
| S12-final-viral-combined_k141_399770\|\|full | 1556 | No | NA | 3 | 0 | 0 | Low-quality | Genome-fragment | 3.36 | AAI-based (high-confidence) | 0 |
| S2-final-viral-combined_k141_102982\|\|lt2gene | 1556 | No | NA | 2 | 1 | 0 | Low-quality | Genome-fragment | 3.02 | AAI-based (high-confidence) | 0 |
| S3-final-viral-combined_k141_80477\|\|full | 2825 | No | NA | 3 | 0 | 2 | Not-determined | Genome-fragment | NA | NA | 0 |
| S2_k141_51878 | 1556 | No | NA | 4 | 0 | 1 | Low-quality | Genome-fragment | 2.15 | AAI-based (medium-confidence) | 0 |
| S3-final-viral-combined_k141_80493\|\|full | 1513 | No | NA | 3 | 0 | 0 | Not-determined | Genome-fragment | NA | NA | 0 |
| S3-final-viral-combined_k141_57455\|\|full | 5740 | No | NA | 6 | 0 | 3 | Not-determined | Genome-fragment | NA | NA | 0 |
| S3-final-viral-combined_k141_63078\|\|full | 1166 | No | NA | 2 | 0 | 1 | Not-determined | Genome-fragment | NA | NA | 0 |
| S1-final-viral-combined_k141_232781\|\|full | 1557 | No | NA | 2 | 0 | 2 | Low-quality | Genome-fragment | 0.43 | AAI-based (high-confidence) | 0 |
| S1-final-viral-combined_k141_333444\|\|full | 1557 | No | NA | 3 | 1 | 0 | Low-quality | Genome-fragment | 37.42 | HMM-based (lower-bound) | 0 |
| S1_k141_221399 | 1557 | No | NA | 3 | 0 | 0 | Low-quality | Genome-fragment | 2.48 | AAI-based (medium-confidence) | 0 |
| S3-final-viral-combined_k141_103118\|\|full | 2852 | No | NA | 4 | 0 | 1 | Not-determined | Genome-fragment | NA | NA | 0 |
| S3-final-viral-combined_k141_74599\|\|full | 1533 | No | NA | 3 | 0 | 1 | Not-determined | Genome-fragment | NA | NA | 0 |
| S3-final-viral-combined_k141_137733\|\|full | 2904 | No | NA | 3 | 0 | 2 | Not-determined | Genome-fragment | NA | NA | 0 |
| S7-final-viral-combined_k141_32821\|\|full | 1557 | No | NA | 4 | 0 | 0 | Low-quality | Genome-fragment | 3.19 | AAI-based (medium-confidence) | 0 |
| S3-final-viral-combined_k141_51575\|\|full | 4403 | No | NA | 4 | 0 | 2 | Not-determined | Genome-fragment | NA | NA | 0 |
| S3-final-viral-combined_k141_63157\|\|full | 3027 | No | NA | 4 | 0 | 4 | Not-determined | Genome-fragment | NA | NA | 0 |
| S3-final-viral-combined_k141_34429\|\|full | 1231 | No | NA | 3 | 0 | 0 | Not-determined | Genome-fragment | NA | NA | 0 |
| S3-final-viral-combined_k141_109160\|\|full | 1668 | No | NA | 3 | 0 | 1 | Not-determined | Genome-fragment | NA | NA | 0 |
| S7-final-viral-combined_k141_243268\|\|full | 1557 | No | NA | 2 | 0 | 2 | Low-quality | Genome-fragment | 0.93 | AAI-based (medium-confidence) | 0 |
| S17-final-viral-combined_k141_178402\|\|full | 1557 | No | NA | 2 | 0 | 1 | Low-quality | Genome-fragment | 0.99 | AAI-based (medium-confidence) | 0 |
| S3-final-viral-combined_k141_103184\|\|full | 2462 | No | NA | 3 | 0 | 1 | Not-determined | Genome-fragment | NA | NA | 0 |
| S3-final-viral-combined_k141_109199\|\|full | 2132 | No | NA | 5 | 0 | 3 | Not-determined | Genome-fragment | NA | NA | 0 |
| S3-final-viral-combined_k141_63238\|\|full | 10611 | No | NA | 10 | 0 | 2 | Not-determined | Genome-fragment | NA | NA | 0 |
| S3-final-viral-combined_k141_109218\|\|full | 1749 | No | NA | 2 | 0 | 0 | Not-determined | Genome-fragment | NA | NA | 0 |
| S3-final-viral-combined_k141_63256\|\|full | 6844 | No | NA | 4 | 0 | 3 | Not-determined | Genome-fragment | NA | NA | 0 |
| S3-final-viral-combined_k141_45814\|\|full | 1024 | No | NA | 3 | 0 | 1 | Not-determined | Genome-fragment | NA | NA | 0 |
| S17-final-viral-combined_k141_11980\|\|full | 1557 | No | NA | 3 | 0 | 0 | Low-quality | Genome-fragment | 3.16 | AAI-based (medium-confidence) | 0 |
| S3-final-viral-combined_k141_45823\|\|full | 2195 | No | NA | 2 | 0 | 2 | Not-determined | Genome-fragment | NA | NA | 0 |
| S3-final-viral-combined_k141_103291\|\|full | 2741 | No | NA | 2 | 0 | 2 | Not-determined | Genome-fragment | NA | NA | 0 |
| S3-final-viral-combined_k141_103293\|\|full | 5131 | No | NA | 8 | 0 | 0 | Not-determined | Genome-fragment | NA | NA | 0 |
| S17-final-viral-combined_k141_167804\|\|full | 1557 | No | NA | 2 | 0 | 0 | Low-quality | Genome-fragment | 2.21 | AAI-based (high-confidence) | 0 |
| S3-final-viral-combined_k141_17186\|\|full | 1172 | No | NA | 2 | 0 | 1 | Not-determined | Genome-fragment | NA | NA | 0 |
| S12-final-viral-combined_k141_240320\|\|full | 1557 | No | NA | 3 | 2 | 0 | Low-quality | Genome-fragment | 3.53 | AAI-based (medium-confidence) | 0 |
| S3-final-viral-combined_k141_45987\|\|full | 2725 | No | NA | 6 | 0 | 0 | Not-determined | Genome-fragment | NA | NA | 0 |
| S3-final-viral-combined_k141_46694\|\|0_partial | 17730 | No | NA | 14 | 0 | 13 | Not-determined | Genome-fragment | NA | NA | 0 |
| S3-final-viral-combined_k141_53130\|\|0_partial | 16672 | No | NA | 16 | 0 | 2 | Not-determined | Genome-fragment | NA | NA | 0 |
| S11-final-viral-combined_k141_41279\|\|full | 1558 | No | NA | 3 | 1 | 0 | Low-quality | Genome-fragment | 0.94 | HMM-based (lower-bound) | 0 |
| S3-final-viral-combined_k141_83057\|\|0_partial | 15074 | No | NA | 13 | 0 | 5 | Not-determined | Genome-fragment | NA | NA | 0 |
| S3-final-viral-combined_k141_106497\|\|full | 1558 | Yes | 333 | 4 | 1 | 1 | Low-quality | Genome-fragment | 0.27 | HMM-based (lower-bound) | 78.63 |
| S3-final-viral-combined_k141_136422\|\|0_partial | 11952 | No | NA | 12 | 0 | 8 | Not-determined | Genome-fragment | NA | NA | 0 |
| S3-final-viral-combined_k141_100652\|\|full | 1558 | No | NA | 2 | 0 | 1 | Low-quality | Genome-fragment | 0.74 | AAI-based (medium-confidence) | 0 |
| S5_k141_73671 | 1558 | No | NA | 3 | 2 | 0 | Low-quality | Genome-fragment | 3.24 | AAI-based (high-confidence) | 0 |
| S4-final-viral-combined_k141_46807\|\|full | 1558 | No | NA | 4 | 1 | 0 | Low-quality | Genome-fragment | 3.45 | AAI-based (medium-confidence) | 0 |
| S8_k141_170563 | 1558 | No | NA | 1 | 0 | 0 | Low-quality | Genome-fragment | 2.94 | AAI-based (high-confidence) | 0 |
| S3-final-viral-combined_k141_6289\|\|lt2gene | 1368 | No | NA | 2 | 0 | 0 | Not-determined | Genome-fragment | NA | NA | 0 |
| S5-final-viral-combined_k141_194404\|\|full | 1559 | No | NA | 2 | 0 | 1 | Low-quality | Genome-fragment | 1.24 | AAI-based (high-confidence) | 0 |
| S3-final-viral-combined_k141_92587\|\|lt2gene | 1412 | No | NA | 1 | 0 | 0 | Not-determined | Genome-fragment | NA | NA | 0 |
| S12-final-viral-combined_k141_123157\|\|full | 1559 | No | NA | 2 | 0 | 1 | Low-quality | Genome-fragment | 1.32 | AAI-based (medium-confidence) | 0 |
| S12-final-viral-combined_k141_114151\|\|full | 1559 | No | NA | 3 | 1 | 0 | Low-quality | Genome-fragment | 3.7 | AAI-based (medium-confidence) | 0 |
| S12-final-viral-combined_k141_291608\|\|lt2gene | 1559 | No | NA | 2 | 1 | 0 | Low-quality | Genome-fragment | 3.11 | AAI-based (high-confidence) | 0 |
| S3-final-viral-combined_k141_52646\|\|lt2gene | 2385 | No | NA | 1 | 0 | 0 | Not-determined | Genome-fragment | NA | NA | 0 |
| S2-final-viral-combined_k141_244811\|\|lt2gene | 1559 | No | NA | 2 | 1 | 0 | Low-quality | Genome-fragment | 2.31 | HMM-based (lower-bound) | 0 |
| S6_k141_18868 | 1559 | No | NA | 6 | 0 | 0 | Low-quality | Genome-fragment | 4.78 | AAI-based (high-confidence) | 0 |
| S11-final-viral-combined_k141_155437\|\|full | 1560 | No | NA | 2 | 1 | 0 | Low-quality | Genome-fragment | 2.9 | HMM-based (lower-bound) | 0 |
| S1-final-viral-combined_k141_171669\|\|full | 1560 | No | NA | 3 | 0 | 0 | Low-quality | Genome-fragment | 2.91 | AAI-based (high-confidence) | 0 |
| S1_k141_152473 | 1560 | No | NA | 1 | 0 | 1 | Low-quality | Genome-fragment | 0.96 | AAI-based (medium-confidence) | 0 |
| S5_k141_178321 | 1560 | No | NA | 1 | 0 | 1 | Low-quality | Genome-fragment | 0.58 | AAI-based (high-confidence) | 0 |
| S3-final-viral-combined_k141_1260\|\|lt2gene | 1255 | No | NA | 1 | 0 | 0 | Not-determined | Genome-fragment | NA | NA | 0 |
| S7_k141_177077 | 1560 | No | NA | 2 | 0 | 1 | Low-quality | Genome-fragment | 2.35 | AAI-based (medium-confidence) | 0 |
| S3-final-viral-combined_k141_93504\|\|lt2gene | 1403 | No | NA | 2 | 0 | 2 | Not-determined | Genome-fragment | NA | NA | 0 |
| S7_k141_182744 | 1560 | No | NA | 3 | 2 | 0 | Low-quality | Genome-fragment | 4.13 | AAI-based (high-confidence) | 0 |
| S9-final-viral-combined_k141_100772\|\|full | 1561 | No | NA | 3 | 0 | 2 | Low-quality | Genome-fragment | 0.93 | AAI-based (medium-confidence) | 0 |
| S3-final-viral-combined_k141_30650\|\|lt2gene | 1163 | No | NA | 1 | 0 | 0 | Not-determined | Genome-fragment | NA | NA | 0 |
| S1-final-viral-combined_k141_338458\|\|full | 1561 | No | NA | 3 | 0 | 0 | Low-quality | Genome-fragment | 4 | AAI-based (high-confidence) | 0 |
| S1_k141_140084 | 1561 | No | NA | 1 | 0 | 1 | Low-quality | Genome-fragment | 0.56 | AAI-based (medium-confidence) | 0 |
| S12-final-viral-combined_k141_84361\|\|lt2gene | 1561 | No | NA | 2 | 1 | 0 | Low-quality | Genome-fragment | 2.1 | HMM-based (lower-bound) | 0 |
| S12-final-viral-combined_k141_221061\|\|lt2gene | 1561 | No | NA | 1 | 1 | 0 | Low-quality | Genome-fragment | 6.92 | AAI-based (high-confidence) | 0 |
| S4_k141_125427 | 1561 | No | NA | 5 | 0 | 0 | Low-quality | Genome-fragment | 1.79 | AAI-based (high-confidence) | 0 |
| S4_k141_27720 | 1561 | No | NA | 1 | 0 | 0 | Low-quality | Genome-fragment | 0.29 | AAI-based (medium-confidence) | 0 |
| S3-final-viral-combined_k141_25515\|\|lt2gene | 1421 | No | NA | 2 | 0 | 1 | Not-determined | Genome-fragment | NA | NA | 0 |
| S11-final-viral-combined_k141_137926\|\|full | 1562 | No | NA | 4 | 1 | 0 | Low-quality | Genome-fragment | 1.42 | AAI-based (medium-confidence) | 0 |
| S3-final-viral-combined_k141_65870\|\|lt2gene | 2322 | No | NA | 2 | 0 | 1 | Not-determined | Genome-fragment | NA | NA | 0 |
| S1_k141_373037 | 1562 | No | NA | 5 | 0 | 2 | Low-quality | Genome-fragment | 2.73 | AAI-based (medium-confidence) | 0 |
| S3-final-viral-combined_k141_16283\|\|full | 1562 | No | NA | 4 | 0 | 0 | Low-quality | Genome-fragment | 3.68 | AAI-based (medium-confidence) | 0 |
| S5-final-viral-combined_k141_4939\|\|full | 1562 | No | NA | 3 | 0 | 0 | Low-quality | Genome-fragment | 2.63 | AAI-based (medium-confidence) | 0 |
| S7-final-viral-combined_k141_233904\|\|full | 1562 | No | NA | 4 | 0 | 0 | Low-quality | Genome-fragment | 0.74 | AAI-based (medium-confidence) | 0 |
| S3-final-viral-combined_k141_112528\|\|lt2gene | 1060 | No | NA | 2 | 0 | 2 | Not-determined | Genome-fragment | NA | NA | 0 |
| S7_k141_81327 | 1562 | No | NA | 3 | 0 | 2 | Low-quality | Genome-fragment | 3.32 | AAI-based (medium-confidence) | 0 |
| S12-final-viral-combined_k141_180386\|\|full | 1562 | No | NA | 3 | 0 | 0 | Low-quality | Genome-fragment | 3.8 | AAI-based (high-confidence) | 0 |
| S8-final-viral-combined_k141_47265\|\|full | 1562 | No | NA | 3 | 0 | 1 | Low-quality | Genome-fragment | 1.3 | AAI-based (medium-confidence) | 0 |
| S3-final-viral-combined_k141_43481\|\|lt2gene | 1169 | No | NA | 1 | 0 | 1 | Not-determined | Genome-fragment | NA | NA | 0 |
| S9-final-viral-combined_k141_19595\|\|full | 1563 | No | NA | 2 | 0 | 0 | Low-quality | Genome-fragment | 1.64 | AAI-based (high-confidence) | 0 |
| S9-final-viral-combined_k141_73709\|\|lt2gene | 1563 | No | NA | 1 | 1 | 0 | Low-quality | Genome-fragment | 3.6 | AAI-based (medium-confidence) | 0 |
| S3-final-viral-combined_k141_118905\|\|lt2gene | 1437 | No | NA | 2 | 0 | 0 | Not-determined | Genome-fragment | NA | NA | 0 |
| S11-final-viral-combined_k141_89341\|\|full | 1563 | No | NA | 2 | 1 | 0 | Low-quality | Genome-fragment | 4.17 | AAI-based (high-confidence) | 0 |
| S7-final-viral-combined_k141_95413\|\|full | 1563 | No | NA | 3 | 0 | 0 | Low-quality | Genome-fragment | 1.52 | AAI-based (medium-confidence) | 0 |
| S3-final-viral-combined_k141_95920\|\|lt2gene | 2607 | No | NA | 2 | 0 | 1 | Not-determined | Genome-fragment | NA | NA | 0 |
| S2-final-viral-combined_k141_225281\|\|full | 1563 | No | NA | 3 | 0 | 0 | Low-quality | Genome-fragment | 2.68 | AAI-based (medium-confidence) | 0 |
| S11-final-viral-combined_k141_50431\|\|full | 1564 | No | NA | 4 | 0 | 0 | Low-quality | Genome-fragment | 3.33 | AAI-based (medium-confidence) | 0 |
| S11-final-viral-combined_k141_37720\|\|full | 1564 | No | NA | 3 | 0 | 0 | Low-quality | Genome-fragment | 2.74 | AAI-based (high-confidence) | 0 |
| S3-final-viral-combined_k141_84623\|\|lt2gene | 1204 | No | NA | 2 | 0 | 0 | Not-determined | Genome-fragment | NA | NA | 0 |
| S1-final-viral-combined_k141_314856\|\|full | 1564 | No | NA | 2 | 0 | 2 | Low-quality | Genome-fragment | 1.46 | AAI-based (medium-confidence) | 0 |
| S3-final-viral-combined_k141_27067\|\|lt2gene | 2507 | No | NA | 2 | 0 | 2 | Not-determined | Genome-fragment | NA | NA | 0 |
| S8-final-viral-combined_k141_116170\|\|full | 1564 | No | NA | 4 | 1 | 0 | Low-quality | Genome-fragment | 3.66 | AAI-based (high-confidence) | 0 |
| S8-final-viral-combined_k141_31856\|\|full | 1564 | No | NA | 4 | 2 | 0 | Low-quality | Genome-fragment | 3.43 | AAI-based (medium-confidence) | 0 |
| S3-final-viral-combined_k141_113636\|\|lt2gene | 1344 | No | NA | 2 | 0 | 0 | Not-determined | Genome-fragment | NA | NA | 0 |
| S3-final-viral-combined_k141_131250\|\|lt2gene | 1005 | No | NA | 2 | 0 | 0 | Not-determined | Genome-fragment | NA | NA | 0 |
| S8-final-viral-combined_k141_147315\|\|lt2gene | 1564 | No | NA | 2 | 1 | 0 | Low-quality | Genome-fragment | 33.23 | AAI-based (high-confidence) | 0 |
| S11-final-viral-combined_k141_304635\|\|full | 1565 | No | NA | 3 | 0 | 1 | Low-quality | Genome-fragment | 1.1 | AAI-based (medium-confidence) | 0 |
| S11-final-viral-combined_k141_326277\|\|full | 1565 | No | NA | 3 | 0 | 1 | Low-quality | Genome-fragment | 2.59 | AAI-based (high-confidence) | 0 |
| S1_k141_249439 | 1565 | No | NA | 4 | 0 | 0 | Low-quality | Genome-fragment | 4.68 | AAI-based (high-confidence) | 0 |
| S3-final-viral-combined_k141_4932\|\|lt2gene | 1148 | No | NA | 1 | 0 | 0 | Not-determined | Genome-fragment | NA | NA | 0 |
| S3-final-viral-combined_k141_125751\|\|lt2gene | 1110 | No | NA | 1 | 0 | 1 | Not-determined | Genome-fragment | NA | NA | 0 |
| S3-final-viral-combined_k141_36537\|\|lt2gene | 1565 | No | NA | 2 | 0 | 1 | Low-quality | Genome-fragment | 0.42 | AAI-based (medium-confidence) | 0 |
| S3-final-viral-combined_k141_91230\|\|lt2gene | 1002 | No | NA | 1 | 0 | 0 | Not-determined | Genome-fragment | NA | NA | 0 |
| S3_k141_19178 | 1565 | No | NA | 2 | 0 | 0 | Low-quality | Genome-fragment | 1.94 | AAI-based (high-confidence) | 0 |
| S7-final-viral-combined_k141_148973\|\|full | 1565 | No | NA | 2 | 2 | 0 | Low-quality | Genome-fragment | 3.76 | AAI-based (high-confidence) | 0 |
| S17-final-viral-combined_k141_200684\|\|full | 1565 | No | NA | 2 | 1 | 0 | Low-quality | Genome-fragment | 1.05 | HMM-based (lower-bound) | 0 |
| S17-final-viral-combined_k141_223554\|\|full | 1565 | No | NA | 3 | 0 | 2 | Low-quality | Genome-fragment | 4.71 | AAI-based (medium-confidence) | 0 |
| S3-final-viral-combined_k141_85632\|\|lt2gene | 3003 | No | NA | 2 | 0 | 0 | Not-determined | Genome-fragment | NA | NA | 0 |
| S12-final-viral-combined_k141_368652\|\|full | 1565 | No | NA | 4 | 1 | 0 | Low-quality | Genome-fragment | 3.58 | AAI-based (high-confidence) | 0 |
| S12-final-viral-combined_k141_51662\|\|full | 1565 | No | NA | 2 | 0 | 2 | Low-quality | Genome-fragment | 1.79 | AAI-based (medium-confidence) | 0 |
| S3-final-viral-combined_k141_33852\|\|lt2gene | 1127 | No | NA | 1 | 0 | 0 | Not-determined | Genome-fragment | NA | NA | 0 |
| S12-final-viral-combined_k141_65512\|\|full | 1565 | No | NA | 2 | 0 | 0 | Low-quality | Genome-fragment | 3.1 | AAI-based (high-confidence) | 0 |
| S12_k141_371448 | 1565 | No | NA | 2 | 0 | 1 | Low-quality | Genome-fragment | 0.39 | AAI-based (medium-confidence) | 0 |
| S8-final-viral-combined_k141_156453\|\|full | 1565 | No | NA | 3 | 1 | 0 | Low-quality | Genome-fragment | 3.34 | AAI-based (high-confidence) | 0 |
| S3-final-viral-combined_k141_45962\|\|lt2gene | 1795 | No | NA | 2 | 0 | 2 | Not-determined | Genome-fragment | NA | NA | 0 |
| S3_k141_86259 | 2004 | No | NA | 1 | 0 | 1 | Not-determined | Genome-fragment | NA | NA | 0 |
| S8-final-viral-combined_k141_210053\|\|lt2gene | 1565 | No | NA | 1 | 0 | 0 | Low-quality | Genome-fragment | 3.52 | AAI-based (medium-confidence) | 0 |
| S3_k141_51769 | 4099 | No | NA | 4 | 0 | 4 | Not-determined | Genome-fragment | NA | NA | 0 |
| S3_k141_126527 | 1550 | No | NA | 1 | 0 | 1 | Not-determined | Genome-fragment | NA | NA | 0 |
| S3_k141_57527 | 1043 | No | NA | 1 | 0 | 0 | Not-determined | Genome-fragment | NA | NA | 0 |
| S3_k141_34551 | 1769 | No | NA | 3 | 0 | 1 | Not-determined | Genome-fragment | NA | NA | 0 |
| S3_k141_109316 | 1173 | No | NA | 2 | 0 | 1 | Not-determined | Genome-fragment | NA | NA | 0 |
| S9-final-viral-combined_k141_75255\|\|full | 1566 | No | NA | 4 | 1 | 0 | Low-quality | Genome-fragment | 4 | AAI-based (high-confidence) | 0 |
| S3_k141_74819 | 1525 | No | NA | 2 | 0 | 1 | Not-determined | Genome-fragment | NA | NA | 0 |
| S3_k141_74833 | 4070 | No | NA | 5 | 0 | 3 | Not-determined | Genome-fragment | NA | NA | 0 |
| S3_k141_11593 | 9989 | No | NA | 10 | 0 | 5 | Not-determined | Genome-fragment | NA | NA | 0 |
| S11-final-viral-combined_k141_54761\|\|full | 1566 | No | NA | 3 | 1 | 0 | Low-quality | Genome-fragment | 3.85 | AAI-based (high-confidence) | 0 |
| S3_k141_97870 | 1578 | No | NA | 2 | 0 | 0 | Not-determined | Genome-fragment | NA | NA | 0 |
| S3_k141_51860 | 1019 | No | NA | 2 | 0 | 0 | Not-determined | Genome-fragment | NA | NA | 0 |
| S3_k141_115166 | 2240 | No | NA | 2 | 0 | 0 | Not-determined | Genome-fragment | NA | NA | 0 |
| S1-final-viral-combined_k141_63465\|\|full | 1566 | No | NA | 3 | 0 | 2 | Low-quality | Genome-fragment | 4.71 | AAI-based (medium-confidence) | 0 |
| S3_k141_69134 | 1511 | No | NA | 1 | 0 | 1 | Not-determined | Genome-fragment | NA | NA | 0 |
| S3_k141_115184 | 1000 | No | NA | 1 | 0 | 0 | Not-determined | Genome-fragment | NA | NA | 0 |
| S3-final-viral-combined_k141_102513\|\|full | 1566 | No | NA | 3 | 0 | 0 | Low-quality | Genome-fragment | 2.92 | AAI-based (high-confidence) | 0 |
| S3_k141_5887 | 3200 | No | NA | 4 | 0 | 1 | Not-determined | Genome-fragment | NA | NA | 0 |
| S3_k141_115210 | 1377 | No | NA | 1 | 0 | 1 | Not-determined | Genome-fragment | NA | NA | 0 |
| S3_k141_92145 | 1062 | No | NA | 2 | 0 | 0 | Not-determined | Genome-fragment | NA | NA | 0 |
| S3_k141_97925 | 1751 | No | NA | 3 | 0 | 2 | Not-determined | Genome-fragment | NA | NA | 0 |
| S5-final-viral-combined_k141_87611\|\|full | 1566 | No | NA | 3 | 2 | 0 | Low-quality | Genome-fragment | 4.16 | AAI-based (high-confidence) | 0 |
| S3_k141_17410 | 4723 | No | NA | 3 | 0 | 2 | Not-determined | Genome-fragment | NA | NA | 0 |
| S3_k141_132474 | 1984 | No | NA | 1 | 0 | 0 | Not-determined | Genome-fragment | NA | NA | 0 |
| S5_k141_189852 | 1566 | No | NA | 3 | 0 | 0 | Low-quality | Genome-fragment | 1.3 | AAI-based (medium-confidence) | 0 |
| S3_k141_109483 | 1211 | No | NA | 2 | 0 | 0 | Not-determined | Genome-fragment | NA | NA | 0 |
| S7-final-viral-combined_k141_162279\|\|lt2gene | 1566 | No | NA | 1 | 0 | 0 | Low-quality | Genome-fragment | 2.91 | AAI-based (medium-confidence) | 0 |
| S3_k141_103719 | 1006 | No | NA | 1 | 0 | 1 | Not-determined | Genome-fragment | NA | NA | 0 |
| S3_k141_5983 | 1897 | No | NA | 2 | 0 | 2 | Not-determined | Genome-fragment | NA | NA | 0 |
| S12-final-viral-combined_k141_377957\|\|full | 1566 | No | NA | 3 | 0 | 1 | Low-quality | Genome-fragment | 2.43 | AAI-based (medium-confidence) | 0 |
| S3_k141_115301 | 1433 | No | NA | 2 | 0 | 1 | Not-determined | Genome-fragment | NA | NA | 0 |
| S3_k141_121038 | 4885 | No | NA | 3 | 0 | 3 | Not-determined | Genome-fragment | NA | NA | 0 |
| S12-final-viral-combined_k141_46328\|\|full | 1566 | No | NA | 3 | 0 | 3 | Low-quality | Genome-fragment | 0.46 | AAI-based (medium-confidence) | 0 |
| S3_k141_52022 | 1631 | No | NA | 3 | 0 | 3 | Not-determined | Genome-fragment | NA | NA | 0 |
| S3_k141_92261 | 1361 | No | NA | 1 | 0 | 1 | Not-determined | Genome-fragment | NA | NA | 0 |
| S3_k141_6033 | 1646 | No | NA | 2 | 0 | 1 | Not-determined | Genome-fragment | NA | NA | 0 |
| S3_k141_34785 | 1338 | No | NA | 1 | 0 | 1 | Not-determined | Genome-fragment | NA | NA | 0 |
| S3_k141_92270 | 1120 | No | NA | 1 | 0 | 0 | Not-determined | Genome-fragment | NA | NA | 0 |
| S12_k141_394057 | 1566 | No | NA | 4 | 0 | 0 | Low-quality | Genome-fragment | 2.72 | AAI-based (high-confidence) | 0 |
| S3_k141_57705 | 3428 | No | NA | 3 | 0 | 0 | Not-determined | Genome-fragment | NA | NA | 0 |
| S3_k141_103795 | 1256 | No | NA | 1 | 0 | 0 | Not-determined | Genome-fragment | NA | NA | 0 |
| S4_k141_146218 | 1566 | No | NA | 2 | 0 | 1 | Low-quality | Genome-fragment | 3.17 | AAI-based (medium-confidence) | 0 |
| S3_k141_52108 | 4798 | No | NA | 4 | 0 | 1 | Not-determined | Genome-fragment | NA | NA | 0 |
| S3_k141_86731 | 1753 | No | NA | 1 | 0 | 0 | Not-determined | Genome-fragment | NA | NA | 0 |
| S3_k141_121098 | 1216 | No | NA | 1 | 0 | 1 | Not-determined | Genome-fragment | NA | NA | 0 |
| S3_k141_109708 | 3557 | No | NA | 2 | 0 | 1 | Not-determined | Genome-fragment | NA | NA | 0 |
| S3_k141_29070 | 1370 | No | NA | 2 | 0 | 1 | Not-determined | Genome-fragment | NA | NA | 0 |
| S6-final-viral-combined_k141_42659\|\|full | 1566 | No | NA | 2 | 0 | 0 | Low-quality | Genome-fragment | 1.95 | AAI-based (high-confidence) | 0 |
| S1_k141_246302 | 1567 | No | NA | 4 | 0 | 1 | Low-quality | Genome-fragment | 4.01 | AAI-based (medium-confidence) | 0 |
| S3_k141_126943 | 1258 | No | NA | 1 | 0 | 0 | Not-determined | Genome-fragment | NA | NA | 0 |
| S3_k141_98089 | 1372 | No | NA | 2 | 0 | 0 | Not-determined | Genome-fragment | NA | NA | 0 |
| S3_k141_40612 | 1868 | No | NA | 1 | 0 | 1 | Not-determined | Genome-fragment | NA | NA | 0 |
| S3_k141_17590 | 1319 | No | NA | 2 | 0 | 0 | Not-determined | Genome-fragment | NA | NA | 0 |
| S3-final-viral-combined_k141_60968\|\|full | 1567 | No | NA | 3 | 0 | 2 | Low-quality | Genome-fragment | 1.18 | AAI-based (medium-confidence) | 0 |
| S3_k141_11838 | 1237 | No | NA | 2 | 0 | 2 | Not-determined | Genome-fragment | NA | NA | 0 |
| S3_k141_69442 | 1068 | No | NA | 1 | 0 | 0 | Not-determined | Genome-fragment | NA | NA | 0 |
| S3_k141_23503 | 1202 | No | NA | 1 | 0 | 0 | Not-determined | Genome-fragment | NA | NA | 0 |
| S5-final-viral-combined_k141_11866\|\|lt2gene | 1567 | No | NA | 2 | 2 | 0 | Low-quality | Genome-fragment | 4.63 | AAI-based (medium-confidence) | 0 |
| S3_k141_103941 | 2904 | No | NA | 1 | 0 | 1 | Not-determined | Genome-fragment | NA | NA | 0 |
| S2_k141_192129 | 1567 | No | NA | 1 | 0 | 1 | Low-quality | Genome-fragment | 1.59 | AAI-based (medium-confidence) | 0 |
| S3_k141_63688 | 1393 | No | NA | 1 | 0 | 0 | Not-determined | Genome-fragment | NA | NA | 0 |
| S3_k141_11882 | 1145 | No | NA | 2 | 0 | 0 | Not-determined | Genome-fragment | NA | NA | 0 |
| S3_k141_5683 | 1568 | No | NA | 6 | 2 | 0 | Low-quality | Genome-fragment | 2.52 | AAI-based (high-confidence) | 0 |
| S3_k141_63704 | 3325 | No | NA | 4 | 0 | 3 | Not-determined | Genome-fragment | NA | NA | 0 |
| S3_k141_52215 | 1638 | No | NA | 2 | 0 | 1 | Not-determined | Genome-fragment | NA | NA | 0 |
| S3_k141_52225 | 1108 | No | NA | 2 | 0 | 1 | Not-determined | Genome-fragment | NA | NA | 0 |
| S3_k141_6283 | 3318 | No | NA | 5 | 0 | 1 | Not-determined | Genome-fragment | NA | NA | 0 |
| S3_k141_92530 | 10075 | No | NA | 8 | 0 | 3 | Not-determined | Genome-fragment | NA | NA | 0 |
| S5-final-viral-combined_k141_30464\|\|full | 1568 | No | NA | 4 | 1 | 0 | Low-quality | Genome-fragment | 3.5 | AAI-based (high-confidence) | 0 |
| S3_k141_29207 | 1054 | No | NA | 1 | 0 | 0 | Not-determined | Genome-fragment | NA | NA | 0 |
| S3_k141_40757 | 1569 | No | NA | 1 | 0 | 1 | Not-determined | Genome-fragment | NA | NA | 0 |
| S3_k141_75231 | 16246 | No | NA | 26 | 0 | 1 | Not-determined | Genome-fragment | NA | NA | 0 |
| S17-final-viral-combined_k141_235632\|\|full | 1568 | No | NA | 2 | 0 | 0 | Low-quality | Genome-fragment | 2.79 | AAI-based (high-confidence) | 0 |
| S3_k141_115604 | 1422 | No | NA | 2 | 0 | 1 | Not-determined | Genome-fragment | NA | NA | 0 |
| S3_k141_627 | 1101 | No | NA | 3 | 0 | 0 | Not-determined | Genome-fragment | NA | NA | 0 |
| S2-final-viral-combined_k141_84876\|\|lt2gene | 1568 | No | NA | 2 | 2 | 0 | Low-quality | Genome-fragment | 3.16 | AAI-based (high-confidence) | 0 |
| S4_k141_149556 | 1568 | No | NA | 5 | 0 | 0 | Low-quality | Genome-fragment | 2.95 | AAI-based (medium-confidence) | 0 |
| S3_k141_132903 | 1455 | No | NA | 2 | 0 | 0 | Not-determined | Genome-fragment | NA | NA | 0 |
| S8-final-viral-combined_k141_3482\|\|full | 1568 | No | NA | 3 | 1 | 0 | Low-quality | Genome-fragment | 2.81 | AAI-based (high-confidence) | 0 |
| S9-final-viral-combined_k141_64108\|\|lt2gene | 1569 | No | NA | 1 | 1 | 0 | Low-quality | Genome-fragment | 3.49 | AAI-based (high-confidence) | 0 |
| S3_k141_6415 | 1785 | No | NA | 2 | 0 | 1 | Not-determined | Genome-fragment | NA | NA | 0 |
| S3_k141_6426 | 1020 | No | NA | 1 | 0 | 1 | Not-determined | Genome-fragment | NA | NA | 0 |
| S3_k141_40886 | 1006 | No | NA | 1 | 0 | 0 | Not-determined | Genome-fragment | NA | NA | 0 |
| S3_k141_12021 | 1762 | No | NA | 2 | 0 | 2 | Not-determined | Genome-fragment | NA | NA | 0 |
| S3_k141_40900 | 1124 | No | NA | 3 | 0 | 2 | Not-determined | Genome-fragment | NA | NA | 0 |
| S3_k141_46718 | 1877 | No | NA | 2 | 0 | 1 | Not-determined | Genome-fragment | NA | NA | 0 |

| | | | | | | | | | | | |
|---|---|---|---|---|---|---|---|---|---|---|---|
| S3_k141_732 | 3069 | No | NA | 2 | 0 | 2 | Not-determined | Genome-fragment | NA | NA | 0 |
| S9_k141_45850 | 1569 | No | NA | 1 | 0 | 1 | Low-quality | Genome-fragment | 3.34 | AAI-based (high-confidence) | 0 |
| S1-final-viral-combined_k141_10704\|\|full | 1569 | No | NA | 2 | 0 | 0 | Low-quality | Genome-fragment | 3.68 | AAI-based (medium-confidence) | 0 |
| S1-final-viral-combined_k141_293288\|\|full | 1569 | No | NA | 5 | 0 | 0 | Low-quality | Genome-fragment | 2.56 | AAI-based (high-confidence) | 0 |
| S7-final-viral-combined_k141_165039\|\|full | 1569 | No | NA | 2 | 0 | 0 | Low-quality | Genome-fragment | 2.92 | AAI-based (medium-confidence) | 0 |
| S3_k141_121422 | 1018 | No | NA | 2 | 0 | 0 | Not-determined | Genome-fragment | NA | NA | 0 |
| S3_k141_133005 | 1016 | No | NA | 1 | 0 | 0 | Not-determined | Genome-fragment | NA | NA | 0 |
| S3_k141_75401 | 1536 | No | NA | 2 | 0 | 1 | Not-determined | Genome-fragment | NA | NA | 0 |
| S3_k141_29348 | 1006 | No | NA | 2 | 0 | 2 | Not-determined | Genome-fragment | NA | NA | 0 |
| S3_k141_98324 | 1366 | No | NA | 2 | 0 | 0 | Not-determined | Genome-fragment | NA | NA | 0 |
| S3_k141_87115 | 4103 | No | NA | 1 | 0 | 0 | Not-determined | Genome-fragment | NA | NA | 0 |
| S2-final-viral-combined_k141_130653\|\|full | 1569 | No | NA | 3 | 0 | 1 | Low-quality | Genome-fragment | 3.6 | AAI-based (high-confidence) | 0 |
| S3_k141_92710 | 1253 | No | NA | 1 | 0 | 1 | Not-determined | Genome-fragment | NA | NA | 0 |
| S3_k141_40988 | 1067 | No | NA | 2 | 0 | 2 | Not-determined | Genome-fragment | NA | NA | 0 |
| S9_k141_86742 | 1570 | No | NA | 2 | 1 | 0 | Low-quality | Genome-fragment | 3.63 | AAI-based (high-confidence) | 0 |
| S9_k141_20395 | 1570 | No | NA | 4 | 0 | 0 | Low-quality | Genome-fragment | 3.85 | AAI-based (medium-confidence) | 0 |
| S3_k141_29425 | 3390 | No | NA | 4 | 0 | 2 | Not-determined | Genome-fragment | NA | NA | 0 |
| S3_k141_92755 | 1131 | No | NA | 1 | 0 | 0 | Not-determined | Genome-fragment | NA | NA | 0 |
| S5-final-viral-combined_k141_30651\|\|full | 1570 | No | NA | 2 | 0 | 0 | Low-quality | Genome-fragment | 0.88 | AAI-based (high-confidence) | 0 |
| S3_k141_64014 | 1940 | No | NA | 2 | 0 | 1 | Not-determined | Genome-fragment | NA | NA | 0 |
| S3_k141_115780 | 3529 | No | NA | 5 | 0 | 4 | Not-determined | Genome-fragment | NA | NA | 0 |
| S3_k141_92774 | 2696 | No | NA | 3 | 0 | 2 | Not-determined | Genome-fragment | NA | NA | 0 |
| S3_k141_81359 | 1757 | No | NA | 2 | 0 | 0 | Not-determined | Genome-fragment | NA | NA | 0 |
| S3_k141_58337 | 1204 | No | NA | 1 | 0 | 1 | Not-determined | Genome-fragment | NA | NA | 0 |
| S3_k141_46906 | 1383 | No | NA | 1 | 0 | 1 | Not-determined | Genome-fragment | NA | NA | 0 |
| S3_k141_18046 | 1154 | No | NA | 1 | 0 | 0 | Not-determined | Genome-fragment | NA | NA | 0 |
| S3_k141_64076 | 1226 | No | NA | 2 | 0 | 1 | Not-determined | Genome-fragment | NA | NA | 0 |
| S3_k141_98436 | 1422 | No | NA | 2 | 0 | 0 | Not-determined | Genome-fragment | NA | NA | 0 |
| S3_k141_69846 | 4971 | No | NA | 6 | 0 | 2 | Not-determined | Genome-fragment | NA | NA | 0 |
| S3_k141_133160 | 3465 | No | NA | 2 | 0 | 2 | Not-determined | Genome-fragment | NA | NA | 0 |
| S3_k141_104372 | 1147 | No | NA | 1 | 0 | 1 | Not-determined | Genome-fragment | NA | NA | 0 |
| S3_k141_75598 | 1110 | No | NA | 1 | 0 | 1 | Not-determined | Genome-fragment | NA | NA | 0 |
| S5-final-viral-combined_k141_157659\|\|full | 1570 | No | NA | 3 | 1 | 0 | Low-quality | Genome-fragment | 2.9 | AAI-based (high-confidence) | 0 |
| S7_k141_225751 | 1570 | No | NA | 2 | 0 | 1 | Low-quality | Genome-fragment | 1.68 | AAI-based (medium-confidence) | 0 |
| S3_k141_121584 | 2204 | No | NA | 2 | 0 | 2 | Not-determined | Genome-fragment | NA | NA | 0 |
| S3_k141_115862 | 2696 | No | NA | 3 | 0 | 3 | Not-determined | Genome-fragment | NA | NA | 0 |
| S3_k141_64149 | 1103 | No | NA | 1 | 0 | 1 | Not-determined | Genome-fragment | NA | NA | 0 |
| S3_k141_127562 | 2331 | No | NA | 2 | 0 | 1 | Not-determined | Genome-fragment | NA | NA | 0 |
| S3_k141_98484 | 1104 | No | NA | 2 | 0 | 2 | Not-determined | Genome-fragment | NA | NA | 0 |
| S3_k141_29552 | 1001 | No | NA | 1 | 0 | 1 | Not-determined | Genome-fragment | NA | NA | 0 |
| S3_k141_104436 | 1212 | No | NA | 2 | 0 | 1 | Not-determined | Genome-fragment | NA | NA | 0 |
| S3_k141_29575 | 2110 | No | NA | 2 | 0 | 0 | Not-determined | Genome-fragment | NA | NA | 0 |
| S3_k141_12247 | 4745 | No | NA | 3 | 0 | 2 | Not-determined | Genome-fragment | NA | NA | 0 |
| S3_k141_23958 | 35821 | No | NA | 51 | 16 | 1 | High-quality | High-quality | 100 | AAI-based (high-confidence) | 0 |
| S3_k141_29610 | 1533 | No | NA | 2 | 0 | 1 | Not-determined | Genome-fragment | NA | NA | 0 |
| S3_k141_29612 | 2467 | No | NA | 2 | 0 | 0 | Not-determined | Genome-fragment | NA | NA | 0 |
| S3_k141_127631 | 2706 | No | NA | 2 | 0 | 0 | Not-determined | Genome-fragment | NA | NA | 0 |
| S3_k141_6777 | 1064 | No | NA | 1 | 0 | 1 | Not-determined | Genome-fragment | NA | NA | 0 |
| S3_k141_64225 | 1306 | No | NA | 1 | 0 | 1 | Not-determined | Genome-fragment | NA | NA | 0 |
| S3_k141_115923 | 1159 | No | NA | 1 | 0 | 1 | Not-determined | Genome-fragment | NA | NA | 0 |
| S6-final-viral-combined_k141_117910\|\|lt2gene | 1570 | No | NA | 1 | 0 | 0 | Low-quality | Genome-fragment | 2.67 | AAI-based (high-confidence) | 0 |
| S8-final-viral-combined_k141_54836\|\|full | 1570 | No | NA | 3 | 2 | 0 | Low-quality | Genome-fragment | 2.26 | HMM-based (lower-bound) | 0 |
| S3_k141_127654 | 1635 | No | NA | 3 | 0 | 3 | Not-determined | Genome-fragment | NA | NA | 0 |
| S3_k141_1094 | 4398 | No | NA | 4 | 0 | 1 | Not-determined | Genome-fragment | NA | NA | 0 |
| S3_k141_64262 | 1387 | No | NA | 1 | 0 | 1 | Not-determined | Genome-fragment | NA | NA | 0 |
| S3_k141_115967 | 2381 | No | NA | 1 | 0 | 1 | Not-determined | Genome-fragment | NA | NA | 0 |
| S3_k141_29652 | 5399 | No | NA | 5 | 0 | 4 | Not-determined | Genome-fragment | NA | NA | 0 |
| S1-final-viral-combined_k141_36556\|\|full | 1571 | No | NA | 3 | 1 | 0 | Low-quality | Genome-fragment | 2.51 | AAI-based (high-confidence) | 0 |
| S3_k141_29660 | 3664 | No | NA | 3 | 0 | 3 | Not-determined | Genome-fragment | NA | NA | 0 |
| S3_k141_52820 | 1572 | No | NA | 3 | 0 | 0 | Not-determined | Genome-fragment | NA | NA | 0 |
| S3_k141_93007 | 1291 | No | NA | 2 | 0 | 0 | Not-determined | Genome-fragment | NA | NA | 0 |
| S5-final-viral-combined_k141_32936\|\|full | 1571 | No | NA | 3 | 0 | 0 | Low-quality | Genome-fragment | 3.31 | AAI-based (medium-confidence) | 0 |
| S3_k141_81594 | 1085 | No | NA | 2 | 0 | 0 | Not-determined | Genome-fragment | NA | NA | 0 |
| S5-final-viral-combined_k141_122754\|\|full | 1571 | Yes | 1091 | 4 | 1 | 1 | Low-quality | Genome-fragment | 2.45 | AAI-based (high-confidence) | 30.55 |
| S3_k141_1167 | 3130 | No | NA | 3 | 0 | 3 | Not-determined | Genome-fragment | NA | NA | 0 |
| S7-final-viral-combined_k141_114092\|\|full | 1571 | No | NA | 3 | 0 | 0 | Low-quality | Genome-fragment | 3.3 | AAI-based (medium-confidence) | 0 |
| S17-final-viral-combined_k141_43559\|\|full | 1571 | No | NA | 3 | 0 | 2 | Low-quality | Genome-fragment | 0.51 | AAI-based (medium-confidence) | 0 |
| S3_k141_24060 | 1177 | No | NA | 2 | 0 | 2 | Not-determined | Genome-fragment | NA | NA | 0 |
| S3_k141_133371 | 1225 | No | NA | 1 | 0 | 0 | Not-determined | Genome-fragment | NA | NA | 0 |
| S12-final-viral-combined_k141_326159\|\|full | 1571 | No | NA | 3 | 1 | 0 | Low-quality | Genome-fragment | 2.33 | HMM-based (lower-bound) | 0 |
| S3_k141_81652 | 1077 | No | NA | 1 | 0 | 1 | Not-determined | Genome-fragment | NA | NA | 0 |
| S4-final-viral-combined_k141_30088\|\|full | 1571 | No | NA | 4 | 0 | 0 | Low-quality | Genome-fragment | 2.94 | AAI-based (high-confidence) | 0 |
| S3_k141_93066 | 1022 | No | NA | 1 | 0 | 1 | Not-determined | Genome-fragment | NA | NA | 0 |
| S8_k141_254010 | 1571 | No | NA | 2 | 1 | 0 | Low-quality | Genome-fragment | 3.83 | AAI-based (high-confidence) | 0 |
| S8_k141_185532 | 1571 | No | NA | 2 | 0 | 1 | Low-quality | Genome-fragment | 3.38 | AAI-based (medium-confidence) | 0 |
| S9-final-viral-combined_k141_100073\|\|full | 1572 | No | NA | 3 | 2 | 0 | Low-quality | Genome-fragment | 2.96 | AAI-based (high-confidence) | 0 |
| S3_k141_127785 | 1126 | No | NA | 2 | 0 | 1 | Not-determined | Genome-fragment | NA | NA | 0 |
| S11-final-viral-combined_k141_191295\|\|full | 1572 | No | NA | 3 | 0 | 1 | Low-quality | Genome-fragment | 5.02 | AAI-based (medium-confidence) | 0 |
| S11_k141_32112 | 1572 | No | NA | 1 | 0 | 1 | Low-quality | Genome-fragment | 2.77 | AAI-based (medium-confidence) | 0 |
| S11_k141_20883 | 1572 | No | NA | 1 | 0 | 1 | Low-quality | Genome-fragment | 0.59 | AAI-based (high-confidence) | 0 |
| S7-final-viral-combined_k141_207044\|\|lt2gene | 1572 | No | NA | 2 | 1 | 0 | Low-quality | Genome-fragment | 3.19 | AAI-based (medium-confidence) | 0 |
| S2-final-viral-combined_k141_199927\|\|full | 1572 | No | NA | 3 | 0 | 0 | Low-quality | Genome-fragment | 2.28 | AAI-based (medium-confidence) | 0 |
| S3_k141_41372 | 1279 | No | NA | 2 | 0 | 0 | Not-determined | Genome-fragment | NA | NA | 0 |
| S3_k141_18294 | 3355 | No | NA | 5 | 0 | 4 | Not-determined | Genome-fragment | NA | NA | 0 |
| S3_k141_41405 | 5120 | No | NA | 5 | 0 | 5 | Not-determined | Genome-fragment | NA | NA | 0 |
| S3_k141_81776 | 1161 | No | NA | 1 | 0 | 1 | Not-determined | Genome-fragment | NA | NA | 0 |
| S3_k141_127911 | 1111 | No | NA | 1 | 0 | 1 | Not-determined | Genome-fragment | NA | NA | 0 |
| S3_k141_24268 | 1116 | No | NA | 1 | 0 | 1 | Not-determined | Genome-fragment | NA | NA | 0 |
| S3_k141_1287 | 2131 | No | NA | 3 | 0 | 1 | Not-determined | Genome-fragment | NA | NA | 0 |
| S3_k141_133549 | 1014 | No | NA | 3 | 0 | 1 | Not-determined | Genome-fragment | NA | NA | 0 |
| S6_k141_134207 | 1572 | No | NA | 3 | 0 | 0 | Low-quality | Genome-fragment | 1.54 | AAI-based (high-confidence) | 0 |
| S3_k141_58779 | 1544 | No | NA | 3 | 0 | 2 | Not-determined | Genome-fragment | NA | NA | 0 |
| S3_k141_64551 | 1186 | No | NA | 1 | 0 | 0 | Not-determined | Genome-fragment | NA | NA | 0 |
| S9-final-viral-combined_k141_109774\|\|full | 1573 | No | NA | 2 | 0 | 1 | Low-quality | Genome-fragment | 1.8 | AAI-based (medium-confidence) | 0 |
| S11-final-viral-combined_k141_16717\|\|full | 1573 | No | NA | 3 | 3 | 0 | Low-quality | Genome-fragment | 3.93 | AAI-based (medium-confidence) | 0 |
| S3_k141_47371 | 2805 | No | NA | 2 | 0 | 0 | Not-determined | Genome-fragment | NA | NA | 0 |
| S3_k141_58798 | 1229 | No | NA | 1 | 0 | 0 | Not-determined | Genome-fragment | NA | NA | 0 |
| S11-final-viral-combined_k141_228759\|\|full | 1573 | No | NA | 2 | 0 | 1 | Low-quality | Genome-fragment | 2.55 | AAI-based (medium-confidence) | 0 |
| S3_k141_98812 | 4551 | No | NA | 5 | 0 | 4 | Not-determined | Genome-fragment | NA | NA | 0 |
| S3_k141_64569 | 3619 | No | NA | 1 | 0 | 0 | Not-determined | Genome-fragment | NA | NA | 0 |
| S1-final-viral-combined_k141_287212\|\|full | 1573 | No | NA | 5 | 0 | 0 | Low-quality | Genome-fragment | 2.92 | AAI-based (high-confidence) | 0 |
| S3_k141_47405 | 1165 | No | NA | 1 | 0 | 0 | Not-determined | Genome-fragment | NA | NA | 0 |
| S3_k141_35781 | 3057 | No | NA | 4 | 0 | 4 | Not-determined | Genome-fragment | NA | NA | 0 |
| S3_k141_122020 | 2062 | No | NA | 1 | 0 | 1 | Not-determined | Genome-fragment | NA | NA | 0 |
| S5_k141_163787 | 1573 | No | NA | 4 | 0 | 0 | Low-quality | Genome-fragment | 0.99 | AAI-based (medium-confidence) | 0 |
| S3_k141_41563 | 1296 | No | NA | 1 | 0 | 1 | Not-determined | Genome-fragment | NA | NA | 0 |
| S3_k141_58893 | 1105 | No | NA | 2 | 0 | 0 | Not-determined | Genome-fragment | NA | NA | 0 |
| S12_k141_389423 | 1573 | No | NA | 1 | 1 | 0 | Low-quality | Genome-fragment | 1.08 | HMM-based (lower-bound) | 0 |
| S2-final-viral-combined_k141_183777\|\|full | 1573 | No | NA | 2 | 0 | 2 | Low-quality | Genome-fragment | 2.98 | AAI-based (high-confidence) | 0 |
| S3_k141_128132 | 1019 | No | NA | 2 | 0 | 0 | Not-determined | Genome-fragment | NA | NA | 0 |
| S3_k141_64653 | 1048 | No | NA | 2 | 0 | 1 | Not-determined | Genome-fragment | NA | NA | 0 |
| S3_k141_128142 | 1692 | No | NA | 1 | 0 | 1 | Not-determined | Genome-fragment | NA | NA | 0 |
| S3_k141_87839 | 1174 | No | NA | 1 | 0 | 0 | Not-determined | Genome-fragment | NA | NA | 0 |
| S3_k141_110682 | 1031 | No | NA | 1 | 0 | 1 | Not-determined | Genome-fragment | NA | NA | 0 |
| S3_k141_122136 | 1059 | No | NA | 1 | 0 | 1 | Not-determined | Genome-fragment | NA | NA | 0 |
| S2-final-viral-combined_k141_153862\|\|full | 1573 | No | NA | 3 | 0 | 1 | Low-quality | Genome-fragment | 3.44 | AAI-based (medium-confidence) | 0 |
| S3_k141_87903 | 1584 | No | NA | 2 | 0 | 1 | Not-determined | Genome-fragment | NA | NA | 0 |
| S4-final-viral-combined_k141_119041\|\|full | 1573 | No | NA | 2 | 0 | 0 | Low-quality | Genome-fragment | 1.88 | AAI-based (medium-confidence) | 0 |
| S3_k141_110746 | 3111 | No | NA | 2 | 0 | 1 | Not-determined | Genome-fragment | NA | NA | 0 |
| S3_k141_104931 | 1083 | No | NA | 2 | 0 | 2 | Not-determined | Genome-fragment | NA | NA | 0 |
| S11-final-viral-combined_k141_214808\|\|full | 1574 | No | NA | 3 | 2 | 0 | Low-quality | Genome-fragment | 1.93 | HMM-based (lower-bound) | 0 |
| S3_k141_59007 | 1123 | No | NA | 2 | 0 | 1 | Not-determined | Genome-fragment | NA | NA | 0 |
| S3_k141_116595 | 1375 | No | NA | 1 | 0 | 1 | Not-determined | Genome-fragment | NA | NA | 0 |
| S3_k141_116623 | 1802 | No | NA | 1 | 0 | 0 | Not-determined | Genome-fragment | NA | NA | 0 |
| S3_k141_76270 | 4544 | No | NA | 5 | 0 | 1 | Not-determined | Genome-fragment | NA | NA | 0 |
| S3_k141_110779 | 1012 | No | NA | 1 | 0 | 0 | Not-determined | Genome-fragment | NA | NA | 0 |
| S3_k141_35979 | 1262 | No | NA | 1 | 0 | 0 | Not-determined | Genome-fragment | NA | NA | 0 |
| S1-final-viral-combined_k141_197653\|\|full | 1574 | No | NA | 3 | 0 | 0 | Low-quality | Genome-fragment | 3.24 | AAI-based (high-confidence) | 0 |
| S3_k141_110802 | 1157 | No | NA | 1 | 0 | 0 | Not-determined | Genome-fragment | NA | NA | 0 |
| S1_k141_44066 | 1574 | No | NA | 1 | 0 | 1 | Low-quality | Genome-fragment | 3.07 | AAI-based (high-confidence) | 0 |
| S3_k141_76324 | 1307 | No | NA | 2 | 0 | 0 | Not-determined | Genome-fragment | NA | NA | 0 |
| S3_k141_133904 | 1932 | No | NA | 2 | 0 | 1 | Not-determined | Genome-fragment | NA | NA | 0 |
| S3_k141_41771 | 1308 | No | NA | 1 | 0 | 1 | Not-determined | Genome-fragment | NA | NA | 0 |
| S3_k141_70563 | 1519 | No | NA | 3 | 0 | 1 | Not-determined | Genome-fragment | NA | NA | 0 |
| S7_k141_29582 | 1574 | No | NA | 2 | 0 | 2 | Low-quality | Genome-fragment | 0.43 | AAI-based (medium-confidence) | 0 |
| S17-final-viral-combined_k141_218889\|\|full | 1574 | No | NA | 3 | 0 | 0 | Low-quality | Genome-fragment | 1.92 | AAI-based (high-confidence) | 0 |
| S3_k141_53447 | 5286 | No | NA | 1 | 0 | 1 | Not-determined | Genome-fragment | NA | NA | 0 |
| S12-final-viral-combined_k141_124531\|\|full | 1574 | No | NA | 4 | 1 | 0 | Low-quality | Genome-fragment | 3.23 | AAI-based (medium-confidence) | 0 |
| S3_k141_30208 | 1021 | No | NA | 3 | 0 | 2 | Not-determined | Genome-fragment | NA | NA | 0 |
| S3_k141_122303 | 1479 | No | NA | 1 | 0 | 1 | Not-determined | Genome-fragment | NA | NA | 0 |
| S3_k141_88046 | 1070 | No | NA | 2 | 0 | 2 | Not-determined | Genome-fragment | NA | NA | 0 |
| S12-final-viral-combined_k141_131825\|\|full | 1574 | No | NA | 4 | 1 | 0 | Low-quality | Genome-fragment | 4.06 | AAI-based (high-confidence) | 0 |
| S3_k141_76357 | 1067 | No | NA | 1 | 0 | 1 | Not-determined | Genome-fragment | NA | NA | 0 |
| S3_k141_30235 | 1655 | No | NA | 2 | 0 | 1 | Not-determined | Genome-fragment | NA | NA | 0 |
| S12-final-viral-combined_k141_268839\|\|lt2gene | 1574 | No | NA | 2 | 1 | 0 | Low-quality | Genome-fragment | 0.84 | HMM-based (lower-bound) | 0 |
| S3_k141_53471 | 1076 | No | NA | 2 | 0 | 1 | Not-determined | Genome-fragment | NA | NA | 0 |
| S3_k141_70649 | 1669 | No | NA | 1 | 0 | 0 | Not-determined | Genome-fragment | NA | NA | 0 |
| S3_k141_59200 | 3176 | No | NA | 1 | 0 | 1 | Not-determined | Genome-fragment | NA | NA | 0 |
| S3_k141_64967 | 1553 | No | NA | 1 | 0 | 1 | Not-determined | Genome-fragment | NA | NA | 0 |
| S3_k141_88083 | 1447 | No | NA | 1 | 0 | 0 | Not-determined | Genome-fragment | NA | NA | 0 |
| S3_k141_110955 | 6631 | No | NA | 3 | 0 | 1 | Not-determined | Genome-fragment | NA | NA | 0 |
| S3_k141_53482 | 1099 | No | NA | 2 | 0 | 0 | Not-determined | Genome-fragment | NA | NA | 0 |
| S2-final-viral-combined_k141_16826\|\|full | 1574 | No | NA | 4 | 0 | 1 | Low-quality | Genome-fragment | 1.07 | AAI-based (high-confidence) | 0 |
| S3_k141_122400 | 1094 | No | NA | 2 | 0 | 2 | Not-determined | Genome-fragment | NA | NA | 0 |
| S3_k141_24699 | 1195 | No | NA | 2 | 0 | 1 | Not-determined | Genome-fragment | NA | NA | 0 |
| S11-final-viral-combined_k141_51704\|\|full | 1575 | No | NA | 3 | 1 | 0 | Low-quality | Genome-fragment | 1.05 | HMM-based (lower-bound) | 0 |
| S5-final-viral-combined_k141_137979\|\|full | 1575 | No | NA | 2 | 0 | 1 | Low-quality | Genome-fragment | 0.84 | AAI-based (medium-confidence) | 0 |
| S17-final-viral-combined_k141_184484\|\|full | 1575 | No | NA | 2 | 0 | 2 | Low-quality | Genome-fragment | 1.89 | AAI-based (medium-confidence) | 0 |
| S3_k141_18874 | 1461 | No | NA | 2 | 0 | 2 | Not-determined | Genome-fragment | NA | NA | 0 |
| S12-final-viral-combined_k141_212597\|\|full | 1575 | No | NA | 3 | 0 | 1 | Low-quality | Genome-fragment | 2.44 | AAI-based (medium-confidence) | 0 |
| S3_k141_116830 | 5745 | No | NA | 7 | 0 | 6 | Not-determined | Genome-fragment | NA | NA | 0 |
| S3_k141_99207 | 1156 | No | NA | 3 | 0 | 0 | Not-determined | Genome-fragment | NA | NA | 0 |
| S3_k141_59247 | 3878 | No | NA | 2 | 0 | 0 | Not-determined | Genome-fragment | NA | NA | 0 |
| S3_k141_128509 | 1203 | No | NA | 2 | 0 | 1 | Not-determined | Genome-fragment | NA | NA | 0 |
| S3_k141_82388 | 1235 | No | NA | 2 | 0 | 0 | Not-determined | Genome-fragment | NA | NA | 0 |
| S3_k141_47863 | 4298 | No | NA | 2 | 0 | 1 | Not-determined | Genome-fragment | NA | NA | 0 |
| S2-final-viral-combined_k141_115107\|\|full | 1575 | No | NA | 4 | 0 | 0 | Low-quality | Genome-fragment | 4.32 | AAI-based (high-confidence) | 0 |
| S3_k141_76503 | 1318 | No | NA | 3 | 0 | 1 | Not-determined | Genome-fragment | NA | NA | 0 |
| S3_k141_30405 | 3030 | No | NA | 5 | 0 | 0 | Not-determined | Genome-fragment | NA | NA | 0 |
| S2-final-viral-combined_k141_84683\|\|full | 1575 | No | NA | 2 | 0 | 0 | Low-quality | Genome-fragment | 3.81 | AAI-based (medium-confidence) | 0 |
| S3_k141_13165 | 2008 | No | NA | 1 | 0 | 1 | Not-determined | Genome-fragment | NA | NA | 0 |
| S2_k141_121428 | 1575 | No | NA | 3 | 0 | 0 | Low-quality | Genome-fragment | 4.77 | AAI-based (medium-confidence) | 0 |
| S3_k141_105345 | 1372 | No | NA | 1 | 0 | 1 | Not-determined | Genome-fragment | NA | NA | 0 |
| S3_k141_93769 | 1065 | No | NA | 2 | 0 | 0 | Not-determined | Genome-fragment | NA | NA | 0 |
| S8-final-viral-combined_k141_113955\|\|full | 1575 | No | NA | 3 | 3 | 0 | Low-quality | Genome-fragment | 3.86 | AAI-based (medium-confidence) | 0 |
| S3_k141_36266 | 1164 | No | NA | 1 | 0 | 0 | Not-determined | Genome-fragment | NA | NA | 0 |
| S8_k141_62038 | 1575 | No | NA | 1 | 0 | 1 | Low-quality | Genome-fragment | 3.9 | AAI-based (high-confidence) | 0 |
| S11-final-viral-combined_k141_31495\|\|full | 1576 | No | NA | 2 | 0 | 0 | Low-quality | Genome-fragment | 1.19 | AAI-based (high-confidence) | 0 |
| S3_k141_65164 | 5042 | No | NA | 4 | 0 | 3 | Not-determined | Genome-fragment | NA | NA | 0 |
| S1-final-viral-combined_k141_114239\|\|full | 1576 | No | NA | 3 | 1 | 0 | Low-quality | Genome-fragment | 2.92 | AAI-based (medium-confidence) | 0 |
| S3_k141_76578 | 1174 | No | NA | 1 | 0 | 1 | Not-determined | Genome-fragment | NA | NA | 0 |
| S3_k141_47921 | 1490 | No | NA | 1 | 0 | 0 | Not-determined | Genome-fragment | NA | NA | 0 |
| S3_k141_105407 | 1482 | No | NA | 2 | 0 | 1 | Not-determined | Genome-fragment | NA | NA | 0 |
| S3_k141_59400 | 3089 | No | NA | 1 | 0 | 1 | Not-determined | Genome-fragment | NA | NA | 0 |
| S1_k141_110502 | 1576 | No | NA | 1 | 1 | 0 | Low-quality | Genome-fragment | 0.99 | HMM-based (lower-bound) | 0 |
| S3_k141_19054 | 1513 | No | NA | 1 | 0 | 1 | Not-determined | Genome-fragment | NA | NA | 0 |
| S3_k141_122648 | 1746 | No | NA | 1 | 0 | 1 | Not-determined | Genome-fragment | NA | NA | 0 |
| S3_k141_37469 | 1576 | No | NA | 1 | 0 | 0 | Low-quality | Genome-fragment | 2.7 | AAI-based (high-confidence) | 0 |
| S3_k141_82530 | 1593 | No | NA | 2 | 0 | 2 | Not-determined | Genome-fragment | NA | NA | 0 |
| S3_k141_76632 | 3236 | No | NA | 3 | 0 | 2 | Not-determined | Genome-fragment | NA | NA | 0 |
| S3_k141_76634 | 1774 | No | NA | 1 | 0 | 1 | Not-determined | Genome-fragment | NA | NA | 0 |
| S3_k141_42153 | 3228 | No | NA | 4 | 0 | 1 | Not-determined | Genome-fragment | NA | NA | 0 |
| S3_k141_128646 | 1410 | No | NA | 1 | 0 | 0 | Not-determined | Genome-fragment | NA | NA | 0 |
| S5-final-viral-combined_k141_162846\|\|full | 1576 | No | NA | 3 | 2 | 0 | Low-quality | Genome-fragment | 3.28 | AAI-based (high-confidence) | 0 |
| S5-final-viral-combined_k141_139079\|\|full | 1576 | No | NA | 3 | 0 | 1 | Low-quality | Genome-fragment | 2.93 | AAI-based (medium-confidence) | 0 |
| S3_k141_111184 | 1123 | No | NA | 1 | 0 | 1 | Not-determined | Genome-fragment | NA | NA | 0 |
| S3_k141_48034 | 1942 | No | NA | 2 | 0 | 0 | Not-determined | Genome-fragment | NA | NA | 0 |
| S3_k141_65284 | 1158 | No | NA | 2 | 0 | 1 | Not-determined | Genome-fragment | NA | NA | 0 |
| S3_k141_128697 | 1065 | No | NA | 3 | 0 | 3 | Not-determined | Genome-fragment | NA | NA | 0 |
| S3_k141_88350 | 1686 | No | NA | 2 | 0 | 2 | Not-determined | Genome-fragment | NA | NA | 0 |
| S3_k141_59503 | 1377 | No | NA | 1 | 0 | 1 | Not-determined | Genome-fragment | NA | NA | 0 |
| S3_k141_2087 | 1124 | No | NA | 2 | 0 | 0 | Not-determined | Genome-fragment | NA | NA | 0 |
| S7-final-viral-combined_k141_129947\|\|full | 1576 | No | NA | 2 | 0 | 1 | Low-quality | Genome-fragment | 1.68 | AAI-based (medium-confidence) | 0 |
| S3_k141_71006 | 3055 | No | NA | 5 | 0 | 4 | Not-determined | Genome-fragment | NA | NA | 0 |
| S17-final-viral-combined_k141_77538\|\|full | 1576 | No | NA | 3 | 0 | 3 | Low-quality | Genome-fragment | 1.2 | AAI-based (medium-confidence) | 0 |
| S12-final-viral-combined_k141_377085\|\|full | 1576 | No | NA | 2 | 0 | 0 | Low-quality | Genome-fragment | 1.09 | AAI-based (medium-confidence) | 0 |
| S3_k141_13416 | 1561 | No | NA | 2 | 0 | 1 | Not-determined | Genome-fragment | NA | NA | 0 |
| S3_k141_105581 | 2110 | No | NA | 2 | 0 | 1 | Not-determined | Genome-fragment | NA | NA | 0 |
| S2-final-viral-combined_k141_145792\|\|full | 1576 | No | NA | 2 | 0 | 1 | Low-quality | Genome-fragment | 0.82 | AAI-based (medium-confidence) | 0 |
| S2-final-viral-combined_k141_178529\|\|full | 1576 | No | NA | 3 | 3 | 0 | Low-quality | Genome-fragment | 3.08 | AAI-based (high-confidence) | 0 |
| S3_k141_88417 | 1460 | No | NA | 1 | 0 | 0 | Not-determined | Genome-fragment | NA | NA | 0 |
| S9_k141_80400 | 1577 | No | NA | 1 | 0 | 0 | Low-quality | Genome-fragment | 3.11 | AAI-based (high-confidence) | 0 |
| S11-final-viral-combined_k141_24585\|\|full | 1577 | No | NA | 2 | 0 | 1 | Low-quality | Genome-fragment | 1.08 | AAI-based (high-confidence) | 0 |
| S5-final-viral-combined_k141_189638\|\|full | 1577 | No | NA | 2 | 0 | 0 | Low-quality | Genome-fragment | 3.72 | AAI-based (medium-confidence) | 0 |
| S5-final-viral-combined_k141_128392\|\|full | 1577 | No | NA | 4 | 1 | 0 | Low-quality | Genome-fragment | 2.58 | AAI-based (medium-confidence) | 0 |
| S3_k141_7764 | 3074 | No | NA | 4 | 0 | 3 | Not-determined | Genome-fragment | NA | NA | 0 |
| S3_k141_48208 | 1886 | No | NA | 4 | 0 | 1 | Not-determined | Genome-fragment | NA | NA | 0 |
| S3_k141_128840 | 1175 | No | NA | 2 | 0 | 1 | Not-determined | Genome-fragment | NA | NA | 0 |
| S3_k141_99564 | 1066 | No | NA | 1 | 0 | 0 | Not-determined | Genome-fragment | NA | NA | 0 |
| S3_k141_59641 | 4084 | No | NA | 4 | 0 | 2 | Not-determined | Genome-fragment | NA | NA | 0 |
| S3_k141_48220 | 1148 | No | NA | 1 | 0 | 0 | Not-determined | Genome-fragment | NA | NA | 0 |
| S3_k141_88547 | 1278 | No | NA | 2 | 0 | 0 | Not-determined | Genome-fragment | NA | NA | 0 |
| S3_k141_94181 | 1021 | No | NA | 3 | 0 | 0 | Not-determined | Genome-fragment | NA | NA | 0 |
| S3_k141_94186 | 4141 | No | NA | 4 | 0 | 3 | Not-determined | Genome-fragment | NA | NA | 0 |
| S5-final-viral-combined_k141_110412\|\|full | 1577 | No | NA | 2 | 0 | 1 | Low-quality | Genome-fragment | 2.29 | AAI-based (medium-confidence) | 0 |
| S3_k141_25220 | 1201 | No | NA | 1 | 0 | 1 | Not-determined | Genome-fragment | NA | NA | 0 |
| S3_k141_99649 | 1226 | No | NA | 1 | 0 | 1 | Not-determined | Genome-fragment | NA | NA | 0 |
| S3_k141_111375 | 1189 | No | NA | 2 | 0 | 0 | Not-determined | Genome-fragment | NA | NA | 0 |
| S3_k141_82810 | 6353 | No | NA | 5 | 0 | 3 | Not-determined | Genome-fragment | NA | NA | 0 |
| S5_k141_10299 | 1577 | No | NA | 1 | 0 | 1 | Low-quality | Genome-fragment | 1.99 | AAI-based (high-confidence) | 0 |
| S17-final-viral-combined_k141_48836\|\|full | 1577 | Yes | 764 | 2 | 1 | 1 | Low-quality | Genome-fragment | 1.63 | AAI-based (medium-confidence) | 51.55 |
| S17-k141_63366 | 1577 | No | NA | 2 | 0 | 1 | Low-quality | Genome-fragment | 1.12 | AAI-based (high-confidence) | 0 |
| S3_k141_19400 | 1011 | No | NA | 1 | 0 | 0 | Not-determined | Genome-fragment | NA | NA | 0 |
| S12_k141_155889 | 1577 | No | NA | 2 | 0 | 1 | Low-quality | Genome-fragment | 1.12 | AAI-based (high-confidence) | 0 |
| S3_k141_30862 | 1023 | No | NA | 1 | 0 | 0 | Not-determined | Genome-fragment | NA | NA | 0 |
| S2-final-viral-combined_k141_108998\|\|full | 1577 | No | NA | 3 | 0 | 2 | Low-quality | Genome-fragment | 3.6 | AAI-based (medium-confidence) | 0 |
| S3_k141_88675 | 1151 | No | NA | 1 | 0 | 1 | Not-determined | Genome-fragment | NA | NA | 0 |
| S3_k141_134626 | 2819 | No | NA | 4 | 0 | 3 | Not-determined | Genome-fragment | NA | NA | 0 |
| S2-final-viral-combined_k141_154822\|\|full | 1577 | No | NA | 4 | 1 | 0 | Low-quality | Genome-fragment | 2.64 | AAI-based (high-confidence) | 0 |
| S3_k141_105784 | 1176 | No | NA | 1 | 0 | 1 | Not-determined | Genome-fragment | NA | NA | 0 |
| S8-final-viral-combined_k141_190742\|\|full | 1577 | No | NA | 4 | 2 | 0 | Low-quality | Genome-fragment | 3.21 | AAI-based (high-confidence) | 0 |
| S1-final-viral-combined_k141_164968\|\|full | 1578 | No | NA | 3 | 0 | 0 | Low-quality | Genome-fragment | 2.92 | AAI-based (high-confidence) | 0 |
| S5-final-viral-combined_k141_9937\|\|full | 1578 | No | NA | 2 | 0 | 2 | Low-quality | Genome-fragment | 2.41 | AAI-based (medium-confidence) | 0 |
| S3_k141_129009 | 4663 | No | NA | 7 | 0 | 4 | Not-determined | Genome-fragment | NA | NA | 0 |
| S17-final-viral-combined_k141_95043\|\|full | 1578 | No | NA | 4 | 1 | 0 | Low-quality | Genome-fragment | 4.04 | AAI-based (high-confidence) | 0 |
| S3_k141_2461 | 1114 | No | NA | 1 | 0 | 1 | Not-determined | Genome-fragment | NA | NA | 0 |
| S3_k141_2464 | 1161 | No | NA | 1 | 0 | 0 | Not-determined | Genome-fragment | NA | NA | 0 |
| S3_k141_71341 | 1170 | No | NA | 4 | 0 | 1 | Not-determined | Genome-fragment | NA | NA | 0 |
| S3_k141_59793 | 1032 | No | NA | 1 | 0 | 1 | Not-determined | Genome-fragment | NA | NA | 0 |
| S3_k141_2480 | 1365 | No | NA | 2 | 0 | 2 | Not-determined | Genome-fragment | NA | NA | 0 |
| S3_k141_36813 | 1035 | No | NA | 1 | 0 | 1 | Not-determined | Genome-fragment | NA | NA | 0 |
| S12_k141_207771 | 1578 | No | NA | 3 | 0 | 0 | Low-quality | Genome-fragment | 1.59 | AAI-based (high-confidence) | 0 |
| S3_k141_88816 | 1851 | No | NA | 3 | 0 | 1 | Not-determined | Genome-fragment | NA | NA | 0 |
| S11-final-viral-combined_k141_241601\|\|full | 1579 | No | NA | 4 | 2 | 0 | Low-quality | Genome-fragment | 2.29 | AAI-based (medium-confidence) | 0 |
| S3_k141_134751 | 10232 | No | NA | 5 | 0 | 3 | Not-determined | Genome-fragment | NA | NA | 0 |
| S3_k141_83024 | 4362 | No | NA | 4 | 0 | 2 | Not-determined | Genome-fragment | NA | NA | 0 |
| S11_k141_219903 | 1579 | No | NA | 3 | 0 | 0 | Low-quality | Genome-fragment | 2.15 | AAI-based (medium-confidence) | 0 |
| S3_k141_88818 | 1508 | No | NA | 2 | 0 | 0 | Not-determined | Genome-fragment | NA | NA | 0 |
| S3_k141_105878 | 1216 | No | NA | 1 | 0 | 1 | Not-determined | Genome-fragment | NA | NA | 0 |
| S3_k141_8065 | 4506 | No | NA | 8 | 0 | 6 | Not-determined | Genome-fragment | NA | NA | 0 |
| S3_k141_99925 | 1042 | No | NA | 2 | 0 | 0 | Not-determined | Genome-fragment | NA | NA | 0 |
| S2_k141_85275 | 1579 | No | NA | 1 | 1 | 0 | Low-quality | Genome-fragment | 0.92 | HMM-based (lower-bound) | 0 |
| S3_k141_111706 | 1059 | No | NA | 1 | 0 | 0 | Not-determined | Genome-fragment | NA | NA | 0 |
| S3_k141_77161 | 5208 | No | NA | 4 | 0 | 4 | Not-determined | Genome-fragment | NA | NA | 0 |
| S3_k141_83097 | 1134 | No | NA | 1 | 0 | 0 | Not-determined | Genome-fragment | NA | NA | 0 |
| S3_k141_94472 | 1132 | No | NA | 3 | 0 | 0 | Not-determined | Genome-fragment | NA | NA | 0 |
| S11-final-viral-combined_k141_126907\|\|full | 1580 | No | NA | 4 | 0 | 0 | Low-quality | Genome-fragment | 2.78 | AAI-based (medium-confidence) | 0 |
| S11_k141_182451 | 1580 | No | NA | 3 | 1 | 0 | Low-quality | Genome-fragment | 2.07 | HMM-based (lower-bound) | 0 |
| S7-final-viral-combined_k141_139510\|\|full | 1580 | No | NA | 4 | 0 | 0 | Low-quality | Genome-fragment | 3.63 | AAI-based (medium-confidence) | 0 |
| S17-final-viral-combined_k141_232699\|\|full | 1580 | No | NA | 2 | 0 | 1 | Low-quality | Genome-fragment | 1.08 | AAI-based (high-confidence) | 0 |
| S3_k141_94494 | 20435 | No | NA | 31 | 14 | 0 | Medium-quality | Genome-fragment | 50.86 | AAI-based (high-confidence) | 0 |
| S3_k141_19691 | 2456 | No | NA | 4 | 0 | 1 | Not-determined | Genome-fragment | NA | NA | 0 |
| S3_k141_48589 | 1309 | No | NA | 2 | 0 | 1 | Not-determined | Genome-fragment | NA | NA | 0 |
| S12-final-viral-combined_k141_87694\|\|full | 1580 | No | NA | 3 | 0 | 1 | Low-quality | Genome-fragment | 1 | AAI-based (high-confidence) | 0 |
| S3_k141_129243 | 4238 | No | NA | 5 | 0 | 1 | Not-determined | Genome-fragment | NA | NA | 0 |
| S12-final-viral-combined_k141_310471\|\|full | 1580 | No | NA | 3 | 3 | 0 | Low-quality | Genome-fragment | 4.91 | AAI-based (high-confidence) | 0 |
| S2-final-viral-combined_k141_14260\|\|full | 1580 | No | NA | 3 | 0 | 0 | Low-quality | Genome-fragment | 3.1 | AAI-based (high-confidence) | 0 |
| S3_k141_88976 | 2780 | No | NA | 3 | 0 | 0 | Not-determined | Genome-fragment | NA | NA | 0 |
| S3_k141_123301 | 1571 | No | NA | 1 | 0 | 1 | Not-determined | Genome-fragment | NA | NA | 0 |
| S3_k141_129258 | 3936 | No | NA | 5 | 0 | 0 | Not-determined | Genome-fragment | NA | NA | 0 |
| S2-final-viral-combined_k141_91384\|\|full | 1580 | No | NA | 5 | 0 | 0 | Low-quality | Genome-fragment | 3.75 | AAI-based (high-confidence) | 0 |
| S2-final-viral-combined_k141_82115\|\|full | 1580 | No | NA | 4 | 1 | 0 | Low-quality | Genome-fragment | 4.19 | AAI-based (high-confidence) | 0 |
| S3_k141_31218 | 1212 | No | NA | 2 | 0 | 1 | Not-determined | Genome-fragment | NA | NA | 0 |
| S9-final-viral-combined_k141_58944\|\|full | 1581 | No | NA | 2 | 0 | 1 | Low-quality | Genome-fragment | 4.13 | AAI-based (medium-confidence) | 0 |
| S3_k141_2717 | 3649 | No | NA | 3 | 0 | 1 | Not-determined | Genome-fragment | NA | NA | 0 |
| S3_k141_111831 | 1194 | No | NA | 1 | 0 | 1 | Not-determined | Genome-fragment | NA | NA | 0 |
| S3_k141_117664 | 1480 | No | NA | 5 | 0 | 5 | Not-determined | Genome-fragment | NA | NA | 0 |
| S3_k141_25608 | 1346 | No | NA | 1 | 0 | 0 | Not-determined | Genome-fragment | NA | NA | 0 |
| S3_k141_19793 | 1037 | No | NA | 1 | 0 | 0 | Not-determined | Genome-fragment | NA | NA | 0 |
| S11-final-viral-combined_k141_7556\|\|full | 1581 | No | NA | 2 | 0 | 1 | Low-quality | Genome-fragment | 1.55 | AAI-based (medium-confidence) | 0 |
| S3_k141_106058 | 2227 | No | NA | 2 | 0 | 1 | Not-determined | Genome-fragment | NA | NA | 0 |
| S3_k141_54310 | 1643 | No | NA | 2 | 0 | 0 | Not-determined | Genome-fragment | NA | NA | 0 |
| S3_k141_94665 | 1147 | No | NA | 2 | 0 | 0 | Not-determined | Genome-fragment | NA | NA | 0 |
| S5-final-viral-combined_k141_137513\|\|lt2gene | 1581 | No | NA | 1 | 1 | 0 | Low-quality | Genome-fragment | 2.54 | HMM-based (lower-bound) | 0 |
| S3_k141_111919 | 6667 | No | NA | 4 | 0 | 1 | Not-determined | Genome-fragment | NA | NA | 0 |
| S3_k141_13964 | 1547 | No | NA | 1 | 0 | 0 | Not-determined | Genome-fragment | NA | NA | 0 |
| S3_k141_77384 | 1281 | No | NA | 1 | 0 | 1 | Not-determined | Genome-fragment | NA | NA | 0 |
| S5_k141_92631 | 1581 | No | NA | 1 | 1 | 0 | Low-quality | Genome-fragment | 3.69 | AAI-based (high-confidence) | 0 |
| S7-final-viral-combined_k141_236851\|\|full | 1581 | No | NA | 4 | 1 | 0 | Low-quality | Genome-fragment | 4.24 | AAI-based (high-confidence) | 0 |
| S3_k141_25730 | 1549 | No | NA | 2 | 0 | 0 | Not-determined | Genome-fragment | NA | NA | 0 |
| S7-final-viral-combined_k141_74071\|\|full | 1581 | No | NA | 3 | 1 | 0 | Low-quality | Genome-fragment | 2.35 | HMM-based (lower-bound) | 0 |
| S3_k141_117746 | 1080 | No | NA | 1 | 0 | 0 | Not-determined | Genome-fragment | NA | NA | 0 |
| S3_k141_83373 | 2129 | No | NA | 1 | 0 | 1 | Not-determined | Genome-fragment | NA | NA | 0 |
| S3_k141_117766 | 1089 | No | NA | 1 | 0 | 0 | Not-determined | Genome-fragment | NA | NA | 0 |
| S3_k141_77417 | 1866 | No | NA | 2 | 0 | 1 | Not-determined | Genome-fragment | NA | NA | 0 |
| S3_k141_100214 | 1776 | No | NA | 1 | 0 | 1 | Not-determined | Genome-fragment | NA | NA | 0 |
| S3_k141_135047 | 1004 | No | NA | 2 | 0 | 2 | Not-determined | Genome-fragment | NA | NA | 0 |
| S3_k141_31426 | 1488 | No | NA | 1 | 0 | 1 | Not-determined | Genome-fragment | NA | NA | 0 |
| S3_k141_2891 | 1125 | No | NA | 2 | 0 | 0 | Not-determined | Genome-fragment | NA | NA | 0 |
| S3_k141_31434 | 1316 | No | NA | 1 | 0 | 1 | Not-determined | Genome-fragment | NA | NA | 0 |
| S7-final-viral-combined_k141_167809\|\|full | 1581 | No | NA | 4 | 0 | 0 | Low-quality | Genome-fragment | 3.35 | AAI-based (medium-confidence) | 0 |
| S12-final-viral-combined_k141_261501\|\|full | 1581 | No | NA | 3 | 0 | 1 | Low-quality | Genome-fragment | 1.05 | AAI-based (high-confidence) | 0 |
| S2-final-viral-combined_k141_138110\|\|full | 1581 | No | NA | 2 | 0 | 1 | Low-quality | Genome-fragment | 1.52 | AAI-based (high-confidence) | 0 |
| S3_k141_31450 | 3328 | No | NA | 2 | 0 | 0 | Not-determined | Genome-fragment | NA | NA | 0 |
| S3_k141_8349 | 1740 | No | NA | 2 | 0 | 0 | Not-determined | Genome-fragment | NA | NA | 0 |
| S3_k141_83422 | 3689 | No | NA | 5 | 0 | 4 | Not-determined | Genome-fragment | NA | NA | 0 |
| S3_k141_65955 | 1067 | No | NA | 3 | 0 | 2 | Not-determined | Genome-fragment | NA | NA | 0 |
| S3_k141_83429 | 1754 | No | NA | 1 | 0 | 0 | Not-determined | Genome-fragment | NA | NA | 0 |
| S3_k141_42941 | 2217 | No | NA | 2 | 0 | 1 | Not-determined | Genome-fragment | NA | NA | 0 |
| S3_k141_83466 | 1047 | No | NA | 1 | 0 | 0 | Not-determined | Genome-fragment | NA | NA | 0 |
| S11-final-viral-combined_k141_323234\|\|full | 1582 | No | NA | 2 | 0 | 1 | Low-quality | Genome-fragment | 1.27 | AAI-based (high-confidence) | 0 |
| S3_k141_37355 | 1797 | No | NA | 3 | 0 | 0 | Not-determined | Genome-fragment | NA | NA | 0 |
| S3_k141_3029 | 1585 | No | NA | 1 | 0 | 0 | Not-determined | Genome-fragment | NA | NA | 0 |
| S11-final-viral-combined_k141_137342\|\|lt2gene | 1582 | No | NA | 2 | 1 | 0 | Low-quality | Genome-fragment | 3.6 | AAI-based (high-confidence) | 0 |
| S3_k141_60340 | 1204 | No | NA | 1 | 0 | 1 | Not-determined | Genome-fragment | NA | NA | 0 |
| S1-final-viral-combined_k141_334552\|\|full | 1582 | No | NA | 3 | 0 | 0 | Low-quality | Genome-fragment | 2.9 | AAI-based (medium-confidence) | 0 |
| S17-final-viral-combined_k141_139703\|\|full | 1582 | No | NA | 4 | 0 | 1 | Low-quality | Genome-fragment | 0.53 | AAI-based (medium-confidence) | 0 |
| S17-final-viral-combined_k141_115802\|\|lt2gene | 1582 | No | NA | 2 | 1 | 0 | Low-quality | Genome-fragment | 3.49 | AAI-based (high-confidence) | 0 |
| S3_k141_20102 | 1032 | No | NA | 2 | 0 | 1 | Not-determined | Genome-fragment | NA | NA | 0 |
| S3_k141_20111 | 1124 | No | NA | 1 | 0 | 0 | Not-determined | Genome-fragment | NA | NA | 0 |
| S3_k141_94944 | 1117 | No | NA | 1 | 0 | 1 | Not-determined | Genome-fragment | NA | NA | 0 |
| S12-final-viral-combined_k141_323779\|\|lt2gene | 1582 | No | NA | 2 | 1 | 0 | Low-quality | Genome-fragment | 2.94 | HMM-based (lower-bound) | 0 |
| S3_k141_48892 | 12703 | No | NA | 11 | 0 | 8 | Not-determined | Genome-fragment | NA | NA | 0 |
| S2_k141_141584 | 1582 | No | NA | 2 | 0 | 0 | Low-quality | Genome-fragment | 2 | AAI-based (medium-confidence) | 0 |
| S3_k141_48897 | 1732 | No | NA | 1 | 0 | 1 | Not-determined | Genome-fragment | NA | NA | 0 |
| S3_k141_14184 | 3945 | No | NA | 7 | 0 | 0 | Not-determined | Genome-fragment | NA | NA | 0 |
| S4_k141_150017 | 1582 | No | NA | 4 | 3 | 0 | Low-quality | Genome-fragment | 1.17 | HMM-based (lower-bound) | 0 |
| S8-final-viral-combined_k141_162761\|\|lt2gene | 1582 | No | NA | 1 | 1 | 0 | Low-quality | Genome-fragment | 3.62 | AAI-based (medium-confidence) | 0 |
| S3_k141_3169 | 1957 | No | NA | 2 | 0 | 2 | Not-determined | Genome-fragment | NA | NA | 0 |
| S1-final-viral-combined_k141_74413\|\|full | 1583 | No | NA | 3 | 0 | 0 | Low-quality | Genome-fragment | 2.85 | AAI-based (medium-confidence) | 0 |
| S3_k141_112191 | 1660 | No | NA | 2 | 0 | 2 | Not-determined | Genome-fragment | NA | NA | 0 |
| S3_k141_89400 | 1057 | No | NA | 1 | 0 | 1 | Not-determined | Genome-fragment | NA | NA | 0 |
| S1-final-viral-combined_k141_389765\|\|lt2gene | 1583 | No | NA | 1 | 1 | 0 | Low-quality | Genome-fragment | 4.22 | AAI-based (medium-confidence) | 0 |
| S3_k141_95036 | 1061 | No | NA | 2 | 0 | 2 | Not-determined | Genome-fragment | NA | NA | 0 |
| S3_k141_100498 | 2213 | No | NA | 3 | 0 | 2 | Not-determined | Genome-fragment | NA | NA | 0 |
| S3_k141_106478 | 1060 | No | NA | 2 | 0 | 1 | Not-determined | Genome-fragment | NA | NA | 0 |
| S17-final-viral-combined_k141_103626\|\|full | 1583 | No | NA | 4 | 0 | 0 | Low-quality | Genome-fragment | 2.6 | AAI-based (medium-confidence) | 0 |
| S3_k141_77643 | 1199 | No | NA | 1 | 0 | 1 | Not-determined | Genome-fragment | NA | NA | 0 |
| S17-k141_153279 | 1583 | No | NA | 1 | 0 | 1 | Low-quality | Genome-fragment | 4.46 | AAI-based (medium-confidence) | 0 |
| S12-final-viral-combined_k141_250632\|\|full | 1583 | No | NA | 2 | 1 | 0 | Low-quality | Genome-fragment | 0.61 | AAI-based (high-confidence) | 0 |
| S3_k141_77663 | 1374 | No | NA | 1 | 0 | 1 | Not-determined | Genome-fragment | NA | NA | 0 |
| S3_k141_100530 | 2545 | No | NA | 1 | 0 | 1 | Not-determined | Genome-fragment | NA | NA | 0 |
| S3_k141_123827 | 1081 | No | NA | 2 | 0 | 2 | Not-determined | Genome-fragment | NA | NA | 0 |
| S3_k141_83679 | 1033 | No | NA | 1 | 0 | 0 | Not-determined | Genome-fragment | NA | NA | 0 |
| S3_k141_31663 | 1397 | No | NA | 1 | 0 | 0 | Not-determined | Genome-fragment | NA | NA | 0 |
| S3_k141_83690 | 1146 | No | NA | 2 | 0 | 1 | Not-determined | Genome-fragment | NA | NA | 0 |
| S4_k141_150726 | 1583 | No | NA | 2 | 0 | 1 | Low-quality | Genome-fragment | 0.44 | AAI-based (high-confidence) | 0 |
| S8_k141_275212 | 1583 | No | NA | 4 | 0 | 0 | Low-quality | Genome-fragment | 2.73 | AAI-based (high-confidence) | 0 |
| S3_k141_60501 | 2717 | No | NA | 3 | 0 | 2 | Not-determined | Genome-fragment | NA | NA | 0 |
| S3_k141_100591 | 1827 | No | NA | 2 | 0 | 0 | Not-determined | Genome-fragment | NA | NA | 0 |
| S3_k141_95128 | 4234 | No | NA | 3 | 0 | 2 | Not-determined | Genome-fragment | NA | NA | 0 |
| S3_k141_83726 | 1393 | No | NA | 4 | 0 | 0 | Not-determined | Genome-fragment | NA | NA | 0 |
| S3_k141_3335 | 2330 | No | NA | 2 | 0 | 1 | Not-determined | Genome-fragment | NA | NA | 0 |
| S3_k141_43263 | 3433 | No | NA | 5 | 0 | 4 | Not-determined | Genome-fragment | NA | NA | 0 |
| S3_k141_118305 | 1018 | No | NA | 2 | 0 | 1 | Not-determined | Genome-fragment | NA | NA | 0 |
| S3_k141_83768 | 1570 | No | NA | 3 | 0 | 0 | Not-determined | Genome-fragment | NA | NA | 0 |
| S3_k141_77822 | 1530 | No | NA | 3 | 0 | 0 | Not-determined | Genome-fragment | NA | NA | 0 |
| S3_k141_112435 | 1052 | No | NA | 1 | 0 | 0 | Not-determined | Genome-fragment | NA | NA | 0 |
| S3_k141_129865 | 1132 | No | NA | 1 | 0 | 0 | Not-determined | Genome-fragment | NA | NA | 0 |
| S3_k141_37694 | 1177 | No | NA | 2 | 0 | 1 | Not-determined | Genome-fragment | NA | NA | 0 |
| S1-final-viral-combined_k141_284505\|\|full | 1584 | No | NA | 3 | 0 | 0 | Low-quality | Genome-fragment | 3.3 | AAI-based (medium-confidence) | 0 |
| S3_k141_8785 | 1199 | No | NA | 1 | 0 | 0 | Not-determined | Genome-fragment | NA | NA | 0 |
| S1-final-viral-combined_k141_86737\|\|full | 1584 | No | NA | 3 | 0 | 0 | Low-quality | Genome-fragment | 1.08 | AAI-based (high-confidence) | 0 |
| S3_k141_95222 | 2030 | No | NA | 1 | 0 | 1 | Not-determined | Genome-fragment | NA | NA | 0 |
| S3_k141_60591 | 1149 | No | NA | 1 | 0 | 0 | Not-determined | Genome-fragment | NA | NA | 0 |
| S1-final-viral-combined_k141_31701\|\|full | 1584 | No | NA | 2 | 0 | 0 | Low-quality | Genome-fragment | 2.06 | AAI-based (high-confidence) | 0 |
| S3_k141_135438 | 1336 | No | NA | 1 | 0 | 1 | Not-determined | Genome-fragment | NA | NA | 0 |
| S17-final-viral-combined_k141_223987\|\|full | 1584 | No | NA | 2 | 0 | 1 | Low-quality | Genome-fragment | 1.97 | AAI-based (high-confidence) | 0 |
| S3_k141_14437 | 1134 | No | NA | 1 | 0 | 1 | Not-determined | Genome-fragment | NA | NA | 0 |
| S3_k141_129941 | 1334 | No | NA | 3 | 0 | 1 | Not-determined | Genome-fragment | NA | NA | 0 |
| S3_k141_72052 | 7425 | No | NA | 5 | 0 | 5 | Not-determined | Genome-fragment | NA | NA | 0 |
| S17-final-viral-combined_k141_57619\|\|full | 1584 | Yes | 1035 | 2 | 1 | 1 | Low-quality | Genome-fragment | 0.57 | HMM-based (lower-bound) | 34.66 |
| S3_k141_31807 | 1048 | No | NA | 2 | 0 | 1 | Not-determined | Genome-fragment | NA | NA | 0 |
| S3_k141_31812 | 1456 | No | NA | 1 | 0 | 1 | Not-determined | Genome-fragment | NA | NA | 0 |
| S3_k141_14509 | 1138 | No | NA | 1 | 0 | 1 | Not-determined | Genome-fragment | NA | NA | 0 |
| S17-final-viral-combined_k141_152772\|\|full | 1584 | No | NA | 4 | 0 | 2 | Low-quality | Genome-fragment | 2.35 | AAI-based (high-confidence) | 0 |
| S3_k141_106685 | 8208 | No | NA | 4 | 0 | 2 | Not-determined | Genome-fragment | NA | NA | 0 |
| S3_k141_8875 | 1224 | No | NA | 3 | 0 | 2 | Not-determined | Genome-fragment | NA | NA | 0 |
| S17-final-viral-combined_k141_171072\|\|lt2gene | 1584 | No | NA | 2 | 2 | 0 | Low-quality | Genome-fragment | 3.49 | AAI-based (high-confidence) | 0 |
| S3_k141_37849 | 1058 | No | NA | 1 | 0 | 1 | Not-determined | Genome-fragment | NA | NA | 0 |
| S3_k141_60687 | 1130 | No | NA | 1 | 0 | 1 | Not-determined | Genome-fragment | NA | NA | 0 |
| S3_k141_83956 | 1043 | No | NA | 5 | 0 | 0 | Not-determined | Genome-fragment | NA | NA | 0 |
| S3_k141_77994 | 1107 | No | NA | 2 | 0 | 1 | Not-determined | Genome-fragment | NA | NA | 0 |
| S3_k141_8936 | 1615 | No | NA | 1 | 0 | 0 | Not-determined | Genome-fragment | NA | NA | 0 |
| S3_k141_26285 | 1009 | No | NA | 1 | 0 | 1 | Not-determined | Genome-fragment | NA | NA | 0 |
| S3_k141_37861 | 1235 | No | NA | 3 | 0 | 0 | Not-determined | Genome-fragment | NA | NA | 0 |
| S3_k141_135602 | 1444 | No | NA | 2 | 0 | 1 | Not-determined | Genome-fragment | NA | NA | 0 |
| S3_k141_26322 | 1206 | No | NA | 1 | 0 | 0 | Not-determined | Genome-fragment | NA | NA | 0 |
| S3_k141_84003 | 4501 | No | NA | 5 | 0 | 2 | Not-determined | Genome-fragment | NA | NA | 0 |
| S3_k141_49225 | 1376 | No | NA | 3 | 0 | 0 | Not-determined | Genome-fragment | NA | NA | 0 |
| S12_k141_323461 | 1584 | No | NA | 2 | 0 | 1 | Low-quality | Genome-fragment | 0.99 | AAI-based (high-confidence) | 0 |
| S3_k141_49232 | 1007 | No | NA | 1 | 0 | 0 | Not-determined | Genome-fragment | NA | NA | 0 |
| S3_k141_8998 | 1082 | No | NA | 2 | 0 | 1 | Not-determined | Genome-fragment | NA | NA | 0 |
| S12_k141_231875 | 1584 | No | NA | 1 | 0 | 1 | Low-quality | Genome-fragment | 3.33 | AAI-based (high-confidence) | 0 |
| S8-final-viral-combined_k141_215803\|\|full | 1584 | No | NA | 3 | 1 | 0 | Low-quality | Genome-fragment | 2.91 | AAI-based (high-confidence) | 0 |
| S3_k141_60796 | 3324 | No | NA | 2 | 0 | 2 | Not-determined | Genome-fragment | NA | NA | 0 |
| S3_k141_100889 | 1674 | No | NA | 3 | 0 | 3 | Not-determined | Genome-fragment | NA | NA | 0 |
| S3_k141_135645 | 1038 | No | NA | 1 | 0 | 0 | Not-determined | Genome-fragment | NA | NA | 0 |
| S3_k141_89904 | 2674 | No | NA | 3 | 0 | 1 | Not-determined | Genome-fragment | NA | NA | 0 |
| S3_k141_124270 | 1400 | No | NA | 1 | 0 | 1 | Not-determined | Genome-fragment | NA | NA | 0 |
| S9-final-viral-combined_k141_51524\|\|full | 1585 | No | NA | 3 | 0 | 0 | Low-quality | Genome-fragment | 2.6 | AAI-based (high-confidence) | 0 |
| S3-final-viral-combined_k141_47421\|\|full | 1585 | No | NA | 3 | 0 | 2 | Low-quality | Genome-fragment | 1.38 | AAI-based (medium-confidence) | 0 |
| S3_k141_124276 | 1485 | No | NA | 3 | 0 | 3 | Not-determined | Genome-fragment | NA | NA | 0 |

| | | | | | | | | | | | |
|---|---|---|---|---|---|---|---|---|---|---|---|
| S3_k141_26428 | 2801 | No | NA | 3 | 0 | 3 | Not-determined | Genome-fragment | NA | NA | 0 |
| S3_k141_49375 | 3228 | No | NA | 4 | 0 | 3 | Not-determined | Genome-fragment | NA | NA | 0 |
| S3_k141_106894 | 3389 | No | NA | 4 | 0 | 1 | Not-determined | Genome-fragment | NA | NA | 0 |
| S5-final-viral-combined_k141_77051\|\|full | 1585 | No | NA | 2 | 1 | 0 | Low-quality | Genome-fragment | 3.71 | AAI-based (high-confidence) | 0 |
| S3_k141_89963 | 1932 | No | NA | 1 | 0 | 1 | Not-determined | Genome-fragment | NA | NA | 0 |
| S3_k141_89973 | 1979 | No | NA | 1 | 0 | 0 | Not-determined | Genome-fragment | NA | NA | 0 |
| S7-final-viral-combined_k141_173052\|\|full | 1585 | No | NA | 3 | 0 | 0 | Low-quality | Genome-fragment | 2.51 | AAI-based (medium-confidence) | 0 |
| S3_k141_78147 | 1064 | No | NA | 1 | 0 | 0 | Not-determined | Genome-fragment | NA | NA | 0 |
| S3_k141_38067 | 1153 | No | NA | 2 | 0 | 1 | Not-determined | Genome-fragment | NA | NA | 0 |
| S3_k141_9084 | 3847 | No | NA | 3 | 0 | 2 | Not-determined | Genome-fragment | NA | NA | 0 |
| S3_k141_38075 | 1512 | No | NA | 2 | 0 | 2 | Not-determined | Genome-fragment | NA | NA | 0 |
| S3_k141_26509 | 2006 | No | NA | 2 | 0 | 1 | Not-determined | Genome-fragment | NA | NA | 0 |
| S3_k141_124338 | 1294 | No | NA | 2 | 0 | 1 | Not-determined | Genome-fragment | NA | NA | 0 |
| S3_k141_100987 | 1189 | No | NA | 2 | 0 | 1 | Not-determined | Genome-fragment | NA | NA | 0 |
| S3_k141_55275 | 3121 | No | NA | 1 | 0 | 0 | Not-determined | Genome-fragment | NA | NA | 0 |
| S17-k141_179680 | 1585 | No | NA | 2 | 0 | 0 | Low-quality | Genome-fragment | 1.86 | AAI-based (high-confidence) | 0 |
| S3_k141_38109 | 1282 | No | NA | 2 | 0 | 1 | Not-determined | Genome-fragment | NA | NA | 0 |
| S3_k141_84119 | 1040 | No | NA | 2 | 0 | 0 | Not-determined | Genome-fragment | NA | NA | 0 |
| S3_k141_84129 | 1990 | No | NA | 2 | 0 | 1 | Not-determined | Genome-fragment | NA | NA | 0 |
| S2-final-viral-combined_k141_150176\|\|full | 1585 | No | NA | 2 | 0 | 1 | Low-quality | Genome-fragment | 1.66 | AAI-based (medium-confidence) | 0 |
| S2-final-viral-combined_k141_96767\|\|lt2gene | 1585 | No | NA | 2 | 1 | 0 | Low-quality | Genome-fragment | 3.61 | AAI-based (high-confidence) | 0 |
| S3_k141_124408 | 1160 | No | NA | 2 | 0 | 0 | Not-determined | Genome-fragment | NA | NA | 0 |
| S3_k141_130309 | 1063 | No | NA | 2 | 0 | 0 | Not-determined | Genome-fragment | NA | NA | 0 |
| S3_k141_95614 | 1993 | No | NA | 2 | 0 | 0 | Not-determined | Genome-fragment | NA | NA | 0 |
| S2_k141_235717 | 1585 | No | NA | 1 | 0 | 1 | Low-quality | Genome-fragment | 0.76 | AAI-based (high-confidence) | 0 |
| S3_k141_106991 | 1005 | No | NA | 1 | 0 | 1 | Not-determined | Genome-fragment | NA | NA | 0 |
| S6_k141_67138 | 1585 | No | NA | 3 | 1 | 0 | Low-quality | Genome-fragment | 2.63 | AAI-based (high-confidence) | 0 |
| S9-final-viral-combined_k141_22181\|\|full | 1586 | No | NA | 4 | 0 | 0 | Low-quality | Genome-fragment | 1.92 | AAI-based (high-confidence) | 0 |
| S3_k141_14836 | 2178 | No | NA | 3 | 0 | 2 | Not-determined | Genome-fragment | NA | NA | 0 |
| S3_k141_32125 | 2050 | No | NA | 2 | 0 | 2 | Not-determined | Genome-fragment | NA | NA | 0 |
| S5-final-viral-combined_k141_175574\|\|full | 1586 | Yes | 717 | 4 | 1 | 1 | Low-quality | Genome-fragment | 1.34 | AAI-based (medium-confidence) | 54.79 |
| S3_k141_66869 | 1139 | No | NA | 1 | 0 | 1 | Not-determined | Genome-fragment | NA | NA | 0 |
| S3_k141_14857 | 1648 | No | NA | 2 | 0 | 2 | Not-determined | Genome-fragment | NA | NA | 0 |
| S3_k141_32148 | 1898 | No | NA | 1 | 0 | 0 | Not-determined | Genome-fragment | NA | NA | 0 |
| S3_k141_72524 | 1079 | No | NA | 1 | 0 | 1 | Not-determined | Genome-fragment | NA | NA | 0 |
| S5-final-viral-combined_k141_199773\|\|lt2gene | 1586 | No | NA | 1 | 1 | 0 | Low-quality | Genome-fragment | 2.96 | AAI-based (medium-confidence) | 0 |
| S3_k141_14886 | 1545 | No | NA | 1 | 0 | 0 | Not-determined | Genome-fragment | NA | NA | 0 |
| S3_k141_61059 | 1444 | No | NA | 3 | 0 | 2 | Not-determined | Genome-fragment | NA | NA | 0 |
| S3_k141_61060 | 1653 | No | NA | 1 | 0 | 1 | Not-determined | Genome-fragment | NA | NA | 0 |
| S3_k141_20892 | 1088 | No | NA | 1 | 0 | 1 | Not-determined | Genome-fragment | NA | NA | 0 |
| S4-final-viral-combined_k141_162101\|\|full | 1586 | No | NA | 4 | 0 | 0 | Low-quality | Genome-fragment | 2.68 | AAI-based (medium-confidence) | 0 |
| S3_k141_84292 | 1290 | No | NA | 1 | 0 | 1 | Not-determined | Genome-fragment | NA | NA | 0 |
| S3_k141_112931 | 1463 | No | NA | 1 | 0 | 1 | Not-determined | Genome-fragment | NA | NA | 0 |
| S3_k141_130467 | 1075 | No | NA | 1 | 0 | 0 | Not-determined | Genome-fragment | NA | NA | 0 |
| S3_k141_124561 | 1816 | No | NA | 2 | 0 | 2 | Not-determined | Genome-fragment | NA | NA | 0 |
| S3_k141_61089 | 2098 | No | NA | 3 | 0 | 3 | Not-determined | Genome-fragment | NA | NA | 0 |
| S4_k141_124600 | 1586 | No | NA | 4 | 0 | 1 | Low-quality | Genome-fragment | 3.01 | AAI-based (medium-confidence) | 0 |
| S3_k141_20919 | 1047 | No | NA | 1 | 0 | 0 | Not-determined | Genome-fragment | NA | NA | 0 |
| S6-final-viral-combined_k141_5934\|\|full | 1586 | No | NA | 2 | 0 | 0 | Low-quality | Genome-fragment | 0.96 | AAI-based (medium-confidence) | 0 |
| S3_k141_49605 | 1451 | No | NA | 1 | 0 | 1 | Not-determined | Genome-fragment | NA | NA | 0 |
| S3_k141_135920 | 1033 | No | NA | 2 | 0 | 0 | Not-determined | Genome-fragment | NA | NA | 0 |
| S3_k141_43782 | 3510 | No | NA | 4 | 0 | 2 | Not-determined | Genome-fragment | NA | NA | 0 |
| S3_k141_135926 | 1914 | No | NA | 1 | 0 | 1 | Not-determined | Genome-fragment | NA | NA | 0 |
| S3_k141_107159 | 1152 | No | NA | 1 | 0 | 1 | Not-determined | Genome-fragment | NA | NA | 0 |
| S3_k141_112974 | 1786 | No | NA | 5 | 0 | 4 | Not-determined | Genome-fragment | NA | NA | 0 |
| S3_k141_118983 | 3388 | No | NA | 2 | 0 | 2 | Not-determined | Genome-fragment | NA | NA | 0 |
| S6_k141_138272 | 1586 | No | NA | 2 | 0 | 0 | Low-quality | Genome-fragment | 1.62 | AAI-based (medium-confidence) | 0 |
| S8-final-viral-combined_k141_253262\|\|full | 1586 | No | NA | 2 | 2 | 0 | Low-quality | Genome-fragment | 4.48 | AAI-based (high-confidence) | 0 |
| S3_k141_84374 | 3876 | No | NA | 4 | 0 | 2 | Not-determined | Genome-fragment | NA | NA | 0 |
| S8_k141_104240 | 1586 | No | NA | 2 | 0 | 0 | Low-quality | Genome-fragment | 1.88 | AAI-based (high-confidence) | 0 |
| S3_k141_113013 | 1299 | No | NA | 4 | 0 | 3 | Not-determined | Genome-fragment | NA | NA | 0 |
| S3_k141_32294 | 1002 | No | NA | 1 | 0 | 1 | Not-determined | Genome-fragment | NA | NA | 0 |
| S5-final-viral-combined_k141_179828\|\|lt2gene | 1587 | No | NA | 1 | 1 | 0 | Low-quality | Genome-fragment | 4.43 | AAI-based (medium-confidence) | 0 |
| S3_k141_9399 | 1342 | No | NA | 1 | 0 | 0 | Not-determined | Genome-fragment | NA | NA | 0 |
| S3_k141_38428 | 2516 | No | NA | 1 | 0 | 1 | Not-determined | Genome-fragment | NA | NA | 0 |
| S3_k141_95841 | 1359 | No | NA | 1 | 0 | 1 | Not-determined | Genome-fragment | NA | NA | 0 |
| S3_k141_15001 | 3802 | No | NA | 3 | 0 | 2 | Not-determined | Genome-fragment | NA | NA | 0 |
| S3_k141_49694 | 1368 | No | NA | 2 | 0 | 1 | Not-determined | Genome-fragment | NA | NA | 0 |
| S3_k141_55615 | 1354 | No | NA | 3 | 0 | 1 | Not-determined | Genome-fragment | NA | NA | 0 |
| S12-final-viral-combined_k141_44704\|\|full | 1587 | No | NA | 2 | 0 | 1 | Low-quality | Genome-fragment | 4.08 | AAI-based (high-confidence) | 0 |
| S3_k141_15049 | 5834 | No | NA | 4 | 0 | 1 | Not-determined | Genome-fragment | NA | NA | 0 |
| S3_k141_90344 | 1112 | No | NA | 2 | 0 | 1 | Not-determined | Genome-fragment | NA | NA | 0 |
| S3_k141_95876 | 1396 | No | NA | 2 | 0 | 1 | Not-determined | Genome-fragment | NA | NA | 0 |
| S3_k141_43953 | 1762 | No | NA | 2 | 0 | 0 | Not-determined | Genome-fragment | NA | NA | 0 |
| S3_k141_26881 | 3076 | No | NA | 3 | 0 | 0 | Not-determined | Genome-fragment | NA | NA | 0 |
| S3_k141_21122 | 6920 | No | NA | 2 | 0 | 2 | Not-determined | Genome-fragment | NA | NA | 0 |
| S3_k141_90386 | 1406 | No | NA | 1 | 0 | 1 | Not-determined | Genome-fragment | NA | NA | 0 |
| S2-final-viral-combined_k141_52608\|\|full | 1587 | No | NA | 3 | 0 | 0 | Low-quality | Genome-fragment | 3.13 | AAI-based (medium-confidence) | 0 |
| S3_k141_4089 | 1005 | No | NA | 1 | 0 | 1 | Not-determined | Genome-fragment | NA | NA | 0 |
| S2-final-viral-combined_k141_196687\|\|full | 1587 | No | NA | 5 | 0 | 0 | Low-quality | Genome-fragment | 1.61 | AAI-based (medium-confidence) | 0 |
| S4-final-viral-combined_k141_12514\|\|full | 1587 | No | NA | 3 | 0 | 0 | Low-quality | Genome-fragment | 4.75 | AAI-based (high-confidence) | 0 |
| S3_k141_15119 | 1043 | No | NA | 2 | 0 | 0 | Not-determined | Genome-fragment | NA | NA | 0 |
| S6-final-viral-combined_k141_102734\|\|full | 1587 | No | NA | 3 | 0 | 1 | Low-quality | Genome-fragment | 1.48 | AAI-based (high-confidence) | 0 |
| S8-final-viral-combined_k141_16125\|\|lt2gene | 1587 | No | NA | 2 | 2 | 0 | Low-quality | Genome-fragment | 2.29 | AAI-based (high-confidence) | 0 |
| S3_k141_101302 | 1072 | No | NA | 1 | 0 | 1 | Not-determined | Genome-fragment | NA | NA | 0 |
| S3_k141_130686 | 1100 | No | NA | 2 | 0 | 0 | Not-determined | Genome-fragment | NA | NA | 0 |
| S1_k141_14768 | 1588 | No | NA | 2 | 0 | 0 | Low-quality | Genome-fragment | 2.46 | AAI-based (high-confidence) | 0 |
| S7_k141_73195 | 1588 | No | NA | 1 | 0 | 1 | Low-quality | Genome-fragment | 1.04 | AAI-based (high-confidence) | 0 |
| S12-final-viral-combined_k141_400843\|\|lt2gene | 1588 | No | NA | 1 | 1 | 0 | Low-quality | Genome-fragment | 3.69 | AAI-based (high-confidence) | 0 |
| S3_k141_107387 | 1353 | No | NA | 2 | 0 | 1 | Not-determined | Genome-fragment | NA | NA | 0 |
| S3_k141_107406 | 1024 | No | NA | 1 | 0 | 1 | Not-determined | Genome-fragment | NA | NA | 0 |
| S3_k141_130716 | 3788 | No | NA | 3 | 0 | 1 | Not-determined | Genome-fragment | NA | NA | 0 |
| S3_k141_32476 | 1103 | No | NA | 2 | 0 | 1 | Not-determined | Genome-fragment | NA | NA | 0 |
| S3_k141_124846 | 1979 | No | NA | 1 | 0 | 0 | Not-determined | Genome-fragment | NA | NA | 0 |
| S12_k141_247168 | 1588 | No | NA | 1 | 0 | 0 | Low-quality | Genome-fragment | 1.6 | AAI-based (medium-confidence) | 0 |
| S3_k141_130736 | 1482 | No | NA | 2 | 0 | 1 | Not-determined | Genome-fragment | NA | NA | 0 |
| S3_k141_61326 | 1078 | No | NA | 2 | 0 | 2 | Not-determined | Genome-fragment | NA | NA | 0 |
| S4_k141_102486 | 1588 | No | NA | 3 | 0 | 0 | Low-quality | Genome-fragment | 3.53 | AAI-based (medium-confidence) | 0 |
| S3_k141_4180 | 2624 | No | NA | 2 | 0 | 1 | Not-determined | Genome-fragment | NA | NA | 0 |
| S3_k141_21266 | 1220 | No | NA | 1 | 0 | 0 | Not-determined | Genome-fragment | NA | NA | 0 |
| S3_k141_21283 | 1392 | No | NA | 2 | 0 | 1 | Not-determined | Genome-fragment | NA | NA | 0 |
| S6-final-viral-combined_k141_88474\|\|full | 1588 | No | NA | 2 | 0 | 1 | Low-quality | Genome-fragment | 1.24 | AAI-based (medium-confidence) | 0 |
| S3_k141_38623 | 1742 | No | NA | 1 | 0 | 1 | Not-determined | Genome-fragment | NA | NA | 0 |
| S3_k141_38626 | 1555 | No | NA | 2 | 0 | 2 | Not-determined | Genome-fragment | NA | NA | 0 |
| S3_k141_136230 | 3357 | No | NA | 2 | 0 | 1 | Not-determined | Genome-fragment | NA | NA | 0 |
| S11-final-viral-combined_k141_168839\|\|full | 1589 | No | NA | 3 | 1 | 0 | Low-quality | Genome-fragment | 3.29 | AAI-based (high-confidence) | 0 |
| S3_k141_49871 | 1436 | No | NA | 2 | 0 | 1 | Not-determined | Genome-fragment | NA | NA | 0 |
| S3_k141_4210 | 1036 | No | NA | 1 | 0 | 0 | Not-determined | Genome-fragment | NA | NA | 0 |
| S3_k141_119354 | 1580 | No | NA | 2 | 0 | 0 | Not-determined | Genome-fragment | NA | NA | 0 |
| S11-final-viral-combined_k141_38858\|\|full | 1589 | No | NA | 3 | 0 | 3 | Low-quality | Genome-fragment | 1.02 | AAI-based (medium-confidence) | 0 |
| S3_k141_78789 | 1037 | No | NA | 1 | 0 | 1 | Not-determined | Genome-fragment | NA | NA | 0 |
| S3_k141_21334 | 1005 | No | NA | 1 | 0 | 1 | Not-determined | Genome-fragment | NA | NA | 0 |
| S3_k141_72905 | 1027 | No | NA | 2 | 0 | 1 | Not-determined | Genome-fragment | NA | NA | 0 |
| S3_k141_15230 | 1132 | No | NA | 2 | 0 | 1 | Not-determined | Genome-fragment | NA | NA | 0 |
| S3_k141_72907 | 1053 | No | NA | 1 | 0 | 0 | Not-determined | Genome-fragment | NA | NA | 0 |
| S3_k141_38720 | 1835 | No | NA | 6 | 0 | 2 | Not-determined | Genome-fragment | NA | NA | 0 |
| S3_k141_44201 | 3678 | No | NA | 4 | 0 | 3 | Not-determined | Genome-fragment | NA | NA | 0 |
| S3_k141_96183 | 1000 | No | NA | 1 | 0 | 1 | Not-determined | Genome-fragment | NA | NA | 0 |
| S3_k141_136359 | 2131 | No | NA | 2 | 0 | 1 | Not-determined | Genome-fragment | NA | NA | 0 |
| S3_k141_119481 | 3160 | No | NA | 1 | 0 | 1 | Not-determined | Genome-fragment | NA | NA | 0 |
| S3_k141_61547 | 1636 | No | NA | 1 | 0 | 0 | Not-determined | Genome-fragment | NA | NA | 0 |
| S3_k141_119495 | 1043 | No | NA | 2 | 0 | 2 | Not-determined | Genome-fragment | NA | NA | 0 |
| S1-final-viral-combined_k141_350963\|\|full | 1589 | No | NA | 2 | 0 | 1 | Low-quality | Genome-fragment | 3.92 | AAI-based (medium-confidence) | 0 |
| S3_k141_9771 | 1155 | No | NA | 2 | 0 | 2 | Not-determined | Genome-fragment | NA | NA | 0 |
| S3_k141_136408 | 1438 | No | NA | 1 | 0 | 1 | Not-determined | Genome-fragment | NA | NA | 0 |
| S1-final-viral-combined_k141_35978\|\|lt2gene | 1589 | No | NA | 1 | 1 | 0 | Low-quality | Genome-fragment | 2.95 | AAI-based (high-confidence) | 0 |
| S5-final-viral-combined_k141_108219\|\|full | 1589 | No | NA | 4 | 0 | 0 | Low-quality | Genome-fragment | 3.74 | AAI-based (high-confidence) | 0 |
| S3_k141_84822 | 1042 | No | NA | 1 | 0 | 1 | Not-determined | Genome-fragment | NA | NA | 0 |
| S3_k141_125123 | 1161 | No | NA | 2 | 0 | 0 | Not-determined | Genome-fragment | NA | NA | 0 |
| S5-final-viral-combined_k141_131875\|\|full | 1589 | No | NA | 2 | 0 | 2 | Low-quality | Genome-fragment | 1.04 | AAI-based (medium-confidence) | 0 |
| S2-final-viral-combined_k141_225887\|\|full | 1589 | No | NA | 2 | 0 | 0 | Low-quality | Genome-fragment | 2.68 | AAI-based (medium-confidence) | 0 |
| S3_k141_125147 | 1739 | No | NA | 1 | 0 | 1 | Not-determined | Genome-fragment | NA | NA | 0 |
| S3_k141_44250 | 1467 | No | NA | 2 | 0 | 1 | Not-determined | Genome-fragment | NA | NA | 0 |
| S2_k141_9487 | 1589 | No | NA | 3 | 0 | 0 | Low-quality | Genome-fragment | 1.91 | AAI-based (high-confidence) | 0 |
| S3_k141_4455 | 3704 | No | NA | 3 | 0 | 3 | Not-determined | Genome-fragment | NA | NA | 0 |
| S3_k141_101649 | 1629 | No | NA | 1 | 0 | 1 | Not-determined | Genome-fragment | NA | NA | 0 |
| S6_k141_37414 | 1589 | No | NA | 5 | 0 | 0 | Low-quality | Genome-fragment | 2.63 | AAI-based (high-confidence) | 0 |
| S3_k141_4469 | 3120 | No | NA | 3 | 0 | 0 | Not-determined | Genome-fragment | NA | NA | 0 |
| S11-final-viral-combined_k141_216220\|\|full | 1590 | No | NA | 2 | 0 | 0 | Low-quality | Genome-fragment | 3.12 | AAI-based (high-confidence) | 0 |
| S3_k141_4477 | 1365 | No | NA | 1 | 0 | 1 | Not-determined | Genome-fragment | NA | NA | 0 |
| S3_k141_131059 | 1042 | No | NA | 1 | 0 | 1 | Not-determined | Genome-fragment | NA | NA | 0 |
| S3_k141_15401 | 6692 | No | NA | 2 | 0 | 1 | Not-determined | Genome-fragment | NA | NA | 0 |
| S12_k141_239390 | 1590 | No | NA | 2 | 0 | 0 | Low-quality | Genome-fragment | 0.73 | AAI-based (high-confidence) | 0 |
| S3_k141_119643 | 1040 | No | NA | 1 | 0 | 0 | Not-determined | Genome-fragment | NA | NA | 0 |
| S3_k141_131082 | 1042 | No | NA | 1 | 0 | 1 | Not-determined | Genome-fragment | NA | NA | 0 |
| S3_k141_101714 | 1230 | No | NA | 1 | 0 | 1 | Not-determined | Genome-fragment | NA | NA | 0 |
| S3_k141_90742 | 1045 | No | NA | 2 | 0 | 2 | Not-determined | Genome-fragment | NA | NA | 0 |
| S3_k141_113633 | 1168 | No | NA | 1 | 0 | 1 | Not-determined | Genome-fragment | NA | NA | 0 |
| S3_k141_9909 | 2409 | No | NA | 3 | 0 | 2 | Not-determined | Genome-fragment | NA | NA | 0 |
| S3_k141_44320 | 1133 | No | NA | 2 | 0 | 1 | Not-determined | Genome-fragment | NA | NA | 0 |
| S4_k141_68299 | 1590 | No | NA | 1 | 0 | 0 | Low-quality | Genome-fragment | 1.63 | AAI-based (high-confidence) | 0 |
| S3_k141_27347 | 1698 | No | NA | 2 | 0 | 1 | Not-determined | Genome-fragment | NA | NA | 0 |
| S3_k141_50155 | 1594 | No | NA | 2 | 0 | 0 | Not-determined | Genome-fragment | NA | NA | 0 |
| S3_k141_90778 | 11861 | No | NA | 9 | 0 | 7 | Not-determined | Genome-fragment | NA | NA | 0 |
| S3_k141_44335 | 1853 | No | NA | 1 | 0 | 1 | Not-determined | Genome-fragment | NA | NA | 0 |
| S3_k141_73148 | 1232 | No | NA | 1 | 0 | 1 | Not-determined | Genome-fragment | NA | NA | 0 |
| S4_k141_96943 | 1590 | No | NA | 1 | 0 | 0 | Low-quality | Genome-fragment | 1.49 | AAI-based (medium-confidence) | 0 |
| S3_k141_119685 | 7847 | No | NA | 5 | 0 | 4 | Not-determined | Genome-fragment | NA | NA | 0 |
| S9-final-viral-combined_k141_54609\|\|full | 1591 | No | NA | 3 | 0 | 0 | Low-quality | Genome-fragment | 1.93 | AAI-based (high-confidence) | 0 |
| S3_k141_85039 | 1042 | No | NA | 3 | 0 | 0 | Not-determined | Genome-fragment | NA | NA | 0 |
| S3_k141_15505 | 1079 | No | NA | 1 | 0 | 1 | Not-determined | Genome-fragment | NA | NA | 0 |
| S9-final-viral-combined_k141_60651\|\|full | 1591 | Yes | 540 | 3 | 1 | 1 | Low-quality | Genome-fragment | 1.12 | AAI-based (medium-confidence) | 66.06 |
| S11-final-viral-combined_k141_329390\|\|full | 1591 | No | NA | 4 | 3 | 0 | Low-quality | Genome-fragment | 4.33 | AAI-based (high-confidence) | 0 |
| S3_k141_27427 | 1043 | No | NA | 1 | 0 | 0 | Not-determined | Genome-fragment | NA | NA | 0 |
| S3_k141_113756 | 1484 | No | NA | 1 | 0 | 1 | Not-determined | Genome-fragment | NA | NA | 0 |
| S3_k141_4619 | 3790 | No | NA | 3 | 0 | 3 | Not-determined | Genome-fragment | NA | NA | 0 |
| S3_k141_44476 | 1155 | No | NA | 1 | 0 | 1 | Not-determined | Genome-fragment | NA | NA | 0 |
| S3_k141_136638 | 1055 | No | NA | 1 | 0 | 0 | Not-determined | Genome-fragment | NA | NA | 0 |
| S3_k141_39100 | 1097 | No | NA | 1 | 0 | 0 | Not-determined | Genome-fragment | NA | NA | 0 |
| S3_k141_4640 | 2334 | No | NA | 3 | 0 | 2 | Not-determined | Genome-fragment | NA | NA | 0 |
| S3_k141_107867 | 1235 | No | NA | 1 | 0 | 0 | Not-determined | Genome-fragment | NA | NA | 0 |
| S3_k141_107868 | 1028 | No | NA | 1 | 0 | 1 | Not-determined | Genome-fragment | NA | NA | 0 |
| S3_k141_10087 | 2166 | No | NA | 1 | 0 | 1 | Not-determined | Genome-fragment | NA | NA | 0 |
| S11-final-viral-combined_k141_302872\|\|lt2gene | 1591 | No | NA | 1 | 1 | 0 | Low-quality | Genome-fragment | 4.05 | AAI-based (high-confidence) | 0 |
| S3_k141_4707 | 1347 | No | NA | 2 | 0 | 2 | Not-determined | Genome-fragment | NA | NA | 0 |
| S3_k141_39158 | 1024 | No | NA | 1 | 0 | 0 | Not-determined | Genome-fragment | NA | NA | 0 |
| S3_k141_67745 | 1117 | No | NA | 1 | 0 | 1 | Not-determined | Genome-fragment | NA | NA | 0 |
| S3_k141_67771 | 1103 | No | NA | 1 | 0 | 1 | Not-determined | Genome-fragment | NA | NA | 0 |
| S1-final-viral-combined_k141_201130\|\|full | 1591 | No | NA | 3 | 0 | 0 | Low-quality | Genome-fragment | 1.02 | AAI-based (high-confidence) | 0 |
| S3_k141_91007 | 1043 | No | NA | 2 | 0 | 1 | Not-determined | Genome-fragment | NA | NA | 0 |
| S1-final-viral-combined_k141_8924\|\|lt2gene | 1591 | No | NA | 2 | 2 | 0 | Low-quality | Genome-fragment | 4.64 | AAI-based (high-confidence) | 0 |
| S3_k141_21846 | 2452 | No | NA | 2 | 0 | 1 | Not-determined | Genome-fragment | NA | NA | 0 |
| S3_k141_4761 | 1885 | No | NA | 1 | 0 | 1 | Not-determined | Genome-fragment | NA | NA | 0 |
| S3_k141_4763 | 1470 | No | NA | 1 | 0 | 1 | Not-determined | Genome-fragment | NA | NA | 0 |
| S3_k141_119915 | 1200 | No | NA | 1 | 0 | 1 | Not-determined | Genome-fragment | NA | NA | 0 |
| S3_k141_107997 | 1014 | No | NA | 1 | 0 | 0 | Not-determined | Genome-fragment | NA | NA | 0 |
| S1_k141_359450 | 1591 | No | NA | 3 | 0 | 1 | Low-quality | Genome-fragment | 1.21 | AAI-based (medium-confidence) | 0 |
| S3_k141_108003 | 1009 | No | NA | 2 | 0 | 0 | Not-determined | Genome-fragment | NA | NA | 0 |
| S5-final-viral-combined_k141_76068\|\|full | 1591 | No | NA | 2 | 1 | 0 | Low-quality | Genome-fragment | 3.54 | HMM-based (lower-bound) | 0 |
| S3_k141_33086 | 1483 | No | NA | 1 | 0 | 1 | Not-determined | Genome-fragment | NA | NA | 0 |
| S3_k141_15727 | 1014 | No | NA | 2 | 0 | 0 | Not-determined | Genome-fragment | NA | NA | 0 |
| S3_k141_108031 | 1899 | No | NA | 1 | 0 | 1 | Not-determined | Genome-fragment | NA | NA | 0 |
| S3_k141_44657 | 3062 | No | NA | 6 | 0 | 0 | Not-determined | Genome-fragment | NA | NA | 0 |
| S5-final-viral-combined_k141_141223\|\|lt2gene | 1591 | No | NA | 2 | 2 | 0 | Low-quality | Genome-fragment | 4.27 | AAI-based (high-confidence) | 0 |
| S3_k141_39288 | 1108 | No | NA | 1 | 0 | 1 | Not-determined | Genome-fragment | NA | NA | 0 |
| S3_k141_21941 | 3176 | No | NA | 3 | 0 | 1 | Not-determined | Genome-fragment | NA | NA | 0 |
| S3_k141_125618 | 1087 | No | NA | 2 | 0 | 0 | Not-determined | Genome-fragment | NA | NA | 0 |
| S3_k141_125633 | 1105 | No | NA | 2 | 0 | 2 | Not-determined | Genome-fragment | NA | NA | 0 |
| S11-final-viral-combined_k141_49290\|\|full | 1592 | No | NA | 4 | 0 | 1 | Low-quality | Genome-fragment | 3.45 | AAI-based (medium-confidence) | 0 |
| S1-final-viral-combined_k141_118405\|\|full | 1592 | No | NA | 4 | 0 | 0 | Low-quality | Genome-fragment | 5.13 | AAI-based (medium-confidence) | 0 |
| S3_k141_96764 | 1035 | No | NA | 2 | 0 | 1 | Not-determined | Genome-fragment | NA | NA | 0 |
| S3_k141_114038 | 1366 | No | NA | 1 | 0 | 1 | Not-determined | Genome-fragment | NA | NA | 0 |
| S3_k141_50489 | 2162 | No | NA | 1 | 0 | 1 | Not-determined | Genome-fragment | NA | NA | 0 |
| S3_k141_73474 | 1183 | No | NA | 3 | 0 | 2 | Not-determined | Genome-fragment | NA | NA | 0 |
| S3-final-viral-combined_k141_88985\|\|full | 1592 | No | NA | 3 | 0 | 1 | Low-quality | Genome-fragment | 0.76 | AAI-based (medium-confidence) | 0 |
| S3_k141_22019 | 1173 | No | NA | 1 | 0 | 1 | Not-determined | Genome-fragment | NA | NA | 0 |
| S3_k141_27814 | 3168 | No | NA | 2 | 0 | 2 | Not-determined | Genome-fragment | NA | NA | 0 |
| S3_k141_15839 | 1015 | No | NA | 1 | 0 | 1 | Not-determined | Genome-fragment | NA | NA | 0 |
| S3_k141_73509 | 1604 | No | NA | 2 | 0 | 2 | Not-determined | Genome-fragment | NA | NA | 0 |
| S3_k141_44774 | 1524 | No | NA | 3 | 0 | 1 | Not-determined | Genome-fragment | NA | NA | 0 |
| S3_k141_62179 | 3027 | No | NA | 4 | 0 | 1 | Not-determined | Genome-fragment | NA | NA | 0 |
| S3_k141_85362 | 9326 | No | NA | 11 | 0 | 2 | Not-determined | Genome-fragment | NA | NA | 0 |
| S3_k141_125747 | 1155 | No | NA | 2 | 0 | 2 | Not-determined | Genome-fragment | NA | NA | 0 |
| S5-final-viral-combined_k141_75692\|\|full | 1592 | No | NA | 4 | 0 | 0 | Low-quality | Genome-fragment | 0.61 | AAI-based (high-confidence) | 0 |
| S3_k141_27884 | 1776 | No | NA | 2 | 0 | 1 | Not-determined | Genome-fragment | NA | NA | 0 |
| S3_k141_136929 | 1282 | No | NA | 1 | 0 | 1 | Not-determined | Genome-fragment | NA | NA | 0 |
| S3_k141_56522 | 1436 | No | NA | 2 | 0 | 1 | Not-determined | Genome-fragment | NA | NA | 0 |
| S3_k141_33291 | 1136 | No | NA | 2 | 0 | 2 | Not-determined | Genome-fragment | NA | NA | 0 |
| S5-final-viral-combined_k141_61615\|\|full | 1592 | No | NA | 3 | 1 | 0 | Low-quality | Genome-fragment | 1.25 | HMM-based (lower-bound) | 0 |
| S3_k141_73620 | 1487 | No | NA | 1 | 0 | 1 | Not-determined | Genome-fragment | NA | NA | 0 |
| S3_k141_136998 | 1010 | No | NA | 1 | 0 | 1 | Not-determined | Genome-fragment | NA | NA | 0 |
| S3_k141_22177 | 1336 | No | NA | 2 | 0 | 1 | Not-determined | Genome-fragment | NA | NA | 0 |
| S3_k141_22184 | 1157 | No | NA | 2 | 0 | 1 | Not-determined | Genome-fragment | NA | NA | 0 |
| S3_k141_10505 | 1326 | No | NA | 1 | 0 | 1 | Not-determined | Genome-fragment | NA | NA | 0 |
| S3_k141_62289 | 2152 | No | NA | 2 | 0 | 1 | Not-determined | Genome-fragment | NA | NA | 0 |
| S3_k141_96938 | 2018 | No | NA | 2 | 0 | 1 | Not-determined | Genome-fragment | NA | NA | 0 |
| S3_k141_33377 | 2041 | No | NA | 5 | 0 | 0 | Not-determined | Genome-fragment | NA | NA | 0 |
| S3_k141_120280 | 1037 | No | NA | 1 | 0 | 1 | Not-determined | Genome-fragment | NA | NA | 0 |
| S3_k141_73649 | 1209 | No | NA | 3 | 0 | 0 | Not-determined | Genome-fragment | NA | NA | 0 |
| S5_k141_57564 | 1592 | No | NA | 3 | 1 | 0 | Low-quality | Genome-fragment | 2.85 | AAI-based (medium-confidence) | 0 |
| S3_k141_33424 | 5120 | No | NA | 6 | 0 | 0 | Not-determined | Genome-fragment | NA | NA | 0 |
| S12_k141_257693 | 1592 | Yes | 440 | 4 | 1 | 1 | Low-quality | Genome-fragment | 1.12 | AAI-based (high-confidence) | 72.36 |
| S3_k141_50705 | 1198 | No | NA | 2 | 0 | 1 | Not-determined | Genome-fragment | NA | NA | 0 |
| S3_k141_62330 | 1976 | No | NA | 3 | 0 | 1 | Not-determined | Genome-fragment | NA | NA | 0 |
| S9-final-viral-combined_k141_106254\|\|full | 1593 | No | NA | 3 | 0 | 1 | Low-quality | Genome-fragment | 0.55 | AAI-based (medium-confidence) | 0 |
| S3_k141_114266 | 3066 | No | NA | 4 | 0 | 2 | Not-determined | Genome-fragment | NA | NA | 0 |
| S3_k141_102274 | 1121 | No | NA | 2 | 0 | 2 | Not-determined | Genome-fragment | NA | NA | 0 |
| S3_k141_39593 | 3384 | No | NA | 3 | 0 | 2 | Not-determined | Genome-fragment | NA | NA | 0 |
| S11-final-viral-combined_k141_255341\|\|full | 1593 | No | NA | 3 | 0 | 1 | Low-quality | Genome-fragment | 2.02 | AAI-based (medium-confidence) | 0 |
| S3_k141_62380 | 1040 | No | NA | 1 | 0 | 1 | Not-determined | Genome-fragment | NA | NA | 0 |
| S11-final-viral-combined_k141_302394\|\|full | 1593 | No | NA | 2 | 0 | 1 | Low-quality | Genome-fragment | 2.46 | AAI-based (medium-confidence) | 0 |
| S3_k141_50759 | 3088 | No | NA | 1 | 0 | 1 | Not-determined | Genome-fragment | NA | NA | 0 |
| S11-final-viral-combined_k141_292904\|\|lt2gene | 1593 | No | NA | 2 | 1 | 0 | Low-quality | Genome-fragment | 3.54 | AAI-based (high-confidence) | 0 |
| S3_k141_28082 | 5300 | No | NA | 2 | 0 | 2 | Not-determined | Genome-fragment | NA | NA | 0 |
| S3_k141_56692 | 8573 | No | NA | 6 | 0 | 2 | Not-determined | Genome-fragment | NA | NA | 0 |
| S3_k141_44982 | 7634 | No | NA | 8 | 0 | 1 | Not-determined | Genome-fragment | NA | NA | 0 |
| S1-final-viral-combined_k141_362628\|\|full | 1593 | No | NA | 3 | 0 | 0 | Low-quality | Genome-fragment | 3.72 | AAI-based (high-confidence) | 0 |
| S3_k141_28116 | 1043 | No | NA | 2 | 0 | 0 | Not-determined | Genome-fragment | NA | NA | 0 |
| S3_k141_97079 | 5303 | No | NA | 5 | 0 | 3 | Not-determined | Genome-fragment | NA | NA | 0 |
| S3_k141_91410 | 2232 | No | NA | 3 | 0 | 2 | Not-determined | Genome-fragment | NA | NA | 0 |
| S3_k141_137119 | 1016 | No | NA | 3 | 0 | 3 | Not-determined | Genome-fragment | NA | NA | 0 |
| S3_k141_79797 | 1353 | No | NA | 1 | 0 | 0 | Not-determined | Genome-fragment | NA | NA | 0 |
| S3_k141_28172 | 1579 | No | NA | 2 | 0 | 2 | Not-determined | Genome-fragment | NA | NA | 0 |
| S3_k141_79814 | 3375 | No | NA | 4 | 0 | 1 | Not-determined | Genome-fragment | NA | NA | 0 |
| S3_k141_79854 | 1154 | No | NA | 1 | 0 | 1 | Not-determined | Genome-fragment | NA | NA | 0 |
| S3_k141_73891 | 4033 | No | NA | 5 | 0 | 1 | Not-determined | Genome-fragment | NA | NA | 0 |
| S1-final-viral-combined_k141_383405\|\|full | 1593 | No | NA | 3 | 0 | 0 | Low-quality | Genome-fragment | 3.47 | AAI-based (high-confidence) | 0 |
| S3_k141_79887 | 1561 | No | NA | 1 | 0 | 1 | Not-determined | Genome-fragment | NA | NA | 0 |
| S3_k141_45164 | 3380 | No | NA | 1 | 0 | 1 | Not-determined | Genome-fragment | NA | NA | 0 |
| S3_k141_45170 | 1024 | No | NA | 1 | 0 | 1 | Not-determined | Genome-fragment | NA | NA | 0 |
| S5-final-viral-combined_k141_1084\|\|full | 1593 | No | NA | 2 | 0 | 2 | Low-quality | Genome-fragment | 0.83 | AAI-based (medium-confidence) | 0 |
| S3_k141_97244 | 2448 | No | NA | 1 | 0 | 1 | Not-determined | Genome-fragment | NA | NA | 0 |
| S5-final-viral-combined_k141_190565\|\|full | 1593 | No | NA | 3 | 0 | 0 | Low-quality | Genome-fragment | 2.73 | AAI-based (high-confidence) | 0 |
| S3_k141_108486 | 6589 | No | NA | 4 | 0 | 4 | Not-determined | Genome-fragment | NA | NA | 0 |
| S3_k141_126267 | 6078 | No | NA | 12 | 0 | 12 | Not-determined | Genome-fragment | NA | NA | 0 |
| S3_k141_114548 | 1170 | No | NA | 4 | 0 | 0 | Not-determined | Genome-fragment | NA | NA | 0 |
| S3_k141_33749 | 1019 | No | NA | 2 | 0 | 1 | Not-determined | Genome-fragment | NA | NA | 0 |
| S3_k141_126335 | 1018 | No | NA | 1 | 0 | 0 | Not-determined | Genome-fragment | NA | NA | 0 |
| S3_k141_22618 | 3923 | No | NA | 3 | 0 | 1 | Not-determined | Genome-fragment | NA | NA | 0 |
| S3_k141_5511 | 1610 | No | NA | 3 | 0 | 3 | Not-determined | Genome-fragment | NA | NA | 0 |
| S7-final-viral-combined_k141_31311\|\|lt2gene | 1593 | No | NA | 2 | 1 | 0 | Low-quality | Genome-fragment | 1.49 | HMM-based (lower-bound) | 0 |
| S7_k141_221573 | 1593 | No | NA | 1 | 0 | 0 | Low-quality | Genome-fragment | 2.76 | AAI-based (high-confidence) | 0 |
| S3_k141_120735 | 1183 | No | NA | 3 | 0 | 2 | Not-determined | Genome-fragment | NA | NA | 0 |
| S3_k141_57032 | 1590 | No | NA | 1 | 0 | 0 | Not-determined | Genome-fragment | NA | NA | 0 |
| S17-final-viral-combined_k141_161528\|\|full | 1593 | No | NA | 2 | 0 | 1 | Low-quality | Genome-fragment | 2.5 | AAI-based (high-confidence) | 0 |
| S3_k141_132057 | 2129 | No | NA | 3 | 0 | 2 | Not-determined | Genome-fragment | NA | NA | 0 |
| S3_k141_85975 | 2706 | No | NA | 3 | 0 | 0 | Not-determined | Genome-fragment | NA | NA | 0 |
| S17-final-viral-combined_k141_161953\|\|full | 1593 | No | NA | 3 | 0 | 2 | Low-quality | Genome-fragment | 1.76 | AAI-based (high-confidence) | 0 |
| S3_k141_137332 | 1003 | No | NA | 1 | 0 | 1 | Not-determined | Genome-fragment | NA | NA | 0 |
| S2_k141_18547 | 1593 | No | NA | 3 | 0 | 0 | Low-quality | Genome-fragment | 1.66 | AAI-based (high-confidence) | 0 |
| S3_k141_74195 | 1189 | No | NA | 2 | 0 | 2 | Not-determined | Genome-fragment | NA | NA | 0 |
| S6-final-viral-combined_k141_42290\|\|full | 1593 | No | NA | 3 | 0 | 1 | Low-quality | Genome-fragment | 1.6 | AAI-based (medium-confidence) | 0 |
| S3_k141_51171 | 1077 | No | NA | 1 | 0 | 0 | Not-determined | Genome-fragment | NA | NA | 0 |
| S3_k141_62757 | 2102 | No | NA | 3 | 0 | 0 | Not-determined | Genome-fragment | NA | NA | 0 |
| S3_k141_126462 | 1575 | No | NA | 4 | 0 | 0 | Not-determined | Genome-fragment | NA | NA | 0 |
| S1-final-viral-combined_k141_161468\|\|full | 1594 | No | NA | 2 | 0 | 2 | Low-quality | Genome-fragment | 1.67 | AAI-based (high-confidence) | 0 |
| S3_k141_68711 | 1129 | No | NA | 1 | 0 | 1 | Not-determined | Genome-fragment | NA | NA | 0 |
| S17-final-viral-combined_k141_169064\|\|full | 1594 | No | NA | 4 | 2 | 0 | Low-quality | Genome-fragment | 2.65 | AAI-based (high-confidence) | 0 |
| S17-k141_52386 | 1594 | No | NA | 3 | 0 | 2 | Low-quality | Genome-fragment | 2.9 | AAI-based (medium-confidence) | 0 |
| S3_k141_74235 | 1614 | No | NA | 2 | 0 | 0 | Not-determined | Genome-fragment | NA | NA | 0 |
| S3_k141_22744 | 48196 | No | NA | 70 | 20 | 0 | High-quality | High-quality | 100 | AAI-based (high-confidence) | 0 |
| S3_k141_102743 | 4355 | No | NA | 2 | 0 | 2 | Not-determined | Genome-fragment | NA | NA | 0 |
| S3_k141_45402 | 3355 | No | NA | 2 | 0 | 0 | Not-determined | Genome-fragment | NA | NA | 0 |
| S12-final-viral-combined_k141_201063\|\|full | 1594 | No | NA | 2 | 0 | 1 | Low-quality | Genome-fragment | 0.46 | AAI-based (high-confidence) | 0 |
| S3_k141_28480 | 1183 | No | NA | 2 | 0 | 1 | Not-determined | Genome-fragment | NA | NA | 0 |
| S3_k141_74318 | 1722 | No | NA | 2 | 0 | 1 | Not-determined | Genome-fragment | NA | NA | 0 |
| S4_k141_166079 | 1594 | No | NA | 1 | 0 | 1 | Low-quality | Genome-fragment | 1.07 | AAI-based (medium-confidence) | 0 |
| S9-final-viral-combined_k141_2256\|\|full | 1595 | No | NA | 4 | 0 | 0 | Low-quality | Genome-fragment | 3.2 | AAI-based (high-confidence) | 0 |
| S3_k141_11141 | 1453 | No | NA | 1 | 0 | 0 | Not-determined | Genome-fragment | NA | NA | 0 |
| S3_k141_51292 | 2255 | No | NA | 2 | 0 | 1 | Not-determined | Genome-fragment | NA | NA | 0 |
| S3_k141_34078 | 1138 | No | NA | 1 | 0 | 1 | Not-determined | Genome-fragment | NA | NA | 0 |
| S11-final-viral-combined_k141_117128\|\|full | 1595 | No | NA | 4 | 1 | 0 | Low-quality | Genome-fragment | 4.45 | AAI-based (medium-confidence) | 0 |
| S3_k141_51323 | 1335 | No | NA | 1 | 0 | 1 | Not-determined | Genome-fragment | NA | NA | 0 |
| S3_k141_68833 | 1414 | No | NA | 1 | 0 | 1 | Not-determined | Genome-fragment | NA | NA | 0 |
| S3_k141_45448 | 5417 | No | NA | 3 | 0 | 2 | Not-determined | Genome-fragment | NA | NA | 0 |
| S3_k141_68841 | 1145 | No | NA | 2 | 0 | 1 | Not-determined | Genome-fragment | NA | NA | 0 |
| S3_k141_68850 | 6921 | No | NA | 6 | 0 | 4 | Not-determined | Genome-fragment | NA | NA | 0 |
| S3_k141_45471 | 2639 | No | NA | 4 | 0 | 2 | Not-determined | Genome-fragment | NA | NA | 0 |
| S11_k141_250961 | 1595 | No | NA | 2 | 0 | 0 | Low-quality | Genome-fragment | 3.47 | AAI-based (high-confidence) | 0 |
| S11_k141_80276 | 1595 | No | NA | 1 | 1 | 0 | Low-quality | Genome-fragment | 2.38 | AAI-based (high-confidence) | 0 |
| S1-final-viral-combined_k141_34223\|\|full | 1595 | No | NA | 3 | 0 | 1 | Low-quality | Genome-fragment | 2.74 | AAI-based (high-confidence) | 0 |
| S1-final-viral-combined_k141_205886\|\|full | 1595 | No | NA | 5 | 1 | 0 | Low-quality | Genome-fragment | 3.59 | AAI-based (medium-confidence) | 0 |
| S3_k141_22930 | 1404 | No | NA | 2 | 0 | 1 | Not-determined | Genome-fragment | NA | NA | 0 |
| S3_k141_62908 | 1297 | No | NA | 2 | 0 | 1 | Not-determined | Genome-fragment | NA | NA | 0 |
| S3_k141_11221 | 3131 | No | NA | 3 | 0 | 1 | Not-determined | Genome-fragment | NA | NA | 0 |
| S3_k141_80321 | 1300 | No | NA | 2 | 0 | 1 | Not-determined | Genome-fragment | NA | NA | 0 |
| S3_k141_11237 | 1514 | No | NA | 1 | 0 | 0 | Not-determined | Genome-fragment | NA | NA | 0 |
| S1-final-viral-combined_k141_386569\|\|full | 1595 | No | NA | 2 | 0 | 0 | Low-quality | Genome-fragment | 3.26 | AAI-based (high-confidence) | 0 |
| S3_k141_16765 | 1002 | No | NA | 1 | 0 | 0 | Not-determined | Genome-fragment | NA | NA | 0 |
| S3_k141_80334 | 1299 | No | NA | 1 | 0 | 1 | Not-determined | Genome-fragment | NA | NA | 0 |
| S3_k141_11249 | 1922 | No | NA | 1 | 0 | 1 | Not-determined | Genome-fragment | NA | NA | 0 |
| S3_k141_22975 | 1861 | No | NA | 2 | 0 | 1 | Not-determined | Genome-fragment | NA | NA | 0 |
| S4-final-viral-combined_k141_99029\|\|lt2gene | 1595 | No | NA | 2 | 1 | 0 | Low-quality | Genome-fragment | 1.57 | AAI-based (high-confidence) | 0 |
| S3_k141_97722 | 2186 | No | NA | 1 | 0 | 1 | Not-determined | Genome-fragment | NA | NA | 0 |
| S4_k141_188 | 1595 | No | NA | 2 | 0 | 1 | Low-quality | Genome-fragment | 1.07 | AAI-based (high-confidence) | 0 |
| S8-final-viral-combined_k141_16714\|\|full | 1595 | No | NA | 2 | 0 | 0 | Low-quality | Genome-fragment | 2.49 | AAI-based (high-confidence) | 0 |
| S3_k141_45555 | 5998 | No | NA | 7 | 0 | 4 | Not-determined | Genome-fragment | NA | NA | 0 |
| S3_k141_80383 | 1302 | No | NA | 2 | 0 | 0 | Not-determined | Genome-fragment | NA | NA | 0 |
| S3_k141_80393 | 3217 | No | NA | 1 | 0 | 1 | Not-determined | Genome-fragment | NA | NA | 0 |
| S3_k141_45577 | 2367 | No | NA | 4 | 0 | 2 | Not-determined | Genome-fragment | NA | NA | 0 |
| S3_k141_103039 | 1454 | No | NA | 2 | 0 | 2 | Not-determined | Genome-fragment | NA | NA | 0 |
| S3_k141_51491 | 1308 | No | NA | 2 | 0 | 1 | Not-determined | Genome-fragment | NA | NA | 0 |
| S11-final-viral-combined_k141_19658\|\|full | 1596 | No | NA | 3 | 2 | 0 | Low-quality | Genome-fragment | 3.7 | AAI-based (high-confidence) | 0 |
| S3_k141_63053 | 1402 | No | NA | 2 | 0 | 2 | Not-determined | Genome-fragment | NA | NA | 0 |
| S3_k141_11378 | 4812 | No | NA | 2 | 0 | 2 | Not-determined | Genome-fragment | NA | NA | 0 |
| S3_k141_80489 | 1136 | No | NA | 1 | 0 | 0 | Not-determined | Genome-fragment | NA | NA | 0 |
| S3_k141_109008 | 1355 | No | NA | 1 | 0 | 1 | Not-determined | Genome-fragment | NA | NA | 0 |
| S3_k141_74581 | 2479 | No | NA | 4 | 0 | 2 | Not-determined | Genome-fragment | NA | NA | 0 |
| S3_k141_63127 | 3127 | No | NA | 3 | 0 | 1 | Not-determined | Genome-fragment | NA | NA | 0 |
| S3_k141_51540 | 1237 | No | NA | 1 | 0 | 1 | Not-determined | Genome-fragment | NA | NA | 0 |
| S3_k141_51565 | 3131 | No | NA | 3 | 0 | 2 | Not-determined | Genome-fragment | NA | NA | 0 |
| S3_k141_137758 | 1139 | No | NA | 1 | 0 | 1 | Not-determined | Genome-fragment | NA | NA | 0 |
| S3_k141_45762 | 1851 | No | NA | 2 | 0 | 1 | Not-determined | Genome-fragment | NA | NA | 0 |
| S3_k141_51632 | 2466 | No | NA | 3 | 0 | 0 | Not-determined | Genome-fragment | NA | NA | 0 |
| S3_k141_35655 | 1596 | No | NA | 2 | 0 | 1 | Low-quality | Genome-fragment | 3.84 | AAI-based (high-confidence) | 0 |
| S7_k141_207367 | 1596 | No | NA | 1 | 0 | 1 | Low-quality | Genome-fragment | 3.44 | AAI-based (high-confidence) | 0 |
| S3_k141_103257 | 1051 | No | NA | 2 | 0 | 2 | Not-determined | Genome-fragment | NA | NA | 0 |
| S12-final-viral-combined_k141_68938\|\|full | 1596 | No | NA | 3 | 0 | 0 | Low-quality | Genome-fragment | 2.67 | AAI-based (high-confidence) | 0 |
| S3_k141_45844 | 2484 | No | NA | 1 | 0 | 1 | Not-determined | Genome-fragment | NA | NA | 0 |
| S3_k141_137877 | 3331 | No | NA | 3 | 0 | 0 | Not-determined | Genome-fragment | NA | NA | 0 |
| S12_k141_110344 | 1596 | No | NA | 3 | 0 | 0 | Low-quality | Genome-fragment | 3.37 | AAI-based (medium-confidence) | 0 |
| S3_k141_137885 | 4589 | No | NA | 4 | 0 | 0 | Not-determined | Genome-fragment | NA | NA | 0 |
| S11-final-viral-combined_k141_256976\|\|full | 1597 | No | NA | 3 | 0 | 0 | Low-quality | Genome-fragment | 2.49 | AAI-based (medium-confidence) | 0 |
| S3_k141_137952 | 1032 | No | NA | 2 | 0 | 0 | Not-determined | Genome-fragment | NA | NA | 0 |
| S3_k141_137953 | 1145 | No | NA | 5 | 0 | 0 | Not-determined | Genome-fragment | NA | NA | 0 |
| S3_k141_45936 | 1001 | No | NA | 2 | 0 | 2 | Not-determined | Genome-fragment | NA | NA | 0 |
| S5-final-viral-combined_k141_53805\|\|full | 2289 | No | NA | 3 | 0 | 0 | Not-determined | Genome-fragment | NA | NA | 0 |
| S5-final-viral-combined_k141_36163\|\|full | 2626 | No | NA | 3 | 0 | 1 | Not-determined | Genome-fragment | NA | NA | 0 |
| S5-final-viral-combined_k141_28935\|\|full | 5594 | No | NA | 9 | 0 | 0 | Not-determined | Genome-fragment | NA | NA | 0 |
| S5-final-viral-combined_k141_181624\|\|full | 6732 | No | NA | 9 | 0 | 0 | Not-determined | Genome-fragment | NA | NA | 0 |
| S5-final-viral-combined_k141_124632\|\|full | 2806 | No | NA | 4 | 0 | 2 | Not-determined | Genome-fragment | NA | NA | 0 |
| S5-final-viral-combined_k141_26061\|\|full | 34223 | No | NA | 53 | 15 | 2 | High-quality | High-quality | 98.24 | AAI-based (high-confidence) | 0 |
| S5-final-viral-combined_k141_213583\|\|full | 3193 | No | NA | 3 | 0 | 0 | Not-determined | Genome-fragment | NA | NA | 0 |
| S1-final-viral-combined_k141_317882\|\|full | 1597 | No | NA | 3 | 0 | 0 | Low-quality | Genome-fragment | 4.37 | AAI-based (medium-confidence) | 0 |
| S3-final-viral-combined_k141_136329\|\|full | 1597 | No | NA | 3 | 0 | 0 | Low-quality | Genome-fragment | 4.22 | AAI-based (high-confidence) | 0 |
| S5-final-viral-combined_k141_169278\|\|full | 1415 | No | NA | 3 | 0 | 0 | Not-determined | Genome-fragment | NA | NA | 0 |
| S5-final-viral-combined_k141_151474\|\|full | 2797 | No | NA | 2 | 0 | 2 | Not-determined | Genome-fragment | NA | NA | 0 |
| S5-final-viral-combined_k141_142555\|\|full | 8633 | No | NA | 7 | 0 | 7 | Not-determined | Genome-fragment | NA | NA | 0 |
| S7-final-viral-combined_k141_185729\|\|full | 1597 | No | NA | 4 | 2 | 0 | Low-quality | Genome-fragment | 3.73 | AAI-based (medium-confidence) | 0 |
| S17-final-viral-combined_k141_49631\|\|full | 1597 | No | NA | 3 | 0 | 0 | Low-quality | Genome-fragment | 1.5 | AAI-based (medium-confidence) | 0 |
| S12_k141_309777 | 1597 | No | NA | 1 | 0 | 0 | Low-quality | Genome-fragment | 0.27 | AAI-based (high-confidence) | 0 |
| S2-final-viral-combined_k141_116459\|\|full | 1597 | No | NA | 1 | 0 | 0 | Low-quality | Genome-fragment | 4.36 | AAI-based (high-confidence) | 0 |
| S5-final-viral-combined_k141_89126\|\|full | 2379 | No | NA | 3 | 0 | 1 | Not-determined | Genome-fragment | NA | NA | 0 |
| S5-final-viral-combined_k141_62414\|\|full | 1526 | No | NA | 3 | 0 | 1 | Not-determined | Genome-fragment | NA | NA | 0 |
| S2_k141_139050 | 1597 | No | NA | 1 | 0 | 0 | Low-quality | Genome-fragment | 1.68 | AAI-based (high-confidence) | 0 |
| S5-final-viral-combined_k141_69\|\|full | 1183 | No | NA | 2 | 0 | 0 | Not-determined | Genome-fragment | NA | NA | 0 |
| S2_k141_140512 | 1597 | No | NA | 2 | 0 | 0 | Low-quality | Genome-fragment | 3.96 | AAI-based (high-confidence) | 0 |
| S8-final-viral-combined_k141_151653\|\|full | 1597 | No | NA | 3 | 0 | 0 | Low-quality | Genome-fragment | 3.6 | AAI-based (medium-confidence) | 0 |
| S8_k141_194757 | 1597 | No | NA | 3 | 2 | 0 | Low-quality | Genome-fragment | 2.25 | AAI-based (high-confidence) | 0 |
| S11-final-viral-combined_k141_59909\|\|full | 1598 | No | NA | 3 | 2 | 0 | Low-quality | Genome-fragment | 3.95 | AAI-based (high-confidence) | 0 |
| S1-final-viral-combined_k141_222287\|\|full | 1598 | No | NA | 3 | 2 | 0 | Low-quality | Genome-fragment | 3.06 | AAI-based (high-confidence) | 0 |
| S1-final-viral-combined_k141_209525\|\|full | 1598 | No | NA | 3 | 1 | 0 | Low-quality | Genome-fragment | 2.71 | AAI-based (medium-confidence) | 0 |
| S7-final-viral-combined_k141_19667\|\|full | 1598 | No | NA | 2 | 2 | 0 | Low-quality | Genome-fragment | 3.3 | AAI-based (high-confidence) | 0 |
| S5-final-viral-combined_k141_196058\|\|full | 2217 | No | NA | 2 | 0 | 2 | Not-determined | Genome-fragment | NA | NA | 0 |
| S12-final-viral-combined_k141_235525\|\|full | 1598 | No | NA | 4 | 0 | 2 | Low-quality | Genome-fragment | 3.8 | AAI-based (high-confidence) | 0 |
| S5-final-viral-combined_k141_71384\|\|full | 5404 | No | NA | 5 | 0 | 5 | Not-determined | Genome-fragment | NA | NA | 0 |
| S2-final-viral-combined_k141_63678\|\|full | 1598 | No | NA | 3 | 0 | 0 | Low-quality | Genome-fragment | 2.99 | AAI-based (high-confidence) | 0 |
| S5-final-viral-combined_k141_115916\|\|full | 7860 | No | NA | 7 | 0 | 6 | Not-determined | Genome-fragment | NA | NA | 0 |
| S5-final-viral-combined_k141_137\|\|full | 3411 | No | NA | 6 | 0 | 0 | Not-determined | Genome-fragment | NA | NA | 0 |
| S5-final-viral-combined_k141_115927\|\|full | 9822 | No | NA | 9 | 0 | 4 | Not-determined | Genome-fragment | NA | NA | 0 |
| S5-final-viral-combined_k141_106988\|\|full | 4438 | No | NA | 6 | 0 | 3 | Not-determined | Genome-fragment | NA | NA | 0 |
| S5-final-viral-combined_k141_107002\|\|full | 1465 | No | NA | 2 | 0 | 1 | Not-determined | Genome-fragment | NA | NA | 0 |
| S5-final-viral-combined_k141_151607\|\|full | 1091 | No | NA | 2 | 0 | 1 | Not-determined | Genome-fragment | NA | NA | 0 |
| S5-final-viral-combined_k141_9091\|\|full | 2562 | No | NA | 4 | 0 | 0 | Not-determined | Genome-fragment | NA | NA | 0 |
| S5-final-viral-combined_k141_173\|\|full | 1027 | No | NA | 3 | 0 | 0 | Not-determined | Genome-fragment | NA | NA | 0 |
| S5-final-viral-combined_k141_107022\|\|full | 2353 | No | NA | 2 | 0 | 2 | Not-determined | Genome-fragment | NA | NA | 0 |
| S5-final-viral-combined_k141_26858\|\|full | 2412 | No | NA | 4 | 0 | 0 | Not-determined | Genome-fragment | NA | NA | 0 |
| S2-final-viral-combined_k141_24948\|\|full | 1598 | No | NA | 3 | 3 | 0 | Low-quality | Genome-fragment | 3.63 | AAI-based (high-confidence) | 0 |
| S8-final-viral-combined_k141_55654\|\|full | 1598 | No | NA | 3 | 1 | 0 | Low-quality | Genome-fragment | 2.85 | AAI-based (high-confidence) | 0 |
| S5-final-viral-combined_k141_133814\|\|full | 20581 | No | NA | 18 | 0 | 14 | Not-determined | Genome-fragment | NA | NA | 0 |
| S1-final-viral-combined_k141_155606\|\|full | 1599 | No | NA | 4 | 0 | 0 | Low-quality | Genome-fragment | 3.97 | AAI-based (medium-confidence) | 0 |
| S5-final-viral-combined_k141_196156\|\|full | 2663 | No | NA | 3 | 0 | 0 | Not-determined | Genome-fragment | NA | NA | 0 |
| S5-final-viral-combined_k141_237\|\|full | 9412 | No | NA | 6 | 0 | 5 | Not-determined | Genome-fragment | NA | NA | 0 |
| S1-final-viral-combined_k141_338499\|\|full | 1599 | No | NA | 3 | 0 | 0 | Low-quality | Genome-fragment | 3.14 | AAI-based (medium-confidence) | 0 |
| S5-final-viral-combined_k141_53567\|\|full | 2786 | No | NA | 4 | 0 | 1 | Not-determined | Genome-fragment | NA | NA | 0 |
| S1_k141_27310 | 1599 | No | NA | 2 | 0 | 2 | Low-quality | Genome-fragment | 0.46 | AAI-based (high-confidence) | 0 |
| S5-final-viral-combined_k141_164418\|\|full | 1599 | No | NA | 2 | 0 | 1 | Low-quality | Genome-fragment | 0.55 | AAI-based (medium-confidence) | 0 |
| S7_k141_129638 | 1599 | No | NA | 2 | 0 | 1 | Low-quality | Genome-fragment | 0.84 | AAI-based (medium-confidence) | 0 |
| S2_k141_162824 | 1599 | No | NA | 1 | 0 | 1 | Low-quality | Genome-fragment | 0.31 | AAI-based (medium-confidence) | 0 |
| S4-final-viral-combined_k141_153881\|\|full | 1599 | No | NA | 3 | 1 | 0 | Low-quality | Genome-fragment | 2.73 | HMM-based (lower-bound) | 0 |
| S6-final-viral-combined_k141_92486\|\|full | 1599 | No | NA | 4 | 0 | 0 | Low-quality | Genome-fragment | 2.98 | AAI-based (high-confidence) | 0 |
| S5-final-viral-combined_k141_62485\|\|full | 3360 | No | NA | 4 | 0 | 2 | Not-determined | Genome-fragment | NA | NA | 0 |
| S6-final-viral-combined_k141_6504\|\|lt2gene | 1599 | No | NA | 1 | 1 | 0 | Low-quality | Genome-fragment | 1.51 | AAI-based (high-confidence) | 0 |

| | | | | | | | | | | | |
|---|---|---|---|---|---|---|---|---|---|---|---|
| S5-final-viral-combined_k141_98163\|\|full | 1126 | No | NA | 2 | 0 | 1 | Not-determined | Genome-fragment | NA | NA | 0 |
| S8_k141_2214 | 1599 | No | NA | 4 | 0 | 0 | Low-quality | Genome-fragment | 3.72 | AAI-based (high-confidence) | 0 |
| S5-final-viral-combined_k141_53622\|\|full | 2678 | No | NA | 2 | 0 | 1 | Not-determined | Genome-fragment | NA | NA | 0 |
| S5-final-viral-combined_k141_107111\|\|full | 8047 | No | NA | 7 | 0 | 5 | Not-determined | Genome-fragment | NA | NA | 0 |
| S5-final-viral-combined_k141_53629\|\|full | 1370 | No | NA | 3 | 0 | 0 | Not-determined | Genome-fragment | NA | NA | 0 |
| S9-final-viral-combined_k141_120991\|\|full | 1600 | No | NA | 3 | 1 | 0 | Low-quality | Genome-fragment | 3.03 | AAI-based (high-confidence) | 0 |
| S5-final-viral-combined_k141_98179\|\|full | 13818 | No | NA | 16 | 0 | 12 | Not-determined | Genome-fragment | NA | NA | 0 |
| S17-final-viral-combined_k141_188704\|\|full | 1600 | No | NA | 3 | 1 | 0 | Low-quality | Genome-fragment | 4.3 | AAI-based (high-confidence) | 0 |
| S17-k141_222018 | 1600 | No | NA | 2 | 0 | 1 | Low-quality | Genome-fragment | 1.19 | AAI-based (medium-confidence) | 0 |
| S9-final-viral-combined_k141_27859\|\|full | 1601 | No | NA | 2 | 0 | 0 | Low-quality | Genome-fragment | 2.64 | AAI-based (high-confidence) | 0 |
| S3-final-viral-combined_k141_70907\|\|full | 1601 | No | NA | 2 | 1 | 0 | Low-quality | Genome-fragment | 3.94 | AAI-based (high-confidence) | 0 |
| S3-final-viral-combined_k141_61537\|\|full | 1601 | No | NA | 4 | 0 | 0 | Low-quality | Genome-fragment | 2.92 | AAI-based (medium-confidence) | 0 |
| S7-final-viral-combined_k141_134212\|\|full | 1601 | No | NA | 2 | 0 | 2 | Low-quality | Genome-fragment | 3.7 | AAI-based (medium-confidence) | 0 |
| S5-final-viral-combined_k141_80372\|\|full | 1804 | No | NA | 3 | 0 | 1 | Not-determined | Genome-fragment | NA | NA | 0 |
| S17-final-viral-combined_k141_141927\|\|full | 1601 | No | NA | 1 | 0 | 1 | Low-quality | Genome-fragment | 4.18 | AAI-based (high-confidence) | 0 |
| S5-final-viral-combined_k141_142683\|\|full | 1676 | No | NA | 2 | 0 | 1 | Not-determined | Genome-fragment | NA | NA | 0 |
| S5-final-viral-combined_k141_53686\|\|full | 3112 | No | NA | 7 | 0 | 0 | Not-determined | Genome-fragment | NA | NA | 0 |
| S17-final-viral-combined_k141_121909\|\|full | 1601 | No | NA | 4 | 1 | 0 | Low-quality | Genome-fragment | 3.01 | AAI-based (medium-confidence) | 0 |
| S12-final-viral-combined_k141_187986\|\|full | 1601 | No | NA | 2 | 1 | 0 | Low-quality | Genome-fragment | 3.63 | AAI-based (high-confidence) | 0 |
| S5-final-viral-combined_k141_142720\|\|full | 2921 | No | NA | 3 | 0 | 0 | Not-determined | Genome-fragment | NA | NA | 0 |
| S5-final-viral-combined_k141_419\|\|full | 1479 | No | NA | 3 | 0 | 1 | Not-determined | Genome-fragment | NA | NA | 0 |
| S5-final-viral-combined_k141_178365\|\|full | 1627 | No | NA | 3 | 0 | 0 | Not-determined | Genome-fragment | NA | NA | 0 |
| S5-final-viral-combined_k141_187241\|\|full | 1606 | No | NA | 3 | 0 | 2 | Not-determined | Genome-fragment | NA | NA | 0 |
| S5-final-viral-combined_k141_124950\|\|full | 2646 | No | NA | 3 | 0 | 0 | Not-determined | Genome-fragment | NA | NA | 0 |
| S5-final-viral-combined_k141_116109\|\|full | 5267 | No | NA | 5 | 0 | 2 | Not-determined | Genome-fragment | NA | NA | 0 |
| S5-final-viral-combined_k141_124965\|\|full | 1260 | No | NA | 3 | 0 | 1 | Not-determined | Genome-fragment | NA | NA | 0 |
| S5-final-viral-combined_k141_71574\|\|full | 2476 | No | NA | 3 | 0 | 0 | Not-determined | Genome-fragment | NA | NA | 0 |
| S5-final-viral-combined_k141_178431\|\|full | 2593 | No | NA | 3 | 0 | 2 | Not-determined | Genome-fragment | NA | NA | 0 |
| S5-final-viral-combined_k141_169349\|\|full | 1173 | No | NA | 2 | 0 | 0 | Not-determined | Genome-fragment | NA | NA | 0 |
| S5-final-viral-combined_k141_124969\|\|full | 6104 | No | NA | 5 | 0 | 1 | Not-determined | Genome-fragment | NA | NA | 0 |
| S5-final-viral-combined_k141_107242\|\|full | 3492 | No | NA | 6 | 0 | 3 | Not-determined | Genome-fragment | NA | NA | 0 |
| S12-final-viral-combined_k141_209607\|\|full | 1601 | No | NA | 6 | 0 | 0 | Low-quality | Genome-fragment | 3.44 | AAI-based (high-confidence) | 0 |
| S5-final-viral-combined_k141_116138\|\|full | 5394 | No | NA | 7 | 0 | 0 | Not-determined | Genome-fragment | NA | NA | 0 |
| S4-final-viral-combined_k141_62445\|\|full | 1601 | No | NA | 3 | 0 | 1 | Low-quality | Genome-fragment | 0.9 | AAI-based (medium-confidence) | 0 |
| S5-final-viral-combined_k141_124987\|\|full | 1496 | No | NA | 3 | 0 | 0 | Not-determined | Genome-fragment | NA | NA | 0 |
| S8-final-viral-combined_k141_268268\|\|full | 1601 | No | NA | 3 | 0 | 0 | Low-quality | Genome-fragment | 3.23 | AAI-based (medium-confidence) | 0 |
| S5-final-viral-combined_k141_116143\|\|full | 2267 | No | NA | 4 | 0 | 1 | Not-determined | Genome-fragment | NA | NA | 0 |
| S5-final-viral-combined_k141_116144\|\|full | 2156 | No | NA | 2 | 0 | 1 | Not-determined | Genome-fragment | NA | NA | 0 |
| S9-final-viral-combined_k141_66492\|\|lt2gene | 1602 | No | NA | 2 | 2 | 0 | Low-quality | Genome-fragment | 3.13 | AAI-based (high-confidence) | 0 |
| S11-final-viral-combined_k141_208039\|\|full | 1602 | No | NA | 4 | 1 | 0 | Low-quality | Genome-fragment | 3.42 | AAI-based (medium-confidence) | 0 |
| S12_k141_173985 | 1602 | No | NA | 2 | 0 | 1 | Low-quality | Genome-fragment | 2.02 | AAI-based (medium-confidence) | 0 |
| S5-final-viral-combined_k141_62591\|\|full | 7507 | No | NA | 11 | 0 | 5 | Not-determined | Genome-fragment | NA | NA | 0 |
| S5-final-viral-combined_k141_53768\|\|full | 1760 | No | NA | 3 | 0 | 0 | Not-determined | Genome-fragment | NA | NA | 0 |
| S4-final-viral-combined_k141_25525\|\|full | 1602 | No | NA | 2 | 0 | 1 | Low-quality | Genome-fragment | 2 | AAI-based (medium-confidence) | 0 |
| S5-final-viral-combined_k141_53786\|\|full | 1148 | No | NA | 2 | 0 | 2 | Not-determined | Genome-fragment | NA | NA | 0 |
| S9-final-viral-combined_k141_42565\|\|full | 1603 | No | NA | 2 | 0 | 1 | Low-quality | Genome-fragment | 2.13 | AAI-based (high-confidence) | 0 |
| S5-final-viral-combined_k141_62606\|\|full | 1201 | No | NA | 2 | 0 | 2 | Not-determined | Genome-fragment | NA | NA | 0 |
| S7_k141_228304 | 1603 | No | NA | 2 | 0 | 1 | Low-quality | Genome-fragment | 0.39 | AAI-based (high-confidence) | 0 |
| S17-final-viral-combined_k141_166973\|\|full | 1603 | No | NA | 2 | 0 | 1 | Low-quality | Genome-fragment | 0.59 | AAI-based (medium-confidence) | 0 |
| S17-final-viral-combined_k141_47025\|\|full | 1603 | No | NA | 3 | 1 | 0 | Low-quality | Genome-fragment | 1.04 | HMM-based (lower-bound) | 0 |
| S12-final-viral-combined_k141_61289\|\|lt2gene | 1603 | No | NA | 2 | 2 | 0 | Low-quality | Genome-fragment | 4.7 | AAI-based (high-confidence) | 0 |
| S12_k141_325124 | 1603 | No | NA | 1 | 0 | 0 | Low-quality | Genome-fragment | 2.53 | AAI-based (high-confidence) | 0 |
| S5-final-viral-combined_k141_205046\|\|full | 34053 | No | NA | 34 | 1 | 18 | High-quality | High-quality | 100 | HMM-based (lower-bound) | 0 |
| S12_k141_26374 | 1603 | No | NA | 1 | 0 | 1 | Low-quality | Genome-fragment | 3.97 | AAI-based (high-confidence) | 0 |
| S5-final-viral-combined_k141_169427\|\|full | 1312 | No | NA | 3 | 0 | 2 | Not-determined | Genome-fragment | NA | NA | 0 |
| S5-final-viral-combined_k141_205049\|\|full | 5605 | No | NA | 8 | 0 | 5 | Not-determined | Genome-fragment | NA | NA | 0 |
| S5-final-viral-combined_k141_62641\|\|full | 2814 | No | NA | 3 | 0 | 3 | Not-determined | Genome-fragment | NA | NA | 0 |
| S2-final-viral-combined_k141_184444\|\|full | 1603 | No | NA | 2 | 0 | 2 | Low-quality | Genome-fragment | 1.34 | AAI-based (high-confidence) | 0 |
| S5-final-viral-combined_k141_205062\|\|full | 11630 | No | NA | 12 | 0 | 7 | Not-determined | Genome-fragment | NA | NA | 0 |
| S5-final-viral-combined_k141_98243\|\|full | 5767 | No | NA | 4 | 0 | 2 | Not-determined | Genome-fragment | NA | NA | 0 |
| S9-final-viral-combined_k141_50158\|\|full | 1604 | No | NA | 4 | 2 | 0 | Low-quality | Genome-fragment | 3.34 | AAI-based (high-confidence) | 0 |
| S9-final-viral-combined_k141_18201\|\|lt2gene | 1604 | No | NA | 2 | 1 | 0 | Low-quality | Genome-fragment | 2.1 | HMM-based (lower-bound) | 0 |
| S5-final-viral-combined_k141_187398\|\|full | 2015 | No | NA | 4 | 0 | 0 | Not-determined | Genome-fragment | NA | NA | 0 |
| S1-final-viral-combined_k141_31274\|\|full | 1604 | No | NA | 4 | 0 | 4 | Low-quality | Genome-fragment | 0.51 | AAI-based (medium-confidence) | 0 |
| S3-final-viral-combined_k141_18954\|\|full | 1604 | No | NA | 3 | 0 | 1 | Low-quality | Genome-fragment | 1 | AAI-based (high-confidence) | 0 |
| S5-final-viral-combined_k141_9406\|\|full | 2497 | No | NA | 4 | 0 | 0 | Not-determined | Genome-fragment | NA | NA | 0 |
| S5-final-viral-combined_k141_183394\|\|full | 1604 | No | NA | 3 | 0 | 1 | Low-quality | Genome-fragment | 2.84 | AAI-based (medium-confidence) | 0 |
| S5-final-viral-combined_k141_9417\|\|full | 2629 | No | NA | 3 | 0 | 1 | Not-determined | Genome-fragment | NA | NA | 0 |
| S17-k141_144298 | 1604 | No | NA | 1 | 0 | 1 | Low-quality | Genome-fragment | 0.65 | AAI-based (medium-confidence) | 0 |
| S12-final-viral-combined_k141_2254\|\|full | 1604 | No | NA | 2 | 0 | 1 | Low-quality | Genome-fragment | 1.93 | AAI-based (high-confidence) | 0 |
| S12-final-viral-combined_k141_258512\|\|full | 1604 | No | NA | 5 | 0 | 0 | Low-quality | Genome-fragment | 5.04 | AAI-based (high-confidence) | 0 |
| S5-final-viral-combined_k141_44736\|\|full | 1402 | No | NA | 2 | 0 | 0 | Not-determined | Genome-fragment | NA | NA | 0 |
| S5-final-viral-combined_k141_9452\|\|full | 2266 | No | NA | 2 | 0 | 2 | Not-determined | Genome-fragment | NA | NA | 0 |
| S5-final-viral-combined_k141_35842\|\|full | 2232 | No | NA | 3 | 0 | 3 | Not-determined | Genome-fragment | NA | NA | 0 |
| S5-final-viral-combined_k141_169496\|\|full | 1220 | No | NA | 2 | 0 | 0 | Not-determined | Genome-fragment | NA | NA | 0 |
| S5-final-viral-combined_k141_187450\|\|full | 1626 | No | NA | 3 | 0 | 0 | Not-determined | Genome-fragment | NA | NA | 0 |
| S12-final-viral-combined_k141_261553\|\|full | 1604 | No | NA | 3 | 1 | 0 | Low-quality | Genome-fragment | 4.41 | HMM-based (lower-bound) | 0 |
| S5-final-viral-combined_k141_9477\|\|full | 2815 | No | NA | 4 | 0 | 1 | Not-determined | Genome-fragment | NA | NA | 0 |
| S12-final-viral-combined_k141_13587\|\|full | 1604 | No | NA | 3 | 0 | 1 | Low-quality | Genome-fragment | 0.83 | AAI-based (medium-confidence) | 0 |
| S5-final-viral-combined_k141_142906\|\|full | 2643 | No | NA | 3 | 0 | 3 | Not-determined | Genome-fragment | NA | NA | 0 |
| S2-final-viral-combined_k141_24619\|\|full | 1604 | No | NA | 3 | 0 | 0 | Low-quality | Genome-fragment | 4.77 | AAI-based (high-confidence) | 0 |
| S5-final-viral-combined_k141_71823\|\|full | 3167 | No | NA | 5 | 0 | 0 | Not-determined | Genome-fragment | NA | NA | 0 |
| S4-final-viral-combined_k141_151061\|\|full | 1604 | No | NA | 3 | 0 | 0 | Low-quality | Genome-fragment | 0.89 | AAI-based (medium-confidence) | 0 |
| S8-final-viral-combined_k141_230909\|\|full | 1604 | No | NA | 4 | 1 | 0 | Low-quality | Genome-fragment | 4.96 | AAI-based (high-confidence) | 0 |
| S11-final-viral-combined_k141_114180\|\|full | 1605 | No | NA | 2 | 0 | 1 | Low-quality | Genome-fragment | 1.6 | AAI-based (medium-confidence) | 0 |
| S5-final-viral-combined_k141_80625\|\|full | 1349 | No | NA | 3 | 0 | 1 | Not-determined | Genome-fragment | NA | NA | 0 |
| S5-final-viral-combined_k141_143002\|\|full | 2995 | No | NA | 4 | 0 | 1 | Not-determined | Genome-fragment | NA | NA | 0 |
| S5-final-viral-combined_k141_44832\|\|full | 3535 | No | NA | 3 | 0 | 1 | Not-determined | Genome-fragment | NA | NA | 0 |
| S2-final-viral-combined_k141_160431\|\|full | 1605 | No | NA | 3 | 0 | 0 | Low-quality | Genome-fragment | 2.74 | AAI-based (medium-confidence) | 0 |
| S9-final-viral-combined_k141_47875\|\|full | 1606 | Yes | 860 | 3 | 1 | 1 | Low-quality | Genome-fragment | 2.12 | AAI-based (high-confidence) | 46.45 |
| S5-final-viral-combined_k141_772\|\|full | 7307 | No | NA | 9 | 0 | 7 | Not-determined | Genome-fragment | NA | NA | 0 |
| S5-final-viral-combined_k141_44898\|\|full | 1142 | No | NA | 3 | 0 | 0 | Not-determined | Genome-fragment | NA | NA | 0 |
| S11-final-viral-combined_k141_167046\|\|full | 1606 | No | NA | 3 | 0 | 0 | Low-quality | Genome-fragment | 3.38 | AAI-based (high-confidence) | 0 |
| S1-final-viral-combined_k141_182015\|\|full | 1606 | No | NA | 3 | 0 | 0 | Low-quality | Genome-fragment | 3.38 | AAI-based (medium-confidence) | 0 |
| S1-final-viral-combined_k141_178754\|\|full | 1606 | No | NA | 3 | 0 | 1 | Low-quality | Genome-fragment | 2.4 | AAI-based (medium-confidence) | 0 |
| S5-final-viral-combined_k141_107415\|\|full | 1584 | No | NA | 3 | 0 | 1 | Not-determined | Genome-fragment | NA | NA | 0 |
| S5-final-viral-combined_k141_169644\|\|full | 1676 | No | NA | 2 | 0 | 2 | Not-determined | Genome-fragment | NA | NA | 0 |
| S1_k141_121029 | 1606 | No | NA | 2 | 0 | 0 | Low-quality | Genome-fragment | 2.14 | AAI-based (medium-confidence) | 0 |
| S5-final-viral-combined_k141_98390\|\|full | 14286 | No | NA | 10 | 0 | 6 | Not-determined | Genome-fragment | NA | NA | 0 |
| S5-final-viral-combined_k141_9583\|\|full | 2594 | No | NA | 3 | 0 | 1 | Not-determined | Genome-fragment | NA | NA | 0 |
| S5_k141_67935 | 1606 | No | NA | 3 | 0 | 2 | Low-quality | Genome-fragment | 3.62 | AAI-based (medium-confidence) | 0 |
| S7_k141_239460 | 1606 | No | NA | 1 | 0 | 0 | Low-quality | Genome-fragment | 0.26 | AAI-based (high-confidence) | 0 |
| S5-final-viral-combined_k141_80730\|\|full | 2469 | No | NA | 3 | 0 | 2 | Not-determined | Genome-fragment | NA | NA | 0 |
| S5-final-viral-combined_k141_143100\|\|full | 1479 | No | NA | 3 | 0 | 2 | Not-determined | Genome-fragment | NA | NA | 0 |
| S5-final-viral-combined_k141_71829\|\|full | 2227 | No | NA | 4 | 0 | 1 | Not-determined | Genome-fragment | NA | NA | 0 |
| S5-final-viral-combined_k141_18356\|\|full | 34910 | Yes | 21165 | 34 | 16 | 8 | Medium-quality | Genome-fragment | 67.13 | AAI-based (medium-confidence) | 39.37 |
| S2-final-viral-combined_k141_120005\|\|full | 1606 | No | NA | 3 | 2 | 0 | Low-quality | Genome-fragment | 4.54 | AAI-based (high-confidence) | 0 |
| S4_k141_14514 | 1606 | No | NA | 3 | 0 | 1 | Low-quality | Genome-fragment | 1.85 | AAI-based (high-confidence) | 0 |
| S5-final-viral-combined_k141_169713\|\|full | 1394 | No | NA | 3 | 0 | 0 | Not-determined | Genome-fragment | NA | NA | 0 |
| S8_k141_239297 | 1606 | No | NA | 2 | 1 | 0 | Low-quality | Genome-fragment | 4.98 | AAI-based (high-confidence) | 0 |
| S5-final-viral-combined_k141_169735\|\|full | 4912 | No | NA | 5 | 0 | 2 | Not-determined | Genome-fragment | NA | NA | 0 |
| S5-final-viral-combined_k141_145367\|\|full | 1607 | No | NA | 4 | 0 | 1 | Low-quality | Genome-fragment | 0.8 | AAI-based (medium-confidence) | 0 |
| S5-final-viral-combined_k141_33324\|\|full | 1607 | No | NA | 5 | 0 | 0 | Low-quality | Genome-fragment | 2.69 | AAI-based (medium-confidence) | 0 |
| S7-final-viral-combined_k141_11997\|\|full | 1607 | No | NA | 4 | 0 | 0 | Low-quality | Genome-fragment | 4.01 | AAI-based (high-confidence) | 0 |
| S7-final-viral-combined_k141_66638\|\|full | 1607 | No | NA | 3 | 0 | 0 | Low-quality | Genome-fragment | 3.93 | AAI-based (medium-confidence) | 0 |
| S5-final-viral-combined_k141_71900\|\|full | 1021 | No | NA | 2 | 0 | 1 | Not-determined | Genome-fragment | NA | NA | 0 |
| S5-final-viral-combined_k141_116289\|\|full | 1681 | No | NA | 2 | 0 | 1 | Not-determined | Genome-fragment | NA | NA | 0 |
| S5-final-viral-combined_k141_116314\|\|full | 1922 | No | NA | 3 | 0 | 2 | Not-determined | Genome-fragment | NA | NA | 0 |
| S5-final-viral-combined_k141_89621\|\|full | 2253 | No | NA | 3 | 0 | 1 | Not-determined | Genome-fragment | NA | NA | 0 |
| S7-final-viral-combined_k141_220414\|\|lt2gene | 1607 | No | NA | 2 | 2 | 0 | Low-quality | Genome-fragment | 2.51 | HMM-based (lower-bound) | 0 |
| S5-final-viral-combined_k141_178485\|\|full | 1499 | No | NA | 3 | 0 | 1 | Not-determined | Genome-fragment | NA | NA | 0 |
| S5-final-viral-combined_k141_205349\|\|full | 7131 | No | NA | 7 | 0 | 6 | Not-determined | Genome-fragment | NA | NA | 0 |
| S5-final-viral-combined_k141_89643\|\|full | 1210 | No | NA | 2 | 0 | 1 | Not-determined | Genome-fragment | NA | NA | 0 |
| S5-final-viral-combined_k141_80788\|\|full | 1533 | No | NA | 3 | 0 | 1 | Not-determined | Genome-fragment | NA | NA | 0 |
| S4-final-viral-combined_k141_107810\|\|full | 1607 | No | NA | 5 | 0 | 0 | Low-quality | Genome-fragment | 2.76 | AAI-based (medium-confidence) | 0 |
| S3_k141_72312 | 1608 | No | NA | 3 | 0 | 0 | Low-quality | Genome-fragment | 0.74 | AAI-based (high-confidence) | 0 |
| S7_k141_61801 | 1608 | No | NA | 1 | 0 | 1 | Low-quality | Genome-fragment | 2.61 | AAI-based (medium-confidence) | 0 |
| S17-final-viral-combined_k141_122285\|\|full | 1608 | No | NA | 3 | 0 | 1 | Low-quality | Genome-fragment | 0.59 | AAI-based (medium-confidence) | 0 |
| S5-final-viral-combined_k141_27141\|\|full | 1053 | No | NA | 3 | 0 | 0 | Not-determined | Genome-fragment | NA | NA | 0 |
| S17-final-viral-combined_k141_119050\|\|full | 1608 | No | NA | 2 | 0 | 2 | Low-quality | Genome-fragment | 0.84 | AAI-based (medium-confidence) | 0 |
| S5-final-viral-combined_k141_98626\|\|full | 1848 | No | NA | 2 | 0 | 0 | Not-determined | Genome-fragment | NA | NA | 0 |
| S5-final-viral-combined_k141_9780\|\|full | 2052 | No | NA | 2 | 0 | 2 | Not-determined | Genome-fragment | NA | NA | 0 |
| S5-final-viral-combined_k141_1057\|\|full | 2172 | No | NA | 4 | 0 | 2 | Not-determined | Genome-fragment | NA | NA | 0 |
| S5-final-viral-combined_k141_89712\|\|full | 1138 | No | NA | 3 | 0 | 1 | Not-determined | Genome-fragment | NA | NA | 0 |
| S12-final-viral-combined_k141_111100\|\|full | 1608 | Yes | 807 | 3 | 1 | 1 | Low-quality | Genome-fragment | 1.16 | AAI-based (high-confidence) | 49.81 |
| S5-final-viral-combined_k141_89727\|\|full | 1350 | No | NA | 3 | 0 | 0 | Not-determined | Genome-fragment | NA | NA | 0 |
| S5-final-viral-combined_k141_169894\|\|full | 5190 | No | NA | 5 | 0 | 4 | Not-determined | Genome-fragment | NA | NA | 0 |
| S2-final-viral-combined_k141_80377\|\|full | 1608 | No | NA | 3 | 1 | 0 | Low-quality | Genome-fragment | 4.22 | AAI-based (high-confidence) | 0 |
| S5-final-viral-combined_k141_107589\|\|full | 2893 | No | NA | 3 | 0 | 0 | Not-determined | Genome-fragment | NA | NA | 0 |
| S2-final-viral-combined_k141_196041\|\|full | 1608 | No | NA | 3 | 0 | 2 | Low-quality | Genome-fragment | 2.25 | AAI-based (high-confidence) | 0 |
| S4_k141_115062 | 1608 | No | NA | 1 | 0 | 1 | Low-quality | Genome-fragment | 0.48 | AAI-based (medium-confidence) | 0 |
| S5-final-viral-combined_k141_9825\|\|full | 1724 | No | NA | 2 | 0 | 2 | Not-determined | Genome-fragment | NA | NA | 0 |
| S5-final-viral-combined_k141_205409\|\|full | 1698 | No | NA | 3 | 0 | 1 | Not-determined | Genome-fragment | NA | NA | 0 |
| S5-final-viral-combined_k141_152066\|\|full | 1435 | No | NA | 3 | 0 | 2 | Not-determined | Genome-fragment | NA | NA | 0 |
| S8-final-viral-combined_k141_237950\|\|full | 1608 | No | NA | 3 | 0 | 0 | Low-quality | Genome-fragment | 2.85 | AAI-based (medium-confidence) | 0 |
| S8_k141_251161 | 1608 | No | NA | 3 | 0 | 0 | Low-quality | Genome-fragment | 3.37 | AAI-based (high-confidence) | 0 |
| S8_k141_261239 | 1608 | No | NA | 2 | 0 | 2 | Low-quality | Genome-fragment | 2.79 | AAI-based (medium-confidence) | 0 |
| S5-final-viral-combined_k141_196767\|\|full | 1234 | No | NA | 2 | 0 | 2 | Not-determined | Genome-fragment | NA | NA | 0 |
| S5-final-viral-combined_k141_169978\|\|full | 5156 | No | NA | 5 | 0 | 1 | Not-determined | Genome-fragment | NA | NA | 0 |
| S5-final-viral-combined_k141_62885\|\|full | 1637 | No | NA | 2 | 0 | 2 | Not-determined | Genome-fragment | NA | NA | 0 |
| S5-final-viral-combined_k141_45124\|\|full | 8462 | No | NA | 9 | 0 | 3 | Not-determined | Genome-fragment | NA | NA | 0 |
| S5-final-viral-combined_k141_36182\|\|full | 1469 | No | NA | 2 | 0 | 1 | Not-determined | Genome-fragment | NA | NA | 0 |
| S5-final-viral-combined_k141_26779\|\|full | 1609 | No | NA | 5 | 2 | 0 | Low-quality | Genome-fragment | 3.5 | AAI-based (high-confidence) | 0 |
| S5-final-viral-combined_k141_67108\|\|full | 1609 | Yes | 656 | 3 | 1 | 1 | Low-quality | Genome-fragment | 0.44 | HMM-based (lower-bound) | 59.23 |
| S17-final-viral-combined_k141_104256\|\|full | 1609 | No | NA | 2 | 0 | 0 | Low-quality | Genome-fragment | 1.71 | AAI-based (high-confidence) | 0 |
| S17-final-viral-combined_k141_218268\|\|full | 1609 | No | NA | 4 | 2 | 0 | Low-quality | Genome-fragment | 4.21 | AAI-based (medium-confidence) | 0 |
| S5-final-viral-combined_k141_36228\|\|full | 1965 | No | NA | 4 | 0 | 1 | Not-determined | Genome-fragment | NA | NA | 0 |
| S5-final-viral-combined_k141_170011\|\|full | 1536 | No | NA | 4 | 0 | 2 | Not-determined | Genome-fragment | NA | NA | 0 |
| S17-final-viral-combined_k141_241362\|\|full | 1609 | No | NA | 2 | 0 | 1 | Low-quality | Genome-fragment | 4.55 | AAI-based (medium-confidence) | 0 |
| S4-final-viral-combined_k141_164359\|\|full | 1609 | No | NA | 4 | 0 | 0 | Low-quality | Genome-fragment | 2.64 | AAI-based (medium-confidence) | 0 |
| S4-final-viral-combined_k141_110617\|\|full | 1609 | No | NA | 4 | 0 | 0 | Low-quality | Genome-fragment | 2.31 | AAI-based (high-confidence) | 0 |
| S5-final-viral-combined_k141_45179\|\|full | 14157 | No | NA | 14 | 0 | 9 | Not-determined | Genome-fragment | NA | NA | 0 |
| S5-final-viral-combined_k141_36254\|\|full | 2420 | No | NA | 3 | 0 | 2 | Not-determined | Genome-fragment | NA | NA | 0 |
| S5-final-viral-combined_k141_205496\|\|full | 2758 | No | NA | 4 | 0 | 0 | Not-determined | Genome-fragment | NA | NA | 0 |
| S8_k141_261462 | 1609 | No | NA | 1 | 0 | 0 | Low-quality | Genome-fragment | 1.64 | AAI-based (medium-confidence) | 0 |
| S5-final-viral-combined_k141_125375\|\|full | 2732 | No | NA | 3 | 0 | 3 | Not-determined | Genome-fragment | NA | NA | 0 |
| S9-final-viral-combined_k141_126669\|\|full | 1610 | No | NA | 2 | 2 | 0 | Low-quality | Genome-fragment | 4.13 | AAI-based (high-confidence) | 0 |
| S5-final-viral-combined_k141_36282\|\|full | 1637 | No | NA | 3 | 0 | 1 | Not-determined | Genome-fragment | NA | NA | 0 |
| S5-final-viral-combined_k141_54116\|\|full | 2015 | No | NA | 3 | 0 | 0 | Not-determined | Genome-fragment | NA | NA | 0 |
| S5-final-viral-combined_k141_1229\|\|full | 1243 | No | NA | 3 | 0 | 0 | Not-determined | Genome-fragment | NA | NA | 0 |
| S5-final-viral-combined_k141_27242\|\|full | 3075 | No | NA | 3 | 0 | 3 | Not-determined | Genome-fragment | NA | NA | 0 |
| S5-final-viral-combined_k141_89990\|\|full | 3088 | No | NA | 2 | 0 | 2 | Not-determined | Genome-fragment | NA | NA | 0 |
| S5-final-viral-combined_k141_107779\|\|full | 7150 | No | NA | 3 | 0 | 3 | Not-determined | Genome-fragment | NA | NA | 0 |
| S5-final-viral-combined_k141_152083\|\|full | 7883 | No | NA | 3 | 0 | 3 | Not-determined | Genome-fragment | NA | NA | 0 |
| S9_k141_80983 | 1610 | No | NA | 2 | 0 | 0 | Low-quality | Genome-fragment | 1.14 | AAI-based (high-confidence) | 0 |
| S11-final-viral-combined_k141_154312\|\|full | 1610 | No | NA | 5 | 2 | 0 | Low-quality | Genome-fragment | 3.77 | AAI-based (medium-confidence) | 0 |
| S5-final-viral-combined_k141_178772\|\|full | 3068 | No | NA | 4 | 0 | 2 | Not-determined | Genome-fragment | NA | NA | 0 |
| S5-final-viral-combined_k141_1291\|\|full | 1916 | No | NA | 3 | 0 | 2 | Not-determined | Genome-fragment | NA | NA | 0 |
| S5-final-viral-combined_k141_18504\|\|full | 1361 | No | NA | 3 | 0 | 2 | Not-determined | Genome-fragment | NA | NA | 0 |
| S5-final-viral-combined_k141_143327\|\|full | 5893 | No | NA | 6 | 0 | 1 | Not-determined | Genome-fragment | NA | NA | 0 |
| S1-final-viral-combined_k141_117926\|\|full | 1610 | No | NA | 2 | 0 | 0 | Low-quality | Genome-fragment | 1.01 | AAI-based (medium-confidence) | 0 |
| S1_k141_311840 | 1610 | No | NA | 1 | 0 | 1 | Low-quality | Genome-fragment | 0.35 | AAI-based (medium-confidence) | 0 |
| S5-final-viral-combined_k141_108039\|\|lt2gene | 1610 | No | NA | 1 | 1 | 0 | Low-quality | Genome-fragment | 1.6 | AAI-based (high-confidence) | 0 |
| S5-final-viral-combined_k141_80873\|\|full | 1148 | No | NA | 3 | 0 | 1 | Not-determined | Genome-fragment | NA | NA | 0 |
| S7-final-viral-combined_k141_13336\|\|full | 1610 | No | NA | 2 | 0 | 2 | Low-quality | Genome-fragment | 0.78 | AAI-based (medium-confidence) | 0 |
| S5-final-viral-combined_k141_80892\|\|full | 1084 | No | NA | 2 | 0 | 2 | Not-determined | Genome-fragment | NA | NA | 0 |
| S12-final-viral-combined_k141_236215\|\|full | 1610 | No | NA | 2 | 0 | 1 | Low-quality | Genome-fragment | 1.64 | AAI-based (medium-confidence) | 0 |
| S12-final-viral-combined_k141_372654\|\|full | 1610 | No | NA | 5 | 0 | 0 | Low-quality | Genome-fragment | 3.98 | AAI-based (medium-confidence) | 0 |
| S5-final-viral-combined_k141_143364\|\|full | 1194 | No | NA | 3 | 0 | 0 | Not-determined | Genome-fragment | NA | NA | 0 |
| S2-final-viral-combined_k141_80608\|\|full | 1610 | No | NA | 4 | 0 | 0 | Low-quality | Genome-fragment | 2.36 | AAI-based (high-confidence) | 0 |
| S5-final-viral-combined_k141_80906\|\|full | 1125 | No | NA | 3 | 0 | 3 | Not-determined | Genome-fragment | NA | NA | 0 |
| S6-final-viral-combined_k141_106419\|\|full | 1610 | No | NA | 3 | 1 | 0 | Low-quality | Genome-fragment | 2.72 | AAI-based (high-confidence) | 0 |
| S5-final-viral-combined_k141_187757\|\|full | 16534 | No | NA | 19 | 0 | 15 | Not-determined | Genome-fragment | NA | NA | 0 |
| S5-final-viral-combined_k141_54284\|\|full | 1581 | No | NA | 3 | 0 | 0 | Not-determined | Genome-fragment | NA | NA | 0 |
| S6-final-viral-combined_k141_50262\|\|full | 1610 | No | NA | 3 | 1 | 0 | Low-quality | Genome-fragment | 1.04 | HMM-based (lower-bound) | 0 |
| S8-final-viral-combined_k141_49663\|\|full | 1610 | No | NA | 1 | 0 | 1 | Low-quality | Genome-fragment | 2.52 | AAI-based (medium-confidence) | 0 |
| S5-final-viral-combined_k141_45351\|\|full | 4413 | No | NA | 5 | 0 | 2 | Not-determined | Genome-fragment | NA | NA | 0 |
| S5-final-viral-combined_k141_161191\|\|full | 185988 | Yes | 33114 | 219 | 21 | 109 | Medium-quality | Genome-fragment | 59.59 | HMM-based (lower-bound) | 82.2 |
| S5-final-viral-combined_k141_1382\|\|full | 1394 | No | NA | 3 | 0 | 1 | Not-determined | Genome-fragment | NA | NA | 0 |
| S11-final-viral-combined_k141_231740\|\|full | 1611 | No | NA | 3 | 1 | 0 | Low-quality | Genome-fragment | 1.32 | AAI-based (high-confidence) | 0 |
| S11-final-viral-combined_k141_68627\|\|full | 1611 | No | NA | 3 | 1 | 0 | Low-quality | Genome-fragment | 2.37 | HMM-based (lower-bound) | 0 |
| S1-final-viral-combined_k141_66424\|\|full | 1611 | No | NA | 3 | 0 | 0 | Low-quality | Genome-fragment | 3.17 | AAI-based (medium-confidence) | 0 |
| S1-final-viral-combined_k141_127209\|\|full | 1611 | No | NA | 4 | 0 | 0 | Low-quality | Genome-fragment | 1.52 | AAI-based (medium-confidence) | 0 |
| S1-final-viral-combined_k141_357601\|\|full | 1611 | No | NA | 2 | 0 | 1 | Low-quality | Genome-fragment | 0.76 | AAI-based (medium-confidence) | 0 |
| S5-final-viral-combined_k141_63072\|\|full | 1940 | No | NA | 2 | 0 | 2 | Not-determined | Genome-fragment | NA | NA | 0 |
| S1-final-viral-combined_k141_316814\|\|lt2gene | 1611 | No | NA | 2 | 1 | 0 | Low-quality | Genome-fragment | 3.37 | AAI-based (high-confidence) | 0 |
| S5-final-viral-combined_k141_143404\|\|full | 3482 | No | NA | 4 | 0 | 1 | Not-determined | Genome-fragment | NA | NA | 0 |
| S5-final-viral-combined_k141_134381\|\|full | 74956 | Yes | 39317 | 102 | 22 | 18 | Medium-quality | Genome-fragment | 83.32 | AAI-based (high-confidence) | 47.55 |
| S5-final-viral-combined_k141_63075\|\|full | 2927 | No | NA | 2 | 0 | 2 | Not-determined | Genome-fragment | NA | NA | 0 |
| S5-final-viral-combined_k141_63081\|\|full | 1330 | No | NA | 4 | 0 | 0 | Not-determined | Genome-fragment | NA | NA | 0 |
| S5-final-viral-combined_k141_45425\|\|full | 1207 | No | NA | 2 | 0 | 1 | Not-determined | Genome-fragment | NA | NA | 0 |
| S5-final-viral-combined_k141_143437\|\|full | 2914 | No | NA | 4 | 0 | 1 | Not-determined | Genome-fragment | NA | NA | 0 |
| S5-final-viral-combined_k141_27494\|\|full | 3706 | No | NA | 6 | 0 | 0 | Not-determined | Genome-fragment | NA | NA | 0 |
| S5-final-viral-combined_k141_178963\|\|full | 2339 | No | NA | 3 | 0 | 2 | Not-determined | Genome-fragment | NA | NA | 0 |
| S5-final-viral-combined_k141_170287\|\|full | 2513 | No | NA | 3 | 0 | 1 | Not-determined | Genome-fragment | NA | NA | 0 |
| S5-final-viral-combined_k141_178998\|\|full | 2325 | No | NA | 3 | 0 | 0 | Not-determined | Genome-fragment | NA | NA | 0 |
| S5-final-viral-combined_k141_45454\|\|full | 1002 | No | NA | 2 | 0 | 0 | Not-determined | Genome-fragment | NA | NA | 0 |
| S1-final-viral-combined_k141_156024\|\|lt2gene | 1611 | No | NA | 2 | 1 | 0 | Low-quality | Genome-fragment | 5.06 | AAI-based (high-confidence) | 0 |
| S5-final-viral-combined_k141_80941\|\|full | 3032 | No | NA | 3 | 0 | 2 | Not-determined | Genome-fragment | NA | NA | 0 |
| S5-final-viral-combined_k141_137867\|\|full | 1611 | No | NA | 4 | 1 | 0 | Low-quality | Genome-fragment | 1.28 | HMM-based (lower-bound) | 0 |
| S5-final-viral-combined_k141_134467\|\|full | 2755 | No | NA | 2 | 0 | 2 | Not-determined | Genome-fragment | NA | NA | 0 |
| S12-final-viral-combined_k141_51936\|\|full | 1611 | No | NA | 4 | 1 | 0 | Low-quality | Genome-fragment | 0.98 | HMM-based (lower-bound) | 0 |
| S5-final-viral-combined_k141_197003\|\|full | 5232 | No | NA | 5 | 0 | 1 | Not-determined | Genome-fragment | NA | NA | 0 |
| S5-final-viral-combined_k141_1525\|\|full | 3934 | No | NA | 4 | 0 | 1 | Not-determined | Genome-fragment | NA | NA | 0 |
| S5-final-viral-combined_k141_1529\|\|full | 2780 | No | NA | 4 | 0 | 3 | Not-determined | Genome-fragment | NA | NA | 0 |
| S5-final-viral-combined_k141_179069\|\|full | 2146 | No | NA | 3 | 0 | 2 | Not-determined | Genome-fragment | NA | NA | 0 |
| S12-final-viral-combined_k141_115794\|\|full | 1611 | No | NA | 4 | 0 | 0 | Low-quality | Genome-fragment | 3.44 | AAI-based (high-confidence) | 0 |
| S4-final-viral-combined_k141_139730\|\|full | 1611 | No | NA | 5 | 1 | 0 | Low-quality | Genome-fragment | 2.2 | AAI-based (high-confidence) | 0 |
| S6-final-viral-combined_k141_104225\|\|full | 1611 | No | NA | 4 | 0 | 1 | Low-quality | Genome-fragment | 1.74 | AAI-based (medium-confidence) | 0 |
| S9-final-viral-combined_k141_39984\|\|full | 1612 | No | NA | 3 | 0 | 1 | Low-quality | Genome-fragment | 3.07 | AAI-based (high-confidence) | 0 |
| S11_k141_218843 | 1612 | No | NA | 1 | 0 | 1 | Low-quality | Genome-fragment | 0.87 | AAI-based (medium-confidence) | 0 |
| S5-final-viral-combined_k141_90277\|\|full | 2411 | No | NA | 2 | 0 | 2 | Not-determined | Genome-fragment | NA | NA | 0 |
| S5-final-viral-combined_k141_108027\|\|full | 1458 | No | NA | 3 | 0 | 2 | Not-determined | Genome-fragment | NA | NA | 0 |
| S5-final-viral-combined_k141_18653\|\|full | 1256 | No | NA | 3 | 0 | 0 | Not-determined | Genome-fragment | NA | NA | 0 |
| S5-final-viral-combined_k141_1578\|\|full | 3307 | No | NA | 4 | 0 | 2 | Not-determined | Genome-fragment | NA | NA | 0 |
| S5-final-viral-combined_k141_54455\|\|full | 1814 | No | NA | 2 | 0 | 1 | Not-determined | Genome-fragment | NA | NA | 0 |
| S5-final-viral-combined_k141_27554\|\|full | 1763 | No | NA | 2 | 0 | 0 | Not-determined | Genome-fragment | NA | NA | 0 |
| S5-final-viral-combined_k141_205701\|\|full | 3285 | No | NA | 3 | 0 | 0 | Not-determined | Genome-fragment | NA | NA | 0 |
| S5-final-viral-combined_k141_205703\|\|full | 1392 | No | NA | 3 | 0 | 1 | Not-determined | Genome-fragment | NA | NA | 0 |
| S1-final-viral-combined_k141_105078\|\|full | 1612 | No | NA | 3 | 0 | 0 | Low-quality | Genome-fragment | 1.11 | AAI-based (medium-confidence) | 0 |
| S5-final-viral-combined_k141_63159\|\|full | 1458 | No | NA | 3 | 0 | 0 | Not-determined | Genome-fragment | NA | NA | 0 |
| S7_k141_185938 | 1612 | No | NA | 2 | 0 | 1 | Low-quality | Genome-fragment | 1.01 | AAI-based (medium-confidence) | 0 |
| S6-final-viral-combined_k141_16512\|\|full | 1612 | No | NA | 3 | 0 | 0 | Low-quality | Genome-fragment | 4.1 | AAI-based (medium-confidence) | 0 |
| S5-final-viral-combined_k141_116805\|\|full | 1981 | No | NA | 2 | 0 | 0 | Not-determined | Genome-fragment | NA | NA | 0 |
| S1-final-viral-combined_k141_82437\|\|full | 1613 | No | NA | 3 | 0 | 2 | Low-quality | Genome-fragment | 4.19 | AAI-based (medium-confidence) | 0 |
| S5-final-viral-combined_k141_116843\|\|full | 5270 | No | NA | 4 | 0 | 3 | Not-determined | Genome-fragment | NA | NA | 0 |
| S5-final-viral-combined_k141_98974\|\|full | 1317 | No | NA | 2 | 0 | 0 | Not-determined | Genome-fragment | NA | NA | 0 |
| S5-final-viral-combined_k141_179203\|\|full | 5314 | No | NA | 4 | 0 | 0 | Not-determined | Genome-fragment | NA | NA | 0 |
| S5-final-viral-combined_k141_187886\|\|full | 1089 | No | NA | 4 | 0 | 0 | Not-determined | Genome-fragment | NA | NA | 0 |
| S5-final-viral-combined_k141_45541\|\|full | 2498 | No | NA | 3 | 0 | 1 | Not-determined | Genome-fragment | NA | NA | 0 |
| S5-final-viral-combined_k141_90315\|\|full | 102707 | Yes | 71984 | 118 | 12 | 25 | High-quality | High-quality | 100 | AAI-based (high-confidence) | 29.91 |
| S5-final-viral-combined_k141_81050\|\|full | 2649 | No | NA | 4 | 0 | 1 | Not-determined | Genome-fragment | NA | NA | 0 |
| S5-final-viral-combined_k141_143597\|\|full | 2114 | No | NA | 2 | 0 | 1 | Not-determined | Genome-fragment | NA | NA | 0 |
| S1-final-viral-combined_k141_111341\|\|full | 1613 | No | NA | 5 | 1 | 0 | Low-quality | Genome-fragment | 4.22 | AAI-based (medium-confidence) | 0 |
| S5-final-viral-combined_k141_143599\|\|full | 1414 | No | NA | 3 | 0 | 2 | Not-determined | Genome-fragment | NA | NA | 0 |
| S5-final-viral-combined_k141_197056\|\|full | 2601 | No | NA | 3 | 0 | 2 | Not-determined | Genome-fragment | NA | NA | 0 |
| S3_k141_61174 | 1613 | No | NA | 3 | 0 | 0 | Low-quality | Genome-fragment | 0.74 | AAI-based (high-confidence) | 0 |
| S7-final-viral-combined_k141_236427\|\|full | 1613 | No | NA | 2 | 0 | 0 | Low-quality | Genome-fragment | 13.58 | AAI-based (medium-confidence) | 0 |
| S7-final-viral-combined_k141_175723\|\|lt2gene | 1613 | No | NA | 2 | 1 | 0 | Low-quality | Genome-fragment | 3.92 | AAI-based (high-confidence) | 0 |
| S17-final-viral-combined_k141_100809\|\|full | 1613 | No | NA | 2 | 0 | 1 | Low-quality | Genome-fragment | 4.21 | AAI-based (medium-confidence) | 0 |
| S17-final-viral-combined_k141_255271\|\|lt2gene | 1613 | No | NA | 2 | 2 | 0 | Low-quality | Genome-fragment | 5.33 | AAI-based (high-confidence) | 0 |
| S5-final-viral-combined_k141_1761\|\|full | 6667 | No | NA | 8 | 0 | 4 | Not-determined | Genome-fragment | NA | NA | 0 |
| S12-final-viral-combined_k141_376196\|\|full | 1613 | No | NA | 2 | 0 | 1 | Low-quality | Genome-fragment | 1.9 | AAI-based (high-confidence) | 0 |
| S12-final-viral-combined_k141_33312\|\|full | 1613 | No | NA | 3 | 0 | 0 | Low-quality | Genome-fragment | 3.85 | AAI-based (medium-confidence) | 0 |
| S12_k141_216787 | 1613 | No | NA | 2 | 0 | 0 | Low-quality | Genome-fragment | 1.18 | AAI-based (high-confidence) | 0 |
| S5-final-viral-combined_k141_197093\|\|full | 3609 | No | NA | 5 | 0 | 1 | Not-determined | Genome-fragment | NA | NA | 0 |
| S5-final-viral-combined_k141_81190\|\|full | 2519 | No | NA | 6 | 0 | 0 | Not-determined | Genome-fragment | NA | NA | 0 |
| S5-final-viral-combined_k141_99051\|\|full | 1945 | No | NA | 2 | 0 | 2 | Not-determined | Genome-fragment | NA | NA | 0 |
| S11-final-viral-combined_k141_165940\|\|full | 1614 | No | NA | 2 | 0 | 0 | Low-quality | Genome-fragment | 3.09 | AAI-based (medium-confidence) | 0 |
| S1-final-viral-combined_k141_56955\|\|full | 1614 | No | NA | 3 | 0 | 0 | Low-quality | Genome-fragment | 3.37 | AAI-based (high-confidence) | 0 |
| S7_k141_114306 | 1614 | No | NA | 3 | 0 | 0 | Low-quality | Genome-fragment | 3.03 | AAI-based (high-confidence) | 0 |
| S5-final-viral-combined_k141_108205\|\|full | 6732 | No | NA | 6 | 0 | 2 | Not-determined | Genome-fragment | NA | NA | 0 |
| S17-final-viral-combined_k141_170059\|\|full | 1614 | No | NA | 2 | 0 | 0 | Low-quality | Genome-fragment | 3.78 | AAI-based (high-confidence) | 0 |
| S5-final-viral-combined_k141_152512\|\|full | 3616 | No | NA | 4 | 0 | 3 | Not-determined | Genome-fragment | NA | NA | 0 |
| S12-final-viral-combined_k141_343801\|\|lt2gene | 1614 | No | NA | 2 | 2 | 0 | Low-quality | Genome-fragment | 2.08 | HMM-based (lower-bound) | 0 |
| S2-final-viral-combined_k141_47781\|\|full | 1614 | No | NA | 3 | 0 | 1 | Low-quality | Genome-fragment | 2.69 | AAI-based (high-confidence) | 0 |
| S6_k141_89835 | 1614 | No | NA | 5 | 1 | 0 | Low-quality | Genome-fragment | 2.68 | AAI-based (high-confidence) | 0 |
| S8-final-viral-combined_k141_27461\|\|full | 1614 | No | NA | 3 | 1 | 0 | Low-quality | Genome-fragment | 1.92 | HMM-based (lower-bound) | 0 |
| S5-final-viral-combined_k141_10168\|\|full | 4713 | No | NA | 4 | 0 | 3 | Not-determined | Genome-fragment | NA | NA | 0 |
| S5-final-viral-combined_k141_81301\|\|full | 1783 | No | NA | 3 | 0 | 0 | Not-determined | Genome-fragment | NA | NA | 0 |
| S5-final-viral-combined_k141_161329\|\|full | 3690 | No | NA | 7 | 0 | 1 | Not-determined | Genome-fragment | NA | NA | 0 |
| S8-final-viral-combined_k141_72959\|\|lt2gene | 1614 | No | NA | 2 | 0 | 0 | Low-quality | Genome-fragment | 2.75 | AAI-based (high-confidence) | 0 |
| S5-final-viral-combined_k141_205873\|\|full | 3171 | No | NA | 3 | 0 | 2 | Not-determined | Genome-fragment | NA | NA | 0 |
| S11_k141_264601 | 1615 | No | NA | 2 | 1 | 0 | Low-quality | Genome-fragment | 4.45 | AAI-based (high-confidence) | 0 |
| S1-final-viral-combined_k141_134992\|\|full | 1615 | No | NA | 5 | 0 | 0 | Low-quality | Genome-fragment | 3.99 | AAI-based (medium-confidence) | 0 |
| S1-final-viral-combined_k141_63190\|\|full | 1615 | No | NA | 5 | 0 | 0 | Low-quality | Genome-fragment | 2.67 | AAI-based (high-confidence) | 0 |
| S7_k141_233094 | 1615 | No | NA | 2 | 0 | 2 | Low-quality | Genome-fragment | 0.75 | AAI-based (medium-confidence) | 0 |
| S12_k141_394396 | 1615 | No | NA | 3 | 0 | 0 | Low-quality | Genome-fragment | 3.39 | AAI-based (medium-confidence) | 0 |
| S5-final-viral-combined_k141_99088\|\|full | 5609 | No | NA | 4 | 0 | 2 | Not-determined | Genome-fragment | NA | NA | 0 |
| S9-final-viral-combined_k141_30925\|\|lt2gene | 1616 | No | NA | 2 | 1 | 0 | Low-quality | Genome-fragment | 2.92 | AAI-based (high-confidence) | 0 |
| S5-final-viral-combined_k141_134743\|\|full | 103549 | Yes | 52229 | 130 | 23 | 27 | Medium-quality | Genome-fragment | 66.11 | HMM-based (lower-bound) | 49.56 |
| S5-final-viral-combined_k141_170493\|\|full | 11226 | No | NA | 17 | 0 | 1 | Not-determined | Genome-fragment | NA | NA | 0 |
| S5-final-viral-combined_k141_108301\|\|full | 1152 | No | NA | 2 | 0 | 1 | Not-determined | Genome-fragment | NA | NA | 0 |
| S11-final-viral-combined_k141_46118\|\|full | 1616 | No | NA | 3 | 1 | 0 | Low-quality | Genome-fragment | 3.39 | AAI-based (high-confidence) | 0 |
| S5-final-viral-combined_k141_134773\|\|full | 1261 | No | NA | 3 | 0 | 0 | Not-determined | Genome-fragment | NA | NA | 0 |
| S5-final-viral-combined_k141_197186\|\|full | 32221 | No | NA | 41 | 1 | 4 | Medium-quality | Genome-fragment | 68.83 | HMM-based (lower-bound) | 0 |
| S5-final-viral-combined_k141_36653\|\|full | 2638 | No | NA | 4 | 0 | 1 | Not-determined | Genome-fragment | NA | NA | 0 |
| S5-final-viral-combined_k141_134832\|\|full | 1446 | No | NA | 3 | 0 | 0 | Not-determined | Genome-fragment | NA | NA | 0 |
| S11-final-viral-combined_k141_258431\|\|full | 1616 | No | NA | 5 | 0 | 0 | Low-quality | Genome-fragment | 3.93 | AAI-based (medium-confidence) | 0 |
| S11-final-viral-combined_k141_203828\|\|full | 1616 | Yes | 881 | 3 | 1 | 1 | Low-quality | Genome-fragment | 1.23 | HMM-based (lower-bound) | 45.48 |
| S5-final-viral-combined_k141_205977\|\|full | 4193 | No | NA | 4 | 0 | 4 | Not-determined | Genome-fragment | NA | NA | 0 |
| S5-final-viral-combined_k141_18774\|\|full | 3340 | No | NA | 2 | 0 | 2 | Not-determined | Genome-fragment | NA | NA | 0 |
| S5-final-viral-combined_k141_45774\|\|full | 2133 | No | NA | 3 | 0 | 2 | Not-determined | Genome-fragment | NA | NA | 0 |
| S1-final-viral-combined_k141_235276\|\|full | 1616 | No | NA | 2 | 0 | 2 | Low-quality | Genome-fragment | 2.7 | AAI-based (medium-confidence) | 0 |
| S5-final-viral-combined_k141_161434\|\|full | 1569 | No | NA | 3 | 0 | 1 | Not-determined | Genome-fragment | NA | NA | 0 |
| S1-final-viral-combined_k141_257827\|\|full | 1616 | No | NA | 3 | 1 | 0 | Low-quality | Genome-fragment | 2.04 | HMM-based (lower-bound) | 0 |
| S5-final-viral-combined_k141_81424\|\|full | 1005 | No | NA | 2 | 0 | 1 | Not-determined | Genome-fragment | NA | NA | 0 |
| S1-final-viral-combined_k141_327975\|\|full | 1616 | No | NA | 5 | 0 | 0 | Low-quality | Genome-fragment | 4.18 | AAI-based (medium-confidence) | 0 |
| S3-final-viral-combined_k141_78005\|\|full | 1616 | No | NA | 2 | 0 | 1 | Low-quality | Genome-fragment | 0.43 | AAI-based (medium-confidence) | 0 |
| S3_k141_133898 | 1616 | No | NA | 1 | 0 | 0 | Low-quality | Genome-fragment | 0.17 | AAI-based (medium-confidence) | 0 |
| S5-final-viral-combined_k141_143896\|\|full | 1388 | No | NA | 2 | 0 | 0 | Not-determined | Genome-fragment | NA | NA | 0 |
| S5-final-viral-combined_k141_117095\|\|full | 26599 | No | NA | 25 | 0 | 12 | Medium-quality | Genome-fragment | 52.6 | AAI-based (medium-confidence) | 0 |
| S5-final-viral-combined_k141_72410\|\|full | 237438 | Yes | 58078 | 252 | 34 | 124 | Complete | High-quality | 100 | Provirus (medium-confidence) | 75.54 |
| S17-final-viral-combined_k141_149353\|\|full | 1616 | No | NA | 2 | 0 | 1 | Low-quality | Genome-fragment | 3.1 | AAI-based (high-confidence) | 0 |
| S5-final-viral-combined_k141_72437\|\|full | 1112 | No | NA | 5 | 0 | 0 | Not-determined | Genome-fragment | NA | NA | 0 |
| S5-final-viral-combined_k141_170708\|\|full | 2070 | No | NA | 4 | 0 | 0 | Not-determined | Genome-fragment | NA | NA | 0 |
| S5-final-viral-combined_k141_90471\|\|full | 16674 | No | NA | 22 | 0 | 5 | Not-determined | Genome-fragment | NA | NA | 0 |
| S17-final-viral-combined_k141_196986\|\|full | 1616 | No | NA | 2 | 0 | 2 | Low-quality | Genome-fragment | 1.38 | AAI-based (medium-confidence) | 0 |
| S17-final-viral-combined_k141_266335\|\|full | 1616 | No | NA | 2 | 0 | 2 | Low-quality | Genome-fragment | 4.06 | AAI-based (medium-confidence) | 0 |
| S17-k141_55676 | 1616 | No | NA | 2 | 0 | 0 | Low-quality | Genome-fragment | 2.1 | AAI-based (high-confidence) | 0 |
| S12-final-viral-combined_k141_127390\|\|full | 1616 | No | NA | 2 | 0 | 0 | Low-quality | Genome-fragment | 2.17 | AAI-based (medium-confidence) | 0 |
| S5-final-viral-combined_k141_170731\|\|full | 1510 | No | NA | 3 | 0 | 1 | Not-determined | Genome-fragment | NA | NA | 0 |
| S12_k141_374118 | 1616 | No | NA | 1 | 0 | 0 | Low-quality | Genome-fragment | 1.28 | AAI-based (medium-confidence) | 0 |
| S12_k141_139898 | 1616 | No | NA | 2 | 0 | 0 | Low-quality | Genome-fragment | 2.7 | AAI-based (high-confidence) | 0 |
| S8-final-viral-combined_k141_148461\|\|full | 1616 | No | NA | 4 | 0 | 0 | Low-quality | Genome-fragment | 3.62 | AAI-based (medium-confidence) | 0 |
| S9-final-viral-combined_k141_86168\|\|full | 1617 | No | NA | 3 | 0 | 1 | Low-quality | Genome-fragment | 1.33 | AAI-based (medium-confidence) | 0 |
| S5-final-viral-combined_k141_170765\|\|full | 3983 | No | NA | 4 | 0 | 1 | Not-determined | Genome-fragment | NA | NA | 0 |
| S5-final-viral-combined_k141_108517\|\|full | 4149 | No | NA | 6 | 0 | 2 | Not-determined | Genome-fragment | NA | NA | 0 |
| S1-final-viral-combined_k141_22841\|\|full | 1617 | No | NA | 4 | 0 | 0 | Low-quality | Genome-fragment | 4.41 | AAI-based (high-confidence) | 0 |
| S1-final-viral-combined_k141_108755\|\|full | 1617 | No | NA | 4 | 1 | 0 | Low-quality | Genome-fragment | 3.77 | AAI-based (high-confidence) | 0 |
| S5-final-viral-combined_k141_1995\|\|full | 4889 | No | NA | 4 | 0 | 4 | Not-determined | Genome-fragment | NA | NA | 0 |
| S12-final-viral-combined_k141_285815\|\|full | 1617 | No | NA | 3 | 1 | 0 | Low-quality | Genome-fragment | 3.53 | AAI-based (high-confidence) | 0 |
| S5-final-viral-combined_k141_179548\|\|full | 2005 | No | NA | 2 | 0 | 2 | Not-determined | Genome-fragment | NA | NA | 0 |
| S5-final-viral-combined_k141_206050\|\|full | 57403 | Yes | 39805 | 61 | 27 | 13 | Medium-quality | Genome-fragment | 79.84 | AAI-based (medium-confidence) | 30.66 |
| S12-final-viral-combined_k141_271964\|\|full | 1617 | No | NA | 2 | 1 | 0 | Low-quality | Genome-fragment | 4.02 | AAI-based (medium-confidence) | 0 |
| S5-final-viral-combined_k141_2043\|\|full | 3569 | No | NA | 5 | 0 | 0 | Not-determined | Genome-fragment | NA | NA | 0 |
| S12-final-viral-combined_k141_81261\|\|full | 1617 | No | NA | 3 | 0 | 1 | Low-quality | Genome-fragment | 0.88 | AAI-based (medium-confidence) | 0 |
| S5-final-viral-combined_k141_197324\|\|full | 1401 | No | NA | 3 | 0 | 0 | Not-determined | Genome-fragment | NA | NA | 0 |
| S2-final-viral-combined_k141_106491\|\|full | 1617 | No | NA | 4 | 0 | 0 | Low-quality | Genome-fragment | 1.31 | AAI-based (medium-confidence) | 0 |
| S9-final-viral-combined_k141_19185\|\|full | 1618 | No | NA | 2 | 0 | 0 | Low-quality | Genome-fragment | 2.7 | AAI-based (high-confidence) | 0 |
| S5-final-viral-combined_k141_72548\|\|full | 1593 | No | NA | 2 | 0 | 0 | Not-determined | Genome-fragment | NA | NA | 0 |
| S5-final-viral-combined_k141_144044\|\|full | 2093 | No | NA | 3 | 0 | 0 | Not-determined | Genome-fragment | NA | NA | 0 |
| S5-final-viral-combined_k141_2088\|\|full | 1492 | No | NA | 2 | 0 | 2 | Not-determined | Genome-fragment | NA | NA | 0 |
| S5-final-viral-combined_k141_197352\|\|full | 11777 | No | NA | 13 | 0 | 4 | Not-determined | Genome-fragment | NA | NA | 0 |
| S11-final-viral-combined_k141_282560\|\|full | 1618 | No | NA | 5 | 3 | 0 | Low-quality | Genome-fragment | 2.6 | AAI-based (high-confidence) | 0 |
| S1-final-viral-combined_k141_9524\|\|full | 1618 | No | NA | 6 | 0 | 0 | Low-quality | Genome-fragment | 2.3 | AAI-based (medium-confidence) | 0 |
| S1-final-viral-combined_k141_96832\|\|full | 1618 | No | NA | 3 | 1 | 0 | Low-quality | Genome-fragment | 29.04 | AAI-based (medium-confidence) | 0 |
| S5-final-viral-combined_k141_179714\|\|full | 6431 | No | NA | 6 | 0 | 6 | Not-determined | Genome-fragment | NA | NA | 0 |
| S5-final-viral-combined_k141_170848\|\|full | 2772 | No | NA | 6 | 0 | 1 | Not-determined | Genome-fragment | NA | NA | 0 |
| S1-final-viral-combined_k141_98143\|\|full | 1618 | No | NA | 2 | 0 | 2 | Low-quality | Genome-fragment | 2.79 | AAI-based (medium-confidence) | 0 |
| S1_k141_150057 | 1618 | No | NA | 2 | 0 | 0 | Low-quality | Genome-fragment | 2.08 | AAI-based (high-confidence) | 0 |
| S5-final-viral-combined_k141_125978\|\|full | 1610 | No | NA | 3 | 0 | 1 | Not-determined | Genome-fragment | NA | NA | 0 |
| S5-final-viral-combined_k141_179761\|\|full | 1883 | No | NA | 5 | 0 | 0 | Not-determined | Genome-fragment | NA | NA | 0 |
| S5-final-viral-combined_k141_90603\|\|full | 1876 | No | NA | 3 | 0 | 1 | Not-determined | Genome-fragment | NA | NA | 0 |
| S5-final-viral-combined_k141_90605\|\|full | 1505 | No | NA | 3 | 0 | 2 | Not-determined | Genome-fragment | NA | NA | 0 |
| S7-final-viral-combined_k141_71324\|\|full | 1618 | No | NA | 3 | 1 | 0 | Low-quality | Genome-fragment | 3.63 | AAI-based (medium-confidence) | 0 |
| S17-final-viral-combined_k141_61992\|\|full | 1618 | No | NA | 3 | 0 | 2 | Low-quality | Genome-fragment | 1.28 | AAI-based (medium-confidence) | 0 |
| S5-final-viral-combined_k141_179830\|\|full | 1040 | No | NA | 2 | 0 | 1 | Not-determined | Genome-fragment | NA | NA | 0 |
| S5-final-viral-combined_k141_63592\|\|full | 3319 | No | NA | 4 | 0 | 0 | Not-determined | Genome-fragment | NA | NA | 0 |
| S2_k141_43188 | 1618 | No | NA | 3 | 0 | 0 | Low-quality | Genome-fragment | 1.12 | AAI-based (medium-confidence) | 0 |
| S5-final-viral-combined_k141_179846\|\|full | 1111 | No | NA | 2 | 0 | 0 | Not-determined | Genome-fragment | NA | NA | 0 |
| S5-final-viral-combined_k141_117249\|\|full | 2790 | No | NA | 3 | 0 | 1 | Not-determined | Genome-fragment | NA | NA | 0 |
| S6-final-viral-combined_k141_124894\|\|full | 1618 | No | NA | 2 | 0 | 1 | Low-quality | Genome-fragment | 2 | AAI-based (high-confidence) | 0 |
| S6-final-viral-combined_k141_58426\|\|full | 1618 | No | NA | 4 | 1 | 0 | Low-quality | Genome-fragment | 3.28 | AAI-based (medium-confidence) | 0 |
| S5-final-viral-combined_k141_81600\|\|full | 2144 | No | NA | 3 | 0 | 3 | Not-determined | Genome-fragment | NA | NA | 0 |
| S5-final-viral-combined_k141_134947\|\|full | 3215 | No | NA | 3 | 0 | 3 | Not-determined | Genome-fragment | NA | NA | 0 |
| S9-final-viral-combined_k141_104815\|\|full | 1619 | No | NA | 5 | 0 | 0 | Low-quality | Genome-fragment | 2.25 | AAI-based (medium-confidence) | 0 |
| S9-final-viral-combined_k141_75718\|\|lt2gene | 1619 | No | NA | 2 | 1 | 0 | Low-quality | Genome-fragment | 3.16 | AAI-based (high-confidence) | 0 |
| S11-final-viral-combined_k141_327243\|\|full | 1619 | No | NA | 3 | 0 | 0 | Low-quality | Genome-fragment | 3.88 | AAI-based (high-confidence) | 0 |
| S1-final-viral-combined_k141_283545\|\|full | 1619 | No | NA | 2 | 0 | 1 | Low-quality | Genome-fragment | 2.49 | AAI-based (medium-confidence) | 0 |
| S5-final-viral-combined_k141_28074\|\|full | 1751 | No | NA | 3 | 0 | 0 | Not-determined | Genome-fragment | NA | NA | 0 |
| S5-final-viral-combined_k141_28081\|\|full | 1127 | No | NA | 2 | 0 | 2 | Not-determined | Genome-fragment | NA | NA | 0 |
| S1-final-viral-combined_k141_194177\|\|full | 1619 | No | NA | 3 | 0 | 0 | Low-quality | Genome-fragment | 4.12 | AAI-based (high-confidence) | 0 |
| S3-final-viral-combined_k141_64361\|\|full | 1619 | No | NA | 3 | 0 | 0 | Low-quality | Genome-fragment | 3.24 | AAI-based (high-confidence) | 0 |
| S5-final-viral-combined_k141_134969\|\|full | 4581 | No | NA | 6 | 0 | 1 | Not-determined | Genome-fragment | NA | NA | 0 |
| S3-final-viral-combined_k141_57910\|\|lt2gene | 1619 | No | NA | 2 | 1 | 0 | Low-quality | Genome-fragment | 2.91 | AAI-based (high-confidence) | 0 |
| S5-final-viral-combined_k141_81637\|\|full | 4410 | No | NA | 6 | 0 | 3 | Not-determined | Genome-fragment | NA | NA | 0 |
| S7-final-viral-combined_k141_157657\|\|full | 1619 | No | NA | 5 | 0 | 0 | Low-quality | Genome-fragment | 3.99 | AAI-based (medium-confidence) | 0 |
| S2-final-viral-combined_k141_223942\|\|full | 1619 | No | NA | 4 | 0 | 0 | Low-quality | Genome-fragment | 1.79 | AAI-based (high-confidence) | 0 |
| S5-final-viral-combined_k141_144235\|\|full | 4021 | No | NA | 4 | 0 | 3 | Not-determined | Genome-fragment | NA | NA | 0 |
| S2-final-viral-combined_k141_39750\|\|full | 1619 | No | NA | 3 | 0 | 0 | Low-quality | Genome-fragment | 3.21 | AAI-based (medium-confidence) | 0 |
| S4-final-viral-combined_k141_103288\|\|full | 1619 | No | NA | 3 | 0 | 1 | Low-quality | Genome-fragment | 3.18 | AAI-based (high-confidence) | 0 |
| S5-final-viral-combined_k141_81666\|\|full | 5280 | No | NA | 8 | 0 | 2 | Not-determined | Genome-fragment | NA | NA | 0 |
| S5-final-viral-combined_k141_134974\|\|full | 35469 | No | NA | 30 | 0 | 20 | Not-determined | Genome-fragment | NA | NA | 0 |
| S5-final-viral-combined_k141_179862\|\|full | 2069 | No | NA | 3 | 0 | 2 | Not-determined | Genome-fragment | NA | NA | 0 |
| S5-final-viral-combined_k141_152902\|\|full | 1015 | No | NA | 2 | 0 | 1 | Not-determined | Genome-fragment | NA | NA | 0 |
| S8-final-viral-combined_k141_171169\|\|full | 1619 | No | NA | 2 | 0 | 1 | Low-quality | Genome-fragment | 1.85 | AAI-based (medium-confidence) | 0 |
| S9-final-viral-combined_k141_118102\|\|full | 1620 | No | NA | 5 | 0 | 0 | Low-quality | Genome-fragment | 2.98 | AAI-based (high-confidence) | 0 |
| S5-final-viral-combined_k141_55055\|\|full | 1194 | No | NA | 3 | 0 | 0 | Not-determined | Genome-fragment | NA | NA | 0 |
| S9-final-viral-combined_k141_9404\|\|full | 1620 | No | NA | 5 | 0 | 0 | Low-quality | Genome-fragment | 3.92 | AAI-based (medium-confidence) | 0 |
| S5-final-viral-combined_k141_126097\|\|full | 1370 | No | NA | 4 | 0 | 0 | Not-determined | Genome-fragment | NA | NA | 0 |
| S5-final-viral-combined_k141_197471\|\|full | 2337 | No | NA | 4 | 0 | 0 | Not-determined | Genome-fragment | NA | NA | 0 |
| S5-final-viral-combined_k141_197477\|\|full | 1731 | No | NA | 2 | 0 | 2 | Not-determined | Genome-fragment | NA | NA | 0 |
| S5-final-viral-combined_k141_171085\|\|full | 1914 | No | NA | 3 | 0 | 1 | Not-determined | Genome-fragment | NA | NA | 0 |
| S1-final-viral-combined_k141_166126\|\|full | 1620 | Yes | 1140 | 3 | 1 | 1 | Low-quality | Genome-fragment | 2.75 | AAI-based (medium-confidence) | 29.63 |
| S1-final-viral-combined_k141_18632\|\|full | 1620 | No | NA | 3 | 0 | 0 | Low-quality | Genome-fragment | 2.08 | AAI-based (medium-confidence) | 0 |
| S5-final-viral-combined_k141_161660\|\|full | 1246 | No | NA | 3 | 0 | 1 | Not-determined | Genome-fragment | NA | NA | 0 |
| S3-final-viral-combined_k141_101830\|\|full | 1620 | No | NA | 3 | 2 | 0 | Low-quality | Genome-fragment | 3.91 | AAI-based (high-confidence) | 0 |
| S17-k141_62846 | 1620 | No | NA | 1 | 0 | 1 | Low-quality | Genome-fragment | 1.49 | AAI-based (medium-confidence) | 0 |
| S12_k141_141057 | 1620 | No | NA | 2 | 0 | 0 | Low-quality | Genome-fragment | 2.71 | AAI-based (high-confidence) | 0 |
| S4-final-viral-combined_k141_111586\|\|full | 1620 | No | NA | 3 | 0 | 0 | Low-quality | Genome-fragment | 3.03 | AAI-based (high-confidence) | 0 |
| S4_k141_88695 | 1620 | No | NA | 3 | 0 | 0 | Low-quality | Genome-fragment | 4.1 | AAI-based (medium-confidence) | 0 |
| S5-final-viral-combined_k141_108659\|\|full | 1259 | No | NA | 2 | 0 | 1 | Not-determined | Genome-fragment | NA | NA | 0 |
| S5-final-viral-combined_k141_10597\|\|full | 2032 | No | NA | 2 | 0 | 0 | Not-determined | Genome-fragment | NA | NA | 0 |
| S9-final-viral-combined_k141_97292\|\|full | 1621 | No | NA | 5 | 4 | 0 | Low-quality | Genome-fragment | 4.62 | AAI-based (medium-confidence) | 0 |
| S5-final-viral-combined_k141_188585\|\|full | 1658 | No | NA | 3 | 0 | 2 | Not-determined | Genome-fragment | NA | NA | 0 |
| S5-final-viral-combined_k141_37055\|\|full | 1632 | No | NA | 3 | 0 | 1 | Not-determined | Genome-fragment | NA | NA | 0 |
| S5-final-viral-combined_k141_126218\|\|full | 11665 | No | NA | 11 | 0 | 9 | Not-determined | Genome-fragment | NA | NA | 0 |
| S5-final-viral-combined_k141_206204\|\|full | 3242 | No | NA | 3 | 0 | 2 | Not-determined | Genome-fragment | NA | NA | 0 |
| S11-final-viral-combined_k141_99670\|\|lt2gene | 1621 | No | NA | 1 | 1 | 0 | Low-quality | Genome-fragment | 2.97 | AAI-based (high-confidence) | 0 |
| S7-final-viral-combined_k141_185032\|\|full | 1621 | No | NA | 3 | 1 | 0 | Low-quality | Genome-fragment | 2.13 | AAI-based (medium-confidence) | 0 |
| S5-final-viral-combined_k141_153040\|\|full | 2453 | No | NA | 2 | 0 | 2 | Not-determined | Genome-fragment | NA | NA | 0 |
| S5-final-viral-combined_k141_37093\|\|full | 2767 | No | NA | 4 | 0 | 3 | Not-determined | Genome-fragment | NA | NA | 0 |

| | | | | | | | | | | | |
|---|---|---|---|---|---|---|---|---|---|---|---|
| S7-final-viral-combined_k141_83185\|\|full | 1621 | No | NA | 4 | 0 | 0 | Low-quality | Genome-fragment | 2.81 | AAI-based (high-confidence) | 0 |
| S5-final-viral-combined_k141_180009\|\|full | 1125 | No | NA | 3 | 0 | 1 | Not-determined | Genome-fragment | NA | NA | 0 |
| S5-final-viral-combined_k141_37094\|\|full | 3537 | No | NA | 4 | 0 | 2 | Not-determined | Genome-fragment | NA | NA | 0 |
| S5-final-viral-combined_k141_72884\|\|full | 1556 | No | NA | 3 | 0 | 1 | Not-determined | Genome-fragment | NA | NA | 0 |
| S17-final-viral-combined_k141_54677\|\|full | 1621 | No | NA | 2 | 0 | 1 | Low-quality | Genome-fragment | 1.02 | AAI-based (high-confidence) | 0 |
| S5-final-viral-combined_k141_19173\|\|full | 1144 | No | NA | 3 | 0 | 1 | Not-determined | Genome-fragment | NA | NA | 0 |
| S4_k141_50136 | 1621 | No | NA | 4 | 0 | 0 | Low-quality | Genome-fragment | 3.53 | AAI-based (high-confidence) | 0 |
| S4_k141_129135 | 1621 | No | NA | 1 | 0 | 1 | Low-quality | Genome-fragment | 0.58 | AAI-based (medium-confidence) | 0 |
| S8_k141_140950 | 1621 | No | NA | 2 | 0 | 2 | Low-quality | Genome-fragment | 3.24 | AAI-based (medium-confidence) | 0 |
| S5-final-viral-combined_k141_99403\|\|full | 2152 | No | NA | 3 | 0 | 0 | Not-determined | Genome-fragment | NA | NA | 0 |
| S5-final-viral-combined_k141_180025\|\|full | 5226 | No | NA | 3 | 0 | 2 | Not-determined | Genome-fragment | NA | NA | 0 |
| S11-final-viral-combined_k141_181763\|\|full | 1622 | No | NA | 3 | 0 | 0 | Low-quality | Genome-fragment | 3.49 | AAI-based (high-confidence) | 0 |
| S5-final-viral-combined_k141_81820\|\|full | 1575 | No | NA | 4 | 0 | 0 | Not-determined | Genome-fragment | NA | NA | 0 |
| S5-final-viral-combined_k141_37141\|\|full | 4116 | No | NA | 3 | 0 | 1 | Not-determined | Genome-fragment | NA | NA | 0 |
| S5-final-viral-combined_k141_161736\|\|full | 2555 | No | NA | 4 | 0 | 1 | Not-determined | Genome-fragment | NA | NA | 0 |
| S5-final-viral-combined_k141_63651\|\|full | 1867 | No | NA | 4 | 0 | 3 | Not-determined | Genome-fragment | NA | NA | 0 |
| S5-final-viral-combined_k141_144416\|\|full | 1715 | No | NA | 2 | 0 | 0 | Not-determined | Genome-fragment | NA | NA | 0 |
| S1_k141_111634 | 1622 | No | NA | 2 | 0 | 2 | Low-quality | Genome-fragment | 1.06 | AAI-based (high-confidence) | 0 |
| S5-final-viral-combined_k141_45379\|\|full | 1622 | No | NA | 2 | 0 | 1 | Low-quality | Genome-fragment | 3.68 | AAI-based (medium-confidence) | 0 |
| S5-final-viral-combined_k141_81138\|\|full | 1622 | No | NA | 2 | 1 | 0 | Low-quality | Genome-fragment | 2.93 | AAI-based (high-confidence) | 0 |
| S5-final-viral-combined_k141_161769\|\|full | 1674 | No | NA | 3 | 0 | 2 | Not-determined | Genome-fragment | NA | NA | 0 |
| S5-final-viral-combined_k141_188639\|\|full | 1049 | No | NA | 2 | 0 | 0 | Not-determined | Genome-fragment | NA | NA | 0 |
| S5-final-viral-combined_k141_90935\|\|full | 10461 | No | NA | 10 | 0 | 1 | Not-determined | Genome-fragment | NA | NA | 0 |
| S5_k141_161663 | 1622 | No | NA | 1 | 0 | 1 | Low-quality | Genome-fragment | 0.19 | AAI-based (medium-confidence) | 0 |
| S5-final-viral-combined_k141_180070\|\|full | 3241 | No | NA | 5 | 0 | 2 | Not-determined | Genome-fragment | NA | NA | 0 |
| S5-final-viral-combined_k141_188661\|\|full | 3203 | No | NA | 6 | 0 | 0 | Not-determined | Genome-fragment | NA | NA | 0 |
| S5-final-viral-combined_k141_171306\|\|full | 2533 | No | NA | 3 | 0 | 1 | Not-determined | Genome-fragment | NA | NA | 0 |
| S5-final-viral-combined_k141_171313\|\|full | 1704 | No | NA | 2 | 0 | 1 | Not-determined | Genome-fragment | NA | NA | 0 |
| S5-final-viral-combined_k141_73026\|\|full | 7265 | No | NA | 12 | 0 | 2 | Not-determined | Genome-fragment | NA | NA | 0 |
| S5-final-viral-combined_k141_90960\|\|full | 2042 | No | NA | 3 | 0 | 3 | Not-determined | Genome-fragment | NA | NA | 0 |
| S7-final-viral-combined_k141_122595\|\|lt2gene | 1622 | No | NA | 1 | 1 | 0 | Low-quality | Genome-fragment | 4.7 | AAI-based (medium-confidence) | 0 |
| S17-k141_264212 | 1622 | No | NA | 1 | 0 | 1 | Low-quality | Genome-fragment | 5.85 | AAI-based (medium-confidence) | 0 |
| S2-final-viral-combined_k141_73739\|\|full | 1622 | No | NA | 2 | 1 | 0 | Low-quality | Genome-fragment | 2.68 | AAI-based (high-confidence) | 0 |
| S5-final-viral-combined_k141_63708\|\|full | 3634 | No | NA | 4 | 0 | 1 | Not-determined | Genome-fragment | NA | NA | 0 |
| S5-final-viral-combined_k141_144463\|\|full | 2444 | No | NA | 3 | 0 | 3 | Not-determined | Genome-fragment | NA | NA | 0 |
| S5-final-viral-combined_k141_180190\|\|full | 1740 | No | NA | 2 | 0 | 2 | Not-determined | Genome-fragment | NA | NA | 0 |
| S5-final-viral-combined_k141_117352\|\|full | 6593 | No | NA | 12 | 1 | 1 | Not-determined | Genome-fragment | NA | NA | 0 |
| S2-final-viral-combined_k141_151227\|\|full | 1622 | No | NA | 2 | 0 | 2 | Low-quality | Genome-fragment | 1.24 | AAI-based (medium-confidence) | 0 |
| S5-final-viral-combined_k141_206321\|\|full | 1924 | No | NA | 4 | 0 | 0 | Not-determined | Genome-fragment | NA | NA | 0 |
| S8_k141_4274 | 1622 | No | NA | 1 | 0 | 0 | Low-quality | Genome-fragment | 3.27 | AAI-based (high-confidence) | 0 |
| S5-final-viral-combined_k141_73106\|\|full | 1918 | No | NA | 3 | 0 | 0 | Not-determined | Genome-fragment | NA | NA | 0 |
| S5-final-viral-combined_k141_144516\|\|full | 3746 | No | NA | 6 | 0 | 4 | Not-determined | Genome-fragment | NA | NA | 0 |
| S9-final-viral-combined_k141_95743\|\|full | 1623 | No | NA | 3 | 0 | 0 | Low-quality | Genome-fragment | 4.55 | AAI-based (medium-confidence) | 0 |
| S5-final-viral-combined_k141_180209\|\|full | 3309 | No | NA | 3 | 0 | 3 | Not-determined | Genome-fragment | NA | NA | 0 |
| S17-final-viral-combined_k141_89000\|\|full | 1623 | No | NA | 3 | 0 | 0 | Low-quality | Genome-fragment | 2.87 | AAI-based (medium-confidence) | 0 |
| S5-final-viral-combined_k141_206341\|\|full | 1693 | No | NA | 3 | 0 | 0 | Not-determined | Genome-fragment | NA | NA | 0 |
| S5-final-viral-combined_k141_206344\|\|full | 1064 | No | NA | 2 | 0 | 1 | Not-determined | Genome-fragment | NA | NA | 0 |
| S8-final-viral-combined_k141_271826\|\|full | 1623 | No | NA | 5 | 0 | 0 | Low-quality | Genome-fragment | 3.79 | AAI-based (high-confidence) | 0 |
| S5-final-viral-combined_k141_99570\|\|full | 1576 | No | NA | 3 | 0 | 0 | Not-determined | Genome-fragment | NA | NA | 0 |
| S11-final-viral-combined_k141_274703\|\|full | 1624 | No | NA | 4 | 0 | 0 | Low-quality | Genome-fragment | 3.39 | AAI-based (high-confidence) | 0 |
| S5-final-viral-combined_k141_73158\|\|full | 1877 | No | NA | 3 | 0 | 0 | Not-determined | Genome-fragment | NA | NA | 0 |
| S1-final-viral-combined_k141_143154\|\|full | 1624 | No | NA | 2 | 0 | 1 | Low-quality | Genome-fragment | 1.53 | AAI-based (high-confidence) | 0 |
| S5-final-viral-combined_k141_144572\|\|full | 2555 | No | NA | 3 | 0 | 2 | Not-determined | Genome-fragment | NA | NA | 0 |
| S5-final-viral-combined_k141_196018\|\|full | 1624 | No | NA | 4 | 1 | 0 | Low-quality | Genome-fragment | 1.4 | HMM-based (lower-bound) | 0 |
| S17-final-viral-combined_k141_80138\|\|full | 1624 | No | NA | 3 | 0 | 0 | Low-quality | Genome-fragment | 2.71 | AAI-based (medium-confidence) | 0 |
| S11-final-viral-combined_k141_194587\|\|full | 1625 | No | NA | 4 | 1 | 0 | Low-quality | Genome-fragment | 3.36 | AAI-based (high-confidence) | 0 |
| S5-final-viral-combined_k141_99623\|\|full | 2429 | No | NA | 6 | 0 | 0 | Not-determined | Genome-fragment | NA | NA | 0 |
| S11-final-viral-combined_k141_140943\|\|lt2gene | 1625 | No | NA | 2 | 2 | 0 | Low-quality | Genome-fragment | 3.34 | AAI-based (high-confidence) | 0 |
| S7-final-viral-combined_k141_121856\|\|full | 1625 | No | NA | 3 | 0 | 0 | Low-quality | Genome-fragment | 1.46 | AAI-based (medium-confidence) | 0 |
| S7-final-viral-combined_k141_29808\|\|full | 1625 | No | NA | 3 | 0 | 0 | Low-quality | Genome-fragment | 2.54 | AAI-based (high-confidence) | 0 |
| S5-final-viral-combined_k141_19295\|\|full | 3475 | No | NA | 5 | 0 | 2 | Not-determined | Genome-fragment | NA | NA | 0 |
| S7_k141_233315 | 1625 | No | NA | 2 | 0 | 0 | Low-quality | Genome-fragment | 2.55 | AAI-based (medium-confidence) | 0 |
| S17-final-viral-combined_k141_59972\|\|full | 1625 | No | NA | 2 | 1 | 0 | Low-quality | Genome-fragment | 4.25 | AAI-based (medium-confidence) | 0 |
| S12-final-viral-combined_k141_99695\|\|lt2gene | 1625 | No | NA | 1 | 1 | 0 | Low-quality | Genome-fragment | 0.63 | HMM-based (lower-bound) | 0 |
| S5-final-viral-combined_k141_197729\|\|full | 1390 | No | NA | 3 | 0 | 1 | Not-determined | Genome-fragment | NA | NA | 0 |
| S5-final-viral-combined_k141_55274\|\|full | 1639 | No | NA | 3 | 0 | 0 | Not-determined | Genome-fragment | NA | NA | 0 |
| S5-final-viral-combined_k141_171536\|\|full | 1589 | No | NA | 3 | 0 | 2 | Not-determined | Genome-fragment | NA | NA | 0 |
| S5-final-viral-combined_k141_82013\|\|full | 2175 | No | NA | 3 | 0 | 2 | Not-determined | Genome-fragment | NA | NA | 0 |
| S5-final-viral-combined_k141_144628\|\|full | 2597 | No | NA | 3 | 0 | 0 | Not-determined | Genome-fragment | NA | NA | 0 |
| S2-final-viral-combined_k141_207526\|\|full | 1625 | No | NA | 2 | 0 | 1 | Low-quality | Genome-fragment | 1.86 | AAI-based (high-confidence) | 0 |
| S4_k141_5855 | 1625 | No | NA | 1 | 0 | 0 | Low-quality | Genome-fragment | 1.65 | AAI-based (medium-confidence) | 0 |
| S8-final-viral-combined_k141_101260\|\|full | 1625 | No | NA | 4 | 2 | 0 | Low-quality | Genome-fragment | 2.36 | HMM-based (lower-bound) | 0 |
| S11-final-viral-combined_k141_254677\|\|full | 1626 | No | NA | 5 | 1 | 0 | Low-quality | Genome-fragment | 3.09 | AAI-based (high-confidence) | 0 |
| S5-final-viral-combined_k141_82052\|\|full | 1174 | No | NA | 3 | 0 | 3 | Not-determined | Genome-fragment | NA | NA | 0 |
| S5-final-viral-combined_k141_188884\|\|full | 4012 | No | NA | 3 | 0 | 0 | Not-determined | Genome-fragment | NA | NA | 0 |
| S11-final-viral-combined_k141_76743\|\|full | 1626 | No | NA | 3 | 1 | 0 | Low-quality | Genome-fragment | 0.98 | HMM-based (lower-bound) | 0 |
| S5-final-viral-combined_k141_117510\|\|full | 3209 | No | NA | 8 | 0 | 0 | Not-determined | Genome-fragment | NA | NA | 0 |
| S1-final-viral-combined_k141_279822\|\|full | 1626 | No | NA | 3 | 0 | 0 | Low-quality | Genome-fragment | 2.77 | AAI-based (high-confidence) | 0 |
| S12-final-viral-combined_k141_34919\|\|full | 1626 | No | NA | 4 | 0 | 1 | Low-quality | Genome-fragment | 1.59 | AAI-based (medium-confidence) | 0 |
| S5-final-viral-combined_k141_73301\|\|full | 20824 | No | NA | 25 | 11 | 0 | Medium-quality | Genome-fragment | 55.48 | AAI-based (high-confidence) | 0 |
| S5-final-viral-combined_k141_171637\|\|full | 1168 | No | NA | 3 | 0 | 1 | Not-determined | Genome-fragment | NA | NA | 0 |
| S5-final-viral-combined_k141_28323\|\|full | 3195 | No | NA | 6 | 0 | 4 | Not-determined | Genome-fragment | NA | NA | 0 |
| S5-final-viral-combined_k141_28331\|\|full | 1346 | No | NA | 1 | 0 | 1 | Not-determined | Genome-fragment | NA | NA | 0 |
| S5-final-viral-combined_k141_63921\|\|full | 4082 | No | NA | 8 | 0 | 0 | Not-determined | Genome-fragment | NA | NA | 0 |
| S12-final-viral-combined_k141_2315\|\|full | 1626 | No | NA | 6 | 1 | 0 | Low-quality | Genome-fragment | 2.89 | AAI-based (high-confidence) | 0 |
| S5-final-viral-combined_k141_2663\|\|full | 1203 | No | NA | 2 | 0 | 1 | Not-determined | Genome-fragment | NA | NA | 0 |
| S2-final-viral-combined_k141_179404\|\|full | 1626 | No | NA | 3 | 0 | 0 | Low-quality | Genome-fragment | 3.71 | AAI-based (medium-confidence) | 0 |
| S6-final-viral-combined_k141_60287\|\|full | 1626 | No | NA | 3 | 0 | 0 | Low-quality | Genome-fragment | 2.29 | AAI-based (medium-confidence) | 0 |
| S6_k141_39834 | 1626 | No | NA | 4 | 0 | 0 | Low-quality | Genome-fragment | 3.02 | AAI-based (high-confidence) | 0 |
| S8-final-viral-combined_k141_72285\|\|full | 1626 | No | NA | 1 | 0 | 0 | Low-quality | Genome-fragment | 2.29 | AAI-based (medium-confidence) | 0 |
| S11-final-viral-combined_k141_58015\|\|lt2gene | 1627 | No | NA | 1 | 1 | 0 | Low-quality | Genome-fragment | 1.06 | AAI-based (high-confidence) | 0 |
| S1-final-viral-combined_k141_186661\|\|full | 1627 | No | NA | 3 | 0 | 0 | Low-quality | Genome-fragment | 4.39 | AAI-based (medium-confidence) | 0 |
| S5-final-viral-combined_k141_114737\|\|full | 1627 | No | NA | 2 | 1 | 0 | Low-quality | Genome-fragment | 3.67 | AAI-based (high-confidence) | 0 |
| S5-final-viral-combined_k141_135373\|\|full | 1869 | No | NA | 2 | 0 | 0 | Not-determined | Genome-fragment | NA | NA | 0 |
| S5-final-viral-combined_k141_114393\|\|lt2gene | 1627 | No | NA | 2 | 2 | 0 | Low-quality | Genome-fragment | 3.56 | AAI-based (high-confidence) | 0 |
| S5-final-viral-combined_k141_197876\|\|full | 1936 | No | NA | 2 | 0 | 1 | Not-determined | Genome-fragment | NA | NA | 0 |
| S5-final-viral-combined_k141_46470\|\|full | 3922 | No | NA | 2 | 0 | 1 | Not-determined | Genome-fragment | NA | NA | 0 |
| S7-final-viral-combined_k141_236380\|\|full | 1627 | No | NA | 2 | 2 | 0 | Low-quality | Genome-fragment | 3.32 | AAI-based (high-confidence) | 0 |
| S5-final-viral-combined_k141_91194\|\|full | 1537 | No | NA | 1 | 0 | 0 | Not-determined | Genome-fragment | NA | NA | 0 |
| S12-final-viral-combined_k141_325312\|\|full | 1627 | No | NA | 2 | 0 | 1 | Low-quality | Genome-fragment | 1.43 | AAI-based (high-confidence) | 0 |
| S2-final-viral-combined_k141_14783\|\|full | 1627 | No | NA | 3 | 1 | 0 | Low-quality | Genome-fragment | 1.87 | HMM-based (lower-bound) | 0 |
| S4_k141_144569 | 1627 | No | NA | 3 | 0 | 1 | Low-quality | Genome-fragment | 0.32 | AAI-based (medium-confidence) | 0 |
| S9-final-viral-combined_k141_84321\|\|full | 1628 | No | NA | 2 | 0 | 1 | Low-quality | Genome-fragment | 0.78 | AAI-based (high-confidence) | 0 |
| S11-final-viral-combined_k141_196688\|\|full | 1628 | No | NA | 3 | 0 | 1 | Low-quality | Genome-fragment | 2.42 | AAI-based (high-confidence) | 0 |
| S5-final-viral-combined_k141_135439\|\|full | 1598 | No | NA | 3 | 0 | 0 | Not-determined | Genome-fragment | NA | NA | 0 |
| S5-final-viral-combined_k141_28378\|\|full | 3246 | No | NA | 5 | 0 | 2 | Not-determined | Genome-fragment | NA | NA | 0 |
| S5-final-viral-combined_k141_55357\|\|full | 3513 | No | NA | 3 | 0 | 3 | Not-determined | Genome-fragment | NA | NA | 0 |
| S5-final-viral-combined_k141_99803\|\|full | 4702 | No | NA | 5 | 0 | 2 | Not-determined | Genome-fragment | NA | NA | 0 |
| S1-final-viral-combined_k141_286412\|\|full | 1628 | No | NA | 2 | 0 | 0 | Low-quality | Genome-fragment | 3.81 | AAI-based (medium-confidence) | 0 |
| S5-final-viral-combined_k141_99804\|\|full | 2390 | No | NA | 2 | 0 | 2 | Not-determined | Genome-fragment | NA | NA | 0 |
| S1_k141_165897 | 1628 | No | NA | 3 | 1 | 0 | Low-quality | Genome-fragment | 2.63 | AAI-based (high-confidence) | 0 |
| S5-final-viral-combined_k141_153406\|\|full | 6829 | No | NA | 6 | 0 | 1 | Not-determined | Genome-fragment | NA | NA | 0 |
| S5-final-viral-combined_k141_144735\|\|full | 1404 | No | NA | 3 | 0 | 0 | Not-determined | Genome-fragment | NA | NA | 0 |
| S5-final-viral-combined_k141_127882\|\|full | 1628 | No | NA | 2 | 0 | 0 | Low-quality | Genome-fragment | 3.78 | AAI-based (high-confidence) | 0 |
| S5-final-viral-combined_k141_2761\|\|full | 6397 | No | NA | 6 | 0 | 0 | Not-determined | Genome-fragment | NA | NA | 0 |
| S5-final-viral-combined_k141_19537\|\|full | 3094 | No | NA | 4 | 0 | 1 | Not-determined | Genome-fragment | NA | NA | 0 |
| S5-final-viral-combined_k141_144773\|\|full | 1103 | No | NA | 3 | 0 | 1 | Not-determined | Genome-fragment | NA | NA | 0 |
| S5-final-viral-combined_k141_64049\|\|full | 1835 | No | NA | 2 | 0 | 1 | Not-determined | Genome-fragment | NA | NA | 0 |
| S17-final-viral-combined_k141_7167\|\|full | 1628 | No | NA | 3 | 0 | 0 | Low-quality | Genome-fragment | 3.4 | AAI-based (medium-confidence) | 0 |
| S5-final-viral-combined_k141_135537\|\|full | 1668 | No | NA | 3 | 0 | 2 | Not-determined | Genome-fragment | NA | NA | 0 |
| S17-final-viral-combined_k141_30720\|\|full | 1628 | No | NA | 3 | 0 | 0 | Low-quality | Genome-fragment | 4.06 | AAI-based (medium-confidence) | 0 |
| S17-final-viral-combined_k141_96378\|\|full | 1628 | No | NA | 3 | 0 | 0 | Low-quality | Genome-fragment | 3.83 | AAI-based (high-confidence) | 0 |
| S5-final-viral-combined_k141_37516\|\|full | 4683 | No | NA | 4 | 0 | 4 | Not-determined | Genome-fragment | NA | NA | 0 |
| S5-final-viral-combined_k141_126710\|\|full | 1266 | No | NA | 3 | 0 | 1 | Not-determined | Genome-fragment | NA | NA | 0 |
| S2_k141_71910 | 1628 | No | NA | 1 | 0 | 1 | Low-quality | Genome-fragment | 0.51 | AAI-based (medium-confidence) | 0 |
| S5-final-viral-combined_k141_126721\|\|full | 4100 | No | NA | 4 | 0 | 0 | Not-determined | Genome-fragment | NA | NA | 0 |
| S8-final-viral-combined_k141_98500\|\|full | 1628 | No | NA | 4 | 1 | 0 | Low-quality | Genome-fragment | 3.2 | AAI-based (medium-confidence) | 0 |
| S9-final-viral-combined_k141_67934\|\|full | 1629 | No | NA | 5 | 3 | 0 | Low-quality | Genome-fragment | 3.7 | AAI-based (high-confidence) | 0 |
| S11-final-viral-combined_k141_279682\|\|full | 1629 | No | NA | 3 | 1 | 0 | Low-quality | Genome-fragment | 4.48 | HMM-based (lower-bound) | 0 |
| S5-final-viral-combined_k141_189106\|\|full | 6710 | No | NA | 6 | 0 | 4 | Not-determined | Genome-fragment | NA | NA | 0 |
| S5-final-viral-combined_k141_206546\|\|full | 1983 | No | NA | 2 | 0 | 0 | Not-determined | Genome-fragment | NA | NA | 0 |
| S11_k141_189941 | 1629 | No | NA | 2 | 0 | 2 | Low-quality | Genome-fragment | 5.43 | AAI-based (high-confidence) | 0 |
| S5-final-viral-combined_k141_46687\|\|full | 1314 | No | NA | 3 | 0 | 0 | Not-determined | Genome-fragment | NA | NA | 0 |
| S1-final-viral-combined_k141_238876\|\|full | 1629 | No | NA | 2 | 0 | 2 | Low-quality | Genome-fragment | 1.87 | AAI-based (medium-confidence) | 0 |
| S5-final-viral-combined_k141_206564\|\|full | 5393 | No | NA | 5 | 0 | 1 | Not-determined | Genome-fragment | NA | NA | 0 |
| S3_k141_78502 | 1629 | No | NA | 3 | 1 | 0 | Low-quality | Genome-fragment | 0.75 | AAI-based (high-confidence) | 0 |
| S5_k141_165622 | 1629 | No | NA | 1 | 0 | 1 | Low-quality | Genome-fragment | 0.4 | AAI-based (high-confidence) | 0 |
| S5-final-viral-combined_k141_2872\|\|full | 2039 | No | NA | 3 | 0 | 1 | Not-determined | Genome-fragment | NA | NA | 0 |
| S5-final-viral-combined_k141_37553\|\|full | 1700 | No | NA | 3 | 0 | 1 | Not-determined | Genome-fragment | NA | NA | 0 |
| S5-final-viral-combined_k141_189164\|\|full | 1095 | No | NA | 1 | 0 | 0 | Not-determined | Genome-fragment | NA | NA | 0 |
| S5-final-viral-combined_k141_126779\|\|full | 1470 | No | NA | 2 | 0 | 0 | Not-determined | Genome-fragment | NA | NA | 0 |
| S7-final-viral-combined_k141_44231\|\|full | 1629 | No | NA | 3 | 0 | 1 | Low-quality | Genome-fragment | 0.68 | AAI-based (medium-confidence) | 0 |
| S7-final-viral-combined_k141_37697\|\|full | 1629 | No | NA | 4 | 2 | 0 | Low-quality | Genome-fragment | 3.81 | AAI-based (medium-confidence) | 0 |
| S5-final-viral-combined_k141_189189\|\|full | 2675 | No | NA | 3 | 0 | 3 | Not-determined | Genome-fragment | NA | NA | 0 |
| S7-final-viral-combined_k141_191360\|\|full | 1629 | No | NA | 5 | 0 | 0 | Low-quality | Genome-fragment | 3.68 | AAI-based (high-confidence) | 0 |
| S5-final-viral-combined_k141_28592\|\|full | 5733 | No | NA | 10 | 0 | 0 | Not-determined | Genome-fragment | NA | NA | 0 |
| S17-final-viral-combined_k141_63806\|\|lt2gene | 1629 | No | NA | 1 | 1 | 0 | Low-quality | Genome-fragment | 3.85 | AAI-based (medium-confidence) | 0 |
| S12-final-viral-combined_k141_284968\|\|full | 1629 | No | NA | 3 | 0 | 0 | Low-quality | Genome-fragment | 3.79 | AAI-based (medium-confidence) | 0 |
| S5-final-viral-combined_k141_37647\|\|full | 1603 | No | NA | 3 | 0 | 1 | Not-determined | Genome-fragment | NA | NA | 0 |
| S5-final-viral-combined_k141_206636\|\|full | 45168 | No | NA | 40 | 0 | 12 | Not-determined | Genome-fragment | NA | NA | 0 |
| S5-final-viral-combined_k141_206642\|\|full | 2283 | No | NA | 5 | 0 | 0 | Not-determined | Genome-fragment | NA | NA | 0 |
| S5-final-viral-combined_k141_108794\|\|full | 177204 | Yes | 49171 | 184 | 24 | 82 | Complete | High-quality | 100 | Provirus (medium-confidence) | 72.25 |
| S5-final-viral-combined_k141_100031\|\|full | 1082 | No | NA | 2 | 0 | 2 | Not-determined | Genome-fragment | NA | NA | 0 |
| S5-final-viral-combined_k141_28626\|\|full | 2003 | No | NA | 4 | 0 | 0 | Not-determined | Genome-fragment | NA | NA | 0 |
| S5-final-viral-combined_k141_162145\|\|full | 7045 | No | NA | 8 | 0 | 4 | Not-determined | Genome-fragment | NA | NA | 0 |
| S5-final-viral-combined_k141_171891\|\|full | 31003 | No | NA | 25 | 0 | 20 | Not-determined | Genome-fragment | NA | NA | 0 |
| S12-final-viral-combined_k141_13758\|\|full | 1629 | No | NA | 2 | 0 | 0 | Low-quality | Genome-fragment | 4.07 | AAI-based (high-confidence) | 0 |
| S11-final-viral-combined_k141_116621\|\|full | 1630 | No | NA | 3 | 0 | 0 | Low-quality | Genome-fragment | 3.16 | AAI-based (high-confidence) | 0 |
| S11_k141_59978 | 1630 | No | NA | 2 | 0 | 2 | Low-quality | Genome-fragment | 0.74 | AAI-based (medium-confidence) | 0 |
| S5-final-viral-combined_k141_100062\|\|full | 2172 | No | NA | 2 | 0 | 2 | Not-determined | Genome-fragment | NA | NA | 0 |
| S5-final-viral-combined_k141_73504\|\|full | 1233 | No | NA | 2 | 0 | 1 | Not-determined | Genome-fragment | NA | NA | 0 |
| S5-final-viral-combined_k141_73513\|\|full | 1070 | No | NA | 3 | 0 | 0 | Not-determined | Genome-fragment | NA | NA | 0 |
| S5-final-viral-combined_k141_100091\|\|full | 2839 | No | NA | 3 | 0 | 0 | Not-determined | Genome-fragment | NA | NA | 0 |
| S5-final-viral-combined_k141_46822\|\|full | 2786 | No | NA | 4 | 0 | 3 | Not-determined | Genome-fragment | NA | NA | 0 |
| S5-final-viral-combined_k141_135569\|\|full | 1691 | No | NA | 2 | 0 | 1 | Not-determined | Genome-fragment | NA | NA | 0 |
| S5-final-viral-combined_k141_198064\|\|full | 2733 | No | NA | 5 | 0 | 1 | Not-determined | Genome-fragment | NA | NA | 0 |
| S5-final-viral-combined_k141_46826\|\|full | 3704 | No | NA | 5 | 0 | 3 | Not-determined | Genome-fragment | NA | NA | 0 |
| S5-final-viral-combined_k141_64147\|\|full | 1159 | No | NA | 2 | 0 | 0 | Not-determined | Genome-fragment | NA | NA | 0 |
| S5-final-viral-combined_k141_47653\|\|full | 1630 | No | NA | 4 | 0 | 0 | Low-quality | Genome-fragment | 3.59 | AAI-based (high-confidence) | 0 |
| S5-final-viral-combined_k141_206740\|\|full | 1333 | No | NA | 2 | 0 | 1 | Not-determined | Genome-fragment | NA | NA | 0 |
| S5-final-viral-combined_k141_46832\|\|full | 2155 | No | NA | 3 | 0 | 2 | Not-determined | Genome-fragment | NA | NA | 0 |
| S17-final-viral-combined_k141_157033\|\|full | 1630 | No | NA | 2 | 0 | 1 | Low-quality | Genome-fragment | 1.11 | AAI-based (high-confidence) | 0 |
| S12-final-viral-combined_k141_253824\|\|full | 1630 | No | NA | 2 | 0 | 2 | Low-quality | Genome-fragment | 1.45 | AAI-based (medium-confidence) | 0 |
| S4-final-viral-combined_k141_79197\|\|full | 1630 | No | NA | 2 | 0 | 0 | Low-quality | Genome-fragment | 1.99 | AAI-based (high-confidence) | 0 |
| S5-final-viral-combined_k141_198100\|\|full | 8247 | No | NA | 8 | 0 | 4 | Not-determined | Genome-fragment | NA | NA | 0 |
| S6_k141_110854 | 1630 | No | NA | 6 | 0 | 0 | Low-quality | Genome-fragment | 5.07 | AAI-based (high-confidence) | 0 |
| S5-final-viral-combined_k141_171987\|\|full | 1143 | No | NA | 4 | 0 | 2 | Not-determined | Genome-fragment | NA | NA | 0 |
| S8-final-viral-combined_k141_3045\|\|full | 1630 | No | NA | 2 | 0 | 1 | Low-quality | Genome-fragment | 3.46 | AAI-based (medium-confidence) | 0 |
| S5-final-viral-combined_k141_55576\|\|full | 3137 | No | NA | 5 | 0 | 0 | Not-determined | Genome-fragment | NA | NA | 0 |
| S8-final-viral-combined_k141_76285\|\|full | 1630 | No | NA | 3 | 1 | 0 | Low-quality | Genome-fragment | 0.89 | HMM-based (lower-bound) | 0 |
| S5-final-viral-combined_k141_37697\|\|full | 2021 | No | NA | 3 | 0 | 1 | Not-determined | Genome-fragment | NA | NA | 0 |
| S8_k141_225811 | 1630 | No | NA | 4 | 3 | 0 | Low-quality | Genome-fragment | 2.69 | AAI-based (high-confidence) | 0 |
| S9-final-viral-combined_k141_104266\|\|full | 1631 | No | NA | 4 | 0 | 2 | Low-quality | Genome-fragment | 1.28 | AAI-based (medium-confidence) | 0 |
| S11-final-viral-combined_k141_202057\|\|full | 1631 | No | NA | 4 | 0 | 0 | Low-quality | Genome-fragment | 1.04 | AAI-based (medium-confidence) | 0 |
| S1-final-viral-combined_k141_87102\|\|full | 1631 | No | NA | 3 | 0 | 1 | Low-quality | Genome-fragment | 1.57 | AAI-based (high-confidence) | 0 |
| S5-final-viral-combined_k141_52291\|\|full | 1631 | No | NA | 2 | 0 | 2 | Low-quality | Genome-fragment | 3.49 | AAI-based (high-confidence) | 0 |
| S5-final-viral-combined_k141_17641\|\|full | 1631 | No | NA | 3 | 1 | 0 | Low-quality | Genome-fragment | 2.47 | HMM-based (lower-bound) | 0 |
| S7-final-viral-combined_k141_86557\|\|full | 1631 | No | NA | 3 | 0 | 0 | Low-quality | Genome-fragment | 3.1 | AAI-based (high-confidence) | 0 |
| S17-k141_106562 | 1631 | No | NA | 3 | 0 | 1 | Low-quality | Genome-fragment | 2.28 | AAI-based (medium-confidence) | 0 |
| S5-final-viral-combined_k141_100184\|\|full | 2214 | No | NA | 2 | 0 | 2 | Not-determined | Genome-fragment | NA | NA | 0 |
| S12_k141_191167 | 1631 | No | NA | 4 | 0 | 0 | Low-quality | Genome-fragment | 2.72 | AAI-based (medium-confidence) | 0 |
| S5-final-viral-combined_k141_28721\|\|full | 1684 | No | NA | 3 | 0 | 2 | Not-determined | Genome-fragment | NA | NA | 0 |
| S5-final-viral-combined_k141_189348\|\|full | 1490 | No | NA | 3 | 0 | 2 | Not-determined | Genome-fragment | NA | NA | 0 |
| S4-final-viral-combined_k141_117655\|\|full | 1631 | No | NA | 2 | 0 | 1 | Low-quality | Genome-fragment | 3.43 | AAI-based (high-confidence) | 0 |
| S5-final-viral-combined_k141_189375\|\|full | 2546 | No | NA | 2 | 0 | 1 | Not-determined | Genome-fragment | NA | NA | 0 |
| S8-final-viral-combined_k141_30067\|\|full | 1631 | No | NA | 3 | 0 | 0 | Low-quality | Genome-fragment | 1.3 | AAI-based (high-confidence) | 0 |
| S9-final-viral-combined_k141_72026\|\|full | 1632 | No | NA | 2 | 0 | 0 | Low-quality | Genome-fragment | 3.23 | AAI-based (high-confidence) | 0 |
| S5-final-viral-combined_k141_100267\|\|full | 1859 | No | NA | 2 | 0 | 0 | Not-determined | Genome-fragment | NA | NA | 0 |
| S3-final-viral-combined_k141_31489\|\|full | 1632 | No | NA | 4 | 0 | 1 | Low-quality | Genome-fragment | 1.29 | AAI-based (medium-confidence) | 0 |
| S5-final-viral-combined_k141_91527\|\|full | 1679 | No | NA | 3 | 0 | 0 | Not-determined | Genome-fragment | NA | NA | 0 |
| S5-final-viral-combined_k141_3003\|\|full | 3578 | No | NA | 4 | 0 | 0 | Not-determined | Genome-fragment | NA | NA | 0 |
| S5-final-viral-combined_k141_144947\|\|full | 1632 | No | NA | 3 | 0 | 1 | Low-quality | Genome-fragment | 1.46 | AAI-based (medium-confidence) | 0 |
| S12_k141_209374 | 1632 | No | NA | 3 | 0 | 0 | Low-quality | Genome-fragment | 1.93 | AAI-based (medium-confidence) | 0 |
| S2-final-viral-combined_k141_68836\|\|full | 1632 | No | NA | 3 | 0 | 1 | Low-quality | Genome-fragment | 0.72 | AAI-based (medium-confidence) | 0 |
| S5-final-viral-combined_k141_73603\|\|full | 2492 | No | NA | 3 | 0 | 3 | Not-determined | Genome-fragment | NA | NA | 0 |
| S4-final-viral-combined_k141_113778\|\|full | 1632 | No | NA | 4 | 2 | 0 | Low-quality | Genome-fragment | 1.96 | AAI-based (high-confidence) | 0 |
| S8-final-viral-combined_k141_27890\|\|full | 1632 | No | NA | 2 | 0 | 0 | Low-quality | Genome-fragment | 2.84 | AAI-based (medium-confidence) | 0 |
| S8-final-viral-combined_k141_248616\|\|full | 1632 | No | NA | 2 | 0 | 0 | Low-quality | Genome-fragment | 1.07 | AAI-based (medium-confidence) | 0 |
| S5-final-viral-combined_k141_3074\|\|full | 2986 | No | NA | 6 | 0 | 5 | Not-determined | Genome-fragment | NA | NA | 0 |
| S5-final-viral-combined_k141_135705\|\|full | 1035 | No | NA | 3 | 0 | 0 | Not-determined | Genome-fragment | NA | NA | 0 |
| S5-final-viral-combined_k141_172035\|\|full | 1773 | No | NA | 2 | 0 | 1 | Not-determined | Genome-fragment | NA | NA | 0 |
| S9-final-viral-combined_k141_121585\|\|full | 1633 | No | NA | 3 | 0 | 0 | Low-quality | Genome-fragment | 0.6 | AAI-based (medium-confidence) | 0 |
| S9-final-viral-combined_k141_90790\|\|full | 1633 | No | NA | 3 | 0 | 0 | Low-quality | Genome-fragment | 2.94 | AAI-based (medium-confidence) | 0 |
| S5-final-viral-combined_k141_28795\|\|full | 1013 | No | NA | 3 | 0 | 0 | Not-determined | Genome-fragment | NA | NA | 0 |
| S11-final-viral-combined_k141_199450\|\|full | 1633 | No | NA | 3 | 0 | 0 | Low-quality | Genome-fragment | 1.23 | AAI-based (high-confidence) | 0 |
| S1-final-viral-combined_k141_292442\|\|full | 1633 | No | NA | 3 | 1 | 0 | Low-quality | Genome-fragment | 3.09 | HMM-based (lower-bound) | 0 |
| S5-final-viral-combined_k141_100415\|\|full | 1264 | No | NA | 4 | 0 | 0 | Not-determined | Genome-fragment | NA | NA | 0 |
| S1-final-viral-combined_k141_131267\|\|full | 1633 | No | NA | 3 | 0 | 0 | Low-quality | Genome-fragment | 3.31 | AAI-based (high-confidence) | 0 |
| S5-final-viral-combined_k141_100436\|\|full | 1678 | No | NA | 4 | 0 | 0 | Not-determined | Genome-fragment | NA | NA | 0 |
| S1_k141_39107 | 1633 | No | NA | 3 | 1 | 0 | Low-quality | Genome-fragment | 4.97 | AAI-based (high-confidence) | 0 |
| S7-final-viral-combined_k141_125230\|\|full | 1633 | No | NA | 3 | 2 | 0 | Low-quality | Genome-fragment | 2.32 | HMM-based (lower-bound) | 0 |
| S5-final-viral-combined_k141_145067\|\|full | 31451 | No | NA | 46 | 13 | 1 | Medium-quality | Genome-fragment | 79.99 | AAI-based (high-confidence) | 0 |
| S5-final-viral-combined_k141_198248\|\|full | 3572 | No | NA | 4 | 0 | 2 | Not-determined | Genome-fragment | NA | NA | 0 |
| S17-k141_225133 | 1633 | No | NA | 1 | 0 | 1 | Low-quality | Genome-fragment | 2.83 | AAI-based (high-confidence) | 0 |
| S5-final-viral-combined_k141_73701\|\|full | 1822 | No | NA | 3 | 0 | 0 | Not-determined | Genome-fragment | NA | NA | 0 |
| S5-final-viral-combined_k141_198278\|\|full | 4397 | No | NA | 3 | 0 | 2 | Not-determined | Genome-fragment | NA | NA | 0 |
| S5-final-viral-combined_k141_127174\|\|full | 4640 | No | NA | 6 | 0 | 4 | Not-determined | Genome-fragment | NA | NA | 0 |
| S5-final-viral-combined_k141_91581\|\|full | 1521 | No | NA | 2 | 0 | 0 | Not-determined | Genome-fragment | NA | NA | 0 |
| S5-final-viral-combined_k141_64496\|\|full | 3162 | No | NA | 2 | 0 | 0 | Not-determined | Genome-fragment | NA | NA | 0 |
| S5-final-viral-combined_k141_135776\|\|full | 1604 | No | NA | 3 | 0 | 1 | Not-determined | Genome-fragment | NA | NA | 0 |
| S2-final-viral-combined_k141_48305\|\|full | 1633 | No | NA | 3 | 0 | 0 | Low-quality | Genome-fragment | 2.13 | AAI-based (medium-confidence) | 0 |
| S8_k141_136281 | 1633 | No | NA | 2 | 0 | 0 | Low-quality | Genome-fragment | 3.58 | AAI-based (high-confidence) | 0 |
| S9_k141_99475 | 1634 | No | NA | 1 | 0 | 1 | Low-quality | Genome-fragment | 1.22 | AAI-based (medium-confidence) | 0 |
| S5-final-viral-combined_k141_198339\|\|full | 3127 | No | NA | 3 | 0 | 2 | Not-determined | Genome-fragment | NA | NA | 0 |
| S5-final-viral-combined_k141_109757\|\|lt2gene | 1634 | No | NA | 2 | 2 | 0 | Low-quality | Genome-fragment | 3.65 | AAI-based (high-confidence) | 0 |
| S5-final-viral-combined_k141_91609\|\|full | 1219 | No | NA | 2 | 0 | 1 | Not-determined | Genome-fragment | NA | NA | 0 |
| S17-final-viral-combined_k141_158877\|\|full | 1634 | No | NA | 5 | 0 | 0 | Low-quality | Genome-fragment | 3.52 | AAI-based (high-confidence) | 0 |
| S2-final-viral-combined_k141_38612\|\|full | 1634 | No | NA | 3 | 2 | 0 | Low-quality | Genome-fragment | 3.88 | AAI-based (high-confidence) | 0 |
| S8-final-viral-combined_k141_14364\|\|full | 1634 | No | NA | 3 | 0 | 1 | Low-quality | Genome-fragment | 2.91 | AAI-based (medium-confidence) | 0 |
| S5-final-viral-combined_k141_117921\|\|full | 2244 | No | NA | 3 | 0 | 2 | Not-determined | Genome-fragment | NA | NA | 0 |
| S5-final-viral-combined_k141_117945\|\|full | 1889 | No | NA | 2 | 0 | 1 | Not-determined | Genome-fragment | NA | NA | 0 |
| S5-final-viral-combined_k141_108949\|\|full | 2820 | No | NA | 3 | 0 | 2 | Not-determined | Genome-fragment | NA | NA | 0 |
| S5-final-viral-combined_k141_145114\|\|full | 4397 | No | NA | 4 | 0 | 1 | Not-determined | Genome-fragment | NA | NA | 0 |
| S11-final-viral-combined_k141_312060\|\|lt2gene | 1635 | No | NA | 1 | 1 | 0 | Low-quality | Genome-fragment | 4.07 | AAI-based (high-confidence) | 0 |
| S5_k141_2350 | 1635 | No | NA | 2 | 0 | 0 | Low-quality | Genome-fragment | 2.94 | AAI-based (medium-confidence) | 0 |
| S5-final-viral-combined_k141_162499\|\|full | 1218 | No | NA | 3 | 0 | 1 | Not-determined | Genome-fragment | NA | NA | 0 |
| S7-final-viral-combined_k141_187427\|\|lt2gene | 1635 | No | NA | 2 | 1 | 0 | Low-quality | Genome-fragment | 4.05 | AAI-based (high-confidence) | 0 |
| S5-final-viral-combined_k141_117991\|\|full | 4854 | No | NA | 6 | 0 | 2 | Not-determined | Genome-fragment | NA | NA | 0 |
| S7-final-viral-combined_k141_88751\|\|lt2gene | 1635 | No | NA | 2 | 2 | 0 | Low-quality | Genome-fragment | 3.66 | AAI-based (high-confidence) | 0 |
| S5-final-viral-combined_k141_118057\|\|full | 3080 | No | NA | 3 | 0 | 2 | Not-determined | Genome-fragment | NA | NA | 0 |
| S17-final-viral-combined_k141_13312\|\|full | 1635 | No | NA | 3 | 0 | 0 | Low-quality | Genome-fragment | 3.89 | AAI-based (medium-confidence) | 0 |
| S5-final-viral-combined_k141_162580\|\|full | 16331 | No | NA | 14 | 0 | 11 | Not-determined | Genome-fragment | NA | NA | 0 |
| S5-final-viral-combined_k141_19708\|\|full | 2792 | No | NA | 4 | 0 | 1 | Not-determined | Genome-fragment | NA | NA | 0 |
| S5-final-viral-combined_k141_64625\|\|full | 15818 | No | NA | 20 | 0 | 14 | Not-determined | Genome-fragment | NA | NA | 0 |
| S12-final-viral-combined_k141_228564\|\|lt2gene | 1635 | No | NA | 2 | 2 | 0 | Low-quality | Genome-fragment | 5.21 | AAI-based (high-confidence) | 0 |
| S8_k141_177947 | 1635 | No | NA | 2 | 0 | 1 | Low-quality | Genome-fragment | 1.9 | AAI-based (medium-confidence) | 0 |
| S9-final-viral-combined_k141_25760\|\|full | 1636 | No | NA | 3 | 2 | 0 | Low-quality | Genome-fragment | 4.14 | AAI-based (medium-confidence) | 0 |
| S5-final-viral-combined_k141_28977\|\|full | 5040 | No | NA | 7 | 0 | 4 | Not-determined | Genome-fragment | NA | NA | 0 |
| S11-final-viral-combined_k141_253857\|\|full | 1636 | No | NA | 3 | 0 | 0 | Low-quality | Genome-fragment | 3.58 | AAI-based (medium-confidence) | 0 |
| S5-final-viral-combined_k141_47147\|\|full | 3184 | No | NA | 4 | 0 | 1 | Not-determined | Genome-fragment | NA | NA | 0 |
| S5-final-viral-combined_k141_145160\|\|full | 5054 | No | NA | 4 | 0 | 0 | Not-determined | Genome-fragment | NA | NA | 0 |
| S5-final-viral-combined_k141_189742\|\|full | 1685 | No | NA | 3 | 0 | 2 | Not-determined | Genome-fragment | NA | NA | 0 |
| S5-final-viral-combined_k141_162620\|\|full | 4130 | No | NA | 4 | 0 | 0 | Not-determined | Genome-fragment | NA | NA | 0 |
| S5-final-viral-combined_k141_135895\|\|full | 1048 | No | NA | 3 | 0 | 0 | Not-determined | Genome-fragment | NA | NA | 0 |
| S5-final-viral-combined_k141_135897\|\|full | 1839 | No | NA | 4 | 0 | 2 | Not-determined | Genome-fragment | NA | NA | 0 |
| S1-final-viral-combined_k141_27533\|\|lt2gene | 1636 | No | NA | 2 | 1 | 0 | Low-quality | Genome-fragment | 2.6 | AAI-based (high-confidence) | 0 |
| S5-final-viral-combined_k141_19732\|\|full | 10738 | No | NA | 3 | 0 | 3 | Not-determined | Genome-fragment | NA | NA | 0 |
| S5-final-viral-combined_k141_180976\|\|full | 1734 | No | NA | 3 | 0 | 1 | Not-determined | Genome-fragment | NA | NA | 0 |
| S5-final-viral-combined_k141_189775\|\|full | 3131 | No | NA | 4 | 0 | 1 | Not-determined | Genome-fragment | NA | NA | 0 |
| S5-final-viral-combined_k141_180981\|\|full | 3244 | No | NA | 4 | 0 | 2 | Not-determined | Genome-fragment | NA | NA | 0 |
| S5-final-viral-combined_k141_64525\|\|full | 1636 | No | NA | 3 | 0 | 0 | Low-quality | Genome-fragment | 2.04 | AAI-based (high-confidence) | 0 |
| S5-final-viral-combined_k141_100722\|\|full | 1232 | No | NA | 3 | 0 | 1 | Not-determined | Genome-fragment | NA | NA | 0 |
| S5-final-viral-combined_k141_138903\|\|full | 1636 | No | NA | 3 | 0 | 0 | Low-quality | Genome-fragment | 1.15 | AAI-based (high-confidence) | 0 |
| S17-final-viral-combined_k141_155369\|\|full | 1636 | No | NA | 3 | 0 | 1 | Low-quality | Genome-fragment | 0.96 | AAI-based (medium-confidence) | 0 |
| S8-final-viral-combined_k141_102708\|\|full | 1636 | No | NA | 3 | 1 | 0 | Low-quality | Genome-fragment | 2.74 | AAI-based (medium-confidence) | 0 |
| S9-final-viral-combined_k141_114935\|\|lt2gene | 1637 | No | NA | 2 | 2 | 0 | Low-quality | Genome-fragment | 4.3 | AAI-based (high-confidence) | 0 |
| S5-final-viral-combined_k141_91715\|\|full | 2977 | No | NA | 3 | 0 | 0 | Not-determined | Genome-fragment | NA | NA | 0 |
| S5-final-viral-combined_k141_3201\|\|full | 33282 | No | NA | 48 | 32 | 0 | Medium-quality | Genome-fragment | 85.21 | AAI-based (medium-confidence) | 0 |
| S5-final-viral-combined_k141_198400\|\|full | 1399 | No | NA | 3 | 0 | 1 | Not-determined | Genome-fragment | NA | NA | 0 |
| S11-final-viral-combined_k141_121710\|\|full | 1637 | No | NA | 4 | 4 | 0 | Low-quality | Genome-fragment | 4.32 | AAI-based (high-confidence) | 0 |
| S1-final-viral-combined_k141_185944\|\|full | 1637 | No | NA | 6 | 3 | 0 | Low-quality | Genome-fragment | 4.2 | AAI-based (high-confidence) | 0 |
| S5-final-viral-combined_k141_181009\|\|full | 1950 | No | NA | 3 | 0 | 1 | Not-determined | Genome-fragment | NA | NA | 0 |
| S5-final-viral-combined_k141_91739\|\|full | 3107 | No | NA | 4 | 0 | 2 | Not-determined | Genome-fragment | NA | NA | 0 |
| S1-final-viral-combined_k141_136945\|\|lt2gene | 1637 | No | NA | 2 | 1 | 0 | Low-quality | Genome-fragment | 0.9 | AAI-based (medium-confidence) | 0 |
| S5-final-viral-combined_k141_91742\|\|full | 1589 | No | NA | 3 | 0 | 1 | Not-determined | Genome-fragment | NA | NA | 0 |
| S1-final-viral-combined_k141_70010\|\|lt2gene | 1637 | No | NA | 2 | 2 | 0 | Low-quality | Genome-fragment | 4.18 | AAI-based (high-confidence) | 0 |
| S5-final-viral-combined_k141_11275\|\|full | 2396 | No | NA | 3 | 0 | 1 | Not-determined | Genome-fragment | NA | NA | 0 |
| S5-final-viral-combined_k141_181026\|\|full | 3497 | No | NA | 2 | 0 | 2 | Not-determined | Genome-fragment | NA | NA | 0 |
| S7_k141_171009 | 1637 | No | NA | 2 | 0 | 0 | Low-quality | Genome-fragment | 2.97 | AAI-based (high-confidence) | 0 |
| S5-final-viral-combined_k141_55675\|\|full | 3891 | No | NA | 5 | 0 | 1 | Not-determined | Genome-fragment | NA | NA | 0 |
| S4-final-viral-combined_k141_140440\|\|full | 1637 | No | NA | 3 | 0 | 3 | Low-quality | Genome-fragment | 1.99 | AAI-based (medium-confidence) | 0 |
| S5-final-viral-combined_k141_91807\|\|full | 3386 | No | NA | 3 | 0 | 2 | Not-determined | Genome-fragment | NA | NA | 0 |
| S5-final-viral-combined_k141_135993\|\|full | 2087 | No | NA | 2 | 0 | 1 | Not-determined | Genome-fragment | NA | NA | 0 |
| S5-final-viral-combined_k141_153827\|\|full | 3331 | No | NA | 4 | 0 | 2 | Not-determined | Genome-fragment | NA | NA | 0 |
| S5-final-viral-combined_k141_37952\|\|full | 1575 | No | NA | 3 | 0 | 0 | Not-determined | Genome-fragment | NA | NA | 0 |
| S11-final-viral-combined_k141_104162\|\|full | 1638 | No | NA | 3 | 1 | 0 | Low-quality | Genome-fragment | 3.54 | AAI-based (medium-confidence) | 0 |
| S11-final-viral-combined_k141_209667\|\|lt2gene | 1638 | No | NA | 2 | 1 | 0 | Low-quality | Genome-fragment | 3.43 | AAI-based (high-confidence) | 0 |
| S5-final-viral-combined_k141_127423\|\|full | 3245 | No | NA | 2 | 0 | 2 | Not-determined | Genome-fragment | NA | NA | 0 |
| S7-final-viral-combined_k141_65429\|\|full | 1638 | No | NA | 2 | 0 | 1 | Low-quality | Genome-fragment | 2.72 | AAI-based (high-confidence) | 0 |
| S7-final-viral-combined_k141_180384\|\|full | 1638 | No | NA | 2 | 0 | 0 | Low-quality | Genome-fragment | 4.96 | AAI-based (medium-confidence) | 0 |
| S17-final-viral-combined_k141_196248\|\|full | 1638 | No | NA | 3 | 0 | 0 | Low-quality | Genome-fragment | 3.03 | AAI-based (high-confidence) | 0 |
| S2-final-viral-combined_k141_228022\|\|full | 1638 | No | NA | 6 | 0 | 0 | Low-quality | Genome-fragment | 3.73 | AAI-based (high-confidence) | 0 |
| S5-final-viral-combined_k141_207291\|\|full | 57653 | Yes | 24305 | 58 | 4 | 9 | Medium-quality | Genome-fragment | 52.91 | AAI-based (high-confidence) | 57.84 |
| S5-final-viral-combined_k141_172483\|\|full | 1717 | No | NA | 2 | 0 | 1 | Not-determined | Genome-fragment | NA | NA | 0 |
| S5-final-viral-combined_k141_47405\|\|full | 1016 | No | NA | 2 | 0 | 1 | Not-determined | Genome-fragment | NA | NA | 0 |
| S5-final-viral-combined_k141_3392\|\|full | 3215 | No | NA | 3 | 0 | 2 | Not-determined | Genome-fragment | NA | NA | 0 |
| S5-final-viral-combined_k141_136148\|\|full | 1503 | No | NA | 2 | 0 | 1 | Not-determined | Genome-fragment | NA | NA | 0 |
| S5-final-viral-combined_k141_73909\|\|full | 2495 | No | NA | 3 | 0 | 0 | Not-determined | Genome-fragment | NA | NA | 0 |
| S5-final-viral-combined_k141_19935\|\|full | 1147 | No | NA | 2 | 0 | 0 | Not-determined | Genome-fragment | NA | NA | 0 |
| S2_k141_124630 | 1638 | No | NA | 2 | 1 | 0 | Low-quality | Genome-fragment | 5.55 | HMM-based (lower-bound) | 0 |
| S9-final-viral-combined_k141_23461\|\|full | 1639 | No | NA | 2 | 0 | 1 | Low-quality | Genome-fragment | 2.12 | AAI-based (medium-confidence) | 0 |
| S5-final-viral-combined_k141_3443\|\|full | 1492 | No | NA | 3 | 0 | 0 | Not-determined | Genome-fragment | NA | NA | 0 |
| S5-final-viral-combined_k141_189934\|\|full | 2938 | No | NA | 8 | 0 | 0 | Not-determined | Genome-fragment | NA | NA | 0 |
| S11-final-viral-combined_k141_65883\|\|full | 1639 | No | NA | 2 | 1 | 0 | Low-quality | Genome-fragment | 2.03 | HMM-based (lower-bound) | 0 |
| S7_k141_6070 | 1639 | No | NA | 1 | 0 | 1 | Low-quality | Genome-fragment | 1.1 | AAI-based (medium-confidence) | 0 |
| S5-final-viral-combined_k141_172539\|\|full | 21226 | No | NA | 29 | 0 | 4 | Not-determined | Genome-fragment | NA | NA | 0 |
| S17-final-viral-combined_k141_19219\|\|lt2gene | 1639 | No | NA | 2 | 2 | 0 | Low-quality | Genome-fragment | 2.66 | AAI-based (high-confidence) | 0 |
| S5-final-viral-combined_k141_127548\|\|full | 1549 | No | NA | 2 | 0 | 2 | Not-determined | Genome-fragment | NA | NA | 0 |
| S5-final-viral-combined_k141_172559\|\|full | 2037 | No | NA | 4 | 0 | 2 | Not-determined | Genome-fragment | NA | NA | 0 |
| S5-final-viral-combined_k141_181161\|\|full | 1864 | No | NA | 4 | 0 | 0 | Not-determined | Genome-fragment | NA | NA | 0 |
| S5-final-viral-combined_k141_136232\|\|full | 2284 | No | NA | 4 | 0 | 1 | Not-determined | Genome-fragment | NA | NA | 0 |
| S2-final-viral-combined_k141_243777\|\|full | 1639 | No | NA | 2 | 0 | 0 | Low-quality | Genome-fragment | 1.32 | AAI-based (medium-confidence) | 0 |
| S5-final-viral-combined_k141_55858\|\|full | 4457 | No | NA | 5 | 0 | 3 | Not-determined | Genome-fragment | NA | NA | 0 |
| S4-final-viral-combined_k141_162879\|\|full | 1639 | No | NA | 5 | 0 | 0 | Low-quality | Genome-fragment | 3.14 | AAI-based (medium-confidence) | 0 |
| S5-final-viral-combined_k141_181194\|\|full | 9505 | No | NA | 13 | 0 | 3 | Not-determined | Genome-fragment | NA | NA | 0 |
| S4-final-viral-combined_k141_83142\|\|lt2gene | 1639 | No | NA | 1 | 0 | 0 | Low-quality | Genome-fragment | 1.63 | AAI-based (medium-confidence) | 0 |
| S5-final-viral-combined_k141_136274\|\|full | 17310 | No | NA | 14 | 0 | 7 | Not-determined | Genome-fragment | NA | NA | 0 |
| S5-final-viral-combined_k141_64783\|\|full | 1114 | No | NA | 2 | 0 | 1 | Not-determined | Genome-fragment | NA | NA | 0 |
| S5-final-viral-combined_k141_181223\|\|full | 3104 | No | NA | 4 | 0 | 0 | Not-determined | Genome-fragment | NA | NA | 0 |
| S5-final-viral-combined_k141_127621\|\|full | 2702 | No | NA | 3 | 0 | 2 | Not-determined | Genome-fragment | NA | NA | 0 |
| S5-final-viral-combined_k141_55913\|\|full | 5562 | No | NA | 7 | 0 | 7 | Not-determined | Genome-fragment | NA | NA | 0 |
| S5-final-viral-combined_k141_162835\|\|full | 9828 | No | NA | 9 | 0 | 5 | Not-determined | Genome-fragment | NA | NA | 0 |
| S5-final-viral-combined_k141_73944\|\|full | 1743 | No | NA | 3 | 0 | 2 | Not-determined | Genome-fragment | NA | NA | 0 |
| S8-final-viral-combined_k141_7271\|\|full | 1639 | No | NA | 2 | 0 | 2 | Low-quality | Genome-fragment | 3.18 | AAI-based (medium-confidence) | 0 |
| S11-final-viral-combined_k141_155786\|\|full | 1640 | No | NA | 3 | 0 | 1 | Low-quality | Genome-fragment | 2.8 | AAI-based (high-confidence) | 0 |
| S5-final-viral-combined_k141_136313\|\|full | 1216 | No | NA | 3 | 0 | 1 | Not-determined | Genome-fragment | NA | NA | 0 |
| S1-final-viral-combined_k141_138561\|\|full | 1640 | No | NA | 4 | 0 | 0 | Low-quality | Genome-fragment | 4.54 | AAI-based (high-confidence) | 0 |
| S5-final-viral-combined_k141_162849\|\|full | 1738 | No | NA | 2 | 0 | 2 | Not-determined | Genome-fragment | NA | NA | 0 |
| S7-final-viral-combined_k141_99605\|\|full | 1640 | No | NA | 4 | 1 | 0 | Low-quality | Genome-fragment | 3.28 | AAI-based (high-confidence) | 0 |
| S5-final-viral-combined_k141_47599\|\|full | 1756 | No | NA | 3 | 0 | 1 | Not-determined | Genome-fragment | NA | NA | 0 |
| S5-final-viral-combined_k141_64866\|\|full | 1028 | No | NA | 3 | 0 | 1 | Not-determined | Genome-fragment | NA | NA | 0 |
| S17-final-viral-combined_k141_58177\|\|full | 1640 | No | NA | 4 | 0 | 0 | Low-quality | Genome-fragment | 1.96 | AAI-based (medium-confidence) | 0 |
| S17-final-viral-combined_k141_262192\|\|full | 1640 | No | NA | 4 | 0 | 2 | Low-quality | Genome-fragment | 0.51 | AAI-based (medium-confidence) | 0 |
| S5-final-viral-combined_k141_38144\|\|full | 1378 | No | NA | 3 | 0 | 0 | Not-determined | Genome-fragment | NA | NA | 0 |
| S12-final-viral-combined_k141_185918\|\|lt2gene | 1640 | No | NA | 2 | 2 | 0 | Low-quality | Genome-fragment | 3.41 | AAI-based (high-confidence) | 0 |
| S5-final-viral-combined_k141_189993\|\|full | 1230 | No | NA | 2 | 0 | 2 | Not-determined | Genome-fragment | NA | NA | 0 |
| S2-final-viral-combined_k141_130722\|\|full | 1640 | No | NA | 2 | 0 | 1 | Low-quality | Genome-fragment | 0.91 | AAI-based (medium-confidence) | 0 |
| S5-final-viral-combined_k141_189998\|\|full | 1182 | No | NA | 3 | 0 | 0 | Not-determined | Genome-fragment | NA | NA | 0 |
| S5-final-viral-combined_k141_64919\|\|full | 1102 | No | NA | 3 | 0 | 1 | Not-determined | Genome-fragment | NA | NA | 0 |
| S5-final-viral-combined_k141_29246\|\|full | 2354 | No | NA | 4 | 0 | 2 | Not-determined | Genome-fragment | NA | NA | 0 |
| S5-final-viral-combined_k141_127702\|\|full | 9200 | No | NA | 15 | 0 | 2 | Not-determined | Genome-fragment | NA | NA | 0 |
| S6-final-viral-combined_k141_47273\|\|full | 1640 | No | NA | 2 | 2 | 0 | Low-quality | Genome-fragment | 4 | AAI-based (medium-confidence) | 0 |
| S1-final-viral-combined_k141_57433\|\|full | 1641 | No | NA | 2 | 0 | 0 | Low-quality | Genome-fragment | 3.05 | AAI-based (high-confidence) | 0 |
| S5-final-viral-combined_k141_38188\|\|full | 5957 | No | NA | 5 | 0 | 5 | Not-determined | Genome-fragment | NA | NA | 0 |
| S5-final-viral-combined_k141_198648\|\|full | 3493 | No | NA | 6 | 0 | 0 | Not-determined | Genome-fragment | NA | NA | 0 |
| S5-final-viral-combined_k141_11430\|\|full | 5601 | No | NA | 6 | 0 | 1 | Not-determined | Genome-fragment | NA | NA | 0 |
| S5-final-viral-combined_k141_38228\|\|full | 1277 | No | NA | 2 | 0 | 2 | Not-determined | Genome-fragment | NA | NA | 0 |
| S5-final-viral-combined_k141_3625\|\|full | 1378 | No | NA | 2 | 0 | 2 | Not-determined | Genome-fragment | NA | NA | 0 |
| S5-final-viral-combined_k141_162980\|\|full | 1203 | No | NA | 3 | 0 | 0 | Not-determined | Genome-fragment | NA | NA | 0 |
| S5-final-viral-combined_k141_172754\|\|full | 5888 | No | NA | 5 | 0 | 1 | Not-determined | Genome-fragment | NA | NA | 0 |
| S5-final-viral-combined_k141_72225\|\|lt2gene | 1641 | No | NA | 2 | 0 | 0 | Low-quality | Genome-fragment | 2.14 | AAI-based (high-confidence) | 0 |
| S5-final-viral-combined_k141_38261\|\|full | 2203 | No | NA | 5 | 0 | 0 | Not-determined | Genome-fragment | NA | NA | 0 |
| S5-final-viral-combined_k141_163011\|\|full | 1039 | No | NA | 3 | 0 | 0 | Not-determined | Genome-fragment | NA | NA | 0 |
| S5-final-viral-combined_k141_3666\|\|full | 3730 | No | NA | 6 | 0 | 1 | Not-determined | Genome-fragment | NA | NA | 0 |
| S17-final-viral-combined_k141_92469\|\|lt2gene | 1641 | No | NA | 1 | 1 | 0 | Low-quality | Genome-fragment | 4.58 | AAI-based (medium-confidence) | 0 |
| S5-final-viral-combined_k141_190116\|\|full | 2561 | No | NA | 3 | 0 | 1 | Not-determined | Genome-fragment | NA | NA | 0 |
| S5-final-viral-combined_k141_207544\|\|full | 1312 | No | NA | 2 | 0 | 0 | Not-determined | Genome-fragment | NA | NA | 0 |
| S5-final-viral-combined_k141_207545\|\|full | 3111 | No | NA | 3 | 0 | 1 | Not-determined | Genome-fragment | NA | NA | 0 |
| S5-final-viral-combined_k141_47798\|\|full | 2588 | No | NA | 3 | 0 | 1 | Not-determined | Genome-fragment | NA | NA | 0 |
| S5-final-viral-combined_k141_181265\|\|full | 169798 | No | NA | 185 | 0 | 38 | Not-determined | Genome-fragment | NA | NA | 0 |
| S12-final-viral-combined_k141_35622\|\|full | 1641 | No | NA | 4 | 2 | 0 | Low-quality | Genome-fragment | 2.62 | AAI-based (high-confidence) | 0 |
| S12-final-viral-combined_k141_240005\|\|full | 1641 | No | NA | 3 | 0 | 1 | Low-quality | Genome-fragment | 0.99 | AAI-based (medium-confidence) | 0 |
| S4-final-viral-combined_k141_42055\|\|full | 1641 | No | NA | 2 | 0 | 0 | Low-quality | Genome-fragment | 3.47 | AAI-based (medium-confidence) | 0 |
| S5-final-viral-combined_k141_109231\|\|full | 5702 | No | NA | 6 | 0 | 1 | Not-determined | Genome-fragment | NA | NA | 0 |
| S8-final-viral-combined_k141_167500\|\|full | 1641 | No | NA | 3 | 1 | 0 | Low-quality | Genome-fragment | 2.75 | AAI-based (medium-confidence) | 0 |
| S5-final-viral-combined_k141_181300\|\|full | 1176 | No | NA | 3 | 0 | 1 | Not-determined | Genome-fragment | NA | NA | 0 |
| S5-final-viral-combined_k141_47841\|\|full | 1368 | No | NA | 3 | 0 | 0 | Not-determined | Genome-fragment | NA | NA | 0 |
| S5-final-viral-combined_k141_190194\|\|full | 1735 | No | NA | 3 | 0 | 1 | Not-determined | Genome-fragment | NA | NA | 0 |
| S5-final-viral-combined_k141_11501\|\|full | 1177 | No | NA | 3 | 0 | 0 | Not-determined | Genome-fragment | NA | NA | 0 |
| S5-final-viral-combined_k141_74132\|\|full | 2874 | No | NA | 4 | 0 | 2 | Not-determined | Genome-fragment | NA | NA | 0 |
| S9-final-viral-combined_k141_92486\|\|full | 1642 | No | NA | 3 | 0 | 0 | Low-quality | Genome-fragment | 2.61 | AAI-based (high-confidence) | 0 |
| S1-final-viral-combined_k141_388155\|\|full | 1642 | No | NA | 2 | 1 | 0 | Low-quality | Genome-fragment | 3.65 | AAI-based (high-confidence) | 0 |
| S5-final-viral-combined_k141_47869\|\|full | 1516 | No | NA | 3 | 0 | 2 | Not-determined | Genome-fragment | NA | NA | 0 |
| S5-final-viral-combined_k141_181336\|\|full | 3124 | No | NA | 3 | 0 | 2 | Not-determined | Genome-fragment | NA | NA | 0 |
| S5-final-viral-combined_k141_66766\|\|full | 1642 | No | NA | 1 | 0 | 0 | Low-quality | Genome-fragment | 2.74 | AAI-based (high-confidence) | 0 |
| S5-final-viral-combined_k141_29272\|\|full | 63967 | No | NA | 70 | 20 | 6 | High-quality | High-quality | 100 | AAI-based (high-confidence) | 0 |
| S5-final-viral-combined_k141_132270\|\|full | 1642 | No | NA | 3 | 0 | 0 | Low-quality | Genome-fragment | 0.93 | AAI-based (high-confidence) | 0 |
| S5-final-viral-combined_k141_97091\|\|full | 1642 | No | NA | 2 | 0 | 1 | Low-quality | Genome-fragment | 0.52 | AAI-based (medium-confidence) | 0 |
| S7-final-viral-combined_k141_35551\|\|full | 1642 | No | NA | 2 | 0 | 1 | Low-quality | Genome-fragment | 1.75 | AAI-based (medium-confidence) | 0 |
| S5-final-viral-combined_k141_74213\|\|full | 2747 | No | NA | 3 | 0 | 1 | Not-determined | Genome-fragment | NA | NA | 0 |
| S17-final-viral-combined_k141_204819\|\|full | 1642 | No | NA | 1 | 0 | 0 | Low-quality | Genome-fragment | 3.94 | AAI-based (medium-confidence) | 0 |
| S5-final-viral-combined_k141_29307\|\|full | 2091 | No | NA | 3 | 0 | 1 | Not-determined | Genome-fragment | NA | NA | 0 |
| S5-final-viral-combined_k141_190295\|\|full | 12056 | No | NA | 16 | 0 | 0 | Not-determined | Genome-fragment | NA | NA | 0 |
| S17-k141_128942 | 1642 | No | NA | 1 | 0 | 1 | Low-quality | Genome-fragment | 0.43 | AAI-based (medium-confidence) | 0 |
| S12-final-viral-combined_k141_334976\|\|full | 1642 | No | NA | 3 | 0 | 1 | Low-quality | Genome-fragment | 2.85 | AAI-based (medium-confidence) | 0 |
| S5-final-viral-combined_k141_207630\|\|full | 4139 | No | NA | 4 | 0 | 2 | Not-determined | Genome-fragment | NA | NA | 0 |
| S12_k141_120409 | 1642 | No | NA | 1 | 0 | 1 | Low-quality | Genome-fragment | 1.95 | AAI-based (high-confidence) | 0 |
| S5-final-viral-combined_k141_101070\|\|full | 2443 | No | NA | 4 | 0 | 0 | Not-determined | Genome-fragment | NA | NA | 0 |
| S4-final-viral-combined_k141_157589\|\|full | 1642 | No | NA | 2 | 0 | 0 | Low-quality | Genome-fragment | 3.72 | AAI-based (high-confidence) | 0 |
| S5-final-viral-combined_k141_154171\|\|full | 2460 | No | NA | 4 | 0 | 3 | Not-determined | Genome-fragment | NA | NA | 0 |
| S1-final-viral-combined_k141_386506\|\|lt2gene | 1643 | No | NA | 1 | 1 | 0 | Low-quality | Genome-fragment | 2.74 | HMM-based (lower-bound) | 0 |
| S5-final-viral-combined_k141_181520\|\|full | 7127 | No | NA | 6 | 0 | 6 | Not-determined | Genome-fragment | NA | NA | 0 |
| S5-final-viral-combined_k141_47992\|\|full | 1108 | No | NA | 2 | 0 | 1 | Not-determined | Genome-fragment | NA | NA | 0 |
| S5-final-viral-combined_k141_48017\|\|full | 2068 | No | NA | 3 | 0 | 0 | Not-determined | Genome-fragment | NA | NA | 0 |
| S5-final-viral-combined_k141_207746\|\|full | 2142 | No | NA | 3 | 0 | 1 | Not-determined | Genome-fragment | NA | NA | 0 |
| S5-final-viral-combined_k141_199172\|\|full | 1643 | No | NA | 3 | 3 | 0 | Low-quality | Genome-fragment | 2.27 | HMM-based (lower-bound) | 0 |
| S5-final-viral-combined_k141_154248\|\|full | 1230 | No | NA | 4 | 0 | 0 | Not-determined | Genome-fragment | NA | NA | 0 |
| S5-final-viral-combined_k141_103943\|\|full | 1643 | No | NA | 3 | 0 | 3 | Low-quality | Genome-fragment | 3.29 | AAI-based (medium-confidence) | 0 |
| S5_k141_197235 | 1643 | No | NA | 1 | 0 | 1 | Low-quality | Genome-fragment | 1.51 | AAI-based (medium-confidence) | 0 |
| S5-final-viral-combined_k141_109454\|\|full | 8037 | No | NA | 11 | 0 | 1 | Not-determined | Genome-fragment | NA | NA | 0 |
| S7-final-viral-combined_k141_64484\|\|full | 1643 | No | NA | 3 | 0 | 2 | Low-quality | Genome-fragment | 0.56 | AAI-based (medium-confidence) | 0 |
| S5-final-viral-combined_k141_92198\|\|full | 1495 | No | NA | 2 | 0 | 0 | Not-determined | Genome-fragment | NA | NA | 0 |
| S5-final-viral-combined_k141_48053\|\|full | 1136 | No | NA | 2 | 0 | 2 | Not-determined | Genome-fragment | NA | NA | 0 |

| | | | | | | | | | | | |
|---|---|---|---|---|---|---|---|---|---|---|---|
| S7-final-viral-combined_k141_18894\|\|full | 1643 | No | NA | 2 | 0 | 0 | Low-quality | Genome-fragment | 2.52 | AAI-based (high-confidence) | 0 |
| S5-final-viral-combined_k141_101218\|\|full | 2771 | No | NA | 3 | 0 | 1 | Not-determined | Genome-fragment | NA | NA | 0 |
| S5-final-viral-combined_k141_190415\|\|full | 4858 | No | NA | 6 | 0 | 4 | Not-determined | Genome-fragment | NA | NA | 0 |
| S17-final-viral-combined_k141_91067\|\|full | 1643 | No | NA | 3 | 0 | 2 | Low-quality | Genome-fragment | 1.08 | AAI-based (medium-confidence) | 0 |
| S17-k141_257018 | 1643 | No | NA | 2 | 0 | 0 | Low-quality | Genome-fragment | 2.15 | AAI-based (high-confidence) | 0 |
| S12-final-viral-combined_k141_8870\|\|full | 1643 | No | NA | 2 | 0 | 1 | Low-quality | Genome-fragment | 1.14 | AAI-based (high-confidence) | 0 |
| S8-final-viral-combined_k141_7245\|\|full | 1643 | No | NA | 4 | 0 | 0 | Low-quality | Genome-fragment | 2.12 | AAI-based (medium-confidence) | 0 |
| S5-final-viral-combined_k141_29396\|\|full | 1276 | No | NA | 3 | 0 | 0 | Not-determined | Genome-fragment | NA | NA | 0 |
| S5-final-viral-combined_k141_109553\|\|full | 2441 | No | NA | 2 | 0 | 1 | Not-determined | Genome-fragment | NA | NA | 0 |
| S1-final-viral-combined_k141_5340\|\|full | 1644 | No | NA | 4 | 0 | 1 | Low-quality | Genome-fragment | 2.96 | AAI-based (medium-confidence) | 0 |
| S5-final-viral-combined_k141_128122\|\|full | 2272 | No | NA | 3 | 0 | 0 | Not-determined | Genome-fragment | NA | NA | 0 |
| S1_k141_323685 | 1644 | No | NA | 2 | 0 | 1 | Low-quality | Genome-fragment | 5.7 | AAI-based (medium-confidence) | 0 |
| S17-final-viral-combined_k141_85515\|\|full | 1644 | No | NA | 5 | 1 | 0 | Low-quality | Genome-fragment | 2.8 | AAI-based (medium-confidence) | 0 |
| S4-final-viral-combined_k141_79997\|\|full | 1644 | No | NA | 6 | 3 | 0 | Low-quality | Genome-fragment | 3.68 | AAI-based (high-confidence) | 0 |
| S5-final-viral-combined_k141_128162\|\|full | 1799 | No | NA | 2 | 0 | 2 | Not-determined | Genome-fragment | NA | NA | 0 |
| S8-final-viral-combined_k141_33450\|\|full | 1644 | No | NA | 6 | 0 | 0 | Low-quality | Genome-fragment | 3.78 | AAI-based (medium-confidence) | 0 |
| S5-final-viral-combined_k141_154358\|\|full | 6667 | No | NA | 5 | 0 | 2 | Not-determined | Genome-fragment | NA | NA | 0 |
| S8-final-viral-combined_k141_216963\|\|full | 1644 | No | NA | 3 | 0 | 1 | Low-quality | Genome-fragment | 1.11 | AAI-based (high-confidence) | 0 |
| S5-final-viral-combined_k141_74353\|\|full | 2432 | No | NA | 3 | 0 | 2 | Not-determined | Genome-fragment | NA | NA | 0 |
| S9-final-viral-combined_k141_5977\|\|full | 1645 | No | NA | 3 | 0 | 0 | Low-quality | Genome-fragment | 2.23 | AAI-based (high-confidence) | 0 |
| S5-final-viral-combined_k141_198810\|\|full | 5153 | No | NA | 4 | 0 | 3 | Not-determined | Genome-fragment | NA | NA | 0 |
| S9-final-viral-combined_k141_37400\|\|full | 1645 | No | NA | 2 | 0 | 2 | Low-quality | Genome-fragment | 0.81 | AAI-based (medium-confidence) | 0 |
| S11-final-viral-combined_k141_284787\|\|full | 1645 | No | NA | 2 | 0 | 0 | Low-quality | Genome-fragment | 2.16 | AAI-based (high-confidence) | 0 |
| S1_k141_4673 | 1645 | No | NA | 4 | 0 | 1 | Low-quality | Genome-fragment | 2.11 | AAI-based (high-confidence) | 0 |
| S5-final-viral-combined_k141_74394\|\|full | 3154 | No | NA | 4 | 0 | 4 | Not-determined | Genome-fragment | NA | NA | 0 |
| S5-final-viral-combined_k141_207812\|\|full | 1379 | No | NA | 2 | 0 | 1 | Not-determined | Genome-fragment | NA | NA | 0 |
| S5-final-viral-combined_k141_172921\|\|full | 1669 | No | NA | 2 | 0 | 0 | Not-determined | Genome-fragment | NA | NA | 0 |
| S5-final-viral-combined_k141_4203\|\|full | 5194 | No | NA | 9 | 0 | 5 | Not-determined | Genome-fragment | NA | NA | 0 |
| S3_k141_133394 | 1645 | No | NA | 2 | 0 | 0 | Low-quality | Genome-fragment | 2.36 | AAI-based (medium-confidence) | 0 |
| S5-final-viral-combined_k141_107676\|\|full | 1645 | No | NA | 2 | 1 | 0 | Low-quality | Genome-fragment | 4.05 | AAI-based (medium-confidence) | 0 |
| S5-final-viral-combined_k141_172930\|\|full | 1426 | No | NA | 2 | 0 | 2 | Not-determined | Genome-fragment | NA | NA | 0 |
| S5-final-viral-combined_k141_4215\|\|full | 1491 | No | NA | 3 | 0 | 0 | Not-determined | Genome-fragment | NA | NA | 0 |
| S5-final-viral-combined_k141_96073\|\|full | 1645 | No | NA | 5 | 2 | 0 | Low-quality | Genome-fragment | 3.69 | AAI-based (high-confidence) | 0 |
| S5-final-viral-combined_k141_65317\|\|full | 3127 | No | NA | 3 | 0 | 0 | Not-determined | Genome-fragment | NA | NA | 0 |
| S5-final-viral-combined_k141_146091\|\|full | 1790 | No | NA | 2 | 0 | 2 | Not-determined | Genome-fragment | NA | NA | 0 |
| S5-final-viral-combined_k141_65323\|\|full | 2101 | No | NA | 3 | 0 | 0 | Not-determined | Genome-fragment | NA | NA | 0 |
| S17-k141_110207 | 1645 | No | NA | 1 | 0 | 1 | Low-quality | Genome-fragment | 0.59 | AAI-based (medium-confidence) | 0 |
| S5-final-viral-combined_k141_4246\|\|full | 2451 | No | NA | 4 | 0 | 1 | Not-determined | Genome-fragment | NA | NA | 0 |
| S12-final-viral-combined_k141_130512\|\|full | 1645 | No | NA | 2 | 0 | 1 | Low-quality | Genome-fragment | 0.47 | AAI-based (high-confidence) | 0 |
| S12_k141_344296 | 1645 | No | NA | 1 | 0 | 1 | Low-quality | Genome-fragment | 0.57 | AAI-based (medium-confidence) | 0 |
| S12_k141_314773 | 1645 | No | NA | 2 | 0 | 0 | Low-quality | Genome-fragment | 0.9 | AAI-based (high-confidence) | 0 |
| S9-final-viral-combined_k141_114689\|\|full | 1646 | No | NA | 4 | 0 | 1 | Low-quality | Genome-fragment | 1.86 | AAI-based (high-confidence) | 0 |
| S5-final-viral-combined_k141_118672\|\|full | 5754 | No | NA | 7 | 0 | 4 | Not-determined | Genome-fragment | NA | NA | 0 |
| S5-final-viral-combined_k141_29447\|\|full | 116107 | Yes | 33613 | 112 | 13 | 46 | Medium-quality | Genome-fragment | 61.36 | HMM-based (lower-bound) | 71.05 |
| S5-final-viral-combined_k141_38514\|\|full | 2286 | No | NA | 4 | 0 | 2 | Not-determined | Genome-fragment | NA | NA | 0 |
| S5-final-viral-combined_k141_118684\|\|full | 1522 | No | NA | 3 | 0 | 0 | Not-determined | Genome-fragment | NA | NA | 0 |
| S5-final-viral-combined_k141_118700\|\|full | 2192 | No | NA | 3 | 0 | 0 | Not-determined | Genome-fragment | NA | NA | 0 |
| S11-final-viral-combined_k141_263833\|\|full | 1646 | No | NA | 3 | 0 | 0 | Low-quality | Genome-fragment | 3.96 | AAI-based (medium-confidence) | 0 |
| S11-final-viral-combined_k141_160646\|\|full | 1646 | No | NA | 2 | 0 | 0 | Low-quality | Genome-fragment | 1.66 | AAI-based (medium-confidence) | 0 |
| S11-final-viral-combined_k141_289399\|\|full | 1646 | No | NA | 4 | 0 | 0 | Low-quality | Genome-fragment | 3.6 | AAI-based (high-confidence) | 0 |
| S5-final-viral-combined_k141_11839\|\|full | 2223 | No | NA | 2 | 0 | 1 | Not-determined | Genome-fragment | NA | NA | 0 |
| S11_k141_126760 | 1646 | No | NA | 3 | 0 | 0 | Low-quality | Genome-fragment | 2.33 | AAI-based (medium-confidence) | 0 |
| S1-final-viral-combined_k141_291299\|\|full | 1646 | No | NA | 2 | 0 | 1 | Low-quality | Genome-fragment | 1.1 | AAI-based (medium-confidence) | 0 |
| S1-final-viral-combined_k141_311613\|\|full | 1646 | No | NA | 4 | 1 | 0 | Low-quality | Genome-fragment | 3.47 | AAI-based (high-confidence) | 0 |
| S5-final-viral-combined_k141_146111\|\|full | 39278 | No | NA | 40 | 0 | 4 | Not-determined | Genome-fragment | NA | NA | 0 |
| S1-final-viral-combined_k141_80688\|\|full | 1646 | No | NA | 3 | 2 | 0 | Low-quality | Genome-fragment | 4.82 | AAI-based (high-confidence) | 0 |
| S3-final-viral-combined_k141_114687\|\|full | 1646 | No | NA | 2 | 0 | 1 | Low-quality | Genome-fragment | 3.53 | AAI-based (high-confidence) | 0 |
| S5-final-viral-combined_k141_182590\|\|full | 1646 | No | NA | 5 | 0 | 0 | Low-quality | Genome-fragment | 3.12 | AAI-based (high-confidence) | 0 |
| S5-final-viral-combined_k141_154442\|\|full | 3780 | No | NA | 6 | 0 | 2 | Not-determined | Genome-fragment | NA | NA | 0 |
| S12-final-viral-combined_k141_129430\|\|full | 1646 | No | NA | 3 | 1 | 0 | Low-quality | Genome-fragment | 1.54 | HMM-based (lower-bound) | 0 |
| S12-final-viral-combined_k141_341091\|\|lt2gene | 1646 | No | NA | 2 | 1 | 0 | Low-quality | Genome-fragment | 6.06 | HMM-based (lower-bound) | 0 |
| S2-final-viral-combined_k141_23515\|\|full | 1646 | No | NA | 2 | 0 | 1 | Low-quality | Genome-fragment | 0.83 | AAI-based (medium-confidence) | 0 |
| S6-final-viral-combined_k141_91735\|\|full | 1646 | No | NA | 6 | 0 | 0 | Low-quality | Genome-fragment | 5.1 | AAI-based (high-confidence) | 0 |
| S5-final-viral-combined_k141_11872\|\|full | 1959 | No | NA | 4 | 0 | 0 | Not-determined | Genome-fragment | NA | NA | 0 |
| S5-final-viral-combined_k141_190536\|\|full | 4706 | No | NA | 6 | 0 | 2 | Not-determined | Genome-fragment | NA | NA | 0 |
| S5-final-viral-combined_k141_38565\|\|full | 3521 | No | NA | 3 | 0 | 1 | Not-determined | Genome-fragment | NA | NA | 0 |
| S6-final-viral-combined_k141_33968\|\|full | 1646 | No | NA | 3 | 1 | 0 | Low-quality | Genome-fragment | 3.63 | AAI-based (high-confidence) | 0 |
| S5-final-viral-combined_k141_65439\|\|full | 4449 | No | NA | 4 | 0 | 1 | Not-determined | Genome-fragment | NA | NA | 0 |
| S11-final-viral-combined_k141_154730\|\|full | 1647 | No | NA | 4 | 0 | 0 | Low-quality | Genome-fragment | 3.77 | AAI-based (medium-confidence) | 0 |
| S11_k141_170813 | 1647 | No | NA | 1 | 0 | 0 | Low-quality | Genome-fragment | 2.19 | AAI-based (high-confidence) | 0 |
| S5-final-viral-combined_k141_83043\|\|full | 1206 | No | NA | 3 | 0 | 1 | Not-determined | Genome-fragment | NA | NA | 0 |
| S5-final-viral-combined_k141_136497\|\|full | 1271 | No | NA | 3 | 0 | 1 | Not-determined | Genome-fragment | NA | NA | 0 |
| S1-final-viral-combined_k141_203196\|\|full | 1647 | No | NA | 6 | 1 | 0 | Low-quality | Genome-fragment | 3.91 | AAI-based (medium-confidence) | 0 |
| S5-final-viral-combined_k141_74610\|\|full | 1038 | No | NA | 2 | 0 | 1 | Not-determined | Genome-fragment | NA | NA | 0 |
| S1-final-viral-combined_k141_334430\|\|full | 1647 | No | NA | 3 | 0 | 0 | Low-quality | Genome-fragment | 4.25 | AAI-based (high-confidence) | 0 |
| S5-final-viral-combined_k141_56188\|\|full | 55731 | No | NA | 50 | 0 | 39 | Not-determined | Genome-fragment | NA | NA | 0 |
| S5-final-viral-combined_k141_136524\|\|full | 1903 | No | NA | 3 | 0 | 2 | Not-determined | Genome-fragment | NA | NA | 0 |
| S5-final-viral-combined_k141_154525\|\|full | 4997 | No | NA | 8 | 0 | 0 | Not-determined | Genome-fragment | NA | NA | 0 |
| S5-final-viral-combined_k141_162395\|\|full | 1647 | No | NA | 4 | 3 | 0 | Low-quality | Genome-fragment | 4.59 | AAI-based (high-confidence) | 0 |
| S7-final-viral-combined_k141_96701\|\|full | 1647 | No | NA | 2 | 0 | 2 | Low-quality | Genome-fragment | 2.81 | AAI-based (high-confidence) | 0 |
| S8-final-viral-combined_k141_229184\|\|full | 1647 | No | NA | 5 | 0 | 0 | Low-quality | Genome-fragment | 3.97 | AAI-based (high-confidence) | 0 |
| S1_k141_230272 | 1648 | No | NA | 2 | 0 | 0 | Low-quality | Genome-fragment | 2.52 | AAI-based (medium-confidence) | 0 |
| S3-final-viral-combined_k141_42417\|\|full | 1648 | No | NA | 3 | 2 | 0 | Low-quality | Genome-fragment | 3.85 | AAI-based (medium-confidence) | 0 |
| S5-final-viral-combined_k141_48302\|\|full | 1895 | No | NA | 3 | 0 | 2 | Not-determined | Genome-fragment | NA | NA | 0 |
| S5-final-viral-combined_k141_207951\|\|full | 1184 | No | NA | 3 | 0 | 2 | Not-determined | Genome-fragment | NA | NA | 0 |
| S5-final-viral-combined_k141_74667\|\|full | 3819 | No | NA | 5 | 0 | 2 | Not-determined | Genome-fragment | NA | NA | 0 |
| S5-final-viral-combined_k141_38634\|\|full | 19486 | No | NA | 16 | 0 | 13 | Not-determined | Genome-fragment | NA | NA | 0 |
| S5-final-viral-combined_k141_74702\|\|full | 6550 | No | NA | 2 | 0 | 1 | Not-determined | Genome-fragment | NA | NA | 0 |
| S5-final-viral-combined_k141_141704\|\|full | 1648 | No | NA | 3 | 0 | 0 | Low-quality | Genome-fragment | 3.07 | AAI-based (medium-confidence) | 0 |
| S12-final-viral-combined_k141_189988\|\|full | 1648 | No | NA | 3 | 0 | 2 | Low-quality | Genome-fragment | 0.88 | AAI-based (medium-confidence) | 0 |
| S2-final-viral-combined_k141_129231\|\|full | 1648 | No | NA | 3 | 0 | 1 | Low-quality | Genome-fragment | 1.86 | AAI-based (medium-confidence) | 0 |
| S5-final-viral-combined_k141_163576\|\|full | 5778 | No | NA | 5 | 0 | 3 | Not-determined | Genome-fragment | NA | NA | 0 |
| S2-final-viral-combined_k141_71266\|\|full | 1648 | No | NA | 3 | 0 | 0 | Low-quality | Genome-fragment | 1.15 | AAI-based (medium-confidence) | 0 |
| S2-final-viral-combined_k141_132846\|\|full | 1648 | No | NA | 4 | 1 | 0 | Low-quality | Genome-fragment | 1.04 | HMM-based (lower-bound) | 0 |
| S5-final-viral-combined_k141_199090\|\|full | 1185 | No | NA | 2 | 0 | 2 | Not-determined | Genome-fragment | NA | NA | 0 |
| S5-final-viral-combined_k141_92463\|\|full | 1569 | No | NA | 2 | 0 | 1 | Not-determined | Genome-fragment | NA | NA | 0 |
| S5-final-viral-combined_k141_92468\|\|full | 1559 | No | NA | 3 | 0 | 1 | Not-determined | Genome-fragment | NA | NA | 0 |
| S5-final-viral-combined_k141_208023\|\|full | 2728 | No | NA | 3 | 0 | 2 | Not-determined | Genome-fragment | NA | NA | 0 |
| S2-final-viral-combined_k141_56916\|\|lt2gene | 1648 | No | NA | 2 | 1 | 0 | Low-quality | Genome-fragment | 3.56 | AAI-based (high-confidence) | 0 |
| S6_k141_135867 | 1648 | No | NA | 2 | 0 | 0 | Low-quality | Genome-fragment | 1.53 | AAI-based (high-confidence) | 0 |
| S5-final-viral-combined_k141_190617\|\|full | 1111 | No | NA | 1 | 0 | 1 | Not-determined | Genome-fragment | NA | NA | 0 |
| S6_k141_2254 | 1648 | No | NA | 2 | 0 | 1 | Low-quality | Genome-fragment | 0.66 | AAI-based (high-confidence) | 0 |
| S5-final-viral-combined_k141_4494\|\|full | 1981 | No | NA | 4 | 0 | 0 | Not-determined | Genome-fragment | NA | NA | 0 |
| S5-final-viral-combined_k141_190623\|\|full | 2716 | No | NA | 4 | 0 | 3 | Not-determined | Genome-fragment | NA | NA | 0 |
| S8-final-viral-combined_k141_135965\|\|lt2gene | 1648 | No | NA | 2 | 1 | 0 | Low-quality | Genome-fragment | 3.92 | AAI-based (medium-confidence) | 0 |
| S5-final-viral-combined_k141_208061\|\|full | 5886 | No | NA | 7 | 0 | 1 | Not-determined | Genome-fragment | NA | NA | 0 |
| S5-final-viral-combined_k141_56317\|\|full | 1246 | No | NA | 4 | 0 | 0 | Not-determined | Genome-fragment | NA | NA | 0 |
| S5-final-viral-combined_k141_4537\|\|full | 1241 | No | NA | 2 | 0 | 2 | Not-determined | Genome-fragment | NA | NA | 0 |
| S5-final-viral-combined_k141_20561\|\|full | 1675 | No | NA | 2 | 0 | 1 | Not-determined | Genome-fragment | NA | NA | 0 |
| S5-final-viral-combined_k141_56338\|\|full | 1705 | No | NA | 4 | 0 | 1 | Not-determined | Genome-fragment | NA | NA | 0 |
| S5-final-viral-combined_k141_4541\|\|full | 4977 | No | NA | 4 | 0 | 4 | Not-determined | Genome-fragment | NA | NA | 0 |
| S5-final-viral-combined_k141_110100\|\|full | 1649 | No | NA | 6 | 1 | 0 | Low-quality | Genome-fragment | 6.1 | AAI-based (high-confidence) | 0 |
| S5-final-viral-combined_k141_208077\|\|full | 3296 | No | NA | 4 | 0 | 3 | Not-determined | Genome-fragment | NA | NA | 0 |
| S7-final-viral-combined_k141_148264\|\|full | 1649 | No | NA | 2 | 0 | 0 | Low-quality | Genome-fragment | 3.15 | AAI-based (medium-confidence) | 0 |
| S12-final-viral-combined_k141_363514\|\|full | 1649 | No | NA | 4 | 0 | 0 | Low-quality | Genome-fragment | 4.28 | AAI-based (high-confidence) | 0 |
| S11-final-viral-combined_k141_330480\|\|full | 1650 | No | NA | 1 | 0 | 1 | Low-quality | Genome-fragment | 2.89 | AAI-based (medium-confidence) | 0 |
| S5-final-viral-combined_k141_4552\|\|full | 15130 | No | NA | 18 | 0 | 14 | Not-determined | Genome-fragment | NA | NA | 0 |
| S5-final-viral-combined_k141_4561\|\|full | 1677 | No | NA | 4 | 0 | 0 | Not-determined | Genome-fragment | NA | NA | 0 |
| S5-final-viral-combined_k141_74864\|\|full | 2113 | No | NA | 3 | 0 | 1 | Not-determined | Genome-fragment | NA | NA | 0 |
| S5-final-viral-combined_k141_101606\|\|full | 2320 | No | NA | 5 | 0 | 0 | Not-determined | Genome-fragment | NA | NA | 0 |
| S5-final-viral-combined_k141_29658\|\|full | 4353 | No | NA | 4 | 0 | 0 | Not-determined | Genome-fragment | NA | NA | 0 |
| S5-final-viral-combined_k141_56367\|\|full | 1165 | No | NA | 2 | 0 | 1 | Not-determined | Genome-fragment | NA | NA | 0 |
| S5-final-viral-combined_k141_163693\|\|full | 6016 | No | NA | 5 | 0 | 4 | Not-determined | Genome-fragment | NA | NA | 0 |
| S1-final-viral-combined_k141_42142\|\|full | 1650 | No | NA | 2 | 0 | 1 | Low-quality | Genome-fragment | 1.44 | AAI-based (medium-confidence) | 0 |
| S3_k141_18248 | 1650 | No | NA | 5 | 1 | 0 | Low-quality | Genome-fragment | 4.16 | AAI-based (high-confidence) | 0 |
| S5-final-viral-combined_k141_56391\|\|full | 2832 | No | NA | 3 | 0 | 0 | Not-determined | Genome-fragment | NA | NA | 0 |
| S5-final-viral-combined_k141_29697\|\|full | 1331 | No | NA | 2 | 0 | 1 | Not-determined | Genome-fragment | NA | NA | 0 |
| S5_k141_50909 | 1650 | No | NA | 1 | 0 | 1 | Low-quality | Genome-fragment | 0.43 | AAI-based (high-confidence) | 0 |
| S5-final-viral-combined_k141_146297\|\|full | 1394 | No | NA | 4 | 0 | 1 | Not-determined | Genome-fragment | NA | NA | 0 |
| S7-final-viral-combined_k141_164832\|\|full | 1650 | No | NA | 5 | 1 | 0 | Low-quality | Genome-fragment | 0.67 | HMM-based (lower-bound) | 0 |
| S5-final-viral-combined_k141_199135\|\|full | 32729 | No | NA | 48 | 16 | 0 | Medium-quality | Genome-fragment | 80.45 | AAI-based (high-confidence) | 0 |
| S7-final-viral-combined_k141_17943\|\|full | 1650 | No | NA | 2 | 0 | 0 | Low-quality | Genome-fragment | 0.72 | AAI-based (medium-confidence) | 0 |
| S12-final-viral-combined_k141_303550\|\|full | 1650 | No | NA | 3 | 0 | 1 | Low-quality | Genome-fragment | 3.17 | AAI-based (high-confidence) | 0 |
| S5-final-viral-combined_k141_154804\|\|full | 4341 | No | NA | 6 | 0 | 0 | Not-determined | Genome-fragment | NA | NA | 0 |
| S12-final-viral-combined_k141_36753\|\|full | 1650 | No | NA | 4 | 0 | 3 | Low-quality | Genome-fragment | 0.54 | AAI-based (medium-confidence) | 0 |
| S3-final-viral-combined_k141_62345\|\|full | 1651 | No | NA | 4 | 3 | 0 | Low-quality | Genome-fragment | 2.76 | AAI-based (high-confidence) | 0 |
| S7-final-viral-combined_k141_26896\|\|full | 1651 | No | NA | 3 | 0 | 0 | Low-quality | Genome-fragment | 2.9 | AAI-based (high-confidence) | 0 |
| S12-final-viral-combined_k141_258339\|\|full | 1651 | No | NA | 3 | 1 | 0 | Low-quality | Genome-fragment | 3.49 | AAI-based (high-confidence) | 0 |
| S5-final-viral-combined_k141_29750\|\|full | 1566 | No | NA | 3 | 0 | 0 | Not-determined | Genome-fragment | NA | NA | 0 |
| S5-final-viral-combined_k141_136845\|\|full | 1567 | No | NA | 3 | 0 | 1 | Not-determined | Genome-fragment | NA | NA | 0 |
| S5-final-viral-combined_k141_173266\|\|full | 5636 | No | NA | 7 | 0 | 7 | Not-determined | Genome-fragment | NA | NA | 0 |
| S2-final-viral-combined_k141_134964\|\|full | 1651 | No | NA | 3 | 2 | 0 | Low-quality | Genome-fragment | 2.99 | AAI-based (medium-confidence) | 0 |
| S5-final-viral-combined_k141_101638\|\|full | 5876 | No | NA | 7 | 0 | 7 | Not-determined | Genome-fragment | NA | NA | 0 |
| S5-final-viral-combined_k141_146354\|\|full | 4350 | No | NA | 8 | 0 | 0 | Not-determined | Genome-fragment | NA | NA | 0 |
| S5-final-viral-combined_k141_199164\|\|full | 2530 | No | NA | 4 | 0 | 2 | Not-determined | Genome-fragment | NA | NA | 0 |
| S11-final-viral-combined_k141_163296\|\|full | 1652 | No | NA | 3 | 0 | 0 | Low-quality | Genome-fragment | 0.84 | AAI-based (high-confidence) | 0 |
| S1-final-viral-combined_k141_106220\|\|full | 1652 | No | NA | 5 | 1 | 0 | Low-quality | Genome-fragment | 4.59 | AAI-based (high-confidence) | 0 |
| S5-final-viral-combined_k141_74945\|\|full | 2036 | No | NA | 2 | 0 | 2 | Not-determined | Genome-fragment | NA | NA | 0 |
| S5-final-viral-combined_k141_173312\|\|full | 7995 | No | NA | 5 | 0 | 4 | Not-determined | Genome-fragment | NA | NA | 0 |
| S5-final-viral-combined_k141_4687\|\|full | 1288 | No | NA | 3 | 0 | 1 | Not-determined | Genome-fragment | NA | NA | 0 |
| S5-final-viral-combined_k141_199195\|\|full | 1723 | No | NA | 3 | 0 | 0 | Not-determined | Genome-fragment | NA | NA | 0 |
| S17-final-viral-combined_k141_223815\|\|lt2gene | 1652 | No | NA | 2 | 0 | 0 | Low-quality | Genome-fragment | 1.92 | AAI-based (medium-confidence) | 0 |
| S5-final-viral-combined_k141_190771\|\|full | 1066 | No | NA | 3 | 0 | 0 | Not-determined | Genome-fragment | NA | NA | 0 |
| S5-final-viral-combined_k141_118985\|\|full | 2334 | No | NA | 4 | 0 | 4 | Not-determined | Genome-fragment | NA | NA | 0 |
| S5-final-viral-combined_k141_101745\|\|full | 1366 | No | NA | 2 | 0 | 1 | Not-determined | Genome-fragment | NA | NA | 0 |
| S1-final-viral-combined_k141_162741\|\|full | 1653 | No | NA | 4 | 2 | 0 | Low-quality | Genome-fragment | 3.56 | AAI-based (high-confidence) | 0 |
| S6-final-viral-combined_k141_123233\|\|full | 1653 | No | NA | 2 | 0 | 2 | Low-quality | Genome-fragment | 0.53 | AAI-based (medium-confidence) | 0 |
| S5-final-viral-combined_k141_92743\|\|full | 3054 | No | NA | 4 | 0 | 1 | Not-determined | Genome-fragment | NA | NA | 0 |
| S5-final-viral-combined_k141_65573\|\|full | 1752 | No | NA | 3 | 0 | 2 | Not-determined | Genome-fragment | NA | NA | 0 |
| S1-final-viral-combined_k141_18149\|\|full | 1654 | No | NA | 4 | 2 | 0 | Low-quality | Genome-fragment | 4.21 | AAI-based (medium-confidence) | 0 |
| S5-final-viral-combined_k141_56493\|\|full | 1934 | No | NA | 4 | 0 | 1 | Not-determined | Genome-fragment | NA | NA | 0 |
| S5-final-viral-combined_k141_20709\|\|full | 1044 | No | NA | 3 | 0 | 0 | Not-determined | Genome-fragment | NA | NA | 0 |
| S5-final-viral-combined_k141_4760\|\|full | 1352 | No | NA | 4 | 0 | 0 | Not-determined | Genome-fragment | NA | NA | 0 |
| S5-final-viral-combined_k141_83370\|\|full | 55883 | Yes | 32897 | 68 | 8 | 13 | Medium-quality | Genome-fragment | 63.84 | AAI-based (high-confidence) | 41.13 |
| S1_k141_76249 | 1654 | No | NA | 5 | 2 | 0 | Low-quality | Genome-fragment | 2.95 | AAI-based (high-confidence) | 0 |
| S5-final-viral-combined_k141_75350\|\|full | 1654 | No | NA | 4 | 1 | 0 | Low-quality | Genome-fragment | 2.8 | HMM-based (lower-bound) | 0 |
| S17-final-viral-combined_k141_144815\|\|full | 1654 | No | NA | 2 | 0 | 1 | Low-quality | Genome-fragment | 2.65 | AAI-based (medium-confidence) | 0 |
| S5-final-viral-combined_k141_38875\|\|full | 1104 | No | NA | 3 | 0 | 1 | Not-determined | Genome-fragment | NA | NA | 0 |
| S17-final-viral-combined_k141_174157\|\|full | 1654 | No | NA | 4 | 1 | 0 | Low-quality | Genome-fragment | 3.32 | AAI-based (high-confidence) | 0 |
| S11-final-viral-combined_k141_212048\|\|full | 1655 | No | NA | 4 | 0 | 0 | Low-quality | Genome-fragment | 3.69 | AAI-based (high-confidence) | 0 |
| S5-final-viral-combined_k141_12295\|\|full | 28602 | No | NA | 42 | 0 | 6 | Not-determined | Genome-fragment | NA | NA | 0 |
| S5-final-viral-combined_k141_83412\|\|full | 29391 | No | NA | 40 | 14 | 1 | Medium-quality | Genome-fragment | 75.69 | AAI-based (medium-confidence) | 0 |
| S5-final-viral-combined_k141_199284\|\|full | 1355 | No | NA | 3 | 0 | 2 | Not-determined | Genome-fragment | NA | NA | 0 |
| S3-final-viral-combined_k141_53780\|\|full | 1655 | No | NA | 6 | 0 | 0 | Low-quality | Genome-fragment | 3.36 | AAI-based (high-confidence) | 0 |
| S5-final-viral-combined_k141_83458\|\|full | 1552 | No | NA | 3 | 0 | 0 | Not-determined | Genome-fragment | NA | NA | 0 |
| S5-final-viral-combined_k141_119091\|\|full | 1596 | No | NA | 2 | 0 | 1 | Not-determined | Genome-fragment | NA | NA | 0 |
| S5-final-viral-combined_k141_4834\|\|full | 3314 | No | NA | 5 | 0 | 1 | Not-determined | Genome-fragment | NA | NA | 0 |
| S3_k141_22540 | 1655 | No | NA | 1 | 0 | 1 | Low-quality | Genome-fragment | 3.52 | AAI-based (medium-confidence) | 0 |
| S17-final-viral-combined_k141_245398\|\|full | 1655 | No | NA | 2 | 0 | 0 | Low-quality | Genome-fragment | 3.2 | AAI-based (medium-confidence) | 0 |
| S12-final-viral-combined_k141_240337\|\|full | 1655 | No | NA | 5 | 1 | 0 | Low-quality | Genome-fragment | 2.69 | AAI-based (high-confidence) | 0 |
| S11-final-viral-combined_k141_245933\|\|full | 1656 | No | NA | 3 | 0 | 0 | Low-quality | Genome-fragment | 3.75 | AAI-based (medium-confidence) | 0 |
| S1-final-viral-combined_k141_161251\|\|full | 1656 | No | NA | 3 | 0 | 0 | Low-quality | Genome-fragment | 1.4 | AAI-based (medium-confidence) | 0 |
| S5-final-viral-combined_k141_65673\|\|full | 3578 | No | NA | 4 | 0 | 3 | Not-determined | Genome-fragment | NA | NA | 0 |
| S1-final-viral-combined_k141_374161\|\|lt2gene | 1656 | No | NA | 2 | 1 | 0 | Low-quality | Genome-fragment | 3.15 | HMM-based (lower-bound) | 0 |
| S5-final-viral-combined_k141_79420\|\|full | 1656 | No | NA | 2 | 0 | 1 | Low-quality | Genome-fragment | 2.34 | AAI-based (high-confidence) | 0 |
| S5-final-viral-combined_k141_173414\|\|full | 9747 | No | NA | 10 | 0 | 4 | Not-determined | Genome-fragment | NA | NA | 0 |
| S17-final-viral-combined_k141_57326\|\|lt2gene | 1656 | No | NA | 2 | 1 | 0 | Low-quality | Genome-fragment | 3.95 | AAI-based (medium-confidence) | 0 |
| S5-final-viral-combined_k141_75140\|\|full | 3031 | No | NA | 3 | 0 | 2 | Not-determined | Genome-fragment | NA | NA | 0 |
| S5-final-viral-combined_k141_146437\|\|full | 3245 | No | NA | 2 | 0 | 2 | Not-determined | Genome-fragment | NA | NA | 0 |
| S5-final-viral-combined_k141_208326\|\|full | 2993 | No | NA | 2 | 0 | 1 | Not-determined | Genome-fragment | NA | NA | 0 |
| S5-final-viral-combined_k141_83541\|\|full | 1551 | No | NA | 3 | 0 | 0 | Not-determined | Genome-fragment | NA | NA | 0 |
| S2_k141_27333 | 1656 | No | NA | 2 | 0 | 0 | Low-quality | Genome-fragment | 3.73 | AAI-based (high-confidence) | 0 |
| S5-final-viral-combined_k141_110057\|\|full | 1868 | No | NA | 5 | 0 | 0 | Not-determined | Genome-fragment | NA | NA | 0 |
| S6-final-viral-combined_k141_7283\|\|full | 1656 | No | NA | 3 | 3 | 0 | Low-quality | Genome-fragment | 2.06 | AAI-based (high-confidence) | 0 |
| S8-final-viral-combined_k141_198976\|\|full | 1656 | No | NA | 2 | 0 | 2 | Low-quality | Genome-fragment | 2.87 | AAI-based (medium-confidence) | 0 |
| S3-final-viral-combined_k141_41644\|\|full | 1657 | No | NA | 3 | 0 | 0 | Low-quality | Genome-fragment | 2.83 | AAI-based (high-confidence) | 0 |
| S5-final-viral-combined_k141_208373\|\|full | 2947 | No | NA | 4 | 0 | 0 | Not-determined | Genome-fragment | NA | NA | 0 |
| S3-final-viral-combined_k141_105633\|\|full | 1657 | No | NA | 2 | 0 | 0 | Low-quality | Genome-fragment | 1.04 | AAI-based (high-confidence) | 0 |
| S5-final-viral-combined_k141_38975\|\|full | 1989 | No | NA | 3 | 0 | 2 | Not-determined | Genome-fragment | NA | NA | 0 |
| S7-final-viral-combined_k141_236563\|\|full | 1657 | No | NA | 3 | 2 | 0 | Low-quality | Genome-fragment | 2.43 | HMM-based (lower-bound) | 0 |
| S5-final-viral-combined_k141_101877\|\|full | 2242 | No | NA | 4 | 0 | 0 | Not-determined | Genome-fragment | NA | NA | 0 |
| S5-final-viral-combined_k141_101880\|\|full | 2024 | No | NA | 3 | 0 | 1 | Not-determined | Genome-fragment | NA | NA | 0 |
| S5-final-viral-combined_k141_163900\|\|full | 1501 | No | NA | 3 | 0 | 2 | Not-determined | Genome-fragment | NA | NA | 0 |
| S12-final-viral-combined_k141_342952\|\|full | 1657 | No | NA | 2 | 0 | 2 | Low-quality | Genome-fragment | 0.53 | AAI-based (medium-confidence) | 0 |
| S5-final-viral-combined_k141_146498\|\|full | 3031 | No | NA | 2 | 0 | 1 | Not-determined | Genome-fragment | NA | NA | 0 |
| S2-final-viral-combined_k141_40852\|\|full | 1657 | No | NA | 2 | 0 | 0 | Low-quality | Genome-fragment | 3.85 | AAI-based (high-confidence) | 0 |
| S5-final-viral-combined_k141_4998\|\|full | 2207 | No | NA | 3 | 0 | 3 | Not-determined | Genome-fragment | NA | NA | 0 |
| S5-final-viral-combined_k141_5021\|\|full | 1939 | No | NA | 3 | 0 | 2 | Not-determined | Genome-fragment | NA | NA | 0 |
| S5-final-viral-combined_k141_29882\|\|full | 5075 | No | NA | 8 | 0 | 1 | Not-determined | Genome-fragment | NA | NA | 0 |
| S5-final-viral-combined_k141_146544\|\|full | 1719 | No | NA | 3 | 0 | 2 | Not-determined | Genome-fragment | NA | NA | 0 |
| S5-final-viral-combined_k141_29897\|\|full | 1448 | No | NA | 2 | 0 | 0 | Not-determined | Genome-fragment | NA | NA | 0 |
| S5-final-viral-combined_k141_83620\|\|full | 1570 | No | NA | 2 | 0 | 0 | Not-determined | Genome-fragment | NA | NA | 0 |
| S5-final-viral-combined_k141_137021\|\|full | 8932 | No | NA | 13 | 0 | 3 | Not-determined | Genome-fragment | NA | NA | 0 |
| S5-final-viral-combined_k141_56781\|\|full | 1636 | No | NA | 2 | 0 | 0 | Not-determined | Genome-fragment | NA | NA | 0 |
| S8-final-viral-combined_k141_274407\|\|lt2gene | 1657 | No | NA | 1 | 1 | 0 | Low-quality | Genome-fragment | 4.62 | AAI-based (high-confidence) | 0 |
| S5-final-viral-combined_k141_56782\|\|full | 1782 | No | NA | 4 | 0 | 2 | Not-determined | Genome-fragment | NA | NA | 0 |
| S9-final-viral-combined_k141_75120\|\|full | 1658 | No | NA | 3 | 0 | 1 | Low-quality | Genome-fragment | 4.83 | AAI-based (high-confidence) | 0 |
| S9-final-viral-combined_k141_30468\|\|full | 1658 | No | NA | 4 | 0 | 0 | Low-quality | Genome-fragment | 4.17 | AAI-based (high-confidence) | 0 |
| S5-final-viral-combined_k141_39039\|\|full | 1446 | No | NA | 3 | 0 | 2 | Not-determined | Genome-fragment | NA | NA | 0 |
| S11_k141_10765 | 1658 | No | NA | 2 | 0 | 0 | Low-quality | Genome-fragment | 2.77 | AAI-based (medium-confidence) | 0 |
| S11_k141_109163 | 1658 | No | NA | 3 | 0 | 1 | Low-quality | Genome-fragment | 4.23 | AAI-based (high-confidence) | 0 |
| S5-final-viral-combined_k141_208457\|\|full | 2457 | No | NA | 4 | 0 | 3 | Not-determined | Genome-fragment | NA | NA | 0 |
| S5-final-viral-combined_k141_48739\|\|full | 4709 | No | NA | 4 | 0 | 3 | Not-determined | Genome-fragment | NA | NA | 0 |
| S5-final-viral-combined_k141_5081\|\|full | 1053 | No | NA | 2 | 0 | 0 | Not-determined | Genome-fragment | NA | NA | 0 |
| S5-final-viral-combined_k141_56879\|\|full | 1431 | No | NA | 2 | 0 | 1 | Not-determined | Genome-fragment | NA | NA | 0 |
| S5-final-viral-combined_k141_191077\|\|full | 3953 | No | NA | 3 | 0 | 2 | Not-determined | Genome-fragment | NA | NA | 0 |
| S5-final-viral-combined_k141_39097\|\|full | 3382 | No | NA | 3 | 0 | 0 | Not-determined | Genome-fragment | NA | NA | 0 |
| S5-final-viral-combined_k141_137100\|\|full | 1776 | No | NA | 2 | 0 | 1 | Not-determined | Genome-fragment | NA | NA | 0 |
| S7-final-viral-combined_k141_2204\|\|full | 1658 | No | NA | 2 | 0 | 0 | Low-quality | Genome-fragment | 9.28 | AAI-based (high-confidence) | 0 |
| S17-final-viral-combined_k141_106947\|\|full | 1658 | No | NA | 3 | 0 | 0 | Low-quality | Genome-fragment | 2.09 | AAI-based (medium-confidence) | 0 |
| S5-final-viral-combined_k141_154937\|\|full | 3410 | No | NA | 5 | 0 | 1 | Not-determined | Genome-fragment | NA | NA | 0 |
| S5-final-viral-combined_k141_199459\|\|full | 1875 | No | NA | 4 | 0 | 0 | Not-determined | Genome-fragment | NA | NA | 0 |
| S12_k141_242187 | 1658 | No | NA | 1 | 0 | 1 | Low-quality | Genome-fragment | 0.4 | AAI-based (medium-confidence) | 0 |
| S5-final-viral-combined_k141_119438\|\|full | 3096 | No | NA | 3 | 0 | 1 | Not-determined | Genome-fragment | NA | NA | 0 |
| S4-final-viral-combined_k141_110997\|\|full | 1658 | No | NA | 2 | 0 | 2 | Low-quality | Genome-fragment | 3.73 | AAI-based (medium-confidence) | 0 |
| S6_k141_28418 | 1658 | No | NA | 5 | 1 | 0 | Low-quality | Genome-fragment | 2.61 | AAI-based (high-confidence) | 0 |
| S5-final-viral-combined_k141_119465\|\|full | 6063 | No | NA | 3 | 0 | 2 | Not-determined | Genome-fragment | NA | NA | 0 |
| S5-final-viral-combined_k141_102167\|\|full | 2813 | No | NA | 4 | 0 | 0 | Not-determined | Genome-fragment | NA | NA | 0 |
| S8-final-viral-combined_k141_96216\|\|full | 1658 | No | NA | 2 | 0 | 2 | Low-quality | Genome-fragment | 2.02 | AAI-based (medium-confidence) | 0 |
| S5-final-viral-combined_k141_83720\|\|full | 1546 | No | NA | 3 | 0 | 1 | Not-determined | Genome-fragment | NA | NA | 0 |
| S5-final-viral-combined_k141_92984\|\|full | 1249 | No | NA | 2 | 0 | 1 | Not-determined | Genome-fragment | NA | NA | 0 |
| S5-final-viral-combined_k141_181970\|\|full | 7557 | No | NA | 2 | 0 | 1 | Not-determined | Genome-fragment | NA | NA | 0 |
| S5-final-viral-combined_k141_66017\|\|full | 32630 | No | NA | 58 | 19 | 0 | Medium-quality | Genome-fragment | 79.35 | AAI-based (medium-confidence) | 0 |
| S8_k141_117090 | 1658 | No | NA | 1 | 0 | 1 | Low-quality | Genome-fragment | 0.45 | AAI-based (medium-confidence) | 0 |
| S8_k141_252220 | 1658 | No | NA | 3 | 1 | 0 | Low-quality | Genome-fragment | 1.6 | AAI-based (high-confidence) | 0 |
| S5-final-viral-combined_k141_128867\|\|full | 1411 | No | NA | 2 | 0 | 1 | Not-determined | Genome-fragment | NA | NA | 0 |
| S5-final-viral-combined_k141_102186\|\|full | 1956 | No | NA | 2 | 0 | 2 | Not-determined | Genome-fragment | NA | NA | 0 |
| S9-final-viral-combined_k141_50977\|\|full | 1659 | No | NA | 3 | 2 | 0 | Low-quality | Genome-fragment | 3.48 | AAI-based (high-confidence) | 0 |
| S5-final-viral-combined_k141_83774\|\|full | 1709 | No | NA | 3 | 0 | 1 | Not-determined | Genome-fragment | NA | NA | 0 |
| S5-final-viral-combined_k141_182017\|\|full | 6939 | No | NA | 9 | 0 | 4 | Not-determined | Genome-fragment | NA | NA | 0 |
| S5-final-viral-combined_k141_93063\|\|full | 7497 | No | NA | 6 | 0 | 4 | Not-determined | Genome-fragment | NA | NA | 0 |
| S9-final-viral-combined_k141_121357\|\|full | 1659 | No | NA | 3 | 2 | 0 | Low-quality | Genome-fragment | 4.13 | AAI-based (high-confidence) | 0 |
| S5-final-viral-combined_k141_199627\|\|full | 2216 | No | NA | 2 | 0 | 0 | Not-determined | Genome-fragment | NA | NA | 0 |
| S5-final-viral-combined_k141_48918\|\|full | 1898 | No | NA | 3 | 0 | 1 | Not-determined | Genome-fragment | NA | NA | 0 |
| S5-final-viral-combined_k141_199634\|\|full | 1801 | No | NA | 2 | 0 | 1 | Not-determined | Genome-fragment | NA | NA | 0 |
| S5-final-viral-combined_k141_29947\|\|full | 3629 | No | NA | 4 | 0 | 1 | Not-determined | Genome-fragment | NA | NA | 0 |
| S11-final-viral-combined_k141_217552\|\|full | 1659 | No | NA | 2 | 0 | 1 | Low-quality | Genome-fragment | 0.75 | AAI-based (high-confidence) | 0 |
| S5-final-viral-combined_k141_199639\|\|full | 1901 | No | NA | 4 | 0 | 0 | Not-determined | Genome-fragment | NA | NA | 0 |
| S5-final-viral-combined_k141_57101\|\|full | 1809 | No | NA | 3 | 0 | 0 | Not-determined | Genome-fragment | NA | NA | 0 |
| S1-final-viral-combined_k141_79320\|\|full | 1659 | No | NA | 6 | 1 | 0 | Low-quality | Genome-fragment | 5.11 | AAI-based (high-confidence) | 0 |
| S5-final-viral-combined_k141_12638\|\|full | 2377 | No | NA | 3 | 0 | 0 | Not-determined | Genome-fragment | NA | NA | 0 |
| S5-final-viral-combined_k141_48953\|\|full | 1765 | No | NA | 2 | 0 | 0 | Not-determined | Genome-fragment | NA | NA | 0 |
| S5-final-viral-combined_k141_128973\|\|full | 1164 | No | NA | 3 | 0 | 1 | Not-determined | Genome-fragment | NA | NA | 0 |
| S7-final-viral-combined_k141_205164\|\|full | 1659 | No | NA | 4 | 0 | 0 | Low-quality | Genome-fragment | 4.36 | AAI-based (high-confidence) | 0 |
| S7-final-viral-combined_k141_240944\|\|full | 1659 | No | NA | 3 | 0 | 0 | Low-quality | Genome-fragment | 3.21 | AAI-based (medium-confidence) | 0 |
| S5-final-viral-combined_k141_146831\|\|full | 2612 | No | NA | 2 | 0 | 1 | Not-determined | Genome-fragment | NA | NA | 0 |
| S5-final-viral-combined_k141_29964\|\|full | 6544 | No | NA | 7 | 0 | 5 | Not-determined | Genome-fragment | NA | NA | 0 |
| S12-final-viral-combined_k141_63727\|\|full | 1659 | No | NA | 3 | 0 | 0 | Low-quality | Genome-fragment | 4.26 | AAI-based (high-confidence) | 0 |
| S12_k141_68216 | 1659 | No | NA | 3 | 0 | 1 | Low-quality | Genome-fragment | 2.17 | AAI-based (medium-confidence) | 0 |
| S5-final-viral-combined_k141_29990\|\|full | 2662 | No | NA | 5 | 0 | 4 | Not-determined | Genome-fragment | NA | NA | 0 |
| S5-final-viral-combined_k141_66097\|\|full | 1739 | No | NA | 2 | 0 | 2 | Not-determined | Genome-fragment | NA | NA | 0 |
| S5-final-viral-combined_k141_21107\|\|full | 2037 | No | NA | 3 | 0 | 1 | Not-determined | Genome-fragment | NA | NA | 0 |
| S5-final-viral-combined_k141_66117\|\|full | 1519 | No | NA | 3 | 0 | 0 | Not-determined | Genome-fragment | NA | NA | 0 |
| S2-final-viral-combined_k141_45357\|\|full | 1659 | No | NA | 4 | 0 | 0 | Low-quality | Genome-fragment | 3.91 | AAI-based (high-confidence) | 0 |
| S11-final-viral-combined_k141_20769\|\|full | 1660 | No | NA | 5 | 1 | 0 | Low-quality | Genome-fragment | 2.58 | AAI-based (high-confidence) | 0 |
| S5-final-viral-combined_k141_137237\|\|full | 2691 | No | NA | 2 | 0 | 1 | Not-determined | Genome-fragment | NA | NA | 0 |
| S1_k141_393719 | 1660 | No | NA | 1 | 0 | 0 | Low-quality | Genome-fragment | 0.73 | AAI-based (medium-confidence) | 0 |
| S17-final-viral-combined_k141_4369\|\|full | 1660 | No | NA | 3 | 0 | 0 | Low-quality | Genome-fragment | 1.54 | AAI-based (high-confidence) | 0 |
| S17-final-viral-combined_k141_96139\|\|full | 1660 | No | NA | 4 | 4 | 0 | Low-quality | Genome-fragment | 4.16 | AAI-based (high-confidence) | 0 |
| S5-final-viral-combined_k141_5391\|\|full | 2180 | No | NA | 2 | 0 | 2 | Not-determined | Genome-fragment | NA | NA | 0 |
| S5-final-viral-combined_k141_110454\|\|full | 1081 | No | NA | 2 | 0 | 0 | Not-determined | Genome-fragment | NA | NA | 0 |
| S17-k141_58460 | 1660 | No | NA | 1 | 0 | 1 | Low-quality | Genome-fragment | 0.72 | AAI-based (medium-confidence) | 0 |
| S5-final-viral-combined_k141_102290\|\|full | 2336 | No | NA | 3 | 0 | 2 | Not-determined | Genome-fragment | NA | NA | 0 |
| S5-final-viral-combined_k141_12807\|\|full | 1382 | No | NA | 3 | 0 | 1 | Not-determined | Genome-fragment | NA | NA | 0 |
| S12-final-viral-combined_k141_124706\|\|full | 1660 | No | NA | 2 | 0 | 1 | Low-quality | Genome-fragment | 2.11 | AAI-based (high-confidence) | 0 |
| S5-final-viral-combined_k141_75560\|\|full | 2515 | No | NA | 3 | 0 | 0 | Not-determined | Genome-fragment | NA | NA | 0 |
| S5-final-viral-combined_k141_57186\|\|full | 4549 | No | NA | 5 | 0 | 2 | Not-determined | Genome-fragment | NA | NA | 0 |
| S12_k141_396478 | 1660 | No | NA | 3 | 0 | 0 | Low-quality | Genome-fragment | 2.64 | AAI-based (high-confidence) | 0 |
| S6-final-viral-combined_k141_123370\|\|full | 1660 | No | NA | 3 | 2 | 0 | Low-quality | Genome-fragment | 3.14 | AAI-based (medium-confidence) | 0 |
| S5-final-viral-combined_k141_49045\|\|full | 1729 | No | NA | 3 | 0 | 1 | Not-determined | Genome-fragment | NA | NA | 0 |
| S5-final-viral-combined_k141_137379\|\|full | 15509 | No | NA | 17 | 0 | 12 | Not-determined | Genome-fragment | NA | NA | 0 |
| S8-final-viral-combined_k141_276313\|\|full | 1660 | No | NA | 3 | 0 | 0 | Low-quality | Genome-fragment | 2.38 | AAI-based (medium-confidence) | 0 |
| S5-final-viral-combined_k141_155177\|\|full | 1584 | No | NA | 2 | 0 | 1 | Not-determined | Genome-fragment | NA | NA | 0 |
| S11-final-viral-combined_k141_153796\|\|full | 1661 | No | NA | 3 | 0 | 0 | Low-quality | Genome-fragment | 2.01 | AAI-based (medium-confidence) | 0 |
| S5-final-viral-combined_k141_93192\|\|full | 1000 | No | NA | 3 | 0 | 0 | Not-determined | Genome-fragment | NA | NA | 0 |
| S7-final-viral-combined_k141_75554\|\|full | 1661 | No | NA | 2 | 0 | 1 | Low-quality | Genome-fragment | 3.19 | AAI-based (medium-confidence) | 0 |
| S5-final-viral-combined_k141_30098\|\|full | 6194 | No | NA | 9 | 0 | 4 | Not-determined | Genome-fragment | NA | NA | 0 |
| S5-final-viral-combined_k141_110551\|\|full | 1232 | No | NA | 4 | 0 | 0 | Not-determined | Genome-fragment | NA | NA | 0 |
| S17-final-viral-combined_k141_186437\|\|full | 1661 | No | NA | 3 | 0 | 2 | Low-quality | Genome-fragment | 0.95 | AAI-based (medium-confidence) | 0 |
| S17-final-viral-combined_k141_144060\|\|full | 1661 | No | NA | 3 | 0 | 2 | Low-quality | Genome-fragment | 2.68 | AAI-based (medium-confidence) | 0 |
| S5-final-viral-combined_k141_66283\|\|full | 2409 | No | NA | 3 | 0 | 0 | Not-determined | Genome-fragment | NA | NA | 0 |
| S5-final-viral-combined_k141_39456\|\|full | 1933 | No | NA | 4 | 0 | 2 | Not-determined | Genome-fragment | NA | NA | 0 |
| S5-final-viral-combined_k141_84061\|\|full | 4428 | No | NA | 5 | 0 | 3 | Not-determined | Genome-fragment | NA | NA | 0 |
| S5-final-viral-combined_k141_146897\|\|full | 1772 | No | NA | 3 | 0 | 1 | Not-determined | Genome-fragment | NA | NA | 0 |
| S17-final-viral-combined_k141_254409\|\|full | 1661 | No | NA | 3 | 0 | 0 | Low-quality | Genome-fragment | 2.27 | AAI-based (high-confidence) | 0 |
| S12-final-viral-combined_k141_103826\|\|full | 1661 | No | NA | 4 | 0 | 0 | Low-quality | Genome-fragment | 5.08 | AAI-based (high-confidence) | 0 |
| S5-final-viral-combined_k141_164301\|\|full | 1858 | No | NA | 3 | 0 | 0 | Not-determined | Genome-fragment | NA | NA | 0 |
| S12-final-viral-combined_k141_307595\|\|full | 1661 | No | NA | 3 | 0 | 0 | Low-quality | Genome-fragment | 0.85 | AAI-based (medium-confidence) | 0 |
| S8_k141_218006 | 1661 | No | NA | 2 | 0 | 0 | Low-quality | Genome-fragment | 1.19 | AAI-based (medium-confidence) | 0 |
| S5-final-viral-combined_k141_137523\|\|full | 4321 | No | NA | 10 | 0 | 1 | Not-determined | Genome-fragment | NA | NA | 0 |
| S11-final-viral-combined_k141_124957\|\|full | 1662 | No | NA | 2 | 0 | 1 | Low-quality | Genome-fragment | 2.67 | AAI-based (medium-confidence) | 0 |
| S11-final-viral-combined_k141_153490\|\|full | 1662 | No | NA | 3 | 1 | 0 | Low-quality | Genome-fragment | 3.67 | AAI-based (medium-confidence) | 0 |
| S5-final-viral-combined_k141_13053\|\|full | 1914 | No | NA | 3 | 0 | 2 | Not-determined | Genome-fragment | NA | NA | 0 |
| S5-final-viral-combined_k141_93265\|\|full | 2252 | No | NA | 2 | 0 | 1 | Not-determined | Genome-fragment | NA | NA | 0 |
| S5-final-viral-combined_k141_159212\|\|full | 1662 | No | NA | 2 | 0 | 0 | Low-quality | Genome-fragment | 1.18 | AAI-based (high-confidence) | 0 |
| S17-final-viral-combined_k141_255778\|\|full | 1662 | No | NA | 3 | 0 | 0 | Low-quality | Genome-fragment | 3.09 | AAI-based (medium-confidence) | 0 |
| S5-final-viral-combined_k141_13076\|\|full | 2171 | No | NA | 3 | 0 | 0 | Not-determined | Genome-fragment | NA | NA | 0 |
| S5-final-viral-combined_k141_173923\|\|full | 7686 | No | NA | 13 | 0 | 0 | Not-determined | Genome-fragment | NA | NA | 0 |
| S5-final-viral-combined_k141_75624\|\|full | 1688 | No | NA | 4 | 0 | 3 | Not-determined | Genome-fragment | NA | NA | 0 |
| S5-final-viral-combined_k141_49082\|\|full | 2934 | No | NA | 3 | 0 | 2 | Not-determined | Genome-fragment | NA | NA | 0 |
| S5-final-viral-combined_k141_30244\|\|full | 5129 | No | NA | 5 | 0 | 1 | Not-determined | Genome-fragment | NA | NA | 0 |
| S5-final-viral-combined_k141_164404\|\|full | 1431 | No | NA | 3 | 0 | 1 | Not-determined | Genome-fragment | NA | NA | 0 |
| S4-final-viral-combined_k141_162724\|\|full | 1662 | No | NA | 2 | 0 | 1 | Low-quality | Genome-fragment | 2.58 | AAI-based (medium-confidence) | 0 |
| S5-final-viral-combined_k141_164409\|\|full | 4964 | No | NA | 7 | 0 | 5 | Not-determined | Genome-fragment | NA | NA | 0 |
| S8-final-viral-combined_k141_207839\|\|lt2gene | 1662 | No | NA | 2 | 1 | 0 | Low-quality | Genome-fragment | 4.41 | AAI-based (medium-confidence) | 0 |
| S9-final-viral-combined_k141_94499\|\|full | 1663 | No | NA | 3 | 0 | 0 | Low-quality | Genome-fragment | 4.38 | AAI-based (high-confidence) | 0 |
| S5-final-viral-combined_k141_208958\|\|full | 2698 | No | NA | 4 | 0 | 0 | Not-determined | Genome-fragment | NA | NA | 0 |
| S11_k141_72220 | 1663 | No | NA | 1 | 0 | 0 | Low-quality | Genome-fragment | 1.1 | AAI-based (medium-confidence) | 0 |
| S11_k141_72256 | 1663 | No | NA | 1 | 0 | 0 | Low-quality | Genome-fragment | 1.1 | AAI-based (medium-confidence) | 0 |
| S5-final-viral-combined_k141_84201\|\|full | 8120 | No | NA | 11 | 0 | 0 | Not-determined | Genome-fragment | NA | NA | 0 |
| S1-final-viral-combined_k141_209523\|\|full | 1663 | No | NA | 5 | 1 | 0 | Low-quality | Genome-fragment | 4.16 | AAI-based (high-confidence) | 0 |
| S1-final-viral-combined_k141_14432\|\|full | 1663 | No | NA | 2 | 0 | 1 | Low-quality | Genome-fragment | 2.61 | AAI-based (high-confidence) | 0 |
| S5_k141_196092 | 1663 | No | NA | 2 | 1 | 0 | Low-quality | Genome-fragment | 4.82 | AAI-based (high-confidence) | 0 |
| S5_k141_77599 | 1663 | No | NA | 1 | 0 | 1 | Low-quality | Genome-fragment | 0.85 | AAI-based (medium-confidence) | 0 |
| S7-final-viral-combined_k141_173915\|\|full | 1663 | No | NA | 2 | 0 | 1 | Low-quality | Genome-fragment | 0.93 | AAI-based (medium-confidence) | 0 |
| S5-final-viral-combined_k141_129198\|\|full | 2355 | No | NA | 3 | 0 | 0 | Not-determined | Genome-fragment | NA | NA | 0 |
| S5-final-viral-combined_k141_5660\|\|full | 6912 | No | NA | 8 | 0 | 7 | Not-determined | Genome-fragment | NA | NA | 0 |
| S5-final-viral-combined_k141_75735\|\|full | 3491 | No | NA | 3 | 0 | 2 | Not-determined | Genome-fragment | NA | NA | 0 |
| S5-final-viral-combined_k141_174088\|\|full | 1403 | No | NA | 3 | 0 | 2 | Not-determined | Genome-fragment | NA | NA | 0 |
| S17-final-viral-combined_k141_209767\|\|full | 1663 | No | NA | 4 | 0 | 1 | Low-quality | Genome-fragment | 1.08 | AAI-based (medium-confidence) | 0 |
| S4-final-viral-combined_k141_88906\|\|full | 1663 | No | NA | 3 | 0 | 1 | Low-quality | Genome-fragment | 1.23 | AAI-based (medium-confidence) | 0 |
| S5-final-viral-combined_k141_93371\|\|full | 1688 | No | NA | 3 | 0 | 1 | Not-determined | Genome-fragment | NA | NA | 0 |
| S5-final-viral-combined_k141_129253\|\|full | 1815 | No | NA | 2 | 0 | 0 | Not-determined | Genome-fragment | NA | NA | 0 |
| S8-final-viral-combined_k141_24556\|\|full | 1663 | No | NA | 3 | 0 | 0 | Low-quality | Genome-fragment | 2.78 | AAI-based (medium-confidence) | 0 |
| S9-final-viral-combined_k141_122429\|\|full | 1664 | No | NA | 2 | 2 | 0 | Low-quality | Genome-fragment | 4.41 | AAI-based (high-confidence) | 0 |
| S5-final-viral-combined_k141_200069\|\|full | 2338 | No | NA | 3 | 0 | 0 | Not-determined | Genome-fragment | NA | NA | 0 |
| S9-final-viral-combined_k141_72830\|\|lt2gene | 1664 | No | NA | 2 | 2 | 0 | Low-quality | Genome-fragment | 5.08 | AAI-based (high-confidence) | 0 |
| S5-final-viral-combined_k141_110880\|\|full | 3106 | No | NA | 4 | 0 | 1 | Not-determined | Genome-fragment | NA | NA | 0 |
| S11-final-viral-combined_k141_223977\|\|full | 1664 | No | NA | 3 | 0 | 1 | Low-quality | Genome-fragment | 1.02 | AAI-based (medium-confidence) | 0 |
| S1-final-viral-combined_k141_206144\|\|full | 1664 | No | NA | 3 | 1 | 0 | Low-quality | Genome-fragment | 3.1 | AAI-based (high-confidence) | 0 |
| S5-final-viral-combined_k141_93398\|\|full | 1823 | No | NA | 4 | 0 | 0 | Not-determined | Genome-fragment | NA | NA | 0 |
| S5-final-viral-combined_k141_5765\|\|full | 2025 | No | NA | 2 | 0 | 0 | Not-determined | Genome-fragment | NA | NA | 0 |
| S5-final-viral-combined_k141_164463\|\|full | 1214 | No | NA | 2 | 0 | 2 | Not-determined | Genome-fragment | NA | NA | 0 |
| S5-final-viral-combined_k141_46069\|\|full | 1664 | No | NA | 3 | 0 | 0 | Low-quality | Genome-fragment | 1.56 | AAI-based (high-confidence) | 0 |
| S5-final-viral-combined_k141_129296\|\|full | 1271 | No | NA | 2 | 0 | 1 | Not-determined | Genome-fragment | NA | NA | 0 |
| S5-final-viral-combined_k141_182293\|\|full | 1643 | No | NA | 2 | 0 | 1 | Not-determined | Genome-fragment | NA | NA | 0 |
| S12_k141_83456 | 1664 | No | NA | 5 | 1 | 0 | Low-quality | Genome-fragment | 5.12 | AAI-based (high-confidence) | 0 |
| S5-final-viral-combined_k141_5798\|\|full | 1788 | No | NA | 1 | 0 | 1 | Not-determined | Genome-fragment | NA | NA | 0 |
| S5-final-viral-combined_k141_84255\|\|full | 4305 | No | NA | 8 | 0 | 1 | Not-determined | Genome-fragment | NA | NA | 0 |
| S2_k141_171449 | 1664 | No | NA | 4 | 1 | 0 | Low-quality | Genome-fragment | 3.17 | AAI-based (medium-confidence) | 0 |
| S4-final-viral-combined_k141_47987\|\|full | 1664 | No | NA | 2 | 0 | 1 | Low-quality | Genome-fragment | 1.19 | AAI-based (high-confidence) | 0 |
| S5-final-viral-combined_k141_182328\|\|full | 1726 | No | NA | 4 | 0 | 1 | Not-determined | Genome-fragment | NA | NA | 0 |
| S5-final-viral-combined_k141_5813\|\|full | 2367 | No | NA | 3 | 0 | 2 | Not-determined | Genome-fragment | NA | NA | 0 |
| S5-final-viral-combined_k141_5845\|\|full | 5108 | No | NA | 6 | 0 | 3 | Not-determined | Genome-fragment | NA | NA | 0 |
| S5-final-viral-combined_k141_147044\|\|full | 1138 | No | NA | 3 | 0 | 1 | Not-determined | Genome-fragment | NA | NA | 0 |
| S5-final-viral-combined_k141_39714\|\|full | 5000 | No | NA | 5 | 0 | 0 | Not-determined | Genome-fragment | NA | NA | 0 |
| S8_k141_30141 | 1664 | No | NA | 1 | 0 | 0 | Low-quality | Genome-fragment | 2.76 | AAI-based (medium-confidence) | 0 |
| S1-final-viral-combined_k141_132719\|\|full | 1665 | No | NA | 3 | 1 | 0 | Low-quality | Genome-fragment | 3.72 | AAI-based (medium-confidence) | 0 |
| S17-final-viral-combined_k141_238467\|\|full | 1665 | No | NA | 3 | 2 | 0 | Low-quality | Genome-fragment | 3.58 | AAI-based (medium-confidence) | 0 |
| S5-final-viral-combined_k141_147106\|\|full | 1333 | No | NA | 3 | 0 | 2 | Not-determined | Genome-fragment | NA | NA | 0 |
| S17-final-viral-combined_k141_31777\|\|full | 1665 | No | NA | 4 | 2 | 0 | Low-quality | Genome-fragment | 2.94 | AAI-based (high-confidence) | 0 |
| S5-final-viral-combined_k141_39767\|\|full | 2200 | No | NA | 3 | 0 | 0 | Not-determined | Genome-fragment | NA | NA | 0 |
| S5-final-viral-combined_k141_191424\|\|full | 1834 | No | NA | 5 | 0 | 1 | Not-determined | Genome-fragment | NA | NA | 0 |
| S5-final-viral-combined_k141_147142\|\|full | 2203 | No | NA | 7 | 0 | 0 | Not-determined | Genome-fragment | NA | NA | 0 |
| S17-k141_188882 | 1665 | No | NA | 2 | 0 | 2 | Low-quality | Genome-fragment | 1.99 | AAI-based (high-confidence) | 0 |
| S5-final-viral-combined_k141_111052\|\|full | 1875 | No | NA | 3 | 0 | 1 | Not-determined | Genome-fragment | NA | NA | 0 |
| S5-final-viral-combined_k141_84402\|\|full | 2207 | No | NA | 3 | 0 | 2 | Not-determined | Genome-fragment | NA | NA | 0 |
| S4_k141_121278 | 1665 | No | NA | 6 | 0 | 0 | Low-quality | Genome-fragment | 11.65 | AAI-based (high-confidence) | 0 |
| S5-final-viral-combined_k141_111070\|\|full | 2019 | No | NA | 2 | 0 | 2 | Not-determined | Genome-fragment | NA | NA | 0 |
| S8-final-viral-combined_k141_129719\|\|full | 1665 | No | NA | 3 | 0 | 0 | Low-quality | Genome-fragment | 5.14 | AAI-based (high-confidence) | 0 |
| S5-final-viral-combined_k141_66662\|\|full | 98874 | Yes | 43806 | 117 | 16 | 44 | High-quality | High-quality | 100 | AAI-based (high-confidence) | 55.7 |
| S11-final-viral-combined_k141_190155\|\|full | 1666 | No | NA | 3 | 2 | 0 | Low-quality | Genome-fragment | 2.78 | AAI-based (medium-confidence) | 0 |
| S5-final-viral-combined_k141_147234\|\|full | 1769 | No | NA | 3 | 0 | 3 | Not-determined | Genome-fragment | NA | NA | 0 |
| S1-final-viral-combined_k141_313471\|\|full | 1666 | No | NA | 5 | 1 | 0 | Low-quality | Genome-fragment | 4.29 | AAI-based (high-confidence) | 0 |
| S1-final-viral-combined_k141_100701\|\|full | 1666 | No | NA | 3 | 0 | 1 | Low-quality | Genome-fragment | 3.91 | AAI-based (medium-confidence) | 0 |
| S5-final-viral-combined_k141_147252\|\|full | 1130 | No | NA | 2 | 0 | 2 | Not-determined | Genome-fragment | NA | NA | 0 |
| S1-final-viral-combined_k141_7045\|\|full | 1666 | No | NA | 2 | 0 | 0 | Low-quality | Genome-fragment | 3.37 | AAI-based (high-confidence) | 0 |
| S7-final-viral-combined_k141_229998\|\|full | 1666 | No | NA | 3 | 1 | 0 | Low-quality | Genome-fragment | 0.13 | HMM-based (lower-bound) | 0 |
| S5-final-viral-combined_k141_39779\|\|full | 1702 | No | NA | 3 | 0 | 1 | Not-determined | Genome-fragment | NA | NA | 0 |
| S5-final-viral-combined_k141_102923\|\|full | 2169 | No | NA | 3 | 0 | 0 | Not-determined | Genome-fragment | NA | NA | 0 |
| S5-final-viral-combined_k141_66714\|\|full | 1327 | No | NA | 3 | 0 | 2 | Not-determined | Genome-fragment | NA | NA | 0 |
| S7_k141_216631 | 1666 | No | NA | 3 | 1 | 0 | Low-quality | Genome-fragment | 3.58 | AAI-based (high-confidence) | 0 |
| S17-final-viral-combined_k141_25260\|\|full | 1666 | No | NA | 2 | 0 | 0 | Low-quality | Genome-fragment | 2.37 | AAI-based (medium-confidence) | 0 |
| S5-final-viral-combined_k141_93599\|\|full | 1051 | No | NA | 3 | 0 | 0 | Not-determined | Genome-fragment | NA | NA | 0 |
| S5-final-viral-combined_k141_66727\|\|full | 2817 | No | NA | 4 | 0 | 1 | Not-determined | Genome-fragment | NA | NA | 0 |
| S17-k141_44159 | 1666 | No | NA | 2 | 1 | 0 | Low-quality | Genome-fragment | 3.77 | HMM-based (lower-bound) | 0 |
| S12_k141_125442 | 1666 | No | NA | 1 | 0 | 1 | Low-quality | Genome-fragment | 0.62 | AAI-based (high-confidence) | 0 |
| S2-final-viral-combined_k141_176415\|\|full | 1666 | No | NA | 3 | 0 | 1 | Low-quality | Genome-fragment | 4.26 | AAI-based (high-confidence) | 0 |
| S5-final-viral-combined_k141_84525\|\|full | 1294 | No | NA | 4 | 0 | 0 | Not-determined | Genome-fragment | NA | NA | 0 |
| S5-final-viral-combined_k141_147370\|\|full | 1592 | No | NA | 1 | 0 | 1 | Not-determined | Genome-fragment | NA | NA | 0 |
| S2-final-viral-combined_k141_130215\|\|full | 1666 | No | NA | 4 | 0 | 2 | Low-quality | Genome-fragment | 1.22 | AAI-based (medium-confidence) | 0 |
| S5-final-viral-combined_k141_147378\|\|full | 1183 | No | NA | 3 | 0 | 1 | Not-determined | Genome-fragment | NA | NA | 0 |
| S5-final-viral-combined_k141_137615\|\|full | 3981 | No | NA | 3 | 0 | 1 | Not-determined | Genome-fragment | NA | NA | 0 |
| S5-final-viral-combined_k141_119948\|\|full | 1963 | No | NA | 2 | 0 | 0 | Not-determined | Genome-fragment | NA | NA | 0 |
| S5-final-viral-combined_k141_119961\|\|full | 2033 | No | NA | 2 | 0 | 0 | Not-determined | Genome-fragment | NA | NA | 0 |
| S8-final-viral-combined_k141_266710\|\|full | 1666 | No | NA | 4 | 1 | 0 | Low-quality | Genome-fragment | 4.03 | AAI-based (high-confidence) | 0 |
| S5-final-viral-combined_k141_30578\|\|full | 1050 | No | NA | 2 | 0 | 1 | Not-determined | Genome-fragment | NA | NA | 0 |
| S5-final-viral-combined_k141_39835\|\|full | 1109 | No | NA | 2 | 0 | 0 | Not-determined | Genome-fragment | NA | NA | 0 |
| S1_k141_17042 | 1667 | No | NA | 2 | 0 | 0 | Low-quality | Genome-fragment | 0.99 | AAI-based (high-confidence) | 0 |
| S7-final-viral-combined_k141_75201\|\|full | 1667 | No | NA | 2 | 0 | 1 | Low-quality | Genome-fragment | 4.02 | AAI-based (medium-confidence) | 0 |
| S7_k141_74296 | 1667 | No | NA | 2 | 0 | 1 | Low-quality | Genome-fragment | 0.8 | AAI-based (high-confidence) | 0 |
| S5-final-viral-combined_k141_93754\|\|full | 1549 | No | NA | 3 | 0 | 1 | Not-determined | Genome-fragment | NA | NA | 0 |
| S5-final-viral-combined_k141_84609\|\|full | 4321 | No | NA | 4 | 0 | 2 | Not-determined | Genome-fragment | NA | NA | 0 |

| | | | | | | | | | | | |
|---|---|---|---|---|---|---|---|---|---|---|---|
| S7_k141_232133 | 1667 | No | NA | 1 | 0 | 0 | Low-quality | Genome-fragment | 1.76 | AAI-based (high-confidence) | 0 |
| S5-final-viral-combined_k141_21370\|\|full | 2402 | No | NA | 5 | 0 | 0 | Not-determined | Genome-fragment | NA | NA | 0 |
| S5-final-viral-combined_k141_129585\|\|full | 2111 | No | NA | 4 | 0 | 1 | Not-determined | Genome-fragment | NA | NA | 0 |
| S5-final-viral-combined_k141_209423\|\|full | 1346 | No | NA | 3 | 0 | 2 | Not-determined | Genome-fragment | NA | NA | 0 |
| S17-k141_157245 | 1667 | No | NA | 6 | 0 | 0 | Low-quality | Genome-fragment | 3.1 | AAI-based (high-confidence) | 0 |
| S5-final-viral-combined_k141_103051\|\|full | 1018 | No | NA | 2 | 0 | 1 | Not-determined | Genome-fragment | NA | NA | 0 |
| S12-final-viral-combined_k141_276296\|\|full | 1667 | No | NA | 3 | 0 | 2 | Low-quality | Genome-fragment | 0.88 | AAI-based (medium-confidence) | 0 |
| S5-final-viral-combined_k141_120052\|\|full | 3465 | No | NA | 4 | 0 | 4 | Not-determined | Genome-fragment | NA | NA | 0 |
| S5-final-viral-combined_k141_111428\|\|full | 2649 | No | NA | 4 | 0 | 3 | Not-determined | Genome-fragment | NA | NA | 0 |
| S12-final-viral-combined_k141_227853\|\|full | 1667 | No | NA | 3 | 0 | 1 | Low-quality | Genome-fragment | 3.65 | AAI-based (medium-confidence) | 0 |
| S2-final-viral-combined_k141_136495\|\|full | 1667 | No | NA | 4 | 1 | 0 | Low-quality | Genome-fragment | 3.48 | AAI-based (high-confidence) | 0 |
| S5-final-viral-combined_k141_182832\|\|full | 1702 | No | NA | 4 | 0 | 1 | Not-determined | Genome-fragment | NA | NA | 0 |
| S2-final-viral-combined_k141_211936\|\|full | 1667 | No | NA | 2 | 0 | 1 | Low-quality | Genome-fragment | 0.93 | AAI-based (medium-confidence) | 0 |
| S2-final-viral-combined_k141_6501\|\|full | 1667 | No | NA | 1 | 0 | 0 | Low-quality | Genome-fragment | 14.03 | AAI-based (medium-confidence) | 0 |
| S5-final-viral-combined_k141_6244\|\|full | 1094 | No | NA | 3 | 0 | 1 | Not-determined | Genome-fragment | NA | NA | 0 |
| S5-final-viral-combined_k141_49277\|\|full | 1702 | No | NA | 2 | 0 | 0 | Not-determined | Genome-fragment | NA | NA | 0 |
| S5-final-viral-combined_k141_191607\|\|full | 1238 | No | NA | 2 | 0 | 0 | Not-determined | Genome-fragment | NA | NA | 0 |
| S2-final-viral-combined_k141_19277\|\|full | 1667 | No | NA | 3 | 0 | 3 | Low-quality | Genome-fragment | 4.35 | AAI-based (medium-confidence) | 0 |
| S5-final-viral-combined_k141_137805\|\|full | 5110 | No | NA | 6 | 0 | 5 | Not-determined | Genome-fragment | NA | NA | 0 |
| S5-final-viral-combined_k141_84762\|\|full | 2140 | No | NA | 3 | 0 | 2 | Not-determined | Genome-fragment | NA | NA | 0 |
| S5-final-viral-combined_k141_182893\|\|full | 1845 | No | NA | 3 | 0 | 0 | Not-determined | Genome-fragment | NA | NA | 0 |
| S2-final-viral-combined_k141_226214\|\|lt2gene | 1667 | No | NA | 2 | 2 | 0 | Low-quality | Genome-fragment | 4.82 | AAI-based (high-confidence) | 0 |
| S5-final-viral-combined_k141_200395\|\|full | 24928 | No | NA | 24 | 0 | 17 | Not-determined | Genome-fragment | NA | NA | 0 |
| S5-final-viral-combined_k141_155691\|\|full | 1127 | No | NA | 3 | 0 | 1 | Not-determined | Genome-fragment | NA | NA | 0 |
| S4-final-viral-combined_k141_162313\|\|full | 1667 | No | NA | 3 | 0 | 1 | Low-quality | Genome-fragment | 3.05 | AAI-based (medium-confidence) | 0 |
| S1-final-viral-combined_k141_37224\|\|full | 1668 | No | NA | 4 | 1 | 0 | Low-quality | Genome-fragment | 2.22 | AAI-based (high-confidence) | 0 |
| S5-final-viral-combined_k141_39970\|\|full | 1524 | No | NA | 2 | 0 | 2 | Not-determined | Genome-fragment | NA | NA | 0 |
| S1-final-viral-combined_k141_238970\|\|lt2gene | 1668 | No | NA | 2 | 2 | 0 | Low-quality | Genome-fragment | 3.55 | AAI-based (high-confidence) | 0 |
| S5-final-viral-combined_k141_6328\|\|full | 1011 | No | NA | 2 | 0 | 0 | Not-determined | Genome-fragment | NA | NA | 0 |
| S5-final-viral-combined_k141_155705\|\|full | 2570 | No | NA | 3 | 0 | 1 | Not-determined | Genome-fragment | NA | NA | 0 |
| S3-final-viral-combined_k141_65239\|\|full | 1668 | No | NA | 2 | 0 | 0 | Low-quality | Genome-fragment | 1.37 | AAI-based (medium-confidence) | 0 |
| S5-final-viral-combined_k141_84818\|\|full | 1138 | No | NA | 2 | 0 | 1 | Not-determined | Genome-fragment | NA | NA | 0 |
| S12-final-viral-combined_k141_375143\|\|full | 1668 | No | NA | 4 | 1 | 0 | Low-quality | Genome-fragment | 5.81 | AAI-based (high-confidence) | 0 |
| S6-final-viral-combined_k141_24269\|\|full | 1668 | No | NA | 3 | 0 | 0 | Low-quality | Genome-fragment | 3.04 | AAI-based (high-confidence) | 0 |
| S6-final-viral-combined_k141_10843\|\|full | 1668 | No | NA | 2 | 0 | 1 | Low-quality | Genome-fragment | 2.12 | AAI-based (high-confidence) | 0 |
| S8-final-viral-combined_k141_248567\|\|full | 1668 | No | NA | 4 | 1 | 0 | Low-quality | Genome-fragment | 5.17 | AAI-based (high-confidence) | 0 |
| S5-final-viral-combined_k141_155747\|\|full | 6031 | No | NA | 6 | 0 | 4 | Not-determined | Genome-fragment | NA | NA | 0 |
| S5-final-viral-combined_k141_13701\|\|full | 2803 | No | NA | 3 | 0 | 1 | Not-determined | Genome-fragment | NA | NA | 0 |
| S5-final-viral-combined_k141_120069\|\|full | 7247 | No | NA | 8 | 0 | 5 | Not-determined | Genome-fragment | NA | NA | 0 |
| S5-final-viral-combined_k141_164798\|\|full | 1082 | No | NA | 2 | 0 | 1 | Not-determined | Genome-fragment | NA | NA | 0 |
| S5-final-viral-combined_k141_164806\|\|full | 2625 | No | NA | 3 | 0 | 3 | Not-determined | Genome-fragment | NA | NA | 0 |
| S8_k141_158186 | 1668 | No | NA | 1 | 0 | 1 | Low-quality | Genome-fragment | 3.45 | AAI-based (medium-confidence) | 0 |
| S9-final-viral-combined_k141_125766\|\|full | 1669 | Yes | 1130 | 4 | 1 | 1 | Low-quality | Genome-fragment | 1.62 | HMM-based (lower-bound) | 32.29 |
| S9_k141_123558 | 1669 | No | NA | 1 | 0 | 0 | Low-quality | Genome-fragment | 2.79 | AAI-based (high-confidence) | 0 |
| S11-final-viral-combined_k141_298146\|\|full | 1669 | No | NA | 4 | 1 | 0 | Low-quality | Genome-fragment | 4.5 | AAI-based (medium-confidence) | 0 |
| S1-final-viral-combined_k141_317605\|\|full | 1669 | No | NA | 2 | 0 | 0 | Low-quality | Genome-fragment | 4.55 | AAI-based (high-confidence) | 0 |
| S5-final-viral-combined_k141_155821\|\|full | 2175 | No | NA | 2 | 0 | 1 | Not-determined | Genome-fragment | NA | NA | 0 |
| S5-final-viral-combined_k141_13805\|\|full | 5760 | No | NA | 9 | 0 | 1 | Not-determined | Genome-fragment | NA | NA | 0 |
| S5-final-viral-combined_k141_40018\|\|full | 2057 | No | NA | 4 | 0 | 1 | Not-determined | Genome-fragment | NA | NA | 0 |
| S5-final-viral-combined_k141_40026\|\|full | 2462 | No | NA | 4 | 0 | 0 | Not-determined | Genome-fragment | NA | NA | 0 |
| S5-final-viral-combined_k141_174460\|\|full | 8125 | No | NA | 7 | 0 | 4 | Not-determined | Genome-fragment | NA | NA | 0 |
| S5-final-viral-combined_k141_40029\|\|full | 1936 | No | NA | 3 | 0 | 3 | Not-determined | Genome-fragment | NA | NA | 0 |
| S5-final-viral-combined_k141_164828\|\|full | 6096 | No | NA | 15 | 0 | 1 | Not-determined | Genome-fragment | NA | NA | 0 |
| S1-final-viral-combined_k141_210227\|\|full | 1669 | No | NA | 2 | 0 | 1 | Low-quality | Genome-fragment | 1.27 | AAI-based (medium-confidence) | 0 |
| S1_k141_140749 | 1669 | No | NA | 1 | 0 | 1 | Low-quality | Genome-fragment | 0.19 | AAI-based (medium-confidence) | 0 |
| S5-final-viral-combined_k141_120116\|\|full | 1326 | No | NA | 3 | 0 | 1 | Not-determined | Genome-fragment | NA | NA | 0 |
| S5-final-viral-combined_k141_40095\|\|full | 1673 | No | NA | 2 | 0 | 1 | Not-determined | Genome-fragment | NA | NA | 0 |
| S5-final-viral-combined_k141_191823\|\|full | 1980 | No | NA | 2 | 0 | 1 | Not-determined | Genome-fragment | NA | NA | 0 |
| S5-final-viral-combined_k141_40114\|\|full | 8453 | No | NA | 10 | 0 | 3 | Not-determined | Genome-fragment | NA | NA | 0 |
| S5-final-viral-combined_k141_111648\|\|full | 42976 | No | NA | 35 | 0 | 24 | Not-determined | Genome-fragment | NA | NA | 0 |
| S5-final-viral-combined_k141_103228\|\|full | 2930 | No | NA | 4 | 0 | 0 | Not-determined | Genome-fragment | NA | NA | 0 |
| S5-final-viral-combined_k141_182955\|\|full | 1408 | No | NA | 3 | 0 | 0 | Not-determined | Genome-fragment | NA | NA | 0 |
| S5-final-viral-combined_k141_13925\|\|full | 3164 | No | NA | 5 | 0 | 1 | Not-determined | Genome-fragment | NA | NA | 0 |
| S3-final-viral-combined_k141_110035\|\|full | 1669 | No | NA | 2 | 0 | 2 | Low-quality | Genome-fragment | 0.57 | AAI-based (medium-confidence) | 0 |
| S5-final-viral-combined_k141_30809\|\|full | 1340 | No | NA | 2 | 0 | 1 | Not-determined | Genome-fragment | NA | NA | 0 |
| S17-final-viral-combined_k141_22650\|\|full | 1669 | No | NA | 4 | 0 | 1 | Low-quality | Genome-fragment | 1.14 | AAI-based (medium-confidence) | 0 |
| S5-final-viral-combined_k141_191866\|\|full | 2351 | No | NA | 4 | 0 | 2 | Not-determined | Genome-fragment | NA | NA | 0 |
| S5-final-viral-combined_k141_84946\|\|full | 1324 | No | NA | 2 | 0 | 1 | Not-determined | Genome-fragment | NA | NA | 0 |
| S17-k141_67953 | 1669 | No | NA | 1 | 0 | 0 | Low-quality | Genome-fragment | 0.42 | AAI-based (high-confidence) | 0 |
| S12-final-viral-combined_k141_236592\|\|full | 1669 | No | NA | 3 | 2 | 0 | Low-quality | Genome-fragment | 4.13 | AAI-based (high-confidence) | 0 |
| S2-final-viral-combined_k141_146262\|\|full | 1669 | No | NA | 4 | 2 | 0 | Low-quality | Genome-fragment | 3.85 | AAI-based (high-confidence) | 0 |
| S5-final-viral-combined_k141_200682\|\|full | 2468 | No | NA | 3 | 0 | 3 | Not-determined | Genome-fragment | NA | NA | 0 |
| S5-final-viral-combined_k141_21614\|\|full | 2622 | No | NA | 5 | 0 | 1 | Not-determined | Genome-fragment | NA | NA | 0 |
| S4-final-viral-combined_k141_28003\|\|full | 1669 | No | NA | 3 | 0 | 0 | Low-quality | Genome-fragment | 4.46 | AAI-based (high-confidence) | 0 |
| S6-final-viral-combined_k141_13726\|\|full | 1669 | No | NA | 4 | 1 | 0 | Low-quality | Genome-fragment | 3.06 | HMM-based (lower-bound) | 0 |
| S5-final-viral-combined_k141_120226\|\|full | 1582 | No | NA | 2 | 0 | 0 | Not-determined | Genome-fragment | NA | NA | 0 |
| S9-final-viral-combined_k141_65965\|\|full | 1670 | No | NA | 5 | 0 | 0 | Low-quality | Genome-fragment | 2.21 | AAI-based (medium-confidence) | 0 |
| S11-final-viral-combined_k141_220599\|\|full | 1670 | No | NA | 5 | 1 | 0 | Low-quality | Genome-fragment | 3.39 | AAI-based (high-confidence) | 0 |
| S5-final-viral-combined_k141_174637\|\|full | 1111 | No | NA | 3 | 0 | 0 | Not-determined | Genome-fragment | NA | NA | 0 |
| S5-final-viral-combined_k141_174638\|\|full | 3066 | No | NA | 5 | 0 | 3 | Not-determined | Genome-fragment | NA | NA | 0 |
| S1-final-viral-combined_k141_314836\|\|full | 1670 | No | NA | 4 | 0 | 0 | Low-quality | Genome-fragment | 3.83 | AAI-based (medium-confidence) | 0 |
| S5-final-viral-combined_k141_76462\|\|full | 1338 | No | NA | 3 | 0 | 0 | Not-determined | Genome-fragment | NA | NA | 0 |
| S3_k141_110076 | 1670 | No | NA | 1 | 0 | 1 | Low-quality | Genome-fragment | 2.76 | AAI-based (medium-confidence) | 0 |
| S5-final-viral-combined_k141_40287\|\|full | 1176 | No | NA | 2 | 0 | 0 | Not-determined | Genome-fragment | NA | NA | 0 |
| S5-final-viral-combined_k141_200166\|\|lt2gene | 1670 | No | NA | 2 | 2 | 0 | Low-quality | Genome-fragment | 3.76 | AAI-based (medium-confidence) | 0 |
| S5-final-viral-combined_k141_120260\|\|full | 1943 | No | NA | 3 | 0 | 1 | Not-determined | Genome-fragment | NA | NA | 0 |
| S7-final-viral-combined_k141_232514\|\|full | 1670 | No | NA | 3 | 2 | 0 | Low-quality | Genome-fragment | 1.95 | HMM-based (lower-bound) | 0 |
| S5-final-viral-combined_k141_30994\|\|full | 2420 | No | NA | 2 | 0 | 2 | Not-determined | Genome-fragment | NA | NA | 0 |
| S17-final-viral-combined_k141_73896\|\|full | 1670 | No | NA | 4 | 0 | 0 | Low-quality | Genome-fragment | 3.86 | AAI-based (high-confidence) | 0 |
| S5-final-viral-combined_k141_120299\|\|full | 1087 | No | NA | 3 | 0 | 0 | Not-determined | Genome-fragment | NA | NA | 0 |
| S5-final-viral-combined_k141_49360\|\|full | 1097 | No | NA | 2 | 0 | 2 | Not-determined | Genome-fragment | NA | NA | 0 |
| S2-final-viral-combined_k141_112333\|\|full | 1670 | Yes | 901 | 4 | 1 | 1 | Low-quality | Genome-fragment | 1.59 | AAI-based (medium-confidence) | 46.05 |
| S9-final-viral-combined_k141_23041\|\|full | 1671 | No | NA | 3 | 0 | 0 | Low-quality | Genome-fragment | 2.88 | AAI-based (high-confidence) | 0 |
| S9-final-viral-combined_k141_88043\|\|lt2gene | 1671 | No | NA | 2 | 2 | 0 | Low-quality | Genome-fragment | 3.8 | AAI-based (medium-confidence) | 0 |
| S1-final-viral-combined_k141_347718\|\|full | 1671 | No | NA | 2 | 0 | 0 | Low-quality | Genome-fragment | 2.9 | AAI-based (high-confidence) | 0 |
| S5-final-viral-combined_k141_49399\|\|full | 2011 | No | NA | 3 | 0 | 3 | Not-determined | Genome-fragment | NA | NA | 0 |
| S1-final-viral-combined_k141_213525\|\|full | 1671 | No | NA | 2 | 0 | 1 | Low-quality | Genome-fragment | 0.48 | AAI-based (high-confidence) | 0 |
| S5-final-viral-combined_k141_22856\|\|full | 1671 | No | NA | 3 | 0 | 2 | Low-quality | Genome-fragment | 2.71 | AAI-based (high-confidence) | 0 |
| S7_k141_118497 | 1671 | No | NA | 3 | 0 | 0 | Low-quality | Genome-fragment | 3.65 | AAI-based (high-confidence) | 0 |
| S5-final-viral-combined_k141_31087\|\|full | 3438 | No | NA | 5 | 0 | 2 | Not-determined | Genome-fragment | NA | NA | 0 |
| S12_k141_307482 | 1671 | No | NA | 4 | 0 | 0 | Low-quality | Genome-fragment | 3.03 | AAI-based (high-confidence) | 0 |
| S5-final-viral-combined_k141_76524\|\|full | 2275 | No | NA | 4 | 0 | 0 | Not-determined | Genome-fragment | NA | NA | 0 |
| S2-final-viral-combined_k141_206329\|\|full | 1671 | No | NA | 4 | 2 | 0 | Low-quality | Genome-fragment | 4.14 | AAI-based (high-confidence) | 0 |
| S5-final-viral-combined_k141_111877\|\|full | 3363 | No | NA | 5 | 0 | 3 | Not-determined | Genome-fragment | NA | NA | 0 |
| S5-final-viral-combined_k141_58302\|\|full | 9341 | No | NA | 7 | 0 | 6 | Not-determined | Genome-fragment | NA | NA | 0 |
| S5-final-viral-combined_k141_49422\|\|full | 2615 | No | NA | 4 | 0 | 1 | Not-determined | Genome-fragment | NA | NA | 0 |
| S5-final-viral-combined_k141_183145\|\|full | 5894 | No | NA | 7 | 0 | 7 | Not-determined | Genome-fragment | NA | NA | 0 |
| S4_k141_20006 | 1671 | No | NA | 3 | 2 | 0 | Low-quality | Genome-fragment | 4.7 | AAI-based (high-confidence) | 0 |
| S5-final-viral-combined_k141_66924\|\|full | 6645 | No | NA | 8 | 0 | 6 | Not-determined | Genome-fragment | NA | NA | 0 |
| S6_k141_67093 | 1671 | No | NA | 3 | 1 | 0 | Low-quality | Genome-fragment | 2.9 | AAI-based (high-confidence) | 0 |
| S5-final-viral-combined_k141_14078\|\|full | 5086 | No | NA | 5 | 0 | 4 | Not-determined | Genome-fragment | NA | NA | 0 |
| S9-final-viral-combined_k141_116102\|\|full | 1672 | No | NA | 3 | 1 | 0 | Low-quality | Genome-fragment | 4.15 | AAI-based (high-confidence) | 0 |
| S1-final-viral-combined_k141_99903\|\|full | 1672 | No | NA | 4 | 0 | 0 | Low-quality | Genome-fragment | 1.24 | AAI-based (medium-confidence) | 0 |
| S3-final-viral-combined_k141_34507\|\|full | 1672 | No | NA | 2 | 1 | 0 | Low-quality | Genome-fragment | 3.06 | AAI-based (high-confidence) | 0 |
| S5-final-viral-combined_k141_14106\|\|full | 1482 | No | NA | 3 | 0 | 1 | Not-determined | Genome-fragment | NA | NA | 0 |
| S12-final-viral-combined_k141_41219\|\|full | 1672 | No | NA | 3 | 0 | 0 | Low-quality | Genome-fragment | 3.71 | AAI-based (medium-confidence) | 0 |
| S5-final-viral-combined_k141_129833\|\|full | 3544 | No | NA | 5 | 0 | 4 | Not-determined | Genome-fragment | NA | NA | 0 |
| S6-final-viral-combined_k141_28020\|\|full | 1672 | Yes | 937 | 2 | 1 | 1 | Low-quality | Genome-fragment | 2.35 | AAI-based (high-confidence) | 43.96 |
| S5-final-viral-combined_k141_165051\|\|full | 2013 | No | NA | 5 | 0 | 4 | Not-determined | Genome-fragment | NA | NA | 0 |
| S5-final-viral-combined_k141_192062\|\|full | 1470 | No | NA | 2 | 0 | 1 | Not-determined | Genome-fragment | NA | NA | 0 |
| S5-final-viral-combined_k141_14151\|\|full | 1997 | No | NA | 3 | 0 | 2 | Not-determined | Genome-fragment | NA | NA | 0 |
| S9-final-viral-combined_k141_60615\|\|lt2gene | 1673 | No | NA | 2 | 2 | 0 | Low-quality | Genome-fragment | 4.1 | AAI-based (high-confidence) | 0 |
| S11-final-viral-combined_k141_235270\|\|full | 1673 | No | NA | 4 | 0 | 0 | Low-quality | Genome-fragment | 2.56 | AAI-based (high-confidence) | 0 |
| S1-final-viral-combined_k141_291193\|\|full | 1673 | No | NA | 3 | 0 | 2 | Low-quality | Genome-fragment | 1.39 | AAI-based (medium-confidence) | 0 |
| S8_k141_65529 | 1673 | No | NA | 6 | 0 | 0 | Low-quality | Genome-fragment | 2.71 | AAI-based (high-confidence) | 0 |
| S5-final-viral-combined_k141_209874\|\|full | 1101 | No | NA | 3 | 0 | 1 | Not-determined | Genome-fragment | NA | NA | 0 |
| S1_k141_390875 | 1674 | No | NA | 1 | 0 | 1 | Low-quality | Genome-fragment | 0.92 | AAI-based (high-confidence) | 0 |
| S3-final-viral-combined_k141_126808\|\|full | 1674 | No | NA | 3 | 0 | 1 | Low-quality | Genome-fragment | 1.72 | AAI-based (medium-confidence) | 0 |
| S5-final-viral-combined_k141_121780\|\|full | 1674 | No | NA | 3 | 0 | 0 | Low-quality | Genome-fragment | 4.89 | AAI-based (high-confidence) | 0 |
| S5-final-viral-combined_k141_165139\|\|full | 6363 | No | NA | 5 | 0 | 3 | Not-determined | Genome-fragment | NA | NA | 0 |
| S7-final-viral-combined_k141_199661\|\|full | 1674 | No | NA | 2 | 0 | 0 | Low-quality | Genome-fragment | 2.27 | AAI-based (high-confidence) | 0 |
| S5-final-viral-combined_k141_209914\|\|full | 1228 | No | NA | 3 | 0 | 1 | Not-determined | Genome-fragment | NA | NA | 0 |
| S5-final-viral-combined_k141_58489\|\|full | 1053 | No | NA | 2 | 0 | 0 | Not-determined | Genome-fragment | NA | NA | 0 |
| S5-final-viral-combined_k141_103384\|\|full | 1147 | No | NA | 1 | 0 | 0 | Not-determined | Genome-fragment | NA | NA | 0 |
| S12_k141_104400 | 1674 | No | NA | 4 | 1 | 0 | Low-quality | Genome-fragment | 3.56 | AAI-based (medium-confidence) | 0 |
| S8-final-viral-combined_k141_190445\|\|full | 1674 | No | NA | 6 | 1 | 0 | Low-quality | Genome-fragment | 0.89 | HMM-based (lower-bound) | 0 |
| S1-final-viral-combined_k141_100303\|\|full | 1675 | No | NA | 3 | 0 | 1 | Low-quality | Genome-fragment | 0.72 | AAI-based (medium-confidence) | 0 |
| S5-final-viral-combined_k141_6876\|\|full | 1582 | No | NA | 3 | 0 | 2 | Not-determined | Genome-fragment | NA | NA | 0 |
| S5-final-viral-combined_k141_49585\|\|full | 5213 | No | NA | 5 | 0 | 4 | Not-determined | Genome-fragment | NA | NA | 0 |
| S1-final-viral-combined_k141_22215\|\|full | 1675 | No | NA | 2 | 0 | 2 | Low-quality | Genome-fragment | 3.14 | AAI-based (medium-confidence) | 0 |
| S3-final-viral-combined_k141_13790\|\|lt2gene | 1675 | No | NA | 2 | 1 | 0 | Low-quality | Genome-fragment | 2.06 | HMM-based (lower-bound) | 0 |
| S5-final-viral-combined_k141_49600\|\|full | 1600 | No | NA | 3 | 0 | 3 | Not-determined | Genome-fragment | NA | NA | 0 |
| S5-final-viral-combined_k141_100734\|\|full | 1675 | No | NA | 3 | 0 | 0 | Low-quality | Genome-fragment | 3.29 | AAI-based (medium-confidence) | 0 |
| S7-final-viral-combined_k141_98860\|\|full | 1675 | No | NA | 2 | 0 | 0 | Low-quality | Genome-fragment | 3.21 | AAI-based (medium-confidence) | 0 |
| S17-final-viral-combined_k141_122710\|\|full | 1675 | No | NA | 3 | 0 | 0 | Low-quality | Genome-fragment | 3.27 | AAI-based (medium-confidence) | 0 |
| S12-final-viral-combined_k141_220708\|\|full | 1675 | No | NA | 3 | 1 | 0 | Low-quality | Genome-fragment | 1.54 | AAI-based (medium-confidence) | 0 |
| S12_k141_280250 | 1675 | No | NA | 1 | 0 | 0 | Low-quality | Genome-fragment | 3.87 | AAI-based (medium-confidence) | 0 |
| S5-final-viral-combined_k141_21981\|\|full | 3296 | No | NA | 3 | 0 | 1 | Not-determined | Genome-fragment | NA | NA | 0 |
| S2_k141_5175 | 1675 | No | NA | 1 | 0 | 1 | Low-quality | Genome-fragment | 0.98 | AAI-based (medium-confidence) | 0 |
| S4-final-viral-combined_k141_32053\|\|full | 1675 | No | NA | 3 | 0 | 2 | Low-quality | Genome-fragment | 2.54 | AAI-based (high-confidence) | 0 |
| S5-final-viral-combined_k141_192128\|\|full | 1796 | No | NA | 3 | 0 | 0 | Not-determined | Genome-fragment | NA | NA | 0 |
| S6_k141_56419 | 1675 | No | NA | 5 | 0 | 0 | Low-quality | Genome-fragment | 3.26 | AAI-based (medium-confidence) | 0 |
| S5-final-viral-combined_k141_31287\|\|full | 2854 | No | NA | 2 | 0 | 2 | Not-determined | Genome-fragment | NA | NA | 0 |
| S5-final-viral-combined_k141_156092\|\|full | 2485 | No | NA | 3 | 0 | 1 | Not-determined | Genome-fragment | NA | NA | 0 |
| S11_k141_59011 | 1676 | No | NA | 1 | 0 | 0 | Low-quality | Genome-fragment | 1.8 | AAI-based (high-confidence) | 0 |
| S5-final-viral-combined_k141_112080\|\|full | 1146 | No | NA | 3 | 0 | 1 | Not-determined | Genome-fragment | NA | NA | 0 |
| S1-final-viral-combined_k141_259675\|\|full | 1676 | No | NA | 3 | 0 | 1 | Low-quality | Genome-fragment | 2.08 | AAI-based (high-confidence) | 0 |
| S5-final-viral-combined_k141_58650\|\|full | 1204 | No | NA | 3 | 0 | 2 | Not-determined | Genome-fragment | NA | NA | 0 |
| S17-final-viral-combined_k141_82307\|\|full | 1676 | No | NA | 2 | 1 | 0 | Low-quality | Genome-fragment | 1.97 | AAI-based (high-confidence) | 0 |
| S5-final-viral-combined_k141_58658\|\|full | 3299 | No | NA | 3 | 0 | 3 | Not-determined | Genome-fragment | NA | NA | 0 |
| S17-final-viral-combined_k141_153649\|\|full | 1676 | Yes | 369 | 3 | 1 | 2 | Low-quality | Genome-fragment | 0.72 | AAI-based (medium-confidence) | 77.98 |
| S2-final-viral-combined_k141_189938\|\|full | 1676 | No | NA | 4 | 1 | 0 | Low-quality | Genome-fragment | 3.23 | AAI-based (high-confidence) | 0 |
| S4-final-viral-combined_k141_39970\|\|full | 1676 | No | NA | 4 | 1 | 0 | Low-quality | Genome-fragment | 4.68 | AAI-based (high-confidence) | 0 |
| S5-final-viral-combined_k141_129972\|\|full | 1994 | No | NA | 2 | 0 | 2 | Not-determined | Genome-fragment | NA | NA | 0 |
| S5-final-viral-combined_k141_40700\|\|full | 1797 | No | NA | 4 | 0 | 2 | Not-determined | Genome-fragment | NA | NA | 0 |
| S5-final-viral-combined_k141_120443\|\|full | 2406 | No | NA | 4 | 0 | 2 | Not-determined | Genome-fragment | NA | NA | 0 |
| S5-final-viral-combined_k141_165230\|\|full | 1149 | No | NA | 2 | 0 | 1 | Not-determined | Genome-fragment | NA | NA | 0 |
| S6-final-viral-combined_k141_89559\|\|full | 1676 | No | NA | 5 | 1 | 0 | Low-quality | Genome-fragment | 4.91 | AAI-based (high-confidence) | 0 |
| S5-final-viral-combined_k141_58696\|\|full | 4133 | No | NA | 7 | 0 | 1 | Not-determined | Genome-fragment | NA | NA | 0 |
| S5-final-viral-combined_k141_130016\|\|full | 6633 | No | NA | 8 | 0 | 3 | Not-determined | Genome-fragment | NA | NA | 0 |
| S11-final-viral-combined_k141_166736\|\|full | 1677 | No | NA | 2 | 0 | 1 | Low-quality | Genome-fragment | 2.43 | AAI-based (medium-confidence) | 0 |
| S5-final-viral-combined_k141_31425\|\|full | 17623 | No | NA | 19 | 0 | 14 | Not-determined | Genome-fragment | NA | NA | 0 |
| S5-final-viral-combined_k141_138406\|\|full | 3133 | No | NA | 4 | 0 | 3 | Not-determined | Genome-fragment | NA | NA | 0 |
| S11-final-viral-combined_k141_138886\|\|full | 1677 | No | NA | 5 | 0 | 0 | Low-quality | Genome-fragment | 3.7 | AAI-based (medium-confidence) | 0 |
| S11-final-viral-combined_k141_62692\|\|full | 1677 | No | NA | 3 | 0 | 1 | Low-quality | Genome-fragment | 0.73 | AAI-based (medium-confidence) | 0 |
| S5-final-viral-combined_k141_138420\|\|full | 2692 | No | NA | 3 | 0 | 1 | Not-determined | Genome-fragment | NA | NA | 0 |
| S5-final-viral-combined_k141_183550\|\|full | 6893 | No | NA | 5 | 0 | 4 | Not-determined | Genome-fragment | NA | NA | 0 |
| S5-final-viral-combined_k141_40859\|\|full | 1766 | No | NA | 2 | 0 | 0 | Not-determined | Genome-fragment | NA | NA | 0 |
| S5-final-viral-combined_k141_4255\|\|full | 1677 | No | NA | 2 | 0 | 2 | Low-quality | Genome-fragment | 2.45 | AAI-based (medium-confidence) | 0 |
| S5-final-viral-combined_k141_165286\|\|full | 1154 | No | NA | 5 | 0 | 0 | Not-determined | Genome-fragment | NA | NA | 0 |
| S5_k141_18793 | 1677 | No | NA | 3 | 0 | 1 | Low-quality | Genome-fragment | 3.71 | AAI-based (high-confidence) | 0 |
| S5-final-viral-combined_k141_174903\|\|full | 2146 | No | NA | 2 | 0 | 2 | Not-determined | Genome-fragment | NA | NA | 0 |
| S7-final-viral-combined_k141_114689\|\|lt2gene | 1677 | No | NA | 2 | 1 | 0 | Low-quality | Genome-fragment | 3.82 | AAI-based (high-confidence) | 0 |
| S5-final-viral-combined_k141_94213\|\|full | 2392 | No | NA | 5 | 0 | 0 | Not-determined | Genome-fragment | NA | NA | 0 |
| S5-final-viral-combined_k141_183626\|\|full | 1546 | No | NA | 2 | 0 | 1 | Not-determined | Genome-fragment | NA | NA | 0 |
| S5-final-viral-combined_k141_138443\|\|full | 3905 | No | NA | 3 | 0 | 2 | Not-determined | Genome-fragment | NA | NA | 0 |
| S5-final-viral-combined_k141_138451\|\|full | 1323 | No | NA | 3 | 0 | 1 | Not-determined | Genome-fragment | NA | NA | 0 |
| S5-final-viral-combined_k141_192308\|\|full | 1917 | No | NA | 2 | 0 | 1 | Not-determined | Genome-fragment | NA | NA | 0 |
| S5-final-viral-combined_k141_138457\|\|full | 1046 | No | NA | 3 | 0 | 1 | Not-determined | Genome-fragment | NA | NA | 0 |
| S17-final-viral-combined_k141_160496\|\|full | 1677 | No | NA | 4 | 2 | 0 | Low-quality | Genome-fragment | 3.96 | AAI-based (high-confidence) | 0 |
| S5-final-viral-combined_k141_130109\|\|full | 1045 | No | NA | 2 | 0 | 0 | Not-determined | Genome-fragment | NA | NA | 0 |
| S17-final-viral-combined_k141_161065\|\|full | 1677 | No | NA | 2 | 0 | 2 | Low-quality | Genome-fragment | 1.26 | AAI-based (high-confidence) | 0 |
| S4-final-viral-combined_k141_156098\|\|full | 1677 | No | NA | 2 | 0 | 2 | Low-quality | Genome-fragment | 0.99 | AAI-based (high-confidence) | 0 |
| S8-final-viral-combined_k141_51869\|\|full | 1677 | No | NA | 3 | 0 | 1 | Low-quality | Genome-fragment | 0.84 | AAI-based (medium-confidence) | 0 |
| S5-final-viral-combined_k141_31466\|\|full | 1130 | No | NA | 3 | 0 | 0 | Not-determined | Genome-fragment | NA | NA | 0 |
| S5-final-viral-combined_k141_130132\|\|full | 1843 | No | NA | 4 | 0 | 0 | Not-determined | Genome-fragment | NA | NA | 0 |
| S5-final-viral-combined_k141_58896\|\|full | 16525 | No | NA | 13 | 0 | 3 | Not-determined | Genome-fragment | NA | NA | 0 |
| S1-final-viral-combined_k141_177849\|\|full | 1678 | No | NA | 3 | 0 | 2 | Low-quality | Genome-fragment | 1.19 | AAI-based (medium-confidence) | 0 |
| S5-final-viral-combined_k141_14342\|\|full | 8164 | No | NA | 6 | 0 | 6 | Not-determined | Genome-fragment | NA | NA | 0 |
| S1_k141_367438 | 1678 | No | NA | 4 | 0 | 0 | Low-quality | Genome-fragment | 3.63 | AAI-based (high-confidence) | 0 |
| S5-final-viral-combined_k141_14360\|\|full | 3058 | No | NA | 5 | 0 | 3 | Not-determined | Genome-fragment | NA | NA | 0 |
| S5-final-viral-combined_k141_112138\|\|full | 1022 | No | NA | 3 | 0 | 1 | Not-determined | Genome-fragment | NA | NA | 0 |
| S3-final-viral-combined_k141_80043\|\|lt2gene | 1678 | No | NA | 1 | 1 | 0 | Low-quality | Genome-fragment | 0.97 | AAI-based (high-confidence) | 0 |
| S7-final-viral-combined_k141_141078\|\|full | 1678 | No | NA | 5 | 1 | 0 | Low-quality | Genome-fragment | 3.71 | AAI-based (high-confidence) | 0 |
| S5-final-viral-combined_k141_103666\|\|full | 2042 | No | NA | 4 | 0 | 0 | Not-determined | Genome-fragment | NA | NA | 0 |
| S5-final-viral-combined_k141_103678\|\|full | 8133 | No | NA | 10 | 0 | 9 | Not-determined | Genome-fragment | NA | NA | 0 |
| S5-final-viral-combined_k141_67259\|\|full | 3226 | No | NA | 3 | 0 | 2 | Not-determined | Genome-fragment | NA | NA | 0 |
| S5-final-viral-combined_k141_31549\|\|full | 3727 | No | NA | 4 | 0 | 0 | Not-determined | Genome-fragment | NA | NA | 0 |
| S5-final-viral-combined_k141_59030\|\|full | 2280 | No | NA | 4 | 0 | 0 | Not-determined | Genome-fragment | NA | NA | 0 |
| S5-final-viral-combined_k141_130269\|\|full | 2857 | No | NA | 3 | 0 | 0 | Not-determined | Genome-fragment | NA | NA | 0 |
| S5-final-viral-combined_k141_130278\|\|full | 2305 | No | NA | 4 | 0 | 0 | Not-determined | Genome-fragment | NA | NA | 0 |
| S5-final-viral-combined_k141_112226\|\|full | 1256 | No | NA | 5 | 0 | 0 | Not-determined | Genome-fragment | NA | NA | 0 |
| S5-final-viral-combined_k141_103740\|\|full | 1181 | No | NA | 2 | 0 | 1 | Not-determined | Genome-fragment | NA | NA | 0 |
| S2-final-viral-combined_k141_125719\|\|full | 1678 | No | NA | 3 | 1 | 0 | Low-quality | Genome-fragment | 3.83 | AAI-based (high-confidence) | 0 |
| S2-final-viral-combined_k141_5225\|\|lt2gene | 1678 | No | NA | 1 | 1 | 0 | Low-quality | Genome-fragment | 2.87 | AAI-based (medium-confidence) | 0 |
| S6-final-viral-combined_k141_20576\|\|full | 1678 | No | NA | 4 | 0 | 2 | Low-quality | Genome-fragment | 2.33 | AAI-based (high-confidence) | 0 |
| S8-final-viral-combined_k141_264778\|\|full | 1678 | No | NA | 3 | 0 | 2 | Low-quality | Genome-fragment | 1.15 | AAI-based (medium-confidence) | 0 |
| S11_k141_312234 | 1679 | No | NA | 3 | 0 | 0 | Low-quality | Genome-fragment | 4.56 | AAI-based (medium-confidence) | 0 |
| S5-final-viral-combined_k141_175060\|\|full | 3271 | No | NA | 3 | 0 | 2 | Not-determined | Genome-fragment | NA | NA | 0 |
| S5-final-viral-combined_k141_41007\|\|full | 4576 | No | NA | 5 | 0 | 1 | Not-determined | Genome-fragment | NA | NA | 0 |
| S5-final-viral-combined_k141_85372\|\|full | 2225 | No | NA | 2 | 0 | 0 | Not-determined | Genome-fragment | NA | NA | 0 |
| S5-final-viral-combined_k141_192521\|\|full | 1707 | No | NA | 3 | 0 | 0 | Not-determined | Genome-fragment | NA | NA | 0 |
| S5-final-viral-combined_k141_94393\|\|full | 1713 | No | NA | 3 | 0 | 1 | Not-determined | Genome-fragment | NA | NA | 0 |
| S5-final-viral-combined_k141_103790\|\|full | 2635 | No | NA | 4 | 0 | 0 | Not-determined | Genome-fragment | NA | NA | 0 |
| S1-final-viral-combined_k141_134168\|\|full | 1679 | No | NA | 2 | 1 | 0 | Low-quality | Genome-fragment | 4 | AAI-based (high-confidence) | 0 |
| S17-final-viral-combined_k141_211420\|\|full | 1679 | No | NA | 3 | 1 | 0 | Low-quality | Genome-fragment | 5.41 | AAI-based (medium-confidence) | 0 |
| S5-final-viral-combined_k141_156266\|\|full | 1309 | No | NA | 2 | 0 | 0 | Not-determined | Genome-fragment | NA | NA | 0 |
| S17-final-viral-combined_k141_12172\|\|full | 1679 | No | NA | 6 | 0 | 0 | Low-quality | Genome-fragment | 3.97 | AAI-based (high-confidence) | 0 |
| S5-final-viral-combined_k141_201136\|\|full | 1023 | No | NA | 4 | 0 | 0 | Not-determined | Genome-fragment | NA | NA | 0 |
| S5-final-viral-combined_k141_22328\|\|full | 7049 | No | NA | 4 | 0 | 2 | Not-determined | Genome-fragment | NA | NA | 0 |
| S5-final-viral-combined_k141_130367\|\|full | 2838 | No | NA | 3 | 0 | 3 | Not-determined | Genome-fragment | NA | NA | 0 |
| S17-final-viral-combined_k141_24181\|\|full | 1679 | No | NA | 3 | 0 | 1 | Low-quality | Genome-fragment | 2.22 | AAI-based (medium-confidence) | 0 |
| S12-final-viral-combined_k141_169185\|\|full | 1679 | No | NA | 3 | 1 | 0 | Low-quality | Genome-fragment | 2.53 | AAI-based (medium-confidence) | 0 |
| S5-final-viral-combined_k141_22342\|\|full | 3189 | No | NA | 2 | 0 | 0 | Not-determined | Genome-fragment | NA | NA | 0 |
| S12-final-viral-combined_k141_190090\|\|full | 1679 | No | NA | 2 | 1 | 0 | Low-quality | Genome-fragment | 3.86 | AAI-based (high-confidence) | 0 |
| S12-final-viral-combined_k141_183173\|\|full | 1679 | No | NA | 2 | 0 | 2 | Low-quality | Genome-fragment | 0.9 | AAI-based (medium-confidence) | 0 |
| S12_k141_112172 | 1679 | No | NA | 2 | 0 | 1 | Low-quality | Genome-fragment | 1.68 | AAI-based (medium-confidence) | 0 |
| S5-final-viral-combined_k141_183734\|\|full | 2354 | No | NA | 3 | 0 | 2 | Not-determined | Genome-fragment | NA | NA | 0 |
| S5-final-viral-combined_k141_156316\|\|full | 3597 | No | NA | 6 | 0 | 0 | Not-determined | Genome-fragment | NA | NA | 0 |
| S2-final-viral-combined_k141_11099\|\|full | 1679 | No | NA | 4 | 0 | 0 | Low-quality | Genome-fragment | 3.94 | AAI-based (high-confidence) | 0 |
| S2-final-viral-combined_k141_158552\|\|full | 1679 | No | NA | 2 | 0 | 0 | Low-quality | Genome-fragment | 4.8 | AAI-based (medium-confidence) | 0 |
| S5-final-viral-combined_k141_147993\|\|full | 18083 | No | NA | 24 | 15 | 0 | Medium-quality | Genome-fragment | 51.07 | AAI-based (high-confidence) | 0 |
| S2-final-viral-combined_k141_180175\|\|full | 1679 | No | NA | 3 | 0 | 1 | Low-quality | Genome-fragment | 1.32 | AAI-based (high-confidence) | 0 |
| S2-final-viral-combined_k141_171691\|\|full | 1679 | No | NA | 4 | 0 | 0 | Low-quality | Genome-fragment | 3.27 | AAI-based (medium-confidence) | 0 |
| S5-final-viral-combined_k141_156329\|\|full | 22989 | No | NA | 35 | 16 | 0 | Medium-quality | Genome-fragment | 59.01 | AAI-based (high-confidence) | 0 |
| S2-final-viral-combined_k141_17532\|\|full | 1679 | No | NA | 4 | 2 | 0 | Low-quality | Genome-fragment | 3.56 | AAI-based (high-confidence) | 0 |
| S5-final-viral-combined_k141_7292\|\|full | 14771 | No | NA | 23 | 13 | 0 | Medium-quality | Genome-fragment | 60.47 | AAI-based (high-confidence) | 0 |
| S2-final-viral-combined_k141_196473\|\|full | 1679 | No | NA | 3 | 0 | 2 | Low-quality | Genome-fragment | 0.91 | AAI-based (medium-confidence) | 0 |
| S5-final-viral-combined_k141_67348\|\|full | 2021 | No | NA | 3 | 0 | 2 | Not-determined | Genome-fragment | NA | NA | 0 |
| S4_k141_22396 | 1679 | No | NA | 3 | 0 | 0 | Low-quality | Genome-fragment | 4.4 | AAI-based (high-confidence) | 0 |
| S5-final-viral-combined_k141_201161\|\|full | 2598 | No | NA | 3 | 0 | 1 | Not-determined | Genome-fragment | NA | NA | 0 |
| S8-final-viral-combined_k141_254486\|\|full | 1679 | No | NA | 3 | 0 | 1 | Low-quality | Genome-fragment | 3 | AAI-based (medium-confidence) | 0 |
| S5-final-viral-combined_k141_183794\|\|full | 2662 | No | NA | 4 | 0 | 0 | Not-determined | Genome-fragment | NA | NA | 0 |
| S5-final-viral-combined_k141_22407\|\|full | 1598 | No | NA | 2 | 0 | 2 | Not-determined | Genome-fragment | NA | NA | 0 |
| S5-final-viral-combined_k141_22408\|\|full | 2287 | No | NA | 4 | 0 | 0 | Not-determined | Genome-fragment | NA | NA | 0 |
| S8-final-viral-combined_k141_249477\|\|full | 1679 | No | NA | 3 | 0 | 0 | Low-quality | Genome-fragment | 3.54 | AAI-based (medium-confidence) | 0 |
| S5-final-viral-combined_k141_59064\|\|full | 117646 | Yes | 31029 | 101 | 7 | 52 | Medium-quality | Genome-fragment | 72.45 | AAI-based (high-confidence) | 73.63 |
| S9-final-viral-combined_k141_49774\|\|full | 1680 | No | NA | 3 | 2 | 0 | Low-quality | Genome-fragment | 4.3 | AAI-based (high-confidence) | 0 |
| S1-final-viral-combined_k141_184157\|\|full | 1680 | No | NA | 5 | 0 | 0 | Low-quality | Genome-fragment | 3.54 | AAI-based (high-confidence) | 0 |
| S1-final-viral-combined_k141_285988\|\|full | 1680 | No | NA | 3 | 0 | 0 | Low-quality | Genome-fragment | 1.01 | AAI-based (high-confidence) | 0 |
| S1-final-viral-combined_k141_354876\|\|full | 1680 | Yes | 773 | 3 | 1 | 1 | Low-quality | Genome-fragment | 0.42 | HMM-based (lower-bound) | 53.99 |
| S1-final-viral-combined_k141_310860\|\|full | 1680 | No | NA | 2 | 0 | 0 | Low-quality | Genome-fragment | 2.67 | AAI-based (medium-confidence) | 0 |
| S5-final-viral-combined_k141_31738\|\|full | 1459 | No | NA | 3 | 0 | 0 | Not-determined | Genome-fragment | NA | NA | 0 |
| S3-final-viral-combined_k141_112807\|\|full | 1680 | No | NA | 6 | 1 | 0 | Low-quality | Genome-fragment | 4.32 | AAI-based (high-confidence) | 0 |
| S3-final-viral-combined_k141_60846\|\|lt2gene | 1680 | No | NA | 1 | 0 | 0 | Low-quality | Genome-fragment | 4.75 | AAI-based (high-confidence) | 0 |
| S5-final-viral-combined_k141_7375\|\|full | 1080 | No | NA | 2 | 0 | 2 | Not-determined | Genome-fragment | NA | NA | 0 |
| S17-final-viral-combined_k141_82688\|\|full | 1680 | No | NA | 3 | 0 | 0 | Low-quality | Genome-fragment | 3.53 | AAI-based (medium-confidence) | 0 |
| S12-final-viral-combined_k141_292685\|\|full | 1680 | No | NA | 2 | 0 | 1 | Low-quality | Genome-fragment | 0.77 | AAI-based (medium-confidence) | 0 |
| S5-final-viral-combined_k141_59092\|\|full | 7061 | No | NA | 5 | 0 | 5 | Not-determined | Genome-fragment | NA | NA | 0 |
| S2-final-viral-combined_k141_13779\|\|full | 1680 | No | NA | 6 | 0 | 0 | Low-quality | Genome-fragment | 3.82 | AAI-based (high-confidence) | 0 |
| S8_k141_227007 | 1680 | No | NA | 5 | 1 | 0 | Low-quality | Genome-fragment | 1.33 | AAI-based (high-confidence) | 0 |
| S5-final-viral-combined_k141_77000\|\|full | 3105 | No | NA | 4 | 0 | 1 | Not-determined | Genome-fragment | NA | NA | 0 |
| S5-final-viral-combined_k141_148170\|\|full | 3258 | No | NA | 4 | 0 | 0 | Not-determined | Genome-fragment | NA | NA | 0 |
| S5-final-viral-combined_k141_183888\|\|full | 4410 | No | NA | 5 | 0 | 2 | Not-determined | Genome-fragment | NA | NA | 0 |
| S9-final-viral-combined_k141_50302\|\|full | 1681 | No | NA | 3 | 0 | 2 | Low-quality | Genome-fragment | 0.95 | AAI-based (medium-confidence) | 0 |
| S1-final-viral-combined_k141_266237\|\|full | 1681 | No | NA | 2 | 0 | 0 | Low-quality | Genome-fragment | 4.36 | AAI-based (medium-confidence) | 0 |
| S3-final-viral-combined_k141_53462\|\|full | 1681 | No | NA | 4 | 1 | 0 | Low-quality | Genome-fragment | 3.13 | AAI-based (high-confidence) | 0 |
| S5-final-viral-combined_k141_50042\|\|full | 1242 | No | NA | 3 | 0 | 0 | Not-determined | Genome-fragment | NA | NA | 0 |
| S5-final-viral-combined_k141_175145\|\|full | 35234 | No | NA | 45 | 23 | 1 | High-quality | High-quality | 93.64 | AAI-based (high-confidence) | 0 |
| S17-final-viral-combined_k141_160405\|\|full | 1681 | No | NA | 2 | 0 | 1 | Low-quality | Genome-fragment | 1.93 | AAI-based (high-confidence) | 0 |
| S17-final-viral-combined_k141_42350\|\|full | 1681 | No | NA | 4 | 1 | 0 | Low-quality | Genome-fragment | 2.44 | AAI-based (high-confidence) | 0 |
| S12-final-viral-combined_k141_327586\|\|full | 1681 | No | NA | 3 | 0 | 0 | Low-quality | Genome-fragment | 2.83 | AAI-based (medium-confidence) | 0 |
| S2-final-viral-combined_k141_241110\|\|full | 1681 | No | NA | 3 | 2 | 0 | Low-quality | Genome-fragment | 3.62 | AAI-based (high-confidence) | 0 |
| S5-final-viral-combined_k141_183935\|\|full | 4411 | No | NA | 5 | 0 | 2 | Not-determined | Genome-fragment | NA | NA | 0 |
| S4-final-viral-combined_k141_100777\|\|full | 1681 | No | NA | 2 | 0 | 2 | Low-quality | Genome-fragment | 2.87 | AAI-based (high-confidence) | 0 |
| S5-final-viral-combined_k141_201316\|\|full | 1625 | No | NA | 2 | 0 | 1 | Not-determined | Genome-fragment | NA | NA | 0 |
| S5-final-viral-combined_k141_156388\|\|full | 2263 | No | NA | 3 | 0 | 0 | Not-determined | Genome-fragment | NA | NA | 0 |
| S5-final-viral-combined_k141_201331\|\|full | 1822 | No | NA | 2 | 0 | 1 | Not-determined | Genome-fragment | NA | NA | 0 |
| S9-final-viral-combined_k141_104734\|\|lt2gene | 1682 | No | NA | 1 | 1 | 0 | Low-quality | Genome-fragment | 4.32 | AAI-based (high-confidence) | 0 |
| S1-final-viral-combined_k141_283393\|\|full | 1682 | No | NA | 2 | 0 | 1 | Low-quality | Genome-fragment | 1.2 | AAI-based (high-confidence) | 0 |
| S1-final-viral-combined_k141_90665\|\|full | 1682 | No | NA | 6 | 0 | 0 | Low-quality | Genome-fragment | 11.3 | AAI-based (high-confidence) | 0 |
| S1-final-viral-combined_k141_241034\|\|full | 1682 | No | NA | 3 | 2 | 0 | Low-quality | Genome-fragment | 4.09 | AAI-based (high-confidence) | 0 |
| S5-final-viral-combined_k141_31859\|\|full | 6903 | No | NA | 11 | 0 | 1 | Not-determined | Genome-fragment | NA | NA | 0 |
| S5-final-viral-combined_k141_67496\|\|full | 2207 | No | NA | 4 | 0 | 1 | Not-determined | Genome-fragment | NA | NA | 0 |
| S5-final-viral-combined_k141_31866\|\|full | 1464 | No | NA | 4 | 0 | 2 | Not-determined | Genome-fragment | NA | NA | 0 |
| S5-final-viral-combined_k141_7484\|\|full | 3737 | No | NA | 4 | 0 | 4 | Not-determined | Genome-fragment | NA | NA | 0 |
| S5-final-viral-combined_k141_156407\|\|full | 1463 | No | NA | 3 | 0 | 0 | Not-determined | Genome-fragment | NA | NA | 0 |
| S5-final-viral-combined_k141_67522\|\|full | 1157 | No | NA | 3 | 0 | 2 | Not-determined | Genome-fragment | NA | NA | 0 |
| S1_k141_129168 | 1682 | No | NA | 2 | 0 | 1 | Low-quality | Genome-fragment | 0.94 | AAI-based (medium-confidence) | 0 |
| S5-final-viral-combined_k141_156442\|\|full | 1637 | No | NA | 3 | 0 | 2 | Not-determined | Genome-fragment | NA | NA | 0 |
| S5-final-viral-combined_k141_104054\|\|full | 2925 | No | NA | 4 | 0 | 0 | Not-determined | Genome-fragment | NA | NA | 0 |
| S5-final-viral-combined_k141_121042\|\|full | 2970 | No | NA | 3 | 0 | 2 | Not-determined | Genome-fragment | NA | NA | 0 |
| S5-final-viral-combined_k141_37383\|\|lt2gene | 1682 | Yes | 1297 | 2 | 1 | 1 | Low-quality | Genome-fragment | 0.37 | HMM-based (lower-bound) | 22.89 |
| S5-final-viral-combined_k141_67561\|\|full | 1451 | No | NA | 3 | 0 | 0 | Not-determined | Genome-fragment | NA | NA | 0 |
| S5-final-viral-combined_k141_22504\|\|full | 1575 | No | NA | 2 | 0 | 1 | Not-determined | Genome-fragment | NA | NA | 0 |
| S17-final-viral-combined_k141_131973\|\|full | 1682 | No | NA | 3 | 2 | 0 | Low-quality | Genome-fragment | 2.77 | HMM-based (lower-bound) | 0 |
| S17-final-viral-combined_k141_151500\|\|lt2gene | 1682 | No | NA | 1 | 1 | 0 | Low-quality | Genome-fragment | 2.65 | AAI-based (high-confidence) | 0 |
| S5-final-viral-combined_k141_94482\|\|full | 59881 | Yes | 51384 | 72 | 4 | 8 | High-quality | High-quality | 100 | AAI-based (medium-confidence) | 14.19 |
| S2-final-viral-combined_k141_67341\|\|full | 1682 | No | NA | 3 | 2 | 0 | Low-quality | Genome-fragment | 5.14 | AAI-based (high-confidence) | 0 |
| S4_k141_132023 | 1682 | No | NA | 1 | 0 | 0 | Low-quality | Genome-fragment | 0.14 | AAI-based (medium-confidence) | 0 |
| S5-final-viral-combined_k141_183940\|\|full | 1814 | No | NA | 2 | 0 | 2 | Not-determined | Genome-fragment | NA | NA | 0 |
| S8_k141_268731 | 1682 | No | NA | 1 | 0 | 0 | Low-quality | Genome-fragment | 1.76 | AAI-based (high-confidence) | 0 |
| S5-final-viral-combined_k141_138888\|\|full | 2464 | No | NA | 3 | 0 | 2 | Not-determined | Genome-fragment | NA | NA | 0 |
| S11-final-viral-combined_k141_84624\|\|full | 1683 | No | NA | 4 | 1 | 0 | Low-quality | Genome-fragment | 1.19 | HMM-based (lower-bound) | 0 |
| S5-final-viral-combined_k141_183973\|\|full | 2437 | No | NA | 3 | 0 | 2 | Not-determined | Genome-fragment | NA | NA | 0 |
| S11-final-viral-combined_k141_103076\|\|full | 1683 | No | NA | 2 | 0 | 0 | Low-quality | Genome-fragment | 2.08 | AAI-based (high-confidence) | 0 |
| S1-final-viral-combined_k141_378865\|\|full | 1683 | No | NA | 3 | 0 | 0 | Low-quality | Genome-fragment | 3.85 | AAI-based (medium-confidence) | 0 |
| S5-final-viral-combined_k141_85727\|\|full | 2206 | No | NA | 3 | 0 | 1 | Not-determined | Genome-fragment | NA | NA | 0 |
| S1_k141_369256 | 1683 | No | NA | 3 | 0 | 0 | Low-quality | Genome-fragment | 2.81 | AAI-based (high-confidence) | 0 |
| S5-final-viral-combined_k141_94495\|\|full | 1016 | No | NA | 2 | 0 | 0 | Not-determined | Genome-fragment | NA | NA | 0 |
| S5-final-viral-combined_k141_112483\|\|full | 1054 | No | NA | 2 | 0 | 1 | Not-determined | Genome-fragment | NA | NA | 0 |
| S5-final-viral-combined_k141_175391\|\|full | 1959 | No | NA | 3 | 0 | 2 | Not-determined | Genome-fragment | NA | NA | 0 |
| S5-final-viral-combined_k141_32050\|\|full | 2370 | No | NA | 3 | 0 | 1 | Not-determined | Genome-fragment | NA | NA | 0 |
| S5-final-viral-combined_k141_50254\|\|full | 1020 | No | NA | 2 | 0 | 0 | Not-determined | Genome-fragment | NA | NA | 0 |
| S5-final-viral-combined_k141_41387\|\|full | 11397 | No | NA | 8 | 0 | 7 | Not-determined | Genome-fragment | NA | NA | 0 |
| S5-final-viral-combined_k141_77233\|\|full | 1254 | No | NA | 1 | 0 | 1 | Not-determined | Genome-fragment | NA | NA | 0 |
| S5-final-viral-combined_k141_87708\|\|full | 1683 | No | NA | 4 | 1 | 0 | Low-quality | Genome-fragment | 3.13 | AAI-based (high-confidence) | 0 |
| S5-final-viral-combined_k141_7525\|\|full | 1205 | No | NA | 3 | 0 | 1 | Not-determined | Genome-fragment | NA | NA | 0 |
| S5-final-viral-combined_k141_14663\|\|full | 2087 | No | NA | 3 | 0 | 0 | Not-determined | Genome-fragment | NA | NA | 0 |
| S5-final-viral-combined_k141_141982\|\|full | 1683 | No | NA | 3 | 3 | 0 | Low-quality | Genome-fragment | 4.27 | AAI-based (high-confidence) | 0 |
| S17-final-viral-combined_k141_234163\|\|full | 1683 | No | NA | 3 | 1 | 0 | Low-quality | Genome-fragment | 1.02 | HMM-based (lower-bound) | 0 |
| S5-final-viral-combined_k141_184036\|\|full | 2302 | No | NA | 3 | 0 | 3 | Not-determined | Genome-fragment | NA | NA | 0 |
| S5-final-viral-combined_k141_112539\|\|full | 2702 | No | NA | 3 | 0 | 2 | Not-determined | Genome-fragment | NA | NA | 0 |
| S5-final-viral-combined_k141_165592\|\|full | 1310 | No | NA | 2 | 0 | 0 | Not-determined | Genome-fragment | NA | NA | 0 |
| S12-final-viral-combined_k141_136219\|\|full | 1683 | No | NA | 3 | 0 | 0 | Low-quality | Genome-fragment | 2.53 | AAI-based (medium-confidence) | 0 |
| S5-final-viral-combined_k141_175449\|\|full | 3389 | No | NA | 4 | 0 | 2 | Not-determined | Genome-fragment | NA | NA | 0 |
| S5-final-viral-combined_k141_130782\|\|full | 1308 | No | NA | 3 | 0 | 0 | Not-determined | Genome-fragment | NA | NA | 0 |
| S5-final-viral-combined_k141_112605\|\|full | 1743 | No | NA | 3 | 0 | 0 | Not-determined | Genome-fragment | NA | NA | 0 |
| S9-final-viral-combined_k141_53399\|\|lt2gene | 1684 | No | NA | 2 | 2 | 0 | Low-quality | Genome-fragment | 4.21 | AAI-based (high-confidence) | 0 |
| S3-final-viral-combined_k141_3699\|\|full | 1684 | No | NA | 4 | 0 | 0 | Low-quality | Genome-fragment | 2.13 | AAI-based (high-confidence) | 0 |
| S3_k141_47154 | 1684 | No | NA | 2 | 1 | 0 | Low-quality | Genome-fragment | 0.78 | AAI-based (high-confidence) | 0 |
| S5-final-viral-combined_k141_14717\|\|full | 14124 | No | NA | 11 | 0 | 10 | Not-determined | Genome-fragment | NA | NA | 0 |
| S5-final-viral-combined_k141_148381\|\|full | 4136 | No | NA | 7 | 0 | 0 | Not-determined | Genome-fragment | NA | NA | 0 |
| S5-final-viral-combined_k141_148385\|\|full | 1619 | No | NA | 2 | 0 | 0 | Not-determined | Genome-fragment | NA | NA | 0 |
| S7-final-viral-combined_k141_237207\|\|full | 1684 | Yes | 1357 | 4 | 1 | 1 | Low-quality | Genome-fragment | 2.47 | HMM-based (lower-bound) | 19.42 |
| S7-final-viral-combined_k141_232575\|\|full | 1684 | No | NA | 2 | 1 | 0 | Low-quality | Genome-fragment | 4.04 | AAI-based (high-confidence) | 0 |
| S17-k141_36168 | 1684 | No | NA | 1 | 0 | 1 | Low-quality | Genome-fragment | 2.58 | AAI-based (medium-confidence) | 0 |
| S5-final-viral-combined_k141_77383\|\|full | 2143 | No | NA | 3 | 0 | 0 | Not-determined | Genome-fragment | NA | NA | 0 |
| S17-k141_182626 | 1684 | No | NA | 4 | 1 | 0 | Low-quality | Genome-fragment | 3.23 | AAI-based (high-confidence) | 0 |
| S12-final-viral-combined_k141_4103\|\|full | 1684 | No | NA | 3 | 0 | 1 | Low-quality | Genome-fragment | 2.13 | AAI-based (medium-confidence) | 0 |
| S4-final-viral-combined_k141_57823\|\|full | 1684 | No | NA | 2 | 0 | 0 | Low-quality | Genome-fragment | 3.77 | AAI-based (medium-confidence) | 0 |
| S11_k141_303751 | 1685 | No | NA | 3 | 1 | 0 | Low-quality | Genome-fragment | 2.96 | AAI-based (high-confidence) | 0 |
| S5-final-viral-combined_k141_165673\|\|full | 2378 | No | NA | 3 | 0 | 2 | Not-determined | Genome-fragment | NA | NA | 0 |
| S5-final-viral-combined_k141_50341\|\|full | 2419 | No | NA | 4 | 0 | 1 | Not-determined | Genome-fragment | NA | NA | 0 |
| S1-final-viral-combined_k141_98797\|\|full | 1685 | No | NA | 3 | 0 | 0 | Low-quality | Genome-fragment | 3.76 | AAI-based (medium-confidence) | 0 |
| S1-final-viral-combined_k141_347614\|\|full | 1685 | No | NA | 2 | 0 | 0 | Low-quality | Genome-fragment | 2.64 | AAI-based (high-confidence) | 0 |
| S5-final-viral-combined_k141_67650\|\|full | 106967 | Yes | 31697 | 90 | 11 | 45 | Medium-quality | Genome-fragment | 51.46 | HMM-based (lower-bound) | 70.37 |
| S5-final-viral-combined_k141_210162\|\|full | 1685 | No | NA | 3 | 0 | 0 | Low-quality | Genome-fragment | 2.86 | AAI-based (medium-confidence) | 0 |
| S5-final-viral-combined_k141_184196\|\|full | 1656 | No | NA | 3 | 0 | 0 | Not-determined | Genome-fragment | NA | NA | 0 |
| S5-final-viral-combined_k141_14842\|\|full | 2637 | No | NA | 4 | 0 | 0 | Not-determined | Genome-fragment | NA | NA | 0 |
| S5-final-viral-combined_k141_50392\|\|full | 1005 | No | NA | 3 | 0 | 2 | Not-determined | Genome-fragment | NA | NA | 0 |
| S17-k141_33192 | 1685 | No | NA | 3 | 0 | 0 | Low-quality | Genome-fragment | 3.03 | AAI-based (medium-confidence) | 0 |
| S2-final-viral-combined_k141_152044\|\|full | 1685 | No | NA | 5 | 0 | 3 | Low-quality | Genome-fragment | 0.55 | AAI-based (medium-confidence) | 0 |
| S5-final-viral-combined_k141_77455\|\|full | 2093 | No | NA | 3 | 0 | 1 | Not-determined | Genome-fragment | NA | NA | 0 |
| S5-final-viral-combined_k141_32196\|\|full | 2760 | No | NA | 5 | 0 | 1 | Not-determined | Genome-fragment | NA | NA | 0 |
| S11-final-viral-combined_k141_319911\|\|full | 1686 | No | NA | 2 | 0 | 0 | Low-quality | Genome-fragment | 0.97 | AAI-based (high-confidence) | 0 |
| S1-final-viral-combined_k141_165393\|\|full | 1686 | No | NA | 2 | 0 | 1 | Low-quality | Genome-fragment | 3.81 | AAI-based (high-confidence) | 0 |
| S5-final-viral-combined_k141_7693\|\|full | 1464 | No | NA | 3 | 0 | 1 | Not-determined | Genome-fragment | NA | NA | 0 |
| S1-final-viral-combined_k141_363829\|\|full | 1686 | No | NA | 3 | 1 | 0 | Low-quality | Genome-fragment | 2.88 | AAI-based (high-confidence) | 0 |
| S5-final-viral-combined_k141_41457\|\|full | 10768 | No | NA | 9 | 0 | 1 | Not-determined | Genome-fragment | NA | NA | 0 |
| S5-final-viral-combined_k141_184297\|\|full | 2041 | No | NA | 5 | 0 | 0 | Not-determined | Genome-fragment | NA | NA | 0 |
| S5-final-viral-combined_k141_32200\|\|full | 2656 | No | NA | 5 | 0 | 1 | Not-determined | Genome-fragment | NA | NA | 0 |
| S1-final-viral-combined_k141_300973\|\|full | 1686 | No | NA | 3 | 0 | 0 | Low-quality | Genome-fragment | 3.54 | AAI-based (medium-confidence) | 0 |
| S5-final-viral-combined_k141_59394\|\|full | 2898 | No | NA | 4 | 0 | 3 | Not-determined | Genome-fragment | NA | NA | 0 |
| S5-final-viral-combined_k141_156737\|\|full | 1293 | No | NA | 3 | 0 | 0 | Not-determined | Genome-fragment | NA | NA | 0 |
| S1-final-viral-combined_k141_73058\|\|lt2gene | 1686 | No | NA | 1 | 1 | 0 | Low-quality | Genome-fragment | 0.71 | HMM-based (lower-bound) | 0 |
| S5-final-viral-combined_k141_184336\|\|full | 3263 | No | NA | 4 | 0 | 0 | Not-determined | Genome-fragment | NA | NA | 0 |
| S5-final-viral-combined_k141_77522\|\|full | 1211 | No | NA | 2 | 0 | 0 | Not-determined | Genome-fragment | NA | NA | 0 |
| S5-final-viral-combined_k141_112779\|\|full | 4579 | No | NA | 7 | 0 | 4 | Not-determined | Genome-fragment | NA | NA | 0 |
| S3_k141_62049 | 1686 | No | NA | 2 | 0 | 2 | Low-quality | Genome-fragment | 1.23 | AAI-based (medium-confidence) | 0 |
| S5-final-viral-combined_k141_22677\|\|full | 1986 | No | NA | 2 | 0 | 2 | Not-determined | Genome-fragment | NA | NA | 0 |
| S5-final-viral-combined_k141_59423\|\|full | 2398 | No | NA | 3 | 0 | 1 | Not-determined | Genome-fragment | NA | NA | 0 |
| S5-final-viral-combined_k141_32263\|\|full | 1259 | No | NA | 3 | 0 | 0 | Not-determined | Genome-fragment | NA | NA | 0 |
| S5-final-viral-combined_k141_93880\|\|full | 1686 | No | NA | 3 | 0 | 3 | Low-quality | Genome-fragment | 3.16 | AAI-based (high-confidence) | 0 |
| S5-final-viral-combined_k141_104557\|\|full | 1686 | No | NA | 3 | 1 | 0 | Low-quality | Genome-fragment | 3.72 | AAI-based (high-confidence) | 0 |
| S5-final-viral-combined_k141_32279\|\|full | 1490 | No | NA | 3 | 0 | 2 | Not-determined | Genome-fragment | NA | NA | 0 |
| S5_k141_26907 | 1686 | No | NA | 4 | 1 | 0 | Low-quality | Genome-fragment | 4.28 | AAI-based (high-confidence) | 0 |
| S17-k141_230875 | 1686 | No | NA | 2 | 0 | 0 | Low-quality | Genome-fragment | 4.61 | AAI-based (high-confidence) | 0 |
| S12-final-viral-combined_k141_327821\|\|full | 1686 | No | NA | 2 | 0 | 0 | Low-quality | Genome-fragment | 0.79 | AAI-based (high-confidence) | 0 |
| S5-final-viral-combined_k141_32289\|\|full | 5476 | No | NA | 7 | 0 | 1 | Not-determined | Genome-fragment | NA | NA | 0 |
| S12-final-viral-combined_k141_366609\|\|full | 1686 | No | NA | 3 | 0 | 1 | Low-quality | Genome-fragment | 3.02 | AAI-based (medium-confidence) | 0 |
| S5-final-viral-combined_k141_59431\|\|full | 25800 | No | NA | 40 | 0 | 6 | Not-determined | Genome-fragment | NA | NA | 0 |
| S12_k141_20242 | 1686 | No | NA | 4 | 0 | 0 | Low-quality | Genome-fragment | 1.31 | AAI-based (high-confidence) | 0 |
| S6_k141_66236 | 1686 | No | NA | 2 | 0 | 1 | Low-quality | Genome-fragment | 3.67 | AAI-based (high-confidence) | 0 |
| S5-final-viral-combined_k141_201612\|\|full | 1903 | No | NA | 3 | 0 | 3 | Not-determined | Genome-fragment | NA | NA | 0 |
| S8-final-viral-combined_k141_223018\|\|full | 1686 | No | NA | 3 | 0 | 0 | Low-quality | Genome-fragment | 2.66 | AAI-based (high-confidence) | 0 |
| S11-final-viral-combined_k141_184189\|\|full | 1687 | No | NA | 3 | 1 | 0 | Low-quality | Genome-fragment | 3.12 | AAI-based (medium-confidence) | 0 |
| S11-final-viral-combined_k141_132230\|\|full | 1687 | No | NA | 2 | 0 | 2 | Low-quality | Genome-fragment | 1.36 | AAI-based (high-confidence) | 0 |
| S5-final-viral-combined_k141_112805\|\|full | 3825 | No | NA | 7 | 0 | 0 | Not-determined | Genome-fragment | NA | NA | 0 |
| S5-final-viral-combined_k141_22746\|\|full | 1708 | No | NA | 4 | 0 | 0 | Not-determined | Genome-fragment | NA | NA | 0 |
| S11-final-viral-combined_k141_61103\|\|lt2gene | 1687 | No | NA | 2 | 2 | 0 | Low-quality | Genome-fragment | 3.71 | AAI-based (high-confidence) | 0 |
| S11_k141_110844 | 1687 | No | NA | 2 | 0 | 0 | Low-quality | Genome-fragment | 1.58 | AAI-based (high-confidence) | 0 |
| S5-final-viral-combined_k141_148626\|\|full | 2972 | No | NA | 3 | 0 | 3 | Not-determined | Genome-fragment | NA | NA | 0 |
| S5-final-viral-combined_k141_32362\|\|full | 1334 | No | NA | 2 | 0 | 0 | Not-determined | Genome-fragment | NA | NA | 0 |
| S11_k141_143888 | 1687 | No | NA | 2 | 0 | 1 | Low-quality | Genome-fragment | 2.03 | AAI-based (high-confidence) | 0 |
| S5-final-viral-combined_k141_14876\|\|full | 1838 | No | NA | 2 | 0 | 0 | Not-determined | Genome-fragment | NA | NA | 0 |
| S5-final-viral-combined_k141_59546\|\|full | 1691 | No | NA | 4 | 0 | 3 | Not-determined | Genome-fragment | NA | NA | 0 |
| S1-final-viral-combined_k141_163600\|\|full | 1687 | No | NA | 4 | 0 | 2 | Low-quality | Genome-fragment | 1.81 | AAI-based (medium-confidence) | 0 |
| S5-final-viral-combined_k141_32393\|\|full | 1158 | No | NA | 2 | 0 | 0 | Not-determined | Genome-fragment | NA | NA | 0 |
| S5-final-viral-combined_k141_86108\|\|full | 1863 | No | NA | 2 | 0 | 0 | Not-determined | Genome-fragment | NA | NA | 0 |
| S3-final-viral-combined_k141_122919\|\|full | 1687 | No | NA | 3 | 0 | 1 | Low-quality | Genome-fragment | 3.35 | AAI-based (medium-confidence) | 0 |
| S5-final-viral-combined_k141_148680\|\|full | 2712 | No | NA | 4 | 0 | 2 | Not-determined | Genome-fragment | NA | NA | 0 |
| S5-final-viral-combined_k141_184548\|\|full | 1772 | No | NA | 4 | 0 | 1 | Not-determined | Genome-fragment | NA | NA | 0 |

| | | | | | | | | | | | | | |
|---|---|---|---|---|---|---|---|---|---|---|---|---|---|
| S17-final-viral-combined_k141_167692\|\|full | 1687 | No | NA | | 3 | 0 | 2 | Low-quality | Genome-fragment | | 0.94 | AAI-based (medium-confidence) | 0 |
| S17-final-viral-combined_k141_53494\|\|full | 1687 | No | NA | | 2 | 0 | 0 | Low-quality | Genome-fragment | | 1.07 | AAI-based (high-confidence) | 0 |
| S5-final-viral-combined_k141_94786\|\|full | 72378 | Yes | | 34416 | 83 | 14 | 25 | Medium-quality | Genome-fragment | | 55.06 | HMM-based (lower-bound) | 52.45 |
| S12-final-viral-combined_k141_275640\|\|lt2gene | 1687 | No | NA | | 1 | 0 | 0 | Low-quality | Genome-fragment | | 5.88 | AAI-based (medium-confidence) | 0 |
| S5-final-viral-combined_k141_59595\|\|full | 1662 | No | NA | | 2 | 0 | 0 | Not-determined | Genome-fragment | NA | | NA | 0 |
| S5-final-viral-combined_k141_148741\|\|full | 1406 | No | NA | | 2 | 0 | 1 | Not-determined | Genome-fragment | NA | | NA | 0 |
| S5-final-viral-combined_k141_210687\|\|full | 2080 | No | NA | | 2 | 0 | 0 | Not-determined | Genome-fragment | NA | | NA | 0 |
| S12-final-viral-combined_k141_113407\|\|lt2gene | 1687 | No | NA | | 1 | 1 | 0 | Low-quality | Genome-fragment | | 3.71 | AAI-based (high-confidence) | 0 |
| S5-final-viral-combined_k141_184608\|\|full | 1416 | No | NA | | 3 | 0 | 3 | Not-determined | Genome-fragment | NA | | NA | 0 |
| S5-final-viral-combined_k141_22839\|\|full | 1436 | No | NA | | 2 | 0 | 1 | Not-determined | Genome-fragment | NA | | NA | 0 |
| S2-final-viral-combined_k141_68880\|\|full | 1687 | No | NA | | 3 | 0 | 0 | Low-quality | Genome-fragment | | 0.95 | AAI-based (medium-confidence) | 0 |
| S4-final-viral-combined_k141_85548\|\|full | 1687 | No | NA | | 2 | 0 | 0 | Low-quality | Genome-fragment | | 1.58 | AAI-based (medium-confidence) | 0 |
| S4-final-viral-combined_k141_59802\|\|full | 1687 | No | NA | | 2 | 0 | 1 | Low-quality | Genome-fragment | | 1.03 | AAI-based (medium-confidence) | 0 |
| S5-final-viral-combined_k141_193234\|\|full | 1042 | No | NA | | 2 | 0 | 0 | Not-determined | Genome-fragment | NA | | NA | 0 |
| S5-final-viral-combined_k141_32437\|\|full | 4307 | No | NA | | 3 | 0 | 3 | Not-determined | Genome-fragment | NA | | NA | 0 |
| S6_k141_41059 | 1687 | No | NA | | 3 | 0 | 1 | Low-quality | Genome-fragment | | 1.46 | AAI-based (medium-confidence) | 0 |
| S5-final-viral-combined_k141_156935\|\|full | 5450 | No | NA | | 5 | 0 | 2 | Not-determined | Genome-fragment | NA | | NA | 0 |
| S8-final-viral-combined_k141_203141\|\|full | 1687 | No | NA | | 3 | 0 | 1 | Low-quality | Genome-fragment | | 0.5 | AAI-based (medium-confidence) | 0 |
| S9-final-viral-combined_k141_93724\|\|full | 1688 | No | NA | | 4 | 0 | 0 | Low-quality | Genome-fragment | | 3.52 | AAI-based (high-confidence) | 0 |
| S11-final-viral-combined_k141_81721\|\|full | 1688 | No | NA | | 3 | 0 | 0 | Low-quality | Genome-fragment | | 0.72 | AAI-based (high-confidence) | 0 |
| S1-final-viral-combined_k141_280797\|\|full | 1688 | No | NA | | 2 | 0 | 2 | Low-quality | Genome-fragment | | 0.59 | AAI-based (medium-confidence) | 0 |
| S17-k141_125584 | 1688 | No | NA | | 3 | 0 | 0 | Low-quality | Genome-fragment | | 3.45 | AAI-based (medium-confidence) | 0 |
| S5-final-viral-combined_k141_166071\|\|full | 6860 | No | NA | | 4 | 0 | 1 | Not-determined | Genome-fragment | NA | | NA | 0 |
| S5-final-viral-combined_k141_201802\|\|full | 1481 | No | NA | | 2 | 0 | 0 | Not-determined | Genome-fragment | NA | | NA | 0 |
| S5-final-viral-combined_k141_86301\|\|full | 6436 | No | NA | | 7 | 0 | 4 | Not-determined | Genome-fragment | NA | | NA | 0 |
| S5-final-viral-combined_k141_86307\|\|full | 5206 | No | NA | | 3 | 0 | 0 | Not-determined | Genome-fragment | NA | | NA | 0 |
| S12-final-viral-combined_k141_363060\|\|lt2gene | 1688 | No | NA | | 1 | 0 | 0 | Low-quality | Genome-fragment | | 3.62 | AAI-based (high-confidence) | 0 |
| S12_k141_281373 | 1688 | No | NA | | 1 | 0 | 0 | Low-quality | Genome-fragment | | 23.86 | AAI-based (medium-confidence) | 0 |
| S4-final-viral-combined_k141_17046\|\|full | 1688 | No | NA | | 3 | 0 | 0 | Low-quality | Genome-fragment | | 3.84 | AAI-based (high-confidence) | 0 |
| S5-final-viral-combined_k141_77751\|\|full | 4511 | No | NA | | 4 | 0 | 1 | Not-determined | Genome-fragment | NA | | NA | 0 |
| S5-final-viral-combined_k141_193295\|\|full | 1635 | No | NA | | 3 | 0 | 2 | Not-determined | Genome-fragment | NA | | NA | 0 |
| S5-final-viral-combined_k141_166114\|\|full | 1091 | No | NA | | 2 | 0 | 0 | Not-determined | Genome-fragment | NA | | NA | 0 |
| S4_k141_15277 | 1688 | No | NA | | 2 | 0 | 0 | Low-quality | Genome-fragment | | 2.99 | AAI-based (high-confidence) | 0 |
| S5-final-viral-combined_k141_86376\|\|full | 1920 | No | NA | | 3 | 0 | 0 | Not-determined | Genome-fragment | NA | | NA | 0 |
| S1-final-viral-combined_k141_111391\|\|full | 1689 | No | NA | | 2 | 0 | 1 | Low-quality | Genome-fragment | | 4.33 | AAI-based (high-confidence) | 0 |
| S5-final-viral-combined_k141_166174\|\|full | 1294 | No | NA | | 2 | 0 | 0 | Not-determined | Genome-fragment | NA | | NA | 0 |
| S5-final-viral-combined_k141_50715\|\|full | 5570 | No | NA | | 6 | 0 | 5 | Not-determined | Genome-fragment | NA | | NA | 0 |
| S5-final-viral-combined_k141_14487\|\|full | 1689 | No | NA | | 3 | 3 | 0 | Low-quality | Genome-fragment | | 4.94 | AAI-based (high-confidence) | 0 |
| S5-final-viral-combined_k141_175940\|\|full | 2245 | No | NA | | 4 | 0 | 1 | Not-determined | Genome-fragment | NA | | NA | 0 |
| S5-final-viral-combined_k141_166186\|\|full | 2067 | No | NA | | 2 | 0 | 0 | Not-determined | Genome-fragment | NA | | NA | 0 |
| S5-final-viral-combined_k141_50733\|\|full | 7596 | No | NA | | 7 | 0 | 6 | Not-determined | Genome-fragment | NA | | NA | 0 |
| S5-final-viral-combined_k141_201933\|\|full | 1484 | No | NA | | 3 | 0 | 2 | Not-determined | Genome-fragment | NA | | NA | 0 |
| S2_k141_99035 | 1689 | No | NA | | 2 | 0 | 0 | Low-quality | Genome-fragment | | 1.07 | AAI-based (high-confidence) | 0 |
| S5-final-viral-combined_k141_50747\|\|full | 1016 | No | NA | | 3 | 0 | 0 | Not-determined | Genome-fragment | NA | | NA | 0 |
| S5-final-viral-combined_k141_175963\|\|full | 4316 | No | NA | | 8 | 0 | 2 | Not-determined | Genome-fragment | NA | | NA | 0 |
| S8-final-viral-combined_k141_97745\|\|full | 1689 | No | NA | | 4 | 1 | 0 | Low-quality | Genome-fragment | | 2.94 | AAI-based (high-confidence) | 0 |
| S5-final-viral-combined_k141_32538\|\|full | 3237 | No | NA | | 4 | 0 | 2 | Not-determined | Genome-fragment | NA | | NA | 0 |
| S5-final-viral-combined_k141_32542\|\|full | 2033 | No | NA | | 4 | 0 | 2 | Not-determined | Genome-fragment | NA | | NA | 0 |
| S5-final-viral-combined_k141_32547\|\|full | 1963 | No | NA | | 3 | 0 | 2 | Not-determined | Genome-fragment | NA | | NA | 0 |
| S1_k141_308365 | 1690 | No | NA | | 6 | 1 | 0 | Low-quality | Genome-fragment | | 4.21 | AAI-based (high-confidence) | 0 |
| S5-final-viral-combined_k141_131454\|\|full | 1771 | No | NA | | 3 | 0 | 3 | Not-determined | Genome-fragment | NA | | NA | 0 |
| S3_k141_76577 | 1690 | No | NA | | 4 | 0 | 0 | Low-quality | Genome-fragment | | 2.67 | AAI-based (high-confidence) | 0 |
| S5-final-viral-combined_k141_15099\|\|full | 1432 | No | NA | | 3 | 0 | 0 | Not-determined | Genome-fragment | NA | | NA | 0 |
| S5-final-viral-combined_k141_201988\|\|full | 1111 | No | NA | | 2 | 0 | 0 | Not-determined | Genome-fragment | NA | | NA | 0 |
| S5-final-viral-combined_k141_32568\|\|full | 1645 | No | NA | | 3 | 0 | 0 | Not-determined | Genome-fragment | NA | | NA | 0 |
| S5-final-viral-combined_k141_113072\|\|full | 2140 | No | NA | | 5 | 0 | 0 | Not-determined | Genome-fragment | NA | | NA | 0 |
| S5-final-viral-combined_k141_91518\|\|full | 1690 | No | NA | | 2 | 0 | 1 | Low-quality | Genome-fragment | | 1.29 | AAI-based (medium-confidence) | 0 |
| S5-final-viral-combined_k141_189735\|\|full | 1690 | No | NA | | 3 | 0 | 1 | Low-quality | Genome-fragment | | 3.77 | AAI-based (high-confidence) | 0 |
| S2-final-viral-combined_k141_21918\|\|full | 1690 | No | NA | | 3 | 1 | 0 | Low-quality | Genome-fragment | | 4.21 | AAI-based (high-confidence) | 0 |
| S2-final-viral-combined_k141_184519\|\|full | 1690 | No | NA | | 2 | 0 | 2 | Low-quality | Genome-fragment | | 3.07 | AAI-based (medium-confidence) | 0 |
| S6-final-viral-combined_k141_115619\|\|lt2gene | 1690 | No | NA | | 2 | 2 | 0 | Low-quality | Genome-fragment | | 5.58 | HMM-based (lower-bound) | 0 |
| S8-final-viral-combined_k141_250551\|\|full | 1690 | No | NA | | 2 | 0 | 1 | Low-quality | Genome-fragment | | 2.93 | AAI-based (medium-confidence) | 0 |
| S5-final-viral-combined_k141_15124\|\|full | 1427 | No | NA | | 3 | 0 | 3 | Not-determined | Genome-fragment | NA | | NA | 0 |
| S5-final-viral-combined_k141_166252\|\|full | 3969 | No | NA | | 4 | 0 | 4 | Not-determined | Genome-fragment | NA | | NA | 0 |
| S8-final-viral-combined_k141_156188\|\|lt2gene | 1690 | No | NA | | 2 | 0 | 0 | Low-quality | Genome-fragment | | 1.1 | AAI-based (high-confidence) | 0 |
| S9-final-viral-combined_k141_40176\|\|full | 1691 | No | NA | | 3 | 0 | 1 | Low-quality | Genome-fragment | | 2.22 | AAI-based (medium-confidence) | 0 |
| S5-final-viral-combined_k141_166271\|\|full | 1314 | No | NA | | 3 | 0 | 1 | Not-determined | Genome-fragment | NA | | NA | 0 |
| S11-final-viral-combined_k141_42751\|\|full | 1691 | No | NA | | 2 | 0 | 0 | Low-quality | Genome-fragment | | 3.52 | AAI-based (medium-confidence) | 0 |
| S1-final-viral-combined_k141_389355\|\|full | 1691 | No | NA | | 7 | 0 | 0 | Low-quality | Genome-fragment | | 3.71 | AAI-based (high-confidence) | 0 |
| S5_k141_122483 | 1691 | No | NA | | 2 | 0 | 1 | Low-quality | Genome-fragment | | 1.6 | AAI-based (medium-confidence) | 0 |
| S7-final-viral-combined_k141_69972\|\|full | 1691 | No | NA | | 2 | 0 | 1 | Low-quality | Genome-fragment | | 1.21 | AAI-based (high-confidence) | 0 |
| S5-final-viral-combined_k141_32593\|\|full | 1394 | No | NA | | 3 | 0 | 1 | Not-determined | Genome-fragment | NA | | NA | 0 |
| S17-final-viral-combined_k141_148866\|\|full | 1691 | No | NA | | 3 | 0 | 1 | Low-quality | Genome-fragment | | 0.51 | AAI-based (medium-confidence) | 0 |
| S5-final-viral-combined_k141_15134\|\|full | 1837 | No | NA | | 4 | 0 | 1 | Not-determined | Genome-fragment | NA | | NA | 0 |
| S5-final-viral-combined_k141_50915\|\|full | 1045 | No | NA | | 2 | 0 | 0 | Not-determined | Genome-fragment | NA | | NA | 0 |
| S5-final-viral-combined_k141_50918\|\|full | 1360 | No | NA | | 4 | 0 | 0 | Not-determined | Genome-fragment | NA | | NA | 0 |
| S5-final-viral-combined_k141_50921\|\|full | 1088 | No | NA | | 2 | 0 | 2 | Not-determined | Genome-fragment | NA | | NA | 0 |
| S12-final-viral-combined_k141_235432\|\|full | 1691 | No | NA | | 4 | 1 | 0 | Low-quality | Genome-fragment | | 4.36 | AAI-based (medium-confidence) | 0 |
| S12-final-viral-combined_k141_288008\|\|full | 1691 | No | NA | | 2 | 0 | 1 | Low-quality | Genome-fragment | | 0.88 | AAI-based (medium-confidence) | 0 |
| S5-final-viral-combined_k141_23106\|\|full | 1674 | No | NA | | 3 | 0 | 0 | Not-determined | Genome-fragment | NA | | NA | 0 |
| S2-final-viral-combined_k141_13935\|\|full | 1691 | No | NA | | 3 | 0 | 0 | Low-quality | Genome-fragment | | 3.67 | AAI-based (medium-confidence) | 0 |
| S2-final-viral-combined_k141_184239\|\|full | 1691 | No | NA | | 5 | 0 | 0 | Low-quality | Genome-fragment | | 2.05 | AAI-based (high-confidence) | 0 |
| S5-final-viral-combined_k141_50955\|\|full | 1958 | No | NA | | 3 | 0 | 1 | Not-determined | Genome-fragment | NA | | NA | 0 |
| S2-final-viral-combined_k141_186226\|\|full | 1691 | No | NA | | 6 | 1 | 0 | Low-quality | Genome-fragment | | 3.35 | AAI-based (high-confidence) | 0 |
| S8-final-viral-combined_k141_187810\|\|lt2gene | 1691 | No | NA | | 2 | 1 | 0 | Low-quality | Genome-fragment | | 4.09 | AAI-based (high-confidence) | 0 |
| S8_k141_76667 | 1691 | No | NA | | 2 | 0 | 2 | Low-quality | Genome-fragment | | 0.28 | AAI-based (high-confidence) | 0 |
| S1-final-viral-combined_k141_49494\|\|full | 1692 | No | NA | | 2 | 0 | 0 | Low-quality | Genome-fragment | | 2.71 | AAI-based (high-confidence) | 0 |
| S1-final-viral-combined_k141_230583\|\|full | 1692 | No | NA | | 3 | 0 | 0 | Low-quality | Genome-fragment | | 1.99 | AAI-based (high-confidence) | 0 |
| S1-final-viral-combined_k141_118478\|\|full | 1692 | No | NA | | 1 | 0 | 0 | Low-quality | Genome-fragment | | 3.84 | AAI-based (medium-confidence) | 0 |
| S5-final-viral-combined_k141_68086\|\|full | 1974 | No | NA | | 3 | 0 | 1 | Not-determined | Genome-fragment | NA | | NA | 0 |
| S5-final-viral-combined_k141_23177\|\|full | 1987 | No | NA | | 4 | 0 | 0 | Not-determined | Genome-fragment | NA | | NA | 0 |
| S5-final-viral-combined_k141_166378\|\|full | 2061 | No | NA | | 3 | 0 | 2 | Not-determined | Genome-fragment | NA | | NA | 0 |
| S5-final-viral-combined_k141_23188\|\|full | 6085 | No | NA | | 12 | 0 | 0 | Not-determined | Genome-fragment | NA | | NA | 0 |
| S1-final-viral-combined_k141_21520\|\|full | 1692 | No | NA | | 5 | 1 | 0 | Low-quality | Genome-fragment | | 4.06 | AAI-based (high-confidence) | 0 |
| S5-final-viral-combined_k141_8153\|\|full | 2497 | No | NA | | 3 | 0 | 1 | Not-determined | Genome-fragment | NA | | NA | 0 |
| S5-final-viral-combined_k141_157409\|\|full | 2231 | No | NA | | 2 | 0 | 1 | Not-determined | Genome-fragment | NA | | NA | 0 |
| S5-final-viral-combined_k141_210959\|\|full | 194243 | Yes | | 61717 | 163 | 7 | 87 | Medium-quality | Genome-fragment | | 85.14 | HMM-based (lower-bound) | 68.23 |
| S5-final-viral-combined_k141_15228\|\|full | 2959 | No | NA | | 4 | 0 | 1 | Not-determined | Genome-fragment | NA | | NA | 0 |
| S5-final-viral-combined_k141_185062\|\|full | 3342 | No | NA | | 4 | 0 | 4 | Not-determined | Genome-fragment | NA | | NA | 0 |
| S5-final-viral-combined_k141_185067\|\|full | 1872 | No | NA | | 3 | 0 | 0 | Not-determined | Genome-fragment | NA | | NA | 0 |
| S5-final-viral-combined_k141_121980\|\|full | 1371 | No | NA | | 3 | 0 | 2 | Not-determined | Genome-fragment | NA | | NA | 0 |
| S1-final-viral-combined_k141_106049\|\|full | 1692 | No | NA | | 4 | 0 | 3 | Low-quality | Genome-fragment | | 1.24 | AAI-based (medium-confidence) | 0 |
| S1_k141_69213 | 1692 | No | NA | | 5 | 1 | 0 | Low-quality | Genome-fragment | | 3.41 | HMM-based (lower-bound) | 0 |
| S2-final-viral-combined_k141_25168\|\|full | 1692 | No | NA | | 3 | 1 | 0 | Low-quality | Genome-fragment | | 0.91 | HMM-based (lower-bound) | 0 |
| S5-final-viral-combined_k141_78162\|\|full | 4991 | No | NA | | 6 | 0 | 4 | Not-determined | Genome-fragment | NA | | NA | 0 |
| S5-final-viral-combined_k141_32764\|\|full | 1004 | No | NA | | 2 | 0 | 1 | Not-determined | Genome-fragment | NA | | NA | 0 |
| S2-final-viral-combined_k141_221803\|\|full | 1692 | No | NA | | 2 | 0 | 1 | Low-quality | Genome-fragment | | 2.36 | AAI-based (high-confidence) | 0 |
| S5-final-viral-combined_k141_104681\|\|full | 7494 | No | NA | | 7 | 0 | 4 | Not-determined | Genome-fragment | NA | | NA | 0 |
| S5-final-viral-combined_k141_185089\|\|full | 6007 | No | NA | | 5 | 0 | 5 | Not-determined | Genome-fragment | NA | | NA | 0 |
| S4_k141_105768 | 1692 | No | NA | | 1 | 0 | 1 | Low-quality | Genome-fragment | | 0.93 | AAI-based (medium-confidence) | 0 |
| S11-final-viral-combined_k141_210447\|\|full | 1693 | No | NA | | 3 | 0 | 0 | Low-quality | Genome-fragment | | 3.13 | AAI-based (high-confidence) | 0 |
| S5-final-viral-combined_k141_193470\|\|full | 1463 | No | NA | | 3 | 0 | 1 | Not-determined | Genome-fragment | NA | | NA | 0 |
| S5-final-viral-combined_k141_104689\|\|full | 3530 | No | NA | | 5 | 0 | 2 | Not-determined | Genome-fragment | NA | | NA | 0 |
| S5-final-viral-combined_k141_149097\|\|full | 1107 | No | NA | | 2 | 0 | 0 | Not-determined | Genome-fragment | NA | | NA | 0 |
| S11_k141_165969 | 1693 | No | NA | | 6 | 2 | 0 | Low-quality | Genome-fragment | | 2.49 | AAI-based (high-confidence) | 0 |
| S1-final-viral-combined_k141_302386\|\|full | 1693 | No | NA | | 8 | 2 | 0 | Low-quality | Genome-fragment | | 2.77 | AAI-based (high-confidence) | 0 |
| S5-final-viral-combined_k141_202101\|\|full | 1383 | No | NA | | 3 | 0 | 1 | Not-determined | Genome-fragment | NA | | NA | 0 |
| S1-final-viral-combined_k141_58543\|\|full | 1693 | No | NA | | 3 | 0 | 0 | Low-quality | Genome-fragment | | 3.93 | AAI-based (high-confidence) | 0 |
| S5-final-viral-combined_k141_122038\|\|full | 5628 | No | NA | | 6 | 0 | 4 | Not-determined | Genome-fragment | NA | | NA | 0 |
| S1_k141_152851 | 1693 | No | NA | | 1 | 0 | 1 | Low-quality | Genome-fragment | | 0.83 | AAI-based (medium-confidence) | 0 |
| S5-final-viral-combined_k141_68268\|\|full | 1029 | No | NA | | 2 | 0 | 1 | Not-determined | Genome-fragment | NA | | NA | 0 |
| S7-final-viral-combined_k141_79721\|\|lt2gene | 1693 | No | NA | | 2 | 2 | 0 | Low-quality | Genome-fragment | | 4 | AAI-based (high-confidence) | 0 |
| S5-final-viral-combined_k141_95083\|\|full | 2234 | No | NA | | 2 | 0 | 2 | Not-determined | Genome-fragment | NA | | NA | 0 |
| S5-final-viral-combined_k141_185138\|\|full | 1896 | No | NA | | 3 | 0 | 0 | Not-determined | Genome-fragment | NA | | NA | 0 |
| S2-final-viral-combined_k141_234655\|\|full | 1693 | No | NA | | 4 | 1 | 0 | Low-quality | Genome-fragment | | 3.54 | AAI-based (medium-confidence) | 0 |
| S5-final-viral-combined_k141_23288\|\|full | 1170 | No | NA | | 2 | 0 | 1 | Not-determined | Genome-fragment | NA | | NA | 0 |
| S5-final-viral-combined_k141_157512\|\|full | 2241 | No | NA | | 3 | 0 | 1 | Not-determined | Genome-fragment | NA | | NA | 0 |
| S5-final-viral-combined_k141_149136\|\|full | 3555 | No | NA | | 4 | 0 | 1 | Not-determined | Genome-fragment | NA | | NA | 0 |
| S5-final-viral-combined_k141_149140\|\|full | 3438 | No | NA | | 6 | 0 | 0 | Not-determined | Genome-fragment | NA | | NA | 0 |
| S5-final-viral-combined_k141_23351\|\|full | 5628 | No | NA | | 5 | 0 | 1 | Not-determined | Genome-fragment | NA | | NA | 0 |
| S5-final-viral-combined_k141_149172\|\|full | 2546 | No | NA | | 3 | 0 | 3 | Not-determined | Genome-fragment | NA | | NA | 0 |
| S6-final-viral-combined_k141_13680\|\|full | 1693 | No | NA | | 2 | 0 | 0 | Low-quality | Genome-fragment | | 3.75 | AAI-based (medium-confidence) | 0 |
| S8-final-viral-combined_k141_107824\|\|full | 1693 | No | NA | | 4 | 0 | 0 | Low-quality | Genome-fragment | | 1.69 | AAI-based (medium-confidence) | 0 |
| S5-final-viral-combined_k141_185141\|\|full | 70563 | No | NA | | 65 | 0 | 14 | Not-determined | Genome-fragment | NA | | NA | 0 |
| S8-final-viral-combined_k141_215848\|\|full | 1693 | No | NA | | 2 | 0 | 1 | Low-quality | Genome-fragment | | 2.69 | AAI-based (high-confidence) | 0 |
| S5-final-viral-combined_k141_41830\|\|full | 1894 | No | NA | | 5 | 0 | 0 | Not-determined | Genome-fragment | NA | | NA | 0 |
| S5-final-viral-combined_k141_139900\|\|full | 3195 | No | NA | | 2 | 0 | 2 | Not-determined | Genome-fragment | NA | | NA | 0 |
| S3-final-viral-combined_k141_70490\|\|full | 1694 | No | NA | | 6 | 0 | 0 | Low-quality | Genome-fragment | | 3.04 | AAI-based (medium-confidence) | 0 |
| S12-final-viral-combined_k141_80527\|\|full | 1694 | No | NA | | 5 | 0 | 0 | Low-quality | Genome-fragment | | 3.57 | AAI-based (high-confidence) | 0 |
| S5-final-viral-combined_k141_193560\|\|full | 4568 | No | NA | | 5 | 0 | 4 | Not-determined | Genome-fragment | NA | | NA | 0 |
| S12_k141_170297 | 1694 | No | NA | | 1 | 0 | 1 | Low-quality | Genome-fragment | | 0.31 | AAI-based (medium-confidence) | 0 |
| S8-final-viral-combined_k141_228519\|\|full | 1694 | No | NA | | 3 | 0 | 1 | Low-quality | Genome-fragment | | 0.47 | AAI-based (medium-confidence) | 0 |
| S5-final-viral-combined_k141_149203\|\|full | 4821 | No | NA | | 4 | 0 | 4 | Not-determined | Genome-fragment | NA | | NA | 0 |
| S5-final-viral-combined_k141_51305\|\|full | 1091 | No | NA | | 2 | 0 | 1 | Not-determined | Genome-fragment | NA | | NA | 0 |
| S5-final-viral-combined_k141_41868\|\|full | 2305 | No | NA | | 2 | 0 | 0 | Not-determined | Genome-fragment | NA | | NA | 0 |
| S11-final-viral-combined_k141_144658\|\|full | 1695 | No | NA | | 3 | 0 | 0 | Low-quality | Genome-fragment | | 3.96 | AAI-based (medium-confidence) | 0 |
| S5-final-viral-combined_k141_95148\|\|full | 5116 | No | NA | | 6 | 0 | 4 | Not-determined | Genome-fragment | NA | | NA | 0 |
| S11-final-viral-combined_k141_217756\|\|full | 1695 | No | NA | | 4 | 0 | 0 | Low-quality | Genome-fragment | | 4.44 | AAI-based (medium-confidence) | 0 |
| S5-final-viral-combined_k141_86846\|\|full | 1713 | No | NA | | 3 | 0 | 1 | Not-determined | Genome-fragment | NA | | NA | 0 |
| S1-final-viral-combined_k141_74441\|\|full | 1695 | No | NA | | 3 | 0 | 0 | Low-quality | Genome-fragment | | 2.95 | AAI-based (medium-confidence) | 0 |
| S5-final-viral-combined_k141_193654\|\|full | 3539 | No | NA | | 2 | 0 | 2 | Not-determined | Genome-fragment | NA | | NA | 0 |
| S5-final-viral-combined_k141_193655\|\|full | 1725 | No | NA | | 3 | 0 | 0 | Not-determined | Genome-fragment | NA | | NA | 0 |
| S5-final-viral-combined_k141_104842\|\|full | 1025 | No | NA | | 2 | 0 | 0 | Not-determined | Genome-fragment | NA | | NA | 0 |
| S5-final-viral-combined_k141_122117\|\|full | 3278 | No | NA | | 4 | 0 | 3 | Not-determined | Genome-fragment | NA | | NA | 0 |
| S5-final-viral-combined_k141_113513\|\|full | 1073 | No | NA | | 3 | 0 | 0 | Not-determined | Genome-fragment | NA | | NA | 0 |
| S5-final-viral-combined_k141_202191\|\|full | 1201 | No | NA | | 2 | 0 | 2 | Not-determined | Genome-fragment | NA | | NA | 0 |
| S1_k141_42460 | 1695 | No | NA | | 3 | 1 | 0 | Low-quality | Genome-fragment | | 4.59 | AAI-based (high-confidence) | 0 |
| S5-final-viral-combined_k141_41950\|\|full | 1953 | No | NA | | 6 | 0 | 0 | Not-determined | Genome-fragment | NA | | NA | 0 |
| S5-final-viral-combined_k141_202208\|\|full | 2855 | No | NA | | 6 | 0 | 3 | Not-determined | Genome-fragment | NA | | NA | 0 |
| S5-final-viral-combined_k141_78673\|\|full | 1695 | No | NA | | 4 | 1 | 0 | Low-quality | Genome-fragment | | 5.13 | AAI-based (medium-confidence) | 0 |
| S12_k141_52544 | 1695 | No | NA | | 3 | 0 | 0 | Low-quality | Genome-fragment | | 3.9 | AAI-based (medium-confidence) | 0 |
| S5-final-viral-combined_k141_51371\|\|full | 1988 | No | NA | | 3 | 0 | 0 | Not-determined | Genome-fragment | NA | | NA | 0 |
| S5-final-viral-combined_k141_166594\|\|full | 1467 | No | NA | | 3 | 0 | 3 | Not-determined | Genome-fragment | NA | | NA | 0 |
| S5-final-viral-combined_k141_166595\|\|full | 1384 | No | NA | | 4 | 0 | 1 | Not-determined | Genome-fragment | NA | | NA | 0 |
| S2_k141_156916 | 1695 | No | NA | | 2 | 0 | 0 | Low-quality | Genome-fragment | | 1.22 | AAI-based (medium-confidence) | 0 |
| S5-final-viral-combined_k141_185242\|\|full | 1505 | No | NA | | 2 | 0 | 1 | Not-determined | Genome-fragment | NA | | NA | 0 |
| S4-final-viral-combined_k141_119712\|\|full | 1695 | No | NA | | 3 | 2 | 0 | Low-quality | Genome-fragment | | 2.76 | AAI-based (medium-confidence) | 0 |
| S5-final-viral-combined_k141_122199\|\|full | 1320 | No | NA | | 3 | 0 | 3 | Not-determined | Genome-fragment | NA | | NA | 0 |
| S4-final-viral-combined_k141_103198\|\|full | 1695 | No | NA | | 4 | 1 | 0 | Low-quality | Genome-fragment | | 2.07 | AAI-based (high-confidence) | 0 |
| S11-final-viral-combined_k141_102388\|\|full | 1696 | No | NA | | 2 | 2 | 0 | Low-quality | Genome-fragment | | 3.18 | HMM-based (lower-bound) | 0 |
| S1-final-viral-combined_k141_97910\|\|full | 1696 | No | NA | | 2 | 0 | 1 | Low-quality | Genome-fragment | | 1.95 | AAI-based (high-confidence) | 0 |
| S5-final-viral-combined_k141_32974\|\|full | 1464 | No | NA | | 3 | 0 | 1 | Not-determined | Genome-fragment | NA | | NA | 0 |
| S5-final-viral-combined_k141_193712\|\|full | 3045 | No | NA | | 6 | 0 | 0 | Not-determined | Genome-fragment | NA | | NA | 0 |
| S5-final-viral-combined_k141_131899\|\|full | 1799 | No | NA | | 3 | 0 | 3 | Not-determined | Genome-fragment | NA | | NA | 0 |
| S5-final-viral-combined_k141_22994\|\|full | 1696 | No | NA | | 3 | 2 | 0 | Low-quality | Genome-fragment | | 3.26 | AAI-based (high-confidence) | 0 |
| S5-final-viral-combined_k141_33006\|\|full | 2233 | No | NA | | 6 | 0 | 0 | Not-determined | Genome-fragment | NA | | NA | 0 |
| S5-final-viral-combined_k141_23159\|\|full | 1696 | No | NA | | 2 | 0 | 1 | Low-quality | Genome-fragment | | 3.29 | AAI-based (high-confidence) | 0 |
| S5-final-viral-combined_k141_104921\|\|full | 1888 | No | NA | | 2 | 0 | 2 | Not-determined | Genome-fragment | NA | | NA | 0 |
| S5-final-viral-combined_k141_122263\|\|full | 2007 | No | NA | | 3 | 0 | 1 | Not-determined | Genome-fragment | NA | | NA | 0 |
| S5-final-viral-combined_k141_33058\|\|full | 1336 | No | NA | | 2 | 0 | 2 | Not-determined | Genome-fragment | NA | | NA | 0 |
| S5-final-viral-combined_k141_51421\|\|full | 3636 | No | NA | | 6 | 0 | 3 | Not-determined | Genome-fragment | NA | | NA | 0 |
| S5-final-viral-combined_k141_60213\|\|full | 1919 | No | NA | | 2 | 0 | 2 | Not-determined | Genome-fragment | NA | | NA | 0 |
| S5-final-viral-combined_k141_68608\|\|full | 1700 | No | NA | | 5 | 0 | 0 | Not-determined | Genome-fragment | NA | | NA | 0 |
| S5-final-viral-combined_k141_34083\|\|full | 1696 | No | NA | | 3 | 1 | 0 | Low-quality | Genome-fragment | | 3.41 | HMM-based (lower-bound) | 0 |
| S5-final-viral-combined_k141_122295\|\|full | 1639 | No | NA | | 2 | 0 | 0 | Not-determined | Genome-fragment | NA | | NA | 0 |
| S5_k141_49741 | 1696 | No | NA | | 2 | 0 | 0 | Low-quality | Genome-fragment | | 3.06 | AAI-based (high-confidence) | 0 |
| S7-final-viral-combined_k141_5322\|\|full | 1696 | No | NA | | 2 | 0 | 2 | Low-quality | Genome-fragment | | 0.93 | AAI-based (medium-confidence) | 0 |
| S17-final-viral-combined_k141_193758\|\|full | 1696 | No | NA | | 3 | 0 | 0 | Low-quality | Genome-fragment | | 2.74 | AAI-based (medium-confidence) | 0 |
| S17-final-viral-combined_k141_92565\|\|full | 1696 | No | NA | | 2 | 0 | 2 | Low-quality | Genome-fragment | | 3.92 | AAI-based (medium-confidence) | 0 |
| S5-final-viral-combined_k141_166780\|\|full | 1081 | No | NA | | 2 | 0 | 1 | Not-determined | Genome-fragment | NA | | NA | 0 |
| S5-final-viral-combined_k141_78434\|\|full | 146444 | No | NA | | 142 | 4 | 25 | Medium-quality | Genome-fragment | | 89.79 | HMM-based (lower-bound) | 0 |
| S17-final-viral-combined_k141_5203\|\|full | 1696 | No | NA | | 3 | 1 | 0 | Low-quality | Genome-fragment | | 2.1 | HMM-based (lower-bound) | 0 |
| S17-k141_88249 | 1696 | No | NA | | 2 | 0 | 0 | Low-quality | Genome-fragment | | 0.7 | AAI-based (high-confidence) | 0 |
| S5-final-viral-combined_k141_105028\|\|full | 2318 | No | NA | | 4 | 0 | 4 | Not-determined | Genome-fragment | NA | | NA | 0 |
| S12-final-viral-combined_k141_43364\|\|full | 1696 | No | NA | | 5 | 2 | 0 | Low-quality | Genome-fragment | | 4.32 | AAI-based (high-confidence) | 0 |
| S5-final-viral-combined_k141_15687\|\|full | 2716 | No | NA | | 6 | 0 | 2 | Not-determined | Genome-fragment | NA | | NA | 0 |
| S2-final-viral-combined_k141_3228\|\|full | 1696 | No | NA | | 4 | 2 | 0 | Low-quality | Genome-fragment | | 3.65 | AAI-based (high-confidence) | 0 |
| S4-final-viral-combined_k141_151269\|\|full | 1696 | No | NA | | 3 | 0 | 0 | Low-quality | Genome-fragment | | 1.08 | AAI-based (high-confidence) | 0 |
| S5-final-viral-combined_k141_23676\|\|full | 1646 | No | NA | | 3 | 0 | 1 | Not-determined | Genome-fragment | NA | | NA | 0 |
| S6-final-viral-combined_k141_9960\|\|full | 1696 | No | NA | | 3 | 0 | 0 | Low-quality | Genome-fragment | | 3.54 | AAI-based (medium-confidence) | 0 |
| S8-final-viral-combined_k141_135995\|\|lt2gene | 1696 | No | NA | | 2 | 1 | 0 | Low-quality | Genome-fragment | | 4.04 | AAI-based (medium-confidence) | 0 |
| S5-final-viral-combined_k141_166890\|\|full | 1642 | No | NA | | 3 | 0 | 1 | Not-determined | Genome-fragment | NA | | NA | 0 |
| S5-final-viral-combined_k141_166901\|\|full | 3867 | No | NA | | 5 | 0 | 5 | Not-determined | Genome-fragment | NA | | NA | 0 |
| S9-final-viral-combined_k141_11148\|\|full | 1697 | No | NA | | 4 | 0 | 0 | Low-quality | Genome-fragment | | 1.81 | AAI-based (medium-confidence) | 0 |
| S5-final-viral-combined_k141_105104\|\|full | 3175 | No | NA | | 4 | 0 | 3 | Not-determined | Genome-fragment | NA | | NA | 0 |
| S11-final-viral-combined_k141_23388\|\|full | 1697 | No | NA | | 2 | 1 | 0 | Low-quality | Genome-fragment | | 3.82 | AAI-based (high-confidence) | 0 |
| S1-final-viral-combined_k141_204207\|\|full | 1697 | No | NA | | 4 | 0 | 0 | Low-quality | Genome-fragment | | 5.09 | AAI-based (high-confidence) | 0 |
| S5-final-viral-combined_k141_78573\|\|full | 2161 | No | NA | | 3 | 0 | 2 | Not-determined | Genome-fragment | NA | | NA | 0 |
| S5-final-viral-combined_k141_60337\|\|full | 8961 | No | NA | | 6 | 0 | 2 | Not-determined | Genome-fragment | NA | | NA | 0 |
| S5-final-viral-combined_k141_146737\|\|full | 1697 | No | NA | | 2 | 0 | 0 | Low-quality | Genome-fragment | | 2 | AAI-based (medium-confidence) | 0 |
| S7-final-viral-combined_k141_161631\|\|full | 1697 | No | NA | | 2 | 0 | 1 | Low-quality | Genome-fragment | | 0.87 | AAI-based (medium-confidence) | 0 |
| S12-final-viral-combined_k141_61176\|\|full | 1697 | No | NA | | 2 | 0 | 0 | Low-quality | Genome-fragment | | 2.15 | AAI-based (high-confidence) | 0 |
| S5-final-viral-combined_k141_211468\|\|full | 2568 | No | NA | | 2 | 0 | 1 | Not-determined | Genome-fragment | NA | | NA | 0 |
| S2-final-viral-combined_k141_105038\|\|full | 1697 | No | NA | | 2 | 1 | 0 | Low-quality | Genome-fragment | | 2.82 | AAI-based (medium-confidence) | 0 |
| S5-final-viral-combined_k141_140300\|\|full | 3651 | No | NA | | 4 | 0 | 3 | Not-determined | Genome-fragment | NA | | NA | 0 |
| S4-final-viral-combined_k141_37661\|\|full | 1697 | No | NA | | 3 | 3 | 0 | Low-quality | Genome-fragment | | 2.73 | AAI-based (high-confidence) | 0 |
| S12_k141_124790 | 1698 | No | NA | | 1 | 0 | 0 | Low-quality | Genome-fragment | | 2.9 | AAI-based (high-confidence) | 0 |
| S5-final-viral-combined_k141_105175\|\|full | 1314 | No | NA | | 3 | 0 | 0 | Not-determined | Genome-fragment | NA | | NA | 0 |
| S2-final-viral-combined_k141_21734\|\|full | 1698 | No | NA | | 3 | 0 | 0 | Low-quality | Genome-fragment | | 5.78 | AAI-based (high-confidence) | 0 |
| S2-final-viral-combined_k141_172945\|\|full | 1698 | No | NA | | 2 | 0 | 0 | Low-quality | Genome-fragment | | 2.14 | AAI-based (high-confidence) | 0 |
| S5-final-viral-combined_k141_105180\|\|full | 1613 | No | NA | | 3 | 0 | 0 | Not-determined | Genome-fragment | NA | | NA | 0 |
| S2-final-viral-combined_k141_117719\|\|lt2gene | 1698 | No | NA | | 2 | 2 | 0 | Low-quality | Genome-fragment | | 4.07 | AAI-based (high-confidence) | 0 |
| S5-final-viral-combined_k141_140330\|\|full | 2069 | No | NA | | 4 | 0 | 1 | Not-determined | Genome-fragment | NA | | NA | 0 |
| S2-final-viral-combined_k141_141096\|\|lt2gene | 1698 | No | NA | | 1 | 1 | 0 | Low-quality | Genome-fragment | | 2.48 | HMM-based (lower-bound) | 0 |
| S5-final-viral-combined_k141_8517\|\|full | 1691 | No | NA | | 2 | 0 | 1 | Not-determined | Genome-fragment | NA | | NA | 0 |
| S4_k141_157432 | 1698 | No | NA | | 1 | 0 | 1 | Low-quality | Genome-fragment | | 2.77 | AAI-based (medium-confidence) | 0 |
| S5-final-viral-combined_k141_122476\|\|full | 1953 | No | NA | | 3 | 0 | 2 | Not-determined | Genome-fragment | NA | | NA | 0 |
| S5-final-viral-combined_k141_105213\|\|full | 1182 | No | NA | | 3 | 0 | 1 | Not-determined | Genome-fragment | NA | | NA | 0 |
| S5-final-viral-combined_k141_166992\|\|full | 1533 | No | NA | | 2 | 0 | 1 | Not-determined | Genome-fragment | NA | | NA | 0 |
| S5-final-viral-combined_k141_122486\|\|full | 1045 | No | NA | | 2 | 0 | 0 | Not-determined | Genome-fragment | NA | | NA | 0 |
| S8-final-viral-combined_k141_98215\|\|full | 1698 | No | NA | | 4 | 1 | 0 | Low-quality | Genome-fragment | | 2.75 | AAI-based (high-confidence) | 0 |
| S11-final-viral-combined_k141_203351\|\|full | 1699 | No | NA | | 3 | 0 | 1 | Low-quality | Genome-fragment | | 2.06 | AAI-based (high-confidence) | 0 |
| S1-final-viral-combined_k141_251703\|\|full | 1699 | No | NA | | 1 | 0 | 1 | Low-quality | Genome-fragment | | 2.79 | AAI-based (medium-confidence) | 0 |
| S1-final-viral-combined_k141_154449\|\|full | 1699 | No | NA | | 2 | 1 | 0 | Low-quality | Genome-fragment | | 2.57 | HMM-based (lower-bound) | 0 |
| S5-final-viral-combined_k141_87267\|\|full | 2137 | No | NA | | 3 | 0 | 1 | Not-determined | Genome-fragment | NA | | NA | 0 |
| S5-final-viral-combined_k141_122510\|\|full | 3782 | No | NA | | 3 | 0 | 3 | Not-determined | Genome-fragment | NA | | NA | 0 |
| S5-final-viral-combined_k141_78692\|\|full | 1892 | No | NA | | 3 | 0 | 0 | Not-determined | Genome-fragment | NA | | NA | 0 |
| S5-final-viral-combined_k141_87288\|\|full | 2204 | No | NA | | 5 | 0 | 0 | Not-determined | Genome-fragment | NA | | NA | 0 |
| S5-final-viral-combined_k141_194125\|\|full | 1959 | No | NA | | 2 | 0 | 0 | Not-determined | Genome-fragment | NA | | NA | 0 |
| S5-final-viral-combined_k141_194126\|\|full | 1036 | No | NA | | 2 | 0 | 1 | Not-determined | Genome-fragment | NA | | NA | 0 |
| S5-final-viral-combined_k141_140366\|\|full | 1494 | No | NA | | 2 | 0 | 0 | Not-determined | Genome-fragment | NA | | NA | 0 |
| S1-final-viral-combined_k141_375253\|\|full | 1699 | No | NA | | 3 | 1 | 0 | Low-quality | Genome-fragment | | 2.85 | AAI-based (high-confidence) | 0 |
| S3-final-viral-combined_k141_9277\|\|full | 1699 | No | NA | | 3 | 0 | 1 | Low-quality | Genome-fragment | | 1.58 | AAI-based (medium-confidence) | 0 |
| S5-final-viral-combined_k141_87311\|\|full | 1184 | No | NA | | 2 | 0 | 1 | Not-determined | Genome-fragment | NA | | NA | 0 |
| S5-final-viral-combined_k141_33297\|\|full | 6286 | No | NA | | 7 | 0 | 3 | Not-determined | Genome-fragment | NA | | NA | 0 |
| S3-final-viral-combined_k141_16610\|\|full | 1699 | No | NA | | 3 | 0 | 1 | Low-quality | Genome-fragment | | 1.17 | AAI-based (medium-confidence) | 0 |
| S12-final-viral-combined_k141_87303\|\|full | 1699 | No | NA | | 3 | 0 | 2 | Low-quality | Genome-fragment | | 2.97 | AAI-based (high-confidence) | 0 |
| S12-final-viral-combined_k141_172251\|\|full | 1699 | No | NA | | 4 | 0 | 1 | Low-quality | Genome-fragment | | 2.57 | AAI-based (medium-confidence) | 0 |
| S4-final-viral-combined_k141_130646\|\|full | 1699 | No | NA | | 3 | 0 | 0 | Low-quality | Genome-fragment | | 3.95 | AAI-based (medium-confidence) | 0 |
| S5-final-viral-combined_k141_122597\|\|full | 1021 | No | NA | | 3 | 0 | 1 | Not-determined | Genome-fragment | NA | | NA | 0 |
| S6_k141_92867 | 1699 | No | NA | | 4 | 1 | 0 | Low-quality | Genome-fragment | | 3.28 | AAI-based (high-confidence) | 0 |
| S5-final-viral-combined_k141_194227\|\|full | 4258 | No | NA | | 7 | 0 | 3 | Not-determined | Genome-fragment | NA | | NA | 0 |
| S5-final-viral-combined_k141_87393\|\|full | 1036 | No | NA | | 3 | 0 | 0 | Not-determined | Genome-fragment | NA | | NA | 0 |
| S11-final-viral-combined_k141_306873\|\|full | 1700 | No | NA | | 4 | 2 | 0 | Low-quality | Genome-fragment | | 3.68 | AAI-based (medium-confidence) | 0 |
| S11-final-viral-combined_k141_176303\|\|full | 1700 | No | NA | | 3 | 0 | 0 | Low-quality | Genome-fragment | | 3.72 | AAI-based (high-confidence) | 0 |
| S1-final-viral-combined_k141_64831\|\|full | 1700 | No | NA | | 2 | 0 | 0 | Low-quality | Genome-fragment | | 3.85 | AAI-based (medium-confidence) | 0 |
| S5-final-viral-combined_k141_68985\|\|full | 1288 | No | NA | | 3 | 0 | 1 | Not-determined | Genome-fragment | NA | | NA | 0 |
| S5-final-viral-combined_k141_23878\|\|full | 5083 | No | NA | | 5 | 0 | 2 | Not-determined | Genome-fragment | NA | | NA | 0 |
| S7_k141_38595 | 1700 | No | NA | | 4 | 2 | 0 | Low-quality | Genome-fragment | | 3.75 | AAI-based (high-confidence) | 0 |
| S17-final-viral-combined_k141_114254\|\|full | 1700 | No | NA | | 2 | 0 | 1 | Low-quality | Genome-fragment | | 1.95 | AAI-based (medium-confidence) | 0 |
| S5-final-viral-combined_k141_185530\|\|full | 1796 | No | NA | | 2 | 0 | 0 | Not-determined | Genome-fragment | NA | | NA | 0 |
| S5-final-viral-combined_k141_42121\|\|full | 2128 | No | NA | | 4 | 0 | 0 | Not-determined | Genome-fragment | NA | | NA | 0 |
| S17-final-viral-combined_k141_193008\|\|full | 1700 | No | NA | | 3 | 0 | 0 | Low-quality | Genome-fragment | | 3.59 | AAI-based (medium-confidence) | 0 |
| S5-final-viral-combined_k141_60650\|\|full | 1882 | No | NA | | 3 | 0 | 0 | Not-determined | Genome-fragment | NA | | NA | 0 |
| S5-final-viral-combined_k141_8760\|\|full | 1765 | No | NA | | 4 | 0 | 1 | Not-determined | Genome-fragment | NA | | NA | 0 |
| S17-final-viral-combined_k141_159885\|\|full | 1700 | No | NA | | 2 | 0 | 1 | Low-quality | Genome-fragment | | 3.19 | AAI-based (medium-confidence) | 0 |
| S5-final-viral-combined_k141_87464\|\|full | 6079 | No | NA | | 3 | 0 | 3 | Not-determined | Genome-fragment | NA | | NA | 0 |
| S17-final-viral-combined_k141_61437\|\|full | 1700 | Yes | | 1001 | 4 | 1 | 1 | Low-quality | Genome-fragment | | 1.44 | HMM-based (lower-bound) | 41.12 |
| S17-final-viral-combined_k141_244453\|\|lt2gene | 1700 | No | NA | | 1 | 1 | 0 | Low-quality | Genome-fragment | | 2.75 | AAI-based (high-confidence) | 0 |
| S2-final-viral-combined_k141_236191\|\|full | 1700 | No | NA | | 3 | 0 | 0 | Low-quality | Genome-fragment | | 1.34 | AAI-based (medium-confidence) | 0 |
| S6-final-viral-combined_k141_10864\|\|full | 1700 | No | NA | | 3 | 0 | 0 | Low-quality | Genome-fragment | | 2.04 | AAI-based (medium-confidence) | 0 |
| S11-final-viral-combined_k141_161278\|\|full | 1701 | No | NA | | 3 | 0 | 0 | Low-quality | Genome-fragment | | 2.3 | AAI-based (medium-confidence) | 0 |
| S11-final-viral-combined_k141_316770\|\|full | 1701 | No | NA | | 5 | 1 | 0 | Low-quality | Genome-fragment | | 4.86 | AAI-based (medium-confidence) | 0 |
| S5-final-viral-combined_k141_33487\|\|full | 2447 | No | NA | | 4 | 0 | 3 | Not-determined | Genome-fragment | NA | | NA | 0 |
| S5-final-viral-combined_k141_149516\|\|full | 3698 | No | NA | | 8 | 0 | 2 | Not-determined | Genome-fragment | NA | | NA | 0 |
| S5-final-viral-combined_k141_149530\|\|full | 4786 | No | NA | | 4 | 0 | 0 | Not-determined | Genome-fragment | NA | | NA | 0 |
| S5-final-viral-combined_k141_15935\|\|full | 1958 | No | NA | | 2 | 0 | 1 | Not-determined | Genome-fragment | NA | | NA | 0 |
| S11_k141_321715 | 1701 | No | NA | | 2 | 0 | 0 | Low-quality | Genome-fragment | | 1.79 | AAI-based (medium-confidence) | 0 |
| S5-final-viral-combined_k141_95763\|\|full | 3826 | No | NA | | 5 | 0 | 0 | Not-determined | Genome-fragment | NA | | NA | 0 |
| S5-final-viral-combined_k141_185558\|\|full | 1275 | No | NA | | 3 | 0 | 0 | Not-determined | Genome-fragment | NA | | NA | 0 |
| S3-final-viral-combined_k141_82027\|\|full | 1701 | No | NA | | 3 | 0 | 2 | Low-quality | Genome-fragment | | 3.55 | AAI-based (medium-confidence) | 0 |
| S5-final-viral-combined_k141_18446\|\|full | 1701 | No | NA | | 3 | 1 | 0 | Low-quality | Genome-fragment | | 3.82 | AAI-based (high-confidence) | 0 |
| S5-final-viral-combined_k141_12875\|\|full | 1701 | No | NA | | 3 | 2 | 0 | Low-quality | Genome-fragment | | 2.81 | AAI-based (high-confidence) | 0 |
| S7-final-viral-combined_k141_171189\|\|full | 1701 | No | NA | | 3 | 2 | 0 | Low-quality | Genome-fragment | | 2.58 | HMM-based (lower-bound) | 0 |
| S7-final-viral-combined_k141_142455\|\|full | 1701 | No | NA | | 3 | 0 | 3 | Low-quality | Genome-fragment | | 1.3 | AAI-based (medium-confidence) | 0 |
| S5-final-viral-combined_k141_51920\|\|full | 5191 | No | NA | | 6 | 0 | 1 | Not-determined | Genome-fragment | NA | | NA | 0 |
| S4-final-viral-combined_k141_81898\|\|full | 1701 | No | NA | | 2 | 0 | 1 | Low-quality | Genome-fragment | | 1.12 | AAI-based (medium-confidence) | 0 |
| S5-final-viral-combined_k141_95841\|\|full | 1489 | No | NA | | 2 | 0 | 2 | Not-determined | Genome-fragment | NA | | NA | 0 |
| S5_k141_99589 | 1702 | No | NA | | 2 | 0 | 0 | Low-quality | Genome-fragment | | 2.64 | AAI-based (medium-confidence) | 0 |
| S5-final-viral-combined_k141_42211\|\|full | 2514 | No | NA | | 3 | 0 | 3 | Not-determined | Genome-fragment | NA | | NA | 0 |
| S5-final-viral-combined_k141_176675\|\|full | 5239 | No | NA | | 3 | 0 | 2 | Not-determined | Genome-fragment | NA | | NA | 0 |
| S5-final-viral-combined_k141_69244\|\|full | 2357 | No | NA | | 4 | 0 | 0 | Not-determined | Genome-fragment | NA | | NA | 0 |
| S7-final-viral-combined_k141_105267\|\|full | 1702 | No | NA | | 3 | 0 | 0 | Low-quality | Genome-fragment | | 2.07 | AAI-based (high-confidence) | 0 |
| S17-k141_11093 | 1702 | No | NA | | 5 | 3 | 0 | Low-quality | Genome-fragment | | 5.9 | AAI-based (high-confidence) | 0 |
| S2-final-viral-combined_k141_63797\|\|full | 1702 | No | NA | | 3 | 1 | 0 | Low-quality | Genome-fragment | | 4.07 | AAI-based (medium-confidence) | 0 |
| S5-final-viral-combined_k141_140651\|\|full | 1668 | No | NA | | 3 | 0 | 2 | Not-determined | Genome-fragment | NA | | NA | 0 |
| S5-final-viral-combined_k141_140662\|\|full | 2416 | No | NA | | 3 | 0 | 1 | Not-determined | Genome-fragment | NA | | NA | 0 |
| S5-final-viral-combined_k141_33669\|\|full | 1245 | No | NA | | 3 | 0 | 0 | Not-determined | Genome-fragment | NA | | NA | 0 |
| S5-final-viral-combined_k141_42258\|\|full | 7236 | No | NA | | 6 | 0 | 0 | Not-determined | Genome-fragment | NA | | NA | 0 |
| S4-final-viral-combined_k141_157658\|\|full | 1702 | No | NA | | 4 | 0 | 0 | Low-quality | Genome-fragment | | 2.8 | AAI-based (high-confidence) | 0 |
| S5-final-viral-combined_k141_122936\|\|full | 1252 | No | NA | | 3 | 0 | 0 | Not-determined | Genome-fragment | NA | | NA | 0 |
| S5-final-viral-combined_k141_52064\|\|full | 1158 | No | NA | | 2 | 0 | 0 | Not-determined | Genome-fragment | NA | | NA | 0 |
| S5-final-viral-combined_k141_24055\|\|full | 2583 | No | NA | | 3 | 0 | 1 | Not-determined | Genome-fragment | NA | | NA | 0 |
| S6-final-viral-combined_k141_104871\|\|full | 1702 | No | NA | | 2 | 0 | 1 | Low-quality | Genome-fragment | | 1.81 | AAI-based (medium-confidence) | 0 |
| S8-final-viral-combined_k141_23252\|\|full | 1702 | No | NA | | 2 | 0 | 1 | Low-quality | Genome-fragment | | 1.81 | AAI-based (medium-confidence) | 0 |
| S8-final-viral-combined_k141_84841\|\|full | 1702 | No | NA | | 3 | 0 | 1 | Low-quality | Genome-fragment | | 3.36 | AAI-based (medium-confidence) | 0 |
| S5-final-viral-combined_k141_202730\|\|full | 1498 | No | NA | | 3 | 0 | 1 | Not-determined | Genome-fragment | NA | | NA | 0 |
| S1-final-viral-combined_k141_313516\|\|full | 1703 | Yes | | 609 | 2 | 1 | 1 | Low-quality | Genome-fragment | | 0.38 | HMM-based (lower-bound) | 64.24 |
| S1-final-viral-combined_k141_149381\|\|full | 1703 | No | NA | | 2 | 1 | 0 | Low-quality | Genome-fragment | | 1.02 | AAI-based (high-confidence) | 0 |
| S5-final-viral-combined_k141_105620\|\|full | 1292 | No | NA | | 3 | 0 | 0 | Not-determined | Genome-fragment | NA | | NA | 0 |
| S1-final-viral-combined_k141_84141\|\|full | 1703 | No | NA | | 5 | 0 | 0 | Low-quality | Genome-fragment | | 0.87 | AAI-based (medium-confidence) | 0 |
| S5-final-viral-combined_k141_105634\|\|full | 1433 | No | NA | | 3 | 0 | 1 | Not-determined | Genome-fragment | NA | | NA | 0 |
| S1-final-viral-combined_k141_393220\|\|full | 1703 | No | NA | | 4 | 2 | 0 | Low-quality | Genome-fragment | | 1.04 | HMM-based (lower-bound) | 0 |
| S5-final-viral-combined_k141_176778\|\|full | 6625 | No | NA | | 7 | 0 | 4 | Not-determined | Genome-fragment | NA | | NA | 0 |
| S1-final-viral-combined_k141_163782\|\|full | 1703 | No | NA | | 5 | 2 | 0 | Low-quality | Genome-fragment | | 2.8 | AAI-based (high-confidence) | 0 |
| S5-final-viral-combined_k141_158204\|\|full | 2413 | No | NA | | 2 | 0 | 2 | Not-determined | Genome-fragment | NA | | NA | 0 |
| S5-final-viral-combined_k141_150420\|\|full | 1703 | No | NA | | 2 | 0 | 2 | Low-quality | Genome-fragment | | 1.6 | AAI-based (medium-confidence) | 0 |
| S5-final-viral-combined_k141_149719\|\|full | 3028 | No | NA | | 5 | 0 | 5 | Not-determined | Genome-fragment | NA | | NA | 0 |
| S17-final-viral-combined_k141_176573\|\|lt2gene | 1703 | No | NA | | 1 | 1 | 0 | Low-quality | Genome-fragment | | 3.16 | AAI-based (medium-confidence) | 0 |
| S5-final-viral-combined_k141_185708\|\|full | 1000 | No | NA | | 3 | 0 | 0 | Not-determined | Genome-fragment | NA | | NA | 0 |
| S12-final-viral-combined_k141_124517\|\|full | 1703 | No | NA | | 5 | 0 | 0 | Low-quality | Genome-fragment | | 4.46 | AAI-based (high-confidence) | 0 |
| S2-final-viral-combined_k141_241825\|\|full | 1703 | No | NA | | 2 | 0 | 1 | Low-quality | Genome-fragment | | 1.39 | AAI-based (high-confidence) | 0 |
| S2_k141_198800 | 1703 | No | NA | | 2 | 0 | 2 | Low-quality | Genome-fragment | | 1.04 | AAI-based (high-confidence) | 0 |
| S8-final-viral-combined_k141_94965\|\|full | 1703 | No | NA | | 2 | 0 | 1 | Low-quality | Genome-fragment | | 0.49 | AAI-based (high-confidence) | 0 |
| S8-final-viral-combined_k141_249448\|\|full | 1703 | No | NA | | 3 | 0 | 0 | Low-quality | Genome-fragment | | 3.53 | AAI-based (medium-confidence) | 0 |
| S5-final-viral-combined_k141_78976\|\|full | 1490 | No | NA | | 4 | 0 | 2 | Not-determined | Genome-fragment | NA | | NA | 0 |
| S11-final-viral-combined_k141_328835\|\|full | 1704 | No | NA | | 2 | 0 | 0 | Low-quality | Genome-fragment | | 2.09 | AAI-based (medium-confidence) | 0 |
| S1_k141_238863 | 1704 | No | NA | | 3 | 0 | 1 | Low-quality | Genome-fragment | | 2.75 | AAI-based (medium-confidence) | 0 |
| S5-final-viral-combined_k141_194551\|\|full | 1203 | No | NA | | 3 | 0 | 0 | Not-determined | Genome-fragment | NA | | NA | 0 |
| S3-final-viral-combined_k141_94841\|\|full | 1704 | No | NA | | 3 | 2 | 0 | Low-quality | Genome-fragment | | 4.47 | AAI-based (high-confidence) | 0 |
| S5-final-viral-combined_k141_156922\|\|full | 1704 | No | NA | | 5 | 0 | 0 | Low-quality | Genome-fragment | | 2.94 | AAI-based (high-confidence) | 0 |
| S5-final-viral-combined_k141_194558\|\|full | 1294 | No | NA | | 3 | 0 | 0 | Not-determined | Genome-fragment | NA | | NA | 0 |
| S5_k141_182329 | 1704 | No | NA | | 5 | 1 | 0 | Low-quality | Genome-fragment | | 3.91 | AAI-based (high-confidence) | 0 |
| S5-final-viral-combined_k141_194573\|\|full | 2733 | No | NA | | 2 | 0 | 1 | Not-determined | Genome-fragment | NA | | NA | 0 |
| S5-final-viral-combined_k141_202878\|\|full | 4908 | No | NA | | 8 | 0 | 3 | Not-determined | Genome-fragment | NA | | NA | 0 |
| S7-final-viral-combined_k141_229803\|\|full | 1704 | No | NA | | 3 | 1 | 0 | Low-quality | Genome-fragment | | 3.22 | HMM-based (lower-bound) | 0 |
| S5-final-viral-combined_k141_149829\|\|full | 2453 | No | NA | | 3 | 0 | 3 | Not-determined | Genome-fragment | NA | | NA | 0 |
| S5-final-viral-combined_k141_149831\|\|full | 1371 | No | NA | | 3 | 0 | 2 | Not-determined | Genome-fragment | NA | | NA | 0 |
| S5-final-viral-combined_k141_79042\|\|full | 2660 | No | NA | | 4 | 0 | 3 | Not-determined | Genome-fragment | NA | | NA | 0 |
| S17-final-viral-combined_k141_45192\|\|full | 1704 | No | NA | | 2 | 2 | 0 | Low-quality | Genome-fragment | | 4.17 | AAI-based (high-confidence) | 0 |
| S5-final-viral-combined_k141_194619\|\|full | 2461 | No | NA | | 3 | 0 | 0 | Not-determined | Genome-fragment | NA | | NA | 0 |
| S5-final-viral-combined_k141_33919\|\|full | 1552 | No | NA | | 3 | 0 | 0 | Not-determined | Genome-fragment | NA | | NA | 0 |
| S5-final-viral-combined_k141_87901\|\|full | 1387 | No | NA | | 2 | 0 | 1 | Not-determined | Genome-fragment | NA | | NA | 0 |
| S5-final-viral-combined_k141_33927\|\|full | 1007 | No | NA | | 3 | 0 | 0 | Not-determined | Genome-fragment | NA | | NA | 0 |
| S12-final-viral-combined_k141_150862\|\|full | 1704 | No | NA | | 3 | 1 | 0 | Low-quality | Genome-fragment | | 3.71 | AAI-based (high-confidence) | 0 |
| S5-final-viral-combined_k141_16036\|\|full | 1150 | No | NA | | 3 | 0 | 1 | Not-determined | Genome-fragment | NA | | NA | 0 |
| S2-final-viral-combined_k141_37686\|\|full | 1704 | No | NA | | 3 | 0 | 2 | Low-quality | Genome-fragment | | 0.57 | AAI-based (medium-confidence) | 0 |
| S5-final-viral-combined_k141_194648\|\|full | 2499 | No | NA | | 3 | 0 | 2 | Not-determined | Genome-fragment | NA | | NA | 0 |
| S5-final-viral-combined_k141_123222\|\|full | 1337 | No | NA | | 2 | 0 | 2 | Not-determined | Genome-fragment | NA | | NA | 0 |
| S2-final-viral-combined_k141_109751\|\|full | 1704 | No | NA | | 1 | 0 | 0 | Low-quality | Genome-fragment | | 5.09 | AAI-based (medium-confidence) | 0 |
| S5-final-viral-combined_k141_177056\|\|full | 1231 | No | NA | | 3 | 0 | 0 | Not-determined | Genome-fragment | NA | | NA | 0 |
| S5-final-viral-combined_k141_167569\|\|full | 2560 | No | NA | | 4 | 0 | 1 | Not-determined | Genome-fragment | NA | | NA | 0 |
| S5-final-viral-combined_k141_194672\|\|full | 2337 | No | NA | | 2 | 0 | 0 | Not-determined | Genome-fragment | NA | | NA | 0 |
| S4-final-viral-combined_k141_77135\|\|full | 1704 | No | NA | | 3 | 2 | 0 | Low-quality | Genome-fragment | | 4.23 | AAI-based (medium-confidence) | 0 |
| S1-final-viral-combined_k141_322854\|\|full | 1705 | No | NA | | 4 | 1 | 0 | Low-quality | Genome-fragment | | 3.85 | AAI-based (high-confidence) | 0 |
| S5-final-viral-combined_k141_24271\|\|full | 8106 | No | NA | | 5 | 0 | 5 | Not-determined | Genome-fragment | NA | | NA | 0 |
| S1-final-viral-combined_k141_146113\|\|full | 1705 | No | NA | | 2 | 0 | 1 | Low-quality | Genome-fragment | | 2.21 | AAI-based (medium-confidence) | 0 |
| S5-final-viral-combined_k141_96080\|\|full | 2567 | No | NA | | 3 | 0 | 0 | Not-determined | Genome-fragment | NA | | NA | 0 |
| S5-final-viral-combined_k141_24288\|\|full | 1441 | No | NA | | 3 | 0 | 2 | Not-determined | Genome-fragment | NA | | NA | 0 |
| S7_k141_51537 | 1705 | No | NA | | 5 | 1 | 0 | Low-quality | Genome-fragment | | 2.18 | AAI-based (high-confidence) | 0 |
| S17-final-viral-combined_k141_121098\|\|full | 1705 | No | NA | | 3 | 1 | 0 | Low-quality | Genome-fragment | | 4.62 | AAI-based (high-confidence) | 0 |
| S5-final-viral-combined_k141_87970\|\|full | 1202 | No | NA | | 2 | 0 | 1 | Not-determined | Genome-fragment | NA | | NA | 0 |
| S12-final-viral-combined_k141_48421\|\|full | 1705 | No | NA | | 4 | 0 | 1 | Low-quality | Genome-fragment | | 3.21 | AAI-based (medium-confidence) | 0 |
| S4_k141_150635 | 1705 | No | NA | | 2 | 0 | 1 | Low-quality | Genome-fragment | | 2.63 | AAI-based (high-confidence) | 0 |
| S6-final-viral-combined_k141_93681\|\|full | 1705 | No | NA | | 2 | 0 | 1 | Low-quality | Genome-fragment | | 1.82 | AAI-based (medium-confidence) | 0 |
| S11-final-viral-combined_k141_306176\|\|full | 1706 | No | NA | | 2 | 0 | 0 | Low-quality | Genome-fragment | | 3.15 | AAI-based (medium-confidence) | 0 |
| S5-final-viral-combined_k141_211970\|\|full | 1024 | No | NA | | 3 | 0 | 0 | Not-determined | Genome-fragment | NA | | NA | 0 |
| S5-final-viral-combined_k141_69357\|\|full | 3486 | No | NA | | 4 | 0 | 3 | Not-determined | Genome-fragment | NA | | NA | 0 |
| S1_k141_262086 | 1706 | No | NA | | 1 | 0 | 1 | Low-quality | Genome-fragment | | 0.75 | AAI-based (medium-confidence) | 0 |
| S1_k141_196242 | 1706 | No | NA | | 2 | 1 | 0 | Low-quality | Genome-fragment | | 1.11 | AAI-based (medium-confidence) | 0 |
| S5-final-viral-combined_k141_123306\|\|full | 4009 | No | NA | | 3 | 0 | 2 | Not-determined | Genome-fragment | NA | | NA | 0 |
| S5-final-viral-combined_k141_167650\|\|full | 3663 | No | NA | | 3 | 0 | 2 | Not-determined | Genome-fragment | NA | | NA | 0 |
| S5-final-viral-combined_k141_130578\|\|lt2gene | 1706 | No | NA | | 1 | 1 | 0 | Low-quality | Genome-fragment | | 4.77 | AAI-based (medium-confidence) | 0 |
| S7-final-viral-combined_k141_239126\|\|lt2gene | 1706 | No | NA | | 2 | 1 | 0 | Low-quality | Genome-fragment | | 4.34 | AAI-based (high-confidence) | 0 |
| S5-final-viral-combined_k141_105757\|\|full | 29322 | No | NA | | 21 | 0 | 11 | Not-determined | Genome-fragment | NA | | NA | 0 |
| S5-final-viral-combined_k141_177143\|\|full | 2092 | No | NA | | 3 | 0 | 0 | Not-determined | Genome-fragment | NA | | NA | 0 |
| S5-final-viral-combined_k141_158459\|\|full | 2897 | No | NA | | 7 | 0 | 1 | Not-determined | Genome-fragment | NA | | NA | 0 |
| S5-final-viral-combined_k141_149962\|\|full | 1367 | No | NA | | 3 | 0 | 2 | Not-determined | Genome-fragment | NA | | NA | 0 |
| S5-final-viral-combined_k141_167690\|\|full | 5460 | No | NA | | 7 | 0 | 1 | Not-determined | Genome-fragment | NA | | NA | 0 |
| S17-final-viral-combined_k141_257609\|\|full | 1706 | No | NA | | 2 | 0 | 1 | Low-quality | Genome-fragment | | 1.17 | AAI-based (medium-confidence) | 0 |
| S12_k141_93500 | 1706 | No | NA | | 3 | 2 | 0 | Low-quality | Genome-fragment | | 4.3 | AAI-based (medium-confidence) | 0 |
| S2-final-viral-combined_k141_190568\|\|full | 1706 | No | NA | | 3 | 0 | 1 | Low-quality | Genome-fragment | | 2.88 | AAI-based (high-confidence) | 0 |
| S2-final-viral-combined_k141_48440\|\|full | 1706 | No | NA | | 3 | 0 | 1 | Low-quality | Genome-fragment | | 0.82 | AAI-based (medium-confidence) | 0 |
| S5-final-viral-combined_k141_123334\|\|full | 4702 | No | NA | | 5 | 0 | 4 | Not-determined | Genome-fragment | NA | | NA | 0 |
| S5-final-viral-combined_k141_212027\|\|full | 1144 | No | NA | | 2 | 0 | 1 | Not-determined | Genome-fragment | NA | | NA | 0 |
| S4_k141_48294 | 1706 | No | NA | | 1 | 0 | 1 | Low-quality | Genome-fragment | | 2.08 | AAI-based (medium-confidence) | 0 |
| S5-final-viral-combined_k141_202965\|\|full | 2401 | No | NA | | 4 | 0 | 4 | Not-determined | Genome-fragment | NA | | NA | 0 |
| S8-final-viral-combined_k141_199477\|\|full | 1706 | No | NA | | 3 | 0 | 0 | Low-quality | Genome-fragment | | 3.42 | AAI-based (high-confidence) | 0 |
| S5-final-viral-combined_k141_79206\|\|full | 2355 | No | NA | | 3 | 0 | 1 | Not-determined | Genome-fragment | NA | | NA | 0 |
| S5-final-viral-combined_k141_42775\|\|full | 2291 | No | NA | | 3 | 0 | 2 | Not-determined | Genome-fragment | NA | | NA | 0 |
| S5-final-viral-combined_k141_16110\|\|full | 1998 | No | NA | | 4 | 0 | 1 | Not-determined | Genome-fragment | NA | | NA | 0 |
| S5-final-viral-combined_k141_185976\|\|full | 1030 | No | NA | | 2 | 0 | 0 | Not-determined | Genome-fragment | NA | | NA | 0 |
| S5-final-viral-combined_k141_185984\|\|full | 1066 | No | NA | | 2 | 0 | 1 | Not-determined | Genome-fragment | NA | | NA | 0 |
| S1-final-viral-combined_k141_254319\|\|full | 1707 | Yes | | 449 | 2 | 1 | 1 | Low-quality | Genome-fragment | | 0.18 | HMM-based (lower-bound) | 73.7 |
| S5-final-viral-combined_k141_16181\|\|full | 1754 | No | NA | | 3 | 0 | 2 | Not-determined | Genome-fragment | NA | | NA | 0 |
| S5-final-viral-combined_k141_185991\|\|full | 1428 | No | NA | | 3 | 0 | 2 | Not-determined | Genome-fragment | NA | | NA | 0 |
| S5-final-viral-combined_k141_114125\|\|full | 7922 | No | NA | | 7 | 0 | 5 | Not-determined | Genome-fragment | NA | | NA | 0 |
| S5-final-viral-combined_k141_16192\|\|full | 2287 | No | NA | | 2 | 0 | 1 | Not-determined | Genome-fragment | NA | | NA | 0 |
| S5-final-viral-combined_k141_79223\|\|full | 1951 | No | NA | | 4 | 0 | 2 | Not-determined | Genome-fragment | NA | | NA | 0 |
| S3-final-viral-combined_k141_34475\|\|full | 1707 | No | NA | | 2 | 0 | 2 | Low-quality | Genome-fragment | | 4.08 | AAI-based (medium-confidence) | 0 |
| S12-final-viral-combined_k141_270579\|\|full | 1707 | No | NA | | 3 | 0 | 0 | Low-quality | Genome-fragment | | 2.84 | AAI-based (medium-confidence) | 0 |
| S5-final-viral-combined_k141_105995\|\|full | 2100 | No | NA | | 3 | 0 | 1 | Not-determined | Genome-fragment | NA | | NA | 0 |
| S2-final-viral-combined_k141_134213\|\|full | 1707 | No | NA | | 3 | 1 | 0 | Low-quality | Genome-fragment | | 3.8 | AAI-based (high-confidence) | 0 |
| S5-final-viral-combined_k141_16235\|\|full | 3802 | No | NA | | 2 | 0 | 0 | Not-determined | Genome-fragment | NA | | NA | 0 |
| S5-final-viral-combined_k141_34172\|\|full | 2786 | No | NA | | 5 | 0 | 3 | Not-determined | Genome-fragment | NA | | NA | 0 |
| S4_k141_42227 | 1707 | No | NA | | 5 | 1 | 0 | Low-quality | Genome-fragment | | 2.37 | AAI-based (high-confidence) | 0 |
| S5-final-viral-combined_k141_167855\|\|full | 3299 | No | NA | | 3 | 0 | 2 | Not-determined | Genome-fragment | NA | | NA | 0 |
| S5-final-viral-combined_k141_24422\|\|full | 2370 | No | NA | | 3 | 0 | 0 | Not-determined | Genome-fragment | NA | | NA | 0 |
| S8-final-viral-combined_k141_2174\|\|1_partial | 1707 | No | NA | | 4 | 0 | 0 | Low-quality | Genome-fragment | | 3.79 | AAI-based (medium-confidence) | 0 |
| S5-final-viral-combined_k141_114191\|\|full | 1794 | No | NA | | 3 | 0 | 0 | Not-determined | Genome-fragment | NA | | NA | 0 |
| S5-final-viral-combined_k141_195069\|\|full | 2028 | No | NA | | 2 | 0 | 2 | Not-determined | Genome-fragment | NA | | NA | 0 |
| S11-final-viral-combined_k141_100718\|\|full | 1708 | No | NA | | 3 | 2 | 0 | Low-quality | Genome-fragment | | 3.16 | AAI-based (medium-confidence) | 0 |
| S5-final-viral-combined_k141_106021\|\|full | 3439 | No | NA | | 4 | 0 | 2 | Not-determined | Genome-fragment | NA | | NA | 0 |

| | | | | | | | | | | | |
|---|---|---|---|---|---|---|---|---|---|---|---|
| S5-final-viral-combined_k141_88204\|\|full | 1376 | No | NA | 2 | 0 | 1 | Not-determined | Genome-fragment | NA | NA | 0 |
| S5-final-viral-combined_k141_114227\|\|full | 2739 | No | NA | 3 | 0 | 2 | Not-determined | Genome-fragment | NA | NA | 0 |
| S11-final-viral-combined_k141_230724\|\|lt2gene | 1708 | No | NA | 1 | 1 | 0 | Low-quality | Genome-fragment | 3.81 | AAI-based (medium-confidence) | 0 |
| S5-final-viral-combined_k141_96447\|\|full | 4907 | No | NA | 5 | 0 | 0 | Not-determined | Genome-fragment | NA | NA | 0 |
| S5-final-viral-combined_k141_16313\|\|full | 2501 | No | NA | 3 | 0 | 3 | Not-determined | Genome-fragment | NA | NA | 0 |
| S5-final-viral-combined_k141_42835\|\|full | 1082 | No | NA | 2 | 0 | 0 | Not-determined | Genome-fragment | NA | NA | 0 |
| S5-final-viral-combined_k141_96453\|\|full | 1442 | No | NA | 2 | 0 | 0 | Not-determined | Genome-fragment | NA | NA | 0 |
| S5-final-viral-combined_k141_106064\|\|full | 1524 | No | NA | 3 | 0 | 1 | Not-determined | Genome-fragment | NA | NA | 0 |
| S5-final-viral-combined_k141_88243\|\|full | 1994 | No | NA | 3 | 0 | 1 | Not-determined | Genome-fragment | NA | NA | 0 |
| S1-final-viral-combined_k141_42505\|\|full | 1708 | No | NA | 3 | 0 | 1 | Low-quality | Genome-fragment | 1.18 | AAI-based (medium-confidence) | 0 |
| S1_k141_13224 | 1708 | No | NA | 3 | 0 | 0 | Low-quality | Genome-fragment | 3.65 | AAI-based (medium-confidence) | 0 |
| S1_k141_394168 | 1708 | No | NA | 4 | 0 | 0 | Low-quality | Genome-fragment | 3.23 | AAI-based (high-confidence) | 0 |
| S5-final-viral-combined_k141_203096\|\|full | 1177 | No | NA | 2 | 0 | 1 | Not-determined | Genome-fragment | NA | NA | 0 |
| S12-final-viral-combined_k141_139311\|\|full | 1708 | No | NA | 4 | 0 | 1 | Low-quality | Genome-fragment | 2.47 | AAI-based (high-confidence) | 0 |
| S5-final-viral-combined_k141_186130\|\|full | 2062 | No | NA | 2 | 0 | 2 | Not-determined | Genome-fragment | NA | NA | 0 |
| S12-final-viral-combined_k141_179434\|\|full | 1708 | No | NA | 3 | 0 | 1 | Low-quality | Genome-fragment | 2.08 | AAI-based (medium-confidence) | 0 |
| S12-final-viral-combined_k141_124015\|\|lt2gene | 1708 | No | NA | 1 | 1 | 0 | Low-quality | Genome-fragment | 2.76 | AAI-based (medium-confidence) | 0 |
| S2-final-viral-combined_k141_104126\|\|full | 1708 | No | NA | 2 | 0 | 1 | Low-quality | Genome-fragment | 2.2 | AAI-based (medium-confidence) | 0 |
| S5-final-viral-combined_k141_114372\|\|full | 2283 | No | NA | 3 | 0 | 1 | Not-determined | Genome-fragment | NA | NA | 0 |
| S8-final-viral-combined_k141_103160\|\|full | 1708 | No | NA | 5 | 1 | 0 | Low-quality | Genome-fragment | 2.81 | AAI-based (medium-confidence) | 0 |
| S1-final-viral-combined_k141_218670\|\|full | 1709 | No | NA | 5 | 2 | 0 | Low-quality | Genome-fragment | 2.79 | AAI-based (high-confidence) | 0 |
| S5_k141_21514 | 1709 | No | NA | 1 | 1 | 0 | Low-quality | Genome-fragment | 0.87 | HMM-based (lower-bound) | 0 |
| S5-final-viral-combined_k141_158758\|\|full | 19544 | No | NA | 17 | 0 | 10 | Not-determined | Genome-fragment | NA | NA | 0 |
| S5-final-viral-combined_k141_123688\|\|full | 2944 | No | NA | 4 | 0 | 1 | Not-determined | Genome-fragment | NA | NA | 0 |
| S5-final-viral-combined_k141_203168\|\|full | 5625 | No | NA | 8 | 0 | 1 | Not-determined | Genome-fragment | NA | NA | 0 |
| S5-final-viral-combined_k141_24694\|\|full | 3508 | No | NA | 5 | 0 | 3 | Not-determined | Genome-fragment | NA | NA | 0 |
| S5-final-viral-combined_k141_123742\|\|full | 4195 | No | NA | 4 | 0 | 3 | Not-determined | Genome-fragment | NA | NA | 0 |
| S5-final-viral-combined_k141_195209\|\|full | 1742 | No | NA | 2 | 0 | 1 | Not-determined | Genome-fragment | NA | NA | 0 |
| S5-final-viral-combined_k141_96664\|\|full | 1677 | No | NA | 3 | 0 | 0 | Not-determined | Genome-fragment | NA | NA | 0 |
| S5-final-viral-combined_k141_88395\|\|full | 13190 | No | NA | 10 | 0 | 3 | Not-determined | Genome-fragment | NA | NA | 0 |
| S5-final-viral-combined_k141_195240\|\|full | 1640 | No | NA | 3 | 0 | 1 | Not-determined | Genome-fragment | NA | NA | 0 |
| S5-final-viral-combined_k141_150331\|\|full | 3689 | No | NA | 7 | 0 | 0 | Not-determined | Genome-fragment | NA | NA | 0 |
| S5-final-viral-combined_k141_52762\|\|full | 1772 | No | NA | 2 | 0 | 0 | Not-determined | Genome-fragment | NA | NA | 0 |
| S4-final-viral-combined_k141_16337\|\|full | 1709 | No | NA | 4 | 1 | 0 | Low-quality | Genome-fragment | 10.32 | AAI-based (high-confidence) | 0 |
| S8-final-viral-combined_k141_123093\|\|full | 1709 | No | NA | 3 | 1 | 0 | Low-quality | Genome-fragment | 4.41 | AAI-based (high-confidence) | 0 |
| S8_k141_223383 | 1709 | No | NA | 6 | 2 | 0 | Low-quality | Genome-fragment | 4.13 | AAI-based (high-confidence) | 0 |
| S5-final-viral-combined_k141_96747\|\|full | 1117 | No | NA | 2 | 0 | 1 | Not-determined | Genome-fragment | NA | NA | 0 |
| S5-final-viral-combined_k141_186223\|\|full | 1585 | No | NA | 2 | 0 | 2 | Not-determined | Genome-fragment | NA | NA | 0 |
| S11-final-viral-combined_k141_46168\|\|full | 1710 | No | NA | 4 | 1 | 0 | Low-quality | Genome-fragment | 2.82 | AAI-based (high-confidence) | 0 |
| S1-final-viral-combined_k141_90409\|\|full | 1710 | No | NA | 4 | 1 | 0 | Low-quality | Genome-fragment | 2.47 | HMM-based (lower-bound) | 0 |
| S17-final-viral-combined_k141_74201\|\|full | 1710 | No | NA | 4 | 2 | 0 | Low-quality | Genome-fragment | 2.65 | AAI-based (high-confidence) | 0 |
| S5-final-viral-combined_k141_150382\|\|full | 5162 | No | NA | 3 | 0 | 3 | Not-determined | Genome-fragment | NA | NA | 0 |
| S5-final-viral-combined_k141_212365\|\|full | 1194 | No | NA | 2 | 0 | 0 | Not-determined | Genome-fragment | NA | NA | 0 |
| S5-final-viral-combined_k141_195284\|\|full | 1748 | No | NA | 3 | 0 | 0 | Not-determined | Genome-fragment | NA | NA | 0 |
| S2-final-viral-combined_k141_119716\|\|lt2gene | 1710 | No | NA | 1 | 1 | 0 | Low-quality | Genome-fragment | 2.85 | HMM-based (lower-bound) | 0 |
| S4_k141_116160 | 1710 | No | NA | 4 | 0 | 1 | Low-quality | Genome-fragment | 2.52 | AAI-based (high-confidence) | 0 |
| S5-final-viral-combined_k141_158917\|\|full | 1576 | No | NA | 3 | 0 | 0 | Not-determined | Genome-fragment | NA | NA | 0 |
| S5-final-viral-combined_k141_158928\|\|full | 3444 | No | NA | 4 | 0 | 0 | Not-determined | Genome-fragment | NA | NA | 0 |
| S8_k141_217207 | 1710 | No | NA | 3 | 0 | 0 | Low-quality | Genome-fragment | 3.69 | AAI-based (high-confidence) | 0 |
| S5-final-viral-combined_k141_168021\|\|full | 3393 | No | NA | 2 | 0 | 2 | Not-determined | Genome-fragment | NA | NA | 0 |
| S8_k141_263368 | 1710 | No | NA | 3 | 0 | 0 | Low-quality | Genome-fragment | 2.77 | AAI-based (high-confidence) | 0 |
| S9-final-viral-combined_k141_101000\|\|full | 1711 | No | NA | 2 | 0 | 1 | Low-quality | Genome-fragment | 1.05 | AAI-based (medium-confidence) | 0 |
| S5-final-viral-combined_k141_123944\|\|full | 1234 | No | NA | 3 | 0 | 0 | Not-determined | Genome-fragment | NA | NA | 0 |
| S11-final-viral-combined_k141_116051\|\|full | 1711 | No | NA | 3 | 0 | 0 | Low-quality | Genome-fragment | 2.98 | AAI-based (high-confidence) | 0 |
| S11_k141_254000 | 1711 | No | NA | 1 | 0 | 1 | Low-quality | Genome-fragment | 1.53 | AAI-based (medium-confidence) | 0 |
| S1-final-viral-combined_k141_116043\|\|full | 1711 | No | NA | 5 | 0 | 0 | Low-quality | Genome-fragment | 6.69 | AAI-based (medium-confidence) | 0 |
| S5-final-viral-combined_k141_123949\|\|full | 2431 | No | NA | 3 | 0 | 2 | Not-determined | Genome-fragment | NA | NA | 0 |
| S3_k141_86156 | 1711 | No | NA | 2 | 0 | 0 | Low-quality | Genome-fragment | 1.67 | AAI-based (medium-confidence) | 0 |
| S7_k141_239773 | 1711 | No | NA | 5 | 1 | 0 | Low-quality | Genome-fragment | 2.3 | HMM-based (lower-bound) | 0 |
| S5-final-viral-combined_k141_16690\|\|full | 5083 | No | NA | 4 | 0 | 3 | Not-determined | Genome-fragment | NA | NA | 0 |
| S5-final-viral-combined_k141_186308\|\|full | 1736 | No | NA | 2 | 0 | 2 | Not-determined | Genome-fragment | NA | NA | 0 |
| S12-final-viral-combined_k141_151985\|\|full | 1711 | No | NA | 5 | 0 | 0 | Low-quality | Genome-fragment | 4.54 | AAI-based (high-confidence) | 0 |
| S12-final-viral-combined_k141_20230\|\|full | 1711 | No | NA | 2 | 0 | 0 | Low-quality | Genome-fragment | 2.68 | AAI-based (high-confidence) | 0 |
| S12_k141_388382 | 1711 | No | NA | 1 | 0 | 0 | Low-quality | Genome-fragment | 1.44 | AAI-based (high-confidence) | 0 |
| S5-final-viral-combined_k141_106394\|\|full | 1400 | No | NA | 2 | 0 | 0 | Not-determined | Genome-fragment | NA | NA | 0 |
| S4-final-viral-combined_k141_71651\|\|full | 1711 | No | NA | 3 | 0 | 1 | Low-quality | Genome-fragment | 4.73 | AAI-based (medium-confidence) | 0 |
| S4_k141_39150 | 1711 | Yes | 1128 | 4 | 1 | 3 | Low-quality | Genome-fragment | 2.34 | AAI-based (medium-confidence) | 34.07 |
| S6-final-viral-combined_k141_32058\|\|full | 1711 | No | NA | 4 | 1 | 0 | Low-quality | Genome-fragment | 3.15 | AAI-based (high-confidence) | 0 |
| S5-final-viral-combined_k141_43072\|\|full | 1840 | No | NA | 2 | 0 | 2 | Not-determined | Genome-fragment | NA | NA | 0 |
| S5-final-viral-combined_k141_141390\|\|full | 2769 | No | NA | 5 | 0 | 0 | Not-determined | Genome-fragment | NA | NA | 0 |
| S5-final-viral-combined_k141_61234\|\|full | 1223 | No | NA | 2 | 0 | 2 | Not-determined | Genome-fragment | NA | NA | 0 |
| S5-final-viral-combined_k141_34562\|\|full | 29164 | No | NA | 31 | 0 | 15 | Not-determined | Genome-fragment | NA | NA | 0 |
| S6-final-viral-combined_k141_27750\|\|full | 1711 | No | NA | 3 | 1 | 0 | Low-quality | Genome-fragment | 3.43 | AAI-based (medium-confidence) | 0 |
| S5-final-viral-combined_k141_79710\|\|full | 4636 | No | NA | 6 | 0 | 4 | Not-determined | Genome-fragment | NA | NA | 0 |
| S9-final-viral-combined_k141_63816\|\|lt2gene | 1712 | No | NA | 1 | 0 | 0 | Low-quality | Genome-fragment | 4.1 | AAI-based (medium-confidence) | 0 |
| S11-final-viral-combined_k141_49548\|\|full | 1712 | No | NA | 3 | 0 | 0 | Low-quality | Genome-fragment | 4.08 | AAI-based (high-confidence) | 0 |
| S11-final-viral-combined_k141_311583\|\|lt2gene | 1712 | No | NA | 1 | 1 | 0 | Low-quality | Genome-fragment | 4.72 | AAI-based (high-confidence) | 0 |
| S1-final-viral-combined_k141_219801\|\|lt2gene | 1712 | No | NA | 2 | 2 | 0 | Low-quality | Genome-fragment | 2.86 | AAI-based (high-confidence) | 0 |
| S5-final-viral-combined_k141_41960\|\|full | 1712 | No | NA | 5 | 3 | 0 | Low-quality | Genome-fragment | 4.43 | AAI-based (medium-confidence) | 0 |
| S5-final-viral-combined_k141_106481\|\|full | 1881 | No | NA | 3 | 0 | 0 | Not-determined | Genome-fragment | NA | NA | 0 |
| S5-final-viral-combined_k141_210438\|\|lt2gene | 1712 | No | NA | 2 | 1 | 0 | Low-quality | Genome-fragment | 1.18 | HMM-based (lower-bound) | 0 |
| S17-final-viral-combined_k141_257006\|\|full | 1712 | No | NA | 2 | 0 | 0 | Low-quality | Genome-fragment | 2.49 | AAI-based (medium-confidence) | 0 |
| S5-final-viral-combined_k141_133140\|\|full | 2171 | No | NA | 3 | 0 | 0 | Not-determined | Genome-fragment | NA | NA | 0 |
| S5-final-viral-combined_k141_97066\|\|full | 4268 | No | NA | 3 | 0 | 2 | Not-determined | Genome-fragment | NA | NA | 0 |
| S5-final-viral-combined_k141_195435\|\|full | 1549 | No | NA | 3 | 0 | 1 | Not-determined | Genome-fragment | NA | NA | 0 |
| S5-final-viral-combined_k141_177496\|\|full | 1254 | No | NA | 3 | 0 | 0 | Not-determined | Genome-fragment | NA | NA | 0 |
| S5-final-viral-combined_k141_168186\|\|full | 2206 | No | NA | 4 | 0 | 0 | Not-determined | Genome-fragment | NA | NA | 0 |
| S12-final-viral-combined_k141_12254\|\|full | 1712 | No | NA | 5 | 0 | 0 | Low-quality | Genome-fragment | 2.7 | AAI-based (high-confidence) | 0 |
| S5-final-viral-combined_k141_133172\|\|full | 3251 | No | NA | 3 | 0 | 3 | Not-determined | Genome-fragment | NA | NA | 0 |
| S2-final-viral-combined_k141_22674\|\|full | 1712 | No | NA | 4 | 0 | 2 | Low-quality | Genome-fragment | 0.58 | AAI-based (medium-confidence) | 0 |
| S5-final-viral-combined_k141_43204\|\|full | 2621 | No | NA | 3 | 0 | 1 | Not-determined | Genome-fragment | NA | NA | 0 |
| S2-final-viral-combined_k141_195014\|\|lt2gene | 1712 | No | NA | 2 | 2 | 0 | Low-quality | Genome-fragment | 4.83 | AAI-based (medium-confidence) | 0 |
| S5-final-viral-combined_k141_43221\|\|full | 2497 | No | NA | 4 | 0 | 1 | Not-determined | Genome-fragment | NA | NA | 0 |
| S5-final-viral-combined_k141_16840\|\|full | 13189 | No | NA | 15 | 0 | 9 | Not-determined | Genome-fragment | NA | NA | 0 |
| S4-final-viral-combined_k141_43329\|\|full | 1712 | No | NA | 2 | 0 | 2 | Low-quality | Genome-fragment | 0.48 | AAI-based (medium-confidence) | 0 |
| S5-final-viral-combined_k141_141438\|\|full | 3746 | No | NA | 5 | 0 | 1 | Not-determined | Genome-fragment | NA | NA | 0 |
| S5-final-viral-combined_k141_79823\|\|full | 1574 | No | NA | 3 | 0 | 0 | Not-determined | Genome-fragment | NA | NA | 0 |
| S6_k141_17184 | 1712 | No | NA | 1 | 0 | 1 | Low-quality | Genome-fragment | 0.9 | AAI-based (medium-confidence) | 0 |
| S5-final-viral-combined_k141_212553\|\|full | 2373 | No | NA | 3 | 0 | 2 | Not-determined | Genome-fragment | NA | NA | 0 |
| S5-final-viral-combined_k141_24966\|\|full | 2171 | No | NA | 3 | 0 | 2 | Not-determined | Genome-fragment | NA | NA | 0 |
| S9-final-viral-combined_k141_12979\|\|full | 1713 | No | NA | 4 | 1 | 0 | Low-quality | Genome-fragment | 4.43 | AAI-based (medium-confidence) | 0 |
| S9-final-viral-combined_k141_68378\|\|full | 1713 | No | NA | 2 | 0 | 0 | Low-quality | Genome-fragment | 4.14 | AAI-based (high-confidence) | 0 |
| S1-final-viral-combined_k141_305434\|\|full | 1713 | No | NA | 4 | 2 | 0 | Low-quality | Genome-fragment | 2.79 | AAI-based (high-confidence) | 0 |
| S5-final-viral-combined_k141_159144\|\|full | 9738 | No | NA | 10 | 0 | 7 | Not-determined | Genome-fragment | NA | NA | 0 |
| S1-final-viral-combined_k141_292743\|\|lt2gene | 1713 | No | NA | 2 | 1 | 0 | Low-quality | Genome-fragment | 4.99 | AAI-based (high-confidence) | 0 |
| S5-final-viral-combined_k141_203520\|\|full | 16250 | No | NA | 18 | 0 | 6 | Not-determined | Genome-fragment | NA | NA | 0 |
| S5-final-viral-combined_k141_139815\|\|full | 1713 | No | NA | 3 | 0 | 0 | Low-quality | Genome-fragment | 4.25 | AAI-based (medium-confidence) | 0 |
| S5-final-viral-combined_k141_203529\|\|full | 2889 | No | NA | 5 | 0 | 2 | Not-determined | Genome-fragment | NA | NA | 0 |
| S5-final-viral-combined_k141_150709\|\|full | 3128 | No | NA | 3 | 0 | 3 | Not-determined | Genome-fragment | NA | NA | 0 |
| S12-final-viral-combined_k141_304540\|\|full | 1713 | No | NA | 3 | 0 | 0 | Low-quality | Genome-fragment | 2.85 | AAI-based (high-confidence) | 0 |
| S12-final-viral-combined_k141_199937\|\|full | 1713 | No | NA | 4 | 2 | 0 | Low-quality | Genome-fragment | 2.78 | AAI-based (high-confidence) | 0 |
| S12-final-viral-combined_k141_132310\|\|lt2gene | 1713 | No | NA | 2 | 1 | 0 | Low-quality | Genome-fragment | 3.85 | AAI-based (medium-confidence) | 0 |
| S5-final-viral-combined_k141_159195\|\|full | 2056 | No | NA | 2 | 0 | 0 | Not-determined | Genome-fragment | NA | NA | 0 |
| S2-final-viral-combined_k141_112222\|\|full | 1713 | No | NA | 3 | 0 | 2 | Low-quality | Genome-fragment | 0.91 | AAI-based (medium-confidence) | 0 |
| S5-final-viral-combined_k141_114701\|\|full | 2526 | No | NA | 2 | 0 | 2 | Not-determined | Genome-fragment | NA | NA | 0 |
| S4-final-viral-combined_k141_116196\|\|full | 1713 | No | NA | 3 | 1 | 0 | Low-quality | Genome-fragment | 1.37 | HMM-based (lower-bound) | 0 |
| S5-final-viral-combined_k141_43288\|\|full | 1923 | No | NA | 3 | 0 | 1 | Not-determined | Genome-fragment | NA | NA | 0 |
| S8-final-viral-combined_k141_158401\|\|full | 1713 | No | NA | 4 | 1 | 0 | Low-quality | Genome-fragment | 2.83 | AAI-based (high-confidence) | 0 |
| S5-final-viral-combined_k141_168346\|\|full | 1985 | No | NA | 3 | 0 | 1 | Not-determined | Genome-fragment | NA | NA | 0 |
| S5-final-viral-combined_k141_114714\|\|full | 2047 | No | NA | 3 | 0 | 2 | Not-determined | Genome-fragment | NA | NA | 0 |
| S5-final-viral-combined_k141_97162\|\|full | 5982 | No | NA | 6 | 0 | 5 | Not-determined | Genome-fragment | NA | NA | 0 |
| S5-final-viral-combined_k141_52962\|\|full | 1828 | No | NA | 5 | 0 | 0 | Not-determined | Genome-fragment | NA | NA | 0 |
| S5-final-viral-combined_k141_212658\|\|full | 1824 | No | NA | 3 | 0 | 1 | Not-determined | Genome-fragment | NA | NA | 0 |
| S5-final-viral-combined_k141_79882\|\|full | 1511 | No | NA | 2 | 0 | 2 | Not-determined | Genome-fragment | NA | NA | 0 |
| S11-final-viral-combined_k141_58746\|\|full | 1714 | No | NA | 3 | 1 | 0 | Low-quality | Genome-fragment | 3.6 | AAI-based (high-confidence) | 0 |
| S5-final-viral-combined_k141_106597\|\|full | 25082 | No | NA | 33 | 13 | 0 | Medium-quality | Genome-fragment | 60.87 | AAI-based (medium-confidence) | 0 |
| S1-final-viral-combined_k141_375959\|\|full | 1714 | No | NA | 5 | 1 | 0 | Low-quality | Genome-fragment | 4.97 | AAI-based (high-confidence) | 0 |
| S5-final-viral-combined_k141_25068\|\|full | 2051 | No | NA | 3 | 0 | 2 | Not-determined | Genome-fragment | NA | NA | 0 |
| S5-final-viral-combined_k141_25075\|\|full | 1759 | No | NA | 3 | 0 | 1 | Not-determined | Genome-fragment | NA | NA | 0 |
| S5-final-viral-combined_k141_124167\|\|full | 1523 | No | NA | 3 | 0 | 1 | Not-determined | Genome-fragment | NA | NA | 0 |
| S5-final-viral-combined_k141_79904\|\|full | 2376 | No | NA | 3 | 0 | 2 | Not-determined | Genome-fragment | NA | NA | 0 |
| S5-final-viral-combined_k141_106635\|\|full | 2451 | No | NA | 3 | 0 | 1 | Not-determined | Genome-fragment | NA | NA | 0 |
| S5-final-viral-combined_k141_141604\|\|full | 5429 | No | NA | 7 | 0 | 4 | Not-determined | Genome-fragment | NA | NA | 0 |
| S5-final-viral-combined_k141_97249\|\|full | 1499 | No | NA | 3 | 0 | 1 | Not-determined | Genome-fragment | NA | NA | 0 |
| S5-final-viral-combined_k141_150798\|\|full | 1597 | No | NA | 3 | 0 | 0 | Not-determined | Genome-fragment | NA | NA | 0 |
| S5-final-viral-combined_k141_70025\|\|full | 3199 | No | NA | 5 | 0 | 1 | Not-determined | Genome-fragment | NA | NA | 0 |
| S5-final-viral-combined_k141_53054\|\|full | 4015 | No | NA | 4 | 0 | 4 | Not-determined | Genome-fragment | NA | NA | 0 |
| S5-final-viral-combined_k141_203673\|\|full | 1672 | No | NA | 3 | 0 | 0 | Not-determined | Genome-fragment | NA | NA | 0 |
| S5-final-viral-combined_k141_141635\|\|full | 6494 | No | NA | 7 | 0 | 4 | Not-determined | Genome-fragment | NA | NA | 0 |
| S1_k141_20434 | 1714 | No | NA | 1 | 0 | 1 | Low-quality | Genome-fragment | 4.83 | AAI-based (medium-confidence) | 0 |
| S17-final-viral-combined_k141_204352\|\|full | 1714 | No | NA | 4 | 0 | 0 | Low-quality | Genome-fragment | 1.85 | AAI-based (medium-confidence) | 0 |
| S5-final-viral-combined_k141_61498\|\|full | 5091 | No | NA | 5 | 0 | 5 | Not-determined | Genome-fragment | NA | NA | 0 |
| S12-final-viral-combined_k141_79985\|\|full | 1714 | No | NA | 4 | 3 | 0 | Low-quality | Genome-fragment | 2.08 | AAI-based (high-confidence) | 0 |
| S2-final-viral-combined_k141_77557\|\|full | 1714 | No | NA | 3 | 0 | 0 | Low-quality | Genome-fragment | 5.07 | AAI-based (high-confidence) | 0 |
| S5-final-viral-combined_k141_88884\|\|full | 3440 | No | NA | 2 | 0 | 2 | Not-determined | Genome-fragment | NA | NA | 0 |
| S4-final-viral-combined_k141_66128\|\|lt2gene | 1714 | No | NA | 2 | 1 | 0 | Low-quality | Genome-fragment | 1.18 | HMM-based (lower-bound) | 0 |
| S5-final-viral-combined_k141_34648\|\|full | 2195 | No | NA | 4 | 0 | 0 | Not-determined | Genome-fragment | NA | NA | 0 |
| S11-final-viral-combined_k141_266938\|\|full | 1715 | No | NA | 4 | 0 | 3 | Low-quality | Genome-fragment | 0.42 | AAI-based (medium-confidence) | 0 |
| S5-final-viral-combined_k141_141680\|\|full | 1026 | No | NA | 2 | 0 | 2 | Not-determined | Genome-fragment | NA | NA | 0 |
| S11-final-viral-combined_k141_200398\|\|full | 1715 | No | NA | 3 | 1 | 0 | Low-quality | Genome-fragment | 3.61 | AAI-based (medium-confidence) | 0 |
| S1-final-viral-combined_k141_232213\|\|full | 1715 | No | NA | 5 | 2 | 0 | Low-quality | Genome-fragment | 4.46 | AAI-based (high-confidence) | 0 |
| S5-final-viral-combined_k141_159341\|\|full | 2724 | No | NA | 5 | 0 | 0 | Not-determined | Genome-fragment | NA | NA | 0 |
| S5-final-viral-combined_k141_124219\|\|full | 7423 | No | NA | 7 | 0 | 6 | Not-determined | Genome-fragment | NA | NA | 0 |
| S5-final-viral-combined_k141_97290\|\|full | 1114 | No | NA | 2 | 0 | 1 | Not-determined | Genome-fragment | NA | NA | 0 |
| S1-final-viral-combined_k141_157669\|\|full | 1715 | No | NA | 4 | 4 | 0 | Low-quality | Genome-fragment | 4.28 | AAI-based (high-confidence) | 0 |
| S3-final-viral-combined_k141_35375\|\|full | 1715 | No | NA | 3 | 0 | 0 | Low-quality | Genome-fragment | 2.42 | AAI-based (medium-confidence) | 0 |
| S5-final-viral-combined_k141_159401\|\|full | 3366 | No | NA | 4 | 0 | 1 | Not-determined | Genome-fragment | NA | NA | 0 |
| S5-final-viral-combined_k141_88938\|\|full | 2488 | No | NA | 3 | 0 | 2 | Not-determined | Genome-fragment | NA | NA | 0 |
| S5-final-viral-combined_k141_61605\|\|full | 1224 | No | NA | 2 | 0 | 1 | Not-determined | Genome-fragment | NA | NA | 0 |
| S3-final-viral-combined_k141_14037\|\|full | 1715 | No | NA | 1 | 0 | 0 | Low-quality | Genome-fragment | 0.5 | AAI-based (medium-confidence) | 0 |
| S5-final-viral-combined_k141_61259\|\|full | 1715 | No | NA | 4 | 0 | 0 | Low-quality | Genome-fragment | 2.8 | AAI-based (high-confidence) | 0 |
| S17-final-viral-combined_k141_246869\|\|full | 1715 | No | NA | 3 | 0 | 1 | Low-quality | Genome-fragment | 1.42 | AAI-based (medium-confidence) | 0 |
| S5-final-viral-combined_k141_70163\|\|full | 2558 | No | NA | 4 | 0 | 3 | Not-determined | Genome-fragment | NA | NA | 0 |
| S5-final-viral-combined_k141_97397\|\|full | 3762 | No | NA | 4 | 0 | 3 | Not-determined | Genome-fragment | NA | NA | 0 |
| S5-final-viral-combined_k141_34740\|\|full | 1233 | No | NA | 3 | 0 | 0 | Not-determined | Genome-fragment | NA | NA | 0 |
| S5-final-viral-combined_k141_124337\|\|full | 1996 | No | NA | 4 | 0 | 1 | Not-determined | Genome-fragment | NA | NA | 0 |
| S12-final-viral-combined_k141_268315\|\|full | 1715 | No | NA | 3 | 2 | 0 | Low-quality | Genome-fragment | 4.41 | AAI-based (high-confidence) | 0 |
| S12-final-viral-combined_k141_154821\|\|full | 1715 | No | NA | 3 | 0 | 0 | Low-quality | Genome-fragment | 1.26 | AAI-based (medium-confidence) | 0 |
| S12-final-viral-combined_k141_93777\|\|lt2gene | 1715 | No | NA | 2 | 1 | 0 | Low-quality | Genome-fragment | 2.86 | AAI-based (high-confidence) | 0 |
| S5-final-viral-combined_k141_70215\|\|full | 2213 | No | NA | 3 | 0 | 0 | Not-determined | Genome-fragment | NA | NA | 0 |
| S5-final-viral-combined_k141_53292\|\|full | 1996 | No | NA | 3 | 0 | 0 | Not-determined | Genome-fragment | NA | NA | 0 |
| S5-final-viral-combined_k141_106759\|\|full | 3032 | No | NA | 3 | 0 | 2 | Not-determined | Genome-fragment | NA | NA | 0 |
| S2-final-viral-combined_k141_105794\|\|full | 1715 | No | NA | 2 | 0 | 2 | Low-quality | Genome-fragment | 2.23 | AAI-based (medium-confidence) | 0 |
| S6-final-viral-combined_k141_99161\|\|full | 1715 | No | NA | 2 | 0 | 1 | Low-quality | Genome-fragment | 1.12 | AAI-based (high-confidence) | 0 |
| S5-final-viral-combined_k141_97497\|\|full | 1707 | No | NA | 2 | 0 | 2 | Not-determined | Genome-fragment | NA | NA | 0 |
| S8-final-viral-combined_k141_246908\|\|full | 1715 | No | NA | 2 | 0 | 0 | Low-quality | Genome-fragment | 3.03 | AAI-based (high-confidence) | 0 |
| S1-final-viral-combined_k141_217068\|\|full | 1716 | No | NA | 3 | 2 | 0 | Low-quality | Genome-fragment | 3.25 | AAI-based (medium-confidence) | 0 |
| S1-final-viral-combined_k141_303326\|\|full | 1716 | No | NA | 2 | 0 | 1 | Low-quality | Genome-fragment | 1.49 | AAI-based (medium-confidence) | 0 |
| S5-final-viral-combined_k141_61730\|\|full | 1596 | No | NA | 3 | 0 | 0 | Not-determined | Genome-fragment | NA | NA | 0 |
| S7-final-viral-combined_k141_82618\|\|full | 1716 | No | NA | 3 | 2 | 0 | Low-quality | Genome-fragment | 4.67 | AAI-based (medium-confidence) | 0 |
| S12-final-viral-combined_k141_89089\|\|full | 1716 | No | NA | 7 | 1 | 0 | Low-quality | Genome-fragment | 4.32 | AAI-based (high-confidence) | 0 |
| S5-final-viral-combined_k141_133518\|\|full | 10021 | No | NA | 12 | 0 | 4 | Not-determined | Genome-fragment | NA | NA | 0 |
| S12-final-viral-combined_k141_32106\|\|full | 1716 | No | NA | 3 | 1 | 0 | Low-quality | Genome-fragment | 4.71 | AAI-based (high-confidence) | 0 |
| S5-final-viral-combined_k141_150977\|\|full | 2211 | No | NA | 3 | 0 | 3 | Not-determined | Genome-fragment | NA | NA | 0 |
| S2-final-viral-combined_k141_116376\|\|full | 1716 | No | NA | 4 | 4 | 0 | Low-quality | Genome-fragment | 3.21 | AAI-based (high-confidence) | 0 |
| S5-final-viral-combined_k141_150995\|\|full | 2149 | No | NA | 2 | 0 | 1 | Not-determined | Genome-fragment | NA | NA | 0 |
| S5-final-viral-combined_k141_150999\|\|full | 1870 | No | NA | 2 | 0 | 0 | Not-determined | Genome-fragment | NA | NA | 0 |
| S2_k141_57825 | 1716 | No | NA | 4 | 0 | 0 | Low-quality | Genome-fragment | 3.55 | AAI-based (medium-confidence) | 0 |
| S4-final-viral-combined_k141_98092\|\|full | 1716 | No | NA | 3 | 0 | 0 | Low-quality | Genome-fragment | 4.11 | AAI-based (high-confidence) | 0 |
| S8-final-viral-combined_k141_187671\|\|full | 1716 | No | NA | 3 | 0 | 1 | Low-quality | Genome-fragment | 3.97 | AAI-based (medium-confidence) | 0 |
| S5-final-viral-combined_k141_168660\|\|full | 4634 | No | NA | 3 | 0 | 1 | Not-determined | Genome-fragment | NA | NA | 0 |
| S5-final-viral-combined_k141_43651\|\|full | 1245 | No | NA | 2 | 0 | 0 | Not-determined | Genome-fragment | NA | NA | 0 |
| S8-final-viral-combined_k141_218454\|\|full | 1716 | No | NA | 6 | 2 | 0 | Low-quality | Genome-fragment | 3.14 | AAI-based (high-confidence) | 0 |
| S5-final-viral-combined_k141_70297\|\|full | 1778 | No | NA | 3 | 0 | 1 | Not-determined | Genome-fragment | NA | NA | 0 |
| S5-final-viral-combined_k141_97636\|\|full | 1699 | No | NA | 3 | 0 | 1 | Not-determined | Genome-fragment | NA | NA | 0 |
| S5-final-viral-combined_k141_97645\|\|full | 3193 | No | NA | 4 | 0 | 3 | Not-determined | Genome-fragment | NA | NA | 0 |
| S9-final-viral-combined_k141_11919\|\|full | 1717 | No | NA | 3 | 0 | 0 | Low-quality | Genome-fragment | 3.5 | AAI-based (medium-confidence) | 0 |
| S11_k141_197710 | 1717 | No | NA | 4 | 0 | 0 | Low-quality | Genome-fragment | 4.07 | AAI-based (high-confidence) | 0 |
| S1_k141_171493 | 1717 | No | NA | 4 | 0 | 0 | Low-quality | Genome-fragment | 1.11 | AAI-based (medium-confidence) | 0 |
| S5-final-viral-combined_k141_198947\|\|full | 1717 | No | NA | 3 | 0 | 0 | Low-quality | Genome-fragment | 2.21 | AAI-based (medium-confidence) | 0 |
| S5-final-viral-combined_k141_97714\|\|full | 1810 | No | NA | 2 | 0 | 0 | Not-determined | Genome-fragment | NA | NA | 0 |
| S5-final-viral-combined_k141_70355\|\|full | 9294 | No | NA | 8 | 0 | 4 | Not-determined | Genome-fragment | NA | NA | 0 |
| S5-final-viral-combined_k141_50868\|\|full | 1717 | No | NA | 3 | 1 | 0 | Low-quality | Genome-fragment | 3.66 | AAI-based (high-confidence) | 0 |
| S17-final-viral-combined_k141_58847\|\|full | 1717 | No | NA | 2 | 0 | 1 | Low-quality | Genome-fragment | 1.09 | AAI-based (medium-confidence) | 0 |
| S5-final-viral-combined_k141_61973\|\|full | 1900 | No | NA | 3 | 0 | 2 | Not-determined | Genome-fragment | NA | NA | 0 |
| S5-final-viral-combined_k141_124605\|\|full | 1525 | No | NA | 3 | 0 | 2 | Not-determined | Genome-fragment | NA | NA | 0 |
| S17-final-viral-combined_k141_87734\|\|full | 1717 | No | NA | 3 | 0 | 1 | Low-quality | Genome-fragment | 2.33 | AAI-based (high-confidence) | 0 |
| S5-final-viral-combined_k141_168751\|\|full | 3545 | No | NA | 5 | 0 | 1 | Not-determined | Genome-fragment | NA | NA | 0 |
| S4_k141_67375 | 1717 | No | NA | 1 | 0 | 1 | Low-quality | Genome-fragment | 2.84 | AAI-based (medium-confidence) | 0 |
| S5-final-viral-combined_k141_115300\|\|full | 4190 | No | NA | 3 | 0 | 1 | Not-determined | Genome-fragment | NA | NA | 0 |
| S5-final-viral-combined_k141_43717\|\|full | 3356 | No | NA | 3 | 0 | 2 | Not-determined | Genome-fragment | NA | NA | 0 |
| S3-final-viral-combined_k141_27682\|\|full | 1718 | No | NA | 4 | 1 | 0 | Low-quality | Genome-fragment | 1.14 | AAI-based (high-confidence) | 0 |
| S3-final-viral-combined_k141_29692\|\|lt2gene | 1718 | No | NA | 2 | 2 | 0 | Low-quality | Genome-fragment | 2.49 | AAI-based (medium-confidence) | 0 |
| S17-k141_39912 | 1718 | No | NA | 1 | 0 | 0 | Low-quality | Genome-fragment | 3.29 | AAI-based (high-confidence) | 0 |
| S5-final-viral-combined_k141_213099\|\|full | 5585 | No | NA | 8 | 0 | 4 | Not-determined | Genome-fragment | NA | NA | 0 |
| S5-final-viral-combined_k141_186775\|\|full | 4932 | No | NA | 4 | 0 | 0 | Not-determined | Genome-fragment | NA | NA | 0 |
| S5-final-viral-combined_k141_177929\|\|full | 2372 | No | NA | 4 | 0 | 1 | Not-determined | Genome-fragment | NA | NA | 0 |
| S5-final-viral-combined_k141_43760\|\|full | 2937 | No | NA | 3 | 0 | 3 | Not-determined | Genome-fragment | NA | NA | 0 |
| S5-final-viral-combined_k141_43778\|\|full | 2111 | No | NA | 2 | 0 | 1 | Not-determined | Genome-fragment | NA | NA | 0 |
| S12-final-viral-combined_k141_155524\|\|full | 1718 | No | NA | 4 | 0 | 0 | Low-quality | Genome-fragment | 4.46 | AAI-based (high-confidence) | 0 |
| S12_k141_273141 | 1718 | No | NA | 4 | 0 | 3 | Low-quality | Genome-fragment | 2.85 | AAI-based (medium-confidence) | 0 |
| S5-final-viral-combined_k141_62005\|\|full | 1747 | No | NA | 2 | 0 | 2 | Not-determined | Genome-fragment | NA | NA | 0 |
| S5-final-viral-combined_k141_159631\|\|full | 2208 | No | NA | 2 | 0 | 1 | Not-determined | Genome-fragment | NA | NA | 0 |
| S5-final-viral-combined_k141_62037\|\|full | 3909 | No | NA | 5 | 0 | 5 | Not-determined | Genome-fragment | NA | NA | 0 |
| S2-final-viral-combined_k141_66656\|\|full | 1718 | No | NA | 3 | 0 | 0 | Low-quality | Genome-fragment | 3.45 | AAI-based (medium-confidence) | 0 |
| S1_k141_336902 | 1719 | No | NA | 2 | 0 | 2 | Low-quality | Genome-fragment | 1.23 | AAI-based (high-confidence) | 0 |
| S7-final-viral-combined_k141_88417\|\|full | 1719 | No | NA | 3 | 1 | 0 | Low-quality | Genome-fragment | 3.91 | AAI-based (high-confidence) | 0 |
| S5-final-viral-combined_k141_178025\|\|full | 1605 | No | NA | 2 | 0 | 1 | Not-determined | Genome-fragment | NA | NA | 0 |
| S5-final-viral-combined_k141_35280\|\|full | 1483 | No | NA | 3 | 0 | 1 | Not-determined | Genome-fragment | NA | NA | 0 |
| S5-final-viral-combined_k141_17329\|\|full | 1046 | No | NA | 2 | 0 | 1 | Not-determined | Genome-fragment | NA | NA | 0 |
| S5-final-viral-combined_k141_70477\|\|full | 7891 | No | NA | 10 | 0 | 2 | Not-determined | Genome-fragment | NA | NA | 0 |
| S5-final-viral-combined_k141_178052\|\|full | 1128 | No | NA | 3 | 0 | 0 | Not-determined | Genome-fragment | NA | NA | 0 |
| S5-final-viral-combined_k141_35318\|\|full | 2098 | No | NA | 3 | 0 | 2 | Not-determined | Genome-fragment | NA | NA | 0 |
| S7_k141_170554 | 1719 | No | NA | 2 | 0 | 0 | Low-quality | Genome-fragment | 1.75 | AAI-based (high-confidence) | 0 |
| S2-final-viral-combined_k141_93396\|\|full | 1719 | No | NA | 2 | 0 | 1 | Low-quality | Genome-fragment | 2.35 | AAI-based (medium-confidence) | 0 |
| S5-final-viral-combined_k141_159809\|\|full | 1836 | No | NA | 4 | 0 | 1 | Not-determined | Genome-fragment | NA | NA | 0 |
| S5-final-viral-combined_k141_17441\|\|full | 1228 | No | NA | 2 | 0 | 1 | Not-determined | Genome-fragment | NA | NA | 0 |
| S5-final-viral-combined_k141_43949\|\|full | 1159 | No | NA | 2 | 0 | 1 | Not-determined | Genome-fragment | NA | NA | 0 |
| S2-final-viral-combined_k141_145816\|\|full | 1719 | No | NA | 1 | 0 | 0 | Low-quality | Genome-fragment | 3.92 | AAI-based (medium-confidence) | 0 |
| S5-final-viral-combined_k141_187031\|\|full | 1701 | No | NA | 3 | 0 | 0 | Not-determined | Genome-fragment | NA | NA | 0 |
| S2-final-viral-combined_k141_162733\|\|full | 1719 | No | NA | 4 | 1 | 0 | Low-quality | Genome-fragment | 2.7 | AAI-based (high-confidence) | 0 |
| S5-final-viral-combined_k141_151416\|\|full | 3588 | No | NA | 5 | 0 | 3 | Not-determined | Genome-fragment | NA | NA | 0 |
| S8-final-viral-combined_k141_121453\|\|full | 1719 | No | NA | 2 | 0 | 1 | Low-quality | Genome-fragment | 2.85 | AAI-based (high-confidence) | 0 |
| S1-final-viral-combined_k141_180742\|\|full | 1720 | No | NA | 3 | 0 | 0 | Low-quality | Genome-fragment | 3.58 | AAI-based (high-confidence) | 0 |
| S5-final-viral-combined_k141_159890\|\|full | 2354 | No | NA | 3 | 0 | 0 | Not-determined | Genome-fragment | NA | NA | 0 |
| S1_k141_230157 | 1720 | No | NA | 5 | 0 | 0 | Low-quality | Genome-fragment | 3.71 | AAI-based (high-confidence) | 0 |
| S5-final-viral-combined_k141_25561\|\|full | 3793 | No | NA | 6 | 0 | 0 | Not-determined | Genome-fragment | NA | NA | 0 |
| S5-final-viral-combined_k141_213174\|\|full | 1697 | No | NA | 2 | 0 | 2 | Not-determined | Genome-fragment | NA | NA | 0 |
| S5-final-viral-combined_k141_17493\|\|full | 3938 | No | NA | 5 | 0 | 1 | Not-determined | Genome-fragment | NA | NA | 0 |
| S5-final-viral-combined_k141_44053\|\|full | 5165 | No | NA | 9 | 0 | 0 | Not-determined | Genome-fragment | NA | NA | 0 |
| S5-final-viral-combined_k141_97925\|\|full | 2928 | No | NA | 6 | 0 | 0 | Not-determined | Genome-fragment | NA | NA | 0 |
| S3-final-viral-combined_k141_123665\|\|full | 1720 | No | NA | 3 | 0 | 0 | Low-quality | Genome-fragment | 3.74 | AAI-based (medium-confidence) | 0 |
| S5-final-viral-combined_k141_94077\|\|full | 1720 | No | NA | 1 | 0 | 1 | Low-quality | Genome-fragment | 2.04 | AAI-based (high-confidence) | 0 |
| S5-final-viral-combined_k141_25590\|\|full | 3110 | No | NA | 5 | 0 | 1 | Not-determined | Genome-fragment | NA | NA | 0 |
| S5-final-viral-combined_k141_97932\|\|full | 2993 | No | NA | 3 | 0 | 2 | Not-determined | Genome-fragment | NA | NA | 0 |
| S7-final-viral-combined_k141_46015\|\|full | 1720 | No | NA | 2 | 0 | 0 | Low-quality | Genome-fragment | 2.8 | AAI-based (medium-confidence) | 0 |
| S5-final-viral-combined_k141_169075\|\|full | 1080 | No | NA | 3 | 0 | 0 | Not-determined | Genome-fragment | NA | NA | 0 |
| S5-final-viral-combined_k141_169084\|\|full | 4285 | No | NA | 4 | 0 | 2 | Not-determined | Genome-fragment | NA | NA | 0 |
| S5-final-viral-combined_k141_160015\|\|full | 4002 | No | NA | 5 | 0 | 0 | Not-determined | Genome-fragment | NA | NA | 0 |
| S2_k141_141589 | 1720 | No | NA | 1 | 0 | 1 | Low-quality | Genome-fragment | 0.63 | AAI-based (high-confidence) | 0 |
| S5-final-viral-combined_k141_115587\|\|full | 3741 | No | NA | 7 | 0 | 0 | Not-determined | Genome-fragment | NA | NA | 0 |
| S5-final-viral-combined_k141_35455\|\|full | 2426 | No | NA | 2 | 0 | 2 | Not-determined | Genome-fragment | NA | NA | 0 |
| S5-final-viral-combined_k141_213249\|\|full | 1919 | No | NA | 3 | 0 | 2 | Not-determined | Genome-fragment | NA | NA | 0 |
| S5-final-viral-combined_k141_160077\|\|full | 1387 | No | NA | 4 | 0 | 1 | Not-determined | Genome-fragment | NA | NA | 0 |
| S5-final-viral-combined_k141_213275\|\|full | 1413 | No | NA | 3 | 0 | 1 | Not-determined | Genome-fragment | NA | NA | 0 |
| S5-final-viral-combined_k141_204394\|\|full | 6658 | No | NA | 6 | 0 | 1 | Not-determined | Genome-fragment | NA | NA | 0 |
| S5-final-viral-combined_k141_142423\|\|full | 4417 | No | NA | 4 | 0 | 1 | Not-determined | Genome-fragment | NA | NA | 0 |
| S5-final-viral-combined_k141_25647\|\|full | 1013 | No | NA | 2 | 0 | 1 | Not-determined | Genome-fragment | NA | NA | 0 |
| S4-final-viral-combined_k141_125409\|\|full | 1720 | No | NA | 2 | 0 | 1 | Low-quality | Genome-fragment | 1.83 | AAI-based (medium-confidence) | 0 |
| S5-final-viral-combined_k141_213287\|\|full | 1185 | No | NA | 3 | 0 | 1 | Not-determined | Genome-fragment | NA | NA | 0 |
| S1-final-viral-combined_k141_141108\|\|full | 1721 | No | NA | 3 | 0 | 0 | Low-quality | Genome-fragment | 2.94 | AAI-based (medium-confidence) | 0 |
| S5-final-viral-combined_k141_204432\|\|full | 3084 | No | NA | 3 | 0 | 1 | Not-determined | Genome-fragment | NA | NA | 0 |
| S5-final-viral-combined_k141_107911\|\|full | 1721 | No | NA | 3 | 0 | 0 | Low-quality | Genome-fragment | 3.14 | AAI-based (high-confidence) | 0 |
| S7-final-viral-combined_k141_66860\|\|full | 1721 | No | NA | 3 | 0 | 1 | Low-quality | Genome-fragment | 1.08 | AAI-based (medium-confidence) | 0 |
| S17-final-viral-combined_k141_89019\|\|full | 1721 | No | NA | 3 | 0 | 0 | Low-quality | Genome-fragment | 4.21 | AAI-based (high-confidence) | 0 |
| S11-final-viral-combined_k141_317505\|\|full | 1722 | No | NA | 2 | 0 | 2 | Low-quality | Genome-fragment | 1.65 | AAI-based (medium-confidence) | 0 |
| S11-final-viral-combined_k141_107940\|\|full | 1722 | No | NA | 3 | 0 | 2 | Low-quality | Genome-fragment | 0.89 | AAI-based (medium-confidence) | 0 |
| S1_k141_27577 | 1722 | No | NA | 2 | 0 | 1 | Low-quality | Genome-fragment | 1.54 | AAI-based (medium-confidence) | 0 |
| S5-final-viral-combined_k141_17673\|\|full | 1298 | No | NA | 3 | 0 | 1 | Not-determined | Genome-fragment | NA | NA | 0 |
| S5-final-viral-combined_k141_25709\|\|full | 1004 | No | NA | 2 | 0 | 0 | Not-determined | Genome-fragment | NA | NA | 0 |
| S5-final-viral-combined_k141_17682\|\|full | 2615 | No | NA | 6 | 0 | 4 | Not-determined | Genome-fragment | NA | NA | 0 |
| S5-final-viral-combined_k141_115728\|\|full | 1206 | No | NA | 3 | 0 | 1 | Not-determined | Genome-fragment | NA | NA | 0 |
| S5-final-viral-combined_k141_142492\|\|full | 1592 | No | NA | 2 | 0 | 0 | Not-determined | Genome-fragment | NA | NA | 0 |
| S3_k141_69993 | 1722 | No | NA | 3 | 1 | 0 | Low-quality | Genome-fragment | 4.08 | AAI-based (high-confidence) | 0 |
| S5-final-viral-combined_k141_35552\|\|full | 12727 | No | NA | 12 | 0 | 4 | Not-determined | Genome-fragment | NA | NA | 0 |
| S5-final-viral-combined_k141_32882\|\|full | 1722 | No | NA | 3 | 0 | 1 | Low-quality | Genome-fragment | 0.9 | AAI-based (medium-confidence) | 0 |
| S5-final-viral-combined_k141_25766\|\|full | 5291 | No | NA | 4 | 0 | 4 | Not-determined | Genome-fragment | NA | NA | 0 |
| S5-final-viral-combined_k141_142512\|\|full | 1937 | No | NA | 2 | 0 | 1 | Not-determined | Genome-fragment | NA | NA | 0 |
| S5-final-viral-combined_k141_25798\|\|full | 1099 | No | NA | 3 | 0 | 1 | Not-determined | Genome-fragment | NA | NA | 0 |
| S5-final-viral-combined_k141_44187\|\|full | 1883 | No | NA | 4 | 0 | 1 | Not-determined | Genome-fragment | NA | NA | 0 |
| S5-final-viral-combined_k141_44191\|\|full | 1454 | No | NA | 2 | 0 | 1 | Not-determined | Genome-fragment | NA | NA | 0 |
| S5-final-viral-combined_k141_115766\|\|full | 1724 | No | NA | 2 | 0 | 1 | Not-determined | Genome-fragment | NA | NA | 0 |
| S5-final-viral-combined_k141_44193\|\|full | 2220 | No | NA | 5 | 0 | 0 | Not-determined | Genome-fragment | NA | NA | 0 |
| S7_k141_121114 | 1722 | No | NA | 2 | 0 | 0 | Low-quality | Genome-fragment | 1.51 | AAI-based (medium-confidence) | 0 |
| S5-final-viral-combined_k141_44227\|\|full | 7022 | No | NA | 6 | 0 | 3 | Not-determined | Genome-fragment | NA | NA | 0 |
| S5-final-viral-combined_k141_17788\|\|full | 2876 | No | NA | 3 | 0 | 2 | Not-determined | Genome-fragment | NA | NA | 0 |
| S5-final-viral-combined_k141_204681\|\|full | 2072 | No | NA | 3 | 0 | 2 | Not-determined | Genome-fragment | NA | NA | 0 |
| S5-final-viral-combined_k141_70908\|\|full | 3904 | No | NA | 3 | 0 | 2 | Not-determined | Genome-fragment | NA | NA | 0 |
| S5-final-viral-combined_k141_204736\|\|full | 2703 | No | NA | 3 | 0 | 2 | Not-determined | Genome-fragment | NA | NA | 0 |
| S12_k141_398392 | 1722 | No | NA | 3 | 0 | 0 | Low-quality | Genome-fragment | 1.24 | AAI-based (medium-confidence) | 0 |
| S5-final-viral-combined_k141_70940\|\|full | 1162 | No | NA | 3 | 0 | 1 | Not-determined | Genome-fragment | NA | NA | 0 |
| S5-final-viral-combined_k141_204782\|\|full | 36604 | No | NA | 56 | 9 | 1 | High-quality | High-quality | 97.09 | AAI-based (high-confidence) | 0 |
| S5-final-viral-combined_k141_62347\|\|full | 2274 | No | NA | 3 | 0 | 2 | Not-determined | Genome-fragment | NA | NA | 0 |
| S5-final-viral-combined_k141_26035\|\|full | 3589 | No | NA | 3 | 0 | 3 | Not-determined | Genome-fragment | NA | NA | 0 |
| S5-final-viral-combined_k141_26058\|\|full | 6753 | No | NA | 6 | 0 | 6 | Not-determined | Genome-fragment | NA | NA | 0 |
| S5-final-viral-combined_k141_204879\|\|full | 1337 | No | NA | 3 | 0 | 0 | Not-determined | Genome-fragment | NA | NA | 0 |
| S5-final-viral-combined_k141_44405\|\|full | 1284 | No | NA | 3 | 0 | 2 | Not-determined | Genome-fragment | NA | NA | 0 |
| S5-final-viral-combined_k141_71033\|\|full | 1274 | No | NA | 3 | 0 | 2 | Not-determined | Genome-fragment | NA | NA | 0 |
| S5-final-viral-combined_k141_44421\|\|full | 1566 | No | NA | 3 | 0 | 3 | Not-determined | Genome-fragment | NA | NA | 0 |
| S5-final-viral-combined_k141_71050\|\|full | 4239 | No | NA | 9 | 0 | 0 | Not-determined | Genome-fragment | NA | NA | 0 |
| S5-final-viral-combined_k141_26120\|\|full | 2096 | No | NA | 5 | 0 | 0 | Not-determined | Genome-fragment | NA | NA | 0 |
| S11-final-viral-combined_k141_181797\|\|lt2gene | 1723 | No | NA | 1 | 1 | 0 | Low-quality | Genome-fragment | 1.8 | HMM-based (lower-bound) | 0 |
| S5-final-viral-combined_k141_71212\|\|full | 1388 | No | NA | 3 | 0 | 1 | Not-determined | Genome-fragment | NA | NA | 0 |
| S5-final-viral-combined_k141_71225\|\|full | 2062 | No | NA | 2 | 0 | 2 | Not-determined | Genome-fragment | NA | NA | 0 |
| S5-final-viral-combined_k141_71249\|\|full | 2983 | No | NA | 3 | 0 | 3 | Not-determined | Genome-fragment | NA | NA | 0 |
| S5-final-viral-combined_k141_71258\|\|full | 1240 | No | NA | 3 | 0 | 2 | Not-determined | Genome-fragment | NA | NA | 0 |
| S11_k141_66588 | 1723 | No | NA | 1 | 0 | 0 | Low-quality | Genome-fragment | 0.93 | AAI-based (medium-confidence) | 0 |
| S5-final-viral-combined_k141_26419\|\|full | 3802 | No | NA | 3 | 0 | 2 | Not-determined | Genome-fragment | NA | NA | 0 |
| S5-final-viral-combined_k141_26503\|\|full | 4608 | No | NA | 5 | 0 | 4 | Not-determined | Genome-fragment | NA | NA | 0 |
| S5-final-viral-combined_k141_26531\|\|full | 2925 | No | NA | 4 | 0 | 2 | Not-determined | Genome-fragment | NA | NA | 0 |
| S5-final-viral-combined_k141_26543\|\|full | 2740 | No | NA | 4 | 0 | 1 | Not-determined | Genome-fragment | NA | NA | 0 |
| S1-final-viral-combined_k141_57130\|\|full | 1723 | No | NA | 2 | 1 | 0 | Low-quality | Genome-fragment | 3.35 | AAI-based (medium-confidence) | 0 |
| S5-final-viral-combined_k141_26647\|\|full | 4137 | No | NA | 4 | 0 | 3 | Not-determined | Genome-fragment | NA | NA | 0 |
| S7-final-viral-combined_k141_43973\|\|full | 1723 | No | NA | 4 | 4 | 0 | Low-quality | Genome-fragment | 2.62 | AAI-based (high-confidence) | 0 |
| S12-final-viral-combined_k141_370921\|\|full | 1723 | No | NA | 4 | 1 | 0 | Low-quality | Genome-fragment | 3.36 | AAI-based (high-confidence) | 0 |
| S2-final-viral-combined_k141_187414\|\|full | 1723 | No | NA | 2 | 0 | 0 | Low-quality | Genome-fragment | 1.48 | AAI-based (high-confidence) | 0 |
| S4-final-viral-combined_k141_67726\|\|full | 1723 | No | NA | 3 | 1 | 0 | Low-quality | Genome-fragment | 2.58 | AAI-based (medium-confidence) | 0 |
| S5-final-viral-combined_k141_196197\|\|0_partial | 12549 | No | NA | 10 | 0 | 9 | Not-determined | Genome-fragment | NA | NA | 0 |
| S9-final-viral-combined_k141_5571\|\|full | 1724 | No | NA | 3 | 2 | 0 | Low-quality | Genome-fragment | 3.6 | AAI-based (medium-confidence) | 0 |
| S5-final-viral-combined_k141_98202\|\|0_partial | 69467 | Yes | 50930 | 59 | 5 | 13 | Medium-quality | Genome-fragment | 84.14 | AAI-based (medium-confidence) | 26.68 |
| S5-final-viral-combined_k141_26971\|\|0_partial | 84424 | Yes | 31361 | 102 | 18 | 51 | Complete | High-quality | 100 | Provirus (high-confidence) | 62.85 |
| S5-final-viral-combined_k141_142873\|\|0_partial | 16880 | No | NA | 10 | 0 | 9 | Not-determined | Genome-fragment | NA | NA | 0 |
| S5-final-viral-combined_k141_154550\|\|full | 1724 | No | NA | 3 | 1 | 0 | Low-quality | Genome-fragment | 2.18 | AAI-based (medium-confidence) | 0 |
| S17-final-viral-combined_k141_246450\|\|full | 1724 | No | NA | 2 | 1 | 0 | Low-quality | Genome-fragment | 4.77 | AAI-based (medium-confidence) | 0 |
| S17-k141_152485 | 1724 | No | NA | 2 | 0 | 0 | Low-quality | Genome-fragment | 1.31 | AAI-based (high-confidence) | 0 |
| S12-final-viral-combined_k141_249861\|\|full | 1724 | No | NA | 2 | 0 | 2 | Low-quality | Genome-fragment | 0.78 | AAI-based (high-confidence) | 0 |
| S5-final-viral-combined_k141_27036\|\|0_partial | 17501 | No | NA | 15 | 0 | 4 | Not-determined | Genome-fragment | NA | NA | 0 |
| S5-final-viral-combined_k141_178432\|\|0_partial | 50459 | No | NA | 60 | 12 | 6 | Medium-quality | Genome-fragment | 60.59 | HMM-based (lower-bound) | 0 |
| S12_k141_329270 | 1724 | No | NA | 3 | 0 | 0 | Low-quality | Genome-fragment | 3.27 | AAI-based (medium-confidence) | 0 |
| S5-final-viral-combined_k141_179241\|\|0_partial | 36095 | Yes | 28068 | 33 | 3 | 7 | Medium-quality | Genome-fragment | 67.37 | AAI-based (medium-confidence) | 22.24 |
| S5-final-viral-combined_k141_161469\|\|0_partial | 20631 | No | NA | 17 | 0 | 12 | Not-determined | Genome-fragment | NA | NA | 0 |
| S9-final-viral-combined_k141_77032\|\|full | 1725 | No | NA | 3 | 1 | 0 | Low-quality | Genome-fragment | 3.53 | AAI-based (high-confidence) | 0 |
| S5-final-viral-combined_k141_152631\|\|0_partial | 17412 | No | NA | 13 | 0 | 7 | Not-determined | Genome-fragment | NA | NA | 0 |
| S5-final-viral-combined_k141_117213\|\|0_partial | 72200 | Yes | 35094 | 81 | 23 | 22 | High-quality | High-quality | 99.25 | AAI-based (high-confidence) | 51.39 |
| S11-final-viral-combined_k141_157020\|\|full | 1725 | No | NA | 4 | 0 | 0 | Low-quality | Genome-fragment | 3 | AAI-based (medium-confidence) | 0 |
| S5-final-viral-combined_k141_36669\|\|full | 1725 | No | NA | 3 | 0 | 1 | Low-quality | Genome-fragment | 4.19 | AAI-based (medium-confidence) | 0 |
| S5-final-viral-combined_k141_51109\|\|full | 1725 | No | NA | 5 | 1 | 0 | Low-quality | Genome-fragment | 3.72 | AAI-based (medium-confidence) | 0 |
| S7_k141_13210 | 1725 | No | NA | 1 | 0 | 1 | Low-quality | Genome-fragment | 1.71 | AAI-based (medium-confidence) | 0 |
| S12-final-viral-combined_k141_155097\|\|full | 1725 | No | NA | 3 | 0 | 3 | Low-quality | Genome-fragment | 0.38 | AAI-based (medium-confidence) | 0 |
| S12-final-viral-combined_k141_199065\|\|full | 1725 | No | NA | 3 | 0 | 0 | Low-quality | Genome-fragment | 2.66 | AAI-based (medium-confidence) | 0 |
| S9-final-viral-combined_k141_37967\|\|full | 1726 | No | NA | 4 | 0 | 0 | Low-quality | Genome-fragment | 3.28 | AAI-based (high-confidence) | 0 |
| S11-final-viral-combined_k141_321561\|\|full | 1726 | No | NA | 2 | 0 | 1 | Low-quality | Genome-fragment | 1.5 | AAI-based (medium-confidence) | 0 |
| S5-final-viral-combined_k141_19663\|\|0_partial | 12851 | No | NA | 8 | 0 | 2 | Not-determined | Genome-fragment | NA | NA | 0 |
| S5-final-viral-combined_k141_162433\|\|0_partial | 10611 | No | NA | 12 | 0 | 1 | Not-determined | Genome-fragment | NA | NA | 0 |
| S1_k141_258005 | 1726 | No | NA | 1 | 0 | 1 | Low-quality | Genome-fragment | 0.15 | AAI-based (medium-confidence) | 0 |
| S17-final-viral-combined_k141_112050\|\|full | 1726 | No | NA | 2 | 0 | 0 | Low-quality | Genome-fragment | 4.05 | AAI-based (high-confidence) | 0 |
| S9-final-viral-combined_k141_27913\|\|full | 1727 | No | NA | 2 | 0 | 1 | Low-quality | Genome-fragment | 1.08 | AAI-based (medium-confidence) | 0 |
| S11-final-viral-combined_k141_49937\|\|full | 1727 | No | NA | 5 | 0 | 1 | Low-quality | Genome-fragment | 5.28 | AAI-based (high-confidence) | 0 |
| S5-final-viral-combined_k141_207323\|\|0_partial | 14551 | No | NA | 16 | 0 | 13 | Not-determined | Genome-fragment | NA | NA | 0 |
| S1-final-viral-combined_k141_268709\|\|full | 1727 | No | NA | 4 | 1 | 0 | Low-quality | Genome-fragment | 4.16 | AAI-based (high-confidence) | 0 |
| S5-final-viral-combined_k141_163165\|\|0_partial | 32777 | Yes | 21719 | 30 | 2 | 6 | High-quality | High-quality | 100 | AAI-based (high-confidence) | 33.74 |
| S5-final-viral-combined_k141_172874\|\|0_partial | 21425 | No | NA | 13 | 0 | 7 | Not-determined | Genome-fragment | NA | NA | 0 |
| S5-final-viral-combined_k141_92221\|\|0_partial | 15119 | No | NA | 8 | 0 | 5 | Not-determined | Genome-fragment | NA | NA | 0 |
| S5-final-viral-combined_k141_48351\|\|0_partial | 16607 | No | NA | 14 | 0 | 5 | Not-determined | Genome-fragment | NA | NA | 0 |
| S3-final-viral-combined_k141_99349\|\|full | 1727 | No | NA | 3 | 0 | 1 | Low-quality | Genome-fragment | 0.39 | AAI-based (medium-confidence) | 0 |
| S3-final-viral-combined_k141_42521\|\|full | 1727 | No | NA | 4 | 0 | 1 | Low-quality | Genome-fragment | 1.03 | AAI-based (medium-confidence) | 0 |
| S5-final-viral-combined_k141_119198\|\|0_partial | 14012 | No | NA | 10 | 0 | 6 | Not-determined | Genome-fragment | NA | NA | 0 |
| S5-final-viral-combined_k141_17310\|\|full | 1727 | No | NA | 3 | 0 | 0 | Low-quality | Genome-fragment | 5.11 | AAI-based (medium-confidence) | 0 |
| S7-final-viral-combined_k141_188756\|\|full | 1727 | No | NA | 2 | 0 | 0 | Low-quality | Genome-fragment | 2.45 | AAI-based (medium-confidence) | 0 |
| S7-final-viral-combined_k141_160513\|\|full | 1727 | No | NA | 4 | 1 | 0 | Low-quality | Genome-fragment | 1.24 | HMM-based (lower-bound) | 0 |
| S2-final-viral-combined_k141_209057\|\|full | 1727 | No | NA | 3 | 0 | 0 | Low-quality | Genome-fragment | 2.88 | AAI-based (high-confidence) | 0 |
| S5-final-viral-combined_k141_173869\|\|0_partial | 14723 | No | NA | 15 | 0 | 3 | Not-determined | Genome-fragment | NA | NA | 0 |
| S5-final-viral-combined_k141_155356\|\|0_partial | 16477 | No | NA | 21 | 0 | 2 | Not-determined | Genome-fragment | NA | NA | 0 |
| S5-final-viral-combined_k141_30475\|\|0_partial | 14679 | No | NA | 7 | 0 | 4 | Not-determined | Genome-fragment | NA | NA | 0 |
| S5-final-viral-combined_k141_39793\|\|0_partial | 2642 | No | NA | 4 | 1 | 0 | High-quality | High-quality | 95.35 | AAI-based (medium-confidence) | 0 |
| S8-final-viral-combined_k141_82373\|\|full | 1727 | No | NA | 3 | 0 | 2 | Low-quality | Genome-fragment | 0.48 | AAI-based (medium-confidence) | 0 |
| S8-final-viral-combined_k141_36540\|\|full | 1727 | No | NA | 2 | 0 | 0 | Low-quality | Genome-fragment | 2.7 | AAI-based (high-confidence) | 0 |
| S1-final-viral-combined_k141_198656\|\|full | 1728 | Yes | 1267 | 4 | 1 | 1 | Low-quality | Genome-fragment | 2.92 | AAI-based (high-confidence) | 26.68 |
| S5-final-viral-combined_k141_21581\|\|0_partial | 10935 | No | NA | 9 | 0 | 6 | Not-determined | Genome-fragment | NA | NA | 0 |
| S1-final-viral-combined_k141_35541\|\|full | 1728 | No | NA | 3 | 1 | 0 | Low-quality | Genome-fragment | 4.09 | AAI-based (high-confidence) | 0 |
| S5-final-viral-combined_k141_183113\|\|0_partial | 29249 | No | NA | 23 | 2 | 9 | Medium-quality | Genome-fragment | 54.11 | HMM-based (lower-bound) | 0 |
| S5-final-viral-combined_k141_200932\|\|0_partial | 11592 | No | NA | 16 | 0 | 2 | Not-determined | Genome-fragment | NA | NA | 0 |
| S1_k141_180956 | 1728 | No | NA | 2 | 0 | 0 | Low-quality | Genome-fragment | 4.4 | AAI-based (high-confidence) | 0 |
| S5-final-viral-combined_k141_156223\|\|0_partial | 13979 | No | NA | 11 | 0 | 2 | Not-determined | Genome-fragment | NA | NA | 0 |
| S3-final-viral-combined_k141_98652\|\|full | 1728 | No | NA | 3 | 0 | 0 | Low-quality | Genome-fragment | 2.7 | AAI-based (medium-confidence) | 0 |
| S5-final-viral-combined_k141_67335\|\|0_partial | 19822 | No | NA | 10 | 0 | 7 | Not-determined | Genome-fragment | NA | NA | 0 |
| S5-final-viral-combined_k141_112407\|\|0_partial | 13968 | No | NA | 11 | 0 | 8 | Not-determined | Genome-fragment | NA | NA | 0 |
| S5-final-viral-combined_k141_130617\|\|0_partial | 34563 | No | NA | 26 | 0 | 20 | Not-determined | Genome-fragment | NA | NA | 0 |
| S7-final-viral-combined_k141_51126\|\|full | 1728 | No | NA | 2 | 0 | 0 | Low-quality | Genome-fragment | 2.96 | AAI-based (high-confidence) | 0 |
| S7-final-viral-combined_k141_117675\|\|full | 1728 | No | NA | 2 | 0 | 1 | Low-quality | Genome-fragment | 1.67 | AAI-based (medium-confidence) | 0 |
| S7-final-viral-combined_k141_106969\|\|full | 1728 | No | NA | 4 | 0 | 0 | Low-quality | Genome-fragment | 4.11 | AAI-based (medium-confidence) | 0 |
| S5-final-viral-combined_k141_15022\|\|0_partial | 21064 | No | NA | 15 | 0 | 12 | Not-determined | Genome-fragment | NA | NA | 0 |
| S5-final-viral-combined_k141_193422\|\|0_partial | 18360 | No | NA | 13 | 0 | 6 | Not-determined | Genome-fragment | NA | NA | 0 |
| S2-final-viral-combined_k141_187621\|\|full | 1728 | No | NA | 3 | 0 | 0 | Low-quality | Genome-fragment | 3.34 | AAI-based (high-confidence) | 0 |
| S9-final-viral-combined_k141_55803\|\|full | 1729 | No | NA | 3 | 0 | 2 | Low-quality | Genome-fragment | 0.6 | AAI-based (medium-confidence) | 0 |
| S7_k141_226673 | 1729 | No | NA | 3 | 0 | 1 | Low-quality | Genome-fragment | 3.44 | AAI-based (medium-confidence) | 0 |
| S2-final-viral-combined_k141_110346\|\|full | 1729 | No | NA | 3 | 0 | 2 | Low-quality | Genome-fragment | 3.23 | AAI-based (medium-confidence) | 0 |
| S5-final-viral-combined_k141_194446\|\|0_partial | 11311 | No | NA | 8 | 0 | 4 | Not-determined | Genome-fragment | NA | NA | 0 |
| S6-final-viral-combined_k141_841\|\|full | 1729 | No | NA | 3 | 0 | 0 | Low-quality | Genome-fragment | 1.59 | AAI-based (high-confidence) | 0 |
| S5-final-viral-combined_k141_211849\|\|0_partial | 17614 | No | NA | 17 | 0 | 13 | Not-determined | Genome-fragment | NA | NA | 0 |
| S5-final-viral-combined_k141_42829\|\|0_partial | 1568 | No | NA | 2 | 0 | 2 | Not-determined | Genome-fragment | NA | NA | 0 |
| S8-final-viral-combined_k141_235003\|\|full | 1729 | No | NA | 6 | 1 | 0 | Low-quality | Genome-fragment | 7.1 | AAI-based (medium-confidence) | 0 |
| S5-final-viral-combined_k141_52741\|\|0_partial | 23075 | No | NA | 33 | 6 | 0 | Medium-quality | Genome-fragment | 68.66 | AAI-based (medium-confidence) | 0 |
| S5-final-viral-combined_k141_167999\|\|0_partial | 11082 | No | NA | 11 | 0 | 9 | Not-determined | Genome-fragment | NA | NA | 0 |
| S5-final-viral-combined_k141_69722\|\|0_partial | 13754 | No | NA | 8 | 0 | 7 | Not-determined | Genome-fragment | NA | NA | 0 |
| S9_k141_126640 | 1730 | No | NA | 1 | 0 | 0 | Low-quality | Genome-fragment | 4.32 | AAI-based (medium-confidence) | 0 |
| S5-final-viral-combined_k141_213019\|\|0_partial | 17367 | No | NA | 10 | 0 | 3 | Not-determined | Genome-fragment | NA | NA | 0 |
| S11-final-viral-combined_k141_296588\|\|full | 1730 | No | NA | 1 | 0 | 1 | Low-quality | Genome-fragment | 0.71 | AAI-based (medium-confidence) | 0 |
| S1_k141_273678 | 1730 | No | NA | 6 | 1 | 0 | Low-quality | Genome-fragment | 3.27 | AAI-based (high-confidence) | 0 |
| S5-final-viral-combined_k141_204892\|\|lt2gene | 1104 | No | NA | 1 | 0 | 0 | Not-determined | Genome-fragment | NA | NA | 0 |
| S17-final-viral-combined_k141_427\|\|full | 1730 | No | NA | 2 | 2 | 0 | Low-quality | Genome-fragment | 3.97 | AAI-based (high-confidence) | 0 |
| S17-final-viral-combined_k141_255298\|\|full | 1730 | No | NA | 2 | 0 | 2 | Low-quality | Genome-fragment | 2.39 | AAI-based (medium-confidence) | 0 |
| S17-k141_40325 | 1730 | No | NA | 2 | 0 | 0 | Low-quality | Genome-fragment | 1.2 | AAI-based (medium-confidence) | 0 |

| | | | | | | | | | | | |
|---|---|---|---|---|---|---|---|---|---|---|---|
| S12-final-viral-combined_k141_221382\|\|full | 1730 | No | NA | 3 | 0 | 1 | Low-quality | Genome-fragment | 0.91 | AAI-based (medium-confidence) | 0 |
| S12_k141_79705 | 1730 | No | NA | 2 | 0 | 2 | Low-quality | Genome-fragment | 1.55 | AAI-based (medium-confidence) | 0 |
| S2_k141_153483 | 1730 | No | NA | 3 | 0 | 3 | Low-quality | Genome-fragment | 2.4 | AAI-based (medium-confidence) | 0 |
| S8_k141_30768 | 1730 | No | NA | 3 | 0 | 0 | Low-quality | Genome-fragment | 3.13 | AAI-based (high-confidence) | 0 |
| S9-final-viral-combined_k141_50268\|\|full | 1731 | No | NA | 3 | 0 | 1 | Low-quality | Genome-fragment | 2.32 | AAI-based (high-confidence) | 0 |
| S11-final-viral-combined_k141_288106\|\|full | 1731 | No | NA | 2 | 0 | 2 | Low-quality | Genome-fragment | 1.03 | AAI-based (high-confidence) | 0 |
| S7-final-viral-combined_k141_130565\|\|full | 1731 | No | NA | 3 | 0 | 0 | Low-quality | Genome-fragment | 9.08 | AAI-based (medium-confidence) | 0 |
| S7_k141_83397 | 1731 | No | NA | 2 | 0 | 0 | Low-quality | Genome-fragment | 1.27 | AAI-based (medium-confidence) | 0 |
| S5-final-viral-combined_k141_143219\|\|lt2gene | 1823 | No | NA | 2 | 0 | 1 | Not-determined | Genome-fragment | NA | NA | 0 |
| S2_k141_127582 | 1731 | No | NA | 2 | 0 | 0 | Low-quality | Genome-fragment | 2.07 | AAI-based (medium-confidence) | 0 |
| S2_k141_116959 | 1731 | No | NA | 6 | 0 | 3 | Low-quality | Genome-fragment | 0.34 | AAI-based (medium-confidence) | 0 |
| S5-final-viral-combined_k141_178561\|\|lt2gene | 1006 | No | NA | 1 | 0 | 0 | Not-determined | Genome-fragment | NA | NA | 0 |
| S5-final-viral-combined_k141_170105\|\|lt2gene | 1565 | No | NA | 1 | 0 | 0 | Not-determined | Genome-fragment | NA | NA | 0 |
| S11-final-viral-combined_k141_75338\|\|full | 1732 | No | NA | 5 | 2 | 0 | Low-quality | Genome-fragment | 2.78 | AAI-based (medium-confidence) | 0 |
| S3-final-viral-combined_k141_49241\|\|lt2gene | 1732 | No | NA | 2 | 1 | 0 | Low-quality | Genome-fragment | 3.36 | AAI-based (high-confidence) | 0 |
| S5-final-viral-combined_k141_9943\|\|lt2gene | 1354 | No | NA | 1 | 0 | 0 | Not-determined | Genome-fragment | NA | NA | 0 |
| S12-final-viral-combined_k141_134575\|\|full | 1732 | No | NA | 3 | 0 | 1 | Low-quality | Genome-fragment | 3.2 | AAI-based (high-confidence) | 0 |
| S12_k141_323838 | 1732 | No | NA | 2 | 0 | 0 | Low-quality | Genome-fragment | 0.93 | AAI-based (high-confidence) | 0 |
| S5-final-viral-combined_k141_98904\|\|lt2gene | 1066 | No | NA | 1 | 0 | 1 | Not-determined | Genome-fragment | NA | NA | 0 |
| S2-final-viral-combined_k141_129963\|\|full | 1732 | No | NA | 2 | 1 | 0 | Low-quality | Genome-fragment | 3.36 | AAI-based (high-confidence) | 0 |
| S4-final-viral-combined_k141_147790\|\|full | 1732 | No | NA | 2 | 0 | 0 | Low-quality | Genome-fragment | 1.27 | AAI-based (high-confidence) | 0 |
| S11-final-viral-combined_k141_189352\|\|full | 1733 | No | NA | 2 | 0 | 2 | Low-quality | Genome-fragment | 0.5 | AAI-based (high-confidence) | 0 |
| S17-final-viral-combined_k141_232815\|\|full | 1733 | No | NA | 3 | 2 | 0 | Low-quality | Genome-fragment | 1.84 | HMM-based (lower-bound) | 0 |
| S17-k141_221813 | 1733 | No | NA | 1 | 0 | 0 | Low-quality | Genome-fragment | 0.15 | AAI-based (medium-confidence) | 0 |
| S12-final-viral-combined_k141_293215\|\|full | 1733 | No | NA | 2 | 0 | 0 | Low-quality | Genome-fragment | 0.95 | AAI-based (medium-confidence) | 0 |
| S12-final-viral-combined_k141_351294\|\|full | 1733 | No | NA | 4 | 0 | 0 | Low-quality | Genome-fragment | 3.39 | AAI-based (high-confidence) | 0 |
| S12-final-viral-combined_k141_224020\|\|lt2gene | 1733 | No | NA | 2 | 1 | 0 | Low-quality | Genome-fragment | 2.27 | HMM-based (lower-bound) | 0 |
| S4-final-viral-combined_k141_97923\|\|full | 1733 | No | NA | 3 | 0 | 0 | Low-quality | Genome-fragment | 2.62 | AAI-based (medium-confidence) | 0 |
| S8-final-viral-combined_k141_214780\|\|full | 1733 | No | NA | 2 | 1 | 0 | Low-quality | Genome-fragment | 4.96 | AAI-based (high-confidence) | 0 |
| S11-final-viral-combined_k141_59061\|\|full | 1734 | No | NA | 7 | 1 | 0 | Low-quality | Genome-fragment | 4.46 | AAI-based (high-confidence) | 0 |
| S1_k141_37428 | 1734 | No | NA | 2 | 0 | 2 | Low-quality | Genome-fragment | 0.88 | AAI-based (medium-confidence) | 0 |
| S3-final-viral-combined_k141_116728\|\|full | 1734 | No | NA | 3 | 1 | 0 | Low-quality | Genome-fragment | 3.02 | AAI-based (medium-confidence) | 0 |
| S5_k141_24636 | 1734 | No | NA | 3 | 0 | 0 | Low-quality | Genome-fragment | 4.59 | AAI-based (high-confidence) | 0 |
| S5-final-viral-combined_k141_72805\|\|lt2gene | 1090 | No | NA | 1 | 0 | 0 | Not-determined | Genome-fragment | NA | NA | 0 |
| S5-final-viral-combined_k141_188612\|\|lt2gene | 1681 | No | NA | 1 | 0 | 1 | Not-determined | Genome-fragment | NA | NA | 0 |
| S5-final-viral-combined_k141_37128\|\|lt2gene | 1501 | No | NA | 1 | 0 | 0 | Not-determined | Genome-fragment | NA | NA | 0 |
| S5-final-viral-combined_k141_117291\|\|lt2gene | 1213 | No | NA | 2 | 0 | 1 | Not-determined | Genome-fragment | NA | NA | 0 |
| S7-final-viral-combined_k141_144551\|\|full | 1734 | No | NA | 2 | 1 | 0 | Low-quality | Genome-fragment | 3.77 | AAI-based (high-confidence) | 0 |
| S7-final-viral-combined_k141_38478\|\|full | 1734 | No | NA | 4 | 2 | 0 | Low-quality | Genome-fragment | 4.29 | AAI-based (high-confidence) | 0 |
| S17-final-viral-combined_k141_201078\|\|full | 1734 | No | NA | 2 | 0 | 2 | Low-quality | Genome-fragment | 0.44 | AAI-based (high-confidence) | 0 |
| S12-final-viral-combined_k141_385023\|\|full | 1734 | No | NA | 4 | 0 | 0 | Low-quality | Genome-fragment | 3.78 | AAI-based (high-confidence) | 0 |
| S2-final-viral-combined_k141_21767\|\|full | 1734 | No | NA | 4 | 0 | 0 | Low-quality | Genome-fragment | 3.14 | AAI-based (medium-confidence) | 0 |
| S11-final-viral-combined_k141_97429\|\|full | 1735 | No | NA | 4 | 1 | 0 | Low-quality | Genome-fragment | 4.32 | HMM-based (lower-bound) | 0 |
| S5-final-viral-combined_k141_171624\|\|lt2gene | 1122 | No | NA | 1 | 0 | 0 | Not-determined | Genome-fragment | NA | NA | 0 |
| S11-final-viral-combined_k141_211701\|\|full | 1735 | No | NA | 3 | 0 | 2 | Low-quality | Genome-fragment | 2.83 | AAI-based (medium-confidence) | 0 |
| S5-final-viral-combined_k141_108761\|\|lt2gene | 1428 | No | NA | 1 | 0 | 0 | Not-determined | Genome-fragment | NA | NA | 0 |
| S11-final-viral-combined_k141_327508\|\|full | 1735 | No | NA | 2 | 0 | 2 | Low-quality | Genome-fragment | 0.41 | AAI-based (medium-confidence) | 0 |
| S7-final-viral-combined_k141_144289\|\|full | 1735 | No | NA | 4 | 1 | 0 | Low-quality | Genome-fragment | 4.46 | AAI-based (medium-confidence) | 0 |
| S7-final-viral-combined_k141_140116\|\|full | 1735 | No | NA | 4 | 0 | 3 | Low-quality | Genome-fragment | 2.45 | AAI-based (medium-confidence) | 0 |
| S5-final-viral-combined_k141_46864\|\|lt2gene | 1492 | No | NA | 2 | 0 | 0 | Not-determined | Genome-fragment | NA | NA | 0 |
| S17-final-viral-combined_k141_100917\|\|full | 1735 | No | NA | 2 | 0 | 2 | Low-quality | Genome-fragment | 3.19 | AAI-based (medium-confidence) | 0 |
| S17-final-viral-combined_k141_124750\|\|full | 1735 | No | NA | 3 | 0 | 2 | Low-quality | Genome-fragment | 1.23 | AAI-based (high-confidence) | 0 |
| S17-final-viral-combined_k141_138428\|\|full | 1735 | No | NA | 3 | 0 | 0 | Low-quality | Genome-fragment | 2.71 | AAI-based (high-confidence) | 0 |
| S8-final-viral-combined_k141_188266\|\|full | 1735 | No | NA | 3 | 2 | 0 | Low-quality | Genome-fragment | 2.78 | AAI-based (medium-confidence) | 0 |
| S5-final-viral-combined_k141_145079\|\|lt2gene | 1722 | No | NA | 1 | 0 | 0 | Not-determined | Genome-fragment | NA | NA | 0 |
| S8_k141_111809 | 1735 | No | NA | 1 | 1 | 0 | Low-quality | Genome-fragment | 0.56 | HMM-based (lower-bound) | 0 |
| S1-final-viral-combined_k141_122069\|\|full | 1736 | No | NA | 5 | 1 | 0 | Low-quality | Genome-fragment | 2.15 | HMM-based (lower-bound) | 0 |
| S1-final-viral-combined_k141_63925\|\|full | 1736 | No | NA | 3 | 0 | 1 | Low-quality | Genome-fragment | 1.05 | AAI-based (medium-confidence) | 0 |
| S1_k141_240105 | 1736 | No | NA | 1 | 0 | 1 | Low-quality | Genome-fragment | 2.95 | AAI-based (high-confidence) | 0 |
| S5-final-viral-combined_k141_172368\|\|lt2gene | 1406 | No | NA | 2 | 0 | 2 | Not-determined | Genome-fragment | NA | NA | 0 |
| S5-final-viral-combined_k141_187857\|\|lt2gene | 1736 | No | NA | 1 | 1 | 0 | Low-quality | Genome-fragment | 4.11 | AAI-based (medium-confidence) | 0 |
| S2-final-viral-combined_k141_57941\|\|full | 1736 | No | NA | 4 | 1 | 0 | Low-quality | Genome-fragment | 2.94 | AAI-based (medium-confidence) | 0 |
| S2-final-viral-combined_k141_143729\|\|full | 1736 | No | NA | 2 | 0 | 1 | Low-quality | Genome-fragment | 1.19 | AAI-based (medium-confidence) | 0 |
| S2_k141_196551 | 1736 | No | NA | 2 | 0 | 2 | Low-quality | Genome-fragment | 2.02 | AAI-based (medium-confidence) | 0 |
| S4-final-viral-combined_k141_141582\|\|full | 1736 | No | NA | 4 | 0 | 0 | Low-quality | Genome-fragment | 3.08 | AAI-based (high-confidence) | 0 |
| S5-final-viral-combined_k141_127601\|\|lt2gene | 1213 | No | NA | 1 | 0 | 0 | Not-determined | Genome-fragment | NA | NA | 0 |
| S5-final-viral-combined_k141_47596\|\|lt2gene | 1822 | No | NA | 2 | 0 | 2 | Not-determined | Genome-fragment | NA | NA | 0 |
| S8-final-viral-combined_k141_94595\|\|full | 1736 | No | NA | 3 | 0 | 0 | Low-quality | Genome-fragment | 1.13 | AAI-based (high-confidence) | 0 |
| S8-final-viral-combined_k141_87578\|\|full | 1736 | No | NA | 2 | 1 | 0 | Low-quality | Genome-fragment | 3.44 | AAI-based (medium-confidence) | 0 |
| S11-final-viral-combined_k141_153833\|\|full | 1737 | No | NA | 3 | 0 | 0 | Low-quality | Genome-fragment | 3.78 | AAI-based (medium-confidence) | 0 |
| S11-final-viral-combined_k141_270585\|\|full | 1737 | No | NA | 2 | 0 | 0 | Low-quality | Genome-fragment | 2.72 | AAI-based (medium-confidence) | 0 |
| S1-final-viral-combined_k141_182040\|\|full | 1737 | No | NA | 3 | 0 | 0 | Low-quality | Genome-fragment | 3.94 | AAI-based (high-confidence) | 0 |
| S5-final-viral-combined_k141_74184\|\|lt2gene | 1474 | No | NA | 1 | 0 | 0 | Not-determined | Genome-fragment | NA | NA | 0 |
| S1-final-viral-combined_k141_388554\|\|full | 1737 | No | NA | 2 | 0 | 0 | Low-quality | Genome-fragment | 2.1 | AAI-based (high-confidence) | 0 |
| S3-final-viral-combined_k141_113265\|\|full | 1737 | No | NA | 2 | 0 | 1 | Low-quality | Genome-fragment | 0.94 | AAI-based (medium-confidence) | 0 |
| S3_k141_47214 | 1737 | No | NA | 2 | 1 | 0 | Low-quality | Genome-fragment | 3.98 | AAI-based (high-confidence) | 0 |
| S17-final-viral-combined_k141_127400\|\|full | 1737 | No | NA | 3 | 0 | 0 | Low-quality | Genome-fragment | 2.13 | AAI-based (medium-confidence) | 0 |
| S5-final-viral-combined_k141_82896\|\|lt2gene | 1096 | No | NA | 1 | 0 | 0 | Not-determined | Genome-fragment | NA | NA | 0 |
| S2-final-viral-combined_k141_98229\|\|full | 1737 | No | NA | 3 | 0 | 2 | Low-quality | Genome-fragment | 2.01 | AAI-based (medium-confidence) | 0 |
| S2-final-viral-combined_k141_67141\|\|full | 1737 | No | NA | 3 | 0 | 0 | Low-quality | Genome-fragment | 2.73 | AAI-based (high-confidence) | 0 |
| S11-final-viral-combined_k141_209033\|\|full | 1738 | No | NA | 3 | 0 | 1 | Low-quality | Genome-fragment | 0.94 | AAI-based (medium-confidence) | 0 |
| S11-final-viral-combined_k141_176329\|\|full | 1738 | No | NA | 4 | 0 | 0 | Low-quality | Genome-fragment | 3.87 | AAI-based (high-confidence) | 0 |
| S1-final-viral-combined_k141_68994\|\|full | 1738 | No | NA | 3 | 0 | 0 | Low-quality | Genome-fragment | 0.92 | AAI-based (medium-confidence) | 0 |
| S1-final-viral-combined_k141_122063\|\|full | 1738 | Yes | 1027 | 3 | 1 | 1 | Low-quality | Genome-fragment | 1.73 | AAI-based (medium-confidence) | 40.91 |
| S5-final-viral-combined_k141_163665\|\|lt2gene | 1044 | No | NA | 1 | 0 | 0 | Not-determined | Genome-fragment | NA | NA | 0 |
| S5-final-viral-combined_k141_29676\|\|lt2gene | 3444 | No | NA | 1 | 0 | 1 | Not-determined | Genome-fragment | NA | NA | 0 |
| S7-final-viral-combined_k141_33219\|\|full | 1738 | No | NA | 5 | 0 | 1 | Low-quality | Genome-fragment | 1.65 | AAI-based (medium-confidence) | 0 |
| S7-final-viral-combined_k141_104839\|\|full | 1738 | No | NA | 3 | 2 | 0 | Low-quality | Genome-fragment | 4.01 | AAI-based (high-confidence) | 0 |
| S5-final-viral-combined_k141_12154\|\|lt2gene | 1350 | No | NA | 1 | 0 | 0 | Not-determined | Genome-fragment | NA | NA | 0 |
| S12-final-viral-combined_k141_205402\|\|full | 1738 | No | NA | 3 | 0 | 0 | Low-quality | Genome-fragment | 3 | AAI-based (high-confidence) | 0 |
| S2-final-viral-combined_k141_47102\|\|full | 1738 | No | NA | 2 | 0 | 1 | Low-quality | Genome-fragment | 0.8 | AAI-based (high-confidence) | 0 |
| S5-final-viral-combined_k141_146545\|\|lt2gene | 1754 | No | NA | 2 | 0 | 0 | Not-determined | Genome-fragment | NA | NA | 0 |
| S6_k141_83090 | 1738 | No | NA | 2 | 0 | 0 | Low-quality | Genome-fragment | 1.84 | AAI-based (high-confidence) | 0 |
| S8-final-viral-combined_k141_243927\|\|full | 1738 | No | NA | 3 | 0 | 0 | Low-quality | Genome-fragment | 4.1 | AAI-based (high-confidence) | 0 |
| S9-final-viral-combined_k141_43540\|\|full | 1739 | No | NA | 3 | 1 | 0 | Low-quality | Genome-fragment | 2.83 | AAI-based (high-confidence) | 0 |
| S11-final-viral-combined_k141_114057\|\|full | 1739 | No | NA | 4 | 2 | 0 | Low-quality | Genome-fragment | 4.68 | AAI-based (high-confidence) | 0 |
| S5-final-viral-combined_k141_119474\|\|lt2gene | 1138 | No | NA | 1 | 0 | 1 | Not-determined | Genome-fragment | NA | NA | 0 |
| S11-final-viral-combined_k141_76364\|\|full | 1739 | No | NA | 3 | 0 | 1 | Low-quality | Genome-fragment | 2.26 | AAI-based (medium-confidence) | 0 |
| S1-final-viral-combined_k141_19306\|\|full | 1739 | No | NA | 4 | 2 | 0 | Low-quality | Genome-fragment | 2.88 | AAI-based (high-confidence) | 0 |
| S1_k141_393605 | 1739 | No | NA | 2 | 0 | 1 | Low-quality | Genome-fragment | 2.1 | AAI-based (high-confidence) | 0 |
| S5-final-viral-combined_k141_174359\|\|full | 1739 | No | NA | 4 | 1 | 0 | Low-quality | Genome-fragment | 4.29 | AAI-based (medium-confidence) | 0 |
| S7-final-viral-combined_k141_107195\|\|full | 1739 | No | NA | 3 | 0 | 1 | Low-quality | Genome-fragment | 2.71 | AAI-based (medium-confidence) | 0 |
| S17-final-viral-combined_k141_236764\|\|full | 1739 | No | NA | 2 | 0 | 1 | Low-quality | Genome-fragment | 0.99 | AAI-based (medium-confidence) | 0 |
| S17-k141_134084 | 1739 | No | NA | 2 | 0 | 2 | Low-quality | Genome-fragment | 0.46 | AAI-based (high-confidence) | 0 |
| S12-final-viral-combined_k141_91452\|\|full | 1739 | No | NA | 2 | 0 | 0 | Low-quality | Genome-fragment | 1.57 | AAI-based (high-confidence) | 0 |
| S12-final-viral-combined_k141_359189\|\|full | 1739 | No | NA | 2 | 0 | 2 | Low-quality | Genome-fragment | 0.5 | AAI-based (medium-confidence) | 0 |
| S4_k141_112142 | 1739 | No | NA | 1 | 0 | 1 | Low-quality | Genome-fragment | 2.73 | AAI-based (high-confidence) | 0 |
| S5-final-viral-combined_k141_57571\|\|lt2gene | 1071 | No | NA | 1 | 0 | 0 | Not-determined | Genome-fragment | NA | NA | 0 |
| S5-final-viral-combined_k141_5857\|\|lt2gene | 1183 | No | NA | 2 | 0 | 0 | Not-determined | Genome-fragment | NA | NA | 0 |
| S6-final-viral-combined_k141_85453\|\|full | 1739 | No | NA | 2 | 0 | 1 | Low-quality | Genome-fragment | 2.85 | AAI-based (medium-confidence) | 0 |
| S5-final-viral-combined_k141_174322\|\|lt2gene | 1476 | No | NA | 2 | 0 | 2 | Not-determined | Genome-fragment | NA | NA | 0 |
| S6_k141_83530 | 1739 | No | NA | 3 | 0 | 3 | Low-quality | Genome-fragment | 1.27 | AAI-based (high-confidence) | 0 |
| S8-final-viral-combined_k141_219544\|\|full | 1739 | No | NA | 3 | 0 | 1 | Low-quality | Genome-fragment | 4.11 | AAI-based (medium-confidence) | 0 |
| S5-final-viral-combined_k141_120239\|\|lt2gene | 1001 | No | NA | 1 | 0 | 0 | Not-determined | Genome-fragment | NA | NA | 0 |
| S5-final-viral-combined_k141_49365\|\|lt2gene | 1600 | No | NA | 1 | 0 | 1 | Not-determined | Genome-fragment | NA | NA | 0 |
| S5-final-viral-combined_k141_156085\|\|lt2gene | 1144 | No | NA | 1 | 0 | 0 | Not-determined | Genome-fragment | NA | NA | 0 |
| S8-final-viral-combined_k141_175110\|\|full | 1739 | No | NA | 3 | 0 | 2 | Low-quality | Genome-fragment | 2.07 | AAI-based (high-confidence) | 0 |
| S11-final-viral-combined_k141_62375\|\|full | 1740 | No | NA | 3 | 1 | 0 | Low-quality | Genome-fragment | 4.75 | AAI-based (high-confidence) | 0 |
| S3-final-viral-combined_k141_6527\|\|full | 1740 | No | NA | 3 | 0 | 1 | Low-quality | Genome-fragment | 2.22 | AAI-based (medium-confidence) | 0 |
| S17-final-viral-combined_k141_127625\|\|full | 1740 | No | NA | 2 | 0 | 1 | Low-quality | Genome-fragment | 2.56 | AAI-based (medium-confidence) | 0 |
| S5-final-viral-combined_k141_31523\|\|lt2gene | 1657 | No | NA | 1 | 0 | 0 | Not-determined | Genome-fragment | NA | NA | 0 |
| S12-final-viral-combined_k141_177548\|\|full | 1740 | No | NA | 2 | 0 | 0 | Low-quality | Genome-fragment | 3.84 | AAI-based (medium-confidence) | 0 |
| S8-final-viral-combined_k141_155092\|\|full | 1740 | Yes | 270 | 3 | 1 | 2 | Low-quality | Genome-fragment | 0.76 | AAI-based (high-confidence) | 84.48 |
| S11-final-viral-combined_k141_258635\|\|full | 1741 | No | NA | 2 | 0 | 2 | Low-quality | Genome-fragment | 0.98 | AAI-based (medium-confidence) | 0 |
| S1-final-viral-combined_k141_323830\|\|full | 1741 | No | NA | 2 | 0 | 1 | Low-quality | Genome-fragment | 0.84 | AAI-based (high-confidence) | 0 |
| S5-final-viral-combined_k141_175360\|\|lt2gene | 1375 | No | NA | 1 | 0 | 0 | Not-determined | Genome-fragment | NA | NA | 0 |
| S1_k141_332191 | 1741 | No | NA | 1 | 0 | 1 | Low-quality | Genome-fragment | 4.91 | AAI-based (medium-confidence) | 0 |
| S5-final-viral-combined_k141_54755\|\|full | 1741 | No | NA | 4 | 1 | 0 | Low-quality | Genome-fragment | 4.02 | AAI-based (medium-confidence) | 0 |
| S8-final-viral-combined_k141_165391\|\|full | 1741 | No | NA | 2 | 0 | 1 | Low-quality | Genome-fragment | 2.28 | AAI-based (medium-confidence) | 0 |
| S11-final-viral-combined_k141_123165\|\|full | 1742 | No | NA | 3 | 1 | 0 | Low-quality | Genome-fragment | 0.14 | HMM-based (lower-bound) | 0 |
| S1-final-viral-combined_k141_234767\|\|full | 1742 | No | NA | 3 | 0 | 0 | Low-quality | Genome-fragment | 4.04 | AAI-based (high-confidence) | 0 |
| S1-final-viral-combined_k141_323952\|\|full | 1742 | No | NA | 2 | 0 | 0 | Low-quality | Genome-fragment | 4.48 | AAI-based (high-confidence) | 0 |
| S1-final-viral-combined_k141_342454\|\|full | 1742 | No | NA | 3 | 0 | 0 | Low-quality | Genome-fragment | 1.95 | AAI-based (medium-confidence) | 0 |
| S5-final-viral-combined_k141_32285\|\|lt2gene | 1772 | No | NA | 1 | 0 | 1 | Not-determined | Genome-fragment | NA | NA | 0 |
| S5_k141_4568 | 1742 | No | NA | 1 | 0 | 1 | Low-quality | Genome-fragment | 0.3 | AAI-based (medium-confidence) | 0 |
| S7_k141_134326 | 1742 | No | NA | 2 | 0 | 0 | Low-quality | Genome-fragment | 3.83 | AAI-based (high-confidence) | 0 |
| S7_k141_91464 | 1742 | No | NA | 2 | 0 | 2 | Low-quality | Genome-fragment | 0.48 | AAI-based (medium-confidence) | 0 |
| S5-final-viral-combined_k141_121411\|\|lt2gene | 1451 | No | NA | 1 | 0 | 0 | Not-determined | Genome-fragment | NA | NA | 0 |
| S17-final-viral-combined_k141_80991\|\|full | 1742 | No | NA | 7 | 1 | 0 | Low-quality | Genome-fragment | 4.13 | AAI-based (high-confidence) | 0 |
| S8_k141_227060 | 1742 | No | NA | 2 | 1 | 0 | Low-quality | Genome-fragment | 2.06 | AAI-based (high-confidence) | 0 |
| S1-final-viral-combined_k141_321985\|\|full | 1743 | No | NA | 2 | 0 | 0 | Low-quality | Genome-fragment | 3.26 | AAI-based (high-confidence) | 0 |
| S7-final-viral-combined_k141_134777\|\|full | 1743 | No | NA | 3 | 3 | 0 | Low-quality | Genome-fragment | 3.83 | AAI-based (high-confidence) | 0 |
| S17-final-viral-combined_k141_36680\|\|full | 1743 | No | NA | 3 | 0 | 0 | Low-quality | Genome-fragment | 4.44 | AAI-based (medium-confidence) | 0 |
| S2-final-viral-combined_k141_94000\|\|full | 1743 | No | NA | 2 | 1 | 0 | Low-quality | Genome-fragment | 3.99 | HMM-based (lower-bound) | 0 |
| S5-final-viral-combined_k141_113230\|\|lt2gene | 3440 | No | NA | 2 | 0 | 0 | Not-determined | Genome-fragment | NA | NA | 0 |
| S5-final-viral-combined_k141_149112\|\|lt2gene | 1216 | No | NA | 1 | 0 | 0 | Not-determined | Genome-fragment | NA | NA | 0 |
| S3-final-viral-combined_k141_88441\|\|full | 1744 | No | NA | 3 | 0 | 0 | Low-quality | Genome-fragment | 2.73 | AAI-based (high-confidence) | 0 |
| S12-final-viral-combined_k141_288621\|\|full | 1744 | No | NA | 2 | 2 | 0 | Low-quality | Genome-fragment | 4.91 | AAI-based (medium-confidence) | 0 |
| S2-final-viral-combined_k141_224117\|\|full | 1744 | No | NA | 3 | 1 | 0 | Low-quality | Genome-fragment | 3.49 | AAI-based (high-confidence) | 0 |
| S5-final-viral-combined_k141_122247\|\|lt2gene | 2424 | No | NA | 1 | 0 | 0 | Not-determined | Genome-fragment | NA | NA | 0 |
| S5-final-viral-combined_k141_23638\|\|lt2gene | 1289 | No | NA | 1 | 0 | 1 | Not-determined | Genome-fragment | NA | NA | 0 |
| S2_k141_150901 | 1744 | No | NA | 2 | 1 | 0 | Low-quality | Genome-fragment | 28.6 | AAI-based (high-confidence) | 0 |
| S11-final-viral-combined_k141_285779\|\|full | 1745 | No | NA | 3 | 0 | 1 | Low-quality | Genome-fragment | 2.62 | AAI-based (medium-confidence) | 0 |
| S5-final-viral-combined_k141_140309\|\|lt2gene | 1209 | No | NA | 2 | 0 | 0 | Not-determined | Genome-fragment | NA | NA | 0 |
| S1-final-viral-combined_k141_377664\|\|full | 1745 | No | NA | 6 | 2 | 0 | Low-quality | Genome-fragment | 4 | AAI-based (high-confidence) | 0 |
| S5-final-viral-combined_k141_122562\|\|lt2gene | 1125 | No | NA | 1 | 0 | 0 | Not-determined | Genome-fragment | NA | NA | 0 |
| S1_k141_114510 | 1745 | No | NA | 1 | 0 | 0 | Low-quality | Genome-fragment | 0.3 | AAI-based (high-confidence) | 0 |
| S7-final-viral-combined_k141_54296\|\|full | 1745 | No | NA | 3 | 2 | 0 | Low-quality | Genome-fragment | 3.4 | AAI-based (high-confidence) | 0 |
| S7-final-viral-combined_k141_75892\|\|full | 1745 | No | NA | 4 | 0 | 0 | Low-quality | Genome-fragment | 2.13 | AAI-based (medium-confidence) | 0 |
| S5-final-viral-combined_k141_149512\|\|lt2gene | 1185 | No | NA | 2 | 0 | 0 | Not-determined | Genome-fragment | NA | NA | 0 |
| S17-final-viral-combined_k141_212711\|\|lt2gene | 1745 | No | NA | 1 | 1 | 0 | Low-quality | Genome-fragment | 1.3 | HMM-based (lower-bound) | 0 |
| S5-final-viral-combined_k141_60777\|\|lt2gene | 1085 | No | NA | 1 | 0 | 0 | Not-determined | Genome-fragment | NA | NA | 0 |
| S4_k141_147275 | 1745 | No | NA | 1 | 0 | 0 | Low-quality | Genome-fragment | 1.73 | AAI-based (medium-confidence) | 0 |
| S8-final-viral-combined_k141_260176\|\|full | 1745 | No | NA | 2 | 2 | 0 | Low-quality | Genome-fragment | 2.26 | AAI-based (high-confidence) | 0 |
| S9-final-viral-combined_k141_57777\|\|full | 1746 | No | NA | 2 | 0 | 2 | Low-quality | Genome-fragment | 2.31 | AAI-based (medium-confidence) | 0 |
| S1-final-viral-combined_k141_365670\|\|full | 1746 | No | NA | 2 | 0 | 0 | Low-quality | Genome-fragment | 1.32 | AAI-based (high-confidence) | 0 |
| S1-final-viral-combined_k141_176149\|\|full | 1746 | No | NA | 3 | 0 | 0 | Low-quality | Genome-fragment | 1.31 | AAI-based (high-confidence) | 0 |
| S5-final-viral-combined_k141_155230\|\|full | 1746 | No | NA | 4 | 2 | 0 | Low-quality | Genome-fragment | 1.96 | AAI-based (high-confidence) | 0 |
| S5-final-viral-combined_k141_42757\|\|lt2gene | 1334 | No | NA | 2 | 0 | 0 | Not-determined | Genome-fragment | NA | NA | 0 |
| S5-final-viral-combined_k141_96762\|\|full | 1746 | No | NA | 4 | 1 | 0 | Low-quality | Genome-fragment | 1.06 | HMM-based (lower-bound) | 0 |
| S2-final-viral-combined_k141_136491\|\|full | 1746 | No | NA | 2 | 0 | 1 | Low-quality | Genome-fragment | 1.2 | AAI-based (high-confidence) | 0 |
| S2-final-viral-combined_k141_19988\|\|lt2gene | 1746 | No | NA | 2 | 2 | 0 | Low-quality | Genome-fragment | 4.31 | AAI-based (high-confidence) | 0 |
| S4-final-viral-combined_k141_157340\|\|full | 1746 | No | NA | 5 | 0 | 0 | Low-quality | Genome-fragment | 3.73 | AAI-based (medium-confidence) | 0 |
| S8-final-viral-combined_k141_98288\|\|full | 1746 | No | NA | 2 | 0 | 0 | Low-quality | Genome-fragment | 3.33 | AAI-based (medium-confidence) | 0 |
| S8_k141_106042 | 1746 | No | NA | 1 | 0 | 1 | Low-quality | Genome-fragment | 3.74 | AAI-based (medium-confidence) | 0 |
| S5-final-viral-combined_k141_90916\|\|full | 1747 | No | NA | 3 | 0 | 0 | Low-quality | Genome-fragment | 2.84 | AAI-based (medium-confidence) | 0 |
| S17-final-viral-combined_k141_146018\|\|full | 1747 | No | NA | 4 | 0 | 0 | Low-quality | Genome-fragment | 3.7 | AAI-based (medium-confidence) | 0 |
| S17-final-viral-combined_k141_88356\|\|full | 1747 | No | NA | 2 | 1 | 0 | Low-quality | Genome-fragment | 4.32 | AAI-based (medium-confidence) | 0 |
| S2-final-viral-combined_k141_244922\|\|full | 1747 | No | NA | 5 | 0 | 0 | Low-quality | Genome-fragment | 1.38 | AAI-based (medium-confidence) | 0 |
| S2_k141_927 | 1747 | No | NA | 3 | 0 | 1 | Low-quality | Genome-fragment | 1.54 | AAI-based (medium-confidence) | 0 |
| S4-final-viral-combined_k141_63016\|\|full | 1747 | No | NA | 2 | 0 | 2 | Low-quality | Genome-fragment | 4.04 | AAI-based (medium-confidence) | 0 |
| S5-final-viral-combined_k141_141718\|\|lt2gene | 1005 | No | NA | 2 | 0 | 1 | Not-determined | Genome-fragment | NA | NA | 0 |
| S5-final-viral-combined_k141_17025\|\|lt2gene | 1778 | No | NA | 2 | 0 | 0 | Not-determined | Genome-fragment | NA | NA | 0 |
| S5-final-viral-combined_k141_177940\|\|lt2gene | 1163 | No | NA | 1 | 0 | 0 | Not-determined | Genome-fragment | NA | NA | 0 |
| S5-final-viral-combined_k141_115428\|\|lt2gene | 1064 | No | NA | 1 | 0 | 0 | Not-determined | Genome-fragment | NA | NA | 0 |
| S4-final-viral-combined_k141_161648\|\|full | 1747 | No | NA | 4 | 0 | 0 | Low-quality | Genome-fragment | 1.76 | AAI-based (high-confidence) | 0 |
| S5-final-viral-combined_k141_62244\|\|lt2gene | 1157 | No | NA | 2 | 0 | 1 | Not-determined | Genome-fragment | NA | NA | 0 |
| S5_k141_44545 | 1385 | No | NA | 2 | 0 | 2 | Not-determined | Genome-fragment | NA | NA | 0 |
| S5_k141_169271 | 3008 | No | NA | 2 | 0 | 2 | Not-determined | Genome-fragment | NA | NA | 0 |
| S5_k141_124747 | 1087 | No | NA | 1 | 0 | 1 | Not-determined | Genome-fragment | NA | NA | 0 |
| S5_k141_124753 | 4086 | No | NA | 3 | 0 | 2 | Not-determined | Genome-fragment | NA | NA | 0 |
| S5_k141_115847 | 6517 | No | NA | 5 | 0 | 3 | Not-determined | Genome-fragment | NA | NA | 0 |
| S8-final-viral-combined_k141_235372\|\|full | 1747 | No | NA | 3 | 1 | 0 | Low-quality | Genome-fragment | 5.92 | HMM-based (lower-bound) | 0 |
| S5_k141_9007 | 1005 | No | NA | 2 | 0 | 2 | Not-determined | Genome-fragment | NA | NA | 0 |
| S1-final-viral-combined_k141_189663\|\|full | 1748 | No | NA | 3 | 0 | 1 | Low-quality | Genome-fragment | 0.85 | AAI-based (medium-confidence) | 0 |
| S9-final-viral-combined_k141_126307\|\|lt2gene | 1749 | No | NA | 2 | 2 | 0 | Low-quality | Genome-fragment | 4.51 | AAI-based (medium-confidence) | 0 |
| S5_k141_124811 | 3941 | No | NA | 3 | 0 | 3 | Not-determined | Genome-fragment | NA | NA | 0 |
| S5_k141_53503 | 1866 | No | NA | 2 | 0 | 2 | Not-determined | Genome-fragment | NA | NA | 0 |
| S5_k141_160468 | 1070 | No | NA | 1 | 0 | 0 | Not-determined | Genome-fragment | NA | NA | 0 |
| S1_k141_53059 | 1749 | No | NA | 1 | 0 | 1 | Low-quality | Genome-fragment | 2.9 | AAI-based (medium-confidence) | 0 |
| S7-final-viral-combined_k141_119150\|\|full | 1749 | No | NA | 4 | 2 | 0 | Low-quality | Genome-fragment | 3.84 | AAI-based (high-confidence) | 0 |
| S5_k141_124859 | 1311 | No | NA | 2 | 0 | 1 | Not-determined | Genome-fragment | NA | NA | 0 |
| S5_k141_9082 | 3180 | No | NA | 2 | 0 | 0 | Not-determined | Genome-fragment | NA | NA | 0 |
| S5_k141_26872 | 1999 | No | NA | 2 | 0 | 0 | Not-determined | Genome-fragment | NA | NA | 0 |
| S17-final-viral-combined_k141_187182\|\|full | 1749 | No | NA | 4 | 0 | 0 | Low-quality | Genome-fragment | 1.92 | AAI-based (high-confidence) | 0 |
| S4-final-viral-combined_k141_41997\|\|full | 1749 | No | NA | 3 | 1 | 0 | Low-quality | Genome-fragment | 1.13 | HMM-based (lower-bound) | 0 |
| S5_k141_107055 | 2559 | No | NA | 1 | 0 | 1 | Not-determined | Genome-fragment | NA | NA | 0 |
| S5_k141_178271 | 1079 | No | NA | 1 | 0 | 1 | Not-determined | Genome-fragment | NA | NA | 0 |
| S5_k141_53621 | 1754 | No | NA | 2 | 0 | 2 | Not-determined | Genome-fragment | NA | NA | 0 |
| S4_k141_52440 | 1749 | No | NA | 2 | 0 | 1 | Low-quality | Genome-fragment | 1.02 | AAI-based (medium-confidence) | 0 |
| S5_k141_80360 | 1124 | No | NA | 1 | 0 | 0 | Not-determined | Genome-fragment | NA | NA | 0 |
| S5_k141_116045 | 6968 | No | NA | 11 | 0 | 2 | Not-determined | Genome-fragment | NA | NA | 0 |
| S5_k141_62583 | 4706 | No | NA | 1 | 0 | 0 | Not-determined | Genome-fragment | NA | NA | 0 |
| S5_k141_178401 | 1070 | No | NA | 1 | 0 | 1 | Not-determined | Genome-fragment | NA | NA | 0 |
| S5_k141_169321 | 6398 | No | NA | 9 | 0 | 3 | Not-determined | Genome-fragment | NA | NA | 0 |
| S5_k141_169347 | 1337 | No | NA | 1 | 0 | 0 | Not-determined | Genome-fragment | NA | NA | 0 |
| S5_k141_169369 | 1011 | No | NA | 1 | 0 | 1 | Not-determined | Genome-fragment | NA | NA | 0 |
| S5_k141_116153 | 1752 | No | NA | 2 | 0 | 1 | Not-determined | Genome-fragment | NA | NA | 0 |
| S8-final-viral-combined_k141_67595\|\|full | 1749 | No | NA | 4 | 1 | 0 | Low-quality | Genome-fragment | 2.38 | HMM-based (lower-bound) | 0 |
| S5_k141_196351 | 1457 | No | NA | 2 | 0 | 1 | Not-determined | Genome-fragment | NA | NA | 0 |
| S5_k141_62598 | 2723 | No | NA | 4 | 0 | 0 | Not-determined | Genome-fragment | NA | NA | 0 |
| S5_k141_187313 | 4845 | No | NA | 8 | 0 | 0 | Not-determined | Genome-fragment | NA | NA | 0 |
| S5_k141_35790 | 3442 | No | NA | 3 | 0 | 3 | Not-determined | Genome-fragment | NA | NA | 0 |
| S9_k141_61823 | 1750 | No | NA | 2 | 0 | 0 | Low-quality | Genome-fragment | 2.01 | AAI-based (high-confidence) | 0 |
| S5_k141_35801 | 2214 | No | NA | 1 | 0 | 0 | Not-determined | Genome-fragment | NA | NA | 0 |
| S5_k141_116227 | 5465 | No | NA | 1 | 0 | 1 | Not-determined | Genome-fragment | NA | NA | 0 |
| S11-final-viral-combined_k141_321787\|\|full | 1750 | No | NA | 4 | 0 | 0 | Low-quality | Genome-fragment | 3.04 | AAI-based (high-confidence) | 0 |
| S5_k141_205074 | 1124 | No | NA | 1 | 0 | 0 | Not-determined | Genome-fragment | NA | NA | 0 |
| S17-k141_59102 | 1750 | No | NA | 2 | 0 | 1 | Low-quality | Genome-fragment | 0.87 | AAI-based (medium-confidence) | 0 |
| S5_k141_9397 | 2862 | No | NA | 2 | 0 | 0 | Not-determined | Genome-fragment | NA | NA | 0 |
| S5_k141_9423 | 3001 | No | NA | 3 | 0 | 1 | Not-determined | Genome-fragment | NA | NA | 0 |
| S5_k141_160816 | 3197 | No | NA | 4 | 0 | 0 | Not-determined | Genome-fragment | NA | NA | 0 |
| S17-k141_141793 | 1750 | No | NA | 1 | 0 | 1 | Low-quality | Genome-fragment | 1.05 | AAI-based (medium-confidence) | 0 |
| S5_k141_89293 | 1240 | No | NA | 1 | 0 | 1 | Not-determined | Genome-fragment | NA | NA | 0 |
| S5_k141_62679 | 29809 | No | NA | 43 | 14 | 1 | High-quality | High-quality | 92.6 | AAI-based (medium-confidence) | 0 |
| S5_k141_196485 | 1807 | No | NA | 1 | 0 | 1 | Not-determined | Genome-fragment | NA | NA | 0 |
| S5_k141_53857 | 1172 | No | NA | 2 | 0 | 2 | Not-determined | Genome-fragment | NA | NA | 0 |
| S5_k141_53872 | 1999 | No | NA | 2 | 0 | 0 | Not-determined | Genome-fragment | NA | NA | 0 |
| S4-final-viral-combined_k141_159586\|\|full | 1750 | No | NA | 2 | 0 | 1 | Low-quality | Genome-fragment | 2.06 | AAI-based (high-confidence) | 0 |
| S6-final-viral-combined_k141_138004\|\|full | 1750 | No | NA | 3 | 2 | 0 | Low-quality | Genome-fragment | 4.89 | AAI-based (high-confidence) | 0 |
| S5_k141_142916 | 1532 | No | NA | 5 | 0 | 5 | Not-determined | Genome-fragment | NA | NA | 0 |
| S5_k141_107314 | 1082 | No | NA | 1 | 0 | 1 | Not-determined | Genome-fragment | NA | NA | 0 |
| S5_k141_62731 | 3450 | No | NA | 2 | 0 | 0 | Not-determined | Genome-fragment | NA | NA | 0 |
| S5_k141_169559 | 1017 | No | NA | 2 | 0 | 1 | Not-determined | Genome-fragment | NA | NA | 0 |
| S5_k141_196532 | 10370 | No | NA | 15 | 0 | 2 | Not-determined | Genome-fragment | NA | NA | 0 |
| S5_k141_151732 | 3580 | No | NA | 5 | 0 | 0 | Not-determined | Genome-fragment | NA | NA | 0 |
| S5_k141_107353 | 1327 | No | NA | 2 | 0 | 1 | Not-determined | Genome-fragment | NA | NA | 0 |
| S5_k141_735 | 17206 | No | NA | 3 | 0 | 2 | Not-determined | Genome-fragment | NA | NA | 0 |
| S8-final-viral-combined_k141_162115\|\|full | 1750 | No | NA | 2 | 0 | 2 | Low-quality | Genome-fragment | 3.55 | AAI-based (medium-confidence) | 0 |
| S5_k141_187568 | 3368 | No | NA | 6 | 0 | 6 | Not-determined | Genome-fragment | NA | NA | 0 |
| S11-final-viral-combined_k141_282358\|\|full | 1751 | No | NA | 2 | 0 | 1 | Low-quality | Genome-fragment | 3.71 | AAI-based (medium-confidence) | 0 |
| S5_k141_9538 | 1198 | No | NA | 1 | 0 | 0 | Not-determined | Genome-fragment | NA | NA | 0 |
| S11-final-viral-combined_k141_168850\|\|lt2gene | 1751 | No | NA | 2 | 1 | 0 | Low-quality | Genome-fragment | 0.97 | HMM-based (lower-bound) | 0 |
| S5_k141_125060 | 1250 | No | NA | 2 | 0 | 1 | Not-determined | Genome-fragment | NA | NA | 0 |
| S1_k141_70033 | 1751 | No | NA | 5 | 0 | 0 | Low-quality | Genome-fragment | 3.31 | AAI-based (high-confidence) | 0 |
| S5_k141_74021 | 1751 | No | NA | 1 | 0 | 0 | Low-quality | Genome-fragment | 3.66 | AAI-based (high-confidence) | 0 |
| S5_k141_36002 | 7797 | No | NA | 8 | 0 | 1 | Not-determined | Genome-fragment | NA | NA | 0 |
| S5_k141_44971 | 1451 | No | NA | 1 | 0 | 1 | Not-determined | Genome-fragment | NA | NA | 0 |
| S5_k141_44972 | 7740 | No | NA | 6 | 0 | 3 | Not-determined | Genome-fragment | NA | NA | 0 |
| S5_k141_841 | 1330 | No | NA | 1 | 0 | 0 | Not-determined | Genome-fragment | NA | NA | 0 |
| S5_k141_842 | 4960 | No | NA | 10 | 0 | 10 | Not-determined | Genome-fragment | NA | NA | 0 |
| S5_k141_54006 | 2935 | No | NA | 3 | 0 | 0 | Not-determined | Genome-fragment | NA | NA | 0 |
| S5_k141_71848 | 1701 | No | NA | 1 | 0 | 1 | Not-determined | Genome-fragment | NA | NA | 0 |
| S7-final-viral-combined_k141_155305\|\|full | 1751 | No | NA | 3 | 0 | 0 | Low-quality | Genome-fragment | 3.49 | AAI-based (medium-confidence) | 0 |
| S5_k141_205298 | 3299 | No | NA | 2 | 0 | 0 | Not-determined | Genome-fragment | NA | NA | 0 |
| S17-final-viral-combined_k141_74855\|\|full | 1751 | No | NA | 3 | 0 | 0 | Low-quality | Genome-fragment | 0.98 | AAI-based (high-confidence) | 0 |
| S5_k141_71922 | 1545 | No | NA | 1 | 0 | 1 | Not-determined | Genome-fragment | NA | NA | 0 |
| S5_k141_957 | 14444 | No | NA | 12 | 0 | 8 | Not-determined | Genome-fragment | NA | NA | 0 |
| S5_k141_196637 | 1907 | No | NA | 1 | 0 | 0 | Not-determined | Genome-fragment | NA | NA | 0 |
| S5_k141_1001 | 1057 | No | NA | 1 | 0 | 1 | Not-determined | Genome-fragment | NA | NA | 0 |
| S5_k141_116366 | 1258 | No | NA | 2 | 0 | 2 | Not-determined | Genome-fragment | NA | NA | 0 |
| S5_k141_1013 | 3587 | No | NA | 4 | 0 | 1 | Not-determined | Genome-fragment | NA | NA | 0 |
| S12-final-viral-combined_k141_101460\|\|full | 1751 | No | NA | 3 | 1 | 0 | Low-quality | Genome-fragment | 1.93 | AAI-based (high-confidence) | 0 |
| S5_k141_125257 | 1698 | No | NA | 1 | 0 | 1 | Not-determined | Genome-fragment | NA | NA | 0 |
| S5_k141_125258 | 1733 | No | NA | 2 | 0 | 1 | Not-determined | Genome-fragment | NA | NA | 0 |
| S5_k141_169869 | 5137 | No | NA | 9 | 0 | 1 | Not-determined | Genome-fragment | NA | NA | 0 |
| S4-final-viral-combined_k141_31172\|\|lt2gene | 1751 | No | NA | 2 | 1 | 0 | Low-quality | Genome-fragment | 1.97 | HMM-based (lower-bound) | 0 |
| S8-final-viral-combined_k141_237195\|\|full | 1751 | No | NA | 4 | 0 | 0 | Low-quality | Genome-fragment | 2.37 | AAI-based (high-confidence) | 0 |
| S5_k141_134254 | 1358 | No | NA | 3 | 0 | 1 | Not-determined | Genome-fragment | NA | NA | 0 |
| S5_k141_54072 | 1898 | No | NA | 1 | 0 | 0 | Not-determined | Genome-fragment | NA | NA | 0 |
| S5_k141_205397 | 1059 | No | NA | 1 | 0 | 0 | Not-determined | Genome-fragment | NA | NA | 0 |
| S11-final-viral-combined_k141_52635\|\|full | 1752 | No | NA | 3 | 2 | 0 | Low-quality | Genome-fragment | 3.86 | AAI-based (high-confidence) | 0 |
| S5_k141_36146 | 1288 | No | NA | 2 | 0 | 1 | Not-determined | Genome-fragment | NA | NA | 0 |
| S5_k141_169929 | 3092 | No | NA | 5 | 0 | 1 | Not-determined | Genome-fragment | NA | NA | 0 |
| S5_k141_45120 | 5586 | No | NA | 5 | 0 | 1 | Not-determined | Genome-fragment | NA | NA | 0 |
| S5_k141_178667 | 5177 | No | NA | 9 | 0 | 0 | Not-determined | Genome-fragment | NA | NA | 0 |
| S7-final-viral-combined_k141_226298\|\|full | 1752 | No | NA | 3 | 0 | 0 | Low-quality | Genome-fragment | 3.76 | AAI-based (medium-confidence) | 0 |
| S7-final-viral-combined_k141_44221\|\|full | 1752 | No | NA | 4 | 0 | 0 | Low-quality | Genome-fragment | 2.8 | AAI-based (high-confidence) | 0 |
| S17-final-viral-combined_k141_177561\|\|full | 1752 | No | NA | 6 | 0 | 0 | Low-quality | Genome-fragment | 3.42 | AAI-based (high-confidence) | 0 |
| S5_k141_178751 | 3483 | No | NA | 6 | 0 | 0 | Not-determined | Genome-fragment | NA | NA | 0 |
| S5_k141_116481 | 1020 | No | NA | 2 | 0 | 2 | Not-determined | Genome-fragment | NA | NA | 0 |
| S12_k141_105456 | 1752 | No | NA | 4 | 1 | 0 | Low-quality | Genome-fragment | 5.46 | AAI-based (high-confidence) | 0 |
| S9-final-viral-combined_k141_81905\|\|full | 1753 | No | NA | 4 | 1 | 0 | Low-quality | Genome-fragment | 2.87 | AAI-based (high-confidence) | 0 |
| S5_k141_205593 | 1331 | No | NA | 1 | 0 | 1 | Not-determined | Genome-fragment | NA | NA | 0 |
| S5_k141_90016 | 1655 | No | NA | 2 | 0 | 1 | Not-determined | Genome-fragment | NA | NA | 0 |
| S1-final-viral-combined_k141_347046\|\|lt2gene | 1753 | No | NA | 2 | 1 | 0 | Low-quality | Genome-fragment | 4.73 | AAI-based (high-confidence) | 0 |
| S5_k141_18510 | 1262 | No | NA | 2 | 0 | 1 | Not-determined | Genome-fragment | NA | NA | 0 |
| S5_k141_152121 | 1411 | No | NA | 3 | 0 | 1 | Not-determined | Genome-fragment | NA | NA | 0 |
| S5_k141_27320 | 1217 | No | NA | 1 | 0 | 1 | Not-determined | Genome-fragment | NA | NA | 0 |
| S5_k141_152161 | 1497 | No | NA | 1 | 0 | 0 | Not-determined | Genome-fragment | NA | NA | 0 |
| S5_k141_62995 | 1261 | No | NA | 1 | 0 | 1 | Not-determined | Genome-fragment | NA | NA | 0 |
| S5_k141_45304 | 3871 | No | NA | 1 | 0 | 1 | Not-determined | Genome-fragment | NA | NA | 0 |
| S5_k141_170231 | 2492 | No | NA | 3 | 0 | 1 | Not-determined | Genome-fragment | NA | NA | 0 |
| S7-final-viral-combined_k141_143543\|\|full | 1753 | No | NA | 2 | 0 | 0 | Low-quality | Genome-fragment | 2.8 | AAI-based (medium-confidence) | 0 |
| S5_k141_27417 | 1125 | No | NA | 1 | 0 | 1 | Not-determined | Genome-fragment | NA | NA | 0 |
| S5_k141_125558 | 1009 | No | NA | 1 | 0 | 0 | Not-determined | Genome-fragment | NA | NA | 0 |
| S7-final-viral-combined_k141_232674\|\|full | 1753 | No | NA | 4 | 3 | 0 | Low-quality | Genome-fragment | 2.22 | AAI-based (high-confidence) | 0 |
| S7-final-viral-combined_k141_60264\|\|full | 1753 | No | NA | 3 | 1 | 0 | Low-quality | Genome-fragment | 2.52 | HMM-based (lower-bound) | 0 |
| S12-final-viral-combined_k141_332959\|\|full | 1753 | No | NA | 3 | 0 | 2 | Low-quality | Genome-fragment | 2.1 | AAI-based (medium-confidence) | 0 |
| S5_k141_27480 | 1390 | No | NA | 3 | 0 | 2 | Not-determined | Genome-fragment | NA | NA | 0 |
| S5_k141_54386 | 1043 | No | NA | 1 | 0 | 1 | Not-determined | Genome-fragment | NA | NA | 0 |
| S5_k141_36406 | 6529 | No | NA | 9 | 0 | 0 | Not-determined | Genome-fragment | NA | NA | 0 |
| S5_k141_45444 | 1089 | No | NA | 1 | 0 | 1 | Not-determined | Genome-fragment | NA | NA | 0 |
| S5_k141_125606 | 1327 | No | NA | 1 | 0 | 1 | Not-determined | Genome-fragment | NA | NA | 0 |
| S5_k141_125631 | 2307 | No | NA | 2 | 0 | 2 | Not-determined | Genome-fragment | NA | NA | 0 |
| S5_k141_18667 | 1157 | No | NA | 1 | 0 | 0 | Not-determined | Genome-fragment | NA | NA | 0 |
| S12-final-viral-combined_k141_99057\|\|full | 1753 | No | NA | 5 | 5 | 0 | Low-quality | Genome-fragment | 4.9 | AAI-based (high-confidence) | 0 |
| S8-final-viral-combined_k141_249245\|\|full | 1753 | No | NA | 2 | 1 | 0 | Low-quality | Genome-fragment | 1.06 | AAI-based (high-confidence) | 0 |
| S5_k141_27570 | 2357 | No | NA | 4 | 0 | 0 | Not-determined | Genome-fragment | NA | NA | 0 |
| S1-final-viral-combined_k141_238647\|\|full | 1754 | No | NA | 4 | 2 | 0 | Low-quality | Genome-fragment | 4.7 | AAI-based (high-confidence) | 0 |
| S5_k141_161304 | 3490 | No | NA | 2 | 0 | 0 | Not-determined | Genome-fragment | NA | NA | 0 |
| S1_k141_335986 | 1754 | No | NA | 2 | 0 | 1 | Low-quality | Genome-fragment | 0.62 | AAI-based (medium-confidence) | 0 |
| S5_k141_212716 | 1754 | No | NA | 3 | 0 | 0 | Low-quality | Genome-fragment | 2.14 | AAI-based (medium-confidence) | 0 |
| S5_k141_152372 | 1068 | No | NA | 1 | 0 | 0 | Not-determined | Genome-fragment | NA | NA | 0 |
| S5_k141_197031 | 1126 | No | NA | 1 | 0 | 0 | Not-determined | Genome-fragment | NA | NA | 0 |
| S12-final-viral-combined_k141_178339\|\|full | 1754 | No | NA | 3 | 0 | 0 | Low-quality | Genome-fragment | 3.89 | AAI-based (medium-confidence) | 0 |
| S5_k141_81054 | 1121 | No | NA | 1 | 0 | 0 | Not-determined | Genome-fragment | NA | NA | 0 |
| S5_k141_10147 | 3844 | No | NA | 1 | 0 | 1 | Not-determined | Genome-fragment | NA | NA | 0 |
| S2-final-viral-combined_k141_23273\|\|full | 1754 | No | NA | 5 | 1 | 0 | Low-quality | Genome-fragment | 4.95 | AAI-based (medium-confidence) | 0 |
| S2-final-viral-combined_k141_79272\|\|full | 1754 | No | NA | 4 | 2 | 0 | Low-quality | Genome-fragment | 2.13 | AAI-based (high-confidence) | 0 |
| S5_k141_1712 | 4080 | No | NA | 8 | 0 | 7 | Not-determined | Genome-fragment | NA | NA | 0 |
| S5_k141_170462 | 1112 | No | NA | 1 | 0 | 0 | Not-determined | Genome-fragment | NA | NA | 0 |
| S5_k141_134673 | 2029 | No | NA | 3 | 0 | 2 | Not-determined | Genome-fragment | NA | NA | 0 |
| S5_k141_125813 | 1407 | No | NA | 1 | 0 | 1 | Not-determined | Genome-fragment | NA | NA | 0 |
| S5_k141_27708 | 1170 | No | NA | 1 | 0 | 1 | Not-determined | Genome-fragment | NA | NA | 0 |
| S5_k141_72288 | 4209 | No | NA | 5 | 0 | 0 | Not-determined | Genome-fragment | NA | NA | 0 |
| S5_k141_27718 | 3326 | No | NA | 5 | 0 | 2 | Not-determined | Genome-fragment | NA | NA | 0 |
| S2-final-viral-combined_k141_220720\|\|lt2gene | 1754 | No | NA | 1 | 1 | 0 | Low-quality | Genome-fragment | 1.3 | HMM-based (lower-bound) | 0 |
| S5_k141_99031 | 3346 | No | NA | 4 | 0 | 2 | Not-determined | Genome-fragment | NA | NA | 0 |
| S5_k141_197104 | 5553 | No | NA | 8 | 0 | 2 | Not-determined | Genome-fragment | NA | NA | 0 |
| S2_k141_92145 | 1754 | No | NA | 2 | 0 | 0 | Low-quality | Genome-fragment | 1.71 | AAI-based (medium-confidence) | 0 |
| S8-final-viral-combined_k141_271423\|\|lt2gene | 1754 | No | NA | 1 | 1 | 0 | Low-quality | Genome-fragment | 5.44 | AAI-based (high-confidence) | 0 |
| S5_k141_197116 | 1237 | No | NA | 1 | 0 | 0 | Not-determined | Genome-fragment | NA | NA | 0 |
| S1-final-viral-combined_k141_143680\|\|full | 1755 | No | NA | 4 | 2 | 0 | Low-quality | Genome-fragment | 4.36 | AAI-based (medium-confidence) | 0 |
| S1-final-viral-combined_k141_113793\|\|full | 1755 | No | NA | 4 | 0 | 1 | Low-quality | Genome-fragment | 2.04 | AAI-based (medium-confidence) | 0 |
| S5_k141_197152 | 1819 | No | NA | 3 | 0 | 2 | Not-determined | Genome-fragment | NA | NA | 0 |
| S5_k141_27851 | 1353 | No | NA | 2 | 0 | 1 | Not-determined | Genome-fragment | NA | NA | 0 |
| S5_k141_45738 | 3184 | No | NA | 4 | 0 | 0 | Not-determined | Genome-fragment | NA | NA | 0 |
| S5_k141_1880 | 1096 | No | NA | 1 | 0 | 1 | Not-determined | Genome-fragment | NA | NA | 0 |
| S7-final-viral-combined_k141_3882\|\|full | 1755 | No | NA | 1 | 0 | 0 | Low-quality | Genome-fragment | 3.53 | AAI-based (medium-confidence) | 0 |
| S5_k141_161439 | 1143 | No | NA | 2 | 0 | 1 | Not-determined | Genome-fragment | NA | NA | 0 |
| S7-final-viral-combined_k141_234079\|\|full | 1755 | No | NA | 2 | 0 | 1 | Low-quality | Genome-fragment | 0.81 | AAI-based (medium-confidence) | 0 |
| S5_k141_72413 | 1190 | No | NA | 1 | 0 | 1 | Not-determined | Genome-fragment | NA | NA | 0 |
| S17-final-viral-combined_k141_9073\|\|full | 1755 | No | NA | 2 | 0 | 2 | Low-quality | Genome-fragment | 1.14 | AAI-based (medium-confidence) | 0 |
| S5_k141_188143 | 1173 | No | NA | 2 | 0 | 2 | Not-determined | Genome-fragment | NA | NA | 0 |
| S5_k141_99197 | 1115 | No | NA | 1 | 0 | 1 | Not-determined | Genome-fragment | NA | NA | 0 |
| S5_k141_188151 | 4188 | No | NA | 9 | 0 | 0 | Not-determined | Genome-fragment | NA | NA | 0 |
| S12-final-viral-combined_k141_15225\|\|full | 1755 | No | NA | 5 | 0 | 2 | Low-quality | Genome-fragment | 3.85 | AAI-based (medium-confidence) | 0 |
| S2-final-viral-combined_k141_191355\|\|full | 1755 | No | NA | 2 | 0 | 0 | Low-quality | Genome-fragment | 3.5 | AAI-based (medium-confidence) | 0 |
| S5_k141_18887 | 1146 | No | NA | 2 | 0 | 0 | Not-determined | Genome-fragment | NA | NA | 0 |
| S5_k141_1951 | 1274 | No | NA | 3 | 0 | 0 | Not-determined | Genome-fragment | NA | NA | 0 |
| S2-final-viral-combined_k141_192434\|\|full | 1755 | No | NA | 3 | 2 | 0 | Low-quality | Genome-fragment | 2.95 | AAI-based (medium-confidence) | 0 |
| S5_k141_108523 | 1180 | No | NA | 1 | 0 | 0 | Not-determined | Genome-fragment | NA | NA | 0 |
| S9_k141_86977 | 1756 | No | NA | 3 | 0 | 0 | Low-quality | Genome-fragment | 4.54 | AAI-based (high-confidence) | 0 |
| S5_k141_179534 | 1082 | No | NA | 1 | 0 | 0 | Not-determined | Genome-fragment | NA | NA | 0 |
| S5_k141_152712 | 1112 | No | NA | 1 | 0 | 1 | Not-determined | Genome-fragment | NA | NA | 0 |
| S5_k141_63461 | 1528 | No | NA | 1 | 0 | 1 | Not-determined | Genome-fragment | NA | NA | 0 |
| S5_k141_10269 | 1385 | No | NA | 3 | 0 | 2 | Not-determined | Genome-fragment | NA | NA | 0 |
| S11-final-viral-combined_k141_281419\|\|full | 1756 | No | NA | 5 | 0 | 0 | Low-quality | Genome-fragment | 3.65 | AAI-based (medium-confidence) | 0 |
| S5_k141_179602 | 1495 | No | NA | 1 | 0 | 1 | Not-determined | Genome-fragment | NA | NA | 0 |
| S5_k141_152755 | 1719 | No | NA | 3 | 0 | 2 | Not-determined | Genome-fragment | NA | NA | 0 |
| S11_k141_39208 | 1756 | No | NA | 5 | 0 | 1 | Low-quality | Genome-fragment | 3.03 | AAI-based (high-confidence) | 0 |
| S1-final-viral-combined_k141_84956\|\|full | 1756 | No | NA | 4 | 0 | 1 | Low-quality | Genome-fragment | 5.26 | AAI-based (high-confidence) | 0 |
| S5_k141_99245 | 1045 | No | NA | 1 | 0 | 1 | Not-determined | Genome-fragment | NA | NA | 0 |
| S5_k141_99249 | 2806 | No | NA | 1 | 0 | 1 | Not-determined | Genome-fragment | NA | NA | 0 |
| S5_k141_81482 | 4799 | No | NA | 3 | 0 | 2 | Not-determined | Genome-fragment | NA | NA | 0 |
| S5_k141_161535 | 1050 | No | NA | 1 | 0 | 1 | Not-determined | Genome-fragment | NA | NA | 0 |
| S5_k141_10349 | 1327 | No | NA | 2 | 0 | 2 | Not-determined | Genome-fragment | NA | NA | 0 |
| S5_k141_81490 | 5792 | No | NA | 1 | 0 | 0 | Not-determined | Genome-fragment | NA | NA | 0 |
| S5_k141_161545 | 8254 | No | NA | 7 | 0 | 2 | Not-determined | Genome-fragment | NA | NA | 0 |
| S5_k141_36881 | 1945 | No | NA | 2 | 0 | 1 | Not-determined | Genome-fragment | NA | NA | 0 |
| S5_k141_170873 | 1156 | No | NA | 1 | 0 | 0 | Not-determined | Genome-fragment | NA | NA | 0 |
| S5_k141_45868 | 1990 | No | NA | 1 | 0 | 1 | Not-determined | Genome-fragment | NA | NA | 0 |
| S5_k141_18958 | 1305 | No | NA | 4 | 0 | 3 | Not-determined | Genome-fragment | NA | NA | 0 |
| S5_k141_18976 | 2038 | No | NA | 1 | 0 | 1 | Not-determined | Genome-fragment | NA | NA | 0 |
| S5_k141_81539 | 3115 | No | NA | 2 | 0 | 2 | Not-determined | Genome-fragment | NA | NA | 0 |
| S5_k141_108562 | 1206 | No | NA | 1 | 0 | 1 | Not-determined | Genome-fragment | NA | NA | 0 |
| S1-final-viral-combined_k141_120584\|\|full | 1756 | No | NA | 2 | 2 | 0 | Low-quality | Genome-fragment | 3.89 | AAI-based (high-confidence) | 0 |
| S5_k141_72670 | 1052 | No | NA | 1 | 0 | 1 | Not-determined | Genome-fragment | NA | NA | 0 |
| S5_k141_72673 | 5789 | No | NA | 9 | 0 | 1 | Not-determined | Genome-fragment | NA | NA | 0 |
| S5_k141_126061 | 2331 | No | NA | 2 | 0 | 2 | Not-determined | Genome-fragment | NA | NA | 0 |
| S5_k141_161588 | 1189 | No | NA | 1 | 0 | 0 | Not-determined | Genome-fragment | NA | NA | 0 |
| S1_k141_151507 | 1756 | No | NA | 2 | 0 | 2 | Low-quality | Genome-fragment | 0.5 | AAI-based (medium-confidence) | 0 |
| S5_k141_72688 | 6718 | No | NA | 7 | 0 | 6 | Not-determined | Genome-fragment | NA | NA | 0 |
| S5_k141_2174 | 3590 | No | NA | 6 | 0 | 1 | Not-determined | Genome-fragment | NA | NA | 0 |
| S5_k141_2187 | 1495 | No | NA | 1 | 0 | 0 | Not-determined | Genome-fragment | NA | NA | 0 |
| S5_k141_170956 | 1254 | No | NA | 2 | 0 | 2 | Not-determined | Genome-fragment | NA | NA | 0 |
| S12-final-viral-combined_k141_170672\|\|full | 1756 | No | NA | 5 | 1 | 0 | Low-quality | Genome-fragment | 1 | HMM-based (lower-bound) | 0 |
| S5_k141_144230 | 1295 | No | NA | 1 | 0 | 1 | Not-determined | Genome-fragment | NA | NA | 0 |
| S5_k141_170972 | 1014 | No | NA | 1 | 0 | 1 | Not-determined | Genome-fragment | NA | NA | 0 |
| S2-final-viral-combined_k141_184174\|\|full | 1756 | No | NA | 2 | 0 | 1 | Low-quality | Genome-fragment | 1.12 | AAI-based (medium-confidence) | 0 |
| S5_k141_144262 | 7519 | No | NA | 7 | 0 | 7 | Not-determined | Genome-fragment | NA | NA | 0 |
| S5_k141_206122 | 3308 | No | NA | 4 | 0 | 0 | Not-determined | Genome-fragment | NA | NA | 0 |
| S5_k141_197474 | 1245 | No | NA | 1 | 0 | 1 | Not-determined | Genome-fragment | NA | NA | 0 |
| S5_k141_144328 | 2876 | No | NA | 2 | 0 | 2 | Not-determined | Genome-fragment | NA | NA | 0 |
| S5_k141_197482 | 1239 | No | NA | 1 | 0 | 1 | Not-determined | Genome-fragment | NA | NA | 0 |
| S2_k141_2972 | 1756 | No | NA | 5 | 0 | 1 | Low-quality | Genome-fragment | 4.5 | AAI-based (medium-confidence) | 0 |
| S5_k141_152953 | 1110 | No | NA | 1 | 0 | 0 | Not-determined | Genome-fragment | NA | NA | 0 |
| S5_k141_171134 | 1052 | No | NA | 4 | 0 | 0 | Not-determined | Genome-fragment | NA | NA | 0 |
| S4-final-viral-combined_k141_112568\|\|full | 1756 | No | NA | 3 | 0 | 0 | Low-quality | Genome-fragment | 2.25 | AAI-based (high-confidence) | 0 |
| S5_k141_2284 | 1968 | No | NA | 2 | 0 | 2 | Not-determined | Genome-fragment | NA | NA | 0 |
| S5_k141_171136 | 3073 | No | NA | 3 | 0 | 0 | Not-determined | Genome-fragment | NA | NA | 0 |
| S5_k141_37036 | 1120 | No | NA | 1 | 0 | 1 | Not-determined | Genome-fragment | NA | NA | 0 |

| | | | | | | | | | | | |
|---|---|---|---|---|---|---|---|---|---|---|---|
| S5_k141_126145 | 1047 | No | NA | 1 | 0 | 1 | Not-determined | Genome-fragment | NA | NA | 0 |
| S5_k141_108678 | 1421 | No | NA | 1 | 0 | 1 | Not-determined | Genome-fragment | NA | NA | 0 |
| S5_k141_99330 | 1802 | No | NA | 1 | 0 | 1 | Not-determined | Genome-fragment | NA | NA | 0 |
| S5_k141_135074 | 4587 | No | NA | 4 | 0 | 2 | Not-determined | Genome-fragment | NA | NA | 0 |
| S8-final-viral-combined_k141_37232\|\|lt2gene | 1756 | No | NA | 1 | 1 | 0 | Low-quality | Genome-fragment | 5.13 | AAI-based (medium-confidence) | 0 |
| S5_k141_10644 | 1178 | No | NA | 2 | 0 | 1 | Not-determined | Genome-fragment | NA | NA | 0 |
| S11-final-viral-combined_k141_314386\|\|full | 1757 | No | NA | 3 | 1 | 0 | Low-quality | Genome-fragment | 3.73 | AAI-based (high-confidence) | 0 |
| S5_k141_37107 | 1475 | No | NA | 4 | 0 | 0 | Not-determined | Genome-fragment | NA | NA | 0 |
| S11_k141_243624 | 1757 | No | NA | 4 | 1 | 0 | Low-quality | Genome-fragment | 1.2 | AAI-based (high-confidence) | 0 |
| S5_k141_161733 | 1340 | No | NA | 3 | 0 | 2 | Not-determined | Genome-fragment | NA | NA | 0 |
| S5_k141_37147 | 8549 | No | NA | 12 | 0 | 1 | Not-determined | Genome-fragment | NA | NA | 0 |
| S5_k141_99456 | 3077 | No | NA | 3 | 0 | 1 | Not-determined | Genome-fragment | NA | NA | 0 |
| S5_k141_37150 | 1225 | No | NA | 1 | 0 | 1 | Not-determined | Genome-fragment | NA | NA | 0 |
| S5_k141_126337 | 1298 | No | NA | 2 | 0 | 1 | Not-determined | Genome-fragment | NA | NA | 0 |
| S5_k141_28176 | 3231 | No | NA | 2 | 0 | 1 | Not-determined | Genome-fragment | NA | NA | 0 |
| S5_k141_161809 | 1030 | No | NA | 1 | 0 | 0 | Not-determined | Genome-fragment | NA | NA | 0 |
| S5_k141_180149 | 1000 | No | NA | 1 | 0 | 1 | Not-determined | Genome-fragment | NA | NA | 0 |
| S5_k141_2448 | 1801 | No | NA | 2 | 0 | 2 | Not-determined | Genome-fragment | NA | NA | 0 |
| S5_k141_73142 | 1353 | No | NA | 1 | 0 | 0 | Not-determined | Genome-fragment | NA | NA | 0 |
| S5_k141_171430 | 1535 | No | NA | 1 | 0 | 1 | Not-determined | Genome-fragment | NA | NA | 0 |
| S5_k141_5051 | 1757 | No | NA | 2 | 0 | 2 | Low-quality | Genome-fragment | 4.43 | AAI-based (medium-confidence) | 0 |
| S7-final-viral-combined_k141_165598\|\|full | 1757 | No | NA | 3 | 0 | 1 | Low-quality | Genome-fragment | 0.77 | AAI-based (medium-confidence) | 0 |
| S7_k141_52534 | 1757 | No | NA | 1 | 1 | 0 | Low-quality | Genome-fragment | 1.84 | AAI-based (high-confidence) | 0 |
| S5_k141_28251 | 7033 | No | NA | 7 | 0 | 1 | Not-determined | Genome-fragment | NA | NA | 0 |
| S5_k141_144584 | 1021 | No | NA | 1 | 0 | 0 | Not-determined | Genome-fragment | NA | NA | 0 |
| S5_k141_206362 | 1198 | No | NA | 2 | 0 | 1 | Not-determined | Genome-fragment | NA | NA | 0 |
| S5_k141_188851 | 5751 | No | NA | 6 | 0 | 4 | Not-determined | Genome-fragment | NA | NA | 0 |
| S17-final-viral-combined_k141_225376\|\|full | 1757 | No | NA | 2 | 0 | 0 | Low-quality | Genome-fragment | 0.99 | AAI-based (medium-confidence) | 0 |
| S5_k141_171526 | 2081 | No | NA | 4 | 0 | 2 | Not-determined | Genome-fragment | NA | NA | 0 |
| S5_k141_10764 | 1229 | No | NA | 1 | 0 | 0 | Not-determined | Genome-fragment | NA | NA | 0 |
| S5_k141_81983 | 1576 | No | NA | 3 | 0 | 2 | Not-determined | Genome-fragment | NA | NA | 0 |
| S5_k141_91051 | 1460 | No | NA | 2 | 0 | 2 | Not-determined | Genome-fragment | NA | NA | 0 |
| S17-final-viral-combined_k141_82370\|\|full | 1757 | No | NA | 3 | 0 | 0 | Low-quality | Genome-fragment | 6.44 | AAI-based (medium-confidence) | 0 |
| S12-final-viral-combined_k141_292028\|\|full | 1757 | No | NA | 2 | 0 | 1 | Low-quality | Genome-fragment | 2.14 | AAI-based (high-confidence) | 0 |
| S5_k141_10769 | 7607 | No | NA | 4 | 0 | 1 | Not-determined | Genome-fragment | NA | NA | 0 |
| S6-final-viral-combined_k141_132973\|\|full | 1757 | No | NA | 3 | 0 | 0 | Low-quality | Genome-fragment | 3.55 | AAI-based (high-confidence) | 0 |
| S5_k141_10783 | 18932 | No | NA | 28 | 1 | 0 | High-quality | High-quality | 100 | AAI-based (medium-confidence) | 0 |
| S11-final-viral-combined_k141_237241\|\|full | 1758 | No | NA | 3 | 0 | 1 | Low-quality | Genome-fragment | 1.85 | AAI-based (medium-confidence) | 0 |
| S5_k141_135296 | 1008 | No | NA | 2 | 0 | 1 | Not-determined | Genome-fragment | NA | NA | 0 |
| S5_k141_2526 | 1061 | No | NA | 1 | 0 | 0 | Not-determined | Genome-fragment | NA | NA | 0 |
| S1-final-viral-combined_k141_168717\|\|full | 1758 | No | NA | 3 | 0 | 0 | Low-quality | Genome-fragment | 2.82 | AAI-based (high-confidence) | 0 |
| S5_k141_63845 | 1564 | No | NA | 1 | 0 | 1 | Not-determined | Genome-fragment | NA | NA | 0 |
| S5_k141_171635 | 2081 | No | NA | 2 | 0 | 0 | Not-determined | Genome-fragment | NA | NA | 0 |
| S5_k141_180354 | 1391 | No | NA | 4 | 0 | 0 | Not-determined | Genome-fragment | NA | NA | 0 |
| S5_k141_28313 | 1244 | No | NA | 2 | 0 | 1 | Not-determined | Genome-fragment | NA | NA | 0 |
| S5_k141_28321 | 2281 | No | NA | 1 | 0 | 0 | Not-determined | Genome-fragment | NA | NA | 0 |
| S5_k141_91154 | 1073 | No | NA | 1 | 0 | 1 | Not-determined | Genome-fragment | NA | NA | 0 |
| S1_k141_201229 | 1758 | No | NA | 1 | 0 | 1 | Low-quality | Genome-fragment | 0.87 | AAI-based (medium-confidence) | 0 |
| S5_k141_28342 | 2259 | No | NA | 3 | 0 | 3 | Not-determined | Genome-fragment | NA | NA | 0 |
| S5_k141_126583 | 3679 | No | NA | 3 | 0 | 3 | Not-determined | Genome-fragment | NA | NA | 0 |
| S5_k141_117557 | 7600 | No | NA | 7 | 0 | 2 | Not-determined | Genome-fragment | NA | NA | 0 |
| S5_k141_2716 | 1137 | No | NA | 1 | 0 | 0 | Not-determined | Genome-fragment | NA | NA | 0 |
| S5_k141_144716 | 2328 | No | NA | 3 | 0 | 2 | Not-determined | Genome-fragment | NA | NA | 0 |
| S5_k141_19512 | 1118 | No | NA | 1 | 0 | 0 | Not-determined | Genome-fragment | NA | NA | 0 |
| S1_k141_219162 | 1758 | No | NA | 1 | 0 | 1 | Low-quality | Genome-fragment | 3.79 | AAI-based (high-confidence) | 0 |
| S5_k141_144768 | 1226 | No | NA | 2 | 0 | 1 | Not-determined | Genome-fragment | NA | NA | 0 |
| S5_k141_108764 | 4077 | No | NA | 5 | 0 | 0 | Not-determined | Genome-fragment | NA | NA | 0 |
| S1_k141_142363 | 1758 | No | NA | 1 | 0 | 0 | Low-quality | Genome-fragment | 0.15 | AAI-based (medium-confidence) | 0 |
| S5_k141_28474 | 3805 | No | NA | 5 | 0 | 4 | Not-determined | Genome-fragment | NA | NA | 0 |
| S3-final-viral-combined_k141_79904\|\|full | 1758 | No | NA | 3 | 0 | 1 | Low-quality | Genome-fragment | 2.44 | AAI-based (medium-confidence) | 0 |
| S7-final-viral-combined_k141_55777\|\|full | 1758 | No | NA | 2 | 0 | 1 | Low-quality | Genome-fragment | 3.45 | AAI-based (medium-confidence) | 0 |
| S5_k141_73427 | 4694 | No | NA | 5 | 0 | 5 | Not-determined | Genome-fragment | NA | NA | 0 |
| S5_k141_144822 | 1038 | No | NA | 2 | 0 | 1 | Not-determined | Genome-fragment | NA | NA | 0 |
| S5_k141_189123 | 3903 | No | NA | 2 | 0 | 1 | Not-determined | Genome-fragment | NA | NA | 0 |
| S5_k141_189141 | 1338 | No | NA | 1 | 0 | 1 | Not-determined | Genome-fragment | NA | NA | 0 |
| S5_k141_99939 | 5469 | No | NA | 11 | 0 | 0 | Not-determined | Genome-fragment | NA | NA | 0 |
| S7-final-viral-combined_k141_211685\|\|full | 1758 | No | NA | 3 | 0 | 0 | Low-quality | Genome-fragment | 2.2 | AAI-based (high-confidence) | 0 |
| S12_k141_312175 | 1758 | No | NA | 2 | 0 | 2 | Low-quality | Genome-fragment | 0.87 | AAI-based (medium-confidence) | 0 |
| S2-final-viral-combined_k141_52356\|\|full | 1758 | No | NA | 3 | 0 | 1 | Low-quality | Genome-fragment | 1.06 | AAI-based (medium-confidence) | 0 |
| S5_k141_197932 | 3419 | No | NA | 4 | 0 | 4 | Not-determined | Genome-fragment | NA | NA | 0 |
| S5_k141_37654 | 1058 | No | NA | 2 | 0 | 1 | Not-determined | Genome-fragment | NA | NA | 0 |
| S8-final-viral-combined_k141_119501\|\|full | 1758 | No | NA | 2 | 0 | 2 | Low-quality | Genome-fragment | 1.16 | AAI-based (medium-confidence) | 0 |
| S5_k141_10842 | 1488 | No | NA | 2 | 0 | 2 | Not-determined | Genome-fragment | NA | NA | 0 |
| S5_k141_37661 | 1280 | No | NA | 1 | 0 | 0 | Not-determined | Genome-fragment | NA | NA | 0 |
| S5_k141_198020 | 1121 | No | NA | 2 | 0 | 1 | Not-determined | Genome-fragment | NA | NA | 0 |
| S11-final-viral-combined_k141_254492\|\|full | 1759 | No | NA | 5 | 0 | 1 | Low-quality | Genome-fragment | 1.31 | AAI-based (medium-confidence) | 0 |
| S17-k141_26337 | 1759 | No | NA | 6 | 0 | 0 | Low-quality | Genome-fragment | 5.42 | AAI-based (high-confidence) | 0 |
| S5_k141_73529 | 14138 | No | NA | 7 | 0 | 4 | Not-determined | Genome-fragment | NA | NA | 0 |
| S17-k141_141329 | 1759 | No | NA | 4 | 0 | 0 | Low-quality | Genome-fragment | 14.8 | AAI-based (high-confidence) | 0 |
| S5_k141_55547 | 4535 | No | NA | 5 | 0 | 2 | Not-determined | Genome-fragment | NA | NA | 0 |
| S5_k141_55553 | 2593 | No | NA | 1 | 0 | 0 | Not-determined | Genome-fragment | NA | NA | 0 |
| S5_k141_126991 | 3041 | No | NA | 4 | 0 | 3 | Not-determined | Genome-fragment | NA | NA | 0 |
| S4_k141_38442 | 1759 | No | NA | 4 | 0 | 0 | Low-quality | Genome-fragment | 0.64 | AAI-based (medium-confidence) | 0 |
| S5_k141_64205 | 1098 | No | NA | 2 | 0 | 1 | Not-determined | Genome-fragment | NA | NA | 0 |
| S5_k141_180568 | 1162 | No | NA | 1 | 0 | 1 | Not-determined | Genome-fragment | NA | NA | 0 |
| S5_k141_153694 | 1039 | No | NA | 2 | 0 | 2 | Not-determined | Genome-fragment | NA | NA | 0 |
| S5_k141_10905 | 3407 | No | NA | 3 | 0 | 2 | Not-determined | Genome-fragment | NA | NA | 0 |
| S5_k141_91467 | 4206 | No | NA | 3 | 0 | 3 | Not-determined | Genome-fragment | NA | NA | 0 |
| S5_k141_46874 | 1504 | No | NA | 1 | 0 | 1 | Not-determined | Genome-fragment | NA | NA | 0 |
| S5_k141_100223 | 1075 | No | NA | 3 | 0 | 0 | Not-determined | Genome-fragment | NA | NA | 0 |
| S5_k141_198189 | 1197 | No | NA | 1 | 0 | 0 | Not-determined | Genome-fragment | NA | NA | 0 |
| S5_k141_189390 | 1062 | No | NA | 2 | 0 | 0 | Not-determined | Genome-fragment | NA | NA | 0 |
| S5_k141_10995 | 1177 | No | NA | 1 | 0 | 1 | Not-determined | Genome-fragment | NA | NA | 0 |
| S5_k141_46912 | 2387 | No | NA | 3 | 0 | 1 | Not-determined | Genome-fragment | NA | NA | 0 |
| S5_k141_19632 | 1577 | No | NA | 2 | 0 | 1 | Not-determined | Genome-fragment | NA | NA | 0 |
| S6-final-viral-combined_k141_111493\|\|lt2gene | 1759 | No | NA | 1 | 1 | 0 | Low-quality | Genome-fragment | 0.97 | HMM-based (lower-bound) | 0 |
| S5_k141_127083 | 1113 | No | NA | 1 | 0 | 0 | Not-determined | Genome-fragment | NA | NA | 0 |
| S5_k141_135662 | 1455 | No | NA | 1 | 0 | 0 | Not-determined | Genome-fragment | NA | NA | 0 |
| S11-final-viral-combined_k141_64945\|\|full | 1760 | No | NA | 2 | 0 | 2 | Low-quality | Genome-fragment | 3.1 | AAI-based (medium-confidence) | 0 |
| S5_k141_135688 | 5451 | No | NA | 3 | 0 | 1 | Not-determined | Genome-fragment | NA | NA | 0 |
| S5_k141_46966 | 1483 | No | NA | 3 | 0 | 0 | Not-determined | Genome-fragment | NA | NA | 0 |
| S5_k141_189522 | 1249 | No | NA | 1 | 0 | 0 | Not-determined | Genome-fragment | NA | NA | 0 |
| S5_k141_64406 | 1058 | No | NA | 1 | 0 | 1 | Not-determined | Genome-fragment | NA | NA | 0 |
| S5_k141_206876 | 3037 | No | NA | 3 | 0 | 1 | Not-determined | Genome-fragment | NA | NA | 0 |
| S5-final-viral-combined_k141_82741\|\|full | 1760 | No | NA | 6 | 1 | 0 | Low-quality | Genome-fragment | 4.81 | AAI-based (high-confidence) | 0 |
| S5_k141_64441 | 1781 | No | NA | 1 | 0 | 0 | Not-determined | Genome-fragment | NA | NA | 0 |
| S17-final-viral-combined_k141_97398\|\|full | 1760 | No | NA | 2 | 0 | 0 | Low-quality | Genome-fragment | 5.51 | AAI-based (high-confidence) | 0 |
| S2-final-viral-combined_k141_162188\|\|lt2gene | 1760 | No | NA | 2 | 1 | 0 | Low-quality | Genome-fragment | 5.13 | AAI-based (high-confidence) | 0 |
| S5_k141_108909 | 1307 | No | NA | 1 | 0 | 1 | Not-determined | Genome-fragment | NA | NA | 0 |
| S5_k141_64474 | 1108 | No | NA | 1 | 0 | 0 | Not-determined | Genome-fragment | NA | NA | 0 |
| S5_k141_145092 | 4471 | No | NA | 6 | 0 | 1 | Not-determined | Genome-fragment | NA | NA | 0 |
| S5_k141_145098 | 1195 | No | NA | 1 | 0 | 0 | Not-determined | Genome-fragment | NA | NA | 0 |
| S5_k141_64595 | 1252 | No | NA | 2 | 0 | 2 | Not-determined | Genome-fragment | NA | NA | 0 |
| S4_k141_18340 | 1760 | No | NA | 3 | 0 | 0 | Low-quality | Genome-fragment | 1.84 | AAI-based (high-confidence) | 0 |
| S6_k141_120284 | 1760 | No | NA | 1 | 0 | 0 | Low-quality | Genome-fragment | 0.9 | AAI-based (high-confidence) | 0 |
| S5_k141_100642 | 1101 | No | NA | 3 | 0 | 0 | Not-determined | Genome-fragment | NA | NA | 0 |
| S5_k141_55641 | 1645 | No | NA | 1 | 0 | 1 | Not-determined | Genome-fragment | NA | NA | 0 |
| S5_k141_207064 | 1084 | No | NA | 2 | 0 | 2 | Not-determined | Genome-fragment | NA | NA | 0 |
| S9_k141_98652 | 1761 | No | NA | 2 | 0 | 2 | Low-quality | Genome-fragment | 1.24 | AAI-based (high-confidence) | 0 |
| S5_k141_82261 | 3524 | No | NA | 3 | 0 | 1 | Not-determined | Genome-fragment | NA | NA | 0 |
| S5_k141_207118 | 1579 | No | NA | 1 | 0 | 0 | Not-determined | Genome-fragment | NA | NA | 0 |
| S5_k141_207124 | 3254 | No | NA | 4 | 0 | 4 | Not-determined | Genome-fragment | NA | NA | 0 |
| S11-final-viral-combined_k141_224021\|\|lt2gene | 1761 | No | NA | 1 | 1 | 0 | Low-quality | Genome-fragment | 3.7 | AAI-based (high-confidence) | 0 |
| S5_k141_19703 | 1619 | No | NA | 2 | 0 | 1 | Not-determined | Genome-fragment | NA | NA | 0 |
| S5_k141_207156 | 4507 | No | NA | 4 | 0 | 4 | Not-determined | Genome-fragment | NA | NA | 0 |
| S5_k141_127294 | 2792 | No | NA | 3 | 0 | 2 | Not-determined | Genome-fragment | NA | NA | 0 |
| S5_k141_145170 | 4776 | No | NA | 3 | 0 | 1 | Not-determined | Genome-fragment | NA | NA | 0 |
| S5_k141_19743 | 1063 | No | NA | 2 | 0 | 0 | Not-determined | Genome-fragment | NA | NA | 0 |
| S5_k141_172299 | 2033 | No | NA | 1 | 0 | 1 | Not-determined | Genome-fragment | NA | NA | 0 |
| S5_k141_172321 | 1059 | No | NA | 2 | 0 | 1 | Not-determined | Genome-fragment | NA | NA | 0 |
| S1-final-viral-combined_k141_230642\|\|full | 1761 | No | NA | 3 | 2 | 0 | Low-quality | Genome-fragment | 4.78 | AAI-based (medium-confidence) | 0 |
| S5_k141_11277 | 1050 | No | NA | 2 | 0 | 0 | Not-determined | Genome-fragment | NA | NA | 0 |
| S5-final-viral-combined_k141_212310\|\|full | 1761 | No | NA | 4 | 2 | 0 | Low-quality | Genome-fragment | 3.92 | AAI-based (high-confidence) | 0 |
| S5_k141_3245 | 1619 | No | NA | 1 | 0 | 0 | Not-determined | Genome-fragment | NA | NA | 0 |
| S5_k141_127324 | 1201 | No | NA | 1 | 0 | 1 | Not-determined | Genome-fragment | NA | NA | 0 |
| S5_k141_181028 | 2984 | No | NA | 1 | 0 | 0 | Not-determined | Genome-fragment | NA | NA | 0 |
| S5_k141_189791 | 3250 | No | NA | 5 | 0 | 3 | Not-determined | Genome-fragment | NA | NA | 0 |
| S17-final-viral-combined_k141_72852\|\|full | 1761 | No | NA | 6 | 0 | 0 | Low-quality | Genome-fragment | 3.85 | AAI-based (medium-confidence) | 0 |
| S5_k141_100797 | 1124 | No | NA | 1 | 0 | 1 | Not-determined | Genome-fragment | NA | NA | 0 |
| S5_k141_55715 | 1307 | No | NA | 1 | 0 | 0 | Not-determined | Genome-fragment | NA | NA | 0 |
| S5_k141_207263 | 1057 | No | NA | 1 | 0 | 1 | Not-determined | Genome-fragment | NA | NA | 0 |
| S5_k141_100825 | 1264 | No | NA | 1 | 0 | 1 | Not-determined | Genome-fragment | NA | NA | 0 |
| S12-final-viral-combined_k141_30163\|\|full | 1761 | No | NA | 4 | 0 | 2 | Low-quality | Genome-fragment | 2.41 | AAI-based (high-confidence) | 0 |
| S5_k141_82415 | 1021 | No | NA | 1 | 0 | 1 | Not-determined | Genome-fragment | NA | NA | 0 |
| S5_k141_64691 | 1112 | No | NA | 1 | 0 | 0 | Not-determined | Genome-fragment | NA | NA | 0 |
| S5_k141_47355 | 1061 | No | NA | 2 | 0 | 2 | Not-determined | Genome-fragment | NA | NA | 0 |
| S4-final-viral-combined_k141_61507\|\|full | 1761 | No | NA | 3 | 0 | 1 | Low-quality | Genome-fragment | 1.34 | AAI-based (medium-confidence) | 0 |
| S5_k141_100843 | 2389 | No | NA | 1 | 0 | 0 | Not-determined | Genome-fragment | NA | NA | 0 |
| S6-final-viral-combined_k141_79170\|\|full | 1761 | No | NA | 3 | 0 | 0 | Low-quality | Genome-fragment | 2.83 | AAI-based (medium-confidence) | 0 |
| S11_k141_109852 | 1762 | No | NA | 2 | 0 | 0 | Low-quality | Genome-fragment | 3.1 | AAI-based (high-confidence) | 0 |
| S12-final-viral-combined_k141_172858\|\|full | 1762 | No | NA | 1 | 1 | 0 | Low-quality | Genome-fragment | 4.02 | AAI-based (medium-confidence) | 0 |
| S5_k141_38069 | 1178 | No | NA | 1 | 0 | 1 | Not-determined | Genome-fragment | NA | NA | 0 |
| S5_k141_73914 | 4532 | No | NA | 1 | 0 | 1 | Not-determined | Genome-fragment | NA | NA | 0 |
| S5_k141_127494 | 4289 | No | NA | 4 | 0 | 0 | Not-determined | Genome-fragment | NA | NA | 0 |
| S5_k141_145366 | 1936 | No | NA | 2 | 0 | 2 | Not-determined | Genome-fragment | NA | NA | 0 |
| S12-final-viral-combined_k141_249310\|\|full | 1762 | No | NA | 2 | 0 | 2 | Low-quality | Genome-fragment | 1.01 | AAI-based (medium-confidence) | 0 |
| S5_k141_198608 | 3221 | No | NA | 2 | 0 | 1 | Not-determined | Genome-fragment | NA | NA | 0 |
| S5_k141_55892 | 2460 | No | NA | 2 | 0 | 1 | Not-determined | Genome-fragment | NA | NA | 0 |
| S5_k141_11331 | 3060 | No | NA | 2 | 0 | 0 | Not-determined | Genome-fragment | NA | NA | 0 |
| S5_k141_162829 | 1535 | No | NA | 2 | 0 | 0 | Not-determined | Genome-fragment | NA | NA | 0 |
| S5_k141_127618 | 3286 | No | NA | 4 | 0 | 0 | Not-determined | Genome-fragment | NA | NA | 0 |
| S5_k141_82518 | 1625 | No | NA | 2 | 0 | 1 | Not-determined | Genome-fragment | NA | NA | 0 |
| S2-final-viral-combined_k141_235354\|\|full | 1762 | No | NA | 3 | 0 | 2 | Low-quality | Genome-fragment | 2.63 | AAI-based (medium-confidence) | 0 |
| S8_k141_167378 | 1762 | No | NA | 2 | 0 | 1 | Low-quality | Genome-fragment | 0.98 | AAI-based (high-confidence) | 0 |
| S3-final-viral-combined_k141_75560\|\|full | 1763 | No | NA | 3 | 1 | 0 | Low-quality | Genome-fragment | 4.63 | AAI-based (high-confidence) | 0 |
| S5_k141_127668 | 1186 | No | NA | 2 | 0 | 1 | Not-determined | Genome-fragment | NA | NA | 0 |
| S5-final-viral-combined_k141_73368\|\|full | 1763 | No | NA | 2 | 0 | 2 | Low-quality | Genome-fragment | 0.41 | AAI-based (medium-confidence) | 0 |
| S7-final-viral-combined_k141_28624\|\|full | 1763 | No | NA | 5 | 1 | 0 | Low-quality | Genome-fragment | 3.57 | AAI-based (medium-confidence) | 0 |
| S2-final-viral-combined_k141_58991\|\|full | 1763 | No | NA | 3 | 0 | 0 | Low-quality | Genome-fragment | 0.72 | AAI-based (medium-confidence) | 0 |
| S5_k141_3677 | 1939 | No | NA | 1 | 0 | 0 | Not-determined | Genome-fragment | NA | NA | 0 |
| S5_k141_20015 | 1053 | No | NA | 1 | 0 | 0 | Not-determined | Genome-fragment | NA | NA | 0 |
| S5_k141_74091 | 1332 | No | NA | 2 | 0 | 1 | Not-determined | Genome-fragment | NA | NA | 0 |
| S5_k141_38295 | 4014 | No | NA | 1 | 0 | 0 | Not-determined | Genome-fragment | NA | NA | 0 |
| S5_k141_11512 | 1116 | No | NA | 1 | 0 | 1 | Not-determined | Genome-fragment | NA | NA | 0 |
| S4-final-viral-combined_k141_146970\|\|full | 1763 | No | NA | 3 | 0 | 1 | Low-quality | Genome-fragment | 2.06 | AAI-based (high-confidence) | 0 |
| S9-final-viral-combined_k141_94183\|\|full | 1764 | No | NA | 3 | 0 | 0 | Low-quality | Genome-fragment | 2.89 | AAI-based (medium-confidence) | 0 |
| S9-final-viral-combined_k141_30540\|\|lt2gene | 1764 | No | NA | 1 | 1 | 0 | Low-quality | Genome-fragment | 4.19 | AAI-based (high-confidence) | 0 |
| S5_k141_198700 | 1204 | No | NA | 1 | 0 | 0 | Not-determined | Genome-fragment | NA | NA | 0 |
| S5_k141_56009 | 1217 | No | NA | 1 | 0 | 0 | Not-determined | Genome-fragment | NA | NA | 0 |
| S5_k141_3797 | 3818 | No | NA | 4 | 0 | 3 | Not-determined | Genome-fragment | NA | NA | 0 |
| S1-final-viral-combined_k141_262853\|\|full | 1764 | No | NA | 3 | 0 | 0 | Low-quality | Genome-fragment | 3.3 | AAI-based (medium-confidence) | 0 |
| S5_k141_20106 | 1066 | No | NA | 2 | 0 | 1 | Not-determined | Genome-fragment | NA | NA | 0 |
| S5_k141_82681 | 1058 | No | NA | 1 | 0 | 1 | Not-determined | Genome-fragment | NA | NA | 0 |
| S5_k141_101064 | 1417 | No | NA | 2 | 0 | 1 | Not-determined | Genome-fragment | NA | NA | 0 |
| S5_k141_181426 | 4999 | No | NA | 4 | 0 | 3 | Not-determined | Genome-fragment | NA | NA | 0 |
| S5_k141_207656 | 1203 | No | NA | 1 | 0 | 0 | Not-determined | Genome-fragment | NA | NA | 0 |
| S5-final-viral-combined_k141_77527\|\|full | 1764 | Yes | 198 | 3 | 1 | 1 | Low-quality | Genome-fragment | 0.47 | AAI-based (high-confidence) | 88.78 |
| S5_k141_142035 | 1764 | No | NA | 3 | 1 | 0 | Low-quality | Genome-fragment | 5.5 | AAI-based (medium-confidence) | 0 |
| S17-final-viral-combined_k141_254747\|\|full | 1764 | No | NA | 3 | 0 | 0 | Low-quality | Genome-fragment | 4.41 | AAI-based (high-confidence) | 0 |
| S12-final-viral-combined_k141_266020\|\|lt2gene | 1764 | No | NA | 2 | 0 | 0 | Low-quality | Genome-fragment | 1.87 | AAI-based (high-confidence) | 0 |
| S12_k141_179845 | 1764 | No | NA | 5 | 1 | 0 | Low-quality | Genome-fragment | 3.51 | AAI-based (high-confidence) | 0 |
| S5_k141_118474 | 1865 | No | NA | 3 | 0 | 2 | Not-determined | Genome-fragment | NA | NA | 0 |
| S5_k141_181543 | 1296 | No | NA | 3 | 0 | 3 | Not-determined | Genome-fragment | NA | NA | 0 |
| S5_k141_118492 | 1250 | No | NA | 3 | 0 | 3 | Not-determined | Genome-fragment | NA | NA | 0 |
| S11-final-viral-combined_k141_47820\|\|full | 1765 | No | NA | 4 | 0 | 3 | Low-quality | Genome-fragment | 4.21 | AAI-based (medium-confidence) | 0 |
| S5_k141_74245 | 1529 | No | NA | 1 | 0 | 0 | Not-determined | Genome-fragment | NA | NA | 0 |
| S1-final-viral-combined_k141_369808\|\|full | 1765 | No | NA | 2 | 0 | 0 | Low-quality | Genome-fragment | 2.53 | AAI-based (medium-confidence) | 0 |
| S5_k141_160405 | 1765 | Yes | 820 | 2 | 1 | 1 | Low-quality | Genome-fragment | 0.53 | AAI-based (medium-confidence) | 53.54 |
| S5_k141_74275 | 1046 | No | NA | 1 | 0 | 0 | Not-determined | Genome-fragment | NA | NA | 0 |
| S5_k141_145884 | 3476 | No | NA | 4 | 0 | 0 | Not-determined | Genome-fragment | NA | NA | 0 |
| S5_k141_38422 | 1316 | No | NA | 1 | 0 | 1 | Not-determined | Genome-fragment | NA | NA | 0 |
| S7_k141_203104 | 1765 | No | NA | 2 | 0 | 1 | Low-quality | Genome-fragment | 1.88 | AAI-based (medium-confidence) | 0 |
| S5_k141_128126 | 1035 | No | NA | 1 | 0 | 1 | Not-determined | Genome-fragment | NA | NA | 0 |
| S12_k141_175953 | 1765 | No | NA | 2 | 0 | 0 | Low-quality | Genome-fragment | 4.01 | AAI-based (high-confidence) | 0 |
| S5_k141_207784 | 1738 | No | NA | 2 | 0 | 1 | Not-determined | Genome-fragment | NA | NA | 0 |
| S5_k141_38431 | 6026 | No | NA | 5 | 0 | 2 | Not-determined | Genome-fragment | NA | NA | 0 |
| S5_k141_145957 | 1392 | No | NA | 2 | 0 | 0 | Not-determined | Genome-fragment | NA | NA | 0 |
| S4-final-viral-combined_k141_17321\|\|full | 1765 | No | NA | 2 | 0 | 1 | Low-quality | Genome-fragment | 1.27 | AAI-based (medium-confidence) | 0 |
| S5_k141_181595 | 1437 | No | NA | 2 | 0 | 0 | Not-determined | Genome-fragment | NA | NA | 0 |
| S1-final-viral-combined_k141_356114\|\|full | 1766 | No | NA | 6 | 3 | 0 | Low-quality | Genome-fragment | 3.85 | AAI-based (high-confidence) | 0 |
| S5_k141_65251 | 2134 | No | NA | 2 | 0 | 2 | Not-determined | Genome-fragment | NA | NA | 0 |
| S1-final-viral-combined_k141_177643\|\|full | 1766 | No | NA | 3 | 1 | 0 | Low-quality | Genome-fragment | 3.23 | AAI-based (medium-confidence) | 0 |
| S5_k141_190479 | 3085 | No | NA | 5 | 0 | 0 | Not-determined | Genome-fragment | NA | NA | 0 |
| S5_k141_172945 | 3484 | No | NA | 3 | 0 | 1 | Not-determined | Genome-fragment | NA | NA | 0 |
| S5-final-viral-combined_k141_136440\|\|full | 1766 | No | NA | 2 | 0 | 1 | Low-quality | Genome-fragment | 1.18 | AAI-based (medium-confidence) | 0 |
| S7-final-viral-combined_k141_4184\|\|full | 1766 | No | NA | 3 | 0 | 2 | Low-quality | Genome-fragment | 0.83 | AAI-based (medium-confidence) | 0 |
| S5_k141_136451 | 1076 | No | NA | 3 | 0 | 2 | Not-determined | Genome-fragment | NA | NA | 0 |
| S5_k141_20417 | 1083 | No | NA | 2 | 0 | 1 | Not-determined | Genome-fragment | NA | NA | 0 |
| S5_k141_4346 | 3029 | No | NA | 2 | 0 | 1 | Not-determined | Genome-fragment | NA | NA | 0 |
| S5_k141_11884 | 1022 | No | NA | 1 | 0 | 0 | Not-determined | Genome-fragment | NA | NA | 0 |
| S5_k141_83030 | 1272 | No | NA | 1 | 0 | 0 | Not-determined | Genome-fragment | NA | NA | 0 |
| S17-final-viral-combined_k141_7348\|\|full | 1766 | No | NA | 3 | 1 | 0 | Low-quality | Genome-fragment | 3.97 | AAI-based (medium-confidence) | 0 |
| S5_k141_29485 | 27661 | No | NA | 31 | 0 | 9 | Not-determined | Genome-fragment | NA | NA | 0 |
| S12-final-viral-combined_k141_83518\|\|full | 1766 | No | NA | 3 | 0 | 0 | Low-quality | Genome-fragment | 2.15 | AAI-based (high-confidence) | 0 |
| S8_k141_22299 | 1766 | No | NA | 6 | 1 | 0 | Low-quality | Genome-fragment | 4.19 | AAI-based (medium-confidence) | 0 |
| S5_k141_154546 | 1297 | No | NA | 1 | 0 | 1 | Not-determined | Genome-fragment | NA | NA | 0 |
| S5_k141_136540 | 1108 | No | NA | 1 | 0 | 1 | Not-determined | Genome-fragment | NA | NA | 0 |
| S5_k141_128413 | 7506 | No | NA | 7 | 0 | 3 | Not-determined | Genome-fragment | NA | NA | 0 |
| S5_k141_173095 | 3565 | No | NA | 3 | 0 | 1 | Not-determined | Genome-fragment | NA | NA | 0 |
| S11-final-viral-combined_k141_252514\|\|full | 1767 | No | NA | 3 | 1 | 0 | Low-quality | Genome-fragment | 4.6 | AAI-based (medium-confidence) | 0 |
| S5_k141_92449 | 9008 | No | NA | 6 | 0 | 5 | Not-determined | Genome-fragment | NA | NA | 0 |
| S17-final-viral-combined_k141_96623\|\|full | 1767 | No | NA | 3 | 0 | 0 | Low-quality | Genome-fragment | 4.37 | AAI-based (medium-confidence) | 0 |
| S17-k141_139193 | 1767 | No | NA | 2 | 0 | 0 | Low-quality | Genome-fragment | 0.71 | AAI-based (medium-confidence) | 0 |
| S5_k141_208039 | 1061 | No | NA | 2 | 0 | 0 | Not-determined | Genome-fragment | NA | NA | 0 |
| S5_k141_48363 | 2236 | No | NA | 1 | 0 | 0 | Not-determined | Genome-fragment | NA | NA | 0 |
| S8-final-viral-combined_k141_88934\|\|full | 1767 | No | NA | 4 | 0 | 0 | Low-quality | Genome-fragment | 4.01 | AAI-based (high-confidence) | 0 |
| S5_k141_190650 | 1163 | No | NA | 4 | 0 | 3 | Not-determined | Genome-fragment | NA | NA | 0 |
| S5_k141_4545 | 1328 | No | NA | 2 | 0 | 2 | Not-determined | Genome-fragment | NA | NA | 0 |
| S5_k141_154666 | 1013 | No | NA | 1 | 0 | 1 | Not-determined | Genome-fragment | NA | NA | 0 |
| S8-final-viral-combined_k141_80226\|\|lt2gene | 1767 | No | NA | 2 | 1 | 0 | Low-quality | Genome-fragment | 3.76 | AAI-based (high-confidence) | 0 |
| S9_k141_64734 | 1768 | No | NA | 1 | 0 | 0 | Low-quality | Genome-fragment | 1.25 | AAI-based (high-confidence) | 0 |
| S5_k141_190680 | 1293 | No | NA | 2 | 0 | 0 | Not-determined | Genome-fragment | NA | NA | 0 |
| S5_k141_163668 | 1289 | No | NA | 1 | 0 | 0 | Not-determined | Genome-fragment | NA | NA | 0 |
| S5_k141_136748 | 1116 | No | NA | 1 | 0 | 1 | Not-determined | Genome-fragment | NA | NA | 0 |
| S11-final-viral-combined_k141_98731\|\|full | 1768 | No | NA | 3 | 0 | 1 | Low-quality | Genome-fragment | 4.06 | AAI-based (high-confidence) | 0 |
| S5_k141_154726 | 1189 | No | NA | 1 | 0 | 0 | Not-determined | Genome-fragment | NA | NA | 0 |
| S1-final-viral-combined_k141_387082\|\|full | 1768 | No | NA | 2 | 0 | 1 | Low-quality | Genome-fragment | 0.48 | AAI-based (medium-confidence) | 0 |
| S5_k141_101624 | 9120 | No | NA | 12 | 0 | 4 | Not-determined | Genome-fragment | NA | NA | 0 |
| S1-final-viral-combined_k141_178516\|\|full | 1768 | No | NA | 2 | 2 | 0 | Low-quality | Genome-fragment | 3.64 | AAI-based (high-confidence) | 0 |
| S3_k141_112707 | 1768 | No | NA | 3 | 1 | 0 | Low-quality | Genome-fragment | 3.39 | AAI-based (high-confidence) | 0 |
| S5_k141_173290 | 1259 | No | NA | 2 | 0 | 1 | Not-determined | Genome-fragment | NA | NA | 0 |
| S5_k141_12178 | 1366 | No | NA | 3 | 0 | 0 | Not-determined | Genome-fragment | NA | NA | 0 |
| S5_k141_92691 | 1041 | No | NA | 1 | 0 | 1 | Not-determined | Genome-fragment | NA | NA | 0 |
| S5_k141_173328 | 1294 | No | NA | 2 | 0 | 2 | Not-determined | Genome-fragment | NA | NA | 0 |
| S5_k141_74956 | 7443 | No | NA | 9 | 0 | 5 | Not-determined | Genome-fragment | NA | NA | 0 |
| S5_k141_199201 | 1060 | No | NA | 1 | 0 | 1 | Not-determined | Genome-fragment | NA | NA | 0 |
| S12-final-viral-combined_k141_132952\|\|full | 1768 | No | NA | 4 | 0 | 4 | Low-quality | Genome-fragment | 0.58 | AAI-based (medium-confidence) | 0 |
| S5_k141_12238 | 2041 | No | NA | 2 | 0 | 2 | Not-determined | Genome-fragment | NA | NA | 0 |
| S5_k141_92763 | 1047 | No | NA | 3 | 0 | 1 | Not-determined | Genome-fragment | NA | NA | 0 |
| S2-final-viral-combined_k141_39440\|\|full | 1768 | No | NA | 4 | 2 | 0 | Low-quality | Genome-fragment | 2.08 | AAI-based (high-confidence) | 0 |
| S6-final-viral-combined_k141_85682\|\|full | 1768 | No | NA | 5 | 1 | 0 | Low-quality | Genome-fragment | 0.98 | HMM-based (lower-bound) | 0 |
| S5_k141_29783 | 1077 | No | NA | 1 | 0 | 1 | Not-determined | Genome-fragment | NA | NA | 0 |
| S8_k141_100816 | 1768 | No | NA | 2 | 0 | 0 | Low-quality | Genome-fragment | 4.08 | AAI-based (high-confidence) | 0 |
| S5_k141_101818 | 1735 | No | NA | 1 | 0 | 1 | Not-determined | Genome-fragment | NA | NA | 0 |
| S5_k141_4862 | 1289 | No | NA | 2 | 0 | 1 | Not-determined | Genome-fragment | NA | NA | 0 |
| S5_k141_146373 | 5747 | No | NA | 6 | 0 | 0 | Not-determined | Genome-fragment | NA | NA | 0 |
| S5-final-viral-combined_k141_57120\|\|full | 1769 | No | NA | 2 | 0 | 0 | Low-quality | Genome-fragment | 2.56 | AAI-based (medium-confidence) | 0 |
| S5_k141_4890 | 1408 | No | NA | 3 | 0 | 1 | Not-determined | Genome-fragment | NA | NA | 0 |
| S5_k141_146441 | 2008 | No | NA | 2 | 0 | 2 | Not-determined | Genome-fragment | NA | NA | 0 |
| S7-final-viral-combined_k141_79409\|\|lt2gene | 1769 | No | NA | 2 | 2 | 0 | Low-quality | Genome-fragment | 4.13 | AAI-based (high-confidence) | 0 |
| S5_k141_101840 | 2551 | No | NA | 5 | 0 | 2 | Not-determined | Genome-fragment | NA | NA | 0 |
| S5_k141_208337 | 1164 | No | NA | 1 | 0 | 1 | Not-determined | Genome-fragment | NA | NA | 0 |
| S4_k141_51935 | 1769 | No | NA | 2 | 0 | 0 | Low-quality | Genome-fragment | 2.95 | AAI-based (high-confidence) | 0 |
| S8_k141_158176 | 1769 | No | NA | 4 | 0 | 0 | Low-quality | Genome-fragment | 2.62 | AAI-based (high-confidence) | 0 |
| S5_k141_163890 | 1229 | No | NA | 1 | 0 | 1 | Not-determined | Genome-fragment | NA | NA | 0 |
| S11-final-viral-combined_k141_183124\|\|full | 1770 | No | NA | 3 | 1 | 0 | Low-quality | Genome-fragment | 5.09 | AAI-based (high-confidence) | 0 |
| S5_k141_12470 | 4112 | No | NA | 1 | 0 | 1 | Not-determined | Genome-fragment | NA | NA | 0 |
| S1-final-viral-combined_k141_361097\|\|full | 1770 | No | NA | 2 | 1 | 0 | Low-quality | Genome-fragment | 4.46 | AAI-based (medium-confidence) | 0 |
| S5_k141_39027 | 1295 | No | NA | 1 | 0 | 1 | Not-determined | Genome-fragment | NA | NA | 0 |
| S5_k141_12483 | 1183 | No | NA | 1 | 0 | 1 | Not-determined | Genome-fragment | NA | NA | 0 |
| S5_k141_75183 | 1546 | No | NA | 2 | 0 | 0 | Not-determined | Genome-fragment | NA | NA | 0 |
| S5_k141_5043 | 1164 | No | NA | 1 | 0 | 1 | Not-determined | Genome-fragment | NA | NA | 0 |
| S5_k141_5050 | 2291 | No | NA | 1 | 0 | 1 | Not-determined | Genome-fragment | NA | NA | 0 |
| S3_k141_83525 | 1770 | No | NA | 1 | 0 | 0 | Low-quality | Genome-fragment | 2.09 | AAI-based (high-confidence) | 0 |
| S5_k141_137032 | 7535 | No | NA | 6 | 0 | 3 | Not-determined | Genome-fragment | NA | NA | 0 |
| S5_k141_191006 | 1081 | No | NA | 1 | 0 | 1 | Not-determined | Genome-fragment | NA | NA | 0 |
| S12-final-viral-combined_k141_337830\|\|full | 1770 | No | NA | 3 | 2 | 0 | Low-quality | Genome-fragment | 3.79 | AAI-based (medium-confidence) | 0 |
| S5_k141_101957 | 2288 | No | NA | 1 | 0 | 0 | Not-determined | Genome-fragment | NA | NA | 0 |
| S5_k141_56862 | 1585 | No | NA | 2 | 0 | 0 | Not-determined | Genome-fragment | NA | NA | 0 |
| S12_k141_313960 | 1770 | No | NA | 2 | 0 | 1 | Low-quality | Genome-fragment | 1.85 | AAI-based (high-confidence) | 0 |
| S2-final-viral-combined_k141_193189\|\|full | 1770 | No | NA | 5 | 1 | 0 | Low-quality | Genome-fragment | 1.19 | HMM-based (lower-bound) | 0 |
| S5_k141_163922 | 1237 | No | NA | 2 | 0 | 0 | Not-determined | Genome-fragment | NA | NA | 0 |
| S4-final-viral-combined_k141_67523\|\|full | 1770 | No | NA | 5 | 0 | 0 | Low-quality | Genome-fragment | 3.53 | AAI-based (high-confidence) | 0 |
| S5_k141_137139 | 5467 | No | NA | 7 | 0 | 2 | Not-determined | Genome-fragment | NA | NA | 0 |
| S5_k141_48778 | 1074 | No | NA | 3 | 0 | 2 | Not-determined | Genome-fragment | NA | NA | 0 |
| S5_k141_56962 | 1458 | No | NA | 3 | 0 | 1 | Not-determined | Genome-fragment | NA | NA | 0 |
| S5_k141_48803 | 3421 | No | NA | 5 | 0 | 5 | Not-determined | Genome-fragment | NA | NA | 0 |
| S5_k141_146734 | 1052 | No | NA | 3 | 0 | 3 | Not-determined | Genome-fragment | NA | NA | 0 |
| S5_k141_146744 | 1075 | No | NA | 1 | 0 | 1 | Not-determined | Genome-fragment | NA | NA | 0 |
| S5_k141_56991 | 1091 | No | NA | 3 | 0 | 0 | Not-determined | Genome-fragment | NA | NA | 0 |
| S5_k141_164032 | 11275 | No | NA | 14 | 10 | 0 | Medium-quality | Genome-fragment | 51.47 | AAI-based (high-confidence) | 0 |
| S1-final-viral-combined_k141_250078\|\|full | 1771 | Yes | 1480 | 3 | 1 | 1 | Low-quality | Genome-fragment | 2.43 | AAI-based (high-confidence) | 16.43 |
| S12-final-viral-combined_k141_97283\|\|full | 1771 | No | NA | 1 | 0 | 0 | Low-quality | Genome-fragment | 1.38 | AAI-based (medium-confidence) | 0 |
| S12-final-viral-combined_k141_401201\|\|full | 1771 | No | NA | 3 | 2 | 0 | Low-quality | Genome-fragment | 0.79 | HMM-based (lower-bound) | 0 |
| S5_k141_102222 | 1463 | No | NA | 2 | 0 | 1 | Not-determined | Genome-fragment | NA | NA | 0 |
| S5_k141_93074 | 2171 | No | NA | 3 | 0 | 2 | Not-determined | Genome-fragment | NA | NA | 0 |
| S2-final-viral-combined_k141_178591\|\|full | 1771 | No | NA | 3 | 0 | 0 | Low-quality | Genome-fragment | 2.58 | AAI-based (medium-confidence) | 0 |
| S5_k141_83857 | 1337 | No | NA | 1 | 0 | 1 | Not-determined | Genome-fragment | NA | NA | 0 |
| S8-final-viral-combined_k141_71656\|\|full | 1771 | No | NA | 2 | 0 | 0 | Low-quality | Genome-fragment | 4.6 | AAI-based (high-confidence) | 0 |
| S11-final-viral-combined_k141_76936\|\|full | 1772 | No | NA | 3 | 0 | 1 | Low-quality | Genome-fragment | 1.06 | AAI-based (medium-confidence) | 0 |
| S5_k141_199704 | 1066 | No | NA | 1 | 0 | 1 | Not-determined | Genome-fragment | NA | NA | 0 |
| S5_k141_75474 | 1659 | No | NA | 1 | 0 | 1 | Not-determined | Genome-fragment | NA | NA | 0 |
| S1-final-viral-combined_k141_87903\|\|full | 1772 | No | NA | 3 | 0 | 1 | Low-quality | Genome-fragment | 0.91 | AAI-based (medium-confidence) | 0 |
| S5_k141_66125 | 8444 | No | NA | 9 | 0 | 1 | Not-determined | Genome-fragment | NA | NA | 0 |
| S1-final-viral-combined_k141_286466\|\|full | 1772 | No | NA | 4 | 1 | 0 | Low-quality | Genome-fragment | 3.05 | AAI-based (high-confidence) | 0 |
| S5_k141_102304 | 1732 | No | NA | 4 | 0 | 3 | Not-determined | Genome-fragment | NA | NA | 0 |
| S5_k141_30018 | 1611 | No | NA | 1 | 0 | 1 | Not-determined | Genome-fragment | NA | NA | 0 |
| S5_k141_110471 | 1285 | No | NA | 2 | 0 | 2 | Not-determined | Genome-fragment | NA | NA | 0 |
| S5_k141_57187 | 3348 | No | NA | 4 | 0 | 1 | Not-determined | Genome-fragment | NA | NA | 0 |
| S5_k141_137359 | 1006 | No | NA | 1 | 0 | 0 | Not-determined | Genome-fragment | NA | NA | 0 |
| S5_k141_57198 | 1012 | No | NA | 1 | 0 | 0 | Not-determined | Genome-fragment | NA | NA | 0 |
| S5_k141_129054 | 6710 | No | NA | 5 | 0 | 1 | Not-determined | Genome-fragment | NA | NA | 0 |
| S5_k141_129085 | 1046 | No | NA | 1 | 0 | 1 | Not-determined | Genome-fragment | NA | NA | 0 |
| S5_k141_102437 | 1262 | No | NA | 2 | 0 | 0 | Not-determined | Genome-fragment | NA | NA | 0 |
| S5_k141_57293 | 1172 | No | NA | 3 | 0 | 0 | Not-determined | Genome-fragment | NA | NA | 0 |
| S5_k141_110553 | 1068 | No | NA | 1 | 0 | 1 | Not-determined | Genome-fragment | NA | NA | 0 |
| S5_k141_66284 | 1486 | No | NA | 2 | 0 | 1 | Not-determined | Genome-fragment | NA | NA | 0 |
| S1_k141_387545 | 1772 | No | NA | 1 | 0 | 1 | Low-quality | Genome-fragment | 0.89 | AAI-based (medium-confidence) | 0 |
| S5_k141_12947 | 1422 | No | NA | 1 | 0 | 1 | Not-determined | Genome-fragment | NA | NA | 0 |
| S5_k141_12964 | 1115 | No | NA | 3 | 0 | 0 | Not-determined | Genome-fragment | NA | NA | 0 |
| S5_k141_30157 | 1296 | No | NA | 1 | 0 | 1 | Not-determined | Genome-fragment | NA | NA | 0 |
| S5_k141_199917 | 1198 | No | NA | 1 | 0 | 1 | Not-determined | Genome-fragment | NA | NA | 0 |
| S5_k141_155324 | 1154 | No | NA | 1 | 0 | 0 | Not-determined | Genome-fragment | NA | NA | 0 |
| S5_k141_199934 | 1174 | No | NA | 1 | 0 | 1 | Not-determined | Genome-fragment | NA | NA | 0 |
| S5_k141_102519 | 7788 | No | NA | 14 | 0 | 0 | Not-determined | Genome-fragment | NA | NA | 0 |
| S3-final-viral-combined_k141_38872\|\|full | 1772 | No | NA | 2 | 0 | 2 | Low-quality | Genome-fragment | 2.91 | AAI-based (medium-confidence) | 0 |
| S5_k141_191193 | 3680 | No | NA | 3 | 0 | 1 | Not-determined | Genome-fragment | NA | NA | 0 |
| S5_k141_84082 | 1233 | No | NA | 2 | 0 | 1 | Not-determined | Genome-fragment | NA | NA | 0 |
| S5_k141_164332 | 1076 | No | NA | 1 | 0 | 0 | Not-determined | Genome-fragment | NA | NA | 0 |
| S5_k141_30201 | 1075 | No | NA | 1 | 0 | 1 | Not-determined | Genome-fragment | NA | NA | 0 |
| S12-final-viral-combined_k141_8166\|\|full | 1772 | No | NA | 5 | 1 | 0 | Low-quality | Genome-fragment | 2.6 | AAI-based (medium-confidence) | 0 |
| S2-final-viral-combined_k141_218935\|\|full | 1772 | No | NA | 3 | 0 | 1 | Low-quality | Genome-fragment | 1.09 | AAI-based (high-confidence) | 0 |
| S4-final-viral-combined_k141_97961\|\|full | 1772 | No | NA | 4 | 2 | 0 | Low-quality | Genome-fragment | 4.31 | AAI-based (medium-confidence) | 0 |
| S5_k141_13092 | 1100 | No | NA | 1 | 0 | 0 | Not-determined | Genome-fragment | NA | NA | 0 |
| S5_k141_75674 | 1176 | No | NA | 1 | 0 | 0 | Not-determined | Genome-fragment | NA | NA | 0 |
| S5_k141_75723 | 1668 | No | NA | 2 | 0 | 2 | Not-determined | Genome-fragment | NA | NA | 0 |
| S5_k141_102689 | 1437 | No | NA | 2 | 0 | 1 | Not-determined | Genome-fragment | NA | NA | 0 |
| S1-final-viral-combined_k141_294595\|\|full | 1773 | No | NA | 4 | 1 | 0 | Low-quality | Genome-fragment | 4.03 | AAI-based (high-confidence) | 0 |
| S5_k141_102751 | 1966 | No | NA | 2 | 0 | 0 | Not-determined | Genome-fragment | NA | NA | 0 |
| S5_k141_5730 | 1038 | No | NA | 2 | 0 | 2 | Not-determined | Genome-fragment | NA | NA | 0 |
| S5_k141_200072 | 1559 | No | NA | 2 | 0 | 1 | Not-determined | Genome-fragment | NA | NA | 0 |
| S5_k141_164452 | 4571 | No | NA | 1 | 0 | 1 | Not-determined | Genome-fragment | NA | NA | 0 |
| S17-final-viral-combined_k141_59858\|\|full | 1773 | No | NA | 2 | 0 | 2 | Low-quality | Genome-fragment | 1.09 | AAI-based (medium-confidence) | 0 |
| S5_k141_209040 | 1795 | No | NA | 1 | 0 | 1 | Not-determined | Genome-fragment | NA | NA | 0 |
| S17-final-viral-combined_k141_63838\|\|full | 1773 | No | NA | 5 | 0 | 0 | Low-quality | Genome-fragment | 4.34 | AAI-based (medium-confidence) | 0 |
| S17-k141_150544 | 1773 | No | NA | 2 | 0 | 0 | Low-quality | Genome-fragment | 0.73 | AAI-based (high-confidence) | 0 |
| S12-final-viral-combined_k141_155196\|\|full | 1773 | No | NA | 2 | 0 | 0 | Low-quality | Genome-fragment | 2.55 | AAI-based (medium-confidence) | 0 |
| S12-final-viral-combined_k141_340617\|\|full | 1773 | No | NA | 3 | 0 | 0 | Low-quality | Genome-fragment | 2.46 | AAI-based (medium-confidence) | 0 |
| S2-final-viral-combined_k141_190856\|\|full | 1773 | No | NA | 5 | 3 | 0 | Low-quality | Genome-fragment | 5.29 | AAI-based (high-confidence) | 0 |
| S5_k141_129368 | 2648 | No | NA | 2 | 0 | 1 | Not-determined | Genome-fragment | NA | NA | 0 |
| S5_k141_110980 | 1260 | No | NA | 1 | 0 | 1 | Not-determined | Genome-fragment | NA | NA | 0 |
| S5_k141_191405 | 1287 | No | NA | 1 | 0 | 1 | Not-determined | Genome-fragment | NA | NA | 0 |
| S5_k141_84350 | 1346 | No | NA | 1 | 0 | 1 | Not-determined | Genome-fragment | NA | NA | 0 |
| S5_k141_57626 | 1771 | No | NA | 1 | 0 | 1 | Not-determined | Genome-fragment | NA | NA | 0 |
| S5_k141_129431 | 1298 | No | NA | 2 | 0 | 2 | Not-determined | Genome-fragment | NA | NA | 0 |
| S5_k141_129432 | 1132 | No | NA | 3 | 0 | 0 | Not-determined | Genome-fragment | NA | NA | 0 |
| S5_k141_147146 | 6196 | No | NA | 1 | 0 | 1 | Not-determined | Genome-fragment | NA | NA | 0 |
| S11-final-viral-combined_k141_314190\|\|full | 1774 | No | NA | 9 | 1 | 0 | Low-quality | Genome-fragment | 4.23 | AAI-based (high-confidence) | 0 |
| S5_k141_6068 | 1056 | No | NA | 2 | 0 | 1 | Not-determined | Genome-fragment | NA | NA | 0 |
| S5_k141_93583 | 4818 | No | NA | 2 | 0 | 0 | Not-determined | Genome-fragment | NA | NA | 0 |
| S5_k141_84499 | 1100 | No | NA | 2 | 0 | 1 | Not-determined | Genome-fragment | NA | NA | 0 |
| S5_k141_111222 | 1590 | No | NA | 3 | 0 | 3 | Not-determined | Genome-fragment | NA | NA | 0 |
| S5_k141_182600 | 1175 | No | NA | 2 | 0 | 1 | Not-determined | Genome-fragment | NA | NA | 0 |
| S5_k141_182620 | 1638 | No | NA | 1 | 0 | 1 | Not-determined | Genome-fragment | NA | NA | 0 |
| S5_k141_200348 | 1389 | No | NA | 1 | 0 | 0 | Not-determined | Genome-fragment | NA | NA | 0 |
| S1_k141_150431 | 1774 | No | NA | 3 | 1 | 0 | Low-quality | Genome-fragment | 2.75 | AAI-based (high-confidence) | 0 |
| S5_k141_182688 | 1300 | No | NA | 2 | 0 | 1 | Not-determined | Genome-fragment | NA | NA | 0 |
| S5_k141_182703 | 1109 | No | NA | 1 | 0 | 1 | Not-determined | Genome-fragment | NA | NA | 0 |
| S7-final-viral-combined_k141_175140\|\|full | 1774 | No | NA | 2 | 0 | 0 | Low-quality | Genome-fragment | 2.02 | AAI-based (high-confidence) | 0 |
| S5_k141_39854 | 1222 | No | NA | 1 | 0 | 1 | Not-determined | Genome-fragment | NA | NA | 0 |
| S5_k141_103031 | 1109 | No | NA | 2 | 0 | 2 | Not-determined | Genome-fragment | NA | NA | 0 |
| S12-final-viral-combined_k141_342921\|\|full | 1774 | No | NA | 6 | 2 | 0 | Low-quality | Genome-fragment | 4.22 | AAI-based (high-confidence) | 0 |
| S5_k141_49270 | 1353 | No | NA | 2 | 0 | 2 | Not-determined | Genome-fragment | NA | NA | 0 |
| S5_k141_13581 | 2773 | No | NA | 2 | 0 | 1 | Not-determined | Genome-fragment | NA | NA | 0 |
| S5_k141_137822 | 1698 | No | NA | 2 | 0 | 1 | Not-determined | Genome-fragment | NA | NA | 0 |
| S5_k141_182902 | 1352 | No | NA | 3 | 0 | 1 | Not-determined | Genome-fragment | NA | NA | 0 |
| S2-final-viral-combined_k141_90541\|\|full | 1774 | No | NA | 4 | 0 | 0 | Low-quality | Genome-fragment | 2.97 | AAI-based (medium-confidence) | 0 |
| S5_k141_137828 | 3422 | No | NA | 1 | 0 | 0 | Not-determined | Genome-fragment | NA | NA | 0 |
| S5_k141_191677 | 1003 | No | NA | 1 | 0 | 1 | Not-determined | Genome-fragment | NA | NA | 0 |
| S5_k141_6338 | 1233 | No | NA | 1 | 0 | 0 | Not-determined | Genome-fragment | NA | NA | 0 |
| S5_k141_49290 | 3774 | No | NA | 4 | 0 | 0 | Not-determined | Genome-fragment | NA | NA | 0 |
| S5_k141_84856 | 1051 | No | NA | 1 | 0 | 1 | Not-determined | Genome-fragment | NA | NA | 0 |
| S5_k141_155753 | 1026 | No | NA | 1 | 0 | 1 | Not-determined | Genome-fragment | NA | NA | 0 |
| S4-final-viral-combined_k141_125861\|\|full | 1774 | No | NA | 3 | 0 | 1 | Low-quality | Genome-fragment | 0.88 | AAI-based (medium-confidence) | 0 |
| S5_k141_49310 | 19132 | No | NA | 28 | 9 | 0 | Medium-quality | Genome-fragment | 53.92 | AAI-based (high-confidence) | 0 |
| S8_k141_244325 | 1774 | No | NA | 5 | 0 | 0 | Low-quality | Genome-fragment | 2.92 | AAI-based (high-confidence) | 0 |
| S5_k141_155803 | 1340 | No | NA | 1 | 0 | 1 | Not-determined | Genome-fragment | NA | NA | 0 |
| S5_k141_200523 | 1732 | No | NA | 1 | 0 | 1 | Not-determined | Genome-fragment | NA | NA | 0 |
| S1-final-viral-combined_k141_234445\|\|full | 1775 | No | NA | 2 | 0 | 1 | Low-quality | Genome-fragment | 1.95 | AAI-based (high-confidence) | 0 |
| S5_k141_6419 | 1060 | No | NA | 1 | 0 | 1 | Not-determined | Genome-fragment | NA | NA | 0 |
| S1-final-viral-combined_k141_63799\|\|full | 1775 | No | NA | 2 | 0 | 2 | Low-quality | Genome-fragment | 1.21 | AAI-based (medium-confidence) | 0 |
| S3-final-viral-combined_k141_105107\|\|full | 1775 | No | NA | 3 | 0 | 0 | Low-quality | Genome-fragment | 5.04 | AAI-based (high-confidence) | 0 |
| S5_k141_200613 | 1107 | No | NA | 2 | 0 | 1 | Not-determined | Genome-fragment | NA | NA | 0 |
| S5_k141_76357 | 2623 | No | NA | 3 | 0 | 1 | Not-determined | Genome-fragment | NA | NA | 0 |
| S5_k141_6457 | 1696 | No | NA | 2 | 0 | 0 | Not-determined | Genome-fragment | NA | NA | 0 |
| S5_k141_155857 | 1016 | No | NA | 1 | 0 | 0 | Not-determined | Genome-fragment | NA | NA | 0 |
| S5-final-viral-combined_k141_142783\|\|full | 1775 | No | NA | 3 | 0 | 1 | Low-quality | Genome-fragment | 3.8 | AAI-based (medium-confidence) | 0 |
| S5_k141_191855 | 1405 | No | NA | 1 | 0 | 0 | Not-determined | Genome-fragment | NA | NA | 0 |
| S5_k141_191892 | 1257 | No | NA | 3 | 0 | 2 | Not-determined | Genome-fragment | NA | NA | 0 |
| S5_k141_58021 | 1146 | No | NA | 1 | 0 | 1 | Not-determined | Genome-fragment | NA | NA | 0 |
| S7_k141_39967 | 1775 | No | NA | 1 | 0 | 1 | Low-quality | Genome-fragment | 3.66 | AAI-based (medium-confidence) | 0 |
| S5_k141_84970 | 1060 | No | NA | 1 | 0 | 0 | Not-determined | Genome-fragment | NA | NA | 0 |
| S5_k141_30896 | 1075 | No | NA | 1 | 0 | 1 | Not-determined | Genome-fragment | NA | NA | 0 |

| | | | | | | | | | | | |
|---|---|---|---|---|---|---|---|---|---|---|---|
| S17-final-viral-combined_k141_214190\|\|full | 1775 | No | NA | 2 | 0 | 1 | Low-quality | Genome-fragment | 1.25 | AAI-based (high-confidence) | 0 |
| S2_k141_174302 | 1775 | No | NA | 3 | 1 | 0 | Low-quality | Genome-fragment | 2.88 | AAI-based (high-confidence) | 0 |
| S5_k141_200784 | 1678 | No | NA | 2 | 0 | 1 | Not-determined | Genome-fragment | NA | NA | 0 |
| S5_k141_209690 | 1597 | No | NA | 1 | 0 | 1 | Not-determined | Genome-fragment | NA | NA | 0 |
| S5_k141_120261 | 1078 | No | NA | 4 | 0 | 4 | Not-determined | Genome-fragment | NA | NA | 0 |
| S5_k141_200879 | 1191 | No | NA | 1 | 0 | 0 | Not-determined | Genome-fragment | NA | NA | 0 |
| S5_k141_200891 | 1302 | No | NA | 1 | 0 | 1 | Not-determined | Genome-fragment | NA | NA | 0 |
| S5_k141_209729 | 1095 | No | NA | 2 | 0 | 1 | Not-determined | Genome-fragment | NA | NA | 0 |
| S5_k141_85099 | 6111 | No | NA | 6 | 0 | 3 | Not-determined | Genome-fragment | NA | NA | 0 |
| S5_k141_209732 | 1222 | No | NA | 3 | 0 | 0 | Not-determined | Genome-fragment | NA | NA | 0 |
| S5_k141_40327 | 7191 | No | NA | 7 | 0 | 3 | Not-determined | Genome-fragment | NA | NA | 0 |
| S5_k141_85138 | 1086 | No | NA | 2 | 0 | 1 | Not-determined | Genome-fragment | NA | NA | 0 |
| S6-final-viral-combined_k141_84085\|\|full | 1775 | No | NA | 3 | 0 | 3 | Low-quality | Genome-fragment | 1.25 | AAI-based (high-confidence) | 0 |
| S8-final-viral-combined_k141_151502\|\|full | 1775 | No | NA | 2 | 0 | 0 | Low-quality | Genome-fragment | 1.67 | AAI-based (medium-confidence) | 0 |
| S5_k141_58226 | 1575 | No | NA | 4 | 0 | 1 | Not-determined | Genome-fragment | NA | NA | 0 |
| S5_k141_58236 | 1023 | No | NA | 2 | 0 | 1 | Not-determined | Genome-fragment | NA | NA | 0 |
| S5_k141_66826 | 1131 | No | NA | 2 | 0 | 1 | Not-determined | Genome-fragment | NA | NA | 0 |
| S5_k141_58269 | 1245 | No | NA | 1 | 0 | 1 | Not-determined | Genome-fragment | NA | NA | 0 |
| S5_k141_164986 | 4105 | No | NA | 4 | 0 | 4 | Not-determined | Genome-fragment | NA | NA | 0 |
| S5_k141_58290 | 1122 | No | NA | 1 | 0 | 0 | Not-determined | Genome-fragment | NA | NA | 0 |
| S5_k141_6618 | 3242 | No | NA | 3 | 0 | 2 | Not-determined | Genome-fragment | NA | NA | 0 |
| S5_k141_49437 | 1344 | No | NA | 2 | 0 | 0 | Not-determined | Genome-fragment | NA | NA | 0 |
| S1-final-viral-combined_k141_238335\|\|full | 1776 | No | NA | 3 | 3 | 0 | Low-quality | Genome-fragment | 5.11 | AAI-based (high-confidence) | 0 |
| S5_k141_129799 | 1083 | No | NA | 2 | 0 | 1 | Not-determined | Genome-fragment | NA | NA | 0 |
| S5_k141_94006 | 1314 | No | NA | 1 | 0 | 1 | Not-determined | Genome-fragment | NA | NA | 0 |
| S5_k141_6686 | 3109 | No | NA | 4 | 0 | 4 | Not-determined | Genome-fragment | NA | NA | 0 |
| S5_k141_6727 | 2203 | No | NA | 3 | 0 | 3 | Not-determined | Genome-fragment | NA | NA | 0 |
| S5_k141_111932 | 1016 | No | NA | 2 | 0 | 1 | Not-determined | Genome-fragment | NA | NA | 0 |
| S5-final-viral-combined_k141_189008\|\|full | 1776 | No | NA | 2 | 0 | 0 | Low-quality | Genome-fragment | 2.33 | AAI-based (high-confidence) | 0 |
| S5_k141_6781 | 1281 | No | NA | 3 | 0 | 3 | Not-determined | Genome-fragment | NA | NA | 0 |
| S5_k141_94098 | 1052 | No | NA | 1 | 0 | 1 | Not-determined | Genome-fragment | NA | NA | 0 |
| S12-final-viral-combined_k141_100913\|\|full | 1776 | No | NA | 4 | 1 | 0 | Low-quality | Genome-fragment | 3.27 | AAI-based (high-confidence) | 0 |
| S2-final-viral-combined_k141_128823\|\|full | 1776 | Yes | 349 | 3 | 1 | 1 | Low-quality | Genome-fragment | 0.73 | HMM-based (lower-bound) | 80.35 |
| S6_k141_54967 | 1776 | No | NA | 3 | 0 | 1 | Low-quality | Genome-fragment | 4.29 | AAI-based (medium-confidence) | 0 |
| S6_k141_40685 | 1776 | No | NA | 1 | 0 | 1 | Low-quality | Genome-fragment | 0.31 | AAI-based (medium-confidence) | 0 |
| S5_k141_40616 | 1185 | No | NA | 1 | 0 | 0 | Not-determined | Genome-fragment | NA | NA | 0 |
| S5_k141_120337 | 1085 | No | NA | 2 | 0 | 0 | Not-determined | Genome-fragment | NA | NA | 0 |
| S5_k141_6891 | 1208 | No | NA | 1 | 0 | 1 | Not-determined | Genome-fragment | NA | NA | 0 |
| S5_k141_21924 | 1105 | No | NA | 2 | 0 | 1 | Not-determined | Genome-fragment | NA | NA | 0 |
| S17-final-viral-combined_k141_213117\|\|full | 1777 | No | NA | 4 | 0 | 0 | Low-quality | Genome-fragment | 3.37 | AAI-based (high-confidence) | 0 |
| S5_k141_76595 | 1024 | No | NA | 1 | 0 | 1 | Not-determined | Genome-fragment | NA | NA | 0 |
| S5_k141_76596 | 1105 | No | NA | 1 | 0 | 1 | Not-determined | Genome-fragment | NA | NA | 0 |
| S12-final-viral-combined_k141_257062\|\|full | 1777 | No | NA | 5 | 0 | 0 | Low-quality | Genome-fragment | 5.48 | AAI-based (high-confidence) | 0 |
| S5_k141_183421 | 3394 | No | NA | 4 | 0 | 1 | Not-determined | Genome-fragment | NA | NA | 0 |
| S5_k141_94172 | 5496 | No | NA | 5 | 0 | 4 | Not-determined | Genome-fragment | NA | NA | 0 |
| S5_k141_22034 | 1005 | No | NA | 1 | 0 | 0 | Not-determined | Genome-fragment | NA | NA | 0 |
| S5_k141_120382 | 7499 | No | NA | 6 | 0 | 4 | Not-determined | Genome-fragment | NA | NA | 0 |
| S12-final-viral-combined_k141_230364\|\|full | 1777 | No | NA | 4 | 0 | 0 | Low-quality | Genome-fragment | 2.89 | AAI-based (medium-confidence) | 0 |
| S5_k141_192202 | 1036 | No | NA | 2 | 0 | 1 | Not-determined | Genome-fragment | NA | NA | 0 |
| S5_k141_49684 | 1209 | No | NA | 3 | 0 | 0 | Not-determined | Genome-fragment | NA | NA | 0 |
| S5_k141_156203 | 1706 | No | NA | 1 | 0 | 0 | Not-determined | Genome-fragment | NA | NA | 0 |
| S5_k141_22134 | 1149 | No | NA | 1 | 0 | 1 | Not-determined | Genome-fragment | NA | NA | 0 |
| S12_k141_218080 | 1777 | No | NA | 3 | 0 | 0 | Low-quality | Genome-fragment | 3.24 | AAI-based (medium-confidence) | 0 |
| S5_k141_40815 | 1069 | No | NA | 1 | 0 | 1 | Not-determined | Genome-fragment | NA | NA | 0 |
| S5_k141_49708 | 2747 | No | NA | 4 | 0 | 3 | Not-determined | Genome-fragment | NA | NA | 0 |
| S12_k141_352051 | 1777 | No | NA | 6 | 0 | 0 | Low-quality | Genome-fragment | 3.4 | AAI-based (high-confidence) | 0 |
| S2-final-viral-combined_k141_66692\|\|lt2gene | 1777 | No | NA | 2 | 2 | 0 | Low-quality | Genome-fragment | 4.16 | AAI-based (high-confidence) | 0 |
| S5_k141_174898 | 1131 | No | NA | 2 | 0 | 2 | Not-determined | Genome-fragment | NA | NA | 0 |
| S5_k141_76724 | 1140 | No | NA | 1 | 0 | 1 | Not-determined | Genome-fragment | NA | NA | 0 |
| S8-final-viral-combined_k141_34817\|\|full | 1777 | No | NA | 5 | 0 | 0 | Low-quality | Genome-fragment | 2.99 | AAI-based (high-confidence) | 0 |
| S8-final-viral-combined_k141_48764\|\|full | 1777 | No | NA | 3 | 0 | 0 | Low-quality | Genome-fragment | 3.59 | AAI-based (medium-confidence) | 0 |
| S5_k141_183642 | 1237 | No | NA | 2 | 0 | 2 | Not-determined | Genome-fragment | NA | NA | 0 |
| S5_k141_130106 | 3122 | No | NA | 2 | 0 | 0 | Not-determined | Genome-fragment | NA | NA | 0 |
| S5_k141_58880 | 2685 | No | NA | 5 | 0 | 1 | Not-determined | Genome-fragment | NA | NA | 0 |
| S5_k141_210203 | 3298 | No | NA | 2 | 0 | 0 | Not-determined | Genome-fragment | NA | NA | 0 |
| S5_k141_147798 | 1032 | No | NA | 2 | 0 | 1 | Not-determined | Genome-fragment | NA | NA | 0 |
| S5_k141_67169 | 39714 | No | NA | 51 | 12 | 2 | High-quality | High-quality | 100 | AAI-based (medium-confidence) | 0 |
| S11-final-viral-combined_k141_247477\|\|full | 1778 | No | NA | 5 | 0 | 0 | Low-quality | Genome-fragment | 3.97 | AAI-based (high-confidence) | 0 |
| S5_k141_94255 | 1020 | No | NA | 2 | 0 | 0 | Not-determined | Genome-fragment | NA | NA | 0 |
| S5_k141_130186 | 1024 | No | NA | 1 | 0 | 0 | Not-determined | Genome-fragment | NA | NA | 0 |
| S5_k141_94280 | 1282 | No | NA | 2 | 0 | 2 | Not-determined | Genome-fragment | NA | NA | 0 |
| S5_k141_22220 | 1096 | No | NA | 3 | 0 | 0 | Not-determined | Genome-fragment | NA | NA | 0 |
| S5_k141_192435 | 1241 | No | NA | 1 | 0 | 1 | Not-determined | Genome-fragment | NA | NA | 0 |
| S5_k141_22226 | 1156 | No | NA | 1 | 0 | 0 | Not-determined | Genome-fragment | NA | NA | 0 |
| S5_k141_138538 | 1222 | No | NA | 2 | 0 | 2 | Not-determined | Genome-fragment | NA | NA | 0 |
| S5_k141_31558 | 3530 | No | NA | 2 | 0 | 0 | Not-determined | Genome-fragment | NA | NA | 0 |
| S5_k141_14398 | 1189 | No | NA | 3 | 0 | 2 | Not-determined | Genome-fragment | NA | NA | 0 |
| S5_k141_130265 | 2370 | No | NA | 2 | 0 | 2 | Not-determined | Genome-fragment | NA | NA | 0 |
| S5_k141_103733 | 1456 | No | NA | 2 | 0 | 1 | Not-determined | Genome-fragment | NA | NA | 0 |
| S5_k141_22250 | 1071 | No | NA | 1 | 0 | 1 | Not-determined | Genome-fragment | NA | NA | 0 |
| S5_k141_175034 | 2748 | No | NA | 4 | 0 | 2 | Not-determined | Genome-fragment | NA | NA | 0 |
| S5_k141_192505 | 1613 | No | NA | 1 | 0 | 0 | Not-determined | Genome-fragment | NA | NA | 0 |
| S5_k141_76829 | 1189 | No | NA | 1 | 0 | 0 | Not-determined | Genome-fragment | NA | NA | 0 |
| S5_k141_94390 | 1127 | No | NA | 1 | 0 | 0 | Not-determined | Genome-fragment | NA | NA | 0 |
| S5_k141_156300 | 4341 | No | NA | 3 | 0 | 3 | Not-determined | Genome-fragment | NA | NA | 0 |
| S7_k141_30341 | 1778 | No | NA | 2 | 0 | 0 | Low-quality | Genome-fragment | 3.2 | AAI-based (medium-confidence) | 0 |
| S5_k141_156330 | 5849 | No | NA | 5 | 0 | 2 | Not-determined | Genome-fragment | NA | NA | 0 |
| S5_k141_148037 | 19695 | No | NA | 19 | 0 | 2 | Not-determined | Genome-fragment | NA | NA | 0 |
| S5_k141_67365 | 1024 | No | NA | 2 | 0 | 1 | Not-determined | Genome-fragment | NA | NA | 0 |
| S5_k141_148059 | 1104 | No | NA | 2 | 0 | 0 | Not-determined | Genome-fragment | NA | NA | 0 |
| S5_k141_201218 | 1179 | No | NA | 1 | 0 | 1 | Not-determined | Genome-fragment | NA | NA | 0 |
| S5_k141_49963 | 1641 | No | NA | 2 | 0 | 0 | Not-determined | Genome-fragment | NA | NA | 0 |
| S17-k141_185500 | 1778 | No | NA | 1 | 0 | 1 | Low-quality | Genome-fragment | 0.47 | AAI-based (medium-confidence) | 0 |
| S5_k141_175143 | 1405 | No | NA | 1 | 0 | 0 | Not-determined | Genome-fragment | NA | NA | 0 |
| S2-final-viral-combined_k141_79886\|\|full | 1778 | No | NA | 4 | 0 | 1 | Low-quality | Genome-fragment | 3.23 | AAI-based (medium-confidence) | 0 |
| S5_k141_103928 | 1883 | No | NA | 2 | 0 | 2 | Not-determined | Genome-fragment | NA | NA | 0 |
| S5_k141_59120 | 1801 | No | NA | 3 | 0 | 2 | Not-determined | Genome-fragment | NA | NA | 0 |
| S5_k141_85650 | 61026 | Yes | 58974 | 69 | 11 | 4 | High-quality | High-quality | 91.2 | AAI-based (high-confidence) | 3.36 |
| S5_k141_175216 | 1348 | No | NA | 1 | 0 | 0 | Not-determined | Genome-fragment | NA | NA | 0 |
| S5_k141_22431 | 1034 | No | NA | 1 | 0 | 0 | Not-determined | Genome-fragment | NA | NA | 0 |
| S5_k141_7477 | 1394 | No | NA | 1 | 0 | 0 | Not-determined | Genome-fragment | NA | NA | 0 |
| S5_k141_77105 | 1322 | No | NA | 1 | 0 | 0 | Not-determined | Genome-fragment | NA | NA | 0 |
| S5_k141_14548 | 2210 | No | NA | 2 | 0 | 1 | Not-determined | Genome-fragment | NA | NA | 0 |
| S5_k141_138839 | 1902 | No | NA | 2 | 0 | 1 | Not-determined | Genome-fragment | NA | NA | 0 |
| S8-final-viral-combined_k141_214187\|\|full | 1778 | No | NA | 2 | 0 | 1 | Low-quality | Genome-fragment | 0.8 | AAI-based (medium-confidence) | 0 |
| S5_k141_41319 | 1132 | No | NA | 1 | 0 | 1 | Not-determined | Genome-fragment | NA | NA | 0 |
| S5_k141_67623 | 1468 | No | NA | 2 | 0 | 0 | Not-determined | Genome-fragment | NA | NA | 0 |
| S5_k141_50225 | 3764 | No | NA | 5 | 0 | 0 | Not-determined | Genome-fragment | NA | NA | 0 |
| S5_k141_165536 | 1047 | No | NA | 1 | 0 | 1 | Not-determined | Genome-fragment | NA | NA | 0 |
| S11_k141_578 | 1779 | No | NA | 2 | 0 | 0 | Low-quality | Genome-fragment | 2.69 | AAI-based (medium-confidence) | 0 |
| S1-final-viral-combined_k141_2800\|\|full | 1779 | Yes | 711 | 3 | 1 | 1 | Low-quality | Genome-fragment | 0.88 | HMM-based (lower-bound) | 60.03 |
| S5_k141_138967 | 1329 | No | NA | 1 | 0 | 1 | Not-determined | Genome-fragment | NA | NA | 0 |
| S5_k141_165585 | 1382 | No | NA | 1 | 0 | 1 | Not-determined | Genome-fragment | NA | NA | 0 |
| S5_k141_67649 | 1002 | No | NA | 3 | 0 | 0 | Not-determined | Genome-fragment | NA | NA | 0 |
| S5_k141_130812 | 1023 | No | NA | 1 | 0 | 1 | Not-determined | Genome-fragment | NA | NA | 0 |
| S1-final-viral-combined_k141_38965\|\|full | 1779 | No | NA | 3 | 1 | 0 | Low-quality | Genome-fragment | 4.73 | AAI-based (high-confidence) | 0 |
| S5_k141_112621 | 2341 | No | NA | 4 | 0 | 0 | Not-determined | Genome-fragment | NA | NA | 0 |
| S12-final-viral-combined_k141_339653\|\|full | 1779 | No | NA | 1 | 0 | 0 | Low-quality | Genome-fragment | 3.92 | AAI-based (medium-confidence) | 0 |
| S5_k141_85796 | 1566 | No | NA | 1 | 0 | 0 | Not-determined | Genome-fragment | NA | NA | 0 |
| S5_k141_32115 | 1252 | No | NA | 1 | 0 | 1 | Not-determined | Genome-fragment | NA | NA | 0 |
| S5_k141_210354 | 1034 | No | NA | 1 | 0 | 0 | Not-determined | Genome-fragment | NA | NA | 0 |
| S5_k141_165656 | 1356 | No | NA | 2 | 0 | 0 | Not-determined | Genome-fragment | NA | NA | 0 |
| S5_k141_192884 | 1041 | No | NA | 2 | 0 | 0 | Not-determined | Genome-fragment | NA | NA | 0 |
| S12-final-viral-combined_k141_391796\|\|full | 1779 | No | NA | 4 | 0 | 0 | Low-quality | Genome-fragment | 3.73 | AAI-based (medium-confidence) | 0 |
| S5_k141_7636 | 3603 | No | NA | 3 | 0 | 1 | Not-determined | Genome-fragment | NA | NA | 0 |
| S5_k141_192913 | 1335 | No | NA | 2 | 0 | 0 | Not-determined | Genome-fragment | NA | NA | 0 |
| S5_k141_184184 | 5980 | No | NA | 9 | 0 | 2 | Not-determined | Genome-fragment | NA | NA | 0 |
| S5_k141_139099 | 3212 | No | NA | 8 | 0 | 1 | Not-determined | Genome-fragment | NA | NA | 0 |
| S5_k141_210415 | 1897 | No | NA | 1 | 0 | 1 | Not-determined | Genome-fragment | NA | NA | 0 |
| S5_k141_184197 | 1383 | No | NA | 1 | 0 | 0 | Not-determined | Genome-fragment | NA | NA | 0 |
| S5_k141_22592 | 1185 | No | NA | 1 | 0 | 1 | Not-determined | Genome-fragment | NA | NA | 0 |
| S5_k141_139124 | 2348 | No | NA | 1 | 0 | 1 | Not-determined | Genome-fragment | NA | NA | 0 |
| S5_k141_192970 | 4143 | No | NA | 6 | 0 | 1 | Not-determined | Genome-fragment | NA | NA | 0 |
| S5_k141_210460 | 20064 | No | NA | 28 | 15 | 0 | Medium-quality | Genome-fragment | 65.41 | AAI-based (medium-confidence) | 0 |
| S5_k141_104174 | 3319 | No | NA | 2 | 0 | 0 | Not-determined | Genome-fragment | NA | NA | 0 |
| S5_k141_121210 | 1739 | No | NA | 1 | 0 | 1 | Not-determined | Genome-fragment | NA | NA | 0 |
| S5_k141_210466 | 1233 | No | NA | 2 | 0 | 0 | Not-determined | Genome-fragment | NA | NA | 0 |
| S5_k141_59372 | 1750 | No | NA | 1 | 0 | 1 | Not-determined | Genome-fragment | NA | NA | 0 |
| S12_k141_206658 | 1779 | No | NA | 3 | 0 | 2 | Low-quality | Genome-fragment | 1.14 | AAI-based (high-confidence) | 0 |
| S5_k141_210477 | 1093 | No | NA | 1 | 0 | 0 | Not-determined | Genome-fragment | NA | NA | 0 |
| S12_k141_192233 | 1779 | No | NA | 3 | 0 | 0 | Low-quality | Genome-fragment | 3.74 | AAI-based (medium-confidence) | 0 |
| S5_k141_139186 | 1216 | No | NA | 1 | 0 | 0 | Not-determined | Genome-fragment | NA | NA | 0 |
| S12_k141_9059 | 1779 | No | NA | 3 | 2 | 0 | Low-quality | Genome-fragment | 4.32 | AAI-based (high-confidence) | 0 |
| S5_k141_184369 | 1721 | No | NA | 1 | 0 | 1 | Not-determined | Genome-fragment | NA | NA | 0 |
| S5_k141_184374 | 1866 | No | NA | 1 | 0 | 0 | Not-determined | Genome-fragment | NA | NA | 0 |
| S5_k141_156791 | 1893 | No | NA | 2 | 0 | 0 | Not-determined | Genome-fragment | NA | NA | 0 |
| S4-final-viral-combined_k141_139669\|\|full | 1779 | No | NA | 3 | 0 | 1 | Low-quality | Genome-fragment | 0.78 | AAI-based (medium-confidence) | 0 |
| S5_k141_94730 | 1216 | No | NA | 1 | 0 | 0 | Not-determined | Genome-fragment | NA | NA | 0 |
| S6-final-viral-combined_k141_76496\|\|full | 1779 | No | NA | 3 | 0 | 1 | Low-quality | Genome-fragment | 3.81 | AAI-based (medium-confidence) | 0 |
| S5_k141_175602 | 7896 | No | NA | 5 | 0 | 2 | Not-determined | Genome-fragment | NA | NA | 0 |
| S5_k141_59510 | 1050 | No | NA | 2 | 0 | 0 | Not-determined | Genome-fragment | NA | NA | 0 |
| S5_k141_104278 | 1330 | No | NA | 1 | 0 | 0 | Not-determined | Genome-fragment | NA | NA | 0 |
| S6-final-viral-combined_k141_122480\|\|full | 1779 | No | NA | 3 | 0 | 0 | Low-quality | Genome-fragment | 3.52 | AAI-based (high-confidence) | 0 |
| S5_k141_86127 | 5233 | No | NA | 6 | 0 | 2 | Not-determined | Genome-fragment | NA | NA | 0 |
| S6_k141_116725 | 1779 | No | NA | 2 | 0 | 1 | Low-quality | Genome-fragment | 1.21 | AAI-based (medium-confidence) | 0 |
| S5_k141_59597 | 3854 | No | NA | 2 | 0 | 1 | Not-determined | Genome-fragment | NA | NA | 0 |
| S8-final-viral-combined_k141_103744\|\|full | 1779 | No | NA | 3 | 0 | 0 | Low-quality | Genome-fragment | 1.51 | AAI-based (medium-confidence) | 0 |
| S8-final-viral-combined_k141_222602\|\|full | 1779 | No | NA | 5 | 1 | 0 | Low-quality | Genome-fragment | 1.42 | AAI-based (high-confidence) | 0 |
| S5_k141_193174 | 1208 | No | NA | 1 | 0 | 1 | Not-determined | Genome-fragment | NA | NA | 0 |
| S5_k141_131245 | 1367 | No | NA | 1 | 0 | 0 | Not-determined | Genome-fragment | NA | NA | 0 |
| S11-final-viral-combined_k141_101443\|\|full | 1780 | No | NA | 2 | 0 | 1 | Low-quality | Genome-fragment | 3.76 | AAI-based (medium-confidence) | 0 |
| S5_k141_86240 | 1223 | No | NA | 1 | 0 | 1 | Not-determined | Genome-fragment | NA | NA | 0 |
| S5_k141_112912 | 1195 | No | NA | 2 | 0 | 1 | Not-determined | Genome-fragment | NA | NA | 0 |
| S5_k141_156948 | 3127 | No | NA | 1 | 0 | 1 | Not-determined | Genome-fragment | NA | NA | 0 |
| S3_k141_127047 | 1780 | No | NA | 3 | 0 | 0 | Low-quality | Genome-fragment | 0.82 | AAI-based (high-confidence) | 0 |
| S5_k141_86269 | 3195 | No | NA | 2 | 0 | 2 | Not-determined | Genome-fragment | NA | NA | 0 |
| S5_k141_50585 | 3373 | No | NA | 1 | 0 | 1 | Not-determined | Genome-fragment | NA | NA | 0 |
| S5_k141_156977 | 1812 | No | NA | 3 | 0 | 2 | Not-determined | Genome-fragment | NA | NA | 0 |
| S5_k141_121485 | 1260 | No | NA | 2 | 0 | 1 | Not-determined | Genome-fragment | NA | NA | 0 |
| S5_k141_121488 | 3279 | No | NA | 4 | 0 | 3 | Not-determined | Genome-fragment | NA | NA | 0 |
| S5_k141_104445 | 1183 | No | NA | 1 | 0 | 1 | Not-determined | Genome-fragment | NA | NA | 0 |
| S5_k141_139508 | 1121 | No | NA | 2 | 0 | 2 | Not-determined | Genome-fragment | NA | NA | 0 |
| S5_k141_157017 | 1566 | No | NA | 2 | 0 | 1 | Not-determined | Genome-fragment | NA | NA | 0 |
| S5_k141_7887 | 6247 | No | NA | 6 | 0 | 6 | Not-determined | Genome-fragment | NA | NA | 0 |
| S2-final-viral-combined_k141_21060\|\|full | 1780 | No | NA | 3 | 0 | 0 | Low-quality | Genome-fragment | 2.78 | AAI-based (medium-confidence) | 0 |
| S5_k141_112946 | 1998 | No | NA | 1 | 0 | 0 | Not-determined | Genome-fragment | NA | NA | 0 |
| S5_k141_112950 | 26449 | No | NA | 21 | 0 | 17 | Not-determined | Genome-fragment | NA | NA | 0 |
| S5_k141_184772 | 1112 | No | NA | 2 | 0 | 2 | Not-determined | Genome-fragment | NA | NA | 0 |
| S6-final-viral-combined_k141_118466\|\|full | 1780 | No | NA | 3 | 1 | 0 | Low-quality | Genome-fragment | 3.23 | AAI-based (high-confidence) | 0 |
| S5_k141_32515 | 1150 | No | NA | 1 | 0 | 1 | Not-determined | Genome-fragment | NA | NA | 0 |
| S5_k141_131367 | 1288 | No | NA | 3 | 0 | 3 | Not-determined | Genome-fragment | NA | NA | 0 |
| S5_k141_148889 | 1118 | No | NA | 1 | 0 | 1 | Not-determined | Genome-fragment | NA | NA | 0 |
| S5_k141_184824 | 3302 | No | NA | 6 | 0 | 1 | Not-determined | Genome-fragment | NA | NA | 0 |
| S5_k141_157133 | 1387 | No | NA | 1 | 0 | 1 | Not-determined | Genome-fragment | NA | NA | 0 |
| S5_k141_131405 | 2062 | No | NA | 1 | 0 | 0 | Not-determined | Genome-fragment | NA | NA | 0 |
| S5_k141_50768 | 2677 | No | NA | 2 | 0 | 0 | Not-determined | Genome-fragment | NA | NA | 0 |
| S5_k141_59830 | 1124 | No | NA | 1 | 0 | 0 | Not-determined | Genome-fragment | NA | NA | 0 |
| S8-final-viral-combined_k141_52432\|\|full | 1780 | No | NA | 4 | 0 | 1 | Low-quality | Genome-fragment | 1.57 | AAI-based (medium-confidence) | 0 |
| S5_k141_94915 | 1163 | No | NA | 1 | 0 | 0 | Not-determined | Genome-fragment | NA | NA | 0 |
| S5_k141_67969 | 4091 | No | NA | 2 | 0 | 0 | Not-determined | Genome-fragment | NA | NA | 0 |
| S5_k141_121745 | 1282 | No | NA | 1 | 0 | 0 | Not-determined | Genome-fragment | NA | NA | 0 |
| S5_k141_131530 | 1044 | No | NA | 1 | 0 | 0 | Not-determined | Genome-fragment | NA | NA | 0 |
| S8-final-viral-combined_k141_168887\|\|full | 1780 | No | NA | 3 | 0 | 0 | Low-quality | Genome-fragment | 4.32 | AAI-based (high-confidence) | 0 |
| S11_k141_235644 | 1781 | No | NA | 4 | 1 | 0 | Low-quality | Genome-fragment | 4.39 | AAI-based (medium-confidence) | 0 |
| S5_k141_184907 | 1026 | No | NA | 1 | 0 | 0 | Not-determined | Genome-fragment | NA | NA | 0 |
| S5_k141_23116 | 1247 | No | NA | 2 | 0 | 0 | Not-determined | Genome-fragment | NA | NA | 0 |
| S5_k141_41679 | 1253 | No | NA | 1 | 0 | 0 | Not-determined | Genome-fragment | NA | NA | 0 |
| S1-final-viral-combined_k141_237555\|\|full | 1781 | No | NA | 3 | 0 | 2 | Low-quality | Genome-fragment | 2.1 | AAI-based (medium-confidence) | 0 |
| S5_k141_157392 | 2075 | No | NA | 1 | 0 | 0 | Not-determined | Genome-fragment | NA | NA | 0 |
| S1_k141_66919 | 1781 | No | NA | 2 | 0 | 1 | Low-quality | Genome-fragment | 4.06 | AAI-based (medium-confidence) | 0 |
| S5_k141_139712 | 1056 | No | NA | 1 | 0 | 1 | Not-determined | Genome-fragment | NA | NA | 0 |
| S5_k141_94965 | 41463 | Yes | 37179 | 61 | 20 | 2 | High-quality | High-quality | 98.56 | AAI-based (high-confidence) | 10.33 |
| S5_k141_51091 | 1291 | No | NA | 1 | 0 | 0 | Not-determined | Genome-fragment | NA | NA | 0 |
| S5_k141_104675 | 1291 | No | NA | 1 | 0 | 0 | Not-determined | Genome-fragment | NA | NA | 0 |
| S5_k141_41714 | 1094 | No | NA | 1 | 0 | 0 | Not-determined | Genome-fragment | NA | NA | 0 |
| S5_k141_193468 | 1117 | No | NA | 1 | 0 | 0 | Not-determined | Genome-fragment | NA | NA | 0 |
| S5_k141_131714 | 11683 | No | NA | 6 | 0 | 4 | Not-determined | Genome-fragment | NA | NA | 0 |
| S1_k141_93039 | 1781 | No | NA | 3 | 0 | 3 | Low-quality | Genome-fragment | 0.59 | AAI-based (high-confidence) | 0 |
| S5_k141_202092 | 1044 | No | NA | 1 | 0 | 1 | Not-determined | Genome-fragment | NA | NA | 0 |
| S5_k141_51151 | 1692 | No | NA | 1 | 0 | 1 | Not-determined | Genome-fragment | NA | NA | 0 |
| S5_k141_185102 | 10757 | No | NA | 6 | 0 | 5 | Not-determined | Genome-fragment | NA | NA | 0 |
| S5_k141_113381 | 1577 | No | NA | 2 | 0 | 1 | Not-determined | Genome-fragment | NA | NA | 0 |
| S5_k141_211058 | 2267 | No | NA | 5 | 0 | 0 | Not-determined | Genome-fragment | NA | NA | 0 |
| S3-final-viral-combined_k141_82369\|\|full | 1781 | No | NA | 3 | 0 | 0 | Low-quality | Genome-fragment | 4.44 | AAI-based (high-confidence) | 0 |
| S5_k141_68299 | 1398 | No | NA | 1 | 0 | 0 | Not-determined | Genome-fragment | NA | NA | 0 |
| S3_k141_75362 | 1781 | No | NA | 4 | 0 | 0 | Low-quality | Genome-fragment | 0.82 | AAI-based (high-confidence) | 0 |
| S5_k141_23298 | 1162 | No | NA | 1 | 0 | 1 | Not-determined | Genome-fragment | NA | NA | 0 |
| S2-final-viral-combined_k141_40200\|\|full | 1781 | No | NA | 4 | 1 | 0 | Low-quality | Genome-fragment | 0.97 | HMM-based (lower-bound) | 0 |
| S5_k141_113390 | 1197 | No | NA | 2 | 0 | 0 | Not-determined | Genome-fragment | NA | NA | 0 |
| S5_k141_193542 | 3394 | No | NA | 6 | 0 | 0 | Not-determined | Genome-fragment | NA | NA | 0 |
| S8-final-viral-combined_k141_143408\|\|full | 1781 | No | NA | 3 | 0 | 0 | Low-quality | Genome-fragment | 2.12 | AAI-based (high-confidence) | 0 |
| S5_k141_41795 | 1198 | No | NA | 2 | 0 | 0 | Not-determined | Genome-fragment | NA | NA | 0 |
| S9-final-viral-combined_k141_109983\|\|full | 1782 | No | NA | 3 | 0 | 0 | Low-quality | Genome-fragment | 4.03 | AAI-based (medium-confidence) | 0 |
| S5_k141_23367 | 1210 | No | NA | 1 | 0 | 1 | Not-determined | Genome-fragment | NA | NA | 0 |
| S5_k141_202138 | 1171 | No | NA | 1 | 0 | 0 | Not-determined | Genome-fragment | NA | NA | 0 |
| S3-final-viral-combined_k141_40371\|\|full | 1782 | No | NA | 3 | 1 | 0 | Low-quality | Genome-fragment | 3.37 | AAI-based (high-confidence) | 0 |
| S5_k141_41811 | 1171 | No | NA | 2 | 0 | 1 | Not-determined | Genome-fragment | NA | NA | 0 |
| S5_k141_139883 | 1429 | No | NA | 2 | 0 | 1 | Not-determined | Genome-fragment | NA | NA | 0 |
| S3_k141_10567 | 1782 | No | NA | 3 | 3 | 0 | Low-quality | Genome-fragment | 3.64 | AAI-based (high-confidence) | 0 |
| S5_k141_51238 | 2175 | No | NA | 2 | 0 | 2 | Not-determined | Genome-fragment | NA | NA | 0 |
| S5_k141_68352 | 4909 | No | NA | 2 | 0 | 0 | Not-determined | Genome-fragment | NA | NA | 0 |
| S4-final-viral-combined_k141_72011\|\|full | 1782 | No | NA | 3 | 0 | 1 | Low-quality | Genome-fragment | 5.69 | AAI-based (high-confidence) | 0 |
| S5_k141_157595 | 1077 | No | NA | 3 | 0 | 0 | Not-determined | Genome-fragment | NA | NA | 0 |
| S5_k141_131793 | 1782 | No | NA | 3 | 0 | 0 | Not-determined | Genome-fragment | NA | NA | 0 |
| S5_k141_131802 | 1510 | No | NA | 2 | 0 | 1 | Not-determined | Genome-fragment | NA | NA | 0 |
| S5_k141_193632 | 2106 | No | NA | 9 | 0 | 0 | Not-determined | Genome-fragment | NA | NA | 0 |
| S5_k141_78374 | 1358 | No | NA | 1 | 0 | 1 | Not-determined | Genome-fragment | NA | NA | 0 |
| S5_k141_113523 | 5828 | No | NA | 6 | 0 | 1 | Not-determined | Genome-fragment | NA | NA | 0 |
| S5_k141_157645 | 1318 | No | NA | 1 | 0 | 1 | Not-determined | Genome-fragment | NA | NA | 0 |
| S8_k141_186408 | 1782 | No | NA | 2 | 0 | 0 | Low-quality | Genome-fragment | 3.94 | AAI-based (high-confidence) | 0 |
| S5_k141_104862 | 1126 | No | NA | 2 | 0 | 2 | Not-determined | Genome-fragment | NA | NA | 0 |
| S11-final-viral-combined_k141_329512\|\|full | 1783 | No | NA | 5 | 1 | 0 | Low-quality | Genome-fragment | 3.57 | AAI-based (high-confidence) | 0 |
| S5_k141_51380 | 3808 | No | NA | 5 | 0 | 2 | Not-determined | Genome-fragment | NA | NA | 0 |
| S5_k141_211164 | 2503 | No | NA | 4 | 0 | 1 | Not-determined | Genome-fragment | NA | NA | 0 |
| S5_k141_51389 | 1175 | No | NA | 2 | 0 | 0 | Not-determined | Genome-fragment | NA | NA | 0 |
| S5_k141_86952 | 1099 | No | NA | 1 | 0 | 1 | Not-determined | Genome-fragment | NA | NA | 0 |
| S5_k141_140072 | 1600 | No | NA | 2 | 0 | 1 | Not-determined | Genome-fragment | NA | NA | 0 |
| S5_k141_104881 | 1207 | No | NA | 3 | 0 | 0 | Not-determined | Genome-fragment | NA | NA | 0 |
| S11-final-viral-combined_k141_248173\|\|lt2gene | 1783 | No | NA | 2 | 1 | 0 | Low-quality | Genome-fragment | 3.39 | HMM-based (lower-bound) | 0 |
| S5_k141_68522 | 1738 | No | NA | 2 | 0 | 0 | Not-determined | Genome-fragment | NA | NA | 0 |
| S5_k141_122230 | 3045 | No | NA | 4 | 0 | 0 | Not-determined | Genome-fragment | NA | NA | 0 |
| S5_k141_211192 | 1490 | No | NA | 2 | 0 | 0 | Not-determined | Genome-fragment | NA | NA | 0 |
| S5_k141_68554 | 1251 | No | NA | 2 | 0 | 2 | Not-determined | Genome-fragment | NA | NA | 0 |
| S1-final-viral-combined_k141_292576\|\|full | 1783 | No | NA | 4 | 0 | 0 | Low-quality | Genome-fragment | 1.18 | AAI-based (medium-confidence) | 0 |
| S5_k141_211255 | 3416 | No | NA | 3 | 0 | 1 | Not-determined | Genome-fragment | NA | NA | 0 |
| S7-final-viral-combined_k141_115575\|\|full | 1783 | No | NA | 3 | 0 | 2 | Low-quality | Genome-fragment | 2.42 | AAI-based (high-confidence) | 0 |
| S5_k141_104943 | 1637 | No | NA | 2 | 0 | 0 | Not-determined | Genome-fragment | NA | NA | 0 |
| S5_k141_193813 | 2748 | No | NA | 5 | 0 | 0 | Not-determined | Genome-fragment | NA | NA | 0 |
| S5_k141_95306 | 1258 | No | NA | 3 | 0 | 0 | Not-determined | Genome-fragment | NA | NA | 0 |
| S5_k141_95309 | 1892 | No | NA | 3 | 0 | 3 | Not-determined | Genome-fragment | NA | NA | 0 |
| S7_k141_215128 | 1783 | No | NA | 2 | 0 | 2 | Low-quality | Genome-fragment | 5.03 | AAI-based (medium-confidence) | 0 |
| S5_k141_157856 | 1462 | No | NA | 3 | 0 | 0 | Not-determined | Genome-fragment | NA | NA | 0 |
| S5_k141_23595 | 1367 | No | NA | 2 | 0 | 1 | Not-determined | Genome-fragment | NA | NA | 0 |
| S5_k141_157859 | 1212 | No | NA | 2 | 0 | 2 | Not-determined | Genome-fragment | NA | NA | 0 |
| S5_k141_60278 | 1946 | No | NA | 1 | 0 | 0 | Not-determined | Genome-fragment | NA | NA | 0 |
| S5_k141_185372 | 5825 | No | NA | 3 | 0 | 2 | Not-determined | Genome-fragment | NA | NA | 0 |
| S17-final-viral-combined_k141_135524\|\|full | 1783 | No | NA | 2 | 0 | 0 | Low-quality | Genome-fragment | 3.16 | AAI-based (medium-confidence) | 0 |
| S5_k141_157926 | 3451 | No | NA | 4 | 0 | 1 | Not-determined | Genome-fragment | NA | NA | 0 |
| S5_k141_15698 | 1197 | No | NA | 1 | 0 | 0 | Not-determined | Genome-fragment | NA | NA | 0 |
| S5_k141_78495 | 1060 | No | NA | 1 | 0 | 0 | Not-determined | Genome-fragment | NA | NA | 0 |
| S5_k141_23653 | 1229 | No | NA | 1 | 0 | 0 | Not-determined | Genome-fragment | NA | NA | 0 |
| S5_k141_157951 | 1858 | No | NA | 2 | 0 | 1 | Not-determined | Genome-fragment | NA | NA | 0 |
| S5_k141_95385 | 1000 | No | NA | 2 | 0 | 2 | Not-determined | Genome-fragment | NA | NA | 0 |
| S5_k141_60326 | 1471 | No | NA | 1 | 0 | 1 | Not-determined | Genome-fragment | NA | NA | 0 |
| S17-final-viral-combined_k141_46295\|\|full | 1783 | No | NA | 3 | 0 | 0 | Low-quality | Genome-fragment | 3.77 | AAI-based (medium-confidence) | 0 |
| S2-final-viral-combined_k141_26245\|\|full | 1783 | No | NA | 3 | 0 | 1 | Low-quality | Genome-fragment | 2.42 | AAI-based (medium-confidence) | 0 |
| S5_k141_105141 | 1244 | No | NA | 1 | 0 | 1 | Not-determined | Genome-fragment | NA | NA | 0 |
| S5_k141_8477 | 1401 | No | NA | 1 | 0 | 0 | Not-determined | Genome-fragment | NA | NA | 0 |
| S8-final-viral-combined_k141_18209\|\|full | 1783 | No | NA | 2 | 1 | 0 | Low-quality | Genome-fragment | 0.64 | HMM-based (lower-bound) | 0 |
| S9-final-viral-combined_k141_70223\|\|full | 1784 | No | NA | 3 | 0 | 0 | Low-quality | Genome-fragment | 3.51 | AAI-based (medium-confidence) | 0 |
| S5_k141_8491 | 1004 | No | NA | 2 | 0 | 1 | Not-determined | Genome-fragment | NA | NA | 0 |
| S5_k141_68903 | 1213 | No | NA | 1 | 0 | 1 | Not-determined | Genome-fragment | NA | NA | 0 |
| S5_k141_78652 | 1059 | No | NA | 3 | 0 | 3 | Not-determined | Genome-fragment | NA | NA | 0 |
| S5_k141_149334 | 10281 | No | NA | 4 | 0 | 1 | Not-determined | Genome-fragment | NA | NA | 0 |
| S7-final-viral-combined_k141_122935\|\|full | 1784 | No | NA | 5 | 2 | 0 | Low-quality | Genome-fragment | 5.26 | AAI-based (high-confidence) | 0 |
| S5_k141_95523 | 3590 | No | NA | 1 | 0 | 0 | Not-determined | Genome-fragment | NA | NA | 0 |
| S5_k141_194101 | 1339 | No | NA | 1 | 0 | 1 | Not-determined | Genome-fragment | NA | NA | 0 |
| S12-final-viral-combined_k141_271114\|\|full | 1784 | No | NA | 3 | 0 | 3 | Low-quality | Genome-fragment | 4.48 | AAI-based (medium-confidence) | 0 |
| S5_k141_122521 | 3040 | No | NA | 1 | 0 | 1 | Not-determined | Genome-fragment | NA | NA | 0 |
| S5_k141_149382 | 4230 | No | NA | 7 | 0 | 1 | Not-determined | Genome-fragment | NA | NA | 0 |
| S5_k141_105244 | 2719 | No | NA | 1 | 0 | 0 | Not-determined | Genome-fragment | NA | NA | 0 |
| S5_k141_149388 | 3435 | No | NA | 5 | 0 | 2 | Not-determined | Genome-fragment | NA | NA | 0 |
| S5_k141_105246 | 7284 | No | NA | 1 | 0 | 1 | Not-determined | Genome-fragment | NA | NA | 0 |
| S5_k141_33275 | 1150 | No | NA | 1 | 0 | 1 | Not-determined | Genome-fragment | NA | NA | 0 |
| S5_k141_78747 | 1087 | No | NA | 1 | 0 | 1 | Not-determined | Genome-fragment | NA | NA | 0 |
| S5_k141_140394 | 1179 | No | NA | 1 | 0 | 1 | Not-determined | Genome-fragment | NA | NA | 0 |
| S5_k141_176517 | 1853 | No | NA | 1 | 0 | 1 | Not-determined | Genome-fragment | NA | NA | 0 |
| S12-final-viral-combined_k141_173505\|\|full | 1784 | No | NA | 2 | 0 | 2 | Low-quality | Genome-fragment | 0.51 | AAI-based (high-confidence) | 0 |
| S5_k141_33362 | 4129 | No | NA | 3 | 0 | 1 | Not-determined | Genome-fragment | NA | NA | 0 |
| S5_k141_149475 | 1084 | No | NA | 1 | 0 | 0 | Not-determined | Genome-fragment | NA | NA | 0 |
| S5_k141_8690 | 1099 | No | NA | 1 | 0 | 1 | Not-determined | Genome-fragment | NA | NA | 0 |
| S5_k141_23872 | 3973 | No | NA | 5 | 0 | 2 | Not-determined | Genome-fragment | NA | NA | 0 |
| S12-final-viral-combined_k141_123595\|\|lt2gene | 1784 | No | NA | 2 | 1 | 0 | Low-quality | Genome-fragment | 3.89 | AAI-based (high-confidence) | 0 |
| S5_k141_149493 | 1054 | No | NA | 1 | 0 | 0 | Not-determined | Genome-fragment | NA | NA | 0 |
| S8-final-viral-combined_k141_8303\|\|full | 1784 | No | NA | 3 | 1 | 0 | Low-quality | Genome-fragment | 0.92 | HMM-based (lower-bound) | 0 |
| S5_k141_68995 | 1949 | No | NA | 1 | 0 | 1 | Not-determined | Genome-fragment | NA | NA | 0 |
| S5_k141_87426 | 1459 | No | NA | 2 | 0 | 0 | Not-determined | Genome-fragment | NA | NA | 0 |
| S9-final-viral-combined_k141_11355\|\|full | 1785 | No | NA | 4 | 2 | 0 | Low-quality | Genome-fragment | 4.25 | AAI-based (medium-confidence) | 0 |
| S5_k141_8756 | 3158 | No | NA | 5 | 0 | 1 | Not-determined | Genome-fragment | NA | NA | 0 |
| S5_k141_60655 | 1024 | No | NA | 2 | 0 | 0 | Not-determined | Genome-fragment | NA | NA | 0 |
| S5_k141_69054 | 3366 | No | NA | 4 | 0 | 1 | Not-determined | Genome-fragment | NA | NA | 0 |
| S5_k141_202510 | 3733 | No | NA | 5 | 0 | 1 | Not-determined | Genome-fragment | NA | NA | 0 |
| S5_k141_33450 | 1677 | No | NA | 1 | 0 | 0 | Not-determined | Genome-fragment | NA | NA | 0 |
| S5_k141_167007 | 1230 | No | NA | 1 | 0 | 1 | Not-determined | Genome-fragment | NA | NA | 0 |
| S5_k141_15901 | 2482 | No | NA | 1 | 0 | 1 | Not-determined | Genome-fragment | NA | NA | 0 |
| S5_k141_132325 | 1347 | No | NA | 2 | 0 | 1 | Not-determined | Genome-fragment | NA | NA | 0 |
| S5_k141_78859 | 1170 | No | NA | 1 | 0 | 1 | Not-determined | Genome-fragment | NA | NA | 0 |
| S5_k141_167062 | 2469 | No | NA | 5 | 0 | 5 | Not-determined | Genome-fragment | NA | NA | 0 |
| S11-final-viral-combined_k141_326963\|\|full | 1785 | No | NA | 3 | 0 | 0 | Low-quality | Genome-fragment | 0.95 | AAI-based (medium-confidence) | 0 |
| S5_k141_15944 | 1157 | No | NA | 1 | 0 | 1 | Not-determined | Genome-fragment | NA | NA | 0 |
| S5_k141_122823 | 1151 | No | NA | 2 | 0 | 0 | Not-determined | Genome-fragment | NA | NA | 0 |
| S3-final-viral-combined_k141_119330\|\|full | 1785 | No | NA | 2 | 0 | 2 | Low-quality | Genome-fragment | 0.71 | AAI-based (medium-confidence) | 0 |
| S5_k141_122875 | 1675 | No | NA | 2 | 0 | 0 | Not-determined | Genome-fragment | NA | NA | 0 |
| S5_k141_140644 | 4950 | No | NA | 2 | 0 | 0 | Not-determined | Genome-fragment | NA | NA | 0 |
| S5-final-viral-combined_k141_79331\|\|full | 1785 | No | NA | 3 | 0 | 1 | Low-quality | Genome-fragment | 4.21 | AAI-based (medium-confidence) | 0 |
| S5_k141_176684 | 1782 | No | NA | 3 | 0 | 1 | Not-determined | Genome-fragment | NA | NA | 0 |
| S5_k141_60912 | 8367 | No | NA | 14 | 0 | 1 | Not-determined | Genome-fragment | NA | NA | 0 |
| S5_k141_132487 | 2775 | No | NA | 2 | 0 | 0 | Not-determined | Genome-fragment | NA | NA | 0 |
| S17-final-viral-combined_k141_38166\|\|full | 1785 | No | NA | 3 | 0 | 0 | Low-quality | Genome-fragment | 3.35 | AAI-based (medium-confidence) | 0 |
| S5_k141_176719 | 1017 | No | NA | 2 | 0 | 2 | Not-determined | Genome-fragment | NA | NA | 0 |
| S5_k141_132511 | 1378 | No | NA | 1 | 0 | 0 | Not-determined | Genome-fragment | NA | NA | 0 |
| S12-final-viral-combined_k141_223527\|\|full | 1785 | No | NA | 3 | 0 | 2 | Low-quality | Genome-fragment | 3.24 | AAI-based (medium-confidence) | 0 |
| S5_k141_95925 | 1136 | No | NA | 3 | 0 | 0 | Not-determined | Genome-fragment | NA | NA | 0 |
| S12-final-viral-combined_k141_316154\|\|full | 1785 | No | NA | 3 | 0 | 1 | Low-quality | Genome-fragment | 0.86 | AAI-based (medium-confidence) | 0 |
| S5_k141_140673 | 13651 | No | NA | 16 | 0 | 6 | Not-determined | Genome-fragment | NA | NA | 0 |
| S12_k141_397452 | 1785 | No | NA | 3 | 0 | 1 | Low-quality | Genome-fragment | 2.25 | AAI-based (medium-confidence) | 0 |
| S5_k141_149647 | 1419 | No | NA | 1 | 0 | 1 | Not-determined | Genome-fragment | NA | NA | 0 |
| S5_k141_185617 | 1553 | No | NA | 2 | 0 | 2 | Not-determined | Genome-fragment | NA | NA | 0 |
| S2-final-viral-combined_k141_58527\|\|full | 1785 | No | NA | 4 | 0 | 0 | Low-quality | Genome-fragment | 4.91 | AAI-based (high-confidence) | 0 |
| S5_k141_42400 | 2759 | No | NA | 5 | 0 | 3 | Not-determined | Genome-fragment | NA | NA | 0 |
| S5_k141_52224 | 1377 | No | NA | 3 | 0 | 1 | Not-determined | Genome-fragment | NA | NA | 0 |
| S6-final-viral-combined_k141_13408\|\|full | 1785 | No | NA | 5 | 2 | 0 | Low-quality | Genome-fragment | 5.05 | AAI-based (medium-confidence) | 0 |
| S5_k141_194503 | 1268 | No | NA | 3 | 0 | 0 | Not-determined | Genome-fragment | NA | NA | 0 |
| S11-final-viral-combined_k141_290247\|\|full | 1786 | No | NA | 3 | 0 | 2 | Low-quality | Genome-fragment | 1.25 | AAI-based (medium-confidence) | 0 |
| S1-final-viral-combined_k141_354887\|\|full | 1786 | No | NA | 2 | 2 | 0 | Low-quality | Genome-fragment | 3.94 | AAI-based (high-confidence) | 0 |
| S5_k141_202880 | 1089 | No | NA | 1 | 0 | 1 | Not-determined | Genome-fragment | NA | NA | 0 |
| S5_k141_202885 | 3946 | No | NA | 7 | 0 | 2 | Not-determined | Genome-fragment | NA | NA | 0 |
| S2-final-viral-combined_k141_11436\|\|full | 1786 | No | NA | 3 | 3 | 0 | Low-quality | Genome-fragment | 4.66 | AAI-based (medium-confidence) | 0 |
| S5_k141_176957 | 1387 | No | NA | 4 | 0 | 2 | Not-determined | Genome-fragment | NA | NA | 0 |
| S2-final-viral-combined_k141_182820\|\|full | 1786 | No | NA | 3 | 1 | 0 | Low-quality | Genome-fragment | 3.8 | AAI-based (medium-confidence) | 0 |
| S5_k141_61020 | 8105 | No | NA | 5 | 0 | 5 | Not-determined | Genome-fragment | NA | NA | 0 |
| S2-final-viral-combined_k141_227820\|\|full | 1786 | No | NA | 2 | 0 | 0 | Low-quality | Genome-fragment | 3.84 | AAI-based (medium-confidence) | 0 |
| S1_k141_186679 | 1787 | No | NA | 1 | 0 | 1 | Low-quality | Genome-fragment | 1.92 | AAI-based (high-confidence) | 0 |
| S12-final-viral-combined_k141_212475\|\|full | 1787 | No | NA | 3 | 0 | 0 | Low-quality | Genome-fragment | 4.19 | AAI-based (high-confidence) | 0 |
| S4-final-viral-combined_k141_53172\|\|full | 1787 | No | NA | 2 | 1 | 0 | Low-quality | Genome-fragment | 3.67 | AAI-based (high-confidence) | 0 |
| S5_k141_149922 | 1250 | No | NA | 1 | 0 | 0 | Not-determined | Genome-fragment | NA | NA | 0 |
| S5_k141_87960 | 1052 | No | NA | 1 | 0 | 1 | Not-determined | Genome-fragment | NA | NA | 0 |
| S5_k141_123281 | 1005 | No | NA | 2 | 0 | 1 | Not-determined | Genome-fragment | NA | NA | 0 |
| S5_k141_177089 | 1579 | No | NA | 2 | 0 | 2 | Not-determined | Genome-fragment | NA | NA | 0 |
| S8-final-viral-combined_k141_28564\|\|full | 1787 | No | NA | 2 | 0 | 1 | Low-quality | Genome-fragment | 3.46 | AAI-based (high-confidence) | 0 |
| S5-final-viral-combined_k141_108519\|\|full | 1788 | No | NA | 3 | 0 | 0 | Low-quality | Genome-fragment | 2.22 | AAI-based (medium-confidence) | 0 |
| S5-final-viral-combined_k141_212672\|\|full | 1788 | No | NA | 4 | 0 | 0 | Low-quality | Genome-fragment | 4.24 | AAI-based (high-confidence) | 0 |
| S5_k141_34031 | 2817 | No | NA | 1 | 0 | 0 | Not-determined | Genome-fragment | NA | NA | 0 |
| S7-final-viral-combined_k141_170584\|\|full | 1788 | No | NA | 2 | 1 | 0 | Low-quality | Genome-fragment | 1.24 | HMM-based (lower-bound) | 0 |
| S5_k141_42660 | 1848 | No | NA | 1 | 0 | 1 | Not-determined | Genome-fragment | NA | NA | 0 |
| S5_k141_177164 | 1096 | No | NA | 1 | 0 | 1 | Not-determined | Genome-fragment | NA | NA | 0 |
| S5_k141_158507 | 1225 | No | NA | 1 | 0 | 1 | Not-determined | Genome-fragment | NA | NA | 0 |
| S7-final-viral-combined_k141_132426\|\|full | 1788 | No | NA | 3 | 0 | 0 | Low-quality | Genome-fragment | 3.55 | AAI-based (medium-confidence) | 0 |
| S5_k141_52566 | 6468 | Yes | 4855 | 7 | 1 | 1 | High-quality | High-quality | 100 | HMM-based (lower-bound) | 24.94 |
| S5_k141_123372 | 1014 | No | NA | 1 | 0 | 0 | Not-determined | Genome-fragment | NA | NA | 0 |
| S5_k141_202966 | 1188 | No | NA | 2 | 0 | 2 | Not-determined | Genome-fragment | NA | NA | 0 |
| S5_k141_88025 | 2926 | No | NA | 2 | 0 | 2 | Not-determined | Genome-fragment | NA | NA | 0 |
| S5_k141_202987 | 1030 | No | NA | 2 | 0 | 0 | Not-determined | Genome-fragment | NA | NA | 0 |
| S5_k141_185947 | 3568 | No | NA | 3 | 0 | 0 | Not-determined | Genome-fragment | NA | NA | 0 |
| S5_k141_203014 | 1039 | No | NA | 1 | 0 | 1 | Not-determined | Genome-fragment | NA | NA | 0 |
| S5_k141_203026 | 1507 | No | NA | 1 | 0 | 1 | Not-determined | Genome-fragment | NA | NA | 0 |
| S5_k141_16143 | 4637 | No | NA | 5 | 0 | 2 | Not-determined | Genome-fragment | NA | NA | 0 |
| S5_k141_16156 | 1086 | No | NA | 3 | 0 | 0 | Not-determined | Genome-fragment | NA | NA | 0 |
| S12_k141_188874 | 1788 | No | NA | 1 | 0 | 1 | Low-quality | Genome-fragment | 1.98 | AAI-based (medium-confidence) | 0 |
| S5_k141_123499 | 3356 | No | NA | 5 | 2 | 0 | Medium-quality | Genome-fragment | 52.73 | AAI-based (high-confidence) | 0 |
| S5_k141_212097 | 1632 | No | NA | 2 | 0 | 0 | Not-determined | Genome-fragment | NA | NA | 0 |
| S5_k141_194967 | 1205 | No | NA | 2 | 0 | 1 | Not-determined | Genome-fragment | NA | NA | 0 |
| S5_k141_150147 | 1302 | No | NA | 2 | 0 | 0 | Not-determined | Genome-fragment | NA | NA | 0 |
| S5_k141_114190 | 1046 | No | NA | 1 | 0 | 1 | Not-determined | Genome-fragment | NA | NA | 0 |
| S5_k141_106046 | 1254 | No | NA | 1 | 0 | 1 | Not-determined | Genome-fragment | NA | NA | 0 |
| S5_k141_42861 | 6942 | No | NA | 12 | 0 | 1 | Not-determined | Genome-fragment | NA | NA | 0 |
| S5_k141_195089 | 1332 | No | NA | 2 | 0 | 2 | Not-determined | Genome-fragment | NA | NA | 0 |
| S5_k141_195106 | 1232 | No | NA | 2 | 0 | 2 | Not-determined | Genome-fragment | NA | NA | 0 |
| S5_k141_16384 | 2018 | No | NA | 2 | 0 | 2 | Not-determined | Genome-fragment | NA | NA | 0 |
| S5_k141_42938 | 3089 | No | NA | 3 | 0 | 0 | Not-determined | Genome-fragment | NA | NA | 0 |
| S4-final-viral-combined_k141_74758\|\|full | 1788 | No | NA | 4 | 0 | 2 | Low-quality | Genome-fragment | 0.88 | AAI-based (medium-confidence) | 0 |
| S5_k141_88336 | 2766 | No | NA | 1 | 0 | 1 | Not-determined | Genome-fragment | NA | NA | 0 |
| S1-final-viral-combined_k141_203347\|\|full | 1789 | No | NA | 3 | 0 | 1 | Low-quality | Genome-fragment | 2.25 | AAI-based (medium-confidence) | 0 |
| S5_k141_123696 | 1614 | No | NA | 1 | 0 | 1 | Not-determined | Genome-fragment | NA | NA | 0 |
| S5_k141_141165 | 9629 | No | NA | 6 | 0 | 6 | Not-determined | Genome-fragment | NA | NA | 0 |
| S5_k141_69683 | 1071 | No | NA | 1 | 0 | 1 | Not-determined | Genome-fragment | NA | NA | 0 |
| S5_k141_203220 | 1450 | No | NA | 1 | 0 | 0 | Not-determined | Genome-fragment | NA | NA | 0 |
| S5_k141_106212 | 3684 | No | NA | 6 | 0 | 2 | Not-determined | Genome-fragment | NA | NA | 0 |
| S5_k141_52784 | 1368 | No | NA | 2 | 0 | 1 | Not-determined | Genome-fragment | NA | NA | 0 |
| S5-final-viral-combined_k141_109944\|\|full | 1789 | No | NA | 2 | 0 | 0 | Low-quality | Genome-fragment | 4.53 | AAI-based (high-confidence) | 0 |
| S5_k141_79545 | 1100 | No | NA | 1 | 0 | 1 | Not-determined | Genome-fragment | NA | NA | 0 |
| S5_k141_96727 | 1306 | No | NA | 1 | 0 | 0 | Not-determined | Genome-fragment | NA | NA | 0 |
| S5_k141_186238 | 3175 | No | NA | 1 | 0 | 1 | Not-determined | Genome-fragment | NA | NA | 0 |
| S5_k141_114481 | 3447 | No | NA | 2 | 0 | 0 | Not-determined | Genome-fragment | NA | NA | 0 |
| S5_k141_150414 | 1095 | No | NA | 1 | 0 | 0 | Not-determined | Genome-fragment | NA | NA | 0 |
| S5_k141_96810 | 1190 | No | NA | 2 | 0 | 0 | Not-determined | Genome-fragment | NA | NA | 0 |
| S5_k141_132960 | 1801 | No | NA | 1 | 0 | 1 | Not-determined | Genome-fragment | NA | NA | 0 |
| S12-final-viral-combined_k141_337768\|\|full | 1789 | No | NA | 3 | 0 | 1 | Low-quality | Genome-fragment | 3.16 | AAI-based (high-confidence) | 0 |
| S5_k141_43010 | 1338 | No | NA | 1 | 0 | 1 | Not-determined | Genome-fragment | NA | NA | 0 |
| S5_k141_150506 | 3048 | No | NA | 3 | 0 | 3 | Not-determined | Genome-fragment | NA | NA | 0 |
| S5_k141_203398 | 1337 | No | NA | 1 | 0 | 0 | Not-determined | Genome-fragment | NA | NA | 0 |
| S5_k141_203402 | 1197 | No | NA | 1 | 0 | 1 | Not-determined | Genome-fragment | NA | NA | 0 |
| S5_k141_133054 | 1362 | No | NA | 2 | 0 | 1 | Not-determined | Genome-fragment | NA | NA | 0 |

| | | | | | | | | | | | |
|---|---|---|---|---|---|---|---|---|---|---|---|
| S5_k141_96929 | 1196 | No | NA | 3 | 0 | 0 | Not-determined | Genome-fragment | NA | NA | 0 |
| S2-final-viral-combined_k141_172157\|\|lt2gene | 1789 | No | NA | 2 | 1 | 0 | Low-quality | Genome-fragment | 2.26 | AAI-based (high-confidence) | 0 |
| S5_k141_195352 | 1561 | No | NA | 2 | 0 | 2 | Not-determined | Genome-fragment | NA | NA | 0 |
| S5_k141_24816 | 1095 | No | NA | 1 | 0 | 1 | Not-determined | Genome-fragment | NA | NA | 0 |
| S5_k141_69772 | 1093 | No | NA | 2 | 0 | 1 | Not-determined | Genome-fragment | NA | NA | 0 |
| S5_k141_168115 | 1064 | No | NA | 2 | 0 | 2 | Not-determined | Genome-fragment | NA | NA | 0 |
| S4_k141_13691 | 1789 | No | NA | 3 | 1 | 0 | Low-quality | Genome-fragment | 3.58 | AAI-based (high-confidence) | 0 |
| S8-final-viral-combined_k141_35746\|\|full | 1789 | No | NA | 2 | 0 | 2 | Low-quality | Genome-fragment | 3.48 | AAI-based (medium-confidence) | 0 |
| S5_k141_133109 | 1052 | No | NA | 2 | 0 | 0 | Not-determined | Genome-fragment | NA | NA | 0 |
| S8-final-viral-combined_k141_155135\|\|full | 1789 | No | NA | 3 | 0 | 0 | Low-quality | Genome-fragment | 1.61 | AAI-based (medium-confidence) | 0 |
| S5_k141_52890 | 1084 | No | NA | 1 | 0 | 0 | Not-determined | Genome-fragment | NA | NA | 0 |
| S5_k141_168160 | 1971 | No | NA | 1 | 0 | 0 | Not-determined | Genome-fragment | NA | NA | 0 |
| S5_k141_195426 | 1020 | No | NA | 1 | 0 | 0 | Not-determined | Genome-fragment | NA | NA | 0 |
| S1-final-viral-combined_k141_42182\|\|full | 1790 | No | NA | 4 | 1 | 0 | Low-quality | Genome-fragment | 2.88 | AAI-based (medium-confidence) | 0 |
| S5_k141_61304 | 1084 | No | NA | 1 | 0 | 1 | Not-determined | Genome-fragment | NA | NA | 0 |
| S5_k141_212488 | 1282 | No | NA | 1 | 0 | 1 | Not-determined | Genome-fragment | NA | NA | 0 |
| S5_k141_212496 | 1041 | No | NA | 2 | 0 | 2 | Not-determined | Genome-fragment | NA | NA | 0 |
| S1-final-viral-combined_k141_227437\|\|full | 1790 | No | NA | 3 | 0 | 1 | Low-quality | Genome-fragment | 0.93 | AAI-based (medium-confidence) | 0 |
| S5_k141_114651 | 1174 | No | NA | 1 | 0 | 1 | Not-determined | Genome-fragment | NA | NA | 0 |
| S5_k141_177505 | 1867 | No | NA | 2 | 0 | 2 | Not-determined | Genome-fragment | NA | NA | 0 |
| S5_k141_159081 | 1011 | No | NA | 1 | 0 | 0 | Not-determined | Genome-fragment | NA | NA | 0 |
| S5_k141_168202 | 1152 | No | NA | 1 | 0 | 0 | Not-determined | Genome-fragment | NA | NA | 0 |
| S5_k141_43208 | 1110 | No | NA | 2 | 0 | 0 | Not-determined | Genome-fragment | NA | NA | 0 |
| S1-final-viral-combined_k141_349214\|\|lt2gene | 1790 | Yes | 1615 | 2 | 1 | 1 | Low-quality | Genome-fragment | 3.47 | AAI-based (high-confidence) | 9.78 |
| S5_k141_141428 | 1021 | No | NA | 1 | 0 | 0 | Not-determined | Genome-fragment | NA | NA | 0 |
| S5_k141_106529 | 2344 | No | NA | 3 | 0 | 2 | Not-determined | Genome-fragment | NA | NA | 0 |
| S5_k141_212596 | 8740 | No | NA | 12 | 6 | 0 | Medium-quality | Genome-fragment | 62.16 | AAI-based (high-confidence) | 0 |
| S5-final-viral-combined_k141_2158\|\|full | 1790 | No | NA | 4 | 0 | 1 | Low-quality | Genome-fragment | 4.59 | AAI-based (medium-confidence) | 0 |
| S5_k141_114705 | 1397 | No | NA | 2 | 0 | 2 | Not-determined | Genome-fragment | NA | NA | 0 |
| S5_k141_124133 | 4555 | No | NA | 2 | 0 | 0 | Not-determined | Genome-fragment | NA | NA | 0 |
| S5_k141_203601 | 1192 | No | NA | 1 | 0 | 1 | Not-determined | Genome-fragment | NA | NA | 0 |
| S5-final-viral-combined_k141_128614\|\|full | 1790 | No | NA | 6 | 1 | 0 | Low-quality | Genome-fragment | 3.23 | AAI-based (medium-confidence) | 0 |
| S12-final-viral-combined_k141_244454\|\|full | 1790 | No | NA | 3 | 0 | 0 | Low-quality | Genome-fragment | 1.38 | AAI-based (medium-confidence) | 0 |
| S5_k141_203622 | 3266 | No | NA | 2 | 0 | 0 | Not-determined | Genome-fragment | NA | NA | 0 |
| S11-final-viral-combined_k141_238818\|\|lt2gene | 1791 | No | NA | 1 | 0 | 0 | Low-quality | Genome-fragment | 2.18 | AAI-based (high-confidence) | 0 |
| S5_k141_195702 | 1077 | No | NA | 1 | 0 | 1 | Not-determined | Genome-fragment | NA | NA | 0 |
| S11_k141_233652 | 1791 | No | NA | 1 | 0 | 1 | Low-quality | Genome-fragment | 0.72 | AAI-based (medium-confidence) | 0 |
| S5_k141_203714 | 1233 | No | NA | 2 | 0 | 1 | Not-determined | Genome-fragment | NA | NA | 0 |
| S5-final-viral-combined_k141_7475\|\|full | 1791 | No | NA | 4 | 2 | 0 | Low-quality | Genome-fragment | 1.2 | HMM-based (lower-bound) | 0 |
| S5_k141_25137 | 1647 | No | NA | 1 | 0 | 1 | Not-determined | Genome-fragment | NA | NA | 0 |
| S5_k141_177613 | 2060 | No | NA | 3 | 0 | 2 | Not-determined | Genome-fragment | NA | NA | 0 |
| S5_k141_53111 | 6683 | No | NA | 6 | 0 | 0 | Not-determined | Genome-fragment | NA | NA | 0 |
| S5_k141_11912 | 1791 | No | NA | 2 | 0 | 0 | Low-quality | Genome-fragment | 4.5 | AAI-based (medium-confidence) | 0 |
| S5_k141_16993 | 3931 | No | NA | 4 | 0 | 0 | Not-determined | Genome-fragment | NA | NA | 0 |
| S5_k141_80016 | 1065 | No | NA | 1 | 0 | 1 | Not-determined | Genome-fragment | NA | NA | 0 |
| S7-final-viral-combined_k141_30127\|\|full | 1791 | No | NA | 4 | 0 | 2 | Low-quality | Genome-fragment | 0.58 | AAI-based (high-confidence) | 0 |
| S5_k141_124300 | 1922 | No | NA | 2 | 0 | 1 | Not-determined | Genome-fragment | NA | NA | 0 |
| S5_k141_61603 | 3027 | No | NA | 2 | 0 | 1 | Not-determined | Genome-fragment | NA | NA | 0 |
| S5_k141_195815 | 1312 | No | NA | 1 | 0 | 1 | Not-determined | Genome-fragment | NA | NA | 0 |
| S5_k141_195823 | 1164 | No | NA | 1 | 0 | 1 | Not-determined | Genome-fragment | NA | NA | 0 |
| S5_k141_115034 | 1073 | No | NA | 2 | 0 | 0 | Not-determined | Genome-fragment | NA | NA | 0 |
| S5_k141_186701 | 1361 | No | NA | 3 | 0 | 0 | Not-determined | Genome-fragment | NA | NA | 0 |
| S5_k141_195864 | 1986 | No | NA | 4 | 0 | 4 | Not-determined | Genome-fragment | NA | NA | 0 |
| S5_k141_177660 | 1257 | No | NA | 2 | 0 | 1 | Not-determined | Genome-fragment | NA | NA | 0 |
| S5_k141_34777 | 1435 | No | NA | 1 | 0 | 1 | Not-determined | Genome-fragment | NA | NA | 0 |
| S17-final-viral-combined_k141_174061\|\|full | 1791 | No | NA | 5 | 1 | 0 | Low-quality | Genome-fragment | 4.1 | AAI-based (medium-confidence) | 0 |
| S5_k141_133486 | 1194 | No | NA | 1 | 0 | 1 | Not-determined | Genome-fragment | NA | NA | 0 |
| S5_k141_53372 | 1308 | No | NA | 1 | 0 | 1 | Not-determined | Genome-fragment | NA | NA | 0 |
| S5_k141_80171 | 1029 | No | NA | 2 | 0 | 1 | Not-determined | Genome-fragment | NA | NA | 0 |
| S5_k141_159463 | 1821 | No | NA | 3 | 0 | 1 | Not-determined | Genome-fragment | NA | NA | 0 |
| S5_k141_115094 | 1116 | No | NA | 2 | 0 | 1 | Not-determined | Genome-fragment | NA | NA | 0 |
| S5_k141_106841 | 1074 | No | NA | 1 | 0 | 1 | Not-determined | Genome-fragment | NA | NA | 0 |
| S5_k141_106862 | 1035 | No | NA | 1 | 0 | 0 | Not-determined | Genome-fragment | NA | NA | 0 |
| S5_k141_43673 | 1122 | No | NA | 2 | 0 | 0 | Not-determined | Genome-fragment | NA | NA | 0 |
| S5_k141_115142 | 1054 | No | NA | 2 | 0 | 1 | Not-determined | Genome-fragment | NA | NA | 0 |
| S5_k141_203920 | 1701 | No | NA | 4 | 0 | 4 | Not-determined | Genome-fragment | NA | NA | 0 |
| S5_k141_70334 | 1463 | No | NA | 3 | 0 | 1 | Not-determined | Genome-fragment | NA | NA | 0 |
| S17-final-viral-combined_k141_120680\|\|full | 1791 | No | NA | 6 | 0 | 0 | Low-quality | Genome-fragment | 13.1 | AAI-based (medium-confidence) | 0 |
| S5_k141_97740 | 2067 | No | NA | 1 | 0 | 1 | Not-determined | Genome-fragment | NA | NA | 0 |
| S5_k141_25341 | 1573 | No | NA | 1 | 0 | 0 | Not-determined | Genome-fragment | NA | NA | 0 |
| S5_k141_141967 | 2040 | No | NA | 2 | 0 | 2 | Not-determined | Genome-fragment | NA | NA | 0 |
| S5_k141_204015 | 1239 | No | NA | 2 | 0 | 2 | Not-determined | Genome-fragment | NA | NA | 0 |
| S5_k141_124622 | 1415 | No | NA | 1 | 0 | 1 | Not-determined | Genome-fragment | NA | NA | 0 |
| S5_k141_177889 | 1182 | No | NA | 1 | 0 | 0 | Not-determined | Genome-fragment | NA | NA | 0 |
| S17-k141_225902 | 1791 | No | NA | 2 | 0 | 0 | Low-quality | Genome-fragment | 3.36 | AAI-based (medium-confidence) | 0 |
| S8-final-viral-combined_k141_224109\|\|full | 1791 | No | NA | 6 | 1 | 0 | Low-quality | Genome-fragment | 4.03 | AAI-based (high-confidence) | 0 |
| S5_k141_70428 | 1086 | No | NA | 1 | 0 | 1 | Not-determined | Genome-fragment | NA | NA | 0 |
| S5_k141_97843 | 3078 | No | NA | 2 | 0 | 2 | Not-determined | Genome-fragment | NA | NA | 0 |
| S5_k141_35230 | 1042 | No | NA | 1 | 0 | 0 | Not-determined | Genome-fragment | NA | NA | 0 |
| S5_k141_97869 | 1069 | No | NA | 1 | 0 | 0 | Not-determined | Genome-fragment | NA | NA | 0 |
| S5_k141_97885 | 1016 | No | NA | 1 | 0 | 0 | Not-determined | Genome-fragment | NA | NA | 0 |
| S5_k141_17308 | 3746 | No | NA | 5 | 0 | 4 | Not-determined | Genome-fragment | NA | NA | 0 |
| S5_k141_97903 | 1211 | No | NA | 1 | 0 | 0 | Not-determined | Genome-fragment | NA | NA | 0 |
| S5_k141_35279 | 1092 | No | NA | 1 | 0 | 0 | Not-determined | Genome-fragment | NA | NA | 0 |
| S5_k141_142112 | 1068 | No | NA | 1 | 0 | 1 | Not-determined | Genome-fragment | NA | NA | 0 |
| S5_k141_43915 | 1432 | No | NA | 1 | 0 | 1 | Not-determined | Genome-fragment | NA | NA | 0 |
| S5_k141_159805 | 1057 | No | NA | 1 | 0 | 0 | Not-determined | Genome-fragment | NA | NA | 0 |
| S5_k141_178157 | 1081 | No | NA | 2 | 0 | 1 | Not-determined | Genome-fragment | NA | NA | 0 |
| S5_k141_115369 | 1106 | No | NA | 1 | 0 | 1 | Not-determined | Genome-fragment | NA | NA | 0 |
| S5_k141_115396 | 1222 | No | NA | 2 | 0 | 2 | Not-determined | Genome-fragment | NA | NA | 0 |
| S5_k141_151362 | 1122 | No | NA | 2 | 0 | 0 | Not-determined | Genome-fragment | NA | NA | 0 |
| S5_k141_168984 | 1948 | No | NA | 5 | 0 | 5 | Not-determined | Genome-fragment | NA | NA | 0 |
| S5_k141_115409 | 1477 | No | NA | 2 | 0 | 2 | Not-determined | Genome-fragment | NA | NA | 0 |
| S5_k141_17470 | 1031 | No | NA | 1 | 0 | 1 | Not-determined | Genome-fragment | NA | NA | 0 |
| S5_k141_187030 | 1679 | No | NA | 5 | 0 | 1 | Not-determined | Genome-fragment | NA | NA | 0 |
| S5_k141_17481 | 1314 | No | NA | 2 | 0 | 1 | Not-determined | Genome-fragment | NA | NA | 0 |
| S5_k141_124719 | 2346 | No | NA | 2 | 0 | 1 | Not-determined | Genome-fragment | NA | NA | 0 |
| S8-final-viral-combined_k141_228550\|\|full | 1791 | No | NA | 2 | 0 | 1 | Low-quality | Genome-fragment | 0.84 | AAI-based (medium-confidence) | 0 |
| S5_k141_70598 | 2567 | No | NA | 4 | 0 | 3 | Not-determined | Genome-fragment | NA | NA | 0 |
| S5_k141_25598 | 1264 | No | NA | 1 | 0 | 0 | Not-determined | Genome-fragment | NA | NA | 0 |
| S5_k141_44083 | 1081 | No | NA | 1 | 0 | 0 | Not-determined | Genome-fragment | NA | NA | 0 |
| S5_k141_115494 | 10940 | No | NA | 7 | 0 | 4 | Not-determined | Genome-fragment | NA | NA | 0 |
| S5_k141_204269 | 1526 | No | NA | 2 | 0 | 0 | Not-determined | Genome-fragment | NA | NA | 0 |
| S1-final-viral-combined_k141_10819\|\|full | 1792 | No | NA | 3 | 0 | 3 | Low-quality | Genome-fragment | 0.62 | AAI-based (high-confidence) | 0 |
| S5_k141_35485 | 3472 | No | NA | 4 | 0 | 1 | Not-determined | Genome-fragment | NA | NA | 0 |
| S3-final-viral-combined_k141_48588\|\|full | 1792 | No | NA | 2 | 0 | 0 | Low-quality | Genome-fragment | 2.22 | AAI-based (medium-confidence) | 0 |
| S5_k141_62316 | 1757 | No | NA | 1 | 0 | 0 | Not-determined | Genome-fragment | NA | NA | 0 |
| S5_k141_44090 | 1105 | No | NA | 1 | 0 | 0 | Not-determined | Genome-fragment | NA | NA | 0 |
| S12-final-viral-combined_k141_93848\|\|full | 1792 | No | NA | 2 | 0 | 0 | Low-quality | Genome-fragment | 3.42 | AAI-based (medium-confidence) | 0 |
| S8-final-viral-combined_k141_266776\|\|full | 1792 | No | NA | 5 | 0 | 0 | Low-quality | Genome-fragment | 5.16 | AAI-based (high-confidence) | 0 |
| S5_k141_44158 | 6918 | No | NA | 7 | 0 | 2 | Not-determined | Genome-fragment | NA | NA | 0 |
| S5_k141_160248 | 1224 | No | NA | 1 | 0 | 0 | Not-determined | Genome-fragment | NA | NA | 0 |
| S5_k141_44205 | 1786 | No | NA | 4 | 0 | 1 | Not-determined | Genome-fragment | NA | NA | 0 |
| S1-final-viral-combined_k141_16840\|\|full | 1793 | No | NA | 4 | 0 | 3 | Low-quality | Genome-fragment | 0.64 | AAI-based (medium-confidence) | 0 |
| S5_k141_44221 | 1348 | No | NA | 2 | 0 | 2 | Not-determined | Genome-fragment | NA | NA | 0 |
| S5_k141_17760 | 1309 | No | NA | 1 | 0 | 1 | Not-determined | Genome-fragment | NA | NA | 0 |
| S5_k141_25885 | 1426 | No | NA | 2 | 0 | 1 | Not-determined | Genome-fragment | NA | NA | 0 |
| S1_k141_47376 | 1793 | No | NA | 1 | 0 | 1 | Low-quality | Genome-fragment | 1.76 | AAI-based (high-confidence) | 0 |
| S5_k141_25988 | 1126 | No | NA | 2 | 0 | 2 | Not-determined | Genome-fragment | NA | NA | 0 |
| S5_k141_70978 | 1120 | No | NA | 2 | 0 | 1 | Not-determined | Genome-fragment | NA | NA | 0 |
| S5_k141_204866 | 1072 | No | NA | 1 | 0 | 0 | Not-determined | Genome-fragment | NA | NA | 0 |
| S5_k141_191179 | 1794 | No | NA | 1 | 0 | 1 | Low-quality | Genome-fragment | 2.89 | AAI-based (medium-confidence) | 0 |
| S5_k141_26201 | 1091 | No | NA | 2 | 0 | 0 | Not-determined | Genome-fragment | NA | NA | 0 |
| S5_k141_71173 | 1760 | No | NA | 2 | 0 | 1 | Not-determined | Genome-fragment | NA | NA | 0 |
| S17-final-viral-combined_k141_83400\|\|full | 1794 | No | NA | 3 | 0 | 2 | Low-quality | Genome-fragment | 1.39 | AAI-based (medium-confidence) | 0 |
| S5_k141_26558 | 2153 | No | NA | 3 | 0 | 2 | Not-determined | Genome-fragment | NA | NA | 0 |
| S5_k141_213586 | 5672 | No | NA | 6 | 0 | 2 | Not-determined | Genome-fragment | NA | NA | 0 |
| S5_k141_213600 | 3628 | No | NA | 4 | 0 | 0 | Not-determined | Genome-fragment | NA | NA | 0 |
| S5_k141_213621 | 5781 | No | NA | 11 | 0 | 0 | Not-determined | Genome-fragment | NA | NA | 0 |
| S17-k141_3644 | 1794 | No | NA | 6 | 1 | 0 | Low-quality | Genome-fragment | 2.93 | AAI-based (high-confidence) | 0 |
| S7-final-viral-combined_k141_83411\|\|full | 7665 | No | NA | 8 | 0 | 6 | Not-determined | Genome-fragment | NA | NA | 0 |
| S7-final-viral-combined_k141_217226\|\|full | 86323 | No | NA | 111 | 16 | 1 | High-quality | High-quality | 98.84 | AAI-based (high-confidence) | 0 |
| S7-final-viral-combined_k141_139683\|\|full | 32722 | No | NA | 51 | 17 | 1 | High-quality | High-quality | 100 | AAI-based (high-confidence) | 0 |
| S7-final-viral-combined_k141_78243\|\|full | 4292 | No | NA | 5 | 0 | 0 | Not-determined | Genome-fragment | NA | NA | 0 |
| S12-final-viral-combined_k141_144183\|\|full | 1794 | No | NA | 4 | 0 | 0 | Low-quality | Genome-fragment | 4.39 | AAI-based (high-confidence) | 0 |
| S2-final-viral-combined_k141_87812\|\|full | 1794 | No | NA | 2 | 0 | 0 | Low-quality | Genome-fragment | 1.13 | AAI-based (high-confidence) | 0 |
| S7-final-viral-combined_k141_61275\|\|full | 1073 | No | NA | 2 | 0 | 2 | Not-determined | Genome-fragment | NA | NA | 0 |
| S7-final-viral-combined_k141_81707\|\|full | 1739 | No | NA | 2 | 0 | 0 | Not-determined | Genome-fragment | NA | NA | 0 |
| S2_k141_209835 | 1794 | No | NA | 2 | 0 | 0 | Low-quality | Genome-fragment | 4.26 | AAI-based (medium-confidence) | 0 |
| S7-final-viral-combined_k141_132769\|\|full | 2148 | No | NA | 3 | 0 | 1 | Not-determined | Genome-fragment | NA | NA | 0 |
| S1_k141_265277 | 1795 | No | NA | 2 | 0 | 2 | Low-quality | Genome-fragment | 0.79 | AAI-based (medium-confidence) | 0 |
| S7-final-viral-combined_k141_173610\|\|full | 1111 | No | NA | 2 | 0 | 1 | Not-determined | Genome-fragment | NA | NA | 0 |
| S7-final-viral-combined_k141_204249\|\|full | 5416 | No | NA | 6 | 0 | 2 | Not-determined | Genome-fragment | NA | NA | 0 |
| S7-final-viral-combined_k141_81712\|\|full | 7247 | No | NA | 7 | 0 | 6 | Not-determined | Genome-fragment | NA | NA | 0 |
| S7-final-viral-combined_k141_153186\|\|full | 1135 | No | NA | 2 | 0 | 2 | Not-determined | Genome-fragment | NA | NA | 0 |
| S7-final-viral-combined_k141_122549\|\|full | 14133 | No | NA | 13 | 0 | 3 | Not-determined | Genome-fragment | NA | NA | 0 |
| S3-final-viral-combined_k141_129973\|\|full | 1795 | No | NA | 2 | 0 | 2 | Low-quality | Genome-fragment | 3.46 | AAI-based (medium-confidence) | 0 |
| S7-final-viral-combined_k141_204252\|\|full | 2338 | No | NA | 2 | 0 | 2 | Not-determined | Genome-fragment | NA | NA | 0 |
| S7-final-viral-combined_k141_51090\|\|full | 1105 | No | NA | 2 | 0 | 1 | Not-determined | Genome-fragment | NA | NA | 0 |
| S7-final-viral-combined_k141_204265\|\|full | 1340 | No | NA | 3 | 0 | 2 | Not-determined | Genome-fragment | NA | NA | 0 |
| S7-final-viral-combined_k141_153205\|\|full | 3574 | No | NA | 6 | 0 | 0 | Not-determined | Genome-fragment | NA | NA | 0 |
| S5-final-viral-combined_k141_178304\|\|full | 1795 | No | NA | 3 | 2 | 0 | Low-quality | Genome-fragment | 4.22 | AAI-based (high-confidence) | 0 |
| S12-final-viral-combined_k141_368708\|\|full | 1795 | No | NA | 3 | 0 | 1 | Low-quality | Genome-fragment | 1.45 | AAI-based (medium-confidence) | 0 |
| S7-final-viral-combined_k141_102157\|\|full | 2469 | No | NA | 2 | 0 | 1 | Not-determined | Genome-fragment | NA | NA | 0 |
| S7-final-viral-combined_k141_112383\|\|full | 1278 | No | NA | 2 | 0 | 0 | Not-determined | Genome-fragment | NA | NA | 0 |
| S7-final-viral-combined_k141_48\|\|full | 1371 | No | NA | 2 | 0 | 0 | Not-determined | Genome-fragment | NA | NA | 0 |
| S7-final-viral-combined_k141_102162\|\|full | 2858 | No | NA | 3 | 0 | 3 | Not-determined | Genome-fragment | NA | NA | 0 |
| S12-final-viral-combined_k141_341762\|\|full | 1795 | No | NA | 3 | 0 | 2 | Low-quality | Genome-fragment | 1.06 | AAI-based (medium-confidence) | 0 |
| S7-final-viral-combined_k141_91934\|\|full | 3762 | No | NA | 5 | 0 | 4 | Not-determined | Genome-fragment | NA | NA | 0 |
| S9-final-viral-combined_k141_83323\|\|full | 1796 | No | NA | 5 | 1 | 0 | Low-quality | Genome-fragment | 3.36 | AAI-based (high-confidence) | 0 |
| S11-final-viral-combined_k141_239446\|\|full | 1796 | No | NA | 3 | 0 | 0 | Low-quality | Genome-fragment | 3.04 | AAI-based (high-confidence) | 0 |
| S7-final-viral-combined_k141_173668\|\|full | 1950 | No | NA | 3 | 0 | 0 | Not-determined | Genome-fragment | NA | NA | 0 |
| S7-final-viral-combined_k141_51135\|\|full | 2605 | No | NA | 3 | 0 | 0 | Not-determined | Genome-fragment | NA | NA | 0 |
| S11_k141_86516 | 1796 | No | NA | 4 | 2 | 0 | Low-quality | Genome-fragment | 4.04 | AAI-based (high-confidence) | 0 |
| S5-final-viral-combined_k141_181155\|\|full | 1796 | No | NA | 4 | 1 | 0 | Low-quality | Genome-fragment | 3.06 | AAI-based (high-confidence) | 0 |
| S7-final-viral-combined_k141_153268\|\|full | 2829 | No | NA | 3 | 0 | 3 | Not-determined | Genome-fragment | NA | NA | 0 |
| S7-final-viral-combined_k141_173675\|\|full | 12302 | No | NA | 12 | 0 | 5 | Not-determined | Genome-fragment | NA | NA | 0 |
| S17-final-viral-combined_k141_235326\|\|full | 1796 | No | NA | 3 | 0 | 0 | Low-quality | Genome-fragment | 5.01 | AAI-based (medium-confidence) | 0 |
| S7-final-viral-combined_k141_112417\|\|full | 2694 | No | NA | 2 | 0 | 0 | Not-determined | Genome-fragment | NA | NA | 0 |
| S17-final-viral-combined_k141_116411\|\|full | 1796 | No | NA | 3 | 0 | 2 | Low-quality | Genome-fragment | 0.5 | AAI-based (medium-confidence) | 0 |
| S7-final-viral-combined_k141_163490\|\|full | 4679 | No | NA | 5 | 0 | 4 | Not-determined | Genome-fragment | NA | NA | 0 |
| S7-final-viral-combined_k141_118\|\|full | 1847 | No | NA | 2 | 0 | 0 | Not-determined | Genome-fragment | NA | NA | 0 |
| S7-final-viral-combined_k141_120\|\|full | 1816 | No | NA | 4 | 0 | 0 | Not-determined | Genome-fragment | NA | NA | 0 |
| S7-final-viral-combined_k141_194085\|\|full | 2894 | No | NA | 4 | 0 | 3 | Not-determined | Genome-fragment | NA | NA | 0 |
| S7-final-viral-combined_k141_214573\|\|full | 1124 | No | NA | 2 | 0 | 2 | Not-determined | Genome-fragment | NA | NA | 0 |
| S7-final-viral-combined_k141_102206\|\|full | 6069 | No | NA | 6 | 0 | 1 | Not-determined | Genome-fragment | NA | NA | 0 |
| S17-k141_142076 | 1796 | No | NA | 2 | 0 | 1 | Low-quality | Genome-fragment | 4.25 | AAI-based (medium-confidence) | 0 |
| S7-final-viral-combined_k141_153325\|\|full | 1352 | No | NA | 2 | 0 | 0 | Not-determined | Genome-fragment | NA | NA | 0 |
| S4-final-viral-combined_k141_26564\|\|full | 1796 | No | NA | 2 | 0 | 1 | Low-quality | Genome-fragment | 1.49 | AAI-based (medium-confidence) | 0 |
| S7-final-viral-combined_k141_40945\|\|full | 2058 | No | NA | 4 | 0 | 1 | Not-determined | Genome-fragment | NA | NA | 0 |
| S11-final-viral-combined_k141_180550\|\|full | 1797 | No | NA | 4 | 3 | 0 | Low-quality | Genome-fragment | 3.74 | AAI-based (high-confidence) | 0 |
| S7-final-viral-combined_k141_51173\|\|full | 2640 | No | NA | 3 | 0 | 2 | Not-determined | Genome-fragment | NA | NA | 0 |
| S3-final-viral-combined_k141_93142\|\|full | 1797 | No | NA | 5 | 1 | 0 | Low-quality | Genome-fragment | 1.14 | HMM-based (lower-bound) | 0 |
| S7-final-viral-combined_k141_224779\|\|full | 1831 | No | NA | 3 | 0 | 3 | Not-determined | Genome-fragment | NA | NA | 0 |
| S3_k141_23093 | 1797 | No | NA | 2 | 0 | 1 | Low-quality | Genome-fragment | 1.28 | AAI-based (high-confidence) | 0 |
| S5-final-viral-combined_k141_83389\|\|full | 1797 | No | NA | 3 | 2 | 0 | Low-quality | Genome-fragment | 3.79 | AAI-based (medium-confidence) | 0 |
| S7-final-viral-combined_k141_112485\|\|full | 3486 | No | NA | 4 | 0 | 1 | Not-determined | Genome-fragment | NA | NA | 0 |
| S7-final-viral-combined_k141_92032\|\|full | 1319 | No | NA | 3 | 0 | 2 | Not-determined | Genome-fragment | NA | NA | 0 |
| S7-final-viral-combined_k141_20484\|\|full | 5740 | No | NA | 5 | 0 | 3 | Not-determined | Genome-fragment | NA | NA | 0 |
| S7-final-viral-combined_k141_102260\|\|full | 1364 | No | NA | 3 | 0 | 2 | Not-determined | Genome-fragment | NA | NA | 0 |
| S7-final-viral-combined_k141_207902\|\|full | 1797 | No | NA | 3 | 0 | 0 | Low-quality | Genome-fragment | 4.35 | AAI-based (high-confidence) | 0 |
| S7_k141_195889 | 1797 | No | NA | 1 | 0 | 1 | Low-quality | Genome-fragment | 0.31 | AAI-based (high-confidence) | 0 |
| S5-final-viral-combined_k141_63859\|\|full | 1798 | No | NA | 2 | 0 | 2 | Low-quality | Genome-fragment | 1.28 | AAI-based (high-confidence) | 0 |
| S7-final-viral-combined_k141_214635\|\|full | 1405 | No | NA | 2 | 0 | 0 | Not-determined | Genome-fragment | NA | NA | 0 |
| S7-final-viral-combined_k141_20515\|\|full | 4707 | No | NA | 7 | 0 | 3 | Not-determined | Genome-fragment | NA | NA | 0 |
| S5-final-viral-combined_k141_52572\|\|full | 1798 | No | NA | 3 | 0 | 0 | Low-quality | Genome-fragment | 3 | AAI-based (medium-confidence) | 0 |
| S7-final-viral-combined_k141_229\|\|full | 4328 | No | NA | 2 | 0 | 2 | Not-determined | Genome-fragment | NA | NA | 0 |
| S12-final-viral-combined_k141_225855\|\|full | 1798 | No | NA | 3 | 0 | 1 | Low-quality | Genome-fragment | 1.21 | AAI-based (medium-confidence) | 0 |
| S7-final-viral-combined_k141_132826\|\|full | 1127 | No | NA | 3 | 0 | 1 | Not-determined | Genome-fragment | NA | NA | 0 |
| S12-final-viral-combined_k141_143387\|\|full | 1798 | No | NA | 3 | 0 | 3 | Low-quality | Genome-fragment | 1.58 | AAI-based (medium-confidence) | 0 |
| S7-final-viral-combined_k141_92069\|\|full | 1129 | No | NA | 3 | 0 | 1 | Not-determined | Genome-fragment | NA | NA | 0 |
| S12_k141_107653 | 1798 | No | NA | 1 | 0 | 1 | Low-quality | Genome-fragment | 0.67 | AAI-based (high-confidence) | 0 |
| S4-final-viral-combined_k141_19054\|\|full | 1798 | No | NA | 2 | 0 | 1 | Low-quality | Genome-fragment | 2.66 | AAI-based (medium-confidence) | 0 |
| S7-final-viral-combined_k141_132842\|\|full | 1529 | No | NA | 2 | 0 | 0 | Not-determined | Genome-fragment | NA | NA | 0 |
| S6-final-viral-combined_k141_129179\|\|full | 1798 | No | NA | 3 | 3 | 0 | Low-quality | Genome-fragment | 2.93 | AAI-based (high-confidence) | 0 |
| S8-final-viral-combined_k141_199028\|\|full | 1798 | No | NA | 2 | 0 | 0 | Low-quality | Genome-fragment | 3.48 | AAI-based (medium-confidence) | 0 |
| S7-final-viral-combined_k141_71592\|\|full | 38751 | Yes | 27062 | 53 | 20 | 8 | Medium-quality | Genome-fragment | 78.69 | AAI-based (high-confidence) | 30.16 |
| S7-final-viral-combined_k141_256\|\|full | 1576 | No | NA | 2 | 0 | 1 | Not-determined | Genome-fragment | NA | NA | 0 |
| S9-final-viral-combined_k141_33481\|\|full | 1799 | No | NA | 2 | 0 | 1 | Low-quality | Genome-fragment | 1.48 | AAI-based (high-confidence) | 0 |
| S7-final-viral-combined_k141_112560\|\|full | 4540 | No | NA | 5 | 0 | 3 | Not-determined | Genome-fragment | NA | NA | 0 |
| S11-final-viral-combined_k141_196395\|\|full | 1799 | No | NA | 2 | 0 | 0 | Low-quality | Genome-fragment | 3.75 | AAI-based (medium-confidence) | 0 |
| S7-final-viral-combined_k141_30793\|\|full | 1263 | No | NA | 3 | 0 | 0 | Not-determined | Genome-fragment | NA | NA | 0 |
| S7-final-viral-combined_k141_235091\|\|full | 1620 | No | NA | 3 | 0 | 3 | Not-determined | Genome-fragment | NA | NA | 0 |
| S11-final-viral-combined_k141_6560\|\|full | 1799 | No | NA | 3 | 0 | 1 | Low-quality | Genome-fragment | 4.07 | AAI-based (high-confidence) | 0 |
| S11-final-viral-combined_k141_50348\|\|full | 1799 | No | NA | 3 | 3 | 0 | Low-quality | Genome-fragment | 4.24 | AAI-based (high-confidence) | 0 |
| S11-final-viral-combined_k141_327609\|\|lt2gene | 1799 | No | NA | 2 | 2 | 0 | Low-quality | Genome-fragment | 3.93 | AAI-based (high-confidence) | 0 |
| S7-final-viral-combined_k141_10331\|\|full | 4641 | No | NA | 4 | 0 | 3 | Not-determined | Genome-fragment | NA | NA | 0 |
| S7-final-viral-combined_k141_214721\|\|full | 8425 | No | NA | 6 | 0 | 5 | Not-determined | Genome-fragment | NA | NA | 0 |
| S7-final-viral-combined_k141_112627\|\|full | 3346 | No | NA | 4 | 0 | 2 | Not-determined | Genome-fragment | NA | NA | 0 |
| S7-final-viral-combined_k141_20568\|\|full | 3193 | No | NA | 2 | 0 | 2 | Not-determined | Genome-fragment | NA | NA | 0 |
| S1-final-viral-combined_k141_52256\|\|full | 1799 | No | NA | 5 | 0 | 2 | Low-quality | Genome-fragment | 3.14 | AAI-based (medium-confidence) | 0 |
| S7-final-viral-combined_k141_122798\|\|full | 2687 | No | NA | 3 | 0 | 0 | Not-determined | Genome-fragment | NA | NA | 0 |
| S7-final-viral-combined_k141_132940\|\|full | 1949 | No | NA | 2 | 0 | 1 | Not-determined | Genome-fragment | NA | NA | 0 |
| S7-final-viral-combined_k141_132952\|\|full | 1463 | No | NA | 3 | 0 | 2 | Not-determined | Genome-fragment | NA | NA | 0 |
| S7-final-viral-combined_k141_204441\|\|full | 4470 | No | NA | 4 | 0 | 3 | Not-determined | Genome-fragment | NA | NA | 0 |
| S7-final-viral-combined_k141_173846\|\|full | 9774 | No | NA | 7 | 0 | 2 | Not-determined | Genome-fragment | NA | NA | 0 |
| S7-final-viral-combined_k141_224954\|\|full | 4180 | No | NA | 7 | 0 | 1 | Not-determined | Genome-fragment | NA | NA | 0 |
| S7-final-viral-combined_k141_194300\|\|full | 1235 | No | NA | 4 | 0 | 0 | Not-determined | Genome-fragment | NA | NA | 0 |
| S17-final-viral-combined_k141_236976\|\|full | 1799 | No | NA | 3 | 0 | 1 | Low-quality | Genome-fragment | 0.61 | AAI-based (medium-confidence) | 0 |
| S7-final-viral-combined_k141_20605\|\|full | 1508 | No | NA | 2 | 0 | 0 | Not-determined | Genome-fragment | NA | NA | 0 |
| S7-final-viral-combined_k141_320\|\|full | 1372 | No | NA | 2 | 0 | 0 | Not-determined | Genome-fragment | NA | NA | 0 |
| S7-final-viral-combined_k141_81801\|\|full | 1280 | No | NA | 2 | 0 | 1 | Not-determined | Genome-fragment | NA | NA | 0 |
| S2-final-viral-combined_k141_137303\|\|full | 1799 | No | NA | 2 | 0 | 2 | Low-quality | Genome-fragment | 0.44 | AAI-based (medium-confidence) | 0 |
| S4-final-viral-combined_k141_67740\|\|full | 1799 | No | NA | 2 | 0 | 2 | Low-quality | Genome-fragment | 2.44 | AAI-based (medium-confidence) | 0 |
| S7-final-viral-combined_k141_153434\|\|full | 1655 | No | NA | 2 | 0 | 1 | Not-determined | Genome-fragment | NA | NA | 0 |
| S11-final-viral-combined_k141_84554\|\|full | 1800 | No | NA | 4 | 1 | 0 | Low-quality | Genome-fragment | 4.47 | AAI-based (high-confidence) | 0 |
| S7-final-viral-combined_k141_235165\|\|full | 31876 | No | NA | 55 | 14 | 0 | High-quality | High-quality | 91.18 | AAI-based (high-confidence) | 0 |
| S7-final-viral-combined_k141_20650\|\|full | 1024 | No | NA | 1 | 0 | 0 | Not-determined | Genome-fragment | NA | NA | 0 |
| S7-final-viral-combined_k141_20654\|\|full | 1935 | No | NA | 3 | 0 | 1 | Not-determined | Genome-fragment | NA | NA | 0 |
| S7-final-viral-combined_k141_194339\|\|full | 1602 | No | NA | 4 | 0 | 1 | Not-determined | Genome-fragment | NA | NA | 0 |
| S7-final-viral-combined_k141_133020\|\|full | 1655 | No | NA | 2 | 0 | 0 | Not-determined | Genome-fragment | NA | NA | 0 |
| S7-final-viral-combined_k141_133025\|\|full | 5175 | No | NA | 5 | 0 | 2 | Not-determined | Genome-fragment | NA | NA | 0 |
| S1-final-viral-combined_k141_352639\|\|full | 1800 | No | NA | 3 | 0 | 0 | Low-quality | Genome-fragment | 2.41 | AAI-based (high-confidence) | 0 |
| S7-final-viral-combined_k141_81917\|\|full | 4016 | No | NA | 6 | 0 | 3 | Not-determined | Genome-fragment | NA | NA | 0 |
| S7_k141_97008 | 1800 | No | NA | 1 | 0 | 0 | Low-quality | Genome-fragment | 3.65 | AAI-based (high-confidence) | 0 |
| S7-final-viral-combined_k141_163830\|\|full | 2850 | No | NA | 4 | 0 | 3 | Not-determined | Genome-fragment | NA | NA | 0 |
| S7-final-viral-combined_k141_61633\|\|full | 1671 | No | NA | 2 | 0 | 0 | Not-determined | Genome-fragment | NA | NA | 0 |
| S7-final-viral-combined_k141_173938\|\|full | 2145 | No | NA | 6 | 0 | 2 | Not-determined | Genome-fragment | NA | NA | 0 |
| S7-final-viral-combined_k141_225068\|\|full | 2019 | No | NA | 3 | 0 | 3 | Not-determined | Genome-fragment | NA | NA | 0 |
| S12-final-viral-combined_k141_229674\|\|full | 1800 | No | NA | 5 | 0 | 0 | Low-quality | Genome-fragment | 2.75 | AAI-based (medium-confidence) | 0 |
| S2_k141_190591 | 1800 | No | NA | 2 | 0 | 2 | Low-quality | Genome-fragment | 1.21 | AAI-based (high-confidence) | 0 |
| S4_k141_136824 | 1800 | No | NA | 7 | 1 | 0 | Low-quality | Genome-fragment | 5.03 | AAI-based (high-confidence) | 0 |
| S5-final-viral-combined_k141_98305\|\|full | 1801 | No | NA | 3 | 0 | 1 | Low-quality | Genome-fragment | 2.45 | AAI-based (medium-confidence) | 0 |
| S5-final-viral-combined_k141_61410\|\|lt2gene | 1801 | No | NA | 1 | 1 | 0 | Low-quality | Genome-fragment | 4.37 | AAI-based (high-confidence) | 0 |
| S7-final-viral-combined_k141_163882\|\|full | 1200 | No | NA | 2 | 0 | 1 | Not-determined | Genome-fragment | NA | NA | 0 |
| S12-final-viral-combined_k141_3581\|\|full | 1801 | No | NA | 3 | 0 | 1 | Low-quality | Genome-fragment | 4.11 | AAI-based (medium-confidence) | 0 |
| S12-final-viral-combined_k141_355814\|\|lt2gene | 1801 | No | NA | 2 | 2 | 0 | Low-quality | Genome-fragment | 4.21 | AAI-based (high-confidence) | 0 |
| S12-final-viral-combined_k141_156206\|\|lt2gene | 1801 | No | NA | 1 | 1 | 0 | Low-quality | Genome-fragment | 5.27 | AAI-based (high-confidence) | 0 |
| S12_k141_121419 | 1801 | No | NA | 3 | 0 | 1 | Low-quality | Genome-fragment | 3.87 | AAI-based (medium-confidence) | 0 |
| S2-final-viral-combined_k141_126217\|\|full | 1801 | No | NA | 3 | 3 | 0 | Low-quality | Genome-fragment | 4.24 | AAI-based (high-confidence) | 0 |
| S4-final-viral-combined_k141_154113\|\|full | 1801 | No | NA | 4 | 0 | 0 | Low-quality | Genome-fragment | 4.34 | AAI-based (medium-confidence) | 0 |
| S6_k141_89643 | 1801 | No | NA | 2 | 2 | 0 | Low-quality | Genome-fragment | 3.52 | AAI-based (medium-confidence) | 0 |
| S7-final-viral-combined_k141_20707\|\|full | 2085 | No | NA | 3 | 0 | 2 | Not-determined | Genome-fragment | NA | NA | 0 |
| S8-final-viral-combined_k141_198030\|\|full | 1801 | No | NA | 3 | 0 | 1 | Low-quality | Genome-fragment | 8.07 | AAI-based (medium-confidence) | 0 |
| S7-final-viral-combined_k141_112861\|\|full | 1820 | No | NA | 3 | 0 | 1 | Not-determined | Genome-fragment | NA | NA | 0 |
| S7-final-viral-combined_k141_163941\|\|full | 1412 | No | NA | 2 | 0 | 1 | Not-determined | Genome-fragment | NA | NA | 0 |
| S1-final-viral-combined_k141_46585\|\|full | 1802 | No | NA | 2 | 0 | 0 | Low-quality | Genome-fragment | 1.72 | AAI-based (medium-confidence) | 0 |
| S7-final-viral-combined_k141_153671\|\|full | 1250 | No | NA | 3 | 0 | 0 | Not-determined | Genome-fragment | NA | NA | 0 |
| S7-final-viral-combined_k141_225176\|\|full | 3390 | No | NA | 5 | 0 | 2 | Not-determined | Genome-fragment | NA | NA | 0 |
| S3-final-viral-combined_k141_92203\|\|full | 1802 | No | NA | 2 | 0 | 0 | Low-quality | Genome-fragment | 2.29 | AAI-based (medium-confidence) | 0 |
| S7-final-viral-combined_k141_82043\|\|full | 2436 | No | NA | 4 | 0 | 3 | Not-determined | Genome-fragment | NA | NA | 0 |
| S5-final-viral-combined_k141_194999\|\|full | 1802 | No | NA | 3 | 0 | 1 | Low-quality | Genome-fragment | 2.02 | AAI-based (high-confidence) | 0 |
| S7-final-viral-combined_k141_204610\|\|full | 4266 | No | NA | 5 | 0 | 2 | Not-determined | Genome-fragment | NA | NA | 0 |
| S7-final-viral-combined_k141_133172\|\|full | 4923 | No | NA | 6 | 0 | 5 | Not-determined | Genome-fragment | NA | NA | 0 |
| S7-final-viral-combined_k141_82062\|\|full | 1235 | No | NA | 2 | 0 | 0 | Not-determined | Genome-fragment | NA | NA | 0 |
| S5-final-viral-combined_k141_145194\|\|lt2gene | 1802 | No | NA | 2 | 1 | 0 | Low-quality | Genome-fragment | 4.65 | AAI-based (high-confidence) | 0 |
| S7-final-viral-combined_k141_102524\|\|full | 4408 | No | NA | 7 | 0 | 1 | Not-determined | Genome-fragment | NA | NA | 0 |
| S7-final-viral-combined_k141_235402\|\|full | 3683 | No | NA | 8 | 0 | 0 | Not-determined | Genome-fragment | NA | NA | 0 |
| S5_k141_100339 | 1802 | No | NA | 2 | 0 | 0 | Low-quality | Genome-fragment | 1.07 | AAI-based (high-confidence) | 0 |
| S7-final-viral-combined_k141_194644\|\|full | 5677 | No | NA | 6 | 0 | 0 | Not-determined | Genome-fragment | NA | NA | 0 |
| S7-final-viral-combined_k141_222785\|\|full | 1802 | No | NA | 3 | 0 | 0 | Low-quality | Genome-fragment | 3.09 | AAI-based (high-confidence) | 0 |
| S7-final-viral-combined_k141_174074\|\|full | 2638 | No | NA | 3 | 0 | 1 | Not-determined | Genome-fragment | NA | NA | 0 |
| S7-final-viral-combined_k141_194656\|\|full | 2800 | No | NA | 3 | 0 | 1 | Not-determined | Genome-fragment | NA | NA | 0 |
| S7-final-viral-combined_k141_225260\|\|full | 15940 | No | NA | 19 | 7 | 0 | High-quality | High-quality | 100 | AAI-based (high-confidence) | 0 |
| S12-final-viral-combined_k141_30931\|\|full | 1802 | No | NA | 2 | 0 | 0 | Low-quality | Genome-fragment | 4.65 | AAI-based (medium-confidence) | 0 |
| S7-final-viral-combined_k141_122944\|\|full | 5428 | No | NA | 7 | 0 | 2 | Not-determined | Genome-fragment | NA | NA | 0 |
| S7-final-viral-combined_k141_31068\|\|full | 27877 | Yes | 18895 | 43 | 7 | 6 | Medium-quality | Genome-fragment | 53.79 | AAI-based (high-confidence) | 32.22 |
| S2-final-viral-combined_k141_162100\|\|full | 1802 | No | NA | 3 | 0 | 2 | Low-quality | Genome-fragment | 0.79 | AAI-based (medium-confidence) | 0 |
| S2_k141_83872 | 1802 | No | NA | 5 | 0 | 0 | Low-quality | Genome-fragment | 2.79 | AAI-based (medium-confidence) | 0 |
| S7-final-viral-combined_k141_71784\|\|full | 3130 | No | NA | 3 | 0 | 0 | Not-determined | Genome-fragment | NA | NA | 0 |
| S9_k141_14273 | 1803 | No | NA | 2 | 0 | 0 | Low-quality | Genome-fragment | 3.24 | AAI-based (medium-confidence) | 0 |
| S7-final-viral-combined_k141_92409\|\|full | 1303 | No | NA | 2 | 0 | 1 | Not-determined | Genome-fragment | NA | NA | 0 |
| S5-final-viral-combined_k141_212258\|\|full | 1803 | No | NA | 3 | 0 | 0 | Low-quality | Genome-fragment | 3.11 | AAI-based (medium-confidence) | 0 |
| S17-final-viral-combined_k141_256045\|\|full | 1803 | No | NA | 3 | 1 | 0 | Low-quality | Genome-fragment | 4.72 | AAI-based (high-confidence) | 0 |
| S7-final-viral-combined_k141_82159\|\|full | 1549 | No | NA | 3 | 0 | 1 | Not-determined | Genome-fragment | NA | NA | 0 |
| S7-final-viral-combined_k141_605\|\|full | 1247 | No | NA | 2 | 0 | 2 | Not-determined | Genome-fragment | NA | NA | 0 |
| S7-final-viral-combined_k141_225326\|\|full | 2542 | No | NA | 3 | 0 | 2 | Not-determined | Genome-fragment | NA | NA | 0 |
| S7-final-viral-combined_k141_20860\|\|full | 2667 | No | NA | 6 | 0 | 0 | Not-determined | Genome-fragment | NA | NA | 0 |
| S12-final-viral-combined_k141_377962\|\|full | 1803 | No | NA | 3 | 0 | 0 | Low-quality | Genome-fragment | 2.57 | AAI-based (medium-confidence) | 0 |
| S7-final-viral-combined_k141_20865\|\|full | 1618 | No | NA | 2 | 0 | 1 | Not-determined | Genome-fragment | NA | NA | 0 |
| S7-final-viral-combined_k141_630\|\|full | 1325 | No | NA | 3 | 0 | 0 | Not-determined | Genome-fragment | NA | NA | 0 |
| S7-final-viral-combined_k141_225344\|\|full | 1790 | No | NA | 2 | 0 | 2 | Not-determined | Genome-fragment | NA | NA | 0 |
| S7-final-viral-combined_k141_61874\|\|full | 1657 | No | NA | 3 | 0 | 1 | Not-determined | Genome-fragment | NA | NA | 0 |
| S7-final-viral-combined_k141_225357\|\|full | 2550 | No | NA | 6 | 0 | 3 | Not-determined | Genome-fragment | NA | NA | 0 |
| S2-final-viral-combined_k141_202552\|\|full | 1803 | No | NA | 3 | 1 | 0 | Low-quality | Genome-fragment | 1.22 | AAI-based (high-confidence) | 0 |
| S7-final-viral-combined_k141_41265\|\|full | 23836 | No | NA | 22 | 0 | 16 | Not-determined | Genome-fragment | NA | NA | 0 |
| S7-final-viral-combined_k141_215082\|\|full | 2483 | No | NA | 2 | 0 | 2 | Not-determined | Genome-fragment | NA | NA | 0 |
| S7-final-viral-combined_k141_681\|\|full | 1369 | No | NA | 4 | 0 | 0 | Not-determined | Genome-fragment | NA | NA | 0 |
| S7-final-viral-combined_k141_102655\|\|full | 2095 | No | NA | 2 | 0 | 2 | Not-determined | Genome-fragment | NA | NA | 0 |
| S7-final-viral-combined_k141_215101\|\|full | 2682 | No | NA | 3 | 0 | 0 | Not-determined | Genome-fragment | NA | NA | 0 |
| S7-final-viral-combined_k141_102666\|\|full | 1079 | No | NA | 2 | 0 | 0 | Not-determined | Genome-fragment | NA | NA | 0 |
| S8-final-viral-combined_k141_159370\|\|full | 1803 | No | NA | 3 | 0 | 0 | Low-quality | Genome-fragment | 2.8 | AAI-based (medium-confidence) | 0 |
| S11-final-viral-combined_k141_230929\|\|full | 1804 | No | NA | 2 | 0 | 2 | Low-quality | Genome-fragment | 0.69 | AAI-based (medium-confidence) | 0 |
| S7-final-viral-combined_k141_194833\|\|full | 8159 | No | NA | 13 | 0 | 3 | Not-determined | Genome-fragment | NA | NA | 0 |
| S7-final-viral-combined_k141_20879\|\|full | 3287 | No | NA | 4 | 0 | 0 | Not-determined | Genome-fragment | NA | NA | 0 |
| S7-final-viral-combined_k141_153937\|\|full | 8940 | No | NA | 10 | 0 | 5 | Not-determined | Genome-fragment | NA | NA | 0 |
| S11-final-viral-combined_k141_54681\|\|full | 1804 | No | NA | 3 | 0 | 2 | Low-quality | Genome-fragment | 5.11 | AAI-based (high-confidence) | 0 |
| S7-final-viral-combined_k141_235567\|\|full | 1677 | No | NA | 2 | 0 | 1 | Not-determined | Genome-fragment | NA | NA | 0 |
| S7-final-viral-combined_k141_51418\|\|full | 1316 | No | NA | 2 | 0 | 0 | Not-determined | Genome-fragment | NA | NA | 0 |
| S7-final-viral-combined_k141_51439\|\|full | 1905 | No | NA | 4 | 0 | 1 | Not-determined | Genome-fragment | NA | NA | 0 |
| S1-final-viral-combined_k141_273233\|\|full | 1804 | No | NA | 3 | 1 | 0 | Low-quality | Genome-fragment | 1.34 | AAI-based (high-confidence) | 0 |
| S1_k141_23140 | 1804 | No | NA | 2 | 0 | 0 | Low-quality | Genome-fragment | 3.41 | AAI-based (high-confidence) | 0 |
| S7-final-viral-combined_k141_153972\|\|full | 1699 | No | NA | 2 | 0 | 2 | Not-determined | Genome-fragment | NA | NA | 0 |
| S1_k141_155854 | 1804 | No | NA | 2 | 0 | 1 | Low-quality | Genome-fragment | 3.51 | AAI-based (high-confidence) | 0 |
| S7-final-viral-combined_k141_235609\|\|full | 2334 | No | NA | 3 | 0 | 1 | Not-determined | Genome-fragment | NA | NA | 0 |
| S3-final-viral-combined_k141_103826\|\|full | 1804 | No | NA | 2 | 0 | 2 | Low-quality | Genome-fragment | 0.81 | AAI-based (medium-confidence) | 0 |
| S7-final-viral-combined_k141_102675\|\|full | 1935 | No | NA | 3 | 0 | 2 | Not-determined | Genome-fragment | NA | NA | 0 |
| S7-final-viral-combined_k141_123085\|\|full | 10061 | No | NA | 11 | 0 | 1 | Medium-quality | Genome-fragment | 70.57 | AAI-based (high-confidence) | 0 |
| S3-final-viral-combined_k141_18544\|\|full | 1804 | No | NA | 4 | 0 | 0 | Low-quality | Genome-fragment | 2.86 | AAI-based (high-confidence) | 0 |
| S5-final-viral-combined_k141_100371\|\|full | 1804 | No | NA | 3 | 2 | 0 | Low-quality | Genome-fragment | 1.04 | AAI-based (high-confidence) | 0 |
| S17-final-viral-combined_k141_219352\|\|full | 1804 | No | NA | 2 | 0 | 0 | Low-quality | Genome-fragment | 2.08 | AAI-based (medium-confidence) | 0 |
| S7-final-viral-combined_k141_82237\|\|full | 3719 | No | NA | 6 | 0 | 2 | Not-determined | Genome-fragment | NA | NA | 0 |
| S7-final-viral-combined_k141_153999\|\|full | 6849 | No | NA | 7 | 0 | 5 | Not-determined | Genome-fragment | NA | NA | 0 |
| S7-final-viral-combined_k141_71973\|\|full | 3184 | No | NA | 2 | 0 | 1 | Not-determined | Genome-fragment | NA | NA | 0 |
| S7-final-viral-combined_k141_164120\|\|full | 3814 | No | NA | 6 | 0 | 2 | Not-determined | Genome-fragment | NA | NA | 0 |
| S7-final-viral-combined_k141_204853\|\|full | 1397 | No | NA | 3 | 0 | 3 | Not-determined | Genome-fragment | NA | NA | 0 |
| S17-final-viral-combined_k141_199825\|\|full | 1804 | No | NA | 4 | 2 | 0 | Low-quality | Genome-fragment | 3.89 | AAI-based (high-confidence) | 0 |
| S7-final-viral-combined_k141_194939\|\|full | 1898 | No | NA | 3 | 0 | 0 | Not-determined | Genome-fragment | NA | NA | 0 |
| S2-final-viral-combined_k141_128443\|\|full | 1804 | No | NA | 2 | 0 | 0 | Low-quality | Genome-fragment | 2.92 | AAI-based (high-confidence) | 0 |
| S7-final-viral-combined_k141_61971\|\|full | 1008 | No | NA | 3 | 0 | 0 | Not-determined | Genome-fragment | NA | NA | 0 |
| S6-final-viral-combined_k141_57322\|\|lt2gene | 1804 | No | NA | 1 | 1 | 0 | Low-quality | Genome-fragment | 2.14 | AAI-based (high-confidence) | 0 |
| S7-final-viral-combined_k141_215272\|\|full | 2360 | No | NA | 3 | 0 | 2 | Not-determined | Genome-fragment | NA | NA | 0 |
| S8-final-viral-combined_k141_186689\|\|full | 1804 | No | NA | 3 | 0 | 0 | Low-quality | Genome-fragment | 3.01 | AAI-based (medium-confidence) | 0 |
| S7-final-viral-combined_k141_72015\|\|full | 8935 | No | NA | 5 | 0 | 4 | Not-determined | Genome-fragment | NA | NA | 0 |
| S7-final-viral-combined_k141_20921\|\|full | 177205 | Yes | 142150 | 206 | 32 | 28 | Medium-quality | Genome-fragment | 81.32 | AAI-based (high-confidence) | 19.78 |
| S7-final-viral-combined_k141_133449\|\|full | 2767 | No | NA | 5 | 0 | 3 | Not-determined | Genome-fragment | NA | NA | 0 |
| S7-final-viral-combined_k141_123227\|\|full | 7346 | No | NA | 10 | 0 | 0 | Not-determined | Genome-fragment | NA | NA | 0 |
| S7-final-viral-combined_k141_72027\|\|full | 1217 | No | NA | 3 | 0 | 1 | Not-determined | Genome-fragment | NA | NA | 0 |
| S7-final-viral-combined_k141_235716\|\|full | 2583 | No | NA | 3 | 0 | 2 | Not-determined | Genome-fragment | NA | NA | 0 |
| S11-final-viral-combined_k141_173092\|\|full | 1805 | No | NA | 2 | 1 | 0 | Low-quality | Genome-fragment | 3.34 | AAI-based (medium-confidence) | 0 |
| S7-final-viral-combined_k141_194979\|\|full | 15543 | No | NA | 17 | 0 | 12 | Not-determined | Genome-fragment | NA | NA | 0 |
| S1-final-viral-combined_k141_36181\|\|full | 1805 | No | NA | 3 | 0 | 3 | Low-quality | Genome-fragment | 0.87 | AAI-based (medium-confidence) | 0 |
| S7-final-viral-combined_k141_72038\|\|full | 4463 | No | NA | 5 | 0 | 2 | Not-determined | Genome-fragment | NA | NA | 0 |
| S5-final-viral-combined_k141_100607\|\|full | 1805 | No | NA | 3 | 0 | 1 | Low-quality | Genome-fragment | 2.76 | AAI-based (medium-confidence) | 0 |
| S7-final-viral-combined_k141_878\|\|full | 2549 | No | NA | 3 | 0 | 2 | Not-determined | Genome-fragment | NA | NA | 0 |
| S17-k141_35251 | 1805 | No | NA | 2 | 0 | 1 | Low-quality | Genome-fragment | 1.18 | AAI-based (high-confidence) | 0 |
| S7-final-viral-combined_k141_174302\|\|full | 4033 | No | NA | 4 | 0 | 2 | Not-determined | Genome-fragment | NA | NA | 0 |
| S7-final-viral-combined_k141_62022\|\|full | 4665 | No | NA | 4 | 0 | 4 | Not-determined | Genome-fragment | NA | NA | 0 |
| S7-final-viral-combined_k141_215344\|\|full | 3203 | No | NA | 5 | 0 | 0 | Not-determined | Genome-fragment | NA | NA | 0 |
| S4-final-viral-combined_k141_136464\|\|full | 1805 | No | NA | 3 | 0 | 1 | Low-quality | Genome-fragment | 1.35 | AAI-based (medium-confidence) | 0 |
| S8-final-viral-combined_k141_207642\|\|full | 1805 | No | NA | 2 | 0 | 0 | Low-quality | Genome-fragment | 2.42 | AAI-based (medium-confidence) | 0 |
| S9-final-viral-combined_k141_70537\|\|full | 1806 | No | NA | 2 | 0 | 1 | Low-quality | Genome-fragment | 0.9 | AAI-based (medium-confidence) | 0 |
| S7-final-viral-combined_k141_225480\|\|full | 1035 | No | NA | 3 | 0 | 1 | Not-determined | Genome-fragment | NA | NA | 0 |
| S9-final-viral-combined_k141_85102\|\|full | 1806 | No | NA | 4 | 3 | 0 | Low-quality | Genome-fragment | 5.31 | AAI-based (high-confidence) | 0 |
| S3_k141_32012 | 1806 | No | NA | 1 | 1 | 0 | Low-quality | Genome-fragment | 4.31 | AAI-based (medium-confidence) | 0 |
| S7-final-viral-combined_k141_10923\|\|full | 9075 | No | NA | 3 | 0 | 1 | Not-determined | Genome-fragment | NA | NA | 0 |
| S12-final-viral-combined_k141_390490\|\|full | 1806 | No | NA | 3 | 0 | 0 | Low-quality | Genome-fragment | 4.03 | AAI-based (medium-confidence) | 0 |
| S7-final-viral-combined_k141_41445\|\|full | 4885 | No | NA | 6 | 0 | 0 | Not-determined | Genome-fragment | NA | NA | 0 |
| S2-final-viral-combined_k141_122223\|\|full | 1806 | No | NA | 2 | 0 | 1 | Low-quality | Genome-fragment | 2.95 | AAI-based (medium-confidence) | 0 |
| S7-final-viral-combined_k141_92556\|\|full | 7718 | No | NA | 9 | 0 | 6 | Not-determined | Genome-fragment | NA | NA | 0 |
| S7-final-viral-combined_k141_20975\|\|full | 1007 | No | NA | 1 | 0 | 1 | Not-determined | Genome-fragment | NA | NA | 0 |
| S7-final-viral-combined_k141_205009\|\|full | 2644 | No | NA | 3 | 0 | 1 | Not-determined | Genome-fragment | NA | NA | 0 |
| S6_k141_81800 | 1806 | No | NA | 2 | 0 | 2 | Low-quality | Genome-fragment | 1.18 | AAI-based (high-confidence) | 0 |
| S11-final-viral-combined_k141_310217\|\|full | 1807 | No | NA | 3 | 3 | 0 | Low-quality | Genome-fragment | 4.31 | AAI-based (high-confidence) | 0 |
| S11-final-viral-combined_k141_118831\|\|full | 1807 | No | NA | 3 | 0 | 0 | Low-quality | Genome-fragment | 3.61 | AAI-based (medium-confidence) | 0 |
| S7-final-viral-combined_k141_205024\|\|full | 5020 | No | NA | 6 | 0 | 5 | Not-determined | Genome-fragment | NA | NA | 0 |
| S7-final-viral-combined_k141_51724\|\|full | 3370 | No | NA | 6 | 0 | 2 | Not-determined | Genome-fragment | NA | NA | 0 |
| S7-final-viral-combined_k141_31423\|\|full | 1307 | No | NA | 2 | 0 | 0 | Not-determined | Genome-fragment | NA | NA | 0 |
| S7-final-viral-combined_k141_31429\|\|full | 2229 | No | NA | 3 | 0 | 0 | Not-determined | Genome-fragment | NA | NA | 0 |
| S1-final-viral-combined_k141_280856\|\|full | 1807 | No | NA | 4 | 2 | 0 | Low-quality | Genome-fragment | 4.09 | AAI-based (high-confidence) | 0 |
| S1-final-viral-combined_k141_279928\|\|lt2gene | 1807 | No | NA | 2 | 1 | 0 | Low-quality | Genome-fragment | 5.41 | AAI-based (high-confidence) | 0 |
| S7-final-viral-combined_k141_51741\|\|full | 5096 | No | NA | 8 | 0 | 4 | Not-determined | Genome-fragment | NA | NA | 0 |
| S7-final-viral-combined_k141_41519\|\|full | 1284 | No | NA | 3 | 0 | 2 | Not-determined | Genome-fragment | NA | NA | 0 |
| S12-final-viral-combined_k141_148908\|\|full | 1807 | No | NA | 3 | 3 | 0 | Low-quality | Genome-fragment | 4.64 | AAI-based (high-confidence) | 0 |
| S6-final-viral-combined_k141_130271\|\|full | 1807 | No | NA | 3 | 0 | 2 | Low-quality | Genome-fragment | 1.29 | AAI-based (medium-confidence) | 0 |
| S7-final-viral-combined_k141_11036\|\|full | 1051 | No | NA | 2 | 0 | 0 | Not-determined | Genome-fragment | NA | NA | 0 |
| S7-final-viral-combined_k141_184632\|\|full | 6503 | No | NA | 6 | 0 | 2 | Not-determined | Genome-fragment | NA | NA | 0 |
| S7-final-viral-combined_k141_235786\|\|full | 2971 | No | NA | 3 | 0 | 0 | Not-determined | Genome-fragment | NA | NA | 0 |
| S7-final-viral-combined_k141_184655\|\|full | 2889 | No | NA | 5 | 0 | 2 | Not-determined | Genome-fragment | NA | NA | 0 |
| S7-final-viral-combined_k141_205084\|\|full | 4118 | No | NA | 8 | 0 | 7 | Not-determined | Genome-fragment | NA | NA | 0 |
| S9-final-viral-combined_k141_5730\|\|full | 1808 | No | NA | 2 | 0 | 1 | Low-quality | Genome-fragment | 1.39 | AAI-based (medium-confidence) | 0 |
| S7-final-viral-combined_k141_11068\|\|full | 1566 | No | NA | 3 | 0 | 1 | Not-determined | Genome-fragment | NA | NA | 0 |
| S7-final-viral-combined_k141_82400\|\|full | 1340 | No | NA | 3 | 0 | 0 | Not-determined | Genome-fragment | NA | NA | 0 |
| S5-final-viral-combined_k141_93385\|\|full | 1808 | No | NA | 4 | 0 | 0 | Low-quality | Genome-fragment | 3.83 | AAI-based (medium-confidence) | 0 |
| S17-final-viral-combined_k141_37798\|\|full | 1808 | No | NA | 4 | 0 | 1 | Low-quality | Genome-fragment | 0.43 | AAI-based (high-confidence) | 0 |
| S2-final-viral-combined_k141_30873\|\|full | 1808 | No | NA | 3 | 0 | 0 | Low-quality | Genome-fragment | 2.73 | AAI-based (high-confidence) | 0 |
| S7-final-viral-combined_k141_235814\|\|full | 5272 | No | NA | 7 | 0 | 0 | Not-determined | Genome-fragment | NA | NA | 0 |
| S7-final-viral-combined_k141_205132\|\|full | 5144 | No | NA | 8 | 0 | 2 | Not-determined | Genome-fragment | NA | NA | 0 |
| S7-final-viral-combined_k141_21075\|\|full | 2371 | No | NA | 4 | 0 | 1 | Not-determined | Genome-fragment | NA | NA | 0 |
| S7-final-viral-combined_k141_82438\|\|full | 2480 | No | NA | 3 | 0 | 1 | Not-determined | Genome-fragment | NA | NA | 0 |
| S4-final-viral-combined_k141_75723\|\|full | 1808 | No | NA | 2 | 0 | 2 | Low-quality | Genome-fragment | 0.92 | AAI-based (medium-confidence) | 0 |
| S7-final-viral-combined_k141_205143\|\|full | 6742 | No | NA | 6 | 0 | 6 | Not-determined | Genome-fragment | NA | NA | 0 |
| S7-final-viral-combined_k141_235835\|\|full | 5412 | No | NA | 7 | 0 | 0 | Not-determined | Genome-fragment | NA | NA | 0 |
| S11-final-viral-combined_k141_136356\|\|full | 1809 | No | NA | 3 | 0 | 0 | Low-quality | Genome-fragment | 3.02 | AAI-based (medium-confidence) | 0 |
| S1-final-viral-combined_k141_42605\|\|full | 1809 | No | NA | 3 | 0 | 0 | Low-quality | Genome-fragment | 3.78 | AAI-based (high-confidence) | 0 |
| S7-final-viral-combined_k141_51866\|\|full | 1649 | No | NA | 2 | 0 | 0 | Not-determined | Genome-fragment | NA | NA | 0 |
| S7-final-viral-combined_k141_235854\|\|full | 4940 | No | NA | 5 | 0 | 0 | Not-determined | Genome-fragment | NA | NA | 0 |
| S7-final-viral-combined_k141_235859\|\|full | 1404 | No | NA | 3 | 0 | 1 | Not-determined | Genome-fragment | NA | NA | 0 |
| S7-final-viral-combined_k141_195218\|\|full | 1841 | No | NA | 5 | 0 | 5 | Not-determined | Genome-fragment | NA | NA | 0 |
| S7-final-viral-combined_k141_143606\|\|full | 1527 | No | NA | 2 | 0 | 1 | Not-determined | Genome-fragment | NA | NA | 0 |
| S3-final-viral-combined_k141_13557\|\|full | 1809 | No | NA | 2 | 0 | 0 | Low-quality | Genome-fragment | 3.78 | AAI-based (high-confidence) | 0 |
| S7-final-viral-combined_k141_143620\|\|full | 2769 | No | NA | 4 | 0 | 0 | Not-determined | Genome-fragment | NA | NA | 0 |
| S7-final-viral-combined_k141_31555\|\|full | 2441 | No | NA | 4 | 0 | 2 | Not-determined | Genome-fragment | NA | NA | 0 |

| | | | | | | | | | | | |
|---|---|---|---|---|---|---|---|---|---|---|---|
| S7-final-viral-combined_k141_225660\|\|full | 1326 | No | NA | 2 | 0 | 1 | Not-determined | Genome-fragment | NA | NA | 0 |
| S7-final-viral-combined_k141_174355\|\|full | 2907 | No | NA | 4 | 0 | 3 | Not-determined | Genome-fragment | NA | NA | 0 |
| S7-final-viral-combined_k141_205197\|\|full | 2829 | No | NA | 5 | 0 | 1 | Not-determined | Genome-fragment | NA | NA | 0 |
| S3_k141_133911 | 1809 | No | NA | 2 | 0 | 0 | Low-quality | Genome-fragment | 0.83 | AAI-based (high-confidence) | 0 |
| S7-final-viral-combined_k141_174378\|\|full | 2715 | No | NA | 3 | 0 | 1 | Not-determined | Genome-fragment | NA | NA | 0 |
| S5-final-viral-combined_k141_92448\|\|full | 1809 | No | NA | 8 | 0 | 0 | Low-quality | Genome-fragment | 4.03 | AAI-based (high-confidence) | 0 |
| S5-final-viral-combined_k141_136943\|\|full | 1809 | No | NA | 4 | 2 | 0 | Low-quality | Genome-fragment | 2.56 | HMM-based (lower-bound) | 0 |
| S7-final-viral-combined_k141_174392\|\|full | 3981 | No | NA | 7 | 0 | 1 | Not-determined | Genome-fragment | NA | NA | 0 |
| S7-final-viral-combined_k141_123499\|\|full | 4003 | No | NA | 6 | 0 | 2 | Not-determined | Genome-fragment | NA | NA | 0 |
| S17-final-viral-combined_k141_27470\|\|full | 1809 | No | NA | 4 | 2 | 0 | Low-quality | Genome-fragment | 5.08 | AAI-based (high-confidence) | 0 |
| S12-final-viral-combined_k141_256819\|\|full | 1809 | No | NA | 3 | 1 | 0 | Low-quality | Genome-fragment | 3.14 | AAI-based (high-confidence) | 0 |
| S7-final-viral-combined_k141_225685\|\|full | 1634 | No | NA | 3 | 0 | 0 | Not-determined | Genome-fragment | NA | NA | 0 |
| S12-final-viral-combined_k141_241177\|\|full | 1809 | No | NA | 6 | 0 | 0 | Low-quality | Genome-fragment | 5.24 | AAI-based (high-confidence) | 0 |
| S7-final-viral-combined_k141_174418\|\|full | 2746 | No | NA | 3 | 0 | 1 | Not-determined | Genome-fragment | NA | NA | 0 |
| S7-final-viral-combined_k141_184872\|\|full | 7850 | No | NA | 6 | 0 | 0 | Not-determined | Genome-fragment | NA | NA | 0 |
| S7-final-viral-combined_k141_133794\|\|full | 2520 | No | NA | 5 | 0 | 0 | Not-determined | Genome-fragment | NA | NA | 0 |
| S7-final-viral-combined_k141_195275\|\|full | 7349 | No | NA | 8 | 0 | 8 | Not-determined | Genome-fragment | NA | NA | 0 |
| S9-final-viral-combined_k141_64290\|\|full | 1810 | No | NA | 6 | 2 | 0 | Low-quality | Genome-fragment | 4.16 | AAI-based (medium-confidence) | 0 |
| S7-final-viral-combined_k141_102833\|\|full | 1238 | No | NA | 2 | 0 | 0 | Not-determined | Genome-fragment | NA | NA | 0 |
| S11-final-viral-combined_k141_286729\|\|full | 1810 | No | NA | 3 | 0 | 0 | Low-quality | Genome-fragment | 1.24 | AAI-based (medium-confidence) | 0 |
| S7-final-viral-combined_k141_41650\|\|full | 2790 | No | NA | 3 | 0 | 1 | Not-determined | Genome-fragment | NA | NA | 0 |
| S7-final-viral-combined_k141_62119\|\|full | 303278 | Yes | 64190 | 313 | 14 | 142 | Medium-quality | Genome-fragment | 57.57 | HMM-based (lower-bound) | 78.83 |
| S1-final-viral-combined_k141_105434\|\|full | 1810 | No | NA | 2 | 0 | 0 | Low-quality | Genome-fragment | 1.13 | AAI-based (high-confidence) | 0 |
| S7-final-viral-combined_k141_123514\|\|full | 1209 | No | NA | 3 | 0 | 2 | Not-determined | Genome-fragment | NA | NA | 0 |
| S7-final-viral-combined_k141_133809\|\|full | 14109 | No | NA | 13 | 0 | 8 | Not-determined | Genome-fragment | NA | NA | 0 |
| S7-final-viral-combined_k141_82603\|\|full | 4059 | No | NA | 5 | 0 | 4 | Not-determined | Genome-fragment | NA | NA | 0 |
| S7-final-viral-combined_k141_62155\|\|full | 4972 | No | NA | 4 | 0 | 4 | Not-determined | Genome-fragment | NA | NA | 0 |
| S7-final-viral-combined_k141_62162\|\|full | 2304 | No | NA | 4 | 0 | 1 | Not-determined | Genome-fragment | NA | NA | 0 |
| S7-final-viral-combined_k141_174489\|\|full | 1122 | No | NA | 2 | 0 | 1 | Not-determined | Genome-fragment | NA | NA | 0 |
| S1-final-viral-combined_k141_190317\|\|full | 1810 | No | NA | 5 | 0 | 0 | Low-quality | Genome-fragment | 1.34 | AAI-based (medium-confidence) | 0 |
| S6-final-viral-combined_k141_135684\|\|full | 1810 | No | NA | 4 | 0 | 0 | Low-quality | Genome-fragment | 4.43 | AAI-based (high-confidence) | 0 |
| S7-final-viral-combined_k141_225756\|\|full | 3832 | No | NA | 4 | 0 | 4 | Not-determined | Genome-fragment | NA | NA | 0 |
| S8-final-viral-combined_k141_266658\|\|full | 1810 | No | NA | 4 | 1 | 0 | Low-quality | Genome-fragment | 4.66 | AAI-based (high-confidence) | 0 |
| S12-final-viral-combined_k141_257775\|\|lt2gene | 1811 | No | NA | 2 | 1 | 0 | Low-quality | Genome-fragment | 4.55 | AAI-based (high-confidence) | 0 |
| S2-final-viral-combined_k141_134069\|\|full | 1811 | No | NA | 4 | 0 | 0 | Low-quality | Genome-fragment | 2.9 | AAI-based (medium-confidence) | 0 |
| S4-final-viral-combined_k141_165110\|\|lt2gene | 1811 | No | NA | 1 | 1 | 0 | Low-quality | Genome-fragment | 1.84 | AAI-based (high-confidence) | 0 |
| S7-final-viral-combined_k141_21159\|\|full | 2637 | No | NA | 4 | 0 | 2 | Not-determined | Genome-fragment | NA | NA | 0 |
| S7-final-viral-combined_k141_102907\|\|full | 4668 | No | NA | 6 | 0 | 5 | Not-determined | Genome-fragment | NA | NA | 0 |
| S7-final-viral-combined_k141_164517\|\|full | 3718 | No | NA | 4 | 0 | 2 | Not-determined | Genome-fragment | NA | NA | 0 |
| S7-final-viral-combined_k141_11306\|\|full | 16792 | No | NA | 15 | 0 | 8 | Not-determined | Genome-fragment | NA | NA | 0 |
| S6-final-viral-combined_k141_34493\|\|full | 1811 | No | NA | 2 | 0 | 1 | Low-quality | Genome-fragment | 3.12 | AAI-based (high-confidence) | 0 |
| S11-final-viral-combined_k141_111758\|\|full | 1812 | No | NA | 3 | 0 | 0 | Low-quality | Genome-fragment | 3.77 | AAI-based (medium-confidence) | 0 |
| S1-final-viral-combined_k141_58588\|\|lt2gene | 1812 | No | NA | 1 | 1 | 0 | Low-quality | Genome-fragment | 3.95 | AAI-based (medium-confidence) | 0 |
| S1_k141_95726 | 1812 | No | NA | 3 | 0 | 2 | Low-quality | Genome-fragment | 0.41 | AAI-based (medium-confidence) | 0 |
| S7-final-viral-combined_k141_21189\|\|full | 2217 | No | NA | 3 | 0 | 2 | Not-determined | Genome-fragment | NA | NA | 0 |
| S1_k141_180813 | 1812 | No | NA | 1 | 0 | 1 | Low-quality | Genome-fragment | 0.49 | AAI-based (high-confidence) | 0 |
| S7-final-viral-combined_k141_205519\|\|full | 2028 | No | NA | 3 | 0 | 3 | Not-determined | Genome-fragment | NA | NA | 0 |
| S1_k141_114291 | 1812 | Yes | 572 | 4 | 1 | 1 | Low-quality | Genome-fragment | 0.94 | AAI-based (high-confidence) | 68.43 |
| S12-final-viral-combined_k141_241401\|\|full | 1812 | No | NA | 3 | 2 | 0 | Low-quality | Genome-fragment | 5.64 | AAI-based (high-confidence) | 0 |
| S7-final-viral-combined_k141_205533\|\|full | 2829 | No | NA | 2 | 0 | 1 | Not-determined | Genome-fragment | NA | NA | 0 |
| S7-final-viral-combined_k141_185010\|\|full | 3099 | No | NA | 2 | 0 | 2 | Not-determined | Genome-fragment | NA | NA | 0 |
| S7-final-viral-combined_k141_31803\|\|full | 4770 | No | NA | 5 | 0 | 1 | Not-determined | Genome-fragment | NA | NA | 0 |
| S7-final-viral-combined_k141_225883\|\|full | 2985 | No | NA | 4 | 0 | 1 | Not-determined | Genome-fragment | NA | NA | 0 |
| S2-final-viral-combined_k141_14989\|\|full | 1812 | No | NA | 3 | 1 | 0 | Low-quality | Genome-fragment | 3.38 | AAI-based (high-confidence) | 0 |
| S4-final-viral-combined_k141_44983\|\|full | 1812 | No | NA | 3 | 0 | 2 | Low-quality | Genome-fragment | 4.09 | AAI-based (medium-confidence) | 0 |
| S7-final-viral-combined_k141_225892\|\|full | 1408 | No | NA | 3 | 0 | 3 | Not-determined | Genome-fragment | NA | NA | 0 |
| S4_k141_42353 | 1812 | No | NA | 3 | 0 | 0 | Low-quality | Genome-fragment | 1.9 | AAI-based (high-confidence) | 0 |
| S7-final-viral-combined_k141_62231\|\|full | 1513 | No | NA | 2 | 0 | 1 | Not-determined | Genome-fragment | NA | NA | 0 |
| S7-final-viral-combined_k141_113636\|\|full | 2352 | No | NA | 3 | 0 | 0 | Not-determined | Genome-fragment | NA | NA | 0 |
| S7-final-viral-combined_k141_143872\|\|full | 2711 | No | NA | 3 | 0 | 2 | Not-determined | Genome-fragment | NA | NA | 0 |
| S7-final-viral-combined_k141_62247\|\|full | 1741 | No | NA | 3 | 0 | 0 | Not-determined | Genome-fragment | NA | NA | 0 |
| S7-final-viral-combined_k141_31822\|\|full | 2903 | No | NA | 4 | 0 | 2 | Not-determined | Genome-fragment | NA | NA | 0 |
| S7-final-viral-combined_k141_143874\|\|full | 2159 | No | NA | 3 | 0 | 2 | Not-determined | Genome-fragment | NA | NA | 0 |
| S6-final-viral-combined_k141_27283\|\|full | 1812 | No | NA | 4 | 0 | 1 | Low-quality | Genome-fragment | 1.07 | AAI-based (medium-confidence) | 0 |
| S8_k141_61519 | 1812 | No | NA | 3 | 1 | 0 | Low-quality | Genome-fragment | 27.72 | AAI-based (high-confidence) | 0 |
| S7-final-viral-combined_k141_185016\|\|full | 41583 | Yes | 30248 | 55 | 12 | 7 | High-quality | High-quality | 93.16 | AAI-based (high-confidence) | 27.26 |
| S7-final-viral-combined_k141_21254\|\|full | 6969 | No | NA | 7 | 0 | 0 | Not-determined | Genome-fragment | NA | NA | 0 |
| S7-final-viral-combined_k141_103052\|\|full | 8843 | No | NA | 11 | 0 | 3 | Not-determined | Genome-fragment | NA | NA | 0 |
| S7-final-viral-combined_k141_185031\|\|full | 1534 | No | NA | 3 | 0 | 1 | Not-determined | Genome-fragment | NA | NA | 0 |
| S5-final-viral-combined_k141_169246\|\|full | 1813 | No | NA | 3 | 1 | 0 | Low-quality | Genome-fragment | 4.01 | AAI-based (medium-confidence) | 0 |
| S7-final-viral-combined_k141_133976\|\|full | 11629 | No | NA | 11 | 0 | 4 | Not-determined | Genome-fragment | NA | NA | 0 |
| S7-final-viral-combined_k141_174707\|\|full | 2630 | No | NA | 2 | 0 | 2 | Not-determined | Genome-fragment | NA | NA | 0 |
| S7-final-viral-combined_k141_41853\|\|full | 2426 | No | NA | 3 | 0 | 2 | Not-determined | Genome-fragment | NA | NA | 0 |
| S7-final-viral-combined_k141_225954\|\|full | 1065 | No | NA | 3 | 0 | 0 | Not-determined | Genome-fragment | NA | NA | 0 |
| S17-k141_246162 | 1813 | No | NA | 3 | 0 | 0 | Low-quality | Genome-fragment | 3.54 | AAI-based (medium-confidence) | 0 |
| S7-final-viral-combined_k141_41875\|\|full | 2172 | No | NA | 3 | 0 | 3 | Not-determined | Genome-fragment | NA | NA | 0 |
| S7-final-viral-combined_k141_31848\|\|full | 3237 | No | NA | 5 | 0 | 3 | Not-determined | Genome-fragment | NA | NA | 0 |
| S7-final-viral-combined_k141_41894\|\|full | 1872 | No | NA | 2 | 0 | 2 | Not-determined | Genome-fragment | NA | NA | 0 |
| S2-final-viral-combined_k141_44268\|\|full | 1813 | No | NA | 3 | 0 | 0 | Low-quality | Genome-fragment | 2.07 | AAI-based (medium-confidence) | 0 |
| S2_k141_149576 | 1813 | No | NA | 4 | 1 | 0 | Low-quality | Genome-fragment | 2.11 | HMM-based (lower-bound) | 0 |
| S7-final-viral-combined_k141_21304\|\|full | 5473 | No | NA | 3 | 0 | 2 | Not-determined | Genome-fragment | NA | NA | 0 |
| S4-final-viral-combined_k141_7651\|\|full | 1813 | No | NA | 3 | 0 | 1 | Low-quality | Genome-fragment | 2.47 | AAI-based (high-confidence) | 0 |
| S7-final-viral-combined_k141_185087\|\|full | 3285 | No | NA | 6 | 0 | 4 | Not-determined | Genome-fragment | NA | NA | 0 |
| S7-final-viral-combined_k141_62329\|\|full | 7504 | No | NA | 7 | 0 | 5 | Not-determined | Genome-fragment | NA | NA | 0 |
| S8-final-viral-combined_k141_230324\|\|full | 1813 | No | NA | 3 | 1 | 0 | Low-quality | Genome-fragment | 3.12 | AAI-based (high-confidence) | 0 |
| S9-final-viral-combined_k141_43472\|\|full | 1814 | No | NA | 2 | 0 | 0 | Low-quality | Genome-fragment | 2.25 | AAI-based (high-confidence) | 0 |
| S1-final-viral-combined_k141_336004\|\|full | 1814 | No | NA | 6 | 5 | 0 | Low-quality | Genome-fragment | 3.39 | AAI-based (high-confidence) | 0 |
| S7-final-viral-combined_k141_195615\|\|full | 2167 | No | NA | 4 | 0 | 1 | Not-determined | Genome-fragment | NA | NA | 0 |
| S7-final-viral-combined_k141_113682\|\|full | 5639 | No | NA | 4 | 0 | 1 | Not-determined | Genome-fragment | NA | NA | 0 |
| S1-final-viral-combined_k141_221630\|\|full | 1814 | No | NA | 2 | 0 | 0 | Low-quality | Genome-fragment | 3.9 | AAI-based (high-confidence) | 0 |
| S5-final-viral-combined_k141_104187\|\|full | 1814 | No | NA | 3 | 0 | 3 | Low-quality | Genome-fragment | 1.79 | AAI-based (medium-confidence) | 0 |
| S7-final-viral-combined_k141_164739\|\|full | 4368 | No | NA | 6 | 0 | 2 | Not-determined | Genome-fragment | NA | NA | 0 |
| S7-final-viral-combined_k141_62358\|\|full | 2092 | No | NA | 2 | 0 | 1 | Not-determined | Genome-fragment | NA | NA | 0 |
| S7-final-viral-combined_k141_31915\|\|full | 1319 | No | NA | 2 | 0 | 1 | Not-determined | Genome-fragment | NA | NA | 0 |
| S7-final-viral-combined_k141_11434\|\|full | 2348 | No | NA | 3 | 0 | 1 | Not-determined | Genome-fragment | NA | NA | 0 |
| S7-final-viral-combined_k141_174821\|\|full | 1967 | No | NA | 2 | 0 | 2 | Not-determined | Genome-fragment | NA | NA | 0 |
| S5_k141_77413 | 1814 | No | NA | 1 | 0 | 1 | Low-quality | Genome-fragment | 1.11 | AAI-based (medium-confidence) | 0 |
| S17-final-viral-combined_k141_15827\|\|full | 1814 | No | NA | 4 | 1 | 0 | Low-quality | Genome-fragment | 1.34 | HMM-based (lower-bound) | 0 |
| S7-final-viral-combined_k141_123797\|\|full | 1114 | No | NA | 2 | 0 | 0 | Not-determined | Genome-fragment | NA | NA | 0 |
| S17-k141_65487 | 1814 | No | NA | 3 | 0 | 0 | Low-quality | Genome-fragment | 2.2 | AAI-based (medium-confidence) | 0 |
| S7-final-viral-combined_k141_215818\|\|full | 2844 | No | NA | 5 | 0 | 0 | Not-determined | Genome-fragment | NA | NA | 0 |
| S7-final-viral-combined_k141_134115\|\|full | 1984 | No | NA | 3 | 0 | 0 | Not-determined | Genome-fragment | NA | NA | 0 |
| S12-final-viral-combined_k141_392955\|\|full | 1814 | No | NA | 3 | 2 | 0 | Low-quality | Genome-fragment | 3.45 | AAI-based (medium-confidence) | 0 |
| S7-final-viral-combined_k141_185182\|\|full | 3573 | No | NA | 3 | 0 | 1 | Not-determined | Genome-fragment | NA | NA | 0 |
| S8_k141_49486 | 1814 | No | NA | 4 | 1 | 0 | Low-quality | Genome-fragment | 2.2 | AAI-based (high-confidence) | 0 |
| S7-final-viral-combined_k141_92820\|\|full | 2203 | No | NA | 4 | 0 | 2 | Not-determined | Genome-fragment | NA | NA | 0 |
| S7-final-viral-combined_k141_52310\|\|full | 2012 | No | NA | 2 | 0 | 1 | Not-determined | Genome-fragment | NA | NA | 0 |
| S7-final-viral-combined_k141_92843\|\|full | 1517 | No | NA | 3 | 0 | 1 | Not-determined | Genome-fragment | NA | NA | 0 |
| S1-final-viral-combined_k141_282745\|\|full | 1815 | No | NA | 3 | 0 | 1 | Low-quality | Genome-fragment | 0.93 | AAI-based (medium-confidence) | 0 |
| S5-final-viral-combined_k141_138131\|\|full | 1815 | No | NA | 4 | 0 | 0 | Low-quality | Genome-fragment | 2.7 | AAI-based (high-confidence) | 0 |
| S7-final-viral-combined_k141_215858\|\|full | 2839 | No | NA | 6 | 0 | 0 | Not-determined | Genome-fragment | NA | NA | 0 |
| S7-final-viral-combined_k141_16213\|\|full | 1815 | No | NA | 5 | 4 | 0 | Low-quality | Genome-fragment | 4.11 | AAI-based (high-confidence) | 0 |
| S7-final-viral-combined_k141_154670\|\|full | 3500 | No | NA | 5 | 0 | 4 | Not-determined | Genome-fragment | NA | NA | 0 |
| S17-final-viral-combined_k141_49649\|\|full | 1815 | No | NA | 3 | 0 | 1 | Low-quality | Genome-fragment | 0.64 | AAI-based (high-confidence) | 0 |
| S12-final-viral-combined_k141_281644\|\|lt2gene | 1815 | No | NA | 2 | 2 | 0 | Low-quality | Genome-fragment | 4.38 | AAI-based (high-confidence) | 0 |
| S7-final-viral-combined_k141_31970\|\|full | 3726 | No | NA | 3 | 0 | 2 | Not-determined | Genome-fragment | NA | NA | 0 |
| S9-final-viral-combined_k141_114902\|\|full | 1816 | No | NA | 2 | 0 | 0 | Low-quality | Genome-fragment | 3.4 | AAI-based (medium-confidence) | 0 |
| S1-final-viral-combined_k141_284540\|\|full | 1816 | No | NA | 3 | 0 | 0 | Low-quality | Genome-fragment | 1.38 | AAI-based (high-confidence) | 0 |
| S7-final-viral-combined_k141_11476\|\|full | 4497 | No | NA | 7 | 0 | 0 | Not-determined | Genome-fragment | NA | NA | 0 |
| S7-final-viral-combined_k141_62455\|\|full | 1271 | No | NA | 3 | 0 | 0 | Not-determined | Genome-fragment | NA | NA | 0 |
| S1-final-viral-combined_k141_236325\|\|full | 1816 | No | NA | 4 | 0 | 1 | Low-quality | Genome-fragment | 1.36 | AAI-based (medium-confidence) | 0 |
| S1-final-viral-combined_k141_236435\|\|full | 1816 | No | NA | 4 | 0 | 0 | Low-quality | Genome-fragment | 4.53 | AAI-based (medium-confidence) | 0 |
| S7-final-viral-combined_k141_215888\|\|full | 2360 | No | NA | 2 | 0 | 0 | Not-determined | Genome-fragment | NA | NA | 0 |
| S5-final-viral-combined_k141_95580\|\|full | 1816 | No | NA | 4 | 1 | 0 | Low-quality | Genome-fragment | 3.03 | AAI-based (high-confidence) | 0 |
| S7-final-viral-combined_k141_123135\|\|full | 1816 | No | NA | 3 | 0 | 0 | Low-quality | Genome-fragment | 3.6 | AAI-based (high-confidence) | 0 |
| S7-final-viral-combined_k141_154718\|\|full | 1498 | No | NA | 3 | 0 | 1 | Not-determined | Genome-fragment | NA | NA | 0 |
| S7-final-viral-combined_k141_62492\|\|full | 5055 | No | NA | 5 | 0 | 4 | Not-determined | Genome-fragment | NA | NA | 0 |
| S7-final-viral-combined_k141_1338\|\|full | 2555 | No | NA | 3 | 0 | 0 | Not-determined | Genome-fragment | NA | NA | 0 |
| S7-final-viral-combined_k141_103239\|\|full | 1470 | No | NA | 3 | 0 | 1 | Not-determined | Genome-fragment | NA | NA | 0 |
| S7-final-viral-combined_k141_157591\|\|full | 1816 | No | NA | 3 | 0 | 0 | Low-quality | Genome-fragment | 2.95 | AAI-based (high-confidence) | 0 |
| S7-final-viral-combined_k141_21386\|\|full | 1327 | No | NA | 3 | 0 | 2 | Not-determined | Genome-fragment | NA | NA | 0 |
| S7-final-viral-combined_k141_41942\|\|full | 6653 | No | NA | 5 | 0 | 4 | Not-determined | Genome-fragment | NA | NA | 0 |
| S7-final-viral-combined_k141_226090\|\|full | 1972 | No | NA | 4 | 0 | 0 | Not-determined | Genome-fragment | NA | NA | 0 |
| S7-final-viral-combined_k141_103252\|\|full | 1712 | No | NA | 3 | 0 | 0 | Not-determined | Genome-fragment | NA | NA | 0 |
| S7-final-viral-combined_k141_215915\|\|full | 3124 | No | NA | 4 | 0 | 0 | Not-determined | Genome-fragment | NA | NA | 0 |
| S17-final-viral-combined_k141_171151\|\|full | 1816 | No | NA | 3 | 0 | 0 | Low-quality | Genome-fragment | 2.84 | AAI-based (high-confidence) | 0 |
| S7-final-viral-combined_k141_164821\|\|full | 2933 | No | NA | 2 | 0 | 0 | Not-determined | Genome-fragment | NA | NA | 0 |
| S7-final-viral-combined_k141_174916\|\|full | 5654 | No | NA | 5 | 0 | 3 | Not-determined | Genome-fragment | NA | NA | 0 |
| S2-final-viral-combined_k141_178214\|\|full | 1816 | No | NA | 3 | 0 | 0 | Low-quality | Genome-fragment | 2.4 | AAI-based (medium-confidence) | 0 |
| S7-final-viral-combined_k141_226112\|\|full | 1066 | No | NA | 2 | 0 | 0 | Not-determined | Genome-fragment | NA | NA | 0 |
| S9-final-viral-combined_k141_87404\|\|lt2gene | 1817 | No | NA | 2 | 1 | 0 | Low-quality | Genome-fragment | 2.38 | HMM-based (lower-bound) | 0 |
| S9-final-viral-combined_k141_4419\|\|lt2gene | 1817 | No | NA | 2 | 1 | 0 | Low-quality | Genome-fragment | 4.71 | AAI-based (medium-confidence) | 0 |
| S11-final-viral-combined_k141_254445\|\|full | 1817 | No | NA | 3 | 2 | 0 | Low-quality | Genome-fragment | 2.23 | HMM-based (lower-bound) | 0 |
| S11-final-viral-combined_k141_275408\|\|full | 1817 | No | NA | 3 | 0 | 0 | Low-quality | Genome-fragment | 2.72 | AAI-based (medium-confidence) | 0 |
| S11_k141_234332 | 1817 | No | NA | 5 | 0 | 0 | Low-quality | Genome-fragment | 4.94 | AAI-based (high-confidence) | 0 |
| S7-final-viral-combined_k141_52355\|\|full | 5458 | No | NA | 7 | 0 | 3 | Not-determined | Genome-fragment | NA | NA | 0 |
| S7-final-viral-combined_k141_154790\|\|full | 2182 | No | NA | 2 | 0 | 1 | Not-determined | Genome-fragment | NA | NA | 0 |
| S1-final-viral-combined_k141_65270\|\|full | 1817 | No | NA | 3 | 0 | 1 | Low-quality | Genome-fragment | 1.82 | AAI-based (medium-confidence) | 0 |
| S7-final-viral-combined_k141_21436\|\|full | 2006 | No | NA | 3 | 0 | 1 | Not-determined | Genome-fragment | NA | NA | 0 |
| S7-final-viral-combined_k141_164890\|\|full | 1246 | No | NA | 3 | 0 | 3 | Not-determined | Genome-fragment | NA | NA | 0 |
| S7-final-viral-combined_k141_226157\|\|full | 1674 | No | NA | 4 | 0 | 1 | Not-determined | Genome-fragment | NA | NA | 0 |
| S7-final-viral-combined_k141_41950\|\|full | 1058 | No | NA | 2 | 0 | 2 | Not-determined | Genome-fragment | NA | NA | 0 |
| S7-final-viral-combined_k141_62551\|\|full | 7084 | No | NA | 6 | 0 | 3 | Not-determined | Genome-fragment | NA | NA | 0 |
| S7-final-viral-combined_k141_226165\|\|full | 2147 | No | NA | 3 | 0 | 1 | Not-determined | Genome-fragment | NA | NA | 0 |
| S7-final-viral-combined_k141_113782\|\|full | 1540 | No | NA | 3 | 0 | 0 | Not-determined | Genome-fragment | NA | NA | 0 |
| S7-final-viral-combined_k141_154810\|\|full | 3521 | No | NA | 5 | 0 | 2 | Not-determined | Genome-fragment | NA | NA | 0 |
| S7-final-viral-combined_k141_72297\|\|full | 1902 | No | NA | 3 | 0 | 2 | Not-determined | Genome-fragment | NA | NA | 0 |
| S7-final-viral-combined_k141_113795\|\|full | 2188 | No | NA | 3 | 0 | 0 | Not-determined | Genome-fragment | NA | NA | 0 |
| S3-final-viral-combined_k141_67105\|\|lt2gene | 1817 | No | NA | 1 | 1 | 0 | Low-quality | Genome-fragment | 1.18 | AAI-based (high-confidence) | 0 |
| S7-final-viral-combined_k141_185418\|\|full | 2318 | No | NA | 2 | 0 | 0 | Not-determined | Genome-fragment | NA | NA | 0 |
| S12-final-viral-combined_k141_389252\|\|full | 1817 | No | NA | 3 | 0 | 2 | Low-quality | Genome-fragment | 3.91 | AAI-based (high-confidence) | 0 |
| S7-final-viral-combined_k141_226195\|\|full | 2311 | No | NA | 3 | 0 | 3 | Not-determined | Genome-fragment | NA | NA | 0 |
| S6-final-viral-combined_k141_8694\|\|full | 1817 | No | NA | 4 | 0 | 1 | Low-quality | Genome-fragment | 3.32 | AAI-based (medium-confidence) | 0 |
| S7-final-viral-combined_k141_174935\|\|full | 60825 | Yes | 21826 | 56 | 12 | 22 | Medium-quality | Genome-fragment | 56.07 | AAI-based (high-confidence) | 64.12 |
| S8_k141_102655 | 1817 | No | NA | 2 | 0 | 0 | Low-quality | Genome-fragment | 2.87 | AAI-based (high-confidence) | 0 |
| S9-final-viral-combined_k141_124874\|\|full | 1818 | No | NA | 5 | 4 | 0 | Low-quality | Genome-fragment | 5.54 | AAI-based (high-confidence) | 0 |
| S7-final-viral-combined_k141_62597\|\|full | 2601 | No | NA | 2 | 0 | 0 | Not-determined | Genome-fragment | NA | NA | 0 |
| S5_k141_168569 | 1818 | No | NA | 1 | 0 | 0 | Low-quality | Genome-fragment | 1.17 | AAI-based (medium-confidence) | 0 |
| S7-final-viral-combined_k141_52449\|\|full | 2725 | No | NA | 5 | 0 | 1 | Not-determined | Genome-fragment | NA | NA | 0 |
| S12-final-viral-combined_k141_188346\|\|full | 1818 | No | NA | 2 | 0 | 0 | Low-quality | Genome-fragment | 4.22 | AAI-based (medium-confidence) | 0 |
| S7-final-viral-combined_k141_92921\|\|full | 10676 | No | NA | 9 | 0 | 5 | Not-determined | Genome-fragment | NA | NA | 0 |
| S7-final-viral-combined_k141_32114\|\|full | 1898 | No | NA | 3 | 0 | 2 | Not-determined | Genome-fragment | NA | NA | 0 |
| S7-final-viral-combined_k141_154852\|\|full | 4768 | No | NA | 2 | 0 | 1 | Not-determined | Genome-fragment | NA | NA | 0 |
| S7-final-viral-combined_k141_52452\|\|full | 2033 | No | NA | 3 | 0 | 2 | Not-determined | Genome-fragment | NA | NA | 0 |
| S12-final-viral-combined_k141_227334\|\|full | 1818 | No | NA | 5 | 1 | 0 | Low-quality | Genome-fragment | 4.01 | HMM-based (lower-bound) | 0 |
| S7-final-viral-combined_k141_92926\|\|full | 2282 | No | NA | 4 | 0 | 0 | Not-determined | Genome-fragment | NA | NA | 0 |
| S7-final-viral-combined_k141_164969\|\|full | 8932 | No | NA | 8 | 0 | 1 | Not-determined | Genome-fragment | NA | NA | 0 |
| S7-final-viral-combined_k141_236361\|\|full | 1502 | No | NA | 3 | 0 | 2 | Not-determined | Genome-fragment | NA | NA | 0 |
| S12-final-viral-combined_k141_264982\|\|full | 1818 | No | NA | 3 | 0 | 1 | Low-quality | Genome-fragment | 0.89 | AAI-based (medium-confidence) | 0 |
| S7-final-viral-combined_k141_11544\|\|full | 2159 | No | NA | 4 | 0 | 0 | Not-determined | Genome-fragment | NA | NA | 0 |
| S7-final-viral-combined_k141_123939\|\|full | 1221 | No | NA | 3 | 0 | 2 | Not-determined | Genome-fragment | NA | NA | 0 |
| S7-final-viral-combined_k141_144112\|\|full | 5217 | No | NA | 5 | 0 | 4 | Not-determined | Genome-fragment | NA | NA | 0 |
| S2-final-viral-combined_k141_60999\|\|full | 1818 | No | NA | 3 | 0 | 0 | Low-quality | Genome-fragment | 2.76 | AAI-based (high-confidence) | 0 |
| S7-final-viral-combined_k141_226248\|\|full | 2000 | No | NA | 3 | 0 | 1 | Not-determined | Genome-fragment | NA | NA | 0 |
| S7-final-viral-combined_k141_117527\|\|full | 1819 | No | NA | 2 | 0 | 1 | Low-quality | Genome-fragment | 0.49 | AAI-based (medium-confidence) | 0 |
| S7-final-viral-combined_k141_121004\|\|full | 1819 | No | NA | 3 | 0 | 0 | Low-quality | Genome-fragment | 1.71 | AAI-based (high-confidence) | 0 |
| S7-final-viral-combined_k141_113896\|\|full | 1678 | No | NA | 2 | 0 | 0 | Not-determined | Genome-fragment | NA | NA | 0 |
| S7-final-viral-combined_k141_82894\|\|full | 2032 | No | NA | 3 | 0 | 1 | Not-determined | Genome-fragment | NA | NA | 0 |
| S2-final-viral-combined_k141_217207\|\|full | 1819 | No | NA | 3 | 0 | 2 | Low-quality | Genome-fragment | 0.5 | AAI-based (medium-confidence) | 0 |
| S7-final-viral-combined_k141_123957\|\|full | 1889 | No | NA | 2 | 0 | 2 | Not-determined | Genome-fragment | NA | NA | 0 |
| S9-final-viral-combined_k141_75058\|\|lt2gene | 1820 | No | NA | 2 | 2 | 0 | Low-quality | Genome-fragment | 4.67 | AAI-based (medium-confidence) | 0 |
| S7-final-viral-combined_k141_134357\|\|full | 5545 | No | NA | 5 | 0 | 1 | Not-determined | Genome-fragment | NA | NA | 0 |
| S7-final-viral-combined_k141_154922\|\|full | 1055 | No | NA | 2 | 0 | 0 | Not-determined | Genome-fragment | NA | NA | 0 |
| S7-final-viral-combined_k141_82905\|\|full | 1613 | No | NA | 3 | 0 | 0 | Not-determined | Genome-fragment | NA | NA | 0 |
| S11-final-viral-combined_k141_153732\|\|full | 1820 | No | NA | 2 | 0 | 0 | Low-quality | Genome-fragment | 1.38 | AAI-based (medium-confidence) | 0 |
| S7-final-viral-combined_k141_62667\|\|full | 1786 | No | NA | 2 | 0 | 0 | Not-determined | Genome-fragment | NA | NA | 0 |
| S11-final-viral-combined_k141_186099\|\|full | 1820 | No | NA | 3 | 3 | 0 | Low-quality | Genome-fragment | 5.7 | AAI-based (high-confidence) | 0 |
| S5-final-viral-combined_k141_10238\|\|lt2gene | 1820 | No | NA | 1 | 1 | 0 | Low-quality | Genome-fragment | 4.03 | AAI-based (medium-confidence) | 0 |
| S17-final-viral-combined_k141_262370\|\|full | 1820 | No | NA | 2 | 0 | 0 | Low-quality | Genome-fragment | 5.35 | AAI-based (high-confidence) | 0 |
| S7-final-viral-combined_k141_1419\|\|full | 1496 | No | NA | 2 | 0 | 2 | Not-determined | Genome-fragment | NA | NA | 0 |
| S7-final-viral-combined_k141_103458\|\|full | 2313 | No | NA | 3 | 0 | 2 | Not-determined | Genome-fragment | NA | NA | 0 |
| S12_k141_231904 | 1820 | No | NA | 3 | 0 | 0 | Low-quality | Genome-fragment | 2.6 | AAI-based (high-confidence) | 0 |
| S7-final-viral-combined_k141_168604\|\|full | 1821 | No | NA | 3 | 2 | 0 | Low-quality | Genome-fragment | 4.4 | AAI-based (high-confidence) | 0 |
| S7-final-viral-combined_k141_82948\|\|full | 1334 | No | NA | 2 | 0 | 2 | Not-determined | Genome-fragment | NA | NA | 0 |
| S7-final-viral-combined_k141_185613\|\|full | 1261 | No | NA | 2 | 0 | 0 | Not-determined | Genome-fragment | NA | NA | 0 |
| S7-final-viral-combined_k141_52598\|\|full | 2110 | No | NA | 3 | 0 | 1 | Not-determined | Genome-fragment | NA | NA | 0 |
| S7-final-viral-combined_k141_123974\|\|full | 2225 | No | NA | 5 | 0 | 0 | Not-determined | Genome-fragment | NA | NA | 0 |
| S7-final-viral-combined_k141_72454\|\|full | 4923 | No | NA | 6 | 0 | 0 | Not-determined | Genome-fragment | NA | NA | 0 |
| S4-final-viral-combined_k141_140469\|\|full | 1821 | No | NA | 5 | 1 | 0 | Low-quality | Genome-fragment | 3.36 | AAI-based (high-confidence) | 0 |
| S8-final-viral-combined_k141_182557\|\|full | 1821 | No | NA | 4 | 1 | 0 | Low-quality | Genome-fragment | 4.54 | AAI-based (high-confidence) | 0 |
| S7-final-viral-combined_k141_236451\|\|full | 6597 | No | NA | 4 | 0 | 2 | Not-determined | Genome-fragment | NA | NA | 0 |
| S9_k141_118286 | 1822 | No | NA | 3 | 2 | 0 | Low-quality | Genome-fragment | 4.37 | AAI-based (high-confidence) | 0 |
| S7-final-viral-combined_k141_122692\|\|full | 1822 | No | NA | 3 | 0 | 0 | Low-quality | Genome-fragment | 3.51 | AAI-based (medium-confidence) | 0 |
| S7-final-viral-combined_k141_154976\|\|full | 3187 | No | NA | 2 | 0 | 2 | Not-determined | Genome-fragment | NA | NA | 0 |
| S7-final-viral-combined_k141_195911\|\|full | 2119 | No | NA | 3 | 0 | 2 | Not-determined | Genome-fragment | NA | NA | 0 |
| S7-final-viral-combined_k141_114008\|\|full | 3265 | No | NA | 3 | 0 | 3 | Not-determined | Genome-fragment | NA | NA | 0 |
| S12-final-viral-combined_k141_186194\|\|full | 1822 | No | NA | 3 | 0 | 1 | Low-quality | Genome-fragment | 3.03 | AAI-based (medium-confidence) | 0 |
| S2_k141_216595 | 1822 | No | NA | 1 | 0 | 0 | Low-quality | Genome-fragment | 1.78 | AAI-based (high-confidence) | 0 |
| S8-final-viral-combined_k141_246017\|\|full | 1822 | No | NA | 6 | 1 | 0 | Low-quality | Genome-fragment | 3.71 | AAI-based (high-confidence) | 0 |
| S7-final-viral-combined_k141_1499\|\|full | 2452 | No | NA | 3 | 0 | 2 | Not-determined | Genome-fragment | NA | NA | 0 |
| S7-final-viral-combined_k141_175098\|\|full | 2531 | No | NA | 4 | 0 | 3 | Not-determined | Genome-fragment | NA | NA | 0 |
| S7-final-viral-combined_k141_124059\|\|full | 1536 | No | NA | 3 | 0 | 2 | Not-determined | Genome-fragment | NA | NA | 0 |
| S9-final-viral-combined_k141_21512\|\|full | 1823 | No | NA | 3 | 1 | 0 | Low-quality | Genome-fragment | 3.13 | HMM-based (lower-bound) | 0 |
| S7-final-viral-combined_k141_92972\|\|full | 3901 | No | NA | 5 | 0 | 4 | Not-determined | Genome-fragment | NA | NA | 0 |
| S7-final-viral-combined_k141_185715\|\|full | 2240 | No | NA | 3 | 0 | 1 | Not-determined | Genome-fragment | NA | NA | 0 |
| S7-final-viral-combined_k141_236536\|\|full | 2056 | No | NA | 3 | 0 | 0 | Not-determined | Genome-fragment | NA | NA | 0 |
| S3_k141_47766 | 1823 | No | NA | 3 | 0 | 2 | Low-quality | Genome-fragment | 1.2 | AAI-based (high-confidence) | 0 |
| S7-final-viral-combined_k141_42177\|\|full | 1925 | No | NA | 2 | 0 | 0 | Not-determined | Genome-fragment | NA | NA | 0 |
| S7_k141_12563 | 1823 | No | NA | 4 | 1 | 0 | Low-quality | Genome-fragment | 2.9 | AAI-based (high-confidence) | 0 |
| S7-final-viral-combined_k141_62796\|\|full | 1424 | No | NA | 3 | 0 | 1 | Not-determined | Genome-fragment | NA | NA | 0 |
| S7_k141_198393 | 1823 | No | NA | 3 | 3 | 0 | Low-quality | Genome-fragment | 4.53 | AAI-based (high-confidence) | 0 |
| S17-final-viral-combined_k141_36743\|\|full | 1823 | No | NA | 4 | 0 | 0 | Low-quality | Genome-fragment | 3.73 | AAI-based (medium-confidence) | 0 |
| S7-final-viral-combined_k141_134452\|\|full | 64594 | No | NA | 67 | 10 | 8 | Medium-quality | Genome-fragment | 89.97 | HMM-based (lower-bound) | 0 |
| S17-final-viral-combined_k141_29143\|\|lt2gene | 1823 | No | NA | 1 | 1 | 0 | Low-quality | Genome-fragment | 3.16 | AAI-based (high-confidence) | 0 |
| S12_k141_19723 | 1823 | No | NA | 2 | 0 | 1 | Low-quality | Genome-fragment | 1.11 | AAI-based (medium-confidence) | 0 |
| S2-final-viral-combined_k141_99668\|\|full | 1823 | No | NA | 2 | 1 | 0 | Low-quality | Genome-fragment | 3.11 | AAI-based (medium-confidence) | 0 |
| S7-final-viral-combined_k141_226420\|\|full | 1931 | No | NA | 3 | 0 | 0 | Not-determined | Genome-fragment | NA | NA | 0 |
| S7-final-viral-combined_k141_32259\|\|full | 23556 | No | NA | 21 | 0 | 8 | Not-determined | Genome-fragment | NA | NA | 0 |
| S7-final-viral-combined_k141_175162\|\|full | 1919 | No | NA | 2 | 0 | 1 | Not-determined | Genome-fragment | NA | NA | 0 |
| S7-final-viral-combined_k141_165064\|\|full | 4653 | No | NA | 5 | 0 | 3 | Not-determined | Genome-fragment | NA | NA | 0 |
| S2_k141_30962 | 1823 | No | NA | 4 | 0 | 2 | Low-quality | Genome-fragment | 2.54 | AAI-based (medium-confidence) | 0 |
| S7-final-viral-combined_k141_175184\|\|full | 1458 | No | NA | 3 | 0 | 0 | Not-determined | Genome-fragment | NA | NA | 0 |
| S8_k141_213437 | 1823 | No | NA | 4 | 2 | 0 | Low-quality | Genome-fragment | 5.37 | AAI-based (high-confidence) | 0 |
| S9_k141_41416 | 1824 | No | NA | 1 | 0 | 1 | Low-quality | Genome-fragment | 0.58 | AAI-based (medium-confidence) | 0 |
| S7-final-viral-combined_k141_1527\|\|full | 4677 | No | NA | 4 | 0 | 4 | Not-determined | Genome-fragment | NA | NA | 0 |
| S7-final-viral-combined_k141_103626\|\|full | 2177 | No | NA | 3 | 0 | 0 | Not-determined | Genome-fragment | NA | NA | 0 |
| S7-final-viral-combined_k141_32311\|\|full | 1353 | No | NA | 3 | 0 | 1 | Not-determined | Genome-fragment | NA | NA | 0 |
| S7-final-viral-combined_k141_165095\|\|full | 1839 | No | NA | 3 | 0 | 1 | Not-determined | Genome-fragment | NA | NA | 0 |
| S7-final-viral-combined_k141_134458\|\|full | 57594 | Yes | 46866 | 79 | 21 | 9 | Medium-quality | Genome-fragment | 80.33 | HMM-based (lower-bound) | 18.63 |
| S11-final-viral-combined_k141_272980\|\|full | 1824 | No | NA | 3 | 1 | 0 | Low-quality | Genome-fragment | 2.53 | AAI-based (high-confidence) | 0 |
| S7-final-viral-combined_k141_93067\|\|full | 2581 | No | NA | 5 | 0 | 0 | Not-determined | Genome-fragment | NA | NA | 0 |
| S11-final-viral-combined_k141_95858\|\|full | 1824 | No | NA | 3 | 0 | 0 | Low-quality | Genome-fragment | 1.76 | AAI-based (high-confidence) | 0 |
| S1-final-viral-combined_k141_111858\|\|full | 1824 | No | NA | 3 | 0 | 2 | Low-quality | Genome-fragment | 0.91 | AAI-based (high-confidence) | 0 |
| S7-final-viral-combined_k141_155056\|\|full | 2647 | No | NA | 2 | 0 | 0 | Not-determined | Genome-fragment | NA | NA | 0 |
| S1-final-viral-combined_k141_31272\|\|full | 1824 | No | NA | 5 | 0 | 0 | Low-quality | Genome-fragment | 3.75 | AAI-based (high-confidence) | 0 |
| S5-final-viral-combined_k141_124613\|\|full | 1824 | No | NA | 4 | 1 | 0 | Low-quality | Genome-fragment | 1.16 | HMM-based (lower-bound) | 0 |
| S7-final-viral-combined_k141_32340\|\|full | 4694 | No | NA | 9 | 0 | 8 | Not-determined | Genome-fragment | NA | NA | 0 |
| S17-final-viral-combined_k141_77663\|\|full | 1824 | No | NA | 4 | 0 | 0 | Low-quality | Genome-fragment | 3.12 | AAI-based (high-confidence) | 0 |
| S7-final-viral-combined_k141_124111\|\|full | 1123 | No | NA | 3 | 0 | 1 | Not-determined | Genome-fragment | NA | NA | 0 |
| S7-final-viral-combined_k141_124115\|\|full | 1526 | No | NA | 4 | 0 | 0 | Not-determined | Genome-fragment | NA | NA | 0 |
| S17-final-viral-combined_k141_31801\|\|full | 1824 | No | NA | 3 | 0 | 0 | Low-quality | Genome-fragment | 4.52 | AAI-based (medium-confidence) | 0 |
| S17-k141_9985 | 1824 | No | NA | 4 | 0 | 0 | Low-quality | Genome-fragment | 1.09 | AAI-based (medium-confidence) | 0 |
| S12-final-viral-combined_k141_314527\|\|full | 1824 | No | NA | 4 | 0 | 2 | Low-quality | Genome-fragment | 3.44 | AAI-based (medium-confidence) | 0 |
| S8-final-viral-combined_k141_151096\|\|full | 1824 | No | NA | 3 | 0 | 1 | Low-quality | Genome-fragment | 1.03 | AAI-based (medium-confidence) | 0 |
| S7-final-viral-combined_k141_32355\|\|full | 2390 | No | NA | 3 | 0 | 1 | Not-determined | Genome-fragment | NA | NA | 0 |
| S8-final-viral-combined_k141_96360\|\|full | 1824 | No | NA | 3 | 0 | 0 | Low-quality | Genome-fragment | 5.28 | AAI-based (high-confidence) | 0 |
| S7-final-viral-combined_k141_124145\|\|full | 1346 | No | NA | 3 | 0 | 0 | Not-determined | Genome-fragment | NA | NA | 0 |
| S8-final-viral-combined_k141_250844\|\|full | 1824 | No | NA | 2 | 0 | 0 | Low-quality | Genome-fragment | 2.22 | AAI-based (high-confidence) | 0 |
| S9_k141_78334 | 1825 | No | NA | 2 | 0 | 1 | Low-quality | Genome-fragment | 3.24 | AAI-based (medium-confidence) | 0 |
| S1-final-viral-combined_k141_97652\|\|full | 1825 | No | NA | 3 | 1 | 0 | Low-quality | Genome-fragment | 3.36 | AAI-based (high-confidence) | 0 |
| S7-final-viral-combined_k141_206166\|\|full | 6242 | No | NA | 9 | 0 | 0 | Not-determined | Genome-fragment | NA | NA | 0 |
| S7-final-viral-combined_k141_83046\|\|full | 4005 | No | NA | 4 | 0 | 4 | Not-determined | Genome-fragment | NA | NA | 0 |
| S7-final-viral-combined_k141_216103\|\|full | 1599 | No | NA | 2 | 0 | 1 | Not-determined | Genome-fragment | NA | NA | 0 |
| S7-final-viral-combined_k141_226532\|\|full | 1262 | No | NA | 3 | 0 | 0 | Not-determined | Genome-fragment | NA | NA | 0 |
| S7-final-viral-combined_k141_206194\|\|full | 2789 | No | NA | 4 | 0 | 2 | Not-determined | Genome-fragment | NA | NA | 0 |
| S1_k141_60074 | 1825 | No | NA | 1 | 0 | 1 | Low-quality | Genome-fragment | 2.21 | AAI-based (medium-confidence) | 0 |
| S7-final-viral-combined_k141_144326\|\|full | 2926 | No | NA | 4 | 0 | 3 | Not-determined | Genome-fragment | NA | NA | 0 |
| S1_k141_344861 | 1825 | No | NA | 3 | 1 | 0 | Low-quality | Genome-fragment | 1.2 | AAI-based (medium-confidence) | 0 |
| S5-final-viral-combined_k141_136473\|\|full | 1825 | No | NA | 3 | 1 | 0 | Low-quality | Genome-fragment | 3.61 | AAI-based (medium-confidence) | 0 |
| S5-final-viral-combined_k141_7815\|\|lt2gene | 1825 | No | NA | 2 | 1 | 0 | Low-quality | Genome-fragment | 4.05 | AAI-based (high-confidence) | 0 |
| S17-final-viral-combined_k141_23700\|\|full | 1825 | No | NA | 4 | 3 | 0 | Low-quality | Genome-fragment | 4.95 | AAI-based (high-confidence) | 0 |
| S7-final-viral-combined_k141_124203\|\|full | 2272 | No | NA | 3 | 0 | 2 | Not-determined | Genome-fragment | NA | NA | 0 |
| S7-final-viral-combined_k141_236680\|\|full | 2707 | No | NA | 4 | 0 | 0 | Not-determined | Genome-fragment | NA | NA | 0 |
| S7-final-viral-combined_k141_124215\|\|full | 2760 | No | NA | 3 | 0 | 2 | Not-determined | Genome-fragment | NA | NA | 0 |
| S17-k141_235906 | 1825 | No | NA | 3 | 2 | 0 | Low-quality | Genome-fragment | 4.51 | AAI-based (medium-confidence) | 0 |
| S7-final-viral-combined_k141_236722\|\|full | 1127 | No | NA | 3 | 0 | 1 | Not-determined | Genome-fragment | NA | NA | 0 |
| S12-final-viral-combined_k141_221828\|\|full | 1825 | No | NA | 3 | 0 | 1 | Low-quality | Genome-fragment | 3.05 | AAI-based (high-confidence) | 0 |
| S12-final-viral-combined_k141_108346\|\|full | 1825 | No | NA | 2 | 0 | 1 | Low-quality | Genome-fragment | 2.64 | AAI-based (medium-confidence) | 0 |
| S7-final-viral-combined_k141_196058\|\|full | 8766 | No | NA | 10 | 0 | 1 | Not-determined | Genome-fragment | NA | NA | 0 |
| S7-final-viral-combined_k141_114294\|\|full | 1807 | No | NA | 3 | 0 | 0 | Not-determined | Genome-fragment | NA | NA | 0 |
| S7-final-viral-combined_k141_72514\|\|full | 1142 | No | NA | 2 | 0 | 0 | Not-determined | Genome-fragment | NA | NA | 0 |
| S7-final-viral-combined_k141_196079\|\|full | 1314 | No | NA | 2 | 0 | 0 | Not-determined | Genome-fragment | NA | NA | 0 |
| S7-final-viral-combined_k141_21661\|\|full | 3250 | No | NA | 3 | 0 | 0 | Not-determined | Genome-fragment | NA | NA | 0 |
| S12-final-viral-combined_k141_130916\|\|full | 1825 | No | NA | 2 | 2 | 0 | Low-quality | Genome-fragment | 2.3 | HMM-based (lower-bound) | 0 |
| S7-final-viral-combined_k141_11700\|\|full | 1761 | No | NA | 3 | 0 | 0 | Not-determined | Genome-fragment | NA | NA | 0 |
| S2-final-viral-combined_k141_132505\|\|lt2gene | 1825 | No | NA | 2 | 1 | 0 | Low-quality | Genome-fragment | 2.62 | AAI-based (medium-confidence) | 0 |
| S7-final-viral-combined_k141_42394\|\|full | 1061 | No | NA | 2 | 0 | 0 | Not-determined | Genome-fragment | NA | NA | 0 |
| S4_k141_21183 | 1825 | No | NA | 1 | 0 | 0 | Low-quality | Genome-fragment | 0.79 | AAI-based (medium-confidence) | 0 |
| S8-final-viral-combined_k141_49910\|\|full | 1825 | No | NA | 4 | 0 | 0 | Low-quality | Genome-fragment | 5.25 | AAI-based (high-confidence) | 0 |
| S8_k141_18358 | 1825 | No | NA | 3 | 0 | 0 | Low-quality | Genome-fragment | 5.53 | AAI-based (medium-confidence) | 0 |
| S7-final-viral-combined_k141_103790\|\|full | 2487 | No | NA | 3 | 0 | 0 | Not-determined | Genome-fragment | NA | NA | 0 |
| S7-final-viral-combined_k141_185924\|\|full | 8573 | No | NA | 9 | 0 | 1 | Not-determined | Genome-fragment | NA | NA | 0 |
| S9_k141_89132 | 1826 | No | NA | 2 | 0 | 0 | Low-quality | Genome-fragment | 4.12 | AAI-based (high-confidence) | 0 |
| S7-final-viral-combined_k141_103808\|\|full | 2241 | No | NA | 3 | 0 | 1 | Not-determined | Genome-fragment | NA | NA | 0 |
| S7-final-viral-combined_k141_124328\|\|full | 1318 | No | NA | 3 | 0 | 1 | Not-determined | Genome-fragment | NA | NA | 0 |
| S7-final-viral-combined_k141_206311\|\|full | 3019 | No | NA | 5 | 0 | 2 | Not-determined | Genome-fragment | NA | NA | 0 |
| S7-final-viral-combined_k141_21693\|\|full | 51935 | No | NA | 60 | 22 | 3 | High-quality | High-quality | 100 | AAI-based (high-confidence) | 0 |
| S7-final-viral-combined_k141_206317\|\|full | 1263 | No | NA | 3 | 0 | 1 | Not-determined | Genome-fragment | NA | NA | 0 |
| S7-final-viral-combined_k141_206324\|\|full | 1175 | No | NA | 3 | 0 | 0 | Not-determined | Genome-fragment | NA | NA | 0 |
| S11-final-viral-combined_k141_263794\|\|full | 1826 | No | NA | 3 | 0 | 2 | Low-quality | Genome-fragment | 1.13 | AAI-based (medium-confidence) | 0 |
| S11-final-viral-combined_k141_198269\|\|full | 1826 | No | NA | 4 | 3 | 0 | Low-quality | Genome-fragment | 4.12 | AAI-based (high-confidence) | 0 |
| S7-final-viral-combined_k141_144388\|\|full | 3000 | No | NA | 3 | 0 | 3 | Not-determined | Genome-fragment | NA | NA | 0 |
| S1-final-viral-combined_k141_260730\|\|full | 1826 | No | NA | 2 | 0 | 1 | Low-quality | Genome-fragment | 1.5 | AAI-based (high-confidence) | 0 |
| S12-final-viral-combined_k141_349017\|\|full | 1826 | No | NA | 1 | 0 | 1 | Low-quality | Genome-fragment | 0.43 | AAI-based (high-confidence) | 0 |
| S7-final-viral-combined_k141_32560\|\|full | 1351 | No | NA | 2 | 0 | 2 | Not-determined | Genome-fragment | NA | NA | 0 |
| S7-final-viral-combined_k141_226704\|\|full | 1106 | No | NA | 2 | 0 | 2 | Not-determined | Genome-fragment | NA | NA | 0 |
| S7-final-viral-combined_k141_114335\|\|full | 3189 | No | NA | 4 | 0 | 3 | Not-determined | Genome-fragment | NA | NA | 0 |
| S7-final-viral-combined_k141_114338\|\|full | 3486 | No | NA | 4 | 0 | 3 | Not-determined | Genome-fragment | NA | NA | 0 |
| S2-final-viral-combined_k141_7444\|\|full | 1826 | No | NA | 3 | 1 | 0 | Low-quality | Genome-fragment | 2.82 | HMM-based (lower-bound) | 0 |
| S4_k141_22621 | 1826 | No | NA | 3 | 1 | 0 | Low-quality | Genome-fragment | 2.96 | AAI-based (high-confidence) | 0 |
| S7-final-viral-combined_k141_83187\|\|full | 2101 | No | NA | 3 | 0 | 2 | Not-determined | Genome-fragment | NA | NA | 0 |
| S7-final-viral-combined_k141_53017\|\|full | 1662 | No | NA | 3 | 0 | 0 | Not-determined | Genome-fragment | NA | NA | 0 |
| S7-final-viral-combined_k141_114379\|\|full | 4770 | No | NA | 3 | 0 | 0 | Not-determined | Genome-fragment | NA | NA | 0 |
| S6-final-viral-combined_k141_24094\|\|full | 1826 | No | NA | 3 | 0 | 1 | Low-quality | Genome-fragment | 1.69 | AAI-based (high-confidence) | 0 |
| S7-final-viral-combined_k141_114395\|\|full | 1193 | No | NA | 3 | 0 | 0 | Not-determined | Genome-fragment | NA | NA | 0 |
| S7-final-viral-combined_k141_21762\|\|full | 1997 | No | NA | 3 | 0 | 2 | Not-determined | Genome-fragment | NA | NA | 0 |
| S7-final-viral-combined_k141_114399\|\|full | 1618 | No | NA | 1 | 0 | 1 | Not-determined | Genome-fragment | NA | NA | 0 |
| S7-final-viral-combined_k141_42522\|\|full | 2992 | No | NA | 3 | 0 | 2 | Not-determined | Genome-fragment | NA | NA | 0 |
| S6-final-viral-combined_k141_56685\|\|full | 1826 | No | NA | 4 | 0 | 2 | Low-quality | Genome-fragment | 5.77 | AAI-based (medium-confidence) | 0 |
| S11-final-viral-combined_k141_286154\|\|full | 1827 | No | NA | 3 | 0 | 0 | Low-quality | Genome-fragment | 2.78 | AAI-based (high-confidence) | 0 |
| S7-final-viral-combined_k141_114429\|\|full | 1195 | No | NA | 3 | 0 | 3 | Not-determined | Genome-fragment | NA | NA | 0 |
| S7-final-viral-combined_k141_1834\|\|full | 3303 | No | NA | 5 | 0 | 2 | Not-determined | Genome-fragment | NA | NA | 0 |
| S5-final-viral-combined_k141_33375\|\|full | 1827 | No | NA | 3 | 0 | 0 | Low-quality | Genome-fragment | 1 | AAI-based (medium-confidence) | 0 |
| S7-final-viral-combined_k141_124462\|\|full | 1661 | No | NA | 3 | 0 | 2 | Not-determined | Genome-fragment | NA | NA | 0 |
| S7-final-viral-combined_k141_155198\|\|full | 2761 | No | NA | 2 | 0 | 2 | Not-determined | Genome-fragment | NA | NA | 0 |
| S12-final-viral-combined_k141_347627\|\|full | 1827 | No | NA | 3 | 0 | 1 | Low-quality | Genome-fragment | 4.98 | AAI-based (high-confidence) | 0 |
| S7-final-viral-combined_k141_72592\|\|full | 1035 | No | NA | 3 | 0 | 0 | Not-determined | Genome-fragment | NA | NA | 0 |
| S7-final-viral-combined_k141_155208\|\|full | 5638 | No | NA | 3 | 0 | 2 | Not-determined | Genome-fragment | NA | NA | 0 |
| S7-final-viral-combined_k141_216272\|\|full | 1245 | No | NA | 3 | 0 | 0 | Not-determined | Genome-fragment | NA | NA | 0 |
| S2_k141_124146 | 1827 | No | NA | 4 | 0 | 1 | Low-quality | Genome-fragment | 4.44 | AAI-based (medium-confidence) | 0 |
| S7-final-viral-combined_k141_104010\|\|full | 6487 | No | NA | 7 | 0 | 1 | Not-determined | Genome-fragment | NA | NA | 0 |
| S7-final-viral-combined_k141_53148\|\|full | 4898 | No | NA | 9 | 0 | 0 | Not-determined | Genome-fragment | NA | NA | 0 |
| S7-final-viral-combined_k141_155247\|\|full | 1073 | No | NA | 3 | 0 | 1 | Not-determined | Genome-fragment | NA | NA | 0 |
| S4-final-viral-combined_k141_121132\|\|full | 1827 | No | NA | 3 | 2 | 0 | Low-quality | Genome-fragment | 5.37 | AAI-based (high-confidence) | 0 |
| S7-final-viral-combined_k141_206514\|\|full | 3369 | No | NA | 4 | 0 | 4 | Not-determined | Genome-fragment | NA | NA | 0 |
| S7-final-viral-combined_k141_124567\|\|full | 3070 | No | NA | 4 | 0 | 1 | Not-determined | Genome-fragment | NA | NA | 0 |
| S7-final-viral-combined_k141_32734\|\|full | 1030 | No | NA | 2 | 0 | 0 | Not-determined | Genome-fragment | NA | NA | 0 |
| S7-final-viral-combined_k141_72627\|\|full | 2062 | No | NA | 2 | 0 | 0 | Not-determined | Genome-fragment | NA | NA | 0 |
| S6-final-viral-combined_k141_13865\|\|full | 1827 | No | NA | 4 | 0 | 0 | Low-quality | Genome-fragment | 3.86 | AAI-based (medium-confidence) | 0 |
| S7-final-viral-combined_k141_155304\|\|full | 1699 | No | NA | 3 | 0 | 2 | Not-determined | Genome-fragment | NA | NA | 0 |
| S8-final-viral-combined_k141_153405\|\|full | 1827 | No | NA | 2 | 0 | 1 | Low-quality | Genome-fragment | 1.53 | AAI-based (high-confidence) | 0 |
| S7-final-viral-combined_k141_104078\|\|full | 2119 | No | NA | 3 | 0 | 0 | Not-determined | Genome-fragment | NA | NA | 0 |
| S8-final-viral-combined_k141_19731\|\|full | 1827 | No | NA | 4 | 1 | 0 | Low-quality | Genome-fragment | 3.62 | AAI-based (medium-confidence) | 0 |
| S9-final-viral-combined_k141_6731\|\|full | 1828 | No | NA | 4 | 1 | 0 | Low-quality | Genome-fragment | 0.96 | HMM-based (lower-bound) | 0 |
| S1-final-viral-combined_k141_359734\|\|full | 1828 | No | NA | 2 | 0 | 1 | Low-quality | Genome-fragment | 1.06 | AAI-based (medium-confidence) | 0 |
| S5_k141_98644 | 1828 | No | NA | 1 | 0 | 0 | Low-quality | Genome-fragment | 3.05 | AAI-based (high-confidence) | 0 |
| S7-final-viral-combined_k141_42685\|\|full | 2642 | No | NA | 5 | 0 | 1 | Not-determined | Genome-fragment | NA | NA | 0 |
| S12-final-viral-combined_k141_367514\|\|full | 1828 | No | NA | 5 | 0 | 0 | Low-quality | Genome-fragment | 2.89 | AAI-based (high-confidence) | 0 |
| S7-final-viral-combined_k141_134692\|\|full | 1186 | No | NA | 3 | 0 | 0 | Not-determined | Genome-fragment | NA | NA | 0 |
| S12_k141_31801 | 1828 | No | NA | 3 | 0 | 1 | Low-quality | Genome-fragment | 1.01 | AAI-based (high-confidence) | 0 |
| S7-final-viral-combined_k141_63112\|\|full | 1008 | No | NA | 2 | 0 | 1 | Not-determined | Genome-fragment | NA | NA | 0 |
| S2-final-viral-combined_k141_187140\|\|full | 1828 | No | NA | 3 | 0 | 3 | Low-quality | Genome-fragment | 3.13 | AAI-based (medium-confidence) | 0 |
| S2_k141_110310 | 1828 | No | NA | 3 | 2 | 0 | Low-quality | Genome-fragment | 2.95 | AAI-based (high-confidence) | 0 |
| S4-final-viral-combined_k141_126140\|\|full | 1828 | No | NA | 4 | 2 | 0 | Low-quality | Genome-fragment | 2.34 | AAI-based (high-confidence) | 0 |
| S7-final-viral-combined_k141_72664\|\|full | 2302 | No | NA | 4 | 0 | 1 | Not-determined | Genome-fragment | NA | NA | 0 |
| S4_k141_47107 | 1828 | No | NA | 2 | 0 | 0 | Low-quality | Genome-fragment | 1.83 | AAI-based (high-confidence) | 0 |
| S7-final-viral-combined_k141_1925\|\|full | 1878 | No | NA | 3 | 0 | 1 | Not-determined | Genome-fragment | NA | NA | 0 |
| S8-final-viral-combined_k141_269106\|\|full | 1828 | No | NA | 3 | 0 | 1 | Low-quality | Genome-fragment | 3.26 | AAI-based (medium-confidence) | 0 |
| S7-final-viral-combined_k141_63124\|\|full | 6484 | No | NA | 10 | 0 | 2 | Not-determined | Genome-fragment | NA | NA | 0 |
| S11-final-viral-combined_k141_237851\|\|full | 1829 | No | NA | 2 | 0 | 1 | Low-quality | Genome-fragment | 2.77 | AAI-based (medium-confidence) | 0 |
| S7-final-viral-combined_k141_155360\|\|full | 2751 | No | NA | 4 | 0 | 0 | Not-determined | Genome-fragment | NA | NA | 0 |
| S7-final-viral-combined_k141_1944\|\|full | 1059 | No | NA | 3 | 0 | 0 | Not-determined | Genome-fragment | NA | NA | 0 |
| S5-final-viral-combined_k141_177437\|\|full | 1829 | No | NA | 2 | 2 | 0 | Low-quality | Genome-fragment | 3.21 | HMM-based (lower-bound) | 0 |
| S7-final-viral-combined_k141_144574\|\|full | 1539 | No | NA | 2 | 0 | 0 | Not-determined | Genome-fragment | NA | NA | 0 |
| S7-final-viral-combined_k141_72762\|\|full | 1829 | No | NA | 2 | 2 | 0 | Low-quality | Genome-fragment | 5.25 | AAI-based (high-confidence) | 0 |
| S12-final-viral-combined_k141_164148\|\|full | 1829 | No | NA | 4 | 1 | 0 | Low-quality | Genome-fragment | 3.39 | AAI-based (medium-confidence) | 0 |
| S6-final-viral-combined_k141_109432\|\|full | 1829 | No | NA | 2 | 0 | 0 | Low-quality | Genome-fragment | 2.22 | AAI-based (medium-confidence) | 0 |
| S11-final-viral-combined_k141_11938\|\|full | 1830 | No | NA | 2 | 0 | 2 | Low-quality | Genome-fragment | 3.16 | AAI-based (medium-confidence) | 0 |
| S1-final-viral-combined_k141_44628\|\|full | 1830 | No | NA | 3 | 0 | 1 | Low-quality | Genome-fragment | 1.52 | AAI-based (medium-confidence) | 0 |
| S1-final-viral-combined_k141_78426\|\|full | 1830 | No | NA | 3 | 1 | 0 | Low-quality | Genome-fragment | 3.75 | AAI-based (high-confidence) | 0 |
| S3-final-viral-combined_k141_17584\|\|full | 1830 | No | NA | 4 | 0 | 0 | Low-quality | Genome-fragment | 3.32 | AAI-based (high-confidence) | 0 |
| S7-final-viral-combined_k141_216438\|\|full | 5657 | No | NA | 5 | 0 | 2 | Not-determined | Genome-fragment | NA | NA | 0 |
| S5-final-viral-combined_k141_66319\|\|full | 1830 | No | NA | 3 | 2 | 0 | Low-quality | Genome-fragment | 5.36 | AAI-based (medium-confidence) | 0 |
| S7-final-viral-combined_k141_144591\|\|full | 3044 | No | NA | 3 | 0 | 3 | Not-determined | Genome-fragment | NA | NA | 0 |
| S17-final-viral-combined_k141_207185\|\|lt2gene | 1830 | No | NA | 1 | 1 | 0 | Low-quality | Genome-fragment | 4.17 | AAI-based (high-confidence) | 0 |
| S7-final-viral-combined_k141_53209\|\|full | 4482 | No | NA | 3 | 0 | 3 | Not-determined | Genome-fragment | NA | NA | 0 |
| S7-final-viral-combined_k141_83366\|\|full | 1232 | No | NA | 2 | 0 | 1 | Not-determined | Genome-fragment | NA | NA | 0 |
| S7-final-viral-combined_k141_165438\|\|full | 3523 | No | NA | 4 | 0 | 3 | Not-determined | Genome-fragment | NA | NA | 0 |
| S7-final-viral-combined_k141_227001\|\|full | 2506 | No | NA | 3 | 0 | 2 | Not-determined | Genome-fragment | NA | NA | 0 |
| S7-final-viral-combined_k141_206657\|\|full | 5769 | No | NA | 10 | 0 | 0 | Not-determined | Genome-fragment | NA | NA | 0 |
| S7-final-viral-combined_k141_114601\|\|full | 1534 | No | NA | 3 | 0 | 0 | Not-determined | Genome-fragment | NA | NA | 0 |
| S7-final-viral-combined_k141_12084\|\|full | 2454 | No | NA | 3 | 0 | 1 | Not-determined | Genome-fragment | NA | NA | 0 |
| S7-final-viral-combined_k141_206670\|\|full | 2232 | No | NA | 3 | 0 | 1 | Not-determined | Genome-fragment | NA | NA | 0 |
| S7-final-viral-combined_k141_72754\|\|full | 2028 | No | NA | 3 | 0 | 1 | Not-determined | Genome-fragment | NA | NA | 0 |
| S7-final-viral-combined_k141_196358\|\|full | 3235 | No | NA | 3 | 0 | 1 | Not-determined | Genome-fragment | NA | NA | 0 |
| S12-final-viral-combined_k141_148930\|\|full | 1830 | No | NA | 2 | 0 | 0 | Low-quality | Genome-fragment | 4.09 | AAI-based (medium-confidence) | 0 |
| S7-final-viral-combined_k141_22029\|\|full | 6634 | No | NA | 7 | 0 | 3 | Not-determined | Genome-fragment | NA | NA | 0 |
| S7-final-viral-combined_k141_144634\|\|full | 1402 | No | NA | 3 | 0 | 1 | Not-determined | Genome-fragment | NA | NA | 0 |
| S2-final-viral-combined_k141_79599\|\|full | 1830 | No | NA | 3 | 1 | 0 | Low-quality | Genome-fragment | 1.12 | HMM-based (lower-bound) | 0 |
| S7-final-viral-combined_k141_104214\|\|full | 1414 | No | NA | 2 | 0 | 2 | Not-determined | Genome-fragment | NA | NA | 0 |
| S8-final-viral-combined_k141_255826\|\|full | 1830 | No | NA | 3 | 0 | 2 | Low-quality | Genome-fragment | 0.73 | AAI-based (medium-confidence) | 0 |
| S7-final-viral-combined_k141_2050\|\|full | 12084 | No | NA | 11 | 0 | 7 | Not-determined | Genome-fragment | NA | NA | 0 |
| S1-final-viral-combined_k141_193101\|\|full | 1831 | No | NA | 3 | 0 | 1 | Low-quality | Genome-fragment | 2.69 | AAI-based (high-confidence) | 0 |
| S7-final-viral-combined_k141_22049\|\|full | 16296 | No | NA | 16 | 0 | 10 | Not-determined | Genome-fragment | NA | NA | 0 |
| S7-final-viral-combined_k141_186225\|\|full | 34162 | No | NA | 33 | 0 | 25 | Not-determined | Genome-fragment | NA | NA | 0 |
| S7-final-viral-combined_k141_2073\|\|full | 2952 | No | NA | 3 | 0 | 2 | Not-determined | Genome-fragment | NA | NA | 0 |
| S7-final-viral-combined_k141_63284\|\|full | 1598 | No | NA | 3 | 0 | 1 | Not-determined | Genome-fragment | NA | NA | 0 |
| S7-final-viral-combined_k141_45599\|\|full | 1831 | No | NA | 6 | 0 | 0 | Low-quality | Genome-fragment | 6.63 | AAI-based (medium-confidence) | 0 |
| S7-final-viral-combined_k141_114637\|\|full | 2091 | No | NA | 2 | 0 | 1 | Not-determined | Genome-fragment | NA | NA | 0 |
| S4-final-viral-combined_k141_13988\|\|lt2gene | 1831 | No | NA | 2 | 1 | 0 | Low-quality | Genome-fragment | 4.77 | AAI-based (high-confidence) | 0 |
| S7-final-viral-combined_k141_155484\|\|full | 7561 | No | NA | 3 | 0 | 2 | Not-determined | Genome-fragment | NA | NA | 0 |
| S7-final-viral-combined_k141_186271\|\|full | 2932 | No | NA | 3 | 0 | 1 | Not-determined | Genome-fragment | NA | NA | 0 |
| S7-final-viral-combined_k141_155501\|\|full | 3809 | No | NA | 5 | 0 | 4 | Not-determined | Genome-fragment | NA | NA | 0 |

| | | | | | | | | | | | |
|---|---|---|---|---|---|---|---|---|---|---|---|
| S7-final-viral-combined_k141_63297\|\|full | 4855 | No | NA | 7 | 0 | 1 | Not-determined | Genome-fragment | NA | NA | 0 |
| S7-final-viral-combined_k141_196453\|\|full | 1330 | No | NA | 2 | 0 | 1 | Not-determined | Genome-fragment | NA | NA | 0 |
| S4_k141_7357 | 1831 | No | NA | 2 | 0 | 1 | Low-quality | Genome-fragment | 3.05 | AAI-based (medium-confidence) | 0 |
| S4_k141_144080 | 1831 | No | NA | 2 | 0 | 0 | Low-quality | Genome-fragment | 3.62 | AAI-based (medium-confidence) | 0 |
| S7-final-viral-combined_k141_63320\|\|full | 1611 | No | NA | 2 | 0 | 1 | Not-determined | Genome-fragment | NA | NA | 0 |
| S7-final-viral-combined_k141_63324\|\|full | 1313 | No | NA | 3 | 0 | 2 | Not-determined | Genome-fragment | NA | NA | 0 |
| S11-final-viral-combined_k141_186300\|\|full | 1832 | No | NA | 2 | 0 | 2 | Low-quality | Genome-fragment | 1.09 | AAI-based (medium-confidence) | 0 |
| S1_k141_98985 | 1832 | No | NA | 1 | 0 | 1 | Low-quality | Genome-fragment | 0.99 | AAI-based (medium-confidence) | 0 |
| S12_k141_174288 | 1832 | No | NA | 3 | 0 | 1 | Low-quality | Genome-fragment | 1.97 | AAI-based (high-confidence) | 0 |
| S12_k141_230147 | 1832 | No | NA | 6 | 2 | 0 | Low-quality | Genome-fragment | 1.92 | AAI-based (high-confidence) | 0 |
| S11_k141_166939 | 1833 | No | NA | 3 | 1 | 0 | Low-quality | Genome-fragment | 2.32 | AAI-based (medium-confidence) | 0 |
| S8-final-viral-combined_k141_215534\|\|full | 1833 | No | NA | 3 | 0 | 0 | Low-quality | Genome-fragment | 5.56 | AAI-based (high-confidence) | 0 |
| S7-final-viral-combined_k141_93424\|\|full | 2135 | No | NA | 3 | 0 | 0 | Not-determined | Genome-fragment | NA | NA | 0 |
| S7-final-viral-combined_k141_2122\|\|full | 50765 | Yes | 31177 | 60 | 20 | 10 | Medium-quality | Genome-fragment | 78.02 | AAI-based (high-confidence) | 38.59 |
| S7-final-viral-combined_k141_206830\|\|full | 1367 | No | NA | 2 | 0 | 1 | Not-determined | Genome-fragment | NA | NA | 0 |
| S9-final-viral-combined_k141_69051\|\|full | 1834 | No | NA | 3 | 0 | 2 | Low-quality | Genome-fragment | 4.24 | AAI-based (medium-confidence) | 0 |
| S12-final-viral-combined_k141_142969\|\|full | 1834 | No | NA | 3 | 0 | 0 | Low-quality | Genome-fragment | 2.94 | AAI-based (medium-confidence) | 0 |
| S4-final-viral-combined_k141_156784\|\|full | 1834 | No | NA | 5 | 2 | 0 | Low-quality | Genome-fragment | 3.77 | AAI-based (high-confidence) | 0 |
| S4_k141_2496 | 1834 | No | NA | 4 | 0 | 0 | Low-quality | Genome-fragment | 3.03 | AAI-based (high-confidence) | 0 |
| S4_k141_144787 | 1834 | No | NA | 3 | 1 | 0 | Low-quality | Genome-fragment | 0.99 | HMM-based (lower-bound) | 0 |
| S7-final-viral-combined_k141_144647\|\|full | 1785 | No | NA | 2 | 0 | 0 | Not-determined | Genome-fragment | NA | NA | 0 |
| S7-final-viral-combined_k141_2151\|\|full | 1063 | No | NA | 3 | 0 | 0 | Not-determined | Genome-fragment | NA | NA | 0 |
| S8-final-viral-combined_k141_275936\|\|full | 1834 | No | NA | 3 | 0 | 1 | Low-quality | Genome-fragment | 2.58 | AAI-based (medium-confidence) | 0 |
| S12-final-viral-combined_k141_234413\|\|full | 1835 | No | NA | 2 | 0 | 1 | Low-quality | Genome-fragment | 3.3 | AAI-based (medium-confidence) | 0 |
| S12-final-viral-combined_k141_187032\|\|full | 1835 | No | NA | 5 | 0 | 0 | Low-quality | Genome-fragment | 4.19 | AAI-based (high-confidence) | 0 |
| S4-final-viral-combined_k141_71258\|\|full | 1835 | No | NA | 3 | 0 | 0 | Low-quality | Genome-fragment | 3.69 | AAI-based (medium-confidence) | 0 |
| S7-final-viral-combined_k141_144674\|\|full | 2841 | No | NA | 3 | 0 | 1 | Not-determined | Genome-fragment | NA | NA | 0 |
| S4_k141_44333 | 1835 | No | NA | 1 | 0 | 1 | Low-quality | Genome-fragment | 3.95 | AAI-based (high-confidence) | 0 |
| S7-final-viral-combined_k141_114672\|\|full | 1943 | No | NA | 2 | 0 | 0 | Not-determined | Genome-fragment | NA | NA | 0 |
| S6-final-viral-combined_k141_140152\|\|full | 1835 | Yes | 725 | 4 | 1 | 1 | Low-quality | Genome-fragment | 0.81 | HMM-based (lower-bound) | 60.49 |
| S6_k141_21192 | 1835 | No | NA | 2 | 1 | 0 | Low-quality | Genome-fragment | 6.29 | HMM-based (lower-bound) | 0 |
| S9-final-viral-combined_k141_34616\|\|full | 1836 | Yes | 881 | 4 | 1 | 1 | Low-quality | Genome-fragment | 1.88 | AAI-based (medium-confidence) | 52.02 |
| S9-final-viral-combined_k141_26687\|\|full | 1836 | No | NA | 3 | 1 | 0 | Low-quality | Genome-fragment | 4.1 | AAI-based (medium-confidence) | 0 |
| S1-final-viral-combined_k141_32956\|\|full | 1836 | No | NA | 4 | 0 | 0 | Low-quality | Genome-fragment | 4.44 | AAI-based (high-confidence) | 0 |
| S7-final-viral-combined_k141_155649\|\|full | 1733 | No | NA | 2 | 0 | 1 | Not-determined | Genome-fragment | NA | NA | 0 |
| S7-final-viral-combined_k141_237247\|\|full | 2642 | No | NA | 3 | 0 | 1 | Not-determined | Genome-fragment | NA | NA | 0 |
| S1-final-viral-combined_k141_7307\|\|full | 1836 | No | NA | 3 | 0 | 2 | Low-quality | Genome-fragment | 2.49 | AAI-based (high-confidence) | 0 |
| S7-final-viral-combined_k141_144746\|\|full | 21391 | No | NA | 19 | 0 | 14 | Not-determined | Genome-fragment | NA | NA | 0 |
| S7-final-viral-combined_k141_144747\|\|full | 1410 | No | NA | 3 | 0 | 1 | Not-determined | Genome-fragment | NA | NA | 0 |
| S7-final-viral-combined_k141_132130\|\|full | 1836 | No | NA | 2 | 0 | 1 | Low-quality | Genome-fragment | 2.29 | AAI-based (medium-confidence) | 0 |
| S17-final-viral-combined_k141_23317\|\|full | 1836 | No | NA | 3 | 2 | 0 | Low-quality | Genome-fragment | 3.6 | AAI-based (high-confidence) | 0 |
| S17-final-viral-combined_k141_129781\|\|full | 1836 | No | NA | 3 | 1 | 0 | Low-quality | Genome-fragment | 2.65 | AAI-based (high-confidence) | 0 |
| S7-final-viral-combined_k141_83578\|\|full | 1310 | No | NA | 2 | 0 | 2 | Not-determined | Genome-fragment | NA | NA | 0 |
| S7-final-viral-combined_k141_216733\|\|full | 2983 | No | NA | 3 | 0 | 2 | Not-determined | Genome-fragment | NA | NA | 0 |
| S7-final-viral-combined_k141_2264\|\|full | 3328 | No | NA | 4 | 0 | 3 | Not-determined | Genome-fragment | NA | NA | 0 |
| S12_k141_211912 | 1836 | No | NA | 1 | 0 | 1 | Low-quality | Genome-fragment | 0.76 | AAI-based (medium-confidence) | 0 |
| S4-final-viral-combined_k141_37571\|\|full | 1836 | No | NA | 3 | 1 | 0 | Low-quality | Genome-fragment | 1.85 | AAI-based (high-confidence) | 0 |
| S7-final-viral-combined_k141_72975\|\|full | 3499 | No | NA | 2 | 0 | 2 | Not-determined | Genome-fragment | NA | NA | 0 |
| S4_k141_160142 | 1836 | No | NA | 1 | 0 | 1 | Low-quality | Genome-fragment | 1 | AAI-based (medium-confidence) | 0 |
| S7-final-viral-combined_k141_206940\|\|full | 3867 | No | NA | 8 | 0 | 6 | Not-determined | Genome-fragment | NA | NA | 0 |
| S8-final-viral-combined_k141_43025\|\|full | 1836 | Yes | 1599 | 3 | 1 | 1 | Low-quality | Genome-fragment | 2.99 | AAI-based (medium-confidence) | 12.91 |
| S1_k141_71906 | 1837 | No | NA | 2 | 0 | 0 | Low-quality | Genome-fragment | 0.8 | AAI-based (medium-confidence) | 0 |
| S7-final-viral-combined_k141_155735\|\|full | 2727 | No | NA | 4 | 0 | 1 | Not-determined | Genome-fragment | NA | NA | 0 |
| S5-final-viral-combined_k141_13679\|\|full | 1837 | No | NA | 4 | 1 | 0 | Low-quality | Genome-fragment | 3.02 | AAI-based (medium-confidence) | 0 |
| S7-final-viral-combined_k141_12375\|\|full | 2747 | No | NA | 4 | 0 | 0 | Not-determined | Genome-fragment | NA | NA | 0 |
| S7-final-viral-combined_k141_224439\|\|full | 1837 | No | NA | 3 | 0 | 1 | Low-quality | Genome-fragment | 3.29 | AAI-based (high-confidence) | 0 |
| S7-final-viral-combined_k141_155754\|\|full | 8940 | No | NA | 7 | 0 | 1 | Not-determined | Genome-fragment | NA | NA | 0 |
| S17-final-viral-combined_k141_68196\|\|full | 1837 | No | NA | 3 | 1 | 0 | Low-quality | Genome-fragment | 3.71 | AAI-based (medium-confidence) | 0 |
| S2-final-viral-combined_k141_171527\|\|full | 1837 | No | NA | 3 | 0 | 0 | Low-quality | Genome-fragment | 2.13 | AAI-based (medium-confidence) | 0 |
| S7-final-viral-combined_k141_144928\|\|full | 1147 | No | NA | 2 | 0 | 0 | Not-determined | Genome-fragment | NA | NA | 0 |
| S7-final-viral-combined_k141_104480\|\|full | 1812 | No | NA | 2 | 0 | 2 | Not-determined | Genome-fragment | NA | NA | 0 |
| S7-final-viral-combined_k141_33047\|\|full | 1300 | No | NA | 4 | 0 | 3 | Not-determined | Genome-fragment | NA | NA | 0 |
| S7-final-viral-combined_k141_63558\|\|full | 1726 | No | NA | 2 | 0 | 0 | Not-determined | Genome-fragment | NA | NA | 0 |
| S7-final-viral-combined_k141_144940\|\|full | 3197 | No | NA | 3 | 0 | 2 | Not-determined | Genome-fragment | NA | NA | 0 |
| S7-final-viral-combined_k141_73048\|\|full | 2293 | No | NA | 4 | 0 | 2 | Not-determined | Genome-fragment | NA | NA | 0 |
| S7-final-viral-combined_k141_216814\|\|full | 12410 | No | NA | 12 | 0 | 2 | Not-determined | Genome-fragment | NA | NA | 0 |
| S7-final-viral-combined_k141_114827\|\|full | 111577 | No | NA | 121 | 0 | 24 | Not-determined | Genome-fragment | NA | NA | 0 |
| S7-final-viral-combined_k141_63582\|\|full | 1724 | No | NA | 4 | 0 | 1 | Not-determined | Genome-fragment | NA | NA | 0 |
| S7-final-viral-combined_k141_114829\|\|full | 1789 | No | NA | 3 | 0 | 2 | Not-determined | Genome-fragment | NA | NA | 0 |
| S8-final-viral-combined_k141_199279\|\|full | 1837 | No | NA | 3 | 0 | 0 | Low-quality | Genome-fragment | 1.73 | AAI-based (high-confidence) | 0 |
| S3-final-viral-combined_k141_117118\|\|full | 1838 | No | NA | 5 | 1 | 0 | Low-quality | Genome-fragment | 4.7 | AAI-based (high-confidence) | 0 |
| S5_k141_75586 | 1838 | No | NA | 4 | 0 | 0 | Low-quality | Genome-fragment | 3.63 | AAI-based (high-confidence) | 0 |
| S7-final-viral-combined_k141_124910\|\|full | 11698 | No | NA | 3 | 0 | 2 | Not-determined | Genome-fragment | NA | NA | 0 |
| S7-final-viral-combined_k141_54943\|\|full | 1838 | No | NA | 3 | 0 | 0 | Low-quality | Genome-fragment | 4.1 | AAI-based (high-confidence) | 0 |
| S7-final-viral-combined_k141_53544\|\|full | 4120 | No | NA | 4 | 0 | 0 | Not-determined | Genome-fragment | NA | NA | 0 |
| S7-final-viral-combined_k141_196841\|\|full | 1578 | No | NA | 2 | 0 | 0 | Not-determined | Genome-fragment | NA | NA | 0 |
| S7-final-viral-combined_k141_237441\|\|full | 10565 | No | NA | 9 | 0 | 5 | Not-determined | Genome-fragment | NA | NA | 0 |
| S7-final-viral-combined_k141_186673\|\|full | 2165 | No | NA | 3 | 0 | 1 | Not-determined | Genome-fragment | NA | NA | 0 |
| S7-final-viral-combined_k141_83782\|\|full | 1732 | No | NA | 3 | 0 | 1 | Not-determined | Genome-fragment | NA | NA | 0 |
| S7-final-viral-combined_k141_210071\|\|full | 1838 | No | NA | 4 | 0 | 0 | Low-quality | Genome-fragment | 2.84 | AAI-based (high-confidence) | 0 |
| S7-final-viral-combined_k141_43161\|\|full | 1816 | No | NA | 2 | 0 | 1 | Not-determined | Genome-fragment | NA | NA | 0 |
| S7_k141_29234 | 1838 | No | NA | 1 | 1 | 0 | Low-quality | Genome-fragment | 4.01 | AAI-based (high-confidence) | 0 |
| S8-final-viral-combined_k141_183673\|\|full | 1838 | No | NA | 4 | 0 | 0 | Low-quality | Genome-fragment | 3.91 | AAI-based (medium-confidence) | 0 |
| S11-final-viral-combined_k141_290523\|\|full | 1839 | No | NA | 3 | 0 | 2 | Low-quality | Genome-fragment | 2.5 | AAI-based (high-confidence) | 0 |
| S7-final-viral-combined_k141_12484\|\|full | 4183 | No | NA | 7 | 0 | 1 | Not-determined | Genome-fragment | NA | NA | 0 |
| S7-final-viral-combined_k141_83809\|\|full | 3842 | No | NA | 4 | 0 | 0 | Not-determined | Genome-fragment | NA | NA | 0 |
| S11-final-viral-combined_k141_73125\|\|full | 1839 | No | NA | 3 | 0 | 0 | Low-quality | Genome-fragment | 4.4 | AAI-based (medium-confidence) | 0 |
| S7-final-viral-combined_k141_186702\|\|full | 2409 | No | NA | 3 | 0 | 1 | Not-determined | Genome-fragment | NA | NA | 0 |
| S7-final-viral-combined_k141_155832\|\|full | 1784 | No | NA | 2 | 0 | 2 | Not-determined | Genome-fragment | NA | NA | 0 |
| S7-final-viral-combined_k141_135049\|\|full | 7902 | No | NA | 8 | 0 | 6 | Not-determined | Genome-fragment | NA | NA | 0 |
| S3-final-viral-combined_k141_12051\|\|full | 1839 | No | NA | 4 | 0 | 0 | Low-quality | Genome-fragment | 1.25 | AAI-based (high-confidence) | 0 |
| S7-final-viral-combined_k141_63735\|\|full | 2431 | No | NA | 4 | 0 | 1 | Not-determined | Genome-fragment | NA | NA | 0 |
| S5-final-viral-combined_k141_73740\|\|full | 1839 | No | NA | 2 | 0 | 0 | Low-quality | Genome-fragment | 3.85 | AAI-based (high-confidence) | 0 |
| S17-final-viral-combined_k141_155169\|\|full | 1839 | No | NA | 3 | 0 | 1 | Low-quality | Genome-fragment | 3.39 | AAI-based (high-confidence) | 0 |
| S17-k141_195720 | 1839 | No | NA | 1 | 0 | 1 | Low-quality | Genome-fragment | 0.99 | AAI-based (medium-confidence) | 0 |
| S2-final-viral-combined_k141_165960\|\|full | 1839 | No | NA | 3 | 1 | 0 | Low-quality | Genome-fragment | 4.34 | AAI-based (high-confidence) | 0 |
| S7-final-viral-combined_k141_237534\|\|full | 1610 | No | NA | 3 | 0 | 2 | Not-determined | Genome-fragment | NA | NA | 0 |
| S7-final-viral-combined_k141_63747\|\|full | 5474 | No | NA | 4 | 0 | 1 | Not-determined | Genome-fragment | NA | NA | 0 |
| S1-final-viral-combined_k141_334150\|\|full | 1840 | No | NA | 3 | 1 | 0 | Low-quality | Genome-fragment | 2.26 | AAI-based (high-confidence) | 0 |
| S1-final-viral-combined_k141_141867\|\|full | 1840 | No | NA | 3 | 1 | 0 | Low-quality | Genome-fragment | 1.12 | HMM-based (lower-bound) | 0 |
| S1-final-viral-combined_k141_142832\|\|full | 1840 | No | NA | 2 | 0 | 1 | Low-quality | Genome-fragment | 1.92 | AAI-based (medium-confidence) | 0 |
| S7-final-viral-combined_k141_83879\|\|full | 4994 | No | NA | 4 | 0 | 3 | Not-determined | Genome-fragment | NA | NA | 0 |
| S1-final-viral-combined_k141_175609\|\|full | 1840 | No | NA | 2 | 1 | 0 | Low-quality | Genome-fragment | 4.52 | AAI-based (high-confidence) | 0 |
| S3-final-viral-combined_k141_30820\|\|full | 1840 | No | NA | 4 | 0 | 0 | Low-quality | Genome-fragment | 1.52 | AAI-based (medium-confidence) | 0 |
| S7-final-viral-combined_k141_2520\|\|full | 2335 | No | NA | 3 | 0 | 1 | Not-determined | Genome-fragment | NA | NA | 0 |
| S7-final-viral-combined_k141_186769\|\|full | 3316 | No | NA | 4 | 0 | 3 | Not-determined | Genome-fragment | NA | NA | 0 |
| S7-final-viral-combined_k141_22411\|\|full | 6763 | No | NA | 6 | 0 | 5 | Not-determined | Genome-fragment | NA | NA | 0 |
| S7-final-viral-combined_k141_165927\|\|full | 1237 | No | NA | 3 | 0 | 2 | Not-determined | Genome-fragment | NA | NA | 0 |
| S7-final-viral-combined_k141_196995\|\|full | 7650 | No | NA | 6 | 0 | 1 | Not-determined | Genome-fragment | NA | NA | 0 |
| S17-final-viral-combined_k141_101007\|\|full | 1840 | No | NA | 2 | 0 | 2 | Low-quality | Genome-fragment | 2.88 | AAI-based (medium-confidence) | 0 |
| S7-final-viral-combined_k141_135121\|\|full | 2679 | No | NA | 4 | 0 | 1 | Not-determined | Genome-fragment | NA | NA | 0 |
| S12_k141_354784 | 1840 | No | NA | 1 | 0 | 0 | Low-quality | Genome-fragment | 3.72 | AAI-based (medium-confidence) | 0 |
| S12_k141_140624 | 1840 | No | NA | 2 | 0 | 0 | Low-quality | Genome-fragment | 5.67 | AAI-based (high-confidence) | 0 |
| S7-final-viral-combined_k141_2596\|\|full | 2707 | No | NA | 3 | 0 | 2 | Not-determined | Genome-fragment | NA | NA | 0 |
| S7-final-viral-combined_k141_165939\|\|full | 5582 | No | NA | 7 | 0 | 3 | Not-determined | Genome-fragment | NA | NA | 0 |
| S7-final-viral-combined_k141_165940\|\|full | 9690 | No | NA | 9 | 0 | 2 | Not-determined | Genome-fragment | NA | NA | 0 |
| S7-final-viral-combined_k141_145080\|\|full | 2292 | No | NA | 4 | 0 | 2 | Not-determined | Genome-fragment | NA | NA | 0 |
| S2-final-viral-combined_k141_55241\|\|full | 1840 | No | NA | 3 | 0 | 0 | Low-quality | Genome-fragment | 3.5 | AAI-based (medium-confidence) | 0 |
| S7-final-viral-combined_k141_114976\|\|full | 3969 | No | NA | 6 | 0 | 0 | Not-determined | Genome-fragment | NA | NA | 0 |
| S7-final-viral-combined_k141_104532\|\|full | 1520 | No | NA | 4 | 0 | 0 | Not-determined | Genome-fragment | NA | NA | 0 |
| S11-final-viral-combined_k141_268335\|\|full | 1841 | No | NA | 3 | 0 | 0 | Low-quality | Genome-fragment | 4.43 | AAI-based (medium-confidence) | 0 |
| S7-final-viral-combined_k141_63867\|\|full | 17572 | No | NA | 19 | 0 | 13 | Not-determined | Genome-fragment | NA | NA | 0 |
| S11-final-viral-combined_k141_119824\|\|full | 1841 | No | NA | 3 | 0 | 1 | Low-quality | Genome-fragment | 2.32 | AAI-based (medium-confidence) | 0 |
| S1-final-viral-combined_k141_135380\|\|full | 1841 | No | NA | 3 | 1 | 0 | Low-quality | Genome-fragment | 6.1 | AAI-based (high-confidence) | 0 |
| S7-final-viral-combined_k141_104579\|\|full | 4673 | No | NA | 5 | 0 | 2 | Not-determined | Genome-fragment | NA | NA | 0 |
| S1-final-viral-combined_k141_250674\|\|lt2gene | 1841 | No | NA | 1 | 1 | 0 | Low-quality | Genome-fragment | 4.46 | AAI-based (high-confidence) | 0 |
| S7-final-viral-combined_k141_104582\|\|full | 2226 | No | NA | 2 | 0 | 2 | Not-determined | Genome-fragment | NA | NA | 0 |
| S7-final-viral-combined_k141_104584\|\|full | 2507 | No | NA | 2 | 0 | 2 | Not-determined | Genome-fragment | NA | NA | 0 |
| S7-final-viral-combined_k141_45332\|\|full | 1841 | No | NA | 4 | 0 | 0 | Low-quality | Genome-fragment | 3.45 | AAI-based (high-confidence) | 0 |
| S17-final-viral-combined_k141_100445\|\|full | 1841 | No | NA | 2 | 2 | 0 | Low-quality | Genome-fragment | 4.47 | AAI-based (high-confidence) | 0 |
| S7-final-viral-combined_k141_135154\|\|full | 50087 | Yes | 29396 | 41 | 4 | 9 | Medium-quality | Genome-fragment | 64.79 | AAI-based (medium-confidence) | 41.31 |
| S17-final-viral-combined_k141_259997\|\|full | 1841 | No | NA | 2 | 0 | 1 | Low-quality | Genome-fragment | 1.18 | AAI-based (medium-confidence) | 0 |
| S2-final-viral-combined_k141_104867\|\|lt2gene | 1841 | No | NA | 1 | 1 | 0 | Low-quality | Genome-fragment | 3.91 | AAI-based (high-confidence) | 0 |
| S7-final-viral-combined_k141_166036\|\|full | 2544 | No | NA | 3 | 0 | 0 | Not-determined | Genome-fragment | NA | NA | 0 |
| S4-final-viral-combined_k141_163409\|\|full | 1841 | No | NA | 1 | 1 | 0 | Low-quality | Genome-fragment | 1.83 | AAI-based (medium-confidence) | 0 |
| S11-final-viral-combined_k141_63956\|\|full | 1842 | No | NA | 2 | 0 | 0 | Low-quality | Genome-fragment | 5.26 | AAI-based (medium-confidence) | 0 |
| S7-final-viral-combined_k141_73312\|\|full | 1113 | No | NA | 2 | 0 | 2 | Not-determined | Genome-fragment | NA | NA | 0 |
| S7-final-viral-combined_k141_93905\|\|full | 1504 | No | NA | 3 | 0 | 0 | Not-determined | Genome-fragment | NA | NA | 0 |
| S1-final-viral-combined_k141_312722\|\|full | 1842 | No | NA | 4 | 0 | 0 | Low-quality | Genome-fragment | 2.13 | AAI-based (medium-confidence) | 0 |
| S7-final-viral-combined_k141_186970\|\|full | 1459 | No | NA | 2 | 0 | 0 | Not-determined | Genome-fragment | NA | NA | 0 |
| S7-final-viral-combined_k141_115103\|\|full | 3745 | No | NA | 3 | 0 | 2 | Not-determined | Genome-fragment | NA | NA | 0 |
| S1_k141_330332 | 1842 | No | NA | 4 | 3 | 0 | Low-quality | Genome-fragment | 3.3 | AAI-based (high-confidence) | 0 |
| S7-final-viral-combined_k141_104666\|\|full | 1076 | No | NA | 2 | 0 | 0 | Not-determined | Genome-fragment | NA | NA | 0 |
| S7-final-viral-combined_k141_104673\|\|full | 1811 | No | NA | 3 | 0 | 2 | Not-determined | Genome-fragment | NA | NA | 0 |
| S7-final-viral-combined_k141_197124\|\|full | 1576 | No | NA | 2 | 0 | 1 | Not-determined | Genome-fragment | NA | NA | 0 |
| S7-final-viral-combined_k141_115119\|\|full | 1845 | No | NA | 3 | 0 | 1 | Not-determined | Genome-fragment | NA | NA | 0 |
| S7-final-viral-combined_k141_175961\|\|full | 2192 | No | NA | 4 | 0 | 4 | Not-determined | Genome-fragment | NA | NA | 0 |
| S7-final-viral-combined_k141_33394\|\|full | 1842 | No | NA | 5 | 2 | 0 | Low-quality | Genome-fragment | 4.64 | AAI-based (high-confidence) | 0 |
| S7-final-viral-combined_k141_237827\|\|full | 1071 | No | NA | 2 | 0 | 1 | Not-determined | Genome-fragment | NA | NA | 0 |
| S8-final-viral-combined_k141_92983\|\|full | 1842 | No | NA | 2 | 0 | 0 | Low-quality | Genome-fragment | 2.97 | AAI-based (medium-confidence) | 0 |
| S7-final-viral-combined_k141_22493\|\|full | 4320 | No | NA | 7 | 0 | 4 | Not-determined | Genome-fragment | NA | NA | 0 |
| S12-final-viral-combined_k141_242329\|\|full | 1843 | No | NA | 6 | 1 | 0 | Low-quality | Genome-fragment | 3.91 | AAI-based (medium-confidence) | 0 |
| S12-final-viral-combined_k141_63546\|\|full | 1843 | No | NA | 4 | 1 | 0 | Low-quality | Genome-fragment | 3.12 | AAI-based (high-confidence) | 0 |
| S7-final-viral-combined_k141_33381\|\|full | 2187 | No | NA | 4 | 0 | 0 | Not-determined | Genome-fragment | NA | NA | 0 |
| S7-final-viral-combined_k141_33387\|\|full | 1277 | No | NA | 3 | 0 | 1 | Not-determined | Genome-fragment | NA | NA | 0 |
| S7-final-viral-combined_k141_125114\|\|full | 1089 | No | NA | 2 | 0 | 0 | Not-determined | Genome-fragment | NA | NA | 0 |
| S2-final-viral-combined_k141_6924\|\|full | 1843 | No | NA | 2 | 0 | 2 | Low-quality | Genome-fragment | 1.3 | AAI-based (medium-confidence) | 0 |
| S7-final-viral-combined_k141_33404\|\|full | 2535 | No | NA | 4 | 0 | 1 | Not-determined | Genome-fragment | NA | NA | 0 |
| S7-final-viral-combined_k141_166135\|\|full | 3542 | No | NA | 5 | 0 | 4 | Not-determined | Genome-fragment | NA | NA | 0 |
| S4_k141_158682 | 1843 | No | NA | 4 | 0 | 1 | Low-quality | Genome-fragment | 1.11 | AAI-based (medium-confidence) | 0 |
| S8-final-viral-combined_k141_102632\|\|lt2gene | 1843 | No | NA | 2 | 1 | 0 | Low-quality | Genome-fragment | 0.15 | HMM-based (lower-bound) | 0 |
| S7-final-viral-combined_k141_237918\|\|full | 3381 | No | NA | 4 | 0 | 3 | Not-determined | Genome-fragment | NA | NA | 0 |
| S1-final-viral-combined_k141_390580\|\|full | 1844 | No | NA | 2 | 0 | 2 | Low-quality | Genome-fragment | 3.37 | AAI-based (medium-confidence) | 0 |
| S7-final-viral-combined_k141_225569\|\|full | 1844 | No | NA | 3 | 0 | 1 | Low-quality | Genome-fragment | 1.92 | AAI-based (high-confidence) | 0 |
| S7-final-viral-combined_k141_227683\|\|full | 2383 | No | NA | 5 | 0 | 0 | Not-determined | Genome-fragment | NA | NA | 0 |
| S7-final-viral-combined_k141_53774\|\|full | 2276 | No | NA | 4 | 0 | 0 | Not-determined | Genome-fragment | NA | NA | 0 |
| S12-final-viral-combined_k141_253665\|\|full | 1844 | No | NA | 2 | 1 | 0 | Low-quality | Genome-fragment | 3.45 | AAI-based (medium-confidence) | 0 |
| S12_k141_362240 | 1844 | No | NA | 3 | 0 | 0 | Low-quality | Genome-fragment | 3.91 | AAI-based (medium-confidence) | 0 |
| S7-final-viral-combined_k141_156234\|\|full | 1191 | No | NA | 2 | 0 | 0 | Not-determined | Genome-fragment | NA | NA | 0 |
| S7-final-viral-combined_k141_84148\|\|full | 2737 | No | NA | 4 | 0 | 1 | Not-determined | Genome-fragment | NA | NA | 0 |
| S4_k141_130277 | 1844 | No | NA | 3 | 3 | 0 | Low-quality | Genome-fragment | 1.06 | AAI-based (high-confidence) | 0 |
| S8_k141_139891 | 1844 | No | NA | 2 | 0 | 0 | Low-quality | Genome-fragment | 2.66 | AAI-based (medium-confidence) | 0 |
| S7-final-viral-combined_k141_237955\|\|full | 2599 | No | NA | 3 | 0 | 0 | Not-determined | Genome-fragment | NA | NA | 0 |
| S7-final-viral-combined_k141_73408\|\|full | 1546 | No | NA | 2 | 0 | 2 | Not-determined | Genome-fragment | NA | NA | 0 |
| S7-final-viral-combined_k141_33479\|\|full | 2997 | No | NA | 2 | 0 | 1 | Not-determined | Genome-fragment | NA | NA | 0 |
| S11_k141_57745 | 1845 | No | NA | 1 | 0 | 0 | Low-quality | Genome-fragment | 3.33 | AAI-based (medium-confidence) | 0 |
| S7-final-viral-combined_k141_141661\|\|full | 1845 | No | NA | 3 | 1 | 0 | Low-quality | Genome-fragment | 2.31 | HMM-based (lower-bound) | 0 |
| S7_k141_69416 | 1845 | No | NA | 3 | 0 | 3 | Low-quality | Genome-fragment | 4.71 | AAI-based (medium-confidence) | 0 |
| S2-final-viral-combined_k141_28213\|\|full | 1845 | No | NA | 4 | 0 | 1 | Low-quality | Genome-fragment | 2.45 | AAI-based (medium-confidence) | 0 |
| S8-final-viral-combined_k141_44209\|\|full | 1845 | No | NA | 4 | 0 | 0 | Low-quality | Genome-fragment | 0.71 | AAI-based (high-confidence) | 0 |
| S7-final-viral-combined_k141_125239\|\|full | 1882 | No | NA | 2 | 0 | 0 | Not-determined | Genome-fragment | NA | NA | 0 |
| S7-final-viral-combined_k141_125245\|\|full | 1935 | No | NA | 2 | 0 | 0 | Not-determined | Genome-fragment | NA | NA | 0 |
| S7-final-viral-combined_k141_73447\|\|full | 4511 | No | NA | 4 | 0 | 2 | Not-determined | Genome-fragment | NA | NA | 0 |
| S7-final-viral-combined_k141_33572\|\|full | 2015 | No | NA | 3 | 0 | 0 | Not-determined | Genome-fragment | NA | NA | 0 |
| S7-final-viral-combined_k141_176154\|\|full | 14792 | No | NA | 15 | 0 | 6 | Not-determined | Genome-fragment | NA | NA | 0 |
| S7-final-viral-combined_k141_217074\|\|full | 1474 | No | NA | 2 | 0 | 2 | Not-determined | Genome-fragment | NA | NA | 0 |
| S7-final-viral-combined_k141_2826\|\|full | 1033 | No | NA | 3 | 0 | 2 | Not-determined | Genome-fragment | NA | NA | 0 |
| S8_k141_123769 | 1845 | No | NA | 4 | 1 | 0 | Low-quality | Genome-fragment | 1.2 | AAI-based (high-confidence) | 0 |
| S7-final-viral-combined_k141_197267\|\|full | 12141 | No | NA | 8 | 0 | 6 | Not-determined | Genome-fragment | NA | NA | 0 |
| S7-final-viral-combined_k141_145313\|\|full | 1495 | No | NA | 3 | 0 | 0 | Not-determined | Genome-fragment | NA | NA | 0 |
| S11_k141_229238 | 1846 | No | NA | 3 | 0 | 0 | Low-quality | Genome-fragment | 1.74 | AAI-based (high-confidence) | 0 |
| S1-final-viral-combined_k141_170590\|\|full | 1846 | No | NA | 4 | 1 | 0 | Low-quality | Genome-fragment | 5.66 | AAI-based (medium-confidence) | 0 |
| S7-final-viral-combined_k141_33613\|\|full | 1255 | No | NA | 3 | 0 | 1 | Not-determined | Genome-fragment | NA | NA | 0 |
| S5_k141_70224 | 1846 | No | NA | 2 | 0 | 0 | Low-quality | Genome-fragment | 2.85 | AAI-based (high-confidence) | 0 |
| S12-final-viral-combined_k141_67593\|\|full | 1846 | No | NA | 2 | 0 | 2 | Low-quality | Genome-fragment | 2.64 | AAI-based (medium-confidence) | 0 |
| S2-final-viral-combined_k141_41291\|\|full | 1846 | No | NA | 3 | 0 | 0 | Low-quality | Genome-fragment | 3.81 | AAI-based (high-confidence) | 0 |
| S7-final-viral-combined_k141_135361\|\|full | 5220 | No | NA | 9 | 0 | 1 | Not-determined | Genome-fragment | NA | NA | 0 |
| S7-final-viral-combined_k141_176215\|\|full | 1255 | No | NA | 3 | 0 | 0 | Not-determined | Genome-fragment | NA | NA | 0 |
| S6-final-viral-combined_k141_46900\|\|full | 1846 | No | NA | 3 | 1 | 0 | Low-quality | Genome-fragment | 4.96 | AAI-based (medium-confidence) | 0 |
| S7-final-viral-combined_k141_166238\|\|full | 3647 | No | NA | 6 | 0 | 0 | Not-determined | Genome-fragment | NA | NA | 0 |
| S9_k141_90828 | 1847 | Yes | 949 | 3 | 1 | 1 | Low-quality | Genome-fragment | 0.73 | AAI-based (medium-confidence) | 48.62 |
| S11-final-viral-combined_k141_163455\|\|full | 1847 | No | NA | 2 | 1 | 0 | Low-quality | Genome-fragment | 2.59 | HMM-based (lower-bound) | 0 |
| S7-final-viral-combined_k141_73532\|\|full | 5993 | No | NA | 5 | 0 | 2 | Not-determined | Genome-fragment | NA | NA | 0 |
| S3-final-viral-combined_k141_19125\|\|full | 1847 | No | NA | 3 | 0 | 1 | Low-quality | Genome-fragment | 1.37 | AAI-based (medium-confidence) | 0 |
| S7-final-viral-combined_k141_156351\|\|full | 21991 | No | NA | 40 | 9 | 1 | Medium-quality | Genome-fragment | 55.52 | AAI-based (high-confidence) | 0 |
| S7-final-viral-combined_k141_94208\|\|full | 5915 | No | NA | 6 | 0 | 3 | Not-determined | Genome-fragment | NA | NA | 0 |
| S7-final-viral-combined_k141_125292\|\|full | 1352 | No | NA | 4 | 0 | 0 | Not-determined | Genome-fragment | NA | NA | 0 |
| S12-final-viral-combined_k141_387022\|\|full | 1847 | No | NA | 4 | 0 | 1 | Low-quality | Genome-fragment | 2.35 | AAI-based (medium-confidence) | 0 |
| S4-final-viral-combined_k141_122879\|\|full | 1847 | No | NA | 4 | 0 | 0 | Low-quality | Genome-fragment | 3.97 | AAI-based (medium-confidence) | 0 |
| S3-final-viral-combined_k141_5293\|\|full | 1848 | No | NA | 3 | 0 | 0 | Low-quality | Genome-fragment | 4.11 | AAI-based (high-confidence) | 0 |
| S7-final-viral-combined_k141_167776\|\|full | 1848 | No | NA | 3 | 0 | 0 | Low-quality | Genome-fragment | 1.52 | AAI-based (medium-confidence) | 0 |
| S7-final-viral-combined_k141_238043\|\|full | 2231 | No | NA | 2 | 0 | 1 | Not-determined | Genome-fragment | NA | NA | 0 |
| S7-final-viral-combined_k141_125311\|\|full | 3169 | No | NA | 3 | 0 | 0 | Not-determined | Genome-fragment | NA | NA | 0 |
| S17-final-viral-combined_k141_106734\|\|lt2gene | 1848 | No | NA | 1 | 1 | 0 | Low-quality | Genome-fragment | 4.78 | AAI-based (medium-confidence) | 0 |
| S7-final-viral-combined_k141_217146\|\|full | 2464 | No | NA | 2 | 0 | 0 | Not-determined | Genome-fragment | NA | NA | 0 |
| S17-k141_79478 | 1848 | No | NA | 1 | 0 | 0 | Low-quality | Genome-fragment | 3.09 | AAI-based (high-confidence) | 0 |
| S7-final-viral-combined_k141_166259\|\|full | 6955 | No | NA | 2 | 0 | 1 | Not-determined | Genome-fragment | NA | NA | 0 |
| S4-final-viral-combined_k141_9470\|\|full | 1848 | Yes | 141 | 5 | 1 | 2 | Low-quality | Genome-fragment | 0.39 | AAI-based (high-confidence) | 92.37 |
| S6-final-viral-combined_k141_50471\|\|full | 1848 | No | NA | 4 | 1 | 0 | Low-quality | Genome-fragment | 3.62 | AAI-based (high-confidence) | 0 |
| S7-final-viral-combined_k141_13115\|\|full | 1478 | No | NA | 4 | 0 | 0 | Not-determined | Genome-fragment | NA | NA | 0 |
| S7-final-viral-combined_k141_104872\|\|full | 31164 | Yes | 27297 | 32 | 3 | 2 | Medium-quality | Genome-fragment | 59.42 | AAI-based (high-confidence) | 12.35 |
| S7-final-viral-combined_k141_94263\|\|full | 5510 | No | NA | 5 | 0 | 3 | Not-determined | Genome-fragment | NA | NA | 0 |
| S9-final-viral-combined_k141_13061\|\|full | 1849 | No | NA | 3 | 0 | 0 | Low-quality | Genome-fragment | 3.51 | AAI-based (high-confidence) | 0 |
| S5-final-viral-combined_k141_133020\|\|full | 1849 | No | NA | 3 | 0 | 0 | Low-quality | Genome-fragment | 3.3 | AAI-based (medium-confidence) | 0 |
| S7-final-viral-combined_k141_197435\|\|full | 1672 | No | NA | 2 | 0 | 1 | Not-determined | Genome-fragment | NA | NA | 0 |
| S7-final-viral-combined_k141_125422\|\|full | 3608 | No | NA | 5 | 0 | 3 | Not-determined | Genome-fragment | NA | NA | 0 |
| S7-final-viral-combined_k141_43726\|\|full | 1540 | No | NA | 3 | 0 | 0 | Not-determined | Genome-fragment | NA | NA | 0 |
| S7-final-viral-combined_k141_104910\|\|full | 1228 | No | NA | 2 | 0 | 2 | Not-determined | Genome-fragment | NA | NA | 0 |
| S7-final-viral-combined_k141_145473\|\|full | 19725 | No | NA | 20 | 0 | 7 | Not-determined | Genome-fragment | NA | NA | 0 |
| S7-final-viral-combined_k141_166345\|\|full | 1723 | No | NA | 3 | 0 | 0 | Not-determined | Genome-fragment | NA | NA | 0 |
| S7-final-viral-combined_k141_125434\|\|full | 3043 | No | NA | 2 | 0 | 2 | Not-determined | Genome-fragment | NA | NA | 0 |
| S7-final-viral-combined_k141_238122\|\|full | 1099 | No | NA | 2 | 0 | 0 | Not-determined | Genome-fragment | NA | NA | 0 |
| S7-final-viral-combined_k141_43747\|\|full | 5479 | No | NA | 7 | 0 | 1 | Not-determined | Genome-fragment | NA | NA | 0 |
| S7-final-viral-combined_k141_145507\|\|full | 1272 | No | NA | 2 | 0 | 1 | Not-determined | Genome-fragment | NA | NA | 0 |
| S5_k141_145898 | 1849 | No | NA | 3 | 1 | 0 | Low-quality | Genome-fragment | 5.02 | AAI-based (high-confidence) | 0 |
| S7-final-viral-combined_k141_166390\|\|full | 5200 | No | NA | 6 | 0 | 1 | Not-determined | Genome-fragment | NA | NA | 0 |
| S7-final-viral-combined_k141_227946\|\|full | 4922 | No | NA | 5 | 0 | 5 | Not-determined | Genome-fragment | NA | NA | 0 |
| S7-final-viral-combined_k141_33719\|\|full | 1174 | No | NA | 2 | 0 | 1 | Not-determined | Genome-fragment | NA | NA | 0 |
| S17-final-viral-combined_k141_64855\|\|full | 1849 | No | NA | 4 | 0 | 2 | Low-quality | Genome-fragment | 3.13 | AAI-based (medium-confidence) | 0 |
| S2_k141_156762 | 1849 | No | NA | 3 | 0 | 0 | Low-quality | Genome-fragment | 3.83 | AAI-based (medium-confidence) | 0 |
| S11_k141_291960 | 1850 | No | NA | 3 | 2 | 0 | Low-quality | Genome-fragment | 3.91 | AAI-based (high-confidence) | 0 |
| S11_k141_79422 | 1850 | No | NA | 3 | 0 | 1 | Low-quality | Genome-fragment | 3.89 | AAI-based (medium-confidence) | 0 |
| S1-final-viral-combined_k141_313140\|\|lt2gene | 1850 | No | NA | 2 | 1 | 0 | Low-quality | Genome-fragment | 3.34 | AAI-based (high-confidence) | 0 |
| S7-final-viral-combined_k141_64111\|\|full | 1962 | No | NA | 3 | 0 | 0 | Not-determined | Genome-fragment | NA | NA | 0 |
| S7-final-viral-combined_k141_29147\|\|full | 1850 | No | NA | 3 | 1 | 0 | Low-quality | Genome-fragment | 0.15 | HMM-based (lower-bound) | 0 |
| S7-final-viral-combined_k141_166419\|\|full | 3451 | No | NA | 3 | 0 | 1 | Not-determined | Genome-fragment | NA | NA | 0 |
| S17-k141_150322 | 1850 | No | NA | 3 | 1 | 0 | Low-quality | Genome-fragment | 3.3 | AAI-based (high-confidence) | 0 |
| S7-final-viral-combined_k141_176375\|\|full | 1519 | No | NA | 3 | 0 | 2 | Not-determined | Genome-fragment | NA | NA | 0 |
| S12-final-viral-combined_k141_50690\|\|lt2gene | 1850 | No | NA | 2 | 2 | 0 | Low-quality | Genome-fragment | 3.91 | AAI-based (high-confidence) | 0 |
| S12_k141_90707 | 1850 | No | NA | 1 | 1 | 0 | Low-quality | Genome-fragment | 1.32 | AAI-based (high-confidence) | 0 |
| S7-final-viral-combined_k141_207882\|\|full | 2996 | No | NA | 2 | 0 | 2 | Not-determined | Genome-fragment | NA | NA | 0 |
| S7-final-viral-combined_k141_54020\|\|full | 1127 | No | NA | 3 | 0 | 1 | Not-determined | Genome-fragment | NA | NA | 0 |
| S7-final-viral-combined_k141_145642\|\|full | 3101 | No | NA | 2 | 0 | 1 | Not-determined | Genome-fragment | NA | NA | 0 |
| S4-final-viral-combined_k141_34662\|\|full | 1850 | No | NA | 6 | 1 | 0 | Low-quality | Genome-fragment | 2.2 | AAI-based (high-confidence) | 0 |
| S7-final-viral-combined_k141_43858\|\|full | 1262 | No | NA | 2 | 0 | 0 | Not-determined | Genome-fragment | NA | NA | 0 |
| S6-final-viral-combined_k141_11083\|\|full | 1850 | No | NA | 1 | 0 | 0 | Low-quality | Genome-fragment | 3.66 | AAI-based (medium-confidence) | 0 |
| S7-final-viral-combined_k141_64141\|\|full | 2246 | No | NA | 2 | 0 | 2 | Not-determined | Genome-fragment | NA | NA | 0 |
| S7-final-viral-combined_k141_238214\|\|full | 1495 | No | NA | 2 | 0 | 2 | Not-determined | Genome-fragment | NA | NA | 0 |
| S8_k141_238296 | 1850 | No | NA | 2 | 0 | 1 | Low-quality | Genome-fragment | 0.32 | AAI-based (high-confidence) | 0 |
| S11-final-viral-combined_k141_262781\|\|full | 1851 | No | NA | 3 | 0 | 0 | Low-quality | Genome-fragment | 3.12 | AAI-based (high-confidence) | 0 |
| S7-final-viral-combined_k141_64156\|\|full | 1004 | No | NA | 2 | 0 | 1 | Not-determined | Genome-fragment | NA | NA | 0 |
| S5-final-viral-combined_k141_15928\|\|full | 1851 | No | NA | 4 | 0 | 0 | Low-quality | Genome-fragment | 3.07 | AAI-based (high-confidence) | 0 |
| S7-final-viral-combined_k141_187276\|\|full | 1640 | No | NA | 4 | 0 | 4 | Not-determined | Genome-fragment | NA | NA | 0 |
| S7-final-viral-combined_k141_200750\|\|full | 1851 | No | NA | 3 | 2 | 0 | Low-quality | Genome-fragment | 2.27 | HMM-based (lower-bound) | 0 |
| S7-final-viral-combined_k141_43915\|\|full | 1718 | No | NA | 3 | 0 | 0 | Not-determined | Genome-fragment | NA | NA | 0 |
| S7-final-viral-combined_k141_43917\|\|full | 2916 | No | NA | 4 | 0 | 0 | Not-determined | Genome-fragment | NA | NA | 0 |
| S7-final-viral-combined_k141_88587\|\|full | 1851 | No | NA | 3 | 0 | 0 | Low-quality | Genome-fragment | 4.44 | AAI-based (high-confidence) | 0 |
| S7-final-viral-combined_k141_54096\|\|full | 2752 | No | NA | 3 | 0 | 2 | Not-determined | Genome-fragment | NA | NA | 0 |
| S2-final-viral-combined_k141_29383\|\|full | 1851 | No | NA | 5 | 5 | 0 | Low-quality | Genome-fragment | 2.94 | AAI-based (high-confidence) | 0 |
| S8_k141_25514 | 1851 | No | NA | 5 | 0 | 0 | Low-quality | Genome-fragment | 3.41 | AAI-based (high-confidence) | 0 |
| S11_k141_98049 | 1852 | No | NA | 2 | 0 | 0 | Low-quality | Genome-fragment | 1.17 | AAI-based (medium-confidence) | 0 |
| S3-final-viral-combined_k141_123706\|\|full | 1852 | No | NA | 4 | 0 | 0 | Low-quality | Genome-fragment | 3.72 | AAI-based (medium-confidence) | 0 |
| S17-final-viral-combined_k141_160122\|\|full | 1852 | No | NA | 4 | 3 | 0 | Low-quality | Genome-fragment | 4.1 | AAI-based (medium-confidence) | 0 |
| S12-final-viral-combined_k141_304828\|\|full | 1852 | No | NA | 3 | 2 | 0 | Low-quality | Genome-fragment | 3.63 | HMM-based (lower-bound) | 0 |
| S7-final-viral-combined_k141_105081\|\|full | 2522 | No | NA | 3 | 0 | 3 | Not-determined | Genome-fragment | NA | NA | 0 |
| S12_k141_128757 | 1852 | No | NA | 1 | 0 | 1 | Low-quality | Genome-fragment | 0.66 | AAI-based (high-confidence) | 0 |
| S2-final-viral-combined_k141_29859\|\|full | 1852 | No | NA | 2 | 1 | 0 | Low-quality | Genome-fragment | 3.22 | AAI-based (high-confidence) | 0 |
| S7-final-viral-combined_k141_166550\|\|full | 1816 | No | NA | 2 | 0 | 0 | Not-determined | Genome-fragment | NA | NA | 0 |
| S7-final-viral-combined_k141_187367\|\|full | 3391 | No | NA | 2 | 0 | 2 | Not-determined | Genome-fragment | NA | NA | 0 |
| S7-final-viral-combined_k141_197682\|\|full | 13129 | No | NA | 9 | 0 | 8 | Not-determined | Genome-fragment | NA | NA | 0 |
| S7-final-viral-combined_k141_238231\|\|full | 83333 | Yes | 32253 | 96 | 14 | 34 | Medium-quality | Genome-fragment | 61.22 | AAI-based (high-confidence) | 61.3 |
| S7-final-viral-combined_k141_64211\|\|full | 3687 | No | NA | 5 | 0 | 1 | Not-determined | Genome-fragment | NA | NA | 0 |
| S7-final-viral-combined_k141_135655\|\|full | 1296 | No | NA | 2 | 0 | 0 | Not-determined | Genome-fragment | NA | NA | 0 |
| S7-final-viral-combined_k141_217230\|\|full | 2874 | No | NA | 5 | 0 | 1 | Not-determined | Genome-fragment | NA | NA | 0 |
| S7-final-viral-combined_k141_33831\|\|full | 3228 | No | NA | 4 | 0 | 2 | Not-determined | Genome-fragment | NA | NA | 0 |
| S7-final-viral-combined_k141_135661\|\|full | 1264 | No | NA | 2 | 0 | 0 | Not-determined | Genome-fragment | NA | NA | 0 |
| S6-final-viral-combined_k141_8378\|\|full | 1852 | No | NA | 3 | 0 | 0 | Low-quality | Genome-fragment | 3.09 | AAI-based (medium-confidence) | 0 |
| S7-final-viral-combined_k141_228154\|\|full | 2073 | No | NA | 3 | 0 | 0 | Not-determined | Genome-fragment | NA | NA | 0 |
| S7-final-viral-combined_k141_43950\|\|full | 57129 | No | NA | 64 | 5 | 10 | High-quality | High-quality | 100 | AAI-based (high-confidence) | 0 |
| S1-final-viral-combined_k141_144320\|\|full | 1853 | No | NA | 3 | 2 | 0 | Low-quality | Genome-fragment | 3.56 | AAI-based (high-confidence) | 0 |
| S7-final-viral-combined_k141_238269\|\|full | 2047 | No | NA | 3 | 0 | 1 | Not-determined | Genome-fragment | NA | NA | 0 |
| S1_k141_354454 | 1853 | No | NA | 1 | 0 | 1 | Low-quality | Genome-fragment | 0.26 | AAI-based (medium-confidence) | 0 |
| S5_k141_201737 | 1853 | No | NA | 1 | 1 | 0 | Low-quality | Genome-fragment | 1.27 | HMM-based (lower-bound) | 0 |
| S7-final-viral-combined_k141_217277\|\|full | 1146 | No | NA | 2 | 0 | 0 | Not-determined | Genome-fragment | NA | NA | 0 |
| S17-final-viral-combined_k141_90069\|\|full | 1853 | No | NA | 2 | 0 | 0 | Low-quality | Genome-fragment | 1.31 | AAI-based (medium-confidence) | 0 |
| S12-final-viral-combined_k141_379515\|\|full | 1853 | No | NA | 4 | 1 | 0 | Low-quality | Genome-fragment | 4.11 | AAI-based (high-confidence) | 0 |
| S7-final-viral-combined_k141_115515\|\|full | 1414 | No | NA | 3 | 0 | 1 | Not-determined | Genome-fragment | NA | NA | 0 |
| S4-final-viral-combined_k141_50577\|\|full | 1853 | No | NA | 4 | 4 | 0 | Low-quality | Genome-fragment | 5.35 | AAI-based (high-confidence) | 0 |
| S11-final-viral-combined_k141_102009\|\|full | 1854 | No | NA | 3 | 1 | 0 | Low-quality | Genome-fragment | 3.67 | AAI-based (high-confidence) | 0 |
| S7-final-viral-combined_k141_197775\|\|full | 1465 | No | NA | 3 | 0 | 1 | Not-determined | Genome-fragment | NA | NA | 0 |
| S7-final-viral-combined_k141_73784\|\|full | 2129 | No | NA | 2 | 0 | 0 | Not-determined | Genome-fragment | NA | NA | 0 |
| S7-final-viral-combined_k141_207979\|\|full | 1062 | No | NA | 3 | 0 | 0 | Not-determined | Genome-fragment | NA | NA | 0 |
| S1-final-viral-combined_k141_321748\|\|full | 1854 | No | NA | 4 | 0 | 1 | Low-quality | Genome-fragment | 1.11 | AAI-based (medium-confidence) | 0 |
| S12-final-viral-combined_k141_353428\|\|full | 1854 | No | NA | 3 | 0 | 1 | Low-quality | Genome-fragment | 1.49 | AAI-based (medium-confidence) | 0 |
| S7-final-viral-combined_k141_115556\|\|full | 1974 | No | NA | 5 | 0 | 2 | Not-determined | Genome-fragment | NA | NA | 0 |
| S12-final-viral-combined_k141_295728\|\|full | 1854 | No | NA | 4 | 0 | 2 | Low-quality | Genome-fragment | 0.61 | AAI-based (medium-confidence) | 0 |
| S7-final-viral-combined_k141_176693\|\|full | 2036 | No | NA | 2 | 0 | 1 | Not-determined | Genome-fragment | NA | NA | 0 |
| S4-final-viral-combined_k141_30157\|\|full | 1854 | No | NA | 4 | 2 | 0 | Low-quality | Genome-fragment | 1.07 | AAI-based (high-confidence) | 0 |
| S7-final-viral-combined_k141_73827\|\|full | 1723 | No | NA | 3 | 0 | 2 | Not-determined | Genome-fragment | NA | NA | 0 |
| S11-final-viral-combined_k141_252625\|\|lt2gene | 1855 | No | NA | 2 | 0 | 0 | Low-quality | Genome-fragment | 4.02 | AAI-based (medium-confidence) | 0 |
| S3-final-viral-combined_k141_23746\|\|full | 1855 | No | NA | 3 | 0 | 3 | Low-quality | Genome-fragment | 3.29 | AAI-based (medium-confidence) | 0 |
| S5_k141_33330 | 1855 | No | NA | 5 | 2 | 0 | Low-quality | Genome-fragment | 4.55 | AAI-based (high-confidence) | 0 |
| S17-final-viral-combined_k141_152393\|\|full | 1855 | No | NA | 3 | 0 | 1 | Low-quality | Genome-fragment | 1.07 | AAI-based (medium-confidence) | 0 |
| S8-final-viral-combined_k141_140447\|\|full | 1855 | No | NA | 4 | 1 | 0 | Low-quality | Genome-fragment | 3.25 | AAI-based (medium-confidence) | 0 |
| S7-final-viral-combined_k141_238357\|\|full | 1122 | No | NA | 2 | 0 | 0 | Not-determined | Genome-fragment | NA | NA | 0 |
| S1-final-viral-combined_k141_224684\|\|0_partial | 1856 | No | NA | 3 | 1 | 0 | Low-quality | Genome-fragment | 32.98 | AAI-based (high-confidence) | 0 |
| S7-final-viral-combined_k141_2492\|\|full | 1856 | No | NA | 3 | 0 | 3 | Low-quality | Genome-fragment | 2.96 | AAI-based (high-confidence) | 0 |
| S2-final-viral-combined_k141_164596\|\|full | 1856 | No | NA | 4 | 2 | 0 | Low-quality | Genome-fragment | 2.46 | HMM-based (lower-bound) | 0 |
| S4_k141_60551 | 1856 | No | NA | 5 | 3 | 0 | Low-quality | Genome-fragment | 10.86 | AAI-based (high-confidence) | 0 |
| S7-final-viral-combined_k141_105281\|\|full | 1469 | No | NA | 2 | 0 | 0 | Not-determined | Genome-fragment | NA | NA | 0 |
| S7-final-viral-combined_k141_176733\|\|full | 2262 | No | NA | 4 | 0 | 2 | Not-determined | Genome-fragment | NA | NA | 0 |
| S4_k141_6850 | 1856 | No | NA | 3 | 0 | 0 | Low-quality | Genome-fragment | 0.94 | AAI-based (medium-confidence) | 0 |
| S7-final-viral-combined_k141_217380\|\|full | 1737 | No | NA | 3 | 0 | 2 | Not-determined | Genome-fragment | NA | NA | 0 |
| S7-final-viral-combined_k141_125815\|\|full | 2300 | No | NA | 3 | 0 | 0 | Not-determined | Genome-fragment | NA | NA | 0 |
| S7-final-viral-combined_k141_217389\|\|full | 3468 | No | NA | 3 | 0 | 3 | Not-determined | Genome-fragment | NA | NA | 0 |
| S7-final-viral-combined_k141_64305\|\|full | 1142 | No | NA | 4 | 0 | 0 | Not-determined | Genome-fragment | NA | NA | 0 |
| S7-final-viral-combined_k141_208121\|\|full | 1024 | No | NA | 2 | 0 | 1 | Not-determined | Genome-fragment | NA | NA | 0 |
| S7-final-viral-combined_k141_84586\|\|full | 9235 | No | NA | 8 | 0 | 3 | Not-determined | Genome-fragment | NA | NA | 0 |
| S7-final-viral-combined_k141_22958\|\|full | 2219 | No | NA | 3 | 0 | 1 | Not-determined | Genome-fragment | NA | NA | 0 |
| S7-final-viral-combined_k141_3208\|\|full | 10106 | No | NA | 18 | 0 | 0 | Not-determined | Genome-fragment | NA | NA | 0 |
| S11-final-viral-combined_k141_319860\|\|full | 1857 | No | NA | 4 | 1 | 0 | Low-quality | Genome-fragment | 4.67 | AAI-based (medium-confidence) | 0 |
| S7-final-viral-combined_k141_135806\|\|full | 1029 | No | NA | 2 | 0 | 1 | Not-determined | Genome-fragment | NA | NA | 0 |
| S11-final-viral-combined_k141_284728\|\|full | 1857 | No | NA | 3 | 1 | 0 | Low-quality | Genome-fragment | 3.54 | AAI-based (medium-confidence) | 0 |
| S7-final-viral-combined_k141_44063\|\|full | 1792 | No | NA | 3 | 0 | 1 | Not-determined | Genome-fragment | NA | NA | 0 |
| S7-final-viral-combined_k141_13513\|\|full | 3278 | No | NA | 1 | 0 | 1 | Not-determined | Genome-fragment | NA | NA | 0 |
| S7-final-viral-combined_k141_145842\|\|full | 8842 | No | NA | 10 | 0 | 2 | Not-determined | Genome-fragment | NA | NA | 0 |
| S7-final-viral-combined_k141_156603\|\|full | 2426 | No | NA | 3 | 0 | 1 | Not-determined | Genome-fragment | NA | NA | 0 |
| S7-final-viral-combined_k141_64348\|\|full | 7613 | No | NA | 6 | 0 | 2 | Not-determined | Genome-fragment | NA | NA | 0 |
| S7-final-viral-combined_k141_187614\|\|full | 2417 | No | NA | 3 | 0 | 1 | Not-determined | Genome-fragment | NA | NA | 0 |
| S7-final-viral-combined_k141_187615\|\|full | 1762 | No | NA | 2 | 0 | 1 | Not-determined | Genome-fragment | NA | NA | 0 |
| S11-final-viral-combined_k141_86951\|\|lt2gene | 1857 | No | NA | 2 | 1 | 0 | Low-quality | Genome-fragment | 3.37 | HMM-based (lower-bound) | 0 |
| S7-final-viral-combined_k141_34112\|\|full | 1807 | No | NA | 2 | 0 | 2 | Not-determined | Genome-fragment | NA | NA | 0 |
| S1-final-viral-combined_k141_216719\|\|0_partial | 1857 | No | NA | 3 | 1 | 0 | Low-quality | Genome-fragment | 35.95 | AAI-based (high-confidence) | 0 |
| S7-final-viral-combined_k141_238377\|\|full | 2774 | No | NA | 3 | 0 | 2 | Not-determined | Genome-fragment | NA | NA | 0 |
| S7-final-viral-combined_k141_68012\|\|full | 1857 | No | NA | 3 | 0 | 0 | Low-quality | Genome-fragment | 2.26 | AAI-based (high-confidence) | 0 |
| S7-final-viral-combined_k141_145882\|\|full | 1162 | No | NA | 3 | 0 | 0 | Not-determined | Genome-fragment | NA | NA | 0 |
| S7-final-viral-combined_k141_187640\|\|full | 1343 | No | NA | 2 | 0 | 2 | Not-determined | Genome-fragment | NA | NA | 0 |
| S7-final-viral-combined_k141_197899\|\|full | 1622 | No | NA | 3 | 0 | 2 | Not-determined | Genome-fragment | NA | NA | 0 |
| S7-final-viral-combined_k141_208201\|\|full | 1624 | No | NA | 3 | 0 | 1 | Not-determined | Genome-fragment | NA | NA | 0 |
| S7-final-viral-combined_k141_54331\|\|full | 1783 | No | NA | 3 | 0 | 1 | Not-determined | Genome-fragment | NA | NA | 0 |
| S7-final-viral-combined_k141_72937\|\|lt2gene | 1857 | No | NA | 2 | 2 | 0 | Low-quality | Genome-fragment | 5.77 | AAI-based (high-confidence) | 0 |
| S2-final-viral-combined_k141_62311\|\|lt2gene | 1857 | No | NA | 2 | 2 | 0 | Low-quality | Genome-fragment | 3.93 | AAI-based (high-confidence) | 0 |
| S7-final-viral-combined_k141_34187\|\|full | 2004 | No | NA | 3 | 0 | 3 | Not-determined | Genome-fragment | NA | NA | 0 |
| S7-final-viral-combined_k141_54341\|\|full | 1504 | No | NA | 3 | 0 | 2 | Not-determined | Genome-fragment | NA | NA | 0 |
| S7-final-viral-combined_k141_228309\|\|full | 2567 | No | NA | 4 | 0 | 3 | Not-determined | Genome-fragment | NA | NA | 0 |
| S7-final-viral-combined_k141_13632\|\|full | 1468 | No | NA | 2 | 0 | 0 | Not-determined | Genome-fragment | NA | NA | 0 |
| S4_k141_61090 | 1857 | No | NA | 1 | 0 | 1 | Low-quality | Genome-fragment | 0.58 | AAI-based (medium-confidence) | 0 |
| S7-final-viral-combined_k141_64465\|\|full | 1687 | No | NA | 3 | 0 | 0 | Not-determined | Genome-fragment | NA | NA | 0 |
| S11-final-viral-combined_k141_88132\|\|full | 1858 | No | NA | 2 | 0 | 0 | Low-quality | Genome-fragment | 6.81 | AAI-based (medium-confidence) | 0 |
| S3_k141_95055 | 1858 | No | NA | 3 | 1 | 0 | Low-quality | Genome-fragment | 4.81 | AAI-based (high-confidence) | 0 |
| S5-final-viral-combined_k141_98189\|\|full | 1858 | No | NA | 3 | 0 | 2 | Low-quality | Genome-fragment | 2.38 | AAI-based (medium-confidence) | 0 |
| S7-final-viral-combined_k141_84688\|\|full | 61878 | Yes | 17213 | 71 | 3 | 21 | Complete | High-quality | 100 | Provirus (high-confidence) | 72.18 |
| S7-final-viral-combined_k141_34214\|\|full | 1701 | No | NA | 2 | 0 | 0 | Not-determined | Genome-fragment | NA | NA | 0 |
| S12-final-viral-combined_k141_388496\|\|full | 1858 | No | NA | 4 | 0 | 0 | Low-quality | Genome-fragment | 2.54 | AAI-based (medium-confidence) | 0 |
| S12-final-viral-combined_k141_391591\|\|full | 1858 | No | NA | 2 | 2 | 0 | Low-quality | Genome-fragment | 3.68 | AAI-based (high-confidence) | 0 |
| S7-final-viral-combined_k141_176907\|\|full | 1860 | No | NA | 5 | 0 | 0 | Not-determined | Genome-fragment | NA | NA | 0 |
| S2-final-viral-combined_k141_51801\|\|full | 1858 | No | NA | 3 | 0 | 0 | Low-quality | Genome-fragment | 2.16 | AAI-based (medium-confidence) | 0 |
| S2-final-viral-combined_k141_244582\|\|full | 1858 | No | NA | 3 | 0 | 0 | Low-quality | Genome-fragment | 0.97 | AAI-based (medium-confidence) | 0 |
| S7-final-viral-combined_k141_34245\|\|full | 1155 | No | NA | 2 | 0 | 1 | Not-determined | Genome-fragment | NA | NA | 0 |
| S7-final-viral-combined_k141_145966\|\|full | 1780 | No | NA | 3 | 0 | 1 | Not-determined | Genome-fragment | NA | NA | 0 |
| S7-final-viral-combined_k141_84708\|\|full | 1761 | No | NA | 3 | 0 | 1 | Not-determined | Genome-fragment | NA | NA | 0 |
| S7-final-viral-combined_k141_135875\|\|full | 1456 | No | NA | 3 | 0 | 1 | Not-determined | Genome-fragment | NA | NA | 0 |
| S7-final-viral-combined_k141_84726\|\|full | 1044 | No | NA | 2 | 0 | 0 | Not-determined | Genome-fragment | NA | NA | 0 |
| S7-final-viral-combined_k141_64564\|\|full | 1664 | No | NA | 2 | 0 | 0 | Not-determined | Genome-fragment | NA | NA | 0 |
| S2-final-viral-combined_k141_71362\|\|full | 1858 | No | NA | 5 | 2 | 0 | Low-quality | Genome-fragment | 2.47 | AAI-based (high-confidence) | 0 |
| S6-final-viral-combined_k141_84623\|\|full | 1858 | No | NA | 2 | 1 | 0 | Low-quality | Genome-fragment | 2.81 | HMM-based (lower-bound) | 0 |
| S7-final-viral-combined_k141_135925\|\|full | 2580 | No | NA | 3 | 0 | 1 | Not-determined | Genome-fragment | NA | NA | 0 |
| S8-final-viral-combined_k141_269407\|\|full | 1858 | No | NA | 3 | 0 | 3 | Low-quality | Genome-fragment | 0.97 | AAI-based (medium-confidence) | 0 |
| S7-final-viral-combined_k141_156817\|\|full | 7669 | No | NA | 7 | 0 | 6 | Not-determined | Genome-fragment | NA | NA | 0 |
| S8-final-viral-combined_k141_178017\|\|full | 1858 | Yes | 675 | 3 | 1 | 1 | Low-quality | Genome-fragment | 0.47 | HMM-based (lower-bound) | 63.67 |
| S7-final-viral-combined_k141_84755\|\|full | 1553 | No | NA | 2 | 0 | 0 | Not-determined | Genome-fragment | NA | NA | 0 |
| S7-final-viral-combined_k141_135953\|\|full | 1057 | No | NA | 2 | 0 | 1 | Not-determined | Genome-fragment | NA | NA | 0 |
| S11_k141_286323 | 1859 | No | NA | 1 | 0 | 1 | Low-quality | Genome-fragment | 0.92 | AAI-based (medium-confidence) | 0 |
| S17-k141_184861 | 1859 | No | NA | 2 | 0 | 0 | Low-quality | Genome-fragment | 1.3 | AAI-based (high-confidence) | 0 |
| S12-final-viral-combined_k141_270472\|\|full | 1859 | No | NA | 2 | 0 | 0 | Low-quality | Genome-fragment | 3.53 | AAI-based (medium-confidence) | 0 |
| S12-final-viral-combined_k141_129833\|\|full | 1859 | No | NA | 5 | 2 | 0 | Low-quality | Genome-fragment | 4.51 | AAI-based (high-confidence) | 0 |
| S2-final-viral-combined_k141_171908\|\|full | 1859 | No | NA | 3 | 2 | 0 | Low-quality | Genome-fragment | 3.49 | AAI-based (high-confidence) | 0 |
| S7-final-viral-combined_k141_94784\|\|full | 5706 | No | NA | 7 | 0 | 0 | Not-determined | Genome-fragment | NA | NA | 0 |
| S6-final-viral-combined_k141_42688\|\|full | 1859 | No | NA | 4 | 0 | 0 | Low-quality | Genome-fragment | 1.59 | AAI-based (medium-confidence) | 0 |
| S7-final-viral-combined_k141_146079\|\|full | 1507 | No | NA | 2 | 0 | 0 | Not-determined | Genome-fragment | NA | NA | 0 |
| S17-final-viral-combined_k141_22577\|\|lt2gene | 1860 | No | NA | 1 | 1 | 0 | Low-quality | Genome-fragment | 3.84 | AAI-based (high-confidence) | 0 |
| S7-final-viral-combined_k141_84793\|\|full | 2406 | No | NA | 3 | 0 | 0 | Not-determined | Genome-fragment | NA | NA | 0 |
| S7-final-viral-combined_k141_64677\|\|full | 10721 | No | NA | 9 | 0 | 9 | Not-determined | Genome-fragment | NA | NA | 0 |
| S8_k141_55435 | 1860 | No | NA | 2 | 0 | 1 | Low-quality | Genome-fragment | 1.14 | AAI-based (medium-confidence) | 0 |
| S7-final-viral-combined_k141_187893\|\|full | 11098 | No | NA | 10 | 0 | 0 | Not-determined | Genome-fragment | NA | NA | 0 |
| S7-final-viral-combined_k141_228448\|\|full | 1271 | No | NA | 4 | 0 | 1 | Not-determined | Genome-fragment | NA | NA | 0 |
| S9-final-viral-combined_k141_14896\|\|full | 1861 | No | NA | 2 | 0 | 1 | Low-quality | Genome-fragment | 1.11 | AAI-based (medium-confidence) | 0 |
| S7-final-viral-combined_k141_13877\|\|full | 2028 | No | NA | 3 | 0 | 1 | Not-determined | Genome-fragment | NA | NA | 0 |
| S7-final-viral-combined_k141_136037\|\|full | 2557 | No | NA | 3 | 0 | 0 | Not-determined | Genome-fragment | NA | NA | 0 |
| S7-final-viral-combined_k141_198124\|\|full | 2290 | No | NA | 3 | 0 | 2 | Not-determined | Genome-fragment | NA | NA | 0 |
| S7-final-viral-combined_k141_105528\|\|full | 8264 | No | NA | 6 | 0 | 4 | Not-determined | Genome-fragment | NA | NA | 0 |
| S7-final-viral-combined_k141_126042\|\|full | 1523 | No | NA | 3 | 0 | 1 | Not-determined | Genome-fragment | NA | NA | 0 |
| S9-final-viral-combined_k141_119171\|\|lt2gene | 1861 | No | NA | 1 | 1 | 0 | Low-quality | Genome-fragment | 1.15 | AAI-based (medium-confidence) | 0 |
| S7-final-viral-combined_k141_126047\|\|full | 2042 | No | NA | 3 | 0 | 1 | Not-determined | Genome-fragment | NA | NA | 0 |
| S7-final-viral-combined_k141_3461\|\|full | 1976 | No | NA | 3 | 0 | 1 | Not-determined | Genome-fragment | NA | NA | 0 |
| S11_k141_70809 | 1861 | No | NA | 1 | 0 | 0 | Low-quality | Genome-fragment | 2.07 | AAI-based (medium-confidence) | 0 |
| S5-final-viral-combined_k141_80337\|\|full | 1861 | No | NA | 3 | 0 | 0 | Low-quality | Genome-fragment | 2.26 | AAI-based (high-confidence) | 0 |
| S17-final-viral-combined_k141_111131\|\|full | 1861 | No | NA | 6 | 2 | 0 | Low-quality | Genome-fragment | 4.48 | AAI-based (high-confidence) | 0 |
| S7-final-viral-combined_k141_217573\|\|full | 5392 | No | NA | 5 | 0 | 2 | Not-determined | Genome-fragment | NA | NA | 0 |
| S7-final-viral-combined_k141_34377\|\|full | 1855 | No | NA | 3 | 0 | 0 | Not-determined | Genome-fragment | NA | NA | 0 |
| S17-final-viral-combined_k141_249608\|\|full | 1861 | No | NA | 3 | 0 | 3 | Low-quality | Genome-fragment | 0.86 | AAI-based (high-confidence) | 0 |
| S7-final-viral-combined_k141_187946\|\|full | 1129 | No | NA | 3 | 0 | 3 | Not-determined | Genome-fragment | NA | NA | 0 |
| S17-k141_155048 | 1861 | No | NA | 2 | 0 | 2 | Low-quality | Genome-fragment | 0.69 | AAI-based (high-confidence) | 0 |

| | | | | | | | | | | | |
|---|---|---|---|---|---|---|---|---|---|---|---|
| S12-final-viral-combined_k141_86952\|\|full | 1861 | No | NA | 3 | 2 | 0 | Low-quality | Genome-fragment | 3.08 | AAI-based (high-confidence) | 0 |
| S7-final-viral-combined_k141_116013\|\|full | 1074 | No | NA | 3 | 0 | 2 | Not-determined | Genome-fragment | NA | NA | 0 |
| S7-final-viral-combined_k141_187957\|\|full | 1068 | No | NA | 2 | 0 | 2 | Not-determined | Genome-fragment | NA | NA | 0 |
| S7-final-viral-combined_k141_187958\|\|full | 3105 | No | NA | 5 | 0 | 3 | Not-determined | Genome-fragment | NA | NA | 0 |
| S12-final-viral-combined_k141_246231\|\|full | 1861 | No | NA | 5 | 4 | 0 | Low-quality | Genome-fragment | 5.38 | AAI-based (high-confidence) | 0 |
| S4-final-viral-combined_k141_51357\|\|full | 1861 | Yes | 294 | 3 | 1 | 1 | Low-quality | Genome-fragment | 0.49 | AAI-based (high-confidence) | 84.2 |
| S7-final-viral-combined_k141_238642\|\|full | 1998 | No | NA | 5 | 0 | 0 | Not-determined | Genome-fragment | NA | NA | 0 |
| S7-final-viral-combined_k141_64730\|\|full | 1721 | No | NA | 3 | 0 | 0 | Not-determined | Genome-fragment | NA | NA | 0 |
| S4-final-viral-combined_k141_137375\|\|full | 1861 | No | NA | 3 | 0 | 2 | Low-quality | Genome-fragment | 2.73 | AAI-based (high-confidence) | 0 |
| S7-final-viral-combined_k141_238678\|\|full | 2093 | No | NA | 3 | 0 | 0 | Not-determined | Genome-fragment | NA | NA | 0 |
| S8_k141_217979 | 1861 | No | NA | 3 | 0 | 0 | Low-quality | Genome-fragment | 2.66 | AAI-based (high-confidence) | 0 |
| S11-final-viral-combined_k141_264513\|\|lt2gene | 1862 | No | NA | 1 | 0 | 0 | Low-quality | Genome-fragment | 5.23 | AAI-based (medium-confidence) | 0 |
| S11_k141_45322 | 1862 | No | NA | 2 | 0 | 1 | Low-quality | Genome-fragment | 1.54 | AAI-based (medium-confidence) | 0 |
| S7-final-viral-combined_k141_34421\|\|full | 1310 | No | NA | 3 | 0 | 0 | Not-determined | Genome-fragment | NA | NA | 0 |
| S7-final-viral-combined_k141_238690\|\|full | 1077 | No | NA | 3 | 0 | 0 | Not-determined | Genome-fragment | NA | NA | 0 |
| S7_k141_6779 | 1862 | No | NA | 2 | 0 | 2 | Low-quality | Genome-fragment | 1.23 | AAI-based (high-confidence) | 0 |
| S7-final-viral-combined_k141_34431\|\|full | 3248 | No | NA | 3 | 0 | 2 | Not-determined | Genome-fragment | NA | NA | 0 |
| S7-final-viral-combined_k141_228579\|\|full | 2010 | No | NA | 3 | 0 | 0 | Not-determined | Genome-fragment | NA | NA | 0 |
| S7-final-viral-combined_k141_44370\|\|full | 1014 | No | NA | 2 | 0 | 1 | Not-determined | Genome-fragment | NA | NA | 0 |
| S7-final-viral-combined_k141_208577\|\|full | 3560 | No | NA | 5 | 0 | 1 | Not-determined | Genome-fragment | NA | NA | 0 |
| S17-k141_58664 | 1862 | No | NA | 2 | 0 | 2 | Low-quality | Genome-fragment | 1.94 | AAI-based (medium-confidence) | 0 |
| S12-final-viral-combined_k141_225901\|\|lt2gene | 1862 | No | NA | 1 | 1 | 0 | Low-quality | Genome-fragment | 3.98 | AAI-based (high-confidence) | 0 |
| S7-final-viral-combined_k141_126166\|\|full | 1634 | No | NA | 3 | 0 | 0 | Not-determined | Genome-fragment | NA | NA | 0 |
| S7-final-viral-combined_k141_146253\|\|full | 3557 | No | NA | 5 | 0 | 0 | Not-determined | Genome-fragment | NA | NA | 0 |
| S4_k141_115556 | 1862 | No | NA | 4 | 0 | 0 | Low-quality | Genome-fragment | 2.95 | AAI-based (high-confidence) | 0 |
| S7-final-viral-combined_k141_177154\|\|full | 3692 | No | NA | 5 | 0 | 4 | Not-determined | Genome-fragment | NA | NA | 0 |
| S7-final-viral-combined_k141_44406\|\|full | 1475 | No | NA | 5 | 0 | 2 | Not-determined | Genome-fragment | NA | NA | 0 |
| S8-final-viral-combined_k141_37636\|\|full | 1862 | No | NA | 3 | 0 | 0 | Low-quality | Genome-fragment | 1.78 | AAI-based (medium-confidence) | 0 |
| S7-final-viral-combined_k141_167246\|\|full | 6456 | No | NA | 6 | 0 | 4 | Not-determined | Genome-fragment | NA | NA | 0 |
| S7-final-viral-combined_k141_44421\|\|full | 3288 | No | NA | 3 | 0 | 2 | Not-determined | Genome-fragment | NA | NA | 0 |
| S7-final-viral-combined_k141_167274\|\|full | 1535 | No | NA | 2 | 0 | 1 | Not-determined | Genome-fragment | NA | NA | 0 |
| S8-final-viral-combined_k141_257330\|\|lt2gene | 1862 | No | NA | 2 | 1 | 0 | Low-quality | Genome-fragment | 29.66 | AAI-based (medium-confidence) | 0 |
| S7-final-viral-combined_k141_116181\|\|full | 3060 | No | NA | 3 | 0 | 2 | Not-determined | Genome-fragment | NA | NA | 0 |
| S7-final-viral-combined_k141_238745\|\|full | 2804 | No | NA | 4 | 0 | 3 | Not-determined | Genome-fragment | NA | NA | 0 |
| S7-final-viral-combined_k141_74356\|\|full | 1387 | No | NA | 3 | 0 | 2 | Not-determined | Genome-fragment | NA | NA | 0 |
| S7-final-viral-combined_k141_85051\|\|full | 2289 | No | NA | 3 | 0 | 0 | Not-determined | Genome-fragment | NA | NA | 0 |
| S1-final-viral-combined_k141_14803\|\|full | 1863 | No | NA | 4 | 0 | 0 | Low-quality | Genome-fragment | 5.28 | AAI-based (high-confidence) | 0 |
| S7-final-viral-combined_k141_198251\|\|full | 1927 | No | NA | 3 | 0 | 2 | Not-determined | Genome-fragment | NA | NA | 0 |
| S7-final-viral-combined_k141_198252\|\|full | 1283 | No | NA | 1 | 0 | 0 | Not-determined | Genome-fragment | NA | NA | 0 |
| S17-final-viral-combined_k141_87459\|\|full | 1863 | No | NA | 2 | 0 | 0 | Low-quality | Genome-fragment | 4.89 | AAI-based (high-confidence) | 0 |
| S7-final-viral-combined_k141_177213\|\|full | 1185 | No | NA | 2 | 0 | 1 | Not-determined | Genome-fragment | NA | NA | 0 |
| S17-final-viral-combined_k141_112349\|\|lt2gene | 1863 | No | NA | 2 | 2 | 0 | Low-quality | Genome-fragment | 4.52 | AAI-based (high-confidence) | 0 |
| S7-final-viral-combined_k141_3669\|\|full | 4619 | No | NA | 4 | 3 | 0 | Medium-quality | Genome-fragment | 72.72 | AAI-based (high-confidence) | 0 |
| S12-final-viral-combined_k141_136331\|\|full | 1863 | No | NA | 4 | 0 | 0 | Low-quality | Genome-fragment | 2.16 | AAI-based (medium-confidence) | 0 |
| S7-final-viral-combined_k141_146359\|\|full | 2837 | No | NA | 3 | 0 | 1 | Not-determined | Genome-fragment | NA | NA | 0 |
| S7-final-viral-combined_k141_208644\|\|full | 2615 | No | NA | 4 | 0 | 0 | Not-determined | Genome-fragment | NA | NA | 0 |
| S2-final-viral-combined_k141_213549\|\|full | 1863 | No | NA | 3 | 2 | 0 | Low-quality | Genome-fragment | 3.04 | HMM-based (lower-bound) | 0 |
| S7-final-viral-combined_k141_126235\|\|full | 4105 | No | NA | 3 | 0 | 0 | Not-determined | Genome-fragment | NA | NA | 0 |
| S7-final-viral-combined_k141_74417\|\|full | 1563 | No | NA | 3 | 0 | 1 | Not-determined | Genome-fragment | NA | NA | 0 |
| S1-final-viral-combined_k141_341080\|\|full | 1864 | No | NA | 2 | 0 | 2 | Low-quality | Genome-fragment | 1.23 | AAI-based (medium-confidence) | 0 |
| S4-final-viral-combined_k141_6646\|\|full | 1864 | No | NA | 3 | 0 | 0 | Low-quality | Genome-fragment | 1.36 | AAI-based (high-confidence) | 0 |
| [illegible] | | | | | | | | | | | |

| | | | | | | | | | | | |
|---|---|---|---|---|---|---|---|---|---|---|---|
| S7-final-viral-combined_k141_4673\|\|full | 3294 | No | NA | 4 | 0 | 0 | Not-determined | Genome-fragment | NA | NA | 0 |
| S7-final-viral-combined_k141_189089\|\|full | 1893 | No | NA | 2 | 0 | 2 | Not-determined | Genome-fragment | NA | NA | 0 |
| S7-final-viral-combined_k141_137398\|\|full | 3212 | No | NA | 3 | 0 | 2 | Not-determined | Genome-fragment | NA | NA | 0 |
| S7-final-viral-combined_k141_168596\|\|full | 1582 | No | NA | 4 | 0 | 1 | Not-determined | Genome-fragment | NA | NA | 0 |
| S5_k141_142601 | 1884 | No | NA | 2 | 1 | 0 | Low-quality | Genome-fragment | 4.38 | AAI-based (high-confidence) | 0 |
| S17-final-viral-combined_k141_175516\|\|full | 1884 | No | NA | 2 | 0 | 0 | Low-quality | Genome-fragment | 3.1 | AAI-based (high-confidence) | 0 |
| S7-final-viral-combined_k141_199535\|\|full | 2159 | No | NA | 3 | 0 | 3 | Not-determined | Genome-fragment | NA | NA | 0 |
| S7-final-viral-combined_k141_75695\|\|full | 1141 | No | NA | 3 | 0 | 2 | Not-determined | Genome-fragment | NA | NA | 0 |
| S12_k141_76083 | 1884 | No | NA | 1 | 0 | 0 | Low-quality | Genome-fragment | 0.17 | AAI-based (medium-confidence) | 0 |
| S7-final-viral-combined_k141_75703\|\|full | 1649 | No | NA | 3 | 0 | 0 | Not-determined | Genome-fragment | NA | NA | 0 |
| S7-final-viral-combined_k141_229767\|\|full | 1909 | No | NA | 3 | 0 | 1 | Not-determined | Genome-fragment | NA | NA | 0 |
| S4-final-viral-combined_k141_43122\|\|full | 1884 | No | NA | 2 | 0 | 2 | Low-quality | Genome-fragment | 4.35 | AAI-based (medium-confidence) | 0 |
| S7-final-viral-combined_k141_189110\|\|full | 9955 | No | NA | 13 | 0 | 0 | Not-determined | Genome-fragment | NA | NA | 0 |
| S7-final-viral-combined_k141_66523\|\|full | 4369 | No | NA | 6 | 0 | 1 | Not-determined | Genome-fragment | NA | NA | 0 |
| S8-final-viral-combined_k141_230540\|\|full | 1884 | No | NA | 3 | 0 | 0 | Low-quality | Genome-fragment | 3.88 | AAI-based (medium-confidence) | 0 |
| S7-final-viral-combined_k141_4769\|\|full | 2674 | No | NA | 4 | 1 | 2 | Not-determined | Genome-fragment | NA | NA | 0 |
| S8-final-viral-combined_k141_254925\|\|full | 1884 | No | NA | 3 | 0 | 0 | Low-quality | Genome-fragment | 3.82 | AAI-based (medium-confidence) | 0 |
| S7-final-viral-combined_k141_45753\|\|full | 2220 | No | NA | 3 | 0 | 2 | Not-determined | Genome-fragment | NA | NA | 0 |
| S7-final-viral-combined_k141_137414\|\|full | 3191 | No | NA | 4 | 0 | 2 | Not-determined | Genome-fragment | NA | NA | 0 |
| S7-final-viral-combined_k141_66567\|\|full | 1312 | No | NA | 2 | 0 | 1 | Not-determined | Genome-fragment | NA | NA | 0 |
| S11-final-viral-combined_k141_327634\|\|full | 1885 | No | NA | 1 | 0 | 1 | Low-quality | Genome-fragment | 2.59 | AAI-based (high-confidence) | 0 |
| S11-final-viral-combined_k141_137334\|\|full | 1885 | No | NA | 2 | 1 | 0 | Low-quality | Genome-fragment | 3.4 | HMM-based (lower-bound) | 0 |
| S1-final-viral-combined_k141_339228\|\|full | 1885 | No | NA | 2 | 0 | 1 | Low-quality | Genome-fragment | 1.11 | AAI-based (medium-confidence) | 0 |
| S7-final-viral-combined_k141_147498\|\|full | 2574 | No | NA | 4 | 0 | 1 | Not-determined | Genome-fragment | NA | NA | 0 |
| S7-final-viral-combined_k141_14898\|\|full | 4461 | No | NA | 6 | 0 | 0 | Not-determined | Genome-fragment | NA | NA | 0 |
| S7-final-viral-combined_k141_229788\|\|full | 6288 | No | NA | 10 | 0 | 2 | Not-determined | Genome-fragment | NA | NA | 0 |
| S7-final-viral-combined_k141_229798\|\|full | 1954 | No | NA | 2 | 0 | 2 | Not-determined | Genome-fragment | NA | NA | 0 |
| S5-final-viral-combined_k141_205194\|\|lt2gene | 1885 | No | NA | 2 | 2 | 0 | Low-quality | Genome-fragment | 5.03 | AAI-based (medium-confidence) | 0 |
| S17-final-viral-combined_k141_174094\|\|full | 1885 | No | NA | 4 | 1 | 0 | Low-quality | Genome-fragment | 3.8 | AAI-based (medium-confidence) | 0 |
| S12-final-viral-combined_k141_169352\|\|full | 1885 | No | NA | 2 | 0 | 0 | Low-quality | Genome-fragment | 3.35 | AAI-based (medium-confidence) | 0 |
| S7-final-viral-combined_k141_127735\|\|full | 1170 | No | NA | 2 | 0 | 0 | Not-determined | Genome-fragment | NA | NA | 0 |
| S7-final-viral-combined_k141_24241\|\|full | 2809 | No | NA | 4 | 0 | 3 | Not-determined | Genome-fragment | NA | NA | 0 |
| S7-final-viral-combined_k141_96499\|\|full | 2505 | No | NA | 3 | 0 | 2 | Not-determined | Genome-fragment | NA | NA | 0 |
| S7-final-viral-combined_k141_199650\|\|full | 1521 | No | NA | 2 | 0 | 0 | Not-determined | Genome-fragment | NA | NA | 0 |
| S12_k141_78212 | 1885 | No | NA | 1 | 1 | 0 | Low-quality | Genome-fragment | 1.01 | AAI-based (high-confidence) | 0 |
| S7-final-viral-combined_k141_96500\|\|full | 3964 | No | NA | 4 | 0 | 3 | Not-determined | Genome-fragment | NA | NA | 0 |
| S3-final-viral-combined_k141_77992\|\|full | 1886 | No | NA | 3 | 0 | 0 | Low-quality | Genome-fragment | 4.4 | AAI-based (high-confidence) | 0 |
| S7-final-viral-combined_k141_96501\|\|full | 1018 | No | NA | 2 | 0 | 1 | Not-determined | Genome-fragment | NA | NA | 0 |
| S7-final-viral-combined_k141_218391\|\|full | 1040 | No | NA | 3 | 0 | 0 | Not-determined | Genome-fragment | NA | NA | 0 |
| S7-final-viral-combined_k141_218394\|\|full | 5302 | No | NA | 3 | 0 | 2 | Not-determined | Genome-fragment | NA | NA | 0 |
| S7-final-viral-combined_k141_151991\|\|full | 1886 | No | NA | 4 | 2 | 0 | Low-quality | Genome-fragment | 4.87 | AAI-based (medium-confidence) | 0 |
| S17-k141_57926 | 1886 | No | NA | 1 | 0 | 1 | Low-quality | Genome-fragment | 0.58 | AAI-based (medium-confidence) | 0 |
| S7-final-viral-combined_k141_137512\|\|full | 4468 | No | NA | 5 | 0 | 0 | Not-determined | Genome-fragment | NA | NA | 0 |
| S17-k141_239564 | 1886 | No | NA | 1 | 0 | 1 | Low-quality | Genome-fragment | 0.9 | AAI-based (medium-confidence) | 0 |
| S7-final-viral-combined_k141_199670\|\|full | 5281 | No | NA | 5 | 0 | 5 | Not-determined | Genome-fragment | NA | NA | 0 |
| S17-k141_239669 | 1886 | No | NA | 1 | 0 | 1 | Low-quality | Genome-fragment | 0.67 | AAI-based (medium-confidence) | 0 |
| S2-final-viral-combined_k141_90777\|\|full | 1886 | No | NA | 4 | 2 | 0 | Low-quality | Genome-fragment | 3.72 | AAI-based (medium-confidence) | 0 |
| S7-final-viral-combined_k141_117605\|\|full | 5343 | No | NA | 4 | 0 | 3 | Not-determined | Genome-fragment | NA | NA | 0 |
| S4-final-viral-combined_k141_143017\|\|full | 1886 | No | NA | 2 | 0 | 1 | Low-quality | Genome-fragment | 3.59 | AAI-based (high-confidence) | 0 |
| S7-final-viral-combined_k141_240327\|\|full | 1326 | No | NA | 3 | 0 | 2 | Not-determined | Genome-fragment | NA | NA | 0 |
| S1-final-viral-combined_k141_55181\|\|full | 1887 | No | NA | 3 | 0 | 0 | Low-quality | Genome-fragment | 1.04 | AAI-based (high-confidence) | 0 |
| S7-final-viral-combined_k141_15027\|\|full | 1895 | No | NA | 3 | 0 | 1 | Not-determined | Genome-fragment | NA | NA | 0 |
| S17-k141_101673 | 1887 | No | NA | 6 | 0 | 0 | Low-quality | Genome-fragment | 4.89 | AAI-based (high-confidence) | 0 |
| S7-final-viral-combined_k141_66698\|\|full | 1161 | No | NA | 3 | 0 | 0 | Not-determined | Genome-fragment | NA | NA | 0 |
| S7-final-viral-combined_k141_75865\|\|full | 1383 | No | NA | 2 | 0 | 0 | Not-determined | Genome-fragment | NA | NA | 0 |
| S7-final-viral-combined_k141_158475\|\|full | 1924 | No | NA | 2 | 0 | 2 | Not-determined | Genome-fragment | NA | NA | 0 |
| S7-final-viral-combined_k141_106774\|\|full | 2668 | No | NA | 3 | 0 | 0 | Not-determined | Genome-fragment | NA | NA | 0 |
| S2_k141_128909 | 1887 | No | NA | 2 | 0 | 1 | Low-quality | Genome-fragment | 1.37 | AAI-based (medium-confidence) | 0 |
| S1-final-viral-combined_k141_22972\|\|full | 1888 | No | NA | 4 | 0 | 0 | Low-quality | Genome-fragment | 1.27 | AAI-based (medium-confidence) | 0 |
| S7-final-viral-combined_k141_189202\|\|full | 3306 | No | NA | 4 | 0 | 1 | Not-determined | Genome-fragment | NA | NA | 0 |
| S1_k141_325103 | 1888 | No | NA | 2 | 0 | 1 | Low-quality | Genome-fragment | 2.64 | AAI-based (medium-confidence) | 0 |
| S7-final-viral-combined_k141_96616\|\|full | 7004 | No | NA | 6 | 0 | 3 | Not-determined | Genome-fragment | NA | NA | 0 |
| S7_k141_30545 | 1888 | No | NA | 2 | 0 | 2 | Low-quality | Genome-fragment | 0.87 | AAI-based (medium-confidence) | 0 |
| S17-final-viral-combined_k141_235977\|\|full | 1888 | No | NA | 3 | 0 | 1 | Low-quality | Genome-fragment | 1.22 | AAI-based (medium-confidence) | 0 |
| S17-k141_42947 | 1888 | No | NA | 2 | 0 | 1 | Low-quality | Genome-fragment | 3.46 | AAI-based (high-confidence) | 0 |
| S1_k141_362982 | 1889 | No | NA | 1 | 0 | 0 | Low-quality | Genome-fragment | 3.6 | AAI-based (high-confidence) | 0 |
| S1_k141_5824 | 1889 | No | NA | 1 | 0 | 0 | Low-quality | Genome-fragment | 1.73 | AAI-based (high-confidence) | 0 |
| S1_k141_220585 | 1889 | No | NA | 1 | 1 | 0 | Low-quality | Genome-fragment | 4.04 | AAI-based (high-confidence) | 0 |
| S2_k141_196876 | 1889 | No | NA | 3 | 3 | 0 | Low-quality | Genome-fragment | 4.6 | AAI-based (high-confidence) | 0 |
| S4-final-viral-combined_k141_22099\|\|full | 1889 | No | NA | 3 | 0 | 0 | Low-quality | Genome-fragment | 1.77 | AAI-based (high-confidence) | 0 |
| S8-final-viral-combined_k141_196541\|\|lt2gene | 1889 | No | NA | 1 | 1 | 0 | Low-quality | Genome-fragment | 4.86 | AAI-based (high-confidence) | 0 |
| S7-final-viral-combined_k141_147643\|\|full | 7683 | No | NA | 6 | 0 | 3 | Not-determined | Genome-fragment | NA | NA | 0 |
| S7-final-viral-combined_k141_96700\|\|full | 8600 | No | NA | 7 | 0 | 3 | Not-determined | Genome-fragment | NA | NA | 0 |
| S7-final-viral-combined_k141_24482\|\|full | 9168 | No | NA | 12 | 0 | 9 | Not-determined | Genome-fragment | NA | NA | 0 |
| S11_k141_14449 | 1890 | No | NA | 4 | 0 | 1 | Low-quality | Genome-fragment | 3.61 | AAI-based (medium-confidence) | 0 |
| S7-final-viral-combined_k141_45818\|\|full | 3023 | No | NA | 5 | 0 | 2 | Not-determined | Genome-fragment | NA | NA | 0 |
| S7-final-viral-combined_k141_189250\|\|full | 34933 | No | NA | 56 | 14 | 0 | High-quality | High-quality | 91.02 | AAI-based (high-confidence) | 0 |
| S7-final-viral-combined_k141_137640\|\|full | 2007 | No | NA | 4 | 0 | 0 | Not-determined | Genome-fragment | NA | NA | 0 |
| S7-final-viral-combined_k141_158548\|\|full | 10699 | No | NA | 10 | 0 | 10 | Not-determined | Genome-fragment | NA | NA | 0 |
| S1-final-viral-combined_k141_108430\|\|full | 1890 | No | NA | 3 | 2 | 0 | Low-quality | Genome-fragment | 4.69 | AAI-based (high-confidence) | 0 |
| S1_k141_205221 | 1890 | No | NA | 1 | 0 | 1 | Low-quality | Genome-fragment | 2.9 | AAI-based (medium-confidence) | 0 |
| S7-final-viral-combined_k141_24503\|\|full | 8811 | No | NA | 5 | 0 | 4 | Not-determined | Genome-fragment | NA | NA | 0 |
| S7-final-viral-combined_k141_35971\|\|full | 1046 | No | NA | 2 | 0 | 0 | Not-determined | Genome-fragment | NA | NA | 0 |
| S7-final-viral-combined_k141_127854\|\|full | 1768 | No | NA | 3 | 0 | 2 | Not-determined | Genome-fragment | NA | NA | 0 |
| S7-final-viral-combined_k141_240421\|\|full | 3323 | No | NA | 4 | 0 | 2 | Not-determined | Genome-fragment | NA | NA | 0 |
| S12-final-viral-combined_k141_361500\|\|full | 1890 | No | NA | 3 | 2 | 0 | Low-quality | Genome-fragment | 4.37 | AAI-based (high-confidence) | 0 |
| S7-final-viral-combined_k141_96750\|\|full | 1739 | No | NA | 1 | 0 | 1 | Not-determined | Genome-fragment | NA | NA | 0 |
| S2_k141_7727 | 1890 | No | NA | 1 | 0 | 1 | Low-quality | Genome-fragment | 3.87 | AAI-based (high-confidence) | 0 |
| S1-final-viral-combined_k141_283370\|\|full | 1891 | No | NA | 2 | 0 | 0 | Low-quality | Genome-fragment | 1.41 | AAI-based (high-confidence) | 0 |
| S1-final-viral-combined_k141_29950\|\|full | 1891 | No | NA | 3 | 0 | 1 | Low-quality | Genome-fragment | 3.27 | AAI-based (high-confidence) | 0 |
| S7-final-viral-combined_k141_216958\|\|full | 1891 | No | NA | 3 | 0 | 0 | Low-quality | Genome-fragment | 3.6 | AAI-based (high-confidence) | 0 |
| S7-final-viral-combined_k141_240448\|\|full | 2950 | No | NA | 4 | 0 | 3 | Not-determined | Genome-fragment | NA | NA | 0 |
| S7-final-viral-combined_k141_117795\|\|full | 1082 | No | NA | 2 | 0 | 0 | Not-determined | Genome-fragment | NA | NA | 0 |
| S12_k141_203604 | 1891 | No | NA | 1 | 0 | 1 | Low-quality | Genome-fragment | 1.98 | AAI-based (medium-confidence) | 0 |
| S7-final-viral-combined_k141_218564\|\|full | 1227 | No | NA | 1 | 0 | 0 | Not-determined | Genome-fragment | NA | NA | 0 |
| S8-final-viral-combined_k141_230915\|\|full | 1891 | No | NA | 2 | 0 | 2 | Low-quality | Genome-fragment | 1.74 | AAI-based (medium-confidence) | 0 |
| S1-final-viral-combined_k141_279078\|\|lt2gene | 1892 | No | NA | 2 | 1 | 0 | Low-quality | Genome-fragment | 4.53 | AAI-based (high-confidence) | 0 |
| S7-final-viral-combined_k141_240469\|\|full | 3274 | No | NA | 5 | 0 | 2 | Not-determined | Genome-fragment | NA | NA | 0 |
| S5_k141_14925 | 1892 | No | NA | 1 | 0 | 1 | Low-quality | Genome-fragment | 3.03 | AAI-based (medium-confidence) | 0 |
| S7-final-viral-combined_k141_45887\|\|full | 1268 | No | NA | 3 | 0 | 0 | Not-determined | Genome-fragment | NA | NA | 0 |
| S7-final-viral-combined_k141_56126\|\|full | 1291 | No | NA | 2 | 0 | 0 | Not-determined | Genome-fragment | NA | NA | 0 |
| S7-final-viral-combined_k141_218595\|\|full | 1450 | No | NA | 3 | 0 | 2 | Not-determined | Genome-fragment | NA | NA | 0 |
| S7-final-viral-combined_k141_66888\|\|full | 2985 | No | NA | 3 | 0 | 0 | Not-determined | Genome-fragment | NA | NA | 0 |
| S7-final-viral-combined_k141_106863\|\|full | 2283 | No | NA | 3 | 0 | 1 | Not-determined | Genome-fragment | NA | NA | 0 |
| S17-final-viral-combined_k141_140950\|\|full | 1892 | No | NA | 2 | 0 | 1 | Low-quality | Genome-fragment | 1.73 | AAI-based (medium-confidence) | 0 |
| S11-final-viral-combined_k141_238230\|\|full | 1893 | No | NA | 2 | 1 | 0 | Low-quality | Genome-fragment | 3.86 | AAI-based (medium-confidence) | 0 |
| S1-final-viral-combined_k141_29271\|\|full | 1893 | No | NA | 2 | 0 | 1 | Low-quality | Genome-fragment | 3.84 | AAI-based (high-confidence) | 0 |
| S7-final-viral-combined_k141_5089\|\|full | 1273 | No | NA | 3 | 0 | 0 | Not-determined | Genome-fragment | NA | NA | 0 |
| S7-final-viral-combined_k141_199970\|\|full | 1906 | No | NA | 2 | 0 | 1 | Not-determined | Genome-fragment | NA | NA | 0 |
| S7-final-viral-combined_k141_15272\|\|full | 4558 | No | NA | 4 | 0 | 2 | Not-determined | Genome-fragment | NA | NA | 0 |
| S7-final-viral-combined_k141_86473\|\|full | 1214 | No | NA | 3 | 0 | 0 | Not-determined | Genome-fragment | NA | NA | 0 |
| S7-final-viral-combined_k141_56204\|\|full | 2017 | No | NA | 2 | 0 | 0 | Not-determined | Genome-fragment | NA | NA | 0 |
| S7-final-viral-combined_k141_45970\|\|full | 2011 | No | NA | 3 | 0 | 1 | Not-determined | Genome-fragment | NA | NA | 0 |
| S7-final-viral-combined_k141_86487\|\|full | 1127 | No | NA | 2 | 0 | 1 | Not-determined | Genome-fragment | NA | NA | 0 |
| S7-final-viral-combined_k141_137807\|\|full | 3381 | No | NA | 3 | 0 | 1 | Not-determined | Genome-fragment | NA | NA | 0 |
| S7-final-viral-combined_k141_56210\|\|full | 2436 | No | NA | 3 | 0 | 2 | Not-determined | Genome-fragment | NA | NA | 0 |
| S1_k141_261501 | 1893 | No | NA | 3 | 0 | 0 | Low-quality | Genome-fragment | 4.13 | AAI-based (high-confidence) | 0 |
| S7-final-viral-combined_k141_96886\|\|full | 1659 | No | NA | 3 | 0 | 2 | Not-determined | Genome-fragment | NA | NA | 0 |
| S7-final-viral-combined_k141_218641\|\|full | 1789 | No | NA | 3 | 0 | 2 | Not-determined | Genome-fragment | NA | NA | 0 |
| S7-final-viral-combined_k141_66915\|\|full | 6209 | No | NA | 6 | 0 | 4 | Not-determined | Genome-fragment | NA | NA | 0 |
| S3-final-viral-combined_k141_68706\|\|full | 1893 | No | NA | 5 | 0 | 4 | Low-quality | Genome-fragment | 0.66 | AAI-based (medium-confidence) | 0 |
| S7-final-viral-combined_k141_74151\|\|full | 1893 | No | NA | 5 | 1 | 0 | Low-quality | Genome-fragment | 2.47 | AAI-based (medium-confidence) | 0 |
| S7-final-viral-combined_k141_189390\|\|full | 1895 | No | NA | 4 | 0 | 0 | Not-determined | Genome-fragment | NA | NA | 0 |
| S17-final-viral-combined_k141_201399\|\|full | 1893 | No | NA | 5 | 1 | 0 | Low-quality | Genome-fragment | 2.78 | AAI-based (medium-confidence) | 0 |
| S12-final-viral-combined_k141_48738\|\|full | 1893 | No | NA | 3 | 0 | 0 | Low-quality | Genome-fragment | 4.75 | AAI-based (medium-confidence) | 0 |
| S6-final-viral-combined_k141_24427\|\|full | 1893 | No | NA | 3 | 0 | 0 | Low-quality | Genome-fragment | 3.66 | AAI-based (high-confidence) | 0 |
| S7-final-viral-combined_k141_218645\|\|full | 18870 | No | NA | 16 | 0 | 7 | Not-determined | Genome-fragment | NA | NA | 0 |
| S8-final-viral-combined_k141_275523\|\|full | 1893 | No | NA | 2 | 0 | 0 | Low-quality | Genome-fragment | 2.28 | AAI-based (high-confidence) | 0 |
| S7-final-viral-combined_k141_127990\|\|full | 3414 | No | NA | 2 | 0 | 0 | Not-determined | Genome-fragment | NA | NA | 0 |
| S7-final-viral-combined_k141_36096\|\|full | 1758 | No | NA | 2 | 0 | 0 | Not-determined | Genome-fragment | NA | NA | 0 |
| S7-final-viral-combined_k141_46006\|\|full | 3181 | No | NA | 2 | 0 | 2 | Not-determined | Genome-fragment | NA | NA | 0 |
| S7-final-viral-combined_k141_179124\|\|full | 1105 | No | NA | 2 | 0 | 2 | Not-determined | Genome-fragment | NA | NA | 0 |
| S11-final-viral-combined_k141_308195\|\|full | 1894 | No | NA | 2 | 0 | 1 | Low-quality | Genome-fragment | 1.27 | AAI-based (medium-confidence) | 0 |
| S11-final-viral-combined_k141_33172\|\|full | 1894 | No | NA | 3 | 2 | 0 | Low-quality | Genome-fragment | 5.13 | AAI-based (high-confidence) | 0 |
| S7-final-viral-combined_k141_128032\|\|full | 1195 | No | NA | 2 | 0 | 1 | Not-determined | Genome-fragment | NA | NA | 0 |
| S7-final-viral-combined_k141_147772\|\|full | 1735 | No | NA | 5 | 0 | 0 | Not-determined | Genome-fragment | NA | NA | 0 |
| S3_k141_74862 | 1894 | No | NA | 4 | 1 | 0 | Low-quality | Genome-fragment | 1.23 | HMM-based (lower-bound) | 0 |
| S12-final-viral-combined_k141_215297\|\|full | 1894 | No | NA | 3 | 2 | 0 | Low-quality | Genome-fragment | 4.21 | AAI-based (medium-confidence) | 0 |
| S7-final-viral-combined_k141_117990\|\|full | 2551 | No | NA | 2 | 0 | 2 | Not-determined | Genome-fragment | NA | NA | 0 |
| S12-final-viral-combined_k141_208917\|\|lt2gene | 1894 | No | NA | 2 | 2 | 0 | Low-quality | Genome-fragment | 4.32 | AAI-based (high-confidence) | 0 |
| S2-final-viral-combined_k141_60773\|\|full | 1894 | No | NA | 6 | 0 | 1 | Low-quality | Genome-fragment | 2.42 | AAI-based (medium-confidence) | 0 |
| S7-final-viral-combined_k141_200043\|\|full | 1607 | No | NA | 3 | 0 | 0 | Not-determined | Genome-fragment | NA | NA | 0 |
| S7-final-viral-combined_k141_5181\|\|full | 4222 | No | NA | 2 | 0 | 0 | Not-determined | Genome-fragment | NA | NA | 0 |
| S7-final-viral-combined_k141_200050\|\|full | 2253 | No | NA | 3 | 0 | 0 | Not-determined | Genome-fragment | NA | NA | 0 |
| S7-final-viral-combined_k141_66997\|\|full | 2868 | No | NA | 3 | 0 | 2 | Not-determined | Genome-fragment | NA | NA | 0 |
| S4-final-viral-combined_k141_142386\|\|full | 1894 | No | NA | 3 | 0 | 2 | Low-quality | Genome-fragment | 3.2 | AAI-based (medium-confidence) | 0 |
| S7-final-viral-combined_k141_137858\|\|full | 50877 | Yes | 27506 | 45 | 3 | 21 | Medium-quality | Genome-fragment | 67.28 | AAI-based (high-confidence) | 45.94 |
| S11-final-viral-combined_k141_219192\|\|full | 1895 | No | NA | 3 | 1 | 0 | Low-quality | Genome-fragment | 1.66 | HMM-based (lower-bound) | 0 |
| S7-final-viral-combined_k141_147822\|\|full | 2700 | No | NA | 4 | 0 | 0 | Not-determined | Genome-fragment | NA | NA | 0 |
| S7-final-viral-combined_k141_24759\|\|full | 6599 | No | NA | 11 | 0 | 4 | Not-determined | Genome-fragment | NA | NA | 0 |
| S1-final-viral-combined_k141_265954\|\|lt2gene | 1895 | No | NA | 1 | 1 | 0 | Low-quality | Genome-fragment | 5.03 | AAI-based (high-confidence) | 0 |
| S7-final-viral-combined_k141_76248\|\|full | 2925 | No | NA | 4 | 0 | 3 | Not-determined | Genome-fragment | NA | NA | 0 |
| S7-final-viral-combined_k141_96940\|\|full | 3818 | No | NA | 3 | 0 | 2 | Not-determined | Genome-fragment | NA | NA | 0 |
| S1_k141_263329 | 1895 | No | NA | 5 | 0 | 0 | Low-quality | Genome-fragment | 5.74 | AAI-based (medium-confidence) | 0 |
| S17-final-viral-combined_k141_192227\|\|full | 1895 | No | NA | 2 | 0 | 1 | Low-quality | Genome-fragment | 1.45 | AAI-based (medium-confidence) | 0 |
| S6_k141_51238 | 1895 | No | NA | 1 | 0 | 0 | Low-quality | Genome-fragment | 1.23 | AAI-based (high-confidence) | 0 |
| S9_k141_89059 | 1896 | No | NA | 2 | 0 | 1 | Low-quality | Genome-fragment | 1.23 | AAI-based (high-confidence) | 0 |
| S7-final-viral-combined_k141_86699\|\|full | 2112 | No | NA | 3 | 0 | 2 | Not-determined | Genome-fragment | NA | NA | 0 |
| S7-final-viral-combined_k141_147939\|\|full | 1591 | No | NA | 2 | 0 | 2 | Not-determined | Genome-fragment | NA | NA | 0 |
| S1_k141_336515 | 1896 | No | NA | 3 | 0 | 2 | Low-quality | Genome-fragment | 1.12 | AAI-based (medium-confidence) | 0 |
| S7-final-viral-combined_k141_118185\|\|full | 1308 | No | NA | 3 | 0 | 0 | Not-determined | Genome-fragment | NA | NA | 0 |
| S7-final-viral-combined_k141_15382\|\|full | 8138 | No | NA | 8 | 0 | 5 | Not-determined | Genome-fragment | NA | NA | 0 |
| S7-final-viral-combined_k141_179296\|\|full | 8242 | No | NA | 14 | 0 | 3 | Not-determined | Genome-fragment | NA | NA | 0 |
| S7-final-viral-combined_k141_179297\|\|full | 1731 | No | NA | 3 | 0 | 1 | Not-determined | Genome-fragment | NA | NA | 0 |
| S7-final-viral-combined_k141_24837\|\|full | 8249 | No | NA | 9 | 0 | 6 | Not-determined | Genome-fragment | NA | NA | 0 |
| S7-final-viral-combined_k141_189622\|\|full | 1575 | No | NA | 3 | 0 | 1 | Not-determined | Genome-fragment | NA | NA | 0 |
| S5_k141_48913 | 1896 | No | NA | 1 | 0 | 1 | Low-quality | Genome-fragment | 0.77 | AAI-based (medium-confidence) | 0 |
| S17-final-viral-combined_k141_120731\|\|full | 1896 | No | NA | 4 | 1 | 0 | Low-quality | Genome-fragment | 1.84 | HMM-based (lower-bound) | 0 |
| S12-final-viral-combined_k141_239601\|\|full | 1896 | No | NA | 4 | 0 | 0 | Low-quality | Genome-fragment | 3.84 | AAI-based (medium-confidence) | 0 |
| S4-final-viral-combined_k141_148607\|\|full | 1896 | No | NA | 1 | 0 | 1 | Low-quality | Genome-fragment | 1.3 | AAI-based (medium-confidence) | 0 |
| S6-final-viral-combined_k141_121398\|\|full | 1896 | No | NA | 3 | 0 | 2 | Low-quality | Genome-fragment | 2.58 | AAI-based (high-confidence) | 0 |
| S7-final-viral-combined_k141_97081\|\|full | 1396 | No | NA | 3 | 0 | 2 | Not-determined | Genome-fragment | NA | NA | 0 |
| S7-final-viral-combined_k141_76434\|\|full | 6449 | No | NA | 9 | 0 | 6 | Not-determined | Genome-fragment | NA | NA | 0 |
| S9-final-viral-combined_k141_62542\|\|full | 1897 | No | NA | 2 | 0 | 2 | Low-quality | Genome-fragment | 2.91 | AAI-based (medium-confidence) | 0 |
| S1_k141_309963 | 1897 | No | NA | 2 | 0 | 0 | Low-quality | Genome-fragment | 1.94 | AAI-based (high-confidence) | 0 |
| S5-final-viral-combined_k141_14966\|\|lt2gene | 1897 | No | NA | 2 | 1 | 0 | Low-quality | Genome-fragment | 1.17 | HMM-based (lower-bound) | 0 |
| S7-final-viral-combined_k141_36346\|\|full | 6311 | No | NA | 4 | 0 | 1 | Not-determined | Genome-fragment | NA | NA | 0 |
| S7-final-viral-combined_k141_169329\|\|full | 3586 | No | NA | 3 | 0 | 0 | Not-determined | Genome-fragment | NA | NA | 0 |
| S7-final-viral-combined_k141_148008\|\|full | 1065 | No | NA | 2 | 0 | 1 | Not-determined | Genome-fragment | NA | NA | 0 |
| S7-final-viral-combined_k141_107239\|\|full | 3372 | No | NA | 5 | 0 | 2 | Not-determined | Genome-fragment | NA | NA | 0 |
| S7-final-viral-combined_k141_97123\|\|full | 3827 | No | NA | 8 | 0 | 4 | Not-determined | Genome-fragment | NA | NA | 0 |
| S7-final-viral-combined_k141_158879\|\|full | 1019 | No | NA | 4 | 0 | 0 | Not-determined | Genome-fragment | NA | NA | 0 |
| S7-final-viral-combined_k141_169360\|\|full | 2572 | No | NA | 3 | 0 | 1 | Not-determined | Genome-fragment | NA | NA | 0 |
| S7-final-viral-combined_k141_118223\|\|full | 2560 | No | NA | 3 | 0 | 0 | Not-determined | Genome-fragment | NA | NA | 0 |
| S17-final-viral-combined_k141_111953\|\|full | 1897 | No | NA | 1 | 0 | 0 | Low-quality | Genome-fragment | 3.11 | AAI-based (medium-confidence) | 0 |
| S7-final-viral-combined_k141_148047\|\|full | 2040 | No | NA | 2 | 0 | 2 | Not-determined | Genome-fragment | NA | NA | 0 |
| S7-final-viral-combined_k141_46302\|\|full | 3922 | No | NA | 5 | 0 | 4 | Not-determined | Genome-fragment | NA | NA | 0 |
| S17-final-viral-combined_k141_213123\|\|full | 1897 | No | NA | 3 | 0 | 0 | Low-quality | Genome-fragment | 0.58 | AAI-based (medium-confidence) | 0 |
| S12-final-viral-combined_k141_255987\|\|full | 1897 | No | NA | 3 | 0 | 2 | Low-quality | Genome-fragment | 2.74 | AAI-based (medium-confidence) | 0 |
| S12_k141_68665 | 1897 | No | NA | 2 | 0 | 1 | Low-quality | Genome-fragment | 3.09 | AAI-based (medium-confidence) | 0 |
| S7-final-viral-combined_k141_36424\|\|full | 3077 | No | NA | 4 | 0 | 2 | Not-determined | Genome-fragment | NA | NA | 0 |
| S7-final-viral-combined_k141_210309\|\|full | 1934 | No | NA | 2 | 0 | 1 | Not-determined | Genome-fragment | NA | NA | 0 |
| S4-final-viral-combined_k141_164686\|\|full | 1897 | No | NA | 6 | 0 | 0 | Low-quality | Genome-fragment | 3.16 | AAI-based (high-confidence) | 0 |
| S7-final-viral-combined_k141_46343\|\|full | 1696 | No | NA | 2 | 0 | 0 | Not-determined | Genome-fragment | NA | NA | 0 |
| S7-final-viral-combined_k141_56462\|\|full | 2534 | No | NA | 6 | 0 | 3 | Not-determined | Genome-fragment | NA | NA | 0 |
| S7-final-viral-combined_k141_97214\|\|full | 2842 | No | NA | 4 | 0 | 0 | Not-determined | Genome-fragment | NA | NA | 0 |
| S8-final-viral-combined_k141_116867\|\|lt2gene | 1897 | No | NA | 1 | 1 | 0 | Low-quality | Genome-fragment | 1.31 | HMM-based (lower-bound) | 0 |
| S7-final-viral-combined_k141_15599\|\|full | 1635 | No | NA | 4 | 0 | 1 | Not-determined | Genome-fragment | NA | NA | 0 |
| S7-final-viral-combined_k141_210323\|\|full | 3508 | No | NA | 4 | 0 | 1 | Not-determined | Genome-fragment | NA | NA | 0 |
| S9-final-viral-combined_k141_100032\|\|full | 1898 | No | NA | 2 | 0 | 1 | Low-quality | Genome-fragment | 0.92 | AAI-based (medium-confidence) | 0 |
| S11-final-viral-combined_k141_82907\|\|full | 1898 | No | NA | 3 | 0 | 0 | Low-quality | Genome-fragment | 3.02 | AAI-based (high-confidence) | 0 |
| S5-final-viral-combined_k141_125933\|\|lt2gene | 1898 | No | NA | 2 | 2 | 0 | Low-quality | Genome-fragment | 4.79 | AAI-based (high-confidence) | 0 |
| S5_k141_82453 | 1898 | No | NA | 2 | 1 | 0 | Low-quality | Genome-fragment | 3.43 | AAI-based (high-confidence) | 0 |
| S7-final-viral-combined_k141_46412\|\|full | 2463 | No | NA | 6 | 0 | 1 | Not-determined | Genome-fragment | NA | NA | 0 |
| S7-final-viral-combined_k141_138150\|\|full | 1467 | No | NA | 2 | 0 | 0 | Not-determined | Genome-fragment | NA | NA | 0 |
| S7-final-viral-combined_k141_237539\|\|full | 1898 | No | NA | 3 | 1 | 0 | Low-quality | Genome-fragment | 2.42 | AAI-based (medium-confidence) | 0 |
| S7-final-viral-combined_k141_230337\|\|full | 1088 | No | NA | 2 | 0 | 1 | Not-determined | Genome-fragment | NA | NA | 0 |
| S7-final-viral-combined_k141_138156\|\|full | 1057 | No | NA | 3 | 0 | 2 | Not-determined | Genome-fragment | NA | NA | 0 |
| S7-final-viral-combined_k141_191938\|\|full | 1898 | No | NA | 3 | 0 | 1 | Low-quality | Genome-fragment | 1.34 | AAI-based (medium-confidence) | 0 |
| S7-final-viral-combined_k141_240728\|\|full | 4222 | No | NA | 8 | 0 | 5 | Not-determined | Genome-fragment | NA | NA | 0 |
| S17-final-viral-combined_k141_77104\|\|full | 1898 | No | NA | 3 | 0 | 2 | Low-quality | Genome-fragment | 1.12 | AAI-based (medium-confidence) | 0 |
| S7-final-viral-combined_k141_107293\|\|full | 3828 | No | NA | 9 | 0 | 1 | Not-determined | Genome-fragment | NA | NA | 0 |
| S12-final-viral-combined_k141_335888\|\|full | 1898 | No | NA | 4 | 1 | 0 | Low-quality | Genome-fragment | 3.65 | AAI-based (medium-confidence) | 0 |
| S12-final-viral-combined_k141_393712\|\|full | 1898 | No | NA | 4 | 0 | 0 | Low-quality | Genome-fragment | 1.58 | AAI-based (medium-confidence) | 0 |
| S7-final-viral-combined_k141_46454\|\|full | 1146 | No | NA | 1 | 0 | 1 | Not-determined | Genome-fragment | NA | NA | 0 |
| S7-final-viral-combined_k141_169375\|\|full | 130069 | No | NA | 125 | 3 | 45 | High-quality | High-quality | 96.12 | HMM-based (lower-bound) | 0 |
| S7-final-viral-combined_k141_86846\|\|full | 2194 | No | NA | 3 | 0 | 2 | Not-determined | Genome-fragment | NA | NA | 0 |
| S7-final-viral-combined_k141_148215\|\|full | 2487 | No | NA | 4 | 0 | 0 | Not-determined | Genome-fragment | NA | NA | 0 |
| S7-final-viral-combined_k141_128274\|\|full | 1525 | No | NA | 3 | 0 | 1 | Not-determined | Genome-fragment | NA | NA | 0 |
| S12-final-viral-combined_k141_211781\|\|lt2gene | 1898 | No | NA | 2 | 1 | 0 | Low-quality | Genome-fragment | 5.66 | AAI-based (high-confidence) | 0 |
| S8-final-viral-combined_k141_176854\|\|full | 1898 | No | NA | 4 | 1 | 0 | Low-quality | Genome-fragment | 4.04 | AAI-based (medium-confidence) | 0 |
| S11-final-viral-combined_k141_297262\|\|full | 1899 | No | NA | 4 | 1 | 0 | Low-quality | Genome-fragment | 4.01 | AAI-based (high-confidence) | 0 |
| S7-final-viral-combined_k141_36605\|\|full | 2230 | No | NA | 2 | 0 | 0 | Not-determined | Genome-fragment | NA | NA | 0 |
| S7-final-viral-combined_k141_189883\|\|full | 17038 | No | NA | 14 | 0 | 5 | Not-determined | Genome-fragment | NA | NA | 0 |
| S11-final-viral-combined_k141_38405\|\|full | 1899 | No | NA | 3 | 0 | 1 | Low-quality | Genome-fragment | 1.34 | AAI-based (high-confidence) | 0 |
| S7-final-viral-combined_k141_15746\|\|full | 3342 | No | NA | 3 | 0 | 2 | Not-determined | Genome-fragment | NA | NA | 0 |
| S1_k141_391283 | 1899 | No | NA | 2 | 0 | 0 | Low-quality | Genome-fragment | 3.55 | AAI-based (medium-confidence) | 0 |
| S7-final-viral-combined_k141_170218\|\|full | 1899 | No | NA | 3 | 2 | 0 | Low-quality | Genome-fragment | 4.96 | AAI-based (high-confidence) | 0 |
| S17-final-viral-combined_k141_248967\|\|lt2gene | 1899 | No | NA | 1 | 1 | 0 | Low-quality | Genome-fragment | 4.55 | AAI-based (medium-confidence) | 0 |
| S7-final-viral-combined_k141_138254\|\|full | 2997 | No | NA | 4 | 0 | 3 | Not-determined | Genome-fragment | NA | NA | 0 |
| S12-final-viral-combined_k141_236657\|\|full | 1899 | No | NA | 5 | 1 | 0 | Low-quality | Genome-fragment | 4.41 | AAI-based (medium-confidence) | 0 |
| S7-final-viral-combined_k141_36639\|\|full | 1021 | No | NA | 3 | 0 | 0 | Not-determined | Genome-fragment | NA | NA | 0 |
| S2-final-viral-combined_k141_65373\|\|full | 1899 | No | NA | 2 | 0 | 0 | Low-quality | Genome-fragment | 3.76 | AAI-based (high-confidence) | 0 |
| S7-final-viral-combined_k141_107418\|\|full | 10304 | No | NA | 17 | 0 | 0 | Not-determined | Genome-fragment | NA | NA | 0 |
| S7-final-viral-combined_k141_138269\|\|full | 1273 | No | NA | 3 | 0 | 0 | Not-determined | Genome-fragment | NA | NA | 0 |
| S7-final-viral-combined_k141_97356\|\|full | 1889 | No | NA | 3 | 0 | 1 | Not-determined | Genome-fragment | NA | NA | 0 |
| S4-final-viral-combined_k141_121491\|\|full | 1899 | No | NA | 3 | 0 | 1 | Low-quality | Genome-fragment | 4.53 | AAI-based (high-confidence) | 0 |
| S4-final-viral-combined_k141_81630\|\|full | 1899 | No | NA | 4 | 1 | 0 | Low-quality | Genome-fragment | 1.91 | AAI-based (high-confidence) | 0 |
| S7-final-viral-combined_k141_148266\|\|full | 9623 | No | NA | 16 | 0 | 1 | Not-determined | Genome-fragment | NA | NA | 0 |
| S7-final-viral-combined_k141_36662\|\|full | 3458 | No | NA | 4 | 0 | 4 | Not-determined | Genome-fragment | NA | NA | 0 |
| S6_k141_88675 | 1899 | No | NA | 2 | 0 | 2 | Low-quality | Genome-fragment | 1.25 | AAI-based (high-confidence) | 0 |
| S7-final-viral-combined_k141_36678\|\|full | 1577 | No | NA | 3 | 0 | 0 | Not-determined | Genome-fragment | NA | NA | 0 |
| S7-final-viral-combined_k141_169479\|\|full | 3191 | No | NA | 5 | 0 | 4 | Not-determined | Genome-fragment | NA | NA | 0 |
| S7-final-viral-combined_k141_86908\|\|full | 3222 | No | NA | 3 | 0 | 3 | Not-determined | Genome-fragment | NA | NA | 0 |
| S7-final-viral-combined_k141_15780\|\|full | 3691 | No | NA | 3 | 0 | 2 | Not-determined | Genome-fragment | NA | NA | 0 |
| S11-final-viral-combined_k141_179205\|\|full | 1900 | No | NA | 3 | 0 | 0 | Low-quality | Genome-fragment | 0.95 | AAI-based (medium-confidence) | 0 |
| S11-final-viral-combined_k141_58024\|\|full | 1900 | No | NA | 2 | 0 | 1 | Low-quality | Genome-fragment | 6.07 | AAI-based (high-confidence) | 0 |
| S11-final-viral-combined_k141_150453\|\|full | 1900 | No | NA | 4 | 0 | 0 | Low-quality | Genome-fragment | 5.12 | AAI-based (high-confidence) | 0 |
| S1-final-viral-combined_k141_58067\|\|full | 1900 | No | NA | 3 | 0 | 2 | Low-quality | Genome-fragment | 2.51 | AAI-based (high-confidence) | 0 |
| S7-final-viral-combined_k141_76673\|\|full | 2783 | No | NA | 4 | 0 | 0 | Not-determined | Genome-fragment | NA | NA | 0 |
| S1-final-viral-combined_k141_262984\|\|full | 1900 | No | NA | 3 | 0 | 2 | Low-quality | Genome-fragment | 3.43 | AAI-based (medium-confidence) | 0 |
| S7-final-viral-combined_k141_189985\|\|full | 2267 | No | NA | 2 | 0 | 2 | Not-determined | Genome-fragment | NA | NA | 0 |
| S7-final-viral-combined_k141_128386\|\|full | 1474 | No | NA | 2 | 0 | 1 | Not-determined | Genome-fragment | NA | NA | 0 |
| S5-final-viral-combined_k141_191622\|\|full | 1900 | No | NA | 3 | 0 | 0 | Low-quality | Genome-fragment | 3.57 | AAI-based (medium-confidence) | 0 |
| S7-final-viral-combined_k141_179571\|\|full | 3389 | No | NA | 4 | 0 | 3 | Not-determined | Genome-fragment | NA | NA | 0 |
| S12-final-viral-combined_k141_87289\|\|full | 1900 | No | NA | 2 | 0 | 1 | Low-quality | Genome-fragment | 1.36 | AAI-based (medium-confidence) | 0 |
| S7-final-viral-combined_k141_25191\|\|full | 1161 | No | NA | 2 | 0 | 0 | Not-determined | Genome-fragment | NA | NA | 0 |
| S6-final-viral-combined_k141_43494\|\|full | 1900 | No | NA | 3 | 2 | 0 | Low-quality | Genome-fragment | 5.74 | AAI-based (medium-confidence) | 0 |
| S7-final-viral-combined_k141_200609\|\|full | 1700 | No | NA | 5 | 0 | 2 | Not-determined | Genome-fragment | NA | NA | 0 |
| S7-final-viral-combined_k141_118425\|\|full | 1686 | No | NA | 2 | 0 | 0 | Not-determined | Genome-fragment | NA | NA | 0 |
| S6-final-viral-combined_k141_31966\|\|full | 1900 | No | NA | 5 | 1 | 0 | Low-quality | Genome-fragment | 5.31 | AAI-based (medium-confidence) | 0 |
| S11-final-viral-combined_k141_299763\|\|full | 1901 | No | NA | 2 | 0 | 0 | Low-quality | Genome-fragment | 3.01 | AAI-based (high-confidence) | 0 |
| S7-final-viral-combined_k141_210613\|\|full | 1333 | No | NA | 3 | 0 | 0 | Not-determined | Genome-fragment | NA | NA | 0 |
| S7-final-viral-combined_k141_179622\|\|full | 1072 | No | NA | 2 | 0 | 0 | Not-determined | Genome-fragment | NA | NA | 0 |
| S7-final-viral-combined_k141_128445\|\|full | 1197 | No | NA | 3 | 0 | 0 | Not-determined | Genome-fragment | NA | NA | 0 |
| S7-final-viral-combined_k141_190073\|\|full | 2503 | No | NA | 3 | 0 | 0 | Not-determined | Genome-fragment | NA | NA | 0 |
| S7-final-viral-combined_k141_36770\|\|full | 2202 | No | NA | 2 | 0 | 0 | Not-determined | Genome-fragment | NA | NA | 0 |
| S7-final-viral-combined_k141_159211\|\|full | 2018 | No | NA | 3 | 0 | 1 | Not-determined | Genome-fragment | NA | NA | 0 |
| S1-final-viral-combined_k141_75659\|\|full | 1901 | No | NA | 3 | 0 | 1 | Low-quality | Genome-fragment | 1.3 | AAI-based (medium-confidence) | 0 |
| S7-final-viral-combined_k141_5625\|\|full | 116688 | Yes | 21872 | 99 | 8 | 43 | Medium-quality | Genome-fragment | 51.07 | AAI-based (high-confidence) | 81.26 |
| S7-final-viral-combined_k141_230519\|\|full | 3168 | No | NA | 7 | 0 | 2 | Not-determined | Genome-fragment | NA | NA | 0 |
| S5-final-viral-combined_k141_19070\|\|full | 1901 | No | NA | 3 | 2 | 0 | Low-quality | Genome-fragment | 4.2 | AAI-based (high-confidence) | 0 |
| S7-final-viral-combined_k141_90\|\|full | 1901 | No | NA | 3 | 0 | 0 | Low-quality | Genome-fragment | 15.1 | AAI-based (medium-confidence) | 0 |
| S7-final-viral-combined_k141_46745\|\|full | 3959 | No | NA | 5 | 0 | 1 | Not-determined | Genome-fragment | NA | NA | 0 |
| S7-final-viral-combined_k141_240982\|\|full | 3410 | No | NA | 4 | 0 | 2 | Not-determined | Genome-fragment | NA | NA | 0 |
| S17-final-viral-combined_k141_162160\|\|lt2gene | 1901 | No | NA | 1 | 0 | 0 | Low-quality | Genome-fragment | 3.43 | AAI-based (medium-confidence) | 0 |
| S7-final-viral-combined_k141_46758\|\|full | 2017 | No | NA | 2 | 0 | 1 | Not-determined | Genome-fragment | NA | NA | 0 |
| S7-final-viral-combined_k141_200700\|\|full | 2616 | No | NA | 2 | 0 | 1 | Not-determined | Genome-fragment | NA | NA | 0 |
| S7-final-viral-combined_k141_128506\|\|full | 1192 | No | NA | 2 | 0 | 1 | Not-determined | Genome-fragment | NA | NA | 0 |
| S2_k141_197410 | 1901 | No | NA | 1 | 0 | 1 | Low-quality | Genome-fragment | 0.16 | AAI-based (medium-confidence) | 0 |
| S7-final-viral-combined_k141_25200\|\|full | 1976 | No | NA | 3 | 0 | 0 | Not-determined | Genome-fragment | NA | NA | 0 |
| S7-final-viral-combined_k141_56810\|\|full | 3356 | No | NA | 4 | 0 | 1 | Not-determined | Genome-fragment | NA | NA | 0 |
| S7-final-viral-combined_k141_128526\|\|full | 1307 | No | NA | 2 | 0 | 0 | Not-determined | Genome-fragment | NA | NA | 0 |
| S11-final-viral-combined_k141_308353\|\|full | 1902 | No | NA | 4 | 0 | 0 | Low-quality | Genome-fragment | 3.74 | AAI-based (medium-confidence) | 0 |
| S7-final-viral-combined_k141_25213\|\|full | 2067 | No | NA | 3 | 0 | 1 | Not-determined | Genome-fragment | NA | NA | 0 |
| S1-final-viral-combined_k141_41859\|\|lt2gene | 1902 | No | NA | 1 | 1 | 0 | Low-quality | Genome-fragment | 3.96 | AAI-based (high-confidence) | 0 |
| S7-final-viral-combined_k141_190216\|\|full | 2106 | No | NA | 3 | 0 | 2 | Not-determined | Genome-fragment | NA | NA | 0 |
| S17-final-viral-combined_k141_104405\|\|full | 1902 | No | NA | 2 | 0 | 1 | Low-quality | Genome-fragment | 0.84 | AAI-based (medium-confidence) | 0 |
| S7-final-viral-combined_k141_190235\|\|full | 5851 | No | NA | 6 | 0 | 1 | Not-determined | Genome-fragment | NA | NA | 0 |
| S7-final-viral-combined_k141_241070\|\|full | 6680 | No | NA | 6 | 0 | 3 | Not-determined | Genome-fragment | NA | NA | 0 |
| S7-final-viral-combined_k141_138469\|\|full | 2481 | No | NA | 3 | 0 | 2 | Not-determined | Genome-fragment | NA | NA | 0 |
| S7-final-viral-combined_k141_25257\|\|full | 5200 | No | NA | 5 | 0 | 2 | Not-determined | Genome-fragment | NA | NA | 0 |
| S17-final-viral-combined_k141_108588\|\|full | 1902 | No | NA | 2 | 0 | 1 | Low-quality | Genome-fragment | 3.64 | AAI-based (high-confidence) | 0 |
| S7-final-viral-combined_k141_67506\|\|full | 2385 | No | NA | 7 | 0 | 0 | Not-determined | Genome-fragment | NA | NA | 0 |
| S7-final-viral-combined_k141_241114\|\|full | 2030 | No | NA | 4 | 0 | 0 | Not-determined | Genome-fragment | NA | NA | 0 |
| S7-final-viral-combined_k141_169747\|\|full | 1823 | No | NA | 2 | 0 | 0 | Not-determined | Genome-fragment | NA | NA | 0 |
| S17-final-viral-combined_k141_65621\|\|full | 1902 | No | NA | 3 | 0 | 3 | Low-quality | Genome-fragment | 0.72 | AAI-based (medium-confidence) | 0 |
| S7-final-viral-combined_k141_159358\|\|full | 1694 | No | NA | 3 | 0 | 1 | Not-determined | Genome-fragment | NA | NA | 0 |
| S7-final-viral-combined_k141_67529\|\|full | 1181 | No | NA | 4 | 0 | 0 | Not-determined | Genome-fragment | NA | NA | 0 |
| S17-final-viral-combined_k141_265501\|\|full | 1902 | No | NA | 6 | 3 | 0 | Low-quality | Genome-fragment | 3.73 | HMM-based (lower-bound) | 0 |
| S7-final-viral-combined_k141_241130\|\|full | 1456 | No | NA | 2 | 0 | 1 | Not-determined | Genome-fragment | NA | NA | 0 |
| S6-final-viral-combined_k141_139569\|\|full | 1902 | No | NA | 5 | 0 | 1 | Low-quality | Genome-fragment | 2.95 | AAI-based (medium-confidence) | 0 |
| S8-final-viral-combined_k141_88442\|\|full | 1902 | No | NA | 2 | 0 | 1 | Low-quality | Genome-fragment | 0.9 | AAI-based (medium-confidence) | 0 |
| S7-final-viral-combined_k141_76831\|\|full | 1566 | No | NA | 3 | 0 | 1 | Not-determined | Genome-fragment | NA | NA | 0 |
| S7-final-viral-combined_k141_118620\|\|full | 3777 | No | NA | 7 | 0 | 0 | Not-determined | Genome-fragment | NA | NA | 0 |
| S7-final-viral-combined_k141_138543\|\|full | 1207 | No | NA | 4 | 0 | 0 | Not-determined | Genome-fragment | NA | NA | 0 |
| S8-final-viral-combined_k141_137259\|\|full | 1902 | No | NA | 4 | 1 | 0 | Low-quality | Genome-fragment | 4.52 | AAI-based (high-confidence) | 0 |
| S11-final-viral-combined_k141_322012\|\|full | 1903 | No | NA | 3 | 0 | 1 | Low-quality | Genome-fragment | 0.86 | AAI-based (high-confidence) | 0 |
| S7-final-viral-combined_k141_138558\|\|full | 1674 | No | NA | 2 | 0 | 0 | Not-determined | Genome-fragment | NA | NA | 0 |
| S11-final-viral-combined_k141_82211\|\|lt2gene | 1903 | No | NA | 2 | 1 | 0 | Low-quality | Genome-fragment | 3.52 | HMM-based (lower-bound) | 0 |
| S7-final-viral-combined_k141_16022\|\|full | 4210 | No | NA | 3 | 0 | 2 | Not-determined | Genome-fragment | NA | NA | 0 |
| S1_k141_337576 | 1903 | No | NA | 1 | 0 | 1 | Low-quality | Genome-fragment | 0.77 | AAI-based (medium-confidence) | 0 |
| S7-final-viral-combined_k141_56910\|\|full | 1072 | No | NA | 3 | 0 | 2 | Not-determined | Genome-fragment | NA | NA | 0 |
| S7-final-viral-combined_k141_179879\|\|full | 2885 | No | NA | 4 | 0 | 4 | Not-determined | Genome-fragment | NA | NA | 0 |
| S3_k141_128347 | 1903 | No | NA | 7 | 0 | 0 | Low-quality | Genome-fragment | 3.31 | AAI-based (high-confidence) | 0 |
| S5-final-viral-combined_k141_179852\|\|full | 1903 | No | NA | 4 | 0 | 0 | Low-quality | Genome-fragment | 3.13 | AAI-based (medium-confidence) | 0 |
| S7-final-viral-combined_k141_241195\|\|full | 1287 | No | NA | 3 | 0 | 1 | Not-determined | Genome-fragment | NA | NA | 0 |
| S5-final-viral-combined_k141_176409\|\|full | 1903 | No | NA | 4 | 1 | 0 | Low-quality | Genome-fragment | 5.76 | AAI-based (medium-confidence) | 0 |
| S17-final-viral-combined_k141_4592\|\|full | 1903 | No | NA | 3 | 0 | 0 | Low-quality | Genome-fragment | 3.31 | AAI-based (high-confidence) | 0 |
| S12-final-viral-combined_k141_263969\|\|full | 1903 | No | NA | 3 | 0 | 2 | Low-quality | Genome-fragment | 2.59 | AAI-based (high-confidence) | 0 |
| S2-final-viral-combined_k141_156165\|\|full | 1903 | No | NA | 2 | 1 | 0 | Low-quality | Genome-fragment | 4.89 | AAI-based (high-confidence) | 0 |
| S7-final-viral-combined_k141_67658\|\|full | 6607 | No | NA | 7 | 0 | 5 | Not-determined | Genome-fragment | NA | NA | 0 |
| S7-final-viral-combined_k141_219169\|\|full | 1405 | No | NA | 3 | 0 | 2 | Not-determined | Genome-fragment | NA | NA | 0 |
| S7-final-viral-combined_k141_241242\|\|full | 2524 | No | NA | 3 | 0 | 2 | Not-determined | Genome-fragment | NA | NA | 0 |
| S8_k141_261092 | 1903 | No | NA | 3 | 0 | 1 | Low-quality | Genome-fragment | 2.29 | AAI-based (high-confidence) | 0 |
| S11-final-viral-combined_k141_317546\|\|full | 1904 | No | NA | 2 | 0 | 1 | Low-quality | Genome-fragment | 1.56 | AAI-based (high-confidence) | 0 |
| S7-final-viral-combined_k141_190281\|\|full | 63064 | Yes | 33724 | 89 | 20 | 14 | High-quality | High-quality | 97.16 | AAI-based (high-confidence) | 46.52 |
| S7-final-viral-combined_k141_210860\|\|full | 2952 | No | NA | 3 | 0 | 3 | Not-determined | Genome-fragment | NA | NA | 0 |
| S7-final-viral-combined_k141_169966\|\|full | 5788 | No | NA | 5 | 0 | 4 | Not-determined | Genome-fragment | NA | NA | 0 |
| S7-final-viral-combined_k141_76886\|\|full | 3389 | No | NA | 3 | 0 | 1 | Not-determined | Genome-fragment | NA | NA | 0 |
| S7-final-viral-combined_k141_118724\|\|full | 1156 | No | NA | 3 | 0 | 0 | Not-determined | Genome-fragment | NA | NA | 0 |
| S11-final-viral-combined_k141_156259\|\|full | 1904 | No | NA | 4 | 1 | 0 | Low-quality | Genome-fragment | 4.11 | AAI-based (high-confidence) | 0 |
| S1-final-viral-combined_k141_116551\|\|full | 1904 | No | NA | 3 | 2 | 0 | Low-quality | Genome-fragment | 1.57 | HMM-based (lower-bound) | 0 |
| S7-final-viral-combined_k141_76896\|\|full | 3679 | No | NA | 6 | 0 | 3 | Not-determined | Genome-fragment | NA | NA | 0 |
| S7-final-viral-combined_k141_219269\|\|full | 2309 | No | NA | 4 | 0 | 1 | Not-determined | Genome-fragment | NA | NA | 0 |
| S1-final-viral-combined_k141_313971\|\|full | 1904 | No | NA | 5 | 1 | 0 | Low-quality | Genome-fragment | 3.2 | AAI-based (high-confidence) | 0 |
| S7-final-viral-combined_k141_190338\|\|full | 11944 | No | NA | 12 | 0 | 9 | Not-determined | Genome-fragment | NA | NA | 0 |
| S7-final-viral-combined_k141_97766\|\|full | 1716 | No | NA | 2 | 0 | 0 | Not-determined | Genome-fragment | NA | NA | 0 |
| S7-final-viral-combined_k141_230782\|\|full | 2905 | No | NA | 4 | 0 | 3 | Not-determined | Genome-fragment | NA | NA | 0 |
| S1-final-viral-combined_k141_155398\|\|full | 1904 | No | NA | 3 | 0 | 0 | Low-quality | Genome-fragment | 4.22 | AAI-based (medium-confidence) | 0 |
| S7-final-viral-combined_k141_97784\|\|full | 2189 | No | NA | 2 | 0 | 2 | Not-determined | Genome-fragment | NA | NA | 0 |
| S7-final-viral-combined_k141_210973\|\|full | 2178 | No | NA | 3 | 0 | 0 | Not-determined | Genome-fragment | NA | NA | 0 |
| S5-final-viral-combined_k141_115971\|\|full | 1904 | No | NA | 4 | 1 | 0 | Low-quality | Genome-fragment | 3.28 | HMM-based (lower-bound) | 0 |
| S5-final-viral-combined_k141_106523\|\|full | 1904 | No | NA | 3 | 2 | 0 | Low-quality | Genome-fragment | 4.17 | AAI-based (high-confidence) | 0 |
| S7-final-viral-combined_k141_134351\|\|full | 1904 | No | NA | 4 | 0 | 0 | Low-quality | Genome-fragment | 3.23 | AAI-based (medium-confidence) | 0 |
| S17-final-viral-combined_k141_255751\|\|full | 1904 | No | NA | 4 | 2 | 0 | Low-quality | Genome-fragment | 3.45 | HMM-based (lower-bound) | 0 |
| S7-final-viral-combined_k141_76994\|\|full | 1107 | No | NA | 3 | 0 | 0 | Not-determined | Genome-fragment | NA | NA | 0 |
| S17-final-viral-combined_k141_151026\|\|full | 1904 | No | NA | 1 | 0 | 1 | Low-quality | Genome-fragment | 1.41 | AAI-based (high-confidence) | 0 |
| S17-final-viral-combined_k141_28249\|\|full | 1904 | No | NA | 3 | 0 | 1 | Low-quality | Genome-fragment | 2.59 | AAI-based (medium-confidence) | 0 |
| S7-final-viral-combined_k141_118827\|\|full | 3354 | No | NA | 5 | 0 | 1 | Not-determined | Genome-fragment | NA | NA | 0 |
| S12-final-viral-combined_k141_169281\|\|full | 1904 | No | NA | 4 | 0 | 1 | Low-quality | Genome-fragment | 1.11 | AAI-based (medium-confidence) | 0 |
| S7-final-viral-combined_k141_97820\|\|full | 1215 | No | NA | 3 | 0 | 0 | Not-determined | Genome-fragment | NA | NA | 0 |
| S7-final-viral-combined_k141_128910\|\|full | 1287 | No | NA | 2 | 0 | 1 | Not-determined | Genome-fragment | NA | NA | 0 |
| S12-final-viral-combined_k141_380868\|\|full | 1904 | No | NA | 3 | 1 | 0 | Low-quality | Genome-fragment | 1.16 | HMM-based (lower-bound) | 0 |
| S7-final-viral-combined_k141_159762\|\|full | 1581 | No | NA | 3 | 0 | 0 | Not-determined | Genome-fragment | NA | NA | 0 |
| S7-final-viral-combined_k141_87473\|\|full | 2868 | No | NA | 4 | 0 | 1 | Not-determined | Genome-fragment | NA | NA | 0 |
| S7-final-viral-combined_k141_180037\|\|full | 1766 | No | NA | 2 | 0 | 1 | Not-determined | Genome-fragment | NA | NA | 0 |
| S7-final-viral-combined_k141_46983\|\|full | 28523 | No | NA | 19 | 0 | 12 | Not-determined | Genome-fragment | NA | NA | 0 |
| S12_k141_103612 | 1904 | No | NA | 1 | 0 | 1 | Low-quality | Genome-fragment | 0.33 | AAI-based (medium-confidence) | 0 |
| S7-final-viral-combined_k141_46985\|\|full | 3638 | No | NA | 3 | 0 | 2 | Not-determined | Genome-fragment | NA | NA | 0 |
| S7-final-viral-combined_k141_148782\|\|full | 1343 | No | NA | 3 | 0 | 0 | Not-determined | Genome-fragment | NA | NA | 0 |
| S7-final-viral-combined_k141_97845\|\|full | 1969 | No | NA | 4 | 0 | 0 | Not-determined | Genome-fragment | NA | NA | 0 |
| S7-final-viral-combined_k141_180058\|\|full | 1922 | No | NA | 3 | 0 | 1 | Not-determined | Genome-fragment | NA | NA | 0 |
| S2-final-viral-combined_k141_38046\|\|full | 1904 | No | NA | 4 | 0 | 2 | Low-quality | Genome-fragment | 3.18 | AAI-based (medium-confidence) | 0 |
| S11-final-viral-combined_k141_119117\|\|full | 1905 | No | NA | 3 | 0 | 2 | Low-quality | Genome-fragment | 0.61 | AAI-based (medium-confidence) | 0 |
| S5-final-viral-combined_k141_190346\|\|full | 1905 | No | NA | 4 | 0 | 0 | Low-quality | Genome-fragment | 4.6 | AAI-based (high-confidence) | 0 |
| S7-final-viral-combined_k141_219420\|\|full | 1004 | No | NA | 2 | 0 | 0 | Not-determined | Genome-fragment | NA | NA | 0 |
| S12-final-viral-combined_k141_255245\|\|full | 1905 | No | NA | 4 | 0 | 0 | Low-quality | Genome-fragment | 4.94 | AAI-based (high-confidence) | 0 |
| S12-final-viral-combined_k141_333544\|\|full | 1905 | No | NA | 2 | 0 | 1 | Low-quality | Genome-fragment | 1.07 | AAI-based (medium-confidence) | 0 |
| S7-final-viral-combined_k141_5990\|\|full | 1708 | No | NA | 2 | 0 | 1 | Not-determined | Genome-fragment | NA | NA | 0 |
| S7-final-viral-combined_k141_148827\|\|full | 1568 | No | NA | 2 | 0 | 2 | Not-determined | Genome-fragment | NA | NA | 0 |
| S7-final-viral-combined_k141_211047\|\|full | 1789 | No | NA | 4 | 0 | 0 | Not-determined | Genome-fragment | NA | NA | 0 |
| S7-final-viral-combined_k141_107703\|\|full | 2037 | No | NA | 3 | 0 | 1 | Not-determined | Genome-fragment | NA | NA | 0 |
| S2-final-viral-combined_k141_167739\|\|full | 1905 | No | NA | 4 | 0 | 2 | Low-quality | Genome-fragment | 0.6 | AAI-based (medium-confidence) | 0 |
| S7-final-viral-combined_k141_241412\|\|full | 2545 | No | NA | 3 | 0 | 1 | Not-determined | Genome-fragment | NA | NA | 0 |
| S7-final-viral-combined_k141_241416\|\|full | 1613 | No | NA | 3 | 0 | 1 | Not-determined | Genome-fragment | NA | NA | 0 |
| S7-final-viral-combined_k141_190594\|\|full | 2504 | No | NA | 3 | 0 | 0 | Not-determined | Genome-fragment | NA | NA | 0 |
| S6-final-viral-combined_k141_95692\|\|full | 1905 | No | NA | 4 | 0 | 0 | Low-quality | Genome-fragment | 4.41 | AAI-based (high-confidence) | 0 |
| S7-final-viral-combined_k141_6032\|\|full | 1556 | No | NA | 2 | 0 | 1 | Not-determined | Genome-fragment | NA | NA | 0 |
| S7-final-viral-combined_k141_37344\|\|full | 3889 | No | NA | 5 | 0 | 1 | Not-determined | Genome-fragment | NA | NA | 0 |
| S7-final-viral-combined_k141_148879\|\|full | 1881 | No | NA | 2 | 0 | 2 | Not-determined | Genome-fragment | NA | NA | 0 |
| S7-final-viral-combined_k141_87554\|\|full | 3523 | No | NA | 6 | 0 | 0 | Not-determined | Genome-fragment | NA | NA | 0 |
| S7-final-viral-combined_k141_148886\|\|full | 1320 | No | NA | 2 | 0 | 0 | Not-determined | Genome-fragment | NA | NA | 0 |
| S7-final-viral-combined_k141_118972\|\|full | 1639 | No | NA | 2 | 0 | 2 | Not-determined | Genome-fragment | NA | NA | 0 |
| S8-final-viral-combined_k141_228966\|\|full | 1905 | No | NA | 2 | 0 | 0 | Low-quality | Genome-fragment | 2.96 | AAI-based (medium-confidence) | 0 |
| S7-final-viral-combined_k141_118998\|\|full | 2032 | No | NA | 3 | 0 | 2 | Not-determined | Genome-fragment | NA | NA | 0 |
| S7-final-viral-combined_k141_87576\|\|full | 1477 | No | NA | 3 | 0 | 2 | Not-determined | Genome-fragment | NA | NA | 0 |
| S8_k141_97296 | 1905 | No | NA | 1 | 0 | 1 | Low-quality | Genome-fragment | 0.5 | AAI-based (high-confidence) | 0 |
| S1-final-viral-combined_k141_144409\|\|full | 1906 | No | NA | 2 | 1 | 0 | Low-quality | Genome-fragment | 2.3 | AAI-based (high-confidence) | 0 |
| S7-final-viral-combined_k141_170263\|\|full | 1276 | No | NA | 4 | 0 | 0 | Not-determined | Genome-fragment | NA | NA | 0 |
| S7-final-viral-combined_k141_138912\|\|full | 1946 | No | NA | 2 | 0 | 1 | Not-determined | Genome-fragment | NA | NA | 0 |
| S1-final-viral-combined_k141_41703\|\|lt2gene | 1906 | Yes | 918 | 2 | 1 | 1 | Low-quality | Genome-fragment | 2.12 | AAI-based (high-confidence) | 51.84 |
| S1_k141_162951 | 1906 | No | NA | 1 | 0 | 1 | Low-quality | Genome-fragment | 1.89 | AAI-based (medium-confidence) | 0 |
| S7-final-viral-combined_k141_129052\|\|full | 1132 | No | NA | 2 | 0 | 1 | Not-determined | Genome-fragment | NA | NA | 0 |
| S7-final-viral-combined_k141_47062\|\|full | 29107 | No | NA | 24 | 0 | 14 | Not-determined | Genome-fragment | NA | NA | 0 |
| S7-final-viral-combined_k141_25764\|\|full | 1754 | No | NA | 3 | 0 | 0 | Not-determined | Genome-fragment | NA | NA | 0 |
| S7-final-viral-combined_k141_68011\|\|full | 1674 | No | NA | 3 | 0 | 2 | Not-determined | Genome-fragment | NA | NA | 0 |
| S7-final-viral-combined_k141_125080\|\|full | 1906 | No | NA | 4 | 1 | 0 | Low-quality | Genome-fragment | 3.75 | HMM-based (lower-bound) | 0 |
| S7-final-viral-combined_k141_119062\|\|full | 1240 | No | NA | 3 | 0 | 1 | Not-determined | Genome-fragment | NA | NA | 0 |
| S7-final-viral-combined_k141_187715\|\|full | 1906 | No | NA | 4 | 3 | 0 | Low-quality | Genome-fragment | 3.91 | AAI-based (high-confidence) | 0 |
| S7-final-viral-combined_k141_190706\|\|full | 1830 | No | NA | 3 | 0 | 1 | Not-determined | Genome-fragment | NA | NA | 0 |
| S12-final-viral-combined_k141_319234\|\|full | 1906 | No | NA | 2 | 0 | 2 | Low-quality | Genome-fragment | 0.14 | AAI-based (medium-confidence) | 0 |
| S7-final-viral-combined_k141_138938\|\|full | 1705 | No | NA | 3 | 0 | 2 | Not-determined | Genome-fragment | NA | NA | 0 |
| S4_k141_2538 | 1906 | No | NA | 2 | 0 | 2 | Low-quality | Genome-fragment | 1.53 | AAI-based (high-confidence) | 0 |
| S7-final-viral-combined_k141_190720\|\|full | 1461 | No | NA | 2 | 0 | 1 | Not-determined | Genome-fragment | NA | NA | 0 |
| S9_k141_96453 | 1907 | No | NA | 1 | 0 | 1 | Low-quality | Genome-fragment | 0.71 | AAI-based (high-confidence) | 0 |
| S1-final-viral-combined_k141_64731\|\|full | 1907 | No | NA | 3 | 0 | 3 | Low-quality | Genome-fragment | 1.14 | AAI-based (medium-confidence) | 0 |
| S7-final-viral-combined_k141_138981\|\|full | 2701 | No | NA | 4 | 0 | 4 | Not-determined | Genome-fragment | NA | NA | 0 |
| S7-final-viral-combined_k141_47186\|\|full | 2035 | No | NA | 3 | 0 | 1 | Not-determined | Genome-fragment | NA | NA | 0 |
| S3-final-viral-combined_k141_76322\|\|full | 1907 | No | NA | 3 | 1 | 0 | Low-quality | Genome-fragment | 5.47 | AAI-based (high-confidence) | 0 |
| S17-final-viral-combined_k141_146640\|\|full | 1907 | No | NA | 3 | 0 | 1 | Low-quality | Genome-fragment | 0.89 | AAI-based (high-confidence) | 0 |
| S7-final-viral-combined_k141_241466\|\|full | 7065 | No | NA | 6 | 0 | 2 | Not-determined | Genome-fragment | NA | NA | 0 |
| S11-final-viral-combined_k141_165293\|\|full | 1908 | No | NA | 3 | 0 | 0 | Low-quality | Genome-fragment | 2.98 | AAI-based (medium-confidence) | 0 |

| | | | | | | | |
|---|---|---|---|---|---|---|---|
| S7-final-viral-combined_k141_231075\|\|full | 1047 | No | NA | 3 | 0 | 3 | [illegible] |
| S7-final-viral-combined_k141_119109\|\|full | 1176 | No | NA | 2 | 0 | 0 | [illegible] |
| S12_k141_182430 | 1908 | No | NA | 7 | 0 | 0 | Low-quality [illegible] |
| S7-final-viral-combined_k141_219501\|\|full | 5125 | No | NA | 4 | 0 | 1 | [illegible] |
| S8-final-viral-combined_k141_153209\|\|full | 1908 | No | NA | 3 | 0 | 0 | Low-quality [illegible] |
| S11-final-viral-combined_k141_29863\|\|full | 1909 | No | NA | 3 | 1 | 0 | Low-quality [illegible] |
| S7-final-viral-combined_k141_180175\|\|full | 2348 | No | NA | 4 | 0 | 1 | [illegible] |
| S11-final-viral-combined_k141_213973\|\|full | 1909 | No | NA | 3 | 3 | 0 | Low-quality [illegible] |
| S7-final-viral-combined_k141_107775\|\|full | 2617 | No | NA | 3 | 0 | 3 | [illegible] |
| S11-final-viral-combined_k141_301769\|\|full | 1909 | No | NA | 2 | 0 | 1 | Low-quality [illegible] |
| S7-final-viral-combined_k141_57516\|\|full | 2511 | No | NA | 3 | 0 | 0 | [illegible] |
| S7-final-viral-combined_k141_77329\|\|full | 2701 | No | NA | 3 | 0 | 3 | [illegible] |
| S7-final-viral-combined_k141_16532\|\|full | 1862 | No | NA | 3 | 0 | 1 | [illegible] |
| S7-final-viral-combined_k141_6201\|\|full | 2416 | No | NA | 4 | 0 | 3 | [illegible] |
| S1-final-viral-combined_k141_285085\|\|full | 1909 | No | NA | 6 | 0 | 0 | Low-quality [illegible] |
| S12_k141_231348 | 1909 | No | NA | 2 | 1 | 0 | Low-quality [illegible] |
| S11-final-viral-combined_k141_97926\|\|full | 1910 | No | NA | 3 | 0 | 0 | Low-quality [illegible] |
| S11-final-viral-combined_k141_301658\|\|full | 1910 | No | NA | 6 | 2 | 0 | Low-quality [illegible] |
| S1_k141_29189 | 1910 | No | NA | 1 | 0 | 1 | Low-quality [illegible] |
| S12_k141_246585 | 1910 | No | NA | 3 | 0 | 0 | Low-quality [illegible] |
| S7-final-viral-combined_k141_6235\|\|full | 1758 | No | NA | 2 | 0 | 1 | [illegible] |
| S7-final-viral-combined_k141_231202\|\|full | 10566 | No | NA | 8 | 0 | 5 | [illegible] |
| S11-final-viral-combined_k141_226549\|\|full | 1911 | No | NA | 3 | 0 | 0 | Low-quality [illegible] |
| S11-final-viral-combined_k141_186329\|\|full | 1911 | No | NA | 3 | 0 | 0 | Low-quality [illegible] |
| S11-final-viral-combined_k141_174777\|\|full | 1911 | No | NA | 2 | 0 | 0 | Low-quality [illegible] |
| S7_k141_6598 | 1911 | No | NA | 3 | 1 | 0 | Low-quality [illegible] |
| S7-final-viral-combined_k141_47278\|\|full | 1265 | No | NA | 4 | 0 | 0 | [illegible] |
| S7-final-viral-combined_k141_139215\|\|full | 1894 | No | NA | 3 | 0 | 3 | [illegible] |
| S7-final-viral-combined_k141_98249\|\|full | 1382 | No | NA | 2 | 0 | 0 | [illegible] |
| S7-final-viral-combined_k141_211277\|\|full | 1243 | No | NA | 2 | 0 | 0 | [illegible] |
| S7-final-viral-combined_k141_16646\|\|full | 1691 | No | NA | 4 | 0 | 1 | [illegible] |
| S4_k141_30944 | 1911 | No | NA | 5 | 0 | 0 | Low-quality [illegible] |
| S9-final-viral-combined_k141_53079\|\|full | 1912 | No | NA | 3 | 0 | 1 | Low-quality [illegible] |
| S7-final-viral-combined_k141_190896\|\|full | 6025 | No | NA | 4 | 0 | 1 | [illegible] |
| S9-final-viral-combined_k141_31973\|\|full | 1912 | No | NA | 3 | 0 | 0 | Low-quality [illegible] |
| S7-final-viral-combined_k141_37604\|\|full | 1808 | No | NA | 3 | 0 | 1 | [illegible] |

[illegible]

| | | | | | | | | | | | |
|---|---|---|---|---|---|---|---|---|---|---|---|
| S7-final-viral-combined_k141_220853\|\|full | 1486 | No | NA | 3 | 0 | 3 | Not-determined | Genome-fragment | NA | NA | 0 |
| S17-final-viral-combined_k141_25954\|\|full | 1935 | No | NA | 4 | 3 | 0 | Low-quality | Genome-fragment | 5.75 | AAI-based (high-confidence) | 0 |
| S9-final-viral-combined_k141_78251\|\|lt2gene | 1936 | No | NA | 1 | 1 | 0 | Low-quality | Genome-fragment | 4.81 | AAI-based (high-confidence) | 0 |
| S7-final-viral-combined_k141_192514\|\|full | 6535 | No | NA | 13 | 0 | 2 | Not-determined | Genome-fragment | NA | NA | 0 |
| S7-final-viral-combined_k141_203124\|\|full | 10655 | No | NA | 8 | 0 | 3 | Not-determined | Genome-fragment | NA | NA | 0 |
| S7-final-viral-combined_k141_182186\|\|full | 1683 | No | NA | 2 | 0 | 1 | Not-determined | Genome-fragment | NA | NA | 0 |
| S11-final-viral-combined_k141_7647\|\|full | 1936 | No | NA | 4 | 1 | 0 | Low-quality | Genome-fragment | 4.45 | AAI-based (medium-confidence) | 0 |
| S7-final-viral-combined_k141_39209\|\|full | 1106 | No | NA | 2 | 0 | 2 | Not-determined | Genome-fragment | NA | NA | 0 |
| S7-final-viral-combined_k141_89306\|\|full | 2391 | No | NA | 5 | 0 | 0 | Not-determined | Genome-fragment | NA | NA | 0 |
| S12-final-viral-combined_k141_126375\|\|full | 1936 | No | NA | 4 | 0 | 1 | Low-quality | Genome-fragment | 2.14 | AAI-based (medium-confidence) | 0 |
| S7-final-viral-combined_k141_220880\|\|full | 5672 | No | NA | 7 | 0 | 4 | Not-determined | Genome-fragment | NA | NA | 0 |
| S7-final-viral-combined_k141_233006\|\|full | 4636 | No | NA | 5 | 0 | 2 | Not-determined | Genome-fragment | NA | NA | 0 |
| S7-final-viral-combined_k141_233007\|\|full | 1577 | No | NA | 2 | 0 | 0 | Not-determined | Genome-fragment | NA | NA | 0 |
| S12-final-viral-combined_k141_250761\|\|full | 1936 | No | NA | 4 | 1 | 0 | Low-quality | Genome-fragment | 3.17 | HMM-based (lower-bound) | 0 |
| S7-final-viral-combined_k141_59394\|\|full | 2901 | No | NA | 2 | 0 | 2 | Not-determined | Genome-fragment | NA | NA | 0 |
| S12-final-viral-combined_k141_63457\|\|lt2gene | 1936 | No | NA | 2 | 1 | 0 | Low-quality | Genome-fragment | 46.52 | HMM-based (lower-bound) | 0 |
| S7-final-viral-combined_k141_100089\|\|full | 1378 | No | NA | 3 | 0 | 0 | Not-determined | Genome-fragment | NA | NA | 0 |
| S2-final-viral-combined_k141_21591\|\|full | 1936 | No | NA | 2 | 0 | 2 | Low-quality | Genome-fragment | 1.55 | AAI-based (high-confidence) | 0 |
| S2-final-viral-combined_k141_120551\|\|lt2gene | 1936 | No | NA | 1 | 1 | 0 | Low-quality | Genome-fragment | 3.21 | HMM-based (lower-bound) | 0 |
| S8_k141_3039 | 1936 | No | NA | 3 | 0 | 3 | Low-quality | Genome-fragment | 0.96 | AAI-based (medium-confidence) | 0 |
| S7-final-viral-combined_k141_59441\|\|full | 1275 | No | NA | 2 | 0 | 1 | Not-determined | Genome-fragment | NA | NA | 0 |
| S7-final-viral-combined_k141_18277\|\|full | 1917 | No | NA | 2 | 0 | 2 | Not-determined | Genome-fragment | NA | NA | 0 |
| S7-final-viral-combined_k141_233051\|\|full | 1473 | No | NA | 2 | 0 | 2 | Not-determined | Genome-fragment | NA | NA | 0 |
| S11-final-viral-combined_k141_289698\|\|lt2gene | 1937 | No | NA | 2 | 1 | 0 | Low-quality | Genome-fragment | 2.54 | HMM-based (lower-bound) | 0 |
| S7-final-viral-combined_k141_18321\|\|full | 1428 | No | NA | 3 | 0 | 0 | Not-determined | Genome-fragment | NA | NA | 0 |
| S7-final-viral-combined_k141_79113\|\|full | 1193 | No | NA | 3 | 0 | 1 | Not-determined | Genome-fragment | NA | NA | 0 |
| S7-final-viral-combined_k141_18326\|\|full | 2068 | No | NA | 4 | 0 | 2 | Not-determined | Genome-fragment | NA | NA | 0 |
| S7-final-viral-combined_k141_59447\|\|full | 1410 | No | NA | 2 | 0 | 0 | Not-determined | Genome-fragment | NA | NA | 0 |
| S1-final-viral-combined_k141_253142\|\|full | 1937 | No | NA | 3 | 0 | 2 | Low-quality | Genome-fragment | 1.3 | AAI-based (medium-confidence) | 0 |
| S7-final-viral-combined_k141_109604\|\|full | 3303 | No | NA | 6 | 0 | 3 | Not-determined | Genome-fragment | NA | NA | 0 |
| S7-final-viral-combined_k141_79159\|\|full | 1739 | No | NA | 3 | 0 | 0 | Not-determined | Genome-fragment | NA | NA | 0 |
| S2-final-viral-combined_k141_12277\|\|full | 1937 | No | NA | 3 | 0 | 0 | Low-quality | Genome-fragment | 2.52 | AAI-based (medium-confidence) | 0 |
| S7-final-viral-combined_k141_182284\|\|full | 1163 | No | NA | 3 | 0 | 1 | Not-determined | Genome-fragment | NA | NA | 0 |
| S7-final-viral-combined_k141_18373\|\|full | 1269 | No | NA | 3 | 0 | 1 | Not-determined | Genome-fragment | NA | NA | 0 |
| S7-final-viral-combined_k141_59492\|\|full | 1756 | No | NA | 3 | 0 | 1 | Not-determined | Genome-fragment | NA | NA | 0 |
| S6_k141_10646 | 1937 | No | NA | 3 | 0 | 2 | Low-quality | Genome-fragment | 1.08 | AAI-based (medium-confidence) | 0 |
| S7-final-viral-combined_k141_150782\|\|full | 3643 | No | NA | 2 | 0 | 1 | Not-determined | Genome-fragment | NA | NA | 0 |
| S1-final-viral-combined_k141_298027\|\|full | 1938 | No | NA | 2 | 0 | 0 | Low-quality | Genome-fragment | 2.95 | AAI-based (medium-confidence) | 0 |
| S7-final-viral-combined_k141_8090\|\|full | 4550 | No | NA | 7 | 0 | 3 | Not-determined | Genome-fragment | NA | NA | 0 |
| S7-final-viral-combined_k141_242650\|\|full | 2401 | No | NA | 5 | 0 | 0 | Not-determined | Genome-fragment | NA | NA | 0 |
| S7-final-viral-combined_k141_242659\|\|full | 1159 | No | NA | 2 | 0 | 0 | Not-determined | Genome-fragment | NA | NA | 0 |
| S7-final-viral-combined_k141_79197\|\|full | 4461 | No | NA | 8 | 0 | 2 | Not-determined | Genome-fragment | NA | NA | 0 |
| S1-final-viral-combined_k141_68663\|\|full | 1938 | No | NA | 4 | 0 | 0 | Low-quality | Genome-fragment | 1.81 | AAI-based (high-confidence) | 0 |
| S1-final-viral-combined_k141_245020\|\|full | 1938 | No | NA | 5 | 0 | 0 | Low-quality | Genome-fragment | 5.03 | AAI-based (high-confidence) | 0 |
| S1_k141_319018 | 1938 | No | NA | 3 | 0 | 1 | Low-quality | Genome-fragment | 3.24 | AAI-based (medium-confidence) | 0 |
| S17-final-viral-combined_k141_90886\|\|full | 1938 | No | NA | 2 | 0 | 1 | Low-quality | Genome-fragment | 2.22 | AAI-based (high-confidence) | 0 |
| S17-final-viral-combined_k141_70260\|\|full | 1938 | No | NA | 3 | 0 | 0 | Low-quality | Genome-fragment | 4.92 | AAI-based (high-confidence) | 0 |
| S7-final-viral-combined_k141_100222\|\|full | 1644 | No | NA | 3 | 0 | 0 | Not-determined | Genome-fragment | NA | NA | 0 |
| S7-final-viral-combined_k141_242714\|\|full | 3769 | No | NA | 3 | 0 | 1 | Not-determined | Genome-fragment | NA | NA | 0 |
| S7-final-viral-combined_k141_120925\|\|full | 2066 | No | NA | 4 | 0 | 1 | Not-determined | Genome-fragment | NA | NA | 0 |
| S7-final-viral-combined_k141_27100\|\|full | 1383 | No | NA | 1 | 0 | 0 | Not-determined | Genome-fragment | NA | NA | 0 |
| S7-final-viral-combined_k141_242742\|\|full | 3138 | No | NA | 4 | 0 | 0 | Not-determined | Genome-fragment | NA | NA | 0 |
| S7-final-viral-combined_k141_109751\|\|full | 4289 | No | NA | 6 | 0 | 0 | Not-determined | Genome-fragment | NA | NA | 0 |
| S17-final-viral-combined_k141_215094\|\|full | 1938 | No | NA | 2 | 0 | 0 | Low-quality | Genome-fragment | 3.03 | AAI-based (high-confidence) | 0 |
| S7-final-viral-combined_k141_162078\|\|full | 2407 | No | NA | 5 | 0 | 1 | Not-determined | Genome-fragment | NA | NA | 0 |
| S2-final-viral-combined_k141_219325\|\|full | 1938 | No | NA | 3 | 2 | 0 | Low-quality | Genome-fragment | 3.17 | HMM-based (lower-bound) | 0 |
| S7-final-viral-combined_k141_120982\|\|full | 1407 | No | NA | 2 | 0 | 1 | Not-determined | Genome-fragment | NA | NA | 0 |
| S7-final-viral-combined_k141_140359\|\|full | 1198 | No | NA | 2 | 0 | 0 | Not-determined | Genome-fragment | NA | NA | 0 |
| S2_k141_213632 | 1938 | No | NA | 3 | 0 | 0 | Low-quality | Genome-fragment | 3.85 | AAI-based (medium-confidence) | 0 |
| S7-final-viral-combined_k141_233270\|\|full | 2037 | No | NA | 3 | 0 | 1 | Not-determined | Genome-fragment | NA | NA | 0 |
| S4_k141_26442 | 1938 | No | NA | 3 | 2 | 0 | Low-quality | Genome-fragment | 1.12 | AAI-based (high-confidence) | 0 |
| S7-final-viral-combined_k141_70027\|\|full | 1449 | No | NA | 2 | 0 | 1 | Not-determined | Genome-fragment | NA | NA | 0 |
| S7-final-viral-combined_k141_70029\|\|full | 6492 | No | NA | 5 | 0 | 2 | Not-determined | Genome-fragment | NA | NA | 0 |
| S8-final-viral-combined_k141_165265\|\|full | 1938 | No | NA | 2 | 0 | 1 | Low-quality | Genome-fragment | 1 | AAI-based (high-confidence) | 0 |
| S11-final-viral-combined_k141_235106\|\|full | 1940 | No | NA | 2 | 0 | 1 | Low-quality | Genome-fragment | 3.41 | AAI-based (medium-confidence) | 0 |
| S7-final-viral-combined_k141_192846\|\|full | 4787 | No | NA | 4 | 0 | 3 | Not-determined | Genome-fragment | NA | NA | 0 |
| S5-final-viral-combined_k141_211975\|\|full | 1940 | No | NA | 3 | 0 | 0 | Low-quality | Genome-fragment | 4.63 | AAI-based (medium-confidence) | 0 |
| S7-final-viral-combined_k141_70052\|\|full | 1042 | No | NA | 3 | 0 | 0 | Not-determined | Genome-fragment | NA | NA | 0 |
| S7-final-viral-combined_k141_140412\|\|full | 6529 | No | NA | 8 | 0 | 5 | Not-determined | Genome-fragment | NA | NA | 0 |
| S7-final-viral-combined_k141_130931\|\|full | 1975 | No | NA | 2 | 0 | 2 | Not-determined | Genome-fragment | NA | NA | 0 |
| S7-final-viral-combined_k141_210481\|\|full | 1940 | No | NA | 3 | 2 | 0 | Low-quality | Genome-fragment | 4.06 | AAI-based (medium-confidence) | 0 |
| S7-final-viral-combined_k141_59656\|\|full | 1629 | No | NA | 3 | 0 | 1 | Not-determined | Genome-fragment | NA | NA | 0 |
| S7-final-viral-combined_k141_140429\|\|full | 2080 | No | NA | 4 | 0 | 0 | Not-determined | Genome-fragment | NA | NA | 0 |
| S7-final-viral-combined_k141_8226\|\|full | 2684 | No | NA | 3 | 0 | 3 | Not-determined | Genome-fragment | NA | NA | 0 |
| S7-final-viral-combined_k141_182495\|\|full | 2296 | No | NA | 6 | 0 | 1 | Not-determined | Genome-fragment | NA | NA | 0 |
| S17-final-viral-combined_k141_892\|\|full | 1940 | No | NA | 4 | 2 | 0 | Low-quality | Genome-fragment | 6.76 | AAI-based (high-confidence) | 0 |
| S12-final-viral-combined_k141_351571\|\|full | 1940 | No | NA | 4 | 1 | 0 | Low-quality | Genome-fragment | 2.55 | HMM-based (lower-bound) | 0 |
| S7-final-viral-combined_k141_121062\|\|full | 29002 | No | NA | 39 | 12 | 0 | Medium-quality | Genome-fragment | 74.07 | AAI-based (high-confidence) | 0 |
| S7-final-viral-combined_k141_59693\|\|full | 1198 | No | NA | 2 | 0 | 0 | Not-determined | Genome-fragment | NA | NA | 0 |
| S7-final-viral-combined_k141_192931\|\|full | 1215 | No | NA | 3 | 0 | 0 | Not-determined | Genome-fragment | NA | NA | 0 |
| S7-final-viral-combined_k141_121093\|\|full | 1148 | No | NA | 3 | 0 | 0 | Not-determined | Genome-fragment | NA | NA | 0 |
| S2_k141_134129 | 1940 | No | NA | 2 | 0 | 2 | Low-quality | Genome-fragment | 0.72 | AAI-based (medium-confidence) | 0 |
| S7-final-viral-combined_k141_89496\|\|full | 10214 | No | NA | 3 | 0 | 2 | Not-determined | Genome-fragment | NA | NA | 0 |
| S7-final-viral-combined_k141_49678\|\|full | 2620 | No | NA | 3 | 0 | 2 | Not-determined | Genome-fragment | NA | NA | 0 |
| S8-final-viral-combined_k141_54178\|\|full | 1940 | No | NA | 3 | 0 | 2 | Low-quality | Genome-fragment | 1.23 | AAI-based (medium-confidence) | 0 |
| S7-final-viral-combined_k141_150918\|\|full | 1098 | No | NA | 2 | 0 | 1 | Not-determined | Genome-fragment | NA | NA | 0 |
| S7-final-viral-combined_k141_27244\|\|full | 1796 | No | NA | 2 | 0 | 1 | Not-determined | Genome-fragment | NA | NA | 0 |
| S9_k141_115407 | 1941 | No | NA | 2 | 0 | 0 | Low-quality | Genome-fragment | 4.59 | AAI-based (medium-confidence) | 0 |
| S7-final-viral-combined_k141_212949\|\|full | 2108 | No | NA | 4 | 0 | 0 | Not-determined | Genome-fragment | NA | NA | 0 |
| S7-final-viral-combined_k141_182554\|\|full | 6181 | No | NA | 5 | 0 | 2 | Not-determined | Genome-fragment | NA | NA | 0 |
| S7-final-viral-combined_k141_192992\|\|full | 1471 | No | NA | 4 | 0 | 0 | Not-determined | Genome-fragment | NA | NA | 0 |
| S7-final-viral-combined_k141_49722\|\|full | 13150 | No | NA | 9 | 0 | 6 | Not-determined | Genome-fragment | NA | NA | 0 |
| S7-final-viral-combined_k141_162167\|\|full | 2468 | No | NA | 6 | 0 | 0 | Not-determined | Genome-fragment | NA | NA | 0 |
| S11-final-viral-combined_k141_281152\|\|full | 1942 | No | NA | 4 | 4 | 0 | Low-quality | Genome-fragment | 4.8 | AAI-based (high-confidence) | 0 |
| S7-final-viral-combined_k141_1690\|\|full | 1942 | No | NA | 2 | 0 | 0 | Low-quality | Genome-fragment | 4.74 | AAI-based (medium-confidence) | 0 |
| S7-final-viral-combined_k141_162187\|\|full | 3462 | No | NA | 3 | 0 | 3 | Not-determined | Genome-fragment | NA | NA | 0 |
| S17-k141_50848 | 1942 | No | NA | 3 | 0 | 0 | Low-quality | Genome-fragment | 3.53 | AAI-based (high-confidence) | 0 |
| S12_k141_367219 | 1942 | No | NA | 1 | 0 | 1 | Low-quality | Genome-fragment | 2.97 | AAI-based (medium-confidence) | 0 |
| S7-final-viral-combined_k141_151009\|\|full | 1791 | No | NA | 2 | 0 | 1 | Not-determined | Genome-fragment | NA | NA | 0 |
| S2-final-viral-combined_k141_232750\|\|full | 1942 | No | NA | 2 | 0 | 0 | Low-quality | Genome-fragment | 3.76 | AAI-based (high-confidence) | 0 |
| S4-final-viral-combined_k141_11866\|\|full | 1942 | No | NA | 5 | 2 | 0 | Low-quality | Genome-fragment | 3.14 | AAI-based (high-confidence) | 0 |
| S7-final-viral-combined_k141_193071\|\|full | 2108 | No | NA | 4 | 0 | 0 | Not-determined | Genome-fragment | NA | NA | 0 |
| S8-final-viral-combined_k141_202121\|\|full | 1942 | No | NA | 4 | 0 | 0 | Low-quality | Genome-fragment | 5.99 | AAI-based (high-confidence) | 0 |
| S7-final-viral-combined_k141_70227\|\|full | 3674 | No | NA | 4 | 0 | 2 | Not-determined | Genome-fragment | NA | NA | 0 |
| S9-final-viral-combined_k141_50064\|\|full | 1943 | No | NA | 3 | 0 | 1 | Low-quality | Genome-fragment | 0.47 | AAI-based (medium-confidence) | 0 |
| S11-final-viral-combined_k141_158100\|\|lt2gene | 1943 | No | NA | 2 | 0 | 0 | Low-quality | Genome-fragment | 3.94 | AAI-based (medium-confidence) | 0 |
| S7-final-viral-combined_k141_100491\|\|full | 2775 | No | NA | 2 | 0 | 2 | Not-determined | Genome-fragment | NA | NA | 0 |
| S1-final-viral-combined_k141_365473\|\|full | 1943 | No | NA | 4 | 1 | 0 | Low-quality | Genome-fragment | 2.79 | HMM-based (lower-bound) | 0 |
| S7-final-viral-combined_k141_233473\|\|full | 1154 | No | NA | 3 | 0 | 1 | Not-determined | Genome-fragment | NA | NA | 0 |
| S7-final-viral-combined_k141_59804\|\|full | 1079 | No | NA | 2 | 0 | 1 | Not-determined | Genome-fragment | NA | NA | 0 |
| S7-final-viral-combined_k141_233475\|\|full | 1888 | No | NA | 3 | 0 | 1 | Not-determined | Genome-fragment | NA | NA | 0 |
| S1-final-viral-combined_k141_109365\|\|full | 1943 | No | NA | 4 | 3 | 0 | Low-quality | Genome-fragment | 4.47 | AAI-based (high-confidence) | 0 |
| S7-final-viral-combined_k141_89604\|\|full | 7495 | No | NA | 6 | 0 | 2 | Not-determined | Genome-fragment | NA | NA | 0 |
| S7-final-viral-combined_k141_70236\|\|full | 1960 | No | NA | 4 | 0 | 3 | Not-determined | Genome-fragment | NA | NA | 0 |
| S7-final-viral-combined_k141_72240\|\|full | 1943 | No | NA | 3 | 0 | 0 | Low-quality | Genome-fragment | 1.27 | AAI-based (high-confidence) | 0 |
| S7-final-viral-combined_k141_213043\|\|full | 2168 | No | NA | 3 | 0 | 1 | Not-determined | Genome-fragment | NA | NA | 0 |
| S7-final-viral-combined_k141_199059\|\|full | 1943 | No | NA | 2 | 0 | 1 | Low-quality | Genome-fragment | 1.73 | AAI-based (high-confidence) | 0 |
| S2-final-viral-combined_k141_160999\|\|full | 1943 | No | NA | 3 | 1 | 0 | Low-quality | Genome-fragment | 3.8 | AAI-based (high-confidence) | 0 |
| S7-final-viral-combined_k141_140657\|\|full | 2652 | No | NA | 4 | 0 | 1 | Not-determined | Genome-fragment | NA | NA | 0 |
| S7-final-viral-combined_k141_100543\|\|full | 2586 | No | NA | 2 | 0 | 2 | Not-determined | Genome-fragment | NA | NA | 0 |
| S2_k141_207062 | 1943 | No | NA | 1 | 0 | 1 | Low-quality | Genome-fragment | 2.95 | AAI-based (medium-confidence) | 0 |
| S7-final-viral-combined_k141_172128\|\|full | 2672 | No | NA | 5 | 0 | 2 | Not-determined | Genome-fragment | NA | NA | 0 |
| S4-final-viral-combined_k141_94097\|\|full | 1943 | No | NA | 2 | 0 | 1 | Low-quality | Genome-fragment | 1.34 | AAI-based (high-confidence) | 0 |
| S8-final-viral-combined_k141_110813\|\|full | 1943 | No | NA | 2 | 0 | 0 | Low-quality | Genome-fragment | 1.58 | AAI-based (medium-confidence) | 0 |
| S7-final-viral-combined_k141_49831\|\|full | 5484 | No | NA | 5 | 0 | 4 | Not-determined | Genome-fragment | NA | NA | 0 |
| S7-final-viral-combined_k141_59832\|\|full | 1915 | No | NA | 4 | 0 | 0 | Not-determined | Genome-fragment | NA | NA | 0 |
| S11-final-viral-combined_k141_149789\|\|full | 1944 | No | NA | 4 | 0 | 2 | Low-quality | Genome-fragment | 2.64 | AAI-based (high-confidence) | 0 |
| S7-final-viral-combined_k141_213104\|\|full | 8269 | No | NA | 4 | 0 | 4 | Not-determined | Genome-fragment | NA | NA | 0 |
| S7-final-viral-combined_k141_110042\|\|full | 2823 | No | NA | 3 | 0 | 1 | Not-determined | Genome-fragment | NA | NA | 0 |
| S11-final-viral-combined_k141_303071\|\|full | 1944 | No | NA | 3 | 0 | 1 | Low-quality | Genome-fragment | 1.53 | AAI-based (high-confidence) | 0 |
| S7-final-viral-combined_k141_233550\|\|full | 16138 | No | NA | 15 | 0 | 9 | Not-determined | Genome-fragment | NA | NA | 0 |
| S1-final-viral-combined_k141_190423\|\|full | 1944 | No | NA | 3 | 0 | 0 | Low-quality | Genome-fragment | 4.43 | AAI-based (medium-confidence) | 0 |
| S7-final-viral-combined_k141_49888\|\|full | 1898 | No | NA | 3 | 0 | 1 | Not-determined | Genome-fragment | NA | NA | 0 |
| S12-final-viral-combined_k141_302939\|\|full | 1944 | No | NA | 3 | 2 | 0 | Low-quality | Genome-fragment | 2.36 | AAI-based (high-confidence) | 0 |
| S12-final-viral-combined_k141_143889\|\|full | 1944 | No | NA | 4 | 1 | 0 | Low-quality | Genome-fragment | 3.08 | AAI-based (high-confidence) | 0 |
| S4-final-viral-combined_k141_85008\|\|full | 1944 | No | NA | 4 | 0 | 0 | Low-quality | Genome-fragment | 3.02 | AAI-based (high-confidence) | 0 |
| S7-final-viral-combined_k141_182657\|\|full | 1093 | No | NA | 3 | 0 | 1 | Not-determined | Genome-fragment | NA | NA | 0 |
| S7-final-viral-combined_k141_162342\|\|full | 3276 | No | NA | 3 | 0 | 1 | Not-determined | Genome-fragment | NA | NA | 0 |
| S8-final-viral-combined_k141_32763\|\|full | 1944 | No | NA | 2 | 0 | 1 | Low-quality | Genome-fragment | 0.54 | AAI-based (high-confidence) | 0 |
| S11_k141_122659 | 1945 | No | NA | 5 | 1 | 0 | Low-quality | Genome-fragment | 4.33 | AAI-based (high-confidence) | 0 |
| S7_k141_163304 | 1945 | No | NA | 7 | 2 | 0 | Low-quality | Genome-fragment | 5.95 | AAI-based (high-confidence) | 0 |
| S12-final-viral-combined_k141_77751\|\|full | 1945 | No | NA | 2 | 0 | 0 | Low-quality | Genome-fragment | 3.75 | AAI-based (medium-confidence) | 0 |
| S2-final-viral-combined_k141_32109\|\|lt2gene | 1945 | No | NA | 2 | 2 | 0 | Low-quality | Genome-fragment | 4.5 | AAI-based (high-confidence) | 0 |
| S4-final-viral-combined_k141_38502\|\|full | 1945 | No | NA | 3 | 1 | 0 | Low-quality | Genome-fragment | 3.93 | AAI-based (medium-confidence) | 0 |
| S8-final-viral-combined_k141_46768\|\|full | 1945 | No | NA | 2 | 0 | 1 | Low-quality | Genome-fragment | 1.15 | AAI-based (medium-confidence) | 0 |
| S7-final-viral-combined_k141_213151\|\|full | 3962 | No | NA | 4 | 0 | 2 | Not-determined | Genome-fragment | NA | NA | 0 |
| S8-final-viral-combined_k141_220497\|\|full | 1945 | No | NA | 3 | 0 | 1 | Low-quality | Genome-fragment | 1.69 | AAI-based (high-confidence) | 0 |
| S7-final-viral-combined_k141_70420\|\|full | 4005 | No | NA | 4 | 0 | 2 | Not-determined | Genome-fragment | NA | NA | 0 |
| S11-final-viral-combined_k141_8356\|\|full | 1946 | No | NA | 4 | 0 | 0 | Low-quality | Genome-fragment | 4.16 | AAI-based (high-confidence) | 0 |
| S7-final-viral-combined_k141_18594\|\|full | 2413 | No | NA | 3 | 0 | 2 | Not-determined | Genome-fragment | NA | NA | 0 |
| S7-final-viral-combined_k141_59934\|\|full | 5142 | No | NA | 5 | 0 | 5 | Not-determined | Genome-fragment | NA | NA | 0 |
| S7-final-viral-combined_k141_79416\|\|full | 7890 | No | NA | 8 | 0 | 4 | Not-determined | Genome-fragment | NA | NA | 0 |
| S7-final-viral-combined_k141_140767\|\|full | 1084 | No | NA | 2 | 0 | 0 | Not-determined | Genome-fragment | NA | NA | 0 |
| S11-final-viral-combined_k141_105378\|\|full | 1946 | No | NA | 4 | 1 | 0 | Low-quality | Genome-fragment | 5.09 | AAI-based (high-confidence) | 0 |
| S11-final-viral-combined_k141_52634\|\|full | 1946 | No | NA | 3 | 0 | 2 | Low-quality | Genome-fragment | 0.83 | AAI-based (medium-confidence) | 0 |
| S1-final-viral-combined_k141_331457\|\|full | 1946 | No | NA | 3 | 0 | 0 | Low-quality | Genome-fragment | 3.41 | AAI-based (medium-confidence) | 0 |
| S7-final-viral-combined_k141_221446\|\|full | 2140 | No | NA | 4 | 0 | 1 | Not-determined | Genome-fragment | NA | NA | 0 |
| S7-final-viral-combined_k141_151233\|\|full | 6937 | No | NA | 7 | 0 | 1 | Not-determined | Genome-fragment | NA | NA | 0 |
| S7-final-viral-combined_k141_27352\|\|full | 1520 | No | NA | 3 | 0 | 1 | Not-determined | Genome-fragment | NA | NA | 0 |
| S7-final-viral-combined_k141_59991\|\|full | 1416 | No | NA | 3 | 0 | 1 | Not-determined | Genome-fragment | NA | NA | 0 |
| S7-final-viral-combined_k141_100602\|\|full | 1014 | No | NA | 2 | 0 | 0 | Not-determined | Genome-fragment | NA | NA | 0 |
| S7-final-viral-combined_k141_110169\|\|full | 7583 | No | NA | 6 | 0 | 2 | Not-determined | Genome-fragment | NA | NA | 0 |
| S7-final-viral-combined_k141_131285\|\|full | 1705 | No | NA | 4 | 0 | 0 | Not-determined | Genome-fragment | NA | NA | 0 |
| S5-final-viral-combined_k141_180583\|\|full | 1946 | No | NA | 2 | 0 | 2 | Low-quality | Genome-fragment | 1.57 | AAI-based (high-confidence) | 0 |
| S5-final-viral-combined_k141_135942\|\|full | 1946 | No | NA | 3 | 1 | 0 | Low-quality | Genome-fragment | 1.73 | AAI-based (medium-confidence) | 0 |
| S7-final-viral-combined_k141_30757\|\|full | 1946 | No | NA | 4 | 1 | 0 | Low-quality | Genome-fragment | 3.12 | AAI-based (medium-confidence) | 0 |
| S7-final-viral-combined_k141_89867\|\|full | 9773 | No | NA | 11 | 0 | 8 | Not-determined | Genome-fragment | NA | NA | 0 |
| S7-final-viral-combined_k141_193340\|\|full | 1101 | No | NA | 3 | 0 | 1 | Not-determined | Genome-fragment | NA | NA | 0 |
| S7-final-viral-combined_k141_89878\|\|full | 5248 | No | NA | 4 | 0 | 3 | Not-determined | Genome-fragment | NA | NA | 0 |
| S7-final-viral-combined_k141_213315\|\|full | 1287 | No | NA | 2 | 0 | 1 | Not-determined | Genome-fragment | NA | NA | 0 |
| S7-final-viral-combined_k141_120303\|\|full | 1946 | No | NA | 2 | 1 | 0 | Low-quality | Genome-fragment | 2.18 | HMM-based (lower-bound) | 0 |
| S7-final-viral-combined_k141_233829\|\|full | 1221 | No | NA | 2 | 0 | 0 | Not-determined | Genome-fragment | NA | NA | 0 |
| S7-final-viral-combined_k141_140812\|\|full | 5417 | No | NA | 6 | 0 | 5 | Not-determined | Genome-fragment | NA | NA | 0 |
| S12-final-viral-combined_k141_149969\|\|full | 1946 | No | NA | 4 | 2 | 0 | Low-quality | Genome-fragment | 4.26 | AAI-based (high-confidence) | 0 |
| S7-final-viral-combined_k141_110254\|\|full | 3338 | No | NA | 4 | 0 | 1 | Not-determined | Genome-fragment | NA | NA | 0 |
| S8-final-viral-combined_k141_116135\|\|full | 1946 | No | NA | 4 | 0 | 0 | Low-quality | Genome-fragment | 3.99 | AAI-based (medium-confidence) | 0 |
| S8-final-viral-combined_k141_264692\|\|full | 1946 | No | NA | 4 | 0 | 0 | Low-quality | Genome-fragment | 1.27 | AAI-based (high-confidence) | 0 |
| S7-final-viral-combined_k141_50153\|\|full | 2172 | No | NA | 3 | 0 | 1 | Not-determined | Genome-fragment | NA | NA | 0 |
| S11-final-viral-combined_k141_185644\|\|full | 1947 | No | NA | 3 | 0 | 0 | Low-quality | Genome-fragment | 2.61 | AAI-based (medium-confidence) | 0 |
| S7-final-viral-combined_k141_60109\|\|full | 4549 | No | NA | 6 | 0 | 5 | Not-determined | Genome-fragment | NA | NA | 0 |
| S11_k141_294577 | 1947 | No | NA | 3 | 0 | 1 | Low-quality | Genome-fragment | 0.72 | AAI-based (medium-confidence) | 0 |
| S1-final-viral-combined_k141_243826\|\|full | 1947 | No | NA | 3 | 0 | 0 | Low-quality | Genome-fragment | 0.88 | AAI-based (medium-confidence) | 0 |
| S7-final-viral-combined_k141_121616\|\|full | 5733 | No | NA | 6 | 0 | 1 | Not-determined | Genome-fragment | NA | NA | 0 |
| S7-final-viral-combined_k141_103144\|\|full | 1947 | No | NA | 4 | 0 | 0 | Low-quality | Genome-fragment | 3.71 | AAI-based (high-confidence) | 0 |
| S7-final-viral-combined_k141_84305\|\|full | 1947 | No | NA | 2 | 0 | 1 | Low-quality | Genome-fragment | 1.42 | AAI-based (high-confidence) | 0 |
| S17-final-viral-combined_k141_69127\|\|full | 1947 | No | NA | 4 | 1 | 0 | Low-quality | Genome-fragment | 3.16 | AAI-based (high-confidence) | 0 |
| S17-final-viral-combined_k141_239624\|\|full | 1947 | No | NA | 3 | 0 | 2 | Low-quality | Genome-fragment | 2.65 | AAI-based (high-confidence) | 0 |
| S7-final-viral-combined_k141_8743\|\|full | 3804 | No | NA | 4 | 0 | 2 | Not-determined | Genome-fragment | NA | NA | 0 |
| S7-final-viral-combined_k141_172424\|\|full | 2874 | No | NA | 4 | 0 | 1 | Not-determined | Genome-fragment | NA | NA | 0 |
| S12-final-viral-combined_k141_340326\|\|full | 1947 | No | NA | 3 | 0 | 0 | Low-quality | Genome-fragment | 3.18 | AAI-based (medium-confidence) | 0 |
| S4-final-viral-combined_k141_1864\|\|full | 1947 | No | NA | 3 | 0 | 0 | Low-quality | Genome-fragment | 6.09 | AAI-based (high-confidence) | 0 |
| S7-final-viral-combined_k141_213393\|\|full | 5118 | No | NA | 5 | 0 | 1 | Not-determined | Genome-fragment | NA | NA | 0 |
| S7-final-viral-combined_k141_151383\|\|full | 1720 | No | NA | 3 | 0 | 2 | Not-determined | Genome-fragment | NA | NA | 0 |
| S4-final-viral-combined_k141_164972\|\|full | 1947 | No | NA | 2 | 0 | 2 | Low-quality | Genome-fragment | 0.52 | AAI-based (medium-confidence) | 0 |
| S7-final-viral-combined_k141_204204\|\|full | 2388 | No | NA | 3 | 0 | 1 | Not-determined | Genome-fragment | NA | NA | 0 |
| S8-final-viral-combined_k141_6389\|\|full | 1947 | No | NA | 4 | 0 | 2 | Low-quality | Genome-fragment | 1.89 | AAI-based (medium-confidence) | 0 |
| S8_k141_266602 | 1947 | No | NA | 2 | 0 | 2 | Low-quality | Genome-fragment | 1.2 | AAI-based (medium-confidence) | 0 |
| S1-final-viral-combined_k141_333984\|\|full | 1948 | Yes | 927 | 4 | 1 | 1 | Low-quality | Genome-fragment | 2.36 | AAI-based (high-confidence) | 52.41 |
| S1_k141_257450 | 1948 | No | NA | 1 | 0 | 1 | Low-quality | Genome-fragment | 0.17 | AAI-based (medium-confidence) | 0 |
| S7-final-viral-combined_k141_221648\|\|full | 3769 | No | NA | 7 | 0 | 3 | Not-determined | Genome-fragment | NA | NA | 0 |
| S7-final-viral-combined_k141_221651\|\|full | 2164 | No | NA | 4 | 0 | 0 | Not-determined | Genome-fragment | NA | NA | 0 |
| S7-final-viral-combined_k141_140867\|\|full | 1524 | No | NA | 2 | 0 | 0 | Not-determined | Genome-fragment | NA | NA | 0 |
| S7-final-viral-combined_k141_110327\|\|full | 2968 | No | NA | 4 | 0 | 4 | Not-determined | Genome-fragment | NA | NA | 0 |
| S7-final-viral-combined_k141_89967\|\|full | 2850 | No | NA | 5 | 0 | 1 | Not-determined | Genome-fragment | NA | NA | 0 |
| S3-final-viral-combined_k141_30643\|\|full | 1948 | No | NA | 2 | 0 | 2 | Low-quality | Genome-fragment | 1.56 | AAI-based (high-confidence) | 0 |
| S7-final-viral-combined_k141_131492\|\|full | 2833 | No | NA | 4 | 0 | 0 | Not-determined | Genome-fragment | NA | NA | 0 |
| S17-final-viral-combined_k141_57578\|\|full | 1948 | No | NA | 3 | 2 | 0 | Low-quality | Genome-fragment | 2.35 | HMM-based (lower-bound) | 0 |
| S7-final-viral-combined_k141_70618\|\|full | 2301 | No | NA | 3 | 0 | 1 | Not-determined | Genome-fragment | NA | NA | 0 |
| S17-final-viral-combined_k141_266257\|\|full | 1948 | No | NA | 2 | 1 | 0 | Low-quality | Genome-fragment | 4.42 | AAI-based (medium-confidence) | 0 |
| S7-final-viral-combined_k141_213432\|\|full | 2216 | No | NA | 3 | 0 | 1 | Not-determined | Genome-fragment | NA | NA | 0 |
| S7-final-viral-combined_k141_243290\|\|full | 2102 | No | NA | 2 | 0 | 2 | Not-determined | Genome-fragment | NA | NA | 0 |
| S7-final-viral-combined_k141_193486\|\|full | 3072 | No | NA | 4 | 0 | 2 | Not-determined | Genome-fragment | NA | NA | 0 |
| S7-final-viral-combined_k141_100812\|\|full | 2489 | No | NA | 3 | 0 | 3 | Not-determined | Genome-fragment | NA | NA | 0 |
| S7-final-viral-combined_k141_121704\|\|full | 2418 | No | NA | 4 | 0 | 1 | Not-determined | Genome-fragment | NA | NA | 0 |
| S7-final-viral-combined_k141_60043\|\|full | 3201 | No | NA | 3 | 0 | 3 | Not-determined | Genome-fragment | NA | NA | 0 |
| S7-final-viral-combined_k141_8771\|\|full | 6795 | No | NA | 8 | 0 | 0 | Not-determined | Genome-fragment | NA | NA | 0 |
| S7-final-viral-combined_k141_40054\|\|full | 1436 | No | NA | 3 | 0 | 0 | Not-determined | Genome-fragment | NA | NA | 0 |
| S7-final-viral-combined_k141_131548\|\|full | 2063 | No | NA | 4 | 0 | 1 | Not-determined | Genome-fragment | NA | NA | 0 |
| S7-final-viral-combined_k141_213451\|\|full | 1357 | No | NA | 3 | 0 | 1 | Not-determined | Genome-fragment | NA | NA | 0 |
| S7-final-viral-combined_k141_40069\|\|full | 1228 | No | NA | 3 | 0 | 2 | Not-determined | Genome-fragment | NA | NA | 0 |
| S7-final-viral-combined_k141_40077\|\|full | 1677 | No | NA | 3 | 0 | 0 | Not-determined | Genome-fragment | NA | NA | 0 |
| S7-final-viral-combined_k141_90035\|\|full | 2458 | No | NA | 3 | 0 | 2 | Not-determined | Genome-fragment | NA | NA | 0 |
| S12-final-viral-combined_k141_82104\|\|lt2gene | 1948 | No | NA | 1 | 1 | 0 | Low-quality | Genome-fragment | 4.16 | HMM-based (lower-bound) | 0 |
| S7-final-viral-combined_k141_213466\|\|full | 3075 | No | NA | 4 | 0 | 1 | Not-determined | Genome-fragment | NA | NA | 0 |
| S12_k141_384283 | 1948 | No | NA | 1 | 1 | 0 | Low-quality | Genome-fragment | 4.31 | AAI-based (high-confidence) | 0 |
| S4-final-viral-combined_k141_55404\|\|full | 1948 | No | NA | 3 | 0 | 3 | Low-quality | Genome-fragment | 3.38 | AAI-based (medium-confidence) | 0 |
| S4_k141_5131 | 1948 | No | NA | 2 | 0 | 1 | Low-quality | Genome-fragment | 0.67 | AAI-based (medium-confidence) | 0 |
| S7-final-viral-combined_k141_172484\|\|full | 3006 | No | NA | 4 | 0 | 3 | Not-determined | Genome-fragment | NA | NA | 0 |
| S11-final-viral-combined_k141_256928\|\|full | 1949 | Yes | 1475 | 3 | 2 | 1 | Low-quality | Genome-fragment | 1.81 | HMM-based (lower-bound) | 24.32 |
| S7-final-viral-combined_k141_172498\|\|full | 3263 | No | NA | 2 | 0 | 1 | Not-determined | Genome-fragment | NA | NA | 0 |
| S7-final-viral-combined_k141_131596\|\|full | 2644 | No | NA | 6 | 0 | 0 | Not-determined | Genome-fragment | NA | NA | 0 |
| S7-final-viral-combined_k141_243361\|\|full | 1059 | No | NA | 2 | 0 | 0 | Not-determined | Genome-fragment | NA | NA | 0 |
| S3-final-viral-combined_k141_9264\|\|full | 1949 | No | NA | 5 | 1 | 0 | Low-quality | Genome-fragment | 4.45 | AAI-based (high-confidence) | 0 |
| S7-final-viral-combined_k141_141006\|\|full | 1607 | No | NA | 2 | 0 | 0 | Not-determined | Genome-fragment | NA | NA | 0 |
| S7_k141_228058 | 1949 | No | NA | 2 | 0 | 1 | Low-quality | Genome-fragment | 4.79 | AAI-based (medium-confidence) | 0 |
| S7-final-viral-combined_k141_79694\|\|full | 1933 | No | NA | 3 | 0 | 2 | Not-determined | Genome-fragment | NA | NA | 0 |
| S7-final-viral-combined_k141_182976\|\|full | 2536 | No | NA | 3 | 0 | 1 | Not-determined | Genome-fragment | NA | NA | 0 |
| S7-final-viral-combined_k141_110434\|\|full | 2651 | No | NA | 4 | 0 | 3 | Not-determined | Genome-fragment | NA | NA | 0 |
| S7-final-viral-combined_k141_213526\|\|full | 5473 | No | NA | 4 | 0 | 2 | Not-determined | Genome-fragment | NA | NA | 0 |
| S7-final-viral-combined_k141_141012\|\|full | 1444 | No | NA | 2 | 0 | 2 | Not-determined | Genome-fragment | NA | NA | 0 |
| S7-final-viral-combined_k141_141024\|\|full | 1019 | No | NA | 2 | 0 | 1 | Not-determined | Genome-fragment | NA | NA | 0 |
| S2-final-viral-combined_k141_151032\|\|full | 1949 | No | NA | 5 | 0 | 0 | Low-quality | Genome-fragment | 1.2 | AAI-based (medium-confidence) | 0 |
| S7-final-viral-combined_k141_151504\|\|full | 1589 | No | NA | 4 | 0 | 1 | Not-determined | Genome-fragment | NA | NA | 0 |
| S7-final-viral-combined_k141_131652\|\|full | 2044 | No | NA | 3 | 0 | 0 | Not-determined | Genome-fragment | NA | NA | 0 |
| S7-final-viral-combined_k141_79714\|\|full | 2815 | No | NA | 3 | 0 | 2 | Not-determined | Genome-fragment | NA | NA | 0 |
| S4-final-viral-combined_k141_10304\|\|full | 1949 | No | NA | 3 | 0 | 0 | Low-quality | Genome-fragment | 3.75 | AAI-based (medium-confidence) | 0 |
| S7-final-viral-combined_k141_40169\|\|full | 10955 | No | NA | 9 | 0 | 6 | Not-determined | Genome-fragment | NA | NA | 0 |
| S1-final-viral-combined_k141_262114\|\|full | 1950 | No | NA | 5 | 2 | 0 | Low-quality | Genome-fragment | 3.34 | AAI-based (high-confidence) | 0 |
| S1_k141_204333 | 1950 | No | NA | 3 | 0 | 2 | Low-quality | Genome-fragment | 0.62 | AAI-based (medium-confidence) | 0 |
| S5-final-viral-combined_k141_198602\|\|full | 1950 | No | NA | 3 | 0 | 0 | Low-quality | Genome-fragment | 1.99 | AAI-based (high-confidence) | 0 |
| S7-final-viral-combined_k141_130965\|\|full | 1950 | No | NA | 4 | 1 | 0 | Low-quality | Genome-fragment | 1.12 | HMM-based (lower-bound) | 0 |
| S7-final-viral-combined_k141_100972\|\|full | 1012 | No | NA | 2 | 0 | 1 | Not-determined | Genome-fragment | NA | NA | 0 |
| S7-final-viral-combined_k141_131685\|\|full | 2917 | No | NA | 4 | 0 | 2 | Not-determined | Genome-fragment | NA | NA | 0 |
| S17-final-viral-combined_k141_56811\|\|full | 1950 | No | NA | 3 | 1 | 0 | Low-quality | Genome-fragment | 3.78 | AAI-based (high-confidence) | 0 |
| S2-final-viral-combined_k141_79224\|\|full | 1950 | No | NA | 4 | 0 | 2 | Low-quality | Genome-fragment | 2.22 | AAI-based (medium-confidence) | 0 |
| S11-final-viral-combined_k141_172081\|\|full | 1951 | No | NA | 3 | 1 | 0 | Low-quality | Genome-fragment | 3.27 | AAI-based (high-confidence) | 0 |
| S7-final-viral-combined_k141_50425\|\|full | 1606 | No | NA | 2 | 0 | 1 | Not-determined | Genome-fragment | NA | NA | 0 |
| S1_k141_192021 | 1951 | No | NA | 3 | 1 | 0 | Low-quality | Genome-fragment | 1.79 | AAI-based (high-confidence) | 0 |
| S7-final-viral-combined_k141_193693\|\|full | 1559 | No | NA | 4 | 0 | 3 | Not-determined | Genome-fragment | NA | NA | 0 |
| S2_k141_132905 | 1951 | No | NA | 1 | 0 | 1 | Low-quality | Genome-fragment | 0.72 | AAI-based (high-confidence) | 0 |
| S7-final-viral-combined_k141_162708\|\|full | 1211 | No | NA | 2 | 0 | 1 | Not-determined | Genome-fragment | NA | NA | 0 |
| S7-final-viral-combined_k141_151593\|\|full | 4386 | No | NA | 5 | 0 | 2 | Not-determined | Genome-fragment | NA | NA | 0 |
| S7-final-viral-combined_k141_221808\|\|full | 2168 | No | NA | 3 | 0 | 0 | Not-determined | Genome-fragment | NA | NA | 0 |
| S6-final-viral-combined_k141_38557\|\|full | 1951 | No | NA | 3 | 2 | 0 | Low-quality | Genome-fragment | 1.17 | HMM-based (lower-bound) | 0 |
| S6_k141_26831 | 1951 | No | NA | 2 | 2 | 0 | Low-quality | Genome-fragment | 2.32 | AAI-based (high-confidence) | 0 |
| S7-final-viral-combined_k141_243484\|\|full | 2699 | No | NA | 2 | 0 | 1 | Not-determined | Genome-fragment | NA | NA | 0 |
| S7-final-viral-combined_k141_221814\|\|full | 9349 | No | NA | 9 | 0 | 6 | Not-determined | Genome-fragment | NA | NA | 0 |
| S7-final-viral-combined_k141_50517\|\|full | 1717 | No | NA | 3 | 0 | 0 | Not-determined | Genome-fragment | NA | NA | 0 |
| S7-final-viral-combined_k141_60353\|\|full | 1397 | No | NA | 2 | 0 | 1 | Not-determined | Genome-fragment | NA | NA | 0 |
| S11-final-viral-combined_k141_185079\|\|full | 1952 | No | NA | 3 | 1 | 0 | Low-quality | Genome-fragment | 2.88 | AAI-based (high-confidence) | 0 |
| S7-final-viral-combined_k141_110634\|\|full | 1347 | No | NA | 3 | 0 | 1 | Not-determined | Genome-fragment | NA | NA | 0 |
| S7-final-viral-combined_k141_121957\|\|full | 1767 | No | NA | 2 | 0 | 1 | Not-determined | Genome-fragment | NA | NA | 0 |
| S11-final-viral-combined_k141_243232\|\|full | 1952 | No | NA | 5 | 1 | 2 | Low-quality | Genome-fragment | 3.07 | AAI-based (medium-confidence) | 0 |
| S11-final-viral-combined_k141_52717\|\|full | 1952 | No | NA | 4 | 0 | 0 | Low-quality | Genome-fragment | 1.27 | AAI-based (high-confidence) | 0 |
| S11-final-viral-combined_k141_214791\|\|lt2gene | 1952 | No | NA | 2 | 1 | 0 | Low-quality | Genome-fragment | 2.85 | HMM-based (lower-bound) | 0 |
| S1-final-viral-combined_k141_350428\|\|lt2gene | 1952 | No | NA | 1 | 1 | 0 | Low-quality | Genome-fragment | 3.47 | AAI-based (high-confidence) | 0 |
| S7-final-viral-combined_k141_60424\|\|full | 3193 | No | NA | 3 | 0 | 1 | Not-determined | Genome-fragment | NA | NA | 0 |
| S7-final-viral-combined_k141_50586\|\|full | 1482 | No | NA | 3 | 0 | 1 | Not-determined | Genome-fragment | NA | NA | 0 |
| S17-final-viral-combined_k141_21983\|\|full | 1952 | No | NA | 3 | 0 | 3 | Low-quality | Genome-fragment | 0.62 | AAI-based (medium-confidence) | 0 |
| S17-k141_150027 | 1952 | No | NA | 4 | 0 | 1 | Low-quality | Genome-fragment | 5.59 | AAI-based (medium-confidence) | 0 |
| S2-final-viral-combined_k141_127345\|\|full | 1952 | No | NA | 6 | 0 | 0 | Low-quality | Genome-fragment | 5.17 | AAI-based (medium-confidence) | 0 |
| S7-final-viral-combined_k141_101073\|\|full | 1948 | No | NA | 3 | 0 | 2 | Not-determined | Genome-fragment | NA | NA | 0 |
| S4_k141_109055 | 1952 | No | NA | 2 | 0 | 0 | Low-quality | Genome-fragment | 3.73 | AAI-based (medium-confidence) | 0 |
| S7-final-viral-combined_k141_18905\|\|full | 3287 | No | NA | 3 | 0 | 3 | Not-determined | Genome-fragment | NA | NA | 0 |
| S7-final-viral-combined_k141_90301\|\|full | 1070 | No | NA | 3 | 0 | 1 | Not-determined | Genome-fragment | NA | NA | 0 |
| S8-final-viral-combined_k141_228107\|\|full | 1952 | No | NA | 3 | 0 | 1 | Low-quality | Genome-fragment | 1.41 | AAI-based (medium-confidence) | 0 |
| S7-final-viral-combined_k141_122002\|\|full | 3895 | No | NA | 4 | 0 | 3 | Not-determined | Genome-fragment | NA | NA | 0 |
| S7-final-viral-combined_k141_234268\|\|full | 17854 | No | NA | 18 | 0 | 6 | Not-determined | Genome-fragment | NA | NA | 0 |
| S7-final-viral-combined_k141_162840\|\|full | 6284 | No | NA | 3 | 0 | 3 | Not-determined | Genome-fragment | NA | NA | 0 |
| S7-final-viral-combined_k141_60484\|\|full | 1519 | No | NA | 4 | 0 | 3 | Not-determined | Genome-fragment | NA | NA | 0 |
| S7-final-viral-combined_k141_8870\|\|full | 1880 | No | NA | 3 | 0 | 2 | Not-determined | Genome-fragment | NA | NA | 0 |
| S11-final-viral-combined_k141_199235\|\|full | 1953 | No | NA | 2 | 0 | 2 | Low-quality | Genome-fragment | 2.61 | AAI-based (medium-confidence) | 0 |
| S7-final-viral-combined_k141_18942\|\|full | 2450 | No | NA | 4 | 0 | 2 | Not-determined | Genome-fragment | NA | NA | 0 |
| S3-final-viral-combined_k141_65782\|\|full | 1953 | No | NA | 2 | 0 | 1 | Low-quality | Genome-fragment | 1.15 | AAI-based (medium-confidence) | 0 |
| S7-final-viral-combined_k141_193893\|\|full | 1515 | No | NA | 2 | 0 | 0 | Not-determined | Genome-fragment | NA | NA | 0 |
| S7-final-viral-combined_k141_183237\|\|full | 2677 | No | NA | 5 | 0 | 1 | Not-determined | Genome-fragment | NA | NA | 0 |
| S7-final-viral-combined_k141_50704\|\|full | 2885 | No | NA | 2 | 0 | 2 | Not-determined | Genome-fragment | NA | NA | 0 |
| S3-final-viral-combined_k141_84953\|\|full | 1953 | No | NA | 2 | 0 | 0 | Low-quality | Genome-fragment | 1.28 | AAI-based (high-confidence) | 0 |
| S7-final-viral-combined_k141_25474\|\|full | 1953 | No | NA | 2 | 2 | 0 | Low-quality | Genome-fragment | 3.19 | HMM-based (lower-bound) | 0 |
| S7-final-viral-combined_k141_213797\|\|full | 1090 | No | NA | 2 | 0 | 2 | Not-determined | Genome-fragment | NA | NA | 0 |
| S17-final-viral-combined_k141_223226\|\|full | 1953 | No | NA | 4 | 1 | 0 | Low-quality | Genome-fragment | 0.79 | HMM-based (lower-bound) | 0 |
| S7-final-viral-combined_k141_18983\|\|full | 3160 | No | NA | 4 | 0 | 2 | Not-determined | Genome-fragment | NA | NA | 0 |
| S7-final-viral-combined_k141_193915\|\|full | 2644 | No | NA | 3 | 0 | 0 | Not-determined | Genome-fragment | NA | NA | 0 |
| S6-final-viral-combined_k141_16968\|\|full | 1953 | No | NA | 4 | 0 | 1 | Low-quality | Genome-fragment | 3.83 | AAI-based (high-confidence) | 0 |
| S8_k141_20882 | 1953 | No | NA | 4 | 0 | 1 | Low-quality | Genome-fragment | 2.51 | AAI-based (medium-confidence) | 0 |
| S7-final-viral-combined_k141_79772\|\|full | 1556 | No | NA | 4 | 0 | 1 | Not-determined | Genome-fragment | NA | NA | 0 |
| S7-final-viral-combined_k141_60527\|\|full | 1924 | No | NA | 2 | 0 | 2 | Not-determined | Genome-fragment | NA | NA | 0 |
| S7-final-viral-combined_k141_193945\|\|full | 5233 | No | NA | 6 | 0 | 2 | Not-determined | Genome-fragment | NA | NA | 0 |
| S11_k141_239559 | 1954 | No | NA | 1 | 0 | 1 | Low-quality | Genome-fragment | 0.72 | AAI-based (high-confidence) | 0 |
| S7-final-viral-combined_k141_183362\|\|full | 1954 | No | NA | 3 | 0 | 1 | Low-quality | Genome-fragment | 1.21 | AAI-based (medium-confidence) | 0 |
| S7-final-viral-combined_k141_8954\|\|full | 4954 | No | NA | 5 | 0 | 3 | Not-determined | Genome-fragment | NA | NA | 0 |
| S4-final-viral-combined_k141_57044\|\|full | 1954 | No | NA | 2 | 0 | 1 | Low-quality | Genome-fragment | 0.22 | AAI-based (medium-confidence) | 0 |
| S7-final-viral-combined_k141_122120\|\|full | 2398 | No | NA | 4 | 0 | 0 | Not-determined | Genome-fragment | NA | NA | 0 |
| S7-final-viral-combined_k141_8959\|\|full | 1260 | No | NA | 3 | 0 | 0 | Not-determined | Genome-fragment | NA | NA | 0 |
| S7-final-viral-combined_k141_19058\|\|full | 5604 | No | NA | 8 | 0 | 0 | Not-determined | Genome-fragment | NA | NA | 0 |
| S1-final-viral-combined_k141_390005\|\|full | 1955 | No | NA | 2 | 0 | 0 | Low-quality | Genome-fragment | 2.9 | AAI-based (high-confidence) | 0 |
| S5_k141_47449 | 1955 | No | NA | 2 | 1 | 0 | Low-quality | Genome-fragment | 1.09 | HMM-based (lower-bound) | 0 |
| S7-final-viral-combined_k141_8982\|\|full | 2873 | No | NA | 4 | 0 | 1 | Not-determined | Genome-fragment | NA | NA | 0 |
| S7-final-viral-combined_k141_50770\|\|full | 2314 | No | NA | 4 | 0 | 0 | Not-determined | Genome-fragment | NA | NA | 0 |
| S7-final-viral-combined_k141_90393\|\|full | 19692 | No | NA | 27 | 9 | 1 | Medium-quality | Genome-fragment | 53.3 | AAI-based (medium-confidence) | 0 |
| S12_k141_34279 | 1955 | No | NA | 3 | 2 | 0 | Low-quality | Genome-fragment | 6.02 | AAI-based (high-confidence) | 0 |
| S2-final-viral-combined_k141_81973\|\|full | 1955 | No | NA | 4 | 0 | 1 | Low-quality | Genome-fragment | 3.45 | AAI-based (high-confidence) | 0 |
| S7-final-viral-combined_k141_183408\|\|full | 3412 | No | NA | 6 | 0 | 4 | Not-determined | Genome-fragment | NA | NA | 0 |
| S7-final-viral-combined_k141_8998\|\|full | 3902 | No | NA | 5 | 0 | 3 | Not-determined | Genome-fragment | NA | NA | 0 |
| S9-final-viral-combined_k141_124633\|\|full | 1956 | No | NA | 6 | 0 | 0 | Low-quality | Genome-fragment | 5.1 | AAI-based (medium-confidence) | 0 |
| S5_k141_156360 | 1956 | No | NA | 4 | 2 | 0 | Low-quality | Genome-fragment | 4.83 | AAI-based (high-confidence) | 0 |
| S7-final-viral-combined_k141_90435\|\|full | 1320 | No | NA | 3 | 0 | 0 | Not-determined | Genome-fragment | NA | NA | 0 |
| S7-final-viral-combined_k141_9009\|\|full | 1680 | No | NA | 3 | 0 | 1 | Not-determined | Genome-fragment | NA | NA | 0 |
| S7-final-viral-combined_k141_183425\|\|full | 1807 | No | NA | 2 | 0 | 1 | Not-determined | Genome-fragment | NA | NA | 0 |
| S17-final-viral-combined_k141_30569\|\|full | 1956 | No | NA | 2 | 0 | 0 | Low-quality | Genome-fragment | 1.23 | AAI-based (medium-confidence) | 0 |
| S8-final-viral-combined_k141_6351\|\|full | 1956 | No | NA | 3 | 0 | 1 | Low-quality | Genome-fragment | 3.93 | AAI-based (medium-confidence) | 0 |
| S7-final-viral-combined_k141_9014\|\|full | 2926 | No | NA | 4 | 0 | 1 | Not-determined | Genome-fragment | NA | NA | 0 |
| S11-final-viral-combined_k141_251695\|\|full | 1957 | No | NA | 3 | 1 | 0 | Low-quality | Genome-fragment | 4.25 | AAI-based (medium-confidence) | 0 |
| S1_k141_185992 | 1957 | No | NA | 1 | 1 | 0 | Low-quality | Genome-fragment | 3.39 | AAI-based (high-confidence) | 0 |
| S7-final-viral-combined_k141_101174\|\|full | 2254 | No | NA | 6 | 0 | 0 | Not-determined | Genome-fragment | NA | NA | 0 |
| S7-final-viral-combined_k141_101186\|\|full | 1804 | No | NA | 3 | 0 | 0 | Not-determined | Genome-fragment | NA | NA | 0 |
| S7-final-viral-combined_k141_183470\|\|full | 3317 | No | NA | 4 | 0 | 3 | Not-determined | Genome-fragment | NA | NA | 0 |
| S7-final-viral-combined_k141_213969\|\|full | 1034 | No | NA | 3 | 0 | 1 | Not-determined | Genome-fragment | NA | NA | 0 |
| S7-final-viral-combined_k141_110974\|\|full | 2216 | No | NA | 3 | 0 | 3 | Not-determined | Genome-fragment | NA | NA | 0 |
| S7-final-viral-combined_k141_155112\|\|full | 1957 | No | NA | 2 | 0 | 0 | Low-quality | Genome-fragment | 1.33 | AAI-based (medium-confidence) | 0 |
| S7-final-viral-combined_k141_213982\|\|full | 4416 | No | NA | 3 | 0 | 3 | Not-determined | Genome-fragment | NA | NA | 0 |
| S7-final-viral-combined_k141_172967\|\|full | 1041 | No | NA | 2 | 0 | 0 | Not-determined | Genome-fragment | NA | NA | 0 |
| S7-final-viral-combined_k141_183501\|\|full | 5580 | No | NA | 6 | 0 | 4 | Not-determined | Genome-fragment | NA | NA | 0 |
| S7_k141_99419 | 1957 | No | NA | 3 | 0 | 0 | Low-quality | Genome-fragment | 2.8 | AAI-based (medium-confidence) | 0 |
| S12-final-viral-combined_k141_393018\|\|full | 1957 | No | NA | 3 | 2 | 0 | Low-quality | Genome-fragment | 4.25 | AAI-based (high-confidence) | 0 |
| S4_k141_145555 | 1957 | No | NA | 3 | 0 | 2 | Low-quality | Genome-fragment | 5 | AAI-based (medium-confidence) | 0 |
| S7-final-viral-combined_k141_71196\|\|full | 2649 | No | NA | 3 | 0 | 0 | Not-determined | Genome-fragment | NA | NA | 0 |
| S7-final-viral-combined_k141_131788\|\|full | 5347 | No | NA | 8 | 0 | 3 | Not-determined | Genome-fragment | NA | NA | 0 |
| S7-final-viral-combined_k141_101246\|\|full | 2181 | No | NA | 3 | 0 | 1 | Not-determined | Genome-fragment | NA | NA | 0 |
| S7-final-viral-combined_k141_151865\|\|full | 2974 | No | NA | 4 | 0 | 0 | Not-determined | Genome-fragment | NA | NA | 0 |
| S7-final-viral-combined_k141_101256\|\|full | 2657 | No | NA | 3 | 0 | 0 | Not-determined | Genome-fragment | NA | NA | 0 |
| S6_k141_54858 | 1957 | No | NA | 4 | 1 | 0 | Low-quality | Genome-fragment | 4.74 | AAI-based (medium-confidence) | 0 |
| S1-final-viral-combined_k141_251926\|\|full | 1958 | Yes | 828 | 3 | 1 | 1 | Low-quality | Genome-fragment | 0.23 | HMM-based (lower-bound) | 57.71 |
| S7-final-viral-combined_k141_163154\|\|full | 1793 | No | NA | 3 | 0 | 1 | Not-determined | Genome-fragment | NA | NA | 0 |
| S7-final-viral-combined_k141_111004\|\|full | 68619 | Yes | 64243 | 101 | 23 | 6 | High-quality | High-quality | 100 | AAI-based (high-confidence) | 6.38 |
| S7-final-viral-combined_k141_111007\|\|full | 5676 | No | NA | 6 | 0 | 1 | Not-determined | Genome-fragment | NA | NA | 0 |
| S7-final-viral-combined_k141_40545\|\|full | 1637 | No | NA | 3 | 0 | 0 | Not-determined | Genome-fragment | NA | NA | 0 |
| S7-final-viral-combined_k141_216542\|\|full | 1958 | No | NA | 3 | 0 | 0 | Low-quality | Genome-fragment | 5.58 | AAI-based (high-confidence) | 0 |
| S7-final-viral-combined_k141_151939\|\|full | 1921 | No | NA | 4 | 0 | 0 | Not-determined | Genome-fragment | NA | NA | 0 |
| S7-final-viral-combined_k141_71300\|\|full | 3236 | No | NA | 4 | 0 | 1 | Not-determined | Genome-fragment | NA | NA | 0 |
| S7-final-viral-combined_k141_80049\|\|full | 3212 | No | NA | 3 | 0 | 1 | Not-determined | Genome-fragment | NA | NA | 0 |
| S7_k141_83265 | 1958 | No | NA | 2 | 0 | 0 | Low-quality | Genome-fragment | 2.94 | AAI-based (medium-confidence) | 0 |
| S7-final-viral-combined_k141_90684\|\|full | 1462 | No | NA | 6 | 0 | 0 | Not-determined | Genome-fragment | NA | NA | 0 |
| S17-final-viral-combined_k141_124411\|\|full | 1958 | No | NA | 4 | 1 | 0 | Low-quality | Genome-fragment | 4.44 | AAI-based (medium-confidence) | 0 |
| S12-final-viral-combined_k141_244480\|\|full | 1958 | No | NA | 2 | 0 | 2 | Low-quality | Genome-fragment | 2.26 | AAI-based (medium-confidence) | 0 |
| S7-final-viral-combined_k141_141384\|\|full | 2462 | No | NA | 3 | 0 | 2 | Not-determined | Genome-fragment | NA | NA | 0 |
| S4-final-viral-combined_k141_136134\|\|full | 1958 | Yes | 666 | 4 | 1 | 1 | Low-quality | Genome-fragment | 1.55 | AAI-based (high-confidence) | 65.99 |
| S7-final-viral-combined_k141_60793\|\|full | 2410 | No | NA | 3 | 0 | 0 | Not-determined | Genome-fragment | NA | NA | 0 |
| S7-final-viral-combined_k141_141398\|\|full | 1817 | No | NA | 2 | 0 | 1 | Not-determined | Genome-fragment | NA | NA | 0 |
| S7-final-viral-combined_k141_183638\|\|full | 1363 | No | NA | 3 | 0 | 0 | Not-determined | Genome-fragment | NA | NA | 0 |
| S7-final-viral-combined_k141_141399\|\|full | 1008 | No | NA | 3 | 0 | 0 | Not-determined | Genome-fragment | NA | NA | 0 |
| S7-final-viral-combined_k141_141409\|\|full | 1117 | No | NA | 3 | 0 | 1 | Not-determined | Genome-fragment | NA | NA | 0 |
| S7-final-viral-combined_k141_152049\|\|full | 6607 | No | NA | 5 | 0 | 4 | Not-determined | Genome-fragment | NA | NA | 0 |
| S9-final-viral-combined_k141_93363\|\|full | 1959 | No | NA | 2 | 0 | 1 | Low-quality | Genome-fragment | 1.35 | AAI-based (high-confidence) | 0 |
| S7-final-viral-combined_k141_19264\|\|full | 1704 | No | NA | 2 | 0 | 0 | Not-determined | Genome-fragment | NA | NA | 0 |
| S11-final-viral-combined_k141_96752\|\|full | 1959 | No | NA | 5 | 1 | 0 | Low-quality | Genome-fragment | 3.3 | AAI-based (high-confidence) | 0 |
| S7-final-viral-combined_k141_90717\|\|full | 7805 | No | NA | 14 | 0 | 2 | Not-determined | Genome-fragment | NA | NA | 0 |
| S11-final-viral-combined_k141_238438\|\|full | 1959 | No | NA | 2 | 0 | 1 | Low-quality | Genome-fragment | 4.19 | AAI-based (medium-confidence) | 0 |
| S1-final-viral-combined_k141_72333\|\|full | 1959 | No | NA | 2 | 0 | 0 | Low-quality | Genome-fragment | 4.48 | AAI-based (high-confidence) | 0 |
| S1-final-viral-combined_k141_10641\|\|full | 1959 | No | NA | 4 | 0 | 3 | Low-quality | Genome-fragment | 0.89 | AAI-based (medium-confidence) | 0 |
| S7-final-viral-combined_k141_111106\|\|full | 6228 | No | NA | 7 | 0 | 6 | Not-determined | Genome-fragment | NA | NA | 0 |
| S7-final-viral-combined_k141_152070\|\|full | 4121 | No | NA | 7 | 0 | 2 | Not-determined | Genome-fragment | NA | NA | 0 |
| S3-final-viral-combined_k141_135450\|\|lt2gene | 1959 | No | NA | 2 | 2 | 0 | Low-quality | Genome-fragment | 5.17 | AAI-based (high-confidence) | 0 |
| S12-final-viral-combined_k141_116649\|\|full | 1959 | No | NA | 3 | 0 | 0 | Low-quality | Genome-fragment | 2.76 | AAI-based (high-confidence) | 0 |
| S7-final-viral-combined_k141_40654\|\|full | 2706 | No | NA | 3 | 0 | 2 | Not-determined | Genome-fragment | NA | NA | 0 |
| S12_k141_193143 | 1959 | No | NA | 1 | 0 | 0 | Low-quality | Genome-fragment | 1.22 | AAI-based (high-confidence) | 0 |
| S9-final-viral-combined_k141_85697\|\|full | 1960 | No | NA | 5 | 4 | 0 | Low-quality | Genome-fragment | 4.86 | AAI-based (high-confidence) | 0 |
| S11-final-viral-combined_k141_95253\|\|full | 1960 | No | NA | 3 | 0 | 1 | Low-quality | Genome-fragment | 2.47 | AAI-based (medium-confidence) | 0 |
| S1-final-viral-combined_k141_218346\|\|full | 1960 | No | NA | 3 | 2 | 0 | Low-quality | Genome-fragment | 4.52 | AAI-based (high-confidence) | 0 |
| S1-final-viral-combined_k141_211068\|\|full | 1960 | No | NA | 6 | 1 | 0 | Low-quality | Genome-fragment | 5.05 | AAI-based (high-confidence) | 0 |
| S7-final-viral-combined_k141_183715\|\|full | 5268 | No | NA | 7 | 0 | 3 | Not-determined | Genome-fragment | NA | NA | 0 |
| S7-final-viral-combined_k141_141473\|\|full | 5288 | No | NA | 6 | 0 | 5 | Not-determined | Genome-fragment | NA | NA | 0 |
| S17-final-viral-combined_k141_129345\|\|full | 1960 | No | NA | 4 | 1 | 0 | Low-quality | Genome-fragment | 4.94 | AAI-based (high-confidence) | 0 |
| S7-final-viral-combined_k141_111152\|\|full | 4592 | No | NA | 3 | 0 | 2 | Not-determined | Genome-fragment | NA | NA | 0 |
| S12-final-viral-combined_k141_136118\|\|full | 1960 | No | NA | 2 | 0 | 0 | Low-quality | Genome-fragment | 0.59 | AAI-based (high-confidence) | 0 |
| S7-final-viral-combined_k141_90728\|\|full | 23541 | No | NA | 28 | 13 | 0 | Medium-quality | Genome-fragment | 58.6 | AAI-based (high-confidence) | 0 |
| S12-final-viral-combined_k141_222231\|\|full | 1960 | No | NA | 6 | 3 | 0 | Low-quality | Genome-fragment | 3.37 | AAI-based (medium-confidence) | 0 |
| S7-final-viral-combined_k141_122336\|\|full | 3266 | No | NA | 3 | 0 | 3 | Not-determined | Genome-fragment | NA | NA | 0 |
| S12-final-viral-combined_k141_96854\|\|full | 1960 | No | NA | 2 | 0 | 2 | Low-quality | Genome-fragment | 0.64 | AAI-based (medium-confidence) | 0 |
| S7-final-viral-combined_k141_183771\|\|full | 2327 | No | NA | 5 | 0 | 0 | Not-determined | Genome-fragment | NA | NA | 0 |
| S7-final-viral-combined_k141_183773\|\|full | 2269 | No | NA | 4 | 0 | 1 | Not-determined | Genome-fragment | NA | NA | 0 |
| S2-final-viral-combined_k141_49731\|\|full | 1960 | Yes | 1436 | 4 | 1 | 1 | Low-quality | Genome-fragment | 2.91 | AAI-based (high-confidence) | 26.73 |
| S7-final-viral-combined_k141_40742\|\|full | 1874 | No | NA | 3 | 0 | 2 | Not-determined | Genome-fragment | NA | NA | 0 |
| S8-final-viral-combined_k141_220744\|\|full | 1960 | No | NA | 6 | 0 | 0 | Low-quality | Genome-fragment | 4.27 | AAI-based (high-confidence) | 0 |
| S7-final-viral-combined_k141_80251\|\|full | 1049 | No | NA | 2 | 0 | 0 | Not-determined | Genome-fragment | NA | NA | 0 |
| S9-final-viral-combined_k141_86762\|\|full | 1961 | No | NA | 3 | 0 | 0 | Low-quality | Genome-fragment | 1.32 | AAI-based (medium-confidence) | 0 |
| S1-final-viral-combined_k141_91843\|\|full | 1961 | No | NA | 3 | 0 | 2 | Low-quality | Genome-fragment | 2.66 | AAI-based (medium-confidence) | 0 |
| S7-final-viral-combined_k141_9489\|\|full | 6497 | No | NA | 6 | 0 | 5 | Not-determined | Genome-fragment | NA | NA | 0 |
| S7-final-viral-combined_k141_222379\|\|full | 2671 | No | NA | 2 | 0 | 1 | Not-determined | Genome-fragment | NA | NA | 0 |
| S7-final-viral-combined_k141_101451\|\|full | 1332 | No | NA | 3 | 0 | 1 | Not-determined | Genome-fragment | NA | NA | 0 |
| S7-final-viral-combined_k141_19521\|\|full | 4029 | No | NA | 8 | 0 | 3 | Not-determined | Genome-fragment | NA | NA | 0 |
| S7-final-viral-combined_k141_141593\|\|full | 3086 | No | NA | 4 | 0 | 1 | Not-determined | Genome-fragment | NA | NA | 0 |
| S7-final-viral-combined_k141_61029\|\|full | 12153 | No | NA | 7 | 0 | 2 | Not-determined | Genome-fragment | NA | NA | 0 |

| | | | | | | | | | | | |
|---|---|---|---|---|---|---|---|---|---|---|---|
| S1-final-viral-combined_k141_289873\|\|full | 1961 | No | NA | 3 | 0 | 2 | Low-quality | Genome-fragment | 1.43 | AAI-based (high-confidence) | 0 |
| S5-final-viral-combined_k141_22188\|\|full | 1961 | No | NA | 4 | 1 | 0 | Low-quality | Genome-fragment | 4.18 | AAI-based (high-confidence) | 0 |
| S7-final-viral-combined_k141_101480\|\|full | 2343 | No | NA | 3 | 0 | 1 | Not-determined | Genome-fragment | NA | NA | 0 |
| S12-final-viral-combined_k141_309582\|\|full | 1961 | No | NA | 2 | 0 | 1 | Low-quality | Genome-fragment | 1.18 | AAI-based (medium-confidence) | 0 |
| S12-final-viral-combined_k141_43241\|\|full | 1961 | No | NA | 4 | 1 | 0 | Low-quality | Genome-fragment | 2.34 | AAI-based (medium-confidence) | 0 |
| S7-final-viral-combined_k141_111235\|\|full | 1285 | No | NA | 2 | 0 | 1 | Not-determined | Genome-fragment | NA | NA | 0 |
| S7-final-viral-combined_k141_50960\|\|full | 1465 | No | NA | 2 | 0 | 1 | Not-determined | Genome-fragment | NA | NA | 0 |
| S7-final-viral-combined_k141_40779\|\|full | 26775 | No | NA | 39 | 8 | 1 | Medium-quality | Genome-fragment | 60.92 | AAI-based (high-confidence) | 0 |
| S7-final-viral-combined_k141_222433\|\|full | 2630 | No | NA | 6 | 0 | 0 | Not-determined | Genome-fragment | NA | NA | 0 |
| S4-final-viral-combined_k141_141423\|\|full | 1961 | No | NA | 4 | 1 | 0 | Low-quality | Genome-fragment | 3.93 | AAI-based (high-confidence) | 0 |
| S7-final-viral-combined_k141_61116\|\|full | 1900 | No | NA | 3 | 0 | 0 | Not-determined | Genome-fragment | NA | NA | 0 |
| S7-final-viral-combined_k141_111264\|\|full | 7452 | No | NA | 9 | 0 | 5 | Not-determined | Genome-fragment | NA | NA | 0 |
| S6_k141_128656 | 1961 | No | NA | 7 | 0 | 0 | Low-quality | Genome-fragment | 2.1 | AAI-based (medium-confidence) | 0 |
| S7-final-viral-combined_k141_141626\|\|full | 1179 | No | NA | 3 | 0 | 1 | Not-determined | Genome-fragment | NA | NA | 0 |
| S7-final-viral-combined_k141_111303\|\|full | 1809 | No | NA | 4 | 0 | 2 | Not-determined | Genome-fragment | NA | NA | 0 |
| S8-final-viral-combined_k141_200194\|\|full | 1961 | No | NA | 7 | 0 | 0 | Low-quality | Genome-fragment | 4.27 | AAI-based (high-confidence) | 0 |
| S7-final-viral-combined_k141_131940\|\|full | 2901 | No | NA | 2 | 0 | 2 | Not-determined | Genome-fragment | NA | NA | 0 |
| S11-final-viral-combined_k141_28625\|\|full | 1962 | No | NA | 3 | 2 | 0 | Low-quality | Genome-fragment | 4.79 | AAI-based (high-confidence) | 0 |
| S7-final-viral-combined_k141_141704\|\|full | 1931 | No | NA | 2 | 0 | 0 | Not-determined | Genome-fragment | NA | NA | 0 |
| S7-final-viral-combined_k141_101581\|\|full | 5434 | No | NA | 5 | 0 | 0 | Not-determined | Genome-fragment | NA | NA | 0 |
| S11-final-viral-combined_k141_294617\|\|full | 1962 | No | NA | 3 | 1 | 0 | Low-quality | Genome-fragment | 2.94 | HMM-based (lower-bound) | 0 |
| S1-final-viral-combined_k141_333769\|\|full | 1962 | No | NA | 3 | 2 | 0 | Low-quality | Genome-fragment | 5.03 | AAI-based (high-confidence) | 0 |
| S7-final-viral-combined_k141_51000\|\|full | 1544 | No | NA | 2 | 0 | 0 | Not-determined | Genome-fragment | NA | NA | 0 |
| S7-final-viral-combined_k141_101595\|\|full | 4770 | No | NA | 3 | 0 | 2 | Not-determined | Genome-fragment | NA | NA | 0 |
| S1-final-viral-combined_k141_81891\|\|lt2gene | 1962 | No | NA | 2 | 1 | 0 | Low-quality | Genome-fragment | 4.69 | AAI-based (high-confidence) | 0 |
| S7-final-viral-combined_k141_222506\|\|full | 1721 | No | NA | 4 | 0 | 2 | Not-determined | Genome-fragment | NA | NA | 0 |
| S17-k141_127870 | 1962 | No | NA | 2 | 0 | 0 | Low-quality | Genome-fragment | 0.9 | AAI-based (medium-confidence) | 0 |
| S6-final-viral-combined_k141_26204\|\|full | 1962 | No | NA | 3 | 2 | 0 | Low-quality | Genome-fragment | 2.33 | AAI-based (high-confidence) | 0 |
| S7-final-viral-combined_k141_131995\|\|full | 1376 | No | NA | 2 | 0 | 2 | Not-determined | Genome-fragment | NA | NA | 0 |
| S8-final-viral-combined_k141_91279\|\|full | 1962 | No | NA | 3 | 1 | 0 | Low-quality | Genome-fragment | 2.65 | AAI-based (high-confidence) | 0 |
| S1-final-viral-combined_k141_331200\|\|full | 1963 | No | NA | 3 | 1 | 0 | Low-quality | Genome-fragment | 2.64 | HMM-based (lower-bound) | 0 |
| S7-final-viral-combined_k141_222573\|\|full | 2223 | No | NA | 4 | 0 | 0 | Not-determined | Genome-fragment | NA | NA | 0 |
| S1-final-viral-combined_k141_381418\|\|full | 1963 | No | NA | 2 | 1 | 0 | Low-quality | Genome-fragment | 4.07 | AAI-based (high-confidence) | 0 |
| S7-final-viral-combined_k141_173503\|\|full | 5852 | No | NA | 9 | 0 | 1 | Not-determined | Genome-fragment | NA | NA | 0 |
| S1-final-viral-combined_k141_337256\|\|full | 1963 | No | NA | 3 | 1 | 0 | Low-quality | Genome-fragment | 4.85 | AAI-based (high-confidence) | 0 |
| S1-final-viral-combined_k141_273006\|\|full | 1963 | No | NA | 3 | 0 | 1 | Low-quality | Genome-fragment | 0.89 | AAI-based (high-confidence) | 0 |
| S1-final-viral-combined_k141_213493\|\|full | 1963 | No | NA | 4 | 0 | 0 | Low-quality | Genome-fragment | 2.5 | AAI-based (medium-confidence) | 0 |
| S5_k141_1414 | 1963 | No | NA | 2 | 0 | 1 | Low-quality | Genome-fragment | 2.09 | AAI-based (medium-confidence) | 0 |
| S17-final-viral-combined_k141_185144\|\|full | 1963 | No | NA | 4 | 2 | 0 | Low-quality | Genome-fragment | 2.95 | AAI-based (medium-confidence) | 0 |
| S7-final-viral-combined_k141_19695\|\|full | 4980 | No | NA | 8 | 0 | 1 | Not-determined | Genome-fragment | NA | NA | 0 |
| S7-final-viral-combined_k141_141763\|\|full | 1194 | No | NA | 3 | 0 | 2 | Not-determined | Genome-fragment | NA | NA | 0 |
| S7-final-viral-combined_k141_173544\|\|full | 2567 | No | NA | 3 | 0 | 1 | Not-determined | Genome-fragment | NA | NA | 0 |
| S7-final-viral-combined_k141_111426\|\|full | 6327 | No | NA | 5 | 0 | 3 | Not-determined | Genome-fragment | NA | NA | 0 |
| S7-final-viral-combined_k141_19707\|\|full | 1040 | No | NA | 2 | 0 | 1 | Not-determined | Genome-fragment | NA | NA | 0 |
| S12-final-viral-combined_k141_118546\|\|full | 1963 | No | NA | 3 | 1 | 0 | Low-quality | Genome-fragment | 4.61 | AAI-based (medium-confidence) | 0 |
| S7-final-viral-combined_k141_111435\|\|full | 1134 | No | NA | 3 | 0 | 3 | Not-determined | Genome-fragment | NA | NA | 0 |
| S7-final-viral-combined_k141_243774\|\|full | 5758 | No | NA | 4 | 0 | 1 | Not-determined | Genome-fragment | NA | NA | 0 |
| S12-final-viral-combined_k141_39766\|\|full | 1963 | No | NA | 2 | 0 | 0 | Low-quality | Genome-fragment | 2.35 | AAI-based (medium-confidence) | 0 |
| S8_k141_16756 | 1963 | No | NA | 5 | 4 | 0 | Low-quality | Genome-fragment | 6.09 | AAI-based (high-confidence) | 0 |
| S7-final-viral-combined_k141_101758\|\|full | 2559 | No | NA | 3 | 0 | 2 | Not-determined | Genome-fragment | NA | NA | 0 |
| S9-final-viral-combined_k141_206\|\|full | 1964 | No | NA | 3 | 0 | 0 | Low-quality | Genome-fragment | 2.12 | AAI-based (high-confidence) | 0 |
| S11_k141_200176 | 1964 | No | NA | 1 | 0 | 1 | Low-quality | Genome-fragment | 5.54 | AAI-based (medium-confidence) | 0 |
| S1-final-viral-combined_k141_238933\|\|full | 1964 | No | NA | 6 | 0 | 0 | Low-quality | Genome-fragment | 5.35 | AAI-based (high-confidence) | 0 |
| S3-final-viral-combined_k141_54522\|\|full | 1964 | No | NA | 3 | 1 | 0 | Low-quality | Genome-fragment | 3.62 | AAI-based (high-confidence) | 0 |
| S7-final-viral-combined_k141_222710\|\|full | 3596 | No | NA | 8 | 0 | 0 | Not-determined | Genome-fragment | NA | NA | 0 |
| S7-final-viral-combined_k141_9849\|\|full | 1103 | No | NA | 3 | 0 | 0 | Not-determined | Genome-fragment | NA | NA | 0 |
| S7-final-viral-combined_k141_238382\|\|full | 1964 | No | NA | 2 | 0 | 1 | Low-quality | Genome-fragment | 1.38 | AAI-based (high-confidence) | 0 |
| S7-final-viral-combined_k141_132200\|\|full | 1205 | No | NA | 3 | 0 | 0 | Not-determined | Genome-fragment | NA | NA | 0 |
| S7-final-viral-combined_k141_132205\|\|full | 3566 | No | NA | 6 | 0 | 4 | Not-determined | Genome-fragment | NA | NA | 0 |
| S12-final-viral-combined_k141_200118\|\|full | 1964 | No | NA | 4 | 0 | 2 | Low-quality | Genome-fragment | 2.26 | AAI-based (medium-confidence) | 0 |
| S11-final-viral-combined_k141_262673\|\|lt2gene | 1965 | No | NA | 1 | 1 | 0 | Low-quality | Genome-fragment | 4.52 | AAI-based (high-confidence) | 0 |
| S1-final-viral-combined_k141_118161\|\|full | 1965 | No | NA | 4 | 0 | 0 | Low-quality | Genome-fragment | 4.21 | AAI-based (medium-confidence) | 0 |
| S7-final-viral-combined_k141_141991\|\|full | 4713 | No | NA | 9 | 0 | 5 | Not-determined | Genome-fragment | NA | NA | 0 |
| S7-final-viral-combined_k141_132255\|\|full | 2761 | No | NA | 4 | 0 | 1 | Not-determined | Genome-fragment | NA | NA | 0 |
| S7-final-viral-combined_k141_28138\|\|full | 30929 | No | NA | 43 | 16 | 1 | Medium-quality | Genome-fragment | 87.91 | AAI-based (high-confidence) | 0 |
| S1-final-viral-combined_k141_61767\|\|full | 1965 | No | NA | 4 | 3 | 0 | Low-quality | Genome-fragment | 3.31 | AAI-based (high-confidence) | 0 |
| S12-final-viral-combined_k141_394925\|\|full | 1965 | No | NA | 4 | 0 | 0 | Low-quality | Genome-fragment | 1.55 | AAI-based (high-confidence) | 0 |
| S2_k141_177130 | 1965 | No | NA | 3 | 0 | 0 | Low-quality | Genome-fragment | 2.06 | AAI-based (high-confidence) | 0 |
| S7-final-viral-combined_k141_101967\|\|full | 8089 | No | NA | 5 | 0 | 5 | Not-determined | Genome-fragment | NA | NA | 0 |
| S8-final-viral-combined_k141_232638\|\|full | 1965 | No | NA | 3 | 0 | 0 | Low-quality | Genome-fragment | 3.61 | AAI-based (medium-confidence) | 0 |
| S7-final-viral-combined_k141_9982\|\|full | 14460 | No | NA | 24 | 0 | 6 | Not-determined | Genome-fragment | NA | NA | 0 |
| S8-final-viral-combined_k141_124828\|\|full | 1965 | No | NA | 3 | 1 | 0 | Low-quality | Genome-fragment | 4.14 | HMM-based (lower-bound) | 0 |
| S7-final-viral-combined_k141_222775\|\|full | 1590 | No | NA | 3 | 0 | 0 | Not-determined | Genome-fragment | NA | NA | 0 |
| S7-final-viral-combined_k141_152644\|\|full | 1915 | No | NA | 2 | 0 | 1 | Not-determined | Genome-fragment | NA | NA | 0 |
| S7-final-viral-combined_k141_71608\|\|full | 1966 | No | NA | 3 | 2 | 0 | Low-quality | Genome-fragment | 2.73 | HMM-based (lower-bound) | 0 |
| S7-final-viral-combined_k141_19976\|\|full | 1374 | No | NA | 2 | 0 | 2 | Not-determined | Genome-fragment | NA | NA | 0 |
| S11-final-viral-combined_k141_198010\|\|full | 1967 | No | NA | 3 | 2 | 0 | Low-quality | Genome-fragment | 5.5 | AAI-based (medium-confidence) | 0 |
| S7-final-viral-combined_k141_243896\|\|full | 4497 | No | NA | 4 | 0 | 2 | Not-determined | Genome-fragment | NA | NA | 0 |
| S7-final-viral-combined_k141_91347\|\|full | 6489 | No | NA | 12 | 0 | 3 | Not-determined | Genome-fragment | NA | NA | 0 |
| S11-final-viral-combined_k141_12454\|\|full | 1967 | No | NA | 3 | 2 | 0 | Low-quality | Genome-fragment | 3.87 | AAI-based (high-confidence) | 0 |
| S3-final-viral-combined_k141_59730\|\|full | 1967 | No | NA | 4 | 0 | 1 | Low-quality | Genome-fragment | 1.12 | AAI-based (medium-confidence) | 0 |
| S7-final-viral-combined_k141_80743\|\|full | 1459 | No | NA | 2 | 0 | 1 | Not-determined | Genome-fragment | NA | NA | 0 |
| S7-final-viral-combined_k141_111872\|\|full | 5800 | No | NA | 7 | 0 | 2 | Not-determined | Genome-fragment | NA | NA | 0 |
| S5-final-viral-combined_k141_52879\|\|full | 1967 | No | NA | 3 | 0 | 0 | Low-quality | Genome-fragment | 3.55 | AAI-based (medium-confidence) | 0 |
| S7-final-viral-combined_k141_91415\|\|full | 2413 | No | NA | 3 | 0 | 1 | Not-determined | Genome-fragment | NA | NA | 0 |
| S5_k141_118438 | 1967 | No | NA | 2 | 0 | 1 | Low-quality | Genome-fragment | 2.01 | AAI-based (high-confidence) | 0 |
| S7-final-viral-combined_k141_132363\|\|full | 1137 | No | NA | 2 | 0 | 0 | Not-determined | Genome-fragment | NA | NA | 0 |
| S7-final-viral-combined_k141_132381\|\|full | 2980 | No | NA | 6 | 0 | 4 | Not-determined | Genome-fragment | NA | NA | 0 |
| S7-final-viral-combined_k141_222913\|\|full | 1860 | No | NA | 2 | 0 | 1 | Not-determined | Genome-fragment | NA | NA | 0 |
| S17-final-viral-combined_k141_18213\|\|full | 1967 | No | NA | 3 | 0 | 2 | Low-quality | Genome-fragment | 0.94 | AAI-based (medium-confidence) | 0 |
| S7-final-viral-combined_k141_80796\|\|full | 5480 | No | NA | 6 | 0 | 1 | Not-determined | Genome-fragment | NA | NA | 0 |
| S7-final-viral-combined_k141_244000\|\|full | 3421 | No | NA | 2 | 0 | 1 | Not-determined | Genome-fragment | NA | NA | 0 |
| S12-final-viral-combined_k141_133044\|\|full | 1967 | No | NA | 4 | 3 | 0 | Low-quality | Genome-fragment | 5.6 | AAI-based (medium-confidence) | 0 |
| S7-final-viral-combined_k141_244016\|\|full | 4697 | No | NA | 4 | 0 | 0 | Not-determined | Genome-fragment | NA | NA | 0 |
| S7-final-viral-combined_k141_222945\|\|full | 8617 | No | NA | 9 | 0 | 1 | Not-determined | Genome-fragment | NA | NA | 0 |
| S7-final-viral-combined_k141_91434\|\|full | 1768 | No | NA | 2 | 0 | 2 | Not-determined | Genome-fragment | NA | NA | 0 |
| S7-final-viral-combined_k141_244049\|\|full | 1210 | No | NA | 2 | 0 | 1 | Not-determined | Genome-fragment | NA | NA | 0 |
| S12-final-viral-combined_k141_247057\|\|lt2gene | 1967 | No | NA | 1 | 1 | 0 | Low-quality | Genome-fragment | 5.46 | AAI-based (high-confidence) | 0 |
| S7-final-viral-combined_k141_152869\|\|full | 1491 | No | NA | 2 | 0 | 0 | Not-determined | Genome-fragment | NA | NA | 0 |
| S7-final-viral-combined_k141_142224\|\|full | 4592 | No | NA | 4 | 0 | 3 | Not-determined | Genome-fragment | NA | NA | 0 |
| S2-final-viral-combined_k141_105049\|\|full | 1967 | No | NA | 2 | 0 | 1 | Low-quality | Genome-fragment | 0.47 | AAI-based (medium-confidence) | 0 |
| S7-final-viral-combined_k141_142228\|\|full | 14512 | No | NA | 7 | 0 | 5 | Not-determined | Genome-fragment | NA | NA | 0 |
| S7-final-viral-combined_k141_20143\|\|full | 3323 | No | NA | 4 | 0 | 4 | Not-determined | Genome-fragment | NA | NA | 0 |
| S7-final-viral-combined_k141_20145\|\|full | 1614 | No | NA | 3 | 0 | 0 | Not-determined | Genome-fragment | NA | NA | 0 |
| S8-final-viral-combined_k141_232282\|\|full | 1967 | No | NA | 2 | 0 | 0 | Low-quality | Genome-fragment | 5.58 | AAI-based (high-confidence) | 0 |
| S1-final-viral-combined_k141_177145\|\|full | 1968 | No | NA | 5 | 1 | 0 | Low-quality | Genome-fragment | 5.58 | AAI-based (high-confidence) | 0 |
| S1_k141_267777 | 1968 | No | NA | 4 | 0 | 0 | Low-quality | Genome-fragment | 5.96 | AAI-based (medium-confidence) | 0 |
| S7-final-viral-combined_k141_80946\|\|full | 1238 | No | NA | 2 | 0 | 2 | Not-determined | Genome-fragment | NA | NA | 0 |
| S7-final-viral-combined_k141_244166\|\|full | 2517 | No | NA | 3 | 0 | 1 | Not-determined | Genome-fragment | NA | NA | 0 |
| S1_k141_114467 | 1968 | No | NA | 4 | 0 | 0 | Low-quality | Genome-fragment | 3.5 | AAI-based (medium-confidence) | 0 |
| S5_k141_206724 | 1968 | No | NA | 5 | 0 | 0 | Low-quality | Genome-fragment | 1.13 | AAI-based (high-confidence) | 0 |
| S7-final-viral-combined_k141_152958\|\|full | 2439 | No | NA | 3 | 0 | 2 | Not-determined | Genome-fragment | NA | NA | 0 |
| S17-final-viral-combined_k141_245271\|\|full | 1968 | No | NA | 4 | 0 | 2 | Low-quality | Genome-fragment | 3.69 | AAI-based (high-confidence) | 0 |
| S17-final-viral-combined_k141_164054\|\|full | 1968 | No | NA | 4 | 0 | 0 | Low-quality | Genome-fragment | 4.72 | AAI-based (medium-confidence) | 0 |
| S7-final-viral-combined_k141_10184\|\|full | 6036 | No | NA | 8 | 0 | 5 | Not-determined | Genome-fragment | NA | NA | 0 |
| S12-final-viral-combined_k141_392085\|\|full | 1968 | No | NA | 3 | 2 | 0 | Low-quality | Genome-fragment | 5 | AAI-based (high-confidence) | 0 |
| S7-final-viral-combined_k141_152991\|\|full | 2053 | No | NA | 3 | 0 | 1 | Not-determined | Genome-fragment | NA | NA | 0 |
| S7-final-viral-combined_k141_153006\|\|full | 2818 | No | NA | 5 | 0 | 0 | Not-determined | Genome-fragment | NA | NA | 0 |
| S7-final-viral-combined_k141_153029\|\|full | 1073 | No | NA | 2 | 0 | 0 | Not-determined | Genome-fragment | NA | NA | 0 |
| S7-final-viral-combined_k141_28709\|\|full | 13955 | No | NA | 12 | 0 | 9 | Not-determined | Genome-fragment | NA | NA | 0 |
| S7-final-viral-combined_k141_223149\|\|full | 1131 | No | NA | 3 | 0 | 2 | Not-determined | Genome-fragment | NA | NA | 0 |
| S4-final-viral-combined_k141_27148\|\|full | 1968 | No | NA | 2 | 0 | 0 | Low-quality | Genome-fragment | 5.31 | AAI-based (medium-confidence) | 0 |
| S11-final-viral-combined_k141_70175\|\|full | 1969 | No | NA | 2 | 0 | 0 | Low-quality | Genome-fragment | 3.53 | AAI-based (medium-confidence) | 0 |
| S7-final-viral-combined_k141_28739\|\|full | 1949 | No | NA | 3 | 0 | 0 | Not-determined | Genome-fragment | NA | NA | 0 |
| S7-final-viral-combined_k141_223165\|\|full | 1428 | No | NA | 4 | 0 | 1 | Not-determined | Genome-fragment | NA | NA | 0 |
| S7-final-viral-combined_k141_244215\|\|full | 1111 | No | NA | 2 | 0 | 0 | Not-determined | Genome-fragment | NA | NA | 0 |
| S7-final-viral-combined_k141_20328\|\|full | 2758 | No | NA | 2 | 0 | 0 | Not-determined | Genome-fragment | NA | NA | 0 |
| S3-final-viral-combined_k141_28998\|\|full | 1969 | No | NA | 2 | 0 | 1 | Low-quality | Genome-fragment | 3.78 | AAI-based (medium-confidence) | 0 |
| S7-final-viral-combined_k141_28834\|\|full | 3665 | No | NA | 4 | 0 | 0 | Not-determined | Genome-fragment | NA | NA | 0 |
| S3-final-viral-combined_k141_12837\|\|full | 1969 | No | NA | 3 | 1 | 0 | Low-quality | Genome-fragment | 2.23 | AAI-based (high-confidence) | 0 |
| S17-final-viral-combined_k141_37093\|\|full | 1969 | No | NA | 3 | 0 | 1 | Low-quality | Genome-fragment | 0.88 | AAI-based (medium-confidence) | 0 |
| S17-final-viral-combined_k141_49916\|\|full | 1969 | No | NA | 2 | 0 | 2 | Low-quality | Genome-fragment | 3.36 | AAI-based (high-confidence) | 0 |
| S7-final-viral-combined_k141_20396\|\|full | 3654 | No | NA | 7 | 0 | 0 | Not-determined | Genome-fragment | NA | NA | 0 |
| S7-final-viral-combined_k141_142501\|\|full | 1796 | No | NA | 3 | 0 | 2 | Not-determined | Genome-fragment | NA | NA | 0 |
| S7-final-viral-combined_k141_91650\|\|full | 1363 | No | NA | 2 | 0 | 1 | Not-determined | Genome-fragment | NA | NA | 0 |
| S7-final-viral-combined_k141_142517\|\|full | 1104 | No | NA | 3 | 0 | 2 | Not-determined | Genome-fragment | NA | NA | 0 |
| S12-final-viral-combined_k141_218151\|\|full | 1969 | No | NA | 3 | 0 | 1 | Low-quality | Genome-fragment | 3.3 | AAI-based (high-confidence) | 0 |
| S7-final-viral-combined_k141_91767\|\|full | 4603 | No | NA | 4 | 0 | 4 | Not-determined | Genome-fragment | NA | NA | 0 |
| S12-final-viral-combined_k141_206464\|\|full | 1969 | Yes | 186 | 3 | 1 | 1 | Low-quality | Genome-fragment | 0.15 | HMM-based (lower-bound) | 90.55 |
| S7-final-viral-combined_k141_244282\|\|full | 2399 | No | NA | 3 | 0 | 2 | Not-determined | Genome-fragment | NA | NA | 0 |
| S7-final-viral-combined_k141_142657\|\|full | 2328 | No | NA | 2 | 0 | 2 | Not-determined | Genome-fragment | NA | NA | 0 |
| S2-final-viral-combined_k141_186269\|\|lt2gene | 1969 | No | NA | 1 | 1 | 0 | Low-quality | Genome-fragment | 4.86 | AAI-based (high-confidence) | 0 |
| S2_k141_199254 | 1969 | No | NA | 2 | 0 | 2 | Low-quality | Genome-fragment | 0.72 | AAI-based (medium-confidence) | 0 |
| S7-final-viral-combined_k141_244332\|\|full | 2873 | No | NA | 4 | 0 | 2 | Not-determined | Genome-fragment | NA | NA | 0 |
| S7-final-viral-combined_k141_142706\|\|full | 1991 | No | NA | 5 | 0 | 0 | Not-determined | Genome-fragment | NA | NA | 0 |
| S8-final-viral-combined_k141_252529\|\|full | 1969 | No | NA | 3 | 1 | 0 | Low-quality | Genome-fragment | 4.79 | AAI-based (high-confidence) | 0 |
| S7-final-viral-combined_k141_29152\|\|full | 1700 | No | NA | 4 | 0 | 0 | Not-determined | Genome-fragment | NA | NA | 0 |
| S1_k141_338349 | 1970 | No | NA | 1 | 0 | 0 | Low-quality | Genome-fragment | 1.79 | AAI-based (high-confidence) | 0 |
| S2-final-viral-combined_k141_164310\|\|full | 1970 | No | NA | 3 | 1 | 0 | Low-quality | Genome-fragment | 1.46 | HMM-based (lower-bound) | 0 |
| S7-final-viral-combined_k141_81330\|\|full | 2505 | No | NA | 4 | 0 | 3 | Not-determined | Genome-fragment | NA | NA | 0 |
| S4_k141_5523 | 1970 | No | NA | 3 | 1 | 0 | Low-quality | Genome-fragment | 5.59 | AAI-based (medium-confidence) | 0 |
| S7-final-viral-combined_k141_142781\|\|full | 3350 | No | NA | 4 | 0 | 3 | Not-determined | Genome-fragment | NA | NA | 0 |
| S7-final-viral-combined_k141_81370\|\|full | 1859 | No | NA | 2 | 0 | 0 | Not-determined | Genome-fragment | NA | NA | 0 |
| S7-final-viral-combined_k141_29211\|\|full | 1023 | No | NA | 3 | 0 | 0 | Not-determined | Genome-fragment | NA | NA | 0 |
| S7-final-viral-combined_k141_91820\|\|full | 1171 | No | NA | 2 | 0 | 1 | Not-determined | Genome-fragment | NA | NA | 0 |
| S7-final-viral-combined_k141_244396\|\|full | 2986 | No | NA | 4 | 0 | 3 | Not-determined | Genome-fragment | NA | NA | 0 |
| S11-final-viral-combined_k141_223739\|\|full | 1971 | No | NA | 3 | 2 | 0 | Low-quality | Genome-fragment | 4.25 | AAI-based (high-confidence) | 0 |
| S1-final-viral-combined_k141_314986\|\|full | 1971 | No | NA | 3 | 1 | 0 | Low-quality | Genome-fragment | 1.75 | HMM-based (lower-bound) | 0 |
| S1-final-viral-combined_k141_335020\|\|full | 1971 | No | NA | 6 | 1 | 0 | Low-quality | Genome-fragment | 3.21 | AAI-based (high-confidence) | 0 |
| S7-final-viral-combined_k141_142945\|\|full | 1386 | No | NA | 3 | 0 | 0 | Not-determined | Genome-fragment | NA | NA | 0 |
| S12-final-viral-combined_k141_191158\|\|full | 1971 | No | NA | 4 | 1 | 0 | Low-quality | Genome-fragment | 2.21 | HMM-based (lower-bound) | 0 |
| S7-final-viral-combined_k141_244512\|\|full | 12658 | No | NA | 10 | 0 | 9 | Not-determined | Genome-fragment | NA | NA | 0 |
| S7-final-viral-combined_k141_223752\|\|full | 1402 | No | NA | 2 | 0 | 1 | Not-determined | Genome-fragment | NA | NA | 0 |
| S7-final-viral-combined_k141_223781\|\|full | 1211 | No | NA | 4 | 0 | 0 | Not-determined | Genome-fragment | NA | NA | 0 |
| S8-final-viral-combined_k141_85211\|\|full | 1971 | No | NA | 5 | 1 | 0 | Low-quality | Genome-fragment | 4.2 | AAI-based (high-confidence) | 0 |
| S7-final-viral-combined_k141_223938\|\|full | 2812 | No | NA | 4 | 0 | 1 | Not-determined | Genome-fragment | NA | NA | 0 |
| S8-final-viral-combined_k141_132977\|\|full | 1971 | No | NA | 3 | 0 | 0 | Low-quality | Genome-fragment | 5.56 | AAI-based (high-confidence) | 0 |
| S7-final-viral-combined_k141_29525\|\|full | 1546 | No | NA | 2 | 0 | 0 | Not-determined | Genome-fragment | NA | NA | 0 |
| S11-final-viral-combined_k141_231550\|\|full | 1972 | No | NA | 4 | 2 | 0 | Low-quality | Genome-fragment | 2.85 | AAI-based (high-confidence) | 0 |
| S1-final-viral-combined_k141_298028\|\|full | 1972 | No | NA | 3 | 0 | 0 | Low-quality | Genome-fragment | 4.15 | AAI-based (medium-confidence) | 0 |
| S1-final-viral-combined_k141_140875\|\|full | 1972 | No | NA | 3 | 0 | 1 | Low-quality | Genome-fragment | 1.73 | AAI-based (medium-confidence) | 0 |
| S7-final-viral-combined_k141_29945\|\|full | 1213 | No | NA | 3 | 0 | 0 | Not-determined | Genome-fragment | NA | NA | 0 |
| S7-final-viral-combined_k141_224199\|\|full | 2113 | No | NA | 3 | 0 | 2 | Not-determined | Genome-fragment | NA | NA | 0 |
| S17-k141_226673 | 1972 | No | NA | 2 | 0 | 1 | Low-quality | Genome-fragment | 2.74 | AAI-based (medium-confidence) | 0 |
| S12-final-viral-combined_k141_92341\|\|full | 1972 | No | NA | 5 | 0 | 0 | Low-quality | Genome-fragment | 5.25 | AAI-based (medium-confidence) | 0 |
| S7-final-viral-combined_k141_244760\|\|full | 3143 | No | NA | 7 | 0 | 1 | Not-determined | Genome-fragment | NA | NA | 0 |
| S2-final-viral-combined_k141_107291\|\|full | 1972 | No | NA | 3 | 0 | 0 | Low-quality | Genome-fragment | 2.88 | AAI-based (medium-confidence) | 0 |
| S7-final-viral-combined_k141_224375\|\|full | 1326 | No | NA | 2 | 0 | 0 | Not-determined | Genome-fragment | NA | NA | 0 |
| S7-final-viral-combined_k141_244765\|\|full | 1818 | No | NA | 2 | 0 | 2 | Not-determined | Genome-fragment | NA | NA | 0 |
| S7-final-viral-combined_k141_224381\|\|full | 11058 | No | NA | 12 | 0 | 2 | Not-determined | Genome-fragment | NA | NA | 0 |
| S4-final-viral-combined_k141_86013\|\|full | 1973 | No | NA | 3 | 2 | 0 | Low-quality | Genome-fragment | 4.63 | AAI-based (high-confidence) | 0 |
| S8-final-viral-combined_k141_150596\|\|full | 1973 | No | NA | 3 | 1 | 0 | Low-quality | Genome-fragment | 1.19 | HMM-based (lower-bound) | 0 |
| S8-final-viral-combined_k141_82084\|\|full | 1973 | No | NA | 3 | 0 | 2 | Low-quality | Genome-fragment | 1.68 | AAI-based (medium-confidence) | 0 |
| S7-final-viral-combined_k141_244803\|\|full | 2926 | No | NA | 3 | 0 | 1 | Not-determined | Genome-fragment | NA | NA | 0 |
| S7-final-viral-combined_k141_224494\|\|full | 2452 | No | NA | 4 | 0 | 2 | Not-determined | Genome-fragment | NA | NA | 0 |
| S7-final-viral-combined_k141_30186\|\|full | 2760 | No | NA | 3 | 0 | 3 | Not-determined | Genome-fragment | NA | NA | 0 |
| S7-final-viral-combined_k141_224523\|\|full | 19119 | No | NA | 31 | 9 | 1 | Medium-quality | Genome-fragment | 50.86 | AAI-based (high-confidence) | 0 |
| S9-final-viral-combined_k141_37253\|\|full | 1974 | No | NA | 3 | 0 | 3 | Low-quality | Genome-fragment | 4.18 | AAI-based (high-confidence) | 0 |
| S7-final-viral-combined_k141_30240\|\|full | 12353 | No | NA | 6 | 0 | 5 | Not-determined | Genome-fragment | NA | NA | 0 |
| S7-final-viral-combined_k141_30276\|\|full | 1956 | No | NA | 2 | 0 | 2 | Not-determined | Genome-fragment | NA | NA | 0 |
| S7-final-viral-combined_k141_30299\|\|full | 3130 | No | NA | 3 | 0 | 3 | Not-determined | Genome-fragment | NA | NA | 0 |
| S7-final-viral-combined_k141_30389\|\|full | 2121 | No | NA | 5 | 0 | 0 | Not-determined | Genome-fragment | NA | NA | 0 |
| S7-final-viral-combined_k141_30463\|\|full | 1187 | No | NA | 3 | 0 | 1 | Not-determined | Genome-fragment | NA | NA | 0 |
| S9_k141_93989 | 1974 | No | NA | 2 | 0 | 1 | Low-quality | Genome-fragment | 0.33 | AAI-based (high-confidence) | 0 |
| S1_k141_229081 | 1974 | No | NA | 4 | 1 | 0 | Low-quality | Genome-fragment | 4.7 | HMM-based (lower-bound) | 0 |
| S7-final-viral-combined_k141_30576\|\|full | 1416 | No | NA | 3 | 0 | 3 | Not-determined | Genome-fragment | NA | NA | 0 |
| S17-k141_138809 | 1974 | No | NA | 1 | 0 | 1 | Low-quality | Genome-fragment | 3.35 | AAI-based (high-confidence) | 0 |
| S7-final-viral-combined_k141_143218\|\|0_partial | 5781 | No | NA | 7 | 1 | 0 | High-quality | High-quality | 100 | HMM-based (lower-bound) | 0 |
| S12-final-viral-combined_k141_340434\|\|full | 1974 | No | NA | 5 | 0 | 0 | Low-quality | Genome-fragment | 5.11 | AAI-based (medium-confidence) | 0 |
| S7-final-viral-combined_k141_184132\|\|0_partial | 12722 | No | NA | 9 | 0 | 6 | Not-determined | Genome-fragment | NA | NA | 0 |
| S7-final-viral-combined_k141_41228\|\|0_partial | 11633 | No | NA | 10 | 0 | 5 | Not-determined | Genome-fragment | NA | NA | 0 |
| S7-final-viral-combined_k141_163980\|\|0_partial | 26788 | No | NA | 35 | 9 | 1 | Medium-quality | Genome-fragment | 75.59 | AAI-based (high-confidence) | 0 |
| S12-final-viral-combined_k141_63002\|\|full | 1974 | No | NA | 4 | 0 | 0 | Low-quality | Genome-fragment | 5.19 | AAI-based (high-confidence) | 0 |
| S7-final-viral-combined_k141_174309\|\|0_partial | 30451 | No | NA | 22 | 0 | 18 | Not-determined | Genome-fragment | NA | NA | 0 |
| S7-final-viral-combined_k141_174309\|\|1_partial | 12056 | No | NA | 16 | 0 | 8 | Not-determined | Genome-fragment | NA | NA | 0 |
| S7-final-viral-combined_k141_72070\|\|0_partial | 55473 | Yes | 41210 | 64 | 3 | 7 | High-quality | High-quality | 92.76 | AAI-based (medium-confidence) | 25.71 |
| S7-final-viral-combined_k141_113668\|\|0_partial | 12615 | No | NA | 11 | 0 | 1 | Not-determined | Genome-fragment | NA | NA | 0 |
| S12_k141_54282 | 1974 | No | NA | 6 | 0 | 0 | Low-quality | Genome-fragment | 0.95 | AAI-based (high-confidence) | 0 |
| S2-final-viral-combined_k141_63005\|\|full | 1974 | No | NA | 2 | 0 | 1 | Low-quality | Genome-fragment | 2.6 | AAI-based (medium-confidence) | 0 |
| S2-final-viral-combined_k141_154779\|\|lt2gene | 1974 | No | NA | 1 | 1 | 0 | Low-quality | Genome-fragment | 1.51 | HMM-based (lower-bound) | 0 |
| S6_k141_120567 | 1974 | No | NA | 4 | 1 | 0 | Low-quality | Genome-fragment | 1.97 | AAI-based (high-confidence) | 0 |
| S7-final-viral-combined_k141_114254\|\|0_partial | 27335 | No | NA | 18 | 0 | 7 | Not-determined | Genome-fragment | NA | NA | 0 |
| S7-final-viral-combined_k141_144442\|\|0_partial | 12026 | No | NA | 14 | 0 | 8 | Not-determined | Genome-fragment | NA | NA | 0 |
| S9-final-viral-combined_k141_124797\|\|full | 1975 | No | NA | 2 | 0 | 1 | Low-quality | Genome-fragment | 1.03 | AAI-based (medium-confidence) | 0 |
| S1-final-viral-combined_k141_52449\|\|full | 1975 | No | NA | 3 | 0 | 2 | Low-quality | Genome-fragment | 0.91 | AAI-based (high-confidence) | 0 |
| S17-final-viral-combined_k141_127402\|\|full | 1975 | No | NA | 4 | 0 | 0 | Low-quality | Genome-fragment | 4.62 | AAI-based (high-confidence) | 0 |
| S17-final-viral-combined_k141_1422\|\|lt2gene | 1975 | No | NA | 2 | 2 | 0 | Low-quality | Genome-fragment | 4.84 | AAI-based (high-confidence) | 0 |
| S1-final-viral-combined_k141_77191\|\|full | 1976 | No | NA | 6 | 1 | 0 | Low-quality | Genome-fragment | 4.46 | AAI-based (medium-confidence) | 0 |
| S5_k141_21700 | 1976 | No | NA | 2 | 0 | 0 | Low-quality | Genome-fragment | 1.42 | AAI-based (high-confidence) | 0 |
| S7-final-viral-combined_k141_237995\|\|0_partial | 14412 | No | NA | 15 | 0 | 6 | Not-determined | Genome-fragment | NA | NA | 0 |
| S7-final-viral-combined_k141_116546\|\|full | 1976 | No | NA | 3 | 0 | 2 | Low-quality | Genome-fragment | 1.41 | AAI-based (high-confidence) | 0 |
| S12-final-viral-combined_k141_310313\|\|full | 1976 | No | NA | 6 | 2 | 0 | Low-quality | Genome-fragment | 3.31 | AAI-based (high-confidence) | 0 |
| S8-final-viral-combined_k141_173674\|\|full | 1976 | No | NA | 3 | 0 | 2 | Low-quality | Genome-fragment | 1.41 | AAI-based (high-confidence) | 0 |
| S8-final-viral-combined_k141_201400\|\|full | 1976 | No | NA | 3 | 0 | 1 | Low-quality | Genome-fragment | 4.05 | AAI-based (high-confidence) | 0 |
| S7-final-viral-combined_k141_136642\|\|0_partial | 24720 | Yes | 16778 | 35 | 7 | 5 | High-quality | High-quality | 99.16 | AAI-based (high-confidence) | 32.13 |
| S7-final-viral-combined_k141_147158\|\|0_partial | 73734 | Yes | 31556 | 85 | 15 | 20 | Medium-quality | Genome-fragment | 78.86 | AAI-based (high-confidence) | 57.2 |
| S7-final-viral-combined_k141_96390\|\|0_partial | 14347 | No | NA | 11 | 0 | 8 | Not-determined | Genome-fragment | NA | NA | 0 |
| S7-final-viral-combined_k141_218379\|\|0_partial | 96077 | Yes | 35165 | 110 | 16 | 42 | Complete | High-quality | 100 | Provirus (high-confidence) | 63.4 |
| S7-final-viral-combined_k141_86413\|\|0_partial | 16113 | No | NA | 19 | 0 | 7 | Not-determined | Genome-fragment | NA | NA | 0 |
| S8-final-viral-combined_k141_125715\|\|full | 1976 | No | NA | 5 | 0 | 0 | Low-quality | Genome-fragment | 3.43 | AAI-based (high-confidence) | 0 |
| S7-final-viral-combined_k141_107679\|\|0_partial | 67608 | Yes | 37921 | 69 | 14 | 18 | Medium-quality | Genome-fragment | 65.46 | HMM-based (lower-bound) | 43.91 |
| S7-final-viral-combined_k141_107679\|\|1_partial | 36183 | Yes | 29250 | 45 | 6 | 8 | Medium-quality | Genome-fragment | 54.76 | AAI-based (high-confidence) | 19.16 |
| S11-final-viral-combined_k141_289325\|\|full | 1977 | No | NA | 3 | 0 | 0 | Low-quality | Genome-fragment | 3.22 | AAI-based (medium-confidence) | 0 |
| S7-final-viral-combined_k141_35099\|\|full | 1977 | No | NA | 3 | 0 | 0 | Low-quality | Genome-fragment | 5.19 | AAI-based (medium-confidence) | 0 |
| S7-final-viral-combined_k141_108165\|\|0_partial | 12878 | No | NA | 8 | 0 | 6 | Not-determined | Genome-fragment | NA | NA | 0 |
| S7-final-viral-combined_k141_211691\|\|0_partial | 23583 | No | NA | 16 | 0 | 10 | Not-determined | Genome-fragment | NA | NA | 0 |
| S2-final-viral-combined_k141_4950\|\|lt2gene | 1977 | No | NA | 1 | 1 | 0 | Low-quality | Genome-fragment | 4.11 | AAI-based (high-confidence) | 0 |
| S7-final-viral-combined_k141_191688\|\|0_partial | 41007 | No | NA | 31 | 0 | 17 | Not-determined | Genome-fragment | NA | NA | 0 |
| S7-final-viral-combined_k141_232465\|\|0_partial | 27193 | No | NA | 25 | 0 | 13 | Not-determined | Genome-fragment | NA | NA | 0 |
| S6-final-viral-combined_k141_57559\|\|full | 1977 | No | NA | 4 | 0 | 0 | Low-quality | Genome-fragment | 1.53 | AAI-based (medium-confidence) | 0 |
| S6_k141_134050 | 1977 | No | NA | 2 | 0 | 1 | Low-quality | Genome-fragment | 2.11 | AAI-based (medium-confidence) | 0 |
| S8-final-viral-combined_k141_109951\|\|lt2gene | 1977 | No | NA | 2 | 2 | 0 | Low-quality | Genome-fragment | 3.66 | HMM-based (lower-bound) | 0 |
| S1-final-viral-combined_k141_376929\|\|full | 1978 | No | NA | 2 | 1 | 0 | Low-quality | Genome-fragment | 2.99 | HMM-based (lower-bound) | 0 |
| S7_k141_96375 | 1978 | No | NA | 3 | 0 | 0 | Low-quality | Genome-fragment | 1.88 | AAI-based (high-confidence) | 0 |
| S7-final-viral-combined_k141_243745\|\|0_partial | 12010 | No | NA | 8 | 0 | 7 | Not-determined | Genome-fragment | NA | NA | 0 |
| S12-final-viral-combined_k141_119504\|\|full | 1978 | No | NA | 3 | 0 | 1 | Low-quality | Genome-fragment | 4.22 | AAI-based (medium-confidence) | 0 |
| S12-final-viral-combined_k141_73507\|\|full | 1978 | No | NA | 4 | 1 | 0 | Low-quality | Genome-fragment | 2.93 | HMM-based (lower-bound) | 0 |
| S12-final-viral-combined_k141_78019\|\|full | 1978 | No | NA | 3 | 1 | 0 | Low-quality | Genome-fragment | 4.43 | AAI-based (medium-confidence) | 0 |
| S7-final-viral-combined_k141_153302\|\|lt2gene | 1052 | No | NA | 2 | 0 | 0 | Not-determined | Genome-fragment | NA | NA | 0 |
| S7-final-viral-combined_k141_122703\|\|lt2gene | 1092 | No | NA | 1 | 0 | 0 | Not-determined | Genome-fragment | NA | NA | 0 |
| S2-final-viral-combined_k141_213345\|\|full | 1978 | No | NA | 2 | 0 | 2 | Low-quality | Genome-fragment | 1 | AAI-based (medium-confidence) | 0 |
| S4-final-viral-combined_k141_11082\|\|full | 1978 | No | NA | 4 | 0 | 3 | Low-quality | Genome-fragment | 0.65 | AAI-based (medium-confidence) | 0 |
| S4_k141_20911 | 1978 | No | NA | 1 | 0 | 1 | Low-quality | Genome-fragment | 0.5 | AAI-based (medium-confidence) | 0 |
| S7-final-viral-combined_k141_82214\|\|lt2gene | 2022 | No | NA | 2 | 0 | 1 | Not-determined | Genome-fragment | NA | NA | 0 |
| S8-final-viral-combined_k141_31179\|\|full | 1978 | No | NA | 5 | 0 | 0 | Low-quality | Genome-fragment | 3.26 | AAI-based (high-confidence) | 0 |
| S11-final-viral-combined_k141_70124\|\|full | 1979 | No | NA | 3 | 0 | 2 | Low-quality | Genome-fragment | 1.22 | AAI-based (medium-confidence) | 0 |
| S11-final-viral-combined_k141_52724\|\|lt2gene | 1979 | No | NA | 2 | 2 | 0 | Low-quality | Genome-fragment | 3.34 | AAI-based (high-confidence) | 0 |
| S1-final-viral-combined_k141_130331\|\|full | 1979 | No | NA | 3 | 0 | 1 | Low-quality | Genome-fragment | 3.41 | AAI-based (medium-confidence) | 0 |
| S7-final-viral-combined_k141_92617\|\|lt2gene | 1262 | No | NA | 1 | 0 | 0 | Not-determined | Genome-fragment | NA | NA | 0 |
| S5_k141_13871 | 1979 | No | NA | 3 | 0 | 0 | Low-quality | Genome-fragment | 3.35 | AAI-based (high-confidence) | 0 |
| S12-final-viral-combined_k141_17178\|\|full | 1979 | No | NA | 4 | 0 | 0 | Low-quality | Genome-fragment | 2.82 | AAI-based (high-confidence) | 0 |
| S7-final-viral-combined_k141_82671\|\|lt2gene | 1170 | No | NA | 1 | 0 | 0 | Not-determined | Genome-fragment | NA | NA | 0 |
| S2-final-viral-combined_k141_140213\|\|full | 1979 | No | NA | 3 | 2 | 0 | Low-quality | Genome-fragment | 4.21 | AAI-based (high-confidence) | 0 |
| S2-final-viral-combined_k141_123836\|\|full | 1979 | No | NA | 6 | 2 | 0 | Low-quality | Genome-fragment | 5.2 | AAI-based (high-confidence) | 0 |
| S2_k141_231006 | 1979 | No | NA | 1 | 0 | 1 | Low-quality | Genome-fragment | 2.12 | AAI-based (medium-confidence) | 0 |
| S11-final-viral-combined_k141_80249\|\|full | 1980 | No | NA | 5 | 3 | 0 | Low-quality | Genome-fragment | 5.01 | AAI-based (medium-confidence) | 0 |
| S11_k141_43229 | 1980 | No | NA | 3 | 2 | 0 | Low-quality | Genome-fragment | 4.33 | AAI-based (high-confidence) | 0 |
| S1-final-viral-combined_k141_89473\|\|full | 1980 | No | NA | 5 | 3 | 0 | Low-quality | Genome-fragment | 5.6 | AAI-based (medium-confidence) | 0 |
| S1-final-viral-combined_k141_123774\|\|full | 1980 | No | NA | 3 | 0 | 1 | Low-quality | Genome-fragment | 1.18 | AAI-based (high-confidence) | 0 |
| S1-final-viral-combined_k141_26072\|\|full | 1980 | No | NA | 3 | 0 | 1 | Low-quality | Genome-fragment | 4.11 | AAI-based (medium-confidence) | 0 |
| S5_k141_205400 | 1980 | No | NA | 1 | 0 | 1 | Low-quality | Genome-fragment | 4.26 | AAI-based (high-confidence) | 0 |
| S12-final-viral-combined_k141_76888\|\|lt2gene | 1980 | No | NA | 1 | 1 | 0 | Low-quality | Genome-fragment | 2.89 | HMM-based (lower-bound) | 0 |
| S12_k141_143414 | 1980 | No | NA | 1 | 0 | 0 | Low-quality | Genome-fragment | 1.44 | AAI-based (high-confidence) | 0 |
| S11-final-viral-combined_k141_168375\|\|full | 1981 | No | NA | 3 | 0 | 2 | Low-quality | Genome-fragment | 1.03 | AAI-based (medium-confidence) | 0 |
| S1-final-viral-combined_k141_276507\|\|full | 1981 | No | NA | 4 | 2 | 0 | Low-quality | Genome-fragment | 5.43 | AAI-based (high-confidence) | 0 |
| S5-final-viral-combined_k141_135702\|\|lt2gene | 1981 | No | NA | 2 | 2 | 0 | Low-quality | Genome-fragment | 4.75 | AAI-based (medium-confidence) | 0 |
| S6-final-viral-combined_k141_30081\|\|full | 1981 | No | NA | 3 | 0 | 1 | Low-quality | Genome-fragment | 6.75 | AAI-based (high-confidence) | 0 |
| S7-final-viral-combined_k141_83747\|\|lt2gene | 1638 | No | NA | 2 | 0 | 1 | Not-determined | Genome-fragment | NA | NA | 0 |
| S6_k141_746 | 1982 | No | NA | 3 | 0 | 2 | Low-quality | Genome-fragment | 1.41 | AAI-based (high-confidence) | 0 |
| S7-final-viral-combined_k141_227491\|\|lt2gene | 1354 | No | NA | 1 | 0 | 0 | Not-determined | Genome-fragment | NA | NA | 0 |
| S8_k141_226277 | 1982 | No | NA | 1 | 0 | 1 | Low-quality | Genome-fragment | 1.48 | AAI-based (medium-confidence) | 0 |
| S17-final-viral-combined_k141_65609\|\|full | 1983 | No | NA | 7 | 0 | 0 | Low-quality | Genome-fragment | 3.68 | AAI-based (medium-confidence) | 0 |
| S11-final-viral-combined_k141_105558\|\|full | 1984 | No | NA | 1 | 0 | 1 | Low-quality | Genome-fragment | 4.21 | AAI-based (medium-confidence) | 0 |
| S7-final-viral-combined_k141_145579\|\|lt2gene | 1358 | No | NA | 1 | 0 | 1 | Not-determined | Genome-fragment | NA | NA | 0 |
| S11-final-viral-combined_k141_31623\|\|lt2gene | 1984 | No | NA | 2 | 2 | 0 | Low-quality | Genome-fragment | 5.19 | AAI-based (medium-confidence) | 0 |
| S1-final-viral-combined_k141_301401\|\|full | 1984 | No | NA | 3 | 0 | 0 | Low-quality | Genome-fragment | 5.54 | AAI-based (medium-confidence) | 0 |
| S7-final-viral-combined_k141_103133\|\|full | 1984 | No | NA | 5 | 2 | 0 | Low-quality | Genome-fragment | 3.58 | AAI-based (high-confidence) | 0 |
| S7-final-viral-combined_k141_44204\|\|lt2gene | 1384 | No | NA | 2 | 0 | 1 | Not-determined | Genome-fragment | NA | NA | 0 |
| S17-final-viral-combined_k141_91219\|\|full | 1984 | No | NA | 4 | 0 | 2 | Low-quality | Genome-fragment | 3.29 | AAI-based (medium-confidence) | 0 |
| S2-final-viral-combined_k141_155413\|\|full | 1984 | No | NA | 4 | 0 | 1 | Low-quality | Genome-fragment | 1.04 | AAI-based (medium-confidence) | 0 |
| S7-final-viral-combined_k141_188446\|\|lt2gene | 1522 | No | NA | 1 | 0 | 0 | Not-determined | Genome-fragment | NA | NA | 0 |
| S8-final-viral-combined_k141_168029\|\|full | 1984 | No | NA | 3 | 0 | 0 | Low-quality | Genome-fragment | 1.81 | AAI-based (high-confidence) | 0 |
| S7-final-viral-combined_k141_423\|\|full | 1985 | No | NA | 3 | 0 | 0 | Low-quality | Genome-fragment | 2.02 | AAI-based (high-confidence) | 0 |
| S7-final-viral-combined_k141_95688\|\|full | 1985 | No | NA | 6 | 0 | 0 | Low-quality | Genome-fragment | 3.58 | AAI-based (high-confidence) | 0 |
| S6_k141_139405 | 1985 | No | NA | 3 | 0 | 3 | Low-quality | Genome-fragment | 0.33 | AAI-based (medium-confidence) | 0 |
| S7-final-viral-combined_k141_157692\|\|lt2gene | 1289 | No | NA | 2 | 0 | 0 | Not-determined | Genome-fragment | NA | NA | 0 |
| S7-final-viral-combined_k141_147075\|\|lt2gene | 2213 | No | NA | 1 | 0 | 0 | Not-determined | Genome-fragment | NA | NA | 0 |
| S9-final-viral-combined_k141_98215\|\|full | 1986 | No | NA | 3 | 2 | 0 | Low-quality | Genome-fragment | 5.23 | AAI-based (high-confidence) | 0 |
| S7-final-viral-combined_k141_96162\|\|lt2gene | 1183 | No | NA | 2 | 0 | 0 | Not-determined | Genome-fragment | NA | NA | 0 |
| S11-final-viral-combined_k141_58014\|\|full | 1986 | No | NA | 3 | 1 | 0 | Low-quality | Genome-fragment | 4.6 | AAI-based (high-confidence) | 0 |
| S1-final-viral-combined_k141_83440\|\|full | 1986 | No | NA | 4 | 0 | 0 | Low-quality | Genome-fragment | 10.32 | AAI-based (high-confidence) | 0 |
| S17-final-viral-combined_k141_130321\|\|full | 1986 | No | NA | 7 | 0 | 0 | Low-quality | Genome-fragment | 5.43 | AAI-based (medium-confidence) | 0 |
| S12-final-viral-combined_k141_175965\|\|full | 1986 | No | NA | 3 | 0 | 1 | Low-quality | Genome-fragment | 3.59 | AAI-based (medium-confidence) | 0 |
| S12-final-viral-combined_k141_365202\|\|full | 1986 | No | NA | 3 | 0 | 0 | Low-quality | Genome-fragment | 5.71 | AAI-based (medium-confidence) | 0 |
| S12-final-viral-combined_k141_351094\|\|full | 1986 | No | NA | 3 | 2 | 0 | Low-quality | Genome-fragment | 4.27 | AAI-based (high-confidence) | 0 |
| S7-final-viral-combined_k141_189219\|\|lt2gene | 5556 | No | NA | 2 | 0 | 2 | Not-determined | Genome-fragment | NA | NA | 0 |
| S7-final-viral-combined_k141_66790\|\|lt2gene | 1415 | No | NA | 1 | 0 | 1 | Not-determined | Genome-fragment | NA | NA | 0 |
| S11-final-viral-combined_k141_146085\|\|full | 1987 | No | NA | 4 | 0 | 0 | Low-quality | Genome-fragment | 3.69 | AAI-based (high-confidence) | 0 |
| S1-final-viral-combined_k141_53356\|\|full | 1987 | No | NA | 4 | 1 | 0 | Low-quality | Genome-fragment | 1.45 | AAI-based (high-confidence) | 0 |
| S5-final-viral-combined_k141_193824\|\|full | 1987 | No | NA | 5 | 0 | 1 | Low-quality | Genome-fragment | 14.86 | AAI-based (medium-confidence) | 0 |
| S17-final-viral-combined_k141_185894\|\|full | 1987 | No | NA | 4 | 0 | 1 | Low-quality | Genome-fragment | 2.55 | AAI-based (medium-confidence) | 0 |
| S12-final-viral-combined_k141_183326\|\|full | 1987 | No | NA | 3 | 1 | 0 | Low-quality | Genome-fragment | 6.93 | AAI-based (high-confidence) | 0 |
| S2-final-viral-combined_k141_220657\|\|lt2gene | 1987 | No | NA | 2 | 2 | 0 | Low-quality | Genome-fragment | 6.98 | HMM-based (lower-bound) | 0 |
| S6_k141_133965 | 1987 | No | NA | 2 | 1 | 0 | Low-quality | Genome-fragment | 32.47 | AAI-based (high-confidence) | 0 |
| S5-final-viral-combined_k141_64198\|\|full | 1988 | No | NA | 4 | 0 | 0 | Low-quality | Genome-fragment | 4.52 | AAI-based (medium-confidence) | 0 |
| S17-final-viral-combined_k141_132870\|\|full | 1988 | No | NA | 4 | 0 | 0 | Low-quality | Genome-fragment | 3.43 | AAI-based (medium-confidence) | 0 |
| S6_k141_52072 | 1988 | No | NA | 1 | 1 | 0 | Low-quality | Genome-fragment | 2.33 | AAI-based (high-confidence) | 0 |
| S1-final-viral-combined_k141_134484\|\|full | 1989 | No | NA | 2 | 0 | 1 | Low-quality | Genome-fragment | 0.96 | AAI-based (medium-confidence) | 0 |
| S7-final-viral-combined_k141_107242\|\|lt2gene | 1121 | No | NA | 1 | 0 | 0 | Not-determined | Genome-fragment | NA | NA | 0 |
| S5-final-viral-combined_k141_451\|\|full | 1989 | No | NA | 4 | 0 | 0 | Low-quality | Genome-fragment | 3.16 | AAI-based (high-confidence) | 0 |
| S7-final-viral-combined_k141_152753\|\|full | 1989 | No | NA | 5 | 1 | 0 | Low-quality | Genome-fragment | 3.88 | AAI-based (high-confidence) | 0 |
| S12-final-viral-combined_k141_293143\|\|full | 1989 | No | NA | 3 | 0 | 2 | Low-quality | Genome-fragment | 3.79 | AAI-based (medium-confidence) | 0 |
| S7-final-viral-combined_k141_67383\|\|lt2gene | 1350 | No | NA | 1 | 0 | 1 | Not-determined | Genome-fragment | NA | NA | 0 |
| S9-final-viral-combined_k141_104477\|\|full | 1990 | No | NA | 4 | 0 | 0 | Low-quality | Genome-fragment | 4.62 | AAI-based (medium-confidence) | 0 |
| S1-final-viral-combined_k141_295109\|\|full | 1990 | No | NA | 2 | 0 | 1 | Low-quality | Genome-fragment | 2.4 | AAI-based (high-confidence) | 0 |
| S1-final-viral-combined_k141_152529\|\|full | 1991 | No | NA | 2 | 0 | 0 | Low-quality | Genome-fragment | 1.94 | AAI-based (medium-confidence) | 0 |
| S1-final-viral-combined_k141_370556\|\|full | 1991 | No | NA | 3 | 1 | 0 | Low-quality | Genome-fragment | 4.23 | AAI-based (medium-confidence) | 0 |
| S5-final-viral-combined_k141_83668\|\|full | 1991 | No | NA | 2 | 0 | 0 | Low-quality | Genome-fragment | 3.46 | AAI-based (medium-confidence) | 0 |
| S7-final-viral-combined_k141_68276\|\|lt2gene | 1273 | No | NA | 2 | 0 | 1 | Not-determined | Genome-fragment | NA | NA | 0 |
| S7-final-viral-combined_k141_129183\|\|lt2gene | 1540 | No | NA | 1 | 0 | 0 | Not-determined | Genome-fragment | NA | NA | 0 |
| S2-final-viral-combined_k141_724\|\|full | 1991 | No | NA | 4 | 1 | 0 | Low-quality | Genome-fragment | 3.36 | AAI-based (medium-confidence) | 0 |
| S4_k141_155005 | 1991 | No | NA | 3 | 0 | 0 | Low-quality | Genome-fragment | 3.16 | AAI-based (high-confidence) | 0 |
| S4_k141_33294 | 1991 | No | NA | 2 | 0 | 0 | Low-quality | Genome-fragment | 4.46 | AAI-based (high-confidence) | 0 |
| S7-final-viral-combined_k141_211419\|\|lt2gene | 1519 | No | NA | 1 | 0 | 0 | Not-determined | Genome-fragment | NA | NA | 0 |
| S6-final-viral-combined_k141_140296\|\|full | 1991 | No | NA | 4 | 2 | 0 | Low-quality | Genome-fragment | 3.29 | AAI-based (high-confidence) | 0 |
| S7-final-viral-combined_k141_98578\|\|lt2gene | 3208 | No | NA | 2 | 0 | 1 | Not-determined | Genome-fragment | NA | NA | 0 |
| S6_k141_22125 | 1991 | No | NA | 3 | 0 | 0 | Low-quality | Genome-fragment | 3.96 | AAI-based (high-confidence) | 0 |
| S9-final-viral-combined_k141_104976\|\|full | 1992 | No | NA | 3 | 0 | 2 | Low-quality | Genome-fragment | 2.48 | AAI-based (medium-confidence) | 0 |
| S11-final-viral-combined_k141_282639\|\|full | 1992 | No | NA | 4 | 1 | 0 | Low-quality | Genome-fragment | 1.29 | HMM-based (lower-bound) | 0 |
| S5-final-viral-combined_k141_23736\|\|full | 1992 | No | NA | 4 | 0 | 0 | Low-quality | Genome-fragment | 4.75 | AAI-based (high-confidence) | 0 |
| S7-final-viral-combined_k141_58361\|\|lt2gene | 1513 | No | NA | 1 | 0 | 1 | Not-determined | Genome-fragment | NA | NA | 0 |
| S2-final-viral-combined_k141_1718\|\|full | 1992 | No | NA | 7 | 0 | 0 | Low-quality | Genome-fragment | 3.29 | AAI-based (high-confidence) | 0 |
| S2-final-viral-combined_k141_178341\|\|full | 1992 | No | NA | 2 | 0 | 2 | Low-quality | Genome-fragment | 1.65 | AAI-based (high-confidence) | 0 |
| S9-final-viral-combined_k141_113241\|\|lt2gene | 1993 | No | NA | 2 | 2 | 0 | Low-quality | Genome-fragment | 4.18 | AAI-based (high-confidence) | 0 |
| S17-k141_62084 | 1993 | No | NA | 1 | 0 | 1 | Low-quality | Genome-fragment | 2.35 | AAI-based (medium-confidence) | 0 |
| S12-final-viral-combined_k141_205350\|\|full | 1993 | No | NA | 2 | 2 | 0 | Low-quality | Genome-fragment | 4.36 | AAI-based (high-confidence) | 0 |
| S4-final-viral-combined_k141_42490\|\|lt2gene | 1993 | No | NA | 1 | 0 | 0 | Low-quality | Genome-fragment | 4.6 | AAI-based (medium-confidence) | 0 |
| S8-final-viral-combined_k141_20559\|\|full | 1993 | No | NA | 3 | 2 | 0 | Low-quality | Genome-fragment | 4.83 | AAI-based (medium-confidence) | 0 |
| S9_k141_126987 | 1994 | No | NA | 4 | 3 | 0 | Low-quality | Genome-fragment | 6.01 | AAI-based (high-confidence) | 0 |
| S11-final-viral-combined_k141_35160\|\|full | 1994 | No | NA | 3 | 0 | 0 | Low-quality | Genome-fragment | 4.02 | AAI-based (high-confidence) | 0 |
| S17-final-viral-combined_k141_158013\|\|full | 1994 | No | NA | 4 | 0 | 0 | Low-quality | Genome-fragment | 3.73 | AAI-based (high-confidence) | 0 |
| S12-final-viral-combined_k141_34862\|\|full | 1994 | No | NA | 5 | 1 | 0 | Low-quality | Genome-fragment | 2.3 | HMM-based (lower-bound) | 0 |
| S12-final-viral-combined_k141_255843\|\|lt2gene | 1994 | No | NA | 2 | 2 | 0 | Low-quality | Genome-fragment | 2.08 | AAI-based (medium-confidence) | 0 |
| S2-final-viral-combined_k141_157818\|\|full | 1994 | No | NA | 2 | 0 | 2 | Low-quality | Genome-fragment | 0.57 | AAI-based (high-confidence) | 0 |
| S2_k141_123084 | 1994 | No | NA | 3 | 0 | 2 | Low-quality | Genome-fragment | 2.05 | AAI-based (medium-confidence) | 0 |
| S7-final-viral-combined_k141_6625\|\|full | 1995 | No | NA | 4 | 0 | 1 | Low-quality | Genome-fragment | 1.42 | AAI-based (medium-confidence) | 0 |
| S7-final-viral-combined_k141_232556\|\|full | 1995 | No | NA | 3 | 0 | 2 | Low-quality | Genome-fragment | 2.89 | AAI-based (medium-confidence) | 0 |
| S12-final-viral-combined_k141_297814\|\|full | 1995 | No | NA | 3 | 0 | 1 | Low-quality | Genome-fragment | 2.55 | AAI-based (medium-confidence) | 0 |
| S12-final-viral-combined_k141_334471\|\|full | 1995 | No | NA | 3 | 0 | 2 | Low-quality | Genome-fragment | 0.99 | AAI-based (medium-confidence) | 0 |
| S7-final-viral-combined_k141_182617\|\|lt2gene | 1268 | No | NA | 2 | 0 | 0 | Not-determined | Genome-fragment | NA | NA | 0 |
| S12_k141_203581 | 1995 | No | NA | 3 | 0 | 2 | Low-quality | Genome-fragment | 1.79 | AAI-based (high-confidence) | 0 |
| S2-final-viral-combined_k141_239400\|\|full | 1995 | No | NA | 4 | 0 | 0 | Low-quality | Genome-fragment | 4.66 | AAI-based (medium-confidence) | 0 |
| S6-final-viral-combined_k141_134397\|\|full | 1995 | No | NA | 3 | 0 | 2 | Low-quality | Genome-fragment | 0.53 | AAI-based (medium-confidence) | 0 |
| S8-final-viral-combined_k141_86931\|\|lt2gene | 1995 | No | NA | 1 | 1 | 0 | Low-quality | Genome-fragment | 3.32 | HMM-based (lower-bound) | 0 |
| S9-final-viral-combined_k141_94238\|\|full | 1996 | No | NA | 4 | 0 | 0 | Low-quality | Genome-fragment | 3.3 | AAI-based (high-confidence) | 0 |
| S11-final-viral-combined_k141_13592\|\|full | 1996 | No | NA | 3 | 0 | 2 | Low-quality | Genome-fragment | 2.56 | AAI-based (medium-confidence) | 0 |
| S7-final-viral-combined_k141_62257\|\|full | 1996 | No | NA | 2 | 0 | 0 | Low-quality | Genome-fragment | 3.72 | AAI-based (medium-confidence) | 0 |
| S17-final-viral-combined_k141_124148\|\|full | 1996 | No | NA | 3 | 0 | 0 | Low-quality | Genome-fragment | 3.73 | AAI-based (medium-confidence) | 0 |
| S12_k141_366298 | 1996 | No | NA | 4 | 2 | 0 | Low-quality | Genome-fragment | 1.09 | AAI-based (high-confidence) | 0 |
| S2-final-viral-combined_k141_192755\|\|full | 1996 | No | NA | 3 | 0 | 2 | Low-quality | Genome-fragment | 3.45 | AAI-based (medium-confidence) | 0 |
| S7-final-viral-combined_k141_19381\|\|lt2gene | 1400 | No | NA | 2 | 0 | 0 | Not-determined | Genome-fragment | NA | NA | 0 |
| S2-final-viral-combined_k141_113401\|\|full | 1996 | No | NA | 3 | 0 | 2 | Low-quality | Genome-fragment | 1.92 | AAI-based (high-confidence) | 0 |
| S2_k141_108193 | 1996 | No | NA | 6 | 1 | 0 | Low-quality | Genome-fragment | 3.09 | AAI-based (high-confidence) | 0 |
| S7-final-viral-combined_k141_50989\|\|lt2gene | 1017 | No | NA | 1 | 0 | 0 | Not-determined | Genome-fragment | NA | NA | 0 |
| S4-final-viral-combined_k141_25262\|\|full | 1996 | No | NA | 3 | 1 | 0 | Low-quality | Genome-fragment | 3.02 | HMM-based (lower-bound) | 0 |
| S7-final-viral-combined_k141_152594\|\|lt2gene | 2201 | No | NA | 1 | 0 | 0 | Not-determined | Genome-fragment | NA | NA | 0 |
| S7-final-viral-combined_k141_91314\|\|lt2gene | 2211 | No | NA | 2 | 0 | 0 | Not-determined | Genome-fragment | NA | NA | 0 |
| S7-final-viral-combined_k141_80959\|\|lt2gene | 1852 | No | NA | 1 | 0 | 0 | Not-determined | Genome-fragment | NA | NA | 0 |
| S8-final-viral-combined_k141_128178\|\|full | 1996 | No | NA | 3 | 0 | 0 | Low-quality | Genome-fragment | 12.9 | AAI-based (high-confidence) | 0 |
| S7_k141_183808 | 1025 | No | NA | 1 | 0 | 1 | Not-determined | Genome-fragment | NA | NA | 0 |
| S8-final-viral-combined_k141_112189\|\|full | 1996 | No | NA | 3 | 0 | 1 | Low-quality | Genome-fragment | 1.15 | AAI-based (medium-confidence) | 0 |
| S7_k141_122547 | 1073 | No | NA | 3 | 0 | 2 | Not-determined | Genome-fragment | NA | NA | 0 |
| S7_k141_163405 | 2731 | No | NA | 1 | 0 | 0 | Not-determined | Genome-fragment | NA | NA | 0 |
| S7_k141_112344 | 5540 | No | NA | 9 | 0 | 0 | Not-determined | Genome-fragment | NA | NA | 0 |
| S7_k141_163418 | 1124 | No | NA | 1 | 0 | 1 | Not-determined | Genome-fragment | NA | NA | 0 |
| S7_k141_30670 | 1965 | No | NA | 4 | 0 | 0 | Not-determined | Genome-fragment | NA | NA | 0 |
| S8-final-viral-combined_k141_201886\|\|lt2gene | 1996 | No | NA | 1 | 0 | 0 | Low-quality | Genome-fragment | 1.31 | AAI-based (high-confidence) | 0 |
| S7_k141_143003 | 3547 | No | NA | 6 | 0 | 2 | Not-determined | Genome-fragment | NA | NA | 0 |
| S7_k141_30678 | 1736 | No | NA | 2 | 0 | 1 | Not-determined | Genome-fragment | NA | NA | 0 |
| S7_k141_153228 | 1030 | No | NA | 1 | 0 | 1 | Not-determined | Genome-fragment | NA | NA | 0 |
| S7_k141_132812 | 1090 | No | NA | 2 | 0 | 0 | Not-determined | Genome-fragment | NA | NA | 0 |
| S7_k141_102180 | 5794 | No | NA | 3 | 0 | 2 | Not-determined | Genome-fragment | NA | NA | 0 |
| S7_k141_30716 | 1025 | No | NA | 2 | 0 | 2 | Not-determined | Genome-fragment | NA | NA | 0 |
| S7_k141_163480 | 1207 | No | NA | 1 | 0 | 1 | Not-determined | Genome-fragment | NA | NA | 0 |
| S11-final-viral-combined_k141_280469\|\|full | 1997 | No | NA | 3 | 0 | 1 | Low-quality | Genome-fragment | 1.49 | AAI-based (high-confidence) | 0 |
| S1_k141_121933 | 1997 | No | NA | 1 | 0 | 1 | Low-quality | Genome-fragment | 0.74 | AAI-based (high-confidence) | 0 |
| S7_k141_214565 | 1522 | No | NA | 2 | 0 | 1 | Not-determined | Genome-fragment | NA | NA | 0 |
| S7_k141_91989 | 1594 | No | NA | 3 | 0 | 3 | Not-determined | Genome-fragment | NA | NA | 0 |
| S7_k141_112441 | 5733 | No | NA | 6 | 0 | 2 | Not-determined | Genome-fragment | NA | NA | 0 |
| S7_k141_65168 | 1997 | No | NA | 3 | 0 | 0 | Low-quality | Genome-fragment | 4.42 | AAI-based (high-confidence) | 0 |
| S7_k141_112444 | 3139 | No | NA | 6 | 0 | 1 | Not-determined | Genome-fragment | NA | NA | 0 |
| S7_k141_20464 | 1269 | No | NA | 2 | 0 | 2 | Not-determined | Genome-fragment | NA | NA | 0 |
| S7_k141_235012 | 4235 | No | NA | 3 | 0 | 2 | Not-determined | Genome-fragment | NA | NA | 0 |
| S7_k141_162 | 4707 | No | NA | 5 | 0 | 4 | Not-determined | Genome-fragment | NA | NA | 0 |
| S17-final-viral-combined_k141_1686\|\|full | 1997 | No | NA | 3 | 0 | 0 | Low-quality | Genome-fragment | 2.41 | AAI-based (high-confidence) | 0 |
| S7_k141_194135 | 1128 | No | NA | 1 | 0 | 0 | Not-determined | Genome-fragment | NA | NA | 0 |
| S12-final-viral-combined_k141_89477\|\|full | 1997 | No | NA | 2 | 0 | 1 | Low-quality | Genome-fragment | 1.72 | AAI-based (medium-confidence) | 0 |
| S7_k141_204332 | 5867 | No | NA | 3 | 0 | 1 | Not-determined | Genome-fragment | NA | NA | 0 |
| S7_k141_112497 | 1005 | No | NA | 2 | 0 | 1 | Not-determined | Genome-fragment | NA | NA | 0 |
| S7_k141_40979 | 1202 | No | NA | 1 | 0 | 1 | Not-determined | Genome-fragment | NA | NA | 0 |
| S7_k141_40985 | 1589 | No | NA | 1 | 0 | 1 | Not-determined | Genome-fragment | NA | NA | 0 |
| S12-final-viral-combined_k141_393616\|\|full | 1997 | No | NA | 4 | 0 | 0 | Low-quality | Genome-fragment | 2.71 | AAI-based (medium-confidence) | 0 |
| S7_k141_20521 | 1532 | No | NA | 1 | 0 | 0 | Not-determined | Genome-fragment | NA | NA | 0 |

| | | | | | | | | | | | | | |
|---|---|---|---|---|---|---|---|---|---|---|---|---|---|
| S7_k141_214653 | 1386 | No | NA | | 2 | 0 | 2 | Not-determined | Genome-fragment | NA | | NA | 0 |
| S7_k141_20533 | 5033 | No | NA | | 2 | 0 | 1 | Not-determined | Genome-fragment | NA | | NA | 0 |
| S7_k141_102307 | 1184 | No | NA | | 1 | 0 | 0 | Not-determined | Genome-fragment | NA | | NA | 0 |
| S7_k141_153386 | 1292 | No | NA | | 1 | 0 | 0 | Not-determined | Genome-fragment | NA | | NA | 0 |
| S7_k141_143216 | 1310 | No | NA | | 3 | 0 | 1 | Not-determined | Genome-fragment | NA | | NA | 0 |
| S7_k141_61465 | 2677 | No | NA | | 5 | 0 | 0 | Not-determined | Genome-fragment | NA | | NA | 0 |
| S7_k141_102323 | 2911 | No | NA | | 2 | 0 | 0 | Not-determined | Genome-fragment | NA | | NA | 0 |
| S7_k141_61483 | 3946 | No | NA | | 1 | 0 | 0 | Not-determined | Genome-fragment | NA | | NA | 0 |
| S6-final-viral-combined_k141_79331\|\|full | 1997 | No | NA | | 2 | 1 | 0 | Low-quality | Genome-fragment | | 3.95 | AAI-based (medium-confidence) | 0 |
| S7_k141_235093 | 1790 | No | NA | | 2 | 0 | 1 | Not-determined | Genome-fragment | NA | | NA | 0 |
| S7_k141_10329 | 3686 | No | NA | | 4 | 0 | 1 | Not-determined | Genome-fragment | NA | | NA | 0 |
| S7_k141_20555 | 5526 | No | NA | | 8 | 0 | 2 | Not-determined | Genome-fragment | NA | | NA | 0 |
| S7_k141_20561 | 2323 | No | NA | | 2 | 0 | 0 | Not-determined | Genome-fragment | NA | | NA | 0 |
| S11-final-viral-combined_k141_314041\|\|full | 1998 | No | NA | | 4 | 0 | 0 | Low-quality | Genome-fragment | | 5.83 | AAI-based (high-confidence) | 0 |
| S11-final-viral-combined_k141_94533\|\|lt2gene | 1998 | No | NA | | 2 | 1 | 0 | Low-quality | Genome-fragment | | 3.51 | AAI-based (high-confidence) | 0 |
| S7_k141_122789 | 3756 | No | NA | | 1 | 0 | 1 | Not-determined | Genome-fragment | NA | | NA | 0 |
| S7_k141_92150 | 1047 | No | NA | | 1 | 0 | 0 | Not-determined | Genome-fragment | NA | | NA | 0 |
| S7_k141_194259 | 2111 | No | NA | | 1 | 0 | 1 | Not-determined | Genome-fragment | NA | | NA | 0 |
| S3-final-viral-combined_k141_83275\|\|full | 1998 | No | NA | | 4 | 0 | 0 | Low-quality | Genome-fragment | | 4.97 | AAI-based (high-confidence) | 0 |
| S7_k141_122832 | 1540 | No | NA | | 1 | 0 | 1 | Not-determined | Genome-fragment | NA | | NA | 0 |
| S7_k141_30869 | 1122 | No | NA | | 1 | 0 | 0 | Not-determined | Genome-fragment | NA | | NA | 0 |
| S5-final-viral-combined_k141_149661\|\|full | 1998 | No | NA | | 4 | 0 | 3 | Low-quality | Genome-fragment | | 2.66 | AAI-based (medium-confidence) | 0 |
| S7_k141_163722 | 1412 | No | NA | | 3 | 0 | 1 | Not-determined | Genome-fragment | NA | | NA | 0 |
| S7_k141_163742 | 1138 | No | NA | | 1 | 0 | 0 | Not-determined | Genome-fragment | NA | | NA | 0 |
| S7_k141_10443 | 1558 | No | NA | | 1 | 0 | 0 | Not-determined | Genome-fragment | NA | | NA | 0 |
| S7_k141_235189 | 2036 | No | NA | | 2 | 0 | 2 | Not-determined | Genome-fragment | NA | | NA | 0 |
| S7_k141_92258 | 1178 | No | NA | | 1 | 0 | 0 | Not-determined | Genome-fragment | NA | | NA | 0 |
| S7_k141_122846 | 27928 | No | NA | | 31 | 0 | 10 | Not-determined | Genome-fragment | NA | | NA | 0 |
| S7_k141_20673 | 4680 | No | NA | | 7 | 0 | 1 | Not-determined | Genome-fragment | NA | | NA | 0 |
| S7-final-viral-combined_k141_33796\|\|full | 1998 | No | NA | | 2 | 0 | 2 | Low-quality | Genome-fragment | | 1.48 | AAI-based (high-confidence) | 0 |
| S7_k141_92305 | 1404 | No | NA | | 2 | 0 | 1 | Not-determined | Genome-fragment | NA | | NA | 0 |
| S7_k141_194410 | 1560 | No | NA | | 2 | 0 | 0 | Not-determined | Genome-fragment | NA | | NA | 0 |
| S7_k141_163832 | 2048 | No | NA | | 3 | 0 | 1 | Not-determined | Genome-fragment | NA | | NA | 0 |
| S12-final-viral-combined_k141_253250\|\|full | 1998 | No | NA | | 4 | 0 | 0 | Low-quality | Genome-fragment | | 2.42 | AAI-based (medium-confidence) | 0 |
| S7_k141_81945 | 3229 | No | NA | | 4 | 0 | 4 | Not-determined | Genome-fragment | NA | | NA | 0 |
| S7_k141_10485 | 1804 | No | NA | | 3 | 0 | 1 | Not-determined | Genome-fragment | NA | | NA | 0 |
| S7_k141_173939 | 3036 | No | NA | | 1 | 0 | 1 | Not-determined | Genome-fragment | NA | | NA | 0 |
| S2_k141_240515 | 1998 | No | NA | | 4 | 0 | 2 | Low-quality | Genome-fragment | | 1.28 | AAI-based (medium-confidence) | 0 |
| S7_k141_143329 | 3822 | No | NA | | 2 | 0 | 2 | Not-determined | Genome-fragment | NA | | NA | 0 |
| S7_k141_194489 | 1092 | No | NA | | 2 | 0 | 1 | Not-determined | Genome-fragment | NA | | NA | 0 |
| S7_k141_163898 | 1816 | No | NA | | 3 | 0 | 0 | Not-determined | Genome-fragment | NA | | NA | 0 |
| S7_k141_10556 | 12514 | No | NA | | 13 | 0 | 11 | Not-determined | Genome-fragment | NA | | NA | 0 |
| S7_k141_225182 | 1009 | No | NA | | 1 | 0 | 0 | Not-determined | Genome-fragment | NA | | NA | 0 |
| S12-final-viral-combined_k141_28905\|\|full | 1999 | No | NA | | 2 | 1 | 0 | Low-quality | Genome-fragment | | 1.52 | AAI-based (medium-confidence) | 0 |
| S7_k141_204611 | 1110 | No | NA | | 2 | 0 | 0 | Not-determined | Genome-fragment | NA | | NA | 0 |
| S7_k141_528 | 1170 | No | NA | | 2 | 0 | 0 | Not-determined | Genome-fragment | NA | | NA | 0 |
| S7_k141_214859 | 1104 | No | NA | | 2 | 0 | 1 | Not-determined | Genome-fragment | NA | | NA | 0 |
| S8_k141_122060 | 1999 | No | NA | | 3 | 0 | 2 | Low-quality | Genome-fragment | | 2.16 | AAI-based (high-confidence) | 0 |
| S7_k141_71719 | 5393 | No | NA | | 2 | 0 | 1 | Not-determined | Genome-fragment | NA | | NA | 0 |
| S7_k141_214875 | 3359 | No | NA | | 4 | 0 | 2 | Not-determined | Genome-fragment | NA | | NA | 0 |
| S7_k141_143480 | 1129 | No | NA | | 1 | 0 | 0 | Not-determined | Genome-fragment | NA | | NA | 0 |
| S7_k141_214886 | 1076 | No | NA | | 1 | 0 | 1 | Not-determined | Genome-fragment | NA | | NA | 0 |
| S7_k141_235426 | 1420 | No | NA | | 2 | 0 | 0 | Not-determined | Genome-fragment | NA | | NA | 0 |
| S7_k141_214898 | 1145 | No | NA | | 1 | 0 | 1 | Not-determined | Genome-fragment | NA | | NA | 0 |
| S7_k141_174068 | 2391 | No | NA | | 3 | 0 | 0 | Not-determined | Genome-fragment | NA | | NA | 0 |
| S3-final-viral-combined_k141_11848\|\|full | 2000 | No | NA | | 4 | 0 | 0 | Low-quality | Genome-fragment | | 3.01 | AAI-based (medium-confidence) | 0 |
| S7_k141_204664 | 4099 | No | NA | | 2 | 0 | 2 | Not-determined | Genome-fragment | NA | | NA | 0 |
| S5-final-viral-combined_k141_115870\|\|full | 2000 | No | NA | | 5 | 0 | 0 | Low-quality | Genome-fragment | | 4.48 | AAI-based (medium-confidence) | 0 |
| S7_k141_92390 | 11420 | No | NA | | 6 | 0 | 2 | Not-determined | Genome-fragment | NA | | NA | 0 |
| S7_k141_61844 | 1151 | No | NA | | 3 | 0 | 0 | Not-determined | Genome-fragment | NA | | NA | 0 |
| S7_k141_184320 | 1301 | No | NA | | 1 | 0 | 1 | Not-determined | Genome-fragment | NA | | NA | 0 |
| S7_k141_10674 | 1951 | No | NA | | 2 | 0 | 0 | Not-determined | Genome-fragment | NA | | NA | 0 |
| S7_k141_174122 | 1819 | No | NA | | 1 | 0 | 1 | Not-determined | Genome-fragment | NA | | NA | 0 |
| S7_k141_10680 | 1918 | No | NA | | 1 | 0 | 1 | Not-determined | Genome-fragment | NA | | NA | 0 |
| S5-final-viral-combined_k141_198391\|\|full | 2000 | No | NA | | 2 | 0 | 1 | Low-quality | Genome-fragment | | 3.61 | AAI-based (medium-confidence) | 0 |
| S5-final-viral-combined_k141_162895\|\|full | 2000 | No | NA | | 2 | 0 | 0 | Low-quality | Genome-fragment | | 9.15 | AAI-based (medium-confidence) | 0 |
| S7_k141_133321 | 1243 | No | NA | | 1 | 0 | 0 | Not-determined | Genome-fragment | NA | | NA | 0 |
| S7_k141_225349 | 1415 | No | NA | | 1 | 0 | 0 | Not-determined | Genome-fragment | NA | | NA | 0 |
| S7_k141_71848 | 3276 | No | NA | | 3 | 0 | 0 | Not-determined | Genome-fragment | NA | | NA | 0 |
| S7_k141_215085 | 1316 | No | NA | | 2 | 0 | 1 | Not-determined | Genome-fragment | NA | | NA | 0 |
| S17-final-viral-combined_k141_110560\|\|full | 2000 | No | NA | | 3 | 0 | 1 | Low-quality | Genome-fragment | | 2.6 | AAI-based (medium-confidence) | 0 |
| S7_k141_706 | 1282 | No | NA | | 1 | 0 | 0 | Not-determined | Genome-fragment | NA | | NA | 0 |
| S7_k141_10756 | 10000 | No | NA | | 7 | 0 | 4 | Not-determined | Genome-fragment | NA | | NA | 0 |
| S17-k141_53522 | 2000 | No | NA | | 5 | 2 | 0 | Low-quality | Genome-fragment | | 4.96 | AAI-based (high-confidence) | 0 |
| S7_k141_71892 | 1445 | No | NA | | 1 | 0 | 0 | Not-determined | Genome-fragment | NA | | NA | 0 |
| S7_k141_235564 | 1292 | No | NA | | 1 | 0 | 1 | Not-determined | Genome-fragment | NA | | NA | 0 |
| S7_k141_82201 | 1723 | No | NA | | 2 | 0 | 1 | Not-determined | Genome-fragment | NA | | NA | 0 |
| S7_k141_31171 | 1034 | No | NA | | 2 | 0 | 0 | Not-determined | Genome-fragment | NA | | NA | 0 |
| S7_k141_71932 | 3638 | No | NA | | 2 | 0 | 1 | Not-determined | Genome-fragment | NA | | NA | 0 |
| S7_k141_194877 | 1525 | No | NA | | 1 | 0 | 0 | Not-determined | Genome-fragment | NA | | NA | 0 |
| S7_k141_102707 | 1021 | No | NA | | 2 | 0 | 1 | Not-determined | Genome-fragment | NA | | NA | 0 |
| S7_k141_164101 | 4786 | No | NA | | 3 | 0 | 3 | Not-determined | Genome-fragment | NA | | NA | 0 |
| S7_k141_113137 | 37141 | No | NA | | 52 | 14 | 0 | High-quality | High-quality | | 100 | AAI-based (medium-confidence) | 0 |
| S7_k141_194924 | 3782 | No | NA | | 4 | 0 | 0 | Not-determined | Genome-fragment | NA | | NA | 0 |
| S7_k141_123136 | 1367 | No | NA | | 2 | 0 | 2 | Not-determined | Genome-fragment | NA | | NA | 0 |
| S6-final-viral-combined_k141_106788\|\|full | 2000 | No | NA | | 3 | 0 | 1 | Low-quality | Genome-fragment | | 2.83 | AAI-based (medium-confidence) | 0 |
| S7_k141_801 | 1220 | No | NA | | 2 | 0 | 0 | Not-determined | Genome-fragment | NA | | NA | 0 |
| S7_k141_184406 | 2055 | No | NA | | 2 | 0 | 1 | Not-determined | Genome-fragment | NA | | NA | 0 |
| S7_k141_815 | 2351 | No | NA | | 4 | 0 | 4 | Not-determined | Genome-fragment | NA | | NA | 0 |
| S7_k141_154061 | 2661 | No | NA | | 4 | 0 | 0 | Not-determined | Genome-fragment | NA | | NA | 0 |
| S1_k141_306933 | 2001 | No | NA | | 5 | 1 | 0 | Low-quality | Genome-fragment | | 1.54 | AAI-based (medium-confidence) | 0 |
| S7_k141_62004 | 1432 | No | NA | | 4 | 0 | 1 | Not-determined | Genome-fragment | NA | | NA | 0 |
| S7_k141_82282 | 3630 | No | NA | | 6 | 0 | 2 | Not-determined | Genome-fragment | NA | | NA | 0 |
| S3_k141_93798 | 2001 | No | NA | | 5 | 1 | 0 | Low-quality | Genome-fragment | | 4.3 | AAI-based (high-confidence) | 0 |
| S12-final-viral-combined_k141_6836\|\|full | 2001 | No | NA | | 4 | 0 | 0 | Low-quality | Genome-fragment | | 3.09 | AAI-based (high-confidence) | 0 |
| S7_k141_31242 | 1175 | No | NA | | 2 | 0 | 1 | Not-determined | Genome-fragment | NA | | NA | 0 |
| S7_k141_51554 | 6033 | No | NA | | 7 | 0 | 1 | Not-determined | Genome-fragment | NA | | NA | 0 |
| S7_k141_194974 | 1111 | No | NA | | 2 | 0 | 1 | Not-determined | Genome-fragment | NA | | NA | 0 |
| S7_k141_113198 | 9381 | No | NA | | 4 | 0 | 2 | Not-determined | Genome-fragment | NA | | NA | 0 |
| S12_k141_397845 | 2001 | No | NA | | 4 | 3 | 0 | Low-quality | Genome-fragment | | 5.71 | AAI-based (high-confidence) | 0 |
| S7_k141_72028 | 1018 | No | NA | | 2 | 0 | 0 | Not-determined | Genome-fragment | NA | | NA | 0 |
| S7_k141_235730 | 3471 | No | NA | | 5 | 0 | 2 | Not-determined | Genome-fragment | NA | | NA | 0 |
| S7_k141_41370 | 2042 | No | NA | | 1 | 0 | 0 | Not-determined | Genome-fragment | NA | | NA | 0 |
| S7_k141_51615 | 1215 | No | NA | | 1 | 0 | 1 | Not-determined | Genome-fragment | NA | | NA | 0 |
| S7_k141_215370 | 1767 | No | NA | | 2 | 0 | 1 | Not-determined | Genome-fragment | NA | | NA | 0 |
| S7_k141_92551 | 1588 | No | NA | | 1 | 0 | 1 | Not-determined | Genome-fragment | NA | | NA | 0 |
| S9_k141_87397 | 2002 | No | NA | | 1 | 0 | 1 | Low-quality | Genome-fragment | | 0.19 | AAI-based (medium-confidence) | 0 |
| S11-final-viral-combined_k141_196050\|\|full | 2002 | No | NA | | 4 | 0 | 1 | Low-quality | Genome-fragment | | 4.19 | AAI-based (medium-confidence) | 0 |
| S7_k141_154219 | 4554 | No | NA | | 8 | 0 | 8 | Not-determined | Genome-fragment | NA | | NA | 0 |
| S7_k141_113290 | 7915 | No | NA | | 3 | 0 | 1 | Not-determined | Genome-fragment | NA | | NA | 0 |
| S7_k141_164259 | 1101 | No | NA | | 2 | 0 | 1 | Not-determined | Genome-fragment | NA | | NA | 0 |
| S7_k141_31379 | 1437 | No | NA | | 1 | 0 | 0 | Not-determined | Genome-fragment | NA | | NA | 0 |
| S7_k141_133572 | 1032 | No | NA | | 1 | 0 | 1 | Not-determined | Genome-fragment | NA | | NA | 0 |
| S7_k141_51719 | 3169 | No | NA | | 5 | 0 | 2 | Not-determined | Genome-fragment | NA | | NA | 0 |
| S7_k141_225513 | 1517 | No | NA | | 2 | 0 | 0 | Not-determined | Genome-fragment | NA | | NA | 0 |
| S11-final-viral-combined_k141_80551\|\|full | 2002 | No | NA | | 4 | 0 | 0 | Low-quality | Genome-fragment | | 1.09 | AAI-based (medium-confidence) | 0 |
| S7_k141_235737 | 2667 | No | NA | | 2 | 0 | 1 | Not-determined | Genome-fragment | NA | | NA | 0 |
| S7_k141_41525 | 7477 | No | NA | | 6 | 0 | 2 | Not-determined | Genome-fragment | NA | | NA | 0 |
| S7_k141_235787 | 1230 | No | NA | | 3 | 0 | 3 | Not-determined | Genome-fragment | NA | | NA | 0 |
| S1-final-viral-combined_k141_36449\|\|full | 2002 | No | NA | | 4 | 2 | 0 | Low-quality | Genome-fragment | | 3.73 | AAI-based (high-confidence) | 0 |
| S5-final-viral-combined_k141_150440\|\|full | 2002 | Yes | | 215 | 4 | 1 | 1 | Low-quality | Genome-fragment | | 0.14 | HMM-based (lower-bound) | 89.26 |
| S7_k141_31481 | 3410 | No | NA | | 3 | 0 | 1 | Not-determined | Genome-fragment | NA | | NA | 0 |
| S7_k141_215462 | 4455 | No | NA | | 5 | 0 | 3 | Not-determined | Genome-fragment | NA | | NA | 0 |
| S7_k141_31511 | 1375 | No | NA | | 2 | 0 | 2 | Not-determined | Genome-fragment | NA | | NA | 0 |
| S7_k141_92679 | 1418 | No | NA | | 1 | 0 | 1 | Not-determined | Genome-fragment | NA | | NA | 0 |
| S7_k141_31518 | 1354 | No | NA | | 1 | 0 | 1 | Not-determined | Genome-fragment | NA | | NA | 0 |
| S5-final-viral-combined_k141_160274\|\|full | 2002 | Yes | | 1322 | 3 | 2 | 1 | Low-quality | Genome-fragment | | 2.99 | AAI-based (high-confidence) | 33.97 |
| S7_k141_154304 | 1066 | No | NA | | 1 | 0 | 0 | Not-determined | Genome-fragment | NA | | NA | 0 |
| S7-final-viral-combined_k141_198832\|\|full | 2002 | No | NA | | 5 | 1 | 0 | Low-quality | Genome-fragment | | 4.03 | AAI-based (medium-confidence) | 0 |
| S7_k141_225558 | 5759 | No | NA | | 4 | 0 | 3 | Not-determined | Genome-fragment | NA | | NA | 0 |
| S7_k141_41575 | 3404 | No | NA | | 2 | 0 | 1 | Not-determined | Genome-fragment | NA | | NA | 0 |
| S7_k141_174331 | 3494 | No | NA | | 4 | 0 | 0 | Not-determined | Genome-fragment | NA | | NA | 0 |
| S2-final-viral-combined_k141_121224\|\|full | 2002 | No | NA | | 4 | 1 | 0 | Low-quality | Genome-fragment | | 2.69 | HMM-based (lower-bound) | 0 |
| S8-final-viral-combined_k141_224129\|\|full | 2002 | No | NA | | 3 | 0 | 1 | Low-quality | Genome-fragment | | 5.47 | AAI-based (high-confidence) | 0 |
| S11-final-viral-combined_k141_189953\|\|full | 2003 | No | NA | | 3 | 2 | 0 | Low-quality | Genome-fragment | | 2.46 | HMM-based (lower-bound) | 0 |
| S7_k141_225611 | 1166 | No | NA | | 3 | 0 | 2 | Not-determined | Genome-fragment | NA | | NA | 0 |
| S7_k141_102740 | 3458 | No | NA | | 4 | 0 | 3 | Not-determined | Genome-fragment | NA | | NA | 0 |
| S7_k141_82481 | 1679 | No | NA | | 3 | 0 | 3 | Not-determined | Genome-fragment | NA | | NA | 0 |
| S5-final-viral-combined_k141_18046\|\|full | 2003 | No | NA | | 2 | 0 | 2 | Low-quality | Genome-fragment | | 3.29 | AAI-based (medium-confidence) | 0 |
| S7_k141_123427 | 3802 | No | NA | | 6 | 0 | 0 | Not-determined | Genome-fragment | NA | | NA | 0 |
| S7_k141_225638 | 1105 | No | NA | | 1 | 0 | 1 | Not-determined | Genome-fragment | NA | | NA | 0 |
| S7_k141_904 | 1307 | No | NA | | 1 | 0 | 0 | Not-determined | Genome-fragment | NA | | NA | 0 |
| S7_k141_225645 | 5073 | No | NA | | 3 | 0 | 3 | Not-determined | Genome-fragment | NA | | NA | 0 |
| S7_k141_205176 | 5005 | No | NA | | 6 | 0 | 2 | Not-determined | Genome-fragment | NA | | NA | 0 |
| S5-final-viral-combined_k141_157278\|\|full | 2003 | No | NA | | 2 | 0 | 1 | Low-quality | Genome-fragment | | 2.41 | AAI-based (high-confidence) | 0 |
| S7_k141_31575 | 3667 | No | NA | | 1 | 0 | 1 | Not-determined | Genome-fragment | NA | | NA | 0 |
| S7_k141_143652 | 1253 | No | NA | | 2 | 0 | 0 | Not-determined | Genome-fragment | NA | | NA | 0 |
| S17-final-viral-combined_k141_56321\|\|full | 2003 | No | NA | | 2 | 0 | 1 | Low-quality | Genome-fragment | | 3.07 | AAI-based (medium-confidence) | 0 |
| S7_k141_133755 | 1121 | No | NA | | 1 | 0 | 0 | Not-determined | Genome-fragment | NA | | NA | 0 |
| S7_k141_133762 | 4566 | No | NA | | 4 | 0 | 3 | Not-determined | Genome-fragment | NA | | NA | 0 |
| S17-final-viral-combined_k141_113239\|\|full | 2003 | No | NA | | 2 | 0 | 1 | Low-quality | Genome-fragment | | 1.76 | AAI-based (medium-confidence) | 0 |
| S7_k141_41668 | 1184 | No | NA | | 3 | 0 | 1 | Not-determined | Genome-fragment | NA | | NA | 0 |
| S12-final-viral-combined_k141_274620\|\|full | 2003 | No | NA | | 2 | 2 | 0 | Low-quality | Genome-fragment | | 5.13 | AAI-based (medium-confidence) | 0 |
| S7_k141_11257 | 2794 | No | NA | | 1 | 0 | 0 | Not-determined | Genome-fragment | NA | | NA | 0 |
| S7_k141_205337 | 1447 | No | NA | | 2 | 0 | 1 | Not-determined | Genome-fragment | NA | | NA | 0 |
| S7_k141_205339 | 1673 | No | NA | | 3 | 0 | 0 | Not-determined | Genome-fragment | NA | | NA | 0 |
| S7_k141_133834 | 1031 | No | NA | | 1 | 0 | 0 | Not-determined | Genome-fragment | NA | | NA | 0 |
| S7_k141_113506 | 2679 | No | NA | | 3 | 0 | 1 | Not-determined | Genome-fragment | NA | | NA | 0 |
| S7_k141_215701 | 2463 | No | NA | | 2 | 0 | 1 | Not-determined | Genome-fragment | NA | | NA | 0 |
| S4_k141_137075 | 2003 | No | NA | | 2 | 0 | 0 | Low-quality | Genome-fragment | | 2.3 | AAI-based (high-confidence) | 0 |
| S7_k141_154489 | 3381 | No | NA | | 2 | 0 | 1 | Not-determined | Genome-fragment | NA | | NA | 0 |
| S7_k141_195355 | 1442 | No | NA | | 2 | 0 | 0 | Not-determined | Genome-fragment | NA | | NA | 0 |
| S11_k141_235674 | 2004 | No | NA | | 3 | 0 | 2 | Low-quality | Genome-fragment | | 0.92 | AAI-based (medium-confidence) | 0 |
| S17-final-viral-combined_k141_227311\|\|full | 2004 | No | NA | | 3 | 0 | 0 | Low-quality | Genome-fragment | | 3.51 | AAI-based (high-confidence) | 0 |
| S17-k141_201425 | 2004 | No | NA | | 4 | 1 | 0 | Low-quality | Genome-fragment | | 1.09 | HMM-based (lower-bound) | 0 |
| S4-final-viral-combined_k141_127185\|\|full | 2004 | No | NA | | 3 | 0 | 2 | Low-quality | Genome-fragment | | 0.93 | AAI-based (high-confidence) | 0 |
| S7_k141_195377 | 1539 | No | NA | | 3 | 0 | 1 | Not-determined | Genome-fragment | NA | | NA | 0 |
| S7_k141_82711 | 1011 | No | NA | | 1 | 0 | 1 | Not-determined | Genome-fragment | NA | | NA | 0 |
| S7_k141_164544 | 2699 | No | NA | | 5 | 0 | 0 | Not-determined | Genome-fragment | NA | | NA | 0 |
| S7_k141_225807 | 1092 | No | NA | | 2 | 0 | 2 | Not-determined | Genome-fragment | NA | | NA | 0 |
| S7_k141_164583 | 3898 | No | NA | | 3 | 0 | 2 | Not-determined | Genome-fragment | NA | | NA | 0 |
| S7_k141_21224 | 1656 | No | NA | | 2 | 0 | 0 | Not-determined | Genome-fragment | NA | | NA | 0 |
| S1-final-viral-combined_k141_370387\|\|full | 2005 | No | NA | | 2 | 2 | 0 | Low-quality | Genome-fragment | | 5.78 | AAI-based (high-confidence) | 0 |
| S7_k141_215729 | 6187 | No | NA | | 3 | 0 | 2 | Not-determined | Genome-fragment | NA | | NA | 0 |
| S7_k141_164634 | 1427 | No | NA | | 1 | 0 | 0 | Not-determined | Genome-fragment | NA | | NA | 0 |
| S7_k141_123692 | 1082 | No | NA | | 1 | 0 | 0 | Not-determined | Genome-fragment | NA | | NA | 0 |
| S7_k141_123693 | 11769 | No | NA | | 8 | 0 | 3 | Not-determined | Genome-fragment | NA | | NA | 0 |
| S7_k141_225903 | 5866 | No | NA | | 3 | 0 | 1 | Not-determined | Genome-fragment | NA | | NA | 0 |
| S7_k141_174676 | 3542 | No | NA | | 4 | 0 | 2 | Not-determined | Genome-fragment | NA | | NA | 0 |
| S7_k141_164672 | 1100 | No | NA | | 1 | 0 | 1 | Not-determined | Genome-fragment | NA | | NA | 0 |
| S7_k141_225908 | 1122 | No | NA | | 1 | 0 | 1 | Not-determined | Genome-fragment | NA | | NA | 0 |
| S7-final-viral-combined_k141_227033\|\|full | 2005 | No | NA | | 2 | 0 | 1 | Low-quality | Genome-fragment | | 1.58 | AAI-based (high-confidence) | 0 |
| S7_k141_225920 | 8884 | No | NA | | 9 | 0 | 2 | Not-determined | Genome-fragment | NA | | NA | 0 |
| S7_k141_11347 | 3686 | No | NA | | 6 | 0 | 0 | Not-determined | Genome-fragment | NA | | NA | 0 |
| S7_k141_154584 | 3720 | No | NA | | 6 | 0 | 2 | Not-determined | Genome-fragment | NA | | NA | 0 |
| S7_k141_92768 | 3201 | No | NA | | 5 | 0 | 5 | Not-determined | Genome-fragment | NA | | NA | 0 |
| S7_k141_1137 | 1120 | No | NA | | 1 | 0 | 1 | Not-determined | Genome-fragment | NA | | NA | 0 |
| S7_k141_205284 | 2005 | No | NA | | 4 | 2 | 0 | Low-quality | Genome-fragment | | 3.96 | AAI-based (high-confidence) | 0 |
| S7_k141_11383 | 3457 | No | NA | | 4 | 0 | 3 | Not-determined | Genome-fragment | NA | | NA | 0 |
| S7_k141_123753 | 4745 | No | NA | | 3 | 0 | 1 | Not-determined | Genome-fragment | NA | | NA | 0 |
| S7_k141_236125 | 1274 | No | NA | | 3 | 0 | 0 | Not-determined | Genome-fragment | NA | | NA | 0 |
| S7_k141_174739 | 3265 | No | NA | | 1 | 0 | 1 | Not-determined | Genome-fragment | NA | | NA | 0 |
| S7_k141_174767 | 3509 | No | NA | | 6 | 0 | 0 | Not-determined | Genome-fragment | NA | | NA | 0 |
| S7_k141_195602 | 6192 | No | NA | | 6 | 0 | 0 | Not-determined | Genome-fragment | NA | | NA | 0 |
| S7_k141_174769 | 5047 | No | NA | | 2 | 0 | 2 | Not-determined | Genome-fragment | NA | | NA | 0 |
| S7_k141_164723 | 3354 | No | NA | | 4 | 0 | 4 | Not-determined | Genome-fragment | NA | | NA | 0 |
| S7_k141_52257 | 2803 | No | NA | | 1 | 0 | 1 | Not-determined | Genome-fragment | NA | | NA | 0 |
| S7_k141_185133 | 3905 | No | NA | | 4 | 0 | 2 | Not-determined | Genome-fragment | NA | | NA | 0 |
| S12-final-viral-combined_k141_294629\|\|full | 2005 | No | NA | | 4 | 2 | 0 | Low-quality | Genome-fragment | | 4.21 | AAI-based (high-confidence) | 0 |
| S7_k141_205744 | 1157 | No | NA | | 2 | 0 | 2 | Not-determined | Genome-fragment | NA | | NA | 0 |
| S7_k141_143942 | 1170 | No | NA | | 1 | 0 | 1 | Not-determined | Genome-fragment | NA | | NA | 0 |
| S12_k141_166904 | 2005 | No | NA | | 3 | 0 | 3 | Low-quality | Genome-fragment | | 0.84 | AAI-based (medium-confidence) | 0 |
| S7_k141_41899 | 1108 | No | NA | | 2 | 0 | 0 | Not-determined | Genome-fragment | NA | | NA | 0 |
| S7_k141_185234 | 1347 | No | NA | | 1 | 0 | 0 | Not-determined | Genome-fragment | NA | | NA | 0 |
| S4-final-viral-combined_k141_102752\|\|full | 2005 | No | NA | | 5 | 0 | 0 | Low-quality | Genome-fragment | | 3.1 | AAI-based (high-confidence) | 0 |
| S6-final-viral-combined_k141_98890\|\|full | 2005 | No | NA | | 2 | 0 | 1 | Low-quality | Genome-fragment | | 1.07 | AAI-based (medium-confidence) | 0 |
| S6_k141_93550 | 2005 | No | NA | | 5 | 2 | 0 | Low-quality | Genome-fragment | | 1.07 | AAI-based (high-confidence) | 0 |
| S7-final-viral-combined_k141_4939\|\|full | 2006 | No | NA | | 2 | 0 | 2 | Low-quality | Genome-fragment | | 3.03 | AAI-based (medium-confidence) | 0 |
| S7_k141_11527 | 1360 | No | NA | | 2 | 0 | 2 | Not-determined | Genome-fragment | NA | | NA | 0 |
| S7_k141_134198 | 1177 | No | NA | | 1 | 0 | 0 | Not-determined | Genome-fragment | NA | | NA | 0 |
| S7_k141_62540 | 1239 | No | NA | | 2 | 0 | 1 | Not-determined | Genome-fragment | NA | | NA | 0 |
| S7_k141_154792 | 1066 | No | NA | | 1 | 0 | 0 | Not-determined | Genome-fragment | NA | | NA | 0 |
| S7_k141_52375 | 1512 | No | NA | | 2 | 0 | 1 | Not-determined | Genome-fragment | NA | | NA | 0 |
| S7_k141_215969 | 1187 | No | NA | | 2 | 0 | 2 | Not-determined | Genome-fragment | NA | | NA | 0 |
| S7_k141_185374 | 1003 | No | NA | | 2 | 0 | 0 | Not-determined | Genome-fragment | NA | | NA | 0 |
| S7_k141_226169 | 1244 | No | NA | | 3 | 0 | 1 | Not-determined | Genome-fragment | NA | | NA | 0 |
| S7_k141_226170 | 1184 | No | NA | | 2 | 0 | 0 | Not-determined | Genome-fragment | NA | | NA | 0 |
| S7_k141_154815 | 3157 | No | NA | | 3 | 0 | 1 | Not-determined | Genome-fragment | NA | | NA | 0 |
| S7_k141_236292 | 1475 | No | NA | | 2 | 0 | 1 | Not-determined | Genome-fragment | NA | | NA | 0 |
| S7_k141_236300 | 1550 | No | NA | | 2 | 0 | 1 | Not-determined | Genome-fragment | NA | | NA | 0 |
| S7-final-viral-combined_k141_98834\|\|full | 2006 | No | NA | | 6 | 0 | 0 | Low-quality | Genome-fragment | | 1.39 | AAI-based (medium-confidence) | 0 |
| S7_k141_226202 | 1064 | No | NA | | 0 | 0 | 0 | Not-determined | Genome-fragment | NA | | NA | 0 |
| S7_k141_226203 | 1533 | No | NA | | 1 | 0 | 1 | Not-determined | Genome-fragment | NA | | NA | 0 |
| S7-final-viral-combined_k141_181104\|\|lt2gene | 2006 | No | NA | | 1 | 1 | 0 | Low-quality | Genome-fragment | | 0.84 | HMM-based (lower-bound) | 0 |
| S7_k141_174946 | 1002 | No | NA | | 1 | 0 | 0 | Not-determined | Genome-fragment | NA | | NA | 0 |
| S12-final-viral-combined_k141_173406\|\|full | 2006 | No | NA | | 3 | 0 | 0 | Low-quality | Genome-fragment | | 4.61 | AAI-based (high-confidence) | 0 |
| S12-final-viral-combined_k141_294861\|\|full | 2006 | No | NA | | 2 | 2 | 0 | Low-quality | Genome-fragment | | 4.23 | AAI-based (high-confidence) | 0 |
| S8-final-viral-combined_k141_264244\|\|full | 2006 | No | NA | | 3 | 1 | 0 | Low-quality | Genome-fragment | | 2.63 | HMM-based (lower-bound) | 0 |
| S7_k141_134342 | 5571 | No | NA | | 9 | 0 | 1 | Not-determined | Genome-fragment | NA | | NA | 0 |
| S7_k141_113847 | 1474 | No | NA | | 3 | 0 | 0 | Not-determined | Genome-fragment | NA | | NA | 0 |
| S7_k141_62615 | 3491 | No | NA | | 8 | 0 | 7 | Not-determined | Genome-fragment | NA | | NA | 0 |
| S9-final-viral-combined_k141_40810\|\|full | 2007 | No | NA | | 4 | 0 | 1 | Low-quality | Genome-fragment | | 2.66 | AAI-based (medium-confidence) | 0 |
| S7_k141_123954 | 21948 | No | NA | | 28 | 10 | 0 | Medium-quality | Genome-fragment | | 76.08 | AAI-based (high-confidence) | 0 |
| S7_k141_42057 | 1042 | No | NA | | 2 | 0 | 1 | Not-determined | Genome-fragment | NA | | NA | 0 |
| S7_k141_62663 | 3448 | No | NA | | 4 | 0 | 3 | Not-determined | Genome-fragment | NA | | NA | 0 |
| S1-final-viral-combined_k141_198034\|\|full | 2007 | No | NA | | 3 | 0 | 1 | Low-quality | Genome-fragment | | 2.85 | AAI-based (medium-confidence) | 0 |
| S7_k141_185577 | 3963 | No | NA | | 2 | 0 | 0 | Not-determined | Genome-fragment | NA | | NA | 0 |
| S7_k141_205889 | 2206 | No | NA | | 3 | 0 | 3 | Not-determined | Genome-fragment | NA | | NA | 0 |
| S7_k141_62688 | 1633 | No | NA | | 3 | 0 | 1 | Not-determined | Genome-fragment | NA | | NA | 0 |
| S7_k141_144216 | 2179 | No | NA | | 1 | 0 | 0 | Not-determined | Genome-fragment | NA | | NA | 0 |
| S7_k141_42087 | 1361 | No | NA | | 1 | 0 | 1 | Not-determined | Genome-fragment | NA | | NA | 0 |
| S7-final-viral-combined_k141_196284\|\|full | 2007 | No | NA | | 6 | 0 | 0 | Low-quality | Genome-fragment | | 3.07 | AAI-based (high-confidence) | 0 |
| S7_k141_1459 | 1517 | No | NA | | 1 | 0 | 1 | Not-determined | Genome-fragment | NA | | NA | 0 |
| S7_k141_175071 | 1354 | No | NA | | 1 | 0 | 0 | Not-determined | Genome-fragment | NA | | NA | 0 |
| S7_k141_226385 | 1765 | No | NA | | 1 | 0 | 0 | Not-determined | Genome-fragment | NA | | NA | 0 |
| S7_k141_124040 | 3249 | No | NA | | 1 | 0 | 0 | Not-determined | Genome-fragment | NA | | NA | 0 |
| S17-k141_111622 | 2007 | No | NA | | 1 | 0 | 1 | Low-quality | Genome-fragment | | 5.66 | AAI-based (medium-confidence) | 0 |
| S7_k141_1497 | 1048 | No | NA | | 1 | 0 | 1 | Not-determined | Genome-fragment | NA | | NA | 0 |
| S7_k141_21530 | 4541 | No | NA | | 7 | 0 | 2 | Not-determined | Genome-fragment | NA | | NA | 0 |
| S7_k141_195964 | 1099 | No | NA | | 2 | 0 | 0 | Not-determined | Genome-fragment | NA | | NA | 0 |
| S7_k141_236522 | 1234 | No | NA | | 1 | 0 | 0 | Not-determined | Genome-fragment | NA | | NA | 0 |
| S7_k141_21540 | 2647 | No | NA | | 3 | 0 | 3 | Not-determined | Genome-fragment | NA | | NA | 0 |
| S7_k141_195986 | 2019 | No | NA | | 1 | 0 | 0 | Not-determined | Genome-fragment | NA | | NA | 0 |
| S7_k141_32251 | 1003 | No | NA | | 2 | 0 | 1 | Not-determined | Genome-fragment | NA | | NA | 0 |
| S7_k141_103552 | 5367 | No | NA | | 5 | 0 | 1 | Not-determined | Genome-fragment | NA | | NA | 0 |
| S7_k141_11612 | 1014 | No | NA | | 2 | 0 | 0 | Not-determined | Genome-fragment | NA | | NA | 0 |
| S7_k141_92987 | 1892 | No | NA | | 2 | 0 | 1 | Not-determined | Genome-fragment | NA | | NA | 0 |
| S7_k141_165060 | 3550 | No | NA | | 2 | 0 | 1 | Not-determined | Genome-fragment | NA | | NA | 0 |
| S7_k141_11627 | 1524 | No | NA | | 1 | 0 | 1 | Not-determined | Genome-fragment | NA | | NA | 0 |
| S7_k141_52738 | 3211 | No | NA | | 3 | 0 | 3 | Not-determined | Genome-fragment | NA | | NA | 0 |
| S8-final-viral-combined_k141_163508\|\|full | 2007 | No | NA | | 4 | 0 | 1 | Low-quality | Genome-fragment | | 1.06 | AAI-based (medium-confidence) | 0 |
| S7_k141_103620 | 3314 | No | NA | | 2 | 0 | 2 | Not-determined | Genome-fragment | NA | | NA | 0 |
| S7_k141_32321 | 1461 | No | NA | | 1 | 0 | 0 | Not-determined | Genome-fragment | NA | | NA | 0 |
| S8-final-viral-combined_k141_119005\|\|full | 2007 | No | NA | | 4 | 0 | 1 | Low-quality | Genome-fragment | | 3.06 | AAI-based (high-confidence) | 0 |
| S7_k141_165147 | 1378 | No | NA | | 3 | 0 | 0 | Not-determined | Genome-fragment | NA | | NA | 0 |
| S7_k141_114208 | 1219 | No | NA | | 2 | 0 | 2 | Not-determined | Genome-fragment | NA | | NA | 0 |
| S7_k141_134519 | 1038 | No | NA | | 2 | 0 | 1 | Not-determined | Genome-fragment | NA | | NA | 0 |
| S8_k141_108030 | 2007 | No | NA | | 3 | 0 | 0 | Low-quality | Genome-fragment | | 3.69 | AAI-based (medium-confidence) | 0 |
| S7_k141_144293 | 2210 | No | NA | | 4 | 0 | 0 | Not-determined | Genome-fragment | NA | | NA | 0 |
| S7_k141_62880 | 3115 | No | NA | | 1 | 0 | 1 | Not-determined | Genome-fragment | NA | | NA | 0 |
| S7_k141_226530 | 2121 | No | NA | | 3 | 0 | 2 | Not-determined | Genome-fragment | NA | | NA | 0 |
| S11-final-viral-combined_k141_151959\|\|full | 2008 | No | NA | | 4 | 1 | 0 | Low-quality | Genome-fragment | | 2.98 | HMM-based (lower-bound) | 0 |
| S7_k141_216107 | 1252 | No | NA | | 2 | 0 | 0 | Not-determined | Genome-fragment | NA | | NA | 0 |
| S7_k141_175223 | 4497 | No | NA | | 3 | 0 | 3 | Not-determined | Genome-fragment | NA | | NA | 0 |
| S7_k141_206225 | 3074 | No | NA | | 2 | 0 | 2 | Not-determined | Genome-fragment | NA | | NA | 0 |
| S7_k141_93128 | 1344 | No | NA | | 3 | 0 | 3 | Not-determined | Genome-fragment | NA | | NA | 0 |
| S7_k141_165249 | 1439 | No | NA | | 2 | 0 | 1 | Not-determined | Genome-fragment | NA | | NA | 0 |
| S7-final-viral-combined_k141_75514\|\|full | 2008 | No | NA | | 2 | 0 | 2 | Low-quality | Genome-fragment | | 0.92 | AAI-based (medium-confidence) | 0 |
| S7_k141_124231 | 6493 | No | NA | | 4 | 0 | 3 | Not-determined | Genome-fragment | NA | | NA | 0 |
| S7_k141_236720 | 1198 | No | NA | | 1 | 0 | 1 | Not-determined | Genome-fragment | NA | | NA | 0 |
| S7_k141_62960 | 2858 | No | NA | | 1 | 0 | 0 | Not-determined | Genome-fragment | NA | | NA | 0 |
| S7_k141_165297 | 1229 | No | NA | | 1 | 0 | 1 | Not-determined | Genome-fragment | NA | | NA | 0 |
| S12-final-viral-combined_k141_168166\|\|full | 2008 | No | NA | | 7 | 0 | 0 | Low-quality | Genome-fragment | | 4.45 | AAI-based (high-confidence) | 0 |
| S12-final-viral-combined_k141_124726\|\|full | 2008 | No | NA | | 3 | 0 | 0 | Low-quality | Genome-fragment | | 1.47 | AAI-based (high-confidence) | 0 |
| S7_k141_236789 | 1367 | No | NA | | 3 | 0 | 0 | Not-determined | Genome-fragment | NA | | NA | 0 |
| S7_k141_185893 | 4323 | No | NA | | 7 | 0 | 0 | Not-determined | Genome-fragment | NA | | NA | 0 |
| S17-final-viral-combined_k141_219374\|\|full | 2009 | No | NA | | 2 | 0 | 0 | Low-quality | Genome-fragment | | 4.64 | AAI-based (high-confidence) | 0 |
| S7_k141_124341 | 2021 | No | NA | | 2 | 0 | 0 | Not-determined | Genome-fragment | NA | | NA | 0 |
| S2-final-viral-combined_k141_130287\|\|full | 2009 | No | NA | | 2 | 0 | 1 | Low-quality | Genome-fragment | | 1.36 | AAI-based (high-confidence) | 0 |
| S7_k141_124350 | 1376 | No | NA | | 1 | 0 | 1 | Not-determined | Genome-fragment | NA | | NA | 0 |
| S9-final-viral-combined_k141_13121\|\|lt2gene | 2010 | No | NA | | 2 | 2 | 0 | Low-quality | Genome-fragment | | 4.45 | AAI-based (high-confidence) | 0 |
| S7_k141_226655 | 1130 | No | NA | | 2 | 0 | 1 | Not-determined | Genome-fragment | NA | | NA | 0 |
| S11-final-viral-combined_k141_194172\|\|full | 2010 | No | NA | | 4 | 1 | 0 | Low-quality | Genome-fragment | | 4.63 | AAI-based (medium-confidence) | 0 |
| S11-final-viral-combined_k141_61014\|\|full | 2010 | No | NA | | 6 | 0 | 0 | Low-quality | Genome-fragment | | 4.41 | AAI-based (high-confidence) | 0 |
| S7_k141_32534 | 1259 | No | NA | | 2 | 0 | 2 | Not-determined | Genome-fragment | NA | | NA | 0 |
| S7_k141_144401 | 6173 | No | NA | | 6 | 0 | 2 | Not-determined | Genome-fragment | NA | | NA | 0 |
| S7_k141_1794 | 1072 | No | NA | | 1 | 0 | 0 | Not-determined | Genome-fragment | NA | | NA | 0 |
| S7_k141_42490 | 7130 | No | NA | | 6 | 0 | 5 | Not-determined | Genome-fragment | NA | | NA | 0 |
| S7_k141_124384 | 1067 | No | NA | | 1 | 0 | 0 | Not-determined | Genome-fragment | NA | | NA | 0 |
| S1-final-viral-combined_k141_289508\|\|full | 2010 | No | NA | | 4 | 0 | 1 | Low-quality | Genome-fragment | | 3.61 | AAI-based (medium-confidence) | 0 |
| S7_k141_83208 | 1381 | No | NA | | 4 | 0 | 4 | Not-determined | Genome-fragment | NA | | NA | 0 |
| S7_k141_236918 | 1265 | No | NA | | 2 | 0 | 2 | Not-determined | Genome-fragment | NA | | NA | 0 |
| S7_k141_185989 | 7296 | No | NA | | 6 | 0 | 6 | Not-determined | Genome-fragment | NA | | NA | 0 |
| S7_k141_42533 | 1258 | No | NA | | 1 | 0 | 0 | Not-determined | Genome-fragment | NA | | NA | 0 |
| S1_k141_376779 | 2010 | No | NA | | 7 | 0 | 1 | Low-quality | Genome-fragment | | 3.05 | AAI-based (high-confidence) | 0 |
| S7-final-viral-combined_k141_221134\|\|full | 2010 | No | NA | | 2 | 0 | 1 | Low-quality | Genome-fragment | | 0.95 | AAI-based (medium-confidence) | 0 |
| S7_k141_226765 | 2706 | No | NA | | 2 | 0 | 1 | Not-determined | Genome-fragment | NA | | NA | 0 |
| S7_k141_144425 | 1016 | No | NA | | 1 | 0 | 0 | Not-determined | Genome-fragment | NA | | NA | 0 |
| S7_k141_21803 | 2453 | No | NA | | 1 | 0 | 0 | Not-determined | Genome-fragment | NA | | NA | 0 |
| S7_k141_42591 | 2173 | No | NA | | 1 | 0 | 0 | Not-determined | Genome-fragment | NA | | NA | 0 |
| S7_k141_206452 | 1002 | No | NA | | 1 | 0 | 0 | Not-determined | Genome-fragment | NA | | NA | 0 |
| S7_k141_196204 | 1070 | No | NA | | 1 | 0 | 0 | Not-determined | Genome-fragment | NA | | NA | 0 |
| S17-final-viral-combined_k141_68064\|\|full | 2010 | No | NA | | 3 | 0 | 0 | Low-quality | Genome-fragment | | 2.03 | AAI-based (medium-confidence) | 0 |
| S7_k141_216305 | 1885 | No | NA | | 3 | 0 | 0 | Not-determined | Genome-fragment | NA | | NA | 0 |
| S7_k141_206500 | 1201 | No | NA | | 1 | 0 | 1 | Not-determined | Genome-fragment | NA | | NA | 0 |
| S7_k141_114456 | 1087 | No | NA | | 3 | 0 | 1 | Not-determined | Genome-fragment | NA | | NA | 0 |
| S7_k141_21871 | 3275 | No | NA | | 7 | 0 | 0 | Not-determined | Genome-fragment | NA | | NA | 0 |
| S7_k141_237073 | 2793 | No | NA | | 4 | 0 | 2 | Not-determined | Genome-fragment | NA | | NA | 0 |
| S17-final-viral-combined_k141_257955\|\|full | 2010 | No | NA | | 7 | 1 | 0 | Low-quality | Genome-fragment | | 2.34 | AAI-based (high-confidence) | 0 |
| S7_k141_186133 | 1135 | No | NA | | 1 | 0 | 0 | Not-determined | Genome-fragment | NA | | NA | 0 |
| S7_k141_114508 | 1801 | No | NA | | 1 | 0 | 0 | Not-determined | Genome-fragment | NA | | NA | 0 |
| S7_k141_1914 | 6790 | No | NA | | 7 | 0 | 5 | Not-determined | Genome-fragment | NA | | NA | 0 |
| S2-final-viral-combined_k141_31180\|\|full | 2010 | No | NA | | 3 | 0 | 0 | Low-quality | Genome-fragment | | 2.14 | AAI-based (medium-confidence) | 0 |
| S4_k141_30296 | 2010 | No | NA | | 1 | 0 | 0 | Low-quality | Genome-fragment | | 2.01 | AAI-based (medium-confidence) | 0 |
| S7_k141_42726 | 3961 | No | NA | | 2 | 0 | 0 | Not-determined | Genome-fragment | NA | | NA | 0 |
| S9-final-viral-combined_k141_13369\|\|full | 2011 | No | NA | | 4 | 0 | 1 | Low-quality | Genome-fragment | | 1.82 | AAI-based (medium-confidence) | 0 |
| S7_k141_32778 | 1174 | No | NA | | 1 | 0 | 1 | Not-determined | Genome-fragment | NA | | NA | 0 |
| S7_k141_114543 | 3388 | No | NA | | 2 | 0 | 1 | Not-determined | Genome-fragment | NA | | NA | 0 |
| S7_k141_175522 | 1252 | No | NA | | 1 | 0 | 0 | Not-determined | Genome-fragment | NA | | NA | 0 |
| S7_k141_226914 | 1653 | No | NA | | 3 | 0 | 2 | Not-determined | Genome-fragment | NA | | NA | 0 |
| S7_k141_196282 | 1154 | No | NA | | 1 | 0 | 1 | Not-determined | Genome-fragment | NA | | NA | 0 |
| S7_k141_226935 | 1616 | No | NA | | 3 | 0 | 1 | Not-determined | Genome-fragment | NA | | NA | 0 |
| S7_k141_144605 | 1120 | No | NA | | 1 | 0 | 1 | Not-determined | Genome-fragment | NA | | NA | 0 |
| S11-final-viral-combined_k141_251130\|\|full | 2011 | No | NA | | 3 | 0 | 0 | Low-quality | Genome-fragment | | 1.63 | AAI-based (medium-confidence) | 0 |
| S7_k141_114565 | 1048 | No | NA | | 2 | 0 | 0 | Not-determined | Genome-fragment | NA | | NA | 0 |
| S7_k141_186187 | 2388 | No | NA | | 1 | 0 | 0 | Not-determined | Genome-fragment | NA | | NA | 0 |
| S7_k141_21981 | 2710 | No | NA | | 1 | 0 | 1 | Not-determined | Genome-fragment | NA | | NA | 0 |
| S1-final-viral-combined_k141_206266\|\|full | 2011 | No | NA | | 2 | 0 | 1 | Low-quality | Genome-fragment | | 4.05 | AAI-based (medium-confidence) | 0 |
| S7_k141_227020 | 2173 | No | NA | | 1 | 0 | 0 | Not-determined | Genome-fragment | NA | | NA | 0 |
| S7_k141_206667 | 1796 | No | NA | | 2 | 0 | 1 | Not-determined | Genome-fragment | NA | | NA | 0 |
| S7_k141_206669 | 1009 | No | NA | | 1 | 0 | 0 | Not-determined | Genome-fragment | NA | | NA | 0 |
| S1-final-viral-combined_k141_27321\|\|full | 2011 | No | NA | | 3 | 0 | 0 | Low-quality | Genome-fragment | | 1.39 | AAI-based (high-confidence) | 0 |
| S7_k141_186205 | 1660 | No | NA | | 2 | 0 | 2 | Not-determined | Genome-fragment | NA | | NA | 0 |
| S7_k141_114615 | 1083 | No | NA | | 1 | 0 | 1 | Not-determined | Genome-fragment | NA | | NA | 0 |
| S7_k141_216487 | 1413 | No | NA | | 1 | 0 | 1 | Not-determined | Genome-fragment | NA | | NA | 0 |
| S7_k141_206697 | 1462 | No | NA | | 1 | 0 | 0 | Not-determined | Genome-fragment | NA | | NA | 0 |
| S7_k141_104221 | 1674 | No | NA | | 3 | 0 | 2 | Not-determined | Genome-fragment | NA | | NA | 0 |
| S7_k141_165493 | 4434 | No | NA | | 7 | 0 | 1 | Not-determined | Genome-fragment | NA | | NA | 0 |
| S7_k141_216509 | 1075 | No | NA | | 1 | 0 | 0 | Not-determined | Genome-fragment | NA | | NA | 0 |
| S12-final-viral-combined_k141_296290\|\|full | 2011 | No | NA | | 4 | 2 | 1 | Low-quality | Genome-fragment | | 1.27 | HMM-based (lower-bound) | 0 |
| S7_k141_227062 | 1042 | No | NA | | 1 | 0 | 0 | Not-determined | Genome-fragment | NA | | NA | 0 |
| S7_k141_175628 | 1153 | No | NA | | 2 | 0 | 2 | Not-determined | Genome-fragment | NA | | NA | 0 |
| S7_k141_124670 | 7821 | No | NA | | 8 | 0 | 2 | Not-determined | Genome-fragment | NA | | NA | 0 |
| S7_k141_227110 | 1187 | No | NA | | 3 | 0 | 0 | Not-determined | Genome-fragment | NA | | NA | 0 |
| S7_k141_165546 | 1305 | No | NA | | 1 | 0 | 1 | Not-determined | Genome-fragment | NA | | NA | 0 |
| S7_k141_206800 | 3475 | No | NA | | 2 | 0 | 1 | Not-determined | Genome-fragment | NA | | NA | 0 |
| S7_k141_165560 | 1144 | No | NA | | 1 | 0 | 1 | Not-determined | Genome-fragment | NA | | NA | 0 |
| S7_k141_63311 | 1147 | No | NA | | 1 | 0 | 1 | Not-determined | Genome-fragment | NA | | NA | 0 |
| S12-final-viral-combined_k141_197661\|\|full | 2012 | No | NA | | 3 | 0 | 1 | Low-quality | Genome-fragment | | 2.42 | AAI-based (high-confidence) | 0 |
| S7_k141_165599 | 1577 | No | NA | | 2 | 0 | 0 | Not-determined | Genome-fragment | NA | | NA | 0 |
| S7_k141_72863 | 1518 | No | NA | | 2 | 0 | 1 | Not-determined | Genome-fragment | NA | | NA | 0 |
| S7_k141_216553 | 1447 | No | NA | | 3 | 0 | 1 | Not-determined | Genome-fragment | NA | | NA | 0 |
| S7_k141_83441 | 7603 | No | NA | | 8 | 0 | 1 | Not-determined | Genome-fragment | NA | | NA | 0 |
| S7_k141_104284 | 1262 | No | NA | | 1 | 0 | 1 | Not-determined | Genome-fragment | NA | | NA | 0 |
| S7_k141_196499 | 1696 | No | NA | | 1 | 0 | 0 | Not-determined | Genome-fragment | NA | | NA | 0 |
| S7_k141_196500 | 1412 | No | NA | | 1 | 0 | 1 | Not-determined | Genome-fragment | NA | | NA | 0 |
| S7_k141_83448 | 4430 | No | NA | | 4 | 0 | 0 | Not-determined | Genome-fragment | NA | | NA | 0 |
| S7_k141_206841 | 1221 | No | NA | | 2 | 0 | 1 | Not-determined | Genome-fragment | NA | | NA | 0 |
| S12_k141_377797 | 2012 | No | NA | | 2 | 0 | 1 | Low-quality | Genome-fragment | | 0.34 | AAI-based (high-confidence) | 0 |
| S7_k141_22188 | 1167 | No | NA | | 2 | 0 | 0 | Not-determined | Genome-fragment | NA | | NA | 0 |
| S7_k141_237194 | 1762 | No | NA | | 1 | 0 | 1 | Not-determined | Genome-fragment | NA | | NA | 0 |
| S4-final-viral-combined_k141_106632\|\|full | 2012 | No | NA | | 3 | 0 | 2 | Low-quality | Genome-fragment | | 3.47 | AAI-based (medium-confidence) | 0 |
| S7_k141_2173 | 5629 | No | NA | | 10 | 0 | 1 | Not-determined | Genome-fragment | NA | | NA | 0 |
| S6-final-viral-combined_k141_82447\|\|full | 2012 | No | NA | | 2 | 0 | 0 | Low-quality | Genome-fragment | | 2.13 | AAI-based (high-confidence) | 0 |
| S7_k141_83466 | 1291 | No | NA | | 1 | 0 | 1 | Not-determined | Genome-fragment | NA | | NA | 0 |
| S7_k141_2200 | 1201 | No | NA | | 3 | 0 | 2 | Not-determined | Genome-fragment | NA | | NA | 0 |
| S7_k141_216674 | 1485 | No | NA | | 2 | 0 | 0 | Not-determined | Genome-fragment | NA | | NA | 0 |
| S7_k141_53338 | 16884 | No | NA | | 27 | 8 | 0 | Medium-quality | Genome-fragment | | 72.45 | AAI-based (medium-confidence) | 0 |
| S7_k141_216675 | 1354 | No | NA | | 2 | 0 | 0 | Not-determined | Genome-fragment | NA | | NA | 0 |
| S7_k141_186427 | 1124 | No | NA | | 2 | 0 | 0 | Not-determined | Genome-fragment | NA | | NA | 0 |
| S7_k141_134970 | 1772 | No | NA | | 2 | 0 | 1 | Not-determined | Genome-fragment | NA | | NA | 0 |
| S7_k141_237236 | 1408 | No | NA | | 2 | 0 | 1 | Not-determined | Genome-fragment | NA | | NA | 0 |
| S7_k141_72924 | 3050 | No | NA | | 5 | 0 | 1 | Not-determined | Genome-fragment | NA | | NA | 0 |
| S7_k141_155640 | 1730 | No | NA | | 1 | 0 | 0 | Not-determined | Genome-fragment | NA | | NA | 0 |
| S7_k141_53382 | 1189 | No | NA | | 1 | 0 | 0 | Not-determined | Genome-fragment | NA | | NA | 0 |
| S7_k141_53426 | 1207 | No | NA | | 1 | 0 | 1 | Not-determined | Genome-fragment | NA | | NA | 0 |
| S7_k141_206904 | 1361 | No | NA | | 1 | 0 | 1 | Not-determined | Genome-fragment | NA | | NA | 0 |
| S7_k141_22302 | 2860 | No | NA | | 1 | 0 | 1 | Not-determined | Genome-fragment | NA | | NA | 0 |
| S7_k141_155680 | 3197 | No | NA | | 3 | 0 | 0 | Not-determined | Genome-fragment | NA | | NA | 0 |
| S7_k141_12316 | 1113 | No | NA | | 1 | 0 | 1 | Not-determined | Genome-fragment | NA | | NA | 0 |
| S7_k141_53445 | 2562 | No | NA | | 3 | 0 | 2 | Not-determined | Genome-fragment | NA | | NA | 0 |
| S7_k141_227292 | 3255 | No | NA | | 4 | 0 | 1 | Not-determined | Genome-fragment | NA | | NA | 0 |
| S7_k141_165760 | 1361 | No | NA | | 4 | 0 | 1 | Not-determined | Genome-fragment | NA | | NA | 0 |
| S7_k141_63541 | 3234 | No | NA | | 4 | 0 | 3 | Not-determined | Genome-fragment | NA | | NA | 0 |
| S7_k141_2306 | 1005 | No | NA | | 1 | 0 | 1 | Not-determined | Genome-fragment | NA | | NA | 0 |
| S7_k141_43059 | 9077 | No | NA | | 11 | 0 | 9 | Not-determined | Genome-fragment | NA | | NA | 0 |
| S11-final-viral-combined_k141_121631\|\|full | 2013 | No | NA | | 3 | 1 | 0 | Low-quality | Genome-fragment | | 1.27 | HMM-based (lower-bound) | 0 |
| S1-final-viral-combined_k141_79258\|\|full | 2013 | No | NA | | 5 | 2 | 0 | Low-quality | Genome-fragment | | 4.76 | AAI-based (high-confidence) | 0 |
| S7_k141_124859 | 1066 | No | NA | | 1 | 0 | 0 | Not-determined | Genome-fragment | NA | | NA | 0 |
| S5-final-viral-combined_k141_37482\|\|full | 2013 | Yes | | 483 | 2 | 1 | 1 | Low-quality | Genome-fragment | | 0.1 | AAI-based (medium-confidence) | 76.01 |
| S7_k141_144942 | 1684 | No | NA | | 1 | 0 | 0 | Not-determined | Genome-fragment | NA | | NA | 0 |
| S5-final-viral-combined_k141_203352\|\|full | 2013 | No | NA | | 5 | 0 | 1 | Low-quality | Genome-fragment | | 4.36 | AAI-based (high-confidence) | 0 |
| S2-final-viral-combined_k141_208778\|\|full | 2013 | No | NA | | 2 | 0 | 1 | Low-quality | Genome-fragment | | 4.12 | AAI-based (high-confidence) | 0 |
| S7_k141_216836 | 2554 | No | NA | | 1 | 0 | 1 | Not-determined | Genome-fragment | NA | | NA | 0 |
| S4-final-viral-combined_k141_101360\|\|full | 2013 | No | NA | | 4 | 2 | 0 | Low-quality | Genome-fragment | | 5.42 | AAI-based (high-confidence) | 0 |
| S7_k141_93692 | 3185 | No | NA | | 2 | 0 | 2 | Not-determined | Genome-fragment | NA | | NA | 0 |
| S7_k141_53525 | 2227 | No | NA | | 3 | 0 | 2 | Not-determined | Genome-fragment | NA | | NA | 0 |
| S7_k141_155782 | 21313 | No | NA | | 15 | 0 | 13 | Not-determined | Genome-fragment | NA | | NA | 0 |
| S7_k141_124913 | 1968 | No | NA | | 1 | 0 | 1 | Not-determined | Genome-fragment | NA | | NA | 0 |
| S7_k141_63637 | 3264 | No | NA | | 4 | 0 | 3 | Not-determined | Genome-fragment | NA | | NA | 0 |
| S7_k141_135022 | 4082 | No | NA | | 6 | 0 | 1 | Not-determined | Genome-fragment | NA | | NA | 0 |
| S7_k141_93725 | 2888 | No | NA | | 2 | 0 | 0 | Not-determined | Genome-fragment | NA | | NA | 0 |
| S4_k141_155997 | 2013 | No | NA | | 1 | 0 | 0 | Low-quality | Genome-fragment | | 2.23 | AAI-based (high-confidence) | 0 |

| | | | | | | | | | | | | |
|---|---|---|---|---|---|---|---|---|---|---|---|---|
| S7_k141_63710 | 4232 | No | NA | | 6 | 0 | 3 | Not-determined | Genome-fragment | NA | NA | 0 |
| S7_k141_114887 | 5066 | No | NA | | 9 | 0 | 0 | Not-determined | Genome-fragment | NA | NA | 0 |
| S7_k141_145014 | 1376 | No | NA | | 2 | 0 | 0 | Not-determined | Genome-fragment | NA | NA | 0 |
| S4_k141_94670 | 2013 | No | NA | | 2 | 0 | 0 | Low-quality | Genome-fragment | 3.36 | AAI-based (high-confidence) | 0 |
| S7_k141_186726 | 2029 | No | NA | | 2 | 0 | 1 | Not-determined | Genome-fragment | NA | NA | 0 |
| S7_k141_63760 | 1852 | No | NA | | 1 | 0 | 0 | Not-determined | Genome-fragment | NA | NA | 0 |
| S7_k141_155915 | 1374 | No | NA | | 1 | 0 | 0 | Not-determined | Genome-fragment | NA | NA | 0 |
| S7_k141_63770 | 1910 | No | NA | | 1 | 0 | 1 | Not-determined | Genome-fragment | NA | NA | 0 |
| S9-final-viral-combined_k141_20809\|\|full | 2014 | No | NA | | 3 | 0 | 1 | Low-quality | Genome-fragment | 1.29 | AAI-based (medium-confidence) | 0 |
| S7_k141_207211 | 1004 | No | NA | | 1 | 0 | 1 | Not-determined | Genome-fragment | NA | NA | 0 |
| S1_k141_221981 | 2014 | No | NA | | 4 | 1 | 0 | Low-quality | Genome-fragment | 2.49 | HMM-based (lower-bound) | 0 |
| S7_k141_84132 | 2014 | No | NA | | 3 | 2 | 0 | Low-quality | Genome-fragment | 5 | AAI-based (medium-confidence) | 0 |
| S7_k141_227570 | 2075 | No | NA | | 2 | 0 | 0 | Not-determined | Genome-fragment | NA | NA | 0 |
| S12-final-viral-combined_k141_82555\|\|lt2gene | 2014 | No | NA | | 1 | 1 | 0 | Low-quality | Genome-fragment | 5.18 | AAI-based (high-confidence) | 0 |
| S7_k141_12640 | 3333 | No | NA | | 4 | 0 | 2 | Not-determined | Genome-fragment | NA | NA | 0 |
| S7_k141_114958 | 1230 | No | NA | | 1 | 0 | 1 | Not-determined | Genome-fragment | NA | NA | 0 |
| S7_k141_175821 | 1520 | No | NA | | 1 | 0 | 0 | Not-determined | Genome-fragment | NA | NA | 0 |
| S7_k141_2608 | 1067 | No | NA | | 1 | 0 | 0 | Not-determined | Genome-fragment | NA | NA | 0 |
| S7_k141_216877 | 1219 | No | NA | | 1 | 0 | 1 | Not-determined | Genome-fragment | NA | NA | 0 |
| S7_k141_83935 | 1155 | No | NA | | 3 | 0 | 0 | Not-determined | Genome-fragment | NA | NA | 0 |
| S7_k141_227620 | 1633 | No | NA | | 1 | 0 | 0 | Not-determined | Genome-fragment | NA | NA | 0 |
| S7_k141_115001 | 1084 | No | NA | | 1 | 0 | 1 | Not-determined | Genome-fragment | NA | NA | 0 |
| S1-final-viral-combined_k141_112272\|\|full | 2015 | No | NA | | 2 | 0 | 1 | Low-quality | Genome-fragment | 1.89 | AAI-based (medium-confidence) | 0 |
| S5-final-viral-combined_k141_70800\|\|full | 2015 | No | NA | | 3 | 2 | 0 | Low-quality | Genome-fragment | 1.09 | HMM-based (lower-bound) | 0 |
| S7_k141_165998 | 1071 | No | NA | | 1 | 0 | 1 | Not-determined | Genome-fragment | NA | NA | 0 |
| S7_k141_2678 | 4747 | No | NA | | 5 | 0 | 2 | Not-determined | Genome-fragment | NA | NA | 0 |
| S7_k141_43413 | 1006 | No | NA | | 1 | 0 | 0 | Not-determined | Genome-fragment | NA | NA | 0 |
| S12-final-viral-combined_k141_251702\|\|full | 2015 | No | NA | | 3 | 2 | 0 | Low-quality | Genome-fragment | 5.37 | AAI-based (high-confidence) | 0 |
| S7_k141_156084 | 1032 | No | NA | | 1 | 0 | 1 | Not-determined | Genome-fragment | NA | NA | 0 |
| S7_k141_135176 | 1100 | No | NA | | 2 | 0 | 1 | Not-determined | Genome-fragment | NA | NA | 0 |
| S7_k141_237746 | 1193 | No | NA | | 2 | 0 | 0 | Not-determined | Genome-fragment | NA | NA | 0 |
| S7_k141_125078 | 1047 | No | NA | | 1 | 0 | 1 | Not-determined | Genome-fragment | NA | NA | 0 |
| S12-final-viral-combined_k141_270984\|\|full | 2015 | No | NA | | 3 | 0 | 1 | Low-quality | Genome-fragment | 2.15 | AAI-based (medium-confidence) | 0 |
| S7_k141_156098 | 1736 | No | NA | | 2 | 0 | 2 | Not-determined | Genome-fragment | NA | NA | 0 |
| S7_k141_84021 | 1275 | No | NA | | 1 | 0 | 0 | Not-determined | Genome-fragment | NA | NA | 0 |
| S7_k141_237768 | 1094 | No | NA | | 1 | 0 | 1 | Not-determined | Genome-fragment | NA | NA | 0 |
| S7_k141_63914 | 1960 | No | NA | | 3 | 0 | 2 | Not-determined | Genome-fragment | NA | NA | 0 |
| S7_k141_207381 | 2663 | No | NA | | 2 | 0 | 0 | Not-determined | Genome-fragment | NA | NA | 0 |
| S7_k141_104654 | 3235 | No | NA | | 3 | 0 | 3 | Not-determined | Genome-fragment | NA | NA | 0 |
| S7_k141_43519 | 5657 | No | NA | | 7 | 0 | 4 | Not-determined | Genome-fragment | NA | NA | 0 |
| S7_k141_63939 | 1240 | No | NA | | 1 | 0 | 0 | Not-determined | Genome-fragment | NA | NA | 0 |
| S7_k141_115144 | 1621 | No | NA | | 2 | 0 | 0 | Not-determined | Genome-fragment | NA | NA | 0 |
| S7_k141_12836 | 1019 | No | NA | | 1 | 0 | 1 | Not-determined | Genome-fragment | NA | NA | 0 |
| S6-final-viral-combined_k141_51965\|\|full | 2015 | No | NA | | 5 | 1 | 0 | Low-quality | Genome-fragment | 2.54 | AAI-based (high-confidence) | 0 |
| S8-final-viral-combined_k141_139032\|\|full | 2015 | No | NA | | 3 | 0 | 0 | Low-quality | Genome-fragment | 4.65 | AAI-based (medium-confidence) | 0 |
| S1-final-viral-combined_k141_232966\|\|full | 2016 | No | NA | | 4 | 1 | 0 | Low-quality | Genome-fragment | 1.89 | HMM-based (lower-bound) | 0 |
| S7-final-viral-combined_k141_148167\|\|full | 2016 | No | NA | | 5 | 1 | 0 | Low-quality | Genome-fragment | 4.33 | AAI-based (high-confidence) | 0 |
| S7_k141_12861 | 1059 | No | NA | | 1 | 0 | 0 | Not-determined | Genome-fragment | NA | NA | 0 |
| S7_k141_227666 | 1286 | No | NA | | 1 | 0 | 0 | Not-determined | Genome-fragment | NA | NA | 0 |
| S9-final-viral-combined_k141_113575\|\|full | 2017 | No | NA | | 4 | 0 | 0 | Low-quality | Genome-fragment | 3 | AAI-based (high-confidence) | 0 |
| S7_k141_43583 | 1844 | No | NA | | 2 | 0 | 0 | Not-determined | Genome-fragment | NA | NA | 0 |
| S11_k141_15069 | 2017 | No | NA | | 3 | 0 | 0 | Low-quality | Genome-fragment | 3.39 | AAI-based (medium-confidence) | 0 |
| S1-final-viral-combined_k141_266985\|\|full | 2017 | No | NA | | 5 | 0 | 0 | Low-quality | Genome-fragment | 3.5 | AAI-based (medium-confidence) | 0 |
| S7_k141_73370 | 4323 | No | NA | | 4 | 0 | 1 | Not-determined | Genome-fragment | NA | NA | 0 |
| S7-final-viral-combined_k141_153611\|\|full | 2017 | No | NA | | 3 | 0 | 1 | Low-quality | Genome-fragment | 4.53 | AAI-based (medium-confidence) | 0 |
| S7_k141_237931 | 1101 | No | NA | | 2 | 0 | 0 | Not-determined | Genome-fragment | NA | NA | 0 |
| S7_k141_166192 | 1295 | No | NA | | 2 | 0 | 0 | Not-determined | Genome-fragment | NA | NA | 0 |
| S7_k141_94029 | 3433 | No | NA | | 7 | 0 | 0 | Not-determined | Genome-fragment | NA | NA | 0 |
| S17-final-viral-combined_k141_54506\|\|full | 2017 | No | NA | | 5 | 0 | 0 | Low-quality | Genome-fragment | 4.04 | AAI-based (high-confidence) | 0 |
| S7_k141_145245 | 7000 | No | NA | | 7 | 0 | 2 | Not-determined | Genome-fragment | NA | NA | 0 |
| S7_k141_33489 | 1422 | No | NA | | 2 | 0 | 1 | Not-determined | Genome-fragment | NA | NA | 0 |
| S7_k141_43678 | 1333 | No | NA | | 2 | 0 | 2 | Not-determined | Genome-fragment | NA | NA | 0 |
| S7_k141_156259 | 1348 | No | NA | | 1 | 0 | 0 | Not-determined | Genome-fragment | NA | NA | 0 |
| S7_k141_2782 | 5413 | No | NA | | 3 | 0 | 0 | Not-determined | Genome-fragment | NA | NA | 0 |
| S2_k141_238533 | 2017 | No | NA | | 3 | 0 | 2 | Low-quality | Genome-fragment | 1.05 | AAI-based (medium-confidence) | 0 |
| S7_k141_176153 | 1383 | No | NA | | 2 | 0 | 0 | Not-determined | Genome-fragment | NA | NA | 0 |
| S7_k141_135291 | 1812 | No | NA | | 2 | 0 | 2 | Not-determined | Genome-fragment | NA | NA | 0 |
| S7_k141_187042 | 1156 | No | NA | | 1 | 0 | 0 | Not-determined | Genome-fragment | NA | NA | 0 |
| S7_k141_197273 | 2339 | No | NA | | 2 | 0 | 1 | Not-determined | Genome-fragment | NA | NA | 0 |
| S7_k141_145317 | 2107 | No | NA | | 2 | 0 | 2 | Not-determined | Genome-fragment | NA | NA | 0 |
| S7_k141_12991 | 1243 | No | NA | | 1 | 0 | 1 | Not-determined | Genome-fragment | NA | NA | 0 |
| S1-final-viral-combined_k141_186662\|\|full | 2018 | Yes | | 570 | 4 | 1 | 1 | Low-quality | Genome-fragment | 0.87 | AAI-based (medium-confidence) | 71.75 |
| S5-final-viral-combined_k141_60429\|\|full | 2018 | No | NA | | 3 | 1 | 0 | Low-quality | Genome-fragment | 4.1 | AAI-based (high-confidence) | 0 |
| S7_k141_166217 | 1242 | No | NA | | 1 | 0 | 1 | Not-determined | Genome-fragment | NA | NA | 0 |
| S17-final-viral-combined_k141_205710\|\|full | 2018 | No | NA | | 2 | 0 | 2 | Low-quality | Genome-fragment | 1.84 | AAI-based (medium-confidence) | 0 |
| S7_k141_176230 | 1259 | No | NA | | 4 | 0 | 1 | Not-determined | Genome-fragment | NA | NA | 0 |
| S7_k141_237999 | 1054 | No | NA | | 1 | 0 | 1 | Not-determined | Genome-fragment | NA | NA | 0 |
| S2-final-viral-combined_k141_86189\|\|full | 2018 | No | NA | | 2 | 0 | 1 | Low-quality | Genome-fragment | 4.06 | AAI-based (high-confidence) | 0 |
| S7_k141_125305 | 1443 | No | NA | | 1 | 0 | 1 | Not-determined | Genome-fragment | NA | NA | 0 |
| S7_k141_115297 | 1606 | No | NA | | 1 | 0 | 1 | Not-determined | Genome-fragment | NA | NA | 0 |
| S7_k141_33677 | 1178 | No | NA | | 2 | 0 | 0 | Not-determined | Genome-fragment | NA | NA | 0 |
| S7_k141_238058 | 2592 | No | NA | | 3 | 0 | 2 | Not-determined | Genome-fragment | NA | NA | 0 |
| S7_k141_22750 | 3877 | No | NA | | 6 | 0 | 2 | Not-determined | Genome-fragment | NA | NA | 0 |
| S4-final-viral-combined_k141_132131\|\|full | 2018 | No | NA | | 2 | 0 | 2 | Low-quality | Genome-fragment | 6.73 | AAI-based (high-confidence) | 0 |
| S7_k141_135443 | 1389 | No | NA | | 2 | 0 | 1 | Not-determined | Genome-fragment | NA | NA | 0 |
| S8-final-viral-combined_k141_134225\|\|full | 2018 | No | NA | | 2 | 0 | 2 | Low-quality | Genome-fragment | 3.87 | AAI-based (medium-confidence) | 0 |
| S7_k141_125364 | 1059 | No | NA | | 1 | 0 | 0 | Not-determined | Genome-fragment | NA | NA | 0 |
| S9-final-viral-combined_k141_18353\|\|full | 2019 | No | NA | | 2 | 0 | 2 | Low-quality | Genome-fragment | 1.54 | AAI-based (high-confidence) | 0 |
| S1_k141_105818 | 2019 | No | NA | | 2 | 0 | 0 | Low-quality | Genome-fragment | 1.94 | AAI-based (medium-confidence) | 0 |
| S17-k141_122739 | 2019 | No | NA | | 2 | 0 | 2 | Low-quality | Genome-fragment | 0.59 | AAI-based (medium-confidence) | 0 |
| S7_k141_135498 | 5533 | No | NA | | 9 | 0 | 1 | Not-determined | Genome-fragment | NA | NA | 0 |
| S7_k141_166355 | 1402 | No | NA | | 1 | 0 | 1 | Not-determined | Genome-fragment | NA | NA | 0 |
| S7_k141_197509 | 4845 | No | NA | | 2 | 0 | 2 | Not-determined | Genome-fragment | NA | NA | 0 |
| S7_k141_104922 | 6946 | No | NA | | 4 | 0 | 0 | Not-determined | Genome-fragment | NA | NA | 0 |
| S12-final-viral-combined_k141_240289\|\|full | 2019 | No | NA | | 3 | 0 | 0 | Low-quality | Genome-fragment | 3.04 | AAI-based (medium-confidence) | 0 |
| S7_k141_207829 | 1550 | No | NA | | 2 | 0 | 1 | Not-determined | Genome-fragment | NA | NA | 0 |
| S7_k141_94362 | 1180 | No | NA | | 2 | 0 | 0 | Not-determined | Genome-fragment | NA | NA | 0 |
| S7_k141_125479 | 3297 | No | NA | | 4 | 0 | 1 | Not-determined | Genome-fragment | NA | NA | 0 |
| S7_k141_145524 | 3873 | No | NA | | 3 | 0 | 2 | Not-determined | Genome-fragment | NA | NA | 0 |
| S12-final-viral-combined_k141_81081\|\|full | 2019 | No | NA | | 3 | 0 | 3 | Low-quality | Genome-fragment | 1.24 | AAI-based (medium-confidence) | 0 |
| S7_k141_2946 | 3328 | No | NA | | 4 | 0 | 2 | Not-determined | Genome-fragment | NA | NA | 0 |
| S12-final-viral-combined_k141_215714\|\|full | 2019 | No | NA | | 4 | 1 | 0 | Low-quality | Genome-fragment | 4.34 | AAI-based (medium-confidence) | 0 |
| S7_k141_187175 | 1298 | No | NA | | 1 | 0 | 0 | Not-determined | Genome-fragment | NA | NA | 0 |
| S12-final-viral-combined_k141_373697\|\|lt2gene | 2019 | No | NA | | 2 | 1 | 0 | Low-quality | Genome-fragment | 2.65 | HMM-based (lower-bound) | 0 |
| S7_k141_207862 | 2894 | No | NA | | 1 | 0 | 1 | Not-determined | Genome-fragment | NA | NA | 0 |
| S7_k141_53996 | 1072 | No | NA | | 1 | 0 | 0 | Not-determined | Genome-fragment | NA | NA | 0 |
| S7_k141_227979 | 1033 | No | NA | | 1 | 0 | 0 | Not-determined | Genome-fragment | NA | NA | 0 |
| S7_k141_115423 | 7868 | No | NA | | 9 | 0 | 2 | Not-determined | Genome-fragment | NA | NA | 0 |
| S7_k141_187273 | 1252 | No | NA | | 2 | 0 | 0 | Not-determined | Genome-fragment | NA | NA | 0 |
| S6-final-viral-combined_k141_113027\|\|full | 2019 | No | NA | | 3 | 0 | 0 | Low-quality | Genome-fragment | 3.99 | AAI-based (medium-confidence) | 0 |
| S7_k141_43927 | 1306 | No | NA | | 1 | 0 | 1 | Not-determined | Genome-fragment | NA | NA | 0 |
| S7_k141_176531 | 1143 | No | NA | | 1 | 0 | 0 | Not-determined | Genome-fragment | NA | NA | 0 |
| S6-final-viral-combined_k141_31536\|\|full | 2019 | No | NA | | 3 | 0 | 1 | Low-quality | Genome-fragment | 0.95 | AAI-based (medium-confidence) | 0 |
| S7_k141_54109 | 4605 | No | NA | | 2 | 0 | 1 | Not-determined | Genome-fragment | NA | NA | 0 |
| S7_k141_156464 | 1307 | No | NA | | 1 | 0 | 1 | Not-determined | Genome-fragment | NA | NA | 0 |
| S7_k141_115472 | 1208 | No | NA | | 1 | 0 | 0 | Not-determined | Genome-fragment | NA | NA | 0 |
| S7_k141_94547 | 1012 | No | NA | | 2 | 0 | 0 | Not-determined | Genome-fragment | NA | NA | 0 |
| S7_k141_13254 | 1826 | No | NA | | 2 | 0 | 1 | Not-determined | Genome-fragment | NA | NA | 0 |
| S7_k141_33829 | 1177 | No | NA | | 2 | 0 | 1 | Not-determined | Genome-fragment | NA | NA | 0 |
| S7_k141_166579 | 1046 | No | NA | | 1 | 0 | 1 | Not-determined | Genome-fragment | NA | NA | 0 |
| S7_k141_187394 | 3835 | No | NA | | 1 | 0 | 0 | Not-determined | Genome-fragment | NA | NA | 0 |
| S7_k141_187396 | 1212 | No | NA | | 1 | 0 | 1 | Not-determined | Genome-fragment | NA | NA | 0 |
| S7_k141_197743 | 2463 | No | NA | | 4 | 0 | 2 | Not-determined | Genome-fragment | NA | NA | 0 |
| S7_k141_13326 | 4145 | No | NA | | 2 | 0 | 2 | Not-determined | Genome-fragment | NA | NA | 0 |
| S7_k141_197751 | 1193 | No | NA | | 1 | 0 | 1 | Not-determined | Genome-fragment | NA | NA | 0 |
| S7_k141_197770 | 1311 | No | NA | | 1 | 0 | 0 | Not-determined | Genome-fragment | NA | NA | 0 |
| S6-final-viral-combined_k141_120696\|\|full | 2019 | No | NA | | 2 | 0 | 0 | Low-quality | Genome-fragment | 4.79 | AAI-based (medium-confidence) | 0 |
| S7_k141_166664 | 1004 | No | NA | | 1 | 0 | 1 | Not-determined | Genome-fragment | NA | NA | 0 |
| S7_k141_197792 | 1411 | No | NA | | 3 | 0 | 0 | Not-determined | Genome-fragment | NA | NA | 0 |
| S7_k141_238315 | 1193 | No | NA | | 2 | 0 | 0 | Not-determined | Genome-fragment | NA | NA | 0 |
| S8-final-viral-combined_k141_159819\|\|full | 2019 | No | NA | | 3 | 0 | 1 | Low-quality | Genome-fragment | 0.52 | AAI-based (medium-confidence) | 0 |
| S7_k141_115550 | 5646 | No | NA | | 3 | 0 | 1 | Not-determined | Genome-fragment | NA | NA | 0 |
| S7-final-viral-combined_k141_57812\|\|full | 2020 | No | NA | | 5 | 3 | 0 | Low-quality | Genome-fragment | 5.21 | AAI-based (high-confidence) | 0 |
| S7_k141_238335 | 4851 | No | NA | | 3 | 0 | 3 | Not-determined | Genome-fragment | NA | NA | 0 |
| S7_k141_22921 | 1061 | No | NA | | 1 | 0 | 1 | Not-determined | Genome-fragment | NA | NA | 0 |
| S7_k141_238342 | 1281 | No | NA | | 1 | 0 | 0 | Not-determined | Genome-fragment | NA | NA | 0 |
| S7_k141_187453 | 5721 | No | NA | | 8 | 0 | 5 | Not-determined | Genome-fragment | NA | NA | 0 |
| S7_k141_64290 | 16852 | No | NA | | 2 | 0 | 1 | Not-determined | Genome-fragment | NA | NA | 0 |
| S7_k141_54172 | 1189 | No | NA | | 1 | 0 | 1 | Not-determined | Genome-fragment | NA | NA | 0 |
| S7_k141_54185 | 2005 | No | NA | | 2 | 0 | 1 | Not-determined | Genome-fragment | NA | NA | 0 |
| S7_k141_3178 | 1563 | No | NA | | 1 | 0 | 0 | Not-determined | Genome-fragment | NA | NA | 0 |
| S7_k141_13490 | 1364 | No | NA | | 1 | 0 | 1 | Not-determined | Genome-fragment | NA | NA | 0 |
| S1-final-viral-combined_k141_59047\|\|full | 2021 | No | NA | | 4 | 1 | 0 | Low-quality | Genome-fragment | 5.28 | AAI-based (high-confidence) | 0 |
| S7_k141_166744 | 1175 | No | NA | | 2 | 0 | 1 | Not-determined | Genome-fragment | NA | NA | 0 |
| S7_k141_208147 | 1127 | No | NA | | 2 | 0 | 0 | Not-determined | Genome-fragment | NA | NA | 0 |
| S7_k141_187543 | 2069 | No | NA | | 2 | 0 | 0 | Not-determined | Genome-fragment | NA | NA | 0 |
| S7_k141_54242 | 3886 | No | NA | | 1 | 0 | 1 | Not-determined | Genome-fragment | NA | NA | 0 |
| S7_k141_73912 | 1311 | No | NA | | 1 | 0 | 1 | Not-determined | Genome-fragment | NA | NA | 0 |
| S7_k141_156578 | 3969 | No | NA | | 2 | 0 | 1 | Not-determined | Genome-fragment | NA | NA | 0 |
| S7_k141_54252 | 1062 | No | NA | | 1 | 0 | 0 | Not-determined | Genome-fragment | NA | NA | 0 |
| S7-final-viral-combined_k141_41915\|\|full | 2021 | No | NA | | 2 | 0 | 1 | Low-quality | Genome-fragment | 3.29 | AAI-based (medium-confidence) | 0 |
| S7_k141_23047 | 1914 | No | NA | | 2 | 0 | 2 | Not-determined | Genome-fragment | NA | NA | 0 |
| S12-final-viral-combined_k141_81110\|\|full | 2021 | No | NA | | 3 | 0 | 3 | Low-quality | Genome-fragment | 1.24 | AAI-based (medium-confidence) | 0 |
| S7_k141_156660 | 1437 | No | NA | | 3 | 0 | 0 | Not-determined | Genome-fragment | NA | NA | 0 |
| S2-final-viral-combined_k141_1049\|\|full | 2021 | No | NA | | 4 | 1 | 0 | Low-quality | Genome-fragment | 1.22 | HMM-based (lower-bound) | 0 |
| S7_k141_197896 | 1061 | No | NA | | 2 | 0 | 2 | Not-determined | Genome-fragment | NA | NA | 0 |
| S7_k141_23064 | 5265 | No | NA | | 2 | 0 | 2 | Not-determined | Genome-fragment | NA | NA | 0 |
| S7_k141_105368 | 3109 | No | NA | | 1 | 0 | 1 | Not-determined | Genome-fragment | NA | NA | 0 |
| S7_k141_176854 | 4060 | No | NA | | 6 | 0 | 4 | Not-determined | Genome-fragment | NA | NA | 0 |
| S11_k141_32628 | 2022 | No | NA | | 5 | 1 | 0 | Low-quality | Genome-fragment | 3.52 | AAI-based (high-confidence) | 0 |
| S1_k141_283207 | 2022 | No | NA | | 1 | 0 | 1 | Low-quality | Genome-fragment | 3.82 | AAI-based (high-confidence) | 0 |
| S7_k141_156711 | 10656 | No | NA | | 18 | 0 | 3 | Not-determined | Genome-fragment | NA | NA | 0 |
| S17-final-viral-combined_k141_126508\|\|full | 2022 | No | NA | | 2 | 0 | 1 | Low-quality | Genome-fragment | 1.91 | AAI-based (high-confidence) | 0 |
| S7_k141_145919 | 1825 | No | NA | | 1 | 0 | 1 | Not-determined | Genome-fragment | NA | NA | 0 |
| S7_k141_217441 | 1205 | No | NA | | 2 | 0 | 1 | Not-determined | Genome-fragment | NA | NA | 0 |
| S7_k141_145939 | 1716 | No | NA | | 1 | 0 | 0 | Not-determined | Genome-fragment | NA | NA | 0 |
| S7_k141_64516 | 1100 | No | NA | | 2 | 0 | 0 | Not-determined | Genome-fragment | NA | NA | 0 |
| S7_k141_228337 | 1429 | No | NA | | 2 | 0 | 2 | Not-determined | Genome-fragment | NA | NA | 0 |
| S7_k141_176909 | 7655 | No | NA | | 8 | 0 | 3 | Not-determined | Genome-fragment | NA | NA | 0 |
| S12-final-viral-combined_k141_110448\|\|full | 2022 | No | NA | | 3 | 0 | 1 | Low-quality | Genome-fragment | 2.59 | AAI-based (medium-confidence) | 0 |
| S7_k141_176912 | 1030 | No | NA | | 1 | 0 | 0 | Not-determined | Genome-fragment | NA | NA | 0 |
| S7_k141_176925 | 4860 | No | NA | | 5 | 0 | 1 | Not-determined | Genome-fragment | NA | NA | 0 |
| S7_k141_84698 | 1607 | No | NA | | 2 | 0 | 2 | Not-determined | Genome-fragment | NA | NA | 0 |
| S7_k141_166924 | 6900 | No | NA | | 8 | 0 | 4 | Not-determined | Genome-fragment | NA | NA | 0 |
| S12_k141_369896 | 2022 | No | NA | | 3 | 0 | 2 | Low-quality | Genome-fragment | 1.23 | AAI-based (high-confidence) | 0 |
| S7_k141_74012 | 2321 | No | NA | | 1 | 0 | 0 | Not-determined | Genome-fragment | NA | NA | 0 |
| S8-final-viral-combined_k141_199183\|\|full | 2022 | No | NA | | 2 | 0 | 0 | Low-quality | Genome-fragment | 4 | AAI-based (high-confidence) | 0 |
| S12-final-viral-combined_k141_359083\|\|full | 2023 | No | NA | | 4 | 0 | 0 | Low-quality | Genome-fragment | 4.67 | AAI-based (medium-confidence) | 0 |
| S7_k141_125980 | 1828 | No | NA | | 2 | 0 | 2 | Not-determined | Genome-fragment | NA | NA | 0 |
| S7_k141_105428 | 3357 | No | NA | | 2 | 0 | 2 | Not-determined | Genome-fragment | NA | NA | 0 |
| S8-final-viral-combined_k141_263034\|\|full | 2023 | Yes | | 1163 | 4 | 1 | 1 | Low-quality | Genome-fragment | 2.07 | AAI-based (medium-confidence) | 42.51 |
| S5-final-viral-combined_k141_136045\|\|full | 2024 | No | NA | | 2 | 0 | 0 | Low-quality | Genome-fragment | 4.94 | AAI-based (medium-confidence) | 0 |
| S7_k141_208414 | 1901 | No | NA | | 1 | 0 | 0 | Not-determined | Genome-fragment | NA | NA | 0 |
| S7_k141_156832 | 2178 | No | NA | | 2 | 0 | 0 | Not-determined | Genome-fragment | NA | NA | 0 |
| S7_k141_44249 | 2557 | No | NA | | 1 | 0 | 1 | Not-determined | Genome-fragment | NA | NA | 0 |
| S7_k141_198108 | 4353 | No | NA | | 5 | 0 | 3 | Not-determined | Genome-fragment | NA | NA | 0 |
| S7_k141_126040 | 3931 | No | NA | | 6 | 0 | 6 | Not-determined | Genome-fragment | NA | NA | 0 |
| S7_k141_228476 | 1262 | No | NA | | 2 | 0 | 1 | Not-determined | Genome-fragment | NA | NA | 0 |
| S12-final-viral-combined_k141_305618\|\|full | 2024 | No | NA | | 6 | 0 | 0 | Low-quality | Genome-fragment | 5.86 | AAI-based (high-confidence) | 0 |
| S7_k141_228507 | 1373 | No | NA | | 2 | 0 | 0 | Not-determined | Genome-fragment | NA | NA | 0 |
| S7_k141_228536 | 1334 | No | NA | | 3 | 0 | 3 | Not-determined | Genome-fragment | NA | NA | 0 |
| S7_k141_187980 | 3142 | No | NA | | 2 | 0 | 0 | Not-determined | Genome-fragment | NA | NA | 0 |
| S12-final-viral-combined_k141_12770\|\|full | 2024 | No | NA | | 3 | 0 | 2 | Low-quality | Genome-fragment | 0.88 | AAI-based (medium-confidence) | 0 |
| S7_k141_238691 | 1423 | No | NA | | 1 | 0 | 1 | Not-determined | Genome-fragment | NA | NA | 0 |
| S4-final-viral-combined_k141_140224\|\|full | 2024 | No | NA | | 4 | 0 | 0 | Low-quality | Genome-fragment | 3.2 | AAI-based (medium-confidence) | 0 |
| S7_k141_177092 | 1277 | No | NA | | 1 | 0 | 0 | Not-determined | Genome-fragment | NA | NA | 0 |
| S4_k141_45879 | 2024 | No | NA | | 1 | 0 | 0 | Low-quality | Genome-fragment | 0.37 | AAI-based (medium-confidence) | 0 |
| S7_k141_3553 | 3874 | No | NA | | 7 | 0 | 7 | Not-determined | Genome-fragment | NA | NA | 0 |
| S7_k141_188042 | 5125 | No | NA | | 6 | 0 | 4 | Not-determined | Genome-fragment | NA | NA | 0 |
| S7_k141_105663 | 1241 | No | NA | | 4 | 0 | 4 | Not-determined | Genome-fragment | NA | NA | 0 |
| S9-final-viral-combined_k141_108725\|\|full | 2025 | No | NA | | 3 | 3 | 0 | Low-quality | Genome-fragment | 4.29 | AAI-based (high-confidence) | 0 |
| S7_k141_116162 | 1028 | No | NA | | 3 | 0 | 2 | Not-determined | Genome-fragment | NA | NA | 0 |
| S11-final-viral-combined_k141_322573\|\|full | 2025 | No | NA | | 3 | 0 | 3 | Low-quality | Genome-fragment | 1.27 | AAI-based (medium-confidence) | 0 |
| S7_k141_146270 | 3659 | No | NA | | 3 | 0 | 1 | Not-determined | Genome-fragment | NA | NA | 0 |
| S7_k141_198217 | 12412 | No | NA | | 15 | 0 | 0 | Not-determined | Genome-fragment | NA | NA | 0 |
| S1-final-viral-combined_k141_23698\|\|full | 2025 | No | NA | | 5 | 1 | 0 | Low-quality | Genome-fragment | 4.79 | AAI-based (medium-confidence) | 0 |
| S7_k141_44459 | 1220 | No | NA | | 1 | 0 | 0 | Not-determined | Genome-fragment | NA | NA | 0 |
| S3_k141_130082 | 2025 | No | NA | | 2 | 0 | 1 | Low-quality | Genome-fragment | 4.36 | AAI-based (high-confidence) | 0 |
| S7_k141_198290 | 1044 | No | NA | | 3 | 0 | 2 | Not-determined | Genome-fragment | NA | NA | 0 |
| S7_k141_146374 | 2531 | No | NA | | 2 | 0 | 2 | Not-determined | Genome-fragment | NA | NA | 0 |
| S7_k141_34554 | 3793 | No | NA | | 1 | 0 | 0 | Not-determined | Genome-fragment | NA | NA | 0 |
| S7_k141_157033 | 1738 | No | NA | | 2 | 0 | 0 | Not-determined | Genome-fragment | NA | NA | 0 |
| S7_k141_177251 | 6189 | No | NA | | 6 | 0 | 2 | Not-determined | Genome-fragment | NA | NA | 0 |
| S7_k141_126288 | 3060 | No | NA | | 3 | 0 | 3 | Not-determined | Genome-fragment | NA | NA | 0 |
| S7-final-viral-combined_k141_230863\|\|full | 2025 | No | NA | | 5 | 0 | 0 | Low-quality | Genome-fragment | 2.8 | AAI-based (medium-confidence) | 0 |
| S7_k141_167385 | 1352 | No | NA | | 1 | 0 | 1 | Not-determined | Genome-fragment | NA | NA | 0 |
| S2-final-viral-combined_k141_123390\|\|full | 2025 | No | NA | | 3 | 0 | 0 | Low-quality | Genome-fragment | 3.57 | AAI-based (medium-confidence) | 0 |
| S7_k141_188215 | 1122 | No | NA | | 1 | 0 | 0 | Not-determined | Genome-fragment | NA | NA | 0 |
| S7_k141_85220 | 1823 | No | NA | | 1 | 0 | 0 | Not-determined | Genome-fragment | NA | NA | 0 |
| S7_k141_34657 | 1037 | No | NA | | 1 | 0 | 0 | Not-determined | Genome-fragment | NA | NA | 0 |
| S7_k141_23447 | 2724 | No | NA | | 1 | 0 | 1 | Not-determined | Genome-fragment | NA | NA | 0 |
| S7_k141_238990 | 1316 | No | NA | | 1 | 0 | 1 | Not-determined | Genome-fragment | NA | NA | 0 |
| S7_k141_95172 | 1249 | No | NA | | 2 | 0 | 2 | Not-determined | Genome-fragment | NA | NA | 0 |
| S7_k141_208817 | 13409 | No | NA | | 12 | 0 | 8 | Not-determined | Genome-fragment | NA | NA | 0 |
| S7_k141_217707 | 1115 | No | NA | | 1 | 0 | 1 | Not-determined | Genome-fragment | NA | NA | 0 |
| S7_k141_95220 | 1015 | No | NA | | 1 | 0 | 1 | Not-determined | Genome-fragment | NA | NA | 0 |
| S7_k141_74694 | 2154 | No | NA | | 1 | 0 | 0 | Not-determined | Genome-fragment | NA | NA | 0 |
| S6-final-viral-combined_k141_113903\|\|full | 2025 | No | NA | | 3 | 0 | 0 | Low-quality | Genome-fragment | 1.15 | AAI-based (medium-confidence) | 0 |
| S7_k141_208825 | 1237 | No | NA | | 3 | 0 | 2 | Not-determined | Genome-fragment | NA | NA | 0 |
| S7_k141_34743 | 1066 | No | NA | | 1 | 0 | 0 | Not-determined | Genome-fragment | NA | NA | 0 |
| S7_k141_95253 | 1250 | No | NA | | 1 | 0 | 0 | Not-determined | Genome-fragment | NA | NA | 0 |
| S7_k141_146590 | 3391 | No | NA | | 5 | 0 | 0 | Not-determined | Genome-fragment | NA | NA | 0 |
| S7_k141_188316 | 2736 | No | NA | | 2 | 0 | 0 | Not-determined | Genome-fragment | NA | NA | 0 |
| S7_k141_146591 | 16112 | No | NA | | 14 | 0 | 8 | Not-determined | Genome-fragment | NA | NA | 0 |
| S7_k141_34805 | 1167 | No | NA | | 1 | 0 | 1 | Not-determined | Genome-fragment | NA | NA | 0 |
| S7_k141_136365 | 3715 | No | NA | | 3 | 0 | 0 | Not-determined | Genome-fragment | NA | NA | 0 |
| S7_k141_54651 | 2121 | No | NA | | 1 | 0 | 0 | Not-determined | Genome-fragment | NA | NA | 0 |
| S7_k141_34809 | 3388 | No | NA | | 3 | 0 | 3 | Not-determined | Genome-fragment | NA | NA | 0 |
| S7_k141_208907 | 4028 | No | NA | | 2 | 0 | 0 | Not-determined | Genome-fragment | NA | NA | 0 |
| S7_k141_198389 | 3261 | No | NA | | 6 | 0 | 1 | Not-determined | Genome-fragment | NA | NA | 0 |
| S6-final-viral-combined_k141_22208\|\|full | 2025 | No | NA | | 3 | 1 | 0 | Low-quality | Genome-fragment | 4.75 | AAI-based (medium-confidence) | 0 |
| S7_k141_44744 | 1616 | No | NA | | 3 | 0 | 0 | Not-determined | Genome-fragment | NA | NA | 0 |
| S1-final-viral-combined_k141_234242\|\|lt2gene | 2026 | No | NA | | 2 | 2 | 0 | Low-quality | Genome-fragment | 5.07 | AAI-based (medium-confidence) | 0 |
| S7_k141_146615 | 5498 | No | NA | | 7 | 0 | 1 | Not-determined | Genome-fragment | NA | NA | 0 |
| S7_k141_105812 | 1113 | No | NA | | 2 | 0 | 2 | Not-determined | Genome-fragment | NA | NA | 0 |
| S7_k141_85358 | 1557 | No | NA | | 2 | 0 | 0 | Not-determined | Genome-fragment | NA | NA | 0 |
| S2-final-viral-combined_k141_211925\|\|full | 2026 | No | NA | | 3 | 0 | 0 | Low-quality | Genome-fragment | 3.85 | AAI-based (medium-confidence) | 0 |
| S7_k141_136441 | 3845 | No | NA | | 3 | 0 | 0 | Not-determined | Genome-fragment | NA | NA | 0 |
| S7_k141_188363 | 1806 | No | NA | | 1 | 0 | 0 | Not-determined | Genome-fragment | NA | NA | 0 |
| S7_k141_54704 | 1007 | No | NA | | 2 | 0 | 1 | Not-determined | Genome-fragment | NA | NA | 0 |
| S7_k141_198446 | 1859 | No | NA | | 3 | 0 | 1 | Not-determined | Genome-fragment | NA | NA | 0 |
| S4-final-viral-combined_k141_2922\|\|full | 2026 | No | NA | | 4 | 0 | 0 | Low-quality | Genome-fragment | 4.63 | AAI-based (high-confidence) | 0 |
| S7_k141_146669 | 2097 | No | NA | | 1 | 0 | 1 | Not-determined | Genome-fragment | NA | NA | 0 |
| S7_k141_157247 | 1520 | No | NA | | 2 | 0 | 1 | Not-determined | Genome-fragment | NA | NA | 0 |
| S7_k141_126595 | 1353 | No | NA | | 1 | 0 | 1 | Not-determined | Genome-fragment | NA | NA | 0 |
| S7_k141_239150 | 2338 | No | NA | | 1 | 0 | 1 | Not-determined | Genome-fragment | NA | NA | 0 |
| S7-final-viral-combined_k141_210738\|\|full | 2027 | No | NA | | 3 | 0 | 2 | Low-quality | Genome-fragment | 2.32 | AAI-based (medium-confidence) | 0 |
| S7_k141_208984 | 1634 | No | NA | | 4 | 0 | 4 | Not-determined | Genome-fragment | NA | NA | 0 |
| S17-final-viral-combined_k141_83987\|\|full | 2027 | No | NA | | 3 | 0 | 0 | Low-quality | Genome-fragment | 3.02 | AAI-based (high-confidence) | 0 |
| S7_k141_34870 | 3558 | No | NA | | 7 | 0 | 2 | Not-determined | Genome-fragment | NA | NA | 0 |
| S7_k141_34872 | 1057 | No | NA | | 2 | 0 | 0 | Not-determined | Genome-fragment | NA | NA | 0 |
| S7_k141_95432 | 1032 | No | NA | | 2 | 0 | 2 | Not-determined | Genome-fragment | NA | NA | 0 |
| S7_k141_188467 | 2707 | No | NA | | 4 | 0 | 1 | Not-determined | Genome-fragment | NA | NA | 0 |
| S12-final-viral-combined_k141_239987\|\|full | 2027 | No | NA | | 4 | 0 | 1 | Low-quality | Genome-fragment | 3.01 | AAI-based (medium-confidence) | 0 |
| S7_k141_188476 | 1028 | No | NA | | 1 | 0 | 1 | Not-determined | Genome-fragment | NA | NA | 0 |
| S2-final-viral-combined_k141_50146\|\|full | 2027 | No | NA | | 3 | 0 | 0 | Low-quality | Genome-fragment | 2.78 | AAI-based (high-confidence) | 0 |
| S7_k141_14129 | 4221 | No | NA | | 5 | 0 | 1 | Not-determined | Genome-fragment | NA | NA | 0 |
| S4-final-viral-combined_k141_143595\|\|full | 2027 | No | NA | | 2 | 0 | 0 | Low-quality | Genome-fragment | 1.33 | AAI-based (high-confidence) | 0 |
| S7_k141_177717 | 3408 | No | NA | | 2 | 0 | 1 | Not-determined | Genome-fragment | NA | NA | 0 |
| S7_k141_85508 | 1319 | No | NA | | 3 | 0 | 1 | Not-determined | Genome-fragment | NA | NA | 0 |
| S7_k141_3874 | 3983 | No | NA | | 2 | 0 | 1 | Not-determined | Genome-fragment | NA | NA | 0 |
| S7_k141_157264 | 15928 | No | NA | | 18 | 5 | 0 | Medium-quality | Genome-fragment | 53.02 | HMM-based (lower-bound) | 0 |
| S7_k141_54852 | 6185 | No | NA | | 5 | 0 | 4 | Not-determined | Genome-fragment | NA | NA | 0 |
| S7_k141_228946 | 1179 | No | NA | | 2 | 0 | 0 | Not-determined | Genome-fragment | NA | NA | 0 |
| S7_k141_177763 | 4402 | No | NA | | 1 | 0 | 0 | Not-determined | Genome-fragment | NA | NA | 0 |
| S7_k141_65358 | 3500 | No | NA | | 4 | 0 | 1 | Not-determined | Genome-fragment | NA | NA | 0 |
| S7_k141_34930 | 6796 | No | NA | | 6 | 0 | 3 | Not-determined | Genome-fragment | NA | NA | 0 |
| S7_k141_177779 | 1006 | No | NA | | 1 | 0 | 0 | Not-determined | Genome-fragment | NA | NA | 0 |
| S7_k141_65389 | 1790 | No | NA | | 2 | 0 | 1 | Not-determined | Genome-fragment | NA | NA | 0 |
| S7_k141_23780 | 1050 | No | NA | | 2 | 0 | 1 | Not-determined | Genome-fragment | NA | NA | 0 |
| S7_k141_23798 | 1313 | No | NA | | 1 | 0 | 1 | Not-determined | Genome-fragment | NA | NA | 0 |
| S9-final-viral-combined_k141_102489\|\|full | 2028 | No | NA | | 3 | 0 | 2 | Low-quality | Genome-fragment | 3.33 | AAI-based (medium-confidence) | 0 |
| S7_k141_45001 | 1042 | No | NA | | 1 | 0 | 0 | Not-determined | Genome-fragment | NA | NA | 0 |
| S7_k141_177829 | 1262 | No | NA | | 3 | 0 | 0 | Not-determined | Genome-fragment | NA | NA | 0 |
| S11-final-viral-combined_k141_117439\|\|full | 2028 | No | NA | | 6 | 0 | 0 | Low-quality | Genome-fragment | 3.38 | AAI-based (medium-confidence) | 0 |
| S7_k141_95561 | 2309 | No | NA | | 2 | 0 | 1 | Not-determined | Genome-fragment | NA | NA | 0 |
| S7_k141_126827 | 2662 | No | NA | | 3 | 0 | 1 | Not-determined | Genome-fragment | NA | NA | 0 |
| S7_k141_23814 | 1119 | No | NA | | 1 | 0 | 1 | Not-determined | Genome-fragment | NA | NA | 0 |
| S7_k141_177865 | 5272 | No | NA | | 2 | 0 | 2 | Not-determined | Genome-fragment | NA | NA | 0 |
| S7_k141_239375 | 1300 | No | NA | | 2 | 0 | 0 | Not-determined | Genome-fragment | NA | NA | 0 |
| S5-final-viral-combined_k141_108907\|\|full | 2028 | No | NA | | 5 | 3 | 0 | Low-quality | Genome-fragment | 4.32 | AAI-based (high-confidence) | 0 |
| S7_k141_106187 | 3394 | No | NA | | 4 | 0 | 3 | Not-determined | Genome-fragment | NA | NA | 0 |
| S7_k141_146845 | 1263 | No | NA | | 1 | 0 | 0 | Not-determined | Genome-fragment | NA | NA | 0 |
| S7_k141_85667 | 1753 | No | NA | | 3 | 0 | 2 | Not-determined | Genome-fragment | NA | NA | 0 |
| S7_k141_85685 | 1213 | No | NA | | 1 | 0 | 1 | Not-determined | Genome-fragment | NA | NA | 0 |
| S7_k141_239426 | 3592 | No | NA | | 6 | 0 | 2 | Not-determined | Genome-fragment | NA | NA | 0 |
| S7_k141_65581 | 2854 | No | NA | | 1 | 0 | 0 | Not-determined | Genome-fragment | NA | NA | 0 |
| S7-final-viral-combined_k141_67262\|\|full | 2028 | No | NA | | 3 | 0 | 1 | Low-quality | Genome-fragment | 1.2 | AAI-based (medium-confidence) | 0 |
| S12-final-viral-combined_k141_115637\|\|full | 2028 | No | NA | | 5 | 2 | 0 | Low-quality | Genome-fragment | 5.43 | AAI-based (high-confidence) | 0 |
| S7_k141_146895 | 1619 | No | NA | | 1 | 0 | 1 | Not-determined | Genome-fragment | NA | NA | 0 |
| S7_k141_177907 | 1119 | No | NA | | 2 | 0 | 1 | Not-determined | Genome-fragment | NA | NA | 0 |
| S7_k141_45105 | 1145 | No | NA | | 1 | 0 | 1 | Not-determined | Genome-fragment | NA | NA | 0 |
| S7_k141_35100 | 2104 | No | NA | | 2 | 0 | 2 | Not-determined | Genome-fragment | NA | NA | 0 |
| S7_k141_177963 | 1001 | No | NA | | 3 | 0 | 0 | Not-determined | Genome-fragment | NA | NA | 0 |
| S7_k141_116736 | 3106 | No | NA | | 3 | 0 | 3 | Not-determined | Genome-fragment | NA | NA | 0 |
| S2-final-viral-combined_k141_199973\|\|full | 2028 | No | NA | | 4 | 3 | 0 | Low-quality | Genome-fragment | 4 | AAI-based (high-confidence) | 0 |
| S7_k141_95675 | 1273 | No | NA | | 2 | 0 | 1 | Not-determined | Genome-fragment | NA | NA | 0 |
| S7_k141_136720 | 1096 | No | NA | | 1 | 0 | 1 | Not-determined | Genome-fragment | NA | NA | 0 |
| S7_k141_85717 | 3762 | No | NA | | 5 | 0 | 1 | Not-determined | Genome-fragment | NA | NA | 0 |
| S7_k141_116757 | 2238 | No | NA | | 2 | 0 | 2 | Not-determined | Genome-fragment | NA | NA | 0 |
| S8-final-viral-combined_k141_43039\|\|full | 2028 | No | NA | | 4 | 1 | 0 | Low-quality | Genome-fragment | 3.25 | AAI-based (medium-confidence) | 0 |
| S7_k141_239597 | 1485 | No | NA | | 2 | 0 | 2 | Not-determined | Genome-fragment | NA | NA | 0 |
| S7_k141_217975 | 1910 | No | NA | | 2 | 0 | 2 | Not-determined | Genome-fragment | NA | NA | 0 |
| S9-final-viral-combined_k141_104056\|\|full | 2029 | No | NA | | 3 | 1 | 0 | Low-quality | Genome-fragment | 3.26 | HMM-based (lower-bound) | 0 |
| S11-final-viral-combined_k141_148488\|\|full | 2029 | No | NA | | 5 | 1 | 0 | Low-quality | Genome-fragment | 4.76 | AAI-based (high-confidence) | 0 |
| S7_k141_239642 | 1434 | No | NA | | 1 | 0 | 1 | Not-determined | Genome-fragment | NA | NA | 0 |
| S7-final-viral-combined_k141_132387\|\|full | 2029 | No | NA | | 3 | 2 | 0 | Low-quality | Genome-fragment | 6.29 | AAI-based (high-confidence) | 0 |
| S2-final-viral-combined_k141_58074\|\|full | 2029 | No | NA | | 3 | 0 | 1 | Low-quality | Genome-fragment | 1.19 | AAI-based (medium-confidence) | 0 |
| S7_k141_95761 | 1217 | No | NA | | 2 | 0 | 1 | Not-determined | Genome-fragment | NA | NA | 0 |
| S7_k141_95767 | 1014 | No | NA | | 1 | 0 | 1 | Not-determined | Genome-fragment | NA | NA | 0 |
| S7_k141_23969 | 1528 | No | NA | | 1 | 0 | 1 | Not-determined | Genome-fragment | NA | NA | 0 |
| S2_k141_155386 | 2029 | No | NA | | 6 | 0 | 0 | Low-quality | Genome-fragment | 3.52 | AAI-based (high-confidence) | 0 |
| S7_k141_188597 | 8498 | No | NA | | 7 | 0 | 6 | Not-determined | Genome-fragment | NA | NA | 0 |
| S7_k141_209360 | 2175 | No | NA | | 2 | 0 | 0 | Not-determined | Genome-fragment | NA | NA | 0 |
| S7_k141_23985 | 3729 | No | NA | | 2 | 0 | 0 | Not-determined | Genome-fragment | NA | NA | 0 |
| S2_k141_45167 | 2029 | No | NA | | 2 | 0 | 2 | Low-quality | Genome-fragment | 5.12 | AAI-based (medium-confidence) | 0 |
| S11-final-viral-combined_k141_224085\|\|full | 2030 | No | NA | | 2 | 1 | 0 | Low-quality | Genome-fragment | 4.03 | AAI-based (high-confidence) | 0 |
| S7_k141_136850 | 1238 | No | NA | | 2 | 0 | 2 | Not-determined | Genome-fragment | NA | NA | 0 |
| S7_k141_14322 | 1852 | No | NA | | 2 | 0 | 2 | Not-determined | Genome-fragment | NA | NA | 0 |
| S7_k141_75155 | 1456 | No | NA | | 2 | 0 | 1 | Not-determined | Genome-fragment | NA | NA | 0 |
| S7_k141_127210 | 3410 | No | NA | | 2 | 0 | 1 | Not-determined | Genome-fragment | NA | NA | 0 |
| S7_k141_14385 | 1139 | No | NA | | 2 | 0 | 0 | Not-determined | Genome-fragment | NA | NA | 0 |
| S11-final-viral-combined_k141_47300\|\|full | 2030 | No | NA | | 3 | 2 | 0 | Low-quality | Genome-fragment | 5.33 | AAI-based (high-confidence) | 0 |
| S7_k141_95836 | 1383 | No | NA | | 1 | 0 | 1 | Not-determined | Genome-fragment | NA | NA | 0 |
| S11-final-viral-combined_k141_219928\|\|full | 2030 | No | NA | | 3 | 0 | 2 | Low-quality | Genome-fragment | 1.28 | AAI-based (medium-confidence) | 0 |
| S12-final-viral-combined_k141_36463\|\|full | 2030 | No | NA | | 3 | 1 | 0 | Low-quality | Genome-fragment | 0.93 | AAI-based (high-confidence) | 0 |
| S7_k141_157784 | 1206 | No | NA | | 1 | 0 | 1 | Not-determined | Genome-fragment | NA | NA | 0 |
| S7_k141_199133 | 1016 | No | NA | | 1 | 0 | 0 | Not-determined | Genome-fragment | NA | NA | 0 |
| S7_k141_199137 | 7570 | No | NA | | 5 | 0 | 1 | Not-determined | Genome-fragment | NA | NA | 0 |
| S7_k141_14460 | 1885 | No | NA | | 2 | 0 | 1 | Not-determined | Genome-fragment | NA | NA | 0 |
| S7_k141_199145 | 4829 | No | NA | | 6 | 0 | 4 | Not-determined | Genome-fragment | NA | NA | 0 |
| S7_k141_85871 | 4503 | No | NA | | 1 | 0 | 0 | Not-determined | Genome-fragment | NA | NA | 0 |
| S7_k141_218143 | 1484 | No | NA | | 1 | 0 | 1 | Not-determined | Genome-fragment | NA | NA | 0 |
| S7_k141_116939 | 1004 | No | NA | | 2 | 0 | 0 | Not-determined | Genome-fragment | NA | NA | 0 |
| S12-final-viral-combined_k141_130713\|\|full | 2030 | No | NA | | 4 | 0 | 0 | Low-quality | Genome-fragment | 4.64 | AAI-based (high-confidence) | 0 |
| S12-final-viral-combined_k141_401094\|\|full | 2030 | No | NA | | 2 | 1 | 0 | Low-quality | Genome-fragment | 4.48 | AAI-based (high-confidence) | 0 |
| S7_k141_178218 | 1231 | No | NA | | 1 | 0 | 1 | Not-determined | Genome-fragment | NA | NA | 0 |
| S7_k141_239833 | 16632 | No | NA | | 28 | 8 | 0 | Medium-quality | Genome-fragment | 75.22 | AAI-based (medium-confidence) | 0 |
| S7_k141_157861 | 1058 | No | NA | | 2 | 0 | 1 | Not-determined | Genome-fragment | NA | NA | 0 |
| S7_k141_127259 | 1604 | No | NA | | 1 | 0 | 1 | Not-determined | Genome-fragment | NA | NA | 0 |
| S7_k141_137008 | 1036 | No | NA | | 1 | 0 | 0 | Not-determined | Genome-fragment | NA | NA | 0 |
| S7_k141_14526 | 1498 | No | NA | | 1 | 0 | 1 | Not-determined | Genome-fragment | NA | NA | 0 |
| S7_k141_147046 | 8784 | No | NA | | 11 | 0 | 9 | Not-determined | Genome-fragment | NA | NA | 0 |
| S7_k141_209584 | 4580 | No | NA | | 3 | 0 | 1 | Not-determined | Genome-fragment | NA | NA | 0 |
| S7_k141_116966 | 6783 | No | NA | | 3 | 0 | 2 | Not-determined | Genome-fragment | NA | NA | 0 |
| S7_k141_35477 | 1151 | No | NA | | 1 | 0 | 0 | Not-determined | Genome-fragment | NA | NA | 0 |
| S7_k141_147072 | 1043 | No | NA | | 1 | 0 | 1 | Not-determined | Genome-fragment | NA | NA | 0 |
| S7_k141_55353 | 1396 | No | NA | | 3 | 0 | 3 | Not-determined | Genome-fragment | NA | NA | 0 |
| S7_k141_85988 | 3621 | No | NA | | 4 | 0 | 2 | Not-determined | Genome-fragment | NA | NA | 0 |
| S12-final-viral-combined_k141_140017\|\|lt2gene | 2030 | No | NA | | 2 | 1 | 0 | Low-quality | Genome-fragment | 1.32 | AAI-based (high-confidence) | 0 |
| S12_k141_235857 | 2030 | No | NA | | 4 | 1 | 0 | Low-quality | Genome-fragment | 1.11 | AAI-based (high-confidence) | 0 |
| S7_k141_55393 | 1191 | No | NA | | 2 | 0 | 0 | Not-determined | Genome-fragment | NA | NA | 0 |
| S2-final-viral-combined_k141_191313\|\|full | 2030 | No | NA | | 3 | 0 | 1 | Low-quality | Genome-fragment | 2.57 | AAI-based (high-confidence) | 0 |
| S7_k141_66025 | 3398 | No | NA | | 2 | 0 | 2 | Not-determined | Genome-fragment | NA | NA | 0 |
| S7-final-viral-combined_k141_131983\|\|full | 2031 | No | NA | | 4 | 1 | 0 | Low-quality | Genome-fragment | 3.99 | AAI-based (medium-confidence) | 0 |
| S17-final-viral-combined_k141_215422\|\|full | 2031 | No | NA | | 2 | 0 | 1 | Low-quality | Genome-fragment | 1.92 | AAI-based (medium-confidence) | 0 |
| S7_k141_209673 | 1532 | No | NA | | 1 | 0 | 0 | Not-determined | Genome-fragment | NA | NA | 0 |
| S7_k141_4473 | 1313 | No | NA | | 1 | 0 | 1 | Not-determined | Genome-fragment | NA | NA | 0 |
| S7_k141_86028 | 3939 | No | NA | | 4 | 0 | 3 | Not-determined | Genome-fragment | NA | NA | 0 |
| S7_k141_158050 | 1005 | No | NA | | 2 | 0 | 0 | Not-determined | Genome-fragment | NA | NA | 0 |
| S7_k141_35586 | 1073 | No | NA | | 1 | 0 | 0 | Not-determined | Genome-fragment | NA | NA | 0 |
| S7_k141_55487 | 1011 | No | NA | | 2 | 0 | 0 | Not-determined | Genome-fragment | NA | NA | 0 |
| S7_k141_66095 | 3001 | No | NA | | 6 | 0 | 0 | Not-determined | Genome-fragment | NA | NA | 0 |
| S12_k141_278491 | 2031 | No | NA | | 2 | 0 | 2 | Low-quality | Genome-fragment | 0.76 | AAI-based (high-confidence) | 0 |
| S7_k141_127415 | 3316 | No | NA | | 3 | 0 | 1 | Not-determined | Genome-fragment | NA | NA | 0 |
| S7_k141_96120 | 1541 | No | NA | | 2 | 0 | 2 | Not-determined | Genome-fragment | NA | NA | 0 |
| S7_k141_4558 | 1046 | No | NA | | 2 | 0 | 0 | Not-determined | Genome-fragment | NA | NA | 0 |
| S7_k141_35699 | 2098 | No | NA | | 3 | 0 | 0 | Not-determined | Genome-fragment | NA | NA | 0 |
| S7_k141_4568 | 3132 | No | NA | | 3 | 0 | 0 | Not-determined | Genome-fragment | NA | NA | 0 |
| S7_k141_199349 | 3610 | No | NA | | 4 | 0 | 4 | Not-determined | Genome-fragment | NA | NA | 0 |
| S7_k141_168332 | 1066 | No | NA | | 2 | 0 | 1 | Not-determined | Genome-fragment | NA | NA | 0 |
| S7_k141_55593 | 1008 | No | NA | | 1 | 0 | 0 | Not-determined | Genome-fragment | NA | NA | 0 |
| S7_k141_240039 | 1066 | No | NA | | 1 | 0 | 1 | Not-determined | Genome-fragment | NA | NA | 0 |
| S7_k141_218294 | 4181 | No | NA | | 4 | 0 | 4 | Not-determined | Genome-fragment | NA | NA | 0 |
| S7_k141_35741 | 3062 | No | NA | | 6 | 0 | 0 | Not-determined | Genome-fragment | NA | NA | 0 |
| S7_k141_106480 | 3961 | No | NA | | 4 | 0 | 0 | Not-determined | Genome-fragment | NA | NA | 0 |
| S7_k141_158193 | 5462 | No | NA | | 7 | 0 | 6 | Not-determined | Genome-fragment | NA | NA | 0 |
| S4-final-viral-combined_k141_159914\|\|full | 2031 | No | NA | | 5 | 0 | 0 | Low-quality | Genome-fragment | 3.14 | AAI-based (medium-confidence) | 0 |
| S7_k141_106522 | 1840 | No | NA | | 1 | 0 | 1 | Not-determined | Genome-fragment | NA | NA | 0 |
| S7_k141_96255 | 1399 | No | NA | | 1 | 0 | 0 | Not-determined | Genome-fragment | NA | NA | 0 |
| S8-final-viral-combined_k141_164834\|\|full | 2031 | Yes | | 920 | 4 | 1 | 1 | Low-quality | Genome-fragment | 0.35 | HMM-based (lower-bound) | 54.7 |
| S7_k141_168444 | 3053 | No | NA | | 6 | 0 | 0 | Not-determined | Genome-fragment | NA | NA | 0 |
| S7_k141_24226 | 1459 | No | NA | | 2 | 0 | 1 | Not-determined | Genome-fragment | NA | NA | 0 |
| S7_k141_96290 | 1146 | No | NA | | 3 | 0 | 2 | Not-determined | Genome-fragment | NA | NA | 0 |
| S7_k141_35852 | 3022 | No | NA | | 3 | 0 | 2 | Not-determined | Genome-fragment | NA | NA | 0 |
| S11-final-viral-combined_k141_69557\|\|lt2gene | 2032 | No | NA | | 1 | 0 | 0 | Low-quality | Genome-fragment | 4.51 | AAI-based (medium-confidence) | 0 |
| S7_k141_168494 | 1249 | No | NA | | 2 | 0 | 1 | Not-determined | Genome-fragment | NA | NA | 0 |
| S7_k141_189000 | 1919 | No | NA | | 3 | 0 | 2 | Not-determined | Genome-fragment | NA | NA | 0 |
| S11_k141_109937 | 2032 | No | NA | | 3 | 1 | 0 | Low-quality | Genome-fragment | 3.44 | AAI-based (high-confidence) | 0 |
| S7_k141_199436 | 3852 | No | NA | | 3 | 0 | 2 | Not-determined | Genome-fragment | NA | NA | 0 |
| S7_k141_106593 | 1759 | No | NA | | 5 | 0 | 4 | Not-determined | Genome-fragment | NA | NA | 0 |
| S7_k141_178759 | 2188 | No | NA | | 2 | 0 | 2 | Not-determined | Genome-fragment | NA | NA | 0 |
| S7_k141_189070 | 3102 | No | NA | | 4 | 0 | 3 | Not-determined | Genome-fragment | NA | NA | 0 |
| S7_k141_240154 | 1471 | No | NA | | 1 | 0 | 1 | Not-determined | Genome-fragment | NA | NA | 0 |
| S1-final-viral-combined_k141_182643\|\|full | 2032 | No | NA | | 3 | 0 | 0 | Low-quality | Genome-fragment | 4.21 | AAI-based (medium-confidence) | 0 |
| S7_k141_137390 | 1314 | No | NA | | 4 | 0 | 1 | Not-determined | Genome-fragment | NA | NA | 0 |
| S7_k141_158352 | 1477 | No | NA | | 1 | 0 | 1 | Not-determined | Genome-fragment | NA | NA | 0 |
| S7_k141_86369 | 3556 | No | NA | | 5 | 0 | 4 | Not-determined | Genome-fragment | NA | NA | 0 |
| S7_k141_14786 | 1081 | No | NA | | 2 | 0 | 1 | Not-determined | Genome-fragment | NA | NA | 0 |
| S7_k141_86386 | 4703 | No | NA | | 9 | 0 | 2 | Not-determined | Genome-fragment | NA | NA | 0 |
| S7_k141_75731 | 1273 | No | NA | | 1 | 0 | 0 | Not-determined | Genome-fragment | NA | NA | 0 |
| S7-final-viral-combined_k141_128275\|\|full | 2032 | No | NA | | 3 | 0 | 1 | Low-quality | Genome-fragment | 1.19 | AAI-based (high-confidence) | 0 |
| S7_k141_168646 | 2193 | No | NA | | 1 | 0 | 1 | Not-determined | Genome-fragment | NA | NA | 0 |

| | | | | | | | | | | | |
|---|---|---|---|---|---|---|---|---|---|---|---|
| S7_k141_147415 | 5202 | No | NA | 4 | 0 | 3 | Not-determined | Genome-fragment | NA | NA | 0 |
| S17-final-viral-combined_k141_47244\|\|full | 2032 | No | NA | 4 | 3 | 0 | Low-quality | Genome-fragment | 5.2 | AAI-based (high-confidence) | 0 |
| S7_k141_147497 | 1590 | No | NA | 2 | 0 | 2 | Not-determined | Genome-fragment | NA | NA | 0 |
| S7_k141_209786 | 3543 | No | NA | 3 | 0 | 3 | Not-determined | Genome-fragment | NA | NA | 0 |
| S7_k141_229778 | 5975 | No | NA | 3 | 0 | 2 | Not-determined | Genome-fragment | NA | NA | 0 |
| S7_k141_147509 | 1419 | No | NA | 3 | 0 | 1 | Not-determined | Genome-fragment | NA | NA | 0 |
| S12-final-viral-combined_k141_366486\|\|full | 2032 | No | NA | 2 | 0 | 2 | Low-quality | Genome-fragment | 2.21 | AAI-based (medium-confidence) | 0 |
| S8-final-viral-combined_k141_237720\|\|full | 2032 | No | NA | 5 | 1 | 0 | Low-quality | Genome-fragment | 5.96 | AAI-based (high-confidence) | 0 |
| S7_k141_199639 | 1701 | No | NA | 5 | 0 | 5 | Not-determined | Genome-fragment | NA | NA | 0 |
| S7_k141_24256 | 3158 | No | NA | 2 | 0 | 2 | Not-determined | Genome-fragment | NA | NA | 0 |
| S7_k141_35932 | 19398 | No | NA | 15 | 0 | 12 | Not-determined | Genome-fragment | NA | NA | 0 |
| S7_k141_56016 | 1049 | No | NA | 1 | 0 | 0 | Not-determined | Genome-fragment | NA | NA | 0 |
| S7_k141_117588 | 1211 | No | NA | 1 | 0 | 0 | Not-determined | Genome-fragment | NA | NA | 0 |
| S11_k141_298394 | 2033 | No | NA | 2 | 0 | 0 | Low-quality | Genome-fragment | 4.6 | AAI-based (high-confidence) | 0 |
| S7_k141_75849 | 1552 | No | NA | 2 | 0 | 2 | Not-determined | Genome-fragment | NA | NA | 0 |
| S7_k141_14998 | 1770 | No | NA | 3 | 0 | 1 | Not-determined | Genome-fragment | NA | NA | 0 |
| S7_k141_178852 | 1612 | No | NA | 1 | 0 | 0 | Not-determined | Genome-fragment | NA | NA | 0 |
| S7_k141_56047 | 1971 | No | NA | 3 | 0 | 1 | Not-determined | Genome-fragment | NA | NA | 0 |
| S7_k141_147599 | 1101 | No | NA | 3 | 0 | 3 | Not-determined | Genome-fragment | NA | NA | 0 |
| S7_k141_229878 | 1393 | No | NA | 2 | 0 | 2 | Not-determined | Genome-fragment | NA | NA | 0 |
| S7_k141_117638 | 1483 | No | NA | 1 | 0 | 0 | Not-determined | Genome-fragment | NA | NA | 0 |
| S7_k141_189177 | 1085 | No | NA | 3 | 0 | 0 | Not-determined | Genome-fragment | NA | NA | 0 |
| S7_k141_24398 | 1617 | No | NA | 2 | 0 | 2 | Not-determined | Genome-fragment | NA | NA | 0 |
| S7_k141_218461 | 1444 | No | NA | 2 | 0 | 1 | Not-determined | Genome-fragment | NA | NA | 0 |
| S7_k141_127801 | 68405 | Yes | 38455 | 83 | 18 | 8 | High-quality | High-quality | 99.6 | AAI-based (high-confidence) | 43.78 |
| S7_k141_229925 | 1095 | No | NA | 1 | 0 | 1 | Not-determined | Genome-fragment | NA | NA | 0 |
| S7_k141_229948 | 1176 | No | NA | 1 | 0 | 0 | Not-determined | Genome-fragment | NA | NA | 0 |
| S7_k141_4945 | 5672 | No | NA | 4 | 0 | 4 | Not-determined | Genome-fragment | NA | NA | 0 |
| S7_k141_158534 | 3224 | No | NA | 3 | 0 | 2 | Not-determined | Genome-fragment | NA | NA | 0 |
| S7_k141_66812 | 22315 | No | NA | 22 | 0 | 5 | Not-determined | Genome-fragment | NA | NA | 0 |
| S5-final-viral-combined_k141_189590\|\|full | 2033 | No | NA | 3 | 2 | 0 | Low-quality | Genome-fragment | 4.57 | AAI-based (high-confidence) | 0 |
| S7-final-viral-combined_k141_228824\|\|full | 2033 | No | NA | 2 | 0 | 2 | Low-quality | Genome-fragment | 3.8 | AAI-based (medium-confidence) | 0 |
| S7_k141_137694 | 3287 | No | NA | 5 | 0 | 5 | Not-determined | Genome-fragment | NA | NA | 0 |
| S7-final-viral-combined_k141_189049\|\|full | 2033 | No | NA | 1 | 0 | 0 | Low-quality | Genome-fragment | 3.5 | AAI-based (medium-confidence) | 0 |
| S7_k141_35999 | 2251 | No | NA | 2 | 0 | 2 | Not-determined | Genome-fragment | NA | NA | 0 |
| S7_k141_240461 | 8570 | No | NA | 8 | 0 | 4 | Not-determined | Genome-fragment | NA | NA | 0 |
| S2-final-viral-combined_k141_230688\|\|full | 2033 | No | NA | 4 | 1 | 0 | Low-quality | Genome-fragment | 0.18 | HMM-based (lower-bound) | 0 |
| S7_k141_45886 | 2017 | No | NA | 1 | 0 | 1 | Not-determined | Genome-fragment | NA | NA | 0 |
| S9-final-viral-combined_k141_53528\|\|full | 2034 | No | NA | 5 | 0 | 1 | Low-quality | Genome-fragment | 5.01 | AAI-based (high-confidence) | 0 |
| S11-final-viral-combined_k141_306978\|\|full | 2034 | No | NA | 3 | 0 | 1 | Low-quality | Genome-fragment | 1.53 | AAI-based (medium-confidence) | 0 |
| S7-final-viral-combined_k141_176191\|\|full | 2034 | No | NA | 2 | 0 | 0 | Low-quality | Genome-fragment | 1.14 | AAI-based (high-confidence) | 0 |
| S7_k141_137754 | 4521 | No | NA | 7 | 0 | 2 | Not-determined | Genome-fragment | NA | NA | 0 |
| S7_k141_147732 | 3010 | No | NA | 5 | 0 | 3 | Not-determined | Genome-fragment | NA | NA | 0 |
| S7_k141_15246 | 1320 | No | NA | 2 | 0 | 1 | Not-determined | Genome-fragment | NA | NA | 0 |
| S7_k141_199943 | 4144 | No | NA | 3 | 0 | 2 | Not-determined | Genome-fragment | NA | NA | 0 |
| S7_k141_137788 | 3808 | No | NA | 3 | 0 | 3 | Not-determined | Genome-fragment | NA | NA | 0 |
| S7_k141_86469 | 1025 | No | NA | 2 | 0 | 0 | Not-determined | Genome-fragment | NA | NA | 0 |
| S7_k141_117900 | 4960 | No | NA | 2 | 0 | 1 | Not-determined | Genome-fragment | NA | NA | 0 |
| S7_k141_45972 | 1448 | No | NA | 1 | 0 | 0 | Not-determined | Genome-fragment | NA | NA | 0 |
| S7_k141_56216 | 1584 | No | NA | 1 | 0 | 0 | Not-determined | Genome-fragment | NA | NA | 0 |
| S7_k141_86494 | 2677 | No | NA | 3 | 0 | 2 | Not-determined | Genome-fragment | NA | NA | 0 |
| S7_k141_117909 | 1065 | No | NA | 3 | 0 | 0 | Not-determined | Genome-fragment | NA | NA | 0 |
| S2-final-viral-combined_k141_75509\|\|full | 2034 | No | NA | 4 | 1 | 0 | Low-quality | Genome-fragment | 3.23 | AAI-based (medium-confidence) | 0 |
| S7_k141_106942 | 1061 | No | NA | 1 | 0 | 0 | Not-determined | Genome-fragment | NA | NA | 0 |
| S7_k141_179125 | 3010 | No | NA | 3 | 0 | 0 | Not-determined | Genome-fragment | NA | NA | 0 |
| S7_k141_117971 | 3345 | No | NA | 4 | 0 | 2 | Not-determined | Genome-fragment | NA | NA | 0 |
| S7_k141_117973 | 1217 | No | NA | 2 | 0 | 1 | Not-determined | Genome-fragment | NA | NA | 0 |
| S7_k141_218663 | 1472 | No | NA | 1 | 0 | 0 | Not-determined | Genome-fragment | NA | NA | 0 |
| S6-final-viral-combined_k141_125246\|\|full | 2034 | No | NA | 2 | 0 | 0 | Low-quality | Genome-fragment | 4.16 | AAI-based (high-confidence) | 0 |
| S7_k141_200046 | 3386 | No | NA | 9 | 0 | 9 | Not-determined | Genome-fragment | NA | NA | 0 |
| S7_k141_118029 | 3597 | No | NA | 3 | 0 | 2 | Not-determined | Genome-fragment | NA | NA | 0 |
| S7_k141_189495 | 3903 | No | NA | 4 | 0 | 3 | Not-determined | Genome-fragment | NA | NA | 0 |
| S7_k141_24742 | 4162 | No | NA | 2 | 0 | 0 | Not-determined | Genome-fragment | NA | NA | 0 |
| S7_k141_118058 | 3285 | No | NA | 3 | 0 | 1 | Not-determined | Genome-fragment | NA | NA | 0 |
| S7_k141_107055 | 1251 | No | NA | 1 | 0 | 1 | Not-determined | Genome-fragment | NA | NA | 0 |
| S7_k141_107060 | 2334 | No | NA | 5 | 0 | 1 | Not-determined | Genome-fragment | NA | NA | 0 |
| S7_k141_76235 | 10432 | No | NA | 9 | 0 | 4 | Not-determined | Genome-fragment | NA | NA | 0 |
| S7_k141_96933 | 1484 | No | NA | 3 | 0 | 0 | Not-determined | Genome-fragment | NA | NA | 0 |
| S7_k141_118074 | 3881 | No | NA | 4 | 0 | 4 | Not-determined | Genome-fragment | NA | NA | 0 |
| S6_k141_33023 | 2034 | No | NA | 1 | 0 | 1 | Low-quality | Genome-fragment | 5.74 | AAI-based (medium-confidence) | 0 |
| S9-final-viral-combined_k141_17436\|\|full | 2035 | No | NA | 3 | 0 | 1 | Low-quality | Genome-fragment | 4.36 | AAI-based (medium-confidence) | 0 |
| S7_k141_179199 | 3603 | No | NA | 4 | 0 | 3 | Not-determined | Genome-fragment | NA | NA | 0 |
| S1-final-viral-combined_k141_353831\|\|lt2gene | 2035 | No | NA | 1 | 1 | 0 | Low-quality | Genome-fragment | 3.69 | AAI-based (medium-confidence) | 0 |
| S7_k141_36183 | 1043 | No | NA | 2 | 0 | 2 | Not-determined | Genome-fragment | NA | NA | 0 |
| S7_k141_67074 | 4769 | No | NA | 1 | 0 | 1 | Not-determined | Genome-fragment | NA | NA | 0 |
| S7_k141_200173 | 2133 | No | NA | 3 | 0 | 1 | Not-determined | Genome-fragment | NA | NA | 0 |
| S1_k141_265939 | 2035 | No | NA | 2 | 0 | 0 | Low-quality | Genome-fragment | 2.4 | AAI-based (high-confidence) | 0 |
| S7_k141_169153 | 1769 | No | NA | 2 | 0 | 1 | Not-determined | Genome-fragment | NA | NA | 0 |
| S7_k141_5268 | 3111 | No | NA | 2 | 0 | 0 | Not-determined | Genome-fragment | NA | NA | 0 |
| S7-final-viral-combined_k141_26575\|\|full | 2035 | No | NA | 3 | 0 | 1 | Low-quality | Genome-fragment | 4.1 | AAI-based (medium-confidence) | 0 |
| S7-final-viral-combined_k141_62901\|\|lt2gene | 2035 | Yes | 629 | 2 | 1 | 1 | Low-quality | Genome-fragment | 1.15 | AAI-based (high-confidence) | 69.09 |
| S7_k141_200190 | 1067 | No | NA | 1 | 0 | 1 | Not-determined | Genome-fragment | NA | NA | 0 |
| S7_k141_97015 | 2022 | No | NA | 2 | 0 | 1 | Not-determined | Genome-fragment | NA | NA | 0 |
| S7_k141_230142 | 2317 | No | NA | 2 | 0 | 0 | Not-determined | Genome-fragment | NA | NA | 0 |
| S7_k141_158786 | 1009 | No | NA | 2 | 0 | 1 | Not-determined | Genome-fragment | NA | NA | 0 |
| S7_k141_76318 | 3167 | No | NA | 2 | 0 | 0 | Not-determined | Genome-fragment | NA | NA | 0 |
| S7_k141_24809 | 1427 | No | NA | 2 | 0 | 1 | Not-determined | Genome-fragment | NA | NA | 0 |
| S7_k141_169244 | 1345 | No | NA | 1 | 0 | 1 | Not-determined | Genome-fragment | NA | NA | 0 |
| S7_k141_189614 | 1146 | No | NA | 1 | 0 | 0 | Not-determined | Genome-fragment | NA | NA | 0 |
| S12-final-viral-combined_k141_73972\|\|full | 2035 | No | NA | 5 | 0 | 0 | Low-quality | Genome-fragment | 32.36 | AAI-based (medium-confidence) | 0 |
| S7_k141_5348 | 14272 | No | NA | 29 | 0 | 28 | Not-determined | Genome-fragment | NA | NA | 0 |
| S7_k141_230213 | 3437 | No | NA | 4 | 0 | 2 | Not-determined | Genome-fragment | NA | NA | 0 |
| S7_k141_36307 | 1367 | No | NA | 1 | 0 | 1 | Not-determined | Genome-fragment | NA | NA | 0 |
| S7_k141_97072 | 1401 | No | NA | 1 | 0 | 1 | Not-determined | Genome-fragment | NA | NA | 0 |
| S2-final-viral-combined_k141_134912\|\|full | 2035 | No | NA | 3 | 0 | 0 | Low-quality | Genome-fragment | 4.18 | AAI-based (medium-confidence) | 0 |
| S7_k141_169309 | 1740 | No | NA | 1 | 0 | 1 | Not-determined | Genome-fragment | NA | NA | 0 |
| S7_k141_67174 | 1430 | No | NA | 1 | 0 | 0 | Not-determined | Genome-fragment | NA | NA | 0 |
| S7_k141_36339 | 1095 | No | NA | 1 | 0 | 0 | Not-determined | Genome-fragment | NA | NA | 0 |
| S1-final-viral-combined_k141_215928\|\|full | 2036 | No | NA | 3 | 1 | 0 | Low-quality | Genome-fragment | 2.73 | AAI-based (medium-confidence) | 0 |
| S7_k141_137975 | 5796 | No | NA | 4 | 0 | 2 | Not-determined | Genome-fragment | NA | NA | 0 |
| S7_k141_107235 | 2495 | No | NA | 4 | 0 | 4 | Not-determined | Genome-fragment | NA | NA | 0 |
| S7_k141_218803 | 2443 | No | NA | 3 | 0 | 1 | Not-determined | Genome-fragment | NA | NA | 0 |
| S7_k141_189685 | 5989 | No | NA | 5 | 0 | 4 | Not-determined | Genome-fragment | NA | NA | 0 |
| S7_k141_56442 | 1217 | No | NA | 2 | 0 | 0 | Not-determined | Genome-fragment | NA | NA | 0 |
| S7_k141_97170 | 6174 | No | NA | 7 | 0 | 4 | Not-determined | Genome-fragment | NA | NA | 0 |
| S7_k141_189715 | 4108 | No | NA | 2 | 0 | 2 | Not-determined | Genome-fragment | NA | NA | 0 |
| S7_k141_240647 | 1337 | No | NA | 4 | 0 | 1 | Not-determined | Genome-fragment | NA | NA | 0 |
| S7_k141_76437 | 81890 | No | NA | 61 | 2 | 6 | High-quality | High-quality | 95.59 | HMM-based (lower-bound) | 0 |
| S7_k141_200340 | 1104 | No | NA | 1 | 0 | 1 | Not-determined | Genome-fragment | NA | NA | 0 |
| S1-final-viral-combined_k141_123536\|\|full | 2036 | No | NA | 3 | 0 | 0 | Low-quality | Genome-fragment | 2.65 | AAI-based (high-confidence) | 0 |
| S3-final-viral-combined_k141_67241\|\|lt2gene | 2036 | No | NA | 1 | 1 | 0 | Low-quality | Genome-fragment | 5.27 | AAI-based (high-confidence) | 0 |
| S7_k141_36462 | 7085 | No | NA | 5 | 0 | 4 | Not-determined | Genome-fragment | NA | NA | 0 |
| S7_k141_118281 | 1414 | No | NA | 1 | 0 | 1 | Not-determined | Genome-fragment | NA | NA | 0 |
| S5_k141_48625 | 2036 | No | NA | 3 | 0 | 1 | Low-quality | Genome-fragment | 2.25 | AAI-based (medium-confidence) | 0 |
| S7_k141_76475 | 1444 | No | NA | 1 | 0 | 0 | Not-determined | Genome-fragment | NA | NA | 0 |
| S7_k141_210338 | 2241 | No | NA | 2 | 0 | 2 | Not-determined | Genome-fragment | NA | NA | 0 |
| S7_k141_107285 | 1405 | No | NA | 3 | 0 | 2 | Not-determined | Genome-fragment | NA | NA | 0 |
| S12-final-viral-combined_k141_9056\|\|full | 2036 | No | NA | 5 | 1 | 0 | Low-quality | Genome-fragment | 4.81 | AAI-based (medium-confidence) | 0 |
| S7_k141_200446 | 5588 | No | NA | 6 | 0 | 4 | Not-determined | Genome-fragment | NA | NA | 0 |
| S2-final-viral-combined_k141_113952\|\|full | 2036 | No | NA | 4 | 3 | 0 | Low-quality | Genome-fragment | 5.52 | AAI-based (medium-confidence) | 0 |
| S7_k141_56564 | 3647 | No | NA | 1 | 0 | 0 | Not-determined | Genome-fragment | NA | NA | 0 |
| S7_k141_36522 | 1097 | No | NA | 1 | 0 | 1 | Not-determined | Genome-fragment | NA | NA | 0 |
| S7_k141_15708 | 1299 | No | NA | 1 | 0 | 0 | Not-determined | Genome-fragment | NA | NA | 0 |
| S7_k141_76566 | 1153 | No | NA | 2 | 0 | 0 | Not-determined | Genome-fragment | NA | NA | 0 |
| S7_k141_189874 | 5597 | No | NA | 6 | 0 | 3 | Not-determined | Genome-fragment | NA | NA | 0 |
| S7_k141_86863 | 1030 | No | NA | 1 | 0 | 0 | Not-determined | Genome-fragment | NA | NA | 0 |
| S2_k141_4361 | 2036 | No | NA | 4 | 1 | 0 | Low-quality | Genome-fragment | 2.01 | AAI-based (high-confidence) | 0 |
| S7_k141_56613 | 1016 | No | NA | 1 | 0 | 0 | Not-determined | Genome-fragment | NA | NA | 0 |
| S7_k141_46507 | 1572 | No | NA | 4 | 0 | 0 | Not-determined | Genome-fragment | NA | NA | 0 |
| S6-final-viral-combined_k141_29031\|\|full | 2036 | No | NA | 2 | 0 | 1 | Low-quality | Genome-fragment | 0.98 | AAI-based (high-confidence) | 0 |
| S7_k141_240801 | 3317 | No | NA | 2 | 0 | 0 | Not-determined | Genome-fragment | NA | NA | 0 |
| S7_k141_169447 | 1574 | No | NA | 1 | 0 | 1 | Not-determined | Genome-fragment | NA | NA | 0 |
| S9-final-viral-combined_k141_10738\|\|full | 2037 | No | NA | 3 | 0 | 0 | Low-quality | Genome-fragment | 3.78 | AAI-based (medium-confidence) | 0 |
| S11-final-viral-combined_k141_146191\|\|full | 2037 | No | NA | 5 | 0 | 0 | Low-quality | Genome-fragment | 5.64 | AAI-based (medium-confidence) | 0 |
| S7_k141_36681 | 1042 | No | NA | 3 | 0 | 0 | Not-determined | Genome-fragment | NA | NA | 0 |
| S7_k141_189943 | 4599 | No | NA | 6 | 0 | 0 | Not-determined | Genome-fragment | NA | NA | 0 |
| S7_k141_118369 | 4987 | No | NA | 3 | 0 | 1 | Not-determined | Genome-fragment | NA | NA | 0 |
| S2-final-viral-combined_k141_453\|\|full | 2037 | No | NA | 2 | 0 | 1 | Low-quality | Genome-fragment | 2.57 | AAI-based (medium-confidence) | 0 |
| S7_k141_46561 | 1159 | No | NA | 1 | 0 | 1 | Not-determined | Genome-fragment | NA | NA | 0 |
| S7_k141_159112 | 3490 | No | NA | 2 | 0 | 1 | Not-determined | Genome-fragment | NA | NA | 0 |
| S7_k141_107490 | 1381 | No | NA | 3 | 0 | 0 | Not-determined | Genome-fragment | NA | NA | 0 |
| S7_k141_159142 | 3574 | No | NA | 8 | 0 | 2 | Not-determined | Genome-fragment | NA | NA | 0 |
| S8-final-viral-combined_k141_178912\|\|full | 2037 | No | NA | 2 | 0 | 2 | Low-quality | Genome-fragment | 1.01 | AAI-based (medium-confidence) | 0 |
| S7_k141_86962 | 1048 | No | NA | 3 | 0 | 0 | Not-determined | Genome-fragment | NA | NA | 0 |
| S7_k141_169522 | 1216 | No | NA | 1 | 0 | 1 | Not-determined | Genome-fragment | NA | NA | 0 |
| S8-final-viral-combined_k141_76790\|\|full | 2037 | No | NA | 3 | 2 | 0 | Low-quality | Genome-fragment | 6.54 | AAI-based (high-confidence) | 0 |
| S7_k141_179596 | 3339 | No | NA | 6 | 0 | 0 | Not-determined | Genome-fragment | NA | NA | 0 |
| S7_k141_210605 | 1151 | No | NA | 3 | 0 | 2 | Not-determined | Genome-fragment | NA | NA | 0 |
| S7_k141_118429 | 9623 | No | NA | 13 | 0 | 2 | Not-determined | Genome-fragment | NA | NA | 0 |
| S8-final-viral-combined_k141_259843\|\|lt2gene | 2037 | No | NA | 2 | 0 | 0 | Low-quality | Genome-fragment | 4.27 | AAI-based (medium-confidence) | 0 |
| S7_k141_190089 | 3458 | No | NA | 2 | 0 | 2 | Not-determined | Genome-fragment | NA | NA | 0 |
| S11-final-viral-combined_k141_322986\|\|full | 2038 | No | NA | 5 | 1 | 0 | Low-quality | Genome-fragment | 4.32 | AAI-based (high-confidence) | 0 |
| S7_k141_138343 | 1011 | No | NA | 1 | 0 | 1 | Not-determined | Genome-fragment | NA | NA | 0 |
| S7_k141_148321 | 1058 | No | NA | 1 | 0 | 1 | Not-determined | Genome-fragment | NA | NA | 0 |
| S1-final-viral-combined_k141_232513\|\|full | 2038 | No | NA | 3 | 2 | 0 | Low-quality | Genome-fragment | 3.79 | AAI-based (high-confidence) | 0 |
| S7_k141_230532 | 5582 | No | NA | 2 | 0 | 1 | Not-determined | Genome-fragment | NA | NA | 0 |
| S7_k141_159250 | 4027 | No | NA | 2 | 0 | 0 | Not-determined | Genome-fragment | NA | NA | 0 |
| S7_k141_97473 | 1569 | No | NA | 2 | 0 | 0 | Not-determined | Genome-fragment | NA | NA | 0 |
| S7_k141_148356 | 1078 | No | NA | 1 | 0 | 1 | Not-determined | Genome-fragment | NA | NA | 0 |
| S7_k141_97478 | 2494 | No | NA | 1 | 0 | 1 | Not-determined | Genome-fragment | NA | NA | 0 |
| S12-final-viral-combined_k141_52990\|\|full | 2038 | No | NA | 2 | 0 | 1 | Low-quality | Genome-fragment | 2.62 | AAI-based (high-confidence) | 0 |
| S7_k141_219009 | 3992 | No | NA | 6 | 0 | 2 | Not-determined | Genome-fragment | NA | NA | 0 |
| S7_k141_190177 | 1595 | No | NA | 3 | 0 | 0 | Not-determined | Genome-fragment | NA | NA | 0 |
| S7_k141_138419 | 1310 | No | NA | 1 | 0 | 0 | Not-determined | Genome-fragment | NA | NA | 0 |
| S7_k141_15904 | 3758 | No | NA | 1 | 0 | 0 | Not-determined | Genome-fragment | NA | NA | 0 |
| S11-final-viral-combined_k141_245670\|\|lt2gene | 2039 | No | NA | 2 | 2 | 0 | Low-quality | Genome-fragment | 4.67 | AAI-based (high-confidence) | 0 |
| S7_k141_87126 | 1136 | No | NA | 3 | 0 | 1 | Not-determined | Genome-fragment | NA | NA | 0 |
| S7_k141_36860 | 1913 | No | NA | 1 | 0 | 0 | Not-determined | Genome-fragment | NA | NA | 0 |
| S7_k141_15916 | 1643 | No | NA | 4 | 0 | 2 | Not-determined | Genome-fragment | NA | NA | 0 |
| S7_k141_138444 | 1352 | No | NA | 2 | 0 | 0 | Not-determined | Genome-fragment | NA | NA | 0 |
| S7_k141_97519 | 5011 | No | NA | 2 | 0 | 2 | Not-determined | Genome-fragment | NA | NA | 0 |
| S7_k141_46797 | 1019 | No | NA | 1 | 0 | 1 | Not-determined | Genome-fragment | NA | NA | 0 |
| S7_k141_179791 | 1805 | No | NA | 7 | 0 | 0 | Not-determined | Genome-fragment | NA | NA | 0 |
| S7_k141_67475 | 1638 | No | NA | 3 | 0 | 2 | Not-determined | Genome-fragment | NA | NA | 0 |
| S7_k141_219046 | 3616 | No | NA | 5 | 0 | 1 | Not-determined | Genome-fragment | NA | NA | 0 |
| S7_k141_15946 | 4388 | No | NA | 4 | 0 | 2 | Not-determined | Genome-fragment | NA | NA | 0 |
| S7_k141_148440 | 9294 | No | NA | 5 | 0 | 1 | Not-determined | Genome-fragment | NA | NA | 0 |
| S1-final-viral-combined_k141_371355\|\|full | 2039 | No | NA | 2 | 0 | 1 | Low-quality | Genome-fragment | 0.73 | AAI-based (medium-confidence) | 0 |
| S7_k141_118607 | 48532 | No | NA | 66 | 21 | 0 | High-quality | High-quality | 100 | AAI-based (high-confidence) | 0 |
| S7_k141_15976 | 1169 | No | NA | 1 | 0 | 0 | Not-determined | Genome-fragment | NA | NA | 0 |
| S7_k141_118622 | 7298 | No | NA | 11 | 0 | 2 | Not-determined | Genome-fragment | NA | NA | 0 |
| S7_k141_128659 | 1019 | No | NA | 1 | 0 | 0 | Not-determined | Genome-fragment | NA | NA | 0 |
| S7_k141_148493 | 1307 | No | NA | 2 | 0 | 0 | Not-determined | Genome-fragment | NA | NA | 0 |
| S7_k141_148505 | 3653 | No | NA | 1 | 0 | 1 | Not-determined | Genome-fragment | NA | NA | 0 |
| S7_k141_37026 | 1150 | No | NA | 2 | 0 | 1 | Not-determined | Genome-fragment | NA | NA | 0 |
| S7_k141_41848 | 2039 | No | NA | 4 | 0 | 0 | Low-quality | Genome-fragment | 3.92 | AAI-based (medium-confidence) | 0 |
| S7_k141_230628 | 1684 | No | NA | 2 | 0 | 2 | Not-determined | Genome-fragment | NA | NA | 0 |
| S7_k141_190270 | 1140 | No | NA | 1 | 0 | 1 | Not-determined | Genome-fragment | NA | NA | 0 |
| S7_k141_159432 | 33004 | No | NA | 50 | 14 | 0 | Medium-quality | Genome-fragment | 67.48 | AAI-based (high-confidence) | 0 |
| S7_k141_230636 | 2902 | No | NA | 1 | 0 | 1 | Not-determined | Genome-fragment | NA | NA | 0 |
| S7_k141_118644 | 2129 | No | NA | 5 | 0 | 0 | Not-determined | Genome-fragment | NA | NA | 0 |
| S7_k141_200751 | 2106 | No | NA | 2 | 0 | 0 | Not-determined | Genome-fragment | NA | NA | 0 |
| S7_k141_210799 | 1036 | No | NA | 2 | 0 | 1 | Not-determined | Genome-fragment | NA | NA | 0 |
| S12-final-viral-combined_k141_396648\|\|full | 2039 | No | NA | 2 | 0 | 2 | Low-quality | Genome-fragment | 3.35 | AAI-based (medium-confidence) | 0 |
| S4_k141_145180 | 2039 | No | NA | 2 | 1 | 0 | Low-quality | Genome-fragment | 2.05 | AAI-based (high-confidence) | 0 |
| S7_k141_16041 | 11402 | No | NA | 16 | 0 | 2 | Not-determined | Genome-fragment | NA | NA | 0 |
| S7_k141_87347 | 4167 | No | NA | 2 | 0 | 0 | Not-determined | Genome-fragment | NA | NA | 0 |
| S7_k141_118674 | 1621 | No | NA | 1 | 0 | 1 | Not-determined | Genome-fragment | NA | NA | 0 |
| S7_k141_56979 | 3288 | No | NA | 4 | 0 | 1 | Not-determined | Genome-fragment | NA | NA | 0 |
| S7_k141_118680 | 1101 | No | NA | 1 | 0 | 1 | Not-determined | Genome-fragment | NA | NA | 0 |
| S11-final-viral-combined_k141_217307\|\|full | 2040 | No | NA | 5 | 1 | 0 | Low-quality | Genome-fragment | 4.78 | AAI-based (medium-confidence) | 0 |
| S7_k141_241266 | 2165 | No | NA | 1 | 0 | 1 | Not-determined | Genome-fragment | NA | NA | 0 |
| S7_k141_5820 | 1655 | No | NA | 1 | 0 | 1 | Not-determined | Genome-fragment | NA | NA | 0 |
| S7_k141_169994 | 1550 | No | NA | 1 | 0 | 1 | Not-determined | Genome-fragment | NA | NA | 0 |
| S7_k141_118728 | 1774 | No | NA | 1 | 0 | 1 | Not-determined | Genome-fragment | NA | NA | 0 |
| S7_k141_159535 | 1569 | No | NA | 2 | 0 | 0 | Not-determined | Genome-fragment | NA | NA | 0 |
| S7_k141_5825 | 1099 | No | NA | 1 | 0 | 1 | Not-determined | Genome-fragment | NA | NA | 0 |
| S7_k141_76889 | 1024 | No | NA | 2 | 0 | 0 | Not-determined | Genome-fragment | NA | NA | 0 |
| S7_k141_148644 | 1488 | No | NA | 2 | 0 | 2 | Not-determined | Genome-fragment | NA | NA | 0 |
| S7_k141_67752 | 3237 | No | NA | 3 | 0 | 3 | Not-determined | Genome-fragment | NA | NA | 0 |
| S7_k141_219300 | 1791 | No | NA | 2 | 0 | 0 | Not-determined | Genome-fragment | NA | NA | 0 |
| S7_k141_230754 | 7706 | No | NA | 11 | 0 | 0 | Not-determined | Genome-fragment | NA | NA | 0 |
| S7_k141_128774 | 1580 | No | NA | 2 | 0 | 0 | Not-determined | Genome-fragment | NA | NA | 0 |
| S4-final-viral-combined_k141_51381\|\|full | 2040 | No | NA | 2 | 0 | 0 | Low-quality | Genome-fragment | 0.77 | AAI-based (high-confidence) | 0 |
| S7_k141_118774 | 1141 | No | NA | 1 | 0 | 0 | Not-determined | Genome-fragment | NA | NA | 0 |
| S7_k141_210965 | 4189 | No | NA | 1 | 0 | 1 | Not-determined | Genome-fragment | NA | NA | 0 |
| S7_k141_190389 | 4378 | No | NA | 2 | 0 | 0 | Not-determined | Genome-fragment | NA | NA | 0 |
| S7_k141_87412 | 1369 | No | NA | 1 | 0 | 1 | Not-determined | Genome-fragment | NA | NA | 0 |
| S4-final-viral-combined_k141_129563\|\|full | 2040 | No | NA | 3 | 0 | 0 | Low-quality | Genome-fragment | 3.63 | AAI-based (high-confidence) | 0 |
| S7_k141_159685 | 3365 | No | NA | 6 | 0 | 5 | Not-determined | Genome-fragment | NA | NA | 0 |
| S7_k141_25490 | 1029 | No | NA | 1 | 0 | 1 | Not-determined | Genome-fragment | NA | NA | 0 |
| S7_k141_148738 | 1138 | No | NA | 2 | 0 | 1 | Not-determined | Genome-fragment | NA | NA | 0 |
| S7_k141_170089 | 1177 | No | NA | 2 | 0 | 2 | Not-determined | Genome-fragment | NA | NA | 0 |
| S7_k141_67871 | 4215 | No | NA | 6 | 0 | 1 | Not-determined | Genome-fragment | NA | NA | 0 |
| S7_k141_148750 | 2117 | No | NA | 3 | 0 | 2 | Not-determined | Genome-fragment | NA | NA | 0 |
| S7_k141_46982 | 2241 | No | NA | 3 | 0 | 2 | Not-determined | Genome-fragment | NA | NA | 0 |
| S7_k141_57204 | 1756 | No | NA | 2 | 0 | 2 | Not-determined | Genome-fragment | NA | NA | 0 |
| S7_k141_170099 | 14213 | No | NA | 7 | 0 | 4 | Not-determined | Genome-fragment | NA | NA | 0 |
| S7_k141_5929 | 1279 | No | NA | 1 | 0 | 1 | Not-determined | Genome-fragment | NA | NA | 0 |
| S7_k141_25574 | 4376 | No | NA | 5 | 0 | 1 | Not-determined | Genome-fragment | NA | NA | 0 |
| S7_k141_211022 | 4463 | No | NA | 3 | 0 | 2 | Not-determined | Genome-fragment | NA | NA | 0 |
| S11-final-viral-combined_k141_278411\|\|full | 2041 | No | NA | 5 | 0 | 1 | Low-quality | Genome-fragment | 1.38 | AAI-based (medium-confidence) | 0 |
| S1_k141_140803 | 2041 | No | NA | 3 | 0 | 0 | Low-quality | Genome-fragment | 2.48 | AAI-based (high-confidence) | 0 |
| S7_k141_138793 | 4512 | No | NA | 3 | 0 | 1 | Not-determined | Genome-fragment | NA | NA | 0 |
| S7_k141_241357 | 1139 | No | NA | 1 | 0 | 0 | Not-determined | Genome-fragment | NA | NA | 0 |
| S17-final-viral-combined_k141_57712\|\|full | 2041 | No | NA | 2 | 0 | 0 | Low-quality | Genome-fragment | 2.33 | AAI-based (medium-confidence) | 0 |
| S7_k141_87512 | 1825 | No | NA | 2 | 0 | 1 | Not-determined | Genome-fragment | NA | NA | 0 |
| S7_k141_241367 | 4172 | No | NA | 5 | 0 | 1 | Not-determined | Genome-fragment | NA | NA | 0 |
| S7_k141_170163 | 3599 | No | NA | 4 | 0 | 3 | Not-determined | Genome-fragment | NA | NA | 0 |
| S17-final-viral-combined_k141_130666\|\|full | 2041 | No | NA | 3 | 0 | 0 | Low-quality | Genome-fragment | 2.92 | AAI-based (high-confidence) | 0 |
| S4-final-viral-combined_k141_54354\|\|full | 2041 | No | NA | 2 | 0 | 0 | Low-quality | Genome-fragment | 3.09 | AAI-based (medium-confidence) | 0 |
| S9-final-viral-combined_k141_79059\|\|full | 2042 | No | NA | 2 | 0 | 1 | Low-quality | Genome-fragment | 0.54 | AAI-based (medium-confidence) | 0 |
| S7_k141_87530 | 1639 | No | NA | 2 | 0 | 1 | Not-determined | Genome-fragment | NA | NA | 0 |
| S7_k141_128982 | 1983 | No | NA | 3 | 0 | 1 | Not-determined | Genome-fragment | NA | NA | 0 |
| S7_k141_230926 | 1363 | No | NA | 1 | 0 | 0 | Not-determined | Genome-fragment | NA | NA | 0 |
| S11-final-viral-combined_k141_119623\|\|full | 2042 | No | NA | 4 | 0 | 1 | Low-quality | Genome-fragment | 1.33 | AAI-based (high-confidence) | 0 |
| S7_k141_190630 | 1639 | No | NA | 2 | 0 | 2 | Not-determined | Genome-fragment | NA | NA | 0 |
| S7_k141_67971 | 1107 | No | NA | 1 | 0 | 1 | Not-determined | Genome-fragment | NA | NA | 0 |
| S7_k141_67982 | 3111 | No | NA | 2 | 0 | 2 | Not-determined | Genome-fragment | NA | NA | 0 |
| S7_k141_67985 | 1232 | No | NA | 2 | 0 | 2 | Not-determined | Genome-fragment | NA | NA | 0 |
| S1-final-viral-combined_k141_350983\|\|full | 2042 | No | NA | 3 | 0 | 1 | Low-quality | Genome-fragment | 2.19 | AAI-based (medium-confidence) | 0 |
| S7_k141_97967 | 3042 | No | NA | 3 | 0 | 3 | Not-determined | Genome-fragment | NA | NA | 0 |
| S7_k141_6040 | 1429 | No | NA | 2 | 0 | 2 | Not-determined | Genome-fragment | NA | NA | 0 |
| S7_k141_87593 | 1514 | No | NA | 3 | 0 | 2 | Not-determined | Genome-fragment | NA | NA | 0 |
| S7_k141_77150 | 1856 | No | NA | 1 | 0 | 1 | Not-determined | Genome-fragment | NA | NA | 0 |
| S7_k141_119042 | 1089 | No | NA | 2 | 0 | 0 | Not-determined | Genome-fragment | NA | NA | 0 |
| S7_k141_211145 | 3801 | No | NA | 2 | 0 | 1 | Not-determined | Genome-fragment | NA | NA | 0 |
| S17-final-viral-combined_k141_58997\|\|full | 2042 | No | NA | 3 | 2 | 0 | Low-quality | Genome-fragment | 5.27 | AAI-based (high-confidence) | 0 |
| S12-final-viral-combined_k141_157041\|\|full | 2042 | No | NA | 3 | 2 | 0 | Low-quality | Genome-fragment | 6.05 | AAI-based (high-confidence) | 0 |
| S7_k141_200936 | 31792 | No | NA | 46 | 13 | 2 | Medium-quality | Genome-fragment | 88.82 | AAI-based (medium-confidence) | 0 |
| S7_k141_16484 | 1294 | No | NA | 1 | 0 | 0 | Not-determined | Genome-fragment | NA | NA | 0 |
| S7_k141_138959 | 1829 | No | NA | 1 | 0 | 1 | Not-determined | Genome-fragment | NA | NA | 0 |
| S7_k141_98050 | 1078 | No | NA | 1 | 0 | 1 | Not-determined | Genome-fragment | NA | NA | 0 |
| S4-final-viral-combined_k141_164421\|\|full | 2042 | Yes | 1502 | 4 | 1 | 1 | Low-quality | Genome-fragment | 3.27 | AAI-based (high-confidence) | 26.44 |
| S7_k141_77233 | 6282 | No | NA | 5 | 0 | 2 | Not-determined | Genome-fragment | NA | NA | 0 |
| S7_k141_231033 | 1313 | No | NA | 2 | 0 | 0 | Not-determined | Genome-fragment | NA | NA | 0 |
| S7_k141_190775 | 1118 | No | NA | 1 | 0 | 0 | Not-determined | Genome-fragment | NA | NA | 0 |
| S7_k141_57482 | 3786 | No | NA | 1 | 0 | 1 | Not-determined | Genome-fragment | NA | NA | 0 |
| S7_k141_201096 | 1025 | No | NA | 1 | 0 | 0 | Not-determined | Genome-fragment | NA | NA | 0 |
| S7_k141_201109 | 1099 | No | NA | 2 | 0 | 0 | Not-determined | Genome-fragment | NA | NA | 0 |
| S7_k141_219549 | 1390 | No | NA | 1 | 0 | 1 | Not-determined | Genome-fragment | NA | NA | 0 |
| S7_k141_77351 | 6483 | No | NA | 3 | 0 | 1 | Not-determined | Genome-fragment | NA | NA | 0 |
| S7_k141_159829 | 4369 | No | NA | 6 | 0 | 3 | Not-determined | Genome-fragment | NA | NA | 0 |
| S7_k141_149047 | 3037 | No | NA | 2 | 0 | 1 | Not-determined | Genome-fragment | NA | NA | 0 |
| S7_k141_57565 | 1384 | No | NA | 2 | 0 | 1 | Not-determined | Genome-fragment | NA | NA | 0 |
| S7_k141_139179 | 1441 | No | NA | 1 | 0 | 1 | Not-determined | Genome-fragment | NA | NA | 0 |
| S7_k141_98222 | 2515 | No | NA | 3 | 0 | 1 | Not-determined | Genome-fragment | NA | NA | 0 |
| S11-final-viral-combined_k141_71279\|\|full | 2043 | No | NA | 4 | 0 | 1 | Low-quality | Genome-fragment | 2.92 | AAI-based (medium-confidence) | 0 |
| S7_k141_57611 | 2260 | No | NA | 2 | 0 | 1 | Not-determined | Genome-fragment | NA | NA | 0 |
| S7_k141_139216 | 1074 | No | NA | 2 | 0 | 2 | Not-determined | Genome-fragment | NA | NA | 0 |
| S7_k141_37546 | 1276 | No | NA | 2 | 0 | 0 | Not-determined | Genome-fragment | NA | NA | 0 |
| S7_k141_98267 | 2622 | No | NA | 3 | 0 | 3 | Not-determined | Genome-fragment | NA | NA | 0 |
| S3_k141_75284 | 2043 | No | NA | 2 | 0 | 0 | Low-quality | Genome-fragment | 1.07 | AAI-based (medium-confidence) | 0 |
| S7_k141_37570 | 1152 | No | NA | 2 | 0 | 0 | Not-determined | Genome-fragment | NA | NA | 0 |
| S7_k141_129177 | 1138 | No | NA | 1 | 0 | 1 | Not-determined | Genome-fragment | NA | NA | 0 |
| S7_k141_6348 | 1602 | No | NA | 2 | 0 | 2 | Not-determined | Genome-fragment | NA | NA | 0 |
| S5-final-viral-combined_k141_79731\|\|full | 2043 | No | NA | 4 | 2 | 0 | Low-quality | Genome-fragment | 3.28 | HMM-based (lower-bound) | 0 |
| S4_k141_62656 | 2043 | No | NA | 2 | 0 | 1 | Low-quality | Genome-fragment | 2.2 | AAI-based (medium-confidence) | 0 |
| S7_k141_16687 | 1095 | No | NA | 3 | 0 | 0 | Not-determined | Genome-fragment | NA | NA | 0 |
| S7_k141_149130 | 1152 | No | NA | 2 | 0 | 1 | Not-determined | Genome-fragment | NA | NA | 0 |
| S7_k141_6385 | 3403 | No | NA | 5 | 0 | 1 | Not-determined | Genome-fragment | NA | NA | 0 |
| S7_k141_170465 | 1378 | No | NA | 2 | 0 | 2 | Not-determined | Genome-fragment | NA | NA | 0 |
| S7_k141_201365 | 3620 | No | NA | 5 | 0 | 3 | Not-determined | Genome-fragment | NA | NA | 0 |
| S7_k141_170539 | 1294 | No | NA | 2 | 0 | 0 | Not-determined | Genome-fragment | NA | NA | 0 |
| S7_k141_190991 | 1133 | No | NA | 1 | 0 | 1 | Not-determined | Genome-fragment | NA | NA | 0 |
| S6-final-viral-combined_k141_75740\|\|full | 2043 | No | NA | 4 | 2 | 0 | Low-quality | Genome-fragment | 5.03 | AAI-based (high-confidence) | 0 |
| S3_k141_86184 | 2044 | No | NA | 1 | 0 | 1 | Low-quality | Genome-fragment | 2.16 | AAI-based (medium-confidence) | 0 |
| S7_k141_160056 | 1020 | No | NA | 1 | 0 | 1 | Not-determined | Genome-fragment | NA | NA | 0 |
| S5-final-viral-combined_k141_210186\|\|full | 2044 | No | NA | 3 | 0 | 2 | Low-quality | Genome-fragment | 0.8 | AAI-based (medium-confidence) | 0 |
| S7_k141_180355 | 1351 | No | NA | 1 | 0 | 0 | Not-determined | Genome-fragment | NA | NA | 0 |
| S7_k141_211335 | 1407 | No | NA | 1 | 0 | 1 | Not-determined | Genome-fragment | NA | NA | 0 |
| S7_k141_191084 | 5799 | No | NA | 4 | 0 | 4 | Not-determined | Genome-fragment | NA | NA | 0 |
| S7_k141_87939 | 4764 | No | NA | 5 | 0 | 5 | Not-determined | Genome-fragment | NA | NA | 0 |
| S7_k141_68491 | 2916 | No | NA | 1 | 0 | 1 | Not-determined | Genome-fragment | NA | NA | 0 |
| S7_k141_232428 | 2044 | No | NA | 2 | 0 | 1 | Low-quality | Genome-fragment | 3.75 | AAI-based (medium-confidence) | 0 |
| S7_k141_26161 | 1705 | No | NA | 2 | 0 | 1 | Not-determined | Genome-fragment | NA | NA | 0 |
| S7_k141_16747 | 4343 | No | NA | 5 | 0 | 2 | Not-determined | Genome-fragment | NA | NA | 0 |
| S7_k141_6526 | 5474 | No | NA | 3 | 0 | 3 | Not-determined | Genome-fragment | NA | NA | 0 |
| S7_k141_191121 | 1303 | No | NA | 1 | 0 | 1 | Not-determined | Genome-fragment | NA | NA | 0 |
| S7_k141_47563 | 1001 | No | NA | 2 | 0 | 1 | Not-determined | Genome-fragment | NA | NA | 0 |
| S7_k141_68567 | 4052 | No | NA | 2 | 0 | 2 | Not-determined | Genome-fragment | NA | NA | 0 |
| S7_k141_98485 | 1327 | No | NA | 1 | 0 | 0 | Not-determined | Genome-fragment | NA | NA | 0 |
| S7_k141_129403 | 7370 | No | NA | 7 | 0 | 3 | Not-determined | Genome-fragment | NA | NA | 0 |
| S7_k141_241759 | 1042 | No | NA | 1 | 0 | 0 | Not-determined | Genome-fragment | NA | NA | 0 |
| S12-final-viral-combined_k141_209466\|\|full | 2044 | No | NA | 4 | 0 | 2 | Low-quality | Genome-fragment | 2.41 | AAI-based (medium-confidence) | 0 |
| S7_k141_149200 | 1178 | No | NA | 1 | 0 | 1 | Not-determined | Genome-fragment | NA | NA | 0 |
| S12_k141_170286 | 2044 | No | NA | 2 | 0 | 0 | Low-quality | Genome-fragment | 4.25 | AAI-based (high-confidence) | 0 |
| S7_k141_77612 | 1316 | No | NA | 1 | 0 | 1 | Not-determined | Genome-fragment | NA | NA | 0 |
| S7_k141_191190 | 2596 | No | NA | 2 | 0 | 1 | Not-determined | Genome-fragment | NA | NA | 0 |
| S7_k141_129448 | 1262 | No | NA | 2 | 0 | 0 | Not-determined | Genome-fragment | NA | NA | 0 |
| S12_k141_49803 | 2044 | No | NA | 5 | 1 | 0 | Low-quality | Genome-fragment | 5.17 | AAI-based (medium-confidence) | 0 |
| S2-final-viral-combined_k141_144945\|\|full | 2044 | No | NA | 2 | 1 | 0 | Low-quality | Genome-fragment | 1.85 | HMM-based (lower-bound) | 0 |
| S7_k141_88081 | 9996 | No | NA | 6 | 0 | 1 | Not-determined | Genome-fragment | NA | NA | 0 |
| S8-final-viral-combined_k141_50984\|\|full | 2044 | No | NA | 2 | 0 | 2 | Low-quality | Genome-fragment | 4.26 | AAI-based (medium-confidence) | 0 |
| S7_k141_88096 | 5676 | No | NA | 7 | 0 | 5 | Not-determined | Genome-fragment | NA | NA | 0 |
| S7_k141_119362 | 1403 | No | NA | 3 | 0 | 2 | Not-determined | Genome-fragment | NA | NA | 0 |
| S7_k141_231478 | 1612 | No | NA | 3 | 0 | 2 | Not-determined | Genome-fragment | NA | NA | 0 |
| S8_k141_132589 | 2044 | No | NA | 3 | 2 | 0 | Low-quality | Genome-fragment | 4.22 | AAI-based (high-confidence) | 0 |
| S7_k141_68611 | 1228 | No | NA | 2 | 0 | 1 | Not-determined | Genome-fragment | NA | NA | 0 |
| S7_k141_26259 | 3701 | No | NA | 6 | 0 | 1 | Not-determined | Genome-fragment | NA | NA | 0 |
| S9-final-viral-combined_k141_53004\|\|lt2gene | 2045 | No | NA | 2 | 2 | 0 | Low-quality | Genome-fragment | 4.93 | AAI-based (high-confidence) | 0 |
| S7_k141_211559 | 1107 | No | NA | 1 | 0 | 0 | Not-determined | Genome-fragment | NA | NA | 0 |
| S7_k141_88222 | 6317 | No | NA | 4 | 0 | 3 | Not-determined | Genome-fragment | NA | NA | 0 |
| S11-final-viral-combined_k141_99353\|\|full | 2045 | No | NA | 2 | 1 | 0 | Low-quality | Genome-fragment | 1.52 | HMM-based (lower-bound) | 0 |
| S11-final-viral-combined_k141_298969\|\|full | 2045 | No | NA | 5 | 0 | 0 | Low-quality | Genome-fragment | 5.52 | AAI-based (medium-confidence) | 0 |
| S7_k141_191299 | 1724 | No | NA | 2 | 0 | 2 | Not-determined | Genome-fragment | NA | NA | 0 |
| S7_k141_68727 | 1807 | No | NA | 2 | 0 | 1 | Not-determined | Genome-fragment | NA | NA | 0 |
| S1_k141_371398 | 2045 | No | NA | 1 | 0 | 1 | Low-quality | Genome-fragment | 0.34 | AAI-based (high-confidence) | 0 |
| S7_k141_26341 | 1768 | No | NA | 2 | 0 | 2 | Not-determined | Genome-fragment | NA | NA | 0 |
| S7_k141_37969 | 3733 | No | NA | 4 | 0 | 4 | Not-determined | Genome-fragment | NA | NA | 0 |
| S7_k141_17042 | 1629 | No | NA | 3 | 0 | 3 | Not-determined | Genome-fragment | NA | NA | 0 |
| S3-final-viral-combined_k141_132374\|\|full | 2045 | No | NA | 3 | 1 | 0 | Low-quality | Genome-fragment | 3.76 | AAI-based (high-confidence) | 0 |
| S17-final-viral-combined_k141_7899\|\|full | 2045 | No | NA | 5 | 4 | 0 | Low-quality | Genome-fragment | 2.48 | AAI-based (high-confidence) | 0 |
| S7_k141_241929 | 1211 | No | NA | 2 | 0 | 0 | Not-determined | Genome-fragment | NA | NA | 0 |
| S7_k141_17081 | 1234 | No | NA | 2 | 0 | 0 | Not-determined | Genome-fragment | NA | NA | 0 |
| S7_k141_160468 | 1233 | No | NA | 1 | 0 | 1 | Not-determined | Genome-fragment | NA | NA | 0 |
| S7_k141_149429 | 1303 | No | NA | 2 | 0 | 1 | Not-determined | Genome-fragment | NA | NA | 0 |
| S7_k141_26405 | 1009 | No | NA | 2 | 0 | 0 | Not-determined | Genome-fragment | NA | NA | 0 |
| S12_k141_352005 | 2045 | No | NA | 1 | 0 | 1 | Low-quality | Genome-fragment | 0.54 | AAI-based (high-confidence) | 0 |
| S7_k141_119550 | 1263 | No | NA | 2 | 0 | 0 | Not-determined | Genome-fragment | NA | NA | 0 |
| S7_k141_38056 | 1017 | No | NA | 1 | 0 | 0 | Not-determined | Genome-fragment | NA | NA | 0 |
| S7_k141_119578 | 1016 | No | NA | 1 | 0 | 0 | Not-determined | Genome-fragment | NA | NA | 0 |
| S11-final-viral-combined_k141_25239\|\|full | 2046 | No | NA | 3 | 1 | 0 | Low-quality | Genome-fragment | 6.26 | AAI-based (high-confidence) | 0 |
| S7_k141_242004 | 1680 | No | NA | 4 | 0 | 2 | Not-determined | Genome-fragment | NA | NA | 0 |
| S5-final-viral-combined_k141_72380\|\|lt2gene | 2046 | No | NA | 2 | 2 | 0 | Low-quality | Genome-fragment | 4.67 | AAI-based (high-confidence) | 0 |
| S7_k141_219936 | 1352 | No | NA | 2 | 0 | 2 | Not-determined | Genome-fragment | NA | NA | 0 |
| S5-final-viral-combined_k141_58959\|\|lt2gene | 2046 | No | NA | 2 | 2 | 0 | Low-quality | Genome-fragment | 4.67 | AAI-based (high-confidence) | 0 |
| S7_k141_17183 | 1234 | No | NA | 1 | 0 | 0 | Not-determined | Genome-fragment | NA | NA | 0 |
| S7_k141_17198 | 1135 | No | NA | 2 | 0 | 0 | Not-determined | Genome-fragment | NA | NA | 0 |
| S7_k141_219948 | 3124 | No | NA | 4 | 0 | 0 | Not-determined | Genome-fragment | NA | NA | 0 |
| S7_k141_17204 | 1867 | No | NA | 3 | 0 | 0 | Not-determined | Genome-fragment | NA | NA | 0 |
| S7_k141_149544 | 1544 | No | NA | 3 | 0 | 0 | Not-determined | Genome-fragment | NA | NA | 0 |
| S7_k141_6958 | 1435 | No | NA | 1 | 0 | 1 | Not-determined | Genome-fragment | NA | NA | 0 |
| S7_k141_38068 | 1551 | No | NA | 2 | 0 | 0 | Not-determined | Genome-fragment | NA | NA | 0 |
| S7_k141_201733 | 1368 | No | NA | 1 | 0 | 0 | Not-determined | Genome-fragment | NA | NA | 0 |
| S9-final-viral-combined_k141_84731\|\|full | 2047 | No | NA | 2 | 0 | 0 | Low-quality | Genome-fragment | 5.55 | AAI-based (medium-confidence) | 0 |
| S11-final-viral-combined_k141_326980\|\|full | 2047 | No | NA | 3 | 0 | 0 | Low-quality | Genome-fragment | 3.31 | AAI-based (high-confidence) | 0 |
| S7_k141_108347 | 1702 | No | NA | 1 | 0 | 1 | Not-determined | Genome-fragment | NA | NA | 0 |
| S7_k141_201817 | 1403 | No | NA | 2 | 0 | 0 | Not-determined | Genome-fragment | NA | NA | 0 |
| S7_k141_220047 | 2014 | No | NA | 2 | 0 | 2 | Not-determined | Genome-fragment | NA | NA | 0 |
| S7_k141_98934 | 3049 | No | NA | 2 | 0 | 2 | Not-determined | Genome-fragment | NA | NA | 0 |
| S7-final-viral-combined_k141_228050\|\|full | 2047 | No | NA | 3 | 0 | 0 | Low-quality | Genome-fragment | 2.49 | AAI-based (high-confidence) | 0 |
| S7_k141_7002 | 1045 | No | NA | 1 | 0 | 0 | Not-determined | Genome-fragment | NA | NA | 0 |
| S7_k141_119824 | 1013 | No | NA | 2 | 0 | 2 | Not-determined | Genome-fragment | NA | NA | 0 |
| S7_k141_139667 | 1111 | No | NA | 1 | 0 | 1 | Not-determined | Genome-fragment | NA | NA | 0 |
| S7_k141_108431 | 1434 | No | NA | 2 | 0 | 0 | Not-determined | Genome-fragment | NA | NA | 0 |
| S7_k141_48067 | 1030 | No | NA | 2 | 0 | 1 | Not-determined | Genome-fragment | NA | NA | 0 |
| S7_k141_242028 | 2047 | No | NA | 4 | 0 | 4 | Low-quality | Genome-fragment | 0.55 | AAI-based (medium-confidence) | 0 |
| S7_k141_7027 | 3539 | No | NA | 5 | 0 | 4 | Not-determined | Genome-fragment | NA | NA | 0 |
| S7_k141_108445 | 1235 | No | NA | 1 | 0 | 0 | Not-determined | Genome-fragment | NA | NA | 0 |
| S7_k141_180943 | 4706 | No | NA | 4 | 0 | 1 | Not-determined | Genome-fragment | NA | NA | 0 |
| S17-final-viral-combined_k141_62178\|\|full | 2047 | No | NA | 3 | 0 | 0 | Low-quality | Genome-fragment | 4.57 | AAI-based (high-confidence) | 0 |
| S17-final-viral-combined_k141_265180\|\|full | 2047 | No | NA | 3 | 0 | 3 | Low-quality | Genome-fragment | 3.74 | AAI-based (medium-confidence) | 0 |
| S12-final-viral-combined_k141_312504\|\|full | 2047 | No | NA | 3 | 0 | 1 | Low-quality | Genome-fragment | 1.3 | AAI-based (medium-confidence) | 0 |
| S7_k141_58300 | 1177 | No | NA | 1 | 0 | 1 | Not-determined | Genome-fragment | NA | NA | 0 |
| S7_k141_220083 | 1125 | No | NA | 1 | 0 | 0 | Not-determined | Genome-fragment | NA | NA | 0 |
| S7_k141_211859 | 3335 | No | NA | 3 | 0 | 1 | Not-determined | Genome-fragment | NA | NA | 0 |
| S7_k141_191656 | 1630 | No | NA | 2 | 0 | 0 | Not-determined | Genome-fragment | NA | NA | 0 |
| S7_k141_48106 | 1563 | No | NA | 3 | 0 | 0 | Not-determined | Genome-fragment | NA | NA | 0 |
| S12_k141_256947 | 2047 | No | NA | 3 | 1 | 0 | Low-quality | Genome-fragment | 1.1 | AAI-based (high-confidence) | 0 |
| S7_k141_231919 | 1805 | No | NA | 1 | 0 | 1 | Not-determined | Genome-fragment | NA | NA | 0 |
| S7_k141_38197 | 1331 | No | NA | 3 | 0 | 3 | Not-determined | Genome-fragment | NA | NA | 0 |
| S7_k141_48126 | 1304 | No | NA | 1 | 0 | 1 | Not-determined | Genome-fragment | NA | NA | 0 |
| S1_k141_23324 | 2048 | No | NA | 3 | 1 | 0 | Low-quality | Genome-fragment | 1.83 | AAI-based (high-confidence) | 0 |
| S7_k141_139671 | 7747 | No | NA | 5 | 0 | 4 | Not-determined | Genome-fragment | NA | NA | 0 |
| S7_k141_181038 | 1805 | No | NA | 2 | 0 | 2 | Not-determined | Genome-fragment | NA | NA | 0 |
| S7_k141_181049 | 3772 | No | NA | 6 | 0 | 6 | Not-determined | Genome-fragment | NA | NA | 0 |
| S7_k141_88469 | 3937 | No | NA | 3 | 0 | 1 | Not-determined | Genome-fragment | NA | NA | 0 |
| S7_k141_26560 | 2524 | No | NA | 3 | 0 | 2 | Not-determined | Genome-fragment | NA | NA | 0 |
| S7_k141_211930 | 8161 | No | NA | 5 | 0 | 0 | Not-determined | Genome-fragment | NA | NA | 0 |
| S2_k141_24217 | 2048 | No | NA | 3 | 0 | 1 | Low-quality | Genome-fragment | 2.28 | AAI-based (medium-confidence) | 0 |
| S7_k141_58393 | 1400 | No | NA | 1 | 0 | 1 | Not-determined | Genome-fragment | NA | NA | 0 |
| S1-final-viral-combined_k141_241222\|\|full | 2049 | No | NA | 5 | 3 | 0 | Low-quality | Genome-fragment | 5.1 | AAI-based (high-confidence) | 0 |
| S7_k141_130000 | 3770 | No | NA | 5 | 0 | 2 | Not-determined | Genome-fragment | NA | NA | 0 |
| S7_k141_88547 | 5309 | No | NA | 4 | 0 | 4 | Not-determined | Genome-fragment | NA | NA | 0 |
| S1_k141_234256 | 2049 | No | NA | 1 | 0 | 0 | Low-quality | Genome-fragment | 2.36 | AAI-based (medium-confidence) | 0 |
| S5-final-viral-combined_k141_97438\|\|full | 2049 | No | NA | 3 | 0 | 3 | Low-quality | Genome-fragment | 0.72 | AAI-based (medium-confidence) | 0 |
| S7_k141_58428 | 11527 | No | NA | 15 | 0 | 9 | Not-determined | Genome-fragment | NA | NA | 0 |
| S7_k141_211969 | 1667 | No | NA | 2 | 0 | 0 | Not-determined | Genome-fragment | NA | NA | 0 |
| S7_k141_181197 | 1604 | No | NA | 2 | 0 | 2 | Not-determined | Genome-fragment | NA | NA | 0 |
| S7_k141_191758 | 6581 | No | NA | 6 | 0 | 4 | Not-determined | Genome-fragment | NA | NA | 0 |
| S7_k141_48292 | 1033 | No | NA | 1 | 0 | 0 | Not-determined | Genome-fragment | NA | NA | 0 |
| S7-final-viral-combined_k141_3895\|\|full | 2049 | No | NA | 3 | 0 | 0 | Low-quality | Genome-fragment | 3.66 | AAI-based (high-confidence) | 0 |

| | | | | | | | | | | | | |
|---|---|---|---|---|---|---|---|---|---|---|---|---|
| S7_k141_191784 | 4509 | No | NA | | 4 | 0 | 3 | Not-determined | Genome-fragment | NA | NA | 0 |
| S2-final-viral-combined_k141_96359\|\|full | 2049 | No | NA | | 3 | 0 | 2 | Low-quality | Genome-fragment | 3.68 | AAI-based (medium-confidence) | 0 |
| S7_k141_26660 | 1175 | No | NA | | 1 | 0 | 1 | Not-determined | Genome-fragment | NA | NA | 0 |
| S2_k141_11083 | 2049 | No | NA | | 1 | 0 | 0 | Low-quality | Genome-fragment | 2.56 | AAI-based (medium-confidence) | 0 |
| S7_k141_139795 | 1434 | No | NA | | 1 | 0 | 0 | Not-determined | Genome-fragment | NA | NA | 0 |
| S6-final-viral-combined_k141_84528\|\|full | 2049 | No | NA | | 3 | 1 | 0 | Low-quality | Genome-fragment | 3.07 | HMM-based (lower-bound) | 0 |
| S7_k141_78238 | 1035 | No | NA | | 2 | 0 | 2 | Not-determined | Genome-fragment | NA | NA | 0 |
| S11-final-viral-combined_k141_230287\|\|full | 2050 | No | NA | | 4 | 1 | 0 | Low-quality | Genome-fragment | 3.75 | AAI-based (high-confidence) | 0 |
| S7_k141_191835 | 1098 | No | NA | | 1 | 0 | 1 | Not-determined | Genome-fragment | NA | NA | 0 |
| S7_k141_99215 | 14829 | No | NA | | 7 | 0 | 5 | Not-determined | Genome-fragment | NA | NA | 0 |
| S7_k141_232174 | 6907 | No | NA | | 8 | 0 | 4 | Not-determined | Genome-fragment | NA | NA | 0 |
| S7_k141_99235 | 3569 | No | NA | | 6 | 0 | 0 | Not-determined | Genome-fragment | NA | NA | 0 |
| S5-final-viral-combined_k141_130796\|\|full | 2050 | No | NA | | 3 | 3 | 0 | Low-quality | Genome-fragment | 4.49 | AAI-based (high-confidence) | 0 |
| S7_k141_232182 | 1109 | No | NA | | 1 | 0 | 1 | Not-determined | Genome-fragment | NA | NA | 0 |
| S7_k141_120169 | 1213 | No | NA | | 2 | 0 | 0 | Not-determined | Genome-fragment | NA | NA | 0 |
| S7_k141_202150 | 1010 | No | NA | | 2 | 0 | 0 | Not-determined | Genome-fragment | NA | NA | 0 |
| S7_k141_149978 | 3790 | No | NA | | 2 | 0 | 2 | Not-determined | Genome-fragment | NA | NA | 0 |
| S7_k141_38400 | 1054 | No | NA | | 1 | 0 | 1 | Not-determined | Genome-fragment | NA | NA | 0 |
| S7_k141_78291 | 1599 | No | NA | | 1 | 0 | 1 | Not-determined | Genome-fragment | NA | NA | 0 |
| S7_k141_38417 | 3198 | No | NA | | 2 | 0 | 2 | Not-determined | Genome-fragment | NA | NA | 0 |
| S7_k141_88748 | 4045 | No | NA | | 4 | 0 | 2 | Not-determined | Genome-fragment | NA | NA | 0 |
| S7_k141_58615 | 1267 | No | NA | | 1 | 0 | 0 | Not-determined | Genome-fragment | NA | NA | 0 |
| S17-final-viral-combined_k141_2345\|\|full | 2050 | No | NA | | 4 | 0 | 0 | Low-quality | Genome-fragment | 3.75 | AAI-based (high-confidence) | 0 |
| S7_k141_69338 | 1506 | No | NA | | 1 | 0 | 1 | Not-determined | Genome-fragment | NA | NA | 0 |
| S7_k141_7254 | 1066 | No | NA | | 1 | 0 | 1 | Not-determined | Genome-fragment | NA | NA | 0 |
| S7_k141_181458 | 1161 | No | NA | | 2 | 0 | 2 | Not-determined | Genome-fragment | NA | NA | 0 |
| S7_k141_48465 | 1016 | No | NA | | 1 | 0 | 1 | Not-determined | Genome-fragment | NA | NA | 0 |
| S17-final-viral-combined_k141_58526\|\|full | 2050 | No | NA | | 6 | 0 | 2 | Low-quality | Genome-fragment | 2.96 | AAI-based (medium-confidence) | 0 |
| S7_k141_202270 | 1698 | No | NA | | 2 | 0 | 0 | Not-determined | Genome-fragment | NA | NA | 0 |
| S7_k141_88801 | 1161 | No | NA | | 2 | 0 | 0 | Not-determined | Genome-fragment | NA | NA | 0 |
| S8-final-viral-combined_k141_147247\|\|lt2gene | 2050 | No | NA | | 1 | 1 | 0 | Low-quality | Genome-fragment | 5.93 | AAI-based (high-confidence) | 0 |
| S7_k141_58677 | 4357 | No | NA | | 2 | 0 | 2 | Not-determined | Genome-fragment | NA | NA | 0 |
| S7_k141_181511 | 1449 | No | NA | | 1 | 0 | 0 | Not-determined | Genome-fragment | NA | NA | 0 |
| S7_k141_78394 | 2196 | No | NA | | 3 | 0 | 0 | Not-determined | Genome-fragment | NA | NA | 0 |
| S7_k141_58723 | 1166 | No | NA | | 1 | 0 | 0 | Not-determined | Genome-fragment | NA | NA | 0 |
| S7_k141_212158 | 1330 | No | NA | | 2 | 0 | 1 | Not-determined | Genome-fragment | NA | NA | 0 |
| S1-final-viral-combined_k141_270009\|\|full | 2051 | No | NA | | 4 | 3 | 0 | Low-quality | Genome-fragment | 4.33 | AAI-based (high-confidence) | 0 |
| S17-final-viral-combined_k141_60660\|\|full | 2051 | No | NA | | 5 | 1 | 0 | Low-quality | Genome-fragment | 4.48 | AAI-based (medium-confidence) | 0 |
| S7_k141_161361 | 2047 | No | NA | | 2 | 0 | 0 | Not-determined | Genome-fragment | NA | NA | 0 |
| S7_k141_212191 | 1271 | No | NA | | 2 | 0 | 0 | Not-determined | Genome-fragment | NA | NA | 0 |
| S7_k141_130299 | 2014 | No | NA | | 1 | 0 | 1 | Not-determined | Genome-fragment | NA | NA | 0 |
| S7_k141_130300 | 1734 | No | NA | | 3 | 0 | 0 | Not-determined | Genome-fragment | NA | NA | 0 |
| S12-final-viral-combined_k141_357226\|\|full | 2051 | No | NA | | 3 | 0 | 1 | Low-quality | Genome-fragment | 3.27 | AAI-based (medium-confidence) | 0 |
| S7_k141_212201 | 1152 | No | NA | | 2 | 0 | 1 | Not-determined | Genome-fragment | NA | NA | 0 |
| S12-final-viral-combined_k141_123803\|\|full | 2051 | No | NA | | 2 | 0 | 0 | Low-quality | Genome-fragment | 1.12 | AAI-based (medium-confidence) | 0 |
| S7_k141_7413 | 2246 | No | NA | | 1 | 0 | 0 | Not-determined | Genome-fragment | NA | NA | 0 |
| S7_k141_130360 | 13539 | No | NA | | 8 | 0 | 7 | Not-determined | Genome-fragment | NA | NA | 0 |
| S2-final-viral-combined_k141_223120\|\|full | 2051 | No | NA | | 3 | 0 | 1 | Low-quality | Genome-fragment | 4.07 | AAI-based (high-confidence) | 0 |
| S7_k141_130362 | 3042 | No | NA | | 2 | 0 | 2 | Not-determined | Genome-fragment | NA | NA | 0 |
| S7_k141_17742 | 3234 | No | NA | | 6 | 0 | 2 | Not-determined | Genome-fragment | NA | NA | 0 |
| S7_k141_192052 | 1113 | No | NA | | 1 | 0 | 0 | Not-determined | Genome-fragment | NA | NA | 0 |
| S2-final-viral-combined_k141_205951\|\|full | 2051 | No | NA | | 6 | 1 | 0 | Low-quality | Genome-fragment | 5.04 | AAI-based (medium-confidence) | 0 |
| S7_k141_108903 | 1455 | No | NA | | 2 | 0 | 0 | Not-determined | Genome-fragment | NA | NA | 0 |
| S7_k141_178967 | 2052 | No | NA | | 1 | 0 | 1 | Low-quality | Genome-fragment | 0.38 | AAI-based (high-confidence) | 0 |
| S7_k141_88931 | 2873 | No | NA | | 1 | 0 | 1 | Not-determined | Genome-fragment | NA | NA | 0 |
| S7_k141_58861 | 2155 | No | NA | | 3 | 0 | 3 | Not-determined | Genome-fragment | NA | NA | 0 |
| S7_k141_181640 | 4393 | No | NA | | 8 | 0 | 8 | Not-determined | Genome-fragment | NA | NA | 0 |
| S7_k141_202479 | 1579 | No | NA | | 2 | 0 | 0 | Not-determined | Genome-fragment | NA | NA | 0 |
| S17-final-viral-combined_k141_7349\|\|full | 2052 | No | NA | | 7 | 2 | 0 | Low-quality | Genome-fragment | 2.5 | HMM-based (lower-bound) | 0 |
| S7_k141_99501 | 3081 | No | NA | | 4 | 0 | 2 | Not-determined | Genome-fragment | NA | NA | 0 |
| S7_k141_38796 | 1383 | No | NA | | 1 | 0 | 0 | Not-determined | Genome-fragment | NA | NA | 0 |
| S12-final-viral-combined_k141_334947\|\|full | 2052 | No | NA | | 3 | 0 | 1 | Low-quality | Genome-fragment | 1.18 | AAI-based (medium-confidence) | 0 |
| S7_k141_69482 | 2324 | No | NA | | 2 | 0 | 1 | Not-determined | Genome-fragment | NA | NA | 0 |
| S7_k141_202545 | 4479 | No | NA | | 4 | 0 | 3 | Not-determined | Genome-fragment | NA | NA | 0 |
| S7_k141_38861 | 1167 | No | NA | | 2 | 0 | 0 | Not-determined | Genome-fragment | NA | NA | 0 |
| S7_k141_109033 | 2089 | No | NA | | 4 | 0 | 1 | Not-determined | Genome-fragment | NA | NA | 0 |
| S7_k141_48761 | 1468 | No | NA | | 6 | 0 | 6 | Not-determined | Genome-fragment | NA | NA | 0 |
| S7_k141_69526 | 1155 | No | NA | | 1 | 0 | 0 | Not-determined | Genome-fragment | NA | NA | 0 |
| S12-final-viral-combined_k141_354280\|\|full | 2052 | No | NA | | 2 | 0 | 2 | Low-quality | Genome-fragment | 0.96 | AAI-based (high-confidence) | 0 |
| S7_k141_161525 | 1140 | No | NA | | 1 | 0 | 1 | Not-determined | Genome-fragment | NA | NA | 0 |
| S7_k141_78659 | 1205 | No | NA | | 2 | 0 | 1 | Not-determined | Genome-fragment | NA | NA | 0 |
| S2-final-viral-combined_k141_68266\|\|full | 2052 | No | NA | | 3 | 0 | 2 | Low-quality | Genome-fragment | 4.9 | AAI-based (medium-confidence) | 0 |
| S7_k141_69545 | 1079 | No | NA | | 2 | 0 | 0 | Not-determined | Genome-fragment | NA | NA | 0 |
| S7_k141_202592 | 1519 | No | NA | | 1 | 0 | 1 | Not-determined | Genome-fragment | NA | NA | 0 |
| S1-final-viral-combined_k141_228648\|\|full | 2053 | No | NA | | 2 | 0 | 2 | Low-quality | Genome-fragment | 2.45 | AAI-based (medium-confidence) | 0 |
| S7_k141_181805 | 1215 | No | NA | | 2 | 0 | 0 | Not-determined | Genome-fragment | NA | NA | 0 |
| S7_k141_130586 | 1515 | No | NA | | 2 | 0 | 2 | Not-determined | Genome-fragment | NA | NA | 0 |
| S5-final-viral-combined_k141_172786\|\|full | 2053 | No | NA | | 6 | 3 | 0 | Low-quality | Genome-fragment | 4.68 | AAI-based (high-confidence) | 0 |
| S7_k141_220555 | 3662 | No | NA | | 4 | 0 | 1 | Not-determined | Genome-fragment | NA | NA | 0 |
| S5_k141_34012 | 2053 | No | NA | | 3 | 1 | 0 | Low-quality | Genome-fragment | 5.11 | AAI-based (medium-confidence) | 0 |
| S7_k141_78721 | 2435 | No | NA | | 1 | 0 | 1 | Not-determined | Genome-fragment | NA | NA | 0 |
| S7-final-viral-combined_k141_78807\|\|full | 2053 | Yes | | 954 | 4 | 1 | 1 | Low-quality | Genome-fragment | 1.65 | AAI-based (high-confidence) | 53.53 |
| S7_k141_48875 | 1313 | No | NA | | 2 | 0 | 1 | Not-determined | Genome-fragment | NA | NA | 0 |
| S7_k141_139959 | 1721 | No | NA | | 2 | 0 | 1 | Not-determined | Genome-fragment | NA | NA | 0 |
| S7_k141_69637 | 1031 | No | NA | | 1 | 0 | 0 | Not-determined | Genome-fragment | NA | NA | 0 |
| S17-final-viral-combined_k141_183309\|\|full | 2053 | No | NA | | 4 | 0 | 0 | Low-quality | Genome-fragment | 5.83 | AAI-based (medium-confidence) | 0 |
| S7_k141_202730 | 1981 | No | NA | | 1 | 0 | 0 | Not-determined | Genome-fragment | NA | NA | 0 |
| S7_k141_181878 | 6675 | No | NA | | 4 | 0 | 1 | Not-determined | Genome-fragment | NA | NA | 0 |
| S7_k141_48956 | 1106 | No | NA | | 1 | 0 | 1 | Not-determined | Genome-fragment | NA | NA | 0 |
| S7_k141_171423 | 1113 | No | NA | | 1 | 0 | 0 | Not-determined | Genome-fragment | NA | NA | 0 |
| S7_k141_232657 | 1630 | No | NA | | 2 | 0 | 0 | Not-determined | Genome-fragment | NA | NA | 0 |
| S7_k141_39036 | 1039 | No | NA | | 1 | 0 | 0 | Not-determined | Genome-fragment | NA | NA | 0 |
| S12-final-viral-combined_k141_270194\|\|full | 2053 | No | NA | | 5 | 1 | 0 | Low-quality | Genome-fragment | 3.96 | AAI-based (medium-confidence) | 0 |
| S7_k141_242462 | 3187 | No | NA | | 4 | 0 | 1 | Not-determined | Genome-fragment | NA | NA | 0 |
| S2-final-viral-combined_k141_130027\|\|full | 2053 | No | NA | | 4 | 0 | 2 | Low-quality | Genome-fragment | 1.53 | AAI-based (high-confidence) | 0 |
| S7_k141_18082 | 3699 | No | NA | | 6 | 0 | 4 | Not-determined | Genome-fragment | NA | NA | 0 |
| S7_k141_130789 | 8513 | No | NA | | 9 | 0 | 8 | Not-determined | Genome-fragment | NA | NA | 0 |
| S7_k141_130790 | 1132 | No | NA | | 2 | 0 | 2 | Not-determined | Genome-fragment | NA | NA | 0 |
| S7_k141_120538 | 1355 | No | NA | | 3 | 0 | 1 | Not-determined | Genome-fragment | NA | NA | 0 |
| S7_k141_242506 | 1222 | No | NA | | 1 | 0 | 0 | Not-determined | Genome-fragment | NA | NA | 0 |
| S7_k141_192344 | 1843 | No | NA | | 1 | 0 | 0 | Not-determined | Genome-fragment | NA | NA | 0 |
| S7_k141_192365 | 1140 | No | NA | | 2 | 0 | 0 | Not-determined | Genome-fragment | NA | NA | 0 |
| S7_k141_232781 | 2231 | No | NA | | 2 | 0 | 2 | Not-determined | Genome-fragment | NA | NA | 0 |
| S4-final-viral-combined_k141_73257\|\|full | 2053 | No | NA | | 4 | 1 | 0 | Low-quality | Genome-fragment | 3.43 | AAI-based (high-confidence) | 0 |
| S7_k141_59267 | 1321 | No | NA | | 1 | 0 | 0 | Not-determined | Genome-fragment | NA | NA | 0 |
| S1-final-viral-combined_k141_123434\|\|full | 2054 | No | NA | | 3 | 3 | 0 | Low-quality | Genome-fragment | 4.92 | AAI-based (high-confidence) | 0 |
| S7_k141_99894 | 1808 | No | NA | | 3 | 0 | 2 | Not-determined | Genome-fragment | NA | NA | 0 |
| S7_k141_150521 | 2888 | No | NA | | 4 | 0 | 2 | Not-determined | Genome-fragment | NA | NA | 0 |
| S7_k141_18162 | 1018 | No | NA | | 1 | 0 | 0 | Not-determined | Genome-fragment | NA | NA | 0 |
| S7_k141_192454 | 1089 | No | NA | | 2 | 0 | 1 | Not-determined | Genome-fragment | NA | NA | 0 |
| S7_k141_232892 | 1106 | No | NA | | 1 | 0 | 0 | Not-determined | Genome-fragment | NA | NA | 0 |
| S7_k141_220788 | 1839 | No | NA | | 3 | 0 | 3 | Not-determined | Genome-fragment | NA | NA | 0 |
| S12-final-viral-combined_k141_293338\|\|full | 2054 | No | NA | | 4 | 1 | 0 | Low-quality | Genome-fragment | 3.77 | HMM-based (lower-bound) | 0 |
| S7_k141_220798 | 1080 | No | NA | | 1 | 0 | 0 | Not-determined | Genome-fragment | NA | NA | 0 |
| S12-final-viral-combined_k141_160933\|\|full | 2054 | No | NA | | 3 | 0 | 1 | Low-quality | Genome-fragment | 1.89 | AAI-based (high-confidence) | 0 |
| S7_k141_39149 | 3075 | No | NA | | 4 | 0 | 1 | Not-determined | Genome-fragment | NA | NA | 0 |
| S7_k141_220835 | 2133 | No | NA | | 5 | 0 | 4 | Not-determined | Genome-fragment | NA | NA | 0 |
| S2-final-viral-combined_k141_164163\|\|full | 2054 | No | NA | | 5 | 1 | 0 | Low-quality | Genome-fragment | 6.08 | AAI-based (high-confidence) | 0 |
| S7_k141_99973 | 1899 | No | NA | | 2 | 0 | 1 | Not-determined | Genome-fragment | NA | NA | 0 |
| S8_k141_74345 | 2054 | No | NA | | 2 | 0 | 2 | Low-quality | Genome-fragment | 1.31 | AAI-based (high-confidence) | 0 |
| S1-final-viral-combined_k141_57756\|\|full | 2055 | No | NA | | 2 | 0 | 2 | Low-quality | Genome-fragment | 3.57 | AAI-based (medium-confidence) | 0 |
| S7_k141_220861 | 2083 | No | NA | | 3 | 0 | 1 | Not-determined | Genome-fragment | NA | NA | 0 |
| S7-final-viral-combined_k141_123922\|\|full | 2055 | No | NA | | 2 | 0 | 0 | Low-quality | Genome-fragment | 4.16 | AAI-based (medium-confidence) | 0 |
| S7-final-viral-combined_k141_118243\|\|full | 2055 | No | NA | | 5 | 1 | 0 | Low-quality | Genome-fragment | 5.26 | AAI-based (high-confidence) | 0 |
| S7_k141_161918 | 1282 | No | NA | | 2 | 0 | 1 | Not-determined | Genome-fragment | NA | NA | 0 |
| S12_k141_25516 | 2055 | No | NA | | 3 | 1 | 0 | Low-quality | Genome-fragment | 4.23 | AAI-based (high-confidence) | 0 |
| S7_k141_150672 | 4678 | No | NA | | 5 | 0 | 0 | Not-determined | Genome-fragment | NA | NA | 0 |
| S7_k141_49237 | 1021 | No | NA | | 1 | 0 | 0 | Not-determined | Genome-fragment | NA | NA | 0 |
| S7_k141_120688 | 1149 | No | NA | | 2 | 0 | 0 | Not-determined | Genome-fragment | NA | NA | 0 |
| S7_k141_192558 | 1405 | No | NA | | 1 | 0 | 1 | Not-determined | Genome-fragment | NA | NA | 0 |
| S7_k141_171722 | 1388 | No | NA | | 2 | 0 | 2 | Not-determined | Genome-fragment | NA | NA | 0 |
| S7_k141_203208 | 1124 | No | NA | | 2 | 0 | 0 | Not-determined | Genome-fragment | NA | NA | 0 |
| S7_k141_182234 | 1891 | No | NA | | 5 | 0 | 5 | Not-determined | Genome-fragment | NA | NA | 0 |
| S7_k141_212777 | 1251 | No | NA | | 2 | 0 | 0 | Not-determined | Genome-fragment | NA | NA | 0 |
| S7_k141_192598 | 1826 | No | NA | | 3 | 0 | 2 | Not-determined | Genome-fragment | NA | NA | 0 |
| S7_k141_18276 | 1826 | No | NA | | 3 | 0 | 0 | Not-determined | Genome-fragment | NA | NA | 0 |
| S7_k141_39291 | 7642 | No | NA | | 4 | 0 | 4 | Not-determined | Genome-fragment | NA | NA | 0 |
| S7_k141_233050 | 1024 | No | NA | | 1 | 0 | 1 | Not-determined | Genome-fragment | NA | NA | 0 |
| S7_k141_203282 | 1094 | No | NA | | 1 | 0 | 1 | Not-determined | Genome-fragment | NA | NA | 0 |
| S7_k141_69898 | 1039 | No | NA | | 1 | 0 | 1 | Not-determined | Genome-fragment | NA | NA | 0 |
| S7_k141_150728 | 1044 | No | NA | | 1 | 0 | 1 | Not-determined | Genome-fragment | NA | NA | 0 |
| S7_k141_69901 | 1992 | No | NA | | 1 | 0 | 1 | Not-determined | Genome-fragment | NA | NA | 0 |
| S2-final-viral-combined_k141_65897\|\|full | 2055 | No | NA | | 2 | 0 | 2 | Low-quality | Genome-fragment | 4.75 | AAI-based (medium-confidence) | 0 |
| S8-final-viral-combined_k141_223975\|\|full | 2055 | No | NA | | 3 | 1 | 0 | Low-quality | Genome-fragment | 5.47 | AAI-based (medium-confidence) | 0 |
| S7_k141_120835 | 1293 | No | NA | | 1 | 0 | 1 | Not-determined | Genome-fragment | NA | NA | 0 |
| S7_k141_242627 | 1071 | No | NA | | 1 | 0 | 1 | Not-determined | Genome-fragment | NA | NA | 0 |
| S7_k141_120843 | 1173 | No | NA | | 5 | 0 | 0 | Not-determined | Genome-fragment | NA | NA | 0 |
| S1_k141_244839 | 2056 | No | NA | | 1 | 0 | 1 | Low-quality | Genome-fragment | 0.39 | AAI-based (medium-confidence) | 0 |
| S7_k141_242661 | 1080 | No | NA | | 2 | 0 | 0 | Not-determined | Genome-fragment | NA | NA | 0 |
| S7_k141_49406 | 1213 | No | NA | | 1 | 0 | 1 | Not-determined | Genome-fragment | NA | NA | 0 |
| S7_k141_27049 | 1081 | No | NA | | 1 | 0 | 1 | Not-determined | Genome-fragment | NA | NA | 0 |
| S7_k141_89418 | 1567 | No | NA | | 3 | 0 | 3 | Not-determined | Genome-fragment | NA | NA | 0 |
| S7_k141_182342 | 2236 | No | NA | | 1 | 0 | 1 | Not-determined | Genome-fragment | NA | NA | 0 |
| S5-final-viral-combined_k141_105010\|\|full | 2056 | No | NA | | 2 | 0 | 0 | Low-quality | Genome-fragment | 4.91 | AAI-based (high-confidence) | 0 |
| S7_k141_89452 | 2731 | No | NA | | 2 | 0 | 1 | Not-determined | Genome-fragment | NA | NA | 0 |
| S7_k141_150798 | 1711 | No | NA | | 2 | 0 | 0 | Not-determined | Genome-fragment | NA | NA | 0 |
| S7_k141_130882 | 1221 | No | NA | | 2 | 0 | 1 | Not-determined | Genome-fragment | NA | NA | 0 |
| S7_k141_39458 | 1148 | No | NA | | 1 | 0 | 1 | Not-determined | Genome-fragment | NA | NA | 0 |
| S7_k141_130887 | 1583 | No | NA | | 1 | 0 | 1 | Not-determined | Genome-fragment | NA | NA | 0 |
| S7_k141_150802 | 1347 | No | NA | | 1 | 0 | 0 | Not-determined | Genome-fragment | NA | NA | 0 |
| S7_k141_79223 | 5332 | No | NA | | 3 | 0 | 1 | Not-determined | Genome-fragment | NA | NA | 0 |
| S7_k141_203452 | 1379 | No | NA | | 1 | 0 | 1 | Not-determined | Genome-fragment | NA | NA | 0 |
| S7_k141_8183 | 4216 | No | NA | | 5 | 0 | 4 | Not-determined | Genome-fragment | NA | NA | 0 |
| S7_k141_49506 | 2068 | No | NA | | 3 | 0 | 0 | Not-determined | Genome-fragment | NA | NA | 0 |
| S7_k141_39502 | 1156 | No | NA | | 1 | 0 | 1 | Not-determined | Genome-fragment | NA | NA | 0 |
| S7-final-viral-combined_k141_232282\|\|full | 2056 | No | NA | | 3 | 0 | 0 | Low-quality | Genome-fragment | 4.94 | AAI-based (high-confidence) | 0 |
| S7_k141_130903 | 1578 | No | NA | | 3 | 0 | 0 | Not-determined | Genome-fragment | NA | NA | 0 |
| S7_k141_8203 | 3036 | No | NA | | 5 | 0 | 3 | Not-determined | Genome-fragment | NA | NA | 0 |
| S7_k141_242853 | 3104 | No | NA | | 4 | 0 | 3 | Not-determined | Genome-fragment | NA | NA | 0 |
| S7_k141_130927 | 8361 | No | NA | | 5 | 0 | 4 | Not-determined | Genome-fragment | NA | NA | 0 |
| S12-final-viral-combined_k141_161015\|\|full | 2056 | No | NA | | 3 | 3 | 0 | Low-quality | Genome-fragment | 5.48 | AAI-based (high-confidence) | 0 |
| S7_k141_171975 | 5868 | No | NA | | 4 | 0 | 2 | Not-determined | Genome-fragment | NA | NA | 0 |
| S7_k141_203588 | 1073 | No | NA | | 1 | 0 | 1 | Not-determined | Genome-fragment | NA | NA | 0 |
| S7_k141_70070 | 1815 | No | NA | | 2 | 0 | 1 | Not-determined | Genome-fragment | NA | NA | 0 |
| S7_k141_100361 | 1043 | No | NA | | 2 | 0 | 2 | Not-determined | Genome-fragment | NA | NA | 0 |
| S7_k141_130956 | 1411 | No | NA | | 1 | 0 | 0 | Not-determined | Genome-fragment | NA | NA | 0 |
| S12_k141_349135 | 2056 | No | NA | | 3 | 1 | 0 | Low-quality | Genome-fragment | 30.12 | AAI-based (medium-confidence) | 0 |
| S7_k141_192953 | 1609 | No | NA | | 3 | 0 | 3 | Not-determined | Genome-fragment | NA | NA | 0 |
| S1-final-viral-combined_k141_326824\|\|full | 2057 | No | NA | | 3 | 0 | 0 | Low-quality | Genome-fragment | 5.27 | AAI-based (medium-confidence) | 0 |
| S7_k141_79268 | 1147 | No | NA | | 2 | 0 | 0 | Not-determined | Genome-fragment | NA | NA | 0 |
| S7_k141_109898 | 1172 | No | NA | | 1 | 0 | 0 | Not-determined | Genome-fragment | NA | NA | 0 |
| S12-final-viral-combined_k141_309749\|\|full | 2057 | No | NA | | 2 | 0 | 1 | Low-quality | Genome-fragment | 1.68 | AAI-based (medium-confidence) | 0 |
| S7_k141_70145 | 1141 | No | NA | | 3 | 0 | 0 | Not-determined | Genome-fragment | NA | NA | 0 |
| S7_k141_172038 | 1319 | No | NA | | 1 | 0 | 0 | Not-determined | Genome-fragment | NA | NA | 0 |
| S7_k141_140570 | 2080 | No | NA | | 3 | 0 | 0 | Not-determined | Genome-fragment | NA | NA | 0 |
| S7_k141_233413 | 1060 | No | NA | | 2 | 0 | 0 | Not-determined | Genome-fragment | NA | NA | 0 |
| S7_k141_59791 | 6198 | No | NA | | 6 | 0 | 2 | Not-determined | Genome-fragment | NA | NA | 0 |
| S7_k141_221280 | 2008 | No | NA | | 2 | 0 | 1 | Not-determined | Genome-fragment | NA | NA | 0 |
| S7_k141_172079 | 1152 | No | NA | | 1 | 0 | 0 | Not-determined | Genome-fragment | NA | NA | 0 |
| S7_k141_39673 | 1147 | No | NA | | 1 | 0 | 1 | Not-determined | Genome-fragment | NA | NA | 0 |
| S12-final-viral-combined_k141_260868\|\|full | 2057 | No | NA | | 3 | 1 | 0 | Low-quality | Genome-fragment | 0.74 | HMM-based (lower-bound) | 0 |
| S7_k141_100478 | 4669 | No | NA | | 4 | 0 | 4 | Not-determined | Genome-fragment | NA | NA | 0 |
| S7_k141_39681 | 1705 | No | NA | | 3 | 0 | 1 | Not-determined | Genome-fragment | NA | NA | 0 |
| S7_k141_49769 | 2691 | No | NA | | 3 | 0 | 0 | Not-determined | Genome-fragment | NA | NA | 0 |
| S7_k141_193085 | 1021 | No | NA | | 2 | 0 | 1 | Not-determined | Genome-fragment | NA | NA | 0 |
| S2-final-viral-combined_k141_234256\|\|full | 2057 | No | NA | | 3 | 0 | 0 | Low-quality | Genome-fragment | 4.21 | AAI-based (high-confidence) | 0 |
| S8-final-viral-combined_k141_98085\|\|full | 2057 | No | NA | | 5 | 1 | 0 | Low-quality | Genome-fragment | 4.06 | AAI-based (high-confidence) | 0 |
| S7_k141_49796 | 1478 | No | NA | | 2 | 0 | 0 | Not-determined | Genome-fragment | NA | NA | 0 |
| S7_k141_233527 | 5041 | No | NA | | 1 | 0 | 1 | Not-determined | Genome-fragment | NA | NA | 0 |
| S7_k141_110020 | 2921 | No | NA | | 2 | 0 | 2 | Not-determined | Genome-fragment | NA | NA | 0 |
| S8-final-viral-combined_k141_29687\|\|full | 2057 | No | NA | | 2 | 0 | 1 | Low-quality | Genome-fragment | 0.92 | AAI-based (medium-confidence) | 0 |
| S7_k141_8489 | 1900 | No | NA | | 3 | 0 | 2 | Not-determined | Genome-fragment | NA | NA | 0 |
| S7_k141_243123 | 1273 | No | NA | | 2 | 0 | 1 | Not-determined | Genome-fragment | NA | NA | 0 |
| S7_k141_89683 | 1307 | No | NA | | 1 | 0 | 1 | Not-determined | Genome-fragment | NA | NA | 0 |
| S7_k141_131193 | 1835 | No | NA | | 2 | 0 | 0 | Not-determined | Genome-fragment | NA | NA | 0 |
| S7_k141_151112 | 28365 | No | NA | | 57 | 13 | 1 | Medium-quality | Genome-fragment | 66.55 | AAI-based (high-confidence) | 0 |
| S7_k141_89719 | 1326 | No | NA | | 3 | 0 | 1 | Not-determined | Genome-fragment | NA | NA | 0 |
| S7_k141_213137 | 1081 | No | NA | | 1 | 0 | 0 | Not-determined | Genome-fragment | NA | NA | 0 |
| S7_k141_193191 | 3219 | No | NA | | 2 | 0 | 0 | Not-determined | Genome-fragment | NA | NA | 0 |
| S7_k141_203893 | 1152 | No | NA | | 2 | 0 | 2 | Not-determined | Genome-fragment | NA | NA | 0 |
| S7_k141_193203 | 1175 | No | NA | | 1 | 0 | 0 | Not-determined | Genome-fragment | NA | NA | 0 |
| S7_k141_121393 | 1248 | No | NA | | 1 | 0 | 1 | Not-determined | Genome-fragment | NA | NA | 0 |
| S7_k141_39793 | 1473 | No | NA | | 1 | 0 | 0 | Not-determined | Genome-fragment | NA | NA | 0 |
| S9-final-viral-combined_k141_87743\|\|full | 2058 | No | NA | | 4 | 1 | 0 | Low-quality | Genome-fragment | 6.68 | AAI-based (high-confidence) | 0 |
| S7_k141_8576 | 3041 | No | NA | | 1 | 0 | 0 | Not-determined | Genome-fragment | NA | NA | 0 |
| S7_k141_213198 | 1912 | No | NA | | 1 | 0 | 1 | Not-determined | Genome-fragment | NA | NA | 0 |
| S7_k141_203944 | 1260 | No | NA | | 2 | 0 | 1 | Not-determined | Genome-fragment | NA | NA | 0 |
| S7_k141_151211 | 1051 | No | NA | | 2 | 0 | 2 | Not-determined | Genome-fragment | NA | NA | 0 |
| S7_k141_50019 | 1190 | No | NA | | 1 | 0 | 1 | Not-determined | Genome-fragment | NA | NA | 0 |
| S7_k141_18602 | 2870 | No | NA | | 3 | 0 | 0 | Not-determined | Genome-fragment | NA | NA | 0 |
| S7_k141_89798 | 1016 | No | NA | | 2 | 0 | 0 | Not-determined | Genome-fragment | NA | NA | 0 |
| S7_k141_79461 | 1099 | No | NA | | 1 | 0 | 1 | Not-determined | Genome-fragment | NA | NA | 0 |
| S7_k141_100606 | 1022 | No | NA | | 1 | 0 | 1 | Not-determined | Genome-fragment | NA | NA | 0 |
| S7_k141_162435 | 13809 | No | NA | | 12 | 0 | 8 | Not-determined | Genome-fragment | NA | NA | 0 |
| S7_k141_221514 | 1161 | No | NA | | 1 | 0 | 0 | Not-determined | Genome-fragment | NA | NA | 0 |
| S7_k141_110182 | 4685 | No | NA | | 6 | 0 | 0 | Not-determined | Genome-fragment | NA | NA | 0 |
| S9_k141_49821 | 2058 | No | NA | | 3 | 0 | 3 | Low-quality | Genome-fragment | 0.76 | AAI-based (medium-confidence) | 0 |
| S7_k141_110202 | 1505 | No | NA | | 1 | 0 | 1 | Not-determined | Genome-fragment | NA | NA | 0 |
| S7_k141_131340 | 1604 | No | NA | | 2 | 0 | 2 | Not-determined | Genome-fragment | NA | NA | 0 |
| S7_k141_233833 | 1051 | No | NA | | 2 | 0 | 0 | Not-determined | Genome-fragment | NA | NA | 0 |
| S7_k141_100658 | 1244 | No | NA | | 2 | 0 | 0 | Not-determined | Genome-fragment | NA | NA | 0 |
| S7_k141_121588 | 3827 | No | NA | | 5 | 0 | 1 | Not-determined | Genome-fragment | NA | NA | 0 |
| S7_k141_172374 | 1041 | No | NA | | 1 | 0 | 0 | Not-determined | Genome-fragment | NA | NA | 0 |
| S7_k141_162536 | 1184 | No | NA | | 1 | 0 | 1 | Not-determined | Genome-fragment | NA | NA | 0 |
| S7_k141_193409 | 1670 | No | NA | | 1 | 0 | 1 | Not-determined | Genome-fragment | NA | NA | 0 |
| S7_k141_131422 | 1055 | No | NA | | 1 | 0 | 1 | Not-determined | Genome-fragment | NA | NA | 0 |
| S7_k141_100700 | 1116 | No | NA | | 1 | 0 | 0 | Not-determined | Genome-fragment | NA | NA | 0 |
| S11-final-viral-combined_k141_57807\|\|full | 2058 | No | NA | | 4 | 1 | 0 | Low-quality | Genome-fragment | 4.06 | AAI-based (medium-confidence) | 0 |
| S7_k141_39949 | 1031 | No | NA | | 1 | 0 | 0 | Not-determined | Genome-fragment | NA | NA | 0 |
| S7_k141_110280 | 1071 | No | NA | | 2 | 0 | 0 | Not-determined | Genome-fragment | NA | NA | 0 |
| S7_k141_39964 | 1313 | No | NA | | 2 | 0 | 1 | Not-determined | Genome-fragment | NA | NA | 0 |
| S5-final-viral-combined_k141_22371\|\|full | 2058 | No | NA | | 2 | 0 | 2 | Low-quality | Genome-fragment | 1.49 | AAI-based (high-confidence) | 0 |
| S7_k141_39969 | 1003 | No | NA | | 1 | 0 | 1 | Not-determined | Genome-fragment | NA | NA | 0 |
| S7_k141_233954 | 1585 | No | NA | | 2 | 0 | 2 | Not-determined | Genome-fragment | NA | NA | 0 |
| S5-final-viral-combined_k141_202409\|\|full | 2058 | No | NA | | 6 | 2 | 0 | Low-quality | Genome-fragment | 5.52 | AAI-based (high-confidence) | 0 |
| S7_k141_151401 | 1313 | No | NA | | 1 | 0 | 1 | Not-determined | Genome-fragment | NA | NA | 0 |
| S7_k141_27440 | 1153 | No | NA | | 2 | 0 | 1 | Not-determined | Genome-fragment | NA | NA | 0 |
| S7_k141_243279 | 1795 | No | NA | | 1 | 0 | 0 | Not-determined | Genome-fragment | NA | NA | 0 |
| S17-final-viral-combined_k141_206059\|\|lt2gene | 2058 | No | NA | | 2 | 1 | 0 | Low-quality | Genome-fragment | 3.45 | HMM-based (lower-bound) | 0 |
| S7_k141_18686 | 3600 | No | NA | | 3 | 0 | 3 | Not-determined | Genome-fragment | NA | NA | 0 |
| S7_k141_121688 | 1221 | No | NA | | 2 | 0 | 2 | Not-determined | Genome-fragment | NA | NA | 0 |
| S17-k141_142391 | 2058 | No | NA | | 1 | 0 | 1 | Low-quality | Genome-fragment | 0.77 | AAI-based (high-confidence) | 0 |
| S7_k141_50289 | 1007 | No | NA | | 1 | 0 | 1 | Not-determined | Genome-fragment | NA | NA | 0 |
| S7_k141_70692 | 1495 | No | NA | | 2 | 0 | 0 | Not-determined | Genome-fragment | NA | NA | 0 |
| S7_k141_79626 | 1980 | No | NA | | 1 | 0 | 0 | Not-determined | Genome-fragment | NA | NA | 0 |
| S8-final-viral-combined_k141_115447\|\|full | 2058 | No | NA | | 3 | 0 | 0 | Low-quality | Genome-fragment | 4.75 | AAI-based (high-confidence) | 0 |
| S7_k141_213486 | 3628 | No | NA | | 2 | 0 | 2 | Not-determined | Genome-fragment | NA | NA | 0 |
| S11-final-viral-combined_k141_259341\|\|full | 2059 | No | NA | | 4 | 0 | 0 | Low-quality | Genome-fragment | 3.75 | AAI-based (medium-confidence) | 0 |
| S1-final-viral-combined_k141_53830\|\|full | 2059 | No | NA | | 2 | 0 | 1 | Low-quality | Genome-fragment | 4.33 | AAI-based (medium-confidence) | 0 |
| S7_k141_162566 | 1152 | No | NA | | 1 | 0 | 1 | Not-determined | Genome-fragment | NA | NA | 0 |
| S7-final-viral-combined_k141_217368\|\|full | 2059 | Yes | | 1483 | 3 | 1 | 1 | Low-quality | Genome-fragment | 4.17 | AAI-based (high-confidence) | 27.97 |
| S7_k141_193584 | 1428 | No | NA | | 3 | 0 | 3 | Not-determined | Genome-fragment | NA | NA | 0 |
| S7_k141_221695 | 1559 | No | NA | | 1 | 0 | 1 | Not-determined | Genome-fragment | NA | NA | 0 |
| S7_k141_50349 | 1017 | No | NA | | 1 | 0 | 0 | Not-determined | Genome-fragment | NA | NA | 0 |
| S7_k141_182986 | 3750 | No | NA | | 4 | 0 | 2 | Not-determined | Genome-fragment | NA | NA | 0 |
| S7_k141_151527 | 3791 | No | NA | | 5 | 0 | 1 | Not-determined | Genome-fragment | NA | NA | 0 |
| S7_k141_243404 | 3297 | No | NA | | 5 | 0 | 2 | Not-determined | Genome-fragment | NA | NA | 0 |
| S2-final-viral-combined_k141_64327\|\|full | 2059 | No | NA | | 4 | 0 | 0 | Low-quality | Genome-fragment | 4.39 | AAI-based (medium-confidence) | 0 |
| S7_k141_70823 | 1079 | No | NA | | 2 | 0 | 0 | Not-determined | Genome-fragment | NA | NA | 0 |
| S7_k141_90136 | 1897 | No | NA | | 2 | 0 | 2 | Not-determined | Genome-fragment | NA | NA | 0 |
| S7_k141_141059 | 1669 | No | NA | | 2 | 0 | 2 | Not-determined | Genome-fragment | NA | NA | 0 |
| S2-final-viral-combined_k141_237298\|\|full | 2059 | No | NA | | 3 | 1 | 0 | Low-quality | Genome-fragment | 1.16 | HMM-based (lower-bound) | 0 |
| S7_k141_70861 | 1443 | No | NA | | 2 | 0 | 1 | Not-determined | Genome-fragment | NA | NA | 0 |
| S6-final-viral-combined_k141_96502\|\|full | 2059 | No | NA | | 3 | 1 | 0 | Low-quality | Genome-fragment | 3.53 | HMM-based (lower-bound) | 0 |
| S7_k141_213645 | 1106 | No | NA | | 1 | 0 | 1 | Not-determined | Genome-fragment | NA | NA | 0 |
| S8_k141_165908 | 2059 | No | NA | | 3 | 1 | 0 | Low-quality | Genome-fragment | 1.21 | HMM-based (lower-bound) | 0 |
| S7_k141_243479 | 1659 | No | NA | | 3 | 0 | 1 | Not-determined | Genome-fragment | NA | NA | 0 |
| S7_k141_70906 | 6602 | No | NA | | 6 | 0 | 6 | Not-determined | Genome-fragment | NA | NA | 0 |
| S1-final-viral-combined_k141_90249\|\|full | 2060 | No | NA | | 3 | 0 | 0 | Low-quality | Genome-fragment | 1.21 | AAI-based (high-confidence) | 0 |
| S17-final-viral-combined_k141_95511\|\|full | 2060 | No | NA | | 4 | 1 | 0 | Low-quality | Genome-fragment | 1.33 | HMM-based (lower-bound) | 0 |
| S7_k141_60341 | 6672 | No | NA | | 6 | 0 | 1 | Not-determined | Genome-fragment | NA | NA | 0 |
| S7_k141_110608 | 1284 | No | NA | | 2 | 0 | 2 | Not-determined | Genome-fragment | NA | NA | 0 |
| S7_k141_60371 | 1173 | No | NA | | 1 | 0 | 0 | Not-determined | Genome-fragment | NA | NA | 0 |
| S7_k141_213732 | 3015 | No | NA | | 5 | 0 | 0 | Not-determined | Genome-fragment | NA | NA | 0 |
| S7_k141_121988 | 1443 | No | NA | | 2 | 0 | 1 | Not-determined | Genome-fragment | NA | NA | 0 |
| S17-final-viral-combined_k141_166246\|\|full | 2060 | No | NA | | 3 | 0 | 1 | Low-quality | Genome-fragment | 1.25 | AAI-based (medium-confidence) | 0 |
| S12-final-viral-combined_k141_250147\|\|full | 2060 | No | NA | | 3 | 0 | 1 | Low-quality | Genome-fragment | 3.13 | AAI-based (high-confidence) | 0 |
| S7_k141_71029 | 9814 | No | NA | | 11 | 0 | 2 | Not-determined | Genome-fragment | NA | NA | 0 |
| S7_k141_18923 | 1992 | No | NA | | 2 | 0 | 1 | Not-determined | Genome-fragment | NA | NA | 0 |
| S7_k141_8864 | 5651 | No | NA | | 6 | 0 | 6 | Not-determined | Genome-fragment | NA | NA | 0 |
| S7_k141_8867 | 1641 | No | NA | | 1 | 0 | 1 | Not-determined | Genome-fragment | NA | NA | 0 |
| S7_k141_122054 | 3740 | No | NA | | 1 | 0 | 1 | Not-determined | Genome-fragment | NA | NA | 0 |
| S11-final-viral-combined_k141_73450\|\|full | 2061 | No | NA | | 5 | 2 | 0 | Low-quality | Genome-fragment | 5.53 | AAI-based (high-confidence) | 0 |
| S7_k141_50718 | 3529 | No | NA | | 1 | 0 | 1 | Not-determined | Genome-fragment | NA | NA | 0 |
| S7_k141_79790 | 5185 | No | NA | | 4 | 0 | 2 | Not-determined | Genome-fragment | NA | NA | 0 |
| S7_k141_222005 | 1408 | No | NA | | 1 | 0 | 0 | Not-determined | Genome-fragment | NA | NA | 0 |
| S7_k141_151635 | 3258 | No | NA | | 9 | 0 | 0 | Not-determined | Genome-fragment | NA | NA | 0 |
| S1-final-viral-combined_k141_202905\|\|full | 2061 | No | NA | | 2 | 0 | 0 | Low-quality | Genome-fragment | 1.13 | AAI-based (medium-confidence) | 0 |
| S7_k141_243676 | 1270 | No | NA | | 1 | 0 | 1 | Not-determined | Genome-fragment | NA | NA | 0 |
| S7_k141_101110 | 3389 | No | NA | | 5 | 0 | 2 | Not-determined | Genome-fragment | NA | NA | 0 |
| S7_k141_101111 | 1220 | No | NA | | 2 | 0 | 1 | Not-determined | Genome-fragment | NA | NA | 0 |
| S7_k141_213884 | 1075 | No | NA | | 1 | 0 | 0 | Not-determined | Genome-fragment | NA | NA | 0 |
| S7_k141_243707 | 1101 | No | NA | | 1 | 0 | 1 | Not-determined | Genome-fragment | NA | NA | 0 |
| S7_k141_50778 | 1707 | No | NA | | 1 | 0 | 1 | Not-determined | Genome-fragment | NA | NA | 0 |
| S7_k141_222064 | 1573 | No | NA | | 1 | 0 | 0 | Not-determined | Genome-fragment | NA | NA | 0 |
| S7_k141_8985 | 12681 | No | NA | | 12 | 0 | 4 | Not-determined | Genome-fragment | NA | NA | 0 |
| S7_k141_162996 | 1873 | No | NA | | 1 | 0 | 1 | Not-determined | Genome-fragment | NA | NA | 0 |
| S7_k141_163001 | 3205 | No | NA | | 2 | 0 | 0 | Not-determined | Genome-fragment | NA | NA | 0 |
| S7_k141_141356 | 1119 | No | NA | | 1 | 0 | 1 | Not-determined | Genome-fragment | NA | NA | 0 |
| S7_k141_101158 | 1165 | No | NA | | 1 | 0 | 0 | Not-determined | Genome-fragment | NA | NA | 0 |
| S7_k141_40371 | 6035 | No | NA | | 6 | 0 | 4 | Not-determined | Genome-fragment | NA | NA | 0 |
| S2_k141_146002 | 2061 | No | NA | | 3 | 0 | 3 | Low-quality | Genome-fragment | 0.76 | AAI-based (medium-confidence) | 0 |
| S6-final-viral-combined_k141_67437\|\|full | 2061 | No | NA | | 4 | 0 | 0 | Low-quality | Genome-fragment | 2.94 | AAI-based (medium-confidence) | 0 |
| S7_k141_90526 | 4673 | No | NA | | 7 | 0 | 0 | Not-determined | Genome-fragment | NA | NA | 0 |
| S17-final-viral-combined_k141_157224\|\|full | 2062 | No | NA | | 1 | 0 | 1 | Low-quality | Genome-fragment | 3.16 | AAI-based (medium-confidence) | 0 |
| S7_k141_79987 | 1794 | No | NA | | 1 | 0 | 1 | Not-determined | Genome-fragment | NA | NA | 0 |
| S7_k141_151851 | 1172 | No | NA | | 2 | 0 | 0 | Not-determined | Genome-fragment | NA | NA | 0 |
| S7_k141_79998 | 2813 | No | NA | | 1 | 0 | 0 | Not-determined | Genome-fragment | NA | NA | 0 |
| S7_k141_214031 | 1861 | No | NA | | 1 | 0 | 0 | Not-determined | Genome-fragment | NA | NA | 0 |
| S7_k141_151942 | 1550 | No | NA | | 1 | 0 | 1 | Not-determined | Genome-fragment | NA | NA | 0 |
| S7_k141_214050 | 1385 | No | NA | | 1 | 0 | 0 | Not-determined | Genome-fragment | NA | NA | 0 |
| S7_k141_9275 | 1477 | No | NA | | 1 | 0 | 0 | Not-determined | Genome-fragment | NA | NA | 0 |
| S7_k141_163221 | 1505 | No | NA | | 2 | 0 | 0 | Not-determined | Genome-fragment | NA | NA | 0 |
| S7_k141_80054 | 1009 | No | NA | | 2 | 0 | 1 | Not-determined | Genome-fragment | NA | NA | 0 |
| S7_k141_152007 | 5883 | No | NA | | 5 | 0 | 3 | Not-determined | Genome-fragment | NA | NA | 0 |
| S7_k141_27703 | 1169 | No | NA | | 1 | 0 | 0 | Not-determined | Genome-fragment | NA | NA | 0 |
| S17-final-viral-combined_k141_264146\|\|full | 2062 | No | NA | | 5 | 4 | 0 | Low-quality | Genome-fragment | 4.8 | AAI-based (high-confidence) | 0 |
| S7_k141_183662 | 1330 | No | NA | | 2 | 0 | 1 | Not-determined | Genome-fragment | NA | NA | 0 |
| S7_k141_60832 | 4096 | No | NA | | 4 | 0 | 4 | Not-determined | Genome-fragment | NA | NA | 0 |
| S4-final-viral-combined_k141_29693\|\|full | 2062 | No | NA | | 3 | 0 | 0 | Low-quality | Genome-fragment | 4.14 | AAI-based (high-confidence) | 0 |
| S9-final-viral-combined_k141_66218\|\|lt2gene | 2063 | No | NA | | 2 | 2 | 0 | Low-quality | Genome-fragment | 2.72 | HMM-based (lower-bound) | 0 |
| S7_k141_40681 | 4700 | No | NA | | 6 | 0 | 6 | Not-determined | Genome-fragment | NA | NA | 0 |
| S7_k141_173169 | 1328 | No | NA | | 2 | 0 | 2 | Not-determined | Genome-fragment | NA | NA | 0 |
| S9_k141_11326 | 2063 | No | NA | | 3 | 1 | 0 | Low-quality | Genome-fragment | 5.2 | AAI-based (high-confidence) | 0 |
| S7_k141_131912 | 7609 | No | NA | | 4 | 0 | 3 | Not-determined | Genome-fragment | NA | NA | 0 |
| S7_k141_27796 | 1646 | No | NA | | 2 | 0 | 1 | Not-determined | Genome-fragment | NA | NA | 0 |
| S7_k141_80184 | 1155 | No | NA | | 1 | 0 | 0 | Not-determined | Genome-fragment | NA | NA | 0 |
| S7_k141_152198 | 2185 | No | NA | | 2 | 0 | 0 | Not-determined | Genome-fragment | NA | NA | 0 |
| S7_k141_141542 | 1354 | No | NA | | 1 | 0 | 1 | Not-determined | Genome-fragment | NA | NA | 0 |
| S7_k141_27820 | 14147 | No | NA | | 12 | 0 | 2 | Not-determined | Genome-fragment | NA | NA | 0 |
| S7_k141_173254 | 1296 | No | NA | | 1 | 0 | 1 | Not-determined | Genome-fragment | NA | NA | 0 |
| S1-final-viral-combined_k141_104476\|\|full | 2063 | No | NA | | 4 | 1 | 0 | Low-quality | Genome-fragment | 3.67 | HMM-based (lower-bound) | 0 |
| S7_k141_9477 | 1494 | No | NA | | 2 | 0 | 1 | Not-determined | Genome-fragment | NA | NA | 0 |
| S7_k141_214115 | 1221 | No | NA | | 2 | 0 | 1 | Not-determined | Genome-fragment | NA | NA | 0 |
| S7_k141_101429 | 1370 | No | NA | | 2 | 0 | 2 | Not-determined | Genome-fragment | NA | NA | 0 |
| S7_k141_80255 | 21021 | No | NA | | 32 | 15 | 0 | Medium-quality | Genome-fragment | 51.45 | AAI-based (high-confidence) | 0 |
| S7_k141_101462 | 1080 | No | NA | | 1 | 0 | 1 | Not-determined | Genome-fragment | NA | NA | 0 |
| S9-final-viral-combined_k141_95563\|\|full | 2064 | No | NA | | 3 | 0 | 2 | Low-quality | Genome-fragment | 2.33 | AAI-based (high-confidence) | 0 |
| S7_k141_152307 | 1209 | No | NA | | 2 | 0 | 2 | Not-determined | Genome-fragment | NA | NA | 0 |
| S7_k141_173295 | 1413 | No | NA | | 2 | 0 | 1 | Not-determined | Genome-fragment | NA | NA | 0 |
| S7_k141_9561 | 1702 | No | NA | | 3 | 0 | 0 | Not-determined | Genome-fragment | NA | NA | 0 |
| S7_k141_19571 | 1500 | No | NA | | 3 | 0 | 0 | Not-determined | Genome-fragment | NA | NA | 0 |
| S11-final-viral-combined_k141_182763\|\|full | 2064 | No | NA | | 6 | 1 | 0 | Low-quality | Genome-fragment | 5.39 | AAI-based (high-confidence) | 0 |
| S7_k141_61082 | 1811 | No | NA | | 2 | 0 | 2 | Not-determined | Genome-fragment | NA | NA | 0 |
| S7_k141_61086 | 1337 | No | NA | | 2 | 0 | 1 | Not-determined | Genome-fragment | NA | NA | 0 |
| S7_k141_234852 | 1450 | No | NA | | 2 | 0 | 2 | Not-determined | Genome-fragment | NA | NA | 0 |
| S7_k141_122462 | 2355 | No | NA | | 3 | 0 | 2 | Not-determined | Genome-fragment | NA | NA | 0 |
| S7_k141_40790 | 2051 | No | NA | | 1 | 0 | 1 | Not-determined | Genome-fragment | NA | NA | 0 |
| S7_k141_214163 | 1761 | No | NA | | 3 | 0 | 2 | Not-determined | Genome-fragment | NA | NA | 0 |
| S7_k141_61112 | 1247 | No | NA | | 2 | 0 | 2 | Not-determined | Genome-fragment | NA | NA | 0 |
| S3-final-viral-combined_k141_109440\|\|lt2gene | 2064 | No | NA | | 2 | 0 | 0 | Low-quality | Genome-fragment | 1.34 | AAI-based (high-confidence) | 0 |
| S7_k141_19609 | 1109 | No | NA | | 1 | 0 | 0 | Not-determined | Genome-fragment | NA | NA | 0 |
| S7_k141_173393 | 3645 | No | NA | | 6 | 0 | 1 | Not-determined | Genome-fragment | NA | NA | 0 |
| S7_k141_27883 | 1232 | No | NA | | 1 | 0 | 1 | Not-determined | Genome-fragment | NA | NA | 0 |
| S7_k141_173423 | 1366 | No | NA | | 1 | 0 | 1 | Not-determined | Genome-fragment | NA | NA | 0 |
| S7_k141_214259 | 4473 | No | NA | | 4 | 0 | 1 | Not-determined | Genome-fragment | NA | NA | 0 |
| S7_k141_9661 | 1409 | No | NA | | 2 | 0 | 0 | Not-determined | Genome-fragment | NA | NA | 0 |
| S7_k141_101594 | 2672 | No | NA | | 2 | 0 | 2 | Not-determined | Genome-fragment | NA | NA | 0 |
| S7_k141_9675 | 1062 | No | NA | | 2 | 0 | 1 | Not-determined | Genome-fragment | NA | NA | 0 |
| S7_k141_141716 | 3492 | No | NA | | 1 | 0 | 0 | Not-determined | Genome-fragment | NA | NA | 0 |
| S7_k141_214270 | 1097 | No | NA | | 1 | 0 | 1 | Not-determined | Genome-fragment | NA | NA | 0 |
| S7_k141_90976 | 4028 | No | NA | | 4 | 0 | 2 | Not-determined | Genome-fragment | NA | NA | 0 |
| S7_k141_9724 | 1099 | No | NA | | 1 | 0 | 1 | Not-determined | Genome-fragment | NA | NA | 0 |
| S7_k141_90988 | 1192 | No | NA | | 1 | 0 | 1 | Not-determined | Genome-fragment | NA | NA | 0 |
| S7-final-viral-combined_k141_140846\|\|full | 2064 | No | NA | | 3 | 0 | 2 | Low-quality | Genome-fragment | 0.6 | AAI-based (high-confidence) | 0 |
| S7_k141_222527 | 1093 | No | NA | | 1 | 0 | 1 | Not-determined | Genome-fragment | NA | NA | 0 |
| S7_k141_101622 | 5999 | No | NA | | 6 | 0 | 6 | Not-determined | Genome-fragment | NA | NA | 0 |
| S7_k141_222537 | 1522 | No | NA | | 3 | 0 | 1 | Not-determined | Genome-fragment | NA | NA | 0 |
| S7_k141_122501 | 1752 | No | NA | | 1 | 0 | 1 | Not-determined | Genome-fragment | NA | NA | 0 |
| S7_k141_91022 | 1460 | No | NA | | 3 | 0 | 3 | Not-determined | Genome-fragment | NA | NA | 0 |
| S17-final-viral-combined_k141_97863\|\|full | 2064 | No | NA | | 4 | 1 | 0 | Low-quality | Genome-fragment | 1.25 | HMM-based (lower-bound) | 0 |
| S7_k141_132071 | 1037 | No | NA | | 1 | 0 | 1 | Not-determined | Genome-fragment | NA | NA | 0 |
| S7_k141_28010 | 1160 | No | NA | | 1 | 0 | 1 | Not-determined | Genome-fragment | NA | NA | 0 |
| S7_k141_91066 | 1824 | No | NA | | 1 | 0 | 0 | Not-determined | Genome-fragment | NA | NA | 0 |
| S4-final-viral-combined_k141_1456\|\|full | 2064 | No | NA | | 3 | 0 | 1 | Low-quality | Genome-fragment | 5.43 | AAI-based (high-confidence) | 0 |
| S7_k141_132131 | 1412 | No | NA | | 3 | 0 | 0 | Not-determined | Genome-fragment | NA | NA | 0 |
| S7-final-viral-combined_k141_102538\|\|full | 2065 | Yes | | 231 | 3 | 1 | 1 | Low-quality | Genome-fragment | 0.07 | HMM-based (lower-bound) | 88.81 |
| S7_k141_111468 | 2087 | No | NA | | 2 | 0 | 2 | Not-determined | Genome-fragment | NA | NA | 0 |
| S7_k141_28037 | 6072 | No | NA | | 4 | 0 | 2 | Not-determined | Genome-fragment | NA | NA | 0 |
| S7-final-viral-combined_k141_202676\|\|full | 2065 | No | NA | | 8 | 0 | 0 | Low-quality | Genome-fragment | 5.29 | AAI-based (high-confidence) | 0 |
| S7_k141_28069 | 2357 | No | NA | | 3 | 0 | 0 | Not-determined | Genome-fragment | NA | NA | 0 |
| S7_k141_80472 | 1169 | No | NA | | 1 | 0 | 0 | Not-determined | Genome-fragment | NA | NA | 0 |
| S7_k141_80518 | 1801 | No | NA | | 2 | 0 | 2 | Not-determined | Genome-fragment | NA | NA | 0 |
| S8-final-viral-combined_k141_74997\|\|full | 2065 | Yes | | 1120 | 4 | 2 | 1 | Low-quality | Genome-fragment | 2.41 | HMM-based (lower-bound) | 45.76 |
| S7_k141_222729 | 1505 | No | NA | | 2 | 0 | 1 | Not-determined | Genome-fragment | NA | NA | 0 |
| S7_k141_101954 | 2275 | No | NA | | 3 | 0 | 2 | Not-determined | Genome-fragment | NA | NA | 0 |
| S7_k141_19939 | 2621 | No | NA | | 3 | 0 | 1 | Not-determined | Genome-fragment | NA | NA | 0 |
| S1-final-viral-combined_k141_258255\|\|full | 2066 | No | NA | | 3 | 0 | 0 | Low-quality | Genome-fragment | 1.06 | AAI-based (medium-confidence) | 0 |
| S7_k141_111679 | 1012 | No | NA | | 1 | 0 | 1 | Not-determined | Genome-fragment | NA | NA | 0 |
| S7_k141_243835 | 1168 | No | NA | | 1 | 0 | 1 | Not-determined | Genome-fragment | NA | NA | 0 |
| S7_k141_222768 | 1149 | No | NA | | 1 | 0 | 1 | Not-determined | Genome-fragment | NA | NA | 0 |
| S7_k141_142082 | 1462 | No | NA | | 1 | 0 | 1 | Not-determined | Genome-fragment | NA | NA | 0 |
| S7_k141_222816 | 1375 | No | NA | | 3 | 0 | 2 | Not-determined | Genome-fragment | NA | NA | 0 |
| S7_k141_142098 | 1077 | No | NA | | 1 | 0 | 1 | Not-determined | Genome-fragment | NA | NA | 0 |
| S7-final-viral-combined_k141_26930\|\|full | 2066 | No | NA | | 2 | 0 | 1 | Low-quality | Genome-fragment | 3.85 | AAI-based (high-confidence) | 0 |
| S7_k141_91374 | 2165 | No | NA | | 2 | 0 | 2 | Not-determined | Genome-fragment | NA | NA | 0 |
| S7_k141_91389 | 1571 | No | NA | | 2 | 0 | 2 | Not-determined | Genome-fragment | NA | NA | 0 |
| S7_k141_152760 | 1451 | No | NA | | 3 | 0 | 2 | Not-determined | Genome-fragment | NA | NA | 0 |
| S7_k141_132370 | 3188 | No | NA | | 3 | 0 | 3 | Not-determined | Genome-fragment | NA | NA | 0 |
| S7_k141_142177 | 1548 | No | NA | | 3 | 0 | 2 | Not-determined | Genome-fragment | NA | NA | 0 |
| S7_k141_152798 | 3137 | No | NA | | 4 | 0 | 1 | Not-determined | Genome-fragment | NA | NA | 0 |
| S7_k141_111899 | 1624 | No | NA | | 2 | 0 | 0 | Not-determined | Genome-fragment | NA | NA | 0 |
| S2-final-viral-combined_k141_242371\|\|full | 2066 | No | NA | | 4 | 2 | 0 | Low-quality | Genome-fragment | 4.83 | AAI-based (high-confidence) | 0 |

| | | | | | | | | | | | |
|---|---|---|---|---|---|---|---|---|---|---|---|
| S7_k141_132533 | 1665 | No | NA | 1 | 0 | 1 | Not-determined | Genome-fragment | NA | NA | 0 |
| S8-final-viral-combined_k141_192242\|\|full | 2066 | No | NA | 3 | 0 | 2 | Low-quality | Genome-fragment | 1.49 | AAI-based (medium-confidence) | 0 |
| S11-final-viral-combined_k141_17221\|\|full | 2067 | No | NA | 3 | 0 | 2 | Low-quality | Genome-fragment | 1.47 | AAI-based (high-confidence) | 0 |
| S1_k141_39094 | 2067 | No | NA | 1 | 0 | 0 | Low-quality | Genome-fragment | 1.62 | AAI-based (medium-confidence) | 0 |
| S17-final-viral-combined_k141_108192\|\|full | 2067 | No | NA | 4 | 1 | 0 | Low-quality | Genome-fragment | 1.22 | HMM-based (lower-bound) | 0 |
| S7_k141_10174 | 2319 | No | NA | 1 | 0 | 0 | Not-determined | Genome-fragment | NA | NA | 0 |
| S7_k141_223109 | 1029 | No | NA | 2 | 0 | 0 | Not-determined | Genome-fragment | NA | NA | 0 |
| S7_k141_132630 | 3749 | No | NA | 3 | 0 | 3 | Not-determined | Genome-fragment | NA | NA | 0 |
| S7_k141_81049 | 1001 | No | NA | 2 | 0 | 2 | Not-determined | Genome-fragment | NA | NA | 0 |
| S7_k141_20233 | 3230 | No | NA | 3 | 0 | 1 | Not-determined | Genome-fragment | NA | NA | 0 |
| S7_k141_20254 | 4325 | No | NA | 2 | 0 | 2 | Not-determined | Genome-fragment | NA | NA | 0 |
| S7_k141_132690 | 1853 | No | NA | 3 | 0 | 1 | Not-determined | Genome-fragment | NA | NA | 0 |
| S17-k141_22004 | 2067 | No | NA | 1 | 0 | 0 | Low-quality | Genome-fragment | 0.18 | AAI-based (medium-confidence) | 0 |
| S11-final-viral-combined_k141_77459\|\|full | 2068 | No | NA | 4 | 1 | 0 | Low-quality | Genome-fragment | 1.34 | HMM-based (lower-bound) | 0 |
| S7_k141_28853 | 1161 | No | NA | 1 | 0 | 1 | Not-determined | Genome-fragment | NA | NA | 0 |
| S7_k141_223263 | 1112 | No | NA | 2 | 0 | 2 | Not-determined | Genome-fragment | NA | NA | 0 |
| S7_k141_91661 | 1100 | No | NA | 1 | 0 | 1 | Not-determined | Genome-fragment | NA | NA | 0 |
| S7_k141_244242 | 1011 | No | NA | 1 | 0 | 1 | Not-determined | Genome-fragment | NA | NA | 0 |
| S7_k141_81221 | 1756 | No | NA | 2 | 0 | 2 | Not-determined | Genome-fragment | NA | NA | 0 |
| S7_k141_29088 | 2328 | No | NA | 2 | 0 | 0 | Not-determined | Genome-fragment | NA | NA | 0 |
| S7_k141_244337 | 1406 | No | NA | 3 | 0 | 3 | Not-determined | Genome-fragment | NA | NA | 0 |
| S7_k141_142746 | 1023 | No | NA | 1 | 0 | 0 | Not-determined | Genome-fragment | NA | NA | 0 |
| S1_k141_142129 | 2068 | No | NA | 1 | 0 | 0 | Low-quality | Genome-fragment | 0.91 | AAI-based (medium-confidence) | 0 |
| S7_k141_223403 | 5150 | No | NA | 10 | 0 | 9 | Not-determined | Genome-fragment | NA | NA | 0 |
| S17-final-viral-combined_k141_53572\|\|full | 2068 | No | NA | 1 | 0 | 0 | Low-quality | Genome-fragment | 2.65 | AAI-based (medium-confidence) | 0 |
| S7_k141_244373 | 8321 | No | NA | 8 | 0 | 6 | Not-determined | Genome-fragment | NA | NA | 0 |
| S7_k141_223421 | 1357 | No | NA | 2 | 0 | 2 | Not-determined | Genome-fragment | NA | NA | 0 |
| S7_k141_223432 | 2049 | No | NA | 1 | 0 | 0 | Not-determined | Genome-fragment | NA | NA | 0 |
| S7_k141_244402 | 1252 | No | NA | 1 | 0 | 1 | Not-determined | Genome-fragment | NA | NA | 0 |
| S7_k141_81467 | 4167 | No | NA | 4 | 0 | 2 | Not-determined | Genome-fragment | NA | NA | 0 |
| S7_k141_142826 | 3930 | No | NA | 4 | 0 | 1 | Not-determined | Genome-fragment | NA | NA | 0 |
| S2-final-viral-combined_k141_42662\|\|full | 2068 | No | NA | 7 | 0 | 0 | Low-quality | Genome-fragment | 2.79 | AAI-based (high-confidence) | 0 |
| S7_k141_29370 | 1207 | No | NA | 1 | 0 | 0 | Not-determined | Genome-fragment | NA | NA | 0 |
| S7_k141_29373 | 1470 | No | NA | 2 | 0 | 1 | Not-determined | Genome-fragment | NA | NA | 0 |
| S7_k141_244425 | 72371 | No | NA | 57 | 7 | 0 | Medium-quality | Genome-fragment | 77.69 | AAI-based (high-confidence) | 0 |
| S7_k141_29389 | 2183 | No | NA | 1 | 0 | 0 | Not-determined | Genome-fragment | NA | NA | 0 |
| S7_k141_244455 | 1037 | No | NA | 2 | 0 | 0 | Not-determined | Genome-fragment | NA | NA | 0 |
| S2_k141_14015 | 2068 | No | NA | 3 | 0 | 2 | Low-quality | Genome-fragment | 1.26 | AAI-based (medium-confidence) | 0 |
| S7_k141_142885 | 1249 | No | NA | 1 | 0 | 0 | Not-determined | Genome-fragment | NA | NA | 0 |
| S7_k141_142895 | 3104 | No | NA | 2 | 0 | 1 | Not-determined | Genome-fragment | NA | NA | 0 |
| S7_k141_142905 | 1254 | No | NA | 2 | 0 | 0 | Not-determined | Genome-fragment | NA | NA | 0 |
| S7_k141_244487 | 4342 | No | NA | 2 | 0 | 1 | Not-determined | Genome-fragment | NA | NA | 0 |
| S7_k141_81559 | 4221 | No | NA | 4 | 0 | 1 | Not-determined | Genome-fragment | NA | NA | 0 |
| S8-final-viral-combined_k141_43065\|\|full | 2068 | No | NA | 3 | 0 | 1 | Low-quality | Genome-fragment | 4.73 | AAI-based (medium-confidence) | 0 |
| S7_k141_223842 | 1391 | No | NA | 2 | 0 | 0 | Not-determined | Genome-fragment | NA | NA | 0 |
| S7_k141_223856 | 1334 | No | NA | 2 | 0 | 1 | Not-determined | Genome-fragment | NA | NA | 0 |
| S8-final-viral-combined_k141_80303\|\|full | 2068 | No | NA | 4 | 3 | 0 | Low-quality | Genome-fragment | 3.41 | AAI-based (high-confidence) | 0 |
| S1-final-viral-combined_k141_100830\|\|full | 2069 | No | NA | 2 | 1 | 0 | Low-quality | Genome-fragment | 5.31 | AAI-based (high-confidence) | 0 |
| S17-final-viral-combined_k141_4582\|\|full | 2069 | No | NA | 5 | 0 | 0 | Low-quality | Genome-fragment | 3.87 | AAI-based (high-confidence) | 0 |
| S7_k141_29716 | 5328 | No | NA | 8 | 0 | 0 | Not-determined | Genome-fragment | NA | NA | 0 |
| S7_k141_244713 | 1577 | No | NA | 3 | 0 | 0 | Not-determined | Genome-fragment | NA | NA | 0 |
| S17-final-viral-combined_k141_84255\|\|full | 2069 | No | NA | 2 | 0 | 1 | Low-quality | Genome-fragment | 0.94 | AAI-based (medium-confidence) | 0 |
| S4-final-viral-combined_k141_11206\|\|full | 2069 | No | NA | 3 | 0 | 2 | Low-quality | Genome-fragment | 2.81 | AAI-based (medium-confidence) | 0 |
| S7_k141_29987 | 4421 | No | NA | 4 | 0 | 3 | Not-determined | Genome-fragment | NA | NA | 0 |
| S7_k141_244773 | 1182 | No | NA | 2 | 0 | 2 | Not-determined | Genome-fragment | NA | NA | 0 |
| S11-final-viral-combined_k141_255543\|\|full | 2070 | No | NA | 3 | 1 | 0 | Low-quality | Genome-fragment | 5.58 | AAI-based (high-confidence) | 0 |
| S7_k141_244797 | 3087 | No | NA | 1 | 0 | 1 | Not-determined | Genome-fragment | NA | NA | 0 |
| S7_k141_30091 | 1322 | No | NA | 1 | 0 | 1 | Not-determined | Genome-fragment | NA | NA | 0 |
| S1-final-viral-combined_k141_329293\|\|full | 2070 | No | NA | 4 | 0 | 2 | Low-quality | Genome-fragment | 2.6 | AAI-based (high-confidence) | 0 |
| S7_k141_30105 | 1040 | No | NA | 1 | 0 | 0 | Not-determined | Genome-fragment | NA | NA | 0 |
| S7_k141_244821 | 3122 | No | NA | 9 | 0 | 0 | Not-determined | Genome-fragment | NA | NA | 0 |
| S7_k141_244866 | 34147 | No | NA | 50 | 12 | 0 | Complete | High-quality | 100 | DTR (high-confidence) | 0 |
| S7-final-viral-combined_k141_209274\|\|full | 2070 | No | NA | 3 | 1 | 0 | Low-quality | Genome-fragment | 1.96 | AAI-based (high-confidence) | 0 |
| S7_k141_30362 | 1153 | No | NA | 2 | 0 | 1 | Not-determined | Genome-fragment | NA | NA | 0 |
| S7_k141_244882 | 3151 | No | NA | 3 | 0 | 0 | Not-determined | Genome-fragment | NA | NA | 0 |
| S7_k141_30446 | 4231 | No | NA | 3 | 0 | 3 | Not-determined | Genome-fragment | NA | NA | 0 |
| S7_k141_244943 | 1428 | No | NA | 1 | 0 | 1 | Not-determined | Genome-fragment | NA | NA | 0 |
| S7_k141_244974 | 3932 | No | NA | 7 | 0 | 0 | Not-determined | Genome-fragment | NA | NA | 0 |
| S12_k141_225066 | 2070 | No | NA | 4 | 0 | 0 | Low-quality | Genome-fragment | 3.35 | AAI-based (medium-confidence) | 0 |
| S2-final-viral-combined_k141_155924\|\|full | 2070 | No | NA | 3 | 2 | 0 | Low-quality | Genome-fragment | 4.57 | AAI-based (high-confidence) | 0 |
| S17-final-viral-combined_k141_167045\|\|full | 6422 | No | NA | 12 | 0 | 2 | Not-determined | Genome-fragment | NA | NA | 0 |
| S17-final-viral-combined_k141_157468\|\|full | 22384 | No | NA | 30 | 3 | 1 | High-quality | High-quality | 100 | AAI-based (high-confidence) | 0 |
| S2-final-viral-combined_k141_162260\|\|full | 2070 | No | NA | 3 | 2 | 0 | Low-quality | Genome-fragment | 4.31 | AAI-based (high-confidence) | 0 |
| S17-final-viral-combined_k141_138473\|\|full | 2732 | No | NA | 6 | 0 | 2 | Not-determined | Genome-fragment | NA | NA | 0 |
| S4-final-viral-combined_k141_16731\|\|full | 2070 | No | NA | 3 | 0 | 2 | Low-quality | Genome-fragment | 2.68 | AAI-based (high-confidence) | 0 |
| S17-final-viral-combined_k141_266513\|\|full | 7267 | No | NA | 8 | 0 | 1 | Not-determined | Genome-fragment | NA | NA | 0 |
| S17-final-viral-combined_k141_266517\|\|full | 2335 | No | NA | 6 | 0 | 0 | Not-determined | Genome-fragment | NA | NA | 0 |
| S17-final-viral-combined_k141_266526\|\|full | 3177 | No | NA | 5 | 0 | 0 | Not-determined | Genome-fragment | NA | NA | 0 |
| S8-final-viral-combined_k141_247870\|\|full | 2070 | No | NA | 5 | 1 | 0 | Low-quality | Genome-fragment | 4.45 | AAI-based (medium-confidence) | 0 |
| S8-final-viral-combined_k141_160009\|\|full | 2070 | No | NA | 3 | 0 | 2 | Low-quality | Genome-fragment | 0.45 | AAI-based (medium-confidence) | 0 |
| S17-final-viral-combined_k141_44464\|\|full | 2278 | No | NA | 2 | 0 | 0 | Not-determined | Genome-fragment | NA | NA | 0 |
| S11-final-viral-combined_k141_147812\|\|full | 2071 | No | NA | 5 | 0 | 0 | Low-quality | Genome-fragment | 5.62 | AAI-based (medium-confidence) | 0 |
| S17-final-viral-combined_k141_244517\|\|full | 3692 | No | NA | 7 | 0 | 7 | Not-determined | Genome-fragment | NA | NA | 0 |
| S5_k141_45792 | 2071 | No | NA | 1 | 0 | 1 | Low-quality | Genome-fragment | 5.84 | AAI-based (medium-confidence) | 0 |
| S17-final-viral-combined_k141_133379\|\|full | 1810 | No | NA | 2 | 0 | 2 | Not-determined | Genome-fragment | NA | NA | 0 |
| S17-final-viral-combined_k141_255625\|\|full | 1978 | No | NA | 3 | 0 | 0 | Not-determined | Genome-fragment | NA | NA | 0 |
| S17-final-viral-combined_k141_200077\|\|full | 2215 | No | NA | 3 | 0 | 2 | Not-determined | Genome-fragment | NA | NA | 0 |
| S17-final-viral-combined_k141_88932\|\|full | 2360 | No | NA | 3 | 0 | 0 | Not-determined | Genome-fragment | NA | NA | 0 |
| S17-final-viral-combined_k141_70088\|\|full | 2071 | No | NA | 4 | 1 | 0 | Low-quality | Genome-fragment | 4.12 | AAI-based (medium-confidence) | 0 |
| S17-final-viral-combined_k141_200089\|\|full | 1296 | No | NA | 3 | 0 | 0 | Not-determined | Genome-fragment | NA | NA | 0 |
| S2-final-viral-combined_k141_1406\|\|full | 2071 | Yes | 1650 | 2 | 1 | 1 | Low-quality | Genome-fragment | 2.75 | HMM-based (lower-bound) | 20.33 |
| S2-final-viral-combined_k141_97892\|\|full | 2071 | No | NA | 4 | 1 | 0 | Low-quality | Genome-fragment | 0.54 | HMM-based (lower-bound) | 0 |
| S17-final-viral-combined_k141_53\|\|full | 3961 | No | NA | 4 | 0 | 3 | Not-determined | Genome-fragment | NA | NA | 0 |
| S17-final-viral-combined_k141_100057\|\|full | 2633 | No | NA | 3 | 0 | 2 | Not-determined | Genome-fragment | NA | NA | 0 |
| S17-final-viral-combined_k141_211195\|\|full | 12650 | No | NA | 12 | 0 | 7 | Not-determined | Genome-fragment | NA | NA | 0 |
| S17-final-viral-combined_k141_100059\|\|full | 2534 | No | NA | 3 | 0 | 1 | Not-determined | Genome-fragment | NA | NA | 0 |
| S11_k141_319223 | 2072 | No | NA | 3 | 3 | 0 | Low-quality | Genome-fragment | 5.68 | AAI-based (high-confidence) | 0 |
| S17-final-viral-combined_k141_200144\|\|full | 10117 | No | NA | 5 | 0 | 2 | Not-determined | Genome-fragment | NA | NA | 0 |
| S1-final-viral-combined_k141_366422\|\|full | 2072 | No | NA | 3 | 0 | 1 | Low-quality | Genome-fragment | 3.34 | AAI-based (medium-confidence) | 0 |
| S7-final-viral-combined_k141_118665\|\|full | 2072 | No | NA | 6 | 2 | 0 | Low-quality | Genome-fragment | 4.79 | AAI-based (high-confidence) | 0 |
| S17-final-viral-combined_k141_33424\|\|full | 6192 | No | NA | 7 | 0 | 0 | Not-determined | Genome-fragment | NA | NA | 0 |
| S17-final-viral-combined_k141_188033\|\|full | 2072 | No | NA | 3 | 0 | 0 | Low-quality | Genome-fragment | 3.97 | AAI-based (medium-confidence) | 0 |
| S17-final-viral-combined_k141_122291\|\|full | 2219 | No | NA | 2 | 0 | 1 | Not-determined | Genome-fragment | NA | NA | 0 |
| S6-final-viral-combined_k141_70921\|\|full | 2072 | No | NA | 2 | 0 | 1 | Low-quality | Genome-fragment | 1.2 | AAI-based (medium-confidence) | 0 |
| S17-final-viral-combined_k141_66729\|\|full | 2420 | No | NA | 2 | 0 | 0 | Not-determined | Genome-fragment | NA | NA | 0 |
| S17-final-viral-combined_k141_88970\|\|full | 1615 | No | NA | 3 | 0 | 1 | Not-determined | Genome-fragment | NA | NA | 0 |
| S17-final-viral-combined_k141_66751\|\|full | 1537 | No | NA | 3 | 0 | 2 | Not-determined | Genome-fragment | NA | NA | 0 |
| S17-final-viral-combined_k141_166775\|\|full | 1393 | No | NA | 2 | 0 | 0 | Not-determined | Genome-fragment | NA | NA | 0 |
| S1-final-viral-combined_k141_335965\|\|full | 2073 | No | NA | 4 | 3 | 0 | Low-quality | Genome-fragment | 4.77 | AAI-based (high-confidence) | 0 |
| S17-final-viral-combined_k141_89875\|\|full | 2073 | No | NA | 2 | 1 | 0 | Low-quality | Genome-fragment | 5.7 | HMM-based (lower-bound) | 0 |
| S17-final-viral-combined_k141_189051\|\|full | 1203 | No | NA | 3 | 0 | 1 | Not-determined | Genome-fragment | NA | NA | 0 |
| S11-final-viral-combined_k141_321430\|\|full | 2074 | No | NA | 2 | 0 | 0 | Low-quality | Genome-fragment | 3.21 | AAI-based (high-confidence) | 0 |
| S17-final-viral-combined_k141_233470\|\|full | 1131 | No | NA | 3 | 0 | 0 | Not-determined | Genome-fragment | NA | NA | 0 |
| S17-final-viral-combined_k141_111245\|\|full | 1475 | No | NA | 3 | 0 | 1 | Not-determined | Genome-fragment | NA | NA | 0 |
| S17-final-viral-combined_k141_222392\|\|full | 1319 | No | NA | 2 | 0 | 0 | Not-determined | Genome-fragment | NA | NA | 0 |
| S3_k141_67785 | 2074 | No | NA | 2 | 0 | 2 | Low-quality | Genome-fragment | 3.77 | AAI-based (medium-confidence) | 0 |
| S7-final-viral-combined_k141_110449\|\|full | 2074 | No | NA | 5 | 0 | 0 | Low-quality | Genome-fragment | 2.03 | AAI-based (medium-confidence) | 0 |
| S17-final-viral-combined_k141_112318\|\|full | 2074 | No | NA | 4 | 1 | 0 | Low-quality | Genome-fragment | 4.81 | AAI-based (medium-confidence) | 0 |
| S12-final-viral-combined_k141_113039\|\|full | 2074 | No | NA | 4 | 2 | 0 | Low-quality | Genome-fragment | 0.96 | AAI-based (high-confidence) | 0 |
| S11-final-viral-combined_k141_188255\|\|full | 2075 | No | NA | 3 | 0 | 2 | Low-quality | Genome-fragment | 1.95 | AAI-based (medium-confidence) | 0 |
| S1-final-viral-combined_k141_144157\|\|full | 2075 | No | NA | 3 | 1 | 0 | Low-quality | Genome-fragment | 1.86 | HMM-based (lower-bound) | 0 |
| S12-final-viral-combined_k141_169242\|\|full | 2075 | No | NA | 5 | 4 | 0 | Low-quality | Genome-fragment | 3.29 | AAI-based (high-confidence) | 0 |
| S12-final-viral-combined_k141_283179\|\|full | 2075 | No | NA | 2 | 0 | 2 | Low-quality | Genome-fragment | 2.39 | AAI-based (medium-confidence) | 0 |
| S17-final-viral-combined_k141_122402\|\|full | 13282 | No | NA | 5 | 0 | 0 | Not-determined | Genome-fragment | NA | NA | 0 |
| S17-final-viral-combined_k141_109\|\|full | 1536 | No | NA | 3 | 0 | 0 | Not-determined | Genome-fragment | NA | NA | 0 |
| S12_k141_191561 | 2075 | No | NA | 2 | 1 | 0 | Low-quality | Genome-fragment | 0.96 | AAI-based (high-confidence) | 0 |
| S17-final-viral-combined_k141_255781\|\|full | 5460 | No | NA | 5 | 0 | 3 | Not-determined | Genome-fragment | NA | NA | 0 |
| S8-final-viral-combined_k141_103247\|\|full | 2075 | No | NA | 2 | 0 | 0 | Low-quality | Genome-fragment | 2.39 | AAI-based (medium-confidence) | 0 |
| S17-final-viral-combined_k141_55680\|\|full | 2054 | No | NA | 2 | 0 | 1 | Not-determined | Genome-fragment | NA | NA | 0 |
| S17-final-viral-combined_k141_222459\|\|full | 2070 | No | NA | 2 | 0 | 1 | Not-determined | Genome-fragment | NA | NA | 0 |
| S17-final-viral-combined_k141_189132\|\|full | 2596 | No | NA | 3 | 0 | 0 | Not-determined | Genome-fragment | NA | NA | 0 |
| S17-final-viral-combined_k141_255794\|\|full | 2133 | No | NA | 3 | 0 | 0 | Not-determined | Genome-fragment | NA | NA | 0 |
| S17-final-viral-combined_k141_258272\|\|lt2gene | 2076 | No | NA | 1 | 1 | 0 | Low-quality | Genome-fragment | 3.27 | AAI-based (high-confidence) | 0 |
| S17-final-viral-combined_k141_189137\|\|full | 3061 | No | NA | 2 | 0 | 1 | Not-determined | Genome-fragment | NA | NA | 0 |
| S17-final-viral-combined_k141_189138\|\|full | 2784 | No | NA | 3 | 0 | 3 | Not-determined | Genome-fragment | NA | NA | 0 |
| S17-final-viral-combined_k141_55728\|\|full | 1114 | No | NA | 1 | 0 | 0 | Not-determined | Genome-fragment | NA | NA | 0 |
| S4_k141_148264 | 2076 | No | NA | 6 | 1 | 0 | Low-quality | Genome-fragment | 6.1 | AAI-based (high-confidence) | 0 |
| S17-final-viral-combined_k141_200335\|\|full | 2018 | No | NA | 3 | 0 | 1 | Not-determined | Genome-fragment | NA | NA | 0 |
| S9-final-viral-combined_k141_69416\|\|full | 2077 | No | NA | 2 | 0 | 2 | Low-quality | Genome-fragment | 2.04 | AAI-based (medium-confidence) | 0 |
| S17-k141_212155 | 2077 | No | NA | 6 | 2 | 0 | Low-quality | Genome-fragment | 5.2 | AAI-based (medium-confidence) | 0 |
| S17-final-viral-combined_k141_211369\|\|full | 1098 | No | NA | 3 | 0 | 1 | Not-determined | Genome-fragment | NA | NA | 0 |
| S17-final-viral-combined_k141_222488\|\|full | 4407 | No | NA | 6 | 0 | 2 | Not-determined | Genome-fragment | NA | NA | 0 |
| S17-final-viral-combined_k141_122443\|\|full | 15181 | No | NA | 12 | 0 | 8 | Not-determined | Genome-fragment | NA | NA | 0 |
| S17-final-viral-combined_k141_222503\|\|full | 2220 | No | NA | 2 | 0 | 1 | Not-determined | Genome-fragment | NA | NA | 0 |
| S12-final-viral-combined_k141_83621\|\|full | 2077 | No | NA | 4 | 0 | 0 | Low-quality | Genome-fragment | 5.39 | AAI-based (high-confidence) | 0 |
| S8-final-viral-combined_k141_22554\|\|full | 2077 | No | NA | 3 | 3 | 0 | Low-quality | Genome-fragment | 5.38 | AAI-based (high-confidence) | 0 |
| S17-final-viral-combined_k141_144770\|\|full | 2687 | No | NA | 6 | 0 | 0 | Not-determined | Genome-fragment | NA | NA | 0 |
| S17-final-viral-combined_k141_155760\|\|full | 2692 | No | NA | 2 | 0 | 0 | Not-determined | Genome-fragment | NA | NA | 0 |
| S17-final-viral-combined_k141_111398\|\|full | 3832 | No | NA | 3 | 0 | 1 | Not-determined | Genome-fragment | NA | NA | 0 |
| S11-final-viral-combined_k141_127292\|\|full | 2078 | No | NA | 3 | 1 | 0 | Low-quality | Genome-fragment | 36.48 | AAI-based (high-confidence) | 0 |
| S17-final-viral-combined_k141_11283\|\|full | 3197 | No | NA | 5 | 0 | 1 | Not-determined | Genome-fragment | NA | NA | 0 |
| S17-final-viral-combined_k141_89084\|\|full | 6710 | No | NA | 6 | 0 | 4 | Not-determined | Genome-fragment | NA | NA | 0 |
| S17-final-viral-combined_k141_178000\|\|full | 2091 | No | NA | 4 | 0 | 2 | Not-determined | Genome-fragment | NA | NA | 0 |
| S11_k141_326853 | 2078 | No | NA | 4 | 1 | 0 | Low-quality | Genome-fragment | 1.63 | AAI-based (high-confidence) | 0 |
| S4-final-viral-combined_k141_62687\|\|lt2gene | 2078 | Yes | 957 | 2 | 1 | 1 | Low-quality | Genome-fragment | 18.62 | AAI-based (high-confidence) | 53.95 |
| S17-final-viral-combined_k141_211423\|\|full | 1121 | No | NA | 2 | 0 | 2 | Not-determined | Genome-fragment | NA | NA | 0 |
| S17-final-viral-combined_k141_255887\|\|full | 4442 | No | NA | 6 | 0 | 5 | Not-determined | Genome-fragment | NA | NA | 0 |
| S5-final-viral-combined_k141_139568\|\|full | 2079 | No | NA | 2 | 0 | 2 | Low-quality | Genome-fragment | 2.47 | AAI-based (medium-confidence) | 0 |
| S17-final-viral-combined_k141_78158\|\|full | 2339 | No | NA | 3 | 0 | 2 | Not-determined | Genome-fragment | NA | NA | 0 |
| S5-final-viral-combined_k141_157969\|\|full | 2079 | No | NA | 3 | 0 | 0 | Low-quality | Genome-fragment | 1.17 | AAI-based (medium-confidence) | 0 |
| S17-final-viral-combined_k141_111403\|\|full | 1289 | No | NA | 3 | 0 | 0 | Not-determined | Genome-fragment | NA | NA | 0 |
| S17-final-viral-combined_k141_111405\|\|full | 2000 | No | NA | 4 | 0 | 3 | Not-determined | Genome-fragment | NA | NA | 0 |
| S17-final-viral-combined_k141_155799\|\|full | 1228 | No | NA | 3 | 0 | 1 | Not-determined | Genome-fragment | NA | NA | 0 |
| S17-final-viral-combined_k141_200386\|\|full | 3041 | No | NA | 2 | 0 | 1 | Not-determined | Genome-fragment | NA | NA | 0 |
| S17-final-viral-combined_k141_218406\|\|full | 2079 | No | NA | 5 | 0 | 0 | Low-quality | Genome-fragment | 4.3 | AAI-based (high-confidence) | 0 |
| S17-k141_223960 | 2079 | No | NA | 4 | 1 | 0 | Low-quality | Genome-fragment | 3.77 | AAI-based (high-confidence) | 0 |
| S17-final-viral-combined_k141_66969\|\|full | 4667 | No | NA | 2 | 0 | 2 | Not-determined | Genome-fragment | NA | NA | 0 |
| S2-final-viral-combined_k141_57705\|\|full | 2079 | No | NA | 4 | 0 | 0 | Low-quality | Genome-fragment | 4.19 | AAI-based (medium-confidence) | 0 |
| S17-final-viral-combined_k141_89129\|\|full | 3470 | No | NA | 3 | 0 | 0 | Not-determined | Genome-fragment | NA | NA | 0 |
| S17-final-viral-combined_k141_233648\|\|full | 1226 | No | NA | 3 | 0 | 0 | Not-determined | Genome-fragment | NA | NA | 0 |
| S11-final-viral-combined_k141_92279\|\|full | 2080 | No | NA | 7 | 0 | 0 | Low-quality | Genome-fragment | 3.09 | AAI-based (high-confidence) | 0 |
| S1-final-viral-combined_k141_150289\|\|full | 2080 | No | NA | 3 | 1 | 0 | Low-quality | Genome-fragment | 1.33 | HMM-based (lower-bound) | 0 |
| S17-final-viral-combined_k141_178014\|\|full | 5132 | No | NA | 3 | 0 | 0 | Not-determined | Genome-fragment | NA | NA | 0 |
| S5-final-viral-combined_k141_4934\|\|full | 2080 | No | NA | 4 | 1 | 0 | Low-quality | Genome-fragment | 2.47 | AAI-based (high-confidence) | 0 |
| S5-final-viral-combined_k141_41177\|\|full | 2080 | No | NA | 5 | 3 | 0 | Low-quality | Genome-fragment | 3.43 | AAI-based (medium-confidence) | 0 |
| S17-final-viral-combined_k141_67037\|\|full | 2014 | No | NA | 3 | 0 | 0 | Not-determined | Genome-fragment | NA | NA | 0 |
| S5_k141_191358 | 2080 | No | NA | 4 | 2 | 0 | Low-quality | Genome-fragment | 5.05 | AAI-based (high-confidence) | 0 |
| S17-final-viral-combined_k141_233705\|\|full | 1195 | No | NA | 2 | 0 | 0 | Not-determined | Genome-fragment | NA | NA | 0 |
| S17-final-viral-combined_k141_67056\|\|full | 3954 | No | NA | 4 | 0 | 2 | Not-determined | Genome-fragment | NA | NA | 0 |
| S17-final-viral-combined_k141_244820\|\|full | 2405 | No | NA | 4 | 0 | 2 | Not-determined | Genome-fragment | NA | NA | 0 |
| S17-final-viral-combined_k141_111461\|\|full | 2932 | No | NA | 3 | 0 | 2 | Not-determined | Genome-fragment | NA | NA | 0 |
| S17-final-viral-combined_k141_303\|\|full | 2182 | No | NA | 3 | 0 | 1 | Not-determined | Genome-fragment | NA | NA | 0 |
| S17-final-viral-combined_k141_244832\|\|full | 1580 | No | NA | 3 | 0 | 2 | Not-determined | Genome-fragment | NA | NA | 0 |
| S17-final-viral-combined_k141_22517\|\|full | 1796 | No | NA | 3 | 0 | 0 | Not-determined | Genome-fragment | NA | NA | 0 |
| S17-final-viral-combined_k141_233742\|\|full | 6322 | No | NA | 4 | 0 | 1 | Not-determined | Genome-fragment | NA | NA | 0 |
| S17-final-viral-combined_k141_222661\|\|full | 2731 | No | NA | 4 | 0 | 3 | Not-determined | Genome-fragment | NA | NA | 0 |
| S12_k141_381437 | 2080 | No | NA | 1 | 1 | 0 | Low-quality | Genome-fragment | 1.12 | AAI-based (high-confidence) | 0 |
| S4-final-viral-combined_k141_55137\|\|full | 2080 | No | NA | 3 | 0 | 2 | Low-quality | Genome-fragment | 1.43 | AAI-based (medium-confidence) | 0 |
| S6-final-viral-combined_k141_70983\|\|full | 2080 | No | NA | 3 | 0 | 2 | Low-quality | Genome-fragment | 1.08 | AAI-based (medium-confidence) | 0 |
| S17-final-viral-combined_k141_89193\|\|full | 1993 | No | NA | 2 | 0 | 0 | Not-determined | Genome-fragment | NA | NA | 0 |
| S17-final-viral-combined_k141_144935\|\|full | 1372 | No | NA | 2 | 0 | 1 | Not-determined | Genome-fragment | NA | NA | 0 |
| S17-final-viral-combined_k141_350\|\|full | 2529 | No | NA | 3 | 0 | 2 | Not-determined | Genome-fragment | NA | NA | 0 |
| S8-final-viral-combined_k141_241635\|\|full | 2080 | No | NA | 4 | 0 | 0 | Low-quality | Genome-fragment | 3.19 | AAI-based (medium-confidence) | 0 |
| S11-final-viral-combined_k141_253447\|\|full | 2081 | No | NA | 5 | 2 | 0 | Low-quality | Genome-fragment | 2.43 | AAI-based (medium-confidence) | 0 |
| S17-final-viral-combined_k141_155873\|\|full | 3382 | No | NA | 5 | 0 | 0 | Not-determined | Genome-fragment | NA | NA | 0 |
| S17-final-viral-combined_k141_78314\|\|full | 2650 | No | NA | 4 | 0 | 0 | Not-determined | Genome-fragment | NA | NA | 0 |
| S11-final-viral-combined_k141_17111\|\|lt2gene | 2081 | No | NA | 2 | 2 | 0 | Low-quality | Genome-fragment | 4.13 | AAI-based (high-confidence) | 0 |
| S17-final-viral-combined_k141_211568\|\|full | 1569 | No | NA | 2 | 0 | 1 | Not-determined | Genome-fragment | NA | NA | 0 |
| S17-final-viral-combined_k141_11387\|\|full | 1117 | No | NA | 3 | 0 | 1 | Not-determined | Genome-fragment | NA | NA | 0 |
| S17-final-viral-combined_k141_222686\|\|full | 4448 | No | NA | 4 | 0 | 2 | Not-determined | Genome-fragment | NA | NA | 0 |
| S1-final-viral-combined_k141_138794\|\|full | 2081 | No | NA | 4 | 0 | 0 | Low-quality | Genome-fragment | 1.99 | AAI-based (high-confidence) | 0 |
| S17-final-viral-combined_k141_233761\|\|full | 44873 | Yes | 36206 | 53 | 3 | 5 | Medium-quality | Genome-fragment | 83.44 | AAI-based (medium-confidence) | 19.31 |
| S17-final-viral-combined_k141_111554\|\|full | 1057 | No | NA | 2 | 0 | 1 | Not-determined | Genome-fragment | NA | NA | 0 |
| S17-final-viral-combined_k141_122637\|\|full | 5798 | No | NA | 9 | 0 | 3 | Not-determined | Genome-fragment | NA | NA | 0 |
| S3-final-viral-combined_k141_87036\|\|full | 2081 | No | NA | 3 | 0 | 2 | Low-quality | Genome-fragment | 1.71 | AAI-based (medium-confidence) | 0 |
| S17-final-viral-combined_k141_111561\|\|full | 1558 | No | NA | 2 | 0 | 1 | Not-determined | Genome-fragment | NA | NA | 0 |
| S17-final-viral-combined_k141_111574\|\|full | 2791 | No | NA | 2 | 0 | 2 | Not-determined | Genome-fragment | NA | NA | 0 |
| S17-final-viral-combined_k141_111583\|\|full | 6768 | No | NA | 5 | 0 | 3 | Not-determined | Genome-fragment | NA | NA | 0 |
| S17-final-viral-combined_k141_89247\|\|full | 3254 | No | NA | 3 | 0 | 1 | Not-determined | Genome-fragment | NA | NA | 0 |
| S7-final-viral-combined_k141_217372\|\|full | 2081 | No | NA | 7 | 0 | 0 | Low-quality | Genome-fragment | 5.57 | AAI-based (medium-confidence) | 0 |
| S7-final-viral-combined_k141_140739\|\|full | 2081 | No | NA | 3 | 1 | 0 | Low-quality | Genome-fragment | 5.3 | AAI-based (high-confidence) | 0 |
| S17-final-viral-combined_k141_157241\|\|full | 2081 | No | NA | 3 | 0 | 0 | Low-quality | Genome-fragment | 3.76 | AAI-based (high-confidence) | 0 |
| S12_k141_87546 | 2081 | No | NA | 2 | 0 | 1 | Low-quality | Genome-fragment | 5.87 | AAI-based (medium-confidence) | 0 |
| S12_k141_273645 | 2081 | No | NA | 1 | 0 | 0 | Low-quality | Genome-fragment | 1.12 | AAI-based (high-confidence) | 0 |
| S6-final-viral-combined_k141_98757\|\|full | 2081 | No | NA | 3 | 0 | 0 | Low-quality | Genome-fragment | 0.52 | AAI-based (medium-confidence) | 0 |
| S17-final-viral-combined_k141_22612\|\|full | 1078 | No | NA | 3 | 0 | 2 | Not-determined | Genome-fragment | NA | NA | 0 |
| S8-final-viral-combined_k141_272972\|\|full | 2081 | No | NA | 3 | 0 | 1 | Low-quality | Genome-fragment | 2.22 | AAI-based (medium-confidence) | 0 |
| S17-final-viral-combined_k141_222788\|\|full | 1905 | No | NA | 3 | 0 | 3 | Not-determined | Genome-fragment | NA | NA | 0 |
| S17-final-viral-combined_k141_133731\|\|full | 7975 | No | NA | 9 | 0 | 0 | Not-determined | Genome-fragment | NA | NA | 0 |
| S17-final-viral-combined_k141_67211\|\|full | 3470 | No | NA | 4 | 0 | 0 | Not-determined | Genome-fragment | NA | NA | 0 |
| S1-final-viral-combined_k141_359141\|\|full | 2082 | No | NA | 4 | 1 | 0 | Low-quality | Genome-fragment | 5.08 | AAI-based (medium-confidence) | 0 |
| S1-final-viral-combined_k141_81584\|\|full | 2082 | No | NA | 4 | 0 | 0 | Low-quality | Genome-fragment | 5.71 | AAI-based (high-confidence) | 0 |
| S17-final-viral-combined_k141_155958\|\|full | 1126 | No | NA | 2 | 0 | 2 | Not-determined | Genome-fragment | NA | NA | 0 |
| S17-final-viral-combined_k141_80\|\|full | 2082 | No | NA | 3 | 0 | 2 | Low-quality | Genome-fragment | 0.59 | AAI-based (high-confidence) | 0 |
| S17-final-viral-combined_k141_200536\|\|full | 1938 | No | NA | 5 | 0 | 0 | Not-determined | Genome-fragment | NA | NA | 0 |
| S17-final-viral-combined_k141_182496\|\|full | 2082 | No | NA | 3 | 0 | 1 | Low-quality | Genome-fragment | 1.46 | AAI-based (high-confidence) | 0 |
| S17-final-viral-combined_k141_188207\|\|full | 2082 | No | NA | 4 | 4 | 0 | Low-quality | Genome-fragment | 4.35 | AAI-based (medium-confidence) | 0 |
| S2-final-viral-combined_k141_94479\|\|full | 2082 | No | NA | 3 | 0 | 1 | Low-quality | Genome-fragment | 1.47 | AAI-based (high-confidence) | 0 |
| S17-final-viral-combined_k141_155965\|\|full | 3064 | No | NA | 3 | 0 | 1 | Not-determined | Genome-fragment | NA | NA | 0 |
| S17-final-viral-combined_k141_56006\|\|full | 96019 | Yes | 79780 | 125 | 29 | 13 | High-quality | High-quality | 100 | AAI-based (high-confidence) | 16.91 |
| S2-final-viral-combined_k141_107976\|\|full | 2082 | Yes | 326 | 3 | 1 | 1 | Low-quality | Genome-fragment | 0.38 | HMM-based (lower-bound) | 84.34 |
| S8_k141_183187 | 2082 | No | NA | 2 | 0 | 2 | Low-quality | Genome-fragment | 0.79 | AAI-based (high-confidence) | 0 |
| S17-final-viral-combined_k141_145085\|\|full | 12732 | No | NA | 12 | 0 | 9 | Not-determined | Genome-fragment | NA | NA | 0 |
| S11-final-viral-combined_k141_252363\|\|full | 2083 | Yes | 1470 | 3 | 1 | 1 | Low-quality | Genome-fragment | 0.74 | HMM-based (lower-bound) | 29.43 |
| S17-final-viral-combined_k141_167032\|\|full | 4423 | No | NA | 5 | 0 | 3 | Not-determined | Genome-fragment | NA | NA | 0 |
| S17-final-viral-combined_k141_486\|\|full | 6055 | No | NA | 6 | 0 | 2 | Not-determined | Genome-fragment | NA | NA | 0 |
| S17-final-viral-combined_k141_78380\|\|full | 4171 | No | NA | 8 | 0 | 0 | Not-determined | Genome-fragment | NA | NA | 0 |
| S17-final-viral-combined_k141_67259\|\|full | 2297 | No | NA | 2 | 0 | 1 | Not-determined | Genome-fragment | NA | NA | 0 |
| S17-final-viral-combined_k141_233799\|\|full | 4634 | No | NA | 3 | 0 | 1 | Not-determined | Genome-fragment | NA | NA | 0 |
| S17-final-viral-combined_k141_167055\|\|full | 1873 | No | NA | 3 | 0 | 0 | Not-determined | Genome-fragment | NA | NA | 0 |
| S11-final-viral-combined_k141_172901\|\|lt2gene | 2083 | No | NA | 1 | 1 | 0 | Low-quality | Genome-fragment | 4.8 | AAI-based (high-confidence) | 0 |
| S1-final-viral-combined_k141_111255\|\|full | 2083 | No | NA | 4 | 0 | 1 | Low-quality | Genome-fragment | 5.75 | AAI-based (high-confidence) | 0 |
| S3_k141_15323 | 2083 | No | NA | 3 | 0 | 1 | Low-quality | Genome-fragment | 4.26 | AAI-based (high-confidence) | 0 |
| S12-final-viral-combined_k141_53886\|\|full | 2083 | No | NA | 3 | 0 | 2 | Low-quality | Genome-fragment | 2.36 | AAI-based (medium-confidence) | 0 |
| S17-final-viral-combined_k141_111697\|\|full | 1148 | No | NA | 3 | 0 | 2 | Not-determined | Genome-fragment | NA | NA | 0 |
| S12-final-viral-combined_k141_15178\|\|full | 2083 | No | NA | 5 | 0 | 0 | Low-quality | Genome-fragment | 0.96 | AAI-based (high-confidence) | 0 |
| S17-final-viral-combined_k141_167090\|\|full | 1173 | No | NA | 2 | 0 | 1 | Not-determined | Genome-fragment | NA | NA | 0 |
| S17-final-viral-combined_k141_33843\|\|full | 1610 | No | NA | 2 | 0 | 1 | Not-determined | Genome-fragment | NA | NA | 0 |
| S2-final-viral-combined_k141_85159\|\|full | 2083 | No | NA | 3 | 0 | 2 | Low-quality | Genome-fragment | 0.56 | AAI-based (medium-confidence) | 0 |
| S8-final-viral-combined_k141_26793\|\|full | 2083 | No | NA | 2 | 1 | 0 | Low-quality | Genome-fragment | 3.24 | AAI-based (medium-confidence) | 0 |
| S8-final-viral-combined_k141_64777\|\|full | 2083 | No | NA | 5 | 4 | 0 | Low-quality | Genome-fragment | 3.6 | HMM-based (lower-bound) | 0 |
| S17-final-viral-combined_k141_200620\|\|full | 1950 | No | NA | 2 | 0 | 1 | Not-determined | Genome-fragment | NA | NA | 0 |
| S17-final-viral-combined_k141_200622\|\|full | 2362 | No | NA | 2 | 0 | 2 | Not-determined | Genome-fragment | NA | NA | 0 |
| S17-final-viral-combined_k141_78476\|\|full | 1699 | No | NA | 2 | 0 | 0 | Not-determined | Genome-fragment | NA | NA | 0 |
| S17-final-viral-combined_k141_222863\|\|full | 1168 | No | NA | 2 | 0 | 0 | Not-determined | Genome-fragment | NA | NA | 0 |
| S17-final-viral-combined_k141_200634\|\|full | 1263 | No | NA | 2 | 0 | 1 | Not-determined | Genome-fragment | NA | NA | 0 |
| S17-final-viral-combined_k141_256074\|\|full | 2161 | No | NA | 2 | 0 | 2 | Not-determined | Genome-fragment | NA | NA | 0 |
| S17-final-viral-combined_k141_178113\|\|full | 31663 | No | NA | 26 | 0 | 14 | Not-determined | Genome-fragment | NA | NA | 0 |
| S17-final-viral-combined_k141_33868\|\|full | 1774 | No | NA | 3 | 0 | 1 | Not-determined | Genome-fragment | NA | NA | 0 |
| S17-final-viral-combined_k141_211718\|\|full | 1793 | No | NA | 2 | 0 | 0 | Not-determined | Genome-fragment | NA | NA | 0 |
| S11-final-viral-combined_k141_173550\|\|lt2gene | 2084 | No | NA | 2 | 2 | 0 | Low-quality | Genome-fragment | 5.43 | AAI-based (high-confidence) | 0 |
| S5-final-viral-combined_k141_31747\|\|full | 2084 | No | NA | 5 | 0 | 0 | Low-quality | Genome-fragment | 4.49 | AAI-based (medium-confidence) | 0 |
| S12-final-viral-combined_k141_367390\|\|full | 2084 | No | NA | 2 | 0 | 2 | Low-quality | Genome-fragment | 1.03 | AAI-based (high-confidence) | 0 |
| S12_k141_103041 | 2084 | No | NA | 7 | 1 | 0 | Low-quality | Genome-fragment | 2.21 | AAI-based (high-confidence) | 0 |
| S4-final-viral-combined_k141_83789\|\|full | 2084 | No | NA | 4 | 0 | 3 | Low-quality | Genome-fragment | 0.75 | AAI-based (medium-confidence) | 0 |
| S4-final-viral-combined_k141_113872\|\|full | 2084 | No | NA | 3 | 0 | 0 | Low-quality | Genome-fragment | 4.42 | AAI-based (high-confidence) | 0 |
| S8-final-viral-combined_k141_171351\|\|full | 2084 | No | NA | 3 | 0 | 2 | Low-quality | Genome-fragment | 1.05 | AAI-based (medium-confidence) | 0 |
| S1-final-viral-combined_k141_57326\|\|full | 2085 | No | NA | 7 | 0 | 0 | Low-quality | Genome-fragment | 3.65 | AAI-based (high-confidence) | 0 |
| S5-final-viral-combined_k141_102541\|\|full | 2085 | No | NA | 3 | 0 | 2 | Low-quality | Genome-fragment | 1.35 | AAI-based (medium-confidence) | 0 |
| S17-final-viral-combined_k141_78507\|\|full | 2529 | No | NA | 4 | 0 | 2 | Not-determined | Genome-fragment | NA | NA | 0 |
| S17-final-viral-combined_k141_256133\|\|full | 1947 | No | NA | 3 | 0 | 0 | Not-determined | Genome-fragment | NA | NA | 0 |
| S17-final-viral-combined_k141_89341\|\|full | 4113 | No | NA | 3 | 0 | 3 | Not-determined | Genome-fragment | NA | NA | 0 |
| S4-final-viral-combined_k141_33102\|\|full | 2085 | No | NA | 4 | 0 | 3 | Low-quality | Genome-fragment | 1.46 | AAI-based (medium-confidence) | 0 |
| S17-final-viral-combined_k141_256143\|\|full | 1400 | No | NA | 2 | 0 | 2 | Not-determined | Genome-fragment | NA | NA | 0 |
| S6-final-viral-combined_k141_72796\|\|full | 2085 | No | NA | 3 | 0 | 1 | Low-quality | Genome-fragment | 1.14 | AAI-based (medium-confidence) | 0 |
| S17-final-viral-combined_k141_101128\|\|full | 2086 | No | NA | 3 | 0 | 2 | Low-quality | Genome-fragment | 1.53 | AAI-based (medium-confidence) | 0 |
| S17-final-viral-combined_k141_178193\|\|full | 4335 | No | NA | 6 | 0 | 5 | Not-determined | Genome-fragment | NA | NA | 0 |
| S17-final-viral-combined_k141_89365\|\|full | 5258 | No | NA | 8 | 0 | 0 | Not-determined | Genome-fragment | NA | NA | 0 |
| S17-final-viral-combined_k141_67414\|\|full | 3316 | No | NA | 3 | 0 | 1 | Not-determined | Genome-fragment | NA | NA | 0 |
| S12-final-viral-combined_k141_323654\|\|full | 2086 | No | NA | 4 | 1 | 0 | Low-quality | Genome-fragment | 5.16 | AAI-based (high-confidence) | 0 |
| S17-final-viral-combined_k141_11542\|\|full | 3267 | No | NA | 3 | 0 | 1 | Not-determined | Genome-fragment | NA | NA | 0 |
| S2-final-viral-combined_k141_188342\|\|full | 2086 | No | NA | 3 | 1 | 0 | Low-quality | Genome-fragment | 5.52 | AAI-based (medium-confidence) | 0 |
| S2-final-viral-combined_k141_220022\|\|full | 2086 | No | NA | 2 | 0 | 0 | Low-quality | Genome-fragment | 4.44 | AAI-based (medium-confidence) | 0 |
| S11-final-viral-combined_k141_256240\|\|full | 2087 | No | NA | 5 | 0 | 0 | Low-quality | Genome-fragment | 2.44 | AAI-based (high-confidence) | 0 |
| S1-final-viral-combined_k141_259646\|\|full | 2087 | No | NA | 3 | 0 | 2 | Low-quality | Genome-fragment | 0.53 | AAI-based (medium-confidence) | 0 |
| S17-final-viral-combined_k141_200730\|\|full | 1479 | No | NA | 3 | 0 | 1 | Not-determined | Genome-fragment | NA | NA | 0 |
| S3_k141_61578 | 2087 | No | NA | 3 | 0 | 0 | Low-quality | Genome-fragment | 0.96 | AAI-based (high-confidence) | 0 |
| S7-final-viral-combined_k141_37685\|\|full | 2087 | No | NA | 1 | 0 | 1 | Low-quality | Genome-fragment | 0.74 | AAI-based (medium-confidence) | 0 |
| S17-final-viral-combined_k141_67443\|\|full | 3250 | No | NA | 6 | 0 | 4 | Not-determined | Genome-fragment | NA | NA | 0 |
| S17-final-viral-combined_k141_122923\|\|full | 1047 | No | NA | 2 | 0 | 1 | Not-determined | Genome-fragment | NA | NA | 0 |
| S17-final-viral-combined_k141_145156\|\|full | 30402 | No | NA | 23 | 0 | 14 | Not-determined | Genome-fragment | NA | NA | 0 |
| S17-final-viral-combined_k141_56061\|\|full | 115134 | No | NA | 96 | 3 | 61 | High-quality | High-quality | 100 | HMM-based (lower-bound) | 0 |
| S17-final-viral-combined_k141_33978\|\|full | 1119 | No | NA | 2 | 0 | 0 | Not-determined | Genome-fragment | NA | NA | 0 |
| S17-final-viral-combined_k141_145166\|\|full | 3534 | No | NA | 6 | 0 | 2 | Not-determined | Genome-fragment | NA | NA | 0 |
| S7_k141_190754 | 2087 | No | NA | 3 | 1 | 0 | Low-quality | Genome-fragment | 4.34 | AAI-based (high-confidence) | 0 |
| S17-final-viral-combined_k141_211838\|\|full | 1901 | No | NA | 3 | 0 | 2 | Not-determined | Genome-fragment | NA | NA | 0 |
| S17-final-viral-combined_k141_200796\|\|full | 1088 | No | NA | 2 | 0 | 1 | Not-determined | Genome-fragment | NA | NA | 0 |
| S12_k141_139745 | 2087 | No | NA | 6 | 1 | 0 | Low-quality | Genome-fragment | 3.31 | AAI-based (high-confidence) | 0 |
| S17-final-viral-combined_k141_256281\|\|full | 1221 | No | NA | 2 | 0 | 0 | Not-determined | Genome-fragment | NA | NA | 0 |
| S17-final-viral-combined_k141_211885\|\|full | 2248 | No | NA | 3 | 0 | 1 | Not-determined | Genome-fragment | NA | NA | 0 |
| S17-final-viral-combined_k141_100509\|\|full | 1376 | No | NA | 2 | 0 | 0 | Not-determined | Genome-fragment | NA | NA | 0 |
| S17-final-viral-combined_k141_178250\|\|full | 2214 | No | NA | 3 | 0 | 1 | Not-determined | Genome-fragment | NA | NA | 0 |
| S11-final-viral-combined_k141_315404\|\|full | 2088 | No | NA | 5 | 0 | 1 | Low-quality | Genome-fragment | 4.78 | AAI-based (high-confidence) | 0 |
| S17-final-viral-combined_k141_660\|\|full | 3637 | No | NA | 4 | 0 | 4 | Not-determined | Genome-fragment | NA | NA | 0 |
| S1-final-viral-combined_k141_209600\|\|full | 2088 | No | NA | 2 | 0 | 1 | Low-quality | Genome-fragment | 0.98 | AAI-based (high-confidence) | 0 |
| S17-final-viral-combined_k141_200836\|\|full | 4307 | No | NA | 5 | 0 | 4 | Not-determined | Genome-fragment | NA | NA | 0 |
| S17-final-viral-combined_k141_156175\|\|full | 1454 | No | NA | 3 | 0 | 0 | Not-determined | Genome-fragment | NA | NA | 0 |
| S5-final-viral-combined_k141_1853\|\|full | 2088 | No | NA | 2 | 0 | 2 | Low-quality | Genome-fragment | 4.21 | AAI-based (medium-confidence) | 0 |
| S7-final-viral-combined_k141_81326\|\|full | 2088 | No | NA | 4 | 0 | 0 | Low-quality | Genome-fragment | 3.38 | AAI-based (high-confidence) | 0 |
| S2-final-viral-combined_k141_1282\|\|full | 2088 | No | NA | 2 | 0 | 2 | Low-quality | Genome-fragment | 3.27 | AAI-based (medium-confidence) | 0 |
| S8-final-viral-combined_k141_203500\|\|full | 2088 | No | NA | 1 | 0 | 0 | Low-quality | Genome-fragment | 1.96 | AAI-based (medium-confidence) | 0 |
| S11_k141_107744 | 2089 | No | NA | 4 | 0 | 0 | Low-quality | Genome-fragment | 3.12 | AAI-based (high-confidence) | 0 |
| S17-final-viral-combined_k141_56142\|\|full | 7487 | No | NA | 8 | 0 | 2 | Not-determined | Genome-fragment | NA | NA | 0 |
| S1-final-viral-combined_k141_35313\|\|full | 2089 | No | NA | 4 | 2 | 0 | Low-quality | Genome-fragment | 4.8 | AAI-based (medium-confidence) | 0 |
| S17-final-viral-combined_k141_56159\|\|full | 2712 | No | NA | 3 | 0 | 0 | Not-determined | Genome-fragment | NA | NA | 0 |
| S1-final-viral-combined_k141_172876\|\|full | 2089 | No | NA | 3 | 3 | 0 | Low-quality | Genome-fragment | 41.28 | AAI-based (high-confidence) | 0 |
| S17-final-viral-combined_k141_189511\|\|full | 20369 | No | NA | 28 | 0 | 10 | Not-determined | Genome-fragment | NA | NA | 0 |
| S17-final-viral-combined_k141_67538\|\|full | 2756 | No | NA | 4 | 0 | 2 | Not-determined | Genome-fragment | NA | NA | 0 |
| S17-final-viral-combined_k141_34004\|\|full | 2711 | No | NA | 3 | 0 | 2 | Not-determined | Genome-fragment | NA | NA | 0 |
| S17-final-viral-combined_k141_89462\|\|full | 1218 | No | NA | 3 | 0 | 2 | Not-determined | Genome-fragment | NA | NA | 0 |
| S17-final-viral-combined_k141_67562\|\|full | 1178 | No | NA | 2 | 0 | 1 | Not-determined | Genome-fragment | NA | NA | 0 |
| S17-final-viral-combined_k141_189540\|\|full | 3160 | No | NA | 4 | 0 | 3 | Not-determined | Genome-fragment | NA | NA | 0 |
| S7-final-viral-combined_k141_165778\|\|full | 2089 | No | NA | 3 | 2 | 0 | Low-quality | Genome-fragment | 4.11 | AAI-based (high-confidence) | 0 |
| S17-final-viral-combined_k141_200874\|\|full | 4456 | No | NA | 7 | 0 | 1 | Not-determined | Genome-fragment | NA | NA | 0 |
| S17-final-viral-combined_k141_241071\|\|full | 2089 | No | NA | 6 | 0 | 0 | Low-quality | Genome-fragment | 4.93 | AAI-based (high-confidence) | 0 |
| S4-final-viral-combined_k141_46518\|\|full | 2089 | No | NA | 6 | 0 | 0 | Low-quality | Genome-fragment | 4.1 | AAI-based (medium-confidence) | 0 |
| S17-final-viral-combined_k141_111978\|\|full | 2862 | No | NA | 4 | 0 | 4 | Not-determined | Genome-fragment | NA | NA | 0 |
| S17-final-viral-combined_k141_256378\|\|full | 2286 | No | NA | 3 | 0 | 0 | Not-determined | Genome-fragment | NA | NA | 0 |
| S17-final-viral-combined_k141_233946\|\|full | 2584 | No | NA | 4 | 0 | 0 | Not-determined | Genome-fragment | NA | NA | 0 |
| S17-final-viral-combined_k141_145284\|\|full | 3205 | No | NA | 4 | 0 | 3 | Not-determined | Genome-fragment | NA | NA | 0 |
| S8-final-viral-combined_k141_214933\|\|full | 2089 | No | NA | 1 | 0 | 0 | Low-quality | Genome-fragment | 7.78 | AAI-based (high-confidence) | 0 |
| S9-final-viral-combined_k141_39778\|\|full | 2090 | No | NA | 5 | 1 | 0 | Low-quality | Genome-fragment | 3.89 | AAI-based (high-confidence) | 0 |
| S17-final-viral-combined_k141_200948\|\|full | 2745 | No | NA | 3 | 0 | 2 | Not-determined | Genome-fragment | NA | NA | 0 |
| S17-final-viral-combined_k141_760\|\|full | 2470 | No | NA | 3 | 0 | 2 | Not-determined | Genome-fragment | NA | NA | 0 |
| S17-final-viral-combined_k141_189597\|\|full | 2139 | No | NA | 3 | 0 | 2 | Not-determined | Genome-fragment | NA | NA | 0 |
| S1_k141_245962 | 2090 | No | NA | 6 | 0 | 0 | Low-quality | Genome-fragment | 3.95 | AAI-based (high-confidence) | 0 |
| S17-final-viral-combined_k141_189605\|\|full | 2254 | No | NA | 4 | 0 | 0 | Not-determined | Genome-fragment | NA | NA | 0 |
| S17-final-viral-combined_k141_112002\|\|full | 1984 | No | NA | 2 | 0 | 1 | Not-determined | Genome-fragment | NA | NA | 0 |
| S4-final-viral-combined_k141_60997\|\|full | 2090 | No | NA | 4 | 2 | 0 | Low-quality | Genome-fragment | 3.83 | HMM-based (lower-bound) | 0 |
| S17-final-viral-combined_k141_78711\|\|full | 1908 | No | NA | 3 | 0 | 0 | Not-determined | Genome-fragment | NA | NA | 0 |
| S17-final-viral-combined_k141_787\|\|full | 1053 | No | NA | 2 | 0 | 1 | Not-determined | Genome-fragment | NA | NA | 0 |
| S17-final-viral-combined_k141_112013\|\|full | 1263 | No | NA | 2 | 0 | 2 | Not-determined | Genome-fragment | NA | NA | 0 |
| S9-final-viral-combined_k141_66740\|\|full | 2091 | No | NA | 2 | 0 | 1 | Low-quality | Genome-fragment | 1.53 | AAI-based (medium-confidence) | 0 |
| S11-final-viral-combined_k141_27620\|\|full | 2091 | No | NA | 4 | 0 | 1 | Low-quality | Genome-fragment | 1.4 | AAI-based (medium-confidence) | 0 |
| S1-final-viral-combined_k141_269247\|\|full | 2091 | No | NA | 2 | 0 | 1 | Low-quality | Genome-fragment | 2.4 | AAI-based (high-confidence) | 0 |
| S17-final-viral-combined_k141_112028\|\|full | 2112 | No | NA | 2 | 0 | 2 | Not-determined | Genome-fragment | NA | NA | 0 |
| S17-final-viral-combined_k141_56263\|\|full | 1938 | No | NA | 3 | 0 | 0 | Not-determined | Genome-fragment | NA | NA | 0 |
| S17-k141_155925 | 2091 | No | NA | 1 | 0 | 0 | Low-quality | Genome-fragment | 1.87 | AAI-based (medium-confidence) | 0 |
| S17-final-viral-combined_k141_78740\|\|full | 2660 | No | NA | 3 | 0 | 0 | Not-determined | Genome-fragment | NA | NA | 0 |
| S17-final-viral-combined_k141_178426\|\|full | 5089 | No | NA | 3 | 0 | 3 | Not-determined | Genome-fragment | NA | NA | 0 |
| S12-final-viral-combined_k141_290319\|\|full | 2091 | No | NA | 4 | 0 | 1 | Low-quality | Genome-fragment | 4.85 | AAI-based (high-confidence) | 0 |
| S2-final-viral-combined_k141_236538\|\|full | 2091 | No | NA | 2 | 0 | 1 | Low-quality | Genome-fragment | 4.89 | AAI-based (medium-confidence) | 0 |
| S4-final-viral-combined_k141_63222\|\|lt2gene | 2091 | No | NA | 1 | 1 | 0 | Low-quality | Genome-fragment | 2.06 | AAI-based (high-confidence) | 0 |
| S17-final-viral-combined_k141_145398\|\|full | 7548 | No | NA | 6 | 0 | 4 | Not-determined | Genome-fragment | NA | NA | 0 |
| S17-final-viral-combined_k141_112085\|\|full | 1236 | No | NA | 3 | 0 | 1 | Not-determined | Genome-fragment | NA | NA | 0 |
| S17-final-viral-combined_k141_201040\|\|full | 1413 | No | NA | 2 | 0 | 2 | Not-determined | Genome-fragment | NA | NA | 0 |
| S6-final-viral-combined_k141_69101\|\|full | 2091 | No | NA | 4 | 0 | 0 | Low-quality | Genome-fragment | 4.89 | AAI-based (high-confidence) | 0 |
| S9-final-viral-combined_k141_37742\|\|full | 2092 | No | NA | 4 | 0 | 0 | Low-quality | Genome-fragment | 3.41 | AAI-based (medium-confidence) | 0 |
| S17-final-viral-combined_k141_178477\|\|full | 1240 | No | NA | 2 | 0 | 2 | Not-determined | Genome-fragment | NA | NA | 0 |
| S9_k141_78280 | 2092 | No | NA | 2 | 0 | 2 | Low-quality | Genome-fragment | 1.42 | AAI-based (medium-confidence) | 0 |
| S11-final-viral-combined_k141_49134\|\|full | 2092 | No | NA | 4 | 1 | 0 | Low-quality | Genome-fragment | 4.87 | AAI-based (high-confidence) | 0 |
| S17-final-viral-combined_k141_212123\|\|full | 1391 | No | NA | 2 | 0 | 2 | Not-determined | Genome-fragment | NA | NA | 0 |
| S17-final-viral-combined_k141_233995\|\|full | 20090 | No | NA | 18 | 0 | 10 | Not-determined | Genome-fragment | NA | NA | 0 |
| S17-final-viral-combined_k141_45035\|\|full | 5737 | No | NA | 9 | 0 | 3 | Not-determined | Genome-fragment | NA | NA | 0 |
| S1-final-viral-combined_k141_352643\|\|full | 2092 | No | NA | 3 | 1 | 0 | Low-quality | Genome-fragment | 3.16 | AAI-based (high-confidence) | 0 |
| S17-final-viral-combined_k141_11787\|\|full | 3110 | No | NA | 5 | 0 | 2 | Not-determined | Genome-fragment | NA | NA | 0 |
| S17-final-viral-combined_k141_154242\|\|full | 2092 | No | NA | 3 | 1 | 0 | Low-quality | Genome-fragment | 3.82 | HMM-based (lower-bound) | 0 |
| S17-final-viral-combined_k141_78825\|\|full | 1356 | No | NA | 3 | 0 | 2 | Not-determined | Genome-fragment | NA | NA | 0 |
| S17-final-viral-combined_k141_56326\|\|full | 5731 | No | NA | 7 | 0 | 4 | Not-determined | Genome-fragment | NA | NA | 0 |
| S17-final-viral-combined_k141_234001\|\|full | 1964 | No | NA | 2 | 0 | 2 | Not-determined | Genome-fragment | NA | NA | 0 |
| S17-final-viral-combined_k141_21890\|\|full | 2092 | No | NA | 5 | 2 | 0 | Low-quality | Genome-fragment | 5.3 | AAI-based (high-confidence) | 0 |
| S17-final-viral-combined_k141_22818\|\|full | 19323 | No | NA | 15 | 0 | 7 | Not-determined | Genome-fragment | NA | NA | 0 |
| S17-final-viral-combined_k141_145461\|\|full | 1350 | No | NA | 3 | 0 | 0 | Not-determined | Genome-fragment | NA | NA | 0 |
| S17-final-viral-combined_k141_45076\|\|full | 2128 | No | NA | 4 | 0 | 4 | Not-determined | Genome-fragment | NA | NA | 0 |
| S17-final-viral-combined_k141_156362\|\|full | 1861 | No | NA | 3 | 0 | 2 | Not-determined | Genome-fragment | NA | NA | 0 |
| S12-final-viral-combined_k141_171633\|\|full | 2092 | No | NA | 4 | 0 | 0 | Low-quality | Genome-fragment | 3.88 | AAI-based (medium-confidence) | 0 |
| S17-final-viral-combined_k141_11850\|\|full | 2018 | No | NA | 2 | 0 | 1 | Not-determined | Genome-fragment | NA | NA | 0 |
| S12-final-viral-combined_k141_38438\|\|full | 2092 | No | NA | 2 | 0 | 1 | Low-quality | Genome-fragment | 1.77 | AAI-based (high-confidence) | 0 |
| S6-final-viral-combined_k141_91044\|\|full | 2092 | No | NA | 4 | 1 | 0 | Low-quality | Genome-fragment | 1.35 | HMM-based (lower-bound) | 0 |
| S17-final-viral-combined_k141_178560\|\|full | 1120 | No | NA | 2 | 0 | 2 | Not-determined | Genome-fragment | NA | NA | 0 |
| S17-final-viral-combined_k141_11855\|\|full | 2018 | No | NA | 2 | 0 | 1 | Not-determined | Genome-fragment | NA | NA | 0 |
| S17-final-viral-combined_k141_45099\|\|full | 3247 | No | NA | 3 | 0 | 3 | Not-determined | Genome-fragment | NA | NA | 0 |
| S17-final-viral-combined_k141_34187\|\|full | 6649 | No | NA | 5 | 0 | 4 | Not-determined | Genome-fragment | NA | NA | 0 |
| S17-final-viral-combined_k141_78886\|\|full | 2797 | No | NA | 3 | 0 | 1 | Not-determined | Genome-fragment | NA | NA | 0 |
| S8-final-viral-combined_k141_88792\|\|full | 2092 | No | NA | 3 | 0 | 0 | Low-quality | Genome-fragment | 1.5 | AAI-based (medium-confidence) | 0 |
| S17-final-viral-combined_k141_78898\|\|full | 2186 | No | NA | 3 | 0 | 2 | Not-determined | Genome-fragment | NA | NA | 0 |
| S9_k141_52605 | 2093 | No | NA | 2 | 0 | 1 | Low-quality | Genome-fragment | 2.33 | AAI-based (medium-confidence) | 0 |
| S17-final-viral-combined_k141_100685\|\|full | 6407 | No | NA | 14 | 0 | 0 | Not-determined | Genome-fragment | NA | NA | 0 |
| S9_k141_128251 | 2093 | No | NA | 2 | 0 | 0 | Low-quality | Genome-fragment | 3.79 | AAI-based (high-confidence) | 0 |
| S1-final-viral-combined_k141_187260\|\|full | 2093 | No | NA | 2 | 0 | 2 | Low-quality | Genome-fragment | 4.84 | AAI-based (medium-confidence) | 0 |
| S17-final-viral-combined_k141_11896\|\|full | 2279 | No | NA | 2 | 0 | 1 | Not-determined | Genome-fragment | NA | NA | 0 |
| S1_k141_17516 | 2093 | No | NA | 1 | 0 | 1 | Low-quality | Genome-fragment | 0.89 | AAI-based (medium-confidence) | 0 |
| S5-final-viral-combined_k141_177049\|\|full | 2093 | No | NA | 4 | 0 | 0 | Low-quality | Genome-fragment | 5.6 | AAI-based (medium-confidence) | 0 |
| S17-final-viral-combined_k141_56476\|\|full | 1485 | No | NA | 3 | 0 | 0 | Not-determined | Genome-fragment | NA | NA | 0 |
| S17-final-viral-combined_k141_67817\|\|full | 1892 | No | NA | 3 | 0 | 1 | Not-determined | Genome-fragment | NA | NA | 0 |
| S7-final-viral-combined_k141_223950\|\|full | 2093 | No | NA | 4 | 0 | 0 | Low-quality | Genome-fragment | 2.54 | AAI-based (high-confidence) | 0 |
| S17-k141_80312 | 2093 | No | NA | 4 | 0 | 3 | Low-quality | Genome-fragment | 1.29 | AAI-based (medium-confidence) | 0 |
| S17-final-viral-combined_k141_256540\|\|full | 2317 | No | NA | 2 | 0 | 2 | Not-determined | Genome-fragment | NA | NA | 0 |
| S17-final-viral-combined_k141_100738\|\|full | 2786 | No | NA | 4 | 0 | 2 | Not-determined | Genome-fragment | NA | NA | 0 |
| S17-final-viral-combined_k141_56483\|\|full | 6678 | No | NA | 7 | 0 | 2 | Not-determined | Genome-fragment | NA | NA | 0 |
| S17-final-viral-combined_k141_156475\|\|full | 2047 | No | NA | 3 | 0 | 3 | Not-determined | Genome-fragment | NA | NA | 0 |
| S17-final-viral-combined_k141_245422\|\|full | 2227 | No | NA | 3 | 0 | 1 | Not-determined | Genome-fragment | NA | NA | 0 |
| S17-final-viral-combined_k141_256547\|\|full | 4724 | No | NA | 5 | 0 | 2 | Not-determined | Genome-fragment | NA | NA | 0 |
| S17-final-viral-combined_k141_100759\|\|full | 3240 | No | NA | 2 | 0 | 2 | Not-determined | Genome-fragment | NA | NA | 0 |
| S12-final-viral-combined_k141_131511\|\|full | 2093 | No | NA | 3 | 0 | 1 | Low-quality | Genome-fragment | 1.45 | AAI-based (medium-confidence) | 0 |
| S12-final-viral-combined_k141_317796\|\|full | 2093 | No | NA | 3 | 3 | 0 | Low-quality | Genome-fragment | 4.71 | AAI-based (high-confidence) | 0 |
| S17-final-viral-combined_k141_79023\|\|full | 1876 | No | NA | 3 | 0 | 1 | Not-determined | Genome-fragment | NA | NA | 0 |
| S17-final-viral-combined_k141_178706\|\|full | 4818 | No | NA | 3 | 0 | 0 | Not-determined | Genome-fragment | NA | NA | 0 |
| S12_k141_319075 | 2093 | No | NA | 4 | 1 | 0 | Low-quality | Genome-fragment | 1.41 | HMM-based (lower-bound) | 0 |
| S2_k141_199531 | 2093 | No | NA | 2 | 0 | 0 | Low-quality | Genome-fragment | 6.92 | AAI-based (high-confidence) | 0 |
| S17-final-viral-combined_k141_201211\|\|full | 3117 | No | NA | 4 | 0 | 3 | Not-determined | Genome-fragment | NA | NA | 0 |
| S17-final-viral-combined_k141_178728\|\|full | 1316 | No | NA | 2 | 0 | 2 | Not-determined | Genome-fragment | NA | NA | 0 |
| S4-final-viral-combined_k141_130767\|\|full | 2093 | No | NA | 2 | 0 | 0 | Low-quality | Genome-fragment | 3.14 | AAI-based (medium-confidence) | 0 |

| | | | | | | | | | | | |
|---|---|---|---|---|---|---|---|---|---|---|---|
| S6-final-viral-combined_k141_67639\|\|full | 2093 | No | NA | 3 | 0 | 3 | Low-quality | Genome-fragment | 0.72 | AAI-based (medium-confidence) | 0 |
| S17-final-viral-combined_k141_256574\|\|full | 1055 | No | NA | 3 | 0 | 1 | Not-determined | Genome-fragment | NA | NA | 0 |
| S9-final-viral-combined_k141_40610\|\|full | 2094 | No | NA | 5 | 0 | 1 | Low-quality | Genome-fragment | 4.12 | AAI-based (high-confidence) | 0 |
| S1-final-viral-combined_k141_26797\|\|full | 2094 | No | NA | 4 | 0 | 0 | Low-quality | Genome-fragment | 5.01 | AAI-based (medium-confidence) | 0 |
| S5-final-viral-combined_k141_139537\|\|full | 2094 | No | NA | 3 | 3 | 0 | Low-quality | Genome-fragment | 5.95 | AAI-based (high-confidence) | 0 |
| S17-final-viral-combined_k141_56555\|\|full | 3895 | No | NA | 3 | 0 | 1 | Not-determined | Genome-fragment | NA | NA | 0 |
| S12-final-viral-combined_k141_378568\|\|full | 2094 | No | NA | 7 | 1 | 0 | Low-quality | Genome-fragment | 6.06 | AAI-based (high-confidence) | 0 |
| S12_k141_126410 | 2094 | No | NA | 2 | 0 | 2 | Low-quality | Genome-fragment | 1.48 | AAI-based (medium-confidence) | 0 |
| S8-final-viral-combined_k141_38328\|\|full | 2094 | No | NA | 4 | 0 | 3 | Low-quality | Genome-fragment | 2.87 | AAI-based (medium-confidence) | 0 |
| S3-final-viral-combined_k141_28341\|\|full | 2095 | No | NA | 3 | 0 | 0 | Low-quality | Genome-fragment | 4.71 | AAI-based (high-confidence) | 0 |
| S5-final-viral-combined_k141_197865\|\|full | 2095 | No | NA | 3 | 1 | 0 | Low-quality | Genome-fragment | 4.73 | AAI-based (medium-confidence) | 0 |
| S17-final-viral-combined_k141_945\|\|full | 1425 | No | NA | 4 | 0 | 2 | Not-determined | Genome-fragment | NA | NA | 0 |
| S17-final-viral-combined_k141_186624\|\|full | 2095 | No | NA | 2 | 0 | 0 | Low-quality | Genome-fragment | 1.1 | AAI-based (medium-confidence) | 0 |
| S17-final-viral-combined_k141_234183\|\|full | 1193 | No | NA | 2 | 0 | 0 | Not-determined | Genome-fragment | NA | NA | 0 |
| S12-final-viral-combined_k141_81054\|\|full | 2095 | No | NA | 4 | 1 | 0 | Low-quality | Genome-fragment | 3.99 | AAI-based (medium-confidence) | 0 |
| S17-final-viral-combined_k141_234185\|\|full | 8040 | No | NA | 6 | 0 | 6 | Not-determined | Genome-fragment | NA | NA | 0 |
| S2_k141_13912 | 2095 | No | NA | 2 | 1 | 0 | Low-quality | Genome-fragment | 35.33 | AAI-based (high-confidence) | 0 |
| S17-final-viral-combined_k141_145641\|\|full | 3526 | No | NA | 4 | 0 | 0 | Not-determined | Genome-fragment | NA | NA | 0 |
| S1-final-viral-combined_k141_371760\|\|full | 2096 | No | NA | 5 | 0 | 0 | Low-quality | Genome-fragment | 5.01 | AAI-based (high-confidence) | 0 |
| S17-final-viral-combined_k141_145643\|\|full | 2790 | No | NA | 3 | 0 | 1 | Not-determined | Genome-fragment | NA | NA | 0 |
| S17-final-viral-combined_k141_234197\|\|full | 6519 | No | NA | 14 | 0 | 1 | Not-determined | Genome-fragment | NA | NA | 0 |
| S2-final-viral-combined_k141_28066\|\|full | 2096 | No | NA | 3 | 0 | 0 | Low-quality | Genome-fragment | 3.98 | AAI-based (high-confidence) | 0 |
| S17-final-viral-combined_k141_12005\|\|full | 2000 | No | NA | 2 | 0 | 0 | Not-determined | Genome-fragment | NA | NA | 0 |
| S4_k141_21374 | 2096 | No | NA | 1 | 0 | 0 | Low-quality | Genome-fragment | 2.06 | AAI-based (high-confidence) | 0 |
| S11-final-viral-combined_k141_28254\|\|full | 2097 | No | NA | 3 | 0 | 0 | Low-quality | Genome-fragment | 3.63 | AAI-based (medium-confidence) | 0 |
| S17-final-viral-combined_k141_123401\|\|full | 2109 | No | NA | 3 | 0 | 3 | Not-determined | Genome-fragment | NA | NA | 0 |
| S17-final-viral-combined_k141_156618\|\|full | 2121 | No | NA | 3 | 0 | 1 | Not-determined | Genome-fragment | NA | NA | 0 |
| S17-final-viral-combined_k141_89722\|\|full | 8443 | No | NA | 7 | 0 | 4 | Not-determined | Genome-fragment | NA | NA | 0 |
| S1-final-viral-combined_k141_266850\|\|full | 2097 | No | NA | 2 | 0 | 1 | Low-quality | Genome-fragment | 3.91 | AAI-based (high-confidence) | 0 |
| S17-final-viral-combined_k141_1027\|\|full | 2989 | No | NA | 4 | 0 | 3 | Not-determined | Genome-fragment | NA | NA | 0 |
| S17-final-viral-combined_k141_212363\|\|full | 4148 | No | NA | 5 | 0 | 1 | Not-determined | Genome-fragment | NA | NA | 0 |
| S5-final-viral-combined_k141_107758\|\|full | 2097 | No | NA | 4 | 1 | 0 | Low-quality | Genome-fragment | 1.27 | HMM-based (lower-bound) | 0 |
| S17-final-viral-combined_k141_34389\|\|full | 3084 | No | NA | 6 | 0 | 0 | Not-determined | Genome-fragment | NA | NA | 0 |
| S7_k141_203604 | 2097 | No | NA | 3 | 1 | 0 | Low-quality | Genome-fragment | 1.94 | HMM-based (lower-bound) | 0 |
| S8-final-viral-combined_k141_79682\|\|full | 2097 | No | NA | 4 | 0 | 0 | Low-quality | Genome-fragment | 2.87 | AAI-based (medium-confidence) | 0 |
| S1_k141_338818 | 2098 | No | NA | 4 | 0 | 0 | Low-quality | Genome-fragment | 3.36 | AAI-based (high-confidence) | 0 |
| S12-final-viral-combined_k141_92901\|\|full | 2098 | No | NA | 5 | 0 | 0 | Low-quality | Genome-fragment | 6.09 | AAI-based (high-confidence) | 0 |
| S17-final-viral-combined_k141_45183\|\|full | 1092 | No | NA | 3 | 0 | 1 | Not-determined | Genome-fragment | NA | NA | 0 |
| S1-final-viral-combined_k141_230082\|\|full | 2099 | No | NA | 3 | 2 | 0 | Low-quality | Genome-fragment | 5.22 | AAI-based (high-confidence) | 0 |
| S5_k141_146090 | 2099 | No | NA | 5 | 4 | 0 | Low-quality | Genome-fragment | 5.16 | AAI-based (high-confidence) | 0 |
| S17-final-viral-combined_k141_111634\|\|full | 2099 | No | NA | 6 | 1 | 0 | Low-quality | Genome-fragment | 5.23 | AAI-based (high-confidence) | 0 |
| S8-final-viral-combined_k141_141687\|\|full | 2099 | No | NA | 4 | 0 | 0 | Low-quality | Genome-fragment | 4.26 | AAI-based (medium-confidence) | 0 |
| S17-final-viral-combined_k141_34436\|\|full | 1094 | No | NA | 3 | 0 | 0 | Not-determined | Genome-fragment | NA | NA | 0 |
| S17-final-viral-combined_k141_223535\|\|full | 7031 | No | NA | 8 | 0 | 5 | Not-determined | Genome-fragment | NA | NA | 0 |
| S17-final-viral-combined_k141_256650\|\|full | 2756 | No | NA | 3 | 0 | 1 | Not-determined | Genome-fragment | NA | NA | 0 |
| S1-final-viral-combined_k141_362547\|\|full | 2100 | No | NA | 3 | 0 | 0 | Low-quality | Genome-fragment | 1.46 | AAI-based (high-confidence) | 0 |
| S3-final-viral-combined_k141_15987\|\|full | 2100 | No | NA | 3 | 0 | 0 | Low-quality | Genome-fragment | 5.88 | AAI-based (high-confidence) | 0 |
| S17-final-viral-combined_k141_145687\|\|full | 2595 | No | NA | 3 | 0 | 3 | Not-determined | Genome-fragment | NA | NA | 0 |
| S12-final-viral-combined_k141_90113\|\|full | 2100 | No | NA | 4 | 0 | 1 | Low-quality | Genome-fragment | 0.84 | AAI-based (medium-confidence) | 0 |
| S12-final-viral-combined_k141_147556\|\|full | 2100 | No | NA | 5 | 0 | 0 | Low-quality | Genome-fragment | 4.79 | AAI-based (medium-confidence) | 0 |
| S17-final-viral-combined_k141_67839\|\|full | 1982 | No | NA | 4 | 0 | 0 | Not-determined | Genome-fragment | NA | NA | 0 |
| S4-final-viral-combined_k141_76036\|\|full | 2100 | No | NA | 2 | 0 | 2 | Low-quality | Genome-fragment | 2.12 | AAI-based (medium-confidence) | 0 |
| S17-final-viral-combined_k141_12076\|\|full | 1531 | No | NA | 4 | 0 | 0 | Not-determined | Genome-fragment | NA | NA | 0 |
| S17-final-viral-combined_k141_112443\|\|full | 2393 | No | NA | 3 | 0 | 0 | Not-determined | Genome-fragment | NA | NA | 0 |
| S1-final-viral-combined_k141_62326\|\|full | 2101 | No | NA | 3 | 0 | 1 | Low-quality | Genome-fragment | 1.01 | AAI-based (medium-confidence) | 0 |
| S17-final-viral-combined_k141_67864\|\|full | 1771 | No | NA | 3 | 0 | 1 | Not-determined | Genome-fragment | NA | NA | 0 |
| S17-final-viral-combined_k141_112446\|\|full | 6414 | No | NA | 6 | 0 | 1 | Not-determined | Genome-fragment | NA | NA | 0 |
| S1-final-viral-combined_k141_376253\|\|full | 2101 | No | NA | 6 | 0 | 0 | Low-quality | Genome-fragment | 3.38 | AAI-based (high-confidence) | 0 |
| S17-final-viral-combined_k141_12087\|\|full | 2265 | No | NA | 3 | 0 | 1 | Not-determined | Genome-fragment | NA | NA | 0 |
| S7-final-viral-combined_k141_158027\|\|full | 2101 | No | NA | 2 | 0 | 2 | Low-quality | Genome-fragment | 1.15 | AAI-based (medium-confidence) | 0 |
| S17-final-viral-combined_k141_212412\|\|full | 1649 | No | NA | 4 | 0 | 0 | Not-determined | Genome-fragment | NA | NA | 0 |
| S17-final-viral-combined_k141_1138\|\|full | 5006 | No | NA | 4 | 0 | 3 | Not-determined | Genome-fragment | NA | NA | 0 |
| S17-final-viral-combined_k141_198071\|\|full | 2101 | No | NA | 3 | 0 | 2 | Low-quality | Genome-fragment | 2.45 | AAI-based (medium-confidence) | 0 |
| S17-final-viral-combined_k141_212417\|\|full | 1514 | No | NA | 2 | 0 | 1 | Not-determined | Genome-fragment | NA | NA | 0 |
| S17-final-viral-combined_k141_89787\|\|full | 3498 | No | NA | 4 | 0 | 3 | Not-determined | Genome-fragment | NA | NA | 0 |
| S17-final-viral-combined_k141_67895\|\|full | 1473 | No | NA | 3 | 0 | 0 | Not-determined | Genome-fragment | NA | NA | 0 |
| S17-final-viral-combined_k141_178974\|\|full | 4848 | No | NA | 5 | 0 | 4 | Not-determined | Genome-fragment | NA | NA | 0 |
| S12_k141_137512 | 2101 | No | NA | 2 | 0 | 0 | Low-quality | Genome-fragment | 2.61 | AAI-based (high-confidence) | 0 |
| S17-final-viral-combined_k141_178984\|\|full | 1778 | No | NA | 2 | 0 | 1 | Not-determined | Genome-fragment | NA | NA | 0 |
| S8-final-viral-combined_k141_23463\|\|full | 2101 | No | NA | 4 | 0 | 1 | Low-quality | Genome-fragment | 4.12 | AAI-based (medium-confidence) | 0 |
| S7-final-viral-combined_k141_54971\|\|full | 2102 | No | NA | 3 | 0 | 1 | Low-quality | Genome-fragment | 2.89 | AAI-based (medium-confidence) | 0 |
| S17-final-viral-combined_k141_79266\|\|full | 2484 | No | NA | 4 | 0 | 3 | Not-determined | Genome-fragment | NA | NA | 0 |
| S7-final-viral-combined_k141_25443\|\|full | 2102 | No | NA | 2 | 0 | 0 | Low-quality | Genome-fragment | 5.63 | AAI-based (medium-confidence) | 0 |
| S12-final-viral-combined_k141_60302\|\|full | 2102 | No | NA | 3 | 0 | 0 | Low-quality | Genome-fragment | 5.45 | AAI-based (high-confidence) | 0 |
| S17-final-viral-combined_k141_23144\|\|full | 29863 | No | NA | 31 | 0 | 19 | Not-determined | Genome-fragment | NA | NA | 0 |
| S2-final-viral-combined_k141_197066\|\|full | 2102 | No | NA | 6 | 2 | 0 | Low-quality | Genome-fragment | 3.45 | AAI-based (high-confidence) | 0 |
| S2-final-viral-combined_k141_37249\|\|full | 2102 | No | NA | 2 | 0 | 1 | Low-quality | Genome-fragment | 2.55 | AAI-based (medium-confidence) | 0 |
| S17-final-viral-combined_k141_12118\|\|full | 1564 | No | NA | 3 | 0 | 1 | Not-determined | Genome-fragment | NA | NA | 0 |
| S8-final-viral-combined_k141_194763\|\|full | 2102 | No | NA | 2 | 1 | 0 | Low-quality | Genome-fragment | 3.54 | AAI-based (high-confidence) | 0 |
| S17-final-viral-combined_k141_34508\|\|full | 1621 | No | NA | 2 | 0 | 2 | Not-determined | Genome-fragment | NA | NA | 0 |
| S1-final-viral-combined_k141_315851\|\|full | 2103 | No | NA | 3 | 2 | 0 | Low-quality | Genome-fragment | 6.94 | HMM-based (lower-bound) | 0 |
| S17-final-viral-combined_k141_67960\|\|full | 3820 | No | NA | 4 | 0 | 3 | Not-determined | Genome-fragment | NA | NA | 0 |
| S17-final-viral-combined_k141_223678\|\|full | 1759 | No | NA | 3 | 0 | 0 | Not-determined | Genome-fragment | NA | NA | 0 |
| S3-final-viral-combined_k141_6389\|\|full | 2103 | No | NA | 2 | 1 | 0 | Low-quality | Genome-fragment | 6.18 | AAI-based (high-confidence) | 0 |
| S17-final-viral-combined_k141_100937\|\|full | 2686 | No | NA | 5 | 0 | 3 | Not-determined | Genome-fragment | NA | NA | 0 |
| S17-final-viral-combined_k141_212470\|\|full | 1616 | No | NA | 2 | 0 | 1 | Not-determined | Genome-fragment | NA | NA | 0 |
| S17-final-viral-combined_k141_123580\|\|full | 3176 | No | NA | 2 | 0 | 2 | Not-determined | Genome-fragment | NA | NA | 0 |
| S17-k141_44577 | 2103 | No | NA | 2 | 0 | 2 | Low-quality | Genome-fragment | 1.18 | AAI-based (high-confidence) | 0 |
| S17-final-viral-combined_k141_212472\|\|full | 2232 | No | NA | 3 | 0 | 0 | Not-determined | Genome-fragment | NA | NA | 0 |
| S17-final-viral-combined_k141_23193\|\|full | 1260 | No | NA | 2 | 0 | 0 | Not-determined | Genome-fragment | NA | NA | 0 |
| S17-final-viral-combined_k141_45357\|\|full | 3471 | No | NA | 5 | 0 | 0 | Not-determined | Genome-fragment | NA | NA | 0 |
| S17-final-viral-combined_k141_223719\|\|full | 1703 | No | NA | 3 | 0 | 2 | Not-determined | Genome-fragment | NA | NA | 0 |
| S17-final-viral-combined_k141_23201\|\|full | 1709 | No | NA | 3 | 0 | 3 | Not-determined | Genome-fragment | NA | NA | 0 |
| S17-final-viral-combined_k141_179084\|\|full | 6455 | No | NA | 9 | 0 | 5 | Not-determined | Genome-fragment | NA | NA | 0 |
| S17-final-viral-combined_k141_223727\|\|full | 5915 | No | NA | 5 | 0 | 4 | Not-determined | Genome-fragment | NA | NA | 0 |
| S17-final-viral-combined_k141_68018\|\|full | 2134 | No | NA | 4 | 0 | 1 | Not-determined | Genome-fragment | NA | NA | 0 |
| S17-final-viral-combined_k141_23213\|\|full | 2269 | No | NA | 3 | 0 | 2 | Not-determined | Genome-fragment | NA | NA | 0 |
| S2-final-viral-combined_k141_174128\|\|full | 2103 | No | NA | 2 | 0 | 1 | Low-quality | Genome-fragment | 6 | AAI-based (medium-confidence) | 0 |
| S17-final-viral-combined_k141_23216\|\|full | 1386 | No | NA | 3 | 0 | 0 | Not-determined | Genome-fragment | NA | NA | 0 |
| S4_k141_66956 | 2103 | No | NA | 2 | 0 | 0 | Low-quality | Genome-fragment | 3.78 | AAI-based (medium-confidence) | 0 |
| S9-final-viral-combined_k141_67917\|\|full | 2104 | No | NA | 2 | 0 | 1 | Low-quality | Genome-fragment | 2.59 | AAI-based (medium-confidence) | 0 |
| S11-final-viral-combined_k141_73398\|\|full | 2104 | No | NA | 5 | 0 | 0 | Low-quality | Genome-fragment | 3.53 | AAI-based (high-confidence) | 0 |
| S17-final-viral-combined_k141_123621\|\|full | 1459 | No | NA | 2 | 0 | 0 | Not-determined | Genome-fragment | NA | NA | 0 |
| S11-final-viral-combined_k141_65465\|\|full | 2104 | No | NA | 2 | 0 | 1 | Low-quality | Genome-fragment | 2.61 | AAI-based (medium-confidence) | 0 |
| S17-final-viral-combined_k141_101010\|\|full | 2829 | No | NA | 3 | 0 | 0 | Not-determined | Genome-fragment | NA | NA | 0 |
| S17-final-viral-combined_k141_212554\|\|full | 4183 | No | NA | 5 | 0 | 4 | Not-determined | Genome-fragment | NA | NA | 0 |
| S11-final-viral-combined_k141_273475\|\|full | 2104 | No | NA | 2 | 0 | 1 | Low-quality | Genome-fragment | 0.75 | AAI-based (medium-confidence) | 0 |
| S17-final-viral-combined_k141_223748\|\|full | 4859 | No | NA | 6 | 1 | 0 | Medium-quality | Genome-fragment | 80.87 | AAI-based (medium-confidence) | 0 |
| S1-final-viral-combined_k141_82328\|\|full | 2104 | No | NA | 3 | 0 | 1 | Low-quality | Genome-fragment | 1.73 | AAI-based (medium-confidence) | 0 |
| S1-final-viral-combined_k141_303839\|\|full | 2104 | No | NA | 5 | 0 | 0 | Low-quality | Genome-fragment | 5.5 | AAI-based (high-confidence) | 0 |
| S5-final-viral-combined_k141_169468\|\|full | 2104 | No | NA | 3 | 2 | 0 | Low-quality | Genome-fragment | 4.02 | AAI-based (high-confidence) | 0 |
| S7-final-viral-combined_k141_115991\|\|full | 2104 | No | NA | 3 | 0 | 0 | Low-quality | Genome-fragment | 3.18 | AAI-based (medium-confidence) | 0 |
| S7-final-viral-combined_k141_119893\|\|full | 2105 | No | NA | 6 | 2 | 0 | Low-quality | Genome-fragment | 3.47 | AAI-based (medium-confidence) | 0 |
| S7-final-viral-combined_k141_40345\|\|full | 2105 | No | NA | 4 | 0 | 3 | Low-quality | Genome-fragment | 0.94 | AAI-based (medium-confidence) | 0 |
| S17-final-viral-combined_k141_34621\|\|full | 2817 | No | NA | 6 | 0 | 0 | Not-determined | Genome-fragment | NA | NA | 0 |
| S17-final-viral-combined_k141_256782\|\|full | 3254 | No | NA | 5 | 0 | 0 | Not-determined | Genome-fragment | NA | NA | 0 |
| S17-final-viral-combined_k141_145887\|\|full | 2401 | No | NA | 2 | 0 | 2 | Not-determined | Genome-fragment | NA | NA | 0 |
| S17-final-viral-combined_k141_223773\|\|full | 7500 | No | NA | 6 | 0 | 5 | Not-determined | Genome-fragment | NA | NA | 0 |
| S17-final-viral-combined_k141_123692\|\|full | 2347 | No | NA | 3 | 0 | 1 | Not-determined | Genome-fragment | NA | NA | 0 |
| S17-final-viral-combined_k141_167857\|\|full | 1184 | No | NA | 3 | 0 | 0 | Not-determined | Genome-fragment | NA | NA | 0 |
| S17-final-viral-combined_k141_68131\|\|full | 2503 | No | NA | 4 | 0 | 1 | Not-determined | Genome-fragment | NA | NA | 0 |
| S17-final-viral-combined_k141_134549\|\|full | 12491 | No | NA | 8 | 0 | 8 | Not-determined | Genome-fragment | NA | NA | 0 |
| S2-final-viral-combined_k141_82256\|\|full | 2105 | No | NA | 2 | 0 | 1 | Low-quality | Genome-fragment | 2.46 | AAI-based (high-confidence) | 0 |
| S17-final-viral-combined_k141_156704\|\|full | 2561 | No | NA | 2 | 0 | 1 | Not-determined | Genome-fragment | NA | NA | 0 |
| S8-final-viral-combined_k141_232362\|\|full | 2105 | No | NA | 4 | 2 | 0 | Low-quality | Genome-fragment | 5.37 | AAI-based (high-confidence) | 0 |
| S9-final-viral-combined_k141_124782\|\|lt2gene | 2106 | No | NA | 2 | 1 | 0 | Low-quality | Genome-fragment | 5.08 | AAI-based (medium-confidence) | 0 |
| S17-final-viral-combined_k141_79491\|\|full | 2272 | No | NA | 4 | 0 | 0 | Not-determined | Genome-fragment | NA | NA | 0 |
| S17-final-viral-combined_k141_89979\|\|full | 1600 | No | NA | 3 | 0 | 0 | Not-determined | Genome-fragment | NA | NA | 0 |
| S5-final-viral-combined_k141_190248\|\|full | 2106 | No | NA | 3 | 0 | 0 | Low-quality | Genome-fragment | 5.89 | AAI-based (medium-confidence) | 0 |
| S2-final-viral-combined_k141_226222\|\|full | 2106 | No | NA | 3 | 2 | 0 | Low-quality | Genome-fragment | 33.55 | AAI-based (high-confidence) | 0 |
| S17-final-viral-combined_k141_123768\|\|full | 1135 | No | NA | 3 | 0 | 1 | Not-determined | Genome-fragment | NA | NA | 0 |
| S17-final-viral-combined_k141_101111\|\|full | 1140 | No | NA | 3 | 0 | 1 | Not-determined | Genome-fragment | NA | NA | 0 |
| S17-final-viral-combined_k141_89994\|\|full | 2329 | No | NA | 4 | 0 | 0 | Not-determined | Genome-fragment | NA | NA | 0 |
| S8_k141_5569 | 2106 | No | NA | 5 | 0 | 0 | Low-quality | Genome-fragment | 3.41 | AAI-based (high-confidence) | 0 |
| S17-final-viral-combined_k141_212670\|\|full | 1030 | No | NA | 2 | 0 | 0 | Not-determined | Genome-fragment | NA | NA | 0 |
| S17-final-viral-combined_k141_201651\|\|full | 6155 | No | NA | 12 | 0 | 0 | Not-determined | Genome-fragment | NA | NA | 0 |
| S9-final-viral-combined_k141_107393\|\|lt2gene | 2107 | No | NA | 2 | 2 | 0 | Low-quality | Genome-fragment | 4.52 | AAI-based (medium-confidence) | 0 |
| S17-final-viral-combined_k141_90005\|\|full | 6562 | No | NA | 8 | 0 | 5 | Not-determined | Genome-fragment | NA | NA | 0 |
| S9_k141_45464 | 2107 | No | NA | 3 | 0 | 1 | Low-quality | Genome-fragment | 3.61 | AAI-based (high-confidence) | 0 |
| S17-final-viral-combined_k141_34624\|\|full | 4548 | No | NA | 8 | 0 | 1 | Not-determined | Genome-fragment | NA | NA | 0 |
| S17-final-viral-combined_k141_45413\|\|full | 3379 | No | NA | 3 | 0 | 2 | Not-determined | Genome-fragment | NA | NA | 0 |
| S11-final-viral-combined_k141_285876\|\|full | 2107 | No | NA | 3 | 0 | 2 | Low-quality | Genome-fragment | 1.01 | AAI-based (medium-confidence) | 0 |
| S11_k141_124222 | 2107 | No | NA | 1 | 0 | 1 | Low-quality | Genome-fragment | 1.23 | AAI-based (medium-confidence) | 0 |
| S12-final-viral-combined_k141_67765\|\|full | 2107 | No | NA | 4 | 1 | 0 | Low-quality | Genome-fragment | 2.69 | HMM-based (lower-bound) | 0 |
| S17-final-viral-combined_k141_212705\|\|full | 1721 | No | NA | 3 | 0 | 0 | Not-determined | Genome-fragment | NA | NA | 0 |
| S17-final-viral-combined_k141_156787\|\|full | 1105 | No | NA | 2 | 0 | 1 | Not-determined | Genome-fragment | NA | NA | 0 |
| S17-final-viral-combined_k141_23338\|\|full | 2925 | No | NA | 7 | 0 | 0 | Not-determined | Genome-fragment | NA | NA | 0 |
| S17-final-viral-combined_k141_79573\|\|full | 1457 | No | NA | 4 | 0 | 0 | Not-determined | Genome-fragment | NA | NA | 0 |
| S17-final-viral-combined_k141_79580\|\|full | 3042 | No | NA | 4 | 0 | 1 | Not-determined | Genome-fragment | NA | NA | 0 |
| S12-final-viral-combined_k141_151775\|\|full | 2107 | No | NA | 3 | 0 | 0 | Low-quality | Genome-fragment | 1.64 | AAI-based (medium-confidence) | 0 |
| S2-final-viral-combined_k141_179583\|\|full | 2107 | No | NA | 3 | 1 | 0 | Low-quality | Genome-fragment | 3.85 | HMM-based (lower-bound) | 0 |
| S17-final-viral-combined_k141_90047\|\|full | 3093 | No | NA | 4 | 0 | 2 | Not-determined | Genome-fragment | NA | NA | 0 |
| S8-final-viral-combined_k141_174315\|\|full | 2107 | No | NA | 2 | 0 | 1 | Low-quality | Genome-fragment | 1.21 | AAI-based (medium-confidence) | 0 |
| S17-final-viral-combined_k141_1373\|\|full | 2141 | No | NA | 4 | 0 | 2 | Not-determined | Genome-fragment | NA | NA | 0 |
| S1-final-viral-combined_k141_215465\|\|full | 2108 | No | NA | 5 | 0 | 0 | Low-quality | Genome-fragment | 4.51 | AAI-based (high-confidence) | 0 |
| S1-final-viral-combined_k141_335152\|\|lt2gene | 2108 | No | NA | 2 | 1 | 0 | Low-quality | Genome-fragment | 4.7 | AAI-based (high-confidence) | 0 |
| S5-final-viral-combined_k141_34014\|\|full | 2108 | No | NA | 3 | 0 | 3 | Low-quality | Genome-fragment | 0.67 | AAI-based (medium-confidence) | 0 |
| S2-final-viral-combined_k141_6864\|\|full | 2108 | No | NA | 6 | 1 | 0 | Low-quality | Genome-fragment | 3.49 | AAI-based (high-confidence) | 0 |
| S17-final-viral-combined_k141_212783\|\|full | 3284 | No | NA | 5 | 0 | 0 | Not-determined | Genome-fragment | NA | NA | 0 |
| S8-final-viral-combined_k141_184839\|\|full | 2108 | No | NA | 6 | 1 | 0 | Low-quality | Genome-fragment | 3.05 | AAI-based (medium-confidence) | 0 |
| S17-final-viral-combined_k141_145970\|\|full | 2071 | No | NA | 3 | 0 | 2 | Not-determined | Genome-fragment | NA | NA | 0 |
| S17-final-viral-combined_k141_45462\|\|full | 2754 | No | NA | 4 | 0 | 1 | Not-determined | Genome-fragment | NA | NA | 0 |
| S9-final-viral-combined_k141_99145\|\|full | 2109 | No | NA | 2 | 0 | 2 | Low-quality | Genome-fragment | 3.43 | AAI-based (medium-confidence) | 0 |
| S17-final-viral-combined_k141_256942\|\|full | 4656 | No | NA | 6 | 0 | 3 | Not-determined | Genome-fragment | NA | NA | 0 |
| S17-final-viral-combined_k141_212836\|\|full | 1438 | No | NA | 2 | 0 | 0 | Not-determined | Genome-fragment | NA | NA | 0 |
| S9-final-viral-combined_k141_69820\|\|lt2gene | 2109 | No | NA | 1 | 1 | 0 | Low-quality | Genome-fragment | 4.81 | AAI-based (medium-confidence) | 0 |
| S17-final-viral-combined_k141_201806\|\|full | 2575 | No | NA | 3 | 0 | 1 | Not-determined | Genome-fragment | NA | NA | 0 |
| S17-final-viral-combined_k141_12288\|\|full | 2031 | No | NA | 4 | 0 | 1 | Not-determined | Genome-fragment | NA | NA | 0 |
| S17-final-viral-combined_k141_201813\|\|full | 2391 | No | NA | 3 | 0 | 2 | Not-determined | Genome-fragment | NA | NA | 0 |
| S11-final-viral-combined_k141_55172\|\|full | 2109 | No | NA | 4 | 1 | 0 | Low-quality | Genome-fragment | 3.1 | HMM-based (lower-bound) | 0 |
| S11_k141_292363 | 2109 | No | NA | 3 | 1 | 0 | Low-quality | Genome-fragment | 4.03 | AAI-based (high-confidence) | 0 |
| S1_k141_257835 | 2109 | No | NA | 2 | 0 | 0 | Low-quality | Genome-fragment | 1.93 | AAI-based (high-confidence) | 0 |
| S17-final-viral-combined_k141_223984\|\|full | 2091 | No | NA | 3 | 0 | 0 | Not-determined | Genome-fragment | NA | NA | 0 |
| S5-final-viral-combined_k141_86936\|\|full | 2109 | No | NA | 3 | 0 | 1 | Low-quality | Genome-fragment | 1.37 | AAI-based (medium-confidence) | 0 |
| S17-final-viral-combined_k141_23378\|\|full | 1518 | No | NA | 3 | 0 | 2 | Not-determined | Genome-fragment | NA | NA | 0 |
| S17-final-viral-combined_k141_79709\|\|full | 19454 | No | NA | 26 | 15 | 0 | Medium-quality | Genome-fragment | 51.21 | AAI-based (medium-confidence) | 0 |
| S17-final-viral-combined_k141_18491\|\|full | 2109 | No | NA | 3 | 0 | 0 | Low-quality | Genome-fragment | 3.34 | AAI-based (high-confidence) | 0 |
| S17-k141_38454 | 2109 | No | NA | 3 | 0 | 0 | Low-quality | Genome-fragment | 5.01 | AAI-based (high-confidence) | 0 |
| S17-final-viral-combined_k141_257015\|\|full | 3315 | No | NA | 4 | 0 | 3 | Not-determined | Genome-fragment | NA | NA | 0 |
| S4-final-viral-combined_k141_152701\|\|full | 2109 | No | NA | 3 | 0 | 3 | Low-quality | Genome-fragment | 0.98 | AAI-based (medium-confidence) | 0 |
| S17-final-viral-combined_k141_257034\|\|full | 1564 | No | NA | 3 | 0 | 1 | Not-determined | Genome-fragment | NA | NA | 0 |
| S17-final-viral-combined_k141_112729\|\|full | 4323 | No | NA | 4 | 0 | 2 | Not-determined | Genome-fragment | NA | NA | 0 |
| S8-final-viral-combined_k141_38151\|\|full | 2109 | No | NA | 6 | 2 | 0 | Low-quality | Genome-fragment | 4.87 | AAI-based (high-confidence) | 0 |
| S8_k141_266875 | 2109 | No | NA | 2 | 0 | 1 | Low-quality | Genome-fragment | 1.01 | AAI-based (high-confidence) | 0 |
| S17-final-viral-combined_k141_123979\|\|full | 3541 | No | NA | 8 | 0 | 0 | Not-determined | Genome-fragment | NA | NA | 0 |
| S9-final-viral-combined_k141_83865\|\|full | 2110 | No | NA | 3 | 1 | 0 | Low-quality | Genome-fragment | 4.61 | AAI-based (high-confidence) | 0 |
| S11-final-viral-combined_k141_246240\|\|lt2gene | 2110 | No | NA | 2 | 1 | 0 | Low-quality | Genome-fragment | 1.3 | HMM-based (lower-bound) | 0 |
| S17-final-viral-combined_k141_257072\|\|full | 1569 | No | NA | 3 | 0 | 1 | Not-determined | Genome-fragment | NA | NA | 0 |
| S17-final-viral-combined_k141_34804\|\|full | 1547 | No | NA | 2 | 0 | 2 | Not-determined | Genome-fragment | NA | NA | 0 |
| S17-final-viral-combined_k141_190365\|\|full | 3937 | No | NA | 2 | 0 | 2 | Not-determined | Genome-fragment | NA | NA | 0 |
| S17-final-viral-combined_k141_146050\|\|full | 40686 | No | NA | 37 | 1 | 13 | Medium-quality | Genome-fragment | 75.56 | AAI-based (medium-confidence) | 0 |
| S1_k141_105294 | 2110 | No | NA | 4 | 0 | 1 | Low-quality | Genome-fragment | 1.48 | AAI-based (high-confidence) | 0 |
| S17-final-viral-combined_k141_201868\|\|full | 1253 | No | NA | 2 | 0 | 1 | Not-determined | Genome-fragment | NA | NA | 0 |
| S17-final-viral-combined_k141_146063\|\|full | 1792 | No | NA | 2 | 0 | 0 | Not-determined | Genome-fragment | NA | NA | 0 |
| S17-final-viral-combined_k141_23465\|\|full | 2618 | No | NA | 5 | 0 | 1 | Not-determined | Genome-fragment | NA | NA | 0 |
| S5-final-viral-combined_k141_30515\|\|full | 2110 | No | NA | 6 | 0 | 0 | Low-quality | Genome-fragment | 4.21 | AAI-based (high-confidence) | 0 |
| S17-final-viral-combined_k141_201904\|\|full | 1641 | No | NA | 4 | 0 | 0 | Not-determined | Genome-fragment | NA | NA | 0 |
| S12-final-viral-combined_k141_308285\|\|full | 2110 | No | NA | 3 | 0 | 0 | Low-quality | Genome-fragment | 2.56 | AAI-based (high-confidence) | 0 |
| S17-final-viral-combined_k141_168111\|\|full | 1462 | No | NA | 3 | 0 | 1 | Not-determined | Genome-fragment | NA | NA | 0 |
| S17-final-viral-combined_k141_146109\|\|full | 5112 | No | NA | 5 | 0 | 3 | Not-determined | Genome-fragment | NA | NA | 0 |
| S17-final-viral-combined_k141_124011\|\|full | 1244 | No | NA | 2 | 0 | 1 | Not-determined | Genome-fragment | NA | NA | 0 |
| S12_k141_323156 | 2110 | No | NA | 5 | 0 | 1 | Low-quality | Genome-fragment | 4.36 | AAI-based (medium-confidence) | 0 |
| S17-final-viral-combined_k141_212884\|\|full | 1344 | No | NA | 3 | 0 | 1 | Not-determined | Genome-fragment | NA | NA | 0 |
| S17-final-viral-combined_k141_34835\|\|full | 1432 | No | NA | 3 | 0 | 1 | Not-determined | Genome-fragment | NA | NA | 0 |
| S17-final-viral-combined_k141_134846\|\|full | 1921 | No | NA | 3 | 0 | 1 | Not-determined | Genome-fragment | NA | NA | 0 |
| S11-final-viral-combined_k141_140481\|\|full | 2111 | No | NA | 7 | 0 | 0 | Low-quality | Genome-fragment | 5.39 | AAI-based (high-confidence) | 0 |
| S17-final-viral-combined_k141_134859\|\|full | 6462 | No | NA | 5 | 0 | 0 | Not-determined | Genome-fragment | NA | NA | 0 |
| S3-final-viral-combined_k141_36758\|\|full | 2111 | No | NA | 3 | 2 | 0 | Low-quality | Genome-fragment | 4.74 | AAI-based (high-confidence) | 0 |
| S3-final-viral-combined_k141_124501\|\|full | 2111 | No | NA | 3 | 0 | 1 | Low-quality | Genome-fragment | 2.45 | AAI-based (high-confidence) | 0 |
| S17-final-viral-combined_k141_257147\|\|full | 15407 | No | NA | 18 | 0 | 1 | Not-determined | Genome-fragment | NA | NA | 0 |
| S17-final-viral-combined_k141_12393\|\|full | 3631 | No | NA | 3 | 0 | 1 | Not-determined | Genome-fragment | NA | NA | 0 |
| S17-final-viral-combined_k141_90155\|\|full | 2337 | No | NA | 3 | 0 | 1 | Not-determined | Genome-fragment | NA | NA | 0 |
| S17-final-viral-combined_k141_179363\|\|full | 18354 | No | NA | 23 | 11 | 0 | Medium-quality | Genome-fragment | 52 | AAI-based (high-confidence) | 0 |
| S17-final-viral-combined_k141_68419\|\|full | 8205 | No | NA | 8 | 0 | 0 | Not-determined | Genome-fragment | NA | NA | 0 |
| S17-final-viral-combined_k141_201943\|\|full | 1732 | No | NA | 4 | 0 | 3 | Not-determined | Genome-fragment | NA | NA | 0 |
| S5-final-viral-combined_k141_58241\|\|full | 2111 | No | NA | 3 | 1 | 0 | Low-quality | Genome-fragment | 3.52 | AAI-based (high-confidence) | 0 |
| S17-final-viral-combined_k141_190496\|\|full | 6091 | No | NA | 5 | 0 | 3 | Not-determined | Genome-fragment | NA | NA | 0 |
| S17-final-viral-combined_k141_23545\|\|full | 1283 | No | NA | 3 | 0 | 1 | Not-determined | Genome-fragment | NA | NA | 0 |
| S7-final-viral-combined_k141_56688\|\|full | 2111 | No | NA | 4 | 0 | 0 | Low-quality | Genome-fragment | 4.46 | AAI-based (medium-confidence) | 0 |
| S17-final-viral-combined_k141_257222\|\|full | 4254 | No | NA | 5 | 0 | 3 | Not-determined | Genome-fragment | NA | NA | 0 |
| S17-final-viral-combined_k141_1617\|\|full | 3654 | No | NA | 3 | 0 | 3 | Not-determined | Genome-fragment | NA | NA | 0 |
| S17-final-viral-combined_k141_146149\|\|full | 4351 | No | NA | 3 | 0 | 2 | Not-determined | Genome-fragment | NA | NA | 0 |
| S17-final-viral-combined_k141_23558\|\|full | 1368 | No | NA | 3 | 0 | 1 | Not-determined | Genome-fragment | NA | NA | 0 |
| S7-final-viral-combined_k141_193198\|\|full | 2111 | No | NA | 3 | 0 | 0 | Low-quality | Genome-fragment | 1.19 | AAI-based (high-confidence) | 0 |
| S2-final-viral-combined_k141_56673\|\|full | 2111 | No | NA | 2 | 0 | 0 | Low-quality | Genome-fragment | 3.96 | AAI-based (medium-confidence) | 0 |
| S17-final-viral-combined_k141_45708\|\|full | 2219 | No | NA | 2 | 0 | 0 | Not-determined | Genome-fragment | NA | NA | 0 |
| S2_k141_101222 | 2111 | No | NA | 2 | 0 | 0 | Low-quality | Genome-fragment | 2.46 | AAI-based (high-confidence) | 0 |
| S4_k141_87211 | 2111 | No | NA | 2 | 0 | 2 | Low-quality | Genome-fragment | 0.94 | AAI-based (medium-confidence) | 0 |
| S17-final-viral-combined_k141_79902\|\|full | 2155 | No | NA | 4 | 0 | 1 | Not-determined | Genome-fragment | NA | NA | 0 |
| S17-final-viral-combined_k141_146178\|\|full | 2951 | No | NA | 3 | 0 | 1 | Not-determined | Genome-fragment | NA | NA | 0 |
| S17-final-viral-combined_k141_1662\|\|full | 3119 | No | NA | 2 | 0 | 2 | Not-determined | Genome-fragment | NA | NA | 0 |
| S17-final-viral-combined_k141_23592\|\|full | 3420 | No | NA | 2 | 0 | 1 | Not-determined | Genome-fragment | NA | NA | 0 |
| S17-final-viral-combined_k141_134960\|\|full | 2111 | No | NA | 6 | 0 | 0 | Not-determined | Genome-fragment | NA | NA | 0 |
| S6_k141_78961 | 2111 | No | NA | 1 | 0 | 1 | Low-quality | Genome-fragment | 0.51 | AAI-based (high-confidence) | 0 |
| S11-final-viral-combined_k141_112409\|\|full | 2112 | No | NA | 5 | 0 | 2 | Low-quality | Genome-fragment | 0.73 | AAI-based (medium-confidence) | 0 |
| S7-final-viral-combined_k141_109244\|\|full | 2112 | No | NA | 4 | 4 | 0 | Low-quality | Genome-fragment | 4.65 | AAI-based (high-confidence) | 0 |
| S17-final-viral-combined_k141_175705\|\|full | 2112 | No | NA | 3 | 1 | 0 | Low-quality | Genome-fragment | 3.06 | AAI-based (high-confidence) | 0 |
| S17-final-viral-combined_k141_202041\|\|full | 1347 | No | NA | 2 | 0 | 2 | Not-determined | Genome-fragment | NA | NA | 0 |
| S8-final-viral-combined_k141_88273\|\|full | 2112 | No | NA | 6 | 2 | 0 | Low-quality | Genome-fragment | 5.16 | AAI-based (medium-confidence) | 0 |
| S17-final-viral-combined_k141_124170\|\|full | 4469 | No | NA | 3 | 0 | 3 | Not-determined | Genome-fragment | NA | NA | 0 |
| S17-final-viral-combined_k141_101445\|\|full | 4428 | No | NA | 2 | 0 | 1 | Not-determined | Genome-fragment | NA | NA | 0 |
| S17-final-viral-combined_k141_168254\|\|full | 2779 | No | NA | 2 | 0 | 2 | Not-determined | Genome-fragment | NA | NA | 0 |
| S1-final-viral-combined_k141_102189\|\|full | 2113 | No | NA | 3 | 0 | 2 | Low-quality | Genome-fragment | 6.8 | AAI-based (high-confidence) | 0 |
| S7-final-viral-combined_k141_172799\|\|full | 2113 | No | NA | 3 | 1 | 0 | Low-quality | Genome-fragment | 5.57 | AAI-based (high-confidence) | 0 |
| S12_k141_350537 | 2113 | No | NA | 3 | 1 | 0 | Low-quality | Genome-fragment | 7.16 | HMM-based (lower-bound) | 0 |
| S6-final-viral-combined_k141_78739\|\|full | 2113 | No | NA | 3 | 0 | 1 | Low-quality | Genome-fragment | 3.73 | AAI-based (medium-confidence) | 0 |
| S17-final-viral-combined_k141_168283\|\|full | 7216 | No | NA | 9 | 0 | 0 | Not-determined | Genome-fragment | NA | NA | 0 |
| S17-final-viral-combined_k141_23688\|\|full | 2034 | No | NA | 5 | 0 | 1 | Not-determined | Genome-fragment | NA | NA | 0 |
| S17-final-viral-combined_k141_213008\|\|full | 2297 | No | NA | 3 | 0 | 3 | Not-determined | Genome-fragment | NA | NA | 0 |
| S5-final-viral-combined_k141_176277\|\|full | 2114 | No | NA | 4 | 1 | 0 | Low-quality | Genome-fragment | 3.38 | AAI-based (medium-confidence) | 0 |
| S17-final-viral-combined_k141_222984\|\|full | 2114 | No | NA | 3 | 2 | 0 | Low-quality | Genome-fragment | 4.42 | AAI-based (medium-confidence) | 0 |
| S6_k141_18168 | 2114 | No | NA | 3 | 0 | 0 | Low-quality | Genome-fragment | 2.23 | AAI-based (high-confidence) | 0 |
| S17-final-viral-combined_k141_113066\|\|full | 1201 | No | NA | 3 | 0 | 0 | Not-determined | Genome-fragment | NA | NA | 0 |
| S17-final-viral-combined_k141_124232\|\|full | 2310 | No | NA | 3 | 0 | 2 | Not-determined | Genome-fragment | NA | NA | 0 |
| S17-final-viral-combined_k141_190683\|\|full | 5531 | No | NA | 5 | 0 | 3 | Not-determined | Genome-fragment | NA | NA | 0 |
| S17-final-viral-combined_k141_190694\|\|full | 17546 | No | NA | 19 | 0 | 12 | Not-determined | Genome-fragment | NA | NA | 0 |
| S17-final-viral-combined_k141_213065\|\|full | 2002 | No | NA | 3 | 0 | 3 | Not-determined | Genome-fragment | NA | NA | 0 |
| S17-final-viral-combined_k141_179572\|\|full | 2875 | No | NA | 4 | 0 | 0 | Not-determined | Genome-fragment | NA | NA | 0 |
| S17-final-viral-combined_k141_224379\|\|full | 1764 | No | NA | 2 | 0 | 2 | Not-determined | Genome-fragment | NA | NA | 0 |
| S17-final-viral-combined_k141_101525\|\|full | 1191 | No | NA | 2 | 0 | 0 | Not-determined | Genome-fragment | NA | NA | 0 |
| S8-final-viral-combined_k141_232953\|\|full | 2114 | No | NA | 5 | 3 | 0 | Low-quality | Genome-fragment | 7.33 | AAI-based (high-confidence) | 0 |
| S17-final-viral-combined_k141_45772\|\|full | 3490 | No | NA | 3 | 0 | 2 | Not-determined | Genome-fragment | NA | NA | 0 |
| S12_k141_398970 | 2115 | No | NA | 2 | 0 | 1 | Low-quality | Genome-fragment | 6.75 | AAI-based (high-confidence) | 0 |
| S17-final-viral-combined_k141_257399\|\|full | 2917 | No | NA | 4 | 0 | 1 | Not-determined | Genome-fragment | NA | NA | 0 |
| S17-final-viral-combined_k141_245912\|\|full | 3582 | No | NA | 4 | 0 | 0 | Not-determined | Genome-fragment | NA | NA | 0 |
| S17-final-viral-combined_k141_79987\|\|full | 1794 | No | NA | 2 | 0 | 2 | Not-determined | Genome-fragment | NA | NA | 0 |
| S17-final-viral-combined_k141_90378\|\|full | 36737 | No | NA | 51 | 23 | 1 | High-quality | High-quality | 100 | AAI-based (high-confidence) | 0 |
| S2-final-viral-combined_k141_184792\|\|full | 2115 | No | NA | 3 | 0 | 0 | Low-quality | Genome-fragment | 5.31 | AAI-based (high-confidence) | 0 |
| S2-final-viral-combined_k141_124048\|\|full | 2115 | No | NA | 3 | 0 | 1 | Low-quality | Genome-fragment | 4.16 | AAI-based (high-confidence) | 0 |
| S17-final-viral-combined_k141_124294\|\|full | 1175 | No | NA | 2 | 0 | 0 | Not-determined | Genome-fragment | NA | NA | 0 |
| S17-final-viral-combined_k141_245938\|\|full | 1233 | No | NA | 3 | 0 | 1 | Not-determined | Genome-fragment | NA | NA | 0 |
| S17-final-viral-combined_k141_79998\|\|full | 2212 | No | NA | 3 | 0 | 2 | Not-determined | Genome-fragment | NA | NA | 0 |
| S17-final-viral-combined_k141_68638\|\|full | 3408 | No | NA | 5 | 0 | 1 | Not-determined | Genome-fragment | NA | NA | 0 |
| S8-final-viral-combined_k141_180221\|\|full | 2115 | No | NA | 2 | 0 | 1 | Low-quality | Genome-fragment | 3.28 | AAI-based (medium-confidence) | 0 |
| S11-final-viral-combined_k141_115978\|\|full | 2116 | No | NA | 3 | 0 | 1 | Low-quality | Genome-fragment | 5.74 | AAI-based (high-confidence) | 0 |
| S17-final-viral-combined_k141_113156\|\|full | 1001 | No | NA | 3 | 0 | 1 | Not-determined | Genome-fragment | NA | NA | 0 |
| S17-final-viral-combined_k141_68679\|\|full | 1855 | No | NA | 4 | 0 | 2 | Not-determined | Genome-fragment | NA | NA | 0 |
| S17-final-viral-combined_k141_190805\|\|full | 3717 | No | NA | 6 | 0 | 3 | Not-determined | Genome-fragment | NA | NA | 0 |
| S17-final-viral-combined_k141_35134\|\|full | 1462 | No | NA | 3 | 0 | 1 | Not-determined | Genome-fragment | NA | NA | 0 |
| S5_k141_122895 | 2116 | Yes | 1435 | 3 | 1 | 1 | Low-quality | Genome-fragment | 12.18 | AAI-based (high-confidence) | 32.18 |
| S17-final-viral-combined_k141_57290\|\|full | 1069 | No | NA | 2 | 0 | 1 | Not-determined | Genome-fragment | NA | NA | 0 |
| S17-final-viral-combined_k141_35142\|\|full | 1074 | No | NA | 2 | 0 | 1 | Not-determined | Genome-fragment | NA | NA | 0 |
| S12-final-viral-combined_k141_21513\|\|full | 2116 | No | NA | 3 | 0 | 2 | Low-quality | Genome-fragment | 1.05 | AAI-based (medium-confidence) | 0 |
| S17-final-viral-combined_k141_1818\|\|full | 2113 | No | NA | 4 | 0 | 1 | Not-determined | Genome-fragment | NA | NA | 0 |
| S17-final-viral-combined_k141_224402\|\|full | 1623 | No | NA | 4 | 0 | 0 | Not-determined | Genome-fragment | NA | NA | 0 |
| S17-final-viral-combined_k141_213194\|\|full | 2230 | No | NA | 4 | 0 | 4 | Not-determined | Genome-fragment | NA | NA | 0 |
| S12-final-viral-combined_k141_260673\|\|full | 2116 | No | NA | 2 | 0 | 1 | Low-quality | Genome-fragment | 1.85 | AAI-based (medium-confidence) | 0 |
| S4_k141_156013 | 2116 | No | NA | 3 | 0 | 2 | Low-quality | Genome-fragment | 0.7 | AAI-based (medium-confidence) | 0 |
| S8-final-viral-combined_k141_236296\|\|full | 2116 | No | NA | 3 | 0 | 0 | Low-quality | Genome-fragment | 3.49 | AAI-based (medium-confidence) | 0 |
| S17-final-viral-combined_k141_213239\|\|full | 2153 | No | NA | 3 | 0 | 1 | Not-determined | Genome-fragment | NA | NA | 0 |
| S8-final-viral-combined_k141_259209\|\|full | 2116 | No | NA | 2 | 0 | 0 | Low-quality | Genome-fragment | 2.43 | AAI-based (medium-confidence) | 0 |
| S17-final-viral-combined_k141_113234\|\|full | 1830 | No | NA | 3 | 0 | 1 | Not-determined | Genome-fragment | NA | NA | 0 |
| S17-final-viral-combined_k141_135279\|\|full | 1012 | No | NA | 4 | 0 | 0 | Not-determined | Genome-fragment | NA | NA | 0 |
| S17-final-viral-combined_k141_80068\|\|full | 1263 | No | NA | 2 | 0 | 2 | Not-determined | Genome-fragment | NA | NA | 0 |
| S17-final-viral-combined_k141_45857\|\|full | 1456 | No | NA | 2 | 0 | 2 | Not-determined | Genome-fragment | NA | NA | 0 |
| S1-final-viral-combined_k141_370367\|\|full | 2117 | Yes | 786 | 2 | 1 | 1 | Low-quality | Genome-fragment | 18.89 | HMM-based (lower-bound) | 62.87 |
| S17-final-viral-combined_k141_168523\|\|full | 1442 | No | NA | 3 | 0 | 2 | Not-determined | Genome-fragment | NA | NA | 0 |
| S3_k141_38893 | 2117 | No | NA | 3 | 0 | 0 | Low-quality | Genome-fragment | 3.8 | AAI-based (medium-confidence) | 0 |
| S17-final-viral-combined_k141_80069\|\|full | 15430 | No | NA | 14 | 0 | 9 | Not-determined | Genome-fragment | NA | NA | 0 |
| S17-final-viral-combined_k141_23836\|\|full | 12888 | No | NA | 16 | 0 | 10 | Not-determined | Genome-fragment | NA | NA | 0 |
| S17-final-viral-combined_k141_221245\|\|full | 2117 | Yes | 1096 | 3 | 1 | 1 | Low-quality | Genome-fragment | 2.7 | AAI-based (high-confidence) | 48.23 |
| S17-final-viral-combined_k141_235013\|\|full | 2151 | No | NA | 3 | 0 | 0 | Not-determined | Genome-fragment | NA | NA | 0 |
| S17-final-viral-combined_k141_213265\|\|full | 1419 | No | NA | 3 | 0 | 1 | Not-determined | Genome-fragment | NA | NA | 0 |
| S17-final-viral-combined_k141_156231\|\|0_partial | 2117 | No | NA | 3 | 1 | 0 | Low-quality | Genome-fragment | 49.29 | AAI-based (medium-confidence) | 0 |
| S17-final-viral-combined_k141_168556\|\|full | 2241 | No | NA | 3 | 0 | 3 | Not-determined | Genome-fragment | NA | NA | 0 |
| S12-final-viral-combined_k141_383671\|\|full | 2117 | No | NA | 2 | 0 | 2 | Low-quality | Genome-fragment | 1.44 | AAI-based (high-confidence) | 0 |
| S2-final-viral-combined_k141_159079\|\|full | 2117 | No | NA | 4 | 0 | 3 | Low-quality | Genome-fragment | 0.67 | AAI-based (medium-confidence) | 0 |
| S1-final-viral-combined_k141_99766\|\|full | 2118 | No | NA | 4 | 0 | 0 | Low-quality | Genome-fragment | 3.71 | AAI-based (medium-confidence) | 0 |
| S17-final-viral-combined_k141_135315\|\|full | 1982 | No | NA | 4 | 0 | 1 | Not-determined | Genome-fragment | NA | NA | 0 |
| S5-final-viral-combined_k141_194723\|\|full | 2118 | No | NA | 4 | 0 | 0 | Low-quality | Genome-fragment | 4.94 | AAI-based (medium-confidence) | 0 |
| S17-final-viral-combined_k141_23864\|\|full | 10356 | No | NA | 11 | 0 | 7 | Not-determined | Genome-fragment | NA | NA | 0 |
| S17-final-viral-combined_k141_90496\|\|full | 7942 | No | NA | 9 | 0 | 9 | Not-determined | Genome-fragment | NA | NA | 0 |
| S17-final-viral-combined_k141_80127\|\|full | 2209 | No | NA | 4 | 0 | 4 | Not-determined | Genome-fragment | NA | NA | 0 |
| S17-final-viral-combined_k141_113311\|\|full | 3074 | No | NA | 4 | 0 | 1 | Not-determined | Genome-fragment | NA | NA | 0 |
| S17-final-viral-combined_k141_1950\|\|full | 2840 | No | NA | 3 | 0 | 2 | Not-determined | Genome-fragment | NA | NA | 0 |
| S5_k141_149127 | 2118 | No | NA | 5 | 4 | 0 | Low-quality | Genome-fragment | 5.25 | AAI-based (high-confidence) | 0 |
| S7-final-viral-combined_k141_41868\|\|full | 2118 | No | NA | 4 | 0 | 1 | Low-quality | Genome-fragment | 1.62 | AAI-based (high-confidence) | 0 |
| S17-final-viral-combined_k141_45893\|\|full | 9258 | No | NA | 12 | 0 | 1 | Not-determined | Genome-fragment | NA | NA | 0 |
| S17-final-viral-combined_k141_202424\|\|full | 1218 | No | NA | 2 | 0 | 2 | Not-determined | Genome-fragment | NA | NA | 0 |
| S7_k141_76682 | 2118 | No | NA | 1 | 0 | 1 | Low-quality | Genome-fragment | 0.91 | AAI-based (medium-confidence) | 0 |
| S17-final-viral-combined_k141_45898\|\|full | 2138 | No | NA | 2 | 0 | 1 | Not-determined | Genome-fragment | NA | NA | 0 |
| S17-final-viral-combined_k141_235038\|\|full | 2100 | No | NA | 4 | 0 | 3 | Not-determined | Genome-fragment | NA | NA | 0 |
| S17-final-viral-combined_k141_101703\|\|full | 1977 | No | NA | 4 | 0 | 1 | Not-determined | Genome-fragment | NA | NA | 0 |
| S17-final-viral-combined_k141_129795\|\|full | 2118 | No | NA | 6 | 1 | 0 | Low-quality | Genome-fragment | 3.92 | AAI-based (high-confidence) | 0 |
| S17-final-viral-combined_k141_90547\|\|full | 1452 | No | NA | 2 | 0 | 1 | Not-determined | Genome-fragment | NA | NA | 0 |
| S11-final-viral-combined_k141_245271\|\|full | 2119 | No | NA | 3 | 0 | 0 | Low-quality | Genome-fragment | 2.37 | AAI-based (medium-confidence) | 0 |
| S17-final-viral-combined_k141_246020\|\|full | 7577 | No | NA | 9 | 0 | 5 | Not-determined | Genome-fragment | NA | NA | 0 |
| S11_k141_33211 | 2119 | No | NA | 1 | 0 | 0 | Low-quality | Genome-fragment | 1.14 | AAI-based (high-confidence) | 0 |
| S17-final-viral-combined_k141_235056\|\|full | 1762 | No | NA | 2 | 0 | 1 | Not-determined | Genome-fragment | NA | NA | 0 |
| S2_k141_1459 | 2119 | No | NA | 3 | 2 | 0 | Low-quality | Genome-fragment | 4.67 | AAI-based (high-confidence) | 0 |
| S4_k141_109016 | 2119 | No | NA | 4 | 0 | 1 | Low-quality | Genome-fragment | 4.43 | AAI-based (medium-confidence) | 0 |
| S11-final-viral-combined_k141_62092\|\|full | 2120 | No | NA | 4 | 3 | 0 | Low-quality | Genome-fragment | 4.77 | AAI-based (medium-confidence) | 0 |
| S11-final-viral-combined_k141_260911\|\|full | 2120 | No | NA | 4 | 2 | 0 | Low-quality | Genome-fragment | 4.61 | AAI-based (high-confidence) | 0 |
| S17-final-viral-combined_k141_224499\|\|full | 1844 | No | NA | 5 | 0 | 0 | Not-determined | Genome-fragment | NA | NA | 0 |
| S17-final-viral-combined_k141_35285\|\|full | 1316 | No | NA | 2 | 0 | 1 | Not-determined | Genome-fragment | NA | NA | 0 |
| S1-final-viral-combined_k141_170761\|\|full | 2120 | No | NA | 5 | 0 | 0 | Low-quality | Genome-fragment | 3.55 | AAI-based (high-confidence) | 0 |
| S17-final-viral-combined_k141_101766\|\|full | 1650 | No | NA | 2 | 0 | 1 | Not-determined | Genome-fragment | NA | NA | 0 |
| S17-final-viral-combined_k141_246070\|\|full | 2262 | No | NA | 4 | 0 | 0 | Not-determined | Genome-fragment | NA | NA | 0 |
| S17-final-viral-combined_k141_179771\|\|full | 43075 | No | NA | 49 | 3 | 3 | High-quality | High-quality | 97.43 | AAI-based (medium-confidence) | 0 |
| S1_k141_40170 | 2120 | No | NA | 3 | 1 | 0 | Low-quality | Genome-fragment | 1.6 | AAI-based (high-confidence) | 0 |
| S17-final-viral-combined_k141_235110\|\|full | 1601 | No | NA | 2 | 0 | 0 | Not-determined | Genome-fragment | NA | NA | 0 |
| S3_k141_16120 | 2120 | No | NA | 6 | 2 | 0 | Low-quality | Genome-fragment | 3.56 | AAI-based (high-confidence) | 0 |
| S12-final-viral-combined_k141_268267\|\|full | 2120 | No | NA | 2 | 0 | 1 | Low-quality | Genome-fragment | 0.95 | AAI-based (medium-confidence) | 0 |
| S6-final-viral-combined_k141_107981\|\|full | 2120 | No | NA | 3 | 0 | 0 | Low-quality | Genome-fragment | 6.5 | AAI-based (high-confidence) | 0 |
| S17-final-viral-combined_k141_2052\|\|full | 1274 | No | NA | 3 | 0 | 1 | Not-determined | Genome-fragment | NA | NA | 0 |
| S8-final-viral-combined_k141_12305\|\|full | 2120 | No | NA | 4 | 1 | 0 | Low-quality | Genome-fragment | 1.37 | HMM-based (lower-bound) | 0 |
| S17-final-viral-combined_k141_157338\|\|full | 1637 | No | NA | 2 | 0 | 1 | Not-determined | Genome-fragment | NA | NA | 0 |
| S8_k141_94469 | 2120 | No | NA | 3 | 1 | 0 | Low-quality | Genome-fragment | 4.66 | AAI-based (high-confidence) | 0 |
| S17-final-viral-combined_k141_57443\|\|full | 2223 | No | NA | 3 | 0 | 3 | Not-determined | Genome-fragment | NA | NA | 0 |
| S1-final-viral-combined_k141_126891\|\|full | 2121 | No | NA | 2 | 0 | 1 | Low-quality | Genome-fragment | 3.56 | AAI-based (high-confidence) | 0 |
| S17-final-viral-combined_k141_90608\|\|full | 1242 | No | NA | 3 | 0 | 2 | Not-determined | Genome-fragment | NA | NA | 0 |
| S7-final-viral-combined_k141_114199\|\|full | 2121 | No | NA | 6 | 1 | 0 | Low-quality | Genome-fragment | 3.28 | AAI-based (high-confidence) | 0 |
| S7-final-viral-combined_k141_89120\|\|full | 2121 | No | NA | 2 | 0 | 2 | Low-quality | Genome-fragment | 0.6 | AAI-based (medium-confidence) | 0 |
| S17-final-viral-combined_k141_257608\|\|full | 3491 | No | NA | 3 | 0 | 2 | Not-determined | Genome-fragment | NA | NA | 0 |
| S17-final-viral-combined_k141_191041\|\|full | 3907 | No | NA | 7 | 0 | 0 | Not-determined | Genome-fragment | NA | NA | 0 |
| S6_k141_13681 | 2121 | No | NA | 3 | 0 | 1 | Low-quality | Genome-fragment | 3.6 | AAI-based (high-confidence) | 0 |
| S17-final-viral-combined_k141_235182\|\|full | 3274 | No | NA | 3 | 0 | 0 | Not-determined | Genome-fragment | NA | NA | 0 |
| S9-final-viral-combined_k141_12576\|\|full | 2122 | No | NA | 1 | 0 | 0 | Low-quality | Genome-fragment | 4.66 | AAI-based (medium-confidence) | 0 |
| S17-final-viral-combined_k141_202572\|\|full | 3606 | No | NA | 5 | 0 | 1 | Not-determined | Genome-fragment | NA | NA | 0 |
| S17-final-viral-combined_k141_146535\|\|full | 2255 | No | NA | 4 | 0 | 2 | Not-determined | Genome-fragment | NA | NA | 0 |
| S17-final-viral-combined_k141_113523\|\|full | 6930 | No | NA | 4 | 0 | 3 | Not-determined | Genome-fragment | NA | NA | 0 |
| S11-final-viral-combined_k141_186710\|\|full | 2122 | No | NA | 3 | 1 | 0 | Low-quality | Genome-fragment | 4.96 | AAI-based (high-confidence) | 0 |
| S17-final-viral-combined_k141_191076\|\|full | 1926 | No | NA | 3 | 0 | 2 | Not-determined | Genome-fragment | NA | NA | 0 |
| S17-final-viral-combined_k141_168787\|\|full | 1063 | No | NA | 2 | 0 | 2 | Not-determined | Genome-fragment | NA | NA | 0 |
| S11_k141_133246 | 2122 | No | NA | 2 | 0 | 1 | Low-quality | Genome-fragment | 5 | AAI-based (high-confidence) | 0 |
| S1_k141_78182 | 2122 | No | NA | 4 | 0 | 4 | Low-quality | Genome-fragment | 1.39 | AAI-based (high-confidence) | 0 |
| S17-final-viral-combined_k141_179882\|\|full | 3572 | No | NA | 3 | 0 | 2 | Not-determined | Genome-fragment | NA | NA | 0 |
| S17-final-viral-combined_k141_146590\|\|full | 2238 | No | NA | 3 | 0 | 3 | Not-determined | Genome-fragment | NA | NA | 0 |
| S17-final-viral-combined_k141_135571\|\|full | 1478 | No | NA | 2 | 0 | 2 | Not-determined | Genome-fragment | NA | NA | 0 |
| S17-final-viral-combined_k141_12641\|\|full | 4859 | No | NA | 4 | 0 | 2 | Not-determined | Genome-fragment | NA | NA | 0 |
| S17-final-viral-combined_k141_57534\|\|full | 1786 | No | NA | 3 | 0 | 0 | Not-determined | Genome-fragment | NA | NA | 0 |
| S17-final-viral-combined_k141_2127\|\|full | 2119 | No | NA | 3 | 0 | 2 | Not-determined | Genome-fragment | NA | NA | 0 |
| S17-final-viral-combined_k141_146600\|\|full | 1999 | No | NA | 3 | 0 | 0 | Not-determined | Genome-fragment | NA | NA | 0 |
| S17-final-viral-combined_k141_68957\|\|full | 1044 | No | NA | 3 | 0 | 0 | Not-determined | Genome-fragment | NA | NA | 0 |
| S17-final-viral-combined_k141_99141\|\|full | 2122 | No | NA | 6 | 1 | 0 | Low-quality | Genome-fragment | 5.54 | AAI-based (medium-confidence) | 0 |
| S17-final-viral-combined_k141_90644\|\|full | 1203 | No | NA | 1 | 0 | 0 | Not-determined | Genome-fragment | NA | NA | 0 |
| S17-final-viral-combined_k141_146613\|\|full | 1508 | No | NA | 3 | 0 | 1 | Not-determined | Genome-fragment | NA | NA | 0 |
| S17-final-viral-combined_k141_246140\|\|full | 2169 | No | NA | 4 | 0 | 0 | Not-determined | Genome-fragment | NA | NA | 0 |
| S12-final-viral-combined_k141_96738\|\|full | 2122 | No | NA | 3 | 0 | 2 | Low-quality | Genome-fragment | 1.3 | AAI-based (medium-confidence) | 0 |
| S17-final-viral-combined_k141_57559\|\|full | 1611 | No | NA | 3 | 0 | 1 | Not-determined | Genome-fragment | NA | NA | 0 |
| S17-final-viral-combined_k141_179911\|\|full | 2848 | No | NA | 4 | 0 | 3 | Not-determined | Genome-fragment | NA | NA | 0 |
| S17-final-viral-combined_k141_35402\|\|full | 1508 | No | NA | 2 | 0 | 1 | Not-determined | Genome-fragment | NA | NA | 0 |
| S7-final-viral-combined_k141_231742\|\|full | 2123 | No | NA | 4 | 0 | 0 | Low-quality | Genome-fragment | 2.65 | AAI-based (high-confidence) | 0 |
| S17-final-viral-combined_k141_191124\|\|full | 2500 | No | NA | 3 | 0 | 1 | Not-determined | Genome-fragment | NA | NA | 0 |
| S17-final-viral-combined_k141_101837\|\|full | 2102 | No | NA | 3 | 0 | 2 | Not-determined | Genome-fragment | NA | NA | 0 |
| S17-final-viral-combined_k141_146406\|\|full | 2123 | No | NA | 3 | 0 | 1 | Low-quality | Genome-fragment | 5.15 | AAI-based (high-confidence) | 0 |
| S12_k141_170157 | 2123 | Yes | 1631 | 4 | 2 | 1 | Low-quality | Genome-fragment | 2 | HMM-based (lower-bound) | 23.17 |
| S9-final-viral-combined_k141_62531\|\|full | 2125 | No | NA | 4 | 0 | 0 | Low-quality | Genome-fragment | 4.01 | AAI-based (medium-confidence) | 0 |
| S2-final-viral-combined_k141_196152\|\|full | 2125 | No | NA | 3 | 0 | 0 | Low-quality | Genome-fragment | 6 | AAI-based (high-confidence) | 0 |
| S4-final-viral-combined_k141_69820\|\|full | 2125 | No | NA | 4 | 0 | 0 | Low-quality | Genome-fragment | 1.18 | AAI-based (high-confidence) | 0 |
| S4-final-viral-combined_k141_138521\|\|full | 2125 | No | NA | 3 | 0 | 0 | Low-quality | Genome-fragment | 3.83 | AAI-based (high-confidence) | 0 |
| S17-final-viral-combined_k141_124531\|\|full | 3337 | No | NA | 3 | 0 | 3 | Not-determined | Genome-fragment | NA | NA | 0 |
| S17-final-viral-combined_k141_168821\|\|full | 6042 | No | NA | 8 | 0 | 6 | Not-determined | Genome-fragment | NA | NA | 0 |
| S8-final-viral-combined_k141_20362\|\|full | 2125 | Yes | 1597 | 5 | 1 | 1 | Low-quality | Genome-fragment | 4.25 | AAI-based (high-confidence) | 24.85 |
| S11-final-viral-combined_k141_204467\|\|full | 2126 | No | NA | 2 | 0 | 0 | Low-quality | Genome-fragment | 1.01 | AAI-based (medium-confidence) | 0 |
| S7-final-viral-combined_k141_198635\|\|full | 2126 | No | NA | 3 | 1 | 0 | Low-quality | Genome-fragment | 2.46 | HMM-based (lower-bound) | 0 |
| S17-final-viral-combined_k141_146671\|\|full | 3605 | No | NA | 4 | 0 | 0 | Not-determined | Genome-fragment | NA | NA | 0 |
| S7-final-viral-combined_k141_244624\|\|full | 2126 | No | NA | 4 | 2 | 0 | Low-quality | Genome-fragment | 4.5 | AAI-based (medium-confidence) | 0 |
| S17-final-viral-combined_k141_12708\|\|full | 1264 | No | NA | 2 | 0 | 1 | Not-determined | Genome-fragment | NA | NA | 0 |
| S12-final-viral-combined_k141_312952\|\|full | 2126 | No | NA | 2 | 0 | 2 | Low-quality | Genome-fragment | 1.71 | AAI-based (high-confidence) | 0 |
| S17-final-viral-combined_k141_213582\|\|full | 4240 | No | NA | 2 | 0 | 2 | Not-determined | Genome-fragment | NA | NA | 0 |
| S12_k141_218282 | 2127 | No | NA | 6 | 0 | 0 | Low-quality | Genome-fragment | 3.33 | AAI-based (medium-confidence) | 0 |
| S11_k141_326684 | 2128 | No | NA | 4 | 3 | 0 | Low-quality | Genome-fragment | 5.26 | AAI-based (high-confidence) | 0 |
| S17-final-viral-combined_k141_257688\|\|full | 2311 | No | NA | 4 | 0 | 1 | Not-determined | Genome-fragment | NA | NA | 0 |
| S17-final-viral-combined_k141_2164\|\|full | 1389 | No | NA | 3 | 0 | 1 | Not-determined | Genome-fragment | NA | NA | 0 |
| S7-final-viral-combined_k141_39707\|\|full | 2128 | No | NA | 3 | 0 | 2 | Low-quality | Genome-fragment | 4.05 | AAI-based (high-confidence) | 0 |
| S17-final-viral-combined_k141_90698\|\|full | 1627 | No | NA | 3 | 0 | 1 | Not-determined | Genome-fragment | NA | NA | 0 |
| S12-final-viral-combined_k141_289567\|\|full | 2128 | No | NA | 5 | 1 | 0 | Low-quality | Genome-fragment | 4.75 | AAI-based (medium-confidence) | 0 |
| S12-final-viral-combined_k141_116163\|\|full | 2128 | No | NA | 3 | 0 | 2 | Low-quality | Genome-fragment | 0.95 | AAI-based (medium-confidence) | 0 |
| S17-final-viral-combined_k141_124611\|\|full | 3534 | No | NA | 3 | 0 | 2 | Not-determined | Genome-fragment | NA | NA | 0 |
| S2_k141_6433 | 2128 | No | NA | 1 | 0 | 1 | Low-quality | Genome-fragment | 2.05 | AAI-based (high-confidence) | 0 |
| S17-final-viral-combined_k141_157501\|\|full | 2951 | No | NA | 3 | 0 | 2 | Not-determined | Genome-fragment | NA | NA | 0 |
| S11-final-viral-combined_k141_57518\|\|full | 2129 | No | NA | 3 | 1 | 0 | Low-quality | Genome-fragment | 2.79 | HMM-based (lower-bound) | 0 |

| | | | | | | | | | | | |
|---|---|---|---|---|---|---|---|---|---|---|---|
| S12-final-viral-combined_k141_124560\|\|full | 2129 | No | NA | 7 | 1 | 0 | Low-quality | Genome-fragment | 3.47 | AAI-based (high-confidence) | 0 |
| S12-final-viral-combined_k141_261145\|\|full | 2129 | No | NA | 5 | 1 | 0 | Low-quality | Genome-fragment | 3.61 | AAI-based (high-confidence) | 0 |
| S17-final-viral-combined_k141_146700\|\|full | 3380 | No | NA | 5 | 0 | 0 | Not-determined | Genome-fragment | NA | NA | 0 |
| S17-final-viral-combined_k141_101903\|\|full | 2754 | No | NA | 2 | 0 | 2 | Not-determined | Genome-fragment | NA | NA | 0 |
| S17-final-viral-combined_k141_24068\|\|full | 3149 | No | NA | 6 | 0 | 3 | Not-determined | Genome-fragment | NA | NA | 0 |
| S17-final-viral-combined_k141_146711\|\|full | 3108 | No | NA | 4 | 0 | 2 | Not-determined | Genome-fragment | NA | NA | 0 |
| S17-final-viral-combined_k141_24079\|\|full | 2092 | No | NA | 4 | 0 | 2 | Not-determined | Genome-fragment | NA | NA | 0 |
| S2-final-viral-combined_k141_34516\|\|full | 2129 | No | NA | 3 | 0 | 0 | Low-quality | Genome-fragment | 2.72 | AAI-based (medium-confidence) | 0 |
| S17-final-viral-combined_k141_191263\|\|full | 4446 | No | NA | 7 | 0 | 4 | Not-determined | Genome-fragment | NA | NA | 0 |
| S17-final-viral-combined_k141_168877\|\|full | 4035 | No | NA | 3 | 0 | 3 | Not-determined | Genome-fragment | NA | NA | 0 |
| S2-final-viral-combined_k141_225923\|\|full | 2129 | No | NA | 5 | 0 | 1 | Low-quality | Genome-fragment | 3.86 | AAI-based (medium-confidence) | 0 |
| S17-final-viral-combined_k141_257743\|\|full | 3753 | No | NA | 2 | 0 | 0 | Not-determined | Genome-fragment | NA | NA | 0 |
| S12-final-viral-combined_k141_379751\|\|full | 2130 | No | NA | 3 | 0 | 0 | Low-quality | Genome-fragment | 2.52 | AAI-based (high-confidence) | 0 |
| S11-final-viral-combined_k141_223325\|\|full | 2131 | No | NA | 3 | 0 | 1 | Low-quality | Genome-fragment | 1.68 | AAI-based (medium-confidence) | 0 |
| S1-final-viral-combined_k141_389008\|\|lt2gene | 2131 | No | NA | 2 | 0 | 0 | Low-quality | Genome-fragment | 2.73 | AAI-based (high-confidence) | 0 |
| S17-final-viral-combined_k141_213646\|\|full | 2575 | No | NA | 4 | 0 | 2 | Not-determined | Genome-fragment | NA | NA | 0 |
| S17-final-viral-combined_k141_113744\|\|full | 1151 | No | NA | 3 | 0 | 1 | Not-determined | Genome-fragment | NA | NA | 0 |
| S17-final-viral-combined_k141_202790\|\|full | 2131 | No | NA | 3 | 1 | 0 | Low-quality | Genome-fragment | 1.56 | HMM-based (lower-bound) | 0 |
| S17-final-viral-combined_k141_35555\|\|full | 2051 | No | NA | 4 | 0 | 1 | Not-determined | Genome-fragment | NA | NA | 0 |
| S8-final-viral-combined_k141_205317\|\|full | 2131 | No | NA | 6 | 0 | 0 | Low-quality | Genome-fragment | 5.31 | AAI-based (high-confidence) | 0 |
| S17-final-viral-combined_k141_202741\|\|full | 1395 | No | NA | 3 | 0 | 2 | Not-determined | Genome-fragment | NA | NA | 0 |
| S1-final-viral-combined_k141_97495\|\|full | 2132 | No | NA | 5 | 2 | 0 | Low-quality | Genome-fragment | 4.5 | AAI-based (high-confidence) | 0 |
| S17-final-viral-combined_k141_135731\|\|full | 1249 | No | NA | 3 | 0 | 0 | Not-determined | Genome-fragment | NA | NA | 0 |
| S17-final-viral-combined_k141_46108\|\|full | 1139 | No | NA | 3 | 0 | 0 | Not-determined | Genome-fragment | NA | NA | 0 |
| S1-final-viral-combined_k141_163945\|\|full | 2132 | No | NA | 5 | 1 | 0 | Low-quality | Genome-fragment | 4.75 | AAI-based (high-confidence) | 0 |
| S17-final-viral-combined_k141_46115\|\|full | 1071 | No | NA | 3 | 0 | 2 | Not-determined | Genome-fragment | NA | NA | 0 |
| S17-final-viral-combined_k141_213668\|\|full | 1337 | No | NA | 3 | 0 | 1 | Not-determined | Genome-fragment | NA | NA | 0 |
| S17-final-viral-combined_k141_235374\|\|full | 3644 | No | NA | 3 | 0 | 2 | Not-determined | Genome-fragment | NA | NA | 0 |
| S17-final-viral-combined_k141_157526\|\|full | 1999 | No | NA | 2 | 0 | 2 | Not-determined | Genome-fragment | NA | NA | 0 |
| S17-final-viral-combined_k141_257777\|\|full | 1509 | No | NA | 4 | 0 | 1 | Not-determined | Genome-fragment | NA | NA | 0 |
| S17-final-viral-combined_k141_235381\|\|full | 2769 | No | NA | 3 | 0 | 1 | Not-determined | Genome-fragment | NA | NA | 0 |
| S17-final-viral-combined_k141_213677\|\|full | 1491 | No | NA | 3 | 0 | 2 | Not-determined | Genome-fragment | NA | NA | 0 |
| S17-final-viral-combined_k141_146753\|\|full | 1228 | No | NA | 3 | 0 | 3 | Not-determined | Genome-fragment | NA | NA | 0 |
| S17-final-viral-combined_k141_202770\|\|full | 2684 | No | NA | 3 | 0 | 0 | Not-determined | Genome-fragment | NA | NA | 0 |
| S17-final-viral-combined_k141_27220\|\|full | 2132 | No | NA | 5 | 0 | 0 | Low-quality | Genome-fragment | 3.07 | AAI-based (medium-confidence) | 0 |
| S17-final-viral-combined_k141_124746\|\|full | 2603 | No | NA | 4 | 0 | 1 | Not-determined | Genome-fragment | NA | NA | 0 |
| S17-final-viral-combined_k141_135767\|\|full | 1786 | No | NA | 3 | 0 | 2 | Not-determined | Genome-fragment | NA | NA | 0 |
| S4_k141_34732 | 2132 | No | NA | 3 | 0 | 3 | Low-quality | Genome-fragment | 0.75 | AAI-based (medium-confidence) | 0 |
| S6-final-viral-combined_k141_89185\|\|full | 2132 | No | NA | 3 | 0 | 3 | Low-quality | Genome-fragment | 1.59 | AAI-based (medium-confidence) | 0 |
| S12_k141_48294 | 2133 | No | NA | 2 | 0 | 1 | Low-quality | Genome-fragment | 2.88 | AAI-based (high-confidence) | 0 |
| S17-final-viral-combined_k141_35604\|\|full | 1387 | No | NA | 2 | 0 | 2 | Not-determined | Genome-fragment | NA | NA | 0 |
| S9-final-viral-combined_k141_118920\|\|full | 2134 | No | NA | 3 | 0 | 1 | Low-quality | Genome-fragment | 1.99 | AAI-based (high-confidence) | 0 |
| S3-final-viral-combined_k141_70230\|\|full | 2134 | No | NA | 4 | 0 | 0 | Low-quality | Genome-fragment | 2.38 | AAI-based (medium-confidence) | 0 |
| S17-final-viral-combined_k141_224722\|\|full | 4838 | No | NA | 7 | 0 | 0 | Not-determined | Genome-fragment | NA | NA | 0 |
| S17-final-viral-combined_k141_90823\|\|full | 2075 | No | NA | 2 | 0 | 1 | Not-determined | Genome-fragment | NA | NA | 0 |
| S12-final-viral-combined_k141_370664\|\|full | 2134 | No | NA | 6 | 0 | 0 | Low-quality | Genome-fragment | 6.62 | AAI-based (high-confidence) | 0 |
| S17-final-viral-combined_k141_224762\|\|full | 9820 | No | NA | 9 | 0 | 2 | Not-determined | Genome-fragment | NA | NA | 0 |
| S17-final-viral-combined_k141_113837\|\|full | 2636 | No | NA | 3 | 0 | 0 | Not-determined | Genome-fragment | NA | NA | 0 |
| S17-final-viral-combined_k141_235451\|\|full | 2620 | No | NA | 3 | 0 | 2 | Not-determined | Genome-fragment | NA | NA | 0 |
| S17-final-viral-combined_k141_2234\|\|full | 70511 | Yes | 55004 | 99 | 22 | 12 | High-quality | High-quality | 100 | AAI-based (high-confidence) | 21.99 |
| S17-final-viral-combined_k141_169026\|\|full | 1847 | No | NA | 2 | 0 | 0 | Not-determined | Genome-fragment | NA | NA | 0 |
| S2_k141_236000 | 2134 | No | NA | 4 | 0 | 0 | Low-quality | Genome-fragment | 3.59 | AAI-based (high-confidence) | 0 |
| S17-final-viral-combined_k141_246314\|\|full | 2129 | No | NA | 3 | 0 | 2 | Not-determined | Genome-fragment | NA | NA | 0 |
| S17-final-viral-combined_k141_191357\|\|full | 1347 | No | NA | 2 | 0 | 1 | Not-determined | Genome-fragment | NA | NA | 0 |
| S17-final-viral-combined_k141_69158\|\|full | 2795 | No | NA | 3 | 0 | 2 | Not-determined | Genome-fragment | NA | NA | 0 |
| S11-final-viral-combined_k141_42956\|\|full | 2135 | No | NA | 3 | 0 | 0 | Low-quality | Genome-fragment | 1.84 | AAI-based (medium-confidence) | 0 |
| S17-final-viral-combined_k141_213724\|\|full | 2500 | No | NA | 4 | 0 | 2 | Not-determined | Genome-fragment | NA | NA | 0 |
| S17-final-viral-combined_k141_146824\|\|full | 2441 | No | NA | 3 | 0 | 2 | Not-determined | Genome-fragment | NA | NA | 0 |
| S11_k141_259751 | 2135 | No | NA | 3 | 0 | 3 | Low-quality | Genome-fragment | 0.86 | AAI-based (medium-confidence) | 0 |
| S17-final-viral-combined_k141_224840\|\|full | 2866 | No | NA | 3 | 0 | 0 | Not-determined | Genome-fragment | NA | NA | 0 |
| S1_k141_371633 | 2135 | No | NA | 3 | 0 | 0 | Low-quality | Genome-fragment | 4.03 | AAI-based (high-confidence) | 0 |
| S17-final-viral-combined_k141_191382\|\|full | 10534 | No | NA | 7 | 0 | 3 | Not-determined | Genome-fragment | NA | NA | 0 |
| S17-final-viral-combined_k141_213743\|\|full | 2891 | No | NA | 3 | 0 | 1 | Not-determined | Genome-fragment | NA | NA | 0 |
| S17-final-viral-combined_k141_257825\|\|full | 1129 | No | NA | 2 | 0 | 1 | Not-determined | Genome-fragment | NA | NA | 0 |
| S17-final-viral-combined_k141_102034\|\|full | 1657 | No | NA | 2 | 0 | 1 | Not-determined | Genome-fragment | NA | NA | 0 |
| S17-final-viral-combined_k141_202847\|\|full | 2238 | No | NA | 2 | 0 | 1 | Not-determined | Genome-fragment | NA | NA | 0 |
| S17-final-viral-combined_k141_201212\|\|full | 2135 | No | NA | 3 | 0 | 3 | Low-quality | Genome-fragment | 0.67 | AAI-based (medium-confidence) | 0 |
| S17-final-viral-combined_k141_180242\|\|full | 1463 | No | NA | 2 | 0 | 1 | Not-determined | Genome-fragment | NA | NA | 0 |
| S17-final-viral-combined_k141_246368\|\|full | 3483 | No | NA | 3 | 0 | 3 | Not-determined | Genome-fragment | NA | NA | 0 |
| S2-final-viral-combined_k141_15148\|\|full | 2135 | No | NA | 6 | 2 | 0 | Low-quality | Genome-fragment | 1.52 | HMM-based (lower-bound) | 0 |
| S17-final-viral-combined_k141_157610\|\|full | 4335 | No | NA | 7 | 0 | 1 | Not-determined | Genome-fragment | NA | NA | 0 |
| S17-final-viral-combined_k141_146850\|\|full | 13505 | Yes | 10278 | 15 | 2 | 2 | Medium-quality | Genome-fragment | 66.51 | AAI-based (medium-confidence) | 23.89 |
| S2-final-viral-combined_k141_243583\|\|full | 2135 | No | NA | 3 | 0 | 2 | Low-quality | Genome-fragment | 2.6 | AAI-based (medium-confidence) | 0 |
| S8-final-viral-combined_k141_88115\|\|full | 2135 | No | NA | 5 | 1 | 0 | Low-quality | Genome-fragment | 1.04 | HMM-based (lower-bound) | 0 |
| S11-final-viral-combined_k141_213229\|\|full | 2136 | No | NA | 3 | 1 | 0 | Low-quality | Genome-fragment | 1.15 | HMM-based (lower-bound) | 0 |
| S17-final-viral-combined_k141_124963\|\|full | 1548 | No | NA | 3 | 0 | 0 | Not-determined | Genome-fragment | NA | NA | 0 |
| S17-final-viral-combined_k141_102080\|\|full | 1857 | No | NA | 3 | 0 | 2 | Not-determined | Genome-fragment | NA | NA | 0 |
| S17-final-viral-combined_k141_57851\|\|full | 14839 | No | NA | 14 | 0 | 13 | Not-determined | Genome-fragment | NA | NA | 0 |
| S17-final-viral-combined_k141_24294\|\|full | 4320 | No | NA | 5 | 0 | 2 | Not-determined | Genome-fragment | NA | NA | 0 |
| S17-final-viral-combined_k141_2283\|\|full | 1656 | No | NA | 2 | 0 | 1 | Not-determined | Genome-fragment | NA | NA | 0 |
| S1-final-viral-combined_k141_53282\|\|full | 2136 | No | NA | 5 | 0 | 0 | Low-quality | Genome-fragment | 3.24 | AAI-based (medium-confidence) | 0 |
| S17-final-viral-combined_k141_90912\|\|full | 1136 | No | NA | 3 | 0 | 0 | Not-determined | Genome-fragment | NA | NA | 0 |
| S17-final-viral-combined_k141_113974\|\|full | 1939 | No | NA | 3 | 0 | 2 | Not-determined | Genome-fragment | NA | NA | 0 |
| S17-final-viral-combined_k141_69263\|\|full | 4482 | No | NA | 4 | 0 | 2 | Not-determined | Genome-fragment | NA | NA | 0 |
| S3-final-viral-combined_k141_60347\|\|full | 2136 | No | NA | 3 | 1 | 0 | Low-quality | Genome-fragment | 3.23 | HMM-based (lower-bound) | 0 |
| S17-final-viral-combined_k141_224927\|\|full | 1332 | No | NA | 3 | 0 | 1 | Not-determined | Genome-fragment | NA | NA | 0 |
| S17-final-viral-combined_k141_113989\|\|full | 4498 | No | NA | 4 | 0 | 2 | Not-determined | Genome-fragment | NA | NA | 0 |
| S17-final-viral-combined_k141_113990\|\|full | 2627 | No | NA | 4 | 0 | 2 | Not-determined | Genome-fragment | NA | NA | 0 |
| S17-final-viral-combined_k141_235587\|\|full | 2350 | No | NA | 2 | 0 | 2 | Not-determined | Genome-fragment | NA | NA | 0 |
| S17-final-viral-combined_k141_90993\|\|full | 2136 | No | NA | 7 | 0 | 0 | Low-quality | Genome-fragment | 4.1 | AAI-based (medium-confidence) | 0 |
| S17-final-viral-combined_k141_12953\|\|full | 1641 | No | NA | 3 | 0 | 2 | Not-determined | Genome-fragment | NA | NA | 0 |
| S17-final-viral-combined_k141_136413\|\|full | 2136 | No | NA | 4 | 2 | 0 | Low-quality | Genome-fragment | 5.05 | AAI-based (high-confidence) | 0 |
| S17-final-viral-combined_k141_46218\|\|full | 1168 | No | NA | 2 | 0 | 0 | Not-determined | Genome-fragment | NA | NA | 0 |
| S12-final-viral-combined_k141_102092\|\|full | 2136 | No | NA | 3 | 0 | 1 | Low-quality | Genome-fragment | 0.61 | AAI-based (medium-confidence) | 0 |
| S2-final-viral-combined_k141_103853\|\|full | 2136 | No | NA | 2 | 0 | 0 | Low-quality | Genome-fragment | 5.31 | AAI-based (medium-confidence) | 0 |
| S2-final-viral-combined_k141_113610\|\|full | 2136 | No | NA | 3 | 0 | 2 | Low-quality | Genome-fragment | 1.3 | AAI-based (high-confidence) | 0 |
| S17-final-viral-combined_k141_246451\|\|full | 6070 | No | NA | 6 | 0 | 5 | Not-determined | Genome-fragment | NA | NA | 0 |
| S5-final-viral-combined_k141_119156\|\|full | 2137 | No | NA | 3 | 0 | 0 | Low-quality | Genome-fragment | 3.5 | AAI-based (high-confidence) | 0 |
| S12-final-viral-combined_k141_170934\|\|full | 2137 | No | NA | 5 | 1 | 0 | Low-quality | Genome-fragment | 2.27 | AAI-based (medium-confidence) | 0 |
| S17-final-viral-combined_k141_35690\|\|full | 2677 | No | NA | 7 | 0 | 0 | Not-determined | Genome-fragment | NA | NA | 0 |
| S17-final-viral-combined_k141_2327\|\|full | 2294 | No | NA | 3 | 0 | 0 | Not-determined | Genome-fragment | NA | NA | 0 |
| S17-final-viral-combined_k141_224988\|\|full | 2018 | No | NA | 4 | 0 | 0 | Not-determined | Genome-fragment | NA | NA | 0 |
| S2-final-viral-combined_k141_736\|\|full | 2137 | No | NA | 2 | 0 | 2 | Low-quality | Genome-fragment | 1.14 | AAI-based (medium-confidence) | 0 |
| S17-final-viral-combined_k141_257999\|\|full | 4265 | No | NA | 4 | 0 | 2 | Not-determined | Genome-fragment | NA | NA | 0 |
| S9-final-viral-combined_k141_96214\|\|full | 2138 | No | NA | 4 | 0 | 0 | Low-quality | Genome-fragment | 3.73 | AAI-based (medium-confidence) | 0 |
| S17-final-viral-combined_k141_28586\|\|full | 2138 | No | NA | 2 | 0 | 0 | Low-quality | Genome-fragment | 2.6 | AAI-based (high-confidence) | 0 |
| S17-final-viral-combined_k141_90990\|\|full | 5745 | No | NA | 4 | 0 | 2 | Not-determined | Genome-fragment | NA | NA | 0 |
| S12-final-viral-combined_k141_236462\|\|full | 2138 | No | NA | 2 | 0 | 1 | Low-quality | Genome-fragment | 2.3 | AAI-based (medium-confidence) | 0 |
| S12_k141_69600 | 2138 | No | NA | 4 | 2 | 0 | Low-quality | Genome-fragment | 3.86 | AAI-based (high-confidence) | 0 |
| S11-final-viral-combined_k141_182862\|\|lt2gene | 2139 | No | NA | 2 | 1 | 0 | Low-quality | Genome-fragment | 4.49 | HMM-based (lower-bound) | 0 |
| S17-final-viral-combined_k141_213887\|\|full | 1164 | No | NA | 3 | 0 | 1 | Not-determined | Genome-fragment | NA | NA | 0 |
| S17-final-viral-combined_k141_135938\|\|full | 1476 | No | NA | 2 | 0 | 2 | Not-determined | Genome-fragment | NA | NA | 0 |
| S12-final-viral-combined_k141_209327\|\|lt2gene | 2139 | No | NA | 1 | 1 | 0 | Low-quality | Genome-fragment | 1.64 | HMM-based (lower-bound) | 0 |
| S2-final-viral-combined_k141_151827\|\|full | 2139 | No | NA | 3 | 0 | 1 | Low-quality | Genome-fragment | 0.56 | AAI-based (medium-confidence) | 0 |
| S17-final-viral-combined_k141_157693\|\|full | 4050 | No | NA | 7 | 0 | 0 | Not-determined | Genome-fragment | NA | NA | 0 |
| S17-final-viral-combined_k141_258027\|\|full | 2077 | No | NA | 2 | 0 | 2 | Not-determined | Genome-fragment | NA | NA | 0 |
| S6-final-viral-combined_k141_51721\|\|full | 2139 | No | NA | 4 | 0 | 1 | Low-quality | Genome-fragment | 2.6 | AAI-based (high-confidence) | 0 |
| S17-final-viral-combined_k141_180309\|\|full | 1693 | No | NA | 3 | 0 | 1 | Not-determined | Genome-fragment | NA | NA | 0 |
| S17-final-viral-combined_k141_191555\|\|full | 6606 | No | NA | 8 | 0 | 3 | Not-determined | Genome-fragment | NA | NA | 0 |
| S9_k141_68922 | 2140 | No | NA | 2 | 0 | 0 | Low-quality | Genome-fragment | 1.51 | AAI-based (medium-confidence) | 0 |
| S17-final-viral-combined_k141_114094\|\|full | 1969 | No | NA | 3 | 0 | 2 | Not-determined | Genome-fragment | NA | NA | 0 |
| S17-final-viral-combined_k141_136009\|\|full | 3437 | No | NA | 3 | 0 | 2 | Not-determined | Genome-fragment | NA | NA | 0 |
| S3-final-viral-combined_k141_92628\|\|full | 2140 | No | NA | 3 | 0 | 2 | Low-quality | Genome-fragment | 1.45 | AAI-based (medium-confidence) | 0 |
| S7_k141_198240 | 2140 | No | NA | 2 | 0 | 1 | Low-quality | Genome-fragment | 3.45 | AAI-based (medium-confidence) | 0 |
| S17-final-viral-combined_k141_136015\|\|full | 2171 | No | NA | 3 | 0 | 2 | Not-determined | Genome-fragment | NA | NA | 0 |
| S4-final-viral-combined_k141_23940\|\|full | 2140 | Yes | 1567 | 3 | 1 | 1 | Low-quality | Genome-fragment | 3.18 | AAI-based (medium-confidence) | 26.78 |
| S17-final-viral-combined_k141_35762\|\|full | 4177 | No | NA | 6 | 0 | 3 | Not-determined | Genome-fragment | NA | NA | 0 |
| S17-final-viral-combined_k141_125057\|\|full | 3463 | No | NA | 4 | 0 | 2 | Not-determined | Genome-fragment | NA | NA | 0 |
| S5-final-viral-combined_k141_88308\|\|full | 2141 | No | NA | 3 | 2 | 0 | Low-quality | Genome-fragment | 3.76 | HMM-based (lower-bound) | 0 |
| S17-final-viral-combined_k141_258086\|\|full | 2088 | No | NA | 2 | 0 | 2 | Not-determined | Genome-fragment | NA | NA | 0 |
| S17-final-viral-combined_k141_114127\|\|full | 4696 | No | NA | 5 | 0 | 2 | Not-determined | Genome-fragment | NA | NA | 0 |
| S17-final-viral-combined_k141_35780\|\|full | 1352 | No | NA | 3 | 0 | 1 | Not-determined | Genome-fragment | NA | NA | 0 |
| S7-final-viral-combined_k141_182320\|\|full | 2141 | No | NA | 6 | 5 | 0 | Low-quality | Genome-fragment | 5.53 | AAI-based (high-confidence) | 0 |
| S17-final-viral-combined_k141_91071\|\|full | 2441 | No | NA | 3 | 0 | 3 | Not-determined | Genome-fragment | NA | NA | 0 |
| S8-final-viral-combined_k141_189230\|\|full | 2141 | No | NA | 5 | 1 | 0 | Low-quality | Genome-fragment | 5.39 | AAI-based (high-confidence) | 0 |
| S17-final-viral-combined_k141_13035\|\|full | 1866 | No | NA | 2 | 0 | 1 | Not-determined | Genome-fragment | NA | NA | 0 |
| S17-final-viral-combined_k141_246521\|\|full | 1370 | No | NA | 2 | 0 | 2 | Not-determined | Genome-fragment | NA | NA | 0 |
| S1-final-viral-combined_k141_99148\|\|full | 2142 | No | NA | 3 | 0 | 1 | Low-quality | Genome-fragment | 0.71 | AAI-based (medium-confidence) | 0 |
| S17-final-viral-combined_k141_39387\|\|full | 2142 | No | NA | 3 | 1 | 0 | Low-quality | Genome-fragment | 3.69 | AAI-based (medium-confidence) | 0 |
| S2-final-viral-combined_k141_110395\|\|full | 2142 | No | NA | 2 | 0 | 1 | Low-quality | Genome-fragment | 1.01 | AAI-based (medium-confidence) | 0 |
| S17-final-viral-combined_k141_258087\|\|full | 25523 | No | NA | 34 | 22 | 0 | Medium-quality | Genome-fragment | 55.71 | AAI-based (high-confidence) | 0 |
| S8-final-viral-combined_k141_184166\|\|full | 2142 | No | NA | 3 | 0 | 0 | Low-quality | Genome-fragment | 4.38 | AAI-based (high-confidence) | 0 |
| S17-final-viral-combined_k141_169290\|\|full | 3640 | No | NA | 9 | 0 | 0 | Not-determined | Genome-fragment | NA | NA | 0 |
| S17-final-viral-combined_k141_80629\|\|full | 2154 | No | NA | 4 | 0 | 3 | Not-determined | Genome-fragment | NA | NA | 0 |
| S17-final-viral-combined_k141_13074\|\|full | 2160 | No | NA | 2 | 0 | 2 | Not-determined | Genome-fragment | NA | NA | 0 |
| S17-final-viral-combined_k141_57957\|\|full | 1743 | No | NA | 3 | 0 | 2 | Not-determined | Genome-fragment | NA | NA | 0 |
| S17-final-viral-combined_k141_213988\|\|full | 2446 | No | NA | 2 | 0 | 0 | Not-determined | Genome-fragment | NA | NA | 0 |
| S17-final-viral-combined_k141_102252\|\|full | 6143 | No | NA | 5 | 0 | 5 | Not-determined | Genome-fragment | NA | NA | 0 |
| S2-final-viral-combined_k141_112065\|\|full | 2143 | No | NA | 2 | 0 | 1 | Low-quality | Genome-fragment | 1.84 | AAI-based (medium-confidence) | 0 |
| S4-final-viral-combined_k141_10239\|\|full | 2143 | No | NA | 3 | 0 | 0 | Low-quality | Genome-fragment | 7.24 | AAI-based (high-confidence) | 0 |
| S3-final-viral-combined_k141_120696\|\|full | 2144 | Yes | 1067 | 3 | 1 | 1 | Low-quality | Genome-fragment | 0.64 | AAI-based (medium-confidence) | 50.23 |
| S17-final-viral-combined_k141_80689\|\|full | 2057 | No | NA | 2 | 0 | 1 | Not-determined | Genome-fragment | NA | NA | 0 |
| S17-final-viral-combined_k141_147166\|\|full | 2255 | No | NA | 2 | 0 | 2 | Not-determined | Genome-fragment | NA | NA | 0 |
| S7-final-viral-combined_k141_35684\|\|full | 2144 | No | NA | 3 | 0 | 2 | Low-quality | Genome-fragment | 3.66 | AAI-based (medium-confidence) | 0 |
| S17-final-viral-combined_k141_191662\|\|full | 1152 | No | NA | 3 | 0 | 0 | Not-determined | Genome-fragment | NA | NA | 0 |
| S7-final-viral-combined_k141_179781\|\|full | 2144 | No | NA | 4 | 0 | 0 | Low-quality | Genome-fragment | 3.46 | AAI-based (medium-confidence) | 0 |
| S17-final-viral-combined_k141_169354\|\|full | 1236 | No | NA | 3 | 0 | 0 | Not-determined | Genome-fragment | NA | NA | 0 |
| S17-final-viral-combined_k141_180420\|\|full | 4283 | No | NA | 4 | 0 | 3 | Not-determined | Genome-fragment | NA | NA | 0 |
| S12_k141_62078 | 2144 | No | NA | 4 | 0 | 0 | Low-quality | Genome-fragment | 1.29 | AAI-based (high-confidence) | 0 |
| S2_k141_92498 | 2144 | No | NA | 2 | 0 | 1 | Low-quality | Genome-fragment | 7.21 | AAI-based (medium-confidence) | 0 |
| S17-final-viral-combined_k141_147191\|\|full | 1845 | No | NA | 3 | 0 | 3 | Not-determined | Genome-fragment | NA | NA | 0 |
| S17-final-viral-combined_k141_13117\|\|full | 1606 | No | NA | 2 | 0 | 2 | Not-determined | Genome-fragment | NA | NA | 0 |
| S17-final-viral-combined_k141_157770\|\|full | 2504 | No | NA | 2 | 0 | 2 | Not-determined | Genome-fragment | NA | NA | 0 |
| S17-final-viral-combined_k141_191680\|\|full | 4008 | No | NA | 4 | 0 | 2 | Not-determined | Genome-fragment | NA | NA | 0 |
| S17-final-viral-combined_k141_136055\|\|full | 5350 | No | NA | 6 | 0 | 5 | Not-determined | Genome-fragment | NA | NA | 0 |
| S17-final-viral-combined_k141_69488\|\|full | 2523 | No | NA | 3 | 0 | 0 | Not-determined | Genome-fragment | NA | NA | 0 |
| S17-final-viral-combined_k141_235838\|\|full | 5853 | No | NA | 6 | 0 | 4 | Not-determined | Genome-fragment | NA | NA | 0 |
| S8-final-viral-combined_k141_208037\|\|full | 2144 | No | NA | 5 | 0 | 0 | Low-quality | Genome-fragment | 3.23 | AAI-based (high-confidence) | 0 |
| S17-final-viral-combined_k141_235851\|\|full | 2518 | No | NA | 2 | 0 | 1 | Not-determined | Genome-fragment | NA | NA | 0 |
| S17-final-viral-combined_k141_258153\|\|full | 1631 | No | NA | 2 | 0 | 1 | Not-determined | Genome-fragment | NA | NA | 0 |
| S17-final-viral-combined_k141_125117\|\|full | 4189 | No | NA | 3 | 0 | 1 | Not-determined | Genome-fragment | NA | NA | 0 |
| S17-final-viral-combined_k141_125118\|\|full | 3315 | No | NA | 4 | 0 | 4 | Not-determined | Genome-fragment | NA | NA | 0 |
| S17-final-viral-combined_k141_214109\|\|full | 6571 | No | NA | 5 | 0 | 4 | Not-determined | Genome-fragment | NA | NA | 0 |
| S17-final-viral-combined_k141_169428\|\|full | 1482 | No | NA | 2 | 0 | 0 | Not-determined | Genome-fragment | NA | NA | 0 |
| S17-final-viral-combined_k141_24403\|\|full | 1207 | No | NA | 2 | 0 | 2 | Not-determined | Genome-fragment | NA | NA | 0 |
| S17-final-viral-combined_k141_136107\|\|full | 2683 | No | NA | 3 | 0 | 0 | Not-determined | Genome-fragment | NA | NA | 0 |
| S17-final-viral-combined_k141_258185\|\|full | 2408 | No | NA | 3 | 0 | 3 | Not-determined | Genome-fragment | NA | NA | 0 |
| S9-final-viral-combined_k141_66153\|\|full | 2145 | No | NA | 6 | 4 | 0 | Low-quality | Genome-fragment | 5.73 | AAI-based (high-confidence) | 0 |
| S17-final-viral-combined_k141_258192\|\|full | 1720 | No | NA | 3 | 0 | 1 | Not-determined | Genome-fragment | NA | NA | 0 |
| S17-final-viral-combined_k141_235889\|\|full | 1366 | No | NA | 3 | 0 | 1 | Not-determined | Genome-fragment | NA | NA | 0 |
| S17-final-viral-combined_k141_102365\|\|full | 1445 | No | NA | 3 | 0 | 1 | Not-determined | Genome-fragment | NA | NA | 0 |
| S17-final-viral-combined_k141_246657\|\|full | 1307 | No | NA | 2 | 0 | 2 | Not-determined | Genome-fragment | NA | NA | 0 |
| S17-final-viral-combined_k141_147245\|\|full | 2740 | No | NA | 5 | 0 | 1 | Not-determined | Genome-fragment | NA | NA | 0 |
| S3-final-viral-combined_k141_90136\|\|full | 2145 | No | NA | 3 | 1 | 0 | Low-quality | Genome-fragment | 5.1 | HMM-based (lower-bound) | 0 |
| S17-final-viral-combined_k141_114388\|\|full | 1237 | No | NA | 3 | 0 | 1 | Not-determined | Genome-fragment | NA | NA | 0 |
| S5-final-viral-combined_k141_70187\|\|full | 2145 | No | NA | 5 | 1 | 0 | Low-quality | Genome-fragment | 4.81 | AAI-based (high-confidence) | 0 |
| S17-final-viral-combined_k141_246671\|\|full | 1291 | No | NA | 3 | 0 | 1 | Not-determined | Genome-fragment | NA | NA | 0 |
| S17-final-viral-combined_k141_180504\|\|full | 1125 | No | NA | 3 | 0 | 1 | Not-determined | Genome-fragment | NA | NA | 0 |
| S17-final-viral-combined_k141_214129\|\|full | 1624 | No | NA | 3 | 0 | 1 | Not-determined | Genome-fragment | NA | NA | 0 |
| S17-final-viral-combined_k141_169492\|\|full | 7673 | No | NA | 10 | 0 | 2 | Not-determined | Genome-fragment | NA | NA | 0 |
| S7-final-viral-combined_k141_31774\|\|full | 2145 | No | NA | 3 | 0 | 2 | Low-quality | Genome-fragment | 1.75 | AAI-based (high-confidence) | 0 |
| S17-final-viral-combined_k141_24459\|\|full | 2161 | No | NA | 2 | 0 | 1 | Not-determined | Genome-fragment | NA | NA | 0 |
| S7-final-viral-combined_k141_239031\|\|full | 2145 | No | NA | 3 | 1 | 0 | Low-quality | Genome-fragment | 5.75 | AAI-based (medium-confidence) | 0 |
| S17-final-viral-combined_k141_214158\|\|full | 2102 | No | NA | 2 | 0 | 2 | Not-determined | Genome-fragment | NA | NA | 0 |
| S17-final-viral-combined_k141_225219\|\|full | 2157 | No | NA | 2 | 0 | 2 | Not-determined | Genome-fragment | NA | NA | 0 |
| S17-final-viral-combined_k141_46406\|\|full | 3756 | No | NA | 4 | 0 | 3 | Not-determined | Genome-fragment | NA | NA | 0 |
| S7-final-viral-combined_k141_224373\|\|full | 2145 | No | NA | 4 | 1 | 0 | Low-quality | Genome-fragment | 3.82 | HMM-based (lower-bound) | 0 |
| S12-final-viral-combined_k141_87756\|\|full | 2145 | No | NA | 4 | 0 | 0 | Low-quality | Genome-fragment | 4.08 | AAI-based (high-confidence) | 0 |
| S1-final-viral-combined_k141_225657\|\|full | 2146 | No | NA | 5 | 0 | 3 | Low-quality | Genome-fragment | 1.77 | AAI-based (medium-confidence) | 0 |
| S17-final-viral-combined_k141_246728\|\|full | 1562 | No | NA | 2 | 0 | 0 | Not-determined | Genome-fragment | NA | NA | 0 |
| S1-final-viral-combined_k141_197214\|\|full | 2146 | No | NA | 9 | 1 | 0 | Low-quality | Genome-fragment | 5.41 | AAI-based (high-confidence) | 0 |
| S12-final-viral-combined_k141_140003\|\|full | 2146 | No | NA | 6 | 1 | 0 | Low-quality | Genome-fragment | 0.99 | AAI-based (high-confidence) | 0 |
| S17-final-viral-combined_k141_35955\|\|full | 4662 | No | NA | 6 | 0 | 2 | Not-determined | Genome-fragment | NA | NA | 0 |
| S17-final-viral-combined_k141_24508\|\|full | 3368 | No | NA | 5 | 0 | 4 | Not-determined | Genome-fragment | NA | NA | 0 |
| S11-final-viral-combined_k141_311381\|\|full | 2147 | No | NA | 2 | 1 | 0 | Low-quality | Genome-fragment | 1.43 | HMM-based (lower-bound) | 0 |
| S17-final-viral-combined_k141_157844\|\|full | 6475 | No | NA | 4 | 0 | 4 | Not-determined | Genome-fragment | NA | NA | 0 |
| S17-final-viral-combined_k141_214239\|\|full | 1052 | No | NA | 2 | 0 | 0 | Not-determined | Genome-fragment | NA | NA | 0 |
| S17-final-viral-combined_k141_80837\|\|full | 1549 | No | NA | 2 | 0 | 1 | Not-determined | Genome-fragment | NA | NA | 0 |
| S5-final-viral-combined_k141_41244\|\|full | 2147 | No | NA | 5 | 0 | 0 | Low-quality | Genome-fragment | 5.52 | AAI-based (medium-confidence) | 0 |
| S17-final-viral-combined_k141_58082\|\|full | 1430 | No | NA | 2 | 0 | 2 | Not-determined | Genome-fragment | NA | NA | 0 |
| S17-final-viral-combined_k141_258267\|\|full | 14318 | No | NA | 16 | 0 | 1 | Not-determined | Genome-fragment | NA | NA | 0 |
| S17-final-viral-combined_k141_180606\|\|full | 1288 | No | NA | 3 | 0 | 1 | Not-determined | Genome-fragment | NA | NA | 0 |
| S17-final-viral-combined_k141_102410\|\|full | 6575 | No | NA | 4 | 0 | 0 | Not-determined | Genome-fragment | NA | NA | 0 |
| S17-final-viral-combined_k141_2599\|\|full | 5245 | No | NA | 7 | 0 | 3 | Not-determined | Genome-fragment | NA | NA | 0 |
| S2_k141_196947 | 2147 | No | NA | 3 | 0 | 0 | Low-quality | Genome-fragment | 3.1 | AAI-based (medium-confidence) | 0 |
| S8_k141_245408 | 2147 | No | NA | 6 | 0 | 0 | Low-quality | Genome-fragment | 6.67 | AAI-based (high-confidence) | 0 |
| S17-final-viral-combined_k141_13308\|\|full | 1576 | No | NA | 3 | 0 | 1 | Not-determined | Genome-fragment | NA | NA | 0 |
| S1-final-viral-combined_k141_264420\|\|full | 2148 | No | NA | 4 | 0 | 0 | Low-quality | Genome-fragment | 4.98 | AAI-based (medium-confidence) | 0 |
| S17-final-viral-combined_k141_157873\|\|full | 3382 | No | NA | 4 | 0 | 2 | Not-determined | Genome-fragment | NA | NA | 0 |
| S1-final-viral-combined_k141_332614\|\|full | 2148 | No | NA | 3 | 0 | 0 | Low-quality | Genome-fragment | 6 | AAI-based (medium-confidence) | 0 |
| S17-final-viral-combined_k141_102431\|\|full | 2075 | No | NA | 3 | 0 | 2 | Not-determined | Genome-fragment | NA | NA | 0 |
| S1-final-viral-combined_k141_174221\|\|full | 2148 | No | NA | 4 | 0 | 3 | Low-quality | Genome-fragment | 0.8 | AAI-based (medium-confidence) | 0 |
| S17-final-viral-combined_k141_58129\|\|full | 1452 | No | NA | 2 | 0 | 2 | Not-determined | Genome-fragment | NA | NA | 0 |
| S17-final-viral-combined_k141_102440\|\|full | 7933 | No | NA | 10 | 0 | 7 | Not-determined | Genome-fragment | NA | NA | 0 |
| S5-final-viral-combined_k141_128974\|\|full | 2148 | No | NA | 3 | 0 | 0 | Low-quality | Genome-fragment | 6.01 | AAI-based (medium-confidence) | 0 |
| S17-final-viral-combined_k141_69994\|\|full | 2148 | No | NA | 8 | 0 | 0 | Low-quality | Genome-fragment | 3.61 | AAI-based (high-confidence) | 0 |
| S12-final-viral-combined_k141_391609\|\|full | 2148 | No | NA | 4 | 0 | 0 | Low-quality | Genome-fragment | 1.61 | AAI-based (medium-confidence) | 0 |
| S17-final-viral-combined_k141_102468\|\|full | 1341 | No | NA | 3 | 0 | 1 | Not-determined | Genome-fragment | NA | NA | 0 |
| S17-final-viral-combined_k141_236035\|\|full | 3123 | No | NA | 3 | 0 | 2 | Not-determined | Genome-fragment | NA | NA | 0 |
| S2-final-viral-combined_k141_68337\|\|full | 2148 | No | NA | 3 | 0 | 0 | Low-quality | Genome-fragment | 5.52 | AAI-based (medium-confidence) | 0 |
| S17-final-viral-combined_k141_36030\|\|full | 2958 | No | NA | 2 | 0 | 2 | Not-determined | Genome-fragment | NA | NA | 0 |
| S1-final-viral-combined_k141_378797\|\|full | 2149 | No | NA | 3 | 1 | 0 | Low-quality | Genome-fragment | 0.19 | HMM-based (lower-bound) | 0 |
| S17-final-viral-combined_k141_236072\|\|full | 1469 | No | NA | 2 | 0 | 0 | Not-determined | Genome-fragment | NA | NA | 0 |
| S17-k141_49085 | 2149 | No | NA | 3 | 0 | 1 | Low-quality | Genome-fragment | 11.96 | AAI-based (high-confidence) | 0 |
| S17-final-viral-combined_k141_136293\|\|full | 2463 | No | NA | 3 | 0 | 2 | Not-determined | Genome-fragment | NA | NA | 0 |
| S17-final-viral-combined_k141_136294\|\|full | 1275 | No | NA | 5 | 0 | 0 | Not-determined | Genome-fragment | NA | NA | 0 |
| S17-final-viral-combined_k141_157954\|\|full | 2105 | No | NA | 2 | 0 | 2 | Not-determined | Genome-fragment | NA | NA | 0 |
| S4-final-viral-combined_k141_156017\|\|full | 2149 | No | NA | 3 | 0 | 1 | Low-quality | Genome-fragment | 1.17 | AAI-based (high-confidence) | 0 |
| S7-final-viral-combined_k141_104301\|\|full | 2150 | No | NA | 3 | 1 | 0 | Low-quality | Genome-fragment | 5.5 | AAI-based (high-confidence) | 0 |
| S12-final-viral-combined_k141_381135\|\|full | 2150 | No | NA | 5 | 0 | 0 | Low-quality | Genome-fragment | 4.26 | AAI-based (medium-confidence) | 0 |
| S12-final-viral-combined_k141_315807\|\|full | 2150 | No | NA | 3 | 0 | 0 | Low-quality | Genome-fragment | 6.88 | AAI-based (high-confidence) | 0 |
| S17-final-viral-combined_k141_114588\|\|full | 1036 | No | NA | 2 | 0 | 1 | Not-determined | Genome-fragment | NA | NA | 0 |
| S17-final-viral-combined_k141_69733\|\|full | 12342 | No | NA | 14 | 0 | 6 | Not-determined | Genome-fragment | NA | NA | 0 |
| S17-final-viral-combined_k141_225356\|\|full | 26833 | No | NA | 37 | 15 | 0 | Medium-quality | Genome-fragment | 53.77 | AAI-based (medium-confidence) | 0 |
| S12-final-viral-combined_k141_115817\|\|full | 2150 | No | NA | 2 | 1 | 0 | Low-quality | Genome-fragment | 5.32 | AAI-based (medium-confidence) | 0 |
| S17-final-viral-combined_k141_13422\|\|full | 1865 | No | NA | 3 | 0 | 1 | Not-determined | Genome-fragment | NA | NA | 0 |
| S17-final-viral-combined_k141_69767\|\|full | 2632 | No | NA | 3 | 0 | 0 | Not-determined | Genome-fragment | NA | NA | 0 |
| S17-final-viral-combined_k141_136355\|\|full | 2805 | No | NA | 3 | 0 | 1 | Not-determined | Genome-fragment | NA | NA | 0 |
| S4-final-viral-combined_k141_48109\|\|full | 2150 | No | NA | 3 | 0 | 2 | Low-quality | Genome-fragment | 1.05 | AAI-based (medium-confidence) | 0 |
| S17-final-viral-combined_k141_69771\|\|full | 1752 | No | NA | 5 | 0 | 0 | Not-determined | Genome-fragment | NA | NA | 0 |
| S17-final-viral-combined_k141_136366\|\|full | 2736 | No | NA | 2 | 0 | 1 | Not-determined | Genome-fragment | NA | NA | 0 |
| S17-final-viral-combined_k141_169595\|\|full | 1981 | No | NA | 3 | 0 | 2 | Not-determined | Genome-fragment | NA | NA | 0 |
| S17-final-viral-combined_k141_2754\|\|full | 2038 | No | NA | 4 | 0 | 0 | Not-determined | Genome-fragment | NA | NA | 0 |
| S8-final-viral-combined_k141_29636\|\|full | 2150 | Yes | 1143 | 2 | 1 | 1 | Low-quality | Genome-fragment | 2.48 | AAI-based (medium-confidence) | 46.84 |
| S8-final-viral-combined_k141_261686\|\|full | 2150 | No | NA | 5 | 3 | 0 | Low-quality | Genome-fragment | 4.9 | AAI-based (high-confidence) | 0 |
| S9_k141_47099 | 2151 | No | NA | 1 | 0 | 1 | Low-quality | Genome-fragment | 3.73 | AAI-based (medium-confidence) | 0 |
| S17-final-viral-combined_k141_69798\|\|full | 1959 | No | NA | 4 | 0 | 0 | Not-determined | Genome-fragment | NA | NA | 0 |
| S17-final-viral-combined_k141_214415\|\|full | 1573 | No | NA | 3 | 0 | 1 | Not-determined | Genome-fragment | NA | NA | 0 |
| S17-final-viral-combined_k141_136389\|\|full | 1807 | No | NA | 3 | 0 | 1 | Not-determined | Genome-fragment | NA | NA | 0 |
| S17-final-viral-combined_k141_102616\|\|full | 2932 | No | NA | 5 | 0 | 1 | Not-determined | Genome-fragment | NA | NA | 0 |
| S17-final-viral-combined_k141_180840\|\|full | 1818 | No | NA | 3 | 0 | 2 | Not-determined | Genome-fragment | NA | NA | 0 |
| S1-final-viral-combined_k141_126938\|\|full | 2151 | No | NA | 3 | 0 | 1 | Low-quality | Genome-fragment | 5.02 | AAI-based (medium-confidence) | 0 |
| S17-final-viral-combined_k141_81076\|\|full | 1663 | No | NA | 2 | 0 | 2 | Not-determined | Genome-fragment | NA | NA | 0 |
| S17-final-viral-combined_k141_102630\|\|full | 1700 | No | NA | 2 | 0 | 0 | Not-determined | Genome-fragment | NA | NA | 0 |
| S1_k141_140618 | 2151 | No | NA | 2 | 0 | 1 | Low-quality | Genome-fragment | 2.56 | AAI-based (high-confidence) | 0 |
| S17-final-viral-combined_k141_81085\|\|full | 2863 | No | NA | 2 | 0 | 1 | Not-determined | Genome-fragment | NA | NA | 0 |
| S17-final-viral-combined_k141_180875\|\|full | 2221 | No | NA | 5 | 0 | 3 | Not-determined | Genome-fragment | NA | NA | 0 |
| S1_k141_387931 | 2151 | No | NA | 4 | 0 | 0 | Low-quality | Genome-fragment | 6.1 | AAI-based (high-confidence) | 0 |
| S17-final-viral-combined_k141_191957\|\|full | 1397 | No | NA | 2 | 0 | 2 | Not-determined | Genome-fragment | NA | NA | 0 |
| S17-final-viral-combined_k141_24687\|\|full | 3122 | No | NA | 3 | 0 | 2 | Not-determined | Genome-fragment | NA | NA | 0 |
| S17-final-viral-combined_k141_102650\|\|full | 3088 | No | NA | 2 | 0 | 2 | Not-determined | Genome-fragment | NA | NA | 0 |
| S3-final-viral-combined_k141_68777\|\|full | 2151 | No | NA | 4 | 0 | 1 | Low-quality | Genome-fragment | 2.91 | AAI-based (high-confidence) | 0 |
| S5-final-viral-combined_k141_182413\|\|full | 2151 | No | NA | 5 | 1 | 0 | Low-quality | Genome-fragment | 3.81 | AAI-based (high-confidence) | 0 |
| S17-final-viral-combined_k141_169661\|\|full | 8603 | No | NA | 10 | 0 | 3 | Not-determined | Genome-fragment | NA | NA | 0 |
| S17-final-viral-combined_k141_24720\|\|full | 1272 | No | NA | 3 | 0 | 0 | Not-determined | Genome-fragment | NA | NA | 0 |
| S17-final-viral-combined_k141_91164\|\|full | 1512 | No | NA | 3 | 0 | 0 | Not-determined | Genome-fragment | NA | NA | 0 |
| S17-final-viral-combined_k141_147452\|\|full | 4533 | No | NA | 1 | 0 | 1 | Not-determined | Genome-fragment | NA | NA | 0 |
| S17-final-viral-combined_k141_102676\|\|full | 3273 | No | NA | 4 | 0 | 3 | Not-determined | Genome-fragment | NA | NA | 0 |
| S17-final-viral-combined_k141_236145\|\|full | 4437 | No | NA | 4 | 0 | 3 | Not-determined | Genome-fragment | NA | NA | 0 |
| S17-final-viral-combined_k141_203434\|\|full | 1989 | No | NA | 3 | 0 | 2 | Not-determined | Genome-fragment | NA | NA | 0 |
| S17-final-viral-combined_k141_225486\|\|full | 2508 | No | NA | 3 | 0 | 2 | Not-determined | Genome-fragment | NA | NA | 0 |
| S7_k141_71171 | 2151 | No | NA | 3 | 1 | 0 | Low-quality | Genome-fragment | 2.18 | AAI-based (high-confidence) | 0 |
| S17-final-viral-combined_k141_147481\|\|full | 2144 | No | NA | 3 | 0 | 0 | Not-determined | Genome-fragment | NA | NA | 0 |
| S12-final-viral-combined_k141_371809\|\|full | 2151 | No | NA | 3 | 0 | 0 | Low-quality | Genome-fragment | 3.85 | AAI-based (medium-confidence) | 0 |
| S17-final-viral-combined_k141_236160\|\|full | 1069 | No | NA | 2 | 0 | 1 | Not-determined | Genome-fragment | NA | NA | 0 |
| S17-final-viral-combined_k141_114730\|\|full | 1519 | No | NA | 2 | 0 | 2 | Not-determined | Genome-fragment | NA | NA | 0 |
| S17-final-viral-combined_k141_246949\|\|full | 3698 | No | NA | 4 | 0 | 1 | Not-determined | Genome-fragment | NA | NA | 0 |
| S17-final-viral-combined_k141_81258\|\|full | 1356 | No | NA | 2 | 0 | 1 | Not-determined | Genome-fragment | NA | NA | 0 |
| S4-final-viral-combined_k141_107120\|\|full | 2151 | No | NA | 2 | 0 | 0 | Low-quality | Genome-fragment | 1.16 | AAI-based (medium-confidence) | 0 |
| S17-final-viral-combined_k141_236204\|\|full | 5673 | No | NA | 7 | 0 | 3 | Not-determined | Genome-fragment | NA | NA | 0 |
| S8-final-viral-combined_k141_96146\|\|full | 2151 | No | NA | 4 | 0 | 1 | Low-quality | Genome-fragment | 4.02 | AAI-based (medium-confidence) | 0 |
| S17-final-viral-combined_k141_81262\|\|full | 9237 | No | NA | 6 | 0 | 1 | Not-determined | Genome-fragment | NA | NA | 0 |
| S8-final-viral-combined_k141_218764\|\|full | 2151 | No | NA | 4 | 0 | 0 | Low-quality | Genome-fragment | 6.06 | AAI-based (medium-confidence) | 0 |
| S9_k141_21613 | 2152 | No | NA | 5 | 4 | 0 | Low-quality | Genome-fragment | 4.12 | AAI-based (medium-confidence) | 0 |
| S17-final-viral-combined_k141_81266\|\|full | 2711 | No | NA | 3 | 0 | 1 | Not-determined | Genome-fragment | NA | NA | 0 |
| S17-final-viral-combined_k141_203503\|\|full | 2489 | No | NA | 2 | 0 | 2 | Not-determined | Genome-fragment | NA | NA | 0 |
| S17-final-viral-combined_k141_246980\|\|full | 1342 | No | NA | 2 | 0 | 0 | Not-determined | Genome-fragment | NA | NA | 0 |
| S17-final-viral-combined_k141_246991\|\|full | 1218 | No | NA | 2 | 0 | 1 | Not-determined | Genome-fragment | NA | NA | 0 |
| S17-final-viral-combined_k141_13588\|\|full | 1364 | No | NA | 3 | 0 | 2 | Not-determined | Genome-fragment | NA | NA | 0 |
| S11-final-viral-combined_k141_111359\|\|full | 2152 | No | NA | 5 | 1 | 0 | Low-quality | Genome-fragment | 1.3 | HMM-based (lower-bound) | 0 |
| S11-final-viral-combined_k141_324371\|\|full | 2152 | No | NA | 5 | 0 | 0 | Low-quality | Genome-fragment | 6.14 | AAI-based (medium-confidence) | 0 |
| S17-final-viral-combined_k141_2892\|\|full | 1235 | No | NA | 2 | 0 | 2 | Not-determined | Genome-fragment | NA | NA | 0 |
| S17-final-viral-combined_k141_81277\|\|full | 1353 | No | NA | 3 | 0 | 1 | Not-determined | Genome-fragment | NA | NA | 0 |
| S17-final-viral-combined_k141_114841\|\|full | 2209 | No | NA | 4 | 0 | 2 | Not-determined | Genome-fragment | NA | NA | 0 |
| S11_k141_191312 | 2152 | No | NA | 4 | 0 | 0 | Low-quality | Genome-fragment | 3.85 | AAI-based (high-confidence) | 0 |
| S11-final-viral-combined_k141_321949\|\|full | 2153 | No | NA | 2 | 0 | 1 | Low-quality | Genome-fragment | 0.98 | AAI-based (high-confidence) | 0 |
| S17-final-viral-combined_k141_258522\|\|full | 3504 | No | NA | 6 | 0 | 3 | Not-determined | Genome-fragment | NA | NA | 0 |
| S12-final-viral-combined_k141_320702\|\|full | 2153 | No | NA | 3 | 0 | 0 | Low-quality | Genome-fragment | 5.63 | AAI-based (high-confidence) | 0 |
| S2-final-viral-combined_k141_59174\|\|full | 2153 | No | NA | 5 | 2 | 0 | Low-quality | Genome-fragment | 3.73 | AAI-based (medium-confidence) | 0 |
| S17-final-viral-combined_k141_258535\|\|full | 1079 | No | NA | 3 | 0 | 2 | Not-determined | Genome-fragment | NA | NA | 0 |
| S4-final-viral-combined_k141_110381\|\|full | 2153 | No | NA | 3 | 0 | 0 | Low-quality | Genome-fragment | 0.77 | AAI-based (medium-confidence) | 0 |
| S5-final-viral-combined_k141_162286\|\|full | 2154 | No | NA | 3 | 0 | 0 | Low-quality | Genome-fragment | 4.92 | AAI-based (medium-confidence) | 0 |
| S17-final-viral-combined_k141_225585\|\|full | 1613 | No | NA | 3 | 0 | 0 | Not-determined | Genome-fragment | NA | NA | 0 |
| S17-final-viral-combined_k141_24859\|\|full | 3354 | No | NA | 6 | 0 | 0 | Not-determined | Genome-fragment | NA | NA | 0 |
| S17-final-viral-combined_k141_114904\|\|full | 1914 | No | NA | 3 | 0 | 1 | Not-determined | Genome-fragment | NA | NA | 0 |
| S17-final-viral-combined_k141_43444\|\|lt2gene | 2154 | No | NA | 1 | 1 | 0 | Low-quality | Genome-fragment | 7.11 | AAI-based (high-confidence) | 0 |
| S12-final-viral-combined_k141_41488\|\|full | 2154 | No | NA | 5 | 0 | 0 | Low-quality | Genome-fragment | 5.69 | AAI-based (high-confidence) | 0 |
| S17-final-viral-combined_k141_169790\|\|full | 2394 | No | NA | 3 | 0 | 1 | Not-determined | Genome-fragment | NA | NA | 0 |
| S17-final-viral-combined_k141_13661\|\|full | 1087 | No | NA | 3 | 0 | 1 | Not-determined | Genome-fragment | NA | NA | 0 |
| S17-final-viral-combined_k141_214648\|\|full | 3198 | No | NA | 5 | 0 | 2 | Not-determined | Genome-fragment | NA | NA | 0 |
| S17-final-viral-combined_k141_69929\|\|full | 1229 | No | NA | 3 | 0 | 1 | Not-determined | Genome-fragment | NA | NA | 0 |
| S17-final-viral-combined_k141_58367\|\|full | 1980 | No | NA | 3 | 0 | 1 | Not-determined | Genome-fragment | NA | NA | 0 |
| S17-final-viral-combined_k141_58368\|\|full | 2964 | No | NA | 3 | 0 | 2 | Not-determined | Genome-fragment | NA | NA | 0 |
| S12-final-viral-combined_k141_262959\|\|full | 2154 | No | NA | 4 | 2 | 0 | Low-quality | Genome-fragment | 2.06 | AAI-based (medium-confidence) | 0 |
| S17-final-viral-combined_k141_203579\|\|full | 2695 | No | NA | 5 | 0 | 2 | Not-determined | Genome-fragment | NA | NA | 0 |
| S17-final-viral-combined_k141_58374\|\|full | 1817 | No | NA | 2 | 0 | 0 | Not-determined | Genome-fragment | NA | NA | 0 |
| S17-final-viral-combined_k141_136622\|\|full | 26553 | No | NA | 40 | 16 | 0 | Medium-quality | Genome-fragment | 67.53 | AAI-based (high-confidence) | 0 |
| S12-final-viral-combined_k141_63905\|\|full | 2154 | No | NA | 3 | 0 | 2 | Low-quality | Genome-fragment | 2.42 | AAI-based (medium-confidence) | 0 |
| S17-final-viral-combined_k141_58384\|\|full | 2483 | No | NA | 4 | 0 | 3 | Not-determined | Genome-fragment | NA | NA | 0 |
| S17-final-viral-combined_k141_158140\|\|full | 1310 | No | NA | 3 | 0 | 1 | Not-determined | Genome-fragment | NA | NA | 0 |
| S17-final-viral-combined_k141_147662\|\|full | 2574 | No | NA | 3 | 0 | 1 | Not-determined | Genome-fragment | NA | NA | 0 |
| S17-final-viral-combined_k141_24923\|\|full | 4067 | No | NA | 3 | 0 | 3 | Not-determined | Genome-fragment | NA | NA | 0 |
| S17-final-viral-combined_k141_114986\|\|full | 1595 | No | NA | 2 | 0 | 1 | Not-determined | Genome-fragment | NA | NA | 0 |
| S12-final-viral-combined_k141_31158\|\|full | 2154 | No | NA | 2 | 0 | 0 | Low-quality | Genome-fragment | 5.88 | AAI-based (high-confidence) | 0 |
| S17-final-viral-combined_k141_3015\|\|full | 5002 | No | NA | 4 | 0 | 3 | Not-determined | Genome-fragment | NA | NA | 0 |
| S12_k141_263753 | 2154 | No | NA | 2 | 0 | 0 | Low-quality | Genome-fragment | 3.79 | AAI-based (high-confidence) | 0 |
| S17-final-viral-combined_k141_203659\|\|full | 1408 | No | NA | 3 | 0 | 2 | Not-determined | Genome-fragment | NA | NA | 0 |
| S2-final-viral-combined_k141_235701\|\|full | 2154 | No | NA | 5 | 1 | 0 | Low-quality | Genome-fragment | 5.56 | AAI-based (medium-confidence) | 0 |
| S17-final-viral-combined_k141_36394\|\|full | 1347 | No | NA | 3 | 0 | 2 | Not-determined | Genome-fragment | NA | NA | 0 |
| S1-final-viral-combined_k141_194852\|\|full | 2155 | No | NA | 4 | 0 | 1 | Low-quality | Genome-fragment | 5.06 | AAI-based (medium-confidence) | 0 |
| S17-final-viral-combined_k141_213357\|\|full | 2155 | No | NA | 5 | 0 | 0 | Low-quality | Genome-fragment | 3.38 | AAI-based (high-confidence) | 0 |
| S12-final-viral-combined_k141_112966\|\|full | 2155 | No | NA | 6 | 3 | 0 | Low-quality | Genome-fragment | 3.07 | AAI-based (high-confidence) | 0 |
| S17-final-viral-combined_k141_70019\|\|full | 1060 | No | NA | 3 | 0 | 0 | Not-determined | Genome-fragment | NA | NA | 0 |
| S17-final-viral-combined_k141_169863\|\|full | 2017 | No | NA | 3 | 0 | 2 | Not-determined | Genome-fragment | NA | NA | 0 |
| S11-final-viral-combined_k141_154535\|\|full | 2156 | No | NA | 4 | 0 | 1 | Low-quality | Genome-fragment | 1.41 | AAI-based (medium-confidence) | 0 |
| S1-final-viral-combined_k141_393177\|\|full | 2156 | No | NA | 4 | 2 | 0 | Low-quality | Genome-fragment | 4.96 | AAI-based (high-confidence) | 0 |
| S17-final-viral-combined_k141_214757\|\|full | 1303 | No | NA | 2 | 0 | 1 | Not-determined | Genome-fragment | NA | NA | 0 |
| S17-final-viral-combined_k141_115050\|\|full | 2631 | No | NA | 2 | 0 | 1 | Not-determined | Genome-fragment | NA | NA | 0 |
| S17-final-viral-combined_k141_3071\|\|full | 5335 | No | NA | 7 | 0 | 4 | Not-determined | Genome-fragment | NA | NA | 0 |
| S7-final-viral-combined_k141_216408\|\|full | 2156 | No | NA | 3 | 0 | 1 | Low-quality | Genome-fragment | 1.04 | AAI-based (medium-confidence) | 0 |
| S17-final-viral-combined_k141_81410\|\|full | 10609 | No | NA | 11 | 0 | 9 | Not-determined | Genome-fragment | NA | NA | 0 |
| S17-final-viral-combined_k141_158684\|\|full | 2156 | No | NA | 3 | 0 | 1 | Low-quality | Genome-fragment | 1.88 | AAI-based (high-confidence) | 0 |
| S2-final-viral-combined_k141_186785\|\|full | 2156 | No | NA | 4 | 0 | 0 | Low-quality | Genome-fragment | 4.43 | AAI-based (medium-confidence) | 0 |
| S17-final-viral-combined_k141_125737\|\|full | 4047 | No | NA | 5 | 0 | 3 | Not-determined | Genome-fragment | NA | NA | 0 |
| S17-final-viral-combined_k141_236396\|\|full | 1703 | No | NA | 3 | 0 | 1 | Not-determined | Genome-fragment | NA | NA | 0 |
| S2-final-viral-combined_k141_224554\|\|full | 2156 | No | NA | 5 | 3 | 0 | Low-quality | Genome-fragment | 6.2 | AAI-based (high-confidence) | 0 |
| S8-final-viral-combined_k141_140320\|\|full | 2156 | No | NA | 5 | 1 | 0 | Low-quality | Genome-fragment | 2.28 | AAI-based (high-confidence) | 0 |
| S8-final-viral-combined_k141_50674\|\|full | 2156 | No | NA | 3 | 0 | 1 | Low-quality | Genome-fragment | 1.14 | AAI-based (medium-confidence) | 0 |
| S12-final-viral-combined_k141_312937\|\|full | 2157 | No | NA | 5 | 4 | 0 | Low-quality | Genome-fragment | 3.36 | AAI-based (high-confidence) | 0 |
| S17-final-viral-combined_k141_46815\|\|full | 2427 | No | NA | 2 | 0 | 2 | Not-determined | Genome-fragment | NA | NA | 0 |
| S17-final-viral-combined_k141_102987\|\|full | 6597 | No | NA | 5 | 0 | 2 | Not-determined | Genome-fragment | NA | NA | 0 |
| S6-final-viral-combined_k141_59505\|\|full | 2157 | No | NA | 3 | 0 | 1 | Low-quality | Genome-fragment | 0.58 | AAI-based (high-confidence) | 0 |
| S11-final-viral-combined_k141_180394\|\|full | 2158 | No | NA | 6 | 0 | 0 | Low-quality | Genome-fragment | 1.61 | AAI-based (high-confidence) | 0 |
| S1-final-viral-combined_k141_6378\|\|full | 2158 | No | NA | 3 | 0 | 1 | Low-quality | Genome-fragment | 1.48 | AAI-based (high-confidence) | 0 |
| S1-final-viral-combined_k141_207919\|\|full | 2158 | No | NA | 3 | 0 | 0 | Low-quality | Genome-fragment | 3.52 | AAI-based (high-confidence) | 0 |
| S17-final-viral-combined_k141_203701\|\|full | 71387 | No | NA | 74 | 1 | 23 | Medium-quality | Genome-fragment | 63.38 | HMM-based (lower-bound) | 0 |
| S7-final-viral-combined_k141_104344\|\|full | 2158 | No | NA | 3 | 0 | 1 | Low-quality | Genome-fragment | 4.84 | AAI-based (high-confidence) | 0 |
| S7-final-viral-combined_k141_222628\|\|full | 2158 | No | NA | 4 | 2 | 0 | Low-quality | Genome-fragment | 2.62 | AAI-based (high-confidence) | 0 |
| S12_k141_384656 | 2158 | No | NA | 1 | 0 | 1 | Low-quality | Genome-fragment | 1.99 | AAI-based (medium-confidence) | 0 |
| S17-final-viral-combined_k141_225744\|\|full | 1463 | No | NA | 2 | 0 | 1 | Not-determined | Genome-fragment | NA | NA | 0 |
| S17-final-viral-combined_k141_169895\|\|full | 1184 | No | NA | 2 | 0 | 1 | Not-determined | Genome-fragment | NA | NA | 0 |
| S17-final-viral-combined_k141_115134\|\|full | 1573 | No | NA | 3 | 0 | 1 | Not-determined | Genome-fragment | NA | NA | 0 |
| S17-final-viral-combined_k141_81521\|\|full | 4554 | No | NA | 6 | 0 | 4 | Not-determined | Genome-fragment | NA | NA | 0 |
| S6-final-viral-combined_k141_96859\|\|full | 2158 | No | NA | 4 | 0 | 0 | Low-quality | Genome-fragment | 3.63 | AAI-based (high-confidence) | 0 |
| S17-final-viral-combined_k141_147836\|\|full | 2198 | No | NA | 3 | 0 | 2 | Not-determined | Genome-fragment | NA | NA | 0 |
| S17-final-viral-combined_k141_103031\|\|full | 4299 | No | NA | 3 | 0 | 2 | Not-determined | Genome-fragment | NA | NA | 0 |
| S9-final-viral-combined_k141_35748\|\|full | 2159 | No | NA | 4 | 2 | 0 | Low-quality | Genome-fragment | 5.4 | AAI-based (medium-confidence) | 0 |
| S3-final-viral-combined_k141_61519\|\|lt2gene | 2159 | No | NA | 1 | 1 | 0 | Low-quality | Genome-fragment | 1.41 | AAI-based (high-confidence) | 0 |
| S17-final-viral-combined_k141_158222\|\|full | 3523 | No | NA | 6 | 0 | 0 | Not-determined | Genome-fragment | NA | NA | 0 |
| S5-final-viral-combined_k141_122001\|\|full | 2159 | No | NA | 4 | 3 | 0 | Low-quality | Genome-fragment | 4.09 | AAI-based (high-confidence) | 0 |
| S17-final-viral-combined_k141_3187\|\|full | 2667 | No | NA | 4 | 0 | 2 | Not-determined | Genome-fragment | NA | NA | 0 |
| S17-final-viral-combined_k141_84302\|\|full | 2159 | No | NA | 3 | 0 | 1 | Low-quality | Genome-fragment | 1.07 | AAI-based (medium-confidence) | 0 |
| S17-final-viral-combined_k141_91426\|\|full | 3503 | No | NA | 3 | 0 | 3 | Not-determined | Genome-fragment | NA | NA | 0 |
| S17-final-viral-combined_k141_36508\|\|full | 1226 | No | NA | 2 | 0 | 2 | Not-determined | Genome-fragment | NA | NA | 0 |
| S17-final-viral-combined_k141_125844\|\|full | 2257 | No | NA | 3 | 0 | 0 | Not-determined | Genome-fragment | NA | NA | 0 |
| S12-final-viral-combined_k141_157529\|\|full | 2159 | No | NA | 4 | 2 | 0 | Low-quality | Genome-fragment | 2.18 | AAI-based (high-confidence) | 0 |
| S17-final-viral-combined_k141_203794\|\|full | 1880 | No | NA | 2 | 0 | 1 | Not-determined | Genome-fragment | NA | NA | 0 |
| S17-final-viral-combined_k141_225790\|\|full | 3558 | No | NA | 8 | 0 | 4 | Not-determined | Genome-fragment | NA | NA | 0 |
| S17-final-viral-combined_k141_136863\|\|full | 2838 | No | NA | 2 | 0 | 2 | Not-determined | Genome-fragment | NA | NA | 0 |
| S17-final-viral-combined_k141_125875\|\|full | 1766 | No | NA | 2 | 0 | 0 | Not-determined | Genome-fragment | NA | NA | 0 |
| S2-final-viral-combined_k141_127985\|\|full | 2159 | No | NA | 4 | 0 | 0 | Low-quality | Genome-fragment | 4.08 | AAI-based (medium-confidence) | 0 |
| S4_k141_107476 | 2159 | No | NA | 2 | 2 | 0 | Low-quality | Genome-fragment | 1.24 | AAI-based (high-confidence) | 0 |
| S8-final-viral-combined_k141_40461\|\|lt2gene | 2159 | No | NA | 2 | 2 | 0 | Low-quality | Genome-fragment | 5.42 | AAI-based (high-confidence) | 0 |
| S11-final-viral-combined_k141_179142\|\|full | 2160 | No | NA | 4 | 0 | 0 | Low-quality | Genome-fragment | 1.85 | AAI-based (medium-confidence) | 0 |
| S1-final-viral-combined_k141_23305\|\|lt2gene | 2160 | No | NA | 2 | 1 | 0 | Low-quality | Genome-fragment | 1.36 | AAI-based (high-confidence) | 0 |
| S17-final-viral-combined_k141_103126\|\|full | 2653 | No | NA | 3 | 0 | 2 | Not-determined | Genome-fragment | NA | NA | 0 |
| S17-final-viral-combined_k141_203826\|\|full | 1700 | No | NA | 2 | 0 | 2 | Not-determined | Genome-fragment | NA | NA | 0 |
| S17-final-viral-combined_k141_13796\|\|full | 1917 | No | NA | 3 | 0 | 0 | Not-determined | Genome-fragment | NA | NA | 0 |
| S17-final-viral-combined_k141_203837\|\|full | 3088 | No | NA | 2 | 0 | 2 | Not-determined | Genome-fragment | NA | NA | 0 |
| S17-final-viral-combined_k141_181421\|\|full | 7718 | No | NA | 9 | 0 | 6 | Not-determined | Genome-fragment | NA | NA | 0 |
| S17-final-viral-combined_k141_192519\|\|full | 2071 | No | NA | 4 | 0 | 1 | Not-determined | Genome-fragment | NA | NA | 0 |
| S5-final-viral-combined_k141_190224\|\|full | 2160 | No | NA | 3 | 0 | 0 | Low-quality | Genome-fragment | 6.03 | AAI-based (medium-confidence) | 0 |
| S5-final-viral-combined_k141_70609\|\|full | 2160 | No | NA | 2 | 1 | 0 | Low-quality | Genome-fragment | 4.31 | AAI-based (medium-confidence) | 0 |
| S7-final-viral-combined_k141_91465\|\|full | 2160 | No | NA | 3 | 3 | 0 | Low-quality | Genome-fragment | 5.64 | AAI-based (medium-confidence) | 0 |
| S17-final-viral-combined_k141_139696\|\|full | 2161 | No | NA | 3 | 2 | 0 | Low-quality | Genome-fragment | 4.89 | AAI-based (high-confidence) | 0 |
| S17-final-viral-combined_k141_81599\|\|full | 11351 | No | NA | 12 | 0 | 6 | Not-determined | Genome-fragment | NA | NA | 0 |
| S17-final-viral-combined_k141_247211\|\|full | 4306 | No | NA | 5 | 0 | 0 | Not-determined | Genome-fragment | NA | NA | 0 |
| S17-final-viral-combined_k141_203868\|\|full | 5083 | No | NA | 5 | 0 | 3 | Not-determined | Genome-fragment | NA | NA | 0 |
| S8-final-viral-combined_k141_94950\|\|full | 2161 | No | NA | 4 | 0 | 0 | Low-quality | Genome-fragment | 5.34 | AAI-based (medium-confidence) | 0 |
| S1-final-viral-combined_k141_381637\|\|full | 2162 | No | NA | 1 | 0 | 0 | Low-quality | Genome-fragment | 4.73 | AAI-based (medium-confidence) | 0 |
| S17-final-viral-combined_k141_170009\|\|full | 1504 | No | NA | 2 | 0 | 2 | Not-determined | Genome-fragment | NA | NA | 0 |
| S5-final-viral-combined_k141_181085\|\|full | 2162 | No | NA | 6 | 2 | 0 | Low-quality | Genome-fragment | 5.04 | AAI-based (high-confidence) | 0 |
| S17-final-viral-combined_k141_167926\|\|full | 2162 | No | NA | 3 | 0 | 3 | Low-quality | Genome-fragment | 1.07 | AAI-based (medium-confidence) | 0 |

| | | | | | | | | | | | |
|---|---|---|---|---|---|---|---|---|---|---|---|
| S12-final-viral-combined_k141_360526||full | 2162 | No | NA | 5 | 0 | 0 | Low-quality | Genome-fragment | 4.15 | AAI-based (high-confidence) | 0 |
| S17-final-viral-combined_k141_36614||full | 1654 | No | NA | 3 | 0 | 1 | Not-determined | Genome-fragment | NA | NA | 0 |
| S12_k141_354730 | 2162 | No | NA | 5 | 1 | 0 | Low-quality | Genome-fragment | 1.18 | AAI-based (high-confidence) | 0 |
| S4-final-viral-combined_k141_35785||full | 2162 | No | NA | 4 | 0 | 0 | Low-quality | Genome-fragment | 1.76 | AAI-based (high-confidence) | 0 |
| S17-final-viral-combined_k141_158354||full | 41783 | No | NA | 55 | 12 | 1 | Medium-quality | Genome-fragment | 64.61 | AAI-based (high-confidence) | 0 |
| S17-final-viral-combined_k141_236583||full | 17497 | No | NA | 15 | 0 | 7 | Not-determined | Genome-fragment | NA | NA | 0 |
| S17-final-viral-combined_k141_36624||full | 2102 | No | NA | 2 | 0 | 1 | Not-determined | Genome-fragment | NA | NA | 0 |
| S17-final-viral-combined_k141_214928||full | 2397 | No | NA | 2 | 0 | 2 | Not-determined | Genome-fragment | NA | NA | 0 |
| S8-final-viral-combined_k141_191159||full | 2162 | No | NA | 4 | 0 | 0 | Low-quality | Genome-fragment | 4.06 | AAI-based (high-confidence) | 0 |
| S17-final-viral-combined_k141_225914||full | 1171 | No | NA | 2 | 0 | 0 | Not-determined | Genome-fragment | NA | NA | 0 |
| S11-final-viral-combined_k141_124092||full | 2163 | No | NA | 5 | 0 | 0 | Low-quality | Genome-fragment | 5.61 | AAI-based (high-confidence) | 0 |
| S17-final-viral-combined_k141_181555||full | 1585 | No | NA | 2 | 0 | 1 | Not-determined | Genome-fragment | NA | NA | 0 |
| S17-final-viral-combined_k141_91559||full | 2050 | No | NA | 3 | 0 | 2 | Not-determined | Genome-fragment | NA | NA | 0 |
| S11-final-viral-combined_k141_18315||full | 2163 | No | NA | 6 | 0 | 1 | Low-quality | Genome-fragment | 4.64 | AAI-based (medium-confidence) | 0 |
| S11-final-viral-combined_k141_149839||lt2gene | 2163 | No | NA | 2 | 2 | 0 | Low-quality | Genome-fragment | 3.54 | AAI-based (high-confidence) | 0 |
| S17-final-viral-combined_k141_91560||full | 4195 | No | NA | 5 | 0 | 0 | Not-determined | Genome-fragment | NA | NA | 0 |
| S17-final-viral-combined_k141_225933||full | 2543 | No | NA | 2 | 0 | 0 | Not-determined | Genome-fragment | NA | NA | 0 |
| S1_k141_168049 | 2163 | No | NA | 1 | 0 | 1 | Low-quality | Genome-fragment | 0.81 | AAI-based (high-confidence) | 0 |
| S17-final-viral-combined_k141_145665||full | 2163 | No | NA | 3 | 0 | 1 | Low-quality | Genome-fragment | 4.24 | AAI-based (high-confidence) | 0 |
| S4_k141_7556 | 2163 | No | NA | 1 | 0 | 0 | Low-quality | Genome-fragment | 0.94 | AAI-based (medium-confidence) | 0 |
| S17-final-viral-combined_k141_192636||full | 2200 | No | NA | 2 | 0 | 1 | Not-determined | Genome-fragment | NA | NA | 0 |
| S4_k141_15012 | 2163 | No | NA | 2 | 0 | 2 | Low-quality | Genome-fragment | 1.65 | AAI-based (medium-confidence) | 0 |
| S6-final-viral-combined_k141_59833||full | 2163 | No | NA | 5 | 1 | 0 | Low-quality | Genome-fragment | 5.25 | AAI-based (medium-confidence) | 0 |
| S11-final-viral-combined_k141_73441||full | 2164 | No | NA | 5 | 2 | 0 | Low-quality | Genome-fragment | 3.33 | AAI-based (high-confidence) | 0 |
| S17-final-viral-combined_k141_192642||full | 5750 | No | NA | 7 | 0 | 6 | Not-determined | Genome-fragment | NA | NA | 0 |
| S1-final-viral-combined_k141_150263||full | 2164 | No | NA | 3 | 0 | 0 | Low-quality | Genome-fragment | 4.12 | AAI-based (medium-confidence) | 0 |
| S17-final-viral-combined_k141_81752||full | 1910 | No | NA | 4 | 0 | 1 | Not-determined | Genome-fragment | NA | NA | 0 |
| S3-final-viral-combined_k141_125099||full | 2164 | No | NA | 5 | 0 | 0 | Low-quality | Genome-fragment | 4.21 | AAI-based (medium-confidence) | 0 |
| S12-final-viral-combined_k141_77913||full | 2164 | No | NA | 3 | 1 | 0 | Low-quality | Genome-fragment | 2.83 | HMM-based (lower-bound) | 0 |
| S17-final-viral-combined_k141_226006||full | 6392 | No | NA | 7 | 0 | 3 | Not-determined | Genome-fragment | NA | NA | 0 |
| S2-final-viral-combined_k141_179018||full | 2164 | No | NA | 3 | 1 | 0 | Low-quality | Genome-fragment | 5.13 | AAI-based (high-confidence) | 0 |
| S17-final-viral-combined_k141_137062||full | 3234 | No | NA | 3 | 0 | 2 | Not-determined | Genome-fragment | NA | NA | 0 |
| S17-final-viral-combined_k141_25313||full | 6851 | No | NA | 8 | 0 | 2 | Not-determined | Genome-fragment | NA | NA | 0 |
| S17-final-viral-combined_k141_126114||full | 3972 | No | NA | 4 | 0 | 4 | Not-determined | Genome-fragment | NA | NA | 0 |
| S2_k141_176397 | 2164 | No | NA | 5 | 0 | 0 | Low-quality | Genome-fragment | 1.12 | AAI-based (high-confidence) | 0 |
| S8-final-viral-combined_k141_244157||full | 2164 | No | NA | 3 | 0 | 1 | Low-quality | Genome-fragment | 2.55 | AAI-based (high-confidence) | 0 |
| S1-final-viral-combined_k141_348199||full | 2165 | No | NA | 2 | 0 | 1 | Low-quality | Genome-fragment | 2.27 | AAI-based (high-confidence) | 0 |
| S17-final-viral-combined_k141_25339||full | 2033 | No | NA | 4 | 0 | 1 | Not-determined | Genome-fragment | NA | NA | 0 |
| S17-k141_161873 | 2165 | No | NA | 3 | 2 | 0 | Low-quality | Genome-fragment | 3.51 | AAI-based (high-confidence) | 0 |
| S17-final-viral-combined_k141_192710||full | 1214 | No | NA | 3 | 0 | 1 | Not-determined | Genome-fragment | NA | NA | 0 |
| S2-final-viral-combined_k141_154395||full | 2165 | Yes | 827 | 2 | 1 | 1 | Low-quality | Genome-fragment | 0.55 | HMM-based (lower-bound) | 61.8 |
| S1_k141_118316 | 2166 | No | NA | 4 | 0 | 1 | Low-quality | Genome-fragment | 3.4 | AAI-based (medium-confidence) | 0 |
| S17-final-viral-combined_k141_236702||full | 1721 | No | NA | 2 | 0 | 2 | Not-determined | Genome-fragment | NA | NA | 0 |
| S17-k141_119097 | 2166 | No | NA | 4 | 2 | 0 | Low-quality | Genome-fragment | 4.98 | AAI-based (high-confidence) | 0 |
| S17-final-viral-combined_k141_25358||full | 1605 | No | NA | 2 | 0 | 2 | Not-determined | Genome-fragment | NA | NA | 0 |
| S2-final-viral-combined_k141_1745||full | 2166 | No | NA | 5 | 2 | 0 | Low-quality | Genome-fragment | 3.95 | AAI-based (medium-confidence) | 0 |
| S17-final-viral-combined_k141_25365||full | 4261 | No | NA | 5 | 0 | 4 | Not-determined | Genome-fragment | NA | NA | 0 |
| S4-final-viral-combined_k141_103586||full | 2166 | No | NA | 3 | 0 | 1 | Low-quality | Genome-fragment | 4.29 | AAI-based (high-confidence) | 0 |
| S11-final-viral-combined_k141_214071||full | 2167 | No | NA | 5 | 1 | 0 | Low-quality | Genome-fragment | 3.3 | HMM-based (lower-bound) | 0 |
| S1-final-viral-combined_k141_232273||full | 2167 | Yes | 862 | 2 | 1 | 1 | Low-quality | Genome-fragment | 1.5 | AAI-based (medium-confidence) | 60.22 |
| S17-final-viral-combined_k141_70450||full | 3552 | No | NA | 4 | 0 | 2 | Not-determined | Genome-fragment | NA | NA | 0 |
| S4-final-viral-combined_k141_39418||lt2gene | 2167 | No | NA | 2 | 1 | 0 | Low-quality | Genome-fragment | 2.94 | AAI-based (high-confidence) | 0 |
| S9-final-viral-combined_k141_83763||full | 2168 | No | NA | 2 | 0 | 2 | Low-quality | Genome-fragment | 1.25 | AAI-based (medium-confidence) | 0 |
| S17-final-viral-combined_k141_36782||full | 1168 | No | NA | 2 | 0 | 2 | Not-determined | Genome-fragment | NA | NA | 0 |
| S9_k141_52596 | 2168 | No | NA | 6 | 0 | 0 | Low-quality | Genome-fragment | 3.62 | AAI-based (medium-confidence) | 0 |
| S11-final-viral-combined_k141_212093||full | 2168 | No | NA | 6 | 0 | 0 | Low-quality | Genome-fragment | 4.83 | AAI-based (high-confidence) | 0 |
| S1-final-viral-combined_k141_34057||full | 2168 | No | NA | 2 | 0 | 2 | Low-quality | Genome-fragment | 1.23 | AAI-based (high-confidence) | 0 |
| S17-final-viral-combined_k141_148196||full | 1266 | No | NA | 3 | 0 | 0 | Not-determined | Genome-fragment | NA | NA | 0 |
| S17-final-viral-combined_k141_70513||full | 1160 | No | NA | 2 | 0 | 1 | Not-determined | Genome-fragment | NA | NA | 0 |
| S1_k141_94798 | 2168 | No | NA | 3 | 1 | 0 | Low-quality | Genome-fragment | 4.3 | AAI-based (high-confidence) | 0 |
| S17-final-viral-combined_k141_192835||full | 1886 | No | NA | 5 | 0 | 1 | Not-determined | Genome-fragment | NA | NA | 0 |
| S17-final-viral-combined_k141_81900||full | 2172 | No | NA | 3 | 0 | 0 | Not-determined | Genome-fragment | NA | NA | 0 |
| S5-final-viral-combined_k141_35659||full | 2168 | No | NA | 7 | 3 | 0 | Low-quality | Genome-fragment | 6.76 | AAI-based (high-confidence) | 0 |
| S17-final-viral-combined_k141_215264||full | 1681 | No | NA | 4 | 0 | 0 | Not-determined | Genome-fragment | NA | NA | 0 |
| S7-final-viral-combined_k141_192776||full | 2168 | No | NA | 3 | 0 | 0 | Low-quality | Genome-fragment | 4.89 | AAI-based (medium-confidence) | 0 |
| S17-final-viral-combined_k141_204122||full | 1376 | No | NA | 2 | 0 | 2 | Not-determined | Genome-fragment | NA | NA | 0 |
| S17-final-viral-combined_k141_215291||full | 2532 | No | NA | 3 | 0 | 0 | Not-determined | Genome-fragment | NA | NA | 0 |
| S17-final-viral-combined_k141_256895||full | 2168 | No | NA | 3 | 1 | 0 | Low-quality | Genome-fragment | 3.72 | AAI-based (high-confidence) | 0 |
| S17-final-viral-combined_k141_91755||full | 6055 | No | NA | 7 | 0 | 3 | Not-determined | Genome-fragment | NA | NA | 0 |
| S17-final-viral-combined_k141_91759||full | 1917 | No | NA | 3 | 0 | 2 | Not-determined | Genome-fragment | NA | NA | 0 |
| S4_k141_71792 | 2168 | No | NA | 3 | 0 | 0 | Low-quality | Genome-fragment | 2.12 | AAI-based (high-confidence) | 0 |
| S17-final-viral-combined_k141_258887||full | 1658 | No | NA | 3 | 0 | 1 | Not-determined | Genome-fragment | NA | NA | 0 |
| S17-final-viral-combined_k141_247494||full | 49880 | Yes | 30128 | 60 | 18 | 9 | Medium-quality | Genome-fragment | 80.49 | AAI-based (high-confidence) | 39.6 |
| S8-final-viral-combined_k141_260340||full | 2168 | No | NA | 3 | 0 | 0 | Low-quality | Genome-fragment | 3.55 | AAI-based (medium-confidence) | 0 |
| S17-final-viral-combined_k141_215333||full | 1163 | No | NA | 2 | 0 | 2 | Not-determined | Genome-fragment | NA | NA | 0 |
| S17-final-viral-combined_k141_226149||full | 1363 | No | NA | 3 | 0 | 0 | Not-determined | Genome-fragment | NA | NA | 0 |
| S17-final-viral-combined_k141_103471||full | 1604 | No | NA | 3 | 0 | 1 | Not-determined | Genome-fragment | NA | NA | 0 |
| S11-final-viral-combined_k141_153081||full | 2169 | No | NA | 3 | 1 | 0 | Low-quality | Genome-fragment | 4.96 | AAI-based (medium-confidence) | 0 |
| S17-final-viral-combined_k141_70605||full | 4849 | No | NA | 4 | 0 | 0 | Not-determined | Genome-fragment | NA | NA | 0 |
| S17-final-viral-combined_k141_247507||full | 2464 | No | NA | 5 | 0 | 2 | Not-determined | Genome-fragment | NA | NA | 0 |
| S17-final-viral-combined_k141_170298||full | 1505 | No | NA | 3 | 0 | 3 | Not-determined | Genome-fragment | NA | NA | 0 |
| S17-final-viral-combined_k141_70624||full | 1087 | No | NA | 2 | 0 | 2 | Not-determined | Genome-fragment | NA | NA | 0 |
| S11-final-viral-combined_k141_224870||full | 2169 | No | NA | 5 | 4 | 0 | Low-quality | Genome-fragment | 4.02 | AAI-based (high-confidence) | 0 |
| S11-final-viral-combined_k141_79246||lt2gene | 2169 | No | NA | 2 | 1 | 0 | Low-quality | Genome-fragment | 4.93 | AAI-based (medium-confidence) | 0 |
| S17-final-viral-combined_k141_58979||full | 2047 | No | NA | 2 | 0 | 2 | Not-determined | Genome-fragment | NA | NA | 0 |
| S1-final-viral-combined_k141_334016||full | 2169 | No | NA | 3 | 0 | 0 | Low-quality | Genome-fragment | 4.69 | AAI-based (high-confidence) | 0 |
| S17-final-viral-combined_k141_103489||full | 8824 | No | NA | 13 | 0 | 2 | Not-determined | Genome-fragment | NA | NA | 0 |
| S1-final-viral-combined_k141_93121||full | 2169 | No | NA | 3 | 2 | 0 | Low-quality | Genome-fragment | 4.81 | AAI-based (high-confidence) | 0 |
| S17-final-viral-combined_k141_158581||full | 1386 | No | NA | 3 | 0 | 1 | Not-determined | Genome-fragment | NA | NA | 0 |
| S17-final-viral-combined_k141_25570||full | 1683 | No | NA | 2 | 0 | 0 | Not-determined | Genome-fragment | NA | NA | 0 |
| S17-final-viral-combined_k141_36805||full | 1869 | No | NA | 3 | 0 | 1 | Not-determined | Genome-fragment | NA | NA | 0 |
| S17-final-viral-combined_k141_126261||full | 1485 | No | NA | 2 | 0 | 0 | Not-determined | Genome-fragment | NA | NA | 0 |
| S3_k141_90413 | 2169 | No | NA | 5 | 0 | 0 | Low-quality | Genome-fragment | 3.48 | AAI-based (high-confidence) | 0 |
| S7-final-viral-combined_k141_111391||full | 2169 | Yes | 1274 | 4 | 1 | 1 | Low-quality | Genome-fragment | 2.14 | AAI-based (medium-confidence) | 41.26 |
| S17-final-viral-combined_k141_36838||full | 3586 | No | NA | 6 | 0 | 0 | Not-determined | Genome-fragment | NA | NA | 0 |
| S17-final-viral-combined_k141_103550||full | 1141 | No | NA | 3 | 0 | 0 | Not-determined | Genome-fragment | NA | NA | 0 |
| S17-final-viral-combined_k141_204206||full | 1967 | No | NA | 4 | 0 | 2 | Not-determined | Genome-fragment | NA | NA | 0 |
| S17-final-viral-combined_k141_203699||full | 2169 | No | NA | 4 | 0 | 2 | Low-quality | Genome-fragment | 6.14 | AAI-based (medium-confidence) | 0 |
| S17-final-viral-combined_k141_192939||full | 3087 | No | NA | 4 | 0 | 4 | Not-determined | Genome-fragment | NA | NA | 0 |
| S17-final-viral-combined_k141_170675||full | 2169 | No | NA | 3 | 0 | 2 | Low-quality | Genome-fragment | 0.6 | AAI-based (high-confidence) | 0 |
| S17-final-viral-combined_k141_49554||full | 2169 | No | NA | 2 | 0 | 1 | Low-quality | Genome-fragment | 3.69 | AAI-based (high-confidence) | 0 |
| S6-final-viral-combined_k141_14217||full | 2169 | No | NA | 3 | 0 | 0 | Low-quality | Genome-fragment | 3.01 | AAI-based (high-confidence) | 0 |
| S17-final-viral-combined_k141_258985||full | 2417 | No | NA | 3 | 0 | 1 | Not-determined | Genome-fragment | NA | NA | 0 |
| S17-final-viral-combined_k141_82086||full | 1699 | No | NA | 2 | 0 | 2 | Not-determined | Genome-fragment | NA | NA | 0 |
| S17-final-viral-combined_k141_226235||full | 2094 | No | NA | 4 | 0 | 1 | Not-determined | Genome-fragment | NA | NA | 0 |
| S17-final-viral-combined_k141_103590||full | 1074 | No | NA | 4 | 0 | 0 | Not-determined | Genome-fragment | NA | NA | 0 |
| S17-final-viral-combined_k141_259001||full | 1916 | No | NA | 2 | 0 | 0 | Not-determined | Genome-fragment | NA | NA | 0 |
| S17-final-viral-combined_k141_158675||full | 2838 | No | NA | 3 | 0 | 1 | Not-determined | Genome-fragment | NA | NA | 0 |
| S17-final-viral-combined_k141_204242||full | 1346 | No | NA | 2 | 0 | 0 | Not-determined | Genome-fragment | NA | NA | 0 |
| S11-final-viral-combined_k141_107251||full | 2170 | No | NA | 5 | 0 | 0 | Low-quality | Genome-fragment | 4.52 | AAI-based (high-confidence) | 0 |
| S17-final-viral-combined_k141_70807||full | 1129 | No | NA | 2 | 0 | 0 | Not-determined | Genome-fragment | NA | NA | 0 |
| S1-final-viral-combined_k141_86525||full | 2170 | No | NA | 3 | 0 | 0 | Low-quality | Genome-fragment | 3.63 | AAI-based (medium-confidence) | 0 |
| S17-final-viral-combined_k141_215481||full | 2545 | No | NA | 5 | 0 | 0 | Not-determined | Genome-fragment | NA | NA | 0 |
| S12-final-viral-combined_k141_19468||full | 2170 | No | NA | 4 | 2 | 0 | Low-quality | Genome-fragment | 2.7 | HMM-based (lower-bound) | 0 |
| S2_k141_130970 | 2170 | No | NA | 3 | 0 | 1 | Low-quality | Genome-fragment | 4.42 | AAI-based (high-confidence) | 0 |
| S4-final-viral-combined_k141_39066||full | 2170 | No | NA | 2 | 0 | 2 | Low-quality | Genome-fragment | 3.4 | AAI-based (medium-confidence) | 0 |
| S9-final-viral-combined_k141_64705||full | 2171 | No | NA | 4 | 3 | 0 | Low-quality | Genome-fragment | 4.73 | AAI-based (high-confidence) | 0 |
| S9-final-viral-combined_k141_38065||full | 2171 | No | NA | 5 | 1 | 0 | Low-quality | Genome-fragment | 3 | AAI-based (medium-confidence) | 0 |
| S17-final-viral-combined_k141_158721||full | 3689 | No | NA | 7 | 0 | 7 | Not-determined | Genome-fragment | NA | NA | 0 |
| S17-final-viral-combined_k141_137299||full | 5258 | No | NA | 5 | 0 | 5 | Not-determined | Genome-fragment | NA | NA | 0 |
| S17-final-viral-combined_k141_115906||full | 1385 | No | NA | 2 | 0 | 2 | Not-determined | Genome-fragment | NA | NA | 0 |
| S7-final-viral-combined_k141_67708||lt2gene | 2171 | No | NA | 2 | 1 | 0 | Low-quality | Genome-fragment | 5.25 | AAI-based (high-confidence) | 0 |
| S17-final-viral-combined_k141_247601||full | 4735 | No | NA | 5 | 0 | 4 | Not-determined | Genome-fragment | NA | NA | 0 |
| S17-final-viral-combined_k141_170447||full | 1777 | No | NA | 3 | 0 | 0 | Not-determined | Genome-fragment | NA | NA | 0 |
| S17-final-viral-combined_k141_137321||full | 2078 | No | NA | 3 | 0 | 0 | Not-determined | Genome-fragment | NA | NA | 0 |
| S12-final-viral-combined_k141_328794||full | 2171 | No | NA | 3 | 0 | 0 | Low-quality | Genome-fragment | 1 | AAI-based (high-confidence) | 0 |
| S17-final-viral-combined_k141_204336||full | 3035 | No | NA | 3 | 0 | 2 | Not-determined | Genome-fragment | NA | NA | 0 |
| S17-final-viral-combined_k141_137335||full | 1133 | No | NA | 3 | 0 | 1 | Not-determined | Genome-fragment | NA | NA | 0 |
| S17-final-viral-combined_k141_204350||full | 1330 | No | NA | 3 | 0 | 0 | Not-determined | Genome-fragment | NA | NA | 0 |
| S12_k141_261183 | 2171 | No | NA | 7 | 0 | 0 | Low-quality | Genome-fragment | 1.19 | AAI-based (high-confidence) | 0 |
| S17-final-viral-combined_k141_182030||full | 4404 | No | NA | 5 | 0 | 1 | Not-determined | Genome-fragment | NA | NA | 0 |
| S4-final-viral-combined_k141_31694||full | 2171 | No | NA | 2 | 1 | 0 | Low-quality | Genome-fragment | 2.12 | AAI-based (high-confidence) | 0 |
| S8-final-viral-combined_k141_107332||full | 2171 | No | NA | 4 | 0 | 0 | Low-quality | Genome-fragment | 1.39 | AAI-based (medium-confidence) | 0 |
| S17-final-viral-combined_k141_115933||full | 14730 | No | NA | 10 | 0 | 5 | Not-determined | Genome-fragment | NA | NA | 0 |
| S17-final-viral-combined_k141_36922||full | 3359 | No | NA | 3 | 0 | 3 | Not-determined | Genome-fragment | NA | NA | 0 |
| S9-final-viral-combined_k141_78931||full | 2172 | No | NA | 3 | 0 | 3 | Low-quality | Genome-fragment | 1.56 | AAI-based (medium-confidence) | 0 |
| S17-final-viral-combined_k141_82191||full | 2789 | No | NA | 3 | 0 | 2 | Not-determined | Genome-fragment | NA | NA | 0 |
| S11-final-viral-combined_k141_85182||full | 2172 | No | NA | 4 | 1 | 0 | Low-quality | Genome-fragment | 2.92 | HMM-based (lower-bound) | 0 |
| S17-final-viral-combined_k141_193086||full | 3240 | No | NA | 6 | 0 | 0 | Not-determined | Genome-fragment | NA | NA | 0 |
| S17-final-viral-combined_k141_25721||full | 1271 | No | NA | 3 | 0 | 1 | Not-determined | Genome-fragment | NA | NA | 0 |
| S17-final-viral-combined_k141_82204||full | 2015 | No | NA | 3 | 0 | 0 | Not-determined | Genome-fragment | NA | NA | 0 |
| S17-final-viral-combined_k141_82212||full | 5699 | No | NA | 4 | 0 | 4 | Not-determined | Genome-fragment | NA | NA | 0 |
| S1-final-viral-combined_k141_356133||full | 2172 | No | NA | 3 | 0 | 0 | Low-quality | Genome-fragment | 7.63 | AAI-based (high-confidence) | 0 |
| S17-final-viral-combined_k141_115972||full | 11473 | No | NA | 6 | 0 | 6 | Not-determined | Genome-fragment | NA | NA | 0 |
| S17-final-viral-combined_k141_247611||full | 6086 | No | NA | 6 | 0 | 5 | Not-determined | Genome-fragment | NA | NA | 0 |
| S1-final-viral-combined_k141_76413||full | 2172 | No | NA | 4 | 1 | 0 | Low-quality | Genome-fragment | 2.84 | HMM-based (lower-bound) | 0 |
| S17-final-viral-combined_k141_158810||full | 1889 | No | NA | 4 | 0 | 0 | Not-determined | Genome-fragment | NA | NA | 0 |
| S12-final-viral-combined_k141_365442||full | 2172 | No | NA | 3 | 0 | 1 | Low-quality | Genome-fragment | 0.62 | AAI-based (high-confidence) | 0 |
| S17-final-viral-combined_k141_193144||full | 2082 | No | NA | 3 | 0 | 2 | Not-determined | Genome-fragment | NA | NA | 0 |
| S8-final-viral-combined_k141_237708||full | 2172 | No | NA | 3 | 0 | 1 | Low-quality | Genome-fragment | 4.5 | AAI-based (medium-confidence) | 0 |
| S17-final-viral-combined_k141_116034||full | 1013 | No | NA | 2 | 0 | 1 | Not-determined | Genome-fragment | NA | NA | 0 |
| S17-final-viral-combined_k141_237002||full | 1058 | No | NA | 3 | 0 | 0 | Not-determined | Genome-fragment | NA | NA | 0 |
| S1-final-viral-combined_k141_3202||full | 2173 | No | NA | 6 | 2 | 0 | Low-quality | Genome-fragment | 5.39 | AAI-based (high-confidence) | 0 |
| S12-final-viral-combined_k141_374762||full | 2173 | No | NA | 3 | 0 | 2 | Low-quality | Genome-fragment | 5.02 | AAI-based (medium-confidence) | 0 |
| S17-final-viral-combined_k141_80152||full | 2174 | No | NA | 4 | 0 | 2 | Low-quality | Genome-fragment | 0.69 | AAI-based (medium-confidence) | 0 |
| S17-final-viral-combined_k141_237008||full | 1224 | No | NA | 1 | 0 | 1 | Not-determined | Genome-fragment | NA | NA | 0 |
| S17-final-viral-combined_k141_193978||full | 2174 | Yes | 1359 | 4 | 1 | 1 | Low-quality | Genome-fragment | 2.49 | HMM-based (lower-bound) | 37.49 |
| S17-final-viral-combined_k141_126411||full | 2021 | No | NA | 3 | 0 | 0 | Not-determined | Genome-fragment | NA | NA | 0 |
| S17-final-viral-combined_k141_70921||full | 29900 | No | NA | 31 | 0 | 19 | Not-determined | Genome-fragment | NA | NA | 0 |
| S9-final-viral-combined_k141_3000||full | 2175 | No | NA | 3 | 1 | 0 | Low-quality | Genome-fragment | 2.6 | AAI-based (high-confidence) | 0 |
| S17-final-viral-combined_k141_237039||full | 1701 | No | NA | 2 | 0 | 1 | Not-determined | Genome-fragment | NA | NA | 0 |
| S1-final-viral-combined_k141_182963||full | 2175 | No | NA | 3 | 0 | 0 | Low-quality | Genome-fragment | 4.94 | AAI-based (high-confidence) | 0 |
| S12-final-viral-combined_k141_56240||full | 2175 | No | NA | 2 | 0 | 1 | Low-quality | Genome-fragment | 4.6 | AAI-based (medium-confidence) | 0 |
| S17-final-viral-combined_k141_70936||full | 1026 | No | NA | 3 | 0 | 1 | Not-determined | Genome-fragment | NA | NA | 0 |
| S12_k141_276172 | 2175 | No | NA | 1 | 0 | 0 | Low-quality | Genome-fragment | 1.17 | AAI-based (high-confidence) | 0 |
| S17-final-viral-combined_k141_3722||full | 6271 | No | NA | 4 | 0 | 1 | Not-determined | Genome-fragment | NA | NA | 0 |
| S17-final-viral-combined_k141_204502||full | 12231 | No | NA | 18 | 0 | 2 | Not-determined | Genome-fragment | NA | NA | 0 |
| S17-final-viral-combined_k141_215651||full | 1706 | No | NA | 2 | 0 | 0 | Not-determined | Genome-fragment | NA | NA | 0 |
| S17-final-viral-combined_k141_126460||full | 4436 | No | NA | 5 | 0 | 4 | Not-determined | Genome-fragment | NA | NA | 0 |
| S17-final-viral-combined_k141_3742||full | 1482 | No | NA | 2 | 0 | 1 | Not-determined | Genome-fragment | NA | NA | 0 |
| S17-final-viral-combined_k141_226537||full | 2484 | No | NA | 3 | 0 | 1 | Not-determined | Genome-fragment | NA | NA | 0 |
| S17-final-viral-combined_k141_59226||full | 1629 | No | NA | 3 | 0 | 2 | Not-determined | Genome-fragment | NA | NA | 0 |
| S17-final-viral-combined_k141_237091||full | 2264 | No | NA | 3 | 0 | 2 | Not-determined | Genome-fragment | NA | NA | 0 |
| S17-final-viral-combined_k141_126494||full | 3101 | No | NA | 3 | 0 | 2 | Not-determined | Genome-fragment | NA | NA | 0 |
| S2_k141_14859 | 2175 | No | NA | 5 | 1 | 0 | Low-quality | Genome-fragment | 3.36 | AAI-based (high-confidence) | 0 |
| S9-final-viral-combined_k141_87497||full | 2176 | No | NA | 5 | 2 | 0 | Low-quality | Genome-fragment | 5.58 | AAI-based (high-confidence) | 0 |
| S17-final-viral-combined_k141_254097||full | 2176 | No | NA | 4 | 2 | 0 | Low-quality | Genome-fragment | 2.62 | HMM-based (lower-bound) | 0 |
| S8-final-viral-combined_k141_19018||full | 2176 | No | NA | 5 | 1 | 0 | Low-quality | Genome-fragment | 3.49 | AAI-based (medium-confidence) | 0 |
| S17-final-viral-combined_k141_70983||full | 27910 | No | NA | 13 | 0 | 5 | Medium-quality | Genome-fragment | 80.49 | AAI-based (medium-confidence) | 0 |
| S17-final-viral-combined_k141_158884||full | 2018 | No | NA | 2 | 0 | 1 | Not-determined | Genome-fragment | NA | NA | 0 |
| S9-final-viral-combined_k141_15532||full | 2177 | No | NA | 5 | 0 | 1 | Low-quality | Genome-fragment | 4.15 | AAI-based (high-confidence) | 0 |
| S17-final-viral-combined_k141_103782||full | 2316 | No | NA | 4 | 0 | 4 | Not-determined | Genome-fragment | NA | NA | 0 |
| S2-final-viral-combined_k141_172130||full | 2177 | No | NA | 1 | 0 | 1 | Low-quality | Genome-fragment | 3.84 | AAI-based (high-confidence) | 0 |
| S17-final-viral-combined_k141_59269||full | 2391 | No | NA | 4 | 0 | 3 | Not-determined | Genome-fragment | NA | NA | 0 |
| S17-final-viral-combined_k141_71002||full | 7057 | No | NA | 6 | 0 | 6 | Not-determined | Genome-fragment | NA | NA | 0 |
| S4-final-viral-combined_k141_43829||full | 2177 | No | NA | 6 | 0 | 0 | Low-quality | Genome-fragment | 2.23 | AAI-based (high-confidence) | 0 |
| S4_k141_63542 | 2177 | No | NA | 4 | 0 | 0 | Low-quality | Genome-fragment | 3.64 | AAI-based (high-confidence) | 0 |
| S17-final-viral-combined_k141_137591||full | 2314 | No | NA | 2 | 0 | 0 | Not-determined | Genome-fragment | NA | NA | 0 |
| S17-final-viral-combined_k141_182290||full | 1660 | No | NA | 3 | 0 | 0 | Not-determined | Genome-fragment | NA | NA | 0 |
| S6-final-viral-combined_k141_120387||full | 2177 | No | NA | 3 | 0 | 0 | Low-quality | Genome-fragment | 2.61 | AAI-based (high-confidence) | 0 |
| S9-final-viral-combined_k141_105543||full | 2178 | No | NA | 3 | 0 | 2 | Low-quality | Genome-fragment | 4.79 | AAI-based (high-confidence) | 0 |
| S17-final-viral-combined_k141_126588||full | 1072 | No | NA | 3 | 0 | 1 | Not-determined | Genome-fragment | NA | NA | 0 |
| S7-final-viral-combined_k141_195391||full | 2178 | No | NA | 4 | 0 | 0 | Low-quality | Genome-fragment | 4.38 | AAI-based (medium-confidence) | 0 |
| S17-final-viral-combined_k141_82436||full | 2403 | No | NA | 3 | 0 | 2 | Not-determined | Genome-fragment | NA | NA | 0 |
| S17-final-viral-combined_k141_126600||full | 8530 | No | NA | 11 | 1 | 3 | High-quality | High-quality | 100 | HMM-based (lower-bound) | 0 |
| S17-final-viral-combined_k141_259111||full | 2722 | No | NA | 3 | 0 | 3 | Not-determined | Genome-fragment | NA | NA | 0 |
| S7-final-viral-combined_k141_75844||full | 2178 | No | NA | 5 | 2 | 0 | Low-quality | Genome-fragment | 5.33 | AAI-based (high-confidence) | 0 |
| S7_k141_150781 | 2178 | No | NA | 3 | 0 | 1 | Low-quality | Genome-fragment | 5.58 | AAI-based (high-confidence) | 0 |
| S17-final-viral-combined_k141_181809||full | 2178 | Yes | 318 | 3 | 1 | 1 | Low-quality | Genome-fragment | 0.17 | HMM-based (lower-bound) | 85.4 |
| S17-final-viral-combined_k141_116230||full | 4158 | No | NA | 4 | 0 | 3 | Not-determined | Genome-fragment | NA | NA | 0 |
| S17-final-viral-combined_k141_249734||full | 2178 | No | NA | 3 | 0 | 1 | Low-quality | Genome-fragment | 1.39 | AAI-based (medium-confidence) | 0 |
| S17-final-viral-combined_k141_71028||full | 1576 | No | NA | 3 | 0 | 1 | Not-determined | Genome-fragment | NA | NA | 0 |
| S17-final-viral-combined_k141_249671||lt2gene | 2178 | No | NA | 1 | 1 | 0 | Low-quality | Genome-fragment | 3.18 | HMM-based (lower-bound) | 0 |
| S17-final-viral-combined_k141_137654||full | 2521 | No | NA | 3 | 0 | 3 | Not-determined | Genome-fragment | NA | NA | 0 |
| S17-final-viral-combined_k141_37171||full | 2462 | No | NA | 3 | 0 | 1 | Not-determined | Genome-fragment | NA | NA | 0 |
| S4_k141_47298 | 2178 | No | NA | 5 | 0 | 0 | Low-quality | Genome-fragment | 2.68 | AAI-based (high-confidence) | 0 |
| S1-final-viral-combined_k141_366412||full | 2179 | No | NA | 3 | 1 | 0 | Low-quality | Genome-fragment | 2.93 | HMM-based (lower-bound) | 0 |
| S17-final-viral-combined_k141_59309||full | 3490 | No | NA | 4 | 0 | 2 | Not-determined | Genome-fragment | NA | NA | 0 |
| S5-final-viral-combined_k141_54081||full | 2179 | No | NA | 6 | 2 | 0 | Low-quality | Genome-fragment | 3.3 | HMM-based (lower-bound) | 0 |
| S17-final-viral-combined_k141_91976||full | 3647 | No | NA | 5 | 0 | 2 | Not-determined | Genome-fragment | NA | NA | 0 |
| S17-final-viral-combined_k141_126612||full | 1665 | No | NA | 2 | 0 | 1 | Not-determined | Genome-fragment | NA | NA | 0 |
| S5-final-viral-combined_k141_152270||full | 2179 | No | NA | 6 | 2 | 0 | Low-quality | Genome-fragment | 3.26 | AAI-based (high-confidence) | 0 |
| S4-final-viral-combined_k141_92014||full | 2179 | No | NA | 3 | 0 | 0 | Low-quality | Genome-fragment | 5.97 | AAI-based (medium-confidence) | 0 |
| S17-final-viral-combined_k141_59314||full | 3270 | No | NA | 4 | 0 | 2 | Not-determined | Genome-fragment | NA | NA | 0 |
| S5-final-viral-combined_k141_127872||full | 2180 | No | NA | 4 | 2 | 0 | Low-quality | Genome-fragment | 6.39 | AAI-based (high-confidence) | 0 |
| S17-final-viral-combined_k141_36790||full | 2180 | No | NA | 2 | 0 | 1 | Low-quality | Genome-fragment | 1.07 | AAI-based (medium-confidence) | 0 |
| S17-final-viral-combined_k141_59332||full | 1652 | No | NA | 2 | 0 | 2 | Not-determined | Genome-fragment | NA | NA | 0 |
| S17-final-viral-combined_k141_237232||full | 5349 | No | NA | 9 | 0 | 2 | Not-determined | Genome-fragment | NA | NA | 0 |
| S17-final-viral-combined_k141_182417||full | 2070 | No | NA | 4 | 0 | 1 | Not-determined | Genome-fragment | NA | NA | 0 |
| S2-final-viral-combined_k141_130934||full | 2180 | No | NA | 6 | 1 | 0 | Low-quality | Genome-fragment | 4.22 | AAI-based (high-confidence) | 0 |
| S17-final-viral-combined_k141_215866||full | 5774 | No | NA | 5 | 0 | 3 | Not-determined | Genome-fragment | NA | NA | 0 |
| S2-final-viral-combined_k141_9144||full | 2180 | No | NA | 4 | 0 | 1 | Low-quality | Genome-fragment | 5.28 | AAI-based (medium-confidence) | 0 |
| S2-final-viral-combined_k141_10194||full | 2180 | No | NA | 3 | 0 | 1 | Low-quality | Genome-fragment | 0.87 | AAI-based (medium-confidence) | 0 |
| S17-final-viral-combined_k141_47100||full | 2137 | No | NA | 3 | 0 | 1 | Not-determined | Genome-fragment | NA | NA | 0 |
| S17-final-viral-combined_k141_71113||full | 2219 | No | NA | 3 | 0 | 2 | Not-determined | Genome-fragment | NA | NA | 0 |
| S17-final-viral-combined_k141_82548||full | 1403 | No | NA | 2 | 0 | 1 | Not-determined | Genome-fragment | NA | NA | 0 |
| S17-final-viral-combined_k141_148737||full | 1397 | No | NA | 3 | 0 | 2 | Not-determined | Genome-fragment | NA | NA | 0 |
| S17-final-viral-combined_k141_14447||full | 1491 | No | NA | 3 | 0 | 0 | Not-determined | Genome-fragment | NA | NA | 0 |
| S17-final-viral-combined_k141_26016||full | 1437 | No | NA | 2 | 0 | 1 | Not-determined | Genome-fragment | NA | NA | 0 |
| S17-final-viral-combined_k141_59387||full | 6092 | No | NA | 8 | 0 | 0 | Not-determined | Genome-fragment | NA | NA | 0 |
| S6_k141_9702 | 2180 | No | NA | 1 | 1 | 0 | Low-quality | Genome-fragment | 2.31 | AAI-based (high-confidence) | 0 |
| S17-final-viral-combined_k141_14476||full | 2490 | No | NA | 4 | 0 | 0 | Not-determined | Genome-fragment | NA | NA | 0 |
| S17-final-viral-combined_k141_193408||full | 1430 | No | NA | 2 | 0 | 1 | Not-determined | Genome-fragment | NA | NA | 0 |
| S11-final-viral-combined_k141_274844||full | 2181 | No | NA | 3 | 1 | 0 | Low-quality | Genome-fragment | 3.4 | HMM-based (lower-bound) | 0 |
| S17-final-viral-combined_k141_47138||full | 1664 | No | NA | 3 | 0 | 0 | Not-determined | Genome-fragment | NA | NA | 0 |
| S17-final-viral-combined_k141_71157||full | 2350 | No | NA | 3 | 0 | 1 | Not-determined | Genome-fragment | NA | NA | 0 |
| S1-final-viral-combined_k141_113420||full | 2181 | No | NA | 3 | 0 | 0 | Low-quality | Genome-fragment | 3.93 | AAI-based (medium-confidence) | 0 |
| S17-final-viral-combined_k141_151241||full | 2181 | No | NA | 4 | 1 | 0 | Low-quality | Genome-fragment | 5.33 | AAI-based (medium-confidence) | 0 |
| S12-final-viral-combined_k141_239759||full | 2181 | No | NA | 7 | 0 | 0 | Low-quality | Genome-fragment | 3.66 | AAI-based (high-confidence) | 0 |
| S12-final-viral-combined_k141_129210||full | 2181 | No | NA | 3 | 0 | 0 | Low-quality | Genome-fragment | 3.69 | AAI-based (medium-confidence) | 0 |
| S17-final-viral-combined_k141_170769||full | 1177 | No | NA | 3 | 0 | 1 | Not-determined | Genome-fragment | NA | NA | 0 |
| S17-final-viral-combined_k141_116382||full | 8503 | No | NA | 5 | 0 | 4 | Not-determined | Genome-fragment | NA | NA | 0 |
| S8-final-viral-combined_k141_66905||full | 2181 | No | NA | 3 | 0 | 3 | Low-quality | Genome-fragment | 6.56 | AAI-based (medium-confidence) | 0 |
| S17-final-viral-combined_k141_226828||full | 8516 | No | NA | 16 | 0 | 1 | Not-determined | Genome-fragment | NA | NA | 0 |
| S17-final-viral-combined_k141_92132||full | 1561 | No | NA | 3 | 0 | 1 | Not-determined | Genome-fragment | NA | NA | 0 |
| S8-final-viral-combined_k141_137121||lt2gene | 2181 | No | NA | 2 | 2 | 0 | Low-quality | Genome-fragment | 5.87 | AAI-based (high-confidence) | 0 |
| S2-final-viral-combined_k141_188514||full | 2182 | No | NA | 3 | 0 | 0 | Low-quality | Genome-fragment | 3.94 | AAI-based (high-confidence) | 0 |
| S8-final-viral-combined_k141_87581||full | 2182 | No | NA | 4 | 2 | 0 | Low-quality | Genome-fragment | 5.07 | AAI-based (high-confidence) | 0 |
| S3-final-viral-combined_k141_65576||full | 2183 | No | NA | 4 | 1 | 0 | Low-quality | Genome-fragment | 1.38 | HMM-based (lower-bound) | 0 |
| S7-final-viral-combined_k141_103020||full | 2183 | No | NA | 5 | 2 | 0 | Low-quality | Genome-fragment | 3.37 | AAI-based (high-confidence) | 0 |
| S1-final-viral-combined_k141_351030||full | 2184 | No | NA | 5 | 3 | 0 | Low-quality | Genome-fragment | 5.66 | AAI-based (high-confidence) | 0 |
| S1-final-viral-combined_k141_261089||full | 2184 | No | NA | 3 | 0 | 1 | Low-quality | Genome-fragment | 0.86 | AAI-based (medium-confidence) | 0 |
| S17-final-viral-combined_k141_71221||full | 1492 | No | NA | 4 | 0 | 1 | Not-determined | Genome-fragment | NA | NA | 0 |
| S17-final-viral-combined_k141_126688||full | 4585 | No | NA | 5 | 0 | 3 | Not-determined | Genome-fragment | NA | NA | 0 |
| S17-final-viral-combined_k141_82687||full | 5457 | No | NA | 3 | 0 | 1 | Not-determined | Genome-fragment | NA | NA | 0 |
| S5-final-viral-combined_k141_166059||full | 2184 | No | NA | 4 | 0 | 0 | Low-quality | Genome-fragment | 5 | AAI-based (medium-confidence) | 0 |
| S7_k141_211150 | 2184 | No | NA | 1 | 0 | 0 | Low-quality | Genome-fragment | 0.4 | AAI-based (medium-confidence) | 0 |
| S17-final-viral-combined_k141_170860||full | 1532 | No | NA | 3 | 0 | 0 | Not-determined | Genome-fragment | NA | NA | 0 |
| S17-final-viral-combined_k141_170865||full | 1166 | No | NA | 3 | 0 | 3 | Not-determined | Genome-fragment | NA | NA | 0 |
| S17-final-viral-combined_k141_237336||full | 1092 | No | NA | 3 | 0 | 1 | Not-determined | Genome-fragment | NA | NA | 0 |
| S17-final-viral-combined_k141_90494||full | 2184 | No | NA | 5 | 2 | 0 | Low-quality | Genome-fragment | 5.2 | AAI-based (medium-confidence) | 0 |
| S17-final-viral-combined_k141_104098||full | 6130 | No | NA | 6 | 0 | 1 | Not-determined | Genome-fragment | NA | NA | 0 |
| S17-final-viral-combined_k141_82737||full | 1523 | No | NA | 2 | 0 | 1 | Not-determined | Genome-fragment | NA | NA | 0 |
| S12-final-viral-combined_k141_192932||full | 2184 | No | NA | 3 | 0 | 2 | Low-quality | Genome-fragment | 1.25 | AAI-based (medium-confidence) | 0 |
| S17-final-viral-combined_k141_226930||full | 5085 | No | NA | 5 | 0 | 5 | Not-determined | Genome-fragment | NA | NA | 0 |
| S2-final-viral-combined_k141_170698||full | 2184 | No | NA | 4 | 1 | 0 | Low-quality | Genome-fragment | 1.38 | HMM-based (lower-bound) | 0 |
| S17-final-viral-combined_k141_148886||full | 4362 | No | NA | 7 | 0 | 0 | Not-determined | Genome-fragment | NA | NA | 0 |
| S8-final-viral-combined_k141_192790||full | 2184 | No | NA | 2 | 0 | 1 | Low-quality | Genome-fragment | 1.05 | AAI-based (medium-confidence) | 0 |
| S4_k141_81160 | 2185 | No | NA | 2 | 0 | 0 | Low-quality | Genome-fragment | 2.25 | AAI-based (high-confidence) | 0 |
| S17-final-viral-combined_k141_82779||full | 1153 | No | NA | 2 | 0 | 1 | Not-determined | Genome-fragment | NA | NA | 0 |
| S17-final-viral-combined_k141_104156||full | 1734 | No | NA | 3 | 0 | 3 | Not-determined | Genome-fragment | NA | NA | 0 |
| S17-final-viral-combined_k141_182580||full | 5257 | No | NA | 7 | 0 | 1 | Not-determined | Genome-fragment | NA | NA | 0 |
| S17-final-viral-combined_k141_237359||full | 5197 | No | NA | 6 | 0 | 3 | Not-determined | Genome-fragment | NA | NA | 0 |
| S1-final-viral-combined_k141_156946||lt2gene | 2186 | No | NA | 2 | 0 | 0 | Low-quality | Genome-fragment | 5.35 | AAI-based (high-confidence) | 0 |
| S17-final-viral-combined_k141_71332||full | 1247 | No | NA | 3 | 0 | 0 | Not-determined | Genome-fragment | NA | NA | 0 |
| S5-final-viral-combined_k141_190324||full | 2186 | No | NA | 2 | 0 | 0 | Low-quality | Genome-fragment | 3.87 | AAI-based (medium-confidence) | 0 |
| S17-final-viral-combined_k141_82797||full | 17582 | No | NA | 17 | 0 | 9 | Not-determined | Genome-fragment | NA | NA | 0 |
| S7-final-viral-combined_k141_215319||lt2gene | 2186 | No | NA | 1 | 0 | 0 | Low-quality | Genome-fragment | 2.85 | AAI-based (high-confidence) | 0 |
| S17-final-viral-combined_k141_216051||full | 1397 | No | NA | 3 | 0 | 1 | Not-determined | Genome-fragment | NA | NA | 0 |
| S17-final-viral-combined_k141_84850||full | 2186 | No | NA | 3 | 0 | 3 | Low-quality | Genome-fragment | 0.64 | AAI-based (medium-confidence) | 0 |
| S17-final-viral-combined_k141_26176||full | 1646 | No | NA | 3 | 0 | 1 | Not-determined | Genome-fragment | NA | NA | 0 |
| S12-final-viral-combined_k141_271017||full | 2186 | No | NA | 3 | 0 | 0 | Low-quality | Genome-fragment | 4.77 | AAI-based (high-confidence) | 0 |
| S17-final-viral-combined_k141_4022||full | 4732 | No | NA | 10 | 0 | 0 | Not-determined | Genome-fragment | NA | NA | 0 |
| S17-final-viral-combined_k141_237391||full | 4895 | No | NA | 8 | 0 | 0 | Not-determined | Genome-fragment | NA | NA | 0 |
| S17-final-viral-combined_k141_4026||full | 3463 | No | NA | 4 | 0 | 3 | Not-determined | Genome-fragment | NA | NA | 0 |
| S12-final-viral-combined_k141_328969||lt2gene | 2186 | No | NA | 2 | 0 | 0 | Low-quality | Genome-fragment | 5.76 | AAI-based (high-confidence) | 0 |
| S17-final-viral-combined_k141_182613||full | 6964 | No | NA | 7 | 0 | 0 | Not-determined | Genome-fragment | NA | NA | 0 |
| S9-final-viral-combined_k141_107930||lt2gene | 2187 | No | NA | 2 | 2 | 0 | Low-quality | Genome-fragment | 5.44 | AAI-based (high-confidence) | 0 |
| S17-final-viral-combined_k141_204734||full | 2306 | No | NA | 3 | 0 | 1 | Not-determined | Genome-fragment | NA | NA | 0 |
| S17-final-viral-combined_k141_207265||full | 2187 | No | NA | 5 | 2 | 0 | Low-quality | Genome-fragment | 5.19 | AAI-based (high-confidence) | 0 |
| S17-final-viral-combined_k141_237416||full | 1509 | No | NA | 3 | 0 | 0 | Not-determined | Genome-fragment | NA | NA | 0 |
| S17-final-viral-combined_k141_237417||full | 3271 | No | NA | 3 | 0 | 1 | Not-determined | Genome-fragment | NA | NA | 0 |
| S2-final-viral-combined_k141_65162||full | 2187 | No | NA | 2 | 1 | 0 | Low-quality | Genome-fragment | 0.78 | HMM-based (lower-bound) | 0 |
| S4-final-viral-combined_k141_115975||full | 2187 | No | NA | 3 | 0 | 2 | Low-quality | Genome-fragment | 0.74 | AAI-based (medium-confidence) | 0 |
| S17-final-viral-combined_k141_182649||full | 1084 | No | NA | 2 | 0 | 0 | Not-determined | Genome-fragment | NA | NA | 0 |
| S11-final-viral-combined_k141_13660||full | 2188 | No | NA | 7 | 0 | 0 | Low-quality | Genome-fragment | 3.46 | AAI-based (high-confidence) | 0 |
| S17-final-viral-combined_k141_126774||full | 66657 | No | NA | 71 | 3 | 11 | High-quality | High-quality | 100 | AAI-based (medium-confidence) | 0 |
| S5-final-viral-combined_k141_141089||full | 2188 | No | NA | 3 | 2 | 0 | Low-quality | Genome-fragment | 4.4 | AAI-based (medium-confidence) | 0 |
| S17-final-viral-combined_k141_126782||full | 1511 | No | NA | 3 | 0 | 2 | Not-determined | Genome-fragment | NA | NA | 0 |
| S17-final-viral-combined_k141_116621||full | 1334 | No | NA | 2 | 0 | 0 | Not-determined | Genome-fragment | NA | NA | 0 |
| S12-final-viral-combined_k141_305627||full | 2188 | No | NA | 7 | 1 | 0 | Low-quality | Genome-fragment | 3.8 | AAI-based (high-confidence) | 0 |
| S6-final-viral-combined_k141_18448||full | 2188 | No | NA | 3 | 0 | 1 | Low-quality | Genome-fragment | 1.64 | AAI-based (high-confidence) | 0 |
| S17-final-viral-combined_k141_14576||full | 2340 | No | NA | 4 | 0 | 1 | Not-determined | Genome-fragment | NA | NA | 0 |
| S8-final-viral-combined_k141_9469||full | 2188 | No | NA | 3 | 0 | 1 | Low-quality | Genome-fragment | 0.5 | AAI-based (medium-confidence) | 0 |
| S17-final-viral-combined_k141_116641||full | 3350 | No | NA | 2 | 0 | 1 | Not-determined | Genome-fragment | NA | NA | 0 |
| S9-final-viral-combined_k141_60878||full | 2189 | No | NA | 4 | 0 | 1 | Low-quality | Genome-fragment | 2.33 | AAI-based (medium-confidence) | 0 |
| S17-final-viral-combined_k141_237474||full | 3144 | No | NA | 5 | 0 | 3 | Not-determined | Genome-fragment | NA | NA | 0 |
| S11-final-viral-combined_k141_128445||full | 2189 | No | NA | 5 | 1 | 0 | Low-quality | Genome-fragment | 5.86 | AAI-based (medium-confidence) | 0 |
| S17-final-viral-combined_k141_248042||full | 1359 | No | NA | 2 | 0 | 0 | Not-determined | Genome-fragment | NA | NA | 0 |
| S17-final-viral-combined_k141_82947||full | 3575 | No | NA | 3 | 0 | 0 | Not-determined | Genome-fragment | NA | NA | 0 |
| S17-final-viral-combined_k141_104279||full | 1287 | No | NA | 2 | 0 | 1 | Not-determined | Genome-fragment | NA | NA | 0 |
| S17-final-viral-combined_k141_171025||full | 1348 | No | NA | 3 | 0 | 0 | Not-determined | Genome-fragment | NA | NA | 0 |
| S17-final-viral-combined_k141_47304||full | 2927 | No | NA | 3 | 0 | 0 | Not-determined | Genome-fragment | NA | NA | 0 |
| S2-final-viral-combined_k141_209596||full | 2189 | No | NA | 2 | 0 | 0 | Low-quality | Genome-fragment | 5.06 | AAI-based (medium-confidence) | 0 |
| S4-final-viral-combined_k141_145190||full | 2189 | No | NA | 5 | 0 | 0 | Low-quality | Genome-fragment | 3.03 | AAI-based (high-confidence) | 0 |
| S1_k141_330017 | 2190 | No | NA | 2 | 0 | 2 | Low-quality | Genome-fragment | 0.97 | AAI-based (medium-confidence) | 0 |
| S17-final-viral-combined_k141_204836||full | 2937 | No | NA | 3 | 0 | 3 | Not-determined | Genome-fragment | NA | NA | 0 |
| S17-final-viral-combined_k141_83019||full | 2124 | No | NA | 4 | 0 | 1 | Not-determined | Genome-fragment | NA | NA | 0 |
| S3-final-viral-combined_k141_36216||full | 2190 | No | NA | 5 | 0 | 0 | Low-quality | Genome-fragment | 1.45 | AAI-based (high-confidence) | 0 |
| S17-final-viral-combined_k141_104313||full | 1447 | No | NA | 2 | 0 | 0 | Not-determined | Genome-fragment | NA | NA | 0 |
| S17-final-viral-combined_k141_59593||full | 7016 | No | NA | 7 | 0 | 4 | Not-determined | Genome-fragment | NA | NA | 0 |
| S17-final-viral-combined_k141_237582||full | 11800 | No | NA | 7 | 0 | 2 | Not-determined | Genome-fragment | NA | NA | 0 |
| S12-final-viral-combined_k141_268939||full | 2190 | No | NA | 3 | 0 | 2 | Low-quality | Genome-fragment | 1.29 | AAI-based (medium-confidence) | 0 |
| S12-final-viral-combined_k141_205933||full | 2190 | No | NA | 3 | 0 | 0 | Low-quality | Genome-fragment | 4.87 | AAI-based (high-confidence) | 0 |
| S17-final-viral-combined_k141_248109||full | 2255 | No | NA | 3 | 0 | 2 | Not-determined | Genome-fragment | NA | NA | 0 |
| S17-final-viral-combined_k141_14661||full | 2982 | No | NA | 3 | 0 | 3 | Not-determined | Genome-fragment | NA | NA | 0 |
| S17-final-viral-combined_k141_204869||full | 11384 | No | NA | 10 | 0 | 8 | Not-determined | Genome-fragment | NA | NA | 0 |
| S17-final-viral-combined_k141_14682||full | 1595 | No | NA | 2 | 0 | 2 | Not-determined | Genome-fragment | NA | NA | 0 |
| S2-final-viral-combined_k141_18787||full | 2190 | No | NA | 3 | 0 | 0 | Low-quality | Genome-fragment | 1.43 | AAI-based (high-confidence) | 0 |
| S3_k141_76712 | 2191 | No | NA | 2 | 0 | 2 | Low-quality | Genome-fragment | 1.25 | AAI-based (high-confidence) | 0 |
| S7_k141_159063 | 2191 | No | NA | 1 | 0 | 1 | Low-quality | Genome-fragment | 4.62 | AAI-based (medium-confidence) | 0 |
| S17-final-viral-combined_k141_59688||full | 2408 | No | NA | 2 | 0 | 1 | Not-determined | Genome-fragment | NA | NA | 0 |
| S17-final-viral-combined_k141_259352||full | 7002 | No | NA | 7 | 0 | 4 | Not-determined | Genome-fragment | NA | NA | 0 |
| S17-final-viral-combined_k141_248153||full | 1048 | No | NA | 2 | 0 | 2 | Not-determined | Genome-fragment | NA | NA | 0 |
| S17-final-viral-combined_k141_149035||full | 1246 | No | NA | 3 | 0 | 0 | Not-determined | Genome-fragment | NA | NA | 0 |
| S12-final-viral-combined_k141_22358||full | 2191 | No | NA | 2 | 0 | 0 | Low-quality | Genome-fragment | 3.3 | AAI-based (medium-confidence) | 0 |
| S17-final-viral-combined_k141_137945||full | 1190 | No | NA | 4 | 0 | 1 | Not-determined | Genome-fragment | NA | NA | 0 |
| S17-final-viral-combined_k141_71454||full | 2821 | No | NA | 4 | 0 | 1 | Not-determined | Genome-fragment | NA | NA | 0 |
| S12_k141_105647 | 2191 | No | NA | 4 | 1 | 0 | Low-quality | Genome-fragment | 2.33 | AAI-based (high-confidence) | 0 |
| S2-final-viral-combined_k141_138999||full | 2191 | No | NA | 2 | 0 | 0 | Low-quality | Genome-fragment | 5.73 | AAI-based (high-confidence) | 0 |
| S17-final-viral-combined_k141_159314||full | 1882 | No | NA | 3 | 0 | 2 | Not-determined | Genome-fragment | NA | NA | 0 |
| S1-final-viral-combined_k141_309347||full | 2192 | No | NA | 5 | 0 | 0 | Low-quality | Genome-fragment | 5.74 | AAI-based (high-confidence) | 0 |
| S7-final-viral-combined_k141_19784||full | 2192 | No | NA | 1 | 0 | 0 | Low-quality | Genome-fragment | 3.42 | AAI-based (medium-confidence) | 0 |
| S8-final-viral-combined_k141_171922||full | 2192 | No | NA | 4 | 2 | 0 | Low-quality | Genome-fragment | 2.7 | AAI-based (high-confidence) | 0 |
| S11-final-viral-combined_k141_206790||full | 2193 | No | NA | 3 | 3 | 0 | Low-quality | Genome-fragment | 3.64 | AAI-based (high-confidence) | 0 |
| S17-final-viral-combined_k141_248251||full | 2228 | No | NA | 3 | 0 | 3 | Not-determined | Genome-fragment | NA | NA | 0 |
| S11-final-viral-combined_k141_196227||full | 2193 | No | NA | 2 | 0 | 1 | Low-quality | Genome-fragment | 1.27 | AAI-based (medium-confidence) | 0 |
| S17-final-viral-combined_k141_182899||full | 3189 | No | NA | 4 | 0 | 3 | Not-determined | Genome-fragment | NA | NA | 0 |
| S1_k141_203268 | 2193 | No | NA | 2 | 0 | 1 | Low-quality | Genome-fragment | 1.01 | AAI-based (medium-confidence) | 0 |
| S17-final-viral-combined_k141_92433||full | 6429 | No | NA | 6 | 0 | 0 | Not-determined | Genome-fragment | NA | NA | 0 |
| S17-final-viral-combined_k141_3186||full | 2193 | No | NA | 3 | 0 | 3 | Low-quality | Genome-fragment | 3.17 | AAI-based (medium-confidence) | 0 |
| S17-final-viral-combined_k141_37654||full | 2432 | No | NA | 2 | 0 | 0 | Not-determined | Genome-fragment | NA | NA | 0 |
| S17-final-viral-combined_k141_216296||full | 1973 | No | NA | 3 | 0 | 2 | Not-determined | Genome-fragment | NA | NA | 0 |
| S17-final-viral-combined_k141_50853||full | 2193 | No | NA | 3 | 0 | 2 | Low-quality | Genome-fragment | 1.08 | AAI-based (medium-confidence) | 0 |
| S12-final-viral-combined_k141_102036||full | 2193 | No | NA | 4 | 0 | 1 | Low-quality | Genome-fragment | 3.74 | AAI-based (medium-confidence) | 0 |
| S17-final-viral-combined_k141_59801||full | 7164 | No | NA | 4 | 0 | 3 | Not-determined | Genome-fragment | NA | NA | 0 |
| S12-final-viral-combined_k141_146017||full | 2193 | No | NA | 2 | 0 | 0 | Low-quality | Genome-fragment | 3.45 | AAI-based (high-confidence) | 0 |
| S17-final-viral-combined_k141_237692||full | 1294 | No | NA | 2 | 0 | 1 | Not-determined | Genome-fragment | NA | NA | 0 |
| S17-final-viral-combined_k141_149168||full | 1930 | No | NA | 2 | 0 | 1 | Not-determined | Genome-fragment | NA | NA | 0 |
| S17-final-viral-combined_k141_227394||full | 1435 | No | NA | 2 | 0 | 1 | Not-determined | Genome-fragment | NA | NA | 0 |
| S2-final-viral-combined_k141_121931||full | 2193 | No | NA | 3 | 0 | 0 | Low-quality | Genome-fragment | 3.96 | AAI-based (high-confidence) | 0 |
| S8-final-viral-combined_k141_247573||full | 2193 | No | NA | 4 | 0 | 1 | Low-quality | Genome-fragment | 1.38 | AAI-based (medium-confidence) | 0 |
| S17-final-viral-combined_k141_193896||full | 1727 | No | NA | 3 | 0 | 1 | Not-determined | Genome-fragment | NA | NA | 0 |
| S5-final-viral-combined_k141_14604||full | 2194 | No | NA | 5 | 1 | 0 | Low-quality | Genome-fragment | 4.32 | AAI-based (high-confidence) | 0 |
| S17-final-viral-combined_k141_47494||full | 3779 | No | NA | 5 | 0 | 0 | Not-determined | Genome-fragment | NA | NA | 0 |
| S5-final-viral-combined_k141_211010||full | 2194 | No | NA | 5 | 0 | 0 | Low-quality | Genome-fragment | 3.74 | AAI-based (medium-confidence) | 0 |
| S17-final-viral-combined_k141_117684||full | 2194 | No | NA | 4 | 1 | 0 | Low-quality | Genome-fragment | 0.89 | HMM-based (lower-bound) | 0 |
| S17-final-viral-combined_k141_37724||full | 2442 | No | NA | 5 | 0 | 1 | Not-determined | Genome-fragment | NA | NA | 0 |
| S2-final-viral-combined_k141_100294||full | 2194 | No | NA | 3 | 0 | 1 | Low-quality | Genome-fragment | 2.34 | AAI-based (medium-confidence) | 0 |
| S17-final-viral-combined_k141_117023||full | 2381 | No | NA | 2 | 0 | 1 | Not-determined | Genome-fragment | NA | NA | 0 |
| S17-final-viral-combined_k141_47495||full | 13375 | No | NA | 15 | 0 | 2 | Not-determined | Genome-fragment | NA | NA | 0 |
| S2-final-viral-combined_k141_101548||full | 2194 | No | NA | 6 | 3 | 0 | Low-quality | Genome-fragment | 5.25 | AAI-based (high-confidence) | 0 |
| S17-final-viral-combined_k141_14811||full | 3738 | No | NA | 5 | 0 | 4 | Not-determined | Genome-fragment | NA | NA | 0 |
| S17-final-viral-combined_k141_26521||full | 7770 | No | NA | 10 | 0 | 6 | Not-determined | Genome-fragment | NA | NA | 0 |
| S17-final-viral-combined_k141_127022||full | 2515 | No | NA | 4 | 0 | 1 | Not-determined | Genome-fragment | NA | NA | 0 |
| S17-final-viral-combined_k141_227491||full | 1282 | No | NA | 2 | 0 | 0 | Not-determined | Genome-fragment | NA | NA | 0 |
| S17-final-viral-combined_k141_47522||full | 1511 | No | NA | 2 | 0 | 1 | Not-determined | Genome-fragment | NA | NA | 0 |
| S17-final-viral-combined_k141_227518||full | 1286 | No | NA | 3 | 0 | 1 | Not-determined | Genome-fragment | NA | NA | 0 |
| S4-final-viral-combined_k141_148887||full | 2194 | Yes | 1591 | 6 | 1 | 1 | Low-quality | Genome-fragment | 3.64 | AAI-based (high-confidence) | 27.48 |
| S4-final-viral-combined_k141_67915||full | 2194 | No | NA | 4 | 0 | 2 | Low-quality | Genome-fragment | 1.55 | AAI-based (medium-confidence) | 0 |
| S17-final-viral-combined_k141_71523||full | 1109 | No | NA | 1 | 0 | 0 | Not-determined | Genome-fragment | NA | NA | 0 |
| S17-final-viral-combined_k141_83278||full | 4083 | No | NA | 5 | 0 | 1 | Not-determined | Genome-fragment | NA | NA | 0 |
| S8-final-viral-combined_k141_185379||full | 2194 | No | NA | 3 | 3 | 0 | Low-quality | Genome-fragment | 5.36 | AAI-based (high-confidence) | 0 |
| S8-final-viral-combined_k141_274599||full | 2194 | No | NA | 3 | 0 | 0 | Low-quality | Genome-fragment | 4.66 | AAI-based (high-confidence) | 0 |
| S12-final-viral-combined_k141_241695||full | 2195 | No | NA | 4 | 1 | 0 | Low-quality | Genome-fragment | 5.73 | AAI-based (high-confidence) | 0 |
| S2_k141_211349 | 2195 | No | NA | 6 | 0 | 0 | Low-quality | Genome-fragment | 5.61 | AAI-based (high-confidence) | 0 |
| S17-final-viral-combined_k141_183064||full | 1057 | No | NA | 2 | 0 | 1 | Not-determined | Genome-fragment | NA | NA | 0 |
| S9_k141_51489 | 2196 | No | NA | 5 | 3 | 0 | Low-quality | Genome-fragment | 4.99 | AAI-based (high-confidence) | 0 |
| S17-final-viral-combined_k141_71565||full | 1888 | No | NA | 3 | 0 | 0 | Not-determined | Genome-fragment | NA | NA | 0 |
| S11-final-viral-combined_k141_49018||full | 2196 | No | NA | 5 | 0 | 1 | Low-quality | Genome-fragment | 3.34 | AAI-based (high-confidence) | 0 |
| S17-final-viral-combined_k141_149322||full | 3316 | No | NA | 3 | 0 | 2 | Not-determined | Genome-fragment | NA | NA | 0 |
| S17-final-viral-combined_k141_205172||full | 2189 | No | NA | 4 | 0 | 2 | Not-determined | Genome-fragment | NA | NA | 0 |
| S17-final-viral-combined_k141_216415||full | 5952 | No | NA | 7 | 0 | 3 | Not-determined | Genome-fragment | NA | NA | 0 |
| S17-final-viral-combined_k141_150739||full | 2196 | No | NA | 3 | 1 | 0 | Low-quality | Genome-fragment | 2.47 | HMM-based (lower-bound) | 0 |
| S12-final-viral-combined_k141_139920||full | 2196 | No | NA | 2 | 0 | 0 | Low-quality | Genome-fragment | 4.81 | AAI-based (high-confidence) | 0 |
| S2-final-viral-combined_k141_146543||full | 2196 | No | NA | 5 | 0 | 0 | Low-quality | Genome-fragment | 4.9 | AAI-based (medium-confidence) | 0 |
| S3-final-viral-combined_k141_92071||full | 2197 | No | NA | 3 | 0 | 1 | Low-quality | Genome-fragment | 3.56 | AAI-based (high-confidence) | 0 |
| S12_k141_7392 | 2197 | No | NA | 1 | 0 | 0 | Low-quality | Genome-fragment | 1.72 | AAI-based (medium-confidence) | 0 |
| S17-final-viral-combined_k141_171305||full | 39241 | No | NA | 56 | 10 | 1 | Medium-quality | Genome-fragment | 62.86 | AAI-based (high-confidence) | 0 |
| S17-final-viral-combined_k141_14867||full | 10119 | No | NA | 11 | 0 | 1 | Not-determined | Genome-fragment | NA | NA | 0 |
| S4_k141_157044 | 2197 | No | NA | 2 | 0 | 2 | Low-quality | Genome-fragment | 0.8 | AAI-based (medium-confidence) | 0 |
| S17-final-viral-combined_k141_248321||full | 127137 | No | NA | 184 | 56 | 16 | High-quality | High-quality | 100 | AAI-based (high-confidence) | 0 |
| S11-final-viral-combined_k141_203151||full | 2198 | No | NA | 3 | 0 | 0 | Low-quality | Genome-fragment | 3.92 | AAI-based (high-confidence) | 0 |
| S4-final-viral-combined_k141_122263||full | 2198 | No | NA | 5 | 1 | 0 | Low-quality | Genome-fragment | 5.32 | AAI-based (high-confidence) | 0 |
| S17-final-viral-combined_k141_127058||full | 1897 | No | NA | 4 | 0 | 3 | Not-determined | Genome-fragment | NA | NA | 0 |
| S9-final-viral-combined_k141_89046||full | 2199 | No | NA | 3 | 0 | 2 | Low-quality | Genome-fragment | 1.2 | AAI-based (medium-confidence) | 0 |
| S11-final-viral-combined_k141_53795||full | 2199 | No | NA | 4 | 1 | 0 | Low-quality | Genome-fragment | 2.54 | HMM-based (lower-bound) | 0 |
| S17-final-viral-combined_k141_248340||full | 1166 | No | NA | 2 | 0 | 2 | Not-determined | Genome-fragment | NA | NA | 0 |
| S2-final-viral-combined_k141_39972||lt2gene | 2199 | No | NA | 1 | 1 | 0 | Low-quality | Genome-fragment | 1.52 | HMM-based (lower-bound) | 0 |
| S17-final-viral-combined_k141_248347||full | 1943 | No | NA | 2 | 0 | 1 | Not-determined | Genome-fragment | NA | NA | 0 |
| S17-final-viral-combined_k141_227643||full | 1915 | No | NA | 2 | 0 | 1 | Not-determined | Genome-fragment | NA | NA | 0 |
| S1-final-viral-combined_k141_187168||full | 2200 | No | NA | 3 | 0 | 1 | Low-quality | Genome-fragment | 2.54 | AAI-based (medium-confidence) | 0 |
| S1_k141_533 | 2200 | No | NA | 7 | 0 | 0 | Low-quality | Genome-fragment | 4.16 | AAI-based (medium-confidence) | 0 |

| | | | | | | | | | | | |
|---|---|---|---|---|---|---|---|---|---|---|---|
| S5_k141_163206 | 2200 | No | NA | 2 | 0 | 1 | Low-quality | Genome-fragment | 5.17 | AAI-based (high-confidence) | 0 |
| S12-final-viral-combined_k141_154100\|\|lt2gene | 2200 | No | NA | 1 | 1 | 0 | Low-quality | Genome-fragment | 1.52 | HMM-based (lower-bound) | 0 |
| S8_k141_250782 | 2200 | No | NA | 3 | 0 | 0 | Low-quality | Genome-fragment | 3.9 | AAI-based (medium-confidence) | 0 |
| S5-final-viral-combined_k141_167023\|\|full | 2201 | No | NA | 4 | 0 | 1 | Low-quality | Genome-fragment | 4.39 | AAI-based (high-confidence) | 0 |
| S7-final-viral-combined_k141_189425\|\|lt2gene | 2201 | No | NA | 2 | 2 | 0 | Low-quality | Genome-fragment | 3.02 | HMM-based (lower-bound) | 0 |
| S17-final-viral-combined_k141_37815\|\|full | 7763 | No | NA | 7 | 0 | 6 | Not-determined | Genome-fragment | NA | NA | 0 |
| S17-final-viral-combined_k141_104601\|\|full | 2173 | No | NA | 4 | 0 | 0 | Not-determined | Genome-fragment | NA | NA | 0 |
| S17-final-viral-combined_k141_83418\|\|full | 2448 | No | NA | 4 | 0 | 0 | Not-determined | Genome-fragment | NA | NA | 0 |
| S17-final-viral-combined_k141_205271\|\|full | 3818 | No | NA | 5 | 0 | 3 | Not-determined | Genome-fragment | NA | NA | 0 |
| S17-final-viral-combined_k141_216543\|\|full | 2425 | No | NA | 3 | 0 | 2 | Not-determined | Genome-fragment | NA | NA | 0 |
| S17-k141_87393 | 2201 | No | NA | 2 | 0 | 2 | Low-quality | Genome-fragment | 1.05 | AAI-based (medium-confidence) | 0 |
| S17-final-viral-combined_k141_216549\|\|full | 3248 | No | NA | 3 | 0 | 0 | Not-determined | Genome-fragment | NA | NA | 0 |
| S17-final-viral-combined_k141_183235\|\|full | 5573 | No | NA | 5 | 0 | 3 | Not-determined | Genome-fragment | NA | NA | 0 |
| S17-final-viral-combined_k141_71684\|\|full | 1893 | No | NA | 3 | 0 | 1 | Not-determined | Genome-fragment | NA | NA | 0 |
| S12-final-viral-combined_k141_2232\|\|full | 2201 | No | NA | 2 | 0 | 0 | Low-quality | Genome-fragment | 3.45 | AAI-based (medium-confidence) | 0 |
| S17-final-viral-combined_k141_71690\|\|full | 1184 | No | NA | 1 | 0 | 1 | Not-determined | Genome-fragment | NA | NA | 0 |
| S17-final-viral-combined_k141_205319\|\|full | 2527 | No | NA | 4 | 0 | 0 | Not-determined | Genome-fragment | NA | NA | 0 |
| S12-final-viral-combined_k141_339030\|\|full | 2201 | No | NA | 3 | 0 | 0 | Low-quality | Genome-fragment | 0.83 | AAI-based (medium-confidence) | 0 |
| S17-final-viral-combined_k141_216594\|\|full | 2625 | No | NA | 4 | 0 | 2 | Not-determined | Genome-fragment | NA | NA | 0 |
| S17-final-viral-combined_k141_193958\|\|full | 101082 | Yes | 35929 | 98 | 15 | 32 | High-quality | High-quality | 100 | AAI-based (medium-confidence) | 64.46 |
| S12-final-viral-combined_k141_346376\|\|full | 2201 | No | NA | 3 | 0 | 1 | Low-quality | Genome-fragment | 2.44 | AAI-based (high-confidence) | 0 |
| S12-final-viral-combined_k141_166769\|\|full | 2201 | No | NA | 2 | 0 | 1 | Low-quality | Genome-fragment | 1.26 | AAI-based (medium-confidence) | 0 |
| S2-final-viral-combined_k141_63787\|\|full | 2201 | No | NA | 3 | 1 | 0 | Low-quality | Genome-fragment | 3.44 | AAI-based (medium-confidence) | 0 |
| S17-final-viral-combined_k141_159685\|\|full | 3627 | No | NA | 3 | 0 | 1 | Not-determined | Genome-fragment | NA | NA | 0 |
| S17-final-viral-combined_k141_92665\|\|full | 2174 | No | NA | 3 | 0 | 1 | Not-determined | Genome-fragment | NA | NA | 0 |
| S17-final-viral-combined_k141_37908\|\|full | 1542 | No | NA | 2 | 0 | 1 | Not-determined | Genome-fragment | NA | NA | 0 |
| S17-final-viral-combined_k141_216630\|\|full | 2223 | No | NA | 3 | 0 | 2 | Not-determined | Genome-fragment | NA | NA | 0 |
| S17-final-viral-combined_k141_237856\|\|full | 4494 | No | NA | 4 | 0 | 1 | Not-determined | Genome-fragment | NA | NA | 0 |
| S17-final-viral-combined_k141_159695\|\|full | 1721 | No | NA | 3 | 0 | 2 | Not-determined | Genome-fragment | NA | NA | 0 |
| S17-final-viral-combined_k141_104657\|\|full | 3049 | No | NA | 3 | 0 | 2 | Not-determined | Genome-fragment | NA | NA | 0 |
| S9-final-viral-combined_k141_123594\|\|full | 2202 | No | NA | 3 | 1 | 0 | Low-quality | Genome-fragment | 4.75 | AAI-based (high-confidence) | 0 |
| S17-final-viral-combined_k141_159706\|\|full | 1089 | No | NA | 3 | 0 | 1 | Not-determined | Genome-fragment | NA | NA | 0 |
| S9-final-viral-combined_k141_127148\|\|full | 2202 | No | NA | 2 | 0 | 2 | Low-quality | Genome-fragment | 2.02 | AAI-based (high-confidence) | 0 |
| S1-final-viral-combined_k141_366982\|\|full | 2202 | No | NA | 6 | 1 | 0 | Low-quality | Genome-fragment | 5.21 | AAI-based (high-confidence) | 0 |
| S17-final-viral-combined_k141_227813\|\|full | 3185 | No | NA | 4 | 0 | 2 | Not-determined | Genome-fragment | NA | NA | 0 |
| S17-final-viral-combined_k141_149479\|\|full | 8489 | No | NA | 11 | 0 | 8 | Not-determined | Genome-fragment | NA | NA | 0 |
| S1-final-viral-combined_k141_113540\|\|full | 2202 | No | NA | 3 | 3 | 0 | Low-quality | Genome-fragment | 4.71 | AAI-based (high-confidence) | 0 |
| S17-final-viral-combined_k141_37969\|\|full | 1646 | No | NA | 2 | 0 | 1 | Not-determined | Genome-fragment | NA | NA | 0 |
| S17-final-viral-combined_k141_47697\|\|full | 1702 | No | NA | 3 | 0 | 2 | Not-determined | Genome-fragment | NA | NA | 0 |
| S17-final-viral-combined_k141_227825\|\|full | 4502 | No | NA | 6 | 0 | 5 | Not-determined | Genome-fragment | NA | NA | 0 |
| S12-final-viral-combined_k141_204964\|\|full | 2202 | No | NA | 3 | 0 | 0 | Low-quality | Genome-fragment | 0.57 | AAI-based (medium-confidence) | 0 |
| S2-final-viral-combined_k141_186736\|\|full | 2202 | No | NA | 4 | 0 | 0 | Low-quality | Genome-fragment | 4.54 | AAI-based (high-confidence) | 0 |
| S2-final-viral-combined_k141_155748\|\|full | 2202 | No | NA | 3 | 2 | 0 | Low-quality | Genome-fragment | 5.05 | AAI-based (high-confidence) | 0 |
| S4-final-viral-combined_k141_138984\|\|full | 2202 | No | NA | 5 | 1 | 0 | Low-quality | Genome-fragment | 3.33 | HMM-based (lower-bound) | 0 |
| S17-final-viral-combined_k141_159738\|\|full | 1543 | No | NA | 3 | 0 | 1 | Not-determined | Genome-fragment | NA | NA | 0 |
| S17-final-viral-combined_k141_205434\|\|full | 1985 | No | NA | 3 | 0 | 0 | Not-determined | Genome-fragment | NA | NA | 0 |
| S9-final-viral-combined_k141_124367\|\|full | 2203 | No | NA | 5 | 2 | 0 | Low-quality | Genome-fragment | 4.86 | AAI-based (high-confidence) | 0 |
| S11-final-viral-combined_k141_194059\|\|full | 2203 | No | NA | 3 | 1 | 0 | Low-quality | Genome-fragment | 1.11 | HMM-based (lower-bound) | 0 |
| S17-final-viral-combined_k141_71783\|\|full | 3751 | No | NA | 4 | 0 | 0 | Not-determined | Genome-fragment | NA | NA | 0 |
| S17-final-viral-combined_k141_26863\|\|full | 2579 | No | NA | 4 | 0 | 3 | Not-determined | Genome-fragment | NA | NA | 0 |
| S17-final-viral-combined_k141_248546\|\|full | 2232 | No | NA | 4 | 0 | 1 | Not-determined | Genome-fragment | NA | NA | 0 |
| S17-final-viral-combined_k141_47737\|\|full | 1185 | No | NA | 1 | 0 | 0 | Not-determined | Genome-fragment | NA | NA | 0 |
| S11-final-viral-combined_k141_282079\|\|full | 2203 | No | NA | 3 | 0 | 2 | Low-quality | Genome-fragment | 1.18 | AAI-based (medium-confidence) | 0 |
| S17-final-viral-combined_k141_51773\|\|full | 2203 | No | NA | 3 | 0 | 0 | Low-quality | Genome-fragment | 3.35 | AAI-based (high-confidence) | 0 |
| S2-final-viral-combined_k141_138401\|\|full | 2203 | No | NA | 5 | 1 | 0 | Low-quality | Genome-fragment | 4.51 | AAI-based (medium-confidence) | 0 |
| S4-final-viral-combined_k141_86666\|\|full | 2203 | No | NA | 4 | 0 | 3 | Low-quality | Genome-fragment | 3.85 | AAI-based (high-confidence) | 0 |
| S17-final-viral-combined_k141_83583\|\|full | 1888 | No | NA | 2 | 0 | 1 | Not-determined | Genome-fragment | NA | NA | 0 |
| S11_k141_322994 | 2204 | No | NA | 7 | 0 | 0 | Low-quality | Genome-fragment | 2.34 | AAI-based (high-confidence) | 0 |
| S17-final-viral-combined_k141_194040\|\|full | 3357 | No | NA | 4 | 0 | 0 | Not-determined | Genome-fragment | NA | NA | 0 |
| S17-final-viral-combined_k141_215055\|\|full | 2204 | No | NA | 4 | 2 | 0 | Low-quality | Genome-fragment | 5.43 | AAI-based (high-confidence) | 0 |
| S17-final-viral-combined_k141_83597\|\|full | 3537 | No | NA | 4 | 0 | 0 | Not-determined | Genome-fragment | NA | NA | 0 |
| S17-final-viral-combined_k141_205462\|\|full | 5437 | No | NA | 10 | 0 | 0 | Not-determined | Genome-fragment | NA | NA | 0 |
| S17-final-viral-combined_k141_127404\|\|full | 9624 | No | NA | 9 | 0 | 7 | Not-determined | Genome-fragment | NA | NA | 0 |
| S12_k141_274637 | 2204 | No | NA | 3 | 0 | 0 | Low-quality | Genome-fragment | 6.32 | AAI-based (medium-confidence) | 0 |
| S17-final-viral-combined_k141_149555\|\|full | 1632 | No | NA | 4 | 0 | 1 | Not-determined | Genome-fragment | NA | NA | 0 |
| S2-final-viral-combined_k141_207287\|\|full | 2204 | No | NA | 3 | 0 | 0 | Low-quality | Genome-fragment | 3.92 | AAI-based (high-confidence) | 0 |
| S17-final-viral-combined_k141_83624\|\|full | 2222 | No | NA | 2 | 0 | 2 | Not-determined | Genome-fragment | NA | NA | 0 |
| S17-final-viral-combined_k141_227935\|\|full | 3802 | No | NA | 5 | 0 | 3 | Not-determined | Genome-fragment | NA | NA | 0 |
| S2-final-viral-combined_k141_231221\|\|full | 2204 | No | NA | 2 | 0 | 2 | Low-quality | Genome-fragment | 3.48 | AAI-based (medium-confidence) | 0 |
| S2-final-viral-combined_k141_244911\|\|full | 2204 | No | NA | 3 | 0 | 2 | Low-quality | Genome-fragment | 2.73 | AAI-based (high-confidence) | 0 |
| S17-final-viral-combined_k141_47824\|\|full | 2474 | No | NA | 3 | 0 | 1 | Not-determined | Genome-fragment | NA | NA | 0 |
| S17-final-viral-combined_k141_227962\|\|full | 2789 | No | NA | 3 | 0 | 2 | Not-determined | Genome-fragment | NA | NA | 0 |
| S4_k141_27080 | 2204 | No | NA | 3 | 2 | 0 | Low-quality | Genome-fragment | 1.27 | AAI-based (high-confidence) | 0 |
| S17-final-viral-combined_k141_227974\|\|full | 1933 | No | NA | 3 | 0 | 3 | Not-determined | Genome-fragment | NA | NA | 0 |
| S17-final-viral-combined_k141_159848\|\|full | 3419 | No | NA | 3 | 0 | 0 | Not-determined | Genome-fragment | NA | NA | 0 |
| S11-final-viral-combined_k141_257065\|\|full | 2205 | No | NA | 5 | 1 | 0 | Low-quality | Genome-fragment | 1.14 | HMM-based (lower-bound) | 0 |
| S17-final-viral-combined_k141_194178\|\|full | 5293 | No | NA | 3 | 0 | 0 | Not-determined | Genome-fragment | NA | NA | 0 |
| S17-final-viral-combined_k141_127503\|\|full | 1619 | No | NA | 3 | 0 | 1 | Not-determined | Genome-fragment | NA | NA | 0 |
| S1-final-viral-combined_k141_300433\|\|full | 2205 | No | NA | 4 | 1 | 0 | Low-quality | Genome-fragment | 3.4 | HMM-based (lower-bound) | 0 |
| S7-final-viral-combined_k141_167741\|\|full | 2205 | No | NA | 4 | 0 | 2 | Low-quality | Genome-fragment | 0.66 | AAI-based (medium-confidence) | 0 |
| S17-final-viral-combined_k141_92788\|\|full | 8364 | No | NA | 7 | 0 | 4 | Not-determined | Genome-fragment | NA | NA | 0 |
| S12-final-viral-combined_k141_181583\|\|full | 2205 | Yes | 1051 | 4 | 1 | 1 | Low-quality | Genome-fragment | 0.84 | HMM-based (lower-bound) | 52.34 |
| S12-final-viral-combined_k141_355897\|\|full | 2206 | No | NA | 4 | 0 | 1 | Low-quality | Genome-fragment | 2.35 | AAI-based (medium-confidence) | 0 |
| S12-final-viral-combined_k141_261221\|\|full | 2206 | No | NA | 5 | 1 | 0 | Low-quality | Genome-fragment | 1.46 | AAI-based (medium-confidence) | 0 |
| S5-final-viral-combined_k141_171725\|\|full | 2207 | No | NA | 4 | 0 | 0 | Low-quality | Genome-fragment | 3.81 | AAI-based (high-confidence) | 0 |
| S12-final-viral-combined_k141_226618\|\|full | 2207 | No | NA | 5 | 2 | 0 | Low-quality | Genome-fragment | 5.83 | AAI-based (high-confidence) | 0 |
| S12-final-viral-combined_k141_180263\|\|full | 2207 | No | NA | 2 | 0 | 1 | Low-quality | Genome-fragment | 4.52 | AAI-based (high-confidence) | 0 |
| S2_k141_9118 | 2207 | No | NA | 2 | 0 | 1 | Low-quality | Genome-fragment | 1.97 | AAI-based (medium-confidence) | 0 |
| S11-final-viral-combined_k141_271124\|\|full | 2208 | No | NA | 3 | 0 | 0 | Low-quality | Genome-fragment | 1.42 | AAI-based (medium-confidence) | 0 |
| S11-final-viral-combined_k141_314653\|\|full | 2208 | No | NA | 3 | 0 | 0 | Low-quality | Genome-fragment | 3.51 | AAI-based (medium-confidence) | 0 |
| S17-final-viral-combined_k141_171744\|\|full | 2352 | No | NA | 3 | 0 | 2 | Not-determined | Genome-fragment | NA | NA | 0 |
| S2-final-viral-combined_k141_147138\|\|full | 2208 | No | NA | 3 | 2 | 0 | Low-quality | Genome-fragment | 5.37 | AAI-based (high-confidence) | 0 |
| S17-final-viral-combined_k141_127551\|\|full | 1014 | No | NA | 2 | 0 | 1 | Not-determined | Genome-fragment | NA | NA | 0 |
| S4-final-viral-combined_k141_95074\|\|full | 2208 | No | NA | 3 | 0 | 0 | Low-quality | Genome-fragment | 4.13 | AAI-based (medium-confidence) | 0 |
| S17-final-viral-combined_k141_194262\|\|full | 1618 | No | NA | 3 | 0 | 1 | Not-determined | Genome-fragment | NA | NA | 0 |
| S17-final-viral-combined_k141_194264\|\|full | 1650 | No | NA | 2 | 0 | 1 | Not-determined | Genome-fragment | NA | NA | 0 |
| S1-final-viral-combined_k141_80967\|\|lt2gene | 2209 | No | NA | 2 | 2 | 0 | Low-quality | Genome-fragment | 3.48 | AAI-based (high-confidence) | 0 |
| S17-final-viral-combined_k141_83764\|\|full | 1513 | No | NA | 2 | 0 | 1 | Not-determined | Genome-fragment | NA | NA | 0 |
| S3-final-viral-combined_k141_108215\|\|full | 2209 | No | NA | 2 | 0 | 1 | Low-quality | Genome-fragment | 2.41 | AAI-based (medium-confidence) | 0 |
| S17-final-viral-combined_k141_150900\|\|full | 2209 | No | NA | 6 | 1 | 0 | Low-quality | Genome-fragment | 5.39 | AAI-based (high-confidence) | 0 |
| S17-final-viral-combined_k141_127564\|\|full | 1870 | No | NA | 2 | 0 | 1 | Not-determined | Genome-fragment | NA | NA | 0 |
| S17-final-viral-combined_k141_92810\|\|full | 40643 | No | NA | 36 | 0 | 30 | Not-determined | Genome-fragment | NA | NA | 0 |
| S17-final-viral-combined_k141_194299\|\|full | 2836 | No | NA | 4 | 0 | 3 | Not-determined | Genome-fragment | NA | NA | 0 |
| S12-final-viral-combined_k141_156882\|\|full | 2209 | No | NA | 3 | 0 | 0 | Low-quality | Genome-fragment | 5.03 | AAI-based (high-confidence) | 0 |
| S17-final-viral-combined_k141_216884\|\|full | 3396 | No | NA | 6 | 0 | 2 | Not-determined | Genome-fragment | NA | NA | 0 |
| S17-final-viral-combined_k141_228015\|\|full | 1607 | No | NA | 3 | 0 | 0 | Not-determined | Genome-fragment | NA | NA | 0 |
| S17-final-viral-combined_k141_92819\|\|full | 1995 | No | NA | 3 | 0 | 1 | Not-determined | Genome-fragment | NA | NA | 0 |
| S17-final-viral-combined_k141_205635\|\|full | 1341 | No | NA | 3 | 0 | 2 | Not-determined | Genome-fragment | NA | NA | 0 |
| S8-final-viral-combined_k141_23348\|\|full | 2209 | No | NA | 3 | 0 | 0 | Low-quality | Genome-fragment | 5.71 | AAI-based (medium-confidence) | 0 |
| S12-final-viral-combined_k141_276925\|\|full | 2210 | No | NA | 3 | 0 | 0 | Low-quality | Genome-fragment | 4.51 | AAI-based (medium-confidence) | 0 |
| S4-final-viral-combined_k141_69975\|\|full | 2210 | No | NA | 3 | 0 | 1 | Low-quality | Genome-fragment | 3.62 | AAI-based (medium-confidence) | 0 |
| S17-final-viral-combined_k141_248708\|\|full | 2679 | No | NA | 3 | 0 | 2 | Not-determined | Genome-fragment | NA | NA | 0 |
| S17-final-viral-combined_k141_47860\|\|full | 55026 | No | NA | 74 | 32 | 0 | Medium-quality | Genome-fragment | 84.62 | AAI-based (high-confidence) | 0 |
| S4_k141_36069 | 2210 | No | NA | 6 | 1 | 0 | Low-quality | Genome-fragment | 2.58 | AAI-based (high-confidence) | 0 |
| S11-final-viral-combined_k141_143000\|\|full | 2211 | No | NA | 3 | 0 | 1 | Low-quality | Genome-fragment | 1.12 | AAI-based (medium-confidence) | 0 |
| S1-final-viral-combined_k141_20695\|\|full | 2211 | No | NA | 4 | 0 | 0 | Low-quality | Genome-fragment | 2.89 | AAI-based (medium-confidence) | 0 |
| S5-final-viral-combined_k141_11598\|\|full | 2211 | No | NA | 3 | 0 | 0 | Low-quality | Genome-fragment | 1.39 | AAI-based (medium-confidence) | 0 |
| S17-final-viral-combined_k141_103084\|\|full | 2211 | Yes | 1399 | 6 | 1 | 1 | Low-quality | Genome-fragment | 2.98 | AAI-based (high-confidence) | 36.73 |
| S17-final-viral-combined_k141_149755\|\|full | 3923 | No | NA | 3 | 0 | 3 | Not-determined | Genome-fragment | NA | NA | 0 |
| S17-final-viral-combined_k141_127628\|\|full | 1025 | No | NA | 2 | 0 | 1 | Not-determined | Genome-fragment | NA | NA | 0 |
| S17-final-viral-combined_k141_47899\|\|full | 1810 | No | NA | 2 | 0 | 1 | Not-determined | Genome-fragment | NA | NA | 0 |
| S17-final-viral-combined_k141_149789\|\|full | 2456 | No | NA | 3 | 0 | 0 | Not-determined | Genome-fragment | NA | NA | 0 |
| S17-final-viral-combined_k141_15291\|\|full | 8254 | No | NA | 7 | 0 | 3 | Not-determined | Genome-fragment | NA | NA | 0 |
| S8-final-viral-combined_k141_246374\|\|full | 2211 | No | NA | 4 | 0 | 2 | Low-quality | Genome-fragment | 1.1 | AAI-based (medium-confidence) | 0 |
| S1_k141_285073 | 2212 | No | NA | 5 | 0 | 0 | Low-quality | Genome-fragment | 3.54 | AAI-based (high-confidence) | 0 |
| S17-final-viral-combined_k141_149802\|\|full | 1415 | No | NA | 2 | 0 | 0 | Not-determined | Genome-fragment | NA | NA | 0 |
| S17-final-viral-combined_k141_15297\|\|full | 4886 | No | NA | 7 | 0 | 0 | Not-determined | Genome-fragment | NA | NA | 0 |
| S17-final-viral-combined_k141_205725\|\|full | 1683 | No | NA | 3 | 0 | 1 | Not-determined | Genome-fragment | NA | NA | 0 |
| S17-final-viral-combined_k141_15299\|\|full | 1247 | No | NA | 3 | 0 | 2 | Not-determined | Genome-fragment | NA | NA | 0 |
| S3-final-viral-combined_k141_88474\|\|full | 2212 | No | NA | 3 | 0 | 0 | Low-quality | Genome-fragment | 3.92 | AAI-based (high-confidence) | 0 |
| S17-final-viral-combined_k141_160051\|\|full | 2965 | No | NA | 5 | 0 | 3 | Not-determined | Genome-fragment | NA | NA | 0 |
| S17-final-viral-combined_k141_259811\|\|full | 69782 | Yes | 48861 | 112 | 21 | 17 | Medium-quality | Genome-fragment | 71.65 | HMM-based (lower-bound) | 29.98 |
| S17-final-viral-combined_k141_228178\|\|full | 1580 | No | NA | 4 | 0 | 0 | Not-determined | Genome-fragment | NA | NA | 0 |
| S11-final-viral-combined_k141_102742\|\|full | 2213 | No | NA | 4 | 1 | 0 | Low-quality | Genome-fragment | 3.3 | HMM-based (lower-bound) | 0 |
| S3-final-viral-combined_k141_125006\|\|full | 2213 | No | NA | 2 | 2 | 0 | Low-quality | Genome-fragment | 0.99 | HMM-based (lower-bound) | 0 |
| S17-final-viral-combined_k141_127688\|\|full | 46043 | No | NA | 64 | 21 | 1 | High-quality | High-quality | 94.05 | AAI-based (high-confidence) | 0 |
| S17-final-viral-combined_k141_27338\|\|full | 2213 | No | NA | 3 | 0 | 0 | Low-quality | Genome-fragment | 3.6 | AAI-based (medium-confidence) | 0 |
| S17-final-viral-combined_k141_174638\|\|full | 2213 | No | NA | 4 | 0 | 3 | Low-quality | Genome-fragment | 0.85 | AAI-based (medium-confidence) | 0 |
| S2_k141_211612 | 2213 | No | NA | 4 | 2 | 0 | Low-quality | Genome-fragment | 8.05 | HMM-based (lower-bound) | 0 |
| S4_k141_43415 | 2213 | No | NA | 1 | 0 | 0 | Low-quality | Genome-fragment | 0.96 | AAI-based (medium-confidence) | 0 |
| S6-final-viral-combined_k141_101333\|\|full | 2213 | No | NA | 4 | 0 | 0 | Low-quality | Genome-fragment | 3.91 | AAI-based (medium-confidence) | 0 |
| S9-final-viral-combined_k141_592\|\|full | 2214 | No | NA | 4 | 0 | 0 | Low-quality | Genome-fragment | 16.78 | AAI-based (medium-confidence) | 0 |
| S1-final-viral-combined_k141_238973\|\|full | 2214 | No | NA | 3 | 0 | 0 | Low-quality | Genome-fragment | 3.88 | AAI-based (high-confidence) | 0 |
| S5-final-viral-combined_k141_132036\|\|full | 2214 | No | NA | 3 | 0 | 0 | Low-quality | Genome-fragment | 4.02 | AAI-based (medium-confidence) | 0 |
| S11_k141_65260 | 2215 | No | NA | 3 | 0 | 3 | Low-quality | Genome-fragment | 1.42 | AAI-based (high-confidence) | 0 |
| S17-final-viral-combined_k141_60524\|\|full | 1064 | No | NA | 3 | 0 | 1 | Not-determined | Genome-fragment | NA | NA | 0 |
| S17-final-viral-combined_k141_117724\|\|full | 1979 | No | NA | 2 | 0 | 1 | Not-determined | Genome-fragment | NA | NA | 0 |
| S17-final-viral-combined_k141_194497\|\|full | 1319 | No | NA | 3 | 0 | 1 | Not-determined | Genome-fragment | NA | NA | 0 |
| S17-final-viral-combined_k141_212980\|\|full | 2215 | No | NA | 2 | 0 | 2 | Low-quality | Genome-fragment | 1.55 | AAI-based (medium-confidence) | 0 |
| S17-final-viral-combined_k141_29385\|\|full | 2215 | No | NA | 2 | 0 | 2 | Low-quality | Genome-fragment | 3.41 | AAI-based (medium-confidence) | 0 |
| S17-final-viral-combined_k141_117737\|\|full | 2157 | No | NA | 4 | 0 | 0 | Not-determined | Genome-fragment | NA | NA | 0 |
| S12-final-viral-combined_k141_9366\|\|full | 2215 | No | NA | 4 | 2 | 0 | Low-quality | Genome-fragment | 2.1 | HMM-based (lower-bound) | 0 |
| S17-final-viral-combined_k141_48016\|\|full | 1907 | No | NA | 3 | 0 | 2 | Not-determined | Genome-fragment | NA | NA | 0 |
| S17-final-viral-combined_k141_238277\|\|full | 2894 | No | NA | 3 | 0 | 3 | Not-determined | Genome-fragment | NA | NA | 0 |
| S17-final-viral-combined_k141_60551\|\|full | 3449 | No | NA | 2 | 0 | 2 | Not-determined | Genome-fragment | NA | NA | 0 |
| S12-final-viral-combined_k141_97490\|\|full | 2215 | No | NA | 4 | 0 | 0 | Low-quality | Genome-fragment | 5.32 | AAI-based (medium-confidence) | 0 |
| S17-final-viral-combined_k141_149916\|\|full | 1557 | No | NA | 3 | 0 | 0 | Not-determined | Genome-fragment | NA | NA | 0 |
| S17-final-viral-combined_k141_205864\|\|full | 2733 | No | NA | 4 | 0 | 1 | Not-determined | Genome-fragment | NA | NA | 0 |
| S17-final-viral-combined_k141_15351\|\|full | 2134 | No | NA | 2 | 0 | 2 | Not-determined | Genome-fragment | NA | NA | 0 |
| S4-final-viral-combined_k141_104120\|\|full | 2215 | No | NA | 2 | 0 | 1 | Low-quality | Genome-fragment | 1.53 | AAI-based (medium-confidence) | 0 |
| S4_k141_156842 | 2216 | No | NA | 1 | 0 | 1 | Low-quality | Genome-fragment | 0.8 | AAI-based (medium-confidence) | 0 |
| S6-final-viral-combined_k141_108515\|\|full | 2216 | No | NA | 4 | 0 | 1 | Low-quality | Genome-fragment | 2.91 | AAI-based (medium-confidence) | 0 |
| S17-final-viral-combined_k141_4934\|\|full | 1854 | No | NA | 3 | 0 | 2 | Not-determined | Genome-fragment | NA | NA | 0 |
| S9-final-viral-combined_k141_108699\|\|full | 2217 | No | NA | 7 | 2 | 0 | Low-quality | Genome-fragment | 5.77 | AAI-based (high-confidence) | 0 |
| S17-final-viral-combined_k141_149959\|\|full | 1451 | No | NA | 3 | 0 | 0 | Not-determined | Genome-fragment | NA | NA | 0 |
| S17-final-viral-combined_k141_72250\|\|full | 4738 | No | NA | 5 | 0 | 4 | Not-determined | Genome-fragment | NA | NA | 0 |
| S11-final-viral-combined_k141_104023\|\|full | 2217 | No | NA | 2 | 0 | 1 | Low-quality | Genome-fragment | 4.21 | AAI-based (medium-confidence) | 0 |
| S17-final-viral-combined_k141_105250\|\|full | 2623 | No | NA | 3 | 0 | 2 | Not-determined | Genome-fragment | NA | NA | 0 |
| S17-final-viral-combined_k141_160214\|\|full | 1466 | No | NA | 2 | 0 | 2 | Not-determined | Genome-fragment | NA | NA | 0 |
| S17-final-viral-combined_k141_60637\|\|full | 5294 | No | NA | 5 | 0 | 4 | Not-determined | Genome-fragment | NA | NA | 0 |
| S11-final-viral-combined_k141_325005\|\|full | 2217 | No | NA | 4 | 0 | 0 | Low-quality | Genome-fragment | 4.76 | AAI-based (high-confidence) | 0 |
| S17-final-viral-combined_k141_194637\|\|full | 17959 | No | NA | 20 | 0 | 14 | Not-determined | Genome-fragment | NA | NA | 0 |
| S17-final-viral-combined_k141_228356\|\|full | 1331 | No | NA | 3 | 0 | 0 | Not-determined | Genome-fragment | NA | NA | 0 |
| S17-final-viral-combined_k141_194653\|\|full | 2322 | No | NA | 3 | 0 | 2 | Not-determined | Genome-fragment | NA | NA | 0 |
| S2_k141_129485 | 2217 | No | NA | 4 | 1 | 0 | Low-quality | Genome-fragment | 2.18 | AAI-based (high-confidence) | 0 |
| S8-final-viral-combined_k141_206059\|\|full | 2217 | No | NA | 5 | 2 | 0 | Low-quality | Genome-fragment | 6.72 | AAI-based (high-confidence) | 0 |
| S9-final-viral-combined_k141_75233\|\|full | 2218 | No | NA | 2 | 0 | 0 | Low-quality | Genome-fragment | 2.73 | AAI-based (medium-confidence) | 0 |
| S17-final-viral-combined_k141_205943\|\|full | 4623 | No | NA | 3 | 0 | 2 | Not-determined | Genome-fragment | NA | NA | 0 |
| S17-final-viral-combined_k141_138984\|\|full | 3598 | No | NA | 3 | 0 | 0 | Not-determined | Genome-fragment | NA | NA | 0 |
| S17-final-viral-combined_k141_194665\|\|full | 1891 | No | NA | 2 | 0 | 2 | Not-determined | Genome-fragment | NA | NA | 0 |
| S11-final-viral-combined_k141_75319\|\|full | 2218 | No | NA | 5 | 1 | 0 | Low-quality | Genome-fragment | 5.02 | AAI-based (high-confidence) | 0 |
| S17-final-viral-combined_k141_172044\|\|full | 2575 | No | NA | 4 | 0 | 3 | Not-determined | Genome-fragment | NA | NA | 0 |
| S17-final-viral-combined_k141_60686\|\|full | 1265 | No | NA | 2 | 0 | 0 | Not-determined | Genome-fragment | NA | NA | 0 |
| S11-final-viral-combined_k141_40657\|\|full | 2218 | No | NA | 3 | 3 | 0 | Low-quality | Genome-fragment | 4.35 | AAI-based (high-confidence) | 0 |
| S17-final-viral-combined_k141_15478\|\|full | 4682 | No | NA | 4 | 0 | 3 | Not-determined | Genome-fragment | NA | NA | 0 |
| S3_k141_24139 | 2218 | No | NA | 4 | 0 | 1 | Low-quality | Genome-fragment | 1.02 | AAI-based (high-confidence) | 0 |
| S1-final-viral-combined_k141_110147\|\|full | 2219 | No | NA | 3 | 0 | 1 | Low-quality | Genome-fragment | 1.72 | AAI-based (high-confidence) | 0 |
| S17-final-viral-combined_k141_183958\|\|full | 1010 | No | NA | 2 | 0 | 1 | Not-determined | Genome-fragment | NA | NA | 0 |
| S17-final-viral-combined_k141_38427\|\|full | 3038 | No | NA | 3 | 0 | 1 | Not-determined | Genome-fragment | NA | NA | 0 |
| S7-final-viral-combined_k141_129560\|\|full | 2219 | No | NA | 4 | 0 | 2 | Low-quality | Genome-fragment | 1.5 | AAI-based (high-confidence) | 0 |
| S8_k141_231345 | 2219 | No | NA | 3 | 0 | 3 | Low-quality | Genome-fragment | 0.66 | AAI-based (medium-confidence) | 0 |
| S17-final-viral-combined_k141_127849\|\|full | 1729 | No | NA | 2 | 0 | 1 | Not-determined | Genome-fragment | NA | NA | 0 |
| S17-final-viral-combined_k141_206029\|\|full | 3376 | No | NA | 3 | 0 | 2 | Not-determined | Genome-fragment | NA | NA | 0 |
| S17-final-viral-combined_k141_217232\|\|full | 4400 | No | NA | 4 | 0 | 3 | Not-determined | Genome-fragment | NA | NA | 0 |
| S9-final-viral-combined_k141_121890\|\|full | 2220 | No | NA | 2 | 0 | 1 | Low-quality | Genome-fragment | 2.93 | AAI-based (high-confidence) | 0 |
| S11-final-viral-combined_k141_99930\|\|full | 2220 | No | NA | 4 | 1 | 0 | Low-quality | Genome-fragment | 5.4 | AAI-based (medium-confidence) | 0 |
| S17-final-viral-combined_k141_260043\|\|full | 3062 | No | NA | 4 | 0 | 2 | Not-determined | Genome-fragment | NA | NA | 0 |
| S11-final-viral-combined_k141_39638\|\|full | 2220 | No | NA | 2 | 2 | 0 | Low-quality | Genome-fragment | 4.91 | AAI-based (medium-confidence) | 0 |
| S17-final-viral-combined_k141_150083\|\|full | 1791 | No | NA | 3 | 0 | 1 | Not-determined | Genome-fragment | NA | NA | 0 |
| S1-final-viral-combined_k141_379035\|\|full | 2220 | No | NA | 2 | 0 | 2 | Low-quality | Genome-fragment | 5.13 | AAI-based (medium-confidence) | 0 |
| S17-final-viral-combined_k141_117949\|\|full | 5301 | No | NA | 8 | 0 | 0 | Not-determined | Genome-fragment | NA | NA | 0 |
| S17-final-viral-combined_k141_194720\|\|full | 13361 | No | NA | 7 | 0 | 5 | Not-determined | Genome-fragment | NA | NA | 0 |
| S2-final-viral-combined_k141_41818\|\|full | 2220 | No | NA | 4 | 2 | 0 | Low-quality | Genome-fragment | 4.97 | AAI-based (high-confidence) | 0 |
| S17-final-viral-combined_k141_183988\|\|full | 2321 | No | NA | 3 | 0 | 1 | Not-determined | Genome-fragment | NA | NA | 0 |
| S1-final-viral-combined_k141_197448\|\|full | 2221 | No | NA | 3 | 3 | 0 | Low-quality | Genome-fragment | 5.7 | AAI-based (high-confidence) | 0 |
| S17-final-viral-combined_k141_194728\|\|full | 1840 | No | NA | 4 | 0 | 1 | Not-determined | Genome-fragment | NA | NA | 0 |
| S17-final-viral-combined_k141_38460\|\|full | 1657 | No | NA | 2 | 0 | 2 | Not-determined | Genome-fragment | NA | NA | 0 |
| S1-final-viral-combined_k141_124153\|\|full | 2221 | No | NA | 2 | 0 | 1 | Low-quality | Genome-fragment | 2.75 | AAI-based (high-confidence) | 0 |
| S17-final-viral-combined_k141_249035\|\|full | 1854 | No | NA | 4 | 0 | 0 | Not-determined | Genome-fragment | NA | NA | 0 |
| S2_k141_152954 | 2221 | No | NA | 1 | 0 | 1 | Low-quality | Genome-fragment | 1.34 | AAI-based (medium-confidence) | 0 |
| S17-final-viral-combined_k141_249036\|\|full | 1652 | No | NA | 2 | 0 | 2 | Not-determined | Genome-fragment | NA | NA | 0 |
| S11_k141_296573 | 2223 | No | NA | 3 | 0 | 1 | Low-quality | Genome-fragment | 7.1 | AAI-based (medium-confidence) | 0 |
| S17-final-viral-combined_k141_38478\|\|full | 2580 | No | NA | 4 | 0 | 1 | Not-determined | Genome-fragment | NA | NA | 0 |
| S17-final-viral-combined_k141_27382\|\|full | 3198 | No | NA | 4 | 0 | 2 | Not-determined | Genome-fragment | NA | NA | 0 |
| S1-final-viral-combined_k141_3559\|\|full | 2223 | No | NA | 3 | 0 | 1 | Low-quality | Genome-fragment | 0.56 | AAI-based (medium-confidence) | 0 |
| S1-final-viral-combined_k141_62410\|\|full | 2223 | No | NA | 2 | 1 | 0 | Low-quality | Genome-fragment | 1.21 | HMM-based (lower-bound) | 0 |
| S3-final-viral-combined_k141_91896\|\|full | 2223 | No | NA | 3 | 2 | 0 | Low-quality | Genome-fragment | 5.56 | AAI-based (medium-confidence) | 0 |
| S17-final-viral-combined_k141_194743\|\|full | 17460 | No | NA | 16 | 0 | 9 | Not-determined | Genome-fragment | NA | NA | 0 |
| S17-final-viral-combined_k141_72437\|\|full | 2497 | No | NA | 3 | 0 | 0 | Not-determined | Genome-fragment | NA | NA | 0 |
| S17-final-viral-combined_k141_249041\|\|full | 10224 | No | NA | 10 | 0 | 6 | Not-determined | Genome-fragment | NA | NA | 0 |
| S17-final-viral-combined_k141_249049\|\|full | 1955 | No | NA | 3 | 0 | 1 | Not-determined | Genome-fragment | NA | NA | 0 |
| S17-final-viral-combined_k141_56620\|\|full | 2223 | No | NA | 3 | 0 | 2 | Low-quality | Genome-fragment | 0.9 | AAI-based (medium-confidence) | 0 |
| S17-final-viral-combined_k141_206096\|\|full | 1702 | No | NA | 3 | 0 | 0 | Not-determined | Genome-fragment | NA | NA | 0 |
| S17-final-viral-combined_k141_93132\|\|full | 2063 | No | NA | 3 | 0 | 2 | Not-determined | Genome-fragment | NA | NA | 0 |
| S17-final-viral-combined_k141_172105\|\|full | 1049 | No | NA | 2 | 0 | 2 | Not-determined | Genome-fragment | NA | NA | 0 |
| S17-final-viral-combined_k141_184050\|\|full | 6028 | No | NA | 5 | 0 | 4 | Not-determined | Genome-fragment | NA | NA | 0 |
| S17-final-viral-combined_k141_48116\|\|full | 1129 | No | NA | 4 | 0 | 0 | Not-determined | Genome-fragment | NA | NA | 0 |
| S17-final-viral-combined_k141_217311\|\|full | 1177 | No | NA | 3 | 0 | 0 | Not-determined | Genome-fragment | NA | NA | 0 |
| S12-final-viral-combined_k141_150847\|\|full | 2223 | No | NA | 3 | 0 | 0 | Low-quality | Genome-fragment | 5.62 | AAI-based (medium-confidence) | 0 |
| S17-final-viral-combined_k141_139169\|\|full | 1617 | No | NA | 5 | 0 | 4 | Not-determined | Genome-fragment | NA | NA | 0 |
| S17-final-viral-combined_k141_48119\|\|full | 14018 | No | NA | 14 | 0 | 10 | Not-determined | Genome-fragment | NA | NA | 0 |
| S17-final-viral-combined_k141_48121\|\|full | 1774 | No | NA | 4 | 0 | 1 | Not-determined | Genome-fragment | NA | NA | 0 |
| S2-final-viral-combined_k141_208842\|\|full | 2223 | No | NA | 4 | 0 | 0 | Low-quality | Genome-fragment | 5.03 | AAI-based (high-confidence) | 0 |
| S2-final-viral-combined_k141_65450\|\|full | 2224 | No | NA | 3 | 0 | 0 | Low-quality | Genome-fragment | 3.81 | AAI-based (high-confidence) | 0 |
| S17-final-viral-combined_k141_206167\|\|full | 1046 | No | NA | 3 | 0 | 1 | Not-determined | Genome-fragment | NA | NA | 0 |
| S1-final-viral-combined_k141_375313\|\|full | 2225 | No | NA | 2 | 0 | 1 | Low-quality | Genome-fragment | 3.99 | AAI-based (medium-confidence) | 0 |
| S1_k141_389677 | 2225 | No | NA | 2 | 0 | 2 | Low-quality | Genome-fragment | 0.97 | AAI-based (medium-confidence) | 0 |
| S12-final-viral-combined_k141_340101\|\|full | 2225 | No | NA | 4 | 0 | 1 | Low-quality | Genome-fragment | 3.01 | AAI-based (high-confidence) | 0 |
| S17-final-viral-combined_k141_84228\|\|full | 3490 | No | NA | 7 | 0 | 2 | Not-determined | Genome-fragment | NA | NA | 0 |
| S17-final-viral-combined_k141_139207\|\|full | 2824 | No | NA | 4 | 0 | 2 | Not-determined | Genome-fragment | NA | NA | 0 |
| S17-final-viral-combined_k141_38594\|\|full | 1278 | No | NA | 3 | 0 | 1 | Not-determined | Genome-fragment | NA | NA | 0 |
| S17-final-viral-combined_k141_194872\|\|full | 2754 | No | NA | 3 | 0 | 3 | Not-determined | Genome-fragment | NA | NA | 0 |
| S12-final-viral-combined_k141_343062\|\|full | 2225 | No | NA | 5 | 2 | 0 | Low-quality | Genome-fragment | 5.46 | AAI-based (medium-confidence) | 0 |
| S17-final-viral-combined_k141_93177\|\|full | 2041 | No | NA | 3 | 0 | 0 | Not-determined | Genome-fragment | NA | NA | 0 |
| S17-final-viral-combined_k141_127960\|\|full | 1449 | No | NA | 3 | 0 | 3 | Not-determined | Genome-fragment | NA | NA | 0 |
| S4-final-viral-combined_k141_157108\|\|full | 2225 | No | NA | 3 | 2 | 0 | Low-quality | Genome-fragment | 1.49 | AAI-based (medium-confidence) | 0 |
| S17-final-viral-combined_k141_160437\|\|full | 16485 | No | NA | 15 | 0 | 13 | Not-determined | Genome-fragment | NA | NA | 0 |
| S8-final-viral-combined_k141_80651\|\|full | 2225 | No | NA | 3 | 0 | 1 | Low-quality | Genome-fragment | 2.59 | AAI-based (high-confidence) | 0 |
| S8-final-viral-combined_k141_63118\|\|full | 2225 | No | NA | 6 | 1 | 0 | Low-quality | Genome-fragment | 3.28 | HMM-based (lower-bound) | 0 |
| S17-final-viral-combined_k141_194893\|\|full | 2515 | No | NA | 4 | 0 | 3 | Not-determined | Genome-fragment | NA | NA | 0 |
| S17-final-viral-combined_k141_84264\|\|full | 4777 | No | NA | 5 | 0 | 5 | Not-determined | Genome-fragment | NA | NA | 0 |
| S17-final-viral-combined_k141_238602\|\|full | 1270 | No | NA | 2 | 0 | 1 | Not-determined | Genome-fragment | NA | NA | 0 |
| S17-final-viral-combined_k141_228583\|\|full | 1701 | No | NA | 3 | 0 | 1 | Not-determined | Genome-fragment | NA | NA | 0 |
| S17-final-viral-combined_k141_238612\|\|full | 1678 | No | NA | 2 | 0 | 1 | Not-determined | Genome-fragment | NA | NA | 0 |
| S17-final-viral-combined_k141_48214\|\|full | 1050 | No | NA | 2 | 0 | 0 | Not-determined | Genome-fragment | NA | NA | 0 |
| S17-final-viral-combined_k141_93219\|\|full | 1121 | No | NA | 3 | 0 | 0 | Not-determined | Genome-fragment | NA | NA | 0 |
| S17-final-viral-combined_k141_15659\|\|full | 1239 | No | NA | 3 | 0 | 0 | Not-determined | Genome-fragment | NA | NA | 0 |
| S5-final-viral-combined_k141_156346\|\|full | 2226 | No | NA | 3 | 0 | 2 | Low-quality | Genome-fragment | 1.09 | AAI-based (medium-confidence) | 0 |
| S7-final-viral-combined_k141_94972\|\|full | 2226 | No | NA | 4 | 0 | 1 | Low-quality | Genome-fragment | 6.48 | AAI-based (medium-confidence) | 0 |
| S12-final-viral-combined_k141_306710\|\|full | 2226 | No | NA | 2 | 0 | 1 | Low-quality | Genome-fragment | 2.17 | AAI-based (medium-confidence) | 0 |
| S17-final-viral-combined_k141_184109\|\|full | 1210 | No | NA | 2 | 0 | 0 | Not-determined | Genome-fragment | NA | NA | 0 |
| S4_k141_56910 | 2226 | No | NA | 1 | 0 | 0 | Low-quality | Genome-fragment | 2.26 | AAI-based (medium-confidence) | 0 |
| S8-final-viral-combined_k141_12093\|\|full | 2226 | No | NA | 2 | 0 | 1 | Low-quality | Genome-fragment | 3.79 | AAI-based (high-confidence) | 0 |
| S11-final-viral-combined_k141_182373\|\|full | 2227 | No | NA | 3 | 0 | 0 | Low-quality | Genome-fragment | 4.65 | AAI-based (high-confidence) | 0 |
| S17-final-viral-combined_k141_118144\|\|full | 5324 | No | NA | 5 | 0 | 5 | Not-determined | Genome-fragment | NA | NA | 0 |
| S1-final-viral-combined_k141_297501\|\|full | 2227 | No | NA | 3 | 0 | 1 | Low-quality | Genome-fragment | 6.51 | AAI-based (medium-confidence) | 0 |
| S3_k141_53929 | 2227 | No | NA | 1 | 0 | 1 | Low-quality | Genome-fragment | 1.07 | AAI-based (medium-confidence) | 0 |
| S17-final-viral-combined_k141_38662\|\|full | 4929 | No | NA | 4 | 0 | 2 | Not-determined | Genome-fragment | NA | NA | 0 |
| S5-final-viral-combined_k141_28199\|\|lt2gene | 2227 | No | NA | 2 | 2 | 0 | Low-quality | Genome-fragment | 6.97 | AAI-based (high-confidence) | 0 |
| S12-final-viral-combined_k141_196770\|\|full | 2227 | No | NA | 3 | 1 | 0 | Low-quality | Genome-fragment | 3.61 | AAI-based (high-confidence) | 0 |
| S17-final-viral-combined_k141_38671\|\|full | 2088 | No | NA | 2 | 0 | 1 | Not-determined | Genome-fragment | NA | NA | 0 |
| S17-final-viral-combined_k141_128066\|\|full | 1357 | No | NA | 2 | 0 | 0 | Not-determined | Genome-fragment | NA | NA | 0 |
| S17-final-viral-combined_k141_72592\|\|full | 1552 | No | NA | 2 | 0 | 1 | Not-determined | Genome-fragment | NA | NA | 0 |
| S17-final-viral-combined_k141_27581\|\|full | 3005 | No | NA | 3 | 0 | 2 | Not-determined | Genome-fragment | NA | NA | 0 |
| S17-final-viral-combined_k141_38686\|\|full | 1526 | No | NA | 3 | 0 | 1 | Not-determined | Genome-fragment | NA | NA | 0 |
| S2-final-viral-combined_k141_154839\|\|full | 2227 | No | NA | 3 | 1 | 0 | Low-quality | Genome-fragment | 5.12 | AAI-based (high-confidence) | 0 |
| S11-final-viral-combined_k141_56923\|\|full | 2228 | No | NA | 2 | 0 | 1 | Low-quality | Genome-fragment | 1.07 | AAI-based (high-confidence) | 0 |
| S17-final-viral-combined_k141_38692\|\|full | 2673 | No | NA | 3 | 0 | 3 | Not-determined | Genome-fragment | NA | NA | 0 |
| S5-final-viral-combined_k141_118234\|\|full | 2228 | No | NA | 3 | 0 | 2 | Low-quality | Genome-fragment | 2.26 | AAI-based (high-confidence) | 0 |
| S17-final-viral-combined_k141_128075\|\|full | 7683 | No | NA | 4 | 0 | 4 | Not-determined | Genome-fragment | NA | NA | 0 |
| S17-final-viral-combined_k141_260238\|\|full | 18403 | No | NA | 20 | 0 | 4 | Not-determined | Genome-fragment | NA | NA | 0 |
| S17-final-viral-combined_k141_15702\|\|full | 2112 | No | NA | 2 | 0 | 2 | Not-determined | Genome-fragment | NA | NA | 0 |
| S17-final-viral-combined_k141_5294\|\|full | 1160 | No | NA | 2 | 0 | 0 | Not-determined | Genome-fragment | NA | NA | 0 |
| S17-final-viral-combined_k141_160569\|\|full | 1193 | No | NA | 2 | 0 | 0 | Not-determined | Genome-fragment | NA | NA | 0 |
| S17-final-viral-combined_k141_228664\|\|full | 1682 | No | NA | 2 | 0 | 1 | Not-determined | Genome-fragment | NA | NA | 0 |
| S17-final-viral-combined_k141_27621\|\|full | 1292 | No | NA | 4 | 0 | 3 | Not-determined | Genome-fragment | NA | NA | 0 |
| S17-final-viral-combined_k141_217540\|\|full | 1230 | No | NA | 3 | 0 | 0 | Not-determined | Genome-fragment | NA | NA | 0 |
| S5-final-viral-combined_k141_105193\|\|full | 2228 | No | NA | 3 | 0 | 0 | Low-quality | Genome-fragment | 1.33 | AAI-based (high-confidence) | 0 |
| S17-final-viral-combined_k141_260253\|\|full | 1115 | No | NA | 2 | 0 | 1 | Not-determined | Genome-fragment | NA | NA | 0 |
| S7-final-viral-combined_k141_132918\|\|full | 2228 | No | NA | 2 | 0 | 2 | Low-quality | Genome-fragment | 2.51 | AAI-based (medium-confidence) | 0 |
| S17-final-viral-combined_k141_184195\|\|full | 2664 | No | NA | 2 | 0 | 0 | Not-determined | Genome-fragment | NA | NA | 0 |
| S17-final-viral-combined_k141_228680\|\|full | 4092 | No | NA | 5 | 0 | 0 | Not-determined | Genome-fragment | NA | NA | 0 |
| S12-final-viral-combined_k141_164559\|\|full | 2228 | No | NA | 2 | 1 | 0 | Low-quality | Genome-fragment | 3.87 | AAI-based (medium-confidence) | 0 |
| S2-final-viral-combined_k141_108166\|\|full | 2228 | No | NA | 6 | 1 | 0 | Low-quality | Genome-fragment | 3.45 | AAI-based (high-confidence) | 0 |
| S6-final-viral-combined_k141_20685\|\|full | 2228 | Yes | 1638 | 4 | 1 | 1 | Low-quality | Genome-fragment | 1.67 | AAI-based (medium-confidence) | 26.48 |
| S17-final-viral-combined_k141_5344\|\|full | 1259 | No | NA | 2 | 0 | 0 | Not-determined | Genome-fragment | NA | NA | 0 |
| S7-final-viral-combined_k141_11235\|\|full | 2229 | No | NA | 5 | 1 | 0 | Low-quality | Genome-fragment | 3.64 | AAI-based (medium-confidence) | 0 |
| S17-final-viral-combined_k141_172266\|\|full | 1902 | No | NA | 3 | 0 | 2 | Not-determined | Genome-fragment | NA | NA | 0 |
| S7-final-viral-combined_k141_94435\|\|full | 2229 | No | NA | 5 | 2 | 0 | Low-quality | Genome-fragment | 2.71 | AAI-based (high-confidence) | 0 |
| S17-final-viral-combined_k141_15761\|\|full | 1011 | No | NA | 2 | 0 | 1 | Not-determined | Genome-fragment | NA | NA | 0 |
| S7-final-viral-combined_k141_233580\|\|full | 2229 | No | NA | 5 | 0 | 1 | Low-quality | Genome-fragment | 12.66 | AAI-based (medium-confidence) | 0 |
| S17-final-viral-combined_k141_128129\|\|full | 2017 | No | NA | 4 | 0 | 2 | Not-determined | Genome-fragment | NA | NA | 0 |
| S2-final-viral-combined_k141_182420\|\|full | 2229 | No | NA | 3 | 0 | 2 | Low-quality | Genome-fragment | 1.43 | AAI-based (medium-confidence) | 0 |
| S8_k141_164156 | 2229 | No | NA | 2 | 0 | 2 | Low-quality | Genome-fragment | 1.39 | AAI-based (medium-confidence) | 0 |
| S17-final-viral-combined_k141_238804\|\|full | 4261 | No | NA | 5 | 0 | 4 | Not-determined | Genome-fragment | NA | NA | 0 |
| S11-final-viral-combined_k141_161431\|\|full | 2230 | No | NA | 3 | 0 | 1 | Low-quality | Genome-fragment | 0.57 | AAI-based (medium-confidence) | 0 |
| S17-final-viral-combined_k141_195105\|\|full | 1823 | No | NA | 4 | 0 | 1 | Not-determined | Genome-fragment | NA | NA | 0 |
| S17-final-viral-combined_k141_48381\|\|full | 1643 | No | NA | 2 | 0 | 2 | Not-determined | Genome-fragment | NA | NA | 0 |
| S1-final-viral-combined_k141_129962\|\|full | 2230 | No | NA | 2 | 0 | 1 | Low-quality | Genome-fragment | 3.4 | AAI-based (high-confidence) | 0 |
| S17-final-viral-combined_k141_5425\|\|full | 8830 | No | NA | 10 | 0 | 4 | Not-determined | Genome-fragment | NA | NA | 0 |
| S17-final-viral-combined_k141_105726\|\|full | 1543 | No | NA | 2 | 0 | 2 | Not-determined | Genome-fragment | NA | NA | 0 |
| S2-final-viral-combined_k141_123987\|\|full | 2230 | No | NA | 2 | 0 | 1 | Low-quality | Genome-fragment | 1.74 | AAI-based (medium-confidence) | 0 |
| S17-final-viral-combined_k141_206531\|\|full | 1253 | No | NA | 2 | 0 | 1 | Not-determined | Genome-fragment | NA | NA | 0 |
| S2-final-viral-combined_k141_246796\|\|full | 2230 | No | NA | 6 | 1 | 0 | Low-quality | Genome-fragment | 6.23 | AAI-based (high-confidence) | 0 |
| S17-final-viral-combined_k141_260336\|\|full | 1596 | No | NA | 2 | 0 | 0 | Not-determined | Genome-fragment | NA | NA | 0 |
| S17-final-viral-combined_k141_160697\|\|full | 3412 | No | NA | 2 | 0 | 2 | Not-determined | Genome-fragment | NA | NA | 0 |
| S17-final-viral-combined_k141_195156\|\|full | 2979 | No | NA | 3 | 0 | 1 | Not-determined | Genome-fragment | NA | NA | 0 |
| S4-final-viral-combined_k141_8280\|\|full | 2230 | No | NA | 2 | 1 | 0 | Low-quality | Genome-fragment | 32.86 | AAI-based (medium-confidence) | 0 |
| S17-final-viral-combined_k141_128232\|\|full | 11539 | No | NA | 16 | 0 | 6 | Not-determined | Genome-fragment | NA | NA | 0 |
| S17-final-viral-combined_k141_72766\|\|full | 1078 | No | NA | 3 | 0 | 0 | Not-determined | Genome-fragment | NA | NA | 0 |
| S9_k141_21177 | 2231 | No | NA | 2 | 2 | 0 | Low-quality | Genome-fragment | 5.54 | AAI-based (high-confidence) | 0 |
| S17-final-viral-combined_k141_195169\|\|full | 1408 | No | NA | 2 | 0 | 2 | Not-determined | Genome-fragment | NA | NA | 0 |
| S17-final-viral-combined_k141_184383\|\|full | 4550 | No | NA | 3 | 0 | 2 | Not-determined | Genome-fragment | NA | NA | 0 |
| S17-final-viral-combined_k141_93398\|\|full | 3009 | No | NA | 6 | 0 | 1 | Not-determined | Genome-fragment | NA | NA | 0 |
| S7-final-viral-combined_k141_1558\|\|full | 2231 | No | NA | 3 | 1 | 0 | Low-quality | Genome-fragment | 3.38 | HMM-based (lower-bound) | 0 |
| S17-final-viral-combined_k141_38772\|\|full | 3589 | No | NA | 3 | 0 | 1 | Not-determined | Genome-fragment | NA | NA | 0 |
| S17-final-viral-combined_k141_260343\|\|full | 46345 | No | NA | 71 | 23 | 1 | High-quality | High-quality | 90.78 | AAI-based (high-confidence) | 0 |
| S17-final-viral-combined_k141_5493\|\|full | 2671 | No | NA | 3 | 0 | 0 | Not-determined | Genome-fragment | NA | NA | 0 |
| S12-final-viral-combined_k141_2274\|\|full | 2231 | No | NA | 4 | 3 | 0 | Low-quality | Genome-fragment | 3.98 | AAI-based (high-confidence) | 0 |
| S12_k141_224849 | 2231 | No | NA | 3 | 0 | 0 | Low-quality | Genome-fragment | 3.18 | AAI-based (medium-confidence) | 0 |
| S1_k141_142187 | 2232 | No | NA | 4 | 2 | 0 | Low-quality | Genome-fragment | 4.65 | AAI-based (high-confidence) | 0 |
| S17-final-viral-combined_k141_195228\|\|full | 1286 | No | NA | 3 | 0 | 0 | Not-determined | Genome-fragment | NA | NA | 0 |
| S1_k141_343422 | 2232 | No | NA | 5 | 3 | 0 | Low-quality | Genome-fragment | 5.3 | AAI-based (high-confidence) | 0 |
| S17-final-viral-combined_k141_72837\|\|full | 38780 | Yes | 34087 | 55 | 16 | 2 | High-quality | High-quality | 96.89 | AAI-based (high-confidence) | 12.1 |
| S17-final-viral-combined_k141_146504\|\|full | 2232 | No | NA | 4 | 0 | 0 | Low-quality | Genome-fragment | 3.79 | AAI-based (medium-confidence) | 0 |
| S4_k141_134809 | 2232 | No | NA | 5 | 0 | 3 | Low-quality | Genome-fragment | 0.66 | AAI-based (medium-confidence) | 0 |
| S8_k141_242711 | 2232 | No | NA | 2 | 0 | 0 | Low-quality | Genome-fragment | 3.51 | AAI-based (high-confidence) | 0 |
| S17-final-viral-combined_k141_128356\|\|full | 2596 | No | NA | 4 | 0 | 3 | Not-determined | Genome-fragment | NA | NA | 0 |
| S17-final-viral-combined_k141_128357\|\|full | 1158 | No | NA | 3 | 0 | 0 | Not-determined | Genome-fragment | NA | NA | 0 |
| S1-final-viral-combined_k141_104347\|\|full | 2233 | No | NA | 2 | 2 | 0 | Low-quality | Genome-fragment | 5.25 | AAI-based (high-confidence) | 0 |
| S17-final-viral-combined_k141_217794\|\|full | 1744 | No | NA | 2 | 0 | 0 | Not-determined | Genome-fragment | NA | NA | 0 |
| S3_k141_8136 | 2233 | No | NA | 3 | 0 | 2 | Low-quality | Genome-fragment | 7.39 | AAI-based (medium-confidence) | 0 |
| S17-k141_69766 | 2233 | No | NA | 4 | 0 | 0 | Low-quality | Genome-fragment | 5.47 | AAI-based (high-confidence) | 0 |
| S17-final-viral-combined_k141_238966\|\|full | 1024 | No | NA | 2 | 0 | 1 | Not-determined | Genome-fragment | NA | NA | 0 |
| S12-final-viral-combined_k141_208524\|\|full | 2233 | No | NA | 4 | 0 | 0 | Low-quality | Genome-fragment | 4.05 | AAI-based (high-confidence) | 0 |
| S17-final-viral-combined_k141_84540\|\|full | 3496 | No | NA | 6 | 0 | 2 | Not-determined | Genome-fragment | NA | NA | 0 |
| S17-final-viral-combined_k141_195277\|\|full | 3264 | No | NA | 4 | 0 | 3 | Not-determined | Genome-fragment | NA | NA | 0 |
| S17-final-viral-combined_k141_195285\|\|full | 1046 | No | NA | 3 | 0 | 1 | Not-determined | Genome-fragment | NA | NA | 0 |
| S17-final-viral-combined_k141_61197\|\|full | 1662 | No | NA | 2 | 0 | 0 | Not-determined | Genome-fragment | NA | NA | 0 |
| S17-final-viral-combined_k141_206691\|\|full | 1432 | No | NA | 3 | 0 | 1 | Not-determined | Genome-fragment | NA | NA | 0 |
| S1_k141_12792 | 2234 | No | NA | 6 | 2 | 0 | Low-quality | Genome-fragment | 4.15 | AAI-based (high-confidence) | 0 |
| S17-final-viral-combined_k141_197090\|\|full | 2234 | No | NA | 2 | 0 | 2 | Low-quality | Genome-fragment | 1.19 | AAI-based (medium-confidence) | 0 |
| S17-final-viral-combined_k141_206700\|\|full | 3935 | No | NA | 3 | 0 | 2 | Not-determined | Genome-fragment | NA | NA | 0 |
| S17-final-viral-combined_k141_150554\|\|full | 2269 | No | NA | 4 | 0 | 3 | Not-determined | Genome-fragment | NA | NA | 0 |
| S2_k141_144027 | 2234 | No | NA | 3 | 0 | 0 | Low-quality | Genome-fragment | 3.51 | AAI-based (high-confidence) | 0 |
| S17-final-viral-combined_k141_184568\|\|full | 2371 | No | NA | 6 | 0 | 0 | Not-determined | Genome-fragment | NA | NA | 0 |
| S17-final-viral-combined_k141_93504\|\|full | 2548 | No | NA | 3 | 0 | 2 | Not-determined | Genome-fragment | NA | NA | 0 |
| S17-final-viral-combined_k141_260469\|\|full | 1389 | No | NA | 2 | 0 | 0 | Not-determined | Genome-fragment | NA | NA | 0 |
| S5-final-viral-combined_k141_172854\|\|full | 2235 | No | NA | 3 | 0 | 1 | Low-quality | Genome-fragment | 1.12 | AAI-based (medium-confidence) | 0 |
| S17-final-viral-combined_k141_195319\|\|full | 2679 | No | NA | 4 | 0 | 2 | Not-determined | Genome-fragment | NA | NA | 0 |
| S17-final-viral-combined_k141_228991\|\|full | 2037 | No | NA | 2 | 0 | 0 | Not-determined | Genome-fragment | NA | NA | 0 |
| S17-final-viral-combined_k141_38903\|\|full | 2136 | No | NA | 2 | 0 | 1 | Not-determined | Genome-fragment | NA | NA | 0 |
| S7-final-viral-combined_k141_195187\|\|full | 2235 | No | NA | 8 | 1 | 0 | Low-quality | Genome-fragment | 5.15 | AAI-based (medium-confidence) | 0 |
| S17-final-viral-combined_k141_15932\|\|full | 1830 | No | NA | 4 | 0 | 0 | Not-determined | Genome-fragment | NA | NA | 0 |
| S17-final-viral-combined_k141_179329\|\|full | 2235 | No | NA | 3 | 3 | 0 | Low-quality | Genome-fragment | 3.17 | AAI-based (high-confidence) | 0 |
| S17-final-viral-combined_k141_195348\|\|full | 1482 | No | NA | 2 | 0 | 0 | Not-determined | Genome-fragment | NA | NA | 0 |
| S2_k141_192319 | 2235 | No | NA | 1 | 0 | 1 | Low-quality | Genome-fragment | 0.59 | AAI-based (high-confidence) | 0 |
| S12-final-viral-combined_k141_211331\|\|full | 2236 | No | NA | 5 | 1 | 0 | Low-quality | Genome-fragment | 4.65 | AAI-based (high-confidence) | 0 |
| S11-final-viral-combined_k141_161971\|\|full | 2237 | No | NA | 6 | 1 | 0 | Low-quality | Genome-fragment | 1.67 | AAI-based (high-confidence) | 0 |
| S11_k141_321168 | 2237 | No | NA | 2 | 0 | 1 | Low-quality | Genome-fragment | 2.29 | AAI-based (medium-confidence) | 0 |
| S5-final-viral-combined_k141_183894\|\|full | 2237 | No | NA | 4 | 0 | 0 | Low-quality | Genome-fragment | 4.86 | AAI-based (medium-confidence) | 0 |
| S17-final-viral-combined_k141_160895\|\|full | 4695 | No | NA | 4 | 0 | 2 | Not-determined | Genome-fragment | NA | NA | 0 |
| S17-final-viral-combined_k141_239082\|\|full | 7432 | No | NA | 2 | 0 | 2 | Not-determined | Genome-fragment | NA | NA | 0 |
| S12-final-viral-combined_k141_397635\|\|full | 2237 | No | NA | 4 | 0 | 1 | Low-quality | Genome-fragment | 1.58 | AAI-based (medium-confidence) | 0 |
| S17-final-viral-combined_k141_38930\|\|full | 1666 | No | NA | 3 | 0 | 1 | Not-determined | Genome-fragment | NA | NA | 0 |
| S2-final-viral-combined_k141_101880\|\|full | 2237 | No | NA | 4 | 0 | 0 | Low-quality | Genome-fragment | 6.32 | AAI-based (medium-confidence) | 0 |
| S17-final-viral-combined_k141_195368\|\|full | 1302 | No | NA | 3 | 0 | 2 | Not-determined | Genome-fragment | NA | NA | 0 |
| S17-final-viral-combined_k141_160915\|\|full | 1323 | No | NA | 2 | 0 | 1 | Not-determined | Genome-fragment | NA | NA | 0 |
| S6-final-viral-combined_k141_74554\|\|full | 2237 | No | NA | 5 | 0 | 1 | Low-quality | Genome-fragment | 4.46 | AAI-based (high-confidence) | 0 |
| S17-final-viral-combined_k141_139759\|\|full | 1559 | No | NA | 3 | 0 | 0 | Not-determined | Genome-fragment | NA | NA | 0 |
| S11_k141_95147 | 2238 | No | NA | 4 | 3 | 0 | Low-quality | Genome-fragment | 3.03 | AAI-based (high-confidence) | 0 |
| S17-final-viral-combined_k141_239103\|\|full | 1820 | No | NA | 3 | 0 | 0 | Not-determined | Genome-fragment | NA | NA | 0 |
| S1-final-viral-combined_k141_318604\|\|full | 2238 | No | NA | 2 | 0 | 1 | Low-quality | Genome-fragment | 4.62 | AAI-based (medium-confidence) | 0 |
| S7-final-viral-combined_k141_126351\|\|full | 2238 | No | NA | 6 | 0 | 1 | Low-quality | Genome-fragment | 5.65 | AAI-based (medium-confidence) | 0 |
| S4_k141_147268 | 2238 | No | NA | 3 | 0 | 0 | Low-quality | Genome-fragment | 2.3 | AAI-based (high-confidence) | 0 |
| S17-final-viral-combined_k141_93597\|\|full | 4660 | No | NA | 5 | 0 | 0 | Not-determined | Genome-fragment | NA | NA | 0 |
| S17-final-viral-combined_k141_93601\|\|full | 1360 | No | NA | 1 | 0 | 1 | Not-determined | Genome-fragment | NA | NA | 0 |
| S17-final-viral-combined_k141_260557\|\|full | 1225 | No | NA | 3 | 0 | 1 | Not-determined | Genome-fragment | NA | NA | 0 |
| S17-final-viral-combined_k141_229111\|\|full | 1465 | No | NA | 3 | 0 | 0 | Not-determined | Genome-fragment | NA | NA | 0 |
| S1-final-viral-combined_k141_79241\|\|full | 2239 | No | NA | 6 | 1 | 0 | Low-quality | Genome-fragment | 5.71 | AAI-based (high-confidence) | 0 |
| S17-final-viral-combined_k141_180062\|\|full | 2239 | No | NA | 3 | 0 | 3 | Low-quality | Genome-fragment | 0.97 | AAI-based (medium-confidence) | 0 |
| S17-final-viral-combined_k141_93625\|\|full | 1973 | No | NA | 3 | 0 | 1 | Not-determined | Genome-fragment | NA | NA | 0 |
| S17-final-viral-combined_k141_27744\|\|full | 2082 | No | NA | 2 | 0 | 1 | Not-determined | Genome-fragment | NA | NA | 0 |
| S17-final-viral-combined_k141_217989\|\|full | 6156 | No | NA | 5 | 0 | 3 | Not-determined | Genome-fragment | NA | NA | 0 |
| S2-final-viral-combined_k141_212204\|\|full | 2239 | No | NA | 2 | 1 | 0 | Low-quality | Genome-fragment | 1.07 | AAI-based (high-confidence) | 0 |
| S17-final-viral-combined_k141_260601\|\|full | 2786 | No | NA | 3 | 0 | 2 | Not-determined | Genome-fragment | NA | NA | 0 |
| S2-final-viral-combined_k141_108043\|\|lt2gene | 2239 | No | NA | 2 | 1 | 0 | Low-quality | Genome-fragment | 0.94 | HMM-based (lower-bound) | 0 |
| S17-final-viral-combined_k141_105932\|\|full | 4597 | No | NA | 4 | 0 | 4 | Not-determined | Genome-fragment | NA | NA | 0 |
| S2-final-viral-combined_k141_7236\|\|lt2gene | 2239 | No | NA | 1 | 1 | 0 | Low-quality | Genome-fragment | 6.87 | AAI-based (high-confidence) | 0 |
| S4-final-viral-combined_k141_41225\|\|full | 2239 | No | NA | 3 | 0 | 0 | Low-quality | Genome-fragment | 3.63 | AAI-based (high-confidence) | 0 |
| S9_k141_108648 | 2240 | No | NA | 4 | 0 | 0 | Low-quality | Genome-fragment | 3.93 | AAI-based (high-confidence) | 0 |
| S17-final-viral-combined_k141_249597\|\|full | 3910 | No | NA | 4 | 0 | 3 | Not-determined | Genome-fragment | NA | NA | 0 |
| S17-final-viral-combined_k141_184765\|\|full | 1953 | No | NA | 3 | 0 | 3 | Not-determined | Genome-fragment | NA | NA | 0 |
| S3_k141_39187 | 2240 | No | NA | 2 | 0 | 2 | Low-quality | Genome-fragment | 3.71 | AAI-based (medium-confidence) | 0 |
| S17-final-viral-combined_k141_229203\|\|full | 2000 | No | NA | 3 | 0 | 2 | Not-determined | Genome-fragment | NA | NA | 0 |
| S17-final-viral-combined_k141_84699\|\|full | 1809 | No | NA | 3 | 0 | 1 | Not-determined | Genome-fragment | NA | NA | 0 |
| S5-final-viral-combined_k141_38543\|\|full | 2240 | Yes | 968 | 4 | 1 | 1 | Low-quality | Genome-fragment | 1.71 | AAI-based (medium-confidence) | 56.79 |
| S17-final-viral-combined_k141_260652\|\|full | 7105 | No | NA | 10 | 0 | 2 | Not-determined | Genome-fragment | NA | NA | 0 |
| S7-final-viral-combined_k141_52680\|\|full | 2240 | No | NA | 2 | 0 | 2 | Low-quality | Genome-fragment | 1.56 | AAI-based (medium-confidence) | 0 |
| S12-final-viral-combined_k141_345529\|\|full | 2240 | No | NA | 4 | 1 | 0 | Low-quality | Genome-fragment | 3.53 | AAI-based (medium-confidence) | 0 |
| S17-final-viral-combined_k141_229230\|\|full | 1653 | No | NA | 5 | 0 | 0 | Not-determined | Genome-fragment | NA | NA | 0 |
| S17-final-viral-combined_k141_128576\|\|full | 1650 | No | NA | 4 | 0 | 0 | Not-determined | Genome-fragment | NA | NA | 0 |
| S1-final-viral-combined_k141_95169\|\|full | 2241 | No | NA | 4 | 1 | 0 | Low-quality | Genome-fragment | 3.59 | HMM-based (lower-bound) | 0 |

| | | | | | | | | | | | |
|---|---|---|---|---|---|---|---|---|---|---|---|
| S7-final-viral-combined_k141_184985\|\|full | 2241 | No | NA | 5 | 1 | 0 | Low-quality | Genome-fragment | 5.24 | AAI-based (medium-confidence) | 0 |
| S17-final-viral-combined_k141_218043\|\|full | 3058 | No | NA | 4 | 0 | 2 | Not-determined | Genome-fragment | NA | NA | 0 |
| S17-final-viral-combined_k141_106024\|\|full | 2328 | No | NA | 3 | 0 | 1 | Not-determined | Genome-fragment | NA | NA | 0 |
| S17-final-viral-combined_k141_128588\|\|full | 1598 | No | NA | 2 | 0 | 1 | Not-determined | Genome-fragment | NA | NA | 0 |
| S2-final-viral-combined_k141_61891\|\|full | 2241 | No | NA | 3 | 0 | 0 | Low-quality | Genome-fragment | 5.69 | AAI-based (medium-confidence) | 0 |
| S4_k141_32550 | 2241 | No | NA | 4 | 1 | 0 | Low-quality | Genome-fragment | 2.37 | AAI-based (high-confidence) | 0 |
| S17-final-viral-combined_k141_48701\|\|full | 2170 | No | NA | 3 | 0 | 1 | Not-determined | Genome-fragment | NA | NA | 0 |
| S17-final-viral-combined_k141_139859\|\|full | 3135 | No | NA | 4 | 0 | 2 | Not-determined | Genome-fragment | NA | NA | 0 |
| S17-final-viral-combined_k141_128605\|\|full | 1900 | No | NA | 3 | 0 | 2 | Not-determined | Genome-fragment | NA | NA | 0 |
| S1_k141_34153 | 2242 | No | NA | 2 | 0 | 0 | Low-quality | Genome-fragment | 2.47 | AAI-based (high-confidence) | 0 |
| S7_k141_180841 | 2242 | No | NA | 4 | 0 | 0 | Low-quality | Genome-fragment | 4.71 | AAI-based (high-confidence) | 0 |
| S17-final-viral-combined_k141_61438\|\|full | 1545 | No | NA | 3 | 0 | 1 | Not-determined | Genome-fragment | NA | NA | 0 |
| S12-final-viral-combined_k141_202043\|\|full | 2242 | No | NA | 4 | 0 | 0 | Low-quality | Genome-fragment | 3.3 | AAI-based (high-confidence) | 0 |
| S17-final-viral-combined_k141_249639\|\|full | 1810 | No | NA | 2 | 0 | 2 | Not-determined | Genome-fragment | NA | NA | 0 |
| S17-final-viral-combined_k141_61454\|\|full | 2951 | No | NA | 5 | 0 | 4 | Not-determined | Genome-fragment | NA | NA | 0 |
| S12-final-viral-combined_k141_307978\|\|full | 2242 | No | NA | 3 | 0 | 2 | Low-quality | Genome-fragment | 4.13 | AAI-based (medium-confidence) | 0 |
| S12_k141_222996 | 2242 | No | NA | 4 | 0 | 0 | Low-quality | Genome-fragment | 4.97 | AAI-based (medium-confidence) | 0 |
| S17-final-viral-combined_k141_139928\|\|full | 1171 | No | NA | 2 | 0 | 0 | Not-determined | Genome-fragment | NA | NA | 0 |
| S8-final-viral-combined_k141_233263\|\|lt2gene | 2242 | No | NA | 1 | 1 | 0 | Low-quality | Genome-fragment | 1.24 | HMM-based (lower-bound) | 0 |
| S7-final-viral-combined_k141_185175\|\|full | 2243 | No | NA | 3 | 3 | 0 | Low-quality | Genome-fragment | 5.87 | AAI-based (high-confidence) | 0 |
| S4-final-viral-combined_k141_141227\|\|full | 2243 | No | NA | 4 | 2 | 0 | Low-quality | Genome-fragment | 1.36 | HMM-based (lower-bound) | 0 |
| S17-final-viral-combined_k141_73143\|\|full | 1106 | No | NA | 2 | 0 | 0 | Not-determined | Genome-fragment | NA | NA | 0 |
| S17-final-viral-combined_k141_218130\|\|full | 5732 | No | NA | 6 | 0 | 3 | Not-determined | Genome-fragment | NA | NA | 0 |
| S17-final-viral-combined_k141_128681\|\|full | 3238 | No | NA | 4 | 0 | 2 | Not-determined | Genome-fragment | NA | NA | 0 |
| S4-final-viral-combined_k141_1988\|\|lt2gene | 2243 | No | NA | 1 | 1 | 0 | Low-quality | Genome-fragment | 3.32 | AAI-based (high-confidence) | 0 |
| S17-final-viral-combined_k141_260744\|\|full | 1346 | No | NA | 2 | 0 | 1 | Not-determined | Genome-fragment | NA | NA | 0 |
| S17-final-viral-combined_k141_207028\|\|full | 1601 | No | NA | 2 | 0 | 1 | Not-determined | Genome-fragment | NA | NA | 0 |
| S8-final-viral-combined_k141_79181\|\|full | 2243 | No | NA | 8 | 1 | 0 | Low-quality | Genome-fragment | 3.88 | AAI-based (high-confidence) | 0 |
| S9-final-viral-combined_k141_55863\|\|full | 2244 | No | NA | 5 | 0 | 2 | Low-quality | Genome-fragment | 1.49 | AAI-based (medium-confidence) | 0 |
| S17-final-viral-combined_k141_260775\|\|full | 2056 | No | NA | 3 | 0 | 2 | Not-determined | Genome-fragment | NA | NA | 0 |
| S9-final-viral-combined_k141_37161\|\|full | 2244 | No | NA | 3 | 0 | 1 | Low-quality | Genome-fragment | 5.46 | AAI-based (high-confidence) | 0 |
| S17-final-viral-combined_k141_84847\|\|full | 9354 | No | NA | 10 | 0 | 6 | Not-determined | Genome-fragment | NA | NA | 0 |
| S17-final-viral-combined_k141_61495\|\|full | 2248 | No | NA | 3 | 0 | 1 | Not-determined | Genome-fragment | NA | NA | 0 |
| S11-final-viral-combined_k141_28067\|\|full | 2244 | No | NA | 4 | 0 | 1 | Low-quality | Genome-fragment | 2.88 | AAI-based (medium-confidence) | 0 |
| S11_k141_146494 | 2244 | No | NA | 4 | 1 | 0 | Low-quality | Genome-fragment | 1.78 | AAI-based (high-confidence) | 0 |
| S11_k141_146538 | 2244 | No | NA | 4 | 1 | 0 | Low-quality | Genome-fragment | 1.78 | AAI-based (high-confidence) | 0 |
| S17-final-viral-combined_k141_118744\|\|full | 4196 | No | NA | 5 | 0 | 2 | Not-determined | Genome-fragment | NA | NA | 0 |
| S17-final-viral-combined_k141_16143\|\|full | 1336 | No | NA | 2 | 0 | 0 | Not-determined | Genome-fragment | NA | NA | 0 |
| S17-final-viral-combined_k141_118747\|\|full | 2079 | No | NA | 5 | 0 | 0 | Not-determined | Genome-fragment | NA | NA | 0 |
| S11_k141_174722 | 2244 | No | NA | 2 | 0 | 1 | Low-quality | Genome-fragment | 1.31 | AAI-based (medium-confidence) | 0 |
| S17-final-viral-combined_k141_139990\|\|full | 2717 | No | NA | 6 | 0 | 0 | Not-determined | Genome-fragment | NA | NA | 0 |
| S7-final-viral-combined_k141_15357\|\|full | 2244 | No | NA | 4 | 0 | 0 | Low-quality | Genome-fragment | 2.74 | AAI-based (high-confidence) | 0 |
| S17-final-viral-combined_k141_207058\|\|full | 6206 | No | NA | 10 | 0 | 2 | Not-determined | Genome-fragment | NA | NA | 0 |
| S17-final-viral-combined_k141_260821\|\|full | 2669 | No | NA | 2 | 0 | 0 | Not-determined | Genome-fragment | NA | NA | 0 |
| S17-final-viral-combined_k141_106127\|\|full | 6206 | No | NA | 5 | 0 | 3 | Not-determined | Genome-fragment | NA | NA | 0 |
| S17-final-viral-combined_k141_229399\|\|full | 1941 | No | NA | 3 | 0 | 0 | Not-determined | Genome-fragment | NA | NA | 0 |
| S12-final-viral-combined_k141_18068\|\|full | 2244 | No | NA | 3 | 1 | 0 | Low-quality | Genome-fragment | 5.86 | AAI-based (medium-confidence) | 0 |
| S12-final-viral-combined_k141_293924\|\|full | 2244 | No | NA | 3 | 0 | 1 | Low-quality | Genome-fragment | 1.41 | AAI-based (medium-confidence) | 0 |
| S17-final-viral-combined_k141_106138\|\|full | 1760 | No | NA | 2 | 0 | 0 | Not-determined | Genome-fragment | NA | NA | 0 |
| S4-final-viral-combined_k141_47273\|\|full | 2245 | No | NA | 3 | 0 | 0 | Low-quality | Genome-fragment | 4.77 | AAI-based (medium-confidence) | 0 |
| S17-final-viral-combined_k141_84903\|\|full | 13748 | No | NA | 17 | 0 | 2 | Not-determined | Genome-fragment | NA | NA | 0 |
| S7-final-viral-combined_k141_8463\|\|full | 2246 | No | NA | 5 | 1 | 0 | Low-quality | Genome-fragment | 1.51 | HMM-based (lower-bound) | 0 |
| S17-final-viral-combined_k141_239357\|\|full | 5455 | No | NA | 5 | 0 | 1 | Not-determined | Genome-fragment | NA | NA | 0 |
| S17-final-viral-combined_k141_218266\|\|full | 5751 | No | NA | 7 | 0 | 1 | Not-determined | Genome-fragment | NA | NA | 0 |
| S17-final-viral-combined_k141_207101\|\|full | 3323 | No | NA | 5 | 0 | 3 | Not-determined | Genome-fragment | NA | NA | 0 |
| S17-final-viral-combined_k141_260864\|\|full | 3076 | No | NA | 5 | 0 | 0 | Not-determined | Genome-fragment | NA | NA | 0 |
| S17-final-viral-combined_k141_243364\|\|full | 2246 | No | NA | 3 | 1 | 0 | Low-quality | Genome-fragment | 1.33 | AAI-based (high-confidence) | 0 |
| S17-final-viral-combined_k141_106175\|\|full | 4243 | No | NA | 3 | 0 | 1 | Not-determined | Genome-fragment | NA | NA | 0 |
| S17-final-viral-combined_k141_207116\|\|full | 1122 | No | NA | 3 | 0 | 0 | Not-determined | Genome-fragment | NA | NA | 0 |
| S12-final-viral-combined_k141_215355\|\|full | 2246 | No | NA | 2 | 0 | 1 | Low-quality | Genome-fragment | 0.74 | AAI-based (medium-confidence) | 0 |
| S8-final-viral-combined_k141_58822\|\|full | 2246 | No | NA | 3 | 0 | 2 | Low-quality | Genome-fragment | 1.06 | AAI-based (high-confidence) | 0 |
| S17-final-viral-combined_k141_172906\|\|full | 2460 | No | NA | 3 | 0 | 2 | Not-determined | Genome-fragment | NA | NA | 0 |
| S17-final-viral-combined_k141_16167\|\|full | 2891 | No | NA | 3 | 0 | 3 | Not-determined | Genome-fragment | NA | NA | 0 |
| S12-final-viral-combined_k141_48073\|\|full | 2247 | No | NA | 5 | 0 | 0 | Low-quality | Genome-fragment | 5.03 | AAI-based (medium-confidence) | 0 |
| S2-final-viral-combined_k141_74703\|\|full | 2247 | No | NA | 3 | 1 | 0 | Low-quality | Genome-fragment | 3.09 | HMM-based (lower-bound) | 0 |
| S17-final-viral-combined_k141_172921\|\|full | 2038 | No | NA | 2 | 0 | 1 | Not-determined | Genome-fragment | NA | NA | 0 |
| S11-final-viral-combined_k141_284340\|\|full | 2248 | No | NA | 5 | 3 | 0 | Low-quality | Genome-fragment | 5.76 | AAI-based (medium-confidence) | 0 |
| S17-final-viral-combined_k141_16188\|\|full | 1687 | No | NA | 3 | 0 | 1 | Not-determined | Genome-fragment | NA | NA | 0 |
| S1-final-viral-combined_k141_263337\|\|full | 2248 | No | NA | 4 | 0 | 4 | Low-quality | Genome-fragment | 1.39 | AAI-based (medium-confidence) | 0 |
| S17-final-viral-combined_k141_239429\|\|full | 1497 | No | NA | 2 | 0 | 2 | Not-determined | Genome-fragment | NA | NA | 0 |
| S17-final-viral-combined_k141_5889\|\|full | 1248 | No | NA | 3 | 0 | 0 | Not-determined | Genome-fragment | NA | NA | 0 |
| S1-final-viral-combined_k141_35017\|\|full | 2248 | No | NA | 2 | 0 | 0 | Low-quality | Genome-fragment | 2.69 | AAI-based (medium-confidence) | 0 |
| S17-final-viral-combined_k141_239432\|\|full | 2419 | No | NA | 4 | 0 | 3 | Not-determined | Genome-fragment | NA | NA | 0 |
| S17-final-viral-combined_k141_39241\|\|full | 1163 | No | NA | 3 | 0 | 1 | Not-determined | Genome-fragment | NA | NA | 0 |
| S17-final-viral-combined_k141_73391\|\|full | 1492 | No | NA | 1 | 0 | 1 | Not-determined | Genome-fragment | NA | NA | 0 |
| S1-final-viral-combined_k141_239540\|\|full | 2248 | Yes | 816 | 3 | 1 | 1 | Low-quality | Genome-fragment | 19.61 | HMM-based (lower-bound) | 63.7 |
| S17-final-viral-combined_k141_239462\|\|full | 3839 | No | NA | 2 | 0 | 2 | Not-determined | Genome-fragment | NA | NA | 0 |
| S7-final-viral-combined_k141_52903\|\|full | 2248 | No | NA | 4 | 0 | 0 | Low-quality | Genome-fragment | 5.04 | AAI-based (medium-confidence) | 0 |
| S7-final-viral-combined_k141_122278\|\|full | 2248 | No | NA | 3 | 0 | 2 | Low-quality | Genome-fragment | 3.94 | AAI-based (medium-confidence) | 0 |
| S17-final-viral-combined_k141_5923\|\|full | 1397 | No | NA | 2 | 0 | 1 | Not-determined | Genome-fragment | NA | NA | 0 |
| S7_k141_111229 | 2248 | No | NA | 1 | 0 | 0 | Low-quality | Genome-fragment | 3.48 | AAI-based (high-confidence) | 0 |
| S17-final-viral-combined_k141_16257\|\|full | 1016 | No | NA | 3 | 0 | 2 | Not-determined | Genome-fragment | NA | NA | 0 |
| S17-final-viral-combined_k141_117295\|\|lt2gene | 2248 | No | NA | 2 | 2 | 0 | Low-quality | Genome-fragment | 5.87 | AAI-based (high-confidence) | 0 |
| S17-k141_121259 | 2248 | No | NA | 5 | 2 | 0 | Low-quality | Genome-fragment | 5.58 | AAI-based (high-confidence) | 0 |
| S17-final-viral-combined_k141_185008\|\|full | 1418 | No | NA | 3 | 0 | 2 | Not-determined | Genome-fragment | NA | NA | 0 |
| S12-final-viral-combined_k141_340956\|\|full | 2248 | No | NA | 3 | 0 | 0 | Low-quality | Genome-fragment | 2.74 | AAI-based (high-confidence) | 0 |
| S17-final-viral-combined_k141_5955\|\|full | 1751 | No | NA | 3 | 0 | 1 | Not-determined | Genome-fragment | NA | NA | 0 |
| S11-final-viral-combined_k141_153033\|\|full | 2249 | No | NA | 4 | 0 | 1 | Low-quality | Genome-fragment | 3.86 | AAI-based (medium-confidence) | 0 |
| S11-final-viral-combined_k141_151050\|\|full | 2249 | No | NA | 3 | 2 | 0 | Low-quality | Genome-fragment | 6.96 | AAI-based (high-confidence) | 0 |
| S17-final-viral-combined_k141_16304\|\|full | 2098 | No | NA | 4 | 0 | 3 | Not-determined | Genome-fragment | NA | NA | 0 |
| S1-final-viral-combined_k141_355095\|\|full | 2249 | No | NA | 3 | 0 | 1 | Low-quality | Genome-fragment | 1.27 | AAI-based (medium-confidence) | 0 |
| S17-final-viral-combined_k141_185049\|\|full | 2108 | No | NA | 2 | 0 | 2 | Not-determined | Genome-fragment | NA | NA | 0 |
| S7-final-viral-combined_k141_244078\|\|full | 2249 | Yes | 1436 | 4 | 1 | 1 | Low-quality | Genome-fragment | 2.63 | HMM-based (lower-bound) | 36.15 |
| S17-final-viral-combined_k141_185058\|\|full | 1530 | No | NA | 4 | 0 | 2 | Not-determined | Genome-fragment | NA | NA | 0 |
| S2-final-viral-combined_k141_125981\|\|full | 2250 | No | NA | 3 | 0 | 1 | Low-quality | Genome-fragment | 1.55 | AAI-based (high-confidence) | 0 |
| S17-final-viral-combined_k141_229556\|\|full | 18658 | No | NA | 33 | 0 | 1 | Not-determined | Genome-fragment | NA | NA | 0 |
| S17-final-viral-combined_k141_195858\|\|full | 1434 | No | NA | 2 | 0 | 1 | Not-determined | Genome-fragment | NA | NA | 0 |
| S11-final-viral-combined_k141_29735\|\|full | 2251 | No | NA | 4 | 1 | 0 | Low-quality | Genome-fragment | 3.13 | AAI-based (medium-confidence) | 0 |
| S17-final-viral-combined_k141_239553\|\|full | 1396 | No | NA | 3 | 0 | 0 | Not-determined | Genome-fragment | NA | NA | 0 |
| S17-final-viral-combined_k141_85094\|\|full | 2973 | No | NA | 3 | 0 | 3 | Not-determined | Genome-fragment | NA | NA | 0 |
| S17-final-viral-combined_k141_16309\|\|full | 20159 | No | NA | 28 | 10 | 0 | Medium-quality | Genome-fragment | 53.78 | AAI-based (medium-confidence) | 0 |
| S7-final-viral-combined_k141_177668\|\|full | 2251 | No | NA | 3 | 1 | 0 | Low-quality | Genome-fragment | 5.31 | AAI-based (high-confidence) | 0 |
| S17-final-viral-combined_k141_140212\|\|full | 1845 | No | NA | 3 | 0 | 2 | Not-determined | Genome-fragment | NA | NA | 0 |
| S17-final-viral-combined_k141_106262\|\|full | 57272 | No | NA | 80 | 24 | 1 | High-quality | High-quality | 100 | AAI-based (high-confidence) | 0 |
| S17-final-viral-combined_k141_207246\|\|full | 7133 | No | NA | 6 | 0 | 2 | Not-determined | Genome-fragment | NA | NA | 0 |
| S17-final-viral-combined_k141_173028\|\|full | 27944 | No | NA | 31 | 0 | 4 | Not-determined | Genome-fragment | NA | NA | 0 |
| S17-final-viral-combined_k141_227002\|\|full | 2251 | No | NA | 3 | 0 | 0 | Low-quality | Genome-fragment | 4.98 | AAI-based (medium-confidence) | 0 |
| S2_k141_5124 | 2251 | No | NA | 3 | 0 | 1 | Low-quality | Genome-fragment | 4.29 | AAI-based (medium-confidence) | 0 |
| S17-final-viral-combined_k141_140227\|\|full | 3666 | No | NA | 4 | 0 | 3 | Not-determined | Genome-fragment | NA | NA | 0 |
| S11-final-viral-combined_k141_89170\|\|full | 2252 | No | NA | 4 | 2 | 0 | Low-quality | Genome-fragment | 2.39 | HMM-based (lower-bound) | 0 |
| S11-final-viral-combined_k141_177546\|\|full | 2252 | No | NA | 3 | 3 | 0 | Low-quality | Genome-fragment | 2.34 | HMM-based (lower-bound) | 0 |
| S11_k141_173759 | 2252 | No | NA | 3 | 2 | 0 | Low-quality | Genome-fragment | 12.81 | AAI-based (high-confidence) | 0 |
| S17-final-viral-combined_k141_185096\|\|full | 2735 | No | NA | 3 | 0 | 1 | Not-determined | Genome-fragment | NA | NA | 0 |
| S17-final-viral-combined_k141_249833\|\|full | 1925 | No | NA | 3 | 0 | 0 | Not-determined | Genome-fragment | NA | NA | 0 |
| S17-final-viral-combined_k141_16340\|\|full | 5948 | No | NA | 11 | 0 | 1 | Not-determined | Genome-fragment | NA | NA | 0 |
| S17-final-viral-combined_k141_161223\|\|full | 2213 | No | NA | 3 | 0 | 2 | Not-determined | Genome-fragment | NA | NA | 0 |
| S17-final-viral-combined_k141_261090\|\|full | 1460 | No | NA | 3 | 0 | 3 | Not-determined | Genome-fragment | NA | NA | 0 |
| S17-final-viral-combined_k141_150947\|\|full | 1039 | No | NA | 3 | 0 | 2 | Not-determined | Genome-fragment | NA | NA | 0 |
| S7-final-viral-combined_k141_1923\|\|full | 2252 | No | NA | 2 | 1 | 0 | Low-quality | Genome-fragment | 1.06 | AAI-based (medium-confidence) | 0 |
| S17-final-viral-combined_k141_39388\|\|full | 1413 | No | NA | 3 | 0 | 0 | Not-determined | Genome-fragment | NA | NA | 0 |
| S17-final-viral-combined_k141_173086\|\|full | 5227 | No | NA | 5 | 0 | 2 | Not-determined | Genome-fragment | NA | NA | 0 |
| S17-final-viral-combined_k141_128829\|\|full | 1354 | No | NA | 2 | 0 | 1 | Not-determined | Genome-fragment | NA | NA | 0 |
| S17-final-viral-combined_k141_124690\|\|full | 2252 | No | NA | 5 | 0 | 0 | Low-quality | Genome-fragment | 5.53 | AAI-based (medium-confidence) | 0 |
| S17-final-viral-combined_k141_261117\|\|full | 2423 | No | NA | 3 | 0 | 0 | Not-determined | Genome-fragment | NA | NA | 0 |
| S17-final-viral-combined_k141_151093\|\|full | 2252 | No | NA | 5 | 0 | 0 | Low-quality | Genome-fragment | 3.35 | AAI-based (medium-confidence) | 0 |
| S2-final-viral-combined_k141_215270\|\|full | 2252 | No | NA | 4 | 0 | 4 | Low-quality | Genome-fragment | 0.73 | AAI-based (medium-confidence) | 0 |
| S17-final-viral-combined_k141_118984\|\|full | 1437 | No | NA | 4 | 0 | 0 | Not-determined | Genome-fragment | NA | NA | 0 |
| S17-final-viral-combined_k141_185157\|\|full | 1893 | No | NA | 3 | 0 | 3 | Not-determined | Genome-fragment | NA | NA | 0 |
| S11-final-viral-combined_k141_107810\|\|full | 2253 | No | NA | 2 | 0 | 2 | Low-quality | Genome-fragment | 4.11 | AAI-based (medium-confidence) | 0 |
| S1-final-viral-combined_k141_249960\|\|full | 2253 | No | NA | 4 | 0 | 0 | Low-quality | Genome-fragment | 14.57 | AAI-based (high-confidence) | 0 |
| S1_k141_215328 | 2253 | No | NA | 1 | 0 | 0 | Low-quality | Genome-fragment | 1.38 | AAI-based (medium-confidence) | 0 |
| S5-final-viral-combined_k141_178769\|\|full | 2253 | No | NA | 2 | 1 | 0 | Low-quality | Genome-fragment | 4.83 | HMM-based (lower-bound) | 0 |
| S12-final-viral-combined_k141_35488\|\|full | 2253 | No | NA | 6 | 1 | 0 | Low-quality | Genome-fragment | 3.73 | AAI-based (medium-confidence) | 0 |
| S17-final-viral-combined_k141_218568\|\|full | 1194 | No | NA | 2 | 0 | 2 | Not-determined | Genome-fragment | NA | NA | 0 |
| S17-final-viral-combined_k141_106311\|\|full | 6700 | No | NA | 6 | 0 | 2 | Not-determined | Genome-fragment | NA | NA | 0 |
| S17-final-viral-combined_k141_28138\|\|full | 1465 | No | NA | 3 | 0 | 0 | Not-determined | Genome-fragment | NA | NA | 0 |
| S17-final-viral-combined_k141_61852\|\|full | 1336 | No | NA | 2 | 0 | 0 | Not-determined | Genome-fragment | NA | NA | 0 |
| S17-final-viral-combined_k141_161284\|\|full | 2617 | No | NA | 2 | 0 | 2 | Not-determined | Genome-fragment | NA | NA | 0 |
| S17-final-viral-combined_k141_28140\|\|full | 2917 | No | NA | 4 | 0 | 2 | Not-determined | Genome-fragment | NA | NA | 0 |
| S4-final-viral-combined_k141_128360\|\|full | 2253 | No | NA | 3 | 1 | 0 | Low-quality | Genome-fragment | 3.79 | AAI-based (medium-confidence) | 0 |
| S17-final-viral-combined_k141_261183\|\|full | 1838 | No | NA | 3 | 0 | 1 | Not-determined | Genome-fragment | NA | NA | 0 |
| S17-final-viral-combined_k141_161302\|\|full | 1915 | No | NA | 2 | 0 | 0 | Not-determined | Genome-fragment | NA | NA | 0 |
| S17-final-viral-combined_k141_207410\|\|full | 6710 | No | NA | 5 | 0 | 5 | Not-determined | Genome-fragment | NA | NA | 0 |
| S9-final-viral-combined_k141_65921\|\|full | 2254 | No | NA | 2 | 1 | 0 | Low-quality | Genome-fragment | 5.21 | AAI-based (high-confidence) | 0 |
| S17-final-viral-combined_k141_196068\|\|full | 2686 | No | NA | 3 | 0 | 1 | Not-determined | Genome-fragment | NA | NA | 0 |
| S17-final-viral-combined_k141_249936\|\|full | 1169 | No | NA | 3 | 0 | 1 | Not-determined | Genome-fragment | NA | NA | 0 |
| S17-final-viral-combined_k141_229686\|\|full | 1324 | No | NA | 3 | 0 | 1 | Not-determined | Genome-fragment | NA | NA | 0 |
| S17-final-viral-combined_k141_39510\|\|full | 1837 | No | NA | 3 | 0 | 0 | Not-determined | Genome-fragment | NA | NA | 0 |
| S9-final-viral-combined_k141_113809\|\|full | 2254 | No | NA | 3 | 1 | 0 | Low-quality | Genome-fragment | 17.63 | AAI-based (medium-confidence) | 0 |
| S17-final-viral-combined_k141_61930\|\|full | 1202 | No | NA | 4 | 0 | 0 | Not-determined | Genome-fragment | NA | NA | 0 |
| S1_k141_344987 | 2254 | No | NA | 4 | 1 | 0 | Low-quality | Genome-fragment | 22.64 | HMM-based (lower-bound) | 0 |
| S17-final-viral-combined_k141_172848\|\|full | 2254 | No | NA | 5 | 0 | 0 | Low-quality | Genome-fragment | 3.68 | AAI-based (medium-confidence) | 0 |
| S6-final-viral-combined_k141_36441\|\|full | 2254 | No | NA | 3 | 0 | 0 | Low-quality | Genome-fragment | 0.9 | AAI-based (medium-confidence) | 0 |
| S17-final-viral-combined_k141_6129\|\|full | 6056 | No | NA | 2 | 0 | 1 | Not-determined | Genome-fragment | NA | NA | 0 |
| S1-final-viral-combined_k141_36355\|\|full | 2255 | No | NA | 3 | 0 | 0 | Low-quality | Genome-fragment | 4.08 | AAI-based (high-confidence) | 0 |
| S1-final-viral-combined_k141_392709\|\|full | 2255 | No | NA | 6 | 2 | 0 | Low-quality | Genome-fragment | 5.68 | AAI-based (high-confidence) | 0 |
| S7-final-viral-combined_k141_199681\|\|full | 2255 | No | NA | 4 | 0 | 3 | Low-quality | Genome-fragment | 1.93 | AAI-based (medium-confidence) | 0 |
| S12-final-viral-combined_k141_257865\|\|full | 2255 | No | NA | 2 | 0 | 2 | Low-quality | Genome-fragment | 2.67 | AAI-based (medium-confidence) | 0 |
| S8-final-viral-combined_k141_83169\|\|full | 2255 | No | NA | 3 | 1 | 0 | Low-quality | Genome-fragment | 4.13 | AAI-based (medium-confidence) | 0 |
| S9-final-viral-combined_k141_92736\|\|full | 2256 | No | NA | 4 | 3 | 0 | Low-quality | Genome-fragment | 4.92 | AAI-based (medium-confidence) | 0 |
| S17-final-viral-combined_k141_229733\|\|full | 1824 | No | NA | 3 | 0 | 2 | Not-determined | Genome-fragment | NA | NA | 0 |
| S9_k141_58468 | 2256 | No | NA | 3 | 0 | 0 | Low-quality | Genome-fragment | 3.77 | AAI-based (high-confidence) | 0 |
| S17-final-viral-combined_k141_218693\|\|full | 3117 | No | NA | 3 | 0 | 0 | Not-determined | Genome-fragment | NA | NA | 0 |
| S11-final-viral-combined_k141_180527\|\|full | 2256 | No | NA | 2 | 0 | 0 | Low-quality | Genome-fragment | 3.05 | AAI-based (high-confidence) | 0 |
| S7-final-viral-combined_k141_63755\|\|full | 2256 | No | NA | 2 | 0 | 0 | Low-quality | Genome-fragment | 1.29 | AAI-based (medium-confidence) | 0 |
| S17-final-viral-combined_k141_173249\|\|full | 15458 | No | NA | 7 | 0 | 6 | Not-determined | Genome-fragment | NA | NA | 0 |
| S17-final-viral-combined_k141_61994\|\|full | 1462 | No | NA | 2 | 0 | 2 | Not-determined | Genome-fragment | NA | NA | 0 |
| S17-final-viral-combined_k141_119083\|\|full | 2800 | No | NA | 3 | 0 | 2 | Not-determined | Genome-fragment | NA | NA | 0 |
| S17-final-viral-combined_k141_84324\|\|full | 2256 | No | NA | 2 | 0 | 1 | Low-quality | Genome-fragment | 0.91 | AAI-based (high-confidence) | 0 |
| S17-k141_176866 | 2256 | No | NA | 5 | 2 | 0 | Low-quality | Genome-fragment | 5.27 | AAI-based (medium-confidence) | 0 |
| S6-final-viral-combined_k141_31639\|\|full | 2256 | No | NA | 3 | 0 | 0 | Low-quality | Genome-fragment | 2.83 | AAI-based (high-confidence) | 0 |
| S17-final-viral-combined_k141_239690\|\|full | 2284 | No | NA | 2 | 0 | 1 | Not-determined | Genome-fragment | NA | NA | 0 |
| S6_k141_33554 | 2256 | No | NA | 2 | 0 | 2 | Low-quality | Genome-fragment | 3.55 | AAI-based (medium-confidence) | 0 |
| S17-final-viral-combined_k141_250014\|\|full | 3170 | No | NA | 5 | 0 | 2 | Not-determined | Genome-fragment | NA | NA | 0 |
| S17-final-viral-combined_k141_16537\|\|full | 21629 | No | NA | 25 | 0 | 8 | Not-determined | Genome-fragment | NA | NA | 0 |
| S17-final-viral-combined_k141_106438\|\|full | 1600 | No | NA | 3 | 0 | 1 | Not-determined | Genome-fragment | NA | NA | 0 |
| S17-final-viral-combined_k141_94015\|\|full | 1513 | No | NA | 2 | 0 | 0 | Not-determined | Genome-fragment | NA | NA | 0 |
| S3-final-viral-combined_k141_129127\|\|full | 2257 | No | NA | 4 | 0 | 0 | Low-quality | Genome-fragment | 4.09 | AAI-based (high-confidence) | 0 |
| S7-final-viral-combined_k141_960\|\|full | 2257 | No | NA | 6 | 0 | 0 | Low-quality | Genome-fragment | 2.81 | AAI-based (medium-confidence) | 0 |
| S17-final-viral-combined_k141_49191\|\|full | 1782 | No | NA | 3 | 0 | 1 | Not-determined | Genome-fragment | NA | NA | 0 |
| S17-final-viral-combined_k141_173283\|\|full | 11557 | No | NA | 11 | 0 | 3 | Not-determined | Genome-fragment | NA | NA | 0 |
| S7-final-viral-combined_k141_83294\|\|full | 2257 | No | NA | 3 | 0 | 0 | Low-quality | Genome-fragment | 5.18 | AAI-based (medium-confidence) | 0 |
| S17-k141_66048 | 2257 | No | NA | 1 | 0 | 1 | Low-quality | Genome-fragment | 0.19 | AAI-based (medium-confidence) | 0 |
| S17-final-viral-combined_k141_106459\|\|full | 2843 | No | NA | 2 | 0 | 2 | Not-determined | Genome-fragment | NA | NA | 0 |
| S2-final-viral-combined_k141_34411\|\|full | 2257 | No | NA | 4 | 0 | 1 | Low-quality | Genome-fragment | 1.55 | AAI-based (high-confidence) | 0 |
| S17-final-viral-combined_k141_239717\|\|full | 1783 | No | NA | 3 | 0 | 3 | Not-determined | Genome-fragment | NA | NA | 0 |
| S17-final-viral-combined_k141_185239\|\|full | 41758 | No | NA | 39 | 1 | 21 | Medium-quality | Genome-fragment | 65.4 | HMM-based (lower-bound) | 0 |
| S17-final-viral-combined_k141_128968\|\|full | 1776 | No | NA | 3 | 0 | 1 | Not-determined | Genome-fragment | NA | NA | 0 |
| S17-final-viral-combined_k141_62067\|\|full | 1104 | No | NA | 3 | 0 | 2 | Not-determined | Genome-fragment | NA | NA | 0 |
| S17-final-viral-combined_k141_151156\|\|full | 1473 | No | NA | 2 | 0 | 2 | Not-determined | Genome-fragment | NA | NA | 0 |
| S2-final-viral-combined_k141_128175\|\|full | 2257 | No | NA | 3 | 0 | 1 | Low-quality | Genome-fragment | 2.5 | AAI-based (medium-confidence) | 0 |
| S17-final-viral-combined_k141_39638\|\|full | 2234 | No | NA | 4 | 0 | 3 | Not-determined | Genome-fragment | NA | NA | 0 |
| S17-final-viral-combined_k141_94064\|\|full | 12139 | No | NA | 9 | 0 | 6 | Not-determined | Genome-fragment | NA | NA | 0 |
| S2-final-viral-combined_k141_112986\|\|full | 2257 | No | NA | 4 | 0 | 0 | Low-quality | Genome-fragment | 6.18 | AAI-based (high-confidence) | 0 |
| S17-final-viral-combined_k141_151179\|\|full | 1411 | No | NA | 2 | 0 | 0 | Not-determined | Genome-fragment | NA | NA | 0 |
| S11-final-viral-combined_k141_181405\|\|full | 2258 | No | NA | 4 | 1 | 0 | Low-quality | Genome-fragment | 5 | AAI-based (high-confidence) | 0 |
| S17-final-viral-combined_k141_85265\|\|full | 2505 | No | NA | 3 | 0 | 1 | Not-determined | Genome-fragment | NA | NA | 0 |
| S17-final-viral-combined_k141_28334\|\|full | 4284 | No | NA | 2 | 0 | 2 | Not-determined | Genome-fragment | NA | NA | 0 |
| S17-k141_197730 | 2258 | No | NA | 3 | 0 | 1 | Low-quality | Genome-fragment | 2.62 | AAI-based (medium-confidence) | 0 |
| S6-final-viral-combined_k141_3674\|\|full | 2258 | No | NA | 2 | 0 | 1 | Low-quality | Genome-fragment | 0.89 | AAI-based (medium-confidence) | 0 |
| S3_k141_55772 | 2259 | No | NA | 2 | 0 | 1 | Low-quality | Genome-fragment | 3.99 | AAI-based (medium-confidence) | 0 |
| S17-final-viral-combined_k141_62122\|\|full | 2022 | No | NA | 2 | 0 | 1 | Not-determined | Genome-fragment | NA | NA | 0 |
| S17-final-viral-combined_k141_229843\|\|full | 2245 | No | NA | 5 | 0 | 4 | Not-determined | Genome-fragment | NA | NA | 0 |
| S5-final-viral-combined_k141_94374\|\|full | 2259 | No | NA | 3 | 0 | 0 | Low-quality | Genome-fragment | 4.58 | AAI-based (medium-confidence) | 0 |
| S17-final-viral-combined_k141_261382\|\|full | 3944 | No | NA | 5 | 0 | 2 | Not-determined | Genome-fragment | NA | NA | 0 |
| S17-final-viral-combined_k141_261386\|\|full | 2478 | No | NA | 2 | 0 | 1 | Not-determined | Genome-fragment | NA | NA | 0 |
| S7-final-viral-combined_k141_119007\|\|full | 2259 | No | NA | 3 | 0 | 3 | Low-quality | Genome-fragment | 1.31 | AAI-based (medium-confidence) | 0 |
| S17-final-viral-combined_k141_39687\|\|full | 24027 | No | NA | 39 | 13 | 0 | Medium-quality | Genome-fragment | 70.37 | AAI-based (high-confidence) | 0 |
| S17-final-viral-combined_k141_151254\|\|full | 2026 | No | NA | 3 | 0 | 0 | Not-determined | Genome-fragment | NA | NA | 0 |
| S17-final-viral-combined_k141_6276\|\|full | 10942 | No | NA | 11 | 0 | 6 | Not-determined | Genome-fragment | NA | NA | 0 |
| S2-final-viral-combined_k141_207286\|\|full | 2259 | No | NA | 3 | 0 | 0 | Low-quality | Genome-fragment | 5.7 | AAI-based (medium-confidence) | 0 |
| S2-final-viral-combined_k141_66044\|\|full | 2259 | No | NA | 3 | 0 | 0 | Low-quality | Genome-fragment | 4.75 | AAI-based (medium-confidence) | 0 |
| S8_k141_232652 | 2259 | No | NA | 3 | 1 | 0 | Low-quality | Genome-fragment | 3.91 | AAI-based (high-confidence) | 0 |
| S11-final-viral-combined_k141_183339\|\|full | 2260 | No | NA | 4 | 0 | 1 | Low-quality | Genome-fragment | 2.43 | AAI-based (medium-confidence) | 0 |
| S11-final-viral-combined_k141_257061\|\|full | 2260 | Yes | 1768 | 3 | 1 | 1 | Low-quality | Genome-fragment | 2.04 | HMM-based (lower-bound) | 21.77 |
| S17-final-viral-combined_k141_106605\|\|full | 2040 | No | NA | 2 | 0 | 1 | Not-determined | Genome-fragment | NA | NA | 0 |
| S11-final-viral-combined_k141_39768\|\|full | 2260 | No | NA | 3 | 0 | 3 | Low-quality | Genome-fragment | 2.71 | AAI-based (medium-confidence) | 0 |
| S17-final-viral-combined_k141_49271\|\|full | 2804 | No | NA | 4 | 0 | 1 | Not-determined | Genome-fragment | NA | NA | 0 |
| S17-final-viral-combined_k141_62199\|\|full | 1510 | No | NA | 4 | 0 | 2 | Not-determined | Genome-fragment | NA | NA | 0 |
| S17-final-viral-combined_k141_129129\|\|full | 2171 | No | NA | 4 | 0 | 2 | Not-determined | Genome-fragment | NA | NA | 0 |
| S3-final-viral-combined_k141_30840\|\|full | 2260 | No | NA | 4 | 1 | 0 | Low-quality | Genome-fragment | 1.52 | HMM-based (lower-bound) | 0 |
| S17-final-viral-combined_k141_62216\|\|full | 1491 | No | NA | 3 | 0 | 1 | Not-determined | Genome-fragment | NA | NA | 0 |
| S17-final-viral-combined_k141_218900\|\|full | 3871 | No | NA | 5 | 0 | 2 | Not-determined | Genome-fragment | NA | NA | 0 |
| S17-final-viral-combined_k141_185427\|\|full | 1660 | No | NA | 3 | 0 | 1 | Not-determined | Genome-fragment | NA | NA | 0 |
| S6-final-viral-combined_k141_32553\|\|full | 2260 | No | NA | 5 | 0 | 0 | Low-quality | Genome-fragment | 4.6 | AAI-based (high-confidence) | 0 |
| S17-final-viral-combined_k141_49295\|\|full | 4889 | No | NA | 4 | 0 | 2 | Not-determined | Genome-fragment | NA | NA | 0 |
| S8-final-viral-combined_k141_140049\|\|full | 2260 | No | NA | 5 | 0 | 1 | Low-quality | Genome-fragment | 4.23 | AAI-based (medium-confidence) | 0 |
| S17-final-viral-combined_k141_85450\|\|full | 1075 | No | NA | 2 | 0 | 0 | Not-determined | Genome-fragment | NA | NA | 0 |
| S5-final-viral-combined_k141_102782\|\|full | 2261 | Yes | 1322 | 4 | 1 | 1 | Low-quality | Genome-fragment | 2.3 | AAI-based (medium-confidence) | 41.53 |
| S2_k141_115974 | 2261 | No | NA | 4 | 0 | 0 | Low-quality | Genome-fragment | 4.77 | AAI-based (high-confidence) | 0 |
| S11-final-viral-combined_k141_232855\|\|full | 2262 | No | NA | 4 | 0 | 1 | Low-quality | Genome-fragment | 4.3 | AAI-based (high-confidence) | 0 |
| S17-final-viral-combined_k141_94158\|\|full | 5005 | No | NA | 6 | 0 | 1 | Not-determined | Genome-fragment | NA | NA | 0 |
| S17-final-viral-combined_k141_106649\|\|full | 1374 | No | NA | 3 | 0 | 3 | Not-determined | Genome-fragment | NA | NA | 0 |
| S17-final-viral-combined_k141_173487\|\|full | 1999 | No | NA | 3 | 0 | 1 | Not-determined | Genome-fragment | NA | NA | 0 |
| S17-final-viral-combined_k141_49317\|\|full | 2760 | No | NA | 3 | 0 | 1 | Not-determined | Genome-fragment | NA | NA | 0 |
| S17-final-viral-combined_k141_239961\|\|full | 3702 | No | NA | 2 | 0 | 1 | Not-determined | Genome-fragment | NA | NA | 0 |
| S17-final-viral-combined_k141_140454\|\|full | 2005 | No | NA | 4 | 0 | 1 | Not-determined | Genome-fragment | NA | NA | 0 |
| S1-final-viral-combined_k141_394592\|\|full | 2262 | No | NA | 4 | 0 | 0 | Low-quality | Genome-fragment | 41.86 | AAI-based (medium-confidence) | 0 |
| S5-final-viral-combined_k141_186998\|\|full | 2262 | No | NA | 2 | 0 | 2 | Low-quality | Genome-fragment | 1.62 | AAI-based (medium-confidence) | 0 |
| S17-final-viral-combined_k141_180660\|\|full | 2262 | No | NA | 4 | 0 | 0 | Low-quality | Genome-fragment | 3.8 | AAI-based (high-confidence) | 0 |
| S4-final-viral-combined_k141_66600\|\|full | 2262 | No | NA | 3 | 0 | 1 | Low-quality | Genome-fragment | 1.55 | AAI-based (medium-confidence) | 0 |
| S11-final-viral-combined_k141_320393\|\|full | 2263 | No | NA | 3 | 1 | 0 | Low-quality | Genome-fragment | 5.18 | AAI-based (high-confidence) | 0 |
| S17-final-viral-combined_k141_73904\|\|full | 3792 | No | NA | 5 | 0 | 3 | Not-determined | Genome-fragment | NA | NA | 0 |
| S17-final-viral-combined_k141_261494\|\|full | 3115 | No | NA | 3 | 0 | 2 | Not-determined | Genome-fragment | NA | NA | 0 |
| S1-final-viral-combined_k141_53277\|\|full | 2263 | No | NA | 5 | 2 | 0 | Low-quality | Genome-fragment | 4.87 | AAI-based (high-confidence) | 0 |
| S1_k141_140950 | 2263 | No | NA | 3 | 0 | 2 | Low-quality | Genome-fragment | 1.64 | AAI-based (high-confidence) | 0 |
| S17-final-viral-combined_k141_218987\|\|full | 2019 | No | NA | 3 | 0 | 2 | Not-determined | Genome-fragment | NA | NA | 0 |
| S17-final-viral-combined_k141_218990\|\|full | 1426 | No | NA | 3 | 0 | 0 | Not-determined | Genome-fragment | NA | NA | 0 |
| S17-final-viral-combined_k141_62277\|\|full | 2792 | No | NA | 5 | 0 | 0 | Not-determined | Genome-fragment | NA | NA | 0 |
| S17-final-viral-combined_k141_119338\|\|full | 1210 | No | NA | 2 | 0 | 0 | Not-determined | Genome-fragment | NA | NA | 0 |
| S17-final-viral-combined_k141_74914\|\|full | 2263 | No | NA | 5 | 4 | 0 | Low-quality | Genome-fragment | 4.37 | AAI-based (medium-confidence) | 0 |
| S8-final-viral-combined_k141_3871\|\|full | 2263 | No | NA | 5 | 2 | 0 | Low-quality | Genome-fragment | 5.27 | AAI-based (high-confidence) | 0 |
| S7-final-viral-combined_k141_37042\|\|full | 2264 | No | NA | 6 | 1 | 0 | Low-quality | Genome-fragment | 3.8 | AAI-based (medium-confidence) | 0 |
| S17-final-viral-combined_k141_230025\|\|full | 16472 | No | NA | 13 | 0 | 10 | Not-determined | Genome-fragment | NA | NA | 0 |
| S17-final-viral-combined_k141_250483\|\|full | 2264 | No | NA | 5 | 0 | 0 | Low-quality | Genome-fragment | 4.11 | AAI-based (high-confidence) | 0 |
| S17-final-viral-combined_k141_40391\|\|full | 2264 | No | NA | 4 | 0 | 3 | Low-quality | Genome-fragment | 5.91 | AAI-based (high-confidence) | 0 |
| S17-final-viral-combined_k141_250242\|\|full | 1085 | No | NA | 2 | 0 | 0 | Not-determined | Genome-fragment | NA | NA | 0 |
| S17-final-viral-combined_k141_119359\|\|full | 1840 | No | NA | 3 | 0 | 0 | Not-determined | Genome-fragment | NA | NA | 0 |
| S17-final-viral-combined_k141_140497\|\|full | 1443 | No | NA | 2 | 0 | 1 | Not-determined | Genome-fragment | NA | NA | 0 |
| S17-final-viral-combined_k141_151448\|\|full | 1295 | No | NA | 3 | 0 | 0 | Not-determined | Genome-fragment | NA | NA | 0 |
| S17-final-viral-combined_k141_252590\|\|full | 2264 | No | NA | 3 | 1 | 0 | Low-quality | Genome-fragment | 6.11 | AAI-based (high-confidence) | 0 |
| S17-final-viral-combined_k141_39792\|\|full | 2338 | No | NA | 2 | 0 | 2 | Not-determined | Genome-fragment | NA | NA | 0 |
| S2-final-viral-combined_k141_7235\|\|full | 2264 | No | NA | 3 | 0 | 1 | Low-quality | Genome-fragment | 2.98 | AAI-based (medium-confidence) | 0 |
| S17-final-viral-combined_k141_196430\|\|full | 1422 | No | NA | 2 | 0 | 2 | Not-determined | Genome-fragment | NA | NA | 0 |
| S2-final-viral-combined_k141_176312\|\|lt2gene | 2264 | No | NA | 2 | 2 | 0 | Low-quality | Genome-fragment | 7.96 | HMM-based (lower-bound) | 0 |
| S6-final-viral-combined_k141_76872\|\|lt2gene | 2264 | No | NA | 1 | 1 | 0 | Low-quality | Genome-fragment | 1.73 | HMM-based (lower-bound) | 0 |
| S17-final-viral-combined_k141_230077\|\|full | 2176 | No | NA | 4 | 0 | 2 | Not-determined | Genome-fragment | NA | NA | 0 |
| S9-final-viral-combined_k141_54400\|\|full | 2265 | No | NA | 4 | 0 | 0 | Low-quality | Genome-fragment | 3.86 | AAI-based (medium-confidence) | 0 |
| S9-final-viral-combined_k141_119555\|\|full | 2265 | No | NA | 6 | 1 | 0 | Low-quality | Genome-fragment | 6.13 | AAI-based (high-confidence) | 0 |
| S17-final-viral-combined_k141_85598\|\|full | 1391 | No | NA | 2 | 0 | 1 | Not-determined | Genome-fragment | NA | NA | 0 |
| S17-final-viral-combined_k141_140517\|\|full | 26302 | No | NA | 38 | 16 | 2 | Medium-quality | Genome-fragment | 55.34 | AAI-based (high-confidence) | 0 |
| S17-final-viral-combined_k141_151483\|\|full | 1560 | No | NA | 2 | 0 | 1 | Not-determined | Genome-fragment | NA | NA | 0 |
| S1-final-viral-combined_k141_138808\|\|full | 2265 | No | NA | 3 | 0 | 3 | Low-quality | Genome-fragment | 1.52 | AAI-based (high-confidence) | 0 |
| S17-final-viral-combined_k141_261595\|\|full | 3068 | No | NA | 4 | 0 | 2 | Not-determined | Genome-fragment | NA | NA | 0 |
| S1-final-viral-combined_k141_145022\|\|full | 2265 | No | NA | 6 | 1 | 0 | Low-quality | Genome-fragment | 5.53 | AAI-based (high-confidence) | 0 |
| S5-final-viral-combined_k141_92205\|\|lt2gene | 2265 | No | NA | 2 | 2 | 0 | Low-quality | Genome-fragment | 4.68 | HMM-based (lower-bound) | 0 |
| S17-final-viral-combined_k141_173643\|\|full | 2022 | No | NA | 2 | 0 | 2 | Not-determined | Genome-fragment | NA | NA | 0 |
| S17-final-viral-combined_k141_4313\|\|full | 2265 | No | NA | 3 | 0 | 0 | Low-quality | Genome-fragment | 3.43 | AAI-based (high-confidence) | 0 |
| S17-final-viral-combined_k141_75683\|\|full | 2265 | No | NA | 4 | 0 | 0 | Low-quality | Genome-fragment | 4.72 | AAI-based (high-confidence) | 0 |
| S17-final-viral-combined_k141_140587\|\|full | 2128 | No | NA | 3 | 0 | 1 | Not-determined | Genome-fragment | NA | NA | 0 |
| S11-final-viral-combined_k141_29406\|\|full | 2266 | No | NA | 3 | 1 | 0 | Low-quality | Genome-fragment | 16.91 | AAI-based (high-confidence) | 0 |
| S3-final-viral-combined_k141_43522\|\|full | 2266 | No | NA | 2 | 0 | 0 | Low-quality | Genome-fragment | 1.46 | AAI-based (high-confidence) | 0 |
| S17-final-viral-combined_k141_83359\|\|full | 2266 | Yes | 927 | 4 | 1 | 1 | Low-quality | Genome-fragment | 1.96 | AAI-based (medium-confidence) | 59.09 |
| S17-final-viral-combined_k141_250349\|\|full | 1543 | No | NA | 3 | 0 | 1 | Not-determined | Genome-fragment | NA | NA | 0 |
| S17-final-viral-combined_k141_83623\|\|full | 2266 | No | NA | 3 | 0 | 2 | Low-quality | Genome-fragment | 1.58 | AAI-based (medium-confidence) | 0 |
| S17-final-viral-combined_k141_119415\|\|full | 2400 | No | NA | 3 | 0 | 2 | Not-determined | Genome-fragment | NA | NA | 0 |
| S6_k141_17484 | 2266 | No | NA | 3 | 1 | 0 | Low-quality | Genome-fragment | 3.37 | AAI-based (high-confidence) | 0 |
| S17-final-viral-combined_k141_173709\|\|full | 3703 | No | NA | 4 | 0 | 3 | Not-determined | Genome-fragment | NA | NA | 0 |
| S17-final-viral-combined_k141_207827\|\|full | 1386 | No | NA | 2 | 0 | 0 | Not-determined | Genome-fragment | NA | NA | 0 |
| S8-final-viral-combined_k141_265298\|\|full | 2266 | No | NA | 6 | 1 | 0 | Low-quality | Genome-fragment | 4.97 | AAI-based (high-confidence) | 0 |
| S8_k141_81125 | 2266 | No | NA | 2 | 1 | 0 | Low-quality | Genome-fragment | 1.62 | AAI-based (high-confidence) | 0 |
| S9-final-viral-combined_k141_123994\|\|full | 2267 | No | NA | 4 | 2 | 0 | Low-quality | Genome-fragment | 5.83 | AAI-based (high-confidence) | 0 |
| S5_k141_199059 | 2267 | No | NA | 1 | 0 | 1 | Low-quality | Genome-fragment | 0.61 | AAI-based (high-confidence) | 0 |
| S17-final-viral-combined_k141_207866\|\|full | 2502 | No | NA | 3 | 0 | 2 | Not-determined | Genome-fragment | NA | NA | 0 |
| S17-final-viral-combined_k141_106809\|\|full | 2810 | No | NA | 3 | 0 | 2 | Not-determined | Genome-fragment | NA | NA | 0 |
| S17-final-viral-combined_k141_185683\|\|full | 2322 | No | NA | 3 | 0 | 2 | Not-determined | Genome-fragment | NA | NA | 0 |
| S12-final-viral-combined_k141_13760\|\|full | 2267 | No | NA | 4 | 1 | 0 | Low-quality | Genome-fragment | 1.46 | HMM-based (lower-bound) | 0 |
| S17-final-viral-combined_k141_106825\|\|full | 2264 | No | NA | 3 | 0 | 0 | Not-determined | Genome-fragment | NA | NA | 0 |
| S17-final-viral-combined_k141_240195\|\|full | 4276 | No | NA | 2 | 0 | 1 | Not-determined | Genome-fragment | NA | NA | 0 |
| S17-final-viral-combined_k141_85713\|\|full | 2267 | No | NA | 2 | 0 | 2 | Not-determined | Genome-fragment | NA | NA | 0 |
| S17-final-viral-combined_k141_250403\|\|full | 1533 | No | NA | 2 | 0 | 2 | Not-determined | Genome-fragment | NA | NA | 0 |
| S17-final-viral-combined_k141_119512\|\|full | 2077 | No | NA | 2 | 0 | 1 | Not-determined | Genome-fragment | NA | NA | 0 |
| S17-final-viral-combined_k141_6542\|\|full | 2171 | No | NA | 3 | 0 | 2 | Not-determined | Genome-fragment | NA | NA | 0 |
| S11-final-viral-combined_k141_224185\|\|full | 2268 | No | NA | 5 | 0 | 0 | Low-quality | Genome-fragment | 3.81 | AAI-based (high-confidence) | 0 |
| S1-final-viral-combined_k141_133968\|\|full | 2268 | No | NA | 5 | 0 | 0 | Low-quality | Genome-fragment | 5.95 | AAI-based (medium-confidence) | 0 |
| S17-final-viral-combined_k141_85731\|\|full | 1179 | No | NA | 3 | 0 | 1 | Not-determined | Genome-fragment | NA | NA | 0 |
| S17-final-viral-combined_k141_6547\|\|full | 1006 | No | NA | 2 | 0 | 1 | Not-determined | Genome-fragment | NA | NA | 0 |
| S17-final-viral-combined_k141_74067\|\|full | 3504 | No | NA | 4 | 0 | 2 | Not-determined | Genome-fragment | NA | NA | 0 |
| S1-final-viral-combined_k141_266575\|\|full | 2268 | No | NA | 5 | 1 | 0 | Low-quality | Genome-fragment | 4 | AAI-based (medium-confidence) | 0 |
| S1-final-viral-combined_k141_332661\|\|full | 2268 | No | NA | 4 | 0 | 3 | Low-quality | Genome-fragment | 1.17 | AAI-based (medium-confidence) | 0 |
| S3-final-viral-combined_k141_65354\|\|full | 2268 | No | NA | 4 | 0 | 0 | Low-quality | Genome-fragment | 1.04 | AAI-based (high-confidence) | 0 |
| S17-final-viral-combined_k141_173796\|\|full | 1340 | No | NA | 2 | 0 | 0 | Not-determined | Genome-fragment | NA | NA | 0 |
| S8-final-viral-combined_k141_214917\|\|full | 2268 | No | NA | 4 | 0 | 4 | Low-quality | Genome-fragment | 1.3 | AAI-based (high-confidence) | 0 |
| S17-final-viral-combined_k141_261758\|\|full | 1700 | No | NA | 5 | 0 | 1 | Not-determined | Genome-fragment | NA | NA | 0 |
| S9-final-viral-combined_k141_71548\|\|full | 2269 | No | NA | 4 | 1 | 0 | Low-quality | Genome-fragment | 1.55 | HMM-based (lower-bound) | 0 |
| S17-final-viral-combined_k141_62539\|\|full | 1827 | No | NA | 2 | 0 | 2 | Not-determined | Genome-fragment | NA | NA | 0 |
| S17-final-viral-combined_k141_85759\|\|full | 1311 | No | NA | 2 | 0 | 0 | Not-determined | Genome-fragment | NA | NA | 0 |
| S17-final-viral-combined_k141_6584\|\|full | 2591 | No | NA | 2 | 0 | 1 | Not-determined | Genome-fragment | NA | NA | 0 |
| S17-final-viral-combined_k141_129553\|\|full | 2782 | No | NA | 3 | 0 | 1 | Not-determined | Genome-fragment | NA | NA | 0 |
| S7-final-viral-combined_k141_80111\|\|full | 2269 | No | NA | 7 | 1 | 0 | Low-quality | Genome-fragment | 2.81 | AAI-based (high-confidence) | 0 |
| S17-final-viral-combined_k141_16904\|\|full | 1047 | No | NA | 4 | 0 | 2 | Not-determined | Genome-fragment | NA | NA | 0 |
| S6-final-viral-combined_k141_5029\|\|full | 2269 | No | NA | 2 | 0 | 1 | Low-quality | Genome-fragment | 1.99 | AAI-based (medium-confidence) | 0 |
| S17-final-viral-combined_k141_49585\|\|full | 1639 | No | NA | 3 | 0 | 0 | Not-determined | Genome-fragment | NA | NA | 0 |
| S11-final-viral-combined_k141_209868\|\|full | 2270 | No | NA | 2 | 0 | 2 | Low-quality | Genome-fragment | 1.61 | AAI-based (medium-confidence) | 0 |
| S17-final-viral-combined_k141_261845\|\|full | 1670 | No | NA | 2 | 0 | 1 | Not-determined | Genome-fragment | NA | NA | 0 |
| S1-final-viral-combined_k141_53870\|\|full | 2270 | No | NA | 3 | 0 | 0 | Low-quality | Genome-fragment | 6.36 | AAI-based (medium-confidence) | 0 |
| S1_k141_161705 | 2270 | No | NA | 2 | 1 | 0 | Low-quality | Genome-fragment | 6.71 | AAI-based (medium-confidence) | 0 |
| S5-final-viral-combined_k141_197230\|\|full | 2270 | No | NA | 3 | 1 | 0 | Low-quality | Genome-fragment | 6.21 | AAI-based (high-confidence) | 0 |
| S17-final-viral-combined_k141_161898\|\|full | 1567 | No | NA | 2 | 0 | 0 | Not-determined | Genome-fragment | NA | NA | 0 |
| S8-final-viral-combined_k141_257262\|\|full | 2270 | No | NA | 3 | 0 | 0 | Low-quality | Genome-fragment | 14.7 | AAI-based (high-confidence) | 0 |
| S17-final-viral-combined_k141_219253\|\|full | 2676 | No | NA | 3 | 0 | 1 | Not-determined | Genome-fragment | NA | NA | 0 |
| S17-final-viral-combined_k141_250536\|\|full | 2058 | No | NA | 4 | 0 | 1 | Not-determined | Genome-fragment | NA | NA | 0 |
| S11-final-viral-combined_k141_318656\|\|full | 2271 | No | NA | 4 | 0 | 0 | Low-quality | Genome-fragment | 2.31 | AAI-based (medium-confidence) | 0 |
| S17-final-viral-combined_k141_261902\|\|full | 1421 | No | NA | 2 | 0 | 0 | Not-determined | Genome-fragment | NA | NA | 0 |
| S17-final-viral-combined_k141_230361\|\|full | 4667 | No | NA | 5 | 0 | 3 | Not-determined | Genome-fragment | NA | NA | 0 |
| S17-final-viral-combined_k141_106940\|\|full | 62680 | Yes | 48510 | 89 | 18 | 9 | Medium-quality | Genome-fragment | 77.06 | AAI-based (high-confidence) | 22.61 |
| S17-final-viral-combined_k141_94511\|\|full | 2556 | No | NA | 3 | 0 | 0 | Not-determined | Genome-fragment | NA | NA | 0 |
| S17-final-viral-combined_k141_261923\|\|full | 3104 | No | NA | 3 | 0 | 3 | Not-determined | Genome-fragment | NA | NA | 0 |
| S17-final-viral-combined_k141_161941\|\|full | 1612 | No | NA | 3 | 0 | 2 | Not-determined | Genome-fragment | NA | NA | 0 |
| S17-final-viral-combined_k141_49661\|\|full | 2347 | No | NA | 5 | 0 | 0 | Not-determined | Genome-fragment | NA | NA | 0 |
| S17-final-viral-combined_k141_85869\|\|full | 2968 | No | NA | 2 | 0 | 2 | Not-determined | Genome-fragment | NA | NA | 0 |
| S11-final-viral-combined_k141_31632\|\|full | 2271 | No | NA | 2 | 0 | 1 | Low-quality | Genome-fragment | 1.5 | AAI-based (medium-confidence) | 0 |
| S17-final-viral-combined_k141_129649\|\|full | 2301 | No | NA | 2 | 0 | 0 | Not-determined | Genome-fragment | NA | NA | 0 |
| S11-final-viral-combined_k141_74837\|\|full | 2271 | No | NA | 5 | 1 | 0 | Low-quality | Genome-fragment | 5.94 | AAI-based (medium-confidence) | 0 |
| S17-final-viral-combined_k141_173978\|\|full | 2958 | No | NA | 5 | 0 | 2 | Not-determined | Genome-fragment | NA | NA | 0 |
| S17-final-viral-combined_k141_129167\|\|full | 2271 | No | NA | 4 | 2 | 0 | Low-quality | Genome-fragment | 4.86 | AAI-based (high-confidence) | 0 |
| S17-k141_167225 | 2271 | No | NA | 3 | 0 | 2 | Low-quality | Genome-fragment | 3 | AAI-based (medium-confidence) | 0 |
| S17-final-viral-combined_k141_40206\|\|full | 3096 | No | NA | 4 | 0 | 2 | Not-determined | Genome-fragment | NA | NA | 0 |
| S12_k141_242069 | 2271 | No | NA | 4 | 1 | 0 | Low-quality | Genome-fragment | 4.48 | AAI-based (high-confidence) | 0 |
| S5_k141_170959 | 2272 | No | NA | 3 | 3 | 0 | Low-quality | Genome-fragment | 3.4 | HMM-based (lower-bound) | 0 |
| S7-final-viral-combined_k141_145577\|\|full | 2272 | No | NA | 4 | 0 | 1 | Low-quality | Genome-fragment | 5.5 | AAI-based (medium-confidence) | 0 |
| S2-final-viral-combined_k141_244728\|\|full | 2272 | No | NA | 3 | 0 | 2 | Low-quality | Genome-fragment | 1.4 | AAI-based (medium-confidence) | 0 |
| S4-final-viral-combined_k141_143403\|\|full | 2272 | No | NA | 4 | 4 | 0 | Low-quality | Genome-fragment | 3.66 | AAI-based (high-confidence) | 0 |
| S8-final-viral-combined_k141_195073\|\|lt2gene | 2272 | No | NA | 2 | 2 | 0 | Low-quality | Genome-fragment | 5.91 | AAI-based (medium-confidence) | 0 |
| S1-final-viral-combined_k141_217618\|\|full | 2273 | No | NA | 4 | 0 | 1 | Low-quality | Genome-fragment | 3.61 | AAI-based (medium-confidence) | 0 |
| S5-final-viral-combined_k141_83069\|\|full | 2273 | No | NA | 3 | 0 | 1 | Low-quality | Genome-fragment | 1.32 | AAI-based (medium-confidence) | 0 |
| S12-final-viral-combined_k141_68509\|\|full | 2273 | No | NA | 4 | 0 | 0 | Low-quality | Genome-fragment | 4 | AAI-based (medium-confidence) | 0 |
| S12-final-viral-combined_k141_112203\|\|full | 2273 | No | NA | 4 | 1 | 0 | Low-quality | Genome-fragment | 1.67 | HMM-based (lower-bound) | 0 |
| S8-final-viral-combined_k141_233042\|\|full | 2273 | No | NA | 7 | 0 | 0 | Low-quality | Genome-fragment | 3.75 | AAI-based (high-confidence) | 0 |
| S8-final-viral-combined_k141_22236\|\|full | 2273 | No | NA | 4 | 0 | 0 | Low-quality | Genome-fragment | 3.69 | AAI-based (medium-confidence) | 0 |
| S4_k141_3280 | 2274 | No | NA | 1 | 0 | 1 | Low-quality | Genome-fragment | 0.9 | AAI-based (medium-confidence) | 0 |
| S8-final-viral-combined_k141_134813\|\|full | 2274 | No | NA | 1 | 0 | 1 | Low-quality | Genome-fragment | 1.42 | AAI-based (medium-confidence) | 0 |
| S17-final-viral-combined_k141_28923\|\|full | 1272 | No | NA | 3 | 0 | 2 | Not-determined | Genome-fragment | NA | NA | 0 |
| S5-final-viral-combined_k141_164901\|\|full | 2275 | No | NA | 4 | 1 | 0 | Low-quality | Genome-fragment | 4.84 | AAI-based (high-confidence) | 0 |
| S17-final-viral-combined_k141_49759\|\|full | 1593 | No | NA | 3 | 0 | 0 | Not-determined | Genome-fragment | NA | NA | 0 |
| S17-final-viral-combined_k141_196171\|\|full | 2275 | No | NA | 3 | 0 | 1 | Low-quality | Genome-fragment | 5.08 | AAI-based (medium-confidence) | 0 |
| S17-final-viral-combined_k141_250594\|\|full | 2121 | No | NA | 3 | 0 | 1 | Not-determined | Genome-fragment | NA | NA | 0 |
| S12-final-viral-combined_k141_57102\|\|full | 2275 | No | NA | 2 | 0 | 0 | Low-quality | Genome-fragment | 3.91 | AAI-based (high-confidence) | 0 |
| S17-final-viral-combined_k141_219423\|\|full | 6216 | No | NA | 6 | 0 | 6 | Not-determined | Genome-fragment | NA | NA | 0 |
| S1-final-viral-combined_k141_337405\|\|full | 2276 | No | NA | 3 | 0 | 1 | Low-quality | Genome-fragment | 5.31 | AAI-based (high-confidence) | 0 |
| S1-final-viral-combined_k141_289113\|\|full | 2276 | No | NA | 2 | 0 | 2 | Low-quality | Genome-fragment | 5.12 | AAI-based (medium-confidence) | 0 |
| S17-final-viral-combined_k141_262044\|\|full | 1600 | No | NA | 3 | 0 | 0 | Not-determined | Genome-fragment | NA | NA | 0 |
| S7-final-viral-combined_k141_89578\|\|full | 2276 | No | NA | 3 | 0 | 0 | Low-quality | Genome-fragment | 5.03 | AAI-based (medium-confidence) | 0 |
| S12-final-viral-combined_k141_376537\|\|full | 2276 | No | NA | 3 | 1 | 0 | Low-quality | Genome-fragment | 3.77 | HMM-based (lower-bound) | 0 |
| S17-final-viral-combined_k141_219461\|\|full | 2402 | No | NA | 3 | 0 | 3 | Not-determined | Genome-fragment | NA | NA | 0 |
| S17-final-viral-combined_k141_94665\|\|full | 1081 | No | NA | 2 | 0 | 1 | Not-determined | Genome-fragment | NA | NA | 0 |
| S17-final-viral-combined_k141_62890\|\|full | 1601 | No | NA | 4 | 0 | 2 | Not-determined | Genome-fragment | NA | NA | 0 |
| S17-final-viral-combined_k141_86053\|\|full | 2858 | No | NA | 3 | 0 | 1 | Not-determined | Genome-fragment | NA | NA | 0 |
| S12-final-viral-combined_k141_228601\|\|full | 2276 | No | NA | 5 | 1 | 0 | Low-quality | Genome-fragment | 7.05 | AAI-based (high-confidence) | 0 |
| S6_k141_81365 | 2276 | No | NA | 6 | 4 | 0 | Low-quality | Genome-fragment | 7.08 | AAI-based (high-confidence) | 0 |
| S5-final-viral-combined_k141_5642\|\|lt2gene | 2277 | No | NA | 2 | 2 | 0 | Low-quality | Genome-fragment | 3.74 | AAI-based (high-confidence) | 0 |
| S17-final-viral-combined_k141_9386\|\|full | 2277 | No | NA | 2 | 0 | 1 | Low-quality | Genome-fragment | 1.53 | AAI-based (medium-confidence) | 0 |
| S17-final-viral-combined_k141_107110\|\|full | 2927 | No | NA | 4 | 0 | 3 | Not-determined | Genome-fragment | NA | NA | 0 |
| S17-final-viral-combined_k141_151933\|\|full | 2419 | No | NA | 3 | 0 | 1 | Not-determined | Genome-fragment | NA | NA | 0 |
| S12_k141_362164 | 2277 | No | NA | 2 | 1 | 0 | Low-quality | Genome-fragment | 2.25 | AAI-based (medium-confidence) | 0 |
| S17-final-viral-combined_k141_107134\|\|full | 2594 | No | NA | 5 | 0 | 0 | Not-determined | Genome-fragment | NA | NA | 0 |
| S17-final-viral-combined_k141_141068\|\|full | 1598 | No | NA | 3 | 0 | 0 | Not-determined | Genome-fragment | NA | NA | 0 |
| S2-final-viral-combined_k141_56363\|\|full | 2277 | No | NA | 2 | 0 | 2 | Low-quality | Genome-fragment | 0.24 | AAI-based (medium-confidence) | 0 |
| S6-final-viral-combined_k141_61796\|\|full | 2277 | No | NA | 3 | 1 | 0 | Low-quality | Genome-fragment | 2.61 | AAI-based (high-confidence) | 0 |
| S17-final-viral-combined_k141_40374\|\|full | 1801 | No | NA | 2 | 0 | 1 | Not-determined | Genome-fragment | NA | NA | 0 |
| S17-final-viral-combined_k141_86107\|\|full | 2671 | No | NA | 3 | 0 | 3 | Not-determined | Genome-fragment | NA | NA | 0 |
| S1-final-viral-combined_k141_187452\|\|full | 2278 | No | NA | 5 | 0 | 0 | Low-quality | Genome-fragment | 5.65 | AAI-based (high-confidence) | 0 |
| S1-final-viral-combined_k141_340649\|\|full | 2278 | No | NA | 2 | 0 | 1 | Low-quality | Genome-fragment | 4.38 | AAI-based (high-confidence) | 0 |
| S2-final-viral-combined_k141_116114\|\|full | 2278 | No | NA | 2 | 0 | 1 | Low-quality | Genome-fragment | 3.65 | AAI-based (high-confidence) | 0 |
| S17-final-viral-combined_k141_119929\|\|full | 1278 | No | NA | 2 | 0 | 1 | Not-determined | Genome-fragment | NA | NA | 0 |
| S4-final-viral-combined_k141_73558\|\|full | 2278 | No | NA | 7 | 0 | 0 | Low-quality | Genome-fragment | 2.91 | AAI-based (high-confidence) | 0 |
| S17-final-viral-combined_k141_162058\|\|full | 15336 | No | NA | 14 | 0 | 4 | Not-determined | Genome-fragment | NA | NA | 0 |
| S1-final-viral-combined_k141_289640\|\|full | 2279 | No | NA | 3 | 0 | 0 | Low-quality | Genome-fragment | 1.69 | AAI-based (high-confidence) | 0 |
| S17-final-viral-combined_k141_162075\|\|full | 1194 | No | NA | 2 | 0 | 0 | Not-determined | Genome-fragment | NA | NA | 0 |
| S17-final-viral-combined_k141_262099\|\|full | 1390 | No | NA | 2 | 0 | 0 | Not-determined | Genome-fragment | NA | NA | 0 |
| S17-final-viral-combined_k141_250695\|\|full | 11161 | No | NA | 10 | 0 | 8 | Not-determined | Genome-fragment | NA | NA | 0 |
| S17-final-viral-combined_k141_219560\|\|full | 2817 | No | NA | 3 | 0 | 1 | Not-determined | Genome-fragment | NA | NA | 0 |
| S3-final-viral-combined_k141_129540\|\|full | 2279 | No | NA | 3 | 0 | 0 | Low-quality | Genome-fragment | 6.14 | AAI-based (medium-confidence) | 0 |
| S4-final-viral-combined_k141_109975\|\|full | 2279 | No | NA | 7 | 2 | 0 | Low-quality | Genome-fragment | 3.76 | AAI-based (high-confidence) | 0 |
| S8-final-viral-combined_k141_180632\|\|full | 2279 | No | NA | 5 | 3 | 0 | Low-quality | Genome-fragment | 2.67 | HMM-based (lower-bound) | 0 |
| S5-final-viral-combined_k141_91210\|\|full | 2280 | No | NA | 2 | 0 | 2 | Low-quality | Genome-fragment | 4.28 | AAI-based (medium-confidence) | 0 |
| S17-final-viral-combined_k141_262151\|\|full | 1174 | No | NA | 3 | 0 | 2 | Not-determined | Genome-fragment | NA | NA | 0 |
| S12_k141_6489 | 2280 | No | NA | 4 | 1 | 0 | Low-quality | Genome-fragment | 5.42 | HMM-based (lower-bound) | 0 |
| S4-final-viral-combined_k141_34820\|\|full | 2280 | No | NA | 2 | 0 | 0 | Low-quality | Genome-fragment | 1.26 | AAI-based (medium-confidence) | 0 |
| S1-final-viral-combined_k141_109733\|\|full | 2281 | No | NA | 3 | 3 | 0 | Low-quality | Genome-fragment | 5.5 | AAI-based (high-confidence) | 0 |
| S5-final-viral-combined_k141_30694\|\|full | 2281 | No | NA | 5 | 3 | 0 | Low-quality | Genome-fragment | 5.06 | AAI-based (medium-confidence) | 0 |
| S17-final-viral-combined_k141_74460\|\|full | 4654 | No | NA | 6 | 0 | 1 | Not-determined | Genome-fragment | NA | NA | 0 |
| S12-final-viral-combined_k141_73483\|\|full | 2281 | No | NA | 3 | 0 | 1 | Low-quality | Genome-fragment | 5.59 | AAI-based (high-confidence) | 0 |

| | | | | | | | | | | | | |
|---|---|---|---|---|---|---|---|---|---|---|---|---|
| S17-final-viral-combined_k141_86171\|\|full | 4822 | No | NA | | 4 | 0 | 3 | Not-determined | Genome-fragment | NA | NA | 0 |
| S17-final-viral-combined_k141_141181\|\|full | 5864 | No | NA | | 11 | 0 | 0 | Not-determined | Genome-fragment | NA | NA | 0 |
| S12-final-viral-combined_k141_45811\|\|full | 2281 | No | NA | | 6 | 0 | 0 | Low-quality | Genome-fragment | 4.23 | AAI-based (high-confidence) | 0 |
| S17-final-viral-combined_k141_141184\|\|full | 2885 | No | NA | | 2 | 0 | 2 | Not-determined | Genome-fragment | NA | NA | 0 |
| S17-final-viral-combined_k141_208385\|\|full | 3572 | No | NA | | 3 | 0 | 1 | Not-determined | Genome-fragment | NA | NA | 0 |
| S17-final-viral-combined_k141_86184\|\|full | 5255 | No | NA | | 8 | 0 | 6 | Not-determined | Genome-fragment | NA | NA | 0 |
| S17-final-viral-combined_k141_29054\|\|full | 3078 | No | NA | | 3 | 0 | 1 | Not-determined | Genome-fragment | NA | NA | 0 |
| S17-final-viral-combined_k141_141199\|\|full | 3381 | No | NA | | 3 | 0 | 3 | Not-determined | Genome-fragment | NA | NA | 0 |
| S12_k141_16670 | 2281 | No | NA | | 2 | 0 | 0 | Low-quality | Genome-fragment | 2.79 | AAI-based (high-confidence) | 0 |
| S2_k141_202083 | 2281 | No | NA | | 3 | 0 | 1 | Low-quality | Genome-fragment | 0.61 | AAI-based (high-confidence) | 0 |
| S17-final-viral-combined_k141_162194\|\|full | 3606 | No | NA | | 3 | 0 | 3 | Not-determined | Genome-fragment | NA | NA | 0 |
| S17-final-viral-combined_k141_174296\|\|full | 4267 | No | NA | | 5 | 0 | 3 | Not-determined | Genome-fragment | NA | NA | 0 |
| S17-final-viral-combined_k141_86207\|\|full | 2256 | No | NA | | 3 | 0 | 1 | Not-determined | Genome-fragment | NA | NA | 0 |
| S4-final-viral-combined_k141_87315\|\|full | 2281 | No | NA | | 3 | 3 | 0 | Low-quality | Genome-fragment | 3.72 | AAI-based (high-confidence) | 0 |
| S1-final-viral-combined_k141_146894\|\|full | 2282 | No | NA | | 3 | 1 | 2 | Low-quality | Genome-fragment | 4.15 | AAI-based (medium-confidence) | 0 |
| S3_k141_126709 | 2282 | No | NA | | 2 | 0 | 1 | Low-quality | Genome-fragment | 3.86 | AAI-based (high-confidence) | 0 |
| S17-final-viral-combined_k141_62993\|\|full | 3522 | No | NA | | 3 | 0 | 0 | Not-determined | Genome-fragment | NA | NA | 0 |
| S17-final-viral-combined_k141_208416\|\|full | 4718 | No | NA | | 5 | 0 | 1 | Not-determined | Genome-fragment | NA | NA | 0 |
| S17-final-viral-combined_k141_120076\|\|full | 9025 | No | NA | | 12 | 0 | 11 | Not-determined | Genome-fragment | NA | NA | 0 |
| S17-final-viral-combined_k141_152658\|\|full | 2282 | No | NA | | 2 | 2 | 0 | Low-quality | Genome-fragment | 3.31 | AAI-based (medium-confidence) | 0 |
| S17-final-viral-combined_k141_86217\|\|full | 6196 | No | NA | | 8 | 0 | 7 | Not-determined | Genome-fragment | NA | NA | 0 |
| S12-final-viral-combined_k141_234655\|\|full | 2282 | No | NA | | 6 | 1 | 0 | Low-quality | Genome-fragment | 4.84 | AAI-based (medium-confidence) | 0 |
| S12-final-viral-combined_k141_329428\|\|full | 2282 | No | NA | | 2 | 0 | 2 | Low-quality | Genome-fragment | 1.1 | AAI-based (medium-confidence) | 0 |
| S17-final-viral-combined_k141_186164\|\|full | 4562 | No | NA | | 4 | 0 | 3 | Not-determined | Genome-fragment | NA | NA | 0 |
| S12-final-viral-combined_k141_11003\|\|full | 2282 | No | NA | | 3 | 1 | 0 | Low-quality | Genome-fragment | 3.33 | HMM-based (lower-bound) | 0 |
| S2-final-viral-combined_k141_35511\|\|full | 2282 | No | NA | | 3 | 2 | 0 | Low-quality | Genome-fragment | 4.95 | AAI-based (high-confidence) | 0 |
| S17-final-viral-combined_k141_74586\|\|full | 2549 | No | NA | | 3 | 0 | 1 | Not-determined | Genome-fragment | NA | NA | 0 |
| S2-final-viral-combined_k141_4822\|\|full | 2282 | No | NA | | 3 | 2 | 0 | Low-quality | Genome-fragment | 4.21 | HMM-based (lower-bound) | 0 |
| S17-final-viral-combined_k141_240539\|\|full | 1928 | No | NA | | 3 | 0 | 2 | Not-determined | Genome-fragment | NA | NA | 0 |
| S17-final-viral-combined_k141_186183\|\|full | 1912 | No | NA | | 3 | 0 | 1 | Not-determined | Genome-fragment | NA | NA | 0 |
| S17-final-viral-combined_k141_86241\|\|full | 1663 | No | NA | | 2 | 0 | 1 | Not-determined | Genome-fragment | NA | NA | 0 |
| [illegible] | [illegible] | [illegible] | [illegible] | [illegible] | [illegible] | [illegible] | [illegible] | [illegible] | [illegible] | [illegible] | [illegible] | [illegible] |

| | | | | | | | | | | | |
|---|---|---|---|---|---|---|---|---|---|---|---|
| S17-final-viral-combined_k141_121537\|\|full | 6670 | No | NA | 4 | 0 | 2 | Not-determined | Genome-fragment | NA | NA | 0 |
| S12-final-viral-combined_k141_13798\|\|full | 2318 | No | NA | 4 | 0 | 2 | Low-quality | Genome-fragment | 0.77 | AAI-based (medium-confidence) | 0 |
| S17-final-viral-combined_k141_76292\|\|full | 1729 | No | NA | 2 | 0 | 1 | Not-determined | Genome-fragment | NA | NA | 0 |
| S17-final-viral-combined_k141_142841\|\|full | 5542 | No | NA | 2 | 0 | 2 | Not-determined | Genome-fragment | NA | NA | 0 |
| S12_k141_169524 | 2318 | No | NA | 4 | 4 | 0 | Low-quality | Genome-fragment | 1.51 | AAI-based (high-confidence) | 0 |
| S17-final-viral-combined_k141_64471\|\|full | 1278 | No | NA | 3 | 0 | 1 | Not-determined | Genome-fragment | NA | NA | 0 |
| S17-final-viral-combined_k141_18692\|\|full | 4803 | No | NA | 7 | 0 | 3 | Not-determined | Genome-fragment | NA | NA | 0 |
| S2-final-viral-combined_k141_247781\|\|full | 2318 | No | NA | 6 | 5 | 0 | Low-quality | Genome-fragment | 5.3 | AAI-based (high-confidence) | 0 |
| S8-final-viral-combined_k141_212274\|\|full | 2318 | No | NA | 5 | 5 | 0 | Low-quality | Genome-fragment | 3.66 | AAI-based (high-confidence) | 0 |
| S17-final-viral-combined_k141_142858\|\|full | 1856 | No | NA | 2 | 0 | 2 | Not-determined | Genome-fragment | NA | NA | 0 |
| S17-final-viral-combined_k141_242067\|\|full | 4101 | No | NA | 7 | 0 | 0 | Not-determined | Genome-fragment | NA | NA | 0 |
| S17-final-viral-combined_k141_51263\|\|full | 2318 | No | NA | 2 | 0 | 2 | Not-determined | Genome-fragment | NA | NA | 0 |
| S17-final-viral-combined_k141_232360\|\|full | 9425 | No | NA | 9 | 0 | 6 | Not-determined | Genome-fragment | NA | NA | 0 |
| S17-final-viral-combined_k141_18707\|\|full | 1657 | No | NA | 2 | 0 | 0 | Not-determined | Genome-fragment | NA | NA | 0 |
| S17-final-viral-combined_k141_163524\|\|full | 2082 | No | NA | 3 | 0 | 1 | Not-determined | Genome-fragment | NA | NA | 0 |
| S11-final-viral-combined_k141_69074\|\|full | 2319 | No | NA | 7 | 0 | 0 | Low-quality | Genome-fragment | 3.8 | AAI-based (medium-confidence) | 0 |
| S17-final-viral-combined_k141_163526\|\|full | 3552 | No | NA | 5 | 0 | 4 | Not-determined | Genome-fragment | NA | NA | 0 |
| S2-final-viral-combined_k141_20179\|\|full | 2319 | No | NA | 2 | 1 | 0 | Low-quality | Genome-fragment | 1.41 | AAI-based (high-confidence) | 0 |
| S17-final-viral-combined_k141_64524\|\|full | 1855 | No | NA | 4 | 0 | 2 | Not-determined | Genome-fragment | NA | NA | 0 |
| S4-final-viral-combined_k141_146037\|\|full | 2319 | No | NA | 3 | 0 | 1 | Low-quality | Genome-fragment | 4.19 | AAI-based (medium-confidence) | 0 |
| S17-final-viral-combined_k141_87902\|\|full | 1668 | No | NA | 2 | 0 | 1 | Not-determined | Genome-fragment | NA | NA | 0 |
| S11-final-viral-combined_k141_28248\|\|full | 2320 | No | NA | 4 | 0 | 1 | Low-quality | Genome-fragment | 0.69 | AAI-based (medium-confidence) | 0 |
| S17-final-viral-combined_k141_142918\|\|full | 8963 | No | NA | 9 | 0 | 7 | Not-determined | Genome-fragment | NA | NA | 0 |
| S1-final-viral-combined_k141_89012\|\|full | 2320 | No | NA | 4 | 1 | 0 | Low-quality | Genome-fragment | 6.44 | AAI-based (high-confidence) | 0 |
| S7-final-viral-combined_k141_236564\|\|full | 2320 | No | NA | 3 | 1 | 0 | Low-quality | Genome-fragment | 3.21 | AAI-based (medium-confidence) | 0 |
| S17-final-viral-combined_k141_76397\|\|full | 2214 | No | NA | 3 | 0 | 2 | Not-determined | Genome-fragment | NA | NA | 0 |
| S17-final-viral-combined_k141_187601\|\|full | 5817 | No | NA | 6 | 0 | 3 | Not-determined | Genome-fragment | NA | NA | 0 |
| S7-final-viral-combined_k141_86229\|\|full | 2320 | No | NA | 4 | 1 | 0 | Low-quality | Genome-fragment | 5.37 | AAI-based (high-confidence) | 0 |
| S17-final-viral-combined_k141_51345\|\|full | 2455 | No | NA | 2 | 0 | 1 | Not-determined | Genome-fragment | NA | NA | 0 |
| S17-final-viral-combined_k141_187616\|\|full | 1477 | No | NA | 5 | 0 | 1 | Not-determined | Genome-fragment | NA | NA | 0 |
| S2-final-viral-combined_k141_81662\|\|full | 2320 | No | NA | 2 | 0 | 1 | Low-quality | Genome-fragment | 1.41 | AAI-based (medium-confidence) | 0 |
| S17-final-viral-combined_k141_96019\|\|full | 1138 | No | NA | 2 | 0 | 2 | Not-determined | Genome-fragment | NA | NA | 0 |
| S3-final-viral-combined_k141_26190\|\|full | 2321 | No | NA | 2 | 0 | 0 | Low-quality | Genome-fragment | 1.63 | AAI-based (high-confidence) | 0 |
| S17-final-viral-combined_k141_232394\|\|full | 2976 | No | NA | 6 | 0 | 2 | Not-determined | Genome-fragment | NA | NA | 0 |
| S17-final-viral-combined_k141_175924\|\|full | 38365 | No | NA | 52 | 16 | 1 | High-quality | High-quality | 100 | AAI-based (medium-confidence) | 0 |
| S17-final-viral-combined_k141_41476\|\|full | 3818 | No | NA | 5 | 0 | 3 | Not-determined | Genome-fragment | NA | NA | 0 |
| S17-final-viral-combined_k141_153606\|\|full | 1747 | No | NA | 3 | 0 | 0 | Not-determined | Genome-fragment | NA | NA | 0 |
| S12-final-viral-combined_k141_141881\|\|full | 2321 | No | NA | 3 | 0 | 0 | Low-quality | Genome-fragment | 5.77 | AAI-based (medium-confidence) | 0 |
| S4-final-viral-combined_k141_31446\|\|full | 2321 | No | NA | 5 | 0 | 0 | Low-quality | Genome-fragment | 5.75 | AAI-based (medium-confidence) | 0 |
| S11-final-viral-combined_k141_80393\|\|full | 2322 | No | NA | 5 | 1 | 0 | Low-quality | Genome-fragment | 6.71 | AAI-based (high-confidence) | 0 |
| S17-final-viral-combined_k141_198899\|\|full | 2401 | No | NA | 3 | 0 | 1 | Not-determined | Genome-fragment | NA | NA | 0 |
| S17-final-viral-combined_k141_209945\|\|full | 1708 | No | NA | 3 | 0 | 0 | Not-determined | Genome-fragment | NA | NA | 0 |
| S1-final-viral-combined_k141_320496\|\|full | 2322 | No | NA | 3 | 0 | 3 | Low-quality | Genome-fragment | 1.18 | AAI-based (medium-confidence) | 0 |
| S1-final-viral-combined_k141_205326\|\|full | 2322 | No | NA | 3 | 2 | 0 | Low-quality | Genome-fragment | 4.18 | HMM-based (lower-bound) | 0 |
| S5-final-viral-combined_k141_195328\|\|full | 2322 | No | NA | 3 | 2 | 0 | Low-quality | Genome-fragment | 5.18 | AAI-based (high-confidence) | 0 |
| S17-final-viral-combined_k141_41511\|\|full | 3037 | No | NA | 4 | 0 | 2 | Not-determined | Genome-fragment | NA | NA | 0 |
| S12-final-viral-combined_k141_313398\|\|full | 2322 | No | NA | 4 | 0 | 2 | Low-quality | Genome-fragment | 3.42 | AAI-based (medium-confidence) | 0 |
| S17-final-viral-combined_k141_153648\|\|full | 4847 | No | NA | 3 | 0 | 3 | Not-determined | Genome-fragment | NA | NA | 0 |
| S9-final-viral-combined_k141_20912\|\|full | 2323 | No | NA | 2 | 0 | 1 | Low-quality | Genome-fragment | 2.18 | AAI-based (medium-confidence) | 0 |
| S11-final-viral-combined_k141_122696\|\|full | 2323 | No | NA | 5 | 0 | 3 | Low-quality | Genome-fragment | 0.92 | AAI-based (medium-confidence) | 0 |
| S1-final-viral-combined_k141_20168\|\|full | 2323 | No | NA | 2 | 2 | 0 | Low-quality | Genome-fragment | 1.76 | AAI-based (medium-confidence) | 0 |
| S8-final-viral-combined_k141_273124\|\|full | 2323 | No | NA | 5 | 1 | 0 | Low-quality | Genome-fragment | 5 | AAI-based (medium-confidence) | 0 |
| S17-final-viral-combined_k141_30609\|\|full | 1126 | No | NA | 2 | 0 | 2 | Not-determined | Genome-fragment | NA | NA | 0 |
| S9-final-viral-combined_k141_44299\|\|full | 2324 | No | NA | 3 | 0 | 1 | Low-quality | Genome-fragment | 1.21 | AAI-based (medium-confidence) | 0 |
| S11-final-viral-combined_k141_41155\|\|full | 2324 | No | NA | 4 | 1 | 0 | Low-quality | Genome-fragment | 3.99 | AAI-based (medium-confidence) | 0 |
| S17-final-viral-combined_k141_221484\|\|full | 3318 | No | NA | 4 | 0 | 4 | Not-determined | Genome-fragment | NA | NA | 0 |
| S17-final-viral-combined_k141_108939\|\|full | 3995 | No | NA | 3 | 0 | 2 | Not-determined | Genome-fragment | NA | NA | 0 |
| S17-final-viral-combined_k141_88049\|\|full | 1022 | No | NA | 2 | 0 | 1 | Not-determined | Genome-fragment | NA | NA | 0 |
| S7-final-viral-combined_k141_209709\|\|full | 2324 | No | NA | 5 | 1 | 0 | Low-quality | Genome-fragment | 4.26 | AAI-based (medium-confidence) | 0 |
| S17-final-viral-combined_k141_143110\|\|full | 1095 | No | NA | 3 | 0 | 1 | Not-determined | Genome-fragment | NA | NA | 0 |
| S17-final-viral-combined_k141_64797\|\|full | 5966 | No | NA | 5 | 0 | 3 | Not-determined | Genome-fragment | NA | NA | 0 |
| S17-final-viral-combined_k141_64812\|\|full | 1084 | No | NA | 2 | 0 | 0 | Not-determined | Genome-fragment | NA | NA | 0 |
| S17-final-viral-combined_k141_131069\|\|full | 1485 | No | NA | 2 | 0 | 1 | Not-determined | Genome-fragment | NA | NA | 0 |
| S12-final-viral-combined_k141_665\|\|full | 2324 | No | NA | 4 | 0 | 0 | Low-quality | Genome-fragment | 4.5 | AAI-based (medium-confidence) | 0 |
| S17-final-viral-combined_k141_18819\|\|full | 1944 | No | NA | 4 | 0 | 0 | Not-determined | Genome-fragment | NA | NA | 0 |
| S12-final-viral-combined_k141_231179\|\|full | 2324 | No | NA | 2 | 0 | 0 | Low-quality | Genome-fragment | 5.58 | AAI-based (high-confidence) | 0 |
| S17-final-viral-combined_k141_131088\|\|full | 6170 | No | NA | 9 | 0 | 1 | Not-determined | Genome-fragment | NA | NA | 0 |
| S2-final-viral-combined_k141_147186\|\|full | 2324 | No | NA | 4 | 0 | 0 | Low-quality | Genome-fragment | 4.95 | AAI-based (medium-confidence) | 0 |
| S17-final-viral-combined_k141_18835\|\|full | 10542 | No | NA | 16 | 0 | 6 | Not-determined | Genome-fragment | NA | NA | 0 |
| S2-final-viral-combined_k141_23974\|\|full | 2324 | No | NA | 6 | 1 | 0 | Low-quality | Genome-fragment | 5.05 | AAI-based (high-confidence) | 0 |
| S8_k141_145341 | 2324 | No | NA | 7 | 1 | 0 | Low-quality | Genome-fragment | 3.81 | AAI-based (high-confidence) | 0 |
| S9_k141_104 | 2325 | No | NA | 2 | 0 | 2 | Low-quality | Genome-fragment | 4.38 | AAI-based (medium-confidence) | 0 |
| S17-final-viral-combined_k141_176075\|\|full | 1408 | No | NA | 3 | 0 | 1 | Not-determined | Genome-fragment | NA | NA | 0 |
| S2_k141_24629 | 2325 | No | NA | 7 | 0 | 0 | Low-quality | Genome-fragment | 2.42 | AAI-based (high-confidence) | 0 |
| S17-final-viral-combined_k141_121843\|\|full | 1014 | No | NA | 3 | 0 | 1 | Not-determined | Genome-fragment | NA | NA | 0 |
| S12-final-viral-combined_k141_128796\|\|full | 2326 | No | NA | 2 | 0 | 0 | Low-quality | Genome-fragment | 3.52 | AAI-based (medium-confidence) | 0 |
| S17-final-viral-combined_k141_163784\|\|full | 2894 | No | NA | 3 | 0 | 0 | Not-determined | Genome-fragment | NA | NA | 0 |
| S17-final-viral-combined_k141_199030\|\|full | 3488 | No | NA | 2 | 0 | 2 | Not-determined | Genome-fragment | NA | NA | 0 |
| S12-final-viral-combined_k141_374386\|\|lt2gene | 2326 | No | NA | 1 | 1 | 0 | Low-quality | Genome-fragment | 5.14 | AAI-based (high-confidence) | 0 |
| S17-final-viral-combined_k141_199036\|\|full | 2258 | No | NA | 2 | 0 | 0 | Not-determined | Genome-fragment | NA | NA | 0 |
| S2-final-viral-combined_k141_50701\|\|full | 2326 | Yes | 1510 | 6 | 1 | 1 | Low-quality | Genome-fragment | 3.41 | AAI-based (high-confidence) | 35.08 |
| S17-final-viral-combined_k141_210112\|\|full | 2065 | No | NA | 3 | 0 | 3 | Not-determined | Genome-fragment | NA | NA | 0 |
| S17-final-viral-combined_k141_96150\|\|full | 4473 | No | NA | 4 | 0 | 3 | Not-determined | Genome-fragment | NA | NA | 0 |
| S17-final-viral-combined_k141_210126\|\|full | 4445 | No | NA | 5 | 0 | 5 | Not-determined | Genome-fragment | NA | NA | 0 |
| S17-final-viral-combined_k141_64870\|\|full | 2079 | No | NA | 2 | 0 | 2 | Not-determined | Genome-fragment | NA | NA | 0 |
| S17-final-viral-combined_k141_143206\|\|full | 1227 | No | NA | 3 | 0 | 0 | Not-determined | Genome-fragment | NA | NA | 0 |
| S17-final-viral-combined_k141_210155\|\|full | 2568 | No | NA | 4 | 0 | 3 | Not-determined | Genome-fragment | NA | NA | 0 |
| S17-final-viral-combined_k141_242341\|\|full | 1744 | No | NA | 4 | 0 | 0 | Not-determined | Genome-fragment | NA | NA | 0 |
| S17-final-viral-combined_k141_88217\|\|full | 1582 | No | NA | 3 | 0 | 1 | Not-determined | Genome-fragment | NA | NA | 0 |
| S17-final-viral-combined_k141_252168\|\|full | 1605 | No | NA | 2 | 0 | 1 | Not-determined | Genome-fragment | NA | NA | 0 |
| S2_k141_134475 | 2326 | No | NA | 6 | 2 | 0 | Low-quality | Genome-fragment | 5.73 | AAI-based (high-confidence) | 0 |
| S8-final-viral-combined_k141_219792\|\|full | 2326 | No | NA | 3 | 0 | 2 | Low-quality | Genome-fragment | 2.28 | AAI-based (medium-confidence) | 0 |
| S17-final-viral-combined_k141_242362\|\|full | 1283 | No | NA | 2 | 0 | 2 | Not-determined | Genome-fragment | NA | NA | 0 |
| S11-final-viral-combined_k141_65268\|\|lt2gene | 2327 | Yes | 1032 | 2 | 1 | 1 | Low-quality | Genome-fragment | 0.71 | HMM-based (lower-bound) | 55.65 |
| S17-final-viral-combined_k141_232684\|\|full | 2758 | No | NA | 3 | 0 | 1 | Not-determined | Genome-fragment | NA | NA | 0 |
| S17-final-viral-combined_k141_88241\|\|full | 2682 | No | NA | 2 | 0 | 0 | Not-determined | Genome-fragment | NA | NA | 0 |
| S17-final-viral-combined_k141_41735\|\|full | 1471 | No | NA | 3 | 0 | 1 | Not-determined | Genome-fragment | NA | NA | 0 |
| S17-k141_110629 | 2327 | No | NA | 3 | 0 | 0 | Low-quality | Genome-fragment | 4.68 | AAI-based (medium-confidence) | 0 |
| S17-final-viral-combined_k141_263710\|\|full | 2066 | No | NA | 2 | 0 | 0 | Not-determined | Genome-fragment | NA | NA | 0 |
| S17-final-viral-combined_k141_221700\|\|full | 2606 | No | NA | 4 | 0 | 2 | Not-determined | Genome-fragment | NA | NA | 0 |
| S12-final-viral-combined_k141_107623\|\|full | 2327 | No | NA | 2 | 0 | 0 | Low-quality | Genome-fragment | 4.25 | AAI-based (high-confidence) | 0 |
| S11-final-viral-combined_k141_189030\|\|full | 2328 | No | NA | 2 | 0 | 2 | Low-quality | Genome-fragment | 1.12 | AAI-based (medium-confidence) | 0 |
| S17-final-viral-combined_k141_232703\|\|full | 1710 | No | NA | 3 | 0 | 1 | Not-determined | Genome-fragment | NA | NA | 0 |
| S17-final-viral-combined_k141_187909\|\|full | 2469 | No | NA | 2 | 0 | 0 | Not-determined | Genome-fragment | NA | NA | 0 |
| S17-final-viral-combined_k141_41768\|\|full | 1683 | No | NA | 3 | 0 | 0 | Not-determined | Genome-fragment | NA | NA | 0 |
| S17-final-viral-combined_k141_143272\|\|full | 1632 | No | NA | 2 | 0 | 1 | Not-determined | Genome-fragment | NA | NA | 0 |
| S11_k141_281688 | 2328 | No | NA | 2 | 0 | 0 | Low-quality | Genome-fragment | 1.77 | AAI-based (high-confidence) | 0 |
| S3_k141_109264 | 2328 | No | NA | 2 | 0 | 1 | Low-quality | Genome-fragment | 3.96 | AAI-based (medium-confidence) | 0 |
| S17-final-viral-combined_k141_232740\|\|full | 4265 | No | NA | 4 | 0 | 4 | Not-determined | Genome-fragment | NA | NA | 0 |
| S3_k141_60225 | 2328 | No | NA | 2 | 0 | 1 | Low-quality | Genome-fragment | 3.53 | AAI-based (medium-confidence) | 0 |
| S17-final-viral-combined_k141_76773\|\|full | 1773 | No | NA | 3 | 0 | 2 | Not-determined | Genome-fragment | NA | NA | 0 |
| S17-final-viral-combined_k141_153950\|\|full | 1695 | No | NA | 3 | 0 | 1 | Not-determined | Genome-fragment | NA | NA | 0 |
| S17-final-viral-combined_k141_163925\|\|full | 1698 | No | NA | 2 | 0 | 2 | Not-determined | Genome-fragment | NA | NA | 0 |
| S17-final-viral-combined_k141_19003\|\|full | 1750 | No | NA | 2 | 0 | 2 | Not-determined | Genome-fragment | NA | NA | 0 |
| S1-final-viral-combined_k141_319466\|\|full | 2329 | No | NA | 5 | 1 | 0 | Low-quality | Genome-fragment | 3.91 | AAI-based (high-confidence) | 0 |
| S17-final-viral-combined_k141_96261\|\|full | 1688 | No | NA | 3 | 0 | 1 | Not-determined | Genome-fragment | NA | NA | 0 |
| S17-final-viral-combined_k141_76790\|\|full | 1490 | No | NA | 2 | 0 | 0 | Not-determined | Genome-fragment | NA | NA | 0 |
| S17-final-viral-combined_k141_199221\|\|full | 5010 | No | NA | 11 | 0 | 0 | Not-determined | Genome-fragment | NA | NA | 0 |
| S17-final-viral-combined_k141_153993\|\|full | 1681 | No | NA | 2 | 0 | 1 | Not-determined | Genome-fragment | NA | NA | 0 |
| S17-final-viral-combined_k141_30857\|\|full | 6439 | No | NA | 10 | 0 | 3 | Not-determined | Genome-fragment | NA | NA | 0 |
| S17-final-viral-combined_k141_199236\|\|full | 4461 | No | NA | 6 | 0 | 6 | Not-determined | Genome-fragment | NA | NA | 0 |
| S7_k141_72176 | 2329 | No | NA | 2 | 0 | 1 | Low-quality | Genome-fragment | 3.65 | AAI-based (medium-confidence) | 0 |
| S12-final-viral-combined_k141_48688\|\|full | 2329 | No | NA | 5 | 1 | 0 | Low-quality | Genome-fragment | 4.73 | HMM-based (lower-bound) | 0 |
| S12-final-viral-combined_k141_49813\|\|full | 2329 | Yes | 1505 | 4 | 1 | 2 | Low-quality | Genome-fragment | 1.14 | AAI-based (medium-confidence) | 35.38 |
| S17-final-viral-combined_k141_176248\|\|full | 2260 | No | NA | 2 | 0 | 1 | Not-determined | Genome-fragment | NA | NA | 0 |
| S17-final-viral-combined_k141_242388\|\|full | 3079 | No | NA | 3 | 0 | 1 | Not-determined | Genome-fragment | NA | NA | 0 |
| S17-final-viral-combined_k141_163950\|\|full | 1188 | No | NA | 3 | 0 | 2 | Not-determined | Genome-fragment | NA | NA | 0 |
| S17-final-viral-combined_k141_232832\|\|full | 3190 | No | NA | 3 | 0 | 1 | Not-determined | Genome-fragment | NA | NA | 0 |
| S1-final-viral-combined_k141_262877\|\|full | 2330 | No | NA | 8 | 0 | 0 | Low-quality | Genome-fragment | 3.76 | AAI-based (high-confidence) | 0 |
| S17-final-viral-combined_k141_199251\|\|full | 1223 | No | NA | 3 | 0 | 1 | Not-determined | Genome-fragment | NA | NA | 0 |
| S5-final-viral-combined_k141_135253\|\|lt2gene | 2330 | No | NA | 1 | 1 | 0 | Low-quality | Genome-fragment | 4.01 | AAI-based (medium-confidence) | 0 |
| S17-final-viral-combined_k141_163963\|\|full | 2668 | No | NA | 3 | 0 | 1 | Not-determined | Genome-fragment | NA | NA | 0 |
| S17-final-viral-combined_k141_131266\|\|full | 1458 | No | NA | 3 | 0 | 3 | Not-determined | Genome-fragment | NA | NA | 0 |
| S17-final-viral-combined_k141_210276\|\|full | 18605 | No | NA | 11 | 0 | 6 | Not-determined | Genome-fragment | NA | NA | 0 |
| S17-final-viral-combined_k141_19025\|\|full | 4032 | No | NA | 3 | 0 | 2 | Not-determined | Genome-fragment | NA | NA | 0 |
| S17-final-viral-combined_k141_163974\|\|full | 3496 | No | NA | 4 | 0 | 2 | Not-determined | Genome-fragment | NA | NA | 0 |
| S17-final-viral-combined_k141_65098\|\|full | 2351 | No | NA | 4 | 0 | 3 | Not-determined | Genome-fragment | NA | NA | 0 |
| S17-final-viral-combined_k141_221862\|\|full | 3397 | No | NA | 4 | 0 | 3 | Not-determined | Genome-fragment | NA | NA | 0 |
| S17-final-viral-combined_k141_242463\|\|full | 1418 | No | NA | 3 | 0 | 1 | Not-determined | Genome-fragment | NA | NA | 0 |
| S17-final-viral-combined_k141_65129\|\|full | 1151 | No | NA | 3 | 0 | 0 | Not-determined | Genome-fragment | NA | NA | 0 |
| S17-final-viral-combined_k141_210309\|\|full | 5252 | No | NA | 6 | 0 | 0 | Not-determined | Genome-fragment | NA | NA | 0 |
| S7-final-viral-combined_k141_204505\|\|full | 2330 | No | NA | 3 | 0 | 2 | Low-quality | Genome-fragment | 1.75 | AAI-based (high-confidence) | 0 |
| S17-final-viral-combined_k141_51685\|\|full | 1292 | No | NA | 2 | 0 | 1 | Not-determined | Genome-fragment | NA | NA | 0 |
| S17-final-viral-combined_k141_80102\|\|full | 2330 | No | NA | 3 | 0 | 3 | Low-quality | Genome-fragment | 1.29 | AAI-based (medium-confidence) | 0 |
| S12-final-viral-combined_k141_321413\|\|full | 2330 | No | NA | 4 | 1 | 0 | Low-quality | Genome-fragment | 1.46 | HMM-based (lower-bound) | 0 |
| S12_k141_126776 | 2330 | No | NA | 1 | 0 | 1 | Low-quality | Genome-fragment | 1.07 | AAI-based (medium-confidence) | 0 |
| S17-final-viral-combined_k141_263850\|\|full | 1353 | No | NA | 3 | 0 | 1 | Not-determined | Genome-fragment | NA | NA | 0 |
| S2-final-viral-combined_k141_147022\|\|0_partial | 2330 | No | NA | 2 | 1 | 0 | Low-quality | Genome-fragment | 6.63 | AAI-based (high-confidence) | 0 |
| S17-final-viral-combined_k141_30924\|\|full | 18303 | No | NA | 10 | 0 | 7 | Not-determined | Genome-fragment | NA | NA | 0 |
| S4-final-viral-combined_k141_44423\|\|full | 2330 | No | NA | 3 | 0 | 2 | Low-quality | Genome-fragment | 6.75 | AAI-based (medium-confidence) | 0 |
| S17-final-viral-combined_k141_263867\|\|full | 1091 | No | NA | 2 | 0 | 2 | Not-determined | Genome-fragment | NA | NA | 0 |
| S11-final-viral-combined_k141_216106\|\|full | 2331 | No | NA | 3 | 0 | 2 | Low-quality | Genome-fragment | 4.16 | AAI-based (medium-confidence) | 0 |
| S17-final-viral-combined_k141_8682\|\|full | 1224 | No | NA | 2 | 0 | 2 | Not-determined | Genome-fragment | NA | NA | 0 |
| S12-final-viral-combined_k141_252602\|\|full | 2331 | No | NA | 4 | 1 | 0 | Low-quality | Genome-fragment | 7.32 | AAI-based (high-confidence) | 0 |
| S11-final-viral-combined_k141_45524\|\|full | 2332 | No | NA | 4 | 0 | 3 | Low-quality | Genome-fragment | 1.48 | AAI-based (medium-confidence) | 0 |
| S17-final-viral-combined_k141_76903\|\|full | 1620 | No | NA | 2 | 0 | 2 | Not-determined | Genome-fragment | NA | NA | 0 |
| S17-final-viral-combined_k141_252371\|\|full | 1457 | No | NA | 3 | 0 | 0 | Not-determined | Genome-fragment | NA | NA | 0 |
| S11-final-viral-combined_k141_294839\|\|full | 2332 | No | NA | 7 | 5 | 0 | Low-quality | Genome-fragment | 6 | AAI-based (high-confidence) | 0 |
| S6-final-viral-combined_k141_94018\|\|full | 2332 | No | NA | 3 | 0 | 1 | Low-quality | Genome-fragment | 4.25 | AAI-based (medium-confidence) | 0 |
| S17-final-viral-combined_k141_242541\|\|full | 2055 | No | NA | 2 | 0 | 2 | Not-determined | Genome-fragment | NA | NA | 0 |
| S11_k141_143394 | 2333 | No | NA | 4 | 0 | 2 | Low-quality | Genome-fragment | 1.07 | AAI-based (medium-confidence) | 0 |
| S17-final-viral-combined_k141_76921\|\|full | 2150 | No | NA | 2 | 0 | 1 | Not-determined | Genome-fragment | NA | NA | 0 |
| S17-final-viral-combined_k141_242550\|\|full | 2459 | No | NA | 4 | 0 | 2 | Not-determined | Genome-fragment | NA | NA | 0 |
| S1-final-viral-combined_k141_85413\|\|full | 2333 | No | NA | 6 | 1 | 0 | Low-quality | Genome-fragment | 3.94 | AAI-based (high-confidence) | 0 |
| S17-final-viral-combined_k141_131405\|\|full | 2663 | No | NA | 5 | 0 | 0 | Not-determined | Genome-fragment | NA | NA | 0 |
| S1-final-viral-combined_k141_343898\|\|full | 2333 | No | NA | 3 | 0 | 1 | Low-quality | Genome-fragment | 4.63 | AAI-based (high-confidence) | 0 |
| S3-final-viral-combined_k141_129181\|\|full | 2333 | No | NA | 3 | 0 | 0 | Low-quality | Genome-fragment | 5.36 | AAI-based (medium-confidence) | 0 |
| S17-final-viral-combined_k141_170078\|\|full | 2333 | No | NA | 2 | 0 | 1 | Low-quality | Genome-fragment | 1.2 | AAI-based (medium-confidence) | 0 |
| S17-final-viral-combined_k141_139720\|\|full | 2333 | No | NA | 3 | 0 | 0 | Low-quality | Genome-fragment | 4.18 | AAI-based (medium-confidence) | 0 |
| S17-final-viral-combined_k141_210470\|\|full | 1481 | No | NA | 2 | 0 | 2 | Not-determined | Genome-fragment | NA | NA | 0 |
| S17-final-viral-combined_k141_233031\|\|full | 1415 | No | NA | 3 | 0 | 0 | Not-determined | Genome-fragment | NA | NA | 0 |
| S4-final-viral-combined_k141_154051\|\|full | 2333 | No | NA | 4 | 1 | 0 | Low-quality | Genome-fragment | 6.21 | AAI-based (medium-confidence) | 0 |
| S17-final-viral-combined_k141_154207\|\|full | 1291 | No | NA | 2 | 0 | 1 | Not-determined | Genome-fragment | NA | NA | 0 |
| S17-final-viral-combined_k141_8791\|\|full | 2292 | No | NA | 3 | 0 | 2 | Not-determined | Genome-fragment | NA | NA | 0 |
| S17-final-viral-combined_k141_31036\|\|full | 1512 | No | NA | 3 | 0 | 0 | Not-determined | Genome-fragment | NA | NA | 0 |
| S17-final-viral-combined_k141_131444\|\|full | 3802 | No | NA | 5 | 0 | 1 | Not-determined | Genome-fragment | NA | NA | 0 |
| S17-final-viral-combined_k141_221990\|\|full | 1654 | No | NA | 2 | 0 | 0 | Not-determined | Genome-fragment | NA | NA | 0 |
| S4-final-viral-combined_k141_29833\|\|full | 2333 | No | NA | 2 | 0 | 0 | Low-quality | Genome-fragment | 1.54 | AAI-based (high-confidence) | 0 |
| S17-final-viral-combined_k141_8795\|\|full | 1241 | No | NA | 2 | 0 | 0 | Not-determined | Genome-fragment | NA | NA | 0 |
| S17-final-viral-combined_k141_88574\|\|full | 4500 | No | NA | 2 | 0 | 1 | Not-determined | Genome-fragment | NA | NA | 0 |
| S3_k141_59314 | 2334 | No | NA | 1 | 0 | 1 | Low-quality | Genome-fragment | 0.62 | AAI-based (high-confidence) | 0 |
| S7-final-viral-combined_k141_64524\|\|full | 2334 | No | NA | 4 | 1 | 0 | Low-quality | Genome-fragment | 4.94 | AAI-based (high-confidence) | 0 |
| S7_k141_114612 | 2334 | No | NA | 1 | 0 | 0 | Low-quality | Genome-fragment | 1.53 | AAI-based (high-confidence) | 0 |
| S17-final-viral-combined_k141_176498\|\|full | 1401 | No | NA | 2 | 0 | 1 | Not-determined | Genome-fragment | NA | NA | 0 |
| S17-final-viral-combined_k141_76988\|\|full | 2462 | No | NA | 3 | 0 | 3 | Not-determined | Genome-fragment | NA | NA | 0 |
| S17-final-viral-combined_k141_109310\|\|full | 2907 | No | NA | 3 | 0 | 3 | Not-determined | Genome-fragment | NA | NA | 0 |
| S17-final-viral-combined_k141_233033\|\|full | 3872 | No | NA | 4 | 0 | 2 | Not-determined | Genome-fragment | NA | NA | 0 |
| S17-final-viral-combined_k141_37775\|\|full | 2334 | No | NA | 7 | 0 | 1 | Low-quality | Genome-fragment | 4.52 | AAI-based (medium-confidence) | 0 |
| S17-final-viral-combined_k141_252454\|\|full | 1485 | No | NA | 2 | 0 | 2 | Not-determined | Genome-fragment | NA | NA | 0 |
| S6-final-viral-combined_k141_11013\|\|full | 2334 | No | NA | 5 | 0 | 0 | Low-quality | Genome-fragment | 0.74 | AAI-based (medium-confidence) | 0 |
| S17-final-viral-combined_k141_109319\|\|full | 2282 | No | NA | 5 | 0 | 4 | Not-determined | Genome-fragment | NA | NA | 0 |
| S17-final-viral-combined_k141_199440\|\|full | 27757 | No | NA | 41 | 10 | 1 | Medium-quality | Genome-fragment | 61.84 | AAI-based (high-confidence) | 0 |
| S9-final-viral-combined_k141_76387\|\|full | 2335 | No | NA | 3 | 0 | 0 | Low-quality | Genome-fragment | 3.06 | AAI-based (high-confidence) | 0 |
| S11-final-viral-combined_k141_23173\|\|full | 2335 | No | NA | 4 | 0 | 2 | Low-quality | Genome-fragment | 1.09 | AAI-based (medium-confidence) | 0 |
| S17-final-viral-combined_k141_8847\|\|full | 1416 | No | NA | 2 | 0 | 2 | Not-determined | Genome-fragment | NA | NA | 0 |
| S17-final-viral-combined_k141_122164\|\|full | 2623 | No | NA | 3 | 0 | 1 | Not-determined | Genome-fragment | NA | NA | 0 |
| S17-final-viral-combined_k141_210567\|\|full | 1185 | No | NA | 3 | 0 | 1 | Not-determined | Genome-fragment | NA | NA | 0 |
| S1-final-viral-combined_k141_2636\|\|full | 2335 | No | NA | 4 | 0 | 2 | Low-quality | Genome-fragment | 2.82 | AAI-based (high-confidence) | 0 |
| S17-final-viral-combined_k141_143636\|\|full | 1230 | No | NA | 4 | 0 | 0 | Not-determined | Genome-fragment | NA | NA | 0 |
| S17-final-viral-combined_k141_222067\|\|full | 5764 | No | NA | 6 | 0 | 3 | Not-determined | Genome-fragment | NA | NA | 0 |
| S17-final-viral-combined_k141_42079\|\|full | 1785 | No | NA | 4 | 0 | 2 | Not-determined | Genome-fragment | NA | NA | 0 |
| S17-final-viral-combined_k141_131504\|\|full | 2087 | No | NA | 2 | 0 | 0 | Not-determined | Genome-fragment | NA | NA | 0 |
| S17-final-viral-combined_k141_164220\|\|full | 1172 | No | NA | 2 | 0 | 1 | Not-determined | Genome-fragment | NA | NA | 0 |
| S17-final-viral-combined_k141_88670\|\|full | 3834 | No | NA | 4 | 0 | 2 | Not-determined | Genome-fragment | NA | NA | 0 |
| S17-final-viral-combined_k141_210606\|\|full | 1226 | No | NA | 3 | 0 | 3 | Not-determined | Genome-fragment | NA | NA | 0 |
| S17-final-viral-combined_k141_222077\|\|full | 8222 | No | NA | 8 | 0 | 6 | Not-determined | Genome-fragment | NA | NA | 0 |
| S17-final-viral-combined_k141_252537\|\|full | 1928 | No | NA | 3 | 0 | 0 | Not-determined | Genome-fragment | NA | NA | 0 |
| S17-final-viral-combined_k141_109394\|\|full | 1202 | No | NA | 1 | 0 | 1 | Not-determined | Genome-fragment | NA | NA | 0 |
| S17-final-viral-combined_k141_131538\|\|full | 3441 | No | NA | 3 | 0 | 2 | Not-determined | Genome-fragment | NA | NA | 0 |
| S17-final-viral-combined_k141_188299\|\|full | 3003 | No | NA | 2 | 0 | 1 | Not-determined | Genome-fragment | NA | NA | 0 |
| S17-final-viral-combined_k141_154356\|\|full | 3197 | No | NA | 3 | 0 | 2 | Not-determined | Genome-fragment | NA | NA | 0 |
| S17-final-viral-combined_k141_88705\|\|full | 1485 | No | NA | 2 | 0 | 1 | Not-determined | Genome-fragment | NA | NA | 0 |
| S7-final-viral-combined_k141_55931\|\|full | 2335 | No | NA | 3 | 1 | 0 | Low-quality | Genome-fragment | 3.09 | HMM-based (lower-bound) | 0 |
| S12-final-viral-combined_k141_282058\|\|full | 2335 | No | NA | 3 | 0 | 2 | Low-quality | Genome-fragment | 1.12 | AAI-based (high-confidence) | 0 |
| S12-final-viral-combined_k141_350666\|\|full | 2335 | No | NA | 4 | 1 | 0 | Low-quality | Genome-fragment | 1.43 | AAI-based (medium-confidence) | 0 |
| S2-final-viral-combined_k141_208078\|\|full | 2335 | No | NA | 3 | 0 | 0 | Low-quality | Genome-fragment | 1.42 | AAI-based (medium-confidence) | 0 |
| S17-final-viral-combined_k141_188330\|\|full | 1120 | No | NA | 3 | 0 | 0 | Not-determined | Genome-fragment | NA | NA | 0 |
| S17-final-viral-combined_k141_199532\|\|full | 1998 | No | NA | 4 | 0 | 3 | Not-determined | Genome-fragment | NA | NA | 0 |
| S17-final-viral-combined_k141_77063\|\|full | 2943 | No | NA | 2 | 0 | 2 | Not-determined | Genome-fragment | NA | NA | 0 |
| S2-final-viral-combined_k141_24712\|\|full | 2335 | No | NA | 3 | 1 | 0 | Low-quality | Genome-fragment | 3.67 | AAI-based (high-confidence) | 0 |
| S17-final-viral-combined_k141_147563\|\|full | 2336 | No | NA | 3 | 0 | 1 | Low-quality | Genome-fragment | 5.36 | AAI-based (high-confidence) | 0 |
| S17-final-viral-combined_k141_210739\|\|full | 1078 | No | NA | 2 | 0 | 2 | Not-determined | Genome-fragment | NA | NA | 0 |
| S17-final-viral-combined_k141_149913\|\|full | 2336 | No | NA | 6 | 1 | 0 | Low-quality | Genome-fragment | 3.62 | AAI-based (medium-confidence) | 0 |
| S17-final-viral-combined_k141_154398\|\|full | 6818 | No | NA | 6 | 0 | 5 | Not-determined | Genome-fragment | NA | NA | 0 |
| S17-final-viral-combined_k141_51778\|\|full | 1708 | No | NA | 4 | 0 | 1 | Not-determined | Genome-fragment | NA | NA | 0 |
| S12-final-viral-combined_k141_77185\|\|full | 2336 | No | NA | 2 | 0 | 0 | Low-quality | Genome-fragment | 3.53 | AAI-based (high-confidence) | 0 |
| S11-final-viral-combined_k141_97828\|\|full | 2337 | No | NA | 4 | 0 | 2 | Low-quality | Genome-fragment | 1.06 | AAI-based (high-confidence) | 0 |
| S17-final-viral-combined_k141_252592\|\|full | 1573 | No | NA | 2 | 0 | 1 | Not-determined | Genome-fragment | NA | NA | 0 |
| S17-final-viral-combined_k141_176691\|\|full | 1199 | No | NA | 2 | 0 | 1 | Not-determined | Genome-fragment | NA | NA | 0 |
| S17-final-viral-combined_k141_19336\|\|full | 1291 | No | NA | 2 | 0 | 2 | Not-determined | Genome-fragment | NA | NA | 0 |
| S7-final-viral-combined_k141_211846\|\|lt2gene | 2337 | No | NA | 2 | 2 | 0 | Low-quality | Genome-fragment | 3.28 | HMM-based (lower-bound) | 0 |
| S17-final-viral-combined_k141_131592\|\|full | 1835 | No | NA | 3 | 0 | 0 | Not-determined | Genome-fragment | NA | NA | 0 |
| S17-final-viral-combined_k141_176695\|\|full | 3112 | No | NA | 4 | 0 | 0 | Not-determined | Genome-fragment | NA | NA | 0 |
| S12-final-viral-combined_k141_48035\|\|lt2gene | 2337 | No | NA | 2 | 1 | 0 | Low-quality | Genome-fragment | 2.27 | HMM-based (lower-bound) | 0 |
| S4_k141_20972 | 2337 | No | NA | 3 | 0 | 2 | Low-quality | Genome-fragment | 1.53 | AAI-based (medium-confidence) | 0 |
| S17-final-viral-combined_k141_233228\|\|full | 4368 | No | NA | 3 | 0 | 2 | Not-determined | Genome-fragment | NA | NA | 0 |
| S17-final-viral-combined_k141_143779\|\|full | 6695 | No | NA | 3 | 0 | 3 | Not-determined | Genome-fragment | NA | NA | 0 |
| S17-final-viral-combined_k141_222197\|\|full | 2010 | No | NA | 4 | 0 | 0 | Not-determined | Genome-fragment | NA | NA | 0 |
| S1-final-viral-combined_k141_255946\|\|full | 2338 | No | NA | 3 | 1 | 0 | Low-quality | Genome-fragment | 3.14 | HMM-based (lower-bound) | 0 |
| S17-final-viral-combined_k141_199615\|\|full | 3347 | No | NA | 4 | 0 | 1 | Not-determined | Genome-fragment | NA | NA | 0 |
| S17-final-viral-combined_k141_9119\|\|full | 1327 | No | NA | 2 | 0 | 2 | Not-determined | Genome-fragment | NA | NA | 0 |
| S17-final-viral-combined_k141_131663\|\|full | 1330 | No | NA | 3 | 0 | 2 | Not-determined | Genome-fragment | NA | NA | 0 |
| S12-final-viral-combined_k141_42924\|\|full | 2338 | No | NA | 2 | 0 | 2 | Low-quality | Genome-fragment | 0.4 | AAI-based (medium-confidence) | 0 |
| S11-final-viral-combined_k141_211685\|\|full | 2339 | No | NA | 4 | 0 | 3 | Low-quality | Genome-fragment | 1.06 | AAI-based (high-confidence) | 0 |
| S17-final-viral-combined_k141_242723\|\|full | 3952 | No | NA | 4 | 0 | 1 | Not-determined | Genome-fragment | NA | NA | 0 |
| S17-final-viral-combined_k141_154496\|\|full | 2507 | No | NA | 4 | 0 | 1 | Not-determined | Genome-fragment | NA | NA | 0 |
| S17-final-viral-combined_k141_154499\|\|full | 3076 | No | NA | 3 | 0 | 1 | Not-determined | Genome-fragment | NA | NA | 0 |
| S17-final-viral-combined_k141_264129\|\|full | 2176 | No | NA | 3 | 0 | 2 | Not-determined | Genome-fragment | NA | NA | 0 |
| S17-final-viral-combined_k141_233299\|\|full | 2773 | No | NA | 3 | 0 | 3 | Not-determined | Genome-fragment | NA | NA | 0 |
| S11-final-viral-combined_k141_295029\|\|full | 2339 | No | NA | 4 | 0 | 0 | Low-quality | Genome-fragment | 4.99 | AAI-based (high-confidence) | 0 |
| S17-final-viral-combined_k141_122221\|\|full | 5250 | No | NA | 5 | 0 | 3 | Not-determined | Genome-fragment | NA | NA | 0 |
| S17-final-viral-combined_k141_27172\|\|full | 2339 | No | NA | 3 | 0 | 0 | Low-quality | Genome-fragment | 5.44 | AAI-based (high-confidence) | 0 |
| S17-final-viral-combined_k141_164346\|\|full | 1547 | No | NA | 3 | 0 | 1 | Not-determined | Genome-fragment | NA | NA | 0 |
| S12-final-viral-combined_k141_25191\|\|full | 2339 | No | NA | 3 | 1 | 0 | Low-quality | Genome-fragment | 6.14 | AAI-based (high-confidence) | 0 |
| S12_k141_286734 | 2339 | No | NA | 2 | 0 | 1 | Low-quality | Genome-fragment | 4.71 | AAI-based (medium-confidence) | 0 |
| S17-final-viral-combined_k141_176816\|\|full | 2424 | No | NA | 3 | 0 | 3 | Not-determined | Genome-fragment | NA | NA | 0 |
| S17-final-viral-combined_k141_131757\|\|full | 1537 | No | NA | 3 | 0 | 2 | Not-determined | Genome-fragment | NA | NA | 0 |
| S1-final-viral-combined_k141_391453\|\|full | 2340 | No | NA | 3 | 3 | 0 | Low-quality | Genome-fragment | 5.03 | AAI-based (high-confidence) | 0 |
| S1_k141_388250 | 2340 | No | NA | 2 | 0 | 1 | Low-quality | Genome-fragment | 3.38 | AAI-based (medium-confidence) | 0 |
| S17-final-viral-combined_k141_65520\|\|full | 1508 | No | NA | 2 | 0 | 0 | Not-determined | Genome-fragment | NA | NA | 0 |
| S3-final-viral-combined_k141_30083\|\|full | 2340 | No | NA | 6 | 2 | 0 | Low-quality | Genome-fragment | 6.05 | AAI-based (high-confidence) | 0 |
| S17-final-viral-combined_k141_131776\|\|full | 3348 | No | NA | 3 | 0 | 2 | Not-determined | Genome-fragment | NA | NA | 0 |
| S17-final-viral-combined_k141_65528\|\|full | 1288 | No | NA | 6 | 0 | 0 | Not-determined | Genome-fragment | NA | NA | 0 |
| S7-final-viral-combined_k141_35819\|\|full | 2340 | No | NA | 3 | 0 | 2 | Low-quality | Genome-fragment | 1.16 | AAI-based (medium-confidence) | 0 |
| S17-final-viral-combined_k141_154555\|\|full | 1088 | No | NA | 3 | 0 | 0 | Not-determined | Genome-fragment | NA | NA | 0 |
| S17-final-viral-combined_k141_222249\|\|full | 3962 | No | NA | 7 | 0 | 0 | Not-determined | Genome-fragment | NA | NA | 0 |
| S17-final-viral-combined_k141_31301\|\|full | 2328 | No | NA | 3 | 0 | 2 | Not-determined | Genome-fragment | NA | NA | 0 |
| S17-final-viral-combined_k141_222255\|\|full | 1079 | No | NA | 2 | 0 | 1 | Not-determined | Genome-fragment | NA | NA | 0 |
| S2-final-viral-combined_k141_170886\|\|full | 2340 | No | NA | 4 | 0 | 2 | Low-quality | Genome-fragment | 6.48 | AAI-based (high-confidence) | 0 |
| S17-final-viral-combined_k141_242828\|\|full | 3582 | No | NA | 4 | 0 | 3 | Not-determined | Genome-fragment | NA | NA | 0 |
| S11-final-viral-combined_k141_324248\|\|full | 2341 | No | NA | 5 | 5 | 0 | Low-quality | Genome-fragment | 4.81 | AAI-based (high-confidence) | 0 |
| S17-final-viral-combined_k141_9220\|\|full | 3321 | No | NA | 4 | 0 | 0 | Not-determined | Genome-fragment | NA | NA | 0 |
| S17-final-viral-combined_k141_51901\|\|full | 1226 | No | NA | 2 | 0 | 2 | Not-determined | Genome-fragment | NA | NA | 0 |
| S17-final-viral-combined_k141_42353\|\|full | 2807 | No | NA | 2 | 0 | 0 | Not-determined | Genome-fragment | NA | NA | 0 |
| S17-final-viral-combined_k141_19487\|\|full | 1402 | No | NA | 2 | 0 | 0 | Not-determined | Genome-fragment | NA | NA | 0 |
| S17-final-viral-combined_k141_65603\|\|full | 2966 | No | NA | 3 | 0 | 2 | Not-determined | Genome-fragment | NA | NA | 0 |
| S3-final-viral-combined_k141_98003\|\|full | 2341 | No | NA | 3 | 0 | 2 | Low-quality | Genome-fragment | 4.42 | AAI-based (medium-confidence) | 0 |
| S17-final-viral-combined_k141_131836\|\|full | 3520 | No | NA | 6 | 0 | 3 | Not-determined | Genome-fragment | NA | NA | 0 |
| S7-final-viral-combined_k141_212490\|\|full | 2341 | No | NA | 2 | 0 | 1 | Low-quality | Genome-fragment | 2.19 | AAI-based (high-confidence) | 0 |
| S17-final-viral-combined_k141_213570\|\|full | 2341 | No | NA | 3 | 0 | 1 | Low-quality | Genome-fragment | 1.03 | AAI-based (medium-confidence) | 0 |
| S17-final-viral-combined_k141_19515\|\|full | 1378 | No | NA | 3 | 0 | 0 | Not-determined | Genome-fragment | NA | NA | 0 |
| S17-final-viral-combined_k141_109700\|\|full | 1801 | No | NA | 4 | 0 | 1 | Not-determined | Genome-fragment | NA | NA | 0 |
| S17-final-viral-combined_k141_143904\|\|full | 5224 | No | NA | 6 | 0 | 6 | Not-determined | Genome-fragment | NA | NA | 0 |
| S17-final-viral-combined_k141_188486\|\|full | 3483 | No | NA | 3 | 0 | 1 | Not-determined | Genome-fragment | NA | NA | 0 |
| S8-final-viral-combined_k141_95267\|\|full | 2341 | No | NA | 3 | 0 | 1 | Low-quality | Genome-fragment | 1.63 | AAI-based (high-confidence) | 0 |
| S17-final-viral-combined_k141_242885\|\|full | 1244 | No | NA | 3 | 0 | 1 | Not-determined | Genome-fragment | NA | NA | 0 |
| S9-final-viral-combined_k141_62601\|\|full | 2342 | No | NA | 3 | 2 | 0 | Low-quality | Genome-fragment | 5.54 | AAI-based (high-confidence) | 0 |
| S17-final-viral-combined_k141_264251\|\|full | 1024 | No | NA | 3 | 0 | 1 | Not-determined | Genome-fragment | NA | NA | 0 |
| S17-final-viral-combined_k141_51997\|\|full | 1155 | No | NA | 1 | 0 | 0 | Not-determined | Genome-fragment | NA | NA | 0 |
| S3_k141_51167 | 2342 | No | NA | 3 | 0 | 0 | Low-quality | Genome-fragment | 2.22 | AAI-based (high-confidence) | 0 |
| S17-final-viral-combined_k141_96837\|\|full | 1037 | No | NA | 2 | 0 | 0 | Not-determined | Genome-fragment | NA | NA | 0 |
| S17-final-viral-combined_k141_242920\|\|full | 2828 | No | NA | 3 | 0 | 3 | Not-determined | Genome-fragment | NA | NA | 0 |
| S7-final-viral-combined_k141_400\|\|full | 2342 | No | NA | 8 | 0 | 0 | Low-quality | Genome-fragment | 4.91 | AAI-based (high-confidence) | 0 |
| S7-final-viral-combined_k141_48896\|\|full | 2342 | No | NA | 3 | 0 | 3 | Low-quality | Genome-fragment | 7.04 | AAI-based (medium-confidence) | 0 |
| S11-final-viral-combined_k141_38900\|\|full | 2343 | No | NA | 4 | 2 | 0 | Low-quality | Genome-fragment | 2.88 | HMM-based (lower-bound) | 0 |
| S17-final-viral-combined_k141_199746\|\|full | 59348 | Yes | 41288 | 64 | 15 | 4 | High-quality | High-quality | 100 | AAI-based (high-confidence) | 30.43 |
| S1-final-viral-combined_k141_105032\|\|full | 2343 | No | NA | 5 | 0 | 0 | Low-quality | Genome-fragment | 2.03 | AAI-based (medium-confidence) | 0 |
| S17-final-viral-combined_k141_177044\|\|full | 1922 | No | NA | 4 | 0 | 1 | Not-determined | Genome-fragment | NA | NA | 0 |
| S17-final-viral-combined_k141_132032\|\|full | 2579 | No | NA | 4 | 0 | 3 | Not-determined | Genome-fragment | NA | NA | 0 |
| S3-final-viral-combined_k141_30527\|\|full | 2343 | No | NA | 4 | 1 | 0 | Low-quality | Genome-fragment | 1.61 | HMM-based (lower-bound) | 0 |
| S17-final-viral-combined_k141_164544\|\|full | 1219 | No | NA | 3 | 0 | 2 | Not-determined | Genome-fragment | NA | NA | 0 |
| S17-final-viral-combined_k141_200355\|\|full | 2343 | No | NA | 2 | 2 | 0 | Low-quality | Genome-fragment | 5.03 | AAI-based (high-confidence) | 0 |
| S17-final-viral-combined_k141_252805\|\|full | 3716 | No | NA | 4 | 0 | 0 | Not-determined | Genome-fragment | NA | NA | 0 |
| S17-final-viral-combined_k141_31511\|\|full | 1941 | No | NA | 3 | 0 | 1 | Not-determined | Genome-fragment | NA | NA | 0 |
| S12-final-viral-combined_k141_227240\|\|full | 2343 | No | NA | 4 | 2 | 0 | Low-quality | Genome-fragment | 5.2 | AAI-based (high-confidence) | 0 |
| S2-final-viral-combined_k141_124426\|\|full | 2343 | No | NA | 4 | 0 | 0 | Low-quality | Genome-fragment | 1.83 | AAI-based (medium-confidence) | 0 |
| S17-final-viral-combined_k141_188597\|\|full | 4080 | No | NA | 3 | 0 | 0 | Not-determined | Genome-fragment | NA | NA | 0 |
| S8-final-viral-combined_k141_256735\|\|full | 2343 | No | NA | 4 | 3 | 0 | Low-quality | Genome-fragment | 4.29 | AAI-based (high-confidence) | 0 |
| S9-final-viral-combined_k141_127690\|\|full | 2344 | No | NA | 3 | 0 | 2 | Low-quality | Genome-fragment | 0.57 | AAI-based (medium-confidence) | 0 |
| S17-final-viral-combined_k141_65785\|\|full | 1629 | No | NA | 4 | 0 | 0 | Not-determined | Genome-fragment | NA | NA | 0 |
| S17-final-viral-combined_k141_211045\|\|full | 1077 | No | NA | 3 | 0 | 1 | Not-determined | Genome-fragment | NA | NA | 0 |
| S1-final-viral-combined_k141_313966\|\|full | 2344 | No | NA | 3 | 0 | 0 | Low-quality | Genome-fragment | 5.22 | AAI-based (medium-confidence) | 0 |
| S12-final-viral-combined_k141_336211\|\|full | 2344 | No | NA | 3 | 0 | 3 | Low-quality | Genome-fragment | 1.08 | AAI-based (medium-confidence) | 0 |
| S2-final-viral-combined_k141_89452\|\|full | 2344 | No | NA | 4 | 2 | 0 | Low-quality | Genome-fragment | 2.88 | HMM-based (lower-bound) | 0 |
| S17-final-viral-combined_k141_264322\|\|full | 10319 | No | NA | 8 | 0 | 0 | Not-determined | Genome-fragment | NA | NA | 0 |
| S9-final-viral-combined_k141_23848\|\|full | 2345 | No | NA | 8 | 2 | 0 | Low-quality | Genome-fragment | 4.37 | AAI-based (high-confidence) | 0 |
| S11-final-viral-combined_k141_193615\|\|full | 2345 | No | NA | 4 | 3 | 0 | Low-quality | Genome-fragment | 3.26 | HMM-based (lower-bound) | 0 |
| S17-final-viral-combined_k141_154784\|\|full | 1755 | No | NA | 2 | 0 | 2 | Not-determined | Genome-fragment | NA | NA | 0 |
| S11-final-viral-combined_k141_4424\|\|full | 2345 | Yes | 998 | 4 | 1 | 1 | Low-quality | Genome-fragment | 1.53 | AAI-based (medium-confidence) | 57.44 |
| S1-final-viral-combined_k141_351782\|\|full | 2345 | No | NA | 3 | 0 | 3 | Low-quality | Genome-fragment | 1.53 | AAI-based (medium-confidence) | 0 |
| S17-final-viral-combined_k141_164655\|\|full | 1801 | No | NA | 3 | 0 | 0 | Not-determined | Genome-fragment | NA | NA | 0 |
| S17-final-viral-combined_k141_19625\|\|full | 3998 | No | NA | 9 | 0 | 2 | Not-determined | Genome-fragment | NA | NA | 0 |
| S5-final-viral-combined_k141_29468\|\|full | 2345 | No | NA | 3 | 0 | 0 | Low-quality | Genome-fragment | 2.59 | AAI-based (high-confidence) | 0 |
| S17-final-viral-combined_k141_31621\|\|full | 4931 | No | NA | 3 | 0 | 3 | Not-determined | Genome-fragment | NA | NA | 0 |
| S2-final-viral-combined_k141_235110\|\|full | 2345 | Yes | 1416 | 2 | 1 | 1 | Low-quality | Genome-fragment | 0.61 | AAI-based (medium-confidence) | 39.62 |
| S4_k141_125094 | 2345 | No | NA | 7 | 4 | 0 | Low-quality | Genome-fragment | 6.79 | AAI-based (high-confidence) | 0 |
| S17-final-viral-combined_k141_9470\|\|full | 1433 | No | NA | 2 | 0 | 1 | Not-determined | Genome-fragment | NA | NA | 0 |
| S17-final-viral-combined_k141_31632\|\|full | 1417 | No | NA | 2 | 0 | 0 | Not-determined | Genome-fragment | NA | NA | 0 |
| S1_k141_365901 | 2346 | No | NA | 1 | 0 | 1 | Low-quality | Genome-fragment | 1.19 | AAI-based (medium-confidence) | 0 |
| S17-final-viral-combined_k141_164683\|\|full | 4657 | No | NA | 4 | 0 | 2 | Not-determined | Genome-fragment | NA | NA | 0 |
| S4-final-viral-combined_k141_70715\|\|full | 2346 | No | NA | 3 | 1 | 0 | Low-quality | Genome-fragment | 5.3 | AAI-based (medium-confidence) | 0 |
| S17-final-viral-combined_k141_252910\|\|full | 1287 | No | NA | 2 | 0 | 1 | Not-determined | Genome-fragment | NA | NA | 0 |
| S17-final-viral-combined_k141_9528\|\|full | 1742 | No | NA | 3 | 0 | 2 | Not-determined | Genome-fragment | NA | NA | 0 |
| S9-final-viral-combined_k141_36764\|\|full | 2347 | No | NA | 2 | 0 | 0 | Low-quality | Genome-fragment | 3.67 | AAI-based (medium-confidence) | 0 |
| S17-final-viral-combined_k141_211141\|\|full | 1374 | No | NA | 2 | 0 | 0 | Not-determined | Genome-fragment | NA | NA | 0 |
| S17-final-viral-combined_k141_154831\|\|full | 3372 | No | NA | 3 | 0 | 0 | Not-determined | Genome-fragment | NA | NA | 0 |
| S1-final-viral-combined_k141_296353\|\|full | 2347 | No | NA | 3 | 0 | 0 | Low-quality | Genome-fragment | 3.78 | AAI-based (medium-confidence) | 0 |
| S1_k141_240407 | 2347 | No | NA | 3 | 0 | 0 | Low-quality | Genome-fragment | 2.03 | AAI-based (high-confidence) | 0 |
| S7-final-viral-combined_k141_170634\|\|full | 2347 | No | NA | 5 | 2 | 0 | Low-quality | Genome-fragment | 4.78 | AAI-based (medium-confidence) | 0 |
| S17-final-viral-combined_k141_154894\|\|full | 2579 | No | NA | 3 | 0 | 2 | Not-determined | Genome-fragment | NA | NA | 0 |
| S17-final-viral-combined_k141_164773\|\|full | 4601 | No | NA | 5 | 0 | 3 | Not-determined | Genome-fragment | NA | NA | 0 |
| S17-final-viral-combined_k141_13280\|\|full | 2347 | No | NA | 2 | 1 | 0 | Low-quality | Genome-fragment | 5 | AAI-based (high-confidence) | 0 |
| S17-final-viral-combined_k141_109863\|\|full | 1923 | No | NA | 2 | 0 | 1 | Not-determined | Genome-fragment | NA | NA | 0 |
| S2-final-viral-combined_k141_168799\|\|full | 2347 | No | NA | 4 | 0 | 1 | Low-quality | Genome-fragment | 4.4 | AAI-based (medium-confidence) | 0 |
| S17-final-viral-combined_k141_31718\|\|full | 2993 | No | NA | 3 | 0 | 3 | Not-determined | Genome-fragment | NA | NA | 0 |
| S17-final-viral-combined_k141_42743\|\|full | 1678 | No | NA | 3 | 0 | 2 | Not-determined | Genome-fragment | NA | NA | 0 |
| S17-final-viral-combined_k141_31727\|\|full | 1182 | No | NA | 3 | 0 | 1 | Not-determined | Genome-fragment | NA | NA | 0 |
| S17-final-viral-combined_k141_164840\|\|full | 1508 | No | NA | 3 | 0 | 1 | Not-determined | Genome-fragment | NA | NA | 0 |
| S4-final-viral-combined_k141_2024\|\|full | 2347 | No | NA | 2 | 0 | 0 | Low-quality | Genome-fragment | 5.34 | AAI-based (high-confidence) | 0 |
| S17-final-viral-combined_k141_199924\|\|full | 2184 | No | NA | 2 | 0 | 1 | Not-determined | Genome-fragment | NA | NA | 0 |
| S17-final-viral-combined_k141_243110\|\|full | 1218 | No | NA | 2 | 0 | 1 | Not-determined | Genome-fragment | NA | NA | 0 |
| S17-final-viral-combined_k141_253044\|\|full | 7763 | No | NA | 6 | 0 | 3 | Not-determined | Genome-fragment | NA | NA | 0 |
| S17-final-viral-combined_k141_243121\|\|full | 2744 | No | NA | 2 | 0 | 0 | Not-determined | Genome-fragment | NA | NA | 0 |
| S17-final-viral-combined_k141_77687\|\|full | 3152 | No | NA | 2 | 0 | 2 | Not-determined | Genome-fragment | NA | NA | 0 |
| S11-final-viral-combined_k141_235152\|\|full | 2348 | No | NA | 4 | 0 | 0 | Low-quality | Genome-fragment | 4.05 | AAI-based (high-confidence) | 0 |
| S1-final-viral-combined_k141_8242\|\|full | 2348 | No | NA | 5 | 2 | 0 | Low-quality | Genome-fragment | 7.01 | AAI-based (high-confidence) | 0 |
| S17-final-viral-combined_k141_132308\|\|full | 1439 | No | NA | 3 | 0 | 2 | Not-determined | Genome-fragment | NA | NA | 0 |
| S17-final-viral-combined_k141_199959\|\|full | 2594 | No | NA | 4 | 0 | 0 | Not-determined | Genome-fragment | NA | NA | 0 |
| S12-final-viral-combined_k141_45875\|\|lt2gene | 2348 | No | NA | 1 | 1 | 0 | Low-quality | Genome-fragment | 5.36 | AAI-based (high-confidence) | 0 |
| S7_k141_239889 | 2349 | No | NA | 2 | 0 | 2 | Low-quality | Genome-fragment | 0.75 | AAI-based (medium-confidence) | 0 |
| S17-final-viral-combined_k141_132331\|\|full | 1297 | No | NA | 3 | 0 | 2 | Not-determined | Genome-fragment | NA | NA | 0 |
| S17-final-viral-combined_k141_66087\|\|full | 7194 | No | NA | 3 | 0 | 1 | Not-determined | Genome-fragment | NA | NA | 0 |
| S17-final-viral-combined_k141_154954\|\|full | 2809 | No | NA | 5 | 0 | 2 | Not-determined | Genome-fragment | NA | NA | 0 |
| S17-final-viral-combined_k141_25890\|\|full | 2349 | No | NA | 3 | 0 | 3 | Low-quality | Genome-fragment | 2.21 | AAI-based (medium-confidence) | 0 |
| S17-final-viral-combined_k141_264520\|\|full | 1794 | No | NA | 3 | 0 | 0 | Not-determined | Genome-fragment | NA | NA | 0 |
| S2-final-viral-combined_k141_145534\|\|lt2gene | 2349 | No | NA | 2 | 1 | 0 | Low-quality | Genome-fragment | 2.09 | HMM-based (lower-bound) | 0 |
| S17-final-viral-combined_k141_177381\|\|full | 3398 | No | NA | 3 | 0 | 1 | Not-determined | Genome-fragment | NA | NA | 0 |
| S8-final-viral-combined_k141_106280\|\|full | 2349 | No | NA | 2 | 0 | 0 | Low-quality | Genome-fragment | 4.71 | AAI-based (medium-confidence) | 0 |
| S17-final-viral-combined_k141_66123\|\|full | 22947 | No | NA | 33 | 21 | 0 | Medium-quality | Genome-fragment | 52.87 | AAI-based (high-confidence) | 0 |
| S11-final-viral-combined_k141_175749\|\|full | 2350 | No | NA | 5 | 0 | 0 | Low-quality | Genome-fragment | 4.27 | AAI-based (medium-confidence) | 0 |
| S17-final-viral-combined_k141_243215\|\|full | 2418 | No | NA | 2 | 0 | 0 | Not-determined | Genome-fragment | NA | NA | 0 |
| S17-final-viral-combined_k141_19814\|\|full | 1035 | No | NA | 4 | 0 | 0 | Not-determined | Genome-fragment | NA | NA | 0 |
| S17-final-viral-combined_k141_110008\|\|full | 1117 | No | NA | 2 | 0 | 0 | Not-determined | Genome-fragment | NA | NA | 0 |
| S3-final-viral-combined_k141_19896\|\|full | 2350 | No | NA | 3 | 0 | 2 | Low-quality | Genome-fragment | 0.56 | AAI-based (medium-confidence) | 0 |
| S7-final-viral-combined_k141_211942\|\|full | 2350 | No | NA | 4 | 0 | 3 | Low-quality | Genome-fragment | 0.8 | AAI-based (medium-confidence) | 0 |
| S17-final-viral-combined_k141_177443\|\|full | 2583 | No | NA | 3 | 0 | 0 | Not-determined | Genome-fragment | NA | NA | 0 |
| S12-final-viral-combined_k141_371914\|\|full | 2350 | No | NA | 4 | 0 | 3 | Low-quality | Genome-fragment | 3.19 | AAI-based (medium-confidence) | 0 |
| S17-final-viral-combined_k141_253163\|\|full | 2539 | No | NA | 3 | 0 | 1 | Not-determined | Genome-fragment | NA | NA | 0 |
| S17-final-viral-combined_k141_19851\|\|full | 2513 | No | NA | 5 | 0 | 0 | Not-determined | Genome-fragment | NA | NA | 0 |
| S17-final-viral-combined_k141_42920\|\|full | 1854 | No | NA | 2 | 0 | 0 | Not-determined | Genome-fragment | NA | NA | 0 |
| S17-final-viral-combined_k141_253191\|\|full | 2478 | No | NA | 2 | 0 | 1 | Not-determined | Genome-fragment | NA | NA | 0 |
| S2-final-viral-combined_k141_175921\|\|full | 2350 | No | NA | 2 | 0 | 1 | Low-quality | Genome-fragment | 3.39 | AAI-based (high-confidence) | 0 |
| S17-final-viral-combined_k141_42932\|\|full | 1653 | No | NA | 2 | 0 | 0 | Not-determined | Genome-fragment | NA | NA | 0 |
| S17-final-viral-combined_k141_31860\|\|full | 2534 | No | NA | 4 | 0 | 2 | Not-determined | Genome-fragment | NA | NA | 0 |
| S8-final-viral-combined_k141_86891\|\|full | 2350 | No | NA | 4 | 1 | 0 | Low-quality | Genome-fragment | 4.97 | AAI-based (medium-confidence) | 0 |
| S9-final-viral-combined_k141_77539\|\|full | 2351 | No | NA | 4 | 0 | 0 | Low-quality | Genome-fragment | 4.11 | AAI-based (high-confidence) | 0 |
| S11-final-viral-combined_k141_328454\|\|full | 2351 | No | NA | 3 | 0 | 0 | Low-quality | Genome-fragment | 0.69 | AAI-based (medium-confidence) | 0 |
| S1-final-viral-combined_k141_161005\|\|full | 2351 | No | NA | 3 | 0 | 2 | Low-quality | Genome-fragment | 0.63 | AAI-based (medium-confidence) | 0 |
| S7-final-viral-combined_k141_194511\|\|full | 2351 | No | NA | 3 | 2 | 0 | Low-quality | Genome-fragment | 5.11 | AAI-based (high-confidence) | 0 |
| S17-final-viral-combined_k141_110133\|\|full | 1159 | No | NA | 3 | 0 | 1 | Not-determined | Genome-fragment | NA | NA | 0 |
| S7-final-viral-combined_k141_214066\|\|full | 2351 | No | NA | 3 | 0 | 1 | Low-quality | Genome-fragment | 0.43 | AAI-based (medium-confidence) | 0 |
| S17-final-viral-combined_k141_160425\|\|full | 2351 | No | NA | 4 | 0 | 1 | Low-quality | Genome-fragment | 4.67 | AAI-based (medium-confidence) | 0 |
| S17-final-viral-combined_k141_264664\|\|full | 1397 | No | NA | 3 | 0 | 1 | Not-determined | Genome-fragment | NA | NA | 0 |
| S17-final-viral-combined_k141_97364\|\|full | 3541 | No | NA | 5 | 0 | 0 | Not-determined | Genome-fragment | NA | NA | 0 |
| S2-final-viral-combined_k141_117652\|\|full | 2351 | No | NA | 8 | 1 | 0 | Low-quality | Genome-fragment | 0.43 | HMM-based (lower-bound) | 0 |
| S17-final-viral-combined_k141_19979\|\|full | 1809 | No | NA | 2 | 0 | 0 | Not-determined | Genome-fragment | NA | NA | 0 |
| S4-final-viral-combined_k141_68618\|\|full | 2351 | No | NA | 4 | 1 | 0 | Low-quality | Genome-fragment | 5.96 | AAI-based (medium-confidence) | 0 |
| S17-final-viral-combined_k141_43028\|\|full | 2641 | No | NA | 4 | 0 | 0 | Not-determined | Genome-fragment | NA | NA | 0 |
| S17-final-viral-combined_k141_97410\|\|full | 1551 | No | NA | 3 | 0 | 1 | Not-determined | Genome-fragment | NA | NA | 0 |
| S17-final-viral-combined_k141_9864\|\|full | 43832 | No | NA | 53 | 21 | 1 | Medium-quality | Genome-fragment | 87.74 | AAI-based (high-confidence) | 0 |
| S17-final-viral-combined_k141_253315\|\|full | 1165 | No | NA | 2 | 0 | 0 | Not-determined | Genome-fragment | NA | NA | 0 |
| S17-final-viral-combined_k141_97424\|\|full | 2355 | No | NA | 4 | 0 | 3 | Not-determined | Genome-fragment | NA | NA | 0 |
| S17-final-viral-combined_k141_9880\|\|full | 1930 | No | NA | 3 | 0 | 3 | Not-determined | Genome-fragment | NA | NA | 0 |
| S17-final-viral-combined_k141_144175\|\|full | 1716 | No | NA | 2 | 0 | 2 | Not-determined | Genome-fragment | NA | NA | 0 |
| S17-final-viral-combined_k141_77760\|\|full | 15866 | No | NA | 15 | 0 | 3 | Not-determined | Genome-fragment | NA | NA | 0 |
| S17-final-viral-combined_k141_165243\|\|full | 7198 | No | NA | 7 | 0 | 4 | Not-determined | Genome-fragment | NA | NA | 0 |
| S17-final-viral-combined_k141_43072\|\|full | 1259 | No | NA | 2 | 0 | 0 | Not-determined | Genome-fragment | NA | NA | 0 |
| S17-final-viral-combined_k141_155076\|\|full | 1237 | No | NA | 1 | 0 | 0 | Not-determined | Genome-fragment | NA | NA | 0 |
| S17-final-viral-combined_k141_155077\|\|full | 2257 | No | NA | 2 | 0 | 1 | Not-determined | Genome-fragment | NA | NA | 0 |
| S6-final-viral-combined_k141_11723\|\|full | 2351 | No | NA | 4 | 0 | 0 | Low-quality | Genome-fragment | 5.65 | AAI-based (high-confidence) | 0 |
| S1-final-viral-combined_k141_337191\|\|full | 2352 | No | NA | 5 | 1 | 0 | Low-quality | Genome-fragment | 1.42 | HMM-based (lower-bound) | 0 |
| S5-final-viral-combined_k141_179816\|\|full | 2352 | No | NA | 3 | 0 | 0 | Low-quality | Genome-fragment | 3.54 | AAI-based (medium-confidence) | 0 |
| S17-final-viral-combined_k141_253354\|\|full | 1559 | No | NA | 2 | 0 | 1 | Not-determined | Genome-fragment | NA | NA | 0 |

| | | | | | | | | | | | |
|---|---|---|---|---|---|---|---|---|---|---|---|
| S17-final-viral-combined_k141_132577\|\|full | 5530 | No | NA | 6 | 0 | 2 | Not-determined | Genome-fragment | NA | NA | 0 |
| S17-final-viral-combined_k141_43180\|\|full | 2744 | No | NA | 3 | 0 | 3 | Not-determined | Genome-fragment | NA | NA | 0 |
| S5-final-viral-combined_k141_81615\|\|full | 2352 | No | NA | 3 | 0 | 2 | Low-quality | Genome-fragment | 1.27 | AAI-based (medium-confidence) | 0 |
| S17-final-viral-combined_k141_43182\|\|full | 1223 | No | NA | 2 | 0 | 2 | Not-determined | Genome-fragment | NA | NA | 0 |
| S17-final-viral-combined_k141_43194\|\|full | 7397 | No | NA | 7 | 0 | 6 | Not-determined | Genome-fragment | NA | NA | 0 |
| S2-final-viral-combined_k141_38356\|\|full | 2352 | No | NA | 5 | 2 | 0 | Low-quality | Genome-fragment | 6.41 | AAI-based (high-confidence) | 0 |
| S1-final-viral-combined_k141_89250\|\|full | 2353 | No | NA | 2 | 0 | 1 | Low-quality | Genome-fragment | 6.14 | AAI-based (medium-confidence) | 0 |
| S17-final-viral-combined_k141_19999\|\|full | 10296 | No | NA | 9 | 0 | 8 | Not-determined | Genome-fragment | NA | NA | 0 |
| S17-final-viral-combined_k141_165334\|\|full | 2011 | No | NA | 5 | 0 | 0 | Not-determined | Genome-fragment | NA | NA | 0 |
| S5-final-viral-combined_k141_14479\|\|full | 2353 | No | NA | 2 | 0 | 0 | Low-quality | Genome-fragment | 5.04 | AAI-based (medium-confidence) | 0 |
| S17-final-viral-combined_k141_253417\|\|full | 1024 | No | NA | 2 | 0 | 1 | Not-determined | Genome-fragment | NA | NA | 0 |
| S5-final-viral-combined_k141_140006\|\|full | 2353 | No | NA | 4 | 0 | 0 | Low-quality | Genome-fragment | 2.66 | AAI-based (high-confidence) | 0 |
| S17-final-viral-combined_k141_66419\|\|full | 2053 | No | NA | 2 | 0 | 2 | Not-determined | Genome-fragment | NA | NA | 0 |
| S17-final-viral-combined_k141_215114\|\|full | 2353 | No | NA | 5 | 4 | 0 | Low-quality | Genome-fragment | 4.71 | AAI-based (high-confidence) | 0 |
| S17-final-viral-combined_k141_43224\|\|full | 6743 | No | NA | 8 | 0 | 5 | Not-determined | Genome-fragment | NA | NA | 0 |
| S11-final-viral-combined_k141_288889\|\|full | 2354 | No | NA | 4 | 0 | 1 | Low-quality | Genome-fragment | 16.68 | AAI-based (medium-confidence) | 0 |
| S17-final-viral-combined_k141_97556\|\|full | 2897 | No | NA | 3 | 0 | 1 | Not-determined | Genome-fragment | NA | NA | 0 |
| S17-final-viral-combined_k141_144284\|\|full | 1400 | No | NA | 2 | 0 | 0 | Not-determined | Genome-fragment | NA | NA | 0 |
| S17-final-viral-combined_k141_155192\|\|full | 4947 | No | NA | 4 | 0 | 2 | Not-determined | Genome-fragment | NA | NA | 0 |
| S17-final-viral-combined_k141_132655\|\|full | 2439 | No | NA | 3 | 0 | 2 | Not-determined | Genome-fragment | NA | NA | 0 |
| S17-final-viral-combined_k141_110306\|\|full | 1034 | No | NA | 3 | 0 | 1 | Not-determined | Genome-fragment | NA | NA | 0 |
| S17-final-viral-combined_k141_155214\|\|full | 1072 | No | NA | 2 | 0 | 2 | Not-determined | Genome-fragment | NA | NA | 0 |
| S17-final-viral-combined_k141_66469\|\|full | 4285 | No | NA | 4 | 0 | 3 | Not-determined | Genome-fragment | NA | NA | 0 |
| S17-k141_103556 | 2354 | No | NA | 5 | 0 | 0 | Low-quality | Genome-fragment | 4.57 | AAI-based (high-confidence) | 0 |
| S17-final-viral-combined_k141_144335\|\|full | 2810 | No | NA | 3 | 0 | 3 | Not-determined | Genome-fragment | NA | NA | 0 |
| S17-final-viral-combined_k141_110337\|\|full | 1089 | No | NA | 2 | 0 | 1 | Not-determined | Genome-fragment | NA | NA | 0 |
| S2-final-viral-combined_k141_48950\|\|full | 2354 | No | NA | 3 | 2 | 0 | Low-quality | Genome-fragment | 6.91 | HMM-based (lower-bound) | 0 |
| S8-final-viral-combined_k141_115821\|\|full | 2354 | No | NA | 4 | 0 | 0 | Low-quality | Genome-fragment | 2.02 | AAI-based (high-confidence) | 0 |
| S17-final-viral-combined_k141_165472\|\|full | 2849 | No | NA | 2 | 0 | 0 | Not-determined | Genome-fragment | NA | NA | 0 |
| S17-final-viral-combined_k141_132744\|\|full | 1811 | No | NA | 3 | 0 | 2 | Not-determined | Genome-fragment | NA | NA | 0 |
| S5-final-viral-combined_k141_34687\|\|full | 2355 | No | NA | 3 | 0 | 1 | Low-quality | Genome-fragment | 0.72 | AAI-based (medium-confidence) | 0 |
| S7-final-viral-combined_k141_173504\|\|full | 2355 | No | NA | 8 | 2 | 0 | Low-quality | Genome-fragment | 3.67 | HMM-based (lower-bound) | 0 |
| S17-final-viral-combined_k141_66512\|\|full | 22094 | No | NA | 19 | 0 | 12 | Not-determined | Genome-fragment | NA | NA | 0 |
| S17-final-viral-combined_k141_165515\|\|full | 1743 | No | NA | 4 | 0 | 0 | Not-determined | Genome-fragment | NA | NA | 0 |
| S17-final-viral-combined_k141_253499\|\|full | 44949 | No | NA | 74 | 13 | 0 | High-quality | High-quality | 99.37 | AAI-based (high-confidence) | 0 |
| S17-final-viral-combined_k141_71281\|\|full | 2355 | No | NA | 3 | 0 | 1 | Low-quality | Genome-fragment | 1.68 | AAI-based (medium-confidence) | 0 |
| S17-final-viral-combined_k141_66522\|\|full | 2417 | No | NA | 4 | 0 | 0 | Not-determined | Genome-fragment | NA | NA | 0 |
| S17-final-viral-combined_k141_43397\|\|full | 1910 | No | NA | 4 | 0 | 0 | Not-determined | Genome-fragment | NA | NA | 0 |
| S1-final-viral-combined_k141_120941\|\|full | 2356 | No | NA | 4 | 0 | 2 | Low-quality | Genome-fragment | 1.19 | AAI-based (medium-confidence) | 0 |
| S17-final-viral-combined_k141_144450\|\|full | 1780 | No | NA | 4 | 0 | 3 | Not-determined | Genome-fragment | NA | NA | 0 |
| S12-final-viral-combined_k141_111442\|\|full | 2356 | No | NA | 5 | 3 | 0 | Low-quality | Genome-fragment | 4.44 | HMM-based (lower-bound) | 0 |
| S12-final-viral-combined_k141_112885\|\|full | 2356 | No | NA | 3 | 2 | 0 | Low-quality | Genome-fragment | 5.79 | AAI-based (high-confidence) | 0 |
| S17-final-viral-combined_k141_97681\|\|full | 1904 | No | NA | 3 | 0 | 1 | Not-determined | Genome-fragment | NA | NA | 0 |
| S17-final-viral-combined_k141_144462\|\|full | 10583 | No | NA | 8 | 0 | 4 | Not-determined | Genome-fragment | NA | NA | 0 |
| S1-final-viral-combined_k141_313630\|\|full | 2357 | No | NA | 3 | 0 | 1 | Low-quality | Genome-fragment | 4.8 | AAI-based (medium-confidence) | 0 |
| S17-final-viral-combined_k141_132809\|\|full | 18407 | No | NA | 9 | 0 | 7 | Not-determined | Genome-fragment | NA | NA | 0 |
| S17-final-viral-combined_k141_97705\|\|full | 5307 | No | NA | 4 | 0 | 1 | Not-determined | Genome-fragment | NA | NA | 0 |
| S17-final-viral-combined_k141_43470\|\|full | 3679 | No | NA | 6 | 0 | 0 | Not-determined | Genome-fragment | NA | NA | 0 |
| S17-final-viral-combined_k141_43481\|\|full | 2828 | No | NA | 3 | 0 | 2 | Not-determined | Genome-fragment | NA | NA | 0 |
| S17-final-viral-combined_k141_265009\|\|full | 1388 | No | NA | 4 | 0 | 0 | Not-determined | Genome-fragment | NA | NA | 0 |
| S17-final-viral-combined_k141_52897\|\|full | 1424 | No | NA | 2 | 0 | 0 | Not-determined | Genome-fragment | NA | NA | 0 |
| S17-final-viral-combined_k141_132833\|\|full | 2302 | No | NA | 3 | 0 | 2 | Not-determined | Genome-fragment | NA | NA | 0 |
| S7-final-viral-combined_k141_59039\|\|full | 2357 | No | NA | 4 | 0 | 1 | Low-quality | Genome-fragment | 3.2 | AAI-based (medium-confidence) | 0 |
| S12-final-viral-combined_k141_258151\|\|full | 2357 | No | NA | 5 | 1 | 0 | Low-quality | Genome-fragment | 4.16 | AAI-based (medium-confidence) | 0 |
| S7-final-viral-combined_k141_117406\|\|full | 2358 | No | NA | 4 | 0 | 1 | Low-quality | Genome-fragment | 0.84 | AAI-based (medium-confidence) | 0 |
| S17-final-viral-combined_k141_43532\|\|full | 1867 | No | NA | 2 | 0 | 0 | Not-determined | Genome-fragment | NA | NA | 0 |
| S17-k141_77700 | 2358 | No | NA | 3 | 0 | 0 | Low-quality | Genome-fragment | 1.33 | AAI-based (medium-confidence) | 0 |
| S2-final-viral-combined_k141_214358\|\|full | 2358 | No | NA | 6 | 1 | 0 | Low-quality | Genome-fragment | 1.59 | HMM-based (lower-bound) | 0 |
| S17-final-viral-combined_k141_10280\|\|full | 1108 | No | NA | 3 | 0 | 1 | Not-determined | Genome-fragment | NA | NA | 0 |
| S9-final-viral-combined_k141_21093\|\|full | 2359 | No | NA | 3 | 0 | 2 | Low-quality | Genome-fragment | 1.22 | AAI-based (medium-confidence) | 0 |
| S17-final-viral-combined_k141_165711\|\|full | 1113 | No | NA | 3 | 0 | 0 | Not-determined | Genome-fragment | NA | NA | 0 |
| S17-final-viral-combined_k141_243733\|\|full | 3759 | No | NA | 3 | 0 | 2 | Not-determined | Genome-fragment | NA | NA | 0 |
| S17-final-viral-combined_k141_10292\|\|full | 1367 | No | NA | 2 | 0 | 2 | Not-determined | Genome-fragment | NA | NA | 0 |
| S11-final-viral-combined_k141_293992\|\|full | 2359 | No | NA | 2 | 0 | 0 | Low-quality | Genome-fragment | 5.24 | AAI-based (medium-confidence) | 0 |
| S1-final-viral-combined_k141_157923\|\|full | 2359 | No | NA | 3 | 0 | 0 | Low-quality | Genome-fragment | 3.68 | AAI-based (medium-confidence) | 0 |
| S17-final-viral-combined_k141_132862\|\|full | 15893 | No | NA | 11 | 0 | 7 | Not-determined | Genome-fragment | NA | NA | 0 |
| S17-final-viral-combined_k141_110518\|\|full | 1942 | No | NA | 3 | 0 | 2 | Not-determined | Genome-fragment | NA | NA | 0 |
| S17-final-viral-combined_k141_155400\|\|full | 1280 | No | NA | 2 | 0 | 2 | Not-determined | Genome-fragment | NA | NA | 0 |
| S1_k141_271026 | 2359 | Yes | 842 | 7 | 1 | 1 | Low-quality | Genome-fragment | 2 | AAI-based (medium-confidence) | 64.31 |
| S12-final-viral-combined_k141_28076\|\|full | 2359 | No | NA | 4 | 1 | 0 | Low-quality | Genome-fragment | 4.23 | AAI-based (medium-confidence) | 0 |
| S17-final-viral-combined_k141_20350\|\|full | 1878 | No | NA | 3 | 0 | 0 | Not-determined | Genome-fragment | NA | NA | 0 |
| S2-final-viral-combined_k141_43799\|\|full | 2359 | No | NA | 7 | 1 | 0 | Low-quality | Genome-fragment | 4.93 | AAI-based (high-confidence) | 0 |
| S2-final-viral-combined_k141_95834\|\|full | 2359 | No | NA | 4 | 0 | 2 | Low-quality | Genome-fragment | 5.18 | AAI-based (medium-confidence) | 0 |
| S8-final-viral-combined_k141_1996\|\|full | 2359 | No | NA | 5 | 2 | 0 | Low-quality | Genome-fragment | 2.97 | HMM-based (lower-bound) | 0 |
| S17-final-viral-combined_k141_10354\|\|full | 2752 | No | NA | 5 | 0 | 0 | Not-determined | Genome-fragment | NA | NA | 0 |
| S1-final-viral-combined_k141_331615\|\|full | 2360 | No | NA | 4 | 0 | 1 | Low-quality | Genome-fragment | 1.1 | AAI-based (medium-confidence) | 0 |
| S11-final-viral-combined_k141_33022\|\|full | 2361 | No | NA | 5 | 2 | 0 | Low-quality | Genome-fragment | 4.19 | AAI-based (high-confidence) | 0 |
| S17-final-viral-combined_k141_132894\|\|full | 3316 | No | NA | 5 | 0 | 2 | Not-determined | Genome-fragment | NA | NA | 0 |
| S17-final-viral-combined_k141_5055\|\|full | 2361 | No | NA | 3 | 2 | 0 | Low-quality | Genome-fragment | 5.6 | AAI-based (medium-confidence) | 0 |
| S17-k141_50145 | 2361 | No | NA | 2 | 0 | 2 | Low-quality | Genome-fragment | 5.62 | AAI-based (high-confidence) | 0 |
| S12_k141_93279 | 2361 | No | NA | 1 | 0 | 1 | Low-quality | Genome-fragment | 6.66 | AAI-based (medium-confidence) | 0 |
| S9-final-viral-combined_k141_6823\|\|full | 2362 | No | NA | 2 | 0 | 2 | Low-quality | Genome-fragment | 0.76 | AAI-based (medium-confidence) | 0 |
| S17-final-viral-combined_k141_243879\|\|full | 2175 | No | NA | 3 | 0 | 1 | Not-determined | Genome-fragment | NA | NA | 0 |
| S17-final-viral-combined_k141_43669\|\|full | 1347 | No | NA | 2 | 0 | 0 | Not-determined | Genome-fragment | NA | NA | 0 |
| S17-final-viral-combined_k141_155507\|\|full | 2548 | No | NA | 4 | 0 | 0 | Not-determined | Genome-fragment | NA | NA | 0 |
| S17-final-viral-combined_k141_43681\|\|full | 2178 | No | NA | 4 | 0 | 4 | Not-determined | Genome-fragment | NA | NA | 0 |
| S17-final-viral-combined_k141_133000\|\|full | 2609 | No | NA | 3 | 0 | 1 | Not-determined | Genome-fragment | NA | NA | 0 |
| S11-final-viral-combined_k141_207298\|\|full | 2362 | No | NA | 3 | 2 | 0 | Low-quality | Genome-fragment | 1.35 | HMM-based (lower-bound) | 0 |
| S1-final-viral-combined_k141_46296\|\|full | 2362 | No | NA | 4 | 2 | 0 | Low-quality | Genome-fragment | 6.95 | AAI-based (high-confidence) | 0 |
| S17-final-viral-combined_k141_165932\|\|full | 3167 | No | NA | 3 | 0 | 1 | Not-determined | Genome-fragment | NA | NA | 0 |
| S17-final-viral-combined_k141_32480\|\|full | 2188 | No | NA | 3 | 0 | 1 | Not-determined | Genome-fragment | NA | NA | 0 |
| S17-final-viral-combined_k141_43786\|\|full | 2462 | No | NA | 3 | 0 | 1 | Not-determined | Genome-fragment | NA | NA | 0 |
| S1-final-viral-combined_k141_55832\|\|full | 2363 | No | NA | 3 | 2 | 0 | Low-quality | Genome-fragment | 2.56 | HMM-based (lower-bound) | 0 |
| S17-final-viral-combined_k141_244027\|\|full | 3971 | No | NA | 7 | 0 | 3 | Not-determined | Genome-fragment | NA | NA | 0 |
| S17-final-viral-combined_k141_97999\|\|full | 2985 | No | NA | 3 | 0 | 0 | Not-determined | Genome-fragment | NA | NA | 0 |
| S17-final-viral-combined_k141_53018\|\|full | 1741 | No | NA | 3 | 0 | 2 | Not-determined | Genome-fragment | NA | NA | 0 |
| S17-final-viral-combined_k141_133090\|\|full | 2053 | No | NA | 3 | 0 | 0 | Not-determined | Genome-fragment | NA | NA | 0 |
| S17-final-viral-combined_k141_165989\|\|full | 1368 | No | NA | 2 | 0 | 2 | Not-determined | Genome-fragment | NA | NA | 0 |
| S17-final-viral-combined_k141_110605\|\|full | 2413 | No | NA | 5 | 0 | 2 | Not-determined | Genome-fragment | NA | NA | 0 |
| S17-final-viral-combined_k141_43813\|\|full | 1260 | No | NA | 2 | 0 | 2 | Not-determined | Genome-fragment | NA | NA | 0 |
| S17-final-viral-combined_k141_244040\|\|full | 1380 | No | NA | 2 | 0 | 1 | Not-determined | Genome-fragment | NA | NA | 0 |
| S12-final-viral-combined_k141_256170\|\|full | 2363 | Yes | 1445 | 3 | 1 | 1 | Low-quality | Genome-fragment | 2.32 | HMM-based (lower-bound) | 38.85 |
| S17-final-viral-combined_k141_166018\|\|full | 1713 | No | NA | 3 | 0 | 3 | Not-determined | Genome-fragment | NA | NA | 0 |
| S6_k141_99729 | 2363 | No | NA | 4 | 1 | 0 | Low-quality | Genome-fragment | 4.68 | AAI-based (high-confidence) | 0 |
| S8-final-viral-combined_k141_23945\|\|full | 2363 | No | NA | 3 | 0 | 0 | Low-quality | Genome-fragment | 1.6 | AAI-based (high-confidence) | 0 |
| S11-final-viral-combined_k141_171219\|\|full | 2364 | No | NA | 3 | 0 | 2 | Low-quality | Genome-fragment | 3.52 | AAI-based (medium-confidence) | 0 |
| S17-final-viral-combined_k141_20678\|\|full | 8463 | No | NA | 6 | 0 | 3 | Not-determined | Genome-fragment | NA | NA | 0 |
| S17-final-viral-combined_k141_133168\|\|full | 1613 | No | NA | 2 | 0 | 2 | Not-determined | Genome-fragment | NA | NA | 0 |
| S17-final-viral-combined_k141_155664\|\|full | 2364 | No | NA | 5 | 1 | 0 | Low-quality | Genome-fragment | 4.22 | AAI-based (high-confidence) | 0 |
| S17-final-viral-combined_k141_133175\|\|full | 1861 | No | NA | 2 | 0 | 1 | Not-determined | Genome-fragment | NA | NA | 0 |
| S12-final-viral-combined_k141_187160\|\|full | 2364 | No | NA | 6 | 0 | 0 | Low-quality | Genome-fragment | 4.71 | AAI-based (high-confidence) | 0 |
| S17-final-viral-combined_k141_254094\|\|full | 2118 | No | NA | 4 | 0 | 1 | Not-determined | Genome-fragment | NA | NA | 0 |
| S12-final-viral-combined_k141_340810\|\|full | 2364 | No | NA | 4 | 0 | 0 | Low-quality | Genome-fragment | 5.08 | AAI-based (high-confidence) | 0 |
| S17-final-viral-combined_k141_43909\|\|full | 1780 | No | NA | 2 | 0 | 1 | Not-determined | Genome-fragment | NA | NA | 0 |
| S4-final-viral-combined_k141_75455\|\|full | 2364 | No | NA | 4 | 2 | 0 | Low-quality | Genome-fragment | 4.83 | AAI-based (high-confidence) | 0 |
| S17-final-viral-combined_k141_110680\|\|full | 3206 | No | NA | 3 | 0 | 1 | Not-determined | Genome-fragment | NA | NA | 0 |
| S17-final-viral-combined_k141_244090\|\|full | 2028 | No | NA | 3 | 0 | 3 | Not-determined | Genome-fragment | NA | NA | 0 |
| S17-final-viral-combined_k141_166124\|\|full | 1659 | No | NA | 5 | 0 | 0 | Not-determined | Genome-fragment | NA | NA | 0 |
| S11-final-viral-combined_k141_291275\|\|full | 2365 | No | NA | 5 | 0 | 0 | Low-quality | Genome-fragment | 6.91 | AAI-based (high-confidence) | 0 |
| S17-final-viral-combined_k141_20694\|\|full | 3076 | No | NA | 6 | 0 | 0 | Not-determined | Genome-fragment | NA | NA | 0 |
| S17-final-viral-combined_k141_43969\|\|full | 4493 | No | NA | 4 | 0 | 2 | Not-determined | Genome-fragment | NA | NA | 0 |
| S5-final-viral-combined_k141_50546\|\|full | 2365 | No | NA | 4 | 1 | 0 | Low-quality | Genome-fragment | 3.8 | AAI-based (medium-confidence) | 0 |
| S17-final-viral-combined_k141_53193\|\|full | 3123 | No | NA | 4 | 0 | 4 | Not-determined | Genome-fragment | NA | NA | 0 |
| S17-final-viral-combined_k141_32685\|\|full | 3009 | No | NA | 4 | 0 | 4 | Not-determined | Genome-fragment | NA | NA | 0 |
| S17-final-viral-combined_k141_20741\|\|full | 6419 | No | NA | 6 | 0 | 4 | Not-determined | Genome-fragment | NA | NA | 0 |
| S7-final-viral-combined_k141_179088\|\|full | 2365 | No | NA | 3 | 0 | 1 | Low-quality | Genome-fragment | 5.66 | AAI-based (medium-confidence) | 0 |
| S17-final-viral-combined_k141_20749\|\|full | 1658 | No | NA | 2 | 0 | 0 | Not-determined | Genome-fragment | NA | NA | 0 |
| S17-final-viral-combined_k141_98256\|\|full | 1264 | No | NA | 2 | 0 | 0 | Not-determined | Genome-fragment | NA | NA | 0 |
| S17-final-viral-combined_k141_44049\|\|full | 3039 | No | NA | 4 | 0 | 0 | Not-determined | Genome-fragment | NA | NA | 0 |
| S17-final-viral-combined_k141_44057\|\|full | 3056 | No | NA | 5 | 0 | 3 | Not-determined | Genome-fragment | NA | NA | 0 |
| S17-final-viral-combined_k141_98288\|\|full | 1680 | No | NA | 3 | 0 | 1 | Not-determined | Genome-fragment | NA | NA | 0 |
| S17-final-viral-combined_k141_166178\|\|full | 2197 | No | NA | 3 | 0 | 3 | Not-determined | Genome-fragment | NA | NA | 0 |
| S4-final-viral-combined_k141_62335\|\|full | 2365 | No | NA | 3 | 0 | 1 | Low-quality | Genome-fragment | 0.32 | AAI-based (medium-confidence) | 0 |
| S17-final-viral-combined_k141_133301\|\|full | 1406 | No | NA | 2 | 0 | 1 | Not-determined | Genome-fragment | NA | NA | 0 |
| S1-final-viral-combined_k141_30315\|\|full | 2366 | No | NA | 3 | 1 | 0 | Low-quality | Genome-fragment | 4.06 | AAI-based (high-confidence) | 0 |
| S7-final-viral-combined_k141_66040\|\|full | 2366 | No | NA | 4 | 0 | 3 | Low-quality | Genome-fragment | 2.64 | AAI-based (high-confidence) | 0 |
| S17-final-viral-combined_k141_32818\|\|full | 1738 | No | NA | 3 | 0 | 0 | Not-determined | Genome-fragment | NA | NA | 0 |
| S12-final-viral-combined_k141_60626\|\|full | 2366 | No | NA | 2 | 1 | 0 | Low-quality | Genome-fragment | 4.4 | AAI-based (medium-confidence) | 0 |
| S4-final-viral-combined_k141_73684\|\|full | 2366 | No | NA | 3 | 0 | 1 | Low-quality | Genome-fragment | 1.51 | AAI-based (medium-confidence) | 0 |
| S17-final-viral-combined_k141_110764\|\|full | 3234 | No | NA | 4 | 0 | 1 | Not-determined | Genome-fragment | NA | NA | 0 |
| S17-final-viral-combined_k141_244286\|\|full | 3729 | No | NA | 2 | 0 | 2 | Not-determined | Genome-fragment | NA | NA | 0 |
| S6-final-viral-combined_k141_136692\|\|full | 2366 | No | NA | 3 | 0 | 1 | Low-quality | Genome-fragment | 2.81 | AAI-based (high-confidence) | 0 |
| S17-final-viral-combined_k141_244306\|\|full | 4881 | No | NA | 7 | 0 | 4 | Not-determined | Genome-fragment | NA | NA | 0 |
| S17-final-viral-combined_k141_244315\|\|full | 1826 | No | NA | 2 | 0 | 0 | Not-determined | Genome-fragment | NA | NA | 0 |
| S17-final-viral-combined_k141_166192\|\|full | 2376 | No | NA | 2 | 0 | 1 | Not-determined | Genome-fragment | NA | NA | 0 |
| S17-final-viral-combined_k141_32886\|\|full | 1339 | No | NA | 3 | 0 | 2 | Not-determined | Genome-fragment | NA | NA | 0 |
| S6-final-viral-combined_k141_40149\|\|full | 2366 | No | NA | 4 | 0 | 1 | Low-quality | Genome-fragment | 1.48 | AAI-based (medium-confidence) | 0 |
| S17-final-viral-combined_k141_265510\|\|full | 1013 | No | NA | 1 | 0 | 0 | Not-determined | Genome-fragment | NA | NA | 0 |
| S17-final-viral-combined_k141_10770\|\|full | 4932 | No | NA | 2 | 0 | 1 | Not-determined | Genome-fragment | NA | NA | 0 |
| S17-final-viral-combined_k141_254285\|\|full | 1373 | No | NA | 3 | 0 | 2 | Not-determined | Genome-fragment | NA | NA | 0 |
| S8-final-viral-combined_k141_135482\|\|full | 2366 | No | NA | 3 | 0 | 2 | Low-quality | Genome-fragment | 0.75 | AAI-based (medium-confidence) | 0 |
| S17-final-viral-combined_k141_98460\|\|full | 1770 | No | NA | 3 | 0 | 2 | Not-determined | Genome-fragment | NA | NA | 0 |
| S17-final-viral-combined_k141_244371\|\|full | 1570 | No | NA | 3 | 0 | 0 | Not-determined | Genome-fragment | NA | NA | 0 |
| S7-final-viral-combined_k141_117002\|\|full | 2367 | No | NA | 4 | 0 | 2 | Low-quality | Genome-fragment | 3.16 | AAI-based (medium-confidence) | 0 |
| S2-final-viral-combined_k141_67360\|\|full | 2367 | No | NA | 4 | 0 | 1 | Low-quality | Genome-fragment | 2.53 | AAI-based (medium-confidence) | 0 |
| S9-final-viral-combined_k141_75046\|\|full | 2368 | No | NA | 2 | 0 | 1 | Low-quality | Genome-fragment | 4.45 | AAI-based (high-confidence) | 0 |
| S5-final-viral-combined_k141_99607\|\|full | 2368 | No | NA | 2 | 0 | 2 | Low-quality | Genome-fragment | 1.6 | AAI-based (medium-confidence) | 0 |
| S17-final-viral-combined_k141_123393\|\|full | 2368 | No | NA | 3 | 2 | 0 | Low-quality | Genome-fragment | 5.27 | AAI-based (medium-confidence) | 0 |
| S17-final-viral-combined_k141_161057\|\|full | 2368 | No | NA | 4 | 0 | 1 | Low-quality | Genome-fragment | 2.88 | AAI-based (high-confidence) | 0 |
| S17-final-viral-combined_k141_21089\|\|full | 11476 | No | NA | 9 | 0 | 9 | Not-determined | Genome-fragment | NA | NA | 0 |
| S17-final-viral-combined_k141_254433\|\|full | 1216 | No | NA | 2 | 0 | 2 | Not-determined | Genome-fragment | NA | NA | 0 |
| S17-final-viral-combined_k141_244432\|\|full | 1174 | No | NA | 2 | 0 | 1 | Not-determined | Genome-fragment | NA | NA | 0 |
| S12-final-viral-combined_k141_335545\|\|full | 2368 | No | NA | 5 | 5 | 0 | Low-quality | Genome-fragment | 5.1 | AAI-based (high-confidence) | 0 |
| S2-final-viral-combined_k141_163927\|\|full | 2368 | No | NA | 4 | 0 | 0 | Low-quality | Genome-fragment | 5.38 | AAI-based (medium-confidence) | 0 |
| S17-final-viral-combined_k141_98567\|\|full | 1085 | No | NA | 2 | 0 | 0 | Not-determined | Genome-fragment | NA | NA | 0 |
| S17-final-viral-combined_k141_33027\|\|full | 1701 | No | NA | 2 | 0 | 2 | Not-determined | Genome-fragment | NA | NA | 0 |
| S17-final-viral-combined_k141_53393\|\|full | 1400 | No | NA | 2 | 0 | 0 | Not-determined | Genome-fragment | NA | NA | 0 |
| S17-final-viral-combined_k141_53414\|\|full | 1184 | No | NA | 2 | 0 | 1 | Not-determined | Genome-fragment | NA | NA | 0 |
| S6_k141_68946 | 2368 | No | NA | 3 | 0 | 2 | Low-quality | Genome-fragment | 1.97 | AAI-based (high-confidence) | 0 |
| S17-final-viral-combined_k141_21215\|\|full | 15744 | No | NA | 15 | 0 | 10 | Not-determined | Genome-fragment | NA | NA | 0 |
| S8-final-viral-combined_k141_219112\|\|lt2gene | 2368 | No | NA | 2 | 2 | 0 | Low-quality | Genome-fragment | 5.2 | AAI-based (high-confidence) | 0 |
| S17-final-viral-combined_k141_33105\|\|full | 1412 | No | NA | 3 | 0 | 3 | Not-determined | Genome-fragment | NA | NA | 0 |
| S17-final-viral-combined_k141_265702\|\|full | 1245 | No | NA | 3 | 0 | 0 | Not-determined | Genome-fragment | NA | NA | 0 |
| S17-final-viral-combined_k141_11041\|\|full | 3112 | No | NA | 2 | 0 | 2 | Not-determined | Genome-fragment | NA | NA | 0 |
| S11-final-viral-combined_k141_182489\|\|full | 2369 | No | NA | 2 | 0 | 2 | Low-quality | Genome-fragment | 1.16 | AAI-based (medium-confidence) | 0 |
| S12-final-viral-combined_k141_99126\|\|full | 2369 | No | NA | 2 | 0 | 0 | Low-quality | Genome-fragment | 1.41 | AAI-based (high-confidence) | 0 |
| S8_k141_126161 | 2369 | No | NA | 3 | 0 | 0 | Low-quality | Genome-fragment | 1.89 | AAI-based (high-confidence) | 0 |
| S17-final-viral-combined_k141_98757\|\|full | 2861 | No | NA | 3 | 0 | 2 | Not-determined | Genome-fragment | NA | NA | 0 |
| S17-final-viral-combined_k141_265756\|\|full | 1178 | No | NA | 3 | 0 | 0 | Not-determined | Genome-fragment | NA | NA | 0 |
| S17-final-viral-combined_k141_265767\|\|full | 1619 | No | NA | 2 | 0 | 1 | Not-determined | Genome-fragment | NA | NA | 0 |
| S1_k141_298129 | 2370 | Yes | 1428 | 6 | 1 | 1 | Low-quality | Genome-fragment | 3.7 | AAI-based (high-confidence) | 39.75 |
| S17-final-viral-combined_k141_21314\|\|full | 1145 | No | NA | 3 | 0 | 1 | Not-determined | Genome-fragment | NA | NA | 0 |
| S17-final-viral-combined_k141_33175\|\|full | 2309 | No | NA | 2 | 0 | 1 | Not-determined | Genome-fragment | NA | NA | 0 |
| S17-final-viral-combined_k141_166391\|\|full | 1210 | No | NA | 3 | 0 | 0 | Not-determined | Genome-fragment | NA | NA | 0 |
| S8-final-viral-combined_k141_116991\|\|full | 2370 | No | NA | 6 | 0 | 0 | Low-quality | Genome-fragment | 3.74 | AAI-based (medium-confidence) | 0 |
| S17-final-viral-combined_k141_254591\|\|full | 2711 | No | NA | 3 | 0 | 3 | Not-determined | Genome-fragment | NA | NA | 0 |
| S17-final-viral-combined_k141_98916\|\|full | 2195 | No | NA | 4 | 0 | 0 | Not-determined | Genome-fragment | NA | NA | 0 |
| S17-final-viral-combined_k141_254627\|\|full | 1068 | No | NA | 2 | 0 | 0 | Not-determined | Genome-fragment | NA | NA | 0 |
| S17-final-viral-combined_k141_21440\|\|full | 3799 | No | NA | 6 | 0 | 0 | Not-determined | Genome-fragment | NA | NA | 0 |
| S17-final-viral-combined_k141_111104\|\|full | 3752 | No | NA | 4 | 0 | 1 | Not-determined | Genome-fragment | NA | NA | 0 |
| S17-final-viral-combined_k141_33293\|\|full | 26635 | No | NA | 20 | 0 | 16 | Not-determined | Genome-fragment | NA | NA | 0 |
| S1-final-viral-combined_k141_68461\|\|full | 2371 | No | NA | 3 | 0 | 0 | Low-quality | Genome-fragment | 2.28 | AAI-based (high-confidence) | 0 |
| S1-final-viral-combined_k141_360846\|\|full | 2371 | Yes | 448 | 4 | 1 | 1 | Low-quality | Genome-fragment | 0.18 | HMM-based (lower-bound) | 81.11 |
| S17-final-viral-combined_k141_266170\|\|full | 1658 | No | NA | 3 | 0 | 2 | Not-determined | Genome-fragment | NA | NA | 0 |
| S17-final-viral-combined_k141_254710\|\|full | 1476 | No | NA | 3 | 0 | 1 | Not-determined | Genome-fragment | NA | NA | 0 |
| S8_k141_297 | 2371 | No | NA | 3 | 0 | 0 | Low-quality | Genome-fragment | 6.54 | AAI-based (high-confidence) | 0 |
| S1-final-viral-combined_k141_155188\|\|full | 2372 | No | NA | 3 | 0 | 0 | Low-quality | Genome-fragment | 1.7 | AAI-based (high-confidence) | 0 |
| S5-final-viral-combined_k141_19304\|\|full | 2372 | No | NA | 3 | 0 | 2 | Low-quality | Genome-fragment | 5.2 | AAI-based (medium-confidence) | 0 |
| S7-final-viral-combined_k141_32554\|\|full | 2372 | No | NA | 4 | 0 | 0 | Low-quality | Genome-fragment | 5.11 | AAI-based (medium-confidence) | 0 |
| S5_k141_25084 | 2373 | No | NA | 4 | 1 | 0 | Low-quality | Genome-fragment | 6.95 | AAI-based (high-confidence) | 0 |
| S12-final-viral-combined_k141_120970\|\|full | 2373 | No | NA | 2 | 0 | 1 | Low-quality | Genome-fragment | 1.53 | AAI-based (medium-confidence) | 0 |
| S17-final-viral-combined_k141_21733\|\|full | 1828 | No | NA | 3 | 0 | 2 | Not-determined | Genome-fragment | NA | NA | 0 |
| S12-final-viral-combined_k141_243938\|\|full | 2373 | No | NA | 3 | 2 | 0 | Low-quality | Genome-fragment | 3.35 | AAI-based (high-confidence) | 0 |
| S17-final-viral-combined_k141_266381\|\|full | 1562 | No | NA | 2 | 0 | 2 | Not-determined | Genome-fragment | NA | NA | 0 |
| S17-final-viral-combined_k141_254877\|\|full | 2562 | No | NA | 5 | 0 | 4 | Not-determined | Genome-fragment | NA | NA | 0 |
| S12-final-viral-combined_k141_316891\|\|full | 2373 | No | NA | 4 | 0 | 3 | Low-quality | Genome-fragment | 1.83 | AAI-based (medium-confidence) | 0 |
| S17-final-viral-combined_k141_254904\|\|full | 2306 | No | NA | 3 | 0 | 1 | Not-determined | Genome-fragment | NA | NA | 0 |
| S4-final-viral-combined_k141_58722\|\|full | 2373 | No | NA | 3 | 0 | 0 | Low-quality | Genome-fragment | 6.15 | AAI-based (high-confidence) | 0 |
| S17-final-viral-combined_k141_99306\|\|full | 4506 | No | NA | 3 | 0 | 3 | Not-determined | Genome-fragment | NA | NA | 0 |
| S1-final-viral-combined_k141_270013\|\|full | 2374 | No | NA | 3 | 0 | 2 | Low-quality | Genome-fragment | 0.69 | AAI-based (medium-confidence) | 0 |
| S1_k141_333993 | 2374 | No | NA | 3 | 0 | 0 | Low-quality | Genome-fragment | 3.51 | AAI-based (medium-confidence) | 0 |
| S17-final-viral-combined_k141_266479\|\|full | 3398 | No | NA | 3 | 0 | 3 | Not-determined | Genome-fragment | NA | NA | 0 |
| S17-final-viral-combined_k141_21908\|\|full | 2486 | No | NA | 3 | 0 | 0 | Not-determined | Genome-fragment | NA | NA | 0 |
| S5-final-viral-combined_k141_55608\|\|full | 2374 | No | NA | 6 | 2 | 0 | Low-quality | Genome-fragment | 5.46 | AAI-based (medium-confidence) | 0 |
| S12-final-viral-combined_k141_169312\|\|full | 2374 | No | NA | 4 | 0 | 0 | Low-quality | Genome-fragment | 5.54 | AAI-based (high-confidence) | 0 |
| S17-final-viral-combined_k141_99522\|\|full | 1725 | No | NA | 3 | 0 | 1 | Not-determined | Genome-fragment | NA | NA | 0 |
| S8-final-viral-combined_k141_123098\|\|full | 2374 | No | NA | 2 | 0 | 2 | Low-quality | Genome-fragment | 1.13 | AAI-based (high-confidence) | 0 |
| S17-final-viral-combined_k141_21986\|\|full | 2086 | No | NA | 4 | 0 | 0 | Not-determined | Genome-fragment | NA | NA | 0 |
| S11-final-viral-combined_k141_184695\|\|full | 2375 | No | NA | 3 | 0 | 0 | Low-quality | Genome-fragment | 2.54 | AAI-based (medium-confidence) | 0 |
| S17-final-viral-combined_k141_54226\|\|full | 1653 | No | NA | 3 | 0 | 3 | Not-determined | Genome-fragment | NA | NA | 0 |
| S17-final-viral-combined_k141_93915\|\|full | 2375 | No | NA | 5 | 1 | 0 | Low-quality | Genome-fragment | 4.08 | AAI-based (high-confidence) | 0 |
| S17-final-viral-combined_k141_99577\|\|full | 2108 | No | NA | 3 | 0 | 0 | Not-determined | Genome-fragment | NA | NA | 0 |
| S17-final-viral-combined_k141_255279\|\|full | 3145 | No | NA | 2 | 0 | 2 | Not-determined | Genome-fragment | NA | NA | 0 |
| S8-final-viral-combined_k141_25814\|\|full | 2375 | No | NA | 4 | 1 | 0 | Low-quality | Genome-fragment | 9.38 | AAI-based (medium-confidence) | 0 |
| S17-final-viral-combined_k141_99687\|\|full | 1711 | No | NA | 3 | 0 | 2 | Not-determined | Genome-fragment | NA | NA | 0 |
| S17-final-viral-combined_k141_99694\|\|full | 2165 | No | NA | 4 | 0 | 0 | Not-determined | Genome-fragment | NA | NA | 0 |
| S1-final-viral-combined_k141_289796\|\|full | 2376 | No | NA | 3 | 0 | 2 | Low-quality | Genome-fragment | 8.17 | AAI-based (medium-confidence) | 0 |
| S3-final-viral-combined_k141_77464\|\|full | 2376 | No | NA | 3 | 0 | 2 | Low-quality | Genome-fragment | 0.99 | AAI-based (medium-confidence) | 0 |
| S5_k141_138856 | 2376 | Yes | 1662 | 3 | 1 | 1 | Low-quality | Genome-fragment | 0.54 | HMM-based (lower-bound) | 30.05 |
| S17-final-viral-combined_k141_54632\|\|full | 2959 | No | NA | 3 | 0 | 2 | Not-determined | Genome-fragment | NA | NA | 0 |
| S12_k141_121782 | 2376 | No | NA | 6 | 0 | 0 | Low-quality | Genome-fragment | 10.09 | AAI-based (medium-confidence) | 0 |
| S17-final-viral-combined_k141_99780\|\|full | 3146 | No | NA | 6 | 0 | 6 | Not-determined | Genome-fragment | NA | NA | 0 |
| S4-final-viral-combined_k141_93251\|\|full | 2376 | No | NA | 2 | 0 | 2 | Low-quality | Genome-fragment | 1.15 | AAI-based (high-confidence) | 0 |
| S17-final-viral-combined_k141_255537\|\|full | 4323 | No | NA | 3 | 0 | 2 | Not-determined | Genome-fragment | NA | NA | 0 |
| S4_k141_137290 | 2376 | No | NA | 2 | 0 | 2 | Low-quality | Genome-fragment | 1.46 | AAI-based (medium-confidence) | 0 |
| S17-final-viral-combined_k141_54743\|\|full | 1008 | No | NA | 2 | 0 | 1 | Not-determined | Genome-fragment | NA | NA | 0 |
| S8-final-viral-combined_k141_12369\|\|full | 2376 | No | NA | 4 | 0 | 0 | Low-quality | Genome-fragment | 3.61 | AAI-based (high-confidence) | 0 |
| S17-final-viral-combined_k141_54873\|\|full | 1391 | No | NA | 3 | 0 | 0 | Not-determined | Genome-fragment | NA | NA | 0 |
| S17-final-viral-combined_k141_99966\|\|full | 1752 | No | NA | 2 | 0 | 1 | Not-determined | Genome-fragment | NA | NA | 0 |
| S17-final-viral-combined_k141_99973\|\|full | 3279 | No | NA | 5 | 0 | 1 | Not-determined | Genome-fragment | NA | NA | 0 |
| S9-final-viral-combined_k141_109001\|\|lt2gene | 2377 | No | NA | 2 | 2 | 0 | Low-quality | Genome-fragment | 4.1 | HMM-based (lower-bound) | 0 |
| S5-final-viral-combined_k141_196567\|\|full | 2377 | No | NA | 6 | 1 | 0 | Low-quality | Genome-fragment | 6.02 | AAI-based (medium-confidence) | 0 |
| S17-final-viral-combined_k141_55239\|\|full | 1072 | No | NA | 3 | 0 | 1 | Not-determined | Genome-fragment | NA | NA | 0 |
| S1-final-viral-combined_k141_332667\|\|full | 2378 | No | NA | 5 | 0 | 0 | Low-quality | Genome-fragment | 2.29 | AAI-based (medium-confidence) | 0 |
| S12-final-viral-combined_k141_92475\|\|full | 2378 | No | NA | 5 | 0 | 0 | Low-quality | Genome-fragment | 6.16 | AAI-based (high-confidence) | 0 |
| S17-final-viral-combined_k141_55550\|\|full | 1232 | No | NA | 2 | 0 | 1 | Not-determined | Genome-fragment | NA | NA | 0 |
| S17-final-viral-combined_k141_55554\|\|full | 1799 | No | NA | 4 | 0 | 1 | Not-determined | Genome-fragment | NA | NA | 0 |
| S12-final-viral-combined_k141_361845\|\|full | 2378 | No | NA | 3 | 0 | 2 | Low-quality | Genome-fragment | 1.96 | AAI-based (high-confidence) | 0 |
| S17-final-viral-combined_k141_22268\|\|0_partial | 11388 | No | NA | 10 | 0 | 2 | Not-determined | Genome-fragment | NA | NA | 0 |
| S12_k141_136889 | 2378 | No | NA | 4 | 1 | 0 | Low-quality | Genome-fragment | 1.09 | AAI-based (high-confidence) | 0 |
| S2_k141_167841 | 2378 | No | NA | 3 | 3 | 0 | Low-quality | Genome-fragment | 2.36 | AAI-based (high-confidence) | 0 |
| S1-final-viral-combined_k141_166054\|\|full | 2379 | No | NA | 3 | 0 | 2 | Low-quality | Genome-fragment | 1.9 | AAI-based (high-confidence) | 0 |
| S1-final-viral-combined_k141_235758\|\|full | 2379 | No | NA | 4 | 0 | 0 | Low-quality | Genome-fragment | 5.65 | AAI-based (high-confidence) | 0 |
| S9-final-viral-combined_k141_20512\|\|full | 2380 | No | NA | 3 | 2 | 0 | Low-quality | Genome-fragment | 6.15 | AAI-based (medium-confidence) | 0 |
| S1-final-viral-combined_k141_378698\|\|full | 2380 | No | NA | 5 | 0 | 0 | Low-quality | Genome-fragment | 5.73 | AAI-based (high-confidence) | 0 |
| S17-final-viral-combined_k141_212874\|\|0_partial | 11807 | No | NA | 8 | 0 | 7 | Not-determined | Genome-fragment | NA | NA | 0 |
| S17-final-viral-combined_k141_203353\|\|0_partial | 14063 | No | NA | 14 | 0 | 9 | Not-determined | Genome-fragment | NA | NA | 0 |
| S5-final-viral-combined_k141_5577\|\|full | 2380 | No | NA | 4 | 0 | 2 | Low-quality | Genome-fragment | 3.23 | AAI-based (high-confidence) | 0 |
| S4_k141_29632 | 2380 | No | NA | 1 | 1 | 0 | Low-quality | Genome-fragment | 3.73 | AAI-based (high-confidence) | 0 |
| S17-final-viral-combined_k141_13918\|\|0_partial | 11421 | No | NA | 8 | 0 | 7 | Not-determined | Genome-fragment | NA | NA | 0 |
| S1-final-viral-combined_k141_82699\|\|full | 2381 | No | NA | 4 | 0 | 1 | Low-quality | Genome-fragment | 4.49 | AAI-based (medium-confidence) | 0 |
| S1-final-viral-combined_k141_1311\|\|full | 2381 | No | NA | 3 | 0 | 0 | Low-quality | Genome-fragment | 5.44 | AAI-based (medium-confidence) | 0 |
| S17-final-viral-combined_k141_249059\|\|0_partial | 16784 | No | NA | 21 | 0 | 4 | Not-determined | Genome-fragment | NA | NA | 0 |
| S2-final-viral-combined_k141_109143\|\|full | 2381 | No | NA | 3 | 0 | 0 | Low-quality | Genome-fragment | 2.82 | AAI-based (medium-confidence) | 0 |
| S1-final-viral-combined_k141_328772\|\|full | 2382 | No | NA | 4 | 0 | 1 | Low-quality | Genome-fragment | 4.82 | AAI-based (medium-confidence) | 0 |
| S17-final-viral-combined_k141_155543\|\|full | 2382 | No | NA | 3 | 0 | 1 | Low-quality | Genome-fragment | 1.16 | AAI-based (medium-confidence) | 0 |
| S17-k141_102308 | 2382 | No | NA | 7 | 0 | 0 | Low-quality | Genome-fragment | 2.89 | AAI-based (high-confidence) | 0 |
| S8-final-viral-combined_k141_78735\|\|full | 2382 | No | NA | 3 | 0 | 0 | Low-quality | Genome-fragment | 4.87 | AAI-based (high-confidence) | 0 |
| S11-final-viral-combined_k141_215378\|\|full | 2383 | No | NA | 3 | 0 | 0 | Low-quality | Genome-fragment | 1.14 | AAI-based (medium-confidence) | 0 |
| S11-final-viral-combined_k141_301057\|\|full | 2383 | No | NA | 5 | 0 | 0 | Low-quality | Genome-fragment | 3.84 | AAI-based (medium-confidence) | 0 |
| S17-final-viral-combined_k141_221567\|\|0_partial | 17406 | No | NA | 17 | 0 | 14 | Not-determined | Genome-fragment | NA | NA | 0 |
| S1-final-viral-combined_k141_386790\|\|full | 2383 | No | NA | 4 | 4 | 0 | Low-quality | Genome-fragment | 5.18 | AAI-based (high-confidence) | 0 |
| S7-final-viral-combined_k141_152624\|\|full | 2383 | No | NA | 4 | 1 | 0 | Low-quality | Genome-fragment | 6.01 | AAI-based (high-confidence) | 0 |
| S17-final-viral-combined_k141_122229\|\|0_partial | 10904 | No | NA | 12 | 0 | 7 | Not-determined | Genome-fragment | NA | NA | 0 |
| S8-final-viral-combined_k141_87853\|\|full | 2383 | No | NA | 3 | 2 | 0 | Low-quality | Genome-fragment | 5.76 | AAI-based (high-confidence) | 0 |
| S17-final-viral-combined_k141_166407\|\|0_partial | 16781 | No | NA | 18 | 0 | 15 | Not-determined | Genome-fragment | NA | NA | 0 |
| S2_k141_201501 | 2384 | No | NA | 4 | 1 | 2 | Low-quality | Genome-fragment | 5.52 | AAI-based (high-confidence) | 0 |
| S17-final-viral-combined_k141_86903\|\|full | 2385 | No | NA | 5 | 1 | 0 | Low-quality | Genome-fragment | 6.26 | AAI-based (medium-confidence) | 0 |
| S17-final-viral-combined_k141_211326\|\|lt2gene | 1309 | No | NA | 1 | 0 | 0 | Not-determined | Genome-fragment | NA | NA | 0 |
| S8-final-viral-combined_k141_104532\|\|full | 2385 | No | NA | 4 | 0 | 0 | Low-quality | Genome-fragment | 15.42 | AAI-based (high-confidence) | 0 |
| S7-final-viral-combined_k141_75492\|\|full | 2386 | No | NA | 2 | 0 | 2 | Low-quality | Genome-fragment | 1.73 | AAI-based (medium-confidence) | 0 |
| S17-final-viral-combined_k141_155785\|\|lt2gene | 1113 | No | NA | 2 | 0 | 1 | Not-determined | Genome-fragment | NA | NA | 0 |
| S12-final-viral-combined_k141_349561\|\|full | 2386 | No | NA | 3 | 2 | 0 | Low-quality | Genome-fragment | 7.7 | AAI-based (high-confidence) | 0 |
| S12_k141_93591 | 2386 | Yes | 1381 | 7 | 1 | 1 | Low-quality | Genome-fragment | 0.75 | AAI-based (high-confidence) | 42.12 |
| S1-final-viral-combined_k141_139789\|\|full | 2387 | No | NA | 3 | 0 | 3 | Low-quality | Genome-fragment | 0.75 | AAI-based (medium-confidence) | 0 |
| S5-final-viral-combined_k141_86134\|\|full | 2387 | No | NA | 4 | 0 | 1 | Low-quality | Genome-fragment | 3.39 | AAI-based (medium-confidence) | 0 |
| S17-final-viral-combined_k141_89332\|\|lt2gene | 1226 | No | NA | 1 | 0 | 0 | Not-determined | Genome-fragment | NA | NA | 0 |
| S17-final-viral-combined_k141_33981\|\|lt2gene | 1047 | No | NA | 1 | 0 | 0 | Not-determined | Genome-fragment | NA | NA | 0 |
| S12-final-viral-combined_k141_302502\|\|full | 2387 | No | NA | 4 | 1 | 0 | Low-quality | Genome-fragment | 4.87 | AAI-based (high-confidence) | 0 |
| S2-final-viral-combined_k141_138869\|\|full | 2387 | No | NA | 7 | 1 | 0 | Low-quality | Genome-fragment | 4.71 | AAI-based (medium-confidence) | 0 |
| S2-final-viral-combined_k141_28309\|\|full | 2387 | No | NA | 6 | 3 | 0 | Low-quality | Genome-fragment | 9.9 | AAI-based (medium-confidence) | 0 |
| S8-final-viral-combined_k141_219854\|\|full | 2387 | No | NA | 3 | 0 | 3 | Low-quality | Genome-fragment | 1.1 | AAI-based (medium-confidence) | 0 |
| S1-final-viral-combined_k141_67061\|\|full | 2388 | No | NA | 2 | 0 | 1 | Low-quality | Genome-fragment | 1.62 | AAI-based (medium-confidence) | 0 |
| S17-final-viral-combined_k141_99543\|\|full | 2388 | No | NA | 4 | 0 | 2 | Low-quality | Genome-fragment | 1.35 | AAI-based (medium-confidence) | 0 |
| S17-final-viral-combined_k141_1274\|\|lt2gene | 1069 | No | NA | 1 | 0 | 0 | Not-determined | Genome-fragment | NA | NA | 0 |
| S12-final-viral-combined_k141_99678\|\|full | 2388 | No | NA | 4 | 3 | 0 | Low-quality | Genome-fragment | 4.77 | AAI-based (medium-confidence) | 0 |
| S4_k141_141336 | 2388 | No | NA | 7 | 1 | 0 | Low-quality | Genome-fragment | 7.05 | AAI-based (high-confidence) | 0 |
| S12_k141_63469 | 2389 | No | NA | 2 | 0 | 0 | Low-quality | Genome-fragment | 5.48 | AAI-based (high-confidence) | 0 |
| S2-final-viral-combined_k141_236123\|\|full | 2389 | No | NA | 7 | 0 | 0 | Low-quality | Genome-fragment | 5.61 | AAI-based (high-confidence) | 0 |
| S11-final-viral-combined_k141_165265\|\|full | 2390 | No | NA | 5 | 1 | 0 | Low-quality | Genome-fragment | 1.45 | HMM-based (lower-bound) | 0 |
| S1-final-viral-combined_k141_206852\|\|full | 2390 | No | NA | 1 | 0 | 1 | Low-quality | Genome-fragment | 1.33 | AAI-based (medium-confidence) | 0 |
| S17-final-viral-combined_k141_23835\|\|lt2gene | 1961 | No | NA | 2 | 0 | 0 | Not-determined | Genome-fragment | NA | NA | 0 |
| S17-final-viral-combined_k141_84130\|\|full | 2390 | No | NA | 4 | 1 | 0 | Low-quality | Genome-fragment | 1.52 | HMM-based (lower-bound) | 0 |
| S17-final-viral-combined_k141_113518\|\|lt2gene | 1254 | No | NA | 1 | 0 | 0 | Not-determined | Genome-fragment | NA | NA | 0 |
| S17-final-viral-combined_k141_35560\|\|lt2gene | 1376 | No | NA | 1 | 0 | 0 | Not-determined | Genome-fragment | NA | NA | 0 |
| S17-final-viral-combined_k141_146765\|\|lt2gene | 1568 | No | NA | 2 | 0 | 0 | Not-determined | Genome-fragment | NA | NA | 0 |
| S17-final-viral-combined_k141_135813\|\|lt2gene | 1652 | No | NA | 2 | 0 | 1 | Not-determined | Genome-fragment | NA | NA | 0 |
| S8-final-viral-combined_k141_118265\|\|full | 2390 | No | NA | 3 | 3 | 0 | Low-quality | Genome-fragment | 5.14 | AAI-based (high-confidence) | 0 |
| S2-final-viral-combined_k141_10680\|\|full | 2391 | No | NA | 5 | 2 | 0 | Low-quality | Genome-fragment | 6.48 | AAI-based (high-confidence) | 0 |
| S8-final-viral-combined_k141_30926\|\|full | 2391 | No | NA | 3 | 0 | 0 | Low-quality | Genome-fragment | 4.28 | AAI-based (medium-confidence) | 0 |
| S17-final-viral-combined_k141_171939\|\|full | 2393 | No | NA | 5 | 0 | 0 | Low-quality | Genome-fragment | 6.23 | AAI-based (high-confidence) | 0 |
| S17-final-viral-combined_k141_246593\|\|lt2gene | 1028 | No | NA | 1 | 0 | 0 | Not-determined | Genome-fragment | NA | NA | 0 |
| S2-final-viral-combined_k141_232608\|\|full | 2393 | No | NA | 3 | 0 | 1 | Low-quality | Genome-fragment | 5.42 | AAI-based (medium-confidence) | 0 |
| S17-final-viral-combined_k141_246612\|\|lt2gene | 1285 | No | NA | 2 | 0 | 0 | Not-determined | Genome-fragment | NA | NA | 0 |
| S5-final-viral-combined_k141_58925\|\|full | 2394 | No | NA | 5 | 0 | 0 | Low-quality | Genome-fragment | 3.59 | AAI-based (medium-confidence) | 0 |
| S12-final-viral-combined_k141_202248\|\|full | 2394 | No | NA | 3 | 2 | 0 | Low-quality | Genome-fragment | 5.27 | AAI-based (high-confidence) | 0 |
| S17-final-viral-combined_k141_58144\|\|lt2gene | 1862 | No | NA | 2 | 0 | 2 | Not-determined | Genome-fragment | NA | NA | 0 |
| S17-final-viral-combined_k141_102499\|\|lt2gene | 1233 | No | NA | 1 | 0 | 1 | Not-determined | Genome-fragment | NA | NA | 0 |
| S17-final-viral-combined_k141_36061\|\|lt2gene | 2155 | No | NA | 1 | 0 | 0 | Not-determined | Genome-fragment | NA | NA | 0 |
| S11-final-viral-combined_k141_72401\|\|full | 2395 | No | NA | 3 | 1 | 0 | Low-quality | Genome-fragment | 4.97 | AAI-based (high-confidence) | 0 |
| S17-final-viral-combined_k141_203421\|\|lt2gene | 1064 | No | NA | 2 | 0 | 1 | Not-determined | Genome-fragment | NA | NA | 0 |
| S5-final-viral-combined_k141_153266\|\|full | 2395 | No | NA | 8 | 1 | 0 | Low-quality | Genome-fragment | 2.68 | AAI-based (high-confidence) | 0 |
| S8-final-viral-combined_k141_178829\|\|full | 2395 | No | NA | 6 | 2 | 0 | Low-quality | Genome-fragment | 4.86 | AAI-based (high-confidence) | 0 |
| S17-final-viral-combined_k141_236282\|\|lt2gene | 1203 | No | NA | 1 | 0 | 0 | Not-determined | Genome-fragment | NA | NA | 0 |
| S11-final-viral-combined_k141_214514\|\|full | 2396 | No | NA | 2 | 0 | 1 | Low-quality | Genome-fragment | 1.42 | AAI-based (high-confidence) | 0 |
| S12-final-viral-combined_k141_231355\|\|full | 2397 | No | NA | 4 | 0 | 0 | Low-quality | Genome-fragment | 5.18 | AAI-based (medium-confidence) | 0 |
| S1-final-viral-combined_k141_1118\|\|full | 2398 | No | NA | 4 | 0 | 1 | Low-quality | Genome-fragment | 2.6 | AAI-based (medium-confidence) | 0 |
| S17-k141_156308 | 2398 | No | NA | 1 | 0 | 0 | Low-quality | Genome-fragment | 3.98 | AAI-based (medium-confidence) | 0 |
| S17-final-viral-combined_k141_115238\|\|lt2gene | 1052 | No | NA | 1 | 0 | 1 | Not-determined | Genome-fragment | NA | NA | 0 |
| S17-final-viral-combined_k141_225887\|\|lt2gene | 1072 | No | NA | 1 | 0 | 0 | Not-determined | Genome-fragment | NA | NA | 0 |
| S12-final-viral-combined_k141_139268\|\|full | 2398 | No | NA | 6 | 0 | 0 | Low-quality | Genome-fragment | 3.58 | AAI-based (medium-confidence) | 0 |
| S17-final-viral-combined_k141_103309\|\|lt2gene | 1665 | No | NA | 2 | 0 | 1 | Not-determined | Genome-fragment | NA | NA | 0 |
| S17-final-viral-combined_k141_236752\|\|lt2gene | 1526 | No | NA | 1 | 0 | 0 | Not-determined | Genome-fragment | NA | NA | 0 |
| S2-final-viral-combined_k141_24995\|\|full | 2398 | No | NA | 5 | 0 | 0 | Low-quality | Genome-fragment | 5.15 | AAI-based (medium-confidence) | 0 |
| S17-final-viral-combined_k141_170433\|\|lt2gene | 1500 | No | NA | 1 | 0 | 0 | Not-determined | Genome-fragment | NA | NA | 0 |
| S8-final-viral-combined_k141_188518\|\|full | 2398 | No | NA | 2 | 0 | 1 | Low-quality | Genome-fragment | 3.38 | AAI-based (high-confidence) | 0 |
| S8_k141_220505 | 2398 | No | NA | 3 | 3 | 0 | Low-quality | Genome-fragment | 7.52 | AAI-based (high-confidence) | 0 |
| S17-final-viral-combined_k141_82313\|\|lt2gene | 1193 | No | NA | 1 | 0 | 0 | Not-determined | Genome-fragment | NA | NA | 0 |
| S17-final-viral-combined_k141_59193\|\|lt2gene | 1219 | No | NA | 1 | 0 | 0 | Not-determined | Genome-fragment | NA | NA | 0 |
| S17-final-viral-combined_k141_215684\|\|lt2gene | 1088 | No | NA | 1 | 0 | 0 | Not-determined | Genome-fragment | NA | NA | 0 |
| S17-final-viral-combined_k141_25848\|\|lt2gene | 2244 | No | NA | 2 | 0 | 0 | Not-determined | Genome-fragment | NA | NA | 0 |
| S17-final-viral-combined_k141_148583\|\|lt2gene | 1162 | No | NA | 1 | 0 | 0 | Not-determined | Genome-fragment | NA | NA | 0 |
| S8-final-viral-combined_k141_71650\|\|full | 2399 | No | NA | 2 | 0 | 1 | Low-quality | Genome-fragment | 1.28 | AAI-based (medium-confidence) | 0 |
| S11-final-viral-combined_k141_133819\|\|full | 2400 | No | NA | 3 | 0 | 3 | Low-quality | Genome-fragment | 1.72 | AAI-based (high-confidence) | 0 |
| S17-final-viral-combined_k141_215754\|\|lt2gene | 1280 | No | NA | 2 | 0 | 0 | Not-determined | Genome-fragment | NA | NA | 0 |
| S7-final-viral-combined_k141_183765\|\|full | 2400 | No | NA | 3 | 2 | 0 | Low-quality | Genome-fragment | 5.39 | AAI-based (medium-confidence) | 0 |
| S7-final-viral-combined_k141_81297\|\|full | 2400 | No | NA | 5 | 0 | 0 | Low-quality | Genome-fragment | 3.89 | AAI-based (high-confidence) | 0 |
| S17-final-viral-combined_k141_207007\|\|full | 2400 | No | NA | 3 | 0 | 0 | Low-quality | Genome-fragment | 6.73 | AAI-based (high-confidence) | 0 |
| S12-final-viral-combined_k141_177679\|\|full | 2400 | No | NA | 4 | 0 | 2 | Low-quality | Genome-fragment | 5.48 | AAI-based (medium-confidence) | 0 |
| S17-final-viral-combined_k141_4271\|\|lt2gene | 1103 | No | NA | 1 | 0 | 0 | Not-determined | Genome-fragment | NA | NA | 0 |
| S12-final-viral-combined_k141_97861\|\|full | 2400 | No | NA | 3 | 0 | 1 | Low-quality | Genome-fragment | 3.22 | AAI-based (medium-confidence) | 0 |
| S6-final-viral-combined_k141_127600\|\|full | 2400 | No | NA | 4 | 3 | 0 | Low-quality | Genome-fragment | 3.19 | HMM-based (lower-bound) | 0 |
| S11-final-viral-combined_k141_292697\|\|full | 2401 | No | NA | 2 | 0 | 0 | Low-quality | Genome-fragment | 1.62 | AAI-based (high-confidence) | 0 |
| S11-final-viral-combined_k141_86154\|\|full | 2402 | No | NA | 2 | 1 | 0 | Low-quality | Genome-fragment | 2.2 | HMM-based (lower-bound) | 0 |
| S11-final-viral-combined_k141_33773\|\|full | 2402 | No | NA | 3 | 0 | 3 | Low-quality | Genome-fragment | 2.74 | AAI-based (medium-confidence) | 0 |
| S17-final-viral-combined_k141_216541\|\|lt2gene | 1062 | No | NA | 1 | 0 | 0 | Not-determined | Genome-fragment | NA | NA | 0 |
| S1-final-viral-combined_k141_150311\|\|full | 2402 | No | NA | 3 | 1 | 0 | Low-quality | Genome-fragment | 5.45 | AAI-based (medium-confidence) | 0 |
| S1-final-viral-combined_k141_258963\|\|full | 2402 | No | NA | 2 | 0 | 1 | Low-quality | Genome-fragment | 2.01 | AAI-based (high-confidence) | 0 |
| S17-final-viral-combined_k141_71796\|\|lt2gene | 1655 | No | NA | 2 | 0 | 2 | Not-determined | Genome-fragment | NA | NA | 0 |
| S12-final-viral-combined_k141_173848\|\|full | 2402 | No | NA | 2 | 0 | 1 | Low-quality | Genome-fragment | 1.71 | AAI-based (medium-confidence) | 0 |
| S2-final-viral-combined_k141_62183\|\|full | 2402 | No | NA | 4 | 0 | 0 | Low-quality | Genome-fragment | 2.79 | AAI-based (medium-confidence) | 0 |
| S17-final-viral-combined_k141_205603\|\|lt2gene | 1424 | No | NA | 1 | 0 | 0 | Not-determined | Genome-fragment | NA | NA | 0 |
| S1-final-viral-combined_k141_387749\|\|full | 2403 | No | NA | 5 | 0 | 1 | Low-quality | Genome-fragment | 0.26 | AAI-based (medium-confidence) | 0 |
| S9-final-viral-combined_k141_69621\|\|full | 2404 | No | NA | 2 | 0 | 1 | Low-quality | Genome-fragment | 2.75 | AAI-based (high-confidence) | 0 |
| S11-final-viral-combined_k141_194753\|\|full | 2404 | No | NA | 7 | 1 | 0 | Low-quality | Genome-fragment | 5.78 | AAI-based (medium-confidence) | 0 |
| S11-final-viral-combined_k141_5604\|\|full | 2404 | No | NA | 4 | 0 | 0 | Low-quality | Genome-fragment | 4.08 | AAI-based (high-confidence) | 0 |
| S1-final-viral-combined_k141_266854\|\|full | 2404 | No | NA | 4 | 0 | 1 | Low-quality | Genome-fragment | 3.51 | AAI-based (medium-confidence) | 0 |
| S7-final-viral-combined_k141_179398\|\|full | 2404 | No | NA | 5 | 0 | 0 | Low-quality | Genome-fragment | 7.63 | AAI-based (medium-confidence) | 0 |
| S7-final-viral-combined_k141_211302\|\|full | 2404 | No | NA | 3 | 0 | 1 | Low-quality | Genome-fragment | 1.9 | AAI-based (high-confidence) | 0 |
| S17-final-viral-combined_k141_145530\|\|full | 2404 | No | NA | 4 | 0 | 1 | Low-quality | Genome-fragment | 5.14 | AAI-based (medium-confidence) | 0 |
| S8-final-viral-combined_k141_2632\|\|full | 2404 | No | NA | 4 | 2 | 0 | Low-quality | Genome-fragment | 4.06 | AAI-based (high-confidence) | 0 |
| S11-final-viral-combined_k141_162465\|\|full | 2405 | No | NA | 3 | 0 | 1 | Low-quality | Genome-fragment | 3.52 | AAI-based (medium-confidence) | 0 |
| S1-final-viral-combined_k141_19109\|\|full | 2405 | No | NA | 2 | 1 | 0 | Low-quality | Genome-fragment | 1.84 | AAI-based (medium-confidence) | 0 |
| S1_k141_152391 | 2405 | No | NA | 2 | 0 | 0 | Low-quality | Genome-fragment | 1.93 | AAI-based (high-confidence) | 0 |
| S5-final-viral-combined_k141_116950\|\|full | 2405 | No | NA | 3 | 0 | 0 | Low-quality | Genome-fragment | 0.84 | AAI-based (medium-confidence) | 0 |
| S17-k141_58064 | 2405 | No | NA | 2 | 0 | 1 | Low-quality | Genome-fragment | 17.17 | AAI-based (medium-confidence) | 0 |
| S6-final-viral-combined_k141_90348\|\|full | 2405 | No | NA | 3 | 0 | 2 | Low-quality | Genome-fragment | 2.32 | AAI-based (medium-confidence) | 0 |
| S9-final-viral-combined_k141_43252\|\|full | 2406 | No | NA | 3 | 0 | 2 | Low-quality | Genome-fragment | 0.94 | AAI-based (medium-confidence) | 0 |
| S7-final-viral-combined_k141_237268\|\|full | 2406 | No | NA | 5 | 0 | 2 | Low-quality | Genome-fragment | 1.64 | AAI-based (medium-confidence) | 0 |
| S7_k141_242278 | 2406 | No | NA | 2 | 0 | 0 | Low-quality | Genome-fragment | 5.62 | AAI-based (medium-confidence) | 0 |
| S17-final-viral-combined_k141_48839\|\|lt2gene | 1462 | No | NA | 1 | 0 | 1 | Not-determined | Genome-fragment | NA | NA | 0 |
| S17-final-viral-combined_k141_239409\|\|lt2gene | 2078 | No | NA | 1 | 0 | 0 | Not-determined | Genome-fragment | NA | NA | 0 |
| S7_k141_40181 | 2406 | No | NA | 1 | 0 | 1 | Low-quality | Genome-fragment | 0.21 | AAI-based (medium-confidence) | 0 |
| S2_k141_199142 | 2406 | No | NA | 7 | 0 | 0 | Low-quality | Genome-fragment | 5.35 | AAI-based (medium-confidence) | 0 |
| S17-final-viral-combined_k141_261137\|\|lt2gene | 1482 | No | NA | 1 | 0 | 0 | Not-determined | Genome-fragment | NA | NA | 0 |
| S8_k141_190401 | 2406 | No | NA | 3 | 0 | 0 | Low-quality | Genome-fragment | 2.87 | AAI-based (medium-confidence) | 0 |
| S17-final-viral-combined_k141_196257\|\|lt2gene | 1563 | No | NA | 2 | 0 | 0 | Not-determined | Genome-fragment | NA | NA | 0 |
| S17-final-viral-combined_k141_119334\|\|lt2gene | 1430 | No | NA | 1 | 0 | 1 | Not-determined | Genome-fragment | NA | NA | 0 |
| S17-final-viral-combined_k141_185501\|\|lt2gene | 1069 | No | NA | 1 | 0 | 0 | Not-determined | Genome-fragment | NA | NA | 0 |
| S1-final-viral-combined_k141_381624\|\|full | 2407 | No | NA | 4 | 0 | 0 | Low-quality | Genome-fragment | 3.31 | AAI-based (high-confidence) | 0 |
| S17-final-viral-combined_k141_80347\|\|full | 2407 | No | NA | 3 | 0 | 0 | Low-quality | Genome-fragment | 3.73 | AAI-based (medium-confidence) | 0 |
| S2-final-viral-combined_k141_20163\|\|full | 2407 | No | NA | 7 | 1 | 0 | Low-quality | Genome-fragment | 5.57 | AAI-based (medium-confidence) | 0 |
| S2-final-viral-combined_k141_134335\|\|full | 2407 | No | NA | 8 | 2 | 0 | Low-quality | Genome-fragment | 4.94 | AAI-based (high-confidence) | 0 |
| S17-final-viral-combined_k141_261870\|\|lt2gene | 1115 | No | NA | 2 | 0 | 0 | Not-determined | Genome-fragment | NA | NA | 0 |
| S17-final-viral-combined_k141_106991\|\|lt2gene | 1057 | No | NA | 1 | 0 | 0 | Not-determined | Genome-fragment | NA | NA | 0 |
| S1_k141_378141 | 2408 | No | NA | 2 | 0 | 2 | Low-quality | Genome-fragment | 0.35 | AAI-based (high-confidence) | 0 |
| S11-final-viral-combined_k141_43411\|\|full | 2409 | No | NA | 5 | 5 | 0 | Low-quality | Genome-fragment | 6.28 | AAI-based (high-confidence) | 0 |
| S7-final-viral-combined_k141_42665\|\|full | 2409 | No | NA | 3 | 0 | 2 | Low-quality | Genome-fragment | 1.93 | AAI-based (high-confidence) | 0 |
| S17-final-viral-combined_k141_197371\|\|lt2gene | 1102 | No | NA | 1 | 0 | 0 | Not-determined | Genome-fragment | NA | NA | 0 |
| S17-final-viral-combined_k141_186588\|\|lt2gene | 2409 | No | NA | 1 | 1 | 0 | Low-quality | Genome-fragment | 5.21 | AAI-based (high-confidence) | 0 |
| S4_k141_140700 | 2409 | No | NA | 2 | 0 | 1 | Low-quality | Genome-fragment | 1.36 | AAI-based (medium-confidence) | 0 |
| S4_k141_71008 | 2409 | No | NA | 4 | 0 | 1 | Low-quality | Genome-fragment | 4.67 | AAI-based (high-confidence) | 0 |
| S8-final-viral-combined_k141_138918\|\|full | 2409 | No | NA | 4 | 0 | 0 | Low-quality | Genome-fragment | 2.99 | AAI-based (high-confidence) | 0 |
| S8-final-viral-combined_k141_47196\|\|full | 2409 | No | NA | 6 | 1 | 0 | Low-quality | Genome-fragment | 5.52 | AAI-based (high-confidence) | 0 |
| S17-final-viral-combined_k141_174990\|\|lt2gene | 1021 | No | NA | 1 | 0 | 1 | Not-determined | Genome-fragment | NA | NA | 0 |
| S11-final-viral-combined_k141_117861\|\|full | 2410 | No | NA | 3 | 0 | 2 | Low-quality | Genome-fragment | 0.63 | AAI-based (medium-confidence) | 0 |
| S17-final-viral-combined_k141_209319\|\|lt2gene | 1200 | No | NA | 2 | 0 | 1 | Not-determined | Genome-fragment | NA | NA | 0 |
| S1_k141_46267 | 2410 | No | NA | 4 | 0 | 3 | Low-quality | Genome-fragment | 5.09 | AAI-based (high-confidence) | 0 |
| S5-final-viral-combined_k141_162361\|\|full | 2410 | No | NA | 4 | 0 | 1 | Low-quality | Genome-fragment | 4.21 | AAI-based (medium-confidence) | 0 |
| S17-final-viral-combined_k141_198301\|\|lt2gene | 1224 | No | NA | 2 | 0 | 0 | Not-determined | Genome-fragment | NA | NA | 0 |
| S2-final-viral-combined_k141_33105\|\|full | 2410 | No | NA | 3 | 0 | 0 | Low-quality | Genome-fragment | 4.31 | AAI-based (high-confidence) | 0 |
| S9_k141_36274 | 2411 | No | NA | 5 | 3 | 0 | Low-quality | Genome-fragment | 5.98 | AAI-based (high-confidence) | 0 |
| S11-final-viral-combined_k141_300272\|\|full | 2411 | No | NA | 2 | 1 | 0 | Low-quality | Genome-fragment | 3.81 | AAI-based (high-confidence) | 0 |

| | | | | | | | | | | | |
|---|---|---|---|---|---|---|---|---|---|---|---|
| S8-final-viral-combined_k141_244456\|\|full | 2411 | No | NA | 3 | 1 | 0 | Low-quality | Genome-fragment | 5.34 | AAI-based (high-confidence) | 0 |
| S17-final-viral-combined_k141_30536\|\|lt2gene | 2174 | No | NA | 2 | 0 | 0 | Not-determined | Genome-fragment | NA | NA | 0 |
| S11-final-viral-combined_k141_316573\|\|full | 2412 | No | NA | 4 | 1 | 0 | Low-quality | Genome-fragment | 1.93 | HMM-based (lower-bound) | 0 |
| S1-final-viral-combined_k141_168728\|\|full | 2412 | No | NA | 2 | 0 | 1 | Low-quality | Genome-fragment | 5.81 | AAI-based (medium-confidence) | 0 |
| S17-final-viral-combined_k141_109095\|\|lt2gene | 1935 | No | NA | 2 | 0 | 0 | Not-determined | Genome-fragment | NA | NA | 0 |
| S17-final-viral-combined_k141_8610\|\|lt2gene | 1250 | No | NA | 1 | 0 | 1 | Not-determined | Genome-fragment | NA | NA | 0 |
| S5-final-viral-combined_k141_55248\|\|full | 2412 | No | NA | 4 | 0 | 1 | Low-quality | Genome-fragment | 0.85 | AAI-based (medium-confidence) | 0 |
| S17-final-viral-combined_k141_8753\|\|lt2gene | 1180 | No | NA | 1 | 0 | 0 | Not-determined | Genome-fragment | NA | NA | 0 |
| S6_k141_107203 | 2412 | No | NA | 5 | 3 | 0 | Low-quality | Genome-fragment | 5.01 | AAI-based (medium-confidence) | 0 |
| S3_k141_24056 | 2413 | No | NA | 4 | 0 | 0 | Low-quality | Genome-fragment | 2.28 | AAI-based (high-confidence) | 0 |
| S5-final-viral-combined_k141_134375\|\|full | 2413 | No | NA | 5 | 5 | 0 | Low-quality | Genome-fragment | 6.03 | AAI-based (high-confidence) | 0 |
| S17-final-viral-combined_k141_9104\|\|lt2gene | 3199 | No | NA | 2 | 0 | 1 | Not-determined | Genome-fragment | NA | NA | 0 |
| S12-final-viral-combined_k141_209695\|\|full | 2413 | No | NA | 4 | 0 | 2 | Low-quality | Genome-fragment | 1.79 | AAI-based (medium-confidence) | 0 |
| S17-final-viral-combined_k141_264240\|\|lt2gene | 1149 | No | NA | 1 | 0 | 0 | Not-determined | Genome-fragment | NA | NA | 0 |
| S17-final-viral-combined_k141_188652\|\|lt2gene | 1015 | No | NA | 2 | 0 | 0 | Not-determined | Genome-fragment | NA | NA | 0 |
| S2-final-viral-combined_k141_54998\|\|full | 2413 | No | NA | 4 | 0 | 0 | Low-quality | Genome-fragment | 1.43 | AAI-based (medium-confidence) | 0 |
| S8-final-viral-combined_k141_244610\|\|full | 2413 | Yes | 742 | 4 | 1 | 1 | Low-quality | Genome-fragment | 0.93 | AAI-based (medium-confidence) | 69.25 |
| S17-final-viral-combined_k141_253243\|\|lt2gene | 1369 | No | NA | 2 | 0 | 1 | Not-determined | Genome-fragment | NA | NA | 0 |
| S17-final-viral-combined_k141_253326\|\|lt2gene | 2042 | No | NA | 2 | 0 | 2 | Not-determined | Genome-fragment | NA | NA | 0 |
| S8-final-viral-combined_k141_9613\|\|full | 2413 | Yes | 742 | 4 | 1 | 1 | Low-quality | Genome-fragment | 0.93 | AAI-based (medium-confidence) | 69.25 |
| S12-final-viral-combined_k141_230686\|\|full | 2414 | No | NA | 6 | 2 | 0 | Low-quality | Genome-fragment | 5.77 | AAI-based (medium-confidence) | 0 |
| S17-final-viral-combined_k141_177777\|\|lt2gene | 1051 | No | NA | 2 | 0 | 0 | Not-determined | Genome-fragment | NA | NA | 0 |
| S17-final-viral-combined_k141_32042\|\|lt2gene | 1283 | No | NA | 2 | 0 | 1 | Not-determined | Genome-fragment | NA | NA | 0 |
| S12_k141_327183 | 2414 | No | NA | 4 | 0 | 0 | Low-quality | Genome-fragment | 1.37 | AAI-based (high-confidence) | 0 |
| S2-final-viral-combined_k141_96616\|\|full | 2414 | No | NA | 3 | 2 | 0 | Low-quality | Genome-fragment | 6.33 | AAI-based (medium-confidence) | 0 |
| S2-final-viral-combined_k141_184754\|\|full | 2414 | No | NA | 2 | 0 | 1 | Low-quality | Genome-fragment | 1.83 | AAI-based (medium-confidence) | 0 |
| S4-final-viral-combined_k141_162184\|\|full | 2414 | No | NA | 5 | 0 | 2 | Low-quality | Genome-fragment | 2.43 | AAI-based (medium-confidence) | 0 |
| S17-final-viral-combined_k141_10511\|\|lt2gene | 1456 | No | NA | 1 | 0 | 1 | Not-determined | Genome-fragment | NA | NA | 0 |
| S7-final-viral-combined_k141_91759\|\|full | 2415 | No | NA | 4 | 0 | 0 | Low-quality | Genome-fragment | 2.88 | AAI-based (medium-confidence) | 0 |
| S17-final-viral-combined_k141_86676\|\|full | 2415 | No | NA | 6 | 2 | 0 | Low-quality | Genome-fragment | 5.83 | AAI-based (high-confidence) | 0 |
| S2-final-viral-combined_k141_111986\|\|full | 2416 | No | NA | 4 | 0 | 1 | Low-quality | Genome-fragment | 3.05 | AAI-based (high-confidence) | 0 |
| S11-final-viral-combined_k141_165927\|\|full | 2417 | No | NA | 4 | 3 | 0 | Low-quality | Genome-fragment | 4.39 | AAI-based (medium-confidence) | 0 |
| S7-final-viral-combined_k141_8752\|\|full | 2417 | No | NA | 6 | 1 | 0 | Low-quality | Genome-fragment | 5.81 | AAI-based (medium-confidence) | 0 |
| S17-k141_55576 | 1585 | No | NA | 2 | 0 | 2 | Not-determined | Genome-fragment | NA | NA | 0 |
| S17-k141_88920 | 1216 | No | NA | 1 | 0 | 0 | Not-determined | Genome-fragment | NA | NA | 0 |
| S17-k141_222270 | 1052 | No | NA | 2 | 0 | 0 | Not-determined | Genome-fragment | NA | NA | 0 |
| S17-k141_111148 | 3197 | No | NA | 5 | 0 | 2 | Not-determined | Genome-fragment | NA | NA | 0 |
| S1-final-viral-combined_k141_89500\|\|full | 2418 | No | NA | 4 | 1 | 0 | Low-quality | Genome-fragment | 6.45 | AAI-based (medium-confidence) | 0 |
| S17-k141_211179 | 1311 | No | NA | 1 | 0 | 1 | Not-determined | Genome-fragment | NA | NA | 0 |
| S17-k141_33391 | 3429 | No | NA | 2 | 0 | 1 | Not-determined | Genome-fragment | NA | NA | 0 |
| S17-k141_255655 | 2525 | No | NA | 1 | 0 | 0 | Not-determined | Genome-fragment | NA | NA | 0 |
| S17-k141_33409 | 1180 | No | NA | 2 | 0 | 1 | Not-determined | Genome-fragment | NA | NA | 0 |
| S17-k141_55608 | 1480 | No | NA | 1 | 0 | 0 | Not-determined | Genome-fragment | NA | NA | 0 |
| S17-k141_222338 | 2060 | No | NA | 2 | 0 | 2 | Not-determined | Genome-fragment | NA | NA | 0 |
| S17-k141_81 | 3194 | No | NA | 4 | 0 | 4 | Not-determined | Genome-fragment | NA | NA | 0 |
| S5-final-viral-combined_k141_113440\|\|lt2gene | 2418 | No | NA | 1 | 1 | 0 | Low-quality | Genome-fragment | 4.58 | HMM-based (lower-bound) | 0 |
| S17-k141_177819 | 1487 | No | NA | 2 | 0 | 2 | Not-determined | Genome-fragment | NA | NA | 0 |
| S11-final-viral-combined_k141_6802\|\|lt2gene | 2419 | No | NA | 2 | 2 | 0 | Low-quality | Genome-fragment | 5.66 | AAI-based (medium-confidence) | 0 |
| S7-final-viral-combined_k141_75494\|\|full | 2419 | No | NA | 5 | 1 | 0 | Low-quality | Genome-fragment | 5.55 | AAI-based (high-confidence) | 0 |
| S17-k141_122331 | 3305 | No | NA | 3 | 0 | 1 | Not-determined | Genome-fragment | NA | NA | 0 |
| S17-k141_144622 | 1092 | No | NA | 2 | 0 | 1 | Not-determined | Genome-fragment | NA | NA | 0 |
| S17-k141_133464 | 1333 | No | NA | 1 | 0 | 0 | Not-determined | Genome-fragment | NA | NA | 0 |
| S17-k141_144647 | 1155 | No | NA | 1 | 0 | 1 | Not-determined | Genome-fragment | NA | NA | 0 |
| S17-final-viral-combined_k141_222056\|\|full | 2419 | No | NA | 4 | 0 | 1 | Low-quality | Genome-fragment | 0.87 | AAI-based (medium-confidence) | 0 |
| S17-k141_11180 | 3812 | No | NA | 4 | 0 | 3 | Not-determined | Genome-fragment | NA | NA | 0 |
| S17-k141_33547 | 2176 | No | NA | 2 | 0 | 1 | Not-determined | Genome-fragment | NA | NA | 0 |
| S12-final-viral-combined_k141_7226\|\|full | 2419 | No | NA | 4 | 0 | 0 | Low-quality | Genome-fragment | 3.8 | AAI-based (medium-confidence) | 0 |
| S17-k141_144699 | 2419 | No | NA | 1 | 0 | 1 | Not-determined | Genome-fragment | NA | NA | 0 |
| S17-k141_144716 | 1351 | No | NA | 1 | 0 | 1 | Not-determined | Genome-fragment | NA | NA | 0 |
| S17-k141_78005 | 1178 | No | NA | 1 | 0 | 1 | Not-determined | Genome-fragment | NA | NA | 0 |
| S17-k141_122424 | 4386 | No | NA | 6 | 0 | 0 | Not-determined | Genome-fragment | NA | NA | 0 |
| S17-k141_100116 | 1004 | No | NA | 1 | 0 | 1 | Not-determined | Genome-fragment | NA | NA | 0 |
| S17-k141_122430 | 4646 | No | NA | 3 | 0 | 1 | Not-determined | Genome-fragment | NA | NA | 0 |
| S4-final-viral-combined_k141_135290\|\|full | 2419 | No | NA | 2 | 0 | 1 | Low-quality | Genome-fragment | 1.38 | AAI-based (medium-confidence) | 0 |
| S17-k141_188 | 1226 | No | NA | 2 | 0 | 1 | Not-determined | Genome-fragment | NA | NA | 0 |
| S17-k141_177942 | 1089 | No | NA | 1 | 0 | 0 | Not-determined | Genome-fragment | NA | NA | 0 |
| S17-k141_222495 | 1638 | No | NA | 3 | 0 | 3 | Not-determined | Genome-fragment | NA | NA | 0 |
| S17-k141_111356 | 7039 | No | NA | 7 | 0 | 2 | Not-determined | Genome-fragment | NA | NA | 0 |
| S1-final-viral-combined_k141_109563\|\|full | 2420 | No | NA | 3 | 0 | 2 | Low-quality | Genome-fragment | 1.1 | AAI-based (medium-confidence) | 0 |
| S17-k141_144760 | 2183 | No | NA | 3 | 0 | 2 | Not-determined | Genome-fragment | NA | NA | 0 |
| S17-k141_78084 | 1979 | No | NA | 3 | 0 | 0 | Not-determined | Genome-fragment | NA | NA | 0 |
| S17-k141_78091 | 1361 | No | NA | 2 | 0 | 1 | Not-determined | Genome-fragment | NA | NA | 0 |
| S17-final-viral-combined_k141_178441\|\|full | 2420 | No | NA | 4 | 0 | 0 | Low-quality | Genome-fragment | 5.4 | AAI-based (medium-confidence) | 0 |
| S17-k141_122487 | 1756 | No | NA | 2 | 0 | 1 | Not-determined | Genome-fragment | NA | NA | 0 |
| S8-final-viral-combined_k141_130631\|\|full | 2420 | No | NA | 5 | 1 | 0 | Low-quality | Genome-fragment | 5.68 | AAI-based (high-confidence) | 0 |
| S17-k141_144788 | 1026 | No | NA | 1 | 0 | 1 | Not-determined | Genome-fragment | NA | NA | 0 |
| S17-k141_33687 | 1002 | No | NA | 1 | 0 | 1 | Not-determined | Genome-fragment | NA | NA | 0 |
| S17-k141_22428 | 1002 | No | NA | 1 | 0 | 1 | Not-determined | Genome-fragment | NA | NA | 0 |
| S12-final-viral-combined_k141_86596\|\|full | 2421 | No | NA | 2 | 0 | 1 | Low-quality | Genome-fragment | 1.53 | AAI-based (medium-confidence) | 0 |
| S17-k141_66941 | 1035 | No | NA | 2 | 0 | 1 | Not-determined | Genome-fragment | NA | NA | 0 |
| S17-k141_33713 | 1078 | No | NA | 1 | 0 | 1 | Not-determined | Genome-fragment | NA | NA | 0 |
| S12_k141_38571 | 2421 | No | NA | 2 | 0 | 1 | Low-quality | Genome-fragment | 0.43 | AAI-based (medium-confidence) | 0 |
| S2-final-viral-combined_k141_210006\|\|full | 2421 | No | NA | 6 | 1 | 0 | Low-quality | Genome-fragment | 3.12 | HMM-based (lower-bound) | 0 |
| S17-k141_222608 | 1840 | No | NA | 2 | 0 | 2 | Not-determined | Genome-fragment | NA | NA | 0 |
| S17-k141_255926 | 1980 | No | NA | 5 | 0 | 0 | Not-determined | Genome-fragment | NA | NA | 0 |
| S17-k141_78189 | 1289 | No | NA | 1 | 0 | 0 | Not-determined | Genome-fragment | NA | NA | 0 |
| S17-k141_100208 | 1815 | No | NA | 3 | 0 | 1 | Not-determined | Genome-fragment | NA | NA | 0 |
| S17-k141_166892 | 4195 | No | NA | 3 | 0 | 3 | Not-determined | Genome-fragment | NA | NA | 0 |
| S17-k141_122542 | 2386 | No | NA | 1 | 0 | 0 | Not-determined | Genome-fragment | NA | NA | 0 |
| S17-k141_122562 | 1701 | No | NA | 4 | 0 | 0 | Not-determined | Genome-fragment | NA | NA | 0 |
| S2-final-viral-combined_k141_86686\|\|full | 2421 | No | NA | 3 | 2 | 0 | Low-quality | Genome-fragment | 6.72 | AAI-based (high-confidence) | 0 |
| S6-final-viral-combined_k141_51782\|\|full | 2421 | No | NA | 4 | 0 | 1 | Low-quality | Genome-fragment | 1.83 | AAI-based (medium-confidence) | 0 |
| S8-final-viral-combined_k141_210978\|\|full | 2421 | No | NA | 6 | 0 | 0 | Low-quality | Genome-fragment | 5.94 | AAI-based (medium-confidence) | 0 |
| S17-k141_233732 | 1219 | No | NA | 2 | 0 | 1 | Not-determined | Genome-fragment | NA | NA | 0 |
| S17-k141_255988 | 1948 | No | NA | 2 | 0 | 2 | Not-determined | Genome-fragment | NA | NA | 0 |
| S17-k141_78258 | 1301 | No | NA | 1 | 0 | 0 | Not-determined | Genome-fragment | NA | NA | 0 |
| S17-k141_166916 | 1194 | No | NA | 1 | 0 | 0 | Not-determined | Genome-fragment | NA | NA | 0 |
| S17-k141_44702 | 3197 | No | NA | 3 | 0 | 2 | Not-determined | Genome-fragment | NA | NA | 0 |
| S11-final-viral-combined_k141_287144\|\|full | 2422 | No | NA | 3 | 1 | 0 | Low-quality | Genome-fragment | 3.87 | AAI-based (medium-confidence) | 0 |
| S17-k141_122645 | 3646 | No | NA | 4 | 0 | 0 | Not-determined | Genome-fragment | NA | NA | 0 |
| S17-k141_11418 | 1619 | No | NA | 2 | 0 | 0 | Not-determined | Genome-fragment | NA | NA | 0 |
| S17-k141_122673 | 1213 | No | NA | 2 | 0 | 1 | Not-determined | Genome-fragment | NA | NA | 0 |
| S17-k141_11438 | 1411 | No | NA | 2 | 0 | 2 | Not-determined | Genome-fragment | NA | NA | 0 |
| S17-k141_166962 | 2923 | No | NA | 3 | 0 | 1 | Not-determined | Genome-fragment | NA | NA | 0 |
| S17-k141_211614 | 1128 | No | NA | 1 | 0 | 1 | Not-determined | Genome-fragment | NA | NA | 0 |
| S5-final-viral-combined_k141_154751\|\|full | 2422 | No | NA | 3 | 0 | 3 | Low-quality | Genome-fragment | 2.27 | AAI-based (medium-confidence) | 0 |
| S17-k141_145035 | 1085 | No | NA | 1 | 0 | 1 | Not-determined | Genome-fragment | NA | NA | 0 |
| S4-final-viral-combined_k141_23794\|\|full | 2422 | No | NA | 6 | 1 | 0 | Low-quality | Genome-fragment | 2.3 | AAI-based (high-confidence) | 0 |
| S8-final-viral-combined_k141_232595\|\|full | 2422 | No | NA | 2 | 0 | 1 | Low-quality | Genome-fragment | 2.83 | AAI-based (high-confidence) | 0 |
| S17-k141_178088 | 1298 | No | NA | 1 | 0 | 1 | Not-determined | Genome-fragment | NA | NA | 0 |
| S17-k141_44774 | 1123 | No | NA | 1 | 0 | 1 | Not-determined | Genome-fragment | NA | NA | 0 |
| S17-k141_111618 | 1884 | No | NA | 3 | 0 | 2 | Not-determined | Genome-fragment | NA | NA | 0 |
| S1-final-viral-combined_k141_298315\|\|full | 2423 | No | NA | 8 | 0 | 0 | Low-quality | Genome-fragment | 5.19 | AAI-based (high-confidence) | 0 |
| S12-final-viral-combined_k141_380203\|\|full | 2423 | No | NA | 4 | 0 | 0 | Low-quality | Genome-fragment | 3.58 | AAI-based (medium-confidence) | 0 |
| S17-k141_111639 | 3002 | No | NA | 2 | 0 | 2 | Not-determined | Genome-fragment | NA | NA | 0 |
| S17-k141_111645 | 3458 | No | NA | 5 | 0 | 5 | Not-determined | Genome-fragment | NA | NA | 0 |
| S17-k141_244998 | 5087 | No | NA | 4 | 0 | 1 | Not-determined | Genome-fragment | NA | NA | 0 |
| S17-k141_33784 | 2009 | No | NA | 2 | 0 | 1 | Not-determined | Genome-fragment | NA | NA | 0 |
| S12-final-viral-combined_k141_47125\|\|full | 2423 | No | NA | 3 | 3 | 0 | Low-quality | Genome-fragment | 5.09 | AAI-based (high-confidence) | 0 |
| S17-k141_122728 | 1041 | No | NA | 2 | 0 | 2 | Not-determined | Genome-fragment | NA | NA | 0 |
| S17-k141_155997 | 1528 | No | NA | 3 | 0 | 1 | Not-determined | Genome-fragment | NA | NA | 0 |
| S17-k141_155999 | 1896 | No | NA | 1 | 0 | 0 | Not-determined | Genome-fragment | NA | NA | 0 |
| S17-k141_211674 | 1149 | No | NA | 2 | 0 | 1 | Not-determined | Genome-fragment | NA | NA | 0 |
| S11-final-viral-combined_k141_105795\|\|full | 2424 | No | NA | 3 | 1 | 0 | Low-quality | Genome-fragment | 4.84 | AAI-based (medium-confidence) | 0 |
| S17-k141_78416 | 1297 | No | NA | 2 | 0 | 0 | Not-determined | Genome-fragment | NA | NA | 0 |
| S11_k141_230377 | 2425 | No | NA | 2 | 0 | 0 | Low-quality | Genome-fragment | 2.58 | AAI-based (high-confidence) | 0 |
| S17-k141_245029 | 1240 | No | NA | 1 | 0 | 0 | Not-determined | Genome-fragment | NA | NA | 0 |
| S17-final-viral-combined_k141_206819\|\|full | 2425 | No | NA | 2 | 0 | 1 | Low-quality | Genome-fragment | 1.72 | AAI-based (medium-confidence) | 0 |
| S2-final-viral-combined_k141_75313\|\|full | 2425 | No | NA | 8 | 5 | 0 | Low-quality | Genome-fragment | 5.69 | AAI-based (high-confidence) | 0 |
| S17-k141_167100 | 3442 | No | NA | 6 | 0 | 3 | Not-determined | Genome-fragment | NA | NA | 0 |
| S17-k141_189332 | 1017 | No | NA | 2 | 0 | 1 | Not-determined | Genome-fragment | NA | NA | 0 |
| S17-k141_167114 | 1621 | No | NA | 5 | 0 | 3 | Not-determined | Genome-fragment | NA | NA | 0 |
| S17-k141_167115 | 1279 | No | NA | 1 | 0 | 1 | Not-determined | Genome-fragment | NA | NA | 0 |
| S7-final-viral-combined_k141_243467\|\|full | 2426 | No | NA | 8 | 2 | 0 | Low-quality | Genome-fragment | 5.89 | AAI-based (high-confidence) | 0 |
| S17-k141_33855 | 1707 | No | NA | 1 | 0 | 1 | Not-determined | Genome-fragment | NA | NA | 0 |
| S17-final-viral-combined_k141_34632\|\|full | 2426 | No | NA | 5 | 2 | 0 | Low-quality | Genome-fragment | 6.99 | AAI-based (high-confidence) | 0 |
| S17-k141_111725 | 1166 | No | NA | 2 | 0 | 2 | Not-determined | Genome-fragment | NA | NA | 0 |
| S17-k141_156061 | 1047 | No | NA | 1 | 0 | 0 | Not-determined | Genome-fragment | NA | NA | 0 |
| S17-k141_133822 | 1673 | No | NA | 1 | 0 | 0 | Not-determined | Genome-fragment | NA | NA | 0 |
| S12-final-viral-combined_k141_241022\|\|full | 2426 | No | NA | 2 | 0 | 2 | Low-quality | Genome-fragment | 1.71 | AAI-based (medium-confidence) | 0 |
| S17-k141_11531 | 3662 | No | NA | 3 | 0 | 3 | Not-determined | Genome-fragment | NA | NA | 0 |
| S17-k141_256117 | 1041 | No | NA | 1 | 0 | 1 | Not-determined | Genome-fragment | NA | NA | 0 |
| S17-k141_67391 | 1827 | No | NA | 2 | 0 | 0 | Not-determined | Genome-fragment | NA | NA | 0 |
| S17-k141_133859 | 1070 | No | NA | 1 | 0 | 1 | Not-determined | Genome-fragment | NA | NA | 0 |
| S17-k141_222909 | 1127 | No | NA | 1 | 0 | 0 | Not-determined | Genome-fragment | NA | NA | 0 |
| S12_k141_401233 | 2426 | No | NA | 3 | 0 | 1 | Low-quality | Genome-fragment | 4.32 | AAI-based (medium-confidence) | 0 |
| S17-k141_67402 | 3346 | No | NA | 2 | 0 | 0 | Not-determined | Genome-fragment | NA | NA | 0 |
| S8_k141_116184 | 2426 | No | NA | 2 | 0 | 2 | Low-quality | Genome-fragment | 0.9 | AAI-based (medium-confidence) | 0 |
| S17-k141_67407 | 3941 | No | NA | 8 | 0 | 1 | Not-determined | Genome-fragment | NA | NA | 0 |
| S17-k141_67408 | 1604 | No | NA | 1 | 0 | 0 | Not-determined | Genome-fragment | NA | NA | 0 |
| S9-final-viral-combined_k141_70044\|\|full | 2427 | No | NA | 4 | 1 | 0 | Low-quality | Genome-fragment | 4.52 | HMM-based (lower-bound) | 0 |
| S1-final-viral-combined_k141_213767\|\|full | 2427 | No | NA | 3 | 1 | 0 | Low-quality | Genome-fragment | 3.67 | HMM-based (lower-bound) | 0 |
| S17-k141_11562 | 1464 | No | NA | 1 | 0 | 1 | Not-determined | Genome-fragment | NA | NA | 0 |
| S17-k141_598 | 1850 | No | NA | 1 | 0 | 1 | Not-determined | Genome-fragment | NA | NA | 0 |
| S17-k141_200729 | 1600 | No | NA | 2 | 0 | 2 | Not-determined | Genome-fragment | NA | NA | 0 |
| S17-final-viral-combined_k141_113892\|\|full | 2427 | No | NA | 4 | 3 | 0 | Low-quality | Genome-fragment | 7.08 | AAI-based (high-confidence) | 0 |
| S17-final-viral-combined_k141_81779\|\|full | 2427 | No | NA | 4 | 0 | 0 | Low-quality | Genome-fragment | 4.54 | AAI-based (medium-confidence) | 0 |
| S2-final-viral-combined_k141_6463\|\|full | 2427 | No | NA | 4 | 0 | 0 | Low-quality | Genome-fragment | 3.99 | AAI-based (high-confidence) | 0 |
| S17-k141_200799 | 1063 | No | NA | 1 | 0 | 1 | Not-determined | Genome-fragment | NA | NA | 0 |
| S17-k141_111881 | 1472 | No | NA | 1 | 0 | 0 | Not-determined | Genome-fragment | NA | NA | 0 |
| S17-k141_89394 | 1186 | No | NA | 1 | 0 | 1 | Not-determined | Genome-fragment | NA | NA | 0 |
| S17-k141_156169 | 1386 | No | NA | 1 | 0 | 0 | Not-determined | Genome-fragment | NA | NA | 0 |
| S17-k141_211884 | 3895 | No | NA | 3 | 0 | 3 | Not-determined | Genome-fragment | NA | NA | 0 |
| S17-k141_100511 | 1457 | No | NA | 1 | 0 | 1 | Not-determined | Genome-fragment | NA | NA | 0 |
| S17-k141_111907 | 1020 | No | NA | 1 | 0 | 0 | Not-determined | Genome-fragment | NA | NA | 0 |
| S17-k141_671 | 1257 | No | NA | 2 | 0 | 0 | Not-determined | Genome-fragment | NA | NA | 0 |
| S2-final-viral-combined_k141_236998\|\|full | 2427 | No | NA | 4 | 0 | 2 | Low-quality | Genome-fragment | 3.11 | AAI-based (high-confidence) | 0 |
| S17-k141_245148 | 14120 | No | NA | 9 | 0 | 2 | Not-determined | Genome-fragment | NA | NA | 0 |
| S4-final-viral-combined_k141_25430\|\|lt2gene | 2427 | No | NA | 1 | 1 | 0 | Low-quality | Genome-fragment | 3.95 | AAI-based (high-confidence) | 0 |
| S17-k141_44906 | 1804 | No | NA | 2 | 0 | 1 | Not-determined | Genome-fragment | NA | NA | 0 |
| S5-final-viral-combined_k141_73010\|\|full | 2428 | No | NA | 3 | 0 | 3 | Low-quality | Genome-fragment | 1.38 | AAI-based (medium-confidence) | 0 |
| S17-k141_189537 | 3474 | No | NA | 4 | 0 | 3 | Not-determined | Genome-fragment | NA | NA | 0 |
| S17-k141_245177 | 2273 | No | NA | 2 | 0 | 1 | Not-determined | Genome-fragment | NA | NA | 0 |
| S17-k141_178323 | 2796 | No | NA | 3 | 0 | 2 | Not-determined | Genome-fragment | NA | NA | 0 |
| S17-k141_11649 | 1091 | No | NA | 2 | 0 | 1 | Not-determined | Genome-fragment | NA | NA | 0 |
| S17-k141_134037 | 6051 | No | NA | 10 | 0 | 1 | Not-determined | Genome-fragment | NA | NA | 0 |
| S17-k141_123043 | 3373 | No | NA | 2 | 0 | 2 | Not-determined | Genome-fragment | NA | NA | 0 |
| S17-k141_145292 | 1167 | No | NA | 1 | 0 | 1 | Not-determined | Genome-fragment | NA | NA | 0 |
| S17-k141_89524 | 1064 | No | NA | 2 | 0 | 1 | Not-determined | Genome-fragment | NA | NA | 0 |
| S17-k141_89530 | 2042 | No | NA | 1 | 0 | 0 | Not-determined | Genome-fragment | NA | NA | 0 |
| S17-k141_56211 | 1039 | No | NA | 1 | 0 | 0 | Not-determined | Genome-fragment | NA | NA | 0 |
| S17-k141_233960 | 1371 | No | NA | 1 | 0 | 1 | Not-determined | Genome-fragment | NA | NA | 0 |
| S17-k141_34122 | 1627 | No | NA | 2 | 0 | 1 | Not-determined | Genome-fragment | NA | NA | 0 |
| S5-final-viral-combined_k141_82549\|\|full | 2428 | Yes | 1122 | 3 | 1 | 1 | Low-quality | Genome-fragment | 0.32 | HMM-based (lower-bound) | 53.79 |
| S17-k141_200982 | 3004 | No | NA | 6 | 0 | 2 | Not-determined | Genome-fragment | NA | NA | 0 |
| S17-k141_112020 | 1472 | No | NA | 1 | 0 | 1 | Not-determined | Genome-fragment | NA | NA | 0 |
| S17-k141_89566 | 6203 | No | NA | 4 | 0 | 4 | Not-determined | Genome-fragment | NA | NA | 0 |
| S17-k141_189647 | 1482 | No | NA | 2 | 0 | 2 | Not-determined | Genome-fragment | NA | NA | 0 |
| S17-k141_56272 | 1379 | No | NA | 2 | 0 | 2 | Not-determined | Genome-fragment | NA | NA | 0 |
| S17-k141_189660 | 1053 | No | NA | 1 | 0 | 0 | Not-determined | Genome-fragment | NA | NA | 0 |
| S11-final-viral-combined_k141_327854\|\|full | 2429 | No | NA | 4 | 0 | 1 | Low-quality | Genome-fragment | 0.74 | AAI-based (medium-confidence) | 0 |
| S17-k141_178440 | 1130 | No | NA | 1 | 0 | 0 | Not-determined | Genome-fragment | NA | NA | 0 |
| S17-k141_201023 | 1582 | No | NA | 2 | 0 | 0 | Not-determined | Genome-fragment | NA | NA | 0 |
| S17-k141_212069 | 1098 | No | NA | 3 | 0 | 2 | Not-determined | Genome-fragment | NA | NA | 0 |
| S17-k141_201024 | 9046 | No | NA | 4 | 0 | 4 | Not-determined | Genome-fragment | NA | NA | 0 |
| S1-final-viral-combined_k141_347842\|\|full | 2429 | No | NA | 6 | 1 | 0 | Low-quality | Genome-fragment | 5.87 | AAI-based (high-confidence) | 0 |
| S17-k141_100635 | 1550 | No | NA | 2 | 0 | 1 | Not-determined | Genome-fragment | NA | NA | 0 |
| S17-k141_67646 | 1978 | No | NA | 1 | 0 | 0 | Not-determined | Genome-fragment | NA | NA | 0 |
| S17-k141_112116 | 1521 | No | NA | 3 | 0 | 1 | Not-determined | Genome-fragment | NA | NA | 0 |
| S17-k141_145418 | 2603 | No | NA | 1 | 0 | 1 | Not-determined | Genome-fragment | NA | NA | 0 |
| S17-k141_11793 | 1055 | No | NA | 2 | 0 | 2 | Not-determined | Genome-fragment | NA | NA | 0 |
| S1-final-viral-combined_k141_76303\|\|full | 2429 | No | NA | 3 | 0 | 0 | Low-quality | Genome-fragment | 5.3 | AAI-based (high-confidence) | 0 |
| S1_k141_47439 | 2429 | No | NA | 3 | 0 | 2 | Low-quality | Genome-fragment | 5.2 | AAI-based (high-confidence) | 0 |
| S17-k141_100662 | 4232 | No | NA | 7 | 0 | 6 | Not-determined | Genome-fragment | NA | NA | 0 |
| S17-k141_234017 | 1140 | No | NA | 1 | 0 | 1 | Not-determined | Genome-fragment | NA | NA | 0 |
| S17-k141_212171 | 3191 | No | NA | 1 | 0 | 0 | Not-determined | Genome-fragment | NA | NA | 0 |
| S7-final-viral-combined_k141_192409\|\|full | 2429 | No | NA | 4 | 0 | 0 | Low-quality | Genome-fragment | 5.65 | AAI-based (high-confidence) | 0 |
| S17-k141_56361 | 1681 | No | NA | 1 | 0 | 1 | Not-determined | Genome-fragment | NA | NA | 0 |
| S7-final-viral-combined_k141_15458\|\|lt2gene | 2429 | No | NA | 2 | 2 | 0 | Low-quality | Genome-fragment | 3.02 | AAI-based (high-confidence) | 0 |
| S1-final-viral-combined_k141_342920\|\|full | 2430 | No | NA | 5 | 1 | 0 | Low-quality | Genome-fragment | 4.79 | AAI-based (high-confidence) | 0 |
| S17-k141_245371 | 1053 | No | NA | 1 | 0 | 0 | Not-determined | Genome-fragment | NA | NA | 0 |
| S17-k141_833 | 8078 | No | NA | 6 | 0 | 2 | Not-determined | Genome-fragment | NA | NA | 0 |
| S17-k141_34184 | 1052 | No | NA | 2 | 0 | 1 | Not-determined | Genome-fragment | NA | NA | 0 |
| S17-k141_178574 | 2146 | No | NA | 2 | 0 | 2 | Not-determined | Genome-fragment | NA | NA | 0 |
| S17-k141_167389 | 1488 | No | NA | 3 | 0 | 0 | Not-determined | Genome-fragment | NA | NA | 0 |
| S17-k141_178583 | 1060 | No | NA | 1 | 0 | 0 | Not-determined | Genome-fragment | NA | NA | 0 |
| S17-k141_189778 | 2308 | No | NA | 1 | 0 | 0 | Not-determined | Genome-fragment | NA | NA | 0 |
| S17-k141_234063 | 1620 | No | NA | 2 | 0 | 2 | Not-determined | Genome-fragment | NA | NA | 0 |
| S17-k141_256504 | 1205 | No | NA | 1 | 0 | 0 | Not-determined | Genome-fragment | NA | NA | 0 |
| S17-k141_167469 | 1396 | No | NA | 1 | 0 | 0 | Not-determined | Genome-fragment | NA | NA | 0 |
| S17-k141_100724 | 3491 | No | NA | 3 | 0 | 2 | Not-determined | Genome-fragment | NA | NA | 0 |
| S17-k141_112183 | 1974 | No | NA | 1 | 0 | 0 | Not-determined | Genome-fragment | NA | NA | 0 |
| S17-k141_234112 | 1214 | No | NA | 1 | 0 | 0 | Not-determined | Genome-fragment | NA | NA | 0 |
| S17-k141_78976 | 3692 | No | NA | 3 | 0 | 2 | Not-determined | Genome-fragment | NA | NA | 0 |
| S17-k141_100730 | 1580 | No | NA | 1 | 1 | 0 | Not-determined | Genome-fragment | NA | NA | 0 |
| S3-final-viral-combined_k141_127282\|\|full | 2430 | No | NA | 10 | 0 | 0 | Low-quality | Genome-fragment | 7.11 | AAI-based (high-confidence) | 0 |
| S17-k141_78999 | 1262 | No | NA | 2 | 0 | 2 | Not-determined | Genome-fragment | NA | NA | 0 |
| S17-k141_876 | 6037 | No | NA | 3 | 0 | 3 | Not-determined | Genome-fragment | NA | NA | 0 |
| S17-k141_100741 | 1116 | No | NA | 2 | 0 | 0 | Not-determined | Genome-fragment | NA | NA | 0 |
| S5-final-viral-combined_k141_596\|\|full | 2430 | No | NA | 3 | 0 | 1 | Low-quality | Genome-fragment | 6.89 | AAI-based (medium-confidence) | 0 |
| S17-k141_56504 | 1275 | No | NA | 4 | 0 | 0 | Not-determined | Genome-fragment | NA | NA | 0 |
| S17-k141_201204 | 1229 | No | NA | 2 | 0 | 2 | Not-determined | Genome-fragment | NA | NA | 0 |
| S17-k141_45130 | 2662 | No | NA | 2 | 0 | 2 | Not-determined | Genome-fragment | NA | NA | 0 |
| S17-k141_56518 | 2185 | No | NA | 2 | 0 | 0 | Not-determined | Genome-fragment | NA | NA | 0 |
| S2_k141_56250 | 2430 | No | NA | 4 | 0 | 0 | Low-quality | Genome-fragment | 1.74 | AAI-based (medium-confidence) | 0 |
| S17-k141_223387 | 5813 | No | NA | 4 | 0 | 3 | Not-determined | Genome-fragment | NA | NA | 0 |
| S17-k141_245460 | 1458 | No | NA | 2 | 0 | 0 | Not-determined | Genome-fragment | NA | NA | 0 |
| S17-k141_178739 | 4360 | No | NA | 4 | 0 | 4 | Not-determined | Genome-fragment | NA | NA | 0 |
| S9-final-viral-combined_k141_83385\|\|full | 2431 | No | NA | 5 | 3 | 0 | Low-quality | Genome-fragment | 6.91 | AAI-based (high-confidence) | 0 |
| S17-k141_123334 | 1214 | No | NA | 2 | 0 | 2 | Not-determined | Genome-fragment | NA | NA | 0 |
| S12-final-viral-combined_k141_135833\|\|full | 2431 | No | NA | 4 | 0 | 0 | Low-quality | Genome-fragment | 2.95 | AAI-based (high-confidence) | 0 |
| S4-final-viral-combined_k141_127794\|\|full | 2431 | No | NA | 2 | 0 | 1 | Low-quality | Genome-fragment | 0.69 | AAI-based (medium-confidence) | 0 |
| S4-final-viral-combined_k141_48319\|\|full | 2431 | No | NA | 5 | 2 | 0 | Low-quality | Genome-fragment | 4.58 | AAI-based (high-confidence) | 0 |
| S17-k141_949 | 8052 | No | NA | 4 | 0 | 0 | Not-determined | Genome-fragment | NA | NA | 0 |
| S17-k141_123367 | 1298 | No | NA | 2 | 0 | 2 | Not-determined | Genome-fragment | NA | NA | 0 |
| S17-k141_201261 | 1399 | No | NA | 2 | 0 | 2 | Not-determined | Genome-fragment | NA | NA | 0 |
| S17-k141_201271 | 1025 | No | NA | 1 | 0 | 1 | Not-determined | Genome-fragment | NA | NA | 0 |
| S17-k141_56627 | 3832 | No | NA | 5 | 0 | 5 | Not-determined | Genome-fragment | NA | NA | 0 |
| S17-k141_1017 | 1291 | No | NA | 1 | 0 | 0 | Not-determined | Genome-fragment | NA | NA | 0 |
| S6_k141_89503 | 2431 | No | NA | 4 | 1 | 0 | Low-quality | Genome-fragment | 7.57 | AAI-based (high-confidence) | 0 |
| S17-k141_89734 | 1265 | No | NA | 2 | 0 | 0 | Not-determined | Genome-fragment | NA | NA | 0 |
| S17-k141_178771 | 3485 | No | NA | 4 | 0 | 2 | Not-determined | Genome-fragment | NA | NA | 0 |
| S17-k141_167615 | 2218 | No | NA | 4 | 0 | 1 | Not-determined | Genome-fragment | NA | NA | 0 |
| S17-k141_112373 | 14552 | No | NA | 10 | 0 | 5 | Not-determined | Genome-fragment | NA | NA | 0 |
| S17-k141_212368 | 10128 | No | NA | 10 | 0 | 8 | Not-determined | Genome-fragment | NA | NA | 0 |
| S17-k141_167628 | 1000 | No | NA | 1 | 0 | 1 | Not-determined | Genome-fragment | NA | NA | 0 |
| S17-k141_189891 | 1125 | No | NA | 3 | 0 | 1 | Not-determined | Genome-fragment | NA | NA | 0 |
| S17-k141_167629 | 4823 | No | NA | 6 | 0 | 5 | Not-determined | Genome-fragment | NA | NA | 0 |
| S17-k141_234239 | 2574 | No | NA | 1 | 0 | 0 | Not-determined | Genome-fragment | NA | NA | 0 |
| S17-k141_1033 | 1455 | No | NA | 1 | 0 | 1 | Not-determined | Genome-fragment | NA | NA | 0 |
| S17-k141_234242 | 1569 | No | NA | 2 | 0 | 0 | Not-determined | Genome-fragment | NA | NA | 0 |
| S17-k141_234243 | 1227 | No | NA | 1 | 0 | 1 | Not-determined | Genome-fragment | NA | NA | 0 |
| S17-k141_23067 | 1346 | No | NA | 1 | 0 | 1 | Not-determined | Genome-fragment | NA | NA | 0 |
| S17-k141_189908 | 1002 | No | NA | 1 | 0 | 1 | Not-determined | Genome-fragment | NA | NA | 0 |
| S17-k141_256638 | 1028 | No | NA | 1 | 0 | 1 | Not-determined | Genome-fragment | NA | NA | 0 |
| S17-k141_56694 | 1065 | No | NA | 1 | 0 | 0 | Not-determined | Genome-fragment | NA | NA | 0 |
| S17-k141_167682 | 1157 | No | NA | 2 | 0 | 2 | Not-determined | Genome-fragment | NA | NA | 0 |
| S17-k141_56699 | 1735 | No | NA | 1 | 0 | 1 | Not-determined | Genome-fragment | NA | NA | 0 |
| S17-k141_123518 | 3285 | No | NA | 2 | 0 | 2 | Not-determined | Genome-fragment | NA | NA | 0 |
| S17-k141_234316 | 1432 | No | NA | 4 | 0 | 0 | Not-determined | Genome-fragment | NA | NA | 0 |
| S17-k141_256684 | 2266 | No | NA | 4 | 0 | 1 | Not-determined | Genome-fragment | NA | NA | 0 |
| S17-k141_145694 | 1105 | No | NA | 1 | 0 | 0 | Not-determined | Genome-fragment | NA | NA | 0 |
| S17-k141_123534 | 1514 | No | NA | 2 | 0 | 2 | Not-determined | Genome-fragment | NA | NA | 0 |
| S17-k141_79240 | 1003 | No | NA | 1 | 0 | 0 | Not-determined | Genome-fragment | NA | NA | 0 |
| S17-k141_201382 | 1242 | No | NA | 3 | 0 | 1 | Not-determined | Genome-fragment | NA | NA | 0 |
| S17-k141_212424 | 7079 | No | NA | 5 | 0 | 4 | Not-determined | Genome-fragment | NA | NA | 0 |
| S17-k141_112478 | 1006 | No | NA | 1 | 0 | 1 | Not-determined | Genome-fragment | NA | NA | 0 |
| S17-k141_167762 | 4912 | No | NA | 2 | 0 | 2 | Not-determined | Genome-fragment | NA | NA | 0 |
| S1_k141_62739 | 2432 | Yes | 1524 | 3 | 2 | 1 | Low-quality | Genome-fragment | 3.48 | AAI-based (high-confidence) | 37.34 |
| S17-k141_167769 | 1157 | No | NA | 2 | 0 | 2 | Not-determined | Genome-fragment | NA | NA | 0 |
| S17-k141_34486 | 1124 | No | NA | 2 | 0 | 0 | Not-determined | Genome-fragment | NA | NA | 0 |
| S5-final-viral-combined_k141_204958\|\|full | 2432 | No | NA | 6 | 1 | 0 | Low-quality | Genome-fragment | 6.82 | AAI-based (high-confidence) | 0 |
| S17-k141_34496 | 1093 | No | NA | 1 | 0 | 1 | Not-determined | Genome-fragment | NA | NA | 0 |
| S7-final-viral-combined_k141_234447\|\|full | 2432 | No | NA | 2 | 0 | 0 | Low-quality | Genome-fragment | 1.53 | AAI-based (high-confidence) | 0 |
| S17-k141_179041 | 3439 | No | NA | 5 | 0 | 1 | Not-determined | Genome-fragment | NA | NA | 0 |
| S12-final-viral-combined_k141_64632\|\|full | 2432 | No | NA | 7 | 0 | 0 | Low-quality | Genome-fragment | 5.3 | AAI-based (medium-confidence) | 0 |
| S17-k141_1179 | 2669 | No | NA | 1 | 0 | 1 | Not-determined | Genome-fragment | NA | NA | 0 |
| S17-k141_79304 | 1456 | No | NA | 2 | 0 | 1 | Not-determined | Genome-fragment | NA | NA | 0 |
| S17-k141_12151 | 1583 | No | NA | 1 | 0 | 1 | Not-determined | Genome-fragment | NA | NA | 0 |
| S9-final-viral-combined_k141_12694\|\|full | 2433 | No | NA | 2 | 0 | 0 | Low-quality | Genome-fragment | 5.25 | AAI-based (high-confidence) | 0 |
| S17-k141_256725 | 2199 | No | NA | 1 | 0 | 1 | Not-determined | Genome-fragment | NA | NA | 0 |
| S17-k141_201436 | 1181 | No | NA | 1 | 0 | 1 | Not-determined | Genome-fragment | NA | NA | 0 |
| S17-k141_134455 | 1315 | No | NA | 1 | 0 | 0 | Not-determined | Genome-fragment | NA | NA | 0 |
| S17-k141_68004 | 1072 | No | NA | 1 | 0 | 1 | Not-determined | Genome-fragment | NA | NA | 0 |
| S17-k141_256740 | 1112 | No | NA | 2 | 0 | 2 | Not-determined | Genome-fragment | NA | NA | 0 |
| S17-k141_134482 | 3754 | No | NA | 4 | 0 | 0 | Not-determined | Genome-fragment | NA | NA | 0 |
| S17-k141_223731 | 4512 | No | NA | 4 | 0 | 0 | Not-determined | Genome-fragment | NA | NA | 0 |
| S17-k141_212528 | 1827 | No | NA | 1 | 0 | 0 | Not-determined | Genome-fragment | NA | NA | 0 |
| S17-k141_68063 | 1198 | No | NA | 1 | 0 | 1 | Not-determined | Genome-fragment | NA | NA | 0 |
| S17-k141_212543 | 1525 | No | NA | 1 | 0 | 1 | Not-determined | Genome-fragment | NA | NA | 0 |
| S17-k141_134534 | 2222 | No | NA | 2 | 0 | 1 | Not-determined | Genome-fragment | NA | NA | 0 |
| S17-k141_23246 | 1120 | No | NA | 2 | 0 | 1 | Not-determined | Genome-fragment | NA | NA | 0 |
| S17-k141_79396 | 1009 | No | NA | 2 | 0 | 2 | Not-determined | Genome-fragment | NA | NA | 0 |
| S17-k141_179110 | 4677 | No | NA | 5 | 0 | 3 | Not-determined | Genome-fragment | NA | NA | 0 |
| S17-k141_145862 | 1288 | No | NA | 2 | 0 | 1 | Not-determined | Genome-fragment | NA | NA | 0 |
| S17-k141_101052 | 1079 | No | NA | 1 | 0 | 1 | Not-determined | Genome-fragment | NA | NA | 0 |
| S17-k141_145886 | 1039 | No | NA | 1 | 0 | 0 | Not-determined | Genome-fragment | NA | NA | 0 |
| S17-k141_89940 | 1057 | No | NA | 1 | 0 | 1 | Not-determined | Genome-fragment | NA | NA | 0 |
| S17-k141_156696 | 1096 | No | NA | 2 | 0 | 0 | Not-determined | Genome-fragment | NA | NA | 0 |
| S17-k141_201551 | 2187 | No | NA | 2 | 0 | 2 | Not-determined | Genome-fragment | NA | NA | 0 |
| S17-k141_1304 | 1798 | No | NA | 1 | 0 | 1 | Not-determined | Genome-fragment | NA | NA | 0 |
| S11-final-viral-combined_k141_60098\|\|full | 2433 | No | NA | 5 | 1 | 0 | Low-quality | Genome-fragment | 1.82 | AAI-based (high-confidence) | 0 |
| S17-k141_179148 | 1499 | No | NA | 1 | 0 | 1 | Not-determined | Genome-fragment | NA | NA | 0 |
| S17-k141_256808 | 1004 | No | NA | 2 | 0 | 0 | Not-determined | Genome-fragment | NA | NA | 0 |
| S17-final-viral-combined_k141_242256\|\|full | 2433 | No | NA | 3 | 0 | 3 | Low-quality | Genome-fragment | 1.2 | AAI-based (medium-confidence) | 0 |
| S17-k141_212634 | 1133 | No | NA | 2 | 0 | 0 | Not-determined | Genome-fragment | NA | NA | 0 |
| S17-k141_245735 | 3459 | No | NA | 6 | 0 | 4 | Not-determined | Genome-fragment | NA | NA | 0 |
| S17-k141_123759 | 2064 | No | NA | 2 | 0 | 2 | Not-determined | Genome-fragment | NA | NA | 0 |
| S17-k141_201613 | 1009 | No | NA | 2 | 0 | 1 | Not-determined | Genome-fragment | NA | NA | 0 |
| S17-k141_223836 | 2481 | No | NA | 4 | 0 | 0 | Not-determined | Genome-fragment | NA | NA | 0 |
| S17-k141_223837 | 6790 | No | NA | 5 | 0 | 0 | Not-determined | Genome-fragment | NA | NA | 0 |
| S17-k141_101133 | 1394 | No | NA | 2 | 0 | 2 | Not-determined | Genome-fragment | NA | NA | 0 |
| S17-k141_45411 | 1002 | No | NA | 1 | 0 | 0 | Not-determined | Genome-fragment | NA | NA | 0 |
| S17-k141_1342 | 3723 | No | NA | 6 | 0 | 0 | Not-determined | Genome-fragment | NA | NA | 0 |
| S17-k141_190207 | 1243 | No | NA | 1 | 0 | 1 | Not-determined | Genome-fragment | NA | NA | 0 |
| S17-k141_201658 | 1364 | No | NA | 2 | 0 | 2 | Not-determined | Genome-fragment | NA | NA | 0 |
| S17-k141_223872 | 1402 | No | NA | 1 | 0 | 1 | Not-determined | Genome-fragment | NA | NA | 0 |
| S17-k141_167892 | 1539 | No | NA | 2 | 0 | 0 | Not-determined | Genome-fragment | NA | NA | 0 |
| S17-k141_212731 | 1099 | No | NA | 1 | 0 | 1 | Not-determined | Genome-fragment | NA | NA | 0 |
| S17-k141_23347 | 1462 | No | NA | 2 | 0 | 2 | Not-determined | Genome-fragment | NA | NA | 0 |
| S17-k141_79592 | 1024 | No | NA | 1 | 0 | 0 | Not-determined | Genome-fragment | NA | NA | 0 |
| S17-k141_34690 | 1838 | No | NA | 1 | 0 | 1 | Not-determined | Genome-fragment | NA | NA | 0 |
| S17-k141_223901 | 4569 | No | NA | 3 | 0 | 0 | Not-determined | Genome-fragment | NA | NA | 0 |
| S17-k141_101194 | 4102 | No | NA | 4 | 0 | 3 | Not-determined | Genome-fragment | NA | NA | 0 |
| S17-k141_201733 | 3352 | No | NA | 2 | 0 | 1 | Not-determined | Genome-fragment | NA | NA | 0 |
| S17-k141_101201 | 1589 | No | NA | 2 | 0 | 2 | Not-determined | Genome-fragment | NA | NA | 0 |
| S17-k141_256927 | 4796 | No | NA | 2 | 0 | 1 | Not-determined | Genome-fragment | NA | NA | 0 |
| S8-final-viral-combined_k141_80885\|\|full | 2434 | No | NA | 5 | 0 | 0 | Low-quality | Genome-fragment | 5.23 | AAI-based (high-confidence) | 0 |
| S17-k141_156883 | 4257 | No | NA | 5 | 0 | 2 | Not-determined | Genome-fragment | NA | NA | 0 |
| S17-k141_79649 | 1126 | No | NA | 1 | 0 | 0 | Not-determined | Genome-fragment | NA | NA | 0 |
| S11-final-viral-combined_k141_64665\|\|full | 2435 | No | NA | 3 | 0 | 2 | Low-quality | Genome-fragment | 0.99 | AAI-based (medium-confidence) | 0 |
| S7-final-viral-combined_k141_114153\|\|full | 2435 | No | NA | 4 | 0 | 0 | Low-quality | Genome-fragment | 2.61 | AAI-based (medium-confidence) | 0 |
| S17-k141_145988 | 1404 | No | NA | 3 | 0 | 1 | Not-determined | Genome-fragment | NA | NA | 0 |
| S17-k141_79675 | 1146 | No | NA | 3 | 0 | 2 | Not-determined | Genome-fragment | NA | NA | 0 |
| S17-k141_90722 | 2435 | No | NA | 9 | 1 | 0 | Low-quality | Genome-fragment | 3.95 | AAI-based (high-confidence) | 0 |
| S17-k141_101234 | 2197 | No | NA | 2 | 0 | 2 | Not-determined | Genome-fragment | NA | NA | 0 |
| S17-k141_45515 | 2574 | No | NA | 2 | 0 | 0 | Not-determined | Genome-fragment | NA | NA | 0 |
| S6-final-viral-combined_k141_132393\|\|full | 2435 | No | NA | 2 | 0 | 2 | Low-quality | Genome-fragment | 1.2 | AAI-based (high-confidence) | 0 |
| S17-k141_257003 | 1693 | No | NA | 1 | 0 | 1 | Not-determined | Genome-fragment | NA | NA | 0 |
| S17-k141_167992 | 5353 | No | NA | 1 | 0 | 1 | Not-determined | Genome-fragment | NA | NA | 0 |
| S17-final-viral-combined_k141_25771\|\|full | 2436 | No | NA | 2 | 0 | 2 | Low-quality | Genome-fragment | 4.13 | AAI-based (medium-confidence) | 0 |
| S17-k141_146044 | 1051 | No | NA | 3 | 0 | 2 | Not-determined | Genome-fragment | NA | NA | 0 |
| S17-k141_56963 | 1074 | No | NA | 1 | 0 | 0 | Not-determined | Genome-fragment | NA | NA | 0 |
| S2-final-viral-combined_k141_79193\|\|full | 2436 | No | NA | 6 | 0 | 1 | Low-quality | Genome-fragment | 2.15 | AAI-based (medium-confidence) | 0 |
| S17-k141_224031 | 1248 | No | NA | 1 | 0 | 1 | Not-determined | Genome-fragment | NA | NA | 0 |
| S17-k141_79755 | 5174 | No | NA | 3 | 0 | 2 | Not-determined | Genome-fragment | NA | NA | 0 |
| S2_k141_226862 | 2436 | No | NA | 4 | 0 | 0 | Low-quality | Genome-fragment | 5.75 | AAI-based (high-confidence) | 0 |
| S17-k141_112753 | 1046 | No | NA | 1 | 0 | 1 | Not-determined | Genome-fragment | NA | NA | 0 |
| S17-k141_12366 | 1468 | No | NA | 2 | 0 | 1 | Not-determined | Genome-fragment | NA | NA | 0 |
| S17-k141_90097 | 4018 | No | NA | 8 | 0 | 8 | Not-determined | Genome-fragment | NA | NA | 0 |
| S4_k141_48942 | 2436 | No | NA | 3 | 0 | 0 | Low-quality | Genome-fragment | 2.98 | AAI-based (medium-confidence) | 0 |
| S17-k141_257123 | 7260 | No | NA | 8 | 0 | 4 | Not-determined | Genome-fragment | NA | NA | 0 |
| S17-k141_190410 | 2357 | No | NA | 2 | 0 | 0 | Not-determined | Genome-fragment | NA | NA | 0 |
| S17-k141_68367 | 1245 | No | NA | 2 | 0 | 2 | Not-determined | Genome-fragment | NA | NA | 0 |
| S17-k141_190413 | 2810 | No | NA | 4 | 0 | 4 | Not-determined | Genome-fragment | NA | NA | 0 |
| S17-k141_179326 | 2804 | No | NA | 3 | 0 | 3 | Not-determined | Genome-fragment | NA | NA | 0 |
| S6-final-viral-combined_k141_26715\|\|full | 2436 | No | NA | 2 | 0 | 1 | Low-quality | Genome-fragment | 1.4 | AAI-based (medium-confidence) | 0 |
| S17-k141_1526 | 1278 | No | NA | 2 | 0 | 0 | Not-determined | Genome-fragment | NA | NA | 0 |
| S17-k141_224107 | 1066 | No | NA | 3 | 0 | 1 | Not-determined | Genome-fragment | NA | NA | 0 |
| S17-k141_179378 | 1183 | No | NA | 2 | 0 | 2 | Not-determined | Genome-fragment | NA | NA | 0 |
| S17-k141_157108 | 1189 | No | NA | 2 | 0 | 0 | Not-determined | Genome-fragment | NA | NA | 0 |
| S17-k141_234745 | 3914 | No | NA | 5 | 0 | 5 | Not-determined | Genome-fragment | NA | NA | 0 |
| S17-k141_112895 | 1057 | No | NA | 1 | 0 | 0 | Not-determined | Genome-fragment | NA | NA | 0 |
| S17-k141_257224 | 4427 | No | NA | 4 | 0 | 2 | Not-determined | Genome-fragment | NA | NA | 0 |
| S17-k141_112898 | 1433 | No | NA | 2 | 0 | 1 | Not-determined | Genome-fragment | NA | NA | 0 |
| S17-k141_1619 | 1413 | No | NA | 1 | 0 | 0 | Not-determined | Genome-fragment | NA | NA | 0 |
| S17-k141_101359 | 4110 | No | NA | 4 | 0 | 3 | Not-determined | Genome-fragment | NA | NA | 0 |
| S17-k141_1624 | 1062 | No | NA | 2 | 0 | 2 | Not-determined | Genome-fragment | NA | NA | 0 |
| S17-k141_190562 | 4347 | No | NA | 6 | 0 | 0 | Not-determined | Genome-fragment | NA | NA | 0 |
| S17-k141_1644 | 3066 | No | NA | 2 | 0 | 2 | Not-determined | Genome-fragment | NA | NA | 0 |
| S17-k141_112949 | 5248 | No | NA | 5 | 0 | 5 | Not-determined | Genome-fragment | NA | NA | 0 |
| S8_k141_240892 | 2436 | No | NA | 5 | 0 | 2 | Low-quality | Genome-fragment | 2.77 | AAI-based (medium-confidence) | 0 |
| S17-k141_190580 | 3590 | No | NA | 3 | 0 | 1 | Not-determined | Genome-fragment | NA | NA | 0 |
| S17-k141_224233 | 1660 | No | NA | 1 | 0 | 1 | Not-determined | Genome-fragment | NA | NA | 0 |
| S17-k141_68544 | 3394 | No | NA | 4 | 0 | 4 | Not-determined | Genome-fragment | NA | NA | 0 |
| S11-final-viral-combined_k141_70307\|\|full | 2437 | Yes | 498 | 3 | 1 | 2 | Low-quality | Genome-fragment | 0.67 | HMM-based (lower-bound) | 79.57 |
| S17-k141_124165 | 1029 | No | NA | 2 | 0 | 1 | Not-determined | Genome-fragment | NA | NA | 0 |
| S17-k141_234843 | 3701 | No | NA | 4 | 0 | 3 | Not-determined | Genome-fragment | NA | NA | 0 |
| S1-final-viral-combined_k141_374231\|\|full | 2437 | No | NA | 5 | 0 | 0 | Low-quality | Genome-fragment | 6.23 | AAI-based (medium-confidence) | 0 |
| S12-final-viral-combined_k141_30890\|\|full | 2437 | No | NA | 3 | 2 | 0 | Low-quality | Genome-fragment | 5.68 | AAI-based (high-confidence) | 0 |
| S17-k141_68586 | 1086 | No | NA | 2 | 0 | 2 | Not-determined | Genome-fragment | NA | NA | 0 |
| S17-k141_1761 | 1214 | No | NA | 1 | 0 | 1 | Not-determined | Genome-fragment | NA | NA | 0 |
| S17-k141_168290 | 1390 | No | NA | 1 | 0 | 1 | Not-determined | Genome-fragment | NA | NA | 0 |
| S17-k141_113069 | 1146 | No | NA | 1 | 0 | 1 | Not-determined | Genome-fragment | NA | NA | 0 |
| S17-k141_202093 | 2134 | No | NA | 2 | 0 | 0 | Not-determined | Genome-fragment | NA | NA | 0 |
| S17-k141_124242 | 1310 | No | NA | 1 | 0 | 1 | Not-determined | Genome-fragment | NA | NA | 0 |
| S5-final-viral-combined_k141_118704\|\|full | 2438 | No | NA | 3 | 2 | 0 | Low-quality | Genome-fragment | 7.11 | AAI-based (high-confidence) | 0 |
| S17-k141_202130 | 4312 | No | NA | 2 | 0 | 2 | Not-determined | Genome-fragment | NA | NA | 0 |
| S17-k141_113075 | 1201 | No | NA | 2 | 0 | 1 | Not-determined | Genome-fragment | NA | NA | 0 |
| S17-k141_257359 | 6395 | No | NA | 6 | 0 | 5 | Not-determined | Genome-fragment | NA | NA | 0 |
| S17-k141_245876 | 1762 | No | NA | 4 | 0 | 0 | Not-determined | Genome-fragment | NA | NA | 0 |
| S17-k141_224366 | 6774 | No | NA | 5 | 0 | 4 | Not-determined | Genome-fragment | NA | NA | 0 |
| S17-k141_190750 | 1548 | No | NA | 1 | 0 | 0 | Not-determined | Genome-fragment | NA | NA | 0 |
| S17-k141_45779 | 1280 | No | NA | 3 | 0 | 3 | Not-determined | Genome-fragment | NA | NA | 0 |
| S17-k141_146277 | 1077 | No | NA | 1 | 0 | 1 | Not-determined | Genome-fragment | NA | NA | 0 |
| S17-k141_257408 | 1253 | No | NA | 1 | 0 | 0 | Not-determined | Genome-fragment | NA | NA | 0 |
| S17-k141_113100 | 1210 | No | NA | 2 | 0 | 1 | Not-determined | Genome-fragment | NA | NA | 0 |
| S17-k141_57242 | 3827 | No | NA | 3 | 0 | 3 | Not-determined | Genome-fragment | NA | NA | 0 |
| S5-final-viral-combined_k141_178008\|\|full | 2438 | No | NA | 5 | 1 | 0 | Low-quality | Genome-fragment | 7.06 | AAI-based (medium-confidence) | 0 |
| S2-final-viral-combined_k141_74206\|\|full | 2438 | No | NA | 4 | 0 | 1 | Low-quality | Genome-fragment | 7.43 | AAI-based (medium-confidence) | 0 |
| S17-k141_113145 | 1571 | No | NA | 1 | 0 | 1 | Not-determined | Genome-fragment | NA | NA | 0 |
| S17-k141_124308 | 22750 | No | NA | 42 | 12 | 0 | Medium-quality | Genome-fragment | 70.13 | AAI-based (high-confidence) | 0 |
| S17-k141_135212 | 1127 | No | NA | 1 | 0 | 1 | Not-determined | Genome-fragment | NA | NA | 0 |

| | | | | | | | | | | | |
|---|---|---|---|---|---|---|---|---|---|---|---|
| S17-k141_190809 | 1683 | No | NA | 2 | 0 | 2 | Not-determined | Genome-fragment | NA | NA | 0 |
| S17-k141_179642 | 2073 | No | NA | 1 | 0 | 0 | Not-determined | Genome-fragment | NA | NA | 0 |
| S17-k141_190826 | 4729 | No | NA | 4 | 0 | 3 | Not-determined | Genome-fragment | NA | NA | 0 |
| S17-k141_245987 | 3179 | No | NA | 4 | 0 | 3 | Not-determined | Genome-fragment | NA | NA | 0 |
| S17-k141_12426 | 1098 | No | NA | 3 | 0 | 2 | Not-determined | Genome-fragment | NA | NA | 0 |
| S17-k141_12435 | 1896 | No | NA | 2 | 0 | 1 | Not-determined | Genome-fragment | NA | NA | 0 |
| S17-k141_124368 | 1586 | No | NA | 1 | 0 | 1 | Not-determined | Genome-fragment | NA | NA | 0 |
| S17-k141_1860 | 3852 | No | NA | 2 | 0 | 2 | Not-determined | Genome-fragment | NA | NA | 0 |
| S6-final-viral-combined_k141_39375\|\|full | 2438 | No | NA | 3 | 0 | 1 | Low-quality | Genome-fragment | 2.47 | AAI-based (medium-confidence) | 0 |
| S17-k141_12474 | 1884 | No | NA | 3 | 0 | 2 | Not-determined | Genome-fragment | NA | NA | 0 |
| S17-k141_213249 | 2365 | No | NA | 3 | 0 | 0 | Not-determined | Genome-fragment | NA | NA | 0 |
| S17-k141_80076 | 3549 | No | NA | 2 | 0 | 2 | Not-determined | Genome-fragment | NA | NA | 0 |
| S17-k141_146364 | 1215 | No | NA | 1 | 0 | 1 | Not-determined | Genome-fragment | NA | NA | 0 |
| S17-k141_90484 | 4909 | No | NA | 3 | 0 | 1 | Not-determined | Genome-fragment | NA | NA | 0 |
| S17-k141_1907 | 1090 | No | NA | 1 | 0 | 1 | Not-determined | Genome-fragment | NA | NA | 0 |
| S17-k141_202366 | 1573 | No | NA | 1 | 0 | 0 | Not-determined | Genome-fragment | NA | NA | 0 |
| S17-k141_68761 | 4050 | No | NA | 4 | 0 | 2 | Not-determined | Genome-fragment | NA | NA | 0 |
| S7-final-viral-combined_k141_13006\|\|full | 2439 | No | NA | 2 | 0 | 0 | Low-quality | Genome-fragment | 3.91 | AAI-based (high-confidence) | 0 |
| S12-final-viral-combined_k141_22480\|\|full | 2439 | No | NA | 4 | 2 | 0 | Low-quality | Genome-fragment | 6.71 | AAI-based (high-confidence) | 0 |
| S12_k141_372279 | 2439 | No | NA | 3 | 0 | 0 | Low-quality | Genome-fragment | 6.49 | AAI-based (high-confidence) | 0 |
| S17-k141_179748 | 1478 | No | NA | 1 | 0 | 1 | Not-determined | Genome-fragment | NA | NA | 0 |
| S6-final-viral-combined_k141_75064\|\|full | 2440 | No | NA | 4 | 0 | 0 | Low-quality | Genome-fragment | 4.41 | AAI-based (medium-confidence) | 0 |
| S17-k141_124445 | 2132 | No | NA | 2 | 0 | 2 | Not-determined | Genome-fragment | NA | NA | 0 |
| S17-k141_235055 | 3796 | No | NA | 6 | 0 | 4 | Not-determined | Genome-fragment | NA | NA | 0 |
| S8-final-viral-combined_k141_43149\|\|full | 2440 | No | NA | 4 | 1 | 0 | Low-quality | Genome-fragment | 3.96 | AAI-based (medium-confidence) | 0 |
| S17-k141_113341 | 1123 | No | NA | 1 | 0 | 1 | Not-determined | Genome-fragment | NA | NA | 0 |
| S8-final-viral-combined_k141_79618\|\|full | 2440 | No | NA | 4 | 0 | 1 | Low-quality | Genome-fragment | 1.44 | AAI-based (high-confidence) | 0 |
| S17-k141_246046 | 1094 | No | NA | 1 | 0 | 0 | Not-determined | Genome-fragment | NA | NA | 0 |
| S17-k141_12525 | 4298 | No | NA | 2 | 0 | 2 | Not-determined | Genome-fragment | NA | NA | 0 |
| S17-k141_168638 | 1284 | No | NA | 2 | 0 | 0 | Not-determined | Genome-fragment | NA | NA | 0 |
| S17-k141_246059 | 1975 | No | NA | 1 | 0 | 0 | Not-determined | Genome-fragment | NA | NA | 0 |
| S11-final-viral-combined_k141_140968\|\|full | 2441 | No | NA | 4 | 0 | 2 | Low-quality | Genome-fragment | 2 | AAI-based (medium-confidence) | 0 |
| S17-k141_23932 | 1185 | No | NA | 2 | 0 | 0 | Not-determined | Genome-fragment | NA | NA | 0 |
| S17-k141_135439 | 3096 | No | NA | 1 | 0 | 1 | Not-determined | Genome-fragment | NA | NA | 0 |
| S17-k141_213401 | 1310 | No | NA | 1 | 0 | 0 | Not-determined | Genome-fragment | NA | NA | 0 |
| S17-k141_35304 | 1848 | No | NA | 3 | 0 | 0 | Not-determined | Genome-fragment | NA | NA | 0 |
| S17-k141_12549 | 3645 | No | NA | 7 | 0 | 5 | Not-determined | Genome-fragment | NA | NA | 0 |
| S17-k141_168704 | 1999 | No | NA | 2 | 0 | 0 | Not-determined | Genome-fragment | NA | NA | 0 |
| S17-k141_57469 | 1082 | No | NA | 2 | 0 | 1 | Not-determined | Genome-fragment | NA | NA | 0 |
| S17-k141_213455 | 3577 | No | NA | 3 | 0 | 3 | Not-determined | Genome-fragment | NA | NA | 0 |
| S17-k141_146525 | 5830 | No | NA | 4 | 0 | 2 | Not-determined | Genome-fragment | NA | NA | 0 |
| S17-k141_179866 | 1375 | No | NA | 1 | 0 | 1 | Not-determined | Genome-fragment | NA | NA | 0 |
| S17-k141_68901 | 1017 | No | NA | 1 | 0 | 1 | Not-determined | Genome-fragment | NA | NA | 0 |
| S17-k141_235214 | 1437 | No | NA | 5 | 0 | 0 | Not-determined | Genome-fragment | NA | NA | 0 |
| S17-k141_146584 | 1107 | No | NA | 1 | 0 | 0 | Not-determined | Genome-fragment | NA | NA | 0 |
| S17-k141_146596 | 1160 | No | NA | 4 | 0 | 3 | Not-determined | Genome-fragment | NA | NA | 0 |
| S11-final-viral-combined_k141_105807\|\|full | 2441 | No | NA | 4 | 0 | 0 | Low-quality | Genome-fragment | 4.78 | AAI-based (medium-confidence) | 0 |
| S17-k141_257633 | 1243 | No | NA | 3 | 0 | 3 | Not-determined | Genome-fragment | NA | NA | 0 |
| S17-k141_68964 | 5702 | No | NA | 15 | 0 | 14 | Not-determined | Genome-fragment | NA | NA | 0 |
| S17-k141_135589 | 1669 | No | NA | 3 | 0 | 2 | Not-determined | Genome-fragment | NA | NA | 0 |
| S17-k141_146627 | 1416 | No | NA | 1 | 0 | 1 | Not-determined | Genome-fragment | NA | NA | 0 |
| S1-final-viral-combined_k141_336399\|\|full | 2441 | No | NA | 2 | 0 | 2 | Low-quality | Genome-fragment | 3.21 | AAI-based (medium-confidence) | 0 |
| S1-final-viral-combined_k141_346383\|\|lt2gene | 2441 | No | NA | 2 | 2 | 0 | Low-quality | Genome-fragment | 5.38 | AAI-based (high-confidence) | 0 |
| S7-final-viral-combined_k141_114773\|\|full | 2441 | No | NA | 3 | 0 | 0 | Low-quality | Genome-fragment | 6.72 | AAI-based (medium-confidence) | 0 |
| S7-final-viral-combined_k141_3235\|\|full | 2441 | No | NA | 4 | 1 | 0 | Low-quality | Genome-fragment | 1.75 | AAI-based (medium-confidence) | 0 |
| S17-k141_113597 | 1796 | No | NA | 2 | 0 | 0 | Not-determined | Genome-fragment | NA | NA | 0 |
| S17-k141_101868 | 1042 | No | NA | 1 | 0 | 0 | Not-determined | Genome-fragment | NA | NA | 0 |
| S17-k141_124555 | 1966 | No | NA | 3 | 0 | 1 | Not-determined | Genome-fragment | NA | NA | 0 |
| S17-k141_124577 | 1812 | No | NA | 2 | 0 | 1 | Not-determined | Genome-fragment | NA | NA | 0 |
| S12-final-viral-combined_k141_1921\|\|full | 2441 | No | NA | 4 | 0 | 1 | Low-quality | Genome-fragment | 4.27 | AAI-based (medium-confidence) | 0 |
| S17-k141_57625 | 1490 | No | NA | 1 | 0 | 1 | Not-determined | Genome-fragment | NA | NA | 0 |
| S17-k141_124587 | 1754 | No | NA | 2 | 0 | 1 | Not-determined | Genome-fragment | NA | NA | 0 |
| S17-k141_80382 | 3778 | No | NA | 2 | 0 | 1 | Not-determined | Genome-fragment | NA | NA | 0 |
| S17-k141_191185 | 1395 | No | NA | 1 | 0 | 1 | Not-determined | Genome-fragment | NA | NA | 0 |
| S17-k141_90685 | 3499 | No | NA | 4 | 0 | 3 | Not-determined | Genome-fragment | NA | NA | 0 |
| S17-k141_235294 | 11140 | No | NA | 8 | 0 | 2 | Not-determined | Genome-fragment | NA | NA | 0 |
| S6_k141_72667 | 2441 | No | NA | 4 | 0 | 2 | Low-quality | Genome-fragment | 0.88 | AAI-based (high-confidence) | 0 |
| S17-k141_224661 | 6757 | No | NA | 10 | 0 | 3 | Not-determined | Genome-fragment | NA | NA | 0 |
| S17-k141_224668 | 1239 | No | NA | 1 | 0 | 0 | Not-determined | Genome-fragment | NA | NA | 0 |
| S17-k141_46023 | 3832 | No | NA | 5 | 0 | 4 | Not-determined | Genome-fragment | NA | NA | 0 |
| S17-k141_235331 | 1076 | No | NA | 2 | 0 | 2 | Not-determined | Genome-fragment | NA | NA | 0 |
| S17-k141_113729 | 1326 | No | NA | 2 | 0 | 0 | Not-determined | Genome-fragment | NA | NA | 0 |
| S17-k141_213641 | 6406 | No | NA | 7 | 0 | 2 | Not-determined | Genome-fragment | NA | NA | 0 |
| S17-k141_46076 | 1264 | No | NA | 2 | 0 | 1 | Not-determined | Genome-fragment | NA | NA | 0 |
| S17-k141_202725 | 3069 | No | NA | 3 | 0 | 2 | Not-determined | Genome-fragment | NA | NA | 0 |
| S17-k141_168893 | 2153 | No | NA | 1 | 0 | 1 | Not-determined | Genome-fragment | NA | NA | 0 |
| S17-k141_124707 | 1016 | No | NA | 1 | 0 | 1 | Not-determined | Genome-fragment | NA | NA | 0 |
| S17-k141_213652 | 5378 | No | NA | 6 | 0 | 2 | Not-determined | Genome-fragment | NA | NA | 0 |
| S17-k141_24119 | 5968 | No | NA | 6 | 0 | 6 | Not-determined | Genome-fragment | NA | NA | 0 |
| S9_k141_75072 | 2442 | No | NA | 3 | 1 | 0 | Low-quality | Genome-fragment | 6.28 | AAI-based (medium-confidence) | 0 |
| S17-k141_157535 | 4085 | No | NA | 3 | 0 | 1 | Not-determined | Genome-fragment | NA | NA | 0 |
| S17-k141_12820 | 4348 | No | NA | 6 | 0 | 2 | Not-determined | Genome-fragment | NA | NA | 0 |
| S3-final-viral-combined_k141_37987\|\|full | 2442 | No | NA | 3 | 0 | 1 | Low-quality | Genome-fragment | 4.99 | AAI-based (medium-confidence) | 0 |
| S17-k141_80499 | 1428 | No | NA | 2 | 0 | 1 | Not-determined | Genome-fragment | NA | NA | 0 |
| S17-k141_168999 | 1339 | No | NA | 2 | 0 | 1 | Not-determined | Genome-fragment | NA | NA | 0 |
| S17-k141_69146 | 1094 | No | NA | 1 | 0 | 0 | Not-determined | Genome-fragment | NA | NA | 0 |
| S17-k141_46176 | 1017 | No | NA | 2 | 0 | 1 | Not-determined | Genome-fragment | NA | NA | 0 |
| S17-k141_113838 | 1434 | No | NA | 2 | 0 | 2 | Not-determined | Genome-fragment | NA | NA | 0 |
| S17-k141_146794 | 2534 | No | NA | 2 | 0 | 0 | Not-determined | Genome-fragment | NA | NA | 0 |
| S17-k141_113846 | 1455 | No | NA | 2 | 0 | 0 | Not-determined | Genome-fragment | NA | NA | 0 |
| S5-final-viral-combined_k141_92884\|\|full | 2442 | No | NA | 3 | 0 | 1 | Low-quality | Genome-fragment | 1.11 | AAI-based (medium-confidence) | 0 |
| S17-k141_224796 | 1015 | No | NA | 1 | 0 | 1 | Not-determined | Genome-fragment | NA | NA | 0 |
| S17-k141_113857 | 1113 | No | NA | 2 | 0 | 0 | Not-determined | Genome-fragment | NA | NA | 0 |
| S17-k141_202824 | 1445 | No | NA | 2 | 0 | 1 | Not-determined | Genome-fragment | NA | NA | 0 |
| S17-k141_124855 | 2175 | No | NA | 1 | 0 | 1 | Not-determined | Genome-fragment | NA | NA | 0 |
| S17-k141_24201 | 10175 | No | NA | 7 | 0 | 4 | Not-determined | Genome-fragment | NA | NA | 0 |
| S17-k141_191378 | 6093 | No | NA | 4 | 0 | 4 | Not-determined | Genome-fragment | NA | NA | 0 |
| S17-k141_180196 | 6305 | No | NA | 8 | 0 | 2 | Not-determined | Genome-fragment | NA | NA | 0 |
| S17-k141_57776 | 2543 | No | NA | 1 | 0 | 1 | Not-determined | Genome-fragment | NA | NA | 0 |
| S17-k141_12848 | 7552 | No | NA | 8 | 0 | 5 | Not-determined | Genome-fragment | NA | NA | 0 |
| S17-k141_12865 | 2008 | No | NA | 2 | 0 | 2 | Not-determined | Genome-fragment | NA | NA | 0 |
| S17-k141_124917 | 1390 | No | NA | 1 | 0 | 1 | Not-determined | Genome-fragment | NA | NA | 0 |
| S17-k141_224873 | 4986 | No | NA | 5 | 0 | 1 | Not-determined | Genome-fragment | NA | NA | 0 |
| S17-k141_113931 | 5110 | No | NA | 7 | 0 | 2 | Not-determined | Genome-fragment | NA | NA | 0 |
| S17-k141_35624 | 1265 | No | NA | 1 | 0 | 1 | Not-determined | Genome-fragment | NA | NA | 0 |
| S17-k141_257871 | 5496 | No | NA | 5 | 0 | 4 | Not-determined | Genome-fragment | NA | NA | 0 |
| S17-k141_90896 | 4217 | No | NA | 6 | 0 | 2 | Not-determined | Genome-fragment | NA | NA | 0 |
| S17-k141_191432 | 1385 | No | NA | 1 | 0 | 1 | Not-determined | Genome-fragment | NA | NA | 0 |
| S17-k141_202889 | 1231 | No | NA | 2 | 0 | 1 | Not-determined | Genome-fragment | NA | NA | 0 |
| S17-k141_235536 | 30415 | No | NA | 17 | 0 | 6 | Medium-quality | Genome-fragment | 87.71 | AAI-based (medium-confidence) | 0 |
| S17-k141_235537 | 1802 | No | NA | 1 | 0 | 0 | Not-determined | Genome-fragment | NA | NA | 0 |
| S17-k141_102077 | 1011 | No | NA | 3 | 0 | 1 | Not-determined | Genome-fragment | NA | NA | 0 |
| S17-k141_24288 | 1374 | No | NA | 3 | 0 | 1 | Not-determined | Genome-fragment | NA | NA | 0 |
| S17-k141_213798 | 3403 | No | NA | 3 | 0 | 3 | Not-determined | Genome-fragment | NA | NA | 0 |
| S17-k141_169121 | 1248 | No | NA | 1 | 0 | 1 | Not-determined | Genome-fragment | NA | NA | 0 |
| S17-k141_213804 | 1502 | No | NA | 2 | 0 | 0 | Not-determined | Genome-fragment | NA | NA | 0 |
| S17-k141_169134 | 8123 | No | NA | 7 | 0 | 5 | Not-determined | Genome-fragment | NA | NA | 0 |
| S5-final-viral-combined_k141_26006\|\|full | 2442 | No | NA | 4 | 2 | 0 | Low-quality | Genome-fragment | 2.71 | AAI-based (high-confidence) | 0 |
| S17-k141_125003 | 1517 | No | NA | 3 | 0 | 3 | Not-determined | Genome-fragment | NA | NA | 0 |
| S17-k141_213833 | 3179 | No | NA | 5 | 0 | 2 | Not-determined | Genome-fragment | NA | NA | 0 |
| S17-k141_114013 | 1135 | No | NA | 4 | 0 | 4 | Not-determined | Genome-fragment | NA | NA | 0 |
| S17-k141_69314 | 4834 | No | NA | 1 | 0 | 0 | Not-determined | Genome-fragment | NA | NA | 0 |
| S17-k141_246455 | 1494 | No | NA | 2 | 0 | 1 | Not-determined | Genome-fragment | NA | NA | 0 |
| S17-k141_169202 | 1123 | No | NA | 2 | 0 | 1 | Not-determined | Genome-fragment | NA | NA | 0 |
| S17-k141_146963 | 1309 | No | NA | 2 | 0 | 0 | Not-determined | Genome-fragment | NA | NA | 0 |
| S17-k141_213862 | 1717 | No | NA | 1 | 0 | 1 | Not-determined | Genome-fragment | NA | NA | 0 |
| S17-k141_157675 | 3398 | No | NA | 2 | 0 | 1 | Not-determined | Genome-fragment | NA | NA | 0 |
| S17-k141_213882 | 3609 | No | NA | 4 | 0 | 3 | Not-determined | Genome-fragment | NA | NA | 0 |
| S17-k141_12987 | 1279 | No | NA | 2 | 0 | 2 | Not-determined | Genome-fragment | NA | NA | 0 |
| S17-k141_135965 | 1016 | No | NA | 1 | 0 | 0 | Not-determined | Genome-fragment | NA | NA | 0 |
| S17-k141_102142 | 1183 | No | NA | 1 | 0 | 1 | Not-determined | Genome-fragment | NA | NA | 0 |
| S17-k141_180302 | 2173 | No | NA | 2 | 0 | 1 | Not-determined | Genome-fragment | NA | NA | 0 |
| S17-k141_203012 | 1370 | No | NA | 2 | 0 | 2 | Not-determined | Genome-fragment | NA | NA | 0 |
| S17-k141_225079 | 1186 | No | NA | 2 | 0 | 2 | Not-determined | Genome-fragment | NA | NA | 0 |
| S17-k141_258074 | 1653 | No | NA | 1 | 0 | 1 | Not-determined | Genome-fragment | NA | NA | 0 |
| S17-k141_213928 | 1819 | No | NA | 1 | 0 | 1 | Not-determined | Genome-fragment | NA | NA | 0 |
| S17-final-viral-combined_k141_104590\|\|full | 2442 | No | NA | 2 | 0 | 2 | Low-quality | Genome-fragment | 2.11 | AAI-based (high-confidence) | 0 |
| S17-k141_235712 | 1202 | No | NA | 2 | 0 | 0 | Not-determined | Genome-fragment | NA | NA | 0 |
| S17-k141_258095 | 1194 | No | NA | 1 | 0 | 0 | Not-determined | Genome-fragment | NA | NA | 0 |
| S17-k141_80625 | 6425 | No | NA | 9 | 0 | 2 | Not-determined | Genome-fragment | NA | NA | 0 |
| S17-k141_114186 | 3140 | No | NA | 2 | 0 | 2 | Not-determined | Genome-fragment | NA | NA | 0 |
| S12-final-viral-combined_k141_176003\|\|full | 2442 | No | NA | 4 | 2 | 0 | Low-quality | Genome-fragment | 4.86 | AAI-based (high-confidence) | 0 |
| S17-k141_180384 | 1059 | No | NA | 1 | 0 | 1 | Not-determined | Genome-fragment | NA | NA | 0 |
| S17-k141_225107 | 7595 | No | NA | 8 | 0 | 6 | Not-determined | Genome-fragment | NA | NA | 0 |
| S17-k141_114216 | 1305 | No | NA | 1 | 0 | 1 | Not-determined | Genome-fragment | NA | NA | 0 |
| S17-k141_203115 | 5110 | No | NA | 5 | 0 | 2 | Not-determined | Genome-fragment | NA | NA | 0 |
| S12-final-viral-combined_k141_217031\|\|full | 2442 | No | NA | 3 | 0 | 2 | Low-quality | Genome-fragment | 1.88 | AAI-based (medium-confidence) | 0 |
| S2-final-viral-combined_k141_231589\|\|full | 2442 | No | NA | 4 | 0 | 2 | Low-quality | Genome-fragment | 1.46 | AAI-based (medium-confidence) | 0 |
| S17-k141_203147 | 1064 | No | NA | 1 | 0 | 1 | Not-determined | Genome-fragment | NA | NA | 0 |
| S17-k141_191668 | 17844 | No | NA | 17 | 0 | 4 | Not-determined | Genome-fragment | NA | NA | 0 |
| S17-k141_157768 | 1050 | No | NA | 2 | 0 | 2 | Not-determined | Genome-fragment | NA | NA | 0 |
| S17-k141_80720 | 1136 | No | NA | 1 | 0 | 1 | Not-determined | Genome-fragment | NA | NA | 0 |
| S17-k141_80722 | 2435 | No | NA | 2 | 0 | 1 | Not-determined | Genome-fragment | NA | NA | 0 |
| S12-final-viral-combined_k141_283803\|\|full | 2443 | No | NA | 4 | 0 | 0 | Low-quality | Genome-fragment | 4.99 | AAI-based (medium-confidence) | 0 |
| S17-k141_180460 | 1228 | No | NA | 1 | 0 | 0 | Not-determined | Genome-fragment | NA | NA | 0 |
| S2_k141_51811 | 2443 | No | NA | 5 | 4 | 0 | Low-quality | Genome-fragment | 5.7 | AAI-based (high-confidence) | 0 |
| S17-k141_2499 | 1872 | No | NA | 1 | 0 | 1 | Not-determined | Genome-fragment | NA | NA | 0 |
| S17-k141_169406 | 4919 | No | NA | 4 | 0 | 1 | Not-determined | Genome-fragment | NA | NA | 0 |
| S17-k141_147211 | 1068 | No | NA | 2 | 0 | 2 | Not-determined | Genome-fragment | NA | NA | 0 |
| S17-k141_114349 | 2647 | No | NA | 3 | 0 | 0 | Not-determined | Genome-fragment | NA | NA | 0 |
| S17-k141_35899 | 1529 | No | NA | 1 | 0 | 0 | Not-determined | Genome-fragment | NA | NA | 0 |
| S17-k141_203214 | 3311 | No | NA | 2 | 0 | 1 | Not-determined | Genome-fragment | NA | NA | 0 |
| S17-k141_125140 | 1104 | No | NA | 1 | 0 | 1 | Not-determined | Genome-fragment | NA | NA | 0 |
| S17-k141_2510 | 4608 | No | NA | 4 | 0 | 2 | Not-determined | Genome-fragment | NA | NA | 0 |
| S8-final-viral-combined_k141_193956\|\|full | 2443 | No | NA | 5 | 2 | 0 | Low-quality | Genome-fragment | 4.47 | AAI-based (high-confidence) | 0 |
| S17-k141_169481 | 4707 | No | NA | 3 | 0 | 2 | Not-determined | Genome-fragment | NA | NA | 0 |
| S17-k141_114393 | 3378 | No | NA | 2 | 0 | 1 | Not-determined | Genome-fragment | NA | NA | 0 |
| S17-k141_246672 | 2849 | No | NA | 1 | 0 | 0 | Not-determined | Genome-fragment | NA | NA | 0 |
| S17-k141_169494 | 1100 | No | NA | 1 | 0 | 1 | Not-determined | Genome-fragment | NA | NA | 0 |
| S17-k141_235920 | 1514 | No | NA | 2 | 0 | 0 | Not-determined | Genome-fragment | NA | NA | 0 |
| S17-k141_147278 | 1230 | No | NA | 1 | 0 | 1 | Not-determined | Genome-fragment | NA | NA | 0 |
| S17-k141_58042 | 1337 | No | NA | 5 | 0 | 0 | Not-determined | Genome-fragment | NA | NA | 0 |
| S11-final-viral-combined_k141_64714\|\|full | 2444 | No | NA | 5 | 0 | 0 | Low-quality | Genome-fragment | 6.59 | AAI-based (high-confidence) | 0 |
| S17-k141_24482 | 1215 | No | NA | 2 | 0 | 2 | Not-determined | Genome-fragment | NA | NA | 0 |
| S17-k141_80819 | 1255 | No | NA | 1 | 0 | 1 | Not-determined | Genome-fragment | NA | NA | 0 |
| S17-k141_147305 | 1292 | No | NA | 2 | 0 | 2 | Not-determined | Genome-fragment | NA | NA | 0 |
| S17-k141_191742 | 1250 | No | NA | 1 | 0 | 1 | Not-determined | Genome-fragment | NA | NA | 0 |
| S12_k141_184114 | 2444 | No | NA | 4 | 2 | 0 | Low-quality | Genome-fragment | 8.79 | HMM-based (lower-bound) | 0 |
| S17-k141_114469 | 7401 | No | NA | 5 | 0 | 5 | Not-determined | Genome-fragment | NA | NA | 0 |
| S6-final-viral-combined_k141_44340\|\|full | 2444 | No | NA | 4 | 0 | 0 | Low-quality | Genome-fragment | 3.85 | AAI-based (high-confidence) | 0 |
| S17-k141_246758 | 3300 | No | NA | 5 | 0 | 3 | Not-determined | Genome-fragment | NA | NA | 0 |
| S9_k141_34557 | 2445 | No | NA | 3 | 3 | 0 | Low-quality | Genome-fragment | 1.6 | HMM-based (lower-bound) | 0 |
| S17-k141_180616 | 2121 | No | NA | 2 | 0 | 2 | Not-determined | Genome-fragment | NA | NA | 0 |
| S17-k141_169565 | 1520 | No | NA | 2 | 0 | 2 | Not-determined | Genome-fragment | NA | NA | 0 |
| S17-k141_191824 | 1527 | No | NA | 2 | 0 | 2 | Not-determined | Genome-fragment | NA | NA | 0 |
| S17-k141_203344 | 2736 | No | NA | 3 | 0 | 2 | Not-determined | Genome-fragment | NA | NA | 0 |
| S17-k141_114515 | 1554 | No | NA | 1 | 0 | 1 | Not-determined | Genome-fragment | NA | NA | 0 |
| S17-k141_46432 | 5736 | No | NA | 9 | 0 | 2 | Not-determined | Genome-fragment | NA | NA | 0 |
| S17-k141_136257 | 1178 | No | NA | 3 | 0 | 1 | Not-determined | Genome-fragment | NA | NA | 0 |
| S17-k141_24561 | 3417 | No | NA | 2 | 0 | 2 | Not-determined | Genome-fragment | NA | NA | 0 |
| S17-k141_180770 | 2270 | No | NA | 1 | 0 | 1 | Not-determined | Genome-fragment | NA | NA | 0 |
| S17-k141_236092 | 1439 | No | NA | 4 | 0 | 1 | Not-determined | Genome-fragment | NA | NA | 0 |
| S1-final-viral-combined_k141_9952\|\|full | 2445 | No | NA | 5 | 2 | 0 | Low-quality | Genome-fragment | 5.14 | AAI-based (medium-confidence) | 0 |
| S12-final-viral-combined_k141_48890\|\|full | 2445 | No | NA | 3 | 0 | 0 | Low-quality | Genome-fragment | 5 | AAI-based (medium-confidence) | 0 |
| S17-k141_136324 | 1057 | No | NA | 2 | 0 | 0 | Not-determined | Genome-fragment | NA | NA | 0 |
| S17-k141_114589 | 1496 | No | NA | 3 | 0 | 0 | Not-determined | Genome-fragment | NA | NA | 0 |
| S17-k141_258352 | 1443 | No | NA | 1 | 0 | 0 | Not-determined | Genome-fragment | NA | NA | 0 |
| S17-k141_191865 | 1273 | No | NA | 1 | 0 | 1 | Not-determined | Genome-fragment | NA | NA | 0 |
| S17-k141_191870 | 1345 | No | NA | 2 | 0 | 2 | Not-determined | Genome-fragment | NA | NA | 0 |
| S17-k141_157988 | 1210 | No | NA | 1 | 0 | 1 | Not-determined | Genome-fragment | NA | NA | 0 |
| S17-k141_225362 | 1011 | No | NA | 2 | 0 | 0 | Not-determined | Genome-fragment | NA | NA | 0 |
| S17-k141_236115 | 11662 | No | NA | 1 | 0 | 0 | Not-determined | Genome-fragment | NA | NA | 0 |
| S2_k141_126898 | 2445 | No | NA | 3 | 0 | 0 | Low-quality | Genome-fragment | 6.2 | AAI-based (medium-confidence) | 0 |
| S8-final-viral-combined_k141_82941\|\|full | 2445 | No | NA | 6 | 2 | 0 | Low-quality | Genome-fragment | 6.1 | AAI-based (high-confidence) | 0 |
| S17-k141_125320 | 1720 | No | NA | 3 | 0 | 2 | Not-determined | Genome-fragment | NA | NA | 0 |
| S9-final-viral-combined_k141_108405\|\|full | 2446 | No | NA | 4 | 0 | 1 | Low-quality | Genome-fragment | 5.13 | AAI-based (medium-confidence) | 0 |
| S17-k141_125338 | 6251 | No | NA | 5 | 0 | 5 | Not-determined | Genome-fragment | NA | NA | 0 |
| S17-k141_225378 | 1560 | No | NA | 1 | 0 | 0 | Not-determined | Genome-fragment | NA | NA | 0 |
| S17-k141_69782 | 1527 | No | NA | 3 | 0 | 2 | Not-determined | Genome-fragment | NA | NA | 0 |
| S17-k141_191905 | 1191 | No | NA | 1 | 0 | 0 | Not-determined | Genome-fragment | NA | NA | 0 |
| S17-k141_158028 | 1115 | No | NA | 1 | 0 | 1 | Not-determined | Genome-fragment | NA | NA | 0 |
| S11-final-viral-combined_k141_289968\|\|full | 2446 | No | NA | 4 | 0 | 1 | Low-quality | Genome-fragment | 4.21 | AAI-based (medium-confidence) | 0 |
| S17-k141_2774 | 1090 | No | NA | 2 | 0 | 1 | Not-determined | Genome-fragment | NA | NA | 0 |
| S17-k141_136399 | 1385 | No | NA | 3 | 0 | 2 | Not-determined | Genome-fragment | NA | NA | 0 |
| S17-k141_180835 | 1095 | No | NA | 2 | 0 | 2 | Not-determined | Genome-fragment | NA | NA | 0 |
| S17-k141_246841 | 3112 | No | NA | 2 | 0 | 1 | Not-determined | Genome-fragment | NA | NA | 0 |
| S17-k141_180836 | 4376 | No | NA | 4 | 0 | 3 | Not-determined | Genome-fragment | NA | NA | 0 |
| S17-k141_114636 | 1564 | No | NA | 1 | 0 | 0 | Not-determined | Genome-fragment | NA | NA | 0 |
| S17-k141_58352 | 6656 | No | NA | 9 | 0 | 4 | Not-determined | Genome-fragment | NA | NA | 0 |
| S8-final-viral-combined_k141_195726\|\|full | 2446 | No | NA | 3 | 0 | 0 | Low-quality | Genome-fragment | 5.66 | AAI-based (high-confidence) | 0 |
| S17-k141_36152 | 1720 | No | NA | 2 | 0 | 0 | Not-determined | Genome-fragment | NA | NA | 0 |
| S11-final-viral-combined_k141_309192\|\|full | 2447 | No | NA | 3 | 0 | 0 | Low-quality | Genome-fragment | 5.92 | AAI-based (medium-confidence) | 0 |
| S11-final-viral-combined_k141_243481\|\|full | 2447 | No | NA | 6 | 1 | 0 | Low-quality | Genome-fragment | 3.22 | AAI-based (high-confidence) | 0 |
| S17-k141_69834 | 1421 | No | NA | 2 | 0 | 2 | Not-determined | Genome-fragment | NA | NA | 0 |
| S1-final-viral-combined_k141_101775\|\|full | 2447 | No | NA | 2 | 0 | 2 | Low-quality | Genome-fragment | 1.41 | AAI-based (medium-confidence) | 0 |
| S7-final-viral-combined_k141_5457\|\|full | 2447 | No | NA | 3 | 0 | 0 | Low-quality | Genome-fragment | 5.39 | AAI-based (medium-confidence) | 0 |
| S17-k141_225497 | 1050 | No | NA | 1 | 0 | 1 | Not-determined | Genome-fragment | NA | NA | 0 |
| S17-k141_81198 | 2183 | No | NA | 2 | 0 | 2 | Not-determined | Genome-fragment | NA | NA | 0 |
| S17-k141_147507 | 1221 | No | NA | 2 | 0 | 0 | Not-determined | Genome-fragment | NA | NA | 0 |
| S17-k141_258460 | 1122 | No | NA | 2 | 0 | 1 | Not-determined | Genome-fragment | NA | NA | 0 |
| S17-final-viral-combined_k141_20347\|\|full | 2447 | No | NA | 6 | 0 | 0 | Low-quality | Genome-fragment | 4.49 | AAI-based (high-confidence) | 0 |
| S17-k141_203490 | 10124 | No | NA | 10 | 0 | 1 | Not-determined | Genome-fragment | NA | NA | 0 |
| S17-k141_81264 | 4924 | No | NA | 3 | 0 | 3 | Not-determined | Genome-fragment | NA | NA | 0 |
| S17-k141_236216 | 1129 | No | NA | 3 | 0 | 1 | Not-determined | Genome-fragment | NA | NA | 0 |
| S17-k141_246987 | 3029 | No | NA | 1 | 0 | 0 | Not-determined | Genome-fragment | NA | NA | 0 |
| S17-k141_236219 | 6982 | No | NA | 7 | 0 | 5 | Not-determined | Genome-fragment | NA | NA | 0 |
| S17-k141_91281 | 3296 | No | NA | 2 | 0 | 2 | Not-determined | Genome-fragment | NA | NA | 0 |
| S17-k141_192044 | 2505 | No | NA | 1 | 0 | 1 | Not-determined | Genome-fragment | NA | NA | 0 |
| S17-k141_258492 | 1150 | No | NA | 1 | 0 | 1 | Not-determined | Genome-fragment | NA | NA | 0 |
| S17-k141_91285 | 1756 | No | NA | 1 | 0 | 1 | Not-determined | Genome-fragment | NA | NA | 0 |
| S17-k141_192053 | 1235 | No | NA | 1 | 0 | 1 | Not-determined | Genome-fragment | NA | NA | 0 |
| S17-k141_236237 | 2762 | No | NA | 4 | 0 | 0 | Not-determined | Genome-fragment | NA | NA | 0 |
| S17-k141_225545 | 1063 | No | NA | 1 | 0 | 0 | Not-determined | Genome-fragment | NA | NA | 0 |
| S17-k141_147578 | 2763 | No | NA | 2 | 0 | 0 | Not-determined | Genome-fragment | NA | NA | 0 |
| S17-k141_114871 | 1756 | No | NA | 1 | 0 | 1 | Not-determined | Genome-fragment | NA | NA | 0 |
| S17-k141_81301 | 5752 | No | NA | 11 | 0 | 1 | Not-determined | Genome-fragment | NA | NA | 0 |
| S17-k141_214620 | 2648 | No | NA | 4 | 0 | 2 | Not-determined | Genome-fragment | NA | NA | 0 |
| S12-final-viral-combined_k141_180671\|\|full | 2447 | No | NA | 4 | 1 | 0 | Low-quality | Genome-fragment | 4.48 | HMM-based (lower-bound) | 0 |
| S17-k141_214630 | 5633 | No | NA | 2 | 0 | 1 | Not-determined | Genome-fragment | NA | NA | 0 |
| S2-final-viral-combined_k141_198995\|\|full | 2447 | No | NA | 4 | 0 | 0 | Low-quality | Genome-fragment | 4.84 | AAI-based (medium-confidence) | 0 |
| S17-k141_69920 | 7412 | No | NA | 7 | 0 | 4 | Not-determined | Genome-fragment | NA | NA | 0 |
| S17-k141_125616 | 1348 | No | NA | 1 | 0 | 1 | Not-determined | Genome-fragment | NA | NA | 0 |
| S17-k141_169816 | 3395 | No | NA | 2 | 0 | 0 | Not-determined | Genome-fragment | NA | NA | 0 |
| S17-final-viral-combined_k141_92112\|\|full | 2448 | No | NA | 3 | 0 | 3 | Low-quality | Genome-fragment | 3.86 | AAI-based (medium-confidence) | 0 |
| S17-k141_258580 | 1171 | No | NA | 2 | 0 | 2 | Not-determined | Genome-fragment | NA | NA | 0 |
| S17-k141_58381 | 1080 | No | NA | 3 | 0 | 2 | Not-determined | Genome-fragment | NA | NA | 0 |
| S17-final-viral-combined_k141_127955\|\|full | 2448 | No | NA | 5 | 1 | 0 | Low-quality | Genome-fragment | 3.59 | HMM-based (lower-bound) | 0 |
| S17-k141_69937 | 1389 | No | NA | 1 | 0 | 0 | Not-determined | Genome-fragment | NA | NA | 0 |
| S17-k141_125703 | 1947 | No | NA | 3 | 0 | 3 | Not-determined | Genome-fragment | NA | NA | 0 |
| S17-k141_81377 | 4195 | No | NA | 4 | 0 | 0 | Not-determined | Genome-fragment | NA | NA | 0 |
| S17-k141_236295 | 1131 | No | NA | 1 | 0 | 0 | Not-determined | Genome-fragment | NA | NA | 0 |
| S17-k141_236297 | 4941 | No | NA | 3 | 0 | 3 | Not-determined | Genome-fragment | NA | NA | 0 |
| S17-k141_192223 | 1209 | No | NA | 4 | 0 | 0 | Not-determined | Genome-fragment | NA | NA | 0 |
| S12-final-viral-combined_k141_169954\|\|lt2gene | 2448 | No | NA | 1 | 0 | 0 | Low-quality | Genome-fragment | 5.12 | AAI-based (medium-confidence) | 0 |
| S17-k141_102923 | 1115 | No | NA | 1 | 0 | 1 | Not-determined | Genome-fragment | NA | NA | 0 |
| S17-k141_236309 | 1057 | No | NA | 1 | 0 | 1 | Not-determined | Genome-fragment | NA | NA | 0 |
| S17-k141_214716 | 2363 | No | NA | 1 | 0 | 1 | Not-determined | Genome-fragment | NA | NA | 0 |
| S17-k141_147696 | 1100 | No | NA | 1 | 0 | 1 | Not-determined | Genome-fragment | NA | NA | 0 |
| S17-k141_181232 | 1566 | No | NA | 1 | 0 | 0 | Not-determined | Genome-fragment | NA | NA | 0 |
| S17-k141_24974 | 1178 | No | NA | 1 | 0 | 1 | Not-determined | Genome-fragment | NA | NA | 0 |
| S2_k141_89525 | 2448 | No | NA | 4 | 1 | 0 | Low-quality | Genome-fragment | 2.55 | AAI-based (high-confidence) | 0 |
| S3-final-viral-combined_k141_35588\|\|full | 2449 | No | NA | 4 | 0 | 0 | Low-quality | Genome-fragment | 4.18 | AAI-based (high-confidence) | 0 |
| S2-final-viral-combined_k141_121663\|\|full | 2449 | No | NA | 5 | 0 | 0 | Low-quality | Genome-fragment | 2.4 | AAI-based (medium-confidence) | 0 |
| S17-k141_125745 | 5264 | No | NA | 5 | 0 | 4 | Not-determined | Genome-fragment | NA | NA | 0 |
| S2-final-viral-combined_k141_9599\|\|full | 2449 | No | NA | 3 | 0 | 1 | Low-quality | Genome-fragment | 1.84 | AAI-based (high-confidence) | 0 |
| S17-k141_70079 | 1070 | No | NA | 1 | 0 | 1 | Not-determined | Genome-fragment | NA | NA | 0 |
| S17-k141_81520 | 1276 | No | NA | 1 | 0 | 1 | Not-determined | Genome-fragment | NA | NA | 0 |
| S17-k141_81527 | 5511 | No | NA | 9 | 0 | 2 | Not-determined | Genome-fragment | NA | NA | 0 |
| S17-k141_225759 | 2148 | No | NA | 2 | 0 | 1 | Not-determined | Genome-fragment | NA | NA | 0 |
| S17-k141_36491 | 5378 | No | NA | 6 | 0 | 6 | Not-determined | Genome-fragment | NA | NA | 0 |
| S5_k141_194704 | 2450 | No | NA | 2 | 1 | 0 | Low-quality | Genome-fragment | 5.51 | AAI-based (high-confidence) | 0 |
| S17-k141_147870 | 1442 | No | NA | 3 | 0 | 0 | Not-determined | Genome-fragment | NA | NA | 0 |
| S17-k141_192435 | 1122 | No | NA | 2 | 0 | 2 | Not-determined | Genome-fragment | NA | NA | 0 |
| S17-k141_147904 | 1757 | No | NA | 2 | 0 | 2 | Not-determined | Genome-fragment | NA | NA | 0 |
| S17-k141_70133 | 3012 | No | NA | 4 | 0 | 2 | Not-determined | Genome-fragment | NA | NA | 0 |
| S17-k141_136878 | 3543 | No | NA | 6 | 0 | 1 | Not-determined | Genome-fragment | NA | NA | 0 |
| S7-final-viral-combined_k141_86528\|\|full | 2450 | No | NA | 4 | 0 | 1 | Low-quality | Genome-fragment | 3.54 | AAI-based (medium-confidence) | 0 |
| S17-k141_158283 | 4769 | No | NA | 6 | 0 | 4 | Not-determined | Genome-fragment | NA | NA | 0 |
| S17-k141_214850 | 1044 | No | NA | 2 | 0 | 2 | Not-determined | Genome-fragment | NA | NA | 0 |
| S17-k141_125885 | 1026 | No | NA | 2 | 0 | 2 | Not-determined | Genome-fragment | NA | NA | 0 |
| S17-k141_136888 | 3737 | No | NA | 4 | 0 | 3 | Not-determined | Genome-fragment | NA | NA | 0 |
| S12-final-viral-combined_k141_197983\|\|full | 2450 | No | NA | 3 | 1 | 0 | Low-quality | Genome-fragment | 2.63 | AAI-based (high-confidence) | 0 |
| S11_k141_267127 | 2451 | No | NA | 1 | 0 | 1 | Low-quality | Genome-fragment | 2.24 | AAI-based (medium-confidence) | 0 |
| S17-k141_258686 | 1569 | No | NA | 1 | 0 | 1 | Not-determined | Genome-fragment | NA | NA | 0 |
| S7-final-viral-combined_k141_64035\|\|full | 2451 | No | NA | 2 | 0 | 0 | Low-quality | Genome-fragment | 1.91 | AAI-based (medium-confidence) | 0 |
| S17-k141_181423 | 2650 | No | NA | 3 | 0 | 2 | Not-determined | Genome-fragment | NA | NA | 0 |
| S17-k141_3287 | 1268 | No | NA | 1 | 0 | 1 | Not-determined | Genome-fragment | NA | NA | 0 |
| S17-k141_147922 | 1126 | No | NA | 1 | 0 | 1 | Not-determined | Genome-fragment | NA | NA | 0 |
| S17-k141_147951 | 1301 | No | NA | 1 | 0 | 0 | Not-determined | Genome-fragment | NA | NA | 0 |
| S17-final-viral-combined_k141_47031\|\|full | 2452 | No | NA | 6 | 0 | 0 | Low-quality | Genome-fragment | 6.08 | AAI-based (medium-confidence) | 0 |
| S17-k141_236595 | 1038 | No | NA | 3 | 0 | 2 | Not-determined | Genome-fragment | NA | NA | 0 |
| S17-k141_25216 | 1189 | No | NA | 2 | 0 | 2 | Not-determined | Genome-fragment | NA | NA | 0 |
| S17-final-viral-combined_k141_5982\|\|full | 2452 | No | NA | 3 | 2 | 0 | Low-quality | Genome-fragment | 6 | AAI-based (high-confidence) | 0 |
| S2-final-viral-combined_k141_210602\|\|full | 2452 | No | NA | 2 | 0 | 0 | Low-quality | Genome-fragment | 5.5 | AAI-based (medium-confidence) | 0 |
| S17-k141_81662 | 2943 | No | NA | 2 | 0 | 0 | Not-determined | Genome-fragment | NA | NA | 0 |
| S1-final-viral-combined_k141_124697\|\|full | 2453 | No | NA | 5 | 1 | 0 | Low-quality | Genome-fragment | 1.55 | HMM-based (lower-bound) | 0 |
| S1-final-viral-combined_k141_369607\|\|full | 2454 | No | NA | 4 | 0 | 0 | Low-quality | Genome-fragment | 4.58 | AAI-based (medium-confidence) | 0 |
| S17-k141_247264 | 8529 | No | NA | 10 | 0 | 3 | Not-determined | Genome-fragment | NA | NA | 0 |
| S17-k141_136991 | 1105 | No | NA | 1 | 0 | 1 | Not-determined | Genome-fragment | NA | NA | 0 |
| S17-k141_247272 | 1193 | No | NA | 1 | 0 | 1 | Not-determined | Genome-fragment | NA | NA | 0 |
| S17-k141_81686 | 1318 | No | NA | 2 | 0 | 1 | Not-determined | Genome-fragment | NA | NA | 0 |
| S17-k141_115314 | 1192 | No | NA | 1 | 0 | 0 | Not-determined | Genome-fragment | NA | NA | 0 |
| S3-final-viral-combined_k141_2079\|\|full | 2454 | No | NA | 6 | 3 | 0 | Low-quality | Genome-fragment | 6.39 | AAI-based (high-confidence) | 0 |
| S17-k141_115321 | 1040 | No | NA | 2 | 0 | 1 | Not-determined | Genome-fragment | NA | NA | 0 |
| S17-k141_158416 | 3011 | No | NA | 4 | 0 | 1 | Not-determined | Genome-fragment | NA | NA | 0 |
| S17-k141_203958 | 2154 | No | NA | 1 | 0 | 1 | Not-determined | Genome-fragment | NA | NA | 0 |
| S12-final-viral-combined_k141_128544\|\|full | 2454 | No | NA | 4 | 1 | 2 | Low-quality | Genome-fragment | 4.06 | AAI-based (medium-confidence) | 0 |
| S17-k141_203977 | 2162 | No | NA | 3 | 0 | 1 | Not-determined | Genome-fragment | NA | NA | 0 |
| S17-k141_225968 | 1139 | No | NA | 2 | 0 | 0 | Not-determined | Genome-fragment | NA | NA | 0 |
| S17-k141_36683 | 2168 | No | NA | 4 | 0 | 3 | Not-determined | Genome-fragment | NA | NA | 0 |
| S4-final-viral-combined_k141_125759\|\|lt2gene | 2454 | No | NA | 2 | 1 | 0 | Low-quality | Genome-fragment | 1.38 | AAI-based (high-confidence) | 0 |
| S17-k141_81731 | 6602 | No | NA | 9 | 0 | 0 | Not-determined | Genome-fragment | NA | NA | 0 |
| S3-final-viral-combined_k141_11444\|\|full | 2455 | No | NA | 6 | 0 | 0 | Low-quality | Genome-fragment | 4.29 | AAI-based (high-confidence) | 0 |
| S4_k141_11211 | 2455 | No | NA | 5 | 1 | 0 | Low-quality | Genome-fragment | 1.35 | AAI-based (high-confidence) | 0 |
| S17-k141_58792 | 3139 | No | NA | 5 | 0 | 2 | Not-determined | Genome-fragment | NA | NA | 0 |
| S2-final-viral-combined_k141_97380\|\|full | 2456 | No | NA | 4 | 1 | 0 | Low-quality | Genome-fragment | 6.05 | AAI-based (medium-confidence) | 0 |
| S17-k141_58804 | 1058 | No | NA | 2 | 0 | 1 | Not-determined | Genome-fragment | NA | NA | 0 |
| S17-k141_192673 | 5347 | No | NA | 10 | 0 | 9 | Not-determined | Genome-fragment | NA | NA | 0 |
| S17-k141_192690 | 2309 | No | NA | 2 | 0 | 1 | Not-determined | Genome-fragment | NA | NA | 0 |
| S17-k141_226017 | 1400 | No | NA | 1 | 0 | 1 | Not-determined | Genome-fragment | NA | NA | 0 |
| S17-k141_236674 | 1763 | No | NA | 1 | 0 | 0 | Not-determined | Genome-fragment | NA | NA | 0 |
| S17-k141_192699 | 1183 | No | NA | 2 | 0 | 2 | Not-determined | Genome-fragment | NA | NA | 0 |
| S17-k141_126120 | 12102 | No | NA | 8 | 0 | 1 | Not-determined | Genome-fragment | NA | NA | 0 |
| S17-k141_226043 | 2710 | No | NA | 2 | 0 | 0 | Not-determined | Genome-fragment | NA | NA | 0 |
| S17-k141_81861 | 1217 | No | NA | 1 | 0 | 1 | Not-determined | Genome-fragment | NA | NA | 0 |
| S17-k141_137104 | 1277 | No | NA | 1 | 0 | 0 | Not-determined | Genome-fragment | NA | NA | 0 |
| S17-k141_36727 | 1702 | No | NA | 1 | 0 | 1 | Not-determined | Genome-fragment | NA | NA | 0 |
| S17-k141_103301 | 2092 | No | NA | 2 | 0 | 2 | Not-determined | Genome-fragment | NA | NA | 0 |
| S17-k141_170215 | 3572 | No | NA | 4 | 0 | 3 | Not-determined | Genome-fragment | NA | NA | 0 |
| S17-k141_192742 | 1377 | No | NA | 2 | 0 | 2 | Not-determined | Genome-fragment | NA | NA | 0 |
| S17-k141_3503 | 1010 | No | NA | 2 | 0 | 0 | Not-determined | Genome-fragment | NA | NA | 0 |
| S17-k141_137141 | 1449 | No | NA | 1 | 0 | 0 | Not-determined | Genome-fragment | NA | NA | 0 |
| S17-k141_36757 | 3682 | No | NA | 4 | 0 | 3 | Not-determined | Genome-fragment | NA | NA | 0 |
| S17-k141_103334 | 1029 | No | NA | 1 | 0 | 0 | Not-determined | Genome-fragment | NA | NA | 0 |
| S17-k141_247460 | 1661 | No | NA | 1 | 0 | 0 | Not-determined | Genome-fragment | NA | NA | 0 |
| S17-k141_148142 | 3769 | No | NA | 3 | 0 | 1 | Not-determined | Genome-fragment | NA | NA | 0 |
| S8_k141_227793 | 2456 | No | NA | 5 | 1 | 0 | Low-quality | Genome-fragment | 1.84 | AAI-based (high-confidence) | 0 |
| S17-k141_3538 | 1562 | No | NA | 2 | 0 | 2 | Not-determined | Genome-fragment | NA | NA | 0 |
| S7-final-viral-combined_k141_90183\|\|full | 2457 | No | NA | 5 | 0 | 0 | Low-quality | Genome-fragment | 4.4 | AAI-based (high-confidence) | 0 |
| S17-k141_70495 | 1120 | No | NA | 3 | 0 | 0 | Not-determined | Genome-fragment | NA | NA | 0 |
| S17-k141_25455 | 1224 | No | NA | 2 | 0 | 1 | Not-determined | Genome-fragment | NA | NA | 0 |
| S7-final-viral-combined_k141_179470\|\|lt2gene | 2457 | No | NA | 1 | 1 | 0 | Low-quality | Genome-fragment | 6.82 | AAI-based (high-confidence) | 0 |
| S17-k141_204094 | 1126 | No | NA | 1 | 0 | 0 | Not-determined | Genome-fragment | NA | NA | 0 |
| S17-k141_115652 | 1177 | No | NA | 1 | 0 | 0 | Not-determined | Genome-fragment | NA | NA | 0 |
| S17-k141_70559 | 1826 | No | NA | 1 | 0 | 1 | Not-determined | Genome-fragment | NA | NA | 0 |
| S17-k141_215296 | 1054 | No | NA | 1 | 0 | 1 | Not-determined | Genome-fragment | NA | NA | 0 |
| S17-k141_148227 | 1922 | No | NA | 1 | 0 | 1 | Not-determined | Genome-fragment | NA | NA | 0 |
| S17-k141_70573 | 1470 | No | NA | 2 | 0 | 1 | Not-determined | Genome-fragment | NA | NA | 0 |
| S17-k141_192861 | 3200 | No | NA | 3 | 0 | 1 | Not-determined | Genome-fragment | NA | NA | 0 |
| S17-k141_170285 | 3228 | No | NA | 4 | 0 | 2 | Not-determined | Genome-fragment | NA | NA | 0 |
| S17-k141_170289 | 1466 | No | NA | 2 | 0 | 0 | Not-determined | Genome-fragment | NA | NA | 0 |
| S17-k141_170294 | 1102 | No | NA | 1 | 0 | 1 | Not-determined | Genome-fragment | NA | NA | 0 |
| S17-k141_70627 | 1503 | No | NA | 1 | 0 | 1 | Not-determined | Genome-fragment | NA | NA | 0 |
| S17-k141_236757 | 1095 | No | NA | 2 | 0 | 2 | Not-determined | Genome-fragment | NA | NA | 0 |
| S17-k141_25525 | 1584 | No | NA | 2 | 0 | 0 | Not-determined | Genome-fragment | NA | NA | 0 |
| S17-k141_103487 | 1100 | No | NA | 2 | 0 | 1 | Not-determined | Genome-fragment | NA | NA | 0 |
| S17-k141_258933 | 1177 | No | NA | 2 | 0 | 0 | Not-determined | Genome-fragment | NA | NA | 0 |
| S17-k141_247545 | 1679 | No | NA | 1 | 0 | 1 | Not-determined | Genome-fragment | NA | NA | 0 |
| S12-final-viral-combined_k141_349617\|\|full | 2457 | No | NA | 3 | 1 | 0 | Low-quality | Genome-fragment | 6.44 | AAI-based (high-confidence) | 0 |
| S17-k141_70676 | 3928 | No | NA | 4 | 0 | 3 | Not-determined | Genome-fragment | NA | NA | 0 |
| S17-k141_81990 | 19438 | No | NA | 14 | 0 | 9 | Not-determined | Genome-fragment | NA | NA | 0 |
| S17-k141_70719 | 1540 | No | NA | 2 | 0 | 0 | Not-determined | Genome-fragment | NA | NA | 0 |
| S17-k141_258967 | 3831 | No | NA | 7 | 0 | 6 | Not-determined | Genome-fragment | NA | NA | 0 |
| S4-final-viral-combined_k141_111316\|\|full | 2457 | No | NA | 4 | 2 | 0 | Low-quality | Genome-fragment | 5.91 | AAI-based (high-confidence) | 0 |
| S17-k141_137201 | 3126 | No | NA | 1 | 0 | 1 | Not-determined | Genome-fragment | NA | NA | 0 |
| S8-final-viral-combined_k141_216552\|\|full | 2457 | No | NA | 3 | 0 | 1 | Low-quality | Genome-fragment | 0.86 | AAI-based (medium-confidence) | 0 |
| S17-k141_247580 | 1276 | No | NA | 3 | 0 | 2 | Not-determined | Genome-fragment | NA | NA | 0 |
| S8-final-viral-combined_k141_203867\|\|full | 2457 | No | NA | 8 | 2 | 0 | Low-quality | Genome-fragment | 4.76 | AAI-based (high-confidence) | 0 |
| S1-final-viral-combined_k141_329266\|\|full | 2458 | No | NA | 4 | 0 | 0 | Low-quality | Genome-fragment | 5.07 | AAI-based (medium-confidence) | 0 |
| S17-k141_36867 | 1629 | No | NA | 1 | 0 | 1 | Not-determined | Genome-fragment | NA | NA | 0 |
| S17-k141_25600 | 1154 | No | NA | 1 | 0 | 1 | Not-determined | Genome-fragment | NA | NA | 0 |
| S17-k141_226261 | 1005 | No | NA | 2 | 0 | 1 | Not-determined | Genome-fragment | NA | NA | 0 |
| S17-k141_193000 | 1399 | No | NA | 3 | 0 | 2 | Not-determined | Genome-fragment | NA | NA | 0 |
| S17-k141_193003 | 1087 | No | NA | 3 | 0 | 0 | Not-determined | Genome-fragment | NA | NA | 0 |
| S17-k141_70819 | 3218 | No | NA | 5 | 0 | 3 | Not-determined | Genome-fragment | NA | NA | 0 |
| S17-k141_70820 | 1070 | No | NA | 1 | 0 | 1 | Not-determined | Genome-fragment | NA | NA | 0 |
| S17-final-viral-combined_k141_46087\|\|full | 2458 | No | NA | 4 | 0 | 0 | Low-quality | Genome-fragment | 6.62 | AAI-based (medium-confidence) | 0 |
| S17-k141_25644 | 1729 | No | NA | 1 | 0 | 1 | Not-determined | Genome-fragment | NA | NA | 0 |
| S17-k141_204277 | 2576 | No | NA | 3 | 0 | 2 | Not-determined | Genome-fragment | NA | NA | 0 |
| S17-k141_14148 | 3405 | No | NA | 2 | 0 | 2 | Not-determined | Genome-fragment | NA | NA | 0 |
| S8-final-viral-combined_k141_70304\|\|full | 2458 | No | NA | 4 | 0 | 0 | Low-quality | Genome-fragment | 1.47 | AAI-based (medium-confidence) | 0 |
| S17-k141_91878 | 1028 | No | NA | 4 | 0 | 1 | Not-determined | Genome-fragment | NA | NA | 0 |
| S8_k141_237035 | 2458 | No | NA | 2 | 1 | 0 | Low-quality | Genome-fragment | 1.89 | HMM-based (lower-bound) | 0 |
| S5-final-viral-combined_k141_151530\|\|full | 2459 | No | NA | 2 | 0 | 1 | Low-quality | Genome-fragment | 1.14 | AAI-based (high-confidence) | 0 |
| S17-k141_14199 | 2186 | No | NA | 1 | 0 | 1 | Not-determined | Genome-fragment | NA | NA | 0 |
| S17-k141_36903 | 21036 | No | NA | 2 | 0 | 2 | Not-determined | Genome-fragment | NA | NA | 0 |
| S5-final-viral-combined_k141_8855\|\|full | 2459 | No | NA | 4 | 1 | 0 | Low-quality | Genome-fragment | 5.18 | AAI-based (medium-confidence) | 0 |
| S17-k141_126327 | 1780 | No | NA | 2 | 0 | 2 | Not-determined | Genome-fragment | NA | NA | 0 |
| S17-k141_91896 | 1185 | No | NA | 1 | 0 | 1 | Not-determined | Genome-fragment | NA | NA | 0 |
| S17-k141_204367 | 2292 | No | NA | 2 | 0 | 2 | Not-determined | Genome-fragment | NA | NA | 0 |
| S17-k141_59101 | 3274 | No | NA | 3 | 0 | 3 | Not-determined | Genome-fragment | NA | NA | 0 |
| S12-final-viral-combined_k141_198556\|\|full | 2459 | No | NA | 3 | 0 | 1 | Low-quality | Genome-fragment | 1.17 | AAI-based (medium-confidence) | 0 |
| S12_k141_398446 | 2459 | No | NA | 5 | 0 | 1 | Low-quality | Genome-fragment | 8.52 | AAI-based (high-confidence) | 0 |
| S11-final-viral-combined_k141_39002\|\|full | 2460 | Yes | 1837 | 3 | 1 | 1 | Low-quality | Genome-fragment | 1.41 | HMM-based (lower-bound) | 25.33 |
| S17-final-viral-combined_k141_56677\|\|full | 2460 | No | NA | 3 | 0 | 0 | Low-quality | Genome-fragment | 5.85 | AAI-based (medium-confidence) | 0 |
| S17-k141_103646 | 1855 | No | NA | 2 | 0 | 2 | Not-determined | Genome-fragment | NA | NA | 0 |
| S17-k141_126346 | 1099 | No | NA | 1 | 0 | 1 | Not-determined | Genome-fragment | NA | NA | 0 |
| S17-k141_14282 | 1151 | No | NA | 1 | 0 | 0 | Not-determined | Genome-fragment | NA | NA | 0 |
| S17-k141_103651 | 1253 | No | NA | 3 | 0 | 2 | Not-determined | Genome-fragment | NA | NA | 0 |
| S17-k141_148486 | 1992 | No | NA | 1 | 0 | 1 | Not-determined | Genome-fragment | NA | NA | 0 |
| S17-final-viral-combined_k141_82120\|\|full | 2460 | No | NA | 3 | 0 | 2 | Low-quality | Genome-fragment | 1.2 | AAI-based (high-confidence) | 0 |
| S17-k141_158828 | 7172 | No | NA | 3 | 0 | 2 | Not-determined | Genome-fragment | NA | NA | 0 |
| S17-k141_82249 | 4451 | No | NA | 5 | 0 | 2 | Not-determined | Genome-fragment | NA | NA | 0 |

| | | | | | | | | | | | |
|---|---|---|---|---|---|---|---|---|---|---|---|
| S17-final-viral-combined_k141_6140\|\|full | 2460 | No | NA | 3 | 0 | 0 | Low-quality | Genome-fragment | 4.86 | AAI-based (medium-confidence) | 0 |
| S17-k141_204456 | 1740 | No | NA | 3 | 0 | 1 | Not-determined | Genome-fragment | NA | NA | 0 |
| S17-k141_116074 | 1077 | No | NA | 1 | 0 | 1 | Not-determined | Genome-fragment | NA | NA | 0 |
| S17-k141_247663 | 1596 | No | NA | 1 | 0 | 1 | Not-determined | Genome-fragment | NA | NA | 0 |
| S17-k141_137472 | 1321 | No | NA | 1 | 0 | 1 | Not-determined | Genome-fragment | NA | NA | 0 |
| S17-k141_148518 | 2165 | No | NA | 1 | 0 | 1 | Not-determined | Genome-fragment | NA | NA | 0 |
| S17-k141_59187 | 6457 | No | NA | 4 | 0 | 4 | Not-determined | Genome-fragment | NA | NA | 0 |
| S12-final-viral-combined_k141_255164\|\|full | 2460 | No | NA | 2 | 0 | 2 | Low-quality | Genome-fragment | 4.73 | AAI-based (medium-confidence) | 0 |
| S2-final-viral-combined_k141_119152\|\|full | 2460 | No | NA | 3 | 0 | 2 | Low-quality | Genome-fragment | 2.39 | AAI-based (medium-confidence) | 0 |
| S17-k141_182193 | 5063 | No | NA | 3 | 0 | 2 | Not-determined | Genome-fragment | NA | NA | 0 |
| S6-final-viral-combined_k141_73953\|\|full | 2460 | No | NA | 4 | 0 | 4 | Low-quality | Genome-fragment | 2.03 | AAI-based (medium-confidence) | 0 |
| S17-k141_182220 | 1278 | No | NA | 1 | 0 | 1 | Not-determined | Genome-fragment | NA | NA | 0 |
| S5-final-viral-combined_k141_58660\|\|full | 2461 | No | NA | 5 | 3 | 0 | Low-quality | Genome-fragment | 5.17 | AAI-based (high-confidence) | 0 |
| S17-k141_137549 | 1127 | No | NA | 1 | 0 | 1 | Not-determined | Genome-fragment | NA | NA | 0 |
| S17-k141_82348 | 6090 | No | NA | 3 | 0 | 0 | Not-determined | Genome-fragment | NA | NA | 0 |
| S17-k141_37065 | 1635 | No | NA | 2 | 0 | 2 | Not-determined | Genome-fragment | NA | NA | 0 |
| S17-k141_148562 | 1327 | No | NA | 2 | 0 | 0 | Not-determined | Genome-fragment | NA | NA | 0 |
| S17-k141_126484 | 7778 | No | NA | 8 | 0 | 6 | Not-determined | Genome-fragment | NA | NA | 0 |
| S17-k141_37072 | 1756 | No | NA | 1 | 0 | 1 | Not-determined | Genome-fragment | NA | NA | 0 |
| S17-k141_215713 | 1037 | No | NA | 2 | 0 | 1 | Not-determined | Genome-fragment | NA | NA | 0 |
| S17-k141_247730 | 1305 | No | NA | 1 | 0 | 1 | Not-determined | Genome-fragment | NA | NA | 0 |
| S17-k141_126496 | 1520 | No | NA | 2 | 0 | 0 | Not-determined | Genome-fragment | NA | NA | 0 |
| S17-k141_126503 | 1116 | No | NA | 1 | 0 | 1 | Not-determined | Genome-fragment | NA | NA | 0 |
| S17-k141_126525 | 4311 | No | NA | 1 | 0 | 1 | Not-determined | Genome-fragment | NA | NA | 0 |
| S17-k141_170615 | 1017 | No | NA | 1 | 0 | 1 | Not-determined | Genome-fragment | NA | NA | 0 |
| S17-k141_182281 | 8336 | No | NA | 11 | 0 | 1 | Not-determined | Genome-fragment | NA | NA | 0 |
| S17-k141_47073 | 1640 | No | NA | 2 | 0 | 1 | Not-determined | Genome-fragment | NA | NA | 0 |
| S17-k141_158929 | 1624 | No | NA | 1 | 0 | 0 | Not-determined | Genome-fragment | NA | NA | 0 |
| S17-k141_37139 | 5057 | No | NA | 3 | 0 | 3 | Not-determined | Genome-fragment | NA | NA | 0 |
| S17-k141_247783 | 1325 | No | NA | 2 | 0 | 0 | Not-determined | Genome-fragment | NA | NA | 0 |
| S17-final-viral-combined_k141_89420\|\|full | 2461 | No | NA | 3 | 0 | 2 | Low-quality | Genome-fragment | 1.43 | AAI-based (high-confidence) | 0 |
| S17-k141_247795 | 4170 | No | NA | 8 | 0 | 8 | Not-determined | Genome-fragment | NA | NA | 0 |
| S17-k141_37173 | 1507 | No | NA | 2 | 0 | 2 | Not-determined | Genome-fragment | NA | NA | 0 |
| S8-final-viral-combined_k141_53056\|\|full | 2461 | No | NA | 6 | 3 | 0 | Low-quality | Genome-fragment | 6.78 | AAI-based (high-confidence) | 0 |
| S17-final-viral-combined_k141_243803\|\|full | 2462 | No | NA | 3 | 0 | 2 | Low-quality | Genome-fragment | 0.78 | AAI-based (medium-confidence) | 0 |
| S17-k141_215854 | 3704 | No | NA | 4 | 0 | 0 | Not-determined | Genome-fragment | NA | NA | 0 |
| S17-k141_116248 | 37748 | No | NA | 56 | 25 | 3 | High-quality | High-quality | 100 | AAI-based (high-confidence) | 0 |
| S17-k141_215855 | 3099 | No | NA | 5 | 0 | 1 | Not-determined | Genome-fragment | NA | NA | 0 |
| S12-final-viral-combined_k141_368806\|\|full | 2462 | No | NA | 3 | 0 | 0 | Low-quality | Genome-fragment | 2.81 | AAI-based (medium-confidence) | 0 |
| S17-k141_82509 | 1117 | No | NA | 2 | 0 | 1 | Not-determined | Genome-fragment | NA | NA | 0 |
| S17-k141_82513 | 2478 | No | NA | 1 | 0 | 0 | Not-determined | Genome-fragment | NA | NA | 0 |
| S17-k141_47113 | 1062 | No | NA | 2 | 0 | 1 | Not-determined | Genome-fragment | NA | NA | 0 |
| S17-k141_237251 | 4899 | No | NA | 3 | 0 | 2 | Not-determined | Genome-fragment | NA | NA | 0 |
| S17-k141_116292 | 1332 | No | NA | 1 | 0 | 1 | Not-determined | Genome-fragment | NA | NA | 0 |
| S17-k141_71135 | 1254 | No | NA | 2 | 0 | 1 | Not-determined | Genome-fragment | NA | NA | 0 |
| S17-k141_82553 | 4136 | No | NA | 4 | 0 | 3 | Not-determined | Genome-fragment | NA | NA | 0 |
| S17-k141_170732 | 2075 | No | NA | 2 | 0 | 1 | Not-determined | Genome-fragment | NA | NA | 0 |
| S12_k141_8902 | 2462 | No | NA | 1 | 0 | 0 | Low-quality | Genome-fragment | 2.27 | AAI-based (medium-confidence) | 0 |
| S1-final-viral-combined_k141_185261\|\|full | 2463 | No | NA | 2 | 0 | 2 | Low-quality | Genome-fragment | 1.39 | AAI-based (medium-confidence) | 0 |
| S17-k141_237314 | 1101 | No | NA | 1 | 0 | 1 | Not-determined | Genome-fragment | NA | NA | 0 |
| S17-k141_82620 | 1275 | No | NA | 1 | 0 | 0 | Not-determined | Genome-fragment | NA | NA | 0 |
| S17-k141_237323 | 1051 | No | NA | 1 | 0 | 1 | Not-determined | Genome-fragment | NA | NA | 0 |
| S17-k141_193470 | 1241 | No | NA | 1 | 0 | 0 | Not-determined | Genome-fragment | NA | NA | 0 |
| S1-final-viral-combined_k141_296151\|\|full | 2463 | No | NA | 4 | 0 | 1 | Low-quality | Genome-fragment | 3.52 | AAI-based (medium-confidence) | 0 |
| S17-k141_37278 | 1075 | No | NA | 2 | 0 | 2 | Not-determined | Genome-fragment | NA | NA | 0 |
| S17-k141_170814 | 1355 | No | NA | 1 | 0 | 0 | Not-determined | Genome-fragment | NA | NA | 0 |
| S17-k141_170826 | 1256 | No | NA | 1 | 0 | 0 | Not-determined | Genome-fragment | NA | NA | 0 |
| S17-k141_237329 | 1728 | No | NA | 2 | 0 | 1 | Not-determined | Genome-fragment | NA | NA | 0 |
| S17-k141_3934 | 1039 | No | NA | 3 | 0 | 0 | Not-determined | Genome-fragment | NA | NA | 0 |
| S17-k141_71229 | 6838 | No | NA | 7 | 0 | 7 | Not-determined | Genome-fragment | NA | NA | 0 |
| S17-k141_3941 | 4177 | No | NA | 4 | 0 | 3 | Not-determined | Genome-fragment | NA | NA | 0 |
| S17-k141_82699 | 1184 | No | NA | 2 | 0 | 1 | Not-determined | Genome-fragment | NA | NA | 0 |
| S17-k141_259176 | 3018 | No | NA | 2 | 0 | 1 | Not-determined | Genome-fragment | NA | NA | 0 |
| S17-k141_159047 | 1344 | No | NA | 2 | 0 | 0 | Not-determined | Genome-fragment | NA | NA | 0 |
| S17-k141_26119 | 3339 | No | NA | 2 | 0 | 2 | Not-determined | Genome-fragment | NA | NA | 0 |
| S17-k141_215981 | 1900 | No | NA | 3 | 0 | 0 | Not-determined | Genome-fragment | NA | NA | 0 |
| S17-k141_137797 | 1885 | No | NA | 3 | 0 | 0 | Not-determined | Genome-fragment | NA | NA | 0 |
| S17-k141_71304 | 1278 | No | NA | 1 | 0 | 1 | Not-determined | Genome-fragment | NA | NA | 0 |
| S17-k141_82792 | 2707 | No | NA | 1 | 0 | 1 | Not-determined | Genome-fragment | NA | NA | 0 |
| S17-k141_193532 | 1092 | No | NA | 2 | 0 | 0 | Not-determined | Genome-fragment | NA | NA | 0 |
| S17-k141_26174 | 1033 | No | NA | 1 | 0 | 1 | Not-determined | Genome-fragment | NA | NA | 0 |
| S17-k141_193547 | 1263 | No | NA | 1 | 0 | 0 | Not-determined | Genome-fragment | NA | NA | 0 |
| S17-k141_247968 | 10900 | No | NA | 14 | 0 | 1 | Not-determined | Genome-fragment | NA | NA | 0 |
| S17-final-viral-combined_k141_254758\|\|full | 2463 | No | NA | 6 | 5 | 0 | Low-quality | Genome-fragment | 6.97 | AAI-based (high-confidence) | 0 |
| S8-final-viral-combined_k141_16712\|\|full | 2463 | No | NA | 3 | 1 | 0 | Low-quality | Genome-fragment | 2.65 | HMM-based (lower-bound) | 0 |
| S1-final-viral-combined_k141_64919\|\|full | 2464 | No | NA | 3 | 0 | 2 | Low-quality | Genome-fragment | 1.81 | AAI-based (high-confidence) | 0 |
| S17-k141_227026 | 4162 | No | NA | 2 | 0 | 2 | Not-determined | Genome-fragment | NA | NA | 0 |
| S3-final-viral-combined_k141_83338\|\|lt2gene | 2464 | No | NA | 2 | 2 | 0 | Low-quality | Genome-fragment | 0.94 | HMM-based (lower-bound) | 0 |
| S4-final-viral-combined_k141_27227\|\|full | 2464 | No | NA | 4 | 0 | 1 | Low-quality | Genome-fragment | 3.23 | AAI-based (medium-confidence) | 0 |
| S17-k141_159150 | 6267 | No | NA | 11 | 0 | 2 | Not-determined | Genome-fragment | NA | NA | 0 |
| S17-k141_104231 | 1258 | No | NA | 1 | 0 | 0 | Not-determined | Genome-fragment | NA | NA | 0 |
| S17-k141_116613 | 1229 | No | NA | 2 | 0 | 1 | Not-determined | Genome-fragment | NA | NA | 0 |
| S17-k141_14573 | 1041 | No | NA | 1 | 0 | 1 | Not-determined | Genome-fragment | NA | NA | 0 |
| S9-final-viral-combined_k141_125026\|\|lt2gene | 2465 | No | NA | 1 | 1 | 0 | Low-quality | Genome-fragment | 6.91 | AAI-based (high-confidence) | 0 |
| S1-final-viral-combined_k141_300736\|\|full | 2465 | No | NA | 3 | 0 | 0 | Low-quality | Genome-fragment | 5.57 | AAI-based (medium-confidence) | 0 |
| S7-final-viral-combined_k141_65629\|\|full | 2465 | No | NA | 4 | 1 | 2 | Low-quality | Genome-fragment | 3.59 | HMM-based (lower-bound) | 0 |
| S17-k141_4125 | 1164 | No | NA | 1 | 0 | 0 | Not-determined | Genome-fragment | NA | NA | 0 |
| S17-k141_204784 | 20084 | No | NA | 24 | 0 | 19 | Not-determined | Genome-fragment | NA | NA | 0 |
| S17-k141_59555 | 1378 | No | NA | 2 | 0 | 2 | Not-determined | Genome-fragment | NA | NA | 0 |
| S17-k141_4155 | 1727 | No | NA | 4 | 0 | 0 | Not-determined | Genome-fragment | NA | NA | 0 |
| S17-k141_237527 | 1033 | No | NA | 2 | 0 | 1 | Not-determined | Genome-fragment | NA | NA | 0 |
| S17-k141_227124 | 1033 | No | NA | 2 | 0 | 0 | Not-determined | Genome-fragment | NA | NA | 0 |
| S17-k141_237536 | 1039 | No | NA | 2 | 0 | 1 | Not-determined | Genome-fragment | NA | NA | 0 |
| S17-k141_237539 | 3761 | No | NA | 4 | 0 | 2 | Not-determined | Genome-fragment | NA | NA | 0 |
| S17-k141_182726 | 1839 | No | NA | 1 | 0 | 1 | Not-determined | Genome-fragment | NA | NA | 0 |
| S17-final-viral-combined_k141_476\|\|full | 2465 | No | NA | 4 | 2 | 0 | Low-quality | Genome-fragment | 5.36 | AAI-based (high-confidence) | 0 |
| S17-k141_26313 | 1066 | No | NA | 2 | 0 | 1 | Not-determined | Genome-fragment | NA | NA | 0 |
| S2-final-viral-combined_k141_29464\|\|full | 2465 | No | NA | 3 | 0 | 1 | Low-quality | Genome-fragment | 2.77 | AAI-based (medium-confidence) | 0 |
| S17-k141_248086 | 3932 | No | NA | 4 | 0 | 1 | Not-determined | Genome-fragment | NA | NA | 0 |
| S8-final-viral-combined_k141_130865\|\|full | 2465 | No | NA | 3 | 0 | 0 | Low-quality | Genome-fragment | 15.9 | AAI-based (high-confidence) | 0 |
| S17-k141_216196 | 3838 | No | NA | 1 | 0 | 1 | Not-determined | Genome-fragment | NA | NA | 0 |
| S17-k141_83041 | 1629 | No | NA | 4 | 0 | 2 | Not-determined | Genome-fragment | NA | NA | 0 |
| S17-k141_14642 | 1167 | No | NA | 2 | 0 | 0 | Not-determined | Genome-fragment | NA | NA | 0 |
| S17-k141_193695 | 3958 | No | NA | 4 | 0 | 3 | Not-determined | Genome-fragment | NA | NA | 0 |
| S8_k141_128847 | 2465 | No | NA | 4 | 0 | 0 | Low-quality | Genome-fragment | 3.23 | AAI-based (medium-confidence) | 0 |
| S17-k141_193701 | 1274 | No | NA | 1 | 0 | 1 | Not-determined | Genome-fragment | NA | NA | 0 |
| S17-k141_259344 | 1328 | No | NA | 2 | 0 | 0 | Not-determined | Genome-fragment | NA | NA | 0 |
| S1-final-viral-combined_k141_234666\|\|full | 2466 | Yes | 568 | 4 | 1 | 1 | Low-quality | Genome-fragment | 1.27 | AAI-based (medium-confidence) | 76.97 |
| S17-k141_193729 | 1358 | No | NA | 3 | 0 | 2 | Not-determined | Genome-fragment | NA | NA | 0 |
| S5-final-viral-combined_k141_104827\|\|full | 2466 | No | NA | 2 | 0 | 0 | Low-quality | Genome-fragment | 5.74 | AAI-based (high-confidence) | 0 |
| S17-k141_237627 | 1423 | No | NA | 2 | 0 | 2 | Not-determined | Genome-fragment | NA | NA | 0 |
| S17-k141_159271 | 1955 | No | NA | 2 | 0 | 0 | Not-determined | Genome-fragment | NA | NA | 0 |
| S17-k141_116795 | 5145 | No | NA | 6 | 0 | 1 | Not-determined | Genome-fragment | NA | NA | 0 |
| S17-k141_26386 | 5054 | No | NA | 7 | 0 | 0 | Not-determined | Genome-fragment | NA | NA | 0 |
| S12_k141_380323 | 2466 | No | NA | 2 | 0 | 2 | Low-quality | Genome-fragment | 1.41 | AAI-based (medium-confidence) | 0 |
| S17-k141_227224 | 1209 | No | NA | 4 | 0 | 0 | Not-determined | Genome-fragment | NA | NA | 0 |
| S17-k141_259365 | 3310 | No | NA | 3 | 0 | 3 | Not-determined | Genome-fragment | NA | NA | 0 |
| S17-k141_149040 | 1120 | No | NA | 1 | 0 | 0 | Not-determined | Genome-fragment | NA | NA | 0 |
| S8-final-viral-combined_k141_148799\|\|full | 2466 | No | NA | 4 | 1 | 0 | Low-quality | Genome-fragment | 2.77 | HMM-based (lower-bound) | 0 |
| S17-k141_137961 | 1341 | No | NA | 1 | 0 | 0 | Not-determined | Genome-fragment | NA | NA | 0 |
| S11-final-viral-combined_k141_185310\|\|full | 2467 | No | NA | 2 | 0 | 1 | Low-quality | Genome-fragment | 1.93 | AAI-based (medium-confidence) | 0 |
| S5-final-viral-combined_k141_49596\|\|full | 2467 | No | NA | 3 | 0 | 2 | Low-quality | Genome-fragment | 1.59 | AAI-based (medium-confidence) | 0 |
| S17-k141_193807 | 1129 | No | NA | 1 | 0 | 1 | Not-determined | Genome-fragment | NA | NA | 0 |
| S17-k141_83064 | 1020 | No | NA | 2 | 0 | 0 | Not-determined | Genome-fragment | NA | NA | 0 |
| S2-final-viral-combined_k141_117840\|\|full | 2467 | No | NA | 3 | 2 | 0 | Low-quality | Genome-fragment | 4.2 | AAI-based (high-confidence) | 0 |
| S17-k141_37607 | 3470 | No | NA | 3 | 0 | 1 | Not-determined | Genome-fragment | NA | NA | 0 |
| S17-k141_216259 | 1244 | No | NA | 1 | 0 | 1 | Not-determined | Genome-fragment | NA | NA | 0 |
| S17-k141_159365 | 1508 | No | NA | 3 | 0 | 3 | Not-determined | Genome-fragment | NA | NA | 0 |
| S17-k141_182900 | 4193 | No | NA | 6 | 0 | 3 | Not-determined | Genome-fragment | NA | NA | 0 |
| S17-k141_149133 | 1574 | No | NA | 1 | 0 | 1 | Not-determined | Genome-fragment | NA | NA | 0 |
| S17-k141_92438 | 1309 | No | NA | 3 | 0 | 0 | Not-determined | Genome-fragment | NA | NA | 0 |
| S17-k141_171172 | 3071 | No | NA | 3 | 0 | 3 | Not-determined | Genome-fragment | NA | NA | 0 |
| S17-k141_92445 | 3245 | No | NA | 3 | 0 | 2 | Not-determined | Genome-fragment | NA | NA | 0 |
| S1-final-viral-combined_k141_305382\|\|full | 2468 | No | NA | 3 | 0 | 0 | Low-quality | Genome-fragment | 2.31 | AAI-based (high-confidence) | 0 |
| S17-k141_149160 | 1210 | No | NA | 1 | 0 | 1 | Not-determined | Genome-fragment | NA | NA | 0 |
| S17-k141_83164 | 1582 | No | NA | 1 | 0 | 0 | Not-determined | Genome-fragment | NA | NA | 0 |
| S17-k141_248288 | 2036 | No | NA | 2 | 0 | 1 | Not-determined | Genome-fragment | NA | NA | 0 |
| S17-k141_193874 | 2051 | No | NA | 2 | 0 | 0 | Not-determined | Genome-fragment | NA | NA | 0 |
| S17-k141_138038 | 3324 | No | NA | 2 | 0 | 1 | Not-determined | Genome-fragment | NA | NA | 0 |
| S17-k141_83207 | 1609 | No | NA | 2 | 0 | 2 | Not-determined | Genome-fragment | NA | NA | 0 |
| S17-k141_37691 | 3395 | No | NA | 3 | 0 | 3 | Not-determined | Genome-fragment | NA | NA | 0 |
| S17-k141_159425 | 2316 | No | NA | 3 | 0 | 2 | Not-determined | Genome-fragment | NA | NA | 0 |
| S17-k141_26461 | 1019 | No | NA | 2 | 0 | 0 | Not-determined | Genome-fragment | NA | NA | 0 |
| S17-k141_59835 | 1054 | No | NA | 2 | 0 | 0 | Not-determined | Genome-fragment | NA | NA | 0 |
| S3-final-viral-combined_k141_104402\|\|full | 2468 | No | NA | 3 | 0 | 1 | Low-quality | Genome-fragment | 2.07 | AAI-based (medium-confidence) | 0 |
| S17-k141_104447 | 3386 | No | NA | 4 | 0 | 2 | Not-determined | Genome-fragment | NA | NA | 0 |
| S17-final-viral-combined_k141_139077\|\|full | 2468 | No | NA | 3 | 0 | 2 | Low-quality | Genome-fragment | 1.77 | AAI-based (high-confidence) | 0 |
| S17-k141_227431 | 4226 | No | NA | 8 | 0 | 4 | Not-determined | Genome-fragment | NA | NA | 0 |
| S17-k141_92474 | 6135 | No | NA | 3 | 0 | 0 | Not-determined | Genome-fragment | NA | NA | 0 |
| S17-k141_59851 | 1514 | No | NA | 3 | 0 | 0 | Not-determined | Genome-fragment | NA | NA | 0 |
| S17-k141_193898 | 1345 | No | NA | 1 | 0 | 0 | Not-determined | Genome-fragment | NA | NA | 0 |
| S17-k141_83236 | 4727 | No | NA | 3 | 0 | 2 | Not-determined | Genome-fragment | NA | NA | 0 |
| S17-k141_183005 | 1279 | No | NA | 2 | 0 | 2 | Not-determined | Genome-fragment | NA | NA | 0 |
| S17-k141_240054 | 2468 | No | NA | 2 | 0 | 0 | Low-quality | Genome-fragment | 3.53 | AAI-based (medium-confidence) | 0 |
| S12-final-viral-combined_k141_120940\|\|full | 2468 | No | NA | 4 | 1 | 0 | Low-quality | Genome-fragment | 7.24 | AAI-based (high-confidence) | 0 |
| S17-k141_183018 | 5487 | No | NA | 8 | 0 | 3 | Not-determined | Genome-fragment | NA | NA | 0 |
| S17-k141_171279 | 1062 | No | NA | 1 | 0 | 1 | Not-determined | Genome-fragment | NA | NA | 0 |
| S17-k141_183024 | 1401 | No | NA | 3 | 0 | 1 | Not-determined | Genome-fragment | NA | NA | 0 |
| S12-final-viral-combined_k141_140198\|\|full | 2468 | No | NA | 2 | 0 | 1 | Low-quality | Genome-fragment | 1.2 | AAI-based (medium-confidence) | 0 |
| S17-k141_237730 | 3499 | No | NA | 4 | 0 | 3 | Not-determined | Genome-fragment | NA | NA | 0 |
| S17-k141_14825 | 1921 | No | NA | 1 | 0 | 1 | Not-determined | Genome-fragment | NA | NA | 0 |
| S17-k141_26539 | 1563 | No | NA | 1 | 0 | 1 | Not-determined | Genome-fragment | NA | NA | 0 |
| S12_k141_241289 | 2468 | No | NA | 2 | 0 | 2 | Low-quality | Genome-fragment | 0.92 | AAI-based (high-confidence) | 0 |
| S17-k141_47532 | 1023 | No | NA | 2 | 0 | 0 | Not-determined | Genome-fragment | NA | NA | 0 |
| S17-k141_205103 | 1875 | No | NA | 2 | 0 | 1 | Not-determined | Genome-fragment | NA | NA | 0 |
| S17-k141_47542 | 1323 | No | NA | 2 | 0 | 1 | Not-determined | Genome-fragment | NA | NA | 0 |
| S17-k141_237762 | 7499 | No | NA | 9 | 0 | 3 | Not-determined | Genome-fragment | NA | NA | 0 |
| S17-k141_205139 | 1235 | No | NA | 2 | 0 | 2 | Not-determined | Genome-fragment | NA | NA | 0 |
| S17-k141_117115 | 1192 | No | NA | 1 | 0 | 1 | Not-determined | Genome-fragment | NA | NA | 0 |
| S8-final-viral-combined_k141_195537\|\|full | 2468 | No | NA | 4 | 0 | 0 | Low-quality | Genome-fragment | 15.95 | AAI-based (high-confidence) | 0 |
| S17-k141_259423 | 1554 | No | NA | 3 | 0 | 0 | Not-determined | Genome-fragment | NA | NA | 0 |
| S11-final-viral-combined_k141_238196\|\|lt2gene | 2469 | No | NA | 1 | 1 | 0 | Low-quality | Genome-fragment | 5.32 | AAI-based (medium-confidence) | 0 |
| S17-k141_216427 | 1375 | No | NA | 2 | 0 | 0 | Not-determined | Genome-fragment | NA | NA | 0 |
| S17-k141_216428 | 9317 | No | NA | 2 | 0 | 2 | Not-determined | Genome-fragment | NA | NA | 0 |
| S17-k141_149339 | 1357 | No | NA | 1 | 0 | 1 | Not-determined | Genome-fragment | NA | NA | 0 |
| S17-k141_237808 | 1109 | No | NA | 1 | 0 | 1 | Not-determined | Genome-fragment | NA | NA | 0 |
| S17-k141_26595 | 3401 | No | NA | 2 | 0 | 1 | Not-determined | Genome-fragment | NA | NA | 0 |
| S17-k141_59956 | 1079 | No | NA | 1 | 0 | 1 | Not-determined | Genome-fragment | NA | NA | 0 |
| S17-k141_183121 | 1059 | No | NA | 1 | 0 | 1 | Not-determined | Genome-fragment | NA | NA | 0 |
| S17-k141_59964 | 1087 | No | NA | 3 | 0 | 1 | Not-determined | Genome-fragment | NA | NA | 0 |
| S17-k141_71615 | 1968 | No | NA | 2 | 0 | 2 | Not-determined | Genome-fragment | NA | NA | 0 |
| S17-k141_127091 | 1333 | No | NA | 2 | 0 | 1 | Not-determined | Genome-fragment | NA | NA | 0 |
| S17-k141_259473 | 1417 | No | NA | 1 | 0 | 0 | Not-determined | Genome-fragment | NA | NA | 0 |
| S2_k141_209200 | 2469 | No | NA | 1 | 0 | 0 | Low-quality | Genome-fragment | 0.21 | AAI-based (medium-confidence) | 0 |
| S17-k141_138215 | 1087 | No | NA | 2 | 0 | 0 | Not-determined | Genome-fragment | NA | NA | 0 |
| S17-k141_248364 | 6363 | No | NA | 5 | 0 | 4 | Not-determined | Genome-fragment | NA | NA | 0 |
| S17-k141_14934 | 1707 | No | NA | 3 | 0 | 0 | Not-determined | Genome-fragment | NA | NA | 0 |
| S17-k141_26677 | 1223 | No | NA | 1 | 0 | 0 | Not-determined | Genome-fragment | NA | NA | 0 |
| S17-k141_259511 | 4011 | No | NA | 3 | 0 | 3 | Not-determined | Genome-fragment | NA | NA | 0 |
| S9_k141_58568 | 2470 | No | NA | 2 | 0 | 0 | Low-quality | Genome-fragment | 5.13 | AAI-based (medium-confidence) | 0 |
| S17-k141_227713 | 1021 | No | NA | 1 | 0 | 1 | Not-determined | Genome-fragment | NA | NA | 0 |
| S17-k141_117293 | 1138 | No | NA | 3 | 0 | 3 | Not-determined | Genome-fragment | NA | NA | 0 |
| S17-k141_37839 | 2453 | No | NA | 2 | 0 | 0 | Not-determined | Genome-fragment | NA | NA | 0 |
| S17-k141_216530 | 2337 | No | NA | 3 | 0 | 3 | Not-determined | Genome-fragment | NA | NA | 0 |
| S17-k141_71640 | 3959 | No | NA | 4 | 0 | 4 | Not-determined | Genome-fragment | NA | NA | 0 |
| S17-k141_127154 | 3092 | No | NA | 3 | 0 | 1 | Not-determined | Genome-fragment | NA | NA | 0 |
| S17-k141_104626 | 1058 | No | NA | 1 | 0 | 1 | Not-determined | Genome-fragment | NA | NA | 0 |
| S17-k141_71652 | 1725 | No | NA | 2 | 0 | 1 | Not-determined | Genome-fragment | NA | NA | 0 |
| S17-k141_259530 | 4552 | No | NA | 6 | 0 | 5 | Not-determined | Genome-fragment | NA | NA | 0 |
| S17-k141_171391 | 1100 | No | NA | 1 | 0 | 0 | Not-determined | Genome-fragment | NA | NA | 0 |
| S17-k141_26750 | 2200 | No | NA | 3 | 0 | 0 | Not-determined | Genome-fragment | NA | NA | 0 |
| S1-final-viral-combined_k141_304182\|\|full | 2470 | No | NA | 2 | 0 | 2 | Low-quality | Genome-fragment | 6.45 | AAI-based (medium-confidence) | 0 |
| S17-k141_259547 | 3639 | No | NA | 3 | 0 | 1 | Not-determined | Genome-fragment | NA | NA | 0 |
| S17-k141_83476 | 1175 | No | NA | 2 | 0 | 1 | Not-determined | Genome-fragment | NA | NA | 0 |
| S17-k141_138316 | 1015 | No | NA | 1 | 0 | 0 | Not-determined | Genome-fragment | NA | NA | 0 |
| S17-k141_248440 | 1035 | No | NA | 1 | 0 | 1 | Not-determined | Genome-fragment | NA | NA | 0 |
| S3-final-viral-combined_k141_52045\|\|full | 2470 | No | NA | 3 | 0 | 2 | Low-quality | Genome-fragment | 1.33 | AAI-based (medium-confidence) | 0 |
| S17-k141_259557 | 1820 | No | NA | 2 | 0 | 2 | Not-determined | Genome-fragment | NA | NA | 0 |
| S17-k141_259564 | 1113 | No | NA | 2 | 0 | 0 | Not-determined | Genome-fragment | NA | NA | 0 |
| S17-k141_193964 | 1127 | No | NA | 3 | 0 | 1 | Not-determined | Genome-fragment | NA | NA | 0 |
| S17-k141_71723 | 1342 | No | NA | 1 | 0 | 1 | Not-determined | Genome-fragment | NA | NA | 0 |
| S17-k141_83506 | 1115 | No | NA | 2 | 0 | 0 | Not-determined | Genome-fragment | NA | NA | 0 |
| S17-k141_149456 | 1889 | No | NA | 1 | 0 | 1 | Not-determined | Genome-fragment | NA | NA | 0 |
| S17-k141_4483 | 2011 | No | NA | 2 | 0 | 1 | Not-determined | Genome-fragment | NA | NA | 0 |
| S17-k141_227771 | 1283 | No | NA | 1 | 0 | 1 | Not-determined | Genome-fragment | NA | NA | 0 |
| S17-k141_92663 | 1092 | No | NA | 1 | 0 | 0 | Not-determined | Genome-fragment | NA | NA | 0 |
| S17-k141_216636 | 1760 | No | NA | 2 | 0 | 2 | Not-determined | Genome-fragment | NA | NA | 0 |
| S17-k141_248474 | 1213 | No | NA | 1 | 0 | 0 | Not-determined | Genome-fragment | NA | NA | 0 |
| S17-k141_205368 | 1782 | No | NA | 1 | 0 | 1 | Not-determined | Genome-fragment | NA | NA | 0 |
| S17-k141_216665 | 1087 | No | NA | 1 | 0 | 0 | Not-determined | Genome-fragment | NA | NA | 0 |
| S17-k141_183322 | 4152 | No | NA | 5 | 0 | 0 | Not-determined | Genome-fragment | NA | NA | 0 |
| S17-k141_104691 | 4677 | No | NA | 4 | 0 | 2 | Not-determined | Genome-fragment | NA | NA | 0 |
| S17-k141_127333 | 1967 | No | NA | 3 | 0 | 2 | Not-determined | Genome-fragment | NA | NA | 0 |
| S17-k141_15027 | 1002 | No | NA | 1 | 0 | 0 | Not-determined | Genome-fragment | NA | NA | 0 |
| S17-k141_183402 | 2140 | No | NA | 1 | 0 | 0 | Not-determined | Genome-fragment | NA | NA | 0 |
| S17-k141_71814 | 1348 | No | NA | 3 | 0 | 1 | Not-determined | Genome-fragment | NA | NA | 0 |
| S17-k141_26916 | 6977 | No | NA | 9 | 0 | 6 | Not-determined | Genome-fragment | NA | NA | 0 |
| S17-k141_83587 | 1396 | No | NA | 1 | 0 | 1 | Not-determined | Genome-fragment | NA | NA | 0 |
| S17-k141_26938 | 1023 | No | NA | 2 | 0 | 0 | Not-determined | Genome-fragment | NA | NA | 0 |
| S2-final-viral-combined_k141_132564\|\|lt2gene | 2470 | No | NA | 2 | 2 | 0 | Low-quality | Genome-fragment | 6.98 | AAI-based (high-confidence) | 0 |
| S17-k141_47792 | 1573 | No | NA | 2 | 0 | 2 | Not-determined | Genome-fragment | NA | NA | 0 |
| S17-k141_117434 | 1087 | No | NA | 2 | 0 | 1 | Not-determined | Genome-fragment | NA | NA | 0 |
| S17-k141_248595 | 3858 | No | NA | 2 | 0 | 2 | Not-determined | Genome-fragment | NA | NA | 0 |
| S17-k141_149568 | 1268 | No | NA | 2 | 0 | 1 | Not-determined | Genome-fragment | NA | NA | 0 |
| S17-k141_194101 | 1246 | No | NA | 1 | 0 | 1 | Not-determined | Genome-fragment | NA | NA | 0 |
| S17-k141_248616 | 1041 | No | NA | 1 | 0 | 1 | Not-determined | Genome-fragment | NA | NA | 0 |
| S17-k141_216774 | 1471 | No | NA | 1 | 0 | 1 | Not-determined | Genome-fragment | NA | NA | 0 |
| S17-k141_4660 | 2583 | No | NA | 3 | 0 | 1 | Not-determined | Genome-fragment | NA | NA | 0 |
| S17-k141_15123 | 3907 | No | NA | 3 | 0 | 3 | Not-determined | Genome-fragment | NA | NA | 0 |
| S17-k141_171662 | 1174 | No | NA | 2 | 0 | 0 | Not-determined | Genome-fragment | NA | NA | 0 |
| S17-k141_205519 | 1051 | No | NA | 2 | 0 | 1 | Not-determined | Genome-fragment | NA | NA | 0 |
| S17-k141_47820 | 5415 | No | NA | 3 | 0 | 0 | Not-determined | Genome-fragment | NA | NA | 0 |
| S17-k141_104811 | 3360 | No | NA | 5 | 0 | 3 | Not-determined | Genome-fragment | NA | NA | 0 |
| S17-k141_159836 | 1385 | No | NA | 1 | 0 | 0 | Not-determined | Genome-fragment | NA | NA | 0 |
| S17-k141_60234 | 1755 | No | NA | 1 | 0 | 1 | Not-determined | Genome-fragment | NA | NA | 0 |
| S17-k141_149622 | 1407 | No | NA | 2 | 0 | 0 | Not-determined | Genome-fragment | NA | NA | 0 |
| S17-k141_205566 | 1592 | No | NA | 1 | 0 | 1 | Not-determined | Genome-fragment | NA | NA | 0 |
| S8_k141_54893 | 2470 | No | NA | 3 | 0 | 0 | Low-quality | Genome-fragment | 3.01 | AAI-based (high-confidence) | 0 |
| S17-k141_216795 | 1309 | No | NA | 2 | 0 | 0 | Not-determined | Genome-fragment | NA | NA | 0 |
| S17-k141_60251 | 1064 | No | NA | 2 | 0 | 0 | Not-determined | Genome-fragment | NA | NA | 0 |
| S17-k141_117443 | 1800 | No | NA | 1 | 0 | 0 | Not-determined | Genome-fragment | NA | NA | 0 |
| S17-k141_83725 | 1026 | No | NA | 1 | 0 | 0 | Not-determined | Genome-fragment | NA | NA | 0 |
| S17-k141_171719 | 1595 | No | NA | 2 | 0 | 2 | Not-determined | Genome-fragment | NA | NA | 0 |
| S17-k141_194199 | 4418 | No | NA | 3 | 0 | 2 | Not-determined | Genome-fragment | NA | NA | 0 |
| S17-k141_83741 | 1197 | No | NA | 2 | 0 | 0 | Not-determined | Genome-fragment | NA | NA | 0 |
| S17-k141_205585 | 4161 | No | NA | 3 | 0 | 0 | Not-determined | Genome-fragment | NA | NA | 0 |
| S1-final-viral-combined_k141_215584\|\|full | 2471 | No | NA | 6 | 1 | 0 | Low-quality | Genome-fragment | 5.56 | AAI-based (high-confidence) | 0 |
| S17-k141_71968 | 1007 | No | NA | 1 | 0 | 1 | Not-determined | Genome-fragment | NA | NA | 0 |
| S17-k141_171751 | 2205 | No | NA | 3 | 0 | 0 | Not-determined | Genome-fragment | NA | NA | 0 |
| S17-k141_149677 | 5699 | No | NA | 5 | 0 | 1 | Not-determined | Genome-fragment | NA | NA | 0 |
| S17-k141_15220 | 1056 | No | NA | 2 | 0 | 1 | Not-determined | Genome-fragment | NA | NA | 0 |
| S17-k141_238073 | 1111 | No | NA | 1 | 0 | 1 | Not-determined | Genome-fragment | NA | NA | 0 |
| S17-k141_117534 | 5511 | No | NA | 5 | 0 | 2 | Not-determined | Genome-fragment | NA | NA | 0 |
| S17-final-viral-combined_k141_68541\|\|full | 2471 | No | NA | 3 | 0 | 1 | Low-quality | Genome-fragment | 6.8 | AAI-based (medium-confidence) | 0 |
| S17-k141_228030 | 1408 | No | NA | 3 | 0 | 2 | Not-determined | Genome-fragment | NA | NA | 0 |
| S17-k141_259762 | 2389 | No | NA | 2 | 0 | 2 | Not-determined | Genome-fragment | NA | NA | 0 |
| S17-k141_228038 | 2030 | No | NA | 3 | 0 | 2 | Not-determined | Genome-fragment | NA | NA | 0 |
| S17-k141_259788 | 1103 | No | NA | 1 | 0 | 1 | Not-determined | Genome-fragment | NA | NA | 0 |
| S17-k141_238123 | 1295 | No | NA | 1 | 0 | 1 | Not-determined | Genome-fragment | NA | NA | 0 |
| S17-k141_38254 | 1362 | No | NA | 2 | 0 | 1 | Not-determined | Genome-fragment | NA | NA | 0 |
| S17-k141_238157 | 1300 | No | NA | 1 | 0 | 1 | Not-determined | Genome-fragment | NA | NA | 0 |
| S17-k141_117623 | 1815 | No | NA | 2 | 0 | 2 | Not-determined | Genome-fragment | NA | NA | 0 |
| S17-k141_183717 | 1157 | No | NA | 2 | 0 | 1 | Not-determined | Genome-fragment | NA | NA | 0 |
| S8-final-viral-combined_k141_168547\|\|full | 2471 | No | NA | 4 | 1 | 0 | Low-quality | Genome-fragment | 6.24 | AAI-based (high-confidence) | 0 |
| S17-k141_15289 | 1084 | No | NA | 1 | 0 | 1 | Not-determined | Genome-fragment | NA | NA | 0 |
| S17-k141_228117 | 1334 | No | NA | 1 | 0 | 0 | Not-determined | Genome-fragment | NA | NA | 0 |
| S11_k141_328240 | 2472 | No | NA | 4 | 0 | 0 | Low-quality | Genome-fragment | 3.49 | AAI-based (high-confidence) | 0 |
| S17-k141_248773 | 8526 | No | NA | 8 | 0 | 4 | Not-determined | Genome-fragment | NA | NA | 0 |
| S2-final-viral-combined_k141_88716\|\|full | 2472 | No | NA | 9 | 0 | 0 | Low-quality | Genome-fragment | 6.67 | AAI-based (medium-confidence) | 0 |
| S17-k141_216951 | 1182 | No | NA | 2 | 0 | 1 | Not-determined | Genome-fragment | NA | NA | 0 |
| S17-k141_60438 | 2890 | No | NA | 2 | 0 | 0 | Not-determined | Genome-fragment | NA | NA | 0 |
| S17-k141_92878 | 1543 | No | NA | 1 | 0 | 0 | Not-determined | Genome-fragment | NA | NA | 0 |
| S17-k141_248833 | 1025 | No | NA | 2 | 0 | 0 | Not-determined | Genome-fragment | NA | NA | 0 |
| S17-k141_183759 | 1313 | No | NA | 1 | 0 | 1 | Not-determined | Genome-fragment | NA | NA | 0 |
| S17-k141_160066 | 3654 | No | NA | 2 | 0 | 0 | Not-determined | Genome-fragment | NA | NA | 0 |
| S2-final-viral-combined_k141_186293\|\|full | 2472 | No | NA | 2 | 1 | 0 | Low-quality | Genome-fragment | 5.37 | AAI-based (medium-confidence) | 0 |
| S17-k141_105092 | 1201 | No | NA | 2 | 0 | 1 | Not-determined | Genome-fragment | NA | NA | 0 |
| S17-k141_38357 | 2886 | No | NA | 5 | 0 | 0 | Not-determined | Genome-fragment | NA | NA | 0 |
| S17-k141_60483 | 2820 | No | NA | 1 | 0 | 0 | Not-determined | Genome-fragment | NA | NA | 0 |
| S17-k141_205790 | 1004 | No | NA | 1 | 0 | 0 | Not-determined | Genome-fragment | NA | NA | 0 |
| S17-k141_4860 | 2992 | No | NA | 3 | 0 | 2 | Not-determined | Genome-fragment | NA | NA | 0 |
| S7-final-viral-combined_k141_214266\|\|full | 2473 | No | NA | 7 | 1 | 0 | Low-quality | Genome-fragment | 1.59 | HMM-based (lower-bound) | 0 |
| S17-k141_105118 | 1413 | No | NA | 2 | 0 | 2 | Not-determined | Genome-fragment | NA | NA | 0 |
| S17-k141_60527 | 1300 | No | NA | 1 | 0 | 1 | Not-determined | Genome-fragment | NA | NA | 0 |
| S17-k141_149873 | 1068 | No | NA | 2 | 0 | 0 | Not-determined | Genome-fragment | NA | NA | 0 |
| S17-k141_160116 | 3642 | No | NA | 3 | 0 | 1 | Not-determined | Genome-fragment | NA | NA | 0 |
| S17-k141_217015 | 2355 | No | NA | 2 | 0 | 2 | Not-determined | Genome-fragment | NA | NA | 0 |
| S17-k141_149889 | 1132 | No | NA | 1 | 0 | 1 | Not-determined | Genome-fragment | NA | NA | 0 |
| S17-k141_238259 | 1767 | No | NA | 2 | 0 | 0 | Not-determined | Genome-fragment | NA | NA | 0 |
| S17-k141_4901 | 4431 | No | NA | 8 | 0 | 2 | Not-determined | Genome-fragment | NA | NA | 0 |
| S17-k141_183852 | 1072 | No | NA | 2 | 0 | 0 | Not-determined | Genome-fragment | NA | NA | 0 |
| S2-final-viral-combined_k141_215216\|\|full | 2473 | No | NA | 3 | 1 | 0 | Low-quality | Genome-fragment | 6.87 | AAI-based (high-confidence) | 0 |
| S17-k141_117781 | 2101 | No | NA | 3 | 0 | 1 | Not-determined | Genome-fragment | NA | NA | 0 |
| S17-k141_60571 | 1101 | No | NA | 3 | 0 | 3 | Not-determined | Genome-fragment | NA | NA | 0 |
| S17-k141_127739 | 2380 | No | NA | 3 | 0 | 3 | Not-determined | Genome-fragment | NA | NA | 0 |
| S17-k141_72233 | 1205 | No | NA | 1 | 0 | 1 | Not-determined | Genome-fragment | NA | NA | 0 |
| S17-k141_60586 | 3066 | No | NA | 2 | 0 | 1 | Not-determined | Genome-fragment | NA | NA | 0 |
| S17-k141_117795 | 7202 | No | NA | 6 | 0 | 1 | Not-determined | Genome-fragment | NA | NA | 0 |
| S17-k141_92949 | 1452 | No | NA | 1 | 0 | 0 | Not-determined | Genome-fragment | NA | NA | 0 |
| S17-k141_84125 | 1271 | No | NA | 1 | 0 | 1 | Not-determined | Genome-fragment | NA | NA | 0 |
| S17-k141_160192 | 1399 | No | NA | 1 | 0 | 0 | Not-determined | Genome-fragment | NA | NA | 0 |
| S7-final-viral-combined_k141_112029\|\|full | 2474 | No | NA | 6 | 5 | 0 | Low-quality | Genome-fragment | 5.3 | AAI-based (high-confidence) | 0 |
| S17-k141_149990 | 1388 | No | NA | 1 | 0 | 1 | Not-determined | Genome-fragment | NA | NA | 0 |
| S17-k141_160210 | 3508 | No | NA | 4 | 0 | 3 | Not-determined | Genome-fragment | NA | NA | 0 |
| S17-k141_27241 | 1354 | No | NA | 2 | 0 | 1 | Not-determined | Genome-fragment | NA | NA | 0 |
| S17-k141_92974 | 3068 | No | NA | 3 | 0 | 3 | Not-determined | Genome-fragment | NA | NA | 0 |
| S5-final-viral-combined_k141_2991\|\|full | 2475 | No | NA | 4 | 0 | 0 | Low-quality | Genome-fragment | 3.39 | AAI-based (medium-confidence) | 0 |
| S17-k141_217145 | 1431 | No | NA | 1 | 0 | 1 | Not-determined | Genome-fragment | NA | NA | 0 |
| S17-k141_217146 | 1307 | No | NA | 1 | 0 | 0 | Not-determined | Genome-fragment | NA | NA | 0 |
| S17-k141_160227 | 1144 | No | NA | 1 | 0 | 1 | Not-determined | Genome-fragment | NA | NA | 0 |
| S17-k141_217164 | 1622 | No | NA | 2 | 0 | 2 | Not-determined | Genome-fragment | NA | NA | 0 |
| S17-final-viral-combined_k141_117931\|\|full | 2475 | No | NA | 3 | 1 | 0 | Low-quality | Genome-fragment | 6.77 | AAI-based (high-confidence) | 0 |
| S17-k141_228368 | 1215 | No | NA | 2 | 0 | 1 | Not-determined | Genome-fragment | NA | NA | 0 |
| S17-final-viral-combined_k141_130651\|\|full | 2475 | No | NA | 3 | 0 | 0 | Low-quality | Genome-fragment | 3.63 | AAI-based (medium-confidence) | 0 |
| S17-k141_183927 | 1292 | No | NA | 2 | 0 | 0 | Not-determined | Genome-fragment | NA | NA | 0 |
| S17-k141_84185 | 1955 | No | NA | 2 | 0 | 2 | Not-determined | Genome-fragment | NA | NA | 0 |
| S17-k141_105285 | 1176 | No | NA | 2 | 0 | 0 | Not-determined | Genome-fragment | NA | NA | 0 |
| S17-k141_72346 | 1753 | No | NA | 2 | 0 | 2 | Not-determined | Genome-fragment | NA | NA | 0 |
| S17-k141_72356 | 1262 | No | NA | 1 | 0 | 1 | Not-determined | Genome-fragment | NA | NA | 0 |
| S17-k141_127826 | 2114 | No | NA | 2 | 0 | 0 | Not-determined | Genome-fragment | NA | NA | 0 |
| S12-final-viral-combined_k141_29761\|\|full | 2475 | No | NA | 2 | 0 | 1 | Low-quality | Genome-fragment | 1.25 | AAI-based (medium-confidence) | 0 |
| S17-k141_183954 | 6012 | No | NA | 6 | 0 | 0 | Not-determined | Genome-fragment | NA | NA | 0 |
| S17-k141_260027 | 4798 | No | NA | 4 | 0 | 1 | Not-determined | Genome-fragment | NA | NA | 0 |
| S17-k141_172092 | 1288 | No | NA | 2 | 0 | 1 | Not-determined | Genome-fragment | NA | NA | 0 |
| S2-final-viral-combined_k141_244489\|\|full | 2475 | No | NA | 3 | 0 | 0 | Low-quality | Genome-fragment | 6.96 | AAI-based (medium-confidence) | 0 |
| S9-final-viral-combined_k141_82615\|\|full | 2476 | No | NA | 4 | 0 | 1 | Low-quality | Genome-fragment | 5.02 | AAI-based (medium-confidence) | 0 |
| S1-final-viral-combined_k141_105476\|\|full | 2476 | No | NA | 4 | 0 | 1 | Low-quality | Genome-fragment | 4.28 | AAI-based (medium-confidence) | 0 |
| S17-k141_5099 | 1169 | No | NA | 2 | 0 | 2 | Not-determined | Genome-fragment | NA | NA | 0 |
| S17-k141_160302 | 1403 | No | NA | 1 | 0 | 1 | Not-determined | Genome-fragment | NA | NA | 0 |
| S17-k141_93062 | 2737 | No | NA | 1 | 0 | 1 | Not-determined | Genome-fragment | NA | NA | 0 |
| S17-k141_15524 | 3082 | No | NA | 2 | 0 | 0 | Not-determined | Genome-fragment | NA | NA | 0 |
| S1_k141_11576 | 2476 | No | NA | 3 | 3 | 0 | Low-quality | Genome-fragment | 4.24 | AAI-based (high-confidence) | 0 |
| S7-final-viral-combined_k141_230685\|\|full | 2476 | No | NA | 5 | 0 | 2 | Low-quality | Genome-fragment | 1.08 | AAI-based (medium-confidence) | 0 |
| S17-k141_206038 | 4297 | No | NA | 3 | 0 | 2 | Not-determined | Genome-fragment | NA | NA | 0 |
| S17-k141_217247 | 3536 | No | NA | 1 | 0 | 0 | Not-determined | Genome-fragment | NA | NA | 0 |
| S17-k141_238452 | 1484 | No | NA | 2 | 0 | 2 | Not-determined | Genome-fragment | NA | NA | 0 |
| S17-k141_105390 | 6073 | No | NA | 4 | 0 | 3 | Not-determined | Genome-fragment | NA | NA | 0 |
| S12-final-viral-combined_k141_381698\|\|full | 2476 | No | NA | 4 | 1 | 0 | Low-quality | Genome-fragment | 3.56 | HMM-based (lower-bound) | 0 |
| S17-k141_38486 | 1001 | No | NA | 2 | 0 | 2 | Not-determined | Genome-fragment | NA | NA | 0 |
| S17-k141_15557 | 2190 | No | NA | 1 | 0 | 1 | Not-determined | Genome-fragment | NA | NA | 0 |
| S17-k141_15569 | 1404 | No | NA | 3 | 0 | 3 | Not-determined | Genome-fragment | NA | NA | 0 |
| S17-k141_139130 | 1166 | No | NA | 3 | 0 | 0 | Not-determined | Genome-fragment | NA | NA | 0 |
| S17-k141_194772 | 1128 | No | NA | 2 | 0 | 0 | Not-determined | Genome-fragment | NA | NA | 0 |
| S17-k141_260127 | 1031 | No | NA | 2 | 0 | 2 | Not-determined | Genome-fragment | NA | NA | 0 |
| S17-k141_127932 | 1013 | No | NA | 2 | 0 | 1 | Not-determined | Genome-fragment | NA | NA | 0 |
| S17-k141_228506 | 2142 | No | NA | 1 | 0 | 1 | Not-determined | Genome-fragment | NA | NA | 0 |
| S17-k141_238518 | 3232 | No | NA | 4 | 0 | 3 | Not-determined | Genome-fragment | NA | NA | 0 |
| S17-k141_93151 | 1795 | No | NA | 1 | 0 | 0 | Not-determined | Genome-fragment | NA | NA | 0 |
| S17-k141_38562 | 1006 | No | NA | 1 | 0 | 1 | Not-determined | Genome-fragment | NA | NA | 0 |
| S17-k141_118093 | 1455 | No | NA | 1 | 0 | 0 | Not-determined | Genome-fragment | NA | NA | 0 |
| S17-k141_260171 | 1688 | No | NA | 1 | 0 | 1 | Not-determined | Genome-fragment | NA | NA | 0 |
| S8-final-viral-combined_k141_106502\|\|full | 2476 | No | NA | 5 | 3 | 0 | Low-quality | Genome-fragment | 3.01 | AAI-based (high-confidence) | 0 |
| S17-k141_38580 | 1236 | No | NA | 1 | 0 | 1 | Not-determined | Genome-fragment | NA | NA | 0 |
| S17-k141_48173 | 1083 | No | NA | 1 | 0 | 1 | Not-determined | Genome-fragment | NA | NA | 0 |
| S17-k141_84233 | 1160 | No | NA | 1 | 0 | 1 | Not-determined | Genome-fragment | NA | NA | 0 |
| S17-k141_60843 | 1209 | No | NA | 1 | 0 | 0 | Not-determined | Genome-fragment | NA | NA | 0 |
| S17-final-viral-combined_k141_58966\|\|full | 2477 | No | NA | 4 | 0 | 1 | Low-quality | Genome-fragment | 3.18 | AAI-based (medium-confidence) | 0 |
| S17-k141_194876 | 2892 | No | NA | 2 | 0 | 2 | Not-determined | Genome-fragment | NA | NA | 0 |
| S17-k141_118121 | 2633 | No | NA | 4 | 0 | 4 | Not-determined | Genome-fragment | NA | NA | 0 |
| S17-k141_194880 | 1003 | No | NA | 1 | 0 | 1 | Not-determined | Genome-fragment | NA | NA | 0 |
| S17-k141_38628 | 2844 | No | NA | 2 | 0 | 1 | Not-determined | Genome-fragment | NA | NA | 0 |
| S17-k141_15623 | 1041 | No | NA | 1 | 0 | 1 | Not-determined | Genome-fragment | NA | NA | 0 |
| S17-k141_206252 | 1329 | No | NA | 2 | 0 | 2 | Not-determined | Genome-fragment | NA | NA | 0 |
| S4_k141_70643 | 2477 | No | NA | 6 | 0 | 0 | Low-quality | Genome-fragment | 6.28 | AAI-based (high-confidence) | 0 |
| S17-k141_249076 | 3778 | No | NA | 4 | 0 | 4 | Not-determined | Genome-fragment | NA | NA | 0 |
| S11-final-viral-combined_k141_293337\|\|full | 2478 | No | NA | 7 | 2 | 0 | Low-quality | Genome-fragment | 3.8 | HMM-based (lower-bound) | 0 |
| S17-k141_118140 | 1030 | No | NA | 1 | 0 | 0 | Not-determined | Genome-fragment | NA | NA | 0 |
| S17-k141_150203 | 2024 | No | NA | 1 | 0 | 1 | Not-determined | Genome-fragment | NA | NA | 0 |
| S17-k141_139285 | 1280 | No | NA | 2 | 0 | 0 | Not-determined | Genome-fragment | NA | NA | 0 |
| S11-final-viral-combined_k141_48117\|\|full | 2478 | No | NA | 4 | 3 | 0 | Low-quality | Genome-fragment | 6.93 | AAI-based (high-confidence) | 0 |
| S7-final-viral-combined_k141_10440\|\|full | 2478 | No | NA | 4 | 0 | 0 | Low-quality | Genome-fragment | 4.36 | AAI-based (high-confidence) | 0 |
| S17-k141_172172 | 1168 | No | NA | 1 | 0 | 0 | Not-determined | Genome-fragment | NA | NA | 0 |
| S17-k141_260224 | 1072 | No | NA | 2 | 0 | 0 | Not-determined | Genome-fragment | NA | NA | 0 |
| S17-k141_206309 | 1114 | No | NA | 1 | 0 | 1 | Not-determined | Genome-fragment | NA | NA | 0 |
| S17-k141_238658 | 1041 | No | NA | 3 | 0 | 0 | Not-determined | Genome-fragment | NA | NA | 0 |
| S17-k141_217476 | 3167 | No | NA | 3 | 0 | 3 | Not-determined | Genome-fragment | NA | NA | 0 |
| S17-k141_60925 | 3431 | No | NA | 2 | 0 | 2 | Not-determined | Genome-fragment | NA | NA | 0 |
| S17-final-viral-combined_k141_157305\|\|full | 2478 | No | NA | 4 | 0 | 0 | Low-quality | Genome-fragment | 3.21 | AAI-based (medium-confidence) | 0 |
| S17-k141_217499 | 1326 | No | NA | 1 | 0 | 1 | Not-determined | Genome-fragment | NA | NA | 0 |
| S17-k141_160554 | 1072 | No | NA | 1 | 0 | 1 | Not-determined | Genome-fragment | NA | NA | 0 |
| S17-k141_238697 | 4697 | No | NA | 5 | 0 | 3 | Not-determined | Genome-fragment | NA | NA | 0 |
| S17-k141_72614 | 2383 | No | NA | 2 | 0 | 2 | Not-determined | Genome-fragment | NA | NA | 0 |
| S17-k141_249150 | 1819 | No | NA | 1 | 0 | 1 | Not-determined | Genome-fragment | NA | NA | 0 |
| S17-k141_184170 | 14093 | No | NA | 12 | 0 | 5 | Not-determined | Genome-fragment | NA | NA | 0 |
| S17-k141_238723 | 2304 | No | NA | 2 | 0 | 2 | Not-determined | Genome-fragment | NA | NA | 0 |
| S17-k141_5320 | 1939 | No | NA | 2 | 0 | 0 | Not-determined | Genome-fragment | NA | NA | 0 |
| S17-k141_206369 | 1773 | No | NA | 1 | 0 | 0 | Not-determined | Genome-fragment | NA | NA | 0 |
| S17-k141_160608 | 1352 | No | NA | 2 | 0 | 2 | Not-determined | Genome-fragment | NA | NA | 0 |
| S17-k141_48308 | 1183 | No | NA | 3 | 0 | 1 | Not-determined | Genome-fragment | NA | NA | 0 |
| S17-k141_48313 | 1486 | No | NA | 2 | 0 | 0 | Not-determined | Genome-fragment | NA | NA | 0 |
| S17-k141_195002 | 2246 | No | NA | 2 | 0 | 2 | Not-determined | Genome-fragment | NA | NA | 0 |
| S17-k141_61016 | 1048 | No | NA | 1 | 0 | 0 | Not-determined | Genome-fragment | NA | NA | 0 |
| S17-k141_139390 | 4061 | No | NA | 5 | 0 | 3 | Not-determined | Genome-fragment | NA | NA | 0 |
| S17-k141_206417 | 1989 | No | NA | 2 | 0 | 2 | Not-determined | Genome-fragment | NA | NA | 0 |
| S17-k141_15722 | 2198 | No | NA | 1 | 0 | 0 | Not-determined | Genome-fragment | NA | NA | 0 |
| S17-k141_217571 | 2472 | No | NA | 3 | 0 | 2 | Not-determined | Genome-fragment | NA | NA | 0 |
| S17-k141_217576 | 1011 | No | NA | 2 | 0 | 1 | Not-determined | Genome-fragment | NA | NA | 0 |
| S17-k141_72700 | 1058 | No | NA | 3 | 0 | 1 | Not-determined | Genome-fragment | NA | NA | 0 |
| S17-k141_48344 | 7008 | No | NA | 5 | 0 | 5 | Not-determined | Genome-fragment | NA | NA | 0 |
| S17-k141_150293 | 1131 | No | NA | 1 | 0 | 1 | Not-determined | Genome-fragment | NA | NA | 0 |
| S17-k141_184269 | 11741 | No | NA | 18 | 0 | 0 | Not-determined | Genome-fragment | NA | NA | 0 |
| S17-k141_206470 | 1000 | No | NA | 1 | 0 | 0 | Not-determined | Genome-fragment | NA | NA | 0 |
| S5-final-viral-combined_k141_66154\|\|full | 2479 | No | NA | 4 | 0 | 1 | Low-quality | Genome-fragment | 3.07 | AAI-based (high-confidence) | 0 |

| | | | | | | | | | | | |
|---|---|---|---|---|---|---|---|---|---|---|---|
| S17-k141_228776 | 3174 | No | NA | 3 | 0 | 3 | Not-determined | Genome-fragment | NA | NA | 0 |
| S17-k141_260317 | 1371 | No | NA | 2 | 0 | 1 | Not-determined | Genome-fragment | NA | NA | 0 |
| S17-k141_150354 | 3394 | No | NA | 4 | 0 | 3 | Not-determined | Genome-fragment | NA | NA | 0 |
| S17-k141_217632 | 1280 | No | NA | 1 | 0 | 1 | Not-determined | Genome-fragment | NA | NA | 0 |
| S17-final-viral-combined_k141_185153\|\|full | 2479 | No | NA | 3 | 0 | 1 | Low-quality | Genome-fragment | 4.31 | AAI-based (medium-confidence) | 0 |
| S17-k141_184318 | 1008 | No | NA | 2 | 0 | 1 | Not-determined | Genome-fragment | NA | NA | 0 |
| S17-k141_61040 | 1150 | No | NA | 2 | 0 | 1 | Not-determined | Genome-fragment | NA | NA | 0 |
| S12-final-viral-combined_k141_35950\|\|full | 2479 | No | NA | 4 | 0 | 2 | Low-quality | Genome-fragment | 1.07 | AAI-based (medium-confidence) | 0 |
| S17-k141_160693 | 1025 | No | NA | 1 | 0 | 1 | Not-determined | Genome-fragment | NA | NA | 0 |
| S17-k141_217663 | 1634 | No | NA | 1 | 0 | 1 | Not-determined | Genome-fragment | NA | NA | 0 |
| S17-k141_105762 | 3039 | No | NA | 3 | 0 | 1 | Not-determined | Genome-fragment | NA | NA | 0 |
| S17-k141_61058 | 4747 | No | NA | 3 | 0 | 3 | Not-determined | Genome-fragment | NA | NA | 0 |
| S12-final-viral-combined_k141_293557\|\|full | 2479 | No | NA | 5 | 3 | 0 | Low-quality | Genome-fragment | 4.45 | AAI-based (medium-confidence) | 0 |
| S8-final-viral-combined_k141_110417\|\|full | 2479 | No | NA | 3 | 0 | 0 | Low-quality | Genome-fragment | 4.88 | AAI-based (high-confidence) | 0 |
| S17-k141_238900 | 1094 | No | NA | 2 | 0 | 1 | Not-determined | Genome-fragment | NA | NA | 0 |
| S17-k141_238901 | 1052 | No | NA | 2 | 0 | 1 | Not-determined | Genome-fragment | NA | NA | 0 |
| S17-k141_128298 | 1424 | No | NA | 2 | 0 | 1 | Not-determined | Genome-fragment | NA | NA | 0 |
| S17-k141_15806 | 1506 | No | NA | 1 | 0 | 1 | Not-determined | Genome-fragment | NA | NA | 0 |
| S17-k141_61099 | 1394 | No | NA | 1 | 0 | 0 | Not-determined | Genome-fragment | NA | NA | 0 |
| S17-k141_184446 | 1190 | No | NA | 2 | 0 | 1 | Not-determined | Genome-fragment | NA | NA | 0 |
| S17-k141_84504 | 4093 | No | NA | 3 | 0 | 0 | Not-determined | Genome-fragment | NA | NA | 0 |
| S17-k141_184492 | 1312 | No | NA | 1 | 0 | 0 | Not-determined | Genome-fragment | NA | NA | 0 |
| S4-final-viral-combined_k141_114301\|\|full | 2480 | Yes | 863 | 4 | 1 | 1 | Low-quality | Genome-fragment | 1.73 | HMM-based (lower-bound) | 65.2 |
| S17-k141_61175 | 1052 | No | NA | 2 | 0 | 0 | Not-determined | Genome-fragment | NA | NA | 0 |
| S17-k141_93481 | 7371 | No | NA | 6 | 0 | 3 | Not-determined | Genome-fragment | NA | NA | 0 |
| S17-k141_217833 | 1007 | No | NA | 1 | 0 | 1 | Not-determined | Genome-fragment | NA | NA | 0 |
| S6-final-viral-combined_k141_11151\|\|full | 2480 | No | NA | 4 | 2 | 0 | Low-quality | Genome-fragment | 5.05 | AAI-based (high-confidence) | 0 |
| S17-k141_118509 | 5493 | No | NA | 3 | 0 | 3 | Not-determined | Genome-fragment | NA | NA | 0 |
| S17-k141_61244 | 1310 | No | NA | 2 | 0 | 1 | Not-determined | Genome-fragment | NA | NA | 0 |
| S17-k141_228972 | 1099 | No | NA | 2 | 0 | 0 | Not-determined | Genome-fragment | NA | NA | 0 |
| S17-k141_206732 | 1226 | No | NA | 3 | 0 | 0 | Not-determined | Genome-fragment | NA | NA | 0 |
| S17-k141_118554 | 2253 | No | NA | 1 | 0 | 1 | Not-determined | Genome-fragment | NA | NA | 0 |
| S17-k141_150604 | 21029 | No | NA | 15 | 0 | 8 | Not-determined | Genome-fragment | NA | NA | 0 |
| S17-k141_105835 | 1382 | No | NA | 2 | 0 | 0 | Not-determined | Genome-fragment | NA | NA | 0 |
| S17-k141_105837 | 1498 | No | NA | 4 | 0 | 0 | Not-determined | Genome-fragment | NA | NA | 0 |
| S17-k141_118592 | 4961 | No | NA | 4 | 0 | 4 | Not-determined | Genome-fragment | NA | NA | 0 |
| S17-k141_38920 | 1056 | No | NA | 1 | 0 | 1 | Not-determined | Genome-fragment | NA | NA | 0 |
| S17-k141_229030 | 3564 | No | NA | 4 | 0 | 3 | Not-determined | Genome-fragment | NA | NA | 0 |
| S17-k141_15957 | 7705 | No | NA | 4 | 0 | 3 | Not-determined | Genome-fragment | NA | NA | 0 |
| S17-k141_61298 | 1443 | No | NA | 1 | 0 | 1 | Not-determined | Genome-fragment | NA | NA | 0 |
| S17-k141_105860 | 2072 | No | NA | 2 | 0 | 1 | Not-determined | Genome-fragment | NA | NA | 0 |
| S17-k141_15990 | 2517 | No | NA | 4 | 0 | 4 | Not-determined | Genome-fragment | NA | NA | 0 |
| S17-k141_105886 | 1730 | No | NA | 1 | 0 | 1 | Not-determined | Genome-fragment | NA | NA | 0 |
| S17-k141_93615 | 5384 | No | NA | 5 | 0 | 1 | Not-determined | Genome-fragment | NA | NA | 0 |
| S17-k141_239123 | 1024 | No | NA | 1 | 0 | 1 | Not-determined | Genome-fragment | NA | NA | 0 |
| S17-k141_118623 | 1029 | No | NA | 1 | 0 | 1 | Not-determined | Genome-fragment | NA | NA | 0 |
| S17-k141_160962 | 1106 | No | NA | 1 | 0 | 0 | Not-determined | Genome-fragment | NA | NA | 0 |
| S17-k141_217993 | 1990 | No | NA | 1 | 0 | 0 | Not-determined | Genome-fragment | NA | NA | 0 |
| S8_k141_176228 | 2480 | No | NA | 2 | 0 | 2 | Low-quality | Genome-fragment | 3.53 | AAI-based (medium-confidence) | 0 |
| S17-k141_172610 | 5588 | No | NA | 5 | 0 | 5 | Not-determined | Genome-fragment | NA | NA | 0 |
| S17-k141_160992 | 2346 | No | NA | 4 | 0 | 1 | Not-determined | Genome-fragment | NA | NA | 0 |
| S17-k141_93666 | 3962 | No | NA | 3 | 0 | 0 | Not-determined | Genome-fragment | NA | NA | 0 |
| S17-k141_260640 | 4286 | No | NA | 5 | 0 | 4 | Not-determined | Genome-fragment | NA | NA | 0 |
| S17-k141_206921 | 3788 | No | NA | 6 | 0 | 2 | Not-determined | Genome-fragment | NA | NA | 0 |
| S1-final-viral-combined_k141_35590\|\|full | 2481 | Yes | 2004 | 6 | 1 | 1 | Low-quality | Genome-fragment | 5.36 | AAI-based (high-confidence) | 19.23 |
| S1-final-viral-combined_k141_105804\|\|full | 2481 | No | NA | 5 | 0 | 0 | Low-quality | Genome-fragment | 6.41 | AAI-based (high-confidence) | 0 |
| S17-k141_172675 | 1093 | No | NA | 1 | 0 | 0 | Not-determined | Genome-fragment | NA | NA | 0 |
| S17-k141_93675 | 7463 | No | NA | 13 | 0 | 3 | Not-determined | Genome-fragment | NA | NA | 0 |
| S1-final-viral-combined_k141_356683\|\|full | 2481 | No | NA | 3 | 2 | 0 | Low-quality | Genome-fragment | 4.73 | AAI-based (medium-confidence) | 0 |
| S17-k141_218031 | 1344 | No | NA | 1 | 0 | 0 | Not-determined | Genome-fragment | NA | NA | 0 |
| S17-k141_260668 | 1193 | No | NA | 2 | 0 | 0 | Not-determined | Genome-fragment | NA | NA | 0 |
| S17-k141_106011 | 1328 | No | NA | 1 | 0 | 1 | Not-determined | Genome-fragment | NA | NA | 0 |
| S17-k141_84749 | 1019 | No | NA | 3 | 0 | 0 | Not-determined | Genome-fragment | NA | NA | 0 |
| S1-final-viral-combined_k141_274450\|\|full | 2481 | No | NA | 3 | 0 | 2 | Low-quality | Genome-fragment | 1.36 | AAI-based (medium-confidence) | 0 |
| S17-k141_128614 | 1817 | No | NA | 1 | 0 | 1 | Not-determined | Genome-fragment | NA | NA | 0 |
| S17-k141_249635 | 1388 | No | NA | 1 | 0 | 0 | Not-determined | Genome-fragment | NA | NA | 0 |
| S17-k141_239232 | 3177 | No | NA | 1 | 0 | 0 | Not-determined | Genome-fragment | NA | NA | 0 |
| S17-k141_195625 | 1117 | No | NA | 2 | 0 | 1 | Not-determined | Genome-fragment | NA | NA | 0 |
| S17-k141_139899 | 2464 | No | NA | 3 | 0 | 2 | Not-determined | Genome-fragment | NA | NA | 0 |
| S17-k141_27822 | 1030 | No | NA | 1 | 0 | 1 | Not-determined | Genome-fragment | NA | NA | 0 |
| S17-k141_48742 | 1060 | No | NA | 2 | 0 | 0 | Not-determined | Genome-fragment | NA | NA | 0 |
| S17-k141_39052 | 1220 | No | NA | 1 | 0 | 1 | Not-determined | Genome-fragment | NA | NA | 0 |
| S17-k141_39060 | 1053 | No | NA | 1 | 0 | 1 | Not-determined | Genome-fragment | NA | NA | 0 |
| S17-k141_139939 | 1106 | No | NA | 3 | 0 | 1 | Not-determined | Genome-fragment | NA | NA | 0 |
| S7-final-viral-combined_k141_18652\|\|full | 2481 | No | NA | 5 | 1 | 1 | Low-quality | Genome-fragment | 1.33 | HMM-based (lower-bound) | 0 |
| S12-final-viral-combined_k141_223969\|\|full | 2481 | No | NA | 6 | 1 | 0 | Low-quality | Genome-fragment | 5.79 | AAI-based (medium-confidence) | 0 |
| S17-k141_172783 | 1049 | No | NA | 1 | 0 | 1 | Not-determined | Genome-fragment | NA | NA | 0 |
| S2_k141_5039 | 2481 | No | NA | 3 | 0 | 1 | Low-quality | Genome-fragment | 1.27 | AAI-based (high-confidence) | 0 |
| S17-k141_249672 | 1360 | No | NA | 3 | 0 | 0 | Not-determined | Genome-fragment | NA | NA | 0 |
| S17-k141_260778 | 1076 | No | NA | 1 | 0 | 0 | Not-determined | Genome-fragment | NA | NA | 0 |
| S17-k141_207039 | 1131 | No | NA | 1 | 0 | 0 | Not-determined | Genome-fragment | NA | NA | 0 |
| S17-k141_84857 | 1546 | No | NA | 2 | 0 | 2 | Not-determined | Genome-fragment | NA | NA | 0 |
| S1-final-viral-combined_k141_225988\|\|full | 2482 | No | NA | 2 | 0 | 2 | Low-quality | Genome-fragment | 1.99 | AAI-based (high-confidence) | 0 |
| S17-k141_218211 | 2049 | No | NA | 3 | 0 | 3 | Not-determined | Genome-fragment | NA | NA | 0 |
| S17-k141_16138 | 2257 | No | NA | 2 | 0 | 1 | Not-determined | Genome-fragment | NA | NA | 0 |
| S17-k141_128694 | 1931 | No | NA | 2 | 0 | 1 | Not-determined | Genome-fragment | NA | NA | 0 |
| S17-k141_39147 | 1626 | No | NA | 1 | 0 | 1 | Not-determined | Genome-fragment | NA | NA | 0 |
| S7-final-viral-combined_k141_178729\|\|full | 2482 | No | NA | 3 | 0 | 1 | Low-quality | Genome-fragment | 6.37 | AAI-based (medium-confidence) | 0 |
| S17-k141_229382 | 1265 | No | NA | 1 | 0 | 1 | Not-determined | Genome-fragment | NA | NA | 0 |
| S7-final-viral-combined_k141_233476\|\|full | 2482 | No | NA | 6 | 1 | 0 | Low-quality | Genome-fragment | 3.48 | HMM-based (lower-bound) | 0 |
| S17-k141_140038 | 2040 | No | NA | 1 | 0 | 0 | Not-determined | Genome-fragment | NA | NA | 0 |
| S17-k141_106139 | 3486 | No | NA | 5 | 0 | 0 | Not-determined | Genome-fragment | NA | NA | 0 |
| S7-final-viral-combined_k141_173345\|\|lt2gene | 2482 | No | NA | 2 | 1 | 0 | Low-quality | Genome-fragment | 5.3 | AAI-based (high-confidence) | 0 |
| S12-final-viral-combined_k141_227238\|\|full | 2482 | No | NA | 2 | 1 | 0 | Low-quality | Genome-fragment | 6.43 | AAI-based (high-confidence) | 0 |
| S17-k141_207073 | 1021 | No | NA | 2 | 0 | 1 | Not-determined | Genome-fragment | NA | NA | 0 |
| S2-final-viral-combined_k141_201973\|\|full | 2482 | No | NA | 4 | 1 | 0 | Low-quality | Genome-fragment | 5.91 | HMM-based (lower-bound) | 0 |
| S17-k141_48857 | 1123 | No | NA | 1 | 0 | 1 | Not-determined | Genome-fragment | NA | NA | 0 |
| S17-k141_118806 | 1435 | No | NA | 1 | 0 | 1 | Not-determined | Genome-fragment | NA | NA | 0 |
| S17-k141_84928 | 1369 | No | NA | 1 | 0 | 1 | Not-determined | Genome-fragment | NA | NA | 0 |
| S17-k141_16150 | 1588 | No | NA | 3 | 0 | 2 | Not-determined | Genome-fragment | NA | NA | 0 |
| S17-k141_218281 | 3085 | No | NA | 3 | 0 | 2 | Not-determined | Genome-fragment | NA | NA | 0 |
| S17-k141_249763 | 1009 | No | NA | 2 | 0 | 1 | Not-determined | Genome-fragment | NA | NA | 0 |
| S17-k141_5864 | 1005 | No | NA | 2 | 0 | 0 | Not-determined | Genome-fragment | NA | NA | 0 |
| S2_k141_210517 | 2482 | No | NA | 4 | 0 | 0 | Low-quality | Genome-fragment | 2.59 | AAI-based (high-confidence) | 0 |
| S17-k141_93786 | 4909 | No | NA | 7 | 0 | 4 | Not-determined | Genome-fragment | NA | NA | 0 |
| S17-k141_106223 | 1010 | No | NA | 1 | 0 | 1 | Not-determined | Genome-fragment | NA | NA | 0 |
| S17-k141_39249 | 1956 | No | NA | 2 | 0 | 1 | Not-determined | Genome-fragment | NA | NA | 0 |
| S9-final-viral-combined_k141_8853\|\|full | 2483 | No | NA | 5 | 3 | 0 | Low-quality | Genome-fragment | 21.1 | AAI-based (high-confidence) | 0 |
| S17-k141_93813 | 6486 | No | NA | 6 | 0 | 2 | Not-determined | Genome-fragment | NA | NA | 0 |
| S17-k141_93817 | 1055 | No | NA | 2 | 0 | 2 | Not-determined | Genome-fragment | NA | NA | 0 |
| S17-k141_195804 | 1042 | No | NA | 3 | 0 | 0 | Not-determined | Genome-fragment | NA | NA | 0 |
| S1-final-viral-combined_k141_282867\|\|full | 2483 | No | NA | 3 | 1 | 0 | Low-quality | Genome-fragment | 5.62 | AAI-based (medium-confidence) | 0 |
| S5-final-viral-combined_k141_63036\|\|full | 2483 | No | NA | 4 | 0 | 4 | Low-quality | Genome-fragment | 0.82 | AAI-based (medium-confidence) | 0 |
| S17-k141_195819 | 1388 | No | NA | 1 | 0 | 1 | Not-determined | Genome-fragment | NA | NA | 0 |
| S17-k141_249785 | 1123 | No | NA | 1 | 0 | 0 | Not-determined | Genome-fragment | NA | NA | 0 |
| S17-k141_140198 | 1344 | No | NA | 1 | 0 | 1 | Not-determined | Genome-fragment | NA | NA | 0 |
| S17-k141_48988 | 3309 | No | NA | 4 | 0 | 2 | Not-determined | Genome-fragment | NA | NA | 0 |
| S17-k141_207255 | 1833 | No | NA | 2 | 0 | 2 | Not-determined | Genome-fragment | NA | NA | 0 |
| S17-k141_229589 | 1347 | No | NA | 1 | 0 | 1 | Not-determined | Genome-fragment | NA | NA | 0 |
| S17-k141_48994 | 3678 | No | NA | 1 | 0 | 0 | Not-determined | Genome-fragment | NA | NA | 0 |
| S5-final-viral-combined_k141_48043\|\|full | 2483 | No | NA | 5 | 1 | 0 | Low-quality | Genome-fragment | 4.43 | AAI-based (medium-confidence) | 0 |
| S17-k141_128787 | 1120 | No | NA | 2 | 0 | 2 | Not-determined | Genome-fragment | NA | NA | 0 |
| S17-k141_150914 | 1476 | No | NA | 1 | 0 | 1 | Not-determined | Genome-fragment | NA | NA | 0 |
| S17-k141_185091 | 4673 | No | NA | 1 | 0 | 1 | Not-determined | Genome-fragment | NA | NA | 0 |
| S17-k141_140253 | 1118 | No | NA | 1 | 0 | 1 | Not-determined | Genome-fragment | NA | NA | 0 |
| S17-k141_261085 | 1502 | No | NA | 2 | 0 | 1 | Not-determined | Genome-fragment | NA | NA | 0 |
| S17-k141_39384 | 1084 | No | NA | 1 | 0 | 1 | Not-determined | Genome-fragment | NA | NA | 0 |
| S5-final-viral-combined_k141_95384\|\|full | 2483 | No | NA | 4 | 0 | 1 | Low-quality | Genome-fragment | 3.93 | AAI-based (high-confidence) | 0 |
| S17-k141_118981 | 1235 | No | NA | 3 | 0 | 3 | Not-determined | Genome-fragment | NA | NA | 0 |
| S17-k141_185146 | 1167 | No | NA | 2 | 0 | 2 | Not-determined | Genome-fragment | NA | NA | 0 |
| S17-k141_93914 | 1672 | No | NA | 1 | 0 | 1 | Not-determined | Genome-fragment | NA | NA | 0 |
| S17-k141_128863 | 1320 | No | NA | 2 | 0 | 0 | Not-determined | Genome-fragment | NA | NA | 0 |
| S17-k141_118991 | 1302 | No | NA | 2 | 0 | 1 | Not-determined | Genome-fragment | NA | NA | 0 |
| S17-k141_6038 | 1046 | No | NA | 2 | 0 | 1 | Not-determined | Genome-fragment | NA | NA | 0 |
| S17-k141_195957 | 3840 | No | NA | 5 | 0 | 5 | Not-determined | Genome-fragment | NA | NA | 0 |
| S17-k141_195958 | 1106 | No | NA | 2 | 0 | 0 | Not-determined | Genome-fragment | NA | NA | 0 |
| S17-k141_16388 | 1510 | No | NA | 1 | 0 | 1 | Not-determined | Genome-fragment | NA | NA | 0 |
| S17-k141_85142 | 1015 | No | NA | 3 | 0 | 1 | Not-determined | Genome-fragment | NA | NA | 0 |
| S17-k141_85143 | 2170 | No | NA | 4 | 0 | 4 | Not-determined | Genome-fragment | NA | NA | 0 |
| S17-k141_229652 | 3361 | No | NA | 4 | 0 | 3 | Not-determined | Genome-fragment | NA | NA | 0 |
| S17-k141_28128 | 1397 | No | NA | 1 | 0 | 0 | Not-determined | Genome-fragment | NA | NA | 0 |
| S17-k141_173169 | 1157 | No | NA | 1 | 0 | 1 | Not-determined | Genome-fragment | NA | NA | 0 |
| S17-k141_28142 | 1195 | No | NA | 4 | 0 | 4 | Not-determined | Genome-fragment | NA | NA | 0 |
| S17-k141_61859 | 2181 | No | NA | 4 | 0 | 4 | Not-determined | Genome-fragment | NA | NA | 0 |
| S17-k141_28154 | 1504 | No | NA | 2 | 0 | 1 | Not-determined | Genome-fragment | NA | NA | 0 |
| S17-k141_207419 | 1556 | No | NA | 1 | 0 | 0 | Not-determined | Genome-fragment | NA | NA | 0 |
| S7-final-viral-combined_k141_99362\|\|full | 2483 | No | NA | 4 | 0 | 2 | Low-quality | Genome-fragment | 3.15 | AAI-based (medium-confidence) | 0 |
| S17-k141_16448 | 9007 | No | NA | 14 | 1 | 1 | Not-determined | Genome-fragment | NA | NA | 0 |
| S17-k141_161348 | 1151 | No | NA | 3 | 0 | 3 | Not-determined | Genome-fragment | NA | NA | 0 |
| S17-k141_6127 | 1943 | No | NA | 1 | 0 | 0 | Not-determined | Genome-fragment | NA | NA | 0 |
| S17-k141_16493 | 3994 | No | NA | 2 | 0 | 1 | Not-determined | Genome-fragment | NA | NA | 0 |
| S17-final-viral-combined_k141_205102\|\|full | 2483 | No | NA | 6 | 1 | 0 | Low-quality | Genome-fragment | 6.44 | AAI-based (high-confidence) | 0 |
| S17-k141_161395 | 1400 | No | NA | 1 | 0 | 1 | Not-determined | Genome-fragment | NA | NA | 0 |
| S17-k141_161397 | 1067 | No | NA | 2 | 0 | 0 | Not-determined | Genome-fragment | NA | NA | 0 |
| S17-k141_261248 | 1077 | No | NA | 1 | 0 | 1 | Not-determined | Genome-fragment | NA | NA | 0 |
| S17-k141_49146 | 3260 | No | NA | 3 | 0 | 1 | Not-determined | Genome-fragment | NA | NA | 0 |
| S17-k141_61989 | 1148 | No | NA | 1 | 0 | 1 | Not-determined | Genome-fragment | NA | NA | 0 |
| S17-k141_161416 | 1386 | No | NA | 2 | 0 | 0 | Not-determined | Genome-fragment | NA | NA | 0 |
| S12-final-viral-combined_k141_319736\|\|full | 2483 | No | NA | 3 | 1 | 0 | Low-quality | Genome-fragment | 5.69 | AAI-based (high-confidence) | 0 |
| S2-final-viral-combined_k141_35196\|\|full | 2483 | No | NA | 4 | 0 | 0 | Low-quality | Genome-fragment | 1.81 | AAI-based (high-confidence) | 0 |
| S2-final-viral-combined_k141_97654\|\|full | 2483 | No | NA | 4 | 1 | 0 | Low-quality | Genome-fragment | 6.59 | AAI-based (high-confidence) | 0 |
| S17-k141_151103 | 1396 | No | NA | 2 | 0 | 0 | Not-determined | Genome-fragment | NA | NA | 0 |
| S2-final-viral-combined_k141_129511\|\|full | 2483 | No | NA | 5 | 0 | 0 | Low-quality | Genome-fragment | 5.93 | AAI-based (medium-confidence) | 0 |
| S17-k141_39591 | 4865 | No | NA | 3 | 0 | 1 | Not-determined | Genome-fragment | NA | NA | 0 |
| S17-k141_128950 | 1377 | No | NA | 2 | 0 | 1 | Not-determined | Genome-fragment | NA | NA | 0 |
| S17-k141_119127 | 7140 | No | NA | 3 | 0 | 0 | Not-determined | Genome-fragment | NA | NA | 0 |
| S17-k141_73685 | 1064 | No | NA | 1 | 0 | 1 | Not-determined | Genome-fragment | NA | NA | 0 |
| S17-k141_239737 | 1058 | No | NA | 1 | 0 | 1 | Not-determined | Genome-fragment | NA | NA | 0 |
| S17-k141_229782 | 1225 | No | NA | 2 | 0 | 0 | Not-determined | Genome-fragment | NA | NA | 0 |
| S4-final-viral-combined_k141_47388\|\|full | 2483 | No | NA | 4 | 0 | 1 | Low-quality | Genome-fragment | 4.16 | AAI-based (high-confidence) | 0 |
| S17-k141_207524 | 1299 | No | NA | 2 | 0 | 2 | Not-determined | Genome-fragment | NA | NA | 0 |
| S17-k141_218753 | 10711 | No | NA | 6 | 0 | 6 | Not-determined | Genome-fragment | NA | NA | 0 |
| S17-k141_94069 | 1567 | No | NA | 2 | 0 | 1 | Not-determined | Genome-fragment | NA | NA | 0 |
| S1-final-viral-combined_k141_284991\|\|full | 2484 | No | NA | 5 | 0 | 2 | Low-quality | Genome-fragment | 2.28 | AAI-based (medium-confidence) | 0 |
| S17-k141_196280 | 1215 | No | NA | 1 | 0 | 1 | Not-determined | Genome-fragment | NA | NA | 0 |
| S1-final-viral-combined_k141_26890\|\|full | 2484 | No | NA | 4 | 0 | 3 | Low-quality | Genome-fragment | 1.29 | AAI-based (medium-confidence) | 0 |
| S17-k141_207532 | 3817 | No | NA | 3 | 0 | 1 | Not-determined | Genome-fragment | NA | NA | 0 |
| S17-final-viral-combined_k141_258330\|\|full | 2484 | No | NA | 5 | 3 | 0 | Low-quality | Genome-fragment | 6.61 | AAI-based (high-confidence) | 0 |
| S17-k141_73723 | 1318 | No | NA | 1 | 0 | 1 | Not-determined | Genome-fragment | NA | NA | 0 |
| S17-k141_229829 | 1040 | No | NA | 2 | 0 | 0 | Not-determined | Genome-fragment | NA | NA | 0 |
| S17-k141_173343 | 1318 | No | NA | 3 | 0 | 1 | Not-determined | Genome-fragment | NA | NA | 0 |
| S17-k141_119180 | 1245 | No | NA | 2 | 0 | 1 | Not-determined | Genome-fragment | NA | NA | 0 |
| S17-k141_85319 | 1052 | No | NA | 2 | 0 | 0 | Not-determined | Genome-fragment | NA | NA | 0 |
| S17-k141_161564 | 1020 | No | NA | 1 | 0 | 1 | Not-determined | Genome-fragment | NA | NA | 0 |
| S17-k141_161566 | 1732 | No | NA | 1 | 0 | 0 | Not-determined | Genome-fragment | NA | NA | 0 |
| S17-k141_73761 | 1013 | No | NA | 1 | 0 | 1 | Not-determined | Genome-fragment | NA | NA | 0 |
| S12-final-viral-combined_k141_251374\|\|full | 2484 | No | NA | 6 | 2 | 0 | Low-quality | Genome-fragment | 5.83 | AAI-based (medium-confidence) | 0 |
| S17-k141_16544 | 2431 | No | NA | 1 | 0 | 0 | Not-determined | Genome-fragment | NA | NA | 0 |
| S17-k141_218814 | 1329 | No | NA | 1 | 0 | 1 | Not-determined | Genome-fragment | NA | NA | 0 |
| S17-k141_28403 | 1070 | No | NA | 1 | 0 | 0 | Not-determined | Genome-fragment | NA | NA | 0 |
| S17-k141_106586 | 1636 | No | NA | 1 | 0 | 0 | Not-determined | Genome-fragment | NA | NA | 0 |
| S17-k141_173400 | 6236 | No | NA | 9 | 0 | 8 | Not-determined | Genome-fragment | NA | NA | 0 |
| S17-k141_16559 | 2861 | No | NA | 2 | 0 | 0 | Not-determined | Genome-fragment | NA | NA | 0 |
| S17-k141_85382 | 3203 | No | NA | 3 | 0 | 2 | Not-determined | Genome-fragment | NA | NA | 0 |
| S17-k141_49256 | 8513 | No | NA | 19 | 0 | 18 | Not-determined | Genome-fragment | NA | NA | 0 |
| S17-k141_239891 | 3516 | No | NA | 3 | 0 | 1 | Not-determined | Genome-fragment | NA | NA | 0 |
| S17-k141_151293 | 1323 | No | NA | 1 | 0 | 1 | Not-determined | Genome-fragment | NA | NA | 0 |
| S17-k141_207600 | 1095 | No | NA | 2 | 0 | 1 | Not-determined | Genome-fragment | NA | NA | 0 |
| S17-k141_39710 | 2397 | No | NA | 2 | 0 | 0 | Not-determined | Genome-fragment | NA | NA | 0 |
| S17-k141_73828 | 1075 | No | NA | 1 | 0 | 0 | Not-determined | Genome-fragment | NA | NA | 0 |
| S17-k141_106618 | 6277 | No | NA | 5 | 0 | 1 | Not-determined | Genome-fragment | NA | NA | 0 |
| S17-k141_151354 | 1099 | No | NA | 1 | 0 | 1 | Not-determined | Genome-fragment | NA | NA | 0 |
| S17-k141_173460 | 1249 | No | NA | 2 | 0 | 0 | Not-determined | Genome-fragment | NA | NA | 0 |
| S17-k141_28466 | 1550 | No | NA | 2 | 0 | 1 | Not-determined | Genome-fragment | NA | NA | 0 |
| S17-k141_85458 | 1034 | No | NA | 2 | 0 | 0 | Not-determined | Genome-fragment | NA | NA | 0 |
| S17-k141_151364 | 1343 | No | NA | 1 | 0 | 1 | Not-determined | Genome-fragment | NA | NA | 0 |
| S17-k141_49307 | 3121 | No | NA | 4 | 0 | 4 | Not-determined | Genome-fragment | NA | NA | 0 |
| S17-k141_229984 | 2560 | No | NA | 2 | 0 | 1 | Not-determined | Genome-fragment | NA | NA | 0 |
| S17-k141_49315 | 3103 | No | NA | 6 | 0 | 0 | Not-determined | Genome-fragment | NA | NA | 0 |
| S12-final-viral-combined_k141_362887\|\|full | 2484 | No | NA | 3 | 0 | 1 | Low-quality | Genome-fragment | 4.24 | AAI-based (high-confidence) | 0 |
| S17-k141_28488 | 1121 | No | NA | 1 | 0 | 0 | Not-determined | Genome-fragment | NA | NA | 0 |
| S17-k141_49326 | 2577 | No | NA | 2 | 0 | 1 | Not-determined | Genome-fragment | NA | NA | 0 |
| S2-final-viral-combined_k141_115095\|\|full | 2485 | No | NA | 7 | 0 | 0 | Low-quality | Genome-fragment | 4.48 | AAI-based (high-confidence) | 0 |
| S17-k141_49352 | 2449 | No | NA | 2 | 0 | 1 | Not-determined | Genome-fragment | NA | NA | 0 |
| S2_k141_78499 | 2485 | No | NA | 3 | 0 | 0 | Low-quality | Genome-fragment | 4.15 | AAI-based (high-confidence) | 0 |
| S17-k141_106671 | 23575 | No | NA | 37 | 21 | 1 | Medium-quality | Genome-fragment | 52.15 | AAI-based (high-confidence) | 0 |
| S17-k141_106677 | 2560 | No | NA | 2 | 0 | 0 | Not-determined | Genome-fragment | NA | NA | 0 |
| S17-k141_73980 | 1722 | No | NA | 2 | 0 | 2 | Not-determined | Genome-fragment | NA | NA | 0 |
| S17-k141_62305 | 2333 | No | NA | 2 | 0 | 1 | Not-determined | Genome-fragment | NA | NA | 0 |
| S17-k141_129265 | 1163 | No | NA | 2 | 0 | 1 | Not-determined | Genome-fragment | NA | NA | 0 |
| S17-k141_28564 | 1136 | No | NA | 3 | 0 | 1 | Not-determined | Genome-fragment | NA | NA | 0 |
| S17-k141_129273 | 3811 | No | NA | 2 | 0 | 0 | Not-determined | Genome-fragment | NA | NA | 0 |
| S8_k141_60282 | 2485 | No | NA | 1 | 0 | 1 | Low-quality | Genome-fragment | 1.28 | AAI-based (medium-confidence) | 0 |
| S17-k141_230051 | 1666 | No | NA | 2 | 0 | 1 | Not-determined | Genome-fragment | NA | NA | 0 |
| S11-final-viral-combined_k141_89701\|\|lt2gene | 2486 | Yes | 1862 | 2 | 1 | 1 | Low-quality | Genome-fragment | 1.01 | HMM-based (lower-bound) | 25.1 |
| S7-final-viral-combined_k141_97860\|\|full | 2486 | No | NA | 5 | 1 | 0 | Low-quality | Genome-fragment | 3.86 | AAI-based (high-confidence) | 0 |
| S17-k141_230086 | 1547 | No | NA | 1 | 0 | 1 | Not-determined | Genome-fragment | NA | NA | 0 |
| S17-k141_219043 | 1322 | No | NA | 2 | 0 | 1 | Not-determined | Genome-fragment | NA | NA | 0 |
| S3-final-viral-combined_k141_127875\|\|full | 2487 | No | NA | 3 | 0 | 0 | Low-quality | Genome-fragment | 4.05 | AAI-based (medium-confidence) | 0 |
| S17-k141_219055 | 1329 | No | NA | 1 | 0 | 1 | Not-determined | Genome-fragment | NA | NA | 0 |
| S17-k141_85616 | 1475 | No | NA | 2 | 0 | 2 | Not-determined | Genome-fragment | NA | NA | 0 |
| S17-k141_219057 | 3174 | No | NA | 4 | 0 | 0 | Not-determined | Genome-fragment | NA | NA | 0 |
| S17-k141_62374 | 2087 | No | NA | 2 | 0 | 2 | Not-determined | Genome-fragment | NA | NA | 0 |
| S17-k141_28595 | 2229 | No | NA | 3 | 0 | 1 | Not-determined | Genome-fragment | NA | NA | 0 |
| S17-k141_28602 | 1216 | No | NA | 1 | 0 | 1 | Not-determined | Genome-fragment | NA | NA | 0 |
| S17-k141_161624 | 4705 | No | NA | 3 | 0 | 3 | Not-determined | Genome-fragment | NA | NA | 0 |
| S17-k141_85679 | 1713 | No | NA | 1 | 0 | 1 | Not-determined | Genome-fragment | NA | NA | 0 |
| S5-final-viral-combined_k141_86182\|\|full | 2487 | No | NA | 3 | 2 | 0 | Low-quality | Genome-fragment | 5.41 | AAI-based (high-confidence) | 0 |
| S17-final-viral-combined_k141_198800\|\|full | 2487 | No | NA | 3 | 1 | 0 | Low-quality | Genome-fragment | 4.79 | AAI-based (high-confidence) | 0 |
| S17-k141_261654 | 2574 | No | NA | 4 | 0 | 3 | Not-determined | Genome-fragment | NA | NA | 0 |
| S4-final-viral-combined_k141_130670\|\|full | 2487 | No | NA | 3 | 0 | 2 | Low-quality | Genome-fragment | 1.2 | AAI-based (medium-confidence) | 0 |
| S11-final-viral-combined_k141_185724\|\|full | 2488 | No | NA | 4 | 1 | 0 | Low-quality | Genome-fragment | 4.81 | AAI-based (high-confidence) | 0 |
| S17-k141_49505 | 1168 | No | NA | 1 | 0 | 1 | Not-determined | Genome-fragment | NA | NA | 0 |
| S17-k141_119436 | 2384 | No | NA | 2 | 0 | 2 | Not-determined | Genome-fragment | NA | NA | 0 |
| S17-k141_16801 | 1005 | No | NA | 1 | 0 | 1 | Not-determined | Genome-fragment | NA | NA | 0 |
| S17-k141_129460 | 2669 | No | NA | 1 | 0 | 0 | Not-determined | Genome-fragment | NA | NA | 0 |
| S6-final-viral-combined_k141_40095\|\|full | 2488 | No | NA | 4 | 2 | 0 | Low-quality | Genome-fragment | 5.14 | HMM-based (lower-bound) | 0 |
| S17-k141_119465 | 1025 | No | NA | 1 | 0 | 1 | Not-determined | Genome-fragment | NA | NA | 0 |
| S17-k141_161740 | 1359 | No | NA | 1 | 0 | 0 | Not-determined | Genome-fragment | NA | NA | 0 |
| S6_k141_34122 | 2488 | No | NA | 6 | 1 | 0 | Low-quality | Genome-fragment | 4.31 | AAI-based (high-confidence) | 0 |
| S17-k141_261731 | 5434 | No | NA | 4 | 0 | 2 | Not-determined | Genome-fragment | NA | NA | 0 |
| S17-k141_106841 | 1707 | No | NA | 1 | 0 | 1 | Not-determined | Genome-fragment | NA | NA | 0 |
| S11-final-viral-combined_k141_8491\|\|lt2gene | 2489 | No | NA | 2 | 1 | 0 | Low-quality | Genome-fragment | 1.54 | HMM-based (lower-bound) | 0 |
| S17-k141_28725 | 2954 | No | NA | 1 | 0 | 0 | Not-determined | Genome-fragment | NA | NA | 0 |
| S17-k141_161770 | 1237 | No | NA | 2 | 0 | 2 | Not-determined | Genome-fragment | NA | NA | 0 |
| S17-k141_6570 | 1334 | No | NA | 2 | 0 | 1 | Not-determined | Genome-fragment | NA | NA | 0 |
| S12-final-viral-combined_k141_338951\|\|full | 2489 | No | NA | 3 | 0 | 2 | Low-quality | Genome-fragment | 4.16 | AAI-based (medium-confidence) | 0 |
| S17-k141_250421 | 1222 | No | NA | 2 | 0 | 1 | Not-determined | Genome-fragment | NA | NA | 0 |
| S12-final-viral-combined_k141_49354\|\|full | 2489 | No | NA | 5 | 0 | 0 | Low-quality | Genome-fragment | 5.32 | AAI-based (high-confidence) | 0 |
| S17-k141_161799 | 1156 | No | NA | 2 | 0 | 2 | Not-determined | Genome-fragment | NA | NA | 0 |
| S17-k141_230238 | 1609 | No | NA | 3 | 0 | 0 | Not-determined | Genome-fragment | NA | NA | 0 |
| S17-k141_40036 | 1468 | No | NA | 1 | 0 | 1 | Not-determined | Genome-fragment | NA | NA | 0 |
| S17-k141_250435 | 5307 | No | NA | 5 | 0 | 2 | Not-determined | Genome-fragment | NA | NA | 0 |
| S8-final-viral-combined_k141_128782\|\|full | 2489 | No | NA | 4 | 4 | 0 | Low-quality | Genome-fragment | 5.43 | AAI-based (high-confidence) | 0 |
| S17-k141_196694 | 1102 | No | NA | 1 | 0 | 0 | Not-determined | Genome-fragment | NA | NA | 0 |
| S3-final-viral-combined_k141_43548\|\|full | 2490 | No | NA | 4 | 2 | 0 | Low-quality | Genome-fragment | 6.71 | AAI-based (high-confidence) | 0 |
| S17-k141_75492 | 2490 | No | NA | 1 | 0 | 1 | Low-quality | Genome-fragment | 0.66 | AAI-based (medium-confidence) | 0 |
| S12-final-viral-combined_k141_152564\|\|full | 2490 | No | NA | 4 | 0 | 0 | Low-quality | Genome-fragment | 6.68 | AAI-based (medium-confidence) | 0 |
| S17-k141_161858 | 1063 | No | NA | 1 | 0 | 0 | Not-determined | Genome-fragment | NA | NA | 0 |
| S17-k141_161859 | 1432 | No | NA | 1 | 0 | 0 | Not-determined | Genome-fragment | NA | NA | 0 |
| S17-k141_28785 | 3205 | No | NA | 2 | 0 | 2 | Not-determined | Genome-fragment | NA | NA | 0 |
| S17-k141_240275 | 5786 | No | NA | 7 | 0 | 7 | Not-determined | Genome-fragment | NA | NA | 0 |
| S17-k141_261823 | 1525 | No | NA | 1 | 0 | 1 | Not-determined | Genome-fragment | NA | NA | 0 |
| S17-k141_74140 | 1067 | No | NA | 1 | 0 | 1 | Not-determined | Genome-fragment | NA | NA | 0 |
| S17-k141_49593 | 1025 | No | NA | 2 | 0 | 0 | Not-determined | Genome-fragment | NA | NA | 0 |
| S17-k141_49595 | 1085 | No | NA | 2 | 0 | 1 | Not-determined | Genome-fragment | NA | NA | 0 |
| S17-k141_16933 | 1960 | No | NA | 4 | 0 | 4 | Not-determined | Genome-fragment | NA | NA | 0 |
| S17-k141_161872 | 1247 | No | NA | 2 | 0 | 1 | Not-determined | Genome-fragment | NA | NA | 0 |
| S6-final-viral-combined_k141_42489\|\|full | 2490 | No | NA | 3 | 0 | 1 | Low-quality | Genome-fragment | 8.35 | AAI-based (high-confidence) | 0 |
| S17-k141_119695 | 1072 | No | NA | 2 | 0 | 1 | Not-determined | Genome-fragment | NA | NA | 0 |
| S17-k141_230345 | 1095 | No | NA | 2 | 0 | 1 | Not-determined | Genome-fragment | NA | NA | 0 |
| S17-k141_119706 | 1130 | No | NA | 1 | 0 | 0 | Not-determined | Genome-fragment | NA | NA | 0 |
| S17-k141_161928 | 1017 | No | NA | 1 | 0 | 0 | Not-determined | Genome-fragment | NA | NA | 0 |
| S17-k141_230371 | 1148 | No | NA | 1 | 0 | 0 | Not-determined | Genome-fragment | NA | NA | 0 |
| S17-k141_240305 | 1602 | No | NA | 3 | 0 | 1 | Not-determined | Genome-fragment | NA | NA | 0 |
| S17-k141_28884 | 1575 | No | NA | 2 | 0 | 1 | Not-determined | Genome-fragment | NA | NA | 0 |
| S17-k141_161956 | 1464 | No | NA | 4 | 0 | 4 | Not-determined | Genome-fragment | NA | NA | 0 |
| S17-k141_173989 | 5744 | No | NA | 6 | 0 | 4 | Not-determined | Genome-fragment | NA | NA | 0 |
| S17-k141_28915 | 1400 | No | NA | 1 | 0 | 0 | Not-determined | Genome-fragment | NA | NA | 0 |
| S17-k141_161971 | 1229 | No | NA | 1 | 0 | 1 | Not-determined | Genome-fragment | NA | NA | 0 |
| S17-k141_261937 | 1729 | No | NA | 2 | 0 | 1 | Not-determined | Genome-fragment | NA | NA | 0 |
| S11_k141_153365 | 2491 | No | NA | 6 | 0 | 0 | Low-quality | Genome-fragment | 2.34 | AAI-based (high-confidence) | 0 |
| S17-k141_219348 | 1400 | No | NA | 1 | 0 | 0 | Not-determined | Genome-fragment | NA | NA | 0 |
| S17-k141_230442 | 1580 | No | NA | 2 | 0 | 2 | Not-determined | Genome-fragment | NA | NA | 0 |
| S17-k141_230446 | 1018 | No | NA | 1 | 0 | 1 | Not-determined | Genome-fragment | NA | NA | 0 |
| S17-k141_140847 | 1179 | No | NA | 2 | 0 | 1 | Not-determined | Genome-fragment | NA | NA | 0 |
| S17-k141_208113 | 1215 | No | NA | 2 | 0 | 1 | Not-determined | Genome-fragment | NA | NA | 0 |
| S17-k141_161992 | 1028 | No | NA | 1 | 0 | 1 | Not-determined | Genome-fragment | NA | NA | 0 |
| S17-k141_119812 | 1081 | No | NA | 3 | 0 | 3 | Not-determined | Genome-fragment | NA | NA | 0 |
| S17-k141_40253 | 1016 | No | NA | 2 | 0 | 0 | Not-determined | Genome-fragment | NA | NA | 0 |
| S17-k141_74272 | 1963 | No | NA | 2 | 0 | 1 | Not-determined | Genome-fragment | NA | NA | 0 |
| S17-k141_208140 | 1317 | No | NA | 1 | 0 | 1 | Not-determined | Genome-fragment | NA | NA | 0 |
| S17-k141_151871 | 1531 | No | NA | 1 | 0 | 1 | Not-determined | Genome-fragment | NA | NA | 0 |
| S17-k141_49750 | 6421 | No | NA | 8 | 0 | 7 | Not-determined | Genome-fragment | NA | NA | 0 |
| S17-k141_17070 | 1075 | No | NA | 2 | 0 | 0 | Not-determined | Genome-fragment | NA | NA | 0 |
| S17-k141_140927 | 1300 | No | NA | 2 | 0 | 1 | Not-determined | Genome-fragment | NA | NA | 0 |
| S1-final-viral-combined_k141_137784\|\|full | 2491 | No | NA | 2 | 0 | 1 | Low-quality | Genome-fragment | 1.33 | AAI-based (medium-confidence) | 0 |
| S17-k141_219414 | 1450 | No | NA | 2 | 0 | 1 | Not-determined | Genome-fragment | NA | NA | 0 |
| S17-k141_262033 | 4065 | No | NA | 3 | 0 | 3 | Not-determined | Genome-fragment | NA | NA | 0 |
| S17-k141_107086 | 9945 | No | NA | 14 | 0 | 14 | Not-determined | Genome-fragment | NA | NA | 0 |
| S17-k141_28938 | 1467 | No | NA | 3 | 0 | 2 | Not-determined | Genome-fragment | NA | NA | 0 |
| S1-final-viral-combined_k141_290543\|\|full | 2491 | No | NA | 3 | 0 | 2 | Low-quality | Genome-fragment | 1.66 | AAI-based (medium-confidence) | 0 |
| S17-k141_230579 | 3099 | No | NA | 3 | 0 | 1 | Not-determined | Genome-fragment | NA | NA | 0 |
| S17-k141_140963 | 1399 | No | NA | 2 | 0 | 2 | Not-determined | Genome-fragment | NA | NA | 0 |
| S3-final-viral-combined_k141_120053\|\|full | 2491 | No | NA | 3 | 0 | 2 | Low-quality | Genome-fragment | 0.73 | AAI-based (medium-confidence) | 0 |
| S7-final-viral-combined_k141_202039\|\|full | 2491 | No | NA | 5 | 0 | 0 | Low-quality | Genome-fragment | 2.05 | AAI-based (medium-confidence) | 0 |
| S17-k141_129767 | 1307 | No | NA | 2 | 0 | 2 | Not-determined | Genome-fragment | NA | NA | 0 |
| S17-k141_196916 | 3305 | No | NA | 3 | 0 | 3 | Not-determined | Genome-fragment | NA | NA | 0 |
| S17-final-viral-combined_k141_242109\|\|full | 2491 | No | NA | 4 | 0 | 0 | Low-quality | Genome-fragment | 3.39 | AAI-based (medium-confidence) | 0 |
| S17-k141_17103 | 11297 | No | NA | 10 | 0 | 1 | Not-determined | Genome-fragment | NA | NA | 0 |
| S17-k141_107114 | 1538 | No | NA | 2 | 0 | 0 | Not-determined | Genome-fragment | NA | NA | 0 |
| S17-k141_74413 | 1190 | No | NA | 2 | 0 | 2 | Not-determined | Genome-fragment | NA | NA | 0 |
| S17-k141_74416 | 1021 | No | NA | 2 | 0 | 2 | Not-determined | Genome-fragment | NA | NA | 0 |
| S17-k141_219529 | 1149 | No | NA | 1 | 0 | 1 | Not-determined | Genome-fragment | NA | NA | 0 |
| S12-final-viral-combined_k141_222692\|\|full | 2491 | No | NA | 3 | 2 | 0 | Low-quality | Genome-fragment | 5.25 | AAI-based (medium-confidence) | 0 |
| S17-k141_230675 | 1128 | No | NA | 1 | 0 | 0 | Not-determined | Genome-fragment | NA | NA | 0 |
| S2-final-viral-combined_k141_241874\|\|full | 2491 | No | NA | 5 | 0 | 1 | Low-quality | Genome-fragment | 5.69 | AAI-based (medium-confidence) | 0 |
| S11_k141_70335 | 2492 | No | NA | 2 | 0 | 2 | Low-quality | Genome-fragment | 0.93 | AAI-based (high-confidence) | 0 |
| S5-final-viral-combined_k141_23874\|\|full | 2492 | No | NA | 5 | 2 | 0 | Low-quality | Genome-fragment | 4.02 | HMM-based (lower-bound) | 0 |
| S17-k141_162071 | 1174 | No | NA | 1 | 0 | 1 | Not-determined | Genome-fragment | NA | NA | 0 |
| S17-k141_162077 | 1066 | No | NA | 1 | 0 | 1 | Not-determined | Genome-fragment | NA | NA | 0 |
| S7-final-viral-combined_k141_154232\|\|full | 2492 | No | NA | 3 | 3 | 0 | Low-quality | Genome-fragment | 6.28 | AAI-based (high-confidence) | 0 |
| S17-k141_262136 | 1462 | No | NA | 2 | 0 | 1 | Not-determined | Genome-fragment | NA | NA | 0 |
| S17-k141_262140 | 6230 | No | NA | 4 | 0 | 3 | Not-determined | Genome-fragment | NA | NA | 0 |
| S17-k141_230781 | 1051 | No | NA | 1 | 0 | 1 | Not-determined | Genome-fragment | NA | NA | 0 |
| S17-k141_86145 | 1236 | No | NA | 1 | 0 | 1 | Not-determined | Genome-fragment | NA | NA | 0 |
| S17-k141_129876 | 2296 | No | NA | 2 | 0 | 1 | Not-determined | Genome-fragment | NA | NA | 0 |
| S17-k141_152015 | 1002 | No | NA | 2 | 0 | 0 | Not-determined | Genome-fragment | NA | NA | 0 |
| S8_k141_275263 | 2492 | No | NA | 5 | 0 | 0 | Low-quality | Genome-fragment | 1.62 | AAI-based (high-confidence) | 0 |
| S17-k141_262152 | 22308 | No | NA | 40 | 0 | 34 | Not-determined | Genome-fragment | NA | NA | 0 |
| S17-k141_86183 | 1416 | No | NA | 2 | 0 | 0 | Not-determined | Genome-fragment | NA | NA | 0 |
| S17-k141_40441 | 8958 | No | NA | 9 | 0 | 8 | Not-determined | Genome-fragment | NA | NA | 0 |
| S17-k141_262166 | 1177 | No | NA | 2 | 0 | 1 | Not-determined | Genome-fragment | NA | NA | 0 |
| S17-k141_230811 | 7052 | No | NA | 5 | 0 | 3 | Not-determined | Genome-fragment | NA | NA | 0 |
| S17-k141_120047 | 1141 | No | NA | 1 | 0 | 1 | Not-determined | Genome-fragment | NA | NA | 0 |
| S17-k141_74539 | 1339 | No | NA | 2 | 0 | 1 | Not-determined | Genome-fragment | NA | NA | 0 |
| S17-k141_174316 | 4832 | No | NA | 3 | 0 | 3 | Not-determined | Genome-fragment | NA | NA | 0 |
| S7-final-viral-combined_k141_187109\|\|full | 2493 | No | NA | 3 | 0 | 2 | Low-quality | Genome-fragment | 1.71 | AAI-based (high-confidence) | 0 |
| S17-k141_262205 | 1538 | No | NA | 1 | 0 | 1 | Not-determined | Genome-fragment | NA | NA | 0 |
| S17-k141_240491 | 1026 | No | NA | 3 | 0 | 3 | Not-determined | Genome-fragment | NA | NA | 0 |
| S17-k141_129966 | 7278 | No | NA | 4 | 0 | 1 | Not-determined | Genome-fragment | NA | NA | 0 |
| S17-k141_208428 | 1359 | No | NA | 1 | 0 | 1 | Not-determined | Genome-fragment | NA | NA | 0 |
| S17-k141_262225 | 3317 | No | NA | 3 | 0 | 3 | Not-determined | Genome-fragment | NA | NA | 0 |
| S17-k141_174362 | 1305 | No | NA | 2 | 0 | 0 | Not-determined | Genome-fragment | NA | NA | 0 |
| S7-final-viral-combined_k141_217905\|\|lt2gene | 2493 | No | NA | 1 | 1 | 0 | Low-quality | Genome-fragment | 1.85 | HMM-based (lower-bound) | 0 |
| S17-k141_17323 | 3329 | No | NA | 1 | 0 | 0 | Not-determined | Genome-fragment | NA | NA | 0 |
| S17-final-viral-combined_k141_222606\|\|full | 2493 | No | NA | 3 | 1 | 0 | Low-quality | Genome-fragment | 5.37 | AAI-based (high-confidence) | 0 |
| S17-k141_250771 | 3632 | No | NA | 3 | 0 | 1 | Not-determined | Genome-fragment | NA | NA | 0 |
| S11-final-viral-combined_k141_229059\|\|full | 2494 | No | NA | 4 | 0 | 2 | Low-quality | Genome-fragment | 2.36 | AAI-based (medium-confidence) | 0 |
| S17-k141_186192 | 1935 | No | NA | 4 | 0 | 4 | Not-determined | Genome-fragment | NA | NA | 0 |
| S2-final-viral-combined_k141_248278\|\|full | 2494 | No | NA | 3 | 0 | 0 | Low-quality | Genome-fragment | 5.35 | AAI-based (high-confidence) | 0 |
| S17-k141_186210 | 1002 | No | NA | 1 | 0 | 1 | Not-determined | Genome-fragment | NA | NA | 0 |
| S17-k141_208488 | 1229 | No | NA | 1 | 0 | 1 | Not-determined | Genome-fragment | NA | NA | 0 |
| S17-k141_29082 | 48174 | No | NA | 69 | 27 | 1 | High-quality | High-quality | 100 | AAI-based (high-confidence) | 0 |
| S17-k141_74656 | 7056 | No | NA | 5 | 0 | 4 | Not-determined | Genome-fragment | NA | NA | 0 |
| S9-final-viral-combined_k141_39556\|\|full | 2495 | No | NA | 5 | 2 | 0 | Low-quality | Genome-fragment | 7.51 | AAI-based (high-confidence) | 0 |
| S17-k141_29090 | 1005 | No | NA | 1 | 0 | 0 | Not-determined | Genome-fragment | NA | NA | 0 |
| S17-k141_29092 | 1615 | No | NA | 2 | 0 | 1 | Not-determined | Genome-fragment | NA | NA | 0 |
| S17-k141_262302 | 1797 | No | NA | 1 | 0 | 0 | Not-determined | Genome-fragment | NA | NA | 0 |
| S17-k141_208562 | 2715 | No | NA | 4 | 0 | 3 | Not-determined | Genome-fragment | NA | NA | 0 |
| S17-k141_219728 | 1581 | No | NA | 1 | 0 | 1 | Not-determined | Genome-fragment | NA | NA | 0 |
| S11-final-viral-combined_k141_83201\|\|full | 2495 | No | NA | 5 | 0 | 0 | Low-quality | Genome-fragment | 8.2 | AAI-based (medium-confidence) | 0 |
| S17-k141_186275 | 1324 | No | NA | 2 | 0 | 1 | Not-determined | Genome-fragment | NA | NA | 0 |
| S17-k141_174512 | 1257 | No | NA | 1 | 0 | 0 | Not-determined | Genome-fragment | NA | NA | 0 |
| S17-k141_230975 | 1203 | No | NA | 1 | 0 | 1 | Not-determined | Genome-fragment | NA | NA | 0 |
| S17-k141_219792 | 1488 | No | NA | 2 | 0 | 1 | Not-determined | Genome-fragment | NA | NA | 0 |
| S11-final-viral-combined_k141_187068\|\|full | 2495 | No | NA | 6 | 0 | 0 | Low-quality | Genome-fragment | 6.17 | AAI-based (medium-confidence) | 0 |
| S17-k141_86417 | 1289 | No | NA | 1 | 0 | 1 | Not-determined | Genome-fragment | NA | NA | 0 |
| S17-k141_231002 | 5135 | No | NA | 3 | 0 | 3 | Not-determined | Genome-fragment | NA | NA | 0 |
| S17-k141_74737 | 1077 | No | NA | 2 | 0 | 1 | Not-determined | Genome-fragment | NA | NA | 0 |
| S17-k141_29162 | 1016 | No | NA | 2 | 0 | 0 | Not-determined | Genome-fragment | NA | NA | 0 |
| S17-k141_152252 | 1833 | No | NA | 2 | 0 | 2 | Not-determined | Genome-fragment | NA | NA | 0 |
| S5-final-viral-combined_k141_61757\|\|full | 2495 | No | NA | 4 | 1 | 0 | Low-quality | Genome-fragment | 5.08 | AAI-based (medium-confidence) | 0 |
| S17-k141_262385 | 1133 | No | NA | 2 | 0 | 1 | Not-determined | Genome-fragment | NA | NA | 0 |
| S17-k141_74766 | 1129 | No | NA | 2 | 0 | 0 | Not-determined | Genome-fragment | NA | NA | 0 |
| S17-k141_17486 | 1365 | No | NA | 2 | 0 | 1 | Not-determined | Genome-fragment | NA | NA | 0 |
| S17-k141_141484 | 3026 | No | NA | 8 | 0 | 8 | Not-determined | Genome-fragment | NA | NA | 0 |
| S7_k141_41712 | 2495 | No | NA | 2 | 0 | 0 | Low-quality | Genome-fragment | 2.37 | AAI-based (high-confidence) | 0 |
| S12-final-viral-combined_k141_286860\|\|full | 2495 | No | NA | 7 | 1 | 0 | Low-quality | Genome-fragment | 4.81 | AAI-based (high-confidence) | 0 |
| S17-k141_74802 | 5917 | No | NA | 3 | 0 | 1 | Not-determined | Genome-fragment | NA | NA | 0 |
| S17-k141_174684 | 1342 | No | NA | 3 | 0 | 2 | Not-determined | Genome-fragment | NA | NA | 0 |
| S17-k141_250950 | 1092 | No | NA | 3 | 0 | 3 | Not-determined | Genome-fragment | NA | NA | 0 |
| S11-final-viral-combined_k141_155290\|\|full | 2496 | No | NA | 8 | 3 | 0 | Low-quality | Genome-fragment | 4.3 | AAI-based (high-confidence) | 0 |
| S17-k141_208772 | 1137 | No | NA | 1 | 0 | 0 | Not-determined | Genome-fragment | NA | NA | 0 |
| S17-k141_174715 | 4147 | No | NA | 5 | 0 | 5 | Not-determined | Genome-fragment | NA | NA | 0 |
| S17-k141_50102 | 5811 | No | NA | 4 | 0 | 3 | Not-determined | Genome-fragment | NA | NA | 0 |
| S8-final-viral-combined_k141_153191\|\|full | 2497 | No | NA | 5 | 1 | 0 | Low-quality | Genome-fragment | 6.14 | AAI-based (high-confidence) | 0 |
| S17-k141_162463 | 2011 | No | NA | 2 | 0 | 2 | Not-determined | Genome-fragment | NA | NA | 0 |
| S17-k141_17549 | 1043 | No | NA | 2 | 0 | 1 | Not-determined | Genome-fragment | NA | NA | 0 |
| S17-k141_162484 | 4116 | No | NA | 3 | 0 | 1 | Not-determined | Genome-fragment | NA | NA | 0 |
| S17-k141_74877 | 3329 | No | NA | 4 | 0 | 3 | Not-determined | Genome-fragment | NA | NA | 0 |
| S17-k141_120310 | 1068 | No | NA | 2 | 0 | 2 | Not-determined | Genome-fragment | NA | NA | 0 |
| S17-k141_208815 | 3267 | No | NA | 2 | 0 | 0 | Not-determined | Genome-fragment | NA | NA | 0 |
| S17-k141_141551 | 1477 | No | NA | 3 | 0 | 1 | Not-determined | Genome-fragment | NA | NA | 0 |
| S11_k141_179336 | 2498 | No | NA | 6 | 1 | 0 | Low-quality | Genome-fragment | 4 | HMM-based (lower-bound) | 0 |
| S17-k141_7000 | 8088 | No | NA | 5 | 0 | 4 | Not-determined | Genome-fragment | NA | NA | 0 |
| S17-k141_120369 | 1339 | No | NA | 1 | 0 | 1 | Not-determined | Genome-fragment | NA | NA | 0 |
| S17-k141_186391 | 13965 | No | NA | 15 | 0 | 1 | Not-determined | Genome-fragment | NA | NA | 0 |
| S17-k141_186395 | 1025 | No | NA | 2 | 0 | 1 | Not-determined | Genome-fragment | NA | NA | 0 |
| S1-final-viral-combined_k141_265445\|\|lt2gene | 2498 | No | NA | 2 | 1 | 0 | Low-quality | Genome-fragment | 4.41 | AAI-based (medium-confidence) | 0 |
| S12-final-viral-combined_k141_157385\|\|full | 2498 | No | NA | 8 | 0 | 0 | Low-quality | Genome-fragment | 4.14 | AAI-based (high-confidence) | 0 |
| S17-k141_240868 | 1053 | No | NA | 2 | 0 | 0 | Not-determined | Genome-fragment | NA | NA | 0 |
| S17-k141_63360 | 1130 | No | NA | 1 | 0 | 1 | Not-determined | Genome-fragment | NA | NA | 0 |

| | | | | | | | | | | | |
|---|---|---|---|---|---|---|---|---|---|---|---|
| S17-k141_120384 | 2052 | No | NA | 3 | 0 | 1 | Not-determined | Genome-fragment | NA | NA | 0 |
| S17-k141_86582 | 2213 | No | NA | 3 | 0 | 3 | Not-determined | Genome-fragment | NA | NA | 0 |
| S17-k141_107626 | 3481 | No | NA | 2 | 0 | 2 | Not-determined | Genome-fragment | NA | NA | 0 |
| S8-final-viral-combined_k141_220612\|\|full | 2498 | No | NA | 4 | 1 | 0 | Low-quality | Genome-fragment | 5.45 | AAI-based (high-confidence) | 0 |
| S17-k141_107631 | 1316 | No | NA | 1 | 0 | 1 | Not-determined | Genome-fragment | NA | NA | 0 |
| S17-k141_197566 | 3601 | No | NA | 4 | 0 | 4 | Not-determined | Genome-fragment | NA | NA | 0 |
| S17-k141_63393 | 1680 | No | NA | 2 | 0 | 0 | Not-determined | Genome-fragment | NA | NA | 0 |
| S17-k141_197605 | 1049 | No | NA | 1 | 0 | 0 | Not-determined | Genome-fragment | NA | NA | 0 |
| S17-k141_220009 | 1306 | No | NA | 2 | 0 | 2 | Not-determined | Genome-fragment | NA | NA | 0 |
| S17-k141_50231 | 3047 | No | NA | 3 | 0 | 0 | Not-determined | Genome-fragment | NA | NA | 0 |
| S17-k141_86650 | 1008 | No | NA | 2 | 0 | 1 | Not-determined | Genome-fragment | NA | NA | 0 |
| S5-final-viral-combined_k141_141550\|\|lt2gene | 2499 | No | NA | 1 | 1 | 0 | Low-quality | Genome-fragment | 1.43 | HMM-based (lower-bound) | 0 |
| S2-final-viral-combined_k141_112956\|\|full | 2499 | No | NA | 2 | 0 | 0 | Low-quality | Genome-fragment | 1.67 | AAI-based (high-confidence) | 0 |
| S17-k141_95060 | 1144 | No | NA | 3 | 0 | 2 | Not-determined | Genome-fragment | NA | NA | 0 |
| S17-k141_141672 | 1288 | No | NA | 2 | 0 | 2 | Not-determined | Genome-fragment | NA | NA | 0 |
| S17-k141_231284 | 1713 | No | NA | 2 | 0 | 0 | Not-determined | Genome-fragment | NA | NA | 0 |
| S17-k141_40493 | 1645 | No | NA | 2 | 0 | 1 | Not-determined | Genome-fragment | NA | NA | 0 |
| S11-final-viral-combined_k141_137063\|\|full | 2500 | No | NA | 3 | 0 | 1 | Low-quality | Genome-fragment | 1.06 | AAI-based (medium-confidence) | 0 |
| S1-final-viral-combined_k141_336982\|\|lt2gene | 2500 | No | NA | 2 | 2 | 0 | Low-quality | Genome-fragment | 5.28 | AAI-based (high-confidence) | 0 |
| S17-k141_262594 | 1412 | No | NA | 2 | 0 | 1 | Not-determined | Genome-fragment | NA | NA | 0 |
| S17-k141_95071 | 1665 | No | NA | 1 | 0 | 1 | Not-determined | Genome-fragment | NA | NA | 0 |
| S17-k141_63432 | 3330 | No | NA | 1 | 0 | 0 | Not-determined | Genome-fragment | NA | NA | 0 |
| S17-k141_95095 | 2160 | No | NA | 5 | 0 | 0 | Not-determined | Genome-fragment | NA | NA | 0 |
| S17-k141_186534 | 1994 | No | NA | 2 | 0 | 2 | Not-determined | Genome-fragment | NA | NA | 0 |
| S17-k141_186548 | 1004 | No | NA | 2 | 0 | 0 | Not-determined | Genome-fragment | NA | NA | 0 |
| S17-k141_7062 | 3731 | No | NA | 4 | 0 | 1 | Not-determined | Genome-fragment | NA | NA | 0 |
| S7-final-viral-combined_k141_244481\|\|full | 2500 | No | NA | 3 | 0 | 0 | Low-quality | Genome-fragment | 4.39 | AAI-based (medium-confidence) | 0 |
| S17-k141_240987 | 3380 | No | NA | 2 | 0 | 1 | Not-determined | Genome-fragment | NA | NA | 0 |
| S17-k141_262618 | 1213 | No | NA | 2 | 0 | 0 | Not-determined | Genome-fragment | NA | NA | 0 |
| S17-k141_152507 | 1010 | No | NA | 2 | 0 | 1 | Not-determined | Genome-fragment | NA | NA | 0 |
| S17-k141_80845 | 2500 | No | NA | 2 | 0 | 2 | Low-quality | Genome-fragment | 5.1 | AAI-based (medium-confidence) | 0 |
| S12-final-viral-combined_k141_83591\|\|full | 2500 | No | NA | 4 | 0 | 2 | Low-quality | Genome-fragment | 1.62 | AAI-based (medium-confidence) | 0 |
| S17-k141_50281 | 14077 | No | NA | 18 | 0 | 0 | Not-determined | Genome-fragment | NA | NA | 0 |
| S17-k141_86723 | 1164 | No | NA | 1 | 0 | 1 | Not-determined | Genome-fragment | NA | NA | 0 |
| S17-k141_231325 | 1209 | No | NA | 1 | 0 | 1 | Not-determined | Genome-fragment | NA | NA | 0 |
| S17-k141_220162 | 1490 | No | NA | 2 | 0 | 1 | Not-determined | Genome-fragment | NA | NA | 0 |
| S8-final-viral-combined_k141_29961\|\|full | 2500 | No | NA | 4 | 2 | 0 | Low-quality | Genome-fragment | 1.58 | HMM-based (lower-bound) | 0 |
| S17-k141_186603 | 1483 | No | NA | 1 | 0 | 1 | Not-determined | Genome-fragment | NA | NA | 0 |
| S1-final-viral-combined_k141_280592\|\|full | 2501 | No | NA | 3 | 0 | 2 | Low-quality | Genome-fragment | 2.47 | AAI-based (medium-confidence) | 0 |
| S17-k141_197713 | 2547 | No | NA | 3 | 0 | 2 | Not-determined | Genome-fragment | NA | NA | 0 |
| S17-k141_231351 | 1904 | No | NA | 2 | 0 | 2 | Not-determined | Genome-fragment | NA | NA | 0 |
| S17-k141_231381 | 13660 | No | NA | 16 | 0 | 3 | Not-determined | Genome-fragment | NA | NA | 0 |
| S17-k141_231401 | 1221 | No | NA | 2 | 0 | 0 | Not-determined | Genome-fragment | NA | NA | 0 |
| S1-final-viral-combined_k141_259641\|\|full | 2501 | No | NA | 4 | 0 | 0 | Low-quality | Genome-fragment | 5.73 | AAI-based (medium-confidence) | 0 |
| S5-final-viral-combined_k141_160464\|\|full | 2501 | No | NA | 3 | 1 | 0 | Low-quality | Genome-fragment | 5.72 | AAI-based (medium-confidence) | 0 |
| S17-k141_63596 | 1427 | No | NA | 3 | 0 | 2 | Not-determined | Genome-fragment | NA | NA | 0 |
| S17-k141_95142 | 1112 | No | NA | 2 | 0 | 0 | Not-determined | Genome-fragment | NA | NA | 0 |
| S17-k141_186691 | 2243 | No | NA | 1 | 0 | 1 | Not-determined | Genome-fragment | NA | NA | 0 |
| S17-k141_251153 | 1096 | No | NA | 2 | 0 | 2 | Not-determined | Genome-fragment | NA | NA | 0 |
| S17-final-viral-combined_k141_147966\|\|full | 2501 | No | NA | 4 | 3 | 0 | Low-quality | Genome-fragment | 5.37 | AAI-based (high-confidence) | 0 |
| S17-k141_162761 | 5869 | No | NA | 5 | 0 | 1 | Not-determined | Genome-fragment | NA | NA | 0 |
| S17-k141_251170 | 1212 | No | NA | 3 | 0 | 1 | Not-determined | Genome-fragment | NA | NA | 0 |
| S17-k141_17835 | 1539 | No | NA | 3 | 0 | 3 | Not-determined | Genome-fragment | NA | NA | 0 |
| S17-k141_75245 | 1047 | No | NA | 2 | 0 | 1 | Not-determined | Genome-fragment | NA | NA | 0 |
| S17-k141_152567 | 1387 | No | NA | 3 | 0 | 0 | Not-determined | Genome-fragment | NA | NA | 0 |
| S17-k141_7269 | 1797 | No | NA | 1 | 0 | 1 | Not-determined | Genome-fragment | NA | NA | 0 |
| S17-k141_209134 | 3880 | No | NA | 5 | 0 | 5 | Not-determined | Genome-fragment | NA | NA | 0 |
| S17-k141_29667 | 1020 | No | NA | 2 | 0 | 2 | Not-determined | Genome-fragment | NA | NA | 0 |
| S17-k141_120670 | 1278 | No | NA | 2 | 0 | 2 | Not-determined | Genome-fragment | NA | NA | 0 |
| S17-k141_40713 | 1440 | No | NA | 3 | 0 | 2 | Not-determined | Genome-fragment | NA | NA | 0 |
| S17-k141_120698 | 1037 | No | NA | 1 | 0 | 1 | Not-determined | Genome-fragment | NA | NA | 0 |
| S17-k141_40738 | 1183 | No | NA | 2 | 0 | 0 | Not-determined | Genome-fragment | NA | NA | 0 |
| S12-final-viral-combined_k141_359350\|\|full | 2501 | No | NA | 2 | 2 | 0 | Low-quality | Genome-fragment | 6.18 | AAI-based (high-confidence) | 0 |
| S17-k141_175029 | 2431 | No | NA | 2 | 0 | 2 | Not-determined | Genome-fragment | NA | NA | 0 |
| S17-k141_220340 | 1389 | No | NA | 2 | 0 | 2 | Not-determined | Genome-fragment | NA | NA | 0 |
| S17-k141_251222 | 1419 | No | NA | 2 | 0 | 0 | Not-determined | Genome-fragment | NA | NA | 0 |
| S17-k141_107851 | 4088 | No | NA | 6 | 0 | 0 | Not-determined | Genome-fragment | NA | NA | 0 |
| S1-final-viral-combined_k141_225538\|\|full | 2502 | No | NA | 2 | 0 | 1 | Low-quality | Genome-fragment | 1.83 | AAI-based (high-confidence) | 0 |
| S8-final-viral-combined_k141_116802\|\|full | 2502 | Yes | 1894 | 6 | 1 | 1 | Low-quality | Genome-fragment | 4.26 | AAI-based (medium-confidence) | 24.3 |
| S17-k141_40756 | 5364 | No | NA | 5 | 0 | 0 | Not-determined | Genome-fragment | NA | NA | 0 |
| S8-final-viral-combined_k141_168831\|\|full | 2502 | No | NA | 6 | 2 | 0 | Low-quality | Genome-fragment | 7.27 | AAI-based (high-confidence) | 0 |
| S17-k141_262684 | 1090 | No | NA | 2 | 0 | 1 | Not-determined | Genome-fragment | NA | NA | 0 |
| S17-k141_17925 | 1462 | No | NA | 1 | 0 | 1 | Not-determined | Genome-fragment | NA | NA | 0 |
| S17-k141_182776 | 2503 | No | NA | 1 | 0 | 1 | Low-quality | Genome-fragment | 1.39 | AAI-based (medium-confidence) | 0 |
| S12-final-viral-combined_k141_398536\|\|full | 2503 | Yes | 1461 | 4 | 1 | 1 | Low-quality | Genome-fragment | 2.99 | HMM-based (lower-bound) | 41.63 |
| S17-k141_75374 | 1006 | No | NA | 1 | 0 | 0 | Not-determined | Genome-fragment | NA | NA | 0 |
| S4-final-viral-combined_k141_39491\|\|full | 2503 | No | NA | 3 | 0 | 1 | Low-quality | Genome-fragment | 3.66 | AAI-based (medium-confidence) | 0 |
| S17-k141_87024 | 4529 | No | NA | 6 | 0 | 3 | Not-determined | Genome-fragment | NA | NA | 0 |
| S17-k141_87038 | 1060 | No | NA | 2 | 0 | 1 | Not-determined | Genome-fragment | NA | NA | 0 |
| S17-k141_141939 | 1681 | No | NA | 1 | 0 | 1 | Not-determined | Genome-fragment | NA | NA | 0 |
| S17-k141_162806 | 1301 | No | NA | 1 | 0 | 1 | Not-determined | Genome-fragment | NA | NA | 0 |
| S17-k141_241338 | 2513 | No | NA | 2 | 0 | 1 | Not-determined | Genome-fragment | NA | NA | 0 |
| S17-k141_75471 | 1374 | No | NA | 1 | 0 | 1 | Not-determined | Genome-fragment | NA | NA | 0 |
| S17-k141_152781 | 2383 | No | NA | 5 | 0 | 2 | Not-determined | Genome-fragment | NA | NA | 0 |
| S4_k141_143841 | 2503 | No | NA | 2 | 0 | 0 | Low-quality | Genome-fragment | 2.53 | AAI-based (high-confidence) | 0 |
| S17-k141_29749 | 1075 | No | NA | 1 | 0 | 1 | Not-determined | Genome-fragment | NA | NA | 0 |
| S8-final-viral-combined_k141_255182\|\|full | 2503 | No | NA | 3 | 0 | 1 | Low-quality | Genome-fragment | 4.11 | AAI-based (high-confidence) | 0 |
| S17-k141_175221 | 1633 | No | NA | 2 | 0 | 0 | Not-determined | Genome-fragment | NA | NA | 0 |
| S17-k141_50547 | 4449 | No | NA | 4 | 0 | 3 | Not-determined | Genome-fragment | NA | NA | 0 |
| S17-k141_198034 | 1245 | No | NA | 2 | 0 | 1 | Not-determined | Genome-fragment | NA | NA | 0 |
| S17-k141_162876 | 1498 | No | NA | 1 | 0 | 1 | Not-determined | Genome-fragment | NA | NA | 0 |
| S7-final-viral-combined_k141_54961\|\|full | 2504 | No | NA | 4 | 1 | 0 | Low-quality | Genome-fragment | 6.78 | AAI-based (high-confidence) | 0 |
| S2-final-viral-combined_k141_57615\|\|full | 2504 | No | NA | 3 | 1 | 0 | Low-quality | Genome-fragment | 1.49 | AAI-based (high-confidence) | 0 |
| S17-k141_262821 | 1135 | No | NA | 1 | 0 | 1 | Not-determined | Genome-fragment | NA | NA | 0 |
| S2-final-viral-combined_k141_215814\|\|full | 2504 | No | NA | 2 | 0 | 2 | Low-quality | Genome-fragment | 1.93 | AAI-based (medium-confidence) | 0 |
| S17-k141_7499 | 1155 | No | NA | 2 | 0 | 2 | Not-determined | Genome-fragment | NA | NA | 0 |
| S1-final-viral-combined_k141_64894\|\|full | 2505 | No | NA | 4 | 0 | 0 | Low-quality | Genome-fragment | 5.35 | AAI-based (medium-confidence) | 0 |
| S17-k141_220610 | 1017 | No | NA | 2 | 0 | 1 | Not-determined | Genome-fragment | NA | NA | 0 |
| S17-k141_198091 | 1024 | No | NA | 3 | 0 | 0 | Not-determined | Genome-fragment | NA | NA | 0 |
| S17-k141_209438 | 3986 | No | NA | 9 | 0 | 9 | Not-determined | Genome-fragment | NA | NA | 0 |
| S17-k141_162939 | 1247 | No | NA | 1 | 0 | 0 | Not-determined | Genome-fragment | NA | NA | 0 |
| S17-k141_142065 | 1001 | No | NA | 2 | 0 | 2 | Not-determined | Genome-fragment | NA | NA | 0 |
| S17-k141_29830 | 1447 | No | NA | 1 | 0 | 0 | Not-determined | Genome-fragment | NA | NA | 0 |
| S17-k141_152857 | 4764 | No | NA | 5 | 0 | 5 | Not-determined | Genome-fragment | NA | NA | 0 |
| S3-final-viral-combined_k141_101341\|\|full | 2505 | No | NA | 4 | 0 | 1 | Low-quality | Genome-fragment | 1.15 | AAI-based (high-confidence) | 0 |
| S7-final-viral-combined_k141_55753\|\|full | 2505 | No | NA | 3 | 0 | 1 | Low-quality | Genome-fragment | 1.73 | AAI-based (high-confidence) | 0 |
| S17-k141_75598 | 3828 | No | NA | 3 | 0 | 3 | Not-determined | Genome-fragment | NA | NA | 0 |
| S12_k141_375701 | 2505 | No | NA | 2 | 0 | 0 | Low-quality | Genome-fragment | 2.6 | AAI-based (high-confidence) | 0 |
| S17-k141_231714 | 1246 | No | NA | 1 | 0 | 0 | Not-determined | Genome-fragment | NA | NA | 0 |
| S17-k141_108027 | 1025 | No | NA | 2 | 0 | 1 | Not-determined | Genome-fragment | NA | NA | 0 |
| S17-k141_186998 | 2185 | No | NA | 3 | 0 | 0 | Not-determined | Genome-fragment | NA | NA | 0 |
| S17-k141_187005 | 1844 | No | NA | 2 | 0 | 2 | Not-determined | Genome-fragment | NA | NA | 0 |
| S17-k141_18073 | 1096 | No | NA | 2 | 0 | 0 | Not-determined | Genome-fragment | NA | NA | 0 |
| S17-k141_50628 | 1241 | No | NA | 1 | 0 | 0 | Not-determined | Genome-fragment | NA | NA | 0 |
| S17-k141_241466 | 1019 | No | NA | 2 | 0 | 1 | Not-determined | Genome-fragment | NA | NA | 0 |
| S17-k141_40939 | 4514 | No | NA | 4 | 0 | 4 | Not-determined | Genome-fragment | NA | NA | 0 |
| S17-k141_7583 | 1284 | No | NA | 1 | 0 | 0 | Not-determined | Genome-fragment | NA | NA | 0 |
| S17-k141_198140 | 5176 | No | NA | 7 | 0 | 0 | Not-determined | Genome-fragment | NA | NA | 0 |
| S1-final-viral-combined_k141_66712\|\|full | 2506 | No | NA | 3 | 0 | 0 | Low-quality | Genome-fragment | 4.07 | AAI-based (medium-confidence) | 0 |
| S17-k141_194614 | 2506 | No | NA | 3 | 1 | 0 | Low-quality | Genome-fragment | 1.73 | AAI-based (medium-confidence) | 0 |
| S17-k141_121050 | 2129 | No | NA | 2 | 0 | 0 | Not-determined | Genome-fragment | NA | NA | 0 |
| S12-final-viral-combined_k141_320877\|\|full | 2506 | No | NA | 5 | 5 | 0 | Low-quality | Genome-fragment | 5.49 | AAI-based (high-confidence) | 0 |
| S17-k141_241502 | 4571 | No | NA | 2 | 0 | 1 | Not-determined | Genome-fragment | NA | NA | 0 |
| S17-k141_29906 | 1192 | No | NA | 1 | 0 | 1 | Not-determined | Genome-fragment | NA | NA | 0 |
| S17-k141_7625 | 3327 | No | NA | 4 | 0 | 1 | Not-determined | Genome-fragment | NA | NA | 0 |
| S17-k141_163050 | 1019 | No | NA | 1 | 0 | 1 | Not-determined | Genome-fragment | NA | NA | 0 |
| S17-k141_18160 | 6480 | No | NA | 6 | 0 | 4 | Not-determined | Genome-fragment | NA | NA | 0 |
| S17-k141_121101 | 1028 | No | NA | 3 | 0 | 2 | Not-determined | Genome-fragment | NA | NA | 0 |
| S17-k141_142204 | 1162 | No | NA | 1 | 0 | 1 | Not-determined | Genome-fragment | NA | NA | 0 |
| S17-k141_95455 | 3162 | No | NA | 2 | 0 | 2 | Not-determined | Genome-fragment | NA | NA | 0 |
| S17-k141_18193 | 2571 | No | NA | 2 | 0 | 2 | Not-determined | Genome-fragment | NA | NA | 0 |
| S17-k141_7666 | 2632 | No | NA | 2 | 0 | 1 | Not-determined | Genome-fragment | NA | NA | 0 |
| S17-k141_153030 | 2787 | No | NA | 2 | 0 | 2 | Not-determined | Genome-fragment | NA | NA | 0 |
| S4_k141_74688 | 2506 | No | NA | 4 | 1 | 0 | Low-quality | Genome-fragment | 2.62 | AAI-based (high-confidence) | 0 |
| S17-k141_153056 | 1851 | No | NA | 2 | 0 | 2 | Not-determined | Genome-fragment | NA | NA | 0 |
| S17-k141_187122 | 1957 | No | NA | 1 | 0 | 0 | Not-determined | Genome-fragment | NA | NA | 0 |
| S17-k141_263009 | 4374 | No | NA | 2 | 0 | 2 | Not-determined | Genome-fragment | NA | NA | 0 |
| S17-k141_95518 | 1389 | No | NA | 1 | 0 | 1 | Not-determined | Genome-fragment | NA | NA | 0 |
| S17-k141_231875 | 4864 | No | NA | 5 | 0 | 3 | Not-determined | Genome-fragment | NA | NA | 0 |
| S6-final-viral-combined_k141_99848\|\|full | 2506 | No | NA | 3 | 0 | 2 | Low-quality | Genome-fragment | 1.21 | AAI-based (medium-confidence) | 0 |
| S1-final-viral-combined_k141_100695\|\|full | 2507 | No | NA | 2 | 1 | 0 | Low-quality | Genome-fragment | 6.99 | AAI-based (medium-confidence) | 0 |
| S17-k141_108229 | 2636 | No | NA | 2 | 0 | 2 | Not-determined | Genome-fragment | NA | NA | 0 |
| S17-final-viral-combined_k141_72173\|\|full | 2507 | No | NA | 7 | 2 | 0 | Low-quality | Genome-fragment | 6.44 | AAI-based (high-confidence) | 0 |
| S17-k141_241612 | 1629 | No | NA | 1 | 0 | 1 | Not-determined | Genome-fragment | NA | NA | 0 |
| S17-k141_18259 | 1292 | No | NA | 4 | 0 | 0 | Not-determined | Genome-fragment | NA | NA | 0 |
| S17-k141_142322 | 1039 | No | NA | 2 | 0 | 0 | Not-determined | Genome-fragment | NA | NA | 0 |
| S17-k141_241656 | 1272 | No | NA | 2 | 0 | 0 | Not-determined | Genome-fragment | NA | NA | 0 |
| S17-k141_209479 | 7129 | No | NA | 8 | 0 | 2 | Not-determined | Genome-fragment | NA | NA | 0 |
| S17-k141_41009 | 1022 | No | NA | 1 | 0 | 1 | Not-determined | Genome-fragment | NA | NA | 0 |
| S17-k141_187188 | 3710 | No | NA | 3 | 0 | 1 | Not-determined | Genome-fragment | NA | NA | 0 |
| S17-k141_7743 | 1621 | No | NA | 1 | 0 | 0 | Not-determined | Genome-fragment | NA | NA | 0 |
| S12-final-viral-combined_k141_22800\|\|full | 2507 | No | NA | 2 | 0 | 0 | Low-quality | Genome-fragment | 6.91 | AAI-based (medium-confidence) | 0 |
| S4-final-viral-combined_k141_1287\|\|full | 2507 | No | NA | 5 | 0 | 0 | Low-quality | Genome-fragment | 4.13 | AAI-based (medium-confidence) | 0 |
| S4-final-viral-combined_k141_89215\|\|full | 2507 | No | NA | 5 | 0 | 0 | Low-quality | Genome-fragment | 3.94 | AAI-based (high-confidence) | 0 |
| S4_k141_144018 | 2507 | No | NA | 4 | 0 | 0 | Low-quality | Genome-fragment | 2.33 | AAI-based (high-confidence) | 0 |
| S17-k141_30046 | 1251 | No | NA | 1 | 0 | 1 | Not-determined | Genome-fragment | NA | NA | 0 |
| S4-final-viral-combined_k141_97471\|\|full | 2508 | No | NA | 4 | 2 | 0 | Low-quality | Genome-fragment | 14.5 | AAI-based (high-confidence) | 0 |
| S17-k141_198286 | 2356 | No | NA | 2 | 0 | 1 | Not-determined | Genome-fragment | NA | NA | 0 |
| S17-k141_175564 | 1463 | No | NA | 1 | 0 | 1 | Not-determined | Genome-fragment | NA | NA | 0 |
| S17-k141_75908 | 1057 | No | NA | 1 | 0 | 1 | Not-determined | Genome-fragment | NA | NA | 0 |
| S17-k141_187272 | 2083 | No | NA | 4 | 0 | 0 | Not-determined | Genome-fragment | NA | NA | 0 |
| S17-k141_263145 | 1369 | No | NA | 1 | 0 | 0 | Not-determined | Genome-fragment | NA | NA | 0 |
| S6-final-viral-combined_k141_68562\|\|full | 2508 | No | NA | 4 | 2 | 0 | Low-quality | Genome-fragment | 7.26 | AAI-based (medium-confidence) | 0 |
| S17-k141_7832 | 5427 | No | NA | 6 | 0 | 3 | Not-determined | Genome-fragment | NA | NA | 0 |
| S8_k141_210455 | 2508 | No | NA | 2 | 0 | 0 | Low-quality | Genome-fragment | 2.2 | AAI-based (medium-confidence) | 0 |
| S17-k141_142469 | 3175 | No | NA | 3 | 0 | 2 | Not-determined | Genome-fragment | NA | NA | 0 |
| S17-k141_187319 | 1397 | No | NA | 2 | 0 | 1 | Not-determined | Genome-fragment | NA | NA | 0 |
| S17-k141_263189 | 1673 | No | NA | 1 | 0 | 1 | Not-determined | Genome-fragment | NA | NA | 0 |
| S17-k141_209593 | 1373 | No | NA | 1 | 0 | 0 | Not-determined | Genome-fragment | NA | NA | 0 |
| S17-k141_163262 | 1113 | No | NA | 1 | 0 | 1 | Not-determined | Genome-fragment | NA | NA | 0 |
| S17-k141_130463 | 4393 | No | NA | 3 | 0 | 3 | Not-determined | Genome-fragment | NA | NA | 0 |
| S17-k141_130465 | 5054 | No | NA | 4 | 0 | 2 | Not-determined | Genome-fragment | NA | NA | 0 |
| S17-k141_30123 | 2562 | No | NA | 4 | 0 | 2 | Not-determined | Genome-fragment | NA | NA | 0 |
| S1_k141_38876 | 2509 | No | NA | 2 | 0 | 2 | Low-quality | Genome-fragment | 0.68 | AAI-based (high-confidence) | 0 |
| S17-k141_263245 | 1477 | No | NA | 2 | 0 | 2 | Not-determined | Genome-fragment | NA | NA | 0 |
| S12-final-viral-combined_k141_29358\|\|full | 2509 | No | NA | 4 | 0 | 0 | Low-quality | Genome-fragment | 1.9 | AAI-based (medium-confidence) | 0 |
| S17-k141_163294 | 2469 | No | NA | 1 | 0 | 1 | Not-determined | Genome-fragment | NA | NA | 0 |
| S2-final-viral-combined_k141_19131\|\|lt2gene | 2509 | No | NA | 1 | 1 | 0 | Low-quality | Genome-fragment | 4.12 | AAI-based (high-confidence) | 0 |
| S17-k141_142523 | 2837 | No | NA | 2 | 0 | 2 | Not-determined | Genome-fragment | NA | NA | 0 |
| S17-k141_163301 | 1127 | No | NA | 1 | 0 | 1 | Not-determined | Genome-fragment | NA | NA | 0 |
| S17-k141_41209 | 2480 | No | NA | 2 | 0 | 2 | Not-determined | Genome-fragment | NA | NA | 0 |
| S8-final-viral-combined_k141_31907\|\|full | 2509 | No | NA | 4 | 0 | 2 | Low-quality | Genome-fragment | 1.49 | AAI-based (medium-confidence) | 0 |
| S17-k141_87627 | 1405 | No | NA | 3 | 0 | 2 | Not-determined | Genome-fragment | NA | NA | 0 |
| S17-k141_142566 | 1140 | No | NA | 1 | 0 | 1 | Not-determined | Genome-fragment | NA | NA | 0 |
| S17-k141_221070 | 6379 | No | NA | 5 | 0 | 4 | Not-determined | Genome-fragment | NA | NA | 0 |
| S17-k141_76036 | 4070 | No | NA | 4 | 0 | 2 | Not-determined | Genome-fragment | NA | NA | 0 |
| S17-k141_108441 | 7830 | No | NA | 4 | 0 | 1 | Not-determined | Genome-fragment | NA | NA | 0 |
| S17-k141_187387 | 1071 | No | NA | 1 | 0 | 0 | Not-determined | Genome-fragment | NA | NA | 0 |
| S17-k141_30209 | 3463 | No | NA | 6 | 0 | 6 | Not-determined | Genome-fragment | NA | NA | 0 |
| S1-final-viral-combined_k141_217493\|\|full | 2510 | No | NA | 4 | 0 | 0 | Low-quality | Genome-fragment | 3.83 | AAI-based (high-confidence) | 0 |
| S17-k141_142612 | 1092 | No | NA | 1 | 0 | 1 | Not-determined | Genome-fragment | NA | NA | 0 |
| S17-k141_50998 | 3039 | No | NA | 4 | 0 | 1 | Not-determined | Genome-fragment | NA | NA | 0 |
| S17-final-viral-combined_k141_103402\|\|full | 2510 | No | NA | 4 | 1 | 0 | Low-quality | Genome-fragment | 7.07 | AAI-based (high-confidence) | 0 |
| S17-k141_198491 | 3881 | No | NA | 3 | 0 | 3 | Not-determined | Genome-fragment | NA | NA | 0 |
| S17-k141_51005 | 1154 | No | NA | 3 | 0 | 0 | Not-determined | Genome-fragment | NA | NA | 0 |
| S12-final-viral-combined_k141_343600\|\|lt2gene | 2510 | No | NA | 1 | 1 | 0 | Low-quality | Genome-fragment | 5.48 | AAI-based (medium-confidence) | 0 |
| S17-k141_87690 | 4611 | No | NA | 6 | 0 | 0 | Not-determined | Genome-fragment | NA | NA | 0 |
| S17-k141_51035 | 1110 | No | NA | 1 | 0 | 1 | Not-determined | Genome-fragment | NA | NA | 0 |
| S17-k141_209729 | 1022 | No | NA | 2 | 0 | 0 | Not-determined | Genome-fragment | NA | NA | 0 |
| S17-k141_18483 | 1211 | No | NA | 1 | 0 | 1 | Not-determined | Genome-fragment | NA | NA | 0 |
| S17-k141_108529 | 1208 | No | NA | 1 | 0 | 1 | Not-determined | Genome-fragment | NA | NA | 0 |
| S17-k141_41279 | 1071 | No | NA | 2 | 0 | 2 | Not-determined | Genome-fragment | NA | NA | 0 |
| S17-k141_142661 | 1182 | No | NA | 2 | 0 | 1 | Not-determined | Genome-fragment | NA | NA | 0 |
| S17-k141_251893 | 1412 | No | NA | 1 | 0 | 0 | Not-determined | Genome-fragment | NA | NA | 0 |
| S17-k141_18513 | 1122 | No | NA | 1 | 0 | 1 | Not-determined | Genome-fragment | NA | NA | 0 |
| S17-k141_209808 | 4183 | No | NA | 4 | 0 | 3 | Not-determined | Genome-fragment | NA | NA | 0 |
| S17-k141_130648 | 1145 | No | NA | 1 | 0 | 1 | Not-determined | Genome-fragment | NA | NA | 0 |
| S17-k141_30317 | 1417 | No | NA | 2 | 0 | 0 | Not-determined | Genome-fragment | NA | NA | 0 |
| S2-final-viral-combined_k141_326\|\|full | 2510 | No | NA | 6 | 0 | 0 | Low-quality | Genome-fragment | 5.03 | AAI-based (high-confidence) | 0 |
| S17-k141_18547 | 2874 | No | NA | 3 | 0 | 3 | Not-determined | Genome-fragment | NA | NA | 0 |
| S17-k141_18548 | 1163 | No | NA | 2 | 0 | 1 | Not-determined | Genome-fragment | NA | NA | 0 |
| S17-k141_8126 | 1020 | No | NA | 1 | 0 | 0 | Not-determined | Genome-fragment | NA | NA | 0 |
| S17-k141_108617 | 14054 | No | NA | 20 | 0 | 8 | Not-determined | Genome-fragment | NA | NA | 0 |
| S11-final-viral-combined_k141_295693\|\|full | 2511 | No | NA | 3 | 0 | 0 | Low-quality | Genome-fragment | 5.77 | AAI-based (medium-confidence) | 0 |
| S17-k141_18559 | 2474 | No | NA | 2 | 0 | 1 | Not-determined | Genome-fragment | NA | NA | 0 |
| S17-k141_76199 | 2330 | No | NA | 2 | 0 | 0 | Not-determined | Genome-fragment | NA | NA | 0 |
| S17-k141_87760 | 1060 | No | NA | 2 | 0 | 2 | Not-determined | Genome-fragment | NA | NA | 0 |
| S17-k141_108645 | 2163 | No | NA | 3 | 0 | 2 | Not-determined | Genome-fragment | NA | NA | 0 |
| S1-final-viral-combined_k141_225815\|\|lt2gene | 2511 | No | NA | 2 | 2 | 0 | Low-quality | Genome-fragment | 4.88 | AAI-based (medium-confidence) | 0 |
| S17-k141_130768 | 1084 | No | NA | 2 | 0 | 1 | Not-determined | Genome-fragment | NA | NA | 0 |
| S5-final-viral-combined_k141_41338\|\|full | 2511 | No | NA | 3 | 0 | 3 | Low-quality | Genome-fragment | 0.87 | AAI-based (medium-confidence) | 0 |
| S17-k141_163439 | 1482 | No | NA | 1 | 0 | 1 | Not-determined | Genome-fragment | NA | NA | 0 |
| S17-k141_130791 | 1076 | No | NA | 2 | 0 | 1 | Not-determined | Genome-fragment | NA | NA | 0 |
| S17-k141_8245 | 2830 | No | NA | 1 | 0 | 1 | Not-determined | Genome-fragment | NA | NA | 0 |
| S17-k141_153515 | 1402 | No | NA | 2 | 0 | 1 | Not-determined | Genome-fragment | NA | NA | 0 |
| S17-k141_18652 | 1141 | No | NA | 2 | 0 | 1 | Not-determined | Genome-fragment | NA | NA | 0 |
| S17-k141_198742 | 3830 | No | NA | 4 | 0 | 4 | Not-determined | Genome-fragment | NA | NA | 0 |
| S17-k141_41372 | 1547 | No | NA | 1 | 0 | 1 | Not-determined | Genome-fragment | NA | NA | 0 |
| S17-k141_41383 | 1594 | No | NA | 1 | 0 | 1 | Not-determined | Genome-fragment | NA | NA | 0 |
| S17-k141_187507 | 2681 | No | NA | 1 | 0 | 1 | Not-determined | Genome-fragment | NA | NA | 0 |
| S17-k141_263384 | 1949 | No | NA | 1 | 0 | 1 | Not-determined | Genome-fragment | NA | NA | 0 |
| S12-final-viral-combined_k141_153560\|\|full | 2511 | No | NA | 8 | 0 | 0 | Low-quality | Genome-fragment | 4.19 | AAI-based (high-confidence) | 0 |
| S17-k141_41392 | 3454 | No | NA | 2 | 0 | 2 | Not-determined | Genome-fragment | NA | NA | 0 |
| S17-k141_153567 | 1591 | No | NA | 1 | 0 | 1 | Not-determined | Genome-fragment | NA | NA | 0 |
| S17-k141_87860 | 1153 | No | NA | 1 | 0 | 0 | Not-determined | Genome-fragment | NA | NA | 0 |
| S17-k141_76326 | 1036 | No | NA | 3 | 0 | 0 | Not-determined | Genome-fragment | NA | NA | 0 |
| S17-k141_221275 | 1255 | No | NA | 1 | 0 | 0 | Not-determined | Genome-fragment | NA | NA | 0 |
| S8-final-viral-combined_k141_12466\|\|full | 2512 | No | NA | 3 | 1 | 0 | Low-quality | Genome-fragment | 4.08 | AAI-based (medium-confidence) | 0 |
| S17-k141_175870 | 3710 | No | NA | 3 | 0 | 1 | Not-determined | Genome-fragment | NA | NA | 0 |
| S17-k141_64529 | 1134 | No | NA | 1 | 0 | 1 | Not-determined | Genome-fragment | NA | NA | 0 |
| S11-final-viral-combined_k141_175192\|\|full | 2514 | No | NA | 3 | 0 | 1 | Low-quality | Genome-fragment | 1.48 | AAI-based (medium-confidence) | 0 |
| S17-k141_108806 | 1137 | No | NA | 2 | 0 | 1 | Not-determined | Genome-fragment | NA | NA | 0 |
| S17-k141_30475 | 1161 | No | NA | 1 | 0 | 1 | Not-determined | Genome-fragment | NA | NA | 0 |
| S17-k141_263429 | 1095 | No | NA | 1 | 0 | 1 | Not-determined | Genome-fragment | NA | NA | 0 |
| S17-k141_87918 | 1243 | No | NA | 1 | 0 | 1 | Not-determined | Genome-fragment | NA | NA | 0 |
| S1-final-viral-combined_k141_215267\|\|full | 2514 | No | NA | 7 | 2 | 0 | Low-quality | Genome-fragment | 3.89 | AAI-based (high-confidence) | 0 |
| S3-final-viral-combined_k141_40663\|\|full | 2514 | No | NA | 5 | 1 | 0 | Low-quality | Genome-fragment | 4.17 | HMM-based (lower-bound) | 0 |
| S17-k141_263483 | 1713 | No | NA | 3 | 0 | 3 | Not-determined | Genome-fragment | NA | NA | 0 |
| S17-k141_142987 | 2672 | No | NA | 2 | 0 | 2 | Not-determined | Genome-fragment | NA | NA | 0 |
| S5-final-viral-combined_k141_101249\|\|full | 2514 | No | NA | 6 | 0 | 0 | Low-quality | Genome-fragment | 3.91 | AAI-based (high-confidence) | 0 |
| S17-k141_51363 | 4383 | No | NA | 5 | 0 | 4 | Not-determined | Genome-fragment | NA | NA | 0 |
| S17-k141_175944 | 1163 | No | NA | 1 | 0 | 1 | Not-determined | Genome-fragment | NA | NA | 0 |
| S5_k141_60193 | 2514 | No | NA | 3 | 0 | 3 | Low-quality | Genome-fragment | 1.62 | AAI-based (high-confidence) | 0 |
| S17-k141_221407 | 2371 | No | NA | 3 | 0 | 2 | Not-determined | Genome-fragment | NA | NA | 0 |
| S17-k141_175958 | 2112 | No | NA | 1 | 0 | 1 | Not-determined | Genome-fragment | NA | NA | 0 |
| S17-k141_187664 | 1138 | No | NA | 2 | 0 | 1 | Not-determined | Genome-fragment | NA | NA | 0 |
| S17-k141_242191 | 1182 | No | NA | 1 | 0 | 1 | Not-determined | Genome-fragment | NA | NA | 0 |
| S17-k141_108895 | 1111 | No | NA | 2 | 0 | 0 | Not-determined | Genome-fragment | NA | NA | 0 |
| S17-k141_130989 | 1811 | No | NA | 4 | 0 | 1 | Not-determined | Genome-fragment | NA | NA | 0 |
| S17-k141_18771 | 1089 | No | NA | 3 | 0 | 1 | Not-determined | Genome-fragment | NA | NA | 0 |
| S17-k141_121739 | 1344 | No | NA | 2 | 0 | 2 | Not-determined | Genome-fragment | NA | NA | 0 |
| S17-k141_64694 | 1058 | No | NA | 3 | 0 | 0 | Not-determined | Genome-fragment | NA | NA | 0 |
| S17-k141_76467 | 1058 | No | NA | 3 | 0 | 1 | Not-determined | Genome-fragment | NA | NA | 0 |
| S17-k141_163658 | 3950 | No | NA | 2 | 0 | 1 | Not-determined | Genome-fragment | NA | NA | 0 |
| S17-k141_108944 | 1052 | No | NA | 2 | 0 | 1 | Not-determined | Genome-fragment | NA | NA | 0 |
| S17-k141_210004 | 1047 | No | NA | 2 | 0 | 2 | Not-determined | Genome-fragment | NA | NA | 0 |
| S17-k141_198983 | 4572 | No | NA | 2 | 0 | 2 | Not-determined | Genome-fragment | NA | NA | 0 |
| S17-k141_232480 | 1896 | No | NA | 2 | 0 | 2 | Not-determined | Genome-fragment | NA | NA | 0 |
| S17-k141_88032 | 8393 | No | NA | 7 | 0 | 4 | Not-determined | Genome-fragment | NA | NA | 0 |
| S17-k141_76518 | 1301 | No | NA | 1 | 0 | 1 | Not-determined | Genome-fragment | NA | NA | 0 |
| S17-k141_143106 | 3143 | No | NA | 5 | 0 | 2 | Not-determined | Genome-fragment | NA | NA | 0 |
| S12-final-viral-combined_k141_184847\|\|full | 2514 | No | NA | 7 | 3 | 0 | Low-quality | Genome-fragment | 6.55 | AAI-based (medium-confidence) | 0 |
| S17-k141_96104 | 2624 | No | NA | 2 | 0 | 0 | Not-determined | Genome-fragment | NA | NA | 0 |
| S12-final-viral-combined_k141_109330\|\|full | 2514 | No | NA | 3 | 1 | 0 | Low-quality | Genome-fragment | 5.83 | AAI-based (medium-confidence) | 0 |
| S2-final-viral-combined_k141_60428\|\|full | 2514 | No | NA | 3 | 0 | 2 | Low-quality | Genome-fragment | 1.84 | AAI-based (high-confidence) | 0 |
| S17-k141_163755 | 1029 | No | NA | 1 | 0 | 1 | Not-determined | Genome-fragment | NA | NA | 0 |
| S17-k141_76577 | 7182 | No | NA | 5 | 0 | 5 | Not-determined | Genome-fragment | NA | NA | 0 |
| S17-k141_252102 | 7107 | No | NA | 6 | 0 | 5 | Not-determined | Genome-fragment | NA | NA | 0 |
| S17-k141_109060 | 2112 | No | NA | 3 | 0 | 3 | Not-determined | Genome-fragment | NA | NA | 0 |
| S8-final-viral-combined_k141_219752\|\|full | 2514 | Yes | 1900 | 5 | 1 | 2 | Low-quality | Genome-fragment | 3.46 | AAI-based (medium-confidence) | 24.42 |
| S17-k141_153860 | 4308 | No | NA | 2 | 0 | 0 | Not-determined | Genome-fragment | NA | NA | 0 |
| S17-k141_221616 | 1038 | No | NA | 2 | 0 | 0 | Not-determined | Genome-fragment | NA | NA | 0 |
| S17-k141_187817 | 2241 | No | NA | 1 | 0 | 1 | Not-determined | Genome-fragment | NA | NA | 0 |
| S1-final-viral-combined_k141_292836\|\|full | 2515 | No | NA | 6 | 1 | 0 | Low-quality | Genome-fragment | 5.89 | AAI-based (high-confidence) | 0 |
| S17-k141_30732 | 1276 | No | NA | 1 | 0 | 1 | Not-determined | Genome-fragment | NA | NA | 0 |
| S6-final-viral-combined_k141_140770\|\|full | 2515 | No | NA | 2 | 1 | 0 | Low-quality | Genome-fragment | 2.02 | HMM-based (lower-bound) | 0 |
| S17-k141_121892 | 3772 | No | NA | 2 | 0 | 2 | Not-determined | Genome-fragment | NA | NA | 0 |
| S17-k141_232626 | 1427 | No | NA | 2 | 0 | 1 | Not-determined | Genome-fragment | NA | NA | 0 |
| S17-k141_187851 | 3353 | No | NA | 4 | 0 | 2 | Not-determined | Genome-fragment | NA | NA | 0 |
| S17-k141_199083 | 1595 | No | NA | 1 | 0 | 1 | Not-determined | Genome-fragment | NA | NA | 0 |
| S17-k141_252165 | 1092 | No | NA | 1 | 0 | 0 | Not-determined | Genome-fragment | NA | NA | 0 |
| S17-k141_143209 | 5242 | No | NA | 4 | 0 | 2 | Not-determined | Genome-fragment | NA | NA | 0 |
| S17-k141_109074 | 7163 | No | NA | 6 | 0 | 3 | Not-determined | Genome-fragment | NA | NA | 0 |
| S17-k141_153888 | 1059 | No | NA | 2 | 0 | 0 | Not-determined | Genome-fragment | NA | NA | 0 |
| S17-k141_153901 | 1131 | No | NA | 2 | 0 | 1 | Not-determined | Genome-fragment | NA | NA | 0 |
| S11-final-viral-combined_k141_179146\|\|full | 2516 | No | NA | 6 | 5 | 0 | Low-quality | Genome-fragment | 6.52 | AAI-based (high-confidence) | 0 |
| S17-k141_76710 | 1328 | No | NA | 2 | 0 | 2 | Not-determined | Genome-fragment | NA | NA | 0 |
| S17-k141_8495 | 2812 | No | NA | 6 | 0 | 0 | Not-determined | Genome-fragment | NA | NA | 0 |
| S9-final-viral-combined_k141_46109\|\|full | 2517 | No | NA | 3 | 3 | 0 | Low-quality | Genome-fragment | 7.54 | AAI-based (high-confidence) | 0 |
| S7-final-viral-combined_k141_61683\|\|full | 2517 | No | NA | 3 | 1 | 0 | Low-quality | Genome-fragment | 1.37 | HMM-based (lower-bound) | 0 |
| S1-final-viral-combined_k141_349464\|\|full | 2518 | No | NA | 3 | 0 | 2 | Low-quality | Genome-fragment | 2.81 | AAI-based (medium-confidence) | 0 |
| S17-k141_88256 | 1568 | No | NA | 2 | 0 | 0 | Not-determined | Genome-fragment | NA | NA | 0 |
| S17-k141_153922 | 2082 | No | NA | 1 | 0 | 1 | Not-determined | Genome-fragment | NA | NA | 0 |
| S17-k141_232713 | 1300 | No | NA | 1 | 0 | 0 | Not-determined | Genome-fragment | NA | NA | 0 |
| S17-final-viral-combined_k141_34611\|\|full | 2518 | No | NA | 5 | 1 | 0 | Low-quality | Genome-fragment | 4.69 | AAI-based (medium-confidence) | 0 |
| S17-k141_30825 | 3134 | No | NA | 2 | 0 | 2 | Not-determined | Genome-fragment | NA | NA | 0 |
| S17-k141_232753 | 1007 | No | NA | 2 | 0 | 0 | Not-determined | Genome-fragment | NA | NA | 0 |
| S17-k141_143302 | 1732 | No | NA | 3 | 0 | 2 | Not-determined | Genome-fragment | NA | NA | 0 |
| S17-k141_41808 | 2049 | No | NA | 3 | 0 | 2 | Not-determined | Genome-fragment | NA | NA | 0 |
| S17-k141_199202 | 1329 | No | NA | 2 | 0 | 2 | Not-determined | Genome-fragment | NA | NA | 0 |
| S17-k141_30838 | 1386 | No | NA | 2 | 0 | 2 | Not-determined | Genome-fragment | NA | NA | 0 |
| S17-k141_143332 | 4380 | No | NA | 4 | 0 | 3 | Not-determined | Genome-fragment | NA | NA | 0 |
| S2_k141_245151 | 2518 | No | NA | 2 | 0 | 1 | Low-quality | Genome-fragment | 2.09 | AAI-based (high-confidence) | 0 |
| S8-final-viral-combined_k141_215258\|\|full | 2518 | No | NA | 5 | 0 | 1 | Low-quality | Genome-fragment | 2.68 | AAI-based (medium-confidence) | 0 |
| S17-k141_65055 | 1190 | No | NA | 2 | 0 | 1 | Not-determined | Genome-fragment | NA | NA | 0 |
| S17-k141_252259 | 1340 | No | NA | 1 | 0 | 1 | Not-determined | Genome-fragment | NA | NA | 0 |
| S3_k141_83623 | 2519 | No | NA | 3 | 0 | 0 | Low-quality | Genome-fragment | 1.64 | AAI-based (high-confidence) | 0 |
| S5-final-viral-combined_k141_98609\|\|full | 2519 | No | NA | 4 | 2 | 0 | Low-quality | Genome-fragment | 6.16 | AAI-based (high-confidence) | 0 |
| S17-k141_30866 | 1881 | No | NA | 1 | 0 | 1 | Not-determined | Genome-fragment | NA | NA | 0 |
| S17-k141_143380 | 3446 | No | NA | 1 | 0 | 1 | Not-determined | Genome-fragment | NA | NA | 0 |
| S17-k141_109161 | 3222 | No | NA | 3 | 0 | 3 | Not-determined | Genome-fragment | NA | NA | 0 |
| S17-k141_76866 | 2671 | No | NA | 2 | 0 | 1 | Not-determined | Genome-fragment | NA | NA | 0 |
| S17-k141_88457 | 1092 | No | NA | 1 | 0 | 0 | Not-determined | Genome-fragment | NA | NA | 0 |
| S17-k141_163996 | 3278 | No | NA | 5 | 0 | 1 | Not-determined | Genome-fragment | NA | NA | 0 |
| S17-k141_232884 | 1067 | No | NA | 1 | 0 | 0 | Not-determined | Genome-fragment | NA | NA | 0 |
| S5-final-viral-combined_k141_58176\|\|full | 2519 | No | NA | 3 | 0 | 0 | Low-quality | Genome-fragment | 6.96 | AAI-based (high-confidence) | 0 |
| S7-final-viral-combined_k141_26571\|\|full | 2519 | No | NA | 2 | 0 | 2 | Low-quality | Genome-fragment | 1.94 | AAI-based (medium-confidence) | 0 |
| S17-k141_30922 | 3085 | No | NA | 3 | 0 | 2 | Not-determined | Genome-fragment | NA | NA | 0 |
| S17-k141_51693 | 2312 | No | NA | 2 | 0 | 0 | Not-determined | Genome-fragment | NA | NA | 0 |
| S17-k141_164023 | 1128 | No | NA | 2 | 0 | 1 | Not-determined | Genome-fragment | NA | NA | 0 |
| S17-k141_199317 | 1747 | No | NA | 1 | 0 | 0 | Not-determined | Genome-fragment | NA | NA | 0 |
| S17-k141_221895 | 2586 | No | NA | 4 | 0 | 1 | Not-determined | Genome-fragment | NA | NA | 0 |
| S17-k141_41882 | 5481 | No | NA | 1 | 0 | 1 | Not-determined | Genome-fragment | NA | NA | 0 |
| S17-k141_232923 | 3492 | No | NA | 3 | 0 | 2 | Not-determined | Genome-fragment | NA | NA | 0 |
| S12-final-viral-combined_k141_365360\|\|full | 2519 | No | NA | 2 | 0 | 2 | Low-quality | Genome-fragment | 0.76 | AAI-based (medium-confidence) | 0 |
| S17-k141_154125 | 1336 | No | NA | 2 | 0 | 2 | Not-determined | Genome-fragment | NA | NA | 0 |
| S17-k141_252359 | 1728 | No | NA | 3 | 0 | 3 | Not-determined | Genome-fragment | NA | NA | 0 |
| S17-k141_19100 | 1082 | No | NA | 1 | 0 | 1 | Not-determined | Genome-fragment | NA | NA | 0 |
| S17-k141_122055 | 1057 | No | NA | 1 | 0 | 1 | Not-determined | Genome-fragment | NA | NA | 0 |
| S17-k141_199358 | 2344 | No | NA | 1 | 0 | 0 | Not-determined | Genome-fragment | NA | NA | 0 |
| S17-k141_19112 | 1184 | No | NA | 2 | 0 | 1 | Not-determined | Genome-fragment | NA | NA | 0 |
| S8_k141_196542 | 2519 | No | NA | 2 | 0 | 0 | Low-quality | Genome-fragment | 4.76 | AAI-based (high-confidence) | 0 |
| S17-k141_76911 | 2566 | No | NA | 3 | 0 | 2 | Not-determined | Genome-fragment | NA | NA | 0 |
| S11-final-viral-combined_k141_118899\|\|full | 2520 | No | NA | 2 | 0 | 1 | Low-quality | Genome-fragment | 5.42 | AAI-based (high-confidence) | 0 |
| S11-final-viral-combined_k141_120303\|\|full | 2520 | No | NA | 3 | 0 | 0 | Low-quality | Genome-fragment | 4.38 | AAI-based (medium-confidence) | 0 |
| S17-k141_176416 | 1100 | No | NA | 3 | 0 | 0 | Not-determined | Genome-fragment | NA | NA | 0 |
| S17-k141_154154 | 1136 | No | NA | 1 | 0 | 0 | Not-determined | Genome-fragment | NA | NA | 0 |
| S1-final-viral-combined_k141_362096\|\|full | 2520 | No | NA | 2 | 1 | 0 | Low-quality | Genome-fragment | 6.18 | AAI-based (high-confidence) | 0 |
| S17-k141_242564 | 1339 | No | NA | 1 | 0 | 1 | Not-determined | Genome-fragment | NA | NA | 0 |
| S17-k141_143502 | 1401 | No | NA | 2 | 0 | 2 | Not-determined | Genome-fragment | NA | NA | 0 |
| S17-k141_242571 | 1334 | No | NA | 2 | 0 | 2 | Not-determined | Genome-fragment | NA | NA | 0 |
| S17-k141_252397 | 1131 | No | NA | 1 | 0 | 0 | Not-determined | Genome-fragment | NA | NA | 0 |
| S17-k141_41942 | 1236 | No | NA | 2 | 0 | 1 | Not-determined | Genome-fragment | NA | NA | 0 |
| S17-k141_122081 | 3313 | No | NA | 6 | 0 | 4 | Not-determined | Genome-fragment | NA | NA | 0 |
| S17-k141_109293 | 2300 | No | NA | 5 | 0 | 2 | Not-determined | Genome-fragment | NA | NA | 0 |
| S17-k141_41991 | 1083 | No | NA | 1 | 0 | 1 | Not-determined | Genome-fragment | NA | NA | 0 |
| S17-k141_176483 | 1536 | No | NA | 1 | 0 | 0 | Not-determined | Genome-fragment | NA | NA | 0 |
| S17-k141_41992 | 1061 | No | NA | 3 | 0 | 2 | Not-determined | Genome-fragment | NA | NA | 0 |
| S17-k141_8811 | 1279 | No | NA | 5 | 0 | 4 | Not-determined | Genome-fragment | NA | NA | 0 |
| S17-k141_188208 | 1058 | No | NA | 1 | 0 | 0 | Not-determined | Genome-fragment | NA | NA | 0 |
| S1-final-viral-combined_k141_111802\|\|full | 2520 | No | NA | 4 | 0 | 0 | Low-quality | Genome-fragment | 5.93 | AAI-based (medium-confidence) | 0 |
| S17-k141_154257 | 1048 | No | NA | 2 | 0 | 1 | Not-determined | Genome-fragment | NA | NA | 0 |
| S17-k141_131476 | 1256 | No | NA | 4 | 0 | 4 | Not-determined | Genome-fragment | NA | NA | 0 |
| S17-k141_65282 | 4258 | No | NA | 4 | 0 | 2 | Not-determined | Genome-fragment | NA | NA | 0 |
| S17-k141_31098 | 1741 | No | NA | 3 | 0 | 0 | Not-determined | Genome-fragment | NA | NA | 0 |
| S1_k141_386913 | 2520 | No | NA | 2 | 0 | 0 | Low-quality | Genome-fragment | 2.74 | AAI-based (medium-confidence) | 0 |
| S17-k141_154274 | 2363 | No | NA | 2 | 0 | 2 | Not-determined | Genome-fragment | NA | NA | 0 |
| S17-k141_176546 | 1047 | No | NA | 1 | 0 | 0 | Not-determined | Genome-fragment | NA | NA | 0 |
| S17-k141_122147 | 2265 | No | NA | 1 | 0 | 0 | Not-determined | Genome-fragment | NA | NA | 0 |
| S17-k141_176555 | 1336 | No | NA | 1 | 0 | 1 | Not-determined | Genome-fragment | NA | NA | 0 |
| S17-k141_176556 | 1266 | No | NA | 1 | 0 | 0 | Not-determined | Genome-fragment | NA | NA | 0 |
| S17-k141_31120 | 1555 | No | NA | 2 | 0 | 0 | Not-determined | Genome-fragment | NA | NA | 0 |
| S17-k141_222066 | 1059 | No | NA | 2 | 0 | 1 | Not-determined | Genome-fragment | NA | NA | 0 |
| S12-final-viral-combined_k141_277068\|\|full | 2520 | No | NA | 3 | 2 | 0 | Low-quality | Genome-fragment | 4.37 | HMM-based (lower-bound) | 0 |
| S17-k141_242644 | 1765 | No | NA | 1 | 0 | 1 | Not-determined | Genome-fragment | NA | NA | 0 |
| S17-k141_263983 | 2053 | No | NA | 3 | 0 | 3 | Not-determined | Genome-fragment | NA | NA | 0 |
| S17-k141_222094 | 2094 | No | NA | 2 | 0 | 2 | Not-determined | Genome-fragment | NA | NA | 0 |
| S17-k141_222095 | 1765 | No | NA | 1 | 0 | 0 | Not-determined | Genome-fragment | NA | NA | 0 |
| S17-k141_31169 | 1844 | No | NA | 2 | 0 | 1 | Not-determined | Genome-fragment | NA | NA | 0 |
| S17-k141_222102 | 5179 | No | NA | 7 | 0 | 1 | Not-determined | Genome-fragment | NA | NA | 0 |
| S17-k141_65376 | 6488 | No | NA | 8 | 0 | 1 | Not-determined | Genome-fragment | NA | NA | 0 |
| S17-k141_109416 | 1270 | No | NA | 2 | 0 | 0 | Not-determined | Genome-fragment | NA | NA | 0 |
| S17-k141_188319 | 6473 | No | NA | 3 | 0 | 3 | Not-determined | Genome-fragment | NA | NA | 0 |
| S17-k141_222143 | 1756 | No | NA | 2 | 0 | 1 | Not-determined | Genome-fragment | NA | NA | 0 |
| S17-k141_77067 | 1358 | No | NA | 2 | 0 | 2 | Not-determined | Genome-fragment | NA | NA | 0 |
| S17-k141_222161 | 1911 | No | NA | 3 | 0 | 2 | Not-determined | Genome-fragment | NA | NA | 0 |
| S17-k141_222165 | 1149 | No | NA | 2 | 0 | 0 | Not-determined | Genome-fragment | NA | NA | 0 |
| S17-k141_143718 | 1087 | No | NA | 1 | 0 | 1 | Not-determined | Genome-fragment | NA | NA | 0 |
| S17-k141_199549 | 1492 | No | NA | 2 | 0 | 2 | Not-determined | Genome-fragment | NA | NA | 0 |
| S17-k141_233198 | 1555 | No | NA | 1 | 0 | 1 | Not-determined | Genome-fragment | NA | NA | 0 |
| S17-k141_96591 | 1275 | No | NA | 1 | 0 | 1 | Not-determined | Genome-fragment | NA | NA | 0 |
| S17-k141_210759 | 2198 | No | NA | 3 | 0 | 1 | Not-determined | Genome-fragment | NA | NA | 0 |
| S17-k141_176676 | 3076 | No | NA | 3 | 0 | 2 | Not-determined | Genome-fragment | NA | NA | 0 |
| S17-k141_65413 | 1058 | No | NA | 2 | 0 | 1 | Not-determined | Genome-fragment | NA | NA | 0 |
| S17-k141_252605 | 1225 | No | NA | 1 | 0 | 1 | Not-determined | Genome-fragment | NA | NA | 0 |
| S17-k141_199589 | 3038 | No | NA | 2 | 0 | 2 | Not-determined | Genome-fragment | NA | NA | 0 |
| S17-k141_19339 | 2044 | No | NA | 3 | 0 | 2 | Not-determined | Genome-fragment | NA | NA | 0 |
| S17-k141_210790 | 1501 | No | NA | 1 | 0 | 0 | Not-determined | Genome-fragment | NA | NA | 0 |
| S17-k141_19346 | 8835 | No | NA | 5 | 0 | 3 | Not-determined | Genome-fragment | NA | NA | 0 |
| S17-k141_31203 | 1009 | No | NA | 1 | 0 | 1 | Not-determined | Genome-fragment | NA | NA | 0 |
| S2-final-viral-combined_k141_125449\|\|full | 2520 | No | NA | 5 | 0 | 1 | Low-quality | Genome-fragment | 5.46 | AAI-based (medium-confidence) | 0 |
| S4-final-viral-combined_k141_147536\|\|full | 2520 | No | NA | 3 | 0 | 0 | Low-quality | Genome-fragment | 4.75 | AAI-based (high-confidence) | 0 |
| S17-k141_96650 | 1461 | No | NA | 2 | 0 | 1 | Not-determined | Genome-fragment | NA | NA | 0 |
| S17-k141_65430 | 1111 | No | NA | 2 | 0 | 2 | Not-determined | Genome-fragment | NA | NA | 0 |
| S17-k141_42245 | 2727 | No | NA | 4 | 0 | 1 | Not-determined | Genome-fragment | NA | NA | 0 |
| S17-k141_210837 | 1773 | No | NA | 2 | 0 | 2 | Not-determined | Genome-fragment | NA | NA | 0 |
| S17-k141_65450 | 1112 | No | NA | 1 | 0 | 0 | Not-determined | Genome-fragment | NA | NA | 0 |
| S8-final-viral-combined_k141_59851\|\|lt2gene | 2520 | No | NA | 2 | 1 | 0 | Low-quality | Genome-fragment | 5.86 | AAI-based (high-confidence) | 0 |
| S17-k141_88801 | 1074 | No | NA | 1 | 0 | 1 | Not-determined | Genome-fragment | NA | NA | 0 |
| S17-k141_154474 | 1071 | No | NA | 1 | 0 | 1 | Not-determined | Genome-fragment | NA | NA | 0 |
| S7-final-viral-combined_k141_159909\|\|full | 2521 | No | NA | 4 | 0 | 2 | Low-quality | Genome-fragment | 5.3 | AAI-based (medium-confidence) | 0 |
| S2_k141_111684 | 2521 | No | NA | 3 | 2 | 0 | Low-quality | Genome-fragment | 7.54 | HMM-based (lower-bound) | 0 |
| S8-final-viral-combined_k141_195778\|\|full | 2521 | No | NA | 3 | 0 | 3 | Low-quality | Genome-fragment | 0.91 | AAI-based (medium-confidence) | 0 |

| | | | | | | | | | | | |
|---|---|---|---|---|---|---|---|---|---|---|---|
| S17-k141_42265 | 1023 | No | NA | 1 | 0 | 1 | Not-determined | Genome-fragment | NA | NA | 0 |
| S17-k141_199673 | 1192 | No | NA | 3 | 0 | 1 | Not-determined | Genome-fragment | NA | NA | 0 |
| S17-k141_77215 | 1319 | No | NA | 1 | 0 | 1 | Not-determined | Genome-fragment | NA | NA | 0 |
| S17-k141_131732 | 1209 | No | NA | 2 | 0 | 1 | Not-determined | Genome-fragment | NA | NA | 0 |
| S17-k141_42285 | 2257 | No | NA | 7 | 0 | 0 | Not-determined | Genome-fragment | NA | NA | 0 |
| S17-k141_242772 | 1322 | No | NA | 2 | 0 | 1 | Not-determined | Genome-fragment | NA | NA | 0 |
| S17-k141_65524 | 3370 | No | NA | 3 | 0 | 1 | Not-determined | Genome-fragment | NA | NA | 0 |
| S17-k141_19432 | 1089 | No | NA | 2 | 0 | 0 | Not-determined | Genome-fragment | NA | NA | 0 |
| S2-final-viral-combined_k141_167397\|\|full | 2522 | No | NA | 6 | 0 | 0 | Low-quality | Genome-fragment | 4.26 | AAI-based (medium-confidence) | 0 |
| S17-k141_131789 | 1704 | No | NA | 2 | 0 | 2 | Not-determined | Genome-fragment | NA | NA | 0 |
| S17-k141_131790 | 2612 | No | NA | 2 | 0 | 2 | Not-determined | Genome-fragment | NA | NA | 0 |
| S2-final-viral-combined_k141_245018\|\|full | 2522 | No | NA | 6 | 1 | 0 | Low-quality | Genome-fragment | 7.14 | AAI-based (high-confidence) | 0 |
| S17-k141_51909 | 1231 | No | NA | 1 | 0 | 0 | Not-determined | Genome-fragment | NA | NA | 0 |
| S6-final-viral-combined_k141_67761\|\|full | 2522 | No | NA | 3 | 0 | 0 | Low-quality | Genome-fragment | 4.44 | AAI-based (medium-confidence) | 0 |
| S17-k141_77298 | 2488 | No | NA | 3 | 0 | 2 | Not-determined | Genome-fragment | NA | NA | 0 |
| S3-final-viral-combined_k141_2322\|\|full | 2523 | No | NA | 4 | 1 | 0 | Low-quality | Genome-fragment | 3.2 | AAI-based (high-confidence) | 0 |
| S17-k141_19524 | 1717 | No | NA | 2 | 0 | 1 | Not-determined | Genome-fragment | NA | NA | 0 |
| S17-k141_31387 | 3402 | No | NA | 5 | 0 | 1 | Not-determined | Genome-fragment | NA | NA | 0 |
| S17-k141_42420 | 1117 | No | NA | 2 | 0 | 0 | Not-determined | Genome-fragment | NA | NA | 0 |
| S17-k141_264238 | 3217 | No | NA | 4 | 0 | 2 | Not-determined | Genome-fragment | NA | NA | 0 |
| S17-k141_143924 | 1624 | No | NA | 3 | 0 | 1 | Not-determined | Genome-fragment | NA | NA | 0 |
| S17-k141_9299 | 1131 | No | NA | 2 | 0 | 2 | Not-determined | Genome-fragment | NA | NA | 0 |
| S17-k141_9302 | 3155 | No | NA | 3 | 0 | 1 | Not-determined | Genome-fragment | NA | NA | 0 |
| S17-k141_242896 | 5813 | No | NA | 3 | 0 | 2 | Not-determined | Genome-fragment | NA | NA | 0 |
| S17-k141_154647 | 1144 | No | NA | 3 | 0 | 1 | Not-determined | Genome-fragment | NA | NA | 0 |
| S17-k141_19576 | 2404 | No | NA | 1 | 0 | 1 | Not-determined | Genome-fragment | NA | NA | 0 |
| S17-final-viral-combined_k141_88082\|\|full | 2523 | No | NA | 2 | 0 | 2 | Low-quality | Genome-fragment | 3.04 | AAI-based (medium-confidence) | 0 |
| S17-k141_51998 | 1056 | No | NA | 3 | 0 | 0 | Not-determined | Genome-fragment | NA | NA | 0 |
| S17-k141_131927 | 1684 | No | NA | 1 | 0 | 0 | Not-determined | Genome-fragment | NA | NA | 0 |
| S17-k141_42458 | 1369 | No | NA | 2 | 0 | 0 | Not-determined | Genome-fragment | NA | NA | 0 |
| S17-k141_131971 | 1325 | No | NA | 1 | 0 | 1 | Not-determined | Genome-fragment | NA | NA | 0 |
| S11-final-viral-combined_k141_159342\|\|full | 2524 | No | NA | 5 | 0 | 0 | Low-quality | Genome-fragment | 6.55 | AAI-based (high-confidence) | 0 |
| S17-k141_164519 | 1536 | No | NA | 1 | 0 | 1 | Not-determined | Genome-fragment | NA | NA | 0 |
| S1-final-viral-combined_k141_337276\|\|full | 2524 | No | NA | 5 | 2 | 0 | Low-quality | Genome-fragment | 4.4 | HMM-based (lower-bound) | 0 |
| S17-k141_164541 | 1536 | No | NA | 1 | 0 | 1 | Not-determined | Genome-fragment | NA | NA | 0 |
| S17-k141_242964 | 1028 | No | NA | 1 | 0 | 1 | Not-determined | Genome-fragment | NA | NA | 0 |
| S17-k141_177064 | 1089 | No | NA | 1 | 0 | 1 | Not-determined | Genome-fragment | NA | NA | 0 |
| S17-k141_65742 | 1040 | No | NA | 2 | 0 | 2 | Not-determined | Genome-fragment | NA | NA | 0 |
| S17-k141_96902 | 1242 | No | NA | 1 | 0 | 1 | Not-determined | Genome-fragment | NA | NA | 0 |
| S5_k141_181776 | 2524 | No | NA | 2 | 0 | 0 | Low-quality | Genome-fragment | 1.6 | AAI-based (high-confidence) | 0 |
| S12-final-viral-combined_k141_122195\|\|full | 2524 | No | NA | 4 | 0 | 0 | Low-quality | Genome-fragment | 4.99 | AAI-based (medium-confidence) | 0 |
| S17-k141_154748 | 1377 | No | NA | 2 | 0 | 1 | Not-determined | Genome-fragment | NA | NA | 0 |
| S17-k141_42570 | 3956 | No | NA | 4 | 0 | 4 | Not-determined | Genome-fragment | NA | NA | 0 |
| S12-final-viral-combined_k141_351061\|\|lt2gene | 2524 | No | NA | 1 | 1 | 0 | Low-quality | Genome-fragment | 4.26 | AAI-based (high-confidence) | 0 |
| S17-k141_243025 | 1051 | No | NA | 3 | 0 | 0 | Not-determined | Genome-fragment | NA | NA | 0 |
| S17-k141_77536 | 1680 | No | NA | 1 | 0 | 1 | Not-determined | Genome-fragment | NA | NA | 0 |
| S17-k141_164642 | 1225 | No | NA | 1 | 0 | 1 | Not-determined | Genome-fragment | NA | NA | 0 |
| S17-k141_264335 | 3737 | No | NA | 4 | 0 | 1 | Not-determined | Genome-fragment | NA | NA | 0 |
| S17-k141_19620 | 1000 | No | NA | 2 | 0 | 2 | Not-determined | Genome-fragment | NA | NA | 0 |
| S17-k141_177148 | 1090 | No | NA | 2 | 0 | 1 | Not-determined | Genome-fragment | NA | NA | 0 |
| S17-k141_31650 | 1184 | No | NA | 1 | 0 | 1 | Not-determined | Genome-fragment | NA | NA | 0 |
| S17-k141_154818 | 1017 | No | NA | 2 | 0 | 1 | Not-determined | Genome-fragment | NA | NA | 0 |
| S17-k141_188677 | 4933 | No | NA | 6 | 0 | 6 | Not-determined | Genome-fragment | NA | NA | 0 |
| S17-k141_164758 | 1135 | No | NA | 3 | 0 | 3 | Not-determined | Genome-fragment | NA | NA | 0 |
| S17-k141_252988 | 1163 | No | NA | 1 | 0 | 0 | Not-determined | Genome-fragment | NA | NA | 0 |
| S17-k141_97051 | 1042 | No | NA | 2 | 0 | 0 | Not-determined | Genome-fragment | NA | NA | 0 |
| S17-k141_19697 | 5360 | No | NA | 4 | 0 | 2 | Not-determined | Genome-fragment | NA | NA | 0 |
| S17-k141_42719 | 4603 | No | NA | 5 | 0 | 5 | Not-determined | Genome-fragment | NA | NA | 0 |
| S17-k141_65958 | 3466 | No | NA | 1 | 0 | 1 | Not-determined | Genome-fragment | NA | NA | 0 |
| S17-k141_19728 | 1026 | No | NA | 2 | 0 | 1 | Not-determined | Genome-fragment | NA | NA | 0 |
| S4-final-viral-combined_k141_29328\|\|full | 2524 | No | NA | 4 | 3 | 0 | Low-quality | Genome-fragment | 5.91 | AAI-based (high-confidence) | 0 |
| S17-k141_253061 | 1033 | No | NA | 1 | 0 | 0 | Not-determined | Genome-fragment | NA | NA | 0 |
| S17-k141_243134 | 1158 | No | NA | 2 | 0 | 1 | Not-determined | Genome-fragment | NA | NA | 0 |
| S4-final-viral-combined_k141_82218\|\|full | 2524 | No | NA | 3 | 0 | 1 | Low-quality | Genome-fragment | 1.83 | AAI-based (high-confidence) | 0 |
| S4-final-viral-combined_k141_124614\|\|full | 2524 | No | NA | 3 | 0 | 0 | Low-quality | Genome-fragment | 2.48 | AAI-based (high-confidence) | 0 |
| S9-final-viral-combined_k141_5294\|\|full | 2525 | No | NA | 7 | 0 | 0 | Low-quality | Genome-fragment | 4.44 | AAI-based (medium-confidence) | 0 |
| S11_k141_125472 | 2525 | No | NA | 2 | 0 | 1 | Low-quality | Genome-fragment | 0.67 | AAI-based (medium-confidence) | 0 |
| S17-k141_253084 | 1077 | No | NA | 2 | 0 | 2 | Not-determined | Genome-fragment | NA | NA | 0 |
| S17-k141_77714 | 4544 | No | NA | 4 | 0 | 2 | Not-determined | Genome-fragment | NA | NA | 0 |
| S17-k141_164892 | 3266 | No | NA | 5 | 0 | 3 | Not-determined | Genome-fragment | NA | NA | 0 |
| S17-k141_109927 | 1209 | No | NA | 1 | 0 | 0 | Not-determined | Genome-fragment | NA | NA | 0 |
| S17-k141_66085 | 1257 | No | NA | 1 | 0 | 0 | Not-determined | Genome-fragment | NA | NA | 0 |
| S17-k141_253099 | 3936 | No | NA | 5 | 0 | 5 | Not-determined | Genome-fragment | NA | NA | 0 |
| S17-k141_9691 | 1502 | No | NA | 2 | 0 | 2 | Not-determined | Genome-fragment | NA | NA | 0 |
| S17-k141_253114 | 1013 | No | NA | 1 | 0 | 1 | Not-determined | Genome-fragment | NA | NA | 0 |
| S17-k141_199985 | 8078 | No | NA | 5 | 0 | 0 | Not-determined | Genome-fragment | NA | NA | 0 |
| S17-k141_42824 | 1632 | No | NA | 2 | 0 | 2 | Not-determined | Genome-fragment | NA | NA | 0 |
| S17-k141_66111 | 1172 | No | NA | 1 | 0 | 1 | Not-determined | Genome-fragment | NA | NA | 0 |
| S17-k141_97188 | 2134 | No | NA | 3 | 0 | 2 | Not-determined | Genome-fragment | NA | NA | 0 |
| S17-k141_177376 | 3496 | No | NA | 6 | 0 | 0 | Not-determined | Genome-fragment | NA | NA | 0 |
| S17-k141_243171 | 2552 | No | NA | 5 | 0 | 5 | Not-determined | Genome-fragment | NA | NA | 0 |
| S17-k141_264518 | 1045 | No | NA | 1 | 0 | 0 | Not-determined | Genome-fragment | NA | NA | 0 |
| S17-k141_264528 | 1127 | No | NA | 1 | 0 | 1 | Not-determined | Genome-fragment | NA | NA | 0 |
| S17-k141_42875 | 5463 | No | NA | 4 | 0 | 0 | Not-determined | Genome-fragment | NA | NA | 0 |
| S17-k141_165027 | 1031 | No | NA | 3 | 0 | 0 | Not-determined | Genome-fragment | NA | NA | 0 |
| S17-k141_243263 | 3241 | No | NA | 2 | 0 | 2 | Not-determined | Genome-fragment | NA | NA | 0 |
| S17-k141_264571 | 4439 | No | NA | 7 | 0 | 0 | Not-determined | Genome-fragment | NA | NA | 0 |
| S7-final-viral-combined_k141_182407\|\|full | 2525 | No | NA | 8 | 1 | 0 | Low-quality | Genome-fragment | 6.6 | AAI-based (high-confidence) | 0 |
| S17-k141_52362 | 1281 | No | NA | 3 | 0 | 1 | Not-determined | Genome-fragment | NA | NA | 0 |
| S12-final-viral-combined_k141_319270\|\|full | 2525 | No | NA | 5 | 1 | 0 | Low-quality | Genome-fragment | 3.01 | HMM-based (lower-bound) | 0 |
| S12-final-viral-combined_k141_137111\|\|full | 2525 | No | NA | 4 | 0 | 2 | Low-quality | Genome-fragment | 1.63 | AAI-based (medium-confidence) | 0 |
| S17-k141_66210 | 6183 | No | NA | 4 | 0 | 3 | Not-determined | Genome-fragment | NA | NA | 0 |
| S17-k141_177504 | 3562 | No | NA | 3 | 0 | 1 | Not-determined | Genome-fragment | NA | NA | 0 |
| S17-k141_132421 | 1009 | No | NA | 2 | 0 | 0 | Not-determined | Genome-fragment | NA | NA | 0 |
| S17-k141_177505 | 1942 | No | NA | 2 | 0 | 2 | Not-determined | Genome-fragment | NA | NA | 0 |
| S17-final-viral-combined_k141_193287\|\|full | 2526 | No | NA | 4 | 0 | 2 | Low-quality | Genome-fragment | 1.41 | AAI-based (medium-confidence) | 0 |
| S17-final-viral-combined_k141_148821\|\|full | 2526 | No | NA | 3 | 0 | 2 | Low-quality | Genome-fragment | 1.35 | AAI-based (medium-confidence) | 0 |
| S17-k141_264634 | 3182 | No | NA | 5 | 0 | 2 | Not-determined | Genome-fragment | NA | NA | 0 |
| S17-k141_19935 | 1042 | No | NA | 1 | 0 | 0 | Not-determined | Genome-fragment | NA | NA | 0 |
| S17-k141_264674 | 1418 | No | NA | 2 | 0 | 0 | Not-determined | Genome-fragment | NA | NA | 0 |
| S17-k141_253279 | 1149 | No | NA | 2 | 0 | 1 | Not-determined | Genome-fragment | NA | NA | 0 |
| S17-k141_43015 | 1124 | No | NA | 2 | 0 | 2 | Not-determined | Genome-fragment | NA | NA | 0 |
| S17-final-viral-combined_k141_15132\|\|full | 2526 | No | NA | 3 | 0 | 1 | Low-quality | Genome-fragment | 1.41 | AAI-based (medium-confidence) | 0 |
| S17-k141_97400 | 1454 | No | NA | 1 | 0 | 1 | Not-determined | Genome-fragment | NA | NA | 0 |
| S17-k141_132511 | 5044 | No | NA | 2 | 0 | 1 | Not-determined | Genome-fragment | NA | NA | 0 |
| S17-k141_43062 | 1116 | No | NA | 2 | 0 | 2 | Not-determined | Genome-fragment | NA | NA | 0 |
| S17-k141_177648 | 1111 | No | NA | 1 | 0 | 0 | Not-determined | Genome-fragment | NA | NA | 0 |
| S17-k141_243339 | 4751 | No | NA | 7 | 0 | 1 | Not-determined | Genome-fragment | NA | NA | 0 |
| S2-final-viral-combined_k141_40678\|\|lt2gene | 2526 | No | NA | 2 | 1 | 0 | Low-quality | Genome-fragment | 6.98 | AAI-based (medium-confidence) | 0 |
| S17-k141_52557 | 1080 | No | NA | 1 | 0 | 1 | Not-determined | Genome-fragment | NA | NA | 0 |
| S17-k141_43119 | 1445 | No | NA | 2 | 0 | 1 | Not-determined | Genome-fragment | NA | NA | 0 |
| S17-k141_132570 | 1000 | No | NA | 1 | 0 | 1 | Not-determined | Genome-fragment | NA | NA | 0 |
| S17-k141_165296 | 1018 | No | NA | 1 | 0 | 0 | Not-determined | Genome-fragment | NA | NA | 0 |
| S9-final-viral-combined_k141_122213\|\|full | 2527 | No | NA | 3 | 0 | 1 | Low-quality | Genome-fragment | 2.36 | AAI-based (medium-confidence) | 0 |
| S17-k141_144251 | 1084 | No | NA | 1 | 0 | 1 | Not-determined | Genome-fragment | NA | NA | 0 |
| S5-final-viral-combined_k141_15165\|\|full | 2527 | No | NA | 3 | 0 | 3 | Low-quality | Genome-fragment | 1.24 | AAI-based (medium-confidence) | 0 |
| S7-final-viral-combined_k141_147385\|\|full | 2527 | No | NA | 4 | 0 | 0 | Low-quality | Genome-fragment | 4.99 | AAI-based (medium-confidence) | 0 |
| S17-k141_52625 | 1347 | No | NA | 1 | 0 | 0 | Not-determined | Genome-fragment | NA | NA | 0 |
| S17-k141_43207 | 3450 | No | NA | 5 | 0 | 1 | Not-determined | Genome-fragment | NA | NA | 0 |
| S17-k141_43211 | 1653 | No | NA | 1 | 0 | 0 | Not-determined | Genome-fragment | NA | NA | 0 |
| S17-k141_144274 | 3703 | No | NA | 3 | 0 | 3 | Not-determined | Genome-fragment | NA | NA | 0 |
| S17-k141_97554 | 2809 | No | NA | 4 | 0 | 1 | Not-determined | Genome-fragment | NA | NA | 0 |
| S17-k141_52657 | 2139 | No | NA | 2 | 0 | 0 | Not-determined | Genome-fragment | NA | NA | 0 |
| S17-final-viral-combined_k141_215550\|\|full | 2527 | No | NA | 3 | 2 | 0 | Low-quality | Genome-fragment | 4.35 | HMM-based (lower-bound) | 0 |
| S17-k141_253438 | 4246 | No | NA | 8 | 0 | 6 | Not-determined | Genome-fragment | NA | NA | 0 |
| S17-final-viral-combined_k141_104243\|\|full | 2527 | Yes | 1740 | 4 | 1 | 1 | Low-quality | Genome-fragment | 1.06 | HMM-based (lower-bound) | 31.14 |
| S17-k141_253476 | 4337 | No | NA | 6 | 0 | 0 | Not-determined | Genome-fragment | NA | NA | 0 |
| S17-final-viral-combined_k141_218132\|\|full | 2527 | Yes | 1843 | 3 | 1 | 2 | Low-quality | Genome-fragment | 1.61 | AAI-based (medium-confidence) | 27.07 |
| S17-k141_243483 | 1286 | No | NA | 2 | 0 | 2 | Not-determined | Genome-fragment | NA | NA | 0 |
| S17-k141_264900 | 3702 | No | NA | 2 | 0 | 2 | Not-determined | Genome-fragment | NA | NA | 0 |
| S17-k141_165510 | 2398 | No | NA | 3 | 0 | 2 | Not-determined | Genome-fragment | NA | NA | 0 |
| S17-k141_110399 | 4117 | No | NA | 3 | 0 | 2 | Not-determined | Genome-fragment | NA | NA | 0 |
| S17-k141_264950 | 1131 | No | NA | 1 | 0 | 1 | Not-determined | Genome-fragment | NA | NA | 0 |
| S17-k141_132796 | 1016 | No | NA | 3 | 0 | 0 | Not-determined | Genome-fragment | NA | NA | 0 |
| S17-k141_97649 | 5442 | No | NA | 8 | 0 | 7 | Not-determined | Genome-fragment | NA | NA | 0 |
| S2-final-viral-combined_k141_202871\|\|full | 2527 | No | NA | 2 | 0 | 0 | Low-quality | Genome-fragment | 5.87 | AAI-based (medium-confidence) | 0 |
| S4_k141_3551 | 2527 | No | NA | 8 | 0 | 0 | Low-quality | Genome-fragment | 6.06 | AAI-based (high-confidence) | 0 |
| S17-k141_20125 | 1248 | No | NA | 2 | 0 | 1 | Not-determined | Genome-fragment | NA | NA | 0 |
| S17-k141_165563 | 1154 | No | NA | 1 | 0 | 1 | Not-determined | Genome-fragment | NA | NA | 0 |
| S17-k141_97673 | 1214 | No | NA | 2 | 0 | 1 | Not-determined | Genome-fragment | NA | NA | 0 |
| S17-k141_52827 | 1239 | No | NA | 3 | 0 | 0 | Not-determined | Genome-fragment | NA | NA | 0 |
| S17-k141_10167 | 4681 | No | NA | 4 | 0 | 2 | Not-determined | Genome-fragment | NA | NA | 0 |
| S17-k141_43450 | 1668 | No | NA | 1 | 0 | 0 | Not-determined | Genome-fragment | NA | NA | 0 |
| S17-k141_97756 | 1508 | No | NA | 2 | 0 | 1 | Not-determined | Genome-fragment | NA | NA | 0 |
| S17-k141_52892 | 1037 | No | NA | 3 | 0 | 0 | Not-determined | Genome-fragment | NA | NA | 0 |
| S8_k141_252674 | 2527 | No | NA | 3 | 3 | 0 | Low-quality | Genome-fragment | 3.26 | AAI-based (high-confidence) | 0 |
| S17-k141_20345 | 1047 | No | NA | 1 | 0 | 1 | Not-determined | Genome-fragment | NA | NA | 0 |
| S17-k141_155422 | 2529 | No | NA | 2 | 0 | 0 | Not-determined | Genome-fragment | NA | NA | 0 |
| S17-k141_165770 | 1469 | No | NA | 1 | 0 | 0 | Not-determined | Genome-fragment | NA | NA | 0 |
| S17-k141_253734 | 1076 | No | NA | 1 | 0 | 1 | Not-determined | Genome-fragment | NA | NA | 0 |
| S17-k141_20404 | 1014 | No | NA | 1 | 0 | 1 | Not-determined | Genome-fragment | NA | NA | 0 |
| S17-k141_243865 | 1131 | No | NA | 2 | 0 | 0 | Not-determined | Genome-fragment | NA | NA | 0 |
| S17-k141_243869 | 2274 | No | NA | 2 | 0 | 1 | Not-determined | Genome-fragment | NA | NA | 0 |
| S9-final-viral-combined_k141_53780\|\|full | 2528 | No | NA | 5 | 0 | 1 | Low-quality | Genome-fragment | 6.63 | AAI-based (medium-confidence) | 0 |
| S17-k141_97896 | 2878 | No | NA | 4 | 0 | 1 | Not-determined | Genome-fragment | NA | NA | 0 |
| S17-k141_20462 | 5304 | No | NA | 6 | 0 | 6 | Not-determined | Genome-fragment | NA | NA | 0 |
| S17-k141_265121 | 2920 | No | NA | 3 | 0 | 1 | Not-determined | Genome-fragment | NA | NA | 0 |
| S17-k141_253829 | 6428 | No | NA | 7 | 0 | 2 | Not-determined | Genome-fragment | NA | NA | 0 |
| S17-k141_20503 | 3032 | No | NA | 2 | 0 | 1 | Not-determined | Genome-fragment | NA | NA | 0 |
| S17-k141_97934 | 3370 | No | NA | 2 | 0 | 1 | Not-determined | Genome-fragment | NA | NA | 0 |
| S17-k141_243910 | 1037 | No | NA | 1 | 0 | 1 | Not-determined | Genome-fragment | NA | NA | 0 |
| S17-k141_155547 | 1149 | No | NA | 1 | 0 | 0 | Not-determined | Genome-fragment | NA | NA | 0 |
| S17-k141_32442 | 1513 | No | NA | 3 | 0 | 1 | Not-determined | Genome-fragment | NA | NA | 0 |
| S3_k141_94704 | 2528 | No | NA | 2 | 1 | 0 | Low-quality | Genome-fragment | 1.16 | AAI-based (high-confidence) | 0 |
| S17-k141_32477 | 1154 | No | NA | 2 | 0 | 0 | Not-determined | Genome-fragment | NA | NA | 0 |
| S5-final-viral-combined_k141_26717\|\|full | 2528 | No | NA | 6 | 0 | 0 | Low-quality | Genome-fragment | 1.45 | AAI-based (high-confidence) | 0 |
| S17-final-viral-combined_k141_204594\|\|full | 2528 | No | NA | 3 | 0 | 0 | Low-quality | Genome-fragment | 6.45 | AAI-based (high-confidence) | 0 |
| S17-k141_133091 | 1224 | No | NA | 1 | 0 | 1 | Not-determined | Genome-fragment | NA | NA | 0 |
| S17-k141_20601 | 1018 | No | NA | 1 | 0 | 0 | Not-determined | Genome-fragment | NA | NA | 0 |
| S17-k141_53032 | 2131 | No | NA | 2 | 0 | 1 | Not-determined | Genome-fragment | NA | NA | 0 |
| S17-k141_265214 | 1016 | No | NA | 1 | 0 | 0 | Not-determined | Genome-fragment | NA | NA | 0 |
| S2-final-viral-combined_k141_228839\|\|full | 2528 | No | NA | 5 | 2 | 0 | Low-quality | Genome-fragment | 4.39 | AAI-based (high-confidence) | 0 |
| S17-k141_265256 | 1343 | No | NA | 1 | 0 | 1 | Not-determined | Genome-fragment | NA | NA | 0 |
| S17-k141_10656 | 3107 | No | NA | 3 | 0 | 3 | Not-determined | Genome-fragment | NA | NA | 0 |
| S17-k141_265275 | 3053 | No | NA | 2 | 0 | 2 | Not-determined | Genome-fragment | NA | NA | 0 |
| S17-k141_166110 | 1272 | No | NA | 1 | 0 | 1 | Not-determined | Genome-fragment | NA | NA | 0 |
| S11-final-viral-combined_k141_52564\|\|full | 2529 | No | NA | 3 | 1 | 0 | Low-quality | Genome-fragment | 6.86 | AAI-based (medium-confidence) | 0 |
| S17-k141_32642 | 4306 | No | NA | 2 | 0 | 2 | Not-determined | Genome-fragment | NA | NA | 0 |
| S17-k141_53161 | 1297 | No | NA | 2 | 0 | 1 | Not-determined | Genome-fragment | NA | NA | 0 |
| S17-k141_32677 | 1551 | No | NA | 2 | 0 | 1 | Not-determined | Genome-fragment | NA | NA | 0 |
| S17-k141_20725 | 10585 | No | NA | 5 | 0 | 4 | Not-determined | Genome-fragment | NA | NA | 0 |
| S17-k141_133230 | 2492 | No | NA | 1 | 0 | 1 | Not-determined | Genome-fragment | NA | NA | 0 |
| S17-k141_20767 | 2336 | No | NA | 2 | 0 | 1 | Not-determined | Genome-fragment | NA | NA | 0 |
| S17-k141_265403 | 1967 | No | NA | 3 | 0 | 2 | Not-determined | Genome-fragment | NA | NA | 0 |
| S17-k141_166162 | 1291 | No | NA | 1 | 0 | 0 | Not-determined | Genome-fragment | NA | NA | 0 |
| S17-k141_265423 | 1172 | No | NA | 2 | 0 | 2 | Not-determined | Genome-fragment | NA | NA | 0 |
| S17-k141_244241 | 2390 | No | NA | 2 | 0 | 1 | Not-determined | Genome-fragment | NA | NA | 0 |
| S17-k141_79713 | 2529 | No | NA | 3 | 0 | 1 | Low-quality | Genome-fragment | 0.29 | AAI-based (high-confidence) | 0 |
| S17-k141_44048 | 1153 | No | NA | 1 | 0 | 1 | Not-determined | Genome-fragment | NA | NA | 0 |
| S17-k141_53240 | 2165 | No | NA | 1 | 0 | 1 | Not-determined | Genome-fragment | NA | NA | 0 |
| S17-k141_32754 | 1022 | No | NA | 1 | 0 | 0 | Not-determined | Genome-fragment | NA | NA | 0 |
| S17-k141_32756 | 1060 | No | NA | 2 | 0 | 0 | Not-determined | Genome-fragment | NA | NA | 0 |
| S2-final-viral-combined_k141_87363\|\|full | 2529 | No | NA | 5 | 1 | 0 | Low-quality | Genome-fragment | 5.89 | AAI-based (high-confidence) | 0 |
| S17-k141_133286 | 3784 | No | NA | 6 | 0 | 6 | Not-determined | Genome-fragment | NA | NA | 0 |
| S17-k141_20823 | 1074 | No | NA | 1 | 0 | 0 | Not-determined | Genome-fragment | NA | NA | 0 |
| S17-k141_53281 | 1795 | No | NA | 1 | 0 | 1 | Not-determined | Genome-fragment | NA | NA | 0 |
| S17-k141_53287 | 1925 | No | NA | 2 | 0 | 1 | Not-determined | Genome-fragment | NA | NA | 0 |
| S17-final-viral-combined_k141_179456\|\|full | 2530 | No | NA | 3 | 0 | 2 | Low-quality | Genome-fragment | 1.57 | AAI-based (medium-confidence) | 0 |
| S17-k141_98338 | 2726 | No | NA | 3 | 0 | 2 | Not-determined | Genome-fragment | NA | NA | 0 |
| S17-k141_265459 | 30938 | No | NA | 31 | 0 | 23 | Not-determined | Genome-fragment | NA | NA | 0 |
| S17-k141_98349 | 1040 | No | NA | 1 | 0 | 0 | Not-determined | Genome-fragment | NA | NA | 0 |
| S17-k141_10726 | 3675 | No | NA | 3 | 0 | 3 | Not-determined | Genome-fragment | NA | NA | 0 |
| S4-final-viral-combined_k141_63059\|\|full | 2530 | No | NA | 3 | 1 | 0 | Low-quality | Genome-fragment | 1.17 | HMM-based (lower-bound) | 0 |
| S17-k141_32864 | 1910 | No | NA | 3 | 0 | 1 | Not-determined | Genome-fragment | NA | NA | 0 |
| S17-k141_133345 | 2650 | No | NA | 2 | 0 | 1 | Not-determined | Genome-fragment | NA | NA | 0 |
| S8-final-viral-combined_k141_33984\|\|full | 2530 | No | NA | 2 | 0 | 1 | Low-quality | Genome-fragment | 1.45 | AAI-based (medium-confidence) | 0 |
| S17-k141_265593 | 1560 | No | NA | 1 | 0 | 0 | Not-determined | Genome-fragment | NA | NA | 0 |
| S17-k141_44267 | 1113 | No | NA | 1 | 0 | 0 | Not-determined | Genome-fragment | NA | NA | 0 |
| S17-k141_44287 | 4763 | No | NA | 10 | 0 | 0 | Not-determined | Genome-fragment | NA | NA | 0 |
| S17-k141_244419 | 1014 | No | NA | 1 | 0 | 1 | Not-determined | Genome-fragment | NA | NA | 0 |
| S17-k141_110843 | 1269 | No | NA | 2 | 0 | 1 | Not-determined | Genome-fragment | NA | NA | 0 |
| S17-k141_32973 | 1369 | No | NA | 1 | 0 | 1 | Not-determined | Genome-fragment | NA | NA | 0 |
| S17-k141_33007 | 2593 | No | NA | 2 | 0 | 2 | Not-determined | Genome-fragment | NA | NA | 0 |
| S17-k141_21109 | 1446 | No | NA | 1 | 0 | 1 | Not-determined | Genome-fragment | NA | NA | 0 |
| S5-final-viral-combined_k141_26926\|\|full | 2531 | No | NA | 4 | 1 | 0 | Low-quality | Genome-fragment | 3.53 | AAI-based (medium-confidence) | 0 |
| S17-k141_53421 | 6712 | No | NA | 4 | 0 | 2 | Not-determined | Genome-fragment | NA | NA | 0 |
| S17-k141_11029 | 1173 | No | NA | 1 | 0 | 0 | Not-determined | Genome-fragment | NA | NA | 0 |
| S17-k141_44436 | 1224 | No | NA | 2 | 0 | 1 | Not-determined | Genome-fragment | NA | NA | 0 |
| S17-k141_265699 | 2177 | No | NA | 2 | 0 | 1 | Not-determined | Genome-fragment | NA | NA | 0 |
| S17-k141_98779 | 3242 | No | NA | 4 | 0 | 1 | Not-determined | Genome-fragment | NA | NA | 0 |
| S17-k141_33140 | 22172 | No | NA | 27 | 0 | 2 | Not-determined | Genome-fragment | NA | NA | 0 |
| S17-k141_265752 | 1079 | No | NA | 2 | 0 | 1 | Not-determined | Genome-fragment | NA | NA | 0 |
| S17-k141_111016 | 2165 | No | NA | 3 | 0 | 0 | Not-determined | Genome-fragment | NA | NA | 0 |
| S17-k141_21300 | 1033 | No | NA | 2 | 0 | 2 | Not-determined | Genome-fragment | NA | NA | 0 |
| S17-k141_11084 | 2675 | No | NA | 1 | 0 | 1 | Not-determined | Genome-fragment | NA | NA | 0 |
| S7-final-viral-combined_k141_124704\|\|full | 2531 | No | NA | 3 | 1 | 0 | Low-quality | Genome-fragment | 3.83 | HMM-based (lower-bound) | 0 |
| S2-final-viral-combined_k141_181995\|\|full | 2531 | No | NA | 3 | 0 | 0 | Low-quality | Genome-fragment | 7.53 | AAI-based (high-confidence) | 0 |
| S17-k141_98828 | 1170 | No | NA | 1 | 0 | 0 | Not-determined | Genome-fragment | NA | NA | 0 |
| S17-k141_33173 | 2029 | No | NA | 4 | 0 | 0 | Not-determined | Genome-fragment | NA | NA | 0 |
| S4-final-viral-combined_k141_58237\|\|full | 2531 | No | NA | 3 | 0 | 0 | Low-quality | Genome-fragment | 3.45 | AAI-based (high-confidence) | 0 |
| S9-final-viral-combined_k141_43866\|\|full | 2532 | No | NA | 6 | 2 | 0 | Low-quality | Genome-fragment | 5.26 | AAI-based (medium-confidence) | 0 |
| S11_k141_179642 | 2532 | No | NA | 5 | 1 | 0 | Low-quality | Genome-fragment | 4.37 | AAI-based (high-confidence) | 0 |
| S17-k141_254587 | 11099 | No | NA | 10 | 0 | 8 | Not-determined | Genome-fragment | NA | NA | 0 |
| S17-k141_265860 | 1024 | No | NA | 1 | 0 | 1 | Not-determined | Genome-fragment | NA | NA | 0 |
| S4-final-viral-combined_k141_119224\|\|full | 2532 | No | NA | 3 | 1 | 0 | Low-quality | Genome-fragment | 5.74 | AAI-based (high-confidence) | 0 |
| S17-k141_53649 | 1613 | No | NA | 1 | 0 | 1 | Not-determined | Genome-fragment | NA | NA | 0 |
| S11-final-viral-combined_k141_55096\|\|full | 2533 | No | NA | 3 | 2 | 0 | Low-quality | Genome-fragment | 6.51 | AAI-based (medium-confidence) | 0 |
| S17-k141_33254 | 1552 | No | NA | 2 | 0 | 2 | Not-determined | Genome-fragment | NA | NA | 0 |
| S3_k141_68863 | 2533 | No | NA | 3 | 0 | 2 | Low-quality | Genome-fragment | 3.05 | AAI-based (medium-confidence) | 0 |
| S17-k141_33264 | 1906 | No | NA | 2 | 0 | 0 | Not-determined | Genome-fragment | NA | NA | 0 |
| S17-k141_125265 | 2533 | No | NA | 3 | 0 | 2 | Low-quality | Genome-fragment | 0.77 | AAI-based (medium-confidence) | 0 |
| S17-k141_21416 | 2289 | No | NA | 3 | 0 | 1 | Not-determined | Genome-fragment | NA | NA | 0 |
| S17-k141_53707 | 1063 | No | NA | 2 | 0 | 1 | Not-determined | Genome-fragment | NA | NA | 0 |
| S17-k141_265966 | 1157 | No | NA | 1 | 0 | 1 | Not-determined | Genome-fragment | NA | NA | 0 |
| S17-k141_21470 | 1013 | No | NA | 1 | 0 | 0 | Not-determined | Genome-fragment | NA | NA | 0 |
| S17-k141_21498 | 1176 | No | NA | 1 | 0 | 1 | Not-determined | Genome-fragment | NA | NA | 0 |
| S17-k141_53802 | 1412 | No | NA | 1 | 0 | 1 | Not-determined | Genome-fragment | NA | NA | 0 |
| S17-k141_266120 | 2195 | No | NA | 2 | 0 | 0 | Not-determined | Genome-fragment | NA | NA | 0 |
| S2-final-viral-combined_k141_52777\|\|full | 2533 | No | NA | 5 | 2 | 0 | Low-quality | Genome-fragment | 4.25 | AAI-based (high-confidence) | 0 |
| S17-k141_21641 | 5689 | No | NA | 6 | 0 | 4 | Not-determined | Genome-fragment | NA | NA | 0 |
| S17-k141_166588 | 1468 | No | NA | 2 | 0 | 0 | Not-determined | Genome-fragment | NA | NA | 0 |
| S17-k141_166601 | 1223 | No | NA | 1 | 0 | 0 | Not-determined | Genome-fragment | NA | NA | 0 |
| S17-k141_266287 | 6161 | No | NA | 7 | 0 | 7 | Not-determined | Genome-fragment | NA | NA | 0 |
| S12-final-viral-combined_k141_289803\|\|full | 2534 | No | NA | 3 | 0 | 3 | Low-quality | Genome-fragment | 1.5 | AAI-based (medium-confidence) | 0 |
| S12-final-viral-combined_k141_255752\|\|full | 2534 | No | NA | 5 | 1 | 0 | Low-quality | Genome-fragment | 7.3 | AAI-based (high-confidence) | 0 |
| S17-k141_53968 | 1253 | No | NA | 3 | 0 | 0 | Not-determined | Genome-fragment | NA | NA | 0 |
| S17-k141_254872 | 6309 | No | NA | 6 | 3 | 0 | High-quality | High-quality | 100 | AAI-based (high-confidence) | 0 |
| S17-k141_21813 | 1002 | No | NA | 1 | 0 | 1 | Not-determined | Genome-fragment | NA | NA | 0 |
| S17-k141_99300 | 1572 | No | NA | 1 | 0 | 1 | Not-determined | Genome-fragment | NA | NA | 0 |
| S17-k141_53992 | 1671 | No | NA | 3 | 0 | 0 | Not-determined | Genome-fragment | NA | NA | 0 |
| S17-k141_21837 | 1121 | No | NA | 3 | 0 | 1 | Not-determined | Genome-fragment | NA | NA | 0 |
| S17-k141_54015 | 1404 | No | NA | 2 | 0 | 1 | Not-determined | Genome-fragment | NA | NA | 0 |
| S17-k141_266448 | 1305 | No | NA | 1 | 0 | 1 | Not-determined | Genome-fragment | NA | NA | 0 |
| S17-k141_99377 | 1485 | No | NA | 1 | 0 | 1 | Not-determined | Genome-fragment | NA | NA | 0 |
| S17-k141_266482 | 3121 | No | NA | 2 | 0 | 1 | Not-determined | Genome-fragment | NA | NA | 0 |
| S17-k141_266507 | 5528 | No | NA | 10 | 0 | 1 | Not-determined | Genome-fragment | NA | NA | 0 |
| S12-final-viral-combined_k141_247553\|\|full | 2534 | No | NA | 4 | 1 | 0 | Low-quality | Genome-fragment | 4.08 | AAI-based (high-confidence) | 0 |
| S17-k141_266511 | 40034 | No | NA | 59 | 44 | 0 | Complete | High-quality | 100 | DTR (high-confidence) | 0 |
| S17-k141_255017 | 1054 | No | NA | 1 | 0 | 1 | Not-determined | Genome-fragment | NA | NA | 0 |
| S17-k141_266523 | 1204 | No | NA | 1 | 0 | 0 | Not-determined | Genome-fragment | NA | NA | 0 |
| S17-k141_266544 | 4526 | No | NA | 8 | 0 | 0 | Not-determined | Genome-fragment | NA | NA | 0 |
| S4_k141_146368 | 2534 | No | NA | 1 | 0 | 0 | Low-quality | Genome-fragment | 2.67 | AAI-based (high-confidence) | 0 |
| S17-k141_22115 | 4616 | No | NA | 3 | 0 | 0 | Not-determined | Genome-fragment | NA | NA | 0 |
| S17-k141_255203 | 2614 | No | NA | 1 | 0 | 1 | Not-determined | Genome-fragment | NA | NA | 0 |
| S17-k141_54414 | 3199 | No | NA | 5 | 0 | 5 | Not-determined | Genome-fragment | NA | NA | 0 |
| S17-k141_54474 | 2887 | No | NA | 1 | 0 | 1 | Not-determined | Genome-fragment | NA | NA | 0 |
| S17-k141_99746 | 1740 | No | NA | 2 | 0 | 1 | Not-determined | Genome-fragment | NA | NA | 0 |
| S1-final-viral-combined_k141_177606\|\|full | 2535 | No | NA | 5 | 1 | 0 | Low-quality | Genome-fragment | 1.7 | HMM-based (lower-bound) | 0 |
| S17-k141_99756 | 4285 | No | NA | 2 | 0 | 1 | Not-determined | Genome-fragment | NA | NA | 0 |
| S17-k141_255494 | 4712 | No | NA | 3 | 0 | 3 | Not-determined | Genome-fragment | NA | NA | 0 |
| S17-k141_99785 | 3065 | No | NA | 6 | 0 | 6 | Not-determined | Genome-fragment | NA | NA | 0 |
| S17-k141_255545 | 4203 | No | NA | 3 | 0 | 2 | Not-determined | Genome-fragment | NA | NA | 0 |
| S17-k141_54784 | 1004 | No | NA | 1 | 0 | 1 | Not-determined | Genome-fragment | NA | NA | 0 |
| S17-k141_54829 | 1199 | No | NA | 2 | 0 | 1 | Not-determined | Genome-fragment | NA | NA | 0 |
| S17-k141_54845 | 1199 | No | NA | 2 | 0 | 1 | Not-determined | Genome-fragment | NA | NA | 0 |
| S17-k141_55118 | 1699 | No | NA | 2 | 0 | 2 | Not-determined | Genome-fragment | NA | NA | 0 |
| S17-k141_55311 | 1917 | No | NA | 1 | 0 | 1 | Not-determined | Genome-fragment | NA | NA | 0 |
| S17-k141_55515 | 2254 | No | NA | 3 | 0 | 1 | Not-determined | Genome-fragment | NA | NA | 0 |
| S12-final-viral-combined_k141_321174\|\|full | 32121 | No | NA | 41 | 14 | 1 | High-quality | High-quality | 98.29 | AAI-based (high-confidence) | 0 |
| S12-final-viral-combined_k141_342900\|\|full | 2133 | No | NA | 3 | 0 | 0 | Not-determined | Genome-fragment | NA | NA | 0 |
| S12-final-viral-combined_k141_383488\|\|full | 1001 | No | NA | 2 | 0 | 0 | Not-determined | Genome-fragment | NA | NA | 0 |
| S12-final-viral-combined_k141_266071\|\|full | 3847 | No | NA | 5 | 0 | 1 | Not-determined | Genome-fragment | NA | NA | 0 |
| S12-final-viral-combined_k141_401374\|\|full | 6103 | No | NA | 5 | 0 | 0 | Not-determined | Genome-fragment | NA | NA | 0 |
| S12-final-viral-combined_k141_401375\|\|full | 5326 | No | NA | 9 | 0 | 1 | Not-determined | Genome-fragment | NA | NA | 0 |
| S12-final-viral-combined_k141_401379\|\|full | 10909 | No | NA | 14 | 0 | 1 | Medium-quality | Genome-fragment | 74.67 | AAI-based (high-confidence) | 0 |
| S12-final-viral-combined_k141_401383\|\|full | 5467 | No | NA | 5 | 2 | 0 | High-quality | High-quality | 99.51 | AAI-based (high-confidence) | 0 |
| S12-final-viral-combined_k141_200877\|\|full | 1536 | No | NA | 2 | 0 | 0 | Not-determined | Genome-fragment | NA | NA | 0 |
| S12-final-viral-combined_k141_385000\|\|full | 3281 | No | NA | 3 | 0 | 2 | Not-determined | Genome-fragment | NA | NA | 0 |
| S12-final-viral-combined_k141_50250\|\|full | 1545 | No | NA | 3 | 0 | 0 | Not-determined | Genome-fragment | NA | NA | 0 |
| S12-final-viral-combined_k141_21\|\|full | 6799 | No | NA | 10 | 0 | 1 | Not-determined | Genome-fragment | NA | NA | 0 |
| S12-final-viral-combined_k141_385009\|\|full | 2545 | No | NA | 3 | 0 | 1 | Not-determined | Genome-fragment | NA | NA | 0 |
| S12-final-viral-combined_k141_83728\|\|full | 6003 | No | NA | 7 | 0 | 3 | Not-determined | Genome-fragment | NA | NA | 0 |
| S6-final-viral-combined_k141_128463\|\|full | 2535 | No | NA | 4 | 0 | 0 | Low-quality | Genome-fragment | 5.9 | AAI-based (high-confidence) | 0 |
| S1-final-viral-combined_k141_244662\|\|full | 2536 | No | NA | 4 | 2 | 0 | Low-quality | Genome-fragment | 5.7 | AAI-based (high-confidence) | 0 |
| S1-final-viral-combined_k141_108769\|\|full | 2537 | No | NA | 4 | 0 | 2 | Low-quality | Genome-fragment | 1.11 | AAI-based (medium-confidence) | 0 |
| S17-final-viral-combined_k141_68100\|\|full | 2537 | No | NA | 4 | 0 | 0 | Low-quality | Genome-fragment | 1.75 | AAI-based (medium-confidence) | 0 |
| S4-final-viral-combined_k141_43602\|\|full | 2537 | No | NA | 4 | 0 | 2 | Low-quality | Genome-fragment | 1.89 | AAI-based (high-confidence) | 0 |
| S12-final-viral-combined_k141_200900\|\|full | 1857 | No | NA | 4 | 0 | 2 | Not-determined | Genome-fragment | NA | NA | 0 |
| S6-final-viral-combined_k141_13438\|\|full | 2537 | Yes | 1271 | 3 | 1 | 2 | Low-quality | Genome-fragment | 0.11 | HMM-based (lower-bound) | 49.9 |
| S12-final-viral-combined_k141_368301\|\|full | 1950 | No | NA | 3 | 0 | 3 | Not-determined | Genome-fragment | NA | NA | 0 |
| S8-final-viral-combined_k141_219128\|\|full | 2537 | No | NA | 6 | 1 | 0 | Low-quality | Genome-fragment | 4.43 | AAI-based (high-confidence) | 0 |
| S12-final-viral-combined_k141_16801\|\|full | 5113 | No | NA | 5 | 0 | 1 | Not-determined | Genome-fragment | NA | NA | 0 |
| S12-final-viral-combined_k141_351543\|\|full | 1023 | No | NA | 3 | 0 | 0 | Not-determined | Genome-fragment | NA | NA | 0 |
| S9-final-viral-combined_k141_9257\|\|full | 2538 | No | NA | 7 | 0 | 0 | Low-quality | Genome-fragment | 12.6 | AAI-based (medium-confidence) | 0 |
| S1-final-viral-combined_k141_334336\|\|full | 2538 | No | NA | 3 | 0 | 2 | Low-quality | Genome-fragment | 1.22 | AAI-based (medium-confidence) | 0 |
| S12-final-viral-combined_k141_368317\|\|full | 3477 | No | NA | 4 | 0 | 0 | Not-determined | Genome-fragment | NA | NA | 0 |
| S1-final-viral-combined_k141_95460\|\|full | 2538 | No | NA | 5 | 2 | 0 | Low-quality | Genome-fragment | 5.84 | AAI-based (medium-confidence) | 0 |
| S12-final-viral-combined_k141_117218\|\|full | 1842 | No | NA | 3 | 0 | 0 | Not-determined | Genome-fragment | NA | NA | 0 |
| S12-final-viral-combined_k141_67021\|\|full | 2574 | No | NA | 3 | 0 | 0 | Not-determined | Genome-fragment | NA | NA | 0 |
| S1-final-viral-combined_k141_48411\|\|full | 2538 | No | NA | 3 | 0 | 2 | Low-quality | Genome-fragment | 1.49 | AAI-based (medium-confidence) | 0 |
| S12-final-viral-combined_k141_251197\|\|full | 1327 | No | NA | 3 | 0 | 1 | Not-determined | Genome-fragment | NA | NA | 0 |
| S12-final-viral-combined_k141_251204\|\|full | 1493 | No | NA | 3 | 0 | 0 | Not-determined | Genome-fragment | NA | NA | 0 |
| S12-final-viral-combined_k141_200985\|\|full | 1155 | No | NA | 3 | 0 | 0 | Not-determined | Genome-fragment | NA | NA | 0 |
| S5-final-viral-combined_k141_180533\|\|full | 2538 | No | NA | 3 | 0 | 1 | Low-quality | Genome-fragment | 6.63 | AAI-based (medium-confidence) | 0 |
| S2_k141_32457 | 2538 | No | NA | 2 | 0 | 1 | Low-quality | Genome-fragment | 0.43 | AAI-based (high-confidence) | 0 |
| S12-final-viral-combined_k141_334806\|\|full | 82744 | Yes | 39697 | 89 | 9 | 37 | High-quality | High-quality | 91.82 | AAI-based (medium-confidence) | 52.02 |
| S4-final-viral-combined_k141_110136\|\|full | 2538 | No | NA | 5 | 3 | 0 | Low-quality | Genome-fragment | 3.91 | AAI-based (high-confidence) | 0 |
| S1-final-viral-combined_k141_30353\|\|full | 2539 | No | NA | 3 | 1 | 0 | Low-quality | Genome-fragment | 6.28 | AAI-based (high-confidence) | 0 |
| S17-final-viral-combined_k141_53072\|\|full | 2539 | No | NA | 2 | 0 | 2 | Low-quality | Genome-fragment | 0.56 | AAI-based (high-confidence) | 0 |
| S12-final-viral-combined_k141_117276\|\|full | 4816 | No | NA | 5 | 0 | 1 | Not-determined | Genome-fragment | NA | NA | 0 |
| S12-final-viral-combined_k141_284745\|\|full | 3231 | No | NA | 3 | 0 | 1 | Not-determined | Genome-fragment | NA | NA | 0 |
| S12-final-viral-combined_k141_175769\|\|full | 2539 | No | NA | 4 | 0 | 2 | Low-quality | Genome-fragment | 6.59 | AAI-based (medium-confidence) | 0 |
| S12-final-viral-combined_k141_83766\|\|full | 1582 | No | NA | 2 | 0 | 2 | Not-determined | Genome-fragment | NA | NA | 0 |
| S12-final-viral-combined_k141_83769\|\|full | 1049 | No | NA | 2 | 0 | 1 | Not-determined | Genome-fragment | NA | NA | 0 |
| S12-final-viral-combined_k141_301426\|\|full | 4655 | No | NA | 3 | 0 | 3 | Not-determined | Genome-fragment | NA | NA | 0 |
| S12-final-viral-combined_k141_117290\|\|full | 4034 | No | NA | 5 | 0 | 0 | Not-determined | Genome-fragment | NA | NA | 0 |
| S12-final-viral-combined_k141_334838\|\|full | 1760 | No | NA | 3 | 0 | 0 | Not-determined | Genome-fragment | NA | NA | 0 |
| S12-final-viral-combined_k141_301435\|\|full | 6336 | No | NA | 11 | 0 | 1 | Not-determined | Genome-fragment | NA | NA | 0 |
| S4_k141_101992 | 2539 | No | NA | 1 | 0 | 1 | Low-quality | Genome-fragment | 1.48 | AAI-based (high-confidence) | 0 |
| S9-final-viral-combined_k141_27191\|\|full | 2540 | No | NA | 2 | 0 | 2 | Low-quality | Genome-fragment | 4.47 | AAI-based (medium-confidence) | 0 |
| S12-final-viral-combined_k141_217795\|\|full | 1662 | No | NA | 2 | 0 | 2 | Not-determined | Genome-fragment | NA | NA | 0 |
| S12-final-viral-combined_k141_284772\|\|full | 13631 | No | NA | 13 | 0 | 11 | Not-determined | Genome-fragment | NA | NA | 0 |
| S12-final-viral-combined_k141_83809\|\|full | 1016 | No | NA | 2 | 0 | 1 | Not-determined | Genome-fragment | NA | NA | 0 |
| S11-final-viral-combined_k141_186120\|\|full | 2540 | No | NA | 4 | 0 | 0 | Low-quality | Genome-fragment | 4.12 | AAI-based (high-confidence) | 0 |
| S12-final-viral-combined_k141_150754\|\|full | 1556 | No | NA | 3 | 0 | 0 | Not-determined | Genome-fragment | NA | NA | 0 |
| S12-final-viral-combined_k141_83825\|\|full | 3550 | No | NA | 3 | 0 | 2 | Not-determined | Genome-fragment | NA | NA | 0 |
| S12-final-viral-combined_k141_301472\|\|full | 2619 | No | NA | 2 | 0 | 1 | Not-determined | Genome-fragment | NA | NA | 0 |
| S5-final-viral-combined_k141_4799\|\|full | 2540 | No | NA | 4 | 1 | 0 | Low-quality | Genome-fragment | 5.28 | AAI-based (high-confidence) | 0 |
| S12-final-viral-combined_k141_150761\|\|full | 2589 | No | NA | 5 | 0 | 0 | Not-determined | Genome-fragment | NA | NA | 0 |
| S2-final-viral-combined_k141_70762\|\|full | 2540 | No | NA | 5 | 2 | 0 | Low-quality | Genome-fragment | 7.16 | AAI-based (high-confidence) | 0 |
| S7-final-viral-combined_k141_201111\|\|full | 2541 | No | NA | 4 | 0 | 0 | Low-quality | Genome-fragment | 5.88 | AAI-based (medium-confidence) | 0 |
| S12-final-viral-combined_k141_301501\|\|full | 4186 | No | NA | 4 | 0 | 3 | Not-determined | Genome-fragment | NA | NA | 0 |
| S12-final-viral-combined_k141_385217\|\|full | 1656 | No | NA | 2 | 0 | 1 | Not-determined | Genome-fragment | NA | NA | 0 |
| S12-final-viral-combined_k141_37768\|\|full | 2541 | No | NA | 3 | 0 | 0 | Low-quality | Genome-fragment | 5.69 | AAI-based (medium-confidence) | 0 |
| S12-final-viral-combined_k141_318216\|\|full | 1376 | No | NA | 4 | 0 | 2 | Not-determined | Genome-fragment | NA | NA | 0 |
| S12-final-viral-combined_k141_117351\|\|full | 2254 | No | NA | 3 | 0 | 0 | Not-determined | Genome-fragment | NA | NA | 0 |
| S12-final-viral-combined_k141_368434\|\|full | 2376 | No | NA | 3 | 0 | 1 | Not-determined | Genome-fragment | NA | NA | 0 |
| S12-final-viral-combined_k141_284843\|\|full | 4886 | No | NA | 6 | 0 | 3 | Not-determined | Genome-fragment | NA | NA | 0 |
| S12-final-viral-combined_k141_368435\|\|full | 3580 | No | NA | 4 | 0 | 4 | Not-determined | Genome-fragment | NA | NA | 0 |
| S12-final-viral-combined_k141_83855\|\|full | 2410 | No | NA | 4 | 0 | 2 | Not-determined | Genome-fragment | NA | NA | 0 |
| S12-final-viral-combined_k141_16917\|\|full | 1339 | No | NA | 1 | 0 | 0 | Not-determined | Genome-fragment | NA | NA | 0 |
| S12-final-viral-combined_k141_184338\|\|full | 50077 | Yes | 38325 | 74 | 20 | 9 | Complete | High-quality | 100 | Provirus (high-confidence) | 23.47 |
| S12_k141_304421 | 2541 | No | NA | 5 | 1 | 0 | Low-quality | Genome-fragment | 7.72 | AAI-based (high-confidence) | 0 |
| S12-final-viral-combined_k141_284847\|\|full | 1674 | No | NA | 6 | 0 | 0 | Not-determined | Genome-fragment | NA | NA | 0 |
| S12-final-viral-combined_k141_184348\|\|full | 1199 | No | NA | 3 | 0 | 2 | Not-determined | Genome-fragment | NA | NA | 0 |
| S2-final-viral-combined_k141_205576\|\|full | 2541 | No | NA | 5 | 1 | 0 | Low-quality | Genome-fragment | 1.45 | HMM-based (lower-bound) | 0 |
| S4_k141_18730 | 2541 | No | NA | 1 | 0 | 0 | Low-quality | Genome-fragment | 2.67 | AAI-based (high-confidence) | 0 |
| S8-final-viral-combined_k141_215500\|\|full | 2541 | No | NA | 3 | 0 | 1 | Low-quality | Genome-fragment | 4.32 | AAI-based (high-confidence) | 0 |
| S12-final-viral-combined_k141_33771\|\|full | 4293 | No | NA | 7 | 0 | 1 | Not-determined | Genome-fragment | NA | NA | 0 |
| S12-final-viral-combined_k141_134144\|\|full | 1814 | No | NA | 2 | 0 | 1 | Not-determined | Genome-fragment | NA | NA | 0 |
| S9-final-viral-combined_k141_109521\|\|full | 2542 | No | NA | 3 | 0 | 1 | Low-quality | Genome-fragment | 1.03 | AAI-based (medium-confidence) | 0 |
| S12-final-viral-combined_k141_318268\|\|full | 2550 | No | NA | 3 | 0 | 1 | Not-determined | Genome-fragment | NA | NA | 0 |
| S1-final-viral-combined_k141_268833\|\|full | 2542 | No | NA | 3 | 0 | 0 | Low-quality | Genome-fragment | 4.68 | AAI-based (high-confidence) | 0 |
| S12-final-viral-combined_k141_234462\|\|full | 2711 | No | NA | 4 | 0 | 3 | Not-determined | Genome-fragment | NA | NA | 0 |
| S12-final-viral-combined_k141_301591\|\|full | 6177 | No | NA | 6 | 0 | 2 | Not-determined | Genome-fragment | NA | NA | 0 |
| S12-final-viral-combined_k141_234470\|\|full | 2162 | No | NA | 5 | 0 | 3 | Not-determined | Genome-fragment | NA | NA | 0 |
| S1-final-viral-combined_k141_194304\|\|full | 2542 | No | NA | 4 | 0 | 0 | Low-quality | Genome-fragment | 6.53 | AAI-based (medium-confidence) | 0 |
| S12-final-viral-combined_k141_67168\|\|full | 2542 | No | NA | 4 | 0 | 0 | Not-determined | Genome-fragment | NA | NA | 0 |
| S12-final-viral-combined_k141_33799\|\|full | 4211 | No | NA | 5 | 0 | 4 | Not-determined | Genome-fragment | NA | NA | 0 |
| S12-final-viral-combined_k141_16988\|\|full | 1785 | No | NA | 4 | 0 | 1 | Not-determined | Genome-fragment | NA | NA | 0 |
| S12-final-viral-combined_k141_368462\|\|full | 55485 | Yes | 51861 | 62 | 23 | 4 | High-quality | High-quality | 100 | AAI-based (high-confidence) | 6.53 |
| S1-final-viral-combined_k141_73113\|\|full | 2543 | No | NA | 4 | 0 | 0 | Low-quality | Genome-fragment | 6.15 | AAI-based (high-confidence) | 0 |
| S12-final-viral-combined_k141_368465\|\|full | 2036 | No | NA | 3 | 0 | 3 | Not-determined | Genome-fragment | NA | NA | 0 |
| S5-final-viral-combined_k141_85510\|\|full | 2543 | No | NA | 8 | 3 | 0 | Low-quality | Genome-fragment | 3.58 | HMM-based (lower-bound) | 0 |
| S12-final-viral-combined_k141_251377\|\|full | 1005 | No | NA | 3 | 0 | 2 | Not-determined | Genome-fragment | NA | NA | 0 |
| S12-final-viral-combined_k141_385270\|\|full | 41955 | Yes | 29371 | 57 | 14 | 4 | Medium-quality | Genome-fragment | 77.49 | AAI-based (high-confidence) | 29.99 |
| S12-final-viral-combined_k141_184376\|\|full | 2165 | No | NA | 3 | 0 | 3 | Not-determined | Genome-fragment | NA | NA | 0 |
| S2-final-viral-combined_k141_83614\|\|full | 2543 | No | NA | 3 | 0 | 3 | Low-quality | Genome-fragment | 3.06 | AAI-based (medium-confidence) | 0 |
| S1-final-viral-combined_k141_37681\|\|full | 2544 | No | NA | 4 | 0 | 0 | Low-quality | Genome-fragment | 6.72 | AAI-based (medium-confidence) | 0 |
| S12-final-viral-combined_k141_117417\|\|full | 1909 | No | NA | 2 | 0 | 0 | Not-determined | Genome-fragment | NA | NA | 0 |
| S12-final-viral-combined_k141_67220\|\|full | 2145 | No | NA | 3 | 0 | 0 | Not-determined | Genome-fragment | NA | NA | 0 |
| S12-final-viral-combined_k141_334907\|\|full | 2168 | No | NA | 4 | 0 | 0 | Not-determined | Genome-fragment | NA | NA | 0 |
| S12-final-viral-combined_k141_301636\|\|full | 8338 | No | NA | 11 | 0 | 3 | Not-determined | Genome-fragment | NA | NA | 0 |
| S5-final-viral-combined_k141_162080\|\|full | 2544 | No | NA | 3 | 0 | 0 | Low-quality | Genome-fragment | 11.12 | AAI-based (medium-confidence) | 0 |
| S12-final-viral-combined_k141_284953\|\|full | 1849 | No | NA | 3 | 0 | 1 | Not-determined | Genome-fragment | NA | NA | 0 |
| S12-final-viral-combined_k141_251392\|\|full | 1446 | No | NA | 3 | 0 | 1 | Not-determined | Genome-fragment | NA | NA | 0 |
| S12-final-viral-combined_k141_116424\|\|full | 2544 | No | NA | 5 | 0 | 0 | Low-quality | Genome-fragment | 3.7 | AAI-based (high-confidence) | 0 |
| S12-final-viral-combined_k141_301677\|\|full | 2713 | No | NA | 3 | 0 | 2 | Not-determined | Genome-fragment | NA | NA | 0 |
| S12-final-viral-combined_k141_67264\|\|full | 3902 | No | NA | 4 | 0 | 3 | Not-determined | Genome-fragment | NA | NA | 0 |
| S12-final-viral-combined_k141_251420\|\|full | 1832 | No | NA | 3 | 0 | 0 | Not-determined | Genome-fragment | NA | NA | 0 |
| S12-final-viral-combined_k141_50556\|\|full | 1417 | No | NA | 2 | 0 | 1 | Not-determined | Genome-fragment | NA | NA | 0 |
| S12-final-viral-combined_k141_234583\|\|full | 8991 | No | NA | 11 | 0 | 9 | Not-determined | Genome-fragment | NA | NA | 0 |
| S12-final-viral-combined_k141_268025\|\|full | 1589 | No | NA | 2 | 0 | 1 | Not-determined | Genome-fragment | NA | NA | 0 |
| S12-final-viral-combined_k141_33881\|\|full | 4470 | No | NA | 10 | 0 | 0 | Not-determined | Genome-fragment | NA | NA | 0 |
| S12-final-viral-combined_k141_33888\|\|full | 6353 | No | NA | 7 | 0 | 2 | Not-determined | Genome-fragment | NA | NA | 0 |
| S12-final-viral-combined_k141_234584\|\|full | 20192 | No | NA | 17 | 0 | 12 | Not-determined | Genome-fragment | NA | NA | 0 |
| S12-final-viral-combined_k141_251449\|\|full | 6099 | No | NA | 6 | 0 | 2 | Not-determined | Genome-fragment | NA | NA | 0 |
| S9-final-viral-combined_k141_3817\|\|full | 2545 | No | NA | 7 | 5 | 0 | Low-quality | Genome-fragment | 5.9 | AAI-based (medium-confidence) | 0 |
| S11-final-viral-combined_k141_42219\|\|full | 2545 | No | NA | 5 | 0 | 0 | Low-quality | Genome-fragment | 6.42 | AAI-based (high-confidence) | 0 |
| S12-final-viral-combined_k141_201125\|\|full | 224193 | Yes | 43348 | 193 | 11 | 102 | Medium-quality | Genome-fragment | 52.63 | HMM-based (lower-bound) | 80.66 |
| S12-final-viral-combined_k141_167761\|\|full | 2851 | No | NA | 2 | 0 | 2 | Not-determined | Genome-fragment | NA | NA | 0 |
| S5-final-viral-combined_k141_45327\|\|full | 2545 | No | NA | 5 | 0 | 0 | Low-quality | Genome-fragment | 2.71 | AAI-based (medium-confidence) | 0 |
| S12-final-viral-combined_k141_318440\|\|full | 1262 | No | NA | 3 | 0 | 1 | Not-determined | Genome-fragment | NA | NA | 0 |

| | | | | | | | | | | | |
|---|---|---|---|---|---|---|---|---|---|---|---|
| S5_k141_193829 | 2545 | No | NA | 2 | 0 | 1 | Low-quality | Genome-fragment | 1.66 | AAI-based (high-confidence) | 0 |
| S12-final-viral-combined_k141_67326||full | 1279 | No | NA | 3 | 0 | 2 | Not-determined | Genome-fragment | NA | NA | 0 |
| S12-final-viral-combined_k141_100749||full | 2314 | No | NA | 4 | 0 | 2 | Not-determined | Genome-fragment | NA | NA | 0 |
| S12-final-viral-combined_k141_334989||full | 1924 | No | NA | 3 | 0 | 1 | Not-determined | Genome-fragment | NA | NA | 0 |
| S12-final-viral-combined_k141_50612||full | 9714 | No | NA | 10 | 0 | 7 | Not-determined | Genome-fragment | NA | NA | 0 |
| S12-final-viral-combined_k141_201138||full | 8995 | No | NA | 8 | 0 | 7 | Not-determined | Genome-fragment | NA | NA | 0 |
| S12-final-viral-combined_k141_201139||full | 2247 | No | NA | 3 | 0 | 2 | Not-determined | Genome-fragment | NA | NA | 0 |
| S7-final-viral-combined_k141_24074||full | 2545 | No | NA | 5 | 2 | 0 | Low-quality | Genome-fragment | 6.24 | HMM-based (lower-bound) | 0 |
| S12-final-viral-combined_k141_268082||full | 3563 | No | NA | 4 | 0 | 1 | Not-determined | Genome-fragment | NA | NA | 0 |
| S12-final-viral-combined_k141_368562||full | 23237 | No | NA | 15 | 0 | 2 | Medium-quality | Genome-fragment | 54.25 | AAI-based (medium-confidence) | 0 |
| S12-final-viral-combined_k141_33984||full | 1781 | No | NA | 3 | 0 | 0 | Not-determined | Genome-fragment | NA | NA | 0 |
| S12-final-viral-combined_k141_318478||full | 1627 | No | NA | 2 | 0 | 1 | Not-determined | Genome-fragment | NA | NA | 0 |
| S12-final-viral-combined_k141_33996||full | 1288 | No | NA | 3 | 0 | 0 | Not-determined | Genome-fragment | NA | NA | 0 |
| S11-final-viral-combined_k141_183928||full | 2546 | No | NA | 4 | 0 | 0 | Low-quality | Genome-fragment | 2.29 | AAI-based (high-confidence) | 0 |
| S12-final-viral-combined_k141_50653||full | 3846 | No | NA | 7 | 0 | 2 | Not-determined | Genome-fragment | NA | NA | 0 |
| S12-final-viral-combined_k141_167831||full | 1759 | No | NA | 2 | 0 | 0 | Not-determined | Genome-fragment | NA | NA | 0 |
| S3-final-viral-combined_k141_133047||lt2gene | 2546 | No | NA | 2 | 0 | 0 | Low-quality | Genome-fragment | 1.17 | AAI-based (high-confidence) | 0 |
| S7-final-viral-combined_k141_174134||full | 2546 | No | NA | 3 | 0 | 0 | Low-quality | Genome-fragment | 5.11 | AAI-based (high-confidence) | 0 |
| S12-final-viral-combined_k141_318505||full | 1167 | No | NA | 3 | 0 | 0 | Not-determined | Genome-fragment | NA | NA | 0 |
| S12-final-viral-combined_k141_368581||full | 1834 | No | NA | 3 | 0 | 0 | Not-determined | Genome-fragment | NA | NA | 0 |
| S12-final-viral-combined_k141_100825||full | 2610 | No | NA | 3 | 0 | 0 | Not-determined | Genome-fragment | NA | NA | 0 |
| S12-final-viral-combined_k141_217996||full | 2712 | No | NA | 3 | 0 | 3 | Not-determined | Genome-fragment | NA | NA | 0 |
| S12-final-viral-combined_k141_268128||full | 6863 | No | NA | 7 | 0 | 4 | Not-determined | Genome-fragment | NA | NA | 0 |
| S12-final-viral-combined_k141_234755||full | 2885 | No | NA | 3 | 0 | 3 | Not-determined | Genome-fragment | NA | NA | 0 |
| S12-final-viral-combined_k141_155781||full | 2546 | No | NA | 3 | 0 | 0 | Low-quality | Genome-fragment | 1.43 | AAI-based (high-confidence) | 0 |
| S12-final-viral-combined_k141_50693||full | 1851 | No | NA | 3 | 0 | 0 | Not-determined | Genome-fragment | NA | NA | 0 |
| S12-final-viral-combined_k141_392568||full | 2546 | No | NA | 2 | 0 | 1 | Low-quality | Genome-fragment | 5.68 | AAI-based (high-confidence) | 0 |
| S12-final-viral-combined_k141_234767||full | 4224 | No | NA | 5 | 0 | 2 | Not-determined | Genome-fragment | NA | NA | 0 |
| S8-final-viral-combined_k141_217377||full | 2546 | No | NA | 3 | 0 | 2 | Low-quality | Genome-fragment | 6.17 | AAI-based (high-confidence) | 0 |
| S12-final-viral-combined_k141_335054||full | 1174 | No | NA | 2 | 0 | 2 | Not-determined | Genome-fragment | NA | NA | 0 |
| S12-final-viral-combined_k141_50745||full | 4445 | No | NA | 4 | 0 | 2 | Not-determined | Genome-fragment | NA | NA | 0 |
| S12-final-viral-combined_k141_385472||full | 1758 | No | NA | 3 | 0 | 2 | Not-determined | Genome-fragment | NA | NA | 0 |
| S5-final-viral-combined_k141_169403||full | 2547 | No | NA | 6 | 1 | 0 | Low-quality | Genome-fragment | 2.86 | AAI-based (high-confidence) | 0 |
| S12-final-viral-combined_k141_201220||full | 2063 | No | NA | 3 | 0 | 1 | Not-determined | Genome-fragment | NA | NA | 0 |
| S12-final-viral-combined_k141_67394||full | 4357 | No | NA | 3 | 0 | 1 | Not-determined | Genome-fragment | NA | NA | 0 |
| S5-final-viral-combined_k141_46896||full | 2547 | No | NA | 4 | 1 | 0 | Low-quality | Genome-fragment | 4.77 | AAI-based (high-confidence) | 0 |
| S12-final-viral-combined_k141_234796||full | 6003 | No | NA | 3 | 0 | 1 | Not-determined | Genome-fragment | NA | NA | 0 |
| S7-final-viral-combined_k141_194825||full | 2547 | No | NA | 8 | 0 | 1 | Low-quality | Genome-fragment | 6.15 | AAI-based (high-confidence) | 0 |
| S12-final-viral-combined_k141_335082||full | 2206 | No | NA | 5 | 0 | 0 | Not-determined | Genome-fragment | NA | NA | 0 |
| S12-final-viral-combined_k141_368885||full | 2547 | No | NA | 4 | 0 | 2 | Low-quality | Genome-fragment | 3.15 | AAI-based (high-confidence) | 0 |
| S12-final-viral-combined_k141_84196||full | 2433 | No | NA | 2 | 0 | 1 | Not-determined | Genome-fragment | NA | NA | 0 |
| S12-final-viral-combined_k141_556||full | 2886 | No | NA | 3 | 0 | 0 | Not-determined | Genome-fragment | NA | NA | 0 |
| S2-final-viral-combined_k141_107467||full | 2547 | No | NA | 2 | 2 | 0 | Low-quality | Genome-fragment | 5.71 | AAI-based (high-confidence) | 0 |
| S12-final-viral-combined_k141_335088||full | 1665 | No | NA | 4 | 0 | 0 | Not-determined | Genome-fragment | NA | NA | 0 |
| S2-final-viral-combined_k141_245682||full | 2547 | No | NA | 4 | 1 | 0 | Low-quality | Genome-fragment | 5.69 | AAI-based (medium-confidence) | 0 |
| S12-final-viral-combined_k141_201275||full | 2934 | No | NA | 3 | 0 | 0 | Not-determined | Genome-fragment | NA | NA | 0 |
| S4_k141_2273 | 2547 | No | NA | 4 | 0 | 0 | Low-quality | Genome-fragment | 15.22 | AAI-based (high-confidence) | 0 |
| S12-final-viral-combined_k141_50792||full | 2211 | No | NA | 3 | 0 | 0 | Not-determined | Genome-fragment | NA | NA | 0 |
| S12-final-viral-combined_k141_218111||full | 2611 | No | NA | 3 | 0 | 2 | Not-determined | Genome-fragment | NA | NA | 0 |
| S12-final-viral-combined_k141_368730||full | 1892 | No | NA | 3 | 0 | 1 | Not-determined | Genome-fragment | NA | NA | 0 |
| S12-final-viral-combined_k141_301863||full | 1259 | No | NA | 3 | 0 | 2 | Not-determined | Genome-fragment | NA | NA | 0 |
| S12-final-viral-combined_k141_285228||full | 9234 | No | NA | 13 | 0 | 1 | Not-determined | Genome-fragment | NA | NA | 0 |
| S12-final-viral-combined_k141_335096||full | 1520 | No | NA | 3 | 0 | 0 | Not-determined | Genome-fragment | NA | NA | 0 |
| S12-final-viral-combined_k141_301868||full | 2844 | No | NA | 3 | 0 | 1 | Not-determined | Genome-fragment | NA | NA | 0 |
| S12-final-viral-combined_k141_617||full | 2611 | No | NA | 3 | 0 | 0 | Not-determined | Genome-fragment | NA | NA | 0 |
| S12-final-viral-combined_k141_285237||full | 2297 | No | NA | 4 | 0 | 3 | Not-determined | Genome-fragment | NA | NA | 0 |
| S12-final-viral-combined_k141_268220||full | 4330 | No | NA | 5 | 0 | 0 | Not-determined | Genome-fragment | NA | NA | 0 |
| S12-final-viral-combined_k141_184608||full | 6415 | No | NA | 7 | 0 | 7 | Not-determined | Genome-fragment | NA | NA | 0 |
| S12-final-viral-combined_k141_100983||full | 1606 | No | NA | 3 | 0 | 2 | Not-determined | Genome-fragment | NA | NA | 0 |
| S12-final-viral-combined_k141_150997||full | 2026 | No | NA | 3 | 0 | 2 | Not-determined | Genome-fragment | NA | NA | 0 |
| S12-final-viral-combined_k141_351767||full | 15789 | No | NA | 16 | 0 | 4 | Not-determined | Genome-fragment | NA | NA | 0 |
| S12-final-viral-combined_k141_234862||full | 1224 | No | NA | 2 | 0 | 0 | Not-determined | Genome-fragment | NA | NA | 0 |
| S12-final-viral-combined_k141_34179||full | 2172 | No | NA | 5 | 0 | 0 | Not-determined | Genome-fragment | NA | NA | 0 |
| S12-final-viral-combined_k141_335103||full | 7382 | No | NA | 10 | 0 | 0 | Not-determined | Genome-fragment | NA | NA | 0 |
| S5_k141_123823 | 2548 | No | NA | 7 | 5 | 0 | Low-quality | Genome-fragment | 6.18 | AAI-based (high-confidence) | 0 |
| S7-final-viral-combined_k141_78567||full | 2548 | No | NA | 5 | 4 | 0 | Low-quality | Genome-fragment | 5.61 | AAI-based (high-confidence) | 0 |
| S17-final-viral-combined_k141_76297||full | 2548 | No | NA | 3 | 0 | 2 | Low-quality | Genome-fragment | 1.7 | AAI-based (high-confidence) | 0 |
| S12_k141_359663 | 2548 | No | NA | 3 | 0 | 0 | Low-quality | Genome-fragment | 1.66 | AAI-based (high-confidence) | 0 |
| S12-final-viral-combined_k141_67493||full | 2745 | No | NA | 5 | 0 | 0 | Not-determined | Genome-fragment | NA | NA | 0 |
| S12-final-viral-combined_k141_234895||full | 3272 | No | NA | 2 | 0 | 2 | Not-determined | Genome-fragment | NA | NA | 0 |
| S12_k141_346211 | 2548 | No | NA | 2 | 0 | 0 | Low-quality | Genome-fragment | 4.6 | AAI-based (medium-confidence) | 0 |
| S12-final-viral-combined_k141_134589||full | 1338 | No | NA | 2 | 0 | 2 | Not-determined | Genome-fragment | NA | NA | 0 |
| S12-final-viral-combined_k141_34217||full | 1696 | No | NA | 2 | 0 | 1 | Not-determined | Genome-fragment | NA | NA | 0 |
| S9-final-viral-combined_k141_101405||full | 2549 | No | NA | 3 | 2 | 0 | Low-quality | Genome-fragment | 4.34 | AAI-based (high-confidence) | 0 |
| S4-final-viral-combined_k141_101094||full | 2549 | No | NA | 7 | 0 | 0 | Low-quality | Genome-fragment | 3.16 | AAI-based (high-confidence) | 0 |
| S8-final-viral-combined_k141_96821||full | 2549 | No | NA | 3 | 2 | 0 | Low-quality | Genome-fragment | 1.93 | AAI-based (high-confidence) | 0 |
| S12-final-viral-combined_k141_201387||full | 1849 | No | NA | 2 | 0 | 2 | Not-determined | Genome-fragment | NA | NA | 0 |
| S12-final-viral-combined_k141_34226||full | 10444 | No | NA | 9 | 0 | 7 | Not-determined | Genome-fragment | NA | NA | 0 |
| S12-final-viral-combined_k141_368824||full | 1512 | No | NA | 2 | 0 | 1 | Not-determined | Genome-fragment | NA | NA | 0 |
| S12-final-viral-combined_k141_34236||full | 1359 | No | NA | 2 | 0 | 1 | Not-determined | Genome-fragment | NA | NA | 0 |
| S12-final-viral-combined_k141_117510||full | 3342 | No | NA | 4 | 0 | 3 | Not-determined | Genome-fragment | NA | NA | 0 |
| S8-final-viral-combined_k141_120990||full | 2549 | No | NA | 3 | 0 | 0 | Low-quality | Genome-fragment | 4.9 | AAI-based (medium-confidence) | 0 |
| S11-final-viral-combined_k141_67879||full | 2550 | No | NA | 4 | 0 | 1 | Low-quality | Genome-fragment | 2.06 | AAI-based (medium-confidence) | 0 |
| S12-final-viral-combined_k141_151054||full | 2822 | No | NA | 4 | 0 | 4 | Not-determined | Genome-fragment | NA | NA | 0 |
| S12-final-viral-combined_k141_151057||full | 2886 | No | NA | 3 | 0 | 2 | Not-determined | Genome-fragment | NA | NA | 0 |
| S12-final-viral-combined_k141_335171||full | 19918 | No | NA | 23 | 0 | 6 | Not-determined | Genome-fragment | NA | NA | 0 |
| S2-final-viral-combined_k141_134426||full | 2550 | No | NA | 3 | 2 | 0 | Low-quality | Genome-fragment | 3.08 | HMM-based (lower-bound) | 0 |
| S12-final-viral-combined_k141_101036||full | 1079 | No | NA | 2 | 0 | 1 | Not-determined | Genome-fragment | NA | NA | 0 |
| S12-final-viral-combined_k141_50905||full | 2940 | No | NA | 5 | 0 | 4 | Not-determined | Genome-fragment | NA | NA | 0 |
| S12-final-viral-combined_k141_117545||full | 2594 | No | NA | 4 | 0 | 1 | Not-determined | Genome-fragment | NA | NA | 0 |
| S12-final-viral-combined_k141_134629||full | 2909 | No | NA | 3 | 0 | 1 | Not-determined | Genome-fragment | NA | NA | 0 |
| S12-final-viral-combined_k141_351847||full | 12931 | No | NA | 15 | 1 | 1 | Medium-quality | Genome-fragment | 61.98 | AAI-based (high-confidence) | 0 |
| S9-final-viral-combined_k141_83138||full | 2551 | No | NA | 3 | 0 | 0 | Low-quality | Genome-fragment | 1.54 | AAI-based (medium-confidence) | 0 |
| S11-final-viral-combined_k141_142885||full | 2551 | No | NA | 3 | 2 | 0 | Low-quality | Genome-fragment | 7.62 | AAI-based (high-confidence) | 0 |
| S12-final-viral-combined_k141_84358||full | 2422 | No | NA | 4 | 0 | 2 | Not-determined | Genome-fragment | NA | NA | 0 |
| S12-final-viral-combined_k141_780||full | 19689 | No | NA | 15 | 0 | 12 | Not-determined | Genome-fragment | NA | NA | 0 |
| S12-final-viral-combined_k141_285466||full | 1468 | No | NA | 4 | 0 | 0 | Not-determined | Genome-fragment | NA | NA | 0 |
| S12-final-viral-combined_k141_184749||full | 1111 | No | NA | 3 | 0 | 0 | Not-determined | Genome-fragment | NA | NA | 0 |
| S12-final-viral-combined_k141_268378||full | 3149 | No | NA | 2 | 0 | 0 | Not-determined | Genome-fragment | NA | NA | 0 |
| S11-final-viral-combined_k141_272097||full | 2551 | No | NA | 4 | 3 | 0 | Low-quality | Genome-fragment | 5.5 | AAI-based (high-confidence) | 0 |
| S17-final-viral-combined_k141_17629||full | 2551 | No | NA | 3 | 0 | 1 | Low-quality | Genome-fragment | 3.04 | AAI-based (medium-confidence) | 0 |
| S12-final-viral-combined_k141_385610||full | 4011 | No | NA | 4 | 0 | 2 | Not-determined | Genome-fragment | NA | NA | 0 |
| S12-final-viral-combined_k141_335208||full | 3210 | No | NA | 3 | 0 | 1 | Not-determined | Genome-fragment | NA | NA | 0 |
| S12-final-viral-combined_k141_360168||full | 2551 | No | NA | 5 | 0 | 1 | Low-quality | Genome-fragment | 1.9 | AAI-based (high-confidence) | 0 |
| S8_k141_139466 | 2551 | No | NA | 2 | 0 | 1 | Low-quality | Genome-fragment | 0.95 | AAI-based (high-confidence) | 0 |
| S5-final-viral-combined_k141_71865||full | 2552 | No | NA | 4 | 1 | 0 | Low-quality | Genome-fragment | 4.71 | AAI-based (medium-confidence) | 0 |
| S12-final-viral-combined_k141_17565||full | 1184 | No | NA | 2 | 0 | 1 | Not-determined | Genome-fragment | NA | NA | 0 |
| S12-final-viral-combined_k141_803||full | 1194 | No | NA | 3 | 0 | 1 | Not-determined | Genome-fragment | NA | NA | 0 |
| S12-final-viral-combined_k141_201556||full | 2317 | No | NA | 3 | 0 | 2 | Not-determined | Genome-fragment | NA | NA | 0 |
| S12-final-viral-combined_k141_318749||full | 4015 | No | NA | 7 | 0 | 3 | Not-determined | Genome-fragment | NA | NA | 0 |
| S12-final-viral-combined_k141_301980||full | 1847 | No | NA | 2 | 0 | 0 | Not-determined | Genome-fragment | NA | NA | 0 |
| S12-final-viral-combined_k141_351936||full | 7284 | No | NA | 10 | 0 | 3 | Not-determined | Genome-fragment | NA | NA | 0 |
| S12-final-viral-combined_k141_285504||full | 1602 | No | NA | 3 | 0 | 1 | Not-determined | Genome-fragment | NA | NA | 0 |
| S5-final-viral-combined_k141_18627||full | 2552 | No | NA | 4 | 0 | 0 | Low-quality | Genome-fragment | 4.92 | AAI-based (high-confidence) | 0 |
| S12-final-viral-combined_k141_287615||full | 2552 | No | NA | 5 | 3 | 0 | Low-quality | Genome-fragment | 5.32 | AAI-based (high-confidence) | 0 |
| S4-final-viral-combined_k141_45269||full | 2552 | No | NA | 3 | 0 | 0 | Low-quality | Genome-fragment | 3.32 | AAI-based (medium-confidence) | 0 |
| S12-final-viral-combined_k141_351942||full | 1379 | No | NA | 3 | 0 | 2 | Not-determined | Genome-fragment | NA | NA | 0 |
| S12-final-viral-combined_k141_84399||full | 8535 | No | NA | 9 | 0 | 6 | Not-determined | Genome-fragment | NA | NA | 0 |
| S12-final-viral-combined_k141_318758||full | 6965 | No | NA | 7 | 0 | 5 | Not-determined | Genome-fragment | NA | NA | 0 |
| S12-final-viral-combined_k141_184806||full | 3469 | No | NA | 4 | 0 | 2 | Not-determined | Genome-fragment | NA | NA | 0 |
| S12-final-viral-combined_k141_151169||full | 1075 | No | NA | 3 | 0 | 0 | Not-determined | Genome-fragment | NA | NA | 0 |
| S12-final-viral-combined_k141_201582||full | 2612 | No | NA | 4 | 0 | 0 | Not-determined | Genome-fragment | NA | NA | 0 |
| S12-final-viral-combined_k141_235047||full | 3309 | No | NA | 4 | 0 | 3 | Not-determined | Genome-fragment | NA | NA | 0 |
| S8-final-viral-combined_k141_11433||full | 2552 | No | NA | 5 | 0 | 0 | Low-quality | Genome-fragment | 4.22 | AAI-based (medium-confidence) | 0 |
| S8-final-viral-combined_k141_109329||lt2gene | 2552 | No | NA | 1 | 1 | 0 | Low-quality | Genome-fragment | 7.4 | AAI-based (high-confidence) | 0 |
| S12-final-viral-combined_k141_268436||full | 1385 | No | NA | 2 | 0 | 1 | Not-determined | Genome-fragment | NA | NA | 0 |
| S9-final-viral-combined_k141_126139||full | 2553 | No | NA | 4 | 1 | 0 | Low-quality | Genome-fragment | 3.76 | HMM-based (lower-bound) | 0 |
| S12-final-viral-combined_k141_201600||full | 6959 | No | NA | 4 | 0 | 2 | Not-determined | Genome-fragment | NA | NA | 0 |
| S2_k141_102086 | 2553 | No | NA | 4 | 0 | 0 | Low-quality | Genome-fragment | 3.1 | AAI-based (medium-confidence) | 0 |
| S9-final-viral-combined_k141_109282||full | 2554 | No | NA | 4 | 0 | 1 | Low-quality | Genome-fragment | 3.98 | AAI-based (medium-confidence) | 0 |
| S5-final-viral-combined_k141_12786||full | 2554 | No | NA | 8 | 1 | 0 | Low-quality | Genome-fragment | 5.29 | AAI-based (high-confidence) | 0 |
| S12-final-viral-combined_k141_84475||full | 2220 | No | NA | 3 | 0 | 1 | Not-determined | Genome-fragment | NA | NA | 0 |
| S12-final-viral-combined_k141_385697||full | 4246 | No | NA | 7 | 0 | 1 | Not-determined | Genome-fragment | NA | NA | 0 |
| S12-final-viral-combined_k141_101110||full | 1006 | No | NA | 3 | 0 | 1 | Not-determined | Genome-fragment | NA | NA | 0 |
| S12-final-viral-combined_k141_34361||full | 1434 | No | NA | 1 | 0 | 0 | Not-determined | Genome-fragment | NA | NA | 0 |
| S12-final-viral-combined_k141_184870||full | 1248 | No | NA | 2 | 0 | 0 | Not-determined | Genome-fragment | NA | NA | 0 |
| S12-final-viral-combined_k141_184872||full | 4622 | No | NA | 3 | 0 | 2 | Not-determined | Genome-fragment | NA | NA | 0 |
| S12-final-viral-combined_k141_264030||full | 2554 | No | NA | 4 | 0 | 3 | Low-quality | Genome-fragment | 2.09 | AAI-based (medium-confidence) | 0 |
| S12-final-viral-combined_k141_134775||full | 1226 | No | NA | 2 | 0 | 1 | Not-determined | Genome-fragment | NA | NA | 0 |
| S12-final-viral-combined_k141_117628||full | 58691 | Yes | 21686 | 65 | 14 | 19 | Medium-quality | Genome-fragment | 67.39 | AAI-based (high-confidence) | 63.05 |
| S12-final-viral-combined_k141_134781||full | 1363 | No | NA | 3 | 0 | 1 | Not-determined | Genome-fragment | NA | NA | 0 |
| S8-final-viral-combined_k141_69903||full | 2554 | No | NA | 4 | 0 | 1 | Low-quality | Genome-fragment | 1.7 | AAI-based (high-confidence) | 0 |
| S12-final-viral-combined_k141_318857||full | 2064 | No | NA | 2 | 0 | 1 | Not-determined | Genome-fragment | NA | NA | 0 |
| S8-final-viral-combined_k141_223549||full | 2554 | No | NA | 4 | 0 | 1 | Low-quality | Genome-fragment | 1.76 | AAI-based (high-confidence) | 0 |
| S12-final-viral-combined_k141_168246||full | 1290 | No | NA | 2 | 0 | 2 | Not-determined | Genome-fragment | NA | NA | 0 |
| S9-final-viral-combined_k141_125036||full | 2555 | No | NA | 2 | 0 | 2 | Low-quality | Genome-fragment | 3.49 | AAI-based (medium-confidence) | 0 |
| S12-final-viral-combined_k141_318871||full | 2388 | No | NA | 2 | 0 | 1 | Not-determined | Genome-fragment | NA | NA | 0 |
| S12-final-viral-combined_k141_34431||full | 2812 | No | NA | 4 | 0 | 1 | Not-determined | Genome-fragment | NA | NA | 0 |
| S11-final-viral-combined_k141_71526||full | 2555 | No | NA | 3 | 0 | 1 | Low-quality | Genome-fragment | 0.58 | AAI-based (medium-confidence) | 0 |
| S12-final-viral-combined_k141_17617||full | 54837 | Yes | 31419 | 70 | 18 | 9 | Medium-quality | Genome-fragment | 70 | AAI-based (high-confidence) | 42.7 |
| S12-final-viral-combined_k141_201683||full | 2287 | No | NA | 3 | 0 | 0 | Not-determined | Genome-fragment | NA | NA | 0 |
| S12-final-viral-combined_k141_184945||full | 2298 | No | NA | 3 | 0 | 0 | Not-determined | Genome-fragment | NA | NA | 0 |
| S12-final-viral-combined_k141_385757||full | 1141 | No | NA | 2 | 0 | 1 | Not-determined | Genome-fragment | NA | NA | 0 |
| S12-final-viral-combined_k141_34457||full | 1415 | No | NA | 2 | 0 | 0 | Not-determined | Genome-fragment | NA | NA | 0 |
| S12-final-viral-combined_k141_285644||full | 1864 | No | NA | 3 | 0 | 2 | Not-determined | Genome-fragment | NA | NA | 0 |
| S12-final-viral-combined_k141_285652||full | 1413 | No | NA | 2 | 0 | 0 | Not-determined | Genome-fragment | NA | NA | 0 |
| S8-final-viral-combined_k141_127985||full | 2555 | No | NA | 7 | 0 | 0 | Low-quality | Genome-fragment | 4.43 | AAI-based (high-confidence) | 0 |
| S9-final-viral-combined_k141_116440||full | 2556 | No | NA | 5 | 0 | 0 | Low-quality | Genome-fragment | 4.62 | AAI-based (medium-confidence) | 0 |
| S11-final-viral-combined_k141_128366||full | 2556 | No | NA | 4 | 3 | 0 | Low-quality | Genome-fragment | 5.9 | AAI-based (high-confidence) | 0 |
| S12-final-viral-combined_k141_117704||full | 1489 | No | NA | 2 | 0 | 1 | Not-determined | Genome-fragment | NA | NA | 0 |
| S12-final-viral-combined_k141_17687||full | 1344 | No | NA | 3 | 0 | 0 | Not-determined | Genome-fragment | NA | NA | 0 |
| S12-final-viral-combined_k141_385802||full | 6687 | No | NA | 9 | 0 | 1 | Not-determined | Genome-fragment | NA | NA | 0 |
| S12-final-viral-combined_k141_101227||full | 2045 | No | NA | 3 | 0 | 2 | Not-determined | Genome-fragment | NA | NA | 0 |
| S12-final-viral-combined_k141_34526||full | 2371 | No | NA | 3 | 0 | 1 | Not-determined | Genome-fragment | NA | NA | 0 |
| S4-final-viral-combined_k141_166412||full | 2556 | No | NA | 2 | 0 | 2 | Low-quality | Genome-fragment | 1.22 | AAI-based (medium-confidence) | 0 |
| S3_k141_15619 | 2557 | No | NA | 3 | 1 | 0 | Low-quality | Genome-fragment | 6.6 | AAI-based (high-confidence) | 0 |
| S12-final-viral-combined_k141_318947||full | 2030 | No | NA | 4 | 0 | 2 | Not-determined | Genome-fragment | NA | NA | 0 |
| S17-final-viral-combined_k141_256800||full | 2557 | No | NA | 3 | 0 | 0 | Low-quality | Genome-fragment | 0.42 | AAI-based (medium-confidence) | 0 |
| S12-final-viral-combined_k141_136846||full | 2557 | No | NA | 4 | 0 | 0 | Low-quality | Genome-fragment | 4.98 | AAI-based (medium-confidence) | 0 |
| S12-final-viral-combined_k141_335362||full | 2185 | No | NA | 5 | 0 | 0 | Not-determined | Genome-fragment | NA | NA | 0 |
| S12-final-viral-combined_k141_51076||full | 120233 | No | NA | 131 | 3 | 20 | Medium-quality | Genome-fragment | 68.52 | HMM-based (lower-bound) | 0 |
| S12-final-viral-combined_k141_335366||full | 4586 | No | NA | 8 | 0 | 1 | Not-determined | Genome-fragment | NA | NA | 0 |
| S12-final-viral-combined_k141_385894||full | 2694 | No | NA | 4 | 0 | 0 | Not-determined | Genome-fragment | NA | NA | 0 |
| S12-final-viral-combined_k141_302234||full | 1971 | No | NA | 4 | 0 | 1 | Not-determined | Genome-fragment | NA | NA | 0 |
| S12_k141_304034 | 2558 | No | NA | 3 | 0 | 1 | Low-quality | Genome-fragment | 3.25 | AAI-based (high-confidence) | 0 |
| S2-final-viral-combined_k141_84514||full | 2558 | No | NA | 5 | 1 | 0 | Low-quality | Genome-fragment | 2.67 | AAI-based (high-confidence) | 0 |
| S9-final-viral-combined_k141_87843||full | 2559 | No | NA | 3 | 0 | 0 | Low-quality | Genome-fragment | 3.32 | AAI-based (medium-confidence) | 0 |
| S12-final-viral-combined_k141_235236||full | 1085 | No | NA | 3 | 0 | 0 | Not-determined | Genome-fragment | NA | NA | 0 |
| S9-final-viral-combined_k141_121235||full | 2559 | No | NA | 5 | 1 | 0 | Low-quality | Genome-fragment | 4.19 | AAI-based (high-confidence) | 0 |
| S12-final-viral-combined_k141_268669||full | 3515 | No | NA | 4 | 0 | 2 | Not-determined | Genome-fragment | NA | NA | 0 |
| S12-final-viral-combined_k141_117743||full | 14230 | No | NA | 14 | 0 | 8 | Not-determined | Genome-fragment | NA | NA | 0 |
| S11-final-viral-combined_k141_36139||full | 2559 | No | NA | 3 | 0 | 3 | Low-quality | Genome-fragment | 3.7 | AAI-based (medium-confidence) | 0 |
| S12-final-viral-combined_k141_285753||full | 16786 | No | NA | 11 | 0 | 4 | Not-determined | Genome-fragment | NA | NA | 0 |
| S12-final-viral-combined_k141_34639||full | 1995 | No | NA | 3 | 0 | 2 | Not-determined | Genome-fragment | NA | NA | 0 |
| S12-final-viral-combined_k141_101305||full | 1556 | No | NA | 2 | 0 | 1 | Not-determined | Genome-fragment | NA | NA | 0 |
| S12-final-viral-combined_k141_285765||full | 2652 | No | NA | 4 | 0 | 2 | Not-determined | Genome-fragment | NA | NA | 0 |
| S12-final-viral-combined_k141_335429||full | 5783 | No | NA | 7 | 0 | 5 | Not-determined | Genome-fragment | NA | NA | 0 |
| S12-final-viral-combined_k141_369144||full | 1915 | No | NA | 3 | 0 | 0 | Not-determined | Genome-fragment | NA | NA | 0 |
| S12-final-viral-combined_k141_285778||full | 1909 | No | NA | 5 | 0 | 1 | Not-determined | Genome-fragment | NA | NA | 0 |
| S1-final-viral-combined_k141_87365||full | 2559 | No | NA | 5 | 0 | 0 | Low-quality | Genome-fragment | 4.03 | AAI-based (high-confidence) | 0 |
| S5-final-viral-combined_k141_92389||full | 2559 | No | NA | 3 | 0 | 3 | Low-quality | Genome-fragment | 1.55 | AAI-based (medium-confidence) | 0 |
| S12-final-viral-combined_k141_385958||full | 1733 | No | NA | 2 | 0 | 2 | Not-determined | Genome-fragment | NA | NA | 0 |
| S12-final-viral-combined_k141_17903||full | 1264 | No | NA | 3 | 0 | 0 | Not-determined | Genome-fragment | NA | NA | 0 |
| S7-final-viral-combined_k141_36067||full | 2559 | No | NA | 4 | 0 | 3 | Low-quality | Genome-fragment | 7.64 | AAI-based (medium-confidence) | 0 |
| S12-final-viral-combined_k141_185086||full | 1504 | No | NA | 2 | 0 | 2 | Not-determined | Genome-fragment | NA | NA | 0 |
| S7-final-viral-combined_k141_48937||full | 2559 | No | NA | 3 | 1 | 0 | Low-quality | Genome-fragment | 1.64 | HMM-based (lower-bound) | 0 |
| S12-final-viral-combined_k141_385991||full | 2392 | No | NA | 5 | 0 | 1 | Not-determined | Genome-fragment | NA | NA | 0 |
| S12-final-viral-combined_k141_385995||full | 2376 | No | NA | 4 | 0 | 0 | Not-determined | Genome-fragment | NA | NA | 0 |
| S2_k141_37816 | 2559 | No | NA | 2 | 0 | 1 | Low-quality | Genome-fragment | 1.14 | AAI-based (medium-confidence) | 0 |
| S12-final-viral-combined_k141_268774||full | 2338 | No | NA | 3 | 0 | 2 | Not-determined | Genome-fragment | NA | NA | 0 |
| S12-final-viral-combined_k141_268779||full | 1712 | No | NA | 5 | 0 | 0 | Not-determined | Genome-fragment | NA | NA | 0 |
| S17-k141_260059 | 2561 | No | NA | 3 | 0 | 0 | Low-quality | Genome-fragment | 6.48 | AAI-based (high-confidence) | 0 |
| S12-final-viral-combined_k141_218512||full | 2719 | No | NA | 4 | 0 | 2 | Not-determined | Genome-fragment | NA | NA | 0 |
| S12-final-viral-combined_k141_268782||full | 8983 | No | NA | 7 | 0 | 3 | Not-determined | Genome-fragment | NA | NA | 0 |
| S12-final-viral-combined_k141_352183||full | 1732 | No | NA | 2 | 0 | 0 | Not-determined | Genome-fragment | NA | NA | 0 |
| S12-final-viral-combined_k141_134875||full | 8667 | No | NA | 8 | 0 | 3 | Not-determined | Genome-fragment | NA | NA | 0 |
| S9-final-viral-combined_k141_43519||full | 2562 | No | NA | 6 | 3 | 0 | Low-quality | Genome-fragment | 6.5 | AAI-based (high-confidence) | 0 |
| S12-final-viral-combined_k141_34742||full | 1135 | No | NA | 2 | 0 | 2 | Not-determined | Genome-fragment | NA | NA | 0 |
| S12-final-viral-combined_k141_17955||full | 10510 | No | NA | 9 | 0 | 8 | Not-determined | Genome-fragment | NA | NA | 0 |
| S12-final-viral-combined_k141_134897||full | 4303 | No | NA | 6 | 0 | 2 | Not-determined | Genome-fragment | NA | NA | 0 |
| S12-final-viral-combined_k141_302330||full | 1677 | No | NA | 2 | 0 | 0 | Not-determined | Genome-fragment | NA | NA | 0 |
| S9_k141_65446 | 2562 | No | NA | 4 | 0 | 0 | Low-quality | Genome-fragment | 4.4 | AAI-based (medium-confidence) | 0 |
| S7-final-viral-combined_k141_124108||full | 2562 | No | NA | 4 | 0 | 1 | Low-quality | Genome-fragment | 2.1 | AAI-based (high-confidence) | 0 |
| S12-final-viral-combined_k141_240863||full | 2562 | No | NA | 4 | 0 | 0 | Low-quality | Genome-fragment | 4.65 | AAI-based (medium-confidence) | 0 |
| S12-final-viral-combined_k141_117824||full | 1729 | No | NA | 3 | 0 | 3 | Not-determined | Genome-fragment | NA | NA | 0 |
| S1-final-viral-combined_k141_156785||full | 2563 | No | NA | 3 | 0 | 3 | Low-quality | Genome-fragment | 3.02 | AAI-based (medium-confidence) | 0 |
| S3-final-viral-combined_k141_27283||full | 2563 | No | NA | 7 | 0 | 0 | Low-quality | Genome-fragment | 4.76 | AAI-based (high-confidence) | 0 |
| S12-final-viral-combined_k141_117841||full | 1322 | No | NA | 3 | 0 | 2 | Not-determined | Genome-fragment | NA | NA | 0 |
| S5-final-viral-combined_k141_118217||full | 2563 | No | NA | 4 | 0 | 2 | Low-quality | Genome-fragment | 1.45 | AAI-based (medium-confidence) | 0 |
| S5_k141_188809 | 2563 | No | NA | 1 | 1 | 0 | Low-quality | Genome-fragment | 1.58 | HMM-based (lower-bound) | 0 |
| S12-final-viral-combined_k141_134926||full | 1864 | No | NA | 2 | 0 | 0 | Not-determined | Genome-fragment | NA | NA | 0 |
| S12-final-viral-combined_k141_319096||full | 1065 | No | NA | 3 | 0 | 0 | Not-determined | Genome-fragment | NA | NA | 0 |
| S12-final-viral-combined_k141_252070||full | 1350 | No | NA | 3 | 0 | 1 | Not-determined | Genome-fragment | NA | NA | 0 |
| S12-final-viral-combined_k141_56950||full | 2563 | No | NA | 3 | 0 | 0 | Low-quality | Genome-fragment | 3.49 | AAI-based (medium-confidence) | 0 |
| S12-final-viral-combined_k141_34773||full | 2655 | No | NA | 2 | 0 | 1 | Not-determined | Genome-fragment | NA | NA | 0 |
| S12_k141_68113 | 2563 | No | NA | 1 | 0 | 1 | Low-quality | Genome-fragment | 1.26 | AAI-based (medium-confidence) | 0 |
| S11_k141_12648 | 2564 | No | NA | 5 | 1 | 0 | Low-quality | Genome-fragment | 1.93 | AAI-based (high-confidence) | 0 |
| S12-final-viral-combined_k141_117866||full | 1376 | No | NA | 2 | 0 | 0 | Not-determined | Genome-fragment | NA | NA | 0 |
| S12-final-viral-combined_k141_252089||full | 4933 | No | NA | 7 | 0 | 2 | Not-determined | Genome-fragment | NA | NA | 0 |
| S12-final-viral-combined_k141_386064||full | 8169 | No | NA | 8 | 0 | 5 | Not-determined | Genome-fragment | NA | NA | 0 |
| S12-final-viral-combined_k141_101422||full | 3117 | No | NA | 3 | 0 | 0 | Not-determined | Genome-fragment | NA | NA | 0 |
| S12-final-viral-combined_k141_285887||full | 3070 | No | NA | 5 | 0 | 2 | Not-determined | Genome-fragment | NA | NA | 0 |
| S1-final-viral-combined_k141_246788||full | 2564 | No | NA | 6 | 0 | 0 | Low-quality | Genome-fragment | 6.16 | AAI-based (high-confidence) | 0 |
| S12-final-viral-combined_k141_204360||full | 2564 | No | NA | 5 | 0 | 0 | Low-quality | Genome-fragment | 1.93 | AAI-based (medium-confidence) | 0 |
| S12-final-viral-combined_k141_386095||full | 3474 | No | NA | 4 | 0 | 4 | Not-determined | Genome-fragment | NA | NA | 0 |
| S12-final-viral-combined_k141_252154||full | 1622 | No | NA | 7 | 0 | 0 | Not-determined | Genome-fragment | NA | NA | 0 |
| S12-final-viral-combined_k141_185206||full | 12841 | No | NA | 20 | 0 | 1 | Not-determined | Genome-fragment | NA | NA | 0 |
| S12_k141_401223 | 2564 | No | NA | 6 | 1 | 0 | Low-quality | Genome-fragment | 1.86 | HMM-based (lower-bound) | 0 |
| S12-final-viral-combined_k141_352313||full | 5990 | No | NA | 8 | 0 | 2 | Not-determined | Genome-fragment | NA | NA | 0 |
| S12-final-viral-combined_k141_352319||full | 2074 | No | NA | 4 | 0 | 0 | Not-determined | Genome-fragment | NA | NA | 0 |
| S1-final-viral-combined_k141_230930||full | 2565 | No | NA | 4 | 0 | 3 | Low-quality | Genome-fragment | 4.43 | AAI-based (medium-confidence) | 0 |
| S17-final-viral-combined_k141_102133||full | 2565 | No | NA | 6 | 0 | 1 | Low-quality | Genome-fragment | 4.42 | AAI-based (high-confidence) | 0 |
| S12-final-viral-combined_k141_1154||full | 1403 | No | NA | 3 | 0 | 2 | Not-determined | Genome-fragment | NA | NA | 0 |
| S2-final-viral-combined_k141_15697||full | 2565 | No | NA | 4 | 2 | 0 | Low-quality | Genome-fragment | 2.85 | HMM-based (lower-bound) | 0 |
| S12-final-viral-combined_k141_302401||full | 2658 | No | NA | 2 | 0 | 2 | Not-determined | Genome-fragment | NA | NA | 0 |
| S2_k141_80876 | 2565 | No | NA | 6 | 0 | 0 | Low-quality | Genome-fragment | 2.49 | AAI-based (high-confidence) | 0 |
| S12-final-viral-combined_k141_185266||full | 2241 | No | NA | 3 | 0 | 1 | Not-determined | Genome-fragment | NA | NA | 0 |
| S12-final-viral-combined_k141_352366||full | 2184 | No | NA | 3 | 0 | 0 | Not-determined | Genome-fragment | NA | NA | 0 |
| S12-final-viral-combined_k141_369324||full | 3286 | No | NA | 4 | 0 | 1 | Not-determined | Genome-fragment | NA | NA | 0 |
| S12-final-viral-combined_k141_117947||full | 1527 | No | NA | 2 | 0 | 0 | Not-determined | Genome-fragment | NA | NA | 0 |
| S12-final-viral-combined_k141_252173||full | 1958 | No | NA | 2 | 0 | 2 | Not-determined | Genome-fragment | NA | NA | 0 |
| S12-final-viral-combined_k141_101476||full | 2735 | No | NA | 3 | 0 | 1 | Not-determined | Genome-fragment | NA | NA | 0 |
| S4-final-viral-combined_k141_103877||full | 2565 | No | NA | 3 | 0 | 2 | Low-quality | Genome-fragment | 1.33 | AAI-based (medium-confidence) | 0 |
| S17-final-viral-combined_k141_62293||full | 2566 | No | NA | 3 | 0 | 0 | Low-quality | Genome-fragment | 3.92 | AAI-based (medium-confidence) | 0 |
| S1-final-viral-combined_k141_278715||full | 2567 | No | NA | 3 | 0 | 3 | Low-quality | Genome-fragment | 3.42 | AAI-based (medium-confidence) | 0 |
| S12-final-viral-combined_k141_157291||full | 2567 | No | NA | 3 | 0 | 0 | Low-quality | Genome-fragment | 5.9 | AAI-based (high-confidence) | 0 |
| S9-final-viral-combined_k141_100267||full | 2568 | No | NA | 2 | 0 | 0 | Low-quality | Genome-fragment | 5.89 | AAI-based (high-confidence) | 0 |
| S7-final-viral-combined_k141_189259||full | 2568 | No | NA | 4 | 0 | 0 | Low-quality | Genome-fragment | 1.85 | AAI-based (medium-confidence) | 0 |
| S12-final-viral-combined_k141_84828||full | 1105 | No | NA | 2 | 0 | 0 | Not-determined | Genome-fragment | NA | NA | 0 |
| S12-final-viral-combined_k141_369412||full | 1146 | No | NA | 2 | 0 | 0 | Not-determined | Genome-fragment | NA | NA | 0 |
| S2-final-viral-combined_k141_53225||full | 2568 | No | NA | 4 | 0 | 0 | Low-quality | Genome-fragment | 5.73 | AAI-based (medium-confidence) | 0 |
| S11-final-viral-combined_k141_108095||full | 2569 | No | NA | 4 | 1 | 0 | Low-quality | Genome-fragment | 1.55 | HMM-based (lower-bound) | 0 |
| S12-final-viral-combined_k141_118009||full | 2466 | No | NA | 3 | 0 | 0 | Not-determined | Genome-fragment | NA | NA | 0 |
| S12-final-viral-combined_k141_18001||full | 1059 | No | NA | 3 | 0 | 0 | Not-determined | Genome-fragment | NA | NA | 0 |
| S12-final-viral-combined_k141_118020||full | 3111 | No | NA | 4 | 0 | 4 | Not-determined | Genome-fragment | NA | NA | 0 |
| S11_k141_258682 | 2569 | No | NA | 1 | 0 | 0 | Low-quality | Genome-fragment | 2.39 | AAI-based (high-confidence) | 0 |
| S12-final-viral-combined_k141_151690||full | 1931 | No | NA | 3 | 0 | 2 | Not-determined | Genome-fragment | NA | NA | 0 |
| S12-final-viral-combined_k141_369441||full | 1631 | No | NA | 3 | 0 | 0 | Not-determined | Genome-fragment | NA | NA | 0 |
| S1-final-viral-combined_k141_357705||full | 2569 | No | NA | 4 | 2 | 0 | Low-quality | Genome-fragment | 5.36 | AAI-based (high-confidence) | 0 |
| S12-final-viral-combined_k141_252259||full | 2737 | No | NA | 2 | 0 | 2 | Not-determined | Genome-fragment | NA | NA | 0 |
| S5-final-viral-combined_k141_114085||full | 2569 | No | NA | 3 | 0 | 1 | Low-quality | Genome-fragment | 3.39 | AAI-based (medium-confidence) | 0 |
| S17-final-viral-combined_k141_264487||full | 2569 | No | NA | 5 | 0 | 0 | Low-quality | Genome-fragment | 1.83 | AAI-based (medium-confidence) | 0 |
| S12_k141_103817 | 2569 | No | NA | 1 | 0 | 1 | Low-quality | Genome-fragment | 5.46 | AAI-based (medium-confidence) | 0 |
| S12-final-viral-combined_k141_302509||full | 2984 | No | NA | 6 | 0 | 2 | Not-determined | Genome-fragment | NA | NA | 0 |
| S12-final-viral-combined_k141_386249||full | 5263 | No | NA | 3 | 0 | 2 | Not-determined | Genome-fragment | NA | NA | 0 |
| S6-final-viral-combined_k141_97286||full | 2569 | No | NA | 4 | 2 | 0 | Low-quality | Genome-fragment | 6.91 | AAI-based (high-confidence) | 0 |
| S11-final-viral-combined_k141_98394||full | 2570 | No | NA | 8 | 0 | 1 | Low-quality | Genome-fragment | 5.67 | AAI-based (medium-confidence) | 0 |
| S3_k141_71088 | 2570 | No | NA | 1 | 0 | 0 | Low-quality | Genome-fragment | 5.73 | AAI-based (high-confidence) | 0 |
| S12-final-viral-combined_k141_151708||full | 2441 | No | NA | 3 | 0 | 1 | Not-determined | Genome-fragment | NA | NA | 0 |
| S12-final-viral-combined_k141_369514||full | 2063 | No | NA | 2 | 0 | 2 | Not-determined | Genome-fragment | NA | NA | 0 |
| S12-final-viral-combined_k141_335661||full | 1101 | No | NA | 2 | 0 | 0 | Not-determined | Genome-fragment | NA | NA | 0 |
| S12-final-viral-combined_k141_84948||full | 1699 | No | NA | 3 | 0 | 3 | Not-determined | Genome-fragment | NA | NA | 0 |
| S8-final-viral-combined_k141_35150||full | 2570 | No | NA | 4 | 1 | 0 | Low-quality | Genome-fragment | 6.76 | AAI-based (medium-confidence) | 0 |
| S5-final-viral-combined_k141_32313||full | 2571 | No | NA | 5 | 1 | 0 | Low-quality | Genome-fragment | 6.75 | AAI-based (medium-confidence) | 0 |
| S12-final-viral-combined_k141_386284||full | 9454 | No | NA | 9 | 0 | 4 | Not-determined | Genome-fragment | NA | NA | 0 |
| S12-final-viral-combined_k141_118091||full | 2569 | No | NA | 5 | 0 | 0 | Not-determined | Genome-fragment | NA | NA | 0 |
| S12-final-viral-combined_k141_285961||full | 1480 | No | NA | 2 | 0 | 0 | Not-determined | Genome-fragment | NA | NA | 0 |
| S12-final-viral-combined_k141_352503||full | 6962 | No | NA | 8 | 0 | 2 | Not-determined | Genome-fragment | NA | NA | 0 |
| S12-final-viral-combined_k141_118097||full | 1401 | No | NA | 2 | 0 | 2 | Not-determined | Genome-fragment | NA | NA | 0 |
| S17-final-viral-combined_k141_241317||full | 2571 | No | NA | 3 | 0 | 1 | Low-quality | Genome-fragment | 1.81 | AAI-based (high-confidence) | 0 |
| S12-final-viral-combined_k141_57726||full | 2571 | No | NA | 4 | 0 | 2 | Low-quality | Genome-fragment | 2.16 | AAI-based (high-confidence) | 0 |
| S12-final-viral-combined_k141_98773||full | 2571 | No | NA | 3 | 0 | 3 | Low-quality | Genome-fragment | 1.51 | AAI-based (medium-confidence) | 0 |
| S6-final-viral-combined_k141_2158||full | 2571 | No | NA | 3 | 0 | 1 | Low-quality | Genome-fragment | 1.96 | AAI-based (high-confidence) | 0 |
| S1-final-viral-combined_k141_12566||full | 2572 | No | NA | 3 | 1 | 0 | Low-quality | Genome-fragment | 4.86 | HMM-based (lower-bound) | 0 |
| S1_k141_134973 | 2572 | No | NA | 4 | 2 | 0 | Low-quality | Genome-fragment | 5.51 | AAI-based (high-confidence) | 0 |
| S2-final-viral-combined_k141_211310||full | 2572 | No | NA | 2 | 0 | 0 | Low-quality | Genome-fragment | 4.25 | AAI-based (medium-confidence) | 0 |
| S12-final-viral-combined_k141_101578||full | 1602 | No | NA | 3 | 0 | 1 | Not-determined | Genome-fragment | NA | NA | 0 |
| S12-final-viral-combined_k141_202063||full | 1553 | No | NA | 4 | 0 | 1 | Not-determined | Genome-fragment | NA | NA | 0 |
| S4-final-viral-combined_k141_86293||full | 2572 | No | NA | 3 | 2 | 0 | Low-quality | Genome-fragment | 15.2 | AAI-based (high-confidence) | 0 |
| S12-final-viral-combined_k141_287350||full | 2573 | No | NA | 3 | 0 | 2 | Low-quality | Genome-fragment | 1.08 | AAI-based (medium-confidence) | 0 |
| S12-final-viral-combined_k141_366473||full | 2573 | No | NA | 6 | 2 | 0 | Low-quality | Genome-fragment | 2.63 | HMM-based (lower-bound) | 0 |
| S2-final-viral-combined_k141_19294||full | 2573 | No | NA | 3 | 0 | 0 | Low-quality | Genome-fragment | 4.9 | AAI-based (high-confidence) | 0 |
| S12-final-viral-combined_k141_101591||full | 1121 | No | NA | 2 | 0 | 1 | Not-determined | Genome-fragment | NA | NA | 0 |
| S12-final-viral-combined_k141_51499||full | 1608 | No | NA | 2 | 0 | 1 | Not-determined | Genome-fragment | NA | NA | 0 |
| S12-final-viral-combined_k141_386371||full | 2502 | No | NA | 4 | 0 | 0 | Not-determined | Genome-fragment | NA | NA | 0 |
| S1-final-viral-combined_k141_48258||lt2gene | 2574 | No | NA | 1 | 0 | 0 | Low-quality | Genome-fragment | 4.45 | AAI-based (medium-confidence) | 0 |
| S12-final-viral-combined_k141_335753||full | 1476 | No | NA | 2 | 0 | 2 | Not-determined | Genome-fragment | NA | NA | 0 |
| S12-final-viral-combined_k141_218798||full | 1475 | No | NA | 4 | 0 | 0 | Not-determined | Genome-fragment | NA | NA | 0 |
| S7-final-viral-combined_k141_120610||full | 2574 | No | NA | 4 | 1 | 0 | Low-quality | Genome-fragment | 0.76 | HMM-based (lower-bound) | 0 |
| S12-final-viral-combined_k141_137190||full | 2574 | No | NA | 3 | 2 | 0 | Low-quality | Genome-fragment | 5.59 | AAI-based (high-confidence) | 0 |
| S12-final-viral-combined_k141_101615||full | 5080 | No | NA | 6 | 0 | 5 | Not-determined | Genome-fragment | NA | NA | 0 |
| S11-final-viral-combined_k141_126460||full | 2575 | No | NA | 4 | 0 | 3 | Low-quality | Genome-fragment | 3.1 | AAI-based (medium-confidence) | 0 |
| S12-final-viral-combined_k141_185466||full | 1118 | No | NA | 2 | 0 | 0 | Not-determined | Genome-fragment | NA | NA | 0 |
| S12-final-viral-combined_k141_168692||full | 3568 | No | NA | 3 | 0 | 1 | Not-determined | Genome-fragment | NA | NA | 0 |
| S12-final-viral-combined_k141_202070||full | 1802 | No | NA | 4 | 0 | 0 | Not-determined | Genome-fragment | NA | NA | 0 |
| S12-final-viral-combined_k141_135136||full | 1309 | No | NA | 2 | 0 | 0 | Not-determined | Genome-fragment | NA | NA | 0 |
| S12-final-viral-combined_k141_68229||full | 3038 | No | NA | 3 | 0 | 2 | Not-determined | Genome-fragment | NA | NA | 0 |
| S1-final-viral-combined_k141_198558||full | 2575 | No | NA | 5 | 2 | 0 | Low-quality | Genome-fragment | 3.63 | AAI-based (high-confidence) | 0 |
| S12-final-viral-combined_k141_252396||full | 1323 | No | NA | 3 | 0 | 0 | Not-determined | Genome-fragment | NA | NA | 0 |
| S7-final-viral-combined_k141_1582||full | 2575 | No | NA | 5 | 0 | 0 | Low-quality | Genome-fragment | 1.89 | AAI-based (medium-confidence) | 0 |
| S17-k141_261651 | 2575 | No | NA | 2 | 0 | 2 | Low-quality | Genome-fragment | 0.62 | AAI-based (medium-confidence) | 0 |
| S4-final-viral-combined_k141_78106||full | 2575 | No | NA | 3 | 0 | 1 | Low-quality | Genome-fragment | 1.18 | AAI-based (high-confidence) | 0 |
| S4-final-viral-combined_k141_88197||full | 2575 | No | NA | 3 | 0 | 2 | Low-quality | Genome-fragment | 2.03 | AAI-based (medium-confidence) | 0 |
| S12-final-viral-combined_k141_34972||full | 1044 | No | NA | 3 | 0 | 2 | Not-determined | Genome-fragment | NA | NA | 0 |
| S12-final-viral-combined_k141_1486||full | 1184 | No | NA | 2 | 0 | 1 | Not-determined | Genome-fragment | NA | NA | 0 |
| S3-final-viral-combined_k141_57461||full | 2576 | No | NA | 2 | 0 | 0 | Low-quality | Genome-fragment | 2.82 | AAI-based (high-confidence) | 0 |
| S7-final-viral-combined_k141_115610||full | 2576 | No | NA | 5 | 0 | 1 | Low-quality | Genome-fragment | 6.01 | AAI-based (medium-confidence) | 0 |
| S12-final-viral-combined_k141_218872||full | 1747 | No | NA | 4 | 0 | 0 | Not-determined | Genome-fragment | NA | NA | 0 |
| S12-final-viral-combined_k141_85057||full | 1678 | No | NA | 3 | 0 | 1 | Not-determined | Genome-fragment | NA | NA | 0 |
| S12-final-viral-combined_k141_335824||full | 8779 | No | NA | 13 | 0 | 0 | Not-determined | Genome-fragment | NA | NA | 0 |
| S12-final-viral-combined_k141_202113||full | 1687 | No | NA | 2 | 0 | 1 | Not-determined | Genome-fragment | NA | NA | 0 |
| S12-final-viral-combined_k141_185580||full | 1409 | No | NA | 2 | 0 | 1 | Not-determined | Genome-fragment | NA | NA | 0 |
| S12-final-viral-combined_k141_88946||full | 2576 | No | NA | 4 | 0 | 0 | Low-quality | Genome-fragment | 2.79 | AAI-based (medium-confidence) | 0 |
| S12-final-viral-combined_k141_335862||full | 1064 | No | NA | 2 | 0 | 0 | Not-determined | Genome-fragment | NA | NA | 0 |
| S12-final-viral-combined_k141_135220||full | 1571 | No | NA | 2 | 0 | 1 | Not-determined | Genome-fragment | NA | NA | 0 |
| S4-final-viral-combined_k141_97029||full | 2576 | No | NA | 5 | 1 | 0 | Low-quality | Genome-fragment | 7.43 | AAI-based (high-confidence) | 0 |
| S8_k141_256176 | 2576 | No | NA | 4 | 0 | 4 | Low-quality | Genome-fragment | 3.41 | AAI-based (medium-confidence) | 0 |
| S7-final-viral-combined_k141_17362||lt2gene | 2577 | No | NA | 2 | 2 | 0 | Low-quality | Genome-fragment | 7.43 | AAI-based (high-confidence) | 0 |
| S12_k141_336640 | 2577 | No | NA | 3 | 0 | 2 | Low-quality | Genome-fragment | 1.25 | AAI-based (medium-confidence) | 0 |
| S12-final-viral-combined_k141_168768||full | 1204 | No | NA | 3 | 0 | 0 | Not-determined | Genome-fragment | NA | NA | 0 |
| S12-final-viral-combined_k141_319427||full | 3426 | No | NA | 8 | 0 | 3 | Not-determined | Genome-fragment | NA | NA | 0 |
| S12-final-viral-combined_k141_302776||full | 3111 | No | NA | 3 | 0 | 0 | Not-determined | Genome-fragment | NA | NA | 0 |
| S12-final-viral-combined_k141_319448||full | 4302 | No | NA | 4 | 0 | 4 | Not-determined | Genome-fragment | NA | NA | 0 |
| S12-final-viral-combined_k141_235706||full | 6869 | No | NA | 6 | 0 | 4 | Not-determined | Genome-fragment | NA | NA | 0 |
| S12-final-viral-combined_k141_151843||full | 128600 | Yes | 54070 | 153 | 30 | 37 | High-quality | High-quality | 100 | AAI-based (high-confidence) | 57.95 |
| S2_k141_164288 | 2577 | No | NA | 2 | 0 | 2 | Low-quality | Genome-fragment | 5.2 | AAI-based (medium-confidence) | 0 |
| S12-final-viral-combined_k141_352653||full | 1307 | No | NA | 1 | 0 | 0 | Not-determined | Genome-fragment | NA | NA | 0 |
| S9-final-viral-combined_k141_28846||full | 2578 | No | NA | 6 | 0 | 0 | Low-quality | Genome-fragment | 7.25 | AAI-based (medium-confidence) | 0 |
| S11-final-viral-combined_k141_202175||full | 2578 | No | NA | 4 | 0 | 0 | Low-quality | Genome-fragment | 5.67 | AAI-based (high-confidence) | 0 |
| S12-final-viral-combined_k141_85178||full | 2582 | No | NA | 4 | 0 | 0 | Not-determined | Genome-fragment | NA | NA | 0 |
| S11-final-viral-combined_k141_203930||full | 2578 | No | NA | 4 | 0 | 2 | Low-quality | Genome-fragment | 3.54 | AAI-based (medium-confidence) | 0 |
| S12-final-viral-combined_k141_1651||full | 1237 | No | NA | 2 | 0 | 1 | Not-determined | Genome-fragment | NA | NA | 0 |
| S12-final-viral-combined_k141_335956||full | 1067 | No | NA | 2 | 0 | 2 | Not-determined | Genome-fragment | NA | NA | 0 |
| S1-final-viral-combined_k141_115184||full | 2578 | No | NA | 5 | 0 | 2 | Low-quality | Genome-fragment | 3.35 | AAI-based (medium-confidence) | 0 |
| S12-final-viral-combined_k141_1667||full | 2853 | No | NA | 6 | 0 | 0 | Not-determined | Genome-fragment | NA | NA | 0 |
| S5-final-viral-combined_k141_77389||full | 2578 | No | NA | 4 | 0 | 2 | Low-quality | Genome-fragment | 4.39 | AAI-based (medium-confidence) | 0 |
| S12-final-viral-combined_k141_101820||full | 7835 | No | NA | 8 | 0 | 3 | Not-determined | Genome-fragment | NA | NA | 0 |
| S12-final-viral-combined_k141_153792||full | 2578 | No | NA | 6 | 2 | 0 | Low-quality | Genome-fragment | 4.64 | AAI-based (high-confidence) | 0 |
| S12-final-viral-combined_k141_191903||full | 2578 | No | NA | 5 | 2 | 0 | Low-quality | Genome-fragment | 4.37 | AAI-based (high-confidence) | 0 |
| S11-final-viral-combined_k141_287278||full | 2579 | No | NA | 4 | 0 | 3 | Low-quality | Genome-fragment | 1.29 | AAI-based (medium-confidence) | 0 |
| S12-final-viral-combined_k141_168868||full | 2467 | No | NA | 3 | 0 | 1 | Not-determined | Genome-fragment | NA | NA | 0 |
| S12-final-viral-combined_k141_168874||full | 2074 | No | NA | 4 | 0 | 3 | Not-determined | Genome-fragment | NA | NA | 0 |
| S12-final-viral-combined_k141_269289||full | 2925 | No | NA | 5 | 0 | 1 | Not-determined | Genome-fragment | NA | NA | 0 |
| S1-final-viral-combined_k141_252029||full | 2579 | No | NA | 4 | 0 | 1 | Low-quality | Genome-fragment | 4.35 | AAI-based (medium-confidence) | 0 |
| S1-final-viral-combined_k141_114162||full | 2579 | No | NA | 2 | 0 | 0 | Low-quality | Genome-fragment | 6.42 | AAI-based (high-confidence) | 0 |
| S1_k141_69200 | 2579 | No | NA | 3 | 1 | 0 | Low-quality | Genome-fragment | 46.59 | AAI-based (high-confidence) | 0 |
| S7-final-viral-combined_k141_22427||full | 2579 | No | NA | 5 | 1 | 0 | Low-quality | Genome-fragment | 4.13 | HMM-based (lower-bound) | 0 |
| S12-final-viral-combined_k141_364489||full | 2579 | No | NA | 3 | 0 | 2 | Low-quality | Genome-fragment | 3.97 | AAI-based (medium-confidence) | 0 |
| S12-final-viral-combined_k141_85259||full | 1564 | No | NA | 3 | 0 | 1 | Not-determined | Genome-fragment | NA | NA | 0 |
| S12-final-viral-combined_k141_302876||full | 24330 | No | NA | 32 | 18 | 0 | Medium-quality | Genome-fragment | 59.7 | AAI-based (high-confidence) | 0 |
| S9_k141_39365 | 2580 | No | NA | 2 | 0 | 2 | Low-quality | Genome-fragment | 0.72 | AAI-based (high-confidence) | 0 |
| S12-final-viral-combined_k141_51779||full | 1546 | No | NA | 3 | 0 | 1 | Not-determined | Genome-fragment | NA | NA | 0 |
| S5-final-viral-combined_k141_212718||full | 2580 | No | NA | 5 | 3 | 0 | Low-quality | Genome-fragment | 3.42 | AAI-based (high-confidence) | 0 |
| S17-final-viral-combined_k141_122572||full | 2580 | No | NA | 4 | 1 | 0 | Low-quality | Genome-fragment | 4.39 | AAI-based (medium-confidence) | 0 |
| S17-final-viral-combined_k141_137588||full | 2580 | No | NA | 5 | 1 | 0 | Low-quality | Genome-fragment | 6.03 | AAI-based (high-confidence) | 0 |
| S12-final-viral-combined_k141_252542||full | 1367 | No | NA | 2 | 0 | 1 | Not-determined | Genome-fragment | NA | NA | 0 |
| S2-final-viral-combined_k141_235897||full | 2580 | No | NA | 3 | 1 | 0 | Low-quality | Genome-fragment | 5.71 | AAI-based (medium-confidence) | 0 |
| S12-final-viral-combined_k141_68434||full | 6911 | No | NA | 8 | 0 | 7 | Not-determined | Genome-fragment | NA | NA | 0 |
| S4-final-viral-combined_k141_75487||full | 2580 | No | NA | 5 | 3 | 0 | Low-quality | Genome-fragment | 6 | AAI-based (high-confidence) | 0 |
| S8_k141_187539 | 2580 | No | NA | 6 | 2 | 0 | Low-quality | Genome-fragment | 1.95 | AAI-based (high-confidence) | 0 |
| S3-final-viral-combined_k141_59589||full | 2581 | No | NA | 3 | 1 | 0 | Low-quality | Genome-fragment | 2.33 | AAI-based (high-confidence) | 0 |
| S12-final-viral-combined_k141_352837||full | 1436 | No | NA | 4 | 0 | 1 | Not-determined | Genome-fragment | NA | NA | 0 |
| S5_k141_126511 | 2581 | No | NA | 4 | 3 | 0 | Low-quality | Genome-fragment | 8.02 | AAI-based (high-confidence) | 0 |
| S12-final-viral-combined_k141_302937||full | 1353 | No | NA | 3 | 0 | 1 | Not-determined | Genome-fragment | NA | NA | 0 |
| S7-final-viral-combined_k141_4450||full | 2581 | No | NA | 4 | 0 | 0 | Low-quality | Genome-fragment | 2.48 | AAI-based (high-confidence) | 0 |
| S7-final-viral-combined_k141_50236||full | 2581 | No | NA | 3 | 0 | 2 | Low-quality | Genome-fragment | 1.23 | AAI-based (medium-confidence) | 0 |
| S12-final-viral-combined_k141_235889||full | 1768 | No | NA | 2 | 0 | 2 | Not-determined | Genome-fragment | NA | NA | 0 |
| S11-final-viral-combined_k141_132575||full | 2582 | No | NA | 2 | 0 | 0 | Low-quality | Genome-fragment | 6.02 | AAI-based (medium-confidence) | 0 |
| S12-final-viral-combined_k141_319671||full | 5945 | No | NA | 9 | 0 | 2 | Not-determined | Genome-fragment | NA | NA | 0 |
| S12-final-viral-combined_k141_101872||full | 1424 | No | NA | 4 | 0 | 1 | Not-determined | Genome-fragment | NA | NA | 0 |
| S12-final-viral-combined_k141_252596||full | 3880 | No | NA | 6 | 0 | 0 | Not-determined | Genome-fragment | NA | NA | 0 |
| S1_k141_241905 | 2582 | No | NA | 7 | 1 | 0 | Low-quality | Genome-fragment | 7.28 | AAI-based (high-confidence) | 0 |
| S12-final-viral-combined_k141_1802||full | 5356 | No | NA | 8 | 0 | 2 | Not-determined | Genome-fragment | NA | NA | 0 |
| S5-final-viral-combined_k141_27682||full | 2582 | No | NA | 7 | 1 | 0 | Low-quality | Genome-fragment | 3.36 | HMM-based (lower-bound) | 0 |
| S5-final-viral-combined_k141_194488||full | 2582 | No | NA | 4 | 0 | 0 | Low-quality | Genome-fragment | 2.51 | AAI-based (medium-confidence) | 0 |
| S12-final-viral-combined_k141_118387||full | 3408 | No | NA | 3 | 0 | 2 | Not-determined | Genome-fragment | NA | NA | 0 |
| S2-final-viral-combined_k141_209667||full | 2582 | No | NA | 4 | 2 | 0 | Low-quality | Genome-fragment | 6.13 | AAI-based (medium-confidence) | 0 |
| S4_k141_155825 | 2582 | No | NA | 2 | 0 | 2 | Low-quality | Genome-fragment | 4.37 | AAI-based (high-confidence) | 0 |
| S12-final-viral-combined_k141_303022||full | 1391 | No | NA | 3 | 0 | 1 | Not-determined | Genome-fragment | NA | NA | 0 |
| S4_k141_122166 | 2582 | No | NA | 5 | 1 | 0 | Low-quality | Genome-fragment | 2.6 | AAI-based (high-confidence) | 0 |
| S1-final-viral-combined_k141_199470||full | 2583 | Yes | 510 | 3 | 1 | 1 | Low-quality | Genome-fragment | 12.26 | HMM-based (lower-bound) | 80.26 |
| S12-final-viral-combined_k141_235945||full | 1717 | No | NA | 3 | 0 | 0 | Not-determined | Genome-fragment | NA | NA | 0 |
| S12-final-viral-combined_k141_101942||full | 1711 | No | NA | 2 | 0 | 0 | Not-determined | Genome-fragment | NA | NA | 0 |
| S12-final-viral-combined_k141_319746||full | 4977 | No | NA | 6 | 0 | 6 | Not-determined | Genome-fragment | NA | NA | 0 |
| S12-final-viral-combined_k141_219180||full | 4714 | No | NA | 4 | 0 | 3 | Not-determined | Genome-fragment | NA | NA | 0 |
| S1-final-viral-combined_k141_339278||full | 2583 | No | NA | 4 | 0 | 1 | Low-quality | Genome-fragment | 2.44 | AAI-based (medium-confidence) | 0 |
| S11-final-viral-combined_k141_297662||full | 2584 | No | NA | 2 | 0 | 0 | Low-quality | Genome-fragment | 2.68 | AAI-based (medium-confidence) | 0 |
| S12-final-viral-combined_k141_169057||full | 2770 | No | NA | 3 | 0 | 2 | Not-determined | Genome-fragment | NA | NA | 0 |
| S12-final-viral-combined_k141_370082||full | 2373 | No | NA | 4 | 0 | 0 | Not-determined | Genome-fragment | NA | NA | 0 |
| S1-final-viral-combined_k141_55930||full | 2584 | No | NA | 3 | 0 | 0 | Low-quality | Genome-fragment | 5.32 | AAI-based (medium-confidence) | 0 |
| S12-final-viral-combined_k141_68518||full | 4083 | No | NA | 4 | 0 | 0 | Not-determined | Genome-fragment | NA | NA | 0 |
| S12-final-viral-combined_k141_18287||full | 98430 | Yes | 32904 | 109 | 17 | 33 | Medium-quality | Genome-fragment | 73.31 | AAI-based (high-confidence) | 66.57 |
| S12-final-viral-combined_k141_85397||full | 1545 | No | NA | 3 | 0 | 2 | Not-determined | Genome-fragment | NA | NA | 0 |

| | | | | | | | | | | | |
|---|---|---|---|---|---|---|---|---|---|---|---|
| S1-final-viral-combined_k141_357156\|\|full | 2584 | No | NA | 1 | 0 | 0 | Low-quality | Genome-fragment | 2.96 | AAI-based (medium-confidence) | 0 |
| S7-final-viral-combined_k141_228771\|\|full | 2584 | No | NA | 3 | 0 | 2 | Low-quality | Genome-fragment | 1.84 | AAI-based (high-confidence) | 0 |
| S12-final-viral-combined_k141_18318\|\|full | 1049 | No | NA | 2 | 0 | 1 | Not-determined | Genome-fragment | NA | NA | 0 |
| S12-final-viral-combined_k141_352905\|\|full | 3932 | No | NA | 5 | 0 | 2 | Not-determined | Genome-fragment | NA | NA | 0 |
| S12-final-viral-combined_k141_102029\|\|full | 1849 | No | NA | 3 | 0 | 2 | Not-determined | Genome-fragment | NA | NA | 0 |
| S12-final-viral-combined_k141_370112\|\|full | 1778 | No | NA | 3 | 0 | 0 | Not-determined | Genome-fragment | NA | NA | 0 |
| S12-final-viral-combined_k141_370117\|\|full | 1071 | No | NA | 3 | 0 | 1 | Not-determined | Genome-fragment | NA | NA | 0 |
| S8-final-viral-combined_k141_208821\|\|full | 2584 | No | NA | 3 | 0 | 2 | Low-quality | Genome-fragment | 1.2 | AAI-based (medium-confidence) | 0 |
| S12-final-viral-combined_k141_118483\|\|full | 1725 | No | NA | 2 | 0 | 2 | Not-determined | Genome-fragment | NA | NA | 0 |
| S12-final-viral-combined_k141_152275\|\|full | 7050 | No | NA | 10 | 0 | 0 | Not-determined | Genome-fragment | NA | NA | 0 |
| S12-final-viral-combined_k141_219219\|\|full | 3551 | No | NA | 4 | 0 | 3 | Not-determined | Genome-fragment | NA | NA | 0 |
| S12-final-viral-combined_k141_202351\|\|full | 3676 | No | NA | 4 | 0 | 1 | Not-determined | Genome-fragment | NA | NA | 0 |
| S8-final-viral-combined_k141_205633\|\|full | 2584 | No | NA | 4 | 0 | 2 | Low-quality | Genome-fragment | 3.57 | AAI-based (medium-confidence) | 0 |
| S12-final-viral-combined_k141_252744\|\|full | 2768 | No | NA | 6 | 0 | 3 | Not-determined | Genome-fragment | NA | NA | 0 |
| S12-final-viral-combined_k141_370127\|\|full | 4477 | No | NA | 3 | 0 | 3 | Not-determined | Genome-fragment | NA | NA | 0 |
| S12-final-viral-combined_k141_269459\|\|full | 2862 | No | NA | 4 | 0 | 1 | Not-determined | Genome-fragment | NA | NA | 0 |
| S12-final-viral-combined_k141_219245\|\|full | 1997 | No | NA | 2 | 0 | 2 | Not-determined | Genome-fragment | NA | NA | 0 |
| S12-final-viral-combined_k141_202371\|\|full | 4970 | No | NA | 3 | 0 | 2 | Not-determined | Genome-fragment | NA | NA | 0 |
| S1-final-viral-combined_k141_209665\|\|full | 2585 | No | NA | 4 | 0 | 1 | Low-quality | Genome-fragment | 5.91 | AAI-based (medium-confidence) | 0 |
| S12-final-viral-combined_k141_236129\|\|full | 2009 | No | NA | 3 | 0 | 0 | Not-determined | Genome-fragment | NA | NA | 0 |
| S7-final-viral-combined_k141_112927\|\|full | 2585 | No | NA | 3 | 0 | 1 | Low-quality | Genome-fragment | 1.84 | AAI-based (medium-confidence) | 0 |
| S12-final-viral-combined_k141_152353\|\|full | 1726 | No | NA | 3 | 0 | 2 | Not-determined | Genome-fragment | NA | NA | 0 |
| S7-final-viral-combined_k141_62583\|\|full | 2585 | No | NA | 6 | 1 | 0 | Low-quality | Genome-fragment | 8.5 | AAI-based (high-confidence) | 0 |
| S11-final-viral-combined_k141_20479\|\|full | 2586 | No | NA | 6 | 4 | 0 | Low-quality | Genome-fragment | 3.96 | HMM-based (lower-bound) | 0 |
| S1-final-viral-combined_k141_68294\|\|full | 2586 | No | NA | 4 | 0 | 0 | Low-quality | Genome-fragment | 5.76 | AAI-based (high-confidence) | 0 |
| S2-final-viral-combined_k141_73512\|\|full | 2586 | No | NA | 2 | 0 | 1 | Low-quality | Genome-fragment | 0.62 | AAI-based (medium-confidence) | 0 |
| S12-final-viral-combined_k141_303251\|\|full | 2664 | No | NA | 4 | 0 | 2 | Not-determined | Genome-fragment | NA | NA | 0 |
| S12-final-viral-combined_k141_219335\|\|full | 2908 | No | NA | 3 | 0 | 1 | Not-determined | Genome-fragment | NA | NA | 0 |
| S12-final-viral-combined_k141_236230\|\|full | 1725 | No | NA | 4 | 0 | 0 | Not-determined | Genome-fragment | NA | NA | 0 |
| S12-final-viral-combined_k141_236231\|\|full | 1334 | No | NA | 3 | 0 | 0 | Not-determined | Genome-fragment | NA | NA | 0 |
| S12-final-viral-combined_k141_18387\|\|full | 1190 | No | NA | 1 | 0 | 1 | Not-determined | Genome-fragment | NA | NA | 0 |
| S12-final-viral-combined_k141_135613\|\|full | 2078 | No | NA | 2 | 0 | 2 | Not-determined | Genome-fragment | NA | NA | 0 |
| S12-final-viral-combined_k141_202426\|\|full | 2355 | No | NA | 3 | 0 | 2 | Not-determined | Genome-fragment | NA | NA | 0 |
| S12-final-viral-combined_k141_35556\|\|full | 3765 | No | NA | 4 | 0 | 3 | Not-determined | Genome-fragment | NA | NA | 0 |
| S12-final-viral-combined_k141_169162\|\|full | 1946 | No | NA | 3 | 0 | 1 | Not-determined | Genome-fragment | NA | NA | 0 |
| S12-final-viral-combined_k141_269492\|\|full | 2601 | No | NA | 5 | 0 | 0 | Not-determined | Genome-fragment | NA | NA | 0 |
| S12-final-viral-combined_k141_353018\|\|full | 5519 | No | NA | 6 | 0 | 1 | Not-determined | Genome-fragment | NA | NA | 0 |
| S6-final-viral-combined_k141_131931\|\|full | 2586 | No | NA | 5 | 1 | 0 | Low-quality | Genome-fragment | 7.71 | AAI-based (high-confidence) | 0 |
| S12-final-viral-combined_k141_52054\|\|full | 1929 | No | NA | 3 | 0 | 3 | Not-determined | Genome-fragment | NA | NA | 0 |
| S12-final-viral-combined_k141_2024\|\|full | 1682 | No | NA | 2 | 0 | 0 | Not-determined | Genome-fragment | NA | NA | 0 |
| S12-final-viral-combined_k141_386896\|\|full | 1154 | No | NA | 3 | 0 | 1 | Not-determined | Genome-fragment | NA | NA | 0 |
| S8_k141_41578 | 2586 | No | NA | 2 | 1 | 0 | Low-quality | Genome-fragment | 4.93 | AAI-based (high-confidence) | 0 |
| S12-final-viral-combined_k141_202482\|\|full | 2200 | No | NA | 3 | 0 | 0 | Not-determined | Genome-fragment | NA | NA | 0 |
| S11-final-viral-combined_k141_255075\|\|full | 2587 | No | NA | 2 | 0 | 2 | Low-quality | Genome-fragment | 2.98 | AAI-based (medium-confidence) | 0 |
| S1-final-viral-combined_k141_314933\|\|full | 2587 | No | NA | 2 | 0 | 1 | Low-quality | Genome-fragment | 1.65 | AAI-based (medium-confidence) | 0 |
| S1-final-viral-combined_k141_255845\|\|full | 2587 | No | NA | 7 | 0 | 0 | Low-quality | Genome-fragment | 6.61 | AAI-based (high-confidence) | 0 |
| S12-final-viral-combined_k141_362846\|\|lt2gene | 2587 | No | NA | 1 | 1 | 0 | Low-quality | Genome-fragment | 4.31 | HMM-based (lower-bound) | 0 |
| S6-final-viral-combined_k141_109910\|\|full | 2587 | No | NA | 2 | 0 | 1 | Low-quality | Genome-fragment | 2.72 | AAI-based (high-confidence) | 0 |
| S12-final-viral-combined_k141_85527\|\|full | 12368 | No | NA | 13 | 0 | 0 | Not-determined | Genome-fragment | NA | NA | 0 |
| S12-final-viral-combined_k141_102098\|\|full | 2226 | No | NA | 4 | 0 | 3 | Not-determined | Genome-fragment | NA | NA | 0 |
| S12-final-viral-combined_k141_85540\|\|full | 1725 | No | NA | 3 | 0 | 2 | Not-determined | Genome-fragment | NA | NA | 0 |
| S8-final-viral-combined_k141_67001\|\|full | 2587 | No | NA | 2 | 0 | 2 | Low-quality | Genome-fragment | 1.8 | AAI-based (medium-confidence) | 0 |
| S8_k141_215861 | 2587 | No | NA | 2 | 0 | 2 | Low-quality | Genome-fragment | 4.98 | AAI-based (medium-confidence) | 0 |
| S12-final-viral-combined_k141_152507\|\|full | 2073 | No | NA | 2 | 0 | 1 | Not-determined | Genome-fragment | NA | NA | 0 |
| S12-final-viral-combined_k141_353098\|\|full | 1561 | No | NA | 4 | 0 | 1 | Not-determined | Genome-fragment | NA | NA | 0 |
| S12-final-viral-combined_k141_353101\|\|full | 1768 | No | NA | 2 | 0 | 2 | Not-determined | Genome-fragment | NA | NA | 0 |
| S12-final-viral-combined_k141_386943\|\|full | 8891 | No | NA | 10 | 0 | 5 | Not-determined | Genome-fragment | NA | NA | 0 |
| S12-final-viral-combined_k141_152515\|\|full | 1082 | No | NA | 1 | 0 | 0 | Not-determined | Genome-fragment | NA | NA | 0 |
| S12-final-viral-combined_k141_52101\|\|full | 24293 | No | NA | 39 | 22 | 0 | Medium-quality | Genome-fragment | 62.44 | AAI-based (medium-confidence) | 0 |
| S1-final-viral-combined_k141_237111\|\|full | 2588 | No | NA | 5 | 1 | 0 | Low-quality | Genome-fragment | 1.94 | AAI-based (high-confidence) | 0 |
| S5-final-viral-combined_k141_205773\|\|full | 2588 | No | NA | 6 | 0 | 0 | Low-quality | Genome-fragment | 1.5 | AAI-based (medium-confidence) | 0 |
| S12-final-viral-combined_k141_118679\|\|full | 1277 | No | NA | 2 | 0 | 0 | Not-determined | Genome-fragment | NA | NA | 0 |
| S12-final-viral-combined_k141_320025\|\|full | 2337 | No | NA | 3 | 0 | 0 | Not-determined | Genome-fragment | NA | NA | 0 |
| S12-final-viral-combined_k141_102131\|\|full | 9281 | No | NA | 11 | 0 | 2 | Not-determined | Genome-fragment | NA | NA | 0 |
| S12-final-viral-combined_k141_320037\|\|full | 3368 | No | NA | 4 | 0 | 0 | Not-determined | Genome-fragment | NA | NA | 0 |
| S12-final-viral-combined_k141_269542\|\|full | 10737 | No | NA | 12 | 0 | 5 | Not-determined | Genome-fragment | NA | NA | 0 |
| S12-final-viral-combined_k141_12179\|\|full | 2588 | No | NA | 3 | 0 | 1 | Low-quality | Genome-fragment | 5.71 | AAI-based (medium-confidence) | 0 |
| S12-final-viral-combined_k141_353150\|\|full | 5505 | No | NA | 10 | 0 | 4 | Not-determined | Genome-fragment | NA | NA | 0 |
| S2-final-viral-combined_k141_190359\|\|full | 2588 | No | NA | 3 | 0 | 0 | Low-quality | Genome-fragment | 3.62 | AAI-based (medium-confidence) | 0 |
| S8-final-viral-combined_k141_83957\|\|full | 2588 | No | NA | 4 | 0 | 2 | Low-quality | Genome-fragment | 4.61 | AAI-based (medium-confidence) | 0 |
| S12-final-viral-combined_k141_299184\|\|full | 2589 | No | NA | 2 | 0 | 0 | Low-quality | Genome-fragment | 3.19 | AAI-based (medium-confidence) | 0 |
| S12-final-viral-combined_k141_219461\|\|full | 1699 | No | NA | 2 | 0 | 0 | Not-determined | Genome-fragment | NA | NA | 0 |
| S2-final-viral-combined_k141_17499\|\|full | 2589 | No | NA | 3 | 0 | 0 | Low-quality | Genome-fragment | 6.6 | AAI-based (high-confidence) | 0 |
| S12-final-viral-combined_k141_386990\|\|full | 4867 | No | NA | 8 | 0 | 3 | Not-determined | Genome-fragment | NA | NA | 0 |
| S12-final-viral-combined_k141_68794\|\|full | 4982 | No | NA | 5 | 0 | 4 | Not-determined | Genome-fragment | NA | NA | 0 |
| S6-final-viral-combined_k141_92584\|\|full | 2589 | No | NA | 4 | 0 | 0 | Low-quality | Genome-fragment | 4.9 | AAI-based (medium-confidence) | 0 |
| S9-final-viral-combined_k141_120932\|\|full | 2590 | No | NA | 6 | 2 | 0 | Low-quality | Genome-fragment | 6.27 | AAI-based (high-confidence) | 0 |
| S12-final-viral-combined_k141_85666\|\|full | 1011 | No | NA | 2 | 0 | 0 | Not-determined | Genome-fragment | NA | NA | 0 |
| [illegible] | | | | | | | | | | | |

| | | | | | | | | | | | |
|---|---|---|---|---|---|---|---|---|---|---|---|
| S12-final-viral-combined_k141_203864\|\|full | 1198 | No | NA | 3 | 0 | 0 | Not-determined | Genome-fragment | NA | NA | 0 |
| S12-final-viral-combined_k141_119887\|\|full | 4280 | No | NA | 5 | 0 | 3 | Not-determined | Genome-fragment | NA | NA | 0 |
| S12-final-viral-combined_k141_170468\|\|full | 1003 | No | NA | 2 | 0 | 1 | Not-determined | Genome-fragment | NA | NA | 0 |
| S12-final-viral-combined_k141_3213\|\|full | 2622 | No | NA | 3 | 0 | 1 | Not-determined | Genome-fragment | NA | NA | 0 |
| S9-final-viral-combined_k141_127484\|\|full | 2636 | No | NA | 4 | 0 | 0 | Low-quality | Genome-fragment | 5.94 | AAI-based (high-confidence) | 0 |
| S12-final-viral-combined_k141_354315\|\|full | 3264 | No | NA | 4 | 0 | 1 | Not-determined | Genome-fragment | NA | NA | 0 |
| S12-final-viral-combined_k141_269508\|\|full | 2636 | Yes | 1197 | 5 | 2 | 1 | Low-quality | Genome-fragment | 3.51 | AAI-based (high-confidence) | 54.59 |
| S12-final-viral-combined_k141_70599\|\|full | 2636 | No | NA | 3 | 0 | 1 | Low-quality | Genome-fragment | 1.52 | AAI-based (medium-confidence) | 0 |
| S12-final-viral-combined_k141_36754\|\|full | 1303 | No | NA | 2 | 0 | 0 | Not-determined | Genome-fragment | NA | NA | 0 |
| S12_k141_190093 | 2636 | No | NA | 5 | 0 | 2 | Low-quality | Genome-fragment | 1.95 | AAI-based (medium-confidence) | 0 |
| S11_k141_94817 | 2637 | No | NA | 6 | 1 | 0 | Low-quality | Genome-fragment | 2.8 | AAI-based (high-confidence) | 0 |
| S12-final-viral-combined_k141_220491\|\|full | 2016 | No | NA | 5 | 0 | 0 | Not-determined | Genome-fragment | NA | NA | 0 |
| S12-final-viral-combined_k141_36771\|\|full | 5066 | No | NA | 5 | 0 | 4 | Not-determined | Genome-fragment | NA | NA | 0 |
| S12-final-viral-combined_k141_354337\|\|full | 18953 | No | NA | 15 | 0 | 12 | Not-determined | Genome-fragment | NA | NA | 0 |
| S12-final-viral-combined_k141_137097\|\|full | 2127 | No | NA | 3 | 0 | 2 | Not-determined | Genome-fragment | NA | NA | 0 |
| S12-final-viral-combined_k141_137109\|\|full | 2497 | No | NA | 2 | 0 | 1 | Not-determined | Genome-fragment | NA | NA | 0 |
| S12-final-viral-combined_k141_103216\|\|full | 1158 | No | NA | 3 | 0 | 1 | Not-determined | Genome-fragment | NA | NA | 0 |
| S1-final-viral-combined_k141_372347\|\|full | 2637 | No | NA | 6 | 1 | 0 | Low-quality | Genome-fragment | 6.79 | AAI-based (high-confidence) | 0 |
| S11-final-viral-combined_k141_262871\|\|full | 2638 | No | NA | 3 | 0 | 1 | Low-quality | Genome-fragment | 1.09 | AAI-based (medium-confidence) | 0 |
| S12-final-viral-combined_k141_3252\|\|full | 2761 | No | NA | 3 | 0 | 1 | Not-determined | Genome-fragment | NA | NA | 0 |
| S12-final-viral-combined_k141_203935\|\|full | 2308 | No | NA | 2 | 0 | 0 | Not-determined | Genome-fragment | NA | NA | 0 |
| S12-final-viral-combined_k141_253753\|\|full | 2586 | No | NA | 4 | 0 | 2 | Not-determined | Genome-fragment | NA | NA | 0 |
| S12-final-viral-combined_k141_220534\|\|full | 1091 | No | NA | 3 | 0 | 1 | Not-determined | Genome-fragment | NA | NA | 0 |
| S12-final-viral-combined_k141_287740\|\|full | 1912 | No | NA | 3 | 0 | 2 | Not-determined | Genome-fragment | NA | NA | 0 |
| S3-final-viral-combined_k141_70145\|\|full | 2638 | No | NA | 4 | 0 | 0 | Low-quality | Genome-fragment | 2.53 | AAI-based (high-confidence) | 0 |
| S12-final-viral-combined_k141_86826\|\|full | 3622 | No | NA | 4 | 0 | 3 | Not-determined | Genome-fragment | NA | NA | 0 |
| S12-final-viral-combined_k141_253773\|\|full | 2875 | No | NA | 4 | 0 | 3 | Not-determined | Genome-fragment | NA | NA | 0 |
| S12-final-viral-combined_k141_220573\|\|full | 1236 | No | NA | 2 | 0 | 1 | Not-determined | Genome-fragment | NA | NA | 0 |
| S12-final-viral-combined_k141_53406\|\|full | 4813 | No | NA | 4 | 0 | 3 | Not-determined | Genome-fragment | NA | NA | 0 |
| S12-final-viral-combined_k141_237333\|\|full | 2721 | No | NA | 4 | 0 | 4 | Not-determined | Genome-fragment | NA | NA | 0 |
| S7-final-viral-combined_k141_148766\|\|full | 2638 | No | NA | 4 | 0 | 0 | Low-quality | Genome-fragment | 2.94 | AAI-based (medium-confidence) | 0 |
| S17-final-viral-combined_k141_37610\|\|full | 2639 | No | NA | 4 | 0 | 0 | Low-quality | Genome-fragment | 6.93 | AAI-based (high-confidence) | 0 |
| S12-final-viral-combined_k141_53417\|\|full | 5323 | No | NA | 4 | 0 | 4 | Not-determined | Genome-fragment | NA | NA | 0 |
| S12-final-viral-combined_k141_388107\|\|full | 6265 | No | NA | 6 | 0 | 4 | Not-determined | Genome-fragment | NA | NA | 0 |
| S5-final-viral-combined_k141_55735\|\|full | 2640 | No | NA | 4 | 0 | 0 | Low-quality | Genome-fragment | 4.8 | AAI-based (high-confidence) | 0 |
| S2-final-viral-combined_k141_207276\|\|full | 2640 | No | NA | 3 | 0 | 2 | Low-quality | Genome-fragment | 5.91 | AAI-based (medium-confidence) | 0 |
| S12-final-viral-combined_k141_86871\|\|full | 2711 | No | NA | 6 | 0 | 1 | Not-determined | Genome-fragment | NA | NA | 0 |
| S12-final-viral-combined_k141_237382\|\|full | 1697 | No | NA | 3 | 0 | 1 | Not-determined | Genome-fragment | NA | NA | 0 |
| S12-final-viral-combined_k141_287800\|\|full | 11530 | No | NA | 9 | 0 | 6 | Not-determined | Genome-fragment | NA | NA | 0 |
| S12-final-viral-combined_k141_153821\|\|full | 1120 | No | NA | 3 | 0 | 0 | Not-determined | Genome-fragment | NA | NA | 0 |
| S12-final-viral-combined_k141_120047\|\|full | 1343 | No | NA | 1 | 0 | 1 | Not-determined | Genome-fragment | NA | NA | 0 |
| S11-final-viral-combined_k141_264978\|\|full | 2641 | Yes | 566 | 4 | 1 | 1 | Low-quality | Genome-fragment | 0.15 | HMM-based (lower-bound) | 78.57 |
| S1-final-viral-combined_k141_49696\|\|full | 2641 | No | NA | 3 | 0 | 2 | Low-quality | Genome-fragment | 1.98 | AAI-based (medium-confidence) | 0 |
| S1-final-viral-combined_k141_150531\|\|full | 2641 | No | NA | 4 | 1 | 0 | Low-quality | Genome-fragment | 5.63 | AAI-based (high-confidence) | 0 |
| S12-final-viral-combined_k141_304787\|\|full | 3779 | No | NA | 3 | 0 | 2 | Not-determined | Genome-fragment | NA | NA | 0 |
| S12-final-viral-combined_k141_206109\|\|full | 2641 | No | NA | 2 | 0 | 0 | Low-quality | Genome-fragment | 1.75 | AAI-based (high-confidence) | 0 |
| S12-final-viral-combined_k141_337466\|\|full | 1063 | No | NA | 3 | 0 | 2 | Not-determined | Genome-fragment | NA | NA | 0 |
| S1-final-viral-combined_k141_278799\|\|full | 2642 | No | NA | 4 | 1 | 0 | Low-quality | Genome-fragment | 1.4 | HMM-based (lower-bound) | 0 |
| S12-final-viral-combined_k141_53511\|\|full | 2009 | No | NA | 3 | 0 | 0 | Not-determined | Genome-fragment | NA | NA | 0 |
| S12-final-viral-combined_k141_120072\|\|full | 1317 | No | NA | 3 | 0 | 0 | Not-determined | Genome-fragment | NA | NA | 0 |
| S1_k141_305650 | 2642 | No | NA | 4 | 0 | 0 | Low-quality | Genome-fragment | 32.07 | AAI-based (medium-confidence) | 0 |
| S5-final-viral-combined_k141_131545\|\|full | 2642 | No | NA | 2 | 0 | 0 | Low-quality | Genome-fragment | 6.5 | AAI-based (medium-confidence) | 0 |
| S17-final-viral-combined_k141_186141\|\|full | 2642 | No | NA | 6 | 0 | 0 | Low-quality | Genome-fragment | 2.71 | AAI-based (medium-confidence) | 0 |
| S12-final-viral-combined_k141_237502\|\|full | 2678 | No | NA | 7 | 0 | 0 | Not-determined | Genome-fragment | NA | NA | 0 |
| S12-final-viral-combined_k141_220648\|\|full | 16404 | No | NA | 16 | 0 | 11 | Not-determined | Genome-fragment | NA | NA | 0 |
| S12-final-viral-combined_k141_354505\|\|full | 2741 | No | NA | 5 | 0 | 0 | Not-determined | Genome-fragment | NA | NA | 0 |
| S12-final-viral-combined_k141_70539\|\|full | 2088 | No | NA | 3 | 0 | 0 | Not-determined | Genome-fragment | NA | NA | 0 |
| S12-final-viral-combined_k141_95732\|\|full | 2642 | No | NA | 5 | 0 | 0 | Low-quality | Genome-fragment | 2.92 | AAI-based (high-confidence) | 0 |
| S1-final-viral-combined_k141_200344\|\|full | 2643 | No | NA | 4 | 0 | 0 | Low-quality | Genome-fragment | 4.5 | AAI-based (medium-confidence) | 0 |
| S12-final-viral-combined_k141_69786\|\|full | 2643 | No | NA | 3 | 1 | 0 | Low-quality | Genome-fragment | 5.57 | HMM-based (lower-bound) | 0 |
| S12-final-viral-combined_k141_170685\|\|full | 1564 | No | NA | 3 | 0 | 1 | Not-determined | Genome-fragment | NA | NA | 0 |
| S12-final-viral-combined_k141_244123\|\|lt2gene | 2643 | No | NA | 1 | 1 | 0 | Low-quality | Genome-fragment | 1.96 | HMM-based (lower-bound) | 0 |
| S12-final-viral-combined_k141_287937\|\|full | 1202 | No | NA | 3 | 0 | 1 | Not-determined | Genome-fragment | NA | NA | 0 |
| S12-final-viral-combined_k141_388206\|\|full | 1133 | No | NA | 1 | 0 | 0 | Not-determined | Genome-fragment | NA | NA | 0 |
| S12-final-viral-combined_k141_270898\|\|full | 1051 | No | NA | 2 | 0 | 0 | Not-determined | Genome-fragment | NA | NA | 0 |
| S6-final-viral-combined_k141_78806\|\|full | 2643 | No | NA | 7 | 1 | 0 | Low-quality | Genome-fragment | 4.81 | AAI-based (medium-confidence) | 0 |
| S12-final-viral-combined_k141_86937\|\|full | 1611 | No | NA | 2 | 0 | 0 | Not-determined | Genome-fragment | NA | NA | 0 |
| S12-final-viral-combined_k141_86940\|\|full | 4138 | No | NA | 3 | 0 | 2 | Not-determined | Genome-fragment | NA | NA | 0 |
| S12-final-viral-combined_k141_287973\|\|full | 1008 | No | NA | 2 | 0 | 0 | Not-determined | Genome-fragment | NA | NA | 0 |
| S6_k141_41535 | 2643 | No | NA | 4 | 1 | 0 | Low-quality | Genome-fragment | 8.2 | AAI-based (high-confidence) | 0 |
| S12-final-viral-combined_k141_287999\|\|full | 2267 | No | NA | 3 | 0 | 1 | Not-determined | Genome-fragment | NA | NA | 0 |
| S8-final-viral-combined_k141_164316\|\|full | 2643 | No | NA | 6 | 4 | 0 | Low-quality | Genome-fragment | 6.73 | AAI-based (medium-confidence) | 0 |
| S1-final-viral-combined_k141_364989\|\|full | 2644 | No | NA | 5 | 0 | 0 | Low-quality | Genome-fragment | 6.88 | AAI-based (high-confidence) | 0 |
| S1-final-viral-combined_k141_122545\|\|full | 2644 | No | NA | 6 | 0 | 0 | Low-quality | Genome-fragment | 4.59 | AAI-based (medium-confidence) | 0 |
| S12-final-viral-combined_k141_288014\|\|full | 5345 | No | NA | 5 | 0 | 3 | Not-determined | Genome-fragment | NA | NA | 0 |
| S5-final-viral-combined_k141_111093\|\|full | 2644 | No | NA | 4 | 3 | 0 | Low-quality | Genome-fragment | 5.05 | AAI-based (medium-confidence) | 0 |
| S12-final-viral-combined_k141_204160\|\|full | 2582 | No | NA | 4 | 0 | 3 | Not-determined | Genome-fragment | NA | NA | 0 |
| S12-final-viral-combined_k141_204184\|\|full | 2061 | No | NA | 2 | 0 | 2 | Not-determined | Genome-fragment | NA | NA | 0 |
| S12-final-viral-combined_k141_3440\|\|full | 9871 | No | NA | 8 | 0 | 5 | Not-determined | Genome-fragment | NA | NA | 0 |
| S12-final-viral-combined_k141_288026\|\|full | 10642 | No | NA | 8 | 0 | 5 | Not-determined | Genome-fragment | NA | NA | 0 |
| S12_k141_11548 | 2644 | No | NA | 3 | 0 | 1 | Low-quality | Genome-fragment | 5.98 | AAI-based (high-confidence) | 0 |
| S12-final-viral-combined_k141_388296\|\|full | 1716 | No | NA | 2 | 0 | 2 | Not-determined | Genome-fragment | NA | NA | 0 |
| S12-final-viral-combined_k141_120133\|\|full | 43578 | Yes | 29566 | 61 | 19 | 12 | Medium-quality | Genome-fragment | 86.79 | AAI-based (high-confidence) | 32.15 |
| S4-final-viral-combined_k141_19331\|\|full | 2644 | No | NA | 2 | 0 | 1 | Low-quality | Genome-fragment | 5.56 | AAI-based (medium-confidence) | 0 |
| S12-final-viral-combined_k141_19780\|\|full | 5050 | No | NA | 7 | 0 | 3 | Not-determined | Genome-fragment | NA | NA | 0 |
| S8-final-viral-combined_k141_228464\|\|full | 2644 | No | NA | 5 | 0 | 0 | Low-quality | Genome-fragment | 5.57 | AAI-based (medium-confidence) | 0 |
| S9-final-viral-combined_k141_103855\|\|full | 2645 | No | NA | 4 | 0 | 1 | Low-quality | Genome-fragment | 1.03 | AAI-based (medium-confidence) | 0 |
| S7-final-viral-combined_k141_215172\|\|full | 2645 | No | NA | 5 | 0 | 1 | Low-quality | Genome-fragment | 3.49 | AAI-based (medium-confidence) | 0 |
| S12-final-viral-combined_k141_137325\|\|full | 1349 | No | NA | 2 | 0 | 0 | Not-determined | Genome-fragment | NA | NA | 0 |
| S12-final-viral-combined_k141_204258\|\|full | 1648 | No | NA | 3 | 0 | 1 | Not-determined | Genome-fragment | NA | NA | 0 |
| S12-final-viral-combined_k141_3499\|\|full | 2341 | No | NA | 3 | 0 | 1 | Not-determined | Genome-fragment | NA | NA | 0 |
| S8-final-viral-combined_k141_272147\|\|full | 2645 | No | NA | 4 | 0 | 2 | Low-quality | Genome-fragment | 3.75 | AAI-based (high-confidence) | 0 |
| S9-final-viral-combined_k141_105817\|\|full | 2646 | No | NA | 3 | 0 | 2 | Low-quality | Genome-fragment | 6.35 | AAI-based (medium-confidence) | 0 |
| S12-final-viral-combined_k141_304942\|\|full | 3395 | No | NA | 6 | 0 | 0 | Not-determined | Genome-fragment | NA | NA | 0 |
| S12-final-viral-combined_k141_354622\|\|full | 3876 | No | NA | 4 | 0 | 3 | Not-determined | Genome-fragment | NA | NA | 0 |
| S12-final-viral-combined_k141_321372\|\|full | 2019 | No | NA | 3 | 0 | 0 | Not-determined | Genome-fragment | NA | NA | 0 |
| S12-final-viral-combined_k141_86985\|\|full | 3807 | No | NA | 6 | 0 | 4 | Not-determined | Genome-fragment | NA | NA | 0 |
| S12-final-viral-combined_k141_103519\|\|full | 3704 | No | NA | 5 | 0 | 3 | Not-determined | Genome-fragment | NA | NA | 0 |
| S1-final-viral-combined_k141_298341\|\|full | 2646 | No | NA | 6 | 0 | 0 | Low-quality | Genome-fragment | 4.66 | AAI-based (medium-confidence) | 0 |
| S12-final-viral-combined_k141_53532\|\|full | 1035 | No | NA | 2 | 0 | 0 | Not-determined | Genome-fragment | NA | NA | 0 |
| S3-final-viral-combined_k141_2372\|\|full | 2646 | No | NA | 5 | 1 | 0 | Low-quality | Genome-fragment | 4.42 | AAI-based (high-confidence) | 0 |
| S17-final-viral-combined_k141_240370\|\|full | 2646 | No | NA | 6 | 1 | 0 | Low-quality | Genome-fragment | 5.92 | AAI-based (medium-confidence) | 0 |
| S9-final-viral-combined_k141_83818\|\|full | 2647 | Yes | 1537 | 5 | 1 | 1 | Low-quality | Genome-fragment | 0.84 | HMM-based (lower-bound) | 41.93 |
| S9-final-viral-combined_k141_62372\|\|full | 2647 | No | NA | 6 | 3 | 0 | Low-quality | Genome-fragment | 7.14 | AAI-based (high-confidence) | 0 |
| S12-final-viral-combined_k141_305029\|\|full | 2790 | No | NA | 2 | 0 | 2 | Not-determined | Genome-fragment | NA | NA | 0 |
| S11-final-viral-combined_k141_166426\|\|full | 2647 | No | NA | 3 | 1 | 0 | Low-quality | Genome-fragment | 4.09 | HMM-based (lower-bound) | 0 |
| S12-final-viral-combined_k141_371845\|\|full | 1024 | No | NA | 2 | 0 | 1 | Not-determined | Genome-fragment | NA | NA | 0 |
| S2-final-viral-combined_k141_131356\|\|full | 2648 | No | NA | 6 | 0 | 3 | Low-quality | Genome-fragment | 3.32 | AAI-based (medium-confidence) | 0 |
| S12-final-viral-combined_k141_103626\|\|full | 1114 | No | NA | 1 | 0 | 0 | Not-determined | Genome-fragment | NA | NA | 0 |
| S6-final-viral-combined_k141_41208\|\|full | 2649 | No | NA | 4 | 0 | 4 | Low-quality | Genome-fragment | 5.65 | AAI-based (high-confidence) | 0 |
| S1-final-viral-combined_k141_3527\|\|full | 2650 | No | NA | 3 | 0 | 0 | Low-quality | Genome-fragment | 7.68 | AAI-based (medium-confidence) | 0 |
| S3-final-viral-combined_k141_93678\|\|full | 2650 | No | NA | 5 | 2 | 0 | Low-quality | Genome-fragment | 7.16 | AAI-based (high-confidence) | 0 |
| S7-final-viral-combined_k141_96669\|\|full | 2650 | No | NA | 3 | 0 | 2 | Low-quality | Genome-fragment | 0.82 | AAI-based (high-confidence) | 0 |
| S7-final-viral-combined_k141_160337\|\|full | 2650 | No | NA | 5 | 1 | 0 | Low-quality | Genome-fragment | 2.52 | AAI-based (medium-confidence) | 0 |
| S7-final-viral-combined_k141_233481\|\|full | 2650 | No | NA | 4 | 0 | 1 | Low-quality | Genome-fragment | 5.26 | AAI-based (high-confidence) | 0 |
| S17-final-viral-combined_k141_84455\|\|full | 2650 | No | NA | 7 | 0 | 0 | Low-quality | Genome-fragment | 5.83 | AAI-based (high-confidence) | 0 |
| S12-final-viral-combined_k141_237715\|\|full | 4994 | No | NA | 4 | 0 | 2 | Not-determined | Genome-fragment | NA | NA | 0 |
| S12-final-viral-combined_k141_3600\|\|full | 10525 | No | NA | 12 | 0 | 2 | Not-determined | Genome-fragment | NA | NA | 0 |
| S12-final-viral-combined_k141_339346\|\|full | 2650 | No | NA | 7 | 4 | 0 | Low-quality | Genome-fragment | 7.36 | AAI-based (medium-confidence) | 0 |
| S11-final-viral-combined_k141_1185\|\|full | 2651 | No | NA | 4 | 0 | 1 | Low-quality | Genome-fragment | 2.18 | AAI-based (medium-confidence) | 0 |
| S1-final-viral-combined_k141_319648\|\|full | 2651 | No | NA | 3 | 0 | 1 | Low-quality | Genome-fragment | 4.37 | AAI-based (high-confidence) | 0 |
| S17-k141_98269 | 2651 | No | NA | 2 | 0 | 0 | Low-quality | Genome-fragment | 5.52 | AAI-based (high-confidence) | 0 |
| S12-final-viral-combined_k141_170975\|\|full | 5771 | No | NA | 8 | 0 | 1 | Not-determined | Genome-fragment | NA | NA | 0 |
| S12-final-viral-combined_k141_103695\|\|full | 3199 | No | NA | 2 | 0 | 2 | Not-determined | Genome-fragment | NA | NA | 0 |
| S12-final-viral-combined_k141_288263\|\|full | 1733 | No | NA | 3 | 0 | 1 | Not-determined | Genome-fragment | NA | NA | 0 |
| S12-final-viral-combined_k141_288287\|\|full | 1412 | No | NA | 2 | 0 | 0 | Not-determined | Genome-fragment | NA | NA | 0 |
| S1-final-viral-combined_k141_323918\|\|full | 2652 | Yes | 1900 | 5 | 1 | 1 | Low-quality | Genome-fragment | 4.24 | AAI-based (medium-confidence) | 28.36 |
| S5-final-viral-combined_k141_18907\|\|full | 2652 | No | NA | 6 | 0 | 0 | Low-quality | Genome-fragment | 6.28 | AAI-based (medium-confidence) | 0 |
| S5-final-viral-combined_k141_162864\|\|full | 2652 | No | NA | 3 | 3 | 0 | Low-quality | Genome-fragment | 4.43 | AAI-based (high-confidence) | 0 |
| S12-final-viral-combined_k141_57475\|\|full | 2652 | No | NA | 4 | 0 | 1 | Low-quality | Genome-fragment | 4.9 | AAI-based (high-confidence) | 0 |
| S4-final-viral-combined_k141_16164\|\|full | 2652 | No | NA | 3 | 0 | 2 | Low-quality | Genome-fragment | 0.84 | AAI-based (medium-confidence) | 0 |
| S5-final-viral-combined_k141_104899\|\|full | 2653 | No | NA | 4 | 0 | 0 | Low-quality | Genome-fragment | 5.38 | AAI-based (medium-confidence) | 0 |
| S12-final-viral-combined_k141_87115\|\|full | 1936 | No | NA | 2 | 0 | 2 | Not-determined | Genome-fragment | NA | NA | 0 |
| S6-final-viral-combined_k141_126817\|\|full | 2653 | No | NA | 5 | 2 | 0 | Low-quality | Genome-fragment | 3.26 | HMM-based (lower-bound) | 0 |
| S3_k141_2425 | 2654 | No | NA | 1 | 0 | 1 | Low-quality | Genome-fragment | 2.41 | AAI-based (medium-confidence) | 0 |
| S12-final-viral-combined_k141_154181\|\|full | 2628 | No | NA | 3 | 0 | 1 | Not-determined | Genome-fragment | NA | NA | 0 |
| S12-final-viral-combined_k141_254150\|\|full | 1079 | No | NA | 3 | 0 | 1 | Not-determined | Genome-fragment | NA | NA | 0 |
| S12-final-viral-combined_k141_271086\|\|full | 5684 | No | NA | 5 | 0 | 4 | Not-determined | Genome-fragment | NA | NA | 0 |
| S12-final-viral-combined_k141_3702\|\|full | 1522 | No | NA | 2 | 0 | 2 | Not-determined | Genome-fragment | NA | NA | 0 |
| S12-final-viral-combined_k141_204381\|\|full | 1252 | No | NA | 2 | 0 | 2 | Not-determined | Genome-fragment | NA | NA | 0 |
| S12-final-viral-combined_k141_53608\|\|full | 2248 | No | NA | 3 | 0 | 2 | Not-determined | Genome-fragment | NA | NA | 0 |
| S7-final-viral-combined_k141_140325\|\|full | 2654 | No | NA | 3 | 1 | 0 | Low-quality | Genome-fragment | 6.69 | AAI-based (medium-confidence) | 0 |
| S12-final-viral-combined_k141_337820\|\|full | 2620 | No | NA | 4 | 0 | 0 | Not-determined | Genome-fragment | NA | NA | 0 |
| S12-final-viral-combined_k141_271113\|\|full | 9708 | No | NA | 9 | 0 | 9 | Not-determined | Genome-fragment | NA | NA | 0 |
| S4-final-viral-combined_k141_85018\|\|full | 2654 | No | NA | 4 | 0 | 3 | Low-quality | Genome-fragment | 1.64 | AAI-based (medium-confidence) | 0 |
| S8_k141_68352 | 2654 | No | NA | 3 | 0 | 2 | Low-quality | Genome-fragment | 1.76 | AAI-based (high-confidence) | 0 |
| S12-final-viral-combined_k141_120220\|\|full | 1170 | No | NA | 3 | 0 | 0 | Not-determined | Genome-fragment | NA | NA | 0 |
| S1-final-viral-combined_k141_59321\|\|full | 2655 | No | NA | 3 | 1 | 0 | Low-quality | Genome-fragment | 7.24 | AAI-based (high-confidence) | 0 |
| S12-final-viral-combined_k141_271126\|\|full | 2633 | No | NA | 2 | 0 | 0 | Not-determined | Genome-fragment | NA | NA | 0 |
| S12-final-viral-combined_k141_220880\|\|full | 1835 | No | NA | 2 | 0 | 2 | Not-determined | Genome-fragment | NA | NA | 0 |
| S12-final-viral-combined_k141_371996\|\|full | 1348 | No | NA | 2 | 0 | 0 | Not-determined | Genome-fragment | NA | NA | 0 |
| S12-final-viral-combined_k141_188086\|\|full | 6673 | No | NA | 5 | 0 | 4 | Not-determined | Genome-fragment | NA | NA | 0 |
| S12-final-viral-combined_k141_305248\|\|full | 1996 | No | NA | 2 | 0 | 0 | Not-determined | Genome-fragment | NA | NA | 0 |
| S12-final-viral-combined_k141_288353\|\|full | 1347 | No | NA | 2 | 0 | 0 | Not-determined | Genome-fragment | NA | NA | 0 |
| S12-final-viral-combined_k141_20114\|\|full | 1155 | No | NA | 3 | 0 | 0 | Not-determined | Genome-fragment | NA | NA | 0 |
| S17-final-viral-combined_k141_97657\|\|full | 2655 | No | NA | 5 | 0 | 0 | Low-quality | Genome-fragment | 3.21 | AAI-based (medium-confidence) | 0 |
| S12-final-viral-combined_k141_288373\|\|full | 2268 | No | NA | 3 | 0 | 1 | Not-determined | Genome-fragment | NA | NA | 0 |
| S12-final-viral-combined_k141_388574\|\|full | 7468 | No | NA | 14 | 0 | 0 | Not-determined | Genome-fragment | NA | NA | 0 |
| S12-final-viral-combined_k141_103806\|\|full | 1440 | No | NA | 2 | 0 | 2 | Not-determined | Genome-fragment | NA | NA | 0 |
| S2_k141_198064 | 2655 | No | NA | 3 | 0 | 3 | Low-quality | Genome-fragment | 5.93 | AAI-based (medium-confidence) | 0 |
| S12-final-viral-combined_k141_372071\|\|full | 1303 | No | NA | 3 | 0 | 1 | Not-determined | Genome-fragment | NA | NA | 0 |
| S12-final-viral-combined_k141_188137\|\|full | 6246 | No | NA | 7 | 0 | 0 | Not-determined | Genome-fragment | NA | NA | 0 |
| S11-final-viral-combined_k141_57035\|\|full | 2656 | No | NA | 5 | 0 | 2 | Low-quality | Genome-fragment | 1.89 | AAI-based (medium-confidence) | 0 |
| S12-final-viral-combined_k141_237865\|\|full | 2625 | No | NA | 3 | 0 | 1 | Not-determined | Genome-fragment | NA | NA | 0 |
| S11-final-viral-combined_k141_210126\|\|full | 2656 | No | NA | 3 | 0 | 1 | Low-quality | Genome-fragment | 2.16 | AAI-based (high-confidence) | 0 |
| S12-final-viral-combined_k141_354794\|\|full | 2042 | No | NA | 3 | 0 | 1 | Not-determined | Genome-fragment | NA | NA | 0 |
| S12-final-viral-combined_k141_288487\|\|full | 2162 | No | NA | 2 | 0 | 1 | Not-determined | Genome-fragment | NA | NA | 0 |
| S12-final-viral-combined_k141_372110\|\|full | 4192 | No | NA | 5 | 0 | 2 | Not-determined | Genome-fragment | NA | NA | 0 |
| S12-final-viral-combined_k141_103856\|\|full | 1484 | No | NA | 3 | 0 | 0 | Not-determined | Genome-fragment | NA | NA | 0 |
| S5-final-viral-combined_k141_93113\|\|full | 2656 | No | NA | 3 | 0 | 2 | Low-quality | Genome-fragment | 1.79 | AAI-based (medium-confidence) | 0 |
| S12-final-viral-combined_k141_3822\|\|full | 3612 | No | NA | 4 | 0 | 3 | Not-determined | Genome-fragment | NA | NA | 0 |
| S7-final-viral-combined_k141_228989\|\|full | 2656 | Yes | 1520 | 3 | 1 | 1 | Low-quality | Genome-fragment | 0.96 | HMM-based (lower-bound) | 42.77 |
| S17-final-viral-combined_k141_260869\|\|full | 2657 | No | NA | 3 | 0 | 2 | Low-quality | Genome-fragment | 4.45 | AAI-based (high-confidence) | 0 |
| S12-final-viral-combined_k141_388678\|\|full | 1789 | No | NA | 3 | 0 | 0 | Not-determined | Genome-fragment | NA | NA | 0 |
| S12-final-viral-combined_k141_20224\|\|full | 1243 | No | NA | 3 | 0 | 0 | Not-determined | Genome-fragment | NA | NA | 0 |
| S1-final-viral-combined_k141_379455\|\|full | 2658 | No | NA | 6 | 1 | 0 | Low-quality | Genome-fragment | 4.66 | AAI-based (high-confidence) | 0 |
| S17-k141_75673 | 2658 | No | NA | 7 | 0 | 0 | Low-quality | Genome-fragment | 5.13 | AAI-based (medium-confidence) | 0 |
| S12_k141_254669 | 2658 | No | NA | 2 | 0 | 2 | Low-quality | Genome-fragment | 0.39 | AAI-based (medium-confidence) | 0 |
| S2-final-viral-combined_k141_17679\|\|full | 2658 | No | NA | 2 | 2 | 0 | Low-quality | Genome-fragment | 4.02 | HMM-based (lower-bound) | 0 |
| S12-final-viral-combined_k141_103866\|\|full | 3743 | No | NA | 2 | 0 | 1 | Not-determined | Genome-fragment | NA | NA | 0 |
| S8_k141_243791 | 2658 | No | NA | 7 | 0 | 0 | Low-quality | Genome-fragment | 2.04 | AAI-based (high-confidence) | 0 |
| S11-final-viral-combined_k141_207949\|\|full | 2659 | No | NA | 3 | 0 | 0 | Low-quality | Genome-fragment | 7.27 | AAI-based (medium-confidence) | 0 |
| S7-final-viral-combined_k141_7942\|\|full | 2659 | No | NA | 2 | 0 | 1 | Low-quality | Genome-fragment | 0.84 | AAI-based (medium-confidence) | 0 |
| S12-final-viral-combined_k141_372155\|\|full | 12012 | No | NA | 13 | 0 | 1 | Not-determined | Genome-fragment | NA | NA | 0 |
| S12-final-viral-combined_k141_71004\|\|full | 1580 | No | NA | 2 | 0 | 1 | Not-determined | Genome-fragment | NA | NA | 0 |
| S17-final-viral-combined_k141_75652\|\|full | 2659 | No | NA | 4 | 2 | 0 | Low-quality | Genome-fragment | 5.68 | AAI-based (medium-confidence) | 0 |
| S2-final-viral-combined_k141_85855\|\|full | 2659 | No | NA | 3 | 0 | 2 | Low-quality | Genome-fragment | 0.76 | AAI-based (medium-confidence) | 0 |
| S12-final-viral-combined_k141_321767\|\|full | 1711 | No | NA | 3 | 0 | 2 | Not-determined | Genome-fragment | NA | NA | 0 |
| S12-final-viral-combined_k141_354884\|\|full | 1595 | No | NA | 2 | 0 | 0 | Not-determined | Genome-fragment | NA | NA | 0 |
| S12-final-viral-combined_k141_388740\|\|full | 1822 | No | NA | 4 | 0 | 3 | Not-determined | Genome-fragment | NA | NA | 0 |
| S9-final-viral-combined_k141_109589\|\|lt2gene | 2660 | No | NA | 2 | 2 | 0 | Low-quality | Genome-fragment | 5.77 | AAI-based (medium-confidence) | 0 |
| S12-final-viral-combined_k141_388751\|\|full | 3453 | No | NA | 3 | 0 | 0 | Not-determined | Genome-fragment | NA | NA | 0 |
| S12-final-viral-combined_k141_321788\|\|full | 5875 | No | NA | 9 | 0 | 1 | Not-determined | Genome-fragment | NA | NA | 0 |
| S11-final-viral-combined_k141_313073\|\|full | 2660 | No | NA | 7 | 1 | 0 | Low-quality | Genome-fragment | 4.2 | AAI-based (high-confidence) | 0 |
| S12-final-viral-combined_k141_103933\|\|full | 1529 | No | NA | 3 | 0 | 1 | Not-determined | Genome-fragment | NA | NA | 0 |
| S1_k141_142250 | 2660 | No | NA | 4 | 0 | 2 | Low-quality | Genome-fragment | 4.51 | AAI-based (high-confidence) | 0 |
| S7-final-viral-combined_k141_14564\|\|full | 2660 | No | NA | 4 | 0 | 1 | Low-quality | Genome-fragment | 1.78 | AAI-based (high-confidence) | 0 |
| S12-final-viral-combined_k141_338091\|\|full | 4725 | No | NA | 5 | 0 | 4 | Not-determined | Genome-fragment | NA | NA | 0 |
| S17-final-viral-combined_k141_55332\|\|full | 2660 | No | NA | 2 | 0 | 2 | Low-quality | Genome-fragment | 8.89 | AAI-based (medium-confidence) | 0 |
| S12-final-viral-combined_k141_238049\|\|full | 1913 | No | NA | 3 | 0 | 3 | Not-determined | Genome-fragment | NA | NA | 0 |
| S12-final-viral-combined_k141_53857\|\|full | 1055 | No | NA | 3 | 0 | 0 | Not-determined | Genome-fragment | NA | NA | 0 |
| S12-final-viral-combined_k141_37407\|\|full | 1659 | No | NA | 2 | 0 | 0 | Not-determined | Genome-fragment | NA | NA | 0 |
| S12-final-viral-combined_k141_338127\|\|full | 2401 | No | NA | 2 | 0 | 2 | Not-determined | Genome-fragment | NA | NA | 0 |
| S12-final-viral-combined_k141_372234\|\|full | 1676 | No | NA | 3 | 0 | 2 | Not-determined | Genome-fragment | NA | NA | 0 |
| S12-final-viral-combined_k141_218832\|\|full | 2660 | No | NA | 5 | 0 | 0 | Low-quality | Genome-fragment | 3.17 | AAI-based (medium-confidence) | 0 |
| S2-final-viral-combined_k141_119266\|\|full | 2660 | No | NA | 1 | 0 | 1 | Low-quality | Genome-fragment | 0.83 | AAI-based (medium-confidence) | 0 |
| S9-final-viral-combined_k141_63871\|\|lt2gene | 2661 | No | NA | 2 | 2 | 0 | Low-quality | Genome-fragment | 7.22 | AAI-based (medium-confidence) | 0 |
| S17-final-viral-combined_k141_163139\|\|full | 2661 | No | NA | 6 | 1 | 0 | Low-quality | Genome-fragment | 4.78 | AAI-based (high-confidence) | 0 |
| S12-final-viral-combined_k141_20412\|\|full | 2112 | No | NA | 3 | 0 | 2 | Not-determined | Genome-fragment | NA | NA | 0 |
| S12-final-viral-combined_k141_221052\|\|full | 1400 | No | NA | 2 | 0 | 1 | Not-determined | Genome-fragment | NA | NA | 0 |
| S12-final-viral-combined_k141_188381\|\|full | 1022 | No | NA | 4 | 0 | 0 | Not-determined | Genome-fragment | NA | NA | 0 |
| S12_k141_84974 | 2661 | No | NA | 4 | 1 | 0 | Low-quality | Genome-fragment | 4.7 | AAI-based (high-confidence) | 0 |
| S12-final-viral-combined_k141_354968\|\|full | 4611 | No | NA | 6 | 0 | 1 | Not-determined | Genome-fragment | NA | NA | 0 |
| S12-final-viral-combined_k141_221066\|\|full | 2924 | No | NA | 3 | 0 | 1 | Not-determined | Genome-fragment | NA | NA | 0 |
| S12-final-viral-combined_k141_321863\|\|full | 4715 | No | NA | 4 | 0 | 0 | Not-determined | Genome-fragment | NA | NA | 0 |
| S6-final-viral-combined_k141_20689\|\|full | 2661 | No | NA | 4 | 0 | 3 | Low-quality | Genome-fragment | 1.31 | AAI-based (medium-confidence) | 0 |
| S6_k141_131828 | 2662 | No | NA | 7 | 0 | 0 | Low-quality | Genome-fragment | 5.64 | AAI-based (medium-confidence) | 0 |
| S12-final-viral-combined_k141_53919\|\|full | 9574 | No | NA | 7 | 0 | 2 | Not-determined | Genome-fragment | NA | NA | 0 |
| S12-final-viral-combined_k141_204538\|\|full | 1300 | No | NA | 3 | 0 | 0 | Not-determined | Genome-fragment | NA | NA | 0 |
| S12-final-viral-combined_k141_171190\|\|full | 1051 | No | NA | 2 | 0 | 0 | Not-determined | Genome-fragment | NA | NA | 0 |
| S9-final-viral-combined_k141_62549\|\|full | 2663 | No | NA | 4 | 4 | 0 | Low-quality | Genome-fragment | 6.08 | AAI-based (high-confidence) | 0 |
| S6-final-viral-combined_k141_108325\|\|full | 2663 | No | NA | 4 | 0 | 0 | Low-quality | Genome-fragment | 6.03 | AAI-based (high-confidence) | 0 |
| S1_k141_20293 | 2664 | No | NA | 3 | 0 | 1 | Low-quality | Genome-fragment | 5.33 | AAI-based (medium-confidence) | 0 |
| S12-final-viral-combined_k141_104014\|\|full | 3711 | No | NA | 8 | 0 | 0 | Not-determined | Genome-fragment | NA | NA | 0 |
| S12-final-viral-combined_k141_53950\|\|full | 1916 | No | NA | 3 | 0 | 2 | Not-determined | Genome-fragment | NA | NA | 0 |
| S12-final-viral-combined_k141_254407\|\|full | 3547 | No | NA | 5 | 0 | 2 | Not-determined | Genome-fragment | NA | NA | 0 |
| S12-final-viral-combined_k141_388909\|\|full | 1669 | No | NA | 2 | 0 | 2 | Not-determined | Genome-fragment | NA | NA | 0 |
| S12-final-viral-combined_k141_221148\|\|full | 3038 | No | NA | 6 | 0 | 0 | Not-determined | Genome-fragment | NA | NA | 0 |
| S12-final-viral-combined_k141_238224\|\|full | 2428 | No | NA | 2 | 0 | 2 | Not-determined | Genome-fragment | NA | NA | 0 |
| S7-final-viral-combined_k141_232128\|\|full | 2664 | No | NA | 4 | 3 | 0 | Low-quality | Genome-fragment | 5.93 | AAI-based (high-confidence) | 0 |
| S12-final-viral-combined_k141_321953\|\|full | 1790 | No | NA | 3 | 0 | 2 | Not-determined | Genome-fragment | NA | NA | 0 |
| S12-final-viral-combined_k141_20486\|\|full | 7055 | No | NA | 7 | 0 | 6 | Not-determined | Genome-fragment | NA | NA | 0 |
| S17-k141_112235 | 2664 | No | NA | 6 | 0 | 4 | Low-quality | Genome-fragment | 1.81 | AAI-based (medium-confidence) | 0 |
| S12-final-viral-combined_k141_355025\|\|full | 2809 | No | NA | 4 | 0 | 1 | Not-determined | Genome-fragment | NA | NA | 0 |
| S12-final-viral-combined_k141_204583\|\|full | 1991 | No | NA | 2 | 0 | 2 | Not-determined | Genome-fragment | NA | NA | 0 |
| S12-final-viral-combined_k141_338233\|\|full | 3000 | No | NA | 2 | 0 | 2 | Not-determined | Genome-fragment | NA | NA | 0 |
| S12-final-viral-combined_k141_20507\|\|full | 2682 | No | NA | 6 | 0 | 5 | Not-determined | Genome-fragment | NA | NA | 0 |
| S12_k141_396392 | 2664 | No | NA | 3 | 0 | 0 | Low-quality | Genome-fragment | 4.69 | AAI-based (high-confidence) | 0 |
| S12-final-viral-combined_k141_338243\|\|full | 3171 | No | NA | 3 | 0 | 2 | Not-determined | Genome-fragment | NA | NA | 0 |
| S11-final-viral-combined_k141_296465\|\|full | 2665 | Yes | 2050 | 4 | 1 | 1 | Low-quality | Genome-fragment | 1.51 | AAI-based (high-confidence) | 23.08 |
| S12-final-viral-combined_k141_305576\|\|full | 1858 | No | NA | 2 | 0 | 0 | Not-determined | Genome-fragment | NA | NA | 0 |
| S1-final-viral-combined_k141_258121\|\|full | 2665 | No | NA | 4 | 0 | 1 | Low-quality | Genome-fragment | 1.95 | AAI-based (high-confidence) | 0 |
| S12-final-viral-combined_k141_271562\|\|full | 1171 | No | NA | 2 | 0 | 1 | Not-determined | Genome-fragment | NA | NA | 0 |
| S12-final-viral-combined_k141_154508\|\|full | 1215 | No | NA | 3 | 0 | 2 | Not-determined | Genome-fragment | NA | NA | 0 |
| S12-final-viral-combined_k141_104098\|\|full | 1445 | No | NA | 3 | 0 | 0 | Not-determined | Genome-fragment | NA | NA | 0 |
| S12-final-viral-combined_k141_210041\|\|full | 2665 | No | NA | 4 | 0 | 1 | Low-quality | Genome-fragment | 4.62 | AAI-based (medium-confidence) | 0 |
| S12-final-viral-combined_k141_54021\|\|full | 2383 | No | NA | 3 | 0 | 2 | Not-determined | Genome-fragment | NA | NA | 0 |
| S12-final-viral-combined_k141_104108\|\|full | 1073 | No | NA | 2 | 0 | 1 | Not-determined | Genome-fragment | NA | NA | 0 |
| S12-final-viral-combined_k141_71235\|\|full | 22756 | No | NA | 28 | 17 | 1 | Medium-quality | Genome-fragment | 64.99 | AAI-based (high-confidence) | 0 |
| S12-final-viral-combined_k141_137917\|\|full | 1060 | No | NA | 3 | 0 | 0 | Not-determined | Genome-fragment | NA | NA | 0 |
| S12-final-viral-combined_k141_221226\|\|full | 2185 | No | NA | 3 | 0 | 2 | Not-determined | Genome-fragment | NA | NA | 0 |
| S12-final-viral-combined_k141_328016\|\|full | 2665 | No | NA | 4 | 0 | 0 | Low-quality | Genome-fragment | 4.82 | AAI-based (high-confidence) | 0 |
| S12-final-viral-combined_k141_338284\|\|full | 3141 | No | NA | 3 | 0 | 1 | Not-determined | Genome-fragment | NA | NA | 0 |
| S12-final-viral-combined_k141_3997\|\|full | 3490 | No | NA | 4 | 0 | 3 | Not-determined | Genome-fragment | NA | NA | 0 |
| S12-final-viral-combined_k141_204676\|\|full | 2486 | No | NA | 3 | 0 | 3 | Not-determined | Genome-fragment | NA | NA | 0 |
| S12-final-viral-combined_k141_316494\|\|full | 2665 | No | NA | 4 | 1 | 0 | Low-quality | Genome-fragment | 6.96 | AAI-based (medium-confidence) | 0 |
| S2-final-viral-combined_k141_190890\|\|full | 2665 | No | NA | 6 | 0 | 0 | Low-quality | Genome-fragment | 5.44 | AAI-based (medium-confidence) | 0 |
| S1_k141_201208 | 2666 | No | NA | 4 | 1 | 0 | Low-quality | Genome-fragment | 5.63 | AAI-based (high-confidence) | 0 |
| S12-final-viral-combined_k141_355068\|\|full | 3221 | No | NA | 3 | 0 | 0 | Not-determined | Genome-fragment | NA | NA | 0 |
| S17-final-viral-combined_k141_172489\|\|full | 2666 | No | NA | 5 | 2 | 0 | Low-quality | Genome-fragment | 6.86 | AAI-based (high-confidence) | 0 |
| S12-final-viral-combined_k141_355072\|\|full | 3262 | No | NA | 8 | 0 | 0 | Not-determined | Genome-fragment | NA | NA | 0 |
| S12-final-viral-combined_k141_312986\|\|full | 2666 | No | NA | 4 | 0 | 4 | Low-quality | Genome-fragment | 3.23 | AAI-based (medium-confidence) | 0 |
| S12-final-viral-combined_k141_288681\|\|full | 1824 | No | NA | 3 | 0 | 0 | Not-determined | Genome-fragment | NA | NA | 0 |
| S12-final-viral-combined_k141_322121\|\|full | 2514 | No | NA | 3 | 0 | 1 | Not-determined | Genome-fragment | NA | NA | 0 |
| S12-final-viral-combined_k141_372497\|\|full | 3827 | No | NA | 5 | 0 | 0 | Not-determined | Genome-fragment | NA | NA | 0 |
| S12-final-viral-combined_k141_154640\|\|full | 4524 | No | NA | 2 | 0 | 2 | Not-determined | Genome-fragment | NA | NA | 0 |
| S2-final-viral-combined_k141_97051\|\|full | 2666 | No | NA | 4 | 1 | 0 | Low-quality | Genome-fragment | 4.89 | HMM-based (lower-bound) | 0 |
| S7-final-viral-combined_k141_88451\|\|full | 2667 | No | NA | 7 | 1 | 0 | Low-quality | Genome-fragment | 7.35 | AAI-based (high-confidence) | 0 |
| S12-final-viral-combined_k141_171396\|\|full | 2156 | No | NA | 3 | 0 | 1 | Not-determined | Genome-fragment | NA | NA | 0 |
| S12-final-viral-combined_k141_271705\|\|full | 6123 | No | NA | 6 | 0 | 4 | Not-determined | Genome-fragment | NA | NA | 0 |
| S12-final-viral-combined_k141_104221\|\|full | 2692 | No | NA | 4 | 0 | 0 | Not-determined | Genome-fragment | NA | NA | 0 |
| S12-final-viral-combined_k141_54173\|\|full | 1397 | No | NA | 2 | 0 | 1 | Not-determined | Genome-fragment | NA | NA | 0 |
| S12-final-viral-combined_k141_275994\|\|full | 2667 | No | NA | 4 | 0 | 1 | Low-quality | Genome-fragment | 6.49 | AAI-based (medium-confidence) | 0 |
| S12-final-viral-combined_k141_305735\|\|full | 3202 | No | NA | 5 | 0 | 2 | Not-determined | Genome-fragment | NA | NA | 0 |
| S12-final-viral-combined_k141_188646\|\|full | 1261 | No | NA | 3 | 0 | 0 | Not-determined | Genome-fragment | NA | NA | 0 |
| S12-final-viral-combined_k141_355162\|\|full | 4807 | No | NA | 6 | 0 | 3 | Not-determined | Genome-fragment | NA | NA | 0 |
| S8-final-viral-combined_k141_770\|\|full | 2667 | No | NA | 6 | 0 | 0 | Low-quality | Genome-fragment | 4.95 | AAI-based (high-confidence) | 0 |
| S9-final-viral-combined_k141_21126\|\|full | 2668 | No | NA | 4 | 1 | 0 | Low-quality | Genome-fragment | 6.43 | AAI-based (high-confidence) | 0 |
| S8_k141_146630 | 2668 | No | NA | 3 | 1 | 0 | Low-quality | Genome-fragment | 2.36 | AAI-based (medium-confidence) | 0 |
| S12-final-viral-combined_k141_221392\|\|full | 2212 | No | NA | 5 | 0 | 0 | Not-determined | Genome-fragment | NA | NA | 0 |
| S12-final-viral-combined_k141_54226\|\|full | 3880 | No | NA | 2 | 0 | 0 | Not-determined | Genome-fragment | NA | NA | 0 |
| S3-final-viral-combined_k141_82905\|\|full | 2669 | No | NA | 2 | 0 | 1 | Low-quality | Genome-fragment | 0.97 | AAI-based (medium-confidence) | 0 |
| S12-final-viral-combined_k141_221414\|\|full | 1367 | No | NA | 3 | 0 | 0 | Not-determined | Genome-fragment | NA | NA | 0 |
| S17-final-viral-combined_k141_204977\|\|full | 2669 | No | NA | 5 | 2 | 0 | Low-quality | Genome-fragment | 5.83 | AAI-based (high-confidence) | 0 |
| S12-final-viral-combined_k141_238402\|\|full | 9732 | No | NA | 9 | 0 | 4 | Not-determined | Genome-fragment | NA | NA | 0 |
| S12-final-viral-combined_k141_134382\|\|full | 2669 | No | NA | 6 | 0 | 0 | Low-quality | Genome-fragment | 6.09 | AAI-based (high-confidence) | 0 |
| S1-final-viral-combined_k141_155203\|\|full | 2670 | No | NA | 2 | 0 | 2 | Low-quality | Genome-fragment | 1.93 | AAI-based (high-confidence) | 0 |
| S12-final-viral-combined_k141_171502\|\|full | 5207 | No | NA | 5 | 0 | 4 | Not-determined | Genome-fragment | NA | NA | 0 |
| S12-final-viral-combined_k141_4138\|\|full | 1634 | No | NA | 3 | 0 | 0 | Not-determined | Genome-fragment | NA | NA | 0 |
| S3_k141_101129 | 2670 | No | NA | 6 | 0 | 0 | Low-quality | Genome-fragment | 4.88 | AAI-based (high-confidence) | 0 |
| S12-final-viral-combined_k141_120783\|\|full | 1590 | No | NA | 3 | 0 | 3 | Not-determined | Genome-fragment | NA | NA | 0 |
| S12-final-viral-combined_k141_271843\|\|full | 2749 | No | NA | 3 | 0 | 2 | Not-determined | Genome-fragment | NA | NA | 0 |
| S12-final-viral-combined_k141_71568\|\|full | 3046 | No | NA | 2 | 0 | 1 | Not-determined | Genome-fragment | NA | NA | 0 |
| S5-final-viral-combined_k141_129479\|\|full | 2670 | No | NA | 3 | 1 | 0 | Low-quality | Genome-fragment | 7.01 | AAI-based (high-confidence) | 0 |
| S12-final-viral-combined_k141_188758\|\|full | 1886 | No | NA | 3 | 0 | 3 | Not-determined | Genome-fragment | NA | NA | 0 |
| S7-final-viral-combined_k141_69537\|\|full | 2670 | No | NA | 4 | 0 | 2 | Low-quality | Genome-fragment | 1.81 | AAI-based (medium-confidence) | 0 |
| S12-final-viral-combined_k141_171529\|\|full | 1173 | No | NA | 3 | 0 | 1 | Not-determined | Genome-fragment | NA | NA | 0 |
| S17-final-viral-combined_k141_7394\|\|full | 2670 | No | NA | 5 | 2 | 0 | Low-quality | Genome-fragment | 4.95 | AAI-based (medium-confidence) | 0 |
| S8_k141_22061 | 2670 | No | NA | 4 | 1 | 0 | Low-quality | Genome-fragment | 2.13 | AAI-based (high-confidence) | 0 |
| S12-final-viral-combined_k141_203755\|\|full | 2671 | No | NA | 5 | 0 | 1 | Low-quality | Genome-fragment | 3.56 | AAI-based (medium-confidence) | 0 |
| S2-final-viral-combined_k141_52499\|\|full | 2671 | No | NA | 3 | 2 | 0 | Low-quality | Genome-fragment | 5.63 | AAI-based (medium-confidence) | 0 |
| S12-final-viral-combined_k141_221490\|\|full | 1195 | No | NA | 2 | 0 | 1 | Not-determined | Genome-fragment | NA | NA | 0 |
| S2_k141_182966 | 2671 | No | NA | 5 | 0 | 0 | Low-quality | Genome-fragment | 4.46 | AAI-based (medium-confidence) | 0 |
| S12-final-viral-combined_k141_188778\|\|full | 5980 | No | NA | 7 | 0 | 4 | Not-determined | Genome-fragment | NA | NA | 0 |
| S8_k141_272364 | 2671 | No | NA | 2 | 0 | 1 | Low-quality | Genome-fragment | 2.39 | AAI-based (medium-confidence) | 0 |
| S11-final-viral-combined_k141_300689\|\|full | 2672 | No | NA | 2 | 1 | 0 | Low-quality | Genome-fragment | 1.35 | HMM-based (lower-bound) | 0 |
| S12-final-viral-combined_k141_238242\|\|full | 2672 | Yes | 1244 | 7 | 1 | 1 | Low-quality | Genome-fragment | 1.92 | HMM-based (lower-bound) | 53.44 |
| S2-final-viral-combined_k141_47640\|\|full | 2672 | No | NA | 7 | 4 | 0 | Low-quality | Genome-fragment | 6.32 | AAI-based (high-confidence) | 0 |
| S12-final-viral-combined_k141_238454\|\|full | 9824 | No | NA | 9 | 0 | 6 | Not-determined | Genome-fragment | NA | NA | 0 |
| S12-final-viral-combined_k141_54265\|\|full | 5238 | No | NA | 5 | 0 | 1 | Not-determined | Genome-fragment | NA | NA | 0 |
| S12-final-viral-combined_k141_322335\|\|full | 2422 | No | NA | 4 | 0 | 1 | Not-determined | Genome-fragment | NA | NA | 0 |
| S8-final-viral-combined_k141_137787\|\|full | 2672 | No | NA | 4 | 0 | 1 | Low-quality | Genome-fragment | 4.72 | AAI-based (medium-confidence) | 0 |
| S11-final-viral-combined_k141_179785\|\|full | 2673 | No | NA | 3 | 2 | 0 | Low-quality | Genome-fragment | 3.88 | AAI-based (high-confidence) | 0 |
| S12-final-viral-combined_k141_104364\|\|full | 2608 | No | NA | 2 | 0 | 0 | Not-determined | Genome-fragment | NA | NA | 0 |
| S12-final-viral-combined_k141_271951\|\|full | 1681 | No | NA | 3 | 0 | 2 | Not-determined | Genome-fragment | NA | NA | 0 |
| S12-final-viral-combined_k141_238475\|\|full | 2465 | No | NA | 2 | 0 | 0 | Not-determined | Genome-fragment | NA | NA | 0 |
| S12-final-viral-combined_k141_54303\|\|full | 1034 | No | NA | 3 | 0 | 1 | Not-determined | Genome-fragment | NA | NA | 0 |
| S12-final-viral-combined_k141_154861\|\|full | 1313 | No | NA | 3 | 0 | 0 | Not-determined | Genome-fragment | NA | NA | 0 |
| S5-final-viral-combined_k141_140102\|\|full | 2673 | No | NA | 2 | 0 | 2 | Low-quality | Genome-fragment | 2.88 | AAI-based (medium-confidence) | 0 |
| S12-final-viral-combined_k141_355358\|\|full | 4686 | No | NA | 3 | 0 | 3 | Not-determined | Genome-fragment | NA | NA | 0 |
| S7-final-viral-combined_k141_236613\|\|full | 2674 | No | NA | 4 | 2 | 0 | Low-quality | Genome-fragment | 6.37 | AAI-based (high-confidence) | 0 |
| S7-final-viral-combined_k141_128737\|\|full | 2674 | No | NA | 3 | 3 | 0 | Low-quality | Genome-fragment | 5.9 | AAI-based (high-confidence) | 0 |
| S12-final-viral-combined_k141_154892\|\|full | 1162 | No | NA | 2 | 0 | 0 | Not-determined | Genome-fragment | NA | NA | 0 |
| S12-final-viral-combined_k141_254836\|\|full | 1304 | No | NA | 2 | 0 | 0 | Not-determined | Genome-fragment | NA | NA | 0 |
| S17-final-viral-combined_k141_201522\|\|full | 2674 | No | NA | 4 | 1 | 0 | Low-quality | Genome-fragment | 3.31 | HMM-based (lower-bound) | 0 |
| S17-final-viral-combined_k141_250629\|\|full | 2675 | No | NA | 6 | 6 | 0 | Low-quality | Genome-fragment | 6.11 | AAI-based (medium-confidence) | 0 |
| S12_k141_208865 | 2675 | No | NA | 5 | 0 | 2 | Low-quality | Genome-fragment | 1.89 | AAI-based (medium-confidence) | 0 |
| S6-final-viral-combined_k141_21255\|\|full | 2675 | No | NA | 3 | 0 | 0 | Low-quality | Genome-fragment | 5 | AAI-based (medium-confidence) | 0 |
| S5-final-viral-combined_k141_156784\|\|full | 2676 | No | NA | 4 | 1 | 0 | Low-quality | Genome-fragment | 4.77 | AAI-based (high-confidence) | 0 |
| S12-final-viral-combined_k141_154924\|\|full | 1739 | No | NA | 2 | 0 | 1 | Not-determined | Genome-fragment | NA | NA | 0 |
| S12-final-viral-combined_k141_221661\|\|full | 1702 | No | NA | 2 | 0 | 0 | Not-determined | Genome-fragment | NA | NA | 0 |
| S12-final-viral-combined_k141_356871\|\|full | 2676 | No | NA | 1 | 1 | 0 | Low-quality | Genome-fragment | 1.85 | HMM-based (lower-bound) | 0 |
| S12-final-viral-combined_k141_154933\|\|full | 2134 | No | NA | 3 | 0 | 2 | Not-determined | Genome-fragment | NA | NA | 0 |
| S1-final-viral-combined_k141_362143\|\|full | 2677 | No | NA | 8 | 0 | 0 | Low-quality | Genome-fragment | 43.38 | AAI-based (medium-confidence) | 0 |
| S5-final-viral-combined_k141_111179\|\|full | 2677 | No | NA | 6 | 1 | 0 | Low-quality | Genome-fragment | 4.65 | AAI-based (high-confidence) | 0 |
| S12-final-viral-combined_k141_19479\|\|full | 2677 | No | NA | 4 | 0 | 2 | Low-quality | Genome-fragment | 4.61 | AAI-based (medium-confidence) | 0 |
| S12-final-viral-combined_k141_205020\|\|full | 4400 | No | NA | 5 | 0 | 3 | Not-determined | Genome-fragment | NA | NA | 0 |
| S12-final-viral-combined_k141_23597\|\|full | 2677 | No | NA | 8 | 4 | 0 | Low-quality | Genome-fragment | 5.98 | AAI-based (medium-confidence) | 0 |
| S12-final-viral-combined_k141_120962\|\|full | 4180 | No | NA | 4 | 0 | 2 | Not-determined | Genome-fragment | NA | NA | 0 |
| S12-final-viral-combined_k141_205028\|\|full | 1133 | No | NA | 3 | 0 | 0 | Not-determined | Genome-fragment | NA | NA | 0 |
| S12-final-viral-combined_k141_138380\|\|full | 6909 | No | NA | 7 | 0 | 1 | Not-determined | Genome-fragment | NA | NA | 0 |
| S12-final-viral-combined_k141_322432\|\|full | 1515 | No | NA | 3 | 0 | 1 | Not-determined | Genome-fragment | NA | NA | 0 |
| S4-final-viral-combined_k141_143647\|\|full | 2677 | No | NA | 8 | 1 | 0 | Low-quality | Genome-fragment | 4.67 | HMM-based (lower-bound) | 0 |
| S11-final-viral-combined_k141_18662\|\|full | 2678 | No | NA | 3 | 0 | 1 | Low-quality | Genome-fragment | 1.21 | AAI-based (medium-confidence) | 0 |
| S12-final-viral-combined_k141_20923\|\|full | 2332 | No | NA | 3 | 0 | 3 | Not-determined | Genome-fragment | NA | NA | 0 |
| S12-final-viral-combined_k141_205050\|\|full | 1835 | No | NA | 4 | 0 | 3 | Not-determined | Genome-fragment | NA | NA | 0 |
| S1-final-viral-combined_k141_18918\|\|full | 2678 | No | NA | 2 | 1 | 0 | Low-quality | Genome-fragment | 4.05 | HMM-based (lower-bound) | 0 |
| S12-final-viral-combined_k141_71784\|\|full | 1224 | No | NA | 3 | 0 | 2 | Not-determined | Genome-fragment | NA | NA | 0 |
| S12-final-viral-combined_k141_306058\|\|full | 2128 | No | NA | 2 | 0 | 0 | Not-determined | Genome-fragment | NA | NA | 0 |
| S12-final-viral-combined_k141_238517\|\|full | 2396 | No | NA | 4 | 0 | 0 | Not-determined | Genome-fragment | NA | NA | 0 |
| S12-final-viral-combined_k141_338777\|\|full | 2939 | No | NA | 3 | 0 | 0 | Not-determined | Genome-fragment | NA | NA | 0 |
| S12-final-viral-combined_k141_120980\|\|full | 1121 | No | NA | 2 | 0 | 1 | Not-determined | Genome-fragment | NA | NA | 0 |
| S12-final-viral-combined_k141_254870\|\|full | 15731 | No | NA | 10 | 0 | 8 | Not-determined | Genome-fragment | NA | NA | 0 |
| S12-final-viral-combined_k141_54403\|\|full | 3884 | No | NA | 4 | 0 | 1 | Not-determined | Genome-fragment | NA | NA | 0 |
| S12-final-viral-combined_k141_238531\|\|full | 3896 | No | NA | 4 | 0 | 0 | Not-determined | Genome-fragment | NA | NA | 0 |
| S7-final-viral-combined_k141_5919\|\|full | 2678 | No | NA | 3 | 0 | 0 | Low-quality | Genome-fragment | 6.99 | AAI-based (high-confidence) | 0 |
| S12-final-viral-combined_k141_37927\|\|full | 1040 | No | NA | 2 | 0 | 1 | Not-determined | Genome-fragment | NA | NA | 0 |
| S12-final-viral-combined_k141_104439\|\|full | 1059 | No | NA | 2 | 0 | 1 | Not-determined | Genome-fragment | NA | NA | 0 |
| S17-final-viral-combined_k141_147679\|\|full | 2678 | No | NA | 7 | 0 | 0 | Low-quality | Genome-fragment | 5.18 | AAI-based (medium-confidence) | 0 |
| S12-final-viral-combined_k141_138441\|\|full | 4784 | No | NA | 5 | 0 | 1 | Not-determined | Genome-fragment | NA | NA | 0 |
| S2_k141_97927 | 2678 | No | NA | 4 | 1 | 0 | Low-quality | Genome-fragment | 42.4 | AAI-based (high-confidence) | 0 |
| S12-final-viral-combined_k141_389390\|\|full | 5474 | No | NA | 7 | 0 | 5 | Not-determined | Genome-fragment | NA | NA | 0 |
| S12-final-viral-combined_k141_21049\|\|full | 1905 | No | NA | 3 | 0 | 1 | Not-determined | Genome-fragment | NA | NA | 0 |
| S12-final-viral-combined_k141_205084\|\|full | 1896 | No | NA | 2 | 0 | 2 | Not-determined | Genome-fragment | NA | NA | 0 |
| S12-final-viral-combined_k141_54451\|\|full | 2571 | No | NA | 6 | 0 | 0 | Not-determined | Genome-fragment | NA | NA | 0 |
| S12-final-viral-combined_k141_104458\|\|full | 1111 | No | NA | 1 | 0 | 0 | Not-determined | Genome-fragment | NA | NA | 0 |
| S12-final-viral-combined_k141_355500\|\|full | 5216 | No | NA | 5 | 0 | 0 | Not-determined | Genome-fragment | NA | NA | 0 |
| S12-final-viral-combined_k141_322545\|\|full | 3511 | No | NA | 4 | 0 | 0 | Not-determined | Genome-fragment | NA | NA | 0 |
| S12-final-viral-combined_k141_171544\|\|full | 1276 | No | NA | 2 | 0 | 1 | Not-determined | Genome-fragment | NA | NA | 0 |
| S12-final-viral-combined_k141_21071\|\|full | 4713 | No | NA | 7 | 0 | 0 | Not-determined | Genome-fragment | NA | NA | 0 |
| S12-final-viral-combined_k141_54478\|\|full | 2691 | No | NA | 4 | 0 | 0 | Not-determined | Genome-fragment | NA | NA | 0 |
| S12-final-viral-combined_k141_154984\|\|full | 4839 | No | NA | 5 | 0 | 2 | Not-determined | Genome-fragment | NA | NA | 0 |
| S4_k141_48940 | 2678 | No | NA | 2 | 1 | 0 | Low-quality | Genome-fragment | 2.63 | AAI-based (high-confidence) | 0 |
| S12-final-viral-combined_k141_189008\|\|full | 3123 | No | NA | 4 | 0 | 1 | Not-determined | Genome-fragment | NA | NA | 0 |
| S12-final-viral-combined_k141_21117\|\|full | 2608 | No | NA | 4 | 0 | 1 | Not-determined | Genome-fragment | NA | NA | 0 |
| S12-final-viral-combined_k141_389414\|\|full | 2154 | No | NA | 2 | 0 | 0 | Not-determined | Genome-fragment | NA | NA | 0 |
| S9_k141_118931 | 2679 | No | NA | 2 | 0 | 1 | Low-quality | Genome-fragment | 1.46 | AAI-based (medium-confidence) | 0 |
| S12-final-viral-combined_k141_87890\|\|full | 1731 | No | NA | 2 | 0 | 1 | Not-determined | Genome-fragment | NA | NA | 0 |
| S12-final-viral-combined_k141_189039\|\|full | 4475 | No | NA | 7 | 0 | 5 | Not-determined | Genome-fragment | NA | NA | 0 |
| S12-final-viral-combined_k141_238663\|\|full | 4064 | No | NA | 4 | 0 | 2 | Not-determined | Genome-fragment | NA | NA | 0 |
| S3-final-viral-combined_k141_135484\|\|full | 2679 | No | NA | 4 | 0 | 3 | Low-quality | Genome-fragment | 0.88 | AAI-based (medium-confidence) | 0 |
| S7-final-viral-combined_k141_108330\|\|full | 2679 | No | NA | 2 | 0 | 1 | Low-quality | Genome-fragment | 1.29 | AAI-based (medium-confidence) | 0 |
| S4-final-viral-combined_k141_21925\|\|full | 2679 | No | NA | 4 | 2 | 0 | Low-quality | Genome-fragment | 7.99 | AAI-based (high-confidence) | 0 |
| S6-final-viral-combined_k141_11675\|\|full | 2679 | No | NA | 4 | 0 | 0 | Low-quality | Genome-fragment | 8.11 | AAI-based (medium-confidence) | 0 |
| S12-final-viral-combined_k141_389469\|\|full | 1593 | No | NA | 2 | 0 | 0 | Not-determined | Genome-fragment | NA | NA | 0 |
| S12-final-viral-combined_k141_306220\|\|full | 1107 | No | NA | 3 | 0 | 0 | Not-determined | Genome-fragment | NA | NA | 0 |
| S12-final-viral-combined_k141_104575\|\|full | 3150 | No | NA | 4 | 0 | 2 | Not-determined | Genome-fragment | NA | NA | 0 |
| S12-final-viral-combined_k141_272171\|\|full | 1485 | No | NA | 3 | 0 | 1 | Not-determined | Genome-fragment | NA | NA | 0 |
| S1-final-viral-combined_k141_355849\|\|full | 2680 | No | NA | 6 | 1 | 0 | Low-quality | Genome-fragment | 6.07 | AAI-based (high-confidence) | 0 |
| S3-final-viral-combined_k141_75588\|\|full | 2680 | No | NA | 2 | 0 | 2 | Low-quality | Genome-fragment | 2.38 | AAI-based (high-confidence) | 0 |
| S12-final-viral-combined_k141_87972\|\|full | 1814 | No | NA | 2 | 0 | 1 | Not-determined | Genome-fragment | NA | NA | 0 |
| S12-final-viral-combined_k141_272225\|\|full | 3296 | No | NA | 6 | 0 | 2 | Not-determined | Genome-fragment | NA | NA | 0 |
| S2-final-viral-combined_k141_143326\|\|full | 2680 | No | NA | 2 | 0 | 1 | Low-quality | Genome-fragment | 0.84 | AAI-based (medium-confidence) | 0 |
| S12-final-viral-combined_k141_289231\|\|full | 16211 | No | NA | 9 | 0 | 3 | Not-determined | Genome-fragment | NA | NA | 0 |
| S12-final-viral-combined_k141_88012\|\|full | 3761 | No | NA | 4 | 0 | 0 | Not-determined | Genome-fragment | NA | NA | 0 |
| S2-final-viral-combined_k141_7837\|\|full | 2680 | No | NA | 5 | 1 | 0 | Low-quality | Genome-fragment | 5.39 | AAI-based (high-confidence) | 0 |
| S12-final-viral-combined_k141_389562\|\|full | 1583 | No | NA | 2 | 0 | 0 | Not-determined | Genome-fragment | NA | NA | 0 |
| S12-final-viral-combined_k141_322640\|\|full | 2346 | No | NA | 5 | 0 | 0 | Not-determined | Genome-fragment | NA | NA | 0 |
| S12-final-viral-combined_k141_289275\|\|full | 1016 | No | NA | 2 | 0 | 2 | Not-determined | Genome-fragment | NA | NA | 0 |
| S7-final-viral-combined_k141_244878\|\|full | 2681 | No | NA | 4 | 0 | 1 | Low-quality | Genome-fragment | 1.42 | AAI-based (medium-confidence) | 0 |
| S12-final-viral-combined_k141_88033\|\|full | 2752 | No | NA | 3 | 0 | 1 | Not-determined | Genome-fragment | NA | NA | 0 |
| S2-final-viral-combined_k141_212138\|\|full | 2681 | No | NA | 6 | 2 | 0 | Low-quality | Genome-fragment | 5.28 | AAI-based (high-confidence) | 0 |
| S2-final-viral-combined_k141_8865\|\|full | 2681 | No | NA | 3 | 1 | 0 | Low-quality | Genome-fragment | 6.05 | AAI-based (high-confidence) | 0 |
| S12-final-viral-combined_k141_171737\|\|full | 1257 | No | NA | 2 | 0 | 0 | Not-determined | Genome-fragment | NA | NA | 0 |
| S12-final-viral-combined_k141_306306\|\|full | 4311 | No | NA | 4 | 0 | 1 | Not-determined | Genome-fragment | NA | NA | 0 |
| S1-final-viral-combined_k141_165479\|\|full | 2682 | No | NA | 2 | 0 | 0 | Low-quality | Genome-fragment | 0.92 | AAI-based (high-confidence) | 0 |
| S12-final-viral-combined_k141_171745\|\|full | 1234 | No | NA | 2 | 0 | 0 | Not-determined | Genome-fragment | NA | NA | 0 |
| S12-final-viral-combined_k141_88074\|\|full | 1358 | No | NA | 2 | 0 | 0 | Not-determined | Genome-fragment | NA | NA | 0 |
| S12-final-viral-combined_k141_289338\|\|full | 2354 | No | NA | 2 | 0 | 2 | Not-determined | Genome-fragment | NA | NA | 0 |
| S1-final-viral-combined_k141_339182\|\|full | 2682 | No | NA | 4 | 4 | 0 | Low-quality | Genome-fragment | 4.08 | HMM-based (lower-bound) | 0 |
| S1_k141_227214 | 2682 | No | NA | 1 | 0 | 1 | Low-quality | Genome-fragment | 0.96 | AAI-based (medium-confidence) | 0 |
| S12-final-viral-combined_k141_104736\|\|full | 2750 | No | NA | 3 | 0 | 1 | Not-determined | Genome-fragment | NA | NA | 0 |
| S11-final-viral-combined_k141_232505\|\|full | 2683 | No | NA | 5 | 2 | 0 | Low-quality | Genome-fragment | 5.71 | AAI-based (high-confidence) | 0 |

| | | | | | | | | | | | |
|---|---|---|---|---|---|---|---|---|---|---|---|
| S17-final-viral-combined_k141_39717\|\|full | 2683 | No | NA | 5 | 0 | 0 | Low-quality | Genome-fragment | 4.43 | AAI-based (medium-confidence) | 0 |
| S12-final-viral-combined_k141_322747\|\|full | 8044 | No | NA | 8 | 0 | 1 | Not-determined | Genome-fragment | NA | NA | 0 |
| S2-final-viral-combined_k141_57222\|\|full | 2683 | No | NA | 6 | 0 | 0 | Low-quality | Genome-fragment | 2.8 | AAI-based (high-confidence) | 0 |
| S12-final-viral-combined_k141_4539\|\|full | 1226 | No | NA | 2 | 0 | 0 | Not-determined | Genome-fragment | NA | NA | 0 |
| S6-final-viral-combined_k141_65998\|\|full | 2683 | Yes | 528 | 3 | 1 | 1 | Low-quality | Genome-fragment | 0.25 | AAI-based (medium-confidence) | 80.32 |
| S12-final-viral-combined_k141_4542\|\|full | 3931 | No | NA | 5 | 0 | 1 | Not-determined | Genome-fragment | NA | NA | 0 |
| S7-final-viral-combined_k141_25023\|\|full | 2684 | No | NA | 4 | 0 | 0 | Low-quality | Genome-fragment | 4.48 | AAI-based (high-confidence) | 0 |
| S12-final-viral-combined_k141_54658\|\|full | 5244 | No | NA | 5 | 0 | 3 | Not-determined | Genome-fragment | NA | NA | 0 |
| S17-k141_111462 | 2684 | No | NA | 5 | 0 | 0 | Low-quality | Genome-fragment | 3.74 | AAI-based (high-confidence) | 0 |
| S12-final-viral-combined_k141_189248\|\|full | 1214 | No | NA | 3 | 0 | 0 | Not-determined | Genome-fragment | NA | NA | 0 |
| S12-final-viral-combined_k141_4544\|\|full | 5080 | No | NA | 4 | 0 | 4 | Not-determined | Genome-fragment | NA | NA | 0 |
| S12-final-viral-combined_k141_238870\|\|full | 1363 | No | NA | 2 | 0 | 1 | Not-determined | Genome-fragment | NA | NA | 0 |
| S2-final-viral-combined_k141_114768\|\|full | 2684 | No | NA | 8 | 0 | 0 | Low-quality | Genome-fragment | 3.46 | AAI-based (high-confidence) | 0 |
| S12-final-viral-combined_k141_21317\|\|full | 1617 | No | NA | 3 | 0 | 3 | Not-determined | Genome-fragment | NA | NA | 0 |
| S12-final-viral-combined_k141_289462\|\|full | 1497 | No | NA | 2 | 0 | 1 | Not-determined | Genome-fragment | NA | NA | 0 |
| S4-final-viral-combined_k141_113621\|\|full | 2684 | No | NA | 4 | 3 | 0 | Low-quality | Genome-fragment | 8.21 | AAI-based (high-confidence) | 0 |
| S8-final-viral-combined_k141_108888\|\|full | 2684 | No | NA | 5 | 1 | 0 | Low-quality | Genome-fragment | 6.64 | AAI-based (high-confidence) | 0 |
| S12-final-viral-combined_k141_322793\|\|full | 1457 | No | NA | 3 | 0 | 0 | Not-determined | Genome-fragment | NA | NA | 0 |
| S12-final-viral-combined_k141_339100\|\|full | 1337 | No | NA | 3 | 0 | 1 | Not-determined | Genome-fragment | NA | NA | 0 |
| S12-final-viral-combined_k141_306437\|\|full | 1474 | No | NA | 2 | 0 | 1 | Not-determined | Genome-fragment | NA | NA | 0 |
| S12-final-viral-combined_k141_272428\|\|full | 4641 | No | NA | 4 | 0 | 3 | Not-determined | Genome-fragment | NA | NA | 0 |
| S12-final-viral-combined_k141_289529\|\|full | 2618 | No | NA | 3 | 0 | 3 | Not-determined | Genome-fragment | NA | NA | 0 |
| S11-final-viral-combined_k141_19588\|\|full | 2685 | No | NA | 3 | 1 | 0 | Low-quality | Genome-fragment | 5.6 | AAI-based (high-confidence) | 0 |
| S11_k141_11422 | 2685 | No | NA | 3 | 0 | 2 | Low-quality | Genome-fragment | 1.08 | AAI-based (high-confidence) | 0 |
| S12-final-viral-combined_k141_221685\|\|full | 2685 | No | NA | 3 | 0 | 2 | Low-quality | Genome-fragment | 6.07 | AAI-based (medium-confidence) | 0 |
| S11-final-viral-combined_k141_262750\|\|full | 2686 | No | NA | 6 | 1 | 0 | Low-quality | Genome-fragment | 2.95 | HMM-based (lower-bound) | 0 |
| S12-final-viral-combined_k141_121243\|\|full | 2512 | No | NA | 3 | 0 | 1 | Not-determined | Genome-fragment | NA | NA | 0 |
| S12-final-viral-combined_k141_355701\|\|full | 1546 | No | NA | 3 | 0 | 2 | Not-determined | Genome-fragment | NA | NA | 0 |
| S1-final-viral-combined_k141_299356\|\|full | 2687 | No | NA | 4 | 0 | 1 | Low-quality | Genome-fragment | 3.83 | AAI-based (high-confidence) | 0 |
| S1-final-viral-combined_k141_334611\|\|full | 2687 | No | NA | 4 | 1 | 0 | Low-quality | Genome-fragment | 2.89 | HMM-based (lower-bound) | 0 |
| S7-final-viral-combined_k141_207855\|\|full | 2687 | No | NA | 5 | 0 | 0 | Low-quality | Genome-fragment | 4.82 | AAI-based (high-confidence) | 0 |
| S12-final-viral-combined_k141_155210\|\|full | 1612 | No | NA | 2 | 0 | 0 | Not-determined | Genome-fragment | NA | NA | 0 |
| S7_k141_133687 | 2687 | No | NA | 6 | 1 | 0 | Low-quality | Genome-fragment | 11.43 | AAI-based (high-confidence) | 0 |
| S12-final-viral-combined_k141_90363\|\|full | 2687 | No | NA | 3 | 3 | 0 | Low-quality | Genome-fragment | 4.57 | AAI-based (high-confidence) | 0 |
| S12-final-viral-combined_k141_155236\|\|full | 3417 | No | NA | 4 | 0 | 2 | Not-determined | Genome-fragment | NA | NA | 0 |
| S12-final-viral-combined_k141_289604\|\|full | 1061 | No | NA | 3 | 0 | 2 | Not-determined | Genome-fragment | NA | NA | 0 |
| S12-final-viral-combined_k141_171915\|\|full | 1383 | No | NA | 3 | 0 | 2 | Not-determined | Genome-fragment | NA | NA | 0 |
| S12-final-viral-combined_k141_62433\|\|full | 2687 | No | NA | 4 | 0 | 1 | Low-quality | Genome-fragment | 3.08 | AAI-based (medium-confidence) | 0 |
| S4-final-viral-combined_k141_25695\|\|full | 2687 | No | NA | 3 | 0 | 2 | Low-quality | Genome-fragment | 1.25 | AAI-based (medium-confidence) | 0 |
| S12-final-viral-combined_k141_322886\|\|full | 1515 | No | NA | 3 | 0 | 0 | Not-determined | Genome-fragment | NA | NA | 0 |
| S12-final-viral-combined_k141_189323\|\|full | 83620 | Yes | 38428 | 71 | 9 | 26 | Medium-quality | Genome-fragment | 89.72 | AAI-based (medium-confidence) | 54.04 |
| S8-final-viral-combined_k141_137167\|\|full | 2687 | No | NA | 4 | 0 | 0 | Low-quality | Genome-fragment | 4.74 | AAI-based (high-confidence) | 0 |
| S11-final-viral-combined_k141_311831\|\|full | 2688 | No | NA | 5 | 2 | 0 | Low-quality | Genome-fragment | 5.74 | AAI-based (medium-confidence) | 0 |
| S12-final-viral-combined_k141_339179\|\|full | 1160 | No | NA | 2 | 0 | 2 | Not-determined | Genome-fragment | NA | NA | 0 |
| S12-final-viral-combined_k141_189357\|\|full | 2605 | No | NA | 3 | 0 | 1 | Not-determined | Genome-fragment | NA | NA | 0 |
| S3-final-viral-combined_k141_61151\|\|full | 2688 | No | NA | 3 | 0 | 1 | Low-quality | Genome-fragment | 4.6 | AAI-based (medium-confidence) | 0 |
| S12-final-viral-combined_k141_38376\|\|full | 5224 | No | NA | 9 | 0 | 1 | Not-determined | Genome-fragment | NA | NA | 0 |
| S12-final-viral-combined_k141_272528\|\|full | 2063 | No | NA | 3 | 0 | 2 | Not-determined | Genome-fragment | NA | NA | 0 |
| S12-final-viral-combined_k141_255327\|\|full | 7435 | No | NA | 4 | 0 | 4 | Not-determined | Genome-fragment | NA | NA | 0 |
| S12-final-viral-combined_k141_389771\|\|full | 1560 | No | NA | 3 | 0 | 1 | Not-determined | Genome-fragment | NA | NA | 0 |
| S12-final-viral-combined_k141_373375\|\|full | 1179 | No | NA | 4 | 0 | 0 | Not-determined | Genome-fragment | NA | NA | 0 |
| S12-final-viral-combined_k141_105003\|\|full | 2056 | No | NA | 2 | 0 | 1 | Not-determined | Genome-fragment | NA | NA | 0 |
| S17-final-viral-combined_k141_122764\|\|full | 2689 | No | NA | 7 | 1 | 0 | Low-quality | Genome-fragment | 1.63 | HMM-based (lower-bound) | 0 |
| S9-final-viral-combined_k141_45840\|\|full | 2690 | No | NA | 2 | 0 | 2 | Low-quality | Genome-fragment | 1.08 | AAI-based (medium-confidence) | 0 |
| S12-final-viral-combined_k141_105041\|\|full | 1603 | No | NA | 2 | 0 | 1 | Not-determined | Genome-fragment | NA | NA | 0 |
| S1_k141_30231 | 2690 | No | NA | 4 | 1 | 0 | Low-quality | Genome-fragment | 5.96 | AAI-based (high-confidence) | 0 |
| S17-final-viral-combined_k141_201203\|\|full | 2690 | No | NA | 3 | 0 | 0 | Low-quality | Genome-fragment | 6.56 | AAI-based (medium-confidence) | 0 |
| S12-final-viral-combined_k141_373418\|\|full | 4225 | No | NA | 4 | 0 | 4 | Not-determined | Genome-fragment | NA | NA | 0 |
| S8-final-viral-combined_k141_202073\|\|full | 2690 | No | NA | 3 | 1 | 0 | Low-quality | Genome-fragment | 3.97 | AAI-based (high-confidence) | 0 |
| S8-final-viral-combined_k141_169312\|\|full | 2690 | No | NA | 8 | 1 | 0 | Low-quality | Genome-fragment | 5.59 | AAI-based (medium-confidence) | 0 |
| S1-final-viral-combined_k141_353876\|\|full | 2691 | No | NA | 3 | 0 | 0 | Low-quality | Genome-fragment | 2.48 | AAI-based (medium-confidence) | 0 |
| S12-final-viral-combined_k141_38465\|\|full | 4034 | No | NA | 5 | 0 | 4 | Not-determined | Genome-fragment | NA | NA | 0 |
| S3-final-viral-combined_k141_136284\|\|full | 2691 | No | NA | 3 | 0 | 2 | Low-quality | Genome-fragment | 2.02 | AAI-based (medium-confidence) | 0 |
| S12-final-viral-combined_k141_155385\|\|full | 3660 | No | NA | 4 | 0 | 3 | Not-determined | Genome-fragment | NA | NA | 0 |
| S12-final-viral-combined_k141_54735\|\|full | 1203 | No | NA | 3 | 0 | 0 | Not-determined | Genome-fragment | NA | NA | 0 |
| S12-final-viral-combined_k141_389813\|\|full | 2123 | No | NA | 3 | 0 | 2 | Not-determined | Genome-fragment | NA | NA | 0 |
| S12-final-viral-combined_k141_33557\|\|full | 2691 | No | NA | 3 | 0 | 2 | Low-quality | Genome-fragment | 1.86 | AAI-based (medium-confidence) | 0 |
| S12-final-viral-combined_k141_105111\|\|full | 5906 | No | NA | 6 | 0 | 5 | Not-determined | Genome-fragment | NA | NA | 0 |
| S2-final-viral-combined_k141_184203\|\|full | 2691 | No | NA | 3 | 0 | 0 | Low-quality | Genome-fragment | 5.98 | AAI-based (medium-confidence) | 0 |
| S12-final-viral-combined_k141_72256\|\|full | 1523 | No | NA | 3 | 0 | 2 | Not-determined | Genome-fragment | NA | NA | 0 |
| S12-final-viral-combined_k141_105165\|\|full | 1052 | No | NA | 2 | 0 | 0 | Not-determined | Genome-fragment | NA | NA | 0 |
| S12-final-viral-combined_k141_389856\|\|full | 1595 | No | NA | 4 | 0 | 2 | Not-determined | Genome-fragment | NA | NA | 0 |
| S12-final-viral-combined_k141_21504\|\|full | 37101 | No | NA | 35 | 16 | 1 | Medium-quality | Genome-fragment | 73.46 | AAI-based (medium-confidence) | 0 |
| S11-final-viral-combined_k141_92821\|\|full | 2692 | No | NA | 5 | 2 | 0 | Low-quality | Genome-fragment | 1.95 | AAI-based (high-confidence) | 0 |
| S12-final-viral-combined_k141_21506\|\|full | 1058 | No | NA | 2 | 0 | 0 | Not-determined | Genome-fragment | NA | NA | 0 |
| S1-final-viral-combined_k141_327726\|\|full | 2692 | No | NA | 7 | 0 | 0 | Low-quality | Genome-fragment | 7.23 | AAI-based (high-confidence) | 0 |
| S2-final-viral-combined_k141_34471\|\|full | 2692 | No | NA | 5 | 0 | 0 | Low-quality | Genome-fragment | 3.55 | AAI-based (high-confidence) | 0 |
| S12-final-viral-combined_k141_323100\|\|full | 2149 | No | NA | 2 | 0 | 2 | Not-determined | Genome-fragment | NA | NA | 0 |
| S12-final-viral-combined_k141_88266\|\|full | 2223 | No | NA | 2 | 0 | 2 | Not-determined | Genome-fragment | NA | NA | 0 |
| S8-final-viral-combined_k141_260529\|\|full | 2692 | No | NA | 3 | 0 | 3 | Low-quality | Genome-fragment | 1.03 | AAI-based (medium-confidence) | 0 |
| S12-final-viral-combined_k141_339379\|\|full | 2583 | No | NA | 3 | 0 | 1 | Not-determined | Genome-fragment | NA | NA | 0 |
| S1-final-viral-combined_k141_173016\|\|full | 2693 | No | NA | 7 | 0 | 0 | Low-quality | Genome-fragment | 6.25 | AAI-based (high-confidence) | 0 |
| S12-final-viral-combined_k141_306622\|\|full | 2144 | No | NA | 3 | 0 | 1 | Not-determined | Genome-fragment | NA | NA | 0 |
| S9-final-viral-combined_k141_51366\|\|full | 2694 | No | NA | 3 | 0 | 0 | Low-quality | Genome-fragment | 5.46 | AAI-based (high-confidence) | 0 |
| S12-final-viral-combined_k141_323121\|\|full | 1187 | No | NA | 2 | 0 | 0 | Not-determined | Genome-fragment | NA | NA | 0 |
| S12-final-viral-combined_k141_301969\|\|full | 2694 | No | NA | 4 | 0 | 1 | Low-quality | Genome-fragment | 4.25 | AAI-based (high-confidence) | 0 |
| S4-final-viral-combined_k141_20923\|\|full | 2694 | No | NA | 5 | 0 | 1 | Low-quality | Genome-fragment | 5.1 | AAI-based (medium-confidence) | 0 |
| S12-final-viral-combined_k141_155504\|\|full | 1679 | No | NA | 3 | 0 | 2 | Not-determined | Genome-fragment | NA | NA | 0 |
| S11-final-viral-combined_k141_253743\|\|full | 2695 | No | NA | 4 | 0 | 2 | Low-quality | Genome-fragment | 1.18 | AAI-based (medium-confidence) | 0 |
| S11-final-viral-combined_k141_61507\|\|full | 2695 | No | NA | 5 | 0 | 0 | Low-quality | Genome-fragment | 5.75 | AAI-based (high-confidence) | 0 |
| S12-final-viral-combined_k141_189593\|\|full | 1206 | No | NA | 3 | 0 | 1 | Not-determined | Genome-fragment | NA | NA | 0 |
| S7-final-viral-combined_k141_141318\|\|full | 2695 | No | NA | 4 | 1 | 0 | Low-quality | Genome-fragment | 6.36 | AAI-based (high-confidence) | 0 |
| S12-final-viral-combined_k141_35140\|\|full | 2695 | No | NA | 6 | 0 | 1 | Low-quality | Genome-fragment | 1.59 | AAI-based (medium-confidence) | 0 |
| S12-final-viral-combined_k141_88330\|\|full | 1532 | No | NA | 4 | 0 | 2 | Not-determined | Genome-fragment | NA | NA | 0 |
| S12-final-viral-combined_k141_38593\|\|full | 6813 | No | NA | 6 | 0 | 4 | Not-determined | Genome-fragment | NA | NA | 0 |
| S12-final-viral-combined_k141_339434\|\|full | 3067 | No | NA | 4 | 0 | 3 | Not-determined | Genome-fragment | NA | NA | 0 |
| S8-final-viral-combined_k141_43407\|\|full | 2695 | No | NA | 3 | 0 | 2 | Low-quality | Genome-fragment | 2.19 | AAI-based (high-confidence) | 0 |
| S12-final-viral-combined_k141_355926\|\|full | 2807 | No | NA | 3 | 0 | 1 | Not-determined | Genome-fragment | NA | NA | 0 |
| S12-final-viral-combined_k141_373579\|\|full | 6180 | No | NA | 6 | 0 | 3 | Not-determined | Genome-fragment | NA | NA | 0 |
| S12-final-viral-combined_k141_355944\|\|full | 4496 | No | NA | 5 | 0 | 1 | Not-determined | Genome-fragment | NA | NA | 0 |
| S11-final-viral-combined_k141_303143\|\|full | 2696 | No | NA | 7 | 0 | 1 | Low-quality | Genome-fragment | 8.52 | AAI-based (medium-confidence) | 0 |
| S11-final-viral-combined_k141_202754\|\|lt2gene | 2696 | No | NA | 2 | 1 | 0 | Low-quality | Genome-fragment | 6.19 | AAI-based (high-confidence) | 0 |
| S12-final-viral-combined_k141_339461\|\|full | 2339 | No | NA | 2 | 0 | 1 | Not-determined | Genome-fragment | NA | NA | 0 |
| S12-final-viral-combined_k141_389988\|\|full | 1387 | No | NA | 2 | 0 | 1 | Not-determined | Genome-fragment | NA | NA | 0 |
| S12-final-viral-combined_k141_355963\|\|full | 1341 | No | NA | 3 | 0 | 1 | Not-determined | Genome-fragment | NA | NA | 0 |
| S12-final-viral-combined_k141_289764\|\|full | 4204 | No | NA | 4 | 0 | 4 | Not-determined | Genome-fragment | NA | NA | 0 |
| S12-final-viral-combined_k141_38653\|\|full | 4956 | No | NA | 6 | 0 | 5 | Not-determined | Genome-fragment | NA | NA | 0 |
| S6-final-viral-combined_k141_42922\|\|full | 2696 | No | NA | 5 | 2 | 0 | Low-quality | Genome-fragment | 7.29 | AAI-based (medium-confidence) | 0 |
| S17-final-viral-combined_k141_119806\|\|full | 2697 | No | NA | 5 | 1 | 0 | Low-quality | Genome-fragment | 5.25 | AAI-based (high-confidence) | 0 |
| S12-final-viral-combined_k141_72400\|\|full | 8976 | No | NA | 6 | 0 | 6 | Not-determined | Genome-fragment | NA | NA | 0 |
| S12-final-viral-combined_k141_189667\|\|full | 2620 | No | NA | 4 | 0 | 1 | Not-determined | Genome-fragment | NA | NA | 0 |
| S12-final-viral-combined_k141_255691\|\|full | 2324 | No | NA | 2 | 0 | 2 | Not-determined | Genome-fragment | NA | NA | 0 |
| S12-final-viral-combined_k141_390053\|\|full | 2946 | No | NA | 3 | 0 | 3 | Not-determined | Genome-fragment | NA | NA | 0 |
| S8-final-viral-combined_k141_74710\|\|full | 2697 | No | NA | 6 | 2 | 0 | Low-quality | Genome-fragment | 3.29 | HMM-based (lower-bound) | 0 |
| S11-final-viral-combined_k141_225274\|\|full | 2698 | No | NA | 7 | 0 | 1 | Low-quality | Genome-fragment | 5.04 | AAI-based (high-confidence) | 0 |
| S1-final-viral-combined_k141_14531\|\|full | 2698 | No | NA | 3 | 0 | 2 | Low-quality | Genome-fragment | 2.01 | AAI-based (medium-confidence) | 0 |
| S12-final-viral-combined_k141_226969\|\|full | 2698 | No | NA | 2 | 0 | 0 | Low-quality | Genome-fragment | 2.08 | AAI-based (high-confidence) | 0 |
| S2-final-viral-combined_k141_21055\|\|full | 2698 | No | NA | 4 | 0 | 1 | Low-quality | Genome-fragment | 2.32 | AAI-based (high-confidence) | 0 |
| S12-final-viral-combined_k141_189720\|\|full | 6432 | No | NA | 6 | 0 | 3 | Not-determined | Genome-fragment | NA | NA | 0 |
| S12-final-viral-combined_k141_72463\|\|full | 1234 | No | NA | 2 | 0 | 2 | Not-determined | Genome-fragment | NA | NA | 0 |
| S1-final-viral-combined_k141_159851\|\|full | 2699 | No | NA | 3 | 0 | 2 | Low-quality | Genome-fragment | 4.23 | AAI-based (high-confidence) | 0 |
| S2-final-viral-combined_k141_39429\|\|full | 2699 | No | NA | 3 | 2 | 0 | Low-quality | Genome-fragment | 42.73 | AAI-based (high-confidence) | 0 |
| S12-final-viral-combined_k141_306798\|\|full | 2050 | No | NA | 3 | 0 | 0 | Not-determined | Genome-fragment | NA | NA | 0 |
| S5-final-viral-combined_k141_200339\|\|full | 2700 | No | NA | 3 | 0 | 0 | Low-quality | Genome-fragment | 1.45 | AAI-based (medium-confidence) | 0 |
| S12-final-viral-combined_k141_235880\|\|full | 2700 | No | NA | 3 | 0 | 2 | Low-quality | Genome-fragment | 3.96 | AAI-based (medium-confidence) | 0 |
| S12-final-viral-combined_k141_121424\|\|full | 2726 | No | NA | 5 | 0 | 1 | Not-determined | Genome-fragment | NA | NA | 0 |
| S12-final-viral-combined_k141_272822\|\|full | 1112 | No | NA | 2 | 0 | 2 | Not-determined | Genome-fragment | NA | NA | 0 |
| S12-final-viral-combined_k141_272828\|\|full | 2607 | No | NA | 3 | 0 | 0 | Not-determined | Genome-fragment | NA | NA | 0 |
| S12-final-viral-combined_k141_21723\|\|full | 1700 | No | NA | 2 | 0 | 1 | Not-determined | Genome-fragment | NA | NA | 0 |
| S12-final-viral-combined_k141_121440\|\|full | 1408 | No | NA | 2 | 0 | 1 | Not-determined | Genome-fragment | NA | NA | 0 |
| S12-final-viral-combined_k141_205586\|\|full | 1392 | No | NA | 3 | 0 | 1 | Not-determined | Genome-fragment | NA | NA | 0 |
| S12-final-viral-combined_k141_54927\|\|full | 1554 | No | NA | 3 | 0 | 1 | Not-determined | Genome-fragment | NA | NA | 0 |
| S12-final-viral-combined_k141_222362\|\|full | 2800 | No | NA | 4 | 0 | 2 | Not-determined | Genome-fragment | NA | NA | 0 |
| S12-final-viral-combined_k141_390174\|\|full | 4923 | No | NA | 5 | 0 | 3 | Not-determined | Genome-fragment | NA | NA | 0 |
| S12-final-viral-combined_k141_339566\|\|full | 1123 | No | NA | 2 | 0 | 1 | Not-determined | Genome-fragment | NA | NA | 0 |
| S12-final-viral-combined_k141_205624\|\|full | 2149 | No | NA | 4 | 0 | 0 | Not-determined | Genome-fragment | NA | NA | 0 |
| S12-final-viral-combined_k141_222405\|\|full | 1503 | No | NA | 3 | 0 | 1 | Not-determined | Genome-fragment | NA | NA | 0 |
| S12-final-viral-combined_k141_239373\|\|full | 2458 | No | NA | 3 | 0 | 1 | Not-determined | Genome-fragment | NA | NA | 0 |
| S12-final-viral-combined_k141_339601\|\|full | 3723 | No | NA | 6 | 0 | 0 | Not-determined | Genome-fragment | NA | NA | 0 |
| S12-final-viral-combined_k141_260503\|\|full | 2700 | No | NA | 2 | 0 | 0 | Low-quality | Genome-fragment | 7.41 | AAI-based (medium-confidence) | 0 |
| S5-final-viral-combined_k141_112282\|\|full | 2701 | No | NA | 2 | 0 | 2 | Low-quality | Genome-fragment | 2.98 | AAI-based (medium-confidence) | 0 |
| S17-k141_265191 | 2701 | No | NA | 3 | 0 | 1 | Low-quality | Genome-fragment | 4.51 | AAI-based (high-confidence) | 0 |
| S12-final-viral-combined_k141_323243\|\|full | 2268 | No | NA | 4 | 0 | 3 | Not-determined | Genome-fragment | NA | NA | 0 |
| S11-final-viral-combined_k141_254995\|\|full | 2702 | Yes | 1429 | 5 | 1 | 1 | Low-quality | Genome-fragment | 1.9 | AAI-based (medium-confidence) | 47.11 |
| S1-final-viral-combined_k141_357734\|\|full | 2702 | No | NA | 6 | 0 | 2 | Low-quality | Genome-fragment | 5.78 | AAI-based (medium-confidence) | 0 |
| S12-final-viral-combined_k141_155702\|\|full | 1748 | No | NA | 2 | 0 | 1 | Not-determined | Genome-fragment | NA | NA | 0 |
| S8-final-viral-combined_k141_96267\|\|full | 2702 | No | NA | 5 | 0 | 0 | Low-quality | Genome-fragment | 6.5 | AAI-based (high-confidence) | 0 |
| S12-final-viral-combined_k141_121494\|\|full | 1586 | No | NA | 2 | 0 | 2 | Not-determined | Genome-fragment | NA | NA | 0 |
| S1-final-viral-combined_k141_332670\|\|full | 2703 | No | NA | 3 | 2 | 0 | Low-quality | Genome-fragment | 3.28 | AAI-based (high-confidence) | 0 |
| S5-final-viral-combined_k141_197034\|\|lt2gene | 2703 | No | NA | 2 | 2 | 0 | Low-quality | Genome-fragment | 3.81 | HMM-based (lower-bound) | 0 |
| S12-final-viral-combined_k141_121521\|\|full | 1735 | No | NA | 2 | 0 | 2 | Not-determined | Genome-fragment | NA | NA | 0 |
| S12-final-viral-combined_k141_307005\|\|full | 7489 | No | NA | 13 | 0 | 2 | Not-determined | Genome-fragment | NA | NA | 0 |
| S11-final-viral-combined_k141_102717\|\|full | 2704 | No | NA | 4 | 0 | 0 | Low-quality | Genome-fragment | 2.55 | AAI-based (high-confidence) | 0 |
| S12-final-viral-combined_k141_121534\|\|full | 1538 | No | NA | 3 | 0 | 0 | Not-determined | Genome-fragment | NA | NA | 0 |
| S12-final-viral-combined_k141_272973\|\|full | 3175 | No | NA | 4 | 0 | 0 | Not-determined | Genome-fragment | NA | NA | 0 |
| S1-final-viral-combined_k141_371137\|\|full | 2704 | No | NA | 7 | 1 | 0 | Low-quality | Genome-fragment | 7.48 | AAI-based (medium-confidence) | 0 |
| S12-final-viral-combined_k141_5012\|\|full | 2219 | No | NA | 2 | 0 | 1 | Not-determined | Genome-fragment | NA | NA | 0 |
| S5-final-viral-combined_k141_125894\|\|lt2gene | 2704 | No | NA | 1 | 1 | 0 | Low-quality | Genome-fragment | 4.27 | HMM-based (lower-bound) | 0 |
| S7-final-viral-combined_k141_102840\|\|full | 2704 | No | NA | 3 | 0 | 1 | Low-quality | Genome-fragment | 1.65 | AAI-based (medium-confidence) | 0 |
| S12-final-viral-combined_k141_121559\|\|full | 1650 | No | NA | 3 | 0 | 1 | Not-determined | Genome-fragment | NA | NA | 0 |
| S12-final-viral-combined_k141_273000\|\|full | 1422 | No | NA | 3 | 0 | 2 | Not-determined | Genome-fragment | NA | NA | 0 |
| S12-final-viral-combined_k141_190848\|\|full | 2704 | No | NA | 3 | 0 | 2 | Low-quality | Genome-fragment | 1.95 | AAI-based (medium-confidence) | 0 |
| S12-final-viral-combined_k141_121567\|\|full | 1258 | No | NA | 3 | 0 | 0 | Not-determined | Genome-fragment | NA | NA | 0 |
| S12-final-viral-combined_k141_222556\|\|full | 1968 | No | NA | 4 | 0 | 0 | Not-determined | Genome-fragment | NA | NA | 0 |
| S12-final-viral-combined_k141_38969\|\|full | 12127 | No | NA | 13 | 0 | 11 | Not-determined | Genome-fragment | NA | NA | 0 |
| S2-final-viral-combined_k141_165492\|\|full | 2704 | No | NA | 4 | 0 | 2 | Low-quality | Genome-fragment | 1.66 | AAI-based (medium-confidence) | 0 |
| S9-final-viral-combined_k141_39618\|\|full | 2705 | No | NA | 3 | 0 | 0 | Low-quality | Genome-fragment | 2.01 | AAI-based (medium-confidence) | 0 |
| S9_k141_54246 | 2705 | No | NA | 1 | 0 | 1 | Low-quality | Genome-fragment | 1.66 | AAI-based (medium-confidence) | 0 |
| S11-final-viral-combined_k141_246629\|\|full | 2705 | No | NA | 2 | 1 | 0 | Low-quality | Genome-fragment | 4.75 | HMM-based (lower-bound) | 0 |
| S11-final-viral-combined_k141_220464\|\|full | 2705 | No | NA | 4 | 0 | 1 | Low-quality | Genome-fragment | 6.12 | AAI-based (high-confidence) | 0 |
| S17-final-viral-combined_k141_239963\|\|full | 2705 | No | NA | 2 | 0 | 1 | Low-quality | Genome-fragment | 1.31 | AAI-based (medium-confidence) | 0 |
| S12-final-viral-combined_k141_222572\|\|full | 1211 | No | NA | 2 | 0 | 1 | Not-determined | Genome-fragment | NA | NA | 0 |
| S12-final-viral-combined_k141_273041\|\|full | 2431 | No | NA | 3 | 0 | 0 | Not-determined | Genome-fragment | NA | NA | 0 |
| S5-final-viral-combined_k141_34509\|\|full | 2706 | No | NA | 4 | 1 | 0 | Low-quality | Genome-fragment | 5.79 | AAI-based (high-confidence) | 0 |
| S12-final-viral-combined_k141_290124\|\|full | 2714 | No | NA | 3 | 0 | 2 | Not-determined | Genome-fragment | NA | NA | 0 |
| S17-k141_246698 | 2706 | No | NA | 2 | 0 | 2 | Low-quality | Genome-fragment | 0.23 | AAI-based (medium-confidence) | 0 |
| S2-final-viral-combined_k141_49817\|\|full | 2706 | No | NA | 4 | 0 | 1 | Low-quality | Genome-fragment | 6.15 | AAI-based (medium-confidence) | 0 |
| S12-final-viral-combined_k141_239433\|\|full | 1467 | No | NA | 2 | 0 | 2 | Not-determined | Genome-fragment | NA | NA | 0 |
| S9-final-viral-combined_k141_104070\|\|full | 2707 | No | NA | 5 | 0 | 0 | Low-quality | Genome-fragment | 21.33 | AAI-based (high-confidence) | 0 |
| S12-final-viral-combined_k141_297276\|\|full | 2707 | No | NA | 2 | 0 | 0 | Low-quality | Genome-fragment | 1.59 | AAI-based (high-confidence) | 0 |
| S12-final-viral-combined_k141_390399\|\|full | 6742 | No | NA | 9 | 0 | 3 | Not-determined | Genome-fragment | NA | NA | 0 |
| S12-final-viral-combined_k141_139209\|\|full | 1103 | No | NA | 2 | 0 | 1 | Not-determined | Genome-fragment | NA | NA | 0 |
| S2-final-viral-combined_k141_2387\|\|full | 2707 | No | NA | 5 | 2 | 0 | Low-quality | Genome-fragment | 5.13 | AAI-based (medium-confidence) | 0 |
| S12-final-viral-combined_k141_105477\|\|full | 1829 | No | NA | 2 | 0 | 1 | Not-determined | Genome-fragment | NA | NA | 0 |
| S12-final-viral-combined_k141_21957\|\|full | 3859 | No | NA | 3 | 0 | 1 | Not-determined | Genome-fragment | NA | NA | 0 |
| S12-final-viral-combined_k141_339679\|\|full | 5488 | No | NA | 7 | 0 | 1 | Not-determined | Genome-fragment | NA | NA | 0 |
| S12-final-viral-combined_k141_139249\|\|full | 2433 | No | NA | 2 | 0 | 1 | Not-determined | Genome-fragment | NA | NA | 0 |
| S12-final-viral-combined_k141_105523\|\|full | 1275 | No | NA | 2 | 0 | 0 | Not-determined | Genome-fragment | NA | NA | 0 |
| S12-final-viral-combined_k141_190066\|\|full | 6745 | No | NA | 6 | 0 | 2 | Not-determined | Genome-fragment | NA | NA | 0 |
| S12-final-viral-combined_k141_5062\|\|full | 4753 | No | NA | 6 | 0 | 0 | Not-determined | Genome-fragment | NA | NA | 0 |
| S12-final-viral-combined_k141_290193\|\|full | 3373 | No | NA | 3 | 0 | 2 | Not-determined | Genome-fragment | NA | NA | 0 |
| S12-final-viral-combined_k141_390469\|\|full | 5215 | No | NA | 4 | 0 | 4 | Not-determined | Genome-fragment | NA | NA | 0 |
| S12-final-viral-combined_k141_39083\|\|full | 3027 | No | NA | 4 | 0 | 3 | Not-determined | Genome-fragment | NA | NA | 0 |
| S2-final-viral-combined_k141_210869\|\|full | 2707 | No | NA | 6 | 1 | 0 | Low-quality | Genome-fragment | 5.15 | AAI-based (medium-confidence) | 0 |
| S9-final-viral-combined_k141_72677\|\|full | 2708 | No | NA | 4 | 1 | 0 | Low-quality | Genome-fragment | 6.95 | AAI-based (medium-confidence) | 0 |
| S1-final-viral-combined_k141_135632\|\|full | 2708 | No | NA | 3 | 0 | 1 | Low-quality | Genome-fragment | 1.24 | AAI-based (medium-confidence) | 0 |
| S12-final-viral-combined_k141_373835\|\|full | 3552 | No | NA | 5 | 0 | 1 | Not-determined | Genome-fragment | NA | NA | 0 |
| S12-final-viral-combined_k141_105568\|\|full | 1358 | No | NA | 3 | 0 | 1 | Not-determined | Genome-fragment | NA | NA | 0 |
| S12-final-viral-combined_k141_339732\|\|full | 8879 | No | NA | 8 | 0 | 6 | Not-determined | Genome-fragment | NA | NA | 0 |
| S12-final-viral-combined_k141_356066\|\|full | 1053 | No | NA | 2 | 0 | 1 | Not-determined | Genome-fragment | NA | NA | 0 |
| S7-final-viral-combined_k141_80307\|\|full | 2708 | No | NA | 5 | 1 | 0 | Low-quality | Genome-fragment | 5.16 | AAI-based (high-confidence) | 0 |
| S17-final-viral-combined_k141_85689\|\|full | 2708 | No | NA | 5 | 1 | 0 | Low-quality | Genome-fragment | 1.52 | HMM-based (lower-bound) | 0 |
| S12-final-viral-combined_k141_72960\|\|full | 2537 | No | NA | 3 | 0 | 2 | Not-determined | Genome-fragment | NA | NA | 0 |
| S12-final-viral-combined_k141_39098\|\|full | 4668 | No | NA | 5 | 0 | 0 | Not-determined | Genome-fragment | NA | NA | 0 |
| S8-final-viral-combined_k141_175441\|\|full | 2708 | No | NA | 2 | 1 | 0 | Low-quality | Genome-fragment | 3.04 | HMM-based (lower-bound) | 0 |
| S5-final-viral-combined_k141_119074\|\|full | 2709 | No | NA | 3 | 0 | 1 | Low-quality | Genome-fragment | 7.68 | AAI-based (high-confidence) | 0 |
| S12-final-viral-combined_k141_190114\|\|full | 36285 | No | NA | 45 | 6 | 4 | Medium-quality | Genome-fragment | 53.01 | HMM-based (lower-bound) | 0 |
| S9-final-viral-combined_k141_47218\|\|full | 2710 | No | NA | 2 | 2 | 0 | Low-quality | Genome-fragment | 7.9 | AAI-based (high-confidence) | 0 |
| S12-final-viral-combined_k141_256135\|\|full | 3530 | No | NA | 6 | 0 | 1 | Not-determined | Genome-fragment | NA | NA | 0 |
| S12-final-viral-combined_k141_172574\|\|full | 1917 | No | NA | 2 | 0 | 1 | Not-determined | Genome-fragment | NA | NA | 0 |
| S3-final-viral-combined_k141_93781\|\|full | 2710 | No | NA | 3 | 0 | 3 | Low-quality | Genome-fragment | 4.07 | AAI-based (medium-confidence) | 0 |
| S11-final-viral-combined_k141_145336\|\|full | 2711 | No | NA | 3 | 0 | 0 | Low-quality | Genome-fragment | 4.21 | AAI-based (high-confidence) | 0 |
| S12-final-viral-combined_k141_239626\|\|full | 1735 | No | NA | 3 | 0 | 2 | Not-determined | Genome-fragment | NA | NA | 0 |
| S12-final-viral-combined_k141_307314\|\|full | 1886 | No | NA | 4 | 0 | 0 | Not-determined | Genome-fragment | NA | NA | 0 |
| S12-final-viral-combined_k141_307331\|\|full | 1078 | No | NA | 2 | 0 | 0 | Not-determined | Genome-fragment | NA | NA | 0 |
| S12-final-viral-combined_k141_22082\|\|full | 1890 | No | NA | 3 | 0 | 2 | Not-determined | Genome-fragment | NA | NA | 0 |
| S12-final-viral-combined_k141_390566\|\|full | 3910 | No | NA | 4 | 0 | 4 | Not-determined | Genome-fragment | NA | NA | 0 |
| S17-final-viral-combined_k141_38219\|\|full | 2712 | No | NA | 4 | 0 | 1 | Low-quality | Genome-fragment | 0.7 | AAI-based (medium-confidence) | 0 |
| S9-final-viral-combined_k141_113629\|\|full | 2713 | No | NA | 5 | 0 | 1 | Low-quality | Genome-fragment | 4.21 | AAI-based (medium-confidence) | 0 |
| S12-final-viral-combined_k141_156057\|\|full | 6339 | No | NA | 9 | 0 | 1 | Not-determined | Genome-fragment | NA | NA | 0 |
| S12-final-viral-combined_k141_139293\|\|full | 2045 | No | NA | 3 | 0 | 3 | Not-determined | Genome-fragment | NA | NA | 0 |
| S1-final-viral-combined_k141_399\|\|full | 2713 | No | NA | 3 | 0 | 2 | Low-quality | Genome-fragment | 1.89 | AAI-based (medium-confidence) | 0 |
| S8-final-viral-combined_k141_187402\|\|full | 2713 | No | NA | 3 | 1 | 0 | Low-quality | Genome-fragment | 2.54 | HMM-based (lower-bound) | 0 |
| S5-final-viral-combined_k141_48601\|\|full | 2714 | No | NA | 6 | 2 | 0 | Low-quality | Genome-fragment | 7.18 | AAI-based (high-confidence) | 0 |
| S12-final-viral-combined_k141_139315\|\|full | 1403 | No | NA | 3 | 0 | 0 | Not-determined | Genome-fragment | NA | NA | 0 |
| S12-final-viral-combined_k141_273269\|\|full | 9494 | No | NA | 6 | 0 | 4 | Not-determined | Genome-fragment | NA | NA | 0 |
| S12-final-viral-combined_k141_222784\|\|full | 1837 | No | NA | 2 | 0 | 0 | Not-determined | Genome-fragment | NA | NA | 0 |
| S12-final-viral-combined_k141_239681\|\|full | 1253 | No | NA | 3 | 0 | 0 | Not-determined | Genome-fragment | NA | NA | 0 |
| S7-final-viral-combined_k141_123991\|\|full | 2714 | No | NA | 3 | 0 | 1 | Low-quality | Genome-fragment | 4.72 | AAI-based (medium-confidence) | 0 |
| S12-final-viral-combined_k141_22134\|\|full | 7666 | No | NA | 3 | 0 | 2 | Not-determined | Genome-fragment | NA | NA | 0 |
| S17-final-viral-combined_k141_216359\|\|full | 2714 | No | NA | 3 | 0 | 0 | Low-quality | Genome-fragment | 5.69 | AAI-based (high-confidence) | 0 |
| S1-final-viral-combined_k141_132379\|\|full | 2715 | No | NA | 3 | 0 | 1 | Low-quality | Genome-fragment | 2.83 | AAI-based (high-confidence) | 0 |
| S8_k141_222360 | 2715 | No | NA | 3 | 0 | 0 | Low-quality | Genome-fragment | 1.97 | AAI-based (high-confidence) | 0 |
| S11-final-viral-combined_k141_52442\|\|full | 2716 | No | NA | 2 | 0 | 1 | Low-quality | Genome-fragment | 0.86 | AAI-based (high-confidence) | 0 |
| S1-final-viral-combined_k141_33725\|\|full | 2716 | No | NA | 5 | 2 | 0 | Low-quality | Genome-fragment | 6.45 | AAI-based (high-confidence) | 0 |
| S1-final-viral-combined_k141_14849\|\|full | 2716 | No | NA | 6 | 0 | 1 | Low-quality | Genome-fragment | 4.13 | AAI-based (high-confidence) | 0 |
| S3-final-viral-combined_k141_112081\|\|full | 2716 | No | NA | 5 | 1 | 0 | Low-quality | Genome-fragment | 4.71 | AAI-based (high-confidence) | 0 |
| S12-final-viral-combined_k141_39106\|\|full | 6621 | No | NA | 8 | 0 | 0 | Not-determined | Genome-fragment | NA | NA | 0 |
| S7-final-viral-combined_k141_87706\|\|full | 2716 | No | NA | 3 | 2 | 0 | Low-quality | Genome-fragment | 5.7 | AAI-based (high-confidence) | 0 |
| S12-final-viral-combined_k141_74742\|\|full | 2716 | No | NA | 5 | 0 | 0 | Low-quality | Genome-fragment | 1.79 | AAI-based (high-confidence) | 0 |
| S12-final-viral-combined_k141_55124\|\|full | 2045 | No | NA | 5 | 0 | 0 | Not-determined | Genome-fragment | NA | NA | 0 |
| S2-final-viral-combined_k141_245876\|\|full | 2716 | No | NA | 4 | 0 | 1 | Low-quality | Genome-fragment | 1.06 | AAI-based (high-confidence) | 0 |
| S1-final-viral-combined_k141_187402\|\|full | 2717 | No | NA | 5 | 0 | 0 | Low-quality | Genome-fragment | 5.99 | AAI-based (high-confidence) | 0 |
| S5_k141_89982 | 2717 | No | NA | 2 | 0 | 0 | Low-quality | Genome-fragment | 3.51 | AAI-based (medium-confidence) | 0 |
| S12-final-viral-combined_k141_256298\|\|full | 7822 | No | NA | 10 | 0 | 1 | Not-determined | Genome-fragment | NA | NA | 0 |
| S12-final-viral-combined_k141_323762\|\|full | 1362 | No | NA | 2 | 0 | 0 | Not-determined | Genome-fragment | NA | NA | 0 |
| S12-final-viral-combined_k141_205979\|\|full | 2940 | No | NA | 3 | 0 | 3 | Not-determined | Genome-fragment | NA | NA | 0 |
| S12-final-viral-combined_k141_307478\|\|full | 4477 | No | NA | 6 | 0 | 3 | Not-determined | Genome-fragment | NA | NA | 0 |
| S12-final-viral-combined_k141_121944\|\|full | 1730 | No | NA | 2 | 0 | 1 | Not-determined | Genome-fragment | NA | NA | 0 |
| S2-final-viral-combined_k141_108174\|\|full | 2717 | No | NA | 4 | 2 | 0 | Low-quality | Genome-fragment | 7.79 | AAI-based (high-confidence) | 0 |
| S4_k141_128331 | 2717 | No | NA | 4 | 0 | 0 | Low-quality | Genome-fragment | 2.85 | AAI-based (high-confidence) | 0 |
| S12-final-viral-combined_k141_256299\|\|full | 53411 | Yes | 46763 | 64 | 12 | 4 | Medium-quality | Genome-fragment | 72.63 | AAI-based (high-confidence) | 12.45 |
| S12-final-viral-combined_k141_121965\|\|full | 2277 | No | NA | 3 | 0 | 1 | Not-determined | Genome-fragment | NA | NA | 0 |
| S12-final-viral-combined_k141_190314\|\|full | 2267 | No | NA | 3 | 0 | 3 | Not-determined | Genome-fragment | NA | NA | 0 |
| S12-final-viral-combined_k141_39251\|\|full | 1414 | No | NA | 3 | 0 | 1 | Not-determined | Genome-fragment | NA | NA | 0 |
| S12-final-viral-combined_k141_156234\|\|full | 46328 | Yes | 30049 | 52 | 17 | 18 | Medium-quality | Genome-fragment | 52.44 | HMM-based (lower-bound) | 35.14 |
| S11-final-viral-combined_k141_181225\|\|full | 2718 | No | NA | 3 | 0 | 0 | Low-quality | Genome-fragment | 4.59 | AAI-based (medium-confidence) | 0 |
| S12-final-viral-combined_k141_55200\|\|full | 1581 | No | NA | 4 | 0 | 0 | Not-determined | Genome-fragment | NA | NA | 0 |
| S12-final-viral-combined_k141_121995\|\|full | 1822 | No | NA | 4 | 0 | 1 | Not-determined | Genome-fragment | NA | NA | 0 |
| S12-final-viral-combined_k141_301330\|\|full | 2718 | No | NA | 4 | 0 | 1 | Low-quality | Genome-fragment | 4.78 | AAI-based (high-confidence) | 0 |
| S12-final-viral-combined_k141_307532\|\|full | 4428 | No | NA | 4 | 0 | 3 | Not-determined | Genome-fragment | NA | NA | 0 |
| S12-final-viral-combined_k141_190370\|\|full | 1150 | No | NA | 3 | 0 | 1 | Not-determined | Genome-fragment | NA | NA | 0 |
| S12-final-viral-combined_k141_223043\|\|full | 20174 | No | NA | 27 | 10 | 0 | Medium-quality | Genome-fragment | 56.36 | AAI-based (medium-confidence) | 0 |
| S12-final-viral-combined_k141_239853\|\|full | 2987 | No | NA | 3 | 0 | 3 | Not-determined | Genome-fragment | NA | NA | 0 |
| S12-final-viral-combined_k141_5272\|\|full | 1880 | No | NA | 4 | 0 | 2 | Not-determined | Genome-fragment | NA | NA | 0 |
| S12-final-viral-combined_k141_55258\|\|full | 2120 | No | NA | 2 | 0 | 2 | Not-determined | Genome-fragment | NA | NA | 0 |
| S2-final-viral-combined_k141_178300\|\|full | 2718 | No | NA | 4 | 2 | 0 | Low-quality | Genome-fragment | 5.56 | AAI-based (high-confidence) | 0 |
| S12-final-viral-combined_k141_239888\|\|full | 2695 | No | NA | 4 | 0 | 0 | Not-determined | Genome-fragment | NA | NA | 0 |
| S12-final-viral-combined_k141_5274\|\|full | 12098 | No | NA | 10 | 0 | 3 | Not-determined | Genome-fragment | NA | NA | 0 |
| S2-final-viral-combined_k141_172956\|\|full | 2718 | No | NA | 3 | 0 | 0 | Low-quality | Genome-fragment | 6.94 | AAI-based (high-confidence) | 0 |
| S12-final-viral-combined_k141_356239\|\|full | 8702 | No | NA | 15 | 0 | 2 | Not-determined | Genome-fragment | NA | NA | 0 |
| S12-final-viral-combined_k141_374031\|\|full | 1080 | No | NA | 2 | 0 | 0 | Not-determined | Genome-fragment | NA | NA | 0 |
| S4-final-viral-combined_k141_130407\|\|full | 2718 | No | NA | 4 | 0 | 0 | Low-quality | Genome-fragment | 6.25 | AAI-based (high-confidence) | 0 |
| S1-final-viral-combined_k141_338624\|\|full | 2719 | No | NA | 3 | 2 | 0 | Low-quality | Genome-fragment | 6.75 | AAI-based (high-confidence) | 0 |
| S12-final-viral-combined_k141_307637\|\|full | 6533 | No | NA | 3 | 0 | 3 | Not-determined | Genome-fragment | NA | NA | 0 |
| S12-final-viral-combined_k141_206092\|\|full | 1949 | No | NA | 2 | 0 | 2 | Not-determined | Genome-fragment | NA | NA | 0 |
| S1-final-viral-combined_k141_15174\|\|full | 2719 | No | NA | 3 | 0 | 2 | Low-quality | Genome-fragment | 1.92 | AAI-based (medium-confidence) | 0 |
| S2-final-viral-combined_k141_28678\|\|full | 2719 | No | NA | 5 | 0 | 2 | Low-quality | Genome-fragment | 2 | AAI-based (high-confidence) | 0 |
| S12-final-viral-combined_k141_105840\|\|full | 2252 | No | NA | 2 | 0 | 1 | Not-determined | Genome-fragment | NA | NA | 0 |
| S12-final-viral-combined_k141_374088\|\|full | 1210 | No | NA | 3 | 0 | 1 | Not-determined | Genome-fragment | NA | NA | 0 |
| S12-final-viral-combined_k141_340076\|\|full | 1671 | No | NA | 3 | 0 | 2 | Not-determined | Genome-fragment | NA | NA | 0 |
| S12-final-viral-combined_k141_323943\|\|full | 55241 | No | NA | 86 | 21 | 2 | High-quality | High-quality | 98.08 | AAI-based (high-confidence) | 0 |
| S12-final-viral-combined_k141_190548\|\|full | 2214 | No | NA | 2 | 0 | 2 | Not-determined | Genome-fragment | NA | NA | 0 |
| S12-final-viral-combined_k141_323944\|\|full | 1183 | No | NA | 2 | 0 | 0 | Not-determined | Genome-fragment | NA | NA | 0 |
| S4-final-viral-combined_k141_3657\|\|full | 2719 | No | NA | 3 | 0 | 0 | Low-quality | Genome-fragment | 5.11 | AAI-based (medium-confidence) | 0 |
| S4-final-viral-combined_k141_27042\|\|full | 2719 | No | NA | 3 | 0 | 3 | Low-quality | Genome-fragment | 2.23 | AAI-based (high-confidence) | 0 |
| S17-final-viral-combined_k141_43221\|\|full | 2720 | No | NA | 3 | 0 | 2 | Low-quality | Genome-fragment | 0.65 | AAI-based (medium-confidence) | 0 |
| S1-final-viral-combined_k141_202082\|\|full | 2721 | No | NA | 2 | 0 | 1 | Low-quality | Genome-fragment | 3.17 | AAI-based (high-confidence) | 0 |
| S12-final-viral-combined_k141_307713\|\|full | 1767 | No | NA | 3 | 0 | 0 | Not-determined | Genome-fragment | NA | NA | 0 |
| S12-final-viral-combined_k141_323984\|\|full | 3479 | No | NA | 3 | 0 | 2 | Not-determined | Genome-fragment | NA | NA | 0 |
| S5-final-viral-combined_k141_6624\|\|full | 2721 | No | NA | 5 | 1 | 0 | Low-quality | Genome-fragment | 1.65 | HMM-based (lower-bound) | 0 |
| S7-final-viral-combined_k141_221213\|\|full | 2721 | No | NA | 6 | 2 | 0 | Low-quality | Genome-fragment | 6.06 | AAI-based (medium-confidence) | 0 |
| S12-final-viral-combined_k141_73199\|\|full | 1454 | No | NA | 3 | 0 | 2 | Not-determined | Genome-fragment | NA | NA | 0 |
| S12-final-viral-combined_k141_206225\|\|full | 2562 | No | NA | 3 | 0 | 3 | Not-determined | Genome-fragment | NA | NA | 0 |
| S12-final-viral-combined_k141_5404\|\|full | 3175 | No | NA | 5 | 0 | 3 | Not-determined | Genome-fragment | NA | NA | 0 |
| S12-final-viral-combined_k141_256515\|\|full | 2356 | No | NA | 2 | 0 | 2 | Not-determined | Genome-fragment | NA | NA | 0 |
| S8-final-viral-combined_k141_266693\|\|full | 2721 | No | NA | 3 | 0 | 3 | Low-quality | Genome-fragment | 0.91 | AAI-based (medium-confidence) | 0 |
| S11-final-viral-combined_k141_89807\|\|full | 2722 | No | NA | 5 | 1 | 0 | Low-quality | Genome-fragment | 4.83 | AAI-based (medium-confidence) | 0 |
| S12-final-viral-combined_k141_39321\|\|full | 2807 | No | NA | 2 | 0 | 1 | Not-determined | Genome-fragment | NA | NA | 0 |
| S1-final-viral-combined_k141_77943\|\|full | 2722 | No | NA | 6 | 4 | 0 | Low-quality | Genome-fragment | 7.78 | AAI-based (high-confidence) | 0 |
| S12-final-viral-combined_k141_122167\|\|full | 4679 | No | NA | 5 | 0 | 5 | Not-determined | Genome-fragment | NA | NA | 0 |
| S4_k141_163400 | 2722 | No | NA | 2 | 0 | 0 | Low-quality | Genome-fragment | 4.03 | AAI-based (medium-confidence) | 0 |
| S5-final-viral-combined_k141_67910\|\|full | 2723 | Yes | 1998 | 4 | 1 | 1 | Low-quality | Genome-fragment | 1.26 | HMM-based (lower-bound) | 26.63 |
| S12-final-viral-combined_k141_240141\|\|full | 3990 | No | NA | 4 | 0 | 2 | Not-determined | Genome-fragment | NA | NA | 0 |
| S12-final-viral-combined_k141_324019\|\|full | 14833 | No | NA | 11 | 0 | 11 | Not-determined | Genome-fragment | NA | NA | 0 |
| S11-final-viral-combined_k141_132032\|\|full | 2724 | No | NA | 7 | 1 | 0 | Low-quality | Genome-fragment | 4.54 | AAI-based (medium-confidence) | 0 |
| S12-final-viral-combined_k141_172997\|\|full | 3683 | No | NA | 4 | 0 | 3 | Not-determined | Genome-fragment | NA | NA | 0 |
| S12-final-viral-combined_k141_39361\|\|full | 3182 | No | NA | 7 | 0 | 2 | Not-determined | Genome-fragment | NA | NA | 0 |
| S1-final-viral-combined_k141_167875\|\|full | 2724 | No | NA | 5 | 1 | 0 | Low-quality | Genome-fragment | 4.9 | AAI-based (high-confidence) | 0 |
| S1-final-viral-combined_k141_319063\|\|full | 2724 | No | NA | 2 | 0 | 2 | Low-quality | Genome-fragment | 1.55 | AAI-based (medium-confidence) | 0 |
| S12-final-viral-combined_k141_105997\|\|full | 1033 | No | NA | 1 | 0 | 0 | Not-determined | Genome-fragment | NA | NA | 0 |
| S12-final-viral-combined_k141_390985\|\|full | 5098 | No | NA | 6 | 2 | 0 | High-quality | High-quality | 95.91 | AAI-based (medium-confidence) | 0 |
| S12-final-viral-combined_k141_173012\|\|full | 1858 | No | NA | 3 | 0 | 2 | Not-determined | Genome-fragment | NA | NA | 0 |
| S12-final-viral-combined_k141_340150\|\|full | 1980 | No | NA | 2 | 0 | 1 | Not-determined | Genome-fragment | NA | NA | 0 |
| S12-final-viral-combined_k141_106021\|\|full | 1432 | No | NA | 2 | 0 | 2 | Not-determined | Genome-fragment | NA | NA | 0 |
| S12-final-viral-combined_k141_256562\|\|full | 1888 | No | NA | 4 | 0 | 0 | Not-determined | Genome-fragment | NA | NA | 0 |
| S12-final-viral-combined_k141_73363\|\|full | 2946 | No | NA | 3 | 0 | 1 | Not-determined | Genome-fragment | NA | NA | 0 |
| S5-final-viral-combined_k141_115194\|\|full | 2724 | No | NA | 3 | 0 | 1 | Low-quality | Genome-fragment | 5.06 | AAI-based (high-confidence) | 0 |
| S12-final-viral-combined_k141_223173\|\|full | 7678 | No | NA | 9 | 0 | 3 | Not-determined | Genome-fragment | NA | NA | 0 |
| S12-final-viral-combined_k141_356426\|\|full | 3564 | No | NA | 4 | 0 | 1 | Not-determined | Genome-fragment | NA | NA | 0 |
| S6-final-viral-combined_k141_94186\|\|full | 2724 | No | NA | 4 | 1 | 0 | Low-quality | Genome-fragment | 3.91 | HMM-based (lower-bound) | 0 |
| S11-final-viral-combined_k141_226604\|\|full | 2726 | No | NA | 3 | 1 | 0 | Low-quality | Genome-fragment | 0.88 | HMM-based (lower-bound) | 0 |
| S12-final-viral-combined_k141_223186\|\|full | 1298 | No | NA | 2 | 0 | 0 | Not-determined | Genome-fragment | NA | NA | 0 |
| S5-final-viral-combined_k141_16694\|\|full | 2726 | No | NA | 5 | 0 | 0 | Low-quality | Genome-fragment | 5.12 | AAI-based (medium-confidence) | 0 |
| S12-final-viral-combined_k141_73384\|\|full | 1893 | No | NA | 3 | 0 | 3 | Not-determined | Genome-fragment | NA | NA | 0 |
| S12-final-viral-combined_k141_240201\|\|full | 1819 | No | NA | 4 | 0 | 0 | Not-determined | Genome-fragment | NA | NA | 0 |
| S12-final-viral-combined_k141_256614\|\|full | 1975 | No | NA | 2 | 0 | 0 | Not-determined | Genome-fragment | NA | NA | 0 |
| S12-final-viral-combined_k141_223202\|\|full | 2228 | No | NA | 4 | 0 | 2 | Not-determined | Genome-fragment | NA | NA | 0 |
| S11-final-viral-combined_k141_218827\|\|full | 2727 | No | NA | 3 | 0 | 1 | Low-quality | Genome-fragment | 2.01 | AAI-based (medium-confidence) | 0 |
| S7-final-viral-combined_k141_1717\|\|full | 2727 | No | NA | 3 | 0 | 0 | Low-quality | Genome-fragment | 4.27 | AAI-based (medium-confidence) | 0 |
| S12-final-viral-combined_k141_5571\|\|full | 1894 | No | NA | 2 | 0 | 1 | Not-determined | Genome-fragment | NA | NA | 0 |
| S12-final-viral-combined_k141_89417\|\|full | 1561 | No | NA | 5 | 0 | 1 | Not-determined | Genome-fragment | NA | NA | 0 |
| S12-final-viral-combined_k141_324087\|\|full | 1267 | No | NA | 3 | 0 | 1 | Not-determined | Genome-fragment | NA | NA | 0 |
| S12-final-viral-combined_k141_374272\|\|full | 1197 | No | NA | 3 | 0 | 0 | Not-determined | Genome-fragment | NA | NA | 0 |
| S17-final-viral-combined_k141_158427\|\|full | 2727 | No | NA | 4 | 0 | 2 | Low-quality | Genome-fragment | 5.06 | AAI-based (medium-confidence) | 0 |
| S12-final-viral-combined_k141_290988\|\|full | 3097 | No | NA | 2 | 0 | 2 | Not-determined | Genome-fragment | NA | NA | 0 |
| S1_k141_130051 | 2728 | No | NA | 4 | 2 | 0 | Low-quality | Genome-fragment | 6.17 | AAI-based (high-confidence) | 0 |
| S12-final-viral-combined_k141_126808\|\|full | 2728 | No | NA | 4 | 1 | 0 | Low-quality | Genome-fragment | 1.28 | HMM-based (lower-bound) | 0 |
| S8-final-viral-combined_k141_117021\|\|full | 2728 | No | NA | 7 | 0 | 0 | Low-quality | Genome-fragment | 8.48 | AAI-based (high-confidence) | 0 |
| S7-final-viral-combined_k141_129131\|\|full | 2729 | No | NA | 5 | 0 | 0 | Low-quality | Genome-fragment | 5.99 | AAI-based (high-confidence) | 0 |
| S12-final-viral-combined_k141_356511\|\|full | 2381 | No | NA | 2 | 0 | 2 | Not-determined | Genome-fragment | NA | NA | 0 |
| S4-final-viral-combined_k141_118833\|\|full | 2729 | No | NA | 4 | 0 | 0 | Low-quality | Genome-fragment | 4.31 | AAI-based (high-confidence) | 0 |
| S12-final-viral-combined_k141_374395\|\|full | 2730 | No | NA | 4 | 1 | 0 | Low-quality | Genome-fragment | 8.2 | AAI-based (high-confidence) | 0 |
| S8-final-viral-combined_k141_22057\|\|full | 2731 | No | NA | 4 | 1 | 0 | Low-quality | Genome-fragment | 46.34 | AAI-based (medium-confidence) | 0 |
| S5-final-viral-combined_k141_178670\|\|full | 2732 | No | NA | 4 | 0 | 0 | Low-quality | Genome-fragment | 4.53 | AAI-based (medium-confidence) | 0 |
| S12-final-viral-combined_k141_307909\|\|full | 1097 | No | NA | 3 | 0 | 2 | Not-determined | Genome-fragment | NA | NA | 0 |
| S12-final-viral-combined_k141_374332\|\|full | 1345 | No | NA | 3 | 0 | 2 | Not-determined | Genome-fragment | NA | NA | 0 |
| S7-final-viral-combined_k141_61397\|\|full | 2732 | No | NA | 3 | 0 | 0 | Low-quality | Genome-fragment | 4.85 | AAI-based (medium-confidence) | 0 |
| S2-final-viral-combined_k141_18817\|\|0_partial | 2732 | Yes | 1435 | 3 | 1 | 1 | Low-quality | Genome-fragment | 34.48 | HMM-based (lower-bound) | 47.47 |
| S12-final-viral-combined_k141_240290\|\|full | 1664 | No | NA | 4 | 0 | 0 | Not-determined | Genome-fragment | NA | NA | 0 |
| S1-final-viral-combined_k141_82924\|\|full | 2733 | No | NA | 4 | 1 | 0 | Low-quality | Genome-fragment | 6.17 | AAI-based (high-confidence) | 0 |
| S12-final-viral-combined_k141_307922\|\|full | 1046 | No | NA | 2 | 0 | 0 | Not-determined | Genome-fragment | NA | NA | 0 |
| S12-final-viral-combined_k141_73574\|\|full | 2086 | No | NA | 3 | 0 | 2 | Not-determined | Genome-fragment | NA | NA | 0 |
| S12-final-viral-combined_k141_58849\|\|full | 2733 | No | NA | 5 | 0 | 1 | Low-quality | Genome-fragment | 6.38 | AAI-based (high-confidence) | 0 |
| S12-final-viral-combined_k141_39658\|\|full | 1490 | No | NA | 3 | 0 | 3 | Not-determined | Genome-fragment | NA | NA | 0 |
| S12-final-viral-combined_k141_307923\|\|full | 2385 | No | NA | 2 | 0 | 0 | Not-determined | Genome-fragment | NA | NA | 0 |
| S12-final-viral-combined_k141_5632\|\|full | 1254 | No | NA | 3 | 0 | 1 | Not-determined | Genome-fragment | NA | NA | 0 |
| S12-final-viral-combined_k141_206517\|\|full | 5204 | No | NA | 9 | 0 | 0 | Not-determined | Genome-fragment | NA | NA | 0 |
| S12-final-viral-combined_k141_356570\|\|full | 1685 | No | NA | 4 | 0 | 2 | Not-determined | Genome-fragment | NA | NA | 0 |
| S12-final-viral-combined_k141_190917\|\|full | 3184 | No | NA | 4 | 0 | 0 | Not-determined | Genome-fragment | NA | NA | 0 |
| S8-final-viral-combined_k141_209789\|\|full | 2733 | No | NA | 3 | 0 | 0 | Low-quality | Genome-fragment | 4.22 | AAI-based (medium-confidence) | 0 |
| S11-final-viral-combined_k141_180827\|\|full | 2734 | No | NA | 3 | 0 | 1 | Low-quality | Genome-fragment | 3.62 | AAI-based (high-confidence) | 0 |
| S7-final-viral-combined_k141_224339\|\|full | 2734 | No | NA | 7 | 0 | 0 | Low-quality | Genome-fragment | 6.09 | AAI-based (high-confidence) | 0 |
| S12-final-viral-combined_k141_46496\|\|full | 2734 | No | NA | 5 | 0 | 1 | Low-quality | Genome-fragment | 1.73 | AAI-based (medium-confidence) | 0 |
| S12-final-viral-combined_k141_391235\|\|full | 1621 | No | NA | 2 | 0 | 0 | Not-determined | Genome-fragment | NA | NA | 0 |
| S12-final-viral-combined_k141_73600\|\|full | 1037 | No | NA | 3 | 0 | 2 | Not-determined | Genome-fragment | NA | NA | 0 |
| S4-final-viral-combined_k141_140280\|\|full | 2734 | No | NA | 3 | 1 | 0 | Low-quality | Genome-fragment | 3.29 | AAI-based (medium-confidence) | 0 |
| S9_k141_33407 | 2735 | No | NA | 3 | 0 | 0 | Low-quality | Genome-fragment | 5.76 | AAI-based (high-confidence) | 0 |
| S12-final-viral-combined_k141_307957\|\|full | 1530 | No | NA | 3 | 0 | 1 | Not-determined | Genome-fragment | NA | NA | 0 |
| S12-final-viral-combined_k141_156785\|\|full | 1931 | No | NA | 5 | 0 | 0 | Not-determined | Genome-fragment | NA | NA | 0 |
| S6-final-viral-combined_k141_4953\|\|full | 2735 | No | NA | 4 | 1 | 0 | Low-quality | Genome-fragment | 3.46 | AAI-based (high-confidence) | 0 |
| S11-final-viral-combined_k141_244293\|\|full | 2736 | No | NA | 4 | 1 | 0 | Low-quality | Genome-fragment | 4.78 | AAI-based (high-confidence) | 0 |
| S1-final-viral-combined_k141_4996\|\|full | 2736 | No | NA | 5 | 0 | 0 | Low-quality | Genome-fragment | 4.43 | AAI-based (medium-confidence) | 0 |
| S1-final-viral-combined_k141_269856\|\|full | 2737 | No | NA | 3 | 0 | 1 | Low-quality | Genome-fragment | 1.3 | AAI-based (medium-confidence) | 0 |
| S12-final-viral-combined_k141_256729\|\|full | 2915 | No | NA | 4 | 0 | 2 | Not-determined | Genome-fragment | NA | NA | 0 |
| S12-final-viral-combined_k141_324302\|\|full | 8177 | No | NA | 10 | 0 | 6 | Not-determined | Genome-fragment | NA | NA | 0 |
| S12-final-viral-combined_k141_223412\|\|full | 1492 | No | NA | 2 | 0 | 1 | Not-determined | Genome-fragment | NA | NA | 0 |
| S3-final-viral-combined_k141_21086\|\|full | 2737 | No | NA | 3 | 0 | 1 | Low-quality | Genome-fragment | 4.8 | AAI-based (high-confidence) | 0 |
| S12-final-viral-combined_k141_307984\|\|full | 1909 | No | NA | 2 | 0 | 1 | Not-determined | Genome-fragment | NA | NA | 0 |
| S12-final-viral-combined_k141_324308\|\|full | 3078 | No | NA | 3 | 0 | 1 | Not-determined | Genome-fragment | NA | NA | 0 |
| S12-final-viral-combined_k141_122499\|\|full | 8579 | No | NA | 6 | 0 | 1 | Not-determined | Genome-fragment | NA | NA | 0 |
| S7-final-viral-combined_k141_28330\|\|full | 2737 | No | NA | 2 | 0 | 2 | Low-quality | Genome-fragment | 0.75 | AAI-based (medium-confidence) | 0 |
| S12-final-viral-combined_k141_256761\|\|full | 4225 | No | NA | 4 | 0 | 2 | Not-determined | Genome-fragment | NA | NA | 0 |
| S5-final-viral-combined_k141_51783\|\|full | 2738 | No | NA | 2 | 1 | 0 | Low-quality | Genome-fragment | 5.92 | AAI-based (high-confidence) | 0 |
| S17-final-viral-combined_k141_113016\|\|full | 2738 | No | NA | 7 | 2 | 0 | Low-quality | Genome-fragment | 6.53 | AAI-based (high-confidence) | 0 |
| S2-final-viral-combined_k141_128739\|\|full | 2738 | No | NA | 8 | 0 | 0 | Low-quality | Genome-fragment | 5.83 | AAI-based (medium-confidence) | 0 |
| S7-final-viral-combined_k141_219734\|\|full | 2739 | No | NA | 4 | 3 | 0 | Low-quality | Genome-fragment | 4.54 | AAI-based (high-confidence) | 0 |
| S2-final-viral-combined_k141_105479\|\|full | 2739 | No | NA | 3 | 0 | 0 | Low-quality | Genome-fragment | 3.7 | AAI-based (high-confidence) | 0 |
| S12-final-viral-combined_k141_55621\|\|full | 3254 | No | NA | 3 | 0 | 2 | Not-determined | Genome-fragment | NA | NA | 0 |
| S12-final-viral-combined_k141_291180\|\|full | 1511 | No | NA | 2 | 0 | 0 | Not-determined | Genome-fragment | NA | NA | 0 |
| S2_k141_233761 | 2739 | No | NA | 3 | 0 | 2 | Low-quality | Genome-fragment | 0.83 | AAI-based (medium-confidence) | 0 |
| S4-final-viral-combined_k141_63968\|\|full | 2739 | No | NA | 2 | 0 | 1 | Low-quality | Genome-fragment | 2.2 | AAI-based (high-confidence) | 0 |
| S12-final-viral-combined_k141_356603\|\|full | 5992 | No | NA | 5 | 0 | 5 | Not-determined | Genome-fragment | NA | NA | 0 |
| S11-final-viral-combined_k141_225930\|\|full | 2740 | No | NA | 4 | 0 | 1 | Low-quality | Genome-fragment | 7.87 | AAI-based (high-confidence) | 0 |
| S2-final-viral-combined_k141_14614\|\|full | 2740 | No | NA | 5 | 0 | 1 | Low-quality | Genome-fragment | 5.55 | AAI-based (medium-confidence) | 0 |
| S8-final-viral-combined_k141_226695\|\|full | 2740 | No | NA | 6 | 5 | 0 | Low-quality | Genome-fragment | 5.4 | AAI-based (high-confidence) | 0 |
| S12-final-viral-combined_k141_89616\|\|full | 4426 | No | NA | 3 | 0 | 3 | Not-determined | Genome-fragment | NA | NA | 0 |
| S12-final-viral-combined_k141_5820\|\|full | 3145 | No | NA | 3 | 0 | 2 | Not-determined | Genome-fragment | NA | NA | 0 |
| S12-final-viral-combined_k141_39848\|\|full | 1561 | No | NA | 3 | 0 | 0 | Not-determined | Genome-fragment | NA | NA | 0 |
| S1-final-viral-combined_k141_374267\|\|full | 2741 | No | NA | 3 | 2 | 0 | Low-quality | Genome-fragment | 1.62 | AAI-based (medium-confidence) | 0 |
| S2_k141_224799 | 2741 | No | NA | 3 | 0 | 3 | Low-quality | Genome-fragment | 3.28 | AAI-based (medium-confidence) | 0 |
| S8-final-viral-combined_k141_5151\|\|full | 2741 | No | NA | 6 | 0 | 0 | Low-quality | Genome-fragment | 7.41 | AAI-based (high-confidence) | 0 |
| S12-final-viral-combined_k141_55678\|\|full | 1691 | No | NA | 3 | 0 | 0 | Not-determined | Genome-fragment | NA | NA | 0 |
| S9-final-viral-combined_k141_6455\|\|full | 2742 | No | NA | 3 | 0 | 3 | Low-quality | Genome-fragment | 1.57 | AAI-based (medium-confidence) | 0 |
| S1-final-viral-combined_k141_144330\|\|full | 2742 | No | NA | 4 | 0 | 3 | Low-quality | Genome-fragment | 0.66 | AAI-based (medium-confidence) | 0 |
| S12-final-viral-combined_k141_308074\|\|full | 1770 | No | NA | 3 | 0 | 1 | Not-determined | Genome-fragment | NA | NA | 0 |
| S11-final-viral-combined_k141_140508\|\|full | 2743 | No | NA | 4 | 0 | 1 | Low-quality | Genome-fragment | 1.73 | AAI-based (medium-confidence) | 0 |
| S12-final-viral-combined_k141_22884\|\|full | 6193 | No | NA | 5 | 0 | 4 | Not-determined | Genome-fragment | NA | NA | 0 |
| S7_k141_59537 | 2743 | No | NA | 6 | 0 | 1 | Low-quality | Genome-fragment | 7.05 | AAI-based (medium-confidence) | 0 |
| S2-final-viral-combined_k141_155737\|\|full | 2743 | No | NA | 4 | 0 | 0 | Low-quality | Genome-fragment | 7.27 | AAI-based (medium-confidence) | 0 |
| S8-final-viral-combined_k141_94178\|\|full | 2743 | No | NA | 7 | 1 | 0 | Low-quality | Genome-fragment | 5.98 | AAI-based (high-confidence) | 0 |
| S12-final-viral-combined_k141_39882\|\|full | 4885 | No | NA | 4 | 0 | 0 | Not-determined | Genome-fragment | NA | NA | 0 |
| S17-final-viral-combined_k141_65518\|\|full | 2744 | No | NA | 3 | 2 | 0 | Low-quality | Genome-fragment | 3.33 | AAI-based (high-confidence) | 0 |

| | | | | | | | | | | | |
|---|---|---|---|---|---|---|---|---|---|---|---|
| S12-final-viral-combined_k141_256826\|\|full | 2018 | No | NA | 3 | 0 | 1 | Not-determined | Genome-fragment | NA | NA | 0 |
| S1-final-viral-combined_k141_282664\|\|full | 2745 | No | NA | 3 | 0 | 2 | Low-quality | Genome-fragment | 1.33 | AAI-based (medium-confidence) | 0 |
| S12-final-viral-combined_k141_106326\|\|full | 1258 | No | NA | 3 | 0 | 2 | Not-determined | Genome-fragment | NA | NA | 0 |
| S12-final-viral-combined_k141_39899\|\|full | 4620 | No | NA | 4 | 0 | 3 | Not-determined | Genome-fragment | NA | NA | 0 |
| S12-final-viral-combined_k141_191072\|\|full | 4826 | No | NA | 2 | 0 | 0 | Not-determined | Genome-fragment | NA | NA | 0 |
| S12-final-viral-combined_k141_223598\|\|full | 2667 | No | NA | 3 | 0 | 2 | Not-determined | Genome-fragment | NA | NA | 0 |
| S12-final-viral-combined_k141_240530\|\|full | 18958 | No | NA | 20 | 0 | 11 | Not-determined | Genome-fragment | NA | NA | 0 |
| S12-final-viral-combined_k141_206736\|\|full | 1957 | No | NA | 2 | 0 | 1 | Not-determined | Genome-fragment | NA | NA | 0 |
| S12-final-viral-combined_k141_206739\|\|full | 1324 | No | NA | 2 | 0 | 2 | Not-determined | Genome-fragment | NA | NA | 0 |
| S12-final-viral-combined_k141_308177\|\|full | 1518 | No | NA | 3 | 0 | 0 | Not-determined | Genome-fragment | NA | NA | 0 |
| S1-final-viral-combined_k141_349738\|\|full | 2745 | No | NA | 3 | 2 | 0 | Low-quality | Genome-fragment | 4.75 | AAI-based (medium-confidence) | 0 |
| S1_k141_220334 | 2745 | No | NA | 4 | 1 | 0 | Low-quality | Genome-fragment | 6.03 | AAI-based (high-confidence) | 0 |
| S12-final-viral-combined_k141_55775\|\|full | 6929 | No | NA | 6 | 0 | 3 | Not-determined | Genome-fragment | NA | NA | 0 |
| S12-final-viral-combined_k141_140159\|\|full | 6641 | No | NA | 12 | 0 | 1 | Not-determined | Genome-fragment | NA | NA | 0 |
| S12-final-viral-combined_k141_5952\|\|full | 2343 | No | NA | 2 | 0 | 2 | Not-determined | Genome-fragment | NA | NA | 0 |
| S7-final-viral-combined_k141_210723\|\|full | 2745 | No | NA | 2 | 0 | 1 | Low-quality | Genome-fragment | 1.74 | AAI-based (medium-confidence) | 0 |
| S17-k141_142240 | 2745 | No | NA | 6 | 3 | 0 | Low-quality | Genome-fragment | 9.52 | AAI-based (high-confidence) | 0 |
| S12-final-viral-combined_k141_73834\|\|full | 3063 | No | NA | 3 | 0 | 1 | Not-determined | Genome-fragment | NA | NA | 0 |
| S12-final-viral-combined_k141_273931\|\|full | 5816 | No | NA | 7 | 0 | 5 | Not-determined | Genome-fragment | NA | NA | 0 |
| S12-final-viral-combined_k141_240577\|\|full | 1106 | No | NA | 2 | 0 | 1 | Not-determined | Genome-fragment | NA | NA | 0 |
| S17-final-viral-combined_k141_44822\|\|full | 2746 | No | NA | 3 | 0 | 3 | Low-quality | Genome-fragment | 1.29 | AAI-based (high-confidence) | 0 |
| S12-final-viral-combined_k141_391435\|\|full | 2147 | No | NA | 4 | 0 | 1 | Not-determined | Genome-fragment | NA | NA | 0 |
| S12-final-viral-combined_k141_39994\|\|full | 5842 | No | NA | 6 | 0 | 3 | Not-determined | Genome-fragment | NA | NA | 0 |
| S12-final-viral-combined_k141_223720\|\|full | 2739 | No | NA | 2 | 0 | 2 | Not-determined | Genome-fragment | NA | NA | 0 |
| S12-final-viral-combined_k141_54331\|\|full | 2746 | No | NA | 2 | 0 | 2 | Low-quality | Genome-fragment | 4.23 | AAI-based (medium-confidence) | 0 |
| S12-final-viral-combined_k141_273989\|\|full | 4600 | No | NA | 3 | 0 | 1 | Not-determined | Genome-fragment | NA | NA | 0 |
| S8-final-viral-combined_k141_151130\|\|full | 2747 | No | NA | 2 | 0 | 1 | Low-quality | Genome-fragment | 2.04 | AAI-based (medium-confidence) | 0 |
| S12-final-viral-combined_k141_308284\|\|full | 5534 | No | NA | 7 | 0 | 3 | Not-determined | Genome-fragment | NA | NA | 0 |
| S11-final-viral-combined_k141_188519\|\|full | 2748 | No | NA | 3 | 2 | 0 | Low-quality | Genome-fragment | 3.38 | HMM-based (lower-bound) | 0 |
| S5-final-viral-combined_k141_34001\|\|full | 2748 | No | NA | 4 | 0 | 1 | Low-quality | Genome-fragment | 0.45 | AAI-based (medium-confidence) | 0 |
| S12-final-viral-combined_k141_374718\|\|full | 2679 | No | NA | 5 | 0 | 3 | Not-determined | Genome-fragment | NA | NA | 0 |
| S12-final-viral-combined_k141_223769\|\|full | 1726 | No | NA | 3 | 0 | 1 | Not-determined | Genome-fragment | NA | NA | 0 |
| S12-final-viral-combined_k141_191198\|\|full | 5280 | No | NA | 5 | 0 | 3 | Not-determined | Genome-fragment | NA | NA | 0 |
| S17-final-viral-combined_k141_258523\|\|full | 2748 | No | NA | 3 | 1 | 0 | Low-quality | Genome-fragment | 6.53 | AAI-based (high-confidence) | 0 |
| S12-final-viral-combined_k141_122762\|\|full | 4950 | No | NA | 5 | 0 | 5 | Not-determined | Genome-fragment | NA | NA | 0 |
| S11-final-viral-combined_k141_292418\|\|full | 2749 | No | NA | 4 | 1 | 0 | Low-quality | Genome-fragment | 4.69 | AAI-based (high-confidence) | 0 |
| S12-final-viral-combined_k141_23063\|\|full | 3020 | No | NA | 6 | 0 | 2 | Not-determined | Genome-fragment | NA | NA | 0 |
| S11_k141_163147 | 2749 | No | NA | 6 | 0 | 0 | Low-quality | Genome-fragment | 5.89 | AAI-based (high-confidence) | 0 |
| S1-final-viral-combined_k141_290976\|\|full | 2749 | No | NA | 4 | 0 | 3 | Low-quality | Genome-fragment | 4.56 | AAI-based (high-confidence) | 0 |
| S7-final-viral-combined_k141_213472\|\|full | 2749 | No | NA | 5 | 2 | 0 | Low-quality | Genome-fragment | 6.74 | AAI-based (high-confidence) | 0 |
| S12-final-viral-combined_k141_55925\|\|full | 1731 | No | NA | 3 | 0 | 2 | Not-determined | Genome-fragment | NA | NA | 0 |
| S12-final-viral-combined_k141_257042\|\|full | 1893 | No | NA | 2 | 0 | 2 | Not-determined | Genome-fragment | NA | NA | 0 |
| S17-final-viral-combined_k141_188074\|\|full | 2749 | No | NA | 3 | 0 | 1 | Low-quality | Genome-fragment | 1.54 | AAI-based (high-confidence) | 0 |
| S12-final-viral-combined_k141_55932\|\|full | 7638 | No | NA | 6 | 0 | 5 | Not-determined | Genome-fragment | NA | NA | 0 |
| S12-final-viral-combined_k141_217436\|\|full | 2749 | No | NA | 5 | 0 | 0 | Low-quality | Genome-fragment | 4.74 | AAI-based (medium-confidence) | 0 |
| S12-final-viral-combined_k141_374812\|\|full | 2564 | No | NA | 3 | 0 | 2 | Not-determined | Genome-fragment | NA | NA | 0 |
| S12-final-viral-combined_k141_240724\|\|full | 1772 | No | NA | 4 | 0 | 0 | Not-determined | Genome-fragment | NA | NA | 0 |
| S1-final-viral-combined_k141_252176\|\|full | 2750 | No | NA | 3 | 0 | 2 | Low-quality | Genome-fragment | 1.24 | AAI-based (medium-confidence) | 0 |
| S7-final-viral-combined_k141_107389\|\|full | 2750 | No | NA | 6 | 1 | 0 | Low-quality | Genome-fragment | 1.85 | HMM-based (lower-bound) | 0 |
| S12-final-viral-combined_k141_23164\|\|full | 1692 | No | NA | 3 | 0 | 3 | Not-determined | Genome-fragment | NA | NA | 0 |
| S4_k141_72058 | 2750 | No | NA | 4 | 3 | 0 | Low-quality | Genome-fragment | 4.53 | AAI-based (high-confidence) | 0 |
| S6-final-viral-combined_k141_16436\|\|full | 2750 | No | NA | 8 | 0 | 0 | Low-quality | Genome-fragment | 4.2 | AAI-based (medium-confidence) | 0 |
| S7-final-viral-combined_k141_228296\|\|full | 2751 | No | NA | 2 | 0 | 1 | Low-quality | Genome-fragment | 1.38 | AAI-based (medium-confidence) | 0 |
| S12-final-viral-combined_k141_23176\|\|full | 3171 | No | NA | 5 | 0 | 0 | Not-determined | Genome-fragment | NA | NA | 0 |
| S12-final-viral-combined_k141_106533\|\|full | 3890 | No | NA | 3 | 0 | 1 | Not-determined | Genome-fragment | NA | NA | 0 |
| S12-final-viral-combined_k141_291590\|\|full | 5092 | No | NA | 5 | 0 | 5 | Not-determined | Genome-fragment | NA | NA | 0 |
| S12-final-viral-combined_k141_56001\|\|full | 1175 | No | NA | 3 | 0 | 0 | Not-determined | Genome-fragment | NA | NA | 0 |
| S12-final-viral-combined_k141_308445\|\|full | 6954 | No | NA | 7 | 0 | 2 | Not-determined | Genome-fragment | NA | NA | 0 |
| S7-final-viral-combined_k141_29307\|\|full | 2751 | No | NA | 3 | 0 | 0 | Low-quality | Genome-fragment | 7.75 | AAI-based (high-confidence) | 0 |
| S12-final-viral-combined_k141_74000\|\|full | 2228 | No | NA | 3 | 0 | 2 | Not-determined | Genome-fragment | NA | NA | 0 |
| S12-final-viral-combined_k141_308460\|\|full | 3255 | No | NA | 4 | 0 | 0 | Not-determined | Genome-fragment | NA | NA | 0 |
| S12-final-viral-combined_k141_106551\|\|full | 7514 | No | NA | 5 | 0 | 2 | Not-determined | Genome-fragment | NA | NA | 0 |
| S12-final-viral-combined_k141_291617\|\|full | 1829 | No | NA | 3 | 0 | 1 | Not-determined | Genome-fragment | NA | NA | 0 |
| S12-final-viral-combined_k141_240796\|\|full | 1536 | No | NA | 2 | 0 | 1 | Not-determined | Genome-fragment | NA | NA | 0 |
| S17-final-viral-combined_k141_210887\|\|full | 2751 | No | NA | 3 | 0 | 1 | Low-quality | Genome-fragment | 2.93 | AAI-based (medium-confidence) | 0 |
| S2-final-viral-combined_k141_73894\|\|full | 2751 | No | NA | 5 | 1 | 0 | Low-quality | Genome-fragment | 6.14 | AAI-based (medium-confidence) | 0 |
| S4-final-viral-combined_k141_56378\|\|full | 2751 | No | NA | 5 | 0 | 0 | Low-quality | Genome-fragment | 3.04 | AAI-based (high-confidence) | 0 |
| S12-final-viral-combined_k141_340568\|\|full | 1025 | No | NA | 4 | 0 | 1 | Not-determined | Genome-fragment | NA | NA | 0 |
| S2-final-viral-combined_k141_5927\|\|full | 2752 | No | NA | 4 | 1 | 0 | Low-quality | Genome-fragment | 1.07 | AAI-based (medium-confidence) | 0 |
| S12-final-viral-combined_k141_291672\|\|full | 3275 | No | NA | 3 | 0 | 2 | Not-determined | Genome-fragment | NA | NA | 0 |
| S12-final-viral-combined_k141_223974\|\|full | 1969 | No | NA | 2 | 0 | 0 | Not-determined | Genome-fragment | NA | NA | 0 |
| S5-final-viral-combined_k141_106763\|\|full | 2753 | No | NA | 3 | 0 | 2 | Low-quality | Genome-fragment | 4.64 | AAI-based (high-confidence) | 0 |
| S17-k141_263028 | 2753 | No | NA | 6 | 0 | 0 | Low-quality | Genome-fragment | 5.38 | AAI-based (high-confidence) | 0 |
| S12-final-viral-combined_k141_223982\|\|full | 8175 | No | NA | 8 | 0 | 7 | Not-determined | Genome-fragment | NA | NA | 0 |
| S2-final-viral-combined_k141_125210\|\|full | 2753 | No | NA | 4 | 0 | 0 | Low-quality | Genome-fragment | 8.11 | AAI-based (high-confidence) | 0 |
| S2-final-viral-combined_k141_33582\|\|full | 2753 | No | NA | 6 | 1 | 0 | Low-quality | Genome-fragment | 4.91 | AAI-based (high-confidence) | 0 |
| S2-final-viral-combined_k141_107335\|\|full | 2753 | No | NA | 4 | 0 | 0 | Low-quality | Genome-fragment | 4.81 | AAI-based (medium-confidence) | 0 |
| S2-final-viral-combined_k141_76653\|\|full | 2753 | No | NA | 6 | 0 | 0 | Low-quality | Genome-fragment | 4.7 | AAI-based (high-confidence) | 0 |
| S12-final-viral-combined_k141_308542\|\|full | 1942 | No | NA | 3 | 0 | 1 | Not-determined | Genome-fragment | NA | NA | 0 |
| S7-final-viral-combined_k141_98641\|\|full | 2754 | No | NA | 6 | 0 | 1 | Low-quality | Genome-fragment | 4.77 | AAI-based (high-confidence) | 0 |
| S2-final-viral-combined_k141_114182\|\|full | 2754 | No | NA | 7 | 0 | 0 | Low-quality | Genome-fragment | 4.59 | AAI-based (high-confidence) | 0 |
| S11-final-viral-combined_k141_276899\|\|full | 2755 | No | NA | 3 | 2 | 0 | Low-quality | Genome-fragment | 6.32 | AAI-based (medium-confidence) | 0 |
| S12-final-viral-combined_k141_324724\|\|full | 1141 | No | NA | 2 | 0 | 2 | Not-determined | Genome-fragment | NA | NA | 0 |
| S1-final-viral-combined_k141_280648\|\|full | 2755 | No | NA | 3 | 0 | 3 | Low-quality | Genome-fragment | 1.64 | AAI-based (medium-confidence) | 0 |
| S12-final-viral-combined_k141_374948\|\|full | 1044 | No | NA | 2 | 0 | 0 | Not-determined | Genome-fragment | NA | NA | 0 |
| S12-final-viral-combined_k141_291701\|\|full | 1921 | No | NA | 3 | 0 | 0 | Not-determined | Genome-fragment | NA | NA | 0 |
| S12-final-viral-combined_k141_224093\|\|full | 2560 | No | NA | 2 | 0 | 2 | Not-determined | Genome-fragment | NA | NA | 0 |
| S8-final-viral-combined_k141_101795\|\|full | 2755 | No | NA | 4 | 0 | 0 | Low-quality | Genome-fragment | 2.58 | AAI-based (medium-confidence) | 0 |
| S12-final-viral-combined_k141_140513\|\|full | 5190 | No | NA | 6 | 0 | 0 | Not-determined | Genome-fragment | NA | NA | 0 |
| S12-final-viral-combined_k141_274273\|\|full | 6973 | No | NA | 6 | 0 | 5 | Not-determined | Genome-fragment | NA | NA | 0 |
| S11-final-viral-combined_k141_227504\|\|full | 2756 | No | NA | 8 | 1 | 0 | Low-quality | Genome-fragment | 5.93 | AAI-based (high-confidence) | 0 |
| S1-final-viral-combined_k141_19446\|\|full | 2756 | No | NA | 3 | 1 | 0 | Low-quality | Genome-fragment | 7.06 | AAI-based (high-confidence) | 0 |
| S3-final-viral-combined_k141_68969\|\|full | 2756 | No | NA | 7 | 0 | 0 | Low-quality | Genome-fragment | 6.43 | AAI-based (high-confidence) | 0 |
| S12-final-viral-combined_k141_106682\|\|full | 3412 | No | NA | 3 | 0 | 0 | Not-determined | Genome-fragment | NA | NA | 0 |
| S5-final-viral-combined_k141_64704\|\|full | 2757 | No | NA | 7 | 0 | 0 | Low-quality | Genome-fragment | 5.02 | AAI-based (medium-confidence) | 0 |
| S7-final-viral-combined_k141_197793\|\|full | 2757 | No | NA | 6 | 0 | 0 | Low-quality | Genome-fragment | 5.25 | AAI-based (high-confidence) | 0 |
| S7-final-viral-combined_k141_28129\|\|full | 2757 | Yes | 224 | 4 | 1 | 1 | Low-quality | Genome-fragment | 0.39 | AAI-based (high-confidence) | 87.89 |
| S2_k141_78029 | 2757 | No | NA | 3 | 0 | 1 | Low-quality | Genome-fragment | 1.16 | AAI-based (medium-confidence) | 0 |
| S12-final-viral-combined_k141_6375\|\|full | 1714 | No | NA | 3 | 0 | 1 | Not-determined | Genome-fragment | NA | NA | 0 |
| S12-final-viral-combined_k141_308618\|\|full | 1893 | No | NA | 5 | 0 | 0 | Not-determined | Genome-fragment | NA | NA | 0 |
| S12-final-viral-combined_k141_140551\|\|full | 6053 | No | NA | 8 | 0 | 4 | Not-determined | Genome-fragment | NA | NA | 0 |
| S12-final-viral-combined_k141_340686\|\|full | 2653 | No | NA | 3 | 0 | 1 | Not-determined | Genome-fragment | NA | NA | 0 |
| S12-final-viral-combined_k141_257323\|\|full | 5351 | No | NA | 5 | 0 | 2 | Not-determined | Genome-fragment | NA | NA | 0 |
| S12_k141_243632 | 2758 | No | NA | 8 | 1 | 0 | Low-quality | Genome-fragment | 1.49 | AAI-based (high-confidence) | 0 |
| S12-final-viral-combined_k141_123056\|\|full | 4928 | No | NA | 6 | 0 | 2 | Not-determined | Genome-fragment | NA | NA | 0 |
| S2-final-viral-combined_k141_210450\|\|full | 2758 | No | NA | 4 | 0 | 0 | Low-quality | Genome-fragment | 5.15 | AAI-based (medium-confidence) | 0 |
| S4-final-viral-combined_k141_50380\|\|full | 2758 | No | NA | 7 | 1 | 0 | Low-quality | Genome-fragment | 7.89 | AAI-based (high-confidence) | 0 |
| S12-final-viral-combined_k141_123060\|\|full | 1521 | No | NA | 3 | 0 | 0 | Not-determined | Genome-fragment | NA | NA | 0 |
| S12-final-viral-combined_k141_56143\|\|full | 2253 | No | NA | 3 | 0 | 2 | Not-determined | Genome-fragment | NA | NA | 0 |
| S12-final-viral-combined_k141_23324\|\|full | 1299 | No | NA | 2 | 0 | 1 | Not-determined | Genome-fragment | NA | NA | 0 |
| S11-final-viral-combined_k141_196584\|\|full | 2759 | No | NA | 4 | 0 | 0 | Low-quality | Genome-fragment | 3.39 | AAI-based (medium-confidence) | 0 |
| S12-final-viral-combined_k141_106754\|\|full | 1921 | No | NA | 4 | 0 | 0 | Not-determined | Genome-fragment | NA | NA | 0 |
| S12-final-viral-combined_k141_257328\|\|full | 6241 | No | NA | 7 | 0 | 1 | Not-determined | Genome-fragment | NA | NA | 0 |
| S11-final-viral-combined_k141_180895\|\|full | 2760 | No | NA | 4 | 0 | 3 | Low-quality | Genome-fragment | 0.8 | AAI-based (medium-confidence) | 0 |
| S12-final-viral-combined_k141_375047\|\|full | 2236 | No | NA | 2 | 0 | 1 | Not-determined | Genome-fragment | NA | NA | 0 |
| S12-final-viral-combined_k141_274362\|\|full | 1026 | No | NA | 3 | 0 | 0 | Not-determined | Genome-fragment | NA | NA | 0 |
| S5-final-viral-combined_k141_152443\|\|full | 2760 | No | NA | 8 | 1 | 0 | Low-quality | Genome-fragment | 4.58 | AAI-based (high-confidence) | 0 |
| S12-final-viral-combined_k141_173874\|\|full | 3234 | No | NA | 4 | 0 | 2 | Not-determined | Genome-fragment | NA | NA | 0 |
| S12-final-viral-combined_k141_224203\|\|full | 5787 | No | NA | 4 | 0 | 2 | Not-determined | Genome-fragment | NA | NA | 0 |
| S12-final-viral-combined_k141_291788\|\|full | 8447 | No | NA | 9 | 0 | 6 | Not-determined | Genome-fragment | NA | NA | 0 |
| S12-final-viral-combined_k141_40380\|\|full | 1493 | No | NA | 2 | 0 | 0 | Not-determined | Genome-fragment | NA | NA | 0 |
| S2-final-viral-combined_k141_14210\|\|full | 2760 | No | NA | 2 | 0 | 0 | Low-quality | Genome-fragment | 7.84 | AAI-based (medium-confidence) | 0 |
| S7-final-viral-combined_k141_221890\|\|full | 2761 | No | NA | 3 | 3 | 0 | Low-quality | Genome-fragment | 5.95 | AAI-based (high-confidence) | 0 |
| S12-final-viral-combined_k141_257395\|\|full | 1619 | No | NA | 3 | 0 | 0 | Not-determined | Genome-fragment | NA | NA | 0 |
| S12-final-viral-combined_k141_90213\|\|full | 7214 | No | NA | 7 | 0 | 3 | Not-determined | Genome-fragment | NA | NA | 0 |
| S12-final-viral-combined_k141_375086\|\|full | 1143 | No | NA | 3 | 0 | 0 | Not-determined | Genome-fragment | NA | NA | 0 |
| S17-final-viral-combined_k141_65558\|\|full | 2761 | No | NA | 4 | 2 | 0 | Low-quality | Genome-fragment | 3.35 | AAI-based (high-confidence) | 0 |
| S1_k141_228102 | 2762 | No | NA | 3 | 0 | 3 | Low-quality | Genome-fragment | 6.49 | AAI-based (high-confidence) | 0 |
| S7-final-viral-combined_k141_143342\|\|full | 2762 | No | NA | 5 | 0 | 0 | Low-quality | Genome-fragment | 2.6 | AAI-based (high-confidence) | 0 |
| S17-k141_118788 | 2762 | No | NA | 3 | 0 | 2 | Low-quality | Genome-fragment | 2.47 | AAI-based (medium-confidence) | 0 |
| S12-final-viral-combined_k141_140920\|\|full | 2762 | No | NA | 5 | 0 | 1 | Low-quality | Genome-fragment | 3.59 | AAI-based (medium-confidence) | 0 |
| S12-final-viral-combined_k141_391913\|\|full | 1972 | No | NA | 3 | 0 | 2 | Not-determined | Genome-fragment | NA | NA | 0 |
| S12-final-viral-combined_k141_257459\|\|full | 1819 | No | NA | 3 | 0 | 3 | Not-determined | Genome-fragment | NA | NA | 0 |
| S12_k141_198833 | 2762 | No | NA | 4 | 0 | 2 | Low-quality | Genome-fragment | 1.33 | AAI-based (medium-confidence) | 0 |
| S12-final-viral-combined_k141_157506\|\|full | 2133 | No | NA | 3 | 0 | 2 | Not-determined | Genome-fragment | NA | NA | 0 |
| S12-final-viral-combined_k141_157512\|\|full | 2544 | No | NA | 3 | 0 | 1 | Not-determined | Genome-fragment | NA | NA | 0 |
| S12-final-viral-combined_k141_274446\|\|full | 6759 | No | NA | 4 | 0 | 4 | Not-determined | Genome-fragment | NA | NA | 0 |
| S12-final-viral-combined_k141_357216\|\|full | 2264 | No | NA | 5 | 0 | 2 | Not-determined | Genome-fragment | NA | NA | 0 |
| S2-final-viral-combined_k141_132867\|\|full | 2762 | No | NA | 8 | 0 | 1 | Low-quality | Genome-fragment | 3.08 | AAI-based (medium-confidence) | 0 |
| S9-final-viral-combined_k141_91869\|\|full | 2763 | No | NA | 4 | 0 | 4 | Low-quality | Genome-fragment | 1.44 | AAI-based (medium-confidence) | 0 |
| S11-final-viral-combined_k141_216637\|\|full | 2763 | No | NA | 3 | 0 | 3 | Low-quality | Genome-fragment | 1.12 | AAI-based (medium-confidence) | 0 |
| S6_k141_127335 | 2764 | No | NA | 7 | 0 | 0 | Low-quality | Genome-fragment | 1.47 | AAI-based (high-confidence) | 0 |
| S12-final-viral-combined_k141_90263\|\|full | 17033 | No | NA | 15 | 0 | 13 | Not-determined | Genome-fragment | NA | NA | 0 |
| S12-final-viral-combined_k141_308751\|\|full | 2814 | No | NA | 3 | 0 | 0 | Not-determined | Genome-fragment | NA | NA | 0 |
| S12-final-viral-combined_k141_23385\|\|full | 2303 | No | NA | 3 | 0 | 0 | Not-determined | Genome-fragment | NA | NA | 0 |
| S8-final-viral-combined_k141_100849\|\|full | 2764 | No | NA | 4 | 0 | 3 | Low-quality | Genome-fragment | 1.92 | AAI-based (medium-confidence) | 0 |
| S12-final-viral-combined_k141_207485\|\|full | 10504 | No | NA | 19 | 0 | 2 | Not-determined | Genome-fragment | NA | NA | 0 |
| S12-final-viral-combined_k141_240827\|\|full | 2361 | No | NA | 3 | 0 | 1 | Not-determined | Genome-fragment | NA | NA | 0 |
| S12-final-viral-combined_k141_257534\|\|full | 2348 | No | NA | 3 | 0 | 0 | Not-determined | Genome-fragment | NA | NA | 0 |
| S11-final-viral-combined_k141_74912\|\|full | 2765 | No | NA | 6 | 2 | 0 | Low-quality | Genome-fragment | 6.59 | AAI-based (high-confidence) | 0 |
| S1-final-viral-combined_k141_379081\|\|full | 2765 | No | NA | 3 | 0 | 3 | Low-quality | Genome-fragment | 1.91 | AAI-based (medium-confidence) | 0 |
| S17-k141_254689 | 2765 | No | NA | 3 | 0 | 0 | Low-quality | Genome-fragment | 5.45 | AAI-based (medium-confidence) | 0 |
| S8-final-viral-combined_k141_174146\|\|full | 2765 | No | NA | 4 | 0 | 1 | Low-quality | Genome-fragment | 4.05 | AAI-based (medium-confidence) | 0 |
| S11-final-viral-combined_k141_14573\|\|full | 2766 | No | NA | 4 | 0 | 2 | Low-quality | Genome-fragment | 1.53 | AAI-based (medium-confidence) | 0 |
| S2_k141_137390 | 2766 | No | NA | 4 | 0 | 0 | Low-quality | Genome-fragment | 6.17 | AAI-based (medium-confidence) | 0 |
| S12-final-viral-combined_k141_191570\|\|full | 26127 | No | NA | 31 | 16 | 0 | Medium-quality | Genome-fragment | 58.02 | AAI-based (high-confidence) | 0 |
| S12-final-viral-combined_k141_340871\|\|full | 3023 | No | NA | 2 | 0 | 0 | Not-determined | Genome-fragment | NA | NA | 0 |
| S6-final-viral-combined_k141_9600\|\|full | 2766 | No | NA | 3 | 0 | 3 | Low-quality | Genome-fragment | 1.27 | AAI-based (medium-confidence) | 0 |
| S12-final-viral-combined_k141_74405\|\|full | 1675 | No | NA | 3 | 0 | 2 | Not-determined | Genome-fragment | NA | NA | 0 |
| S12-final-viral-combined_k141_191584\|\|full | 1723 | No | NA | 2 | 0 | 0 | Not-determined | Genome-fragment | NA | NA | 0 |
| S9-final-viral-combined_k141_18377\|\|full | 2767 | No | NA | 3 | 0 | 0 | Low-quality | Genome-fragment | 5.57 | AAI-based (medium-confidence) | 0 |
| S12-final-viral-combined_k141_140715\|\|full | 2354 | No | NA | 3 | 0 | 0 | Not-determined | Genome-fragment | NA | NA | 0 |
| S5-final-viral-combined_k141_192195\|\|full | 2767 | No | NA | 5 | 2 | 0 | Low-quality | Genome-fragment | 6.39 | AAI-based (high-confidence) | 0 |
| S5-final-viral-combined_k141_32588\|\|full | 2767 | No | NA | 4 | 1 | 0 | Low-quality | Genome-fragment | 7.03 | AAI-based (high-confidence) | 0 |
| S11-final-viral-combined_k141_109952\|\|full | 2768 | No | NA | 4 | 0 | 0 | Low-quality | Genome-fragment | 4.79 | AAI-based (high-confidence) | 0 |
| S12-final-viral-combined_k141_90354\|\|full | 1717 | No | NA | 4 | 0 | 1 | Not-determined | Genome-fragment | NA | NA | 0 |
| S12-final-viral-combined_k141_340935\|\|full | 1372 | No | NA | 2 | 0 | 2 | Not-determined | Genome-fragment | NA | NA | 0 |
| S5-final-viral-combined_k141_37582\|\|full | 2768 | No | NA | 3 | 1 | 0 | Low-quality | Genome-fragment | 4.79 | HMM-based (lower-bound) | 0 |
| S12-final-viral-combined_k141_6547\|\|full | 1058 | No | NA | 2 | 0 | 0 | Not-determined | Genome-fragment | NA | NA | 0 |
| S12-final-viral-combined_k141_325007\|\|full | 2048 | No | NA | 3 | 0 | 1 | Not-determined | Genome-fragment | NA | NA | 0 |
| S12-final-viral-combined_k141_207599\|\|full | 1815 | No | NA | 3 | 0 | 0 | Not-determined | Genome-fragment | NA | NA | 0 |
| S12-final-viral-combined_k141_240932\|\|full | 2954 | No | NA | 4 | 0 | 3 | Not-determined | Genome-fragment | NA | NA | 0 |
| S12-final-viral-combined_k141_392063\|\|full | 1796 | No | NA | 3 | 0 | 1 | Not-determined | Genome-fragment | NA | NA | 0 |
| S12-final-viral-combined_k141_90382\|\|full | 3362 | No | NA | 3 | 0 | 2 | Not-determined | Genome-fragment | NA | NA | 0 |
| S6_k141_105721 | 2768 | Yes | 971 | 3 | 1 | 1 | Low-quality | Genome-fragment | 1.64 | AAI-based (medium-confidence) | 64.92 |
| S12-final-viral-combined_k141_140766\|\|full | 6981 | No | NA | 7 | 0 | 3 | Not-determined | Genome-fragment | NA | NA | 0 |
| S11-final-viral-combined_k141_50742\|\|full | 2769 | No | NA | 4 | 1 | 0 | Low-quality | Genome-fragment | 6.04 | AAI-based (high-confidence) | 0 |
| S12-final-viral-combined_k141_6591\|\|full | 1066 | No | NA | 2 | 0 | 2 | Not-determined | Genome-fragment | NA | NA | 0 |
| S12-final-viral-combined_k141_123310\|\|full | 1967 | No | NA | 4 | 0 | 2 | Not-determined | Genome-fragment | NA | NA | 0 |
| S1-final-viral-combined_k141_318859\|\|full | 2769 | No | NA | 5 | 0 | 2 | Low-quality | Genome-fragment | 2.95 | AAI-based (medium-confidence) | 0 |
| S12-final-viral-combined_k141_274608\|\|full | 3263 | No | NA | 5 | 0 | 3 | Not-determined | Genome-fragment | NA | NA | 0 |
| S17-final-viral-combined_k141_87525\|\|full | 2769 | No | NA | 4 | 2 | 0 | Low-quality | Genome-fragment | 6.59 | AAI-based (medium-confidence) | 0 |
| S4-final-viral-combined_k141_38479\|\|full | 2769 | No | NA | 3 | 0 | 0 | Low-quality | Genome-fragment | 4.73 | AAI-based (medium-confidence) | 0 |
| S12-final-viral-combined_k141_123322\|\|full | 6543 | No | NA | 5 | 0 | 1 | Not-determined | Genome-fragment | NA | NA | 0 |
| S12-final-viral-combined_k141_325061\|\|full | 2418 | No | NA | 2 | 0 | 0 | Not-determined | Genome-fragment | NA | NA | 0 |
| S8-final-viral-combined_k141_93325\|\|full | 2769 | No | NA | 2 | 0 | 2 | Low-quality | Genome-fragment | 5.06 | AAI-based (medium-confidence) | 0 |
| S11-final-viral-combined_k141_133135\|\|full | 2770 | No | NA | 3 | 0 | 2 | Low-quality | Genome-fragment | 2.24 | AAI-based (high-confidence) | 0 |
| S12-final-viral-combined_k141_224383\|\|full | 1262 | No | NA | 3 | 0 | 0 | Not-determined | Genome-fragment | NA | NA | 0 |
| S1-final-viral-combined_k141_14151\|\|full | 2770 | No | NA | 3 | 0 | 0 | Low-quality | Genome-fragment | 1.81 | AAI-based (high-confidence) | 0 |
| S6-final-viral-combined_k141_41577\|\|full | 2770 | No | NA | 9 | 1 | 0 | Low-quality | Genome-fragment | 4.6 | AAI-based (high-confidence) | 0 |
| S8_k141_122760 | 2770 | No | NA | 2 | 1 | 0 | Low-quality | Genome-fragment | 4.61 | AAI-based (medium-confidence) | 0 |
| S12-final-viral-combined_k141_157773\|\|full | 2069 | No | NA | 3 | 0 | 1 | Not-determined | Genome-fragment | NA | NA | 0 |
| S12-final-viral-combined_k141_257661\|\|full | 2162 | No | NA | 2 | 0 | 1 | Not-determined | Genome-fragment | NA | NA | 0 |
| S4-final-viral-combined_k141_1019\|\|full | 2771 | No | NA | 5 | 2 | 0 | Low-quality | Genome-fragment | 6.74 | AAI-based (medium-confidence) | 0 |
| S11-final-viral-combined_k141_303262\|\|full | 2772 | No | NA | 6 | 6 | 0 | Low-quality | Genome-fragment | 4.58 | HMM-based (lower-bound) | 0 |
| S11-final-viral-combined_k141_49449\|\|full | 2772 | Yes | 1601 | 4 | 2 | 1 | Low-quality | Genome-fragment | 3.34 | AAI-based (high-confidence) | 42.24 |
| S17-final-viral-combined_k141_67373\|\|full | 2772 | No | NA | 3 | 3 | 0 | Low-quality | Genome-fragment | 5.98 | AAI-based (high-confidence) | 0 |
| S17-final-viral-combined_k141_2347\|\|full | 2772 | No | NA | 5 | 1 | 0 | Low-quality | Genome-fragment | 3.87 | AAI-based (medium-confidence) | 0 |
| S12-final-viral-combined_k141_23502\|\|full | 6568 | No | NA | 4 | 0 | 2 | Not-determined | Genome-fragment | NA | NA | 0 |
| S12-final-viral-combined_k141_257685\|\|full | 1848 | No | NA | 5 | 0 | 0 | Not-determined | Genome-fragment | NA | NA | 0 |
| S12-final-viral-combined_k141_274723\|\|full | 2325 | No | NA | 3 | 0 | 0 | Not-determined | Genome-fragment | NA | NA | 0 |
| S12-final-viral-combined_k141_23509\|\|full | 1819 | No | NA | 2 | 0 | 0 | Not-determined | Genome-fragment | NA | NA | 0 |
| S12-final-viral-combined_k141_7739\|\|full | 2773 | No | NA | 3 | 0 | 1 | Low-quality | Genome-fragment | 2.53 | AAI-based (medium-confidence) | 0 |
| S12-final-viral-combined_k141_157796\|\|full | 9787 | No | NA | 15 | 0 | 8 | Not-determined | Genome-fragment | NA | NA | 0 |
| S12-final-viral-combined_k141_176426\|\|full | 2773 | No | NA | 4 | 0 | 1 | Low-quality | Genome-fragment | 6.99 | AAI-based (high-confidence) | 0 |
| S12-final-viral-combined_k141_257705\|\|full | 5244 | No | NA | 4 | 0 | 2 | Not-determined | Genome-fragment | NA | NA | 0 |
| S12-final-viral-combined_k141_375276\|\|full | 3390 | No | NA | 3 | 0 | 2 | Not-determined | Genome-fragment | NA | NA | 0 |
| S12-final-viral-combined_k141_207721\|\|full | 2047 | No | NA | 3 | 0 | 1 | Not-determined | Genome-fragment | NA | NA | 0 |
| S17-final-viral-combined_k141_11924\|\|full | 2774 | No | NA | 6 | 2 | 0 | Low-quality | Genome-fragment | 6.01 | AAI-based (high-confidence) | 0 |
| S12-final-viral-combined_k141_375292\|\|full | 10212 | No | NA | 6 | 0 | 1 | Not-determined | Genome-fragment | NA | NA | 0 |
| S12-final-viral-combined_k141_207743\|\|full | 1855 | No | NA | 3 | 0 | 2 | Not-determined | Genome-fragment | NA | NA | 0 |
| S12-final-viral-combined_k141_191753\|\|full | 2692 | No | NA | 3 | 0 | 0 | Not-determined | Genome-fragment | NA | NA | 0 |
| S12-final-viral-combined_k141_274766\|\|full | 4453 | No | NA | 6 | 0 | 3 | Not-determined | Genome-fragment | NA | NA | 0 |
| S12-final-viral-combined_k141_81198\|\|full | 2774 | No | NA | 5 | 0 | 4 | Low-quality | Genome-fragment | 1.71 | AAI-based (medium-confidence) | 0 |
| S12-final-viral-combined_k141_357490\|\|full | 9321 | No | NA | 10 | 0 | 1 | Not-determined | Genome-fragment | NA | NA | 0 |
| S2_k141_94002 | 2774 | No | NA | 2 | 0 | 1 | Low-quality | Genome-fragment | 1.37 | AAI-based (medium-confidence) | 0 |
| S12-final-viral-combined_k141_325185\|\|full | 2453 | No | NA | 3 | 0 | 0 | Not-determined | Genome-fragment | NA | NA | 0 |
| S12-final-viral-combined_k141_224480\|\|full | 2241 | No | NA | 4 | 0 | 1 | Not-determined | Genome-fragment | NA | NA | 0 |
| S1_k141_77626 | 2775 | No | NA | 2 | 0 | 2 | Low-quality | Genome-fragment | 0.67 | AAI-based (medium-confidence) | 0 |
| S12-final-viral-combined_k141_23517\|\|full | 75931 | Yes | 42420 | 111 | 19 | 24 | High-quality | High-quality | 100 | AAI-based (high-confidence) | 44.13 |
| S2-final-viral-combined_k141_105590\|\|full | 2775 | No | NA | 3 | 0 | 2 | Low-quality | Genome-fragment | 2.48 | AAI-based (medium-confidence) | 0 |
| S12-final-viral-combined_k141_56477\|\|full | 1202 | No | NA | 2 | 0 | 1 | Not-determined | Genome-fragment | NA | NA | 0 |
| S12-final-viral-combined_k141_56484\|\|full | 1654 | No | NA | 3 | 0 | 0 | Not-determined | Genome-fragment | NA | NA | 0 |
| S12-final-viral-combined_k141_90523\|\|full | 1153 | No | NA | 3 | 0 | 1 | Not-determined | Genome-fragment | NA | NA | 0 |
| S12-final-viral-combined_k141_56495\|\|full | 2489 | No | NA | 3 | 0 | 3 | Not-determined | Genome-fragment | NA | NA | 0 |
| S12-final-viral-combined_k141_157915\|\|full | 4902 | No | NA | 4 | 0 | 3 | Not-determined | Genome-fragment | NA | NA | 0 |
| S12-final-viral-combined_k141_375343\|\|full | 52960 | Yes | 29654 | 60 | 10 | 14 | Medium-quality | Genome-fragment | 53.26 | AAI-based (high-confidence) | 44.01 |
| S5-final-viral-combined_k141_126253\|\|full | 2776 | No | NA | 3 | 1 | 0 | Low-quality | Genome-fragment | 7.29 | AAI-based (medium-confidence) | 0 |
| S9_k141_93735 | 2777 | No | NA | 6 | 6 | 0 | Low-quality | Genome-fragment | 8.45 | AAI-based (high-confidence) | 0 |
| S12-final-viral-combined_k141_107135\|\|full | 2079 | No | NA | 3 | 0 | 1 | Not-determined | Genome-fragment | NA | NA | 0 |
| S12-final-viral-combined_k141_40641\|\|full | 1417 | No | NA | 2 | 0 | 0 | Not-determined | Genome-fragment | NA | NA | 0 |
| S12-final-viral-combined_k141_107153\|\|full | 1242 | No | NA | 2 | 0 | 1 | Not-determined | Genome-fragment | NA | NA | 0 |
| S12-final-viral-combined_k141_375359\|\|full | 12265 | No | NA | 26 | 0 | 0 | Not-determined | Genome-fragment | NA | NA | 0 |
| S12-final-viral-combined_k141_375361\|\|full | 3099 | No | NA | 3 | 0 | 1 | Not-determined | Genome-fragment | NA | NA | 0 |
| S12-final-viral-combined_k141_158276\|\|full | 2777 | No | NA | 8 | 0 | 0 | Low-quality | Genome-fragment | 4.73 | AAI-based (high-confidence) | 0 |
| S12-final-viral-combined_k141_341137\|\|full | 8130 | No | NA | 11 | 0 | 2 | Not-determined | Genome-fragment | NA | NA | 0 |
| S12_k141_264792 | 2777 | No | NA | 4 | 0 | 0 | Low-quality | Genome-fragment | 1.52 | AAI-based (high-confidence) | 0 |
| S3_k141_134099 | 2778 | No | NA | 4 | 3 | 0 | Low-quality | Genome-fragment | 4.4 | AAI-based (high-confidence) | 0 |
| S12-final-viral-combined_k141_392269\|\|full | 2164 | No | NA | 3 | 0 | 0 | Not-determined | Genome-fragment | NA | NA | 0 |
| S12-final-viral-combined_k141_375379\|\|full | 1916 | No | NA | 3 | 0 | 1 | Not-determined | Genome-fragment | NA | NA | 0 |
| S12-final-viral-combined_k141_309039\|\|full | 1442 | No | NA | 3 | 0 | 0 | Not-determined | Genome-fragment | NA | NA | 0 |
| S5-final-viral-combined_k141_173261\|\|full | 2779 | No | NA | 7 | 5 | 0 | Low-quality | Genome-fragment | 7.46 | AAI-based (high-confidence) | 0 |
| S12_k141_25580 | 2779 | No | NA | 7 | 0 | 1 | Low-quality | Genome-fragment | 3.47 | AAI-based (high-confidence) | 0 |
| S12-final-viral-combined_k141_23614\|\|full | 1163 | No | NA | 2 | 0 | 0 | Not-determined | Genome-fragment | NA | NA | 0 |
| S17-k141_136393 | 2780 | No | NA | 2 | 0 | 1 | Low-quality | Genome-fragment | 19.85 | AAI-based (medium-confidence) | 0 |
| S12-final-viral-combined_k141_23641\|\|full | 1629 | No | NA | 3 | 0 | 0 | Not-determined | Genome-fragment | NA | NA | 0 |
| S12-final-viral-combined_k141_157955\|\|full | 30578 | No | NA | 21 | 0 | 3 | Not-determined | Genome-fragment | NA | NA | 0 |
| S12-final-viral-combined_k141_157969\|\|full | 1746 | No | NA | 2 | 0 | 2 | Not-determined | Genome-fragment | NA | NA | 0 |
| S8-final-viral-combined_k141_94116\|\|full | 2780 | No | NA | 8 | 0 | 0 | Low-quality | Genome-fragment | 6.08 | AAI-based (high-confidence) | 0 |
| S12-final-viral-combined_k141_123588\|\|full | 2475 | No | NA | 4 | 0 | 0 | Not-determined | Genome-fragment | NA | NA | 0 |
| S5-final-viral-combined_k141_55814\|\|full | 2781 | No | NA | 8 | 5 | 0 | Low-quality | Genome-fragment | 4.24 | AAI-based (high-confidence) | 0 |
| S12-final-viral-combined_k141_392364\|\|full | 4729 | No | NA | 5 | 0 | 5 | Not-determined | Genome-fragment | NA | NA | 0 |
| S17-final-viral-combined_k141_110434\|\|full | 2781 | No | NA | 2 | 2 | 0 | Low-quality | Genome-fragment | 6.85 | AAI-based (medium-confidence) | 0 |
| S2_k141_109257 | 2781 | No | NA | 2 | 0 | 0 | Low-quality | Genome-fragment | 4.62 | AAI-based (high-confidence) | 0 |
| S12-final-viral-combined_k141_56622\|\|full | 1207 | No | NA | 2 | 0 | 0 | Not-determined | Genome-fragment | NA | NA | 0 |
| S12-final-viral-combined_k141_158017\|\|full | 1262 | No | NA | 2 | 0 | 1 | Not-determined | Genome-fragment | NA | NA | 0 |
| S11-final-viral-combined_k141_314324\|\|full | 2782 | No | NA | 6 | 0 | 0 | Low-quality | Genome-fragment | 5.39 | AAI-based (medium-confidence) | 0 |
| S12-final-viral-combined_k141_257960\|\|full | 10194 | No | NA | 9 | 0 | 6 | Not-determined | Genome-fragment | NA | NA | 0 |
| S12-final-viral-combined_k141_325354\|\|full | 2590 | No | NA | 4 | 0 | 2 | Not-determined | Genome-fragment | NA | NA | 0 |
| S5-final-viral-combined_k141_70060\|\|full | 2782 | No | NA | 4 | 0 | 0 | Low-quality | Genome-fragment | 6.83 | AAI-based (medium-confidence) | 0 |
| S2_k141_230012 | 2782 | No | NA | 3 | 0 | 2 | Low-quality | Genome-fragment | 0.53 | AAI-based (medium-confidence) | 0 |
| S12-final-viral-combined_k141_375532\|\|full | 1603 | No | NA | 3 | 0 | 1 | Not-determined | Genome-fragment | NA | NA | 0 |
| S12-final-viral-combined_k141_357682\|\|full | 4454 | No | NA | 7 | 0 | 0 | Not-determined | Genome-fragment | NA | NA | 0 |
| S12-final-viral-combined_k141_309126\|\|full | 4069 | No | NA | 5 | 0 | 1 | Not-determined | Genome-fragment | NA | NA | 0 |
| S12-final-viral-combined_k141_309135\|\|full | 2920 | No | NA | 6 | 0 | 0 | Not-determined | Genome-fragment | NA | NA | 0 |
| S12-final-viral-combined_k141_241387\|\|full | 1148 | No | NA | 2 | 0 | 2 | Not-determined | Genome-fragment | NA | NA | 0 |
| S12-final-viral-combined_k141_6941\|\|full | 1622 | No | NA | 2 | 0 | 0 | Not-determined | Genome-fragment | NA | NA | 0 |
| S8-final-viral-combined_k141_58542\|\|full | 2782 | No | NA | 6 | 0 | 0 | Low-quality | Genome-fragment | 2.94 | AAI-based (medium-confidence) | 0 |
| S12-final-viral-combined_k141_158068\|\|full | 1661 | No | NA | 3 | 0 | 1 | Not-determined | Genome-fragment | NA | NA | 0 |
| S11-final-viral-combined_k141_134925\|\|full | 2783 | No | NA | 4 | 0 | 3 | Low-quality | Genome-fragment | 6.05 | AAI-based (medium-confidence) | 0 |
| S1_k141_183559 | 2783 | No | NA | 4 | 1 | 0 | Low-quality | Genome-fragment | 4.8 | AAI-based (high-confidence) | 0 |
| S12-final-viral-combined_k141_56734\|\|full | 7023 | No | NA | 6 | 0 | 5 | Not-determined | Genome-fragment | NA | NA | 0 |
| S12-final-viral-combined_k141_325407\|\|full | 1934 | No | NA | 3 | 0 | 0 | Not-determined | Genome-fragment | NA | NA | 0 |
| S12-final-viral-combined_k141_224751\|\|full | 1728 | No | NA | 2 | 0 | 2 | Not-determined | Genome-fragment | NA | NA | 0 |
| S12-final-viral-combined_k141_6948\|\|full | 1627 | No | NA | 3 | 0 | 0 | Not-determined | Genome-fragment | NA | NA | 0 |
| S12-final-viral-combined_k141_141079\|\|full | 1773 | No | NA | 3 | 0 | 0 | Not-determined | Genome-fragment | NA | NA | 0 |
| S3_k141_53117 | 2783 | No | NA | 2 | 0 | 0 | Low-quality | Genome-fragment | 1.28 | AAI-based (high-confidence) | 0 |
| S12-final-viral-combined_k141_174358\|\|full | 1082 | No | NA | 2 | 0 | 1 | Not-determined | Genome-fragment | NA | NA | 0 |
| S11-final-viral-combined_k141_66376\|\|full | 2784 | No | NA | 3 | 1 | 0 | Low-quality | Genome-fragment | 1.32 | AAI-based (medium-confidence) | 0 |
| S3-final-viral-combined_k141_117963\|\|full | 2784 | No | NA | 3 | 0 | 2 | Low-quality | Genome-fragment | 4.05 | AAI-based (medium-confidence) | 0 |
| S12-final-viral-combined_k141_325418\|\|full | 34704 | No | NA | 36 | 9 | 0 | Medium-quality | Genome-fragment | 71.85 | AAI-based (high-confidence) | 0 |
| S12-final-viral-combined_k141_341324\|\|full | 2086 | No | NA | 3 | 0 | 2 | Not-determined | Genome-fragment | NA | NA | 0 |
| S12-final-viral-combined_k141_192017\|\|full | 4766 | No | NA | 4 | 0 | 4 | Not-determined | Genome-fragment | NA | NA | 0 |
| S12-final-viral-combined_k141_309184\|\|full | 1202 | No | NA | 3 | 0 | 0 | Not-determined | Genome-fragment | NA | NA | 0 |
| S12-final-viral-combined_k141_193531\|\|full | 2784 | No | NA | 4 | 1 | 0 | Low-quality | Genome-fragment | 6.93 | AAI-based (high-confidence) | 0 |
| S12-final-viral-combined_k141_192021\|\|full | 1592 | No | NA | 3 | 0 | 3 | Not-determined | Genome-fragment | NA | NA | 0 |
| S12-final-viral-combined_k141_192023\|\|full | 2329 | No | NA | 3 | 0 | 2 | Not-determined | Genome-fragment | NA | NA | 0 |
| S12-final-viral-combined_k141_309188\|\|full | 3401 | No | NA | 5 | 0 | 3 | Not-determined | Genome-fragment | NA | NA | 0 |
| S12-final-viral-combined_k141_241513\|\|full | 2770 | No | NA | 5 | 0 | 0 | Not-determined | Genome-fragment | NA | NA | 0 |
| S12-final-viral-combined_k141_341336\|\|full | 2444 | No | NA | 2 | 0 | 1 | Not-determined | Genome-fragment | NA | NA | 0 |
| S12-final-viral-combined_k141_40848\|\|full | 1984 | No | NA | 3 | 0 | 2 | Not-determined | Genome-fragment | NA | NA | 0 |
| S12-final-viral-combined_k141_341347\|\|full | 1682 | No | NA | 2 | 0 | 1 | Not-determined | Genome-fragment | NA | NA | 0 |
| S12-final-viral-combined_k141_141143\|\|full | 1989 | No | NA | 3 | 0 | 2 | Not-determined | Genome-fragment | NA | NA | 0 |
| S12-final-viral-combined_k141_192038\|\|full | 1974 | No | NA | 2 | 0 | 2 | Not-determined | Genome-fragment | NA | NA | 0 |
| S4_k141_87387 | 2784 | No | NA | 3 | 0 | 3 | Low-quality | Genome-fragment | 1.58 | AAI-based (medium-confidence) | 0 |
| S12-final-viral-combined_k141_23861\|\|full | 10779 | No | NA | 15 | 0 | 1 | Not-determined | Genome-fragment | NA | NA | 0 |
| S12-final-viral-combined_k141_375700\|\|full | 4116 | No | NA | 4 | 0 | 3 | Not-determined | Genome-fragment | NA | NA | 0 |
| S12-final-viral-combined_k141_23875\|\|full | 5658 | No | NA | 5 | 0 | 4 | Not-determined | Genome-fragment | NA | NA | 0 |
| S11-final-viral-combined_k141_109695\|\|full | 2785 | No | NA | 3 | 2 | 0 | Low-quality | Genome-fragment | 6.18 | AAI-based (high-confidence) | 0 |
| S11-final-viral-combined_k141_192444\|\|full | 2785 | No | NA | 3 | 3 | 0 | Low-quality | Genome-fragment | 7.26 | AAI-based (high-confidence) | 0 |
| S1-final-viral-combined_k141_374958\|\|full | 2785 | No | NA | 3 | 1 | 0 | Low-quality | Genome-fragment | 0.79 | HMM-based (lower-bound) | 0 |
| S12-final-viral-combined_k141_341380\|\|full | 3298 | No | NA | 4 | 0 | 0 | Not-determined | Genome-fragment | NA | NA | 0 |
| S12-final-viral-combined_k141_309237\|\|full | 2925 | No | NA | 6 | 0 | 0 | Not-determined | Genome-fragment | NA | NA | 0 |
| S12-final-viral-combined_k141_107354\|\|full | 4049 | No | NA | 4 | 0 | 0 | Not-determined | Genome-fragment | NA | NA | 0 |
| S1-final-viral-combined_k141_343398\|\|full | 2785 | No | NA | 3 | 0 | 2 | Low-quality | Genome-fragment | 0.89 | AAI-based (medium-confidence) | 0 |
| S12-final-viral-combined_k141_107356\|\|full | 1583 | No | NA | 3 | 0 | 1 | Not-determined | Genome-fragment | NA | NA | 0 |
| S5-final-viral-combined_k141_57164\|\|lt2gene | 2785 | No | NA | 2 | 2 | 0 | Low-quality | Genome-fragment | 6.98 | AAI-based (high-confidence) | 0 |
| S12-final-viral-combined_k141_104011\|\|full | 2785 | No | NA | 4 | 1 | 0 | Low-quality | Genome-fragment | 1.29 | HMM-based (lower-bound) | 0 |
| S12-final-viral-combined_k141_207952\|\|full | 1954 | No | NA | 3 | 0 | 2 | Not-determined | Genome-fragment | NA | NA | 0 |
| S1-final-viral-combined_k141_390276\|\|full | 2786 | Yes | 286 | 5 | 1 | 2 | Low-quality | Genome-fragment | 0.22 | HMM-based (lower-bound) | 89.73 |
| S8-final-viral-combined_k141_248385\|\|full | 2786 | No | NA | 4 | 0 | 2 | Low-quality | Genome-fragment | 7.26 | AAI-based (high-confidence) | 0 |
| S12-final-viral-combined_k141_207977\|\|full | 1712 | No | NA | 2 | 0 | 2 | Not-determined | Genome-fragment | NA | NA | 0 |
| S12-final-viral-combined_k141_158246\|\|full | 2318 | No | NA | 3 | 0 | 0 | Not-determined | Genome-fragment | NA | NA | 0 |
| S3-final-viral-combined_k141_26678\|\|full | 2787 | No | NA | 5 | 1 | 0 | Low-quality | Genome-fragment | 7.31 | AAI-based (high-confidence) | 0 |
| S12-final-viral-combined_k141_309365\|\|full | 1571 | No | NA | 3 | 0 | 0 | Not-determined | Genome-fragment | NA | NA | 0 |
| S12-final-viral-combined_k141_241559\|\|full | 2847 | No | NA | 4 | 0 | 3 | Not-determined | Genome-fragment | NA | NA | 0 |
| S3-final-viral-combined_k141_137528\|\|full | 2787 | No | NA | 6 | 0 | 0 | Low-quality | Genome-fragment | 3.9 | AAI-based (medium-confidence) | 0 |
| S12-final-viral-combined_k141_208007\|\|full | 1334 | No | NA | 3 | 0 | 0 | Not-determined | Genome-fragment | NA | NA | 0 |
| S12-final-viral-combined_k141_325547\|\|full | 8891 | No | NA | 9 | 0 | 6 | Not-determined | Genome-fragment | NA | NA | 0 |
| S12-final-viral-combined_k141_238144\|\|full | 2787 | No | NA | 3 | 0 | 0 | Low-quality | Genome-fragment | 3.18 | AAI-based (medium-confidence) | 0 |
| S12-final-viral-combined_k141_258130\|\|full | 2021 | No | NA | 3 | 0 | 2 | Not-determined | Genome-fragment | NA | NA | 0 |
| S12-final-viral-combined_k141_305852\|\|full | 2787 | No | NA | 2 | 0 | 1 | Low-quality | Genome-fragment | 4.61 | AAI-based (medium-confidence) | 0 |
| S11-final-viral-combined_k141_46553\|\|full | 2788 | No | NA | 3 | 0 | 1 | Low-quality | Genome-fragment | 4.71 | AAI-based (high-confidence) | 0 |
| S2-final-viral-combined_k141_177980\|\|full | 2788 | No | NA | 3 | 1 | 0 | Low-quality | Genome-fragment | 1.51 | HMM-based (lower-bound) | 0 |
| S12-final-viral-combined_k141_192144\|\|full | 2465 | No | NA | 3 | 0 | 1 | Not-determined | Genome-fragment | NA | NA | 0 |
| S12-final-viral-combined_k141_341469\|\|full | 4099 | No | NA | 5 | 0 | 0 | Not-determined | Genome-fragment | NA | NA | 0 |
| S1-final-viral-combined_k141_105650\|\|full | 2789 | Yes | 1289 | 6 | 1 | 1 | Low-quality | Genome-fragment | 3.34 | AAI-based (high-confidence) | 53.78 |
| S12-final-viral-combined_k141_392591\|\|full | 2061 | No | NA | 2 | 0 | 0 | Not-determined | Genome-fragment | NA | NA | 0 |
| S12-final-viral-combined_k141_56943\|\|full | 1029 | No | NA | 3 | 0 | 0 | Not-determined | Genome-fragment | NA | NA | 0 |
| S12-final-viral-combined_k141_7035\|\|full | 5833 | No | NA | 5 | 0 | 4 | Not-determined | Genome-fragment | NA | NA | 0 |
| S7-final-viral-combined_k141_240142\|\|full | 2789 | No | NA | 5 | 0 | 0 | Low-quality | Genome-fragment | 5.87 | AAI-based (medium-confidence) | 0 |
| S12-final-viral-combined_k141_357980\|\|full | 3101 | No | NA | 4 | 0 | 2 | Not-determined | Genome-fragment | NA | NA | 0 |
| S12-final-viral-combined_k141_7038\|\|full | 3696 | No | NA | 3 | 0 | 1 | Not-determined | Genome-fragment | NA | NA | 0 |
| S12-final-viral-combined_k141_174546\|\|full | 1674 | No | NA | 4 | 0 | 0 | Not-determined | Genome-fragment | NA | NA | 0 |
| S6-final-viral-combined_k141_122927\|\|full | 2789 | No | NA | 4 | 0 | 1 | Low-quality | Genome-fragment | 1.34 | AAI-based (medium-confidence) | 0 |
| S9-final-viral-combined_k141_50415\|\|full | 2790 | No | NA | 3 | 0 | 0 | Low-quality | Genome-fragment | 3.01 | AAI-based (high-confidence) | 0 |
| S5-final-viral-combined_k141_186501\|\|full | 2791 | Yes | 2088 | 4 | 1 | 1 | Low-quality | Genome-fragment | 1.31 | HMM-based (lower-bound) | 25.19 |
| S12-final-viral-combined_k141_56953\|\|full | 2333 | No | NA | 3 | 0 | 1 | Not-determined | Genome-fragment | NA | NA | 0 |
| S12-final-viral-combined_k141_7053\|\|full | 4275 | No | NA | 8 | 0 | 2 | Not-determined | Genome-fragment | NA | NA | 0 |
| S7-final-viral-combined_k141_62438\|\|full | 2791 | No | NA | 4 | 0 | 3 | Low-quality | Genome-fragment | 2.28 | AAI-based (high-confidence) | 0 |
| S2-final-viral-combined_k141_157626\|\|full | 2791 | No | NA | 3 | 0 | 2 | Low-quality | Genome-fragment | 1.94 | AAI-based (medium-confidence) | 0 |
| S12-final-viral-combined_k141_275301\|\|full | 2852 | No | NA | 3 | 0 | 1 | Not-determined | Genome-fragment | NA | NA | 0 |
| S12-final-viral-combined_k141_56986\|\|full | 2360 | No | NA | 3 | 0 | 2 | Not-determined | Genome-fragment | NA | NA | 0 |
| S12-final-viral-combined_k141_375876\|\|full | 1866 | No | NA | 2 | 0 | 0 | Not-determined | Genome-fragment | NA | NA | 0 |
| S12-final-viral-combined_k141_309525\|\|full | 7748 | No | NA | 8 | 0 | 7 | Not-determined | Genome-fragment | NA | NA | 0 |
| S12-final-viral-combined_k141_292370\|\|full | 1758 | No | NA | 4 | 0 | 0 | Not-determined | Genome-fragment | NA | NA | 0 |
| S12-final-viral-combined_k141_7068\|\|full | 4083 | No | NA | 2 | 0 | 2 | Not-determined | Genome-fragment | NA | NA | 0 |
| S12-final-viral-combined_k141_158445\|\|full | 1676 | No | NA | 2 | 0 | 2 | Not-determined | Genome-fragment | NA | NA | 0 |
| S12-final-viral-combined_k141_392657\|\|full | 1257 | No | NA | 2 | 0 | 1 | Not-determined | Genome-fragment | NA | NA | 0 |
| S5-final-viral-combined_k141_117729\|\|full | 2792 | No | NA | 5 | 0 | 0 | Low-quality | Genome-fragment | 6.64 | AAI-based (medium-confidence) | 0 |
| S12-final-viral-combined_k141_341576\|\|full | 2166 | No | NA | 3 | 0 | 3 | Not-determined | Genome-fragment | NA | NA | 0 |
| S12-final-viral-combined_k141_375911\|\|full | 1426 | No | NA | 2 | 0 | 2 | Not-determined | Genome-fragment | NA | NA | 0 |
| S12-final-viral-combined_k141_292405\|\|full | 2842 | No | NA | 3 | 0 | 0 | Not-determined | Genome-fragment | NA | NA | 0 |
| S17-k141_241280 | 2792 | No | NA | 3 | 0 | 2 | Low-quality | Genome-fragment | 3.66 | AAI-based (medium-confidence) | 0 |
| S12-final-viral-combined_k141_341578\|\|full | 6019 | No | NA | 4 | 0 | 3 | Not-determined | Genome-fragment | NA | NA | 0 |
| S12-final-viral-combined_k141_84630\|\|full | 2792 | No | NA | 4 | 0 | 0 | Low-quality | Genome-fragment | 4.58 | AAI-based (medium-confidence) | 0 |
| S12-final-viral-combined_k141_41160\|\|full | 22653 | No | NA | 29 | 8 | 1 | Medium-quality | Genome-fragment | 56.12 | AAI-based (medium-confidence) | 0 |
| S17-final-viral-combined_k141_142809\|\|full | 2793 | No | NA | 3 | 0 | 0 | Low-quality | Genome-fragment | 2.49 | AAI-based (high-confidence) | 0 |
| S12-final-viral-combined_k141_325718\|\|full | 4099 | No | NA | 3 | 0 | 3 | Not-determined | Genome-fragment | NA | NA | 0 |
| S12-final-viral-combined_k141_141368\|\|full | 2271 | No | NA | 3 | 0 | 1 | Not-determined | Genome-fragment | NA | NA | 0 |
| S12-final-viral-combined_k141_341629\|\|full | 1788 | No | NA | 4 | 0 | 0 | Not-determined | Genome-fragment | NA | NA | 0 |
| S12-final-viral-combined_k141_44435\|\|full | 2793 | No | NA | 4 | 1 | 0 | Low-quality | Genome-fragment | 3.48 | AAI-based (high-confidence) | 0 |
| S11-final-viral-combined_k141_256995\|\|full | 2794 | No | NA | 4 | 2 | 0 | Low-quality | Genome-fragment | 3.39 | AAI-based (high-confidence) | 0 |
| S12-final-viral-combined_k141_7174\|\|full | 25484 | No | NA | 33 | 18 | 0 | Medium-quality | Genome-fragment | 53.42 | AAI-based (high-confidence) | 0 |
| S12-final-viral-combined_k141_309621\|\|full | 3094 | No | NA | 2 | 0 | 1 | Not-determined | Genome-fragment | NA | NA | 0 |
| S12-final-viral-combined_k141_192314\|\|full | 9097 | No | NA | 7 | 0 | 6 | Not-determined | Genome-fragment | NA | NA | 0 |
| S12-final-viral-combined_k141_225052\|\|full | 6903 | No | NA | 10 | 0 | 3 | Not-determined | Genome-fragment | NA | NA | 0 |
| S1-final-viral-combined_k141_366627\|\|full | 2794 | No | NA | 5 | 0 | 0 | Low-quality | Genome-fragment | 18.09 | AAI-based (high-confidence) | 0 |
| S12-final-viral-combined_k141_292496\|\|full | 4123 | No | NA | 2 | 0 | 1 | Not-determined | Genome-fragment | NA | NA | 0 |
| S12-final-viral-combined_k141_174714\|\|full | 1892 | No | NA | 4 | 0 | 0 | Not-determined | Genome-fragment | NA | NA | 0 |
| S12-final-viral-combined_k141_174716\|\|full | 1501 | No | NA | 2 | 0 | 0 | Not-determined | Genome-fragment | NA | NA | 0 |
| S7-final-viral-combined_k141_42766\|\|full | 2794 | No | NA | 5 | 0 | 1 | Low-quality | Genome-fragment | 7.46 | AAI-based (high-confidence) | 0 |
| S12-final-viral-combined_k141_90819\|\|full | 1368 | No | NA | 2 | 0 | 1 | Not-determined | Genome-fragment | NA | NA | 0 |
| S12-final-viral-combined_k141_131772\|\|full | 2794 | No | NA | 4 | 0 | 0 | Low-quality | Genome-fragment | 1.59 | AAI-based (medium-confidence) | 0 |
| S8-final-viral-combined_k141_208348\|\|full | 2794 | Yes | 1876 | 3 | 1 | 1 | Low-quality | Genome-fragment | 4.27 | AAI-based (medium-confidence) | 32.86 |
| S9-final-viral-combined_k141_115184\|\|full | 2795 | No | NA | 6 | 5 | 0 | Low-quality | Genome-fragment | 6.04 | AAI-based (high-confidence) | 0 |
| S12-final-viral-combined_k141_24254\|\|full | 1596 | No | NA | 2 | 0 | 1 | Not-determined | Genome-fragment | NA | NA | 0 |
| S1-final-viral-combined_k141_15076\|\|full | 2795 | No | NA | 4 | 3 | 0 | Low-quality | Genome-fragment | 4.43 | AAI-based (high-confidence) | 0 |
| S12-final-viral-combined_k141_376009\|\|full | 1237 | No | NA | 2 | 0 | 1 | Not-determined | Genome-fragment | NA | NA | 0 |
| S12-final-viral-combined_k141_225092\|\|full | 2468 | No | NA | 2 | 0 | 0 | Not-determined | Genome-fragment | NA | NA | 0 |
| S12-final-viral-combined_k141_358010\|\|full | 57083 | Yes | 52856 | 77 | 11 | 4 | High-quality | High-quality | 100 | AAI-based (medium-confidence) | 7.41 |
| S2-final-viral-combined_k141_199781\|\|full | 2796 | No | NA | 5 | 2 | 0 | Low-quality | Genome-fragment | 6.56 | AAI-based (high-confidence) | 0 |
| S12-final-viral-combined_k141_341757\|\|full | 3191 | No | NA | 4 | 0 | 2 | Not-determined | Genome-fragment | NA | NA | 0 |
| S2-final-viral-combined_k141_113184\|\|full | 2796 | No | NA | 4 | 0 | 3 | Low-quality | Genome-fragment | 0.74 | AAI-based (medium-confidence) | 0 |
| S6-final-viral-combined_k141_140344\|\|full | 2796 | No | NA | 3 | 0 | 3 | Low-quality | Genome-fragment | 5.34 | AAI-based (medium-confidence) | 0 |
| S11-final-viral-combined_k141_48556\|\|full | 2797 | No | NA | 6 | 2 | 0 | Low-quality | Genome-fragment | 6.96 | AAI-based (medium-confidence) | 0 |
| S4_k141_32645 | 2797 | No | NA | 4 | 0 | 0 | Low-quality | Genome-fragment | 5.86 | AAI-based (medium-confidence) | 0 |
| S12-final-viral-combined_k141_392791\|\|full | 4568 | No | NA | 6 | 0 | 0 | Not-determined | Genome-fragment | NA | NA | 0 |
| S12-final-viral-combined_k141_225176\|\|full | 1833 | No | NA | 3 | 0 | 1 | Not-determined | Genome-fragment | NA | NA | 0 |
| S12-final-viral-combined_k141_90895\|\|full | 3508 | No | NA | 5 | 0 | 1 | Not-determined | Genome-fragment | NA | NA | 0 |
| S11-final-viral-combined_k141_119910\|\|full | 2798 | No | NA | 5 | 3 | 0 | Low-quality | Genome-fragment | 3.61 | HMM-based (lower-bound) | 0 |
| S1-final-viral-combined_k141_285138\|\|full | 2798 | No | NA | 9 | 0 | 0 | Low-quality | Genome-fragment | 7.4 | AAI-based (high-confidence) | 0 |
| S12-final-viral-combined_k141_57237\|\|full | 2031 | No | NA | 2 | 0 | 1 | Not-determined | Genome-fragment | NA | NA | 0 |
| S12-final-viral-combined_k141_210901\|\|full | 2798 | No | NA | 5 | 0 | 0 | Low-quality | Genome-fragment | 3.37 | AAI-based (medium-confidence) | 0 |
| S12-final-viral-combined_k141_141590\|\|full | 1009 | No | NA | 2 | 0 | 1 | Not-determined | Genome-fragment | NA | NA | 0 |
| S12-final-viral-combined_k141_141602\|\|full | 2719 | No | NA | 3 | 0 | 2 | Not-determined | Genome-fragment | NA | NA | 0 |
| S2-final-viral-combined_k141_214163\|\|full | 2798 | No | NA | 11 | 1 | 0 | Low-quality | Genome-fragment | 6.62 | AAI-based (high-confidence) | 0 |
| S12-final-viral-combined_k141_7337\|\|full | 2454 | No | NA | 2 | 0 | 1 | Not-determined | Genome-fragment | NA | NA | 0 |
| S4-final-viral-combined_k141_29636\|\|full | 2798 | Yes | 1095 | 4 | 1 | 1 | Low-quality | Genome-fragment | 0.7 | HMM-based (lower-bound) | 60.86 |
| S11-final-viral-combined_k141_184869\|\|full | 2799 | No | NA | 5 | 2 | 0 | Low-quality | Genome-fragment | 6 | AAI-based (high-confidence) | 0 |
| S12-final-viral-combined_k141_141624\|\|full | 6035 | No | NA | 9 | 0 | 1 | Not-determined | Genome-fragment | NA | NA | 0 |
| S4-final-viral-combined_k141_46794\|\|full | 2799 | No | NA | 3 | 0 | 2 | Low-quality | Genome-fragment | 5.91 | AAI-based (medium-confidence) | 0 |
| S12-final-viral-combined_k141_225264\|\|full | 2285 | No | NA | 2 | 0 | 2 | Not-determined | Genome-fragment | NA | NA | 0 |
| S1-final-viral-combined_k141_131786\|\|full | 2800 | No | NA | 3 | 0 | 1 | Low-quality | Genome-fragment | 6.03 | AAI-based (high-confidence) | 0 |
| S12-final-viral-combined_k141_192506\|\|full | 11301 | No | NA | 8 | 0 | 5 | Not-determined | Genome-fragment | NA | NA | 0 |
| S12-final-viral-combined_k141_242052\|\|full | 1351 | No | NA | 2 | 0 | 1 | Not-determined | Genome-fragment | NA | NA | 0 |
| S12-final-viral-combined_k141_57334\|\|full | 1113 | No | NA | 2 | 0 | 1 | Not-determined | Genome-fragment | NA | NA | 0 |
| S1-final-viral-combined_k141_73034\|\|full | 2800 | No | NA | 3 | 0 | 3 | Low-quality | Genome-fragment | 2.47 | AAI-based (medium-confidence) | 0 |
| S3-final-viral-combined_k141_25636\|\|full | 2800 | No | NA | 7 | 2 | 0 | Low-quality | Genome-fragment | 4.83 | AAI-based (high-confidence) | 0 |
| S12-final-viral-combined_k141_225333\|\|full | 1346 | No | NA | 2 | 0 | 1 | Not-determined | Genome-fragment | NA | NA | 0 |
| S12-final-viral-combined_k141_174831\|\|full | 2082 | No | NA | 3 | 0 | 1 | Not-determined | Genome-fragment | NA | NA | 0 |
| S12-final-viral-combined_k141_56376\|\|full | 2800 | No | NA | 3 | 0 | 0 | Low-quality | Genome-fragment | 6.23 | AAI-based (high-confidence) | 0 |
| S2_k141_201818 | 2800 | No | NA | 6 | 0 | 1 | Low-quality | Genome-fragment | 6.84 | AAI-based (medium-confidence) | 0 |

| | | | | | | | | | | | |
|---|---|---|---|---|---|---|---|---|---|---|---|
| S12-final-viral-combined_k141_107738\|\|full | 2266 | No | NA | 4 | 0 | 0 | Not-determined | Genome-fragment | NA | NA | 0 |
| S12-final-viral-combined_k141_225395\|\|full | 1209 | No | NA | 4 | 0 | 0 | Not-determined | Genome-fragment | NA | NA | 0 |
| S12-final-viral-combined_k141_275743\|\|full | 2817 | No | NA | 3 | 0 | 2 | Not-determined | Genome-fragment | NA | NA | 0 |
| S9-final-viral-combined_k141_89958\|\|full | 2801 | Yes | 2249 | 5 | 1 | 1 | Low-quality | Genome-fragment | 4.12 | AAI-based (medium-confidence) | 19.71 |
| S11-final-viral-combined_k141_230749\|\|lt2gene | 2801 | No | NA | 2 | 1 | 0 | Low-quality | Genome-fragment | 6.2 | AAI-based (medium-confidence) | 0 |
| S12-final-viral-combined_k141_376177\|\|full | 2001 | No | NA | 3 | 0 | 3 | Not-determined | Genome-fragment | NA | NA | 0 |
| S12-final-viral-combined_k141_7541\|\|full | 1851 | No | NA | 2 | 0 | 0 | Not-determined | Genome-fragment | NA | NA | 0 |
| S17-final-viral-combined_k141_211514\|\|full | 2803 | No | NA | 4 | 0 | 0 | Low-quality | Genome-fragment | 5.38 | AAI-based (medium-confidence) | 0 |
| S12-final-viral-combined_k141_158722\|\|full | 1484 | No | NA | 2 | 0 | 1 | Not-determined | Genome-fragment | NA | NA | 0 |
| S2-final-viral-combined_k141_159729\|\|full | 2803 | No | NA | 6 | 1 | 0 | Low-quality | Genome-fragment | 1.78 | HMM-based (lower-bound) | 0 |
| S12-final-viral-combined_k141_57449\|\|full | 3025 | No | NA | 3 | 0 | 1 | Not-determined | Genome-fragment | NA | NA | 0 |
| S12-final-viral-combined_k141_57471\|\|full | 3778 | No | NA | 5 | 0 | 0 | Not-determined | Genome-fragment | NA | NA | 0 |
| S1-final-viral-combined_k141_151256\|\|full | 2804 | No | NA | 3 | 0 | 1 | Low-quality | Genome-fragment | 1.76 | AAI-based (medium-confidence) | 0 |
| S2-final-viral-combined_k141_203588\|\|full | 2804 | No | NA | 5 | 0 | 0 | Low-quality | Genome-fragment | 3.51 | AAI-based (medium-confidence) | 0 |
| S12-final-viral-combined_k141_124131\|\|full | 2308 | No | NA | 3 | 0 | 3 | Not-determined | Genome-fragment | NA | NA | 0 |
| S12-final-viral-combined_k141_24433\|\|full | 1347 | No | NA | 3 | 0 | 2 | Not-determined | Genome-fragment | NA | NA | 0 |
| S12-final-viral-combined_k141_158772\|\|full | 5808 | No | NA | 4 | 0 | 2 | Not-determined | Genome-fragment | NA | NA | 0 |
| S12-final-viral-combined_k141_376237\|\|full | 6458 | No | NA | 5 | 0 | 2 | Not-determined | Genome-fragment | NA | NA | 0 |
| S12-final-viral-combined_k141_7620\|\|full | 1398 | No | NA | 5 | 0 | 0 | Not-determined | Genome-fragment | NA | NA | 0 |
| S7-final-viral-combined_k141_12023\|\|full | 2805 | No | NA | 3 | 0 | 1 | Low-quality | Genome-fragment | 4.64 | AAI-based (medium-confidence) | 0 |
| S7-final-viral-combined_k141_178445\|\|full | 2805 | No | NA | 3 | 1 | 0 | Low-quality | Genome-fragment | 3.15 | HMM-based (lower-bound) | 0 |
| S17-final-viral-combined_k141_16830\|\|full | 2805 | No | NA | 10 | 0 | 0 | Low-quality | Genome-fragment | 6.8 | AAI-based (high-confidence) | 0 |
| S12-final-viral-combined_k141_376272\|\|full | 1124 | No | NA | 2 | 0 | 1 | Not-determined | Genome-fragment | NA | NA | 0 |
| S12-final-viral-combined_k141_75176\|\|full | 2394 | No | NA | 3 | 0 | 1 | Not-determined | Genome-fragment | NA | NA | 0 |
| S2-final-viral-combined_k141_10633\|\|full | 2805 | No | NA | 6 | 0 | 0 | Low-quality | Genome-fragment | 4.26 | AAI-based (high-confidence) | 0 |
| S12-final-viral-combined_k141_242218\|\|full | 1172 | No | NA | 2 | 0 | 1 | Not-determined | Genome-fragment | NA | NA | 0 |
| S12-final-viral-combined_k141_41530\|\|full | 1431 | No | NA | 3 | 0 | 2 | Not-determined | Genome-fragment | NA | NA | 0 |
| S12-final-viral-combined_k141_75191\|\|full | 4143 | No | NA | 4 | 0 | 4 | Not-determined | Genome-fragment | NA | NA | 0 |
| S12-final-viral-combined_k141_225537\|\|full | 5267 | No | NA | 8 | 0 | 4 | Not-determined | Genome-fragment | NA | NA | 0 |
| S12-final-viral-combined_k141_7666\|\|full | 1604 | No | NA | 3 | 0 | 2 | Not-determined | Genome-fragment | NA | NA | 0 |
| S11-final-viral-combined_k141_71662\|\|full | 2806 | No | NA | 4 | 0 | 0 | Low-quality | Genome-fragment | 1.8 | AAI-based (medium-confidence) | 0 |
| S12-final-viral-combined_k141_7668\|\|full | 2085 | No | NA | 2 | 0 | 1 | Not-determined | Genome-fragment | NA | NA | 0 |
| S11_k141_289779 | 2806 | No | NA | 4 | 4 | 0 | Low-quality | Genome-fragment | 6.44 | AAI-based (high-confidence) | 0 |
| S12-final-viral-combined_k141_24521\|\|full | 1621 | No | NA | 4 | 0 | 0 | Not-determined | Genome-fragment | NA | NA | 0 |
| S12-final-viral-combined_k141_342095\|\|full | 2947 | No | NA | 3 | 0 | 3 | Not-determined | Genome-fragment | NA | NA | 0 |
| S12-final-viral-combined_k141_242274\|\|full | 2029 | No | NA | 2 | 0 | 1 | Not-determined | Genome-fragment | NA | NA | 0 |
| S7-final-viral-combined_k141_78001\|\|full | 2806 | No | NA | 4 | 0 | 3 | Low-quality | Genome-fragment | 2.25 | AAI-based (medium-confidence) | 0 |
| S2-final-viral-combined_k141_163935\|\|full | 2806 | No | NA | 6 | 2 | 0 | Low-quality | Genome-fragment | 6.47 | AAI-based (high-confidence) | 0 |
| S4-final-viral-combined_k141_6281\|\|full | 2806 | No | NA | 3 | 0 | 1 | Low-quality | Genome-fragment | 0.82 | AAI-based (medium-confidence) | 0 |
| S12-final-viral-combined_k141_342134\|\|full | 1057 | No | NA | 3 | 0 | 2 | Not-determined | Genome-fragment | NA | NA | 0 |
| S2-final-viral-combined_k141_44026\|\|full | 2807 | No | NA | 4 | 0 | 3 | Low-quality | Genome-fragment | 4.77 | AAI-based (high-confidence) | 0 |
| S8-final-viral-combined_k141_216926\|\|full | 2807 | No | NA | 4 | 0 | 0 | Low-quality | Genome-fragment | 7.33 | AAI-based (medium-confidence) | 0 |
| S1-final-viral-combined_k141_346687\|\|full | 2808 | No | NA | 4 | 2 | 0 | Low-quality | Genome-fragment | 6.7 | AAI-based (high-confidence) | 0 |
| S12-final-viral-combined_k141_7722\|\|full | 2434 | No | NA | 3 | 0 | 2 | Not-determined | Genome-fragment | NA | NA | 0 |
| S5-final-viral-combined_k141_80100\|\|full | 2808 | No | NA | 5 | 0 | 0 | Low-quality | Genome-fragment | 9.71 | AAI-based (medium-confidence) | 0 |
| S12-final-viral-combined_k141_358269\|\|full | 5159 | No | NA | 6 | 0 | 0 | Not-determined | Genome-fragment | NA | NA | 0 |
| S12-final-viral-combined_k141_158878\|\|full | 3045 | No | NA | 6 | 0 | 3 | Not-determined | Genome-fragment | NA | NA | 0 |
| S6-final-viral-combined_k141_42814\|\|full | 2808 | No | NA | 2 | 0 | 2 | Low-quality | Genome-fragment | 2.84 | AAI-based (medium-confidence) | 0 |
| S11-final-viral-combined_k141_326657\|\|full | 2809 | No | NA | 3 | 0 | 3 | Low-quality | Genome-fragment | 3.29 | AAI-based (high-confidence) | 0 |
| S1_k141_342989 | 2809 | No | NA | 1 | 0 | 1 | Low-quality | Genome-fragment | 2.53 | AAI-based (medium-confidence) | 0 |
| S12-final-viral-combined_k141_24590\|\|full | 1946 | No | NA | 3 | 0 | 1 | Not-determined | Genome-fragment | NA | NA | 0 |
| S3-final-viral-combined_k141_28261\|\|full | 2809 | No | NA | 3 | 0 | 0 | Low-quality | Genome-fragment | 3.18 | AAI-based (medium-confidence) | 0 |
| S12-final-viral-combined_k141_358303\|\|full | 2470 | No | NA | 2 | 0 | 2 | Not-determined | Genome-fragment | NA | NA | 0 |
| S12-final-viral-combined_k141_158892\|\|full | 2691 | No | NA | 2 | 0 | 1 | Not-determined | Genome-fragment | NA | NA | 0 |
| S12-final-viral-combined_k141_91175\|\|full | 3242 | No | NA | 3 | 0 | 3 | Not-determined | Genome-fragment | NA | NA | 0 |
| S2-final-viral-combined_k141_18335\|\|full | 2809 | No | NA | 6 | 1 | 0 | Low-quality | Genome-fragment | 5.91 | AAI-based (high-confidence) | 0 |
| S8-final-viral-combined_k141_232121\|\|full | 2809 | No | NA | 2 | 1 | 0 | Low-quality | Genome-fragment | 2.16 | HMM-based (lower-bound) | 0 |
| S12-final-viral-combined_k141_358315\|\|full | 4855 | No | NA | 4 | 0 | 2 | Not-determined | Genome-fragment | NA | NA | 0 |
| S3-final-viral-combined_k141_16369\|\|full | 2810 | No | NA | 5 | 0 | 0 | Low-quality | Genome-fragment | 6.04 | AAI-based (high-confidence) | 0 |
| S5-final-viral-combined_k141_71638\|\|full | 2810 | No | NA | 2 | 2 | 0 | Low-quality | Genome-fragment | 7.43 | AAI-based (high-confidence) | 0 |
| S5-final-viral-combined_k141_171584\|\|full | 2810 | No | NA | 8 | 3 | 0 | Low-quality | Genome-fragment | 6.66 | AAI-based (high-confidence) | 0 |
| S12-final-viral-combined_k141_7773\|\|full | 1011 | No | NA | 2 | 0 | 0 | Not-determined | Genome-fragment | NA | NA | 0 |
| S12-final-viral-combined_k141_75351\|\|full | 1241 | No | NA | 3 | 0 | 0 | Not-determined | Genome-fragment | NA | NA | 0 |
| S5-final-viral-combined_k141_80700\|\|full | 2811 | No | NA | 3 | 0 | 1 | Low-quality | Genome-fragment | 1.28 | AAI-based (medium-confidence) | 0 |
| S12-final-viral-combined_k141_111170\|\|full | 2811 | No | NA | 6 | 1 | 0 | Low-quality | Genome-fragment | 2.01 | HMM-based (lower-bound) | 0 |
| S12-final-viral-combined_k141_258818\|\|full | 2632 | No | NA | 3 | 0 | 2 | Not-determined | Genome-fragment | NA | NA | 0 |
| S1_k141_116384 | 2812 | No | NA | 6 | 2 | 0 | Low-quality | Genome-fragment | 6.55 | AAI-based (high-confidence) | 0 |
| S12-final-viral-combined_k141_75367\|\|full | 4911 | No | NA | 6 | 0 | 2 | Not-determined | Genome-fragment | NA | NA | 0 |
| S12-final-viral-combined_k141_276054\|\|full | 1196 | No | NA | 2 | 0 | 0 | Not-determined | Genome-fragment | NA | NA | 0 |
| S12-final-viral-combined_k141_75379\|\|full | 1042 | No | NA | 2 | 0 | 0 | Not-determined | Genome-fragment | NA | NA | 0 |
| S5-final-viral-combined_k141_1537\|\|full | 2812 | No | NA | 4 | 0 | 1 | Low-quality | Genome-fragment | 6.5 | AAI-based (medium-confidence) | 0 |
| S12-final-viral-combined_k141_41697\|\|full | 3943 | No | NA | 4 | 0 | 1 | Not-determined | Genome-fragment | NA | NA | 0 |
| S12-final-viral-combined_k141_24666\|\|full | 3724 | No | NA | 3 | 0 | 1 | Not-determined | Genome-fragment | NA | NA | 0 |
| S12-final-viral-combined_k141_376326\|\|full | 46498 | No | NA | 56 | 21 | 0 | Medium-quality | Genome-fragment | 78.92 | AAI-based (medium-confidence) | 0 |
| S12-final-viral-combined_k141_142052\|\|full | 1350 | No | NA | 4 | 0 | 0 | Not-determined | Genome-fragment | NA | NA | 0 |
| S7-final-viral-combined_k141_133607\|\|full | 2812 | No | NA | 4 | 0 | 0 | Low-quality | Genome-fragment | 3.51 | AAI-based (high-confidence) | 0 |
| S17-k141_3095 | 2812 | No | NA | 6 | 0 | 1 | Low-quality | Genome-fragment | 4.81 | AAI-based (medium-confidence) | 0 |
| S12-final-viral-combined_k141_376348\|\|full | 1980 | No | NA | 4 | 0 | 0 | Not-determined | Genome-fragment | NA | NA | 0 |
| S12-final-viral-combined_k141_142083\|\|full | 2432 | No | NA | 3 | 0 | 0 | Not-determined | Genome-fragment | NA | NA | 0 |
| S12-final-viral-combined_k141_91231\|\|full | 49609 | No | NA | 37 | 3 | 3 | Medium-quality | Genome-fragment | 78.28 | AAI-based (high-confidence) | 0 |
| S12-final-viral-combined_k141_173720\|\|full | 2812 | No | NA | 3 | 0 | 1 | Low-quality | Genome-fragment | 5.98 | AAI-based (medium-confidence) | 0 |
| S12-final-viral-combined_k141_127543\|\|full | 2812 | No | NA | 4 | 0 | 1 | Low-quality | Genome-fragment | 2.68 | AAI-based (medium-confidence) | 0 |
| S12-final-viral-combined_k141_107942\|\|full | 30890 | No | NA | 37 | 23 | 0 | Medium-quality | Genome-fragment | 51.21 | AAI-based (high-confidence) | 0 |
| S12-final-viral-combined_k141_159015\|\|full | 7414 | No | NA | 7 | 0 | 7 | Not-determined | Genome-fragment | NA | NA | 0 |
| S12-final-viral-combined_k141_297529\|\|full | 2812 | No | NA | 3 | 0 | 1 | Low-quality | Genome-fragment | 3.32 | AAI-based (medium-confidence) | 0 |
| S6-final-viral-combined_k141_125878\|\|full | 2813 | No | NA | 4 | 0 | 1 | Low-quality | Genome-fragment | 1.76 | AAI-based (medium-confidence) | 0 |
| S12-final-viral-combined_k141_293286\|\|full | 1493 | No | NA | 3 | 0 | 1 | Not-determined | Genome-fragment | NA | NA | 0 |
| S12-final-viral-combined_k141_393277\|\|full | 6529 | No | NA | 7 | 0 | 1 | Not-determined | Genome-fragment | NA | NA | 0 |
| S12-final-viral-combined_k141_393282\|\|full | 1135 | No | NA | 2 | 0 | 1 | Not-determined | Genome-fragment | NA | NA | 0 |
| S12-final-viral-combined_k141_376377\|\|full | 6114 | No | NA | 2 | 0 | 2 | Not-determined | Genome-fragment | NA | NA | 0 |
| S12-final-viral-combined_k141_366161\|\|full | 2814 | No | NA | 3 | 0 | 1 | Low-quality | Genome-fragment | 6.5 | AAI-based (high-confidence) | 0 |
| S8-final-viral-combined_k141_63937\|\|full | 2814 | No | NA | 2 | 0 | 1 | Low-quality | Genome-fragment | 0.9 | AAI-based (high-confidence) | 0 |
| S12-final-viral-combined_k141_192997\|\|full | 2417 | No | NA | 3 | 0 | 1 | Not-determined | Genome-fragment | NA | NA | 0 |
| S9-final-viral-combined_k141_65678\|\|full | 2815 | No | NA | 5 | 1 | 0 | Low-quality | Genome-fragment | 5.47 | AAI-based (high-confidence) | 0 |
| S11-final-viral-combined_k141_163285\|\|full | 2815 | No | NA | 2 | 0 | 2 | Low-quality | Genome-fragment | 1.29 | AAI-based (medium-confidence) | 0 |
| S12-final-viral-combined_k141_193023\|\|full | 1576 | No | NA | 2 | 0 | 1 | Not-determined | Genome-fragment | NA | NA | 0 |
| S12-final-viral-combined_k141_91284\|\|full | 1703 | No | NA | 3 | 0 | 1 | Not-determined | Genome-fragment | NA | NA | 0 |
| S11-final-viral-combined_k141_287534\|\|full | 2815 | No | NA | 3 | 2 | 0 | Low-quality | Genome-fragment | 4.44 | HMM-based (lower-bound) | 0 |
| S1-final-viral-combined_k141_291219\|\|full | 2815 | No | NA | 3 | 0 | 1 | Low-quality | Genome-fragment | 3.82 | AAI-based (medium-confidence) | 0 |
| S17-final-viral-combined_k141_134948\|\|full | 2815 | No | NA | 6 | 0 | 1 | Low-quality | Genome-fragment | 5.37 | AAI-based (medium-confidence) | 0 |
| S12-final-viral-combined_k141_124392\|\|full | 4736 | No | NA | 8 | 0 | 0 | Not-determined | Genome-fragment | NA | NA | 0 |
| S12_k141_274512 | 2815 | No | NA | 3 | 3 | 0 | Low-quality | Genome-fragment | 7.51 | AAI-based (high-confidence) | 0 |
| S12-final-viral-combined_k141_75502\|\|full | 2150 | No | NA | 3 | 0 | 0 | Not-determined | Genome-fragment | NA | NA | 0 |
| S9-final-viral-combined_k141_70318\|\|full | 2816 | No | NA | 6 | 1 | 0 | Low-quality | Genome-fragment | 5.26 | AAI-based (medium-confidence) | 0 |
| S1-final-viral-combined_k141_382632\|\|full | 2816 | No | NA | 3 | 0 | 1 | Low-quality | Genome-fragment | 1.79 | AAI-based (high-confidence) | 0 |
| S12-final-viral-combined_k141_276249\|\|full | 1891 | No | NA | 6 | 0 | 3 | Not-determined | Genome-fragment | NA | NA | 0 |
| S17-final-viral-combined_k141_161742\|\|full | 2816 | No | NA | 4 | 0 | 0 | Low-quality | Genome-fragment | 49.73 | AAI-based (medium-confidence) | 0 |
| S12-final-viral-combined_k141_392609\|\|full | 2816 | No | NA | 3 | 0 | 2 | Low-quality | Genome-fragment | 3.41 | AAI-based (medium-confidence) | 0 |
| S12-final-viral-combined_k141_276263\|\|full | 1705 | No | NA | 2 | 0 | 1 | Not-determined | Genome-fragment | NA | NA | 0 |
| S12-final-viral-combined_k141_342430\|\|full | 4522 | No | NA | 4 | 0 | 4 | Not-determined | Genome-fragment | NA | NA | 0 |
| S11-final-viral-combined_k141_126093\|\|full | 2817 | No | NA | 3 | 0 | 0 | Low-quality | Genome-fragment | 6.18 | AAI-based (medium-confidence) | 0 |
| S1_k141_165445 | 2817 | No | NA | 1 | 0 | 1 | Low-quality | Genome-fragment | 6.07 | AAI-based (high-confidence) | 0 |
| S12-final-viral-combined_k141_225818\|\|full | 3298 | No | NA | 3 | 0 | 1 | Not-determined | Genome-fragment | NA | NA | 0 |
| S12-final-viral-combined_k141_376483\|\|full | 2259 | No | NA | 4 | 0 | 0 | Not-determined | Genome-fragment | NA | NA | 0 |
| S12-final-viral-combined_k141_124442\|\|full | 1954 | No | NA | 4 | 0 | 2 | Not-determined | Genome-fragment | NA | NA | 0 |
| S1_k141_355195 | 2817 | No | NA | 5 | 1 | 0 | Low-quality | Genome-fragment | 3.8 | HMM-based (lower-bound) | 0 |
| S12-final-viral-combined_k141_342471\|\|full | 3078 | No | NA | 4 | 0 | 3 | Not-determined | Genome-fragment | NA | NA | 0 |
| S12-final-viral-combined_k141_276295\|\|full | 24242 | No | NA | 12 | 0 | 7 | Not-determined | Genome-fragment | NA | NA | 0 |
| S5-final-viral-combined_k141_190237\|\|full | 2817 | No | NA | 5 | 0 | 0 | Low-quality | Genome-fragment | 3.14 | AAI-based (medium-confidence) | 0 |
| S12-final-viral-combined_k141_293354\|\|full | 40065 | Yes | 37843 | 58 | 13 | 2 | Medium-quality | Genome-fragment | 74.45 | AAI-based (high-confidence) | 5.55 |
| S12-final-viral-combined_k141_376497\|\|full | 9149 | No | NA | 11 | 0 | 2 | Not-determined | Genome-fragment | NA | NA | 0 |
| S17-final-viral-combined_k141_223560\|\|full | 2817 | No | NA | 3 | 0 | 1 | Low-quality | Genome-fragment | 1.42 | AAI-based (medium-confidence) | 0 |
| S12-final-viral-combined_k141_24956\|\|full | 1800 | No | NA | 3 | 0 | 2 | Not-determined | Genome-fragment | NA | NA | 0 |
| S12-final-viral-combined_k141_259057\|\|full | 3555 | No | NA | 3 | 0 | 1 | Not-determined | Genome-fragment | NA | NA | 0 |
| S17-final-viral-combined_k141_49179\|\|full | 2817 | Yes | 1134 | 3 | 1 | 1 | Low-quality | Genome-fragment | 22.13 | AAI-based (medium-confidence) | 59.74 |
| S12-final-viral-combined_k141_326390\|\|full | 1220 | No | NA | 2 | 0 | 1 | Not-determined | Genome-fragment | NA | NA | 0 |
| S12-final-viral-combined_k141_142256\|\|full | 1320 | No | NA | 4 | 0 | 0 | Not-determined | Genome-fragment | NA | NA | 0 |
| S1-final-viral-combined_k141_362745\|\|lt2gene | 2818 | No | NA | 1 | 1 | 0 | Low-quality | Genome-fragment | 5.84 | AAI-based (high-confidence) | 0 |
| S17-final-viral-combined_k141_169246\|\|full | 2818 | No | NA | 4 | 0 | 1 | Low-quality | Genome-fragment | 2.96 | AAI-based (high-confidence) | 0 |
| S17-final-viral-combined_k141_32272\|\|full | 2818 | No | NA | 3 | 0 | 0 | Low-quality | Genome-fragment | 0.24 | AAI-based (medium-confidence) | 0 |
| S12-final-viral-combined_k141_242557\|\|full | 1022 | No | NA | 2 | 0 | 0 | Not-determined | Genome-fragment | NA | NA | 0 |
| S17-final-viral-combined_k141_105417\|\|lt2gene | 2818 | No | NA | 1 | 1 | 0 | Low-quality | Genome-fragment | 4.67 | HMM-based (lower-bound) | 0 |
| S12-final-viral-combined_k141_342521\|\|full | 1259 | No | NA | 2 | 0 | 1 | Not-determined | Genome-fragment | NA | NA | 0 |
| S11-final-viral-combined_k141_129834\|\|full | 2819 | No | NA | 3 | 0 | 1 | Low-quality | Genome-fragment | 5.04 | AAI-based (high-confidence) | 0 |
| S12-final-viral-combined_k141_108149\|\|full | 2888 | No | NA | 3 | 0 | 3 | Not-determined | Genome-fragment | NA | NA | 0 |
| S12-final-viral-combined_k141_57958\|\|full | 3191 | No | NA | 3 | 0 | 3 | Not-determined | Genome-fragment | NA | NA | 0 |
| S12-final-viral-combined_k141_142273\|\|full | 1913 | No | NA | 2 | 0 | 0 | Not-determined | Genome-fragment | NA | NA | 0 |
| S12-final-viral-combined_k141_193271\|\|full | 1483 | No | NA | 3 | 0 | 1 | Not-determined | Genome-fragment | NA | NA | 0 |
| S12-final-viral-combined_k141_342557\|\|full | 1940 | No | NA | 2 | 0 | 1 | Not-determined | Genome-fragment | NA | NA | 0 |
| S12-final-viral-combined_k141_358560\|\|full | 2441 | No | NA | 3 | 0 | 0 | Not-determined | Genome-fragment | NA | NA | 0 |
| S11-final-viral-combined_k141_20658\|\|full | 2819 | No | NA | 3 | 1 | 0 | Low-quality | Genome-fragment | 6.79 | AAI-based (high-confidence) | 0 |
| S3-final-viral-combined_k141_43443\|\|full | 2820 | No | NA | 3 | 1 | 0 | Low-quality | Genome-fragment | 6.68 | AAI-based (high-confidence) | 0 |
| S2-final-viral-combined_k141_110475\|\|full | 2820 | No | NA | 6 | 0 | 0 | Low-quality | Genome-fragment | 5.43 | AAI-based (high-confidence) | 0 |
| S12-final-viral-combined_k141_276398\|\|full | 1941 | No | NA | 4 | 0 | 0 | Not-determined | Genome-fragment | NA | NA | 0 |
| S12-final-viral-combined_k141_242616\|\|full | 1045 | No | NA | 2 | 0 | 1 | Not-determined | Genome-fragment | NA | NA | 0 |
| S12-final-viral-combined_k141_310517\|\|full | 2707 | No | NA | 4 | 0 | 3 | Not-determined | Genome-fragment | NA | NA | 0 |
| S12-final-viral-combined_k141_208881\|\|full | 5803 | No | NA | 7 | 0 | 5 | Not-determined | Genome-fragment | NA | NA | 0 |
| S12-final-viral-combined_k141_342581\|\|full | 2099 | No | NA | 2 | 0 | 0 | Not-determined | Genome-fragment | NA | NA | 0 |
| S8_k141_102398 | 2820 | No | NA | 3 | 0 | 2 | Low-quality | Genome-fragment | 4.76 | AAI-based (high-confidence) | 0 |
| S11-final-viral-combined_k141_113904\|\|full | 2821 | No | NA | 7 | 1 | 0 | Low-quality | Genome-fragment | 4.31 | AAI-based (medium-confidence) | 0 |
| S5-final-viral-combined_k141_140037\|\|full | 2821 | No | NA | 6 | 2 | 0 | Low-quality | Genome-fragment | 6.82 | AAI-based (high-confidence) | 0 |
| S12-final-viral-combined_k141_242652\|\|full | 1437 | No | NA | 2 | 0 | 0 | Not-determined | Genome-fragment | NA | NA | 0 |
| S7-final-viral-combined_k141_199686\|\|full | 2821 | No | NA | 5 | 1 | 0 | Low-quality | Genome-fragment | 12.39 | AAI-based (high-confidence) | 0 |
| S12-final-viral-combined_k141_326505\|\|full | 3826 | No | NA | 2 | 0 | 2 | Not-determined | Genome-fragment | NA | NA | 0 |
| S12-final-viral-combined_k141_42015\|\|full | 2844 | No | NA | 5 | 0 | 4 | Not-determined | Genome-fragment | NA | NA | 0 |
| S17-final-viral-combined_k141_83744\|\|full | 2821 | No | NA | 4 | 0 | 2 | Low-quality | Genome-fragment | 4.4 | AAI-based (medium-confidence) | 0 |
| S11-final-viral-combined_k141_98900\|\|full | 2822 | No | NA | 6 | 2 | 0 | Low-quality | Genome-fragment | 5.83 | AAI-based (high-confidence) | 0 |
| S17-final-viral-combined_k141_141277\|\|full | 2823 | No | NA | 4 | 1 | 0 | Low-quality | Genome-fragment | 7.56 | AAI-based (high-confidence) | 0 |
| S17-final-viral-combined_k141_164152\|\|full | 2823 | No | NA | 4 | 0 | 3 | Low-quality | Genome-fragment | 2.36 | AAI-based (medium-confidence) | 0 |
| S12-final-viral-combined_k141_175446\|\|full | 1168 | No | NA | 3 | 0 | 1 | Not-determined | Genome-fragment | NA | NA | 0 |
| S9-final-viral-combined_k141_38477\|\|full | 2824 | No | NA | 3 | 0 | 3 | Low-quality | Genome-fragment | 1.41 | AAI-based (medium-confidence) | 0 |
| S9-final-viral-combined_k141_98957\|\|full | 2825 | No | NA | 5 | 3 | 0 | Low-quality | Genome-fragment | 4.31 | HMM-based (lower-bound) | 0 |
| S12-final-viral-combined_k141_42038\|\|full | 1334 | No | NA | 2 | 0 | 0 | Not-determined | Genome-fragment | NA | NA | 0 |
| S12-final-viral-combined_k141_326543\|\|full | 1121 | No | NA | 2 | 0 | 2 | Not-determined | Genome-fragment | NA | NA | 0 |
| S12-final-viral-combined_k141_124594\|\|full | 2035 | No | NA | 2 | 0 | 1 | Not-determined | Genome-fragment | NA | NA | 0 |
| S12-final-viral-combined_k141_91470\|\|full | 1295 | No | NA | 2 | 0 | 0 | Not-determined | Genome-fragment | NA | NA | 0 |
| S11-final-viral-combined_k141_193193\|\|full | 2825 | No | NA | 5 | 0 | 0 | Low-quality | Genome-fragment | 1.84 | AAI-based (high-confidence) | 0 |
| S12-final-viral-combined_k141_226056\|\|full | 3970 | No | NA | 4 | 0 | 3 | Not-determined | Genome-fragment | NA | NA | 0 |
| S12-final-viral-combined_k141_326559\|\|full | 1537 | No | NA | 3 | 0 | 1 | Not-determined | Genome-fragment | NA | NA | 0 |
| S12-final-viral-combined_k141_259192\|\|full | 1712 | No | NA | 3 | 0 | 1 | Not-determined | Genome-fragment | NA | NA | 0 |
| S11-final-viral-combined_k141_185932\|\|full | 2825 | No | NA | 3 | 3 | 0 | Low-quality | Genome-fragment | 4.65 | AAI-based (high-confidence) | 0 |
| S17-final-viral-combined_k141_110615\|\|full | 2825 | No | NA | 6 | 2 | 0 | Low-quality | Genome-fragment | 2.75 | HMM-based (lower-bound) | 0 |
| S1-final-viral-combined_k141_231184\|\|full | 2826 | No | NA | 6 | 2 | 0 | Low-quality | Genome-fragment | 3.52 | AAI-based (high-confidence) | 0 |
| S12-final-viral-combined_k141_58097\|\|full | 1150 | No | NA | 2 | 0 | 1 | Not-determined | Genome-fragment | NA | NA | 0 |
| S1-final-viral-combined_k141_366652\|\|full | 2826 | No | NA | 1 | 0 | 1 | Low-quality | Genome-fragment | 1.44 | AAI-based (medium-confidence) | 0 |
| S12-final-viral-combined_k141_209030\|\|full | 1003 | No | NA | 2 | 0 | 1 | Not-determined | Genome-fragment | NA | NA | 0 |
| S12-final-viral-combined_k141_342710\|\|full | 1198 | No | NA | 2 | 0 | 2 | Not-determined | Genome-fragment | NA | NA | 0 |
| S12-final-viral-combined_k141_226129\|\|full | 1856 | No | NA | 2 | 0 | 2 | Not-determined | Genome-fragment | NA | NA | 0 |
| S12-final-viral-combined_k141_197442\|\|full | 2826 | No | NA | 4 | 0 | 1 | Low-quality | Genome-fragment | 0.68 | AAI-based (medium-confidence) | 0 |
| S1-final-viral-combined_k141_209580\|\|full | 2827 | No | NA | 7 | 3 | 0 | Low-quality | Genome-fragment | 4.7 | AAI-based (high-confidence) | 0 |
| S5-final-viral-combined_k141_50861\|\|full | 2827 | No | NA | 3 | 1 | 0 | Low-quality | Genome-fragment | 2.44 | HMM-based (lower-bound) | 0 |
| S12-final-viral-combined_k141_293637\|\|full | 3885 | No | NA | 3 | 0 | 2 | Not-determined | Genome-fragment | NA | NA | 0 |
| S12-final-viral-combined_k141_193460\|\|full | 1270 | No | NA | 2 | 0 | 1 | Not-determined | Genome-fragment | NA | NA | 0 |
| S5-final-viral-combined_k141_193491\|\|full | 2827 | No | NA | 4 | 2 | 1 | Low-quality | Genome-fragment | 6.01 | AAI-based (high-confidence) | 0 |
| S7_k141_216895 | 2827 | No | NA | 5 | 3 | 0 | Low-quality | Genome-fragment | 6.32 | AAI-based (high-confidence) | 0 |
| S1-final-viral-combined_k141_66882\|\|full | 2828 | No | NA | 3 | 0 | 1 | Low-quality | Genome-fragment | 5.17 | AAI-based (medium-confidence) | 0 |
| S17-final-viral-combined_k141_66376\|\|lt2gene | 2828 | No | NA | 1 | 1 | 0 | Low-quality | Genome-fragment | 5.46 | AAI-based (medium-confidence) | 0 |
| S12-final-viral-combined_k141_253439\|\|full | 2828 | No | NA | 3 | 0 | 1 | Low-quality | Genome-fragment | 2.97 | AAI-based (medium-confidence) | 0 |
| S12-final-viral-combined_k141_54656\|\|full | 2828 | No | NA | 3 | 0 | 3 | Low-quality | Genome-fragment | 8.5 | AAI-based (medium-confidence) | 0 |
| S11_k141_178652 | 2829 | No | NA | 4 | 2 | 0 | Low-quality | Genome-fragment | 10.3 | HMM-based (lower-bound) | 0 |
| S12-final-viral-combined_k141_293682\|\|full | 1123 | No | NA | 2 | 0 | 1 | Not-determined | Genome-fragment | NA | NA | 0 |
| S12-final-viral-combined_k141_142570\|\|full | 2171 | No | NA | 3 | 0 | 1 | Not-determined | Genome-fragment | NA | NA | 0 |
| S17-final-viral-combined_k141_146168\|\|full | 2829 | No | NA | 4 | 2 | 0 | Low-quality | Genome-fragment | 7.05 | AAI-based (high-confidence) | 0 |
| S8-final-viral-combined_k141_129280\|\|full | 2829 | No | NA | 3 | 0 | 0 | Low-quality | Genome-fragment | 5.66 | AAI-based (medium-confidence) | 0 |
| S8-final-viral-combined_k141_272158\|\|full | 2829 | No | NA | 6 | 1 | 0 | Low-quality | Genome-fragment | 4.71 | AAI-based (medium-confidence) | 0 |
| S11-final-viral-combined_k141_284601\|\|full | 2830 | No | NA | 3 | 0 | 2 | Low-quality | Genome-fragment | 0.43 | AAI-based (high-confidence) | 0 |
| S12-final-viral-combined_k141_276582\|\|full | 12185 | No | NA | 5 | 0 | 1 | Not-determined | Genome-fragment | NA | NA | 0 |
| S12-final-viral-combined_k141_108375\|\|full | 2955 | No | NA | 3 | 0 | 2 | Not-determined | Genome-fragment | NA | NA | 0 |
| S1-final-viral-combined_k141_217082\|\|full | 2830 | No | NA | 2 | 2 | 0 | Low-quality | Genome-fragment | 4.65 | AAI-based (high-confidence) | 0 |
| S12-final-viral-combined_k141_75733\|\|full | 5432 | No | NA | 7 | 1 | 0 | High-quality | High-quality | 98.25 | AAI-based (high-confidence) | 0 |
| S12-final-viral-combined_k141_326765\|\|full | 3879 | No | NA | 3 | 0 | 1 | Not-determined | Genome-fragment | NA | NA | 0 |
| S4_k141_23138 | 2830 | No | NA | 9 | 0 | 0 | Low-quality | Genome-fragment | 17.09 | AAI-based (high-confidence) | 0 |
| S12-final-viral-combined_k141_276607\|\|full | 1442 | No | NA | 3 | 0 | 1 | Not-determined | Genome-fragment | NA | NA | 0 |
| S12-final-viral-combined_k141_124792\|\|full | 1749 | No | NA | 3 | 0 | 1 | Not-determined | Genome-fragment | NA | NA | 0 |
| S12-final-viral-combined_k141_326784\|\|full | 2508 | No | NA | 4 | 0 | 0 | Not-determined | Genome-fragment | NA | NA | 0 |
| S12-final-viral-combined_k141_175610\|\|full | 5266 | No | NA | 6 | 0 | 2 | Not-determined | Genome-fragment | NA | NA | 0 |
| S12-final-viral-combined_k141_58247\|\|full | 6679 | No | NA | 5 | 0 | 3 | Not-determined | Genome-fragment | NA | NA | 0 |
| S6-final-viral-combined_k141_74882\|\|full | 2830 | No | NA | 3 | 2 | 0 | Low-quality | Genome-fragment | 10.3 | HMM-based (lower-bound) | 0 |
| S12-final-viral-combined_k141_276628\|\|full | 2928 | No | NA | 6 | 0 | 1 | Not-determined | Genome-fragment | NA | NA | 0 |
| S12-final-viral-combined_k141_159339\|\|full | 2388 | No | NA | 4 | 0 | 0 | Not-determined | Genome-fragment | NA | NA | 0 |
| S8-final-viral-combined_k141_218666\|\|full | 2830 | No | NA | 5 | 0 | 2 | Low-quality | Genome-fragment | 3.63 | AAI-based (medium-confidence) | 0 |
| S12-final-viral-combined_k141_259320\|\|full | 1786 | No | NA | 2 | 0 | 0 | Not-determined | Genome-fragment | NA | NA | 0 |
| S8_k141_26665 | 2831 | No | NA | 2 | 1 | 0 | Low-quality | Genome-fragment | 2.39 | AAI-based (high-confidence) | 0 |
| S12-final-viral-combined_k141_393703\|\|full | 11842 | No | NA | 9 | 1 | 2 | High-quality | High-quality | 100 | HMM-based (lower-bound) | 0 |
| S12-final-viral-combined_k141_226239\|\|full | 1433 | No | NA | 2 | 0 | 1 | Not-determined | Genome-fragment | NA | NA | 0 |
| S12-final-viral-combined_k141_259346\|\|full | 1359 | No | NA | 4 | 0 | 0 | Not-determined | Genome-fragment | NA | NA | 0 |
| S11-final-viral-combined_k141_160703\|\|full | 2832 | No | NA | 3 | 0 | 0 | Low-quality | Genome-fragment | 6.37 | AAI-based (high-confidence) | 0 |
| S1_k141_94197 | 2832 | No | NA | 4 | 1 | 0 | Low-quality | Genome-fragment | 6 | AAI-based (high-confidence) | 0 |
| S12-final-viral-combined_k141_242798\|\|full | 3762 | No | NA | 5 | 0 | 2 | Not-determined | Genome-fragment | NA | NA | 0 |
| S12-final-viral-combined_k141_259368\|\|full | 1506 | No | NA | 2 | 0 | 1 | Not-determined | Genome-fragment | NA | NA | 0 |
| S12-final-viral-combined_k141_108486\|\|full | 5617 | No | NA | 6 | 0 | 3 | Not-determined | Genome-fragment | NA | NA | 0 |
| S12-final-viral-combined_k141_58319\|\|full | 3732 | No | NA | 3 | 0 | 0 | Not-determined | Genome-fragment | NA | NA | 0 |
| S12-final-viral-combined_k141_376807\|\|full | 1264 | No | NA | 2 | 0 | 1 | Not-determined | Genome-fragment | NA | NA | 0 |
| S7-final-viral-combined_k141_40333\|\|full | 2832 | Yes | 1782 | 5 | 1 | 1 | Low-quality | Genome-fragment | 1.26 | AAI-based (high-confidence) | 37.08 |
| S12-final-viral-combined_k141_358797\|\|full | 41816 | Yes | 27363 | 42 | 9 | 14 | Medium-quality | Genome-fragment | 56.81 | AAI-based (medium-confidence) | 34.56 |
| S12-final-viral-combined_k141_91698\|\|full | 4318 | No | NA | 4 | 0 | 2 | Not-determined | Genome-fragment | NA | NA | 0 |
| S4_k141_27668 | 2832 | No | NA | 6 | 0 | 0 | Low-quality | Genome-fragment | 5.09 | AAI-based (medium-confidence) | 0 |
| S12-final-viral-combined_k141_208921\|\|full | 2833 | No | NA | 3 | 0 | 1 | Low-quality | Genome-fragment | 3.5 | AAI-based (high-confidence) | 0 |
| S12-final-viral-combined_k141_42376\|\|full | 2466 | No | NA | 3 | 0 | 3 | Not-determined | Genome-fragment | NA | NA | 0 |
| S12-final-viral-combined_k141_226356\|\|full | 3408 | No | NA | 3 | 0 | 2 | Not-determined | Genome-fragment | NA | NA | 0 |
| S11-final-viral-combined_k141_107884\|\|full | 2834 | No | NA | 3 | 0 | 2 | Low-quality | Genome-fragment | 1.01 | AAI-based (high-confidence) | 0 |
| S12-final-viral-combined_k141_242912\|\|full | 3295 | No | NA | 2 | 0 | 0 | Not-determined | Genome-fragment | NA | NA | 0 |
| S7-final-viral-combined_k141_199801\|\|full | 2834 | No | NA | 3 | 0 | 1 | Low-quality | Genome-fragment | 5.23 | AAI-based (high-confidence) | 0 |
| S12-final-viral-combined_k141_124966\|\|full | 3535 | No | NA | 5 | 0 | 1 | Not-determined | Genome-fragment | NA | NA | 0 |
| S2-final-viral-combined_k141_86823\|\|full | 2834 | No | NA | 2 | 0 | 1 | Low-quality | Genome-fragment | 0.94 | AAI-based (medium-confidence) | 0 |
| S12-final-viral-combined_k141_276734\|\|full | 1267 | No | NA | 2 | 0 | 1 | Not-determined | Genome-fragment | NA | NA | 0 |
| S12-final-viral-combined_k141_242952\|\|full | 1008 | No | NA | 3 | 0 | 0 | Not-determined | Genome-fragment | NA | NA | 0 |
| S8_k141_132391 | 2834 | No | NA | 5 | 3 | 0 | Low-quality | Genome-fragment | 4.59 | AAI-based (high-confidence) | 0 |
| S11-final-viral-combined_k141_263184\|\|full | 2835 | No | NA | 3 | 2 | 0 | Low-quality | Genome-fragment | 6.2 | AAI-based (high-confidence) | 0 |
| S12-final-viral-combined_k141_376865\|\|full | 1724 | No | NA | 2 | 0 | 2 | Not-determined | Genome-fragment | NA | NA | 0 |
| S12-final-viral-combined_k141_293892\|\|full | 2036 | No | NA | 4 | 0 | 3 | Not-determined | Genome-fragment | NA | NA | 0 |
| S11-final-viral-combined_k141_209862\|\|full | 2835 | No | NA | 4 | 0 | 2 | Low-quality | Genome-fragment | 2.61 | AAI-based (medium-confidence) | 0 |
| S2-final-viral-combined_k141_197947\|\|full | 2835 | No | NA | 4 | 0 | 0 | Low-quality | Genome-fragment | 5.14 | AAI-based (medium-confidence) | 0 |
| S12-final-viral-combined_k141_125009\|\|full | 2047 | No | NA | 3 | 0 | 0 | Not-determined | Genome-fragment | NA | NA | 0 |
| S2-final-viral-combined_k141_182771\|\|full | 2835 | No | NA | 6 | 0 | 0 | Low-quality | Genome-fragment | 5.53 | AAI-based (medium-confidence) | 0 |
| S8-final-viral-combined_k141_272718\|\|full | 2835 | No | NA | 3 | 1 | 0 | Low-quality | Genome-fragment | 5.34 | AAI-based (high-confidence) | 0 |
| S3-final-viral-combined_k141_20137\|\|full | 2836 | No | NA | 4 | 0 | 0 | Low-quality | Genome-fragment | 4.85 | AAI-based (high-confidence) | 0 |
| S12-final-viral-combined_k141_293929\|\|full | 4052 | No | NA | 3 | 0 | 2 | Not-determined | Genome-fragment | NA | NA | 0 |
| S7-final-viral-combined_k141_82909\|\|full | 2836 | No | NA | 3 | 0 | 0 | Low-quality | Genome-fragment | 5.23 | AAI-based (medium-confidence) | 0 |
| S12-final-viral-combined_k141_209293\|\|full | 1341 | No | NA | 3 | 0 | 0 | Not-determined | Genome-fragment | NA | NA | 0 |
| S12-final-viral-combined_k141_58436\|\|full | 1419 | No | NA | 4 | 0 | 0 | Not-determined | Genome-fragment | NA | NA | 0 |
| S1-final-viral-combined_k141_40941\|\|full | 2837 | No | NA | 4 | 1 | 0 | Low-quality | Genome-fragment | 6.55 | AAI-based (medium-confidence) | 0 |
| S12-final-viral-combined_k141_358941\|\|full | 1201 | No | NA | 3 | 0 | 1 | Not-determined | Genome-fragment | NA | NA | 0 |
| S12-final-viral-combined_k141_159568\|\|full | 1564 | No | NA | 2 | 0 | 1 | Not-determined | Genome-fragment | NA | NA | 0 |
| S4_k141_6399 | 2837 | No | NA | 3 | 0 | 2 | Low-quality | Genome-fragment | 0.75 | AAI-based (high-confidence) | 0 |
| S12-final-viral-combined_k141_108651\|\|full | 1501 | No | NA | 2 | 0 | 0 | Not-determined | Genome-fragment | NA | NA | 0 |
| S12-final-viral-combined_k141_91840\|\|full | 1472 | No | NA | 3 | 0 | 1 | Not-determined | Genome-fragment | NA | NA | 0 |
| S12-final-viral-combined_k141_25459\|\|full | 47975 | No | NA | 36 | 2 | 20 | High-quality | High-quality | 100 | HMM-based (lower-bound) | 0 |
| S5-final-viral-combined_k141_19109\|\|full | 2838 | No | NA | 4 | 3 | 0 | Low-quality | Genome-fragment | 6.33 | AAI-based (high-confidence) | 0 |
| S9-final-viral-combined_k141_12584\|\|full | 2839 | No | NA | 3 | 0 | 0 | Low-quality | Genome-fragment | 5.57 | AAI-based (medium-confidence) | 0 |
| S12-final-viral-combined_k141_76064\|\|full | 1497 | No | NA | 2 | 0 | 0 | Not-determined | Genome-fragment | NA | NA | 0 |
| S12-final-viral-combined_k141_293992\|\|full | 1236 | No | NA | 2 | 0 | 0 | Not-determined | Genome-fragment | NA | NA | 0 |
| S12-final-viral-combined_k141_58516\|\|full | 3004 | No | NA | 4 | 0 | 0 | Not-determined | Genome-fragment | NA | NA | 0 |
| S12-final-viral-combined_k141_209386\|\|full | 3613 | No | NA | 7 | 0 | 1 | Not-determined | Genome-fragment | NA | NA | 0 |
| S12-final-viral-combined_k141_294022\|\|full | 3826 | No | NA | 3 | 0 | 2 | Not-determined | Genome-fragment | NA | NA | 0 |
| S12-final-viral-combined_k141_393973\|\|full | 4855 | No | NA | 4 | 0 | 4 | Not-determined | Genome-fragment | NA | NA | 0 |
| S12-final-viral-combined_k141_193690\|\|full | 5948 | No | NA | 5 | 0 | 0 | Not-determined | Genome-fragment | NA | NA | 0 |
| S12-final-viral-combined_k141_359037\|\|full | 1544 | No | NA | 2 | 0 | 1 | Not-determined | Genome-fragment | NA | NA | 0 |
| S11-final-viral-combined_k141_273453\|\|full | 2839 | No | NA | 5 | 0 | 1 | Low-quality | Genome-fragment | 2.36 | AAI-based (medium-confidence) | 0 |
| S12-final-viral-combined_k141_142957\|\|full | 1911 | No | NA | 3 | 0 | 2 | Not-determined | Genome-fragment | NA | NA | 0 |
| S17-k141_15555 | 2839 | No | NA | 2 | 0 | 0 | Low-quality | Genome-fragment | 6.02 | AAI-based (medium-confidence) | 0 |
| S12-final-viral-combined_k141_226633\|\|full | 1239 | No | NA | 2 | 0 | 1 | Not-determined | Genome-fragment | NA | NA | 0 |
| S12_k141_53455 | 2839 | No | NA | 7 | 0 | 0 | Low-quality | Genome-fragment | 4.49 | AAI-based (medium-confidence) | 0 |
| S4-final-viral-combined_k141_21590\|\|full | 2839 | No | NA | 3 | 0 | 1 | Low-quality | Genome-fragment | 0.9 | AAI-based (high-confidence) | 0 |
| S12-final-viral-combined_k141_175940\|\|full | 5763 | No | NA | 6 | 0 | 4 | Not-determined | Genome-fragment | NA | NA | 0 |
| S1-final-viral-combined_k141_180273\|\|full | 2840 | No | NA | 4 | 0 | 2 | Low-quality | Genome-fragment | 1.54 | AAI-based (medium-confidence) | 0 |
| S3-final-viral-combined_k141_123415\|\|full | 2840 | No | NA | 3 | 0 | 0 | Low-quality | Genome-fragment | 3.35 | AAI-based (medium-confidence) | 0 |
| S17-final-viral-combined_k141_73251\|\|full | 2840 | No | NA | 5 | 0 | 0 | Low-quality | Genome-fragment | 5.63 | AAI-based (medium-confidence) | 0 |
| S12-final-viral-combined_k141_343081\|\|full | 1396 | No | NA | 3 | 0 | 0 | Not-determined | Genome-fragment | NA | NA | 0 |
| S12-final-viral-combined_k141_125230\|\|full | 4943 | No | NA | 6 | 0 | 4 | Not-determined | Genome-fragment | NA | NA | 0 |
| S6-final-viral-combined_k141_100160\|\|full | 2840 | No | NA | 8 | 0 | 0 | Low-quality | Genome-fragment | 8.03 | AAI-based (medium-confidence) | 0 |
| S12-final-viral-combined_k141_294056\|\|full | 1552 | No | NA | 3 | 0 | 0 | Not-determined | Genome-fragment | NA | NA | 0 |
| S11_k141_299650 | 2841 | No | NA | 6 | 0 | 0 | Low-quality | Genome-fragment | 4.76 | AAI-based (high-confidence) | 0 |
| S12-final-viral-combined_k141_159627\|\|full | 1958 | No | NA | 3 | 0 | 1 | Not-determined | Genome-fragment | NA | NA | 0 |
| S12-final-viral-combined_k141_25573\|\|full | 3344 | No | NA | 3 | 0 | 1 | Not-determined | Genome-fragment | NA | NA | 0 |
| S12-final-viral-combined_k141_176000\|\|full | 1020 | No | NA | 2 | 0 | 2 | Not-determined | Genome-fragment | NA | NA | 0 |
| S12-final-viral-combined_k141_209493\|\|full | 1200 | No | NA | 2 | 0 | 1 | Not-determined | Genome-fragment | NA | NA | 0 |
| S12-final-viral-combined_k141_42589\|\|full | 3562 | No | NA | 5 | 0 | 1 | Not-determined | Genome-fragment | NA | NA | 0 |
| S1-final-viral-combined_k141_53368\|\|full | 2841 | No | NA | 6 | 0 | 0 | Low-quality | Genome-fragment | 7.28 | AAI-based (high-confidence) | 0 |
| S12-final-viral-combined_k141_108839\|\|full | 2297 | No | NA | 4 | 0 | 1 | Not-determined | Genome-fragment | NA | NA | 0 |
| S7-final-viral-combined_k141_51742\|\|full | 2841 | No | NA | 4 | 0 | 4 | Low-quality | Genome-fragment | 0.8 | AAI-based (medium-confidence) | 0 |
| S12-final-viral-combined_k141_209515\|\|full | 1515 | No | NA | 3 | 0 | 1 | Not-determined | Genome-fragment | NA | NA | 0 |
| S7-final-viral-combined_k141_217797\|\|full | 2841 | No | NA | 3 | 0 | 3 | Low-quality | Genome-fragment | 5.16 | AAI-based (medium-confidence) | 0 |
| S12-final-viral-combined_k141_76219\|\|full | 1422 | No | NA | 3 | 0 | 0 | Not-determined | Genome-fragment | NA | NA | 0 |
| S6-final-viral-combined_k141_12575\|\|full | 2841 | No | NA | 5 | 0 | 4 | Low-quality | Genome-fragment | 2.22 | AAI-based (medium-confidence) | 0 |
| S12-final-viral-combined_k141_343179\|\|full | 1493 | No | NA | 3 | 0 | 1 | Not-determined | Genome-fragment | NA | NA | 0 |
| S12-final-viral-combined_k141_343188\|\|full | 2938 | No | NA | 3 | 0 | 0 | Not-determined | Genome-fragment | NA | NA | 0 |
| S12-final-viral-combined_k141_143074\|\|full | 2028 | No | NA | 3 | 0 | 0 | Not-determined | Genome-fragment | NA | NA | 0 |
| S12-final-viral-combined_k141_343196\|\|full | 1201 | No | NA | 3 | 0 | 1 | Not-determined | Genome-fragment | NA | NA | 0 |
| S1-final-viral-combined_k141_200140\|\|full | 2842 | No | NA | 3 | 0 | 2 | Low-quality | Genome-fragment | 0.85 | AAI-based (medium-confidence) | 0 |
| S3-final-viral-combined_k141_1339\|\|full | 2842 | No | NA | 3 | 0 | 3 | Low-quality | Genome-fragment | 1.39 | AAI-based (medium-confidence) | 0 |
| S12-final-viral-combined_k141_310274\|\|full | 2842 | No | NA | 4 | 3 | 0 | Low-quality | Genome-fragment | 4.4 | AAI-based (high-confidence) | 0 |
| S12-final-viral-combined_k141_394038\|\|full | 2004 | No | NA | 2 | 0 | 2 | Not-determined | Genome-fragment | NA | NA | 0 |
| S2-final-viral-combined_k141_189775\|\|full | 2842 | No | NA | 3 | 0 | 0 | Low-quality | Genome-fragment | 6.43 | AAI-based (medium-confidence) | 0 |
| S2-final-viral-combined_k141_231626\|\|full | 2842 | No | NA | 3 | 1 | 0 | Low-quality | Genome-fragment | 5.44 | AAI-based (medium-confidence) | 0 |
| S2_k141_151826 | 2843 | No | NA | 3 | 0 | 1 | Low-quality | Genome-fragment | 2.78 | AAI-based (high-confidence) | 0 |
| S12-final-viral-combined_k141_193910\|\|full | 1916 | No | NA | 2 | 0 | 0 | Not-determined | Genome-fragment | NA | NA | 0 |
| S12-final-viral-combined_k141_226810\|\|full | 1210 | No | NA | 2 | 0 | 0 | Not-determined | Genome-fragment | NA | NA | 0 |
| S4-final-viral-combined_k141_156476\|\|full | 2843 | No | NA | 3 | 3 | 0 | Low-quality | Genome-fragment | 7.62 | AAI-based (high-confidence) | 0 |
| S12-final-viral-combined_k141_343240\|\|full | 2063 | No | NA | 3 | 0 | 2 | Not-determined | Genome-fragment | NA | NA | 0 |
| S1-final-viral-combined_k141_242105\|\|full | 2844 | No | NA | 6 | 0 | 0 | Low-quality | Genome-fragment | 6.42 | AAI-based (high-confidence) | 0 |
| S12-final-viral-combined_k141_377116\|\|full | 1855 | No | NA | 3 | 0 | 2 | Not-determined | Genome-fragment | NA | NA | 0 |
| S12-final-viral-combined_k141_96148\|\|full | 2844 | No | NA | 2 | 0 | 1 | Low-quality | Genome-fragment | 0.84 | AAI-based (medium-confidence) | 0 |
| S12-final-viral-combined_k141_176070\|\|full | 3831 | No | NA | 4 | 0 | 2 | Not-determined | Genome-fragment | NA | NA | 0 |
| S12-final-viral-combined_k141_226847\|\|full | 3773 | No | NA | 3 | 0 | 2 | Not-determined | Genome-fragment | NA | NA | 0 |
| S12-final-viral-combined_k141_394111\|\|full | 7714 | No | NA | 3 | 0 | 3 | Not-determined | Genome-fragment | NA | NA | 0 |
| S12-final-viral-combined_k141_400782\|\|full | 2844 | No | NA | 2 | 0 | 0 | Low-quality | Genome-fragment | 7.64 | AAI-based (high-confidence) | 0 |
| S12-final-viral-combined_k141_58715\|\|full | 1692 | No | NA | 3 | 0 | 1 | Not-determined | Genome-fragment | NA | NA | 0 |
| S12-final-viral-combined_k141_8742\|\|full | 1065 | No | NA | 3 | 0 | 1 | Not-determined | Genome-fragment | NA | NA | 0 |
| S12-final-viral-combined_k141_359111\|\|full | 5840 | No | NA | 6 | 0 | 4 | Not-determined | Genome-fragment | NA | NA | 0 |
| S12-final-viral-combined_k141_209717\|\|full | 2749 | No | NA | 2 | 0 | 1 | Not-determined | Genome-fragment | NA | NA | 0 |
| S12-final-viral-combined_k141_8762\|\|full | 3610 | No | NA | 3 | 0 | 2 | Not-determined | Genome-fragment | NA | NA | 0 |
| S17-final-viral-combined_k141_138120\|\|full | 2845 | No | NA | 2 | 0 | 1 | Low-quality | Genome-fragment | 1.96 | AAI-based (medium-confidence) | 0 |
| S12-final-viral-combined_k141_42716\|\|full | 2056 | No | NA | 3 | 0 | 2 | Not-determined | Genome-fragment | NA | NA | 0 |
| S12-final-viral-combined_k141_294248\|\|full | 1475 | No | NA | 3 | 0 | 0 | Not-determined | Genome-fragment | NA | NA | 0 |
| S17-final-viral-combined_k141_164026\|\|full | 2845 | No | NA | 2 | 0 | 1 | Low-quality | Genome-fragment | 3.29 | AAI-based (high-confidence) | 0 |
| S12-final-viral-combined_k141_76440\|\|full | 2038 | No | NA | 2 | 0 | 2 | Not-determined | Genome-fragment | NA | NA | 0 |
| S1-final-viral-combined_k141_724\|\|full | 2846 | No | NA | 4 | 0 | 2 | Low-quality | Genome-fragment | 2.18 | AAI-based (high-confidence) | 0 |
| S12-final-viral-combined_k141_76444\|\|full | 4586 | No | NA | 4 | 0 | 4 | Not-determined | Genome-fragment | NA | NA | 0 |
| S12-final-viral-combined_k141_92094\|\|full | 10072 | No | NA | 11 | 0 | 9 | Not-determined | Genome-fragment | NA | NA | 0 |
| S12-final-viral-combined_k141_194098\|\|full | 1112 | No | NA | 3 | 0 | 0 | Not-determined | Genome-fragment | NA | NA | 0 |
| S12-final-viral-combined_k141_311290\|\|full | 1661 | No | NA | 2 | 0 | 1 | Not-determined | Genome-fragment | NA | NA | 0 |
| S1-final-viral-combined_k141_262573\|\|full | 2846 | No | NA | 6 | 1 | 0 | Low-quality | Genome-fragment | 4.84 | AAI-based (high-confidence) | 0 |
| S12-final-viral-combined_k141_125327\|\|full | 10730 | No | NA | 10 | 0 | 2 | Not-determined | Genome-fragment | NA | NA | 0 |
| S1-final-viral-combined_k141_229473\|\|lt2gene | 2846 | No | NA | 2 | 1 | 0 | Low-quality | Genome-fragment | 4.16 | HMM-based (lower-bound) | 0 |
| S12-final-viral-combined_k141_113192\|\|full | 2846 | No | NA | 5 | 1 | 0 | Low-quality | Genome-fragment | 1.54 | HMM-based (lower-bound) | 0 |
| S12-final-viral-combined_k141_377256\|\|full | 3856 | No | NA | 3 | 0 | 2 | Not-determined | Genome-fragment | NA | NA | 0 |
| S12-final-viral-combined_k141_226902\|\|full | 1415 | No | NA | 2 | 0 | 1 | Not-determined | Genome-fragment | NA | NA | 0 |
| S4-final-viral-combined_k141_165978\|\|full | 2846 | No | NA | 4 | 1 | 0 | Low-quality | Genome-fragment | 3.19 | HMM-based (lower-bound) | 0 |
| S11-final-viral-combined_k141_274935\|\|full | 2848 | No | NA | 4 | 0 | 0 | Low-quality | Genome-fragment | 7.86 | AAI-based (medium-confidence) | 0 |
| S12-final-viral-combined_k141_185034\|\|full | 2848 | No | NA | 5 | 3 | 0 | Low-quality | Genome-fragment | 5.46 | AAI-based (medium-confidence) | 0 |
| S2_k141_62366 | 2848 | No | NA | 4 | 1 | 0 | Low-quality | Genome-fragment | 4.65 | AAI-based (high-confidence) | 0 |
| S12-final-viral-combined_k141_311328\|\|full | 1205 | No | NA | 2 | 0 | 0 | Not-determined | Genome-fragment | NA | NA | 0 |
| S12-final-viral-combined_k141_277105\|\|full | 2729 | No | NA | 4 | 0 | 0 | Not-determined | Genome-fragment | NA | NA | 0 |
| S12-final-viral-combined_k141_277106\|\|full | 1072 | No | NA | 3 | 0 | 0 | Not-determined | Genome-fragment | NA | NA | 0 |
| S12-final-viral-combined_k141_394234\|\|full | 2983 | No | NA | 4 | 0 | 0 | Not-determined | Genome-fragment | NA | NA | 0 |
| S12-final-viral-combined_k141_243365\|\|full | 2268 | No | NA | 3 | 0 | 0 | Not-determined | Genome-fragment | NA | NA | 0 |
| S12-final-viral-combined_k141_259713\|\|full | 3090 | No | NA | 7 | 0 | 0 | Not-determined | Genome-fragment | NA | NA | 0 |
| S1-final-viral-combined_k141_304481\|\|full | 2849 | No | NA | 8 | 2 | 0 | Low-quality | Genome-fragment | 7.15 | AAI-based (high-confidence) | 0 |
| S1-final-viral-combined_k141_243092\|\|full | 2849 | No | NA | 2 | 0 | 2 | Low-quality | Genome-fragment | 2.51 | AAI-based (medium-confidence) | 0 |
| S12-final-viral-combined_k141_109126\|\|full | 2752 | No | NA | 5 | 0 | 2 | Not-determined | Genome-fragment | NA | NA | 0 |
| S12-final-viral-combined_k141_159899\|\|full | 4469 | No | NA | 3 | 0 | 0 | Not-determined | Genome-fragment | NA | NA | 0 |
| S12-final-viral-combined_k141_327518\|\|full | 1534 | No | NA | 4 | 0 | 0 | Not-determined | Genome-fragment | NA | NA | 0 |
| S12-final-viral-combined_k141_42852\|\|full | 1212 | No | NA | 2 | 0 | 0 | Not-determined | Genome-fragment | NA | NA | 0 |
| S12-final-viral-combined_k141_109130\|\|full | 5345 | No | NA | 6 | 0 | 0 | Not-determined | Genome-fragment | NA | NA | 0 |
| S7-final-viral-combined_k141_91921\|\|full | 2849 | No | NA | 3 | 0 | 0 | Low-quality | Genome-fragment | 2.15 | AAI-based (high-confidence) | 0 |
| S12-final-viral-combined_k141_8887\|\|full | 5957 | No | NA | 2 | 0 | 2 | Not-determined | Genome-fragment | NA | NA | 0 |
| S17-final-viral-combined_k141_12359\|\|full | 2849 | No | NA | 5 | 2 | 0 | Low-quality | Genome-fragment | 7.44 | AAI-based (high-confidence) | 0 |
| S12-final-viral-combined_k141_226964\|\|full | 1562 | No | NA | 2 | 0 | 1 | Not-determined | Genome-fragment | NA | NA | 0 |
| S12-final-viral-combined_k141_324296\|\|full | 2849 | No | NA | 5 | 1 | 0 | Low-quality | Genome-fragment | 7.73 | AAI-based (high-confidence) | 0 |
| S12-final-viral-combined_k141_378601\|\|full | 2849 | No | NA | 4 | 0 | 0 | Low-quality | Genome-fragment | 3.98 | AAI-based (medium-confidence) | 0 |
| S12-final-viral-combined_k141_327554\|\|full | 2285 | No | NA | 3 | 0 | 0 | Not-determined | Genome-fragment | NA | NA | 0 |
| S1-final-viral-combined_k141_142613\|\|full | 2851 | No | NA | 3 | 1 | 0 | Low-quality | Genome-fragment | 6.86 | AAI-based (high-confidence) | 0 |
| S1-final-viral-combined_k141_365739\|\|full | 2852 | No | NA | 6 | 1 | 0 | Low-quality | Genome-fragment | 1.73 | HMM-based (lower-bound) | 0 |
| S12-final-viral-combined_k141_42869\|\|full | 2389 | No | NA | 2 | 0 | 1 | Not-determined | Genome-fragment | NA | NA | 0 |
| S12-final-viral-combined_k141_58867\|\|full | 3118 | No | NA | 4 | 0 | 3 | Not-determined | Genome-fragment | NA | NA | 0 |
| S12-final-viral-combined_k141_311386\|\|full | 1428 | No | NA | 2 | 0 | 2 | Not-determined | Genome-fragment | NA | NA | 0 |
| S12-final-viral-combined_k141_42878\|\|full | 6728 | No | NA | 10 | 0 | 3 | Not-determined | Genome-fragment | NA | NA | 0 |
| S12-final-viral-combined_k141_377339\|\|full | 2628 | No | NA | 3 | 0 | 1 | Not-determined | Genome-fragment | NA | NA | 0 |
| S12-final-viral-combined_k141_227015\|\|full | 1042 | No | NA | 2 | 0 | 1 | Not-determined | Genome-fragment | NA | NA | 0 |
| S1-final-viral-combined_k141_194669\|\|full | 2852 | No | NA | 7 | 0 | 0 | Low-quality | Genome-fragment | 2.12 | AAI-based (high-confidence) | 0 |
| S1-final-viral-combined_k141_195529\|\|full | 2852 | No | NA | 1 | 0 | 1 | Low-quality | Genome-fragment | 1.44 | AAI-based (medium-confidence) | 0 |
| S12-final-viral-combined_k141_343480\|\|full | 11984 | No | NA | 15 | 0 | 14 | Not-determined | Genome-fragment | NA | NA | 0 |
| S12-final-viral-combined_k141_343484\|\|full | 1179 | No | NA | 3 | 0 | 1 | Not-determined | Genome-fragment | NA | NA | 0 |
| S12-final-viral-combined_k141_76613\|\|full | 1508 | No | NA | 4 | 0 | 0 | Not-determined | Genome-fragment | NA | NA | 0 |
| S12-final-viral-combined_k141_135978\|\|full | 2852 | No | NA | 3 | 0 | 0 | Low-quality | Genome-fragment | 7.33 | AAI-based (high-confidence) | 0 |
| S12-final-viral-combined_k141_343487\|\|full | 4446 | No | NA | 7 | 0 | 5 | Not-determined | Genome-fragment | NA | NA | 0 |
| S12-final-viral-combined_k141_42925\|\|full | 2585 | No | NA | 5 | 0 | 1 | Not-determined | Genome-fragment | NA | NA | 0 |
| S5-final-viral-combined_k141_107049\|\|full | 2853 | No | NA | 3 | 0 | 3 | Low-quality | Genome-fragment | 1.37 | AAI-based (medium-confidence) | 0 |
| S12-final-viral-combined_k141_359190\|\|full | 64793 | Yes | 27741 | 82 | 19 | 26 | Medium-quality | Genome-fragment | 58.55 | AAI-based (high-confidence) | 57.19 |
| S17-final-viral-combined_k141_200530\|\|full | 2854 | No | NA | 5 | 1 | 0 | Low-quality | Genome-fragment | 1.82 | HMM-based (lower-bound) | 0 |
| S8-final-viral-combined_k141_82005\|\|full | 2854 | No | NA | 4 | 2 | 0 | Low-quality | Genome-fragment | 6 | AAI-based (medium-confidence) | 0 |
| S12-final-viral-combined_k141_209881\|\|full | 10045 | No | NA | 9 | 0 | 3 | Not-determined | Genome-fragment | NA | NA | 0 |
| S12-final-viral-combined_k141_311452\|\|full | 1165 | No | NA | 2 | 0 | 2 | Not-determined | Genome-fragment | NA | NA | 0 |
| S12-final-viral-combined_k141_259783\|\|full | 45415 | No | NA | 66 | 15 | 0 | High-quality | High-quality | 97.98 | AAI-based (high-confidence) | 0 |
| S8-final-viral-combined_k141_217791\|\|full | 2854 | No | NA | 4 | 0 | 3 | Low-quality | Genome-fragment | 0.99 | AAI-based (medium-confidence) | 0 |
| S12-final-viral-combined_k141_109216\|\|full | 2802 | No | NA | 3 | 0 | 2 | Not-determined | Genome-fragment | NA | NA | 0 |
| S12-final-viral-combined_k141_125337\|\|full | 2836 | No | NA | 3 | 0 | 2 | Not-determined | Genome-fragment | NA | NA | 0 |
| S8_k141_58232 | 2854 | No | NA | 4 | 0 | 0 | Low-quality | Genome-fragment | 3.02 | AAI-based (high-confidence) | 0 |
| S12-final-viral-combined_k141_38141\|\|full | 2855 | No | NA | 3 | 0 | 3 | Low-quality | Genome-fragment | 2.22 | AAI-based (high-confidence) | 0 |
| S12-final-viral-combined_k141_259806\|\|full | 1220 | No | NA | 3 | 0 | 1 | Not-determined | Genome-fragment | NA | NA | 0 |
| S12-final-viral-combined_k141_377424\|\|full | 5747 | No | NA | 2 | 0 | 2 | Not-determined | Genome-fragment | NA | NA | 0 |
| S12-final-viral-combined_k141_8965\|\|full | 7910 | No | NA | 8 | 0 | 7 | Not-determined | Genome-fragment | NA | NA | 0 |
| S2-final-viral-combined_k141_187356\|\|full | 2855 | No | NA | 2 | 0 | 2 | Low-quality | Genome-fragment | 0.69 | AAI-based (medium-confidence) | 0 |
| S12-final-viral-combined_k141_58922\|\|full | 2457 | No | NA | 3 | 0 | 1 | Not-determined | Genome-fragment | NA | NA | 0 |
| S8_k141_232309 | 2855 | No | NA | 7 | 1 | 0 | Low-quality | Genome-fragment | 1.92 | HMM-based (lower-bound) | 0 |
| S12-final-viral-combined_k141_125364\|\|full | 3550 | No | NA | 4 | 0 | 1 | Not-determined | Genome-fragment | NA | NA | 0 |
| S7_k141_235089 | 2856 | No | NA | 3 | 0 | 0 | Low-quality | Genome-fragment | 4.77 | AAI-based (high-confidence) | 0 |
| S12-final-viral-combined_k141_343556\|\|full | 2120 | No | NA | 2 | 0 | 2 | Not-determined | Genome-fragment | NA | NA | 0 |
| S12-final-viral-combined_k141_311480\|\|full | 3413 | No | NA | 2 | 0 | 2 | Not-determined | Genome-fragment | NA | NA | 0 |
| S12-final-viral-combined_k141_294376\|\|full | 2229 | No | NA | 3 | 0 | 2 | Not-determined | Genome-fragment | NA | NA | 0 |
| S12-final-viral-combined_k141_243482\|\|full | 1201 | No | NA | 2 | 0 | 0 | Not-determined | Genome-fragment | NA | NA | 0 |
| S12-final-viral-combined_k141_176569\|\|full | 2856 | No | NA | 5 | 0 | 1 | Low-quality | Genome-fragment | 5.09 | AAI-based (high-confidence) | 0 |
| S12-final-viral-combined_k141_311519\|\|full | 1003 | No | NA | 3 | 0 | 1 | Not-determined | Genome-fragment | NA | NA | 0 |
| S12-final-viral-combined_k141_209981\|\|full | 1136 | No | NA | 2 | 0 | 0 | Not-determined | Genome-fragment | NA | NA | 0 |
| S11-final-viral-combined_k141_174205\|\|lt2gene | 2857 | No | NA | 2 | 0 | 0 | Low-quality | Genome-fragment | 4.77 | AAI-based (high-confidence) | 0 |
| S12-final-viral-combined_k141_76689\|\|full | 5369 | No | NA | 3 | 0 | 2 | Not-determined | Genome-fragment | NA | NA | 0 |
| S12-final-viral-combined_k141_343604\|\|full | 1879 | No | NA | 3 | 0 | 2 | Not-determined | Genome-fragment | NA | NA | 0 |
| S1-final-viral-combined_k141_337423\|\|full | 2857 | No | NA | 5 | 0 | 1 | Low-quality | Genome-fragment | 3.04 | AAI-based (medium-confidence) | 0 |
| S12-final-viral-combined_k141_209997\|\|full | 1114 | No | NA | 2 | 0 | 1 | Not-determined | Genome-fragment | NA | NA | 0 |
| S12-final-viral-combined_k141_176353\|\|full | 1309 | No | NA | 2 | 0 | 0 | Not-determined | Genome-fragment | NA | NA | 0 |
| S2-final-viral-combined_k141_220599\|\|full | 2857 | No | NA | 4 | 2 | 0 | Low-quality | Genome-fragment | 4.66 | HMM-based (lower-bound) | 0 |
| S12-final-viral-combined_k141_109291\|\|full | 1252 | No | NA | 2 | 0 | 0 | Not-determined | Genome-fragment | NA | NA | 0 |

| | | | | | | | | | | | |
|---|---|---|---|---|---|---|---|---|---|---|---|
| S12-final-viral-combined_k141_9046\|\|full | 4357 | No | NA | 3 | 0 | 2 | Not-determined | Genome-fragment | NA | NA | 0 |
| S11-final-viral-combined_k141_146153\|\|full | 2858 | No | NA | 6 | 3 | 0 | Low-quality | Genome-fragment | 7.78 | AAI-based (high-confidence) | 0 |
| S12-final-viral-combined_k141_327726\|\|full | 3297 | No | NA | 3 | 0 | 2 | Not-determined | Genome-fragment | NA | NA | 0 |
| S12-final-viral-combined_k141_125425\|\|full | 2646 | No | NA | 3 | 0 | 2 | Not-determined | Genome-fragment | NA | NA | 0 |
| S1-final-viral-combined_k141_271801\|\|full | 2858 | No | NA | 3 | 0 | 0 | Low-quality | Genome-fragment | 7.2 | AAI-based (medium-confidence) | 0 |
| S12-final-viral-combined_k141_210039\|\|full | 11138 | No | NA | 9 | 0 | 5 | Not-determined | Genome-fragment | NA | NA | 0 |
| S1-final-viral-combined_k141_355216\|\|full | 2858 | No | NA | 5 | 0 | 0 | Low-quality | Genome-fragment | 6.14 | AAI-based (medium-confidence) | 0 |
| S11-final-viral-combined_k141_310765\|\|full | 2859 | No | NA | 7 | 1 | 0 | Low-quality | Genome-fragment | 4.99 | AAI-based (high-confidence) | 0 |
| S12-final-viral-combined_k141_9065\|\|full | 1282 | No | NA | 2 | 0 | 2 | Not-determined | Genome-fragment | NA | NA | 0 |
| S12-final-viral-combined_k141_9071\|\|full | 5352 | No | NA | 4 | 0 | 4 | Not-determined | Genome-fragment | NA | NA | 0 |
| S12-final-viral-combined_k141_227205\|\|full | 1038 | No | NA | 3 | 0 | 1 | Not-determined | Genome-fragment | NA | NA | 0 |
| S12-final-viral-combined_k141_259850\|\|full | 2147 | No | NA | 2 | 0 | 1 | Not-determined | Genome-fragment | NA | NA | 0 |
| S12-final-viral-combined_k141_75458\|\|full | 2859 | No | NA | 3 | 2 | 0 | Low-quality | Genome-fragment | 4.64 | AAI-based (medium-confidence) | 0 |
| S4-final-viral-combined_k141_61999\|\|full | 2859 | No | NA | 4 | 0 | 2 | Low-quality | Genome-fragment | 1.41 | AAI-based (medium-confidence) | 0 |
| S1-final-viral-combined_k141_382093\|\|full | 2860 | No | NA | 3 | 2 | 0 | Low-quality | Genome-fragment | 6.59 | AAI-based (high-confidence) | 0 |
| S12-final-viral-combined_k141_243608\|\|full | 2450 | No | NA | 3 | 0 | 3 | Not-determined | Genome-fragment | NA | NA | 0 |
| S12-final-viral-combined_k141_43044\|\|full | 1609 | No | NA | 3 | 0 | 0 | Not-determined | Genome-fragment | NA | NA | 0 |
| S2-final-viral-combined_k141_41161\|\|full | 2861 | No | NA | 6 | 1 | 0 | Low-quality | Genome-fragment | 2.37 | HMM-based (lower-bound) | 0 |
| S12-final-viral-combined_k141_176443\|\|full | 1051 | No | NA | 3 | 0 | 2 | Not-determined | Genome-fragment | NA | NA | 0 |
| S6-final-viral-combined_k141_20065\|\|full | 2861 | No | NA | 8 | 3 | 0 | Low-quality | Genome-fragment | 7.55 | AAI-based (high-confidence) | 0 |
| S8-final-viral-combined_k141_255410\|\|full | 2861 | No | NA | 7 | 3 | 0 | Low-quality | Genome-fragment | 7.03 | AAI-based (high-confidence) | 0 |
| S17-final-viral-combined_k141_166894\|\|full | 2862 | No | NA | 3 | 0 | 0 | Low-quality | Genome-fragment | 6.41 | AAI-based (medium-confidence) | 0 |
| S12-final-viral-combined_k141_110683\|\|full | 2862 | No | NA | 6 | 2 | 0 | Low-quality | Genome-fragment | 4.17 | AAI-based (high-confidence) | 0 |
| S12-final-viral-combined_k141_59003\|\|full | 1074 | No | NA | 1 | 0 | 0 | Not-determined | Genome-fragment | NA | NA | 0 |
| S2-final-viral-combined_k141_164179\|\|full | 2862 | No | NA | 7 | 1 | 0 | Low-quality | Genome-fragment | 5.28 | AAI-based (high-confidence) | 0 |
| S12-final-viral-combined_k141_160214\|\|full | 2853 | No | NA | 3 | 0 | 2 | Not-determined | Genome-fragment | NA | NA | 0 |
| S12-final-viral-combined_k141_227293\|\|full | 1447 | No | NA | 3 | 0 | 0 | Not-determined | Genome-fragment | NA | NA | 0 |
| S12-final-viral-combined_k141_294500\|\|full | 2268 | No | NA | 3 | 0 | 0 | Not-determined | Genome-fragment | NA | NA | 0 |
| S12-final-viral-combined_k141_176564\|\|full | 1303 | No | NA | 4 | 0 | 1 | Not-determined | Genome-fragment | NA | NA | 0 |
| S4_k141_13355 | 2862 | No | NA | 3 | 0 | 1 | Low-quality | Genome-fragment | 0.76 | AAI-based (high-confidence) | 0 |
| S12-final-viral-combined_k141_311738\|\|full | 4230 | No | NA | 4 | 0 | 0 | Not-determined | Genome-fragment | NA | NA | 0 |
| S5-final-viral-combined_k141_80244\|\|full | 2863 | No | NA | 6 | 0 | 0 | Low-quality | Genome-fragment | 6.43 | AAI-based (medium-confidence) | 0 |
| S12-final-viral-combined_k141_59079\|\|full | 1090 | No | NA | 3 | 0 | 1 | Not-determined | Genome-fragment | NA | NA | 0 |
| S12-final-viral-combined_k141_160255\|\|full | 1842 | No | NA | 3 | 0 | 1 | Not-determined | Genome-fragment | NA | NA | 0 |
| S12-final-viral-combined_k141_92525\|\|full | 1258 | No | NA | 3 | 0 | 0 | Not-determined | Genome-fragment | NA | NA | 0 |
| S12-final-viral-combined_k141_277420\|\|full | 4788 | No | NA | 3 | 0 | 3 | Not-determined | Genome-fragment | NA | NA | 0 |
| S11-final-viral-combined_k141_308316\|\|full | 2864 | No | NA | 2 | 0 | 1 | Low-quality | Genome-fragment | 1.53 | AAI-based (high-confidence) | 0 |
| S12-final-viral-combined_k141_109515\|\|full | 1305 | No | NA | 3 | 0 | 0 | Not-determined | Genome-fragment | NA | NA | 0 |
| S12-final-viral-combined_k141_59121\|\|full | 2375 | No | NA | 3 | 0 | 1 | Not-determined | Genome-fragment | NA | NA | 0 |
| S12-final-viral-combined_k141_343870\|\|full | 3722 | No | NA | 3 | 0 | 1 | Not-determined | Genome-fragment | NA | NA | 0 |
| S12-final-viral-combined_k141_294595\|\|full | 5012 | No | NA | 4 | 0 | 3 | Not-determined | Genome-fragment | NA | NA | 0 |
| S12-final-viral-combined_k141_26156\|\|full | 2257 | No | NA | 3 | 0 | 1 | Not-determined | Genome-fragment | NA | NA | 0 |
| S12-final-viral-combined_k141_227420\|\|full | 1426 | No | NA | 3 | 0 | 0 | Not-determined | Genome-fragment | NA | NA | 0 |
| S12-final-viral-combined_k141_294625\|\|full | 1136 | No | NA | 2 | 0 | 1 | Not-determined | Genome-fragment | NA | NA | 0 |
| S12-final-viral-combined_k141_386973\|\|full | 2864 | No | NA | 5 | 0 | 0 | Low-quality | Genome-fragment | 3.71 | AAI-based (medium-confidence) | 0 |
| S12-final-viral-combined_k141_59179\|\|full | 1598 | No | NA | 2 | 0 | 1 | Not-determined | Genome-fragment | NA | NA | 0 |
| S12-final-viral-combined_k141_314230\|\|full | 2864 | No | NA | 6 | 0 | 0 | Low-quality | Genome-fragment | 5.58 | AAI-based (medium-confidence) | 0 |
| S12-final-viral-combined_k141_76911\|\|full | 1296 | No | NA | 1 | 0 | 0 | Not-determined | Genome-fragment | NA | NA | 0 |
| S8-final-viral-combined_k141_212835\|\|full | 2864 | No | NA | 6 | 2 | 0 | Low-quality | Genome-fragment | 6.75 | AAI-based (high-confidence) | 0 |
| S7-final-viral-combined_k141_199651\|\|full | 2865 | No | NA | 5 | 0 | 1 | Low-quality | Genome-fragment | 2.77 | AAI-based (medium-confidence) | 0 |
| S12-final-viral-combined_k141_260021\|\|full | 1165 | No | NA | 2 | 0 | 0 | Not-determined | Genome-fragment | NA | NA | 0 |
| S12-final-viral-combined_k141_176644\|\|full | 2008 | No | NA | 3 | 0 | 1 | Not-determined | Genome-fragment | NA | NA | 0 |
| S12-final-viral-combined_k141_26209\|\|full | 2771 | No | NA | 4 | 0 | 2 | Not-determined | Genome-fragment | NA | NA | 0 |
| S8-final-viral-combined_k141_246790\|\|full | 2865 | No | NA | 2 | 0 | 2 | Low-quality | Genome-fragment | 7.48 | AAI-based (medium-confidence) | 0 |
| S11-final-viral-combined_k141_285875\|\|full | 2866 | No | NA | 2 | 0 | 2 | Low-quality | Genome-fragment | 3.32 | AAI-based (high-confidence) | 0 |
| S12-final-viral-combined_k141_210270\|\|full | 1369 | No | NA | 2 | 0 | 0 | Not-determined | Genome-fragment | NA | NA | 0 |
| S17-final-viral-combined_k141_195378\|\|full | 2866 | No | NA | 3 | 0 | 0 | Low-quality | Genome-fragment | 1.89 | AAI-based (high-confidence) | 0 |
| S2-final-viral-combined_k141_42317\|\|full | 2866 | No | NA | 4 | 1 | 0 | Low-quality | Genome-fragment | 4.34 | HMM-based (lower-bound) | 0 |
| S12-final-viral-combined_k141_194465\|\|full | 6796 | No | NA | 10 | 0 | 0 | Not-determined | Genome-fragment | NA | NA | 0 |
| S8-final-viral-combined_k141_130098\|\|full | 2866 | No | NA | 2 | 0 | 0 | Low-quality | Genome-fragment | 0.82 | AAI-based (medium-confidence) | 0 |
| S12-final-viral-combined_k141_359610\|\|full | 1291 | No | NA | 3 | 0 | 0 | Not-determined | Genome-fragment | NA | NA | 0 |
| S5-final-viral-combined_k141_115242\|\|full | 2867 | No | NA | 3 | 0 | 1 | Low-quality | Genome-fragment | 2.61 | AAI-based (high-confidence) | 0 |
| S8-final-viral-combined_k141_245999\|\|full | 2867 | No | NA | 2 | 0 | 2 | Low-quality | Genome-fragment | 4.91 | AAI-based (high-confidence) | 0 |
| S12-final-viral-combined_k141_260036\|\|full | 32189 | No | NA | 30 | 4 | 0 | Medium-quality | Genome-fragment | 54.91 | AAI-based (high-confidence) | 0 |
| S12-final-viral-combined_k141_9168\|\|full | 2132 | No | NA | 4 | 0 | 3 | Not-determined | Genome-fragment | NA | NA | 0 |
| S12-final-viral-combined_k141_311914\|\|full | 1895 | No | NA | 3 | 0 | 3 | Not-determined | Genome-fragment | NA | NA | 0 |
| S11-final-viral-combined_k141_184681\|\|full | 2868 | No | NA | 3 | 0 | 0 | Low-quality | Genome-fragment | 7.12 | AAI-based (medium-confidence) | 0 |
| S3-final-viral-combined_k141_96686\|\|full | 2868 | No | NA | 6 | 1 | 0 | Low-quality | Genome-fragment | 6.61 | AAI-based (medium-confidence) | 0 |
| S5-final-viral-combined_k141_194407\|\|full | 2869 | Yes | 1404 | 5 | 1 | 1 | Low-quality | Genome-fragment | 1.57 | AAI-based (high-confidence) | 51.06 |
| S12-final-viral-combined_k141_43365\|\|full | 2270 | No | NA | 3 | 0 | 0 | Not-determined | Genome-fragment | NA | NA | 0 |
| S12-final-viral-combined_k141_125581\|\|full | 1652 | No | NA | 4 | 0 | 0 | Not-determined | Genome-fragment | NA | NA | 0 |
| S7-final-viral-combined_k141_61686\|\|full | 2869 | No | NA | 4 | 2 | 0 | Low-quality | Genome-fragment | 6.47 | AAI-based (medium-confidence) | 0 |
| S12-final-viral-combined_k141_311955\|\|full | 10977 | No | NA | 11 | 0 | 0 | Not-determined | Genome-fragment | NA | NA | 0 |
| S12-final-viral-combined_k141_109607\|\|full | 1135 | No | NA | 2 | 0 | 1 | Not-determined | Genome-fragment | NA | NA | 0 |
| S12-final-viral-combined_k141_160755\|\|full | 2869 | No | NA | 3 | 2 | 0 | Low-quality | Genome-fragment | 6.67 | AAI-based (medium-confidence) | 0 |
| S12-final-viral-combined_k141_260119\|\|full | 3193 | No | NA | 3 | 0 | 0 | Not-determined | Genome-fragment | NA | NA | 0 |
| S12-final-viral-combined_k141_277632\|\|full | 1228 | No | NA | 2 | 0 | 1 | Not-determined | Genome-fragment | NA | NA | 0 |
| S12-final-viral-combined_k141_344112\|\|full | 1599 | No | NA | 4 | 0 | 1 | Not-determined | Genome-fragment | NA | NA | 0 |
| S12-final-viral-combined_k141_92672\|\|full | 1396 | No | NA | 2 | 0 | 0 | Not-determined | Genome-fragment | NA | NA | 0 |
| S12_k141_358180 | 2869 | No | NA | 8 | 2 | 0 | Low-quality | Genome-fragment | 6.5 | AAI-based (high-confidence) | 0 |
| S12-final-viral-combined_k141_109639\|\|full | 1628 | No | NA | 3 | 0 | 0 | Not-determined | Genome-fragment | NA | NA | 0 |
| S17-final-viral-combined_k141_219544\|\|full | 2870 | No | NA | 3 | 0 | 2 | Low-quality | Genome-fragment | 1.6 | AAI-based (medium-confidence) | 0 |
| S12-final-viral-combined_k141_260206\|\|full | 2870 | No | NA | 3 | 0 | 2 | Low-quality | Genome-fragment | 1.39 | AAI-based (medium-confidence) | 0 |
| S12-final-viral-combined_k141_176790\|\|full | 1356 | No | NA | 3 | 0 | 1 | Not-determined | Genome-fragment | NA | NA | 0 |
| S8-final-viral-combined_k141_37056\|\|full | 2870 | No | NA | 6 | 4 | 0 | Low-quality | Genome-fragment | 6.33 | AAI-based (high-confidence) | 0 |
| S12-final-viral-combined_k141_294804\|\|full | 1002 | No | NA | 3 | 0 | 0 | Not-determined | Genome-fragment | NA | NA | 0 |
| S12-final-viral-combined_k141_327845\|\|full | 2216 | No | NA | 2 | 0 | 0 | Not-determined | Genome-fragment | NA | NA | 0 |
| S12-final-viral-combined_k141_194581\|\|full | 1895 | No | NA | 2 | 0 | 2 | Not-determined | Genome-fragment | NA | NA | 0 |
| S7_k141_113390 | 2871 | No | NA | 4 | 0 | 2 | Low-quality | Genome-fragment | 0.67 | AAI-based (medium-confidence) | 0 |
| S12-final-viral-combined_k141_194590\|\|full | 12460 | No | NA | 18 | 0 | 9 | Not-determined | Genome-fragment | NA | NA | 0 |
| S6-final-viral-combined_k141_137884\|\|full | 2871 | No | NA | 3 | 0 | 2 | Low-quality | Genome-fragment | 2.06 | AAI-based (medium-confidence) | 0 |
| S12-final-viral-combined_k141_359687\|\|full | 6663 | No | NA | 6 | 0 | 1 | Not-determined | Genome-fragment | NA | NA | 0 |
| S8-final-viral-combined_k141_119838\|\|full | 2871 | No | NA | 2 | 0 | 2 | Low-quality | Genome-fragment | 2.81 | AAI-based (medium-confidence) | 0 |
| S12-final-viral-combined_k141_92715\|\|full | 1430 | No | NA | 2 | 0 | 0 | Not-determined | Genome-fragment | NA | NA | 0 |
| S12-final-viral-combined_k141_143328\|\|full | 1109 | No | NA | 2 | 0 | 1 | Not-determined | Genome-fragment | NA | NA | 0 |
| S1-final-viral-combined_k141_358989\|\|full | 2872 | No | NA | 7 | 6 | 0 | Low-quality | Genome-fragment | 6.72 | AAI-based (high-confidence) | 0 |
| S12-final-viral-combined_k141_359698\|\|full | 1500 | No | NA | 3 | 0 | 0 | Not-determined | Genome-fragment | NA | NA | 0 |
| S8_k141_124722 | 2872 | No | NA | 5 | 0 | 0 | Low-quality | Genome-fragment | 3.02 | AAI-based (high-confidence) | 0 |
| S5-final-viral-combined_k141_6227\|\|full | 2873 | No | NA | 6 | 1 | 0 | Low-quality | Genome-fragment | 1.93 | HMM-based (lower-bound) | 0 |
| S12-final-viral-combined_k141_59377\|\|full | 2273 | No | NA | 3 | 0 | 2 | Not-determined | Genome-fragment | NA | NA | 0 |
| S12-final-viral-combined_k141_109701\|\|full | 2067 | No | NA | 6 | 0 | 1 | Not-determined | Genome-fragment | NA | NA | 0 |
| S12-final-viral-combined_k141_43513\|\|full | 5381 | No | NA | 8 | 0 | 0 | Not-determined | Genome-fragment | NA | NA | 0 |
| S12-final-viral-combined_k141_312103\|\|full | 1873 | No | NA | 3 | 0 | 1 | Not-determined | Genome-fragment | NA | NA | 0 |
| S2_k141_128825 | 2873 | No | NA | 5 | 1 | 0 | Low-quality | Genome-fragment | 1.87 | AAI-based (medium-confidence) | 0 |
| S12-final-viral-combined_k141_109744\|\|full | 2130 | No | NA | 3 | 0 | 1 | Not-determined | Genome-fragment | NA | NA | 0 |
| S12-final-viral-combined_k141_143360\|\|full | 3818 | No | NA | 3 | 0 | 3 | Not-determined | Genome-fragment | NA | NA | 0 |
| S12-final-viral-combined_k141_61247\|\|full | 2874 | No | NA | 3 | 0 | 3 | Low-quality | Genome-fragment | 8.64 | AAI-based (medium-confidence) | 0 |
| S12-final-viral-combined_k141_77259\|\|full | 1173 | No | NA | 3 | 0 | 0 | Not-determined | Genome-fragment | NA | NA | 0 |
| S12-final-viral-combined_k141_347163\|\|full | 2874 | No | NA | 4 | 0 | 1 | Low-quality | Genome-fragment | 2.29 | AAI-based (medium-confidence) | 0 |
| S8-final-viral-combined_k141_25192\|\|full | 2874 | No | NA | 6 | 0 | 1 | Low-quality | Genome-fragment | 4.15 | AAI-based (medium-confidence) | 0 |
| S12-final-viral-combined_k141_312162\|\|full | 2185 | No | NA | 3 | 0 | 2 | Not-determined | Genome-fragment | NA | NA | 0 |
| S11-final-viral-combined_k141_298984\|\|full | 2875 | No | NA | 5 | 5 | 0 | Low-quality | Genome-fragment | 7.11 | AAI-based (high-confidence) | 0 |
| S3-final-viral-combined_k141_50624\|\|full | 2875 | No | NA | 3 | 0 | 0 | Low-quality | Genome-fragment | 8.21 | AAI-based (high-confidence) | 0 |
| S2-final-viral-combined_k141_53671\|\|full | 2875 | No | NA | 5 | 0 | 0 | Low-quality | Genome-fragment | 5.54 | AAI-based (medium-confidence) | 0 |
| S12-final-viral-combined_k141_26505\|\|full | 2370 | No | NA | 4 | 0 | 1 | Not-determined | Genome-fragment | NA | NA | 0 |
| S12-final-viral-combined_k141_160581\|\|full | 1610 | No | NA | 3 | 0 | 2 | Not-determined | Genome-fragment | NA | NA | 0 |
| S12-final-viral-combined_k141_227764\|\|full | 2299 | No | NA | 3 | 0 | 3 | Not-determined | Genome-fragment | NA | NA | 0 |
| S12-final-viral-combined_k141_294961\|\|full | 2716 | No | NA | 3 | 0 | 1 | Not-determined | Genome-fragment | NA | NA | 0 |
| S12-final-viral-combined_k141_344250\|\|full | 2131 | No | NA | 3 | 0 | 2 | Not-determined | Genome-fragment | NA | NA | 0 |
| S9-final-viral-combined_k141_42364\|\|full | 2876 | No | NA | 9 | 2 | 0 | Low-quality | Genome-fragment | 7.51 | AAI-based (high-confidence) | 0 |
| S11-final-viral-combined_k141_98086\|\|full | 2876 | No | NA | 5 | 1 | 0 | Low-quality | Genome-fragment | 2.33 | HMM-based (lower-bound) | 0 |
| S12-final-viral-combined_k141_312216\|\|full | 2009 | No | NA | 2 | 0 | 1 | Not-determined | Genome-fragment | NA | NA | 0 |
| S7-final-viral-combined_k141_195846\|\|full | 2876 | No | NA | 5 | 0 | 0 | Low-quality | Genome-fragment | 2.13 | AAI-based (high-confidence) | 0 |
| S8-final-viral-combined_k141_40045\|\|full | 2876 | No | NA | 4 | 0 | 0 | Low-quality | Genome-fragment | 1.87 | AAI-based (high-confidence) | 0 |
| S12-final-viral-combined_k141_24526\|\|full | 2877 | Yes | 516 | 3 | 1 | 1 | Low-quality | Genome-fragment | 0.27 | AAI-based (medium-confidence) | 82.06 |
| S12-final-viral-combined_k141_125795\|\|full | 3109 | No | NA | 3 | 0 | 2 | Not-determined | Genome-fragment | NA | NA | 0 |
| S12-final-viral-combined_k141_344303\|\|full | 1231 | No | NA | 3 | 0 | 2 | Not-determined | Genome-fragment | NA | NA | 0 |
| S12-final-viral-combined_k141_247524\|\|full | 2877 | No | NA | 5 | 0 | 2 | Low-quality | Genome-fragment | 3.27 | AAI-based (medium-confidence) | 0 |
| S2-final-viral-combined_k141_147651\|\|full | 2877 | No | NA | 5 | 0 | 3 | Low-quality | Genome-fragment | 4.65 | AAI-based (high-confidence) | 0 |
| S11-final-viral-combined_k141_264927\|\|full | 2878 | No | NA | 3 | 1 | 0 | Low-quality | Genome-fragment | 7.65 | AAI-based (medium-confidence) | 0 |
| S12-final-viral-combined_k141_328080\|\|full | 6392 | No | NA | 4 | 0 | 3 | Not-determined | Genome-fragment | NA | NA | 0 |
| S12-final-viral-combined_k141_312310\|\|full | 4726 | No | NA | 4 | 0 | 3 | Not-determined | Genome-fragment | NA | NA | 0 |
| S1-final-viral-combined_k141_196806\|\|full | 2878 | No | NA | 8 | 5 | 0 | Low-quality | Genome-fragment | 9.02 | AAI-based (high-confidence) | 0 |
| S12-final-viral-combined_k141_260373\|\|full | 1901 | No | NA | 3 | 0 | 2 | Not-determined | Genome-fragment | NA | NA | 0 |
| S11-final-viral-combined_k141_250315\|\|full | 2879 | No | NA | 3 | 0 | 0 | Low-quality | Genome-fragment | 5.79 | AAI-based (medium-confidence) | 0 |
| S12-final-viral-combined_k141_295072\|\|full | 1287 | No | NA | 3 | 0 | 1 | Not-determined | Genome-fragment | NA | NA | 0 |
| S12-final-viral-combined_k141_394752\|\|full | 2195 | No | NA | 2 | 0 | 2 | Not-determined | Genome-fragment | NA | NA | 0 |
| S12-final-viral-combined_k141_74553\|\|full | 2879 | No | NA | 3 | 1 | 0 | Low-quality | Genome-fragment | 6.87 | AAI-based (medium-confidence) | 0 |
| S12-final-viral-combined_k141_148018\|\|full | 2879 | No | NA | 4 | 0 | 1 | Low-quality | Genome-fragment | 7.89 | AAI-based (medium-confidence) | 0 |
| S12-final-viral-combined_k141_9434\|\|full | 3208 | No | NA | 3 | 0 | 3 | Not-determined | Genome-fragment | NA | NA | 0 |
| S9-final-viral-combined_k141_123341\|\|full | 2880 | Yes | 246 | 4 | 1 | 1 | Low-quality | Genome-fragment | 0.07 | HMM-based (lower-bound) | 91.46 |
| S12-final-viral-combined_k141_328103\|\|full | 6026 | No | NA | 6 | 0 | 2 | Not-determined | Genome-fragment | NA | NA | 0 |
| S12-final-viral-combined_k141_18570\|\|full | 2880 | No | NA | 5 | 0 | 3 | Low-quality | Genome-fragment | 3.27 | AAI-based (high-confidence) | 0 |
| S12-final-viral-combined_k141_92952\|\|full | 3539 | No | NA | 5 | 0 | 4 | Not-determined | Genome-fragment | NA | NA | 0 |
| S12-final-viral-combined_k141_26691\|\|full | 4319 | No | NA | 4 | 0 | 1 | Not-determined | Genome-fragment | NA | NA | 0 |
| S2-final-viral-combined_k141_130040\|\|full | 2880 | No | NA | 3 | 0 | 2 | Low-quality | Genome-fragment | 0.69 | AAI-based (high-confidence) | 0 |
| S12-final-viral-combined_k141_243926\|\|full | 3679 | No | NA | 4 | 0 | 4 | Not-determined | Genome-fragment | NA | NA | 0 |
| S1-final-viral-combined_k141_28427\|\|full | 2881 | No | NA | 3 | 0 | 2 | Low-quality | Genome-fragment | 8.18 | AAI-based (medium-confidence) | 0 |
| S2_k141_138200 | 2881 | No | NA | 3 | 1 | 0 | Low-quality | Genome-fragment | 1.99 | HMM-based (lower-bound) | 0 |
| S11-final-viral-combined_k141_287706\|\|full | 2882 | No | NA | 5 | 0 | 0 | Low-quality | Genome-fragment | 5.36 | AAI-based (high-confidence) | 0 |
| S1-final-viral-combined_k141_121446\|\|full | 2882 | No | NA | 3 | 0 | 2 | Low-quality | Genome-fragment | 2.65 | AAI-based (high-confidence) | 0 |
| S17-k141_193561 | 2882 | No | NA | 3 | 0 | 3 | Low-quality | Genome-fragment | 1.4 | AAI-based (medium-confidence) | 0 |
| S8-final-viral-combined_k141_140062\|\|full | 2882 | No | NA | 3 | 0 | 0 | Low-quality | Genome-fragment | 6.51 | AAI-based (medium-confidence) | 0 |
| S12-final-viral-combined_k141_125940\|\|full | 1715 | No | NA | 4 | 0 | 0 | Not-determined | Genome-fragment | NA | NA | 0 |
| S9-final-viral-combined_k141_57869\|\|full | 2883 | No | NA | 3 | 0 | 2 | Low-quality | Genome-fragment | 1.04 | AAI-based (medium-confidence) | 0 |
| S12-final-viral-combined_k141_125951\|\|full | 2928 | No | NA | 3 | 0 | 1 | Not-determined | Genome-fragment | NA | NA | 0 |
| S5-final-viral-combined_k141_101917\|\|full | 2883 | No | NA | 8 | 0 | 1 | Low-quality | Genome-fragment | 4.56 | AAI-based (medium-confidence) | 0 |
| S12-final-viral-combined_k141_43858\|\|full | 2725 | No | NA | 4 | 0 | 2 | Not-determined | Genome-fragment | NA | NA | 0 |
| S12-final-viral-combined_k141_59730\|\|full | 3981 | No | NA | 6 | 0 | 0 | Not-determined | Genome-fragment | NA | NA | 0 |
| S12-final-viral-combined_k141_247493\|\|full | 2883 | No | NA | 4 | 0 | 4 | Low-quality | Genome-fragment | 1.03 | AAI-based (medium-confidence) | 0 |
| S12-final-viral-combined_k141_243986\|\|full | 1954 | No | NA | 4 | 0 | 0 | Not-determined | Genome-fragment | NA | NA | 0 |
| S12-final-viral-combined_k141_59763\|\|full | 5181 | No | NA | 7 | 0 | 6 | Not-determined | Genome-fragment | NA | NA | 0 |
| S12-final-viral-combined_k141_77583\|\|full | 1794 | No | NA | 2 | 0 | 1 | Not-determined | Genome-fragment | NA | NA | 0 |
| S12-final-viral-combined_k141_260478\|\|full | 1855 | No | NA | 2 | 0 | 0 | Not-determined | Genome-fragment | NA | NA | 0 |
| S2_k141_221565 | 2883 | No | NA | 3 | 0 | 0 | Low-quality | Genome-fragment | 4.82 | AAI-based (high-confidence) | 0 |
| S12-final-viral-combined_k141_9603\|\|full | 1527 | No | NA | 2 | 0 | 1 | Not-determined | Genome-fragment | NA | NA | 0 |
| S11-final-viral-combined_k141_188923\|\|full | 2884 | No | NA | 2 | 2 | 0 | Low-quality | Genome-fragment | 3.72 | HMM-based (lower-bound) | 0 |
| S12-final-viral-combined_k141_77603\|\|full | 1690 | No | NA | 3 | 0 | 0 | Not-determined | Genome-fragment | NA | NA | 0 |
| S7_k141_122241 | 2884 | No | NA | 1 | 0 | 0 | Low-quality | Genome-fragment | 2.7 | AAI-based (high-confidence) | 0 |
| S8-final-viral-combined_k141_104959\|\|full | 2884 | No | NA | 8 | 2 | 0 | Low-quality | Genome-fragment | 5.23 | AAI-based (high-confidence) | 0 |
| S12-final-viral-combined_k141_210663\|\|full | 3318 | No | NA | 3 | 0 | 0 | Not-determined | Genome-fragment | NA | NA | 0 |
| S7-final-viral-combined_k141_53818\|\|full | 2885 | No | NA | 3 | 0 | 2 | Low-quality | Genome-fragment | 0.33 | AAI-based (medium-confidence) | 0 |
| S12-final-viral-combined_k141_378280\|\|full | 1191 | No | NA | 3 | 0 | 1 | Not-determined | Genome-fragment | NA | NA | 0 |
| S12-final-viral-combined_k141_278014\|\|full | 2845 | No | NA | 4 | 0 | 2 | Not-determined | Genome-fragment | NA | NA | 0 |
| S8_k141_119456 | 2885 | No | NA | 5 | 0 | 3 | Low-quality | Genome-fragment | 1.89 | AAI-based (high-confidence) | 0 |
| S12-final-viral-combined_k141_26805\|\|full | 6140 | No | NA | 6 | 0 | 2 | Not-determined | Genome-fragment | NA | NA | 0 |
| S11-final-viral-combined_k141_9009\|\|full | 2886 | No | NA | 4 | 0 | 4 | Low-quality | Genome-fragment | 1.99 | AAI-based (medium-confidence) | 0 |
| S12-final-viral-combined_k141_210669\|\|full | 8273 | No | NA | 9 | 0 | 3 | Not-determined | Genome-fragment | NA | NA | 0 |
| S2_k141_158226 | 2886 | No | NA | 2 | 0 | 2 | Low-quality | Genome-fragment | 0.44 | AAI-based (medium-confidence) | 0 |
| S12-final-viral-combined_k141_260518\|\|full | 1552 | No | NA | 3 | 0 | 0 | Not-determined | Genome-fragment | NA | NA | 0 |
| S12-final-viral-combined_k141_26813\|\|full | 1877 | No | NA | 3 | 0 | 1 | Not-determined | Genome-fragment | NA | NA | 0 |
| S12-final-viral-combined_k141_278028\|\|full | 2017 | No | NA | 3 | 0 | 3 | Not-determined | Genome-fragment | NA | NA | 0 |
| S12-final-viral-combined_k141_328262\|\|full | 1592 | No | NA | 2 | 0 | 0 | Not-determined | Genome-fragment | NA | NA | 0 |
| S12-final-viral-combined_k141_143565\|\|full | 2633 | No | NA | 4 | 0 | 3 | Not-determined | Genome-fragment | NA | NA | 0 |
| S12-final-viral-combined_k141_59856\|\|full | 10289 | No | NA | 8 | 0 | 5 | Not-determined | Genome-fragment | NA | NA | 0 |
| S12-final-viral-combined_k141_394942\|\|full | 11493 | No | NA | 11 | 0 | 6 | Not-determined | Genome-fragment | NA | NA | 0 |
| S12-final-viral-combined_k141_328298\|\|full | 1034 | No | NA | 3 | 0 | 0 | Not-determined | Genome-fragment | NA | NA | 0 |
| S12-final-viral-combined_k141_44023\|\|full | 1119 | No | NA | 2 | 0 | 0 | Not-determined | Genome-fragment | NA | NA | 0 |
| S12-final-viral-combined_k141_344543\|\|full | 1340 | No | NA | 4 | 0 | 2 | Not-determined | Genome-fragment | NA | NA | 0 |
| S12-final-viral-combined_k141_295290\|\|full | 1492 | No | NA | 2 | 0 | 0 | Not-determined | Genome-fragment | NA | NA | 0 |
| S12-final-viral-combined_k141_378367\|\|full | 1333 | No | NA | 3 | 0 | 1 | Not-determined | Genome-fragment | NA | NA | 0 |
| S12-final-viral-combined_k141_378369\|\|full | 4140 | No | NA | 4 | 0 | 2 | Not-determined | Genome-fragment | NA | NA | 0 |
| S12-final-viral-combined_k141_312602\|\|full | 2949 | No | NA | 4 | 0 | 0 | Not-determined | Genome-fragment | NA | NA | 0 |
| S12-final-viral-combined_k141_44083\|\|full | 1132 | No | NA | 2 | 0 | 1 | Not-determined | Genome-fragment | NA | NA | 0 |
| S8-final-viral-combined_k141_152707\|\|full | 2886 | Yes | 1378 | 4 | 2 | 1 | Low-quality | Genome-fragment | 2.28 | AAI-based (high-confidence) | 52.25 |
| S12-final-viral-combined_k141_160908\|\|full | 1054 | No | NA | 3 | 0 | 0 | Not-determined | Genome-fragment | NA | NA | 0 |
| S12-final-viral-combined_k141_17796\|\|full | 2887 | No | NA | 2 | 0 | 2 | Low-quality | Genome-fragment | 1.58 | AAI-based (medium-confidence) | 0 |
| S12-final-viral-combined_k141_59953\|\|full | 2373 | No | NA | 3 | 0 | 2 | Not-determined | Genome-fragment | NA | NA | 0 |
| S12-final-viral-combined_k141_210809\|\|full | 2850 | No | NA | 3 | 0 | 1 | Not-determined | Genome-fragment | NA | NA | 0 |
| S11-final-viral-combined_k141_172027\|\|lt2gene | 2888 | No | NA | 2 | 1 | 0 | Low-quality | Genome-fragment | 7.53 | AAI-based (high-confidence) | 0 |
| S12-final-viral-combined_k141_160917\|\|full | 3245 | No | NA | 4 | 0 | 2 | Not-determined | Genome-fragment | NA | NA | 0 |
| S1-final-viral-combined_k141_233643\|\|full | 2888 | No | NA | 4 | 0 | 0 | Low-quality | Genome-fragment | 3.58 | AAI-based (medium-confidence) | 0 |
| S3-final-viral-combined_k141_54440\|\|full | 2888 | No | NA | 5 | 1 | 0 | Low-quality | Genome-fragment | 7.56 | AAI-based (high-confidence) | 0 |
| S12-final-viral-combined_k141_378423\|\|full | 1848 | No | NA | 2 | 0 | 0 | Not-determined | Genome-fragment | NA | NA | 0 |
| S12-final-viral-combined_k141_328335\|\|full | 1987 | No | NA | 5 | 0 | 0 | Not-determined | Genome-fragment | NA | NA | 0 |
| S17-final-viral-combined_k141_181617\|\|full | 2888 | No | NA | 4 | 0 | 3 | Low-quality | Genome-fragment | 1.31 | AAI-based (medium-confidence) | 0 |
| S12-final-viral-combined_k141_44136\|\|full | 6721 | No | NA | 9 | 0 | 4 | Not-determined | Genome-fragment | NA | NA | 0 |
| S12-final-viral-combined_k141_328356\|\|full | 6616 | No | NA | 8 | 0 | 4 | Not-determined | Genome-fragment | NA | NA | 0 |
| S1-final-viral-combined_k141_96718\|\|full | 2889 | No | NA | 4 | 0 | 0 | Low-quality | Genome-fragment | 2.76 | AAI-based (medium-confidence) | 0 |
| S2-final-viral-combined_k141_241588\|\|full | 2889 | No | NA | 5 | 1 | 0 | Low-quality | Genome-fragment | 1.94 | HMM-based (lower-bound) | 0 |
| S12-final-viral-combined_k141_177313\|\|full | 1700 | No | NA | 2 | 0 | 2 | Not-determined | Genome-fragment | NA | NA | 0 |
| S2-final-viral-combined_k141_100335\|\|full | 2889 | No | NA | 5 | 1 | 2 | Low-quality | Genome-fragment | 2.15 | AAI-based (medium-confidence) | 0 |
| S11-final-viral-combined_k141_8513\|\|full | 2890 | No | NA | 3 | 0 | 0 | Low-quality | Genome-fragment | 7.28 | AAI-based (high-confidence) | 0 |
| S12-final-viral-combined_k141_77726\|\|full | 4128 | No | NA | 4 | 0 | 1 | Not-determined | Genome-fragment | NA | NA | 0 |
| S12-final-viral-combined_k141_109997\|\|full | 1061 | No | NA | 3 | 0 | 0 | Not-determined | Genome-fragment | NA | NA | 0 |
| S5-final-viral-combined_k141_201163\|\|full | 2890 | No | NA | 4 | 0 | 0 | Low-quality | Genome-fragment | 6.62 | AAI-based (medium-confidence) | 0 |
| S17-final-viral-combined_k141_13938\|\|full | 2890 | No | NA | 3 | 0 | 1 | Low-quality | Genome-fragment | 5.08 | AAI-based (medium-confidence) | 0 |
| S12-final-viral-combined_k141_44191\|\|full | 1816 | No | NA | 3 | 0 | 1 | Not-determined | Genome-fragment | NA | NA | 0 |
| S8-final-viral-combined_k141_18319\|\|full | 2890 | No | NA | 4 | 0 | 2 | Low-quality | Genome-fragment | 2.41 | AAI-based (high-confidence) | 0 |
| S1-final-viral-combined_k141_52637\|\|full | 2891 | No | NA | 5 | 0 | 0 | Low-quality | Genome-fragment | 6.4 | AAI-based (medium-confidence) | 0 |
| S1-final-viral-combined_k141_109001\|\|full | 2891 | No | NA | 2 | 0 | 2 | Low-quality | Genome-fragment | 1.87 | AAI-based (medium-confidence) | 0 |
| S6-final-viral-combined_k141_18127\|\|full | 2891 | No | NA | 4 | 0 | 2 | Low-quality | Genome-fragment | 3.59 | AAI-based (medium-confidence) | 0 |
| S5-final-viral-combined_k141_203097\|\|full | 2892 | No | NA | 6 | 2 | 0 | Low-quality | Genome-fragment | 3.21 | AAI-based (high-confidence) | 0 |
| S12-final-viral-combined_k141_312704\|\|full | 1738 | No | NA | 2 | 0 | 2 | Not-determined | Genome-fragment | NA | NA | 0 |
| S12-final-viral-combined_k141_228207\|\|full | 8641 | No | NA | 10 | 0 | 5 | Not-determined | Genome-fragment | NA | NA | 0 |
| S12-final-viral-combined_k141_161018\|\|full | 5833 | No | NA | 10 | 0 | 0 | Not-determined | Genome-fragment | NA | NA | 0 |
| S5-final-viral-combined_k141_26412\|\|full | 2892 | No | NA | 4 | 1 | 0 | Low-quality | Genome-fragment | 6.6 | AAI-based (medium-confidence) | 0 |
| S12-final-viral-combined_k141_60031\|\|full | 1764 | No | NA | 2 | 0 | 2 | Not-determined | Genome-fragment | NA | NA | 0 |
| S17-final-viral-combined_k141_14838\|\|full | 2893 | No | NA | 5 | 0 | 0 | Low-quality | Genome-fragment | 8.13 | AAI-based (high-confidence) | 0 |
| S12-final-viral-combined_k141_177344\|\|full | 2288 | No | NA | 3 | 0 | 2 | Not-determined | Genome-fragment | NA | NA | 0 |
| S1-final-viral-combined_k141_214619\|\|full | 2894 | Yes | 2567 | 6 | 2 | 1 | Low-quality | Genome-fragment | 7.38 | AAI-based (high-confidence) | 11.3 |
| S5-final-viral-combined_k141_136922\|\|full | 2894 | Yes | 2293 | 6 | 1 | 1 | Low-quality | Genome-fragment | 5.38 | AAI-based (medium-confidence) | 20.77 |
| S12-final-viral-combined_k141_26995\|\|full | 8478 | No | NA | 8 | 0 | 7 | Not-determined | Genome-fragment | NA | NA | 0 |
| S12-final-viral-combined_k141_395157\|\|full | 5720 | No | NA | 9 | 0 | 4 | Not-determined | Genome-fragment | NA | NA | 0 |
| S12-final-viral-combined_k141_395159\|\|full | 2437 | No | NA | 3 | 0 | 1 | Not-determined | Genome-fragment | NA | NA | 0 |
| S12-final-viral-combined_k141_328444\|\|full | 1271 | No | NA | 3 | 0 | 0 | Not-determined | Genome-fragment | NA | NA | 0 |
| S17-final-viral-combined_k141_46301\|\|full | 2894 | No | NA | 4 | 2 | 0 | Low-quality | Genome-fragment | 6.45 | AAI-based (medium-confidence) | 0 |
| S12-final-viral-combined_k141_60088\|\|full | 1204 | No | NA | 1 | 0 | 0 | Not-determined | Genome-fragment | NA | NA | 0 |
| S12-final-viral-combined_k141_77770\|\|full | 1815 | No | NA | 3 | 0 | 1 | Not-determined | Genome-fragment | NA | NA | 0 |
| S9-final-viral-combined_k141_56813\|\|full | 2895 | No | NA | 3 | 0 | 0 | Low-quality | Genome-fragment | 7 | AAI-based (medium-confidence) | 0 |
| S12-final-viral-combined_k141_244257\|\|full | 1863 | No | NA | 3 | 0 | 0 | Not-determined | Genome-fragment | NA | NA | 0 |
| S12-final-viral-combined_k141_210942\|\|full | 18527 | No | NA | 28 | 2 | 1 | High-quality | High-quality | 100 | AAI-based (high-confidence) | 0 |
| S11-final-viral-combined_k141_112515\|\|full | 2895 | No | NA | 5 | 4 | 0 | Low-quality | Genome-fragment | 6.28 | AAI-based (high-confidence) | 0 |
| S2-final-viral-combined_k141_247413\|\|full | 2895 | No | NA | 3 | 0 | 3 | Low-quality | Genome-fragment | 1.84 | AAI-based (medium-confidence) | 0 |
| S12-final-viral-combined_k141_328499\|\|full | 1171 | No | NA | 3 | 0 | 2 | Not-determined | Genome-fragment | NA | NA | 0 |
| S12-final-viral-combined_k141_344806\|\|full | 1416 | No | NA | 3 | 0 | 0 | Not-determined | Genome-fragment | NA | NA | 0 |
| S4-final-viral-combined_k141_107369\|\|full | 2895 | No | NA | 2 | 0 | 2 | Low-quality | Genome-fragment | 0.92 | AAI-based (medium-confidence) | 0 |
| S1_k141_118737 | 2896 | Yes | 2192 | 4 | 1 | 1 | Low-quality | Genome-fragment | 2.58 | AAI-based (high-confidence) | 24.31 |
| S3_k141_76027 | 2896 | No | NA | 3 | 1 | 0 | Low-quality | Genome-fragment | 1.33 | AAI-based (high-confidence) | 0 |
| S11-final-viral-combined_k141_244481\|\|full | 2897 | No | NA | 8 | 1 | 0 | Low-quality | Genome-fragment | 6.56 | AAI-based (high-confidence) | 0 |
| S17-final-viral-combined_k141_177502\|\|full | 2897 | No | NA | 4 | 0 | 0 | Low-quality | Genome-fragment | 4.57 | AAI-based (medium-confidence) | 0 |
| S12-final-viral-combined_k141_9787\|\|full | 2037 | No | NA | 3 | 0 | 3 | Not-determined | Genome-fragment | NA | NA | 0 |
| S17-final-viral-combined_k141_254911\|\|full | 2897 | Yes | 1127 | 6 | 1 | 1 | Low-quality | Genome-fragment | 2.63 | AAI-based (high-confidence) | 61.1 |
| S12-final-viral-combined_k141_278391\|\|full | 2325 | No | NA | 2 | 0 | 2 | Not-determined | Genome-fragment | NA | NA | 0 |
| S12-final-viral-combined_k141_278392\|\|full | 1351 | No | NA | 2 | 0 | 2 | Not-determined | Genome-fragment | NA | NA | 0 |
| S12-final-viral-combined_k141_389187\|\|full | 2898 | No | NA | 5 | 2 | 0 | Low-quality | Genome-fragment | 5.03 | HMM-based (lower-bound) | 0 |
| S12-final-viral-combined_k141_27039\|\|full | 2367 | No | NA | 4 | 0 | 2 | Not-determined | Genome-fragment | NA | NA | 0 |
| S12-final-viral-combined_k141_395277\|\|full | 1035 | No | NA | 3 | 0 | 0 | Not-determined | Genome-fragment | NA | NA | 0 |
| S12-final-viral-combined_k141_211005\|\|full | 4551 | No | NA | 9 | 0 | 2 | Not-determined | Genome-fragment | NA | NA | 0 |
| S12-final-viral-combined_k141_195050\|\|full | 3329 | No | NA | 3 | 0 | 3 | Not-determined | Genome-fragment | NA | NA | 0 |
| S12-final-viral-combined_k141_395333\|\|full | 1071 | No | NA | 3 | 0 | 1 | Not-determined | Genome-fragment | NA | NA | 0 |
| S12-final-viral-combined_k141_195094\|\|full | 2167 | No | NA | 3 | 0 | 1 | Not-determined | Genome-fragment | NA | NA | 0 |
| S12-final-viral-combined_k141_328554\|\|full | 43415 | Yes | 24021 | 48 | 5 | 5 | Medium-quality | Genome-fragment | 50.97 | AAI-based (high-confidence) | 44.67 |
| S11-final-viral-combined_k141_216113\|\|full | 2899 | No | NA | 2 | 0 | 0 | Low-quality | Genome-fragment | 0.96 | AAI-based (medium-confidence) | 0 |
| S12-final-viral-combined_k141_126272\|\|full | 1239 | No | NA | 2 | 0 | 0 | Not-determined | Genome-fragment | NA | NA | 0 |
| S7-final-viral-combined_k141_110426\|\|full | 2899 | No | NA | 10 | 2 | 0 | Low-quality | Genome-fragment | 7.4 | AAI-based (high-confidence) | 0 |
| S12-final-viral-combined_k141_328566\|\|full | 2329 | No | NA | 3 | 0 | 0 | Not-determined | Genome-fragment | NA | NA | 0 |
| S12-final-viral-combined_k141_93479\|\|full | 7045 | No | NA | 5 | 0 | 4 | Not-determined | Genome-fragment | NA | NA | 0 |
| S11-final-viral-combined_k141_112228\|\|full | 2900 | No | NA | 3 | 0 | 0 | Low-quality | Genome-fragment | 5.64 | AAI-based (medium-confidence) | 0 |
| S12-final-viral-combined_k141_228454\|\|full | 2816 | No | NA | 4 | 0 | 0 | Not-determined | Genome-fragment | NA | NA | 0 |
| S1-final-viral-combined_k141_55278\|\|full | 2900 | No | NA | 5 | 0 | 1 | Low-quality | Genome-fragment | 6.15 | AAI-based (medium-confidence) | 0 |
| S6-final-viral-combined_k141_42088\|\|full | 2900 | No | NA | 4 | 0 | 1 | Low-quality | Genome-fragment | 3.77 | AAI-based (medium-confidence) | 0 |
| S12-final-viral-combined_k141_77922\|\|full | 1979 | No | NA | 3 | 0 | 2 | Not-determined | Genome-fragment | NA | NA | 0 |
| S12-final-viral-combined_k141_378746\|\|full | 1194 | No | NA | 2 | 0 | 1 | Not-determined | Genome-fragment | NA | NA | 0 |
| S12-final-viral-combined_k141_44453\|\|full | 3934 | No | NA | 4 | 0 | 2 | Not-determined | Genome-fragment | NA | NA | 0 |
| S8-final-viral-combined_k141_130462\|\|full | 2900 | No | NA | 5 | 0 | 0 | Low-quality | Genome-fragment | 1.47 | AAI-based (medium-confidence) | 0 |
| S5-final-viral-combined_k141_28347\|\|full | 2901 | No | NA | 4 | 0 | 0 | Low-quality | Genome-fragment | 7.06 | AAI-based (high-confidence) | 0 |
| S12-final-viral-combined_k141_77942\|\|full | 1944 | No | NA | 3 | 0 | 1 | Not-determined | Genome-fragment | NA | NA | 0 |
| S12-final-viral-combined_k141_177525\|\|full | 2720 | No | NA | 3 | 0 | 1 | Not-determined | Genome-fragment | NA | NA | 0 |
| S12-final-viral-combined_k141_312860\|\|full | 1697 | No | NA | 2 | 0 | 1 | Not-determined | Genome-fragment | NA | NA | 0 |
| S12-final-viral-combined_k141_93534\|\|full | 2266 | No | NA | 2 | 0 | 1 | Not-determined | Genome-fragment | NA | NA | 0 |
| S12-final-viral-combined_k141_60358\|\|full | 6460 | No | NA | 4 | 0 | 2 | Not-determined | Genome-fragment | NA | NA | 0 |
| S12-final-viral-combined_k141_355014\|\|full | 2901 | No | NA | 5 | 0 | 0 | Low-quality | Genome-fragment | 5.6 | AAI-based (high-confidence) | 0 |
| S4-final-viral-combined_k141_4595\|\|full | 2901 | No | NA | 3 | 0 | 1 | Low-quality | Genome-fragment | 1.07 | AAI-based (medium-confidence) | 0 |
| S12-final-viral-combined_k141_161180\|\|full | 2800 | No | NA | 3 | 0 | 0 | Not-determined | Genome-fragment | NA | NA | 0 |
| S4_k141_6639 | 2901 | No | NA | 4 | 0 | 2 | Low-quality | Genome-fragment | 1.43 | AAI-based (medium-confidence) | 0 |
| S12-final-viral-combined_k141_77990\|\|full | 1163 | No | NA | 3 | 0 | 0 | Not-determined | Genome-fragment | NA | NA | 0 |
| S9_k141_85704 | 2902 | No | NA | 1 | 0 | 1 | Low-quality | Genome-fragment | 1.56 | AAI-based (medium-confidence) | 0 |
| S12-final-viral-combined_k141_161194\|\|full | 1007 | No | NA | 2 | 0 | 0 | Not-determined | Genome-fragment | NA | NA | 0 |
| S12-final-viral-combined_k141_195200\|\|full | 5017 | No | NA | 2 | 0 | 2 | Not-determined | Genome-fragment | NA | NA | 0 |
| S11-final-viral-combined_k141_136747\|\|full | 2902 | No | NA | 5 | 3 | 0 | Low-quality | Genome-fragment | 5.51 | AAI-based (high-confidence) | 0 |
| S12-final-viral-combined_k141_195209\|\|full | 1848 | No | NA | 4 | 0 | 0 | Not-determined | Genome-fragment | NA | NA | 0 |
| S12-final-viral-combined_k141_244463\|\|full | 1522 | No | NA | 3 | 0 | 0 | Not-determined | Genome-fragment | NA | NA | 0 |
| S5-final-viral-combined_k141_125347\|\|full | 2902 | No | NA | 4 | 0 | 1 | Low-quality | Genome-fragment | 4.76 | AAI-based (high-confidence) | 0 |
| S1-final-viral-combined_k141_85645\|\|full | 2903 | No | NA | 4 | 0 | 0 | Low-quality | Genome-fragment | 5.6 | AAI-based (medium-confidence) | 0 |
| S12-final-viral-combined_k141_9971\|\|full | 10461 | No | NA | 8 | 0 | 5 | Not-determined | Genome-fragment | NA | NA | 0 |
| S17-final-viral-combined_k141_233803\|\|full | 2903 | No | NA | 6 | 0 | 2 | Low-quality | Genome-fragment | 5.74 | AAI-based (high-confidence) | 0 |
| S12-final-viral-combined_k141_211176\|\|full | 2732 | No | NA | 4 | 0 | 3 | Not-determined | Genome-fragment | NA | NA | 0 |
| S17-final-viral-combined_k141_156553\|\|full | 2904 | No | NA | 6 | 1 | 0 | Low-quality | Genome-fragment | 1.3 | HMM-based (lower-bound) | 0 |
| S9-final-viral-combined_k141_53378\|\|full | 2905 | No | NA | 4 | 0 | 0 | Low-quality | Genome-fragment | 5.31 | AAI-based (high-confidence) | 0 |
| S12-final-viral-combined_k141_27161\|\|full | 13742 | No | NA | 15 | 0 | 2 | Not-determined | Genome-fragment | NA | NA | 0 |
| S12-final-viral-combined_k141_278630\|\|full | 2044 | No | NA | 3 | 0 | 1 | Not-determined | Genome-fragment | NA | NA | 0 |
| S1-final-viral-combined_k141_379453\|\|full | 2905 | No | NA | 4 | 0 | 4 | Low-quality | Genome-fragment | 0.7 | AAI-based (medium-confidence) | 0 |
| S12-final-viral-combined_k141_110283\|\|full | 2329 | No | NA | 4 | 0 | 0 | Not-determined | Genome-fragment | NA | NA | 0 |
| S12-final-viral-combined_k141_360282\|\|full | 6545 | No | NA | 4 | 0 | 3 | Not-determined | Genome-fragment | NA | NA | 0 |
| S12-final-viral-combined_k141_261048\|\|full | 5846 | No | NA | 4 | 0 | 2 | Not-determined | Genome-fragment | NA | NA | 0 |
| S12-final-viral-combined_k141_281225\|\|full | 2905 | No | NA | 3 | 0 | 1 | Low-quality | Genome-fragment | 5.54 | AAI-based (high-confidence) | 0 |
| S12-final-viral-combined_k141_167325\|\|full | 2905 | No | NA | 3 | 1 | 0 | Low-quality | Genome-fragment | 3.08 | AAI-based (medium-confidence) | 0 |
| S12-final-viral-combined_k141_328599\|\|full | 8287 | No | NA | 7 | 0 | 2 | Not-determined | Genome-fragment | NA | NA | 0 |
| S12-final-viral-combined_k141_195279\|\|full | 5990 | No | NA | 6 | 0 | 5 | Not-determined | Genome-fragment | NA | NA | 0 |
| S1-final-viral-combined_k141_291361\|\|full | 2906 | No | NA | 4 | 0 | 2 | Low-quality | Genome-fragment | 7.59 | AAI-based (medium-confidence) | 0 |
| S5-final-viral-combined_k141_146992\|\|full | 2906 | No | NA | 4 | 3 | 0 | Low-quality | Genome-fragment | 7.93 | AAI-based (high-confidence) | 0 |
| S12-final-viral-combined_k141_157802\|\|full | 2906 | No | NA | 2 | 0 | 2 | Low-quality | Genome-fragment | 0.93 | AAI-based (medium-confidence) | 0 |
| S12-final-viral-combined_k141_261121\|\|full | 1026 | No | NA | 4 | 0 | 1 | Not-determined | Genome-fragment | NA | NA | 0 |
| S12-final-viral-combined_k141_295543\|\|full | 1354 | No | NA | 2 | 0 | 0 | Not-determined | Genome-fragment | NA | NA | 0 |
| S1-final-viral-combined_k141_86\|\|full | 2907 | No | NA | 3 | 0 | 2 | Low-quality | Genome-fragment | 1.03 | AAI-based (high-confidence) | 0 |
| S1_k141_8648 | 2907 | No | NA | 7 | 1 | 0 | Low-quality | Genome-fragment | 4.43 | AAI-based (high-confidence) | 0 |
| S12-final-viral-combined_k141_328678\|\|full | 2011 | No | NA | 3 | 0 | 2 | Not-determined | Genome-fragment | NA | NA | 0 |
| S2-final-viral-combined_k141_1309\|\|full | 2907 | No | NA | 4 | 2 | 0 | Low-quality | Genome-fragment | 8.67 | AAI-based (high-confidence) | 0 |
| S12-final-viral-combined_k141_78098\|\|full | 9323 | No | NA | 11 | 0 | 4 | Not-determined | Genome-fragment | NA | NA | 0 |
| S17-final-viral-combined_k141_145733\|\|full | 2908 | No | NA | 8 | 0 | 0 | Low-quality | Genome-fragment | 2.34 | AAI-based (medium-confidence) | 0 |
| S1_k141_82837 | 2909 | No | NA | 3 | 1 | 0 | Low-quality | Genome-fragment | 7.3 | AAI-based (high-confidence) | 0 |
| S12-final-viral-combined_k141_345119\|\|full | 1321 | No | NA | 2 | 0 | 0 | Not-determined | Genome-fragment | NA | NA | 0 |
| S1-final-viral-combined_k141_75696\|\|full | 2910 | No | NA | 4 | 3 | 0 | Low-quality | Genome-fragment | 5.44 | AAI-based (high-confidence) | 0 |
| S12-final-viral-combined_k141_139337\|\|full | 2910 | No | NA | 5 | 3 | 0 | Low-quality | Genome-fragment | 1.41 | HMM-based (lower-bound) | 0 |
| S12-final-viral-combined_k141_144221\|\|full | 2499 | No | NA | 4 | 0 | 3 | Not-determined | Genome-fragment | NA | NA | 0 |
| S12-final-viral-combined_k141_93714\|\|full | 3913 | No | NA | 3 | 0 | 1 | Not-determined | Genome-fragment | NA | NA | 0 |
| S12-final-viral-combined_k141_261211\|\|full | 1957 | No | NA | 2 | 0 | 1 | Not-determined | Genome-fragment | NA | NA | 0 |
| S12_k141_359256 | 2910 | No | NA | 4 | 0 | 0 | Low-quality | Genome-fragment | 6.64 | AAI-based (high-confidence) | 0 |
| S12-final-viral-combined_k141_78165\|\|full | 1566 | No | NA | 4 | 0 | 1 | Not-determined | Genome-fragment | NA | NA | 0 |
| S12-final-viral-combined_k141_244637\|\|full | 1620 | No | NA | 3 | 0 | 1 | Not-determined | Genome-fragment | NA | NA | 0 |
| S12-final-viral-combined_k141_10077\|\|full | 4210 | No | NA | 3 | 0 | 2 | Not-determined | Genome-fragment | NA | NA | 0 |
| S5-final-viral-combined_k141_166331\|\|full | 2911 | No | NA | 3 | 0 | 2 | Low-quality | Genome-fragment | 3.6 | AAI-based (medium-confidence) | 0 |
| S12-final-viral-combined_k141_194391\|\|full | 2911 | Yes | 2575 | 4 | 2 | 1 | Low-quality | Genome-fragment | 6.16 | AAI-based (high-confidence) | 11.54 |
| S11_k141_206429 | 2912 | No | NA | 2 | 0 | 2 | Low-quality | Genome-fragment | 0.44 | AAI-based (medium-confidence) | 0 |
| S12-final-viral-combined_k141_278868\|\|full | 2000 | No | NA | 2 | 0 | 0 | Not-determined | Genome-fragment | NA | NA | 0 |
| S12-final-viral-combined_k141_313039\|\|full | 2281 | No | NA | 3 | 0 | 2 | Not-determined | Genome-fragment | NA | NA | 0 |
| S12-final-viral-combined_k141_360460\|\|full | 3090 | No | NA | 2 | 0 | 2 | Not-determined | Genome-fragment | NA | NA | 0 |
| S1-final-viral-combined_k141_8796\|\|full | 2912 | No | NA | 4 | 0 | 2 | Low-quality | Genome-fragment | 2.09 | AAI-based (medium-confidence) | 0 |
| S12-final-viral-combined_k141_27374\|\|full | 8831 | No | NA | 11 | 0 | 2 | Not-determined | Genome-fragment | NA | NA | 0 |
| S12-final-viral-combined_k141_395579\|\|full | 1693 | No | NA | 2 | 0 | 0 | Not-determined | Genome-fragment | NA | NA | 0 |
| S12-final-viral-combined_k141_27392\|\|full | 1318 | No | NA | 3 | 0 | 0 | Not-determined | Genome-fragment | NA | NA | 0 |
| S2-final-viral-combined_k141_47115\|\|full | 2912 | No | NA | 3 | 3 | 0 | Low-quality | Genome-fragment | 5.43 | AAI-based (high-confidence) | 0 |
| S4-final-viral-combined_k141_6344\|\|full | 2912 | No | NA | 3 | 0 | 1 | Low-quality | Genome-fragment | 7.11 | AAI-based (high-confidence) | 0 |
| S12-final-viral-combined_k141_211464\|\|full | 1693 | No | NA | 4 | 0 | 0 | Not-determined | Genome-fragment | NA | NA | 0 |
| S12-final-viral-combined_k141_379063\|\|full | 1120 | No | NA | 2 | 0 | 0 | Not-determined | Genome-fragment | NA | NA | 0 |
| S4_k141_48165 | 2913 | No | NA | 2 | 0 | 0 | Low-quality | Genome-fragment | 3.08 | AAI-based (high-confidence) | 0 |
| S12-final-viral-combined_k141_161487\|\|full | 3929 | No | NA | 5 | 0 | 2 | Not-determined | Genome-fragment | NA | NA | 0 |
| S12-final-viral-combined_k141_60572\|\|full | 9401 | No | NA | 9 | 0 | 7 | Not-determined | Genome-fragment | NA | NA | 0 |
| S12-final-viral-combined_k141_27430\|\|full | 1586 | No | NA | 3 | 0 | 1 | Not-determined | Genome-fragment | NA | NA | 0 |
| S9-final-viral-combined_k141_98184\|\|full | 2914 | No | NA | 4 | 0 | 2 | Low-quality | Genome-fragment | 4.02 | AAI-based (medium-confidence) | 0 |
| S12-final-viral-combined_k141_10137\|\|full | 2756 | No | NA | 3 | 0 | 1 | Not-determined | Genome-fragment | NA | NA | 0 |
| S12-final-viral-combined_k141_44745\|\|full | 2212 | No | NA | 3 | 0 | 2 | Not-determined | Genome-fragment | NA | NA | 0 |
| S17-final-viral-combined_k141_74072\|\|full | 2914 | No | NA | 4 | 0 | 2 | Low-quality | Genome-fragment | 1.27 | AAI-based (medium-confidence) | 0 |
| S4-final-viral-combined_k141_97566\|\|full | 2915 | No | NA | 4 | 0 | 1 | Low-quality | Genome-fragment | 4.59 | AAI-based (medium-confidence) | 0 |
| S12-final-viral-combined_k141_144247\|\|full | 1402 | No | NA | 4 | 0 | 2 | Not-determined | Genome-fragment | NA | NA | 0 |
| S6-final-viral-combined_k141_6446\|\|full | 2915 | No | NA | 5 | 0 | 1 | Low-quality | Genome-fragment | 4.2 | AAI-based (medium-confidence) | 0 |
| S5-final-viral-combined_k141_3899\|\|full | 2916 | No | NA | 6 | 1 | 0 | Low-quality | Genome-fragment | 5.01 | AAI-based (medium-confidence) | 0 |
| S12-final-viral-combined_k141_228802\|\|full | 1955 | No | NA | 4 | 0 | 1 | Not-determined | Genome-fragment | NA | NA | 0 |
| S12-final-viral-combined_k141_244786\|\|full | 1198 | No | NA | 2 | 0 | 2 | Not-determined | Genome-fragment | NA | NA | 0 |
| S8-final-viral-combined_k141_1749\|\|full | 2916 | No | NA | 6 | 0 | 1 | Low-quality | Genome-fragment | 7.72 | AAI-based (medium-confidence) | 0 |
| S7-final-viral-combined_k141_226368\|\|full | 2917 | No | NA | 6 | 0 | 0 | Low-quality | Genome-fragment | 4.2 | AAI-based (medium-confidence) | 0 |
| S12-final-viral-combined_k141_335099\|\|0_partial | 2917 | Yes | 1426 | 4 | 1 | 1 | Low-quality | Genome-fragment | 34.27 | HMM-based (lower-bound) | 51.11 |
| S12-final-viral-combined_k141_328858\|\|full | 1549 | No | NA | 2 | 0 | 1 | Not-determined | Genome-fragment | NA | NA | 0 |
| S12-final-viral-combined_k141_144286\|\|full | 2650 | No | NA | 3 | 0 | 3 | Not-determined | Genome-fragment | NA | NA | 0 |
| S12-final-viral-combined_k141_177881\|\|full | 1104 | No | NA | 3 | 0 | 0 | Not-determined | Genome-fragment | NA | NA | 0 |
| S1-final-viral-combined_k141_167149\|\|full | 2918 | No | NA | 2 | 0 | 2 | Low-quality | Genome-fragment | 4.98 | AAI-based (high-confidence) | 0 |
| S12-final-viral-combined_k141_144293\|\|full | 4186 | No | NA | 7 | 0 | 2 | Not-determined | Genome-fragment | NA | NA | 0 |
| S12-final-viral-combined_k141_78344\|\|full | 2835 | No | NA | 5 | 0 | 2 | Not-determined | Genome-fragment | NA | NA | 0 |
| S12-final-viral-combined_k141_328884\|\|full | 1013 | No | NA | 2 | 0 | 2 | Not-determined | Genome-fragment | NA | NA | 0 |
| S12-final-viral-combined_k141_379169\|\|full | 1386 | No | NA | 3 | 0 | 1 | Not-determined | Genome-fragment | NA | NA | 0 |
| S12-final-viral-combined_k141_10194\|\|full | 2098 | No | NA | 3 | 0 | 1 | Not-determined | Genome-fragment | NA | NA | 0 |
| S1-final-viral-combined_k141_391112\|\|full | 2918 | No | NA | 3 | 0 | 2 | Low-quality | Genome-fragment | 0.7 | AAI-based (medium-confidence) | 0 |
| S5-final-viral-combined_k141_211462\|\|full | 2918 | No | NA | 4 | 0 | 3 | Low-quality | Genome-fragment | 3.29 | AAI-based (medium-confidence) | 0 |
| S12-final-viral-combined_k141_244844\|\|full | 1468 | No | NA | 3 | 0 | 0 | Not-determined | Genome-fragment | NA | NA | 0 |
| S12-final-viral-combined_k141_244855\|\|full | 3438 | No | NA | 3 | 0 | 0 | Not-determined | Genome-fragment | NA | NA | 0 |
| S12-final-viral-combined_k141_93884\|\|full | 2209 | No | NA | 3 | 0 | 2 | Not-determined | Genome-fragment | NA | NA | 0 |
| S12-final-viral-combined_k141_27573\|\|full | 1324 | No | NA | 3 | 0 | 0 | Not-determined | Genome-fragment | NA | NA | 0 |
| S12-final-viral-combined_k141_211613\|\|full | 8560 | No | NA | 11 | 0 | 0 | Not-determined | Genome-fragment | NA | NA | 0 |
| S17-final-viral-combined_k141_259398\|\|full | 2919 | No | NA | 6 | 0 | 0 | Low-quality | Genome-fragment | 7.91 | AAI-based (high-confidence) | 0 |
| S12-final-viral-combined_k141_78431\|\|full | 1032 | No | NA | 3 | 0 | 0 | Not-determined | Genome-fragment | NA | NA | 0 |
| S8_k141_45772 | 2919 | No | NA | 4 | 1 | 0 | Low-quality | Genome-fragment | 3.89 | AAI-based (high-confidence) | 0 |
| S17-final-viral-combined_k141_65836\|\|full | 2920 | No | NA | 5 | 0 | 1 | Low-quality | Genome-fragment | 5.55 | AAI-based (high-confidence) | 0 |
| S2-final-viral-combined_k141_195284\|\|full | 2920 | No | NA | 5 | 1 | 0 | Low-quality | Genome-fragment | 5.21 | AAI-based (high-confidence) | 0 |
| S7-final-viral-combined_k141_30649\|\|full | 2921 | No | NA | 7 | 1 | 0 | Low-quality | Genome-fragment | 6.42 | AAI-based (high-confidence) | 0 |
| S12-final-viral-combined_k141_93939\|\|full | 3777 | No | NA | 7 | 0 | 0 | Not-determined | Genome-fragment | NA | NA | 0 |
| S12-final-viral-combined_k141_395714\|\|full | 1020 | No | NA | 3 | 0 | 0 | Not-determined | Genome-fragment | NA | NA | 0 |
| S8-final-viral-combined_k141_230095\|\|full | 2921 | No | NA | 3 | 1 | 0 | Low-quality | Genome-fragment | 3.68 | HMM-based (lower-bound) | 0 |
| S5-final-viral-combined_k141_135028\|\|full | 2922 | No | NA | 4 | 0 | 0 | Low-quality | Genome-fragment | 7.46 | AAI-based (high-confidence) | 0 |
| S12-final-viral-combined_k141_44844\|\|full | 2003 | No | NA | 4 | 0 | 0 | Not-determined | Genome-fragment | NA | NA | 0 |
| S2-final-viral-combined_k141_218932\|\|full | 2922 | Yes | 2203 | 4 | 1 | 1 | Low-quality | Genome-fragment | 1.48 | HMM-based (lower-bound) | 24.61 |
| S12-final-viral-combined_k141_211679\|\|full | 4202 | No | NA | 7 | 0 | 5 | Not-determined | Genome-fragment | NA | NA | 0 |
| S12-final-viral-combined_k141_211685\|\|full | 1006 | No | NA | 2 | 0 | 0 | Not-determined | Genome-fragment | NA | NA | 0 |
| S9-final-viral-combined_k141_64007\|\|full | 2923 | No | NA | 7 | 3 | 0 | Low-quality | Genome-fragment | 6.51 | AAI-based (high-confidence) | 0 |
| S6-final-viral-combined_k141_29467\|\|0_partial | 2923 | Yes | 1423 | 3 | 1 | 1 | Low-quality | Genome-fragment | 6.37 | AAI-based (medium-confidence) | 51.32 |
| S11-final-viral-combined_k141_273577\|\|full | 2924 | Yes | 2362 | 5 | 1 | 1 | Low-quality | Genome-fragment | 1.5 | HMM-based (lower-bound) | 19.22 |
| S12-final-viral-combined_k141_126751\|\|full | 1220 | No | NA | 3 | 0 | 2 | Not-determined | Genome-fragment | NA | NA | 0 |
| S12-final-viral-combined_k141_345467\|\|full | 1976 | No | NA | 4 | 0 | 1 | Not-determined | Genome-fragment | NA | NA | 0 |
| S9-final-viral-combined_k141_97169\|\|full | 2925 | No | NA | 4 | 0 | 4 | Low-quality | Genome-fragment | 0.9 | AAI-based (medium-confidence) | 0 |
| S12-final-viral-combined_k141_126794\|\|full | 1558 | No | NA | 3 | 0 | 0 | Not-determined | Genome-fragment | NA | NA | 0 |
| S12_k141_206981 | 2925 | No | NA | 5 | 1 | 0 | Low-quality | Genome-fragment | 5.01 | AAI-based (high-confidence) | 0 |
| S12-final-viral-combined_k141_153689\|\|full | 2926 | No | NA | 6 | 2 | 0 | Low-quality | Genome-fragment | 5.11 | AAI-based (medium-confidence) | 0 |
| S12-final-viral-combined_k141_295863\|\|full | 8616 | No | NA | 3 | 0 | 1 | Not-determined | Genome-fragment | NA | NA | 0 |
| S2-final-viral-combined_k141_127550\|\|lt2gene | 2926 | No | NA | 1 | 1 | 0 | Low-quality | Genome-fragment | 6.13 | AAI-based (high-confidence) | 0 |
| S5-final-viral-combined_k141_68222\|\|full | 2927 | No | NA | 6 | 1 | 0 | Low-quality | Genome-fragment | 2.09 | HMM-based (lower-bound) | 0 |
| S12-final-viral-combined_k141_379406\|\|full | 2366 | No | NA | 4 | 0 | 2 | Not-determined | Genome-fragment | NA | NA | 0 |
| S1-final-viral-combined_k141_240048\|\|full | 2928 | No | NA | 4 | 3 | 0 | Low-quality | Genome-fragment | 8.41 | AAI-based (high-confidence) | 0 |
| S12-final-viral-combined_k141_78559\|\|full | 6177 | No | NA | 7 | 0 | 4 | Not-determined | Genome-fragment | NA | NA | 0 |
| S1_k141_209530 | 2928 | No | NA | 2 | 0 | 2 | Low-quality | Genome-fragment | 0.46 | AAI-based (medium-confidence) | 0 |
| S12-final-viral-combined_k141_360733\|\|full | 2644 | No | NA | 3 | 0 | 3 | Not-determined | Genome-fragment | NA | NA | 0 |
| S12-final-viral-combined_k141_10359\|\|full | 3510 | No | NA | 4 | 0 | 2 | Not-determined | Genome-fragment | NA | NA | 0 |
| S12-final-viral-combined_k141_211788\|\|full | 12974 | No | NA | 15 | 0 | 9 | Not-determined | Genome-fragment | NA | NA | 0 |
| S12-final-viral-combined_k141_27708\|\|full | 1348 | No | NA | 2 | 0 | 0 | Not-determined | Genome-fragment | NA | NA | 0 |

| | | | | | | | | | | | |
|---|---|---|---|---|---|---|---|---|---|---|---|
| S7-final-viral-combined_k141_68829\|\|full | 2928 | No | NA | 5 | 0 | 1 | Low-quality | Genome-fragment | 3.82 | AAI-based (medium-confidence) | 0 |
| S12-final-viral-combined_k141_377108\|\|full | 2928 | No | NA | 4 | 0 | 0 | Low-quality | Genome-fragment | 6.92 | AAI-based (high-confidence) | 0 |
| S1-final-viral-combined_k141_391551\|\|full | 2929 | No | NA | 5 | 0 | 0 | Low-quality | Genome-fragment | 5.6 | AAI-based (medium-confidence) | 0 |
| S7-final-viral-combined_k141_8801\|\|full | 2929 | No | NA | 6 | 0 | 0 | Low-quality | Genome-fragment | 5.34 | AAI-based (medium-confidence) | 0 |
| S12-final-viral-combined_k141_178007\|\|full | 1003 | No | NA | 2 | 0 | 0 | Not-determined | Genome-fragment | NA | NA | 0 |
| S12-final-viral-combined_k141_229152\|\|full | 1520 | No | NA | 2 | 0 | 1 | Not-determined | Genome-fragment | NA | NA | 0 |
| S12-final-viral-combined_k141_10395\|\|full | 1396 | No | NA | 3 | 0 | 0 | Not-determined | Genome-fragment | NA | NA | 0 |
| S11-final-viral-combined_k141_143621\|\|full | 2930 | No | NA | 5 | 0 | 3 | Low-quality | Genome-fragment | 1.42 | AAI-based (medium-confidence) | 0 |
| S12-final-viral-combined_k141_126912\|\|full | 2040 | No | NA | 3 | 0 | 1 | Not-determined | Genome-fragment | NA | NA | 0 |
| S12-final-viral-combined_k141_295914\|\|full | 1329 | No | NA | 3 | 0 | 2 | Not-determined | Genome-fragment | NA | NA | 0 |
| S7-final-viral-combined_k141_191727\|\|full | 2931 | No | NA | 3 | 0 | 0 | Low-quality | Genome-fragment | 5.33 | AAI-based (medium-confidence) | 0 |
| S12-final-viral-combined_k141_279262\|\|full | 1892 | No | NA | 3 | 0 | 2 | Not-determined | Genome-fragment | NA | NA | 0 |
| S12-final-viral-combined_k141_261587\|\|full | 6696 | No | NA | 10 | 0 | 0 | Not-determined | Genome-fragment | NA | NA | 0 |
| S8-final-viral-combined_k141_134605\|\|full | 2931 | No | NA | 3 | 1 | 0 | Low-quality | Genome-fragment | 5.88 | AAI-based (high-confidence) | 0 |
| S17-final-viral-combined_k141_6698\|\|full | 2932 | No | NA | 3 | 0 | 1 | Low-quality | Genome-fragment | 1.84 | AAI-based (high-confidence) | 0 |
| S12-final-viral-combined_k141_395892\|\|full | 1119 | No | NA | 3 | 0 | 0 | Not-determined | Genome-fragment | NA | NA | 0 |
| S12-final-viral-combined_k141_261620\|\|full | 2319 | No | NA | 5 | 0 | 1 | Not-determined | Genome-fragment | NA | NA | 0 |
| S12-final-viral-combined_k141_45020\|\|full | 1754 | No | NA | 3 | 0 | 0 | Not-determined | Genome-fragment | NA | NA | 0 |
| S12-final-viral-combined_k141_295965\|\|full | 1528 | No | NA | 3 | 0 | 0 | Not-determined | Genome-fragment | NA | NA | 0 |
| S12-final-viral-combined_k141_313383\|\|full | 3814 | No | NA | 3 | 0 | 3 | Not-determined | Genome-fragment | NA | NA | 0 |
| S8-final-viral-combined_k141_58953\|\|full | 2932 | No | NA | 3 | 1 | 0 | Low-quality | Genome-fragment | 5.65 | HMM-based (lower-bound) | 0 |
| S12-final-viral-combined_k141_45042\|\|full | 3284 | No | NA | 5 | 0 | 2 | Not-determined | Genome-fragment | NA | NA | 0 |
| S2-final-viral-combined_k141_116825\|\|full | 2933 | No | NA | 3 | 0 | 2 | Low-quality | Genome-fragment | 5.61 | AAI-based (medium-confidence) | 0 |
| S2_k141_229433 | 2933 | No | NA | 3 | 0 | 1 | Low-quality | Genome-fragment | 2.08 | AAI-based (medium-confidence) | 0 |
| S12-final-viral-combined_k141_261688\|\|full | 1407 | No | NA | 1 | 0 | 0 | Not-determined | Genome-fragment | NA | NA | 0 |
| S8-final-viral-combined_k141_87389\|\|full | 2933 | No | NA | 7 | 1 | 0 | Low-quality | Genome-fragment | 5.14 | AAI-based (high-confidence) | 0 |
| S12-final-viral-combined_k141_60846\|\|full | 1875 | No | NA | 3 | 0 | 2 | Not-determined | Genome-fragment | NA | NA | 0 |
| S12-final-viral-combined_k141_161688\|\|full | 4079 | No | NA | 5 | 0 | 1 | Not-determined | Genome-fragment | NA | NA | 0 |
| S12-final-viral-combined_k141_10495\|\|full | 2871 | No | NA | 3 | 0 | 1 | Not-determined | Genome-fragment | NA | NA | 0 |
| S12-final-viral-combined_k141_329217\|\|full | 3829 | No | NA | 6 | 0 | 0 | Not-determined | Genome-fragment | NA | NA | 0 |
| S17-final-viral-combined_k141_131205\|\|full | 2934 | No | NA | 3 | 1 | 0 | Low-quality | Genome-fragment | 3.94 | HMM-based (lower-bound) | 0 |
| S4-final-viral-combined_k141_46327\|\|full | 2934 | No | NA | 5 | 0 | 0 | Low-quality | Genome-fragment | 4.33 | AAI-based (high-confidence) | 0 |
| S12-final-viral-combined_k141_229310\|\|full | 5999 | No | NA | 9 | 0 | 3 | Not-determined | Genome-fragment | NA | NA | 0 |
| S12-final-viral-combined_k141_78676\|\|full | 1120 | No | NA | 2 | 0 | 1 | Not-determined | Genome-fragment | NA | NA | 0 |
| S12-final-viral-combined_k141_345597\|\|full | 1407 | No | NA | 2 | 0 | 1 | Not-determined | Genome-fragment | NA | NA | 0 |
| S12-final-viral-combined_k141_396034\|\|full | 3595 | No | NA | 2 | 0 | 2 | Not-determined | Genome-fragment | NA | NA | 0 |
| S12-final-viral-combined_k141_78710\|\|full | 5170 | No | NA | 4 | 0 | 1 | Not-determined | Genome-fragment | NA | NA | 0 |
| S12-final-viral-combined_k141_245206\|\|full | 15190 | No | NA | 18 | 0 | 4 | Not-determined | Genome-fragment | NA | NA | 0 |
| S12-final-viral-combined_k141_360922\|\|full | 2517 | No | NA | 2 | 0 | 1 | Not-determined | Genome-fragment | NA | NA | 0 |
| S8-final-viral-combined_k141_151071\|\|full | 2934 | No | NA | 4 | 0 | 0 | Low-quality | Genome-fragment | 4.71 | AAI-based (medium-confidence) | 0 |
| S12-final-viral-combined_k141_329242\|\|full | 50647 | No | NA | 66 | 4 | 7 | High-quality | High-quality | 100 | AAI-based (high-confidence) | 0 |
| S12-final-viral-combined_k141_178134\|\|full | 1672 | No | NA | 3 | 0 | 3 | Not-determined | Genome-fragment | NA | NA | 0 |
| S12-final-viral-combined_k141_195784\|\|full | 1574 | No | NA | 3 | 0 | 0 | Not-determined | Genome-fragment | NA | NA | 0 |
| S8-final-viral-combined_k141_120889\|\|full | 2936 | No | NA | 3 | 2 | 0 | Low-quality | Genome-fragment | 5.18 | HMM-based (lower-bound) | 0 |
| S12-final-viral-combined_k141_60965\|\|full | 4134 | No | NA | 5 | 0 | 4 | Not-determined | Genome-fragment | NA | NA | 0 |
| S12-final-viral-combined_k141_110835\|\|full | 2065 | No | NA | 2 | 0 | 0 | Not-determined | Genome-fragment | NA | NA | 0 |
| S12-final-viral-combined_k141_195808\|\|full | 1537 | No | NA | 3 | 0 | 1 | Not-determined | Genome-fragment | NA | NA | 0 |
| S12-final-viral-combined_k141_313557\|\|full | 1175 | No | NA | 4 | 0 | 0 | Not-determined | Genome-fragment | NA | NA | 0 |
| S12-final-viral-combined_k141_345669\|\|full | 5382 | No | NA | 5 | 0 | 3 | Not-determined | Genome-fragment | NA | NA | 0 |
| S12-final-viral-combined_k141_45215\|\|full | 1059 | No | NA | 2 | 0 | 1 | Not-determined | Genome-fragment | NA | NA | 0 |
| S1-final-viral-combined_k141_120663\|\|full | 2937 | No | NA | 5 | 1 | 0 | Low-quality | Genome-fragment | 5.38 | AAI-based (medium-confidence) | 0 |
| S12-final-viral-combined_k141_127160\|\|full | 2466 | No | NA | 3 | 0 | 3 | Not-determined | Genome-fragment | NA | NA | 0 |
| S12-final-viral-combined_k141_261850\|\|full | 7370 | No | NA | 5 | 0 | 5 | Not-determined | Genome-fragment | NA | NA | 0 |
| S12-final-viral-combined_k141_61007\|\|full | 2342 | No | NA | 4 | 0 | 2 | Not-determined | Genome-fragment | NA | NA | 0 |
| S12-final-viral-combined_k141_379769\|\|full | 1532 | No | NA | 3 | 0 | 1 | Not-determined | Genome-fragment | NA | NA | 0 |
| S12-final-viral-combined_k141_127171\|\|full | 2452 | No | NA | 3 | 0 | 0 | Not-determined | Genome-fragment | NA | NA | 0 |
| S12-final-viral-combined_k141_379774\|\|full | 2321 | No | NA | 4 | 0 | 3 | Not-determined | Genome-fragment | NA | NA | 0 |
| S12-final-viral-combined_k141_212050\|\|full | 1617 | No | NA | 3 | 0 | 2 | Not-determined | Genome-fragment | NA | NA | 0 |
| S12-final-viral-combined_k141_127185\|\|full | 2263 | No | NA | 4 | 0 | 0 | Not-determined | Genome-fragment | NA | NA | 0 |
| S7-final-viral-combined_k141_8789\|\|full | 2937 | Yes | 561 | 4 | 1 | 1 | Low-quality | Genome-fragment | 0.34 | HMM-based (lower-bound) | 80.9 |
| S12-final-viral-combined_k141_127196\|\|full | 9025 | No | NA | 6 | 0 | 3 | Not-determined | Genome-fragment | NA | NA | 0 |
| S17-final-viral-combined_k141_91634\|\|full | 2937 | No | NA | 3 | 0 | 1 | Low-quality | Genome-fragment | 6.75 | AAI-based (medium-confidence) | 0 |
| S12-final-viral-combined_k141_195915\|\|full | 1366 | No | NA | 2 | 0 | 0 | Not-determined | Genome-fragment | NA | NA | 0 |
| S12-final-viral-combined_k141_78852\|\|full | 2612 | No | NA | 3 | 0 | 2 | Not-determined | Genome-fragment | NA | NA | 0 |
| S12-final-viral-combined_k141_127209\|\|full | 1622 | No | NA | 3 | 0 | 0 | Not-determined | Genome-fragment | NA | NA | 0 |
| S12-final-viral-combined_k141_195917\|\|full | 1679 | No | NA | 3 | 0 | 2 | Not-determined | Genome-fragment | NA | NA | 0 |
| S12-final-viral-combined_k141_229516\|\|full | 1913 | No | NA | 4 | 0 | 0 | Not-determined | Genome-fragment | NA | NA | 0 |
| S12-final-viral-combined_k141_279537\|\|full | 7284 | No | NA | 9 | 0 | 8 | Not-determined | Genome-fragment | NA | NA | 0 |
| S17-final-viral-combined_k141_61717\|\|full | 2937 | No | NA | 3 | 3 | 0 | Low-quality | Genome-fragment | 6.84 | AAI-based (medium-confidence) | 0 |
| S12-final-viral-combined_k141_61059\|\|full | 3882 | No | NA | 5 | 0 | 4 | Not-determined | Genome-fragment | NA | NA | 0 |
| S4-final-viral-combined_k141_136867\|\|full | 2937 | No | NA | 5 | 1 | 0 | Low-quality | Genome-fragment | 6.95 | AAI-based (medium-confidence) | 0 |
| S12-final-viral-combined_k141_27953\|\|full | 2054 | No | NA | 3 | 0 | 2 | Not-determined | Genome-fragment | NA | NA | 0 |
| S12-final-viral-combined_k141_229558\|\|full | 3929 | No | NA | 4 | 0 | 4 | Not-determined | Genome-fragment | NA | NA | 0 |
| S12-final-viral-combined_k141_78901\|\|full | 6434 | No | NA | 8 | 0 | 5 | Not-determined | Genome-fragment | NA | NA | 0 |
| S12-final-viral-combined_k141_379860\|\|full | 6970 | No | NA | 8 | 0 | 6 | Not-determined | Genome-fragment | NA | NA | 0 |
| S12-final-viral-combined_k141_178296\|\|full | 1620 | No | NA | 3 | 0 | 1 | Not-determined | Genome-fragment | NA | NA | 0 |
| S1-final-viral-combined_k141_13183\|\|full | 2938 | No | NA | 4 | 0 | 2 | Low-quality | Genome-fragment | 3.45 | AAI-based (high-confidence) | 0 |
| S12-final-viral-combined_k141_396215\|\|full | 2614 | No | NA | 4 | 0 | 1 | Not-determined | Genome-fragment | NA | NA | 0 |
| S12-final-viral-combined_k141_341582\|\|full | 2938 | No | NA | 6 | 0 | 1 | Low-quality | Genome-fragment | 8.61 | AAI-based (high-confidence) | 0 |
| S12-final-viral-combined_k141_45302\|\|full | 7017 | No | NA | 10 | 0 | 0 | Not-determined | Genome-fragment | NA | NA | 0 |
| S4-final-viral-combined_k141_69601\|\|full | 2938 | No | NA | 3 | 3 | 0 | Low-quality | Genome-fragment | 1.69 | AAI-based (high-confidence) | 0 |
| S12-final-viral-combined_k141_229611\|\|full | 1474 | No | NA | 3 | 0 | 0 | Not-determined | Genome-fragment | NA | NA | 0 |
| S12-final-viral-combined_k141_127332\|\|full | 1520 | No | NA | 3 | 0 | 1 | Not-determined | Genome-fragment | NA | NA | 0 |
| S12-final-viral-combined_k141_161957\|\|full | 4386 | No | NA | 3 | 0 | 2 | Not-determined | Genome-fragment | NA | NA | 0 |
| S2-final-viral-combined_k141_92333\|\|full | 2939 | No | NA | 6 | 1 | 0 | Low-quality | Genome-fragment | 8 | AAI-based (high-confidence) | 0 |
| S5-final-viral-combined_k141_6360\|\|full | 2940 | No | NA | 3 | 0 | 0 | Low-quality | Genome-fragment | 3.03 | AAI-based (high-confidence) | 0 |
| S12-final-viral-combined_k141_396269\|\|full | 1859 | No | NA | 3 | 0 | 0 | Not-determined | Genome-fragment | NA | NA | 0 |
| S12-final-viral-combined_k141_379916\|\|full | 3608 | No | NA | 5 | 0 | 1 | Not-determined | Genome-fragment | NA | NA | 0 |
| S12-final-viral-combined_k141_329359\|\|full | 2360 | No | NA | 2 | 0 | 2 | Not-determined | Genome-fragment | NA | NA | 0 |
| S12-final-viral-combined_k141_325137\|\|full | 2940 | No | NA | 3 | 2 | 0 | Low-quality | Genome-fragment | 7.36 | AAI-based (high-confidence) | 0 |
| S12-final-viral-combined_k141_313781\|\|full | 1178 | No | NA | 2 | 0 | 1 | Not-determined | Genome-fragment | NA | NA | 0 |
| S12-final-viral-combined_k141_162016\|\|full | 1240 | No | NA | 3 | 0 | 2 | Not-determined | Genome-fragment | NA | NA | 0 |
| S9-final-viral-combined_k141_100477\|\|full | 2941 | No | NA | 7 | 3 | 0 | Low-quality | Genome-fragment | 7.45 | AAI-based (medium-confidence) | 0 |
| S5-final-viral-combined_k141_9815\|\|full | 2941 | Yes | 502 | 6 | 1 | 4 | Low-quality | Genome-fragment | 0.87 | HMM-based (lower-bound) | 82.93 |
| S12-final-viral-combined_k141_78935\|\|full | 1715 | No | NA | 6 | 0 | 0 | Not-determined | Genome-fragment | NA | NA | 0 |
| S5-final-viral-combined_k141_172205\|\|lt2gene | 2941 | No | NA | 2 | 2 | 0 | Low-quality | Genome-fragment | 5.17 | AAI-based (medium-confidence) | 0 |
| S4-final-viral-combined_k141_11726\|\|full | 2941 | Yes | 1947 | 5 | 1 | 1 | Low-quality | Genome-fragment | 1.23 | HMM-based (lower-bound) | 33.8 |
| S12-final-viral-combined_k141_94412\|\|full | 1536 | No | NA | 4 | 0 | 0 | Not-determined | Genome-fragment | NA | NA | 0 |
| S7-final-viral-combined_k141_129496\|\|full | 2943 | No | NA | 3 | 1 | 0 | Low-quality | Genome-fragment | 1.92 | AAI-based (high-confidence) | 0 |
| S17-k141_22600 | 2943 | No | NA | 4 | 0 | 1 | Low-quality | Genome-fragment | 3.24 | AAI-based (medium-confidence) | 0 |
| S12-final-viral-combined_k141_144693\|\|full | 8448 | No | NA | 7 | 0 | 6 | Not-determined | Genome-fragment | NA | NA | 0 |
| S8-final-viral-combined_k141_236032\|\|full | 2943 | No | NA | 3 | 1 | 0 | Low-quality | Genome-fragment | 5.24 | HMM-based (lower-bound) | 0 |
| S12-final-viral-combined_k141_178429\|\|full | 2381 | No | NA | 3 | 0 | 2 | Not-determined | Genome-fragment | NA | NA | 0 |
| S12-final-viral-combined_k141_10790\|\|full | 2324 | No | NA | 3 | 0 | 1 | Not-determined | Genome-fragment | NA | NA | 0 |
| S8_k141_109011 | 2943 | No | NA | 6 | 0 | 0 | Low-quality | Genome-fragment | 5.34 | AAI-based (high-confidence) | 0 |
| S1-final-viral-combined_k141_117861\|\|full | 2944 | No | NA | 3 | 0 | 0 | Low-quality | Genome-fragment | 3.92 | AAI-based (medium-confidence) | 0 |
| S4_k141_128142 | 2944 | No | NA | 5 | 0 | 2 | Low-quality | Genome-fragment | 5.86 | AAI-based (medium-confidence) | 0 |
| S2_k141_234550 | 2945 | No | NA | 4 | 0 | 1 | Low-quality | Genome-fragment | 6.53 | AAI-based (high-confidence) | 0 |
| S11-final-viral-combined_k141_46779\|\|full | 2947 | No | NA | 3 | 0 | 0 | Low-quality | Genome-fragment | 2.1 | AAI-based (high-confidence) | 0 |
| S12-final-viral-combined_k141_212237\|\|full | 2397 | No | NA | 4 | 0 | 2 | Not-determined | Genome-fragment | NA | NA | 0 |
| S7_k141_104822 | 2947 | No | NA | 3 | 3 | 0 | Low-quality | Genome-fragment | 4.94 | AAI-based (high-confidence) | 0 |
| S17-final-viral-combined_k141_240176\|\|full | 2947 | No | NA | 4 | 0 | 0 | Low-quality | Genome-fragment | 3.58 | AAI-based (high-confidence) | 0 |
| S12-final-viral-combined_k141_61258\|\|full | 1457 | No | NA | 3 | 0 | 1 | Not-determined | Genome-fragment | NA | NA | 0 |
| S12-final-viral-combined_k141_229712\|\|full | 4026 | No | NA | 5 | 0 | 0 | Not-determined | Genome-fragment | NA | NA | 0 |
| S17-final-viral-combined_k141_86345\|\|full | 2947 | No | NA | 2 | 0 | 1 | Low-quality | Genome-fragment | 1.48 | AAI-based (medium-confidence) | 0 |
| S12-final-viral-combined_k141_261974\|\|full | 1985 | No | NA | 3 | 0 | 1 | Not-determined | Genome-fragment | NA | NA | 0 |
| S2_k141_241190 | 2947 | Yes | 997 | 3 | 1 | 1 | Low-quality | Genome-fragment | 0.61 | HMM-based (lower-bound) | 66.17 |
| S6-final-viral-combined_k141_46245\|\|full | 2947 | Yes | 1800 | 5 | 1 | 2 | Low-quality | Genome-fragment | 3.29 | AAI-based (medium-confidence) | 38.92 |
| S11-final-viral-combined_k141_96946\|\|full | 2948 | No | NA | 6 | 1 | 0 | Low-quality | Genome-fragment | 1.78 | HMM-based (lower-bound) | 0 |
| S12-final-viral-combined_k141_299965\|\|full | 2948 | No | NA | 7 | 1 | 0 | Low-quality | Genome-fragment | 7.22 | AAI-based (high-confidence) | 0 |
| S12-final-viral-combined_k141_229745\|\|full | 1154 | No | NA | 4 | 0 | 0 | Not-determined | Genome-fragment | NA | NA | 0 |
| S12-final-viral-combined_k141_396396\|\|full | 2699 | No | NA | 7 | 0 | 0 | Not-determined | Genome-fragment | NA | NA | 0 |
| S12-final-viral-combined_k141_229750\|\|full | 2190 | No | NA | 3 | 0 | 1 | Not-determined | Genome-fragment | NA | NA | 0 |
| S8-final-viral-combined_k141_63343\|\|full | 2948 | No | NA | 9 | 0 | 1 | Low-quality | Genome-fragment | 6.77 | AAI-based (medium-confidence) | 0 |
| S12-final-viral-combined_k141_345948\|\|full | 33538 | Yes | 28220 | 49 | 18 | 4 | Medium-quality | Genome-fragment | 81.24 | AAI-based (high-confidence) | 15.86 |
| S8-final-viral-combined_k141_258267\|\|full | 2949 | No | NA | 3 | 0 | 3 | Low-quality | Genome-fragment | 1.69 | AAI-based (medium-confidence) | 0 |
| S12-final-viral-combined_k141_162155\|\|full | 1490 | No | NA | 3 | 0 | 2 | Not-determined | Genome-fragment | NA | NA | 0 |
| S12-final-viral-combined_k141_10831\|\|full | 1138 | No | NA | 3 | 0 | 1 | Not-determined | Genome-fragment | NA | NA | 0 |
| S17-final-viral-combined_k141_77559\|\|full | 2950 | No | NA | 7 | 2 | 0 | Low-quality | Genome-fragment | 6.37 | AAI-based (medium-confidence) | 0 |
| S2_k141_189206 | 2950 | No | NA | 5 | 2 | 0 | Low-quality | Genome-fragment | 6.56 | AAI-based (medium-confidence) | 0 |
| S1-final-viral-combined_k141_100469\|\|full | 2951 | No | NA | 4 | 0 | 0 | Low-quality | Genome-fragment | 6.63 | AAI-based (medium-confidence) | 0 |
| S12-final-viral-combined_k141_94590\|\|full | 1462 | No | NA | 1 | 0 | 0 | Not-determined | Genome-fragment | NA | NA | 0 |
| S12-final-viral-combined_k141_10854\|\|full | 1886 | No | NA | 3 | 0 | 2 | Not-determined | Genome-fragment | NA | NA | 0 |
| S7-final-viral-combined_k141_209608\|\|full | 2951 | No | NA | 2 | 0 | 0 | Low-quality | Genome-fragment | 5.95 | AAI-based (high-confidence) | 0 |
| S12-final-viral-combined_k141_212317\|\|full | 2213 | No | NA | 4 | 0 | 1 | Not-determined | Genome-fragment | NA | NA | 0 |
| S12-final-viral-combined_k141_127577\|\|full | 1775 | No | NA | 2 | 0 | 0 | Not-determined | Genome-fragment | NA | NA | 0 |
| S12-final-viral-combined_k141_245666\|\|full | 1350 | No | NA | 3 | 0 | 0 | Not-determined | Genome-fragment | NA | NA | 0 |
| S12-final-viral-combined_k141_262126\|\|full | 2008 | No | NA | 3 | 0 | 2 | Not-determined | Genome-fragment | NA | NA | 0 |
| S9_k141_43404 | 2952 | No | NA | 10 | 0 | 0 | Low-quality | Genome-fragment | 6.02 | AAI-based (medium-confidence) | 0 |
| S12-final-viral-combined_k141_127601\|\|full | 2883 | No | NA | 2 | 0 | 1 | Not-determined | Genome-fragment | NA | NA | 0 |
| S12-final-viral-combined_k141_178584\|\|full | 1497 | No | NA | 2 | 0 | 0 | Not-determined | Genome-fragment | NA | NA | 0 |
| S1_k141_130302 | 2952 | No | NA | 6 | 1 | 0 | Low-quality | Genome-fragment | 1.32 | HMM-based (lower-bound) | 0 |
| S12-final-viral-combined_k141_262144\|\|full | 2003 | No | NA | 3 | 0 | 0 | Not-determined | Genome-fragment | NA | NA | 0 |
| S11-final-viral-combined_k141_218910\|\|full | 2953 | No | NA | 4 | 2 | 0 | Low-quality | Genome-fragment | 6.47 | AAI-based (high-confidence) | 0 |
| S12-final-viral-combined_k141_178608\|\|full | 1811 | No | NA | 3 | 0 | 1 | Not-determined | Genome-fragment | NA | NA | 0 |
| S12-final-viral-combined_k141_262155\|\|full | 6562 | No | NA | 8 | 0 | 8 | Not-determined | Genome-fragment | NA | NA | 0 |
| S12-final-viral-combined_k141_313953\|\|full | 61118 | Yes | 52155 | 72 | 20 | 7 | High-quality | High-quality | 100 | AAI-based (medium-confidence) | 14.67 |
| S12-final-viral-combined_k141_262159\|\|full | 1923 | No | NA | 3 | 0 | 2 | Not-determined | Genome-fragment | NA | NA | 0 |
| S1-final-viral-combined_k141_166078\|\|full | 2953 | No | NA | 3 | 0 | 1 | Low-quality | Genome-fragment | 11.4 | AAI-based (high-confidence) | 0 |
| S1-final-viral-combined_k141_206478\|\|full | 2953 | No | NA | 3 | 0 | 2 | Low-quality | Genome-fragment | 2.09 | AAI-based (medium-confidence) | 0 |
| S12-final-viral-combined_k141_127638\|\|full | 1070 | No | NA | 2 | 0 | 1 | Not-determined | Genome-fragment | NA | NA | 0 |
| S2-final-viral-combined_k141_33073\|\|full | 2953 | No | NA | 6 | 3 | 0 | Low-quality | Genome-fragment | 9.6 | AAI-based (medium-confidence) | 0 |
| S12-final-viral-combined_k141_329584\|\|full | 8565 | No | NA | 8 | 0 | 3 | Not-determined | Genome-fragment | NA | NA | 0 |
| S12-final-viral-combined_k141_45633\|\|full | 1912 | No | NA | 2 | 0 | 2 | Not-determined | Genome-fragment | NA | NA | 0 |
| S12-final-viral-combined_k141_127688\|\|full | 1325 | No | NA | 3 | 0 | 2 | Not-determined | Genome-fragment | NA | NA | 0 |
| S12-final-viral-combined_k141_396599\|\|full | 4361 | No | NA | 5 | 0 | 2 | Not-determined | Genome-fragment | NA | NA | 0 |
| S12-final-viral-combined_k141_178704\|\|full | 1905 | No | NA | 4 | 0 | 0 | Not-determined | Genome-fragment | NA | NA | 0 |
| S3-final-viral-combined_k141_54251\|\|full | 2954 | No | NA | 3 | 0 | 1 | Low-quality | Genome-fragment | 4.57 | AAI-based (high-confidence) | 0 |
| S12-final-viral-combined_k141_279968\|\|full | 2595 | No | NA | 3 | 0 | 1 | Not-determined | Genome-fragment | NA | NA | 0 |
| S17-final-viral-combined_k141_156178\|\|full | 2954 | No | NA | 5 | 3 | 0 | Low-quality | Genome-fragment | 5.69 | AAI-based (high-confidence) | 0 |
| S12-final-viral-combined_k141_119021\|\|full | 2954 | No | NA | 7 | 1 | 0 | Low-quality | Genome-fragment | 4.14 | HMM-based (lower-bound) | 0 |
| S12-final-viral-combined_k141_245755\|\|full | 29036 | No | NA | 45 | 19 | 0 | Medium-quality | Genome-fragment | 88.69 | AAI-based (high-confidence) | 0 |
| S12-final-viral-combined_k141_144938\|\|full | 2384 | No | NA | 4 | 0 | 3 | Not-determined | Genome-fragment | NA | NA | 0 |
| S12-final-viral-combined_k141_103579\|\|full | 2954 | Yes | 1228 | 3 | 1 | 1 | Low-quality | Genome-fragment | 0.77 | HMM-based (lower-bound) | 58.43 |
| S12_k141_350225 | 2954 | No | NA | 2 | 0 | 2 | Low-quality | Genome-fragment | 3.7 | AAI-based (medium-confidence) | 0 |
| S12-final-viral-combined_k141_162365\|\|full | 2781 | No | NA | 4 | 0 | 1 | Not-determined | Genome-fragment | NA | NA | 0 |
| S4-final-viral-combined_k141_138105\|\|full | 2954 | No | NA | 3 | 0 | 2 | Low-quality | Genome-fragment | 0.96 | AAI-based (medium-confidence) | 0 |
| S12-final-viral-combined_k141_212506\|\|full | 4160 | No | NA | 6 | 0 | 3 | Not-determined | Genome-fragment | NA | NA | 0 |
| S8_k141_250112 | 2954 | No | NA | 6 | 0 | 1 | Low-quality | Genome-fragment | 2.41 | AAI-based (high-confidence) | 0 |
| S3-final-viral-combined_k141_42715\|\|full | 2955 | No | NA | 2 | 0 | 1 | Low-quality | Genome-fragment | 6.58 | AAI-based (high-confidence) | 0 |
| S9_k141_83055 | 2957 | No | NA | 2 | 0 | 2 | Low-quality | Genome-fragment | 0.79 | AAI-based (high-confidence) | 0 |
| S12-final-viral-combined_k141_296512\|\|full | 2165 | No | NA | 2 | 0 | 2 | Not-determined | Genome-fragment | NA | NA | 0 |
| S12-final-viral-combined_k141_262291\|\|full | 3218 | No | NA | 4 | 0 | 4 | Not-determined | Genome-fragment | NA | NA | 0 |
| S11-final-viral-combined_k141_207026\|\|full | 2957 | No | NA | 3 | 0 | 1 | Low-quality | Genome-fragment | 3.43 | AAI-based (medium-confidence) | 0 |
| S11-final-viral-combined_k141_109524\|\|full | 2957 | No | NA | 5 | 0 | 1 | Low-quality | Genome-fragment | 5.5 | AAI-based (medium-confidence) | 0 |
| S1-final-viral-combined_k141_239826\|\|full | 2957 | No | NA | 3 | 0 | 1 | Low-quality | Genome-fragment | 1.31 | AAI-based (medium-confidence) | 0 |
| S12-final-viral-combined_k141_245863\|\|full | 3167 | No | NA | 4 | 0 | 1 | Not-determined | Genome-fragment | NA | NA | 0 |
| S12-final-viral-combined_k141_11089\|\|full | 1087 | No | NA | 2 | 0 | 1 | Not-determined | Genome-fragment | NA | NA | 0 |
| S2-final-viral-combined_k141_82062\|\|full | 2959 | No | NA | 3 | 0 | 0 | Low-quality | Genome-fragment | 6.51 | AAI-based (high-confidence) | 0 |
| S9-final-viral-combined_k141_47976\|\|full | 2960 | No | NA | 3 | 1 | 0 | Low-quality | Genome-fragment | 2.33 | AAI-based (medium-confidence) | 0 |
| S11-final-viral-combined_k141_69880\|\|full | 2961 | No | NA | 4 | 1 | 0 | Low-quality | Genome-fragment | 6.25 | AAI-based (high-confidence) | 0 |
| S12-final-viral-combined_k141_280051\|\|full | 2396 | No | NA | 3 | 0 | 1 | Not-determined | Genome-fragment | NA | NA | 0 |
| S12-final-viral-combined_k141_314112\|\|full | 3833 | No | NA | 5 | 0 | 2 | Not-determined | Genome-fragment | NA | NA | 0 |
| S12-final-viral-combined_k141_178836\|\|full | 1831 | No | NA | 3 | 0 | 2 | Not-determined | Genome-fragment | NA | NA | 0 |
| S12-final-viral-combined_k141_262380\|\|full | 2169 | No | NA | 3 | 0 | 2 | Not-determined | Genome-fragment | NA | NA | 0 |
| S1-final-viral-combined_k141_76201\|\|full | 2961 | No | NA | 3 | 0 | 3 | Low-quality | Genome-fragment | 4.91 | AAI-based (medium-confidence) | 0 |
| S12-final-viral-combined_k141_196360\|\|full | 3899 | No | NA | 4 | 0 | 0 | Not-determined | Genome-fragment | NA | NA | 0 |
| S12-final-viral-combined_k141_380294\|\|full | 3808 | No | NA | 6 | 0 | 3 | Not-determined | Genome-fragment | NA | NA | 0 |
| S12_k141_54364 | 2961 | No | NA | 7 | 1 | 0 | Low-quality | Genome-fragment | 4.05 | AAI-based (medium-confidence) | 0 |
| S12-final-viral-combined_k141_361604\|\|full | 3926 | No | NA | 7 | 0 | 1 | Not-determined | Genome-fragment | NA | NA | 0 |
| S11-final-viral-combined_k141_1884\|\|full | 2962 | No | NA | 3 | 0 | 0 | Low-quality | Genome-fragment | 5.19 | AAI-based (high-confidence) | 0 |
| S12-final-viral-combined_k141_396844\|\|full | 5941 | No | NA | 5 | 0 | 3 | Not-determined | Genome-fragment | NA | NA | 0 |
| S12-final-viral-combined_k141_380319\|\|full | 2041 | No | NA | 2 | 0 | 2 | Not-determined | Genome-fragment | NA | NA | 0 |
| S12-final-viral-combined_k141_162535\|\|full | 4958 | No | NA | 9 | 0 | 3 | Not-determined | Genome-fragment | NA | NA | 0 |
| S17-final-viral-combined_k141_233056\|\|full | 2962 | No | NA | 2 | 1 | 0 | Low-quality | Genome-fragment | 4.05 | HMM-based (lower-bound) | 0 |
| S12-final-viral-combined_k141_28185\|\|full | 1201 | No | NA | 2 | 0 | 2 | Not-determined | Genome-fragment | NA | NA | 0 |
| S12-final-viral-combined_k141_229984\|\|full | 2057 | No | NA | 4 | 0 | 1 | Not-determined | Genome-fragment | NA | NA | 0 |
| S12-final-viral-combined_k141_329804\|\|full | 2410 | No | NA | 3 | 0 | 2 | Not-determined | Genome-fragment | NA | NA | 0 |
| S1_k141_212188 | 2963 | No | NA | 4 | 0 | 2 | Low-quality | Genome-fragment | 1.05 | AAI-based (medium-confidence) | 0 |
| S12-final-viral-combined_k141_28205\|\|full | 1893 | No | NA | 4 | 0 | 1 | Not-determined | Genome-fragment | NA | NA | 0 |
| S2-final-viral-combined_k141_195936\|\|full | 2963 | No | NA | 4 | 0 | 3 | Low-quality | Genome-fragment | 3.29 | AAI-based (medium-confidence) | 0 |
| S12-final-viral-combined_k141_127919\|\|full | 2832 | No | NA | 6 | 0 | 1 | Not-determined | Genome-fragment | NA | NA | 0 |
| S6_k141_66476 | 2963 | No | NA | 2 | 0 | 1 | Low-quality | Genome-fragment | 5.49 | AAI-based (high-confidence) | 0 |
| S12-final-viral-combined_k141_61595\|\|full | 2498 | No | NA | 3 | 0 | 1 | Not-determined | Genome-fragment | NA | NA | 0 |
| S12-final-viral-combined_k141_145209\|\|full | 1300 | No | NA | 5 | 0 | 0 | Not-determined | Genome-fragment | NA | NA | 0 |
| S9-final-viral-combined_k141_92358\|\|full | 2964 | No | NA | 5 | 1 | 0 | Low-quality | Genome-fragment | 7.1 | AAI-based (high-confidence) | 0 |
| S12-final-viral-combined_k141_380381\|\|full | 7273 | No | NA | 4 | 0 | 3 | Not-determined | Genome-fragment | NA | NA | 0 |
| S12-final-viral-combined_k141_178995\|\|full | 1832 | No | NA | 2 | 0 | 2 | Not-determined | Genome-fragment | NA | NA | 0 |
| S12-final-viral-combined_k141_230048\|\|full | 1091 | No | NA | 2 | 0 | 1 | Not-determined | Genome-fragment | NA | NA | 0 |
| S12_k141_354722 | 2964 | No | NA | 2 | 0 | 1 | Low-quality | Genome-fragment | 2.65 | AAI-based (medium-confidence) | 0 |
| S12-final-viral-combined_k141_230071\|\|full | 1490 | No | NA | 3 | 0 | 2 | Not-determined | Genome-fragment | NA | NA | 0 |
| S12-final-viral-combined_k141_11293\|\|full | 3370 | No | NA | 4 | 0 | 2 | Not-determined | Genome-fragment | NA | NA | 0 |
| S12-final-viral-combined_k141_212781\|\|full | 12269 | No | NA | 9 | 0 | 8 | Not-determined | Genome-fragment | NA | NA | 0 |
| S12-final-viral-combined_k141_61684\|\|full | 3505 | No | NA | 3 | 0 | 3 | Not-determined | Genome-fragment | NA | NA | 0 |
| S2-final-viral-combined_k141_4045\|\|full | 2964 | No | NA | 2 | 0 | 2 | Low-quality | Genome-fragment | 7.74 | AAI-based (medium-confidence) | 0 |
| S12-final-viral-combined_k141_28314\|\|full | 4089 | No | NA | 4 | 0 | 1 | Not-determined | Genome-fragment | NA | NA | 0 |
| S12-final-viral-combined_k141_262615\|\|full | 2725 | No | NA | 2 | 0 | 2 | Not-determined | Genome-fragment | NA | NA | 0 |
| S6-final-viral-combined_k141_113118\|\|full | 2964 | No | NA | 4 | 0 | 3 | Low-quality | Genome-fragment | 2.27 | AAI-based (high-confidence) | 0 |
| S11-final-viral-combined_k141_268434\|\|full | 2965 | No | NA | 4 | 3 | 0 | Low-quality | Genome-fragment | 7.14 | AAI-based (medium-confidence) | 0 |
| S12-final-viral-combined_k141_280303\|\|full | 2556 | No | NA | 2 | 0 | 1 | Not-determined | Genome-fragment | NA | NA | 0 |
| S12-final-viral-combined_k141_230171\|\|full | 1626 | No | NA | 3 | 0 | 0 | Not-determined | Genome-fragment | NA | NA | 0 |
| S12-final-viral-combined_k141_45973\|\|full | 1425 | No | NA | 3 | 0 | 0 | Not-determined | Genome-fragment | NA | NA | 0 |
| S12-final-viral-combined_k141_397001\|\|full | 1006 | No | NA | 2 | 0 | 1 | Not-determined | Genome-fragment | NA | NA | 0 |
| S12-final-viral-combined_k141_230189\|\|full | 1114 | No | NA | 2 | 0 | 1 | Not-determined | Genome-fragment | NA | NA | 0 |
| S12-final-viral-combined_k141_246021\|\|full | 1503 | No | NA | 2 | 0 | 1 | Not-determined | Genome-fragment | NA | NA | 0 |
| S12-final-viral-combined_k141_111524\|\|full | 106106 | Yes | 43797 | 80 | 5 | 38 | Medium-quality | Genome-fragment | 72.35 | AAI-based (medium-confidence) | 58.72 |
| S12-final-viral-combined_k141_79345\|\|full | 3323 | No | NA | 4 | 0 | 2 | Not-determined | Genome-fragment | NA | NA | 0 |
| S12-final-viral-combined_k141_95141\|\|full | 11312 | No | NA | 9 | 0 | 7 | Not-determined | Genome-fragment | NA | NA | 0 |
| S1-final-viral-combined_k141_245362\|\|full | 2965 | No | NA | 4 | 0 | 1 | Low-quality | Genome-fragment | 3.58 | AAI-based (medium-confidence) | 0 |
| S12-final-viral-combined_k141_212855\|\|full | 6773 | No | NA | 12 | 0 | 0 | Not-determined | Genome-fragment | NA | NA | 0 |
| S11_k141_90645 | 2966 | No | NA | 5 | 1 | 0 | Low-quality | Genome-fragment | 6.5 | AAI-based (high-confidence) | 0 |
| S5-final-viral-combined_k141_136404\|\|full | 2966 | Yes | 962 | 6 | 1 | 1 | Low-quality | Genome-fragment | 2.54 | AAI-based (medium-confidence) | 67.57 |
| S12-final-viral-combined_k141_262696\|\|full | 1386 | No | NA | 3 | 0 | 2 | Not-determined | Genome-fragment | NA | NA | 0 |
| S12_k141_174799 | 2966 | No | NA | 3 | 0 | 1 | Low-quality | Genome-fragment | 5.32 | AAI-based (high-confidence) | 0 |
| S12-final-viral-combined_k141_380516\|\|full | 4971 | No | NA | 3 | 0 | 2 | Not-determined | Genome-fragment | NA | NA | 0 |
| S12-final-viral-combined_k141_111575\|\|full | 2620 | No | NA | 3 | 0 | 2 | Not-determined | Genome-fragment | NA | NA | 0 |
| S12-final-viral-combined_k141_314318\|\|full | 1735 | No | NA | 2 | 0 | 1 | Not-determined | Genome-fragment | NA | NA | 0 |
| S12-final-viral-combined_k141_397059\|\|full | 2699 | No | NA | 3 | 0 | 2 | Not-determined | Genome-fragment | NA | NA | 0 |
| S12-final-viral-combined_k141_179126\|\|full | 4867 | No | NA | 8 | 0 | 0 | Not-determined | Genome-fragment | NA | NA | 0 |
| S8-final-viral-combined_k141_239910\|\|full | 2966 | No | NA | 4 | 1 | 0 | Low-quality | Genome-fragment | 4.49 | HMM-based (lower-bound) | 0 |
| S12-final-viral-combined_k141_380528\|\|full | 9786 | No | NA | 10 | 0 | 5 | Not-determined | Genome-fragment | NA | NA | 0 |
| S11-final-viral-combined_k141_92938\|\|full | 2967 | No | NA | 4 | 1 | 0 | Low-quality | Genome-fragment | 6.99 | AAI-based (medium-confidence) | 0 |
| S3-final-viral-combined_k141_44915\|\|full | 2967 | No | NA | 5 | 1 | 0 | Low-quality | Genome-fragment | 4.76 | HMM-based (lower-bound) | 0 |
| S12-final-viral-combined_k141_46099\|\|full | 3037 | No | NA | 4 | 0 | 1 | Not-determined | Genome-fragment | NA | NA | 0 |
| S12-final-viral-combined_k141_262727\|\|full | 15826 | No | NA | 13 | 0 | 9 | Not-determined | Genome-fragment | NA | NA | 0 |
| S12-final-viral-combined_k141_128239\|\|full | 2388 | No | NA | 3 | 0 | 1 | Not-determined | Genome-fragment | NA | NA | 0 |
| S12-final-viral-combined_k141_330037\|\|full | 1469 | No | NA | 4 | 0 | 0 | Not-determined | Genome-fragment | NA | NA | 0 |
| S2-final-viral-combined_k141_10791\|\|full | 2967 | No | NA | 4 | 2 | 0 | Low-quality | Genome-fragment | 1.5 | AAI-based (high-confidence) | 0 |
| S12-final-viral-combined_k141_246092\|\|full | 2298 | No | NA | 3 | 0 | 1 | Not-determined | Genome-fragment | NA | NA | 0 |
| S12-final-viral-combined_k141_28485\|\|full | 2214 | No | NA | 3 | 0 | 1 | Not-determined | Genome-fragment | NA | NA | 0 |
| S1-final-viral-combined_k141_184241\|\|full | 2968 | No | NA | 4 | 1 | 0 | Low-quality | Genome-fragment | 7.25 | AAI-based (high-confidence) | 0 |
| S12-final-viral-combined_k141_314375\|\|full | 1623 | No | NA | 2 | 0 | 0 | Not-determined | Genome-fragment | NA | NA | 0 |
| S12-final-viral-combined_k141_380594\|\|full | 8122 | No | NA | 7 | 0 | 4 | Not-determined | Genome-fragment | NA | NA | 0 |
| S17-final-viral-combined_k141_188998\|\|full | 2968 | Yes | 1410 | 5 | 1 | 1 | Low-quality | Genome-fragment | 2.85 | AAI-based (medium-confidence) | 52.49 |
| S12-final-viral-combined_k141_262789\|\|full | 1010 | No | NA | 2 | 0 | 2 | Not-determined | Genome-fragment | NA | NA | 0 |
| S8-final-viral-combined_k141_258904\|\|full | 2968 | No | NA | 3 | 2 | 0 | Low-quality | Genome-fragment | 4.81 | AAI-based (medium-confidence) | 0 |
| S9-final-viral-combined_k141_120578\|\|full | 2969 | Yes | 1219 | 6 | 2 | 1 | Low-quality | Genome-fragment | 3.09 | AAI-based (high-confidence) | 58.94 |
| S12-final-viral-combined_k141_346606\|\|full | 2412 | No | NA | 3 | 0 | 2 | Not-determined | Genome-fragment | NA | NA | 0 |
| S9-final-viral-combined_k141_89465\|\|full | 2969 | No | NA | 6 | 0 | 0 | Low-quality | Genome-fragment | 7.45 | AAI-based (high-confidence) | 0 |
| S17-final-viral-combined_k141_97011\|\|full | 2970 | No | NA | 4 | 2 | 0 | Low-quality | Genome-fragment | 1.71 | AAI-based (medium-confidence) | 0 |
| S9-final-viral-combined_k141_48140\|\|full | 2971 | No | NA | 4 | 0 | 2 | Low-quality | Genome-fragment | 0.85 | AAI-based (medium-confidence) | 0 |
| S17-final-viral-combined_k141_147740\|\|full | 2971 | No | NA | 7 | 0 | 0 | Low-quality | Genome-fragment | 3.31 | AAI-based (medium-confidence) | 0 |
| S12-final-viral-combined_k141_145498\|\|full | 5502 | No | NA | 6 | 0 | 3 | Not-determined | Genome-fragment | NA | NA | 0 |
| S12-final-viral-combined_k141_111671\|\|full | 3432 | No | NA | 4 | 0 | 3 | Not-determined | Genome-fragment | NA | NA | 0 |
| S2-final-viral-combined_k141_7232\|\|full | 2971 | No | NA | 5 | 0 | 0 | Low-quality | Genome-fragment | 5.98 | AAI-based (medium-confidence) | 0 |
| S12-final-viral-combined_k141_145503\|\|full | 4612 | No | NA | 7 | 0 | 4 | Not-determined | Genome-fragment | NA | NA | 0 |
| S1_k141_337647 | 2972 | No | NA | 2 | 0 | 2 | Low-quality | Genome-fragment | 1.1 | AAI-based (high-confidence) | 0 |
| S12-final-viral-combined_k141_361893\|\|full | 3878 | No | NA | 5 | 0 | 2 | Not-determined | Genome-fragment | NA | NA | 0 |
| S12-final-viral-combined_k141_262844\|\|full | 1511 | No | NA | 4 | 0 | 0 | Not-determined | Genome-fragment | NA | NA | 0 |
| S12-final-viral-combined_k141_128350\|\|full | 1697 | No | NA | 3 | 0 | 0 | Not-determined | Genome-fragment | NA | NA | 0 |
| S12-final-viral-combined_k141_397191\|\|full | 1108 | No | NA | 2 | 0 | 1 | Not-determined | Genome-fragment | NA | NA | 0 |
| S1-final-viral-combined_k141_213804\|\|full | 2974 | No | NA | 5 | 0 | 2 | Low-quality | Genome-fragment | 6.02 | AAI-based (high-confidence) | 0 |
| S12-final-viral-combined_k141_297012\|\|full | 2856 | No | NA | 4 | 0 | 1 | Not-determined | Genome-fragment | NA | NA | 0 |
| S5-final-viral-combined_k141_133403\|\|full | 2974 | No | NA | 3 | 3 | 0 | Low-quality | Genome-fragment | 7.78 | AAI-based (high-confidence) | 0 |
| S12-final-viral-combined_k141_145590\|\|full | 1579 | No | NA | 3 | 0 | 0 | Not-determined | Genome-fragment | NA | NA | 0 |
| S12-final-viral-combined_k141_346748\|\|full | 3131 | No | NA | 3 | 0 | 0 | Not-determined | Genome-fragment | NA | NA | 0 |
| S7-final-viral-combined_k141_204478\|\|full | 2974 | Yes | 441 | 5 | 1 | 2 | Low-quality | Genome-fragment | 0.34 | AAI-based (high-confidence) | 85.17 |
| S12_k141_101456 | 2974 | No | NA | 3 | 1 | 0 | Low-quality | Genome-fragment | 7.16 | AAI-based (high-confidence) | 0 |
| S12-final-viral-combined_k141_62031\|\|full | 1196 | No | NA | 2 | 0 | 1 | Not-determined | Genome-fragment | NA | NA | 0 |
| S12-final-viral-combined_k141_128429\|\|full | 1980 | No | NA | 2 | 0 | 1 | Not-determined | Genome-fragment | NA | NA | 0 |
| S12-final-viral-combined_k141_46312\|\|full | 1725 | No | NA | 3 | 0 | 0 | Not-determined | Genome-fragment | NA | NA | 0 |
| S8-final-viral-combined_k141_183717\|\|full | 2974 | No | NA | 4 | 0 | 2 | Low-quality | Genome-fragment | 0.84 | AAI-based (medium-confidence) | 0 |
| S9-final-viral-combined_k141_40274\|\|full | 2975 | No | NA | 3 | 0 | 0 | Low-quality | Genome-fragment | 7.41 | AAI-based (high-confidence) | 0 |
| S1-final-viral-combined_k141_215089\|\|full | 2976 | No | NA | 3 | 0 | 2 | Low-quality | Genome-fragment | 6.21 | AAI-based (medium-confidence) | 0 |
| S12-final-viral-combined_k141_330246\|\|full | 1001 | No | NA | 2 | 0 | 1 | Not-determined | Genome-fragment | NA | NA | 0 |
| S12-final-viral-combined_k141_79615\|\|full | 1543 | No | NA | 4 | 0 | 0 | Not-determined | Genome-fragment | NA | NA | 0 |
| S12-final-viral-combined_k141_213104\|\|full | 1774 | No | NA | 2 | 0 | 1 | Not-determined | Genome-fragment | NA | NA | 0 |
| S12-final-viral-combined_k141_397241\|\|full | 1736 | No | NA | 5 | 0 | 1 | Not-determined | Genome-fragment | NA | NA | 0 |
| S12-final-viral-combined_k141_246289\|\|full | 1242 | No | NA | 3 | 0 | 2 | Not-determined | Genome-fragment | NA | NA | 0 |
| S1-final-viral-combined_k141_209982\|\|full | 2976 | No | NA | 4 | 0 | 0 | Low-quality | Genome-fragment | 1.11 | AAI-based (medium-confidence) | 0 |
| S1-final-viral-combined_k141_343282\|\|full | 2976 | No | NA | 4 | 0 | 3 | Low-quality | Genome-fragment | 1.25 | AAI-based (medium-confidence) | 0 |
| S12-final-viral-combined_k141_48886\|\|full | 2976 | No | NA | 7 | 1 | 0 | Low-quality | Genome-fragment | 2.37 | HMM-based (lower-bound) | 0 |
| S12-final-viral-combined_k141_330276\|\|full | 3533 | No | NA | 4 | 0 | 3 | Not-determined | Genome-fragment | NA | NA | 0 |
| S2-final-viral-combined_k141_111776\|\|full | 2976 | No | NA | 3 | 0 | 1 | Low-quality | Genome-fragment | 1.99 | AAI-based (medium-confidence) | 0 |
| S12-final-viral-combined_k141_297095\|\|full | 2888 | No | NA | 3 | 0 | 0 | Not-determined | Genome-fragment | NA | NA | 0 |
| S7_k141_12538 | 2977 | No | NA | 9 | 1 | 0 | Low-quality | Genome-fragment | 4.71 | AAI-based (high-confidence) | 0 |
| S8-final-viral-combined_k141_114151\|\|full | 2977 | No | NA | 4 | 0 | 4 | Low-quality | Genome-fragment | 1.01 | AAI-based (medium-confidence) | 0 |
| S12-final-viral-combined_k141_230330\|\|full | 1674 | No | NA | 3 | 0 | 1 | Not-determined | Genome-fragment | NA | NA | 0 |
| S9-final-viral-combined_k141_17003\|\|full | 2978 | No | NA | 5 | 2 | 0 | Low-quality | Genome-fragment | 5.93 | AAI-based (high-confidence) | 0 |
| S12-final-viral-combined_k141_111834\|\|full | 1820 | No | NA | 3 | 0 | 2 | Not-determined | Genome-fragment | NA | NA | 0 |
| S12-final-viral-combined_k141_95463\|\|full | 11698 | No | NA | 10 | 0 | 4 | Not-determined | Genome-fragment | NA | NA | 0 |
| S11-final-viral-combined_k141_56874\|\|full | 2978 | Yes | 1433 | 4 | 1 | 1 | Low-quality | Genome-fragment | 2.99 | AAI-based (medium-confidence) | 51.88 |
| S12-final-viral-combined_k141_179403\|\|full | 1811 | No | NA | 4 | 0 | 0 | Not-determined | Genome-fragment | NA | NA | 0 |
| S12-final-viral-combined_k141_111855\|\|full | 1494 | No | NA | 3 | 0 | 0 | Not-determined | Genome-fragment | NA | NA | 0 |
| S11_k141_260580 | 2978 | No | NA | 2 | 0 | 2 | Low-quality | Genome-fragment | 1.28 | AAI-based (medium-confidence) | 0 |
| S12-final-viral-combined_k141_196598\|\|full | 4203 | No | NA | 4 | 0 | 3 | Not-determined | Genome-fragment | NA | NA | 0 |
| S12-final-viral-combined_k141_79694\|\|full | 2559 | No | NA | 3 | 0 | 1 | Not-determined | Genome-fragment | NA | NA | 0 |
| S12-final-viral-combined_k141_128573\|\|full | 1092 | No | NA | 2 | 0 | 0 | Not-determined | Genome-fragment | NA | NA | 0 |
| S1-final-viral-combined_k141_99311\|\|full | 2978 | No | NA | 2 | 0 | 1 | Low-quality | Genome-fragment | 2.31 | AAI-based (medium-confidence) | 0 |
| S12-final-viral-combined_k141_280691\|\|full | 1191 | No | NA | 2 | 0 | 0 | Not-determined | Genome-fragment | NA | NA | 0 |
| S12-final-viral-combined_k141_263061\|\|full | 5836 | No | NA | 5 | 0 | 2 | Not-determined | Genome-fragment | NA | NA | 0 |
| S12-final-viral-combined_k141_62191\|\|full | 1578 | No | NA | 3 | 0 | 0 | Not-determined | Genome-fragment | NA | NA | 0 |
| S12-final-viral-combined_k141_362034\|\|full | 4787 | No | NA | 10 | 0 | 3 | Not-determined | Genome-fragment | NA | NA | 0 |
| S12-final-viral-combined_k141_255951\|\|full | 2978 | No | NA | 3 | 2 | 0 | Low-quality | Genome-fragment | 5.99 | HMM-based (lower-bound) | 0 |
| S2-final-viral-combined_k141_107444\|\|full | 2978 | No | NA | 4 | 0 | 1 | Low-quality | Genome-fragment | 6.31 | AAI-based (medium-confidence) | 0 |
| S9-final-viral-combined_k141_10970\|\|full | 2979 | No | NA | 2 | 0 | 2 | Low-quality | Genome-fragment | 7.78 | AAI-based (high-confidence) | 0 |
| S12-final-viral-combined_k141_196623\|\|full | 1072 | No | NA | 2 | 0 | 2 | Not-determined | Genome-fragment | NA | NA | 0 |
| S12-final-viral-combined_k141_11666\|\|full | 1057 | No | NA | 3 | 0 | 0 | Not-determined | Genome-fragment | NA | NA | 0 |
| S11-final-viral-combined_k141_196346\|\|full | 2979 | No | NA | 4 | 0 | 0 | Low-quality | Genome-fragment | 3.77 | AAI-based (medium-confidence) | 0 |
| S12-final-viral-combined_k141_79737\|\|full | 2653 | No | NA | 4 | 0 | 4 | Not-determined | Genome-fragment | NA | NA | 0 |
| S17-final-viral-combined_k141_116546\|\|full | 2979 | No | NA | 6 | 1 | 0 | Low-quality | Genome-fragment | 6.97 | AAI-based (high-confidence) | 0 |
| S12-final-viral-combined_k141_40470\|\|full | 2979 | No | NA | 3 | 0 | 2 | Low-quality | Genome-fragment | 5.93 | AAI-based (medium-confidence) | 0 |
| S12-final-viral-combined_k141_345635\|\|full | 2979 | No | NA | 4 | 0 | 2 | Low-quality | Genome-fragment | 4.05 | AAI-based (medium-confidence) | 0 |
| S8-final-viral-combined_k141_50172\|\|full | 2979 | No | NA | 7 | 1 | 0 | Low-quality | Genome-fragment | 7.6 | AAI-based (high-confidence) | 0 |
| S12-final-viral-combined_k141_246376\|\|full | 1240 | No | NA | 2 | 0 | 1 | Not-determined | Genome-fragment | NA | NA | 0 |
| S12-final-viral-combined_k141_95538\|\|full | 4947 | No | NA | 9 | 0 | 8 | Not-determined | Genome-fragment | NA | NA | 0 |
| S12-final-viral-combined_k141_179533\|\|full | 1455 | No | NA | 2 | 0 | 1 | Not-determined | Genome-fragment | NA | NA | 0 |
| S12-final-viral-combined_k141_213271\|\|full | 2393 | No | NA | 4 | 0 | 3 | Not-determined | Genome-fragment | NA | NA | 0 |
| S12-final-viral-combined_k141_95554\|\|full | 1551 | No | NA | 3 | 0 | 3 | Not-determined | Genome-fragment | NA | NA | 0 |
| S12-final-viral-combined_k141_380787\|\|full | 2185 | No | NA | 3 | 0 | 0 | Not-determined | Genome-fragment | NA | NA | 0 |
| S12-final-viral-combined_k141_179570\|\|full | 1389 | No | NA | 2 | 0 | 0 | Not-determined | Genome-fragment | NA | NA | 0 |
| S12-final-viral-combined_k141_173591\|\|full | 2980 | No | NA | 5 | 0 | 0 | Low-quality | Genome-fragment | 5.96 | AAI-based (medium-confidence) | 0 |
| S12-final-viral-combined_k141_62254\|\|full | 2551 | No | NA | 5 | 0 | 3 | Not-determined | Genome-fragment | NA | NA | 0 |
| S17-final-viral-combined_k141_215611\|\|full | 2981 | No | NA | 4 | 0 | 2 | Low-quality | Genome-fragment | 2.27 | AAI-based (high-confidence) | 0 |
| S1-final-viral-combined_k141_370587\|\|full | 2983 | No | NA | 4 | 0 | 2 | Low-quality | Genome-fragment | 1.3 | AAI-based (medium-confidence) | 0 |
| S12-final-viral-combined_k141_280821\|\|full | 2594 | No | NA | 3 | 0 | 0 | Not-determined | Genome-fragment | NA | NA | 0 |
| S12-final-viral-combined_k141_280824\|\|full | 1066 | No | NA | 2 | 0 | 2 | Not-determined | Genome-fragment | NA | NA | 0 |
| S12-final-viral-combined_k141_179626\|\|full | 2948 | No | NA | 6 | 0 | 0 | Not-determined | Genome-fragment | NA | NA | 0 |
| S4_k141_148152 | 2983 | No | NA | 6 | 1 | 0 | Low-quality | Genome-fragment | 4.73 | AAI-based (high-confidence) | 0 |
| S8-final-viral-combined_k141_230165\|\|full | 2983 | No | NA | 4 | 1 | 0 | Low-quality | Genome-fragment | 3.35 | HMM-based (lower-bound) | 0 |
| S12-final-viral-combined_k141_280850\|\|full | 1487 | No | NA | 3 | 0 | 2 | Not-determined | Genome-fragment | NA | NA | 0 |
| S4-final-viral-combined_k141_21317\|\|full | 2984 | No | NA | 3 | 0 | 2 | Low-quality | Genome-fragment | 1.84 | AAI-based (medium-confidence) | 0 |
| S12-final-viral-combined_k141_196785\|\|full | 1773 | No | NA | 3 | 0 | 0 | Not-determined | Genome-fragment | NA | NA | 0 |
| S12-final-viral-combined_k141_362132\|\|full | 1785 | No | NA | 3 | 0 | 1 | Not-determined | Genome-fragment | NA | NA | 0 |
| S4-final-viral-combined_k141_166360\|\|full | 2984 | No | NA | 3 | 0 | 1 | Low-quality | Genome-fragment | 6.44 | AAI-based (medium-confidence) | 0 |
| S12-final-viral-combined_k141_297278\|\|full | 2070 | No | NA | 3 | 0 | 1 | Not-determined | Genome-fragment | NA | NA | 0 |
| S1-final-viral-combined_k141_7027\|\|full | 2985 | No | NA | 5 | 0 | 0 | Low-quality | Genome-fragment | 4.98 | AAI-based (medium-confidence) | 0 |
| S12-final-viral-combined_k141_230479\|\|full | 1420 | No | NA | 2 | 0 | 0 | Not-determined | Genome-fragment | NA | NA | 0 |
| S12-final-viral-combined_k141_79899\|\|full | 3251 | No | NA | 4 | 0 | 0 | Not-determined | Genome-fragment | NA | NA | 0 |
| S12-final-viral-combined_k141_314686\|\|full | 12627 | No | NA | 14 | 0 | 0 | Not-determined | Genome-fragment | NA | NA | 0 |
| S12-final-viral-combined_k141_297311\|\|full | 1149 | No | NA | 2 | 0 | 1 | Not-determined | Genome-fragment | NA | NA | 0 |
| S12-final-viral-combined_k141_111991\|\|full | 7472 | No | NA | 7 | 0 | 5 | Not-determined | Genome-fragment | NA | NA | 0 |
| S12-final-viral-combined_k141_397406\|\|full | 1492 | No | NA | 2 | 0 | 2 | Not-determined | Genome-fragment | NA | NA | 0 |
| S12-final-viral-combined_k141_263219\|\|full | 5375 | No | NA | 4 | 0 | 4 | Not-determined | Genome-fragment | NA | NA | 0 |
| S12-final-viral-combined_k141_196869\|\|full | 2277 | No | NA | 3 | 0 | 2 | Not-determined | Genome-fragment | NA | NA | 0 |
| S7-final-viral-combined_k141_4669\|\|full | 2985 | No | NA | 6 | 0 | 4 | Low-quality | Genome-fragment | 2.11 | AAI-based (medium-confidence) | 0 |
| S12-final-viral-combined_k141_79981\|\|full | 1587 | No | NA | 2 | 0 | 0 | Not-determined | Genome-fragment | NA | NA | 0 |
| S4-final-viral-combined_k141_145266\|\|full | 2985 | No | NA | 6 | 0 | 0 | Low-quality | Genome-fragment | 8.8 | AAI-based (medium-confidence) | 0 |
| S9-final-viral-combined_k141_104094\|\|full | 2986 | No | NA | 3 | 0 | 3 | Low-quality | Genome-fragment | 3.09 | AAI-based (medium-confidence) | 0 |
| S8_k141_253009 | 2986 | No | NA | 9 | 0 | 0 | Low-quality | Genome-fragment | 5.64 | AAI-based (high-confidence) | 0 |
| S2-final-viral-combined_k141_192841\|\|full | 2987 | Yes | 2170 | 5 | 3 | 1 | Low-quality | Genome-fragment | 5.48 | AAI-based (high-confidence) | 27.35 |
| S4-final-viral-combined_k141_28444\|\|full | 2987 | No | NA | 4 | 0 | 1 | Low-quality | Genome-fragment | 5.79 | AAI-based (medium-confidence) | 0 |
| S12-final-viral-combined_k141_28842\|\|full | 1936 | No | NA | 2 | 0 | 2 | Not-determined | Genome-fragment | NA | NA | 0 |
| S12-final-viral-combined_k141_95616\|\|full | 4207 | No | NA | 5 | 0 | 0 | Not-determined | Genome-fragment | NA | NA | 0 |
| S12-final-viral-combined_k141_380902\|\|full | 1178 | No | NA | 2 | 0 | 0 | Not-determined | Genome-fragment | NA | NA | 0 |
| S12-final-viral-combined_k141_213484\|\|full | 1915 | No | NA | 3 | 0 | 2 | Not-determined | Genome-fragment | NA | NA | 0 |
| S3-final-viral-combined_k141_83356\|\|full | 2988 | No | NA | 5 | 2 | 0 | Low-quality | Genome-fragment | 4.8 | AAI-based (high-confidence) | 0 |
| S12-final-viral-combined_k141_230592\|\|full | 1585 | No | NA | 3 | 0 | 3 | Not-determined | Genome-fragment | NA | NA | 0 |
| S12-final-viral-combined_k141_46690\|\|full | 1601 | No | NA | 3 | 0 | 1 | Not-determined | Genome-fragment | NA | NA | 0 |
| S12-final-viral-combined_k141_196936\|\|full | 2833 | No | NA | 5 | 0 | 0 | Not-determined | Genome-fragment | NA | NA | 0 |
| S12-final-viral-combined_k141_397440\|\|full | 2030 | No | NA | 2 | 0 | 2 | Not-determined | Genome-fragment | NA | NA | 0 |
| S12-final-viral-combined_k141_230595\|\|full | 3337 | No | NA | 3 | 0 | 1 | Not-determined | Genome-fragment | NA | NA | 0 |
| S12-final-viral-combined_k141_128830\|\|full | 1295 | No | NA | 2 | 0 | 1 | Not-determined | Genome-fragment | NA | NA | 0 |
| S12-final-viral-combined_k141_230599\|\|full | 7345 | No | NA | 6 | 0 | 3 | Not-determined | Genome-fragment | NA | NA | 0 |
| S12-final-viral-combined_k141_246532\|\|full | 1039 | No | NA | 2 | 0 | 2 | Not-determined | Genome-fragment | NA | NA | 0 |
| S8-final-viral-combined_k141_174579\|\|full | 2988 | No | NA | 3 | 0 | 1 | Low-quality | Genome-fragment | 1 | AAI-based (medium-confidence) | 0 |
| S12-final-viral-combined_k141_112127\|\|full | 2091 | No | NA | 4 | 0 | 0 | Not-determined | Genome-fragment | NA | NA | 0 |
| S12-final-viral-combined_k141_80064\|\|full | 5976 | No | NA | 8 | 0 | 0 | Not-determined | Genome-fragment | NA | NA | 0 |
| S11-final-viral-combined_k141_62071\|\|full | 2989 | No | NA | 6 | 1 | 0 | Low-quality | Genome-fragment | 5.97 | AAI-based (medium-confidence) | 0 |
| S7-final-viral-combined_k141_148799\|\|full | 2989 | No | NA | 2 | 0 | 1 | Low-quality | Genome-fragment | 1.5 | AAI-based (medium-confidence) | 0 |
| S12-final-viral-combined_k141_297465\|\|full | 2433 | No | NA | 3 | 0 | 2 | Not-determined | Genome-fragment | NA | NA | 0 |
| S12-final-viral-combined_k141_230665\|\|full | 1797 | No | NA | 3 | 0 | 0 | Not-determined | Genome-fragment | NA | NA | 0 |
| S12-final-viral-combined_k141_128883\|\|full | 1182 | No | NA | 3 | 0 | 0 | Not-determined | Genome-fragment | NA | NA | 0 |
| S5-final-viral-combined_k141_20689\|\|full | 2990 | No | NA | 4 | 0 | 2 | Low-quality | Genome-fragment | 2.28 | AAI-based (medium-confidence) | 0 |
| S1_k141_240967 | 2991 | No | NA | 5 | 1 | 0 | Low-quality | Genome-fragment | 1.62 | HMM-based (lower-bound) | 0 |
| S12-final-viral-combined_k141_263319\|\|full | 7658 | No | NA | 9 | 0 | 1 | Not-determined | Genome-fragment | NA | NA | 0 |
| S12-final-viral-combined_k141_46785\|\|full | 2492 | No | NA | 4 | 0 | 1 | Not-determined | Genome-fragment | NA | NA | 0 |
| S5-final-viral-combined_k141_18875\|\|full | 2991 | No | NA | 3 | 0 | 0 | Low-quality | Genome-fragment | 5.84 | AAI-based (high-confidence) | 0 |
| S11-final-viral-combined_k141_208094\|\|full | 2992 | No | NA | 9 | 0 | 0 | Low-quality | Genome-fragment | 5.01 | AAI-based (high-confidence) | 0 |
| S7-final-viral-combined_k141_12645\|\|full | 2992 | No | NA | 3 | 0 | 1 | Low-quality | Genome-fragment | 2.47 | AAI-based (medium-confidence) | 0 |
| S12-final-viral-combined_k141_151655\|\|full | 2992 | No | NA | 4 | 0 | 0 | Low-quality | Genome-fragment | 6.24 | AAI-based (high-confidence) | 0 |
| S12-final-viral-combined_k141_80132\|\|full | 1005 | No | NA | 2 | 0 | 0 | Not-determined | Genome-fragment | NA | NA | 0 |
| S4-final-viral-combined_k141_139830\|\|full | 2992 | No | NA | 3 | 0 | 2 | Low-quality | Genome-fragment | 3.49 | AAI-based (high-confidence) | 0 |
| S11-final-viral-combined_k141_210602\|\|full | 2994 | No | NA | 4 | 1 | 0 | Low-quality | Genome-fragment | 4.66 | HMM-based (lower-bound) | 0 |
| S12-final-viral-combined_k141_330636\|\|full | 2060 | No | NA | 3 | 0 | 3 | Not-determined | Genome-fragment | NA | NA | 0 |
| S12-final-viral-combined_k141_246687\|\|full | 1717 | No | NA | 3 | 0 | 3 | Not-determined | Genome-fragment | NA | NA | 0 |
| S12-final-viral-combined_k141_330641\|\|full | 1081 | No | NA | 2 | 0 | 0 | Not-determined | Genome-fragment | NA | NA | 0 |
| S12-final-viral-combined_k141_80163\|\|full | 2105 | No | NA | 2 | 0 | 2 | Not-determined | Genome-fragment | NA | NA | 0 |
| S12-final-viral-combined_k141_80174\|\|full | 1072 | No | NA | 3 | 0 | 0 | Not-determined | Genome-fragment | NA | NA | 0 |
| S12-final-viral-combined_k141_397571\|\|full | 1228 | No | NA | 2 | 0 | 1 | Not-determined | Genome-fragment | NA | NA | 0 |
| S12-final-viral-combined_k141_180019\|\|full | 1264 | No | NA | 2 | 0 | 1 | Not-determined | Genome-fragment | NA | NA | 0 |
| S1-final-viral-combined_k141_221154\|\|full | 2994 | No | NA | 6 | 0 | 3 | Low-quality | Genome-fragment | 2.67 | AAI-based (medium-confidence) | 0 |
| S12-final-viral-combined_k141_95834\|\|full | 2679 | No | NA | 4 | 0 | 3 | Not-determined | Genome-fragment | NA | NA | 0 |
| S4_k141_92384 | 2994 | No | NA | 3 | 0 | 2 | Low-quality | Genome-fragment | 2.5 | AAI-based (high-confidence) | 0 |
| S12-final-viral-combined_k141_112250\|\|full | 1349 | No | NA | 2 | 0 | 0 | Not-determined | Genome-fragment | NA | NA | 0 |
| S12-final-viral-combined_k141_397624\|\|full | 6204 | No | NA | 4 | 0 | 3 | Not-determined | Genome-fragment | NA | NA | 0 |
| S7-final-viral-combined_k141_117351\|\|lt2gene | 2995 | No | NA | 2 | 1 | 0 | Low-quality | Genome-fragment | 5.36 | AAI-based (high-confidence) | 0 |
| S12-final-viral-combined_k141_163413\|\|full | 1158 | No | NA | 3 | 0 | 0 | Not-determined | Genome-fragment | NA | NA | 0 |
| S12-final-viral-combined_k141_95850\|\|full | 3129 | No | NA | 3 | 0 | 0 | Not-determined | Genome-fragment | NA | NA | 0 |
| S12-final-viral-combined_k141_149666\|\|full | 2995 | No | NA | 8 | 0 | 0 | Low-quality | Genome-fragment | 4.54 | AAI-based (high-confidence) | 0 |
| S12-final-viral-combined_k141_397641\|\|full | 1602 | No | NA | 4 | 0 | 0 | Not-determined | Genome-fragment | NA | NA | 0 |
| S11-final-viral-combined_k141_290683\|\|full | 2996 | No | NA | 7 | 1 | 0 | Low-quality | Genome-fragment | 7.58 | AAI-based (medium-confidence) | 0 |
| S1-final-viral-combined_k141_48419\|\|full | 2996 | No | NA | 6 | 2 | 0 | Low-quality | Genome-fragment | 7.7 | AAI-based (medium-confidence) | 0 |

| | | | | | | | | | | | |
|---|---|---|---|---|---|---|---|---|---|---|---|
| S12-final-viral-combined_k141_163428\|\|full | 4387 | No | NA | 4 | 0 | 3 | Not-determined | Genome-fragment | NA | NA | 0 |
| S12-final-viral-combined_k141_381084\|\|full | 2593 | No | NA | 5 | 0 | 0 | Not-determined | Genome-fragment | NA | NA | 0 |
| S12-final-viral-combined_k141_213666\|\|full | 1329 | No | NA | 3 | 0 | 0 | Not-determined | Genome-fragment | NA | NA | 0 |
| S12-final-viral-combined_k141_163430\|\|full | 1937 | No | NA | 3 | 0 | 2 | Not-determined | Genome-fragment | NA | NA | 0 |
| S12-final-viral-combined_k141_263377\|\|full | 59167 | No | NA | 85 | 21 | 1 | High-quality | High-quality | 97.94 | AAI-based (high-confidence) | 0 |
| S12-final-viral-combined_k141_146165\|\|full | 2578 | No | NA | 3 | 0 | 3 | Not-determined | Genome-fragment | NA | NA | 0 |
| S12-final-viral-combined_k141_29078\|\|full | 1107 | No | NA | 3 | 0 | 1 | Not-determined | Genome-fragment | NA | NA | 0 |
| S8-final-viral-combined_k141_14823\|\|full | 2997 | No | NA | 3 | 0 | 1 | Low-quality | Genome-fragment | 1.38 | AAI-based (medium-confidence) | 0 |
| S11-final-viral-combined_k141_255017\|\|full | 2998 | No | NA | 7 | 1 | 0 | Low-quality | Genome-fragment | 8.16 | AAI-based (high-confidence) | 0 |
| S12-final-viral-combined_k141_12161\|\|full | 2700 | No | NA | 4 | 0 | 0 | Not-determined | Genome-fragment | NA | NA | 0 |
| S7_k141_115390 | 2999 | No | NA | 1 | 0 | 0 | Low-quality | Genome-fragment | 4.97 | AAI-based (medium-confidence) | 0 |
| S7_k141_237373 | 3000 | No | NA | 3 | 0 | 0 | Low-quality | Genome-fragment | 3.42 | AAI-based (medium-confidence) | 0 |
| S12-final-viral-combined_k141_62718\|\|full | 1659 | No | NA | 2 | 0 | 2 | Not-determined | Genome-fragment | NA | NA | 0 |
| S12-final-viral-combined_k141_213726\|\|full | 1682 | No | NA | 5 | 0 | 0 | Not-determined | Genome-fragment | NA | NA | 0 |
| S12_k141_140254 | 3000 | No | NA | 4 | 2 | 0 | Low-quality | Genome-fragment | 8 | AAI-based (high-confidence) | 0 |
| S12-final-viral-combined_k141_95951\|\|full | 1101 | No | NA | 2 | 0 | 1 | Not-determined | Genome-fragment | NA | NA | 0 |
| S12-final-viral-combined_k141_263457\|\|full | 1218 | No | NA | 3 | 0 | 2 | Not-determined | Genome-fragment | NA | NA | 0 |
| S12-final-viral-combined_k141_297545\|\|full | 135267 | Yes | 52934 | 131 | 15 | 50 | Medium-quality | Genome-fragment | 66.71 | HMM-based (lower-bound) | 60.87 |
| S12-final-viral-combined_k141_213759\|\|full | 6027 | No | NA | 7 | 0 | 5 | Not-determined | Genome-fragment | NA | NA | 0 |
| S8-final-viral-combined_k141_251499\|\|full | 3000 | No | NA | 4 | 0 | 1 | Low-quality | Genome-fragment | 4.66 | AAI-based (medium-confidence) | 0 |
| S12-final-viral-combined_k141_146283\|\|full | 5356 | No | NA | 5 | 0 | 2 | Not-determined | Genome-fragment | NA | NA | 0 |
| S1-final-viral-combined_k141_237312\|\|full | 3001 | No | NA | 5 | 0 | 1 | Low-quality | Genome-fragment | 4.94 | AAI-based (medium-confidence) | 0 |
| S12-final-viral-combined_k141_146289\|\|full | 2841 | No | NA | 4 | 0 | 0 | Not-determined | Genome-fragment | NA | NA | 0 |
| S12-final-viral-combined_k141_112345\|\|full | 1969 | No | NA | 2 | 0 | 2 | Not-determined | Genome-fragment | NA | NA | 0 |
| S12-final-viral-combined_k141_297578\|\|full | 1080 | No | NA | 2 | 0 | 0 | Not-determined | Genome-fragment | NA | NA | 0 |
| S17-final-viral-combined_k141_164489\|\|full | 3001 | No | NA | 3 | 0 | 2 | Low-quality | Genome-fragment | 0.69 | AAI-based (high-confidence) | 0 |
| S12-final-viral-combined_k141_397759\|\|full | 1726 | No | NA | 3 | 0 | 0 | Not-determined | Genome-fragment | NA | NA | 0 |
| S11-final-viral-combined_k141_185782\|\|full | 3002 | No | NA | 6 | 0 | 0 | Low-quality | Genome-fragment | 6.47 | AAI-based (high-confidence) | 0 |
| S6_k141_112786 | 3002 | No | NA | 6 | 0 | 0 | Low-quality | Genome-fragment | 3.2 | AAI-based (medium-confidence) | 0 |
| S12-final-viral-combined_k141_129156\|\|full | 2602 | No | NA | 3 | 0 | 1 | Not-determined | Genome-fragment | NA | NA | 0 |
| S12-final-viral-combined_k141_281375\|\|full | 3606 | No | NA | 3 | 0 | 2 | Not-determined | Genome-fragment | NA | NA | 0 |
| S5-final-viral-combined_k141_83639\|\|lt2gene | 3003 | No | NA | 2 | 1 | 0 | Low-quality | Genome-fragment | 7.22 | AAI-based (high-confidence) | 0 |
| S12-final-viral-combined_k141_62784\|\|full | 1603 | No | NA | 3 | 0 | 1 | Not-determined | Genome-fragment | NA | NA | 0 |
| S5_k141_87521 | 3003 | No | NA | 3 | 0 | 0 | Low-quality | Genome-fragment | 2.92 | AAI-based (high-confidence) | 0 |
| S12-final-viral-combined_k141_246855\|\|full | 1971 | No | NA | 3 | 0 | 1 | Not-determined | Genome-fragment | NA | NA | 0 |
| S2_k141_168441 | 3003 | No | NA | 4 | 0 | 0 | Low-quality | Genome-fragment | 6.48 | AAI-based (medium-confidence) | 0 |
| S1-final-viral-combined_k141_102706\|\|full | 3004 | No | NA | 6 | 3 | 0 | Low-quality | Genome-fragment | 4.92 | AAI-based (high-confidence) | 0 |
| S12-final-viral-combined_k141_230907\|\|full | 1494 | No | NA | 3 | 0 | 1 | Not-determined | Genome-fragment | NA | NA | 0 |
| S1_k141_384257 | 3004 | No | NA | 4 | 0 | 0 | Low-quality | Genome-fragment | 13.07 | AAI-based (medium-confidence) | 0 |
| S12-final-viral-combined_k141_180229\|\|full | 1193 | No | NA | 3 | 0 | 1 | Not-determined | Genome-fragment | NA | NA | 0 |
| S5-final-viral-combined_k141_208581\|\|full | 3004 | No | NA | 8 | 1 | 0 | Low-quality | Genome-fragment | 4.77 | AAI-based (high-confidence) | 0 |
| S12-final-viral-combined_k141_197274\|\|full | 5050 | No | NA | 8 | 0 | 1 | Not-determined | Genome-fragment | NA | NA | 0 |
| S12-final-viral-combined_k141_246893\|\|full | 2790 | No | NA | 2 | 0 | 1 | Not-determined | Genome-fragment | NA | NA | 0 |
| S7-final-viral-combined_k141_53584\|\|full | 3004 | No | NA | 3 | 1 | 0 | Low-quality | Genome-fragment | 2.84 | AAI-based (high-confidence) | 0 |
| S12-final-viral-combined_k141_129194\|\|full | 1236 | No | NA | 3 | 0 | 1 | Not-determined | Genome-fragment | NA | NA | 0 |
| S12-final-viral-combined_k141_281458\|\|full | 1581 | No | NA | 4 | 0 | 0 | Not-determined | Genome-fragment | NA | NA | 0 |
| S8_k141_252385 | 3004 | No | NA | 2 | 0 | 2 | Low-quality | Genome-fragment | 5.12 | AAI-based (medium-confidence) | 0 |
| S9-final-viral-combined_k141_100813\|\|full | 3005 | No | NA | 3 | 0 | 1 | Low-quality | Genome-fragment | 5.41 | AAI-based (high-confidence) | 0 |
| S12-final-viral-combined_k141_362562\|\|full | 5419 | No | NA | 3 | 0 | 3 | Not-determined | Genome-fragment | NA | NA | 0 |
| S1-final-viral-combined_k141_11881\|\|full | 3005 | No | NA | 7 | 1 | 0 | Low-quality | Genome-fragment | 5.59 | AAI-based (high-confidence) | 0 |
| S7_k141_26847 | 3005 | No | NA | 1 | 0 | 1 | Low-quality | Genome-fragment | 2.71 | AAI-based (medium-confidence) | 0 |
| S11-final-viral-combined_k141_328156\|\|full | 3006 | No | NA | 5 | 1 | 0 | Low-quality | Genome-fragment | 5.99 | AAI-based (high-confidence) | 0 |
| S12-final-viral-combined_k141_263564\|\|full | 1104 | No | NA | 2 | 0 | 1 | Not-determined | Genome-fragment | NA | NA | 0 |
| S12-final-viral-combined_k141_197314\|\|full | 2118 | No | NA | 3 | 0 | 2 | Not-determined | Genome-fragment | NA | NA | 0 |
| S11_k141_54529 | 3006 | No | NA | 4 | 0 | 2 | Low-quality | Genome-fragment | 1.97 | AAI-based (medium-confidence) | 0 |
| S3-final-viral-combined_k141_38873\|\|full | 3006 | No | NA | 4 | 2 | 0 | Low-quality | Genome-fragment | 7.86 | AAI-based (high-confidence) | 0 |
| S12-final-viral-combined_k141_49121\|\|full | 3006 | No | NA | 4 | 3 | 0 | Low-quality | Genome-fragment | 7.61 | AAI-based (medium-confidence) | 0 |
| S12-final-viral-combined_k141_29313\|\|full | 3117 | No | NA | 2 | 0 | 1 | Not-determined | Genome-fragment | NA | NA | 0 |
| S12-final-viral-combined_k141_146410\|\|full | 1409 | No | NA | 4 | 0 | 1 | Not-determined | Genome-fragment | NA | NA | 0 |
| S2-final-viral-combined_k141_81542\|\|full | 3006 | No | NA | 3 | 0 | 2 | Low-quality | Genome-fragment | 0.87 | AAI-based (medium-confidence) | 0 |
| S12-final-viral-combined_k141_12404\|\|full | 1234 | No | NA | 2 | 0 | 2 | Not-determined | Genome-fragment | NA | NA | 0 |
| S12-final-viral-combined_k141_281512\|\|full | 2041 | No | NA | 3 | 0 | 3 | Not-determined | Genome-fragment | NA | NA | 0 |
| S12-final-viral-combined_k141_347759\|\|full | 1932 | No | NA | 3 | 0 | 1 | Not-determined | Genome-fragment | NA | NA | 0 |
| S12-final-viral-combined_k141_12427\|\|full | 2270 | No | NA | 3 | 0 | 2 | Not-determined | Genome-fragment | NA | NA | 0 |
| S1_k141_376155 | 3007 | No | NA | 3 | 0 | 2 | Low-quality | Genome-fragment | 2.83 | AAI-based (medium-confidence) | 0 |
| S12-final-viral-combined_k141_180358\|\|full | 2147 | No | NA | 4 | 0 | 3 | Not-determined | Genome-fragment | NA | NA | 0 |
| S12-final-viral-combined_k141_231085\|\|full | 3222 | No | NA | 5 | 0 | 0 | Not-determined | Genome-fragment | NA | NA | 0 |
| S12-final-viral-combined_k141_12446\|\|full | 1460 | No | NA | 3 | 0 | 1 | Not-determined | Genome-fragment | NA | NA | 0 |
| S12-final-viral-combined_k141_231102\|\|full | 1216 | No | NA | 3 | 0 | 0 | Not-determined | Genome-fragment | NA | NA | 0 |
| S17-final-viral-combined_k141_236398\|\|full | 3007 | No | NA | 6 | 0 | 0 | Low-quality | Genome-fragment | 6.57 | AAI-based (medium-confidence) | 0 |
| S2-final-viral-combined_k141_126628\|\|full | 3007 | No | NA | 3 | 0 | 1 | Low-quality | Genome-fragment | 2.13 | AAI-based (high-confidence) | 0 |
| S12-final-viral-combined_k141_62886\|\|full | 4537 | No | NA | 4 | 0 | 2 | Not-determined | Genome-fragment | NA | NA | 0 |
| S12-final-viral-combined_k141_197428\|\|full | 1620 | No | NA | 2 | 0 | 2 | Not-determined | Genome-fragment | NA | NA | 0 |
| S12-final-viral-combined_k141_163821\|\|full | 5388 | No | NA | 5 | 0 | 2 | Not-determined | Genome-fragment | NA | NA | 0 |
| S8-final-viral-combined_k141_37125\|\|0_partial | 3007 | No | NA | 4 | 1 | 0 | Low-quality | Genome-fragment | 6.22 | AAI-based (medium-confidence) | 0 |
| S12-final-viral-combined_k141_80457\|\|full | 3729 | No | NA | 4 | 0 | 3 | Not-determined | Genome-fragment | NA | NA | 0 |
| S12-final-viral-combined_k141_197436\|\|full | 1162 | No | NA | 3 | 0 | 1 | Not-determined | Genome-fragment | NA | NA | 0 |
| S12-final-viral-combined_k141_281615\|\|full | 1012 | No | NA | 2 | 0 | 0 | Not-determined | Genome-fragment | NA | NA | 0 |
| S12-final-viral-combined_k141_381385\|\|full | 3272 | No | NA | 3 | 0 | 1 | Not-determined | Genome-fragment | NA | NA | 0 |
| S12-final-viral-combined_k141_112535\|\|full | 1810 | No | NA | 2 | 0 | 1 | Not-determined | Genome-fragment | NA | NA | 0 |
| S8_k141_28603 | 3007 | No | NA | 2 | 0 | 2 | Low-quality | Genome-fragment | 0.81 | AAI-based (high-confidence) | 0 |
| S12-final-viral-combined_k141_129352\|\|full | 10804 | No | NA | 11 | 0 | 6 | Not-determined | Genome-fragment | NA | NA | 0 |
| S12-final-viral-combined_k141_231154\|\|full | 1040 | No | NA | 3 | 0 | 1 | Not-determined | Genome-fragment | NA | NA | 0 |
| S5_k141_184591 | 3009 | No | NA | 7 | 2 | 0 | Low-quality | Genome-fragment | 3.92 | HMM-based (lower-bound) | 0 |
| S12-final-viral-combined_k141_163847\|\|full | 2200 | No | NA | 2 | 0 | 2 | Not-determined | Genome-fragment | NA | NA | 0 |
| S12-final-viral-combined_k141_397951\|\|full | 1812 | No | NA | 3 | 0 | 0 | Not-determined | Genome-fragment | NA | NA | 0 |
| S12-final-viral-combined_k141_365561\|\|full | 3009 | No | NA | 3 | 0 | 0 | Low-quality | Genome-fragment | 4.78 | AAI-based (medium-confidence) | 0 |
| S12-final-viral-combined_k141_331143\|\|full | 7504 | No | NA | 8 | 0 | 4 | Not-determined | Genome-fragment | NA | NA | 0 |
| S8-final-viral-combined_k141_73966\|\|full | 3009 | No | NA | 4 | 0 | 3 | Low-quality | Genome-fragment | 2.53 | AAI-based (medium-confidence) | 0 |
| S8-final-viral-combined_k141_157208\|\|full | 3009 | No | NA | 4 | 0 | 2 | Low-quality | Genome-fragment | 4.5 | AAI-based (medium-confidence) | 0 |
| S5-final-viral-combined_k141_204459\|\|full | 3010 | No | NA | 4 | 1 | 0 | Low-quality | Genome-fragment | 7.71 | AAI-based (medium-confidence) | 0 |
| S12-final-viral-combined_k141_397960\|\|full | 1277 | No | NA | 2 | 0 | 0 | Not-determined | Genome-fragment | NA | NA | 0 |
| S2-final-viral-combined_k141_248228\|\|full | 3010 | No | NA | 2 | 0 | 1 | Low-quality | Genome-fragment | 1.28 | AAI-based (high-confidence) | 0 |
| S12-final-viral-combined_k141_362773\|\|full | 2062 | No | NA | 3 | 0 | 1 | Not-determined | Genome-fragment | NA | NA | 0 |
| S11-final-viral-combined_k141_212975\|\|full | 3011 | No | NA | 6 | 0 | 0 | Low-quality | Genome-fragment | 8.15 | AAI-based (high-confidence) | 0 |
| S12-final-viral-combined_k141_331154\|\|full | 1203 | No | NA | 2 | 0 | 1 | Not-determined | Genome-fragment | NA | NA | 0 |
| S12-final-viral-combined_k141_62948\|\|full | 10597 | No | NA | 10 | 0 | 5 | Not-determined | Genome-fragment | NA | NA | 0 |
| S12-final-viral-combined_k141_12539\|\|full | 1446 | No | NA | 3 | 0 | 2 | Not-determined | Genome-fragment | NA | NA | 0 |
| S3_k141_25854 | 3011 | No | NA | 5 | 1 | 0 | Low-quality | Genome-fragment | 7.57 | AAI-based (high-confidence) | 0 |
| S12-final-viral-combined_k141_112654\|\|full | 1714 | No | NA | 3 | 0 | 0 | Not-determined | Genome-fragment | NA | NA | 0 |
| S12-final-viral-combined_k141_315408\|\|full | 1323 | No | NA | 3 | 0 | 0 | Not-determined | Genome-fragment | NA | NA | 0 |
| S17-final-viral-combined_k141_155012\|\|full | 3011 | No | NA | 3 | 0 | 3 | Low-quality | Genome-fragment | 1.27 | AAI-based (medium-confidence) | 0 |
| S8_k141_4896 | 3011 | No | NA | 6 | 0 | 0 | Low-quality | Genome-fragment | 6.58 | AAI-based (high-confidence) | 0 |
| S12-final-viral-combined_k141_129420\|\|full | 1343 | No | NA | 2 | 0 | 1 | Not-determined | Genome-fragment | NA | NA | 0 |
| S12-final-viral-combined_k141_214051\|\|full | 1750 | No | NA | 3 | 0 | 1 | Not-determined | Genome-fragment | NA | NA | 0 |
| S3-final-viral-combined_k141_62421\|\|full | 3012 | No | NA | 3 | 0 | 2 | Low-quality | Genome-fragment | 1.55 | AAI-based (medium-confidence) | 0 |
| S12-final-viral-combined_k141_231250\|\|full | 1698 | No | NA | 3 | 0 | 1 | Not-determined | Genome-fragment | NA | NA | 0 |
| S7-final-viral-combined_k141_117345\|\|full | 3012 | No | NA | 5 | 0 | 0 | Low-quality | Genome-fragment | 7.05 | AAI-based (medium-confidence) | 0 |
| S12-final-viral-combined_k141_163946\|\|full | 1201 | No | NA | 3 | 0 | 2 | Not-determined | Genome-fragment | NA | NA | 0 |
| S12-final-viral-combined_k141_281744\|\|full | 2616 | No | NA | 2 | 0 | 2 | Not-determined | Genome-fragment | NA | NA | 0 |
| S12-final-viral-combined_k141_297804\|\|full | 4791 | No | NA | 4 | 0 | 2 | Not-determined | Genome-fragment | NA | NA | 0 |
| S12-final-viral-combined_k141_231263\|\|full | 1433 | No | NA | 3 | 0 | 1 | Not-determined | Genome-fragment | NA | NA | 0 |
| S12-final-viral-combined_k141_163962\|\|full | 2098 | No | NA | 3 | 0 | 2 | Not-determined | Genome-fragment | NA | NA | 0 |
| S12-final-viral-combined_k141_398009\|\|full | 2008 | No | NA | 2 | 0 | 1 | Not-determined | Genome-fragment | NA | NA | 0 |
| S4_k141_50998 | 3012 | No | NA | 2 | 0 | 1 | Low-quality | Genome-fragment | 0.96 | AAI-based (medium-confidence) | 0 |
| S12-final-viral-combined_k141_197519\|\|full | 2954 | No | NA | 3 | 0 | 2 | Not-determined | Genome-fragment | NA | NA | 0 |
| S5_k141_77347 | 3013 | No | NA | 5 | 0 | 0 | Low-quality | Genome-fragment | 6.27 | AAI-based (medium-confidence) | 0 |
| S12-final-viral-combined_k141_347964\|\|full | 1078 | No | NA | 3 | 0 | 1 | Not-determined | Genome-fragment | NA | NA | 0 |
| S12-final-viral-combined_k141_362844\|\|full | 2324 | No | NA | 2 | 0 | 2 | Not-determined | Genome-fragment | NA | NA | 0 |
| S17-final-viral-combined_k141_147736\|\|full | 3013 | No | NA | 3 | 0 | 3 | Low-quality | Genome-fragment | 4.99 | AAI-based (medium-confidence) | 0 |
| S12-final-viral-combined_k141_388665\|\|full | 3013 | No | NA | 4 | 0 | 0 | Low-quality | Genome-fragment | 4.95 | AAI-based (high-confidence) | 0 |
| S12-final-viral-combined_k141_180499\|\|full | 1680 | No | NA | 3 | 0 | 2 | Not-determined | Genome-fragment | NA | NA | 0 |
| S12-final-viral-combined_k141_129498\|\|full | 2633 | No | NA | 3 | 0 | 2 | Not-determined | Genome-fragment | NA | NA | 0 |
| S12-final-viral-combined_k141_297839\|\|full | 2136 | No | NA | 2 | 0 | 2 | Not-determined | Genome-fragment | NA | NA | 0 |
| S12-final-viral-combined_k141_362863\|\|full | 2193 | No | NA | 4 | 0 | 1 | Not-determined | Genome-fragment | NA | NA | 0 |
| S12-final-viral-combined_k141_12610\|\|full | 1684 | No | NA | 3 | 0 | 1 | Not-determined | Genome-fragment | NA | NA | 0 |
| S9-final-viral-combined_k141_96950\|\|full | 3014 | No | NA | 7 | 4 | 0 | Low-quality | Genome-fragment | 6.96 | AAI-based (high-confidence) | 0 |
| S12-final-viral-combined_k141_398055\|\|full | 1255 | No | NA | 2 | 0 | 2 | Not-determined | Genome-fragment | NA | NA | 0 |
| S17-k141_133662 | 3014 | No | NA | 5 | 1 | 0 | Low-quality | Genome-fragment | 4.81 | AAI-based (high-confidence) | 0 |
| S12-final-viral-combined_k141_263735\|\|full | 1450 | No | NA | 5 | 0 | 1 | Not-determined | Genome-fragment | NA | NA | 0 |
| S2_k141_171248 | 3015 | No | NA | 6 | 4 | 0 | Low-quality | Genome-fragment | 6.88 | AAI-based (high-confidence) | 0 |
| S12-final-viral-combined_k141_29582\|\|full | 4085 | No | NA | 6 | 0 | 0 | Not-determined | Genome-fragment | NA | NA | 0 |
| S12-final-viral-combined_k141_80630\|\|full | 2109 | No | NA | 3 | 0 | 2 | Not-determined | Genome-fragment | NA | NA | 0 |
| S12-final-viral-combined_k141_197614\|\|full | 1288 | No | NA | 3 | 0 | 0 | Not-determined | Genome-fragment | NA | NA | 0 |
| S11-final-viral-combined_k141_98127\|\|full | 3016 | No | NA | 3 | 0 | 0 | Low-quality | Genome-fragment | 2.95 | AAI-based (high-confidence) | 0 |
| S5-final-viral-combined_k141_160883\|\|full | 3016 | No | NA | 7 | 5 | 0 | Low-quality | Genome-fragment | 6.99 | AAI-based (medium-confidence) | 0 |
| S12-final-viral-combined_k141_129554\|\|full | 27558 | No | NA | 39 | 15 | 0 | Medium-quality | Genome-fragment | 56.85 | AAI-based (high-confidence) | 0 |
| S8-final-viral-combined_k141_84940\|\|full | 3016 | No | NA | 3 | 0 | 2 | Low-quality | Genome-fragment | 2.17 | AAI-based (medium-confidence) | 0 |
| S12-final-viral-combined_k141_80663\|\|full | 1738 | No | NA | 3 | 0 | 0 | Not-determined | Genome-fragment | NA | NA | 0 |
| S12-final-viral-combined_k141_331337\|\|full | 2014 | No | NA | 4 | 0 | 1 | Not-determined | Genome-fragment | NA | NA | 0 |
| S11-final-viral-combined_k141_116899\|\|full | 3017 | No | NA | 3 | 0 | 3 | Low-quality | Genome-fragment | 4.72 | AAI-based (medium-confidence) | 0 |
| S12-final-viral-combined_k141_96484\|\|full | 6645 | No | NA | 7 | 0 | 6 | Not-determined | Genome-fragment | NA | NA | 0 |
| S1-final-viral-combined_k141_217574\|\|full | 3017 | No | NA | 5 | 0 | 1 | Low-quality | Genome-fragment | 3.21 | AAI-based (medium-confidence) | 0 |
| S1_k141_210877 | 3018 | No | NA | 2 | 0 | 1 | Low-quality | Genome-fragment | 1.29 | AAI-based (medium-confidence) | 0 |
| S7_k141_192382 | 3018 | No | NA | 3 | 0 | 1 | Low-quality | Genome-fragment | 21.55 | AAI-based (medium-confidence) | 0 |
| S17-final-viral-combined_k141_239089\|\|full | 3018 | No | NA | 3 | 0 | 0 | Low-quality | Genome-fragment | 1.45 | AAI-based (medium-confidence) | 0 |
| S12-final-viral-combined_k141_231434\|\|full | 1608 | No | NA | 2 | 0 | 0 | Not-determined | Genome-fragment | NA | NA | 0 |
| S2-final-viral-combined_k141_75606\|\|full | 3018 | No | NA | 4 | 0 | 0 | Low-quality | Genome-fragment | 4.74 | AAI-based (medium-confidence) | 0 |
| S1-final-viral-combined_k141_321800\|\|full | 3019 | No | NA | 5 | 0 | 2 | Low-quality | Genome-fragment | 4.34 | AAI-based (medium-confidence) | 0 |
| S12-final-viral-combined_k141_247304\|\|full | 1069 | No | NA | 2 | 0 | 1 | Not-determined | Genome-fragment | NA | NA | 0 |
| S7-final-viral-combined_k141_27748\|\|full | 3019 | No | NA | 5 | 0 | 0 | Low-quality | Genome-fragment | 2.07 | AAI-based (medium-confidence) | 0 |
| S12-final-viral-combined_k141_63020\|\|full | 1484 | No | NA | 2 | 0 | 0 | Not-determined | Genome-fragment | NA | NA | 0 |
| S12-final-viral-combined_k141_231447\|\|full | 3127 | No | NA | 4 | 0 | 0 | Not-determined | Genome-fragment | NA | NA | 0 |
| S12-final-viral-combined_k141_63023\|\|full | 2311 | No | NA | 3 | 0 | 3 | Not-determined | Genome-fragment | NA | NA | 0 |
| S12-final-viral-combined_k141_214160\|\|full | 1077 | No | NA | 2 | 0 | 1 | Not-determined | Genome-fragment | NA | NA | 0 |
| S12-final-viral-combined_k141_12668\|\|full | 2930 | No | NA | 5 | 0 | 1 | Not-determined | Genome-fragment | NA | NA | 0 |
| S12-final-viral-combined_k141_381741\|\|full | 3138 | No | NA | 5 | 0 | 2 | Not-determined | Genome-fragment | NA | NA | 0 |
| S12-final-viral-combined_k141_297960\|\|full | 2456 | No | NA | 4 | 0 | 1 | Not-determined | Genome-fragment | NA | NA | 0 |
| S12-final-viral-combined_k141_348107\|\|full | 10689 | No | NA | 9 | 0 | 9 | Not-determined | Genome-fragment | NA | NA | 0 |
| S12-final-viral-combined_k141_29699\|\|full | 1111 | No | NA | 2 | 0 | 2 | Not-determined | Genome-fragment | NA | NA | 0 |
| S12-final-viral-combined_k141_214198\|\|full | 1348 | No | NA | 4 | 0 | 0 | Not-determined | Genome-fragment | NA | NA | 0 |
| S12-final-viral-combined_k141_29704\|\|full | 3158 | No | NA | 3 | 0 | 2 | Not-determined | Genome-fragment | NA | NA | 0 |
| S12-final-viral-combined_k141_96566\|\|full | 1485 | No | NA | 3 | 0 | 3 | Not-determined | Genome-fragment | NA | NA | 0 |
| S1-final-viral-combined_k141_303925\|\|full | 3020 | No | NA | 5 | 0 | 3 | Low-quality | Genome-fragment | 1.76 | AAI-based (medium-confidence) | 0 |
| S12-final-viral-combined_k141_331426\|\|full | 1005 | No | NA | 2 | 0 | 1 | Not-determined | Genome-fragment | NA | NA | 0 |
| S12-final-viral-combined_k141_348123\|\|full | 3679 | No | NA | 4 | 0 | 3 | Not-determined | Genome-fragment | NA | NA | 0 |
| S12-final-viral-combined_k141_180641\|\|full | 2513 | No | NA | 2 | 0 | 1 | Not-determined | Genome-fragment | NA | NA | 0 |
| S1-final-viral-combined_k141_221700\|\|full | 3020 | No | NA | 2 | 0 | 2 | Low-quality | Genome-fragment | 1.37 | AAI-based (medium-confidence) | 0 |
| S3_k141_46956 | 3020 | No | NA | 3 | 0 | 1 | Low-quality | Genome-fragment | 3.83 | AAI-based (medium-confidence) | 0 |
| S12-final-viral-combined_k141_47635\|\|full | 2529 | No | NA | 2 | 0 | 2 | Not-determined | Genome-fragment | NA | NA | 0 |
| S17-k141_220874 | 3020 | No | NA | 3 | 0 | 1 | Low-quality | Genome-fragment | 0.73 | AAI-based (medium-confidence) | 0 |
| S11-final-viral-combined_k141_298642\|\|full | 3021 | No | NA | 3 | 1 | 2 | Low-quality | Genome-fragment | 1.15 | HMM-based (lower-bound) | 0 |
| S12-final-viral-combined_k141_63056\|\|full | 35070 | No | NA | 41 | 8 | 3 | Medium-quality | Genome-fragment | 62.07 | AAI-based (medium-confidence) | 0 |
| S12-final-viral-combined_k141_363069\|\|full | 1486 | No | NA | 3 | 0 | 0 | Not-determined | Genome-fragment | NA | NA | 0 |
| S12-final-viral-combined_k141_164241\|\|full | 6213 | No | NA | 4 | 0 | 3 | Not-determined | Genome-fragment | NA | NA | 0 |
| S12-final-viral-combined_k141_263961\|\|full | 2757 | No | NA | 4 | 0 | 2 | Not-determined | Genome-fragment | NA | NA | 0 |
| S5-final-viral-combined_k141_154739\|\|full | 3021 | Yes | 354 | 4 | 1 | 3 | Low-quality | Genome-fragment | 0.16 | HMM-based (lower-bound) | 88.28 |
| S17-k141_155194 | 3021 | No | NA | 4 | 0 | 2 | Low-quality | Genome-fragment | 6.35 | AAI-based (medium-confidence) | 0 |
| S12-final-viral-combined_k141_164253\|\|full | 3318 | No | NA | 5 | 0 | 1 | Not-determined | Genome-fragment | NA | NA | 0 |
| S11-final-viral-combined_k141_185655\|\|full | 3022 | No | NA | 3 | 0 | 2 | Low-quality | Genome-fragment | 1.13 | AAI-based (medium-confidence) | 0 |
| S12-final-viral-combined_k141_231552\|\|full | 2049 | No | NA | 4 | 0 | 2 | Not-determined | Genome-fragment | NA | NA | 0 |
| S12-final-viral-combined_k141_146830\|\|full | 1876 | No | NA | 2 | 0 | 1 | Not-determined | Genome-fragment | NA | NA | 0 |
| S5_k141_180047 | 3022 | No | NA | 2 | 0 | 0 | Low-quality | Genome-fragment | 4 | AAI-based (high-confidence) | 0 |
| S12-final-viral-combined_k141_214254\|\|full | 2772 | No | NA | 5 | 0 | 0 | Not-determined | Genome-fragment | NA | NA | 0 |
| S12-final-viral-combined_k141_315618\|\|full | 3217 | No | NA | 2 | 0 | 1 | Not-determined | Genome-fragment | NA | NA | 0 |
| S12-final-viral-combined_k141_47707\|\|full | 1804 | No | NA | 3 | 0 | 1 | Not-determined | Genome-fragment | NA | NA | 0 |
| S12-final-viral-combined_k141_348202\|\|full | 1520 | No | NA | 3 | 0 | 1 | Not-determined | Genome-fragment | NA | NA | 0 |
| S12-final-viral-combined_k141_263989\|\|full | 1068 | No | NA | 2 | 0 | 1 | Not-determined | Genome-fragment | NA | NA | 0 |
| S12-final-viral-combined_k141_282143\|\|full | 3022 | No | NA | 3 | 0 | 1 | Low-quality | Genome-fragment | 6.4 | AAI-based (medium-confidence) | 0 |
| S2-final-viral-combined_k141_239384\|\|full | 3022 | Yes | 1948 | 6 | 1 | 1 | Low-quality | Genome-fragment | 2.46 | AAI-based (medium-confidence) | 35.54 |
| S17-final-viral-combined_k141_100484\|\|full | 3023 | No | NA | 3 | 0 | 3 | Low-quality | Genome-fragment | 1.31 | AAI-based (medium-confidence) | 0 |
| S17-k141_236933 | 3023 | No | NA | 3 | 0 | 0 | Low-quality | Genome-fragment | 7.11 | AAI-based (medium-confidence) | 0 |
| S17-k141_19140 | 3023 | No | NA | 4 | 1 | 0 | Low-quality | Genome-fragment | 7.65 | AAI-based (high-confidence) | 0 |
| S8-final-viral-combined_k141_273786\|\|full | 3023 | No | NA | 3 | 0 | 1 | Low-quality | Genome-fragment | 1.37 | AAI-based (medium-confidence) | 0 |
| S12-final-viral-combined_k141_12789\|\|full | 2903 | No | NA | 5 | 0 | 1 | Not-determined | Genome-fragment | NA | NA | 0 |
| S12-final-viral-combined_k141_80810\|\|full | 2168 | No | NA | 2 | 0 | 1 | Not-determined | Genome-fragment | NA | NA | 0 |
| S12-final-viral-combined_k141_247459\|\|full | 2308 | No | NA | 3 | 0 | 2 | Not-determined | Genome-fragment | NA | NA | 0 |
| S12-final-viral-combined_k141_282077\|\|full | 2557 | No | NA | 2 | 0 | 1 | Not-determined | Genome-fragment | NA | NA | 0 |
| S1_k141_149386 | 3025 | No | NA | 4 | 1 | 0 | Low-quality | Genome-fragment | 1.36 | HMM-based (lower-bound) | 0 |
| S12-final-viral-combined_k141_129740\|\|full | 2707 | No | NA | 2 | 0 | 2 | Not-determined | Genome-fragment | NA | NA | 0 |
| S3-final-viral-combined_k141_58212\|\|full | 3025 | No | NA | 5 | 0 | 2 | Low-quality | Genome-fragment | 7.86 | AAI-based (medium-confidence) | 0 |
| S12-final-viral-combined_k141_146863\|\|full | 5194 | No | NA | 6 | 0 | 0 | Not-determined | Genome-fragment | NA | NA | 0 |
| S12-final-viral-combined_k141_112981\|\|full | 2263 | No | NA | 3 | 0 | 1 | Not-determined | Genome-fragment | NA | NA | 0 |
| S12-final-viral-combined_k141_164334\|\|full | 1461 | No | NA | 2 | 0 | 1 | Not-determined | Genome-fragment | NA | NA | 0 |
| S12-final-viral-combined_k141_197822\|\|full | 4921 | No | NA | 6 | 0 | 2 | Not-determined | Genome-fragment | NA | NA | 0 |
| S12-final-viral-combined_k141_164336\|\|full | 1298 | No | NA | 3 | 0 | 1 | Not-determined | Genome-fragment | NA | NA | 0 |
| S5-final-viral-combined_k141_189959\|\|lt2gene | 3025 | No | NA | 2 | 2 | 0 | Low-quality | Genome-fragment | 1.47 | HMM-based (lower-bound) | 0 |
| S12-final-viral-combined_k141_282104\|\|full | 4356 | No | NA | 5 | 0 | 3 | Not-determined | Genome-fragment | NA | NA | 0 |
| S12-final-viral-combined_k141_197834\|\|full | 3431 | No | NA | 3 | 0 | 1 | Not-determined | Genome-fragment | NA | NA | 0 |
| S12-final-viral-combined_k141_12814\|\|full | 2218 | No | NA | 3 | 0 | 1 | Not-determined | Genome-fragment | NA | NA | 0 |
| S12-final-viral-combined_k141_381872\|\|full | 13772 | No | NA | 19 | 0 | 3 | Not-determined | Genome-fragment | NA | NA | 0 |
| S12-final-viral-combined_k141_298079\|\|full | 2480 | No | NA | 2 | 0 | 2 | Not-determined | Genome-fragment | NA | NA | 0 |
| S8-final-viral-combined_k141_177778\|\|full | 3025 | Yes | 1751 | 3 | 1 | 1 | Low-quality | Genome-fragment | 4.08 | AAI-based (medium-confidence) | 42.12 |
| S9_k141_56516 | 3026 | Yes | 777 | 5 | 1 | 1 | Low-quality | Genome-fragment | 1.46 | AAI-based (high-confidence) | 74.32 |
| S12-final-viral-combined_k141_180831\|\|full | 1604 | No | NA | 3 | 0 | 2 | Not-determined | Genome-fragment | NA | NA | 0 |
| S9_k141_127788 | 3026 | No | NA | 5 | 2 | 0 | Low-quality | Genome-fragment | 7.62 | AAI-based (high-confidence) | 0 |
| S3-final-viral-combined_k141_115846\|\|full | 3026 | No | NA | 3 | 0 | 2 | Low-quality | Genome-fragment | 0.58 | AAI-based (medium-confidence) | 0 |
| S12-final-viral-combined_k141_214351\|\|full | 1025 | No | NA | 2 | 0 | 0 | Not-determined | Genome-fragment | NA | NA | 0 |
| S12-final-viral-combined_k141_113038\|\|full | 1637 | No | NA | 3 | 0 | 1 | Not-determined | Genome-fragment | NA | NA | 0 |
| S17-final-viral-combined_k141_169129\|\|full | 3026 | No | NA | 2 | 0 | 2 | Low-quality | Genome-fragment | 2.89 | AAI-based (medium-confidence) | 0 |
| S17-final-viral-combined_k141_41517\|\|full | 3026 | No | NA | 3 | 2 | 0 | Low-quality | Genome-fragment | 8.29 | AAI-based (medium-confidence) | 0 |
| S12-final-viral-combined_k141_363169\|\|full | 4728 | No | NA | 6 | 0 | 0 | Not-determined | Genome-fragment | NA | NA | 0 |
| S12-final-viral-combined_k141_315758\|\|full | 2023 | No | NA | 3 | 0 | 1 | Not-determined | Genome-fragment | NA | NA | 0 |
| S17-k141_86449 | 3026 | No | NA | 3 | 1 | 0 | Low-quality | Genome-fragment | 6.53 | AAI-based (high-confidence) | 0 |
| S12-final-viral-combined_k141_113078\|\|full | 1705 | No | NA | 3 | 0 | 0 | Not-determined | Genome-fragment | NA | NA | 0 |
| S12-final-viral-combined_k141_197918\|\|full | 2789 | No | NA | 4 | 0 | 2 | Not-determined | Genome-fragment | NA | NA | 0 |
| S12-final-viral-combined_k141_231691\|\|full | 1019 | No | NA | 2 | 0 | 0 | Not-determined | Genome-fragment | NA | NA | 0 |
| S12-final-viral-combined_k141_247576\|\|full | 1600 | No | NA | 2 | 0 | 0 | Not-determined | Genome-fragment | NA | NA | 0 |
| S12-final-viral-combined_k141_264074\|\|full | 1231 | No | NA | 3 | 0 | 1 | Not-determined | Genome-fragment | NA | NA | 0 |
| S12_k141_16557 | 3026 | No | NA | 5 | 0 | 1 | Low-quality | Genome-fragment | 5.9 | AAI-based (medium-confidence) | 0 |
| S12-final-viral-combined_k141_63322\|\|full | 3565 | No | NA | 2 | 0 | 1 | Not-determined | Genome-fragment | NA | NA | 0 |
| S6_k141_116762 | 3027 | No | NA | 3 | 0 | 2 | Low-quality | Genome-fragment | 0.43 | AAI-based (medium-confidence) | 0 |
| S1-final-viral-combined_k141_131500\|\|full | 3028 | No | NA | 3 | 0 | 3 | Low-quality | Genome-fragment | 1.47 | AAI-based (medium-confidence) | 0 |
| S12-final-viral-combined_k141_363245\|\|full | 1800 | No | NA | 3 | 0 | 0 | Not-determined | Genome-fragment | NA | NA | 0 |
| S12-final-viral-combined_k141_214422\|\|full | 3679 | No | NA | 4 | 0 | 2 | Not-determined | Genome-fragment | NA | NA | 0 |
| S12-final-viral-combined_k141_96781\|\|full | 3501 | No | NA | 5 | 0 | 0 | Not-determined | Genome-fragment | NA | NA | 0 |
| S12-final-viral-combined_k141_197946\|\|full | 9220 | No | NA | 9 | 0 | 7 | Not-determined | Genome-fragment | NA | NA | 0 |
| S12-final-viral-combined_k141_264077\|\|full | 32216 | No | NA | 53 | 8 | 1 | Medium-quality | Genome-fragment | 60.94 | AAI-based (high-confidence) | 0 |
| S12-final-viral-combined_k141_247647\|\|full | 2174 | No | NA | 2 | 0 | 0 | Not-determined | Genome-fragment | NA | NA | 0 |
| S12-final-viral-combined_k141_146944\|\|full | 1193 | No | NA | 3 | 0 | 1 | Not-determined | Genome-fragment | NA | NA | 0 |
| S12-final-viral-combined_k141_315852\|\|full | 1472 | No | NA | 2 | 0 | 1 | Not-determined | Genome-fragment | NA | NA | 0 |
| S12-final-viral-combined_k141_180924\|\|full | 7751 | No | NA | 10 | 0 | 6 | Not-determined | Genome-fragment | NA | NA | 0 |
| S7-final-viral-combined_k141_152066\|\|full | 3028 | No | NA | 4 | 2 | 0 | Low-quality | Genome-fragment | 5.52 | AAI-based (high-confidence) | 0 |
| S12-final-viral-combined_k141_47819\|\|full | 3896 | No | NA | 5 | 0 | 2 | Not-determined | Genome-fragment | NA | NA | 0 |
| S2_k141_175743 | 3028 | No | NA | 1 | 0 | 1 | Low-quality | Genome-fragment | 2.97 | AAI-based (medium-confidence) | 0 |
| S12-final-viral-combined_k141_363282\|\|full | 3112 | No | NA | 4 | 0 | 2 | Not-determined | Genome-fragment | NA | NA | 0 |
| S6_k141_98410 | 3028 | No | NA | 4 | 1 | 0 | Low-quality | Genome-fragment | 3.07 | AAI-based (high-confidence) | 0 |
| S8-final-viral-combined_k141_256488\|\|full | 3028 | No | NA | 5 | 0 | 1 | Low-quality | Genome-fragment | 7.69 | AAI-based (medium-confidence) | 0 |
| S1-final-viral-combined_k141_273238\|\|full | 3029 | No | NA | 4 | 0 | 1 | Low-quality | Genome-fragment | 3.57 | AAI-based (medium-confidence) | 0 |
| S1_k141_73152 | 3029 | No | NA | 4 | 1 | 0 | Low-quality | Genome-fragment | 3.61 | HMM-based (lower-bound) | 0 |
| S12-final-viral-combined_k141_180947\|\|full | 2725 | No | NA | 5 | 0 | 3 | Not-determined | Genome-fragment | NA | NA | 0 |
| S12-final-viral-combined_k141_231802\|\|full | 1896 | No | NA | 2 | 0 | 1 | Not-determined | Genome-fragment | NA | NA | 0 |
| S12-final-viral-combined_k141_198000\|\|full | 3081 | No | NA | 5 | 0 | 5 | Not-determined | Genome-fragment | NA | NA | 0 |
| S7-final-viral-combined_k141_243623\|\|full | 3029 | No | NA | 5 | 0 | 0 | Low-quality | Genome-fragment | 3.76 | AAI-based (high-confidence) | 0 |
| S12-final-viral-combined_k141_164553\|\|full | 1684 | No | NA | 3 | 0 | 1 | Not-determined | Genome-fragment | NA | NA | 0 |
| S4-final-viral-combined_k141_40140\|\|full | 3029 | No | NA | 3 | 0 | 3 | Low-quality | Genome-fragment | 2.09 | AAI-based (medium-confidence) | 0 |
| S8-final-viral-combined_k141_207261\|\|full | 3029 | No | NA | 5 | 0 | 0 | Low-quality | Genome-fragment | 8.38 | AAI-based (high-confidence) | 0 |
| S12-final-viral-combined_k141_282330\|\|full | 1326 | No | NA | 3 | 0 | 0 | Not-determined | Genome-fragment | NA | NA | 0 |
| S9-final-viral-combined_k141_43537\|\|full | 3030 | No | NA | 6 | 0 | 0 | Low-quality | Genome-fragment | 7.59 | AAI-based (medium-confidence) | 0 |
| S12-final-viral-combined_k141_129799\|\|full | 86035 | Yes | 43504 | 102 | 19 | 24 | Complete | High-quality | 100 | Provirus (medium-confidence) | 49.43 |
| S12-final-viral-combined_k141_96857\|\|full | 2210 | No | NA | 5 | 0 | 0 | Not-determined | Genome-fragment | NA | NA | 0 |
| S1-final-viral-combined_k141_153822\|\|full | 3030 | No | NA | 5 | 0 | 3 | Low-quality | Genome-fragment | 6.88 | AAI-based (medium-confidence) | 0 |
| S1-final-viral-combined_k141_23000\|\|full | 3030 | No | NA | 3 | 0 | 2 | Low-quality | Genome-fragment | 2.12 | AAI-based (medium-confidence) | 0 |
| S1-final-viral-combined_k141_336393\|\|full | 3030 | No | NA | 2 | 0 | 2 | Low-quality | Genome-fragment | 2.76 | AAI-based (medium-confidence) | 0 |
| S12-final-viral-combined_k141_13003\|\|full | 1264 | No | NA | 3 | 0 | 0 | Not-determined | Genome-fragment | NA | NA | 0 |
| S3-final-viral-combined_k141_66894\|\|full | 3030 | No | NA | 4 | 1 | 0 | Low-quality | Genome-fragment | 9.12 | AAI-based (high-confidence) | 0 |
| S12-final-viral-combined_k141_315917\|\|full | 2540 | No | NA | 2 | 0 | 2 | Not-determined | Genome-fragment | NA | NA | 0 |
| S12-final-viral-combined_k141_181030\|\|full | 4725 | No | NA | 6 | 0 | 2 | Not-determined | Genome-fragment | NA | NA | 0 |
| S12_k141_253156 | 3030 | No | NA | 7 | 5 | 0 | Low-quality | Genome-fragment | 9.34 | AAI-based (high-confidence) | 0 |
| S17-final-viral-combined_k141_124415\|\|full | 3031 | No | NA | 4 | 0 | 0 | Low-quality | Genome-fragment | 2.61 | AAI-based (medium-confidence) | 0 |
| S12-final-viral-combined_k141_331792\|\|full | 3604 | No | NA | 4 | 0 | 0 | Not-determined | Genome-fragment | NA | NA | 0 |
| S4_k141_28899 | 3031 | No | NA | 3 | 0 | 3 | Low-quality | Genome-fragment | 7.12 | AAI-based (medium-confidence) | 0 |
| S12-final-viral-combined_k141_47887\|\|full | 1435 | No | NA | 5 | 0 | 0 | Not-determined | Genome-fragment | NA | NA | 0 |
| S12-final-viral-combined_k141_381999\|\|full | 3653 | No | NA | 3 | 0 | 2 | Not-determined | Genome-fragment | NA | NA | 0 |
| S12-final-viral-combined_k141_47896\|\|full | 2578 | No | NA | 3 | 0 | 1 | Not-determined | Genome-fragment | NA | NA | 0 |
| S4_k141_98576 | 3031 | No | NA | 3 | 0 | 2 | Low-quality | Genome-fragment | 0.86 | AAI-based (medium-confidence) | 0 |
| S12-final-viral-combined_k141_363325\|\|full | 1383 | No | NA | 4 | 0 | 0 | Not-determined | Genome-fragment | NA | NA | 0 |
| S8-final-viral-combined_k141_54404\|\|full | 3031 | Yes | 1419 | 5 | 1 | 1 | Low-quality | Genome-fragment | 3.24 | AAI-based (medium-confidence) | 53.18 |
| S12-final-viral-combined_k141_198096\|\|full | 1927 | No | NA | 6 | 0 | 1 | Not-determined | Genome-fragment | NA | NA | 0 |
| S12-final-viral-combined_k141_363344\|\|full | 2974 | No | NA | 2 | 0 | 2 | Not-determined | Genome-fragment | NA | NA | 0 |
| S9-final-viral-combined_k141_83417\|\|full | 3032 | No | NA | 4 | 0 | 3 | Low-quality | Genome-fragment | 5.03 | AAI-based (high-confidence) | 0 |
| S12-final-viral-combined_k141_298258\|\|full | 3034 | No | NA | 3 | 0 | 2 | Not-determined | Genome-fragment | NA | NA | 0 |
| S12-final-viral-combined_k141_81020\|\|full | 1422 | No | NA | 2 | 0 | 2 | Not-determined | Genome-fragment | NA | NA | 0 |
| S12-final-viral-combined_k141_30120\|\|full | 8868 | No | NA | 10 | 0 | 1 | Not-determined | Genome-fragment | NA | NA | 0 |
| S12-final-viral-combined_k141_181120\|\|full | 5663 | No | NA | 6 | 0 | 2 | Not-determined | Genome-fragment | NA | NA | 0 |
| S12-final-viral-combined_k141_96959\|\|full | 1926 | No | NA | 3 | 0 | 0 | Not-determined | Genome-fragment | NA | NA | 0 |
| S12-final-viral-combined_k141_247836\|\|full | 3086 | No | NA | 5 | 0 | 0 | Not-determined | Genome-fragment | NA | NA | 0 |
| S11-final-viral-combined_k141_179046\|\|full | 3032 | No | NA | 2 | 1 | 0 | Low-quality | Genome-fragment | 6.62 | AAI-based (medium-confidence) | 0 |
| S12-final-viral-combined_k141_13125\|\|full | 1287 | No | NA | 2 | 0 | 0 | Not-determined | Genome-fragment | NA | NA | 0 |
| S1-final-viral-combined_k141_40195\|\|full | 3032 | No | NA | 8 | 4 | 0 | Low-quality | Genome-fragment | 5.64 | AAI-based (high-confidence) | 0 |
| S1_k141_290738 | 3032 | No | NA | 5 | 1 | 0 | Low-quality | Genome-fragment | 1.25 | HMM-based (lower-bound) | 0 |
| S12-final-viral-combined_k141_181144\|\|full | 1765 | No | NA | 4 | 0 | 1 | Not-determined | Genome-fragment | NA | NA | 0 |
| S12-final-viral-combined_k141_382063\|\|full | 3483 | No | NA | 4 | 0 | 4 | Not-determined | Genome-fragment | NA | NA | 0 |
| S12-final-viral-combined_k141_30146\|\|full | 3443 | No | NA | 5 | 0 | 0 | Not-determined | Genome-fragment | NA | NA | 0 |
| S12-final-viral-combined_k141_13157\|\|full | 4198 | No | NA | 3 | 0 | 2 | Not-determined | Genome-fragment | NA | NA | 0 |
| S12-final-viral-combined_k141_398335\|\|full | 1066 | No | NA | 2 | 0 | 1 | Not-determined | Genome-fragment | NA | NA | 0 |
| S11_k141_291830 | 3033 | No | NA | 4 | 2 | 0 | Low-quality | Genome-fragment | 10.34 | HMM-based (lower-bound) | 0 |
| S12-final-viral-combined_k141_63514\|\|full | 1071 | No | NA | 2 | 0 | 1 | Not-determined | Genome-fragment | NA | NA | 0 |
| S1-final-viral-combined_k141_382999\|\|full | 3033 | No | NA | 6 | 2 | 0 | Low-quality | Genome-fragment | 3.7 | HMM-based (lower-bound) | 0 |
| S12-final-viral-combined_k141_298329\|\|full | 14352 | No | NA | 15 | 0 | 10 | Not-determined | Genome-fragment | NA | NA | 0 |
| S1_k141_165188 | 3033 | No | NA | 5 | 0 | 0 | Low-quality | Genome-fragment | 4.72 | AAI-based (high-confidence) | 0 |
| S12-final-viral-combined_k141_13181\|\|full | 10830 | No | NA | 6 | 0 | 0 | Not-determined | Genome-fragment | NA | NA | 0 |
| S5-final-viral-combined_k141_106309\|\|full | 3033 | No | NA | 3 | 0 | 2 | Low-quality | Genome-fragment | 3.73 | AAI-based (medium-confidence) | 0 |
| S7_k141_196797 | 3033 | No | NA | 3 | 0 | 0 | Low-quality | Genome-fragment | 0.83 | AAI-based (medium-confidence) | 0 |
| S12-final-viral-combined_k141_81142\|\|full | 1779 | No | NA | 3 | 0 | 1 | Not-determined | Genome-fragment | NA | NA | 0 |
| S12-final-viral-combined_k141_214783\|\|full | 4648 | No | NA | 7 | 0 | 1 | Not-determined | Genome-fragment | NA | NA | 0 |
| S9-final-viral-combined_k141_46146\|\|full | 3034 | No | NA | 2 | 0 | 0 | Low-quality | Genome-fragment | 8.28 | AAI-based (high-confidence) | 0 |
| S12-final-viral-combined_k141_13212\|\|full | 1555 | No | NA | 3 | 0 | 0 | Not-determined | Genome-fragment | NA | NA | 0 |
| S12-final-viral-combined_k141_81159\|\|full | 2718 | No | NA | 5 | 0 | 2 | Not-determined | Genome-fragment | NA | NA | 0 |
| S12-final-viral-combined_k141_63577\|\|full | 1547 | No | NA | 2 | 0 | 2 | Not-determined | Genome-fragment | NA | NA | 0 |
| S11-final-viral-combined_k141_69706\|\|full | 3034 | No | NA | 4 | 3 | 0 | Low-quality | Genome-fragment | 2 | HMM-based (lower-bound) | 0 |
| S12-final-viral-combined_k141_331983\|\|full | 1727 | No | NA | 2 | 0 | 2 | Not-determined | Genome-fragment | NA | NA | 0 |
| S12-final-viral-combined_k141_363436\|\|full | 6051 | No | NA | 6 | 0 | 6 | Not-determined | Genome-fragment | NA | NA | 0 |
| S11-final-viral-combined_k141_94051\|\|full | 3034 | Yes | 1911 | 2 | 1 | 1 | Low-quality | Genome-fragment | 0.91 | AAI-based (medium-confidence) | 37.01 |
| S12-final-viral-combined_k141_113423\|\|full | 1185 | No | NA | 3 | 0 | 1 | Not-determined | Genome-fragment | NA | NA | 0 |
| S11_k141_287352 | 3034 | No | NA | 7 | 1 | 0 | Low-quality | Genome-fragment | 10.4 | HMM-based (lower-bound) | 0 |
| S4-final-viral-combined_k141_109994\|\|full | 3034 | No | NA | 3 | 0 | 0 | Low-quality | Genome-fragment | 7.39 | AAI-based (medium-confidence) | 0 |
| S12-final-viral-combined_k141_247953\|\|full | 3298 | No | NA | 5 | 0 | 1 | Not-determined | Genome-fragment | NA | NA | 0 |
| S12-final-viral-combined_k141_181244\|\|full | 1560 | No | NA | 2 | 0 | 0 | Not-determined | Genome-fragment | NA | NA | 0 |
| S12-final-viral-combined_k141_398447\|\|full | 1292 | No | NA | 3 | 0 | 1 | Not-determined | Genome-fragment | NA | NA | 0 |
| S12-final-viral-combined_k141_264309\|\|full | 1757 | No | NA | 3 | 0 | 1 | Not-determined | Genome-fragment | NA | NA | 0 |
| S9_k141_80582 | 3035 | No | NA | 5 | 5 | 0 | Low-quality | Genome-fragment | 6.47 | AAI-based (high-confidence) | 0 |
| S1_k141_129374 | 3035 | No | NA | 2 | 0 | 1 | Low-quality | Genome-fragment | 7.17 | AAI-based (medium-confidence) | 0 |
| S12-final-viral-combined_k141_30184\|\|full | 6524 | No | NA | 7 | 0 | 4 | Not-determined | Genome-fragment | NA | NA | 0 |
| S12-final-viral-combined_k141_198224\|\|full | 1742 | No | NA | 4 | 0 | 1 | Not-determined | Genome-fragment | NA | NA | 0 |
| S12-final-viral-combined_k141_63599\|\|full | 4468 | No | NA | 2 | 0 | 2 | Not-determined | Genome-fragment | NA | NA | 0 |
| S12-final-viral-combined_k141_332026\|\|full | 2532 | No | NA | 4 | 0 | 1 | Not-determined | Genome-fragment | NA | NA | 0 |
| S12-final-viral-combined_k141_363475\|\|full | 2146 | No | NA | 3 | 0 | 2 | Not-determined | Genome-fragment | NA | NA | 0 |
| S12-final-viral-combined_k141_181294\|\|full | 1794 | No | NA | 4 | 0 | 0 | Not-determined | Genome-fragment | NA | NA | 0 |
| S12-final-viral-combined_k141_398476\|\|full | 1144 | No | NA | 2 | 0 | 0 | Not-determined | Genome-fragment | NA | NA | 0 |
| S2-final-viral-combined_k141_8088\|\|full | 3035 | No | NA | 7 | 0 | 0 | Low-quality | Genome-fragment | 4.92 | AAI-based (medium-confidence) | 0 |
| S2_k141_113438 | 3035 | No | NA | 2 | 2 | 0 | Low-quality | Genome-fragment | 10.01 | HMM-based (lower-bound) | 0 |
| S12-final-viral-combined_k141_48142\|\|full | 1058 | No | NA | 2 | 0 | 1 | Not-determined | Genome-fragment | NA | NA | 0 |
| S6-final-viral-combined_k141_110981\|\|full | 3035 | No | NA | 2 | 0 | 2 | Low-quality | Genome-fragment | 1.61 | AAI-based (medium-confidence) | 0 |
| S12-final-viral-combined_k141_147163\|\|full | 3976 | No | NA | 7 | 0 | 3 | Not-determined | Genome-fragment | NA | NA | 0 |
| S12-final-viral-combined_k141_21897\|\|full | 3036 | No | NA | 4 | 0 | 2 | Low-quality | Genome-fragment | 4.18 | AAI-based (medium-confidence) | 0 |
| S2-final-viral-combined_k141_96861\|\|full | 3036 | No | NA | 3 | 0 | 0 | Low-quality | Genome-fragment | 3.31 | AAI-based (medium-confidence) | 0 |
| S12-final-viral-combined_k141_232106\|\|full | 1504 | No | NA | 3 | 0 | 0 | Not-determined | Genome-fragment | NA | NA | 0 |
| S12-final-viral-combined_k141_348660\|\|full | 2809 | No | NA | 5 | 0 | 1 | Not-determined | Genome-fragment | NA | NA | 0 |
| S12-final-viral-combined_k141_48145\|\|full | 7282 | No | NA | 5 | 0 | 1 | Not-determined | Genome-fragment | NA | NA | 0 |
| S12-final-viral-combined_k141_63649\|\|full | 13148 | No | NA | 13 | 1 | 1 | Medium-quality | Genome-fragment | 84.79 | AAI-based (high-confidence) | 0 |
| S12-final-viral-combined_k141_164711\|\|full | 1070 | No | NA | 2 | 0 | 1 | Not-determined | Genome-fragment | NA | NA | 0 |
| S12-final-viral-combined_k141_316224\|\|full | 1178 | No | NA | 2 | 0 | 2 | Not-determined | Genome-fragment | NA | NA | 0 |
| S6-final-viral-combined_k141_135357\|\|full | 3036 | No | NA | 2 | 0 | 1 | Low-quality | Genome-fragment | 4.87 | AAI-based (medium-confidence) | 0 |
| S12-final-viral-combined_k141_316226\|\|full | 2851 | No | NA | 3 | 0 | 0 | Not-determined | Genome-fragment | NA | NA | 0 |
| S1-final-viral-combined_k141_326497\|\|full | 3037 | No | NA | 3 | 0 | 2 | Low-quality | Genome-fragment | 6.2 | AAI-based (high-confidence) | 0 |
| S12-final-viral-combined_k141_214902\|\|full | 2673 | No | NA | 4 | 0 | 1 | Not-determined | Genome-fragment | NA | NA | 0 |
| S12-final-viral-combined_k141_250505\|\|full | 3037 | No | NA | 2 | 0 | 0 | Low-quality | Genome-fragment | 8.31 | AAI-based (medium-confidence) | 0 |
| S12-final-viral-combined_k141_214940\|\|full | 1776 | No | NA | 3 | 0 | 0 | Not-determined | Genome-fragment | NA | NA | 0 |
| S12_k141_140912 | 3037 | No | NA | 3 | 0 | 2 | Low-quality | Genome-fragment | 0.99 | AAI-based (medium-confidence) | 0 |
| S12-final-viral-combined_k141_113546\|\|full | 6996 | No | NA | 5 | 0 | 2 | Not-determined | Genome-fragment | NA | NA | 0 |
| S2-final-viral-combined_k141_207876\|\|full | 3037 | No | NA | 4 | 0 | 3 | Low-quality | Genome-fragment | 1.47 | AAI-based (medium-confidence) | 0 |
| S8-final-viral-combined_k141_141859\|\|full | 3037 | No | NA | 5 | 2 | 0 | Low-quality | Genome-fragment | 5.36 | HMM-based (lower-bound) | 0 |
| S12-final-viral-combined_k141_332125\|\|full | 2651 | No | NA | 4 | 0 | 4 | Not-determined | Genome-fragment | NA | NA | 0 |
| S12-final-viral-combined_k141_181434\|\|full | 1843 | No | NA | 4 | 0 | 2 | Not-determined | Genome-fragment | NA | NA | 0 |
| S9_k141_56766 | 3038 | No | NA | 3 | 0 | 3 | Low-quality | Genome-fragment | 1.14 | AAI-based (high-confidence) | 0 |
| S3_k141_135787 | 3038 | No | NA | 2 | 0 | 2 | Low-quality | Genome-fragment | 1.06 | AAI-based (high-confidence) | 0 |
| S12-final-viral-combined_k141_363561\|\|full | 9583 | Yes | 7172 | 9 | 1 | 1 | Not-determined | Genome-fragment | NA | NA | 25.16 |
| S12-final-viral-combined_k141_282594\|\|full | 93773 | Yes | 40043 | 92 | 6 | 28 | Medium-quality | Genome-fragment | 52.76 | HMM-based (lower-bound) | 57.3 |
| S12-final-viral-combined_k141_81290\|\|full | 3921 | No | NA | 8 | 0 | 1 | Not-determined | Genome-fragment | NA | NA | 0 |
| S7-final-viral-combined_k141_60704\|\|full | 3038 | No | NA | 3 | 0 | 1 | Low-quality | Genome-fragment | 2.14 | AAI-based (high-confidence) | 0 |
| S12-final-viral-combined_k141_147318\|\|full | 1753 | No | NA | 1 | 0 | 0 | Not-determined | Genome-fragment | NA | NA | 0 |
| S12-final-viral-combined_k141_298595\|\|full | 5072 | No | NA | 4 | 0 | 2 | Not-determined | Genome-fragment | NA | NA | 0 |
| S12-final-viral-combined_k141_113647\|\|full | 1736 | No | NA | 3 | 0 | 0 | Not-determined | Genome-fragment | NA | NA | 0 |
| S12-final-viral-combined_k141_130241\|\|full | 1033 | No | NA | 2 | 0 | 0 | Not-determined | Genome-fragment | NA | NA | 0 |
| S2-final-viral-combined_k141_210010\|\|full | 3038 | No | NA | 5 | 0 | 3 | Low-quality | Genome-fragment | 8.54 | AAI-based (medium-confidence) | 0 |
| S12-final-viral-combined_k141_332192\|\|full | 1991 | No | NA | 2 | 0 | 0 | Not-determined | Genome-fragment | NA | NA | 0 |
| S12-final-viral-combined_k141_48259\|\|full | 7925 | No | NA | 8 | 0 | 4 | Not-determined | Genome-fragment | NA | NA | 0 |
| S12-final-viral-combined_k141_332195\|\|full | 3248 | No | NA | 5 | 0 | 1 | Not-determined | Genome-fragment | NA | NA | 0 |
| S9_k141_76540 | 3039 | No | NA | 10 | 1 | 0 | Low-quality | Genome-fragment | 7.1 | AAI-based (high-confidence) | 0 |
| S1-final-viral-combined_k141_221894\|\|full | 3039 | No | NA | 3 | 2 | 0 | Low-quality | Genome-fragment | 3.15 | HMM-based (lower-bound) | 0 |
| S12-final-viral-combined_k141_130280\|\|full | 2702 | No | NA | 4 | 0 | 2 | Not-determined | Genome-fragment | NA | NA | 0 |
| S12-final-viral-combined_k141_97265\|\|full | 2971 | No | NA | 4 | 0 | 3 | Not-determined | Genome-fragment | NA | NA | 0 |
| S12-final-viral-combined_k141_13530\|\|full | 1038 | No | NA | 2 | 0 | 0 | Not-determined | Genome-fragment | NA | NA | 0 |
| S12-final-viral-combined_k141_298686\|\|full | 3811 | No | NA | 6 | 0 | 4 | Not-determined | Genome-fragment | NA | NA | 0 |
| S1_k141_24755 | 3039 | No | NA | 2 | 0 | 2 | Low-quality | Genome-fragment | 7.23 | AAI-based (high-confidence) | 0 |
| S12-final-viral-combined_k141_264425\|\|full | 46657 | No | NA | 68 | 13 | 1 | High-quality | High-quality | 100 | AAI-based (high-confidence) | 0 |
| S12-final-viral-combined_k141_316430\|\|full | 1243 | No | NA | 2 | 0 | 1 | Not-determined | Genome-fragment | NA | NA | 0 |
| S12-final-viral-combined_k141_164916\|\|full | 1934 | No | NA | 2 | 0 | 1 | Not-determined | Genome-fragment | NA | NA | 0 |
| S12-final-viral-combined_k141_264441\|\|full | 1316 | No | NA | 2 | 0 | 2 | Not-determined | Genome-fragment | NA | NA | 0 |
| S12-final-viral-combined_k141_181525\|\|full | 1024 | No | NA | 2 | 0 | 0 | Not-determined | Genome-fragment | NA | NA | 0 |
| S3_k141_70888 | 3039 | No | NA | 2 | 0 | 1 | Low-quality | Genome-fragment | 5.92 | AAI-based (medium-confidence) | 0 |
| S17-final-viral-combined_k141_140907\|\|full | 3039 | No | NA | 3 | 0 | 0 | Low-quality | Genome-fragment | 7.02 | AAI-based (high-confidence) | 0 |
| S12-final-viral-combined_k141_50420\|\|full | 3039 | No | NA | 4 | 0 | 2 | Low-quality | Genome-fragment | 1.43 | AAI-based (medium-confidence) | 0 |
| S12-final-viral-combined_k141_363701\|\|full | 2239 | No | NA | 3 | 0 | 1 | Not-determined | Genome-fragment | NA | NA | 0 |
| S12-final-viral-combined_k141_298753\|\|full | 1659 | No | NA | 3 | 0 | 0 | Not-determined | Genome-fragment | NA | NA | 0 |
| S12-final-viral-combined_k141_164972\|\|full | 1715 | No | NA | 2 | 0 | 0 | Not-determined | Genome-fragment | NA | NA | 0 |
| S8-final-viral-combined_k141_55332\|\|full | 3039 | No | NA | 4 | 0 | 1 | Low-quality | Genome-fragment | 3.24 | AAI-based (medium-confidence) | 0 |
| S3-final-viral-combined_k141_63491\|\|full | 3040 | No | NA | 3 | 0 | 3 | Low-quality | Genome-fragment | 1.53 | AAI-based (medium-confidence) | 0 |
| S12-final-viral-combined_k141_113756\|\|full | 3492 | No | NA | 3 | 0 | 0 | Not-determined | Genome-fragment | NA | NA | 0 |
| S12-final-viral-combined_k141_332298\|\|full | 2001 | No | NA | 4 | 0 | 1 | Not-determined | Genome-fragment | NA | NA | 0 |
| S12-final-viral-combined_k141_363714\|\|full | 3634 | No | NA | 4 | 0 | 1 | Not-determined | Genome-fragment | NA | NA | 0 |
| S17-final-viral-combined_k141_47675\|\|full | 3040 | Yes | 2019 | 4 | 2 | 1 | Low-quality | Genome-fragment | 7.04 | AAI-based (high-confidence) | 33.59 |
| S12-final-viral-combined_k141_97359\|\|full | 3751 | No | NA | 6 | 0 | 2 | Not-determined | Genome-fragment | NA | NA | 0 |
| S3_k141_630 | 3041 | No | NA | 5 | 0 | 1 | Low-quality | Genome-fragment | 5.66 | AAI-based (medium-confidence) | 0 |
| S12-final-viral-combined_k141_186497\|\|full | 3041 | Yes | 1168 | 4 | 1 | 1 | Low-quality | Genome-fragment | 2.87 | AAI-based (medium-confidence) | 61.59 |
| S2-final-viral-combined_k141_72828\|\|full | 3041 | No | NA | 3 | 0 | 0 | Low-quality | Genome-fragment | 6.9 | AAI-based (medium-confidence) | 0 |
| S11-final-viral-combined_k141_324092\|\|full | 3042 | No | NA | 3 | 0 | 3 | Low-quality | Genome-fragment | 1.74 | AAI-based (medium-confidence) | 0 |
| S1-final-viral-combined_k141_278162\|\|full | 3042 | No | NA | 4 | 0 | 0 | Low-quality | Genome-fragment | 2.49 | AAI-based (medium-confidence) | 0 |
| S1_k141_209319 | 3042 | No | NA | 4 | 3 | 0 | Low-quality | Genome-fragment | 7.68 | AAI-based (high-confidence) | 0 |
| S12-final-viral-combined_k141_316505\|\|full | 5744 | No | NA | 7 | 0 | 4 | Not-determined | Genome-fragment | NA | NA | 0 |
| S12-final-viral-combined_k141_348967\|\|full | 3923 | No | NA | 4 | 0 | 3 | Not-determined | Genome-fragment | NA | NA | 0 |
| S7-final-viral-combined_k141_157570\|\|full | 3042 | No | NA | 3 | 0 | 2 | Low-quality | Genome-fragment | 5.18 | AAI-based (high-confidence) | 0 |
| S12-final-viral-combined_k141_398676\|\|full | 2270 | No | NA | 4 | 0 | 2 | Not-determined | Genome-fragment | NA | NA | 0 |
| S4-final-viral-combined_k141_80275\|\|full | 3042 | No | NA | 5 | 1 | 0 | Low-quality | Genome-fragment | 6.72 | AAI-based (high-confidence) | 0 |
| S7-final-viral-combined_k141_79970\|\|full | 3043 | No | NA | 3 | 0 | 1 | Low-quality | Genome-fragment | 3.2 | AAI-based (high-confidence) | 0 |
| S12-final-viral-combined_k141_282825\|\|full | 1303 | No | NA | 2 | 0 | 0 | Not-determined | Genome-fragment | NA | NA | 0 |
| S12-final-viral-combined_k141_147453\|\|full | 1385 | No | NA | 2 | 0 | 1 | Not-determined | Genome-fragment | NA | NA | 0 |
| S12-final-viral-combined_k141_63890\|\|full | 2266 | No | NA | 4 | 0 | 0 | Not-determined | Genome-fragment | NA | NA | 0 |
| S12_k141_167233 | 3043 | No | NA | 6 | 0 | 0 | Low-quality | Genome-fragment | 7.73 | AAI-based (medium-confidence) | 0 |
| S12-final-viral-combined_k141_63900\|\|full | 1252 | No | NA | 3 | 0 | 1 | Not-determined | Genome-fragment | NA | NA | 0 |
| S12-final-viral-combined_k141_147467\|\|full | 3095 | No | NA | 4 | 0 | 3 | Not-determined | Genome-fragment | NA | NA | 0 |
| S8-final-viral-combined_k141_275540\|\|full | 3043 | No | NA | 4 | 0 | 0 | Low-quality | Genome-fragment | 8.98 | AAI-based (high-confidence) | 0 |

| | | | | | | | | | | | |
|---|---|---|---|---|---|---|---|---|---|---|---|
| S12-final-viral-combined_k141_48446\|\|full | 6461 | No | NA | 5 | 0 | 3 | Not-determined | Genome-fragment | NA | NA | 0 |
| S12-final-viral-combined_k141_63932\|\|full | 1662 | No | NA | 2 | 0 | 1 | Not-determined | Genome-fragment | NA | NA | 0 |
| S12-final-viral-combined_k141_232454\|\|full | 1793 | No | NA | 2 | 0 | 0 | Not-determined | Genome-fragment | NA | NA | 0 |
| S12-final-viral-combined_k141_198603\|\|full | 3550 | No | NA | 4 | 0 | 4 | Not-determined | Genome-fragment | NA | NA | 0 |
| S12-final-viral-combined_k141_97440\|\|full | 1955 | No | NA | 1 | 0 | 1 | Not-determined | Genome-fragment | NA | NA | 0 |
| S12-final-viral-combined_k141_282842\|\|full | 2254 | No | NA | 2 | 0 | 2 | Not-determined | Genome-fragment | NA | NA | 0 |
| S9-final-viral-combined_k141_50920\|\|full | 3044 | No | NA | 4 | 0 | 3 | Low-quality | Genome-fragment | 5.05 | AAI-based (medium-confidence) | 0 |
| S12-final-viral-combined_k141_232466\|\|full | 1481 | No | NA | 2 | 0 | 1 | Not-determined | Genome-fragment | NA | NA | 0 |
| S12-final-viral-combined_k141_398705\|\|full | 1311 | No | NA | 3 | 0 | 0 | Not-determined | Genome-fragment | NA | NA | 0 |
| S9-final-viral-combined_k141_34244\|\|full | 3044 | No | NA | 3 | 1 | 0 | Low-quality | Genome-fragment | 8.11 | AAI-based (medium-confidence) | 0 |
| S12-final-viral-combined_k141_248343\|\|full | 1929 | No | NA | 2 | 0 | 1 | Not-determined | Genome-fragment | NA | NA | 0 |
| S11-final-viral-combined_k141_209946\|\|full | 3044 | No | NA | 4 | 1 | 0 | Low-quality | Genome-fragment | 4.61 | HMM-based (lower-bound) | 0 |
| S12-final-viral-combined_k141_130456\|\|full | 3001 | No | NA | 6 | 0 | 0 | Not-determined | Genome-fragment | NA | NA | 0 |
| S12-final-viral-combined_k141_30523\|\|full | 1413 | No | NA | 3 | 0 | 0 | Not-determined | Genome-fragment | NA | NA | 0 |
| S1-final-viral-combined_k141_353133\|\|full | 3044 | No | NA | 4 | 0 | 3 | Low-quality | Genome-fragment | 0.6 | AAI-based (medium-confidence) | 0 |
| S12-final-viral-combined_k141_332480\|\|full | 1346 | No | NA | 2 | 0 | 0 | Not-determined | Genome-fragment | NA | NA | 0 |
| S12-final-viral-combined_k141_30524\|\|full | 8060 | No | NA | 9 | 0 | 1 | Not-determined | Genome-fragment | NA | NA | 0 |
| S4-final-viral-combined_k141_6622\|\|full | 3044 | No | NA | 3 | 0 | 2 | Low-quality | Genome-fragment | 0.86 | AAI-based (high-confidence) | 0 |
| S1-final-viral-combined_k141_20001\|\|full | 3045 | No | NA | 6 | 0 | 0 | Low-quality | Genome-fragment | 5.29 | AAI-based (high-confidence) | 0 |
| S1-final-viral-combined_k141_301327\|\|full | 3045 | No | NA | 2 | 0 | 1 | Low-quality | Genome-fragment | 4.26 | AAI-based (high-confidence) | 0 |
| S5-final-viral-combined_k141_10407\|\|full | 3045 | No | NA | 4 | 0 | 1 | Low-quality | Genome-fragment | 1.43 | AAI-based (medium-confidence) | 0 |
| S17-final-viral-combined_k141_16547\|\|full | 3045 | No | NA | 6 | 2 | 0 | Low-quality | Genome-fragment | 7.34 | AAI-based (high-confidence) | 0 |
| S17-final-viral-combined_k141_174054\|\|full | 3046 | No | NA | 4 | 0 | 1 | Low-quality | Genome-fragment | 4.07 | AAI-based (medium-confidence) | 0 |
| S17-final-viral-combined_k141_98756\|\|full | 3046 | No | NA | 4 | 0 | 2 | Low-quality | Genome-fragment | 1.64 | AAI-based (medium-confidence) | 0 |
| S4-final-viral-combined_k141_59135\|\|full | 3046 | No | NA | 6 | 0 | 0 | Low-quality | Genome-fragment | 2.02 | AAI-based (high-confidence) | 0 |
| S1_k141_13823 | 3047 | No | NA | 5 | 0 | 0 | Low-quality | Genome-fragment | 5.01 | AAI-based (high-confidence) | 0 |
| S12-final-viral-combined_k141_30559\|\|full | 5005 | No | NA | 7 | 1 | 1 | Not-determined | Genome-fragment | NA | NA | 0 |
| S12-final-viral-combined_k141_165054\|\|full | 1649 | No | NA | 2 | 0 | 2 | Not-determined | Genome-fragment | NA | NA | 0 |
| S12-final-viral-combined_k141_282985\|\|full | 4681 | No | NA | 9 | 0 | 0 | Not-determined | Genome-fragment | NA | NA | 0 |
| S12-final-viral-combined_k141_298949\|\|full | 3192 | No | NA | 3 | 0 | 1 | Not-determined | Genome-fragment | NA | NA | 0 |
| S5_k141_154321 | 3047 | No | NA | 3 | 0 | 3 | Low-quality | Genome-fragment | 1.15 | AAI-based (high-confidence) | 0 |
| S12-final-viral-combined_k141_97586\|\|full | 3376 | No | NA | 4 | 0 | 3 | Not-determined | Genome-fragment | NA | NA | 0 |
| S4-final-viral-combined_k141_45560\|\|full | 3047 | No | NA | 4 | 0 | 2 | Low-quality | Genome-fragment | 2.59 | AAI-based (medium-confidence) | 0 |
| S17-final-viral-combined_k141_13767\|\|full | 3048 | No | NA | 5 | 2 | 0 | Low-quality | Genome-fragment | 8.18 | AAI-based (high-confidence) | 0 |
| S12-final-viral-combined_k141_97588\|\|full | 3059 | No | NA | 3 | 0 | 0 | Not-determined | Genome-fragment | NA | NA | 0 |
| S12-final-viral-combined_k141_181744\|\|full | 1268 | No | NA | 2 | 0 | 1 | Not-determined | Genome-fragment | NA | NA | 0 |
| S12-final-viral-combined_k141_113992\|\|full | 3350 | No | NA | 4 | 0 | 2 | Not-determined | Genome-fragment | NA | NA | 0 |
| S12-final-viral-combined_k141_30585\|\|full | 1707 | No | NA | 3 | 0 | 2 | Not-determined | Genome-fragment | NA | NA | 0 |
| S12-final-viral-combined_k141_64022\|\|full | 3211 | No | NA | 4 | 0 | 2 | Not-determined | Genome-fragment | NA | NA | 0 |
| S11-final-viral-combined_k141_278828\|\|full | 3049 | No | NA | 6 | 5 | 0 | Low-quality | Genome-fragment | 6.45 | AAI-based (medium-confidence) | 0 |
| S11_k141_133717 | 3049 | No | NA | 4 | 0 | 3 | Low-quality | Genome-fragment | 1.12 | AAI-based (medium-confidence) | 0 |
| S12-final-viral-combined_k141_298996\|\|full | 2251 | No | NA | 5 | 0 | 0 | Not-determined | Genome-fragment | NA | NA | 0 |
| S12-final-viral-combined_k141_147568\|\|full | 4362 | No | NA | 6 | 0 | 3 | Not-determined | Genome-fragment | NA | NA | 0 |
| S12-final-viral-combined_k141_97624\|\|full | 1986 | No | NA | 3 | 0 | 1 | Not-determined | Genome-fragment | NA | NA | 0 |
| S12-final-viral-combined_k141_332610\|\|full | 1109 | No | NA | 2 | 0 | 1 | Not-determined | Genome-fragment | NA | NA | 0 |
| S12-final-viral-combined_k141_13859\|\|full | 1516 | No | NA | 2 | 0 | 1 | Not-determined | Genome-fragment | NA | NA | 0 |
| S12-final-viral-combined_k141_48540\|\|full | 2324 | No | NA | 6 | 0 | 2 | Not-determined | Genome-fragment | NA | NA | 0 |
| S12-final-viral-combined_k141_30624\|\|full | 7186 | No | NA | 4 | 0 | 1 | Not-determined | Genome-fragment | NA | NA | 0 |
| S12-final-viral-combined_k141_212856\|\|full | 3049 | No | NA | 2 | 0 | 2 | Low-quality | Genome-fragment | 0.51 | AAI-based (medium-confidence) | 0 |
| S2-final-viral-combined_k141_15070\|\|full | 3049 | No | NA | 3 | 0 | 2 | Low-quality | Genome-fragment | 1.79 | AAI-based (medium-confidence) | 0 |
| S12_k141_357369 | 3050 | No | NA | 5 | 0 | 0 | Low-quality | Genome-fragment | 1.42 | AAI-based (high-confidence) | 0 |
| S12-final-viral-combined_k141_198803\|\|full | 3019 | No | NA | 3 | 0 | 3 | Not-determined | Genome-fragment | NA | NA | 0 |
| S12-final-viral-combined_k141_81657\|\|full | 3224 | No | NA | 3 | 0 | 0 | Not-determined | Genome-fragment | NA | NA | 0 |
| S12-final-viral-combined_k141_81677\|\|full | 2029 | No | NA | 3 | 0 | 2 | Not-determined | Genome-fragment | NA | NA | 0 |
| S12-final-viral-combined_k141_332684\|\|full | 2909 | No | NA | 3 | 0 | 1 | Not-determined | Genome-fragment | NA | NA | 0 |
| S12-final-viral-combined_k141_299088\|\|full | 1732 | No | NA | 4 | 0 | 0 | Not-determined | Genome-fragment | NA | NA | 0 |
| S11-final-viral-combined_k141_123925\|\|full | 3051 | No | NA | 5 | 0 | 0 | Low-quality | Genome-fragment | 6.86 | AAI-based (medium-confidence) | 0 |
| S12-final-viral-combined_k141_316810\|\|full | 1127 | No | NA | 3 | 0 | 0 | Not-determined | Genome-fragment | NA | NA | 0 |
| S12-final-viral-combined_k141_232737\|\|full | 1844 | No | NA | 3 | 0 | 1 | Not-determined | Genome-fragment | NA | NA | 0 |
| S12-final-viral-combined_k141_215504\|\|full | 1875 | No | NA | 3 | 0 | 1 | Not-determined | Genome-fragment | NA | NA | 0 |
| S12-final-viral-combined_k141_114116\|\|full | 1878 | No | NA | 2 | 0 | 1 | Not-determined | Genome-fragment | NA | NA | 0 |
| S12-final-viral-combined_k141_181890\|\|full | 1967 | No | NA | 2 | 0 | 0 | Not-determined | Genome-fragment | NA | NA | 0 |
| S5-final-viral-combined_k141_171418\|\|full | 3051 | No | NA | 4 | 0 | 0 | Low-quality | Genome-fragment | 1.97 | AAI-based (medium-confidence) | 0 |
| S2-final-viral-combined_k141_5772\|\|full | 3051 | No | NA | 6 | 0 | 0 | Low-quality | Genome-fragment | 8.35 | AAI-based (medium-confidence) | 0 |
| S2_k141_82545 | 3051 | No | NA | 1 | 0 | 1 | Low-quality | Genome-fragment | 1.64 | AAI-based (medium-confidence) | 0 |
| S11-final-viral-combined_k141_217901\|\|full | 3052 | No | NA | 6 | 2 | 0 | Low-quality | Genome-fragment | 1.81 | HMM-based (lower-bound) | 0 |
| S12-final-viral-combined_k141_81782\|\|full | 1189 | No | NA | 2 | 0 | 1 | Not-determined | Genome-fragment | NA | NA | 0 |
| S12-final-viral-combined_k141_81784\|\|full | 6778 | No | NA | 4 | 0 | 4 | Not-determined | Genome-fragment | NA | NA | 0 |
| S12-final-viral-combined_k141_198880\|\|full | 1062 | No | NA | 3 | 0 | 1 | Not-determined | Genome-fragment | NA | NA | 0 |
| S12-final-viral-combined_k141_198895\|\|full | 2006 | No | NA | 2 | 0 | 0 | Not-determined | Genome-fragment | NA | NA | 0 |
| S11-final-viral-combined_k141_316162\|\|full | 3052 | No | NA | 3 | 2 | 0 | Low-quality | Genome-fragment | 7.74 | AAI-based (high-confidence) | 0 |
| S12-final-viral-combined_k141_147628\|\|full | 79515 | Yes | 60141 | 101 | 20 | 13 | Complete | High-quality | 100 | Provirus (high-confidence) | 24.37 |
| S1_k141_348471 | 3052 | No | NA | 3 | 0 | 0 | Low-quality | Genome-fragment | 6.41 | AAI-based (medium-confidence) | 0 |
| S12-final-viral-combined_k141_48623\|\|full | 2439 | No | NA | 2 | 0 | 1 | Not-determined | Genome-fragment | NA | NA | 0 |
| S12-final-viral-combined_k141_97759\|\|full | 1323 | No | NA | 3 | 0 | 2 | Not-determined | Genome-fragment | NA | NA | 0 |
| S12-final-viral-combined_k141_81813\|\|full | 1822 | No | NA | 4 | 0 | 1 | Not-determined | Genome-fragment | NA | NA | 0 |
| S12-final-viral-combined_k141_349295\|\|full | 3767 | No | NA | 4 | 0 | 2 | Not-determined | Genome-fragment | NA | NA | 0 |
| S12-final-viral-combined_k141_299234\|\|full | 1534 | No | NA | 3 | 0 | 2 | Not-determined | Genome-fragment | NA | NA | 0 |
| S4_k141_32415 | 3052 | No | NA | 4 | 2 | 0 | Low-quality | Genome-fragment | 1.76 | AAI-based (high-confidence) | 0 |
| S12-final-viral-combined_k141_114191\|\|full | 2307 | No | NA | 3 | 0 | 1 | Not-determined | Genome-fragment | NA | NA | 0 |
| S12-final-viral-combined_k141_215585\|\|full | 1181 | No | NA | 2 | 0 | 1 | Not-determined | Genome-fragment | NA | NA | 0 |
| S12-final-viral-combined_k141_349316\|\|full | 2841 | No | NA | 3 | 0 | 1 | Not-determined | Genome-fragment | NA | NA | 0 |
| S12-final-viral-combined_k141_264795\|\|full | 1088 | No | NA | 2 | 0 | 1 | Not-determined | Genome-fragment | NA | NA | 0 |
| S12-final-viral-combined_k141_48648\|\|full | 11195 | No | NA | 16 | 0 | 1 | Medium-quality | Genome-fragment | 73.71 | AAI-based (high-confidence) | 0 |
| S7-final-viral-combined_k141_163035\|\|full | 3053 | No | NA | 5 | 0 | 1 | Low-quality | Genome-fragment | 8.46 | AAI-based (medium-confidence) | 0 |
| S12-final-viral-combined_k141_398950\|\|full | 7644 | No | NA | 8 | 0 | 1 | Not-determined | Genome-fragment | NA | NA | 0 |
| S1_k141_111051 | 3054 | No | NA | 4 | 0 | 0 | Low-quality | Genome-fragment | 3.71 | AAI-based (high-confidence) | 0 |
| S12-final-viral-combined_k141_130836\|\|full | 4745 | No | NA | 9 | 0 | 3 | Not-determined | Genome-fragment | NA | NA | 0 |
| S12-final-viral-combined_k141_382723\|\|full | 1992 | No | NA | 3 | 0 | 0 | Not-determined | Genome-fragment | NA | NA | 0 |
| S12-final-viral-combined_k141_283334\|\|full | 2730 | No | NA | 3 | 0 | 2 | Not-determined | Genome-fragment | NA | NA | 0 |
| S17-final-viral-combined_k141_256575\|\|full | 3054 | No | NA | 4 | 0 | 1 | Low-quality | Genome-fragment | 2.89 | AAI-based (medium-confidence) | 0 |
| S12-final-viral-combined_k141_64161\|\|full | 1878 | No | NA | 3 | 0 | 1 | Not-determined | Genome-fragment | NA | NA | 0 |
| S12-final-viral-combined_k141_264845\|\|full | 19738 | No | NA | 22 | 1 | 12 | High-quality | High-quality | 100 | AAI-based (medium-confidence) | 0 |
| S5-final-viral-combined_k141_50347\|\|full | 3055 | No | NA | 5 | 2 | 0 | Low-quality | Genome-fragment | 4.97 | AAI-based (high-confidence) | 0 |
| S12-final-viral-combined_k141_182084\|\|full | 3904 | No | NA | 3 | 0 | 2 | Not-determined | Genome-fragment | NA | NA | 0 |
| S12-final-viral-combined_k141_48726\|\|full | 2015 | No | NA | 4 | 0 | 2 | Not-determined | Genome-fragment | NA | NA | 0 |
| S7-final-viral-combined_k141_104121\|\|full | 3055 | No | NA | 4 | 0 | 3 | Low-quality | Genome-fragment | 3.62 | AAI-based (medium-confidence) | 0 |
| S12-final-viral-combined_k141_104793\|\|full | 3055 | No | NA | 7 | 0 | 1 | Low-quality | Genome-fragment | 3.26 | AAI-based (medium-confidence) | 0 |
| S12-final-viral-combined_k141_264890\|\|full | 1670 | No | NA | 3 | 0 | 1 | Not-determined | Genome-fragment | NA | NA | 0 |
| S12-final-viral-combined_k141_96679\|\|full | 3055 | No | NA | 8 | 0 | 7 | Low-quality | Genome-fragment | 1.29 | AAI-based (medium-confidence) | 0 |
| S12-final-viral-combined_k141_97929\|\|full | 3837 | No | NA | 5 | 0 | 0 | Not-determined | Genome-fragment | NA | NA | 0 |
| S12-final-viral-combined_k141_81942\|\|full | 1382 | No | NA | 3 | 0 | 3 | Not-determined | Genome-fragment | NA | NA | 0 |
| S12-final-viral-combined_k141_97935\|\|full | 3653 | No | NA | 4 | 0 | 2 | Not-determined | Genome-fragment | NA | NA | 0 |
| S11-final-viral-combined_k141_96760\|\|full | 3056 | No | NA | 2 | 0 | 0 | Low-quality | Genome-fragment | 4.6 | AAI-based (high-confidence) | 0 |
| S12-final-viral-combined_k141_349406\|\|full | 1692 | No | NA | 3 | 0 | 2 | Not-determined | Genome-fragment | NA | NA | 0 |
| S11_k141_308750 | 3056 | No | NA | 5 | 3 | 0 | Low-quality | Genome-fragment | 7.48 | AAI-based (high-confidence) | 0 |
| S12-final-viral-combined_k141_97949\|\|full | 2863 | No | NA | 3 | 0 | 1 | Not-determined | Genome-fragment | NA | NA | 0 |
| S12-final-viral-combined_k141_299293\|\|full | 74597 | Yes | 46591 | 86 | 15 | 17 | Complete | High-quality | 100 | Provirus (high-confidence) | 37.54 |
| S12-final-viral-combined_k141_264943\|\|full | 1225 | No | NA | 2 | 0 | 0 | Not-determined | Genome-fragment | NA | NA | 0 |
| S12-final-viral-combined_k141_349413\|\|full | 5480 | No | NA | 2 | 0 | 2 | Not-determined | Genome-fragment | NA | NA | 0 |
| S12-final-viral-combined_k141_299297\|\|full | 1149 | No | NA | 2 | 0 | 0 | Not-determined | Genome-fragment | NA | NA | 0 |
| S12-final-viral-combined_k141_81957\|\|full | 9870 | No | NA | 9 | 0 | 4 | Not-determined | Genome-fragment | NA | NA | 0 |
| S12-final-viral-combined_k141_64291\|\|full | 4071 | No | NA | 4 | 0 | 1 | Not-determined | Genome-fragment | NA | NA | 0 |
| S17-final-viral-combined_k141_8872\|\|lt2gene | 3056 | No | NA | 2 | 1 | 0 | Low-quality | Genome-fragment | 8.21 | AAI-based (medium-confidence) | 0 |
| S2-final-viral-combined_k141_162219\|\|full | 3056 | No | NA | 2 | 0 | 1 | Low-quality | Genome-fragment | 1.53 | AAI-based (medium-confidence) | 0 |
| S11_k141_45569 | 3058 | No | NA | 3 | 0 | 0 | Low-quality | Genome-fragment | 5.48 | AAI-based (medium-confidence) | 0 |
| S17-final-viral-combined_k141_37291\|\|full | 3058 | Yes | 997 | 7 | 1 | 2 | Low-quality | Genome-fragment | 0.54 | HMM-based (lower-bound) | 67.4 |
| S17-final-viral-combined_k141_140490\|\|full | 3058 | No | NA | 5 | 0 | 1 | Low-quality | Genome-fragment | 2.77 | AAI-based (medium-confidence) | 0 |
| S7-final-viral-combined_k141_93841\|\|full | 3059 | No | NA | 6 | 0 | 0 | Low-quality | Genome-fragment | 5.31 | AAI-based (high-confidence) | 0 |
| S12-final-viral-combined_k141_30862\|\|full | 1295 | No | NA | 2 | 0 | 2 | Not-determined | Genome-fragment | NA | NA | 0 |
| S2_k141_248154 | 3059 | No | NA | 2 | 0 | 2 | Low-quality | Genome-fragment | 0.49 | AAI-based (medium-confidence) | 0 |
| S12-final-viral-combined_k141_332983\|\|full | 7009 | No | NA | 6 | 0 | 3 | Not-determined | Genome-fragment | NA | NA | 0 |
| S12-final-viral-combined_k141_332987\|\|full | 2634 | No | NA | 4 | 0 | 3 | Not-determined | Genome-fragment | NA | NA | 0 |
| S12-final-viral-combined_k141_232946\|\|full | 2450 | No | NA | 3 | 0 | 2 | Not-determined | Genome-fragment | NA | NA | 0 |
| S12-final-viral-combined_k141_265034\|\|full | 4969 | No | NA | 5 | 0 | 3 | Not-determined | Genome-fragment | NA | NA | 0 |
| S12-final-viral-combined_k141_182235\|\|full | 1687 | No | NA | 3 | 0 | 0 | Not-determined | Genome-fragment | NA | NA | 0 |
| S12_k141_253268 | 3060 | No | NA | 7 | 2 | 0 | Low-quality | Genome-fragment | 7.67 | AAI-based (medium-confidence) | 0 |
| S12-final-viral-combined_k141_130990\|\|full | 1360 | No | NA | 3 | 0 | 0 | Not-determined | Genome-fragment | NA | NA | 0 |
| S6-final-viral-combined_k141_24766\|\|full | 3060 | No | NA | 2 | 0 | 1 | Low-quality | Genome-fragment | 3.71 | AAI-based (high-confidence) | 0 |
| S12-final-viral-combined_k141_248714\|\|full | 6055 | No | NA | 9 | 0 | 2 | Not-determined | Genome-fragment | NA | NA | 0 |
| S2_k141_3658 | 3061 | No | NA | 10 | 5 | 0 | Low-quality | Genome-fragment | 6.42 | AAI-based (high-confidence) | 0 |
| S4_k141_74160 | 3061 | No | NA | 3 | 0 | 0 | Low-quality | Genome-fragment | 12.29 | AAI-based (medium-confidence) | 0 |
| S4_k141_144845 | 3061 | No | NA | 2 | 0 | 2 | Low-quality | Genome-fragment | 7.28 | AAI-based (high-confidence) | 0 |
| S8-final-viral-combined_k141_78173\|\|full | 3061 | No | NA | 7 | 2 | 0 | Low-quality | Genome-fragment | 4.47 | AAI-based (medium-confidence) | 0 |
| S11-final-viral-combined_k141_165584\|\|full | 3062 | No | NA | 6 | 6 | 0 | Low-quality | Genome-fragment | 6.54 | AAI-based (high-confidence) | 0 |
| S12-final-viral-combined_k141_233003\|\|full | 1519 | No | NA | 4 | 0 | 2 | Not-determined | Genome-fragment | NA | NA | 0 |
| S12-final-viral-combined_k141_215733\|\|full | 1033 | No | NA | 2 | 0 | 1 | Not-determined | Genome-fragment | NA | NA | 0 |
| S1-final-viral-combined_k141_109260\|\|full | 3062 | No | NA | 2 | 0 | 2 | Low-quality | Genome-fragment | 4.72 | AAI-based (medium-confidence) | 0 |
| S17-k141_108097 | 3062 | No | NA | 4 | 0 | 0 | Low-quality | Genome-fragment | 3.49 | AAI-based (medium-confidence) | 0 |
| S2-final-viral-combined_k141_151957\|\|full | 3062 | No | NA | 3 | 0 | 0 | Low-quality | Genome-fragment | 2.55 | AAI-based (medium-confidence) | 0 |
| S12-final-viral-combined_k141_64419\|\|full | 1010 | No | NA | 2 | 0 | 0 | Not-determined | Genome-fragment | NA | NA | 0 |
| S8_k141_69768 | 3062 | No | NA | 4 | 1 | 0 | Low-quality | Genome-fragment | 4.1 | AAI-based (high-confidence) | 0 |
| S12-final-viral-combined_k141_165500\|\|full | 1114 | No | NA | 2 | 0 | 1 | Not-determined | Genome-fragment | NA | NA | 0 |
| S12-final-viral-combined_k141_233025\|\|full | 4987 | No | NA | 2 | 0 | 1 | Not-determined | Genome-fragment | NA | NA | 0 |
| S12-final-viral-combined_k141_283580\|\|full | 2552 | No | NA | 4 | 0 | 4 | Not-determined | Genome-fragment | NA | NA | 0 |
| S12-final-viral-combined_k141_182297\|\|full | 1802 | No | NA | 2 | 0 | 2 | Not-determined | Genome-fragment | NA | NA | 0 |
| S12-final-viral-combined_k141_131083\|\|full | 2396 | No | NA | 5 | 0 | 1 | Not-determined | Genome-fragment | NA | NA | 0 |
| S12-final-viral-combined_k141_349544\|\|full | 2503 | No | NA | 3 | 0 | 1 | Not-determined | Genome-fragment | NA | NA | 0 |
| S5_k141_18535 | 3063 | No | NA | 3 | 2 | 0 | Low-quality | Genome-fragment | 8.02 | AAI-based (medium-confidence) | 0 |
| S12-final-viral-combined_k141_165522\|\|full | 2914 | No | NA | 4 | 0 | 2 | Not-determined | Genome-fragment | NA | NA | 0 |
| S12-final-viral-combined_k141_382858\|\|full | 1690 | No | NA | 3 | 0 | 1 | Not-determined | Genome-fragment | NA | NA | 0 |
| S12-final-viral-combined_k141_215786\|\|full | 1832 | No | NA | 3 | 0 | 1 | Not-determined | Genome-fragment | NA | NA | 0 |
| S1-final-viral-combined_k141_187472\|\|full | 3064 | No | NA | 4 | 1 | 0 | Low-quality | Genome-fragment | 3.61 | AAI-based (medium-confidence) | 0 |
| S4-final-viral-combined_k141_93396\|\|full | 3064 | No | NA | 6 | 0 | 0 | Low-quality | Genome-fragment | 4.74 | AAI-based (high-confidence) | 0 |
| S12-final-viral-combined_k141_317220\|\|full | 1954 | No | NA | 3 | 0 | 1 | Not-determined | Genome-fragment | NA | NA | 0 |
| S12-final-viral-combined_k141_98046\|\|full | 89205 | Yes | 39193 | 77 | 21 | 24 | Medium-quality | Genome-fragment | 61.36 | AAI-based (high-confidence) | 56.06 |
| S11-final-viral-combined_k141_291053\|\|full | 3066 | No | NA | 4 | 0 | 1 | Low-quality | Genome-fragment | 9.18 | AAI-based (high-confidence) | 0 |
| S4-final-viral-combined_k141_66663\|\|full | 3066 | No | NA | 4 | 0 | 1 | Low-quality | Genome-fragment | 1.93 | AAI-based (medium-confidence) | 0 |
| S12-final-viral-combined_k141_148021\|\|full | 1776 | No | NA | 3 | 0 | 0 | Not-determined | Genome-fragment | NA | NA | 0 |
| S5_k141_185889 | 3067 | No | NA | 7 | 7 | 0 | Low-quality | Genome-fragment | 7.54 | AAI-based (high-confidence) | 0 |
| S17-final-viral-combined_k141_59438\|\|full | 3067 | No | NA | 5 | 0 | 1 | Low-quality | Genome-fragment | 5.23 | AAI-based (medium-confidence) | 0 |
| S12-final-viral-combined_k141_165617\|\|full | 2211 | No | NA | 3 | 0 | 3 | Not-determined | Genome-fragment | NA | NA | 0 |
| S17-final-viral-combined_k141_127863\|\|full | 3067 | No | NA | 4 | 0 | 2 | Low-quality | Genome-fragment | 2.44 | AAI-based (high-confidence) | 0 |
| S12_k141_270280 | 3067 | No | NA | 6 | 0 | 4 | Low-quality | Genome-fragment | 6.56 | AAI-based (medium-confidence) | 0 |
| S2_k141_207823 | 3067 | No | NA | 3 | 0 | 2 | Low-quality | Genome-fragment | 5 | AAI-based (medium-confidence) | 0 |
| S12-final-viral-combined_k141_64596\|\|full | 3725 | No | NA | 4 | 0 | 2 | Not-determined | Genome-fragment | NA | NA | 0 |
| S12-final-viral-combined_k141_131226\|\|full | 4468 | No | NA | 8 | 0 | 1 | Not-determined | Genome-fragment | NA | NA | 0 |
| S12-final-viral-combined_k141_382953\|\|full | 3041 | No | NA | 4 | 0 | 2 | Not-determined | Genome-fragment | NA | NA | 0 |
| S12-final-viral-combined_k141_215927\|\|full | 2690 | No | NA | 3 | 0 | 0 | Not-determined | Genome-fragment | NA | NA | 0 |
| S4-final-viral-combined_k141_47790\|\|full | 3067 | No | NA | 2 | 0 | 0 | Low-quality | Genome-fragment | 2.18 | AAI-based (medium-confidence) | 0 |
| S12-final-viral-combined_k141_299467\|\|full | 2211 | No | NA | 3 | 0 | 2 | Not-determined | Genome-fragment | NA | NA | 0 |
| S12-final-viral-combined_k141_114579\|\|full | 1817 | No | NA | 3 | 0 | 1 | Not-determined | Genome-fragment | NA | NA | 0 |
| S12-final-viral-combined_k141_182456\|\|full | 4870 | No | NA | 5 | 0 | 3 | Not-determined | Genome-fragment | NA | NA | 0 |
| S12-final-viral-combined_k141_148114\|\|full | 1747 | No | NA | 2 | 0 | 1 | Not-determined | Genome-fragment | NA | NA | 0 |
| S12-final-viral-combined_k141_399191\|\|full | 1021 | No | NA | 2 | 0 | 0 | Not-determined | Genome-fragment | NA | NA | 0 |
| S4_k141_63903 | 3067 | No | NA | 2 | 0 | 0 | Low-quality | Genome-fragment | 3.19 | AAI-based (medium-confidence) | 0 |
| S4_k141_48135 | 3067 | No | NA | 3 | 0 | 1 | Low-quality | Genome-fragment | 10.69 | AAI-based (high-confidence) | 0 |
| S12-final-viral-combined_k141_299519\|\|full | 2124 | No | NA | 2 | 0 | 1 | Not-determined | Genome-fragment | NA | NA | 0 |
| S1-final-viral-combined_k141_244081\|\|full | 3068 | No | NA | 3 | 0 | 1 | Low-quality | Genome-fragment | 2.78 | AAI-based (medium-confidence) | 0 |
| S4-final-viral-combined_k141_136853\|\|full | 3068 | Yes | 1680 | 2 | 1 | 1 | Low-quality | Genome-fragment | 0.71 | HMM-based (lower-bound) | 45.24 |
| S1-final-viral-combined_k141_286050\|\|full | 3069 | No | NA | 9 | 1 | 0 | Low-quality | Genome-fragment | 9.19 | HMM-based (lower-bound) | 0 |
| S12-final-viral-combined_k141_399208\|\|full | 3078 | No | NA | 4 | 0 | 3 | Not-determined | Genome-fragment | NA | NA | 0 |
| S12-final-viral-combined_k141_165717\|\|full | 1223 | No | NA | 2 | 0 | 1 | Not-determined | Genome-fragment | NA | NA | 0 |
| S1-final-viral-combined_k141_387362\|\|full | 3069 | No | NA | 6 | 0 | 3 | Low-quality | Genome-fragment | 2.52 | AAI-based (medium-confidence) | 0 |
| S12-final-viral-combined_k141_199260\|\|full | 2516 | No | NA | 4 | 0 | 1 | Not-determined | Genome-fragment | NA | NA | 0 |
| S1_k141_364714 | 3069 | No | NA | 4 | 1 | 0 | Low-quality | Genome-fragment | 2.19 | AAI-based (medium-confidence) | 0 |
| S12-final-viral-combined_k141_283709\|\|full | 2469 | No | NA | 2 | 0 | 2 | Not-determined | Genome-fragment | NA | NA | 0 |
| S12-final-viral-combined_k141_283716\|\|full | 1160 | No | NA | 3 | 0 | 1 | Not-determined | Genome-fragment | NA | NA | 0 |
| S12-final-viral-combined_k141_98189\|\|full | 8462 | No | NA | 10 | 0 | 7 | Not-determined | Genome-fragment | NA | NA | 0 |
| S12-final-viral-combined_k141_216026\|\|full | 1506 | No | NA | 3 | 0 | 1 | Not-determined | Genome-fragment | NA | NA | 0 |
| S12-final-viral-combined_k141_31241\|\|full | 8199 | No | NA | 8 | 0 | 7 | Not-determined | Genome-fragment | NA | NA | 0 |
| S12-final-viral-combined_k141_165742\|\|full | 2084 | No | NA | 3 | 0 | 0 | Not-determined | Genome-fragment | NA | NA | 0 |
| S12-final-viral-combined_k141_399299\|\|full | 1232 | No | NA | 2 | 0 | 0 | Not-determined | Genome-fragment | NA | NA | 0 |
| S1_k141_197079 | 3069 | No | NA | 1 | 0 | 1 | Low-quality | Genome-fragment | 3.23 | AAI-based (medium-confidence) | 0 |
| S12_k141_318757 | 3069 | No | NA | 3 | 0 | 1 | Low-quality | Genome-fragment | 5.69 | AAI-based (high-confidence) | 0 |
| S12-final-viral-combined_k141_31270\|\|full | 1163 | No | NA | 4 | 0 | 0 | Not-determined | Genome-fragment | NA | NA | 0 |
| S2-final-viral-combined_k141_155952\|\|full | 3069 | No | NA | 4 | 3 | 1 | Low-quality | Genome-fragment | 6.5 | AAI-based (high-confidence) | 0 |
| S12-final-viral-combined_k141_131446\|\|full | 1924 | No | NA | 2 | 0 | 1 | Not-determined | Genome-fragment | NA | NA | 0 |
| S8-final-viral-combined_k141_107078\|\|full | 3069 | No | NA | 4 | 0 | 0 | Low-quality | Genome-fragment | 5.85 | AAI-based (medium-confidence) | 0 |
| S12-final-viral-combined_k141_216123\|\|full | 1128 | No | NA | 3 | 0 | 0 | Not-determined | Genome-fragment | NA | NA | 0 |
| S8-final-viral-combined_k141_249735\|\|full | 3069 | No | NA | 6 | 2 | 0 | Low-quality | Genome-fragment | 7.09 | AAI-based (medium-confidence) | 0 |
| S9_k141_25580 | 3070 | No | NA | 5 | 0 | 1 | Low-quality | Genome-fragment | 3.62 | AAI-based (high-confidence) | 0 |
| S7_k141_189327 | 3070 | No | NA | 4 | 1 | 0 | Low-quality | Genome-fragment | 6.88 | AAI-based (high-confidence) | 0 |
| S12-final-viral-combined_k141_299628\|\|full | 1226 | No | NA | 3 | 0 | 1 | Not-determined | Genome-fragment | NA | NA | 0 |
| S12-final-viral-combined_k141_216165\|\|full | 2100 | No | NA | 3 | 0 | 0 | Not-determined | Genome-fragment | NA | NA | 0 |
| S2-final-viral-combined_k141_75495\|\|full | 3070 | No | NA | 3 | 0 | 1 | Low-quality | Genome-fragment | 4.99 | AAI-based (high-confidence) | 0 |
| S12-final-viral-combined_k141_349875\|\|full | 2098 | No | NA | 3 | 0 | 2 | Not-determined | Genome-fragment | NA | NA | 0 |
| S12-final-viral-combined_k141_49167\|\|full | 2705 | No | NA | 3 | 0 | 1 | Not-determined | Genome-fragment | NA | NA | 0 |
| S12-final-viral-combined_k141_98378\|\|full | 4022 | No | NA | 3 | 0 | 2 | Not-determined | Genome-fragment | NA | NA | 0 |
| S12-final-viral-combined_k141_148383\|\|full | 1279 | No | NA | 3 | 0 | 0 | Not-determined | Genome-fragment | NA | NA | 0 |
| S12-final-viral-combined_k141_349928\|\|full | 1126 | No | NA | 3 | 0 | 0 | Not-determined | Genome-fragment | NA | NA | 0 |
| S12-final-viral-combined_k141_49219\|\|full | 3252 | No | NA | 2 | 0 | 0 | Not-determined | Genome-fragment | NA | NA | 0 |
| S12-final-viral-combined_k141_399487\|\|full | 1570 | No | NA | 2 | 0 | 1 | Not-determined | Genome-fragment | NA | NA | 0 |
| S12-final-viral-combined_k141_233510\|\|full | 1231 | No | NA | 2 | 0 | 1 | Not-determined | Genome-fragment | NA | NA | 0 |
| S11-final-viral-combined_k141_163945\|\|full | 3071 | No | NA | 3 | 0 | 2 | Low-quality | Genome-fragment | 1.68 | AAI-based (medium-confidence) | 0 |
| S12-final-viral-combined_k141_82530\|\|full | 1954 | No | NA | 5 | 0 | 0 | Not-determined | Genome-fragment | NA | NA | 0 |
| S11_k141_119113 | 3072 | No | NA | 5 | 1 | 0 | Low-quality | Genome-fragment | 2.31 | AAI-based (high-confidence) | 0 |
| S7-final-viral-combined_k141_31890\|\|full | 3072 | No | NA | 7 | 1 | 0 | Low-quality | Genome-fragment | 8.05 | AAI-based (medium-confidence) | 0 |
| S12-final-viral-combined_k141_182671\|\|full | 1414 | No | NA | 2 | 0 | 0 | Not-determined | Genome-fragment | NA | NA | 0 |
| S12-final-viral-combined_k141_199460\|\|full | 1325 | No | NA | 2 | 0 | 1 | Not-determined | Genome-fragment | NA | NA | 0 |
| S12-final-viral-combined_k141_249198\|\|full | 1210 | No | NA | 2 | 0 | 1 | Not-determined | Genome-fragment | NA | NA | 0 |
| S12-final-viral-combined_k141_199476\|\|full | 1235 | No | NA | 2 | 0 | 1 | Not-determined | Genome-fragment | NA | NA | 0 |
| S7-final-viral-combined_k141_176199\|\|full | 3072 | No | NA | 4 | 0 | 1 | Low-quality | Genome-fragment | 3.63 | AAI-based (medium-confidence) | 0 |
| S12-final-viral-combined_k141_14735\|\|full | 2278 | No | NA | 3 | 0 | 1 | Not-determined | Genome-fragment | NA | NA | 0 |
| S12-final-viral-combined_k141_148470\|\|full | 1121 | No | NA | 2 | 0 | 1 | Not-determined | Genome-fragment | NA | NA | 0 |
| S12-final-viral-combined_k141_199485\|\|full | 5462 | No | NA | 5 | 0 | 2 | Not-determined | Genome-fragment | NA | NA | 0 |
| S12-final-viral-combined_k141_165925\|\|full | 1883 | No | NA | 5 | 0 | 0 | Not-determined | Genome-fragment | NA | NA | 0 |
| S6-final-viral-combined_k141_106585\|\|full | 3072 | No | NA | 3 | 0 | 2 | Low-quality | Genome-fragment | 2.04 | AAI-based (medium-confidence) | 0 |
| S3_k141_85840 | 3073 | No | NA | 4 | 0 | 1 | Low-quality | Genome-fragment | 5.47 | AAI-based (medium-confidence) | 0 |
| S7_k141_18653 | 3073 | No | NA | 2 | 0 | 2 | Low-quality | Genome-fragment | 9.24 | AAI-based (medium-confidence) | 0 |
| S1-final-viral-combined_k141_38244\|\|full | 3074 | No | NA | 3 | 1 | 0 | Low-quality | Genome-fragment | 8.33 | AAI-based (high-confidence) | 0 |
| S12-final-viral-combined_k141_165948\|\|full | 1723 | No | NA | 3 | 0 | 3 | Not-determined | Genome-fragment | NA | NA | 0 |
| S1_k141_8417 | 3074 | No | NA | 9 | 2 | 0 | Low-quality | Genome-fragment | 2.27 | AAI-based (high-confidence) | 0 |
| S3_k141_128792 | 3074 | No | NA | 5 | 3 | 0 | Low-quality | Genome-fragment | 8.04 | AAI-based (medium-confidence) | 0 |
| S11_k141_303559 | 3075 | No | NA | 3 | 3 | 0 | Low-quality | Genome-fragment | 7.17 | AAI-based (medium-confidence) | 0 |
| S17-final-viral-combined_k141_140628\|\|full | 3075 | No | NA | 4 | 1 | 0 | Low-quality | Genome-fragment | 7 | AAI-based (medium-confidence) | 0 |
| S12-final-viral-combined_k141_182784\|\|full | 2635 | No | NA | 4 | 0 | 3 | Not-determined | Genome-fragment | NA | NA | 0 |
| S12-final-viral-combined_k141_49343\|\|full | 1776 | No | NA | 3 | 0 | 0 | Not-determined | Genome-fragment | NA | NA | 0 |
| S12-final-viral-combined_k141_165975\|\|full | 3435 | No | NA | 3 | 0 | 2 | Not-determined | Genome-fragment | NA | NA | 0 |
| S12-final-viral-combined_k141_364955\|\|full | 3386 | No | NA | 3 | 0 | 2 | Not-determined | Genome-fragment | NA | NA | 0 |
| S12-final-viral-combined_k141_260260\|\|full | 3075 | No | NA | 5 | 2 | 0 | Low-quality | Genome-fragment | 8.37 | AAI-based (high-confidence) | 0 |
| S12-final-viral-combined_k141_82644\|\|full | 4968 | No | NA | 6 | 0 | 1 | Not-determined | Genome-fragment | NA | NA | 0 |
| S12-final-viral-combined_k141_399567\|\|full | 2212 | No | NA | 2 | 0 | 1 | Not-determined | Genome-fragment | NA | NA | 0 |
| S2-final-viral-combined_k141_11256\|\|full | 3075 | No | NA | 3 | 0 | 2 | Low-quality | Genome-fragment | 8.03 | AAI-based (medium-confidence) | 0 |
| S2_k141_161745 | 3075 | No | NA | 9 | 0 | 0 | Low-quality | Genome-fragment | 2.35 | AAI-based (high-confidence) | 0 |
| S12-final-viral-combined_k141_148583\|\|full | 2949 | No | NA | 3 | 0 | 2 | Not-determined | Genome-fragment | NA | NA | 0 |
| S12-final-viral-combined_k141_131616\|\|full | 4946 | No | NA | 10 | 0 | 0 | Not-determined | Genome-fragment | NA | NA | 0 |
| S12-final-viral-combined_k141_49408\|\|full | 4184 | No | NA | 5 | 0 | 0 | Not-determined | Genome-fragment | NA | NA | 0 |
| S8_k141_25585 | 3076 | No | NA | 5 | 3 | 0 | Low-quality | Genome-fragment | 9.57 | AAI-based (high-confidence) | 0 |
| S17-k141_72562 | 3077 | No | NA | 4 | 0 | 0 | Low-quality | Genome-fragment | 4.17 | AAI-based (high-confidence) | 0 |
| S12-final-viral-combined_k141_284006\|\|full | 3799 | No | NA | 4 | 0 | 4 | Not-determined | Genome-fragment | NA | NA | 0 |
| S12-final-viral-combined_k141_383236\|\|full | 2584 | No | NA | 2 | 0 | 1 | Not-determined | Genome-fragment | NA | NA | 0 |
| S17-k141_28336 | 3077 | No | NA | 4 | 2 | 0 | Low-quality | Genome-fragment | 4.99 | AAI-based (high-confidence) | 0 |
| S8_k141_270098 | 3077 | No | NA | 5 | 4 | 0 | Low-quality | Genome-fragment | 9 | AAI-based (high-confidence) | 0 |
| S9-final-viral-combined_k141_74363\|\|full | 3078 | No | NA | 3 | 0 | 1 | Low-quality | Genome-fragment | 3.47 | AAI-based (medium-confidence) | 0 |
| S12-final-viral-combined_k141_365054\|\|full | 3326 | No | NA | 3 | 0 | 1 | Not-determined | Genome-fragment | NA | NA | 0 |
| S12-final-viral-combined_k141_333615\|\|full | 1614 | No | NA | 2 | 0 | 2 | Not-determined | Genome-fragment | NA | NA | 0 |
| S9_k141_77388 | 3078 | No | NA | 4 | 0 | 0 | Low-quality | Genome-fragment | 5.26 | AAI-based (high-confidence) | 0 |
| S1-final-viral-combined_k141_213590\|\|full | 3078 | No | NA | 3 | 0 | 2 | Low-quality | Genome-fragment | 3.2 | AAI-based (medium-confidence) | 0 |
| S3-final-viral-combined_k141_121309\|\|full | 3078 | No | NA | 2 | 0 | 0 | Low-quality | Genome-fragment | 5.16 | AAI-based (high-confidence) | 0 |
| S12-final-viral-combined_k141_299872\|\|full | 30946 | No | NA | 48 | 13 | 0 | Medium-quality | Genome-fragment | 86.46 | AAI-based (medium-confidence) | 0 |
| S12-final-viral-combined_k141_350150\|\|full | 2970 | No | NA | 3 | 0 | 1 | Not-determined | Genome-fragment | NA | NA | 0 |
| S12-final-viral-combined_k141_31662\|\|full | 1748 | No | NA | 2 | 0 | 1 | Not-determined | Genome-fragment | NA | NA | 0 |
| S12-final-viral-combined_k141_333671\|\|full | 1748 | No | NA | 3 | 0 | 0 | Not-determined | Genome-fragment | NA | NA | 0 |
| S12-final-viral-combined_k141_65253\|\|full | 2565 | No | NA | 4 | 0 | 1 | Not-determined | Genome-fragment | NA | NA | 0 |
| S12-final-viral-combined_k141_65269\|\|full | 2603 | No | NA | 5 | 0 | 1 | Not-determined | Genome-fragment | NA | NA | 0 |
| S12-final-viral-combined_k141_65273\|\|full | 1168 | No | NA | 3 | 0 | 2 | Not-determined | Genome-fragment | NA | NA | 0 |
| S12-final-viral-combined_k141_98647\|\|full | 2046 | No | NA | 2 | 0 | 0 | Not-determined | Genome-fragment | NA | NA | 0 |
| S3_k141_36013 | 3078 | No | NA | 4 | 0 | 1 | Low-quality | Genome-fragment | 5.63 | AAI-based (medium-confidence) | 0 |
| S11-final-viral-combined_k141_300134\|\|full | 3079 | No | NA | 4 | 3 | 0 | Low-quality | Genome-fragment | 8.02 | AAI-based (medium-confidence) | 0 |
| S12-final-viral-combined_k141_249425\|\|full | 2818 | No | NA | 4 | 0 | 1 | Not-determined | Genome-fragment | NA | NA | 0 |
| S8-final-viral-combined_k141_83129\|\|full | 3079 | No | NA | 3 | 0 | 0 | Low-quality | Genome-fragment | 7.75 | AAI-based (high-confidence) | 0 |
| S12-final-viral-combined_k141_148723\|\|full | 1552 | No | NA | 2 | 0 | 2 | Not-determined | Genome-fragment | NA | NA | 0 |
| S12-final-viral-combined_k141_98666\|\|full | 6018 | No | NA | 3 | 0 | 2 | Not-determined | Genome-fragment | NA | NA | 0 |
| S12-final-viral-combined_k141_365165\|\|full | 3167 | No | NA | 4 | 0 | 1 | Not-determined | Genome-fragment | NA | NA | 0 |
| S12-final-viral-combined_k141_82820\|\|full | 2071 | No | NA | 3 | 0 | 2 | Not-determined | Genome-fragment | NA | NA | 0 |
| S17-k141_58626 | 3080 | No | NA | 2 | 0 | 2 | Low-quality | Genome-fragment | 3.51 | AAI-based (medium-confidence) | 0 |
| S12-final-viral-combined_k141_219989\|\|full | 3080 | Yes | 1568 | 5 | 1 | 1 | Low-quality | Genome-fragment | 2.88 | HMM-based (lower-bound) | 49.09 |
| S2-final-viral-combined_k141_199638\|\|full | 3080 | No | NA | 2 | 0 | 0 | Low-quality | Genome-fragment | 4.83 | AAI-based (medium-confidence) | 0 |
| S12-final-viral-combined_k141_114981\|\|full | 2381 | No | NA | 2 | 0 | 0 | Not-determined | Genome-fragment | NA | NA | 0 |
| S12-final-viral-combined_k141_350169\|\|full | 1203 | No | NA | 3 | 0 | 0 | Not-determined | Genome-fragment | NA | NA | 0 |
| S8_k141_132075 | 3080 | No | NA | 4 | 1 | 0 | Low-quality | Genome-fragment | 2.32 | AAI-based (high-confidence) | 0 |
| S12-final-viral-combined_k141_115004\|\|full | 2039 | No | NA | 2 | 0 | 1 | Not-determined | Genome-fragment | NA | NA | 0 |
| S1-final-viral-combined_k141_285390\|\|full | 3081 | No | NA | 4 | 0 | 3 | Low-quality | Genome-fragment | 2.56 | AAI-based (medium-confidence) | 0 |
| S12-final-viral-combined_k141_49594\|\|full | 6585 | No | NA | 5 | 0 | 1 | Not-determined | Genome-fragment | NA | NA | 0 |
| S12-final-viral-combined_k141_14856\|\|full | 15255 | No | NA | 17 | 0 | 6 | Not-determined | Genome-fragment | NA | NA | 0 |
| S12-final-viral-combined_k141_82869\|\|full | 1087 | No | NA | 2 | 0 | 1 | Not-determined | Genome-fragment | NA | NA | 0 |
| S12-final-viral-combined_k141_148741\|\|full | 41121 | Yes | 37297 | 52 | 7 | 4 | High-quality | High-quality | 94.73 | AAI-based (high-confidence) | 9.3 |
| S7_k141_95658 | 3081 | No | NA | 6 | 0 | 1 | Low-quality | Genome-fragment | 5.94 | AAI-based (high-confidence) | 0 |
| S17-k141_193606 | 3081 | No | NA | 4 | 0 | 3 | Low-quality | Genome-fragment | 1.15 | AAI-based (medium-confidence) | 0 |
| S12-final-viral-combined_k141_98767\|\|full | 4582 | No | NA | 6 | 0 | 4 | Not-determined | Genome-fragment | NA | NA | 0 |
| S12_k141_233335 | 3081 | No | NA | 5 | 2 | 0 | Low-quality | Genome-fragment | 6.23 | AAI-based (medium-confidence) | 0 |
| S12-final-viral-combined_k141_233937\|\|full | 1532 | No | NA | 3 | 0 | 1 | Not-determined | Genome-fragment | NA | NA | 0 |
| S12-final-viral-combined_k141_333793\|\|full | 2373 | No | NA | 3 | 0 | 0 | Not-determined | Genome-fragment | NA | NA | 0 |
| S12-final-viral-combined_k141_333803\|\|full | 2192 | No | NA | 3 | 0 | 2 | Not-determined | Genome-fragment | NA | NA | 0 |
| S4_k141_128264 | 3081 | No | NA | 5 | 0 | 0 | Low-quality | Genome-fragment | 1.98 | AAI-based (high-confidence) | 0 |
| S17-final-viral-combined_k141_6136\|\|full | 3082 | No | NA | 4 | 0 | 1 | Low-quality | Genome-fragment | 5.73 | AAI-based (medium-confidence) | 0 |
| S12-final-viral-combined_k141_31761\|\|full | 40932 | Yes | 36180 | 62 | 14 | 7 | High-quality | High-quality | 100 | AAI-based (high-confidence) | 11.61 |
| S12-final-viral-combined_k141_350226\|\|full | 2579 | No | NA | 2 | 0 | 2 | Not-determined | Genome-fragment | NA | NA | 0 |
| S12-final-viral-combined_k141_249514\|\|full | 1235 | No | NA | 3 | 0 | 2 | Not-determined | Genome-fragment | NA | NA | 0 |
| S12_k141_232654 | 3082 | No | NA | 2 | 0 | 2 | Low-quality | Genome-fragment | 0.95 | AAI-based (medium-confidence) | 0 |
| S12-final-viral-combined_k141_249516\|\|full | 4518 | No | NA | 4 | 0 | 2 | Not-determined | Genome-fragment | NA | NA | 0 |
| S12-final-viral-combined_k141_82942\|\|full | 1483 | No | NA | 3 | 0 | 3 | Not-determined | Genome-fragment | NA | NA | 0 |
| S5-final-viral-combined_k141_167643\|\|full | 3083 | No | NA | 4 | 1 | 0 | Low-quality | Genome-fragment | 0.49 | HMM-based (lower-bound) | 0 |
| S12-final-viral-combined_k141_350257\|\|full | 1733 | No | NA | 2 | 0 | 1 | Not-determined | Genome-fragment | NA | NA | 0 |
| S17-final-viral-combined_k141_53460\|\|full | 3083 | No | NA | 3 | 0 | 2 | Low-quality | Genome-fragment | 3.7 | AAI-based (medium-confidence) | 0 |
| S12-final-viral-combined_k141_169931\|\|full | 3083 | No | NA | 5 | 2 | 1 | Low-quality | Genome-fragment | 6.12 | AAI-based (medium-confidence) | 0 |
| S8-final-viral-combined_k141_125445\|\|full | 3083 | No | NA | 2 | 1 | 0 | Low-quality | Genome-fragment | 7.56 | AAI-based (high-confidence) | 0 |
| S12-final-viral-combined_k141_148792\|\|full | 1144 | No | NA | 2 | 0 | 1 | Not-determined | Genome-fragment | NA | NA | 0 |
| S5-final-viral-combined_k141_188693\|\|full | 3084 | No | NA | 7 | 1 | 0 | Low-quality | Genome-fragment | 2.32 | AAI-based (medium-confidence) | 0 |
| S5-final-viral-combined_k141_134047\|\|full | 3085 | No | NA | 3 | 0 | 2 | Low-quality | Genome-fragment | 3.65 | AAI-based (medium-confidence) | 0 |
| S17-k141_43791 | 3085 | No | NA | 8 | 1 | 0 | Low-quality | Genome-fragment | 3.75 | AAI-based (high-confidence) | 0 |
| S12-final-viral-combined_k141_383463\|\|full | 1609 | No | NA | 3 | 0 | 0 | Not-determined | Genome-fragment | NA | NA | 0 |
| S12-final-viral-combined_k141_199948\|\|full | 2967 | No | NA | 6 | 0 | 0 | Not-determined | Genome-fragment | NA | NA | 0 |
| S12_k141_20452 | 3085 | Yes | 586 | 4 | 1 | 2 | Low-quality | Genome-fragment | 0.53 | HMM-based (lower-bound) | 81 |
| S4-final-viral-combined_k141_136536\|\|full | 3085 | No | NA | 4 | 1 | 0 | Low-quality | Genome-fragment | 7.3 | AAI-based (high-confidence) | 0 |
| S12-final-viral-combined_k141_333867\|\|full | 1075 | No | NA | 2 | 0 | 1 | Not-determined | Genome-fragment | NA | NA | 0 |
| S12-final-viral-combined_k141_365317\|\|full | 5229 | No | NA | 7 | 0 | 4 | Not-determined | Genome-fragment | NA | NA | 0 |
| S12-final-viral-combined_k141_365323\|\|full | 1433 | No | NA | 2 | 0 | 2 | Not-determined | Genome-fragment | NA | NA | 0 |
| S12-final-viral-combined_k141_350343\|\|full | 5760 | No | NA | 4 | 0 | 2 | Not-determined | Genome-fragment | NA | NA | 0 |
| S12-final-viral-combined_k141_15016\|\|full | 1056 | No | NA | 3 | 0 | 0 | Not-determined | Genome-fragment | NA | NA | 0 |
| S1-final-viral-combined_k141_133954\|\|full | 3086 | No | NA | 7 | 2 | 0 | Low-quality | Genome-fragment | 6.73 | AAI-based (high-confidence) | 0 |
| S7-final-viral-combined_k141_170149\|\|full | 3086 | No | NA | 3 | 2 | 0 | Low-quality | Genome-fragment | 3.75 | HMM-based (lower-bound) | 0 |
| S12-final-viral-combined_k141_131947\|\|full | 2777 | No | NA | 2 | 0 | 2 | Not-determined | Genome-fragment | NA | NA | 0 |
| S7_k141_99046 | 3086 | No | NA | 6 | 4 | 0 | Low-quality | Genome-fragment | 7.6 | AAI-based (high-confidence) | 0 |
| S3_k141_53282 | 3087 | No | NA | 2 | 1 | 0 | Low-quality | Genome-fragment | 4.93 | AAI-based (high-confidence) | 0 |
| S6_k141_84172 | 3087 | No | NA | 4 | 0 | 2 | Low-quality | Genome-fragment | 2.13 | AAI-based (high-confidence) | 0 |
| S8_k141_236488 | 3087 | No | NA | 6 | 0 | 0 | Low-quality | Genome-fragment | 2.47 | AAI-based (high-confidence) | 0 |
| S12-final-viral-combined_k141_98904\|\|full | 3346 | No | NA | 4 | 0 | 2 | Not-determined | Genome-fragment | NA | NA | 0 |
| S12-final-viral-combined_k141_365380\|\|full | 1793 | No | NA | 3 | 0 | 1 | Not-determined | Genome-fragment | NA | NA | 0 |
| S12-final-viral-combined_k141_65468\|\|full | 4532 | No | NA | 8 | 0 | 0 | Not-determined | Genome-fragment | NA | NA | 0 |
| S12-final-viral-combined_k141_115143\|\|full | 5246 | No | NA | 6 | 0 | 5 | Not-determined | Genome-fragment | NA | NA | 0 |
| S1-final-viral-combined_k141_265559\|\|lt2gene | 3088 | No | NA | 2 | 2 | 0 | Low-quality | Genome-fragment | 4.78 | HMM-based (lower-bound) | 0 |
| S4_k141_107922 | 3088 | No | NA | 2 | 0 | 1 | Low-quality | Genome-fragment | 4.88 | AAI-based (high-confidence) | 0 |
| S6-final-viral-combined_k141_73461\|\|full | 3088 | No | NA | 4 | 0 | 2 | Low-quality | Genome-fragment | 6.86 | AAI-based (medium-confidence) | 0 |
| S12-final-viral-combined_k141_199962\|\|full | 2241 | No | NA | 3 | 0 | 3 | Not-determined | Genome-fragment | NA | NA | 0 |
| S12-final-viral-combined_k141_216850\|\|full | 2943 | No | NA | 5 | 0 | 2 | Not-determined | Genome-fragment | NA | NA | 0 |
| S8-final-viral-combined_k141_228389\|\|full | 3088 | No | NA | 6 | 0 | 0 | Low-quality | Genome-fragment | 4.22 | AAI-based (medium-confidence) | 0 |
| S9-final-viral-combined_k141_11076\|\|full | 3090 | No | NA | 3 | 0 | 1 | Low-quality | Genome-fragment | 0.43 | AAI-based (medium-confidence) | 0 |
| S12-final-viral-combined_k141_42770\|\|full | 3090 | No | NA | 4 | 0 | 3 | Low-quality | Genome-fragment | 3.77 | AAI-based (medium-confidence) | 0 |
| S12-final-viral-combined_k141_199989\|\|full | 2018 | No | NA | 2 | 0 | 1 | Not-determined | Genome-fragment | NA | NA | 0 |
| S12-final-viral-combined_k141_234097\|\|full | 1847 | No | NA | 3 | 0 | 2 | Not-determined | Genome-fragment | NA | NA | 0 |
| S12_k141_174005 | 3090 | No | NA | 3 | 0 | 1 | Low-quality | Genome-fragment | 2.86 | AAI-based (medium-confidence) | 0 |
| S12-final-viral-combined_k141_199996\|\|full | 1683 | No | NA | 3 | 0 | 0 | Not-determined | Genome-fragment | NA | NA | 0 |
| S12-final-viral-combined_k141_166359\|\|full | 1629 | No | NA | 3 | 0 | 1 | Not-determined | Genome-fragment | NA | NA | 0 |
| S8-final-viral-combined_k141_197865\|\|full | 3090 | No | NA | 4 | 0 | 0 | Low-quality | Genome-fragment | 2.53 | AAI-based (high-confidence) | 0 |
| S12-final-viral-combined_k141_265696\|\|full | 1945 | No | NA | 2 | 0 | 1 | Not-determined | Genome-fragment | NA | NA | 0 |
| S12-final-viral-combined_k141_31975\|\|full | 1428 | No | NA | 2 | 0 | 0 | Not-determined | Genome-fragment | NA | NA | 0 |
| S12-final-viral-combined_k141_350458\|\|full | 4035 | No | NA | 5 | 0 | 1 | Not-determined | Genome-fragment | NA | NA | 0 |
| S8-final-viral-combined_k141_31023\|\|full | 3090 | No | NA | 3 | 0 | 0 | Low-quality | Genome-fragment | 6.26 | AAI-based (medium-confidence) | 0 |
| S12-final-viral-combined_k141_83129\|\|full | 2080 | No | NA | 3 | 0 | 2 | Not-determined | Genome-fragment | NA | NA | 0 |
| S12-final-viral-combined_k141_115233\|\|full | 1684 | No | NA | 3 | 0 | 1 | Not-determined | Genome-fragment | NA | NA | 0 |
| S12-final-viral-combined_k141_317966\|\|full | 4161 | No | NA | 5 | 0 | 5 | Not-determined | Genome-fragment | NA | NA | 0 |
| S12-final-viral-combined_k141_115236\|\|full | 1286 | No | NA | 2 | 0 | 1 | Not-determined | Genome-fragment | NA | NA | 0 |
| S9-final-viral-combined_k141_8243\|\|full | 3091 | No | NA | 3 | 0 | 3 | Low-quality | Genome-fragment | 2.29 | AAI-based (medium-confidence) | 0 |
| S1-final-viral-combined_k141_345966\|\|full | 3091 | No | NA | 4 | 0 | 4 | Low-quality | Genome-fragment | 2.51 | AAI-based (medium-confidence) | 0 |
| S7-final-viral-combined_k141_176840\|\|full | 3091 | No | NA | 3 | 0 | 2 | Low-quality | Genome-fragment | 1.39 | AAI-based (medium-confidence) | 0 |
| S12-final-viral-combined_k141_383614\|\|full | 1247 | No | NA | 2 | 0 | 1 | Not-determined | Genome-fragment | NA | NA | 0 |
| S12-final-viral-combined_k141_15233\|\|full | 3088 | No | NA | 3 | 0 | 0 | Not-determined | Genome-fragment | NA | NA | 0 |
| S12-final-viral-combined_k141_98966\|\|full | 1271 | No | NA | 2 | 0 | 0 | Not-determined | Genome-fragment | NA | NA | 0 |
| S12-final-viral-combined_k141_83188\|\|full | 1286 | No | NA | 3 | 0 | 0 | Not-determined | Genome-fragment | NA | NA | 0 |
| S12-final-viral-combined_k141_166417\|\|full | 1485 | No | NA | 2 | 0 | 1 | Not-determined | Genome-fragment | NA | NA | 0 |
| S12-final-viral-combined_k141_234200\|\|full | 2293 | No | NA | 4 | 0 | 1 | Not-determined | Genome-fragment | NA | NA | 0 |
| S8-final-viral-combined_k141_266419\|\|full | 3091 | No | NA | 6 | 2 | 0 | Low-quality | Genome-fragment | 8.59 | AAI-based (medium-confidence) | 0 |
| S1-final-viral-combined_k141_375923\|\|full | 3092 | No | NA | 4 | 1 | 0 | Low-quality | Genome-fragment | 2.38 | HMM-based (lower-bound) | 0 |
| S7_k141_93445 | 3092 | No | NA | 1 | 0 | 1 | Low-quality | Genome-fragment | 6.66 | AAI-based (high-confidence) | 0 |
| S17-k141_71843 | 3092 | No | NA | 4 | 1 | 0 | Low-quality | Genome-fragment | 0.25 | HMM-based (lower-bound) | 0 |
| S12-final-viral-combined_k141_166464\|\|full | 1034 | No | NA | 4 | 0 | 0 | Not-determined | Genome-fragment | NA | NA | 0 |
| S4_k141_109411 | 3092 | No | NA | 3 | 0 | 0 | Low-quality | Genome-fragment | 5.11 | AAI-based (medium-confidence) | 0 |
| S8_k141_93486 | 3092 | No | NA | 7 | 1 | 0 | Low-quality | Genome-fragment | 7.43 | AAI-based (high-confidence) | 0 |
| S12-final-viral-combined_k141_234239\|\|full | 3086 | No | NA | 3 | 0 | 2 | Not-determined | Genome-fragment | NA | NA | 0 |
| S12-final-viral-combined_k141_399981\|\|full | 2280 | No | NA | 5 | 0 | 0 | Not-determined | Genome-fragment | NA | NA | 0 |
| S12-final-viral-combined_k141_166477\|\|full | 1288 | No | NA | 2 | 0 | 0 | Not-determined | Genome-fragment | NA | NA | 0 |
| S9-final-viral-combined_k141_59260\|\|full | 3093 | No | NA | 3 | 0 | 0 | Low-quality | Genome-fragment | 6.58 | AAI-based (medium-confidence) | 0 |
| S11-final-viral-combined_k141_241532\|\|full | 3093 | No | NA | 4 | 0 | 0 | Low-quality | Genome-fragment | 4.38 | AAI-based (medium-confidence) | 0 |
| S1-final-viral-combined_k141_160635\|\|full | 3093 | No | NA | 3 | 0 | 1 | Low-quality | Genome-fragment | 5.51 | AAI-based (high-confidence) | 0 |
| S12-final-viral-combined_k141_15357\|\|full | 1110 | No | NA | 3 | 0 | 0 | Not-determined | Genome-fragment | NA | NA | 0 |
| S5-final-viral-combined_k141_52561\|\|full | 3093 | No | NA | 5 | 0 | 1 | Low-quality | Genome-fragment | 3.59 | AAI-based (medium-confidence) | 0 |
| S12-final-viral-combined_k141_300361\|\|full | 1810 | No | NA | 3 | 0 | 2 | Not-determined | Genome-fragment | NA | NA | 0 |
| S17-k141_91181 | 3093 | No | NA | 3 | 0 | 2 | Low-quality | Genome-fragment | 1.49 | AAI-based (medium-confidence) | 0 |
| S12-final-viral-combined_k141_132177\|\|full | 3198 | No | NA | 3 | 0 | 2 | Not-determined | Genome-fragment | NA | NA | 0 |
| S12-final-viral-combined_k141_136292\|\|full | 3093 | No | NA | 3 | 1 | 0 | Low-quality | Genome-fragment | 7.21 | AAI-based (medium-confidence) | 0 |
| S4-final-viral-combined_k141_77551\|\|full | 3093 | No | NA | 2 | 0 | 1 | Low-quality | Genome-fragment | 4.4 | AAI-based (medium-confidence) | 0 |
| S12-final-viral-combined_k141_183461\|\|full | 1878 | No | NA | 3 | 0 | 1 | Not-determined | Genome-fragment | NA | NA | 0 |
| S9-final-viral-combined_k141_86755\|\|full | 3094 | No | NA | 3 | 0 | 1 | Low-quality | Genome-fragment | 2.88 | AAI-based (medium-confidence) | 0 |
| S7-final-viral-combined_k141_202007\|\|full | 3094 | No | NA | 5 | 0 | 0 | Low-quality | Genome-fragment | 5.45 | AAI-based (high-confidence) | 0 |
| S12-final-viral-combined_k141_83366\|\|full | 1439 | No | NA | 2 | 0 | 2 | Not-determined | Genome-fragment | NA | NA | 0 |
| S12-final-viral-combined_k141_249787\|\|full | 16419 | No | NA | 14 | 0 | 10 | Not-determined | Genome-fragment | NA | NA | 0 |
| S12-final-viral-combined_k141_350620\|\|full | 2158 | No | NA | 3 | 0 | 1 | Not-determined | Genome-fragment | NA | NA | 0 |
| S12-final-viral-combined_k141_234325\|\|full | 1532 | No | NA | 2 | 0 | 1 | Not-determined | Genome-fragment | NA | NA | 0 |
| S12-final-viral-combined_k141_65755\|\|full | 3287 | No | NA | 4 | 0 | 0 | Not-determined | Genome-fragment | NA | NA | 0 |
| S12-final-viral-combined_k141_183494\|\|full | 3913 | No | NA | 5 | 0 | 5 | Not-determined | Genome-fragment | NA | NA | 0 |
| S2-final-viral-combined_k141_103858\|\|full | 3094 | No | NA | 4 | 1 | 0 | Low-quality | Genome-fragment | 2.37 | HMM-based (lower-bound) | 0 |
| S12-final-viral-combined_k141_265966\|\|full | 4476 | No | NA | 7 | 0 | 1 | Not-determined | Genome-fragment | NA | NA | 0 |
| S12-final-viral-combined_k141_50100\|\|full | 3695 | No | NA | 4 | 0 | 3 | Not-determined | Genome-fragment | NA | NA | 0 |
| S12_k141_188934 | 3095 | No | NA | 4 | 0 | 0 | Low-quality | Genome-fragment | 3.11 | AAI-based (high-confidence) | 0 |
| S12-final-viral-combined_k141_15480\|\|full | 2338 | No | NA | 4 | 0 | 3 | Not-determined | Genome-fragment | NA | NA | 0 |
| S1_k141_343240 | 3096 | No | NA | 4 | 0 | 0 | Low-quality | Genome-fragment | 6.83 | AAI-based (high-confidence) | 0 |
| S1-final-viral-combined_k141_178551\|\|full | 3097 | No | NA | 6 | 3 | 0 | Low-quality | Genome-fragment | 4.98 | AAI-based (high-confidence) | 0 |
| S12-final-viral-combined_k141_200245\|\|full | 2107 | No | NA | 3 | 0 | 0 | Not-determined | Genome-fragment | NA | NA | 0 |
| S12-final-viral-combined_k141_99239\|\|full | 1031 | No | NA | 3 | 0 | 1 | Not-determined | Genome-fragment | NA | NA | 0 |
| S12_k141_260153 | 3097 | No | NA | 6 | 0 | 2 | Low-quality | Genome-fragment | 7.18 | AAI-based (medium-confidence) | 0 |
| S12-final-viral-combined_k141_266011\|\|full | 3740 | No | NA | 6 | 0 | 2 | Not-determined | Genome-fragment | NA | NA | 0 |
| S2-final-viral-combined_k141_238125\|\|full | 3097 | No | NA | 3 | 0 | 1 | Low-quality | Genome-fragment | 1.4 | AAI-based (medium-confidence) | 0 |
| S12-final-viral-combined_k141_383822\|\|full | 3046 | No | NA | 4 | 0 | 1 | Not-determined | Genome-fragment | NA | NA | 0 |
| S7-final-viral-combined_k141_240486\|\|full | 3098 | No | NA | 5 | 0 | 2 | Low-quality | Genome-fragment | 1.06 | AAI-based (medium-confidence) | 0 |
| S12-final-viral-combined_k141_217152\|\|full | 2215 | No | NA | 3 | 0 | 1 | Not-determined | Genome-fragment | NA | NA | 0 |
| S12-final-viral-combined_k141_334290\|\|full | 1376 | No | NA | 3 | 0 | 1 | Not-determined | Genome-fragment | NA | NA | 0 |
| S17-k141_167540 | 3098 | No | NA | 7 | 1 | 0 | Low-quality | Genome-fragment | 1.19 | HMM-based (lower-bound) | 0 |
| S12-final-viral-combined_k141_50197\|\|full | 1785 | No | NA | 2 | 0 | 0 | Not-determined | Genome-fragment | NA | NA | 0 |
| S12-final-viral-combined_k141_350735\|\|full | 2800 | No | NA | 3 | 0 | 2 | Not-determined | Genome-fragment | NA | NA | 0 |
| S12_k141_20173 | 3098 | No | NA | 10 | 2 | 0 | Low-quality | Genome-fragment | 8.12 | AAI-based (high-confidence) | 0 |
| S12-final-viral-combined_k141_183652\|\|full | 2416 | No | NA | 3 | 0 | 3 | Not-determined | Genome-fragment | NA | NA | 0 |

| | | | | | | | | | | | |
|---|---|---|---|---|---|---|---|---|---|---|---|
| S8_k141_133679 | 3098 | No | NA | 6 | 0 | 2 | Low-quality | Genome-fragment | 8.17 | AAI-based (medium-confidence) | 0 |
| S12-final-viral-combined_k141_65847\|\|full | 2898 | No | NA | 6 | 0 | 0 | Not-determined | Genome-fragment | NA | NA | 0 |
| S11-final-viral-combined_k141_302379\|\|full | 3099 | No | NA | 4 | 1 | 0 | Low-quality | Genome-fragment | 6.94 | AAI-based (medium-confidence) | 0 |
| S12-final-viral-combined_k141_365539\|\|full | 1573 | No | NA | 2 | 0 | 1 | Not-determined | Genome-fragment | NA | NA | 0 |
| S8-final-viral-combined_k141_207262\|\|full | 3100 | No | NA | 8 | 0 | 0 | Low-quality | Genome-fragment | 9.71 | AAI-based (high-confidence) | 0 |
| S11_k141_9061 | 3101 | No | NA | 3 | 0 | 0 | Low-quality | Genome-fragment | 3.1 | AAI-based (high-confidence) | 0 |
| S12-final-viral-combined_k141_166694\|\|full | 2337 | No | NA | 2 | 0 | 0 | Not-determined | Genome-fragment | NA | NA | 0 |
| S12-final-viral-combined_k141_115590\|\|full | 1141 | No | NA | 2 | 0 | 1 | Not-determined | Genome-fragment | NA | NA | 0 |
| S7-final-viral-combined_k141_4737\|\|full | 3101 | No | NA | 4 | 0 | 4 | Low-quality | Genome-fragment | 1.5 | AAI-based (medium-confidence) | 0 |
| S12-final-viral-combined_k141_15537\|\|full | 1600 | No | NA | 2 | 0 | 0 | Not-determined | Genome-fragment | NA | NA | 0 |
| S12-final-viral-combined_k141_99347\|\|full | 1831 | No | NA | 3 | 0 | 0 | Not-determined | Genome-fragment | NA | NA | 0 |
| S12-final-viral-combined_k141_115607\|\|full | 1141 | No | NA | 2 | 0 | 1 | Not-determined | Genome-fragment | NA | NA | 0 |
| S12-final-viral-combined_k141_200398\|\|full | 2633 | No | NA | 3 | 0 | 0 | Not-determined | Genome-fragment | NA | NA | 0 |
| S2-final-viral-combined_k141_245789\|\|full | 3101 | No | NA | 6 | 6 | 0 | Low-quality | Genome-fragment | 6.5 | AAI-based (high-confidence) | 0 |
| S12-final-viral-combined_k141_32289\|\|full | 2784 | No | NA | 3 | 0 | 3 | Not-determined | Genome-fragment | NA | NA | 0 |
| S11-final-viral-combined_k141_225999\|\|full | 3102 | No | NA | 4 | 1 | 0 | Low-quality | Genome-fragment | 1.26 | HMM-based (lower-bound) | 0 |
| S1_k141_4305 | 3103 | No | NA | 3 | 0 | 2 | Low-quality | Genome-fragment | 8.04 | AAI-based (high-confidence) | 0 |
| S12-final-viral-combined_k141_217232\|\|full | 2137 | No | NA | 2 | 0 | 1 | Not-determined | Genome-fragment | NA | NA | 0 |
| S12-final-viral-combined_k141_166770\|\|full | 2352 | No | NA | 3 | 0 | 1 | Not-determined | Genome-fragment | NA | NA | 0 |
| S1-final-viral-combined_k141_133074\|\|full | 3104 | No | NA | 3 | 0 | 1 | Low-quality | Genome-fragment | 1.16 | AAI-based (medium-confidence) | 0 |
| S1_k141_220138 | 3104 | No | NA | 4 | 0 | 2 | Low-quality | Genome-fragment | 5.08 | AAI-based (medium-confidence) | 0 |
| S7-final-viral-combined_k141_9550\|\|full | 3104 | No | NA | 4 | 0 | 1 | Low-quality | Genome-fragment | 4.76 | AAI-based (medium-confidence) | 0 |
| S12-final-viral-combined_k141_32343\|\|full | 2572 | No | NA | 3 | 0 | 0 | Not-determined | Genome-fragment | NA | NA | 0 |
| S12-final-viral-combined_k141_246160\|\|full | 3104 | No | NA | 4 | 0 | 1 | Low-quality | Genome-fragment | 2.53 | AAI-based (high-confidence) | 0 |
| S12-final-viral-combined_k141_249951\|\|full | 1064 | No | NA | 3 | 0 | 1 | Not-determined | Genome-fragment | NA | NA | 0 |
| S12-final-viral-combined_k141_149283\|\|full | 1713 | No | NA | 3 | 0 | 1 | Not-determined | Genome-fragment | NA | NA | 0 |
| S12-final-viral-combined_k141_115692\|\|full | 1939 | No | NA | 2 | 0 | 1 | Not-determined | Genome-fragment | NA | NA | 0 |
| S8-final-viral-combined_k141_160554\|\|full | 3104 | No | NA | 8 | 0 | 0 | Low-quality | Genome-fragment | 5.65 | AAI-based (high-confidence) | 0 |
| S1-final-viral-combined_k141_315164\|\|full | 3105 | No | NA | 4 | 0 | 1 | Low-quality | Genome-fragment | 7.29 | AAI-based (medium-confidence) | 0 |
| S12-final-viral-combined_k141_217319\|\|full | 6002 | No | NA | 6 | 0 | 3 | Not-determined | Genome-fragment | NA | NA | 0 |
| S1-final-viral-combined_k141_8450\|\|full | 3105 | No | NA | 2 | 0 | 0 | Low-quality | Genome-fragment | 1.88 | AAI-based (high-confidence) | 0 |
| S12-final-viral-combined_k141_300679\|\|full | 5001 | No | NA | 5 | 0 | 3 | Not-determined | Genome-fragment | NA | NA | 0 |
| S12-final-viral-combined_k141_183803\|\|full | 1299 | No | NA | 3 | 0 | 1 | Not-determined | Genome-fragment | NA | NA | 0 |
| S12-final-viral-combined_k141_250023\|\|full | 1310 | No | NA | 3 | 0 | 0 | Not-determined | Genome-fragment | NA | NA | 0 |
| S12-final-viral-combined_k141_15683\|\|full | 3209 | No | NA | 2 | 0 | 1 | Not-determined | Genome-fragment | NA | NA | 0 |
| S1_k141_173531 | 3105 | No | NA | 3 | 0 | 1 | Low-quality | Genome-fragment | 3.75 | AAI-based (high-confidence) | 0 |
| S12-final-viral-combined_k141_66088\|\|full | 3885 | No | NA | 3 | 0 | 3 | Not-determined | Genome-fragment | NA | NA | 0 |
| S17-k141_119172 | 3105 | No | NA | 6 | 0 | 1 | Low-quality | Genome-fragment | 6.85 | AAI-based (medium-confidence) | 0 |
| S6_k141_96157 | 3105 | No | NA | 4 | 1 | 0 | Low-quality | Genome-fragment | 5.33 | AAI-based (high-confidence) | 0 |
| S12-final-viral-combined_k141_132581\|\|full | 1355 | No | NA | 2 | 0 | 1 | Not-determined | Genome-fragment | NA | NA | 0 |
| S12-final-viral-combined_k141_115777\|\|full | 10045 | No | NA | 17 | 0 | 0 | Not-determined | Genome-fragment | NA | NA | 0 |
| S12-final-viral-combined_k141_200563\|\|full | 10572 | No | NA | 9 | 0 | 6 | Not-determined | Genome-fragment | NA | NA | 0 |
| S12-final-viral-combined_k141_300710\|\|full | 1412 | No | NA | 2 | 0 | 0 | Not-determined | Genome-fragment | NA | NA | 0 |
| S9_k141_49282 | 3106 | No | NA | 3 | 2 | 0 | Low-quality | Genome-fragment | 7.73 | AAI-based (high-confidence) | 0 |
| S12-final-viral-combined_k141_334532\|\|full | 5146 | No | NA | 5 | 0 | 3 | Not-determined | Genome-fragment | NA | NA | 0 |
| S12-final-viral-combined_k141_132598\|\|full | 2122 | No | NA | 2 | 0 | 1 | Not-determined | Genome-fragment | NA | NA | 0 |
| S1-final-viral-combined_k141_166385\|\|full | 3106 | No | NA | 6 | 0 | 4 | Low-quality | Genome-fragment | 4.52 | AAI-based (medium-confidence) | 0 |
| S12-final-viral-combined_k141_365771\|\|full | 1093 | No | NA | 3 | 0 | 2 | Not-determined | Genome-fragment | NA | NA | 0 |
| S3-final-viral-combined_k141_83200\|\|full | 3106 | No | NA | 3 | 0 | 0 | Low-quality | Genome-fragment | 7.34 | AAI-based (medium-confidence) | 0 |
| S12-final-viral-combined_k141_250105\|\|full | 2679 | No | NA | 3 | 0 | 3 | Not-determined | Genome-fragment | NA | NA | 0 |
| S3-final-viral-combined_k141_67163\|\|full | 3106 | No | NA | 3 | 0 | 2 | Low-quality | Genome-fragment | 3.27 | AAI-based (medium-confidence) | 0 |
| S12-final-viral-combined_k141_351016\|\|full | 3426 | No | NA | 5 | 0 | 3 | Not-determined | Genome-fragment | NA | NA | 0 |
| S12-final-viral-combined_k141_167034\|\|full | 1573 | No | NA | 3 | 0 | 2 | Not-determined | Genome-fragment | NA | NA | 0 |
| S12-final-viral-combined_k141_183068\|\|full | 3106 | No | NA | 2 | 0 | 2 | Low-quality | Genome-fragment | 1.58 | AAI-based (medium-confidence) | 0 |
| S12-final-viral-combined_k141_99626\|\|full | 1523 | No | NA | 2 | 0 | 0 | Not-determined | Genome-fragment | NA | NA | 0 |
| S12_k141_238054 | 3106 | No | NA | 3 | 0 | 0 | Low-quality | Genome-fragment | 4.39 | AAI-based (medium-confidence) | 0 |
| S2_k141_197641 | 3107 | No | NA | 2 | 0 | 2 | Low-quality | Genome-fragment | 0.6 | AAI-based (medium-confidence) | 0 |
| S2_k141_7957 | 3107 | No | NA | 3 | 0 | 1 | Low-quality | Genome-fragment | 3.31 | AAI-based (medium-confidence) | 0 |
| S3-final-viral-combined_k141_136862\|\|lt2gene | 3108 | No | NA | 1 | 0 | 0 | Low-quality | Genome-fragment | 2.03 | AAI-based (high-confidence) | 0 |
| S12_k141_61535 | 3108 | No | NA | 4 | 0 | 0 | Low-quality | Genome-fragment | 6.49 | AAI-based (medium-confidence) | 0 |
| S9-final-viral-combined_k141_99246\|\|full | 3109 | No | NA | 6 | 0 | 0 | Low-quality | Genome-fragment | 6.99 | AAI-based (high-confidence) | 0 |
| S11-final-viral-combined_k141_111652\|\|full | 3109 | Yes | 993 | 5 | 1 | 1 | Low-quality | Genome-fragment | 0.46 | HMM-based (lower-bound) | 68.06 |
| S12-final-viral-combined_k141_149427\|\|full | 2208 | No | NA | 3 | 0 | 2 | Not-determined | Genome-fragment | NA | NA | 0 |
| S12-final-viral-combined_k141_32644\|\|full | 13102 | No | NA | 13 | 0 | 7 | Not-determined | Genome-fragment | NA | NA | 0 |
| S12-final-viral-combined_k141_351122\|\|full | 1803 | No | NA | 2 | 0 | 0 | Not-determined | Genome-fragment | NA | NA | 0 |
| S12-final-viral-combined_k141_99697\|\|full | 2306 | No | NA | 4 | 0 | 2 | Not-determined | Genome-fragment | NA | NA | 0 |
| S12-final-viral-combined_k141_183957\|\|full | 1639 | No | NA | 3 | 0 | 0 | Not-determined | Genome-fragment | NA | NA | 0 |
| S4-final-viral-combined_k141_92371\|\|full | 3109 | No | NA | 5 | 0 | 3 | Low-quality | Genome-fragment | 3.57 | AAI-based (high-confidence) | 0 |
| S12-final-viral-combined_k141_400125\|\|full | 4391 | No | NA | 2 | 0 | 2 | Not-determined | Genome-fragment | NA | NA | 0 |
| S12-final-viral-combined_k141_167146\|\|full | 1799 | No | NA | 2 | 0 | 0 | Not-determined | Genome-fragment | NA | NA | 0 |
| S17-final-viral-combined_k141_226757\|\|full | 3110 | No | NA | 6 | 1 | 0 | Low-quality | Genome-fragment | 1.11 | HMM-based (lower-bound) | 0 |
| S12-final-viral-combined_k141_217509\|\|full | 6472 | No | NA | 7 | 0 | 6 | Not-determined | Genome-fragment | NA | NA | 0 |
| S12-final-viral-combined_k141_15837\|\|full | 2450 | No | NA | 5 | 0 | 0 | Not-determined | Genome-fragment | NA | NA | 0 |
| S12-final-viral-combined_k141_132751\|\|full | 4615 | No | NA | 3 | 0 | 3 | Not-determined | Genome-fragment | NA | NA | 0 |
| S12-final-viral-combined_k141_99704\|\|full | 32142 | No | NA | 40 | 24 | 0 | Medium-quality | Genome-fragment | 67.85 | AAI-based (high-confidence) | 0 |
| S9-final-viral-combined_k141_35129\|\|full | 3111 | No | NA | 3 | 0 | 3 | Low-quality | Genome-fragment | 2.23 | AAI-based (medium-confidence) | 0 |
| S12-final-viral-combined_k141_15865\|\|full | 2056 | No | NA | 3 | 0 | 2 | Not-determined | Genome-fragment | NA | NA | 0 |
| S12-final-viral-combined_k141_32707\|\|full | 1283 | No | NA | 2 | 0 | 1 | Not-determined | Genome-fragment | NA | NA | 0 |
| S1-final-viral-combined_k141_284487\|\|full | 3111 | No | NA | 3 | 0 | 3 | Low-quality | Genome-fragment | 3.57 | AAI-based (medium-confidence) | 0 |
| S12-final-viral-combined_k141_116027\|\|full | 10048 | No | NA | 8 | 0 | 6 | Not-determined | Genome-fragment | NA | NA | 0 |
| S12-final-viral-combined_k141_66402\|\|full | 5537 | No | NA | 8 | 0 | 0 | Not-determined | Genome-fragment | NA | NA | 0 |
| S12-final-viral-combined_k141_66403\|\|full | 1251 | No | NA | 2 | 0 | 0 | Not-determined | Genome-fragment | NA | NA | 0 |
| S11-final-viral-combined_k141_287690\|\|full | 3112 | No | NA | 4 | 1 | 0 | Low-quality | Genome-fragment | 5.68 | HMM-based (lower-bound) | 0 |
| S12-final-viral-combined_k141_334700\|\|full | 1093 | No | NA | 2 | 0 | 1 | Not-determined | Genome-fragment | NA | NA | 0 |
| S12-final-viral-combined_k141_116050\|\|full | 2330 | No | NA | 4 | 0 | 2 | Not-determined | Genome-fragment | NA | NA | 0 |
| S17-final-viral-combined_k141_229010\|\|full | 3112 | No | NA | 6 | 1 | 0 | Low-quality | Genome-fragment | 5.87 | AAI-based (high-confidence) | 0 |
| S12-final-viral-combined_k141_32764\|\|full | 2432 | No | NA | 4 | 0 | 1 | Not-determined | Genome-fragment | NA | NA | 0 |
| S12-final-viral-combined_k141_384315\|\|full | 3615 | No | NA | 5 | 0 | 2 | Not-determined | Genome-fragment | NA | NA | 0 |
| S12-final-viral-combined_k141_366045\|\|full | 1292 | No | NA | 3 | 0 | 0 | Not-determined | Genome-fragment | NA | NA | 0 |
| S11-final-viral-combined_k141_125908\|\|full | 3113 | No | NA | 5 | 4 | 0 | Low-quality | Genome-fragment | 6.84 | AAI-based (high-confidence) | 0 |
| S11_k141_191820 | 3113 | No | NA | 5 | 0 | 2 | Low-quality | Genome-fragment | 1.06 | AAI-based (medium-confidence) | 0 |
| S8_k141_150166 | 3113 | No | NA | 7 | 0 | 0 | Low-quality | Genome-fragment | 3.59 | AAI-based (high-confidence) | 0 |
| S12-final-viral-combined_k141_334734\|\|full | 2236 | No | NA | 3 | 0 | 0 | Not-determined | Genome-fragment | NA | NA | 0 |
| S12-final-viral-combined_k141_149677\|\|full | 1459 | No | NA | 3 | 0 | 2 | Not-determined | Genome-fragment | NA | NA | 0 |
| S9_k141_50689 | 3114 | No | NA | 2 | 0 | 2 | Low-quality | Genome-fragment | 3.88 | AAI-based (medium-confidence) | 0 |
| S7-final-viral-combined_k141_28640\|\|full | 3114 | No | NA | 4 | 3 | 0 | Low-quality | Genome-fragment | 6.67 | AAI-based (high-confidence) | 0 |
| S17-k141_65196 | 3114 | No | NA | 4 | 0 | 0 | Low-quality | Genome-fragment | 3.56 | AAI-based (medium-confidence) | 0 |
| S1-final-viral-combined_k141_114773\|\|full | 3115 | No | NA | 4 | 0 | 1 | Low-quality | Genome-fragment | 2.24 | AAI-based (medium-confidence) | 0 |
| S12-final-viral-combined_k141_334757\|\|full | 4789 | No | NA | 6 | 0 | 4 | Not-determined | Genome-fragment | NA | NA | 0 |
| S12-final-viral-combined_k141_99868\|\|full | 3260 | No | NA | 3 | 0 | 2 | Not-determined | Genome-fragment | NA | NA | 0 |
| S12-final-viral-combined_k141_384391\|\|full | 3240 | No | NA | 3 | 0 | 1 | Not-determined | Genome-fragment | NA | NA | 0 |
| S12-final-viral-combined_k141_149714\|\|full | 2610 | No | NA | 4 | 0 | 1 | Not-determined | Genome-fragment | NA | NA | 0 |
| S12_k141_155308 | 3115 | No | NA | 8 | 0 | 0 | Low-quality | Genome-fragment | 3.14 | AAI-based (high-confidence) | 0 |
| S2-final-viral-combined_k141_69715\|\|full | 3115 | No | NA | 4 | 0 | 2 | Low-quality | Genome-fragment | 2.38 | AAI-based (medium-confidence) | 0 |
| S8-final-viral-combined_k141_20674\|\|full | 3115 | No | NA | 4 | 1 | 0 | Low-quality | Genome-fragment | 5.38 | AAI-based (high-confidence) | 0 |
| S8_k141_6282 | 3115 | No | NA | 1 | 0 | 1 | Low-quality | Genome-fragment | 1.68 | AAI-based (medium-confidence) | 0 |
| S1-final-viral-combined_k141_151131\|\|full | 3116 | No | NA | 3 | 0 | 0 | Low-quality | Genome-fragment | 2.4 | AAI-based (medium-confidence) | 0 |
| S7_k141_114524 | 3116 | No | NA | 4 | 0 | 0 | Low-quality | Genome-fragment | 5.22 | AAI-based (medium-confidence) | 0 |
| S6_k141_102968 | 3116 | No | NA | 4 | 0 | 3 | Low-quality | Genome-fragment | 2.25 | AAI-based (medium-confidence) | 0 |
| S9-final-viral-combined_k141_70947\|\|full | 3117 | No | NA | 4 | 0 | 2 | Low-quality | Genome-fragment | 3.89 | AAI-based (high-confidence) | 0 |
| S12-final-viral-combined_k141_86007\|\|full | 3117 | No | NA | 4 | 1 | 0 | Low-quality | Genome-fragment | 2.31 | AAI-based (high-confidence) | 0 |
| S12-final-viral-combined_k141_366171\|\|full | 3561 | No | NA | 5 | 0 | 0 | Not-determined | Genome-fragment | NA | NA | 0 |
| S12-final-viral-combined_k141_132907\|\|full | 41330 | No | NA | 61 | 30 | 0 | High-quality | High-quality | 90.9 | AAI-based (high-confidence) | 0 |
| S12-final-viral-combined_k141_366173\|\|full | 2954 | No | NA | 5 | 0 | 2 | Not-determined | Genome-fragment | NA | NA | 0 |
| S12-final-viral-combined_k141_351312\|\|full | 1575 | No | NA | 2 | 0 | 0 | Not-determined | Genome-fragment | NA | NA | 0 |
| S7-final-viral-combined_k141_182599\|\|full | 3118 | No | NA | 3 | 0 | 0 | Low-quality | Genome-fragment | 5.56 | AAI-based (medium-confidence) | 0 |
| S17-final-viral-combined_k141_43557\|\|full | 3118 | No | NA | 6 | 0 | 0 | Low-quality | Genome-fragment | 5.66 | AAI-based (high-confidence) | 0 |
| S12-final-viral-combined_k141_149756\|\|full | 1429 | No | NA | 2 | 0 | 0 | Not-determined | Genome-fragment | NA | NA | 0 |
| S2-final-viral-combined_k141_57090\|\|full | 3118 | No | NA | 7 | 0 | 0 | Low-quality | Genome-fragment | 5.88 | AAI-based (medium-confidence) | 0 |
| S12-final-viral-combined_k141_66622\|\|full | 2525 | No | NA | 4 | 0 | 0 | Not-determined | Genome-fragment | NA | NA | 0 |
| S12-final-viral-combined_k141_384460\|\|full | 1027 | No | NA | 2 | 0 | 1 | Not-determined | Genome-fragment | NA | NA | 0 |
| S9_k141_118003 | 3119 | No | NA | 5 | 1 | 0 | Low-quality | Genome-fragment | 5.8 | AAI-based (high-confidence) | 0 |
| S12-final-viral-combined_k141_132974\|\|full | 2935 | No | NA | 2 | 0 | 2 | Not-determined | Genome-fragment | NA | NA | 0 |
| S1-final-viral-combined_k141_363234\|\|full | 3119 | No | NA | 6 | 4 | 0 | Low-quality | Genome-fragment | 5.53 | HMM-based (lower-bound) | 0 |
| S2-final-viral-combined_k141_131633\|\|full | 3119 | No | NA | 4 | 0 | 3 | Low-quality | Genome-fragment | 0.92 | AAI-based (high-confidence) | 0 |
| S9-final-viral-combined_k141_113210\|\|full | 3120 | No | NA | 4 | 0 | 2 | Low-quality | Genome-fragment | 0.92 | AAI-based (medium-confidence) | 0 |
| S12-final-viral-combined_k141_266567\|\|full | 1403 | No | NA | 2 | 0 | 1 | Not-determined | Genome-fragment | NA | NA | 0 |
| S12-final-viral-combined_k141_384520\|\|full | 4218 | No | NA | 5 | 0 | 3 | Not-determined | Genome-fragment | NA | NA | 0 |
| S11-final-viral-combined_k141_11761\|\|full | 3120 | No | NA | 6 | 6 | 0 | Low-quality | Genome-fragment | 6.68 | AAI-based (high-confidence) | 0 |
| S1-final-viral-combined_k141_276808\|\|full | 3120 | No | NA | 3 | 0 | 1 | Low-quality | Genome-fragment | 7.14 | AAI-based (high-confidence) | 0 |
| S5_k141_118665 | 3120 | No | NA | 3 | 0 | 0 | Low-quality | Genome-fragment | 3.32 | AAI-based (medium-confidence) | 0 |
| S2-final-viral-combined_k141_32050\|\|full | 3120 | No | NA | 3 | 1 | 0 | Low-quality | Genome-fragment | 3.7 | HMM-based (lower-bound) | 0 |
| S2_k141_180204 | 3120 | No | NA | 3 | 0 | 1 | Low-quality | Genome-fragment | 4.89 | AAI-based (medium-confidence) | 0 |
| S12-final-viral-combined_k141_250551\|\|full | 41741 | No | NA | 45 | 5 | 2 | High-quality | High-quality | 97.46 | AAI-based (high-confidence) | 0 |
| S8_k141_255148 | 3120 | No | NA | 3 | 0 | 2 | Low-quality | Genome-fragment | 1.55 | AAI-based (medium-confidence) | 0 |
| S12-final-viral-combined_k141_66749\|\|full | 1223 | No | NA | 2 | 0 | 0 | Not-determined | Genome-fragment | NA | NA | 0 |
| S1-final-viral-combined_k141_383983\|\|full | 3121 | No | NA | 2 | 1 | 0 | Low-quality | Genome-fragment | 1.69 | HMM-based (lower-bound) | 0 |
| S12-final-viral-combined_k141_100046\|\|full | 1215 | No | NA | 3 | 0 | 1 | Not-determined | Genome-fragment | NA | NA | 0 |
| S1_k141_330699 | 3121 | No | NA | 4 | 0 | 1 | Low-quality | Genome-fragment | 5.16 | AAI-based (medium-confidence) | 0 |
| S17-final-viral-combined_k141_155963\|\|full | 3121 | No | NA | 5 | 2 | 0 | Low-quality | Genome-fragment | 7.4 | AAI-based (high-confidence) | 0 |
| S12-final-viral-combined_k141_351414\|\|full | 2120 | No | NA | 4 | 0 | 0 | Not-determined | Genome-fragment | NA | NA | 0 |
| S12-final-viral-combined_k141_16344\|\|full | 4442 | No | NA | 5 | 0 | 3 | Not-determined | Genome-fragment | NA | NA | 0 |
| S12-final-viral-combined_k141_316726\|\|full | 3121 | No | NA | 7 | 0 | 0 | Low-quality | Genome-fragment | 3.48 | AAI-based (medium-confidence) | 0 |
| S11-final-viral-combined_k141_102994\|\|full | 3122 | Yes | 2026 | 6 | 1 | 1 | Low-quality | Genome-fragment | 4.63 | AAI-based (medium-confidence) | 35.11 |
| S12-final-viral-combined_k141_266672\|\|full | 1410 | No | NA | 2 | 0 | 1 | Not-determined | Genome-fragment | NA | NA | 0 |
| S12-final-viral-combined_k141_66812\|\|full | 4356 | No | NA | 5 | 0 | 0 | Not-determined | Genome-fragment | NA | NA | 0 |
| S3-final-viral-combined_k141_131056\|\|full | 3122 | No | NA | 3 | 0 | 0 | Low-quality | Genome-fragment | 2.19 | AAI-based (high-confidence) | 0 |
| S12-final-viral-combined_k141_133180\|\|full | 1525 | No | NA | 3 | 0 | 1 | Not-determined | Genome-fragment | NA | NA | 0 |
| S12-final-viral-combined_k141_133187\|\|full | 3429 | No | NA | 6 | 0 | 4 | Not-determined | Genome-fragment | NA | NA | 0 |
| S12-final-viral-combined_k141_266681\|\|full | 1268 | No | NA | 2 | 0 | 0 | Not-determined | Genome-fragment | NA | NA | 0 |
| S12-final-viral-combined_k141_133215\|\|full | 1861 | No | NA | 2 | 0 | 2 | Not-determined | Genome-fragment | NA | NA | 0 |
| S12-final-viral-combined_k141_400553\|\|full | 1743 | No | NA | 4 | 0 | 0 | Not-determined | Genome-fragment | NA | NA | 0 |
| S12-final-viral-combined_k141_100130\|\|full | 1013 | No | NA | 2 | 0 | 1 | Not-determined | Genome-fragment | NA | NA | 0 |
| S12-final-viral-combined_k141_250647\|\|full | 1533 | No | NA | 2 | 0 | 0 | Not-determined | Genome-fragment | NA | NA | 0 |
| S5-final-viral-combined_k141_14477\|\|full | 3122 | No | NA | 3 | 2 | 0 | Low-quality | Genome-fragment | 5.24 | AAI-based (high-confidence) | 0 |
| S9-final-viral-combined_k141_55180\|\|full | 3123 | No | NA | 6 | 0 | 0 | Low-quality | Genome-fragment | 5.34 | AAI-based (medium-confidence) | 0 |
| S17-final-viral-combined_k141_89747\|\|full | 3123 | No | NA | 7 | 7 | 0 | Low-quality | Genome-fragment | 9.14 | AAI-based (high-confidence) | 0 |
| S4_k141_112261 | 3123 | No | NA | 3 | 0 | 0 | Low-quality | Genome-fragment | 5.22 | AAI-based (high-confidence) | 0 |
| S8-final-viral-combined_k141_144162\|\|full | 3123 | No | NA | 5 | 0 | 2 | Low-quality | Genome-fragment | 2.31 | AAI-based (high-confidence) | 0 |
| S12-final-viral-combined_k141_100195\|\|full | 3291 | No | NA | 4 | 0 | 1 | Not-determined | Genome-fragment | NA | NA | 0 |
| S12-final-viral-combined_k141_301284\|\|full | 3010 | No | NA | 4 | 0 | 0 | Not-determined | Genome-fragment | NA | NA | 0 |
| S12-final-viral-combined_k141_400590\|\|full | 2681 | No | NA | 3 | 0 | 3 | Not-determined | Genome-fragment | NA | NA | 0 |
| S12-final-viral-combined_k141_33129\|\|full | 1211 | No | NA | 1 | 0 | 0 | Not-determined | Genome-fragment | NA | NA | 0 |
| S12-final-viral-combined_k141_301290\|\|full | 3028 | No | NA | 4 | 0 | 0 | Not-determined | Genome-fragment | NA | NA | 0 |
| S17-final-viral-combined_k141_80617\|\|full | 3124 | Yes | 998 | 4 | 1 | 2 | Low-quality | Genome-fragment | 1.89 | AAI-based (medium-confidence) | 68.05 |
| S17-final-viral-combined_k141_92220\|\|full | 3124 | No | NA | 5 | 2 | 0 | Low-quality | Genome-fragment | 9.09 | AAI-based (high-confidence) | 0 |
| S12-final-viral-combined_k141_116465\|\|full | 3242 | No | NA | 6 | 0 | 0 | Not-determined | Genome-fragment | NA | NA | 0 |
| S9-final-viral-combined_k141_62622\|\|full | 3125 | No | NA | 3 | 0 | 1 | Low-quality | Genome-fragment | 0.94 | AAI-based (medium-confidence) | 0 |
| S12-final-viral-combined_k141_400622\|\|full | 1114 | No | NA | 3 | 0 | 0 | Not-determined | Genome-fragment | NA | NA | 0 |
| S12-final-viral-combined_k141_33193\|\|full | 1782 | No | NA | 3 | 0 | 2 | Not-determined | Genome-fragment | NA | NA | 0 |
| S12-final-viral-combined_k141_250658\|\|full | 3297 | No | NA | 2 | 0 | 1 | Not-determined | Genome-fragment | NA | NA | 0 |
| S12-final-viral-combined_k141_250671\|\|full | 2250 | No | NA | 5 | 0 | 1 | Not-determined | Genome-fragment | NA | NA | 0 |
| S11-final-viral-combined_k141_234816\|\|lt2gene | 3125 | No | NA | 2 | 2 | 0 | Low-quality | Genome-fragment | 7.22 | AAI-based (high-confidence) | 0 |
| S11_k141_69232 | 3125 | No | NA | 8 | 2 | 0 | Low-quality | Genome-fragment | 7.21 | AAI-based (high-confidence) | 0 |
| S12-final-viral-combined_k141_250741\|\|full | 11977 | No | NA | 10 | 0 | 6 | Not-determined | Genome-fragment | NA | NA | 0 |
| S11_k141_154113 | 3125 | No | NA | 1 | 0 | 1 | Low-quality | Genome-fragment | 1.68 | AAI-based (medium-confidence) | 0 |
| S1_k141_368104 | 3125 | No | NA | 3 | 0 | 2 | Low-quality | Genome-fragment | 0.43 | AAI-based (medium-confidence) | 0 |
| S12_k141_228030 | 3125 | No | NA | 3 | 0 | 0 | Low-quality | Genome-fragment | 2.85 | AAI-based (medium-confidence) | 0 |
| S2-final-viral-combined_k141_88994\|\|full | 3125 | No | NA | 6 | 4 | 0 | Low-quality | Genome-fragment | 5.4 | HMM-based (lower-bound) | 0 |
| S12-final-viral-combined_k141_384694\|\|full | 2554 | No | NA | 4 | 0 | 0 | Not-determined | Genome-fragment | NA | NA | 0 |
| S2_k141_193076 | 3125 | No | NA | 3 | 0 | 0 | Low-quality | Genome-fragment | 0.94 | AAI-based (medium-confidence) | 0 |
| S12-final-viral-combined_k141_16623\|\|full | 2519 | No | NA | 5 | 0 | 0 | Not-determined | Genome-fragment | NA | NA | 0 |
| S8_k141_266286 | 3125 | Yes | 1415 | 4 | 1 | 1 | Low-quality | Genome-fragment | 0.95 | HMM-based (lower-bound) | 54.72 |
| S17-final-viral-combined_k141_230004\|\|full | 3126 | No | NA | 3 | 2 | 0 | Low-quality | Genome-fragment | 6.35 | HMM-based (lower-bound) | 0 |
| S12-final-viral-combined_k141_133461\|\|full | 2997 | No | NA | 4 | 0 | 1 | Not-determined | Genome-fragment | NA | NA | 0 |
| S12-final-viral-combined_k141_366743\|\|full | 2822 | No | NA | 5 | 0 | 0 | Not-determined | Genome-fragment | NA | NA | 0 |
| S12-final-viral-combined_k141_250828\|\|full | 2220 | No | NA | 4 | 0 | 2 | Not-determined | Genome-fragment | NA | NA | 0 |
| S12-final-viral-combined_k141_384780\|\|full | 1206 | No | NA | 2 | 0 | 0 | Not-determined | Genome-fragment | NA | NA | 0 |
| S1-final-viral-combined_k141_378628\|\|full | 3127 | No | NA | 3 | 2 | 0 | Low-quality | Genome-fragment | 1.94 | HMM-based (lower-bound) | 0 |
| S12-final-viral-combined_k141_384796\|\|full | 1543 | No | NA | 3 | 0 | 0 | Not-determined | Genome-fragment | NA | NA | 0 |
| S9-final-viral-combined_k141_11298\|\|full | 3128 | No | NA | 7 | 3 | 0 | Low-quality | Genome-fragment | 8.4 | AAI-based (high-confidence) | 0 |
| S12-final-viral-combined_k141_366762\|\|full | 3199 | No | NA | 3 | 0 | 2 | Not-determined | Genome-fragment | NA | NA | 0 |
| S12-final-viral-combined_k141_150361\|\|full | 2939 | No | NA | 3 | 0 | 2 | Not-determined | Genome-fragment | NA | NA | 0 |
| S12-final-viral-combined_k141_150392\|\|full | 1095 | No | NA | 2 | 0 | 2 | Not-determined | Genome-fragment | NA | NA | 0 |
| S17-final-viral-combined_k141_43305\|\|full | 3129 | No | NA | 3 | 0 | 2 | Low-quality | Genome-fragment | 0.99 | AAI-based (medium-confidence) | 0 |
| S12-final-viral-combined_k141_366812\|\|full | 3216 | No | NA | 3 | 0 | 2 | Not-determined | Genome-fragment | NA | NA | 0 |
| S12-final-viral-combined_k141_250924\|\|full | 1306 | No | NA | 3 | 0 | 0 | Not-determined | Genome-fragment | NA | NA | 0 |
| S12-final-viral-combined_k141_400863\|\|full | 1779 | No | NA | 4 | 0 | 2 | Not-determined | Genome-fragment | NA | NA | 0 |
| S4_k141_81231 | 3129 | No | NA | 3 | 3 | 0 | Low-quality | Genome-fragment | 8.25 | AAI-based (high-confidence) | 0 |
| S12-final-viral-combined_k141_116827\|\|full | 5519 | No | NA | 4 | 0 | 4 | Not-determined | Genome-fragment | NA | NA | 0 |
| S7-final-viral-combined_k141_208002\|\|full | 3130 | No | NA | 3 | 0 | 2 | Low-quality | Genome-fragment | 3.97 | AAI-based (medium-confidence) | 0 |
| S12-final-viral-combined_k141_170058\|\|full | 3131 | No | NA | 4 | 2 | 0 | Low-quality | Genome-fragment | 7.54 | AAI-based (medium-confidence) | 0 |
| S12-final-viral-combined_k141_267237\|\|full | 1075 | No | NA | 3 | 0 | 1 | Not-determined | Genome-fragment | NA | NA | 0 |
| S12-final-viral-combined_k141_251007\|\|full | 3296 | No | NA | 3 | 0 | 2 | Not-determined | Genome-fragment | NA | NA | 0 |
| S12-final-viral-combined_k141_296771\|\|full | 3131 | No | NA | 3 | 0 | 1 | Low-quality | Genome-fragment | 5.19 | AAI-based (medium-confidence) | 0 |
| S4-final-viral-combined_k141_121732\|\|full | 3131 | No | NA | 3 | 0 | 2 | Low-quality | Genome-fragment | 3.76 | AAI-based (medium-confidence) | 0 |
| S8-final-viral-combined_k141_70200\|\|full | 3131 | No | NA | 7 | 3 | 0 | Low-quality | Genome-fragment | 8.87 | AAI-based (high-confidence) | 0 |
| S12-final-viral-combined_k141_150507\|\|full | 1221 | No | NA | 2 | 0 | 1 | Not-determined | Genome-fragment | NA | NA | 0 |
| S8_k141_161256 | 3131 | Yes | 2571 | 4 | 1 | 1 | Low-quality | Genome-fragment | 3.89 | HMM-based (lower-bound) | 17.89 |
| S12-final-viral-combined_k141_367020\|\|full | 1177 | No | NA | 4 | 0 | 1 | Not-determined | Genome-fragment | NA | NA | 0 |
| S12-final-viral-combined_k141_116925\|\|full | 3206 | No | NA | 5 | 0 | 2 | Not-determined | Genome-fragment | NA | NA | 0 |
| S12-final-viral-combined_k141_400981\|\|full | 2630 | No | NA | 4 | 0 | 0 | Not-determined | Genome-fragment | NA | NA | 0 |
| S12-final-viral-combined_k141_400988\|\|full | 1050 | No | NA | 2 | 0 | 1 | Not-determined | Genome-fragment | NA | NA | 0 |
| S12-final-viral-combined_k141_267339\|\|full | 13918 | No | NA | 10 | 0 | 3 | Not-determined | Genome-fragment | NA | NA | 0 |
| S9-final-viral-combined_k141_29356\|\|full | 3132 | No | NA | 5 | 0 | 0 | Low-quality | Genome-fragment | 4.86 | AAI-based (high-confidence) | 0 |
| S12-final-viral-combined_k141_117007\|\|full | 1575 | No | NA | 3 | 0 | 1 | Not-determined | Genome-fragment | NA | NA | 0 |
| S12-final-viral-combined_k141_150632\|\|full | 2687 | No | NA | 4 | 0 | 1 | Not-determined | Genome-fragment | NA | NA | 0 |
| S12-final-viral-combined_k141_267377\|\|full | 1951 | No | NA | 2 | 0 | 0 | Not-determined | Genome-fragment | NA | NA | 0 |
| S12-final-viral-combined_k141_117056\|\|full | 5039 | No | NA | 4 | 0 | 2 | Not-determined | Genome-fragment | NA | NA | 0 |
| S11-final-viral-combined_k141_304529\|\|full | 3133 | No | NA | 4 | 3 | 0 | Low-quality | Genome-fragment | 4.34 | HMM-based (lower-bound) | 0 |
| S7-final-viral-combined_k141_33352\|\|full | 3133 | No | NA | 4 | 0 | 0 | Low-quality | Genome-fragment | 2.33 | AAI-based (high-confidence) | 0 |
| S12-final-viral-combined_k141_117113\|\|full | 1136 | No | NA | 2 | 0 | 1 | Not-determined | Genome-fragment | NA | NA | 0 |
| S12-final-viral-combined_k141_267423\|\|full | 1717 | No | NA | 2 | 0 | 0 | Not-determined | Genome-fragment | NA | NA | 0 |
| S17-final-viral-combined_k141_192416\|\|full | 3133 | No | NA | 5 | 2 | 0 | Low-quality | Genome-fragment | 3.86 | AAI-based (high-confidence) | 0 |
| S12-final-viral-combined_k141_133759\|\|full | 2689 | No | NA | 4 | 0 | 3 | Not-determined | Genome-fragment | NA | NA | 0 |
| S12_k141_182334 | 3133 | No | NA | 2 | 0 | 2 | Low-quality | Genome-fragment | 7.45 | AAI-based (high-confidence) | 0 |
| S2_k141_237319 | 3133 | No | NA | 7 | 2 | 0 | Low-quality | Genome-fragment | 3.26 | AAI-based (high-confidence) | 0 |
| S12-final-viral-combined_k141_133789\|\|full | 1051 | No | NA | 3 | 0 | 2 | Not-determined | Genome-fragment | NA | NA | 0 |
| S12-final-viral-combined_k141_401196\|\|full | 5239 | No | NA | 11 | 0 | 1 | Not-determined | Genome-fragment | NA | NA | 0 |
| S2_k141_245586 | 3133 | Yes | 2351 | 7 | 2 | 1 | Low-quality | Genome-fragment | 6.34 | AAI-based (high-confidence) | 24.96 |
| S12-final-viral-combined_k141_401222\|\|full | 1395 | No | NA | 2 | 0 | 0 | Not-determined | Genome-fragment | NA | NA | 0 |
| S7-final-viral-combined_k141_42140\|\|full | 3134 | No | NA | 7 | 0 | 0 | Low-quality | Genome-fragment | 5.55 | AAI-based (medium-confidence) | 0 |
| S12-final-viral-combined_k141_401278\|\|full | 6251 | No | NA | 3 | 0 | 2 | Not-determined | Genome-fragment | NA | NA | 0 |
| S12-final-viral-combined_k141_267531\|\|full | 1499 | No | NA | 4 | 0 | 1 | Not-determined | Genome-fragment | NA | NA | 0 |
| S7-final-viral-combined_k141_201858\|\|full | 3134 | No | NA | 5 | 3 | 0 | Low-quality | Genome-fragment | 8.05 | AAI-based (medium-confidence) | 0 |
| S17-final-viral-combined_k141_201536\|\|full | 3134 | No | NA | 8 | 2 | 0 | Low-quality | Genome-fragment | 8.43 | AAI-based (high-confidence) | 0 |
| S12-final-viral-combined_k141_401346\|\|full | 3482 | No | NA | 5 | 0 | 1 | Not-determined | Genome-fragment | NA | NA | 0 |
| S12-final-viral-combined_k141_401348\|\|full | 2958 | No | NA | 3 | 0 | 0 | Not-determined | Genome-fragment | NA | NA | 0 |
| S17-final-viral-combined_k141_192348\|\|full | 3134 | No | NA | 5 | 2 | 0 | Low-quality | Genome-fragment | 3.69 | AAI-based (high-confidence) | 0 |
| S12-final-viral-combined_k141_367585\|\|full | 1103 | No | NA | 3 | 0 | 0 | Not-determined | Genome-fragment | NA | NA | 0 |
| S12-final-viral-combined_k141_367590\|\|full | 1727 | No | NA | 3 | 0 | 0 | Not-determined | Genome-fragment | NA | NA | 0 |
| S6-final-viral-combined_k141_25729\|\|full | 3134 | No | NA | 4 | 0 | 2 | Low-quality | Genome-fragment | 2.43 | AAI-based (high-confidence) | 0 |
| S12-final-viral-combined_k141_367632\|\|full | 1294 | No | NA | 3 | 0 | 0 | Not-determined | Genome-fragment | NA | NA | 0 |
| S8-final-viral-combined_k141_52743\|\|lt2gene | 3134 | No | NA | 1 | 1 | 0 | Low-quality | Genome-fragment | 3.05 | HMM-based (lower-bound) | 0 |
| S12-final-viral-combined_k141_367827\|\|full | 5837 | No | NA | 5 | 0 | 4 | Not-determined | Genome-fragment | NA | NA | 0 |
| S12-final-viral-combined_k141_267809\|\|full | 1652 | No | NA | 3 | 0 | 1 | Not-determined | Genome-fragment | NA | NA | 0 |
| S12-final-viral-combined_k141_368190\|\|full | 112724 | Yes | 52158 | 118 | 13 | 26 | Complete | High-quality | 100 | Provirus (medium-confidence) | 53.73 |
| S12-final-viral-combined_k141_368216\|\|full | 1502 | No | NA | 2 | 0 | 0 | Not-determined | Genome-fragment | NA | NA | 0 |
| S1_k141_172255 | 3135 | No | NA | 2 | 1 | 0 | Low-quality | Genome-fragment | 6.73 | AAI-based (high-confidence) | 0 |
| S12-final-viral-combined_k141_234454\|\|0_partial | 25769 | No | NA | 20 | 0 | 9 | Not-determined | Genome-fragment | NA | NA | 0 |
| S12-final-viral-combined_k141_334917\|\|0_partial | 16813 | No | NA | 14 | 0 | 4 | Not-determined | Genome-fragment | NA | NA | 0 |
| S12_k141_53474 | 3135 | No | NA | 5 | 1 | 0 | Low-quality | Genome-fragment | 2.52 | HMM-based (lower-bound) | 0 |
| S12-final-viral-combined_k141_117447\|\|0_partial | 4229 | No | NA | 5 | 1 | 0 | Medium-quality | Genome-fragment | 51 | AAI-based (medium-confidence) | 0 |
| S2-final-viral-combined_k141_77766\|\|full | 3135 | No | NA | 4 | 0 | 1 | Low-quality | Genome-fragment | 3.92 | AAI-based (high-confidence) | 0 |
| S5-final-viral-combined_k141_173343\|\|full | 3136 | No | NA | 6 | 1 | 0 | Low-quality | Genome-fragment | 1.65 | HMM-based (lower-bound) | 0 |
| S1-final-viral-combined_k141_148693\|\|full | 3137 | Yes | 2306 | 5 | 1 | 1 | Low-quality | Genome-fragment | 4.25 | AAI-based (high-confidence) | 26.49 |
| S12-final-viral-combined_k141_168259\|\|0_partial | 40030 | Yes | 30653 | 51 | 6 | 8 | High-quality | High-quality | 91.95 | AAI-based (high-confidence) | 23.42 |
| S12-final-viral-combined_k141_84788\|\|0_partial | 22085 | No | NA | 24 | 0 | 6 | Not-determined | Genome-fragment | NA | NA | 0 |
| S5-final-viral-combined_k141_188342\|\|full | 3137 | No | NA | 5 | 0 | 0 | Low-quality | Genome-fragment | 2.52 | AAI-based (high-confidence) | 0 |
| S1_k141_528 | 3138 | No | NA | 5 | 1 | 0 | Low-quality | Genome-fragment | 4.75 | HMM-based (lower-bound) | 0 |
| S12-final-viral-combined_k141_185931\|\|0_partial | 13114 | No | NA | 17 | 0 | 5 | Not-determined | Genome-fragment | NA | NA | 0 |
| S5-final-viral-combined_k141_124006\|\|full | 3138 | No | NA | 9 | 3 | 0 | Low-quality | Genome-fragment | 3.52 | AAI-based (high-confidence) | 0 |
| S7-final-viral-combined_k141_222241\|\|full | 3138 | No | NA | 5 | 0 | 2 | Low-quality | Genome-fragment | 5.51 | AAI-based (medium-confidence) | 0 |
| S12-final-viral-combined_k141_36018\|\|0_partial | 17461 | No | NA | 14 | 0 | 10 | Not-determined | Genome-fragment | NA | NA | 0 |
| S7-final-viral-combined_k141_173507\|\|full | 3138 | No | NA | 4 | 0 | 3 | Low-quality | Genome-fragment | 7.28 | AAI-based (medium-confidence) | 0 |
| S5_k141_156954 | 3139 | Yes | 1599 | 5 | 1 | 2 | Low-quality | Genome-fragment | 4.92 | AAI-based (high-confidence) | 49.06 |
| S12-final-viral-combined_k141_220029\|\|0_partial | 10676 | No | NA | 8 | 0 | 7 | Not-determined | Genome-fragment | NA | NA | 0 |
| S12-final-viral-combined_k141_270839\|\|0_partial | 54725 | Yes | 34624 | 63 | 6 | 17 | Complete | High-quality | 100 | Provirus (high-confidence) | 36.73 |
| S12-final-viral-combined_k141_120125\|\|0_partial | 12078 | No | NA | 15 | 0 | 11 | Not-determined | Genome-fragment | NA | NA | 0 |
| S12-final-viral-combined_k141_87166\|\|0_partial | 10772 | No | NA | 7 | 0 | 6 | Not-determined | Genome-fragment | NA | NA | 0 |
| S2-final-viral-combined_k141_175574\|\|full | 3139 | No | NA | 4 | 0 | 3 | Low-quality | Genome-fragment | 1.6 | AAI-based (medium-confidence) | 0 |
| S12-final-viral-combined_k141_138432\|\|0_partial | 11277 | No | NA | 11 | 0 | 4 | Not-determined | Genome-fragment | NA | NA | 0 |
| S11-final-viral-combined_k141_239134\|\|full | 3140 | No | NA | 6 | 0 | 0 | Low-quality | Genome-fragment | 7.78 | AAI-based (medium-confidence) | 0 |
| S17-final-viral-combined_k141_157967\|\|full | 3140 | No | NA | 5 | 0 | 0 | Low-quality | Genome-fragment | 6.18 | AAI-based (medium-confidence) | 0 |
| S12-final-viral-combined_k141_4752\|\|0_partial | 13259 | No | NA | 10 | 0 | 7 | Not-determined | Genome-fragment | NA | NA | 0 |
| S12-final-viral-combined_k141_373648\|\|0_partial | 21152 | No | NA | 8 | 0 | 4 | Not-determined | Genome-fragment | NA | NA | 0 |
| S12-final-viral-combined_k141_323658\|\|0_partial | 11952 | No | NA | 10 | 0 | 10 | Not-determined | Genome-fragment | NA | NA | 0 |
| S12-final-viral-combined_k141_223051\|\|0_partial | 14781 | No | NA | 18 | 0 | 4 | Not-determined | Genome-fragment | NA | NA | 0 |
| S17-final-viral-combined_k141_99075\|\|full | 3140 | No | NA | 3 | 1 | 0 | Low-quality | Genome-fragment | 19.34 | AAI-based (high-confidence) | 0 |
| S12-final-viral-combined_k141_157167\|\|0_partial | 3883 | No | NA | 5 | 1 | 0 | High-quality | High-quality | 93.31 | HMM-based (lower-bound) | 0 |
| S7_k141_238291 | 3141 | No | NA | 2 | 0 | 0 | Low-quality | Genome-fragment | 1.66 | AAI-based (medium-confidence) | 0 |
| S9-final-viral-combined_k141_31567\|\|full | 3142 | No | NA | 8 | 2 | 0 | Low-quality | Genome-fragment | 6.94 | AAI-based (high-confidence) | 0 |
| S1-final-viral-combined_k141_43000\|\|full | 3142 | No | NA | 5 | 0 | 1 | Low-quality | Genome-fragment | 2.16 | AAI-based (medium-confidence) | 0 |
| S8-final-viral-combined_k141_85754\|\|full | 3142 | Yes | 282 | 4 | 1 | 3 | Low-quality | Genome-fragment | 0.54 | HMM-based (lower-bound) | 91.02 |
| S17-final-viral-combined_k141_231246\|\|full | 3143 | No | NA | 5 | 0 | 2 | Low-quality | Genome-fragment | 5.05 | AAI-based (medium-confidence) | 0 |
| S12-final-viral-combined_k141_141157\|\|0_partial | 12794 | No | NA | 11 | 0 | 2 | Not-determined | Genome-fragment | NA | NA | 0 |
| S12_k141_302331 | 3143 | No | NA | 4 | 0 | 1 | Low-quality | Genome-fragment | 1.34 | AAI-based (medium-confidence) | 0 |
| S17-final-viral-combined_k141_29659\|\|full | 3144 | No | NA | 6 | 4 | 0 | Low-quality | Genome-fragment | 6.53 | AAI-based (medium-confidence) | 0 |
| S8-final-viral-combined_k141_267887\|\|full | 3144 | No | NA | 5 | 0 | 1 | Low-quality | Genome-fragment | 4.26 | AAI-based (medium-confidence) | 0 |
| S11-final-viral-combined_k141_128218\|\|full | 3145 | No | NA | 4 | 0 | 1 | Low-quality | Genome-fragment | 2.09 | AAI-based (medium-confidence) | 0 |
| S1-final-viral-combined_k141_152109\|\|full | 3145 | No | NA | 6 | 1 | 0 | Low-quality | Genome-fragment | 1.99 | HMM-based (lower-bound) | 0 |
| S7_k141_30503 | 3145 | Yes | 2027 | 3 | 1 | 1 | Low-quality | Genome-fragment | 3.07 | HMM-based (lower-bound) | 35.55 |
| S12-final-viral-combined_k141_312983\|\|0_partial | 11027 | No | NA | 11 | 0 | 8 | Not-determined | Genome-fragment | NA | NA | 0 |
| S8-final-viral-combined_k141_168056\|\|full | 3145 | No | NA | 4 | 0 | 2 | Low-quality | Genome-fragment | 4.35 | AAI-based (medium-confidence) | 0 |
| S1-final-viral-combined_k141_368679\|\|full | 3146 | No | NA | 5 | 2 | 0 | Low-quality | Genome-fragment | 4.55 | HMM-based (lower-bound) | 0 |
| S12-final-viral-combined_k141_162415\|\|0_partial | 10884 | No | NA | 15 | 0 | 11 | Not-determined | Genome-fragment | NA | NA | 0 |
| S1-final-viral-combined_k141_172615\|\|full | 3146 | No | NA | 3 | 0 | 3 | Low-quality | Genome-fragment | 2.4 | AAI-based (medium-confidence) | 0 |
| S12-final-viral-combined_k141_112207\|\|0_partial | 15004 | No | NA | 13 | 0 | 10 | Not-determined | Genome-fragment | NA | NA | 0 |
| S12-final-viral-combined_k141_112382\|\|0_partial | 13708 | No | NA | 12 | 0 | 3 | Not-determined | Genome-fragment | NA | NA | 0 |
| S8-final-viral-combined_k141_187369\|\|full | 3146 | No | NA | 7 | 2 | 0 | Low-quality | Genome-fragment | 7.06 | AAI-based (medium-confidence) | 0 |
| S2-final-viral-combined_k141_232128\|\|full | 3147 | No | NA | 4 | 1 | 0 | Low-quality | Genome-fragment | 4.95 | AAI-based (high-confidence) | 0 |
| S8-final-viral-combined_k141_88440\|\|full | 3147 | No | NA | 3 | 1 | 0 | Low-quality | Genome-fragment | 6.59 | AAI-based (high-confidence) | 0 |
| S12-final-viral-combined_k141_164656\|\|0_partial | 24358 | No | NA | 25 | 0 | 7 | Medium-quality | Genome-fragment | 69.41 | AAI-based (medium-confidence) | 0 |
| S7_k141_2875 | 3148 | No | NA | 2 | 0 | 2 | Low-quality | Genome-fragment | 1.42 | AAI-based (high-confidence) | 0 |
| S4_k141_63394 | 3148 | No | NA | 3 | 0 | 1 | Low-quality | Genome-fragment | 3.69 | AAI-based (high-confidence) | 0 |
| S9_k141_4527 | 3149 | No | NA | 3 | 3 | 0 | Low-quality | Genome-fragment | 9.49 | AAI-based (high-confidence) | 0 |
| S12-final-viral-combined_k141_133608\|\|0_partial | 3151 | No | NA | 3 | 1 | 0 | Medium-quality | Genome-fragment | 62.56 | AAI-based (medium-confidence) | 0 |
| S7_k141_54313 | 3149 | No | NA | 1 | 0 | 1 | Low-quality | Genome-fragment | 5.65 | AAI-based (high-confidence) | 0 |
| S1-final-viral-combined_k141_94560\|\|full | 3150 | No | NA | 6 | 5 | 0 | Low-quality | Genome-fragment | 6.78 | AAI-based (medium-confidence) | 0 |
| S12_k141_168303 | 3151 | No | NA | 6 | 1 | 0 | Low-quality | Genome-fragment | 5.36 | AAI-based (high-confidence) | 0 |
| S1-final-viral-combined_k141_214507\|\|full | 3152 | No | NA | 4 | 1 | 0 | Low-quality | Genome-fragment | 3.37 | AAI-based (medium-confidence) | 0 |
| S7-final-viral-combined_k141_85940\|\|full | 3152 | No | NA | 5 | 1 | 0 | Low-quality | Genome-fragment | 1.79 | HMM-based (lower-bound) | 0 |
| S12-final-viral-combined_k141_251364\|\|lt2gene | 1595 | No | NA | 1 | 0 | 0 | Not-determined | Genome-fragment | NA | NA | 0 |
| S17-k141_78509 | 3152 | No | NA | 3 | 0 | 2 | Low-quality | Genome-fragment | 0.84 | AAI-based (high-confidence) | 0 |
| S12-final-viral-combined_k141_84075\|\|lt2gene | 1408 | No | NA | 2 | 0 | 1 | Not-determined | Genome-fragment | NA | NA | 0 |
| S4-final-viral-combined_k141_156689\|\|full | 3152 | No | NA | 3 | 0 | 1 | Low-quality | Genome-fragment | 0.93 | AAI-based (high-confidence) | 0 |
| S4_k141_42049 | 3152 | No | NA | 2 | 0 | 1 | Low-quality | Genome-fragment | 0.98 | AAI-based (medium-confidence) | 0 |
| S12-final-viral-combined_k141_100851\|\|lt2gene | 1069 | No | NA | 1 | 0 | 0 | Not-determined | Genome-fragment | NA | NA | 0 |
| S8_k141_115790 | 3152 | Yes | 2493 | 2 | 1 | 1 | Low-quality | Genome-fragment | 1.98 | AAI-based (high-confidence) | 20.91 |
| S8_k141_123722 | 3152 | No | NA | 1 | 0 | 0 | Low-quality | Genome-fragment | 3.28 | AAI-based (medium-confidence) | 0 |
| S11_k141_172206 | 3153 | No | NA | 4 | 0 | 1 | Low-quality | Genome-fragment | 7.74 | AAI-based (high-confidence) | 0 |
| S12-final-viral-combined_k141_335335\|\|lt2gene | 1585 | No | NA | 2 | 0 | 0 | Not-determined | Genome-fragment | NA | NA | 0 |
| S12-final-viral-combined_k141_354704\|\|full | 3153 | Yes | 1898 | 4 | 2 | 1 | Low-quality | Genome-fragment | 3.87 | HMM-based (lower-bound) | 39.8 |
| S8-final-viral-combined_k141_12787\|\|full | 3153 | No | NA | 4 | 4 | 0 | Low-quality | Genome-fragment | 7.84 | AAI-based (high-confidence) | 0 |
| S11-final-viral-combined_k141_322835\|\|full | 3154 | No | NA | 4 | 1 | 0 | Low-quality | Genome-fragment | 2.01 | HMM-based (lower-bound) | 0 |
| S17-final-viral-combined_k141_180774\|\|full | 3154 | No | NA | 6 | 2 | 0 | Low-quality | Genome-fragment | 6.65 | AAI-based (medium-confidence) | 0 |
| S12-final-viral-combined_k141_135067\|\|lt2gene | 1171 | No | NA | 1 | 0 | 0 | Not-determined | Genome-fragment | NA | NA | 0 |
| S12-final-viral-combined_k141_1508\|\|lt2gene | 1413 | No | NA | 2 | 0 | 0 | Not-determined | Genome-fragment | NA | NA | 0 |
| S12-final-viral-combined_k141_369780\|\|lt2gene | 1163 | No | NA | 1 | 0 | 1 | Not-determined | Genome-fragment | NA | NA | 0 |
| S12-final-viral-combined_k141_355954\|\|full | 3154 | No | NA | 6 | 0 | 1 | Low-quality | Genome-fragment | 6.77 | AAI-based (medium-confidence) | 0 |
| S12_k141_115148 | 3155 | No | NA | 5 | 0 | 0 | Low-quality | Genome-fragment | 4.09 | AAI-based (medium-confidence) | 0 |
| S6-final-viral-combined_k141_57050\|\|full | 3155 | No | NA | 2 | 0 | 1 | Low-quality | Genome-fragment | 1.42 | AAI-based (medium-confidence) | 0 |
| S12-final-viral-combined_k141_152472\|\|lt2gene | 3503 | No | NA | 2 | 0 | 0 | Not-determined | Genome-fragment | NA | NA | 0 |
| S12-final-viral-combined_k141_319986\|\|lt2gene | 1373 | No | NA | 1 | 0 | 0 | Not-determined | Genome-fragment | NA | NA | 0 |
| S2-final-viral-combined_k141_102643\|\|full | 3156 | No | NA | 3 | 2 | 0 | Low-quality | Genome-fragment | 12.48 | AAI-based (medium-confidence) | 0 |
| S1-final-viral-combined_k141_185341\|\|full | 3157 | No | NA | 6 | 0 | 0 | Low-quality | Genome-fragment | 7.51 | AAI-based (medium-confidence) | 0 |
| S17-final-viral-combined_k141_141473\|\|full | 3157 | No | NA | 6 | 0 | 1 | Low-quality | Genome-fragment | 5.3 | AAI-based (high-confidence) | 0 |
| S8_k141_231842 | 3157 | No | NA | 2 | 0 | 1 | Low-quality | Genome-fragment | 6.71 | AAI-based (medium-confidence) | 0 |
| S11-final-viral-combined_k141_323154\|\|full | 3158 | No | NA | 3 | 1 | 0 | Low-quality | Genome-fragment | 7.79 | AAI-based (high-confidence) | 0 |
| S1_k141_298426 | 3158 | No | NA | 5 | 0 | 0 | Low-quality | Genome-fragment | 6.08 | AAI-based (medium-confidence) | 0 |
| S7-final-viral-combined_k141_233347\|\|full | 3158 | No | NA | 5 | 1 | 0 | Low-quality | Genome-fragment | 8.55 | AAI-based (medium-confidence) | 0 |
| S8_k141_23724 | 3158 | No | NA | 4 | 2 | 0 | Low-quality | Genome-fragment | 7.94 | AAI-based (medium-confidence) | 0 |
| S11_k141_280237 | 3159 | No | NA | 2 | 0 | 0 | Low-quality | Genome-fragment | 3.96 | AAI-based (medium-confidence) | 0 |
| S7_k141_30101 | 3159 | No | NA | 5 | 0 | 3 | Low-quality | Genome-fragment | 7.43 | AAI-based (high-confidence) | 0 |
| S12-final-viral-combined_k141_369842\|\|full | 3159 | No | NA | 5 | 1 | 0 | Low-quality | Genome-fragment | 7.07 | AAI-based (high-confidence) | 0 |
| S12_k141_91319 | 3159 | No | NA | 6 | 0 | 0 | Low-quality | Genome-fragment | 5.28 | AAI-based (high-confidence) | 0 |
| S3-final-viral-combined_k141_7862\|\|full | 3160 | No | NA | 2 | 0 | 2 | Low-quality | Genome-fragment | 1.06 | AAI-based (high-confidence) | 0 |
| S3_k141_57253 | 3160 | No | NA | 5 | 0 | 0 | Low-quality | Genome-fragment | 2.24 | AAI-based (medium-confidence) | 0 |
| S12-final-viral-combined_k141_96093\|\|full | 3160 | No | NA | 8 | 2 | 0 | Low-quality | Genome-fragment | 5.66 | AAI-based (high-confidence) | 0 |
| S5-final-viral-combined_k141_117838\|\|full | 3161 | No | NA | 4 | 0 | 3 | Low-quality | Genome-fragment | 1.72 | AAI-based (medium-confidence) | 0 |
| S5_k141_14629 | 3161 | No | NA | 6 | 0 | 2 | Low-quality | Genome-fragment | 1.63 | AAI-based (medium-confidence) | 0 |
| S5_k141_69797 | 3161 | No | NA | 6 | 6 | 0 | Low-quality | Genome-fragment | 7.64 | AAI-based (high-confidence) | 0 |
| S12-final-viral-combined_k141_105090\|\|full | 3161 | No | NA | 4 | 4 | 0 | Low-quality | Genome-fragment | 8.42 | AAI-based (high-confidence) | 0 |
| S2_k141_26530 | 3161 | No | NA | 3 | 0 | 0 | Low-quality | Genome-fragment | 2.26 | AAI-based (high-confidence) | 0 |
| S12-final-viral-combined_k141_170635\|\|lt2gene | 1155 | No | NA | 1 | 0 | 0 | Not-determined | Genome-fragment | NA | NA | 0 |
| S6_k141_81839 | 3161 | No | NA | 5 | 0 | 3 | Low-quality | Genome-fragment | 3.88 | AAI-based (medium-confidence) | 0 |
| S8_k141_122851 | 3161 | No | NA | 7 | 1 | 0 | Low-quality | Genome-fragment | 4.2 | AAI-based (high-confidence) | 0 |
| S12-final-viral-combined_k141_321344\|\|lt2gene | 1130 | No | NA | 1 | 0 | 0 | Not-determined | Genome-fragment | NA | NA | 0 |
| S9_k141_62472 | 3162 | No | NA | 4 | 4 | 0 | Low-quality | Genome-fragment | 8.2 | AAI-based (high-confidence) | 0 |
| S12-final-viral-combined_k141_137352\|\|lt2gene | 1417 | No | NA | 1 | 0 | 0 | Not-determined | Genome-fragment | NA | NA | 0 |
| S12-final-viral-combined_k141_388404\|\|lt2gene | 1020 | No | NA | 1 | 0 | 1 | Not-determined | Genome-fragment | NA | NA | 0 |
| S11-final-viral-combined_k141_107876\|\|full | 3162 | No | NA | 4 | 0 | 2 | Low-quality | Genome-fragment | 10.1 | AAI-based (medium-confidence) | 0 |
| S3-final-viral-combined_k141_29468\|\|full | 3162 | No | NA | 5 | 0 | 1 | Low-quality | Genome-fragment | 4.22 | AAI-based (medium-confidence) | 0 |
| S5-final-viral-combined_k141_12796\|\|full | 3162 | No | NA | 2 | 0 | 2 | Low-quality | Genome-fragment | 1.13 | AAI-based (high-confidence) | 0 |
| S7_k141_22515 | 3162 | No | NA | 1 | 0 | 0 | Low-quality | Genome-fragment | 7.26 | AAI-based (high-confidence) | 0 |
| S12_k141_138703 | 3162 | No | NA | 2 | 1 | 0 | Low-quality | Genome-fragment | 1.77 | HMM-based (lower-bound) | 0 |
| S3-final-viral-combined_k141_23530\|\|full | 3163 | No | NA | 3 | 0 | 2 | Low-quality | Genome-fragment | 0.9 | AAI-based (high-confidence) | 0 |
| S12-final-viral-combined_k141_120407\|\|lt2gene | 1140 | No | NA | 1 | 0 | 0 | Not-determined | Genome-fragment | NA | NA | 0 |
| S12-final-viral-combined_k141_372248\|\|lt2gene | 1007 | No | NA | 2 | 0 | 0 | Not-determined | Genome-fragment | NA | NA | 0 |
| S3-final-viral-combined_k141_13225\|\|full | 3163 | No | NA | 7 | 1 | 0 | Low-quality | Genome-fragment | 1.8 | HMM-based (lower-bound) | 0 |
| S17-final-viral-combined_k141_71090\|\|full | 3163 | No | NA | 2 | 0 | 1 | Low-quality | Genome-fragment | 0.83 | AAI-based (medium-confidence) | 0 |
| S12-final-viral-combined_k141_290518\|\|full | 3163 | No | NA | 7 | 0 | 0 | Low-quality | Genome-fragment | 5.09 | AAI-based (high-confidence) | 0 |
| S12-final-viral-combined_k141_54041\|\|lt2gene | 2158 | No | NA | 1 | 0 | 1 | Not-determined | Genome-fragment | NA | NA | 0 |
| S8_k141_38394 | 3164 | No | NA | 2 | 0 | 1 | Low-quality | Genome-fragment | 4.22 | AAI-based (medium-confidence) | 0 |
| S3_k141_9611 | 3165 | No | NA | 2 | 1 | 0 | Low-quality | Genome-fragment | 1.88 | AAI-based (high-confidence) | 0 |
| S12-final-viral-combined_k141_137479\|\|full | 3165 | No | NA | 2 | 0 | 1 | Low-quality | Genome-fragment | 6.26 | AAI-based (medium-confidence) | 0 |
| S12-final-viral-combined_k141_88080\|\|lt2gene | 1049 | No | NA | 1 | 0 | 1 | Not-determined | Genome-fragment | NA | NA | 0 |
| S12-final-viral-combined_k141_243120\|\|full | 3165 | No | NA | 7 | 1 | 0 | Low-quality | Genome-fragment | 6.31 | AAI-based (medium-confidence) | 0 |
| S12-final-viral-combined_k141_234116\|\|full | 3165 | No | NA | 10 | 1 | 0 | Low-quality | Genome-fragment | 6.95 | AAI-based (medium-confidence) | 0 |
| S12-final-viral-combined_k141_4597\|\|lt2gene | 1429 | No | NA | 1 | 0 | 0 | Not-determined | Genome-fragment | NA | NA | 0 |
| S2_k141_198203 | 3165 | No | NA | 6 | 0 | 3 | Low-quality | Genome-fragment | 10.52 | AAI-based (medium-confidence) | 0 |
| S7_k141_131717 | 3166 | No | NA | 3 | 0 | 2 | Low-quality | Genome-fragment | 1.18 | AAI-based (high-confidence) | 0 |
| S17-final-viral-combined_k141_25943\|\|full | 3166 | No | NA | 5 | 0 | 1 | Low-quality | Genome-fragment | 2.12 | AAI-based (high-confidence) | 0 |
| S12_k141_321245 | 3166 | No | NA | 3 | 2 | 0 | Low-quality | Genome-fragment | 5.35 | AAI-based (high-confidence) | 0 |
| S12_k141_318767 | 3167 | No | NA | 3 | 0 | 0 | Low-quality | Genome-fragment | 6.39 | AAI-based (medium-confidence) | 0 |
| S8-final-viral-combined_k141_102371\|\|full | 3167 | No | NA | 4 | 3 | 0 | Low-quality | Genome-fragment | 8.04 | AAI-based (high-confidence) | 0 |
| S5_k141_211099 | 3168 | No | NA | 2 | 0 | 1 | Low-quality | Genome-fragment | 2.85 | AAI-based (medium-confidence) | 0 |
| S12_k141_323234 | 3168 | No | NA | 3 | 1 | 0 | Low-quality | Genome-fragment | 1.82 | AAI-based (high-confidence) | 0 |
| S9-final-viral-combined_k141_56403\|\|full | 3169 | No | NA | 3 | 0 | 2 | Low-quality | Genome-fragment | 3.06 | AAI-based (medium-confidence) | 0 |
| S5_k141_102036 | 3169 | No | NA | 4 | 1 | 0 | Low-quality | Genome-fragment | 8.47 | AAI-based (medium-confidence) | 0 |
| S12-final-viral-combined_k141_121935\|\|lt2gene | 1275 | No | NA | 2 | 0 | 0 | Not-determined | Genome-fragment | NA | NA | 0 |
| S2_k141_121106 | 3169 | No | NA | 2 | 0 | 0 | Low-quality | Genome-fragment | 4.08 | AAI-based (medium-confidence) | 0 |
| S11-final-viral-combined_k141_96396\|\|full | 3170 | No | NA | 6 | 0 | 3 | Low-quality | Genome-fragment | 1.53 | AAI-based (medium-confidence) | 0 |
| S5-final-viral-combined_k141_115529\|\|full | 3170 | No | NA | 5 | 4 | 0 | Low-quality | Genome-fragment | 8.75 | AAI-based (high-confidence) | 0 |
| S12_k141_78288 | 3170 | No | NA | 3 | 0 | 3 | Low-quality | Genome-fragment | 1.18 | AAI-based (medium-confidence) | 0 |
| S6-final-viral-combined_k141_111148\|\|full | 3170 | No | NA | 5 | 0 | 1 | Low-quality | Genome-fragment | 5.81 | AAI-based (medium-confidence) | 0 |
| S17-final-viral-combined_k141_131850\|\|full | 3171 | No | NA | 4 | 0 | 2 | Low-quality | Genome-fragment | 4.31 | AAI-based (high-confidence) | 0 |
| S12-final-viral-combined_k141_185754\|\|full | 3171 | No | NA | 5 | 0 | 1 | Low-quality | Genome-fragment | 3.38 | AAI-based (high-confidence) | 0 |
| S2_k141_240285 | 3171 | No | NA | 3 | 0 | 3 | Low-quality | Genome-fragment | 1.16 | AAI-based (high-confidence) | 0 |
| S8_k141_207885 | 3171 | No | NA | 4 | 0 | 0 | Low-quality | Genome-fragment | 8.15 | AAI-based (medium-confidence) | 0 |
| S12-final-viral-combined_k141_380265\|\|full | 3172 | No | NA | 3 | 1 | 0 | Low-quality | Genome-fragment | 7.66 | AAI-based (high-confidence) | 0 |
| S1-final-viral-combined_k141_21366\|\|full | 3173 | No | NA | 3 | 1 | 0 | Low-quality | Genome-fragment | 4.8 | HMM-based (lower-bound) | 0 |
| S12-final-viral-combined_k141_156934\|\|lt2gene | 2066 | No | NA | 1 | 0 | 0 | Not-determined | Genome-fragment | NA | NA | 0 |
| S2-final-viral-combined_k141_48831\|\|full | 3173 | No | NA | 7 | 2 | 0 | Low-quality | Genome-fragment | 6.88 | AAI-based (high-confidence) | 0 |
| S12-final-viral-combined_k141_73914\|\|lt2gene | 1701 | No | NA | 2 | 0 | 0 | Not-determined | Genome-fragment | NA | NA | 0 |
| S12-final-viral-combined_k141_40056\|\|lt2gene | 1554 | No | NA | 2 | 0 | 1 | Not-determined | Genome-fragment | NA | NA | 0 |
| S12-final-viral-combined_k141_106501\|\|lt2gene | 1976 | No | NA | 1 | 0 | 1 | Not-determined | Genome-fragment | NA | NA | 0 |
| S12-final-viral-combined_k141_274143\|\|lt2gene | 1412 | No | NA | 1 | 0 | 0 | Not-determined | Genome-fragment | NA | NA | 0 |
| S5-final-viral-combined_k141_156067\|\|full | 3174 | No | NA | 3 | 0 | 0 | Low-quality | Genome-fragment | 8.36 | AAI-based (high-confidence) | 0 |
| S12-final-viral-combined_k141_3963\|\|full | 3174 | No | NA | 4 | 1 | 0 | Low-quality | Genome-fragment | 8.14 | AAI-based (medium-confidence) | 0 |
| S4_k141_161877 | 3174 | No | NA | 9 | 1 | 0 | Low-quality | Genome-fragment | 5.12 | AAI-based (high-confidence) | 0 |
| S2_k141_202482 | 3175 | No | NA | 3 | 0 | 1 | Low-quality | Genome-fragment | 1.32 | AAI-based (medium-confidence) | 0 |
| S11_k141_258242 | 3176 | No | NA | 6 | 0 | 0 | Low-quality | Genome-fragment | 6.23 | AAI-based (high-confidence) | 0 |

| | | | | | | | | | | | |
|---|---|---|---|---|---|---|---|---|---|---|---|
| S12-final-viral-combined_k141_140494\|\|lt2gene | 3189 | No | NA | 1 | 0 | 0 | Not-determined | Genome-fragment | NA | NA | 0 |
| S1-final-viral-combined_k141_112091\|\|full | 3176 | No | NA | 4 | 0 | 1 | Low-quality | Genome-fragment | 5.84 | AAI-based (medium-confidence) | 0 |
| S12-final-viral-combined_k141_207614\|\|lt2gene | 1470 | No | NA | 1 | 0 | 0 | Not-determined | Genome-fragment | NA | NA | 0 |
| S1_k141_82765 | 3176 | No | NA | 3 | 0 | 2 | Low-quality | Genome-fragment | 1.75 | AAI-based (high-confidence) | 0 |
| S12-final-viral-combined_k141_6656\|\|lt2gene | 1025 | No | NA | 1 | 0 | 0 | Not-determined | Genome-fragment | NA | NA | 0 |
| S5-final-viral-combined_k141_82625\|\|full | 3176 | No | NA | 2 | 0 | 2 | Low-quality | Genome-fragment | 1.29 | AAI-based (medium-confidence) | 0 |
| S5_k141_31160 | 3176 | Yes | 1343 | 3 | 1 | 1 | Low-quality | Genome-fragment | 0.62 | HMM-based (lower-bound) | 57.71 |
| S12_k141_311994 | 3176 | No | NA | 3 | 0 | 1 | Low-quality | Genome-fragment | 6.07 | AAI-based (high-confidence) | 0 |
| S17-k141_26252 | 3177 | No | NA | 5 | 3 | 0 | Low-quality | Genome-fragment | 7.53 | AAI-based (high-confidence) | 0 |
| S2-final-viral-combined_k141_16835\|\|full | 3177 | No | NA | 5 | 0 | 1 | Low-quality | Genome-fragment | 3.38 | AAI-based (high-confidence) | 0 |
| S8-final-viral-combined_k141_83167\|\|full | 3177 | No | NA | 5 | 0 | 1 | Low-quality | Genome-fragment | 2.42 | AAI-based (high-confidence) | 0 |
| S9-final-viral-combined_k141_59156\|\|full | 3179 | No | NA | 8 | 0 | 2 | Low-quality | Genome-fragment | 6.37 | AAI-based (medium-confidence) | 0 |
| S12-final-viral-combined_k141_375598\|\|lt2gene | 1571 | No | NA | 1 | 0 | 1 | Not-determined | Genome-fragment | NA | NA | 0 |
| S9-final-viral-combined_k141_82389\|\|full | 3179 | No | NA | 4 | 0 | 1 | Low-quality | Genome-fragment | 4.23 | AAI-based (medium-confidence) | 0 |
| S1-final-viral-combined_k141_246677\|\|full | 3179 | No | NA | 2 | 0 | 1 | Low-quality | Genome-fragment | 3.1 | AAI-based (medium-confidence) | 0 |
| S1_k141_320103 | 3179 | No | NA | 9 | 0 | 0 | Low-quality | Genome-fragment | 5.24 | AAI-based (high-confidence) | 0 |
| S3-final-viral-combined_k141_15745\|\|full | 3179 | No | NA | 6 | 4 | 0 | Low-quality | Genome-fragment | 5.26 | AAI-based (high-confidence) | 0 |
| S12-final-viral-combined_k141_325575\|\|lt2gene | 1484 | No | NA | 1 | 0 | 0 | Not-determined | Genome-fragment | NA | NA | 0 |
| S12-final-viral-combined_k141_258218\|\|lt2gene | 1950 | No | NA | 1 | 0 | 0 | Not-determined | Genome-fragment | NA | NA | 0 |
| S17-final-viral-combined_k141_86531\|\|full | 3179 | No | NA | 4 | 1 | 0 | Low-quality | Genome-fragment | 2.18 | HMM-based (lower-bound) | 0 |
| S12-final-viral-combined_k141_24097\|\|lt2gene | 2096 | No | NA | 1 | 0 | 1 | Not-determined | Genome-fragment | NA | NA | 0 |
| S7-final-viral-combined_k141_28998\|\|full | 3180 | No | NA | 3 | 0 | 0 | Low-quality | Genome-fragment | 3.24 | AAI-based (high-confidence) | 0 |
| S8_k141_109932 | 3180 | No | NA | 8 | 2 | 0 | Low-quality | Genome-fragment | 7.81 | AAI-based (medium-confidence) | 0 |
| S8-final-viral-combined_k141_242302\|\|full | 3181 | No | NA | 6 | 0 | 0 | Low-quality | Genome-fragment | 7.36 | AAI-based (medium-confidence) | 0 |
| S3-final-viral-combined_k141_21786\|\|full | 3182 | No | NA | 7 | 4 | 0 | Low-quality | Genome-fragment | 9.12 | AAI-based (high-confidence) | 0 |
| S7-final-viral-combined_k141_90881\|\|full | 3182 | No | NA | 4 | 0 | 3 | Low-quality | Genome-fragment | 4.18 | AAI-based (medium-confidence) | 0 |
| S12-final-viral-combined_k141_41484\|\|lt2gene | 1315 | No | NA | 1 | 0 | 1 | Not-determined | Genome-fragment | NA | NA | 0 |
| S12-final-viral-combined_k141_255397\|\|full | 3182 | No | NA | 6 | 0 | 0 | Low-quality | Genome-fragment | 7.09 | AAI-based (high-confidence) | 0 |
| S12-final-viral-combined_k141_358423\|\|lt2gene | 1419 | No | NA | 1 | 0 | 0 | Not-determined | Genome-fragment | NA | NA | 0 |
| S1-final-viral-combined_k141_181420\|\|full | 3183 | Yes | 2302 | 4 | 1 | 1 | Low-quality | Genome-fragment | 1.42 | HMM-based (lower-bound) | 27.68 |
| S12-final-viral-combined_k141_208775\|\|lt2gene | 3396 | No | NA | 1 | 0 | 1 | Not-determined | Genome-fragment | NA | NA | 0 |
| S1_k141_343190 | 3183 | No | NA | 4 | 0 | 4 | Low-quality | Genome-fragment | 0.82 | AAI-based (medium-confidence) | 0 |
| S2-final-viral-combined_k141_239696\|\|full | 3183 | No | NA | 4 | 0 | 0 | Low-quality | Genome-fragment | 5.28 | AAI-based (medium-confidence) | 0 |
| S1-final-viral-combined_k141_226848\|\|full | 3185 | No | NA | 2 | 0 | 1 | Low-quality | Genome-fragment | 4.94 | AAI-based (medium-confidence) | 0 |
| S12-final-viral-combined_k141_142365\|\|lt2gene | 1644 | No | NA | 2 | 0 | 0 | Not-determined | Genome-fragment | NA | NA | 0 |
| S12-final-viral-combined_k141_108522\|\|lt2gene | 1079 | No | NA | 1 | 0 | 0 | Not-determined | Genome-fragment | NA | NA | 0 |
| S1_k141_370285 | 3185 | No | NA | 7 | 5 | 0 | Low-quality | Genome-fragment | 8.04 | AAI-based (high-confidence) | 0 |
| S7_k141_159482 | 3185 | No | NA | 5 | 0 | 0 | Low-quality | Genome-fragment | 5.06 | AAI-based (high-confidence) | 0 |
| S12-final-viral-combined_k141_74476\|\|full | 3185 | No | NA | 2 | 0 | 1 | Low-quality | Genome-fragment | 6.3 | AAI-based (medium-confidence) | 0 |
| S12-final-viral-combined_k141_276791\|\|lt2gene | 1194 | No | NA | 1 | 0 | 0 | Not-determined | Genome-fragment | NA | NA | 0 |
| S12-final-viral-combined_k141_42623\|\|lt2gene | 1295 | No | NA | 1 | 0 | 0 | Not-determined | Genome-fragment | NA | NA | 0 |
| S6-final-viral-combined_k141_24254\|\|full | 3185 | No | NA | 5 | 0 | 0 | Low-quality | Genome-fragment | 8.36 | AAI-based (high-confidence) | 0 |
| S12-final-viral-combined_k141_25744\|\|lt2gene | 1543 | No | NA | 2 | 0 | 2 | Not-determined | Genome-fragment | NA | NA | 0 |
| S12_k141_203713 | 3186 | Yes | 549 | 4 | 1 | 2 | Low-quality | Genome-fragment | 0.68 | HMM-based (lower-bound) | 82.77 |
| S12-final-viral-combined_k141_311331\|\|lt2gene | 1274 | No | NA | 2 | 0 | 0 | Not-determined | Genome-fragment | NA | NA | 0 |
| S12_k141_120373 | 3186 | No | NA | 2 | 0 | 2 | Low-quality | Genome-fragment | 2.16 | AAI-based (medium-confidence) | 0 |
| S8_k141_220193 | 3186 | No | NA | 5 | 0 | 3 | Low-quality | Genome-fragment | 3.88 | AAI-based (medium-confidence) | 0 |
| S12-final-viral-combined_k141_42972\|\|lt2gene | 1224 | No | NA | 1 | 0 | 0 | Not-determined | Genome-fragment | NA | NA | 0 |
| S8_k141_202741 | 3186 | Yes | 729 | 3 | 1 | 1 | Low-quality | Genome-fragment | 0.33 | HMM-based (lower-bound) | 77.12 |
| S12-final-viral-combined_k141_243596\|\|lt2gene | 1392 | No | NA | 1 | 0 | 0 | Not-determined | Genome-fragment | NA | NA | 0 |
| S11-final-viral-combined_k141_302243\|\|full | 3188 | No | NA | 2 | 0 | 0 | Low-quality | Genome-fragment | 2.39 | AAI-based (medium-confidence) | 0 |
| S7-final-viral-combined_k141_126021\|\|full | 3190 | No | NA | 5 | 2 | 0 | Low-quality | Genome-fragment | 5 | HMM-based (lower-bound) | 0 |
| S1-final-viral-combined_k141_267982\|\|full | 3191 | No | NA | 4 | 0 | 3 | Low-quality | Genome-fragment | 1.54 | AAI-based (medium-confidence) | 0 |
| S1-final-viral-combined_k141_128806\|\|full | 3191 | No | NA | 4 | 0 | 3 | Low-quality | Genome-fragment | 5.99 | AAI-based (medium-confidence) | 0 |
| S12-final-viral-combined_k141_9176\|\|lt2gene | 1341 | No | NA | 1 | 0 | 1 | Not-determined | Genome-fragment | NA | NA | 0 |
| S1-final-viral-combined_k141_341729\|\|full | 3191 | No | NA | 7 | 0 | 0 | Low-quality | Genome-fragment | 6.61 | AAI-based (medium-confidence) | 0 |
| S12-final-viral-combined_k141_160512\|\|lt2gene | 1135 | No | NA | 2 | 0 | 0 | Not-determined | Genome-fragment | NA | NA | 0 |
| S12-final-viral-combined_k141_74091\|\|full | 3191 | No | NA | 3 | 3 | 0 | Low-quality | Genome-fragment | 7.13 | AAI-based (high-confidence) | 0 |
| S11-final-viral-combined_k141_303354\|\|full | 3192 | No | NA | 5 | 3 | 0 | Low-quality | Genome-fragment | 5.28 | AAI-based (high-confidence) | 0 |
| S1_k141_371977 | 3192 | No | NA | 4 | 0 | 3 | Low-quality | Genome-fragment | 6.98 | AAI-based (medium-confidence) | 0 |
| S12-final-viral-combined_k141_92965\|\|lt2gene | 2475 | No | NA | 1 | 0 | 0 | Not-determined | Genome-fragment | NA | NA | 0 |
| S12-final-viral-combined_k141_243976\|\|lt2gene | 1390 | No | NA | 2 | 0 | 1 | Not-determined | Genome-fragment | NA | NA | 0 |
| S17-final-viral-combined_k141_26999\|\|full | 3192 | No | NA | 5 | 0 | 0 | Low-quality | Genome-fragment | 5.92 | AAI-based (medium-confidence) | 0 |
| S12-final-viral-combined_k141_77666\|\|lt2gene | 2016 | No | NA | 2 | 0 | 0 | Not-determined | Genome-fragment | NA | NA | 0 |
| S17-final-viral-combined_k141_56682\|\|full | 3193 | No | NA | 3 | 0 | 1 | Low-quality | Genome-fragment | 6.77 | AAI-based (medium-confidence) | 0 |
| S12-final-viral-combined_k141_177264\|\|lt2gene | 1036 | No | NA | 1 | 0 | 0 | Not-determined | Genome-fragment | NA | NA | 0 |
| S4-final-viral-combined_k141_140017\|\|full | 3194 | No | NA | 4 | 0 | 3 | Low-quality | Genome-fragment | 1.44 | AAI-based (medium-confidence) | 0 |
| S12-final-viral-combined_k141_44213\|\|lt2gene | 1324 | No | NA | 2 | 0 | 0 | Not-determined | Genome-fragment | NA | NA | 0 |
| S12-final-viral-combined_k141_395165\|\|lt2gene | 1310 | No | NA | 1 | 0 | 0 | Not-determined | Genome-fragment | NA | NA | 0 |
| S1-final-viral-combined_k141_211281\|\|full | 3195 | No | NA | 3 | 0 | 1 | Low-quality | Genome-fragment | 3.18 | AAI-based (medium-confidence) | 0 |
| S1-final-viral-combined_k141_13874\|\|full | 3195 | No | NA | 3 | 0 | 1 | Low-quality | Genome-fragment | 7.52 | AAI-based (medium-confidence) | 0 |
| S12-final-viral-combined_k141_295407\|\|lt2gene | 1802 | No | NA | 1 | 0 | 0 | Not-determined | Genome-fragment | NA | NA | 0 |
| S12-final-viral-combined_k141_119547\|\|full | 3195 | No | NA | 4 | 0 | 4 | Low-quality | Genome-fragment | 1.02 | AAI-based (medium-confidence) | 0 |
| S12-final-viral-combined_k141_211134\|\|lt2gene | 1750 | No | NA | 1 | 0 | 0 | Not-determined | Genome-fragment | NA | NA | 0 |
| S12-final-viral-combined_k141_115262\|\|lt2gene | 3195 | No | NA | 1 | 1 | 0 | Low-quality | Genome-fragment | 6.81 | AAI-based (high-confidence) | 0 |
| S12-final-viral-combined_k141_295648\|\|lt2gene | 1280 | No | NA | 2 | 0 | 1 | Not-determined | Genome-fragment | NA | NA | 0 |
| S11-final-viral-combined_k141_202212\|\|full | 3196 | No | NA | 7 | 2 | 0 | Low-quality | Genome-fragment | 4.95 | HMM-based (lower-bound) | 0 |
| S1-final-viral-combined_k141_205918\|\|full | 3196 | No | NA | 4 | 2 | 0 | Low-quality | Genome-fragment | 8.55 | AAI-based (medium-confidence) | 0 |
| S9-final-viral-combined_k141_126119\|\|full | 3197 | No | NA | 4 | 0 | 3 | Low-quality | Genome-fragment | 3.74 | AAI-based (medium-confidence) | 0 |
| S12-final-viral-combined_k141_395592\|\|lt2gene | 1015 | No | NA | 2 | 0 | 0 | Not-determined | Genome-fragment | NA | NA | 0 |
| S1_k141_99533 | 3197 | No | NA | 6 | 0 | 0 | Low-quality | Genome-fragment | 7.54 | AAI-based (medium-confidence) | 0 |
| S2_k141_120242 | 3197 | No | NA | 7 | 0 | 0 | Low-quality | Genome-fragment | 4.4 | AAI-based (medium-confidence) | 0 |
| S1-final-viral-combined_k141_311831\|\|full | 3198 | No | NA | 4 | 1 | 0 | Low-quality | Genome-fragment | 5.81 | AAI-based (medium-confidence) | 0 |
| S3_k141_94901 | 3198 | No | NA | 8 | 1 | 0 | Low-quality | Genome-fragment | 5.79 | AAI-based (high-confidence) | 0 |
| S7_k141_91120 | 3198 | No | NA | 5 | 0 | 4 | Low-quality | Genome-fragment | 3.48 | AAI-based (medium-confidence) | 0 |
| S12-final-viral-combined_k141_44944\|\|lt2gene | 1453 | No | NA | 1 | 0 | 0 | Not-determined | Genome-fragment | NA | NA | 0 |
| S2-final-viral-combined_k141_207191\|\|full | 3198 | No | NA | 7 | 0 | 0 | Low-quality | Genome-fragment | 6.47 | AAI-based (medium-confidence) | 0 |
| S4-final-viral-combined_k141_105507\|\|full | 3198 | No | NA | 3 | 0 | 2 | Low-quality | Genome-fragment | 3.76 | AAI-based (medium-confidence) | 0 |
| S6-final-viral-combined_k141_629\|\|full | 3198 | No | NA | 4 | 0 | 1 | Low-quality | Genome-fragment | 5.84 | AAI-based (medium-confidence) | 0 |
| S3_k141_109191 | 3199 | No | NA | 1 | 0 | 0 | Low-quality | Genome-fragment | 2.73 | AAI-based (medium-confidence) | 0 |
| S17-final-viral-combined_k141_51277\|\|full | 3199 | No | NA | 5 | 1 | 2 | Low-quality | Genome-fragment | 5.65 | AAI-based (high-confidence) | 0 |
| S2-final-viral-combined_k141_104200\|\|full | 3199 | No | NA | 3 | 0 | 3 | Low-quality | Genome-fragment | 2.59 | AAI-based (high-confidence) | 0 |
| S8_k141_110686 | 3199 | No | NA | 1 | 0 | 1 | Low-quality | Genome-fragment | 1.72 | AAI-based (medium-confidence) | 0 |
| S12-final-viral-combined_k141_178214\|\|lt2gene | 1100 | No | NA | 2 | 0 | 0 | Not-determined | Genome-fragment | NA | NA | 0 |
| S12-final-viral-combined_k141_10619\|\|lt2gene | 1447 | No | NA | 1 | 0 | 0 | Not-determined | Genome-fragment | NA | NA | 0 |
| S1-final-viral-combined_k141_207856\|\|full | 3200 | No | NA | 5 | 0 | 0 | Low-quality | Genome-fragment | 8.62 | AAI-based (medium-confidence) | 0 |
| S5-final-viral-combined_k141_33215\|\|full | 3200 | No | NA | 3 | 3 | 0 | Low-quality | Genome-fragment | 8.01 | AAI-based (high-confidence) | 0 |
| S1-final-viral-combined_k141_167466\|\|full | 3201 | No | NA | 4 | 1 | 0 | Low-quality | Genome-fragment | 2.04 | HMM-based (lower-bound) | 0 |
| S3_k141_5095 | 3201 | No | NA | 6 | 2 | 0 | Low-quality | Genome-fragment | 5.3 | AAI-based (high-confidence) | 0 |
| S12-final-viral-combined_k141_45688\|\|lt2gene | 2024 | No | NA | 1 | 0 | 0 | Not-determined | Genome-fragment | NA | NA | 0 |
| S12-final-viral-combined_k141_296509\|\|lt2gene | 1073 | No | NA | 1 | 0 | 0 | Not-determined | Genome-fragment | NA | NA | 0 |
| S12_k141_215106 | 3201 | No | NA | 6 | 3 | 0 | Low-quality | Genome-fragment | 5.79 | AAI-based (high-confidence) | 0 |
| S2_k141_174802 | 3201 | No | NA | 5 | 2 | 0 | Low-quality | Genome-fragment | 3.65 | AAI-based (medium-confidence) | 0 |
| S3-final-viral-combined_k141_112327\|\|full | 3202 | No | NA | 3 | 0 | 2 | Low-quality | Genome-fragment | 1.39 | AAI-based (medium-confidence) | 0 |
| S3_k141_114964 | 3202 | No | NA | 1 | 0 | 1 | Low-quality | Genome-fragment | 3.01 | AAI-based (medium-confidence) | 0 |
| S7-final-viral-combined_k141_84272\|\|full | 3202 | No | NA | 3 | 0 | 1 | Low-quality | Genome-fragment | 1.79 | AAI-based (medium-confidence) | 0 |
| S12-final-viral-combined_k141_212807\|\|lt2gene | 1049 | No | NA | 1 | 0 | 0 | Not-determined | Genome-fragment | NA | NA | 0 |
| S12-final-viral-combined_k141_11477\|\|lt2gene | 1113 | No | NA | 1 | 0 | 0 | Not-determined | Genome-fragment | NA | NA | 0 |
| S17-final-viral-combined_k141_264523\|\|full | 3202 | No | NA | 4 | 1 | 0 | Low-quality | Genome-fragment | 8.68 | AAI-based (medium-confidence) | 0 |
| S8-final-viral-combined_k141_97652\|\|full | 3202 | No | NA | 3 | 0 | 2 | Low-quality | Genome-fragment | 0.81 | AAI-based (medium-confidence) | 0 |
| S5-final-viral-combined_k141_29846\|\|full | 3203 | No | NA | 6 | 5 | 0 | Low-quality | Genome-fragment | 7.08 | AAI-based (medium-confidence) | 0 |
| S12-final-viral-combined_k141_314756\|\|lt2gene | 1716 | No | NA | 1 | 0 | 1 | Not-determined | Genome-fragment | NA | NA | 0 |
| S12-final-viral-combined_k141_397559\|\|lt2gene | 1214 | No | NA | 1 | 0 | 0 | Not-determined | Genome-fragment | NA | NA | 0 |
| S12-final-viral-combined_k141_362483\|\|lt2gene | 1532 | No | NA | 2 | 0 | 0 | Not-determined | Genome-fragment | NA | NA | 0 |
| S8-final-viral-combined_k141_176166\|\|full | 3204 | No | NA | 3 | 0 | 0 | Low-quality | Genome-fragment | 1.8 | AAI-based (medium-confidence) | 0 |
| S5-final-viral-combined_k141_188288\|\|full | 3205 | No | NA | 3 | 0 | 2 | Low-quality | Genome-fragment | 1.15 | AAI-based (medium-confidence) | 0 |
| S12-final-viral-combined_k141_23244\|\|full | 3205 | No | NA | 6 | 0 | 0 | Low-quality | Genome-fragment | 7.43 | AAI-based (medium-confidence) | 0 |
| S12-final-viral-combined_k141_397837\|\|lt2gene | 1462 | No | NA | 1 | 0 | 0 | Not-determined | Genome-fragment | NA | NA | 0 |
| S2-final-viral-combined_k141_160960\|\|full | 3205 | No | NA | 3 | 0 | 1 | Low-quality | Genome-fragment | 1.02 | AAI-based (medium-confidence) | 0 |
| S12-final-viral-combined_k141_388007\|\|full | 3206 | No | NA | 3 | 2 | 0 | Low-quality | Genome-fragment | 3.1 | HMM-based (lower-bound) | 0 |
| S2-final-viral-combined_k141_18214\|\|full | 3206 | Yes | 2303 | 6 | 1 | 1 | Low-quality | Genome-fragment | 4.56 | HMM-based (lower-bound) | 28.17 |
| S17-final-viral-combined_k141_26175\|\|full | 3207 | No | NA | 6 | 0 | 0 | Low-quality | Genome-fragment | 5.25 | AAI-based (medium-confidence) | 0 |
| S4-final-viral-combined_k141_33934\|\|full | 3207 | No | NA | 4 | 2 | 0 | Low-quality | Genome-fragment | 8.43 | AAI-based (high-confidence) | 0 |
| S1-final-viral-combined_k141_149462\|\|full | 3208 | No | NA | 3 | 0 | 0 | Low-quality | Genome-fragment | 6.23 | AAI-based (medium-confidence) | 0 |
| S9-final-viral-combined_k141_45801\|\|full | 3209 | No | NA | 6 | 0 | 1 | Low-quality | Genome-fragment | 3.42 | AAI-based (medium-confidence) | 0 |
| S11-final-viral-combined_k141_45538\|\|full | 3209 | No | NA | 4 | 0 | 2 | Low-quality | Genome-fragment | 8.63 | AAI-based (medium-confidence) | 0 |
| S2-final-viral-combined_k141_246632\|\|full | 3209 | No | NA | 3 | 0 | 0 | Low-quality | Genome-fragment | 20.69 | AAI-based (high-confidence) | 0 |
| S9_k141_23304 | 3210 | No | NA | 2 | 1 | 0 | Low-quality | Genome-fragment | 7.09 | AAI-based (high-confidence) | 0 |
| S12-final-viral-combined_k141_135982\|\|full | 3210 | No | NA | 2 | 0 | 1 | Low-quality | Genome-fragment | 1.07 | AAI-based (medium-confidence) | 0 |
| S1_k141_579 | 3211 | No | NA | 3 | 0 | 0 | Low-quality | Genome-fragment | 6.61 | AAI-based (high-confidence) | 0 |
| S12-final-viral-combined_k141_113256\|\|lt2gene | 1032 | No | NA | 2 | 0 | 0 | Not-determined | Genome-fragment | NA | NA | 0 |
| S5-final-viral-combined_k141_38613\|\|full | 3211 | No | NA | 5 | 3 | 0 | Low-quality | Genome-fragment | 8.6 | AAI-based (high-confidence) | 0 |
| S17-final-viral-combined_k141_26376\|\|full | 3211 | No | NA | 4 | 0 | 1 | Low-quality | Genome-fragment | 2.31 | AAI-based (medium-confidence) | 0 |
| S1-final-viral-combined_k141_357579\|\|full | 3213 | No | NA | 6 | 0 | 2 | Low-quality | Genome-fragment | 2.88 | AAI-based (high-confidence) | 0 |
| S17-k141_41051 | 3213 | No | NA | 5 | 3 | 0 | Low-quality | Genome-fragment | 7.94 | AAI-based (high-confidence) | 0 |
| S12-final-viral-combined_k141_298456\|\|lt2gene | 1288 | No | NA | 1 | 0 | 0 | Not-determined | Genome-fragment | NA | NA | 0 |
| S12-final-viral-combined_k141_141755\|\|full | 3213 | No | NA | 6 | 4 | 0 | Low-quality | Genome-fragment | 6.08 | HMM-based (lower-bound) | 0 |
| S2-final-viral-combined_k141_110035\|\|full | 3213 | No | NA | 6 | 1 | 0 | Low-quality | Genome-fragment | 11.01 | HMM-based (lower-bound) | 0 |
| S4-final-viral-combined_k141_63447\|\|full | 3213 | No | NA | 5 | 0 | 3 | Low-quality | Genome-fragment | 1.97 | AAI-based (medium-confidence) | 0 |
| S12-final-viral-combined_k141_232298\|\|lt2gene | 1081 | No | NA | 2 | 0 | 1 | Not-determined | Genome-fragment | NA | NA | 0 |
| S7-final-viral-combined_k141_125640\|\|full | 3214 | No | NA | 4 | 0 | 0 | Low-quality | Genome-fragment | 8.28 | AAI-based (medium-confidence) | 0 |
| S12-final-viral-combined_k141_11658\|\|full | 3214 | No | NA | 3 | 0 | 0 | Low-quality | Genome-fragment | 1.57 | AAI-based (medium-confidence) | 0 |
| S12-final-viral-combined_k141_147499\|\|lt2gene | 1862 | No | NA | 1 | 0 | 0 | Not-determined | Genome-fragment | NA | NA | 0 |
| S12_k141_287692 | 3214 | No | NA | 5 | 0 | 0 | Low-quality | Genome-fragment | 2.58 | AAI-based (high-confidence) | 0 |
| S1_k141_314202 | 3215 | No | NA | 3 | 0 | 1 | Low-quality | Genome-fragment | 2.03 | AAI-based (high-confidence) | 0 |
| S5-final-viral-combined_k141_200369\|\|full | 3215 | No | NA | 11 | 2 | 0 | Low-quality | Genome-fragment | 5.77 | AAI-based (high-confidence) | 0 |
| S12-final-viral-combined_k141_31215\|\|lt2gene | 1002 | No | NA | 1 | 0 | 0 | Not-determined | Genome-fragment | NA | NA | 0 |
| S12-final-viral-combined_k141_82343\|\|lt2gene | 1291 | No | NA | 1 | 0 | 0 | Not-determined | Genome-fragment | NA | NA | 0 |
| S2_k141_80011 | 3215 | No | NA | 3 | 0 | 3 | Low-quality | Genome-fragment | 1.08 | AAI-based (high-confidence) | 0 |
| S12-final-viral-combined_k141_349893\|\|lt2gene | 1955 | No | NA | 2 | 0 | 0 | Not-determined | Genome-fragment | NA | NA | 0 |
| S3-final-viral-combined_k141_103334\|\|full | 3216 | No | NA | 4 | 4 | 0 | Low-quality | Genome-fragment | 5.88 | AAI-based (high-confidence) | 0 |
| S5-final-viral-combined_k141_75231\|\|full | 3216 | No | NA | 3 | 0 | 1 | Low-quality | Genome-fragment | 6.92 | AAI-based (high-confidence) | 0 |
| S7-final-viral-combined_k141_96771\|\|full | 3216 | No | NA | 8 | 2 | 0 | Low-quality | Genome-fragment | 9.45 | AAI-based (high-confidence) | 0 |
| S17-final-viral-combined_k141_35045\|\|full | 3216 | No | NA | 4 | 2 | 0 | Low-quality | Genome-fragment | 2 | HMM-based (lower-bound) | 0 |
| S12_k141_277198 | 3216 | No | NA | 5 | 2 | 0 | Low-quality | Genome-fragment | 2.4 | AAI-based (high-confidence) | 0 |
| S12-final-viral-combined_k141_216660\|\|lt2gene | 1305 | No | NA | 1 | 0 | 0 | Not-determined | Genome-fragment | NA | NA | 0 |
| S12-final-viral-combined_k141_284257\|\|lt2gene | 1712 | No | NA | 2 | 0 | 0 | Not-determined | Genome-fragment | NA | NA | 0 |
| S12-final-viral-combined_k141_317884\|\|lt2gene | 1714 | No | NA | 1 | 0 | 0 | Not-determined | Genome-fragment | NA | NA | 0 |
| S4-final-viral-combined_k141_134640\|\|full | 3217 | No | NA | 3 | 0 | 0 | Low-quality | Genome-fragment | 5.99 | AAI-based (high-confidence) | 0 |
| S7-final-viral-combined_k141_155516\|\|full | 3218 | No | NA | 4 | 0 | 1 | Low-quality | Genome-fragment | 1.54 | AAI-based (high-confidence) | 0 |
| S12-final-viral-combined_k141_98960\|\|lt2gene | 1315 | No | NA | 1 | 0 | 0 | Not-determined | Genome-fragment | NA | NA | 0 |
| S8_k141_256648 | 3218 | Yes | 349 | 2 | 1 | 1 | Low-quality | Genome-fragment | 0.12 | HMM-based (lower-bound) | 89.15 |
| S12-final-viral-combined_k141_132105\|\|lt2gene | 1029 | No | NA | 1 | 0 | 0 | Not-determined | Genome-fragment | NA | NA | 0 |
| S12-final-viral-combined_k141_399957\|\|lt2gene | 1087 | No | NA | 2 | 0 | 0 | Not-determined | Genome-fragment | NA | NA | 0 |
| S9_k141_74080 | 3219 | No | NA | 3 | 0 | 3 | Low-quality | Genome-fragment | 7.56 | AAI-based (medium-confidence) | 0 |
| S1-final-viral-combined_k141_107001\|\|full | 3219 | No | NA | 3 | 0 | 0 | Low-quality | Genome-fragment | 1.64 | AAI-based (medium-confidence) | 0 |
| S1_k141_2290 | 3219 | No | NA | 4 | 0 | 1 | Low-quality | Genome-fragment | 8.37 | AAI-based (high-confidence) | 0 |
| S12-final-viral-combined_k141_149018\|\|lt2gene | 1272 | No | NA | 1 | 0 | 0 | Not-determined | Genome-fragment | NA | NA | 0 |
| S5-final-viral-combined_k141_79713\|\|full | 3219 | No | NA | 3 | 3 | 0 | Low-quality | Genome-fragment | 9.03 | AAI-based (high-confidence) | 0 |
| S7_k141_166395 | 3219 | No | NA | 7 | 0 | 0 | Low-quality | Genome-fragment | 5.62 | AAI-based (high-confidence) | 0 |
| S12-final-viral-combined_k141_56578\|\|full | 3219 | No | NA | 2 | 0 | 2 | Low-quality | Genome-fragment | 1.91 | AAI-based (high-confidence) | 0 |
| S2_k141_28021 | 3219 | No | NA | 1 | 0 | 0 | Low-quality | Genome-fragment | 7.54 | AAI-based (medium-confidence) | 0 |
| S12_k141_392957 | 3220 | No | NA | 3 | 0 | 0 | Low-quality | Genome-fragment | 3.18 | AAI-based (high-confidence) | 0 |
| S1_k141_365635 | 3221 | No | NA | 4 | 0 | 1 | Low-quality | Genome-fragment | 4.66 | AAI-based (medium-confidence) | 0 |
| S12-final-viral-combined_k141_99899\|\|lt2gene | 1338 | No | NA | 1 | 0 | 0 | Not-determined | Genome-fragment | NA | NA | 0 |
| S12-final-viral-combined_k141_384421\|\|lt2gene | 1753 | No | NA | 2 | 0 | 1 | Not-determined | Genome-fragment | NA | NA | 0 |
| S5-final-viral-combined_k141_196613\|\|full | 3221 | No | NA | 5 | 0 | 1 | Low-quality | Genome-fragment | 8.07 | AAI-based (medium-confidence) | 0 |
| S17-final-viral-combined_k141_171333\|\|full | 3221 | No | NA | 4 | 0 | 2 | Low-quality | Genome-fragment | 2.45 | AAI-based (high-confidence) | 0 |
| S12-final-viral-combined_k141_297057\|\|full | 3221 | No | NA | 4 | 0 | 2 | Low-quality | Genome-fragment | 3.59 | AAI-based (medium-confidence) | 0 |
| S1-final-viral-combined_k141_190010\|\|full | 3222 | No | NA | 6 | 0 | 0 | Low-quality | Genome-fragment | 5.45 | AAI-based (high-confidence) | 0 |
| S17-final-viral-combined_k141_241588\|\|lt2gene | 3222 | No | NA | 1 | 1 | 0 | Low-quality | Genome-fragment | 6.61 | AAI-based (high-confidence) | 0 |
| S12-final-viral-combined_k141_255858\|\|full | 3222 | No | NA | 6 | 3 | 0 | Low-quality | Genome-fragment | 4.36 | HMM-based (lower-bound) | 0 |
| S12-final-viral-combined_k141_367516\|\|lt2gene | 1016 | No | NA | 1 | 0 | 1 | Not-determined | Genome-fragment | NA | NA | 0 |
| S6-final-viral-combined_k141_138583\|\|full | 3222 | No | NA | 6 | 2 | 0 | Low-quality | Genome-fragment | 6.27 | AAI-based (medium-confidence) | 0 |
| S12_k141_217609 | 1014 | No | NA | 3 | 0 | 0 | Not-determined | Genome-fragment | NA | NA | 0 |
| S12_k141_16745 | 1531 | No | NA | 1 | 0 | 1 | Not-determined | Genome-fragment | NA | NA | 0 |
| S12_k141_83709 | 1584 | No | NA | 2 | 0 | 0 | Not-determined | Genome-fragment | NA | NA | 0 |
| S12_k141_234364 | 1003 | No | NA | 2 | 0 | 1 | Not-determined | Genome-fragment | NA | NA | 0 |
| S12_k141_83713 | 1538 | No | NA | 3 | 0 | 3 | Not-determined | Genome-fragment | NA | NA | 0 |
| S12_k141_117184 | 45814 | No | NA | 33 | 0 | 19 | Not-determined | Genome-fragment | NA | NA | 0 |
| S12_k141_117196 | 3776 | No | NA | 6 | 0 | 3 | Not-determined | Genome-fragment | NA | NA | 0 |
| S12_k141_251143 | 1828 | No | NA | 2 | 0 | 2 | Not-determined | Genome-fragment | NA | NA | 0 |
| S12_k141_301366 | 1037 | No | NA | 1 | 0 | 1 | Not-determined | Genome-fragment | NA | NA | 0 |
| S12_k141_301367 | 2245 | No | NA | 1 | 0 | 0 | Not-determined | Genome-fragment | NA | NA | 0 |
| S6-final-viral-combined_k141_16246\|\|full | 3222 | No | NA | 7 | 1 | 0 | Low-quality | Genome-fragment | 4.9 | HMM-based (lower-bound) | 0 |
| S12_k141_67008 | 3970 | No | NA | 4 | 0 | 2 | Not-determined | Genome-fragment | NA | NA | 0 |
| S12_k141_301383 | 1665 | No | NA | 4 | 0 | 0 | Not-determined | Genome-fragment | NA | NA | 0 |
| S12_k141_318084 | 2057 | No | NA | 2 | 0 | 1 | Not-determined | Genome-fragment | NA | NA | 0 |
| S12_k141_167503 | 1108 | No | NA | 3 | 0 | 1 | Not-determined | Genome-fragment | NA | NA | 0 |
| S12_k141_301394 | 1561 | No | NA | 4 | 0 | 4 | Not-determined | Genome-fragment | NA | NA | 0 |
| S12_k141_134004 | 4003 | No | NA | 2 | 0 | 1 | Not-determined | Genome-fragment | NA | NA | 0 |
| S12_k141_368352 | 1117 | No | NA | 2 | 0 | 0 | Not-determined | Genome-fragment | NA | NA | 0 |
| S12_k141_334807 | 1448 | No | NA | 1 | 0 | 1 | Not-determined | Genome-fragment | NA | NA | 0 |
| S12_k141_134017 | 1005 | No | NA | 2 | 0 | 2 | Not-determined | Genome-fragment | NA | NA | 0 |
| S12_k141_16833 | 3240 | No | NA | 2 | 0 | 0 | Not-determined | Genome-fragment | NA | NA | 0 |
| S12_k141_167 | 1907 | No | NA | 2 | 0 | 0 | Not-determined | Genome-fragment | NA | NA | 0 |
| S12_k141_184276 | 30698 | No | NA | 44 | 18 | 1 | High-quality | High-quality | 95.99 | AAI-based (high-confidence) | 0 |
| S12_k141_217794 | 7498 | No | NA | 3 | 0 | 3 | Not-determined | Genome-fragment | NA | NA | 0 |
| S12_k141_351619 | 1298 | No | NA | 2 | 0 | 1 | Not-determined | Genome-fragment | NA | NA | 0 |
| S12_k141_351622 | 4636 | No | NA | 7 | 0 | 0 | Not-determined | Genome-fragment | NA | NA | 0 |
| S12_k141_201107 | 3155 | No | NA | 3 | 0 | 2 | Not-determined | Genome-fragment | NA | NA | 0 |
| S12_k141_301478 | 1151 | No | NA | 1 | 0 | 1 | Not-determined | Genome-fragment | NA | NA | 0 |
| S12_k141_50445 | 1566 | No | NA | 2 | 0 | 2 | Not-determined | Genome-fragment | NA | NA | 0 |
| S12_k141_284828 | 1214 | No | NA | 1 | 0 | 0 | Not-determined | Genome-fragment | NA | NA | 0 |
| S12_k141_318246 | 1844 | No | NA | 4 | 0 | 0 | Not-determined | Genome-fragment | NA | NA | 0 |
| S12_k141_351653 | 28451 | No | NA | 36 | 0 | 13 | Not-determined | Genome-fragment | NA | NA | 0 |
| S12_k141_301576 | 1176 | No | NA | 1 | 0 | 0 | Not-determined | Genome-fragment | NA | NA | 0 |
| S5-final-viral-combined_k141_172358\|\|full | 3223 | No | NA | 4 | 2 | 0 | Low-quality | Genome-fragment | 7.09 | AAI-based (high-confidence) | 0 |
| S17-final-viral-combined_k141_234546\|\|full | 3223 | No | NA | 5 | 1 | 0 | Low-quality | Genome-fragment | 4.88 | HMM-based (lower-bound) | 0 |
| S12_k141_117385 | 1330 | No | NA | 2 | 0 | 1 | Not-determined | Genome-fragment | NA | NA | 0 |
| S12_k141_334897 | 1298 | No | NA | 4 | 0 | 4 | Not-determined | Genome-fragment | NA | NA | 0 |
| S12_k141_368473 | 1351 | No | NA | 1 | 0 | 1 | Not-determined | Genome-fragment | NA | NA | 0 |
| S12_k141_67216 | 1227 | No | NA | 2 | 0 | 0 | Not-determined | Genome-fragment | NA | NA | 0 |
| S12_k141_334902 | 4424 | No | NA | 1 | 0 | 1 | Not-determined | Genome-fragment | NA | NA | 0 |
| S12_k141_385271 | 15581 | No | NA | 32 | 0 | 29 | Not-determined | Genome-fragment | NA | NA | 0 |
| S12_k141_100688 | 3128 | No | NA | 1 | 0 | 1 | Not-determined | Genome-fragment | NA | NA | 0 |
| S12_k141_67243 | 1420 | No | NA | 1 | 0 | 1 | Not-determined | Genome-fragment | NA | NA | 0 |
| S12_k141_267999 | 1183 | No | NA | 1 | 0 | 1 | Not-determined | Genome-fragment | NA | NA | 0 |
| S12_k141_351710 | 1319 | No | NA | 1 | 0 | 1 | Not-determined | Genome-fragment | NA | NA | 0 |
| S2-final-viral-combined_k141_158119\|\|full | 3223 | No | NA | 8 | 0 | 0 | Low-quality | Genome-fragment | 8.57 | AAI-based (medium-confidence) | 0 |
| S3_k141_87486 | 3224 | No | NA | 6 | 0 | 0 | Low-quality | Genome-fragment | 3.05 | AAI-based (high-confidence) | 0 |
| S12_k141_234576 | 1048 | No | NA | 1 | 0 | 0 | Not-determined | Genome-fragment | NA | NA | 0 |
| S12_k141_100730 | 5102 | No | NA | 6 | 0 | 5 | Not-determined | Genome-fragment | NA | NA | 0 |
| S11-final-viral-combined_k141_116212\|\|full | 3225 | No | NA | 8 | 4 | 0 | Low-quality | Genome-fragment | 8.97 | AAI-based (high-confidence) | 0 |
| S12_k141_301712 | 2959 | No | NA | 2 | 0 | 2 | Not-determined | Genome-fragment | NA | NA | 0 |
| S12_k141_318401 | 3022 | No | NA | 4 | 0 | 0 | Not-determined | Genome-fragment | NA | NA | 0 |
| S12_k141_268053 | 1859 | No | NA | 2 | 0 | 1 | Not-determined | Genome-fragment | NA | NA | 0 |
| S1-final-viral-combined_k141_25136\|\|full | 3225 | No | NA | 4 | 0 | 0 | Low-quality | Genome-fragment | 2.21 | AAI-based (medium-confidence) | 0 |
| S12_k141_67308 | 1933 | No | NA | 3 | 0 | 1 | Not-determined | Genome-fragment | NA | NA | 0 |
| S12_k141_334943 | 2140 | No | NA | 1 | 0 | 0 | Not-determined | Genome-fragment | NA | NA | 0 |
| S12_k141_334957 | 2747 | No | NA | 5 | 0 | 5 | Not-determined | Genome-fragment | NA | NA | 0 |
| S12_k141_318428 | 1213 | No | NA | 2 | 0 | 0 | Not-determined | Genome-fragment | NA | NA | 0 |
| S1-final-viral-combined_k141_157791\|\|full | 3225 | No | NA | 4 | 0 | 3 | Low-quality | Genome-fragment | 1.02 | AAI-based (medium-confidence) | 0 |
| S7-final-viral-combined_k141_12554\|\|full | 3225 | Yes | 2361 | 5 | 2 | 1 | Low-quality | Genome-fragment | 2.9 | HMM-based (lower-bound) | 26.79 |
| S12_k141_134294 | 1358 | No | NA | 1 | 0 | 1 | Not-determined | Genome-fragment | NA | NA | 0 |
| S12_k141_167780 | 1367 | No | NA | 1 | 0 | 0 | Not-determined | Genome-fragment | NA | NA | 0 |
| S12_k141_50619 | 1029 | No | NA | 1 | 0 | 0 | Not-determined | Genome-fragment | NA | NA | 0 |
| S12_k141_318456 | 1213 | No | NA | 1 | 0 | 0 | Not-determined | Genome-fragment | NA | NA | 0 |
| S12_k141_201146 | 1497 | No | NA | 2 | 0 | 1 | Not-determined | Genome-fragment | NA | NA | 0 |
| S12_k141_268091 | 1796 | No | NA | 1 | 0 | 0 | Not-determined | Genome-fragment | NA | NA | 0 |
| S12_k141_17160 | 2468 | No | NA | 5 | 0 | 1 | Not-determined | Genome-fragment | NA | NA | 0 |
| S12_k141_50633 | 8375 | No | NA | 3 | 0 | 0 | Not-determined | Genome-fragment | NA | NA | 0 |
| S12_k141_33998 | 1300 | No | NA | 1 | 0 | 0 | Not-determined | Genome-fragment | NA | NA | 0 |
| S12_k141_34023 | 1028 | No | NA | 2 | 0 | 2 | Not-determined | Genome-fragment | NA | NA | 0 |
| S12_k141_234725 | 1268 | No | NA | 2 | 0 | 1 | Not-determined | Genome-fragment | NA | NA | 0 |
| S5-final-viral-combined_k141_161547\|\|full | 3227 | No | NA | 3 | 0 | 0 | Low-quality | Genome-fragment | 3.36 | AAI-based (high-confidence) | 0 |
| S12_k141_471 | 7288 | No | NA | 5 | 0 | 1 | Not-determined | Genome-fragment | NA | NA | 0 |
| S12_k141_217997 | 5874 | No | NA | 4 | 0 | 2 | Not-determined | Genome-fragment | NA | NA | 0 |
| S12_k141_268134 | 1305 | No | NA | 2 | 0 | 1 | Not-determined | Genome-fragment | NA | NA | 0 |
| S12_k141_234749 | 1175 | No | NA | 1 | 0 | 1 | Not-determined | Genome-fragment | NA | NA | 0 |
| S17-final-viral-combined_k141_111257\|\|full | 3228 | No | NA | 4 | 0 | 2 | Low-quality | Genome-fragment | 2.35 | AAI-based (medium-confidence) | 0 |
| S12_k141_17242 | 3671 | No | NA | 9 | 0 | 8 | Not-determined | Genome-fragment | NA | NA | 0 |
| S12_k141_50712 | 7040 | No | NA | 7 | 0 | 0 | Not-determined | Genome-fragment | NA | NA | 0 |
| S12_k141_285183 | 2929 | No | NA | 1 | 0 | 0 | Not-determined | Genome-fragment | NA | NA | 0 |
| S12_k141_50714 | 2884 | No | NA | 3 | 0 | 3 | Not-determined | Genome-fragment | NA | NA | 0 |
| S12_k141_67390 | 1017 | No | NA | 1 | 0 | 0 | Not-determined | Genome-fragment | NA | NA | 0 |
| S12_k141_351747 | 4005 | No | NA | 1 | 0 | 0 | Not-determined | Genome-fragment | NA | NA | 0 |
| S12_k141_318600 | 1185 | No | NA | 1 | 0 | 0 | Not-determined | Genome-fragment | NA | NA | 0 |
| S12_k141_60072 | 3228 | No | NA | 4 | 1 | 0 | Low-quality | Genome-fragment | 8.79 | AAI-based (high-confidence) | 0 |
| S12_k141_318618 | 2449 | No | NA | 2 | 0 | 1 | Not-determined | Genome-fragment | NA | NA | 0 |
| S12_k141_50772 | 1614 | No | NA | 1 | 0 | 1 | Not-determined | Genome-fragment | NA | NA | 0 |
| S17-final-viral-combined_k141_20713\|\|full | 3229 | No | NA | 6 | 0 | 1 | Low-quality | Genome-fragment | 6.58 | AAI-based (medium-confidence) | 0 |
| S12_k141_318651 | 6133 | No | NA | 5 | 0 | 5 | Not-determined | Genome-fragment | NA | NA | 0 |
| S12_k141_318659 | 1247 | No | NA | 1 | 0 | 1 | Not-determined | Genome-fragment | NA | NA | 0 |
| S12_k141_368713 | 2764 | No | NA | 2 | 0 | 2 | Not-determined | Genome-fragment | NA | NA | 0 |
| S1-final-viral-combined_k141_392693\|\|full | 3230 | No | NA | 3 | 0 | 2 | Low-quality | Genome-fragment | 2.24 | AAI-based (high-confidence) | 0 |
| S12_k141_201304 | 1196 | No | NA | 1 | 0 | 0 | Not-determined | Genome-fragment | NA | NA | 0 |
| S12_k141_234838 | 5005 | No | NA | 7 | 0 | 0 | Not-determined | Genome-fragment | NA | NA | 0 |
| S12_k141_285256 | 2214 | No | NA | 1 | 0 | 1 | Not-determined | Genome-fragment | NA | NA | 0 |
| S12_k141_100979 | 8392 | No | NA | 6 | 0 | 4 | Not-determined | Genome-fragment | NA | NA | 0 |
| S12_k141_285264 | 1670 | No | NA | 2 | 0 | 2 | Not-determined | Genome-fragment | NA | NA | 0 |
| S12_k141_285281 | 1857 | No | NA | 2 | 0 | 1 | Not-determined | Genome-fragment | NA | NA | 0 |
| S7-final-viral-combined_k141_175740\|\|full | 3230 | Yes | 720 | 4 | 1 | 3 | Low-quality | Genome-fragment | 0.31 | HMM-based (lower-bound) | 77.71 |
| S12_k141_285298 | 1186 | No | NA | 1 | 0 | 0 | Not-determined | Genome-fragment | NA | NA | 0 |
| S12_k141_682 | 5257 | No | NA | 5 | 0 | 2 | Not-determined | Genome-fragment | NA | NA | 0 |
| S12_k141_67502 | 1004 | No | NA | 2 | 0 | 0 | Not-determined | Genome-fragment | NA | NA | 0 |
| S12_k141_50871 | 1214 | No | NA | 2 | 0 | 0 | Not-determined | Genome-fragment | NA | NA | 0 |
| S12_k141_335137 | 5322 | No | NA | 6 | 0 | 6 | Not-determined | Genome-fragment | NA | NA | 0 |
| S7_k141_223725 | 3230 | No | NA | 3 | 0 | 3 | Low-quality | Genome-fragment | 1.01 | AAI-based (medium-confidence) | 0 |
| S5-final-viral-combined_k141_132138\|\|full | 3231 | No | NA | 3 | 0 | 1 | Low-quality | Genome-fragment | 20.86 | AAI-based (medium-confidence) | 0 |
| S12_k141_218132 | 1267 | No | NA | 1 | 0 | 1 | Not-determined | Genome-fragment | NA | NA | 0 |
| S12_k141_117504 | 1382 | No | NA | 1 | 0 | 1 | Not-determined | Genome-fragment | NA | NA | 0 |
| S12_k141_17412 | 4301 | No | NA | 5 | 0 | 1 | Not-determined | Genome-fragment | NA | NA | 0 |
| S7_k141_43109 | 3231 | No | NA | 2 | 0 | 0 | Low-quality | Genome-fragment | 11.2 | AAI-based (medium-confidence) | 0 |
| S12_k141_34238 | 1333 | No | NA | 1 | 0 | 1 | Not-determined | Genome-fragment | NA | NA | 0 |
| S12_k141_117514 | 1158 | No | NA | 1 | 0 | 1 | Not-determined | Genome-fragment | NA | NA | 0 |
| S12_k141_251676 | 4919 | No | NA | 4 | 0 | 4 | Not-determined | Genome-fragment | NA | NA | 0 |
| S12_k141_117537 | 6035 | No | NA | 5 | 0 | 4 | Not-determined | Genome-fragment | NA | NA | 0 |
| S12_k141_117542 | 1248 | No | NA | 4 | 0 | 0 | Not-determined | Genome-fragment | NA | NA | 0 |
| S12_k141_17504 | 3048 | No | NA | 4 | 0 | 1 | Not-determined | Genome-fragment | NA | NA | 0 |
| S12_k141_117577 | 2198 | No | NA | 2 | 0 | 0 | Not-determined | Genome-fragment | NA | NA | 0 |
| S12-final-viral-combined_k141_122876\|\|full | 3231 | No | NA | 4 | 0 | 2 | Low-quality | Genome-fragment | 9.24 | AAI-based (medium-confidence) | 0 |
| S12_k141_134671 | 1021 | No | NA | 1 | 0 | 1 | Not-determined | Genome-fragment | NA | NA | 0 |
| S12_k141_268342 | 3415 | No | NA | 5 | 0 | 0 | Not-determined | Genome-fragment | NA | NA | 0 |
| S12_k141_334728 | 3231 | No | NA | 1 | 0 | 0 | Low-quality | Genome-fragment | 4.79 | AAI-based (medium-confidence) | 0 |
| S8_k141_145278 | 3231 | No | NA | 1 | 0 | 1 | Low-quality | Genome-fragment | 3.81 | AAI-based (medium-confidence) | 0 |
| S12_k141_268359 | 1018 | No | NA | 1 | 0 | 0 | Not-determined | Genome-fragment | NA | NA | 0 |
| S12_k141_117610 | 1309 | No | NA | 2 | 0 | 1 | Not-determined | Genome-fragment | NA | NA | 0 |
| S12_k141_151119 | 1666 | No | NA | 1 | 0 | 0 | Not-determined | Genome-fragment | NA | NA | 0 |
| S12_k141_335203 | 20405 | No | NA | 24 | 0 | 7 | Not-determined | Genome-fragment | NA | NA | 0 |
| S12_k141_84380 | 3723 | No | NA | 1 | 0 | 1 | Not-determined | Genome-fragment | NA | NA | 0 |
| S12_k141_182590 | 3232 | No | NA | 3 | 0 | 2 | Low-quality | Genome-fragment | 1.5 | AAI-based (high-confidence) | 0 |
| S5-final-viral-combined_k141_172580\|\|full | 3233 | No | NA | 7 | 0 | 2 | Low-quality | Genome-fragment | 5.74 | AAI-based (medium-confidence) | 0 |
| S12_k141_385613 | 1222 | No | NA | 1 | 0 | 1 | Not-determined | Genome-fragment | NA | NA | 0 |
| S12_k141_134694 | 1374 | No | NA | 1 | 0 | 0 | Not-determined | Genome-fragment | NA | NA | 0 |
| S12-final-viral-combined_k141_248341\|\|full | 3233 | No | NA | 6 | 0 | 1 | Low-quality | Genome-fragment | 7.98 | AAI-based (medium-confidence) | 0 |
| S12_k141_201563 | 2163 | No | NA | 2 | 0 | 1 | Not-determined | Genome-fragment | NA | NA | 0 |
| S9-final-viral-combined_k141_76002\|\|lt2gene | 3234 | No | NA | 1 | 1 | 0 | Low-quality | Genome-fragment | 2.24 | HMM-based (lower-bound) | 0 |
| S17-final-viral-combined_k141_38838\|\|full | 3234 | No | NA | 8 | 0 | 0 | Low-quality | Genome-fragment | 6.05 | AAI-based (high-confidence) | 0 |
| S11-final-viral-combined_k141_295430\|\|full | 3235 | No | NA | 4 | 2 | 0 | Low-quality | Genome-fragment | 10.88 | AAI-based (high-confidence) | 0 |
| S12_k141_218255 | 1220 | No | NA | 1 | 0 | 0 | Not-determined | Genome-fragment | NA | NA | 0 |
| S12_k141_335241 | 1812 | No | NA | 4 | 0 | 1 | Not-determined | Genome-fragment | NA | NA | 0 |
| S12_k141_151225 | 1159 | No | NA | 1 | 0 | 1 | Not-determined | Genome-fragment | NA | NA | 0 |
| S6_k141_64331 | 3235 | No | NA | 3 | 0 | 0 | Low-quality | Genome-fragment | 4.22 | AAI-based (medium-confidence) | 0 |
| S12_k141_201619 | 7315 | Yes | 1865 | 8 | 1 | 5 | Not-determined | Genome-fragment | NA | NA | 74.5 |
| S12_k141_34369 | 5173 | No | NA | 12 | 0 | 0 | Not-determined | Genome-fragment | NA | NA | 0 |
| S7-final-viral-combined_k141_198537\|\|full | 3236 | No | NA | 5 | 2 | 0 | Low-quality | Genome-fragment | 3.04 | AAI-based (high-confidence) | 0 |
| S12_k141_368977 | 1732 | No | NA | 2 | 0 | 1 | Not-determined | Genome-fragment | NA | NA | 0 |
| S12_k141_34398 | 3263 | No | NA | 6 | 0 | 2 | Not-determined | Genome-fragment | NA | NA | 0 |
| S12_k141_51065 | 4759 | No | NA | 4 | 0 | 1 | Not-determined | Genome-fragment | NA | NA | 0 |
| S2_k141_24253 | 3236 | No | NA | 4 | 0 | 2 | Low-quality | Genome-fragment | 1.66 | AAI-based (medium-confidence) | 0 |
| S2-final-viral-combined_k141_145207\|\|full | 3237 | No | NA | 2 | 0 | 2 | Low-quality | Genome-fragment | 1.03 | AAI-based (medium-confidence) | 0 |
| S12_k141_302103 | 3013 | No | NA | 5 | 0 | 5 | Not-determined | Genome-fragment | NA | NA | 0 |
| S12_k141_235125 | 23589 | No | NA | 16 | 1 | 0 | Medium-quality | Genome-fragment | 55.38 | AAI-based (high-confidence) | 0 |
| S12_k141_201673 | 5153 | No | NA | 4 | 0 | 1 | Not-determined | Genome-fragment | NA | NA | 0 |
| S12_k141_101153 | 2254 | No | NA | 3 | 0 | 1 | Not-determined | Genome-fragment | NA | NA | 0 |
| S12_k141_268553 | 1428 | No | NA | 3 | 0 | 0 | Not-determined | Genome-fragment | NA | NA | 0 |
| S2_k141_114224 | 3237 | Yes | 2189 | 5 | 1 | 1 | Low-quality | Genome-fragment | 3.92 | AAI-based (high-confidence) | 32.38 |
| S12_k141_369055 | 1946 | No | NA | 2 | 0 | 2 | Not-determined | Genome-fragment | NA | NA | 0 |
| S12_k141_101207 | 2638 | No | NA | 2 | 0 | 2 | Not-determined | Genome-fragment | NA | NA | 0 |
| S8-final-viral-combined_k141_111114\|\|full | 3237 | No | NA | 3 | 0 | 1 | Low-quality | Genome-fragment | 6.2 | AAI-based (medium-confidence) | 0 |
| S11-final-viral-combined_k141_248146\|\|full | 3238 | No | NA | 5 | 0 | 1 | Low-quality | Genome-fragment | 1.59 | AAI-based (medium-confidence) | 0 |
| S7-final-viral-combined_k141_34963\|\|full | 3238 | No | NA | 7 | 0 | 1 | Low-quality | Genome-fragment | 8.9 | AAI-based (medium-confidence) | 0 |
| S12-final-viral-combined_k141_47180\|\|full | 3238 | No | NA | 6 | 0 | 0 | Low-quality | Genome-fragment | 7.03 | AAI-based (medium-confidence) | 0 |
| S12_k141_285662 | 2744 | No | NA | 1 | 0 | 1 | Not-determined | Genome-fragment | NA | NA | 0 |
| S17-k141_170969 | 3239 | No | NA | 3 | 0 | 1 | Low-quality | Genome-fragment | 2.75 | AAI-based (high-confidence) | 0 |
| S12_k141_352070 | 1274 | No | NA | 1 | 0 | 1 | Not-determined | Genome-fragment | NA | NA | 0 |
| S12_k141_201706 | 2184 | No | NA | 2 | 0 | 1 | Not-determined | Genome-fragment | NA | NA | 0 |
| S12_k141_17717 | 4321 | No | NA | 6 | 0 | 2 | Not-determined | Genome-fragment | NA | NA | 0 |
| S12_k141_251853 | 1291 | No | NA | 4 | 0 | 0 | Not-determined | Genome-fragment | NA | NA | 0 |
| S12_k141_285727 | 1522 | No | NA | 1 | 0 | 1 | Not-determined | Genome-fragment | NA | NA | 0 |
| S12_k141_352087 | 1232 | No | NA | 1 | 0 | 0 | Not-determined | Genome-fragment | NA | NA | 0 |
| S8-final-viral-combined_k141_105589\|\|full | 3239 | No | NA | 8 | 1 | 0 | Low-quality | Genome-fragment | 8.72 | AAI-based (high-confidence) | 0 |
| S12_k141_302224 | 2598 | No | NA | 1 | 0 | 1 | Not-determined | Genome-fragment | NA | NA | 0 |
| S5_k141_209903 | 3240 | No | NA | 6 | 1 | 0 | Low-quality | Genome-fragment | 5.43 | AAI-based (medium-confidence) | 0 |
| S12_k141_235221 | 3545 | No | NA | 10 | 0 | 9 | Not-determined | Genome-fragment | NA | NA | 0 |
| S12_k141_268651 | 2278 | No | NA | 1 | 0 | 0 | Not-determined | Genome-fragment | NA | NA | 0 |
| S17-k141_103752 | 3240 | No | NA | 6 | 1 | 0 | Low-quality | Genome-fragment | 5.03 | AAI-based (high-confidence) | 0 |
| S8_k141_71716 | 3241 | No | NA | 4 | 1 | 0 | Low-quality | Genome-fragment | 3.95 | AAI-based (high-confidence) | 0 |
| S7-final-viral-combined_k141_163551\|\|full | 3242 | No | NA | 3 | 0 | 2 | Low-quality | Genome-fragment | 10.9 | AAI-based (medium-confidence) | 0 |
| S2-final-viral-combined_k141_71295\|\|full | 3242 | No | NA | 4 | 2 | 0 | Low-quality | Genome-fragment | 8.83 | AAI-based (high-confidence) | 0 |
| S12_k141_268753 | 2761 | No | NA | 2 | 0 | 1 | Not-determined | Genome-fragment | NA | NA | 0 |
| S12_k141_268764 | 1100 | No | NA | 2 | 0 | 0 | Not-determined | Genome-fragment | NA | NA | 0 |
| S12_k141_134864 | 5842 | No | NA | 7 | 0 | 1 | Not-determined | Genome-fragment | NA | NA | 0 |
| S12_k141_268771 | 2261 | No | NA | 2 | 0 | 0 | Not-determined | Genome-fragment | NA | NA | 0 |
| S12_k141_34724 | 8596 | No | NA | 11 | 0 | 9 | Not-determined | Genome-fragment | NA | NA | 0 |
| S12_k141_168338 | 3322 | No | NA | 1 | 0 | 0 | Not-determined | Genome-fragment | NA | NA | 0 |
| S12_k141_101370 | 15744 | No | NA | 15 | 0 | 5 | Not-determined | Genome-fragment | NA | NA | 0 |
| S12_k141_17950 | 11870 | No | NA | 19 | 7 | 0 | Medium-quality | Genome-fragment | 89.5 | AAI-based (high-confidence) | 0 |
| S2-final-viral-combined_k141_225691\|\|full | 3242 | No | NA | 5 | 1 | 0 | Low-quality | Genome-fragment | 2.66 | HMM-based (lower-bound) | 0 |
| S12_k141_201818 | 1093 | No | NA | 1 | 0 | 1 | Not-determined | Genome-fragment | NA | NA | 0 |
| S12_k141_335526 | 1012 | No | NA | 2 | 0 | 0 | Not-determined | Genome-fragment | NA | NA | 0 |
| S11-final-viral-combined_k141_247407\|\|full | 3243 | No | NA | 6 | 0 | 0 | Low-quality | Genome-fragment | 6.98 | AAI-based (high-confidence) | 0 |
| S12_k141_67919 | 1157 | No | NA | 1 | 0 | 1 | Not-determined | Genome-fragment | NA | NA | 0 |
| S12_k141_201837 | 1141 | No | NA | 1 | 0 | 0 | Not-determined | Genome-fragment | NA | NA | 0 |
| S2-final-viral-combined_k141_129338\|\|full | 3243 | No | NA | 6 | 0 | 0 | Low-quality | Genome-fragment | 3.91 | AAI-based (high-confidence) | 0 |
| S12_k141_335549 | 1092 | No | NA | 1 | 0 | 0 | Not-determined | Genome-fragment | NA | NA | 0 |
| S12_k141_268820 | 10662 | No | NA | 11 | 0 | 8 | Not-determined | Genome-fragment | NA | NA | 0 |
| S12_k141_34767 | 3028 | No | NA | 8 | 0 | 8 | Not-determined | Genome-fragment | NA | NA | 0 |
| S12_k141_369162 | 1494 | No | NA | 1 | 0 | 1 | Not-determined | Genome-fragment | NA | NA | 0 |
| S12_k141_352255 | 1157 | No | NA | 1 | 0 | 0 | Not-determined | Genome-fragment | NA | NA | 0 |
| S12_k141_134937 | 3108 | No | NA | 3 | 0 | 3 | Not-determined | Genome-fragment | NA | NA | 0 |
| S6_k141_40182 | 3243 | No | NA | 4 | 0 | 0 | Low-quality | Genome-fragment | 3.43 | AAI-based (high-confidence) | 0 |
| S12_k141_201878 | 2041 | No | NA | 1 | 0 | 1 | Not-determined | Genome-fragment | NA | NA | 0 |
| S12_k141_185197 | 1415 | No | NA | 1 | 0 | 1 | Not-determined | Genome-fragment | NA | NA | 0 |
| S12_k141_101427 | 7039 | No | NA | 6 | 0 | 3 | Not-determined | Genome-fragment | NA | NA | 0 |
| S12_k141_335612 | 5661 | No | NA | 7 | 0 | 4 | Not-determined | Genome-fragment | NA | NA | 0 |
| S12_k141_84776 | 3391 | No | NA | 2 | 0 | 0 | Not-determined | Genome-fragment | NA | NA | 0 |
| S8_k141_224085 | 3243 | No | NA | 7 | 5 | 0 | Low-quality | Genome-fragment | 10.08 | AAI-based (high-confidence) | 0 |
| S12_k141_101447 | 1950 | No | NA | 1 | 0 | 1 | Not-determined | Genome-fragment | NA | NA | 0 |
| S8_k141_120545 | 3243 | No | NA | 3 | 1 | 0 | Low-quality | Genome-fragment | 2.43 | AAI-based (high-confidence) | 0 |
| S12_k141_168427 | 1729 | No | NA | 3 | 0 | 1 | Not-determined | Genome-fragment | NA | NA | 0 |
| S12_k141_34835 | 1009 | No | NA | 1 | 0 | 0 | Not-determined | Genome-fragment | NA | NA | 0 |
| S12_k141_352336 | 2001 | No | NA | 2 | 0 | 1 | Not-determined | Genome-fragment | NA | NA | 0 |
| S12_k141_1193 | 2571 | No | NA | 1 | 0 | 1 | Not-determined | Genome-fragment | NA | NA | 0 |
| S12_k141_302419 | 3415 | No | NA | 2 | 0 | 0 | Not-determined | Genome-fragment | NA | NA | 0 |
| S12_k141_117951 | 1344 | No | NA | 1 | 0 | 1 | Not-determined | Genome-fragment | NA | NA | 0 |
| S12_k141_101474 | 3255 | No | NA | 3 | 0 | 0 | Not-determined | Genome-fragment | NA | NA | 0 |
| S11-final-viral-combined_k141_266398\|\|full | 3244 | No | NA | 4 | 0 | 2 | Low-quality | Genome-fragment | 1.73 | AAI-based (medium-confidence) | 0 |
| S12_k141_319215 | 1631 | No | NA | 1 | 0 | 0 | Not-determined | Genome-fragment | NA | NA | 0 |
| S12_k141_268929 | 1207 | No | NA | 3 | 0 | 0 | Not-determined | Genome-fragment | NA | NA | 0 |
| S12_k141_1208 | 1278 | No | NA | 1 | 0 | 0 | Not-determined | Genome-fragment | NA | NA | 0 |
| S12-final-viral-combined_k141_252401\|\|full | 3244 | No | NA | 7 | 1 | 0 | Low-quality | Genome-fragment | 4.76 | HMM-based (lower-bound) | 0 |
| S12-final-viral-combined_k141_27496\|\|full | 3244 | No | NA | 6 | 0 | 0 | Low-quality | Genome-fragment | 7.62 | AAI-based (medium-confidence) | 0 |
| S12_k141_319242 | 1429 | No | NA | 1 | 0 | 0 | Not-determined | Genome-fragment | NA | NA | 0 |
| S12_k141_51347 | 1250 | No | NA | 1 | 0 | 1 | Not-determined | Genome-fragment | NA | NA | 0 |
| S12_k141_117981 | 1820 | No | NA | 1 | 0 | 1 | Not-determined | Genome-fragment | NA | NA | 0 |
| S12_k141_84833 | 2683 | No | NA | 6 | 0 | 0 | Not-determined | Genome-fragment | NA | NA | 0 |
| S12_k141_51374 | 4681 | No | NA | 4 | 0 | 1 | Not-determined | Genome-fragment | NA | NA | 0 |
| S12_k141_51376 | 1074 | No | NA | 1 | 0 | 1 | Not-determined | Genome-fragment | NA | NA | 0 |
| S2_k141_43409 | 3244 | No | NA | 5 | 0 | 2 | Low-quality | Genome-fragment | 1.49 | AAI-based (medium-confidence) | 0 |
| S12_k141_51423 | 1137 | No | NA | 1 | 0 | 1 | Not-determined | Genome-fragment | NA | NA | 0 |
| S12_k141_68068 | 2324 | No | NA | 3 | 0 | 2 | Not-determined | Genome-fragment | NA | NA | 0 |

| | | | | | | | | | | | |
|---|---|---|---|---|---|---|---|---|---|---|---|
| S12_k141_18055 | 1154 | No | NA | 1 | 0 | 0 | Not-determined | Genome-fragment | NA | NA | 0 |
| S12-final-viral-combined_k141_329791\|\|full | 3245 | No | NA | 6 | 0 | 0 | Low-quality | Genome-fragment | 7.59 | AAI-based (medium-confidence) | 0 |
| S12_k141_118053 | 3027 | No | NA | 2 | 0 | 2 | Not-determined | Genome-fragment | NA | NA | 0 |
| S12_k141_168543 | 1242 | No | NA | 1 | 0 | 1 | Not-determined | Genome-fragment | NA | NA | 0 |
| S4_k141_35748 | 3245 | No | NA | 2 | 0 | 2 | Low-quality | Genome-fragment | 0.62 | AAI-based (medium-confidence) | 0 |
| S12_k141_185387 | 1068 | No | NA | 1 | 0 | 1 | Not-determined | Genome-fragment | NA | NA | 0 |
| S12_k141_269001 | 6049 | No | NA | 6 | 0 | 6 | Not-determined | Genome-fragment | NA | NA | 0 |
| S11_k141_123858 | 3246 | No | NA | 5 | 4 | 0 | Low-quality | Genome-fragment | 7.93 | AAI-based (high-confidence) | 0 |
| S12_k141_369537 | 4213 | No | NA | 7 | 0 | 6 | Not-determined | Genome-fragment | NA | NA | 0 |
| S12_k141_202017 | 3667 | No | NA | 5 | 0 | 0 | Not-determined | Genome-fragment | NA | NA | 0 |
| S12_k141_51478 | 6878 | No | NA | 9 | 0 | 5 | Not-determined | Genome-fragment | NA | NA | 0 |
| S12-final-viral-combined_k141_155410\|\|full | 3247 | No | NA | 3 | 0 | 2 | Low-quality | Genome-fragment | 1.14 | AAI-based (medium-confidence) | 0 |
| S12_k141_285969 | 1278 | No | NA | 1 | 0 | 0 | Not-determined | Genome-fragment | NA | NA | 0 |
| S12_k141_302588 | 1085 | No | NA | 1 | 0 | 1 | Not-determined | Genome-fragment | NA | NA | 0 |
| S12_k141_319300 | 1070 | No | NA | 1 | 0 | 0 | Not-determined | Genome-fragment | NA | NA | 0 |
| S12_k141_135073 | 1123 | No | NA | 2 | 0 | 1 | Not-determined | Genome-fragment | NA | NA | 0 |
| S12_k141_352538 | 1225 | No | NA | 2 | 0 | 0 | Not-determined | Genome-fragment | NA | NA | 0 |
| S12_k141_202052 | 4652 | No | NA | 2 | 0 | 2 | Not-determined | Genome-fragment | NA | NA | 0 |
| S12_k141_68176 | 2135 | No | NA | 1 | 0 | 0 | Not-determined | Genome-fragment | NA | NA | 0 |
| S12_k141_319344 | 1616 | No | NA | 2 | 0 | 0 | Not-determined | Genome-fragment | NA | NA | 0 |
| S12_k141_1361 | 1211 | No | NA | 1 | 0 | 1 | Not-determined | Genome-fragment | NA | NA | 0 |
| S12_k141_151796 | 2538 | No | NA | 1 | 0 | 0 | Not-determined | Genome-fragment | NA | NA | 0 |
| S12_k141_286005 | 1336 | No | NA | 2 | 0 | 0 | Not-determined | Genome-fragment | NA | NA | 0 |
| S12_k141_85009 | 14995 | No | NA | 16 | 0 | 9 | Not-determined | Genome-fragment | NA | NA | 0 |
| S9-final-viral-combined_k141_126804\|\|full | 3249 | No | NA | 5 | 0 | 1 | Low-quality | Genome-fragment | 6.8 | AAI-based (high-confidence) | 0 |
| S12_k141_335776 | 1505 | No | NA | 2 | 0 | 0 | Not-determined | Genome-fragment | NA | NA | 0 |
| S12_k141_34929 | 3687 | No | NA | 2 | 0 | 1 | Not-determined | Genome-fragment | NA | NA | 0 |
| S12_k141_335781 | 1000 | No | NA | 1 | 0 | 1 | Not-determined | Genome-fragment | NA | NA | 0 |
| S9_k141_59086 | 3249 | No | NA | 5 | 0 | 1 | Low-quality | Genome-fragment | 3.46 | AAI-based (medium-confidence) | 0 |
| S12_k141_168693 | 2346 | No | NA | 1 | 0 | 1 | Not-determined | Genome-fragment | NA | NA | 0 |
| S12_k141_269130 | 1120 | No | NA | 1 | 0 | 0 | Not-determined | Genome-fragment | NA | NA | 0 |
| S12_k141_335797 | 1656 | No | NA | 1 | 0 | 1 | Not-determined | Genome-fragment | NA | NA | 0 |
| S12_k141_18223 | 1213 | No | NA | 2 | 0 | 0 | Not-determined | Genome-fragment | NA | NA | 0 |
| S17-final-viral-combined_k141_8118\|\|full | 3249 | No | NA | 2 | 0 | 1 | Low-quality | Genome-fragment | 2.59 | AAI-based (medium-confidence) | 0 |
| S12_k141_135167 | 1425 | No | NA | 1 | 0 | 0 | Not-determined | Genome-fragment | NA | NA | 0 |
| S12_k141_286045 | 5043 | No | NA | 3 | 0 | 1 | Not-determined | Genome-fragment | NA | NA | 0 |
| S2-final-viral-combined_k141_186421\|\|full | 3249 | No | NA | 5 | 1 | 2 | Low-quality | Genome-fragment | 7.89 | HMM-based (lower-bound) | 0 |
| S2-final-viral-combined_k141_23219\|\|full | 3249 | No | NA | 3 | 0 | 2 | Low-quality | Genome-fragment | 1.62 | AAI-based (medium-confidence) | 0 |
| S12_k141_185545 | 3028 | No | NA | 6 | 0 | 1 | Not-determined | Genome-fragment | NA | NA | 0 |
| S12_k141_218877 | 2208 | No | NA | 2 | 0 | 0 | Not-determined | Genome-fragment | NA | NA | 0 |
| S12_k141_1510 | 1578 | No | NA | 2 | 0 | 1 | Not-determined | Genome-fragment | NA | NA | 0 |
| S12_k141_218883 | 2454 | No | NA | 4 | 0 | 2 | Not-determined | Genome-fragment | NA | NA | 0 |
| S12_k141_202104 | 1453 | No | NA | 2 | 0 | 0 | Not-determined | Genome-fragment | NA | NA | 0 |
| S2-final-viral-combined_k141_171011\|\|full | 3249 | No | NA | 5 | 0 | 0 | Low-quality | Genome-fragment | 6.71 | AAI-based (medium-confidence) | 0 |
| S12_k141_335833 | 1533 | No | NA | 1 | 0 | 1 | Not-determined | Genome-fragment | NA | NA | 0 |
| S12_k141_218891 | 6999 | No | NA | 6 | 0 | 1 | Not-determined | Genome-fragment | NA | NA | 0 |
| S12_k141_386424 | 4502 | No | NA | 4 | 0 | 2 | Not-determined | Genome-fragment | NA | NA | 0 |
| S2-final-viral-combined_k141_195339\|\|full | 3249 | No | NA | 6 | 1 | 0 | Low-quality | Genome-fragment | 7.85 | AAI-based (high-confidence) | 0 |
| S12_k141_68253 | 1166 | No | NA | 2 | 0 | 2 | Not-determined | Genome-fragment | NA | NA | 0 |
| S12_k141_185603 | 1570 | No | NA | 2 | 0 | 0 | Not-determined | Genome-fragment | NA | NA | 0 |
| S12_k141_386457 | 1085 | No | NA | 1 | 0 | 1 | Not-determined | Genome-fragment | NA | NA | 0 |
| S12_k141_235677 | 1087 | No | NA | 2 | 0 | 0 | Not-determined | Genome-fragment | NA | NA | 0 |
| S12_k141_35052 | 1227 | No | NA | 3 | 0 | 0 | Not-determined | Genome-fragment | NA | NA | 0 |
| S12_k141_118217 | 1556 | No | NA | 2 | 0 | 0 | Not-determined | Genome-fragment | NA | NA | 0 |
| S12_k141_185673 | 6014 | No | NA | 1 | 0 | 1 | Not-determined | Genome-fragment | NA | NA | 0 |
| S12_k141_60295 | 3250 | No | NA | 1 | 0 | 1 | Low-quality | Genome-fragment | 2.92 | AAI-based (medium-confidence) | 0 |
| S12_k141_101766 | 2263 | No | NA | 2 | 0 | 1 | Not-determined | Genome-fragment | NA | NA | 0 |
| S12_k141_135291 | 35092 | No | NA | 36 | 1 | 11 | Medium-quality | Genome-fragment | 50.05 | AAI-based (medium-confidence) | 0 |
| S12_k141_151863 | 1141 | No | NA | 2 | 0 | 0 | Not-determined | Genome-fragment | NA | NA | 0 |
| S12_k141_1638 | 4484 | No | NA | 4 | 0 | 3 | Not-determined | Genome-fragment | NA | NA | 0 |
| S12_k141_222930 | 3251 | No | NA | 7 | 0 | 2 | Low-quality | Genome-fragment | 7.7 | AAI-based (medium-confidence) | 0 |
| S12_k141_286188 | 2114 | No | NA | 2 | 0 | 0 | Not-determined | Genome-fragment | NA | NA | 0 |
| S12_k141_352667 | 5722 | No | NA | 4 | 0 | 2 | Not-determined | Genome-fragment | NA | NA | 0 |
| S12_k141_1670 | 3215 | No | NA | 3 | 0 | 1 | Not-determined | Genome-fragment | NA | NA | 0 |
| S12_k141_85206 | 2499 | No | NA | 6 | 0 | 1 | Not-determined | Genome-fragment | NA | NA | 0 |
| S12_k141_1675 | 1763 | No | NA | 1 | 0 | 0 | Not-determined | Genome-fragment | NA | NA | 0 |
| S12_k141_235772 | 1445 | No | NA | 1 | 0 | 1 | Not-determined | Genome-fragment | NA | NA | 0 |
| S12_k141_51684 | 3683 | No | NA | 2 | 0 | 2 | Not-determined | Genome-fragment | NA | NA | 0 |
| S12_k141_319474 | 1214 | No | NA | 3 | 0 | 1 | Not-determined | Genome-fragment | NA | NA | 0 |
| S12_k141_386558 | 1740 | No | NA | 2 | 0 | 0 | Not-determined | Genome-fragment | NA | NA | 0 |
| S12_k141_85225 | 1350 | No | NA | 2 | 0 | 0 | Not-determined | Genome-fragment | NA | NA | 0 |
| S12_k141_269283 | 1708 | No | NA | 3 | 0 | 3 | Not-determined | Genome-fragment | NA | NA | 0 |
| S12_k141_101852 | 1209 | No | NA | 5 | 0 | 3 | Not-determined | Genome-fragment | NA | NA | 0 |
| S12_k141_168913 | 1354 | No | NA | 1 | 0 | 1 | Not-determined | Genome-fragment | NA | NA | 0 |
| S12_k141_269316 | 13979 | No | NA | 8 | 0 | 6 | Not-determined | Genome-fragment | NA | NA | 0 |
| S8-final-viral-combined_k141_147277\|\|full | 3251 | No | NA | 6 | 1 | 0 | Low-quality | Genome-fragment | 19.48 | AAI-based (medium-confidence) | 0 |
| S12_k141_269329 | 3186 | No | NA | 5 | 0 | 3 | Not-determined | Genome-fragment | NA | NA | 0 |
| S12_k141_386576 | 3065 | No | NA | 4 | 0 | 1 | Not-determined | Genome-fragment | NA | NA | 0 |
| S7-final-viral-combined_k141_211380\|\|full | 3252 | No | NA | 5 | 0 | 1 | Low-quality | Genome-fragment | 7.59 | AAI-based (medium-confidence) | 0 |
| S12_k141_386586 | 1211 | No | NA | 1 | 0 | 0 | Not-determined | Genome-fragment | NA | NA | 0 |
| S12_k141_118288 | 3608 | No | NA | 5 | 0 | 3 | Not-determined | Genome-fragment | NA | NA | 0 |
| S12_k141_51772 | 3752 | No | NA | 7 | 0 | 7 | Not-determined | Genome-fragment | NA | NA | 0 |
| S7_k141_123878 | 3252 | No | NA | 4 | 0 | 0 | Low-quality | Genome-fragment | 10.05 | AAI-based (high-confidence) | 0 |
| S12_k141_68422 | 1508 | No | NA | 2 | 0 | 0 | Not-determined | Genome-fragment | NA | NA | 0 |
| S12_k141_168965 | 2884 | No | NA | 3 | 0 | 2 | Not-determined | Genome-fragment | NA | NA | 0 |
| S12_k141_369945 | 2149 | No | NA | 2 | 0 | 2 | Not-determined | Genome-fragment | NA | NA | 0 |
| S12_k141_1786 | 7601 | No | NA | 7 | 0 | 1 | Not-determined | Genome-fragment | NA | NA | 0 |
| S12_k141_252584 | 3028 | No | NA | 1 | 0 | 0 | Not-determined | Genome-fragment | NA | NA | 0 |
| S12_k141_336019 | 1164 | No | NA | 2 | 0 | 0 | Not-determined | Genome-fragment | NA | NA | 0 |
| S12_k141_369991 | 2564 | No | NA | 3 | 0 | 2 | Not-determined | Genome-fragment | NA | NA | 0 |
| S12_k141_302972 | 1071 | No | NA | 1 | 0 | 1 | Not-determined | Genome-fragment | NA | NA | 0 |
| S12_k141_302993 | 1071 | No | NA | 1 | 0 | 1 | Not-determined | Genome-fragment | NA | NA | 0 |
| S12_k141_51826 | 11676 | No | NA | 7 | 0 | 2 | Not-determined | Genome-fragment | NA | NA | 0 |
| S12_k141_319710 | 6227 | No | NA | 2 | 0 | 0 | Not-determined | Genome-fragment | NA | NA | 0 |
| S12_k141_51845 | 1445 | No | NA | 1 | 0 | 1 | Not-determined | Genome-fragment | NA | NA | 0 |
| S12_k141_1849 | 3288 | No | NA | 4 | 0 | 0 | Not-determined | Genome-fragment | NA | NA | 0 |
| S17-final-viral-combined_k141_92730\|\|full | 3252 | No | NA | 7 | 0 | 3 | Low-quality | Genome-fragment | 6.93 | AAI-based (medium-confidence) | 0 |
| S12_k141_219167 | 1086 | No | NA | 1 | 0 | 0 | Not-determined | Genome-fragment | NA | NA | 0 |
| S12_k141_352846 | 67870 | No | NA | 78 | 5 | 0 | High-quality | High-quality | 98.66 | AAI-based (high-confidence) | 0 |
| S1_k141_149751 | 3253 | No | NA | 4 | 0 | 0 | Low-quality | Genome-fragment | 4.95 | AAI-based (medium-confidence) | 0 |
| S12_k141_1871 | 1040 | No | NA | 1 | 0 | 1 | Not-determined | Genome-fragment | NA | NA | 0 |
| S12_k141_386692 | 40153 | No | NA | 53 | 8 | 1 | Complete | High-quality | 100 | DTR (high-confidence) | 0 |
| S12_k141_51854 | 1263 | No | NA | 1 | 0 | 0 | Not-determined | Genome-fragment | NA | NA | 0 |
| S12_k141_370050 | 2120 | No | NA | 4 | 0 | 2 | Not-determined | Genome-fragment | NA | NA | 0 |
| S12_k141_169042 | 4320 | No | NA | 2 | 0 | 2 | Not-determined | Genome-fragment | NA | NA | 0 |
| S12_k141_151046 | 3255 | No | NA | 5 | 0 | 0 | Low-quality | Genome-fragment | 6.88 | AAI-based (medium-confidence) | 0 |
| S12_k141_386704 | 2798 | No | NA | 4 | 0 | 3 | Not-determined | Genome-fragment | NA | NA | 0 |
| S8_k141_6190 | 3255 | No | NA | 2 | 0 | 2 | Low-quality | Genome-fragment | 0.74 | AAI-based (medium-confidence) | 0 |
| S8_k141_43671 | 3255 | No | NA | 3 | 0 | 1 | Low-quality | Genome-fragment | 7 | AAI-based (medium-confidence) | 0 |
| S12_k141_286341 | 3426 | No | NA | 1 | 0 | 1 | Not-determined | Genome-fragment | NA | NA | 0 |
| S12_k141_286348 | 1334 | No | NA | 1 | 0 | 1 | Not-determined | Genome-fragment | NA | NA | 0 |
| S12_k141_169087 | 1508 | No | NA | 2 | 0 | 2 | Not-determined | Genome-fragment | NA | NA | 0 |
| S12_k141_85409 | 1183 | No | NA | 2 | 0 | 0 | Not-determined | Genome-fragment | NA | NA | 0 |
| S12_k141_51890 | 1167 | No | NA | 1 | 0 | 1 | Not-determined | Genome-fragment | NA | NA | 0 |
| S12_k141_102048 | 1134 | No | NA | 1 | 0 | 1 | Not-determined | Genome-fragment | NA | NA | 0 |
| S12_k141_352918 | 1451 | No | NA | 2 | 0 | 1 | Not-determined | Genome-fragment | NA | NA | 0 |
| S12_k141_35450 | 1502 | No | NA | 2 | 0 | 2 | Not-determined | Genome-fragment | NA | NA | 0 |
| S12_k141_269439 | 5518 | No | NA | 6 | 0 | 2 | Not-determined | Genome-fragment | NA | NA | 0 |
| S3-final-viral-combined_k141_99377\|\|full | 3256 | No | NA | 6 | 0 | 1 | Low-quality | Genome-fragment | 5.44 | AAI-based (medium-confidence) | 0 |
| S12_k141_35453 | 1513 | No | NA | 2 | 0 | 1 | Not-determined | Genome-fragment | NA | NA | 0 |
| S12_k141_102071 | 1054 | No | NA | 1 | 0 | 0 | Not-determined | Genome-fragment | NA | NA | 0 |
| S12_k141_252746 | 1122 | No | NA | 2 | 0 | 2 | Not-determined | Genome-fragment | NA | NA | 0 |
| S12_k141_152304 | 3837 | No | NA | 5 | 0 | 0 | Not-determined | Genome-fragment | NA | NA | 0 |
| S12_k141_386795 | 1193 | No | NA | 1 | 0 | 1 | Not-determined | Genome-fragment | NA | NA | 0 |
| S17-final-viral-combined_k141_35846\|\|full | 3256 | No | NA | 3 | 0 | 2 | Low-quality | Genome-fragment | 7.89 | AAI-based (medium-confidence) | 0 |
| S12_k141_68615 | 3850 | No | NA | 6 | 0 | 2 | Not-determined | Genome-fragment | NA | NA | 0 |
| S12_k141_336176 | 1023 | No | NA | 1 | 0 | 0 | Not-determined | Genome-fragment | NA | NA | 0 |
| S12_k141_236192 | 1275 | No | NA | 2 | 0 | 1 | Not-determined | Genome-fragment | NA | NA | 0 |
| S2_k141_54886 | 3256 | No | NA | 5 | 0 | 1 | Low-quality | Genome-fragment | 7.37 | AAI-based (high-confidence) | 0 |
| S12_k141_118532 | 6869 | No | NA | 11 | 0 | 4 | Not-determined | Genome-fragment | NA | NA | 0 |
| S12_k141_303254 | 1035 | No | NA | 1 | 0 | 0 | Not-determined | Genome-fragment | NA | NA | 0 |
| S12_k141_303262 | 1305 | No | NA | 1 | 0 | 0 | Not-determined | Genome-fragment | NA | NA | 0 |
| S12_k141_386860 | 1105 | No | NA | 1 | 0 | 1 | Not-determined | Genome-fragment | NA | NA | 0 |
| S12_k141_169151 | 1536 | No | NA | 1 | 0 | 1 | Not-determined | Genome-fragment | NA | NA | 0 |
| S1-final-viral-combined_k141_91687\|\|full | 3257 | No | NA | 5 | 0 | 1 | Low-quality | Genome-fragment | 7.28 | AAI-based (medium-confidence) | 0 |
| S12_k141_219390 | 1122 | No | NA | 1 | 0 | 1 | Not-determined | Genome-fragment | NA | NA | 0 |
| S12_k141_286577 | 3361 | No | NA | 4 | 0 | 1 | Not-determined | Genome-fragment | NA | NA | 0 |
| S12_k141_303333 | 2732 | No | NA | 2 | 0 | 2 | Not-determined | Genome-fragment | NA | NA | 0 |
| S12_k141_303341 | 3120 | No | NA | 5 | 0 | 1 | Not-determined | Genome-fragment | NA | NA | 0 |
| S12_k141_202461 | 1600 | No | NA | 1 | 0 | 1 | Not-determined | Genome-fragment | NA | NA | 0 |
| S4_k141_96371 | 3257 | No | NA | 6 | 2 | 0 | Low-quality | Genome-fragment | 3.31 | AAI-based (high-confidence) | 0 |
| S12_k141_286599 | 1817 | No | NA | 2 | 0 | 1 | Not-determined | Genome-fragment | NA | NA | 0 |
| S12_k141_68668 | 1615 | No | NA | 3 | 0 | 0 | Not-determined | Genome-fragment | NA | NA | 0 |
| S12_k141_85533 | 1374 | No | NA | 2 | 0 | 1 | Not-determined | Genome-fragment | NA | NA | 0 |
| S12_k141_319984 | 1037 | No | NA | 1 | 0 | 0 | Not-determined | Genome-fragment | NA | NA | 0 |
| S9_k141_53203 | 3258 | No | NA | 4 | 0 | 1 | Low-quality | Genome-fragment | 0.86 | AAI-based (medium-confidence) | 0 |
| S12_k141_336229 | 1247 | No | NA | 2 | 0 | 2 | Not-determined | Genome-fragment | NA | NA | 0 |
| S12_k141_303370 | 10384 | No | NA | 13 | 0 | 2 | Not-determined | Genome-fragment | NA | NA | 0 |
| S12_k141_319993 | 3329 | No | NA | 2 | 0 | 2 | Not-determined | Genome-fragment | NA | NA | 0 |
| S12_k141_68746 | 1466 | No | NA | 1 | 0 | 1 | Not-determined | Genome-fragment | NA | NA | 0 |
| S12_k141_386949 | 1222 | No | NA | 1 | 0 | 1 | Not-determined | Genome-fragment | NA | NA | 0 |
| S11_k141_245860 | 3258 | Yes | 2620 | 6 | 1 | 1 | Low-quality | Genome-fragment | 5.66 | AAI-based (medium-confidence) | 19.58 |
| S12_k141_186086 | 2093 | No | NA | 1 | 0 | 1 | Not-determined | Genome-fragment | NA | NA | 0 |
| S12_k141_353127 | 1612 | No | NA | 1 | 0 | 1 | Not-determined | Genome-fragment | NA | NA | 0 |
| S12_k141_85599 | 1044 | No | NA | 1 | 0 | 0 | Not-determined | Genome-fragment | NA | NA | 0 |
| S12_k141_85609 | 1107 | No | NA | 1 | 0 | 1 | Not-determined | Genome-fragment | NA | NA | 0 |
| S1-final-viral-combined_k141_24492\|\|full | 3258 | No | NA | 3 | 0 | 1 | Low-quality | Genome-fragment | 7.1 | AAI-based (medium-confidence) | 0 |
| S7-final-viral-combined_k141_217392\|\|lt2gene | 3258 | No | NA | 2 | 1 | 0 | Low-quality | Genome-fragment | 5.94 | AAI-based (high-confidence) | 0 |
| S12_k141_320059 | 3095 | No | NA | 7 | 0 | 3 | Not-determined | Genome-fragment | NA | NA | 0 |
| S12_k141_320061 | 1252 | No | NA | 1 | 0 | 0 | Not-determined | Genome-fragment | NA | NA | 0 |
| S12_k141_303436 | 1146 | No | NA | 2 | 0 | 0 | Not-determined | Genome-fragment | NA | NA | 0 |
| S12_k141_85641 | 1559 | No | NA | 2 | 0 | 1 | Not-determined | Genome-fragment | NA | NA | 0 |
| S2-final-viral-combined_k141_234743\|\|full | 3258 | No | NA | 6 | 0 | 0 | Low-quality | Genome-fragment | 6.09 | AAI-based (high-confidence) | 0 |
| S8-final-viral-combined_k141_61239\|\|full | 3259 | No | NA | 3 | 1 | 0 | Low-quality | Genome-fragment | 2.06 | HMM-based (lower-bound) | 0 |
| S12_k141_370389 | 3273 | No | NA | 4 | 0 | 0 | Not-determined | Genome-fragment | NA | NA | 0 |
| S12_k141_35709 | 2127 | No | NA | 2 | 0 | 0 | Not-determined | Genome-fragment | NA | NA | 0 |
| S12_k141_269572 | 1016 | No | NA | 2 | 0 | 1 | Not-determined | Genome-fragment | NA | NA | 0 |
| S12_k141_68832 | 4316 | No | NA | 3 | 0 | 1 | Not-determined | Genome-fragment | NA | NA | 0 |
| S12_k141_202596 | 1219 | No | NA | 3 | 0 | 0 | Not-determined | Genome-fragment | NA | NA | 0 |
| S3-final-viral-combined_k141_41575\|\|full | 3260 | No | NA | 9 | 0 | 0 | Low-quality | Genome-fragment | 7.56 | AAI-based (high-confidence) | 0 |
| S12_k141_353256 | 1002 | No | NA | 2 | 0 | 0 | Not-determined | Genome-fragment | NA | NA | 0 |
| S12_k141_169266 | 1113 | No | NA | 1 | 0 | 0 | Not-determined | Genome-fragment | NA | NA | 0 |
| S12_k141_2181 | 3914 | No | NA | 5 | 0 | 1 | Not-determined | Genome-fragment | NA | NA | 0 |
| S5_k141_8711 | 3260 | No | NA | 6 | 0 | 1 | Low-quality | Genome-fragment | 5.45 | AAI-based (high-confidence) | 0 |
| S12_k141_286743 | 3905 | No | NA | 5 | 0 | 1 | Not-determined | Genome-fragment | NA | NA | 0 |
| S12_k141_102300 | 1156 | No | NA | 2 | 0 | 1 | Not-determined | Genome-fragment | NA | NA | 0 |
| S2-final-viral-combined_k141_198048\|\|full | 3260 | No | NA | 4 | 0 | 1 | Low-quality | Genome-fragment | 4.2 | AAI-based (medium-confidence) | 0 |
| S17-final-viral-combined_k141_61625\|\|full | 3261 | No | NA | 5 | 0 | 1 | Low-quality | Genome-fragment | 3.96 | AAI-based (medium-confidence) | 0 |
| S12_k141_169364 | 1046 | No | NA | 2 | 0 | 0 | Not-determined | Genome-fragment | NA | NA | 0 |
| S12_k141_2228 | 3805 | No | NA | 3 | 0 | 2 | Not-determined | Genome-fragment | NA | NA | 0 |
| S12_k141_387147 | 4127 | No | NA | 4 | 0 | 0 | Not-determined | Genome-fragment | NA | NA | 0 |
| S12_k141_286813 | 1017 | No | NA | 2 | 0 | 1 | Not-determined | Genome-fragment | NA | NA | 0 |
| S12_k141_269758 | 3483 | No | NA | 4 | 0 | 3 | Not-determined | Genome-fragment | NA | NA | 0 |
| S12_k141_253013 | 3205 | No | NA | 4 | 0 | 3 | Not-determined | Genome-fragment | NA | NA | 0 |
| S12_k141_102346 | 1108 | No | NA | 2 | 0 | 0 | Not-determined | Genome-fragment | NA | NA | 0 |
| S12_k141_68978 | 2782 | No | NA | 2 | 0 | 2 | Not-determined | Genome-fragment | NA | NA | 0 |
| S12_k141_269762 | 8317 | No | NA | 9 | 0 | 9 | Not-determined | Genome-fragment | NA | NA | 0 |
| S12_k141_118958 | 1700 | No | NA | 2 | 0 | 1 | Not-determined | Genome-fragment | NA | NA | 0 |
| S12_k141_303635 | 1547 | No | NA | 1 | 0 | 1 | Not-determined | Genome-fragment | NA | NA | 0 |
| S12-final-viral-combined_k141_136636\|\|full | 3261 | No | NA | 1 | 0 | 0 | Low-quality | Genome-fragment | 3.53 | AAI-based (high-confidence) | 0 |
| S12_k141_387194 | 1882 | No | NA | 2 | 0 | 0 | Not-determined | Genome-fragment | NA | NA | 0 |
| S12_k141_2308 | 4153 | No | NA | 3 | 0 | 3 | Not-determined | Genome-fragment | NA | NA | 0 |
| S12_k141_186413 | 1070 | No | NA | 1 | 0 | 1 | Not-determined | Genome-fragment | NA | NA | 0 |
| S12_k141_364927 | 3261 | No | NA | 10 | 1 | 0 | Low-quality | Genome-fragment | 5.81 | AAI-based (high-confidence) | 0 |
| S2_k141_129766 | 3261 | No | NA | 6 | 0 | 1 | Low-quality | Genome-fragment | 2.99 | AAI-based (medium-confidence) | 0 |
| S12_k141_2318 | 1064 | No | NA | 2 | 0 | 0 | Not-determined | Genome-fragment | NA | NA | 0 |
| S11-final-viral-combined_k141_210964\|\|full | 3262 | No | NA | 5 | 3 | 0 | Low-quality | Genome-fragment | 2.64 | HMM-based (lower-bound) | 0 |
| S12_k141_102429 | 1911 | No | NA | 2 | 0 | 0 | Not-determined | Genome-fragment | NA | NA | 0 |
| S12_k141_52423 | 1119 | No | NA | 2 | 0 | 1 | Not-determined | Genome-fragment | NA | NA | 0 |
| S1-final-viral-combined_k141_78639\|\|full | 3262 | No | NA | 4 | 0 | 1 | Low-quality | Genome-fragment | 7.47 | AAI-based (medium-confidence) | 0 |
| S12_k141_2338 | 1824 | No | NA | 3 | 0 | 0 | Not-determined | Genome-fragment | NA | NA | 0 |
| S12_k141_236304 | 5552 | No | NA | 5 | 0 | 1 | Not-determined | Genome-fragment | NA | NA | 0 |
| S12_k141_119026 | 1384 | No | NA | 1 | 0 | 0 | Not-determined | Genome-fragment | NA | NA | 0 |
| S12_k141_320300 | 3187 | No | NA | 3 | 0 | 3 | Not-determined | Genome-fragment | NA | NA | 0 |
| S12_k141_202803 | 42999 | No | NA | 48 | 13 | 0 | High-quality | High-quality | 100 | AAI-based (high-confidence) | 0 |
| S12_k141_286909 | 2237 | No | NA | 1 | 0 | 1 | Not-determined | Genome-fragment | NA | NA | 0 |
| S12_k141_2347 | 5073 | No | NA | 4 | 0 | 2 | Not-determined | Genome-fragment | NA | NA | 0 |
| S12_k141_202816 | 1061 | No | NA | 3 | 0 | 0 | Not-determined | Genome-fragment | NA | NA | 0 |
| S12_k141_152893 | 1134 | No | NA | 1 | 0 | 1 | Not-determined | Genome-fragment | NA | NA | 0 |
| S12_k141_136023 | 15739 | No | NA | 12 | 0 | 1 | Not-determined | Genome-fragment | NA | NA | 0 |
| S12_k141_186537 | 4379 | No | NA | 4 | 0 | 4 | Not-determined | Genome-fragment | NA | NA | 0 |
| S12_k141_69163 | 1050 | No | NA | 2 | 0 | 1 | Not-determined | Genome-fragment | NA | NA | 0 |
| S12_k141_336564 | 4247 | No | NA | 3 | 0 | 1 | Not-determined | Genome-fragment | NA | NA | 0 |
| S17-final-viral-combined_k141_220311\|\|full | 3262 | No | NA | 5 | 2 | 0 | Low-quality | Genome-fragment | 3.93 | HMM-based (lower-bound) | 0 |
| S12_k141_269908 | 14275 | No | NA | 16 | 0 | 3 | Not-determined | Genome-fragment | NA | NA | 0 |
| S12_k141_52513 | 2244 | No | NA | 1 | 0 | 1 | Not-determined | Genome-fragment | NA | NA | 0 |
| S9-final-viral-combined_k141_106860\|\|full | 3264 | No | NA | 4 | 0 | 2 | Low-quality | Genome-fragment | 1.13 | AAI-based (medium-confidence) | 0 |
| S5-final-viral-combined_k141_65963\|\|full | 3264 | No | NA | 4 | 0 | 1 | Low-quality | Genome-fragment | 2.34 | AAI-based (medium-confidence) | 0 |
| S12_k141_353524 | 1333 | No | NA | 1 | 0 | 1 | Not-determined | Genome-fragment | NA | NA | 0 |
| S12_k141_320377 | 1473 | No | NA | 1 | 0 | 1 | Not-determined | Genome-fragment | NA | NA | 0 |
| S12_k141_85957 | 1318 | No | NA | 2 | 0 | 1 | Not-determined | Genome-fragment | NA | NA | 0 |
| S12_k141_69210 | 3415 | No | NA | 2 | 0 | 2 | Not-determined | Genome-fragment | NA | NA | 0 |
| S12-final-viral-combined_k141_210605\|\|full | 3264 | No | NA | 3 | 3 | 0 | Low-quality | Genome-fragment | 7.92 | AAI-based (high-confidence) | 0 |
| S2_k141_85751 | 3264 | No | NA | 4 | 2 | 0 | Low-quality | Genome-fragment | 9.11 | AAI-based (high-confidence) | 0 |
| S12_k141_36005 | 1057 | No | NA | 2 | 0 | 1 | Not-determined | Genome-fragment | NA | NA | 0 |
| S12_k141_269978 | 1614 | No | NA | 2 | 0 | 1 | Not-determined | Genome-fragment | NA | NA | 0 |
| S12_k141_52589 | 3466 | No | NA | 7 | 0 | 0 | Not-determined | Genome-fragment | NA | NA | 0 |
| S12_k141_253187 | 1044 | No | NA | 1 | 0 | 0 | Not-determined | Genome-fragment | NA | NA | 0 |
| S12_k141_136169 | 4111 | No | NA | 6 | 0 | 0 | Not-determined | Genome-fragment | NA | NA | 0 |
| S12_k141_320463 | 1070 | No | NA | 1 | 0 | 1 | Not-determined | Genome-fragment | NA | NA | 0 |
| S12_k141_52625 | 1555 | No | NA | 3 | 0 | 2 | Not-determined | Genome-fragment | NA | NA | 0 |
| S4_k141_14780 | 3264 | No | NA | 3 | 0 | 2 | Low-quality | Genome-fragment | 1.36 | AAI-based (medium-confidence) | 0 |
| S11_k141_188008 | 3265 | No | NA | 4 | 2 | 0 | Low-quality | Genome-fragment | 5.14 | AAI-based (medium-confidence) | 0 |
| S12_k141_387338 | 1357 | No | NA | 1 | 0 | 0 | Not-determined | Genome-fragment | NA | NA | 0 |
| S12_k141_69371 | 1410 | No | NA | 2 | 0 | 1 | Not-determined | Genome-fragment | NA | NA | 0 |
| S12_k141_69376 | 1858 | No | NA | 3 | 0 | 0 | Not-determined | Genome-fragment | NA | NA | 0 |
| S11_k141_11796 | 3265 | No | NA | 6 | 3 | 0 | Low-quality | Genome-fragment | 6.92 | AAI-based (high-confidence) | 0 |
| S4-final-viral-combined_k141_51751\|\|full | 3265 | No | NA | 2 | 0 | 1 | Low-quality | Genome-fragment | 1.35 | AAI-based (medium-confidence) | 0 |
| S8-final-viral-combined_k141_144193\|\|full | 3265 | No | NA | 5 | 0 | 3 | Low-quality | Genome-fragment | 4.26 | AAI-based (medium-confidence) | 0 |
| S12_k141_336638 | 1210 | No | NA | 3 | 0 | 1 | Not-determined | Genome-fragment | NA | NA | 0 |
| S12_k141_287119 | 1609 | No | NA | 2 | 0 | 2 | Not-determined | Genome-fragment | NA | NA | 0 |
| S9-final-viral-combined_k141_11894\|\|lt2gene | 3267 | No | NA | 1 | 1 | 0 | Low-quality | Genome-fragment | 2.26 | HMM-based (lower-bound) | 0 |
| S12_k141_353715 | 1013 | No | NA | 2 | 0 | 0 | Not-determined | Genome-fragment | NA | NA | 0 |
| S12_k141_219746 | 1180 | No | NA | 1 | 0 | 1 | Not-determined | Genome-fragment | NA | NA | 0 |
| S12_k141_36057 | 7481 | No | NA | 10 | 0 | 7 | Not-determined | Genome-fragment | NA | NA | 0 |
| S12_k141_153135 | 1513 | No | NA | 1 | 0 | 0 | Not-determined | Genome-fragment | NA | NA | 0 |
| S12_k141_18813 | 1153 | No | NA | 1 | 0 | 0 | Not-determined | Genome-fragment | NA | NA | 0 |
| S12_k141_304010 | 1037 | No | NA | 2 | 0 | 1 | Not-determined | Genome-fragment | NA | NA | 0 |
| S12_k141_18850 | 2049 | No | NA | 3 | 0 | 3 | Not-determined | Genome-fragment | NA | NA | 0 |
| S8-final-viral-combined_k141_36715\|\|full | 3267 | No | NA | 4 | 0 | 3 | Low-quality | Genome-fragment | 1.01 | AAI-based (high-confidence) | 0 |
| S8_k141_271660 | 3267 | No | NA | 6 | 0 | 0 | Low-quality | Genome-fragment | 7.71 | AAI-based (medium-confidence) | 0 |
| S12_k141_387427 | 1435 | No | NA | 3 | 0 | 0 | Not-determined | Genome-fragment | NA | NA | 0 |
| S12_k141_169732 | 3195 | No | NA | 3 | 0 | 3 | Not-determined | Genome-fragment | NA | NA | 0 |
| S12_k141_52806 | 1665 | No | NA | 1 | 0 | 0 | Not-determined | Genome-fragment | NA | NA | 0 |
| S12_k141_370926 | 2353 | No | NA | 4 | 0 | 2 | Not-determined | Genome-fragment | NA | NA | 0 |
| S5-final-viral-combined_k141_208886\|\|full | 3268 | No | NA | 5 | 1 | 0 | Low-quality | Genome-fragment | 8.98 | HMM-based (lower-bound) | 0 |
| S12_k141_270017 | 1197 | No | NA | 1 | 0 | 1 | Not-determined | Genome-fragment | NA | NA | 0 |
| S12_k141_387454 | 3467 | No | NA | 3 | 0 | 2 | Not-determined | Genome-fragment | NA | NA | 0 |
| S12_k141_304089 | 3005 | No | NA | 4 | 0 | 2 | Not-determined | Genome-fragment | NA | NA | 0 |
| S12_k141_119245 | 3281 | No | NA | 6 | 0 | 0 | Not-determined | Genome-fragment | NA | NA | 0 |
| S1-final-viral-combined_k141_163982\|\|full | 3269 | Yes | 1779 | 5 | 1 | 1 | Low-quality | Genome-fragment | 42.75 | HMM-based (lower-bound) | 45.58 |
| S12_k141_2579 | 2740 | No | NA | 3 | 0 | 1 | Not-determined | Genome-fragment | NA | NA | 0 |
| S12_k141_304118 | 1042 | No | NA | 2 | 0 | 1 | Not-determined | Genome-fragment | NA | NA | 0 |
| S8-final-viral-combined_k141_65280\|\|full | 3269 | No | NA | 3 | 0 | 1 | Low-quality | Genome-fragment | 1.64 | AAI-based (medium-confidence) | 0 |
| S12_k141_203168 | 1455 | No | NA | 1 | 0 | 1 | Not-determined | Genome-fragment | NA | NA | 0 |
| S12_k141_353879 | 1292 | No | NA | 1 | 0 | 1 | Not-determined | Genome-fragment | NA | NA | 0 |
| S12_k141_387537 | 1654 | No | NA | 2 | 0 | 1 | Not-determined | Genome-fragment | NA | NA | 0 |
| S12_k141_86190 | 1086 | No | NA | 1 | 0 | 1 | Not-determined | Genome-fragment | NA | NA | 0 |
| S12_k141_387558 | 2315 | No | NA | 1 | 0 | 0 | Not-determined | Genome-fragment | NA | NA | 0 |
| S11-final-viral-combined_k141_245152\|\|full | 3270 | No | NA | 4 | 1 | 0 | Low-quality | Genome-fragment | 7.24 | AAI-based (high-confidence) | 0 |
| S12_k141_86208 | 1129 | No | NA | 1 | 0 | 0 | Not-determined | Genome-fragment | NA | NA | 0 |
| S12_k141_387572 | 1443 | No | NA | 2 | 0 | 2 | Not-determined | Genome-fragment | NA | NA | 0 |
| S12_k141_119302 | 1577 | No | NA | 1 | 0 | 0 | Not-determined | Genome-fragment | NA | NA | 0 |
| S5-final-viral-combined_k141_148491\|\|full | 3270 | No | NA | 5 | 3 | 0 | Low-quality | Genome-fragment | 5.06 | HMM-based (lower-bound) | 0 |
| S11-final-viral-combined_k141_10611\|\|full | 3271 | No | NA | 4 | 0 | 1 | Low-quality | Genome-fragment | 1.73 | AAI-based (medium-confidence) | 0 |
| S12_k141_253414 | 3463 | No | NA | 3 | 0 | 2 | Not-determined | Genome-fragment | NA | NA | 0 |
| S12_k141_69756 | 1922 | No | NA | 3 | 0 | 1 | Not-determined | Genome-fragment | NA | NA | 0 |
| S12_k141_153395 | 1423 | No | NA | 1 | 0 | 1 | Not-determined | Genome-fragment | NA | NA | 0 |
| S12_k141_69808 | 1223 | No | NA | 1 | 0 | 1 | Not-determined | Genome-fragment | NA | NA | 0 |
| S12_k141_86284 | 2100 | No | NA | 1 | 0 | 1 | Not-determined | Genome-fragment | NA | NA | 0 |
| S12-final-viral-combined_k141_86967\|\|full | 3271 | No | NA | 8 | 1 | 0 | Low-quality | Genome-fragment | 6.99 | AAI-based (medium-confidence) | 0 |
| S12_k141_169960 | 3841 | No | NA | 3 | 0 | 2 | Not-determined | Genome-fragment | NA | NA | 0 |
| S12_k141_2741 | 1373 | No | NA | 1 | 0 | 1 | Not-determined | Genome-fragment | NA | NA | 0 |
| S12_k141_287372 | 1404 | No | NA | 2 | 0 | 0 | Not-determined | Genome-fragment | NA | NA | 0 |
| S12_k141_354100 | 1660 | No | NA | 5 | 0 | 0 | Not-determined | Genome-fragment | NA | NA | 0 |
| S12_k141_287385 | 1103 | No | NA | 1 | 0 | 1 | Not-determined | Genome-fragment | NA | NA | 0 |
| S7_k141_111989 | 3273 | No | NA | 3 | 0 | 0 | Low-quality | Genome-fragment | 4.98 | AAI-based (medium-confidence) | 0 |
| S17-final-viral-combined_k141_17823\|\|full | 3274 | No | NA | 4 | 3 | 0 | Low-quality | Genome-fragment | 7.8 | AAI-based (high-confidence) | 0 |
| S12_k141_270255 | 1341 | No | NA | 1 | 0 | 1 | Not-determined | Genome-fragment | NA | NA | 0 |
| S12_k141_169990 | 1280 | No | NA | 1 | 0 | 0 | Not-determined | Genome-fragment | NA | NA | 0 |
| S12_k141_219969 | 1349 | No | NA | 2 | 0 | 1 | Not-determined | Genome-fragment | NA | NA | 0 |
| S12_k141_153441 | 1163 | No | NA | 2 | 0 | 1 | Not-determined | Genome-fragment | NA | NA | 0 |
| S1-final-viral-combined_k141_178650\|\|full | 3275 | No | NA | 3 | 3 | 0 | Low-quality | Genome-fragment | 7.05 | AAI-based (high-confidence) | 0 |
| S12_k141_336780 | 9397 | No | NA | 16 | 1 | 1 | Medium-quality | Genome-fragment | 50.86 | AAI-based (high-confidence) | 0 |
| S12_k141_18935 | 6895 | No | NA | 7 | 0 | 7 | Not-determined | Genome-fragment | NA | NA | 0 |
| S12_k141_371198 | 1882 | No | NA | 1 | 0 | 0 | Not-determined | Genome-fragment | NA | NA | 0 |
| S12_k141_18966 | 3397 | No | NA | 5 | 0 | 1 | Not-determined | Genome-fragment | NA | NA | 0 |
| S12_k141_102902 | 1394 | No | NA | 2 | 0 | 0 | Not-determined | Genome-fragment | NA | NA | 0 |
| S12_k141_203415 | 2334 | No | NA | 1 | 0 | 0 | Not-determined | Genome-fragment | NA | NA | 0 |
| S1_k141_145547 | 3275 | No | NA | 1 | 0 | 1 | Low-quality | Genome-fragment | 0.28 | AAI-based (medium-confidence) | 0 |
| S12_k141_19013 | 1315 | No | NA | 2 | 0 | 0 | Not-determined | Genome-fragment | NA | NA | 0 |
| S12_k141_86396 | 1028 | No | NA | 1 | 0 | 1 | Not-determined | Genome-fragment | NA | NA | 0 |
| S12_k141_119592 | 1016 | No | NA | 1 | 0 | 0 | Not-determined | Genome-fragment | NA | NA | 0 |
| S12_k141_336871 | 3604 | No | NA | 10 | 0 | 0 | Not-determined | Genome-fragment | NA | NA | 0 |
| S12_k141_320820 | 1365 | No | NA | 1 | 0 | 0 | Not-determined | Genome-fragment | NA | NA | 0 |
| S4_k141_126707 | 3275 | No | NA | 4 | 0 | 0 | Low-quality | Genome-fragment | 3.92 | AAI-based (high-confidence) | 0 |
| S12_k141_320835 | 3097 | No | NA | 4 | 0 | 3 | Not-determined | Genome-fragment | NA | NA | 0 |
| S12_k141_136675 | 1158 | No | NA | 3 | 0 | 2 | Not-determined | Genome-fragment | NA | NA | 0 |
| S17-final-viral-combined_k141_191307\|\|full | 3276 | No | NA | 4 | 0 | 0 | Low-quality | Genome-fragment | 5.26 | AAI-based (medium-confidence) | 0 |
| S12_k141_220055 | 1484 | No | NA | 2 | 0 | 0 | Not-determined | Genome-fragment | NA | NA | 0 |
| S12_k141_387766 | 1938 | No | NA | 2 | 0 | 0 | Not-determined | Genome-fragment | NA | NA | 0 |
| S12-final-viral-combined_k141_153634\|\|full | 3276 | Yes | 2213 | 6 | 1 | 1 | Low-quality | Genome-fragment | 2.82 | AAI-based (medium-confidence) | 32.45 |
| S4-final-viral-combined_k141_150442\|\|full | 3276 | No | NA | 5 | 0 | 1 | Low-quality | Genome-fragment | 2.73 | AAI-based (high-confidence) | 0 |
| S12_k141_270459 | 11480 | No | NA | 13 | 0 | 13 | Not-determined | Genome-fragment | NA | NA | 0 |
| S12_k141_203568 | 2236 | No | NA | 2 | 0 | 1 | Not-determined | Genome-fragment | NA | NA | 0 |
| S3-final-viral-combined_k141_44502\|\|full | 3277 | No | NA | 3 | 0 | 3 | Low-quality | Genome-fragment | 8.55 | AAI-based (medium-confidence) | 0 |
| S12_k141_136746 | 1128 | No | NA | 1 | 0 | 0 | Not-determined | Genome-fragment | NA | NA | 0 |
| S12_k141_136747 | 3848 | No | NA | 5 | 0 | 4 | Not-determined | Genome-fragment | NA | NA | 0 |
| S8-final-viral-combined_k141_234686\|\|full | 3277 | No | NA | 3 | 0 | 2 | Low-quality | Genome-fragment | 1.92 | AAI-based (medium-confidence) | 0 |
| S12_k141_2957 | 1203 | No | NA | 2 | 0 | 1 | Not-determined | Genome-fragment | NA | NA | 0 |
| S9_k141_29301 | 3278 | No | NA | 2 | 0 | 0 | Low-quality | Genome-fragment | 3.43 | AAI-based (high-confidence) | 0 |
| S11_k141_316883 | 3279 | No | NA | 9 | 1 | 0 | Low-quality | Genome-fragment | 2.43 | HMM-based (lower-bound) | 0 |
| S1_k141_361083 | 3279 | No | NA | 7 | 0 | 0 | Low-quality | Genome-fragment | 5.42 | AAI-based (medium-confidence) | 0 |
| S12_k141_220139 | 4356 | No | NA | 7 | 0 | 0 | Not-determined | Genome-fragment | NA | NA | 0 |
| S8-final-viral-combined_k141_67818\|\|full | 3279 | No | NA | 6 | 0 | 2 | Low-quality | Genome-fragment | 2.22 | AAI-based (medium-confidence) | 0 |
| S9_k141_100169 | 3280 | No | NA | 6 | 4 | 0 | Low-quality | Genome-fragment | 6.86 | AAI-based (high-confidence) | 0 |
| S11-final-viral-combined_k141_128843\|\|full | 3280 | No | NA | 5 | 0 | 1 | Low-quality | Genome-fragment | 4.22 | AAI-based (high-confidence) | 0 |
| S12_k141_3004 | 1003 | No | NA | 1 | 0 | 1 | Not-determined | Genome-fragment | NA | NA | 0 |
| S12_k141_136811 | 1333 | No | NA | 1 | 0 | 0 | Not-determined | Genome-fragment | NA | NA | 0 |
| S12_k141_19145 | 1743 | No | NA | 2 | 0 | 1 | Not-determined | Genome-fragment | NA | NA | 0 |
| S12_k141_3017 | 2867 | No | NA | 1 | 0 | 0 | Not-determined | Genome-fragment | NA | NA | 0 |
| S12_k141_53106 | 6859 | No | NA | 6 | 0 | 4 | Not-determined | Genome-fragment | NA | NA | 0 |
| S12_k141_320896 | 3914 | No | NA | 4 | 0 | 1 | Not-determined | Genome-fragment | NA | NA | 0 |
| S12_k141_320898 | 6436 | No | NA | 6 | 0 | 6 | Not-determined | Genome-fragment | NA | NA | 0 |
| S12_k141_320908 | 1009 | No | NA | 1 | 0 | 0 | Not-determined | Genome-fragment | NA | NA | 0 |
| S12_k141_187329 | 1707 | No | NA | 4 | 0 | 1 | Not-determined | Genome-fragment | NA | NA | 0 |
| S12_k141_220234 | 1034 | No | NA | 1 | 0 | 1 | Not-determined | Genome-fragment | NA | NA | 0 |
| S12_k141_287611 | 3648 | No | NA | 4 | 0 | 4 | Not-determined | Genome-fragment | NA | NA | 0 |
| S12_k141_86573 | 3808 | No | NA | 4 | 0 | 3 | Not-determined | Genome-fragment | NA | NA | 0 |
| S12_k141_119816 | 28082 | No | NA | 35 | 0 | 7 | Not-determined | Genome-fragment | NA | NA | 0 |
| S12-final-viral-combined_k141_329347\|\|lt2gene | 3280 | No | NA | 1 | 1 | 0 | Low-quality | Genome-fragment | 7.12 | AAI-based (medium-confidence) | 0 |
| S12-final-viral-combined_k141_179048\|\|lt2gene | 3280 | No | NA | 2 | 2 | 0 | Low-quality | Genome-fragment | 4.49 | HMM-based (lower-bound) | 0 |
| S12_k141_289003 | 3280 | No | NA | 2 | 0 | 1 | Low-quality | Genome-fragment | 1.26 | AAI-based (medium-confidence) | 0 |
| S9_k141_116108 | 3282 | No | NA | 3 | 0 | 2 | Low-quality | Genome-fragment | 2.93 | AAI-based (medium-confidence) | 0 |
| S11-final-viral-combined_k141_191476\|\|full | 3282 | No | NA | 6 | 2 | 0 | Low-quality | Genome-fragment | 7.55 | AAI-based (high-confidence) | 0 |
| S12_k141_270631 | 5979 | No | NA | 5 | 0 | 1 | Not-determined | Genome-fragment | NA | NA | 0 |
| S5-final-viral-combined_k141_182535\|\|full | 3282 | No | NA | 6 | 1 | 0 | Low-quality | Genome-fragment | 7.21 | AAI-based (medium-confidence) | 0 |
| S12_k141_19262 | 1178 | No | NA | 3 | 0 | 2 | Not-determined | Genome-fragment | NA | NA | 0 |
| S12_k141_187393 | 1451 | No | NA | 1 | 0 | 1 | Not-determined | Genome-fragment | NA | NA | 0 |
| S1-final-viral-combined_k141_41561\|\|full | 3283 | No | NA | 8 | 0 | 0 | Low-quality | Genome-fragment | 5.27 | AAI-based (high-confidence) | 0 |
| S2-final-viral-combined_k141_197167\|\|full | 3283 | No | NA | 4 | 0 | 0 | Low-quality | Genome-fragment | 5.99 | AAI-based (medium-confidence) | 0 |
| S12_k141_3082 | 1189 | No | NA | 2 | 0 | 1 | Not-determined | Genome-fragment | NA | NA | 0 |
| S2-final-viral-combined_k141_96323\|\|full | 3283 | No | NA | 5 | 0 | 1 | Low-quality | Genome-fragment | 5.82 | AAI-based (high-confidence) | 0 |
| S7-final-viral-combined_k141_226147\|\|full | 3284 | No | NA | 3 | 0 | 1 | Low-quality | Genome-fragment | 4.17 | AAI-based (medium-confidence) | 0 |
| S12_k141_371470 | 1058 | No | NA | 2 | 0 | 0 | Not-determined | Genome-fragment | NA | NA | 0 |
| S4_k141_151366 | 3284 | No | NA | 3 | 0 | 0 | Low-quality | Genome-fragment | 6.33 | AAI-based (medium-confidence) | 0 |
| S12_k141_136968 | 5179 | No | NA | 6 | 0 | 0 | Not-determined | Genome-fragment | NA | NA | 0 |
| S12_k141_270687 | 1512 | No | NA | 1 | 0 | 0 | Not-determined | Genome-fragment | NA | NA | 0 |
| S12_k141_86661 | 4868 | No | NA | 3 | 0 | 1 | Not-determined | Genome-fragment | NA | NA | 0 |
| S11-final-viral-combined_k141_151801\|\|full | 3286 | No | NA | 3 | 1 | 0 | Low-quality | Genome-fragment | 6.1 | AAI-based (medium-confidence) | 0 |
| S12_k141_3122 | 6689 | No | NA | 4 | 0 | 2 | Not-determined | Genome-fragment | NA | NA | 0 |
| S12_k141_320982 | 5071 | No | NA | 3 | 0 | 3 | Not-determined | Genome-fragment | NA | NA | 0 |
| S12_k141_70277 | 3427 | No | NA | 3 | 0 | 0 | Not-determined | Genome-fragment | NA | NA | 0 |
| S12_k141_354271 | 3650 | No | NA | 1 | 0 | 0 | Not-determined | Genome-fragment | NA | NA | 0 |
| S12_k141_304658 | 1030 | No | NA | 2 | 0 | 2 | Not-determined | Genome-fragment | NA | NA | 0 |
| S12_k141_314460 | 3286 | No | NA | 3 | 0 | 2 | Low-quality | Genome-fragment | 1.56 | AAI-based (medium-confidence) | 0 |
| S8_k141_173713 | 3287 | No | NA | 3 | 0 | 2 | Low-quality | Genome-fragment | 1.84 | AAI-based (medium-confidence) | 0 |
| S12-final-viral-combined_k141_259553\|\|full | 3288 | No | NA | 4 | 3 | 0 | Low-quality | Genome-fragment | 8.34 | AAI-based (high-confidence) | 0 |
| S12_k141_337286 | 2117 | No | NA | 2 | 0 | 0 | Not-determined | Genome-fragment | NA | NA | 0 |
| S12_k141_270809 | 2884 | No | NA | 2 | 0 | 0 | Not-determined | Genome-fragment | NA | NA | 0 |
| S12-final-viral-combined_k141_307256\|\|full | 3289 | No | NA | 4 | 0 | 0 | Low-quality | Genome-fragment | 4.5 | AAI-based (medium-confidence) | 0 |
| S12_k141_203884 | 1118 | No | NA | 1 | 0 | 1 | Not-determined | Genome-fragment | NA | NA | 0 |
| S11-final-viral-combined_k141_11547\|\|full | 3290 | No | NA | 4 | 1 | 0 | Low-quality | Genome-fragment | 3.08 | HMM-based (lower-bound) | 0 |
| S1-final-viral-combined_k141_77878\|\|full | 3290 | No | NA | 5 | 1 | 0 | Low-quality | Genome-fragment | 10.91 | AAI-based (high-confidence) | 0 |
| S12_k141_170558 | 1024 | No | NA | 1 | 0 | 1 | Not-determined | Genome-fragment | NA | NA | 0 |
| S12_k141_304707 | 1049 | No | NA | 2 | 0 | 0 | Not-determined | Genome-fragment | NA | NA | 0 |
| S12_k141_19441 | 1157 | No | NA | 2 | 0 | 2 | Not-determined | Genome-fragment | NA | NA | 0 |
| S12_k141_253768 | 1338 | No | NA | 2 | 0 | 0 | Not-determined | Genome-fragment | NA | NA | 0 |
| S12_k141_321084 | 1414 | No | NA | 1 | 0 | 1 | Not-determined | Genome-fragment | NA | NA | 0 |
| S12_k141_321096 | 1217 | No | NA | 2 | 0 | 1 | Not-determined | Genome-fragment | NA | NA | 0 |
| S12_k141_371612 | 1186 | No | NA | 1 | 0 | 0 | Not-determined | Genome-fragment | NA | NA | 0 |
| S12_k141_371614 | 1275 | No | NA | 1 | 0 | 1 | Not-determined | Genome-fragment | NA | NA | 0 |
| S12_k141_371616 | 1353 | No | NA | 1 | 0 | 1 | Not-determined | Genome-fragment | NA | NA | 0 |
| S12_k141_137140 | 3674 | No | NA | 7 | 0 | 3 | Not-determined | Genome-fragment | NA | NA | 0 |
| S1-final-viral-combined_k141_49100\|\|full | 3290 | Yes | 1891 | 6 | 2 | 1 | Low-quality | Genome-fragment | 4.03 | AAI-based (high-confidence) | 42.52 |
| S12_k141_388119 | 9342 | No | NA | 20 | 0 | 19 | Not-determined | Genome-fragment | NA | NA | 0 |
| S12-final-viral-combined_k141_224104\|\|full | 3290 | No | NA | 7 | 3 | 0 | Low-quality | Genome-fragment | 8.62 | AAI-based (medium-confidence) | 0 |
| S12_k141_354447 | 5457 | No | NA | 7 | 0 | 2 | Not-determined | Genome-fragment | NA | NA | 0 |
| S12_k141_187647 | 3570 | No | NA | 4 | 0 | 2 | Not-determined | Genome-fragment | NA | NA | 0 |
| S2_k141_138271 | 3290 | No | NA | 7 | 1 | 0 | Low-quality | Genome-fragment | 1.73 | HMM-based (lower-bound) | 0 |
| S8_k141_21669 | 3290 | No | NA | 3 | 0 | 0 | Low-quality | Genome-fragment | 4.86 | AAI-based (medium-confidence) | 0 |
| S1-final-viral-combined_k141_352634\|\|full | 3291 | No | NA | 6 | 3 | 0 | Low-quality | Genome-fragment | 11.47 | AAI-based (high-confidence) | 0 |
| S5-final-viral-combined_k141_144576\|\|full | 3291 | No | NA | 4 | 0 | 0 | Low-quality | Genome-fragment | 7.78 | AAI-based (medium-confidence) | 0 |
| S12_k141_321194 | 1149 | No | NA | 2 | 0 | 2 | Not-determined | Genome-fragment | NA | NA | 0 |
| S12_k141_321216 | 4603 | No | NA | 4 | 0 | 1 | Not-determined | Genome-fragment | NA | NA | 0 |
| S12_k141_153892 | 1205 | No | NA | 1 | 0 | 0 | Not-determined | Genome-fragment | NA | NA | 0 |
| S12_k141_253885 | 1173 | No | NA | 2 | 0 | 2 | Not-determined | Genome-fragment | NA | NA | 0 |
| S5_k141_59078 | 3291 | No | NA | 2 | 0 | 2 | Low-quality | Genome-fragment | 5.04 | AAI-based (medium-confidence) | 0 |
| S7-final-viral-combined_k141_153353\|\|full | 3291 | No | NA | 10 | 1 | 0 | Low-quality | Genome-fragment | 6.28 | AAI-based (high-confidence) | 0 |
| S1_k141_229783 | 3292 | No | NA | 4 | 0 | 1 | Low-quality | Genome-fragment | 4.34 | AAI-based (medium-confidence) | 0 |
| S12_k141_304889 | 3750 | No | NA | 3 | 0 | 0 | Not-determined | Genome-fragment | NA | NA | 0 |
| S12_k141_253903 | 1627 | No | NA | 3 | 0 | 1 | Not-determined | Genome-fragment | NA | NA | 0 |
| S17-final-viral-combined_k141_173472\|\|full | 3292 | No | NA | 10 | 1 | 0 | Low-quality | Genome-fragment | 8.69 | AAI-based (high-confidence) | 0 |
| S12_k141_270903 | 2755 | No | NA | 2 | 0 | 2 | Not-determined | Genome-fragment | NA | NA | 0 |
| S12_k141_103422 | 4827 | No | NA | 9 | 0 | 8 | Not-determined | Genome-fragment | NA | NA | 0 |
| S12_k141_204119 | 4831 | No | NA | 8 | 0 | 1 | Not-determined | Genome-fragment | NA | NA | 0 |
| S12_k141_70575 | 3321 | No | NA | 2 | 0 | 0 | Not-determined | Genome-fragment | NA | NA | 0 |
| S11-final-viral-combined_k141_80280\|\|full | 3293 | Yes | 2683 | 6 | 1 | 1 | Low-quality | Genome-fragment | 5.26 | AAI-based (high-confidence) | 18.52 |
| S12_k141_19725 | 3255 | No | NA | 5 | 0 | 0 | Not-determined | Genome-fragment | NA | NA | 0 |
| S12_k141_170756 | 1068 | No | NA | 2 | 0 | 0 | Not-determined | Genome-fragment | NA | NA | 0 |
| S12_k141_187837 | 3315 | No | NA | 2 | 0 | 2 | Not-determined | Genome-fragment | NA | NA | 0 |
| S1-final-viral-combined_k141_136081\|\|full | 3293 | No | NA | 4 | 0 | 2 | Low-quality | Genome-fragment | 6.34 | AAI-based (medium-confidence) | 0 |
| S12_k141_3452 | 1013 | No | NA | 1 | 0 | 0 | Not-determined | Genome-fragment | NA | NA | 0 |

| | | | | | | | | | | | |
|---|---|---|---|---|---|---|---|---|---|---|---|
| S12_k141_170781 | 1159 | No | NA | 1 | 0 | 1 | Not-determined | Genome-fragment | NA | NA | 0 |
| S3-final-viral-combined_k141_7109\|\|full | 3293 | No | NA | 7 | 2 | 0 | Low-quality | Genome-fragment | 9.51 | AAI-based (high-confidence) | 0 |
| S12_k141_354565 | 8175 | No | NA | 1 | 0 | 1 | Not-determined | Genome-fragment | NA | NA | 0 |
| S12_k141_253985 | 4648 | No | NA | 4 | 0 | 3 | Not-determined | Genome-fragment | NA | NA | 0 |
| S12_k141_321355 | 4763 | No | NA | 4 | 0 | 2 | Not-determined | Genome-fragment | NA | NA | 0 |
| S12_k141_70699 | 5102 | No | NA | 2 | 0 | 1 | Not-determined | Genome-fragment | NA | NA | 0 |
| S12_k141_86962 | 1519 | No | NA | 2 | 0 | 0 | Not-determined | Genome-fragment | NA | NA | 0 |
| S12_k141_304939 | 7103 | No | NA | 6 | 0 | 4 | Not-determined | Genome-fragment | NA | NA | 0 |
| S12_k141_270930 | 4935 | No | NA | 3 | 0 | 1 | Not-determined | Genome-fragment | NA | NA | 0 |
| S12_k141_170823 | 1716 | No | NA | 1 | 0 | 1 | Not-determined | Genome-fragment | NA | NA | 0 |
| S12_k141_187928 | 1198 | No | NA | 1 | 0 | 0 | Not-determined | Genome-fragment | NA | NA | 0 |
| S5-final-viral-combined_k141_161271\|\|full | 3293 | No | NA | 5 | 1 | 0 | Low-quality | Genome-fragment | 4.98 | HMM-based (lower-bound) | 0 |
| S7_k141_175807 | 3293 | No | NA | 3 | 0 | 1 | Low-quality | Genome-fragment | 4.3 | AAI-based (high-confidence) | 0 |
| S17-final-viral-combined_k141_224651\|\|full | 3293 | No | NA | 8 | 0 | 0 | Low-quality | Genome-fragment | 4 | AAI-based (high-confidence) | 0 |
| S12_k141_154030 | 9087 | No | NA | 7 | 0 | 7 | Not-determined | Genome-fragment | NA | NA | 0 |
| S12_k141_388388 | 3876 | No | NA | 5 | 0 | 3 | Not-determined | Genome-fragment | NA | NA | 0 |
| S4-final-viral-combined_k141_28635\|\|full | 3293 | No | NA | 3 | 0 | 3 | Low-quality | Genome-fragment | 5.6 | AAI-based (high-confidence) | 0 |
| S12_k141_187990 | 3686 | No | NA | 4 | 0 | 3 | Not-determined | Genome-fragment | NA | NA | 0 |
| S1-final-viral-combined_k141_365932\|\|full | 3294 | No | NA | 7 | 0 | 3 | Low-quality | Genome-fragment | 7.87 | AAI-based (medium-confidence) | 0 |
| S12_k141_305013 | 2519 | No | NA | 1 | 0 | 1 | Not-determined | Genome-fragment | NA | NA | 0 |
| S12_k141_19872 | 1921 | No | NA | 2 | 0 | 2 | Not-determined | Genome-fragment | NA | NA | 0 |
| S12_k141_154044 | 1113 | No | NA | 1 | 0 | 0 | Not-determined | Genome-fragment | NA | NA | 0 |
| S12_k141_220752 | 1312 | No | NA | 2 | 0 | 1 | Not-determined | Genome-fragment | NA | NA | 0 |
| S12_k141_270979 | 1225 | No | NA | 1 | 0 | 0 | Not-determined | Genome-fragment | NA | NA | 0 |
| S12_k141_137351 | 1316 | No | NA | 2 | 0 | 1 | Not-determined | Genome-fragment | NA | NA | 0 |
| S12_k141_37095 | 11686 | No | NA | 6 | 0 | 4 | Not-determined | Genome-fragment | NA | NA | 0 |
| S5_k141_167355 | 3294 | No | NA | 4 | 0 | 3 | Low-quality | Genome-fragment | 1.06 | AAI-based (medium-confidence) | 0 |
| S12_k141_70762 | 3050 | No | NA | 2 | 0 | 1 | Not-determined | Genome-fragment | NA | NA | 0 |
| S12_k141_37126 | 1646 | No | NA | 1 | 0 | 0 | Not-determined | Genome-fragment | NA | NA | 0 |
| S12_k141_53561 | 1280 | No | NA | 4 | 0 | 0 | Not-determined | Genome-fragment | NA | NA | 0 |
| S12_k141_3599 | 1232 | No | NA | 1 | 0 | 1 | Not-determined | Genome-fragment | NA | NA | 0 |
| S12_k141_37172 | 1536 | No | NA | 1 | 0 | 1 | Not-determined | Genome-fragment | NA | NA | 0 |
| S12_k141_288235 | 1421 | No | NA | 6 | 0 | 0 | Not-determined | Genome-fragment | NA | NA | 0 |
| S6-final-viral-combined_k141_42017\|\|full | 3294 | No | NA | 3 | 0 | 3 | Low-quality | Genome-fragment | 1.59 | AAI-based (medium-confidence) | 0 |
| S12_k141_288257 | 4268 | No | NA | 4 | 0 | 1 | Not-determined | Genome-fragment | NA | NA | 0 |
| S12_k141_103708 | 2083 | No | NA | 2 | 0 | 0 | Not-determined | Genome-fragment | NA | NA | 0 |
| S12_k141_271034 | 2926 | No | NA | 2 | 0 | 2 | Not-determined | Genome-fragment | NA | NA | 0 |
| S12_k141_3653 | 1095 | No | NA | 1 | 0 | 0 | Not-determined | Genome-fragment | NA | NA | 0 |
| S12_k141_237753 | 1478 | No | NA | 2 | 0 | 2 | Not-determined | Genome-fragment | NA | NA | 0 |
| S12_k141_137441 | 5079 | No | NA | 3 | 0 | 3 | Not-determined | Genome-fragment | NA | NA | 0 |
| S12_k141_70854 | 3133 | No | NA | 2 | 0 | 1 | Not-determined | Genome-fragment | NA | NA | 0 |
| S12_k141_321528 | 1072 | No | NA | 1 | 0 | 1 | Not-determined | Genome-fragment | NA | NA | 0 |
| S12_k141_3687 | 1469 | No | NA | 1 | 0 | 0 | Not-determined | Genome-fragment | NA | NA | 0 |
| S12_k141_271078 | 3467 | No | NA | 3 | 0 | 1 | Not-determined | Genome-fragment | NA | NA | 0 |
| S12_k141_120170 | 2156 | No | NA | 1 | 0 | 0 | Not-determined | Genome-fragment | NA | NA | 0 |
| S3_k141_29974 | 3295 | No | NA | 1 | 0 | 1 | Low-quality | Genome-fragment | 5.44 | AAI-based (high-confidence) | 0 |
| S7-final-viral-combined_k141_108004\|\|full | 3295 | No | NA | 4 | 2 | 0 | Low-quality | Genome-fragment | 7.25 | AAI-based (high-confidence) | 0 |
| S12_k141_3703 | 6804 | No | NA | 5 | 0 | 2 | Not-determined | Genome-fragment | NA | NA | 0 |
| S12_k141_220858 | 1551 | No | NA | 2 | 0 | 0 | Not-determined | Genome-fragment | NA | NA | 0 |
| S12_k141_20061 | 1084 | No | NA | 1 | 0 | 0 | Not-determined | Genome-fragment | NA | NA | 0 |
| S17-final-viral-combined_k141_83307\|\|full | 3295 | No | NA | 6 | 1 | 0 | Low-quality | Genome-fragment | 2.1 | HMM-based (lower-bound) | 0 |
| S12_k141_305218 | 5153 | No | NA | 3 | 0 | 0 | Not-determined | Genome-fragment | NA | NA | 0 |
| S12_k141_120223 | 1155 | No | NA | 3 | 0 | 3 | Not-determined | Genome-fragment | NA | NA | 0 |
| S4-final-viral-combined_k141_14212\|\|full | 3295 | No | NA | 4 | 0 | 3 | Low-quality | Genome-fragment | 2.31 | AAI-based (medium-confidence) | 0 |
| S5-final-viral-combined_k141_160778\|\|full | 3297 | Yes | 700 | 5 | 1 | 2 | Low-quality | Genome-fragment | 0.53 | AAI-based (high-confidence) | 78.77 |
| S12_k141_288366 | 1025 | No | NA | 1 | 0 | 0 | Not-determined | Genome-fragment | NA | NA | 0 |
| S7-final-viral-combined_k141_179564\|\|full | 3298 | No | NA | 4 | 0 | 0 | Low-quality | Genome-fragment | 8.43 | AAI-based (high-confidence) | 0 |
| S12_k141_372034 | 1479 | No | NA | 2 | 0 | 2 | Not-determined | Genome-fragment | NA | NA | 0 |
| S12_k141_220917 | 1492 | No | NA | 2 | 0 | 1 | Not-determined | Genome-fragment | NA | NA | 0 |
| S12_k141_137532 | 4106 | No | NA | 2 | 0 | 1 | Not-determined | Genome-fragment | NA | NA | 0 |
| S12_k141_3764 | 2375 | No | NA | 4 | 0 | 2 | Not-determined | Genome-fragment | NA | NA | 0 |
| S12_k141_67773 | 3298 | No | NA | 4 | 0 | 1 | Low-quality | Genome-fragment | 3.83 | AAI-based (medium-confidence) | 0 |
| S12_k141_37281 | 3113 | No | NA | 3 | 0 | 2 | Not-determined | Genome-fragment | NA | NA | 0 |
| S2-final-viral-combined_k141_90952\|\|full | 3298 | Yes | 2720 | 4 | 2 | 1 | Low-quality | Genome-fragment | 4.21 | AAI-based (high-confidence) | 17.53 |
| S12_k141_254232 | 3194 | No | NA | 6 | 0 | 0 | Not-determined | Genome-fragment | NA | NA | 0 |
| S8_k141_203434 | 3298 | No | NA | 3 | 0 | 1 | Low-quality | Genome-fragment | 6.09 | AAI-based (high-confidence) | 0 |
| S12_k141_271183 | 1009 | No | NA | 1 | 0 | 0 | Not-determined | Genome-fragment | NA | NA | 0 |
| S12_k141_337922 | 2640 | No | NA | 1 | 0 | 0 | Not-determined | Genome-fragment | NA | NA | 0 |
| S12_k141_288468 | 1062 | No | NA | 2 | 0 | 2 | Not-determined | Genome-fragment | NA | NA | 0 |
| S12_k141_271203 | 1160 | No | NA | 3 | 0 | 2 | Not-determined | Genome-fragment | NA | NA | 0 |
| S12_k141_103844 | 1735 | No | NA | 2 | 0 | 0 | Not-determined | Genome-fragment | NA | NA | 0 |
| S12-final-viral-combined_k141_243477\|\|full | 3299 | No | NA | 5 | 0 | 0 | Low-quality | Genome-fragment | 2.37 | AAI-based (medium-confidence) | 0 |
| S12_k141_271242 | 1124 | No | NA | 1 | 0 | 0 | Not-determined | Genome-fragment | NA | NA | 0 |
| S12_k141_237945 | 1168 | No | NA | 2 | 0 | 2 | Not-determined | Genome-fragment | NA | NA | 0 |
| S12_k141_388698 | 4026 | No | NA | 4 | 0 | 3 | Not-determined | Genome-fragment | NA | NA | 0 |
| S11_k141_81322 | 3300 | Yes | 2859 | 6 | 2 | 1 | Low-quality | Genome-fragment | 5.01 | AAI-based (high-confidence) | 13.36 |
| S12_k141_237952 | 2233 | No | NA | 2 | 0 | 1 | Not-determined | Genome-fragment | NA | NA | 0 |
| S12_k141_305408 | 1111 | No | NA | 1 | 0 | 0 | Not-determined | Genome-fragment | NA | NA | 0 |
| S12_k141_305411 | 4496 | No | NA | 2 | 0 | 2 | Not-determined | Genome-fragment | NA | NA | 0 |
| S12_k141_221009 | 1573 | No | NA | 2 | 0 | 0 | Not-determined | Genome-fragment | NA | NA | 0 |
| S12_k141_120296 | 1934 | No | NA | 3 | 0 | 1 | Not-determined | Genome-fragment | NA | NA | 0 |
| S12_k141_305422 | 1472 | No | NA | 3 | 0 | 1 | Not-determined | Genome-fragment | NA | NA | 0 |
| S17-final-viral-combined_k141_215974\|\|full | 3300 | Yes | 3039 | 4 | 1 | 1 | Low-quality | Genome-fragment | 3.41 | HMM-based (lower-bound) | 7.91 |
| S12_k141_71013 | 1184 | No | NA | 2 | 0 | 2 | Not-determined | Genome-fragment | NA | NA | 0 |
| S12_k141_120317 | 1337 | No | NA | 2 | 0 | 1 | Not-determined | Genome-fragment | NA | NA | 0 |
| S12_k141_271316 | 3542 | No | NA | 3 | 0 | 3 | Not-determined | Genome-fragment | NA | NA | 0 |
| S12_k141_20320 | 1198 | No | NA | 2 | 0 | 1 | Not-determined | Genome-fragment | NA | NA | 0 |
| S8-final-viral-combined_k141_85842\|\|full | 3300 | No | NA | 8 | 2 | 0 | Low-quality | Genome-fragment | 7.82 | AAI-based (high-confidence) | 0 |
| S12_k141_350656 | 3301 | No | NA | 11 | 2 | 0 | Low-quality | Genome-fragment | 10.08 | AAI-based (high-confidence) | 0 |
| S1_k141_101175 | 3302 | No | NA | 3 | 0 | 3 | Low-quality | Genome-fragment | 2.62 | AAI-based (medium-confidence) | 0 |
| S12_k141_137729 | 1573 | No | NA | 1 | 0 | 0 | Not-determined | Genome-fragment | NA | NA | 0 |
| S8-final-viral-combined_k141_273217\|\|full | 3302 | No | NA | 5 | 1 | 0 | Low-quality | Genome-fragment | 4.41 | AAI-based (medium-confidence) | 0 |
| S11-final-viral-combined_k141_224508\|\|full | 3303 | No | NA | 5 | 0 | 3 | Low-quality | Genome-fragment | 1.53 | AAI-based (medium-confidence) | 0 |
| S12_k141_37402 | 1009 | No | NA | 2 | 0 | 0 | Not-determined | Genome-fragment | NA | NA | 0 |
| S12_k141_53872 | 1177 | No | NA | 2 | 0 | 0 | Not-determined | Genome-fragment | NA | NA | 0 |
| S12_k141_3893 | 3229 | No | NA | 2 | 0 | 2 | Not-determined | Genome-fragment | NA | NA | 0 |
| S12_k141_53885 | 1626 | No | NA | 1 | 0 | 0 | Not-determined | Genome-fragment | NA | NA | 0 |
| S12_k141_71074 | 5287 | No | NA | 6 | 0 | 6 | Not-determined | Genome-fragment | NA | NA | 0 |
| S12_k141_171144 | 2001 | No | NA | 1 | 0 | 0 | Not-determined | Genome-fragment | NA | NA | 0 |
| S12_k141_354958 | 3035 | No | NA | 4 | 0 | 0 | Not-determined | Genome-fragment | NA | NA | 0 |
| S1-final-viral-combined_k141_291380\|\|full | 3303 | No | NA | 5 | 0 | 0 | Low-quality | Genome-fragment | 7.19 | AAI-based (medium-confidence) | 0 |
| S12_k141_20417 | 4678 | No | NA | 2 | 0 | 2 | Not-determined | Genome-fragment | NA | NA | 0 |
| S12_k141_221060 | 1039 | No | NA | 3 | 0 | 2 | Not-determined | Genome-fragment | NA | NA | 0 |
| S12_k141_388860 | 2241 | No | NA | 3 | 0 | 1 | Not-determined | Genome-fragment | NA | NA | 0 |
| S12_k141_372266 | 1086 | No | NA | 3 | 0 | 2 | Not-determined | Genome-fragment | NA | NA | 0 |
| S12_k141_305533 | 2698 | No | NA | 1 | 0 | 0 | Not-determined | Genome-fragment | NA | NA | 0 |
| S12_k141_355006 | 1126 | No | NA | 2 | 0 | 2 | Not-determined | Genome-fragment | NA | NA | 0 |
| S9_k141_93207 | 3304 | No | NA | 6 | 3 | 0 | Low-quality | Genome-fragment | 5.79 | HMM-based (lower-bound) | 0 |
| S12_k141_355013 | 1246 | No | NA | 1 | 0 | 1 | Not-determined | Genome-fragment | NA | NA | 0 |
| S8-final-viral-combined_k141_45310\|\|full | 3304 | No | NA | 6 | 0 | 0 | Low-quality | Genome-fragment | 5.62 | AAI-based (medium-confidence) | 0 |
| S12_k141_171207 | 6471 | No | NA | 8 | 0 | 2 | Not-determined | Genome-fragment | NA | NA | 0 |
| S12_k141_254423 | 1177 | No | NA | 2 | 0 | 1 | Not-determined | Genome-fragment | NA | NA | 0 |
| S8_k141_270783 | 3304 | No | NA | 6 | 0 | 0 | Low-quality | Genome-fragment | 6.6 | AAI-based (medium-confidence) | 0 |
| S12_k141_171230 | 1109 | No | NA | 1 | 0 | 0 | Not-determined | Genome-fragment | NA | NA | 0 |
| S7-final-viral-combined_k141_184416\|\|full | 3305 | No | NA | 3 | 0 | 0 | Low-quality | Genome-fragment | 4.01 | AAI-based (high-confidence) | 0 |
| S12_k141_120476 | 1180 | No | NA | 2 | 0 | 0 | Not-determined | Genome-fragment | NA | NA | 0 |
| S12_k141_120477 | 1351 | No | NA | 1 | 0 | 0 | Not-determined | Genome-fragment | NA | NA | 0 |
| S12_k141_288596 | 1265 | No | NA | 1 | 0 | 1 | Not-determined | Genome-fragment | NA | NA | 0 |
| S17-final-viral-combined_k141_90382\|\|full | 3305 | No | NA | 7 | 1 | 0 | Low-quality | Genome-fragment | 5.88 | HMM-based (lower-bound) | 0 |
| S12_k141_204588 | 1338 | No | NA | 1 | 0 | 0 | Not-determined | Genome-fragment | NA | NA | 0 |
| S12_k141_20510 | 1288 | No | NA | 1 | 0 | 1 | Not-determined | Genome-fragment | NA | NA | 0 |
| S12_k141_137885 | 1133 | No | NA | 1 | 0 | 1 | Not-determined | Genome-fragment | NA | NA | 0 |
| S12_k141_71210 | 1215 | No | NA | 2 | 0 | 1 | Not-determined | Genome-fragment | NA | NA | 0 |
| S12_k141_20532 | 3301 | No | NA | 3 | 0 | 2 | Not-determined | Genome-fragment | NA | NA | 0 |
| S12_k141_137901 | 1847 | No | NA | 3 | 0 | 0 | Not-determined | Genome-fragment | NA | NA | 0 |
| S12-final-viral-combined_k141_281370\|\|full | 3305 | Yes | 630 | 4 | 1 | 2 | Low-quality | Genome-fragment | 0.36 | HMM-based (lower-bound) | 80.94 |
| S12_k141_20569 | 1192 | No | NA | 2 | 0 | 1 | Not-determined | Genome-fragment | NA | NA | 0 |
| S12_k141_154511 | 3531 | No | NA | 7 | 0 | 3 | Not-determined | Genome-fragment | NA | NA | 0 |
| S12_k141_188511 | 1122 | No | NA | 2 | 0 | 1 | Not-determined | Genome-fragment | NA | NA | 0 |
| S12_k141_254504 | 1521 | No | NA | 1 | 0 | 1 | Not-determined | Genome-fragment | NA | NA | 0 |
| S12_k141_3996 | 1137 | No | NA | 2 | 0 | 0 | Not-determined | Genome-fragment | NA | NA | 0 |
| S12_k141_154555 | 1258 | No | NA | 4 | 0 | 0 | Not-determined | Genome-fragment | NA | NA | 0 |
| S12_k141_254530 | 1070 | No | NA | 2 | 0 | 0 | Not-determined | Genome-fragment | NA | NA | 0 |
| S12_k141_322089 | 1104 | No | NA | 1 | 0 | 1 | Not-determined | Genome-fragment | NA | NA | 0 |
| S12_k141_271637 | 1383 | No | NA | 1 | 0 | 0 | Not-determined | Genome-fragment | NA | NA | 0 |
| S12_k141_71295 | 10829 | No | NA | 14 | 0 | 3 | Not-determined | Genome-fragment | NA | NA | 0 |
| S12_k141_120636 | 1097 | No | NA | 1 | 0 | 0 | Not-determined | Genome-fragment | NA | NA | 0 |
| S12_k141_104198 | 2630 | No | NA | 1 | 0 | 0 | Not-determined | Genome-fragment | NA | NA | 0 |
| S12_k141_188576 | 5969 | No | NA | 11 | 0 | 1 | Not-determined | Genome-fragment | NA | NA | 0 |
| S8-final-viral-combined_k141_106254\|\|full | 3305 | No | NA | 5 | 0 | 5 | Low-quality | Genome-fragment | 1.23 | AAI-based (medium-confidence) | 0 |
| S12_k141_271691 | 2559 | No | NA | 1 | 0 | 0 | Not-determined | Genome-fragment | NA | NA | 0 |
| S12_k141_171390 | 4281 | No | NA | 3 | 0 | 2 | Not-determined | Genome-fragment | NA | NA | 0 |
| S12_k141_71362 | 5716 | No | NA | 1 | 0 | 1 | Not-determined | Genome-fragment | NA | NA | 0 |
| S9-final-viral-combined_k141_19005\|\|full | 3306 | No | NA | 4 | 0 | 1 | Low-quality | Genome-fragment | 5.31 | AAI-based (medium-confidence) | 0 |
| S12_k141_104223 | 1766 | No | NA | 1 | 0 | 0 | Not-determined | Genome-fragment | NA | NA | 0 |
| S8-final-viral-combined_k141_161928\|\|full | 3306 | No | NA | 3 | 0 | 1 | Low-quality | Genome-fragment | 1.64 | AAI-based (medium-confidence) | 0 |
| S12_k141_389023 | 3713 | No | NA | 3 | 0 | 3 | Not-determined | Genome-fragment | NA | NA | 0 |
| S5-final-viral-combined_k141_210046\|\|full | 3308 | No | NA | 6 | 2 | 0 | Low-quality | Genome-fragment | 7.4 | AAI-based (medium-confidence) | 0 |
| S12_k141_204755 | 1149 | No | NA | 1 | 0 | 1 | Not-determined | Genome-fragment | NA | NA | 0 |
| S12_k141_204767 | 1988 | No | NA | 2 | 0 | 1 | Not-determined | Genome-fragment | NA | NA | 0 |
| S12_k141_355173 | 1338 | No | NA | 3 | 0 | 1 | Not-determined | Genome-fragment | NA | NA | 0 |
| S12_k141_254638 | 1005 | No | NA | 1 | 0 | 0 | Not-determined | Genome-fragment | NA | NA | 0 |
| S12_k141_389079 | 5066 | No | NA | 7 | 0 | 1 | Not-determined | Genome-fragment | NA | NA | 0 |
| S12_k141_305750 | 1653 | No | NA | 3 | 0 | 0 | Not-determined | Genome-fragment | NA | NA | 0 |
| S12_k141_138115 | 1845 | No | NA | 2 | 0 | 2 | Not-determined | Genome-fragment | NA | NA | 0 |
| S12-final-viral-combined_k141_68410\|\|full | 3308 | No | NA | 7 | 1 | 0 | Low-quality | Genome-fragment | 2.02 | HMM-based (lower-bound) | 0 |
| S12_k141_389107 | 1280 | No | NA | 1 | 0 | 0 | Not-determined | Genome-fragment | NA | NA | 0 |
| S12-final-viral-combined_k141_371792\|\|full | 3309 | No | NA | 6 | 0 | 0 | Low-quality | Genome-fragment | 6.77 | AAI-based (medium-confidence) | 0 |
| S12_k141_288747 | 1558 | No | NA | 3 | 0 | 0 | Not-determined | Genome-fragment | NA | NA | 0 |
| S12_k141_188741 | 1359 | No | NA | 1 | 0 | 1 | Not-determined | Genome-fragment | NA | NA | 0 |
| S12_k141_305762 | 1134 | No | NA | 3 | 0 | 2 | Not-determined | Genome-fragment | NA | NA | 0 |
| S12_k141_254711 | 3695 | No | NA | 5 | 0 | 3 | Not-determined | Genome-fragment | NA | NA | 0 |
| S12_k141_71567 | 7371 | No | NA | 3 | 0 | 0 | Not-determined | Genome-fragment | NA | NA | 0 |
| S12_k141_120796 | 4592 | No | NA | 3 | 0 | 1 | Not-determined | Genome-fragment | NA | NA | 0 |
| S12_k141_35754 | 3309 | No | NA | 4 | 0 | 1 | Low-quality | Genome-fragment | 1.96 | AAI-based (medium-confidence) | 0 |
| S12_k141_221513 | 1524 | No | NA | 2 | 0 | 2 | Not-determined | Genome-fragment | NA | NA | 0 |
| S4_k141_127624 | 3309 | No | NA | 4 | 0 | 0 | Low-quality | Genome-fragment | 6.34 | AAI-based (medium-confidence) | 0 |
| S12_k141_221518 | 2075 | No | NA | 1 | 0 | 0 | Not-determined | Genome-fragment | NA | NA | 0 |
| S1_k141_102902 | 3310 | Yes | 1868 | 4 | 1 | 1 | Low-quality | Genome-fragment | 3.39 | HMM-based (lower-bound) | 43.56 |
| S12_k141_372682 | 1021 | No | NA | 2 | 0 | 0 | Not-determined | Genome-fragment | NA | NA | 0 |
| S12_k141_138278 | 2111 | No | NA | 1 | 0 | 1 | Not-determined | Genome-fragment | NA | NA | 0 |
| S7-final-viral-combined_k141_85113\|\|full | 3310 | No | NA | 3 | 2 | 0 | Low-quality | Genome-fragment | 4.66 | AAI-based (medium-confidence) | 0 |
| S12_k141_71660 | 3088 | No | NA | 2 | 0 | 0 | Not-determined | Genome-fragment | NA | NA | 0 |
| S12_k141_20790 | 2118 | No | NA | 2 | 0 | 0 | Not-determined | Genome-fragment | NA | NA | 0 |
| S12_k141_254810 | 3660 | No | NA | 2 | 0 | 2 | Not-determined | Genome-fragment | NA | NA | 0 |
| S5-final-viral-combined_k141_59565\|\|full | 3311 | No | NA | 3 | 0 | 2 | Low-quality | Genome-fragment | 0.34 | AAI-based (medium-confidence) | 0 |
| S12_k141_254815 | 1079 | No | NA | 1 | 0 | 0 | Not-determined | Genome-fragment | NA | NA | 0 |
| S8-final-viral-combined_k141_38918\|\|full | 3311 | No | NA | 5 | 1 | 0 | Low-quality | Genome-fragment | 2.09 | HMM-based (lower-bound) | 0 |
| S12_k141_288903 | 3151 | No | NA | 3 | 0 | 1 | Not-determined | Genome-fragment | NA | NA | 0 |
| S12_k141_188855 | 1424 | No | NA | 2 | 0 | 1 | Not-determined | Genome-fragment | NA | NA | 0 |
| S12_k141_87679 | 1288 | No | NA | 2 | 0 | 0 | Not-determined | Genome-fragment | NA | NA | 0 |
| S12_k141_54300 | 1041 | No | NA | 1 | 0 | 0 | Not-determined | Genome-fragment | NA | NA | 0 |
| S17-final-viral-combined_k141_111858\|\|full | 3312 | No | NA | 11 | 3 | 0 | Low-quality | Genome-fragment | 8.64 | AAI-based (medium-confidence) | 0 |
| S12_k141_4178 | 1015 | No | NA | 3 | 0 | 3 | Not-determined | Genome-fragment | NA | NA | 0 |
| S12_k141_54314 | 1065 | No | NA | 2 | 0 | 1 | Not-determined | Genome-fragment | NA | NA | 0 |
| S12_k141_204954 | 1124 | No | NA | 1 | 0 | 0 | Not-determined | Genome-fragment | NA | NA | 0 |
| S4-final-viral-combined_k141_151509\|\|full | 3312 | No | NA | 3 | 0 | 2 | Low-quality | Genome-fragment | 8.14 | AAI-based (medium-confidence) | 0 |
| S1-final-viral-combined_k141_105504\|\|full | 3313 | No | NA | 4 | 0 | 2 | Low-quality | Genome-fragment | 2.04 | AAI-based (medium-confidence) | 0 |
| S12_k141_389265 | 5309 | No | NA | 4 | 0 | 1 | Not-determined | Genome-fragment | NA | NA | 0 |
| S12_k141_87735 | 5317 | No | NA | 3 | 0 | 2 | Not-determined | Genome-fragment | NA | NA | 0 |
| S1-final-viral-combined_k141_12877\|\|full | 3313 | No | NA | 4 | 0 | 0 | Low-quality | Genome-fragment | 9.46 | AAI-based (medium-confidence) | 0 |
| S12_k141_372801 | 1019 | No | NA | 1 | 0 | 1 | Not-determined | Genome-fragment | NA | NA | 0 |
| S12_k141_221693 | 5063 | No | NA | 4 | 0 | 4 | Not-determined | Genome-fragment | NA | NA | 0 |
| S12_k141_54354 | 1238 | No | NA | 1 | 0 | 1 | Not-determined | Genome-fragment | NA | NA | 0 |
| S11-final-viral-combined_k141_296448\|\|full | 3314 | No | NA | 3 | 0 | 3 | Low-quality | Genome-fragment | 7.02 | AAI-based (medium-confidence) | 0 |
| S12_k141_154970 | 11765 | No | NA | 12 | 0 | 0 | Not-determined | Genome-fragment | NA | NA | 0 |
| S12_k141_338771 | 2374 | No | NA | 2 | 0 | 1 | Not-determined | Genome-fragment | NA | NA | 0 |
| S12_k141_54394 | 3139 | No | NA | 6 | 0 | 1 | Not-determined | Genome-fragment | NA | NA | 0 |
| S12_k141_4284 | 3873 | No | NA | 2 | 0 | 1 | Not-determined | Genome-fragment | NA | NA | 0 |
| S12_k141_288983 | 2693 | No | NA | 1 | 0 | 0 | Not-determined | Genome-fragment | NA | NA | 0 |
| S1-final-viral-combined_k141_13979\|\|full | 3314 | No | NA | 11 | 1 | 0 | Low-quality | Genome-fragment | 6.74 | AAI-based (high-confidence) | 0 |
| S12_k141_87806 | 2410 | No | NA | 3 | 0 | 1 | Not-determined | Genome-fragment | NA | NA | 0 |
| S5_k141_33143 | 3314 | No | NA | 4 | 0 | 1 | Low-quality | Genome-fragment | 2.03 | AAI-based (medium-confidence) | 0 |
| S12_k141_37935 | 5463 | No | NA | 4 | 0 | 0 | Not-determined | Genome-fragment | NA | NA | 0 |
| S1_k141_53739 | 3315 | No | NA | 1 | 0 | 0 | Low-quality | Genome-fragment | 7.86 | AAI-based (high-confidence) | 0 |
| S12_k141_37950 | 1312 | No | NA | 2 | 0 | 1 | Not-determined | Genome-fragment | NA | NA | 0 |
| S12_k141_4299 | 1240 | No | NA | 3 | 0 | 0 | Not-determined | Genome-fragment | NA | NA | 0 |
| S12_k141_221821 | 1216 | No | NA | 1 | 0 | 1 | Not-determined | Genome-fragment | NA | NA | 0 |
| S12_k141_138453 | 1139 | No | NA | 2 | 0 | 0 | Not-determined | Genome-fragment | NA | NA | 0 |
| S12_k141_171531 | 1674 | No | NA | 1 | 0 | 1 | Not-determined | Genome-fragment | NA | NA | 0 |
| S12_k141_322554 | 3563 | No | NA | 2 | 0 | 2 | Not-determined | Genome-fragment | NA | NA | 0 |
| S9_k141_103955 | 3316 | No | NA | 4 | 0 | 0 | Low-quality | Genome-fragment | 5.31 | AAI-based (high-confidence) | 0 |
| S2-final-viral-combined_k141_66557\|\|full | 3316 | No | NA | 4 | 2 | 0 | Low-quality | Genome-fragment | 7.73 | AAI-based (medium-confidence) | 0 |
| S12_k141_289088 | 1256 | No | NA | 2 | 0 | 0 | Not-determined | Genome-fragment | NA | NA | 0 |
| S12_k141_171563 | 3349 | No | NA | 2 | 0 | 0 | Not-determined | Genome-fragment | NA | NA | 0 |
| S12_k141_205114 | 1120 | No | NA | 2 | 0 | 1 | Not-determined | Genome-fragment | NA | NA | 0 |
| S12_k141_372949 | 1263 | No | NA | 2 | 0 | 1 | Not-determined | Genome-fragment | NA | NA | 0 |
| S2-final-viral-combined_k141_213214\|\|full | 3317 | No | NA | 3 | 0 | 3 | Low-quality | Genome-fragment | 0.31 | AAI-based (medium-confidence) | 0 |
| S12_k141_4362 | 1862 | No | NA | 3 | 0 | 1 | Not-determined | Genome-fragment | NA | NA | 0 |
| S2-final-viral-combined_k141_213916\|\|full | 3317 | No | NA | 4 | 1 | 0 | Low-quality | Genome-fragment | 3.72 | HMM-based (lower-bound) | 0 |
| S12_k141_221892 | 1059 | No | NA | 1 | 0 | 1 | Not-determined | Genome-fragment | NA | NA | 0 |
| S12_k141_306204 | 2765 | No | NA | 4 | 0 | 1 | Not-determined | Genome-fragment | NA | NA | 0 |
| S2-final-viral-combined_k141_8216\|\|full | 3317 | No | NA | 4 | 0 | 0 | Low-quality | Genome-fragment | 10.14 | AAI-based (medium-confidence) | 0 |
| S12_k141_255023 | 1318 | No | NA | 2 | 0 | 0 | Not-determined | Genome-fragment | NA | NA | 0 |
| S12_k141_205150 | 3016 | No | NA | 3 | 0 | 3 | Not-determined | Genome-fragment | NA | NA | 0 |
| S12_k141_272178 | 1022 | No | NA | 1 | 0 | 0 | Not-determined | Genome-fragment | NA | NA | 0 |
| S12_k141_255041 | 4187 | No | NA | 2 | 0 | 2 | Not-determined | Genome-fragment | NA | NA | 0 |
| S12_k141_289197 | 2444 | No | NA | 1 | 0 | 0 | Not-determined | Genome-fragment | NA | NA | 0 |
| S12_k141_138598 | 2216 | No | NA | 3 | 0 | 1 | Not-determined | Genome-fragment | NA | NA | 0 |
| S12_k141_138599 | 1479 | No | NA | 2 | 0 | 1 | Not-determined | Genome-fragment | NA | NA | 0 |
| S4-final-viral-combined_k141_126705\|\|full | 3317 | Yes | 1566 | 4 | 1 | 1 | Low-quality | Genome-fragment | 2.23 | HMM-based (lower-bound) | 52.79 |
| S12_k141_338939 | 1046 | No | NA | 1 | 0 | 0 | Not-determined | Genome-fragment | NA | NA | 0 |
| S8-final-viral-combined_k141_12361\|\|full | 3318 | No | NA | 4 | 0 | 2 | Low-quality | Genome-fragment | 3.94 | AAI-based (high-confidence) | 0 |
| S12_k141_322600 | 1307 | No | NA | 3 | 0 | 2 | Not-determined | Genome-fragment | NA | NA | 0 |
| S1_k141_164399 | 3319 | No | NA | 7 | 0 | 0 | Low-quality | Genome-fragment | 6.88 | AAI-based (high-confidence) | 0 |
| S12_k141_189181 | 1226 | No | NA | 1 | 0 | 1 | Not-determined | Genome-fragment | NA | NA | 0 |
| S12_k141_84867 | 3319 | Yes | 649 | 5 | 1 | 1 | Low-quality | Genome-fragment | 0.7 | AAI-based (high-confidence) | 80.45 |
| S12_k141_238774 | 1076 | No | NA | 1 | 0 | 1 | Not-determined | Genome-fragment | NA | NA | 0 |
| S12_k141_104697 | 8408 | No | NA | 9 | 0 | 2 | Not-determined | Genome-fragment | NA | NA | 0 |
| S12_k141_322661 | 1401 | No | NA | 3 | 0 | 1 | Not-determined | Genome-fragment | NA | NA | 0 |
| S12_k141_272308 | 2336 | No | NA | 3 | 0 | 1 | Not-determined | Genome-fragment | NA | NA | 0 |
| S12_k141_238797 | 1041 | No | NA | 1 | 0 | 0 | Not-determined | Genome-fragment | NA | NA | 0 |
| S11-final-viral-combined_k141_271652\|\|full | 3320 | No | NA | 4 | 3 | 0 | Low-quality | Genome-fragment | 7.02 | AAI-based (high-confidence) | 0 |
| S12_k141_71942 | 2184 | No | NA | 4 | 0 | 0 | Not-determined | Genome-fragment | NA | NA | 0 |
| S1-final-viral-combined_k141_132092\|\|full | 3321 | No | NA | 4 | 0 | 2 | Low-quality | Genome-fragment | 6.89 | AAI-based (medium-confidence) | 0 |
| S12_k141_21261 | 1479 | No | NA | 3 | 0 | 1 | Not-determined | Genome-fragment | NA | NA | 0 |
| S12_k141_171743 | 1722 | No | NA | 1 | 0 | 1 | Not-determined | Genome-fragment | NA | NA | 0 |
| S12_k141_322703 | 1166 | No | NA | 1 | 0 | 0 | Not-determined | Genome-fragment | NA | NA | 0 |
| S12_k141_88088 | 1545 | No | NA | 2 | 0 | 0 | Not-determined | Genome-fragment | NA | NA | 0 |
| S12_k141_88091 | 1504 | No | NA | 1 | 0 | 1 | Not-determined | Genome-fragment | NA | NA | 0 |
| S12_k141_138744 | 1170 | No | NA | 1 | 0 | 1 | Not-determined | Genome-fragment | NA | NA | 0 |
| S12_k141_171769 | 3987 | No | NA | 1 | 0 | 1 | Not-determined | Genome-fragment | NA | NA | 0 |
| S7-final-viral-combined_k141_177871\|\|full | 3321 | No | NA | 3 | 0 | 2 | Low-quality | Genome-fragment | 1.91 | AAI-based (medium-confidence) | 0 |
| S12_k141_54595 | 3104 | No | NA | 3 | 0 | 0 | Not-determined | Genome-fragment | NA | NA | 0 |
| S12_k141_71981 | 8191 | No | NA | 6 | 0 | 3 | Not-determined | Genome-fragment | NA | NA | 0 |
| S12_k141_104771 | 5312 | No | NA | 2 | 0 | 2 | Not-determined | Genome-fragment | NA | NA | 0 |
| S12_k141_389666 | 1924 | No | NA | 3 | 0 | 1 | Not-determined | Genome-fragment | NA | NA | 0 |
| S12-final-viral-combined_k141_189992\|\|full | 3321 | No | NA | 6 | 0 | 0 | Low-quality | Genome-fragment | 10.38 | AAI-based (high-confidence) | 0 |
| S12_k141_289420 | 1210 | No | NA | 1 | 0 | 1 | Not-determined | Genome-fragment | NA | NA | 0 |
| S12_k141_4543 | 4124 | No | NA | 7 | 0 | 0 | Not-determined | Genome-fragment | NA | NA | 0 |
| S4-final-viral-combined_k141_55706\|\|full | 3321 | No | NA | 3 | 0 | 2 | Low-quality | Genome-fragment | 5.2 | AAI-based (high-confidence) | 0 |
| S12_k141_189246 | 1322 | No | NA | 2 | 0 | 0 | Not-determined | Genome-fragment | NA | NA | 0 |
| S12_k141_38193 | 1218 | No | NA | 1 | 0 | 1 | Not-determined | Genome-fragment | NA | NA | 0 |
| S12_k141_121151 | 1117 | No | NA | 2 | 0 | 2 | Not-determined | Genome-fragment | NA | NA | 0 |
| S12_k141_306399 | 1404 | No | NA | 2 | 0 | 0 | Not-determined | Genome-fragment | NA | NA | 0 |
| S11-final-viral-combined_k141_227427\|\|full | 3322 | No | NA | 4 | 0 | 0 | Low-quality | Genome-fragment | 5.89 | AAI-based (medium-confidence) | 0 |
| S12_k141_38220 | 2160 | No | NA | 3 | 0 | 2 | Not-determined | Genome-fragment | NA | NA | 0 |
| S12_k141_38223 | 1503 | No | NA | 2 | 0 | 1 | Not-determined | Genome-fragment | NA | NA | 0 |
| S12_k141_205346 | 1867 | No | NA | 1 | 0 | 0 | Not-determined | Genome-fragment | NA | NA | 0 |
| S12_k141_339104 | 1763 | No | NA | 4 | 0 | 3 | Not-determined | Genome-fragment | NA | NA | 0 |
| S12_k141_21347 | 3087 | No | NA | 6 | 0 | 2 | Not-determined | Genome-fragment | NA | NA | 0 |
| S12_k141_189314 | 1092 | No | NA | 1 | 0 | 0 | Not-determined | Genome-fragment | NA | NA | 0 |
| S12_k141_38245 | 1015 | No | NA | 1 | 0 | 0 | Not-determined | Genome-fragment | NA | NA | 0 |
| S12_k141_221990 | 1370 | No | NA | 2 | 0 | 2 | Not-determined | Genome-fragment | NA | NA | 0 |
| S12_k141_238923 | 1919 | No | NA | 2 | 0 | 1 | Not-determined | Genome-fragment | NA | NA | 0 |
| S12_k141_21358 | 14258 | No | NA | 11 | 0 | 7 | Not-determined | Genome-fragment | NA | NA | 0 |
| S12_k141_289540 | 1768 | No | NA | 1 | 0 | 0 | Not-determined | Genome-fragment | NA | NA | 0 |
| S12_k141_289548 | 1122 | No | NA | 2 | 0 | 0 | Not-determined | Genome-fragment | NA | NA | 0 |
| S12_k141_355700 | 1168 | No | NA | 1 | 0 | 0 | Not-determined | Genome-fragment | NA | NA | 0 |
| S12_k141_155188 | 4510 | No | NA | 4 | 0 | 1 | Not-determined | Genome-fragment | NA | NA | 0 |
| S17-final-viral-combined_k141_229212\|\|full | 3322 | No | NA | 3 | 3 | 0 | Low-quality | Genome-fragment | 9.63 | AAI-based (high-confidence) | 0 |
| S12_k141_238970 | 2036 | No | NA | 2 | 0 | 1 | Not-determined | Genome-fragment | NA | NA | 0 |
| S12_k141_98757 | 3322 | No | NA | 3 | 3 | 0 | Low-quality | Genome-fragment | 8.69 | AAI-based (high-confidence) | 0 |
| S12_k141_272489 | 3184 | No | NA | 4 | 0 | 3 | Not-determined | Genome-fragment | NA | NA | 0 |
| S4-final-viral-combined_k141_146589\|\|full | 3322 | No | NA | 7 | 0 | 0 | Low-quality | Genome-fragment | 3.07 | AAI-based (medium-confidence) | 0 |
| S3_k141_13859 | 3323 | No | NA | 11 | 2 | 0 | Low-quality | Genome-fragment | 5.75 | AAI-based (high-confidence) | 0 |
| S12_k141_72136 | 2441 | No | NA | 3 | 0 | 0 | Not-determined | Genome-fragment | NA | NA | 0 |
| S12_k141_239012 | 4683 | No | NA | 11 | 0 | 3 | Not-determined | Genome-fragment | NA | NA | 0 |
| S12_k141_373345 | 1069 | No | NA | 1 | 0 | 1 | Not-determined | Genome-fragment | NA | NA | 0 |
| S12_k141_339185 | 1248 | No | NA | 2 | 0 | 0 | Not-determined | Genome-fragment | NA | NA | 0 |
| S17-final-viral-combined_k141_147925\|\|full | 3323 | Yes | 2595 | 5 | 1 | 1 | Low-quality | Genome-fragment | 1.63 | HMM-based (lower-bound) | 21.91 |
| S12_k141_389767 | 5271 | No | NA | 3 | 0 | 0 | Not-determined | Genome-fragment | NA | NA | 0 |
| S2-final-viral-combined_k141_185082\|\|full | 3323 | No | NA | 6 | 0 | 0 | Low-quality | Genome-fragment | 7.3 | AAI-based (high-confidence) | 0 |
| S12_k141_373399 | 1325 | No | NA | 2 | 0 | 2 | Not-determined | Genome-fragment | NA | NA | 0 |
| S12_k141_138921 | 2558 | No | NA | 2 | 0 | 0 | Not-determined | Genome-fragment | NA | NA | 0 |
| S12_k141_255409 | 12821 | No | NA | 11 | 0 | 1 | Not-determined | Genome-fragment | NA | NA | 0 |
| S12_k141_306515 | 1731 | No | NA | 2 | 0 | 0 | Not-determined | Genome-fragment | NA | NA | 0 |
| S12_k141_323024 | 1630 | No | NA | 3 | 0 | 0 | Not-determined | Genome-fragment | NA | NA | 0 |
| S12_k141_121301 | 1249 | No | NA | 2 | 0 | 0 | Not-determined | Genome-fragment | NA | NA | 0 |
| S12_k141_38501 | 2230 | No | NA | 3 | 0 | 1 | Not-determined | Genome-fragment | NA | NA | 0 |
| S12_k141_105140 | 1367 | No | NA | 2 | 0 | 1 | Not-determined | Genome-fragment | NA | NA | 0 |
| S12_k141_155415 | 1330 | No | NA | 1 | 0 | 1 | Not-determined | Genome-fragment | NA | NA | 0 |
| S12_k141_272637 | 10142 | No | NA | 1 | 0 | 0 | Not-determined | Genome-fragment | NA | NA | 0 |
| S12_k141_255472 | 1235 | No | NA | 1 | 0 | 1 | Not-determined | Genome-fragment | NA | NA | 0 |
| S12_k141_54752 | 1615 | No | NA | 2 | 0 | 1 | Not-determined | Genome-fragment | NA | NA | 0 |
| S12_k141_355889 | 1098 | No | NA | 1 | 0 | 1 | Not-determined | Genome-fragment | NA | NA | 0 |
| S12_k141_339373 | 1747 | No | NA | 2 | 0 | 0 | Not-determined | Genome-fragment | NA | NA | 0 |
| S11-final-viral-combined_k141_196505\|\|full | 3324 | No | NA | 5 | 0 | 3 | Low-quality | Genome-fragment | 2.54 | AAI-based (medium-confidence) | 0 |
| S2-final-viral-combined_k141_79562\|\|full | 3325 | No | NA | 3 | 0 | 2 | Low-quality | Genome-fragment | 3.06 | AAI-based (medium-confidence) | 0 |
| S12_k141_355902 | 1071 | No | NA | 1 | 0 | 1 | Not-determined | Genome-fragment | NA | NA | 0 |
| S4_k141_1954 | 3325 | No | NA | 3 | 0 | 1 | Low-quality | Genome-fragment | 5.3 | AAI-based (medium-confidence) | 0 |
| S12_k141_72313 | 1028 | No | NA | 1 | 0 | 0 | Not-determined | Genome-fragment | NA | NA | 0 |
| S1-final-viral-combined_k141_69353\|\|full | 3326 | No | NA | 3 | 0 | 2 | Low-quality | Genome-fragment | 1.57 | AAI-based (medium-confidence) | 0 |
| S12_k141_4686 | 2190 | No | NA | 3 | 0 | 2 | Not-determined | Genome-fragment | NA | NA | 0 |
| S12_k141_155523 | 1920 | No | NA | 2 | 0 | 0 | Not-determined | Genome-fragment | NA | NA | 0 |
| S1_k141_135140 | 3326 | No | NA | 1 | 0 | 1 | Low-quality | Genome-fragment | 1.79 | AAI-based (medium-confidence) | 0 |
| S12_k141_389922 | 1550 | No | NA | 2 | 0 | 0 | Not-determined | Genome-fragment | NA | NA | 0 |
| S12_k141_355935 | 1754 | No | NA | 2 | 0 | 2 | Not-determined | Genome-fragment | NA | NA | 0 |
| S12_k141_88406 | 1002 | No | NA | 2 | 0 | 1 | Not-determined | Genome-fragment | NA | NA | 0 |
| S1_k141_335678 | 3326 | No | NA | 8 | 1 | 0 | Low-quality | Genome-fragment | 2.43 | AAI-based (high-confidence) | 0 |
| S12_k141_189664 | 1677 | No | NA | 1 | 0 | 1 | Not-determined | Genome-fragment | NA | NA | 0 |
| S12_k141_339471 | 3544 | No | NA | 2 | 0 | 2 | Not-determined | Genome-fragment | NA | NA | 0 |
| S5-final-viral-combined_k141_28335\|\|full | 3326 | No | NA | 5 | 0 | 4 | Low-quality | Genome-fragment | 3.12 | AAI-based (medium-confidence) | 0 |
| S12_k141_390023 | 2881 | No | NA | 3 | 0 | 2 | Not-determined | Genome-fragment | NA | NA | 0 |
| S12_k141_38674 | 1011 | No | NA | 1 | 0 | 1 | Not-determined | Genome-fragment | NA | NA | 0 |
| S12_k141_54893 | 4734 | No | NA | 3 | 0 | 3 | Not-determined | Genome-fragment | NA | NA | 0 |
| S5-final-viral-combined_k141_8092\|\|full | 3326 | Yes | 792 | 3 | 1 | 1 | Low-quality | Genome-fragment | 1.5 | AAI-based (medium-confidence) | 76.19 |
| S12_k141_339486 | 3478 | No | NA | 5 | 0 | 1 | Not-determined | Genome-fragment | NA | NA | 0 |
| S12_k141_390054 | 3512 | No | NA | 4 | 0 | 1 | Not-determined | Genome-fragment | NA | NA | 0 |
| S12_k141_272751 | 1093 | No | NA | 1 | 0 | 1 | Not-determined | Genome-fragment | NA | NA | 0 |
| S12_k141_139072 | 6835 | No | NA | 6 | 0 | 1 | Not-determined | Genome-fragment | NA | NA | 0 |
| S12_k141_172101 | 2202 | No | NA | 2 | 0 | 2 | Not-determined | Genome-fragment | NA | NA | 0 |
| S12_k141_239264 | 1016 | No | NA | 1 | 0 | 1 | Not-determined | Genome-fragment | NA | NA | 0 |
| S12_k141_38717 | 2193 | No | NA | 2 | 0 | 1 | Not-determined | Genome-fragment | NA | NA | 0 |
| S12_k141_88495 | 2585 | No | NA | 2 | 0 | 0 | Not-determined | Genome-fragment | NA | NA | 0 |
| S17-k141_202635 | 3327 | No | NA | 3 | 0 | 0 | Low-quality | Genome-fragment | 3.08 | AAI-based (high-confidence) | 0 |
| S4_k141_115111 | 3327 | No | NA | 6 | 0 | 0 | Low-quality | Genome-fragment | 3.22 | AAI-based (high-confidence) | 0 |
| S12_k141_38752 | 1125 | No | NA | 4 | 0 | 0 | Not-determined | Genome-fragment | NA | NA | 0 |
| S12_k141_205562 | 1132 | No | NA | 1 | 0 | 0 | Not-determined | Genome-fragment | NA | NA | 0 |
| S12_k141_373671 | 1010 | No | NA | 1 | 0 | 0 | Not-determined | Genome-fragment | NA | NA | 0 |
| S12_k141_239281 | 1260 | No | NA | 1 | 0 | 0 | Not-determined | Genome-fragment | NA | NA | 0 |
| S3-final-viral-combined_k141_56008\|\|full | 3328 | No | NA | 4 | 0 | 0 | Low-quality | Genome-fragment | 1.8 | AAI-based (medium-confidence) | 0 |
| S12_k141_390142 | 1781 | No | NA | 1 | 0 | 1 | Not-determined | Genome-fragment | NA | NA | 0 |
| S11-final-viral-combined_k141_235683\|\|full | 3329 | No | NA | 2 | 0 | 2 | Low-quality | Genome-fragment | 3.11 | AAI-based (medium-confidence) | 0 |
| S12_k141_306819 | 1170 | No | NA | 2 | 0 | 1 | Not-determined | Genome-fragment | NA | NA | 0 |
| S8-final-viral-combined_k141_222927\|\|full | 3329 | No | NA | 6 | 0 | 1 | Low-quality | Genome-fragment | 6.64 | AAI-based (medium-confidence) | 0 |
| S12_k141_239314 | 1178 | No | NA | 1 | 0 | 1 | Not-determined | Genome-fragment | NA | NA | 0 |
| S12_k141_72516 | 2695 | No | NA | 5 | 0 | 3 | Not-determined | Genome-fragment | NA | NA | 0 |
| S12_k141_121452 | 7044 | No | NA | 6 | 0 | 6 | Not-determined | Genome-fragment | NA | NA | 0 |
| S12_k141_306854 | 3128 | No | NA | 7 | 0 | 1 | Not-determined | Genome-fragment | NA | NA | 0 |
| S12_k141_390169 | 3906 | No | NA | 5 | 0 | 1 | Not-determined | Genome-fragment | NA | NA | 0 |
| S12_k141_390978 | 3330 | No | NA | 6 | 0 | 0 | Low-quality | Genome-fragment | 11.86 | AAI-based (medium-confidence) | 0 |
| S2_k141_90687 | 3330 | No | NA | 2 | 0 | 2 | Low-quality | Genome-fragment | 2.06 | AAI-based (medium-confidence) | 0 |
| S4-final-viral-combined_k141_74123\|\|full | 3330 | No | NA | 9 | 0 | 0 | Low-quality | Genome-fragment | 6.72 | AAI-based (medium-confidence) | 0 |
| S12_k141_306920 | 3918 | No | NA | 5 | 0 | 3 | Not-determined | Genome-fragment | NA | NA | 0 |
| S11-final-viral-combined_k141_192319\|\|full | 3332 | Yes | 2601 | 4 | 2 | 1 | Low-quality | Genome-fragment | 4.3 | AAI-based (high-confidence) | 21.94 |
| S12_k141_356053 | 1180 | No | NA | 1 | 0 | 1 | Not-determined | Genome-fragment | NA | NA | 0 |
| S12_k141_21758 | 2423 | No | NA | 1 | 0 | 0 | Not-determined | Genome-fragment | NA | NA | 0 |
| S12_k141_390219 | 2787 | No | NA | 1 | 0 | 0 | Not-determined | Genome-fragment | NA | NA | 0 |
| S12_k141_272914 | 1352 | No | NA | 3 | 0 | 3 | Not-determined | Genome-fragment | NA | NA | 0 |
| S12_k141_390228 | 1176 | No | NA | 1 | 0 | 0 | Not-determined | Genome-fragment | NA | NA | 0 |
| S12_k141_390240 | 1491 | No | NA | 2 | 0 | 1 | Not-determined | Genome-fragment | NA | NA | 0 |
| S12_k141_255873 | 1160 | No | NA | 1 | 0 | 1 | Not-determined | Genome-fragment | NA | NA | 0 |
| S12_k141_88621 | 5854 | No | NA | 4 | 0 | 3 | Not-determined | Genome-fragment | NA | NA | 0 |
| S12_k141_4980 | 3172 | No | NA | 3 | 0 | 3 | Not-determined | Genome-fragment | NA | NA | 0 |
| S12_k141_4999 | 1181 | No | NA | 2 | 0 | 0 | Not-determined | Genome-fragment | NA | NA | 0 |
| S12_k141_272969 | 13227 | No | NA | 6 | 0 | 5 | Not-determined | Genome-fragment | NA | NA | 0 |
| S12_k141_222501 | 1939 | No | NA | 1 | 0 | 0 | Not-determined | Genome-fragment | NA | NA | 0 |
| S11_k141_11976 | 3332 | No | NA | 6 | 3 | 0 | Low-quality | Genome-fragment | 8.68 | AAI-based (high-confidence) | 0 |
| S12_k141_307062 | 3912 | No | NA | 1 | 0 | 1 | Not-determined | Genome-fragment | NA | NA | 0 |
| S12_k141_21868 | 1137 | No | NA | 1 | 0 | 1 | Not-determined | Genome-fragment | NA | NA | 0 |
| S12_k141_273017 | 1000 | No | NA | 1 | 0 | 0 | Not-determined | Genome-fragment | NA | NA | 0 |
| S12_k141_273035 | 1003 | No | NA | 1 | 0 | 0 | Not-determined | Genome-fragment | NA | NA | 0 |
| S12_k141_189975 | 1065 | No | NA | 1 | 0 | 1 | Not-determined | Genome-fragment | NA | NA | 0 |
| S12_k141_323338 | 1588 | No | NA | 2 | 0 | 1 | Not-determined | Genome-fragment | NA | NA | 0 |
| S12_k141_307155 | 3045 | No | NA | 4 | 0 | 2 | Not-determined | Genome-fragment | NA | NA | 0 |
| S12_k141_390386 | 1139 | No | NA | 2 | 0 | 2 | Not-determined | Genome-fragment | NA | NA | 0 |
| S1-final-viral-combined_k141_148963\|\|full | 3332 | No | NA | 4 | 0 | 0 | Low-quality | Genome-fragment | 6.25 | AAI-based (medium-confidence) | 0 |
| S12_k141_239436 | 1143 | No | NA | 1 | 0 | 1 | Not-determined | Genome-fragment | NA | NA | 0 |
| S12_k141_39033 | 1225 | No | NA | 1 | 0 | 1 | Not-determined | Genome-fragment | NA | NA | 0 |
| S12_k141_373775 | 14211 | No | NA | 23 | 2 | 1 | High-quality | High-quality | 97.66 | AAI-based (high-confidence) | 0 |
| S12_k141_323394 | 1003 | No | NA | 1 | 0 | 0 | Not-determined | Genome-fragment | NA | NA | 0 |
| S12_k141_155888 | 3105 | No | NA | 5 | 0 | 0 | Not-determined | Genome-fragment | NA | NA | 0 |
| S1_k141_358238 | 3332 | No | NA | 3 | 0 | 2 | Low-quality | Genome-fragment | 0.37 | AAI-based (medium-confidence) | 0 |
| S12_k141_39071 | 6507 | No | NA | 8 | 0 | 3 | Not-determined | Genome-fragment | NA | NA | 0 |
| S17-k141_63763 | 3332 | No | NA | 2 | 0 | 2 | Low-quality | Genome-fragment | 3.3 | AAI-based (medium-confidence) | 0 |
| S12_k141_105532 | 3314 | No | NA | 2 | 0 | 1 | Not-determined | Genome-fragment | NA | NA | 0 |
| S12_k141_290208 | 1405 | No | NA | 2 | 0 | 2 | Not-determined | Genome-fragment | NA | NA | 0 |
| S1_k141_177585 | 3333 | No | NA | 3 | 1 | 0 | Low-quality | Genome-fragment | 5.26 | HMM-based (lower-bound) | 0 |
| S12_k141_88820 | 1426 | No | NA | 2 | 0 | 1 | Not-determined | Genome-fragment | NA | NA | 0 |
| S2-final-viral-combined_k141_8273\|\|full | 3333 | No | NA | 3 | 0 | 3 | Low-quality | Genome-fragment | 7.36 | AAI-based (medium-confidence) | 0 |
| S6_k141_140139 | 3333 | No | NA | 5 | 0 | 0 | Low-quality | Genome-fragment | 8.18 | AAI-based (medium-confidence) | 0 |
| S12_k141_256070 | 1671 | No | NA | 2 | 0 | 0 | Not-determined | Genome-fragment | NA | NA | 0 |
| S12_k141_356079 | 1126 | No | NA | 1 | 0 | 1 | Not-determined | Genome-fragment | NA | NA | 0 |
| S12_k141_105590 | 2892 | No | NA | 3 | 0 | 3 | Not-determined | Genome-fragment | NA | NA | 0 |
| S12_k141_356084 | 1388 | No | NA | 3 | 0 | 0 | Not-determined | Genome-fragment | NA | NA | 0 |
| S12_k141_323475 | 2343 | No | NA | 1 | 0 | 1 | Not-determined | Genome-fragment | NA | NA | 0 |
| S12-final-viral-combined_k141_114503\|\|full | 3334 | No | NA | 5 | 4 | 0 | Low-quality | Genome-fragment | 7.8 | AAI-based (medium-confidence) | 0 |
| S12_k141_307254 | 3847 | No | NA | 8 | 0 | 8 | Not-determined | Genome-fragment | NA | NA | 0 |
| S12_k141_373898 | 1699 | No | NA | 2 | 0 | 1 | Not-determined | Genome-fragment | NA | NA | 0 |
| S7-final-viral-combined_k141_226036\|\|full | 3335 | No | NA | 6 | 0 | 0 | Low-quality | Genome-fragment | 5.44 | AAI-based (high-confidence) | 0 |
| S1_k141_23558 | 3338 | No | NA | 10 | 2 | 0 | Low-quality | Genome-fragment | 8.58 | AAI-based (high-confidence) | 0 |
| S12_k141_139284 | 4119 | No | NA | 7 | 0 | 0 | Not-determined | Genome-fragment | NA | NA | 0 |
| S12_k141_205816 | 7077 | No | NA | 6 | 0 | 2 | Not-determined | Genome-fragment | NA | NA | 0 |
| S9-final-viral-combined_k141_81554\|\|full | 3339 | No | NA | 5 | 0 | 2 | Low-quality | Genome-fragment | 2.71 | AAI-based (high-confidence) | 0 |
| S12_k141_273227 | 6963 | No | NA | 4 | 0 | 3 | Not-determined | Genome-fragment | NA | NA | 0 |
| S7-final-viral-combined_k141_61355\|\|full | 3339 | No | NA | 5 | 1 | 0 | Low-quality | Genome-fragment | 3.84 | HMM-based (lower-bound) | 0 |
| S12_k141_290357 | 1367 | No | NA | 1 | 0 | 0 | Not-determined | Genome-fragment | NA | NA | 0 |
| S12_k141_323588 | 5942 | No | NA | 1 | 0 | 1 | Not-determined | Genome-fragment | NA | NA | 0 |
| S12_k141_156073 | 1002 | No | NA | 1 | 0 | 0 | Not-determined | Genome-fragment | NA | NA | 0 |
| S12_k141_121787 | 8398 | No | NA | 5 | 0 | 4 | Not-determined | Genome-fragment | NA | NA | 0 |
| S7_k141_164505 | 3339 | No | NA | 2 | 0 | 2 | Low-quality | Genome-fragment | 0.92 | AAI-based (medium-confidence) | 0 |
| S12_k141_307338 | 1448 | No | NA | 2 | 0 | 2 | Not-determined | Genome-fragment | NA | NA | 0 |

| | | | | | | | | | | | |
|---|---|---|---|---|---|---|---|---|---|---|---|
| S12_k141_390589 | 1913 | No | NA | 2 | 0 | 1 | Not-determined | Genome-fragment | NA | NA | 0 |
| S12_k141_205867 | 1042 | No | NA | 1 | 0 | 0 | Not-determined | Genome-fragment | NA | NA | 0 |
| S12_k141_205871 | 3574 | No | NA | 2 | 0 | 1 | Not-determined | Genome-fragment | NA | NA | 0 |
| S12_k141_88933 | 4059 | No | NA | 4 | 0 | 2 | Not-determined | Genome-fragment | NA | NA | 0 |
| S12_k141_222810 | 1043 | No | NA | 1 | 0 | 1 | Not-determined | Genome-fragment | NA | NA | 0 |
| S12_k141_307388 | 1304 | No | NA | 2 | 0 | 0 | Not-determined | Genome-fragment | NA | NA | 0 |
| S12_k141_390635 | 1444 | No | NA | 1 | 0 | 0 | Not-determined | Genome-fragment | NA | NA | 0 |
| S12_k141_390649 | 1214 | No | NA | 2 | 0 | 1 | Not-determined | Genome-fragment | NA | NA | 0 |
| S12_k141_55044 | 1283 | No | NA | 1 | 0 | 0 | Not-determined | Genome-fragment | NA | NA | 0 |
| S12_k141_307410 | 2028 | No | NA | 1 | 0 | 1 | Not-determined | Genome-fragment | NA | NA | 0 |
| S12_k141_134453 | 3339 | No | NA | 4 | 1 | 0 | Low-quality | Genome-fragment | 9.51 | AAI-based (medium-confidence) | 0 |
| S12_k141_390687 | 1015 | No | NA | 2 | 0 | 1 | Not-determined | Genome-fragment | NA | NA | 0 |
| S12_k141_39147 | 1242 | No | NA | 1 | 0 | 1 | Not-determined | Genome-fragment | NA | NA | 0 |
| S12_k141_55097 | 1775 | No | NA | 1 | 0 | 1 | Not-determined | Genome-fragment | NA | NA | 0 |
| S12_k141_256291 | 1147 | No | NA | 2 | 0 | 1 | Not-determined | Genome-fragment | NA | NA | 0 |
| S12_k141_156198 | 4264 | No | NA | 3 | 0 | 2 | Not-determined | Genome-fragment | NA | NA | 0 |
| S11-final-viral-combined_k141_209135\|\|full | 3340 | No | NA | 4 | 1 | 0 | Low-quality | Genome-fragment | 6.71 | AAI-based (high-confidence) | 0 |
| S12_k141_39182 | 1106 | No | NA | 1 | 0 | 1 | Not-determined | Genome-fragment | NA | NA | 0 |
| S12_k141_121921 | 1058 | No | NA | 1 | 0 | 1 | Not-determined | Genome-fragment | NA | NA | 0 |
| S11-final-viral-combined_k141_204350\|\|full | 3340 | No | NA | 6 | 0 | 0 | Low-quality | Genome-fragment | 8.28 | AAI-based (high-confidence) | 0 |
| S5-final-viral-combined_k141_186913\|\|full | 3340 | No | NA | 7 | 0 | 1 | Low-quality | Genome-fragment | 5.59 | AAI-based (high-confidence) | 0 |
| S12_k141_323787 | 1800 | No | NA | 1 | 0 | 1 | Not-determined | Genome-fragment | NA | NA | 0 |
| S12_k141_5190 | 1411 | No | NA | 1 | 0 | 0 | Not-determined | Genome-fragment | NA | NA | 0 |
| S5_k141_142011 | 3340 | No | NA | 4 | 0 | 0 | Low-quality | Genome-fragment | 6.43 | AAI-based (medium-confidence) | 0 |
| S12_k141_5206 | 6774 | No | NA | 6 | 0 | 6 | Not-determined | Genome-fragment | NA | NA | 0 |
| S12_k141_55184 | 2142 | No | NA | 2 | 0 | 2 | Not-determined | Genome-fragment | NA | NA | 0 |
| S12_k141_139483 | 2586 | No | NA | 1 | 0 | 1 | Not-determined | Genome-fragment | NA | NA | 0 |
| S12_k141_55206 | 1410 | No | NA | 2 | 0 | 2 | Not-determined | Genome-fragment | NA | NA | 0 |
| S7_k141_14842 | 3340 | No | NA | 4 | 0 | 0 | Low-quality | Genome-fragment | 6.36 | AAI-based (medium-confidence) | 0 |
| S4_k141_160823 | 3340 | No | NA | 7 | 0 | 0 | Low-quality | Genome-fragment | 10.29 | AAI-based (high-confidence) | 0 |
| S12_k141_39288 | 1028 | No | NA | 1 | 0 | 0 | Not-determined | Genome-fragment | NA | NA | 0 |
| S12_k141_390779 | 8486 | No | NA | 5 | 0 | 3 | Not-determined | Genome-fragment | NA | NA | 0 |
| S12_k141_256324 | 1043 | No | NA | 1 | 0 | 0 | Not-determined | Genome-fragment | NA | NA | 0 |
| S12_k141_256326 | 1577 | No | NA | 1 | 0 | 1 | Not-determined | Genome-fragment | NA | NA | 0 |
| S12_k141_55217 | 1279 | No | NA | 1 | 0 | 1 | Not-determined | Genome-fragment | NA | NA | 0 |
| S12_k141_390789 | 1222 | No | NA | 2 | 0 | 0 | Not-determined | Genome-fragment | NA | NA | 0 |
| S12_k141_390790 | 1508 | No | NA | 1 | 0 | 0 | Not-determined | Genome-fragment | NA | NA | 0 |
| S12_k141_273437 | 1222 | No | NA | 2 | 0 | 1 | Not-determined | Genome-fragment | NA | NA | 0 |
| S12_k141_105716 | 1401 | No | NA | 2 | 0 | 1 | Not-determined | Genome-fragment | NA | NA | 0 |
| S8-final-viral-combined_k141_22615\|\|full | 3340 | Yes | 2604 | 7 | 3 | 1 | Low-quality | Genome-fragment | 6.88 | AAI-based (high-confidence) | 22.04 |
| S12_k141_374007 | 1036 | No | NA | 1 | 0 | 1 | Not-determined | Genome-fragment | NA | NA | 0 |
| S12_k141_390843 | 1239 | No | NA | 3 | 0 | 0 | Not-determined | Genome-fragment | NA | NA | 0 |
| S11-final-viral-combined_k141_208002\|\|full | 3341 | No | NA | 5 | 0 | 0 | Low-quality | Genome-fragment | 6.99 | AAI-based (high-confidence) | 0 |
| S12_k141_55259 | 3224 | No | NA | 1 | 0 | 1 | Not-determined | Genome-fragment | NA | NA | 0 |
| S12_k141_223053 | 1178 | No | NA | 1 | 0 | 0 | Not-determined | Genome-fragment | NA | NA | 0 |
| S12_k141_206028 | 3203 | No | NA | 2 | 0 | 2 | Not-determined | Genome-fragment | NA | NA | 0 |
| S12_k141_105771 | 1202 | No | NA | 2 | 0 | 2 | Not-determined | Genome-fragment | NA | NA | 0 |
| S12_k141_139537 | 1623 | No | NA | 2 | 0 | 1 | Not-determined | Genome-fragment | NA | NA | 0 |
| S12_k141_256380 | 12505 | No | NA | 24 | 0 | 23 | Not-determined | Genome-fragment | NA | NA | 0 |
| S12_k141_156330 | 2457 | No | NA | 3 | 0 | 2 | Not-determined | Genome-fragment | NA | NA | 0 |
| S12_k141_190463 | 1037 | No | NA | 1 | 0 | 0 | Not-determined | Genome-fragment | NA | NA | 0 |
| S12_k141_105778 | 1643 | No | NA | 2 | 0 | 1 | Not-determined | Genome-fragment | NA | NA | 0 |
| S11_k141_296593 | 3341 | No | NA | 2 | 0 | 2 | Low-quality | Genome-fragment | 7.19 | AAI-based (high-confidence) | 0 |
| S12_k141_273504 | 1397 | No | NA | 1 | 0 | 1 | Not-determined | Genome-fragment | NA | NA | 0 |
| S12_k141_55294 | 1343 | No | NA | 2 | 0 | 1 | Not-determined | Genome-fragment | NA | NA | 0 |
| S12_k141_239926 | 1473 | No | NA | 3 | 0 | 0 | Not-determined | Genome-fragment | NA | NA | 0 |
| S12_k141_89168 | 1276 | No | NA | 1 | 0 | 1 | Not-determined | Genome-fragment | NA | NA | 0 |
| S12_k141_55300 | 2165 | No | NA | 1 | 0 | 1 | Not-determined | Genome-fragment | NA | NA | 0 |
| S12_k141_307638 | 1419 | No | NA | 2 | 0 | 0 | Not-determined | Genome-fragment | NA | NA | 0 |
| S7-final-viral-combined_k141_63740\|\|full | 3341 | No | NA | 4 | 0 | 1 | Low-quality | Genome-fragment | 5.95 | AAI-based (medium-confidence) | 0 |
| S12_k141_139604 | 1129 | No | NA | 2 | 0 | 1 | Not-determined | Genome-fragment | NA | NA | 0 |
| S7-final-viral-combined_k141_173177\|\|full | 3341 | No | NA | 2 | 0 | 0 | Low-quality | Genome-fragment | 1.69 | AAI-based (medium-confidence) | 0 |
| S12_k141_356329 | 1693 | No | NA | 1 | 0 | 1 | Not-determined | Genome-fragment | NA | NA | 0 |
| S12_k141_73169 | 1194 | No | NA | 1 | 0 | 1 | Not-determined | Genome-fragment | NA | NA | 0 |
| S12_k141_374101 | 1615 | No | NA | 3 | 0 | 1 | Not-determined | Genome-fragment | NA | NA | 0 |
| S11-final-viral-combined_k141_96961\|\|full | 3342 | No | NA | 5 | 0 | 1 | Low-quality | Genome-fragment | 2.32 | AAI-based (medium-confidence) | 0 |
| S12_k141_374114 | 1616 | No | NA | 3 | 0 | 0 | Not-determined | Genome-fragment | NA | NA | 0 |
| S7_k141_62079 | 3342 | No | NA | 4 | 0 | 3 | Low-quality | Genome-fragment | 6.01 | AAI-based (medium-confidence) | 0 |
| S12_k141_139659 | 7870 | No | NA | 4 | 0 | 3 | Not-determined | Genome-fragment | NA | NA | 0 |
| S12_k141_374124 | 1641 | No | NA | 2 | 0 | 1 | Not-determined | Genome-fragment | NA | NA | 0 |
| S7-final-viral-combined_k141_56109\|\|full | 3344 | No | NA | 8 | 1 | 4 | Low-quality | Genome-fragment | 6.7 | HMM-based (lower-bound) | 0 |
| S9-final-viral-combined_k141_109571\|\|full | 3345 | No | NA | 4 | 0 | 0 | Low-quality | Genome-fragment | 5.09 | AAI-based (high-confidence) | 0 |
| S17-k141_240350 | 3346 | No | NA | 3 | 0 | 2 | Low-quality | Genome-fragment | 5.18 | AAI-based (medium-confidence) | 0 |
| S1_k141_318563 | 3347 | No | NA | 6 | 0 | 0 | Low-quality | Genome-fragment | 2.98 | AAI-based (high-confidence) | 0 |
| S5-final-viral-combined_k141_80491\|\|full | 3347 | No | NA | 7 | 3 | 0 | Low-quality | Genome-fragment | 7.5 | AAI-based (high-confidence) | 0 |
| S12_k141_374189 | 2655 | No | NA | 1 | 0 | 1 | Not-determined | Genome-fragment | NA | NA | 0 |
| S12_k141_89257 | 1987 | No | NA | 4 | 0 | 1 | Not-determined | Genome-fragment | NA | NA | 0 |
| S17-k141_67039 | 3347 | No | NA | 4 | 0 | 4 | Low-quality | Genome-fragment | 1.23 | AAI-based (medium-confidence) | 0 |
| S12_k141_39311 | 1683 | No | NA | 2 | 0 | 0 | Not-determined | Genome-fragment | NA | NA | 0 |
| S12_k141_256533 | 1007 | No | NA | 2 | 0 | 1 | Not-determined | Genome-fragment | NA | NA | 0 |
| S12_k141_223082 | 1317 | No | NA | 3 | 0 | 0 | Not-determined | Genome-fragment | NA | NA | 0 |
| S12_k141_290764 | 3655 | No | NA | 1 | 0 | 1 | Not-determined | Genome-fragment | NA | NA | 0 |
| S12_k141_273642 | 3431 | No | NA | 1 | 0 | 0 | Not-determined | Genome-fragment | NA | NA | 0 |
| S2-final-viral-combined_k141_242247\|\|full | 3347 | No | NA | 2 | 0 | 1 | Low-quality | Genome-fragment | 1.28 | AAI-based (medium-confidence) | 0 |
| S12_k141_223098 | 1807 | No | NA | 1 | 0 | 0 | Not-determined | Genome-fragment | NA | NA | 0 |
| S4-final-viral-combined_k141_113762\|\|full | 3347 | No | NA | 2 | 0 | 1 | Low-quality | Genome-fragment | 3.65 | AAI-based (high-confidence) | 0 |
| S12_k141_390955 | 1034 | No | NA | 1 | 0 | 0 | Not-determined | Genome-fragment | NA | NA | 0 |
| S12_k141_190711 | 10526 | No | NA | 7 | 0 | 0 | Not-determined | Genome-fragment | NA | NA | 0 |
| S6-final-viral-combined_k141_126383\|\|full | 3347 | Yes | 1487 | 3 | 1 | 1 | Low-quality | Genome-fragment | 0.83 | HMM-based (lower-bound) | 55.57 |
| S12_k141_173000 | 1081 | No | NA | 2 | 0 | 1 | Not-determined | Genome-fragment | NA | NA | 0 |
| S12_k141_39369 | 1013 | No | NA | 2 | 0 | 0 | Not-determined | Genome-fragment | NA | NA | 0 |
| S12_k141_273683 | 2449 | No | NA | 3 | 0 | 1 | Not-determined | Genome-fragment | NA | NA | 0 |
| S12_k141_5468 | 5547 | No | NA | 3 | 0 | 3 | Not-determined | Genome-fragment | NA | NA | 0 |
| S12_k141_273692 | 3245 | No | NA | 2 | 0 | 2 | Not-determined | Genome-fragment | NA | NA | 0 |
| S12_k141_122249 | 3431 | No | NA | 4 | 0 | 1 | Not-determined | Genome-fragment | NA | NA | 0 |
| S12_k141_391001 | 1203 | No | NA | 1 | 0 | 0 | Not-determined | Genome-fragment | NA | NA | 0 |
| S12_k141_206290 | 1477 | No | NA | 2 | 0 | 0 | Not-determined | Genome-fragment | NA | NA | 0 |
| S12_k141_290867 | 1110 | No | NA | 2 | 0 | 1 | Not-determined | Genome-fragment | NA | NA | 0 |
| S12_k141_156556 | 1639 | No | NA | 2 | 0 | 2 | Not-determined | Genome-fragment | NA | NA | 0 |
| S3-final-viral-combined_k141_89637\|\|full | 3348 | No | NA | 6 | 2 | 0 | Low-quality | Genome-fragment | 8.14 | AAI-based (high-confidence) | 0 |
| S12_k141_223143 | 1138 | No | NA | 1 | 0 | 1 | Not-determined | Genome-fragment | NA | NA | 0 |
| S12_k141_340187 | 1185 | No | NA | 1 | 0 | 0 | Not-determined | Genome-fragment | NA | NA | 0 |
| S12_k141_190757 | 1145 | No | NA | 1 | 0 | 1 | Not-determined | Genome-fragment | NA | NA | 0 |
| S1_k141_313629 | 3349 | No | NA | 3 | 0 | 2 | Low-quality | Genome-fragment | 1.55 | AAI-based (medium-confidence) | 0 |
| S12_k141_122288 | 1694 | No | NA | 2 | 0 | 1 | Not-determined | Genome-fragment | NA | NA | 0 |
| S12_k141_5514 | 1049 | No | NA | 1 | 0 | 1 | Not-determined | Genome-fragment | NA | NA | 0 |
| S12_k141_5516 | 6122 | No | NA | 8 | 0 | 4 | Not-determined | Genome-fragment | NA | NA | 0 |
| S12-final-viral-combined_k141_205415\|\|full | 3349 | No | NA | 7 | 0 | 0 | Low-quality | Genome-fragment | 8.59 | AAI-based (high-confidence) | 0 |
| S12_k141_89377 | 3429 | No | NA | 3 | 0 | 1 | Not-determined | Genome-fragment | NA | NA | 0 |
| S12_k141_206366 | 1707 | No | NA | 2 | 0 | 2 | Not-determined | Genome-fragment | NA | NA | 0 |
| S12_k141_89411 | 1018 | No | NA | 1 | 0 | 1 | Not-determined | Genome-fragment | NA | NA | 0 |
| S12_k141_356467 | 1576 | No | NA | 3 | 0 | 1 | Not-determined | Genome-fragment | NA | NA | 0 |
| S12_k141_89423 | 1027 | No | NA | 1 | 0 | 1 | Not-determined | Genome-fragment | NA | NA | 0 |
| S12_k141_223235 | 3957 | No | NA | 5 | 1 | 0 | High-quality | High-quality | 100 | AAI-based (medium-confidence) | 0 |
| S12_k141_39539 | 1250 | No | NA | 2 | 0 | 1 | Not-determined | Genome-fragment | NA | NA | 0 |
| S12_k141_206405 | 4209 | No | NA | 3 | 0 | 3 | Not-determined | Genome-fragment | NA | NA | 0 |
| S12_k141_5587 | 4352 | No | NA | 5 | 0 | 4 | Not-determined | Genome-fragment | NA | NA | 0 |
| S12_k141_223249 | 2509 | No | NA | 2 | 0 | 1 | Not-determined | Genome-fragment | NA | NA | 0 |
| S12_k141_156655 | 2341 | No | NA | 2 | 0 | 1 | Not-determined | Genome-fragment | NA | NA | 0 |
| S12_k141_5594 | 1579 | No | NA | 1 | 0 | 1 | Not-determined | Genome-fragment | NA | NA | 0 |
| S12_k141_73489 | 1034 | No | NA | 2 | 0 | 0 | Not-determined | Genome-fragment | NA | NA | 0 |
| S2-final-viral-combined_k141_50746\|\|full | 3350 | No | NA | 5 | 0 | 3 | Low-quality | Genome-fragment | 0.98 | AAI-based (medium-confidence) | 0 |
| S12_k141_22716 | 3302 | No | NA | 3 | 0 | 3 | Not-determined | Genome-fragment | NA | NA | 0 |
| S12_k141_206475 | 2269 | No | NA | 3 | 0 | 0 | Not-determined | Genome-fragment | NA | NA | 0 |
| S12_k141_55480 | 3418 | No | NA | 4 | 0 | 2 | Not-determined | Genome-fragment | NA | NA | 0 |
| S12_k141_307877 | 1442 | No | NA | 2 | 0 | 2 | Not-determined | Genome-fragment | NA | NA | 0 |
| S12_k141_356521 | 1015 | No | NA | 1 | 0 | 1 | Not-determined | Genome-fragment | NA | NA | 0 |
| S12_k141_22737 | 1434 | No | NA | 1 | 0 | 1 | Not-determined | Genome-fragment | NA | NA | 0 |
| S12_k141_173134 | 1165 | No | NA | 1 | 0 | 0 | Not-determined | Genome-fragment | NA | NA | 0 |
| S12_k141_156690 | 1135 | No | NA | 2 | 0 | 1 | Not-determined | Genome-fragment | NA | NA | 0 |
| S12_k141_324220 | 1077 | No | NA | 3 | 0 | 2 | Not-determined | Genome-fragment | NA | NA | 0 |
| S17-final-viral-combined_k141_78447\|\|full | 3351 | No | NA | 4 | 3 | 0 | Low-quality | Genome-fragment | 8.61 | AAI-based (medium-confidence) | 0 |
| S12_k141_106186 | 1509 | No | NA | 2 | 0 | 1 | Not-determined | Genome-fragment | NA | NA | 0 |
| S17-final-viral-combined_k141_151661\|\|full | 3351 | No | NA | 6 | 0 | 1 | Low-quality | Genome-fragment | 7.94 | AAI-based (high-confidence) | 0 |
| S12_k141_106189 | 1024 | No | NA | 1 | 0 | 0 | Not-determined | Genome-fragment | NA | NA | 0 |
| S12_k141_256644 | 1577 | No | NA | 1 | 0 | 1 | Not-determined | Genome-fragment | NA | NA | 0 |
| S12_k141_374362 | 1985 | No | NA | 2 | 0 | 0 | Not-determined | Genome-fragment | NA | NA | 0 |
| S17-k141_90348 | 3351 | No | NA | 1 | 1 | 0 | Low-quality | Genome-fragment | 3.56 | HMM-based (lower-bound) | 0 |
| S12_k141_122425 | 1377 | No | NA | 2 | 0 | 0 | Not-determined | Genome-fragment | NA | NA | 0 |
| S12_k141_240303 | 2978 | No | NA | 3 | 0 | 0 | Not-determined | Genome-fragment | NA | NA | 0 |
| S12_k141_307933 | 1291 | No | NA | 1 | 0 | 1 | Not-determined | Genome-fragment | NA | NA | 0 |
| S12_k141_240312 | 1114 | No | NA | 2 | 0 | 1 | Not-determined | Genome-fragment | NA | NA | 0 |
| S12_k141_122432 | 5460 | No | NA | 4 | 0 | 3 | Not-determined | Genome-fragment | NA | NA | 0 |
| S12_k141_22330 | 3351 | No | NA | 2 | 0 | 2 | Low-quality | Genome-fragment | 9.45 | AAI-based (medium-confidence) | 0 |
| S12_k141_391234 | 1648 | No | NA | 3 | 0 | 2 | Not-determined | Genome-fragment | NA | NA | 0 |
| S12_k141_55556 | 1117 | No | NA | 1 | 0 | 1 | Not-determined | Genome-fragment | NA | NA | 0 |
| S12_k141_324293 | 1531 | No | NA | 1 | 0 | 0 | Not-determined | Genome-fragment | NA | NA | 0 |
| S12_k141_55572 | 1292 | No | NA | 2 | 0 | 2 | Not-determined | Genome-fragment | NA | NA | 0 |
| S12_k141_122490 | 1794 | No | NA | 3 | 0 | 0 | Not-determined | Genome-fragment | NA | NA | 0 |
| S12_k141_173248 | 1615 | No | NA | 1 | 0 | 1 | Not-determined | Genome-fragment | NA | NA | 0 |
| S2-final-viral-combined_k141_159874\|\|full | 3352 | No | NA | 4 | 0 | 2 | Low-quality | Genome-fragment | 1.85 | AAI-based (medium-confidence) | 0 |
| S11-final-viral-combined_k141_45324\|\|full | 3353 | No | NA | 6 | 3 | 0 | Low-quality | Genome-fragment | 4.45 | HMM-based (lower-bound) | 0 |
| S12_k141_55611 | 4802 | No | NA | 6 | 0 | 3 | Not-determined | Genome-fragment | NA | NA | 0 |
| S12_k141_256782 | 1057 | No | NA | 1 | 0 | 1 | Not-determined | Genome-fragment | NA | NA | 0 |
| S12_k141_223458 | 4481 | No | NA | 4 | 0 | 2 | Not-determined | Genome-fragment | NA | NA | 0 |
| S12_k141_240419 | 2091 | No | NA | 1 | 0 | 1 | Not-determined | Genome-fragment | NA | NA | 0 |
| S12_k141_156849 | 3873 | No | NA | 4 | 0 | 1 | Not-determined | Genome-fragment | NA | NA | 0 |
| S1-final-viral-combined_k141_27202\|\|full | 3353 | No | NA | 4 | 0 | 3 | Low-quality | Genome-fragment | 0.96 | AAI-based (medium-confidence) | 0 |
| S12_k141_5799 | 1049 | No | NA | 1 | 0 | 0 | Not-determined | Genome-fragment | NA | NA | 0 |
| S12_k141_89612 | 1252 | No | NA | 1 | 0 | 1 | Not-determined | Genome-fragment | NA | NA | 0 |
| S12-final-viral-combined_k141_349808\|\|full | 3353 | No | NA | 5 | 0 | 0 | Low-quality | Genome-fragment | 5.42 | AAI-based (medium-confidence) | 0 |
| S12_k141_55672 | 2343 | No | NA | 2 | 0 | 1 | Not-determined | Genome-fragment | NA | NA | 0 |
| S12_k141_291258 | 8014 | No | NA | 5 | 0 | 2 | Not-determined | Genome-fragment | NA | NA | 0 |
| S1-final-viral-combined_k141_254135\|\|full | 3354 | No | NA | 3 | 0 | 2 | Low-quality | Genome-fragment | 1.51 | AAI-based (high-confidence) | 0 |
| S11-final-viral-combined_k141_157611\|\|full | 3356 | No | NA | 3 | 0 | 0 | Low-quality | Genome-fragment | 5.06 | AAI-based (medium-confidence) | 0 |
| S12_k141_291281 | 1400 | No | NA | 2 | 0 | 1 | Not-determined | Genome-fragment | NA | NA | 0 |
| S12_k141_122642 | 1752 | No | NA | 2 | 0 | 1 | Not-determined | Genome-fragment | NA | NA | 0 |
| S12_k141_308104 | 3128 | No | NA | 3 | 0 | 3 | Not-determined | Genome-fragment | NA | NA | 0 |
| S3-final-viral-combined_k141_61142\|\|full | 3356 | No | NA | 6 | 1 | 0 | Low-quality | Genome-fragment | 21.72 | AAI-based (high-confidence) | 0 |
| S12_k141_89685 | 2106 | No | NA | 1 | 0 | 1 | Not-determined | Genome-fragment | NA | NA | 0 |
| S12_k141_391336 | 41654 | No | NA | 43 | 0 | 9 | Not-determined | Genome-fragment | NA | NA | 0 |
| S12_k141_122669 | 1255 | No | NA | 1 | 0 | 0 | Not-determined | Genome-fragment | NA | NA | 0 |
| S12_k141_5876 | 3778 | No | NA | 1 | 0 | 1 | Not-determined | Genome-fragment | NA | NA | 0 |
| S2-final-viral-combined_k141_13200\|\|full | 3356 | No | NA | 9 | 1 | 0 | Low-quality | Genome-fragment | 3.9 | AAI-based (medium-confidence) | 0 |
| S12_k141_22953 | 3282 | No | NA | 2 | 0 | 2 | Not-determined | Genome-fragment | NA | NA | 0 |
| S12_k141_356711 | 1179 | No | NA | 1 | 0 | 0 | Not-determined | Genome-fragment | NA | NA | 0 |
| S12_k141_308201 | 1364 | No | NA | 1 | 0 | 0 | Not-determined | Genome-fragment | NA | NA | 0 |
| S12_k141_256922 | 1112 | No | NA | 1 | 0 | 0 | Not-determined | Genome-fragment | NA | NA | 0 |
| S12_k141_391426 | 2079 | No | NA | 2 | 0 | 0 | Not-determined | Genome-fragment | NA | NA | 0 |
| S12_k141_5966 | 2612 | No | NA | 2 | 0 | 2 | Not-determined | Genome-fragment | NA | NA | 0 |
| S4-final-viral-combined_k141_41039\|\|full | 3356 | Yes | 1911 | 4 | 1 | 1 | Low-quality | Genome-fragment | 2.53 | HMM-based (lower-bound) | 43.06 |
| S12_k141_256952 | 1055 | No | NA | 1 | 0 | 1 | Not-determined | Genome-fragment | NA | NA | 0 |
| S12_k141_340348 | 2570 | No | NA | 3 | 0 | 2 | Not-determined | Genome-fragment | NA | NA | 0 |
| S8_k141_216736 | 3356 | No | NA | 5 | 0 | 4 | Low-quality | Genome-fragment | 2.61 | AAI-based (medium-confidence) | 0 |
| S12_k141_273987 | 1695 | No | NA | 4 | 0 | 0 | Not-determined | Genome-fragment | NA | NA | 0 |
| S12_k141_106401 | 3422 | No | NA | 3 | 0 | 1 | Not-determined | Genome-fragment | NA | NA | 0 |
| S12_k141_391455 | 1149 | No | NA | 1 | 0 | 1 | Not-determined | Genome-fragment | NA | NA | 0 |
| S12_k141_106412 | 1025 | No | NA | 1 | 0 | 1 | Not-determined | Genome-fragment | NA | NA | 0 |
| S12_k141_356798 | 1290 | No | NA | 1 | 0 | 0 | Not-determined | Genome-fragment | NA | NA | 0 |
| S4_k141_147206 | 3357 | No | NA | 2 | 0 | 2 | Low-quality | Genome-fragment | 6.85 | AAI-based (medium-confidence) | 0 |
| S12_k141_73881 | 1376 | No | NA | 2 | 0 | 0 | Not-determined | Genome-fragment | NA | NA | 0 |
| S12_k141_356816 | 2593 | No | NA | 1 | 0 | 0 | Not-determined | Genome-fragment | NA | NA | 0 |
| S8-final-viral-combined_k141_140974\|\|full | 3357 | Yes | 1388 | 5 | 1 | 2 | Low-quality | Genome-fragment | 0.72 | HMM-based (lower-bound) | 58.65 |
| S12_k141_271882 | 3358 | No | NA | 6 | 2 | 0 | Low-quality | Genome-fragment | 10.17 | AAI-based (high-confidence) | 0 |
| S12_k141_374725 | 1205 | No | NA | 2 | 0 | 0 | Not-determined | Genome-fragment | NA | NA | 0 |
| S2-final-viral-combined_k141_131149\|\|full | 3358 | No | NA | 4 | 1 | 0 | Low-quality | Genome-fragment | 7.92 | HMM-based (lower-bound) | 0 |
| S12_k141_173608 | 21222 | No | NA | 21 | 0 | 16 | Not-determined | Genome-fragment | NA | NA | 0 |
| S12_k141_257004 | 3836 | No | NA | 5 | 0 | 1 | Not-determined | Genome-fragment | NA | NA | 0 |
| S12_k141_324563 | 1042 | No | NA | 1 | 0 | 1 | Not-determined | Genome-fragment | NA | NA | 0 |
| S12_k141_6077 | 1074 | No | NA | 2 | 0 | 1 | Not-determined | Genome-fragment | NA | NA | 0 |
| S12_k141_340437 | 1386 | No | NA | 1 | 0 | 0 | Not-determined | Genome-fragment | NA | NA | 0 |
| S12_k141_324569 | 8406 | No | NA | 7 | 0 | 2 | Not-determined | Genome-fragment | NA | NA | 0 |
| S12_k141_191239 | 1124 | No | NA | 1 | 0 | 1 | Not-determined | Genome-fragment | NA | NA | 0 |
| S11-final-viral-combined_k141_209501\|\|full | 3359 | No | NA | 4 | 2 | 0 | Low-quality | Genome-fragment | 5.83 | HMM-based (lower-bound) | 0 |
| S12_k141_206962 | 3897 | No | NA | 5 | 0 | 3 | Not-determined | Genome-fragment | NA | NA | 0 |
| S12_k141_73923 | 14119 | No | NA | 13 | 0 | 9 | Not-determined | Genome-fragment | NA | NA | 0 |
| S12_k141_191264 | 1121 | No | NA | 3 | 0 | 2 | Not-determined | Genome-fragment | NA | NA | 0 |
| S12_k141_122809 | 1159 | No | NA | 2 | 0 | 1 | Not-determined | Genome-fragment | NA | NA | 0 |
| S17-final-viral-combined_k141_78377\|\|full | 3359 | No | NA | 7 | 6 | 0 | Low-quality | Genome-fragment | 4.08 | AAI-based (high-confidence) | 0 |
| S12_k141_173617 | 3110 | No | NA | 4 | 0 | 2 | Not-determined | Genome-fragment | NA | NA | 0 |
| S12_k141_40088 | 2455 | No | NA | 3 | 0 | 3 | Not-determined | Genome-fragment | NA | NA | 0 |
| S12_k141_122839 | 1521 | No | NA | 1 | 0 | 1 | Not-determined | Genome-fragment | NA | NA | 0 |
| S12-final-viral-combined_k141_314679\|\|full | 3360 | Yes | 1574 | 3 | 2 | 1 | Low-quality | Genome-fragment | 5.49 | AAI-based (high-confidence) | 53.15 |
| S12_k141_118726 | 3360 | No | NA | 4 | 2 | 0 | Low-quality | Genome-fragment | 6.02 | AAI-based (high-confidence) | 0 |
| S12_k141_207006 | 1089 | No | NA | 1 | 0 | 0 | Not-determined | Genome-fragment | NA | NA | 0 |
| S11-final-viral-combined_k141_308448\|\|full | 3361 | No | NA | 2 | 0 | 1 | Low-quality | Genome-fragment | 3.16 | AAI-based (medium-confidence) | 0 |
| S12_k141_157140 | 3638 | No | NA | 5 | 0 | 2 | Not-determined | Genome-fragment | NA | NA | 0 |
| S17-final-viral-combined_k141_89666\|\|full | 3361 | No | NA | 12 | 2 | 0 | Low-quality | Genome-fragment | 7.8 | AAI-based (medium-confidence) | 0 |
| S12_k141_140397 | 1351 | No | NA | 2 | 0 | 0 | Not-determined | Genome-fragment | NA | NA | 0 |
| S12_k141_89931 | 3096 | No | NA | 3 | 0 | 3 | Not-determined | Genome-fragment | NA | NA | 0 |
| S12_k141_223907 | 1048 | No | NA | 3 | 0 | 0 | Not-determined | Genome-fragment | NA | NA | 0 |
| S12_k141_6158 | 3661 | No | NA | 5 | 0 | 3 | Not-determined | Genome-fragment | NA | NA | 0 |
| S12_k141_240789 | 1428 | No | NA | 2 | 0 | 0 | Not-determined | Genome-fragment | NA | NA | 0 |
| S9_k141_86593 | 3362 | No | NA | 2 | 1 | 0 | Low-quality | Genome-fragment | 9.19 | AAI-based (high-confidence) | 0 |
| S12_k141_122897 | 1030 | No | NA | 1 | 0 | 1 | Not-determined | Genome-fragment | NA | NA | 0 |
| S12_k141_308493 | 1062 | No | NA | 1 | 0 | 0 | Not-determined | Genome-fragment | NA | NA | 0 |
| S12_k141_223965 | 3552 | No | NA | 2 | 0 | 2 | Not-determined | Genome-fragment | NA | NA | 0 |
| S12_k141_274202 | 1046 | No | NA | 1 | 0 | 1 | Not-determined | Genome-fragment | NA | NA | 0 |
| S2-final-viral-combined_k141_172827\|\|full | 3362 | No | NA | 4 | 0 | 0 | Low-quality | Genome-fragment | 7.24 | AAI-based (medium-confidence) | 0 |
| S9_k141_12154 | 3363 | No | NA | 5 | 3 | 0 | Low-quality | Genome-fragment | 8.35 | AAI-based (medium-confidence) | 0 |
| S12_k141_340619 | 1013 | No | NA | 2 | 0 | 2 | Not-determined | Genome-fragment | NA | NA | 0 |
| S11-final-viral-combined_k141_226613\|\|full | 3363 | No | NA | 10 | 0 | 0 | Low-quality | Genome-fragment | 8.29 | AAI-based (high-confidence) | 0 |
| S9-final-viral-combined_k141_36222\|\|full | 3364 | No | NA | 5 | 0 | 1 | Low-quality | Genome-fragment | 4.16 | AAI-based (medium-confidence) | 0 |
| S12_k141_391699 | 1041 | No | NA | 1 | 0 | 0 | Not-determined | Genome-fragment | NA | NA | 0 |
| S12_k141_207152 | 3176 | No | NA | 3 | 0 | 1 | Not-determined | Genome-fragment | NA | NA | 0 |
| S12_k141_207154 | 1585 | No | NA | 1 | 0 | 1 | Not-determined | Genome-fragment | NA | NA | 0 |
| S12_k141_224040 | 1755 | No | NA | 1 | 0 | 0 | Not-determined | Genome-fragment | NA | NA | 0 |
| S11-final-viral-combined_k141_307756\|\|full | 3364 | No | NA | 9 | 4 | 0 | Low-quality | Genome-fragment | 5.78 | AAI-based (high-confidence) | 0 |
| S12_k141_257265 | 1183 | No | NA | 2 | 0 | 2 | Not-determined | Genome-fragment | NA | NA | 0 |
| S12_k141_6277 | 5134 | No | NA | 3 | 0 | 1 | Not-determined | Genome-fragment | NA | NA | 0 |
| S5-final-viral-combined_k141_105097\|\|full | 3364 | No | NA | 5 | 0 | 4 | Low-quality | Genome-fragment | 2.35 | AAI-based (medium-confidence) | 0 |
| S12_k141_224095 | 3835 | No | NA | 2 | 0 | 0 | Not-determined | Genome-fragment | NA | NA | 0 |
| S12_k141_157270 | 8538 | No | NA | 9 | 0 | 8 | Not-determined | Genome-fragment | NA | NA | 0 |
| S12_k141_40223 | 1200 | No | NA | 1 | 0 | 1 | Not-determined | Genome-fragment | NA | NA | 0 |
| S12_k141_340665 | 1462 | No | NA | 2 | 0 | 1 | Not-determined | Genome-fragment | NA | NA | 0 |
| S12_k141_74154 | 1542 | No | NA | 3 | 0 | 0 | Not-determined | Genome-fragment | NA | NA | 0 |
| S12_k141_6353 | 1481 | No | NA | 1 | 0 | 0 | Not-determined | Genome-fragment | NA | NA | 0 |
| S12_k141_324825 | 1256 | No | NA | 2 | 0 | 1 | Not-determined | Genome-fragment | NA | NA | 0 |
| S9-final-viral-combined_k141_78014\|\|full | 3365 | No | NA | 6 | 0 | 2 | Low-quality | Genome-fragment | 5.93 | AAI-based (high-confidence) | 0 |
| S12_k141_207240 | 3121 | No | NA | 1 | 0 | 0 | Not-determined | Genome-fragment | NA | NA | 0 |
| S12_k141_257306 | 3559 | No | NA | 2 | 0 | 2 | Not-determined | Genome-fragment | NA | NA | 0 |
| S12_k141_40277 | 1080 | No | NA | 1 | 0 | 0 | Not-determined | Genome-fragment | NA | NA | 0 |
| S8_k141_188879 | 3365 | No | NA | 5 | 0 | 0 | Low-quality | Genome-fragment | 5.95 | AAI-based (medium-confidence) | 0 |
| S12_k141_140549 | 3936 | No | NA | 8 | 0 | 1 | Not-determined | Genome-fragment | NA | NA | 0 |
| S12_k141_123040 | 1065 | No | NA | 2 | 0 | 0 | Not-determined | Genome-fragment | NA | NA | 0 |
| S12_k141_106711 | 1257 | No | NA | 1 | 0 | 1 | Not-determined | Genome-fragment | NA | NA | 0 |
| S12_k141_90134 | 1055 | No | NA | 2 | 0 | 0 | Not-determined | Genome-fragment | NA | NA | 0 |
| S1_k141_266011 | 3366 | No | NA | 4 | 0 | 1 | Low-quality | Genome-fragment | 11.67 | AAI-based (medium-confidence) | 0 |
| S12_k141_274336 | 1361 | No | NA | 2 | 0 | 1 | Not-determined | Genome-fragment | NA | NA | 0 |
| S12_k141_375035 | 1621 | No | NA | 2 | 0 | 2 | Not-determined | Genome-fragment | NA | NA | 0 |
| S12_k141_207323 | 5357 | No | NA | 5 | 0 | 5 | Not-determined | Genome-fragment | NA | NA | 0 |
| S12_k141_90159 | 6100 | No | NA | 1 | 0 | 0 | Not-determined | Genome-fragment | NA | NA | 0 |
| S12_k141_391841 | 1029 | No | NA | 1 | 0 | 1 | Not-determined | Genome-fragment | NA | NA | 0 |
| S12_k141_6452 | 2433 | No | NA | 2 | 0 | 1 | Not-determined | Genome-fragment | NA | NA | 0 |
| S7-final-viral-combined_k141_96391\|\|full | 3366 | No | NA | 3 | 0 | 2 | Low-quality | Genome-fragment | 1.02 | AAI-based (medium-confidence) | 0 |
| S12_k141_257374 | 7852 | No | NA | 9 | 0 | 5 | Not-determined | Genome-fragment | NA | NA | 0 |
| S12_k141_375080 | 1022 | No | NA | 1 | 0 | 1 | Not-determined | Genome-fragment | NA | NA | 0 |
| S12_k141_140579 | 3315 | No | NA | 5 | 0 | 4 | Not-determined | Genome-fragment | NA | NA | 0 |
| S12_k141_40394 | 2659 | No | NA | 5 | 0 | 1 | Not-determined | Genome-fragment | NA | NA | 0 |
| S3_k141_35260 | 3367 | Yes | 1229 | 4 | 1 | 1 | Low-quality | Genome-fragment | 1.86 | HMM-based (lower-bound) | 63.5 |
| S5-final-viral-combined_k141_339\|\|full | 3367 | No | NA | 5 | 1 | 1 | Low-quality | Genome-fragment | 6.74 | HMM-based (lower-bound) | 0 |
| S12_k141_157460 | 3202 | No | NA | 2 | 0 | 0 | Not-determined | Genome-fragment | NA | NA | 0 |
| S2-final-viral-combined_k141_199406\|\|full | 3367 | No | NA | 7 | 1 | 0 | Low-quality | Genome-fragment | 10 | AAI-based (high-confidence) | 0 |
| S12_k141_157473 | 1574 | No | NA | 2 | 0 | 1 | Not-determined | Genome-fragment | NA | NA | 0 |
| S12_k141_291815 | 1222 | No | NA | 1 | 0 | 0 | Not-determined | Genome-fragment | NA | NA | 0 |
| S12_k141_157477 | 4442 | No | NA | 4 | 0 | 0 | Not-determined | Genome-fragment | NA | NA | 0 |
| S12_k141_191485 | 9647 | No | NA | 7 | 0 | 4 | Not-determined | Genome-fragment | NA | NA | 0 |
| S2_k141_105147 | 3367 | No | NA | 2 | 0 | 0 | Low-quality | Genome-fragment | 3.34 | AAI-based (medium-confidence) | 0 |
| S12_k141_207417 | 1085 | No | NA | 1 | 0 | 0 | Not-determined | Genome-fragment | NA | NA | 0 |
| S8-final-viral-combined_k141_78589\|\|full | 3367 | No | NA | 3 | 3 | 0 | Low-quality | Genome-fragment | 8.27 | AAI-based (high-confidence) | 0 |
| S5_k141_82001 | 3368 | No | NA | 2 | 0 | 2 | Low-quality | Genome-fragment | 0.85 | AAI-based (high-confidence) | 0 |
| S12_k141_274423 | 1249 | No | NA | 1 | 0 | 1 | Not-determined | Genome-fragment | NA | NA | 0 |
| S12_k141_106838 | 3261 | No | NA | 9 | 0 | 2 | Not-determined | Genome-fragment | NA | NA | 0 |
| S12_k141_191510 | 1476 | No | NA | 4 | 0 | 0 | Not-determined | Genome-fragment | NA | NA | 0 |
| S12_k141_90260 | 1351 | No | NA | 2 | 0 | 2 | Not-determined | Genome-fragment | NA | NA | 0 |
| S12_k141_173914 | 2324 | No | NA | 1 | 0 | 1 | Not-determined | Genome-fragment | NA | NA | 0 |
| S12_k141_375159 | 1210 | No | NA | 2 | 0 | 0 | Not-determined | Genome-fragment | NA | NA | 0 |
| S12_k141_74333 | 1018 | No | NA | 2 | 0 | 0 | Not-determined | Genome-fragment | NA | NA | 0 |
| S11-final-viral-combined_k141_212804\|\|full | 3369 | No | NA | 9 | 0 | 0 | Low-quality | Genome-fragment | 6.88 | AAI-based (medium-confidence) | 0 |
| S12_k141_291887 | 2585 | No | NA | 6 | 0 | 1 | Not-determined | Genome-fragment | NA | NA | 0 |
| S1-final-viral-combined_k141_3363\|\|full | 3369 | No | NA | 4 | 0 | 2 | Low-quality | Genome-fragment | 7.37 | AAI-based (medium-confidence) | 0 |
| S12_k141_257552 | 1333 | No | NA | 2 | 0 | 0 | Not-determined | Genome-fragment | NA | NA | 0 |
| S1-final-viral-combined_k141_76650\|\|full | 3369 | No | NA | 4 | 1 | 0 | Low-quality | Genome-fragment | 10.26 | AAI-based (high-confidence) | 0 |
| S12_k141_90330 | 1854 | No | NA | 1 | 0 | 1 | Not-determined | Genome-fragment | NA | NA | 0 |
| S3-final-viral-combined_k141_81591\|\|full | 3369 | Yes | 1538 | 5 | 1 | 1 | Low-quality | Genome-fragment | 2.35 | AAI-based (medium-confidence) | 54.35 |
| S12_k141_391999 | 1669 | No | NA | 2 | 0 | 0 | Not-determined | Genome-fragment | NA | NA | 0 |
| S3-final-viral-combined_k141_2539\|\|full | 3369 | No | NA | 3 | 0 | 3 | Low-quality | Genome-fragment | 0.94 | AAI-based (high-confidence) | 0 |
| S12_k141_207544 | 1641 | No | NA | 1 | 0 | 0 | Not-determined | Genome-fragment | NA | NA | 0 |
| S5_k141_27214 | 3369 | No | NA | 6 | 0 | 0 | Low-quality | Genome-fragment | 7.14 | AAI-based (high-confidence) | 0 |
| S12_k141_56329 | 1474 | No | NA | 1 | 0 | 0 | Not-determined | Genome-fragment | NA | NA | 0 |
| S7_k141_44415 | 3369 | No | NA | 4 | 0 | 0 | Low-quality | Genome-fragment | 16 | AAI-based (high-confidence) | 0 |
| S12_k141_357330 | 1298 | No | NA | 2 | 0 | 1 | Not-determined | Genome-fragment | NA | NA | 0 |
| S12_k141_324994 | 3890 | No | NA | 2 | 0 | 1 | Not-determined | Genome-fragment | NA | NA | 0 |
| S12_k141_207571 | 3409 | No | NA | 4 | 0 | 4 | Not-determined | Genome-fragment | NA | NA | 0 |
| S12_k141_40552 | 3731 | No | NA | 4 | 0 | 1 | Not-determined | Genome-fragment | NA | NA | 0 |
| S12_k141_240927 | 1851 | No | NA | 2 | 0 | 2 | Not-determined | Genome-fragment | NA | NA | 0 |
| S12_k141_140751 | 6391 | No | NA | 5 | 0 | 4 | Not-determined | Genome-fragment | NA | NA | 0 |
| S17-final-viral-combined_k141_6586\|\|lt2gene | 3369 | No | NA | 1 | 1 | 0 | Low-quality | Genome-fragment | 8.91 | AAI-based (high-confidence) | 0 |
| S12_k141_106947 | 3334 | No | NA | 1 | 0 | 1 | Not-determined | Genome-fragment | NA | NA | 0 |
| S12_k141_40568 | 1366 | No | NA | 1 | 0 | 1 | Not-determined | Genome-fragment | NA | NA | 0 |
| S12_k141_340947 | 1295 | No | NA | 2 | 0 | 2 | Not-determined | Genome-fragment | NA | NA | 0 |
| S12_k141_157667 | 2405 | No | NA | 1 | 0 | 1 | Not-determined | Genome-fragment | NA | NA | 0 |
| S12_k141_174095 | 1041 | No | NA | 1 | 0 | 1 | Not-determined | Genome-fragment | NA | NA | 0 |
| S11_k141_153239 | 3371 | No | NA | 5 | 0 | 4 | Low-quality | Genome-fragment | 1.25 | AAI-based (medium-confidence) | 0 |
| S12_k141_274598 | 2150 | No | NA | 2 | 0 | 1 | Not-determined | Genome-fragment | NA | NA | 0 |
| S12_k141_90400 | 10601 | No | NA | 6 | 0 | 4 | Not-determined | Genome-fragment | NA | NA | 0 |
| S3-final-viral-combined_k141_82376\|\|full | 3372 | No | NA | 4 | 0 | 2 | Low-quality | Genome-fragment | 2.24 | AAI-based (medium-confidence) | 0 |
| S6_k141_59820 | 3372 | No | NA | 5 | 0 | 4 | Low-quality | Genome-fragment | 2.05 | AAI-based (medium-confidence) | 0 |
| S6-final-viral-combined_k141_81283\|\|full | 3373 | No | NA | 6 | 0 | 2 | Low-quality | Genome-fragment | 1.92 | AAI-based (high-confidence) | 0 |
| S12_k141_274616 | 3459 | No | NA | 2 | 0 | 1 | Not-determined | Genome-fragment | NA | NA | 0 |
| S8_k141_231632 | 3373 | No | NA | 4 | 0 | 3 | Low-quality | Genome-fragment | 5.68 | AAI-based (medium-confidence) | 0 |
| S11-final-viral-combined_k141_47927\|\|full | 3374 | No | NA | 5 | 0 | 0 | Low-quality | Genome-fragment | 28.25 | AAI-based (high-confidence) | 0 |
| S17-final-viral-combined_k141_14950\|\|full | 3374 | No | NA | 4 | 1 | 0 | Low-quality | Genome-fragment | 5.11 | HMM-based (lower-bound) | 0 |
| S12_k141_106988 | 1265 | No | NA | 1 | 0 | 1 | Not-determined | Genome-fragment | NA | NA | 0 |
| S12_k141_265650 | 3374 | No | NA | 2 | 0 | 2 | Low-quality | Genome-fragment | 1.82 | AAI-based (medium-confidence) | 0 |
| S2-final-viral-combined_k141_45243\|\|full | 3374 | No | NA | 3 | 1 | 0 | Low-quality | Genome-fragment | 9.09 | AAI-based (high-confidence) | 0 |
| S12_k141_274654 | 1204 | No | NA | 2 | 0 | 1 | Not-determined | Genome-fragment | NA | NA | 0 |
| S1-final-viral-combined_k141_172801\|\|full | 3375 | No | NA | 3 | 0 | 3 | Low-quality | Genome-fragment | 1.91 | AAI-based (medium-confidence) | 0 |
| S12_k141_308946 | 3278 | No | NA | 2 | 0 | 1 | Not-determined | Genome-fragment | NA | NA | 0 |
| S12_k141_292015 | 2034 | No | NA | 1 | 0 | 1 | Not-determined | Genome-fragment | NA | NA | 0 |
| S12-final-viral-combined_k141_334664\|\|full | 3375 | No | NA | 3 | 0 | 3 | Low-quality | Genome-fragment | 1.69 | AAI-based (medium-confidence) | 0 |
| S12_k141_292021 | 1334 | No | NA | 2 | 0 | 2 | Not-determined | Genome-fragment | NA | NA | 0 |
| S9-final-viral-combined_k141_90144\|\|full | 3377 | No | NA | 6 | 3 | 0 | Low-quality | Genome-fragment | 7.03 | AAI-based (high-confidence) | 0 |
| S12_k141_6645 | 1685 | No | NA | 2 | 0 | 1 | Not-determined | Genome-fragment | NA | NA | 0 |
| S1-final-viral-combined_k141_65480\|\|full | 3377 | No | NA | 4 | 1 | 0 | Low-quality | Genome-fragment | 8.06 | AAI-based (high-confidence) | 0 |
| S12_k141_123350 | 8783 | No | NA | 7 | 0 | 2 | Not-determined | Genome-fragment | NA | NA | 0 |
| S12_k141_308976 | 1196 | No | NA | 2 | 0 | 1 | Not-determined | Genome-fragment | NA | NA | 0 |
| S17-final-viral-combined_k141_210687\|\|full | 3377 | No | NA | 7 | 0 | 0 | Low-quality | Genome-fragment | 26.1 | AAI-based (high-confidence) | 0 |
| S12_k141_257715 | 1768 | No | NA | 2 | 0 | 2 | Not-determined | Genome-fragment | NA | NA | 0 |
| S12_k141_309003 | 1109 | No | NA | 2 | 0 | 1 | Not-determined | Genome-fragment | NA | NA | 0 |
| S12_k141_107049 | 2360 | No | NA | 2 | 0 | 1 | Not-determined | Genome-fragment | NA | NA | 0 |
| S12_k141_309024 | 3333 | No | NA | 3 | 0 | 2 | Not-determined | Genome-fragment | NA | NA | 0 |
| S12_k141_56413 | 1203 | No | NA | 1 | 0 | 1 | Not-determined | Genome-fragment | NA | NA | 0 |
| S12_k141_140849 | 1274 | No | NA | 1 | 0 | 1 | Not-determined | Genome-fragment | NA | NA | 0 |
| S12_k141_292101 | 1045 | No | NA | 1 | 0 | 0 | Not-determined | Genome-fragment | NA | NA | 0 |
| S8_k141_213935 | 3377 | No | NA | 4 | 1 | 0 | Low-quality | Genome-fragment | 1.63 | AAI-based (medium-confidence) | 0 |
| S12_k141_375338 | 3323 | No | NA | 3 | 0 | 2 | Not-determined | Genome-fragment | NA | NA | 0 |
| S7-final-viral-combined_k141_221549\|\|full | 3378 | No | NA | 6 | 2 | 0 | Low-quality | Genome-fragment | 7.44 | AAI-based (high-confidence) | 0 |
| S12_k141_292178 | 2094 | No | NA | 3 | 0 | 0 | Not-determined | Genome-fragment | NA | NA | 0 |
| S12_k141_325196 | 33104 | No | NA | 49 | 0 | 43 | Not-determined | Genome-fragment | NA | NA | 0 |
| S12-final-viral-combined_k141_352399\|\|full | 3378 | No | NA | 5 | 0 | 1 | Low-quality | Genome-fragment | 3.57 | AAI-based (high-confidence) | 0 |
| S12_k141_224521 | 3865 | No | NA | 1 | 0 | 0 | Not-determined | Genome-fragment | NA | NA | 0 |
| S12_k141_257861 | 1311 | No | NA | 2 | 0 | 2 | Not-determined | Genome-fragment | NA | NA | 0 |
| S9-final-viral-combined_k141_106874\|\|full | 3379 | No | NA | 5 | 0 | 1 | Low-quality | Genome-fragment | 9.35 | AAI-based (high-confidence) | 0 |
| S12_k141_90560 | 4484 | No | NA | 4 | 0 | 2 | Not-determined | Genome-fragment | NA | NA | 0 |
| S9_k141_111592 | 3379 | No | NA | 4 | 0 | 3 | Low-quality | Genome-fragment | 3.17 | AAI-based (medium-confidence) | 0 |
| S12_k141_325254 | 1326 | No | NA | 1 | 0 | 1 | Not-determined | Genome-fragment | NA | NA | 0 |
| S12_k141_40682 | 1053 | No | NA | 1 | 0 | 0 | Not-determined | Genome-fragment | NA | NA | 0 |
| S12_k141_375382 | 4383 | No | NA | 3 | 0 | 2 | Not-determined | Genome-fragment | NA | NA | 0 |
| S12_k141_325278 | 1013 | No | NA | 1 | 0 | 0 | Not-determined | Genome-fragment | NA | NA | 0 |
| S12_k141_392296 | 3369 | No | NA | 2 | 0 | 2 | Not-determined | Genome-fragment | NA | NA | 0 |
| S12_k141_274871 | 9412 | No | NA | 6 | 0 | 4 | Not-determined | Genome-fragment | NA | NA | 0 |
| S12_k141_123557 | 3776 | No | NA | 4 | 0 | 2 | Not-determined | Genome-fragment | NA | NA | 0 |
| S12_k141_224617 | 1066 | No | NA | 2 | 0 | 2 | Not-determined | Genome-fragment | NA | NA | 0 |
| S12_k141_325296 | 1504 | No | NA | 3 | 0 | 2 | Not-determined | Genome-fragment | NA | NA | 0 |
| S12_k141_325298 | 1317 | No | NA | 1 | 0 | 0 | Not-determined | Genome-fragment | NA | NA | 0 |
| S12_k141_6816 | 1315 | No | NA | 1 | 0 | 1 | Not-determined | Genome-fragment | NA | NA | 0 |
| S17-final-viral-combined_k141_9676\|\|full | 3379 | No | NA | 8 | 1 | 1 | Low-quality | Genome-fragment | 8.95 | AAI-based (high-confidence) | 0 |
| S12_k141_158006 | 7821 | No | NA | 1 | 0 | 1 | Not-determined | Genome-fragment | NA | NA | 0 |
| S12_k141_140960 | 1236 | No | NA | 1 | 0 | 1 | Not-determined | Genome-fragment | NA | NA | 0 |
| S12_k141_191889 | 1350 | No | NA | 3 | 0 | 1 | Not-determined | Genome-fragment | NA | NA | 0 |
| S12_k141_309105 | 1034 | No | NA | 1 | 0 | 1 | Not-determined | Genome-fragment | NA | NA | 0 |
| S12_k141_56629 | 1296 | No | NA | 1 | 0 | 0 | Not-determined | Genome-fragment | NA | NA | 0 |
| S17-k141_40755 | 3379 | No | NA | 5 | 0 | 0 | Low-quality | Genome-fragment | 8.01 | AAI-based (high-confidence) | 0 |
| S12_k141_375520 | 1391 | No | NA | 2 | 0 | 2 | Not-determined | Genome-fragment | NA | NA | 0 |
| S11-final-viral-combined_k141_253202\|\|full | 3380 | No | NA | 6 | 0 | 3 | Low-quality | Genome-fragment | 3.23 | AAI-based (medium-confidence) | 0 |
| S12_k141_6902 | 1059 | No | NA | 1 | 0 | 1 | Not-determined | Genome-fragment | NA | NA | 0 |
| S12-final-viral-combined_k141_232386\|\|full | 3380 | No | NA | 4 | 0 | 3 | Low-quality | Genome-fragment | 1.78 | AAI-based (medium-confidence) | 0 |
| S12_k141_224706 | 6653 | No | NA | 14 | 0 | 0 | Not-determined | Genome-fragment | NA | NA | 0 |
| S12_k141_141035 | 1194 | No | NA | 2 | 0 | 1 | Not-determined | Genome-fragment | NA | NA | 0 |
| S12_k141_56694 | 3362 | No | NA | 2 | 0 | 2 | Not-determined | Genome-fragment | NA | NA | 0 |
| S12_k141_241388 | 1070 | No | NA | 1 | 0 | 1 | Not-determined | Genome-fragment | NA | NA | 0 |
| S12_k141_191928 | 1007 | No | NA | 2 | 0 | 0 | Not-determined | Genome-fragment | NA | NA | 0 |
| S12_k141_6935 | 1140 | No | NA | 4 | 0 | 0 | Not-determined | Genome-fragment | NA | NA | 0 |
| S9-final-viral-combined_k141_89018\|\|full | 3381 | Yes | 2273 | 4 | 1 | 1 | Low-quality | Genome-fragment | 1.15 | HMM-based (lower-bound) | 32.77 |
| S12_k141_392438 | 1373 | No | NA | 3 | 0 | 2 | Not-determined | Genome-fragment | NA | NA | 0 |
| S12_k141_123724 | 1428 | No | NA | 1 | 0 | 1 | Not-determined | Genome-fragment | NA | NA | 0 |
| S12_k141_207881 | 1360 | No | NA | 1 | 0 | 1 | Not-determined | Genome-fragment | NA | NA | 0 |
| S12_k141_74688 | 3461 | No | NA | 7 | 0 | 4 | Not-determined | Genome-fragment | NA | NA | 0 |
| S1_k141_106138 | 3381 | No | NA | 4 | 0 | 3 | Low-quality | Genome-fragment | 7.4 | AAI-based (medium-confidence) | 0 |
| S12_k141_6957 | 13249 | No | NA | 25 | 0 | 24 | Not-determined | Genome-fragment | NA | NA | 0 |
| S17-k141_120196 | 3381 | No | NA | 5 | 3 | 0 | Low-quality | Genome-fragment | 9.08 | AAI-based (medium-confidence) | 0 |
| S12_k141_357777 | 2308 | No | NA | 4 | 0 | 2 | Not-determined | Genome-fragment | NA | NA | 0 |
| S12_k141_90571 | 1169 | No | NA | 1 | 0 | 1 | Not-determined | Genome-fragment | NA | NA | 0 |
| S12_k141_325423 | 1193 | No | NA | 2 | 0 | 2 | Not-determined | Genome-fragment | NA | NA | 0 |
| S12_k141_141109 | 4948 | No | NA | 13 | 0 | 13 | Not-determined | Genome-fragment | NA | NA | 0 |
| S12_k141_241504 | 1365 | No | NA | 1 | 0 | 1 | Not-determined | Genome-fragment | NA | NA | 0 |
| S12_k141_141116 | 1067 | No | NA | 1 | 0 | 1 | Not-determined | Genome-fragment | NA | NA | 0 |
| S12_k141_74763 | 1563 | No | NA | 1 | 0 | 0 | Not-determined | Genome-fragment | NA | NA | 0 |
| S12_k141_275049 | 1573 | No | NA | 3 | 0 | 2 | Not-determined | Genome-fragment | NA | NA | 0 |
| S12_k141_141132 | 1302 | No | NA | 2 | 0 | 2 | Not-determined | Genome-fragment | NA | NA | 0 |
| S12_k141_307997 | 3381 | No | NA | 4 | 0 | 0 | Low-quality | Genome-fragment | 5.06 | AAI-based (medium-confidence) | 0 |
| S12_k141_158163 | 2024 | No | NA | 3 | 0 | 0 | Not-determined | Genome-fragment | NA | NA | 0 |
| S12_k141_174414 | 1976 | No | NA | 3 | 0 | 3 | Not-determined | Genome-fragment | NA | NA | 0 |
| S6_k141_108151 | 3381 | No | NA | 3 | 3 | 0 | Low-quality | Genome-fragment | 8.17 | AAI-based (medium-confidence) | 0 |
| S8_k141_76328 | 3382 | No | NA | 3 | 0 | 3 | Low-quality | Genome-fragment | 3.65 | AAI-based (medium-confidence) | 0 |
| S12_k141_224854 | 7698 | No | NA | 14 | 0 | 0 | Not-determined | Genome-fragment | NA | NA | 0 |
| S12_k141_56840 | 1017 | No | NA | 1 | 0 | 1 | Not-determined | Genome-fragment | NA | NA | 0 |
| S12_k141_275125 | 1394 | No | NA | 1 | 0 | 0 | Not-determined | Genome-fragment | NA | NA | 0 |
| S12_k141_325468 | 1634 | No | NA | 2 | 0 | 0 | Not-determined | Genome-fragment | NA | NA | 0 |
| S1_k141_276180 | 3383 | No | NA | 5 | 0 | 1 | Low-quality | Genome-fragment | 3.01 | AAI-based (high-confidence) | 0 |
| S12_k141_392475 | 6343 | No | NA | 5 | 0 | 4 | Not-determined | Genome-fragment | NA | NA | 0 |
| S12_k141_23926 | 1677 | No | NA | 5 | 0 | 4 | Not-determined | Genome-fragment | NA | NA | 0 |

| | | | | | | | | | | | |
|---|---|---|---|---|---|---|---|---|---|---|---|
| S12_k141_56880 | 7814 | No | NA | 7 | 0 | 3 | Not-determined | Genome-fragment | NA | NA | 0 |
| S12_k141_357897 | 1078 | No | NA | 2 | 0 | 2 | Not-determined | Genome-fragment | NA | NA | 0 |
| S1_k141_161293 | 3383 | No | NA | 7 | 0 | 0 | Low-quality | Genome-fragment | 7.31 | AAI-based (high-confidence) | 0 |
| S12_k141_341447 | 1746 | No | NA | 1 | 0 | 1 | Not-determined | Genome-fragment | NA | NA | 0 |
| S12_k141_357923 | 1638 | No | NA | 2 | 0 | 1 | Not-determined | Genome-fragment | NA | NA | 0 |
| S12_k141_107443 | 1417 | No | NA | 1 | 0 | 0 | Not-determined | Genome-fragment | NA | NA | 0 |
| S12_k141_325542 | 1356 | No | NA | 1 | 0 | 1 | Not-determined | Genome-fragment | NA | NA | 0 |
| S12_k141_392597 | 2656 | No | NA | 5 | 0 | 0 | Not-determined | Genome-fragment | NA | NA | 0 |
| S12_k141_309432 | 1012 | No | NA | 3 | 0 | 0 | Not-determined | Genome-fragment | NA | NA | 0 |
| S5-final-viral-combined_k141_172356\|\|full | 3383 | No | NA | 5 | 3 | 0 | Low-quality | Genome-fragment | 4.85 | HMM-based (lower-bound) | 0 |
| S12_k141_41054 | 4096 | No | NA | 5 | 0 | 2 | Not-determined | Genome-fragment | NA | NA | 0 |
| S12_k141_141260 | 1588 | No | NA | 3 | 0 | 2 | Not-determined | Genome-fragment | NA | NA | 0 |
| S12_k141_174575 | 1641 | No | NA | 3 | 0 | 1 | Not-determined | Genome-fragment | NA | NA | 0 |
| S12_k141_90684 | 4489 | No | NA | 4 | 0 | 0 | Not-determined | Genome-fragment | NA | NA | 0 |
| S5-final-viral-combined_k141_168820\|\|full | 3383 | No | NA | 3 | 0 | 1 | Low-quality | Genome-fragment | 0.99 | AAI-based (medium-confidence) | 0 |
| S12_k141_258219 | 3462 | No | NA | 3 | 0 | 0 | Not-determined | Genome-fragment | NA | NA | 0 |
| S7_k141_244589 | 3383 | Yes | 2036 | 5 | 1 | 1 | Low-quality | Genome-fragment | 2.57 | AAI-based (medium-confidence) | 39.82 |
| S12_k141_341552 | 3570 | No | NA | 1 | 0 | 0 | Not-determined | Genome-fragment | NA | NA | 0 |
| S12_k141_375880 | 2381 | No | NA | 2 | 0 | 0 | Not-determined | Genome-fragment | NA | NA | 0 |
| S12_k141_258249 | 1113 | No | NA | 1 | 0 | 0 | Not-determined | Genome-fragment | NA | NA | 0 |
| S12-final-viral-combined_k141_99880\|\|full | 3384 | No | NA | 6 | 1 | 0 | Low-quality | Genome-fragment | 10.47 | AAI-based (medium-confidence) | 0 |
| S12_k141_375884 | 2364 | No | NA | 4 | 0 | 3 | Not-determined | Genome-fragment | NA | NA | 0 |
| S12_k141_375892 | 2504 | No | NA | 3 | 0 | 0 | Not-determined | Genome-fragment | NA | NA | 0 |
| S12-final-viral-combined_k141_301129\|\|full | 3384 | No | NA | 4 | 0 | 3 | Low-quality | Genome-fragment | 2.79 | AAI-based (medium-confidence) | 0 |
| S12_k141_375903 | 1568 | No | NA | 1 | 0 | 0 | Not-determined | Genome-fragment | NA | NA | 0 |
| S9-final-viral-combined_k141_26272\|\|full | 3385 | No | NA | 4 | 0 | 0 | Low-quality | Genome-fragment | 5.34 | AAI-based (high-confidence) | 0 |
| S12-final-viral-combined_k141_213644\|\|full | 3386 | Yes | 2126 | 6 | 1 | 1 | Low-quality | Genome-fragment | 2.91 | HMM-based (lower-bound) | 37.21 |
| S12_k141_158476 | 1022 | No | NA | 2 | 0 | 1 | Not-determined | Genome-fragment | NA | NA | 0 |
| S12_k141_123897 | 2185 | No | NA | 1 | 0 | 0 | Not-determined | Genome-fragment | NA | NA | 0 |
| S12_k141_74987 | 4724 | No | NA | 5 | 0 | 4 | Not-determined | Genome-fragment | NA | NA | 0 |
| S12_k141_325743 | 1434 | No | NA | 1 | 0 | 0 | Not-determined | Genome-fragment | NA | NA | 0 |
| S12_k141_341642 | 1031 | No | NA | 1 | 0 | 1 | Not-determined | Genome-fragment | NA | NA | 0 |
| S12_k141_57089 | 3436 | No | NA | 1 | 0 | 0 | Not-determined | Genome-fragment | NA | NA | 0 |
| S8-final-viral-combined_k141_51393\|\|full | 3386 | No | NA | 3 | 0 | 0 | Low-quality | Genome-fragment | 9.2 | AAI-based (medium-confidence) | 0 |
| S12_k141_90788 | 7016 | No | NA | 8 | 0 | 6 | Not-determined | Genome-fragment | NA | NA | 0 |
| S12_k141_375959 | 1480 | No | NA | 2 | 0 | 0 | Not-determined | Genome-fragment | NA | NA | 0 |
| [illegible] | [illegible] | [illegible] | [illegible] | [illegible] | [illegible] | [illegible] | [illegible] | [illegible] | [illegible] | [illegible] | [illegible] |

| | | | | | | | | | | | |
|---|---|---|---|---|---|---|---|---|---|---|---|
| S17-final-viral-combined_k141_180545\|\|full | 3425 | No | NA | 4 | 1 | 0 | Low-quality | Genome-fragment | 2.26 | AAI-based (medium-confidence) | 0 |
| S12_k141_303957 | 3425 | No | NA | 4 | 1 | 0 | Low-quality | Genome-fragment | 3.3 | AAI-based (high-confidence) | 0 |
| S12_k141_210330 | 1388 | No | NA | 2 | 0 | 1 | Not-determined | Genome-fragment | NA | NA | 0 |
| S12_k141_59292 | 1516 | No | NA | 1 | 0 | 1 | Not-determined | Genome-fragment | NA | NA | 0 |
| S12_k141_77050 | 1496 | No | NA | 2 | 0 | 2 | Not-determined | Genome-fragment | NA | NA | 0 |
| S12_k141_9211 | 1395 | No | NA | 1 | 0 | 1 | Not-determined | Genome-fragment | NA | NA | 0 |
| S12_k141_125606 | 1270 | No | NA | 1 | 0 | 1 | Not-determined | Genome-fragment | NA | NA | 0 |
| S12_k141_210346 | 1165 | No | NA | 2 | 0 | 0 | Not-determined | Genome-fragment | NA | NA | 0 |
| S4_k141_149808 | 3425 | No | NA | 7 | 1 | 0 | Low-quality | Genome-fragment | 4.92 | HMM-based (lower-bound) | 0 |
| S12_k141_377890 | 1188 | No | NA | 2 | 0 | 1 | Not-determined | Genome-fragment | NA | NA | 0 |
| S2_k141_64325 | 3426 | No | NA | 1 | 0 | 1 | Low-quality | Genome-fragment | 5.21 | AAI-based (medium-confidence) | 0 |
| S11-final-viral-combined_k141_199136\|\|full | 3427 | No | NA | 6 | 1 | 0 | Low-quality | Genome-fragment | 3.63 | AAI-based (high-confidence) | 0 |
| S12_k141_125640 | 1323 | No | NA | 2 | 0 | 0 | Not-determined | Genome-fragment | NA | NA | 0 |
| S17-k141_31281 | 3427 | No | NA | 4 | 0 | 4 | Low-quality | Genome-fragment | 1.27 | AAI-based (high-confidence) | 0 |
| S12_k141_344155 | 2587 | No | NA | 3 | 0 | 3 | Not-determined | Genome-fragment | NA | NA | 0 |
| S12_k141_394473 | 3715 | No | NA | 5 | 0 | 4 | Not-determined | Genome-fragment | NA | NA | 0 |
| S12_k141_176791 | 1243 | No | NA | 1 | 0 | 1 | Not-determined | Genome-fragment | NA | NA | 0 |
| S12-final-viral-combined_k141_252276\|\|full | 3427 | No | NA | 4 | 0 | 3 | Low-quality | Genome-fragment | 4.83 | AAI-based (medium-confidence) | 0 |
| S12_k141_312037 | 1012 | No | NA | 1 | 0 | 0 | Not-determined | Genome-fragment | NA | NA | 0 |
| S12_k141_160521 | 1177 | No | NA | 1 | 0 | 0 | Not-determined | Genome-fragment | NA | NA | 0 |
| S8-final-viral-combined_k141_148152\|\|full | 3427 | No | NA | 2 | 0 | 2 | Low-quality | Genome-fragment | 1.88 | AAI-based (medium-confidence) | 0 |
| S3-final-viral-combined_k141_61852\|\|full | 3428 | No | NA | 6 | 0 | 0 | Low-quality | Genome-fragment | 8.21 | AAI-based (medium-confidence) | 0 |
| S12_k141_176824 | 1588 | No | NA | 1 | 0 | 1 | Not-determined | Genome-fragment | NA | NA | 0 |
| S2-final-viral-combined_k141_160697\|\|full | 3428 | No | NA | 3 | 2 | 0 | Low-quality | Genome-fragment | 11.31 | HMM-based (lower-bound) | 0 |
| S12_k141_227658 | 1211 | No | NA | 1 | 0 | 0 | Not-determined | Genome-fragment | NA | NA | 0 |
| S12_k141_92710 | 11468 | No | NA | 13 | 0 | 7 | Not-determined | Genome-fragment | NA | NA | 0 |
| S3-final-viral-combined_k141_103732\|\|full | 3429 | No | NA | 2 | 0 | 0 | Low-quality | Genome-fragment | 6.53 | AAI-based (medium-confidence) | 0 |
| S12_k141_77187 | 1164 | No | NA | 2 | 0 | 2 | Not-determined | Genome-fragment | NA | NA | 0 |
| S12_k141_359697 | 1629 | No | NA | 3 | 0 | 0 | Not-determined | Genome-fragment | NA | NA | 0 |
| S12_k141_394519 | 1137 | No | NA | 2 | 0 | 1 | Not-determined | Genome-fragment | NA | NA | 0 |
| S3-final-viral-combined_k141_72185\|\|full | 3429 | No | NA | 5 | 0 | 5 | Low-quality | Genome-fragment | 0.82 | AAI-based (medium-confidence) | 0 |
| S8-final-viral-combined_k141_276049\|\|full | 3429 | No | NA | 6 | 2 | 0 | Low-quality | Genome-fragment | 6.01 | HMM-based (lower-bound) | 0 |
| S12_k141_294859 | 13605 | No | NA | 8 | 0 | 3 | Not-determined | Genome-fragment | NA | NA | 0 |
| S12_k141_109731 | 4247 | No | NA | 4 | 0 | 3 | Not-determined | Genome-fragment | NA | NA | 0 |
| S12_k141_227697 | 1083 | No | NA | 1 | 0 | 0 | Not-determined | Genome-fragment | NA | NA | 0 |
| S12_k141_26459 | 3295 | No | NA | 2 | 0 | 2 | Not-determined | Genome-fragment | NA | NA | 0 |
| S12_k141_59415 | 1883 | No | NA | 2 | 0 | 0 | Not-determined | Genome-fragment | NA | NA | 0 |
| S8_k141_130384 | 3429 | No | NA | 5 | 0 | 4 | Low-quality | Genome-fragment | 1.33 | AAI-based (medium-confidence) | 0 |
| S11-final-viral-combined_k141_140224\|\|full | 3430 | No | NA | 3 | 1 | 0 | Low-quality | Genome-fragment | 5.97 | AAI-based (medium-confidence) | 0 |
| S12_k141_312180 | 1406 | No | NA | 1 | 0 | 1 | Not-determined | Genome-fragment | NA | NA | 0 |
| S12_k141_394634 | 1083 | No | NA | 2 | 0 | 0 | Not-determined | Genome-fragment | NA | NA | 0 |
| S5_k141_180003 | 3430 | No | NA | 3 | 0 | 2 | Low-quality | Genome-fragment | 0.89 | AAI-based (medium-confidence) | 0 |
| S12_k141_176943 | 1545 | No | NA | 1 | 0 | 0 | Not-determined | Genome-fragment | NA | NA | 0 |
| S2_k141_116853 | 3430 | No | NA | 4 | 0 | 1 | Low-quality | Genome-fragment | 3.57 | AAI-based (high-confidence) | 0 |
| S12_k141_59486 | 6087 | No | NA | 2 | 0 | 1 | Not-determined | Genome-fragment | NA | NA | 0 |
| S12_k141_160651 | 3272 | No | NA | 4 | 0 | 4 | Not-determined | Genome-fragment | NA | NA | 0 |
| S11_k141_72198 | 3431 | No | NA | 11 | 0 | 0 | Low-quality | Genome-fragment | 7.64 | AAI-based (medium-confidence) | 0 |
| S12_k141_59524 | 1169 | No | NA | 1 | 0 | 0 | Not-determined | Genome-fragment | NA | NA | 0 |
| S12_k141_295018 | 6084 | No | NA | 7 | 0 | 4 | Not-determined | Genome-fragment | NA | NA | 0 |
| S12_k141_160659 | 2447 | No | NA | 2 | 0 | 2 | Not-determined | Genome-fragment | NA | NA | 0 |
| S2_k141_204260 | 3431 | No | NA | 4 | 0 | 0 | Low-quality | Genome-fragment | 3.19 | AAI-based (high-confidence) | 0 |
| S1-final-viral-combined_k141_258917\|\|full | 3433 | No | NA | 6 | 0 | 0 | Low-quality | Genome-fragment | 7.34 | AAI-based (medium-confidence) | 0 |
| S12_k141_143415 | 1312 | No | NA | 2 | 0 | 2 | Not-determined | Genome-fragment | NA | NA | 0 |
| S12_k141_160676 | 3184 | No | NA | 3 | 0 | 1 | Not-determined | Genome-fragment | NA | NA | 0 |
| S12_k141_77377 | 1061 | No | NA | 1 | 0 | 0 | Not-determined | Genome-fragment | NA | NA | 0 |
| S12_k141_9400 | 1359 | No | NA | 2 | 0 | 0 | Not-determined | Genome-fragment | NA | NA | 0 |
| S12_k141_260359 | 3390 | No | NA | 7 | 0 | 6 | Not-determined | Genome-fragment | NA | NA | 0 |
| S12_k141_394741 | 10156 | No | NA | 12 | 0 | 8 | Not-determined | Genome-fragment | NA | NA | 0 |
| S12_k141_394758 | 1229 | No | NA | 1 | 0 | 1 | Not-determined | Genome-fragment | NA | NA | 0 |
| S12_k141_277895 | 1742 | No | NA | 2 | 0 | 1 | Not-determined | Genome-fragment | NA | NA | 0 |
| S12_k141_92950 | 1393 | No | NA | 1 | 0 | 1 | Not-determined | Genome-fragment | NA | NA | 0 |
| S12_k141_378242 | 1516 | No | NA | 1 | 0 | 0 | Not-determined | Genome-fragment | NA | NA | 0 |
| S12_k141_59626 | 1027 | No | NA | 1 | 0 | 1 | Not-determined | Genome-fragment | NA | NA | 0 |
| S12_k141_312379 | 1176 | No | NA | 1 | 0 | 1 | Not-determined | Genome-fragment | NA | NA | 0 |
| S9_k141_82440 | 3434 | No | NA | 7 | 1 | 0 | Low-quality | Genome-fragment | 7.21 | AAI-based (high-confidence) | 0 |
| S12_k141_59637 | 2959 | No | NA | 1 | 0 | 0 | Not-determined | Genome-fragment | NA | NA | 0 |
| S12_k141_109916 | 1524 | No | NA | 2 | 0 | 0 | Not-determined | Genome-fragment | NA | NA | 0 |
| S12_k141_394788 | 2489 | No | NA | 2 | 0 | 2 | Not-determined | Genome-fragment | NA | NA | 0 |
| S12_k141_26718 | 1489 | No | NA | 3 | 0 | 3 | Not-determined | Genome-fragment | NA | NA | 0 |
| S11_k141_304922 | 3434 | No | NA | 4 | 0 | 0 | Low-quality | Genome-fragment | 2.04 | AAI-based (medium-confidence) | 0 |
| S12_k141_160753 | 1637 | No | NA | 1 | 0 | 0 | Not-determined | Genome-fragment | NA | NA | 0 |
| S12_k141_109950 | 1024 | No | NA | 1 | 0 | 1 | Not-determined | Genome-fragment | NA | NA | 0 |
| S12_k141_143512 | 2220 | No | NA | 4 | 0 | 0 | Not-determined | Genome-fragment | NA | NA | 0 |
| S8-final-viral-combined_k141_255297\|\|full | 3434 | No | NA | 4 | 1 | 0 | Low-quality | Genome-fragment | 2.46 | HMM-based (lower-bound) | 0 |
| S12_k141_9511 | 1177 | No | NA | 1 | 0 | 1 | Not-determined | Genome-fragment | NA | NA | 0 |
| S12_k141_160772 | 1673 | No | NA | 3 | 0 | 1 | Not-determined | Genome-fragment | NA | NA | 0 |
| S12_k141_394814 | 1124 | No | NA | 1 | 0 | 0 | Not-determined | Genome-fragment | NA | NA | 0 |
| S9-final-viral-combined_k141_114452\|\|full | 3435 | No | NA | 4 | 0 | 0 | Low-quality | Genome-fragment | 5.74 | AAI-based (medium-confidence) | 0 |
| S1-final-viral-combined_k141_56493\|\|full | 3435 | No | NA | 4 | 3 | 0 | Low-quality | Genome-fragment | 4.12 | HMM-based (lower-bound) | 0 |
| S12_k141_77542 | 1061 | No | NA | 3 | 0 | 0 | Not-determined | Genome-fragment | NA | NA | 0 |
| S12_k141_344442 | 17444 | No | NA | 3 | 0 | 0 | Not-determined | Genome-fragment | NA | NA | 0 |
| S12_k141_125955 | 6224 | No | NA | 5 | 0 | 5 | Not-determined | Genome-fragment | NA | NA | 0 |
| S12_k141_394847 | 2243 | No | NA | 1 | 0 | 0 | Not-determined | Genome-fragment | NA | NA | 0 |
| S12_k141_210646 | 1052 | No | NA | 1 | 0 | 0 | Not-determined | Genome-fragment | NA | NA | 0 |
| S12_k141_59787 | 1042 | No | NA | 1 | 0 | 1 | Not-determined | Genome-fragment | NA | NA | 0 |
| S12_k141_277989 | 1154 | No | NA | 3 | 0 | 0 | Not-determined | Genome-fragment | NA | NA | 0 |
| S5-final-viral-combined_k141_212556\|\|full | 3435 | No | NA | 7 | 0 | 0 | Low-quality | Genome-fragment | 9.08 | AAI-based (high-confidence) | 0 |
| S12_k141_394898 | 1246 | No | NA | 2 | 0 | 2 | Not-determined | Genome-fragment | NA | NA | 0 |
| S12_k141_344486 | 1512 | No | NA | 2 | 0 | 1 | Not-determined | Genome-fragment | NA | NA | 0 |
| S12_k141_244029 | 1355 | No | NA | 3 | 0 | 1 | Not-determined | Genome-fragment | NA | NA | 0 |
| S12_k141_394921 | 1027 | No | NA | 1 | 0 | 0 | Not-determined | Genome-fragment | NA | NA | 0 |
| S12_k141_24243 | 3435 | No | NA | 7 | 2 | 0 | Low-quality | Genome-fragment | 7.63 | AAI-based (medium-confidence) | 0 |
| S9-final-viral-combined_k141_102429\|\|full | 3436 | No | NA | 7 | 0 | 1 | Low-quality | Genome-fragment | 7.97 | AAI-based (medium-confidence) | 0 |
| S12_k141_9636 | 3495 | No | NA | 1 | 0 | 0 | Not-determined | Genome-fragment | NA | NA | 0 |
| S12_k141_344514 | 1340 | No | NA | 1 | 0 | 1 | Not-determined | Genome-fragment | NA | NA | 0 |
| S12_k141_360112 | 1045 | No | NA | 1 | 0 | 1 | Not-determined | Genome-fragment | NA | NA | 0 |
| S12_k141_44024 | 1049 | No | NA | 3 | 0 | 2 | Not-determined | Genome-fragment | NA | NA | 0 |
| S9-final-viral-combined_k141_73918\|\|full | 3436 | No | NA | 9 | 4 | 0 | Low-quality | Genome-fragment | 8.51 | AAI-based (high-confidence) | 0 |
| S12_k141_260565 | 9033 | No | NA | 5 | 0 | 3 | Not-determined | Genome-fragment | NA | NA | 0 |
| S12_k141_210730 | 1102 | No | NA | 2 | 0 | 1 | Not-determined | Genome-fragment | NA | NA | 0 |
| S12_k141_360146 | 3022 | No | NA | 2 | 0 | 1 | Not-determined | Genome-fragment | NA | NA | 0 |
| S12_k141_44049 | 1444 | No | NA | 1 | 0 | 1 | Not-determined | Genome-fragment | NA | NA | 0 |
| S12_k141_44057 | 1178 | No | NA | 1 | 0 | 0 | Not-determined | Genome-fragment | NA | NA | 0 |
| S12_k141_59926 | 3366 | No | NA | 4 | 0 | 2 | Not-determined | Genome-fragment | NA | NA | 0 |
| S12_k141_160894 | 3726 | No | NA | 2 | 0 | 2 | Not-determined | Genome-fragment | NA | NA | 0 |
| S12_k141_394981 | 1399 | No | NA | 3 | 0 | 0 | Not-determined | Genome-fragment | NA | NA | 0 |
| S12_k141_210802 | 1488 | No | NA | 1 | 0 | 0 | Not-determined | Genome-fragment | NA | NA | 0 |
| S12_k141_260623 | 1610 | No | NA | 6 | 0 | 0 | Not-determined | Genome-fragment | NA | NA | 0 |
| S12_k141_344557 | 1077 | No | NA | 1 | 0 | 1 | Not-determined | Genome-fragment | NA | NA | 0 |
| S12_k141_59948 | 7204 | No | NA | 9 | 0 | 2 | Not-determined | Genome-fragment | NA | NA | 0 |
| S12_k141_278072 | 5018 | No | NA | 7 | 0 | 1 | Not-determined | Genome-fragment | NA | NA | 0 |
| S12_k141_210808 | 1021 | No | NA | 1 | 0 | 1 | Not-determined | Genome-fragment | NA | NA | 0 |
| S12_k141_177242 | 1545 | No | NA | 3 | 0 | 1 | Not-determined | Genome-fragment | NA | NA | 0 |
| S5_k141_103265 | 3436 | Yes | 1229 | 4 | 1 | 1 | Low-quality | Genome-fragment | 0.72 | HMM-based (lower-bound) | 64.23 |
| S7_k141_207241 | 3436 | No | NA | 4 | 1 | 0 | Low-quality | Genome-fragment | 7.22 | AAI-based (medium-confidence) | 0 |
| S12_k141_244145 | 8378 | No | NA | 9 | 0 | 9 | Not-determined | Genome-fragment | NA | NA | 0 |
| S12_k141_295355 | 1388 | No | NA | 2 | 0 | 2 | Not-determined | Genome-fragment | NA | NA | 0 |
| S12_k141_109985 | 6205 | No | NA | 6 | 0 | 2 | Not-determined | Genome-fragment | NA | NA | 0 |
| S12_k141_160982 | 1287 | No | NA | 1 | 0 | 1 | Not-determined | Genome-fragment | NA | NA | 0 |
| S12_k141_278122 | 1264 | No | NA | 2 | 0 | 1 | Not-determined | Genome-fragment | NA | NA | 0 |
| S12_k141_395076 | 1154 | No | NA | 2 | 0 | 1 | Not-determined | Genome-fragment | NA | NA | 0 |
| S12_k141_210867 | 1564 | No | NA | 1 | 0 | 0 | Not-determined | Genome-fragment | NA | NA | 0 |
| S6-final-viral-combined_k141_138435\|\|full | 3436 | No | NA | 7 | 2 | 0 | Low-quality | Genome-fragment | 3.56 | AAI-based (medium-confidence) | 0 |
| S12_k141_395082 | 1047 | No | NA | 1 | 0 | 0 | Not-determined | Genome-fragment | NA | NA | 0 |
| S12_k141_278169 | 1172 | No | NA | 3 | 0 | 1 | Not-determined | Genome-fragment | NA | NA | 0 |
| S1-final-viral-combined_k141_240533\|\|full | 3437 | No | NA | 9 | 1 | 0 | Low-quality | Genome-fragment | 9.1 | AAI-based (high-confidence) | 0 |
| S12_k141_194918 | 2404 | No | NA | 2 | 0 | 0 | Not-determined | Genome-fragment | NA | NA | 0 |
| S5-final-viral-combined_k141_18479\|\|full | 3437 | No | NA | 5 | 1 | 0 | Low-quality | Genome-fragment | 2.36 | HMM-based (lower-bound) | 0 |
| S12_k141_278220 | 1206 | No | NA | 1 | 0 | 0 | Not-determined | Genome-fragment | NA | NA | 0 |
| S12_k141_210927 | 1136 | No | NA | 1 | 0 | 1 | Not-determined | Genome-fragment | NA | NA | 0 |
| S12_k141_378581 | 1850 | No | NA | 1 | 0 | 0 | Not-determined | Genome-fragment | NA | NA | 0 |
| S12-final-viral-combined_k141_180988\|\|full | 3437 | No | NA | 4 | 0 | 2 | Low-quality | Genome-fragment | 1.46 | AAI-based (medium-confidence) | 0 |
| S12_k141_328482 | 1526 | No | NA | 2 | 0 | 1 | Not-determined | Genome-fragment | NA | NA | 0 |
| S12_k141_60124 | 3561 | No | NA | 2 | 0 | 2 | Not-determined | Genome-fragment | NA | NA | 0 |
| S12_k141_244295 | 7935 | No | NA | 6 | 0 | 3 | Not-determined | Genome-fragment | NA | NA | 0 |
| S12_k141_77837 | 1197 | No | NA | 2 | 0 | 2 | Not-determined | Genome-fragment | NA | NA | 0 |
| S6-final-viral-combined_k141_45485\|\|full | 3437 | No | NA | 4 | 0 | 1 | Low-quality | Genome-fragment | 7.41 | AAI-based (high-confidence) | 0 |
| S12_k141_210954 | 1867 | No | NA | 2 | 0 | 2 | Not-determined | Genome-fragment | NA | NA | 0 |
| S12_k141_60159 | 1008 | No | NA | 1 | 0 | 1 | Not-determined | Genome-fragment | NA | NA | 0 |
| S12_k141_110025 | 3137 | No | NA | 2 | 0 | 1 | Not-determined | Genome-fragment | NA | NA | 0 |
| S12_k141_395244 | 8405 | No | NA | 8 | 0 | 5 | Not-determined | Genome-fragment | NA | NA | 0 |
| S12_k141_195025 | 2607 | No | NA | 1 | 0 | 1 | Not-determined | Genome-fragment | NA | NA | 0 |
| S12_k141_228334 | 6318 | No | NA | 5 | 0 | 3 | Not-determined | Genome-fragment | NA | NA | 0 |
| S12_k141_27049 | 1229 | No | NA | 4 | 0 | 2 | Not-determined | Genome-fragment | NA | NA | 0 |
| S12_k141_126250 | 1123 | No | NA | 2 | 0 | 0 | Not-determined | Genome-fragment | NA | NA | 0 |
| S12_k141_195072 | 1216 | No | NA | 1 | 0 | 1 | Not-determined | Genome-fragment | NA | NA | 0 |
| S12_k141_211035 | 3283 | No | NA | 3 | 0 | 2 | Not-determined | Genome-fragment | NA | NA | 0 |
| S12_k141_110101 | 1125 | No | NA | 1 | 0 | 1 | Not-determined | Genome-fragment | NA | NA | 0 |
| S12_k141_177442 | 5617 | No | NA | 1 | 0 | 1 | Not-determined | Genome-fragment | NA | NA | 0 |
| S12_k141_195097 | 1264 | No | NA | 1 | 0 | 0 | Not-determined | Genome-fragment | NA | NA | 0 |
| S12_k141_126261 | 9719 | No | NA | 10 | 0 | 2 | Not-determined | Genome-fragment | NA | NA | 0 |
| S12_k141_126271 | 1019 | No | NA | 2 | 0 | 1 | Not-determined | Genome-fragment | NA | NA | 0 |
| S12_k141_143870 | 3288 | No | NA | 1 | 0 | 0 | Not-determined | Genome-fragment | NA | NA | 0 |
| S5-final-viral-combined_k141_71990\|\|lt2gene | 3438 | No | NA | 2 | 2 | 0 | Low-quality | Genome-fragment | 2.13 | HMM-based (lower-bound) | 0 |
| S7-final-viral-combined_k141_188798\|\|full | 3438 | No | NA | 3 | 0 | 2 | Low-quality | Genome-fragment | 5.82 | AAI-based (medium-confidence) | 0 |
| S12-final-viral-combined_k141_351401\|\|full | 3438 | No | NA | 5 | 2 | 0 | Low-quality | Genome-fragment | 3.9 | AAI-based (medium-confidence) | 0 |
| S12_k141_370502 | 3440 | No | NA | 5 | 0 | 0 | Low-quality | Genome-fragment | 2.12 | AAI-based (high-confidence) | 0 |
| S12_k141_278514 | 1252 | No | NA | 1 | 0 | 0 | Not-determined | Genome-fragment | NA | NA | 0 |
| S12_k141_228460 | 1501 | No | NA | 2 | 0 | 1 | Not-determined | Genome-fragment | NA | NA | 0 |
| S12_k141_126305 | 4258 | No | NA | 3 | 0 | 1 | Not-determined | Genome-fragment | NA | NA | 0 |
| S12_k141_27079 | 1315 | No | NA | 2 | 0 | 2 | Not-determined | Genome-fragment | NA | NA | 0 |
| S12_k141_344900 | 4745 | No | NA | 2 | 0 | 2 | Not-determined | Genome-fragment | NA | NA | 0 |
| S12_k141_278554 | 1835 | No | NA | 1 | 0 | 0 | Not-determined | Genome-fragment | NA | NA | 0 |
| S5-final-viral-combined_k141_189071\|\|full | 3441 | No | NA | 6 | 0 | 2 | Low-quality | Genome-fragment | 5.7 | AAI-based (medium-confidence) | 0 |
| S12_k141_44529 | 7548 | No | NA | 3 | 0 | 2 | Not-determined | Genome-fragment | NA | NA | 0 |
| S12_k141_60382 | 2594 | No | NA | 1 | 0 | 1 | Not-determined | Genome-fragment | NA | NA | 0 |
| S12_k141_143990 | 1279 | No | NA | 3 | 0 | 0 | Not-determined | Genome-fragment | NA | NA | 0 |
| S12_k141_211128 | 1063 | No | NA | 2 | 0 | 2 | Not-determined | Genome-fragment | NA | NA | 0 |
| S5_k141_146378 | 3441 | No | NA | 3 | 0 | 3 | Low-quality | Genome-fragment | 7.21 | AAI-based (medium-confidence) | 0 |
| S12_k141_195228 | 1186 | No | NA | 1 | 0 | 1 | Not-determined | Genome-fragment | NA | NA | 0 |
| S12_k141_360242 | 1482 | No | NA | 3 | 0 | 2 | Not-determined | Genome-fragment | NA | NA | 0 |
| S12_k141_360249 | 1257 | No | NA | 1 | 0 | 1 | Not-determined | Genome-fragment | NA | NA | 0 |
| S12_k141_211164 | 1176 | No | NA | 1 | 0 | 1 | Not-determined | Genome-fragment | NA | NA | 0 |
| S12_k141_244483 | 4229 | No | NA | 4 | 0 | 2 | Not-determined | Genome-fragment | NA | NA | 0 |
| S17-k141_49763 | 3441 | No | NA | 5 | 2 | 0 | Low-quality | Genome-fragment | 13.61 | AAI-based (high-confidence) | 0 |
| S12_k141_378851 | 3359 | No | NA | 5 | 0 | 2 | Not-determined | Genome-fragment | NA | NA | 0 |
| S12_k141_161239 | 25372 | Yes | 19782 | 38 | 16 | 4 | Medium-quality | Genome-fragment | 54.65 | AAI-based (high-confidence) | 22.03 |
| S12_k141_161248 | 1240 | No | NA | 1 | 0 | 0 | Not-determined | Genome-fragment | NA | NA | 0 |
| S12_k141_9981 | 1027 | No | NA | 1 | 0 | 1 | Not-determined | Genome-fragment | NA | NA | 0 |
| S12_k141_9989 | 1528 | No | NA | 2 | 0 | 2 | Not-determined | Genome-fragment | NA | NA | 0 |
| S12_k141_261075 | 1444 | No | NA | 3 | 0 | 2 | Not-determined | Genome-fragment | NA | NA | 0 |
| S2_k141_21384 | 3442 | No | NA | 6 | 2 | 0 | Low-quality | Genome-fragment | 2.37 | AAI-based (high-confidence) | 0 |
| S12_k141_328621 | 1092 | No | NA | 1 | 0 | 0 | Not-determined | Genome-fragment | NA | NA | 0 |
| S12_k141_195292 | 1175 | No | NA | 1 | 0 | 1 | Not-determined | Genome-fragment | NA | NA | 0 |
| S12_k141_244579 | 1004 | No | NA | 1 | 0 | 0 | Not-determined | Genome-fragment | NA | NA | 0 |
| S12_k141_244581 | 1407 | No | NA | 1 | 0 | 0 | Not-determined | Genome-fragment | NA | NA | 0 |
| S12_k141_93629 | 1078 | No | NA | 1 | 0 | 1 | Not-determined | Genome-fragment | NA | NA | 0 |
| S12_k141_44587 | 1146 | No | NA | 2 | 0 | 2 | Not-determined | Genome-fragment | NA | NA | 0 |
| S12_k141_261128 | 1883 | No | NA | 1 | 0 | 0 | Not-determined | Genome-fragment | NA | NA | 0 |
| S12_k141_278713 | 1110 | No | NA | 1 | 0 | 0 | Not-determined | Genome-fragment | NA | NA | 0 |
| S8-final-viral-combined_k141_232066\|\|full | 3442 | No | NA | 4 | 1 | 0 | Low-quality | Genome-fragment | 2.23 | AAI-based (high-confidence) | 0 |
| S17-k141_169983 | 3443 | Yes | 2498 | 4 | 1 | 1 | Low-quality | Genome-fragment | 0.74 | HMM-based (lower-bound) | 27.45 |
| S12_k141_161330 | 1330 | No | NA | 2 | 0 | 2 | Not-determined | Genome-fragment | NA | NA | 0 |
| S12_k141_60495 | 7847 | No | NA | 7 | 0 | 5 | Not-determined | Genome-fragment | NA | NA | 0 |
| S12_k141_278741 | 3092 | No | NA | 2 | 0 | 2 | Not-determined | Genome-fragment | NA | NA | 0 |
| S12_k141_295590 | 2359 | No | NA | 2 | 0 | 2 | Not-determined | Genome-fragment | NA | NA | 0 |
| S12_k141_219815 | 3443 | No | NA | 4 | 2 | 0 | Low-quality | Genome-fragment | 10.71 | AAI-based (high-confidence) | 0 |
| S12_k141_161364 | 1462 | No | NA | 2 | 0 | 1 | Not-determined | Genome-fragment | NA | NA | 0 |
| S12_k141_360368 | 1617 | No | NA | 1 | 0 | 1 | Not-determined | Genome-fragment | NA | NA | 0 |
| S4-final-viral-combined_k141_3729\|\|full | 3443 | No | NA | 3 | 1 | 0 | Low-quality | Genome-fragment | 6.08 | AAI-based (high-confidence) | 0 |
| S12_k141_228694 | 3447 | No | NA | 3 | 0 | 2 | Not-determined | Genome-fragment | NA | NA | 0 |
| S17-final-viral-combined_k141_75443\|\|full | 3445 | No | NA | 6 | 1 | 0 | Low-quality | Genome-fragment | 8.34 | AAI-based (high-confidence) | 0 |
| S12_k141_161385 | 1065 | No | NA | 1 | 0 | 0 | Not-determined | Genome-fragment | NA | NA | 0 |
| S12_k141_345152 | 2694 | No | NA | 2 | 0 | 2 | Not-determined | Genome-fragment | NA | NA | 0 |
| S12_k141_360398 | 1255 | No | NA | 3 | 0 | 0 | Not-determined | Genome-fragment | NA | NA | 0 |
| S12_k141_278805 | 1698 | No | NA | 1 | 0 | 1 | Not-determined | Genome-fragment | NA | NA | 0 |
| S8-final-viral-combined_k141_114833\|\|full | 3445 | No | NA | 3 | 0 | 2 | Low-quality | Genome-fragment | 6.61 | AAI-based (medium-confidence) | 0 |
| S12_k141_295614 | 2110 | No | NA | 1 | 0 | 1 | Not-determined | Genome-fragment | NA | NA | 0 |
| S12_k141_295626 | 1497 | No | NA | 2 | 0 | 0 | Not-determined | Genome-fragment | NA | NA | 0 |
| S17-final-viral-combined_k141_28922\|\|full | 3446 | No | NA | 5 | 2 | 0 | Low-quality | Genome-fragment | 9.96 | AAI-based (high-confidence) | 0 |
| S12_k141_278856 | 4079 | No | NA | 2 | 0 | 1 | Not-determined | Genome-fragment | NA | NA | 0 |
| S9-final-viral-combined_k141_44063\|\|full | 3447 | No | NA | 5 | 0 | 0 | Low-quality | Genome-fragment | 8.33 | AAI-based (medium-confidence) | 0 |
| S12_k141_395536 | 1074 | No | NA | 1 | 0 | 1 | Not-determined | Genome-fragment | NA | NA | 0 |
| S2_k141_201367 | 3447 | No | NA | 5 | 0 | 0 | Low-quality | Genome-fragment | 3.77 | AAI-based (medium-confidence) | 0 |
| S12_k141_295645 | 3133 | No | NA | 2 | 0 | 1 | Not-determined | Genome-fragment | NA | NA | 0 |
| S12_k141_161466 | 10084 | No | NA | 7 | 0 | 4 | Not-determined | Genome-fragment | NA | NA | 0 |
| S12_k141_110441 | 1059 | No | NA | 1 | 0 | 0 | Not-determined | Genome-fragment | NA | NA | 0 |
| S12_k141_295671 | 1440 | No | NA | 3 | 0 | 3 | Not-determined | Genome-fragment | NA | NA | 0 |
| S17-final-viral-combined_k141_259627\|\|full | 3448 | No | NA | 4 | 2 | 0 | Low-quality | Genome-fragment | 10.09 | AAI-based (high-confidence) | 0 |
| S12_k141_110452 | 1063 | No | NA | 1 | 0 | 1 | Not-determined | Genome-fragment | NA | NA | 0 |
| S12_k141_278943 | 4554 | No | NA | 3 | 0 | 2 | Not-determined | Genome-fragment | NA | NA | 0 |
| S12_k141_78296 | 1613 | No | NA | 3 | 0 | 0 | Not-determined | Genome-fragment | NA | NA | 0 |
| S12_k141_244773 | 1475 | No | NA | 4 | 0 | 3 | Not-determined | Genome-fragment | NA | NA | 0 |
| S12_k141_177855 | 1354 | No | NA | 3 | 0 | 0 | Not-determined | Genome-fragment | NA | NA | 0 |
| S12_k141_328849 | 1239 | No | NA | 2 | 0 | 1 | Not-determined | Genome-fragment | NA | NA | 0 |
| S2-final-viral-combined_k141_32731\|\|full | 3448 | No | NA | 6 | 1 | 0 | Low-quality | Genome-fragment | 4.39 | HMM-based (lower-bound) | 0 |
| S12_k141_395617 | 2144 | No | NA | 2 | 0 | 2 | Not-determined | Genome-fragment | NA | NA | 0 |
| S12_k141_379141 | 1692 | No | NA | 1 | 0 | 0 | Not-determined | Genome-fragment | NA | NA | 0 |
| S4-final-viral-combined_k141_132708\|\|full | 3448 | No | NA | 3 | 0 | 3 | Low-quality | Genome-fragment | 6.9 | AAI-based (medium-confidence) | 0 |
| S12_k141_279002 | 1602 | No | NA | 2 | 0 | 1 | Not-determined | Genome-fragment | NA | NA | 0 |
| S12_k141_177914 | 1125 | No | NA | 3 | 0 | 3 | Not-determined | Genome-fragment | NA | NA | 0 |
| S12_k141_126654 | 1697 | No | NA | 2 | 0 | 2 | Not-determined | Genome-fragment | NA | NA | 0 |
| S12_k141_27534 | 1333 | No | NA | 1 | 0 | 1 | Not-determined | Genome-fragment | NA | NA | 0 |
| S5_k141_185495 | 3449 | No | NA | 10 | 2 | 0 | Low-quality | Genome-fragment | 8.51 | AAI-based (high-confidence) | 0 |
| S17-k141_140056 | 3449 | No | NA | 4 | 0 | 3 | Low-quality | Genome-fragment | 1.42 | AAI-based (high-confidence) | 0 |
| S12_k141_126667 | 10829 | No | NA | 9 | 0 | 5 | Not-determined | Genome-fragment | NA | NA | 0 |
| S12_k141_345327 | 1002 | No | NA | 1 | 0 | 1 | Not-determined | Genome-fragment | NA | NA | 0 |
| S12_k141_78410 | 2162 | No | NA | 3 | 0 | 2 | Not-determined | Genome-fragment | NA | NA | 0 |
| S12_k141_244883 | 1074 | No | NA | 1 | 0 | 0 | Not-determined | Genome-fragment | NA | NA | 0 |
| S12_k141_10216 | 1001 | No | NA | 2 | 0 | 0 | Not-determined | Genome-fragment | NA | NA | 0 |
| S12_k141_379227 | 1503 | No | NA | 2 | 0 | 0 | Not-determined | Genome-fragment | NA | NA | 0 |
| S12_k141_10217 | 13478 | No | NA | 16 | 0 | 11 | Not-determined | Genome-fragment | NA | NA | 0 |
| S12-final-viral-combined_k141_204217\|\|full | 3450 | No | NA | 5 | 1 | 0 | Low-quality | Genome-fragment | 2.36 | HMM-based (lower-bound) | 0 |
| S12_k141_345382 | 2248 | No | NA | 4 | 0 | 3 | Not-determined | Genome-fragment | NA | NA | 0 |
| S12_k141_228938 | 1077 | No | NA | 1 | 0 | 1 | Not-determined | Genome-fragment | NA | NA | 0 |
| S12_k141_360647 | 1279 | No | NA | 2 | 0 | 1 | Not-determined | Genome-fragment | NA | NA | 0 |
| S12_k141_10241 | 1322 | No | NA | 1 | 0 | 1 | Not-determined | Genome-fragment | NA | NA | 0 |
| S12_k141_44817 | 34968 | No | NA | 59 | 10 | 0 | Medium-quality | Genome-fragment | 76.17 | AAI-based (high-confidence) | 0 |
| S12_k141_211667 | 1190 | No | NA | 2 | 0 | 2 | Not-determined | Genome-fragment | NA | NA | 0 |
| S12_k141_93954 | 1766 | No | NA | 1 | 0 | 1 | Not-determined | Genome-fragment | NA | NA | 0 |
| S12-final-viral-combined_k141_191002\|\|full | 3450 | No | NA | 5 | 1 | 0 | Low-quality | Genome-fragment | 7.13 | AAI-based (medium-confidence) | 0 |
| S12_k141_110574 | 5631 | No | NA | 6 | 0 | 5 | Not-determined | Genome-fragment | NA | NA | 0 |
| S12_k141_313257 | 6112 | No | NA | 6 | 0 | 2 | Not-determined | Genome-fragment | NA | NA | 0 |
| S12_k141_126766 | 1043 | No | NA | 1 | 0 | 1 | Not-determined | Genome-fragment | NA | NA | 0 |
| S12_k141_261445 | 1084 | No | NA | 2 | 0 | 0 | Not-determined | Genome-fragment | NA | NA | 0 |
| S2-final-viral-combined_k141_82146\|\|full | 3452 | No | NA | 5 | 3 | 0 | Low-quality | Genome-fragment | 7.48 | AAI-based (high-confidence) | 0 |
| S12_k141_395776 | 4468 | No | NA | 4 | 0 | 2 | Not-determined | Genome-fragment | NA | NA | 0 |
| S12_k141_78526 | 3768 | No | NA | 2 | 0 | 2 | Not-determined | Genome-fragment | NA | NA | 0 |
| S12_k141_360682 | 2069 | No | NA | 1 | 0 | 0 | Not-determined | Genome-fragment | NA | NA | 0 |
| S12_k141_379385 | 1128 | No | NA | 1 | 0 | 0 | Not-determined | Genome-fragment | NA | NA | 0 |
| S12_k141_211735 | 1101 | No | NA | 1 | 0 | 1 | Not-determined | Genome-fragment | NA | NA | 0 |
| S12_k141_261492 | 1152 | No | NA | 1 | 0 | 0 | Not-determined | Genome-fragment | NA | NA | 0 |
| S12_k141_211761 | 3505 | No | NA | 3 | 0 | 3 | Not-determined | Genome-fragment | NA | NA | 0 |
| S12_k141_161638 | 1092 | No | NA | 3 | 0 | 0 | Not-determined | Genome-fragment | NA | NA | 0 |
| S12_k141_211771 | 1011 | No | NA | 2 | 0 | 1 | Not-determined | Genome-fragment | NA | NA | 0 |
| S12_k141_229105 | 1211 | No | NA | 2 | 0 | 0 | Not-determined | Genome-fragment | NA | NA | 0 |
| S12_k141_379454 | 1033 | No | NA | 1 | 0 | 1 | Not-determined | Genome-fragment | NA | NA | 0 |
| S12_k141_195519 | 1158 | No | NA | 4 | 0 | 3 | Not-determined | Genome-fragment | NA | NA | 0 |
| S12_k141_279254 | 1050 | No | NA | 1 | 0 | 0 | Not-determined | Genome-fragment | NA | NA | 0 |
| S12_k141_27740 | 1486 | No | NA | 2 | 0 | 1 | Not-determined | Genome-fragment | NA | NA | 0 |
| S7-final-viral-combined_k141_170460\|\|full | 3453 | No | NA | 5 | 1 | 0 | Low-quality | Genome-fragment | 9.24 | AAI-based (high-confidence) | 0 |
| S4-final-viral-combined_k141_88864\|\|full | 3453 | No | NA | 5 | 0 | 0 | Low-quality | Genome-fragment | 4.13 | AAI-based (high-confidence) | 0 |
| S12_k141_379504 | 1547 | No | NA | 1 | 0 | 1 | Not-determined | Genome-fragment | NA | NA | 0 |
| S12_k141_110692 | 1149 | No | NA | 2 | 0 | 0 | Not-determined | Genome-fragment | NA | NA | 0 |
| S12_k141_261600 | 1226 | No | NA | 2 | 0 | 0 | Not-determined | Genome-fragment | NA | NA | 0 |
| S12_k141_245080 | 1228 | No | NA | 1 | 0 | 1 | Not-determined | Genome-fragment | NA | NA | 0 |
| S12_k141_279281 | 1013 | No | NA | 1 | 0 | 1 | Not-determined | Genome-fragment | NA | NA | 0 |
| S12_k141_360794 | 4749 | No | NA | 2 | 0 | 2 | Not-determined | Genome-fragment | NA | NA | 0 |
| S12_k141_178060 | 1098 | No | NA | 2 | 0 | 1 | Not-determined | Genome-fragment | NA | NA | 0 |
| S12_k141_360805 | 9708 | No | NA | 20 | 0 | 6 | Not-determined | Genome-fragment | NA | NA | 0 |
| S12_k141_360808 | 1291 | No | NA | 1 | 0 | 1 | Not-determined | Genome-fragment | NA | NA | 0 |
| S12_k141_229277 | 1006 | No | NA | 1 | 0 | 1 | Not-determined | Genome-fragment | NA | NA | 0 |
| S11-final-viral-combined_k141_18600\|\|full | 3454 | No | NA | 4 | 0 | 3 | Low-quality | Genome-fragment | 1.14 | AAI-based (medium-confidence) | 0 |
| S12_k141_45078 | 5071 | No | NA | 3 | 0 | 1 | Not-determined | Genome-fragment | NA | NA | 0 |
| S7_k141_175086 | 3456 | No | NA | 1 | 0 | 1 | Low-quality | Genome-fragment | 1.86 | AAI-based (medium-confidence) | 0 |
| S12_k141_144393 | 1071 | No | NA | 1 | 0 | 0 | Not-determined | Genome-fragment | NA | NA | 0 |
| S12_k141_261718 | 1379 | No | NA | 1 | 0 | 1 | Not-determined | Genome-fragment | NA | NA | 0 |
| S12-final-viral-combined_k141_142586\|\|full | 3456 | No | NA | 4 | 0 | 2 | Low-quality | Genome-fragment | 7.2 | AAI-based (medium-confidence) | 0 |
| S12_k141_45095 | 1254 | No | NA | 1 | 0 | 1 | Not-determined | Genome-fragment | NA | NA | 0 |
| S12-final-viral-combined_k141_378779\|\|full | 3456 | No | NA | 2 | 0 | 2 | Low-quality | Genome-fragment | 1.4 | AAI-based (medium-confidence) | 0 |
| S12_k141_144427 | 1546 | No | NA | 2 | 0 | 1 | Not-determined | Genome-fragment | NA | NA | 0 |
| S12_k141_36119 | 3456 | No | NA | 1 | 0 | 0 | Low-quality | Genome-fragment | 5.73 | AAI-based (medium-confidence) | 0 |
| S12_k141_94218 | 1032 | No | NA | 1 | 0 | 0 | Not-determined | Genome-fragment | NA | NA | 0 |
| S12_k141_45144 | 1109 | No | NA | 1 | 0 | 1 | Not-determined | Genome-fragment | NA | NA | 0 |
| S12_k141_60894 | 1265 | No | NA | 1 | 0 | 0 | Not-determined | Genome-fragment | NA | NA | 0 |
| S5-final-viral-combined_k141_129140\|\|full | 3457 | No | NA | 4 | 0 | 3 | Low-quality | Genome-fragment | 3.87 | AAI-based (medium-confidence) | 0 |
| S12_k141_127038 | 10930 | No | NA | 6 | 0 | 5 | Not-determined | Genome-fragment | NA | NA | 0 |
| S8-final-viral-combined_k141_51977\|\|full | 3457 | No | NA | 7 | 0 | 0 | Low-quality | Genome-fragment | 4.37 | AAI-based (medium-confidence) | 0 |
| S12_k141_161756 | 2061 | No | NA | 2 | 0 | 1 | Not-determined | Genome-fragment | NA | NA | 0 |
| S8_k141_216577 | 3457 | No | NA | 10 | 1 | 0 | Low-quality | Genome-fragment | 3.61 | AAI-based (high-confidence) | 0 |
| S12_k141_396055 | 1006 | No | NA | 1 | 0 | 1 | Not-determined | Genome-fragment | NA | NA | 0 |
| S12_k141_211941 | 1206 | No | NA | 1 | 0 | 0 | Not-determined | Genome-fragment | NA | NA | 0 |
| S12_k141_45196 | 3118 | No | NA | 3 | 0 | 1 | Not-determined | Genome-fragment | NA | NA | 0 |
| S12_k141_127111 | 2132 | No | NA | 4 | 0 | 1 | Not-determined | Genome-fragment | NA | NA | 0 |
| S12_k141_345637 | 1069 | No | NA | 1 | 0 | 1 | Not-determined | Genome-fragment | NA | NA | 0 |
| S12_k141_279453 | 1196 | No | NA | 1 | 0 | 1 | Not-determined | Genome-fragment | NA | NA | 0 |
| S12_k141_127134 | 1149 | No | NA | 1 | 0 | 1 | Not-determined | Genome-fragment | NA | NA | 0 |
| S7-final-viral-combined_k141_237716\|\|full | 3458 | No | NA | 3 | 0 | 1 | Low-quality | Genome-fragment | 3.77 | AAI-based (medium-confidence) | 0 |
| S12_k141_212018 | 3466 | No | NA | 4 | 0 | 4 | Not-determined | Genome-fragment | NA | NA | 0 |
| S7-final-viral-combined_k141_190802\|\|full | 3458 | No | NA | 6 | 0 | 0 | Low-quality | Genome-fragment | 8.59 | AAI-based (high-confidence) | 0 |
| S12_k141_195842 | 1127 | No | NA | 2 | 0 | 0 | Not-determined | Genome-fragment | NA | NA | 0 |
| S12_k141_396139 | 3856 | No | NA | 8 | 0 | 7 | Not-determined | Genome-fragment | NA | NA | 0 |
| S12_k141_94290 | 1142 | No | NA | 2 | 0 | 2 | Not-determined | Genome-fragment | NA | NA | 0 |
| S12_k141_212075 | 3649 | No | NA | 7 | 0 | 0 | Not-determined | Genome-fragment | NA | NA | 0 |
| S12_k141_212079 | 2185 | No | NA | 2 | 0 | 2 | Not-determined | Genome-fragment | NA | NA | 0 |
| S12_k141_78917 | 1890 | No | NA | 2 | 0 | 0 | Not-determined | Genome-fragment | NA | NA | 0 |
| S12_k141_94348 | 1564 | No | NA | 2 | 0 | 2 | Not-determined | Genome-fragment | NA | NA | 0 |
| S7-final-viral-combined_k141_243564\|\|full | 3458 | No | NA | 5 | 2 | 0 | Low-quality | Genome-fragment | 9.86 | AAI-based (high-confidence) | 0 |
| S7_k141_162194 | 3458 | No | NA | 4 | 0 | 0 | Low-quality | Genome-fragment | 4.68 | AAI-based (medium-confidence) | 0 |
| S12_k141_144626 | 1168 | No | NA | 1 | 0 | 1 | Not-determined | Genome-fragment | NA | NA | 0 |
| S12_k141_10665 | 1277 | No | NA | 1 | 0 | 0 | Not-determined | Genome-fragment | NA | NA | 0 |
| S12_k141_296125 | 1573 | No | NA | 2 | 0 | 2 | Not-determined | Genome-fragment | NA | NA | 0 |
| S12_k141_45313 | 3480 | No | NA | 3 | 0 | 1 | Not-determined | Genome-fragment | NA | NA | 0 |
| S2-final-viral-combined_k141_96348\|\|full | 3458 | No | NA | 9 | 0 | 3 | Low-quality | Genome-fragment | 6.2 | AAI-based (medium-confidence) | 0 |
| S12_k141_196005 | 3483 | No | NA | 1 | 0 | 0 | Not-determined | Genome-fragment | NA | NA | 0 |
| S12_k141_245405 | 1088 | No | NA | 2 | 0 | 2 | Not-determined | Genome-fragment | NA | NA | 0 |
| S12_k141_245440 | 2014 | No | NA | 1 | 0 | 0 | Not-determined | Genome-fragment | NA | NA | 0 |
| S12_k141_396305 | 2950 | No | NA | 3 | 0 | 1 | Not-determined | Genome-fragment | NA | NA | 0 |
| S2_k141_45071 | 3458 | No | NA | 3 | 0 | 2 | Low-quality | Genome-fragment | 4.31 | AAI-based (medium-confidence) | 0 |
| S12_k141_127423 | 1652 | No | NA | 3 | 0 | 1 | Not-determined | Genome-fragment | NA | NA | 0 |
| S12_k141_396347 | 1719 | No | NA | 1 | 0 | 0 | Not-determined | Genome-fragment | NA | NA | 0 |
| S1_k141_229423 | 3459 | No | NA | 6 | 0 | 1 | Low-quality | Genome-fragment | 9.07 | AAI-based (medium-confidence) | 0 |
| S12_k141_229713 | 1172 | No | NA | 1 | 0 | 1 | Not-determined | Genome-fragment | NA | NA | 0 |
| S12_k141_127469 | 1682 | No | NA | 1 | 0 | 0 | Not-determined | Genome-fragment | NA | NA | 0 |
| S12_k141_61269 | 5244 | No | NA | 6 | 0 | 3 | Not-determined | Genome-fragment | NA | NA | 0 |
| S5_k141_195397 | 3459 | No | NA | 4 | 1 | 0 | Low-quality | Genome-fragment | 3.91 | AAI-based (high-confidence) | 0 |
| S12_k141_313917 | 1282 | No | NA | 1 | 0 | 1 | Not-determined | Genome-fragment | NA | NA | 0 |
| S12_k141_111070 | 3413 | No | NA | 4 | 0 | 2 | Not-determined | Genome-fragment | NA | NA | 0 |
| S7-final-viral-combined_k141_114146\|\|full | 3459 | No | NA | 6 | 0 | 0 | Low-quality | Genome-fragment | 6.47 | AAI-based (high-confidence) | 0 |
| S12_k141_245580 | 2242 | No | NA | 1 | 0 | 1 | Not-determined | Genome-fragment | NA | NA | 0 |
| S12_k141_345954 | 2595 | No | NA | 2 | 0 | 0 | Not-determined | Genome-fragment | NA | NA | 0 |
| S12_k141_345960 | 1380 | No | NA | 2 | 0 | 2 | Not-determined | Genome-fragment | NA | NA | 0 |
| S12_k141_396435 | 2159 | No | NA | 1 | 0 | 0 | Not-determined | Genome-fragment | NA | NA | 0 |
| S12_k141_79007 | 3015 | No | NA | 2 | 0 | 1 | Not-determined | Genome-fragment | NA | NA | 0 |
| S2-final-viral-combined_k141_131656\|\|full | 3459 | No | NA | 5 | 2 | 0 | Low-quality | Genome-fragment | 6.76 | AAI-based (high-confidence) | 0 |
| S12_k141_162189 | 6173 | No | NA | 4 | 0 | 0 | Not-determined | Genome-fragment | NA | NA | 0 |
| S12_k141_345999 | 2857 | No | NA | 3 | 0 | 0 | Not-determined | Genome-fragment | NA | NA | 0 |
| S11-final-viral-combined_k141_79898\|\|full | 3460 | No | NA | 4 | 0 | 1 | Low-quality | Genome-fragment | 2.05 | AAI-based (medium-confidence) | 0 |
| S12_k141_61316 | 1106 | No | NA | 2 | 0 | 0 | Not-determined | Genome-fragment | NA | NA | 0 |
| S12_k141_262103 | 4712 | No | NA | 1 | 0 | 1 | Not-determined | Genome-fragment | NA | NA | 0 |
| S12_k141_346092 | 3450 | No | NA | 4 | 0 | 3 | Not-determined | Genome-fragment | NA | NA | 0 |
| S12_k141_127608 | 1024 | No | NA | 2 | 0 | 2 | Not-determined | Genome-fragment | NA | NA | 0 |
| S2-final-viral-combined_k141_192331\|\|full | 3460 | No | NA | 7 | 1 | 0 | Low-quality | Genome-fragment | 6.58 | AAI-based (medium-confidence) | 0 |
| S12_k141_10908 | 3915 | No | NA | 3 | 0 | 2 | Not-determined | Genome-fragment | NA | NA | 0 |
| S12_k141_245724 | 1397 | No | NA | 1 | 0 | 1 | Not-determined | Genome-fragment | NA | NA | 0 |
| S6_k141_48562 | 3460 | No | NA | 8 | 1 | 0 | Low-quality | Genome-fragment | 10.36 | AAI-based (high-confidence) | 0 |
| S12-final-viral-combined_k141_157295\|\|full | 3462 | No | NA | 6 | 0 | 0 | Low-quality | Genome-fragment | 6.94 | AAI-based (medium-confidence) | 0 |
| S12_k141_196224 | 1069 | No | NA | 2 | 0 | 1 | Not-determined | Genome-fragment | NA | NA | 0 |
| S12_k141_61401 | 5468 | No | NA | 2 | 0 | 2 | Not-determined | Genome-fragment | NA | NA | 0 |
| S12_k141_380115 | 6005 | No | NA | 3 | 0 | 3 | Not-determined | Genome-fragment | NA | NA | 0 |
| S8_k141_69678 | 3462 | No | NA | 2 | 2 | 0 | Low-quality | Genome-fragment | 5.74 | AAI-based (high-confidence) | 0 |
| S12_k141_178677 | 1376 | No | NA | 1 | 0 | 0 | Not-determined | Genome-fragment | NA | NA | 0 |
| S9-final-viral-combined_k141_60288\|\|full | 3463 | Yes | 1706 | 7 | 1 | 1 | Low-quality | Genome-fragment | 2.11 | HMM-based (lower-bound) | 50.74 |
| S12_k141_10992 | 1024 | No | NA | 2 | 0 | 0 | Not-determined | Genome-fragment | NA | NA | 0 |
| S7-final-viral-combined_k141_79760\|\|full | 3463 | No | NA | 4 | 0 | 1 | Low-quality | Genome-fragment | 7.08 | AAI-based (high-confidence) | 0 |
| S3-final-viral-combined_k141_120835\|\|full | 3464 | No | NA | 8 | 4 | 0 | Low-quality | Genome-fragment | 9.16 | AAI-based (high-confidence) | 0 |
| S1_k141_274652 | 3465 | Yes | 1968 | 7 | 1 | 2 | Low-quality | Genome-fragment | 3.28 | AAI-based (medium-confidence) | 43.2 |
| S2_k141_85479 | 3465 | No | NA | 3 | 0 | 0 | Low-quality | Genome-fragment | 5.05 | AAI-based (high-confidence) | 0 |
| S12_k141_296477 | 1258 | No | NA | 3 | 0 | 0 | Not-determined | Genome-fragment | NA | NA | 0 |
| S12_k141_380179 | 1805 | No | NA | 1 | 0 | 1 | Not-determined | Genome-fragment | NA | NA | 0 |
| S12_k141_144950 | 1463 | No | NA | 5 | 0 | 3 | Not-determined | Genome-fragment | NA | NA | 0 |
| S12_k141_11028 | 1516 | No | NA | 2 | 0 | 2 | Not-determined | Genome-fragment | NA | NA | 0 |
| S12_k141_111269 | 1264 | No | NA | 1 | 0 | 1 | Not-determined | Genome-fragment | NA | NA | 0 |
| S12_k141_346180 | 2070 | No | NA | 4 | 0 | 0 | Not-determined | Genome-fragment | NA | NA | 0 |
| S12_k141_162403 | 3558 | No | NA | 3 | 0 | 1 | Not-determined | Genome-fragment | NA | NA | 0 |
| S12_k141_346201 | 1534 | No | NA | 2 | 0 | 1 | Not-determined | Genome-fragment | NA | NA | 0 |
| S11-final-viral-combined_k141_246810\|\|full | 3467 | No | NA | 3 | 1 | 0 | Low-quality | Genome-fragment | 2.58 | HMM-based (lower-bound) | 0 |
| S17-final-viral-combined_k141_224051\|\|full | 3468 | No | NA | 6 | 0 | 0 | Low-quality | Genome-fragment | 8.04 | AAI-based (high-confidence) | 0 |
| S12_k141_145014 | 1203 | No | NA | 1 | 0 | 1 | Not-determined | Genome-fragment | NA | NA | 0 |
| S12_k141_212562 | 1404 | No | NA | 1 | 0 | 1 | Not-determined | Genome-fragment | NA | NA | 0 |
| S4-final-viral-combined_k141_158011\|\|full | 3468 | No | NA | 2 | 0 | 2 | Low-quality | Genome-fragment | 3.1 | AAI-based (medium-confidence) | 0 |
| S4_k141_144431 | 3468 | No | NA | 3 | 0 | 3 | Low-quality | Genome-fragment | 1.72 | AAI-based (medium-confidence) | 0 |
| S12_k141_296529 | 2097 | No | NA | 1 | 0 | 0 | Not-determined | Genome-fragment | NA | NA | 0 |
| S12_k141_145046 | 1822 | No | NA | 3 | 0 | 0 | Not-determined | Genome-fragment | NA | NA | 0 |
| S12_k141_380227 | 1053 | No | NA | 2 | 0 | 1 | Not-determined | Genome-fragment | NA | NA | 0 |
| S12_k141_361516 | 1097 | No | NA | 2 | 0 | 1 | Not-determined | Genome-fragment | NA | NA | 0 |
| S12_k141_94795 | 1021 | No | NA | 1 | 0 | 1 | Not-determined | Genome-fragment | NA | NA | 0 |
| S9-final-viral-combined_k141_82458\|\|lt2gene | 3469 | No | NA | 2 | 2 | 0 | Low-quality | Genome-fragment | 8.63 | AAI-based (medium-confidence) | 0 |
| S12_k141_111370 | 3014 | No | NA | 4 | 0 | 0 | Not-determined | Genome-fragment | NA | NA | 0 |
| S12_k141_280040 | 7414 | No | NA | 10 | 0 | 2 | Not-determined | Genome-fragment | NA | NA | 0 |
| S12-final-viral-combined_k141_12228\|\|full | 3469 | No | NA | 5 | 0 | 0 | Low-quality | Genome-fragment | 7.97 | AAI-based (medium-confidence) | 0 |
| S4_k141_10110 | 3469 | No | NA | 3 | 0 | 1 | Low-quality | Genome-fragment | 6.65 | AAI-based (medium-confidence) | 0 |
| S12_k141_45727 | 5609 | No | NA | 6 | 0 | 5 | Not-determined | Genome-fragment | NA | NA | 0 |
| S12_k141_346258 | 4814 | No | NA | 2 | 0 | 2 | Not-determined | Genome-fragment | NA | NA | 0 |
| S12_k141_196342 | 1003 | No | NA | 1 | 0 | 0 | Not-determined | Genome-fragment | NA | NA | 0 |
| S4_k141_25923 | 3469 | No | NA | 4 | 0 | 2 | Low-quality | Genome-fragment | 19.31 | AAI-based (high-confidence) | 0 |
| S12_k141_280094 | 3388 | No | NA | 4 | 0 | 2 | Not-determined | Genome-fragment | NA | NA | 0 |
| S12_k141_94828 | 1076 | No | NA | 1 | 0 | 0 | Not-determined | Genome-fragment | NA | NA | 0 |
| S12_k141_45763 | 1298 | No | NA | 2 | 0 | 1 | Not-determined | Genome-fragment | NA | NA | 0 |
| S12_k141_162519 | 1026 | No | NA | 1 | 0 | 0 | Not-determined | Genome-fragment | NA | NA | 0 |
| S12_k141_111457 | 1279 | No | NA | 2 | 0 | 1 | Not-determined | Genome-fragment | NA | NA | 0 |
| S5_k141_36641 | 3470 | No | NA | 5 | 4 | 0 | Low-quality | Genome-fragment | 10.86 | AAI-based (high-confidence) | 0 |
| S12_k141_289576 | 3470 | No | NA | 4 | 4 | 0 | Low-quality | Genome-fragment | 8.2 | AAI-based (high-confidence) | 0 |
| S12_k141_111461 | 4358 | No | NA | 4 | 0 | 3 | Not-determined | Genome-fragment | NA | NA | 0 |
| S12_k141_296603 | 3368 | No | NA | 2 | 0 | 2 | Not-determined | Genome-fragment | NA | NA | 0 |
| S8-final-viral-combined_k141_245434\|\|full | 3470 | No | NA | 3 | 0 | 2 | Low-quality | Genome-fragment | 2.8 | AAI-based (medium-confidence) | 0 |
| S12_k141_396859 | 3757 | No | NA | 2 | 0 | 1 | Not-determined | Genome-fragment | NA | NA | 0 |
| S9-final-viral-combined_k141_84816\|\|full | 3471 | No | NA | 4 | 1 | 0 | Low-quality | Genome-fragment | 24.54 | AAI-based (medium-confidence) | 0 |
| S12_k141_396875 | 2484 | No | NA | 1 | 0 | 1 | Not-determined | Genome-fragment | NA | NA | 0 |
| S12_k141_79300 | 1543 | No | NA | 2 | 0 | 2 | Not-determined | Genome-fragment | NA | NA | 0 |
| S12_k141_262501 | 1047 | No | NA | 1 | 0 | 0 | Not-determined | Genome-fragment | NA | NA | 0 |
| S12_k141_346363 | 1919 | No | NA | 3 | 0 | 0 | Not-determined | Genome-fragment | NA | NA | 0 |
| S12_k141_296628 | 1009 | No | NA | 3 | 0 | 1 | Not-determined | Genome-fragment | NA | NA | 0 |

| | | | | | | | | | | | |
|---|---|---|---|---|---|---|---|---|---|---|---|
| S5-final-viral-combined_k141_196663\|\|full | 3471 | No | NA | 9 | 0 | 0 | Low-quality | Genome-fragment | 5.16 | AAI-based (medium-confidence) | 0 |
| S12-final-viral-combined_k141_43696\|\|full | 3471 | No | NA | 2 | 0 | 2 | Low-quality | Genome-fragment | 4.24 | AAI-based (medium-confidence) | 0 |
| S12_k141_212728 | 1521 | No | NA | 1 | 0 | 0 | Not-determined | Genome-fragment | NA | NA | 0 |
| S12_k141_245929 | 1407 | No | NA | 2 | 0 | 1 | Not-determined | Genome-fragment | NA | NA | 0 |
| S12_k141_262550 | 1120 | No | NA | 1 | 0 | 1 | Not-determined | Genome-fragment | NA | NA | 0 |
| S12_k141_127988 | 1011 | No | NA | 1 | 0 | 1 | Not-determined | Genome-fragment | NA | NA | 0 |
| S11-final-viral-combined_k141_63951\|\|full | 3472 | No | NA | 4 | 0 | 0 | Low-quality | Genome-fragment | 6.76 | AAI-based (medium-confidence) | 0 |
| S5-final-viral-combined_k141_171176\|\|full | 3472 | No | NA | 8 | 2 | 0 | Low-quality | Genome-fragment | 3.67 | HMM-based (lower-bound) | 0 |
| S12_k141_296714 | 1270 | No | NA | 2 | 0 | 1 | Not-determined | Genome-fragment | NA | NA | 0 |
| S12_k141_296736 | 1018 | No | NA | 1 | 0 | 0 | Not-determined | Genome-fragment | NA | NA | 0 |
| S12_k141_95026 | 1070 | No | NA | 1 | 0 | 1 | Not-determined | Genome-fragment | NA | NA | 0 |
| S12_k141_262602 | 1031 | No | NA | 4 | 0 | 0 | Not-determined | Genome-fragment | NA | NA | 0 |
| S12_k141_179067 | 1147 | No | NA | 2 | 0 | 0 | Not-determined | Genome-fragment | NA | NA | 0 |
| S12_k141_230128 | 3477 | No | NA | 3 | 0 | 1 | Not-determined | Genome-fragment | NA | NA | 0 |
| S2-final-viral-combined_k141_164749\|\|full | 3472 | No | NA | 3 | 0 | 3 | Low-quality | Genome-fragment | 4.14 | AAI-based (medium-confidence) | 0 |
| S12_k141_28307 | 1843 | No | NA | 2 | 0 | 2 | Not-determined | Genome-fragment | NA | NA | 0 |
| S12_k141_230145 | 1070 | No | NA | 2 | 0 | 1 | Not-determined | Genome-fragment | NA | NA | 0 |
| S8_k141_122355 | 3472 | No | NA | 3 | 0 | 1 | Low-quality | Genome-fragment | 7.47 | AAI-based (high-confidence) | 0 |
| S12_k141_11300 | 55721 | No | NA | 57 | 0 | 13 | Not-determined | Genome-fragment | NA | NA | 0 |
| S12_k141_212830 | 1188 | No | NA | 2 | 0 | 1 | Not-determined | Genome-fragment | NA | NA | 0 |
| S12_k141_280299 | 1018 | No | NA | 1 | 0 | 1 | Not-determined | Genome-fragment | NA | NA | 0 |
| S12_k141_45951 | 1003 | No | NA | 2 | 0 | 1 | Not-determined | Genome-fragment | NA | NA | 0 |
| S12_k141_45955 | 3615 | No | NA | 5 | 0 | 1 | Not-determined | Genome-fragment | NA | NA | 0 |
| S12_k141_11319 | 1259 | No | NA | 2 | 0 | 0 | Not-determined | Genome-fragment | NA | NA | 0 |
| S12_k141_61748 | 1842 | No | NA | 1 | 0 | 0 | Not-determined | Genome-fragment | NA | NA | 0 |
| S12_k141_230195 | 3489 | No | NA | 5 | 0 | 0 | Not-determined | Genome-fragment | NA | NA | 0 |
| S12_k141_262678 | 1210 | No | NA | 2 | 0 | 0 | Not-determined | Genome-fragment | NA | NA | 0 |
| S12_k141_230218 | 1014 | No | NA | 2 | 0 | 0 | Not-determined | Genome-fragment | NA | NA | 0 |
| S12_k141_296834 | 1062 | No | NA | 1 | 0 | 1 | Not-determined | Genome-fragment | NA | NA | 0 |
| S12_k141_329939 | 2146 | No | NA | 2 | 0 | 1 | Not-determined | Genome-fragment | NA | NA | 0 |
| S12_k141_128138 | 1365 | No | NA | 1 | 0 | 0 | Not-determined | Genome-fragment | NA | NA | 0 |
| S12_k141_296852 | 1395 | No | NA | 2 | 0 | 0 | Not-determined | Genome-fragment | NA | NA | 0 |
| S8-final-viral-combined_k141_52202\|\|full | 3473 | No | NA | 4 | 0 | 2 | Low-quality | Genome-fragment | 2.4 | AAI-based (high-confidence) | 0 |
| S1-final-viral-combined_k141_363128\|\|full | 3474 | No | NA | 5 | 0 | 0 | Low-quality | Genome-fragment | 6.98 | AAI-based (medium-confidence) | 0 |
| S12_k141_128185 | 1894 | No | NA | 1 | 0 | 0 | Not-determined | Genome-fragment | NA | NA | 0 |
| S12_k141_330009 | 1192 | No | NA | 1 | 0 | 1 | Not-determined | Genome-fragment | NA | NA | 0 |
| S12_k141_196457 | 1017 | No | NA | 2 | 0 | 0 | Not-determined | Genome-fragment | NA | NA | 0 |
| S12_k141_79410 | 11566 | No | NA | 9 | 0 | 9 | Not-determined | Genome-fragment | NA | NA | 0 |
| S12_k141_46100 | 2381 | No | NA | 3 | 0 | 1 | Not-determined | Genome-fragment | NA | NA | 0 |
| S12_k141_145424 | 21494 | No | NA | 18 | 0 | 16 | Not-determined | Genome-fragment | NA | NA | 0 |
| S12_k141_380569 | 1143 | No | NA | 1 | 0 | 1 | Not-determined | Genome-fragment | NA | NA | 0 |
| S1-final-viral-combined_k141_131600\|\|full | 3474 | No | NA | 6 | 1 | 0 | Low-quality | Genome-fragment | 2.2 | HMM-based (lower-bound) | 0 |
| S12_k141_196503 | 3285 | No | NA | 3 | 0 | 2 | Not-determined | Genome-fragment | NA | NA | 0 |
| S12_k141_162739 | 1920 | No | NA | 3 | 0 | 0 | Not-determined | Genome-fragment | NA | NA | 0 |
| S12_k141_280425 | 1808 | No | NA | 1 | 0 | 1 | Not-determined | Genome-fragment | NA | NA | 0 |
| S12_k141_79456 | 1779 | No | NA | 2 | 0 | 1 | Not-determined | Genome-fragment | NA | NA | 0 |
| S12_k141_280446 | 1409 | No | NA | 1 | 0 | 0 | Not-determined | Genome-fragment | NA | NA | 0 |
| S12_k141_330087 | 1085 | No | NA | 1 | 0 | 0 | Not-determined | Genome-fragment | NA | NA | 0 |
| S12_k141_46162 | 1005 | No | NA | 2 | 0 | 0 | Not-determined | Genome-fragment | NA | NA | 0 |
| S12_k141_330109 | 1339 | No | NA | 1 | 0 | 1 | Not-determined | Genome-fragment | NA | NA | 0 |
| S12_k141_179244 | 2142 | No | NA | 5 | 0 | 2 | Not-determined | Genome-fragment | NA | NA | 0 |
| S8-final-viral-combined_k141_45371\|\|full | 3474 | No | NA | 5 | 1 | 0 | Low-quality | Genome-fragment | 2.35 | HMM-based (lower-bound) | 0 |
| S12_k141_280486 | 1601 | No | NA | 1 | 0 | 0 | Not-determined | Genome-fragment | NA | NA | 0 |
| S12_k141_79543 | 2090 | No | NA | 2 | 0 | 1 | Not-determined | Genome-fragment | NA | NA | 0 |
| S12_k141_61963 | 8069 | No | NA | 3 | 0 | 3 | Not-determined | Genome-fragment | NA | NA | 0 |
| S11-final-viral-combined_k141_23122\|\|full | 3476 | No | NA | 8 | 1 | 0 | Low-quality | Genome-fragment | 6.64 | HMM-based (lower-bound) | 0 |
| S12_k141_361917 | 1132 | No | NA | 1 | 0 | 0 | Not-determined | Genome-fragment | NA | NA | 0 |
| S12_k141_46222 | 1087 | No | NA | 2 | 0 | 0 | Not-determined | Genome-fragment | NA | NA | 0 |
| S12_k141_297009 | 2004 | No | NA | 2 | 0 | 1 | Not-determined | Genome-fragment | NA | NA | 0 |
| S12_k141_145550 | 2076 | No | NA | 1 | 0 | 1 | Not-determined | Genome-fragment | NA | NA | 0 |
| S11-final-viral-combined_k141_107797\|\|full | 3476 | No | NA | 5 | 0 | 2 | Low-quality | Genome-fragment | 3.17 | AAI-based (medium-confidence) | 0 |
| S5-final-viral-combined_k141_147723\|\|full | 3476 | No | NA | 4 | 1 | 0 | Low-quality | Genome-fragment | 2.38 | HMM-based (lower-bound) | 0 |
| S12_k141_262894 | 4449 | No | NA | 4 | 0 | 4 | Not-determined | Genome-fragment | NA | NA | 0 |
| S12_k141_314494 | 1337 | No | NA | 1 | 0 | 0 | Not-determined | Genome-fragment | NA | NA | 0 |
| S6-final-viral-combined_k141_88710\|\|full | 3476 | No | NA | 5 | 0 | 0 | Low-quality | Genome-fragment | 9.08 | AAI-based (medium-confidence) | 0 |
| S12_k141_46291 | 1003 | No | NA | 2 | 0 | 1 | Not-determined | Genome-fragment | NA | NA | 0 |
| S8-final-viral-combined_k141_164489\|\|full | 3476 | No | NA | 9 | 3 | 0 | Low-quality | Genome-fragment | 8.36 | AAI-based (high-confidence) | 0 |
| S12_k141_297063 | 1350 | No | NA | 2 | 0 | 2 | Not-determined | Genome-fragment | NA | NA | 0 |
| S12_k141_130412 | 3477 | No | NA | 3 | 0 | 0 | Low-quality | Genome-fragment | 6.68 | AAI-based (high-confidence) | 0 |
| S8-final-viral-combined_k141_109360\|\|full | 3477 | No | NA | 5 | 0 | 1 | Low-quality | Genome-fragment | 1.69 | AAI-based (medium-confidence) | 0 |
| S12_k141_95416 | 3220 | No | NA | 2 | 0 | 1 | Not-determined | Genome-fragment | NA | NA | 0 |
| S12_k141_145661 | 1016 | No | NA | 1 | 0 | 1 | Not-determined | Genome-fragment | NA | NA | 0 |
| S12_k141_213107 | 1040 | No | NA | 2 | 0 | 0 | Not-determined | Genome-fragment | NA | NA | 0 |
| S12_k141_95442 | 1632 | No | NA | 2 | 0 | 1 | Not-determined | Genome-fragment | NA | NA | 0 |
| S12_k141_62103 | 5658 | No | NA | 6 | 0 | 3 | Not-determined | Genome-fragment | NA | NA | 0 |
| S12_k141_179388 | 45584 | No | NA | 64 | 15 | 0 | High-quality | High-quality | 99.32 | AAI-based (high-confidence) | 0 |
| S12_k141_361977 | 1194 | No | NA | 2 | 0 | 1 | Not-determined | Genome-fragment | NA | NA | 0 |
| S12_k141_314576 | 3127 | No | NA | 3 | 0 | 3 | Not-determined | Genome-fragment | NA | NA | 0 |
| S12_k141_380704 | 1150 | No | NA | 2 | 0 | 0 | Not-determined | Genome-fragment | NA | NA | 0 |
| S1-final-viral-combined_k141_341146\|\|full | 3478 | No | NA | 9 | 0 | 1 | Low-quality | Genome-fragment | 25.88 | AAI-based (high-confidence) | 0 |
| S12_k141_314580 | 4790 | No | NA | 5 | 0 | 0 | Not-determined | Genome-fragment | NA | NA | 0 |
| S12_k141_346869 | 1094 | No | NA | 1 | 0 | 1 | Not-determined | Genome-fragment | NA | NA | 0 |
| S12_k141_362011 | 2882 | No | NA | 1 | 0 | 1 | Not-determined | Genome-fragment | NA | NA | 0 |
| S5_k141_175936 | 3478 | Yes | 902 | 6 | 1 | 1 | Low-quality | Genome-fragment | 1.06 | AAI-based (medium-confidence) | 74.07 |
| S12-final-viral-combined_k141_170642\|\|full | 3478 | No | NA | 7 | 1 | 0 | Low-quality | Genome-fragment | 7.47 | AAI-based (medium-confidence) | 0 |
| S12_k141_263052 | 1105 | No | NA | 1 | 0 | 1 | Not-determined | Genome-fragment | NA | NA | 0 |
| S4_k141_164644 | 3478 | No | NA | 3 | 0 | 2 | Low-quality | Genome-fragment | 7.77 | AAI-based (medium-confidence) | 0 |
| S12_k141_11643 | 1910 | No | NA | 1 | 0 | 1 | Not-determined | Genome-fragment | NA | NA | 0 |
| S12_k141_359968 | 3479 | No | NA | 2 | 0 | 2 | Low-quality | Genome-fragment | 1.78 | AAI-based (medium-confidence) | 0 |
| S12_k141_145743 | 1756 | No | NA | 2 | 0 | 1 | Not-determined | Genome-fragment | NA | NA | 0 |
| S12_k141_397261 | 22820 | No | NA | 22 | 0 | 5 | Not-determined | Genome-fragment | NA | NA | 0 |
| S12_k141_95514 | 1278 | No | NA | 2 | 0 | 0 | Not-determined | Genome-fragment | NA | NA | 0 |
| S12_k141_62203 | 5500 | No | NA | 5 | 0 | 2 | Not-determined | Genome-fragment | NA | NA | 0 |
| S12_k141_179481 | 1200 | No | NA | 1 | 0 | 0 | Not-determined | Genome-fragment | NA | NA | 0 |
| S12_k141_346945 | 1177 | No | NA | 2 | 0 | 0 | Not-determined | Genome-fragment | NA | NA | 0 |
| S12_k141_330351 | 1016 | No | NA | 2 | 0 | 1 | Not-determined | Genome-fragment | NA | NA | 0 |
| S12_k141_230402 | 1076 | No | NA | 1 | 0 | 1 | Not-determined | Genome-fragment | NA | NA | 0 |
| S12_k141_196679 | 1025 | No | NA | 2 | 0 | 1 | Not-determined | Genome-fragment | NA | NA | 0 |
| S12_k141_179524 | 1322 | No | NA | 1 | 0 | 1 | Not-determined | Genome-fragment | NA | NA | 0 |
| S7-final-viral-combined_k141_107229\|\|full | 3480 | No | NA | 2 | 0 | 2 | Low-quality | Genome-fragment | 2.52 | AAI-based (high-confidence) | 0 |
| S12_k141_62232 | 2281 | No | NA | 5 | 0 | 5 | Not-determined | Genome-fragment | NA | NA | 0 |
| S12_k141_397282 | 3202 | No | NA | 4 | 0 | 1 | Not-determined | Genome-fragment | NA | NA | 0 |
| S12_k141_46514 | 2528 | No | NA | 3 | 0 | 3 | Not-determined | Genome-fragment | NA | NA | 0 |
| S12_k141_380779 | 4060 | No | NA | 3 | 0 | 1 | Not-determined | Genome-fragment | NA | NA | 0 |
| S12_k141_79814 | 1557 | No | NA | 3 | 0 | 1 | Not-determined | Genome-fragment | NA | NA | 0 |
| S12_k141_280794 | 2137 | No | NA | 1 | 0 | 1 | Not-determined | Genome-fragment | NA | NA | 0 |
| S12_k141_111902 | 1451 | No | NA | 1 | 0 | 0 | Not-determined | Genome-fragment | NA | NA | 0 |
| S12_k141_155582 | 3481 | No | NA | 7 | 0 | 0 | Low-quality | Genome-fragment | 7.55 | AAI-based (high-confidence) | 0 |
| S12_k141_62264 | 1104 | No | NA | 1 | 0 | 0 | Not-determined | Genome-fragment | NA | NA | 0 |
| S12_k141_196751 | 1238 | No | NA | 2 | 0 | 0 | Not-determined | Genome-fragment | NA | NA | 0 |
| S12_k141_297559 | 3481 | No | NA | 5 | 0 | 2 | Low-quality | Genome-fragment | 5.98 | AAI-based (medium-confidence) | 0 |
| S12_k141_79867 | 1862 | No | NA | 4 | 0 | 1 | Not-determined | Genome-fragment | NA | NA | 0 |
| S12_k141_380832 | 1050 | No | NA | 1 | 0 | 0 | Not-determined | Genome-fragment | NA | NA | 0 |
| S17-final-viral-combined_k141_37510\|\|full | 3483 | Yes | 2756 | 7 | 2 | 1 | Low-quality | Genome-fragment | 4.29 | AAI-based (high-confidence) | 20.87 |
| S12_k141_46574 | 1315 | No | NA | 1 | 0 | 1 | Not-determined | Genome-fragment | NA | NA | 0 |
| S12_k141_179642 | 1799 | No | NA | 1 | 0 | 1 | Not-determined | Genome-fragment | NA | NA | 0 |
| S12_k141_79894 | 3417 | No | NA | 1 | 0 | 1 | Not-determined | Genome-fragment | NA | NA | 0 |
| S12_k141_263186 | 1061 | No | NA | 3 | 0 | 1 | Not-determined | Genome-fragment | NA | NA | 0 |
| S12_k141_11801 | 3045 | No | NA | 2 | 0 | 2 | Not-determined | Genome-fragment | NA | NA | 0 |
| S12_k141_314707 | 1125 | No | NA | 2 | 0 | 1 | Not-determined | Genome-fragment | NA | NA | 0 |
| S12_k141_397369 | 2978 | No | NA | 4 | 0 | 3 | Not-determined | Genome-fragment | NA | NA | 0 |
| S12_k141_213422 | 1847 | No | NA | 2 | 0 | 1 | Not-determined | Genome-fragment | NA | NA | 0 |
| S1-final-viral-combined_k141_378608\|\|full | 3484 | No | NA | 7 | 0 | 1 | Low-quality | Genome-fragment | 7.62 | AAI-based (medium-confidence) | 0 |
| S5_k141_164925 | 3484 | No | NA | 10 | 7 | 0 | Low-quality | Genome-fragment | 15.9 | AAI-based (high-confidence) | 0 |
| S12_k141_145876 | 2000 | No | NA | 4 | 0 | 2 | Not-determined | Genome-fragment | NA | NA | 0 |
| S1_k141_72421 | 3485 | No | NA | 7 | 1 | 0 | Low-quality | Genome-fragment | 8.78 | AAI-based (high-confidence) | 0 |
| S6-final-viral-combined_k141_89406\|\|full | 3485 | No | NA | 5 | 1 | 0 | Low-quality | Genome-fragment | 4.14 | AAI-based (high-confidence) | 0 |
| S12_k141_145905 | 1216 | No | NA | 1 | 0 | 1 | Not-determined | Genome-fragment | NA | NA | 0 |
| S7_k141_44374 | 3486 | Yes | 2659 | 3 | 1 | 1 | Low-quality | Genome-fragment | 5.4 | AAI-based (medium-confidence) | 23.72 |
| S12_k141_280956 | 4675 | No | NA | 5 | 0 | 0 | Not-determined | Genome-fragment | NA | NA | 0 |
| S12_k141_196924 | 1347 | No | NA | 3 | 0 | 3 | Not-determined | Genome-fragment | NA | NA | 0 |
| S17-k141_216420 | 3486 | No | NA | 3 | 1 | 0 | Low-quality | Genome-fragment | 2.59 | HMM-based (lower-bound) | 0 |
| S12_k141_280963 | 1318 | No | NA | 2 | 0 | 2 | Not-determined | Genome-fragment | NA | NA | 0 |
| S12-final-viral-combined_k141_75682\|\|full | 3486 | No | NA | 9 | 1 | 0 | Low-quality | Genome-fragment | 4.36 | AAI-based (medium-confidence) | 0 |
| S4-final-viral-combined_k141_9728\|\|full | 3486 | No | NA | 6 | 0 | 1 | Low-quality | Genome-fragment | 7.02 | AAI-based (high-confidence) | 0 |
| S9-final-viral-combined_k141_74458\|\|full | 3487 | Yes | 1617 | 3 | 1 | 2 | Low-quality | Genome-fragment | 0.79 | AAI-based (high-confidence) | 53.63 |
| S11-final-viral-combined_k141_156992\|\|full | 3487 | No | NA | 2 | 2 | 0 | Low-quality | Genome-fragment | 7.95 | AAI-based (high-confidence) | 0 |
| S12_k141_281013 | 9649 | No | NA | 13 | 0 | 1 | Not-determined | Genome-fragment | NA | NA | 0 |
| S12_k141_263287 | 1541 | No | NA | 1 | 0 | 0 | Not-determined | Genome-fragment | NA | NA | 0 |
| S12_k141_263290 | 4058 | No | NA | 4 | 0 | 0 | Not-determined | Genome-fragment | NA | NA | 0 |
| S12_k141_397477 | 1017 | No | NA | 2 | 0 | 0 | Not-determined | Genome-fragment | NA | NA | 0 |
| S12_k141_347313 | 1472 | No | NA | 1 | 0 | 1 | Not-determined | Genome-fragment | NA | NA | 0 |
| S5-final-viral-combined_k141_93762\|\|full | 3488 | No | NA | 6 | 1 | 0 | Low-quality | Genome-fragment | 5.49 | AAI-based (high-confidence) | 0 |
| S12_k141_297467 | 1630 | No | NA | 2 | 0 | 1 | Not-determined | Genome-fragment | NA | NA | 0 |
| S12_k141_11962 | 3469 | No | NA | 2 | 0 | 1 | Not-determined | Genome-fragment | NA | NA | 0 |
| S7_k141_137740 | 3488 | No | NA | 2 | 0 | 2 | Low-quality | Genome-fragment | 1.65 | AAI-based (medium-confidence) | 0 |
| S12-final-viral-combined_k141_102157\|\|full | 3488 | No | NA | 2 | 0 | 2 | Low-quality | Genome-fragment | 2.13 | AAI-based (medium-confidence) | 0 |
| S12_k141_347337 | 3333 | No | NA | 6 | 0 | 1 | Not-determined | Genome-fragment | NA | NA | 0 |
| S12_k141_95700 | 1837 | No | NA | 2 | 0 | 0 | Not-determined | Genome-fragment | NA | NA | 0 |
| S12_k141_347347 | 3071 | No | NA | 3 | 0 | 0 | Not-determined | Genome-fragment | NA | NA | 0 |
| S12_k141_314882 | 1072 | No | NA | 2 | 0 | 0 | Not-determined | Genome-fragment | NA | NA | 0 |
| S11-final-viral-combined_k141_243513\|\|full | 3489 | No | NA | 3 | 0 | 3 | Low-quality | Genome-fragment | 2.15 | AAI-based (medium-confidence) | 0 |
| S12_k141_362394 | 2422 | No | NA | 3 | 0 | 0 | Not-determined | Genome-fragment | NA | NA | 0 |
| S12_k141_28952 | 4586 | No | NA | 6 | 0 | 5 | Not-determined | Genome-fragment | NA | NA | 0 |
| S1_k141_293423 | 3490 | No | NA | 3 | 0 | 2 | Low-quality | Genome-fragment | 1.69 | AAI-based (medium-confidence) | 0 |
| S12_k141_128911 | 1052 | No | NA | 1 | 0 | 0 | Not-determined | Genome-fragment | NA | NA | 0 |
| S12-final-viral-combined_k141_75237\|\|full | 3490 | No | NA | 9 | 2 | 0 | Low-quality | Genome-fragment | 9.62 | AAI-based (high-confidence) | 0 |
| S12_k141_197033 | 1209 | No | NA | 2 | 0 | 0 | Not-determined | Genome-fragment | NA | NA | 0 |
| S12_k141_397534 | 2006 | No | NA | 3 | 0 | 1 | Not-determined | Genome-fragment | NA | NA | 0 |
| S12_k141_347429 | 1302 | No | NA | 2 | 0 | 1 | Not-determined | Genome-fragment | NA | NA | 0 |
| S12_k141_281147 | 1146 | No | NA | 1 | 0 | 1 | Not-determined | Genome-fragment | NA | NA | 0 |
| S12_k141_128926 | 2124 | No | NA | 2 | 0 | 1 | Not-determined | Genome-fragment | NA | NA | 0 |
| S12_k141_180047 | 3877 | No | NA | 5 | 0 | 5 | Not-determined | Genome-fragment | NA | NA | 0 |
| S12_k141_128943 | 1164 | No | NA | 1 | 0 | 0 | Not-determined | Genome-fragment | NA | NA | 0 |
| S12_k141_381057 | 1057 | No | NA | 2 | 0 | 1 | Not-determined | Genome-fragment | NA | NA | 0 |
| S12_k141_112259 | 2065 | No | NA | 1 | 0 | 0 | Not-determined | Genome-fragment | NA | NA | 0 |
| S12_k141_62658 | 1409 | No | NA | 2 | 0 | 2 | Not-determined | Genome-fragment | NA | NA | 0 |
| S12_k141_397659 | 1107 | No | NA | 1 | 0 | 1 | Not-determined | Genome-fragment | NA | NA | 0 |
| S12_k141_246754 | 1305 | No | NA | 1 | 0 | 1 | Not-determined | Genome-fragment | NA | NA | 0 |
| S12_k141_180130 | 1224 | No | NA | 1 | 0 | 1 | Not-determined | Genome-fragment | NA | NA | 0 |
| S12_k141_213706 | 4020 | No | NA | 4 | 0 | 0 | Not-determined | Genome-fragment | NA | NA | 0 |
| S12_k141_62719 | 1356 | No | NA | 2 | 0 | 1 | Not-determined | Genome-fragment | NA | NA | 0 |
| S12_k141_29105 | 1162 | No | NA | 1 | 0 | 1 | Not-determined | Genome-fragment | NA | NA | 0 |
| S12_k141_146249 | 1006 | No | NA | 1 | 0 | 0 | Not-determined | Genome-fragment | NA | NA | 0 |
| S12_k141_263441 | 2322 | No | NA | 2 | 0 | 1 | Not-determined | Genome-fragment | NA | NA | 0 |
| S12_k141_180149 | 7935 | No | NA | 8 | 0 | 2 | Not-determined | Genome-fragment | NA | NA | 0 |
| S12_k141_29147 | 3299 | No | NA | 3 | 0 | 0 | Not-determined | Genome-fragment | NA | NA | 0 |
| S7-final-viral-combined_k141_236010\|\|full | 3491 | No | NA | 3 | 0 | 3 | Low-quality | Genome-fragment | 1.86 | AAI-based (medium-confidence) | 0 |
| S12_k141_95987 | 1403 | No | NA | 3 | 0 | 3 | Not-determined | Genome-fragment | NA | NA | 0 |
| S12_k141_315144 | 1657 | No | NA | 1 | 0 | 1 | Not-determined | Genome-fragment | NA | NA | 0 |
| S12_k141_281327 | 4861 | No | NA | 2 | 0 | 2 | Not-determined | Genome-fragment | NA | NA | 0 |
| S12_k141_315165 | 1248 | No | NA | 2 | 0 | 0 | Not-determined | Genome-fragment | NA | NA | 0 |
| S12_k141_263487 | 3840 | No | NA | 2 | 0 | 2 | Not-determined | Genome-fragment | NA | NA | 0 |
| S12_k141_146310 | 1082 | No | NA | 1 | 0 | 1 | Not-determined | Genome-fragment | NA | NA | 0 |
| S12_k141_80304 | 1746 | No | NA | 2 | 0 | 1 | Not-determined | Genome-fragment | NA | NA | 0 |
| S12_k141_12204 | 6819 | No | NA | 7 | 0 | 3 | Not-determined | Genome-fragment | NA | NA | 0 |
| S12_k141_230861 | 1461 | No | NA | 1 | 0 | 1 | Not-determined | Genome-fragment | NA | NA | 0 |
| S12_k141_47064 | 1217 | No | NA | 1 | 0 | 1 | Not-determined | Genome-fragment | NA | NA | 0 |
| S12-final-viral-combined_k141_377776\|\|full | 3491 | No | NA | 3 | 2 | 0 | Low-quality | Genome-fragment | 7.65 | AAI-based (high-confidence) | 0 |
| S12_k141_315203 | 1758 | No | NA | 1 | 0 | 1 | Not-determined | Genome-fragment | NA | NA | 0 |
| S12_k141_381194 | 1164 | No | NA | 1 | 0 | 1 | Not-determined | Genome-fragment | NA | NA | 0 |
| S12_k141_263532 | 2983 | No | NA | 4 | 0 | 1 | Not-determined | Genome-fragment | NA | NA | 0 |
| S12_k141_12239 | 3011 | No | NA | 4 | 0 | 1 | Not-determined | Genome-fragment | NA | NA | 0 |
| S12_k141_29216 | 4468 | No | NA | 2 | 0 | 0 | Not-determined | Genome-fragment | NA | NA | 0 |
| S7-final-viral-combined_k141_119820\|\|full | 3492 | No | NA | 3 | 0 | 3 | Low-quality | Genome-fragment | 2.04 | AAI-based (medium-confidence) | 0 |
| S12_k141_297652 | 2522 | No | NA | 2 | 0 | 2 | Not-determined | Genome-fragment | NA | NA | 0 |
| S5-final-viral-combined_k141_69601\|\|full | 3493 | No | NA | 7 | 4 | 0 | Low-quality | Genome-fragment | 9.27 | AAI-based (medium-confidence) | 0 |
| S12_k141_397827 | 1314 | No | NA | 3 | 0 | 3 | Not-determined | Genome-fragment | NA | NA | 0 |
| S12_k141_246897 | 1151 | No | NA | 1 | 0 | 1 | Not-determined | Genome-fragment | NA | NA | 0 |
| S8_k141_242103 | 3493 | No | NA | 2 | 1 | 0 | Low-quality | Genome-fragment | 6.34 | AAI-based (high-confidence) | 0 |
| S12_k141_180258 | 1137 | No | NA | 2 | 0 | 1 | Not-determined | Genome-fragment | NA | NA | 0 |
| S12_k141_230966 | 1099 | No | NA | 2 | 0 | 2 | Not-determined | Genome-fragment | NA | NA | 0 |
| S12_k141_347683 | 1163 | No | NA | 1 | 0 | 0 | Not-determined | Genome-fragment | NA | NA | 0 |
| S12_k141_96120 | 1005 | No | NA | 2 | 0 | 2 | Not-determined | Genome-fragment | NA | NA | 0 |
| S12_k141_347702 | 1646 | No | NA | 2 | 0 | 2 | Not-determined | Genome-fragment | NA | NA | 0 |
| S12_k141_347714 | 1210 | No | NA | 2 | 0 | 0 | Not-determined | Genome-fragment | NA | NA | 0 |
| S12_k141_163715 | 1790 | No | NA | 1 | 0 | 1 | Not-determined | Genome-fragment | NA | NA | 0 |
| S12_k141_180306 | 1816 | No | NA | 1 | 0 | 0 | Not-determined | Genome-fragment | NA | NA | 0 |
| S12_k141_163741 | 1791 | No | NA | 2 | 0 | 1 | Not-determined | Genome-fragment | NA | NA | 0 |
| S12_k141_231034 | 3025 | No | NA | 2 | 0 | 1 | Not-determined | Genome-fragment | NA | NA | 0 |
| S12_k141_96186 | 1285 | No | NA | 2 | 0 | 1 | Not-determined | Genome-fragment | NA | NA | 0 |
| S5_k141_53973 | 3494 | No | NA | 8 | 0 | 2 | Low-quality | Genome-fragment | 2.27 | AAI-based (high-confidence) | 0 |
| S12_k141_347739 | 4416 | No | NA | 5 | 0 | 3 | Not-determined | Genome-fragment | NA | NA | 0 |
| S12_k141_197362 | 2504 | No | NA | 3 | 0 | 0 | Not-determined | Genome-fragment | NA | NA | 0 |
| S12_k141_315368 | 3460 | No | NA | 4 | 0 | 4 | Not-determined | Genome-fragment | NA | NA | 0 |
| S12_k141_281520 | 1986 | No | NA | 1 | 0 | 0 | Not-determined | Genome-fragment | NA | NA | 0 |
| S9-final-viral-combined_k141_55574\|\|full | 3495 | No | NA | 3 | 0 | 2 | Low-quality | Genome-fragment | 4.79 | AAI-based (medium-confidence) | 0 |
| S12_k141_96212 | 1163 | No | NA | 1 | 0 | 1 | Not-determined | Genome-fragment | NA | NA | 0 |
| S12_k141_80418 | 1704 | No | NA | 1 | 0 | 1 | Not-determined | Genome-fragment | NA | NA | 0 |
| S12_k141_80421 | 1207 | No | NA | 2 | 0 | 2 | Not-determined | Genome-fragment | NA | NA | 0 |
| S12_k141_231096 | 2859 | No | NA | 2 | 0 | 2 | Not-determined | Genome-fragment | NA | NA | 0 |
| S12_k141_96225 | 3309 | No | NA | 3 | 0 | 3 | Not-determined | Genome-fragment | NA | NA | 0 |
| S12_k141_247016 | 2594 | No | NA | 1 | 0 | 0 | Not-determined | Genome-fragment | NA | NA | 0 |
| S12_k141_347811 | 2049 | No | NA | 1 | 0 | 0 | Not-determined | Genome-fragment | NA | NA | 0 |
| S12_k141_96238 | 1595 | No | NA | 2 | 0 | 1 | Not-determined | Genome-fragment | NA | NA | 0 |
| S12_k141_347820 | 1012 | No | NA | 1 | 0 | 1 | Not-determined | Genome-fragment | NA | NA | 0 |
| S12_k141_96250 | 1702 | No | NA | 2 | 0 | 2 | Not-determined | Genome-fragment | NA | NA | 0 |
| S12_k141_12464 | 1285 | No | NA | 1 | 0 | 1 | Not-determined | Genome-fragment | NA | NA | 0 |
| S2_k141_168482 | 3495 | No | NA | 5 | 0 | 2 | Low-quality | Genome-fragment | 28.24 | AAI-based (medium-confidence) | 0 |
| S12_k141_180375 | 1729 | No | NA | 1 | 0 | 1 | Not-determined | Genome-fragment | NA | NA | 0 |
| S12_k141_331106 | 2368 | No | NA | 2 | 0 | 1 | Not-determined | Genome-fragment | NA | NA | 0 |
| S12_k141_231150 | 1545 | No | NA | 2 | 0 | 1 | Not-determined | Genome-fragment | NA | NA | 0 |
| S12_k141_397937 | 1015 | No | NA | 2 | 0 | 1 | Not-determined | Genome-fragment | NA | NA | 0 |
| S2_k141_57282 | 3495 | No | NA | 3 | 0 | 3 | Low-quality | Genome-fragment | 1.61 | AAI-based (high-confidence) | 0 |
| S8_k141_164020 | 3495 | No | NA | 5 | 0 | 0 | Low-quality | Genome-fragment | 3.64 | AAI-based (high-confidence) | 0 |
| S12_k141_213980 | 1187 | No | NA | 2 | 0 | 0 | Not-determined | Genome-fragment | NA | NA | 0 |
| S12_k141_397958 | 1190 | No | NA | 2 | 0 | 2 | Not-determined | Genome-fragment | NA | NA | 0 |
| S12_k141_231194 | 1334 | No | NA | 1 | 0 | 1 | Not-determined | Genome-fragment | NA | NA | 0 |
| S12_k141_29375 | 1835 | No | NA | 1 | 0 | 1 | Not-determined | Genome-fragment | NA | NA | 0 |
| S12_k141_62950 | 1330 | No | NA | 2 | 0 | 0 | Not-determined | Genome-fragment | NA | NA | 0 |
| S12_k141_96368 | 1044 | No | NA | 1 | 0 | 1 | Not-determined | Genome-fragment | NA | NA | 0 |
| S12_k141_347898 | 1116 | No | NA | 2 | 0 | 0 | Not-determined | Genome-fragment | NA | NA | 0 |
| S12_k141_331180 | 1106 | No | NA | 2 | 0 | 2 | Not-determined | Genome-fragment | NA | NA | 0 |
| S8_k141_51097 | 3495 | No | NA | 3 | 0 | 0 | Low-quality | Genome-fragment | 6.16 | AAI-based (high-confidence) | 0 |
| S12_k141_163923 | 1021 | No | NA | 1 | 0 | 0 | Not-determined | Genome-fragment | NA | NA | 0 |
| S12_k141_315424 | 1377 | No | NA | 1 | 0 | 1 | Not-determined | Genome-fragment | NA | NA | 0 |
| S1-final-viral-combined_k141_207439\|\|full | 3497 | No | NA | 5 | 3 | 0 | Low-quality | Genome-fragment | 7.94 | AAI-based (high-confidence) | 0 |
| S12_k141_146621 | 1461 | No | NA | 2 | 0 | 0 | Not-determined | Genome-fragment | NA | NA | 0 |
| S12_k141_129447 | 6334 | No | NA | 3 | 0 | 2 | Not-determined | Genome-fragment | NA | NA | 0 |
| S12_k141_146633 | 1200 | No | NA | 2 | 0 | 0 | Not-determined | Genome-fragment | NA | NA | 0 |
| S17-k141_175274 | 3497 | No | NA | 6 | 0 | 0 | Low-quality | Genome-fragment | 9.92 | AAI-based (medium-confidence) | 0 |
| S12_k141_231283 | 1119 | No | NA | 1 | 0 | 0 | Not-determined | Genome-fragment | NA | NA | 0 |
| S12_k141_381549 | 2375 | No | NA | 1 | 0 | 1 | Not-determined | Genome-fragment | NA | NA | 0 |
| S12_k141_47392 | 1739 | No | NA | 1 | 0 | 1 | Not-determined | Genome-fragment | NA | NA | 0 |
| S12_k141_29483 | 1729 | No | NA | 3 | 0 | 0 | Not-determined | Genome-fragment | NA | NA | 0 |
| S12_k141_263687 | 1513 | No | NA | 3 | 0 | 2 | Not-determined | Genome-fragment | NA | NA | 0 |
| S12_k141_297844 | 1204 | No | NA | 1 | 0 | 0 | Not-determined | Genome-fragment | NA | NA | 0 |
| S12_k141_197568 | 3689 | No | NA | 5 | 0 | 1 | Not-determined | Genome-fragment | NA | NA | 0 |
| S12_k141_47441 | 2265 | No | NA | 1 | 0 | 1 | Not-determined | Genome-fragment | NA | NA | 0 |
| S12_k141_146673 | 4674 | No | NA | 2 | 0 | 2 | Not-determined | Genome-fragment | NA | NA | 0 |
| S12_k141_281843 | 1903 | No | NA | 3 | 0 | 3 | Not-determined | Genome-fragment | NA | NA | 0 |
| S6_k141_52657 | 3497 | No | NA | 3 | 0 | 1 | Low-quality | Genome-fragment | 7.53 | AAI-based (high-confidence) | 0 |
| S12_k141_281846 | 2682 | No | NA | 4 | 0 | 2 | Not-determined | Genome-fragment | NA | NA | 0 |
| S9-final-viral-combined_k141_93650\|\|full | 3498 | No | NA | 4 | 0 | 0 | Low-quality | Genome-fragment | 7.91 | AAI-based (high-confidence) | 0 |
| S12_k141_214084 | 1248 | No | NA | 2 | 0 | 0 | Not-determined | Genome-fragment | NA | NA | 0 |
| S12_k141_263798 | 1930 | No | NA | 2 | 0 | 0 | Not-determined | Genome-fragment | NA | NA | 0 |
| S12_k141_96466 | 7632 | No | NA | 11 | 0 | 1 | Not-determined | Genome-fragment | NA | NA | 0 |
| S12_k141_197636 | 1221 | No | NA | 1 | 0 | 0 | Not-determined | Genome-fragment | NA | NA | 0 |
| S12_k141_62978 | 1526 | No | NA | 1 | 0 | 1 | Not-determined | Genome-fragment | NA | NA | 0 |
| S7_k141_83792 | 3499 | No | NA | 3 | 0 | 3 | Low-quality | Genome-fragment | 4.5 | AAI-based (medium-confidence) | 0 |
| S12_k141_112855 | 3524 | No | NA | 1 | 0 | 1 | Not-determined | Genome-fragment | NA | NA | 0 |
| S12_k141_214121 | 4189 | No | NA | 5 | 0 | 2 | Not-determined | Genome-fragment | NA | NA | 0 |
| S12_k141_112872 | 4275 | No | NA | 1 | 0 | 0 | Not-determined | Genome-fragment | NA | NA | 0 |
| S12_k141_197660 | 6193 | No | NA | 6 | 0 | 5 | Not-determined | Genome-fragment | NA | NA | 0 |
| S12_k141_331376 | 1018 | No | NA | 1 | 0 | 1 | Not-determined | Genome-fragment | NA | NA | 0 |
| S12_k141_12658 | 58622 | Yes | 46331 | 72 | 13 | 6 | High-quality | High-quality | 91.18 | AAI-based (high-confidence) | 20.97 |
| S12_k141_197678 | 1032 | No | NA | 2 | 0 | 1 | Not-determined | Genome-fragment | NA | NA | 0 |
| S12_k141_29673 | 1501 | No | NA | 2 | 0 | 1 | Not-determined | Genome-fragment | NA | NA | 0 |
| S12_k141_129584 | 1173 | No | NA | 2 | 0 | 0 | Not-determined | Genome-fragment | NA | NA | 0 |
| S12_k141_247321 | 1627 | No | NA | 2 | 0 | 1 | Not-determined | Genome-fragment | NA | NA | 0 |
| S12_k141_297949 | 4581 | No | NA | 2 | 0 | 1 | Not-determined | Genome-fragment | NA | NA | 0 |
| S12_k141_47595 | 1807 | No | NA | 2 | 0 | 2 | Not-determined | Genome-fragment | NA | NA | 0 |
| S12_k141_281953 | 2077 | No | NA | 1 | 0 | 1 | Not-determined | Genome-fragment | NA | NA | 0 |
| S8-final-viral-combined_k141_4389\|\|full | 3499 | No | NA | 7 | 0 | 0 | Low-quality | Genome-fragment | 6.63 | AAI-based (high-confidence) | 0 |
| S12_k141_214210 | 3319 | No | NA | 5 | 0 | 1 | Not-determined | Genome-fragment | NA | NA | 0 |
| S11_k141_235888 | 3500 | No | NA | 6 | 2 | 0 | Low-quality | Genome-fragment | 7.67 | AAI-based (high-confidence) | 0 |
| S12_k141_247346 | 1261 | No | NA | 2 | 0 | 0 | Not-determined | Genome-fragment | NA | NA | 0 |
| S2-final-viral-combined_k141_207961\|\|full | 3500 | No | NA | 7 | 0 | 0 | Low-quality | Genome-fragment | 9.36 | AAI-based (high-confidence) | 0 |
| S12_k141_214235 | 1440 | No | NA | 2 | 0 | 1 | Not-determined | Genome-fragment | NA | NA | 0 |
| S12_k141_129641 | 6040 | No | NA | 8 | 0 | 2 | Not-determined | Genome-fragment | NA | NA | 0 |
| S12_k141_298005 | 6123 | No | NA | 9 | 0 | 0 | Not-determined | Genome-fragment | NA | NA | 0 |
| S8_k141_110259 | 3500 | No | NA | 4 | 0 | 4 | Low-quality | Genome-fragment | 1.29 | AAI-based (high-confidence) | 0 |
| S12_k141_164248 | 5255 | No | NA | 2 | 0 | 2 | Not-determined | Genome-fragment | NA | NA | 0 |
| S12_k141_307679 | 3501 | No | NA | 8 | 3 | 0 | Low-quality | Genome-fragment | 5.78 | AAI-based (high-confidence) | 0 |
| S12_k141_214249 | 1117 | No | NA | 1 | 0 | 0 | Not-determined | Genome-fragment | NA | NA | 0 |
| S12_k141_47708 | 1439 | No | NA | 1 | 0 | 1 | Not-determined | Genome-fragment | NA | NA | 0 |
| S12_k141_348207 | 1062 | No | NA | 1 | 0 | 0 | Not-determined | Genome-fragment | NA | NA | 0 |
| S12_k141_96681 | 4984 | No | NA | 4 | 0 | 3 | Not-determined | Genome-fragment | NA | NA | 0 |
| S12_k141_282065 | 3049 | No | NA | 2 | 0 | 2 | Not-determined | Genome-fragment | NA | NA | 0 |
| S12_k141_381831 | 1502 | No | NA | 4 | 0 | 0 | Not-determined | Genome-fragment | NA | NA | 0 |
| S12_k141_282090 | 1630 | No | NA | 2 | 0 | 2 | Not-determined | Genome-fragment | NA | NA | 0 |
| S12_k141_348258 | 1555 | No | NA | 3 | 0 | 1 | Not-determined | Genome-fragment | NA | NA | 0 |
| S4-final-viral-combined_k141_50393\|\|full | 3501 | No | NA | 6 | 0 | 2 | Low-quality | Genome-fragment | 7.43 | AAI-based (medium-confidence) | 0 |
| S12_k141_129784 | 1719 | No | NA | 3 | 0 | 0 | Not-determined | Genome-fragment | NA | NA | 0 |
| S12_k141_282117 | 1675 | No | NA | 2 | 0 | 1 | Not-determined | Genome-fragment | NA | NA | 0 |
| S1-final-viral-combined_k141_326470\|\|full | 3502 | No | NA | 4 | 0 | 4 | Low-quality | Genome-fragment | 4.5 | AAI-based (medium-confidence) | 0 |
| S12_k141_247513 | 1464 | No | NA | 2 | 0 | 0 | Not-determined | Genome-fragment | NA | NA | 0 |
| S12_k141_197850 | 1417 | No | NA | 1 | 0 | 1 | Not-determined | Genome-fragment | NA | NA | 0 |
| S12_k141_29810 | 1167 | No | NA | 2 | 0 | 2 | Not-determined | Genome-fragment | NA | NA | 0 |
| S12_k141_298175 | 1003 | No | NA | 1 | 0 | 1 | Not-determined | Genome-fragment | NA | NA | 0 |
| S12_k141_298104 | 1588 | No | NA | 2 | 0 | 1 | Not-determined | Genome-fragment | NA | NA | 0 |
| S12_k141_113089 | 4474 | No | NA | 3 | 0 | 2 | Not-determined | Genome-fragment | NA | NA | 0 |
| S3-final-viral-combined_k141_57670\|\|full | 3502 | No | NA | 4 | 0 | 1 | Low-quality | Genome-fragment | 4.44 | AAI-based (high-confidence) | 0 |
| S12_k141_113098 | 1082 | No | NA | 1 | 0 | 1 | Not-determined | Genome-fragment | NA | NA | 0 |
| S12_k141_96764 | 3011 | No | NA | 4 | 0 | 1 | Not-determined | Genome-fragment | NA | NA | 0 |
| S12_k141_29849 | 1073 | No | NA | 2 | 0 | 1 | Not-determined | Genome-fragment | NA | NA | 0 |
| S12_k141_398210 | 1041 | No | NA | 2 | 0 | 2 | Not-determined | Genome-fragment | NA | NA | 0 |
| S12_k141_247571 | 3039 | No | NA | 2 | 0 | 2 | Not-determined | Genome-fragment | NA | NA | 0 |
| S12_k141_231696 | 2656 | No | NA | 2 | 0 | 1 | Not-determined | Genome-fragment | NA | NA | 0 |
| S12_k141_282204 | 1537 | No | NA | 2 | 0 | 1 | Not-determined | Genome-fragment | NA | NA | 0 |
| S12-final-viral-combined_k141_363884\|\|full | 3502 | No | NA | 8 | 1 | 0 | Low-quality | Genome-fragment | 4.72 | AAI-based (high-confidence) | 0 |
| S12_k141_247637 | 1197 | No | NA | 1 | 0 | 1 | Not-determined | Genome-fragment | NA | NA | 0 |
| S12_k141_113159 | 1382 | No | NA | 2 | 0 | 1 | Not-determined | Genome-fragment | NA | NA | 0 |
| S12_k141_197952 | 3385 | No | NA | 9 | 0 | 0 | Not-determined | Genome-fragment | NA | NA | 0 |
| S12_k141_146945 | 1083 | No | NA | 1 | 0 | 0 | Not-determined | Genome-fragment | NA | NA | 0 |
| S11-final-viral-combined_k141_329871\|\|full | 3503 | No | NA | 4 | 3 | 0 | Low-quality | Genome-fragment | 7.89 | AAI-based (high-confidence) | 0 |
| S12_k141_180926 | 1341 | No | NA | 1 | 0 | 0 | Not-determined | Genome-fragment | NA | NA | 0 |
| S12_k141_146958 | 1042 | No | NA | 1 | 0 | 0 | Not-determined | Genome-fragment | NA | NA | 0 |
| S12_k141_63350 | 1203 | No | NA | 1 | 0 | 0 | Not-determined | Genome-fragment | NA | NA | 0 |
| S12_k141_282284 | 1135 | No | NA | 1 | 0 | 0 | Not-determined | Genome-fragment | NA | NA | 0 |
| S12_k141_180953 | 3509 | No | NA | 2 | 0 | 1 | Not-determined | Genome-fragment | NA | NA | 0 |
| S12_k141_315900 | 1150 | No | NA | 1 | 0 | 0 | Not-determined | Genome-fragment | NA | NA | 0 |
| S12_k141_2909 | 3503 | No | NA | 7 | 0 | 0 | Low-quality | Genome-fragment | 7.8 | AAI-based (medium-confidence) | 0 |
| S17-final-viral-combined_k141_83105\|\|full | 3504 | No | NA | 4 | 1 | 0 | Low-quality | Genome-fragment | 4.64 | HMM-based (lower-bound) | 0 |
| S12_k141_348467 | 3238 | No | NA | 3 | 0 | 3 | Not-determined | Genome-fragment | NA | NA | 0 |
| S12_k141_264141 | 1151 | No | NA | 1 | 0 | 0 | Not-determined | Genome-fragment | NA | NA | 0 |
| S11-final-viral-combined_k141_81950\|\|full | 3505 | No | NA | 7 | 1 | 0 | Low-quality | Genome-fragment | 4.96 | AAI-based (high-confidence) | 0 |
| S12_k141_13018 | 1141 | No | NA | 2 | 0 | 1 | Not-determined | Genome-fragment | NA | NA | 0 |
| S12_k141_164586 | 2575 | No | NA | 3 | 0 | 1 | Not-determined | Genome-fragment | NA | NA | 0 |
| S17-final-viral-combined_k141_57980\|\|full | 3505 | No | NA | 6 | 0 | 0 | Low-quality | Genome-fragment | 7.42 | AAI-based (medium-confidence) | 0 |
| S12_k141_381987 | 1095 | No | NA | 3 | 0 | 0 | Not-determined | Genome-fragment | NA | NA | 0 |
| S17-k141_6953 | 3505 | No | NA | 5 | 0 | 0 | Low-quality | Genome-fragment | 8.39 | AAI-based (medium-confidence) | 0 |
| S12_k141_129857 | 2317 | No | NA | 2 | 0 | 2 | Not-determined | Genome-fragment | NA | NA | 0 |
| S12_k141_147034 | 2101 | No | NA | 1 | 0 | 1 | Not-determined | Genome-fragment | NA | NA | 0 |
| S8_k141_149573 | 3505 | No | NA | 5 | 1 | 0 | Low-quality | Genome-fragment | 4.42 | HMM-based (lower-bound) | 0 |
| S12_k141_298254 | 1434 | No | NA | 2 | 0 | 1 | Not-determined | Genome-fragment | NA | NA | 0 |
| S7-final-viral-combined_k141_103195\|\|full | 3506 | No | NA | 3 | 0 | 3 | Low-quality | Genome-fragment | 2.88 | AAI-based (medium-confidence) | 0 |
| S12-final-viral-combined_k141_220740\|\|full | 3506 | No | NA | 4 | 0 | 4 | Low-quality | Genome-fragment | 1.79 | AAI-based (medium-confidence) | 0 |
| S12_k141_214675 | 1445 | No | NA | 1 | 0 | 1 | Not-determined | Genome-fragment | NA | NA | 0 |
| S8-final-viral-combined_k141_237255\|\|full | 3506 | No | NA | 7 | 4 | 0 | Low-quality | Genome-fragment | 5.08 | HMM-based (lower-bound) | 0 |
| S12_k141_398287 | 6485 | No | NA | 3 | 0 | 3 | Not-determined | Genome-fragment | NA | NA | 0 |
| S12_k141_214697 | 4816 | No | NA | 7 | 0 | 6 | Not-determined | Genome-fragment | NA | NA | 0 |
| S1_k141_4547 | 3507 | No | NA | 5 | 1 | 0 | Low-quality | Genome-fragment | 2.9 | AAI-based (high-confidence) | 0 |
| S7_k141_93412 | 3507 | No | NA | 4 | 0 | 0 | Low-quality | Genome-fragment | 4.28 | AAI-based (medium-confidence) | 0 |
| S12_k141_13135 | 1051 | No | NA | 1 | 0 | 0 | Not-determined | Genome-fragment | NA | NA | 0 |
| S12_k141_398301 | 1329 | No | NA | 3 | 0 | 2 | Not-determined | Genome-fragment | NA | NA | 0 |
| S12_k141_81082 | 3856 | No | NA | 3 | 0 | 1 | Not-determined | Genome-fragment | NA | NA | 0 |
| S12_k141_398326 | 1243 | No | NA | 2 | 0 | 0 | Not-determined | Genome-fragment | NA | NA | 0 |
| S3-final-viral-combined_k141_9263\|\|full | 3508 | Yes | 1326 | 3 | 1 | 1 | Low-quality | Genome-fragment | 2.36 | AAI-based (medium-confidence) | 62.2 |
| S12_k141_331927 | 1280 | No | NA | 2 | 0 | 2 | Not-determined | Genome-fragment | NA | NA | 0 |
| S12_k141_298334 | 1415 | No | NA | 2 | 0 | 0 | Not-determined | Genome-fragment | NA | NA | 0 |
| S12_k141_48020 | 7977 | No | NA | 16 | 0 | 15 | Not-determined | Genome-fragment | NA | NA | 0 |
| S12_k141_81144 | 3082 | No | NA | 4 | 0 | 2 | Not-determined | Genome-fragment | NA | NA | 0 |
| S12_k141_157266 | 3508 | No | NA | 4 | 0 | 1 | Low-quality | Genome-fragment | 3.64 | AAI-based (medium-confidence) | 0 |
| S1-final-viral-combined_k141_271545\|\|full | 3509 | No | NA | 4 | 0 | 3 | Low-quality | Genome-fragment | 2.44 | AAI-based (high-confidence) | 0 |
| S1_k141_200308 | 3509 | No | NA | 4 | 4 | 0 | Low-quality | Genome-fragment | 5.7 | AAI-based (high-confidence) | 0 |
| S12_k141_363426 | 1680 | No | NA | 3 | 0 | 1 | Not-determined | Genome-fragment | NA | NA | 0 |
| S12_k141_48066 | 1817 | No | NA | 2 | 0 | 0 | Not-determined | Genome-fragment | NA | NA | 0 |
| S11-final-viral-combined_k141_99387\|\|full | 3510 | No | NA | 3 | 0 | 3 | Low-quality | Genome-fragment | 1.56 | AAI-based (medium-confidence) | 0 |
| S1-final-viral-combined_k141_308053\|\|full | 3510 | No | NA | 4 | 0 | 0 | Low-quality | Genome-fragment | 5.72 | AAI-based (medium-confidence) | 0 |
| S12_k141_282577 | 1004 | No | NA | 2 | 0 | 0 | Not-determined | Genome-fragment | NA | NA | 0 |
| S12_k141_81208 | 1836 | No | NA | 3 | 0 | 0 | Not-determined | Genome-fragment | NA | NA | 0 |
| S12_k141_81238 | 1651 | No | NA | 1 | 0 | 0 | Not-determined | Genome-fragment | NA | NA | 0 |
| S12_k141_348655 | 1762 | No | NA | 1 | 0 | 1 | Not-determined | Genome-fragment | NA | NA | 0 |
| S5-final-viral-combined_k141_61617\|\|full | 3510 | No | NA | 5 | 2 | 0 | Low-quality | Genome-fragment | 13.96 | AAI-based (high-confidence) | 0 |
| S12_k141_198259 | 1010 | No | NA | 2 | 0 | 1 | Not-determined | Genome-fragment | NA | NA | 0 |
| S6_k141_25388 | 3510 | No | NA | 6 | 1 | 0 | Low-quality | Genome-fragment | 3.78 | AAI-based (high-confidence) | 0 |
| S12_k141_13328 | 9651 | No | NA | 4 | 0 | 3 | Not-determined | Genome-fragment | NA | NA | 0 |
| S12_k141_198288 | 3316 | No | NA | 1 | 0 | 1 | Not-determined | Genome-fragment | NA | NA | 0 |
| S12_k141_113541 | 3776 | No | NA | 1 | 0 | 1 | Not-determined | Genome-fragment | NA | NA | 0 |
| S11-final-viral-combined_k141_8135\|\|lt2gene | 3511 | No | NA | 2 | 1 | 0 | Low-quality | Genome-fragment | 9.69 | AAI-based (medium-confidence) | 0 |
| S12_k141_97192 | 1201 | No | NA | 2 | 0 | 1 | Not-determined | Genome-fragment | NA | NA | 0 |
| S12_k141_214977 | 1041 | No | NA | 2 | 0 | 2 | Not-determined | Genome-fragment | NA | NA | 0 |
| S12_k141_363557 | 1394 | No | NA | 1 | 0 | 1 | Not-determined | Genome-fragment | NA | NA | 0 |
| S12_k141_81286 | 1750 | No | NA | 4 | 0 | 4 | Not-determined | Genome-fragment | NA | NA | 0 |
| S12_k141_382280 | 3415 | No | NA | 4 | 0 | 1 | Not-determined | Genome-fragment | NA | NA | 0 |
| S12_k141_164822 | 1002 | No | NA | 1 | 0 | 1 | Not-determined | Genome-fragment | NA | NA | 0 |
| S2_k141_21354 | 3511 | No | NA | 2 | 0 | 1 | Low-quality | Genome-fragment | 5.85 | AAI-based (medium-confidence) | 0 |
| S12_k141_48219 | 3025 | No | NA | 2 | 0 | 2 | Not-determined | Genome-fragment | NA | NA | 0 |
| S12_k141_382329 | 1461 | No | NA | 3 | 0 | 2 | Not-determined | Genome-fragment | NA | NA | 0 |
| S12_k141_248158 | 1546 | No | NA | 1 | 0 | 1 | Not-determined | Genome-fragment | NA | NA | 0 |
| S12_k141_48240 | 1087 | No | NA | 1 | 0 | 0 | Not-determined | Genome-fragment | NA | NA | 0 |
| S12_k141_81344 | 1266 | No | NA | 1 | 0 | 1 | Not-determined | Genome-fragment | NA | NA | 0 |
| S12_k141_282664 | 1088 | No | NA | 2 | 0 | 1 | Not-determined | Genome-fragment | NA | NA | 0 |
| S12_k141_81383 | 1048 | No | NA | 1 | 0 | 1 | Not-determined | Genome-fragment | NA | NA | 0 |
| S12_k141_363617 | 2617 | No | NA | 1 | 0 | 1 | Not-determined | Genome-fragment | NA | NA | 0 |
| S8-final-viral-combined_k141_161032\|\|full | 3511 | No | NA | 4 | 0 | 1 | Low-quality | Genome-fragment | 6.27 | AAI-based (high-confidence) | 0 |
| S9-final-viral-combined_k141_59476\|\|full | 3512 | No | NA | 4 | 0 | 2 | Low-quality | Genome-fragment | 7.48 | AAI-based (medium-confidence) | 0 |
| S12_k141_363634 | 1283 | No | NA | 1 | 0 | 0 | Not-determined | Genome-fragment | NA | NA | 0 |
| S12_k141_398578 | 1004 | No | NA | 1 | 0 | 0 | Not-determined | Genome-fragment | NA | NA | 0 |
| S12_k141_48303 | 1366 | No | NA | 2 | 0 | 2 | Not-determined | Genome-fragment | NA | NA | 0 |
| S12_k141_147362 | 1588 | No | NA | 3 | 0 | 2 | Not-determined | Genome-fragment | NA | NA | 0 |
| S12_k141_198475 | 1302 | No | NA | 2 | 0 | 2 | Not-determined | Genome-fragment | NA | NA | 0 |
| S12_k141_48320 | 1360 | No | NA | 1 | 0 | 1 | Not-determined | Genome-fragment | NA | NA | 0 |
| S12_k141_48324 | 1167 | No | NA | 3 | 0 | 3 | Not-determined | Genome-fragment | NA | NA | 0 |
| S4-final-viral-combined_k141_104799\|\|full | 3512 | No | NA | 3 | 0 | 3 | Low-quality | Genome-fragment | 4.26 | AAI-based (medium-confidence) | 0 |
| S12_k141_13562 | 3259 | No | NA | 3 | 0 | 2 | Not-determined | Genome-fragment | NA | NA | 0 |
| S12_k141_164925 | 3133 | No | NA | 3 | 0 | 2 | Not-determined | Genome-fragment | NA | NA | 0 |
| S12_k141_84007 | 3513 | No | NA | 4 | 0 | 1 | Low-quality | Genome-fragment | 8.51 | AAI-based (medium-confidence) | 0 |
| S12_k141_298727 | 1180 | No | NA | 2 | 0 | 1 | Not-determined | Genome-fragment | NA | NA | 0 |
| S12_k141_382401 | 6044 | No | NA | 3 | 0 | 2 | Not-determined | Genome-fragment | NA | NA | 0 |
| S2_k141_248328 | 3513 | No | NA | 4 | 0 | 0 | Low-quality | Genome-fragment | 3.03 | AAI-based (high-confidence) | 0 |
| S12_k141_198506 | 1653 | No | NA | 3 | 0 | 3 | Not-determined | Genome-fragment | NA | NA | 0 |

| | | | | | | | | | | | |
|---|---|---|---|---|---|---|---|---|---|---|---|
| S12_k141_147385 | 1284 | No | NA | 1 | 0 | 1 | Not-determined | Genome-fragment | NA | NA | 0 |
| S12_k141_282743 | 1090 | No | NA | 2 | 0 | 2 | Not-determined | Genome-fragment | NA | NA | 0 |
| S12_k141_97326 | 4572 | No | NA | 4 | 0 | 3 | Not-determined | Genome-fragment | NA | NA | 0 |
| S12_k141_215108 | 3887 | No | NA | 2 | 0 | 2 | Not-determined | Genome-fragment | NA | NA | 0 |
| S12_k141_232333 | 1939 | No | NA | 1 | 0 | 1 | Not-determined | Genome-fragment | NA | NA | 0 |
| S12_k141_298774 | 1522 | No | NA | 1 | 0 | 0 | Not-determined | Genome-fragment | NA | NA | 0 |
| S12_k141_316493 | 2561 | No | NA | 4 | 0 | 2 | Not-determined | Genome-fragment | NA | NA | 0 |
| S12_k141_282775 | 5476 | No | NA | 2 | 0 | 2 | Not-determined | Genome-fragment | NA | NA | 0 |
| S12_k141_248295 | 1010 | No | NA | 2 | 0 | 1 | Not-determined | Genome-fragment | NA | NA | 0 |
| S12_k141_48406 | 1192 | No | NA | 3 | 0 | 1 | Not-determined | Genome-fragment | NA | NA | 0 |
| S12_k141_382449 | 12399 | No | NA | 9 | 0 | 8 | Not-determined | Genome-fragment | NA | NA | 0 |
| S12_k141_215139 | 3990 | No | NA | 4 | 0 | 4 | Not-determined | Genome-fragment | NA | NA | 0 |
| S12_k141_97371 | 1660 | No | NA | 1 | 0 | 1 | Not-determined | Genome-fragment | NA | NA | 0 |
| S12_k141_63875 | 3951 | No | NA | 7 | 0 | 6 | Not-determined | Genome-fragment | NA | NA | 0 |
| S17-final-viral-combined_k141_100922\|\|full | 3514 | Yes | 996 | 5 | 1 | 2 | Low-quality | Genome-fragment | 0.63 | HMM-based (lower-bound) | 71.66 |
| S12_k141_398687 | 7060 | No | NA | 7 | 0 | 5 | Not-determined | Genome-fragment | NA | NA | 0 |
| S12_k141_215211 | 3483 | No | NA | 4 | 0 | 3 | Not-determined | Genome-fragment | NA | NA | 0 |
| S12_k141_48448 | 1705 | No | NA | 1 | 0 | 1 | Not-determined | Genome-fragment | NA | NA | 0 |
| S12_k141_316568 | 1669 | No | NA | 1 | 0 | 0 | Not-determined | Genome-fragment | NA | NA | 0 |
| S17-k141_86651 | 3514 | No | NA | 6 | 0 | 1 | Low-quality | Genome-fragment | 4.2 | AAI-based (medium-confidence) | 0 |
| S12_k141_48461 | 1233 | No | NA | 2 | 0 | 1 | Not-determined | Genome-fragment | NA | NA | 0 |
| S12_k141_298840 | 1611 | No | NA | 2 | 0 | 0 | Not-determined | Genome-fragment | NA | NA | 0 |
| S12_k141_13695 | 1430 | No | NA | 1 | 0 | 0 | Not-determined | Genome-fragment | NA | NA | 0 |
| S12_k141_316595 | 3009 | No | NA | 4 | 0 | 4 | Not-determined | Genome-fragment | NA | NA | 0 |
| S12_k141_198646 | 1699 | No | NA | 2 | 0 | 1 | Not-determined | Genome-fragment | NA | NA | 0 |
| S9_k141_37252 | 3515 | No | NA | 3 | 0 | 0 | Low-quality | Genome-fragment | 8.02 | AAI-based (medium-confidence) | 0 |
| S12_k141_63971 | 1163 | No | NA | 1 | 0 | 1 | Not-determined | Genome-fragment | NA | NA | 0 |
| S12_k141_130494 | 1237 | No | NA | 4 | 0 | 0 | Not-determined | Genome-fragment | NA | NA | 0 |
| S12_k141_215308 | 2255 | No | NA | 2 | 0 | 2 | Not-determined | Genome-fragment | NA | NA | 0 |
| S12_k141_232547 | 1015 | No | NA | 1 | 0 | 0 | Not-determined | Genome-fragment | NA | NA | 0 |
| S11-final-viral-combined_k141_50045\|\|full | 3515 | No | NA | 3 | 0 | 2 | Low-quality | Genome-fragment | 25.09 | AAI-based (medium-confidence) | 0 |
| S12-final-viral-combined_k141_315951\|\|full | 3515 | No | NA | 4 | 0 | 0 | Low-quality | Genome-fragment | 7.87 | AAI-based (medium-confidence) | 0 |
| S4_k141_128212 | 3515 | No | NA | 2 | 0 | 1 | Low-quality | Genome-fragment | 0.64 | AAI-based (medium-confidence) | 0 |
| S12_k141_349055 | 1143 | No | NA | 2 | 0 | 1 | Not-determined | Genome-fragment | NA | NA | 0 |
| S12_k141_398795 | 1477 | No | NA | 1 | 0 | 0 | Not-determined | Genome-fragment | NA | NA | 0 |
| S6-final-viral-combined_k141_95243\|\|full | 3515 | No | NA | 5 | 0 | 2 | Low-quality | Genome-fragment | 3.78 | AAI-based (medium-confidence) | 0 |
| S12_k141_13805 | 1281 | No | NA | 1 | 0 | 0 | Not-determined | Genome-fragment | NA | NA | 0 |
| S12_k141_232595 | 1187 | No | NA | 2 | 0 | 0 | Not-determined | Genome-fragment | NA | NA | 0 |
| S12_k141_30588 | 1564 | No | NA | 2 | 0 | 0 | Not-determined | Genome-fragment | NA | NA | 0 |
| S12_k141_165090 | 1419 | No | NA | 2 | 0 | 1 | Not-determined | Genome-fragment | NA | NA | 0 |
| S12_k141_215396 | 2136 | No | NA | 4 | 0 | 1 | Not-determined | Genome-fragment | NA | NA | 0 |
| S12_k141_81629 | 4265 | No | NA | 4 | 0 | 0 | Not-determined | Genome-fragment | NA | NA | 0 |
| S12_k141_283063 | 6190 | No | NA | 14 | 0 | 1 | Not-determined | Genome-fragment | NA | NA | 0 |
| S7-final-viral-combined_k141_166169\|\|full | 3516 | No | NA | 5 | 0 | 2 | Low-quality | Genome-fragment | 4.79 | AAI-based (high-confidence) | 0 |
| S1-final-viral-combined_k141_215135\|\|full | 3517 | No | NA | 7 | 0 | 1 | Low-quality | Genome-fragment | 5.72 | AAI-based (high-confidence) | 0 |
| S17-final-viral-combined_k141_218247\|\|full | 3519 | No | NA | 6 | 2 | 0 | Low-quality | Genome-fragment | 6.6 | AAI-based (medium-confidence) | 0 |
| S12_k141_264675 | 1070 | No | NA | 1 | 0 | 1 | Not-determined | Genome-fragment | NA | NA | 0 |
| S12_k141_97660 | 4240 | No | NA | 9 | 0 | 6 | Not-determined | Genome-fragment | NA | NA | 0 |
| S12-final-viral-combined_k141_293752\|\|full | 3519 | No | NA | 9 | 1 | 0 | Low-quality | Genome-fragment | 8.7 | AAI-based (high-confidence) | 0 |
| S12_k141_64106 | 1144 | No | NA | 2 | 0 | 1 | Not-determined | Genome-fragment | NA | NA | 0 |
| S12-final-viral-combined_k141_246114\|\|full | 3519 | No | NA | 5 | 3 | 0 | Low-quality | Genome-fragment | 7.71 | AAI-based (high-confidence) | 0 |
| S9-final-viral-combined_k141_121091\|\|full | 3520 | No | NA | 4 | 0 | 0 | Low-quality | Genome-fragment | 5.47 | AAI-based (medium-confidence) | 0 |
| S12_k141_130688 | 1910 | No | NA | 2 | 0 | 2 | Not-determined | Genome-fragment | NA | NA | 0 |
| S12_k141_198847 | 1167 | No | NA | 2 | 0 | 0 | Not-determined | Genome-fragment | NA | NA | 0 |
| S12_k141_81759 | 2148 | No | NA | 2 | 0 | 0 | Not-determined | Genome-fragment | NA | NA | 0 |
| S12_k141_232745 | 1122 | No | NA | 1 | 0 | 1 | Not-determined | Genome-fragment | NA | NA | 0 |
| S9_k141_12809 | 3520 | No | NA | 7 | 2 | 1 | Low-quality | Genome-fragment | 8.73 | AAI-based (high-confidence) | 0 |
| S12_k141_398884 | 1048 | No | NA | 1 | 0 | 0 | Not-determined | Genome-fragment | NA | NA | 0 |
| S12_k141_48587 | 2146 | No | NA | 2 | 0 | 1 | Not-determined | Genome-fragment | NA | NA | 0 |
| S5-final-viral-combined_k141_140230\|\|full | 3520 | No | NA | 9 | 3 | 0 | Low-quality | Genome-fragment | 9.83 | AAI-based (high-confidence) | 0 |
| S12_k141_13973 | 3177 | No | NA | 4 | 0 | 1 | Not-determined | Genome-fragment | NA | NA | 0 |
| S12_k141_232762 | 2970 | No | NA | 2 | 0 | 2 | Not-determined | Genome-fragment | NA | NA | 0 |
| S12_k141_299171 | 7814 | No | NA | 5 | 0 | 2 | Not-determined | Genome-fragment | NA | NA | 0 |
| S12_k141_30722 | 1712 | No | NA | 2 | 0 | 0 | Not-determined | Genome-fragment | NA | NA | 0 |
| S12_k141_283215 | 1463 | No | NA | 1 | 0 | 0 | Not-determined | Genome-fragment | NA | NA | 0 |
| S12_k141_81801 | 1279 | No | NA | 1 | 0 | 1 | Not-determined | Genome-fragment | NA | NA | 0 |
| S12_k141_81831 | 2148 | No | NA | 2 | 0 | 1 | Not-determined | Genome-fragment | NA | NA | 0 |
| S12_k141_349304 | 1029 | No | NA | 1 | 0 | 1 | Not-determined | Genome-fragment | NA | NA | 0 |
| S4-final-viral-combined_k141_96200\|\|full | 3520 | No | NA | 5 | 0 | 1 | Low-quality | Genome-fragment | 8.18 | AAI-based (medium-confidence) | 0 |
| S6-final-viral-combined_k141_72578\|\|full | 3520 | No | NA | 3 | 0 | 2 | Low-quality | Genome-fragment | 1.67 | AAI-based (medium-confidence) | 0 |
| S6-final-viral-combined_k141_98512\|\|full | 3521 | No | NA | 5 | 1 | 0 | Low-quality | Genome-fragment | 2.19 | HMM-based (lower-bound) | 0 |
| S12_k141_48719 | 1079 | No | NA | 1 | 0 | 0 | Not-determined | Genome-fragment | NA | NA | 0 |
| S12_k141_81899 | 1440 | No | NA | 2 | 0 | 2 | Not-determined | Genome-fragment | NA | NA | 0 |
| S5-final-viral-combined_k141_141753\|\|full | 3522 | No | NA | 9 | 9 | 0 | Low-quality | Genome-fragment | 5.75 | AAI-based (high-confidence) | 0 |
| S5_k141_164362 | 3523 | Yes | 906 | 4 | 1 | 2 | Low-quality | Genome-fragment | 1.95 | AAI-based (medium-confidence) | 74.28 |
| S12_k141_64180 | 1159 | No | NA | 1 | 0 | 1 | Not-determined | Genome-fragment | NA | NA | 0 |
| S12_k141_64182 | 5389 | No | NA | 4 | 0 | 2 | Not-determined | Genome-fragment | NA | NA | 0 |
| S12_k141_198982 | 9769 | No | NA | 7 | 0 | 1 | Not-determined | Genome-fragment | NA | NA | 0 |
| S12_k141_81927 | 3035 | No | NA | 3 | 0 | 2 | Not-determined | Genome-fragment | NA | NA | 0 |
| S12_k141_48756 | 4056 | No | NA | 7 | 0 | 0 | Not-determined | Genome-fragment | NA | NA | 0 |
| S12_k141_165304 | 1111 | No | NA | 2 | 0 | 0 | Not-determined | Genome-fragment | NA | NA | 0 |
| S8-final-viral-combined_k141_46679\|\|full | 3523 | No | NA | 7 | 1 | 0 | Low-quality | Genome-fragment | 8.01 | AAI-based (high-confidence) | 0 |
| S12_k141_332895 | 1052 | No | NA | 1 | 0 | 1 | Not-determined | Genome-fragment | NA | NA | 0 |
| S12_k141_364230 | 3360 | No | NA | 4 | 0 | 1 | Not-determined | Genome-fragment | NA | NA | 0 |
| S12_k141_165341 | 1121 | No | NA | 3 | 0 | 1 | Not-determined | Genome-fragment | NA | NA | 0 |
| S12_k141_48794 | 1455 | No | NA | 2 | 0 | 2 | Not-determined | Genome-fragment | NA | NA | 0 |
| S12_k141_30823 | 1465 | No | NA | 1 | 0 | 1 | Not-determined | Genome-fragment | NA | NA | 0 |
| S12_k141_349434 | 1958 | No | NA | 1 | 0 | 0 | Not-determined | Genome-fragment | NA | NA | 0 |
| S11-final-viral-combined_k141_245452\|\|full | 3524 | No | NA | 2 | 0 | 1 | Low-quality | Genome-fragment | 2.44 | AAI-based (high-confidence) | 0 |
| S12_k141_199050 | 4039 | No | NA | 7 | 0 | 1 | Not-determined | Genome-fragment | NA | NA | 0 |
| S12_k141_349469 | 1034 | No | NA | 1 | 0 | 1 | Not-determined | Genome-fragment | NA | NA | 0 |
| S12_k141_48837 | 2187 | No | NA | 1 | 0 | 1 | Not-determined | Genome-fragment | NA | NA | 0 |
| S17-final-viral-combined_k141_2919\|\|full | 3524 | No | NA | 3 | 3 | 0 | Low-quality | Genome-fragment | 7.48 | AAI-based (high-confidence) | 0 |
| S12_k141_14191 | 1298 | No | NA | 1 | 0 | 0 | Not-determined | Genome-fragment | NA | NA | 0 |
| S12_k141_232938 | 1356 | No | NA | 2 | 0 | 2 | Not-determined | Genome-fragment | NA | NA | 0 |
| S12_k141_264992 | 1398 | No | NA | 3 | 0 | 0 | Not-determined | Genome-fragment | NA | NA | 0 |
| S12_k141_165402 | 3245 | No | NA | 3 | 0 | 2 | Not-determined | Genome-fragment | NA | NA | 0 |
| S12_k141_14256 | 1443 | No | NA | 1 | 0 | 0 | Not-determined | Genome-fragment | NA | NA | 0 |
| S12_k141_283541 | 1459 | No | NA | 1 | 0 | 0 | Not-determined | Genome-fragment | NA | NA | 0 |
| S12_k141_382818 | 2514 | No | NA | 2 | 0 | 2 | Not-determined | Genome-fragment | NA | NA | 0 |
| S12_k141_199142 | 1167 | No | NA | 1 | 0 | 1 | Not-determined | Genome-fragment | NA | NA | 0 |
| S12_k141_48909 | 1057 | No | NA | 1 | 0 | 0 | Not-determined | Genome-fragment | NA | NA | 0 |
| S12_k141_233022 | 1168 | No | NA | 1 | 0 | 1 | Not-determined | Genome-fragment | NA | NA | 0 |
| S12_k141_248763 | 1515 | No | NA | 3 | 0 | 1 | Not-determined | Genome-fragment | NA | NA | 0 |
| S12_k141_48918 | 1543 | No | NA | 4 | 0 | 0 | Not-determined | Genome-fragment | NA | NA | 0 |
| S12_k141_82169 | 3496 | No | NA | 1 | 0 | 1 | Not-determined | Genome-fragment | NA | NA | 0 |
| S12_k141_333064 | 1770 | No | NA | 2 | 0 | 2 | Not-determined | Genome-fragment | NA | NA | 0 |
| S12_k141_165505 | 1278 | No | NA | 2 | 0 | 1 | Not-determined | Genome-fragment | NA | NA | 0 |
| S12_k141_233037 | 1019 | No | NA | 1 | 0 | 1 | Not-determined | Genome-fragment | NA | NA | 0 |
| S12_k141_114392 | 4911 | No | NA | 6 | 0 | 0 | Not-determined | Genome-fragment | NA | NA | 0 |
| S12_k141_199148 | 1048 | No | NA | 1 | 0 | 0 | Not-determined | Genome-fragment | NA | NA | 0 |
| S12_k141_333083 | 1251 | No | NA | 1 | 0 | 0 | Not-determined | Genome-fragment | NA | NA | 0 |
| S12_k141_333087 | 3976 | No | NA | 4 | 0 | 3 | Not-determined | Genome-fragment | NA | NA | 0 |
| S12_k141_165531 | 2916 | No | NA | 2 | 0 | 1 | Not-determined | Genome-fragment | NA | NA | 0 |
| S12_k141_31007 | 5171 | No | NA | 5 | 0 | 1 | Not-determined | Genome-fragment | NA | NA | 0 |
| S12_k141_333096 | 3705 | No | NA | 5 | 0 | 0 | Not-determined | Genome-fragment | NA | NA | 0 |
| S5_k141_90342 | 3525 | No | NA | 3 | 0 | 1 | Low-quality | Genome-fragment | 5.92 | AAI-based (medium-confidence) | 0 |
| S12_k141_215793 | 1042 | No | NA | 1 | 0 | 1 | Not-determined | Genome-fragment | NA | NA | 0 |
| S12_k141_31026 | 1087 | No | NA | 1 | 0 | 1 | Not-determined | Genome-fragment | NA | NA | 0 |
| S12_k141_131142 | 2672 | No | NA | 2 | 0 | 2 | Not-determined | Genome-fragment | NA | NA | 0 |
| S12_k141_182396 | 3035 | No | NA | 3 | 0 | 2 | Not-determined | Genome-fragment | NA | NA | 0 |
| S8-final-viral-combined_k141_66042\|\|full | 3525 | No | NA | 3 | 0 | 1 | Low-quality | Genome-fragment | 2.4 | AAI-based (medium-confidence) | 0 |
| S12_k141_98086 | 5522 | No | NA | 4 | 0 | 3 | Not-determined | Genome-fragment | NA | NA | 0 |
| S1-final-viral-combined_k141_56934\|\|full | 3526 | No | NA | 4 | 0 | 0 | Low-quality | Genome-fragment | 2.1 | AAI-based (high-confidence) | 0 |
| S12_k141_98092 | 1094 | No | NA | 2 | 0 | 1 | Not-determined | Genome-fragment | NA | NA | 0 |
| S12_k141_14463 | 1183 | No | NA | 4 | 0 | 0 | Not-determined | Genome-fragment | NA | NA | 0 |
| S12_k141_248852 | 2300 | No | NA | 2 | 0 | 2 | Not-determined | Genome-fragment | NA | NA | 0 |
| S12_k141_114566 | 1201 | No | NA | 1 | 0 | 0 | Not-determined | Genome-fragment | NA | NA | 0 |
| S12_k141_82295 | 7754 | No | NA | 4 | 0 | 3 | Not-determined | Genome-fragment | NA | NA | 0 |
| S12_k141_114610 | 1043 | No | NA | 2 | 0 | 2 | Not-determined | Genome-fragment | NA | NA | 0 |
| S12_k141_25475 | 3526 | No | NA | 9 | 2 | 0 | Low-quality | Genome-fragment | 2.63 | AAI-based (high-confidence) | 0 |
| S12_k141_283697 | 1029 | No | NA | 1 | 0 | 1 | Not-determined | Genome-fragment | NA | NA | 0 |
| S12_k141_248911 | 1077 | No | NA | 1 | 0 | 0 | Not-determined | Genome-fragment | NA | NA | 0 |
| S12_k141_31195 | 1010 | No | NA | 1 | 0 | 1 | Not-determined | Genome-fragment | NA | NA | 0 |
| S2-final-viral-combined_k141_200155\|\|full | 3526 | No | NA | 2 | 0 | 1 | Low-quality | Genome-fragment | 2.22 | AAI-based (medium-confidence) | 0 |
| S12_k141_364639 | 1932 | No | NA | 2 | 0 | 1 | Not-determined | Genome-fragment | NA | NA | 0 |
| S12_k141_216043 | 1284 | No | NA | 1 | 0 | 1 | Not-determined | Genome-fragment | NA | NA | 0 |
| S12_k141_364650 | 1044 | No | NA | 2 | 0 | 1 | Not-determined | Genome-fragment | NA | NA | 0 |
| S12_k141_349787 | 1297 | No | NA | 1 | 0 | 0 | Not-determined | Genome-fragment | NA | NA | 0 |
| S12_k141_49070 | 3131 | No | NA | 3 | 0 | 3 | Not-determined | Genome-fragment | NA | NA | 0 |
| S12_k141_248990 | 1032 | No | NA | 2 | 0 | 0 | Not-determined | Genome-fragment | NA | NA | 0 |
| S12_k141_349795 | 3803 | No | NA | 4 | 0 | 2 | Not-determined | Genome-fragment | NA | NA | 0 |
| S12_k141_216070 | 1085 | No | NA | 1 | 0 | 0 | Not-determined | Genome-fragment | NA | NA | 0 |
| S12_k141_148279 | 1937 | No | NA | 1 | 0 | 1 | Not-determined | Genome-fragment | NA | NA | 0 |
| S12_k141_265193 | 1639 | No | NA | 1 | 0 | 1 | Not-determined | Genome-fragment | NA | NA | 0 |
| S12_k141_317469 | 1071 | No | NA | 1 | 0 | 1 | Not-determined | Genome-fragment | NA | NA | 0 |
| S12_k141_317479 | 1125 | No | NA | 3 | 0 | 0 | Not-determined | Genome-fragment | NA | NA | 0 |
| S12_k141_199359 | 1467 | No | NA | 2 | 0 | 1 | Not-determined | Genome-fragment | NA | NA | 0 |
| S12_k141_364756 | 1222 | No | NA | 1 | 0 | 0 | Not-determined | Genome-fragment | NA | NA | 0 |
| S12_k141_216133 | 1679 | No | NA | 1 | 0 | 1 | Not-determined | Genome-fragment | NA | NA | 0 |
| S12_k141_64853 | 1451 | No | NA | 1 | 0 | 1 | Not-determined | Genome-fragment | NA | NA | 0 |
| S2-final-viral-combined_k141_143245\|\|full | 3526 | No | NA | 5 | 0 | 3 | Low-quality | Genome-fragment | 1.09 | AAI-based (medium-confidence) | 0 |
| S12_k141_364776 | 2179 | No | NA | 3 | 0 | 2 | Not-determined | Genome-fragment | NA | NA | 0 |
| S12_k141_216151 | 1289 | No | NA | 1 | 0 | 1 | Not-determined | Genome-fragment | NA | NA | 0 |
| S12_k141_49142 | 6061 | No | NA | 10 | 0 | 6 | Not-determined | Genome-fragment | NA | NA | 0 |
| S12_k141_14638 | 1846 | No | NA | 2 | 0 | 0 | Not-determined | Genome-fragment | NA | NA | 0 |
| S12_k141_283831 | 1499 | No | NA | 2 | 0 | 2 | Not-determined | Genome-fragment | NA | NA | 0 |
| S9_k141_91393 | 3527 | No | NA | 6 | 0 | 0 | Low-quality | Genome-fragment | 10.85 | AAI-based (medium-confidence) | 0 |
| S7-final-viral-combined_k141_226500\|\|full | 3527 | No | NA | 6 | 1 | 0 | Low-quality | Genome-fragment | 3.43 | AAI-based (high-confidence) | 0 |
| S12_k141_299685 | 1085 | No | NA | 2 | 0 | 2 | Not-determined | Genome-fragment | NA | NA | 0 |
| S12_k141_364842 | 1573 | No | NA | 1 | 0 | 0 | Not-determined | Genome-fragment | NA | NA | 0 |
| S17-final-viral-combined_k141_222001\|\|full | 3527 | No | NA | 2 | 0 | 2 | Low-quality | Genome-fragment | 3.66 | AAI-based (medium-confidence) | 0 |
| S12_k141_64920 | 1353 | No | NA | 1 | 0 | 1 | Not-determined | Genome-fragment | NA | NA | 0 |
| S11-final-viral-combined_k141_278168\|\|full | 3528 | No | NA | 5 | 3 | 0 | Low-quality | Genome-fragment | 9.56 | AAI-based (medium-confidence) | 0 |
| S12_k141_216232 | 2129 | No | NA | 2 | 0 | 2 | Not-determined | Genome-fragment | NA | NA | 0 |
| S1-final-viral-combined_k141_314356\|\|full | 3528 | No | NA | 10 | 1 | 0 | Low-quality | Genome-fragment | 5.81 | AAI-based (high-confidence) | 0 |
| S12_k141_399480 | 1079 | No | NA | 1 | 0 | 1 | Not-determined | Genome-fragment | NA | NA | 0 |
| S5-final-viral-combined_k141_6608\|\|full | 3528 | No | NA | 5 | 0 | 0 | Low-quality | Genome-fragment | 5.21 | AAI-based (medium-confidence) | 0 |
| S12_k141_82540 | 1056 | No | NA | 2 | 0 | 0 | Not-determined | Genome-fragment | NA | NA | 0 |
| S7-final-viral-combined_k141_72695\|\|full | 3528 | No | NA | 8 | 0 | 0 | Low-quality | Genome-fragment | 8.54 | AAI-based (high-confidence) | 0 |
| S12_k141_148436 | 4762 | No | NA | 4 | 0 | 4 | Not-determined | Genome-fragment | NA | NA | 0 |
| S12_k141_49253 | 1401 | No | NA | 1 | 0 | 1 | Not-determined | Genome-fragment | NA | NA | 0 |
| S12_k141_265340 | 1043 | No | NA | 2 | 0 | 2 | Not-determined | Genome-fragment | NA | NA | 0 |
| S12_k141_148440 | 3354 | No | NA | 4 | 0 | 0 | Not-determined | Genome-fragment | NA | NA | 0 |
| S12_k141_31463 | 2194 | No | NA | 3 | 0 | 0 | Not-determined | Genome-fragment | NA | NA | 0 |
| S12_k141_265350 | 2294 | No | NA | 1 | 0 | 1 | Not-determined | Genome-fragment | NA | NA | 0 |
| S12_k141_31478 | 1367 | No | NA | 2 | 0 | 0 | Not-determined | Genome-fragment | NA | NA | 0 |
| S12_k141_249218 | 1372 | No | NA | 2 | 0 | 1 | Not-determined | Genome-fragment | NA | NA | 0 |
| S12_k141_98428 | 1012 | No | NA | 1 | 0 | 1 | Not-determined | Genome-fragment | NA | NA | 0 |
| S7_k141_87798 | 3528 | No | NA | 3 | 0 | 2 | Low-quality | Genome-fragment | 8.9 | AAI-based (medium-confidence) | 0 |
| S12_k141_216319 | 1194 | No | NA | 1 | 0 | 0 | Not-determined | Genome-fragment | NA | NA | 0 |
| S12_k141_98468 | 1495 | No | NA | 2 | 0 | 1 | Not-determined | Genome-fragment | NA | NA | 0 |
| S12_k141_199535 | 1034 | No | NA | 2 | 0 | 0 | Not-determined | Genome-fragment | NA | NA | 0 |
| S12_k141_14783 | 1863 | No | NA | 3 | 0 | 1 | Not-determined | Genome-fragment | NA | NA | 0 |
| S12_k141_165944 | 6743 | No | NA | 7 | 0 | 6 | Not-determined | Genome-fragment | NA | NA | 0 |
| S12_k141_383147 | 4781 | No | NA | 4 | 0 | 2 | Not-determined | Genome-fragment | NA | NA | 0 |
| S12_k141_165947 | 1682 | No | NA | 1 | 0 | 0 | Not-determined | Genome-fragment | NA | NA | 0 |
| S12_k141_206100 | 3528 | No | NA | 4 | 0 | 0 | Low-quality | Genome-fragment | 4.83 | AAI-based (medium-confidence) | 0 |
| S12_k141_98490 | 1355 | No | NA | 1 | 0 | 0 | Not-determined | Genome-fragment | NA | NA | 0 |
| S2-final-viral-combined_k141_10659\|\|full | 3528 | No | NA | 2 | 0 | 2 | Low-quality | Genome-fragment | 3.91 | AAI-based (medium-confidence) | 0 |
| S12_k141_265373 | 1058 | No | NA | 1 | 0 | 0 | Not-determined | Genome-fragment | NA | NA | 0 |
| S12_k141_333490 | 1665 | No | NA | 1 | 0 | 1 | Not-determined | Genome-fragment | NA | NA | 0 |
| S12_k141_399536 | 3097 | No | NA | 3 | 0 | 2 | Not-determined | Genome-fragment | NA | NA | 0 |
| S12_k141_317638 | 3431 | No | NA | 7 | 0 | 5 | Not-determined | Genome-fragment | NA | NA | 0 |
| S12_k141_65105 | 1180 | No | NA | 1 | 0 | 1 | Not-determined | Genome-fragment | NA | NA | 0 |
| S1-final-viral-combined_k141_223319\|\|full | 3529 | No | NA | 3 | 0 | 0 | Low-quality | Genome-fragment | 4.92 | AAI-based (medium-confidence) | 0 |
| S12_k141_114805 | 1013 | No | NA | 2 | 0 | 0 | Not-determined | Genome-fragment | NA | NA | 0 |
| S12_k141_399564 | 1162 | No | NA | 2 | 0 | 1 | Not-determined | Genome-fragment | NA | NA | 0 |
| S12_k141_317671 | 1163 | No | NA | 2 | 0 | 1 | Not-determined | Genome-fragment | NA | NA | 0 |
| S12_k141_399573 | 1862 | No | NA | 2 | 0 | 2 | Not-determined | Genome-fragment | NA | NA | 0 |
| S12_k141_148580 | 2092 | No | NA | 2 | 0 | 1 | Not-determined | Genome-fragment | NA | NA | 0 |
| S12_k141_283993 | 1292 | No | NA | 2 | 0 | 0 | Not-determined | Genome-fragment | NA | NA | 0 |
| S1_k141_232989 | 3529 | No | NA | 6 | 1 | 0 | Low-quality | Genome-fragment | 6.74 | AAI-based (high-confidence) | 0 |
| S17-final-viral-combined_k141_132727\|\|full | 3529 | No | NA | 2 | 0 | 2 | Low-quality | Genome-fragment | 2.03 | AAI-based (medium-confidence) | 0 |
| S12_k141_299847 | 1330 | No | NA | 2 | 0 | 1 | Not-determined | Genome-fragment | NA | NA | 0 |
| S12_k141_82699 | 1734 | No | NA | 5 | 0 | 0 | Not-determined | Genome-fragment | NA | NA | 0 |
| S12_k141_284022 | 1367 | No | NA | 2 | 0 | 0 | Not-determined | Genome-fragment | NA | NA | 0 |
| S12_k141_114877 | 1759 | No | NA | 3 | 0 | 2 | Not-determined | Genome-fragment | NA | NA | 0 |
| S2-final-viral-combined_k141_32547\|\|full | 3529 | No | NA | 6 | 2 | 0 | Low-quality | Genome-fragment | 4.79 | HMM-based (lower-bound) | 0 |
| S12_k141_114880 | 1206 | No | NA | 1 | 0 | 0 | Not-determined | Genome-fragment | NA | NA | 0 |
| S12_k141_233738 | 4572 | No | NA | 5 | 0 | 0 | Not-determined | Genome-fragment | NA | NA | 0 |
| S12_k141_82747 | 3132 | No | NA | 5 | 0 | 2 | Not-determined | Genome-fragment | NA | NA | 0 |
| S12_k141_31621 | 1149 | No | NA | 2 | 0 | 0 | Not-determined | Genome-fragment | NA | NA | 0 |
| S12_k141_31675 | 1117 | No | NA | 1 | 0 | 1 | Not-determined | Genome-fragment | NA | NA | 0 |
| S12_k141_284088 | 1084 | No | NA | 1 | 0 | 0 | Not-determined | Genome-fragment | NA | NA | 0 |
| S12_k141_284095 | 1954 | No | NA | 1 | 0 | 1 | Not-determined | Genome-fragment | NA | NA | 0 |
| S12_k141_333692 | 1255 | No | NA | 1 | 0 | 1 | Not-determined | Genome-fragment | NA | NA | 0 |
| S12_k141_14823 | 1417 | No | NA | 2 | 0 | 0 | Not-determined | Genome-fragment | NA | NA | 0 |
| S11-final-viral-combined_k141_249591\|\|full | 3530 | No | NA | 5 | 0 | 0 | Low-quality | Genome-fragment | 9.46 | AAI-based (medium-confidence) | 0 |
| S12_k141_299939 | 1460 | No | NA | 3 | 0 | 2 | Not-determined | Genome-fragment | NA | NA | 0 |
| S12_k141_98675 | 1094 | No | NA | 1 | 0 | 1 | Not-determined | Genome-fragment | NA | NA | 0 |
| S17-k141_129557 | 3530 | No | NA | 3 | 0 | 0 | Low-quality | Genome-fragment | 5.41 | AAI-based (medium-confidence) | 0 |
| S8-final-viral-combined_k141_167570\|\|full | 3530 | No | NA | 6 | 0 | 1 | Low-quality | Genome-fragment | 8.62 | AAI-based (high-confidence) | 0 |
| S12_k141_233853 | 4599 | No | NA | 2 | 0 | 1 | Not-determined | Genome-fragment | NA | NA | 0 |
| S12_k141_98685 | 2317 | No | NA | 3 | 0 | 3 | Not-determined | Genome-fragment | NA | NA | 0 |
| S12_k141_216597 | 1072 | No | NA | 1 | 0 | 0 | Not-determined | Genome-fragment | NA | NA | 0 |
| S12_k141_199823 | 1073 | No | NA | 3 | 0 | 3 | Not-determined | Genome-fragment | NA | NA | 0 |
| S17-final-viral-combined_k141_214803\|\|full | 3532 | No | NA | 9 | 2 | 0 | Low-quality | Genome-fragment | 9.57 | AAI-based (medium-confidence) | 0 |
| S12_k141_216605 | 1188 | No | NA | 1 | 0 | 1 | Not-determined | Genome-fragment | NA | NA | 0 |
| S12_k141_166105 | 8341 | No | NA | 12 | 0 | 11 | Not-determined | Genome-fragment | NA | NA | 0 |
| S12_k141_216632 | 1238 | No | NA | 1 | 0 | 0 | Not-determined | Genome-fragment | NA | NA | 0 |
| S12_k141_98751 | 1403 | No | NA | 2 | 0 | 0 | Not-determined | Genome-fragment | NA | NA | 0 |
| S12_k141_399704 | 1024 | No | NA | 1 | 0 | 1 | Not-determined | Genome-fragment | NA | NA | 0 |
| S12-final-viral-combined_k141_181179\|\|full | 3533 | No | NA | 6 | 1 | 0 | Low-quality | Genome-fragment | 9.64 | AAI-based (medium-confidence) | 0 |
| S12_k141_383381 | 1535 | No | NA | 2 | 0 | 2 | Not-determined | Genome-fragment | NA | NA | 0 |
| S12_k141_131798 | 1172 | No | NA | 2 | 0 | 1 | Not-determined | Genome-fragment | NA | NA | 0 |
| S12_k141_383387 | 2287 | No | NA | 2 | 0 | 0 | Not-determined | Genome-fragment | NA | NA | 0 |
| S12_k141_333802 | 1438 | No | NA | 2 | 0 | 0 | Not-determined | Genome-fragment | NA | NA | 0 |
| S12_k141_284157 | 4566 | No | NA | 4 | 0 | 3 | Not-determined | Genome-fragment | NA | NA | 0 |
| S17-k141_207129 | 3534 | No | NA | 1 | 0 | 1 | Low-quality | Genome-fragment | 1.6 | AAI-based (medium-confidence) | 0 |
| S12_k141_249509 | 4921 | No | NA | 5 | 0 | 4 | Not-determined | Genome-fragment | NA | NA | 0 |
| S12_k141_279989 | 3534 | No | NA | 1 | 0 | 0 | Low-quality | Genome-fragment | 2.77 | AAI-based (medium-confidence) | 0 |
| S8-final-viral-combined_k141_188103\|\|0_partial | 3534 | Yes | 2038 | 4 | 1 | 1 | Low-quality | Genome-fragment | 48.97 | HMM-based (lower-bound) | 42.33 |
| S12_k141_131821 | 1027 | No | NA | 1 | 0 | 0 | Not-determined | Genome-fragment | NA | NA | 0 |
| S12_k141_199898 | 1493 | No | NA | 3 | 0 | 1 | Not-determined | Genome-fragment | NA | NA | 0 |
| S12_k141_49667 | 1254 | No | NA | 1 | 0 | 1 | Not-determined | Genome-fragment | NA | NA | 0 |
| S1_k141_98448 | 3537 | No | NA | 4 | 1 | 0 | Low-quality | Genome-fragment | 8.02 | AAI-based (high-confidence) | 0 |
| S12_k141_365251 | 1957 | No | NA | 1 | 0 | 1 | Not-determined | Genome-fragment | NA | NA | 0 |
| S12_k141_216700 | 1442 | No | NA | 1 | 0 | 1 | Not-determined | Genome-fragment | NA | NA | 0 |
| S12_k141_49673 | 2343 | No | NA | 2 | 0 | 1 | Not-determined | Genome-fragment | NA | NA | 0 |
| S12_k141_31783 | 1806 | No | NA | 2 | 0 | 1 | Not-determined | Genome-fragment | NA | NA | 0 |
| S12_k141_333848 | 1255 | No | NA | 1 | 0 | 0 | Not-determined | Genome-fragment | NA | NA | 0 |
| S12_k141_49690 | 1529 | No | NA | 1 | 0 | 1 | Not-determined | Genome-fragment | NA | NA | 0 |
| S9-final-viral-combined_k141_7173\|\|full | 3538 | No | NA | 2 | 0 | 2 | Low-quality | Genome-fragment | 4.87 | AAI-based (medium-confidence) | 0 |
| S5-final-viral-combined_k141_73744\|\|full | 3538 | No | NA | 6 | 4 | 0 | Low-quality | Genome-fragment | 8.71 | AAI-based (high-confidence) | 0 |
| S12_k141_365281 | 1065 | No | NA | 2 | 0 | 1 | Not-determined | Genome-fragment | NA | NA | 0 |
| S12_k141_216740 | 1028 | No | NA | 2 | 0 | 2 | Not-determined | Genome-fragment | NA | NA | 0 |
| S12_k141_49707 | 1010 | No | NA | 1 | 0 | 0 | Not-determined | Genome-fragment | NA | NA | 0 |
| S12_k141_98831 | 1370 | No | NA | 1 | 0 | 0 | Not-determined | Genome-fragment | NA | NA | 0 |
| S4-final-viral-combined_k141_43190\|\|full | 3538 | No | NA | 5 | 1 | 0 | Low-quality | Genome-fragment | 7.85 | AAI-based (medium-confidence) | 0 |
| S1-final-viral-combined_k141_233138\|\|full | 3539 | No | NA | 9 | 2 | 0 | Low-quality | Genome-fragment | 5.18 | HMM-based (lower-bound) | 0 |
| S12_k141_350318 | 1745 | No | NA | 2 | 0 | 0 | Not-determined | Genome-fragment | NA | NA | 0 |
| S12_k141_14983 | 1529 | No | NA | 1 | 0 | 1 | Not-determined | Genome-fragment | NA | NA | 0 |
| S12_k141_131903 | 3380 | No | NA | 3 | 0 | 3 | Not-determined | Genome-fragment | NA | NA | 0 |
| S12_k141_49765 | 1146 | No | NA | 1 | 0 | 1 | Not-determined | Genome-fragment | NA | NA | 0 |
| S12-final-viral-combined_k141_330785\|\|full | 3539 | No | NA | 5 | 1 | 0 | Low-quality | Genome-fragment | 2.64 | AAI-based (high-confidence) | 0 |
| S2-final-viral-combined_k141_153569\|\|full | 3539 | No | NA | 4 | 0 | 0 | Low-quality | Genome-fragment | 5.35 | AAI-based (medium-confidence) | 0 |
| S12_k141_49794 | 1076 | No | NA | 2 | 0 | 1 | Not-determined | Genome-fragment | NA | NA | 0 |
| S12_k141_300168 | 1037 | No | NA | 1 | 0 | 0 | Not-determined | Genome-fragment | NA | NA | 0 |
| S12_k141_83038 | 1844 | No | NA | 2 | 0 | 2 | Not-determined | Genome-fragment | NA | NA | 0 |
| S12_k141_166296 | 1186 | No | NA | 2 | 0 | 2 | Not-determined | Genome-fragment | NA | NA | 0 |
| S11-final-viral-combined_k141_88124\|\|full | 3540 | No | NA | 3 | 1 | 0 | Low-quality | Genome-fragment | 2.52 | AAI-based (medium-confidence) | 0 |
| S2-final-viral-combined_k141_229308\|\|full | 3540 | No | NA | 4 | 0 | 2 | Low-quality | Genome-fragment | 1.81 | AAI-based (medium-confidence) | 0 |
| S12_k141_166309 | 1443 | No | NA | 3 | 0 | 3 | Not-determined | Genome-fragment | NA | NA | 0 |
| S12_k141_216819 | 1220 | No | NA | 1 | 0 | 0 | Not-determined | Genome-fragment | NA | NA | 0 |
| S12_k141_284258 | 1905 | No | NA | 3 | 0 | 3 | Not-determined | Genome-fragment | NA | NA | 0 |
| S12_k141_65478 | 1010 | No | NA | 1 | 0 | 0 | Not-determined | Genome-fragment | NA | NA | 0 |
| S12_k141_183171 | 1038 | No | NA | 1 | 0 | 1 | Not-determined | Genome-fragment | NA | NA | 0 |
| S8-final-viral-combined_k141_206685\|\|full | 3540 | No | NA | 3 | 2 | 0 | Low-quality | Genome-fragment | 7.88 | AAI-based (high-confidence) | 0 |
| S12_k141_284293 | 1212 | No | NA | 1 | 0 | 0 | Not-determined | Genome-fragment | NA | NA | 0 |
| S12_k141_317910 | 1152 | No | NA | 1 | 0 | 1 | Not-determined | Genome-fragment | NA | NA | 0 |
| S12_k141_31950 | 1014 | No | NA | 2 | 0 | 1 | Not-determined | Genome-fragment | NA | NA | 0 |
| S12_k141_115215 | 1051 | No | NA | 2 | 0 | 0 | Not-determined | Genome-fragment | NA | NA | 0 |
| S9-final-viral-combined_k141_97492\|\|full | 3541 | No | NA | 4 | 0 | 2 | Low-quality | Genome-fragment | 2.53 | AAI-based (high-confidence) | 0 |
| S12_k141_115218 | 4889 | No | NA | 1 | 0 | 1 | Not-determined | Genome-fragment | NA | NA | 0 |
| S12_k141_31977 | 1137 | No | NA | 1 | 0 | 0 | Not-determined | Genome-fragment | NA | NA | 0 |
| S12_k141_166364 | 3281 | No | NA | 1 | 0 | 0 | Not-determined | Genome-fragment | NA | NA | 0 |
| S12_k141_31988 | 1271 | No | NA | 1 | 0 | 0 | Not-determined | Genome-fragment | NA | NA | 0 |
| S12_k141_284353 | 1052 | No | NA | 1 | 0 | 1 | Not-determined | Genome-fragment | NA | NA | 0 |
| S9_k141_52496 | 3541 | No | NA | 3 | 0 | 2 | Low-quality | Genome-fragment | 1.47 | AAI-based (medium-confidence) | 0 |
| S12_k141_65573 | 1302 | No | NA | 3 | 0 | 1 | Not-determined | Genome-fragment | NA | NA | 0 |
| S12_k141_65582 | 1178 | No | NA | 1 | 0 | 1 | Not-determined | Genome-fragment | NA | NA | 0 |
| S7_k141_228390 | 3541 | No | NA | 5 | 0 | 3 | Low-quality | Genome-fragment | 1.66 | AAI-based (high-confidence) | 0 |
| S12_k141_399932 | 1179 | No | NA | 1 | 0 | 1 | Not-determined | Genome-fragment | NA | NA | 0 |
| S12_k141_15232 | 1017 | No | NA | 1 | 0 | 1 | Not-determined | Genome-fragment | NA | NA | 0 |
| S12_k141_383613 | 2455 | No | NA | 2 | 0 | 2 | Not-determined | Genome-fragment | NA | NA | 0 |
| S12_k141_200063 | 1564 | No | NA | 1 | 0 | 0 | Not-determined | Genome-fragment | NA | NA | 0 |
| S17-final-viral-combined_k141_198687\|\|full | 3541 | No | NA | 2 | 0 | 2 | Low-quality | Genome-fragment | 1.78 | AAI-based (medium-confidence) | 0 |
| S12_k141_99008 | 1079 | No | NA | 3 | 0 | 3 | Not-determined | Genome-fragment | NA | NA | 0 |
| S12_k141_265799 | 1005 | No | NA | 2 | 0 | 1 | Not-determined | Genome-fragment | NA | NA | 0 |
| S12_k141_115319 | 1711 | No | NA | 3 | 0 | 0 | Not-determined | Genome-fragment | NA | NA | 0 |
| S12_k141_284483 | 1467 | No | NA | 1 | 0 | 1 | Not-determined | Genome-fragment | NA | NA | 0 |
| S17-final-viral-combined_k141_53060\|\|full | 3541 | No | NA | 2 | 0 | 1 | Low-quality | Genome-fragment | 1.8 | AAI-based (medium-confidence) | 0 |
| S12_k141_265812 | 2204 | No | NA | 1 | 0 | 0 | Not-determined | Genome-fragment | NA | NA | 0 |
| S12_k141_265814 | 1300 | No | NA | 1 | 0 | 1 | Not-determined | Genome-fragment | NA | NA | 0 |
| S12_k141_350539 | 2834 | No | NA | 2 | 0 | 2 | Not-determined | Genome-fragment | NA | NA | 0 |
| S12_k141_334112 | 11259 | No | NA | 6 | 0 | 6 | Not-determined | Genome-fragment | NA | NA | 0 |
| S12_k141_300324 | 12930 | No | NA | 12 | 0 | 8 | Not-determined | Genome-fragment | NA | NA | 0 |
| S12_k141_300330 | 1069 | No | NA | 2 | 0 | 1 | Not-determined | Genome-fragment | NA | NA | 0 |
| S12_k141_115392 | 2045 | No | NA | 2 | 0 | 1 | Not-determined | Genome-fragment | NA | NA | 0 |
| S12_k141_65653 | 1509 | No | NA | 3 | 0 | 1 | Not-determined | Genome-fragment | NA | NA | 0 |
| S12_k141_132171 | 1584 | No | NA | 5 | 0 | 1 | Not-determined | Genome-fragment | NA | NA | 0 |
| S12_k141_334150 | 1079 | No | NA | 1 | 0 | 0 | Not-determined | Genome-fragment | NA | NA | 0 |
| S12_k141_166575 | 2035 | No | NA | 1 | 0 | 1 | Not-determined | Genome-fragment | NA | NA | 0 |
| S12_k141_65668 | 1085 | No | NA | 1 | 0 | 1 | Not-determined | Genome-fragment | NA | NA | 0 |
| S12_k141_15395 | 3485 | No | NA | 2 | 0 | 0 | Not-determined | Genome-fragment | NA | NA | 0 |
| S12-final-viral-combined_k141_66958\|\|full | 3541 | No | NA | 4 | 1 | 0 | Low-quality | Genome-fragment | 6.4 | AAI-based (medium-confidence) | 0 |
| S3_k141_120199 | 3542 | No | NA | 6 | 0 | 1 | Low-quality | Genome-fragment | 4.22 | AAI-based (medium-confidence) | 0 |
| S12_k141_132193 | 1072 | No | NA | 2 | 0 | 1 | Not-determined | Genome-fragment | NA | NA | 0 |
| S12_k141_234315 | 1396 | No | NA | 1 | 0 | 1 | Not-determined | Genome-fragment | NA | NA | 0 |
| S12_k141_15452 | 2841 | No | NA | 2 | 0 | 1 | Not-determined | Genome-fragment | NA | NA | 0 |
| S12_k141_265968 | 3454 | No | NA | 2 | 0 | 2 | Not-determined | Genome-fragment | NA | NA | 0 |
| S12_k141_50108 | 2881 | No | NA | 3 | 0 | 1 | Not-determined | Genome-fragment | NA | NA | 0 |
| S12_k141_65780 | 3245 | No | NA | 2 | 0 | 1 | Not-determined | Genome-fragment | NA | NA | 0 |
| S5-final-viral-combined_k141_6562\|\|full | 3542 | No | NA | 5 | 0 | 0 | Low-quality | Genome-fragment | 5.39 | AAI-based (medium-confidence) | 0 |
| S12_k141_249848 | 2539 | No | NA | 3 | 0 | 0 | Not-determined | Genome-fragment | NA | NA | 0 |
| S12_k141_99205 | 1286 | No | NA | 1 | 0 | 0 | Not-determined | Genome-fragment | NA | NA | 0 |
| S12_k141_265987 | 1095 | No | NA | 2 | 0 | 2 | Not-determined | Genome-fragment | NA | NA | 0 |
| S6_k141_134674 | 3542 | No | NA | 3 | 0 | 2 | Low-quality | Genome-fragment | 0.46 | AAI-based (medium-confidence) | 0 |
| S12_k141_183584 | 1087 | No | NA | 3 | 0 | 1 | Not-determined | Genome-fragment | NA | NA | 0 |
| S12_k141_200266 | 1347 | No | NA | 2 | 0 | 1 | Not-determined | Genome-fragment | NA | NA | 0 |
| S12_k141_300461 | 1331 | No | NA | 1 | 0 | 1 | Not-determined | Genome-fragment | NA | NA | 0 |
| S11_k141_95355 | 3543 | No | NA | 7 | 0 | 0 | Low-quality | Genome-fragment | 3.43 | AAI-based (medium-confidence) | 0 |
| S12_k141_149041 | 1055 | No | NA | 1 | 0 | 1 | Not-determined | Genome-fragment | NA | NA | 0 |
| S12_k141_340984 | 3543 | No | NA | 2 | 0 | 1 | Low-quality | Genome-fragment | 1.75 | AAI-based (medium-confidence) | 0 |
| S12_k141_300476 | 1310 | No | NA | 1 | 0 | 0 | Not-determined | Genome-fragment | NA | NA | 0 |
| S12_k141_83497 | 1761 | No | NA | 2 | 0 | 1 | Not-determined | Genome-fragment | NA | NA | 0 |
| S12_k141_300494 | 1008 | No | NA | 2 | 0 | 2 | Not-determined | Genome-fragment | NA | NA | 0 |
| S12_k141_383851 | 22736 | No | NA | 23 | 1 | 3 | Medium-quality | Genome-fragment | 55.69 | AAI-based (medium-confidence) | 0 |
| S12_k141_99315 | 1052 | No | NA | 1 | 0 | 0 | Not-determined | Genome-fragment | NA | NA | 0 |
| S12_k141_365550 | 2699 | No | NA | 3 | 0 | 2 | Not-determined | Genome-fragment | NA | NA | 0 |
| S12_k141_32259 | 1000 | No | NA | 1 | 0 | 0 | Not-determined | Genome-fragment | NA | NA | 0 |
| S12_k141_249906 | 1699 | No | NA | 1 | 0 | 0 | Not-determined | Genome-fragment | NA | NA | 0 |
| S12_k141_65889 | 1478 | No | NA | 1 | 0 | 1 | Not-determined | Genome-fragment | NA | NA | 0 |
| S12_k141_149138 | 1118 | No | NA | 2 | 0 | 0 | Not-determined | Genome-fragment | NA | NA | 0 |
| S12_k141_115614 | 1104 | No | NA | 1 | 0 | 1 | Not-determined | Genome-fragment | NA | NA | 0 |
| S12_k141_149154 | 1974 | No | NA | 1 | 0 | 0 | Not-determined | Genome-fragment | NA | NA | 0 |
| S12_k141_249925 | 1378 | No | NA | 2 | 0 | 0 | Not-determined | Genome-fragment | NA | NA | 0 |
| S12_k141_166757 | 1134 | No | NA | 1 | 0 | 0 | Not-determined | Genome-fragment | NA | NA | 0 |
| S1_k141_209301 | 3544 | No | NA | 2 | 0 | 1 | Low-quality | Genome-fragment | 7.25 | AAI-based (medium-confidence) | 0 |
| S12_k141_300562 | 2002 | No | NA | 2 | 0 | 2 | Not-determined | Genome-fragment | NA | NA | 0 |
| S12_k141_83593 | 4189 | No | NA | 5 | 0 | 1 | Not-determined | Genome-fragment | NA | NA | 0 |
| S12_k141_217242 | 1317 | No | NA | 1 | 0 | 0 | Not-determined | Genome-fragment | NA | NA | 0 |
| S12_k141_99409 | 1178 | No | NA | 4 | 1 | 0 | Not-determined | Genome-fragment | NA | NA | 0 |
| S2-final-viral-combined_k141_114313\|\|full | 3544 | No | NA | 3 | 0 | 2 | Low-quality | Genome-fragment | 5.85 | AAI-based (high-confidence) | 0 |
| S12_k141_200445 | 1061 | No | NA | 2 | 0 | 1 | Not-determined | Genome-fragment | NA | NA | 0 |
| S12_k141_183741 | 1697 | No | NA | 1 | 0 | 1 | Not-determined | Genome-fragment | NA | NA | 0 |
| S4-final-viral-combined_k141_134603\|\|full | 3544 | No | NA | 7 | 0 | 1 | Low-quality | Genome-fragment | 6.15 | AAI-based (high-confidence) | 0 |
| S12_k141_266155 | 1092 | No | NA | 1 | 0 | 0 | Not-determined | Genome-fragment | NA | NA | 0 |
| S12_k141_350881 | 3671 | No | NA | 4 | 0 | 4 | Not-determined | Genome-fragment | NA | NA | 0 |
| S12_k141_99449 | 3926 | No | NA | 3 | 0 | 1 | Not-determined | Genome-fragment | NA | NA | 0 |
| S12_k141_249993 | 1097 | No | NA | 1 | 0 | 0 | Not-determined | Genome-fragment | NA | NA | 0 |
| S12_k141_132504 | 1064 | No | NA | 1 | 0 | 0 | Not-determined | Genome-fragment | NA | NA | 0 |
| S8_k141_208923 | 3544 | No | NA | 5 | 1 | 0 | Low-quality | Genome-fragment | 4.76 | HMM-based (lower-bound) | 0 |
| S12_k141_32418 | 8231 | No | NA | 7 | 0 | 2 | Not-determined | Genome-fragment | NA | NA | 0 |
| S11_k141_117634 | 3545 | No | NA | 5 | 2 | 0 | Low-quality | Genome-fragment | 2.23 | HMM-based (lower-bound) | 0 |
| S12_k141_365673 | 2243 | No | NA | 2 | 0 | 1 | Not-determined | Genome-fragment | NA | NA | 0 |
| S11-final-viral-combined_k141_194475\|\|full | 3546 | No | NA | 6 | 0 | 3 | Low-quality | Genome-fragment | 17.32 | AAI-based (medium-confidence) | 0 |
| S12_k141_266212 | 1057 | No | NA | 2 | 0 | 2 | Not-determined | Genome-fragment | NA | NA | 0 |
| S12_k141_217327 | 1063 | No | NA | 1 | 0 | 1 | Not-determined | Genome-fragment | NA | NA | 0 |
| S11-final-viral-combined_k141_288706\|\|full | 3546 | No | NA | 4 | 0 | 3 | Low-quality | Genome-fragment | 2.13 | AAI-based (medium-confidence) | 0 |
| S7-final-viral-combined_k141_236237\|\|full | 3546 | No | NA | 4 | 0 | 1 | Low-quality | Genome-fragment | 7.57 | AAI-based (medium-confidence) | 0 |
| S12_k141_99515 | 1275 | No | NA | 3 | 0 | 0 | Not-determined | Genome-fragment | NA | NA | 0 |
| S12_k141_115727 | 4824 | No | NA | 7 | 0 | 2 | Not-determined | Genome-fragment | NA | NA | 0 |
| S7_k141_147198 | 3546 | No | NA | 4 | 0 | 0 | Low-quality | Genome-fragment | 4.66 | AAI-based (medium-confidence) | 0 |
| S12_k141_334474 | 1100 | No | NA | 1 | 0 | 0 | Not-determined | Genome-fragment | NA | NA | 0 |
| S12_k141_99524 | 1040 | No | NA | 2 | 0 | 2 | Not-determined | Genome-fragment | NA | NA | 0 |
| S12_k141_166937 | 1106 | No | NA | 1 | 0 | 1 | Not-determined | Genome-fragment | NA | NA | 0 |
| S12_k141_15692 | 1807 | No | NA | 1 | 0 | 1 | Not-determined | Genome-fragment | NA | NA | 0 |
| S12_k141_32467 | 2109 | No | NA | 3 | 0 | 2 | Not-determined | Genome-fragment | NA | NA | 0 |
| S12_k141_334511 | 2168 | No | NA | 2 | 0 | 2 | Not-determined | Genome-fragment | NA | NA | 0 |
| S12_k141_365735 | 1217 | No | NA | 2 | 0 | 1 | Not-determined | Genome-fragment | NA | NA | 0 |
| S12_k141_384079 | 1425 | No | NA | 2 | 0 | 1 | Not-determined | Genome-fragment | NA | NA | 0 |
| S12_k141_149368 | 3535 | No | NA | 6 | 0 | 0 | Not-determined | Genome-fragment | NA | NA | 0 |
| S12_k141_384087 | 1113 | No | NA | 1 | 0 | 1 | Not-determined | Genome-fragment | NA | NA | 0 |
| S12_k141_15700 | 46190 | No | NA | 51 | 6 | 0 | Medium-quality | Genome-fragment | 64.59 | AAI-based (high-confidence) | 0 |
| S12_k141_32555 | 1157 | No | NA | 2 | 0 | 0 | Not-determined | Genome-fragment | NA | NA | 0 |
| S12_k141_66182 | 1286 | No | NA | 1 | 0 | 0 | Not-determined | Genome-fragment | NA | NA | 0 |
| S2_k141_64188 | 3547 | No | NA | 3 | 0 | 2 | Low-quality | Genome-fragment | 0.54 | AAI-based (high-confidence) | 0 |
| S12_k141_400061 | 1132 | No | NA | 1 | 0 | 1 | Not-determined | Genome-fragment | NA | NA | 0 |
| S12_k141_66232 | 1421 | No | NA | 2 | 0 | 0 | Not-determined | Genome-fragment | NA | NA | 0 |
| S12_k141_300857 | 2066 | No | NA | 3 | 0 | 1 | Not-determined | Genome-fragment | NA | NA | 0 |
| S9-final-viral-combined_k141_123189\|\|full | 3548 | No | NA | 4 | 0 | 1 | Low-quality | Genome-fragment | 8.35 | AAI-based (high-confidence) | 0 |
| S12_k141_149439 | 1426 | No | NA | 3 | 0 | 0 | Not-determined | Genome-fragment | NA | NA | 0 |
| S12-final-viral-combined_k141_289260\|\|full | 3548 | No | NA | 5 | 1 | 0 | Low-quality | Genome-fragment | 8.24 | AAI-based (medium-confidence) | 0 |
| S8-final-viral-combined_k141_1785\|\|full | 3548 | No | NA | 2 | 0 | 2 | Low-quality | Genome-fragment | 2.13 | AAI-based (medium-confidence) | 0 |
| S12_k141_400130 | 1149 | No | NA | 1 | 0 | 1 | Not-determined | Genome-fragment | NA | NA | 0 |
| S12_k141_384198 | 3969 | No | NA | 1 | 0 | 1 | Not-determined | Genome-fragment | NA | NA | 0 |
| S12_k141_334668 | 3454 | No | NA | 5 | 0 | 3 | Not-determined | Genome-fragment | NA | NA | 0 |
| S12_k141_365942 | 2539 | No | NA | 2 | 0 | 0 | Not-determined | Genome-fragment | NA | NA | 0 |
| S5-final-viral-combined_k141_161844\|\|full | 3549 | No | NA | 4 | 2 | 0 | Low-quality | Genome-fragment | 4.31 | HMM-based (lower-bound) | 0 |
| S9_k141_103568 | 3550 | No | NA | 3 | 3 | 0 | Low-quality | Genome-fragment | 7.61 | AAI-based (high-confidence) | 0 |
| S12_k141_149561 | 1736 | No | NA | 3 | 0 | 3 | Not-determined | Genome-fragment | NA | NA | 0 |
| S12_k141_250334 | 1452 | No | NA | 1 | 0 | 1 | Not-determined | Genome-fragment | NA | NA | 0 |
| S12_k141_132779 | 2289 | No | NA | 2 | 0 | 1 | Not-determined | Genome-fragment | NA | NA | 0 |
| S12_k141_116039 | 1131 | No | NA | 2 | 0 | 0 | Not-determined | Genome-fragment | NA | NA | 0 |
| S17-final-viral-combined_k141_45377\|\|full | 3550 | No | NA | 4 | 1 | 0 | Low-quality | Genome-fragment | 5.1 | AAI-based (medium-confidence) | 0 |
| S12_k141_184014 | 1040 | No | NA | 3 | 0 | 2 | Not-determined | Genome-fragment | NA | NA | 0 |
| S12_k141_266489 | 1291 | No | NA | 1 | 0 | 1 | Not-determined | Genome-fragment | NA | NA | 0 |
| S12_k141_149587 | 1250 | No | NA | 1 | 0 | 0 | Not-determined | Genome-fragment | NA | NA | 0 |
| S2-final-viral-combined_k141_3899\|\|full | 3550 | No | NA | 2 | 0 | 2 | Low-quality | Genome-fragment | 4 | AAI-based (medium-confidence) | 0 |
| S12_k141_400211 | 1870 | No | NA | 2 | 0 | 0 | Not-determined | Genome-fragment | NA | NA | 0 |
| S6-final-viral-combined_k141_47485\|\|full | 3550 | No | NA | 6 | 3 | 0 | Low-quality | Genome-fragment | 2.98 | HMM-based (lower-bound) | 0 |

| | | | | | | | | | | | |
|---|---|---|---|---|---|---|---|---|---|---|---|
| S12_k141_300983 | 1565 | No | NA | 1 | 0 | 1 | Not-determined | Genome-fragment | NA | NA | 0 |
| S11_k141_259789 | 3551 | No | NA | 3 | 0 | 1 | Low-quality | Genome-fragment | 4.54 | AAI-based (medium-confidence) | 0 |
| S12-final-viral-combined_k141_154136\|\|full | 3551 | No | NA | 3 | 3 | 0 | Low-quality | Genome-fragment | 8.36 | AAI-based (high-confidence) | 0 |
| S12_k141_167256 | 6883 | No | NA | 6 | 0 | 6 | Not-determined | Genome-fragment | NA | NA | 0 |
| S12_k141_149694 | 1181 | No | NA | 2 | 0 | 0 | Not-determined | Genome-fragment | NA | NA | 0 |
| S12_k141_66514 | 2161 | No | NA | 2 | 0 | 2 | Not-determined | Genome-fragment | NA | NA | 0 |
| S12_k141_334771 | 1119 | No | NA | 1 | 0 | 1 | Not-determined | Genome-fragment | NA | NA | 0 |
| S12_k141_384396 | 2030 | No | NA | 4 | 0 | 1 | Not-determined | Genome-fragment | NA | NA | 0 |
| S12_k141_132918 | 1608 | No | NA | 1 | 0 | 1 | Not-determined | Genome-fragment | NA | NA | 0 |
| S12_k141_301091 | 1199 | No | NA | 1 | 0 | 0 | Not-determined | Genome-fragment | NA | NA | 0 |
| S12_k141_167330 | 1454 | No | NA | 3 | 0 | 1 | Not-determined | Genome-fragment | NA | NA | 0 |
| S12_k141_99923 | 1087 | No | NA | 2 | 0 | 2 | Not-determined | Genome-fragment | NA | NA | 0 |
| S12_k141_99933 | 3990 | No | NA | 2 | 0 | 2 | Not-determined | Genome-fragment | NA | NA | 0 |
| S1-final-viral-combined_k141_216435\|\|full | 3552 | No | NA | 2 | 0 | 0 | Low-quality | Genome-fragment | 6.01 | AAI-based (high-confidence) | 0 |
| S12_k141_384492 | 3833 | No | NA | 2 | 0 | 0 | Not-determined | Genome-fragment | NA | NA | 0 |
| S12_k141_16189 | 4273 | No | NA | 2 | 0 | 0 | Not-determined | Genome-fragment | NA | NA | 0 |
| S12_k141_133021 | 1514 | No | NA | 2 | 0 | 0 | Not-determined | Genome-fragment | NA | NA | 0 |
| S12_k141_384513 | 1097 | No | NA | 1 | 0 | 0 | Not-determined | Genome-fragment | NA | NA | 0 |
| S12-final-viral-combined_k141_72673\|\|full | 3552 | No | NA | 5 | 1 | 0 | Low-quality | Genome-fragment | 7.11 | HMM-based (lower-bound) | 0 |
| S2_k141_94716 | 3552 | No | NA | 1 | 0 | 1 | Low-quality | Genome-fragment | 6.37 | AAI-based (high-confidence) | 0 |
| S12_k141_400422 | 4031 | No | NA | 6 | 0 | 1 | Not-determined | Genome-fragment | NA | NA | 0 |
| S12_k141_16249 | 2091 | No | NA | 1 | 0 | 0 | Not-determined | Genome-fragment | NA | NA | 0 |
| S12_k141_351399 | 1266 | No | NA | 1 | 0 | 0 | Not-determined | Genome-fragment | NA | NA | 0 |
| S12_k141_250561 | 2115 | No | NA | 3 | 0 | 1 | Not-determined | Genome-fragment | NA | NA | 0 |
| S12_k141_301179 | 3227 | No | NA | 2 | 0 | 2 | Not-determined | Genome-fragment | NA | NA | 0 |
| S12_k141_366323 | 1330 | No | NA | 2 | 0 | 0 | Not-determined | Genome-fragment | NA | NA | 0 |
| S11-final-viral-combined_k141_157054\|\|full | 3553 | No | NA | 9 | 1 | 0 | Low-quality | Genome-fragment | 8.33 | AAI-based (high-confidence) | 0 |
| S12_k141_250602 | 4145 | No | NA | 1 | 0 | 0 | Not-determined | Genome-fragment | NA | NA | 0 |
| S12_k141_400506 | 1269 | No | NA | 2 | 0 | 0 | Not-determined | Genome-fragment | NA | NA | 0 |
| S1_k141_239271 | 3554 | No | NA | 5 | 1 | 0 | Low-quality | Genome-fragment | 7.71 | AAI-based (high-confidence) | 0 |
| S12_k141_133206 | 1575 | No | NA | 3 | 0 | 2 | Not-determined | Genome-fragment | NA | NA | 0 |
| S12_k141_116396 | 2822 | No | NA | 2 | 0 | 0 | Not-determined | Genome-fragment | NA | NA | 0 |
| S12_k141_66825 | 1065 | No | NA | 1 | 0 | 1 | Not-determined | Genome-fragment | NA | NA | 0 |
| S12_k141_149964 | 7169 | No | NA | 8 | 0 | 4 | Not-determined | Genome-fragment | NA | NA | 0 |
| S12_k141_366452 | 4179 | No | NA | 3 | 0 | 0 | Not-determined | Genome-fragment | NA | NA | 0 |
| S12_k141_100155 | 1045 | No | NA | 2 | 0 | 0 | Not-determined | Genome-fragment | NA | NA | 0 |
| S12_k141_266739 | 1276 | No | NA | 2 | 0 | 1 | Not-determined | Genome-fragment | NA | NA | 0 |
| S12_k141_33115 | 1169 | No | NA | 3 | 0 | 0 | Not-determined | Genome-fragment | NA | NA | 0 |
| S12_k141_100201 | 1451 | No | NA | 2 | 0 | 0 | Not-determined | Genome-fragment | NA | NA | 0 |
| S5-final-viral-combined_k141_48135\|\|full | 3554 | Yes | 2806 | 5 | 1 | 1 | Low-quality | Genome-fragment | 1.59 | HMM-based (lower-bound) | 21.05 |
| S7-final-viral-combined_k141_233589\|\|full | 3554 | No | NA | 4 | 0 | 1 | Low-quality | Genome-fragment | 5.52 | AAI-based (high-confidence) | 0 |
| S12_k141_366551 | 1279 | No | NA | 1 | 0 | 0 | Not-determined | Genome-fragment | NA | NA | 0 |
| S12_k141_366554 | 3944 | No | NA | 4 | 0 | 3 | Not-determined | Genome-fragment | NA | NA | 0 |
| S12_k141_400619 | 1342 | No | NA | 2 | 0 | 2 | Not-determined | Genome-fragment | NA | NA | 0 |
| S12_k141_16483 | 5981 | No | NA | 6 | 0 | 3 | Not-determined | Genome-fragment | NA | NA | 0 |
| S12_k141_133285 | 3103 | No | NA | 4 | 0 | 3 | Not-determined | Genome-fragment | NA | NA | 0 |
| S12-final-viral-combined_k141_290833\|\|full | 3554 | No | NA | 4 | 0 | 0 | Low-quality | Genome-fragment | 7.21 | AAI-based (medium-confidence) | 0 |
| S12_k141_60891 | 3554 | No | NA | 4 | 0 | 0 | Low-quality | Genome-fragment | 5.44 | AAI-based (high-confidence) | 0 |
| S6-final-viral-combined_k141_131181\|\|full | 3554 | No | NA | 8 | 1 | 0 | Low-quality | Genome-fragment | 2.15 | HMM-based (lower-bound) | 0 |
| S12_k141_266928 | 1002 | No | NA | 1 | 0 | 1 | Not-determined | Genome-fragment | NA | NA | 0 |
| S12_k141_16594 | 6382 | No | NA | 6 | 0 | 2 | Not-determined | Genome-fragment | NA | NA | 0 |
| S7-final-viral-combined_k141_145582\|\|full | 3556 | No | NA | 4 | 3 | 0 | Low-quality | Genome-fragment | 7.69 | AAI-based (high-confidence) | 0 |
| S12_k141_33292 | 1198 | No | NA | 1 | 0 | 1 | Not-determined | Genome-fragment | NA | NA | 0 |
| S12_k141_116644 | 5096 | No | NA | 5 | 0 | 3 | Not-determined | Genome-fragment | NA | NA | 0 |
| S12_k141_266993 | 4174 | No | NA | 3 | 0 | 0 | Not-determined | Genome-fragment | NA | NA | 0 |
| S12_k141_266998 | 1615 | No | NA | 4 | 0 | 0 | Not-determined | Genome-fragment | NA | NA | 0 |
| S5_k141_90019 | 3557 | No | NA | 7 | 4 | 0 | Low-quality | Genome-fragment | 8.51 | AAI-based (high-confidence) | 0 |
| S12_k141_400760 | 1063 | No | NA | 2 | 0 | 1 | Not-determined | Genome-fragment | NA | NA | 0 |
| S12_k141_16716 | 4781 | No | NA | 7 | 0 | 6 | Not-determined | Genome-fragment | NA | NA | 0 |
| S12_k141_400775 | 2030 | No | NA | 2 | 0 | 0 | Not-determined | Genome-fragment | NA | NA | 0 |
| S12_k141_384805 | 1842 | No | NA | 4 | 0 | 1 | Not-determined | Genome-fragment | NA | NA | 0 |
| S12_k141_250877 | 3378 | No | NA | 3 | 0 | 1 | Not-determined | Genome-fragment | NA | NA | 0 |
| S12_k141_150362 | 1616 | No | NA | 2 | 0 | 1 | Not-determined | Genome-fragment | NA | NA | 0 |
| S12_k141_267097 | 5750 | No | NA | 4 | 0 | 1 | Not-determined | Genome-fragment | NA | NA | 0 |
| S12_k141_116812 | 1199 | No | NA | 1 | 0 | 1 | Not-determined | Genome-fragment | NA | NA | 0 |
| S12_k141_384908 | 4111 | No | NA | 4 | 0 | 3 | Not-determined | Genome-fragment | NA | NA | 0 |
| S12-final-viral-combined_k141_31466\|\|full | 3558 | No | NA | 6 | 0 | 0 | Low-quality | Genome-fragment | 5.13 | AAI-based (medium-confidence) | 0 |
| S12_k141_367008 | 1223 | No | NA | 1 | 0 | 1 | Not-determined | Genome-fragment | NA | NA | 0 |
| S2_k141_10651 | 3558 | No | NA | 4 | 1 | 0 | Low-quality | Genome-fragment | 7.43 | AAI-based (medium-confidence) | 0 |
| S12_k141_367044 | 1208 | No | NA | 1 | 0 | 0 | Not-determined | Genome-fragment | NA | NA | 0 |
| S12_k141_367082 | 2801 | No | NA | 2 | 0 | 2 | Not-determined | Genome-fragment | NA | NA | 0 |
| S12_k141_401004 | 1030 | No | NA | 1 | 0 | 0 | Not-determined | Genome-fragment | NA | NA | 0 |
| S12_k141_133643 | 3212 | No | NA | 3 | 0 | 1 | Not-determined | Genome-fragment | NA | NA | 0 |
| S12_k141_401080 | 1634 | No | NA | 1 | 0 | 1 | Not-determined | Genome-fragment | NA | NA | 0 |
| S8-final-viral-combined_k141_97165\|\|full | 3558 | No | NA | 3 | 0 | 1 | Low-quality | Genome-fragment | 17.5 | AAI-based (medium-confidence) | 0 |
| S12_k141_367247 | 4880 | No | NA | 3 | 0 | 3 | Not-determined | Genome-fragment | NA | NA | 0 |
| S12_k141_267450 | 3450 | No | NA | 6 | 0 | 0 | Not-determined | Genome-fragment | NA | NA | 0 |
| S12_k141_367276 | 1035 | No | NA | 2 | 0 | 1 | Not-determined | Genome-fragment | NA | NA | 0 |
| S12_k141_367306 | 4217 | No | NA | 2 | 0 | 2 | Not-determined | Genome-fragment | NA | NA | 0 |
| S17-final-viral-combined_k141_1541\|\|full | 3559 | No | NA | 5 | 0 | 2 | Low-quality | Genome-fragment | 2.18 | AAI-based (medium-confidence) | 0 |
| S2-final-viral-combined_k141_45594\|\|full | 3559 | No | NA | 4 | 0 | 1 | Low-quality | Genome-fragment | 6.4 | AAI-based (high-confidence) | 0 |
| S12_k141_367350 | 1173 | No | NA | 1 | 0 | 0 | Not-determined | Genome-fragment | NA | NA | 0 |
| S12_k141_367407 | 6084 | No | NA | 5 | 0 | 1 | Not-determined | Genome-fragment | NA | NA | 0 |
| S12_k141_267512 | 8796 | No | NA | 10 | 0 | 3 | Not-determined | Genome-fragment | NA | NA | 0 |
| S12_k141_267515 | 3189 | No | NA | 3 | 0 | 3 | Not-determined | Genome-fragment | NA | NA | 0 |
| S12_k141_401283 | 6581 | No | NA | 2 | 0 | 1 | Not-determined | Genome-fragment | NA | NA | 0 |
| S12_k141_267538 | 4255 | No | NA | 7 | 0 | 1 | Not-determined | Genome-fragment | NA | NA | 0 |
| S12_k141_267620 | 9550 | No | NA | 5 | 0 | 0 | Not-determined | Genome-fragment | NA | NA | 0 |
| S12_k141_401366 | 45090 | No | NA | 66 | 14 | 0 | Complete | High-quality | 100 | DTR (high-confidence) | 0 |
| S12_k141_267680 | 3219 | No | NA | 5 | 0 | 1 | Not-determined | Genome-fragment | NA | NA | 0 |
| S12_k141_401372 | 87910 | No | NA | 111 | 17 | 4 | Complete | High-quality | 100 | DTR (high-confidence) | 0 |
| S12_k141_367796 | 1159 | No | NA | 2 | 0 | 1 | Not-determined | Genome-fragment | NA | NA | 0 |
| S12_k141_401381 | 4542 | No | NA | 8 | 0 | 1 | Not-determined | Genome-fragment | NA | NA | 0 |
| S12_k141_401422 | 2043 | No | NA | 3 | 0 | 0 | Not-determined | Genome-fragment | NA | NA | 0 |
| S12_k141_401423 | 3605 | No | NA | 5 | 0 | 0 | Not-determined | Genome-fragment | NA | NA | 0 |
| S12_k141_367960 | 1140 | No | NA | 1 | 0 | 1 | Not-determined | Genome-fragment | NA | NA | 0 |
| S12_k141_401480 | 1062 | No | NA | 1 | 0 | 0 | Not-determined | Genome-fragment | NA | NA | 0 |
| S1_k141_85911 | 3560 | No | NA | 5 | 0 | 2 | Low-quality | Genome-fragment | 7.1 | AAI-based (medium-confidence) | 0 |
| S12_k141_368136 | 1004 | No | NA | 2 | 0 | 1 | Not-determined | Genome-fragment | NA | NA | 0 |
| S2-final-viral-combined_k141_135289\|\|full | 2191 | No | NA | 2 | 0 | 0 | Not-determined | Genome-fragment | NA | NA | 0 |
| S2-final-viral-combined_k141_207832\|\|full | 46322 | No | NA | 70 | 19 | 0 | High-quality | High-quality | 98.24 | AAI-based (high-confidence) | 0 |
| S2-final-viral-combined_k141_125246\|\|full | 12671 | No | NA | 18 | 0 | 4 | Not-determined | Genome-fragment | NA | NA | 0 |
| S2-final-viral-combined_k141_177256\|\|full | 3366 | No | NA | 4 | 0 | 0 | Not-determined | Genome-fragment | NA | NA | 0 |
| S2-final-viral-combined_k141_114705\|\|full | 3560 | No | NA | 2 | 0 | 2 | Low-quality | Genome-fragment | 2.82 | AAI-based (medium-confidence) | 0 |
| S2-final-viral-combined_k141_15783\|\|full | 7543 | No | NA | 3 | 0 | 2 | Not-determined | Genome-fragment | NA | NA | 0 |
| S2-final-viral-combined_k141_248318\|\|full | 24915 | No | NA | 40 | 16 | 0 | High-quality | High-quality | 98.49 | AAI-based (high-confidence) | 0 |
| S2-final-viral-combined_k141_248329\|\|full | 31650 | No | NA | 46 | 17 | 0 | High-quality | High-quality | 100 | AAI-based (medium-confidence) | 0 |
| S2-final-viral-combined_k141_248339\|\|full | 2859 | No | NA | 4 | 0 | 0 | Not-determined | Genome-fragment | NA | NA | 0 |
| S2-final-viral-combined_k141_113920\|\|full | 1043 | No | NA | 2 | 0 | 1 | Not-determined | Genome-fragment | NA | NA | 0 |
| S2-final-viral-combined_k141_31082\|\|full | 1697 | No | NA | 3 | 0 | 1 | Not-determined | Genome-fragment | NA | NA | 0 |
| S2-final-viral-combined_k141_31083\|\|full | 1656 | No | NA | 3 | 0 | 0 | Not-determined | Genome-fragment | NA | NA | 0 |
| S2-final-viral-combined_k141_196775\|\|full | 1667 | No | NA | 4 | 0 | 0 | Not-determined | Genome-fragment | NA | NA | 0 |
| S2-final-viral-combined_k141_72505\|\|full | 5330 | No | NA | 10 | 0 | 0 | Not-determined | Genome-fragment | NA | NA | 0 |
| S2-final-viral-combined_k141_134641\|\|full | 1162 | No | NA | 2 | 0 | 0 | Not-determined | Genome-fragment | NA | NA | 0 |
| S1_k141_353380 | 3561 | No | NA | 4 | 0 | 1 | Low-quality | Genome-fragment | 6.69 | AAI-based (high-confidence) | 0 |
| S2-final-viral-combined_k141_72507\|\|full | 1503 | No | NA | 3 | 0 | 3 | Not-determined | Genome-fragment | NA | NA | 0 |
| S4-final-viral-combined_k141_2897\|\|full | 3561 | No | NA | 3 | 0 | 2 | Low-quality | Genome-fragment | 3.59 | AAI-based (medium-confidence) | 0 |
| S11-final-viral-combined_k141_301407\|\|full | 3562 | No | NA | 4 | 0 | 1 | Low-quality | Genome-fragment | 5.34 | AAI-based (medium-confidence) | 0 |
| S2-final-viral-combined_k141_134646\|\|full | 6100 | No | NA | 6 | 0 | 3 | Not-determined | Genome-fragment | NA | NA | 0 |
| S2-final-viral-combined_k141_207148\|\|full | 1651 | No | NA | 2 | 0 | 1 | Not-determined | Genome-fragment | NA | NA | 0 |
| S11_k141_82014 | 3562 | No | NA | 2 | 0 | 2 | Low-quality | Genome-fragment | 1.92 | AAI-based (high-confidence) | 0 |
| S2-final-viral-combined_k141_31084\|\|full | 31856 | No | NA | 47 | 14 | 1 | Medium-quality | Genome-fragment | 51.91 | AAI-based (high-confidence) | 0 |
| S2-final-viral-combined_k141_207156\|\|full | 4597 | No | NA | 3 | 0 | 1 | Not-determined | Genome-fragment | NA | NA | 0 |
| S12-final-viral-combined_k141_151748\|\|full | 3562 | No | NA | 3 | 0 | 2 | Low-quality | Genome-fragment | 2.15 | AAI-based (medium-confidence) | 0 |
| S2-final-viral-combined_k141_113951\|\|full | 2562 | No | NA | 2 | 0 | 1 | Not-determined | Genome-fragment | NA | NA | 0 |
| S4-final-viral-combined_k141_37737\|\|full | 3562 | No | NA | 4 | 0 | 0 | Low-quality | Genome-fragment | 8.33 | AAI-based (medium-confidence) | 0 |
| S2-final-viral-combined_k141_82879\|\|full | 1629 | No | NA | 3 | 0 | 1 | Not-determined | Genome-fragment | NA | NA | 0 |
| S2-final-viral-combined_k141_227855\|\|full | 2241 | No | NA | 3 | 0 | 1 | Not-determined | Genome-fragment | NA | NA | 0 |
| S2-final-viral-combined_k141_176111\|\|full | 2391 | No | NA | 2 | 0 | 1 | Not-determined | Genome-fragment | NA | NA | 0 |
| S2_k141_221289 | 3563 | No | NA | 3 | 0 | 3 | Low-quality | Genome-fragment | 8.99 | AAI-based (medium-confidence) | 0 |
| S2-final-viral-combined_k141_196783\|\|full | 53725 | Yes | 39647 | 70 | 22 | 8 | Complete | High-quality | 100 | Provirus (medium-confidence) | 26.2 |
| S2-final-viral-combined_k141_134695\|\|full | 2483 | No | NA | 5 | 0 | 2 | Not-determined | Genome-fragment | NA | NA | 0 |
| S2-final-viral-combined_k141_176138\|\|full | 6520 | No | NA | 7 | 0 | 0 | Not-determined | Genome-fragment | NA | NA | 0 |
| S11-final-viral-combined_k141_237807\|\|full | 3564 | No | NA | 6 | 0 | 0 | Low-quality | Genome-fragment | 8.9 | AAI-based (high-confidence) | 0 |
| S11_k141_321605 | 3564 | No | NA | 3 | 1 | 0 | Low-quality | Genome-fragment | 7.66 | AAI-based (high-confidence) | 0 |
| S2-final-viral-combined_k141_165747\|\|full | 2413 | No | NA | 3 | 0 | 1 | Not-determined | Genome-fragment | NA | NA | 0 |
| S2-final-viral-combined_k141_186481\|\|full | 6676 | No | NA | 9 | 0 | 3 | Not-determined | Genome-fragment | NA | NA | 0 |
| S2-final-viral-combined_k141_41559\|\|full | 3130 | No | NA | 5 | 0 | 1 | Not-determined | Genome-fragment | NA | NA | 0 |
| S2-final-viral-combined_k141_51857\|\|full | 1018 | No | NA | 3 | 0 | 0 | Not-determined | Genome-fragment | NA | NA | 0 |
| S17-final-viral-combined_k141_235297\|\|full | 3564 | No | NA | 5 | 0 | 0 | Low-quality | Genome-fragment | 15.5 | AAI-based (medium-confidence) | 0 |
| S2-final-viral-combined_k141_196818\|\|full | 2045 | No | NA | 4 | 0 | 0 | Not-determined | Genome-fragment | NA | NA | 0 |
| S6_k141_12552 | 3564 | No | NA | 2 | 0 | 1 | Low-quality | Genome-fragment | 1.15 | AAI-based (medium-confidence) | 0 |
| S2-final-viral-combined_k141_176176\|\|full | 1665 | No | NA | 2 | 0 | 1 | Not-determined | Genome-fragment | NA | NA | 0 |
| S2-final-viral-combined_k141_113980\|\|full | 1326 | No | NA | 2 | 0 | 0 | Not-determined | Genome-fragment | NA | NA | 0 |
| S2-final-viral-combined_k141_10444\|\|full | 1760 | No | NA | 2 | 0 | 2 | Not-determined | Genome-fragment | NA | NA | 0 |
| S6_k141_1321 | 3564 | No | NA | 5 | 3 | 0 | Low-quality | Genome-fragment | 5.77 | AAI-based (high-confidence) | 0 |
| S2-final-viral-combined_k141_134776\|\|full | 2418 | No | NA | 2 | 0 | 1 | Not-determined | Genome-fragment | NA | NA | 0 |
| S2-final-viral-combined_k141_124\|\|full | 1583 | No | NA | 2 | 0 | 1 | Not-determined | Genome-fragment | NA | NA | 0 |
| S2-final-viral-combined_k141_49352\|\|full | 3565 | No | NA | 4 | 2 | 0 | Low-quality | Genome-fragment | 4.27 | HMM-based (lower-bound) | 0 |
| S1_k141_349800 | 3566 | No | NA | 3 | 0 | 1 | Low-quality | Genome-fragment | 3.55 | AAI-based (medium-confidence) | 0 |
| S2-final-viral-combined_k141_81016\|\|full | 3566 | No | NA | 4 | 3 | 0 | Low-quality | Genome-fragment | 4.63 | AAI-based (high-confidence) | 0 |
| S2-final-viral-combined_k141_185270\|\|full | 3566 | No | NA | 3 | 0 | 1 | Low-quality | Genome-fragment | 8.63 | AAI-based (medium-confidence) | 0 |
| S8-final-viral-combined_k141_156727\|\|full | 3566 | No | NA | 4 | 0 | 0 | Low-quality | Genome-fragment | 2.75 | AAI-based (medium-confidence) | 0 |
| S11-final-viral-combined_k141_47614\|\|full | 3568 | No | NA | 10 | 1 | 0 | Low-quality | Genome-fragment | 7.81 | AAI-based (high-confidence) | 0 |
| S2-final-viral-combined_k141_130\|\|full | 1768 | No | NA | 2 | 0 | 1 | Not-determined | Genome-fragment | NA | NA | 0 |
| S11_k141_184124 | 3568 | No | NA | 4 | 1 | 0 | Low-quality | Genome-fragment | 5.83 | AAI-based (medium-confidence) | 0 |
| S1-final-viral-combined_k141_189176\|\|full | 3568 | No | NA | 9 | 3 | 0 | Low-quality | Genome-fragment | 6.32 | AAI-based (high-confidence) | 0 |
| S12_k141_369496 | 3568 | No | NA | 4 | 0 | 4 | Low-quality | Genome-fragment | 11.8 | AAI-based (high-confidence) | 0 |
| S1-final-viral-combined_k141_207362\|\|full | 3570 | No | NA | 6 | 0 | 2 | Low-quality | Genome-fragment | 1.62 | AAI-based (medium-confidence) | 0 |
| S2-final-viral-combined_k141_165832\|\|full | 3752 | No | NA | 5 | 0 | 2 | Not-determined | Genome-fragment | NA | NA | 0 |
| S2-final-viral-combined_k141_41626\|\|full | 2764 | No | NA | 2 | 0 | 2 | Not-determined | Genome-fragment | NA | NA | 0 |
| S2-final-viral-combined_k141_150\|\|full | 4946 | No | NA | 5 | 0 | 0 | Not-determined | Genome-fragment | NA | NA | 0 |
| S2-final-viral-combined_k141_137385\|\|full | 3570 | No | NA | 6 | 0 | 3 | Low-quality | Genome-fragment | 3.63 | AAI-based (high-confidence) | 0 |
| S2-final-viral-combined_k141_31196\|\|full | 1790 | No | NA | 2 | 0 | 2 | Not-determined | Genome-fragment | NA | NA | 0 |
| S2-final-viral-combined_k141_155495\|\|full | 3169 | No | NA | 3 | 0 | 3 | Not-determined | Genome-fragment | NA | NA | 0 |
| S2-final-viral-combined_k141_31212\|\|full | 4469 | No | NA | 4 | 0 | 1 | Not-determined | Genome-fragment | NA | NA | 0 |
| S2-final-viral-combined_k141_165833\|\|full | 30167 | No | NA | 40 | 13 | 1 | Medium-quality | Genome-fragment | 84.28 | AAI-based (high-confidence) | 0 |
| S2-final-viral-combined_k141_155497\|\|full | 6490 | No | NA | 5 | 0 | 4 | Not-determined | Genome-fragment | NA | NA | 0 |
| S11_k141_30458 | 3571 | No | NA | 7 | 3 | 0 | Low-quality | Genome-fragment | 8.06 | AAI-based (high-confidence) | 0 |
| S2-final-viral-combined_k141_93339\|\|full | 1910 | No | NA | 4 | 0 | 1 | Not-determined | Genome-fragment | NA | NA | 0 |
| S2-final-viral-combined_k141_72568\|\|full | 2569 | No | NA | 4 | 0 | 1 | Not-determined | Genome-fragment | NA | NA | 0 |
| S2-final-viral-combined_k141_62281\|\|full | 5483 | No | NA | 5 | 0 | 1 | Not-determined | Genome-fragment | NA | NA | 0 |
| S2-final-viral-combined_k141_83023\|\|full | 1572 | No | NA | 2 | 0 | 2 | Not-determined | Genome-fragment | NA | NA | 0 |
| S7-final-viral-combined_k141_146613\|\|full | 3573 | No | NA | 12 | 0 | 0 | Low-quality | Genome-fragment | 10.27 | AAI-based (high-confidence) | 0 |
| S7_k141_242824 | 3573 | No | NA | 3 | 0 | 0 | Low-quality | Genome-fragment | 3.76 | AAI-based (medium-confidence) | 0 |
| S17-final-viral-combined_k141_54638\|\|full | 3573 | No | NA | 4 | 0 | 3 | Low-quality | Genome-fragment | 5.08 | AAI-based (medium-confidence) | 0 |
| S17-k141_213355 | 3573 | No | NA | 9 | 2 | 0 | Low-quality | Genome-fragment | 9.16 | AAI-based (high-confidence) | 0 |
| S11-final-viral-combined_k141_75404\|\|full | 3574 | No | NA | 4 | 1 | 0 | Low-quality | Genome-fragment | 4.22 | AAI-based (high-confidence) | 0 |
| S2-final-viral-combined_k141_196964\|\|full | 2534 | No | NA | 3 | 0 | 3 | Not-determined | Genome-fragment | NA | NA | 0 |
| S2-final-viral-combined_k141_41682\|\|full | 1751 | No | NA | 2 | 0 | 0 | Not-determined | Genome-fragment | NA | NA | 0 |
| S2-final-viral-combined_k141_114055\|\|full | 12876 | No | NA | 8 | 0 | 8 | Not-determined | Genome-fragment | NA | NA | 0 |
| S2-final-viral-combined_k141_155523\|\|full | 3463 | No | NA | 3 | 0 | 2 | Not-determined | Genome-fragment | NA | NA | 0 |
| S2-final-viral-combined_k141_238345\|\|full | 3756 | No | NA | 6 | 0 | 1 | Not-determined | Genome-fragment | NA | NA | 0 |
| S2-final-viral-combined_k141_72603\|\|full | 4621 | No | NA | 4 | 0 | 4 | Not-determined | Genome-fragment | NA | NA | 0 |
| S12-final-viral-combined_k141_65457\|\|full | 3574 | No | NA | 4 | 0 | 2 | Low-quality | Genome-fragment | 1.7 | AAI-based (medium-confidence) | 0 |
| S8-final-viral-combined_k141_275574\|\|full | 3574 | No | NA | 7 | 0 | 1 | Low-quality | Genome-fragment | 9.26 | AAI-based (high-confidence) | 0 |
| S17-final-viral-combined_k141_2033\|\|full | 3575 | No | NA | 6 | 0 | 0 | Low-quality | Genome-fragment | 11.88 | AAI-based (medium-confidence) | 0 |
| S2-final-viral-combined_k141_41729\|\|full | 1541 | No | NA | 2 | 0 | 0 | Not-determined | Genome-fragment | NA | NA | 0 |
| S2-final-viral-combined_k141_176376\|\|full | 41888 | No | NA | 59 | 20 | 1 | Medium-quality | Genome-fragment | 62.84 | AAI-based (high-confidence) | 0 |
| S9-final-viral-combined_k141_125750\|\|full | 3576 | No | NA | 3 | 0 | 2 | Low-quality | Genome-fragment | 1.55 | AAI-based (medium-confidence) | 0 |
| S2-final-viral-combined_k141_228051\|\|full | 1490 | No | NA | 1 | 0 | 0 | Not-determined | Genome-fragment | NA | NA | 0 |
| S2-final-viral-combined_k141_243\|\|full | 1716 | No | NA | 3 | 0 | 0 | Not-determined | Genome-fragment | NA | NA | 0 |
| S5-final-viral-combined_k141_100111\|\|full | 3576 | No | NA | 6 | 0 | 1 | Low-quality | Genome-fragment | 4.5 | AAI-based (medium-confidence) | 0 |
| S2-final-viral-combined_k141_114071\|\|full | 2570 | No | NA | 3 | 0 | 2 | Not-determined | Genome-fragment | NA | NA | 0 |
| S12-final-viral-combined_k141_4400\|\|full | 3576 | Yes | 3276 | 6 | 3 | 1 | Low-quality | Genome-fragment | 6.69 | AAI-based (medium-confidence) | 8.39 |
| S2-final-viral-combined_k141_134893\|\|full | 2021 | No | NA | 3 | 0 | 0 | Not-determined | Genome-fragment | NA | NA | 0 |
| S2-final-viral-combined_k141_41774\|\|full | 2319 | No | NA | 3 | 0 | 2 | Not-determined | Genome-fragment | NA | NA | 0 |
| S2_k141_172368 | 3576 | No | NA | 1 | 0 | 1 | Low-quality | Genome-fragment | 7.71 | AAI-based (high-confidence) | 0 |
| S7-final-viral-combined_k141_206994\|\|full | 3577 | No | NA | 2 | 0 | 2 | Low-quality | Genome-fragment | 4.76 | AAI-based (medium-confidence) | 0 |
| S2-final-viral-combined_k141_31352\|\|full | 4190 | No | NA | 4 | 0 | 2 | Not-determined | Genome-fragment | NA | NA | 0 |
| S8-final-viral-combined_k141_117192\|\|full | 3578 | No | NA | 3 | 0 | 0 | Low-quality | Genome-fragment | 2.69 | AAI-based (high-confidence) | 0 |
| S2-final-viral-combined_k141_217740\|\|full | 6062 | No | NA | 5 | 0 | 1 | Not-determined | Genome-fragment | NA | NA | 0 |
| S8-final-viral-combined_k141_45502\|\|full | 3578 | No | NA | 5 | 4 | 0 | Low-quality | Genome-fragment | 4.23 | AAI-based (medium-confidence) | 0 |
| S2-final-viral-combined_k141_217742\|\|full | 1743 | No | NA | 3 | 0 | 0 | Not-determined | Genome-fragment | NA | NA | 0 |
| S2-final-viral-combined_k141_228148\|\|full | 2495 | No | NA | 2 | 0 | 2 | Not-determined | Genome-fragment | NA | NA | 0 |
| S2-final-viral-combined_k141_298\|\|full | 1828 | No | NA | 3 | 0 | 1 | Not-determined | Genome-fragment | NA | NA | 0 |
| S11-final-viral-combined_k141_111704\|\|full | 3579 | No | NA | 4 | 3 | 0 | Low-quality | Genome-fragment | 7.69 | AAI-based (high-confidence) | 0 |
| S1_k141_322949 | 3579 | No | NA | 4 | 0 | 4 | Low-quality | Genome-fragment | 1.33 | AAI-based (high-confidence) | 0 |
| S17-k141_65453 | 3579 | No | NA | 4 | 0 | 1 | Low-quality | Genome-fragment | 1.78 | AAI-based (medium-confidence) | 0 |
| S2-final-viral-combined_k141_72674\|\|full | 4042 | No | NA | 8 | 0 | 0 | Not-determined | Genome-fragment | NA | NA | 0 |
| S2_k141_149354 | 3579 | No | NA | 8 | 1 | 0 | Low-quality | Genome-fragment | 6.42 | AAI-based (medium-confidence) | 0 |
| S2-final-viral-combined_k141_31398\|\|full | 6369 | No | NA | 7 | 0 | 0 | Not-determined | Genome-fragment | NA | NA | 0 |
| S8-final-viral-combined_k141_51590\|\|full | 3579 | Yes | 2065 | 8 | 1 | 1 | Low-quality | Genome-fragment | 0.54 | HMM-based (lower-bound) | 42.3 |
| S9-final-viral-combined_k141_71234\|\|full | 3580 | No | NA | 4 | 0 | 0 | Low-quality | Genome-fragment | 6.52 | AAI-based (medium-confidence) | 0 |
| S17-final-viral-combined_k141_200493\|\|full | 3580 | No | NA | 5 | 0 | 1 | Low-quality | Genome-fragment | 8.07 | AAI-based (medium-confidence) | 0 |
| S2-final-viral-combined_k141_83111\|\|full | 1103 | No | NA | 2 | 0 | 0 | Not-determined | Genome-fragment | NA | NA | 0 |
| S2-final-viral-combined_k141_37461\|\|full | 3580 | No | NA | 3 | 0 | 0 | Low-quality | Genome-fragment | 5.86 | AAI-based (medium-confidence) | 0 |
| S5-final-viral-combined_k141_54884\|\|full | 3581 | No | NA | 3 | 0 | 1 | Low-quality | Genome-fragment | 1.65 | AAI-based (medium-confidence) | 0 |
| S2-final-viral-combined_k141_217769\|\|full | 25156 | No | NA | 23 | 0 | 14 | Not-determined | Genome-fragment | NA | NA | 0 |
| S2-final-viral-combined_k141_133879\|\|full | 3581 | No | NA | 2 | 0 | 1 | Low-quality | Genome-fragment | 2.27 | AAI-based (high-confidence) | 0 |
| S2-final-viral-combined_k141_103956\|\|full | 1688 | No | NA | 2 | 0 | 1 | Not-determined | Genome-fragment | NA | NA | 0 |
| S2-final-viral-combined_k141_41861\|\|full | 2871 | No | NA | 5 | 0 | 2 | Not-determined | Genome-fragment | NA | NA | 0 |
| S2-final-viral-combined_k141_93497\|\|full | 3704 | No | NA | 4 | 0 | 3 | Not-determined | Genome-fragment | NA | NA | 0 |
| S11_k141_18395 | 3582 | No | NA | 8 | 1 | 1 | Low-quality | Genome-fragment | 6.08 | AAI-based (medium-confidence) | 0 |
| S7-final-viral-combined_k141_97955\|\|full | 3582 | Yes | 2868 | 6 | 1 | 1 | Low-quality | Genome-fragment | 3.69 | AAI-based (high-confidence) | 19.93 |
| S2-final-viral-combined_k141_238439\|\|full | 5716 | No | NA | 3 | 0 | 1 | Not-determined | Genome-fragment | NA | NA | 0 |
| S2-final-viral-combined_k141_155626\|\|full | 2231 | No | NA | 3 | 0 | 2 | Not-determined | Genome-fragment | NA | NA | 0 |
| S2-final-viral-combined_k141_134979\|\|full | 4791 | No | NA | 4 | 0 | 2 | Not-determined | Genome-fragment | NA | NA | 0 |
| S17-k141_4207 | 3582 | No | NA | 3 | 0 | 2 | Low-quality | Genome-fragment | 0.95 | AAI-based (high-confidence) | 0 |
| S1-final-viral-combined_k141_55395\|\|full | 3583 | No | NA | 3 | 0 | 3 | Low-quality | Genome-fragment | 1.71 | AAI-based (medium-confidence) | 0 |
| S2-final-viral-combined_k141_186737\|\|full | 2181 | No | NA | 3 | 0 | 1 | Not-determined | Genome-fragment | NA | NA | 0 |
| S2-final-viral-combined_k141_124554\|\|full | 2289 | No | NA | 2 | 0 | 1 | Not-determined | Genome-fragment | NA | NA | 0 |
| S2-final-viral-combined_k141_62377\|\|full | 60903 | No | NA | 99 | 33 | 0 | High-quality | High-quality | 100 | AAI-based (high-confidence) | 0 |
| S2-final-viral-combined_k141_93509\|\|full | 1756 | No | NA | 3 | 0 | 0 | Not-determined | Genome-fragment | NA | NA | 0 |
| S2-final-viral-combined_k141_228178\|\|full | 6900 | No | NA | 5 | 0 | 2 | Not-determined | Genome-fragment | NA | NA | 0 |
| S2-final-viral-combined_k141_228180\|\|full | 3971 | No | NA | 7 | 0 | 4 | Not-determined | Genome-fragment | NA | NA | 0 |
| S2-final-viral-combined_k141_124575\|\|full | 1014 | No | NA | 3 | 0 | 1 | Not-determined | Genome-fragment | NA | NA | 0 |
| S2-final-viral-combined_k141_10564\|\|full | 2224 | No | NA | 2 | 0 | 1 | Not-determined | Genome-fragment | NA | NA | 0 |
| S1-final-viral-combined_k141_6495\|\|full | 3583 | No | NA | 3 | 0 | 3 | Low-quality | Genome-fragment | 1.69 | AAI-based (medium-confidence) | 0 |
| S2-final-viral-combined_k141_62416\|\|full | 1200 | No | NA | 4 | 0 | 0 | Not-determined | Genome-fragment | NA | NA | 0 |
| S2-final-viral-combined_k141_4087\|\|full | 3583 | No | NA | 8 | 0 | 0 | Low-quality | Genome-fragment | 10.51 | AAI-based (high-confidence) | 0 |
| S2-final-viral-combined_k141_10576\|\|full | 1066 | No | NA | 2 | 0 | 0 | Not-determined | Genome-fragment | NA | NA | 0 |
| S2-final-viral-combined_k141_41914\|\|full | 9150 | No | NA | 10 | 0 | 1 | Not-determined | Genome-fragment | NA | NA | 0 |
| S3-final-viral-combined_k141_114706\|\|full | 3584 | No | NA | 8 | 1 | 0 | Low-quality | Genome-fragment | 3.74 | AAI-based (medium-confidence) | 0 |
| S7-final-viral-combined_k141_153332\|\|full | 3584 | No | NA | 3 | 0 | 2 | Low-quality | Genome-fragment | 5.77 | AAI-based (high-confidence) | 0 |
| S2-final-viral-combined_k141_72725\|\|full | 29807 | No | NA | 47 | 6 | 0 | Medium-quality | Genome-fragment | 80.47 | AAI-based (high-confidence) | 0 |
| S17-final-viral-combined_k141_257940\|\|full | 3584 | No | NA | 5 | 1 | 0 | Low-quality | Genome-fragment | 9 | AAI-based (high-confidence) | 0 |
| S12-final-viral-combined_k141_351400\|\|full | 3584 | No | NA | 5 | 0 | 0 | Low-quality | Genome-fragment | 7.92 | AAI-based (high-confidence) | 0 |
| S2-final-viral-combined_k141_168949\|\|full | 3584 | No | NA | 2 | 0 | 2 | Low-quality | Genome-fragment | 3.2 | AAI-based (medium-confidence) | 0 |
| S3-final-viral-combined_k141_59956\|\|full | 3585 | No | NA | 4 | 3 | 0 | Low-quality | Genome-fragment | 9.04 | AAI-based (high-confidence) | 0 |
| S2-final-viral-combined_k141_354\|\|full | 1208 | No | NA | 3 | 0 | 1 | Not-determined | Genome-fragment | NA | NA | 0 |
| S2-final-viral-combined_k141_135014\|\|full | 30532 | No | NA | 34 | 16 | 2 | Medium-quality | Genome-fragment | 67.23 | AAI-based (medium-confidence) | 0 |
| S2-final-viral-combined_k141_135024\|\|full | 1953 | No | NA | 3 | 0 | 0 | Not-determined | Genome-fragment | NA | NA | 0 |
| S2-final-viral-combined_k141_10608\|\|full | 2434 | No | NA | 3 | 0 | 0 | Not-determined | Genome-fragment | NA | NA | 0 |
| S2-final-viral-combined_k141_72774\|\|full | 1812 | No | NA | 4 | 0 | 1 | Not-determined | Genome-fragment | NA | NA | 0 |
| S2-final-viral-combined_k141_114138\|\|full | 1989 | No | NA | 3 | 0 | 0 | Not-determined | Genome-fragment | NA | NA | 0 |
| S7-final-viral-combined_k141_202248\|\|full | 3585 | No | NA | 4 | 3 | 0 | Low-quality | Genome-fragment | 8.93 | AAI-based (high-confidence) | 0 |
| S8_k141_163655 | 3585 | No | NA | 4 | 0 | 3 | Low-quality | Genome-fragment | 1.04 | AAI-based (medium-confidence) | 0 |
| S11-final-viral-combined_k141_202266\|\|full | 3586 | No | NA | 8 | 2 | 0 | Low-quality | Genome-fragment | 6.95 | AAI-based (medium-confidence) | 0 |
| S3_k141_65362 | 3586 | No | NA | 2 | 0 | 1 | Low-quality | Genome-fragment | 4.54 | AAI-based (high-confidence) | 0 |
| S17-final-viral-combined_k141_212435\|\|full | 3586 | No | NA | 2 | 0 | 2 | Low-quality | Genome-fragment | 6.99 | AAI-based (medium-confidence) | 0 |
| S2-final-viral-combined_k141_135056\|\|full | 1448 | No | NA | 3 | 0 | 2 | Not-determined | Genome-fragment | NA | NA | 0 |
| S11_k141_242203 | 3587 | No | NA | 3 | 0 | 2 | Low-quality | Genome-fragment | 0.97 | AAI-based (high-confidence) | 0 |
| S7-final-viral-combined_k141_191471\|\|full | 3587 | No | NA | 6 | 1 | 0 | Low-quality | Genome-fragment | 7.95 | AAI-based (high-confidence) | 0 |
| S2-final-viral-combined_k141_238538\|\|full | 1507 | No | NA | 3 | 0 | 2 | Not-determined | Genome-fragment | NA | NA | 0 |
| S2-final-viral-combined_k141_176516\|\|full | 2021 | No | NA | 2 | 0 | 2 | Not-determined | Genome-fragment | NA | NA | 0 |
| S8-final-viral-combined_k141_88922\|\|full | 3587 | No | NA | 2 | 0 | 2 | Low-quality | Genome-fragment | 3.52 | AAI-based (medium-confidence) | 0 |
| S1-final-viral-combined_k141_288044\|\|full | 3588 | Yes | 375 | 3 | 1 | 1 | Low-quality | Genome-fragment | 0.24 | HMM-based (lower-bound) | 89.55 |
| S2-final-viral-combined_k141_124700\|\|full | 1278 | No | NA | 3 | 0 | 0 | Not-determined | Genome-fragment | NA | NA | 0 |
| S12-final-viral-combined_k141_18258\|\|full | 3588 | No | NA | 4 | 0 | 2 | Low-quality | Genome-fragment | 0.4 | AAI-based (medium-confidence) | 0 |
| S2-final-viral-combined_k141_124701\|\|full | 4463 | No | NA | 5 | 0 | 2 | Not-determined | Genome-fragment | NA | NA | 0 |
| S3-final-viral-combined_k141_26268\|\|full | 3589 | No | NA | 3 | 0 | 2 | Low-quality | Genome-fragment | 0.37 | AAI-based (medium-confidence) | 0 |
| S2-final-viral-combined_k141_83184\|\|full | 1731 | No | NA | 3 | 0 | 2 | Not-determined | Genome-fragment | NA | NA | 0 |
| S5-final-viral-combined_k141_175320\|\|full | 3589 | No | NA | 4 | 0 | 1 | Low-quality | Genome-fragment | 6.29 | AAI-based (high-confidence) | 0 |
| S2-final-viral-combined_k141_72847\|\|full | 5001 | No | NA | 8 | 0 | 0 | Not-determined | Genome-fragment | NA | NA | 0 |
| S12-final-viral-combined_k141_181609\|\|full | 3590 | Yes | 1955 | 3 | 1 | 1 | Low-quality | Genome-fragment | 1.63 | AAI-based (high-confidence) | 45.54 |
| S7-final-viral-combined_k141_169246\|\|lt2gene | 3591 | No | NA | 1 | 1 | 0 | Low-quality | Genome-fragment | 7.38 | AAI-based (high-confidence) | 0 |
| S8-final-viral-combined_k141_941\|\|full | 3591 | No | NA | 4 | 0 | 4 | Low-quality | Genome-fragment | 2.13 | AAI-based (medium-confidence) | 0 |
| S2-final-viral-combined_k141_83203\|\|full | 6302 | No | NA | 6 | 0 | 2 | Not-determined | Genome-fragment | NA | NA | 0 |
| S8-final-viral-combined_k141_82897\|\|full | 3591 | No | NA | 5 | 0 | 0 | Low-quality | Genome-fragment | 7.82 | AAI-based (high-confidence) | 0 |
| S2-final-viral-combined_k141_10634\|\|full | 1729 | No | NA | 2 | 0 | 1 | Not-determined | Genome-fragment | NA | NA | 0 |
| S2-final-viral-combined_k141_83225\|\|full | 1443 | No | NA | 3 | 0 | 0 | Not-determined | Genome-fragment | NA | NA | 0 |
| S9-final-viral-combined_k141_48449\|\|full | 3592 | Yes | 2143 | 6 | 1 | 1 | Low-quality | Genome-fragment | 4.31 | AAI-based (medium-confidence) | 40.34 |
| S2-final-viral-combined_k141_155769\|\|full | 6690 | No | NA | 10 | 0 | 2 | Not-determined | Genome-fragment | NA | NA | 0 |
| S2-final-viral-combined_k141_83254\|\|full | 1160 | No | NA | 2 | 0 | 1 | Not-determined | Genome-fragment | NA | NA | 0 |
| S2-final-viral-combined_k141_145428\|\|full | 9162 | No | NA | 7 | 0 | 6 | Not-determined | Genome-fragment | NA | NA | 0 |
| S2-final-viral-combined_k141_62519\|\|full | 85237 | Yes | 34105 | 94 | 19 | 33 | Medium-quality | Genome-fragment | 59.25 | AAI-based (medium-confidence) | 59.99 |
| S1_k141_153526 | 3592 | No | NA | 9 | 3 | 0 | Low-quality | Genome-fragment | 6.15 | AAI-based (high-confidence) | 0 |
| S2-final-viral-combined_k141_207441\|\|full | 15540 | No | NA | 18 | 0 | 13 | Not-determined | Genome-fragment | NA | NA | 0 |
| S2-final-viral-combined_k141_31489\|\|full | 3275 | No | NA | 8 | 0 | 0 | Not-determined | Genome-fragment | NA | NA | 0 |
| S2-final-viral-combined_k141_42126\|\|full | 2744 | No | NA | 4 | 0 | 2 | Not-determined | Genome-fragment | NA | NA | 0 |
| S5-final-viral-combined_k141_183967\|\|full | 3592 | No | NA | 3 | 0 | 0 | Low-quality | Genome-fragment | 4.18 | AAI-based (medium-confidence) | 0 |
| S2-final-viral-combined_k141_114239\|\|full | 2296 | No | NA | 4 | 0 | 0 | Not-determined | Genome-fragment | NA | NA | 0 |
| S17-final-viral-combined_k141_201332\|\|full | 3592 | No | NA | 3 | 0 | 0 | Low-quality | Genome-fragment | 7.23 | AAI-based (high-confidence) | 0 |
| S4-final-viral-combined_k141_50377\|\|full | 3592 | No | NA | 2 | 0 | 2 | Low-quality | Genome-fragment | 2.69 | AAI-based (medium-confidence) | 0 |
| S7-final-viral-combined_k141_101630\|\|full | 3593 | No | NA | 3 | 0 | 2 | Low-quality | Genome-fragment | 1.12 | AAI-based (medium-confidence) | 0 |
| S2_k141_49153 | 3593 | No | NA | 6 | 1 | 0 | Low-quality | Genome-fragment | 17.64 | AAI-based (medium-confidence) | 0 |
| S2_k141_237698 | 3593 | Yes | 793 | 2 | 1 | 1 | Low-quality | Genome-fragment | 0.42 | HMM-based (lower-bound) | 77.93 |
| S1-final-viral-combined_k141_99312\|\|full | 3594 | No | NA | 6 | 2 | 0 | Low-quality | Genome-fragment | 5.96 | AAI-based (high-confidence) | 0 |
| S2-final-viral-combined_k141_93682\|\|full | 7839 | No | NA | 5 | 0 | 3 | Not-determined | Genome-fragment | NA | NA | 0 |
| S12-final-viral-combined_k141_231109\|\|full | 3594 | No | NA | 7 | 1 | 0 | Low-quality | Genome-fragment | 7.9 | AAI-based (high-confidence) | 0 |
| S2-final-viral-combined_k141_238687\|\|full | 1062 | No | NA | 2 | 0 | 0 | Not-determined | Genome-fragment | NA | NA | 0 |
| S4-final-viral-combined_k141_64557\|\|full | 3594 | Yes | 1743 | 5 | 1 | 1 | Low-quality | Genome-fragment | 2.07 | HMM-based (lower-bound) | 51.5 |
| S2-final-viral-combined_k141_470\|\|full | 4996 | No | NA | 5 | 0 | 3 | Not-determined | Genome-fragment | NA | NA | 0 |
| S4_k141_97457 | 3594 | No | NA | 5 | 0 | 0 | Low-quality | Genome-fragment | 7.3 | AAI-based (high-confidence) | 0 |
| S2-final-viral-combined_k141_72949\|\|full | 7957 | No | NA | 7 | 0 | 6 | Not-determined | Genome-fragment | NA | NA | 0 |
| S2-final-viral-combined_k141_135139\|\|full | 116446 | Yes | 34141 | 86 | 6 | 38 | Medium-quality | Genome-fragment | 61.89 | HMM-based (lower-bound) | 70.68 |
| S2-final-viral-combined_k141_186930\|\|full | 4141 | No | NA | 3 | 0 | 2 | Not-determined | Genome-fragment | NA | NA | 0 |
| S2-final-viral-combined_k141_72950\|\|full | 1038 | No | NA | 2 | 0 | 0 | Not-determined | Genome-fragment | NA | NA | 0 |
| S2-final-viral-combined_k141_114260\|\|full | 1982 | No | NA | 3 | 0 | 3 | Not-determined | Genome-fragment | NA | NA | 0 |
| S11-final-viral-combined_k141_194075\|\|full | 3595 | No | NA | 10 | 0 | 0 | Low-quality | Genome-fragment | 11.16 | AAI-based (high-confidence) | 0 |
| S1-final-viral-combined_k141_309110\|\|full | 3595 | No | NA | 3 | 0 | 1 | Low-quality | Genome-fragment | 7.81 | AAI-based (medium-confidence) | 0 |
| S2-final-viral-combined_k141_197015\|\|full | 10008 | No | NA | 10 | 0 | 5 | Not-determined | Genome-fragment | NA | NA | 0 |
| S2-final-viral-combined_k141_166098\|\|full | 1054 | No | NA | 2 | 0 | 0 | Not-determined | Genome-fragment | NA | NA | 0 |
| S2-final-viral-combined_k141_42138\|\|full | 8567 | No | NA | 14 | 0 | 0 | Not-determined | Genome-fragment | NA | NA | 0 |
| S3-final-viral-combined_k141_85621\|\|full | 3595 | No | NA | 5 | 0 | 1 | Low-quality | Genome-fragment | 1.95 | AAI-based (medium-confidence) | 0 |
| S2-final-viral-combined_k141_155834\|\|full | 2763 | No | NA | 3 | 0 | 0 | Not-determined | Genome-fragment | NA | NA | 0 |
| S2-final-viral-combined_k141_124775\|\|full | 3451 | No | NA | 2 | 0 | 1 | Not-determined | Genome-fragment | NA | NA | 0 |
| S2-final-viral-combined_k141_42156\|\|full | 1819 | No | NA | 3 | 0 | 2 | Not-determined | Genome-fragment | NA | NA | 0 |
| S2-final-viral-combined_k141_218002\|\|full | 76226 | Yes | 33528 | 89 | 17 | 30 | Medium-quality | Genome-fragment | 58.75 | HMM-based (lower-bound) | 56.02 |
| S5_k141_99176 | 3595 | No | NA | 8 | 0 | 0 | Low-quality | Genome-fragment | 8.4 | AAI-based (medium-confidence) | 0 |
| S2-final-viral-combined_k141_72991\|\|full | 2987 | No | NA | 3 | 0 | 2 | Not-determined | Genome-fragment | NA | NA | 0 |
| S2-final-viral-combined_k141_52213\|\|full | 1569 | No | NA | 3 | 0 | 2 | Not-determined | Genome-fragment | NA | NA | 0 |
| S2-final-viral-combined_k141_10778\|\|full | 5910 | No | NA | 8 | 0 | 1 | Not-determined | Genome-fragment | NA | NA | 0 |
| S9_k141_9891 | 3596 | No | NA | 5 | 0 | 2 | Low-quality | Genome-fragment | 6.01 | AAI-based (medium-confidence) | 0 |
| S1_k141_46305 | 3596 | No | NA | 5 | 1 | 0 | Low-quality | Genome-fragment | 5.48 | AAI-based (high-confidence) | 0 |
| S9-final-viral-combined_k141_118531\|\|full | 3597 | Yes | 2868 | 7 | 1 | 1 | Low-quality | Genome-fragment | 6.92 | AAI-based (high-confidence) | 20.27 |
| S2-final-viral-combined_k141_135154\|\|full | 66430 | Yes | 23131 | 91 | 12 | 47 | Medium-quality | Genome-fragment | 55.05 | AAI-based (high-confidence) | 65.18 |
| S2-final-viral-combined_k141_42204\|\|full | 1597 | No | NA | 2 | 0 | 0 | Not-determined | Genome-fragment | NA | NA | 0 |
| S2-final-viral-combined_k141_197043\|\|full | 1820 | No | NA | 3 | 0 | 1 | Not-determined | Genome-fragment | NA | NA | 0 |
| S2-final-viral-combined_k141_207584\|\|full | 2168 | No | NA | 3 | 0 | 1 | Not-determined | Genome-fragment | NA | NA | 0 |
| S2-final-viral-combined_k141_83324\|\|full | 1951 | No | NA | 3 | 0 | 0 | Not-determined | Genome-fragment | NA | NA | 0 |
| S2-final-viral-combined_k141_73061\|\|full | 2938 | No | NA | 4 | 0 | 2 | Not-determined | Genome-fragment | NA | NA | 0 |
| S2-final-viral-combined_k141_52266\|\|full | 1855 | No | NA | 2 | 0 | 1 | Not-determined | Genome-fragment | NA | NA | 0 |
| S3-final-viral-combined_k141_134168\|\|full | 3597 | No | NA | 4 | 0 | 0 | Low-quality | Genome-fragment | 2.84 | AAI-based (high-confidence) | 0 |
| S7-final-viral-combined_k141_206439\|\|full | 3597 | No | NA | 7 | 0 | 0 | Low-quality | Genome-fragment | 7.87 | AAI-based (high-confidence) | 0 |
| S2-final-viral-combined_k141_52284\|\|full | 1555 | No | NA | 2 | 0 | 1 | Not-determined | Genome-fragment | NA | NA | 0 |
| S2-final-viral-combined_k141_62615\|\|full | 1726 | No | NA | 3 | 0 | 1 | Not-determined | Genome-fragment | NA | NA | 0 |
| S11-final-viral-combined_k141_135994\|\|full | 3598 | No | NA | 2 | 0 | 1 | Low-quality | Genome-fragment | 9.66 | AAI-based (high-confidence) | 0 |
| S2-final-viral-combined_k141_124921\|\|full | 7446 | No | NA | 9 | 0 | 3 | Not-determined | Genome-fragment | NA | NA | 0 |
| S2-final-viral-combined_k141_42307\|\|full | 1390 | No | NA | 2 | 0 | 2 | Not-determined | Genome-fragment | NA | NA | 0 |
| S2-final-viral-combined_k141_572\|\|full | 1750 | No | NA | 3 | 0 | 2 | Not-determined | Genome-fragment | NA | NA | 0 |
| S11_k141_102396 | 3598 | No | NA | 4 | 2 | 0 | Low-quality | Genome-fragment | 7.75 | AAI-based (high-confidence) | 0 |
| S2-final-viral-combined_k141_31693\|\|full | 2787 | No | NA | 3 | 0 | 2 | Not-determined | Genome-fragment | NA | NA | 0 |
| S2-final-viral-combined_k141_21267\|\|full | 1007 | No | NA | 2 | 0 | 1 | Not-determined | Genome-fragment | NA | NA | 0 |
| S2-final-viral-combined_k141_155997\|\|full | 2376 | No | NA | 4 | 0 | 0 | Not-determined | Genome-fragment | NA | NA | 0 |
| S2-final-viral-combined_k141_73132\|\|full | 1941 | No | NA | 3 | 0 | 1 | Not-determined | Genome-fragment | NA | NA | 0 |
| S2-final-viral-combined_k141_176676\|\|full | 1366 | No | NA | 3 | 0 | 0 | Not-determined | Genome-fragment | NA | NA | 0 |
| S2-final-viral-combined_k141_166175\|\|full | 1641 | No | NA | 3 | 0 | 0 | Not-determined | Genome-fragment | NA | NA | 0 |
| S2-final-viral-combined_k141_42359\|\|full | 8767 | No | NA | 7 | 0 | 7 | Not-determined | Genome-fragment | NA | NA | 0 |
| S2-final-viral-combined_k141_156015\|\|full | 2760 | No | NA | 4 | 0 | 1 | Not-determined | Genome-fragment | NA | NA | 0 |
| S3_k141_124445 | 3598 | No | NA | 2 | 0 | 2 | Low-quality | Genome-fragment | 2.06 | AAI-based (medium-confidence) | 0 |
| S4-final-viral-combined_k141_18363\|\|full | 3598 | No | NA | 7 | 2 | 0 | Low-quality | Genome-fragment | 6.38 | HMM-based (lower-bound) | 0 |
| S2-final-viral-combined_k141_156017\|\|full | 2084 | No | NA | 3 | 0 | 3 | Not-determined | Genome-fragment | NA | NA | 0 |
| S8-final-viral-combined_k141_33138\|\|full | 3598 | No | NA | 3 | 3 | 0 | Low-quality | Genome-fragment | 8.97 | AAI-based (high-confidence) | 0 |
| S17-k141_127772 | 3599 | No | NA | 4 | 0 | 0 | Low-quality | Genome-fragment | 5.76 | AAI-based (medium-confidence) | 0 |
| S2-final-viral-combined_k141_187086\|\|full | 3421 | No | NA | 2 | 0 | 1 | Not-determined | Genome-fragment | NA | NA | 0 |
| S2-final-viral-combined_k141_135228\|\|full | 4064 | No | NA | 6 | 0 | 4 | Not-determined | Genome-fragment | NA | NA | 0 |
| S12-final-viral-combined_k141_34777\|\|full | 3599 | No | NA | 6 | 2 | 0 | Low-quality | Genome-fragment | 6.99 | AAI-based (high-confidence) | 0 |
| S11_k141_287145 | 3600 | No | NA | 8 | 2 | 0 | Low-quality | Genome-fragment | 8.33 | AAI-based (high-confidence) | 0 |
| S2-final-viral-combined_k141_104192\|\|full | 6659 | No | NA | 9 | 0 | 4 | Not-determined | Genome-fragment | NA | NA | 0 |
| S2-final-viral-combined_k141_207696\|\|full | 6676 | No | NA | 7 | 0 | 2 | Not-determined | Genome-fragment | NA | NA | 0 |
| S7-final-viral-combined_k141_18453\|\|full | 3600 | No | NA | 4 | 3 | 0 | Low-quality | Genome-fragment | 7.1 | AAI-based (high-confidence) | 0 |
| S2-final-viral-combined_k141_228451\|\|full | 6118 | No | NA | 4 | 0 | 3 | Not-determined | Genome-fragment | NA | NA | 0 |
| S2-final-viral-combined_k141_166234\|\|full | 3130 | No | NA | 6 | 0 | 1 | Not-determined | Genome-fragment | NA | NA | 0 |
| S2-final-viral-combined_k141_135284\|\|full | 1307 | No | NA | 2 | 0 | 1 | Not-determined | Genome-fragment | NA | NA | 0 |
| S2-final-viral-combined_k141_10849\|\|full | 1845 | No | NA | 2 | 0 | 1 | Not-determined | Genome-fragment | NA | NA | 0 |
| S12_k141_384605 | 3600 | No | NA | 3 | 0 | 3 | Low-quality | Genome-fragment | 2.92 | AAI-based (high-confidence) | 0 |
| S1-final-viral-combined_k141_117161\|\|full | 3601 | No | NA | 3 | 0 | 0 | Low-quality | Genome-fragment | 5.48 | AAI-based (medium-confidence) | 0 |
| S2-final-viral-combined_k141_238929\|\|full | 42118 | No | NA | 52 | 19 | 1 | High-quality | High-quality | 92.62 | AAI-based (medium-confidence) | 0 |
| S2-final-viral-combined_k141_176760\|\|full | 2997 | No | NA | 3 | 0 | 0 | Not-determined | Genome-fragment | NA | NA | 0 |
| S2-final-viral-combined_k141_73186\|\|full | 1653 | No | NA | 2 | 0 | 2 | Not-determined | Genome-fragment | NA | NA | 0 |
| S2-final-viral-combined_k141_93800\|\|full | 2087 | No | NA | 3 | 0 | 0 | Not-determined | Genome-fragment | NA | NA | 0 |
| S2-final-viral-combined_k141_197149\|\|full | 1046 | No | NA | 2 | 0 | 1 | Not-determined | Genome-fragment | NA | NA | 0 |
| S1-final-viral-combined_k141_171494\|\|full | 3601 | No | NA | 4 | 1 | 2 | Low-quality | Genome-fragment | 3.87 | AAI-based (medium-confidence) | 0 |
| S2-final-viral-combined_k141_154318\|\|full | 3602 | No | NA | 2 | 1 | 0 | Low-quality | Genome-fragment | 4.2 | AAI-based (high-confidence) | 0 |
| S2-final-viral-combined_k141_112572\|\|full | 3602 | No | NA | 8 | 1 | 0 | Low-quality | Genome-fragment | 8.59 | AAI-based (high-confidence) | 0 |
| S8-final-viral-combined_k141_45812\|\|full | 3602 | No | NA | 4 | 0 | 2 | Low-quality | Genome-fragment | 6.65 | AAI-based (medium-confidence) | 0 |
| S2-final-viral-combined_k141_705\|\|full | 1941 | No | NA | 4 | 0 | 3 | Not-determined | Genome-fragment | NA | NA | 0 |
| S11-final-viral-combined_k141_291762\|\|full | 3603 | No | NA | 3 | 1 | 0 | Low-quality | Genome-fragment | 9.17 | AAI-based (medium-confidence) | 0 |
| S5-final-viral-combined_k141_171127\|\|full | 3603 | Yes | 1346 | 6 | 1 | 2 | Low-quality | Genome-fragment | 1.93 | AAI-based (medium-confidence) | 62.64 |
| S2-final-viral-combined_k141_83423\|\|full | 6155 | No | NA | 6 | 0 | 4 | Not-determined | Genome-fragment | NA | NA | 0 |
| S2-final-viral-combined_k141_165546\|\|full | 3603 | No | NA | 6 | 0 | 1 | Low-quality | Genome-fragment | 1.53 | AAI-based (medium-confidence) | 0 |
| S2-final-viral-combined_k141_83433\|\|full | 4080 | No | NA | 4 | 0 | 2 | Not-determined | Genome-fragment | NA | NA | 0 |
| S6-final-viral-combined_k141_130668\|\|full | 3603 | No | NA | 3 | 0 | 2 | Low-quality | Genome-fragment | 4.26 | AAI-based (medium-confidence) | 0 |
| S11-final-viral-combined_k141_83882\|\|full | 3604 | No | NA | 3 | 3 | 0 | Low-quality | Genome-fragment | 7.65 | AAI-based (high-confidence) | 0 |
| S2-final-viral-combined_k141_31803\|\|full | 10268 | No | NA | 9 | 0 | 7 | Not-determined | Genome-fragment | NA | NA | 0 |
| S2-final-viral-combined_k141_166358\|\|full | 1151 | No | NA | 2 | 0 | 1 | Not-determined | Genome-fragment | NA | NA | 0 |
| S2-final-viral-combined_k141_228587\|\|full | 1703 | No | NA | 2 | 0 | 1 | Not-determined | Genome-fragment | NA | NA | 0 |
| S11-final-viral-combined_k141_129352\|\|full | 3604 | No | NA | 6 | 2 | 0 | Low-quality | Genome-fragment | 8.31 | AAI-based (high-confidence) | 0 |
| S2-final-viral-combined_k141_104277\|\|full | 1691 | No | NA | 4 | 0 | 3 | Not-determined | Genome-fragment | NA | NA | 0 |
| S11-final-viral-combined_k141_161046\|\|full | 3604 | No | NA | 2 | 0 | 1 | Low-quality | Genome-fragment | 1.68 | AAI-based (medium-confidence) | 0 |
| S2-final-viral-combined_k141_114351\|\|full | 1929 | No | NA | 3 | 0 | 2 | Not-determined | Genome-fragment | NA | NA | 0 |
| S1-final-viral-combined_k141_389588\|\|full | 3604 | No | NA | 3 | 3 | 0 | Low-quality | Genome-fragment | 10.27 | AAI-based (high-confidence) | 0 |
| S2-final-viral-combined_k141_42586\|\|full | 3335 | No | NA | 2 | 0 | 1 | Not-determined | Genome-fragment | NA | NA | 0 |
| S2-final-viral-combined_k141_114372\|\|full | 2015 | No | NA | 2 | 0 | 1 | Not-determined | Genome-fragment | NA | NA | 0 |
| S1-final-viral-combined_k141_159083\|\|full | 3606 | No | NA | 4 | 1 | 0 | Low-quality | Genome-fragment | 6.69 | AAI-based (high-confidence) | 0 |
| S2-final-viral-combined_k141_11070\|\|full | 2967 | No | NA | 4 | 0 | 1 | Not-determined | Genome-fragment | NA | NA | 0 |
| S2-final-viral-combined_k141_187180\|\|full | 1278 | No | NA | 2 | 0 | 0 | Not-determined | Genome-fragment | NA | NA | 0 |
| S2-final-viral-combined_k141_176886\|\|full | 2961 | No | NA | 3 | 0 | 2 | Not-determined | Genome-fragment | NA | NA | 0 |
| S11-final-viral-combined_k141_147905\|\|full | 3607 | No | NA | 5 | 0 | 3 | Low-quality | Genome-fragment | 1.69 | AAI-based (medium-confidence) | 0 |
| S2-final-viral-combined_k141_166415\|\|full | 1789 | No | NA | 3 | 0 | 0 | Not-determined | Genome-fragment | NA | NA | 0 |
| S11-final-viral-combined_k141_283223\|\|full | 3608 | No | NA | 7 | 3 | 0 | Low-quality | Genome-fragment | 9.25 | AAI-based (high-confidence) | 0 |
| S2-final-viral-combined_k141_208405\|\|full | 3608 | No | NA | 4 | 0 | 1 | Low-quality | Genome-fragment | 6.94 | AAI-based (medium-confidence) | 0 |
| S2-final-viral-combined_k141_104308\|\|full | 2020 | No | NA | 3 | 0 | 1 | Not-determined | Genome-fragment | NA | NA | 0 |
| S2-final-viral-combined_k141_187211\|\|full | 1710 | No | NA | 2 | 0 | 0 | Not-determined | Genome-fragment | NA | NA | 0 |
| S2-final-viral-combined_k141_228710\|\|full | 2105 | No | NA | 6 | 0 | 0 | Not-determined | Genome-fragment | NA | NA | 0 |
| S8-final-viral-combined_k141_166286\|\|full | 3608 | No | NA | 7 | 0 | 0 | Low-quality | Genome-fragment | 6.12 | AAI-based (high-confidence) | 0 |
| S2-final-viral-combined_k141_73231\|\|full | 4139 | No | NA | 8 | 0 | 2 | Not-determined | Genome-fragment | NA | NA | 0 |
| S2-final-viral-combined_k141_145813\|\|full | 2768 | No | NA | 3 | 0 | 0 | Not-determined | Genome-fragment | NA | NA | 0 |
| S1-final-viral-combined_k141_2113\|\|full | 3610 | No | NA | 3 | 0 | 2 | Low-quality | Genome-fragment | 3.16 | AAI-based (medium-confidence) | 0 |
| S2-final-viral-combined_k141_197236\|\|full | 1749 | No | NA | 3 | 0 | 1 | Not-determined | Genome-fragment | NA | NA | 0 |
| S8_k141_76908 | 3610 | No | NA | 3 | 0 | 2 | Low-quality | Genome-fragment | 6.24 | AAI-based (high-confidence) | 0 |

| | | | | | | | | | | | |
|---|---|---|---|---|---|---|---|---|---|---|---|
| S8_k141_125374 | 3610 | No | NA | 2 | 0 | 2 | Low-quality | Genome-fragment | 1.94 | AAI-based (medium-confidence) | 0 |
| S1-final-viral-combined_k141_169950\|\|full | 3611 | No | NA | 4 | 0 | 1 | Low-quality | Genome-fragment | 2.18 | AAI-based (high-confidence) | 0 |
| S2-final-viral-combined_k141_114448\|\|full | 2795 | No | NA | 3 | 0 | 1 | Not-determined | Genome-fragment | NA | NA | 0 |
| S2-final-viral-combined_k141_156134\|\|full | 1376 | No | NA | 2 | 0 | 2 | Not-determined | Genome-fragment | NA | NA | 0 |
| S4-final-viral-combined_k141_149457\|\|full | 3611 | No | NA | 5 | 0 | 3 | Low-quality | Genome-fragment | 8.69 | AAI-based (medium-confidence) | 0 |
| S2-final-viral-combined_k141_114465\|\|full | 1933 | No | NA | 4 | 0 | 1 | Not-determined | Genome-fragment | NA | NA | 0 |
| S8-final-viral-combined_k141_10276\|\|full | 3611 | No | NA | 4 | 0 | 1 | Low-quality | Genome-fragment | 3.78 | AAI-based (medium-confidence) | 0 |
| S2-final-viral-combined_k141_246159\|\|full | 3612 | No | NA | 4 | 0 | 0 | Low-quality | Genome-fragment | 2.57 | AAI-based (high-confidence) | 0 |
| S1-final-viral-combined_k141_314889\|\|full | 3613 | No | NA | 3 | 0 | 2 | Low-quality | Genome-fragment | 2.31 | AAI-based (medium-confidence) | 0 |
| S2-final-viral-combined_k141_93845\|\|full | 56715 | Yes | 29207 | 64 | 16 | 18 | Medium-quality | Genome-fragment | 56.12 | AAI-based (medium-confidence) | 48.5 |
| S7-final-viral-combined_k141_227826\|\|full | 3613 | No | NA | 4 | 0 | 2 | Low-quality | Genome-fragment | 2.6 | AAI-based (medium-confidence) | 0 |
| S2-final-viral-combined_k141_52473\|\|full | 1785 | No | NA | 3 | 0 | 1 | Not-determined | Genome-fragment | NA | NA | 0 |
| S12-final-viral-combined_k141_87171\|\|full | 3613 | No | NA | 3 | 0 | 0 | Low-quality | Genome-fragment | 2.76 | AAI-based (medium-confidence) | 0 |
| S2-final-viral-combined_k141_218169\|\|full | 2161 | No | NA | 4 | 0 | 2 | Not-determined | Genome-fragment | NA | NA | 0 |
| S2-final-viral-combined_k141_104355\|\|full | 2860 | No | NA | 3 | 0 | 0 | Not-determined | Genome-fragment | NA | NA | 0 |
| S2-final-viral-combined_k141_73300\|\|full | 1630 | No | NA | 3 | 0 | 1 | Not-determined | Genome-fragment | NA | NA | 0 |
| S2-final-viral-combined_k141_62727\|\|full | 1737 | No | NA | 2 | 0 | 1 | Not-determined | Genome-fragment | NA | NA | 0 |
| S4-final-viral-combined_k141_33225\|\|full | 3613 | No | NA | 7 | 3 | 0 | Low-quality | Genome-fragment | 8.41 | AAI-based (high-confidence) | 0 |
| S2-final-viral-combined_k141_218200\|\|full | 4848 | No | NA | 2 | 0 | 2 | Not-determined | Genome-fragment | NA | NA | 0 |
| S2-final-viral-combined_k141_176970\|\|full | 1260 | No | NA | 3 | 0 | 2 | Not-determined | Genome-fragment | NA | NA | 0 |
| S11-final-viral-combined_k141_167524\|\|full | 3614 | No | NA | 3 | 0 | 2 | Low-quality | Genome-fragment | 5.79 | AAI-based (high-confidence) | 0 |
| S2-final-viral-combined_k141_145913\|\|full | 1495 | No | NA | 4 | 0 | 1 | Not-determined | Genome-fragment | NA | NA | 0 |
| S3_k141_44345 | 3614 | No | NA | 1 | 0 | 0 | Low-quality | Genome-fragment | 3.67 | AAI-based (medium-confidence) | 0 |
| S2-final-viral-combined_k141_83595\|\|full | 2830 | No | NA | 3 | 0 | 3 | Not-determined | Genome-fragment | NA | NA | 0 |
| S2-final-viral-combined_k141_166507\|\|full | 9213 | No | NA | 12 | 0 | 5 | Not-determined | Genome-fragment | NA | NA | 0 |
| S2-final-viral-combined_k141_218221\|\|full | 2474 | No | NA | 2 | 0 | 1 | Not-determined | Genome-fragment | NA | NA | 0 |
| S2-final-viral-combined_k141_104387\|\|full | 2319 | No | NA | 3 | 0 | 1 | Not-determined | Genome-fragment | NA | NA | 0 |
| S17-k141_2151 | 3614 | No | NA | 4 | 0 | 2 | Low-quality | Genome-fragment | 1.83 | AAI-based (medium-confidence) | 0 |
| S4_k141_59819 | 3614 | No | NA | 3 | 0 | 0 | Low-quality | Genome-fragment | 8.03 | AAI-based (medium-confidence) | 0 |
| S2-final-viral-combined_k141_135364\|\|full | 4185 | No | NA | 3 | 0 | 1 | Not-determined | Genome-fragment | NA | NA | 0 |
| S2-final-viral-combined_k141_11146\|\|full | 4329 | No | NA | 5 | 0 | 4 | Not-determined | Genome-fragment | NA | NA | 0 |
| S5-final-viral-combined_k141_197338\|\|full | 3615 | No | NA | 3 | 0 | 2 | Low-quality | Genome-fragment | 5.78 | AAI-based (medium-confidence) | 0 |
| S2-final-viral-combined_k141_228847\|\|full | 1355 | No | NA | 2 | 0 | 1 | Not-determined | Genome-fragment | NA | NA | 0 |
| S17-final-viral-combined_k141_194274\|\|full | 3615 | No | NA | 3 | 3 | 0 | Low-quality | Genome-fragment | 8.57 | AAI-based (high-confidence) | 0 |
| S17-final-viral-combined_k141_95173\|\|full | 3615 | No | NA | 7 | 1 | 0 | Low-quality | Genome-fragment | 5.2 | AAI-based (high-confidence) | 0 |
| S2-final-viral-combined_k141_228871\|\|full | 1324 | No | NA | 3 | 0 | 2 | Not-determined | Genome-fragment | NA | NA | 0 |
| S2-final-viral-combined_k141_125155\|\|full | 5689 | No | NA | 6 | 0 | 4 | Not-determined | Genome-fragment | NA | NA | 0 |
| S2-final-viral-combined_k141_11168\|\|full | 1633 | No | NA | 2 | 0 | 1 | Not-determined | Genome-fragment | NA | NA | 0 |
| S2-final-viral-combined_k141_135386\|\|full | 8152 | No | NA | 6 | 0 | 5 | Not-determined | Genome-fragment | NA | NA | 0 |
| S6-final-viral-combined_k141_64398\|\|full | 3615 | No | NA | 2 | 0 | 1 | Low-quality | Genome-fragment | 1.82 | AAI-based (medium-confidence) | 0 |
| S11_k141_179762 | 3616 | No | NA | 2 | 0 | 1 | Low-quality | Genome-fragment | 1.83 | AAI-based (medium-confidence) | 0 |
| S2-final-viral-combined_k141_73327\|\|full | 9812 | No | NA | 13 | 1 | 4 | High-quality | High-quality | 100 | HMM-based (lower-bound) | 0 |
| S2-final-viral-combined_k141_62783\|\|full | 5462 | No | NA | 7 | 0 | 1 | Not-determined | Genome-fragment | NA | NA | 0 |
| S17-final-viral-combined_k141_258315\|\|full | 3616 | No | NA | 4 | 2 | 0 | Low-quality | Genome-fragment | 8.68 | AAI-based (high-confidence) | 0 |
| S2-final-viral-combined_k141_166564\|\|full | 1146 | No | NA | 3 | 0 | 0 | Not-determined | Genome-fragment | NA | NA | 0 |
| S2-final-viral-combined_k141_145936\|\|full | 2105 | No | NA | 4 | 0 | 0 | Not-determined | Genome-fragment | NA | NA | 0 |
| S2-final-viral-combined_k141_104459\|\|full | 1242 | No | NA | 2 | 0 | 1 | Not-determined | Genome-fragment | NA | NA | 0 |
| S2-final-viral-combined_k141_73345\|\|full | 3577 | No | NA | 2 | 0 | 2 | Not-determined | Genome-fragment | NA | NA | 0 |
| S8-final-viral-combined_k141_247043\|\|full | 3616 | Yes | 3096 | 5 | 2 | 1 | Low-quality | Genome-fragment | 4.31 | HMM-based (lower-bound) | 14.28 |
| S2-final-viral-combined_k141_11204\|\|full | 1096 | No | NA | 2 | 0 | 1 | Not-determined | Genome-fragment | NA | NA | 0 |
| S2-final-viral-combined_k141_228916\|\|full | 3815 | No | NA | 5 | 0 | 2 | Not-determined | Genome-fragment | NA | NA | 0 |
| S2-final-viral-combined_k141_228917\|\|full | 1268 | No | NA | 3 | 0 | 0 | Not-determined | Genome-fragment | NA | NA | 0 |
| S8-final-viral-combined_k141_87847\|\|full | 3616 | No | NA | 4 | 0 | 0 | Low-quality | Genome-fragment | 5.84 | AAI-based (medium-confidence) | 0 |
| S2-final-viral-combined_k141_42769\|\|full | 2702 | No | NA | 4 | 0 | 0 | Not-determined | Genome-fragment | NA | NA | 0 |
| S2-final-viral-combined_k141_83696\|\|full | 1520 | No | NA | 2 | 0 | 1 | Not-determined | Genome-fragment | NA | NA | 0 |
| S2-final-viral-combined_k141_83698\|\|full | 1412 | No | NA | 3 | 0 | 0 | Not-determined | Genome-fragment | NA | NA | 0 |
| S2-final-viral-combined_k141_145952\|\|full | 1631 | No | NA | 2 | 0 | 1 | Not-determined | Genome-fragment | NA | NA | 0 |
| S2-final-viral-combined_k141_238956\|\|full | 1199 | No | NA | 3 | 0 | 0 | Not-determined | Genome-fragment | NA | NA | 0 |
| S2-final-viral-combined_k141_145960\|\|full | 1099 | No | NA | 2 | 0 | 0 | Not-determined | Genome-fragment | NA | NA | 0 |
| S2-final-viral-combined_k141_83705\|\|full | 8622 | No | NA | 12 | 0 | 6 | Not-determined | Genome-fragment | NA | NA | 0 |
| S8_k141_176093 | 3617 | No | NA | 4 | 0 | 2 | Low-quality | Genome-fragment | 0.98 | AAI-based (high-confidence) | 0 |
| S2-final-viral-combined_k141_52568\|\|full | 49409 | Yes | 36225 | 63 | 8 | 10 | Complete | High-quality | 100 | Provirus (medium-confidence) | 26.68 |
| S2-final-viral-combined_k141_114609\|\|full | 6196 | No | NA | 3 | 0 | 1 | Not-determined | Genome-fragment | NA | NA | 0 |
| S7-final-viral-combined_k141_141573\|\|full | 3618 | No | NA | 2 | 0 | 1 | Low-quality | Genome-fragment | 1.45 | AAI-based (medium-confidence) | 0 |
| S1_k141_13812 | 3619 | No | NA | 2 | 0 | 0 | Low-quality | Genome-fragment | 4.18 | AAI-based (medium-confidence) | 0 |
| S5-final-viral-combined_k141_100048\|\|full | 3619 | No | NA | 2 | 0 | 2 | Low-quality | Genome-fragment | 8.08 | AAI-based (medium-confidence) | 0 |
| S2-final-viral-combined_k141_125228\|\|full | 4171 | No | NA | 5 | 0 | 3 | Not-determined | Genome-fragment | NA | NA | 0 |
| S2-final-viral-combined_k141_238981\|\|full | 2099 | No | NA | 3 | 0 | 3 | Not-determined | Genome-fragment | NA | NA | 0 |
| S11-final-viral-combined_k141_266568\|\|full | 3620 | No | NA | 6 | 0 | 1 | Low-quality | Genome-fragment | 4.4 | AAI-based (high-confidence) | 0 |
| S11_k141_233787 | 3620 | No | NA | 6 | 2 | 0 | Low-quality | Genome-fragment | 7.7 | AAI-based (high-confidence) | 0 |
| S2-final-viral-combined_k141_228940\|\|full | 1133 | No | NA | 3 | 0 | 0 | Not-determined | Genome-fragment | NA | NA | 0 |
| S1-final-viral-combined_k141_55781\|\|full | 3620 | No | NA | 3 | 3 | 0 | Low-quality | Genome-fragment | 8.73 | AAI-based (medium-confidence) | 0 |
| S4_k141_7728 | 3620 | No | NA | 7 | 2 | 0 | Low-quality | Genome-fragment | 2.09 | AAI-based (high-confidence) | 0 |
| S2-final-viral-combined_k141_166595\|\|full | 1643 | No | NA | 2 | 0 | 2 | Not-determined | Genome-fragment | NA | NA | 0 |
| S2-final-viral-combined_k141_62824\|\|full | 2942 | No | NA | 4 | 0 | 3 | Not-determined | Genome-fragment | NA | NA | 0 |
| S2-final-viral-combined_k141_239023\|\|full | 1618 | No | NA | 2 | 0 | 0 | Not-determined | Genome-fragment | NA | NA | 0 |
| S1-final-viral-combined_k141_332944\|\|full | 3621 | No | NA | 3 | 0 | 2 | Low-quality | Genome-fragment | 2.69 | AAI-based (medium-confidence) | 0 |
| S2-final-viral-combined_k141_197350\|\|full | 1892 | No | NA | 3 | 0 | 0 | Not-determined | Genome-fragment | NA | NA | 0 |
| S17-final-viral-combined_k141_9363\|\|full | 3621 | No | NA | 4 | 0 | 0 | Low-quality | Genome-fragment | 3.63 | AAI-based (medium-confidence) | 0 |
| S11-final-viral-combined_k141_211141\|\|full | 3622 | Yes | 1421 | 5 | 1 | 1 | Low-quality | Genome-fragment | 34.15 | HMM-based (lower-bound) | 60.77 |
| S1-final-viral-combined_k141_102816\|\|full | 3623 | No | NA | 2 | 0 | 2 | Low-quality | Genome-fragment | 7.54 | AAI-based (medium-confidence) | 0 |
| S2-final-viral-combined_k141_125294\|\|full | 1055 | No | NA | 3 | 0 | 1 | Not-determined | Genome-fragment | NA | NA | 0 |
| S2-final-viral-combined_k141_218355\|\|full | 1054 | No | NA | 2 | 0 | 0 | Not-determined | Genome-fragment | NA | NA | 0 |
| S12_k141_279257 | 3625 | No | NA | 4 | 0 | 1 | Low-quality | Genome-fragment | 1.78 | AAI-based (medium-confidence) | 0 |
| S2-final-viral-combined_k141_52646\|\|full | 3705 | No | NA | 4 | 0 | 3 | Not-determined | Genome-fragment | NA | NA | 0 |
| S2-final-viral-combined_k141_32027\|\|full | 8289 | No | NA | 16 | 0 | 3 | Not-determined | Genome-fragment | NA | NA | 0 |
| S2-final-viral-combined_k141_208008\|\|full | 3479 | No | NA | 3 | 0 | 1 | Not-determined | Genome-fragment | NA | NA | 0 |
| S1-final-viral-combined_k141_238726\|\|full | 3626 | No | NA | 5 | 0 | 0 | Low-quality | Genome-fragment | 8.02 | AAI-based (medium-confidence) | 0 |
| S5-final-viral-combined_k141_141914\|\|full | 3626 | No | NA | 4 | 1 | 0 | Low-quality | Genome-fragment | 5.3 | HMM-based (lower-bound) | 0 |
| S12-final-viral-combined_k141_331090\|\|full | 3626 | Yes | 1752 | 6 | 1 | 3 | Low-quality | Genome-fragment | 1.57 | AAI-based (medium-confidence) | 51.68 |
| S4_k141_101708 | 3626 | No | NA | 3 | 0 | 2 | Low-quality | Genome-fragment | 2.07 | AAI-based (medium-confidence) | 0 |
| S2-final-viral-combined_k141_73404\|\|full | 1018 | No | NA | 3 | 0 | 0 | Not-determined | Genome-fragment | NA | NA | 0 |
| S2-final-viral-combined_k141_73410\|\|full | 2135 | No | NA | 3 | 0 | 1 | Not-determined | Genome-fragment | NA | NA | 0 |
| S9_k141_124043 | 3627 | No | NA | 4 | 0 | 1 | Low-quality | Genome-fragment | 4.5 | AAI-based (medium-confidence) | 0 |
| S12_k141_42404 | 3627 | No | NA | 2 | 0 | 1 | Low-quality | Genome-fragment | 7.7 | AAI-based (medium-confidence) | 0 |
| S2-final-viral-combined_k141_166665\|\|full | 46255 | Yes | 30719 | 63 | 18 | 6 | Medium-quality | Genome-fragment | 89.88 | AAI-based (medium-confidence) | 33.59 |
| S4-final-viral-combined_k141_78806\|\|full | 3627 | No | NA | 5 | 0 | 2 | Low-quality | Genome-fragment | 8.5 | AAI-based (high-confidence) | 0 |
| S1-final-viral-combined_k141_101310\|\|full | 3628 | No | NA | 4 | 0 | 0 | Low-quality | Genome-fragment | 3.28 | AAI-based (medium-confidence) | 0 |
| S17-final-viral-combined_k141_221545\|\|full | 3628 | No | NA | 7 | 0 | 1 | Low-quality | Genome-fragment | 4.17 | AAI-based (high-confidence) | 0 |
| S12-final-viral-combined_k141_117063\|\|full | 3628 | No | NA | 5 | 3 | 0 | Low-quality | Genome-fragment | 7.27 | AAI-based (medium-confidence) | 0 |
| S1-final-viral-combined_k141_240744\|\|full | 3629 | No | NA | 2 | 0 | 2 | Low-quality | Genome-fragment | 0.87 | AAI-based (medium-confidence) | 0 |
| S2-final-viral-combined_k141_11372\|\|full | 1398 | No | NA | 1 | 0 | 1 | Not-determined | Genome-fragment | NA | NA | 0 |
| S12_k141_205431 | 3629 | No | NA | 3 | 0 | 3 | Low-quality | Genome-fragment | 1.93 | AAI-based (medium-confidence) | 0 |
| S9-final-viral-combined_k141_94597\|\|full | 3630 | No | NA | 3 | 0 | 2 | Low-quality | Genome-fragment | 3.54 | AAI-based (medium-confidence) | 0 |
| S2-final-viral-combined_k141_1005\|\|full | 1696 | No | NA | 2 | 0 | 2 | Not-determined | Genome-fragment | NA | NA | 0 |
| S2-final-viral-combined_k141_62839\|\|full | 145503 | No | NA | 190 | 36 | 5 | High-quality | High-quality | 100 | AAI-based (high-confidence) | 0 |
| S2-final-viral-combined_k141_1018\|\|full | 1033 | No | NA | 3 | 0 | 0 | Not-determined | Genome-fragment | NA | NA | 0 |
| S2-final-viral-combined_k141_166703\|\|full | 2481 | No | NA | 2 | 0 | 1 | Not-determined | Genome-fragment | NA | NA | 0 |
| S9_k141_55538 | 3630 | No | NA | 5 | 0 | 0 | Low-quality | Genome-fragment | 10.61 | AAI-based (high-confidence) | 0 |
| S11-final-viral-combined_k141_99553\|\|full | 3630 | No | NA | 3 | 0 | 3 | Low-quality | Genome-fragment | 2.45 | AAI-based (medium-confidence) | 0 |
| S11-final-viral-combined_k141_127130\|\|full | 3630 | No | NA | 9 | 0 | 0 | Low-quality | Genome-fragment | 8.48 | AAI-based (high-confidence) | 0 |
| S7-final-viral-combined_k141_12233\|\|full | 3630 | No | NA | 6 | 0 | 0 | Low-quality | Genome-fragment | 8.65 | AAI-based (high-confidence) | 0 |
| S12-final-viral-combined_k141_187469\|\|full | 3630 | No | NA | 5 | 0 | 3 | Low-quality | Genome-fragment | 2.44 | AAI-based (high-confidence) | 0 |
| S2-final-viral-combined_k141_156420\|\|full | 11985 | No | NA | 11 | 0 | 2 | Not-determined | Genome-fragment | NA | NA | 0 |
| S11-final-viral-combined_k141_182618\|\|full | 3631 | No | NA | 6 | 1 | 0 | Low-quality | Genome-fragment | 5.98 | AAI-based (medium-confidence) | 0 |
| S2-final-viral-combined_k141_73542\|\|full | 5109 | No | NA | 4 | 0 | 1 | Not-determined | Genome-fragment | NA | NA | 0 |
| S2-final-viral-combined_k141_146104\|\|full | 14197 | No | NA | 13 | 0 | 2 | Not-determined | Genome-fragment | NA | NA | 0 |
| S2-final-viral-combined_k141_229085\|\|full | 7986 | No | NA | 9 | 0 | 6 | Not-determined | Genome-fragment | NA | NA | 0 |
| S2-final-viral-combined_k141_11465\|\|full | 4732 | No | NA | 5 | 0 | 4 | Not-determined | Genome-fragment | NA | NA | 0 |
| S2-final-viral-combined_k141_239083\|\|full | 5896 | No | NA | 5 | 0 | 3 | Not-determined | Genome-fragment | NA | NA | 0 |
| S1-final-viral-combined_k141_8560\|\|full | 3631 | No | NA | 3 | 0 | 0 | Low-quality | Genome-fragment | 6.99 | AAI-based (medium-confidence) | 0 |
| S2-final-viral-combined_k141_83918\|\|full | 2781 | No | NA | 3 | 0 | 2 | Not-determined | Genome-fragment | NA | NA | 0 |
| S2-final-viral-combined_k141_146139\|\|full | 1750 | No | NA | 2 | 0 | 0 | Not-determined | Genome-fragment | NA | NA | 0 |
| S2-final-viral-combined_k141_52691\|\|full | 2189 | No | NA | 2 | 0 | 0 | Not-determined | Genome-fragment | NA | NA | 0 |
| S2-final-viral-combined_k141_43000\|\|full | 1225 | No | NA | 3 | 0 | 1 | Not-determined | Genome-fragment | NA | NA | 0 |
| S8_k141_120244 | 3631 | No | NA | 4 | 0 | 0 | Low-quality | Genome-fragment | 5.21 | AAI-based (medium-confidence) | 0 |
| S11_k141_139484 | 3632 | No | NA | 5 | 1 | 0 | Low-quality | Genome-fragment | 30.41 | AAI-based (high-confidence) | 0 |
| S2-final-viral-combined_k141_229099\|\|full | 29644 | No | NA | 23 | 0 | 13 | Not-determined | Genome-fragment | NA | NA | 0 |
| S2-final-viral-combined_k141_229101\|\|full | 6429 | No | NA | 5 | 0 | 4 | Not-determined | Genome-fragment | NA | NA | 0 |
| S2-final-viral-combined_k141_114846\|\|full | 2664 | No | NA | 5 | 0 | 2 | Not-determined | Genome-fragment | NA | NA | 0 |
| S1-final-viral-combined_k141_314917\|\|full | 3632 | No | NA | 3 | 0 | 3 | Low-quality | Genome-fragment | 1.77 | AAI-based (medium-confidence) | 0 |
| S2-final-viral-combined_k141_62908\|\|full | 2668 | No | NA | 3 | 0 | 2 | Not-determined | Genome-fragment | NA | NA | 0 |
| S2-final-viral-combined_k141_73629\|\|full | 1264 | No | NA | 3 | 0 | 0 | Not-determined | Genome-fragment | NA | NA | 0 |
| S2-final-viral-combined_k141_135615\|\|full | 3880 | No | NA | 3 | 0 | 1 | Not-determined | Genome-fragment | NA | NA | 0 |
| S2-final-viral-combined_k141_177379\|\|full | 2258 | No | NA | 3 | 0 | 0 | Not-determined | Genome-fragment | NA | NA | 0 |
| S2-final-viral-combined_k141_177382\|\|full | 4095 | No | NA | 7 | 0 | 0 | Not-determined | Genome-fragment | NA | NA | 0 |
| S2-final-viral-combined_k141_166817\|\|full | 2066 | No | NA | 3 | 0 | 1 | Not-determined | Genome-fragment | NA | NA | 0 |
| S2-final-viral-combined_k141_229120\|\|full | 1263 | No | NA | 1 | 0 | 0 | Not-determined | Genome-fragment | NA | NA | 0 |
| S8-final-viral-combined_k141_275061\|\|full | 3632 | No | NA | 4 | 0 | 3 | Low-quality | Genome-fragment | 12.2 | AAI-based (medium-confidence) | 0 |
| S11-final-viral-combined_k141_118857\|\|full | 3633 | No | NA | 8 | 0 | 0 | Low-quality | Genome-fragment | 5.39 | AAI-based (high-confidence) | 0 |
| S5-final-viral-combined_k141_103771\|\|full | 3633 | No | NA | 3 | 0 | 2 | Low-quality | Genome-fragment | 1.83 | AAI-based (medium-confidence) | 0 |
| S2-final-viral-combined_k141_166825\|\|full | 5842 | No | NA | 5 | 0 | 3 | Not-determined | Genome-fragment | NA | NA | 0 |
| S5-final-viral-combined_k141_53333\|\|full | 3633 | No | NA | 6 | 0 | 0 | Low-quality | Genome-fragment | 6.57 | AAI-based (high-confidence) | 0 |
| S2-final-viral-combined_k141_166835\|\|full | 3398 | No | NA | 3 | 0 | 0 | Not-determined | Genome-fragment | NA | NA | 0 |
| S12-final-viral-combined_k141_292248\|\|full | 3633 | No | NA | 6 | 0 | 4 | Low-quality | Genome-fragment | 6.64 | AAI-based (medium-confidence) | 0 |
| S2-final-viral-combined_k141_146215\|\|full | 4695 | No | NA | 4 | 0 | 2 | Not-determined | Genome-fragment | NA | NA | 0 |
| S2-final-viral-combined_k141_135640\|\|full | 2980 | No | NA | 4 | 0 | 1 | Not-determined | Genome-fragment | NA | NA | 0 |
| S2_k141_116154 | 3633 | No | NA | 3 | 0 | 0 | Low-quality | Genome-fragment | 13.26 | AAI-based (high-confidence) | 0 |
| S2-final-viral-combined_k141_32127\|\|full | 1830 | No | NA | 3 | 0 | 0 | Not-determined | Genome-fragment | NA | NA | 0 |
| S2-final-viral-combined_k141_104716\|\|full | 1860 | No | NA | 2 | 0 | 1 | Not-determined | Genome-fragment | NA | NA | 0 |
| S2-final-viral-combined_k141_125430\|\|full | 4626 | No | NA | 3 | 0 | 2 | Not-determined | Genome-fragment | NA | NA | 0 |
| S2-final-viral-combined_k141_32132\|\|full | 3338 | No | NA | 6 | 0 | 3 | Not-determined | Genome-fragment | NA | NA | 0 |
| S2-final-viral-combined_k141_32135\|\|full | 2284 | No | NA | 2 | 0 | 1 | Not-determined | Genome-fragment | NA | NA | 0 |
| S2-final-viral-combined_k141_156480\|\|full | 1199 | No | NA | 2 | 0 | 1 | Not-determined | Genome-fragment | NA | NA | 0 |
| S2-final-viral-combined_k141_208192\|\|full | 1334 | No | NA | 2 | 0 | 0 | Not-determined | Genome-fragment | NA | NA | 0 |
| S2-final-viral-combined_k141_84063\|\|full | 6593 | No | NA | 6 | 0 | 4 | Not-determined | Genome-fragment | NA | NA | 0 |
| S2-final-viral-combined_k141_208204\|\|full | 1978 | No | NA | 3 | 0 | 2 | Not-determined | Genome-fragment | NA | NA | 0 |
| S1-final-viral-combined_k141_267815\|\|full | 3635 | No | NA | 6 | 0 | 2 | Low-quality | Genome-fragment | 8.4 | AAI-based (medium-confidence) | 0 |
| S7-final-viral-combined_k141_85068\|\|full | 3635 | No | NA | 3 | 0 | 2 | Low-quality | Genome-fragment | 4.39 | AAI-based (high-confidence) | 0 |
| S2-final-viral-combined_k141_94221\|\|full | 2064 | No | NA | 3 | 0 | 2 | Not-determined | Genome-fragment | NA | NA | 0 |
| S11-final-viral-combined_k141_244775\|\|full | 3636 | No | NA | 4 | 4 | 0 | Low-quality | Genome-fragment | 5.89 | AAI-based (high-confidence) | 0 |
| S2-final-viral-combined_k141_146270\|\|full | 1483 | No | NA | 3 | 0 | 0 | Not-determined | Genome-fragment | NA | NA | 0 |
| S2-final-viral-combined_k141_146275\|\|full | 2265 | No | NA | 3 | 0 | 2 | Not-determined | Genome-fragment | NA | NA | 0 |
| S2-final-viral-combined_k141_52813\|\|full | 1019 | No | NA | 2 | 0 | 1 | Not-determined | Genome-fragment | NA | NA | 0 |
| S2-final-viral-combined_k141_177407\|\|full | 6776 | No | NA | 4 | 0 | 1 | Not-determined | Genome-fragment | NA | NA | 0 |
| S3-final-viral-combined_k141_77483\|\|full | 3636 | No | NA | 4 | 0 | 1 | Low-quality | Genome-fragment | 6.32 | AAI-based (medium-confidence) | 0 |
| S2-final-viral-combined_k141_125489\|\|full | 2386 | No | NA | 3 | 0 | 2 | Not-determined | Genome-fragment | NA | NA | 0 |
| S2-final-viral-combined_k141_84096\|\|full | 1178 | No | NA | 3 | 0 | 1 | Not-determined | Genome-fragment | NA | NA | 0 |
| S2-final-viral-combined_k141_43140\|\|full | 3092 | No | NA | 3 | 0 | 1 | Not-determined | Genome-fragment | NA | NA | 0 |
| S5-final-viral-combined_k141_177688\|\|full | 3636 | No | NA | 3 | 0 | 0 | Low-quality | Genome-fragment | 5 | AAI-based (medium-confidence) | 0 |
| S2-final-viral-combined_k141_73770\|\|full | 5111 | No | NA | 8 | 0 | 0 | Not-determined | Genome-fragment | NA | NA | 0 |
| S7-final-viral-combined_k141_49838\|\|full | 3636 | No | NA | 2 | 0 | 1 | Low-quality | Genome-fragment | 10.12 | AAI-based (medium-confidence) | 0 |
| S4-final-viral-combined_k141_64133\|\|full | 3636 | No | NA | 5 | 0 | 0 | Low-quality | Genome-fragment | 3.71 | AAI-based (high-confidence) | 0 |
| S2-final-viral-combined_k141_73780\|\|full | 1719 | No | NA | 4 | 0 | 0 | Not-determined | Genome-fragment | NA | NA | 0 |
| S11_k141_64486 | 3637 | No | NA | 3 | 0 | 1 | Low-quality | Genome-fragment | 8.75 | AAI-based (high-confidence) | 0 |
| S1-final-viral-combined_k141_174621\|\|full | 3637 | No | NA | 6 | 1 | 0 | Low-quality | Genome-fragment | 7.88 | AAI-based (high-confidence) | 0 |
| S2-final-viral-combined_k141_177471\|\|full | 3752 | No | NA | 6 | 0 | 2 | Not-determined | Genome-fragment | NA | NA | 0 |
| S7-final-viral-combined_k141_161180\|\|full | 3637 | No | NA | 3 | 0 | 2 | Low-quality | Genome-fragment | 7.55 | AAI-based (medium-confidence) | 0 |
| S2-final-viral-combined_k141_1205\|\|full | 5426 | No | NA | 5 | 0 | 3 | Not-determined | Genome-fragment | NA | NA | 0 |
| S4_k141_134429 | 3637 | No | NA | 4 | 0 | 1 | Low-quality | Genome-fragment | 5.38 | AAI-based (medium-confidence) | 0 |
| S1-final-viral-combined_k141_106584\|\|full | 3638 | No | NA | 4 | 4 | 0 | Low-quality | Genome-fragment | 8.38 | AAI-based (high-confidence) | 0 |
| S2-final-viral-combined_k141_197577\|\|full | 64386 | No | NA | 78 | 9 | 1 | Medium-quality | Genome-fragment | 82.7 | AAI-based (high-confidence) | 0 |
| S2-final-viral-combined_k141_166968\|\|full | 5336 | No | NA | 6 | 0 | 5 | Not-determined | Genome-fragment | NA | NA | 0 |
| S2-final-viral-combined_k141_1222\|\|full | 3909 | No | NA | 7 | 0 | 0 | Not-determined | Genome-fragment | NA | NA | 0 |
| S2-final-viral-combined_k141_177509\|\|full | 1024 | No | NA | 2 | 0 | 0 | Not-determined | Genome-fragment | NA | NA | 0 |
| S7-final-viral-combined_k141_187713\|\|full | 3638 | No | NA | 4 | 2 | 0 | Low-quality | Genome-fragment | 6.32 | HMM-based (lower-bound) | 0 |
| S2-final-viral-combined_k141_1252\|\|full | 6289 | No | NA | 7 | 0 | 5 | Not-determined | Genome-fragment | NA | NA | 0 |
| S2-final-viral-combined_k141_135722\|\|full | 1502 | No | NA | 2 | 0 | 1 | Not-determined | Genome-fragment | NA | NA | 0 |
| S2-final-viral-combined_k141_135750\|\|full | 4493 | No | NA | 6 | 0 | 1 | Not-determined | Genome-fragment | NA | NA | 0 |
| S2-final-viral-combined_k141_73720\|\|full | 3638 | No | NA | 7 | 0 | 3 | Low-quality | Genome-fragment | 6.25 | AAI-based (high-confidence) | 0 |
| S2-final-viral-combined_k141_125523\|\|full | 2967 | No | NA | 3 | 0 | 3 | Not-determined | Genome-fragment | NA | NA | 0 |
| S2-final-viral-combined_k141_135755\|\|full | 3640 | No | NA | 4 | 0 | 2 | Not-determined | Genome-fragment | NA | NA | 0 |
| S2-final-viral-combined_k141_229242\|\|full | 1117 | No | NA | 3 | 0 | 1 | Not-determined | Genome-fragment | NA | NA | 0 |
| S8_k141_257322 | 3638 | No | NA | 6 | 0 | 3 | Low-quality | Genome-fragment | 9.29 | AAI-based (medium-confidence) | 0 |
| S9_k141_99376 | 3639 | No | NA | 5 | 3 | 0 | Low-quality | Genome-fragment | 7.66 | AAI-based (medium-confidence) | 0 |
| S2-final-viral-combined_k141_63013\|\|full | 1892 | No | NA | 4 | 0 | 1 | Not-determined | Genome-fragment | NA | NA | 0 |
| S12_k141_101967 | 3639 | Yes | 2679 | 6 | 1 | 1 | Low-quality | Genome-fragment | 4.91 | AAI-based (high-confidence) | 26.38 |
| S2-final-viral-combined_k141_229248\|\|full | 2266 | No | NA | 2 | 0 | 1 | Not-determined | Genome-fragment | NA | NA | 0 |
| S2_k141_214084 | 3640 | No | NA | 5 | 0 | 1 | Low-quality | Genome-fragment | 5.19 | AAI-based (medium-confidence) | 0 |
| S2-final-viral-combined_k141_208294\|\|full | 1221 | No | NA | 3 | 0 | 2 | Not-determined | Genome-fragment | NA | NA | 0 |
| S8_k141_26162 | 3640 | No | NA | 5 | 0 | 2 | Low-quality | Genome-fragment | 3.42 | AAI-based (medium-confidence) | 0 |
| S9_k141_54932 | 3641 | No | NA | 7 | 0 | 2 | Low-quality | Genome-fragment | 5.7 | AAI-based (medium-confidence) | 0 |
| S2-final-viral-combined_k141_208318\|\|full | 3701 | No | NA | 4 | 0 | 2 | Not-determined | Genome-fragment | NA | NA | 0 |
| S2-final-viral-combined_k141_197668\|\|full | 3607 | No | NA | 5 | 0 | 2 | Not-determined | Genome-fragment | NA | NA | 0 |
| S2-final-viral-combined_k141_229294\|\|full | 2757 | No | NA | 4 | 0 | 3 | Not-determined | Genome-fragment | NA | NA | 0 |
| S1_k141_3811 | 3641 | No | NA | 1 | 0 | 0 | Low-quality | Genome-fragment | 8.75 | AAI-based (high-confidence) | 0 |
| S2-final-viral-combined_k141_167119\|\|full | 10786 | No | NA | 8 | 0 | 4 | Not-determined | Genome-fragment | NA | NA | 0 |
| S17-final-viral-combined_k141_124595\|\|full | 3641 | No | NA | 3 | 0 | 1 | Low-quality | Genome-fragment | 1.15 | AAI-based (medium-confidence) | 0 |
| S2-final-viral-combined_k141_43341\|\|full | 32512 | No | NA | 30 | 0 | 21 | Not-determined | Genome-fragment | NA | NA | 0 |
| S17-final-viral-combined_k141_5879\|\|full | 3641 | No | NA | 5 | 0 | 3 | Low-quality | Genome-fragment | 4.04 | AAI-based (medium-confidence) | 0 |
| S2-final-viral-combined_k141_11741\|\|full | 6210 | No | NA | 11 | 0 | 1 | Not-determined | Genome-fragment | NA | NA | 0 |
| S2-final-viral-combined_k141_150185\|\|full | 3641 | No | NA | 3 | 0 | 1 | Low-quality | Genome-fragment | 2.02 | AAI-based (medium-confidence) | 0 |
| S2-final-viral-combined_k141_63035\|\|full | 1659 | No | NA | 2 | 0 | 0 | Not-determined | Genome-fragment | NA | NA | 0 |
| S9-final-viral-combined_k141_11519\|\|full | 3642 | No | NA | 3 | 0 | 0 | Low-quality | Genome-fragment | 4.38 | AAI-based (medium-confidence) | 0 |
| S2-final-viral-combined_k141_94424\|\|full | 1069 | No | NA | 2 | 0 | 0 | Not-determined | Genome-fragment | NA | NA | 0 |
| S2-final-viral-combined_k141_229315\|\|full | 1279 | No | NA | 2 | 0 | 2 | Not-determined | Genome-fragment | NA | NA | 0 |
| S2-final-viral-combined_k141_11746\|\|full | 3450 | No | NA | 3 | 0 | 0 | Not-determined | Genome-fragment | NA | NA | 0 |
| S1_k141_391923 | 3642 | Yes | 2060 | 6 | 1 | 2 | Low-quality | Genome-fragment | 5.3 | AAI-based (high-confidence) | 43.44 |
| S2-final-viral-combined_k141_104871\|\|full | 6429 | No | NA | 8 | 0 | 4 | Not-determined | Genome-fragment | NA | NA | 0 |
| S2-final-viral-combined_k141_94435\|\|full | 1072 | No | NA | 3 | 0 | 0 | Not-determined | Genome-fragment | NA | NA | 0 |
| S2-final-viral-combined_k141_114961\|\|full | 2498 | No | NA | 8 | 0 | 0 | Not-determined | Genome-fragment | NA | NA | 0 |
| S2-final-viral-combined_k141_197689\|\|full | 1325 | No | NA | 4 | 0 | 0 | Not-determined | Genome-fragment | NA | NA | 0 |
| S2-final-viral-combined_k141_125573\|\|full | 2571 | No | NA | 3 | 0 | 2 | Not-determined | Genome-fragment | NA | NA | 0 |
| S2-final-viral-combined_k141_239411\|\|full | 1978 | No | NA | 2 | 0 | 1 | Not-determined | Genome-fragment | NA | NA | 0 |
| S2-final-viral-combined_k141_208360\|\|full | 11368 | No | NA | 10 | 0 | 5 | Not-determined | Genome-fragment | NA | NA | 0 |
| S1_k141_343445 | 3642 | No | NA | 5 | 1 | 0 | Low-quality | Genome-fragment | 7.44 | AAI-based (high-confidence) | 0 |
| S2-final-viral-combined_k141_146447\|\|full | 1234 | No | NA | 3 | 0 | 1 | Not-determined | Genome-fragment | NA | NA | 0 |
| S2-final-viral-combined_k141_94477\|\|full | 1653 | No | NA | 4 | 0 | 1 | Not-determined | Genome-fragment | NA | NA | 0 |
| S17-final-viral-combined_k141_255563\|\|full | 3643 | No | NA | 4 | 0 | 3 | Low-quality | Genome-fragment | 4.15 | AAI-based (medium-confidence) | 0 |
| S2-final-viral-combined_k141_11828\|\|full | 1535 | No | NA | 3 | 0 | 2 | Not-determined | Genome-fragment | NA | NA | 0 |
| S12-final-viral-combined_k141_259206\|\|full | 3643 | No | NA | 3 | 0 | 3 | Low-quality | Genome-fragment | 3.14 | AAI-based (medium-confidence) | 0 |
| S1-final-viral-combined_k141_257356\|\|full | 3644 | No | NA | 5 | 3 | 0 | Low-quality | Genome-fragment | 8.47 | AAI-based (medium-confidence) | 0 |
| S2-final-viral-combined_k141_63118\|\|full | 3791 | No | NA | 3 | 0 | 3 | Not-determined | Genome-fragment | NA | NA | 0 |
| S5-final-viral-combined_k141_201264\|\|full | 3645 | No | NA | 2 | 0 | 0 | Low-quality | Genome-fragment | 6.81 | AAI-based (medium-confidence) | 0 |
| S17-final-viral-combined_k141_15892\|\|full | 3646 | No | NA | 3 | 0 | 2 | Low-quality | Genome-fragment | 1.07 | AAI-based (medium-confidence) | 0 |
| S2-final-viral-combined_k141_63137\|\|full | 6267 | No | NA | 9 | 0 | 4 | Not-determined | Genome-fragment | NA | NA | 0 |
| S17-final-viral-combined_k141_149839\|\|full | 3647 | No | NA | 7 | 4 | 0 | Low-quality | Genome-fragment | 9.19 | AAI-based (high-confidence) | 0 |
| S2-final-viral-combined_k141_104892\|\|full | 8376 | No | NA | 8 | 0 | 6 | Not-determined | Genome-fragment | NA | NA | 0 |
| S17-final-viral-combined_k141_242773\|\|full | 3647 | No | NA | 3 | 0 | 3 | Low-quality | Genome-fragment | 1.07 | AAI-based (medium-confidence) | 0 |
| S2-final-viral-combined_k141_146492\|\|full | 4047 | No | NA | 4 | 0 | 0 | Not-determined | Genome-fragment | NA | NA | 0 |
| S2-final-viral-combined_k141_94510\|\|full | 1871 | No | NA | 3 | 0 | 1 | Not-determined | Genome-fragment | NA | NA | 0 |
| S2-final-viral-combined_k141_239433\|\|full | 6373 | No | NA | 8 | 0 | 4 | Not-determined | Genome-fragment | NA | NA | 0 |
| S2-final-viral-combined_k141_1351\|\|full | 2397 | No | NA | 3 | 0 | 0 | Not-determined | Genome-fragment | NA | NA | 0 |
| S2-final-viral-combined_k141_146496\|\|full | 6892 | No | NA | 5 | 0 | 2 | Not-determined | Genome-fragment | NA | NA | 0 |
| S2-final-viral-combined_k141_63150\|\|full | 5483 | No | NA | 4 | 0 | 3 | Not-determined | Genome-fragment | NA | NA | 0 |
| S2-final-viral-combined_k141_167271\|\|full | 4646 | No | NA | 7 | 0 | 0 | Not-determined | Genome-fragment | NA | NA | 0 |
| S2-final-viral-combined_k141_115001\|\|full | 1501 | No | NA | 3 | 0 | 0 | Not-determined | Genome-fragment | NA | NA | 0 |
| S2-final-viral-combined_k141_84325\|\|full | 2273 | No | NA | 3 | 0 | 2 | Not-determined | Genome-fragment | NA | NA | 0 |
| S2-final-viral-combined_k141_73944\|\|full | 4712 | No | NA | 9 | 0 | 0 | Not-determined | Genome-fragment | NA | NA | 0 |
| S2-final-viral-combined_k141_187687\|\|full | 3112 | No | NA | 4 | 0 | 3 | Not-determined | Genome-fragment | NA | NA | 0 |
| S2-final-viral-combined_k141_135925\|\|full | 1640 | No | NA | 2 | 0 | 1 | Not-determined | Genome-fragment | NA | NA | 0 |
| S11_k141_141581 | 3648 | No | NA | 6 | 2 | 0 | Low-quality | Genome-fragment | 8 | AAI-based (high-confidence) | 0 |
| S2-final-viral-combined_k141_218675\|\|full | 8830 | No | NA | 8 | 0 | 3 | Not-determined | Genome-fragment | NA | NA | 0 |
| S2-final-viral-combined_k141_43459\|\|full | 1513 | No | NA | 2 | 0 | 0 | Not-determined | Genome-fragment | NA | NA | 0 |
| S2-final-viral-combined_k141_218682\|\|full | 7047 | No | NA | 7 | 0 | 3 | Not-determined | Genome-fragment | NA | NA | 0 |
| S2-final-viral-combined_k141_73982\|\|full | 2792 | No | NA | 3 | 0 | 2 | Not-determined | Genome-fragment | NA | NA | 0 |
| S2-final-viral-combined_k141_197753\|\|full | 1395 | No | NA | 2 | 0 | 1 | Not-determined | Genome-fragment | NA | NA | 0 |
| S3-final-viral-combined_k141_62207\|\|full | 3648 | No | NA | 9 | 1 | 0 | Low-quality | Genome-fragment | 4.04 | AAI-based (high-confidence) | 0 |
| S2-final-viral-combined_k141_137951\|\|full | 3648 | No | NA | 3 | 0 | 1 | Low-quality | Genome-fragment | 10.24 | AAI-based (medium-confidence) | 0 |
| S2-final-viral-combined_k141_167341\|\|full | 4618 | No | NA | 7 | 0 | 1 | Not-determined | Genome-fragment | NA | NA | 0 |
| S2-final-viral-combined_k141_156647\|\|full | 2491 | No | NA | 3 | 0 | 2 | Not-determined | Genome-fragment | NA | NA | 0 |
| S2-final-viral-combined_k141_43516\|\|full | 2523 | No | NA | 4 | 0 | 1 | Not-determined | Genome-fragment | NA | NA | 0 |
| S7-final-viral-combined_k141_73413\|\|full | 3649 | No | NA | 6 | 2 | 0 | Low-quality | Genome-fragment | 9.82 | AAI-based (medium-confidence) | 0 |
| S2-final-viral-combined_k141_197773\|\|full | 3029 | No | NA | 4 | 0 | 1 | Not-determined | Genome-fragment | NA | NA | 0 |
| S2-final-viral-combined_k141_43533\|\|full | 1485 | No | NA | 3 | 0 | 0 | Not-determined | Genome-fragment | NA | NA | 0 |
| S2-final-viral-combined_k141_115057\|\|full | 3112 | No | NA | 7 | 0 | 1 | Not-determined | Genome-fragment | NA | NA | 0 |
| S2-final-viral-combined_k141_167373\|\|full | 1953 | No | NA | 3 | 0 | 2 | Not-determined | Genome-fragment | NA | NA | 0 |
| S2-final-viral-combined_k141_167374\|\|full | 1209 | No | NA | 3 | 0 | 1 | Not-determined | Genome-fragment | NA | NA | 0 |
| S2-final-viral-combined_k141_43548\|\|full | 1317 | No | NA | 3 | 0 | 1 | Not-determined | Genome-fragment | NA | NA | 0 |
| S12_k141_24898 | 3649 | No | NA | 4 | 0 | 2 | Low-quality | Genome-fragment | 19.98 | AAI-based (medium-confidence) | 0 |
| S2-final-viral-combined_k141_146573\|\|full | 1028 | No | NA | 3 | 0 | 1 | Not-determined | Genome-fragment | NA | NA | 0 |
| S2-final-viral-combined_k141_43564\|\|full | 2806 | No | NA | 3 | 0 | 3 | Not-determined | Genome-fragment | NA | NA | 0 |
| S2-final-viral-combined_k141_167401\|\|full | 4664 | No | NA | 8 | 0 | 1 | Not-determined | Genome-fragment | NA | NA | 0 |
| S2-final-viral-combined_k141_43568\|\|full | 1707 | No | NA | 2 | 0 | 1 | Not-determined | Genome-fragment | NA | NA | 0 |
| S2-final-viral-combined_k141_1393\|\|full | 6032 | No | NA | 4 | 0 | 4 | Not-determined | Genome-fragment | NA | NA | 0 |
| S2-final-viral-combined_k141_84390\|\|full | 1205 | No | NA | 2 | 0 | 0 | Not-determined | Genome-fragment | NA | NA | 0 |
| S2-final-viral-combined_k141_84393\|\|full | 1046 | No | NA | 3 | 0 | 0 | Not-determined | Genome-fragment | NA | NA | 0 |
| S2-final-viral-combined_k141_167408\|\|full | 2033 | No | NA | 3 | 0 | 2 | Not-determined | Genome-fragment | NA | NA | 0 |
| S2-final-viral-combined_k141_105003\|\|full | 1544 | No | NA | 3 | 0 | 0 | Not-determined | Genome-fragment | NA | NA | 0 |
| S2-final-viral-combined_k141_22311\|\|full | 1515 | No | NA | 3 | 0 | 3 | Not-determined | Genome-fragment | NA | NA | 0 |
| S2-final-viral-combined_k141_115092\|\|full | 2447 | No | NA | 4 | 0 | 2 | Not-determined | Genome-fragment | NA | NA | 0 |
| S2_k141_137512 | 3649 | No | NA | 3 | 0 | 0 | Low-quality | Genome-fragment | 1.59 | AAI-based (medium-confidence) | 0 |
| S2-final-viral-combined_k141_43591\|\|full | 1551 | No | NA | 3 | 0 | 2 | Not-determined | Genome-fragment | NA | NA | 0 |
| S4-final-viral-combined_k141_32761\|\|full | 3649 | Yes | 1160 | 4 | 1 | 3 | Low-quality | Genome-fragment | 0.42 | HMM-based (lower-bound) | 68.21 |
| S2-final-viral-combined_k141_197827\|\|full | 1569 | No | NA | 3 | 0 | 1 | Not-determined | Genome-fragment | NA | NA | 0 |
| S2-final-viral-combined_k141_1410\|\|full | 5136 | No | NA | 2 | 0 | 1 | Not-determined | Genome-fragment | NA | NA | 0 |
| S11-final-viral-combined_k141_328877\|\|full | 3650 | No | NA | 3 | 0 | 2 | Low-quality | Genome-fragment | 4.05 | AAI-based (medium-confidence) | 0 |
| S2-final-viral-combined_k141_94664\|\|full | 2629 | No | NA | 2 | 0 | 2 | Not-determined | Genome-fragment | NA | NA | 0 |
| S17-final-viral-combined_k141_156994\|\|full | 3650 | No | NA | 5 | 1 | 0 | Low-quality | Genome-fragment | 8.09 | AAI-based (medium-confidence) | 0 |
| S2-final-viral-combined_k141_187770\|\|full | 18550 | No | NA | 20 | 0 | 14 | Not-determined | Genome-fragment | NA | NA | 0 |
| S17-final-viral-combined_k141_13670\|\|full | 3651 | No | NA | 5 | 2 | 0 | Low-quality | Genome-fragment | 3.86 | HMM-based (lower-bound) | 0 |
| S2-final-viral-combined_k141_187775\|\|full | 2207 | No | NA | 4 | 0 | 0 | Not-determined | Genome-fragment | NA | NA | 0 |
| S2-final-viral-combined_k141_135983\|\|full | 8574 | No | NA | 10 | 0 | 0 | Not-determined | Genome-fragment | NA | NA | 0 |
| S2-final-viral-combined_k141_53044\|\|full | 1707 | No | NA | 3 | 0 | 2 | Not-determined | Genome-fragment | NA | NA | 0 |
| S2-final-viral-combined_k141_125678\|\|full | 2306 | No | NA | 3 | 0 | 0 | Not-determined | Genome-fragment | NA | NA | 0 |
| S2-final-viral-combined_k141_63390\|\|full | 2799 | No | NA | 2 | 0 | 1 | Not-determined | Genome-fragment | NA | NA | 0 |
| S17-k141_167586 | 3651 | No | NA | 3 | 0 | 2 | Low-quality | Genome-fragment | 0.96 | AAI-based (high-confidence) | 0 |
| S2-final-viral-combined_k141_94685\|\|full | 3332 | No | NA | 4 | 0 | 1 | Not-determined | Genome-fragment | NA | NA | 0 |
| S1_k141_102346 | 3652 | No | NA | 5 | 0 | 4 | Low-quality | Genome-fragment | 7.27 | AAI-based (medium-confidence) | 0 |
| S2-final-viral-combined_k141_136002\|\|full | 1969 | No | NA | 3 | 0 | 0 | Not-determined | Genome-fragment | NA | NA | 0 |
| S2-final-viral-combined_k141_1449\|\|full | 5210 | No | NA | 3 | 0 | 2 | Not-determined | Genome-fragment | NA | NA | 0 |
| S2-final-viral-combined_k141_167514\|\|full | 1352 | No | NA | 2 | 0 | 1 | Not-determined | Genome-fragment | NA | NA | 0 |
| S2-final-viral-combined_k141_94696\|\|full | 1486 | No | NA | 2 | 0 | 2 | Not-determined | Genome-fragment | NA | NA | 0 |
| S2-final-viral-combined_k141_84498\|\|full | 4480 | No | NA | 4 | 0 | 1 | Not-determined | Genome-fragment | NA | NA | 0 |
| S2-final-viral-combined_k141_84508\|\|full | 1121 | No | NA | 2 | 0 | 1 | Not-determined | Genome-fragment | NA | NA | 0 |
| S2-final-viral-combined_k141_156727\|\|full | 1271 | No | NA | 2 | 0 | 0 | Not-determined | Genome-fragment | NA | NA | 0 |
| S2-final-viral-combined_k141_214863\|\|full | 3652 | No | NA | 5 | 1 | 0 | Low-quality | Genome-fragment | 7.61 | AAI-based (high-confidence) | 0 |
| S2_k141_98810 | 3652 | No | NA | 7 | 2 | 0 | Low-quality | Genome-fragment | 8.09 | HMM-based (lower-bound) | 0 |
| S2-final-viral-combined_k141_208559\|\|full | 68162 | No | NA | 60 | 2 | 36 | Medium-quality | Genome-fragment | 88.92 | HMM-based (lower-bound) | 0 |
| S2-final-viral-combined_k141_208583\|\|full | 2605 | No | NA | 3 | 0 | 2 | Not-determined | Genome-fragment | NA | NA | 0 |
| S2-final-viral-combined_k141_239502\|\|full | 5118 | No | NA | 6 | 0 | 3 | Not-determined | Genome-fragment | NA | NA | 0 |
| S6-final-viral-combined_k141_43423\|\|full | 3652 | No | NA | 3 | 1 | 0 | Low-quality | Genome-fragment | 7.42 | HMM-based (lower-bound) | 0 |
| S2-final-viral-combined_k141_115247\|\|full | 3721 | No | NA | 3 | 0 | 2 | Not-determined | Genome-fragment | NA | NA | 0 |
| S7-final-viral-combined_k141_110340\|\|full | 3653 | No | NA | 9 | 0 | 0 | Low-quality | Genome-fragment | 8.68 | AAI-based (medium-confidence) | 0 |
| S2-final-viral-combined_k141_53171\|\|full | 10462 | No | NA | 10 | 0 | 7 | Not-determined | Genome-fragment | NA | NA | 0 |
| S2-final-viral-combined_k141_11991\|\|full | 9902 | No | NA | 12 | 0 | 8 | Not-determined | Genome-fragment | NA | NA | 0 |
| S2-final-viral-combined_k141_187909\|\|full | 3262 | No | NA | 3 | 0 | 0 | Not-determined | Genome-fragment | NA | NA | 0 |
| S2-final-viral-combined_k141_86561\|\|full | 3654 | No | NA | 3 | 0 | 3 | Low-quality | Genome-fragment | 2.07 | AAI-based (medium-confidence) | 0 |
| S2-final-viral-combined_k141_146713\|\|full | 4895 | No | NA | 3 | 0 | 2 | Not-determined | Genome-fragment | NA | NA | 0 |
| S9_k141_68040 | 3655 | No | NA | 4 | 0 | 1 | Low-quality | Genome-fragment | 2.47 | AAI-based (high-confidence) | 0 |
| S2-final-viral-combined_k141_12003\|\|full | 1031 | No | NA | 3 | 0 | 1 | Not-determined | Genome-fragment | NA | NA | 0 |
| S2-final-viral-combined_k141_1465\|\|full | 3428 | No | NA | 2 | 0 | 2 | Not-determined | Genome-fragment | NA | NA | 0 |
| S2-final-viral-combined_k141_105064\|\|full | 10608 | No | NA | 10 | 0 | 6 | Not-determined | Genome-fragment | NA | NA | 0 |
| S2-final-viral-combined_k141_53216\|\|full | 7057 | No | NA | 6 | 0 | 3 | Not-determined | Genome-fragment | NA | NA | 0 |
| S6_k141_28884 | 3655 | No | NA | 2 | 0 | 1 | Low-quality | Genome-fragment | 3.33 | AAI-based (high-confidence) | 0 |
| S2-final-viral-combined_k141_177885\|\|full | 1463 | No | NA | 3 | 0 | 0 | Not-determined | Genome-fragment | NA | NA | 0 |
| S8-final-viral-combined_k141_34380\|\|full | 3655 | No | NA | 6 | 0 | 0 | Low-quality | Genome-fragment | 2.99 | AAI-based (medium-confidence) | 0 |
| S2-final-viral-combined_k141_105093\|\|full | 1191 | No | NA | 2 | 0 | 0 | Not-determined | Genome-fragment | NA | NA | 0 |
| S2-final-viral-combined_k141_197894\|\|full | 1062 | No | NA | 2 | 0 | 2 | Not-determined | Genome-fragment | NA | NA | 0 |
| S2-final-viral-combined_k141_115255\|\|full | 1921 | No | NA | 2 | 0 | 0 | Not-determined | Genome-fragment | NA | NA | 0 |
| S2-final-viral-combined_k141_63487\|\|full | 3712 | No | NA | 5 | 0 | 0 | Not-determined | Genome-fragment | NA | NA | 0 |
| S9_k141_31754 | 3656 | No | NA | 1 | 0 | 1 | Low-quality | Genome-fragment | 3.96 | AAI-based (high-confidence) | 0 |
| S2-final-viral-combined_k141_229529\|\|full | 113248 | Yes | 59461 | 143 | 14 | 30 | Complete | High-quality | 100 | Provirus (high-confidence) | 47.49 |
| S2-final-viral-combined_k141_94788\|\|full | 5179 | No | NA | 5 | 0 | 4 | Not-determined | Genome-fragment | NA | NA | 0 |
| S1-final-viral-combined_k141_262514\|\|full | 3656 | No | NA | 3 | 0 | 3 | Low-quality | Genome-fragment | 1 | AAI-based (medium-confidence) | 0 |
| S2-final-viral-combined_k141_43767\|\|full | 3108 | No | NA | 4 | 0 | 0 | Not-determined | Genome-fragment | NA | NA | 0 |
| S12_k141_252735 | 3656 | No | NA | 3 | 0 | 0 | Low-quality | Genome-fragment | 4.27 | AAI-based (medium-confidence) | 0 |
| S2-final-viral-combined_k141_146761\|\|full | 1782 | No | NA | 3 | 0 | 1 | Not-determined | Genome-fragment | NA | NA | 0 |
| S2-final-viral-combined_k141_1514\|\|full | 4867 | No | NA | 4 | 0 | 0 | Not-determined | Genome-fragment | NA | NA | 0 |
| S2-final-viral-combined_k141_177911\|\|full | 1224 | No | NA | 2 | 0 | 0 | Not-determined | Genome-fragment | NA | NA | 0 |
| S2-final-viral-combined_k141_162064\|\|full | 3656 | No | NA | 7 | 0 | 0 | Low-quality | Genome-fragment | 6.75 | AAI-based (high-confidence) | 0 |
| S2-final-viral-combined_k141_177940\|\|full | 2713 | No | NA | 2 | 0 | 1 | Not-determined | Genome-fragment | NA | NA | 0 |
| S4_k141_145917 | 3656 | No | NA | 9 | 3 | 0 | Low-quality | Genome-fragment | 9.83 | AAI-based (high-confidence) | 0 |
| S2-final-viral-combined_k141_239621\|\|full | 2446 | No | NA | 3 | 0 | 1 | Not-determined | Genome-fragment | NA | NA | 0 |
| S2-final-viral-combined_k141_247242\|\|full | 3657 | No | NA | 9 | 0 | 0 | Low-quality | Genome-fragment | 6.25 | AAI-based (high-confidence) | 0 |
| S2-final-viral-combined_k141_12029\|\|full | 7841 | No | NA | 9 | 0 | 5 | Not-determined | Genome-fragment | NA | NA | 0 |
| S2-final-viral-combined_k141_43822\|\|full | 1564 | No | NA | 2 | 0 | 1 | Not-determined | Genome-fragment | NA | NA | 0 |
| S6-final-viral-combined_k141_133390\|\|full | 3658 | No | NA | 3 | 0 | 1 | Low-quality | Genome-fragment | 3.21 | AAI-based (medium-confidence) | 0 |
| S2-final-viral-combined_k141_156851\|\|full | 3271 | No | NA | 4 | 0 | 1 | Not-determined | Genome-fragment | NA | NA | 0 |
| S2-final-viral-combined_k141_156853\|\|full | 4308 | No | NA | 3 | 0 | 1 | Not-determined | Genome-fragment | NA | NA | 0 |
| S2-final-viral-combined_k141_125828\|\|full | 2342 | No | NA | 4 | 0 | 0 | Not-determined | Genome-fragment | NA | NA | 0 |
| S8_k141_229254 | 3658 | No | NA | 2 | 0 | 0 | Low-quality | Genome-fragment | 4.33 | AAI-based (medium-confidence) | 0 |
| S2-final-viral-combined_k141_177981\|\|full | 1825 | No | NA | 3 | 0 | 1 | Not-determined | Genome-fragment | NA | NA | 0 |
| S2-final-viral-combined_k141_115352\|\|full | 3640 | No | NA | 6 | 0 | 0 | Medium-quality | Genome-fragment | 82.92 | AAI-based (medium-confidence) | 0 |
| S1-final-viral-combined_k141_205552\|\|full | 3659 | No | NA | 2 | 0 | 2 | Low-quality | Genome-fragment | 4.35 | AAI-based (medium-confidence) | 0 |
| S12-final-viral-combined_k141_143780\|\|full | 3659 | No | NA | 4 | 0 | 1 | Low-quality | Genome-fragment | 3.88 | AAI-based (medium-confidence) | 0 |
| S2-final-viral-combined_k141_32509\|\|full | 5140 | No | NA | 5 | 0 | 5 | Not-determined | Genome-fragment | NA | NA | 0 |
| S2-final-viral-combined_k141_22437\|\|full | 2461 | No | NA | 4 | 0 | 0 | Not-determined | Genome-fragment | NA | NA | 0 |
| S2_k141_169142 | 3659 | No | NA | 3 | 0 | 0 | Low-quality | Genome-fragment | 4.29 | AAI-based (medium-confidence) | 0 |
| S11_k141_101516 | 3660 | No | NA | 4 | 0 | 1 | Low-quality | Genome-fragment | 9.06 | AAI-based (medium-confidence) | 0 |
| S17-final-viral-combined_k141_120914\|\|full | 3660 | No | NA | 6 | 2 | 0 | Low-quality | Genome-fragment | 10.2 | AAI-based (medium-confidence) | 0 |
| S2-final-viral-combined_k141_125886\|\|full | 6718 | No | NA | 5 | 0 | 2 | Not-determined | Genome-fragment | NA | NA | 0 |
| S11-final-viral-combined_k141_197402\|\|full | 3661 | No | NA | 3 | 0 | 2 | Low-quality | Genome-fragment | 8.18 | AAI-based (medium-confidence) | 0 |
| S2-final-viral-combined_k141_1555\|\|full | 4036 | No | NA | 4 | 0 | 0 | Not-determined | Genome-fragment | NA | NA | 0 |
| S2-final-viral-combined_k141_125901\|\|full | 1879 | No | NA | 2 | 0 | 2 | Not-determined | Genome-fragment | NA | NA | 0 |
| S2-final-viral-combined_k141_105170\|\|full | 1108 | No | NA | 6 | 0 | 1 | Not-determined | Genome-fragment | NA | NA | 0 |
| S17-final-viral-combined_k141_172922\|\|full | 3661 | No | NA | 7 | 5 | 0 | Low-quality | Genome-fragment | 9.86 | AAI-based (high-confidence) | 0 |
| S2-final-viral-combined_k141_84608\|\|full | 129176 | Yes | 44483 | 97 | 20 | 31 | Medium-quality | Genome-fragment | 78.65 | HMM-based (lower-bound) | 65.56 |
| S2-final-viral-combined_k141_1574\|\|full | 5785 | No | NA | 5 | 0 | 3 | Not-determined | Genome-fragment | NA | NA | 0 |
| S2-final-viral-combined_k141_32597\|\|full | 11770 | No | NA | 15 | 0 | 5 | Not-determined | Genome-fragment | NA | NA | 0 |
| S2-final-viral-combined_k141_94877\|\|full | 1063 | No | NA | 2 | 0 | 2 | Not-determined | Genome-fragment | NA | NA | 0 |
| S2-final-viral-combined_k141_43953\|\|full | 8763 | No | NA | 8 | 0 | 7 | Not-determined | Genome-fragment | NA | NA | 0 |
| S17-k141_46338 | 3661 | No | NA | 6 | 0 | 0 | Low-quality | Genome-fragment | 6.95 | AAI-based (high-confidence) | 0 |
| S4_k141_43879 | 3661 | No | NA | 4 | 0 | 0 | Low-quality | Genome-fragment | 6.6 | AAI-based (high-confidence) | 0 |
| S8_k141_170405 | 3661 | No | NA | 9 | 1 | 1 | Low-quality | Genome-fragment | 8.11 | AAI-based (high-confidence) | 0 |
| S2-final-viral-combined_k141_32620\|\|full | 14433 | No | NA | 13 | 0 | 10 | Not-determined | Genome-fragment | NA | NA | 0 |
| S11-final-viral-combined_k141_230617\|\|full | 3662 | No | NA | 5 | 4 | 0 | Low-quality | Genome-fragment | 6.39 | AAI-based (high-confidence) | 0 |
| S2-final-viral-combined_k141_125947\|\|full | 2157 | No | NA | 3 | 0 | 1 | Not-determined | Genome-fragment | NA | NA | 0 |
| S2-final-viral-combined_k141_208723\|\|full | 3662 | No | NA | 5 | 0 | 2 | Low-quality | Genome-fragment | 4.57 | AAI-based (medium-confidence) | 0 |
| S2-final-viral-combined_k141_218969\|\|full | 2885 | No | NA | 3 | 0 | 0 | Not-determined | Genome-fragment | NA | NA | 0 |
| S2-final-viral-combined_k141_188592\|\|full | 3662 | No | NA | 6 | 0 | 0 | Low-quality | Genome-fragment | 7.88 | AAI-based (medium-confidence) | 0 |
| S2-final-viral-combined_k141_125948\|\|full | 5281 | No | NA | 7 | 0 | 1 | Not-determined | Genome-fragment | NA | NA | 0 |
| S4-final-viral-combined_k141_127515\|\|full | 3662 | No | NA | 5 | 1 | 0 | Low-quality | Genome-fragment | 7.51 | HMM-based (lower-bound) | 0 |
| S2-final-viral-combined_k141_136283\|\|full | 4846 | No | NA | 5 | 0 | 1 | Not-determined | Genome-fragment | NA | NA | 0 |
| S2-final-viral-combined_k141_105215\|\|full | 2953 | No | NA | 5 | 0 | 0 | Not-determined | Genome-fragment | NA | NA | 0 |
| S8-final-viral-combined_k141_35168\|\|full | 3662 | No | NA | 4 | 0 | 4 | Low-quality | Genome-fragment | 4.68 | AAI-based (medium-confidence) | 0 |
| S7-final-viral-combined_k141_151557\|\|full | 3664 | No | NA | 4 | 0 | 2 | Low-quality | Genome-fragment | 7.78 | AAI-based (medium-confidence) | 0 |
| S2-final-viral-combined_k141_1661\|\|full | 1120 | No | NA | 3 | 0 | 2 | Not-determined | Genome-fragment | NA | NA | 0 |
| S6_k141_52102 | 3664 | No | NA | 5 | 4 | 0 | Low-quality | Genome-fragment | 6.24 | AAI-based (high-confidence) | 0 |
| S2-final-viral-combined_k141_94965\|\|full | 3261 | No | NA | 4 | 0 | 2 | Not-determined | Genome-fragment | NA | NA | 0 |
| S5-final-viral-combined_k141_117726\|\|lt2gene | 3665 | No | NA | 2 | 2 | 0 | Low-quality | Genome-fragment | 7.72 | AAI-based (high-confidence) | 0 |
| S2-final-viral-combined_k141_178079\|\|full | 1066 | No | NA | 3 | 0 | 0 | Not-determined | Genome-fragment | NA | NA | 0 |
| S2_k141_138396 | 3666 | No | NA | 2 | 0 | 1 | Low-quality | Genome-fragment | 2.14 | AAI-based (medium-confidence) | 0 |
| S11-final-viral-combined_k141_194195\|\|full | 3667 | No | NA | 4 | 0 | 3 | Low-quality | Genome-fragment | 1.14 | AAI-based (medium-confidence) | 0 |
| S3-final-viral-combined_k141_34957\|\|full | 3667 | No | NA | 4 | 0 | 0 | Low-quality | Genome-fragment | 3.42 | AAI-based (medium-confidence) | 0 |
| S2-final-viral-combined_k141_22579\|\|full | 5495 | No | NA | 5 | 0 | 4 | Not-determined | Genome-fragment | NA | NA | 0 |
| S11-final-viral-combined_k141_302736\|\|full | 3668 | No | NA | 3 | 0 | 1 | Low-quality | Genome-fragment | 2.56 | AAI-based (medium-confidence) | 0 |
| S4_k141_30851 | 3668 | No | NA | 5 | 0 | 2 | Low-quality | Genome-fragment | 3.29 | AAI-based (medium-confidence) | 0 |
| S2-final-viral-combined_k141_239774\|\|full | 1348 | No | NA | 3 | 0 | 2 | Not-determined | Genome-fragment | NA | NA | 0 |
| S9-final-viral-combined_k141_106974\|\|full | 3669 | No | NA | 5 | 1 | 0 | Low-quality | Genome-fragment | 5.55 | HMM-based (lower-bound) | 0 |
| S2-final-viral-combined_k141_44053\|\|full | 1700 | No | NA | 4 | 0 | 0 | Not-determined | Genome-fragment | NA | NA | 0 |
| S11_k141_312794 | 3670 | No | NA | 6 | 1 | 0 | Low-quality | Genome-fragment | 7.76 | AAI-based (high-confidence) | 0 |
| S7-final-viral-combined_k141_79304\|\|full | 3670 | No | NA | 8 | 0 | 0 | Low-quality | Genome-fragment | 8.31 | AAI-based (high-confidence) | 0 |
| S2-final-viral-combined_k141_198050\|\|full | 1512 | No | NA | 3 | 0 | 2 | Not-determined | Genome-fragment | NA | NA | 0 |
| S17-k141_109881 | 3670 | No | NA | 3 | 0 | 2 | Low-quality | Genome-fragment | 8.39 | AAI-based (medium-confidence) | 0 |
| S17-final-viral-combined_k141_78984\|\|full | 3671 | No | NA | 4 | 0 | 3 | Low-quality | Genome-fragment | 3.01 | AAI-based (high-confidence) | 0 |
| S12_k141_262180 | 3671 | No | NA | 3 | 0 | 2 | Low-quality | Genome-fragment | 6.21 | AAI-based (high-confidence) | 0 |
| S8-final-viral-combined_k141_231213\|\|full | 3671 | No | NA | 3 | 0 | 3 | Low-quality | Genome-fragment | 2.06 | AAI-based (medium-confidence) | 0 |
| S8-final-viral-combined_k141_191582\|\|full | 3671 | No | NA | 9 | 1 | 0 | Low-quality | Genome-fragment | 9.92 | AAI-based (medium-confidence) | 0 |
| S9_k141_105686 | 3672 | No | NA | 3 | 0 | 3 | Low-quality | Genome-fragment | 3.03 | AAI-based (medium-confidence) | 0 |
| S2-final-viral-combined_k141_229724\|\|full | 1240 | No | NA | 2 | 0 | 2 | Not-determined | Genome-fragment | NA | NA | 0 |
| S2-final-viral-combined_k141_22625\|\|full | 15970 | No | NA | 17 | 0 | 1 | Not-determined | Genome-fragment | NA | NA | 0 |
| S12_k141_350000 | 3672 | No | NA | 3 | 0 | 3 | Low-quality | Genome-fragment | 2.98 | AAI-based (medium-confidence) | 0 |

| | | | | | | | | | | | |
|---|---|---|---|---|---|---|---|---|---|---|---|
| S1_k141_296730 | 3673 | No | NA | 6 | 4 | 0 | Low-quality | Genome-fragment | 5.94 | AAI-based (medium-confidence) | 0 |
| S7_k141_186544 | 3673 | No | NA | 3 | 0 | 2 | Low-quality | Genome-fragment | 1.17 | AAI-based (medium-confidence) | 0 |
| S17-k141_228823 | 3673 | No | NA | 3 | 0 | 0 | Low-quality | Genome-fragment | 6.99 | AAI-based (medium-confidence) | 0 |
| S2-final-viral-combined_k141_178216\|\|full | 1216 | No | NA | 3 | 0 | 2 | Not-determined | Genome-fragment | NA | NA | 0 |
| S2-final-viral-combined_k141_227625\|\|full | 3673 | No | NA | 5 | 1 | 0 | Low-quality | Genome-fragment | 4.86 | HMM-based (lower-bound) | 0 |
| S2-final-viral-combined_k141_208743\|\|full | 1658 | No | NA | 2 | 0 | 2 | Not-determined | Genome-fragment | NA | NA | 0 |
| S1-final-viral-combined_k141_121284\|\|full | 3674 | No | NA | 3 | 0 | 2 | Low-quality | Genome-fragment | 1.99 | AAI-based (medium-confidence) | 0 |
| S2-final-viral-combined_k141_115492\|\|full | 2326 | No | NA | 3 | 0 | 2 | Not-determined | Genome-fragment | NA | NA | 0 |
| S7-final-viral-combined_k141_941\|\|full | 3674 | No | NA | 4 | 0 | 1 | Low-quality | Genome-fragment | 3.09 | AAI-based (medium-confidence) | 0 |
| S6-final-viral-combined_k141_58768\|\|full | 3674 | No | NA | 3 | 0 | 3 | Low-quality | Genome-fragment | 1.12 | AAI-based (medium-confidence) | 0 |
| S2-final-viral-combined_k141_178223\|\|full | 1951 | No | NA | 4 | 0 | 2 | Not-determined | Genome-fragment | NA | NA | 0 |
| S7-final-viral-combined_k141_174293\|\|full | 3675 | No | NA | 5 | 0 | 0 | Low-quality | Genome-fragment | 9.06 | AAI-based (medium-confidence) | 0 |
| S7-final-viral-combined_k141_92885\|\|full | 3675 | No | NA | 5 | 0 | 0 | Low-quality | Genome-fragment | 5.6 | AAI-based (high-confidence) | 0 |
| S17-k141_204933 | 3675 | No | NA | 2 | 0 | 1 | Low-quality | Genome-fragment | 6.61 | AAI-based (medium-confidence) | 0 |
| S2-final-viral-combined_k141_239859\|\|full | 2825 | No | NA | 3 | 0 | 0 | Not-determined | Genome-fragment | NA | NA | 0 |
| S2-final-viral-combined_k141_63791\|\|full | 2558 | No | NA | 3 | 0 | 1 | Not-determined | Genome-fragment | NA | NA | 0 |
| S4-final-viral-combined_k141_75526\|\|full | 3675 | No | NA | 3 | 0 | 3 | Low-quality | Genome-fragment | 4.36 | AAI-based (high-confidence) | 0 |
| S11-final-viral-combined_k141_317598\|\|full | 3676 | No | NA | 5 | 4 | 0 | Low-quality | Genome-fragment | 8.44 | AAI-based (high-confidence) | 0 |
| S11-final-viral-combined_k141_192190\|\|full | 3676 | No | NA | 6 | 3 | 0 | Low-quality | Genome-fragment | 9.88 | AAI-based (high-confidence) | 0 |
| S5-final-viral-combined_k141_96849\|\|full | 3676 | No | NA | 4 | 0 | 2 | Low-quality | Genome-fragment | 6.27 | AAI-based (high-confidence) | 0 |
| S11_k141_191172 | 3677 | No | NA | 6 | 2 | 0 | Low-quality | Genome-fragment | 6.06 | AAI-based (high-confidence) | 0 |
| S1_k141_216264 | 3677 | No | NA | 3 | 0 | 1 | Low-quality | Genome-fragment | 4.28 | AAI-based (high-confidence) | 0 |
| S2-final-viral-combined_k141_198099\|\|full | 3579 | No | NA | 4 | 0 | 1 | Not-determined | Genome-fragment | NA | NA | 0 |
| S5_k141_130088 | 3678 | No | NA | 12 | 0 | 0 | Low-quality | Genome-fragment | 9.46 | AAI-based (high-confidence) | 0 |
| S2-final-viral-combined_k141_84716\|\|full | 5351 | No | NA | 6 | 0 | 1 | Not-determined | Genome-fragment | NA | NA | 0 |
| S2-final-viral-combined_k141_84723\|\|full | 2690 | No | NA | 3 | 0 | 2 | Not-determined | Genome-fragment | NA | NA | 0 |
| S2-final-viral-combined_k141_84725\|\|full | 3309 | No | NA | 5 | 0 | 0 | Not-determined | Genome-fragment | NA | NA | 0 |
| S2-final-viral-combined_k141_198686\|\|full | 3678 | No | NA | 5 | 2 | 0 | Low-quality | Genome-fragment | 6.7 | AAI-based (medium-confidence) | 0 |
| S2-final-viral-combined_k141_115577\|\|full | 1020 | No | NA | 2 | 0 | 0 | Not-determined | Genome-fragment | NA | NA | 0 |
| S2-final-viral-combined_k141_126044\|\|full | 13865 | No | NA | 11 | 0 | 9 | Not-determined | Genome-fragment | NA | NA | 0 |
| S2-final-viral-combined_k141_198109\|\|full | 1799 | No | NA | 3 | 0 | 1 | Not-determined | Genome-fragment | NA | NA | 0 |
| S4-final-viral-combined_k141_42159\|\|full | 3678 | No | NA | 4 | 0 | 3 | Low-quality | Genome-fragment | 2.42 | AAI-based (high-confidence) | 0 |
| S8-final-viral-combined_k141_197875\|\|full | 3678 | No | NA | 4 | 1 | 0 | Low-quality | Genome-fragment | 3.96 | HMM-based (lower-bound) | 0 |
| S9-final-viral-combined_k141_92496\|\|full | 3679 | No | NA | 5 | 0 | 1 | Low-quality | Genome-fragment | 1.15 | AAI-based (medium-confidence) | 0 |
| S2-final-viral-combined_k141_115606\|\|full | 6815 | No | NA | 6 | 0 | 3 | Not-determined | Genome-fragment | NA | NA | 0 |
| S8-final-viral-combined_k141_102600\|\|full | 3679 | No | NA | 3 | 2 | 0 | Low-quality | Genome-fragment | 8.71 | AAI-based (medium-confidence) | 0 |
| S2-final-viral-combined_k141_126071\|\|full | 1803 | No | NA | 3 | 0 | 2 | Not-determined | Genome-fragment | NA | NA | 0 |
| S2-final-viral-combined_k141_74183\|\|full | 1901 | No | NA | 3 | 0 | 0 | Not-determined | Genome-fragment | NA | NA | 0 |
| S2-final-viral-combined_k141_115627\|\|full | 1598 | No | NA | 3 | 0 | 0 | Not-determined | Genome-fragment | NA | NA | 0 |
| S2-final-viral-combined_k141_74189\|\|full | 1321 | No | NA | 3 | 0 | 0 | Not-determined | Genome-fragment | NA | NA | 0 |
| S4-final-viral-combined_k141_51653\|\|full | 3680 | No | NA | 3 | 0 | 2 | Low-quality | Genome-fragment | 1.93 | AAI-based (medium-confidence) | 0 |
| S2-final-viral-combined_k141_53511\|\|full | 2219 | No | NA | 3 | 0 | 1 | Not-determined | Genome-fragment | NA | NA | 0 |
| S8_k141_32178 | 3680 | No | NA | 3 | 3 | 0 | Low-quality | Genome-fragment | 5.96 | AAI-based (high-confidence) | 0 |
| S2-final-viral-combined_k141_198142\|\|full | 3409 | No | NA | 3 | 0 | 0 | Not-determined | Genome-fragment | NA | NA | 0 |
| S4-final-viral-combined_k141_46987\|\|full | 3681 | No | NA | 3 | 0 | 3 | Low-quality | Genome-fragment | 4.43 | AAI-based (medium-confidence) | 0 |
| S2-final-viral-combined_k141_178339\|\|full | 4134 | No | NA | 3 | 0 | 0 | Not-determined | Genome-fragment | NA | NA | 0 |
| S5-final-viral-combined_k141_196460\|\|full | 3683 | No | NA | 3 | 0 | 0 | Low-quality | Genome-fragment | 9.4 | AAI-based (high-confidence) | 0 |
| S2-final-viral-combined_k141_167879\|\|full | 1274 | No | NA | 2 | 0 | 1 | Not-determined | Genome-fragment | NA | NA | 0 |
| S2-final-viral-combined_k141_105472\|\|full | 2072 | No | NA | 2 | 0 | 1 | Not-determined | Genome-fragment | NA | NA | 0 |
| S5-final-viral-combined_k141_134138\|\|full | 3683 | No | NA | 5 | 0 | 4 | Low-quality | Genome-fragment | 1.54 | AAI-based (medium-confidence) | 0 |
| S5-final-viral-combined_k141_201369\|\|full | 3683 | No | NA | 4 | 2 | 0 | Low-quality | Genome-fragment | 6.23 | AAI-based (high-confidence) | 0 |
| S2-final-viral-combined_k141_22693\|\|full | 2490 | No | NA | 3 | 0 | 0 | Not-determined | Genome-fragment | NA | NA | 0 |
| S2-final-viral-combined_k141_208826\|\|full | 5746 | No | NA | 5 | 0 | 3 | Not-determined | Genome-fragment | NA | NA | 0 |
| S2_k141_137952 | 3683 | No | NA | 10 | 2 | 0 | Low-quality | Genome-fragment | 7.41 | AAI-based (medium-confidence) | 0 |
| S2-final-viral-combined_k141_198179\|\|full | 1125 | No | NA | 3 | 0 | 1 | Not-determined | Genome-fragment | NA | NA | 0 |
| S6_k141_36056 | 3683 | No | NA | 3 | 1 | 0 | Low-quality | Genome-fragment | 3.61 | AAI-based (high-confidence) | 0 |
| S9-final-viral-combined_k141_93726\|\|full | 3684 | No | NA | 2 | 0 | 0 | Low-quality | Genome-fragment | 2.72 | AAI-based (medium-confidence) | 0 |
| S11_k141_204587 | 3684 | No | NA | 5 | 3 | 0 | Low-quality | Genome-fragment | 12.99 | HMM-based (lower-bound) | 0 |
| S3_k141_100561 | 3684 | No | NA | 2 | 0 | 2 | Low-quality | Genome-fragment | 1.5 | AAI-based (medium-confidence) | 0 |
| S2-final-viral-combined_k141_63865\|\|full | 2924 | No | NA | 3 | 0 | 2 | Not-determined | Genome-fragment | NA | NA | 0 |
| S2-final-viral-combined_k141_74276\|\|full | 2887 | No | NA | 5 | 0 | 1 | Not-determined | Genome-fragment | NA | NA | 0 |
| S12_k141_145030 | 3686 | No | NA | 1 | 0 | 0 | Low-quality | Genome-fragment | 3.62 | AAI-based (medium-confidence) | 0 |
| S1-final-viral-combined_k141_185011\|\|full | 3687 | No | NA | 4 | 0 | 4 | Low-quality | Genome-fragment | 4.94 | AAI-based (medium-confidence) | 0 |
| S2-final-viral-combined_k141_178381\|\|full | 13819 | No | NA | 23 | 0 | 3 | Not-determined | Genome-fragment | NA | NA | 0 |
| S2-final-viral-combined_k141_128367\|\|full | 3687 | No | NA | 4 | 0 | 3 | Low-quality | Genome-fragment | 2.48 | AAI-based (medium-confidence) | 0 |
| S2-final-viral-combined_k141_219102\|\|full | 5166 | No | NA | 6 | 0 | 2 | Not-determined | Genome-fragment | NA | NA | 0 |
| S2-final-viral-combined_k141_126134\|\|full | 19616 | No | NA | 35 | 9 | 1 | Medium-quality | Genome-fragment | 52.51 | AAI-based (medium-confidence) | 0 |
| S4_k141_133165 | 3687 | No | NA | 3 | 0 | 1 | Low-quality | Genome-fragment | 2.35 | AAI-based (high-confidence) | 0 |
| S6-final-viral-combined_k141_99514\|\|full | 3687 | No | NA | 4 | 0 | 0 | Low-quality | Genome-fragment | 5.81 | AAI-based (high-confidence) | 0 |
| S2-final-viral-combined_k141_53612\|\|full | 1802 | No | NA | 2 | 0 | 1 | Not-determined | Genome-fragment | NA | NA | 0 |
| S2-final-viral-combined_k141_84863\|\|full | 2249 | No | NA | 5 | 0 | 1 | Not-determined | Genome-fragment | NA | NA | 0 |
| S2-final-viral-combined_k141_53648\|\|full | 1490 | No | NA | 4 | 0 | 0 | Not-determined | Genome-fragment | NA | NA | 0 |
| S2-final-viral-combined_k141_146901\|\|full | 1313 | No | NA | 2 | 0 | 2 | Not-determined | Genome-fragment | NA | NA | 0 |
| S3_k141_65649 | 3688 | No | NA | 3 | 0 | 0 | Low-quality | Genome-fragment | 5.66 | AAI-based (medium-confidence) | 0 |
| S2-final-viral-combined_k141_239953\|\|full | 1443 | No | NA | 4 | 0 | 0 | Not-determined | Genome-fragment | NA | NA | 0 |
| S2-final-viral-combined_k141_157159\|\|full | 2653 | No | NA | 6 | 0 | 0 | Not-determined | Genome-fragment | NA | NA | 0 |
| S2-final-viral-combined_k141_22758\|\|full | 2558 | No | NA | 3 | 0 | 2 | Not-determined | Genome-fragment | NA | NA | 0 |
| S7_k141_55211 | 3688 | No | NA | 3 | 0 | 1 | Low-quality | Genome-fragment | 7.42 | AAI-based (medium-confidence) | 0 |
| S17-final-viral-combined_k141_75971\|\|full | 3688 | No | NA | 3 | 0 | 1 | Low-quality | Genome-fragment | 7.13 | AAI-based (medium-confidence) | 0 |
| S11-final-viral-combined_k141_142327\|\|full | 3689 | No | NA | 6 | 0 | 1 | Low-quality | Genome-fragment | 8.17 | AAI-based (high-confidence) | 0 |
| S2-final-viral-combined_k141_22763\|\|full | 7234 | No | NA | 8 | 0 | 2 | Not-determined | Genome-fragment | NA | NA | 0 |
| S12-final-viral-combined_k141_306574\|\|full | 3689 | No | NA | 6 | 3 | 0 | Low-quality | Genome-fragment | 8.78 | AAI-based (high-confidence) | 0 |
| S12-final-viral-combined_k141_90565\|\|full | 3689 | No | NA | 3 | 0 | 1 | Low-quality | Genome-fragment | 3.69 | AAI-based (medium-confidence) | 0 |
| S2-final-viral-combined_k141_126210\|\|full | 1181 | No | NA | 3 | 0 | 2 | Not-determined | Genome-fragment | NA | NA | 0 |
| S2-final-viral-combined_k141_44385\|\|full | 2241 | No | NA | 3 | 0 | 3 | Not-determined | Genome-fragment | NA | NA | 0 |
| S12-final-viral-combined_k141_12449\|\|full | 3689 | No | NA | 2 | 0 | 1 | Low-quality | Genome-fragment | 1.91 | AAI-based (medium-confidence) | 0 |
| S2-final-viral-combined_k141_53736\|\|full | 1703 | No | NA | 3 | 0 | 1 | Not-determined | Genome-fragment | NA | NA | 0 |
| S2-final-viral-combined_k141_178447\|\|full | 2096 | No | NA | 3 | 0 | 1 | Not-determined | Genome-fragment | NA | NA | 0 |
| S4-final-viral-combined_k141_4295\|\|full | 3689 | No | NA | 3 | 0 | 1 | Low-quality | Genome-fragment | 5.24 | AAI-based (medium-confidence) | 0 |
| S2-final-viral-combined_k141_133374\|\|full | 3691 | No | NA | 2 | 0 | 1 | Low-quality | Genome-fragment | 3.96 | AAI-based (medium-confidence) | 0 |
| S17-k141_60244 | 3692 | No | NA | 4 | 0 | 1 | Low-quality | Genome-fragment | 7.22 | AAI-based (medium-confidence) | 0 |
| S2-final-viral-combined_k141_188317\|\|full | 3926 | No | NA | 4 | 0 | 4 | Not-determined | Genome-fragment | NA | NA | 0 |
| S2-final-viral-combined_k141_2029\|\|full | 31399 | No | NA | 28 | 0 | 18 | Not-determined | Genome-fragment | NA | NA | 0 |
| S2-final-viral-combined_k141_170154\|\|full | 3692 | No | NA | 3 | 0 | 1 | Low-quality | Genome-fragment | 1.85 | AAI-based (medium-confidence) | 0 |
| S2-final-viral-combined_k141_64021\|\|full | 5175 | No | NA | 7 | 0 | 4 | Not-determined | Genome-fragment | NA | NA | 0 |
| S2-final-viral-combined_k141_146942\|\|full | 5784 | No | NA | 10 | 0 | 3 | Not-determined | Genome-fragment | NA | NA | 0 |
| S11-final-viral-combined_k141_210912\|\|full | 3693 | No | NA | 6 | 1 | 0 | Low-quality | Genome-fragment | 8.8 | AAI-based (high-confidence) | 0 |
| S2-final-viral-combined_k141_44488\|\|full | 1318 | No | NA | 2 | 0 | 1 | Not-determined | Genome-fragment | NA | NA | 0 |
| S11-final-viral-combined_k141_13162\|\|full | 3693 | No | NA | 6 | 1 | 0 | Low-quality | Genome-fragment | 6.13 | AAI-based (high-confidence) | 0 |
| S17-k141_76814 | 3693 | No | NA | 7 | 1 | 0 | Low-quality | Genome-fragment | 9.33 | AAI-based (medium-confidence) | 0 |
| S2-final-viral-combined_k141_84969\|\|full | 6635 | No | NA | 6 | 0 | 3 | Not-determined | Genome-fragment | NA | NA | 0 |
| S2-final-viral-combined_k141_188360\|\|full | 1662 | No | NA | 4 | 0 | 2 | Not-determined | Genome-fragment | NA | NA | 0 |
| S2-final-viral-combined_k141_53804\|\|full | 2888 | No | NA | 4 | 0 | 2 | Not-determined | Genome-fragment | NA | NA | 0 |
| S2-final-viral-combined_k141_74387\|\|full | 60095 | Yes | 37755 | 72 | 20 | 16 | Complete | High-quality | 100 | Provirus (high-confidence) | 37.17 |
| S2-final-viral-combined_k141_105689\|\|full | 3376 | No | NA | 2 | 0 | 1 | Not-determined | Genome-fragment | NA | NA | 0 |
| S2-final-viral-combined_k141_32777\|\|full | 1271 | No | NA | 3 | 0 | 3 | Not-determined | Genome-fragment | NA | NA | 0 |
| S2-final-viral-combined_k141_95249\|\|full | 2862 | No | NA | 4 | 0 | 0 | Not-determined | Genome-fragment | NA | NA | 0 |
| S6-final-viral-combined_k141_102573\|\|full | 3693 | No | NA | 3 | 0 | 1 | Low-quality | Genome-fragment | 1.29 | AAI-based (medium-confidence) | 0 |
| S2-final-viral-combined_k141_167926\|\|full | 1683 | No | NA | 2 | 0 | 1 | Not-determined | Genome-fragment | NA | NA | 0 |
| S2-final-viral-combined_k141_198264\|\|full | 2060 | No | NA | 3 | 0 | 2 | Not-determined | Genome-fragment | NA | NA | 0 |
| S8_k141_58020 | 3693 | No | NA | 4 | 0 | 3 | Low-quality | Genome-fragment | 3.9 | AAI-based (high-confidence) | 0 |
| S1-final-viral-combined_k141_282339\|\|full | 3694 | No | NA | 5 | 0 | 0 | Low-quality | Genome-fragment | 8.96 | AAI-based (medium-confidence) | 0 |
| S3-final-viral-combined_k141_2387\|\|full | 3694 | No | NA | 5 | 2 | 0 | Low-quality | Genome-fragment | 9.28 | AAI-based (high-confidence) | 0 |
| S2-final-viral-combined_k141_126259\|\|full | 3797 | No | NA | 4 | 0 | 1 | Not-determined | Genome-fragment | NA | NA | 0 |
| S4-final-viral-combined_k141_58293\|\|full | 3694 | No | NA | 3 | 0 | 3 | Low-quality | Genome-fragment | 1.77 | AAI-based (medium-confidence) | 0 |
| S2-final-viral-combined_k141_22839\|\|full | 7345 | No | NA | 6 | 0 | 3 | Not-determined | Genome-fragment | NA | NA | 0 |
| S2-final-viral-combined_k141_157319\|\|full | 2901 | No | NA | 2 | 0 | 1 | Not-determined | Genome-fragment | NA | NA | 0 |
| S2-final-viral-combined_k141_136795\|\|full | 6399 | No | NA | 6 | 0 | 4 | Not-determined | Genome-fragment | NA | NA | 0 |
| S2-final-viral-combined_k141_85059\|\|full | 2127 | No | NA | 3 | 0 | 2 | Not-determined | Genome-fragment | NA | NA | 0 |
| S1-final-viral-combined_k141_345613\|\|full | 3695 | No | NA | 8 | 3 | 0 | Low-quality | Genome-fragment | 10 | AAI-based (high-confidence) | 0 |
| S2-final-viral-combined_k141_22869\|\|full | 3095 | No | NA | 2 | 0 | 2 | Not-determined | Genome-fragment | NA | NA | 0 |
| S5_k141_36894 | 3695 | No | NA | 11 | 2 | 0 | Low-quality | Genome-fragment | 6.76 | AAI-based (high-confidence) | 0 |
| S2-final-viral-combined_k141_105771\|\|full | 2155 | No | NA | 3 | 0 | 0 | Not-determined | Genome-fragment | NA | NA | 0 |
| S2-final-viral-combined_k141_178546\|\|full | 3004 | No | NA | 4 | 0 | 0 | Not-determined | Genome-fragment | NA | NA | 0 |
| S2-final-viral-combined_k141_105775\|\|full | 1637 | No | NA | 3 | 0 | 0 | Not-determined | Genome-fragment | NA | NA | 0 |
| S2-final-viral-combined_k141_230135\|\|full | 5317 | No | NA | 5 | 0 | 1 | Not-determined | Genome-fragment | NA | NA | 0 |
| S1-final-viral-combined_k141_348653\|\|full | 3696 | No | NA | 4 | 0 | 1 | Low-quality | Genome-fragment | 9.18 | AAI-based (high-confidence) | 0 |
| S2-final-viral-combined_k141_64152\|\|full | 2751 | No | NA | 3 | 0 | 3 | Not-determined | Genome-fragment | NA | NA | 0 |
| S2-final-viral-combined_k141_95330\|\|full | 1619 | No | NA | 2 | 0 | 1 | Not-determined | Genome-fragment | NA | NA | 0 |
| S2-final-viral-combined_k141_146948\|\|full | 1375 | No | NA | 3 | 0 | 0 | Not-determined | Genome-fragment | NA | NA | 0 |
| S1_k141_278753 | 3696 | No | NA | 5 | 4 | 0 | Low-quality | Genome-fragment | 6.36 | HMM-based (lower-bound) | 0 |
| S7-final-viral-combined_k141_147833\|\|full | 3697 | No | NA | 4 | 0 | 4 | Low-quality | Genome-fragment | 2.48 | AAI-based (medium-confidence) | 0 |
| S12_k141_223525 | 3697 | No | NA | 2 | 0 | 1 | Low-quality | Genome-fragment | 3.83 | AAI-based (medium-confidence) | 0 |
| S8_k141_128221 | 3697 | No | NA | 3 | 1 | 0 | Low-quality | Genome-fragment | 3.44 | AAI-based (high-confidence) | 0 |
| S2-final-viral-combined_k141_95357\|\|full | 1811 | No | NA | 3 | 0 | 2 | Not-determined | Genome-fragment | NA | NA | 0 |
| S2-final-viral-combined_k141_230197\|\|full | 2183 | No | NA | 2 | 0 | 2 | Not-determined | Genome-fragment | NA | NA | 0 |
| S11-final-viral-combined_k141_227385\|\|full | 3699 | No | NA | 4 | 3 | 0 | Low-quality | Genome-fragment | 7.57 | AAI-based (high-confidence) | 0 |
| S2-final-viral-combined_k141_136860\|\|full | 9800 | No | NA | 6 | 0 | 2 | Not-determined | Genome-fragment | NA | NA | 0 |
| S1-final-viral-combined_k141_375368\|\|full | 3699 | No | NA | 3 | 0 | 2 | Low-quality | Genome-fragment | 2.71 | AAI-based (high-confidence) | 0 |
| S2-final-viral-combined_k141_95378\|\|full | 4101 | No | NA | 3 | 0 | 1 | Not-determined | Genome-fragment | NA | NA | 0 |
| S17-k141_163430 | 3699 | No | NA | 4 | 0 | 4 | Low-quality | Genome-fragment | 1.24 | AAI-based (medium-confidence) | 0 |
| S2-final-viral-combined_k141_240154\|\|full | 2009 | No | NA | 3 | 0 | 0 | Not-determined | Genome-fragment | NA | NA | 0 |
| S2-final-viral-combined_k141_44570\|\|full | 2714 | No | NA | 3 | 0 | 1 | Not-determined | Genome-fragment | NA | NA | 0 |
| S2-final-viral-combined_k141_240161\|\|full | 6025 | No | NA | 5 | 0 | 3 | Not-determined | Genome-fragment | NA | NA | 0 |
| S4_k141_113591 | 3699 | No | NA | 5 | 1 | 0 | Low-quality | Genome-fragment | 3.57 | AAI-based (high-confidence) | 0 |
| S2-final-viral-combined_k141_230263\|\|full | 1117 | No | NA | 3 | 0 | 1 | Not-determined | Genome-fragment | NA | NA | 0 |
| S2-final-viral-combined_k141_188458\|\|full | 3150 | No | NA | 3 | 0 | 2 | Not-determined | Genome-fragment | NA | NA | 0 |
| S2-final-viral-combined_k141_157411\|\|full | 1233 | No | NA | 4 | 0 | 0 | Not-determined | Genome-fragment | NA | NA | 0 |
| S1-final-viral-combined_k141_85425\|\|full | 3700 | No | NA | 5 | 0 | 5 | Low-quality | Genome-fragment | 2.88 | AAI-based (medium-confidence) | 0 |
| S2-final-viral-combined_k141_12331\|\|full | 5691 | No | NA | 8 | 0 | 4 | Not-determined | Genome-fragment | NA | NA | 0 |
| S2-final-viral-combined_k141_230270\|\|full | 3175 | No | NA | 4 | 0 | 0 | Not-determined | Genome-fragment | NA | NA | 0 |
| S2-final-viral-combined_k141_157422\|\|full | 1629 | No | NA | 3 | 0 | 1 | Not-determined | Genome-fragment | NA | NA | 0 |
| S2-final-viral-combined_k141_64241\|\|full | 1769 | No | NA | 3 | 0 | 0 | Not-determined | Genome-fragment | NA | NA | 0 |
| S9-final-viral-combined_k141_20029\|\|full | 3701 | No | NA | 3 | 0 | 1 | Low-quality | Genome-fragment | 2.37 | AAI-based (high-confidence) | 0 |
| S2-final-viral-combined_k141_230282\|\|full | 3813 | No | NA | 4 | 1 | 0 | Medium-quality | Genome-fragment | 85.27 | AAI-based (medium-confidence) | 0 |
| S12-final-viral-combined_k141_216588\|\|full | 3701 | No | NA | 9 | 0 | 0 | Low-quality | Genome-fragment | 6.21 | AAI-based (high-confidence) | 0 |
| S2-final-viral-combined_k141_155050\|\|full | 3701 | No | NA | 9 | 0 | 0 | Low-quality | Genome-fragment | 9.78 | AAI-based (medium-confidence) | 0 |
| S5_k141_205706 | 3702 | No | NA | 5 | 0 | 2 | Low-quality | Genome-fragment | 6.18 | AAI-based (medium-confidence) | 0 |
| S2-final-viral-combined_k141_44627\|\|full | 8315 | No | NA | 5 | 0 | 5 | Not-determined | Genome-fragment | NA | NA | 0 |
| S2-final-viral-combined_k141_240214\|\|full | 1964 | No | NA | 3 | 0 | 1 | Not-determined | Genome-fragment | NA | NA | 0 |
| S1-final-viral-combined_k141_330288\|\|full | 3703 | No | NA | 5 | 0 | 0 | Low-quality | Genome-fragment | 2.76 | AAI-based (high-confidence) | 0 |
| S7_k141_184 | 3703 | No | NA | 3 | 0 | 0 | Low-quality | Genome-fragment | 4.29 | AAI-based (medium-confidence) | 0 |
| S2-final-viral-combined_k141_22869\|\|full | 1469 | No | NA | 3 | 0 | 2 | Not-determined | Genome-fragment | NA | NA | 0 |
| S2-final-viral-combined_k141_147003\|\|full | 1400 | No | NA | 3 | 0 | 1 | Not-determined | Genome-fragment | NA | NA | 0 |
| S9-final-viral-combined_k141_31862\|\|full | 3704 | No | NA | 2 | 0 | 2 | Low-quality | Genome-fragment | 2.03 | AAI-based (medium-confidence) | 0 |
| S11-final-viral-combined_k141_30153\|\|full | 3704 | No | NA | 3 | 0 | 3 | Low-quality | Genome-fragment | 8.27 | AAI-based (medium-confidence) | 0 |
| S2-final-viral-combined_k141_53986\|\|full | 1506 | No | NA | 4 | 0 | 2 | Not-determined | Genome-fragment | NA | NA | 0 |
| S2-final-viral-combined_k141_137000\|\|full | 4506 | No | NA | 5 | 0 | 1 | Not-determined | Genome-fragment | NA | NA | 0 |
| S1-final-viral-combined_k141_151720\|\|full | 3704 | No | NA | 4 | 2 | 0 | Low-quality | Genome-fragment | 8.86 | AAI-based (high-confidence) | 0 |
| S2-final-viral-combined_k141_230341\|\|full | 4460 | No | NA | 5 | 0 | 3 | Not-determined | Genome-fragment | NA | NA | 0 |
| S5-final-viral-combined_k141_107036\|\|full | 3704 | No | NA | 8 | 0 | 0 | Low-quality | Genome-fragment | 6.77 | AAI-based (high-confidence) | 0 |
| S2-final-viral-combined_k141_137024\|\|full | 6410 | No | NA | 5 | 0 | 4 | Not-determined | Genome-fragment | NA | NA | 0 |
| S5-final-viral-combined_k141_185862\|\|full | 3704 | No | NA | 7 | 6 | 0 | Low-quality | Genome-fragment | 8.09 | AAI-based (high-confidence) | 0 |
| S17-final-viral-combined_k141_152455\|\|full | 3704 | No | NA | 3 | 0 | 3 | Low-quality | Genome-fragment | 2.77 | AAI-based (medium-confidence) | 0 |
| S12-final-viral-combined_k141_245430\|\|full | 3704 | No | NA | 3 | 0 | 3 | Low-quality | Genome-fragment | 2.17 | AAI-based (high-confidence) | 0 |
| S2-final-viral-combined_k141_168182\|\|full | 2838 | No | NA | 3 | 0 | 0 | Not-determined | Genome-fragment | NA | NA | 0 |
| S2-final-viral-combined_k141_126440\|\|full | 23248 | No | NA | 31 | 10 | 0 | Medium-quality | Genome-fragment | 59.14 | AAI-based (medium-confidence) | 0 |
| S2-final-viral-combined_k141_230359\|\|full | 2661 | No | NA | 6 | 0 | 3 | Not-determined | Genome-fragment | NA | NA | 0 |
| S2-final-viral-combined_k141_126447\|\|full | 1469 | No | NA | 1 | 0 | 0 | Not-determined | Genome-fragment | NA | NA | 0 |
| S2-final-viral-combined_k141_230369\|\|full | 2267 | No | NA | 3 | 0 | 2 | Not-determined | Genome-fragment | NA | NA | 0 |
| S2-final-viral-combined_k141_85214\|\|full | 5643 | No | NA | 4 | 0 | 4 | Not-determined | Genome-fragment | NA | NA | 0 |
| S2-final-viral-combined_k141_230370\|\|full | 2220 | No | NA | 3 | 0 | 2 | Not-determined | Genome-fragment | NA | NA | 0 |
| S2-final-viral-combined_k141_188570\|\|full | 3056 | No | NA | 4 | 0 | 0 | Not-determined | Genome-fragment | NA | NA | 0 |
| S2-final-viral-combined_k141_126452\|\|full | 16461 | No | NA | 16 | 0 | 11 | Not-determined | Genome-fragment | NA | NA | 0 |
| S2-final-viral-combined_k141_32950\|\|full | 2510 | No | NA | 5 | 0 | 0 | Not-determined | Genome-fragment | NA | NA | 0 |
| S2-final-viral-combined_k141_230378\|\|full | 6076 | No | NA | 11 | 0 | 0 | Not-determined | Genome-fragment | NA | NA | 0 |
| S2-final-viral-combined_k141_105906\|\|full | 2541 | No | NA | 4 | 0 | 0 | Not-determined | Genome-fragment | NA | NA | 0 |
| S2-final-viral-combined_k141_44670\|\|full | 1195 | No | NA | 3 | 0 | 0 | Not-determined | Genome-fragment | NA | NA | 0 |
| S12_k141_79714 | 3705 | No | NA | 3 | 0 | 3 | Low-quality | Genome-fragment | 0.57 | AAI-based (medium-confidence) | 0 |
| S2-final-viral-combined_k141_99378\|\|full | 3705 | No | NA | 3 | 0 | 3 | Low-quality | Genome-fragment | 1.78 | AAI-based (medium-confidence) | 0 |
| S7-final-viral-combined_k141_207680\|\|full | 3707 | No | NA | 7 | 6 | 0 | Low-quality | Genome-fragment | 7.49 | AAI-based (medium-confidence) | 0 |
| S2-final-viral-combined_k141_157509\|\|full | 1384 | No | NA | 2 | 0 | 0 | Not-determined | Genome-fragment | NA | NA | 0 |
| S2-final-viral-combined_k141_2159\|\|full | 1054 | No | NA | 2 | 0 | 1 | Not-determined | Genome-fragment | NA | NA | 0 |
| S2-final-viral-combined_k141_44692\|\|full | 1935 | No | NA | 4 | 0 | 4 | Not-determined | Genome-fragment | NA | NA | 0 |
| S2-final-viral-combined_k141_219340\|\|full | 1753 | No | NA | 3 | 0 | 2 | Not-determined | Genome-fragment | NA | NA | 0 |
| S2-final-viral-combined_k141_105951\|\|full | 2306 | No | NA | 4 | 0 | 2 | Not-determined | Genome-fragment | NA | NA | 0 |
| S2-final-viral-combined_k141_64312\|\|full | 1577 | No | NA | 3 | 0 | 1 | Not-determined | Genome-fragment | NA | NA | 0 |
| S17-final-viral-combined_k141_257798\|\|full | 3707 | No | NA | 5 | 3 | 0 | Low-quality | Genome-fragment | 9.84 | AAI-based (high-confidence) | 0 |
| S2-final-viral-combined_k141_168221\|\|full | 2059 | No | NA | 2 | 0 | 0 | Not-determined | Genome-fragment | NA | NA | 0 |
| S12_k141_218707 | 3707 | No | NA | 3 | 0 | 0 | Low-quality | Genome-fragment | 9.22 | AAI-based (medium-confidence) | 0 |
| S7-final-viral-combined_k141_1665\|\|full | 3708 | No | NA | 3 | 0 | 3 | Low-quality | Genome-fragment | 5.04 | AAI-based (medium-confidence) | 0 |
| S17-final-viral-combined_k141_44142\|\|full | 3708 | No | NA | 7 | 4 | 0 | Low-quality | Genome-fragment | 9.77 | AAI-based (high-confidence) | 0 |
| S2-final-viral-combined_k141_240261\|\|full | 2149 | No | NA | 3 | 0 | 1 | Not-determined | Genome-fragment | NA | NA | 0 |
| S2-final-viral-combined_k141_44731\|\|full | 1516 | No | NA | 3 | 0 | 2 | Not-determined | Genome-fragment | NA | NA | 0 |
| S2-final-viral-combined_k141_64342\|\|full | 4468 | No | NA | 6 | 0 | 0 | Not-determined | Genome-fragment | NA | NA | 0 |
| S2-final-viral-combined_k141_2179\|\|full | 2624 | No | NA | 3 | 0 | 2 | Not-determined | Genome-fragment | NA | NA | 0 |
| S2-final-viral-combined_k141_198450\|\|full | 2292 | No | NA | 2 | 0 | 1 | Not-determined | Genome-fragment | NA | NA | 0 |
| S2-final-viral-combined_k141_240262\|\|full | 23789 | No | NA | 25 | 0 | 6 | Not-determined | Genome-fragment | NA | NA | 0 |
| S2-final-viral-combined_k141_126564\|\|full | 1013 | No | NA | 2 | 0 | 0 | Not-determined | Genome-fragment | NA | NA | 0 |
| S2-final-viral-combined_k141_95614\|\|full | 6211 | No | NA | 14 | 0 | 0 | Not-determined | Genome-fragment | NA | NA | 0 |
| S2-final-viral-combined_k141_168275\|\|full | 1345 | No | NA | 2 | 0 | 1 | Not-determined | Genome-fragment | NA | NA | 0 |
| S2-final-viral-combined_k141_230460\|\|full | 13823 | No | NA | 14 | 0 | 2 | Not-determined | Genome-fragment | NA | NA | 0 |
| S2-final-viral-combined_k141_23111\|\|full | 4832 | No | NA | 3 | 0 | 3 | Not-determined | Genome-fragment | NA | NA | 0 |
| S2-final-viral-combined_k141_54125\|\|full | 2903 | No | NA | 4 | 0 | 3 | Not-determined | Genome-fragment | NA | NA | 0 |
| S1_k141_104359 | 3709 | Yes | 1545 | 5 | 1 | 2 | Low-quality | Genome-fragment | 1.91 | HMM-based (lower-bound) | 58.34 |
| S7-final-viral-combined_k141_223110\|\|full | 3709 | No | NA | 2 | 0 | 1 | Low-quality | Genome-fragment | 2.33 | AAI-based (high-confidence) | 0 |
| S2-final-viral-combined_k141_54153\|\|full | 4796 | No | NA | 9 | 0 | 1 | Not-determined | Genome-fragment | NA | NA | 0 |
| S2-final-viral-combined_k141_116047\|\|full | 5236 | No | NA | 9 | 0 | 1 | Not-determined | Genome-fragment | NA | NA | 0 |
| S2-final-viral-combined_k141_44829\|\|full | 1433 | No | NA | 3 | 0 | 1 | Not-determined | Genome-fragment | NA | NA | 0 |
| S6-final-viral-combined_k141_114133\|\|full | 3709 | No | NA | 4 | 0 | 2 | Low-quality | Genome-fragment | 1.08 | AAI-based (medium-confidence) | 0 |
| S2-final-viral-combined_k141_147065\|\|full | 156271 | No | NA | 131 | 6 | 69 | High-quality | High-quality | 100 | HMM-based (lower-bound) | 0 |
| S8-final-viral-combined_k141_76254\|\|full | 3709 | No | NA | 3 | 0 | 2 | Low-quality | Genome-fragment | 3.31 | AAI-based (medium-confidence) | 0 |
| S2-final-viral-combined_k141_116057\|\|full | 1229 | No | NA | 3 | 0 | 0 | Not-determined | Genome-fragment | NA | NA | 0 |
| S11-final-viral-combined_k141_154493\|\|full | 3710 | No | NA | 7 | 0 | 1 | Low-quality | Genome-fragment | 9.62 | AAI-based (high-confidence) | 0 |
| S2-final-viral-combined_k141_137119\|\|full | 96227 | Yes | 51652 | 130 | 19 | 17 | High-quality | High-quality | 100 | AAI-based (high-confidence) | 46.32 |
| S2-final-viral-combined_k141_137121\|\|full | 3035 | No | NA | 3 | 0 | 3 | Not-determined | Genome-fragment | NA | NA | 0 |
| S2-final-viral-combined_k141_188718\|\|full | 2564 | No | NA | 3 | 0 | 0 | Not-determined | Genome-fragment | NA | NA | 0 |
| S1_k141_288732 | 3710 | No | NA | 3 | 0 | 1 | Low-quality | Genome-fragment | 6.35 | AAI-based (high-confidence) | 0 |
| S2-final-viral-combined_k141_137132\|\|full | 1230 | No | NA | 2 | 0 | 0 | Not-determined | Genome-fragment | NA | NA | 0 |
| S7-final-viral-combined_k141_205398\|\|full | 3710 | No | NA | 5 | 0 | 2 | Low-quality | Genome-fragment | 1.89 | AAI-based (medium-confidence) | 0 |
| S2-final-viral-combined_k141_168351\|\|full | 6387 | No | NA | 9 | 0 | 1 | Not-determined | Genome-fragment | NA | NA | 0 |
| S2-final-viral-combined_k141_44869\|\|full | 2063 | No | NA | 2 | 0 | 0 | Not-determined | Genome-fragment | NA | NA | 0 |
| S17-final-viral-combined_k141_202684\|\|full | 3710 | No | NA | 3 | 0 | 2 | Low-quality | Genome-fragment | 8.42 | AAI-based (medium-confidence) | 0 |
| S2-final-viral-combined_k141_209211\|\|full | 2982 | No | NA | 2 | 0 | 2 | Not-determined | Genome-fragment | NA | NA | 0 |
| S2-final-viral-combined_k141_126611\|\|full | 10792 | No | NA | 10 | 0 | 4 | Not-determined | Genome-fragment | NA | NA | 0 |
| S12_k141_63444 | 3710 | No | NA | 1 | 0 | 1 | Low-quality | Genome-fragment | 3.04 | AAI-based (medium-confidence) | 0 |
| S8-final-viral-combined_k141_41597\|\|full | 3710 | No | NA | 3 | 0 | 2 | Low-quality | Genome-fragment | 1.71 | AAI-based (medium-confidence) | 0 |
| S7_k141_77868 | 3711 | No | NA | 6 | 1 | 0 | Low-quality | Genome-fragment | 3.6 | AAI-based (high-confidence) | 0 |
| S2-final-viral-combined_k141_85381\|\|full | 43616 | Yes | 27079 | 47 | 15 | 9 | Medium-quality | Genome-fragment | 56.47 | AAI-based (medium-confidence) | 37.91 |
| S17-final-viral-combined_k141_110958\|\|full | 3711 | No | NA | 6 | 0 | 0 | Low-quality | Genome-fragment | 3.32 | AAI-based (high-confidence) | 0 |
| S2-final-viral-combined_k141_116090\|\|full | 2045 | No | NA | 2 | 0 | 2 | Not-determined | Genome-fragment | NA | NA | 0 |
| S2-final-viral-combined_k141_126664\|\|full | 5829 | No | NA | 8 | 0 | 2 | Not-determined | Genome-fragment | NA | NA | 0 |
| S2-final-viral-combined_k141_12604\|\|full | 8257 | No | NA | 6 | 0 | 2 | Not-determined | Genome-fragment | NA | NA | 0 |
| S12-final-viral-combined_k141_130654\|\|full | 3711 | No | NA | 4 | 0 | 1 | Low-quality | Genome-fragment | 8.42 | AAI-based (medium-confidence) | 0 |
| S2-final-viral-combined_k141_157648\|\|full | 3272 | No | NA | 2 | 0 | 2 | Not-determined | Genome-fragment | NA | NA | 0 |
| S2-final-viral-combined_k141_188794\|\|full | 1677 | No | NA | 4 | 0 | 0 | Not-determined | Genome-fragment | NA | NA | 0 |
| S2-final-viral-combined_k141_12619\|\|full | 4253 | No | NA | 3 | 0 | 2 | Not-determined | Genome-fragment | NA | NA | 0 |
| S2-final-viral-combined_k141_147159\|\|full | 1573 | No | NA | 3 | 0 | 3 | Not-determined | Genome-fragment | NA | NA | 0 |
| S2-final-viral-combined_k141_219527\|\|full | 11248 | No | NA | 13 | 0 | 4 | Not-determined | Genome-fragment | NA | NA | 0 |
| S2-final-viral-combined_k141_209294\|\|full | 2896 | No | NA | 3 | 0 | 2 | Not-determined | Genome-fragment | NA | NA | 0 |
| S2-final-viral-combined_k141_178657\|\|full | 1229 | No | NA | 3 | 0 | 1 | Not-determined | Genome-fragment | NA | NA | 0 |
| S1-final-viral-combined_k141_201859\|\|full | 3712 | No | NA | 9 | 5 | 0 | Low-quality | Genome-fragment | 10.54 | AAI-based (high-confidence) | 0 |
| S2-final-viral-combined_k141_23162\|\|full | 7303 | No | NA | 7 | 0 | 7 | Not-determined | Genome-fragment | NA | NA | 0 |
| S2-final-viral-combined_k141_44945\|\|full | 4549 | No | NA | 5 | 0 | 3 | Not-determined | Genome-fragment | NA | NA | 0 |
| S1_k141_67226 | 3712 | No | NA | 7 | 0 | 0 | Low-quality | Genome-fragment | 7.84 | AAI-based (high-confidence) | 0 |
| S2-final-viral-combined_k141_12658\|\|full | 1436 | No | NA | 2 | 0 | 0 | Not-determined | Genome-fragment | NA | NA | 0 |
| S2-final-viral-combined_k141_54252\|\|full | 3186 | No | NA | 3 | 0 | 1 | Not-determined | Genome-fragment | NA | NA | 0 |
| S2-final-viral-combined_k141_219559\|\|full | 1695 | No | NA | 4 | 0 | 0 | Not-determined | Genome-fragment | NA | NA | 0 |
| S7_k141_153097 | 3712 | No | NA | 5 | 1 | 0 | Low-quality | Genome-fragment | 6.89 | AAI-based (high-confidence) | 0 |
| S12_k141_170141 | 3712 | No | NA | 5 | 0 | 2 | Low-quality | Genome-fragment | 3.69 | AAI-based (medium-confidence) | 0 |
| S8-final-viral-combined_k141_114901\|\|full | 3712 | No | NA | 3 | 0 | 3 | Low-quality | Genome-fragment | 1.19 | AAI-based (medium-confidence) | 0 |
| S2-final-viral-combined_k141_54297\|\|full | 2237 | No | NA | 3 | 0 | 0 | Not-determined | Genome-fragment | NA | NA | 0 |
| S8_k141_274740 | 3712 | No | NA | 8 | 4 | 0 | Low-quality | Genome-fragment | 10.18 | AAI-based (medium-confidence) | 0 |
| S2-final-viral-combined_k141_44984\|\|full | 1192 | No | NA | 2 | 0 | 1 | Not-determined | Genome-fragment | NA | NA | 0 |
| S3-final-viral-combined_k141_38285\|\|full | 3713 | No | NA | 3 | 0 | 2 | Low-quality | Genome-fragment | 2.04 | AAI-based (medium-confidence) | 0 |
| S2-final-viral-combined_k141_54301\|\|full | 1694 | No | NA | 3 | 0 | 1 | Not-determined | Genome-fragment | NA | NA | 0 |
| S2-final-viral-combined_k141_209367\|\|full | 1580 | No | NA | 2 | 0 | 2 | Not-determined | Genome-fragment | NA | NA | 0 |
| S5_k141_102801 | 3713 | No | NA | 3 | 1 | 0 | Low-quality | Genome-fragment | 1.86 | HMM-based (lower-bound) | 0 |
| S2-final-viral-combined_k141_64388\|\|full | 9697 | No | NA | 17 | 0 | 1 | Not-determined | Genome-fragment | NA | NA | 0 |
| S12_k141_165246 | 3713 | No | NA | 5 | 2 | 0 | Low-quality | Genome-fragment | 9.47 | AAI-based (medium-confidence) | 0 |
| S2-final-viral-combined_k141_54331\|\|full | 2007 | No | NA | 5 | 0 | 0 | Not-determined | Genome-fragment | NA | NA | 0 |
| S11-final-viral-combined_k141_306442\|\|full | 3714 | No | NA | 9 | 0 | 0 | Low-quality | Genome-fragment | 7.04 | AAI-based (high-confidence) | 0 |
| S2-final-viral-combined_k141_240385\|\|full | 2485 | No | NA | 2 | 0 | 0 | Not-determined | Genome-fragment | NA | NA | 0 |
| S2-final-viral-combined_k141_230652\|\|full | 2335 | No | NA | 5 | 0 | 1 | Not-determined | Genome-fragment | NA | NA | 0 |
| S2-final-viral-combined_k141_178742\|\|full | 1532 | No | NA | 3 | 0 | 1 | Not-determined | Genome-fragment | NA | NA | 0 |
| S7_k141_232508 | 3714 | No | NA | 11 | 3 | 1 | Low-quality | Genome-fragment | 8.64 | AAI-based (high-confidence) | 0 |
| S2-final-viral-combined_k141_240394\|\|full | 2232 | No | NA | 4 | 0 | 1 | Not-determined | Genome-fragment | NA | NA | 0 |
| S2-final-viral-combined_k141_209441\|\|full | 1045 | No | NA | 3 | 0 | 0 | Not-determined | Genome-fragment | NA | NA | 0 |
| S2-final-viral-combined_k141_157794\|\|full | 8276 | No | NA | 11 | 0 | 4 | Not-determined | Genome-fragment | NA | NA | 0 |
| S2-final-viral-combined_k141_230685\|\|full | 1568 | No | NA | 3 | 0 | 2 | Not-determined | Genome-fragment | NA | NA | 0 |
| S2-final-viral-combined_k141_23254\|\|full | 2285 | No | NA | 4 | 0 | 4 | Not-determined | Genome-fragment | NA | NA | 0 |
| S9-final-viral-combined_k141_79640\|\|full | 3715 | No | NA | 5 | 0 | 1 | Low-quality | Genome-fragment | 16.77 | AAI-based (medium-confidence) | 0 |
| S2-final-viral-combined_k141_168524\|\|full | 1500 | No | NA | 2 | 0 | 2 | Not-determined | Genome-fragment | NA | NA | 0 |
| S1-final-viral-combined_k141_42968\|\|full | 3715 | No | NA | 6 | 0 | 1 | Low-quality | Genome-fragment | 8.6 | AAI-based (high-confidence) | 0 |
| S5-final-viral-combined_k141_112708\|\|full | 3715 | No | NA | 5 | 1 | 0 | Low-quality | Genome-fragment | 4.17 | HMM-based (lower-bound) | 0 |
| S2-final-viral-combined_k141_85490\|\|full | 2259 | No | NA | 3 | 0 | 2 | Not-determined | Genome-fragment | NA | NA | 0 |
| S2-final-viral-combined_k141_12754\|\|full | 2290 | No | NA | 3 | 0 | 2 | Not-determined | Genome-fragment | NA | NA | 0 |
| S11-final-viral-combined_k141_164548\|\|full | 3716 | No | NA | 5 | 0 | 0 | Low-quality | Genome-fragment | 3.53 | AAI-based (medium-confidence) | 0 |
| S1-final-viral-combined_k141_354637\|\|full | 3716 | Yes | 1728 | 5 | 1 | 1 | Low-quality | Genome-fragment | 1.09 | HMM-based (lower-bound) | 53.5 |
| S2-final-viral-combined_k141_23281\|\|full | 2356 | No | NA | 2 | 0 | 2 | Not-determined | Genome-fragment | NA | NA | 0 |
| S2-final-viral-combined_k141_33220\|\|full | 2145 | No | NA | 3 | 0 | 3 | Not-determined | Genome-fragment | NA | NA | 0 |
| S2-final-viral-combined_k141_147277\|\|full | 1941 | No | NA | 4 | 0 | 0 | Not-determined | Genome-fragment | NA | NA | 0 |
| S6_k141_30981 | 3716 | No | NA | 2 | 0 | 0 | Low-quality | Genome-fragment | 6.36 | AAI-based (high-confidence) | 0 |
| S9_k141_71368 | 3717 | No | NA | 2 | 0 | 1 | Low-quality | Genome-fragment | 6.13 | AAI-based (medium-confidence) | 0 |
| S2-final-viral-combined_k141_198663\|\|full | 10086 | No | NA | 12 | 0 | 10 | Not-determined | Genome-fragment | NA | NA | 0 |
| S1-final-viral-combined_k141_84309\|\|full | 3718 | No | NA | 7 | 2 | 0 | Low-quality | Genome-fragment | 6.57 | AAI-based (high-confidence) | 0 |
| S2-final-viral-combined_k141_33253\|\|full | 2933 | No | NA | 3 | 0 | 0 | Not-determined | Genome-fragment | NA | NA | 0 |
| S2-final-viral-combined_k141_23321\|\|full | 3795 | No | NA | 4 | 0 | 2 | Not-determined | Genome-fragment | NA | NA | 0 |
| S7_k141_54705 | 3719 | No | NA | 3 | 0 | 2 | Low-quality | Genome-fragment | 0.88 | AAI-based (medium-confidence) | 0 |
| S2-final-viral-combined_k141_85528\|\|full | 9967 | No | NA | 20 | 0 | 6 | Not-determined | Genome-fragment | NA | NA | 0 |
| S2-final-viral-combined_k141_198680\|\|full | 4427 | No | NA | 4 | 0 | 1 | Not-determined | Genome-fragment | NA | NA | 0 |
| S2-final-viral-combined_k141_178844\|\|full | 3630 | No | NA | 5 | 0 | 4 | Not-determined | Genome-fragment | NA | NA | 0 |
| S2-final-viral-combined_k141_126699\|\|full | 1673 | No | NA | 3 | 0 | 1 | Not-determined | Genome-fragment | NA | NA | 0 |
| S12-final-viral-combined_k141_287910\|\|full | 3719 | No | NA | 6 | 0 | 5 | Low-quality | Genome-fragment | 1.76 | AAI-based (medium-confidence) | 0 |
| S1-final-viral-combined_k141_109061\|\|full | 3720 | No | NA | 3 | 0 | 3 | Low-quality | Genome-fragment | 2.26 | AAI-based (medium-confidence) | 0 |
| S2-final-viral-combined_k141_45090\|\|full | 44627 | No | NA | 34 | 0 | 27 | Not-determined | Genome-fragment | NA | NA | 0 |
| S2-final-viral-combined_k141_126715\|\|full | 5399 | No | NA | 5 | 0 | 5 | Not-determined | Genome-fragment | NA | NA | 0 |
| S2-final-viral-combined_k141_2391\|\|full | 5780 | No | NA | 5 | 0 | 4 | Not-determined | Genome-fragment | NA | NA | 0 |
| S11-final-viral-combined_k141_59005\|\|full | 3721 | Yes | 2565 | 4 | 1 | 1 | Low-quality | Genome-fragment | 5.56 | AAI-based (medium-confidence) | 31.07 |
| S12-final-viral-combined_k141_337999\|\|full | 3722 | No | NA | 3 | 0 | 2 | Low-quality | Genome-fragment | 3.03 | AAI-based (high-confidence) | 0 |
| S2-final-viral-combined_k141_178886\|\|full | 1146 | No | NA | 4 | 0 | 2 | Not-determined | Genome-fragment | NA | NA | 0 |
| S2-final-viral-combined_k141_12850\|\|full | 2418 | No | NA | 3 | 0 | 2 | Not-determined | Genome-fragment | NA | NA | 0 |
| S2-final-viral-combined_k141_63690\|\|full | 3722 | No | NA | 3 | 0 | 2 | Low-quality | Genome-fragment | 1.88 | AAI-based (medium-confidence) | 0 |
| S2-final-viral-combined_k141_2402\|\|full | 4034 | No | NA | 6 | 0 | 1 | Not-determined | Genome-fragment | NA | NA | 0 |
| S2-final-viral-combined_k141_137280\|\|full | 7115 | No | NA | 9 | 0 | 7 | Not-determined | Genome-fragment | NA | NA | 0 |
| S2-final-viral-combined_k141_137289\|\|full | 1205 | No | NA | 3 | 0 | 0 | Not-determined | Genome-fragment | NA | NA | 0 |
| S2-final-viral-combined_k141_64481\|\|full | 5363 | No | NA | 4 | 0 | 3 | Not-determined | Genome-fragment | NA | NA | 0 |
| S2_k141_118679 | 3722 | No | NA | 4 | 1 | 0 | Low-quality | Genome-fragment | 3.88 | AAI-based (high-confidence) | 0 |
| S2-final-viral-combined_k141_64492\|\|full | 1074 | No | NA | 3 | 0 | 0 | Not-determined | Genome-fragment | NA | NA | 0 |
| S2-final-viral-combined_k141_116221\|\|full | 90374 | Yes | 78965 | 126 | 36 | 22 | High-quality | High-quality | 94.24 | HMM-based (lower-bound) | 12.62 |
| S7-final-viral-combined_k141_55830\|\|full | 3723 | No | NA | 4 | 0 | 3 | Low-quality | Genome-fragment | 3.66 | AAI-based (medium-confidence) | 0 |
| S2-final-viral-combined_k141_132494\|\|full | 3723 | No | NA | 6 | 0 | 0 | Low-quality | Genome-fragment | 2.28 | AAI-based (high-confidence) | 0 |
| S5-final-viral-combined_k141_163427\|\|full | 3724 | No | NA | 4 | 3 | 0 | Low-quality | Genome-fragment | 10.42 | AAI-based (high-confidence) | 0 |
| S1-final-viral-combined_k141_252977\|\|full | 3725 | No | NA | 5 | 0 | 0 | Low-quality | Genome-fragment | 12.57 | AAI-based (medium-confidence) | 0 |
| S1_k141_357341 | 3725 | No | NA | 3 | 0 | 3 | Low-quality | Genome-fragment | 1.38 | AAI-based (high-confidence) | 0 |
| S7_k141_3283 | 3725 | No | NA | 2 | 0 | 1 | Low-quality | Genome-fragment | 7.12 | AAI-based (medium-confidence) | 0 |
| S2-final-viral-combined_k141_240562\|\|full | 3372 | No | NA | 4 | 0 | 2 | Not-determined | Genome-fragment | NA | NA | 0 |
| S2-final-viral-combined_k141_198713\|\|full | 6283 | No | NA | 6 | 0 | 2 | Not-determined | Genome-fragment | NA | NA | 0 |
| S2-final-viral-combined_k141_177155\|\|full | 3725 | No | NA | 4 | 0 | 2 | Low-quality | Genome-fragment | 3.04 | AAI-based (medium-confidence) | 0 |
| S2-final-viral-combined_k141_85654\|\|full | 1339 | No | NA | 3 | 0 | 1 | Not-determined | Genome-fragment | NA | NA | 0 |
| S2-final-viral-combined_k141_95987\|\|full | 10749 | No | NA | 21 | 1 | 2 | High-quality | High-quality | 100 | AAI-based (high-confidence) | 0 |
| S2-final-viral-combined_k141_54571\|\|full | 2896 | No | NA | 3 | 0 | 2 | Not-determined | Genome-fragment | NA | NA | 0 |
| S2-final-viral-combined_k141_95991\|\|full | 1724 | No | NA | 4 | 0 | 0 | Not-determined | Genome-fragment | NA | NA | 0 |
| S11-final-viral-combined_k141_13584\|\|full | 3727 | No | NA | 9 | 1 | 0 | Low-quality | Genome-fragment | 3.95 | HMM-based (lower-bound) | 0 |
| S2-final-viral-combined_k141_23395\|\|full | 3743 | No | NA | 5 | 0 | 4 | Not-determined | Genome-fragment | NA | NA | 0 |
| S3_k141_43567 | 3728 | No | NA | 6 | 1 | 0 | Low-quality | Genome-fragment | 2.47 | AAI-based (high-confidence) | 0 |
| S2-final-viral-combined_k141_23404\|\|full | 2248 | No | NA | 3 | 0 | 1 | Not-determined | Genome-fragment | NA | NA | 0 |
| S2-final-viral-combined_k141_136275\|\|full | 3728 | No | NA | 4 | 2 | 0 | Low-quality | Genome-fragment | 8.84 | AAI-based (medium-confidence) | 0 |
| S2-final-viral-combined_k141_178960\|\|full | 3311 | No | NA | 5 | 0 | 3 | Not-determined | Genome-fragment | NA | NA | 0 |
| S2_k141_63293 | 3728 | No | NA | 9 | 1 | 0 | Low-quality | Genome-fragment | 4.31 | HMM-based (lower-bound) | 0 |
| S9-final-viral-combined_k141_48293\|\|full | 3729 | No | NA | 3 | 0 | 2 | Low-quality | Genome-fragment | 2.59 | AAI-based (medium-confidence) | 0 |
| S11-final-viral-combined_k141_298567\|\|full | 3729 | No | NA | 5 | 0 | 1 | Low-quality | Genome-fragment | 4.62 | AAI-based (medium-confidence) | 0 |
| S2-final-viral-combined_k141_2481\|\|full | 1112 | No | NA | 2 | 0 | 0 | Not-determined | Genome-fragment | NA | NA | 0 |
| S2-final-viral-combined_k141_33454\|\|full | 2254 | No | NA | 3 | 0 | 1 | Not-determined | Genome-fragment | NA | NA | 0 |
| S2-final-viral-combined_k141_137400\|\|full | 2270 | No | NA | 3 | 0 | 2 | Not-determined | Genome-fragment | NA | NA | 0 |
| S2-final-viral-combined_k141_2484\|\|full | 1834 | No | NA | 3 | 0 | 0 | Not-determined | Genome-fragment | NA | NA | 0 |
| S2-final-viral-combined_k141_126823\|\|full | 3882 | No | NA | 8 | 0 | 1 | Not-determined | Genome-fragment | NA | NA | 0 |
| S1-final-viral-combined_k141_362286\|\|full | 3729 | No | NA | 7 | 0 | 2 | Low-quality | Genome-fragment | 7.82 | AAI-based (medium-confidence) | 0 |
| S2-final-viral-combined_k141_189073\|\|full | 5001 | No | NA | 8 | 0 | 1 | Not-determined | Genome-fragment | NA | NA | 0 |
| S2-final-viral-combined_k141_126840\|\|full | 1942 | No | NA | 4 | 0 | 3 | Not-determined | Genome-fragment | NA | NA | 0 |
| S2-final-viral-combined_k141_147415\|\|full | 2529 | No | NA | 3 | 0 | 1 | Not-determined | Genome-fragment | NA | NA | 0 |
| S17-final-viral-combined_k141_86201\|\|full | 3729 | Yes | 2744 | 7 | 2 | 1 | Low-quality | Genome-fragment | 4.28 | HMM-based (lower-bound) | 26.41 |
| S2-final-viral-combined_k141_240606\|\|full | 4922 | No | NA | 6 | 0 | 3 | Not-determined | Genome-fragment | NA | NA | 0 |
| S2-final-viral-combined_k141_179020\|\|full | 3165 | No | NA | 5 | 0 | 0 | Not-determined | Genome-fragment | NA | NA | 0 |
| S2-final-viral-combined_k141_64630\|\|full | 2379 | No | NA | 3 | 0 | 1 | Not-determined | Genome-fragment | NA | NA | 0 |
| S12_k141_47935 | 3729 | No | NA | 9 | 3 | 0 | Low-quality | Genome-fragment | 9.94 | AAI-based (high-confidence) | 0 |
| S2-final-viral-combined_k141_179022\|\|full | 2736 | No | NA | 4 | 0 | 0 | Not-determined | Genome-fragment | NA | NA | 0 |
| S2-final-viral-combined_k141_157934\|\|full | 96819 | Yes | 33133 | 111 | 14 | 49 | Complete | High-quality | 100 | Provirus (high-confidence) | 65.78 |
| S2-final-viral-combined_k141_147438\|\|full | 8243 | No | NA | 9 | 0 | 7 | Not-determined | Genome-fragment | NA | NA | 0 |
| S2-final-viral-combined_k141_198826\|\|full | 2658 | No | NA | 2 | 0 | 2 | Not-determined | Genome-fragment | NA | NA | 0 |
| S4_k141_1791 | 3729 | No | NA | 3 | 0 | 2 | Low-quality | Genome-fragment | 1.99 | AAI-based (medium-confidence) | 0 |
| S11-final-viral-combined_k141_92953\|\|full | 3730 | No | NA | 3 | 0 | 1 | Low-quality | Genome-fragment | 1.13 | AAI-based (medium-confidence) | 0 |
| S2-final-viral-combined_k141_106223\|\|full | 5360 | No | NA | 5 | 0 | 4 | Not-determined | Genome-fragment | NA | NA | 0 |
| S8-final-viral-combined_k141_69340\|\|full | 3730 | No | NA | 5 | 1 | 0 | Low-quality | Genome-fragment | 5.82 | HMM-based (lower-bound) | 0 |
| S2-final-viral-combined_k141_12966\|\|full | 1220 | No | NA | 2 | 0 | 1 | Not-determined | Genome-fragment | NA | NA | 0 |
| S2-final-viral-combined_k141_2540\|\|full | 2715 | No | NA | 3 | 0 | 3 | Not-determined | Genome-fragment | NA | NA | 0 |
| S8_k141_197909 | 3731 | No | NA | 9 | 2 | 0 | Low-quality | Genome-fragment | 8.09 | AAI-based (high-confidence) | 0 |
| S2-final-viral-combined_k141_168708\|\|full | 4575 | No | NA | 8 | 0 | 2 | Not-determined | Genome-fragment | NA | NA | 0 |
| S1-final-viral-combined_k141_2755\|\|full | 3732 | No | NA | 5 | 0 | 4 | Low-quality | Genome-fragment | 1.95 | AAI-based (medium-confidence) | 0 |
| S2-final-viral-combined_k141_106238\|\|full | 2865 | No | NA | 2 | 0 | 0 | Not-determined | Genome-fragment | NA | NA | 0 |
| S5_k141_100312 | 3732 | No | NA | 7 | 2 | 0 | Low-quality | Genome-fragment | 5.08 | AAI-based (medium-confidence) | 0 |
| S8-final-viral-combined_k141_70101\|\|full | 3732 | No | NA | 3 | 0 | 2 | Low-quality | Genome-fragment | 3.77 | AAI-based (medium-confidence) | 0 |
| S2-final-viral-combined_k141_168733\|\|full | 1621 | No | NA | 3 | 0 | 0 | Not-determined | Genome-fragment | NA | NA | 0 |
| S2-final-viral-combined_k141_64668\|\|full | 2419 | No | NA | 2 | 0 | 1 | Not-determined | Genome-fragment | NA | NA | 0 |
| S8-final-viral-combined_k141_233058\|\|full | 3732 | No | NA | 3 | 0 | 2 | Low-quality | Genome-fragment | 3.99 | AAI-based (medium-confidence) | 0 |
| S5-final-viral-combined_k141_139612\|\|full | 3733 | No | NA | 6 | 2 | 0 | Low-quality | Genome-fragment | 7.01 | AAI-based (high-confidence) | 0 |
| S2-final-viral-combined_k141_116332\|\|full | 2711 | No | NA | 5 | 0 | 2 | Not-determined | Genome-fragment | NA | NA | 0 |
| S2-final-viral-combined_k141_106259\|\|full | 2088 | No | NA | 2 | 0 | 1 | Not-determined | Genome-fragment | NA | NA | 0 |
| S2-final-viral-combined_k141_209630\|\|full | 3450 | No | NA | 7 | 0 | 3 | Not-determined | Genome-fragment | NA | NA | 0 |
| S2-final-viral-combined_k141_157996\|\|full | 1644 | No | NA | 3 | 0 | 2 | Not-determined | Genome-fragment | NA | NA | 0 |
| S17-k141_210410 | 3733 | No | NA | 3 | 1 | 0 | Low-quality | Genome-fragment | 5.65 | HMM-based (lower-bound) | 0 |
| S8_k141_96573 | 3733 | Yes | 2077 | 4 | 1 | 1 | Low-quality | Genome-fragment | 5.09 | AAI-based (medium-confidence) | 44.36 |
| S1-final-viral-combined_k141_276014\|\|full | 3734 | No | NA | 3 | 0 | 3 | Low-quality | Genome-fragment | 2.8 | AAI-based (medium-confidence) | 0 |
| S1-final-viral-combined_k141_200210\|\|full | 3735 | No | NA | 4 | 0 | 0 | Low-quality | Genome-fragment | 7 | AAI-based (medium-confidence) | 0 |
| S17-final-viral-combined_k141_204149\|\|full | 3735 | No | NA | 4 | 3 | 0 | Low-quality | Genome-fragment | 5.47 | AAI-based (high-confidence) | 0 |
| S2-final-viral-combined_k141_106275\|\|full | 5425 | No | NA | 2 | 0 | 0 | Not-determined | Genome-fragment | NA | NA | 0 |
| S2-final-viral-combined_k141_96169\|\|full | 1922 | No | NA | 2 | 0 | 2 | Not-determined | Genome-fragment | NA | NA | 0 |
| S2-final-viral-combined_k141_202794\|\|full | 3735 | No | NA | 4 | 0 | 1 | Low-quality | Genome-fragment | 9.37 | AAI-based (medium-confidence) | 0 |
| S2-final-viral-combined_k141_240641\|\|full | 1187 | No | NA | 2 | 0 | 1 | Not-determined | Genome-fragment | NA | NA | 0 |
| S2-final-viral-combined_k141_54682\|\|full | 5512 | No | NA | 4 | 0 | 4 | Not-determined | Genome-fragment | NA | NA | 0 |
| S4_k141_71916 | 3735 | No | NA | 5 | 2 | 0 | Low-quality | Genome-fragment | 6.85 | AAI-based (high-confidence) | 0 |
| S2-final-viral-combined_k141_33631\|\|full | 2026 | No | NA | 3 | 0 | 2 | Not-determined | Genome-fragment | NA | NA | 0 |
| S2-final-viral-combined_k141_179069\|\|full | 2307 | No | NA | 4 | 0 | 0 | Not-determined | Genome-fragment | NA | NA | 0 |
| S2-final-viral-combined_k141_64692\|\|full | 44384 | No | NA | 80 | 20 | 3 | High-quality | High-quality | 100 | AAI-based (high-confidence) | 0 |
| S2-final-viral-combined_k141_179072\|\|full | 4798 | No | NA | 5 | 0 | 2 | Not-determined | Genome-fragment | NA | NA | 0 |
| S2-final-viral-combined_k141_85820\|\|full | 28702 | No | NA | 23 | 0 | 5 | Not-determined | Genome-fragment | NA | NA | 0 |
| S2-final-viral-combined_k141_179079\|\|full | 2558 | No | NA | 5 | 0 | 4 | Not-determined | Genome-fragment | NA | NA | 0 |
| S6-final-viral-combined_k141_136070\|\|full | 3735 | No | NA | 3 | 0 | 3 | Low-quality | Genome-fragment | 5.88 | AAI-based (medium-confidence) | 0 |
| S2-final-viral-combined_k141_33655\|\|full | 2881 | No | NA | 3 | 0 | 2 | Not-determined | Genome-fragment | NA | NA | 0 |
| S7-final-viral-combined_k141_20334\|\|full | 3736 | No | NA | 6 | 0 | 0 | Low-quality | Genome-fragment | 9.08 | AAI-based (high-confidence) | 0 |
| S2-final-viral-combined_k141_209654\|\|full | 2228 | No | NA | 3 | 0 | 2 | Not-determined | Genome-fragment | NA | NA | 0 |
| S2-final-viral-combined_k141_168820\|\|full | 5118 | No | NA | 6 | 0 | 4 | Not-determined | Genome-fragment | NA | NA | 0 |
| S12_k141_169984 | 3736 | No | NA | 2 | 0 | 1 | Low-quality | Genome-fragment | 3.39 | AAI-based (medium-confidence) | 0 |
| S2-final-viral-combined_k141_75089\|\|full | 3093 | No | NA | 2 | 0 | 1 | Not-determined | Genome-fragment | NA | NA | 0 |
| S2-final-viral-combined_k141_147578\|\|full | 3886 | No | NA | 7 | 0 | 0 | Not-determined | Genome-fragment | NA | NA | 0 |
| S1-final-viral-combined_k141_240763\|\|full | 3737 | Yes | 1164 | 3 | 1 | 2 | Low-quality | Genome-fragment | 2.73 | AAI-based (high-confidence) | 68.85 |
| S1-final-viral-combined_k141_13635\|\|full | 3737 | No | NA | 7 | 2 | 0 | Low-quality | Genome-fragment | 10.34 | AAI-based (high-confidence) | 0 |
| S2-final-viral-combined_k141_240670\|\|full | 19696 | No | NA | 12 | 0 | 4 | Not-determined | Genome-fragment | NA | NA | 0 |
| S2-final-viral-combined_k141_75094\|\|full | 4288 | No | NA | 3 | 0 | 0 | Not-determined | Genome-fragment | NA | NA | 0 |
| S2-final-viral-combined_k141_85859\|\|full | 1169 | No | NA | 2 | 0 | 1 | Not-determined | Genome-fragment | NA | NA | 0 |
| S2-final-viral-combined_k141_179141\|\|full | 1666 | No | NA | 4 | 0 | 1 | Not-determined | Genome-fragment | NA | NA | 0 |
| S2-final-viral-combined_k141_23550\|\|full | 5684 | No | NA | 7 | 0 | 7 | Not-determined | Genome-fragment | NA | NA | 0 |
| S2-final-viral-combined_k141_230913\|\|full | 3558 | No | NA | 5 | 0 | 2 | Not-determined | Genome-fragment | NA | NA | 0 |

| | | | | | | | | | | | |
|---|---|---|---|---|---|---|---|---|---|---|---|
| S2-final-viral-combined_k141_189327\|\|full | 4289 | No | NA | 3 | 0 | 2 | Not-determined | Genome-fragment | NA | NA | 0 |
| S3-final-viral-combined_k141_63889\|\|full | 3737 | No | NA | 3 | 1 | 0 | Low-quality | Genome-fragment | 7.57 | AAI-based (medium-confidence) | 0 |
| S2-final-viral-combined_k141_96299\|\|full | 1191 | No | NA | 3 | 0 | 0 | Not-determined | Genome-fragment | NA | NA | 0 |
| S2-final-viral-combined_k141_23559\|\|full | 1357 | No | NA | 2 | 0 | 0 | Not-determined | Genome-fragment | NA | NA | 0 |
| S2-final-viral-combined_k141_106249\|\|full | 3737 | No | NA | 5 | 0 | 0 | Low-quality | Genome-fragment | 2.48 | AAI-based (medium-confidence) | 0 |
| S11-final-viral-combined_k141_143699\|\|full | 3738 | No | NA | 7 | 0 | 1 | Low-quality | Genome-fragment | 7.48 | AAI-based (medium-confidence) | 0 |
| S2-final-viral-combined_k141_179184\|\|full | 1471 | No | NA | 3 | 0 | 0 | Not-determined | Genome-fragment | NA | NA | 0 |
| S2-final-viral-combined_k141_2624\|\|full | 1223 | No | NA | 3 | 0 | 0 | Not-determined | Genome-fragment | NA | NA | 0 |
| S2-final-viral-combined_k141_168890\|\|full | 4610 | No | NA | 3 | 0 | 2 | Not-determined | Genome-fragment | NA | NA | 0 |
| S11_k141_33379 | 3738 | No | NA | 7 | 0 | 0 | Low-quality | Genome-fragment | 8.03 | AAI-based (high-confidence) | 0 |
| S1-final-viral-combined_k141_238376\|\|full | 3739 | No | NA | 5 | 3 | 0 | Low-quality | Genome-fragment | 10.9 | AAI-based (high-confidence) | 0 |
| S12-final-viral-combined_k141_382032\|\|full | 3739 | No | NA | 9 | 0 | 0 | Low-quality | Genome-fragment | 5.99 | AAI-based (high-confidence) | 0 |
| S2-final-viral-combined_k141_127080\|\|full | 25447 | No | NA | 47 | 1 | 1 | Medium-quality | Genome-fragment | 61.63 | AAI-based (high-confidence) | 0 |
| S7-final-viral-combined_k141_218457\|\|full | 3740 | No | NA | 3 | 0 | 2 | Low-quality | Genome-fragment | 1.24 | AAI-based (medium-confidence) | 0 |
| S2-final-viral-combined_k141_33758\|\|full | 2322 | No | NA | 4 | 0 | 0 | Not-determined | Genome-fragment | NA | NA | 0 |
| S2-final-viral-combined_k141_34016\|\|full | 3741 | No | NA | 3 | 0 | 2 | Low-quality | Genome-fragment | 8.9 | AAI-based (high-confidence) | 0 |
| S2-final-viral-combined_k141_158124\|\|full | 1287 | No | NA | 3 | 0 | 1 | Not-determined | Genome-fragment | NA | NA | 0 |
| S4-final-viral-combined_k141_139951\|\|full | 3741 | No | NA | 4 | 0 | 2 | Low-quality | Genome-fragment | 12.51 | AAI-based (high-confidence) | 0 |
| S8-final-viral-combined_k141_256831\|\|full | 3741 | No | NA | 6 | 1 | 0 | Low-quality | Genome-fragment | 9.62 | AAI-based (high-confidence) | 0 |
| S8_k141_58200 | 3741 | No | NA | 9 | 1 | 0 | Low-quality | Genome-fragment | 7.18 | AAI-based (high-confidence) | 0 |
| S2-final-viral-combined_k141_106388\|\|full | 1582 | No | NA | 3 | 0 | 0 | Not-determined | Genome-fragment | NA | NA | 0 |
| S2_k141_167563 | 3742 | No | NA | 2 | 0 | 0 | Low-quality | Genome-fragment | 5.63 | AAI-based (medium-confidence) | 0 |
| S2-final-viral-combined_k141_147708\|\|full | 5475 | No | NA | 5 | 0 | 5 | Not-determined | Genome-fragment | NA | NA | 0 |
| S4-final-viral-combined_k141_6619\|\|full | 3742 | No | NA | 10 | 0 | 0 | Low-quality | Genome-fragment | 8.34 | AAI-based (high-confidence) | 0 |
| S2-final-viral-combined_k141_198911\|\|full | 1641 | No | NA | 3 | 0 | 0 | Not-determined | Genome-fragment | NA | NA | 0 |
| S17-final-viral-combined_k141_38939\|\|full | 3744 | No | NA | 6 | 1 | 2 | Low-quality | Genome-fragment | 8.13 | AAI-based (high-confidence) | 0 |
| S2-final-viral-combined_k141_143034\|\|full | 3744 | No | NA | 5 | 0 | 5 | Low-quality | Genome-fragment | 1.7 | AAI-based (medium-confidence) | 0 |
| S2-final-viral-combined_k141_127090\|\|full | 1433 | No | NA | 2 | 0 | 1 | Not-determined | Genome-fragment | NA | NA | 0 |
| S2-final-viral-combined_k141_179272\|\|full | 1937 | No | NA | 2 | 0 | 1 | Not-determined | Genome-fragment | NA | NA | 0 |
| S2-final-viral-combined_k141_127096\|\|full | 2551 | No | NA | 3 | 0 | 2 | Not-determined | Genome-fragment | NA | NA | 0 |
| S2-final-viral-combined_k141_13147\|\|full | 2514 | No | NA | 5 | 0 | 3 | Not-determined | Genome-fragment | NA | NA | 0 |
| S4_k141_67396 | 3744 | No | NA | 1 | 0 | 0 | Low-quality | Genome-fragment | 3.8 | AAI-based (high-confidence) | 0 |
| S11-final-viral-combined_k141_157470\|\|full | 3745 | No | NA | 4 | 0 | 3 | Low-quality | Genome-fragment | 1.81 | AAI-based (medium-confidence) | 0 |
| S11-final-viral-combined_k141_156268\|\|lt2gene | 3745 | No | NA | 1 | 1 | 0 | Low-quality | Genome-fragment | 6.69 | AAI-based (high-confidence) | 0 |
| S1-final-viral-combined_k141_386248\|\|full | 3745 | No | NA | 3 | 0 | 3 | Low-quality | Genome-fragment | 1.79 | AAI-based (medium-confidence) | 0 |
| S2-final-viral-combined_k141_54850\|\|full | 1094 | No | NA | 3 | 0 | 2 | Not-determined | Genome-fragment | NA | NA | 0 |
| S7_k141_14205 | 3745 | Yes | 3620 | 6 | 1 | 1 | Low-quality | Genome-fragment | 2.34 | HMM-based (lower-bound) | 3.34 |
| S8-final-viral-combined_k141_9804\|\|full | 3745 | No | NA | 6 | 2 | 0 | Low-quality | Genome-fragment | 10.97 | AAI-based (high-confidence) | 0 |
| S2-final-viral-combined_k141_198922\|\|full | 1435 | No | NA | 2 | 0 | 0 | Not-determined | Genome-fragment | NA | NA | 0 |
| S1-final-viral-combined_k141_199638\|\|full | 3746 | No | NA | 3 | 0 | 0 | Low-quality | Genome-fragment | 6.41 | AAI-based (medium-confidence) | 0 |
| S2-final-viral-combined_k141_158155\|\|full | 2918 | No | NA | 3 | 0 | 0 | Not-determined | Genome-fragment | NA | NA | 0 |
| S2-final-viral-combined_k141_158174\|\|full | 3039 | No | NA | 3 | 0 | 2 | Not-determined | Genome-fragment | NA | NA | 0 |
| S4_k141_117539 | 3746 | No | NA | 7 | 2 | 0 | Low-quality | Genome-fragment | 5.77 | AAI-based (high-confidence) | 0 |
| S11-final-viral-combined_k141_84140\|\|full | 3747 | No | NA | 5 | 2 | 0 | Low-quality | Genome-fragment | 10.11 | AAI-based (high-confidence) | 0 |
| S11-final-viral-combined_k141_202714\|\|full | 3747 | No | NA | 3 | 0 | 0 | Low-quality | Genome-fragment | 9.38 | AAI-based (high-confidence) | 0 |
| S2-final-viral-combined_k141_127208\|\|full | 6433 | No | NA | 5 | 0 | 1 | Not-determined | Genome-fragment | NA | NA | 0 |
| S2-final-viral-combined_k141_13225\|\|full | 3339 | No | NA | 5 | 0 | 0 | Not-determined | Genome-fragment | NA | NA | 0 |
| S5-final-viral-combined_k141_181319\|\|full | 3747 | No | NA | 3 | 0 | 0 | Low-quality | Genome-fragment | 3.4 | AAI-based (high-confidence) | 0 |
| S6_k141_53698 | 3747 | No | NA | 8 | 1 | 0 | Low-quality | Genome-fragment | 2 | AAI-based (high-confidence) | 0 |
| S2-final-viral-combined_k141_106423\|\|full | 2395 | No | NA | 3 | 0 | 2 | Not-determined | Genome-fragment | NA | NA | 0 |
| S2-final-viral-combined_k141_198975\|\|full | 5849 | No | NA | 6 | 0 | 0 | Not-determined | Genome-fragment | NA | NA | 0 |
| S11-final-viral-combined_k141_86731\|\|full | 3748 | No | NA | 4 | 0 | 1 | Low-quality | Genome-fragment | 1.17 | AAI-based (medium-confidence) | 0 |
| S1-final-viral-combined_k141_156117\|\|full | 3749 | No | NA | 1 | 0 | 1 | Low-quality | Genome-fragment | 13.07 | AAI-based (medium-confidence) | 0 |
| S12_k141_210000 | 3750 | No | NA | 3 | 1 | 0 | Low-quality | Genome-fragment | 2.05 | AAI-based (high-confidence) | 0 |
| S2-final-viral-combined_k141_231027\|\|full | 11280 | No | NA | 12 | 0 | 4 | Not-determined | Genome-fragment | NA | NA | 0 |
| S2-final-viral-combined_k141_198987\|\|full | 2466 | No | NA | 2 | 0 | 0 | Not-determined | Genome-fragment | NA | NA | 0 |
| S2-final-viral-combined_k141_2738\|\|full | 2793 | No | NA | 3 | 0 | 3 | Not-determined | Genome-fragment | NA | NA | 0 |
| S2-final-viral-combined_k141_33953\|\|full | 1993 | No | NA | 3 | 0 | 1 | Not-determined | Genome-fragment | NA | NA | 0 |
| S2-final-viral-combined_k141_106435\|\|full | 1717 | No | NA | 2 | 0 | 2 | Not-determined | Genome-fragment | NA | NA | 0 |
| S2-final-viral-combined_k141_39779\|\|full | 3750 | No | NA | 4 | 0 | 1 | Low-quality | Genome-fragment | 8.61 | AAI-based (high-confidence) | 0 |
| S2-final-viral-combined_k141_158224\|\|full | 3749 | No | NA | 4 | 0 | 0 | Not-determined | Genome-fragment | NA | NA | 0 |
| S12-final-viral-combined_k141_103719\|\|full | 3751 | No | NA | 4 | 0 | 2 | Low-quality | Genome-fragment | 4.52 | AAI-based (medium-confidence) | 0 |
| S2-final-viral-combined_k141_240765\|\|full | 1069 | No | NA | 2 | 0 | 0 | Not-determined | Genome-fragment | NA | NA | 0 |
| S2-final-viral-combined_k141_137697\|\|full | 1507 | No | NA | 3 | 0 | 1 | Not-determined | Genome-fragment | NA | NA | 0 |
| S2-final-viral-combined_k141_240771\|\|full | 2509 | No | NA | 5 | 0 | 0 | Not-determined | Genome-fragment | NA | NA | 0 |
| S2-final-viral-combined_k141_116462\|\|full | 5537 | No | NA | 6 | 0 | 0 | Not-determined | Genome-fragment | NA | NA | 0 |
| S2-final-viral-combined_k141_189557\|\|full | 3856 | No | NA | 3 | 0 | 3 | Not-determined | Genome-fragment | NA | NA | 0 |
| S2-final-viral-combined_k141_175033\|\|full | 3751 | No | NA | 4 | 0 | 2 | Low-quality | Genome-fragment | 1.72 | AAI-based (medium-confidence) | 0 |
| S12-final-viral-combined_k141_202595\|\|full | 3752 | Yes | 985 | 3 | 1 | 2 | Low-quality | Genome-fragment | 0.57 | HMM-based (lower-bound) | 73.75 |
| S2-final-viral-combined_k141_137784\|\|full | 2448 | No | NA | 4 | 0 | 2 | Not-determined | Genome-fragment | NA | NA | 0 |
| S2-final-viral-combined_k141_2813\|\|full | 10208 | No | NA | 12 | 0 | 4 | Not-determined | Genome-fragment | NA | NA | 0 |
| S2-final-viral-combined_k141_158292\|\|full | 4671 | No | NA | 6 | 0 | 0 | Not-determined | Genome-fragment | NA | NA | 0 |
| S2-final-viral-combined_k141_139409\|\|full | 3752 | No | NA | 3 | 0 | 3 | Low-quality | Genome-fragment | 4.01 | AAI-based (medium-confidence) | 0 |
| S12-final-viral-combined_k141_282912\|\|full | 3753 | No | NA | 5 | 0 | 0 | Low-quality | Genome-fragment | 5.06 | AAI-based (high-confidence) | 0 |
| S2-final-viral-combined_k141_231103\|\|full | 2946 | No | NA | 6 | 0 | 0 | Not-determined | Genome-fragment | NA | NA | 0 |
| S2-final-viral-combined_k141_127304\|\|full | 3655 | No | NA | 4 | 0 | 2 | Not-determined | Genome-fragment | NA | NA | 0 |
| S2-final-viral-combined_k141_96545\|\|full | 5921 | No | NA | 5 | 0 | 0 | Not-determined | Genome-fragment | NA | NA | 0 |
| S2-final-viral-combined_k141_86090\|\|full | 4673 | No | NA | 4 | 0 | 2 | Not-determined | Genome-fragment | NA | NA | 0 |
| S2-final-viral-combined_k141_189634\|\|full | 1315 | No | NA | 3 | 0 | 0 | Not-determined | Genome-fragment | NA | NA | 0 |
| S2-final-viral-combined_k141_96554\|\|full | 5174 | No | NA | 6 | 0 | 1 | Not-determined | Genome-fragment | NA | NA | 0 |
| S2_k141_155408 | 3753 | No | NA | 2 | 0 | 2 | Low-quality | Genome-fragment | 0.32 | AAI-based (medium-confidence) | 0 |
| S8_k141_71346 | 3753 | No | NA | 10 | 2 | 0 | Low-quality | Genome-fragment | 11.56 | AAI-based (high-confidence) | 0 |
| S12-final-viral-combined_k141_270748\|\|full | 3755 | No | NA | 6 | 0 | 5 | Low-quality | Genome-fragment | 2.87 | AAI-based (medium-confidence) | 0 |
| S2-final-viral-combined_k141_116545\|\|full | 1681 | No | NA | 2 | 0 | 1 | Not-determined | Genome-fragment | NA | NA | 0 |
| S12_k141_271905 | 3755 | No | NA | 7 | 3 | 0 | Low-quality | Genome-fragment | 7.84 | AAI-based (high-confidence) | 0 |
| S8-final-viral-combined_k141_267727\|\|full | 3757 | No | NA | 3 | 0 | 3 | Low-quality | Genome-fragment | 3.15 | AAI-based (medium-confidence) | 0 |
| S11-final-viral-combined_k141_12531\|\|full | 3758 | No | NA | 2 | 0 | 1 | Low-quality | Genome-fragment | 1.23 | AAI-based (high-confidence) | 0 |
| S11_k141_21073 | 3758 | No | NA | 6 | 3 | 0 | Low-quality | Genome-fragment | 8.2 | AAI-based (high-confidence) | 0 |
| S1-final-viral-combined_k141_10996\|\|full | 3758 | No | NA | 11 | 0 | 0 | Low-quality | Genome-fragment | 9.64 | AAI-based (medium-confidence) | 0 |
| S2-final-viral-combined_k141_2881\|\|full | 4401 | No | NA | 2 | 0 | 1 | Not-determined | Genome-fragment | NA | NA | 0 |
| S2-final-viral-combined_k141_127382\|\|full | 1501 | No | NA | 3 | 0 | 0 | Not-determined | Genome-fragment | NA | NA | 0 |
| S11-final-viral-combined_k141_323077\|\|full | 3759 | No | NA | 4 | 0 | 1 | Low-quality | Genome-fragment | 1.92 | AAI-based (medium-confidence) | 0 |
| S11_k141_255564 | 3759 | No | NA | 2 | 0 | 1 | Low-quality | Genome-fragment | 7.99 | AAI-based (high-confidence) | 0 |
| S1-final-viral-combined_k141_260691\|\|full | 3759 | No | NA | 3 | 0 | 1 | Low-quality | Genome-fragment | 6.13 | AAI-based (medium-confidence) | 0 |
| S8-final-viral-combined_k141_2443\|\|full | 3759 | No | NA | 3 | 0 | 1 | Low-quality | Genome-fragment | 2.11 | AAI-based (medium-confidence) | 0 |
| S2-final-viral-combined_k141_219987\|\|full | 1630 | No | NA | 4 | 0 | 1 | Not-determined | Genome-fragment | NA | NA | 0 |
| S2-final-viral-combined_k141_2910\|\|full | 9311 | No | NA | 11 | 0 | 2 | Not-determined | Genome-fragment | NA | NA | 0 |
| S2-final-viral-combined_k141_179506\|\|full | 6880 | No | NA | 5 | 0 | 4 | Not-determined | Genome-fragment | NA | NA | 0 |
| S2-final-viral-combined_k141_219995\|\|full | 1156 | No | NA | 3 | 0 | 0 | Not-determined | Genome-fragment | NA | NA | 0 |
| S2-final-viral-combined_k141_2913\|\|full | 1718 | No | NA | 2 | 0 | 2 | Not-determined | Genome-fragment | NA | NA | 0 |
| S2-final-viral-combined_k141_75177\|\|full | 1386 | No | NA | 3 | 0 | 1 | Not-determined | Genome-fragment | NA | NA | 0 |
| S2-final-viral-combined_k141_209943\|\|full | 1026 | No | NA | 2 | 0 | 1 | Not-determined | Genome-fragment | NA | NA | 0 |
| S2-final-viral-combined_k141_199123\|\|full | 4590 | No | NA | 4 | 0 | 3 | Not-determined | Genome-fragment | NA | NA | 0 |
| S2-final-viral-combined_k141_147885\|\|full | 1586 | No | NA | 3 | 0 | 0 | Not-determined | Genome-fragment | NA | NA | 0 |
| S8_k141_34025 | 3759 | No | NA | 3 | 0 | 2 | Low-quality | Genome-fragment | 3.79 | AAI-based (high-confidence) | 0 |
| S2-final-viral-combined_k141_116628\|\|full | 36543 | No | NA | 33 | 0 | 25 | Not-determined | Genome-fragment | NA | NA | 0 |
| S12-final-viral-combined_k141_77085\|\|full | 3760 | No | NA | 4 | 0 | 0 | Low-quality | Genome-fragment | 8.52 | AAI-based (high-confidence) | 0 |
| S2-final-viral-combined_k141_86151\|\|full | 5300 | No | NA | 5 | 0 | 1 | Not-determined | Genome-fragment | NA | NA | 0 |
| S2-final-viral-combined_k141_127443\|\|full | 1713 | No | NA | 3 | 0 | 0 | Not-determined | Genome-fragment | NA | NA | 0 |
| S2-final-viral-combined_k141_116642\|\|full | 4750 | No | NA | 6 | 0 | 1 | Not-determined | Genome-fragment | NA | NA | 0 |
| S2-final-viral-combined_k141_75203\|\|full | 14256 | No | NA | 16 | 0 | 10 | Not-determined | Genome-fragment | NA | NA | 0 |
| S2-final-viral-combined_k141_54926\|\|full | 2510 | No | NA | 3 | 0 | 1 | Not-determined | Genome-fragment | NA | NA | 0 |
| S1-final-viral-combined_k141_167884\|\|full | 3761 | Yes | 2376 | 7 | 2 | 1 | Low-quality | Genome-fragment | 4.54 | AAI-based (medium-confidence) | 36.83 |
| S2-final-viral-combined_k141_13246\|\|full | 240958 | Yes | 37802 | 261 | 19 | 148 | Medium-quality | Genome-fragment | 64.53 | HMM-based (lower-bound) | 84.31 |
| S3-final-viral-combined_k141_14541\|\|full | 3761 | No | NA | 3 | 0 | 3 | Low-quality | Genome-fragment | 8.54 | AAI-based (medium-confidence) | 0 |
| S2-final-viral-combined_k141_65024\|\|full | 1614 | No | NA | 3 | 0 | 1 | Not-determined | Genome-fragment | NA | NA | 0 |
| S17-k141_259368 | 3761 | No | NA | 11 | 1 | 0 | Low-quality | Genome-fragment | 4.57 | AAI-based (high-confidence) | 0 |
| S2-final-viral-combined_k141_127472\|\|full | 1412 | No | NA | 4 | 0 | 0 | Not-determined | Genome-fragment | NA | NA | 0 |
| S17-final-viral-combined_k141_172154\|\|full | 3762 | No | NA | 5 | 0 | 2 | Low-quality | Genome-fragment | 4.33 | AAI-based (high-confidence) | 0 |
| S12-final-viral-combined_k141_320715\|\|full | 3762 | No | NA | 3 | 0 | 2 | Low-quality | Genome-fragment | 5.12 | AAI-based (medium-confidence) | 0 |
| S4-final-viral-combined_k141_50682\|\|full | 3762 | No | NA | 6 | 0 | 1 | Low-quality | Genome-fragment | 6.27 | AAI-based (medium-confidence) | 0 |
| S8-final-viral-combined_k141_127867\|\|full | 3762 | No | NA | 5 | 0 | 2 | Low-quality | Genome-fragment | 1.24 | AAI-based (medium-confidence) | 0 |
| S1-final-viral-combined_k141_389969\|\|full | 3763 | No | NA | 6 | 0 | 0 | Low-quality | Genome-fragment | 8.84 | AAI-based (medium-confidence) | 0 |
| S3_k141_137878 | 3763 | No | NA | 5 | 0 | 0 | Low-quality | Genome-fragment | 4.81 | AAI-based (medium-confidence) | 0 |
| S12-final-viral-combined_k141_175869\|\|full | 3763 | No | NA | 6 | 1 | 1 | Low-quality | Genome-fragment | 3.92 | AAI-based (medium-confidence) | 0 |
| S2-final-viral-combined_k141_77207\|\|full | 3763 | No | NA | 4 | 0 | 2 | Low-quality | Genome-fragment | 2.5 | AAI-based (medium-confidence) | 0 |
| S9_k141_63471 | 3764 | No | NA | 4 | 0 | 1 | Low-quality | Genome-fragment | 4.11 | AAI-based (medium-confidence) | 0 |
| S2-final-viral-combined_k141_138022\|\|full | 3096 | No | NA | 5 | 0 | 4 | Not-determined | Genome-fragment | NA | NA | 0 |
| S2-final-viral-combined_k141_45747\|\|full | 3109 | No | NA | 3 | 0 | 0 | Not-determined | Genome-fragment | NA | NA | 0 |
| S11_k141_137052 | 3764 | No | NA | 4 | 0 | 3 | Low-quality | Genome-fragment | 3.78 | AAI-based (medium-confidence) | 0 |
| S2-final-viral-combined_k141_179539\|\|full | 4476 | No | NA | 5 | 0 | 2 | Not-determined | Genome-fragment | NA | NA | 0 |
| S2-final-viral-combined_k141_75262\|\|full | 1300 | No | NA | 3 | 0 | 0 | Not-determined | Genome-fragment | NA | NA | 0 |
| S2-final-viral-combined_k141_147965\|\|full | 2674 | No | NA | 4 | 0 | 1 | Not-determined | Genome-fragment | NA | NA | 0 |
| S5-final-viral-combined_k141_73334\|\|full | 3764 | No | NA | 5 | 0 | 2 | Low-quality | Genome-fragment | 3.1 | AAI-based (medium-confidence) | 0 |
| S17-final-viral-combined_k141_150547\|\|full | 3764 | No | NA | 8 | 2 | 0 | Low-quality | Genome-fragment | 7.27 | AAI-based (high-confidence) | 0 |
| S2-final-viral-combined_k141_169367\|\|full | 2143 | No | NA | 4 | 0 | 0 | Not-determined | Genome-fragment | NA | NA | 0 |
| S2-final-viral-combined_k141_199156\|\|full | 1942 | No | NA | 3 | 0 | 1 | Not-determined | Genome-fragment | NA | NA | 0 |
| S2-final-viral-combined_k141_109730\|\|full | 3764 | No | NA | 3 | 0 | 1 | Low-quality | Genome-fragment | 4.43 | AAI-based (medium-confidence) | 0 |
| S2-final-viral-combined_k141_189858\|\|full | 1418 | No | NA | 2 | 0 | 0 | Not-determined | Genome-fragment | NA | NA | 0 |
| S2-final-viral-combined_k141_148016\|\|full | 2831 | No | NA | 5 | 0 | 0 | Not-determined | Genome-fragment | NA | NA | 0 |
| S5-final-viral-combined_k141_77539\|\|full | 3765 | No | NA | 6 | 2 | 0 | Low-quality | Genome-fragment | 6.74 | AAI-based (medium-confidence) | 0 |
| S2-final-viral-combined_k141_106721\|\|full | 1352 | No | NA | 3 | 0 | 0 | Not-determined | Genome-fragment | NA | NA | 0 |
| S2-final-viral-combined_k141_13295\|\|full | 4778 | No | NA | 7 | 0 | 0 | Not-determined | Genome-fragment | NA | NA | 0 |
| S2-final-viral-combined_k141_86263\|\|full | 1375 | No | NA | 2 | 0 | 0 | Not-determined | Genome-fragment | NA | NA | 0 |
| S7-final-viral-combined_k141_57831\|\|full | 3766 | No | NA | 5 | 0 | 1 | Low-quality | Genome-fragment | 2.93 | AAI-based (high-confidence) | 0 |
| S2-final-viral-combined_k141_5327\|\|full | 3766 | No | NA | 7 | 2 | 0 | Low-quality | Genome-fragment | 8.25 | AAI-based (medium-confidence) | 0 |
| S1-final-viral-combined_k141_337184\|\|full | 3767 | No | NA | 6 | 6 | 0 | Low-quality | Genome-fragment | 8.25 | AAI-based (high-confidence) | 0 |
| S2-final-viral-combined_k141_231313\|\|full | 1343 | No | NA | 3 | 0 | 1 | Not-determined | Genome-fragment | NA | NA | 0 |
| S2-final-viral-combined_k141_210115\|\|full | 1639 | No | NA | 3 | 0 | 1 | Not-determined | Genome-fragment | NA | NA | 0 |
| S2-final-viral-combined_k141_231320\|\|full | 3491 | No | NA | 5 | 0 | 4 | Not-determined | Genome-fragment | NA | NA | 0 |
| S2-final-viral-combined_k141_211682\|\|full | 3767 | No | NA | 3 | 2 | 0 | Low-quality | Genome-fragment | 11.14 | HMM-based (lower-bound) | 0 |
| S2-final-viral-combined_k141_179675\|\|full | 2547 | No | NA | 3 | 0 | 0 | Not-determined | Genome-fragment | NA | NA | 0 |
| S2-final-viral-combined_k141_231353\|\|full | 2423 | No | NA | 3 | 0 | 2 | Not-determined | Genome-fragment | NA | NA | 0 |
| S2-final-viral-combined_k141_231356\|\|full | 2489 | No | NA | 3 | 0 | 2 | Not-determined | Genome-fragment | NA | NA | 0 |
| S4-final-viral-combined_k141_48774\|\|full | 3767 | No | NA | 5 | 0 | 1 | Low-quality | Genome-fragment | 5.86 | AAI-based (medium-confidence) | 0 |
| S2-final-viral-combined_k141_96717\|\|full | 1185 | No | NA | 2 | 0 | 0 | Not-determined | Genome-fragment | NA | NA | 0 |
| S11_k141_166086 | 3768 | No | NA | 10 | 2 | 0 | Low-quality | Genome-fragment | 4.58 | AAI-based (high-confidence) | 0 |
| S3-final-viral-combined_k141_1540\|\|full | 3768 | No | NA | 7 | 2 | 1 | Low-quality | Genome-fragment | 6.01 | AAI-based (high-confidence) | 0 |
| S2-final-viral-combined_k141_45927\|\|full | 3038 | No | NA | 4 | 0 | 2 | Not-determined | Genome-fragment | NA | NA | 0 |
| S2-final-viral-combined_k141_148103\|\|full | 1527 | No | NA | 3 | 0 | 2 | Not-determined | Genome-fragment | NA | NA | 0 |
| S5_k141_191346 | 3768 | No | NA | 3 | 0 | 0 | Low-quality | Genome-fragment | 6.25 | AAI-based (medium-confidence) | 0 |
| S17-final-viral-combined_k141_220279\|\|full | 3768 | No | NA | 4 | 0 | 1 | Low-quality | Genome-fragment | 6.02 | AAI-based (medium-confidence) | 0 |
| S2-final-viral-combined_k141_148123\|\|full | 1591 | No | NA | 2 | 0 | 1 | Not-determined | Genome-fragment | NA | NA | 0 |
| S2-final-viral-combined_k141_65281\|\|full | 2241 | No | NA | 3 | 0 | 1 | Not-determined | Genome-fragment | NA | NA | 0 |
| S2-final-viral-combined_k141_230035\|\|full | 3768 | No | NA | 6 | 5 | 0 | Low-quality | Genome-fragment | 8.67 | AAI-based (high-confidence) | 0 |
| S2-final-viral-combined_k141_106843\|\|full | 2368 | No | NA | 2 | 0 | 2 | Not-determined | Genome-fragment | NA | NA | 0 |
| S2-final-viral-combined_k141_210253\|\|full | 2707 | No | NA | 3 | 0 | 2 | Not-determined | Genome-fragment | NA | NA | 0 |
| S1_k141_37110 | 3770 | No | NA | 3 | 0 | 0 | Low-quality | Genome-fragment | 5.23 | AAI-based (high-confidence) | 0 |
| S2-final-viral-combined_k141_138130\|\|full | 1535 | No | NA | 2 | 0 | 2 | Not-determined | Genome-fragment | NA | NA | 0 |
| S2-final-viral-combined_k141_45992\|\|full | 3517 | No | NA | 5 | 0 | 0 | Not-determined | Genome-fragment | NA | NA | 0 |
| S2-final-viral-combined_k141_231416\|\|full | 3180 | No | NA | 2 | 0 | 2 | Not-determined | Genome-fragment | NA | NA | 0 |
| S4-final-viral-combined_k141_155739\|\|full | 3770 | No | NA | 8 | 1 | 0 | Low-quality | Genome-fragment | 6.94 | AAI-based (high-confidence) | 0 |
| S2-final-viral-combined_k141_138159\|\|full | 1242 | No | NA | 2 | 0 | 0 | Not-determined | Genome-fragment | NA | NA | 0 |
| S2-final-viral-combined_k141_158518\|\|full | 3166 | No | NA | 5 | 0 | 0 | Not-determined | Genome-fragment | NA | NA | 0 |
| S2-final-viral-combined_k141_138171\|\|full | 2626 | No | NA | 3 | 0 | 2 | Not-determined | Genome-fragment | NA | NA | 0 |
| S8_k141_8740 | 3770 | No | NA | 5 | 1 | 0 | Low-quality | Genome-fragment | 2.38 | HMM-based (lower-bound) | 0 |
| S2-final-viral-combined_k141_116858\|\|full | 2197 | No | NA | 3 | 0 | 0 | Not-determined | Genome-fragment | NA | NA | 0 |
| S2-final-viral-combined_k141_158524\|\|full | 4268 | No | NA | 6 | 0 | 2 | Not-determined | Genome-fragment | NA | NA | 0 |
| S8-final-viral-combined_k141_210736\|\|full | 3771 | No | NA | 3 | 0 | 0 | Low-quality | Genome-fragment | 2.06 | AAI-based (medium-confidence) | 0 |
| S2-final-viral-combined_k141_13432\|\|full | 4855 | No | NA | 3 | 0 | 3 | Not-determined | Genome-fragment | NA | NA | 0 |
| S2-final-viral-combined_k141_210302\|\|full | 1841 | No | NA | 2 | 0 | 1 | Not-determined | Genome-fragment | NA | NA | 0 |
| S2-final-viral-combined_k141_116869\|\|full | 2679 | No | NA | 3 | 0 | 2 | Not-determined | Genome-fragment | NA | NA | 0 |
| S9-final-viral-combined_k141_59008\|\|full | 3773 | Yes | 2029 | 7 | 1 | 1 | Low-quality | Genome-fragment | 2.51 | HMM-based (lower-bound) | 46.22 |
| S9-final-viral-combined_k141_94849\|\|full | 3773 | No | NA | 2 | 0 | 2 | Low-quality | Genome-fragment | 2.22 | AAI-based (medium-confidence) | 0 |
| S2-final-viral-combined_k141_116885\|\|full | 1957 | No | NA | 3 | 0 | 1 | Not-determined | Genome-fragment | NA | NA | 0 |
| S2-final-viral-combined_k141_34377\|\|full | 2130 | No | NA | 5 | 0 | 1 | Not-determined | Genome-fragment | NA | NA | 0 |
| S6_k141_6862 | 3773 | No | NA | 7 | 0 | 0 | Low-quality | Genome-fragment | 3.96 | AAI-based (high-confidence) | 0 |
| S1-final-viral-combined_k141_33854\|\|full | 3774 | No | NA | 5 | 1 | 0 | Low-quality | Genome-fragment | 6.98 | AAI-based (high-confidence) | 0 |
| S2-final-viral-combined_k141_46035\|\|full | 1777 | No | NA | 2 | 0 | 2 | Not-determined | Genome-fragment | NA | NA | 0 |
| S8-final-viral-combined_k141_82355\|\|full | 3776 | No | NA | 3 | 2 | 0 | Low-quality | Genome-fragment | 24.31 | AAI-based (high-confidence) | 0 |
| S2-final-viral-combined_k141_127635\|\|full | 3521 | No | NA | 4 | 0 | 2 | Not-determined | Genome-fragment | NA | NA | 0 |
| S7-final-viral-combined_k141_30548\|\|full | 3777 | No | NA | 5 | 1 | 0 | Low-quality | Genome-fragment | 2.21 | HMM-based (lower-bound) | 0 |
| S2-final-viral-combined_k141_86427\|\|full | 2719 | No | NA | 3 | 0 | 0 | Not-determined | Genome-fragment | NA | NA | 0 |
| S17-k141_17871 | 3777 | No | NA | 7 | 0 | 0 | Low-quality | Genome-fragment | 5.37 | AAI-based (high-confidence) | 0 |
| S2-final-viral-combined_k141_158148\|\|full | 3777 | No | NA | 4 | 0 | 3 | Low-quality | Genome-fragment | 4.47 | AAI-based (high-confidence) | 0 |
| S8_k141_15539 | 3777 | No | NA | 2 | 0 | 0 | Low-quality | Genome-fragment | 4.25 | AAI-based (medium-confidence) | 0 |
| S2-final-viral-combined_k141_241134\|\|full | 2567 | No | NA | 3 | 0 | 0 | Not-determined | Genome-fragment | NA | NA | 0 |
| S2-final-viral-combined_k141_148151\|\|full | 4712 | No | NA | 3 | 0 | 2 | Not-determined | Genome-fragment | NA | NA | 0 |
| S1-final-viral-combined_k141_339577\|\|full | 3778 | No | NA | 3 | 0 | 2 | Low-quality | Genome-fragment | 4.01 | AAI-based (medium-confidence) | 0 |
| S2-final-viral-combined_k141_75508\|\|full | 2142 | No | NA | 3 | 0 | 3 | Not-determined | Genome-fragment | NA | NA | 0 |
| S2-final-viral-combined_k141_46079\|\|full | 7766 | No | NA | 8 | 0 | 3 | Not-determined | Genome-fragment | NA | NA | 0 |
| S12-final-viral-combined_k141_143636\|\|full | 3778 | No | NA | 7 | 5 | 0 | Low-quality | Genome-fragment | 9.54 | AAI-based (medium-confidence) | 0 |
| S11-final-viral-combined_k141_284514\|\|full | 3779 | Yes | 2419 | 3 | 1 | 1 | Low-quality | Genome-fragment | 5.15 | AAI-based (high-confidence) | 35.99 |
| S2-final-viral-combined_k141_189984\|\|full | 1618 | No | NA | 2 | 0 | 0 | Not-determined | Genome-fragment | NA | NA | 0 |
| S2-final-viral-combined_k141_13466\|\|full | 4637 | No | NA | 4 | 0 | 4 | Not-determined | Genome-fragment | NA | NA | 0 |
| S2-final-viral-combined_k141_46113\|\|full | 1245 | No | NA | 4 | 0 | 1 | Not-determined | Genome-fragment | NA | NA | 0 |
| S5-final-viral-combined_k141_96078\|\|full | 3780 | No | NA | 4 | 0 | 0 | Low-quality | Genome-fragment | 2.74 | AAI-based (medium-confidence) | 0 |
| S2-final-viral-combined_k141_220051\|\|full | 1661 | No | NA | 2 | 0 | 0 | Not-determined | Genome-fragment | NA | NA | 0 |
| S7_k141_173637 | 3780 | No | NA | 6 | 0 | 0 | Low-quality | Genome-fragment | 10.66 | AAI-based (high-confidence) | 0 |
| S2-final-viral-combined_k141_96806\|\|full | 1448 | No | NA | 3 | 0 | 1 | Not-determined | Genome-fragment | NA | NA | 0 |
| S6_k141_124004 | 3780 | No | NA | 7 | 0 | 0 | Low-quality | Genome-fragment | 9.96 | AAI-based (high-confidence) | 0 |
| S2-final-viral-combined_k141_127654\|\|full | 1529 | No | NA | 2 | 0 | 1 | Not-determined | Genome-fragment | NA | NA | 0 |
| S2-final-viral-combined_k141_158619\|\|full | 4822 | No | NA | 7 | 0 | 2 | Not-determined | Genome-fragment | NA | NA | 0 |
| S7-final-viral-combined_k141_115057\|\|full | 3781 | No | NA | 4 | 0 | 0 | Low-quality | Genome-fragment | 3.9 | AAI-based (medium-confidence) | 0 |
| S2-final-viral-combined_k141_3284\|\|full | 3146 | No | NA | 4 | 0 | 4 | Not-determined | Genome-fragment | NA | NA | 0 |
| S8_k141_41054 | 3781 | No | NA | 7 | 3 | 0 | Low-quality | Genome-fragment | 8.42 | AAI-based (high-confidence) | 0 |
| S9-final-viral-combined_k141_38974\|\|full | 3782 | No | NA | 2 | 0 | 2 | Low-quality | Genome-fragment | 1.18 | AAI-based (medium-confidence) | 0 |
| S2-final-viral-combined_k141_3286\|\|full | 2954 | No | NA | 3 | 0 | 1 | Not-determined | Genome-fragment | NA | NA | 0 |
| S9-final-viral-combined_k141_127435\|\|full | 3782 | No | NA | 6 | 4 | 0 | Low-quality | Genome-fragment | 7.86 | AAI-based (high-confidence) | 0 |
| S2-final-viral-combined_k141_210423\|\|full | 2058 | No | NA | 4 | 0 | 0 | Not-determined | Genome-fragment | NA | NA | 0 |
| S17-k141_113325 | 3783 | No | NA | 3 | 0 | 1 | Low-quality | Genome-fragment | 13.19 | AAI-based (high-confidence) | 0 |
| S2-final-viral-combined_k141_96834\|\|full | 2589 | No | NA | 4 | 0 | 3 | Not-determined | Genome-fragment | NA | NA | 0 |
| S3_k141_69560 | 3784 | No | NA | 7 | 0 | 1 | Low-quality | Genome-fragment | 7.11 | AAI-based (medium-confidence) | 0 |
| S12-final-viral-combined_k141_109218\|\|full | 3784 | No | NA | 4 | 1 | 0 | Low-quality | Genome-fragment | 5.73 | HMM-based (lower-bound) | 0 |
| S2-final-viral-combined_k141_152630\|\|full | 3784 | No | NA | 4 | 2 | 0 | Low-quality | Genome-fragment | 11.11 | AAI-based (medium-confidence) | 0 |
| S2-final-viral-combined_k141_86532\|\|full | 2631 | No | NA | 3 | 0 | 1 | Not-determined | Genome-fragment | NA | NA | 0 |
| S2_k141_87482 | 3784 | No | NA | 6 | 2 | 0 | Low-quality | Genome-fragment | 9.64 | AAI-based (high-confidence) | 0 |
| S4_k141_149757 | 3784 | No | NA | 2 | 0 | 1 | Low-quality | Genome-fragment | 11.39 | AAI-based (high-confidence) | 0 |
| S8-final-viral-combined_k141_113798\|\|full | 3784 | No | NA | 3 | 0 | 3 | Low-quality | Genome-fragment | 0.54 | AAI-based (medium-confidence) | 0 |
| S2-final-viral-combined_k141_13591\|\|full | 1560 | No | NA | 3 | 0 | 0 | Not-determined | Genome-fragment | NA | NA | 0 |
| S2-final-viral-combined_k141_55174\|\|full | 1028 | No | NA | 3 | 0 | 1 | Not-determined | Genome-fragment | NA | NA | 0 |
| S2-final-viral-combined_k141_107034\|\|full | 1470 | No | NA | 2 | 0 | 0 | Not-determined | Genome-fragment | NA | NA | 0 |
| S11_k141_311803 | 3785 | No | NA | 7 | 0 | 0 | Low-quality | Genome-fragment | 6.57 | AAI-based (high-confidence) | 0 |
| S2-final-viral-combined_k141_148279\|\|full | 1087 | No | NA | 2 | 0 | 1 | Not-determined | Genome-fragment | NA | NA | 0 |
| S17-final-viral-combined_k141_166076\|\|full | 3786 | No | NA | 6 | 2 | 0 | Low-quality | Genome-fragment | 7.37 | AAI-based (high-confidence) | 0 |
| S17-k141_203657 | 3786 | No | NA | 11 | 0 | 0 | Low-quality | Genome-fragment | 6.4 | AAI-based (medium-confidence) | 0 |
| S4-final-viral-combined_k141_8739\|\|full | 3786 | No | NA | 4 | 0 | 2 | Low-quality | Genome-fragment | 2.25 | AAI-based (medium-confidence) | 0 |
| S8-final-viral-combined_k141_90250\|\|full | 3786 | No | NA | 4 | 0 | 0 | Low-quality | Genome-fragment | 6.51 | AAI-based (high-confidence) | 0 |
| S17-final-viral-combined_k141_151985\|\|full | 3787 | No | NA | 4 | 0 | 1 | Low-quality | Genome-fragment | 8.24 | AAI-based (high-confidence) | 0 |
| S2-final-viral-combined_k141_138218\|\|full | 1008 | No | NA | 2 | 0 | 0 | Not-determined | Genome-fragment | NA | NA | 0 |
| S2-final-viral-combined_k141_199384\|\|full | 17330 | No | NA | 27 | 12 | 1 | Medium-quality | Genome-fragment | 54.9 | AAI-based (medium-confidence) | 0 |
| S4-final-viral-combined_k141_71184\|\|full | 3787 | No | NA | 2 | 0 | 2 | Low-quality | Genome-fragment | 3.37 | AAI-based (medium-confidence) | 0 |
| S2-final-viral-combined_k141_96874\|\|full | 2458 | No | NA | 4 | 0 | 1 | Not-determined | Genome-fragment | NA | NA | 0 |
| S12_k141_225076 | 3788 | No | NA | 5 | 0 | 1 | Low-quality | Genome-fragment | 6.33 | AAI-based (medium-confidence) | 0 |
| S2-final-viral-combined_k141_96892\|\|full | 1428 | No | NA | 2 | 0 | 1 | Not-determined | Genome-fragment | NA | NA | 0 |
| S2-final-viral-combined_k141_241238\|\|full | 2193 | No | NA | 3 | 0 | 0 | Not-determined | Genome-fragment | NA | NA | 0 |
| S8_k141_232740 | 3788 | No | NA | 2 | 0 | 0 | Low-quality | Genome-fragment | 2.31 | AAI-based (high-confidence) | 0 |
| S2-final-viral-combined_k141_169746\|\|full | 1789 | No | NA | 3 | 0 | 0 | Not-determined | Genome-fragment | NA | NA | 0 |
| S3-final-viral-combined_k141_23381\|\|full | 3790 | No | NA | 4 | 0 | 0 | Low-quality | Genome-fragment | 6.23 | AAI-based (high-confidence) | 0 |
| S12-final-viral-combined_k141_355626\|\|full | 3790 | No | NA | 9 | 5 | 0 | Low-quality | Genome-fragment | 10.47 | AAI-based (high-confidence) | 0 |
| S2-final-viral-combined_k141_117194\|\|full | 8207 | No | NA | 7 | 0 | 7 | Not-determined | Genome-fragment | NA | NA | 0 |
| S2-final-viral-combined_k141_231716\|\|full | 4140 | No | NA | 4 | 0 | 1 | Not-determined | Genome-fragment | NA | NA | 0 |
| S2-final-viral-combined_k141_75648\|\|full | 5368 | No | NA | 4 | 0 | 2 | Not-determined | Genome-fragment | NA | NA | 0 |
| S2-final-viral-combined_k141_190130\|\|full | 3392 | No | NA | 2 | 0 | 0 | Not-determined | Genome-fragment | NA | NA | 0 |
| S1_k141_167749 | 3791 | No | NA | 10 | 1 | 0 | Low-quality | Genome-fragment | 5.87 | AAI-based (high-confidence) | 0 |
| S2-final-viral-combined_k141_54503\|\|full | 3791 | No | NA | 5 | 1 | 0 | Low-quality | Genome-fragment | 2.45 | HMM-based (lower-bound) | 0 |
| S1-final-viral-combined_k141_134649\|\|full | 3792 | No | NA | 10 | 1 | 0 | Low-quality | Genome-fragment | 6.22 | AAI-based (high-confidence) | 0 |
| S8-final-viral-combined_k141_233416\|\|full | 3792 | No | NA | 5 | 0 | 0 | Low-quality | Genome-fragment | 5.67 | AAI-based (medium-confidence) | 0 |
| S2-final-viral-combined_k141_13704\|\|full | 1469 | No | NA | 3 | 0 | 1 | Not-determined | Genome-fragment | NA | NA | 0 |
| S2-final-viral-combined_k141_220191\|\|full | 2728 | No | NA | 2 | 0 | 2 | Not-determined | Genome-fragment | NA | NA | 0 |
| S2-final-viral-combined_k141_190170\|\|full | 2371 | No | NA | 4 | 0 | 0 | Not-determined | Genome-fragment | NA | NA | 0 |
| S2-final-viral-combined_k141_148424\|\|full | 1291 | No | NA | 3 | 0 | 1 | Not-determined | Genome-fragment | NA | NA | 0 |
| S2-final-viral-combined_k141_46269\|\|full | 1564 | No | NA | 3 | 0 | 1 | Not-determined | Genome-fragment | NA | NA | 0 |
| S2-final-viral-combined_k141_148431\|\|full | 2157 | No | NA | 2 | 0 | 2 | Not-determined | Genome-fragment | NA | NA | 0 |
| S2-final-viral-combined_k141_158709\|\|full | 1871 | No | NA | 2 | 0 | 2 | Not-determined | Genome-fragment | NA | NA | 0 |
| S2-final-viral-combined_k141_86640\|\|full | 3103 | No | NA | 4 | 0 | 2 | Not-determined | Genome-fragment | NA | NA | 0 |
| S2-final-viral-combined_k141_97011\|\|full | 1010 | No | NA | 2 | 0 | 0 | Not-determined | Genome-fragment | NA | NA | 0 |
| S2-final-viral-combined_k141_3403\|\|full | 6814 | No | NA | 4 | 0 | 2 | Not-determined | Genome-fragment | NA | NA | 0 |
| S2-final-viral-combined_k141_117253\|\|full | 3698 | No | NA | 5 | 0 | 1 | Not-determined | Genome-fragment | NA | NA | 0 |
| S11-final-viral-combined_k141_270445\|\|full | 3793 | No | NA | 7 | 0 | 0 | Low-quality | Genome-fragment | 4.97 | AAI-based (medium-confidence) | 0 |
| S2-final-viral-combined_k141_75724\|\|full | 1264 | No | NA | 2 | 0 | 0 | Not-determined | Genome-fragment | NA | NA | 0 |
| S2-final-viral-combined_k141_231788\|\|full | 2169 | No | NA | 3 | 0 | 2 | Not-determined | Genome-fragment | NA | NA | 0 |
| S2-final-viral-combined_k141_148481\|\|full | 1368 | No | NA | 2 | 0 | 1 | Not-determined | Genome-fragment | NA | NA | 0 |
| S2-final-viral-combined_k141_55307\|\|full | 4286 | No | NA | 5 | 0 | 1 | Not-determined | Genome-fragment | NA | NA | 0 |
| S2-final-viral-combined_k141_148494\|\|full | 6836 | No | NA | 7 | 0 | 1 | Not-determined | Genome-fragment | NA | NA | 0 |
| S2-final-viral-combined_k141_46354\|\|full | 1174 | No | NA | 3 | 0 | 1 | Not-determined | Genome-fragment | NA | NA | 0 |
| S2-final-viral-combined_k141_65640\|\|full | 4512 | No | NA | 10 | 0 | 0 | Not-determined | Genome-fragment | NA | NA | 0 |
| S2-final-viral-combined_k141_199489\|\|full | 4464 | No | NA | 5 | 0 | 3 | Not-determined | Genome-fragment | NA | NA | 0 |
| S2-final-viral-combined_k141_86674\|\|full | 1707 | No | NA | 4 | 0 | 0 | Not-determined | Genome-fragment | NA | NA | 0 |
| S1-final-viral-combined_k141_70607\|\|full | 3793 | No | NA | 3 | 0 | 2 | Low-quality | Genome-fragment | 1.98 | AAI-based (medium-confidence) | 0 |
| S2-final-viral-combined_k141_169863\|\|full | 1377 | No | NA | 3 | 0 | 2 | Not-determined | Genome-fragment | NA | NA | 0 |
| S2-final-viral-combined_k141_3459\|\|full | 5294 | No | NA | 5 | 0 | 4 | Not-determined | Genome-fragment | NA | NA | 0 |
| S7-final-viral-combined_k141_6204\|\|full | 3793 | No | NA | 6 | 0 | 3 | Low-quality | Genome-fragment | 6.11 | AAI-based (medium-confidence) | 0 |
| S2-final-viral-combined_k141_46378\|\|full | 5843 | No | NA | 6 | 0 | 4 | Not-determined | Genome-fragment | NA | NA | 0 |
| S4-final-viral-combined_k141_113497\|\|full | 3793 | No | NA | 4 | 0 | 3 | Low-quality | Genome-fragment | 1.26 | AAI-based (medium-confidence) | 0 |
| S2-final-viral-combined_k141_117277\|\|full | 2729 | No | NA | 4 | 0 | 3 | Not-determined | Genome-fragment | NA | NA | 0 |
| S2-final-viral-combined_k141_231862\|\|full | 1237 | No | NA | 3 | 0 | 1 | Not-determined | Genome-fragment | NA | NA | 0 |
| S2-final-viral-combined_k141_190257\|\|full | 3438 | No | NA | 4 | 0 | 1 | Not-determined | Genome-fragment | NA | NA | 0 |
| S2-final-viral-combined_k141_3475\|\|full | 1376 | No | NA | 2 | 0 | 1 | Not-determined | Genome-fragment | NA | NA | 0 |
| S2-final-viral-combined_k141_241334\|\|full | 2043 | No | NA | 2 | 0 | 2 | Not-determined | Genome-fragment | NA | NA | 0 |
| S2-final-viral-combined_k141_86682\|\|full | 3258 | No | NA | 4 | 0 | 3 | Not-determined | Genome-fragment | NA | NA | 0 |
| S2-final-viral-combined_k141_210576\|\|full | 4598 | No | NA | 5 | 0 | 4 | Not-determined | Genome-fragment | NA | NA | 0 |
| S6_k141_117080 | 3793 | No | NA | 11 | 0 | 0 | Low-quality | Genome-fragment | 6.44 | AAI-based (high-confidence) | 0 |
| S2-final-viral-combined_k141_3476\|\|full | 2669 | No | NA | 2 | 0 | 0 | Not-determined | Genome-fragment | NA | NA | 0 |
| S11-final-viral-combined_k141_144071\|\|full | 3794 | No | NA | 9 | 2 | 0 | Low-quality | Genome-fragment | 7.91 | AAI-based (high-confidence) | 0 |
| S2-final-viral-combined_k141_65692\|\|full | 2432 | No | NA | 3 | 0 | 2 | Not-determined | Genome-fragment | NA | NA | 0 |
| S17-final-viral-combined_k141_93453\|\|full | 3794 | No | NA | 5 | 0 | 2 | Low-quality | Genome-fragment | 3.37 | AAI-based (high-confidence) | 0 |
| S12_k141_103488 | 3794 | No | NA | 2 | 0 | 1 | Low-quality | Genome-fragment | 3.73 | AAI-based (medium-confidence) | 0 |
| S2-final-viral-combined_k141_190273\|\|full | 1958 | No | NA | 3 | 0 | 0 | Not-determined | Genome-fragment | NA | NA | 0 |
| S9-final-viral-combined_k141_51966\|\|full | 3796 | No | NA | 5 | 0 | 1 | Low-quality | Genome-fragment | 8.99 | AAI-based (medium-confidence) | 0 |
| S2-final-viral-combined_k141_55337\|\|full | 1055 | No | NA | 2 | 0 | 2 | Not-determined | Genome-fragment | NA | NA | 0 |
| S2-final-viral-combined_k141_75836\|\|full | 1799 | No | NA | 2 | 0 | 1 | Not-determined | Genome-fragment | NA | NA | 0 |
| S2-final-viral-combined_k141_3541\|\|full | 2385 | No | NA | 2 | 0 | 1 | Not-determined | Genome-fragment | NA | NA | 0 |
| S2-final-viral-combined_k141_86723\|\|full | 13443 | No | NA | 19 | 0 | 0 | Not-determined | Genome-fragment | NA | NA | 0 |
| S2-final-viral-combined_k141_190324\|\|full | 2083 | No | NA | 3 | 0 | 1 | Not-determined | Genome-fragment | NA | NA | 0 |
| S2-final-viral-combined_k141_210642\|\|full | 3063 | No | NA | 2 | 0 | 1 | Not-determined | Genome-fragment | NA | NA | 0 |
| S1-final-viral-combined_k141_156075\|\|full | 3797 | No | NA | 4 | 0 | 1 | Low-quality | Genome-fragment | 5.41 | AAI-based (medium-confidence) | 0 |
| S2-final-viral-combined_k141_117431\|\|full | 9997 | No | NA | 13 | 0 | 1 | Not-determined | Genome-fragment | NA | NA | 0 |
| S11-final-viral-combined_k141_109096\|\|full | 3798 | No | NA | 6 | 0 | 0 | Low-quality | Genome-fragment | 6.65 | AAI-based (high-confidence) | 0 |
| S2-final-viral-combined_k141_180257\|\|full | 2775 | No | NA | 5 | 0 | 2 | Not-determined | Genome-fragment | NA | NA | 0 |
| S2-final-viral-combined_k141_241384\|\|full | 6558 | No | NA | 4 | 0 | 2 | Not-determined | Genome-fragment | NA | NA | 0 |
| S2-final-viral-combined_k141_138460\|\|full | 1162 | No | NA | 3 | 0 | 1 | Not-determined | Genome-fragment | NA | NA | 0 |
| S2-final-viral-combined_k141_3564\|\|full | 4424 | No | NA | 6 | 0 | 1 | Not-determined | Genome-fragment | NA | NA | 0 |
| S2-final-viral-combined_k141_231965\|\|full | 1288 | No | NA | 3 | 0 | 1 | Not-determined | Genome-fragment | NA | NA | 0 |
| S12-final-viral-combined_k141_352224\|\|full | 3798 | No | NA | 11 | 2 | 0 | Low-quality | Genome-fragment | 5.1 | HMM-based (lower-bound) | 0 |
| S2-final-viral-combined_k141_180302\|\|full | 7146 | No | NA | 7 | 0 | 5 | Not-determined | Genome-fragment | NA | NA | 0 |
| S2-final-viral-combined_k141_180321\|\|full | 1139 | No | NA | 3 | 0 | 0 | Not-determined | Genome-fragment | NA | NA | 0 |
| S2-final-viral-combined_k141_55495\|\|full | 17732 | No | NA | 24 | 11 | 0 | Medium-quality | Genome-fragment | 52.38 | AAI-based (medium-confidence) | 0 |
| S2-final-viral-combined_k141_13921\|\|full | 3018 | No | NA | 3 | 0 | 3 | Not-determined | Genome-fragment | NA | NA | 0 |
| S2-final-viral-combined_k141_97178\|\|full | 1242 | No | NA | 3 | 0 | 3 | Not-determined | Genome-fragment | NA | NA | 0 |
| S2-final-viral-combined_k141_180322\|\|full | 3494 | No | NA | 3 | 0 | 0 | Not-determined | Genome-fragment | NA | NA | 0 |
| S2_k141_164961 | 3798 | No | NA | 5 | 2 | 0 | Low-quality | Genome-fragment | 8.4 | AAI-based (medium-confidence) | 0 |
| S2-final-viral-combined_k141_170018\|\|full | 7429 | No | NA | 4 | 0 | 3 | Not-determined | Genome-fragment | NA | NA | 0 |
| S2-final-viral-combined_k141_158821\|\|full | 29638 | Yes | 25251 | 41 | 13 | 2 | Medium-quality | Genome-fragment | 74.04 | AAI-based (high-confidence) | 14.8 |
| S2-final-viral-combined_k141_107332\|\|full | 1331 | No | NA | 2 | 0 | 1 | Not-determined | Genome-fragment | NA | NA | 0 |
| S1-final-viral-combined_k141_153292\|\|full | 3799 | No | NA | 8 | 1 | 0 | Low-quality | Genome-fragment | 10.6 | AAI-based (high-confidence) | 0 |
| S2-final-viral-combined_k141_138542\|\|full | 5392 | No | NA | 6 | 0 | 3 | Not-determined | Genome-fragment | NA | NA | 0 |
| S7-final-viral-combined_k141_105603\|\|full | 3799 | No | NA | 8 | 5 | 0 | Low-quality | Genome-fragment | 11.11 | AAI-based (medium-confidence) | 0 |
| S2-final-viral-combined_k141_220326\|\|full | 1441 | No | NA | 2 | 0 | 2 | Not-determined | Genome-fragment | NA | NA | 0 |
| S2-final-viral-combined_k141_138568\|\|full | 2903 | No | NA | 4 | 0 | 0 | Not-determined | Genome-fragment | NA | NA | 0 |
| S11-final-viral-combined_k141_215662\|\|full | 3800 | No | NA | 7 | 1 | 0 | Low-quality | Genome-fragment | 6.61 | HMM-based (lower-bound) | 0 |
| S2-final-viral-combined_k141_107369\|\|full | 2017 | No | NA | 2 | 0 | 1 | Not-determined | Genome-fragment | NA | NA | 0 |
| S1-final-viral-combined_k141_309613\|\|full | 3800 | No | NA | 3 | 0 | 3 | Low-quality | Genome-fragment | 0.55 | AAI-based (medium-confidence) | 0 |
| S2-final-viral-combined_k141_138570\|\|full | 3105 | No | NA | 4 | 0 | 2 | Not-determined | Genome-fragment | NA | NA | 0 |
| S2-final-viral-combined_k141_14008\|\|full | 1524 | No | NA | 3 | 0 | 1 | Not-determined | Genome-fragment | NA | NA | 0 |
| S2-final-viral-combined_k141_107396\|\|full | 3105 | No | NA | 4 | 0 | 2 | Not-determined | Genome-fragment | NA | NA | 0 |
| S12-final-viral-combined_k141_187314\|\|full | 3800 | No | NA | 3 | 0 | 3 | Low-quality | Genome-fragment | 1.69 | AAI-based (medium-confidence) | 0 |
| S2-final-viral-combined_k141_34875\|\|full | 2215 | No | NA | 2 | 0 | 0 | Not-determined | Genome-fragment | NA | NA | 0 |
| S2-final-viral-combined_k141_117532\|\|full | 1287 | No | NA | 3 | 0 | 1 | Not-determined | Genome-fragment | NA | NA | 0 |
| S12-final-viral-combined_k141_94376\|\|full | 3800 | No | NA | 8 | 5 | 0 | Low-quality | Genome-fragment | 8.94 | AAI-based (medium-confidence) | 0 |
| S2-final-viral-combined_k141_158849\|\|full | 1182 | No | NA | 2 | 0 | 0 | Not-determined | Genome-fragment | NA | NA | 0 |
| S12-final-viral-combined_k141_183212\|\|full | 3800 | No | NA | 3 | 0 | 2 | Low-quality | Genome-fragment | 4.57 | AAI-based (medium-confidence) | 0 |
| S2_k141_85743 | 3800 | No | NA | 4 | 0 | 0 | Low-quality | Genome-fragment | 4.87 | AAI-based (medium-confidence) | 0 |
| S2-final-viral-combined_k141_55659\|\|full | 2156 | No | NA | 3 | 0 | 0 | Not-determined | Genome-fragment | NA | NA | 0 |
| S4_k141_123259 | 3800 | No | NA | 4 | 1 | 0 | Low-quality | Genome-fragment | 3.79 | AAI-based (high-confidence) | 0 |
| S8_k141_2537 | 3801 | No | NA | 8 | 0 | 0 | Low-quality | Genome-fragment | 8.35 | AAI-based (medium-confidence) | 0 |
| S9-final-viral-combined_k141_81163\|\|full | 3802 | No | NA | 5 | 0 | 1 | Low-quality | Genome-fragment | 6.2 | AAI-based (medium-confidence) | 0 |
| S2-final-viral-combined_k141_107474\|\|full | 2764 | No | NA | 2 | 0 | 1 | Not-determined | Genome-fragment | NA | NA | 0 |
| S11-final-viral-combined_k141_73534\|\|full | 3802 | No | NA | 3 | 0 | 2 | Low-quality | Genome-fragment | 2.49 | AAI-based (medium-confidence) | 0 |
| S2-final-viral-combined_k141_158882\|\|full | 1853 | No | NA | 3 | 0 | 2 | Not-determined | Genome-fragment | NA | NA | 0 |
| S2-final-viral-combined_k141_127729\|\|full | 1949 | No | NA | 3 | 0 | 0 | Not-determined | Genome-fragment | NA | NA | 0 |
| S2-final-viral-combined_k141_46614\|\|full | 1978 | No | NA | 3 | 0 | 0 | Not-determined | Genome-fragment | NA | NA | 0 |
| S2-final-viral-combined_k141_220407\|\|full | 6665 | No | NA | 5 | 0 | 4 | Not-determined | Genome-fragment | NA | NA | 0 |
| S2-final-viral-combined_k141_107496\|\|full | 4551 | No | NA | 3 | 0 | 1 | Not-determined | Genome-fragment | NA | NA | 0 |
| S2-final-viral-combined_k141_158897\|\|full | 6999 | No | NA | 5 | 0 | 4 | Not-determined | Genome-fragment | NA | NA | 0 |
| S7-final-viral-combined_k141_170797\|\|full | 3802 | No | NA | 5 | 0 | 1 | Low-quality | Genome-fragment | 7.94 | AAI-based (medium-confidence) | 0 |
| S2-final-viral-combined_k141_107498\|\|full | 2189 | No | NA | 3 | 0 | 1 | Not-determined | Genome-fragment | NA | NA | 0 |
| S2-final-viral-combined_k141_24448\|\|full | 1980 | No | NA | 3 | 0 | 2 | Not-determined | Genome-fragment | NA | NA | 0 |
| S17-k141_146465 | 3803 | No | NA | 3 | 0 | 2 | Low-quality | Genome-fragment | 8.13 | AAI-based (medium-confidence) | 0 |
| S2-final-viral-combined_k141_76036\|\|full | 1382 | No | NA | 2 | 0 | 0 | Not-determined | Genome-fragment | NA | NA | 0 |
| S12-final-viral-combined_k141_1249\|\|full | 3803 | No | NA | 4 | 0 | 1 | Low-quality | Genome-fragment | 2.57 | AAI-based (medium-confidence) | 0 |
| S1_k141_36273 | 3804 | Yes | 2319 | 5 | 1 | 1 | Low-quality | Genome-fragment | 2.98 | AAI-based (high-confidence) | 39.04 |
| S2-final-viral-combined_k141_117614\|\|full | 2536 | No | NA | 3 | 0 | 0 | Not-determined | Genome-fragment | NA | NA | 0 |
| S12_k141_97554 | 3804 | No | NA | 5 | 0 | 4 | Low-quality | Genome-fragment | 6.29 | AAI-based (medium-confidence) | 0 |
| S2-final-viral-combined_k141_152163\|\|full | 3804 | No | NA | 4 | 0 | 2 | Low-quality | Genome-fragment | 2.13 | AAI-based (medium-confidence) | 0 |
| S2-final-viral-combined_k141_76071\|\|full | 1521 | No | NA | 3 | 0 | 1 | Not-determined | Genome-fragment | NA | NA | 0 |
| S2-final-viral-combined_k141_86970\|\|full | 3145 | No | NA | 5 | 0 | 0 | Not-determined | Genome-fragment | NA | NA | 0 |
| S2-final-viral-combined_k141_210822\|\|full | 92842 | No | NA | 91 | 1 | 22 | High-quality | High-quality | 100 | AAI-based (medium-confidence) | 0 |
| S6_k141_57574 | 3804 | No | NA | 4 | 0 | 2 | Low-quality | Genome-fragment | 2.61 | AAI-based (high-confidence) | 0 |
| S2-final-viral-combined_k141_158941\|\|full | 1791 | No | NA | 3 | 0 | 2 | Not-determined | Genome-fragment | NA | NA | 0 |
| S9_k141_93513 | 3805 | No | NA | 3 | 1 | 0 | Low-quality | Genome-fragment | 7.02 | AAI-based (high-confidence) | 0 |
| S2-final-viral-combined_k141_24506\|\|full | 1642 | No | NA | 4 | 0 | 0 | Not-determined | Genome-fragment | NA | NA | 0 |
| S2-final-viral-combined_k141_220494\|\|full | 2322 | No | NA | 4 | 0 | 0 | Not-determined | Genome-fragment | NA | NA | 0 |
| S2-final-viral-combined_k141_232081\|\|full | 2884 | No | NA | 3 | 0 | 2 | Not-determined | Genome-fragment | NA | NA | 0 |
| S5-final-viral-combined_k141_38699\|\|full | 3805 | No | NA | 4 | 0 | 2 | Low-quality | Genome-fragment | 2.65 | AAI-based (high-confidence) | 0 |
| S2-final-viral-combined_k141_24530\|\|full | 1860 | No | NA | 4 | 0 | 1 | Not-determined | Genome-fragment | NA | NA | 0 |
| S2-final-viral-combined_k141_87089\|\|full | 2025 | No | NA | 4 | 0 | 0 | Not-determined | Genome-fragment | NA | NA | 0 |
| S4-final-viral-combined_k141_42052\|\|full | 3805 | No | NA | 3 | 0 | 3 | Low-quality | Genome-fragment | 1.26 | AAI-based (medium-confidence) | 0 |
| S2-final-viral-combined_k141_65910\|\|full | 3111 | No | NA | 4 | 0 | 0 | Not-determined | Genome-fragment | NA | NA | 0 |
| S2-final-viral-combined_k141_190624\|\|full | 26624 | No | NA | 35 | 10 | 1 | Medium-quality | Genome-fragment | 64.41 | AAI-based (medium-confidence) | 0 |
| S17-final-viral-combined_k141_99430\|\|full | 3806 | No | NA | 8 | 1 | 0 | Low-quality | Genome-fragment | 9.35 | AAI-based (medium-confidence) | 0 |
| S2-final-viral-combined_k141_76165\|\|full | 1533 | No | NA | 4 | 0 | 2 | Not-determined | Genome-fragment | NA | NA | 0 |
| S2-final-viral-combined_k141_97320\|\|full | 1648 | No | NA | 3 | 0 | 2 | Not-determined | Genome-fragment | NA | NA | 0 |
| S2-final-viral-combined_k141_190645\|\|full | 1577 | No | NA | 3 | 0 | 1 | Not-determined | Genome-fragment | NA | NA | 0 |
| S2-final-viral-combined_k141_199567\|\|full | 1626 | No | NA | 3 | 0 | 1 | Not-determined | Genome-fragment | NA | NA | 0 |
| S2-final-viral-combined_k141_46761\|\|full | 37782 | No | NA | 36 | 0 | 12 | Not-determined | Genome-fragment | NA | NA | 0 |
| S2-final-viral-combined_k141_65917\|\|full | 3404 | No | NA | 4 | 0 | 2 | Not-determined | Genome-fragment | NA | NA | 0 |
| S6-final-viral-combined_k141_41236\|\|full | 3806 | No | NA | 4 | 0 | 3 | Low-quality | Genome-fragment | 3.57 | AAI-based (medium-confidence) | 0 |
| S2-final-viral-combined_k141_65919\|\|full | 11761 | No | NA | 16 | 0 | 3 | Not-determined | Genome-fragment | NA | NA | 0 |
| S2-final-viral-combined_k141_159057\|\|full | 1293 | No | NA | 3 | 0 | 0 | Not-determined | Genome-fragment | NA | NA | 0 |
| S2-final-viral-combined_k141_199582\|\|full | 8065 | No | NA | 6 | 0 | 5 | Not-determined | Genome-fragment | NA | NA | 0 |
| S1-final-viral-combined_k141_4950\|\|full | 3807 | No | NA | 3 | 0 | 3 | Low-quality | Genome-fragment | 7.53 | AAI-based (medium-confidence) | 0 |
| S7_k141_197402 | 3807 | No | NA | 5 | 0 | 1 | Low-quality | Genome-fragment | 6.36 | AAI-based (high-confidence) | 0 |
| S2-final-viral-combined_k141_24549\|\|full | 65681 | Yes | 25345 | 81 | 14 | 35 | Medium-quality | Genome-fragment | 63.84 | AAI-based (high-confidence) | 61.41 |
| S2-final-viral-combined_k141_3745\|\|full | 5955 | No | NA | 8 | 0 | 6 | Not-determined | Genome-fragment | NA | NA | 0 |
| S2-final-viral-combined_k141_220539\|\|full | 9651 | No | NA | 9 | 0 | 7 | Not-determined | Genome-fragment | NA | NA | 0 |
| S17-final-viral-combined_k141_250936\|\|full | 3807 | No | NA | 3 | 0 | 1 | Low-quality | Genome-fragment | 8.69 | AAI-based (medium-confidence) | 0 |
| S1-final-viral-combined_k141_63574\|\|full | 3808 | No | NA | 6 | 0 | 1 | Low-quality | Genome-fragment | 5.74 | AAI-based (high-confidence) | 0 |
| S2-final-viral-combined_k141_35153\|\|full | 1293 | No | NA | 2 | 0 | 1 | Not-determined | Genome-fragment | NA | NA | 0 |
| S2-final-viral-combined_k141_127900\|\|full | 3435 | No | NA | 3 | 0 | 2 | Not-determined | Genome-fragment | NA | NA | 0 |
| S7-final-viral-combined_k141_99822\|\|full | 3808 | No | NA | 7 | 2 | 0 | Low-quality | Genome-fragment | 6.59 | AAI-based (medium-confidence) | 0 |
| S2-final-viral-combined_k141_55860\|\|full | 2014 | No | NA | 4 | 0 | 1 | Not-determined | Genome-fragment | NA | NA | 0 |
| S2-final-viral-combined_k141_138875\|\|full | 2467 | No | NA | 3 | 0 | 3 | Not-determined | Genome-fragment | NA | NA | 0 |
| S11_k141_204358 | 3809 | No | NA | 6 | 0 | 3 | Low-quality | Genome-fragment | 6.36 | AAI-based (medium-confidence) | 0 |
| S17-final-viral-combined_k141_103988\|\|full | 3809 | No | NA | 2 | 0 | 0 | Low-quality | Genome-fragment | 1.25 | AAI-based (medium-confidence) | 0 |
| S8-final-viral-combined_k141_261213\|\|full | 3809 | No | NA | 4 | 0 | 0 | Low-quality | Genome-fragment | 4.34 | AAI-based (medium-confidence) | 0 |
| S2-final-viral-combined_k141_87150\|\|full | 7018 | No | NA | 7 | 0 | 5 | Not-determined | Genome-fragment | NA | NA | 0 |
| S2-final-viral-combined_k141_107668\|\|full | 1831 | No | NA | 2 | 0 | 2 | Not-determined | Genome-fragment | NA | NA | 0 |
| S2-final-viral-combined_k141_14278\|\|full | 2825 | No | NA | 6 | 0 | 1 | Not-determined | Genome-fragment | NA | NA | 0 |
| S2-final-viral-combined_k141_232212\|\|full | 4339 | No | NA | 7 | 0 | 1 | Not-determined | Genome-fragment | NA | NA | 0 |
| S2-final-viral-combined_k141_46858\|\|full | 4492 | No | NA | 5 | 0 | 1 | Not-determined | Genome-fragment | NA | NA | 0 |
| S2-final-viral-combined_k141_220567\|\|full | 2315 | No | NA | 4 | 0 | 1 | Not-determined | Genome-fragment | NA | NA | 0 |
| S1-final-viral-combined_k141_43666\|\|full | 3810 | Yes | 2133 | 4 | 1 | 1 | Low-quality | Genome-fragment | 2.72 | AAI-based (high-confidence) | 44.02 |

| | | | | | | | | | | | |
|---|---|---|---|---|---|---|---|---|---|---|---|
| S3-final-viral-combined_k141_48566\|\|full | 3810 | No | NA | 7 | 0 | 0 | Low-quality | Genome-fragment | 2.52 | AAI-based (high-confidence) | 0 |
| S5-final-viral-combined_k141_123026\|\|full | 3810 | No | NA | 6 | 4 | 0 | Low-quality | Genome-fragment | 9.78 | AAI-based (high-confidence) | 0 |
| S8-final-viral-combined_k141_160557\|\|full | 3810 | No | NA | 4 | 0 | 1 | Low-quality | Genome-fragment | 0.35 | AAI-based (medium-confidence) | 0 |
| S2-final-viral-combined_k141_46880\|\|full | 1140 | No | NA | 2 | 0 | 0 | Not-determined | Genome-fragment | NA | NA | 0 |
| S11-final-viral-combined_k141_37151\|\|full | 3812 | No | NA | 7 | 1 | 0 | Low-quality | Genome-fragment | 7.62 | AAI-based (medium-confidence) | 0 |
| S1-final-viral-combined_k141_99120\|\|full | 3812 | No | NA | 3 | 0 | 3 | Low-quality | Genome-fragment | 2.43 | AAI-based (medium-confidence) | 0 |
| S2-final-viral-combined_k141_149022\|\|full | 1507 | No | NA | 3 | 0 | 1 | Not-determined | Genome-fragment | NA | NA | 0 |
| S1-final-viral-combined_k141_102198\|\|full | 3813 | No | NA | 3 | 0 | 1 | Low-quality | Genome-fragment | 1.28 | AAI-based (medium-confidence) | 0 |
| S2-final-viral-combined_k141_107738\|\|full | 2183 | No | NA | 3 | 0 | 0 | Not-determined | Genome-fragment | NA | NA | 0 |
| S2-final-viral-combined_k141_180640\|\|full | 30412 | No | NA | 46 | 15 | 0 | Medium-quality | Genome-fragment | 61.73 | AAI-based (high-confidence) | 0 |
| S17-final-viral-combined_k141_244869\|\|full | 3813 | No | NA | 2 | 0 | 2 | Low-quality | Genome-fragment | 1.37 | AAI-based (medium-confidence) | 0 |
| S2-final-viral-combined_k141_232282\|\|full | 5874 | No | NA | 9 | 0 | 7 | Not-determined | Genome-fragment | NA | NA | 0 |
| S12-final-viral-combined_k141_174159\|\|full | 3813 | No | NA | 3 | 0 | 2 | Low-quality | Genome-fragment | 2.47 | AAI-based (medium-confidence) | 0 |
| S4-final-viral-combined_k141_164401\|\|full | 3813 | No | NA | 4 | 0 | 0 | Low-quality | Genome-fragment | 9.1 | AAI-based (high-confidence) | 0 |
| S7-final-viral-combined_k141_63003\|\|full | 3814 | No | NA | 5 | 0 | 4 | Low-quality | Genome-fragment | 2.12 | AAI-based (medium-confidence) | 0 |
| S2-final-viral-combined_k141_46955\|\|full | 1248 | No | NA | 3 | 0 | 0 | Not-determined | Genome-fragment | NA | NA | 0 |
| S17-final-viral-combined_k141_103203\|\|full | 3815 | No | NA | 10 | 4 | 0 | Low-quality | Genome-fragment | 9.2 | AAI-based (high-confidence) | 0 |
| S1-final-viral-combined_k141_230433\|\|full | 3816 | No | NA | 5 | 4 | 0 | Low-quality | Genome-fragment | 8.06 | AAI-based (high-confidence) | 0 |
| S2-final-viral-combined_k141_14360\|\|full | 6004 | No | NA | 4 | 0 | 3 | Not-determined | Genome-fragment | NA | NA | 0 |
| S2-final-viral-combined_k141_87191\|\|full | 6588 | No | NA | 9 | 0 | 6 | Not-determined | Genome-fragment | NA | NA | 0 |
| S2-final-viral-combined_k141_220664\|\|full | 2882 | No | NA | 4 | 0 | 2 | Not-determined | Genome-fragment | NA | NA | 0 |
| S1-final-viral-combined_k141_257376\|\|full | 3816 | No | NA | 5 | 0 | 0 | Low-quality | Genome-fragment | 6.86 | AAI-based (medium-confidence) | 0 |
| S1_k141_148975 | 3816 | No | NA | 4 | 0 | 0 | Low-quality | Genome-fragment | 8.5 | AAI-based (medium-confidence) | 0 |
| S2-final-viral-combined_k141_138944\|\|full | 4234 | No | NA | 2 | 0 | 1 | Not-determined | Genome-fragment | NA | NA | 0 |
| S7_k141_232595 | 3817 | No | NA | 2 | 0 | 2 | Low-quality | Genome-fragment | 4.56 | AAI-based (medium-confidence) | 0 |
| S11_k141_10643 | 3818 | No | NA | 1 | 0 | 1 | Low-quality | Genome-fragment | 0.33 | AAI-based (medium-confidence) | 0 |
| S1_k141_59103 | 3818 | No | NA | 9 | 3 | 0 | Low-quality | Genome-fragment | 9.69 | AAI-based (high-confidence) | 0 |
| S2-final-viral-combined_k141_66073\|\|full | 1489 | No | NA | 3 | 0 | 0 | Not-determined | Genome-fragment | NA | NA | 0 |
| S2-final-viral-combined_k141_117904\|\|full | 3598 | No | NA | 3 | 0 | 1 | Not-determined | Genome-fragment | NA | NA | 0 |
| S2-final-viral-combined_k141_87266\|\|full | 1916 | No | NA | 2 | 0 | 1 | Not-determined | Genome-fragment | NA | NA | 0 |
| S2-final-viral-combined_k141_138991\|\|full | 2629 | No | NA | 5 | 0 | 0 | Not-determined | Genome-fragment | NA | NA | 0 |
| S5-final-viral-combined_k141_178183\|\|full | 3818 | Yes | 2395 | 4 | 1 | 1 | Low-quality | Genome-fragment | 2.69 | HMM-based (lower-bound) | 37.27 |
| S2-final-viral-combined_k141_24691\|\|full | 1086 | No | NA | 2 | 0 | 0 | Not-determined | Genome-fragment | NA | NA | 0 |
| S2-final-viral-combined_k141_47045\|\|full | 3444 | No | NA | 4 | 0 | 4 | Not-determined | Genome-fragment | NA | NA | 0 |
| S2-final-viral-combined_k141_138870\|\|full | 3818 | No | NA | 7 | 1 | 0 | Low-quality | Genome-fragment | 7.65 | HMM-based (lower-bound) | 0 |
| S2-final-viral-combined_k141_199763\|\|full | 1330 | No | NA | 3 | 0 | 1 | Not-determined | Genome-fragment | NA | NA | 0 |
| S2-final-viral-combined_k141_97558\|\|full | 4850 | No | NA | 4 | 0 | 3 | Not-determined | Genome-fragment | NA | NA | 0 |
| S2-final-viral-combined_k141_159229\|\|full | 1335 | No | NA | 4 | 0 | 0 | Not-determined | Genome-fragment | NA | NA | 0 |
| S12-final-viral-combined_k141_249051\|\|full | 3820 | No | NA | 6 | 0 | 0 | Low-quality | Genome-fragment | 6.06 | AAI-based (high-confidence) | 0 |
| S2-final-viral-combined_k141_127951\|\|full | 10795 | No | NA | 11 | 0 | 6 | Not-determined | Genome-fragment | NA | NA | 0 |
| S11-final-viral-combined_k141_240883\|\|full | 3821 | No | NA | 3 | 0 | 3 | Low-quality | Genome-fragment | 1.37 | AAI-based (medium-confidence) | 0 |
| S2-final-viral-combined_k141_127956\|\|full | 5247 | No | NA | 6 | 0 | 0 | Not-determined | Genome-fragment | NA | NA | 0 |
| S1-final-viral-combined_k141_280401\|\|full | 3821 | No | NA | 4 | 0 | 3 | Low-quality | Genome-fragment | 3.11 | AAI-based (medium-confidence) | 0 |
| S2-final-viral-combined_k141_170180\|\|full | 2648 | No | NA | 3 | 0 | 1 | Not-determined | Genome-fragment | NA | NA | 0 |
| S17-final-viral-combined_k141_173024\|\|full | 3821 | No | NA | 5 | 0 | 3 | Low-quality | Genome-fragment | 2.71 | AAI-based (high-confidence) | 0 |
| S11_k141_278976 | 3822 | No | NA | 3 | 0 | 0 | Low-quality | Genome-fragment | 8.83 | AAI-based (high-confidence) | 0 |
| S2-final-viral-combined_k141_180762\|\|full | 2016 | No | NA | 3 | 0 | 0 | Not-determined | Genome-fragment | NA | NA | 0 |
| S7-final-viral-combined_k141_207952\|\|full | 3822 | No | NA | 4 | 0 | 2 | Low-quality | Genome-fragment | 1.78 | AAI-based (medium-confidence) | 0 |
| S2-final-viral-combined_k141_66124\|\|full | 4632 | No | NA | 5 | 0 | 3 | Not-determined | Genome-fragment | NA | NA | 0 |
| S2_k141_836 | 3823 | No | NA | 2 | 0 | 2 | Low-quality | Genome-fragment | 6.29 | AAI-based (high-confidence) | 0 |
| S2-final-viral-combined_k141_117964\|\|full | 1247 | No | NA | 3 | 0 | 1 | Not-determined | Genome-fragment | NA | NA | 0 |
| S2-final-viral-combined_k141_139036\|\|full | 2157 | No | NA | 3 | 0 | 2 | Not-determined | Genome-fragment | NA | NA | 0 |
| S2-final-viral-combined_k141_180785\|\|full | 3205 | No | NA | 2 | 0 | 2 | Not-determined | Genome-fragment | NA | NA | 0 |
| S2-final-viral-combined_k141_180791\|\|full | 4651 | No | NA | 6 | 0 | 3 | Not-determined | Genome-fragment | NA | NA | 0 |
| S9_k141_16730 | 3824 | No | NA | 3 | 0 | 3 | Low-quality | Genome-fragment | 1.37 | AAI-based (medium-confidence) | 0 |
| S2-final-viral-combined_k141_118024\|\|full | 1500 | No | NA | 2 | 0 | 1 | Not-determined | Genome-fragment | NA | NA | 0 |
| S2-final-viral-combined_k141_87390\|\|full | 1187 | No | NA | 3 | 0 | 0 | Not-determined | Genome-fragment | NA | NA | 0 |
| S2-final-viral-combined_k141_107826\|\|full | 4959 | No | NA | 6 | 0 | 4 | Not-determined | Genome-fragment | NA | NA | 0 |
| S2-final-viral-combined_k141_210966\|\|full | 1351 | No | NA | 2 | 0 | 1 | Not-determined | Genome-fragment | NA | NA | 0 |
| S2-final-viral-combined_k141_35389\|\|full | 1553 | No | NA | 2 | 0 | 1 | Not-determined | Genome-fragment | NA | NA | 0 |
| S2-final-viral-combined_k141_38933\|\|full | 3824 | No | NA | 7 | 2 | 0 | Low-quality | Genome-fragment | 6.45 | AAI-based (medium-confidence) | 0 |
| S2-final-viral-combined_k141_24812\|\|full | 1188 | No | NA | 3 | 0 | 1 | Not-determined | Genome-fragment | NA | NA | 0 |
| S2-final-viral-combined_k141_35409\|\|full | 1774 | No | NA | 3 | 0 | 0 | Not-determined | Genome-fragment | NA | NA | 0 |
| S2-final-viral-combined_k141_241696\|\|full | 2786 | No | NA | 3 | 0 | 0 | Not-determined | Genome-fragment | NA | NA | 0 |
| S8-final-viral-combined_k141_177418\|\|full | 3824 | No | NA | 6 | 0 | 0 | Low-quality | Genome-fragment | 5.99 | AAI-based (medium-confidence) | 0 |
| S2-final-viral-combined_k141_199903\|\|full | 3587 | No | NA | 7 | 0 | 2 | Not-determined | Genome-fragment | NA | NA | 0 |
| S5-final-viral-combined_k141_104159\|\|full | 3825 | No | NA | 9 | 0 | 0 | Low-quality | Genome-fragment | 4.84 | AAI-based (medium-confidence) | 0 |
| S2-final-viral-combined_k141_118078\|\|full | 1950 | No | NA | 4 | 0 | 0 | Not-determined | Genome-fragment | NA | NA | 0 |
| S4_k141_31382 | 3825 | No | NA | 9 | 4 | 0 | Low-quality | Genome-fragment | 9.11 | AAI-based (high-confidence) | 0 |
| S11-final-viral-combined_k141_51749\|\|full | 3826 | No | NA | 6 | 0 | 1 | Low-quality | Genome-fragment | 7.9 | AAI-based (medium-confidence) | 0 |
| S17-final-viral-combined_k141_77172\|\|full | 3826 | No | NA | 3 | 0 | 3 | Low-quality | Genome-fragment | 7.2 | AAI-based (medium-confidence) | 0 |
| S2-final-viral-combined_k141_87445\|\|full | 1205 | No | NA | 2 | 0 | 1 | Not-determined | Genome-fragment | NA | NA | 0 |
| S12-final-viral-combined_k141_179401\|\|full | 3826 | No | NA | 2 | 0 | 0 | Low-quality | Genome-fragment | 24.56 | AAI-based (high-confidence) | 0 |
| S8-final-viral-combined_k141_210048\|\|full | 3826 | No | NA | 4 | 0 | 1 | Low-quality | Genome-fragment | 2.72 | AAI-based (medium-confidence) | 0 |
| S2-final-viral-combined_k141_35474\|\|full | 2320 | No | NA | 3 | 0 | 0 | Not-determined | Genome-fragment | NA | NA | 0 |
| S3_k141_26151 | 3827 | No | NA | 6 | 0 | 1 | Low-quality | Genome-fragment | 8.13 | AAI-based (medium-confidence) | 0 |
| S2-final-viral-combined_k141_170351\|\|full | 1648 | No | NA | 3 | 0 | 0 | Not-determined | Genome-fragment | NA | NA | 0 |
| S2-final-viral-combined_k141_211040\|\|full | 1478 | No | NA | 2 | 0 | 2 | Not-determined | Genome-fragment | NA | NA | 0 |
| S7-final-viral-combined_k141_12612\|\|full | 3827 | No | NA | 5 | 1 | 0 | Low-quality | Genome-fragment | 4.82 | AAI-based (medium-confidence) | 0 |
| S2-final-viral-combined_k141_118114\|\|full | 2285 | No | NA | 3 | 0 | 2 | Not-determined | Genome-fragment | NA | NA | 0 |
| S1-final-viral-combined_k141_205553\|\|full | 3828 | No | NA | 3 | 1 | 0 | Low-quality | Genome-fragment | 11.69 | AAI-based (medium-confidence) | 0 |
| S2-final-viral-combined_k141_97678\|\|full | 5530 | No | NA | 8 | 0 | 0 | Not-determined | Genome-fragment | NA | NA | 0 |
| S2-final-viral-combined_k141_170387\|\|full | 1233 | No | NA | 2 | 0 | 1 | Not-determined | Genome-fragment | NA | NA | 0 |
| S2-final-viral-combined_k141_107883\|\|full | 1248 | No | NA | 4 | 0 | 0 | Not-determined | Genome-fragment | NA | NA | 0 |
| S2-final-viral-combined_k141_35498\|\|full | 1760 | No | NA | 2 | 0 | 2 | Not-determined | Genome-fragment | NA | NA | 0 |
| S2-final-viral-combined_k141_170404\|\|full | 1720 | No | NA | 5 | 0 | 0 | Not-determined | Genome-fragment | NA | NA | 0 |
| S2-final-viral-combined_k141_56276\|\|full | 1881 | No | NA | 3 | 0 | 2 | Not-determined | Genome-fragment | NA | NA | 0 |
| S7-final-viral-combined_k141_145933\|\|full | 3828 | No | NA | 6 | 0 | 3 | Low-quality | Genome-fragment | 2.92 | AAI-based (high-confidence) | 0 |
| S2-final-viral-combined_k141_97696\|\|full | 3044 | No | NA | 6 | 0 | 1 | Not-determined | Genome-fragment | NA | NA | 0 |
| S12-final-viral-combined_k141_54255\|\|full | 3828 | No | NA | 8 | 3 | 0 | Low-quality | Genome-fragment | 9.74 | AAI-based (medium-confidence) | 0 |
| S4_k141_54974 | 3828 | No | NA | 4 | 0 | 4 | Low-quality | Genome-fragment | 1.64 | AAI-based (high-confidence) | 0 |
| S2-final-viral-combined_k141_97708\|\|full | 1403 | No | NA | 2 | 0 | 2 | Not-determined | Genome-fragment | NA | NA | 0 |
| S1-final-viral-combined_k141_308049\|\|full | 3829 | No | NA | 4 | 0 | 2 | Low-quality | Genome-fragment | 6.43 | AAI-based (medium-confidence) | 0 |
| S2-final-viral-combined_k141_146095\|\|full | 3829 | No | NA | 7 | 0 | 0 | Low-quality | Genome-fragment | 7.53 | AAI-based (high-confidence) | 0 |
| S2-final-viral-combined_k141_56322\|\|full | 1899 | No | NA | 4 | 0 | 1 | Not-determined | Genome-fragment | NA | NA | 0 |
| S2-final-viral-combined_k141_244920\|\|full | 3829 | No | NA | 3 | 0 | 3 | Low-quality | Genome-fragment | 1.37 | AAI-based (medium-confidence) | 0 |
| S2-final-viral-combined_k141_181005\|\|full | 1270 | No | NA | 3 | 0 | 1 | Not-determined | Genome-fragment | NA | NA | 0 |
| S2-final-viral-combined_k141_35567\|\|full | 2007 | No | NA | 3 | 0 | 0 | Not-determined | Genome-fragment | NA | NA | 0 |
| S8_k141_168738 | 3829 | No | NA | 9 | 3 | 0 | Low-quality | Genome-fragment | 8.73 | AAI-based (medium-confidence) | 0 |
| S8_k141_91870 | 3829 | No | NA | 6 | 2 | 0 | Low-quality | Genome-fragment | 5.26 | AAI-based (high-confidence) | 0 |
| S11-final-viral-combined_k141_327041\|\|full | 3830 | No | NA | 4 | 0 | 0 | Low-quality | Genome-fragment | 7.7 | AAI-based (medium-confidence) | 0 |
| S2-final-viral-combined_k141_190897\|\|full | 1657 | No | NA | 3 | 0 | 2 | Not-determined | Genome-fragment | NA | NA | 0 |
| S2-final-viral-combined_k141_87581\|\|full | 2266 | No | NA | 3 | 0 | 1 | Not-determined | Genome-fragment | NA | NA | 0 |
| S2-final-viral-combined_k141_220926\|\|full | 27225 | No | NA | 24 | 0 | 17 | Not-determined | Genome-fragment | NA | NA | 0 |
| S2-final-viral-combined_k141_181066\|\|full | 2426 | No | NA | 3 | 0 | 2 | Not-determined | Genome-fragment | NA | NA | 0 |
| S2-final-viral-combined_k141_87607\|\|full | 3064 | No | NA | 5 | 0 | 2 | Not-determined | Genome-fragment | NA | NA | 0 |
| S2-final-viral-combined_k141_3948\|\|full | 1741 | No | NA | 3 | 0 | 1 | Not-determined | Genome-fragment | NA | NA | 0 |
| S2-final-viral-combined_k141_181075\|\|full | 5375 | No | NA | 5 | 0 | 5 | Not-determined | Genome-fragment | NA | NA | 0 |
| S1-final-viral-combined_k141_252357\|\|full | 3830 | No | NA | 3 | 0 | 3 | Low-quality | Genome-fragment | 2.88 | AAI-based (medium-confidence) | 0 |
| S17-final-viral-combined_k141_221568\|\|full | 3830 | No | NA | 5 | 0 | 0 | Low-quality | Genome-fragment | 2.7 | AAI-based (medium-confidence) | 0 |
| S2-final-viral-combined_k141_149183\|\|full | 5110 | No | NA | 5 | 0 | 3 | Not-determined | Genome-fragment | NA | NA | 0 |
| S12-final-viral-combined_k141_211219\|\|full | 3830 | No | NA | 3 | 2 | 0 | Low-quality | Genome-fragment | 8.36 | AAI-based (high-confidence) | 0 |
| S2-final-viral-combined_k141_159359\|\|full | 1751 | No | NA | 2 | 0 | 2 | Not-determined | Genome-fragment | NA | NA | 0 |
| S2-final-viral-combined_k141_47312\|\|full | 4893 | No | NA | 4 | 0 | 4 | Not-determined | Genome-fragment | NA | NA | 0 |
| S2-final-viral-combined_k141_190938\|\|full | 33381 | No | NA | 26 | 0 | 12 | Not-determined | Genome-fragment | NA | NA | 0 |
| S2-final-viral-combined_k141_169395\|\|full | 3830 | No | NA | 3 | 0 | 2 | Low-quality | Genome-fragment | 5.39 | AAI-based (medium-confidence) | 0 |
| S11_k141_105072 | 3832 | No | NA | 3 | 3 | 0 | Low-quality | Genome-fragment | 6.62 | AAI-based (high-confidence) | 0 |
| S9_k141_47343 | 3833 | No | NA | 5 | 0 | 4 | Low-quality | Genome-fragment | 1.06 | AAI-based (medium-confidence) | 0 |
| S11-final-viral-combined_k141_325922\|\|full | 3833 | No | NA | 6 | 4 | 0 | Low-quality | Genome-fragment | 11.17 | AAI-based (medium-confidence) | 0 |
| S2-final-viral-combined_k141_87662\|\|full | 2017 | No | NA | 3 | 0 | 2 | Not-determined | Genome-fragment | NA | NA | 0 |
| S2-final-viral-combined_k141_159408\|\|full | 1678 | No | NA | 3 | 0 | 2 | Not-determined | Genome-fragment | NA | NA | 0 |
| S2-final-viral-combined_k141_159410\|\|full | 1227 | No | NA | 2 | 0 | 1 | Not-determined | Genome-fragment | NA | NA | 0 |
| S2-final-viral-combined_k141_128072\|\|full | 5839 | No | NA | 5 | 0 | 3 | Not-determined | Genome-fragment | NA | NA | 0 |
| S3-final-viral-combined_k141_99737\|\|full | 3833 | No | NA | 6 | 0 | 0 | Low-quality | Genome-fragment | 5.97 | AAI-based (medium-confidence) | 0 |
| S11-final-viral-combined_k141_160511\|\|full | 3834 | No | NA | 8 | 5 | 0 | Low-quality | Genome-fragment | 10.3 | AAI-based (high-confidence) | 0 |
| S2-final-viral-combined_k141_87696\|\|full | 2321 | No | NA | 3 | 0 | 3 | Not-determined | Genome-fragment | NA | NA | 0 |
| S2-final-viral-combined_k141_149262\|\|full | 1736 | No | NA | 2 | 0 | 2 | Not-determined | Genome-fragment | NA | NA | 0 |
| S2-final-viral-combined_k141_159460\|\|full | 3055 | No | NA | 4 | 0 | 1 | Not-determined | Genome-fragment | NA | NA | 0 |
| S2-final-viral-combined_k141_200107\|\|full | 2594 | No | NA | 4 | 0 | 0 | Not-determined | Genome-fragment | NA | NA | 0 |
| S5-final-viral-combined_k141_13756\|\|full | 3834 | No | NA | 4 | 0 | 1 | Low-quality | Genome-fragment | 8.61 | AAI-based (medium-confidence) | 0 |
| S7-final-viral-combined_k141_61381\|\|full | 3834 | No | NA | 7 | 2 | 0 | Low-quality | Genome-fragment | 9.46 | AAI-based (high-confidence) | 0 |
| S2-final-viral-combined_k141_139331\|\|full | 1476 | No | NA | 3 | 0 | 0 | Not-determined | Genome-fragment | NA | NA | 0 |
| S2-final-viral-combined_k141_56532\|\|full | 1564 | No | NA | 2 | 0 | 1 | Not-determined | Genome-fragment | NA | NA | 0 |
| S2-final-viral-combined_k141_149280\|\|full | 2579 | No | NA | 3 | 0 | 3 | Not-determined | Genome-fragment | NA | NA | 0 |
| S1_k141_108538 | 3835 | No | NA | 9 | 1 | 0 | Low-quality | Genome-fragment | 8.48 | AAI-based (medium-confidence) | 0 |
| S2-final-viral-combined_k141_159488\|\|full | 2532 | No | NA | 2 | 0 | 1 | Not-determined | Genome-fragment | NA | NA | 0 |
| S17-final-viral-combined_k141_123342\|\|full | 3835 | No | NA | 4 | 0 | 2 | Low-quality | Genome-fragment | 6 | AAI-based (medium-confidence) | 0 |
| S2-final-viral-combined_k141_181154\|\|full | 4079 | No | NA | 5 | 0 | 4 | Not-determined | Genome-fragment | NA | NA | 0 |
| S2-final-viral-combined_k141_211188\|\|full | 3633 | No | NA | 5 | 0 | 0 | Not-determined | Genome-fragment | NA | NA | 0 |
| S12-final-viral-combined_k141_21234\|\|full | 3835 | No | NA | 3 | 0 | 2 | Low-quality | Genome-fragment | 4.08 | AAI-based (medium-confidence) | 0 |
| S5-final-viral-combined_k141_24574\|\|full | 3836 | No | NA | 3 | 0 | 2 | Low-quality | Genome-fragment | 7.38 | AAI-based (high-confidence) | 0 |
| S17-k141_1721 | 3836 | No | NA | 5 | 0 | 0 | Low-quality | Genome-fragment | 9.95 | AAI-based (medium-confidence) | 0 |
| S2-final-viral-combined_k141_97908\|\|full | 2233 | No | NA | 3 | 0 | 3 | Not-determined | Genome-fragment | NA | NA | 0 |
| S2-final-viral-combined_k141_14626\|\|full | 1277 | No | NA | 3 | 0 | 1 | Not-determined | Genome-fragment | NA | NA | 0 |
| S2-final-viral-combined_k141_4086\|\|full | 2100 | No | NA | 2 | 0 | 1 | Not-determined | Genome-fragment | NA | NA | 0 |
| S2-final-viral-combined_k141_220960\|\|full | 1560 | No | NA | 2 | 0 | 1 | Not-determined | Genome-fragment | NA | NA | 0 |
| S2-final-viral-combined_k141_139377\|\|full | 3456 | No | NA | 3 | 0 | 1 | Not-determined | Genome-fragment | NA | NA | 0 |
| S11-final-viral-combined_k141_3330\|\|full | 3837 | No | NA | 5 | 3 | 0 | Low-quality | Genome-fragment | 8.12 | AAI-based (high-confidence) | 0 |
| S2-final-viral-combined_k141_76821\|\|full | 1035 | No | NA | 3 | 0 | 0 | Not-determined | Genome-fragment | NA | NA | 0 |
| S1-final-viral-combined_k141_97742\|\|full | 3837 | No | NA | 4 | 1 | 0 | Low-quality | Genome-fragment | 2.36 | HMM-based (lower-bound) | 0 |
| S12-final-viral-combined_k141_119783\|\|full | 3837 | No | NA | 3 | 0 | 2 | Low-quality | Genome-fragment | 1.93 | AAI-based (medium-confidence) | 0 |
| S2-final-viral-combined_k141_128171\|\|full | 2091 | No | NA | 4 | 0 | 0 | Not-determined | Genome-fragment | NA | NA | 0 |
| S2-final-viral-combined_k141_76837\|\|full | 1919 | No | NA | 3 | 0 | 2 | Not-determined | Genome-fragment | NA | NA | 0 |
| S7-final-viral-combined_k141_97622\|\|full | 3838 | No | NA | 6 | 0 | 0 | Low-quality | Genome-fragment | 5.84 | AAI-based (high-confidence) | 0 |
| S12-final-viral-combined_k141_126106\|\|full | 3838 | No | NA | 5 | 2 | 0 | Low-quality | Genome-fragment | 6.89 | AAI-based (high-confidence) | 0 |
| S2-final-viral-combined_k141_220983\|\|full | 4058 | No | NA | 7 | 0 | 0 | Not-determined | Genome-fragment | NA | NA | 0 |
| S9_k141_116999 | 3839 | No | NA | 3 | 0 | 2 | Low-quality | Genome-fragment | 3.44 | AAI-based (medium-confidence) | 0 |
| S2-final-viral-combined_k141_191046\|\|full | 2087 | No | NA | 3 | 0 | 0 | Not-determined | Genome-fragment | NA | NA | 0 |
| S2-final-viral-combined_k141_24976\|\|full | 2479 | No | NA | 4 | 0 | 0 | Not-determined | Genome-fragment | NA | NA | 0 |
| S2_k141_201102 | 3841 | No | NA | 4 | 0 | 4 | Low-quality | Genome-fragment | 2 | AAI-based (medium-confidence) | 0 |
| S2-final-viral-combined_k141_66535\|\|full | 1898 | No | NA | 3 | 0 | 1 | Not-determined | Genome-fragment | NA | NA | 0 |
| S2-final-viral-combined_k141_47481\|\|full | 1675 | No | NA | 2 | 0 | 0 | Not-determined | Genome-fragment | NA | NA | 0 |
| S2-final-viral-combined_k141_211253\|\|full | 3815 | No | NA | 3 | 0 | 2 | Not-determined | Genome-fragment | NA | NA | 0 |
| S2-final-viral-combined_k141_221012\|\|full | 2026 | No | NA | 4 | 0 | 0 | Not-determined | Genome-fragment | NA | NA | 0 |
| S2_k141_101339 | 3841 | No | NA | 12 | 1 | 0 | Low-quality | Genome-fragment | 2.43 | AAI-based (high-confidence) | 0 |
| S11-final-viral-combined_k141_28530\|\|full | 3842 | No | NA | 3 | 0 | 0 | Low-quality | Genome-fragment | 8.46 | AAI-based (medium-confidence) | 0 |
| S2-final-viral-combined_k141_211280\|\|full | 1673 | No | NA | 2 | 0 | 2 | Not-determined | Genome-fragment | NA | NA | 0 |
| S2-final-viral-combined_k141_108144\|\|full | 2624 | No | NA | 3 | 0 | 2 | Not-determined | Genome-fragment | NA | NA | 0 |
| S1-final-viral-combined_k141_335664\|\|full | 3842 | No | NA | 2 | 0 | 0 | Low-quality | Genome-fragment | 8.69 | AAI-based (high-confidence) | 0 |
| S1-final-viral-combined_k141_237828\|\|full | 3842 | No | NA | 5 | 0 | 3 | Low-quality | Genome-fragment | 3.16 | AAI-based (medium-confidence) | 0 |
| S3_k141_119947 | 3842 | No | NA | 6 | 0 | 0 | Low-quality | Genome-fragment | 1.77 | AAI-based (high-confidence) | 0 |
| S2-final-viral-combined_k141_76883\|\|full | 1187 | No | NA | 3 | 0 | 1 | Not-determined | Genome-fragment | NA | NA | 0 |
| S17-k141_179426 | 3842 | No | NA | 2 | 0 | 1 | Low-quality | Genome-fragment | 8.26 | AAI-based (high-confidence) | 0 |
| S2_k141_17511 | 3843 | No | NA | 4 | 0 | 2 | Low-quality | Genome-fragment | 0.54 | AAI-based (high-confidence) | 0 |
| S2-final-viral-combined_k141_211299\|\|full | 1483 | No | NA | 3 | 0 | 2 | Not-determined | Genome-fragment | NA | NA | 0 |
| S17-final-viral-combined_k141_92222\|\|full | 3845 | No | NA | 4 | 0 | 2 | Low-quality | Genome-fragment | 0.45 | AAI-based (medium-confidence) | 0 |
| S2-final-viral-combined_k141_56566\|\|full | 5405 | No | NA | 7 | 0 | 4 | Not-determined | Genome-fragment | NA | NA | 0 |
| S2-final-viral-combined_k141_145339\|\|full | 3845 | No | NA | 3 | 0 | 0 | Low-quality | Genome-fragment | 11.64 | AAI-based (high-confidence) | 0 |
| S2-final-viral-combined_k141_66596\|\|full | 1491 | No | NA | 2 | 0 | 0 | Not-determined | Genome-fragment | NA | NA | 0 |
| S2-final-viral-combined_k141_221044\|\|full | 15701 | No | NA | 15 | 0 | 10 | Not-determined | Genome-fragment | NA | NA | 0 |
| S8_k141_90744 | 3845 | No | NA | 6 | 0 | 0 | Low-quality | Genome-fragment | 9.38 | AAI-based (medium-confidence) | 0 |
| S9-final-viral-combined_k141_66239\|\|full | 3846 | No | NA | 3 | 0 | 0 | Low-quality | Genome-fragment | 8.02 | AAI-based (medium-confidence) | 0 |
| S9-final-viral-combined_k141_85111\|\|full | 3846 | No | NA | 5 | 0 | 0 | Low-quality | Genome-fragment | 7.67 | AAI-based (medium-confidence) | 0 |
| S2-final-viral-combined_k141_66631\|\|full | 1914 | No | NA | 4 | 0 | 1 | Not-determined | Genome-fragment | NA | NA | 0 |
| S1-final-viral-combined_k141_149018\|\|full | 3846 | No | NA | 8 | 0 | 0 | Low-quality | Genome-fragment | 9.89 | AAI-based (medium-confidence) | 0 |
| S2-final-viral-combined_k141_241885\|\|full | 1978 | No | NA | 2 | 0 | 0 | Not-determined | Genome-fragment | NA | NA | 0 |
| S3-final-viral-combined_k141_29882\|\|lt2gene | 3846 | No | NA | 1 | 1 | 0 | Low-quality | Genome-fragment | 2.66 | HMM-based (lower-bound) | 0 |
| S1-final-viral-combined_k141_315003\|\|full | 3847 | No | NA | 5 | 0 | 0 | Low-quality | Genome-fragment | 1.11 | AAI-based (medium-confidence) | 0 |
| S8-final-viral-combined_k141_101464\|\|full | 3847 | No | NA | 6 | 1 | 0 | Low-quality | Genome-fragment | 28.38 | AAI-based (medium-confidence) | 0 |
| S2-final-viral-combined_k141_25014\|\|full | 1239 | No | NA | 2 | 0 | 0 | Not-determined | Genome-fragment | NA | NA | 0 |
| S9-final-viral-combined_k141_49423\|\|full | 3848 | No | NA | 6 | 4 | 0 | Low-quality | Genome-fragment | 10.31 | AAI-based (high-confidence) | 0 |
| S11_k141_124225 | 3848 | No | NA | 3 | 0 | 1 | Low-quality | Genome-fragment | 5.28 | AAI-based (high-confidence) | 0 |
| S1-final-viral-combined_k141_281269\|\|full | 3848 | No | NA | 5 | 1 | 0 | Low-quality | Genome-fragment | 9.99 | AAI-based (high-confidence) | 0 |
| S2-final-viral-combined_k141_200190\|\|full | 2406 | No | NA | 2 | 0 | 2 | Not-determined | Genome-fragment | NA | NA | 0 |
| S2-final-viral-combined_k141_25030\|\|full | 1477 | No | NA | 3 | 0 | 2 | Not-determined | Genome-fragment | NA | NA | 0 |
| S2-final-viral-combined_k141_76991\|\|full | 1148 | No | NA | 2 | 0 | 0 | Not-determined | Genome-fragment | NA | NA | 0 |
| S2-final-viral-combined_k141_77008\|\|full | 1129 | No | NA | 3 | 0 | 0 | Not-determined | Genome-fragment | NA | NA | 0 |
| S3-final-viral-combined_k141_1718\|\|full | 3848 | No | NA | 4 | 0 | 0 | Low-quality | Genome-fragment | 3.61 | AAI-based (medium-confidence) | 0 |
| S2-final-viral-combined_k141_35909\|\|full | 2188 | No | NA | 3 | 0 | 3 | Not-determined | Genome-fragment | NA | NA | 0 |
| S2-final-viral-combined_k141_181388\|\|full | 1556 | No | NA | 3 | 0 | 1 | Not-determined | Genome-fragment | NA | NA | 0 |
| S2-final-viral-combined_k141_139486\|\|full | 2219 | No | NA | 2 | 0 | 0 | Not-determined | Genome-fragment | NA | NA | 0 |
| S2-final-viral-combined_k141_170845\|\|full | 48753 | Yes | 33515 | 50 | 17 | 9 | Medium-quality | Genome-fragment | 70.53 | AAI-based (medium-confidence) | 31.26 |
| S2-final-viral-combined_k141_221109\|\|full | 1869 | No | NA | 3 | 0 | 1 | Not-determined | Genome-fragment | NA | NA | 0 |
| S5-final-viral-combined_k141_99011\|\|full | 3848 | No | NA | 6 | 0 | 1 | Low-quality | Genome-fragment | 2.93 | AAI-based (medium-confidence) | 0 |
| S2-final-viral-combined_k141_66704\|\|full | 1075 | No | NA | 2 | 0 | 2 | Not-determined | Genome-fragment | NA | NA | 0 |
| S2-final-viral-combined_k141_25059\|\|full | 2191 | No | NA | 5 | 0 | 1 | Not-determined | Genome-fragment | NA | NA | 0 |
| S2-final-viral-combined_k141_170875\|\|full | 3441 | No | NA | 4 | 0 | 0 | Not-determined | Genome-fragment | NA | NA | 0 |
| S2-final-viral-combined_k141_47618\|\|full | 9289 | No | NA | 7 | 0 | 7 | Not-determined | Genome-fragment | NA | NA | 0 |
| S2-final-viral-combined_k141_159664\|\|full | 3979 | No | NA | 6 | 0 | 1 | Not-determined | Genome-fragment | NA | NA | 0 |
| S17-final-viral-combined_k141_240892\|\|full | 3848 | No | NA | 2 | 0 | 2 | Low-quality | Genome-fragment | 1.3 | AAI-based (medium-confidence) | 0 |
| S2-final-viral-combined_k141_149527\|\|full | 1721 | No | NA | 5 | 0 | 5 | Not-determined | Genome-fragment | NA | NA | 0 |
| S2-final-viral-combined_k141_56710\|\|full | 4961 | No | NA | 4 | 0 | 4 | Not-determined | Genome-fragment | NA | NA | 0 |
| S12_k141_360585 | 3848 | No | NA | 3 | 0 | 1 | Low-quality | Genome-fragment | 0.94 | AAI-based (medium-confidence) | 0 |
| S2-final-viral-combined_k141_66729\|\|full | 1507 | No | NA | 2 | 0 | 1 | Not-determined | Genome-fragment | NA | NA | 0 |
| S2-final-viral-combined_k141_118606\|\|full | 2978 | No | NA | 3 | 0 | 1 | Not-determined | Genome-fragment | NA | NA | 0 |
| S2-final-viral-combined_k141_4259\|\|full | 2650 | No | NA | 9 | 0 | 0 | Not-determined | Genome-fragment | NA | NA | 0 |
| S2-final-viral-combined_k141_159670\|\|full | 36742 | Yes | 24122 | 50 | 16 | 7 | Medium-quality | Genome-fragment | 64.72 | AAI-based (high-confidence) | 34.35 |
| S2-final-viral-combined_k141_56750\|\|full | 2425 | No | NA | 3 | 0 | 1 | Not-determined | Genome-fragment | NA | NA | 0 |
| S2-final-viral-combined_k141_149542\|\|full | 3533 | No | NA | 4 | 0 | 1 | Not-determined | Genome-fragment | NA | NA | 0 |
| S2-final-viral-combined_k141_98219\|\|full | 1269 | No | NA | 2 | 0 | 1 | Not-determined | Genome-fragment | NA | NA | 0 |
| S2-final-viral-combined_k141_87905\|\|full | 2518 | No | NA | 2 | 0 | 1 | Not-determined | Genome-fragment | NA | NA | 0 |
| S12_k141_313871 | 3849 | No | NA | 5 | 0 | 1 | Low-quality | Genome-fragment | 5.95 | AAI-based (medium-confidence) | 0 |
| S4_k141_65119 | 3849 | No | NA | 2 | 0 | 1 | Low-quality | Genome-fragment | 2.42 | AAI-based (medium-confidence) | 0 |
| S2-final-viral-combined_k141_36021\|\|full | 1185 | No | NA | 2 | 0 | 1 | Not-determined | Genome-fragment | NA | NA | 0 |
| S7_k141_119389 | 3850 | No | NA | 4 | 0 | 4 | Low-quality | Genome-fragment | 0.93 | AAI-based (medium-confidence) | 0 |
| S2-final-viral-combined_k141_200273\|\|full | 3167 | No | NA | 2 | 0 | 1 | Not-determined | Genome-fragment | NA | NA | 0 |
| S2-final-viral-combined_k141_191238\|\|full | 1808 | No | NA | 2 | 0 | 1 | Not-determined | Genome-fragment | NA | NA | 0 |
| S2-final-viral-combined_k141_211366\|\|full | 1521 | No | NA | 2 | 0 | 2 | Not-determined | Genome-fragment | NA | NA | 0 |
| S2-final-viral-combined_k141_146330\|\|full | 3850 | No | NA | 3 | 0 | 2 | Low-quality | Genome-fragment | 1.37 | AAI-based (medium-confidence) | 0 |
| S4_k141_1652 | 3851 | No | NA | 2 | 0 | 2 | Low-quality | Genome-fragment | 0.67 | AAI-based (medium-confidence) | 0 |
| S3_k141_26014 | 3852 | No | NA | 3 | 2 | 0 | Low-quality | Genome-fragment | 10.13 | AAI-based (high-confidence) | 0 |
| S8_k141_219084 | 3852 | No | NA | 9 | 1 | 0 | Low-quality | Genome-fragment | 4.45 | HMM-based (lower-bound) | 0 |
| S3-final-viral-combined_k141_70034\|\|full | 3853 | No | NA | 6 | 1 | 0 | Low-quality | Genome-fragment | 27.02 | AAI-based (high-confidence) | 0 |
| S2-final-viral-combined_k141_200308\|\|full | 1726 | No | NA | 3 | 0 | 1 | Not-determined | Genome-fragment | NA | NA | 0 |
| S2-final-viral-combined_k141_232760\|\|full | 2614 | No | NA | 2 | 0 | 2 | Not-determined | Genome-fragment | NA | NA | 0 |
| S2-final-viral-combined_k141_191255\|\|full | 1202 | No | NA | 2 | 0 | 1 | Not-determined | Genome-fragment | NA | NA | 0 |
| S2-final-viral-combined_k141_139615\|\|full | 1921 | No | NA | 3 | 0 | 2 | Not-determined | Genome-fragment | NA | NA | 0 |
| S2-final-viral-combined_k141_98266\|\|full | 5349 | No | NA | 9 | 0 | 0 | Not-determined | Genome-fragment | NA | NA | 0 |
| S2-final-viral-combined_k141_181391\|\|full | 142071 | Yes | 34174 | 167 | 12 | 102 | Medium-quality | Genome-fragment | 66.2 | HMM-based (lower-bound) | 75.95 |
| S4-final-viral-combined_k141_29538\|\|full | 3853 | No | NA | 3 | 0 | 3 | Low-quality | Genome-fragment | 11.59 | AAI-based (medium-confidence) | 0 |
| S8-final-viral-combined_k141_128083\|\|full | 3853 | No | NA | 4 | 0 | 1 | Low-quality | Genome-fragment | 2.63 | AAI-based (high-confidence) | 0 |
| S2-final-viral-combined_k141_128381\|\|full | 20658 | No | NA | 21 | 0 | 15 | Not-determined | Genome-fragment | NA | NA | 0 |
| S4-final-viral-combined_k141_56345\|\|full | 3855 | No | NA | 6 | 0 | 0 | Low-quality | Genome-fragment | 4.19 | AAI-based (high-confidence) | 0 |
| S2-final-viral-combined_k141_159752\|\|full | 2372 | No | NA | 2 | 0 | 1 | Not-determined | Genome-fragment | NA | NA | 0 |
| S5_k141_153520 | 3856 | No | NA | 5 | 0 | 0 | Low-quality | Genome-fragment | 7.8 | AAI-based (medium-confidence) | 0 |
| S2-final-viral-combined_k141_159774\|\|full | 2089 | No | NA | 3 | 0 | 2 | Not-determined | Genome-fragment | NA | NA | 0 |
| S2-final-viral-combined_k141_139667\|\|full | 2978 | No | NA | 3 | 0 | 0 | Not-determined | Genome-fragment | NA | NA | 0 |
| S2-final-viral-combined_k141_221256\|\|full | 4154 | No | NA | 6 | 0 | 1 | Not-determined | Genome-fragment | NA | NA | 0 |
| S2-final-viral-combined_k141_149648\|\|full | 15803 | No | NA | 16 | 0 | 12 | Not-determined | Genome-fragment | NA | NA | 0 |
| S17-k141_259837 | 3856 | No | NA | 3 | 3 | 0 | Low-quality | Genome-fragment | 5.18 | AAI-based (high-confidence) | 0 |
| S1-final-viral-combined_k141_18942\|\|full | 3857 | No | NA | 6 | 0 | 3 | Low-quality | Genome-fragment | 4.11 | AAI-based (medium-confidence) | 0 |
| S7_k141_144451 | 3857 | No | NA | 4 | 0 | 3 | Low-quality | Genome-fragment | 2.7 | AAI-based (medium-confidence) | 0 |
| S2-final-viral-combined_k141_85772\|\|full | 3857 | No | NA | 5 | 3 | 0 | Low-quality | Genome-fragment | 13.11 | AAI-based (high-confidence) | 0 |
| S2-final-viral-combined_k141_77255\|\|full | 3857 | No | NA | 5 | 3 | 0 | Low-quality | Genome-fragment | 9.65 | AAI-based (medium-confidence) | 0 |
| S2-final-viral-combined_k141_66769\|\|full | 1447 | No | NA | 4 | 0 | 1 | Not-determined | Genome-fragment | NA | NA | 0 |
| S12-final-viral-combined_k141_193317\|\|full | 3860 | No | NA | 5 | 0 | 0 | Low-quality | Genome-fragment | 5.88 | AAI-based (high-confidence) | 0 |
| S2-final-viral-combined_k141_102873\|\|full | 3860 | No | NA | 8 | 2 | 0 | Low-quality | Genome-fragment | 6.98 | AAI-based (high-confidence) | 0 |
| S7-final-viral-combined_k141_32458\|\|full | 3861 | No | NA | 8 | 4 | 0 | Low-quality | Genome-fragment | 9.58 | AAI-based (high-confidence) | 0 |
| S2-final-viral-combined_k141_139768\|\|full | 4381 | No | NA | 6 | 0 | 2 | Not-determined | Genome-fragment | NA | NA | 0 |
| S2-final-viral-combined_k141_159835\|\|full | 2518 | No | NA | 3 | 0 | 1 | Not-determined | Genome-fragment | NA | NA | 0 |
| S2-final-viral-combined_k141_181464\|\|full | 2653 | No | NA | 5 | 0 | 2 | Not-determined | Genome-fragment | NA | NA | 0 |
| S2-final-viral-combined_k141_1768\|\|full | 3862 | No | NA | 8 | 0 | 1 | Low-quality | Genome-fragment | 6.87 | AAI-based (medium-confidence) | 0 |
| S5-final-viral-combined_k141_160665\|\|full | 3864 | No | NA | 11 | 0 | 0 | Low-quality | Genome-fragment | 10.44 | AAI-based (high-confidence) | 0 |
| S2_k141_187505 | 3864 | No | NA | 5 | 0 | 0 | Low-quality | Genome-fragment | 5.96 | AAI-based (high-confidence) | 0 |
| S9-final-viral-combined_k141_5657\|\|full | 3865 | No | NA | 3 | 0 | 3 | Low-quality | Genome-fragment | 3.99 | AAI-based (medium-confidence) | 0 |
| S11-final-viral-combined_k141_110603\|\|full | 3865 | No | NA | 9 | 0 | 1 | Low-quality | Genome-fragment | 5.71 | AAI-based (high-confidence) | 0 |
| S2-final-viral-combined_k141_149708\|\|full | 2294 | No | NA | 4 | 0 | 1 | Not-determined | Genome-fragment | NA | NA | 0 |
| S2-final-viral-combined_k141_4366\|\|full | 2513 | No | NA | 4 | 0 | 3 | Not-determined | Genome-fragment | NA | NA | 0 |
| S2-final-viral-combined_k141_211561\|\|full | 1002 | No | NA | 2 | 0 | 1 | Not-determined | Genome-fragment | NA | NA | 0 |
| S1-final-viral-combined_k141_131908\|\|full | 3866 | No | NA | 8 | 0 | 1 | Low-quality | Genome-fragment | 5.7 | AAI-based (high-confidence) | 0 |
| S5-final-viral-combined_k141_34210\|\|full | 3866 | No | NA | 4 | 4 | 0 | Low-quality | Genome-fragment | 9.63 | AAI-based (high-confidence) | 0 |
| S2-final-viral-combined_k141_77288\|\|full | 1756 | No | NA | 3 | 0 | 1 | Not-determined | Genome-fragment | NA | NA | 0 |
| S2-final-viral-combined_k141_15017\|\|full | 6572 | No | NA | 3 | 0 | 2 | Not-determined | Genome-fragment | NA | NA | 0 |
| S2-final-viral-combined_k141_139835\|\|full | 3735 | No | NA | 6 | 0 | 3 | Not-determined | Genome-fragment | NA | NA | 0 |
| S2-final-viral-combined_k141_242080\|\|full | 1440 | No | NA | 2 | 0 | 0 | Not-determined | Genome-fragment | NA | NA | 0 |
| S2-final-viral-combined_k141_149769\|\|full | 1346 | No | NA | 2 | 0 | 0 | Not-determined | Genome-fragment | NA | NA | 0 |
| S17-final-viral-combined_k141_158807\|\|full | 3866 | No | NA | 6 | 4 | 0 | Low-quality | Genome-fragment | 9.2 | AAI-based (medium-confidence) | 0 |
| S2-final-viral-combined_k141_211577\|\|full | 2685 | No | NA | 5 | 0 | 1 | Not-determined | Genome-fragment | NA | NA | 0 |
| S2-final-viral-combined_k141_205090\|\|full | 3866 | No | NA | 3 | 0 | 2 | Low-quality | Genome-fragment | 0.92 | AAI-based (medium-confidence) | 0 |
| S2-final-viral-combined_k141_200390\|\|full | 90161 | Yes | 37148 | 105 | 17 | 26 | Complete | High-quality | 100 | Provirus (medium-confidence) | 58.8 |
| S1-final-viral-combined_k141_230541\|\|full | 3867 | No | NA | 5 | 0 | 3 | Low-quality | Genome-fragment | 3.11 | AAI-based (high-confidence) | 0 |
| S2-final-viral-combined_k141_25356\|\|full | 6455 | No | NA | 7 | 0 | 3 | Not-determined | Genome-fragment | NA | NA | 0 |
| S1-final-viral-combined_k141_341466\|\|full | 3867 | No | NA | 4 | 2 | 0 | Low-quality | Genome-fragment | 10.32 | AAI-based (medium-confidence) | 0 |
| S4-final-viral-combined_k141_12595\|\|full | 3868 | No | NA | 8 | 2 | 0 | Low-quality | Genome-fragment | 2.23 | AAI-based (high-confidence) | 0 |
| S2-final-viral-combined_k141_139881\|\|full | 1644 | No | NA | 4 | 0 | 0 | Not-determined | Genome-fragment | NA | NA | 0 |
| S2-final-viral-combined_k141_221491\|\|full | 6769 | No | NA | 4 | 0 | 2 | Not-determined | Genome-fragment | NA | NA | 0 |
| S12-final-viral-combined_k141_390553\|\|full | 3869 | No | NA | 6 | 4 | 0 | Low-quality | Genome-fragment | 8.92 | AAI-based (high-confidence) | 0 |
| S2-final-viral-combined_k141_77370\|\|full | 1061 | No | NA | 2 | 0 | 1 | Not-determined | Genome-fragment | NA | NA | 0 |
| S2-final-viral-combined_k141_77376\|\|full | 4111 | No | NA | 8 | 0 | 1 | Not-determined | Genome-fragment | NA | NA | 0 |
| S1-final-viral-combined_k141_383458\|\|full | 3870 | No | NA | 6 | 0 | 1 | Low-quality | Genome-fragment | 6.8 | AAI-based (high-confidence) | 0 |
| S2-final-viral-combined_k141_98523\|\|full | 10338 | No | NA | 12 | 0 | 6 | Not-determined | Genome-fragment | NA | NA | 0 |
| S5-final-viral-combined_k141_33077\|\|full | 3870 | No | NA | 8 | 7 | 0 | Low-quality | Genome-fragment | 10.79 | AAI-based (high-confidence) | 0 |
| S2-final-viral-combined_k141_47947\|\|full | 2580 | No | NA | 7 | 0 | 1 | Not-determined | Genome-fragment | NA | NA | 0 |
| S2-final-viral-combined_k141_98531\|\|full | 1152 | No | NA | 2 | 0 | 0 | Not-determined | Genome-fragment | NA | NA | 0 |
| S2-final-viral-combined_k141_15140\|\|full | 1817 | No | NA | 2 | 0 | 1 | Not-determined | Genome-fragment | NA | NA | 0 |
| S17-final-viral-combined_k141_3832\|\|full | 3870 | No | NA | 5 | 0 | 4 | Low-quality | Genome-fragment | 2.25 | AAI-based (medium-confidence) | 0 |
| S12-final-viral-combined_k141_188215\|\|full | 3870 | No | NA | 5 | 4 | 0 | Low-quality | Genome-fragment | 11.75 | AAI-based (high-confidence) | 0 |
| S2-final-viral-combined_k141_118817\|\|full | 1342 | No | NA | 2 | 0 | 0 | Not-determined | Genome-fragment | NA | NA | 0 |
| S2-final-viral-combined_k141_139949\|\|full | 1504 | No | NA | 2 | 0 | 1 | Not-determined | Genome-fragment | NA | NA | 0 |
| S2-final-viral-combined_k141_200464\|\|full | 3010 | No | NA | 3 | 0 | 3 | Not-determined | Genome-fragment | NA | NA | 0 |
| S2-final-viral-combined_k141_36327\|\|full | 8240 | No | NA | 7 | 0 | 4 | Not-determined | Genome-fragment | NA | NA | 0 |
| S2-final-viral-combined_k141_47978\|\|full | 1323 | No | NA | 2 | 0 | 0 | Not-determined | Genome-fragment | NA | NA | 0 |
| S9-final-viral-combined_k141_77130\|\|full | 3871 | No | NA | 5 | 0 | 2 | Low-quality | Genome-fragment | 1.69 | AAI-based (medium-confidence) | 0 |
| S2-final-viral-combined_k141_88034\|\|full | 1077 | No | NA | 2 | 0 | 0 | Not-determined | Genome-fragment | NA | NA | 0 |
| S2-final-viral-combined_k141_36343\|\|full | 3428 | No | NA | 5 | 0 | 2 | Not-determined | Genome-fragment | NA | NA | 0 |
| S2-final-viral-combined_k141_191337\|\|full | 1597 | No | NA | 3 | 0 | 0 | Not-determined | Genome-fragment | NA | NA | 0 |
| S2-final-viral-combined_k141_66891\|\|full | 2332 | No | NA | 3 | 0 | 0 | Not-determined | Genome-fragment | NA | NA | 0 |
| S2-final-viral-combined_k141_47984\|\|full | 3800 | No | NA | 6 | 0 | 1 | Not-determined | Genome-fragment | NA | NA | 0 |
| S1-final-viral-combined_k141_114071\|\|full | 3871 | No | NA | 4 | 0 | 1 | Low-quality | Genome-fragment | 2.65 | AAI-based (high-confidence) | 0 |
| S2-final-viral-combined_k141_15218\|\|full | 2921 | No | NA | 5 | 0 | 0 | Not-determined | Genome-fragment | NA | NA | 0 |
| S2-final-viral-combined_k141_149857\|\|full | 2715 | No | NA | 3 | 0 | 1 | Not-determined | Genome-fragment | NA | NA | 0 |
| S2-final-viral-combined_k141_211717\|\|full | 1498 | No | NA | 2 | 0 | 0 | Not-determined | Genome-fragment | NA | NA | 0 |
| S2-final-viral-combined_k141_48029\|\|full | 4337 | No | NA | 2 | 0 | 1 | Not-determined | Genome-fragment | NA | NA | 0 |
| S2-final-viral-combined_k141_57001\|\|full | 1534 | No | NA | 2 | 0 | 0 | Not-determined | Genome-fragment | NA | NA | 0 |
| S5_k141_16038 | 3871 | Yes | 1587 | 4 | 1 | 1 | Low-quality | Genome-fragment | 1.47 | AAI-based (medium-confidence) | 59 |
| S9-final-viral-combined_k141_86696\|\|full | 3872 | Yes | 2649 | 7 | 1 | 1 | Low-quality | Genome-fragment | 6.38 | AAI-based (high-confidence) | 31.59 |
| S2-final-viral-combined_k141_200504\|\|full | 1903 | No | NA | 2 | 0 | 0 | Not-determined | Genome-fragment | NA | NA | 0 |
| S9-final-viral-combined_k141_68348\|\|full | 3872 | No | NA | 4 | 0 | 0 | Low-quality | Genome-fragment | 7.6 | AAI-based (high-confidence) | 0 |
| S2-final-viral-combined_k141_57036\|\|full | 3124 | No | NA | 4 | 0 | 4 | Not-determined | Genome-fragment | NA | NA | 0 |
| S2-final-viral-combined_k141_200508\|\|full | 5137 | No | NA | 3 | 0 | 3 | Not-determined | Genome-fragment | NA | NA | 0 |
| S2-final-viral-combined_k141_233107\|\|full | 16186 | No | NA | 16 | 0 | 5 | Medium-quality | Genome-fragment | 52.08 | AAI-based (high-confidence) | 0 |
| S11_k141_212423 | 3872 | No | NA | 4 | 0 | 1 | Low-quality | Genome-fragment | 2.2 | AAI-based (medium-confidence) | 0 |
| S2-final-viral-combined_k141_98670\|\|full | 2197 | No | NA | 3 | 0 | 0 | Not-determined | Genome-fragment | NA | NA | 0 |
| S2-final-viral-combined_k141_233111\|\|full | 1026 | No | NA | 3 | 0 | 0 | Not-determined | Genome-fragment | NA | NA | 0 |
| S17-final-viral-combined_k141_167908\|\|full | 3874 | No | NA | 3 | 0 | 3 | Low-quality | Genome-fragment | 6.89 | AAI-based (medium-confidence) | 0 |
| S2-final-viral-combined_k141_200524\|\|full | 2559 | No | NA | 3 | 0 | 1 | Not-determined | Genome-fragment | NA | NA | 0 |
| S2-final-viral-combined_k141_67014\|\|full | 1734 | No | NA | 2 | 0 | 1 | Not-determined | Genome-fragment | NA | NA | 0 |
| S2-final-viral-combined_k141_57065\|\|full | 1940 | No | NA | 2 | 0 | 2 | Not-determined | Genome-fragment | NA | NA | 0 |
| S2-final-viral-combined_k141_98683\|\|full | 1623 | No | NA | 4 | 0 | 1 | Not-determined | Genome-fragment | NA | NA | 0 |
| S2-final-viral-combined_k141_191396\|\|full | 4334 | No | NA | 5 | 0 | 1 | Not-determined | Genome-fragment | NA | NA | 0 |
| S2-final-viral-combined_k141_200527\|\|full | 1416 | No | NA | 2 | 0 | 2 | Not-determined | Genome-fragment | NA | NA | 0 |
| S2-final-viral-combined_k141_181598\|\|full | 1799 | No | NA | 3 | 0 | 0 | Not-determined | Genome-fragment | NA | NA | 0 |
| S12-final-viral-combined_k141_168808\|\|full | 3874 | No | NA | 6 | 1 | 0 | Low-quality | Genome-fragment | 9.04 | AAI-based (high-confidence) | 0 |
| S2-final-viral-combined_k141_191428\|\|full | 4206 | No | NA | 4 | 0 | 1 | Not-determined | Genome-fragment | NA | NA | 0 |
| S8-final-viral-combined_k141_233567\|\|full | 3874 | No | NA | 6 | 0 | 0 | Low-quality | Genome-fragment | 8.62 | AAI-based (medium-confidence) | 0 |
| S9_k141_73702 | 3875 | No | NA | 5 | 0 | 5 | Low-quality | Genome-fragment | 1.45 | AAI-based (high-confidence) | 0 |
| S11_k141_60769 | 3875 | No | NA | 2 | 0 | 2 | Low-quality | Genome-fragment | 0.62 | AAI-based (high-confidence) | 0 |
| S2-final-viral-combined_k141_221554\|\|full | 1317 | No | NA | 2 | 0 | 1 | Not-determined | Genome-fragment | NA | NA | 0 |
| S2-final-viral-combined_k141_24494\|\|full | 3875 | No | NA | 4 | 0 | 1 | Low-quality | Genome-fragment | 4.91 | AAI-based (high-confidence) | 0 |
| S4-final-viral-combined_k141_133956\|\|full | 3875 | No | NA | 5 | 0 | 4 | Low-quality | Genome-fragment | 5.94 | AAI-based (medium-confidence) | 0 |
| S2-final-viral-combined_k141_25482\|\|full | 1589 | No | NA | 3 | 0 | 1 | Not-determined | Genome-fragment | NA | NA | 0 |
| S5-final-viral-combined_k141_137151\|\|full | 3876 | No | NA | 6 | 0 | 0 | Low-quality | Genome-fragment | 8.79 | AAI-based (medium-confidence) | 0 |
| S2-final-viral-combined_k141_233219\|\|full | 7892 | No | NA | 10 | 0 | 2 | Not-determined | Genome-fragment | NA | NA | 0 |
| S2-final-viral-combined_k141_221584\|\|full | 2283 | No | NA | 4 | 0 | 1 | Not-determined | Genome-fragment | NA | NA | 0 |
| S17-k141_164878 | 3876 | Yes | 3094 | 5 | 1 | 1 | Low-quality | Genome-fragment | 8.17 | AAI-based (high-confidence) | 20.18 |
| S2-final-viral-combined_k141_25487\|\|full | 2185 | No | NA | 2 | 0 | 0 | Not-determined | Genome-fragment | NA | NA | 0 |
| S12-final-viral-combined_k141_239813\|\|full | 3876 | No | NA | 13 | 0 | 0 | Low-quality | Genome-fragment | 10.27 | AAI-based (high-confidence) | 0 |
| S1-final-viral-combined_k141_179639\|\|full | 3877 | Yes | 2681 | 9 | 2 | 1 | Low-quality | Genome-fragment | 6.71 | AAI-based (high-confidence) | 30.85 |
| S2-final-viral-combined_k141_57212\|\|full | 1698 | No | NA | 3 | 0 | 0 | Not-determined | Genome-fragment | NA | NA | 0 |
| S3_k141_66733 | 3877 | No | NA | 2 | 0 | 1 | Low-quality | Genome-fragment | 0.33 | AAI-based (medium-confidence) | 0 |
| S5_k141_120358 | 3877 | No | NA | 8 | 1 | 0 | Low-quality | Genome-fragment | 4.31 | AAI-based (high-confidence) | 0 |
| S12_k141_187868 | 3877 | No | NA | 3 | 0 | 1 | Low-quality | Genome-fragment | 2.62 | AAI-based (medium-confidence) | 0 |
| S1-final-viral-combined_k141_238111\|\|full | 3878 | No | NA | 3 | 0 | 2 | Low-quality | Genome-fragment | 1.22 | AAI-based (medium-confidence) | 0 |
| S2-final-viral-combined_k141_88223\|\|full | 18499 | No | NA | 21 | 0 | 15 | Not-determined | Genome-fragment | NA | NA | 0 |
| S2-final-viral-combined_k141_88241\|\|full | 3159 | No | NA | 5 | 0 | 1 | Not-determined | Genome-fragment | NA | NA | 0 |
| S12-final-viral-combined_k141_67564\|\|full | 3878 | No | NA | 6 | 1 | 2 | Low-quality | Genome-fragment | 2.46 | HMM-based (lower-bound) | 0 |
| S2-final-viral-combined_k141_233304\|\|full | 2483 | No | NA | 4 | 0 | 2 | Not-determined | Genome-fragment | NA | NA | 0 |
| S2-final-viral-combined_k141_191507\|\|full | 1633 | No | NA | 3 | 0 | 0 | Not-determined | Genome-fragment | NA | NA | 0 |
| S2-final-viral-combined_k141_242402\|\|full | 6128 | No | NA | 5 | 0 | 4 | Not-determined | Genome-fragment | NA | NA | 0 |
| S2-final-viral-combined_k141_181711\|\|full | 4911 | No | NA | 6 | 0 | 4 | Not-determined | Genome-fragment | NA | NA | 0 |
| S1-final-viral-combined_k141_97264\|\|full | 3880 | No | NA | 4 | 0 | 1 | Low-quality | Genome-fragment | 6.09 | AAI-based (medium-confidence) | 0 |
| S12-final-viral-combined_k141_169558\|\|full | 3880 | No | NA | 3 | 0 | 3 | Low-quality | Genome-fragment | 2.51 | AAI-based (medium-confidence) | 0 |
| S2-final-viral-combined_k141_36626\|\|full | 2382 | No | NA | 3 | 0 | 1 | Not-determined | Genome-fragment | NA | NA | 0 |
| S2-final-viral-combined_k141_108417\|\|full | 5441 | No | NA | 4 | 0 | 1 | Not-determined | Genome-fragment | NA | NA | 0 |
| S8_k141_126269 | 3880 | No | NA | 4 | 1 | 0 | Low-quality | Genome-fragment | 8.55 | HMM-based (lower-bound) | 0 |
| S17-final-viral-combined_k141_163594\|\|full | 3882 | No | NA | 3 | 0 | 2 | Low-quality | Genome-fragment | 1.33 | AAI-based (medium-confidence) | 0 |
| S1-final-viral-combined_k141_382161\|\|full | 3883 | No | NA | 6 | 0 | 1 | Low-quality | Genome-fragment | 6.15 | AAI-based (medium-confidence) | 0 |
| S11-final-viral-combined_k141_175625\|\|full | 3884 | No | NA | 4 | 0 | 2 | Low-quality | Genome-fragment | 6.96 | AAI-based (high-confidence) | 0 |
| S7_k141_175559 | 3885 | No | NA | 3 | 0 | 2 | Low-quality | Genome-fragment | 7.08 | AAI-based (medium-confidence) | 0 |
| S4-final-viral-combined_k141_136614\|\|full | 3885 | No | NA | 3 | 0 | 2 | Low-quality | Genome-fragment | 2.73 | AAI-based (medium-confidence) | 0 |
| S17-k141_36161 | 3886 | No | NA | 1 | 0 | 1 | Low-quality | Genome-fragment | 1.76 | AAI-based (medium-confidence) | 0 |
| S2-final-viral-combined_k141_48227\|\|full | 2514 | No | NA | 4 | 0 | 1 | Not-determined | Genome-fragment | NA | NA | 0 |
| S2-final-viral-combined_k141_181780\|\|full | 7684 | No | NA | 10 | 0 | 4 | Not-determined | Genome-fragment | NA | NA | 0 |
| S2-final-viral-combined_k141_119120\|\|full | 1517 | No | NA | 2 | 0 | 0 | Not-determined | Genome-fragment | NA | NA | 0 |
| S2-final-viral-combined_k141_242461\|\|full | 2616 | No | NA | 2 | 0 | 0 | Not-determined | Genome-fragment | NA | NA | 0 |
| S2-final-viral-combined_k141_4466\|\|full | 21009 | No | NA | 22 | 0 | 14 | Not-determined | Genome-fragment | NA | NA | 0 |
| S2-final-viral-combined_k141_67221\|\|full | 2219 | No | NA | 2 | 0 | 2 | Not-determined | Genome-fragment | NA | NA | 0 |
| S2-final-viral-combined_k141_83896\|\|full | 3886 | No | NA | 3 | 0 | 2 | Low-quality | Genome-fragment | 5.09 | AAI-based (medium-confidence) | 0 |
| S8_k141_255552 | 3886 | No | NA | 2 | 0 | 2 | Low-quality | Genome-fragment | 1.69 | AAI-based (medium-confidence) | 0 |
| S2-final-viral-combined_k141_119146\|\|full | 4357 | No | NA | 5 | 0 | 1 | Not-determined | Genome-fragment | NA | NA | 0 |
| S5_k141_161239 | 3887 | Yes | 2322 | 5 | 1 | 1 | Low-quality | Genome-fragment | 6.27 | HMM-based (lower-bound) | 40.26 |
| S2-final-viral-combined_k141_77700\|\|full | 1831 | No | NA | 4 | 0 | 2 | Not-determined | Genome-fragment | NA | NA | 0 |
| S2-final-viral-combined_k141_119156\|\|full | 3516 | No | NA | 3 | 0 | 1 | Not-determined | Genome-fragment | NA | NA | 0 |
| S7-final-viral-combined_k141_59407\|\|full | 3887 | No | NA | 8 | 0 | 1 | Low-quality | Genome-fragment | 10.43 | AAI-based (medium-confidence) | 0 |
| S2-final-viral-combined_k141_67256\|\|full | 3017 | No | NA | 5 | 0 | 1 | Not-determined | Genome-fragment | NA | NA | 0 |
| S2-final-viral-combined_k141_57293\|\|full | 47253 | No | NA | 48 | 3 | 23 | High-quality | High-quality | 100 | AAI-based (high-confidence) | 0 |
| S17-final-viral-combined_k141_64848\|\|full | 3887 | No | NA | 3 | 0 | 3 | Low-quality | Genome-fragment | 4.2 | AAI-based (medium-confidence) | 0 |
| S2-final-viral-combined_k141_221713\|\|full | 14765 | No | NA | 17 | 0 | 2 | Not-determined | Genome-fragment | NA | NA | 0 |
| S2-final-viral-combined_k141_242504\|\|full | 2032 | No | NA | 3 | 0 | 0 | Not-determined | Genome-fragment | NA | NA | 0 |
| S9_k141_37725 | 3888 | No | NA | 4 | 0 | 4 | Low-quality | Genome-fragment | 1.44 | AAI-based (high-confidence) | 0 |
| S1-final-viral-combined_k141_277382\|\|full | 3888 | No | NA | 8 | 7 | 0 | Low-quality | Genome-fragment | 8.87 | AAI-based (high-confidence) | 0 |
| S7-final-viral-combined_k141_180495\|\|full | 3889 | No | NA | 5 | 1 | 0 | Low-quality | Genome-fragment | 8.64 | HMM-based (lower-bound) | 0 |
| S12-final-viral-combined_k141_215334\|\|full | 3889 | No | NA | 3 | 0 | 0 | Low-quality | Genome-fragment | 10.01 | AAI-based (high-confidence) | 0 |
| S1-final-viral-combined_k141_48284\|\|full | 3890 | No | NA | 5 | 0 | 0 | Low-quality | Genome-fragment | 10.66 | AAI-based (medium-confidence) | 0 |
| S2-final-viral-combined_k141_191613\|\|full | 79617 | Yes | 22352 | 97 | 7 | 38 | Medium-quality | Genome-fragment | 52.41 | AAI-based (high-confidence) | 71.93 |
| S3_k141_57895 | 3891 | No | NA | 2 | 0 | 1 | Low-quality | Genome-fragment | 1.93 | AAI-based (medium-confidence) | 0 |
| S1-final-viral-combined_k141_16569\|\|full | 3892 | No | NA | 5 | 0 | 1 | Low-quality | Genome-fragment | 8.31 | AAI-based (medium-confidence) | 0 |
| S2-final-viral-combined_k141_212041\|\|full | 1815 | No | NA | 2 | 0 | 0 | Not-determined | Genome-fragment | NA | NA | 0 |
| S1_k141_362899 | 3892 | No | NA | 5 | 0 | 3 | Low-quality | Genome-fragment | 2.17 | AAI-based (medium-confidence) | 0 |
| S17-final-viral-combined_k141_226938\|\|full | 3892 | No | NA | 4 | 1 | 0 | Low-quality | Genome-fragment | 9.1 | AAI-based (high-confidence) | 0 |
| S2-final-viral-combined_k141_233528\|\|full | 2269 | No | NA | 4 | 0 | 1 | Not-determined | Genome-fragment | NA | NA | 0 |
| S8-final-viral-combined_k141_94646\|\|full | 3892 | No | NA | 4 | 3 | 0 | Low-quality | Genome-fragment | 8.5 | AAI-based (high-confidence) | 0 |
| S2-final-viral-combined_k141_191703\|\|full | 1133 | No | NA | 3 | 0 | 1 | Not-determined | Genome-fragment | NA | NA | 0 |
| S2-final-viral-combined_k141_200862\|\|full | 2445 | No | NA | 3 | 0 | 0 | Not-determined | Genome-fragment | NA | NA | 0 |
| S2-final-viral-combined_k141_128702\|\|full | 2116 | No | NA | 4 | 0 | 3 | Not-determined | Genome-fragment | NA | NA | 0 |
| S2-final-viral-combined_k141_140183\|\|full | 2344 | No | NA | 3 | 0 | 0 | Not-determined | Genome-fragment | NA | NA | 0 |
| S2-final-viral-combined_k141_200867\|\|full | 1180 | No | NA | 3 | 0 | 0 | Not-determined | Genome-fragment | NA | NA | 0 |
| S2-final-viral-combined_k141_200871\|\|full | 1488 | No | NA | 2 | 0 | 2 | Not-determined | Genome-fragment | NA | NA | 0 |
| S2-final-viral-combined_k141_140195\|\|full | 4939 | No | NA | 4 | 0 | 4 | Not-determined | Genome-fragment | NA | NA | 0 |
| S2-final-viral-combined_k141_160257\|\|full | 1859 | No | NA | 3 | 0 | 0 | Not-determined | Genome-fragment | NA | NA | 0 |
| S2-final-viral-combined_k141_108483\|\|full | 5632 | No | NA | 6 | 0 | 2 | Not-determined | Genome-fragment | NA | NA | 0 |
| S2-final-viral-combined_k141_36866\|\|full | 3910 | No | NA | 5 | 0 | 0 | Not-determined | Genome-fragment | NA | NA | 0 |
| S2-final-viral-combined_k141_98888\|\|full | 107937 | Yes | 61979 | 134 | 33 | 35 | High-quality | High-quality | 100 | AAI-based (high-confidence) | 42.58 |

| | | | | | | | | | | | |
|---|---|---|---|---|---|---|---|---|---|---|---|
| S2-final-viral-combined_k141_181849\|\|full | 57130 | Yes | 33154 | 57 | 7 | 13 | Complete | High-quality | 100 | Provirus (medium-confidence) | 41.97 |
| S1-final-viral-combined_k141_324691\|\|full | 3893 | No | NA | 5 | 0 | 4 | Low-quality | Genome-fragment | 2.68 | AAI-based (high-confidence) | 0 |
| S2-final-viral-combined_k141_140218\|\|full | 6496 | No | NA | 6 | 0 | 5 | Not-determined | Genome-fragment | NA | NA | 0 |
| S2-final-viral-combined_k141_212116\|\|full | 1183 | No | NA | 2 | 0 | 2 | Not-determined | Genome-fragment | NA | NA | 0 |
| S3-final-viral-combined_k141_28363\|\|full | 3893 | No | NA | 5 | 0 | 3 | Low-quality | Genome-fragment | 3.13 | AAI-based (medium-confidence) | 0 |
| S3_k141_84543 | 3893 | No | NA | 4 | 2 | 0 | Low-quality | Genome-fragment | 7.43 | AAI-based (high-confidence) | 0 |
| S2-final-viral-combined_k141_108518\|\|full | 7863 | No | NA | 6 | 0 | 6 | Not-determined | Genome-fragment | NA | NA | 0 |
| S1_k141_140523 | 3894 | No | NA | 2 | 0 | 1 | Low-quality | Genome-fragment | 1.22 | AAI-based (medium-confidence) | 0 |
| S5_k141_145841 | 3894 | No | NA | 3 | 0 | 1 | Low-quality | Genome-fragment | 1.57 | AAI-based (medium-confidence) | 0 |
| S7-final-viral-combined_k141_141468\|\|full | 3895 | No | NA | 9 | 0 | 0 | Low-quality | Genome-fragment | 9.58 | AAI-based (high-confidence) | 0 |
| S17-k141_211228 | 3895 | No | NA | 2 | 0 | 0 | Low-quality | Genome-fragment | 4.11 | AAI-based (medium-confidence) | 0 |
| S2-final-viral-combined_k141_57504\|\|full | 1014 | No | NA | 3 | 0 | 1 | Not-determined | Genome-fragment | NA | NA | 0 |
| S2-final-viral-combined_k141_233610\|\|full | 2894 | No | NA | 3 | 0 | 1 | Not-determined | Genome-fragment | NA | NA | 0 |
| S2-final-viral-combined_k141_77969\|\|full | 3049 | No | NA | 3 | 0 | 2 | Not-determined | Genome-fragment | NA | NA | 0 |
| S2-final-viral-combined_k141_212124\|\|full | 1307 | No | NA | 2 | 0 | 1 | Not-determined | Genome-fragment | NA | NA | 0 |
| S2-final-viral-combined_k141_171685\|\|full | 2110 | No | NA | 3 | 0 | 1 | Not-determined | Genome-fragment | NA | NA | 0 |
| S2-final-viral-combined_k141_77979\|\|full | 2504 | No | NA | 3 | 0 | 1 | Not-determined | Genome-fragment | NA | NA | 0 |
| S2-final-viral-combined_k141_119408\|\|full | 1873 | No | NA | 3 | 0 | 1 | Not-determined | Genome-fragment | NA | NA | 0 |
| S1-final-viral-combined_k141_114832\|\|full | 3896 | Yes | 2541 | 8 | 1 | 1 | Low-quality | Genome-fragment | 5.93 | AAI-based (high-confidence) | 34.78 |
| S17-final-viral-combined_k141_48698\|\|full | 3896 | No | NA | 5 | 0 | 1 | Low-quality | Genome-fragment | 9.03 | AAI-based (medium-confidence) | 0 |
| S9-final-viral-combined_k141_48570\|\|full | 3898 | No | NA | 6 | 3 | 0 | Low-quality | Genome-fragment | 8.48 | AAI-based (medium-confidence) | 0 |
| S12_k141_61966 | 3898 | No | NA | 3 | 0 | 2 | Low-quality | Genome-fragment | 2.15 | AAI-based (medium-confidence) | 0 |
| S2-final-viral-combined_k141_119423\|\|full | 1103 | No | NA | 2 | 0 | 0 | Not-determined | Genome-fragment | NA | NA | 0 |
| S2-final-viral-combined_k141_77988\|\|full | 2201 | No | NA | 3 | 0 | 2 | Not-determined | Genome-fragment | NA | NA | 0 |
| S11-final-viral-combined_k141_105554\|\|full | 3900 | No | NA | 5 | 1 | 0 | Low-quality | Genome-fragment | 10.16 | AAI-based (medium-confidence) | 0 |
| S2-final-viral-combined_k141_171731\|\|full | 2262 | No | NA | 3 | 0 | 0 | Not-determined | Genome-fragment | NA | NA | 0 |
| S2-final-viral-combined_k141_15708\|\|full | 1131 | No | NA | 2 | 0 | 2 | Not-determined | Genome-fragment | NA | NA | 0 |
| S1-final-viral-combined_k141_111587\|\|full | 3900 | No | NA | 9 | 1 | 0 | Low-quality | Genome-fragment | 26.26 | AAI-based (high-confidence) | 0 |
| S17-k141_38652 | 3900 | No | NA | 4 | 0 | 1 | Low-quality | Genome-fragment | 12.45 | AAI-based (medium-confidence) | 0 |
| S2-final-viral-combined_k141_108597\|\|full | 1768 | No | NA | 3 | 0 | 0 | Not-determined | Genome-fragment | NA | NA | 0 |
| S2-final-viral-combined_k141_171746\|\|full | 1334 | No | NA | 2 | 0 | 1 | Not-determined | Genome-fragment | NA | NA | 0 |
| S2-final-viral-combined_k141_191753\|\|full | 1057 | No | NA | 3 | 0 | 1 | Not-determined | Genome-fragment | NA | NA | 0 |
| S2-final-viral-combined_k141_88649\|\|full | 1280 | No | NA | 4 | 0 | 0 | Not-determined | Genome-fragment | NA | NA | 0 |
| S2-final-viral-combined_k141_191761\|\|full | 2061 | No | NA | 2 | 0 | 1 | Not-determined | Genome-fragment | NA | NA | 0 |
| S2-final-viral-combined_k141_119479\|\|full | 1179 | No | NA | 2 | 0 | 0 | Not-determined | Genome-fragment | NA | NA | 0 |
| S2-final-viral-combined_k141_128756\|\|full | 3194 | No | NA | 3 | 0 | 3 | Not-determined | Genome-fragment | NA | NA | 0 |
| S8-final-viral-combined_k141_209496\|\|full | 3900 | No | NA | 10 | 4 | 0 | Low-quality | Genome-fragment | 10.82 | AAI-based (high-confidence) | 0 |
| S2-final-viral-combined_k141_160276\|\|full | 2219 | No | NA | 2 | 0 | 2 | Not-determined | Genome-fragment | NA | NA | 0 |
| S2-final-viral-combined_k141_201017\|\|full | 3291 | No | NA | 3 | 0 | 2 | Not-determined | Genome-fragment | NA | NA | 0 |
| S2-final-viral-combined_k141_150394\|\|full | 1707 | No | NA | 3 | 0 | 0 | Not-determined | Genome-fragment | NA | NA | 0 |
| S2-final-viral-combined_k141_150414\|\|full | 1616 | No | NA | 2 | 0 | 2 | Not-determined | Genome-fragment | NA | NA | 0 |
| S2-final-viral-combined_k141_88675\|\|full | 1005 | No | NA | 3 | 0 | 0 | Not-determined | Genome-fragment | NA | NA | 0 |
| S4-final-viral-combined_k141_75191\|\|full | 3901 | No | NA | 5 | 0 | 0 | Low-quality | Genome-fragment | 4.13 | AAI-based (high-confidence) | 0 |
| S2-final-viral-combined_k141_233689\|\|full | 1522 | No | NA | 2 | 0 | 0 | Not-determined | Genome-fragment | NA | NA | 0 |
| S2-final-viral-combined_k141_4638\|\|full | 2486 | No | NA | 3 | 0 | 0 | Not-determined | Genome-fragment | NA | NA | 0 |
| S2-final-viral-combined_k141_221846\|\|full | 1476 | No | NA | 2 | 0 | 0 | Not-determined | Genome-fragment | NA | NA | 0 |
| S1-final-viral-combined_k141_285722\|\|full | 3902 | No | NA | 8 | 1 | 0 | Low-quality | Genome-fragment | 9.78 | AAI-based (high-confidence) | 0 |
| S5-final-viral-combined_k141_52179\|\|full | 3902 | No | NA | 4 | 4 | 0 | Low-quality | Genome-fragment | 9.61 | AAI-based (high-confidence) | 0 |
| S11-final-viral-combined_k141_325666\|\|full | 3903 | No | NA | 5 | 1 | 0 | Low-quality | Genome-fragment | 8.89 | AAI-based (high-confidence) | 0 |
| S2-final-viral-combined_k141_37241\|\|full | 3903 | No | NA | 5 | 1 | 0 | Low-quality | Genome-fragment | 8.23 | AAI-based (high-confidence) | 0 |
| S2-final-viral-combined_k141_67354\|\|full | 33027 | No | NA | 24 | 0 | 16 | Not-determined | Genome-fragment | NA | NA | 0 |
| S1-final-viral-combined_k141_165148\|\|full | 3904 | No | NA | 5 | 0 | 4 | Low-quality | Genome-fragment | 2.38 | AAI-based (medium-confidence) | 0 |
| S2-final-viral-combined_k141_78153\|\|full | 2250 | No | NA | 3 | 0 | 2 | Not-determined | Genome-fragment | NA | NA | 0 |
| S2-final-viral-combined_k141_48531\|\|full | 3065 | No | NA | 4 | 0 | 4 | Not-determined | Genome-fragment | NA | NA | 0 |
| S12-final-viral-combined_k141_350639\|\|full | 3904 | No | NA | 8 | 2 | 0 | Low-quality | Genome-fragment | 10.03 | AAI-based (high-confidence) | 0 |
| S2-final-viral-combined_k141_108667\|\|full | 1097 | No | NA | 2 | 0 | 1 | Not-determined | Genome-fragment | NA | NA | 0 |
| S9-final-viral-combined_k141_40233\|\|full | 3905 | No | NA | 3 | 0 | 1 | Low-quality | Genome-fragment | 2.03 | AAI-based (medium-confidence) | 0 |
| S11-final-viral-combined_k141_7294\|\|full | 3905 | No | NA | 2 | 0 | 2 | Low-quality | Genome-fragment | 2.27 | AAI-based (medium-confidence) | 0 |
| S3_k141_83034 | 3905 | No | NA | 6 | 2 | 0 | Low-quality | Genome-fragment | 6.24 | AAI-based (high-confidence) | 0 |
| S2-final-viral-combined_k141_201110\|\|full | 3616 | No | NA | 7 | 0 | 0 | Not-determined | Genome-fragment | NA | NA | 0 |
| S2-final-viral-combined_k141_171889\|\|full | 1700 | No | NA | 3 | 0 | 1 | Not-determined | Genome-fragment | NA | NA | 0 |
| S2-final-viral-combined_k141_221963\|\|full | 1726 | No | NA | 2 | 0 | 2 | Not-determined | Genome-fragment | NA | NA | 0 |
| S2-final-viral-combined_k141_140471\|\|full | 1586 | No | NA | 4 | 0 | 0 | Not-determined | Genome-fragment | NA | NA | 0 |
| S12-final-viral-combined_k141_72493\|\|full | 3905 | No | NA | 3 | 0 | 3 | Low-quality | Genome-fragment | 1.31 | AAI-based (medium-confidence) | 0 |
| S2-final-viral-combined_k141_48617\|\|full | 2521 | No | NA | 3 | 0 | 2 | Not-determined | Genome-fragment | NA | NA | 0 |
| S2-final-viral-combined_k141_119623\|\|full | 1034 | No | NA | 2 | 0 | 0 | Not-determined | Genome-fragment | NA | NA | 0 |
| S2-final-viral-combined_k141_140475\|\|full | 2855 | No | NA | 3 | 0 | 2 | Not-determined | Genome-fragment | NA | NA | 0 |
| S2-final-viral-combined_k141_233822\|\|full | 1254 | No | NA | 3 | 0 | 0 | Not-determined | Genome-fragment | NA | NA | 0 |
| S12-final-viral-combined_k141_231792\|\|full | 3905 | No | NA | 4 | 0 | 3 | Low-quality | Genome-fragment | 1.81 | AAI-based (medium-confidence) | 0 |
| S2-final-viral-combined_k141_4688\|\|full | 1123 | No | NA | 2 | 0 | 2 | Not-determined | Genome-fragment | NA | NA | 0 |
| S2-final-viral-combined_k141_233832\|\|full | 2027 | No | NA | 2 | 0 | 2 | Not-determined | Genome-fragment | NA | NA | 0 |
| S2-final-viral-combined_k141_242767\|\|full | 1719 | No | NA | 2 | 0 | 1 | Not-determined | Genome-fragment | NA | NA | 0 |
| S12_k141_187374 | 3905 | No | NA | 1 | 0 | 1 | Low-quality | Genome-fragment | 3.95 | AAI-based (medium-confidence) | 0 |
| S12_k141_131500 | 3905 | No | NA | 3 | 0 | 1 | Low-quality | Genome-fragment | 5.99 | AAI-based (medium-confidence) | 0 |
| S2-final-viral-combined_k141_182029\|\|full | 1378 | No | NA | 2 | 0 | 1 | Not-determined | Genome-fragment | NA | NA | 0 |
| S11_k141_43251 | 3906 | Yes | 3331 | 10 | 6 | 1 | Low-quality | Genome-fragment | 7.32 | AAI-based (high-confidence) | 14.72 |
| S4_k141_152999 | 3906 | No | NA | 7 | 1 | 0 | Low-quality | Genome-fragment | 4.09 | AAI-based (high-confidence) | 0 |
| S2-final-viral-combined_k141_48661\|\|full | 3829 | No | NA | 5 | 0 | 1 | Medium-quality | Genome-fragment | 79.05 | AAI-based (medium-confidence) | 0 |
| S2-final-viral-combined_k141_67419\|\|full | 3820 | No | NA | 4 | 0 | 1 | Not-determined | Genome-fragment | NA | NA | 0 |
| S2-final-viral-combined_k141_67426\|\|full | 1087 | No | NA | 2 | 0 | 0 | Not-determined | Genome-fragment | NA | NA | 0 |
| S2-final-viral-combined_k141_78253\|\|full | 1129 | No | NA | 3 | 0 | 1 | Not-determined | Genome-fragment | NA | NA | 0 |
| S8_k141_225113 | 3906 | Yes | 3360 | 8 | 1 | 1 | Low-quality | Genome-fragment | 8.76 | AAI-based (high-confidence) | 13.98 |
| S2-final-viral-combined_k141_48682\|\|full | 4904 | No | NA | 5 | 0 | 5 | Not-determined | Genome-fragment | NA | NA | 0 |
| S2-final-viral-combined_k141_78262\|\|full | 2071 | No | NA | 2 | 0 | 1 | Not-determined | Genome-fragment | NA | NA | 0 |
| S2-final-viral-combined_k141_57643\|\|full | 2558 | No | NA | 2 | 0 | 1 | Not-determined | Genome-fragment | NA | NA | 0 |
| S12-final-viral-combined_k141_108031\|\|full | 3908 | No | NA | 4 | 0 | 3 | Low-quality | Genome-fragment | 6.34 | AAI-based (medium-confidence) | 0 |
| S2-final-viral-combined_k141_78276\|\|full | 1051 | No | NA | 3 | 0 | 0 | Not-determined | Genome-fragment | NA | NA | 0 |
| S2-final-viral-combined_k141_191961\|\|full | 1168 | No | NA | 4 | 0 | 0 | Not-determined | Genome-fragment | NA | NA | 0 |
| S2-final-viral-combined_k141_212262\|\|full | 1835 | No | NA | 3 | 0 | 0 | Not-determined | Genome-fragment | NA | NA | 0 |
| S2-final-viral-combined_k141_78289\|\|full | 3954 | No | NA | 4 | 0 | 2 | Not-determined | Genome-fragment | NA | NA | 0 |
| S2-final-viral-combined_k141_15948\|\|full | 1885 | No | NA | 3 | 0 | 3 | Not-determined | Genome-fragment | NA | NA | 0 |
| S2-final-viral-combined_k141_57661\|\|full | 1130 | No | NA | 3 | 0 | 1 | Not-determined | Genome-fragment | NA | NA | 0 |
| S2-final-viral-combined_k141_201205\|\|full | 30077 | No | NA | 39 | 3 | 0 | High-quality | High-quality | 90.45 | AAI-based (high-confidence) | 0 |
| S2_k141_214798 | 3908 | No | NA | 2 | 0 | 2 | Low-quality | Genome-fragment | 4.39 | AAI-based (medium-confidence) | 0 |
| S2-final-viral-combined_k141_191972\|\|full | 2043 | No | NA | 3 | 0 | 0 | Not-determined | Genome-fragment | NA | NA | 0 |
| S2-final-viral-combined_k141_172078\|\|full | 3802 | No | NA | 3 | 0 | 0 | Not-determined | Genome-fragment | NA | NA | 0 |
| S2-final-viral-combined_k141_242876\|\|full | 3935 | No | NA | 3 | 0 | 3 | Not-determined | Genome-fragment | NA | NA | 0 |
| S7-final-viral-combined_k141_61653\|\|full | 3911 | No | NA | 4 | 0 | 4 | Low-quality | Genome-fragment | 1.3 | AAI-based (medium-confidence) | 0 |
| S8_k141_60716 | 3911 | No | NA | 8 | 1 | 0 | Low-quality | Genome-fragment | 4.2 | AAI-based (high-confidence) | 0 |
| S3-final-viral-combined_k141_75905\|\|full | 3912 | No | NA | 3 | 0 | 2 | Low-quality | Genome-fragment | 1.73 | AAI-based (medium-confidence) | 0 |
| S2-final-viral-combined_k141_201227\|\|full | 1357 | No | NA | 2 | 0 | 2 | Not-determined | Genome-fragment | NA | NA | 0 |
| S2-final-viral-combined_k141_25547\|\|full | 8336 | No | NA | 17 | 0 | 0 | Not-determined | Genome-fragment | NA | NA | 0 |
| S2-final-viral-combined_k141_48786\|\|full | 2425 | No | NA | 2 | 0 | 2 | Not-determined | Genome-fragment | NA | NA | 0 |
| S2-final-viral-combined_k141_150644\|\|full | 1064 | No | NA | 3 | 0 | 1 | Not-determined | Genome-fragment | NA | NA | 0 |
| S2-final-viral-combined_k141_88875\|\|full | 3355 | No | NA | 4 | 0 | 1 | Not-determined | Genome-fragment | NA | NA | 0 |
| S2-final-viral-combined_k141_201236\|\|full | 1873 | No | NA | 3 | 0 | 0 | Not-determined | Genome-fragment | NA | NA | 0 |
| S2-final-viral-combined_k141_150658\|\|full | 1075 | No | NA | 3 | 0 | 0 | Not-determined | Genome-fragment | NA | NA | 0 |
| S2_k141_238038 | 3912 | No | NA | 10 | 0 | 2 | Low-quality | Genome-fragment | 7.51 | AAI-based (medium-confidence) | 0 |
| S3-final-viral-combined_k141_116919\|\|full | 3913 | Yes | 3604 | 7 | 2 | 1 | Low-quality | Genome-fragment | 9.69 | AAI-based (high-confidence) | 7.9 |
| S2-final-viral-combined_k141_172092\|\|full | 8266 | No | NA | 4 | 0 | 3 | Not-determined | Genome-fragment | NA | NA | 0 |
| S2-final-viral-combined_k141_16036\|\|full | 1166 | No | NA | 2 | 0 | 2 | Not-determined | Genome-fragment | NA | NA | 0 |
| S5-final-viral-combined_k141_181983\|\|full | 3913 | Yes | 2609 | 8 | 2 | 1 | Low-quality | Genome-fragment | 6.1 | AAI-based (medium-confidence) | 33.32 |
| S3_k141_47739 | 3914 | No | NA | 9 | 2 | 0 | Low-quality | Genome-fragment | 10.13 | AAI-based (high-confidence) | 0 |
| S12-final-viral-combined_k141_359207\|\|full | 3914 | No | NA | 3 | 0 | 3 | Low-quality | Genome-fragment | 3.02 | AAI-based (medium-confidence) | 0 |
| S2-final-viral-combined_k141_78386\|\|full | 2200 | No | NA | 2 | 0 | 1 | Not-determined | Genome-fragment | NA | NA | 0 |
| S4-final-viral-combined_k141_152156\|\|full | 3914 | No | NA | 4 | 0 | 2 | Low-quality | Genome-fragment | 2.31 | AAI-based (medium-confidence) | 0 |
| S2-final-viral-combined_k141_140641\|\|full | 1881 | No | NA | 3 | 0 | 1 | Not-determined | Genome-fragment | NA | NA | 0 |
| S2-final-viral-combined_k141_119807\|\|full | 1132 | No | NA | 2 | 0 | 0 | Not-determined | Genome-fragment | NA | NA | 0 |
| S2_k141_129814 | 3915 | No | NA | 15 | 1 | 0 | Low-quality | Genome-fragment | 2.17 | AAI-based (high-confidence) | 0 |
| S2-final-viral-combined_k141_233936\|\|full | 1588 | No | NA | 2 | 0 | 0 | Not-determined | Genome-fragment | NA | NA | 0 |
| S7-final-viral-combined_k141_48183\|\|full | 3916 | No | NA | 4 | 0 | 2 | Low-quality | Genome-fragment | 6.44 | AAI-based (medium-confidence) | 0 |
| S17-k141_68643 | 3916 | No | NA | 3 | 0 | 2 | Low-quality | Genome-fragment | 0.67 | AAI-based (high-confidence) | 0 |
| S8-final-viral-combined_k141_33584\|\|full | 3916 | No | NA | 6 | 0 | 2 | Low-quality | Genome-fragment | 2.06 | AAI-based (high-confidence) | 0 |
| S2-final-viral-combined_k141_4772\|\|full | 2460 | No | NA | 4 | 0 | 0 | Not-determined | Genome-fragment | NA | NA | 0 |
| S8_k141_261063 | 3917 | No | NA | 3 | 0 | 1 | Low-quality | Genome-fragment | 6.32 | AAI-based (medium-confidence) | 0 |
| S7-final-viral-combined_k141_52642\|\|full | 3918 | No | NA | 7 | 6 | 0 | Low-quality | Genome-fragment | 7.99 | AAI-based (high-confidence) | 0 |
| S17-k141_67927 | 3918 | Yes | 1605 | 5 | 1 | 1 | Low-quality | Genome-fragment | 2.72 | AAI-based (medium-confidence) | 59.04 |
| S2-final-viral-combined_k141_37171\|\|full | 2392 | No | NA | 3 | 0 | 0 | Not-determined | Genome-fragment | NA | NA | 0 |
| S2-final-viral-combined_k141_48914\|\|full | 2786 | No | NA | 3 | 0 | 1 | Not-determined | Genome-fragment | NA | NA | 0 |
| S2-final-viral-combined_k141_48929\|\|full | 1276 | No | NA | 2 | 0 | 2 | Not-determined | Genome-fragment | NA | NA | 0 |
| S2-final-viral-combined_k141_140700\|\|full | 1761 | No | NA | 2 | 0 | 0 | Not-determined | Genome-fragment | NA | NA | 0 |
| S2-final-viral-combined_k141_108908\|\|full | 2891 | No | NA | 4 | 0 | 1 | Not-determined | Genome-fragment | NA | NA | 0 |
| S2-final-viral-combined_k141_88973\|\|full | 3823 | No | NA | 4 | 0 | 3 | Not-determined | Genome-fragment | NA | NA | 0 |
| S12-final-viral-combined_k141_348571\|\|full | 3919 | No | NA | 4 | 1 | 0 | Low-quality | Genome-fragment | 3.05 | AAI-based (medium-confidence) | 0 |
| S2-final-viral-combined_k141_208664\|\|full | 3919 | No | NA | 3 | 0 | 3 | Low-quality | Genome-fragment | 0.85 | AAI-based (medium-confidence) | 0 |
| S2-final-viral-combined_k141_150760\|\|full | 1017 | No | NA | 1 | 0 | 0 | Not-determined | Genome-fragment | NA | NA | 0 |
| S2-final-viral-combined_k141_99265\|\|full | 1499 | No | NA | 2 | 0 | 0 | Not-determined | Genome-fragment | NA | NA | 0 |
| S2-final-viral-combined_k141_160507\|\|full | 2174 | No | NA | 4 | 0 | 2 | Not-determined | Genome-fragment | NA | NA | 0 |
| S2-final-viral-combined_k141_78449\|\|full | 10483 | No | NA | 10 | 0 | 9 | Not-determined | Genome-fragment | NA | NA | 0 |
| S12_k141_204037 | 3920 | No | NA | 5 | 0 | 1 | Low-quality | Genome-fragment | 4.17 | AAI-based (medium-confidence) | 0 |
| S2-final-viral-combined_k141_196781\|\|full | 3921 | No | NA | 8 | 6 | 0 | Low-quality | Genome-fragment | 8.36 | AAI-based (medium-confidence) | 0 |
| S2-final-viral-combined_k141_160535\|\|full | 1513 | No | NA | 2 | 0 | 1 | Not-determined | Genome-fragment | NA | NA | 0 |
| S2-final-viral-combined_k141_99311\|\|full | 1694 | No | NA | 3 | 0 | 1 | Not-determined | Genome-fragment | NA | NA | 0 |
| S2-final-viral-combined_k141_172207\|\|full | 2584 | No | NA | 3 | 0 | 0 | Not-determined | Genome-fragment | NA | NA | 0 |
| S2-final-viral-combined_k141_160544\|\|full | 1513 | No | NA | 2 | 0 | 1 | Not-determined | Genome-fragment | NA | NA | 0 |
| S2-final-viral-combined_k141_108971\|\|full | 1745 | No | NA | 3 | 0 | 0 | Not-determined | Genome-fragment | NA | NA | 0 |
| S11_k141_222322 | 3924 | No | NA | 5 | 3 | 0 | Low-quality | Genome-fragment | 9.58 | AAI-based (high-confidence) | 0 |
| S2-final-viral-combined_k141_212307\|\|full | 2773 | No | NA | 4 | 0 | 1 | Not-determined | Genome-fragment | NA | NA | 0 |
| S2-final-viral-combined_k141_108980\|\|full | 3463 | No | NA | 3 | 0 | 2 | Not-determined | Genome-fragment | NA | NA | 0 |
| S1-final-viral-combined_k141_22934\|\|full | 3924 | No | NA | 6 | 0 | 2 | Low-quality | Genome-fragment | 4.51 | AAI-based (medium-confidence) | 0 |
| S2-final-viral-combined_k141_244902\|\|full | 3924 | No | NA | 11 | 0 | 1 | Low-quality | Genome-fragment | 10.84 | AAI-based (high-confidence) | 0 |
| S2-final-viral-combined_k141_182326\|\|full | 1274 | No | NA | 4 | 0 | 0 | Not-determined | Genome-fragment | NA | NA | 0 |
| S2-final-viral-combined_k141_37244\|\|full | 2761 | No | NA | 3 | 0 | 3 | Not-determined | Genome-fragment | NA | NA | 0 |
| S2-final-viral-combined_k141_37246\|\|full | 2559 | No | NA | 4 | 0 | 3 | Not-determined | Genome-fragment | NA | NA | 0 |
| S2-final-viral-combined_k141_37247\|\|full | 3087 | No | NA | 2 | 0 | 1 | Not-determined | Genome-fragment | NA | NA | 0 |
| S9-final-viral-combined_k141_68057\|\|full | 3926 | No | NA | 3 | 0 | 1 | Low-quality | Genome-fragment | 13.68 | AAI-based (medium-confidence) | 0 |
| S5-final-viral-combined_k141_492\|\|full | 3926 | No | NA | 4 | 0 | 0 | Low-quality | Genome-fragment | 44.22 | AAI-based (high-confidence) | 0 |
| S2-final-viral-combined_k141_212352\|\|full | 6650 | No | NA | 4 | 0 | 3 | Not-determined | Genome-fragment | NA | NA | 0 |
| S2-final-viral-combined_k141_109014\|\|full | 2791 | No | NA | 3 | 0 | 0 | Not-determined | Genome-fragment | NA | NA | 0 |
| S2-final-viral-combined_k141_150874\|\|full | 1283 | No | NA | 4 | 0 | 1 | Not-determined | Genome-fragment | NA | NA | 0 |
| S2_k141_134758 | 3926 | No | NA | 6 | 0 | 1 | Low-quality | Genome-fragment | 6.52 | AAI-based (medium-confidence) | 0 |
| S2-final-viral-combined_k141_201349\|\|full | 2865 | No | NA | 3 | 0 | 1 | Not-determined | Genome-fragment | NA | NA | 0 |
| S2-final-viral-combined_k141_140864\|\|full | 2330 | No | NA | 3 | 0 | 2 | Not-determined | Genome-fragment | NA | NA | 0 |
| S2-final-viral-combined_k141_99371\|\|full | 12023 | No | NA | 10 | 0 | 9 | Not-determined | Genome-fragment | NA | NA | 0 |
| S2-final-viral-combined_k141_16127\|\|full | 1833 | No | NA | 2 | 0 | 1 | Not-determined | Genome-fragment | NA | NA | 0 |
| S2-final-viral-combined_k141_89127\|\|full | 2343 | No | NA | 3 | 0 | 0 | Not-determined | Genome-fragment | NA | NA | 0 |
| S7-final-viral-combined_k141_114432\|\|full | 3927 | No | NA | 9 | 1 | 0 | Low-quality | Genome-fragment | 8.65 | AAI-based (high-confidence) | 0 |
| S12-final-viral-combined_k141_379591\|\|full | 3927 | No | NA | 3 | 0 | 2 | Low-quality | Genome-fragment | 4.68 | AAI-based (medium-confidence) | 0 |
| S1-final-viral-combined_k141_376022\|\|full | 3928 | No | NA | 6 | 0 | 1 | Low-quality | Genome-fragment | 3.49 | AAI-based (high-confidence) | 0 |
| S2-final-viral-combined_k141_57858\|\|full | 1918 | No | NA | 3 | 0 | 2 | Not-determined | Genome-fragment | NA | NA | 0 |
| S2-final-viral-combined_k141_16152\|\|full | 3085 | No | NA | 4 | 0 | 3 | Not-determined | Genome-fragment | NA | NA | 0 |
| S2-final-viral-combined_k141_49073\|\|full | 7931 | No | NA | 12 | 0 | 8 | Not-determined | Genome-fragment | NA | NA | 0 |
| S2-final-viral-combined_k141_172306\|\|full | 2372 | No | NA | 3 | 0 | 3 | Not-determined | Genome-fragment | NA | NA | 0 |
| S1-final-viral-combined_k141_354987\|\|full | 3929 | No | NA | 3 | 0 | 2 | Low-quality | Genome-fragment | 1.73 | AAI-based (medium-confidence) | 0 |
| S2-final-viral-combined_k141_109123\|\|full | 1646 | No | NA | 3 | 0 | 1 | Not-determined | Genome-fragment | NA | NA | 0 |
| S2-final-viral-combined_k141_25606\|\|full | 1869 | No | NA | 3 | 0 | 0 | Not-determined | Genome-fragment | NA | NA | 0 |
| S2-final-viral-combined_k141_150934\|\|full | 2472 | No | NA | 2 | 0 | 1 | Not-determined | Genome-fragment | NA | NA | 0 |
| S7_k141_93684 | 3930 | No | NA | 6 | 1 | 0 | Low-quality | Genome-fragment | 11.12 | AAI-based (high-confidence) | 0 |
| S17-final-viral-combined_k141_201609\|\|full | 3930 | Yes | 1628 | 5 | 1 | 1 | Low-quality | Genome-fragment | 1.32 | AAI-based (medium-confidence) | 58.58 |
| S2-final-viral-combined_k141_192167\|\|full | 1901 | No | NA | 2 | 0 | 2 | Not-determined | Genome-fragment | NA | NA | 0 |
| S2-final-viral-combined_k141_170486\|\|full | 3930 | No | NA | 3 | 0 | 2 | Low-quality | Genome-fragment | 3.26 | AAI-based (medium-confidence) | 0 |
| S2-final-viral-combined_k141_120028\|\|full | 5178 | No | NA | 3 | 0 | 3 | Not-determined | Genome-fragment | NA | NA | 0 |
| S3-final-viral-combined_k141_122955\|\|full | 3931 | No | NA | 5 | 0 | 0 | Low-quality | Genome-fragment | 6.82 | AAI-based (high-confidence) | 0 |
| S17-k141_112294 | 3931 | No | NA | 7 | 1 | 0 | Low-quality | Genome-fragment | 5.33 | AAI-based (high-confidence) | 0 |
| S2-final-viral-combined_k141_78578\|\|full | 32898 | No | NA | 26 | 0 | 18 | Not-determined | Genome-fragment | NA | NA | 0 |
| S6_k141_125554 | 3932 | No | NA | 4 | 0 | 3 | Low-quality | Genome-fragment | 0.76 | AAI-based (medium-confidence) | 0 |
| S2-final-viral-combined_k141_25646\|\|full | 11166 | No | NA | 16 | 0 | 3 | Not-determined | Genome-fragment | NA | NA | 0 |
| S2-final-viral-combined_k141_172422\|\|full | 1293 | No | NA | 3 | 0 | 0 | Not-determined | Genome-fragment | NA | NA | 0 |
| S8-final-viral-combined_k141_153683\|\|full | 3932 | No | NA | 8 | 1 | 0 | Low-quality | Genome-fragment | 4.32 | HMM-based (lower-bound) | 0 |
| S2-final-viral-combined_k141_99446\|\|full | 43104 | Yes | 18481 | 49 | 18 | 15 | Medium-quality | Genome-fragment | 51.83 | AAI-based (high-confidence) | 57.12 |
| S2-final-viral-combined_k141_57914\|\|full | 13451 | No | NA | 12 | 0 | 11 | Not-determined | Genome-fragment | NA | NA | 0 |
| S5-final-viral-combined_k141_7478\|\|lt2gene | 3933 | No | NA | 1 | 1 | 0 | Low-quality | Genome-fragment | 10.66 | AAI-based (medium-confidence) | 0 |
| S1-final-viral-combined_k141_65362\|\|full | 3937 | No | NA | 4 | 0 | 2 | Low-quality | Genome-fragment | 1.99 | AAI-based (medium-confidence) | 0 |
| S2-final-viral-combined_k141_192212\|\|full | 1971 | No | NA | 2 | 0 | 2 | Not-determined | Genome-fragment | NA | NA | 0 |
| S2-final-viral-combined_k141_120091\|\|full | 1986 | No | NA | 3 | 0 | 1 | Not-determined | Genome-fragment | NA | NA | 0 |
| S12-final-viral-combined_k141_125960\|\|full | 3937 | No | NA | 3 | 0 | 3 | Low-quality | Genome-fragment | 1.63 | AAI-based (medium-confidence) | 0 |
| S12_k141_170699 | 3937 | No | NA | 6 | 0 | 1 | Low-quality | Genome-fragment | 7.81 | AAI-based (medium-confidence) | 0 |
| S2-final-viral-combined_k141_172481\|\|full | 3764 | No | NA | 4 | 0 | 2 | Not-determined | Genome-fragment | NA | NA | 0 |
| S2-final-viral-combined_k141_14310\|\|full | 3937 | No | NA | 13 | 1 | 0 | Low-quality | Genome-fragment | 9.59 | AAI-based (high-confidence) | 0 |
| S6-final-viral-combined_k141_37573\|\|full | 3937 | No | NA | 4 | 0 | 2 | Low-quality | Genome-fragment | 3.15 | AAI-based (medium-confidence) | 0 |
| S2-final-viral-combined_k141_243050\|\|full | 1792 | No | NA | 3 | 0 | 0 | Not-determined | Genome-fragment | NA | NA | 0 |
| S2-final-viral-combined_k141_99461\|\|full | 1100 | No | NA | 3 | 0 | 1 | Not-determined | Genome-fragment | NA | NA | 0 |
| S8-final-viral-combined_k141_235053\|\|full | 3937 | No | NA | 4 | 0 | 0 | Low-quality | Genome-fragment | 7.98 | AAI-based (medium-confidence) | 0 |
| S2-final-viral-combined_k141_243091\|\|full | 2760 | No | NA | 2 | 0 | 1 | Not-determined | Genome-fragment | NA | NA | 0 |
| S2-final-viral-combined_k141_120171\|\|full | 7252 | No | NA | 6 | 0 | 3 | Not-determined | Genome-fragment | NA | NA | 0 |
| S2-final-viral-combined_k141_129085\|\|full | 1182 | No | NA | 2 | 0 | 1 | Not-determined | Genome-fragment | NA | NA | 0 |
| S2-final-viral-combined_k141_120181\|\|full | 2504 | No | NA | 2 | 0 | 1 | Not-determined | Genome-fragment | NA | NA | 0 |
| S1-final-viral-combined_k141_329924\|\|full | 3940 | Yes | 2614 | 9 | 2 | 1 | Low-quality | Genome-fragment | 6.6 | AAI-based (medium-confidence) | 33.65 |
| S2-final-viral-combined_k141_49291\|\|full | 6783 | No | NA | 9 | 0 | 4 | Not-determined | Genome-fragment | NA | NA | 0 |
| S17-final-viral-combined_k141_206105\|\|full | 3940 | No | NA | 9 | 2 | 0 | Low-quality | Genome-fragment | 5.58 | HMM-based (lower-bound) | 0 |
| S2-final-viral-combined_k141_172584\|\|full | 1255 | No | NA | 2 | 0 | 1 | Not-determined | Genome-fragment | NA | NA | 0 |
| S17-k141_250448 | 3941 | No | NA | 8 | 6 | 0 | Low-quality | Genome-fragment | 12.99 | AAI-based (high-confidence) | 0 |
| S2-final-viral-combined_k141_120197\|\|full | 1895 | No | NA | 2 | 0 | 2 | Not-determined | Genome-fragment | NA | NA | 0 |
| S2-final-viral-combined_k141_25727\|\|full | 57878 | Yes | 24597 | 79 | 15 | 23 | Medium-quality | Genome-fragment | 66.52 | AAI-based (high-confidence) | 57.5 |
| S2-final-viral-combined_k141_160777\|\|full | 1514 | No | NA | 2 | 0 | 2 | Not-determined | Genome-fragment | NA | NA | 0 |
| S4-final-viral-combined_k141_93375\|\|full | 3941 | No | NA | 5 | 0 | 0 | Low-quality | Genome-fragment | 10.45 | AAI-based (high-confidence) | 0 |
| S2-final-viral-combined_k141_16288\|\|full | 7141 | No | NA | 5 | 0 | 4 | Not-determined | Genome-fragment | NA | NA | 0 |
| S2-final-viral-combined_k141_99542\|\|full | 1822 | No | NA | 3 | 0 | 0 | Not-determined | Genome-fragment | NA | NA | 0 |
| S2-final-viral-combined_k141_109217\|\|full | 1568 | No | NA | 2 | 0 | 0 | Not-determined | Genome-fragment | NA | NA | 0 |
| S8-final-viral-combined_k141_98983\|\|full | 3941 | No | NA | 5 | 0 | 0 | Low-quality | Genome-fragment | 7.21 | AAI-based (medium-confidence) | 0 |
| S11-final-viral-combined_k141_84739\|\|full | 3942 | No | NA | 7 | 0 | 3 | Low-quality | Genome-fragment | 1.62 | AAI-based (medium-confidence) | 0 |
| S2-final-viral-combined_k141_109238\|\|full | 1493 | No | NA | 4 | 0 | 0 | Not-determined | Genome-fragment | NA | NA | 0 |
| S2-final-viral-combined_k141_58013\|\|full | 2209 | No | NA | 4 | 0 | 3 | Not-determined | Genome-fragment | NA | NA | 0 |
| S2-final-viral-combined_k141_109245\|\|full | 6484 | No | NA | 9 | 0 | 2 | Not-determined | Genome-fragment | NA | NA | 0 |
| S2-final-viral-combined_k141_16312\|\|full | 1941 | No | NA | 3 | 0 | 1 | Not-determined | Genome-fragment | NA | NA | 0 |
| S2-final-viral-combined_k141_109249\|\|full | 1438 | No | NA | 3 | 0 | 1 | Not-determined | Genome-fragment | NA | NA | 0 |
| S11-final-viral-combined_k141_142552\|\|full | 3943 | No | NA | 6 | 0 | 4 | Low-quality | Genome-fragment | 2.27 | AAI-based (medium-confidence) | 0 |
| S2-final-viral-combined_k141_78668\|\|full | 2360 | No | NA | 3 | 0 | 2 | Not-determined | Genome-fragment | NA | NA | 0 |
| S2-final-viral-combined_k141_89393\|\|full | 1124 | No | NA | 2 | 0 | 2 | Not-determined | Genome-fragment | NA | NA | 0 |
| S12-final-viral-combined_k141_134507\|\|full | 3943 | No | NA | 7 | 0 | 0 | Low-quality | Genome-fragment | 7.02 | AAI-based (medium-confidence) | 0 |
| S2-final-viral-combined_k141_78670\|\|full | 33299 | No | NA | 49 | 8 | 2 | Medium-quality | Genome-fragment | 72.69 | AAI-based (high-confidence) | 0 |
| S2-final-viral-combined_k141_201632\|\|full | 1165 | No | NA | 3 | 0 | 1 | Not-determined | Genome-fragment | NA | NA | 0 |
| S2-final-viral-combined_k141_89419\|\|full | 2311 | No | NA | 4 | 0 | 3 | Not-determined | Genome-fragment | NA | NA | 0 |
| S1_k141_27445 | 3944 | No | NA | 5 | 0 | 1 | Low-quality | Genome-fragment | 10.89 | AAI-based (medium-confidence) | 0 |
| S2-final-viral-combined_k141_4884\|\|full | 4561 | No | NA | 4 | 0 | 3 | Not-determined | Genome-fragment | NA | NA | 0 |
| S2-final-viral-combined_k141_67984\|\|full | 8671 | No | NA | 6 | 0 | 5 | Not-determined | Genome-fragment | NA | NA | 0 |
| S2-final-viral-combined_k141_172729\|\|full | 6270 | No | NA | 5 | 0 | 5 | Not-determined | Genome-fragment | NA | NA | 0 |
| S6_k141_76398 | 3945 | No | NA | 9 | 1 | 0 | Low-quality | Genome-fragment | 9.39 | HMM-based (lower-bound) | 0 |
| S8-final-viral-combined_k141_150949\|\|full | 3945 | Yes | 2193 | 6 | 1 | 1 | Low-quality | Genome-fragment | 3.55 | AAI-based (high-confidence) | 44.41 |
| S2-final-viral-combined_k141_243234\|\|full | 1675 | No | NA | 2 | 0 | 2 | Not-determined | Genome-fragment | NA | NA | 0 |
| S7_k141_224652 | 3946 | No | NA | 6 | 0 | 0 | Low-quality | Genome-fragment | 6.59 | AAI-based (high-confidence) | 0 |
| S2-final-viral-combined_k141_4894\|\|full | 4783 | No | NA | 5 | 0 | 0 | Not-determined | Genome-fragment | NA | NA | 0 |
| S2-final-viral-combined_k141_49408\|\|full | 1718 | No | NA | 2 | 0 | 0 | Not-determined | Genome-fragment | NA | NA | 0 |
| S2-final-viral-combined_k141_68003\|\|full | 1075 | No | NA | 3 | 0 | 0 | Not-determined | Genome-fragment | NA | NA | 0 |
| S17-k141_226374 | 3946 | No | NA | 3 | 0 | 1 | Low-quality | Genome-fragment | 7.51 | AAI-based (medium-confidence) | 0 |
| S2-final-viral-combined_k141_222185\|\|full | 1086 | No | NA | 3 | 0 | 1 | Not-determined | Genome-fragment | NA | NA | 0 |
| S11-final-viral-combined_k141_174859\|\|lt2gene | 3947 | No | NA | 2 | 1 | 0 | Low-quality | Genome-fragment | 6.85 | HMM-based (lower-bound) | 0 |
| S3-final-viral-combined_k141_134727\|\|full | 3947 | No | NA | 5 | 1 | 0 | Low-quality | Genome-fragment | 7.22 | AAI-based (high-confidence) | 0 |
| S2-final-viral-combined_k141_16389\|\|full | 1958 | No | NA | 2 | 0 | 1 | Not-determined | Genome-fragment | NA | NA | 0 |
| S7-final-viral-combined_k141_201074\|\|full | 3947 | No | NA | 2 | 0 | 0 | Low-quality | Genome-fragment | 1.76 | AAI-based (medium-confidence) | 0 |
| S2-final-viral-combined_k141_68033\|\|full | 5590 | No | NA | 7 | 0 | 1 | Not-determined | Genome-fragment | NA | NA | 0 |
| S2-final-viral-combined_k141_201691\|\|full | 5415 | No | NA | 2 | 0 | 2 | Not-determined | Genome-fragment | NA | NA | 0 |
| S2-final-viral-combined_k141_234407\|\|full | 1306 | No | NA | 2 | 0 | 1 | Not-determined | Genome-fragment | NA | NA | 0 |
| S8-final-viral-combined_k141_35288\|\|full | 3947 | No | NA | 4 | 1 | 0 | Low-quality | Genome-fragment | 2.65 | HMM-based (lower-bound) | 0 |
| S9-final-viral-combined_k141_75877\|\|full | 3949 | No | NA | 4 | 2 | 0 | Low-quality | Genome-fragment | 8.41 | AAI-based (high-confidence) | 0 |
| S1-final-viral-combined_k141_382092\|\|full | 3949 | No | NA | 3 | 0 | 0 | Low-quality | Genome-fragment | 25.51 | AAI-based (high-confidence) | 0 |
| S1_k141_9033 | 3949 | No | NA | 5 | 0 | 0 | Low-quality | Genome-fragment | 9.42 | AAI-based (high-confidence) | 0 |
| S2-final-viral-combined_k141_68058\|\|full | 2094 | No | NA | 3 | 0 | 0 | Not-determined | Genome-fragment | NA | NA | 0 |
| S2-final-viral-combined_k141_58121\|\|full | 27276 | No | NA | 31 | 0 | 16 | Medium-quality | Genome-fragment | 62.68 | AAI-based (medium-confidence) | 0 |
| S2-final-viral-combined_k141_172851\|\|full | 1810 | No | NA | 3 | 0 | 3 | Not-determined | Genome-fragment | NA | NA | 0 |
| S2-final-viral-combined_k141_192398\|\|full | 5201 | No | NA | 4 | 0 | 1 | Not-determined | Genome-fragment | NA | NA | 0 |
| S2-final-viral-combined_k141_49506\|\|full | 1820 | No | NA | 3 | 0 | 0 | Not-determined | Genome-fragment | NA | NA | 0 |
| S17-final-viral-combined_k141_255759\|\|full | 3949 | No | NA | 2 | 0 | 1 | Low-quality | Genome-fragment | 3.7 | AAI-based (medium-confidence) | 0 |
| S9_k141_78576 | 3950 | No | NA | 4 | 0 | 2 | Low-quality | Genome-fragment | 1.41 | AAI-based (medium-confidence) | 0 |
| S17-k141_110513 | 3950 | No | NA | 6 | 0 | 1 | Low-quality | Genome-fragment | 6.7 | AAI-based (medium-confidence) | 0 |
| S12_k141_185715 | 3950 | No | NA | 1 | 0 | 1 | Low-quality | Genome-fragment | 3.59 | AAI-based (high-confidence) | 0 |
| S2-final-viral-combined_k141_192418\|\|full | 2729 | No | NA | 3 | 0 | 2 | Not-determined | Genome-fragment | NA | NA | 0 |
| S2-final-viral-combined_k141_25914\|\|full | 2371 | No | NA | 2 | 0 | 1 | Not-determined | Genome-fragment | NA | NA | 0 |
| S2-final-viral-combined_k141_172892\|\|full | 1703 | No | NA | 2 | 0 | 1 | Not-determined | Genome-fragment | NA | NA | 0 |
| S2-final-viral-combined_k141_89509\|\|full | 4677 | No | NA | 6 | 0 | 6 | Not-determined | Genome-fragment | NA | NA | 0 |
| S2-final-viral-combined_k141_140996\|\|full | 243800 | Yes | 52444 | 279 | 31 | 125 | Complete | High-quality | 100 | Provirus (medium-confidence) | 78.49 |
| S2-final-viral-combined_k141_4925\|\|full | 1511 | No | NA | 2 | 0 | 1 | Not-determined | Genome-fragment | NA | NA | 0 |
| S2-final-viral-combined_k141_151373\|\|full | 1691 | No | NA | 2 | 0 | 1 | Not-determined | Genome-fragment | NA | NA | 0 |
| S2-final-viral-combined_k141_172914\|\|full | 2094 | No | NA | 2 | 0 | 1 | Not-determined | Genome-fragment | NA | NA | 0 |
| S4_k141_6681 | 3950 | No | NA | 7 | 0 | 2 | Low-quality | Genome-fragment | 4.24 | AAI-based (medium-confidence) | 0 |
| S2-final-viral-combined_k141_25941\|\|full | 2276 | No | NA | 3 | 0 | 0 | Not-determined | Genome-fragment | NA | NA | 0 |
| S12_k141_265338 | 3951 | No | NA | 1 | 0 | 1 | Low-quality | Genome-fragment | 2.13 | AAI-based (medium-confidence) | 0 |
| S11-final-viral-combined_k141_89377\|\|full | 3952 | Yes | 1229 | 4 | 1 | 2 | Low-quality | Genome-fragment | 2.81 | AAI-based (medium-confidence) | 68.9 |
| S5-final-viral-combined_k141_197043\|\|full | 3952 | No | NA | 5 | 0 | 1 | Low-quality | Genome-fragment | 4.81 | AAI-based (medium-confidence) | 0 |
| S2-final-viral-combined_k141_160969\|\|full | 10453 | No | NA | 5 | 0 | 3 | Not-determined | Genome-fragment | NA | NA | 0 |
| S7_k141_196585 | 3953 | No | NA | 2 | 0 | 2 | Low-quality | Genome-fragment | 6.54 | AAI-based (medium-confidence) | 0 |
| S2-final-viral-combined_k141_99775\|\|full | 1357 | No | NA | 4 | 0 | 2 | Not-determined | Genome-fragment | NA | NA | 0 |
| S2-final-viral-combined_k141_120333\|\|full | 2164 | No | NA | 5 | 0 | 0 | Not-determined | Genome-fragment | NA | NA | 0 |
| S2-final-viral-combined_k141_58210\|\|full | 2367 | No | NA | 6 | 0 | 0 | Not-determined | Genome-fragment | NA | NA | 0 |
| S12-final-viral-combined_k141_279970\|\|full | 3953 | No | NA | 8 | 3 | 0 | Low-quality | Genome-fragment | 9.69 | AAI-based (high-confidence) | 0 |
| S2-final-viral-combined_k141_89572\|\|full | 1934 | No | NA | 2 | 0 | 1 | Not-determined | Genome-fragment | NA | NA | 0 |
| S2-final-viral-combined_k141_120359\|\|full | 2316 | No | NA | 3 | 0 | 1 | Not-determined | Genome-fragment | NA | NA | 0 |
| S2-final-viral-combined_k141_141077\|\|full | 1975 | No | NA | 4 | 0 | 1 | Not-determined | Genome-fragment | NA | NA | 0 |
| S2-final-viral-combined_k141_243322\|\|full | 1723 | No | NA | 2 | 0 | 1 | Not-determined | Genome-fragment | NA | NA | 0 |
| S2-final-viral-combined_k141_234563\|\|full | 1323 | No | NA | 3 | 0 | 1 | Not-determined | Genome-fragment | NA | NA | 0 |
| S2-final-viral-combined_k141_58232\|\|full | 7679 | No | NA | 5 | 0 | 3 | Not-determined | Genome-fragment | NA | NA | 0 |
| S6_k141_131685 | 3953 | No | NA | 5 | 0 | 0 | Low-quality | Genome-fragment | 5.91 | AAI-based (high-confidence) | 0 |
| S2-final-viral-combined_k141_109462\|\|full | 1209 | No | NA | 5 | 0 | 1 | Not-determined | Genome-fragment | NA | NA | 0 |
| S1_k141_7909 | 3954 | No | NA | 5 | 0 | 0 | Low-quality | Genome-fragment | 4.71 | AAI-based (medium-confidence) | 0 |
| S2-final-viral-combined_k141_192509\|\|full | 1489 | No | NA | 3 | 0 | 1 | Not-determined | Genome-fragment | NA | NA | 0 |
| S2-final-viral-combined_k141_173086\|\|full | 2540 | No | NA | 4 | 0 | 0 | Not-determined | Genome-fragment | NA | NA | 0 |
| S5-final-viral-combined_k141_192570\|\|full | 3955 | No | NA | 11 | 3 | 0 | Low-quality | Genome-fragment | 3.24 | HMM-based (lower-bound) | 0 |
| S4_k141_70887 | 3955 | No | NA | 3 | 0 | 2 | Low-quality | Genome-fragment | 1.69 | AAI-based (medium-confidence) | 0 |
| S2-final-viral-combined_k141_5015\|\|full | 1478 | No | NA | 2 | 0 | 0 | Not-determined | Genome-fragment | NA | NA | 0 |
| S1-final-viral-combined_k141_4164\|\|full | 3956 | No | NA | 6 | 4 | 0 | Low-quality | Genome-fragment | 9.78 | AAI-based (high-confidence) | 0 |
| S5-final-viral-combined_k141_179394\|\|full | 3956 | No | NA | 7 | 1 | 0 | Low-quality | Genome-fragment | 9.77 | AAI-based (high-confidence) | 0 |
| S2-final-viral-combined_k141_5024\|\|full | 1163 | No | NA | 3 | 0 | 0 | Not-determined | Genome-fragment | NA | NA | 0 |
| S2-final-viral-combined_k141_68198\|\|full | 9320 | No | NA | 9 | 0 | 6 | Not-determined | Genome-fragment | NA | NA | 0 |
| S2-final-viral-combined_k141_156648\|\|full | 3956 | No | NA | 4 | 2 | 0 | Low-quality | Genome-fragment | 19.81 | AAI-based (high-confidence) | 0 |
| S2-final-viral-combined_k141_129371\|\|full | 2399 | No | NA | 3 | 0 | 0 | Not-determined | Genome-fragment | NA | NA | 0 |
| S11_k141_326764 | 3957 | No | NA | 7 | 6 | 0 | Low-quality | Genome-fragment | 10.94 | AAI-based (high-confidence) | 0 |
| S12-final-viral-combined_k141_328801\|\|full | 3957 | No | NA | 3 | 0 | 2 | Low-quality | Genome-fragment | 5.25 | AAI-based (medium-confidence) | 0 |
| S17-final-viral-combined_k141_91392\|\|full | 3958 | No | NA | 9 | 0 | 0 | Low-quality | Genome-fragment | 6.63 | AAI-based (high-confidence) | 0 |
| S2-final-viral-combined_k141_161060\|\|full | 2151 | No | NA | 4 | 0 | 0 | Not-determined | Genome-fragment | NA | NA | 0 |
| S8_k141_6710 | 3958 | No | NA | 5 | 0 | 1 | Low-quality | Genome-fragment | 6.53 | AAI-based (medium-confidence) | 0 |
| S2-final-viral-combined_k141_49652\|\|full | 2682 | No | NA | 2 | 0 | 2 | Not-determined | Genome-fragment | NA | NA | 0 |
| S2-final-viral-combined_k141_99890\|\|full | 35924 | No | NA | 22 | 0 | 17 | Not-determined | Genome-fragment | NA | NA | 0 |
| S2-final-viral-combined_k141_99910\|\|full | 1231 | No | NA | 3 | 0 | 0 | Not-determined | Genome-fragment | NA | NA | 0 |
| S2-final-viral-combined_k141_182801\|\|full | 2407 | No | NA | 4 | 0 | 0 | Not-determined | Genome-fragment | NA | NA | 0 |
| S2-final-viral-combined_k141_120457\|\|full | 3872 | No | NA | 4 | 0 | 3 | Not-determined | Genome-fragment | NA | NA | 0 |
| S1_k141_233150 | 3959 | No | NA | 6 | 0 | 1 | Low-quality | Genome-fragment | 5.86 | AAI-based (medium-confidence) | 0 |
| S17-k141_147973 | 3959 | No | NA | 5 | 1 | 0 | Low-quality | Genome-fragment | 9.56 | AAI-based (medium-confidence) | 0 |
| S2-final-viral-combined_k141_234725\|\|full | 4475 | No | NA | 2 | 0 | 1 | Not-determined | Genome-fragment | NA | NA | 0 |
| S7-final-viral-combined_k141_102064\|\|full | 3960 | No | NA | 5 | 0 | 1 | Low-quality | Genome-fragment | 2.6 | AAI-based (high-confidence) | 0 |
| S4_k141_72890 | 3960 | No | NA | 1 | 0 | 1 | Low-quality | Genome-fragment | 7.21 | AAI-based (medium-confidence) | 0 |
| S1-final-viral-combined_k141_220987\|\|full | 3961 | Yes | 2063 | 7 | 1 | 1 | Low-quality | Genome-fragment | 1.88 | HMM-based (lower-bound) | 47.92 |
| S2-final-viral-combined_k141_49732\|\|full | 1245 | No | NA | 2 | 0 | 0 | Not-determined | Genome-fragment | NA | NA | 0 |
| S2-final-viral-combined_k141_99957\|\|full | 1704 | No | NA | 3 | 0 | 1 | Not-determined | Genome-fragment | NA | NA | 0 |
| S2-final-viral-combined_k141_129488\|\|full | 9261 | No | NA | 8 | 0 | 5 | Not-determined | Genome-fragment | NA | NA | 0 |
| S2-final-viral-combined_k141_243366\|\|full | 1154 | No | NA | 3 | 0 | 2 | Not-determined | Genome-fragment | NA | NA | 0 |
| S2-final-viral-combined_k141_234780\|\|full | 3950 | No | NA | 7 | 0 | 0 | Not-determined | Genome-fragment | NA | NA | 0 |
| S2-final-viral-combined_k141_173195\|\|full | 1194 | No | NA | 2 | 0 | 1 | Not-determined | Genome-fragment | NA | NA | 0 |
| S17-k141_187345 | 3961 | No | NA | 4 | 0 | 0 | Low-quality | Genome-fragment | 6.86 | AAI-based (medium-confidence) | 0 |
| S12_k141_335424 | 3961 | Yes | 2612 | 3 | 1 | 1 | Low-quality | Genome-fragment | 6.57 | AAI-based (high-confidence) | 34.06 |
| S2-final-viral-combined_k141_120569\|\|full | 2266 | No | NA | 2 | 0 | 2 | Not-determined | Genome-fragment | NA | NA | 0 |
| S2-final-viral-combined_k141_89734\|\|full | 1469 | No | NA | 3 | 0 | 0 | Not-determined | Genome-fragment | NA | NA | 0 |
| S2-final-viral-combined_k141_195714\|\|full | 3961 | No | NA | 6 | 0 | 0 | Low-quality | Genome-fragment | 10.1 | AAI-based (medium-confidence) | 0 |
| S7_k141_151612 | 3963 | No | NA | 4 | 1 | 0 | Low-quality | Genome-fragment | 9.45 | AAI-based (high-confidence) | 0 |
| S2-final-viral-combined_k141_120603\|\|full | 1794 | No | NA | 4 | 0 | 0 | Not-determined | Genome-fragment | NA | NA | 0 |
| S2-final-viral-combined_k141_219315\|\|full | 3963 | Yes | 2060 | 4 | 1 | 1 | Low-quality | Genome-fragment | 5.03 | AAI-based (high-confidence) | 48.02 |
| S2-final-viral-combined_k141_89742\|\|full | 24414 | No | NA | 36 | 15 | 0 | Medium-quality | Genome-fragment | 63.53 | AAI-based (high-confidence) | 0 |
| S2-final-viral-combined_k141_49828\|\|full | 1429 | No | NA | 2 | 0 | 1 | Not-determined | Genome-fragment | NA | NA | 0 |
| S2-final-viral-combined_k141_182935\|\|full | 1843 | No | NA | 3 | 0 | 1 | Not-determined | Genome-fragment | NA | NA | 0 |
| S8_k141_154873 | 3964 | No | NA | 9 | 3 | 1 | Low-quality | Genome-fragment | 12.18 | AAI-based (high-confidence) | 0 |
| S5-final-viral-combined_k141_92477\|\|full | 3965 | No | NA | 4 | 0 | 2 | Low-quality | Genome-fragment | 1.26 | AAI-based (medium-confidence) | 0 |
| S12-final-viral-combined_k141_168141\|\|full | 3965 | No | NA | 6 | 1 | 0 | Low-quality | Genome-fragment | 4.99 | HMM-based (lower-bound) | 0 |
| S1-final-viral-combined_k141_40900\|\|full | 3966 | No | NA | 10 | 2 | 0 | Low-quality | Genome-fragment | 10.74 | AAI-based (high-confidence) | 0 |
| S1_k141_51893 | 3966 | No | NA | 6 | 0 | 0 | Low-quality | Genome-fragment | 6.62 | AAI-based (high-confidence) | 0 |
| S2-final-viral-combined_k141_182958\|\|full | 1504 | No | NA | 2 | 0 | 0 | Not-determined | Genome-fragment | NA | NA | 0 |
| S2-final-viral-combined_k141_202025\|\|full | 1889 | No | NA | 3 | 0 | 0 | Not-determined | Genome-fragment | NA | NA | 0 |
| S8-final-viral-combined_k141_3670\|\|full | 3966 | Yes | 2724 | 7 | 4 | 1 | Low-quality | Genome-fragment | 5.56 | AAI-based (high-confidence) | 31.32 |
| S2-final-viral-combined_k141_58366\|\|full | 4421 | No | NA | 4 | 0 | 2 | Not-determined | Genome-fragment | NA | NA | 0 |
| S2-final-viral-combined_k141_89806\|\|full | 2261 | No | NA | 4 | 0 | 0 | Not-determined | Genome-fragment | NA | NA | 0 |
| S2-final-viral-combined_k141_173283\|\|full | 4107 | No | NA | 4 | 0 | 1 | Not-determined | Genome-fragment | NA | NA | 0 |
| S2-final-viral-combined_k141_89814\|\|full | 2762 | No | NA | 3 | 0 | 3 | Not-determined | Genome-fragment | NA | NA | 0 |
| S8_k141_4046 | 3966 | No | NA | 6 | 2 | 0 | Low-quality | Genome-fragment | 6.43 | AAI-based (high-confidence) | 0 |
| S2-final-viral-combined_k141_222535\|\|full | 38886 | No | NA | 45 | 2 | 2 | Medium-quality | Genome-fragment | 56.54 | HMM-based (lower-bound) | 0 |
| S2-final-viral-combined_k141_89825\|\|full | 3158 | No | NA | 3 | 0 | 1 | Not-determined | Genome-fragment | NA | NA | 0 |
| S2-final-viral-combined_k141_65109\|\|full | 3967 | No | NA | 7 | 2 | 0 | Low-quality | Genome-fragment | 9.34 | AAI-based (high-confidence) | 0 |
| S2-final-viral-combined_k141_109655\|\|full | 1928 | No | NA | 3 | 0 | 0 | Not-determined | Genome-fragment | NA | NA | 0 |
| S2-final-viral-combined_k141_68331\|\|full | 2761 | No | NA | 5 | 0 | 0 | Not-determined | Genome-fragment | NA | NA | 0 |
| S2-final-viral-combined_k141_222556\|\|full | 3716 | No | NA | 6 | 0 | 1 | Not-determined | Genome-fragment | NA | NA | 0 |
| S3-final-viral-combined_k141_104495\|\|lt2gene | 3968 | No | NA | 1 | 1 | 0 | Low-quality | Genome-fragment | 6.34 | AAI-based (high-confidence) | 0 |
| S2-final-viral-combined_k141_109666\|\|full | 1600 | No | NA | 3 | 0 | 1 | Not-determined | Genome-fragment | NA | NA | 0 |
| S2_k141_178298 | 3969 | No | NA | 8 | 2 | 0 | Low-quality | Genome-fragment | 3.97 | AAI-based (medium-confidence) | 0 |
| S4_k141_75306 | 3969 | No | NA | 4 | 1 | 0 | Low-quality | Genome-fragment | 3.58 | AAI-based (high-confidence) | 0 |
| S2-final-viral-combined_k141_141268\|\|full | 1329 | No | NA | 2 | 0 | 2 | Not-determined | Genome-fragment | NA | NA | 0 |
| S2-final-viral-combined_k141_58482\|\|full | 11359 | No | NA | 12 | 0 | 7 | Not-determined | Genome-fragment | NA | NA | 0 |
| S2-final-viral-combined_k141_151818\|\|full | 2893 | No | NA | 5 | 0 | 2 | Not-determined | Genome-fragment | NA | NA | 0 |
| S4_k141_102708 | 3969 | No | NA | 13 | 2 | 0 | Low-quality | Genome-fragment | 5.03 | AAI-based (medium-confidence) | 0 |
| S2-final-viral-combined_k141_173426\|\|full | 1757 | No | NA | 3 | 0 | 0 | Not-determined | Genome-fragment | NA | NA | 0 |
| S2-final-viral-combined_k141_151847\|\|full | 1312 | No | NA | 4 | 0 | 0 | Not-determined | Genome-fragment | NA | NA | 0 |
| S9-final-viral-combined_k141_77243\|\|full | 3970 | No | NA | 4 | 1 | 0 | Low-quality | Genome-fragment | 11.03 | AAI-based (medium-confidence) | 0 |
| S3-final-viral-combined_k141_30245\|\|full | 3970 | No | NA | 3 | 0 | 3 | Low-quality | Genome-fragment | 2.34 | AAI-based (medium-confidence) | 0 |
| S2-final-viral-combined_k141_173446\|\|full | 5384 | No | NA | 11 | 0 | 1 | Not-determined | Genome-fragment | NA | NA | 0 |
| S2-final-viral-combined_k141_17005\|\|full | 3311 | No | NA | 4 | 0 | 3 | Not-determined | Genome-fragment | NA | NA | 0 |
| S17-final-viral-combined_k141_81496\|\|full | 3970 | No | NA | 5 | 0 | 5 | Low-quality | Genome-fragment | 2.76 | AAI-based (high-confidence) | 0 |
| S8-final-viral-combined_k141_175622\|\|full | 3970 | No | NA | 3 | 0 | 3 | Low-quality | Genome-fragment | 2.8 | AAI-based (medium-confidence) | 0 |
| S12_k141_288124 | 3971 | No | NA | 3 | 0 | 2 | Low-quality | Genome-fragment | 5.82 | AAI-based (medium-confidence) | 0 |
| S2-final-viral-combined_k141_234897\|\|full | 1937 | No | NA | 3 | 0 | 3 | Not-determined | Genome-fragment | NA | NA | 0 |
| S2-final-viral-combined_k141_173490\|\|full | 2373 | No | NA | 4 | 0 | 2 | Not-determined | Genome-fragment | NA | NA | 0 |
| S2-final-viral-combined_k141_26136\|\|full | 2578 | No | NA | 3 | 0 | 1 | Not-determined | Genome-fragment | NA | NA | 0 |
| S8-final-viral-combined_k141_6451\|\|full | 3971 | No | NA | 6 | 1 | 0 | Low-quality | Genome-fragment | 4.86 | HMM-based (lower-bound) | 0 |
| S2-final-viral-combined_k141_243479\|\|full | 1877 | No | NA | 3 | 0 | 0 | Not-determined | Genome-fragment | NA | NA | 0 |
| S2-final-viral-combined_k141_202117\|\|full | 5713 | No | NA | 4 | 0 | 2 | Not-determined | Genome-fragment | NA | NA | 0 |
| S2-final-viral-combined_k141_68414\|\|full | 3566 | No | NA | 3 | 0 | 2 | Not-determined | Genome-fragment | NA | NA | 0 |
| S2-final-viral-combined_k141_202136\|\|full | 2279 | No | NA | 3 | 0 | 1 | Not-determined | Genome-fragment | NA | NA | 0 |
| S2-final-viral-combined_k141_17046\|\|full | 5515 | No | NA | 10 | 0 | 0 | Not-determined | Genome-fragment | NA | NA | 0 |
| S2-final-viral-combined_k141_37916\|\|full | 1522 | No | NA | 2 | 0 | 1 | Not-determined | Genome-fragment | NA | NA | 0 |
| S2-final-viral-combined_k141_213082\|\|full | 1652 | No | NA | 3 | 0 | 0 | Not-determined | Genome-fragment | NA | NA | 0 |
| S1_k141_185976 | 3972 | No | NA | 4 | 0 | 0 | Low-quality | Genome-fragment | 3.7 | AAI-based (medium-confidence) | 0 |
| S2-final-viral-combined_k141_100192\|\|full | 14501 | No | NA | 22 | 0 | 3 | Not-determined | Genome-fragment | NA | NA | 0 |
| S17-final-viral-combined_k141_248283\|\|full | 3972 | No | NA | 4 | 0 | 1 | Low-quality | Genome-fragment | 2.87 | AAI-based (high-confidence) | 0 |
| S2-final-viral-combined_k141_202192\|\|full | 2338 | No | NA | 3 | 0 | 2 | Not-determined | Genome-fragment | NA | NA | 0 |
| S2-final-viral-combined_k141_151917\|\|full | 1123 | No | NA | 2 | 0 | 1 | Not-determined | Genome-fragment | NA | NA | 0 |
| S2-final-viral-combined_k141_213111\|\|full | 6280 | No | NA | 5 | 0 | 3 | Not-determined | Genome-fragment | NA | NA | 0 |
| S17-final-viral-combined_k141_83329\|\|full | 3972 | Yes | 1674 | 7 | 2 | 1 | Low-quality | Genome-fragment | 3.84 | AAI-based (high-confidence) | 57.85 |
| S2-final-viral-combined_k141_17102\|\|full | 2560 | No | NA | 3 | 0 | 2 | Not-determined | Genome-fragment | NA | NA | 0 |
| S17-final-viral-combined_k141_97765\|\|full | 3973 | No | NA | 4 | 0 | 2 | Low-quality | Genome-fragment | 3.33 | AAI-based (medium-confidence) | 0 |
| S12-final-viral-combined_k141_21786\|\|full | 3973 | No | NA | 4 | 3 | 0 | Low-quality | Genome-fragment | 9.07 | AAI-based (high-confidence) | 0 |
| S12_k141_5180 | 3973 | No | NA | 3 | 0 | 1 | Low-quality | Genome-fragment | 5.87 | AAI-based (high-confidence) | 0 |
| S2-final-viral-combined_k141_141431\|\|full | 1156 | No | NA | 3 | 0 | 0 | Not-determined | Genome-fragment | NA | NA | 0 |
| S2-final-viral-combined_k141_202244\|\|full | 3298 | No | NA | 6 | 0 | 1 | Not-determined | Genome-fragment | NA | NA | 0 |
| S2-final-viral-combined_k141_58563\|\|full | 53508 | Yes | 27398 | 65 | 24 | 17 | Medium-quality | Genome-fragment | 59.82 | AAI-based (high-confidence) | 48.8 |
| S2-final-viral-combined_k141_17154\|\|full | 2406 | No | NA | 2 | 0 | 2 | Not-determined | Genome-fragment | NA | NA | 0 |
| S2-final-viral-combined_k141_141457\|\|full | 1108 | No | NA | 3 | 0 | 0 | Not-determined | Genome-fragment | NA | NA | 0 |
| S4-final-viral-combined_k141_105020\|\|full | 3973 | No | NA | 6 | 1 | 0 | Low-quality | Genome-fragment | 2.45 | HMM-based (lower-bound) | 0 |
| S2-final-viral-combined_k141_141468\|\|full | 1104 | No | NA | 3 | 0 | 0 | Not-determined | Genome-fragment | NA | NA | 0 |
| S11-final-viral-combined_k141_222263\|\|full | 3974 | No | NA | 10 | 3 | 0 | Low-quality | Genome-fragment | 7.21 | AAI-based (high-confidence) | 0 |
| S17-final-viral-combined_k141_219373\|\|full | 3975 | No | NA | 10 | 1 | 0 | Low-quality | Genome-fragment | 31.3 | AAI-based (high-confidence) | 0 |
| S8-final-viral-combined_k141_272160\|\|full | 3975 | No | NA | 5 | 1 | 0 | Low-quality | Genome-fragment | 6.21 | HMM-based (lower-bound) | 0 |
| S12-final-viral-combined_k141_386199\|\|full | 3976 | No | NA | 7 | 2 | 0 | Low-quality | Genome-fragment | 4.23 | HMM-based (lower-bound) | 0 |
| S2-final-viral-combined_k141_120873\|\|full | 1512 | No | NA | 3 | 0 | 0 | Not-determined | Genome-fragment | NA | NA | 0 |
| S7_k141_39188 | 3977 | No | NA | 4 | 0 | 2 | Low-quality | Genome-fragment | 2.71 | AAI-based (medium-confidence) | 0 |
| S2-final-viral-combined_k141_202265\|\|full | 1268 | No | NA | 2 | 0 | 0 | Not-determined | Genome-fragment | NA | NA | 0 |
| S3-final-viral-combined_k141_71425\|\|full | 3978 | No | NA | 5 | 1 | 0 | Low-quality | Genome-fragment | 9.56 | AAI-based (high-confidence) | 0 |
| S2-final-viral-combined_k141_79120\|\|full | 5446 | No | NA | 5 | 0 | 2 | Not-determined | Genome-fragment | NA | NA | 0 |
| S2-final-viral-combined_k141_68542\|\|full | 1871 | No | NA | 2 | 0 | 0 | Not-determined | Genome-fragment | NA | NA | 0 |

| | | | | | | | | | | | |
|---|---|---|---|---|---|---|---|---|---|---|---|
| S2-final-viral-combined_k141_152012\|\|full | 2136 | No | NA | 4 | 0 | 1 | Not-determined | Genome-fragment | NA | NA | 0 |
| S2-final-viral-combined_k141_90141\|\|full | 1555 | No | NA | 3 | 0 | 0 | Not-determined | Genome-fragment | NA | NA | 0 |
| S2-final-viral-combined_k141_243647\|\|full | 6549 | No | NA | 5 | 0 | 4 | Not-determined | Genome-fragment | NA | NA | 0 |
| S2-final-viral-combined_k141_235071\|\|full | 4167 | No | NA | 4 | 0 | 2 | Not-determined | Genome-fragment | NA | NA | 0 |
| S2-final-viral-combined_k141_50216\|\|full | 3323 | No | NA | 4 | 0 | 4 | Not-determined | Genome-fragment | NA | NA | 0 |
| S4-final-viral-combined_k141_103485\|\|full | 3978 | No | NA | 3 | 0 | 2 | Low-quality | Genome-fragment | 2.98 | AAI-based (medium-confidence) | 0 |
| S2-final-viral-combined_k141_243669\|\|full | 6097 | No | NA | 6 | 0 | 5 | Not-determined | Genome-fragment | NA | NA | 0 |
| S4_k141_36293 | 3978 | No | NA | 4 | 0 | 4 | Low-quality | Genome-fragment | 4.05 | AAI-based (medium-confidence) | 0 |
| S6-final-viral-combined_k141_115664\|\|full | 3978 | No | NA | 5 | 0 | 0 | Low-quality | Genome-fragment | 9.31 | AAI-based (medium-confidence) | 0 |
| S1-final-viral-combined_k141_356411\|\|full | 3980 | No | NA | 3 | 0 | 3 | Low-quality | Genome-fragment | 0.38 | AAI-based (medium-confidence) | 0 |
| S2-final-viral-combined_k141_68607\|\|full | 1934 | No | NA | 2 | 0 | 1 | Not-determined | Genome-fragment | NA | NA | 0 |
| S1-final-viral-combined_k141_338965\|\|full | 3980 | No | NA | 6 | 0 | 0 | Low-quality | Genome-fragment | 10.23 | AAI-based (medium-confidence) | 0 |
| S1_k141_55360 | 3980 | No | NA | 5 | 0 | 1 | Low-quality | Genome-fragment | 5.6 | AAI-based (medium-confidence) | 0 |
| S2-final-viral-combined_k141_64924\|\|full | 3980 | No | NA | 2 | 0 | 1 | Low-quality | Genome-fragment | 1.32 | AAI-based (high-confidence) | 0 |
| S2-final-viral-combined_k141_68620\|\|full | 1046 | No | NA | 2 | 0 | 0 | Not-determined | Genome-fragment | NA | NA | 0 |
| S2-final-viral-combined_k141_100339\|\|full | 4202 | No | NA | 3 | 0 | 3 | Not-determined | Genome-fragment | NA | NA | 0 |
| S2-final-viral-combined_k141_68625\|\|full | 3929 | No | NA | 5 | 0 | 3 | Not-determined | Genome-fragment | NA | NA | 0 |
| S11_k141_293129 | 3981 | No | NA | 9 | 0 | 0 | Low-quality | Genome-fragment | 5.14 | AAI-based (high-confidence) | 0 |
| S12-final-viral-combined_k141_342029\|\|full | 3981 | No | NA | 3 | 0 | 2 | Low-quality | Genome-fragment | 7.38 | AAI-based (medium-confidence) | 0 |
| S2_k141_240613 | 3982 | No | NA | 1 | 0 | 0 | Low-quality | Genome-fragment | 9.9 | AAI-based (medium-confidence) | 0 |
| S2-final-viral-combined_k141_100348\|\|full | 2025 | No | NA | 3 | 0 | 1 | Not-determined | Genome-fragment | NA | NA | 0 |
| S2-final-viral-combined_k141_213219\|\|full | 2779 | No | NA | 4 | 0 | 2 | Not-determined | Genome-fragment | NA | NA | 0 |
| S2-final-viral-combined_k141_38201\|\|full | 2106 | No | NA | 4 | 0 | 0 | Not-determined | Genome-fragment | NA | NA | 0 |
| S2_k141_192275 | 3983 | No | NA | 8 | 2 | 0 | Low-quality | Genome-fragment | 8.35 | AAI-based (medium-confidence) | 0 |
| S7-final-viral-combined_k141_147737\|\|full | 3984 | No | NA | 5 | 1 | 0 | Low-quality | Genome-fragment | 2.87 | HMM-based (lower-bound) | 0 |
| S11-final-viral-combined_k141_189912\|\|full | 3985 | No | NA | 5 | 0 | 1 | Low-quality | Genome-fragment | 9.25 | AAI-based (medium-confidence) | 0 |
| S2-final-viral-combined_k141_100397\|\|full | 1896 | No | NA | 4 | 0 | 1 | Not-determined | Genome-fragment | NA | NA | 0 |
| S2-final-viral-combined_k141_202429\|\|full | 1053 | No | NA | 2 | 0 | 1 | Not-determined | Genome-fragment | NA | NA | 0 |
| S2-final-viral-combined_k141_183196\|\|full | 1497 | No | NA | 1 | 0 | 0 | Not-determined | Genome-fragment | NA | NA | 0 |
| S2-final-viral-combined_k141_50366\|\|full | 1387 | No | NA | 2 | 0 | 1 | Not-determined | Genome-fragment | NA | NA | 0 |
| S2-final-viral-combined_k141_159684\|\|full | 3985 | Yes | 2348 | 6 | 1 | 1 | Low-quality | Genome-fragment | 1.49 | HMM-based (lower-bound) | 41.08 |
| S2-final-viral-combined_k141_90297\|\|full | 3345 | No | NA | 3 | 0 | 2 | Not-determined | Genome-fragment | NA | NA | 0 |
| S2-final-viral-combined_k141_50378\|\|full | 2762 | No | NA | 4 | 0 | 1 | Not-determined | Genome-fragment | NA | NA | 0 |
| S2-final-viral-combined_k141_68718\|\|full | 1250 | No | NA | 2 | 0 | 0 | Not-determined | Genome-fragment | NA | NA | 0 |
| S2-final-viral-combined_k141_129885\|\|full | 2180 | No | NA | 2 | 0 | 2 | Not-determined | Genome-fragment | NA | NA | 0 |
| S17-k141_74080 | 3986 | No | NA | 3 | 0 | 3 | Low-quality | Genome-fragment | 2.26 | AAI-based (medium-confidence) | 0 |
| S2-final-viral-combined_k141_193048\|\|full | 2063 | No | NA | 3 | 0 | 1 | Not-determined | Genome-fragment | NA | NA | 0 |
| S2-final-viral-combined_k141_50419\|\|full | 3039 | No | NA | 4 | 0 | 3 | Not-determined | Genome-fragment | NA | NA | 0 |
| S2-final-viral-combined_k141_50424\|\|full | 1933 | No | NA | 4 | 0 | 1 | Not-determined | Genome-fragment | NA | NA | 0 |
| S12-final-viral-combined_k141_69884\|\|full | 3986 | No | NA | 7 | 0 | 0 | Low-quality | Genome-fragment | 8.12 | AAI-based (high-confidence) | 0 |
| S2-final-viral-combined_k141_173770\|\|full | 1885 | No | NA | 3 | 0 | 2 | Not-determined | Genome-fragment | NA | NA | 0 |
| S2-final-viral-combined_k141_193074\|\|full | 2456 | No | NA | 3 | 0 | 3 | Not-determined | Genome-fragment | NA | NA | 0 |
| S11-final-viral-combined_k141_41031\|\|full | 3987 | No | NA | 3 | 1 | 0 | Low-quality | Genome-fragment | 5.78 | AAI-based (high-confidence) | 0 |
| S7_k141_240588 | 3987 | No | NA | 7 | 0 | 0 | Low-quality | Genome-fragment | 9.4 | AAI-based (high-confidence) | 0 |
| S17-final-viral-combined_k141_182180\|\|full | 3987 | No | NA | 7 | 0 | 2 | Low-quality | Genome-fragment | 7.48 | AAI-based (medium-confidence) | 0 |
| S2-final-viral-combined_k141_129954\|\|full | 3251 | No | NA | 2 | 0 | 1 | Not-determined | Genome-fragment | NA | NA | 0 |
| S2-final-viral-combined_k141_235239\|\|full | 3357 | No | NA | 3 | 0 | 1 | Not-determined | Genome-fragment | NA | NA | 0 |
| S2-final-viral-combined_k141_152148\|\|full | 2499 | No | NA | 5 | 0 | 1 | Not-determined | Genome-fragment | NA | NA | 0 |
| S2-final-viral-combined_k141_235248\|\|full | 2447 | No | NA | 2 | 0 | 2 | Not-determined | Genome-fragment | NA | NA | 0 |
| S12_k141_1977 | 3987 | Yes | 468 | 6 | 1 | 4 | Low-quality | Genome-fragment | 0.67 | HMM-based (lower-bound) | 88.26 |
| S2-final-viral-combined_k141_50492\|\|full | 1321 | No | NA | 2 | 0 | 1 | Not-determined | Genome-fragment | NA | NA | 0 |
| S2-final-viral-combined_k141_161610\|\|full | 2002 | No | NA | 3 | 0 | 3 | Not-determined | Genome-fragment | NA | NA | 0 |
| S2-final-viral-combined_k141_335\|\|full | 3987 | No | NA | 6 | 0 | 2 | Low-quality | Genome-fragment | 4.54 | AAI-based (medium-confidence) | 0 |
| S2-final-viral-combined_k141_109824\|\|full | 1962 | No | NA | 2 | 0 | 0 | Not-determined | Genome-fragment | NA | NA | 0 |
| S17-final-viral-combined_k141_3436\|\|full | 3988 | No | NA | 7 | 0 | 1 | Low-quality | Genome-fragment | 6.19 | AAI-based (medium-confidence) | 0 |
| S2-final-viral-combined_k141_245516\|\|full | 3988 | No | NA | 4 | 2 | 1 | Low-quality | Genome-fragment | 7.3 | AAI-based (high-confidence) | 0 |
| S17-k141_186319 | 3989 | No | NA | 2 | 0 | 1 | Low-quality | Genome-fragment | 0.34 | AAI-based (medium-confidence) | 0 |
| S12-final-viral-combined_k141_119540\|\|full | 3989 | No | NA | 8 | 1 | 0 | Low-quality | Genome-fragment | 8.69 | AAI-based (high-confidence) | 0 |
| S2_k141_136474 | 3989 | No | NA | 7 | 0 | 0 | Low-quality | Genome-fragment | 4.16 | AAI-based (high-confidence) | 0 |
| S2-final-viral-combined_k141_17292\|\|full | 1288 | No | NA | 2 | 0 | 0 | Not-determined | Genome-fragment | NA | NA | 0 |
| S6-final-viral-combined_k141_80629\|\|full | 3989 | No | NA | 4 | 0 | 1 | Low-quality | Genome-fragment | 4.95 | AAI-based (medium-confidence) | 0 |
| S2-final-viral-combined_k141_161651\|\|full | 2703 | No | NA | 2 | 0 | 0 | Not-determined | Genome-fragment | NA | NA | 0 |
| S9-final-viral-combined_k141_61625\|\|full | 3990 | No | NA | 2 | 0 | 1 | Low-quality | Genome-fragment | 3.87 | AAI-based (medium-confidence) | 0 |
| S2-final-viral-combined_k141_100507\|\|full | 4040 | No | NA | 5 | 0 | 2 | Not-determined | Genome-fragment | NA | NA | 0 |
| S1-final-viral-combined_k141_310218\|\|full | 3990 | No | NA | 4 | 0 | 3 | Low-quality | Genome-fragment | 2.05 | AAI-based (medium-confidence) | 0 |
| S2-final-viral-combined_k141_68918\|\|full | 1489 | No | NA | 2 | 0 | 1 | Not-determined | Genome-fragment | NA | NA | 0 |
| S2-final-viral-combined_k141_38404\|\|full | 3327 | No | NA | 4 | 0 | 3 | Not-determined | Genome-fragment | NA | NA | 0 |
| S2-final-viral-combined_k141_213451\|\|full | 2415 | No | NA | 3 | 0 | 2 | Not-determined | Genome-fragment | NA | NA | 0 |
| S17-final-viral-combined_k141_26300\|\|full | 3990 | No | NA | 4 | 1 | 0 | Low-quality | Genome-fragment | 10.13 | AAI-based (high-confidence) | 0 |
| S4-final-viral-combined_k141_108998\|\|full | 3990 | No | NA | 5 | 0 | 0 | Low-quality | Genome-fragment | 6.36 | AAI-based (medium-confidence) | 0 |
| S2-final-viral-combined_k141_100543\|\|full | 1264 | No | NA | 3 | 0 | 1 | Not-determined | Genome-fragment | NA | NA | 0 |
| S8-final-viral-combined_k141_239082\|\|full | 3990 | No | NA | 2 | 0 | 2 | Low-quality | Genome-fragment | 2.08 | AAI-based (medium-confidence) | 0 |
| S2-final-viral-combined_k141_38449\|\|full | 2420 | No | NA | 2 | 0 | 1 | Not-determined | Genome-fragment | NA | NA | 0 |
| S2-final-viral-combined_k141_121206\|\|full | 1068 | No | NA | 3 | 0 | 0 | Not-determined | Genome-fragment | NA | NA | 0 |
| S1-final-viral-combined_k141_251832\|\|full | 3991 | Yes | 1397 | 6 | 1 | 2 | Low-quality | Genome-fragment | 0.76 | HMM-based (lower-bound) | 65 |
| S2-final-viral-combined_k141_130049\|\|full | 1078 | No | NA | 3 | 0 | 0 | Not-determined | Genome-fragment | NA | NA | 0 |
| S3-final-viral-combined_k141_101290\|\|full | 3991 | Yes | 1784 | 5 | 1 | 1 | Low-quality | Genome-fragment | 3.51 | AAI-based (medium-confidence) | 55.3 |
| S2-final-viral-combined_k141_152318\|\|full | 1348 | No | NA | 2 | 0 | 1 | Not-determined | Genome-fragment | NA | NA | 0 |
| S11_k141_41901 | 3992 | No | NA | 5 | 0 | 4 | Low-quality | Genome-fragment | 2.17 | AAI-based (medium-confidence) | 0 |
| S2-final-viral-combined_k141_235355\|\|full | 2737 | No | NA | 2 | 0 | 2 | Not-determined | Genome-fragment | NA | NA | 0 |
| S2-final-viral-combined_k141_130071\|\|full | 4827 | No | NA | 5 | 0 | 4 | Not-determined | Genome-fragment | NA | NA | 0 |
| S1-final-viral-combined_k141_187756\|\|full | 3992 | Yes | 3266 | 4 | 1 | 1 | Low-quality | Genome-fragment | 6.69 | AAI-based (medium-confidence) | 18.19 |
| S2-final-viral-combined_k141_121237\|\|full | 1008 | No | NA | 3 | 0 | 0 | Not-determined | Genome-fragment | NA | NA | 0 |
| S2-final-viral-combined_k141_173945\|\|full | 4061 | No | NA | 2 | 0 | 0 | Not-determined | Genome-fragment | NA | NA | 0 |
| S2-final-viral-combined_k141_58822\|\|full | 10205 | No | NA | 11 | 0 | 8 | Not-determined | Genome-fragment | NA | NA | 0 |
| S2-final-viral-combined_k141_58823\|\|full | 2222 | No | NA | 2 | 0 | 2 | Not-determined | Genome-fragment | NA | NA | 0 |
| S2-final-viral-combined_k141_26468\|\|full | 3222 | No | NA | 4 | 0 | 2 | Not-determined | Genome-fragment | NA | NA | 0 |
| S1_k141_167543 | 3994 | No | NA | 10 | 1 | 0 | Low-quality | Genome-fragment | 9.39 | AAI-based (high-confidence) | 0 |
| S3-final-viral-combined_k141_14319\|\|full | 3994 | No | NA | 7 | 0 | 0 | Low-quality | Genome-fragment | 8.86 | AAI-based (high-confidence) | 0 |
| S2-final-viral-combined_k141_235434\|\|full | 4119 | No | NA | 4 | 0 | 0 | Not-determined | Genome-fragment | NA | NA | 0 |
| S12-final-viral-combined_k141_70768\|\|full | 3994 | Yes | 2583 | 5 | 1 | 1 | Low-quality | Genome-fragment | 2.62 | AAI-based (high-confidence) | 35.33 |
| S6-final-viral-combined_k141_100106\|\|full | 3994 | No | NA | 4 | 0 | 3 | Low-quality | Genome-fragment | 2.98 | AAI-based (medium-confidence) | 0 |
| S4-final-viral-combined_k141_126893\|\|full | 3995 | No | NA | 3 | 0 | 3 | Low-quality | Genome-fragment | 9.06 | AAI-based (medium-confidence) | 0 |
| S4_k141_160307 | 3995 | No | NA | 5 | 0 | 0 | Low-quality | Genome-fragment | 8.95 | AAI-based (high-confidence) | 0 |
| S7-final-viral-combined_k141_236251\|\|full | 3996 | Yes | 2209 | 4 | 1 | 1 | Low-quality | Genome-fragment | 4.57 | AAI-based (medium-confidence) | 44.72 |
| S2-final-viral-combined_k141_50722\|\|full | 5745 | No | NA | 9 | 0 | 1 | Not-determined | Genome-fragment | NA | NA | 0 |
| S2-final-viral-combined_k141_213543\|\|full | 2395 | No | NA | 3 | 0 | 1 | Not-determined | Genome-fragment | NA | NA | 0 |
| S17-final-viral-combined_k141_161467\|\|full | 3996 | No | NA | 3 | 0 | 1 | Low-quality | Genome-fragment | 0.46 | AAI-based (medium-confidence) | 0 |
| S2-final-viral-combined_k141_141865\|\|full | 3192 | No | NA | 4 | 0 | 4 | Not-determined | Genome-fragment | NA | NA | 0 |
| S2-final-viral-combined_k141_79567\|\|full | 12997 | No | NA | 10 | 0 | 8 | Not-determined | Genome-fragment | NA | NA | 0 |
| S11-final-viral-combined_k141_226159\|\|full | 3998 | No | NA | 3 | 2 | 0 | Low-quality | Genome-fragment | 10.04 | AAI-based (high-confidence) | 0 |
| S9-final-viral-combined_k141_93043\|\|full | 3999 | No | NA | 4 | 0 | 3 | Low-quality | Genome-fragment | 1.65 | AAI-based (medium-confidence) | 0 |
| S2-final-viral-combined_k141_17496\|\|full | 4256 | No | NA | 4 | 0 | 2 | Not-determined | Genome-fragment | NA | NA | 0 |
| S2-final-viral-combined_k141_174080\|\|full | 1796 | No | NA | 2 | 0 | 2 | Not-determined | Genome-fragment | NA | NA | 0 |
| S11-final-viral-combined_k141_243218\|\|full | 3999 | No | NA | 9 | 0 | 0 | Low-quality | Genome-fragment | 5.99 | AAI-based (medium-confidence) | 0 |
| S1-final-viral-combined_k141_317472\|\|full | 3999 | No | NA | 4 | 0 | 1 | Low-quality | Genome-fragment | 6.44 | AAI-based (medium-confidence) | 0 |
| S3-final-viral-combined_k141_4556\|\|full | 3999 | No | NA | 5 | 0 | 0 | Low-quality | Genome-fragment | 5.88 | AAI-based (medium-confidence) | 0 |
| S3_k141_71002 | 4001 | No | NA | 3 | 0 | 1 | Low-quality | Genome-fragment | 2.05 | AAI-based (medium-confidence) | 0 |
| S2-final-viral-combined_k141_90501\|\|full | 1201 | No | NA | 2 | 0 | 1 | Not-determined | Genome-fragment | NA | NA | 0 |
| S2-final-viral-combined_k141_243886\|\|full | 1460 | No | NA | 2 | 0 | 1 | Not-determined | Genome-fragment | NA | NA | 0 |
| S2-final-viral-combined_k141_235503\|\|full | 1647 | No | NA | 3 | 0 | 2 | Not-determined | Genome-fragment | NA | NA | 0 |
| S2-final-viral-combined_k141_58920\|\|full | 1799 | No | NA | 3 | 0 | 1 | Not-determined | Genome-fragment | NA | NA | 0 |
| S2-final-viral-combined_k141_174123\|\|full | 1174 | No | NA | 3 | 0 | 0 | Not-determined | Genome-fragment | NA | NA | 0 |
| S17-k141_627 | 4001 | No | NA | 3 | 0 | 1 | Low-quality | Genome-fragment | 13.87 | AAI-based (high-confidence) | 0 |
| S2-final-viral-combined_k141_188776\|\|full | 4003 | No | NA | 4 | 0 | 3 | Low-quality | Genome-fragment | 8.16 | AAI-based (medium-confidence) | 0 |
| S2-final-viral-combined_k141_202627\|\|full | 8554 | No | NA | 10 | 0 | 2 | Not-determined | Genome-fragment | NA | NA | 0 |
| S2-final-viral-combined_k141_213573\|\|full | 11741 | No | NA | 13 | 0 | 8 | Not-determined | Genome-fragment | NA | NA | 0 |
| S2-final-viral-combined_k141_121391\|\|full | 1984 | No | NA | 2 | 0 | 2 | Not-determined | Genome-fragment | NA | NA | 0 |
| S2-final-viral-combined_k141_223159\|\|full | 1775 | No | NA | 2 | 0 | 2 | Not-determined | Genome-fragment | NA | NA | 0 |
| S2-final-viral-combined_k141_79579\|\|full | 1872 | No | NA | 2 | 0 | 0 | Not-determined | Genome-fragment | NA | NA | 0 |
| S12_k141_220399 | 4003 | No | NA | 5 | 1 | 0 | Low-quality | Genome-fragment | 10.11 | AAI-based (high-confidence) | 0 |
| S7-final-viral-combined_k141_244459\|\|full | 4004 | No | NA | 3 | 0 | 2 | Low-quality | Genome-fragment | 2.2 | AAI-based (medium-confidence) | 0 |
| S2-final-viral-combined_k141_213603\|\|full | 1087 | No | NA | 2 | 0 | 0 | Not-determined | Genome-fragment | NA | NA | 0 |
| S4-final-viral-combined_k141_37778\|\|full | 4004 | No | NA | 5 | 0 | 0 | Low-quality | Genome-fragment | 3.4 | AAI-based (high-confidence) | 0 |
| S1-final-viral-combined_k141_373295\|\|full | 4005 | No | NA | 10 | 0 | 1 | Low-quality | Genome-fragment | 6.11 | AAI-based (medium-confidence) | 0 |
| S2-final-viral-combined_k141_38694\|\|full | 1818 | No | NA | 3 | 0 | 0 | Not-determined | Genome-fragment | NA | NA | 0 |
| S2-final-viral-combined_k141_141971\|\|full | 1715 | No | NA | 3 | 0 | 1 | Not-determined | Genome-fragment | NA | NA | 0 |
| S2-final-viral-combined_k141_100845\|\|full | 1475 | No | NA | 2 | 0 | 0 | Not-determined | Genome-fragment | NA | NA | 0 |
| S4-final-viral-combined_k141_139600\|\|full | 4005 | No | NA | 4 | 0 | 3 | Low-quality | Genome-fragment | 4.7 | AAI-based (medium-confidence) | 0 |
| S11-final-viral-combined_k141_163738\|\|full | 4007 | No | NA | 4 | 0 | 1 | Low-quality | Genome-fragment | 2.85 | AAI-based (medium-confidence) | 0 |
| S2-final-viral-combined_k141_50882\|\|full | 11772 | No | NA | 15 | 0 | 8 | Not-determined | Genome-fragment | NA | NA | 0 |
| S1-final-viral-combined_k141_295817\|\|full | 4007 | No | NA | 3 | 0 | 0 | Low-quality | Genome-fragment | 2.1 | AAI-based (medium-confidence) | 0 |
| S2-final-viral-combined_k141_110080\|\|full | 3630 | No | NA | 4 | 0 | 1 | Not-determined | Genome-fragment | NA | NA | 0 |
| S2-final-viral-combined_k141_90557\|\|full | 1882 | No | NA | 4 | 0 | 2 | Not-determined | Genome-fragment | NA | NA | 0 |
| S2-final-viral-combined_k141_152508\|\|full | 4720 | No | NA | 4 | 0 | 2 | Not-determined | Genome-fragment | NA | NA | 0 |
| S2-final-viral-combined_k141_174208\|\|full | 1912 | No | NA | 3 | 0 | 2 | Not-determined | Genome-fragment | NA | NA | 0 |
| S2-final-viral-combined_k141_5721\|\|full | 1109 | No | NA | 2 | 0 | 0 | Not-determined | Genome-fragment | NA | NA | 0 |
| S2-final-viral-combined_k141_193425\|\|full | 2448 | No | NA | 3 | 0 | 2 | Not-determined | Genome-fragment | NA | NA | 0 |
| S2-final-viral-combined_k141_90574\|\|full | 3036 | No | NA | 3 | 0 | 0 | Not-determined | Genome-fragment | NA | NA | 0 |
| S3_k141_55502 | 4007 | No | NA | 1 | 0 | 1 | Low-quality | Genome-fragment | 1.84 | AAI-based (high-confidence) | 0 |
| S2-final-viral-combined_k141_223240\|\|full | 1843 | No | NA | 3 | 0 | 2 | Not-determined | Genome-fragment | NA | NA | 0 |
| S2-final-viral-combined_k141_193460\|\|full | 2896 | No | NA | 5 | 0 | 0 | Not-determined | Genome-fragment | NA | NA | 0 |
| S2-final-viral-combined_k141_17628\|\|full | 1115 | No | NA | 2 | 0 | 1 | Not-determined | Genome-fragment | NA | NA | 0 |
| S2-final-viral-combined_k141_235633\|\|full | 7783 | No | NA | 11 | 0 | 9 | Not-determined | Genome-fragment | NA | NA | 0 |
| S2-final-viral-combined_k141_38802\|\|full | 1153 | No | NA | 2 | 0 | 1 | Not-determined | Genome-fragment | NA | NA | 0 |
| S2-final-viral-combined_k141_183459\|\|full | 3277 | No | NA | 6 | 0 | 2 | Not-determined | Genome-fragment | NA | NA | 0 |
| S9-final-viral-combined_k141_118479\|\|lt2gene | 4008 | No | NA | 2 | 2 | 0 | Low-quality | Genome-fragment | 9.94 | AAI-based (high-confidence) | 0 |
| S2-final-viral-combined_k141_161996\|\|full | 4008 | No | NA | 4 | 0 | 3 | Low-quality | Genome-fragment | 7.21 | AAI-based (medium-confidence) | 0 |
| S7_k141_125962 | 4009 | No | NA | 3 | 0 | 2 | Low-quality | Genome-fragment | 4.8 | AAI-based (medium-confidence) | 0 |
| S17-final-viral-combined_k141_145626\|\|full | 4009 | No | NA | 6 | 0 | 1 | Low-quality | Genome-fragment | 7.46 | AAI-based (medium-confidence) | 0 |
| S8-final-viral-combined_k141_44273\|\|full | 4009 | No | NA | 4 | 0 | 2 | Low-quality | Genome-fragment | 1.21 | AAI-based (medium-confidence) | 0 |
| S2-final-viral-combined_k141_243989\|\|full | 28377 | No | NA | 30 | 0 | 3 | Not-determined | Genome-fragment | NA | NA | 0 |
| S2-final-viral-combined_k141_174241\|\|full | 61290 | Yes | 44828 | 64 | 22 | 3 | Medium-quality | Genome-fragment | 89.69 | AAI-based (medium-confidence) | 26.86 |
| S2-final-viral-combined_k141_59042\|\|full | 1047 | No | NA | 3 | 0 | 0 | Not-determined | Genome-fragment | NA | NA | 0 |
| S2-final-viral-combined_k141_121463\|\|full | 1800 | No | NA | 2 | 0 | 1 | Not-determined | Genome-fragment | NA | NA | 0 |
| S2-final-viral-combined_k141_121465\|\|full | 1382 | No | NA | 2 | 0 | 0 | Not-determined | Genome-fragment | NA | NA | 0 |
| S7-final-viral-combined_k141_90243\|\|full | 4010 | No | NA | 3 | 0 | 0 | Low-quality | Genome-fragment | 4.76 | AAI-based (high-confidence) | 0 |
| S2-final-viral-combined_k141_244004\|\|full | 3120 | No | NA | 4 | 0 | 2 | Not-determined | Genome-fragment | NA | NA | 0 |
| S17-k141_264585 | 4010 | No | NA | 7 | 0 | 0 | Low-quality | Genome-fragment | 6.22 | AAI-based (medium-confidence) | 0 |
| S2-final-viral-combined_k141_174272\|\|full | 2274 | No | NA | 4 | 0 | 1 | Not-determined | Genome-fragment | NA | NA | 0 |
| S4-final-viral-combined_k141_110092\|\|full | 4010 | No | NA | 9 | 3 | 0 | Low-quality | Genome-fragment | 6.56 | AAI-based (high-confidence) | 0 |
| S2-final-viral-combined_k141_152605\|\|full | 2794 | No | NA | 2 | 0 | 0 | Not-determined | Genome-fragment | NA | NA | 0 |
| S2-final-viral-combined_k141_183504\|\|full | 6935 | No | NA | 9 | 0 | 1 | Not-determined | Genome-fragment | NA | NA | 0 |
| S4_k141_57247 | 4010 | No | NA | 5 | 0 | 2 | Low-quality | Genome-fragment | 3.04 | AAI-based (medium-confidence) | 0 |
| S2-final-viral-combined_k141_100951\|\|full | 1222 | No | NA | 3 | 0 | 0 | Not-determined | Genome-fragment | NA | NA | 0 |
| S5_k141_208254 | 4011 | No | NA | 5 | 1 | 0 | Low-quality | Genome-fragment | 11.02 | HMM-based (lower-bound) | 0 |
| S2-final-viral-combined_k141_59071\|\|full | 1179 | No | NA | 2 | 0 | 1 | Not-determined | Genome-fragment | NA | NA | 0 |
| S2-final-viral-combined_k141_100954\|\|full | 2698 | No | NA | 3 | 0 | 2 | Not-determined | Genome-fragment | NA | NA | 0 |
| S2-final-viral-combined_k141_121503\|\|full | 1040 | No | NA | 2 | 0 | 1 | Not-determined | Genome-fragment | NA | NA | 0 |
| S2-final-viral-combined_k141_183511\|\|full | 4127 | No | NA | 6 | 0 | 4 | Not-determined | Genome-fragment | NA | NA | 0 |
| S8_k141_148349 | 4011 | No | NA | 1 | 0 | 0 | Low-quality | Genome-fragment | 10.17 | AAI-based (medium-confidence) | 0 |
| S2-final-viral-combined_k141_213776\|\|full | 5450 | No | NA | 4 | 0 | 3 | Not-determined | Genome-fragment | NA | NA | 0 |
| S2-final-viral-combined_k141_79707\|\|full | 4270 | No | NA | 4 | 0 | 2 | Not-determined | Genome-fragment | NA | NA | 0 |
| S2_k141_177484 | 4012 | No | NA | 5 | 0 | 2 | Low-quality | Genome-fragment | 6.26 | AAI-based (medium-confidence) | 0 |
| S2-final-viral-combined_k141_193557\|\|full | 5470 | No | NA | 6 | 0 | 0 | Not-determined | Genome-fragment | NA | NA | 0 |
| S2-final-viral-combined_k141_79732\|\|full | 1701 | No | NA | 2 | 0 | 2 | Not-determined | Genome-fragment | NA | NA | 0 |
| S11_k141_283306 | 4014 | No | NA | 6 | 1 | 0 | Low-quality | Genome-fragment | 10.3 | AAI-based (high-confidence) | 0 |
| S2-final-viral-combined_k141_152666\|\|full | 1275 | No | NA | 3 | 0 | 0 | Not-determined | Genome-fragment | NA | NA | 0 |
| S2-final-viral-combined_k141_223362\|\|full | 3640 | No | NA | 3 | 0 | 2 | Not-determined | Genome-fragment | NA | NA | 0 |
| S5_k141_107381 | 4014 | No | NA | 5 | 5 | 0 | Low-quality | Genome-fragment | 2.19 | HMM-based (lower-bound) | 0 |
| S5-final-viral-combined_k141_162090\|\|full | 4015 | Yes | 3226 | 6 | 3 | 1 | Low-quality | Genome-fragment | 8.29 | AAI-based (high-confidence) | 19.65 |
| S2-final-viral-combined_k141_235809\|\|full | 3268 | No | NA | 3 | 0 | 1 | Not-determined | Genome-fragment | NA | NA | 0 |
| S2-final-viral-combined_k141_69282\|\|full | 1792 | No | NA | 3 | 0 | 0 | Not-determined | Genome-fragment | NA | NA | 0 |
| S2-final-viral-combined_k141_51011\|\|full | 3148 | No | NA | 3 | 0 | 2 | Not-determined | Genome-fragment | NA | NA | 0 |
| S7-final-viral-combined_k141_161936\|\|full | 4015 | No | NA | 4 | 3 | 0 | Low-quality | Genome-fragment | 9.56 | AAI-based (high-confidence) | 0 |
| S2-final-viral-combined_k141_17703\|\|full | 2835 | No | NA | 4 | 0 | 1 | Not-determined | Genome-fragment | NA | NA | 0 |
| S2-final-viral-combined_k141_235828\|\|full | 4062 | No | NA | 3 | 0 | 2 | Not-determined | Genome-fragment | NA | NA | 0 |
| S2-final-viral-combined_k141_100992\|\|full | 3786 | No | NA | 3 | 0 | 3 | Not-determined | Genome-fragment | NA | NA | 0 |
| S2-final-viral-combined_k141_79788\|\|full | 2951 | No | NA | 3 | 0 | 2 | Not-determined | Genome-fragment | NA | NA | 0 |
| S4-final-viral-combined_k141_39737\|\|full | 4016 | No | NA | 6 | 1 | 0 | Low-quality | Genome-fragment | 2.07 | HMM-based (lower-bound) | 0 |
| S7-final-viral-combined_k141_11473\|\|full | 4017 | No | NA | 4 | 0 | 2 | Low-quality | Genome-fragment | 7.95 | AAI-based (high-confidence) | 0 |
| S7-final-viral-combined_k141_66091\|\|full | 4017 | No | NA | 5 | 0 | 0 | Low-quality | Genome-fragment | 3.54 | AAI-based (medium-confidence) | 0 |
| S2-final-viral-combined_k141_223445\|\|full | 2074 | No | NA | 3 | 0 | 1 | Not-determined | Genome-fragment | NA | NA | 0 |
| S2-final-viral-combined_k141_110215\|\|full | 78637 | Yes | 66879 | 107 | 21 | 12 | High-quality | High-quality | 100 | AAI-based (medium-confidence) | 14.95 |
| S2-final-viral-combined_k141_17746\|\|full | 5179 | No | NA | 4 | 0 | 3 | Not-determined | Genome-fragment | NA | NA | 0 |
| S2-final-viral-combined_k141_213824\|\|full | 7279 | No | NA | 7 | 0 | 6 | Not-determined | Genome-fragment | NA | NA | 0 |
| S2-final-viral-combined_k141_235874\|\|full | 3519 | No | NA | 6 | 0 | 0 | Not-determined | Genome-fragment | NA | NA | 0 |
| S2-final-viral-combined_k141_79838\|\|full | 5317 | No | NA | 7 | 0 | 4 | Not-determined | Genome-fragment | NA | NA | 0 |
| S2-final-viral-combined_k141_69362\|\|full | 6690 | No | NA | 5 | 0 | 3 | Not-determined | Genome-fragment | NA | NA | 0 |
| S2-final-viral-combined_k141_38922\|\|full | 3902 | No | NA | 5 | 0 | 3 | Not-determined | Genome-fragment | NA | NA | 0 |
| S1-final-viral-combined_k141_167080\|\|full | 4018 | No | NA | 6 | 3 | 0 | Low-quality | Genome-fragment | 10.3 | AAI-based (high-confidence) | 0 |
| S2-final-viral-combined_k141_235901\|\|full | 1549 | No | NA | 3 | 0 | 2 | Not-determined | Genome-fragment | NA | NA | 0 |
| S2-final-viral-combined_k141_69390\|\|full | 2720 | No | NA | 2 | 0 | 1 | Not-determined | Genome-fragment | NA | NA | 0 |
| S2-final-viral-combined_k141_69396\|\|full | 1004 | No | NA | 2 | 0 | 1 | Not-determined | Genome-fragment | NA | NA | 0 |
| S8-final-viral-combined_k141_161990\|\|full | 4020 | No | NA | 5 | 1 | 0 | Low-quality | Genome-fragment | 2.9 | HMM-based (lower-bound) | 0 |
| S2-final-viral-combined_k141_5791\|\|full | 143252 | Yes | 30986 | 138 | 14 | 69 | Medium-quality | Genome-fragment | 59.4 | HMM-based (lower-bound) | 78.37 |
| S2-final-viral-combined_k141_193717\|\|full | 1345 | No | NA | 2 | 0 | 0 | Not-determined | Genome-fragment | NA | NA | 0 |
| S2-final-viral-combined_k141_235927\|\|full | 2411 | No | NA | 3 | 0 | 0 | Not-determined | Genome-fragment | NA | NA | 0 |
| S2-final-viral-combined_k141_244177\|\|full | 3479 | No | NA | 3 | 0 | 0 | Not-determined | Genome-fragment | NA | NA | 0 |
| S9_k141_63772 | 4023 | No | NA | 10 | 0 | 0 | Low-quality | Genome-fragment | 9.99 | AAI-based (high-confidence) | 0 |
| S2-final-viral-combined_k141_193719\|\|full | 10475 | No | NA | 11 | 0 | 2 | Not-determined | Genome-fragment | NA | NA | 0 |
| S5-final-viral-combined_k141_85112\|\|full | 4023 | No | NA | 5 | 0 | 4 | Low-quality | Genome-fragment | 1.79 | AAI-based (medium-confidence) | 0 |
| S2-final-viral-combined_k141_161940\|\|full | 2243 | No | NA | 4 | 0 | 1 | Not-determined | Genome-fragment | NA | NA | 0 |
| S2-final-viral-combined_k141_235953\|\|full | 5087 | No | NA | 9 | 0 | 1 | Not-determined | Genome-fragment | NA | NA | 0 |
| S17-final-viral-combined_k141_37257\|\|full | 4023 | No | NA | 7 | 2 | 0 | Low-quality | Genome-fragment | 10.08 | AAI-based (high-confidence) | 0 |
| S2-final-viral-combined_k141_69474\|\|full | 1063 | No | NA | 2 | 0 | 1 | Not-determined | Genome-fragment | NA | NA | 0 |
| S2-final-viral-combined_k141_193748\|\|full | 3020 | No | NA | 2 | 0 | 0 | Not-determined | Genome-fragment | NA | NA | 0 |
| S2-final-viral-combined_k141_223527\|\|full | 5479 | No | NA | 5 | 0 | 5 | Not-determined | Genome-fragment | NA | NA | 0 |
| S11-final-viral-combined_k141_121447\|\|full | 4024 | No | NA | 10 | 0 | 1 | Low-quality | Genome-fragment | 5.35 | AAI-based (medium-confidence) | 0 |
| S2-final-viral-combined_k141_203031\|\|full | 3641 | No | NA | 5 | 0 | 1 | Not-determined | Genome-fragment | NA | NA | 0 |
| S2-final-viral-combined_k141_90670\|\|full | 2055 | No | NA | 2 | 0 | 2 | Not-determined | Genome-fragment | NA | NA | 0 |
| S2-final-viral-combined_k141_26642\|\|full | 23822 | No | NA | 22 | 0 | 16 | Not-determined | Genome-fragment | NA | NA | 0 |
| S2-final-viral-combined_k141_79863\|\|full | 3259 | No | NA | 2 | 0 | 0 | Not-determined | Genome-fragment | NA | NA | 0 |
| S2-final-viral-combined_k141_152757\|\|full | 2267 | No | NA | 3 | 0 | 1 | Not-determined | Genome-fragment | NA | NA | 0 |
| S12_k141_371834 | 4024 | No | NA | 6 | 0 | 5 | Low-quality | Genome-fragment | 1.49 | AAI-based (medium-confidence) | 0 |
| S8_k141_5883 | 4025 | No | NA | 3 | 0 | 0 | Low-quality | Genome-fragment | 3.27 | AAI-based (high-confidence) | 0 |
| S2-final-viral-combined_k141_236030\|\|full | 4001 | No | NA | 5 | 0 | 0 | Not-determined | Genome-fragment | NA | NA | 0 |
| S5-final-viral-combined_k141_160666\|\|full | 4026 | No | NA | 3 | 3 | 0 | Low-quality | Genome-fragment | 8.67 | AAI-based (high-confidence) | 0 |
| S2-final-viral-combined_k141_193830\|\|full | 2739 | No | NA | 3 | 0 | 2 | Not-determined | Genome-fragment | NA | NA | 0 |
| S12_k141_24807 | 4026 | No | NA | 2 | 0 | 2 | Low-quality | Genome-fragment | 2.03 | AAI-based (medium-confidence) | 0 |
| S2-final-viral-combined_k141_142260\|\|full | 7282 | No | NA | 4 | 0 | 2 | Not-determined | Genome-fragment | NA | NA | 0 |
| S2-final-viral-combined_k141_236061\|\|full | 7797 | No | NA | 10 | 0 | 6 | Not-determined | Genome-fragment | NA | NA | 0 |
| S2-final-viral-combined_k141_38538\|\|full | 4026 | No | NA | 5 | 2 | 0 | Low-quality | Genome-fragment | 12.19 | AAI-based (high-confidence) | 0 |
| S2-final-viral-combined_k141_130454\|\|full | 3649 | No | NA | 5 | 0 | 3 | Not-determined | Genome-fragment | NA | NA | 0 |
| S2-final-viral-combined_k141_236065\|\|full | 2747 | No | NA | 3 | 0 | 0 | Not-determined | Genome-fragment | NA | NA | 0 |
| S2-final-viral-combined_k141_51227\|\|full | 1296 | No | NA | 3 | 0 | 0 | Not-determined | Genome-fragment | NA | NA | 0 |
| S2-final-viral-combined_k141_39079\|\|full | 4295 | No | NA | 6 | 0 | 2 | Not-determined | Genome-fragment | NA | NA | 0 |
| S2-final-viral-combined_k141_130464\|\|full | 3368 | No | NA | 3 | 0 | 1 | Not-determined | Genome-fragment | NA | NA | 0 |
| S8_k141_24268 | 4026 | No | NA | 7 | 0 | 1 | Low-quality | Genome-fragment | 6.29 | AAI-based (medium-confidence) | 0 |
| S11-final-viral-combined_k141_253245\|\|full | 4027 | No | NA | 6 | 1 | 0 | Low-quality | Genome-fragment | 7.48 | AAI-based (high-confidence) | 0 |
| S7_k141_175287 | 4027 | No | NA | 3 | 0 | 3 | Low-quality | Genome-fragment | 1.74 | AAI-based (medium-confidence) | 0 |
| S2-final-viral-combined_k141_39099\|\|full | 2422 | No | NA | 3 | 0 | 0 | Not-determined | Genome-fragment | NA | NA | 0 |
| S17-final-viral-combined_k141_228639\|\|full | 4027 | No | NA | 3 | 0 | 0 | Low-quality | Genome-fragment | 4.89 | AAI-based (high-confidence) | 0 |
| S2-final-viral-combined_k141_130507\|\|full | 3662 | No | NA | 3 | 0 | 2 | Not-determined | Genome-fragment | NA | NA | 0 |
| S2-final-viral-combined_k141_90764\|\|full | 2142 | No | NA | 2 | 0 | 0 | Not-determined | Genome-fragment | NA | NA | 0 |
| S2-final-viral-combined_k141_51271\|\|full | 3473 | No | NA | 3 | 0 | 3 | Not-determined | Genome-fragment | NA | NA | 0 |
| S2-final-viral-combined_k141_59414\|\|full | 4101 | No | NA | 7 | 0 | 5 | Not-determined | Genome-fragment | NA | NA | 0 |
| S17-final-viral-combined_k141_250348\|\|full | 4027 | No | NA | 5 | 4 | 0 | Low-quality | Genome-fragment | 12.45 | AAI-based (high-confidence) | 0 |
| S2-final-viral-combined_k141_90768\|\|full | 8702 | No | NA | 12 | 0 | 1 | Not-determined | Genome-fragment | NA | NA | 0 |
| S5-final-viral-combined_k141_85750\|\|full | 4028 | No | NA | 4 | 0 | 0 | Low-quality | Genome-fragment | 2.85 | AAI-based (medium-confidence) | 0 |
| S2-final-viral-combined_k141_174491\|\|full | 1877 | No | NA | 4 | 0 | 1 | Not-determined | Genome-fragment | NA | NA | 0 |
| S2-final-viral-combined_k141_183818\|\|full | 6078 | No | NA | 6 | 0 | 2 | Not-determined | Genome-fragment | NA | NA | 0 |
| S2-final-viral-combined_k141_174502\|\|full | 4173 | No | NA | 7 | 0 | 5 | Not-determined | Genome-fragment | NA | NA | 0 |
| S2-final-viral-combined_k141_236081\|\|full | 2393 | No | NA | 3 | 0 | 1 | Not-determined | Genome-fragment | NA | NA | 0 |
| S2-final-viral-combined_k141_183859\|\|full | 4484 | No | NA | 6 | 0 | 2 | Not-determined | Genome-fragment | NA | NA | 0 |
| S2-final-viral-combined_k141_152960\|\|full | 1002 | No | NA | 3 | 0 | 0 | Not-determined | Genome-fragment | NA | NA | 0 |
| S2-final-viral-combined_k141_152968\|\|full | 2342 | No | NA | 2 | 0 | 0 | Not-determined | Genome-fragment | NA | NA | 0 |
| S2-final-viral-combined_k141_79966\|\|full | 1182 | No | NA | 2 | 0 | 0 | Not-determined | Genome-fragment | NA | NA | 0 |
| S17-final-viral-combined_k141_135631\|\|full | 4028 | Yes | 3668 | 10 | 2 | 1 | Low-quality | Genome-fragment | 6.86 | AAI-based (high-confidence) | 8.94 |
| S1-final-viral-combined_k141_79706\|\|full | 4029 | Yes | 829 | 8 | 1 | 3 | Low-quality | Genome-fragment | 0.57 | HMM-based (lower-bound) | 79.42 |
| S12-final-viral-combined_k141_384243\|\|full | 4029 | No | NA | 2 | 0 | 2 | Low-quality | Genome-fragment | 1.39 | AAI-based (medium-confidence) | 0 |
| S2-final-viral-combined_k141_51330\|\|full | 4137 | No | NA | 3 | 0 | 3 | Not-determined | Genome-fragment | NA | NA | 0 |
| S2-final-viral-combined_k141_223720\|\|full | 1949 | No | NA | 2 | 0 | 0 | Not-determined | Genome-fragment | NA | NA | 0 |
| S6-final-viral-combined_k141_125932\|\|full | 4029 | Yes | 1675 | 5 | 1 | 2 | Low-quality | Genome-fragment | 1.93 | HMM-based (lower-bound) | 58.43 |
| S17-final-viral-combined_k141_88455\|\|lt2gene | 4030 | No | NA | 2 | 2 | 0 | Low-quality | Genome-fragment | 9.72 | AAI-based (high-confidence) | 0 |
| S2-final-viral-combined_k141_214108\|\|full | 1516 | No | NA | 3 | 0 | 1 | Not-determined | Genome-fragment | NA | NA | 0 |
| S2_k141_213095 | 4030 | No | NA | 6 | 0 | 2 | Low-quality | Genome-fragment | 4.8 | AAI-based (medium-confidence) | 0 |
| S17-final-viral-combined_k141_26381\|\|full | 4033 | No | NA | 4 | 0 | 3 | Low-quality | Genome-fragment | 5.48 | AAI-based (high-confidence) | 0 |
| S2-final-viral-combined_k141_203180\|\|full | 1604 | No | NA | 3 | 0 | 2 | Not-determined | Genome-fragment | NA | NA | 0 |
| S1-final-viral-combined_k141_68832\|\|full | 4034 | Yes | 1355 | 5 | 1 | 1 | Low-quality | Genome-fragment | 1.71 | AAI-based (medium-confidence) | 66.41 |
| S12-final-viral-combined_k141_394058\|\|full | 4035 | Yes | 2711 | 5 | 1 | 1 | Low-quality | Genome-fragment | 4.06 | AAI-based (medium-confidence) | 32.81 |
| S12_k141_183236 | 4035 | No | NA | 8 | 2 | 0 | Low-quality | Genome-fragment | 6.03 | AAI-based (high-confidence) | 0 |
| S6-final-viral-combined_k141_57048\|\|full | 4035 | No | NA | 5 | 2 | 0 | Low-quality | Genome-fragment | 10.01 | AAI-based (medium-confidence) | 0 |
| S11-final-viral-combined_k141_308541\|\|full | 4036 | No | NA | 7 | 0 | 0 | Low-quality | Genome-fragment | 4.17 | AAI-based (medium-confidence) | 0 |
| S2-final-viral-combined_k141_244305\|\|full | 2649 | No | NA | 3 | 0 | 1 | Not-determined | Genome-fragment | NA | NA | 0 |
| S2-final-viral-combined_k141_17988\|\|full | 2180 | No | NA | 3 | 0 | 2 | Not-determined | Genome-fragment | NA | NA | 0 |
| S5_k141_49524 | 4037 | No | NA | 6 | 0 | 0 | Low-quality | Genome-fragment | 6.74 | AAI-based (high-confidence) | 0 |
| S17-final-viral-combined_k141_2511\|\|full | 4037 | Yes | 2244 | 6 | 2 | 2 | Low-quality | Genome-fragment | 5.25 | AAI-based (high-confidence) | 44.41 |
| S12-final-viral-combined_k141_317141\|\|full | 4037 | No | NA | 3 | 0 | 2 | Low-quality | Genome-fragment | 0.39 | AAI-based (medium-confidence) | 0 |
| S12_k141_97051 | 4037 | No | NA | 3 | 0 | 1 | Low-quality | Genome-fragment | 5.9 | AAI-based (medium-confidence) | 0 |
| S4_k141_9054 | 4037 | No | NA | 6 | 0 | 1 | Low-quality | Genome-fragment | 6.25 | AAI-based (high-confidence) | 0 |
| S2-final-viral-combined_k141_101360\|\|full | 1899 | No | NA | 3 | 0 | 2 | Not-determined | Genome-fragment | NA | NA | 0 |
| S2-final-viral-combined_k141_203245\|\|full | 5087 | No | NA | 7 | 0 | 3 | Not-determined | Genome-fragment | NA | NA | 0 |
| S6_k141_17795 | 4037 | No | NA | 7 | 2 | 0 | Low-quality | Genome-fragment | 8.28 | AAI-based (medium-confidence) | 0 |
| S8-final-viral-combined_k141_264592\|\|full | 4037 | No | NA | 2 | 0 | 2 | Low-quality | Genome-fragment | 9.02 | AAI-based (medium-confidence) | 0 |
| S2-final-viral-combined_k141_26718\|\|full | 8395 | No | NA | 6 | 0 | 1 | Not-determined | Genome-fragment | NA | NA | 0 |
| S12-final-viral-combined_k141_213782\|\|full | 4038 | No | NA | 4 | 0 | 4 | Low-quality | Genome-fragment | 1.07 | AAI-based (medium-confidence) | 0 |
| S2-final-viral-combined_k141_162091\|\|full | 5089 | No | NA | 3 | 0 | 0 | Not-determined | Genome-fragment | NA | NA | 0 |
| S9-final-viral-combined_k141_44248\|\|full | 4039 | No | NA | 5 | 0 | 1 | Low-quality | Genome-fragment | 1.15 | AAI-based (medium-confidence) | 0 |
| S1-final-viral-combined_k141_229640\|\|full | 4039 | No | NA | 4 | 0 | 1 | Low-quality | Genome-fragment | 3.81 | AAI-based (medium-confidence) | 0 |
| S2-final-viral-combined_k141_110548\|\|full | 1261 | No | NA | 3 | 0 | 0 | Not-determined | Genome-fragment | NA | NA | 0 |
| S2-final-viral-combined_k141_184039\|\|full | 5336 | No | NA | 5 | 0 | 0 | Not-determined | Genome-fragment | NA | NA | 0 |
| S17-k141_234669 | 4039 | No | NA | 6 | 1 | 0 | Low-quality | Genome-fragment | 4.81 | HMM-based (lower-bound) | 0 |
| S2-final-viral-combined_k141_110549\|\|full | 3491 | No | NA | 5 | 0 | 1 | Not-determined | Genome-fragment | NA | NA | 0 |
| S12-final-viral-combined_k141_321017\|\|full | 4039 | No | NA | 5 | 0 | 3 | Low-quality | Genome-fragment | 8.68 | AAI-based (high-confidence) | 0 |
| S2-final-viral-combined_k141_6063\|\|full | 2336 | No | NA | 4 | 0 | 0 | Not-determined | Genome-fragment | NA | NA | 0 |
| S2-final-viral-combined_k141_18055\|\|full | 2207 | No | NA | 3 | 0 | 0 | Not-determined | Genome-fragment | NA | NA | 0 |
| S2-final-viral-combined_k141_236201\|\|full | 2374 | No | NA | 2 | 0 | 1 | Not-determined | Genome-fragment | NA | NA | 0 |
| S17-final-viral-combined_k141_28068\|\|full | 4040 | No | NA | 6 | 0 | 0 | Low-quality | Genome-fragment | 3.13 | AAI-based (medium-confidence) | 0 |
| S1_k141_195455 | 4042 | No | NA | 3 | 0 | 1 | Low-quality | Genome-fragment | 6.91 | AAI-based (high-confidence) | 0 |
| S12-final-viral-combined_k141_151862\|\|full | 4042 | No | NA | 5 | 3 | 0 | Low-quality | Genome-fragment | 8.6 | AAI-based (high-confidence) | 0 |
| S2-final-viral-combined_k141_110592\|\|full | 1488 | No | NA | 3 | 0 | 2 | Not-determined | Genome-fragment | NA | NA | 0 |
| S2-final-viral-combined_k141_5014\|\|full | 4042 | Yes | 3030 | 4 | 1 | 1 | Low-quality | Genome-fragment | 1.82 | AAI-based (high-confidence) | 25.04 |
| S2-final-viral-combined_k141_184119\|\|full | 2728 | No | NA | 4 | 0 | 0 | Not-determined | Genome-fragment | NA | NA | 0 |
| S2-final-viral-combined_k141_121890\|\|full | 1327 | No | NA | 2 | 0 | 1 | Not-determined | Genome-fragment | NA | NA | 0 |
| S2-final-viral-combined_k141_214222\|\|full | 1420 | No | NA | 4 | 0 | 1 | Not-determined | Genome-fragment | NA | NA | 0 |
| S1-final-viral-combined_k141_345092\|\|full | 4044 | No | NA | 8 | 4 | 0 | Low-quality | Genome-fragment | 12.67 | AAI-based (high-confidence) | 0 |
| S1_k141_388587 | 4044 | No | NA | 9 | 1 | 0 | Low-quality | Genome-fragment | 11.49 | AAI-based (high-confidence) | 0 |
| S6-final-viral-combined_k141_54916\|\|full | 4045 | No | NA | 8 | 1 | 3 | Low-quality | Genome-fragment | 9.25 | AAI-based (high-confidence) | 0 |
| S2-final-viral-combined_k141_51446\|\|full | 8622 | No | NA | 13 | 3 | 0 | Medium-quality | Genome-fragment | 64.37 | AAI-based (medium-confidence) | 0 |
| S6-final-viral-combined_k141_139467\|\|full | 4045 | No | NA | 4 | 0 | 2 | Low-quality | Genome-fragment | 2.27 | AAI-based (medium-confidence) | 0 |
| S2-final-viral-combined_k141_51451\|\|full | 1270 | No | NA | 3 | 0 | 1 | Not-determined | Genome-fragment | NA | NA | 0 |
| S2-final-viral-combined_k141_142403\|\|full | 2336 | No | NA | 2 | 0 | 0 | Not-determined | Genome-fragment | NA | NA | 0 |
| S12-final-viral-combined_k141_80269\|\|full | 4046 | No | NA | 4 | 0 | 3 | Low-quality | Genome-fragment | 0.61 | AAI-based (medium-confidence) | 0 |
| S9-final-viral-combined_k141_83339\|\|full | 4047 | No | NA | 4 | 0 | 3 | Low-quality | Genome-fragment | 3.94 | AAI-based (medium-confidence) | 0 |
| S2-final-viral-combined_k141_174793\|\|full | 1297 | No | NA | 3 | 0 | 2 | Not-determined | Genome-fragment | NA | NA | 0 |
| S2-final-viral-combined_k141_110619\|\|full | 1393 | No | NA | 3 | 0 | 1 | Not-determined | Genome-fragment | NA | NA | 0 |
| S2-final-viral-combined_k141_214275\|\|full | 1177 | No | NA | 3 | 0 | 1 | Not-determined | Genome-fragment | NA | NA | 0 |
| S2-final-viral-combined_k141_184384\|\|full | 4047 | No | NA | 6 | 1 | 0 | Low-quality | Genome-fragment | 5.01 | HMM-based (lower-bound) | 0 |
| S2_k141_50603 | 4050 | No | NA | 2 | 0 | 2 | Low-quality | Genome-fragment | 2.69 | AAI-based (medium-confidence) | 0 |
| S2-final-viral-combined_k141_214294\|\|full | 1036 | No | NA | 3 | 0 | 0 | Not-determined | Genome-fragment | NA | NA | 0 |
| S2-final-viral-combined_k141_236292\|\|full | 1657 | No | NA | 3 | 0 | 0 | Not-determined | Genome-fragment | NA | NA | 0 |
| S4_k141_104441 | 4050 | No | NA | 3 | 0 | 0 | Low-quality | Genome-fragment | 6.82 | AAI-based (medium-confidence) | 0 |
| S2-final-viral-combined_k141_130761\|\|full | 1775 | No | NA | 3 | 0 | 1 | Not-determined | Genome-fragment | NA | NA | 0 |
| S9-final-viral-combined_k141_41241\|\|full | 4051 | No | NA | 10 | 2 | 0 | Low-quality | Genome-fragment | 8.94 | AAI-based (medium-confidence) | 0 |
| S1-final-viral-combined_k141_56592\|\|full | 4051 | No | NA | 8 | 5 | 0 | Low-quality | Genome-fragment | 9.55 | AAI-based (medium-confidence) | 0 |
| S11-final-viral-combined_k141_260540\|\|full | 4052 | No | NA | 3 | 0 | 2 | Low-quality | Genome-fragment | 2.46 | AAI-based (medium-confidence) | 0 |
| S12-final-viral-combined_k141_66382\|\|lt2gene | 4052 | No | NA | 1 | 1 | 0 | Low-quality | Genome-fragment | 2.2 | HMM-based (lower-bound) | 0 |
| S2-final-viral-combined_k141_104682\|\|full | 4052 | Yes | 3363 | 6 | 1 | 1 | Low-quality | Genome-fragment | 0.95 | HMM-based (lower-bound) | 17 |
| S2-final-viral-combined_k141_236319\|\|full | 10139 | No | NA | 7 | 0 | 2 | Not-determined | Genome-fragment | NA | NA | 0 |
| S2-final-viral-combined_k141_6205\|\|full | 1336 | No | NA | 2 | 0 | 0 | Not-determined | Genome-fragment | NA | NA | 0 |
| S11_k141_96702 | 4053 | No | NA | 5 | 0 | 1 | Low-quality | Genome-fragment | 12.49 | AAI-based (high-confidence) | 0 |
| S2-final-viral-combined_k141_6220\|\|full | 6280 | No | NA | 4 | 0 | 0 | Not-determined | Genome-fragment | NA | NA | 0 |
| S2-final-viral-combined_k141_194183\|\|full | 2265 | No | NA | 2 | 0 | 2 | Not-determined | Genome-fragment | NA | NA | 0 |
| S2-final-viral-combined_k141_39529\|\|full | 2557 | No | NA | 3 | 0 | 2 | Not-determined | Genome-fragment | NA | NA | 0 |
| S1-final-viral-combined_k141_159744\|\|full | 4053 | No | NA | 5 | 0 | 3 | Low-quality | Genome-fragment | 9.37 | AAI-based (medium-confidence) | 0 |
| S2-final-viral-combined_k141_130874\|\|full | 2961 | No | NA | 4 | 0 | 3 | Not-determined | Genome-fragment | NA | NA | 0 |
| S8-final-viral-combined_k141_27999\|\|full | 4054 | No | NA | 6 | 0 | 2 | Low-quality | Genome-fragment | 2.15 | AAI-based (medium-confidence) | 0 |
| S2-final-viral-combined_k141_223922\|\|full | 1157 | No | NA | 2 | 0 | 2 | Not-determined | Genome-fragment | NA | NA | 0 |
| S2-final-viral-combined_k141_130888\|\|full | 2602 | No | NA | 3 | 0 | 1 | Not-determined | Genome-fragment | NA | NA | 0 |
| S2-final-viral-combined_k141_194228\|\|full | 1699 | No | NA | 2 | 0 | 2 | Not-determined | Genome-fragment | NA | NA | 0 |
| S11-final-viral-combined_k141_253838\|\|full | 4055 | No | NA | 7 | 2 | 1 | Low-quality | Genome-fragment | 8.65 | AAI-based (high-confidence) | 0 |
| S1-final-viral-combined_k141_139442\|\|full | 4055 | No | NA | 7 | 3 | 0 | Low-quality | Genome-fragment | 6.82 | AAI-based (high-confidence) | 0 |
| S2-final-viral-combined_k141_184409\|\|full | 2294 | No | NA | 3 | 0 | 0 | Not-determined | Genome-fragment | NA | NA | 0 |
| S2-final-viral-combined_k141_223937\|\|full | 1773 | No | NA | 3 | 0 | 2 | Not-determined | Genome-fragment | NA | NA | 0 |
| S2-final-viral-combined_k141_236400\|\|full | 2973 | No | NA | 9 | 0 | 0 | Not-determined | Genome-fragment | NA | NA | 0 |
| S12-final-viral-combined_k141_142017\|\|full | 4056 | No | NA | 5 | 0 | 2 | Low-quality | Genome-fragment | 2.79 | AAI-based (high-confidence) | 0 |
| S2-final-viral-combined_k141_51577\|\|full | 3151 | No | NA | 3 | 0 | 3 | Not-determined | Genome-fragment | NA | NA | 0 |
| S7-final-viral-combined_k141_237481\|\|full | 4057 | No | NA | 3 | 0 | 3 | Low-quality | Genome-fragment | 2.05 | AAI-based (medium-confidence) | 0 |
| S2-final-viral-combined_k141_59767\|\|full | 1320 | No | NA | 2 | 0 | 0 | Not-determined | Genome-fragment | NA | NA | 0 |
| S17-final-viral-combined_k141_248903\|\|full | 4057 | No | NA | 5 | 0 | 2 | Low-quality | Genome-fragment | 8.83 | AAI-based (medium-confidence) | 0 |
| S2-final-viral-combined_k141_91177\|\|full | 3067 | No | NA | 3 | 0 | 3 | Not-determined | Genome-fragment | NA | NA | 0 |
| S2-final-viral-combined_k141_51586\|\|full | 9411 | No | NA | 16 | 0 | 0 | Not-determined | Genome-fragment | NA | NA | 0 |
| S2-final-viral-combined_k141_80179\|\|full | 3928 | No | NA | 4 | 0 | 2 | Not-determined | Genome-fragment | NA | NA | 0 |
| S2-final-viral-combined_k141_91189\|\|full | 1714 | No | NA | 3 | 0 | 1 | Not-determined | Genome-fragment | NA | NA | 0 |
| S2-final-viral-combined_k141_184430\|\|full | 15780 | No | NA | 17 | 0 | 10 | Not-determined | Genome-fragment | NA | NA | 0 |
| S2-final-viral-combined_k141_168888\|\|full | 4058 | No | NA | 7 | 2 | 0 | Low-quality | Genome-fragment | 9.58 | AAI-based (high-confidence) | 0 |
| S2-final-viral-combined_k141_80204\|\|full | 2007 | No | NA | 3 | 0 | 2 | Not-determined | Genome-fragment | NA | NA | 0 |
| S2-final-viral-combined_k141_69977\|\|full | 2456 | No | NA | 2 | 0 | 2 | Not-determined | Genome-fragment | NA | NA | 0 |
| S1-final-viral-combined_k141_74342\|\|full | 4059 | No | NA | 4 | 1 | 0 | Low-quality | Genome-fragment | 9.25 | AAI-based (high-confidence) | 0 |
| S2-final-viral-combined_k141_6301\|\|full | 1141 | No | NA | 2 | 0 | 1 | Not-determined | Genome-fragment | NA | NA | 0 |
| S2_k141_244030 | 4059 | No | NA | 5 | 5 | 0 | Low-quality | Genome-fragment | 9.61 | AAI-based (high-confidence) | 0 |
| S2-final-viral-combined_k141_27032\|\|full | 1691 | No | NA | 2 | 0 | 2 | Not-determined | Genome-fragment | NA | NA | 0 |
| S11_k141_236885 | 4060 | No | NA | 5 | 1 | 0 | Low-quality | Genome-fragment | 10.34 | AAI-based (medium-confidence) | 0 |
| S2-final-viral-combined_k141_51604\|\|full | 17451 | No | NA | 18 | 0 | 12 | Not-determined | Genome-fragment | NA | NA | 0 |
| S2-final-viral-combined_k141_6311\|\|full | 1126 | No | NA | 2 | 0 | 1 | Not-determined | Genome-fragment | NA | NA | 0 |
| S2-final-viral-combined_k141_18347\|\|full | 2096 | No | NA | 3 | 0 | 1 | Not-determined | Genome-fragment | NA | NA | 0 |
| S1-final-viral-combined_k141_66520\|\|full | 4060 | No | NA | 6 | 1 | 0 | Low-quality | Genome-fragment | 2.93 | AAI-based (high-confidence) | 0 |
| S2-final-viral-combined_k141_142650\|\|full | 24175 | No | NA | 15 | 0 | 4 | Not-determined | Genome-fragment | NA | NA | 0 |
| S11-final-viral-combined_k141_155445\|\|full | 4061 | No | NA | 7 | 2 | 0 | Low-quality | Genome-fragment | 9.3 | AAI-based (medium-confidence) | 0 |
| S2-final-viral-combined_k141_18361\|\|full | 1264 | No | NA | 2 | 0 | 2 | Not-determined | Genome-fragment | NA | NA | 0 |
| S2-final-viral-combined_k141_142661\|\|full | 5527 | No | NA | 8 | 0 | 2 | Not-determined | Genome-fragment | NA | NA | 0 |
| S2-final-viral-combined_k141_110778\|\|full | 2099 | No | NA | 3 | 0 | 2 | Not-determined | Genome-fragment | NA | NA | 0 |
| S2-final-viral-combined_k141_70047\|\|full | 1396 | No | NA | 2 | 0 | 0 | Not-determined | Genome-fragment | NA | NA | 0 |
| S2-final-viral-combined_k141_70060\|\|full | 2709 | No | NA | 3 | 0 | 2 | Not-determined | Genome-fragment | NA | NA | 0 |
| S2-final-viral-combined_k141_162396\|\|full | 1881 | No | NA | 3 | 0 | 1 | Not-determined | Genome-fragment | NA | NA | 0 |
| S2-final-viral-combined_k141_101751\|\|full | 2846 | No | NA | 4 | 0 | 1 | Not-determined | Genome-fragment | NA | NA | 0 |
| S1_k141_186326 | 4061 | No | NA | 10 | 0 | 0 | Low-quality | Genome-fragment | 11.35 | AAI-based (high-confidence) | 0 |
| S12-final-viral-combined_k141_257400\|\|full | 4061 | No | NA | 3 | 0 | 0 | Low-quality | Genome-fragment | 4.08 | AAI-based (medium-confidence) | 0 |
| S2-final-viral-combined_k141_93974\|\|full | 4062 | No | NA | 3 | 0 | 1 | Low-quality | Genome-fragment | 3.34 | AAI-based (medium-confidence) | 0 |
| S2-final-viral-combined_k141_236489\|\|full | 5863 | No | NA | 5 | 0 | 3 | Not-determined | Genome-fragment | NA | NA | 0 |
| S11_k141_204910 | 4063 | No | NA | 4 | 1 | 0 | Low-quality | Genome-fragment | 9.19 | AAI-based (high-confidence) | 0 |
| S2-final-viral-combined_k141_194352\|\|full | 2410 | No | NA | 2 | 0 | 2 | Not-determined | Genome-fragment | NA | NA | 0 |
| S2-final-viral-combined_k141_80279\|\|full | 1483 | No | NA | 3 | 0 | 1 | Not-determined | Genome-fragment | NA | NA | 0 |
| S2-final-viral-combined_k141_224016\|\|full | 1082 | No | NA | 3 | 0 | 1 | Not-determined | Genome-fragment | NA | NA | 0 |
| S3_k141_1214 | 4063 | No | NA | 9 | 7 | 0 | Low-quality | Genome-fragment | 11.09 | AAI-based (high-confidence) | 0 |
| S2-final-viral-combined_k141_80298\|\|full | 1759 | No | NA | 2 | 0 | 2 | Not-determined | Genome-fragment | NA | NA | 0 |
| S1-final-viral-combined_k141_124486\|\|full | 4064 | No | NA | 3 | 0 | 2 | Low-quality | Genome-fragment | 2.94 | AAI-based (medium-confidence) | 0 |
| S2-final-viral-combined_k141_122248\|\|full | 5029 | No | NA | 6 | 0 | 1 | Not-determined | Genome-fragment | NA | NA | 0 |
| S2-final-viral-combined_k141_39752\|\|full | 2110 | No | NA | 4 | 0 | 0 | Not-determined | Genome-fragment | NA | NA | 0 |
| S4_k141_140003 | 4065 | No | NA | 4 | 0 | 1 | Low-quality | Genome-fragment | 4.84 | AAI-based (high-confidence) | 0 |
| S8-final-viral-combined_k141_130411\|\|full | 4065 | No | NA | 3 | 0 | 2 | Low-quality | Genome-fragment | 4.64 | AAI-based (medium-confidence) | 0 |
| S12_k141_182631 | 4067 | No | NA | 9 | 1 | 0 | Low-quality | Genome-fragment | 6.25 | AAI-based (high-confidence) | 0 |
| S2-final-viral-combined_k141_224043\|\|full | 7151 | No | NA | 5 | 0 | 4 | Not-determined | Genome-fragment | NA | NA | 0 |
| S1_k141_49560 | 4068 | No | NA | 4 | 4 | 0 | Low-quality | Genome-fragment | 10.09 | AAI-based (high-confidence) | 0 |
| S2-final-viral-combined_k141_194389\|\|full | 1845 | No | NA | 2 | 0 | 1 | Not-determined | Genome-fragment | NA | NA | 0 |
| S2-final-viral-combined_k141_110853\|\|full | 1066 | No | NA | 3 | 0 | 0 | Not-determined | Genome-fragment | NA | NA | 0 |
| S2-final-viral-combined_k141_91308\|\|full | 4478 | No | NA | 7 | 0 | 6 | Not-determined | Genome-fragment | NA | NA | 0 |
| S2-final-viral-combined_k141_6360\|\|full | 13088 | No | NA | 5 | 0 | 3 | Not-determined | Genome-fragment | NA | NA | 0 |
| S2-final-viral-combined_k141_122300\|\|full | 2860 | No | NA | 5 | 0 | 0 | Not-determined | Genome-fragment | NA | NA | 0 |
| S7-final-viral-combined_k141_225489\|\|full | 4069 | No | NA | 3 | 0 | 3 | Low-quality | Genome-fragment | 2.85 | AAI-based (medium-confidence) | 0 |
| S2-final-viral-combined_k141_91311\|\|full | 1318 | No | NA | 4 | 0 | 1 | Not-determined | Genome-fragment | NA | NA | 0 |
| S2-final-viral-combined_k141_122332\|\|full | 2670 | No | NA | 3 | 0 | 3 | Not-determined | Genome-fragment | NA | NA | 0 |
| S2-final-viral-combined_k141_101826\|\|full | 62201 | Yes | 26132 | 76 | 19 | 16 | Complete | High-quality | 100 | Provirus (medium-confidence) | 57.99 |
| S2_k141_118805 | 4069 | No | NA | 8 | 1 | 0 | Low-quality | Genome-fragment | 10.1 | AAI-based (medium-confidence) | 0 |
| S2-final-viral-combined_k141_101829\|\|full | 3193 | No | NA | 3 | 0 | 0 | Not-determined | Genome-fragment | NA | NA | 0 |
| S6_k141_16444 | 4069 | No | NA | 3 | 0 | 3 | Low-quality | Genome-fragment | 9.68 | AAI-based (high-confidence) | 0 |
| S2-final-viral-combined_k141_175187\|\|full | 2545 | No | NA | 3 | 0 | 3 | Not-determined | Genome-fragment | NA | NA | 0 |
| S2-final-viral-combined_k141_162517\|\|full | 2749 | No | NA | 3 | 0 | 2 | Not-determined | Genome-fragment | NA | NA | 0 |
| S2-final-viral-combined_k141_236594\|\|full | 1615 | No | NA | 3 | 0 | 0 | Not-determined | Genome-fragment | NA | NA | 0 |
| S7_k141_179225 | 4071 | No | NA | 5 | 0 | 4 | Low-quality | Genome-fragment | 0.98 | AAI-based (medium-confidence) | 0 |
| S2-final-viral-combined_k141_214463\|\|full | 1564 | No | NA | 2 | 0 | 1 | Not-determined | Genome-fragment | NA | NA | 0 |
| S6-final-viral-combined_k141_80653\|\|full | 4071 | No | NA | 5 | 0 | 4 | Low-quality | Genome-fragment | 2.34 | AAI-based (medium-confidence) | 0 |

| | | | | | | | | | | | |
|---|---|---|---|---|---|---|---|---|---|---|---|
| S2-final-viral-combined_k141_122342\|\|full | 2261 | No | NA | 3 | 0 | 1 | Not-determined | Genome-fragment | NA | NA | 0 |
| S11-final-viral-combined_k141_89231\|\|full | 4072 | No | NA | 3 | 3 | 0 | Low-quality | Genome-fragment | 4.95 | AAI-based (high-confidence) | 0 |
| S2-final-viral-combined_k141_80368\|\|full | 2119 | No | NA | 3 | 0 | 1 | Not-determined | Genome-fragment | NA | NA | 0 |
| S1-final-viral-combined_k141_170988\|\|full | 4072 | No | NA | 3 | 0 | 3 | Low-quality | Genome-fragment | 4.22 | AAI-based (high-confidence) | 0 |
| S3-final-viral-combined_k141_3523\|\|full | 4072 | No | NA | 3 | 0 | 0 | Low-quality | Genome-fragment | 4.42 | AAI-based (medium-confidence) | 0 |
| S2-final-viral-combined_k141_175235\|\|full | 8000 | No | NA | 9 | 0 | 6 | Not-determined | Genome-fragment | NA | NA | 0 |
| S2-final-viral-combined_k141_122357\|\|full | 5838 | No | NA | 4 | 0 | 2 | Not-determined | Genome-fragment | NA | NA | 0 |
| S11_k141_78703 | 4073 | No | NA | 2 | 0 | 0 | Low-quality | Genome-fragment | 4.31 | AAI-based (high-confidence) | 0 |
| S7-final-viral-combined_k141_50765\|\|full | 4074 | No | NA | 9 | 3 | 0 | Low-quality | Genome-fragment | 7.58 | AAI-based (high-confidence) | 0 |
| S17-final-viral-combined_k141_232205\|\|full | 4075 | No | NA | 5 | 0 | 2 | Low-quality | Genome-fragment | 1.87 | AAI-based (medium-confidence) | 0 |
| S2-final-viral-combined_k141_194580\|\|full | 2741 | No | NA | 3 | 0 | 1 | Not-determined | Genome-fragment | NA | NA | 0 |
| S2-final-viral-combined_k141_59985\|\|full | 2001 | No | NA | 3 | 0 | 0 | Not-determined | Genome-fragment | NA | NA | 0 |
| S2-final-viral-combined_k141_110975\|\|full | 1034 | No | NA | 3 | 0 | 0 | Not-determined | Genome-fragment | NA | NA | 0 |
| S2-final-viral-combined_k141_70283\|\|full | 1488 | No | NA | 1 | 0 | 0 | Not-determined | Genome-fragment | NA | NA | 0 |
| S2-final-viral-combined_k141_39944\|\|full | 2325 | No | NA | 3 | 0 | 2 | Not-determined | Genome-fragment | NA | NA | 0 |
| S8-final-viral-combined_k141_58355\|\|full | 4075 | No | NA | 5 | 0 | 2 | Low-quality | Genome-fragment | 4.34 | AAI-based (medium-confidence) | 0 |
| S2-final-viral-combined_k141_110998\|\|full | 1605 | No | NA | 2 | 0 | 1 | Not-determined | Genome-fragment | NA | NA | 0 |
| S2-final-viral-combined_k141_111007\|\|full | 5274 | No | NA | 4 | 0 | 2 | Not-determined | Genome-fragment | NA | NA | 0 |
| S17-final-viral-combined_k141_66718\|\|full | 4076 | No | NA | 4 | 0 | 2 | Low-quality | Genome-fragment | 4.62 | AAI-based (medium-confidence) | 0 |
| S12-final-viral-combined_k141_316301\|\|full | 4076 | No | NA | 4 | 0 | 2 | Low-quality | Genome-fragment | 2.53 | AAI-based (medium-confidence) | 0 |
| S2-final-viral-combined_k141_70297\|\|full | 3145 | No | NA | 5 | 0 | 0 | Not-determined | Genome-fragment | NA | NA | 0 |
| S11_k141_330202 | 4077 | No | NA | 7 | 1 | 0 | Low-quality | Genome-fragment | 6.17 | HMM-based (lower-bound) | 0 |
| S2-final-viral-combined_k141_27188\|\|full | 2744 | No | NA | 3 | 0 | 0 | Not-determined | Genome-fragment | NA | NA | 0 |
| S2-final-viral-combined_k141_122435\|\|full | 4585 | No | NA | 6 | 0 | 2 | Not-determined | Genome-fragment | NA | NA | 0 |
| S2-final-viral-combined_k141_40028\|\|full | 4892 | No | NA | 4 | 0 | 3 | Not-determined | Genome-fragment | NA | NA | 0 |
| S2-final-viral-combined_k141_101920\|\|full | 1601 | No | NA | 2 | 0 | 1 | Not-determined | Genome-fragment | NA | NA | 0 |
| S2-final-viral-combined_k141_101927\|\|full | 4055 | No | NA | 3 | 0 | 1 | Not-determined | Genome-fragment | NA | NA | 0 |
| S2-final-viral-combined_k141_60077\|\|full | 2428 | No | NA | 2 | 0 | 0 | Not-determined | Genome-fragment | NA | NA | 0 |
| S3_k141_54557 | 4077 | No | NA | 3 | 0 | 0 | Low-quality | Genome-fragment | 10.34 | AAI-based (medium-confidence) | 0 |
| S2-final-viral-combined_k141_70352\|\|full | 3719 | No | NA | 6 | 0 | 0 | Not-determined | Genome-fragment | NA | NA | 0 |
| S2-final-viral-combined_k141_40040\|\|full | 1575 | No | NA | 3 | 0 | 2 | Not-determined | Genome-fragment | NA | NA | 0 |
| S2-final-viral-combined_k141_40042\|\|full | 5383 | No | NA | 3 | 0 | 0 | Not-determined | Genome-fragment | NA | NA | 0 |
| S2-final-viral-combined_k141_214637\|\|full | 2012 | No | NA | 3 | 0 | 2 | Not-determined | Genome-fragment | NA | NA | 0 |
| S6-final-viral-combined_k141_42788\|\|full | 4078 | No | NA | 9 | 0 | 0 | Low-quality | Genome-fragment | 7.88 | AAI-based (high-confidence) | 0 |
| S2-final-viral-combined_k141_214639\|\|full | 1614 | No | NA | 2 | 0 | 1 | Not-determined | Genome-fragment | NA | NA | 0 |
| S7-final-viral-combined_k141_153593\|\|full | 4079 | No | NA | 5 | 0 | 4 | Low-quality | Genome-fragment | 5.13 | AAI-based (medium-confidence) | 0 |
| S2-final-viral-combined_k141_91563\|\|full | 21509 | No | NA | 25 | 0 | 17 | Not-determined | Genome-fragment | NA | NA | 0 |
| S12-final-viral-combined_k141_147773\|\|full | 4079 | No | NA | 3 | 1 | 0 | Low-quality | Genome-fragment | 9.59 | AAI-based (high-confidence) | 0 |
| S2-final-viral-combined_k141_122457\|\|full | 56154 | Yes | 22785 | 67 | 21 | 24 | Medium-quality | Genome-fragment | 52.39 | AAI-based (high-confidence) | 59.42 |
| S2-final-viral-combined_k141_91582\|\|full | 5063 | No | NA | 5 | 0 | 0 | Not-determined | Genome-fragment | NA | NA | 0 |
| S2-final-viral-combined_k141_6439\|\|full | 3778 | No | NA | 5 | 0 | 4 | Not-determined | Genome-fragment | NA | NA | 0 |
| S6-final-viral-combined_k141_120656\|\|full | 4079 | No | NA | 5 | 0 | 4 | Low-quality | Genome-fragment | 1.94 | AAI-based (medium-confidence) | 0 |
| S2-final-viral-combined_k141_40090\|\|full | 3941 | No | NA | 6 | 0 | 0 | Not-determined | Genome-fragment | NA | NA | 0 |
| S2-final-viral-combined_k141_27325\|\|full | 2943 | No | NA | 4 | 0 | 0 | Not-determined | Genome-fragment | NA | NA | 0 |
| S2-final-viral-combined_k141_175363\|\|full | 3970 | No | NA | 5 | 0 | 2 | Not-determined | Genome-fragment | NA | NA | 0 |
| S11-final-viral-combined_k141_257596\|\|full | 4080 | Yes | 3415 | 8 | 1 | 1 | Low-quality | Genome-fragment | 7.97 | AAI-based (high-confidence) | 16.3 |
| S2-final-viral-combined_k141_88097\|\|full | 4081 | No | NA | 6 | 2 | 0 | Low-quality | Genome-fragment | 8.69 | AAI-based (high-confidence) | 0 |
| S2-final-viral-combined_k141_194774\|\|full | 6657 | No | NA | 6 | 0 | 2 | Not-determined | Genome-fragment | NA | NA | 0 |
| S1_k141_70132 | 4082 | No | NA | 4 | 2 | 0 | Low-quality | Genome-fragment | 8.85 | AAI-based (high-confidence) | 0 |
| S2-final-viral-combined_k141_143041\|\|full | 1455 | No | NA | 4 | 0 | 0 | Not-determined | Genome-fragment | NA | NA | 0 |
| S17-final-viral-combined_k141_206298\|\|full | 4082 | Yes | 1401 | 6 | 1 | 1 | Low-quality | Genome-fragment | 3.89 | AAI-based (high-confidence) | 65.68 |
| S2-final-viral-combined_k141_6494\|\|full | 1624 | No | NA | 4 | 0 | 0 | Not-determined | Genome-fragment | NA | NA | 0 |
| S7-final-viral-combined_k141_209123\|\|full | 4083 | No | NA | 4 | 0 | 2 | Low-quality | Genome-fragment | 6.6 | AAI-based (medium-confidence) | 0 |
| S9-final-viral-combined_k141_77661\|\|full | 4084 | No | NA | 8 | 3 | 0 | Low-quality | Genome-fragment | 7.54 | AAI-based (high-confidence) | 0 |
| S2-final-viral-combined_k141_102085\|\|full | 3394 | No | NA | 4 | 0 | 3 | Not-determined | Genome-fragment | NA | NA | 0 |
| S1_k141_35301 | 4084 | No | NA | 7 | 0 | 0 | Low-quality | Genome-fragment | 7.02 | AAI-based (medium-confidence) | 0 |
| S1_k141_352445 | 4085 | Yes | 3227 | 9 | 1 | 1 | Low-quality | Genome-fragment | 7.62 | AAI-based (high-confidence) | 21 |
| S2-final-viral-combined_k141_236791\|\|full | 1057 | No | NA | 2 | 0 | 0 | Not-determined | Genome-fragment | NA | NA | 0 |
| S2-final-viral-combined_k141_162632\|\|full | 2351 | No | NA | 2 | 0 | 1 | Not-determined | Genome-fragment | NA | NA | 0 |
| S2-final-viral-combined_k141_80578\|\|full | 2045 | No | NA | 2 | 0 | 1 | Not-determined | Genome-fragment | NA | NA | 0 |
| S2-final-viral-combined_k141_194843\|\|full | 6611 | No | NA | 9 | 0 | 5 | Not-determined | Genome-fragment | NA | NA | 0 |
| S17-final-viral-combined_k141_95819\|\|full | 4085 | No | NA | 5 | 0 | 1 | Low-quality | Genome-fragment | 9.89 | AAI-based (high-confidence) | 0 |
| S17-k141_253540 | 4085 | No | NA | 5 | 0 | 2 | Low-quality | Genome-fragment | 6.03 | AAI-based (medium-confidence) | 0 |
| S12-final-viral-combined_k141_379842\|\|full | 4085 | No | NA | 3 | 0 | 2 | Low-quality | Genome-fragment | 5.27 | AAI-based (medium-confidence) | 0 |
| S2-final-viral-combined_k141_40189\|\|full | 2312 | No | NA | 3 | 0 | 3 | Not-determined | Genome-fragment | NA | NA | 0 |
| S8-final-viral-combined_k141_269059\|\|full | 4085 | No | NA | 4 | 0 | 1 | Low-quality | Genome-fragment | 1.28 | AAI-based (medium-confidence) | 0 |
| S5_k141_194502 | 4086 | No | NA | 5 | 2 | 0 | Low-quality | Genome-fragment | 2.36 | AAI-based (high-confidence) | 0 |
| S2-final-viral-combined_k141_131152\|\|full | 3527 | No | NA | 5 | 0 | 1 | Not-determined | Genome-fragment | NA | NA | 0 |
| S12_k141_388053 | 4086 | No | NA | 3 | 0 | 2 | Low-quality | Genome-fragment | 1.45 | AAI-based (medium-confidence) | 0 |
| S2-final-viral-combined_k141_18806\|\|full | 1828 | No | NA | 2 | 0 | 1 | Not-determined | Genome-fragment | NA | NA | 0 |
| S2-final-viral-combined_k141_91772\|\|full | 1802 | No | NA | 5 | 0 | 0 | Not-determined | Genome-fragment | NA | NA | 0 |
| S2_k141_181654 | 4086 | No | NA | 3 | 0 | 1 | Low-quality | Genome-fragment | 5.75 | AAI-based (medium-confidence) | 0 |
| S8-final-viral-combined_k141_9410\|\|full | 4086 | No | NA | 4 | 0 | 3 | Low-quality | Genome-fragment | 1.93 | AAI-based (medium-confidence) | 0 |
| S2-final-viral-combined_k141_236827\|\|full | 2955 | No | NA | 3 | 0 | 3 | Not-determined | Genome-fragment | NA | NA | 0 |
| S2-final-viral-combined_k141_60307\|\|full | 2183 | No | NA | 4 | 0 | 1 | Not-determined | Genome-fragment | NA | NA | 0 |
| S2-final-viral-combined_k141_122546\|\|full | 1968 | No | NA | 2 | 0 | 1 | Not-determined | Genome-fragment | NA | NA | 0 |
| S2-final-viral-combined_k141_214807\|\|full | 2651 | No | NA | 3 | 0 | 1 | Not-determined | Genome-fragment | NA | NA | 0 |
| S2-final-viral-combined_k141_184743\|\|full | 1109 | No | NA | 2 | 0 | 0 | Not-determined | Genome-fragment | NA | NA | 0 |
| S2-final-viral-combined_k141_184748\|\|full | 2905 | No | NA | 5 | 0 | 1 | Not-determined | Genome-fragment | NA | NA | 0 |
| S2-final-viral-combined_k141_102182\|\|full | 2863 | No | NA | 4 | 0 | 0 | Not-determined | Genome-fragment | NA | NA | 0 |
| S4_k141_143421 | 4087 | No | NA | 6 | 0 | 1 | Low-quality | Genome-fragment | 4.47 | AAI-based (medium-confidence) | 0 |
| S2-final-viral-combined_k141_236866\|\|full | 1890 | No | NA | 4 | 0 | 0 | Not-determined | Genome-fragment | NA | NA | 0 |
| S11-final-viral-combined_k141_77973\|\|full | 4088 | No | NA | 4 | 1 | 0 | Low-quality | Genome-fragment | 3.27 | HMM-based (lower-bound) | 0 |
| S2-final-viral-combined_k141_80623\|\|full | 1124 | No | NA | 3 | 0 | 1 | Not-determined | Genome-fragment | NA | NA | 0 |
| S2-final-viral-combined_k141_60337\|\|full | 1094 | No | NA | 2 | 0 | 0 | Not-determined | Genome-fragment | NA | NA | 0 |
| S2-final-viral-combined_k141_70521\|\|full | 1361 | No | NA | 3 | 0 | 0 | Not-determined | Genome-fragment | NA | NA | 0 |
| S2-final-viral-combined_k141_102211\|\|full | 1146 | No | NA | 3 | 0 | 1 | Not-determined | Genome-fragment | NA | NA | 0 |
| S2-final-viral-combined_k141_102223\|\|full | 2096 | No | NA | 3 | 0 | 2 | Not-determined | Genome-fragment | NA | NA | 0 |
| S3-final-viral-combined_k141_115299\|\|full | 4088 | No | NA | 3 | 0 | 2 | Low-quality | Genome-fragment | 3.02 | AAI-based (medium-confidence) | 0 |
| S2-final-viral-combined_k141_6567\|\|full | 2647 | No | NA | 5 | 0 | 2 | Not-determined | Genome-fragment | NA | NA | 0 |
| S2-final-viral-combined_k141_111133\|\|full | 2064 | No | NA | 2 | 0 | 1 | Not-determined | Genome-fragment | NA | NA | 0 |
| S5-final-viral-combined_k141_192548\|\|full | 4090 | No | NA | 6 | 0 | 0 | Low-quality | Genome-fragment | 5.41 | AAI-based (medium-confidence) | 0 |
| S12-final-viral-combined_k141_121898\|\|full | 4090 | No | NA | 5 | 0 | 1 | Low-quality | Genome-fragment | 4.35 | AAI-based (medium-confidence) | 0 |
| S2-final-viral-combined_k141_203546\|\|full | 1575 | No | NA | 4 | 0 | 1 | Not-determined | Genome-fragment | NA | NA | 0 |
| S2-final-viral-combined_k141_80653\|\|full | 5724 | No | NA | 5 | 0 | 4 | Not-determined | Genome-fragment | NA | NA | 0 |
| S2-final-viral-combined_k141_102282\|\|full | 2003 | No | NA | 4 | 0 | 1 | Not-determined | Genome-fragment | NA | NA | 0 |
| S5-final-viral-combined_k141_81920\|\|full | 4091 | No | NA | 5 | 0 | 1 | Low-quality | Genome-fragment | 8.55 | AAI-based (medium-confidence) | 0 |
| S2-final-viral-combined_k141_111267\|\|full | 4091 | No | NA | 4 | 0 | 3 | Low-quality | Genome-fragment | 4.45 | AAI-based (medium-confidence) | 0 |
| S2-final-viral-combined_k141_244564\|\|full | 71440 | No | NA | 80 | 21 | 5 | High-quality | High-quality | 100 | AAI-based (high-confidence) | 0 |
| S2-final-viral-combined_k141_236945\|\|full | 10710 | No | NA | 7 | 0 | 6 | Not-determined | Genome-fragment | NA | NA | 0 |
| S2-final-viral-combined_k141_44964\|\|full | 4092 | No | NA | 4 | 0 | 1 | Low-quality | Genome-fragment | 2.06 | AAI-based (medium-confidence) | 0 |
| S2-final-viral-combined_k141_91904\|\|full | 1824 | No | NA | 3 | 0 | 2 | Not-determined | Genome-fragment | NA | NA | 0 |
| S8-final-viral-combined_k141_68074\|\|full | 4092 | No | NA | 2 | 0 | 1 | Low-quality | Genome-fragment | 0.85 | AAI-based (medium-confidence) | 0 |
| S2-final-viral-combined_k141_70606\|\|full | 3568 | No | NA | 3 | 0 | 0 | Not-determined | Genome-fragment | NA | NA | 0 |
| S2-final-viral-combined_k141_224387\|\|full | 3837 | No | NA | 3 | 0 | 1 | Not-determined | Genome-fragment | NA | NA | 0 |
| S2-final-viral-combined_k141_18823\|\|full | 3316 | No | NA | 6 | 0 | 3 | Not-determined | Genome-fragment | NA | NA | 0 |
| S1-final-viral-combined_k141_371391\|\|full | 4093 | No | NA | 10 | 2 | 0 | Low-quality | Genome-fragment | 10.55 | AAI-based (high-confidence) | 0 |
| S17-final-viral-combined_k141_82762\|\|lt2gene | 4093 | No | NA | 2 | 1 | 0 | Low-quality | Genome-fragment | 6.15 | AAI-based (high-confidence) | 0 |
| S12-final-viral-combined_k141_166687\|\|full | 4093 | No | NA | 3 | 0 | 2 | Low-quality | Genome-fragment | 2.85 | AAI-based (medium-confidence) | 0 |
| S2-final-viral-combined_k141_111177\|\|full | 1447 | No | NA | 3 | 0 | 2 | Not-determined | Genome-fragment | NA | NA | 0 |
| S2-final-viral-combined_k141_224399\|\|full | 2247 | No | NA | 2 | 0 | 0 | Not-determined | Genome-fragment | NA | NA | 0 |
| S2-final-viral-combined_k141_111181\|\|full | 1442 | No | NA | 2 | 0 | 2 | Not-determined | Genome-fragment | NA | NA | 0 |
| S2-final-viral-combined_k141_35262\|\|full | 4094 | Yes | 3568 | 8 | 1 | 1 | Low-quality | Genome-fragment | 2.25 | HMM-based (lower-bound) | 12.85 |
| S2-final-viral-combined_k141_40324\|\|full | 3120 | No | NA | 4 | 0 | 3 | Not-determined | Genome-fragment | NA | NA | 0 |
| S2-final-viral-combined_k141_203575\|\|full | 3294 | No | NA | 3 | 0 | 2 | Not-determined | Genome-fragment | NA | NA | 0 |
| S2-final-viral-combined_k141_244603\|\|full | 1941 | No | NA | 3 | 0 | 2 | Not-determined | Genome-fragment | NA | NA | 0 |
| S2-final-viral-combined_k141_40325\|\|full | 1222 | No | NA | 2 | 0 | 0 | Not-determined | Genome-fragment | NA | NA | 0 |
| S12-final-viral-combined_k141_60155\|\|full | 4095 | No | NA | 9 | 1 | 0 | Low-quality | Genome-fragment | 6.47 | AAI-based (high-confidence) | 0 |
| S2-final-viral-combined_k141_118502\|\|full | 4095 | No | NA | 9 | 0 | 2 | Low-quality | Genome-fragment | 7.7 | AAI-based (medium-confidence) | 0 |
| S5-final-viral-combined_k141_113367\|\|full | 4096 | No | NA | 4 | 0 | 1 | Low-quality | Genome-fragment | 5.07 | AAI-based (high-confidence) | 0 |
| S2-final-viral-combined_k141_27438\|\|full | 2629 | No | NA | 4 | 0 | 0 | Not-determined | Genome-fragment | NA | NA | 0 |
| S2-final-viral-combined_k141_214926\|\|full | 2107 | No | NA | 3 | 0 | 2 | Not-determined | Genome-fragment | NA | NA | 0 |
| S7_k141_97908 | 4096 | No | NA | 11 | 1 | 0 | Low-quality | Genome-fragment | 9.51 | AAI-based (high-confidence) | 0 |
| S2-final-viral-combined_k141_131305\|\|full | 1202 | No | NA | 2 | 0 | 0 | Not-determined | Genome-fragment | NA | NA | 0 |
| S2-final-viral-combined_k141_162801\|\|full | 2167 | No | NA | 3 | 0 | 0 | Not-determined | Genome-fragment | NA | NA | 0 |
| S2-final-viral-combined_k141_6605\|\|full | 80208 | No | NA | 90 | 5 | 19 | High-quality | High-quality | 100 | AAI-based (high-confidence) | 0 |
| S12-final-viral-combined_k141_121792\|\|full | 4096 | No | NA | 7 | 0 | 0 | Low-quality | Genome-fragment | 7.93 | AAI-based (medium-confidence) | 0 |
| S2-final-viral-combined_k141_214950\|\|full | 2890 | No | NA | 4 | 0 | 2 | Not-determined | Genome-fragment | NA | NA | 0 |
| S1-final-viral-combined_k141_367920\|\|full | 4097 | No | NA | 4 | 0 | 4 | Low-quality | Genome-fragment | 1.35 | AAI-based (medium-confidence) | 0 |
| S5_k141_41801 | 4097 | No | NA | 10 | 1 | 0 | Low-quality | Genome-fragment | 4.59 | AAI-based (high-confidence) | 0 |
| S2-final-viral-combined_k141_91979\|\|full | 2547 | No | NA | 3 | 0 | 2 | Not-determined | Genome-fragment | NA | NA | 0 |
| S2-final-viral-combined_k141_244676\|\|full | 5142 | No | NA | 7 | 0 | 5 | Not-determined | Genome-fragment | NA | NA | 0 |
| S2-final-viral-combined_k141_162826\|\|full | 1965 | No | NA | 3 | 0 | 0 | Not-determined | Genome-fragment | NA | NA | 0 |
| S2-final-viral-combined_k141_91982\|\|full | 2144 | No | NA | 2 | 0 | 0 | Not-determined | Genome-fragment | NA | NA | 0 |
| S2-final-viral-combined_k141_6629\|\|full | 1403 | No | NA | 3 | 0 | 1 | Not-determined | Genome-fragment | NA | NA | 0 |
| S2-final-viral-combined_k141_27469\|\|full | 2243 | No | NA | 5 | 0 | 2 | Not-determined | Genome-fragment | NA | NA | 0 |
| S2-final-viral-combined_k141_70726\|\|full | 2715 | No | NA | 2 | 0 | 1 | Not-determined | Genome-fragment | NA | NA | 0 |
| S2-final-viral-combined_k141_214970\|\|full | 1454 | No | NA | 3 | 0 | 1 | Not-determined | Genome-fragment | NA | NA | 0 |
| S2-final-viral-combined_k141_122674\|\|full | 1100 | No | NA | 2 | 0 | 1 | Not-determined | Genome-fragment | NA | NA | 0 |
| S2-final-viral-combined_k141_70754\|\|full | 4187 | No | NA | 5 | 0 | 0 | Not-determined | Genome-fragment | NA | NA | 0 |
| S17-k141_265936 | 4097 | No | NA | 3 | 0 | 2 | Low-quality | Genome-fragment | 2.18 | AAI-based (medium-confidence) | 0 |
| S2_k141_92744 | 4097 | No | NA | 4 | 3 | 0 | Low-quality | Genome-fragment | 2.26 | AAI-based (high-confidence) | 0 |
| S2-final-viral-combined_k141_122677\|\|full | 6233 | No | NA | 9 | 0 | 3 | Not-determined | Genome-fragment | NA | NA | 0 |
| S7-final-viral-combined_k141_216126\|\|full | 4098 | No | NA | 4 | 0 | 3 | Low-quality | Genome-fragment | 4.83 | AAI-based (high-confidence) | 0 |
| S2-final-viral-combined_k141_162854\|\|full | 1660 | No | NA | 3 | 0 | 1 | Not-determined | Genome-fragment | NA | NA | 0 |
| S2-final-viral-combined_k141_60600\|\|full | 4715 | No | NA | 6 | 0 | 6 | Not-determined | Genome-fragment | NA | NA | 0 |
| S17-final-viral-combined_k141_138690\|\|full | 4099 | No | NA | 3 | 0 | 2 | Low-quality | Genome-fragment | 2.84 | AAI-based (medium-confidence) | 0 |
| S1-final-viral-combined_k141_57149\|\|full | 4100 | No | NA | 7 | 1 | 0 | Low-quality | Genome-fragment | 4.98 | AAI-based (high-confidence) | 0 |
| S1-final-viral-combined_k141_173406\|\|full | 4100 | No | NA | 7 | 1 | 0 | Low-quality | Genome-fragment | 10.27 | AAI-based (medium-confidence) | 0 |
| S2-final-viral-combined_k141_153532\|\|full | 2173 | No | NA | 3 | 0 | 1 | Not-determined | Genome-fragment | NA | NA | 0 |
| S2-final-viral-combined_k141_215015\|\|full | 6379 | No | NA | 9 | 0 | 6 | Not-determined | Genome-fragment | NA | NA | 0 |
| S1-final-viral-combined_k141_294563\|\|full | 4100 | Yes | 3845 | 9 | 1 | 1 | Low-quality | Genome-fragment | 12.05 | AAI-based (high-confidence) | 6.22 |
| S2-final-viral-combined_k141_40418\|\|full | 4262 | No | NA | 5 | 0 | 0 | Not-determined | Genome-fragment | NA | NA | 0 |
| S7_k141_66849 | 4102 | No | NA | 7 | 1 | 0 | Low-quality | Genome-fragment | 6.62 | HMM-based (lower-bound) | 0 |
| S2-final-viral-combined_k141_224525\|\|full | 9233 | No | NA | 14 | 0 | 4 | Not-determined | Genome-fragment | NA | NA | 0 |
| S2-final-viral-combined_k141_155302\|\|full | 4103 | No | NA | 5 | 1 | 0 | Low-quality | Genome-fragment | 11.27 | AAI-based (medium-confidence) | 0 |
| S2-final-viral-combined_k141_192816\|\|full | 4104 | No | NA | 6 | 1 | 0 | Low-quality | Genome-fragment | 2.76 | AAI-based (high-confidence) | 0 |
| S2-final-viral-combined_k141_40851\|\|full | 4106 | No | NA | 5 | 0 | 0 | Low-quality | Genome-fragment | 9.51 | AAI-based (medium-confidence) | 0 |
| S2-final-viral-combined_k141_237147\|\|full | 1665 | No | NA | 3 | 0 | 3 | Not-determined | Genome-fragment | NA | NA | 0 |
| S7-final-viral-combined_k141_189782\|\|full | 4108 | No | NA | 3 | 0 | 3 | Low-quality | Genome-fragment | 1.3 | AAI-based (medium-confidence) | 0 |
| S2-final-viral-combined_k141_60641\|\|full | 1408 | No | NA | 2 | 0 | 1 | Not-determined | Genome-fragment | NA | NA | 0 |
| S2-final-viral-combined_k141_6685\|\|full | 2611 | No | NA | 3 | 0 | 2 | Not-determined | Genome-fragment | NA | NA | 0 |
| S2_k141_20743 | 4109 | No | NA | 6 | 0 | 0 | Low-quality | Genome-fragment | 8.04 | AAI-based (medium-confidence) | 0 |
| S2-final-viral-combined_k141_215058\|\|full | 1909 | No | NA | 2 | 0 | 1 | Not-determined | Genome-fragment | NA | NA | 0 |
| S2-final-viral-combined_k141_244754\|\|full | 1157 | No | NA | 2 | 0 | 0 | Not-determined | Genome-fragment | NA | NA | 0 |
| S2-final-viral-combined_k141_92129\|\|full | 12732 | No | NA | 11 | 0 | 2 | Not-determined | Genome-fragment | NA | NA | 0 |
| S2-final-viral-combined_k141_49136\|\|full | 4112 | No | NA | 11 | 1 | 0 | Low-quality | Genome-fragment | 6.37 | AAI-based (medium-confidence) | 0 |
| S2-final-viral-combined_k141_244765\|\|full | 1197 | No | NA | 2 | 0 | 1 | Not-determined | Genome-fragment | NA | NA | 0 |
| S2-final-viral-combined_k141_131442\|\|full | 3986 | No | NA | 11 | 0 | 0 | Not-determined | Genome-fragment | NA | NA | 0 |
| S9_k141_100567 | 4113 | Yes | 738 | 5 | 1 | 1 | Low-quality | Genome-fragment | 0.46 | HMM-based (lower-bound) | 82.06 |
| S2-final-viral-combined_k141_122798\|\|full | 1515 | No | NA | 2 | 0 | 0 | Not-determined | Genome-fragment | NA | NA | 0 |
| S3_k141_107322 | 4113 | No | NA | 5 | 1 | 0 | Low-quality | Genome-fragment | 5.21 | AAI-based (high-confidence) | 0 |
| S7_k141_184657 | 4113 | No | NA | 13 | 2 | 0 | Low-quality | Genome-fragment | 7.8 | AAI-based (high-confidence) | 0 |
| S2-final-viral-combined_k141_203716\|\|full | 2399 | No | NA | 4 | 0 | 1 | Not-determined | Genome-fragment | NA | NA | 0 |
| S2-final-viral-combined_k141_27524\|\|full | 2876 | No | NA | 4 | 0 | 2 | Not-determined | Genome-fragment | NA | NA | 0 |
| S2-final-viral-combined_k141_102403\|\|full | 1017 | No | NA | 2 | 0 | 0 | Not-determined | Genome-fragment | NA | NA | 0 |
| S2-final-viral-combined_k141_153649\|\|full | 5766 | No | NA | 5 | 0 | 5 | Not-determined | Genome-fragment | NA | NA | 0 |
| S12-final-viral-combined_k141_217002\|\|full | 4113 | No | NA | 5 | 0 | 4 | Low-quality | Genome-fragment | 1.13 | AAI-based (medium-confidence) | 0 |
| S2-final-viral-combined_k141_60715\|\|full | 6825 | No | NA | 5 | 0 | 4 | Not-determined | Genome-fragment | NA | NA | 0 |
| S2-final-viral-combined_k141_162923\|\|full | 1337 | No | NA | 4 | 0 | 0 | Not-determined | Genome-fragment | NA | NA | 0 |
| S2-final-viral-combined_k141_111343\|\|full | 1137 | No | NA | 2 | 0 | 0 | Not-determined | Genome-fragment | NA | NA | 0 |
| S2-final-viral-combined_k141_237260\|\|full | 1975 | No | NA | 4 | 0 | 3 | Not-determined | Genome-fragment | NA | NA | 0 |
| S9_k141_49477 | 4114 | No | NA | 6 | 4 | 0 | Low-quality | Genome-fragment | 11.02 | AAI-based (high-confidence) | 0 |
| S2-final-viral-combined_k141_215185\|\|full | 1172 | No | NA | 3 | 0 | 0 | Not-determined | Genome-fragment | NA | NA | 0 |
| S2-final-viral-combined_k141_122847\|\|full | 3289 | No | NA | 5 | 0 | 0 | Not-determined | Genome-fragment | NA | NA | 0 |
| S5-final-viral-combined_k141_89492\|\|full | 4114 | No | NA | 3 | 0 | 1 | Low-quality | Genome-fragment | 14.34 | AAI-based (medium-confidence) | 0 |
| S12-final-viral-combined_k141_364582\|\|full | 4114 | No | NA | 5 | 4 | 0 | Low-quality | Genome-fragment | 9.65 | AAI-based (medium-confidence) | 0 |
| S2-final-viral-combined_k141_200366\|\|full | 4114 | Yes | 1330 | 4 | 1 | 2 | Low-quality | Genome-fragment | 1.91 | AAI-based (medium-confidence) | 67.67 |
| S1-final-viral-combined_k141_61104\|\|full | 4115 | No | NA | 4 | 4 | 0 | Low-quality | Genome-fragment | 9.17 | AAI-based (high-confidence) | 0 |
| S1_k141_16705 | 4115 | No | NA | 4 | 0 | 3 | Low-quality | Genome-fragment | 9.79 | AAI-based (high-confidence) | 0 |
| S2-final-viral-combined_k141_215206\|\|full | 1394 | No | NA | 3 | 0 | 1 | Not-determined | Genome-fragment | NA | NA | 0 |
| S2-final-viral-combined_k141_19018\|\|full | 2939 | No | NA | 3 | 0 | 1 | Not-determined | Genome-fragment | NA | NA | 0 |
| S2-final-viral-combined_k141_80910\|\|full | 2181 | No | NA | 3 | 0 | 2 | Not-determined | Genome-fragment | NA | NA | 0 |
| S2-final-viral-combined_k141_195046\|\|full | 1941 | No | NA | 3 | 0 | 1 | Not-determined | Genome-fragment | NA | NA | 0 |
| S2-final-viral-combined_k141_122869\|\|full | 1047 | No | NA | 3 | 0 | 0 | Not-determined | Genome-fragment | NA | NA | 0 |
| S5-final-viral-combined_k141_52943\|\|full | 4115 | No | NA | 5 | 0 | 1 | Low-quality | Genome-fragment | 11.02 | AAI-based (high-confidence) | 0 |
| S1-final-viral-combined_k141_257408\|\|full | 4116 | No | NA | 5 | 0 | 0 | Low-quality | Genome-fragment | 3.87 | AAI-based (medium-confidence) | 0 |
| S2-final-viral-combined_k141_175830\|\|full | 1475 | No | NA | 4 | 0 | 0 | Not-determined | Genome-fragment | NA | NA | 0 |
| S2-final-viral-combined_k141_131480\|\|full | 9921 | No | NA | 3 | 0 | 2 | Not-determined | Genome-fragment | NA | NA | 0 |
| S2-final-viral-combined_k141_184987\|\|full | 1527 | No | NA | 2 | 0 | 1 | Not-determined | Genome-fragment | NA | NA | 0 |
| S2-final-viral-combined_k141_143546\|\|full | 7173 | No | NA | 4 | 0 | 3 | Not-determined | Genome-fragment | NA | NA | 0 |
| S2-final-viral-combined_k141_215267\|\|full | 3347 | No | NA | 4 | 0 | 1 | Not-determined | Genome-fragment | NA | NA | 0 |
| S2-final-viral-combined_k141_80987\|\|full | 1377 | No | NA | 3 | 0 | 2 | Not-determined | Genome-fragment | NA | NA | 0 |
| S17-final-viral-combined_k141_213958\|\|full | 4118 | No | NA | 3 | 0 | 1 | Low-quality | Genome-fragment | 2.77 | AAI-based (high-confidence) | 0 |
| S7-final-viral-combined_k141_30497\|\|full | 4119 | No | NA | 4 | 0 | 1 | Low-quality | Genome-fragment | 0.95 | AAI-based (high-confidence) | 0 |
| S2-final-viral-combined_k141_40579\|\|full | 1335 | No | NA | 2 | 0 | 0 | Not-determined | Genome-fragment | NA | NA | 0 |
| S12-final-viral-combined_k141_337087\|\|full | 4119 | No | NA | 3 | 0 | 3 | Low-quality | Genome-fragment | 1.28 | AAI-based (medium-confidence) | 0 |
| S2-final-viral-combined_k141_40593\|\|full | 1004 | No | NA | 3 | 0 | 2 | Not-determined | Genome-fragment | NA | NA | 0 |
| S4_k141_17766 | 4119 | No | NA | 3 | 0 | 1 | Low-quality | Genome-fragment | 4.8 | AAI-based (medium-confidence) | 0 |
| S12-final-viral-combined_k141_218731\|\|full | 4120 | No | NA | 7 | 0 | 1 | Low-quality | Genome-fragment | 3.05 | AAI-based (medium-confidence) | 0 |
| S2-final-viral-combined_k141_131519\|\|full | 1039 | No | NA | 3 | 0 | 1 | Not-determined | Genome-fragment | NA | NA | 0 |
| S2_k141_153048 | 4120 | No | NA | 8 | 0 | 0 | Low-quality | Genome-fragment | 6.71 | AAI-based (medium-confidence) | 0 |
| S2-final-viral-combined_k141_143591\|\|full | 1396 | No | NA | 2 | 0 | 1 | Not-determined | Genome-fragment | NA | NA | 0 |
| S2-final-viral-combined_k141_244858\|\|full | 1283 | No | NA | 3 | 0 | 1 | Not-determined | Genome-fragment | NA | NA | 0 |
| S2-final-viral-combined_k141_19155\|\|full | 1411 | No | NA | 2 | 0 | 1 | Not-determined | Genome-fragment | NA | NA | 0 |
| S2-final-viral-combined_k141_203864\|\|full | 4048 | No | NA | 4 | 0 | 0 | Not-determined | Genome-fragment | NA | NA | 0 |
| S2-final-viral-combined_k141_131561\|\|full | 1137 | No | NA | 2 | 0 | 1 | Not-determined | Genome-fragment | NA | NA | 0 |
| S2-final-viral-combined_k141_185038\|\|full | 6288 | No | NA | 3 | 0 | 3 | Not-determined | Genome-fragment | NA | NA | 0 |
| S8-final-viral-combined_k141_20386\|\|full | 4120 | No | NA | 5 | 0 | 3 | Low-quality | Genome-fragment | 3.09 | AAI-based (medium-confidence) | 0 |
| S2-final-viral-combined_k141_185042\|\|full | 2092 | No | NA | 2 | 0 | 1 | Not-determined | Genome-fragment | NA | NA | 0 |
| S17-final-viral-combined_k141_196813\|\|full | 4121 | No | NA | 4 | 0 | 1 | Low-quality | Genome-fragment | 2.75 | AAI-based (high-confidence) | 0 |
| S17-k141_254618 | 4121 | No | NA | 7 | 2 | 0 | Low-quality | Genome-fragment | 1.89 | HMM-based (lower-bound) | 0 |
| S2-final-viral-combined_k141_175888\|\|full | 3362 | No | NA | 5 | 0 | 0 | Not-determined | Genome-fragment | NA | NA | 0 |
| S2-final-viral-combined_k141_237454\|\|full | 1411 | No | NA | 2 | 0 | 0 | Not-determined | Genome-fragment | NA | NA | 0 |
| S12_k141_376867 | 4122 | No | NA | 2 | 0 | 0 | Low-quality | Genome-fragment | 18.17 | AAI-based (medium-confidence) | 0 |
| S2-final-viral-combined_k141_102666\|\|full | 2019 | No | NA | 3 | 0 | 1 | Not-determined | Genome-fragment | NA | NA | 0 |
| S2-final-viral-combined_k141_173439\|\|full | 4122 | No | NA | 4 | 3 | 0 | Low-quality | Genome-fragment | 8.64 | AAI-based (medium-confidence) | 0 |
| S12-final-viral-combined_k141_385975\|\|full | 4123 | No | NA | 6 | 1 | 0 | Low-quality | Genome-fragment | 6.79 | AAI-based (medium-confidence) | 0 |
| S2-final-viral-combined_k141_175922\|\|full | 1045 | No | NA | 3 | 0 | 1 | Not-determined | Genome-fragment | NA | NA | 0 |
| S2-final-viral-combined_k141_215377\|\|full | 2093 | No | NA | 4 | 0 | 0 | Not-determined | Genome-fragment | NA | NA | 0 |
| S2-final-viral-combined_k141_111492\|\|full | 1648 | No | NA | 2 | 0 | 2 | Not-determined | Genome-fragment | NA | NA | 0 |
| S6-final-viral-combined_k141_115867\|\|full | 4124 | No | NA | 4 | 0 | 0 | Low-quality | Genome-fragment | 3.1 | AAI-based (high-confidence) | 0 |
| S2-final-viral-combined_k141_215393\|\|full | 1155 | No | NA | 2 | 0 | 0 | Not-determined | Genome-fragment | NA | NA | 0 |
| S2-final-viral-combined_k141_162987\|\|full | 1884 | No | NA | 2 | 0 | 1 | Not-determined | Genome-fragment | NA | NA | 0 |
| S2-final-viral-combined_k141_215396\|\|full | 1382 | No | NA | 3 | 0 | 2 | Not-determined | Genome-fragment | NA | NA | 0 |
| S2-final-viral-combined_k141_19215\|\|full | 1872 | No | NA | 3 | 0 | 1 | Not-determined | Genome-fragment | NA | NA | 0 |
| S2-final-viral-combined_k141_71035\|\|full | 1609 | No | NA | 2 | 0 | 0 | Not-determined | Genome-fragment | NA | NA | 0 |
| S2-final-viral-combined_k141_195237\|\|full | 2177 | No | NA | 4 | 0 | 1 | Not-determined | Genome-fragment | NA | NA | 0 |
| S2-final-viral-combined_k141_19230\|\|full | 6071 | No | NA | 11 | 0 | 1 | Not-determined | Genome-fragment | NA | NA | 0 |
| S17-final-viral-combined_k141_234984\|\|full | 4125 | No | NA | 5 | 1 | 0 | Low-quality | Genome-fragment | 9.47 | AAI-based (high-confidence) | 0 |
| S2_k141_83144 | 4127 | No | NA | 4 | 1 | 0 | Low-quality | Genome-fragment | 7.49 | AAI-based (high-confidence) | 0 |
| S2-final-viral-combined_k141_81124\|\|full | 1360 | No | NA | 2 | 0 | 0 | Not-determined | Genome-fragment | NA | NA | 0 |
| S2-final-viral-combined_k141_185122\|\|full | 1648 | No | NA | 2 | 0 | 1 | Not-determined | Genome-fragment | NA | NA | 0 |
| S11-final-viral-combined_k141_285822\|\|full | 4128 | No | NA | 10 | 0 | 0 | Low-quality | Genome-fragment | 5.84 | AAI-based (high-confidence) | 0 |
| S8_k141_115051 | 4129 | No | NA | 7 | 0 | 1 | Low-quality | Genome-fragment | 6.39 | AAI-based (medium-confidence) | 0 |
| S1-final-viral-combined_k141_138328\|\|full | 4131 | No | NA | 5 | 0 | 2 | Low-quality | Genome-fragment | 2.85 | AAI-based (high-confidence) | 0 |
| S4-final-viral-combined_k141_109177\|\|full | 4131 | No | NA | 8 | 0 | 1 | Low-quality | Genome-fragment | 12.11 | AAI-based (high-confidence) | 0 |
| S2-final-viral-combined_k141_19266\|\|full | 3653 | No | NA | 3 | 0 | 2 | Not-determined | Genome-fragment | NA | NA | 0 |
| S5-final-viral-combined_k141_145349\|\|full | 4132 | No | NA | 5 | 0 | 4 | Low-quality | Genome-fragment | 12.51 | AAI-based (high-confidence) | 0 |
| S2-final-viral-combined_k141_102750\|\|full | 3198 | No | NA | 6 | 0 | 0 | Not-determined | Genome-fragment | NA | NA | 0 |
| S8_k141_157917 | 4132 | No | NA | 7 | 0 | 0 | Low-quality | Genome-fragment | 8.08 | AAI-based (high-confidence) | 0 |
| S12-final-viral-combined_k141_364413\|\|full | 4134 | No | NA | 3 | 3 | 0 | Low-quality | Genome-fragment | 10.56 | AAI-based (high-confidence) | 0 |
| S12_k141_325046 | 4134 | No | NA | 6 | 3 | 0 | Low-quality | Genome-fragment | 11.12 | AAI-based (high-confidence) | 0 |
| S2-final-viral-combined_k141_153938\|\|full | 2587 | No | NA | 3 | 0 | 2 | Not-determined | Genome-fragment | NA | NA | 0 |
| S2-final-viral-combined_k141_204011\|\|full | 1379 | No | NA | 2 | 0 | 1 | Not-determined | Genome-fragment | NA | NA | 0 |
| S2-final-viral-combined_k141_71124\|\|full | 1975 | No | NA | 4 | 0 | 0 | Not-determined | Genome-fragment | NA | NA | 0 |
| S12-final-viral-combined_k141_376660\|\|full | 4135 | No | NA | 7 | 1 | 0 | Low-quality | Genome-fragment | 7.3 | AAI-based (high-confidence) | 0 |
| S2-final-viral-combined_k141_198338\|\|full | 4135 | No | NA | 12 | 1 | 0 | Low-quality | Genome-fragment | 3.06 | HMM-based (lower-bound) | 0 |
| S7-final-viral-combined_k141_169723\|\|full | 4137 | No | NA | 3 | 0 | 1 | Low-quality | Genome-fragment | 2.07 | AAI-based (medium-confidence) | 0 |
| S2-final-viral-combined_k141_195330\|\|full | 2439 | No | NA | 3 | 0 | 1 | Not-determined | Genome-fragment | NA | NA | 0 |
| S8_k141_30367 | 4137 | No | NA | 11 | 2 | 0 | Low-quality | Genome-fragment | 11.73 | AAI-based (high-confidence) | 0 |
| S9-final-viral-combined_k141_101956\|\|full | 4138 | No | NA | 10 | 1 | 0 | Low-quality | Genome-fragment | 8.6 | AAI-based (high-confidence) | 0 |
| S7-final-viral-combined_k141_4851\|\|full | 4138 | No | NA | 6 | 2 | 0 | Low-quality | Genome-fragment | 9.99 | AAI-based (high-confidence) | 0 |
| S6_k141_44614 | 4139 | No | NA | 3 | 0 | 3 | Low-quality | Genome-fragment | 2.88 | AAI-based (medium-confidence) | 0 |
| S2-final-viral-combined_k141_131584\|\|full | 4098 | No | NA | 4 | 0 | 3 | Not-determined | Genome-fragment | NA | NA | 0 |
| S2-final-viral-combined_k141_215492\|\|full | 1360 | No | NA | 2 | 0 | 0 | Not-determined | Genome-fragment | NA | NA | 0 |
| S9_k141_123787 | 4140 | No | NA | 3 | 0 | 0 | Low-quality | Genome-fragment | 5.46 | AAI-based (medium-confidence) | 0 |
| S2-final-viral-combined_k141_163106\|\|full | 2629 | No | NA | 4 | 0 | 0 | Not-determined | Genome-fragment | NA | NA | 0 |
| S2-final-viral-combined_k141_153997\|\|full | 1151 | No | NA | 4 | 0 | 0 | Not-determined | Genome-fragment | NA | NA | 0 |
| S2-final-viral-combined_k141_224787\|\|full | 2533 | No | NA | 3 | 0 | 0 | Not-determined | Genome-fragment | NA | NA | 0 |
| S1-final-viral-combined_k141_347869\|\|full | 4140 | No | NA | 5 | 4 | 0 | Low-quality | Genome-fragment | 10.13 | AAI-based (medium-confidence) | 0 |
| S5-final-viral-combined_k141_94693\|\|full | 4141 | No | NA | 5 | 4 | 0 | Low-quality | Genome-fragment | 9.33 | AAI-based (high-confidence) | 0 |
| S7-final-viral-combined_k141_126782\|\|full | 4142 | No | NA | 5 | 0 | 1 | Low-quality | Genome-fragment | 7.63 | AAI-based (medium-confidence) | 0 |
| S2-final-viral-combined_k141_102832\|\|full | 1658 | No | NA | 3 | 0 | 0 | Not-determined | Genome-fragment | NA | NA | 0 |
| S2_k141_98926 | 4142 | No | NA | 2 | 0 | 1 | Low-quality | Genome-fragment | 2.6 | AAI-based (high-confidence) | 0 |
| S1-final-viral-combined_k141_68775\|\|full | 4143 | Yes | 3373 | 8 | 2 | 1 | Low-quality | Genome-fragment | 8.76 | AAI-based (high-confidence) | 18.59 |
| S2-final-viral-combined_k141_92394\|\|full | 1309 | No | NA | 2 | 0 | 2 | Not-determined | Genome-fragment | NA | NA | 0 |
| S2-final-viral-combined_k141_27737\|\|full | 1087 | No | NA | 4 | 0 | 0 | Not-determined | Genome-fragment | NA | NA | 0 |
| S2-final-viral-combined_k141_176001\|\|full | 1278 | No | NA | 2 | 0 | 0 | Not-determined | Genome-fragment | NA | NA | 0 |
| S3-final-viral-combined_k141_77671\|\|full | 4143 | No | NA | 5 | 0 | 0 | Low-quality | Genome-fragment | 1.91 | AAI-based (high-confidence) | 0 |
| S7-final-viral-combined_k141_64705\|\|full | 4145 | No | NA | 5 | 1 | 0 | Low-quality | Genome-fragment | 8.76 | AAI-based (medium-confidence) | 0 |
| S12_k141_174703 | 4145 | Yes | 2195 | 6 | 2 | 2 | Low-quality | Genome-fragment | 5.6 | AAI-based (medium-confidence) | 47.04 |
| S2-final-viral-combined_k141_81349\|\|full | 1154 | No | NA | 2 | 0 | 2 | Not-determined | Genome-fragment | NA | NA | 0 |
| S2-final-viral-combined_k141_163222\|\|full | 4445 | No | NA | 4 | 0 | 2 | Not-determined | Genome-fragment | NA | NA | 0 |
| S11-final-viral-combined_k141_219770\|\|full | 4146 | No | NA | 4 | 0 | 3 | Low-quality | Genome-fragment | 0.46 | AAI-based (medium-confidence) | 0 |
| S17-k141_41686 | 4146 | No | NA | 2 | 0 | 2 | Low-quality | Genome-fragment | 13.82 | AAI-based (high-confidence) | 0 |
| S12_k141_115948 | 4146 | No | NA | 4 | 0 | 2 | Low-quality | Genome-fragment | 1.72 | AAI-based (medium-confidence) | 0 |
| S17-final-viral-combined_k141_81918\|\|full | 4147 | No | NA | 5 | 0 | 1 | Low-quality | Genome-fragment | 5.97 | AAI-based (medium-confidence) | 0 |
| S12-final-viral-combined_k141_399843\|\|full | 4147 | No | NA | 8 | 7 | 0 | Low-quality | Genome-fragment | 6.4 | AAI-based (high-confidence) | 0 |
| S2-final-viral-combined_k141_245027\|\|full | 5665 | No | NA | 5 | 0 | 3 | Not-determined | Genome-fragment | NA | NA | 0 |
| S2-final-viral-combined_k141_185336\|\|full | 1795 | No | NA | 3 | 0 | 0 | Not-determined | Genome-fragment | NA | NA | 0 |
| S8_k141_177750 | 4147 | No | NA | 1 | 0 | 0 | Low-quality | Genome-fragment | 6.38 | AAI-based (medium-confidence) | 0 |
| S11-final-viral-combined_k141_167715\|\|full | 4149 | No | NA | 3 | 0 | 2 | Low-quality | Genome-fragment | 5.88 | AAI-based (medium-confidence) | 0 |
| S2-final-viral-combined_k141_61142\|\|full | 2582 | No | NA | 3 | 0 | 3 | Not-determined | Genome-fragment | NA | NA | 0 |
| S17-final-viral-combined_k141_190898\|\|full | 4149 | No | NA | 8 | 0 | 0 | Low-quality | Genome-fragment | 7.77 | AAI-based (high-confidence) | 0 |
| S1-final-viral-combined_k141_5694\|\|full | 4150 | No | NA | 4 | 0 | 0 | Low-quality | Genome-fragment | 11 | AAI-based (high-confidence) | 0 |
| S2-final-viral-combined_k141_131753\|\|full | 1439 | No | NA | 3 | 0 | 0 | Not-determined | Genome-fragment | NA | NA | 0 |
| S2-final-viral-combined_k141_40905\|\|full | 2099 | No | NA | 2 | 0 | 1 | Not-determined | Genome-fragment | NA | NA | 0 |
| S2-final-viral-combined_k141_61207\|\|full | 1141 | No | NA | 3 | 0 | 1 | Not-determined | Genome-fragment | NA | NA | 0 |
| S2-final-viral-combined_k141_154102\|\|full | 4907 | No | NA | 5 | 0 | 0 | Not-determined | Genome-fragment | NA | NA | 0 |
| S5-final-viral-combined_k141_147422\|\|full | 4150 | No | NA | 4 | 2 | 0 | Low-quality | Genome-fragment | 8.54 | AAI-based (high-confidence) | 0 |
| S2-final-viral-combined_k141_195655\|\|full | 1019 | No | NA | 2 | 0 | 0 | Not-determined | Genome-fragment | NA | NA | 0 |
| S2-final-viral-combined_k141_92460\|\|full | 1614 | No | NA | 2 | 0 | 1 | Not-determined | Genome-fragment | NA | NA | 0 |
| S7-final-viral-combined_k141_204242\|\|full | 4150 | No | NA | 4 | 0 | 4 | Low-quality | Genome-fragment | 5.78 | AAI-based (medium-confidence) | 0 |
| S2-final-viral-combined_k141_185452\|\|full | 3337 | No | NA | 4 | 0 | 2 | Not-determined | Genome-fragment | NA | NA | 0 |
| S2-final-viral-combined_k141_245157\|\|full | 3476 | No | NA | 4 | 0 | 1 | Not-determined | Genome-fragment | NA | NA | 0 |
| S2-final-viral-combined_k141_215686\|\|full | 2084 | No | NA | 3 | 0 | 2 | Not-determined | Genome-fragment | NA | NA | 0 |
| S2-final-viral-combined_k141_61268\|\|full | 1163 | No | NA | 3 | 0 | 0 | Not-determined | Genome-fragment | NA | NA | 0 |
| S2-final-viral-combined_k141_111810\|\|full | 2792 | No | NA | 3 | 0 | 1 | Not-determined | Genome-fragment | NA | NA | 0 |
| S17-final-viral-combined_k141_158947\|\|full | 4151 | No | NA | 3 | 0 | 2 | Low-quality | Genome-fragment | 1.07 | AAI-based (medium-confidence) | 0 |
| S2-final-viral-combined_k141_19429\|\|full | 6279 | No | NA | 7 | 0 | 1 | Not-determined | Genome-fragment | NA | NA | 0 |
| S7-final-viral-combined_k141_121178\|\|full | 4153 | No | NA | 10 | 1 | 0 | Low-quality | Genome-fragment | 7.91 | HMM-based (lower-bound) | 0 |
| S11_k141_68215 | 4154 | No | NA | 7 | 0 | 0 | Low-quality | Genome-fragment | 4.04 | AAI-based (high-confidence) | 0 |
| S2-final-viral-combined_k141_185470\|\|full | 1562 | No | NA | 2 | 0 | 2 | Not-determined | Genome-fragment | NA | NA | 0 |
| S2-final-viral-combined_k141_154174\|\|full | 1015 | No | NA | 2 | 0 | 1 | Not-determined | Genome-fragment | NA | NA | 0 |
| S2-final-viral-combined_k141_203609\|\|full | 4154 | No | NA | 3 | 0 | 2 | Low-quality | Genome-fragment | 4.32 | AAI-based (medium-confidence) | 0 |
| S2-final-viral-combined_k141_154182\|\|full | 1951 | No | NA | 3 | 0 | 3 | Not-determined | Genome-fragment | NA | NA | 0 |
| S2-final-viral-combined_k141_195740\|\|full | 12345 | No | NA | 16 | 0 | 8 | Not-determined | Genome-fragment | NA | NA | 0 |
| S2-final-viral-combined_k141_123198\|\|full | 1812 | No | NA | 4 | 0 | 0 | Not-determined | Genome-fragment | NA | NA | 0 |
| S17-k141_2447 | 4155 | No | NA | 3 | 0 | 0 | Low-quality | Genome-fragment | 6.3 | AAI-based (medium-confidence) | 0 |
| S2-final-viral-combined_k141_19485\|\|full | 7136 | No | NA | 7 | 0 | 1 | Not-determined | Genome-fragment | NA | NA | 0 |
| S2-final-viral-combined_k141_195756\|\|full | 1099 | No | NA | 3 | 0 | 0 | Not-determined | Genome-fragment | NA | NA | 0 |
| S2-final-viral-combined_k141_11636\|\|full | 4156 | No | NA | 5 | 1 | 0 | Low-quality | Genome-fragment | 6.29 | HMM-based (lower-bound) | 0 |
| S2-final-viral-combined_k141_215808\|\|full | 5201 | No | NA | 3 | 0 | 1 | Not-determined | Genome-fragment | NA | NA | 0 |
| S2-final-viral-combined_k141_19544\|\|full | 8697 | No | NA | 9 | 0 | 5 | Not-determined | Genome-fragment | NA | NA | 0 |
| S2-final-viral-combined_k141_195818\|\|full | 1021 | No | NA | 2 | 0 | 1 | Not-determined | Genome-fragment | NA | NA | 0 |
| S9-final-viral-combined_k141_93102\|\|full | 4157 | No | NA | 4 | 0 | 2 | Low-quality | Genome-fragment | 1.39 | AAI-based (medium-confidence) | 0 |
| S2-final-viral-combined_k141_143956\|\|full | 5138 | No | NA | 7 | 0 | 1 | Not-determined | Genome-fragment | NA | NA | 0 |
| S2-final-viral-combined_k141_19571\|\|full | 1025 | No | NA | 3 | 0 | 0 | Not-determined | Genome-fragment | NA | NA | 0 |
| S2-final-viral-combined_k141_19573\|\|full | 2287 | No | NA | 4 | 0 | 0 | Not-determined | Genome-fragment | NA | NA | 0 |
| S3-final-viral-combined_k141_71687\|\|full | 4157 | No | NA | 6 | 0 | 4 | Low-quality | Genome-fragment | 2.53 | AAI-based (medium-confidence) | 0 |
| S2-final-viral-combined_k141_131901\|\|full | 5226 | No | NA | 4 | 0 | 2 | Not-determined | Genome-fragment | NA | NA | 0 |
| S2-final-viral-combined_k141_19579\|\|full | 2291 | No | NA | 3 | 0 | 1 | Not-determined | Genome-fragment | NA | NA | 0 |
| S2-final-viral-combined_k141_131909\|\|full | 6215 | No | NA | 4 | 0 | 1 | Not-determined | Genome-fragment | NA | NA | 0 |
| S3-final-viral-combined_k141_22117\|\|full | 4157 | No | NA | 6 | 3 | 0 | Low-quality | Genome-fragment | 7.66 | AAI-based (high-confidence) | 0 |
| S2-final-viral-combined_k141_7155\|\|full | 2181 | No | NA | 3 | 0 | 0 | Not-determined | Genome-fragment | NA | NA | 0 |
| S12_k141_26267 | 4157 | No | NA | 2 | 0 | 2 | Low-quality | Genome-fragment | 2.42 | AAI-based (high-confidence) | 0 |
| S2-final-viral-combined_k141_103120\|\|full | 2566 | No | NA | 4 | 0 | 1 | Not-determined | Genome-fragment | NA | NA | 0 |
| S2-final-viral-combined_k141_7178\|\|full | 7380 | No | NA | 12 | 0 | 1 | Not-determined | Genome-fragment | NA | NA | 0 |
| S11_k141_213952 | 4159 | No | NA | 4 | 0 | 0 | Low-quality | Genome-fragment | 7.32 | AAI-based (medium-confidence) | 0 |
| S2-final-viral-combined_k141_131953\|\|full | 5057 | No | NA | 10 | 0 | 0 | Not-determined | Genome-fragment | NA | NA | 0 |
| S1-final-viral-combined_k141_178615\|\|full | 4161 | No | NA | 4 | 0 | 3 | Low-quality | Genome-fragment | 2.31 | AAI-based (medium-confidence) | 0 |
| S7_k141_52436 | 4161 | No | NA | 3 | 0 | 2 | Low-quality | Genome-fragment | 10.72 | AAI-based (high-confidence) | 0 |
| S17-k141_120429 | 4162 | No | NA | 8 | 1 | 3 | Low-quality | Genome-fragment | 5.06 | AAI-based (high-confidence) | 0 |
| S2-final-viral-combined_k141_27994\|\|full | 11367 | No | NA | 11 | 0 | 7 | Not-determined | Genome-fragment | NA | NA | 0 |
| S2-final-viral-combined_k141_204291\|\|full | 2527 | No | NA | 3 | 0 | 1 | Not-determined | Genome-fragment | NA | NA | 0 |
| S17-final-viral-combined_k141_212052\|\|full | 4163 | No | NA | 4 | 0 | 0 | Low-quality | Genome-fragment | 3.64 | AAI-based (medium-confidence) | 0 |
| S4_k141_107295 | 4163 | No | NA | 6 | 0 | 1 | Low-quality | Genome-fragment | 7.72 | AAI-based (medium-confidence) | 0 |
| S2-final-viral-combined_k141_28022\|\|full | 1591 | No | NA | 2 | 0 | 0 | Not-determined | Genome-fragment | NA | NA | 0 |
| S12-final-viral-combined_k141_303350\|\|full | 4166 | No | NA | 3 | 0 | 3 | Low-quality | Genome-fragment | 3.36 | AAI-based (medium-confidence) | 0 |
| S11-final-viral-combined_k141_290482\|\|full | 4167 | No | NA | 5 | 2 | 0 | Low-quality | Genome-fragment | 8.68 | AAI-based (high-confidence) | 0 |
| S5_k141_83878 | 4167 | No | NA | 6 | 1 | 0 | Low-quality | Genome-fragment | 2.26 | HMM-based (lower-bound) | 0 |
| S2-final-viral-combined_k141_41125\|\|full | 3002 | No | NA | 3 | 0 | 0 | Not-determined | Genome-fragment | NA | NA | 0 |
| S2-final-viral-combined_k141_81744\|\|full | 1518 | No | NA | 3 | 0 | 1 | Not-determined | Genome-fragment | NA | NA | 0 |
| S7-final-viral-combined_k141_89938\|\|full | 4168 | No | NA | 5 | 2 | 0 | Low-quality | Genome-fragment | 9.71 | AAI-based (medium-confidence) | 0 |
| S2-final-viral-combined_k141_19692\|\|full | 2753 | No | NA | 5 | 0 | 3 | Not-determined | Genome-fragment | NA | NA | 0 |
| S17-final-viral-combined_k141_4514\|\|full | 4169 | No | NA | 4 | 0 | 3 | Low-quality | Genome-fragment | 3.35 | AAI-based (medium-confidence) | 0 |
| S2-final-viral-combined_k141_185494\|\|full | 234793 | Yes | 30808 | 236 | 23 | 159 | Medium-quality | Genome-fragment | 65.16 | HMM-based (lower-bound) | 86.88 |
| S2-final-viral-combined_k141_195944\|\|full | 2435 | No | NA | 2 | 0 | 2 | Not-determined | Genome-fragment | NA | NA | 0 |
| S9_k141_45397 | 4170 | No | NA | 5 | 2 | 0 | Low-quality | Genome-fragment | 8.68 | AAI-based (high-confidence) | 0 |
| S1_k141_334914 | 4170 | No | NA | 1 | 0 | 1 | Low-quality | Genome-fragment | 2.09 | AAI-based (medium-confidence) | 0 |
| S2-final-viral-combined_k141_61484\|\|full | 1260 | No | NA | 2 | 0 | 2 | Not-determined | Genome-fragment | NA | NA | 0 |
| S2-final-viral-combined_k141_144188\|\|full | 6099 | No | NA | 7 | 0 | 2 | Not-determined | Genome-fragment | NA | NA | 0 |
| S1-final-viral-combined_k141_391062\|\|full | 4171 | No | NA | 5 | 4 | 0 | Low-quality | Genome-fragment | 10.64 | AAI-based (high-confidence) | 0 |
| S8-final-viral-combined_k141_216580\|\|full | 4174 | No | NA | 4 | 3 | 0 | Low-quality | Genome-fragment | 10.37 | AAI-based (high-confidence) | 0 |
| S2-final-viral-combined_k141_185529\|\|full | 3363 | No | NA | 3 | 0 | 2 | Not-determined | Genome-fragment | NA | NA | 0 |
| S2-final-viral-combined_k141_111888\|\|full | 2253 | No | NA | 5 | 0 | 1 | Not-determined | Genome-fragment | NA | NA | 0 |
| S2-final-viral-combined_k141_225300\|\|full | 2224 | No | NA | 2 | 0 | 2 | Not-determined | Genome-fragment | NA | NA | 0 |
| S8-final-viral-combined_k141_242753\|\|full | 4175 | No | NA | 6 | 0 | 4 | Low-quality | Genome-fragment | 6.54 | AAI-based (medium-confidence) | 0 |
| S17-final-viral-combined_k141_43746\|\|full | 4176 | No | NA | 4 | 4 | 0 | Low-quality | Genome-fragment | 9.17 | AAI-based (high-confidence) | 0 |
| S2-final-viral-combined_k141_185563\|\|full | 4832 | No | NA | 5 | 0 | 3 | Not-determined | Genome-fragment | NA | NA | 0 |
| S1-final-viral-combined_k141_261163\|\|full | 4177 | No | NA | 5 | 0 | 2 | Low-quality | Genome-fragment | 2.18 | AAI-based (medium-confidence) | 0 |
| S5-final-viral-combined_k141_40149\|\|full | 4177 | No | NA | 5 | 0 | 4 | Low-quality | Genome-fragment | 9.65 | AAI-based (medium-confidence) | 0 |
| S12-final-viral-combined_k141_317976\|\|full | 4177 | No | NA | 5 | 3 | 1 | Low-quality | Genome-fragment | 9.77 | AAI-based (medium-confidence) | 0 |
| S9-final-viral-combined_k141_72200\|\|full | 4178 | No | NA | 4 | 0 | 2 | Low-quality | Genome-fragment | 4.77 | AAI-based (medium-confidence) | 0 |
| S4_k141_133399 | 4178 | No | NA | 9 | 0 | 1 | Low-quality | Genome-fragment | 7.11 | AAI-based (medium-confidence) | 0 |
| S2-final-viral-combined_k141_237909\|\|full | 2209 | No | NA | 2 | 0 | 2 | Not-determined | Genome-fragment | NA | NA | 0 |
| S2-final-viral-combined_k141_7293\|\|full | 1066 | No | NA | 3 | 0 | 1 | Not-determined | Genome-fragment | NA | NA | 0 |
| S2-final-viral-combined_k141_215886\|\|full | 6275 | No | NA | 6 | 0 | 6 | Not-determined | Genome-fragment | NA | NA | 0 |
| S2-final-viral-combined_k141_154544\|\|full | 1755 | No | NA | 3 | 0 | 2 | Not-determined | Genome-fragment | NA | NA | 0 |
| S5_k141_1655 | 4179 | No | NA | 3 | 0 | 0 | Low-quality | Genome-fragment | 5.31 | AAI-based (medium-confidence) | 0 |
| S17-k141_249075 | 4179 | No | NA | 8 | 0 | 0 | Low-quality | Genome-fragment | 5.95 | AAI-based (high-confidence) | 0 |
| S11-final-viral-combined_k141_82198\|\|full | 4180 | No | NA | 5 | 2 | 0 | Low-quality | Genome-fragment | 8.98 | AAI-based (medium-confidence) | 0 |
| S2-final-viral-combined_k141_112005\|\|full | 3543 | No | NA | 4 | 0 | 3 | Not-determined | Genome-fragment | NA | NA | 0 |
| S1_k141_282919 | 4181 | No | NA | 4 | 0 | 1 | Low-quality | Genome-fragment | 11.33 | AAI-based (medium-confidence) | 0 |
| S2-final-viral-combined_k141_92796\|\|full | 1178 | No | NA | 2 | 0 | 1 | Not-determined | Genome-fragment | NA | NA | 0 |
| S3-final-viral-combined_k141_84993\|\|full | 4182 | No | NA | 8 | 3 | 0 | Low-quality | Genome-fragment | 8.54 | AAI-based (medium-confidence) | 0 |
| S17-final-viral-combined_k141_215070\|\|full | 4183 | No | NA | 2 | 0 | 1 | Low-quality | Genome-fragment | 6.97 | AAI-based (medium-confidence) | 0 |
| S2-final-viral-combined_k141_204382\|\|full | 1367 | No | NA | 3 | 0 | 0 | Not-determined | Genome-fragment | NA | NA | 0 |
| S2-final-viral-combined_k141_144334\|\|full | 4497 | No | NA | 6 | 0 | 1 | Not-determined | Genome-fragment | NA | NA | 0 |
| S2_k141_76159 | 4183 | No | NA | 4 | 0 | 0 | Low-quality | Genome-fragment | 9.37 | AAI-based (medium-confidence) | 0 |
| S9-final-viral-combined_k141_120812\|\|full | 4184 | No | NA | 3 | 0 | 2 | Low-quality | Genome-fragment | 4.88 | AAI-based (medium-confidence) | 0 |
| S2-final-viral-combined_k141_185668\|\|full | 4189 | No | NA | 3 | 0 | 2 | Not-determined | Genome-fragment | NA | NA | 0 |
| S7_k141_48069 | 4184 | No | NA | 8 | 0 | 1 | Low-quality | Genome-fragment | 4.84 | AAI-based (medium-confidence) | 0 |
| S12-final-viral-combined_k141_304571\|\|full | 4186 | No | NA | 8 | 2 | 0 | Low-quality | Genome-fragment | 10.38 | AAI-based (high-confidence) | 0 |
| S2-final-viral-combined_k141_28319\|\|full | 1837 | No | NA | 2 | 0 | 0 | Not-determined | Genome-fragment | NA | NA | 0 |
| S12-final-viral-combined_k141_188429\|\|full | 4186 | No | NA | 7 | 3 | 0 | Low-quality | Genome-fragment | 9.68 | AAI-based (high-confidence) | 0 |
| S2-final-viral-combined_k141_87688\|\|full | 4187 | No | NA | 3 | 0 | 1 | Low-quality | Genome-fragment | 2.1 | AAI-based (medium-confidence) | 0 |
| S5-final-viral-combined_k141_22926\|\|full | 4188 | No | NA | 7 | 1 | 0 | Low-quality | Genome-fragment | 4.42 | AAI-based (medium-confidence) | 0 |
| S2-final-viral-combined_k141_217053\|\|full | 4189 | No | NA | 7 | 0 | 1 | Low-quality | Genome-fragment | 7.76 | AAI-based (medium-confidence) | 0 |
| S4_k141_123017 | 4189 | No | NA | 2 | 0 | 1 | Low-quality | Genome-fragment | 2.06 | AAI-based (medium-confidence) | 0 |
| S2-final-viral-combined_k141_112071\|\|full | 3644 | No | NA | 4 | 0 | 2 | Not-determined | Genome-fragment | NA | NA | 0 |
| S2-final-viral-combined_k141_154687\|\|full | 1671 | No | NA | 3 | 0 | 1 | Not-determined | Genome-fragment | NA | NA | 0 |
| S2-final-viral-combined_k141_28345\|\|full | 2357 | No | NA | 2 | 0 | 1 | Not-determined | Genome-fragment | NA | NA | 0 |
| S2-final-viral-combined_k141_81965\|\|full | 1047 | No | NA | 2 | 0 | 1 | Not-determined | Genome-fragment | NA | NA | 0 |
| S1_k141_198891 | 4190 | No | NA | 4 | 0 | 4 | Low-quality | Genome-fragment | 1.16 | AAI-based (medium-confidence) | 0 |
| S2-final-viral-combined_k141_111820\|\|full | 4191 | No | NA | 6 | 1 | 0 | Low-quality | Genome-fragment | 2.67 | HMM-based (lower-bound) | 0 |
| S2-final-viral-combined_k141_71926\|\|full | 2390 | No | NA | 3 | 0 | 0 | Not-determined | Genome-fragment | NA | NA | 0 |
| S11-final-viral-combined_k141_310904\|\|full | 4192 | No | NA | 3 | 0 | 1 | Low-quality | Genome-fragment | 4.85 | AAI-based (medium-confidence) | 0 |
| S2-final-viral-combined_k141_61668\|\|full | 1014 | No | NA | 3 | 0 | 2 | Not-determined | Genome-fragment | NA | NA | 0 |
| S12-final-viral-combined_k141_169282\|\|full | 4192 | No | NA | 3 | 0 | 3 | Low-quality | Genome-fragment | 2.26 | AAI-based (medium-confidence) | 0 |
| S6-final-viral-combined_k141_26999\|\|full | 4193 | No | NA | 4 | 0 | 1 | Low-quality | Genome-fragment | 0.49 | AAI-based (medium-confidence) | 0 |
| S2-final-viral-combined_k141_123517\|\|full | 2188 | No | NA | 7 | 0 | 0 | Not-determined | Genome-fragment | NA | NA | 0 |

| | | | | | | | | | | | |
|---|---|---|---|---|---|---|---|---|---|---|---|
| S2-final-viral-combined_k141_41354\|\|full | 2129 | No | NA | 4 | 0 | 0 | Not-determined | Genome-fragment | NA | NA | 0 |
| S12-final-viral-combined_k141_110188\|\|full | 4194 | No | NA | 6 | 0 | 0 | Low-quality | Genome-fragment | 3.77 | AAI-based (medium-confidence) | 0 |
| S2-final-viral-combined_k141_19982\|\|full | 1231 | No | NA | 3 | 0 | 0 | Not-determined | Genome-fragment | NA | NA | 0 |
| S2-final-viral-combined_k141_123531\|\|full | 2269 | No | NA | 3 | 0 | 1 | Not-determined | Genome-fragment | NA | NA | 0 |
| S12_k141_12991 | 4194 | No | NA | 13 | 0 | 0 | Low-quality | Genome-fragment | 6.91 | AAI-based (high-confidence) | 0 |
| S2-final-viral-combined_k141_238082\|\|full | 6763 | No | NA | 5 | 0 | 2 | Not-determined | Genome-fragment | NA | NA | 0 |
| S2-final-viral-combined_k141_163780\|\|full | 5179 | No | NA | 9 | 0 | 0 | Not-determined | Genome-fragment | NA | NA | 0 |
| S2-final-viral-combined_k141_28422\|\|full | 3341 | No | NA | 3 | 0 | 1 | Not-determined | Genome-fragment | NA | NA | 0 |
| S2-final-viral-combined_k141_185838\|\|full | 1012 | No | NA | 2 | 0 | 0 | Not-determined | Genome-fragment | NA | NA | 0 |
| S2-final-viral-combined_k141_72010\|\|full | 2178 | No | NA | 2 | 0 | 2 | Not-determined | Genome-fragment | NA | NA | 0 |
| S2-final-viral-combined_k141_82057\|\|full | 1835 | No | NA | 3 | 0 | 2 | Not-determined | Genome-fragment | NA | NA | 0 |
| S2-final-viral-combined_k141_185867\|\|full | 1012 | No | NA | 2 | 0 | 2 | Not-determined | Genome-fragment | NA | NA | 0 |
| S1-final-viral-combined_k141_362619\|\|full | 4195 | No | NA | 8 | 0 | 0 | Low-quality | Genome-fragment | 9.69 | AAI-based (high-confidence) | 0 |
| S2-final-viral-combined_k141_82059\|\|full | 1224 | No | NA | 2 | 0 | 2 | Not-determined | Genome-fragment | NA | NA | 0 |
| S7-final-viral-combined_k141_178511\|\|full | 4195 | No | NA | 9 | 1 | 0 | Low-quality | Genome-fragment | 6.81 | AAI-based (high-confidence) | 0 |
| S2-final-viral-combined_k141_188155\|\|full | 4196 | No | NA | 6 | 0 | 0 | Low-quality | Genome-fragment | 7.83 | AAI-based (high-confidence) | 0 |
| S3-final-viral-combined_k141_101919\|\|full | 4197 | No | NA | 3 | 0 | 1 | Low-quality | Genome-fragment | 1.33 | AAI-based (medium-confidence) | 0 |
| S7_k141_207454 | 4197 | No | NA | 2 | 0 | 1 | Low-quality | Genome-fragment | 4.08 | AAI-based (high-confidence) | 0 |
| S2-final-viral-combined_k141_61792\|\|full | 1744 | No | NA | 3 | 0 | 0 | Not-determined | Genome-fragment | NA | NA | 0 |
| S2-final-viral-combined_k141_7570\|\|full | 2109 | No | NA | 2 | 0 | 0 | Not-determined | Genome-fragment | NA | NA | 0 |
| S2-final-viral-combined_k141_61802\|\|full | 1556 | No | NA | 3 | 0 | 1 | Not-determined | Genome-fragment | NA | NA | 0 |
| S17-final-viral-combined_k141_151902\|\|full | 4198 | No | NA | 4 | 0 | 1 | Low-quality | Genome-fragment | 4.32 | AAI-based (high-confidence) | 0 |
| S2-final-viral-combined_k141_238105\|\|full | 1201 | No | NA | 2 | 0 | 1 | Not-determined | Genome-fragment | NA | NA | 0 |
| S2-final-viral-combined_k141_7586\|\|full | 2096 | No | NA | 4 | 0 | 1 | Not-determined | Genome-fragment | NA | NA | 0 |
| S2-final-viral-combined_k141_103367\|\|full | 1742 | No | NA | 4 | 0 | 1 | Not-determined | Genome-fragment | NA | NA | 0 |
| S2-final-viral-combined_k141_119706\|\|full | 4198 | No | NA | 13 | 3 | 0 | Low-quality | Genome-fragment | 7.88 | AAI-based (high-confidence) | 0 |
| S2-final-viral-combined_k141_132311\|\|full | 1525 | No | NA | 2 | 0 | 2 | Not-determined | Genome-fragment | NA | NA | 0 |
| S12-final-viral-combined_k141_344553\|\|full | 4199 | No | NA | 4 | 3 | 0 | Low-quality | Genome-fragment | 6.47 | HMM-based (lower-bound) | 0 |
| S2-final-viral-combined_k141_216248\|\|full | 2141 | No | NA | 4 | 0 | 3 | Not-determined | Genome-fragment | NA | NA | 0 |
| S2-final-viral-combined_k141_124746\|\|full | 4200 | No | NA | 5 | 3 | 0 | Low-quality | Genome-fragment | 14.81 | HMM-based (lower-bound) | 0 |
| S4_k141_77235 | 4201 | No | NA | 3 | 0 | 3 | Low-quality | Genome-fragment | 0.61 | AAI-based (medium-confidence) | 0 |
| S11-final-viral-combined_k141_80014\|\|full | 4203 | No | NA | 6 | 2 | 0 | Low-quality | Genome-fragment | 11.78 | AAI-based (medium-confidence) | 0 |
| S2-final-viral-combined_k141_112229\|\|full | 1703 | No | NA | 2 | 0 | 0 | Not-determined | Genome-fragment | NA | NA | 0 |
| S17-final-viral-combined_k141_71045\|\|full | 4203 | No | NA | 3 | 0 | 2 | Low-quality | Genome-fragment | 1.4 | AAI-based (medium-confidence) | 0 |
| S2-final-viral-combined_k141_144534\|\|full | 1566 | No | NA | 2 | 0 | 1 | Not-determined | Genome-fragment | NA | NA | 0 |
| S11-final-viral-combined_k141_300027\|\|full | 4204 | No | NA | 4 | 0 | 3 | Low-quality | Genome-fragment | 2.37 | AAI-based (medium-confidence) | 0 |
| S17-final-viral-combined_k141_257221\|\|full | 4204 | No | NA | 3 | 0 | 2 | Low-quality | Genome-fragment | 1.92 | AAI-based (medium-confidence) | 0 |
| S17-final-viral-combined_k141_263886\|\|full | 4205 | No | NA | 6 | 0 | 2 | Low-quality | Genome-fragment | 4.8 | AAI-based (medium-confidence) | 0 |
| S2_k141_18580 | 4205 | No | NA | 3 | 0 | 3 | Low-quality | Genome-fragment | 1.19 | AAI-based (medium-confidence) | 0 |
| S2-final-viral-combined_k141_225668\|\|full | 4835 | No | NA | 7 | 0 | 0 | Not-determined | Genome-fragment | NA | NA | 0 |
| S2-final-viral-combined_k141_123728\|\|full | 1843 | No | NA | 3 | 0 | 0 | Not-determined | Genome-fragment | NA | NA | 0 |
| S2-final-viral-combined_k141_7669\|\|full | 1889 | No | NA | 2 | 0 | 0 | Not-determined | Genome-fragment | NA | NA | 0 |
| S2-final-viral-combined_k141_82252\|\|full | 2490 | No | NA | 5 | 0 | 0 | Not-determined | Genome-fragment | NA | NA | 0 |
| S17-final-viral-combined_k141_18940\|\|full | 4207 | No | NA | 3 | 3 | 0 | Low-quality | Genome-fragment | 8.08 | AAI-based (high-confidence) | 0 |
| S2-final-viral-combined_k141_7683\|\|full | 1577 | No | NA | 2 | 0 | 1 | Not-determined | Genome-fragment | NA | NA | 0 |
| S2-final-viral-combined_k141_144616\|\|full | 5182 | No | NA | 5 | 0 | 2 | Not-determined | Genome-fragment | NA | NA | 0 |
| S2_k141_207854 | 4207 | No | NA | 2 | 0 | 2 | Low-quality | Genome-fragment | 6.97 | AAI-based (medium-confidence) | 0 |
| S5-final-viral-combined_k141_31238\|\|full | 4208 | No | NA | 5 | 0 | 0 | Low-quality | Genome-fragment | 2.66 | AAI-based (medium-confidence) | 0 |
| S2-final-viral-combined_k141_103443\|\|full | 1149 | No | NA | 3 | 0 | 1 | Not-determined | Genome-fragment | NA | NA | 0 |
| S5-final-viral-combined_k141_96888\|\|full | 4208 | No | NA | 4 | 0 | 1 | Low-quality | Genome-fragment | 2.79 | AAI-based (high-confidence) | 0 |
| S2-final-viral-combined_k141_7714\|\|full | 3030 | No | NA | 3 | 0 | 0 | Not-determined | Genome-fragment | NA | NA | 0 |
| S6_k141_137430 | 4208 | No | NA | 3 | 0 | 3 | Low-quality | Genome-fragment | 1.67 | AAI-based (medium-confidence) | 0 |
| S1_k141_347713 | 4209 | No | NA | 1 | 0 | 0 | Low-quality | Genome-fragment | 2.6 | AAI-based (medium-confidence) | 0 |
| S2-final-viral-combined_k141_216304\|\|full | 2026 | No | NA | 2 | 0 | 2 | Not-determined | Genome-fragment | NA | NA | 0 |
| S2-final-viral-combined_k141_245445\|\|full | 1112 | No | NA | 2 | 0 | 1 | Not-determined | Genome-fragment | NA | NA | 0 |
| S17-final-viral-combined_k141_167086\|\|full | 4210 | No | NA | 3 | 0 | 2 | Low-quality | Genome-fragment | 6.03 | AAI-based (medium-confidence) | 0 |
| S17-final-viral-combined_k141_131244\|\|full | 4210 | No | NA | 4 | 0 | 4 | Low-quality | Genome-fragment | 1.88 | AAI-based (medium-confidence) | 0 |
| S6-final-viral-combined_k141_86188\|\|full | 4210 | No | NA | 3 | 0 | 3 | Low-quality | Genome-fragment | 2 | AAI-based (medium-confidence) | 0 |
| S3-final-viral-combined_k141_93252\|\|full | 4211 | Yes | 438 | 5 | 1 | 2 | Low-quality | Genome-fragment | 0.28 | HMM-based (lower-bound) | 89.6 |
| S2-final-viral-combined_k141_112393\|\|full | 1619 | No | NA | 3 | 0 | 0 | Not-determined | Genome-fragment | NA | NA | 0 |
| S2_k141_146120 | 4211 | No | NA | 2 | 0 | 1 | Low-quality | Genome-fragment | 14.68 | AAI-based (medium-confidence) | 0 |
| S5-final-viral-combined_k141_23433\|\|full | 4212 | No | NA | 9 | 1 | 0 | Low-quality | Genome-fragment | 10.34 | AAI-based (high-confidence) | 0 |
| S2-final-viral-combined_k141_132565\|\|full | 2474 | No | NA | 2 | 0 | 2 | Not-determined | Genome-fragment | NA | NA | 0 |
| S2-final-viral-combined_k141_103504\|\|full | 4993 | No | NA | 6 | 0 | 3 | Not-determined | Genome-fragment | NA | NA | 0 |
| S12_k141_38550 | 4212 | No | NA | 5 | 0 | 0 | Low-quality | Genome-fragment | 9 | AAI-based (medium-confidence) | 0 |
| S2-final-viral-combined_k141_132570\|\|full | 1869 | No | NA | 3 | 0 | 0 | Not-determined | Genome-fragment | NA | NA | 0 |
| S2-final-viral-combined_k141_216331\|\|full | 2242 | No | NA | 3 | 0 | 2 | Not-determined | Genome-fragment | NA | NA | 0 |
| S2-final-viral-combined_k141_186092\|\|full | 1982 | No | NA | 2 | 0 | 2 | Not-determined | Genome-fragment | NA | NA | 0 |
| S2-final-viral-combined_k141_196500\|\|full | 1118 | No | NA | 3 | 0 | 1 | Not-determined | Genome-fragment | NA | NA | 0 |
| S2-final-viral-combined_k141_186112\|\|full | 2387 | No | NA | 4 | 0 | 1 | Not-determined | Genome-fragment | NA | NA | 0 |
| S2-final-viral-combined_k141_93115\|\|full | 1001 | No | NA | 2 | 0 | 1 | Not-determined | Genome-fragment | NA | NA | 0 |
| S2-final-viral-combined_k141_196549\|\|full | 2510 | No | NA | 5 | 0 | 2 | Not-determined | Genome-fragment | NA | NA | 0 |
| S17-final-viral-combined_k141_244577\|\|full | 4213 | No | NA | 7 | 1 | 0 | Low-quality | Genome-fragment | 3.04 | HMM-based (lower-bound) | 0 |
| S2-final-viral-combined_k141_196574\|\|full | 3273 | No | NA | 5 | 0 | 2 | Not-determined | Genome-fragment | NA | NA | 0 |
| S17-final-viral-combined_k141_199379\|\|full | 4213 | No | NA | 6 | 0 | 1 | Low-quality | Genome-fragment | 12.87 | AAI-based (medium-confidence) | 0 |
| S6-final-viral-combined_k141_138089\|\|full | 4213 | No | NA | 8 | 3 | 0 | Low-quality | Genome-fragment | 4.06 | HMM-based (lower-bound) | 0 |
| S2-final-viral-combined_k141_20176\|\|full | 1041 | No | NA | 2 | 0 | 2 | Not-determined | Genome-fragment | NA | NA | 0 |
| S2-final-viral-combined_k141_82408\|\|full | 6604 | No | NA | 4 | 0 | 1 | Not-determined | Genome-fragment | NA | NA | 0 |
| S2-final-viral-combined_k141_196599\|\|full | 1103 | No | NA | 2 | 0 | 0 | Not-determined | Genome-fragment | NA | NA | 0 |
| S7_k141_129045 | 4214 | No | NA | 8 | 1 | 0 | Low-quality | Genome-fragment | 2.29 | HMM-based (lower-bound) | 0 |
| S1_k141_154305 | 4215 | No | NA | 1 | 0 | 0 | Low-quality | Genome-fragment | 4.96 | AAI-based (medium-confidence) | 0 |
| S5-final-viral-combined_k141_171816\|\|full | 4216 | No | NA | 5 | 0 | 2 | Low-quality | Genome-fragment | 8.25 | AAI-based (high-confidence) | 0 |
| S2-final-viral-combined_k141_62002\|\|full | 3307 | No | NA | 5 | 0 | 4 | Not-determined | Genome-fragment | NA | NA | 0 |
| S2-final-viral-combined_k141_132681\|\|full | 1094 | No | NA | 2 | 0 | 1 | Not-determined | Genome-fragment | NA | NA | 0 |
| S2-final-viral-combined_k141_62014\|\|full | 1393 | No | NA | 2 | 0 | 1 | Not-determined | Genome-fragment | NA | NA | 0 |
| S2-final-viral-combined_k141_204808\|\|full | 2519 | No | NA | 4 | 0 | 0 | Not-determined | Genome-fragment | NA | NA | 0 |
| S2-final-viral-combined_k141_245564\|\|full | 1260 | No | NA | 2 | 0 | 2 | Not-determined | Genome-fragment | NA | NA | 0 |
| S12_k141_134676 | 4217 | No | NA | 4 | 0 | 1 | Low-quality | Genome-fragment | 6.48 | AAI-based (medium-confidence) | 0 |
| S4-final-viral-combined_k141_122507\|\|full | 4217 | No | NA | 3 | 0 | 3 | Low-quality | Genome-fragment | 0.68 | AAI-based (high-confidence) | 0 |
| S2-final-viral-combined_k141_144742\|\|full | 4477 | No | NA | 6 | 0 | 0 | Not-determined | Genome-fragment | NA | NA | 0 |
| S2-final-viral-combined_k141_204829\|\|full | 2838 | No | NA | 3 | 0 | 2 | Not-determined | Genome-fragment | NA | NA | 0 |
| S2-final-viral-combined_k141_164110\|\|full | 4452 | No | NA | 5 | 0 | 3 | Not-determined | Genome-fragment | NA | NA | 0 |
| S5-final-viral-combined_k141_28044\|\|full | 4218 | No | NA | 3 | 0 | 2 | Low-quality | Genome-fragment | 3.78 | AAI-based (medium-confidence) | 0 |
| S2-final-viral-combined_k141_20260\|\|full | 2208 | No | NA | 2 | 0 | 0 | Not-determined | Genome-fragment | NA | NA | 0 |
| S1-final-viral-combined_k141_78167\|\|full | 4219 | No | NA | 5 | 0 | 0 | Low-quality | Genome-fragment | 2.25 | AAI-based (medium-confidence) | 0 |
| S2-final-viral-combined_k141_103553\|\|full | 1218 | No | NA | 2 | 0 | 0 | Not-determined | Genome-fragment | NA | NA | 0 |
| S3-final-viral-combined_k141_1526\|\|full | 4220 | No | NA | 2 | 0 | 1 | Low-quality | Genome-fragment | 5.87 | AAI-based (medium-confidence) | 0 |
| S2-final-viral-combined_k141_141200\|\|full | 4220 | No | NA | 5 | 0 | 2 | Low-quality | Genome-fragment | 2.9 | AAI-based (medium-confidence) | 0 |
| S2-final-viral-combined_k141_93143\|\|full | 11977 | No | NA | 13 | 0 | 6 | Not-determined | Genome-fragment | NA | NA | 0 |
| S2-final-viral-combined_k141_132842\|\|full | 1090 | No | NA | 3 | 0 | 0 | Not-determined | Genome-fragment | NA | NA | 0 |
| S2-final-viral-combined_k141_93148\|\|full | 2581 | No | NA | 4 | 0 | 0 | Not-determined | Genome-fragment | NA | NA | 0 |
| S4_k141_8613 | 4221 | Yes | 2113 | 5 | 1 | 2 | Low-quality | Genome-fragment | 2.62 | HMM-based (lower-bound) | 49.94 |
| S2-final-viral-combined_k141_155173\|\|full | 1256 | No | NA | 3 | 0 | 0 | Not-determined | Genome-fragment | NA | NA | 0 |
| S11_k141_243932 | 4222 | No | NA | 3 | 0 | 1 | Low-quality | Genome-fragment | 2.98 | AAI-based (high-confidence) | 0 |
| S1_k141_269942 | 4222 | No | NA | 3 | 0 | 1 | Low-quality | Genome-fragment | 9.08 | AAI-based (high-confidence) | 0 |
| S2-final-viral-combined_k141_196708\|\|full | 1315 | No | NA | 2 | 0 | 0 | Not-determined | Genome-fragment | NA | NA | 0 |
| S12-final-viral-combined_k141_254747\|\|full | 4223 | No | NA | 5 | 0 | 3 | Low-quality | Genome-fragment | 1.29 | AAI-based (medium-confidence) | 0 |
| S17-final-viral-combined_k141_9468\|\|full | 4224 | No | NA | 6 | 1 | 0 | Low-quality | Genome-fragment | 9.92 | AAI-based (medium-confidence) | 0 |
| S11_k141_32929 | 4225 | No | NA | 10 | 1 | 0 | Low-quality | Genome-fragment | 3.18 | AAI-based (high-confidence) | 0 |
| S2-final-viral-combined_k141_132901\|\|full | 2268 | No | NA | 4 | 0 | 1 | Not-determined | Genome-fragment | NA | NA | 0 |
| S17-final-viral-combined_k141_244073\|\|full | 4225 | No | NA | 4 | 0 | 2 | Low-quality | Genome-fragment | 1.17 | AAI-based (medium-confidence) | 0 |
| S11-final-viral-combined_k141_166600\|\|full | 4226 | Yes | 2903 | 9 | 5 | 1 | Low-quality | Genome-fragment | 6.81 | AAI-based (high-confidence) | 31.31 |
| S2-final-viral-combined_k141_245680\|\|full | 7487 | No | NA | 6 | 0 | 5 | Not-determined | Genome-fragment | NA | NA | 0 |
| S4-final-viral-combined_k141_134762\|\|full | 4226 | No | NA | 5 | 0 | 1 | Low-quality | Genome-fragment | 11.85 | AAI-based (high-confidence) | 0 |
| S9-final-viral-combined_k141_84448\|\|full | 4227 | No | NA | 11 | 2 | 0 | Low-quality | Genome-fragment | 10.29 | AAI-based (medium-confidence) | 0 |
| S4-final-viral-combined_k141_15055\|\|full | 4227 | Yes | 3102 | 7 | 3 | 1 | Low-quality | Genome-fragment | 6.64 | AAI-based (high-confidence) | 26.61 |
| S2-final-viral-combined_k141_186248\|\|full | 2430 | No | NA | 2 | 0 | 2 | Not-determined | Genome-fragment | NA | NA | 0 |
| S2-final-viral-combined_k141_196729\|\|full | 3961 | No | NA | 7 | 0 | 3 | Not-determined | Genome-fragment | NA | NA | 0 |
| S2-final-viral-combined_k141_88844\|\|full | 4229 | No | NA | 8 | 2 | 0 | Low-quality | Genome-fragment | 22.98 | AAI-based (high-confidence) | 0 |
| S2-final-viral-combined_k141_132966\|\|full | 4461 | No | NA | 11 | 0 | 0 | Medium-quality | Genome-fragment | 69.54 | AAI-based (high-confidence) | 0 |
| S2-final-viral-combined_k141_144798\|\|full | 5750 | No | NA | 13 | 0 | 1 | Not-determined | Genome-fragment | NA | NA | 0 |
| S2-final-viral-combined_k141_245706\|\|full | 16662 | No | NA | 18 | 0 | 18 | Not-determined | Genome-fragment | NA | NA | 0 |
| S2-final-viral-combined_k141_123972\|\|full | 4311 | No | NA | 4 | 0 | 4 | Not-determined | Genome-fragment | NA | NA | 0 |
| S9_k141_28690 | 4230 | No | NA | 8 | 6 | 0 | Low-quality | Genome-fragment | 10.69 | AAI-based (high-confidence) | 0 |
| S4-final-viral-combined_k141_113901\|\|full | 4230 | Yes | 2772 | 3 | 1 | 1 | Low-quality | Genome-fragment | 4.01 | AAI-based (medium-confidence) | 34.47 |
| S2-final-viral-combined_k141_196754\|\|full | 5447 | No | NA | 6 | 0 | 2 | Not-determined | Genome-fragment | NA | NA | 0 |
| S2-final-viral-combined_k141_112577\|\|full | 1341 | No | NA | 2 | 0 | 0 | Not-determined | Genome-fragment | NA | NA | 0 |
| S8-final-viral-combined_k141_124192\|\|full | 4230 | Yes | 2784 | 5 | 2 | 1 | Low-quality | Genome-fragment | 1.65 | HMM-based (lower-bound) | 34.18 |
| S2-final-viral-combined_k141_245718\|\|full | 3594 | No | NA | 2 | 0 | 1 | Not-determined | Genome-fragment | NA | NA | 0 |
| S2-final-viral-combined_k141_123993\|\|full | 6366 | No | NA | 3 | 0 | 3 | Not-determined | Genome-fragment | NA | NA | 0 |
| S2-final-viral-combined_k141_20439\|\|full | 43600 | No | NA | 38 | 0 | 25 | Not-determined | Genome-fragment | NA | NA | 0 |
| S4_k141_68072 | 4231 | No | NA | 1 | 0 | 0 | Low-quality | Genome-fragment | 4.39 | AAI-based (medium-confidence) | 0 |
| S2-final-viral-combined_k141_186349\|\|full | 1846 | No | NA | 2 | 0 | 2 | Not-determined | Genome-fragment | NA | NA | 0 |
| S2-final-viral-combined_k141_225908\|\|full | 195686 | Yes | 34900 | 203 | 17 | 108 | Medium-quality | Genome-fragment | 51.14 | HMM-based (lower-bound) | 82.17 |
| S2-final-viral-combined_k141_82535\|\|full | 2344 | No | NA | 5 | 0 | 1 | Not-determined | Genome-fragment | NA | NA | 0 |
| S8-final-viral-combined_k141_164630\|\|full | 4231 | No | NA | 9 | 0 | 0 | Low-quality | Genome-fragment | 6.35 | AAI-based (high-confidence) | 0 |
| S2-final-viral-combined_k141_225925\|\|full | 2356 | No | NA | 4 | 0 | 1 | Not-determined | Genome-fragment | NA | NA | 0 |
| S2-final-viral-combined_k141_112634\|\|full | 3955 | No | NA | 3 | 0 | 3 | Not-determined | Genome-fragment | NA | NA | 0 |
| S2-final-viral-combined_k141_216747\|\|full | 7337 | No | NA | 4 | 0 | 2 | Not-determined | Genome-fragment | NA | NA | 0 |
| S6-final-viral-combined_k141_92365\|\|full | 4232 | No | NA | 4 | 0 | 3 | Low-quality | Genome-fragment | 1.2 | AAI-based (high-confidence) | 0 |
| S2-final-viral-combined_k141_112677\|\|full | 4176 | No | NA | 6 | 0 | 4 | Not-determined | Genome-fragment | NA | NA | 0 |
| S2-final-viral-combined_k141_112684\|\|full | 1868 | No | NA | 3 | 0 | 0 | Not-determined | Genome-fragment | NA | NA | 0 |
| S2-final-viral-combined_k141_216773\|\|full | 1072 | No | NA | 2 | 0 | 1 | Not-determined | Genome-fragment | NA | NA | 0 |
| S2-final-viral-combined_k141_216787\|\|full | 1996 | No | NA | 2 | 0 | 1 | Not-determined | Genome-fragment | NA | NA | 0 |
| S2-final-viral-combined_k141_216841\|\|full | 1636 | No | NA | 2 | 0 | 2 | Not-determined | Genome-fragment | NA | NA | 0 |
| S6-final-viral-combined_k141_27675\|\|full | 4232 | No | NA | 3 | 0 | 1 | Low-quality | Genome-fragment | 4.14 | AAI-based (medium-confidence) | 0 |
| S11_k141_9590 | 4233 | No | NA | 3 | 0 | 1 | Low-quality | Genome-fragment | 2.67 | AAI-based (medium-confidence) | 0 |
| S1_k141_288013 | 4233 | No | NA | 4 | 0 | 2 | Low-quality | Genome-fragment | 4.64 | AAI-based (high-confidence) | 0 |
| S2-final-viral-combined_k141_164275\|\|full | 1891 | No | NA | 5 | 0 | 0 | Not-determined | Genome-fragment | NA | NA | 0 |
| S2-final-viral-combined_k141_216889\|\|full | 5684 | No | NA | 7 | 0 | 0 | Not-determined | Genome-fragment | NA | NA | 0 |
| S2-final-viral-combined_k141_64430\|\|full | 4233 | No | NA | 5 | 0 | 0 | Low-quality | Genome-fragment | 7.37 | AAI-based (medium-confidence) | 0 |
| S2-final-viral-combined_k141_112759\|\|full | 1542 | No | NA | 2 | 0 | 2 | Not-determined | Genome-fragment | NA | NA | 0 |
| S2-final-viral-combined_k141_82646\|\|full | 1827 | No | NA | 2 | 0 | 0 | Not-determined | Genome-fragment | NA | NA | 0 |
| S12-final-viral-combined_k141_196766\|\|full | 4234 | No | NA | 9 | 1 | 0 | Low-quality | Genome-fragment | 9.33 | AAI-based (high-confidence) | 0 |
| S1-final-viral-combined_k141_9727\|\|full | 4238 | No | NA | 4 | 0 | 2 | Low-quality | Genome-fragment | 3.72 | AAI-based (medium-confidence) | 0 |
| S3_k141_18597 | 4238 | No | NA | 5 | 0 | 1 | Low-quality | Genome-fragment | 9.89 | AAI-based (high-confidence) | 0 |
| S2-final-viral-combined_k141_28958\|\|full | 1731 | No | NA | 3 | 0 | 0 | Not-determined | Genome-fragment | NA | NA | 0 |
| S8-final-viral-combined_k141_220991\|\|full | 4238 | No | NA | 5 | 0 | 1 | Low-quality | Genome-fragment | 6.37 | AAI-based (medium-confidence) | 0 |
| S2-final-viral-combined_k141_226022\|\|full | 1038 | No | NA | 3 | 0 | 0 | Not-determined | Genome-fragment | NA | NA | 0 |
| S11-final-viral-combined_k141_179179\|\|full | 4239 | No | NA | 5 | 3 | 0 | Low-quality | Genome-fragment | 9.33 | AAI-based (high-confidence) | 0 |
| S3-final-viral-combined_k141_23580\|\|full | 4240 | No | NA | 3 | 0 | 3 | Low-quality | Genome-fragment | 3.03 | AAI-based (high-confidence) | 0 |
| S5-final-viral-combined_k141_27948\|\|full | 4240 | No | NA | 7 | 0 | 1 | Low-quality | Genome-fragment | 7.49 | AAI-based (medium-confidence) | 0 |
| S2-final-viral-combined_k141_216934\|\|full | 2949 | No | NA | 4 | 0 | 1 | Not-determined | Genome-fragment | NA | NA | 0 |
| S2-final-viral-combined_k141_112795\|\|full | 1148 | No | NA | 2 | 0 | 0 | Not-determined | Genome-fragment | NA | NA | 0 |
| S2-final-viral-combined_k141_112809\|\|full | 1271 | No | NA | 2 | 0 | 1 | Not-determined | Genome-fragment | NA | NA | 0 |
| S2-final-viral-combined_k141_226117\|\|full | 3400 | No | NA | 2 | 0 | 1 | Not-determined | Genome-fragment | NA | NA | 0 |
| S7-final-viral-combined_k141_196798\|\|full | 4240 | No | NA | 4 | 2 | 0 | Low-quality | Genome-fragment | 6.7 | HMM-based (lower-bound) | 0 |
| S8-final-viral-combined_k141_2840\|\|full | 4240 | No | NA | 4 | 0 | 1 | Low-quality | Genome-fragment | 0.49 | AAI-based (medium-confidence) | 0 |
| S2-final-viral-combined_k141_132996\|\|full | 3325 | No | NA | 3 | 0 | 3 | Not-determined | Genome-fragment | NA | NA | 0 |
| S2-final-viral-combined_k141_164434\|\|full | 2987 | No | NA | 3 | 0 | 3 | Not-determined | Genome-fragment | NA | NA | 0 |
| S2-final-viral-combined_k141_7770\|\|full | 2460 | No | NA | 5 | 0 | 0 | Not-determined | Genome-fragment | NA | NA | 0 |
| S11-final-viral-combined_k141_153340\|\|full | 4241 | No | NA | 3 | 0 | 2 | Low-quality | Genome-fragment | 4.58 | AAI-based (medium-confidence) | 0 |
| S11-final-viral-combined_k141_198518\|\|full | 4241 | No | NA | 7 | 0 | 1 | Low-quality | Genome-fragment | 9.41 | AAI-based (medium-confidence) | 0 |
| S2-final-viral-combined_k141_124270\|\|full | 1712 | No | NA | 3 | 0 | 1 | Not-determined | Genome-fragment | NA | NA | 0 |
| S7-final-viral-combined_k141_145998\|\|full | 4241 | Yes | 2909 | 4 | 1 | 1 | Low-quality | Genome-fragment | 4.18 | HMM-based (lower-bound) | 31.41 |
| S12_k141_301583 | 4241 | No | NA | 5 | 0 | 0 | Low-quality | Genome-fragment | 16.02 | AAI-based (high-confidence) | 0 |
| S6-final-viral-combined_k141_130033\|\|full | 4241 | No | NA | 4 | 0 | 3 | Low-quality | Genome-fragment | 1.97 | AAI-based (medium-confidence) | 0 |
| S2-final-viral-combined_k141_7839\|\|full | 1755 | No | NA | 2 | 0 | 0 | Not-determined | Genome-fragment | NA | NA | 0 |
| S2-final-viral-combined_k141_29160\|\|full | 1519 | No | NA | 3 | 0 | 1 | Not-determined | Genome-fragment | NA | NA | 0 |
| S2-final-viral-combined_k141_226332\|\|full | 1414 | No | NA | 3 | 0 | 1 | Not-determined | Genome-fragment | NA | NA | 0 |
| S3-final-viral-combined_k141_22449\|\|full | 4243 | No | NA | 2 | 0 | 0 | Low-quality | Genome-fragment | 7.59 | AAI-based (medium-confidence) | 0 |
| S2-final-viral-combined_k141_217157\|\|full | 1292 | No | NA | 2 | 0 | 0 | Not-determined | Genome-fragment | NA | NA | 0 |
| S2-final-viral-combined_k141_205387\|\|full | 1361 | No | NA | 2 | 0 | 2 | Not-determined | Genome-fragment | NA | NA | 0 |
| S2-final-viral-combined_k141_217162\|\|full | 1038 | No | NA | 2 | 0 | 0 | Not-determined | Genome-fragment | NA | NA | 0 |
| S2-final-viral-combined_k141_217174\|\|full | 1725 | No | NA | 3 | 0 | 0 | Not-determined | Genome-fragment | NA | NA | 0 |
| S4-final-viral-combined_k141_26854\|\|full | 4243 | No | NA | 14 | 1 | 0 | Low-quality | Genome-fragment | 11.43 | AAI-based (high-confidence) | 0 |
| S4_k141_29291 | 4243 | No | NA | 3 | 0 | 3 | Low-quality | Genome-fragment | 0.73 | AAI-based (high-confidence) | 0 |
| S4_k141_58829 | 4243 | Yes | 602 | 3 | 1 | 2 | Low-quality | Genome-fragment | 1.3 | AAI-based (high-confidence) | 85.81 |
| S2-final-viral-combined_k141_217189\|\|full | 1253 | No | NA | 2 | 0 | 1 | Not-determined | Genome-fragment | NA | NA | 0 |
| S2-final-viral-combined_k141_7919\|\|full | 3211 | No | NA | 2 | 0 | 1 | Not-determined | Genome-fragment | NA | NA | 0 |
| S2-final-viral-combined_k141_226455\|\|full | 1976 | No | NA | 3 | 0 | 2 | Not-determined | Genome-fragment | NA | NA | 0 |
| S8_k141_37108 | 4243 | No | NA | 7 | 1 | 0 | Low-quality | Genome-fragment | 11.67 | HMM-based (lower-bound) | 0 |
| S2-final-viral-combined_k141_29221\|\|full | 2788 | No | NA | 6 | 0 | 3 | Not-determined | Genome-fragment | NA | NA | 0 |
| S2-final-viral-combined_k141_217219\|\|full | 1820 | No | NA | 3 | 0 | 2 | Not-determined | Genome-fragment | NA | NA | 0 |
| S2-final-viral-combined_k141_164677\|\|full | 5108 | No | NA | 5 | 0 | 3 | Not-determined | Genome-fragment | NA | NA | 0 |
| S2-final-viral-combined_k141_29229\|\|full | 4801 | No | NA | 6 | 0 | 0 | Not-determined | Genome-fragment | NA | NA | 0 |
| S2-final-viral-combined_k141_29241\|\|full | 1624 | No | NA | 3 | 0 | 0 | Not-determined | Genome-fragment | NA | NA | 0 |
| S9_k141_10606 | 4244 | No | NA | 6 | 4 | 0 | Low-quality | Genome-fragment | 10.27 | AAI-based (high-confidence) | 0 |
| S2-final-viral-combined_k141_164738\|\|full | 2832 | No | NA | 3 | 0 | 3 | Not-determined | Genome-fragment | NA | NA | 0 |
| S4-final-viral-combined_k141_17033\|\|full | 4244 | No | NA | 8 | 2 | 0 | Low-quality | Genome-fragment | 11.27 | AAI-based (high-confidence) | 0 |
| S1-final-viral-combined_k141_213012\|\|full | 4246 | No | NA | 6 | 3 | 0 | Low-quality | Genome-fragment | 7.3 | HMM-based (lower-bound) | 0 |
| S2-final-viral-combined_k141_226508\|\|full | 2794 | No | NA | 4 | 0 | 0 | Not-determined | Genome-fragment | NA | NA | 0 |
| S2-final-viral-combined_k141_113129\|\|full | 1002 | No | NA | 3 | 0 | 0 | Not-determined | Genome-fragment | NA | NA | 0 |
| S2-final-viral-combined_k141_113146\|\|full | 1032 | No | NA | 3 | 0 | 0 | Not-determined | Genome-fragment | NA | NA | 0 |
| S2-final-viral-combined_k141_205507\|\|full | 2439 | No | NA | 3 | 0 | 0 | Not-determined | Genome-fragment | NA | NA | 0 |
| S12-final-viral-combined_k141_358572\|\|full | 4246 | No | NA | 7 | 0 | 7 | Low-quality | Genome-fragment | 2.26 | AAI-based (medium-confidence) | 0 |
| S12-final-viral-combined_k141_399616\|\|full | 4246 | No | NA | 8 | 6 | 0 | Low-quality | Genome-fragment | 6.77 | AAI-based (high-confidence) | 0 |
| S8-final-viral-combined_k141_118353\|\|full | 4246 | No | NA | 7 | 2 | 0 | Low-quality | Genome-fragment | 11.4 | AAI-based (high-confidence) | 0 |
| S2-final-viral-combined_k141_205538\|\|full | 7256 | No | NA | 8 | 0 | 6 | Not-determined | Genome-fragment | NA | NA | 0 |
| S1-final-viral-combined_k141_389605\|\|full | 4247 | No | NA | 4 | 0 | 4 | Low-quality | Genome-fragment | 5.11 | AAI-based (medium-confidence) | 0 |
| S7-final-viral-combined_k141_55176\|\|full | 4247 | No | NA | 3 | 0 | 1 | Low-quality | Genome-fragment | 3.1 | AAI-based (high-confidence) | 0 |
| S4-final-viral-combined_k141_132144\|\|lt2gene | 4247 | No | NA | 2 | 0 | 0 | Low-quality | Genome-fragment | 4.12 | AAI-based (medium-confidence) | 0 |
| S2-final-viral-combined_k141_164859\|\|full | 1070 | No | NA | 2 | 0 | 2 | Not-determined | Genome-fragment | NA | NA | 0 |
| S11_k141_124077 | 4248 | No | NA | 7 | 0 | 0 | Low-quality | Genome-fragment | 8.11 | AAI-based (medium-confidence) | 0 |
| S2-final-viral-combined_k141_217364\|\|full | 1782 | No | NA | 3 | 0 | 2 | Not-determined | Genome-fragment | NA | NA | 0 |
| S2-final-viral-combined_k141_113279\|\|full | 1365 | No | NA | 2 | 0 | 1 | Not-determined | Genome-fragment | NA | NA | 0 |
| S7_k141_204707 | 4248 | No | NA | 4 | 0 | 0 | Low-quality | Genome-fragment | 3.99 | AAI-based (high-confidence) | 0 |
| S17-k141_162669 | 4249 | No | NA | 6 | 1 | 0 | Low-quality | Genome-fragment | 10.71 | AAI-based (high-confidence) | 0 |
| S11-final-viral-combined_k141_93237\|\|full | 4250 | No | NA | 7 | 1 | 0 | Low-quality | Genome-fragment | 2.57 | HMM-based (lower-bound) | 0 |
| S1_k141_2265 | 4250 | No | NA | 3 | 0 | 3 | Low-quality | Genome-fragment | 2.76 | AAI-based (medium-confidence) | 0 |
| S2-final-viral-combined_k141_246243\|\|full | 1454 | No | NA | 3 | 0 | 2 | Not-determined | Genome-fragment | NA | NA | 0 |
| S2-final-viral-combined_k141_29399\|\|full | 11429 | No | NA | 9 | 0 | 3 | Not-determined | Genome-fragment | NA | NA | 0 |
| S9-final-viral-combined_k141_74532\|\|full | 4252 | No | NA | 5 | 0 | 1 | Low-quality | Genome-fragment | 2.08 | AAI-based (medium-confidence) | 0 |
| S2-final-viral-combined_k141_226780\|\|full | 2424 | No | NA | 3 | 0 | 1 | Not-determined | Genome-fragment | NA | NA | 0 |
| S8-final-viral-combined_k141_155563\|\|full | 4253 | No | NA | 6 | 1 | 0 | Low-quality | Genome-fragment | 7.96 | AAI-based (medium-confidence) | 0 |
| S2-final-viral-combined_k141_205720\|\|full | 1373 | No | NA | 2 | 0 | 2 | Not-determined | Genome-fragment | NA | NA | 0 |
| S17-final-viral-combined_k141_199786\|\|full | 4254 | No | NA | 4 | 0 | 4 | Low-quality | Genome-fragment | 3.11 | AAI-based (medium-confidence) | 0 |
| S11-final-viral-combined_k141_37681\|\|full | 4257 | No | NA | 6 | 4 | 0 | Low-quality | Genome-fragment | 9.48 | AAI-based (high-confidence) | 0 |
| S2-final-viral-combined_k141_29476\|\|full | 1257 | No | NA | 3 | 0 | 0 | Not-determined | Genome-fragment | NA | NA | 0 |
| S2-final-viral-combined_k141_205725\|\|full | 2943 | No | NA | 3 | 0 | 1 | Not-determined | Genome-fragment | NA | NA | 0 |
| S2-final-viral-combined_k141_205727\|\|full | 5575 | No | NA | 3 | 0 | 3 | Not-determined | Genome-fragment | NA | NA | 0 |
| S2-final-viral-combined_k141_8112\|\|full | 1126 | No | NA | 2 | 0 | 0 | Not-determined | Genome-fragment | NA | NA | 0 |
| S11_k141_4704 | 4257 | No | NA | 2 | 0 | 1 | Low-quality | Genome-fragment | 9.17 | AAI-based (high-confidence) | 0 |
| S8-final-viral-combined_k141_209841\|\|full | 4257 | No | NA | 12 | 1 | 0 | Low-quality | Genome-fragment | 6.9 | AAI-based (high-confidence) | 0 |
| S2-final-viral-combined_k141_29561\|\|full | 3042 | No | NA | 4 | 0 | 4 | Not-determined | Genome-fragment | NA | NA | 0 |
| S2-final-viral-combined_k141_246322\|\|full | 1058 | No | NA | 3 | 0 | 0 | Not-determined | Genome-fragment | NA | NA | 0 |
| S9-final-viral-combined_k141_126819\|\|full | 4258 | Yes | 3151 | 5 | 1 | 1 | Low-quality | Genome-fragment | 5.64 | AAI-based (high-confidence) | 26 |
| S2-final-viral-combined_k141_226896\|\|full | 2165 | No | NA | 3 | 0 | 0 | Not-determined | Genome-fragment | NA | NA | 0 |
| S2-final-viral-combined_k141_165169\|\|full | 2274 | No | NA | 3 | 0 | 0 | Not-determined | Genome-fragment | NA | NA | 0 |
| S2-final-viral-combined_k141_165171\|\|full | 2595 | No | NA | 4 | 0 | 3 | Not-determined | Genome-fragment | NA | NA | 0 |
| S2-final-viral-combined_k141_8232\|\|full | 4905 | No | NA | 3 | 0 | 1 | Not-determined | Genome-fragment | NA | NA | 0 |
| S11-final-viral-combined_k141_271586\|\|full | 4258 | No | NA | 7 | 0 | 2 | Low-quality | Genome-fragment | 7.54 | AAI-based (medium-confidence) | 0 |
| S7_k141_206191 | 4258 | No | NA | 4 | 0 | 1 | Low-quality | Genome-fragment | 2.27 | AAI-based (medium-confidence) | 0 |
| S2-final-viral-combined_k141_205807\|\|full | 2861 | No | NA | 4 | 0 | 3 | Not-determined | Genome-fragment | NA | NA | 0 |
| S2-final-viral-combined_k141_113432\|\|full | 1750 | No | NA | 3 | 0 | 2 | Not-determined | Genome-fragment | NA | NA | 0 |
| S7_k141_207819 | 4258 | No | NA | 5 | 4 | 0 | Low-quality | Genome-fragment | 7.08 | AAI-based (high-confidence) | 0 |
| S2-final-viral-combined_k141_226978\|\|full | 1024 | No | NA | 2 | 0 | 0 | Not-determined | Genome-fragment | NA | NA | 0 |
| S2-final-viral-combined_k141_113463\|\|full | 1920 | No | NA | 3 | 0 | 1 | Not-determined | Genome-fragment | NA | NA | 0 |
| S17-final-viral-combined_k141_200726\|\|full | 4258 | No | NA | 5 | 0 | 0 | Low-quality | Genome-fragment | 5.15 | AAI-based (medium-confidence) | 0 |
| S2-final-viral-combined_k141_113475\|\|full | 2141 | No | NA | 4 | 0 | 0 | Not-determined | Genome-fragment | NA | NA | 0 |
| S12-final-viral-combined_k141_158265\|\|full | 4258 | No | NA | 8 | 0 | 1 | Low-quality | Genome-fragment | 5.54 | AAI-based (medium-confidence) | 0 |
| S9_k141_108578 | 4260 | No | NA | 6 | 4 | 0 | Low-quality | Genome-fragment | 9.13 | AAI-based (high-confidence) | 0 |
| S2-final-viral-combined_k141_113486\|\|full | 1386 | No | NA | 2 | 0 | 1 | Not-determined | Genome-fragment | NA | NA | 0 |
| S2-final-viral-combined_k141_227056\|\|full | 1251 | No | NA | 2 | 0 | 1 | Not-determined | Genome-fragment | NA | NA | 0 |
| S2-final-viral-combined_k141_246423\|\|full | 1380 | No | NA | 3 | 0 | 0 | Not-determined | Genome-fragment | NA | NA | 0 |
| S2-final-viral-combined_k141_227064\|\|full | 1333 | No | NA | 2 | 0 | 1 | Not-determined | Genome-fragment | NA | NA | 0 |
| S11_k141_282779 | 4260 | No | NA | 6 | 0 | 0 | Low-quality | Genome-fragment | 6.36 | AAI-based (high-confidence) | 0 |
| S5-final-viral-combined_k141_90407\|\|full | 4260 | No | NA | 2 | 0 | 1 | Low-quality | Genome-fragment | 0.84 | AAI-based (medium-confidence) | 0 |
| S12-final-viral-combined_k141_335106\|\|full | 4260 | Yes | 667 | 7 | 1 | 2 | Low-quality | Genome-fragment | 1.41 | AAI-based (high-confidence) | 84.34 |
| S2-final-viral-combined_k141_113521\|\|full | 5296 | No | NA | 5 | 0 | 4 | Not-determined | Genome-fragment | NA | NA | 0 |
| S2-final-viral-combined_k141_227099\|\|full | 1719 | No | NA | 2 | 0 | 1 | Not-determined | Genome-fragment | NA | NA | 0 |
| S2-final-viral-combined_k141_165394\|\|full | 3098 | No | NA | 2 | 0 | 2 | Not-determined | Genome-fragment | NA | NA | 0 |
| S2-final-viral-combined_k141_113535\|\|full | 2434 | No | NA | 3 | 0 | 2 | Not-determined | Genome-fragment | NA | NA | 0 |
| S2-final-viral-combined_k141_29690\|\|full | 1979 | No | NA | 3 | 0 | 1 | Not-determined | Genome-fragment | NA | NA | 0 |
| S2-final-viral-combined_k141_133643\|\|full | 2793 | No | NA | 4 | 0 | 0 | Not-determined | Genome-fragment | NA | NA | 0 |
| S2-final-viral-combined_k141_205997\|\|full | 1439 | No | NA | 3 | 0 | 2 | Not-determined | Genome-fragment | NA | NA | 0 |
| S2-final-viral-combined_k141_29714\|\|full | 1217 | No | NA | 3 | 0 | 1 | Not-determined | Genome-fragment | NA | NA | 0 |
| S2-final-viral-combined_k141_227127\|\|full | 1579 | No | NA | 3 | 0 | 0 | Not-determined | Genome-fragment | NA | NA | 0 |
| S2-final-viral-combined_k141_246480\|\|full | 30382 | No | NA | 46 | 17 | 0 | Medium-quality | Genome-fragment | 66.75 | AAI-based (high-confidence) | 0 |
| S2-final-viral-combined_k141_8340\|\|full | 2424 | No | NA | 3 | 0 | 3 | Not-determined | Genome-fragment | NA | NA | 0 |
| S12_k141_338199 | 4260 | No | NA | 5 | 0 | 3 | Low-quality | Genome-fragment | 1.59 | AAI-based (medium-confidence) | 0 |
| S2-final-viral-combined_k141_165483\|\|full | 4724 | No | NA | 6 | 0 | 4 | Not-determined | Genome-fragment | NA | NA | 0 |
| S2-final-viral-combined_k141_10277\|\|full | 4260 | No | NA | 4 | 0 | 3 | Low-quality | Genome-fragment | 1.47 | AAI-based (medium-confidence) | 0 |
| S2-final-viral-combined_k141_165503\|\|full | 3371 | No | NA | 2 | 0 | 2 | Not-determined | Genome-fragment | NA | NA | 0 |
| S2-final-viral-combined_k141_113619\|\|full | 49583 | No | NA | 71 | 15 | 2 | High-quality | High-quality | 90.89 | AAI-based (high-confidence) | 0 |
| S2-final-viral-combined_k141_8388\|\|full | 1099 | No | NA | 2 | 0 | 1 | Not-determined | Genome-fragment | NA | NA | 0 |
| S11-final-viral-combined_k141_43404\|\|full | 4261 | No | NA | 5 | 0 | 5 | Low-quality | Genome-fragment | 4.2 | AAI-based (medium-confidence) | 0 |
| S12_k141_129279 | 4262 | No | NA | 6 | 1 | 0 | Low-quality | Genome-fragment | 10.69 | AAI-based (medium-confidence) | 0 |
| S2-final-viral-combined_k141_227244\|\|full | 1228 | No | NA | 3 | 0 | 1 | Not-determined | Genome-fragment | NA | NA | 0 |
| S9-final-viral-combined_k141_22762\|\|full | 4264 | No | NA | 5 | 0 | 2 | Low-quality | Genome-fragment | 1.5 | AAI-based (medium-confidence) | 0 |
| S2-final-viral-combined_k141_133746\|\|full | 3275 | No | NA | 7 | 0 | 1 | Not-determined | Genome-fragment | NA | NA | 0 |
| S17-k141_59632 | 4264 | No | NA | 3 | 0 | 2 | Low-quality | Genome-fragment | 0.37 | AAI-based (medium-confidence) | 0 |
| S2-final-viral-combined_k141_227313\|\|full | 8866 | No | NA | 9 | 0 | 6 | Not-determined | Genome-fragment | NA | NA | 0 |
| S2-final-viral-combined_k141_227317\|\|full | 2074 | No | NA | 3 | 0 | 2 | Not-determined | Genome-fragment | NA | NA | 0 |
| S9_k141_49002 | 4265 | No | NA | 4 | 0 | 2 | Low-quality | Genome-fragment | 7.93 | AAI-based (medium-confidence) | 0 |
| S2-final-viral-combined_k141_29862\|\|full | 1720 | No | NA | 2 | 0 | 0 | Not-determined | Genome-fragment | NA | NA | 0 |
| S2-final-viral-combined_k141_206168\|\|full | 3301 | No | NA | 2 | 0 | 2 | Not-determined | Genome-fragment | NA | NA | 0 |
| S2-final-viral-combined_k141_227333\|\|full | 5756 | No | NA | 4 | 0 | 1 | Not-determined | Genome-fragment | NA | NA | 0 |
| S2-final-viral-combined_k141_8451\|\|full | 4876 | No | NA | 4 | 0 | 2 | Not-determined | Genome-fragment | NA | NA | 0 |
| S9_k141_30737 | 4265 | No | NA | 7 | 0 | 1 | Low-quality | Genome-fragment | 5.34 | AAI-based (medium-confidence) | 0 |
| S2-final-viral-combined_k141_133834\|\|full | 2429 | No | NA | 5 | 0 | 0 | Not-determined | Genome-fragment | NA | NA | 0 |
| S2-final-viral-combined_k141_206191\|\|full | 2773 | No | NA | 5 | 0 | 4 | Not-determined | Genome-fragment | NA | NA | 0 |
| S2-final-viral-combined_k141_133861\|\|full | 3239 | No | NA | 3 | 0 | 1 | Not-determined | Genome-fragment | NA | NA | 0 |
| S7_k141_5574 | 4266 | No | NA | 4 | 0 | 0 | Low-quality | Genome-fragment | 4.19 | AAI-based (high-confidence) | 0 |
| S2-final-viral-combined_k141_113741\|\|full | 3714 | No | NA | 7 | 0 | 0 | Not-determined | Genome-fragment | NA | NA | 0 |
| S9-final-viral-combined_k141_45200\|\|full | 4268 | No | NA | 5 | 0 | 0 | Low-quality | Genome-fragment | 9.97 | AAI-based (high-confidence) | 0 |
| S12-final-viral-combined_k141_335527\|\|full | 4268 | No | NA | 5 | 0 | 2 | Low-quality | Genome-fragment | 8.28 | AAI-based (medium-confidence) | 0 |
| S2-final-viral-combined_k141_133918\|\|full | 3639 | No | NA | 2 | 0 | 0 | Not-determined | Genome-fragment | NA | NA | 0 |
| S2-final-viral-combined_k141_133924\|\|full | 2234 | No | NA | 3 | 0 | 2 | Not-determined | Genome-fragment | NA | NA | 0 |
| S5-final-viral-combined_k141_37281\|\|full | 4272 | No | NA | 7 | 4 | 0 | Low-quality | Genome-fragment | 8.62 | AAI-based (high-confidence) | 0 |
| S2-final-viral-combined_k141_8504\|\|full | 3257 | No | NA | 3 | 0 | 3 | Not-determined | Genome-fragment | NA | NA | 0 |
| S2-final-viral-combined_k141_133959\|\|full | 1753 | No | NA | 3 | 0 | 3 | Not-determined | Genome-fragment | NA | NA | 0 |
| S7-final-viral-combined_k141_226473\|\|full | 4272 | Yes | 2143 | 7 | 3 | 1 | Low-quality | Genome-fragment | 6.06 | AAI-based (high-confidence) | 49.84 |
| S12_k141_195654 | 4274 | No | NA | 3 | 0 | 2 | Low-quality | Genome-fragment | 1.17 | AAI-based (high-confidence) | 0 |
| S9-final-viral-combined_k141_441\|\|full | 4275 | No | NA | 4 | 4 | 0 | Low-quality | Genome-fragment | 9.22 | AAI-based (high-confidence) | 0 |
| S11_k141_293399 | 4275 | No | NA | 4 | 0 | 1 | Low-quality | Genome-fragment | 4.14 | AAI-based (medium-confidence) | 0 |
| S2-final-viral-combined_k141_8582\|\|full | 1131 | No | NA | 1 | 0 | 1 | Not-determined | Genome-fragment | NA | NA | 0 |
| S17-final-viral-combined_k141_119635\|\|full | 4275 | No | NA | 7 | 2 | 0 | Low-quality | Genome-fragment | 9.98 | AAI-based (medium-confidence) | 0 |
| S2-final-viral-combined_k141_30245\|\|full | 3994 | No | NA | 3 | 0 | 3 | Not-determined | Genome-fragment | NA | NA | 0 |
| S11-final-viral-combined_k141_17685\|\|full | 4276 | No | NA | 3 | 0 | 2 | Low-quality | Genome-fragment | 2.15 | AAI-based (medium-confidence) | 0 |
| S7-final-viral-combined_k141_72078\|\|full | 4276 | No | NA | 5 | 0 | 2 | Low-quality | Genome-fragment | 3.37 | AAI-based (high-confidence) | 0 |
| S2-final-viral-combined_k141_30299\|\|full | 1041 | No | NA | 3 | 0 | 0 | Not-determined | Genome-fragment | NA | NA | 0 |
| S9-final-viral-combined_k141_14751\|\|full | 4278 | No | NA | 7 | 2 | 0 | Low-quality | Genome-fragment | 11.69 | AAI-based (medium-confidence) | 0 |
| S2-final-viral-combined_k141_30319\|\|full | 2415 | No | NA | 3 | 0 | 1 | Not-determined | Genome-fragment | NA | NA | 0 |
| S2-final-viral-combined_k141_134319\|\|full | 2862 | No | NA | 3 | 0 | 3 | Not-determined | Genome-fragment | NA | NA | 0 |
| S11-final-viral-combined_k141_292602\|\|full | 4278 | No | NA | 6 | 1 | 0 | Low-quality | Genome-fragment | 9.78 | AAI-based (medium-confidence) | 0 |
| S11-final-viral-combined_k141_316386\|\|full | 4278 | No | NA | 6 | 5 | 0 | Low-quality | Genome-fragment | 9.52 | AAI-based (medium-confidence) | 0 |
| S12-final-viral-combined_k141_309275\|\|full | 4279 | Yes | 502 | 7 | 2 | 2 | Low-quality | Genome-fragment | 0.79 | AAI-based (high-confidence) | 88.27 |
| S2-final-viral-combined_k141_30391\|\|full | 1794 | No | NA | 3 | 0 | 2 | Not-determined | Genome-fragment | NA | NA | 0 |
| S2-final-viral-combined_k141_206540\|\|full | 23376 | No | NA | 18 | 0 | 12 | Not-determined | Genome-fragment | NA | NA | 0 |
| S8-final-viral-combined_k141_68105\|\|full | 4279 | No | NA | 3 | 1 | 0 | Low-quality | Genome-fragment | 8.01 | AAI-based (high-confidence) | 0 |
| S2-final-viral-combined_k141_247105\|\|full | 4299 | No | NA | 5 | 0 | 4 | Not-determined | Genome-fragment | NA | NA | 0 |
| S12-final-viral-combined_k141_120983\|\|full | 4280 | No | NA | 9 | 2 | 0 | Low-quality | Genome-fragment | 12.99 | AAI-based (high-confidence) | 0 |
| S11-final-viral-combined_k141_250437\|\|full | 4281 | No | NA | 4 | 0 | 4 | Low-quality | Genome-fragment | 5.45 | AAI-based (medium-confidence) | 0 |
| S2-final-viral-combined_k141_30454\|\|full | 1176 | No | NA | 2 | 0 | 1 | Not-determined | Genome-fragment | NA | NA | 0 |
| S4-final-viral-combined_k141_98994\|\|full | 4282 | No | NA | 5 | 1 | 0 | Low-quality | Genome-fragment | 6.69 | HMM-based (lower-bound) | 0 |
| S2-final-viral-combined_k141_227741\|\|full | 1563 | No | NA | 2 | 0 | 1 | Not-determined | Genome-fragment | NA | NA | 0 |
| S11_k141_234728 | 4284 | No | NA | 4 | 3 | 0 | Low-quality | Genome-fragment | 7.4 | AAI-based (high-confidence) | 0 |
| S1-final-viral-combined_k141_315180\|\|full | 4284 | No | NA | 8 | 2 | 0 | Low-quality | Genome-fragment | 9.59 | AAI-based (high-confidence) | 0 |
| S2-final-viral-combined_k141_134488\|\|full | 23097 | No | NA | 21 | 0 | 16 | Not-determined | Genome-fragment | NA | NA | 0 |
| S2-final-viral-combined_k141_30577\|\|full | 2026 | No | NA | 2 | 0 | 0 | Not-determined | Genome-fragment | NA | NA | 0 |
| S2-final-viral-combined_k141_206698\|\|full | 3858 | No | NA | 3 | 0 | 1 | Not-determined | Genome-fragment | NA | NA | 0 |
| S2-final-viral-combined_k141_8687\|\|full | 1329 | No | NA | 2 | 0 | 2 | Not-determined | Genome-fragment | NA | NA | 0 |
| S2-final-viral-combined_k141_227800\|\|full | 36179 | Yes | 21073 | 46 | 11 | 6 | Medium-quality | Genome-fragment | 58.22 | AAI-based (medium-confidence) | 41.75 |
| S12_k141_362088 | 4284 | No | NA | 5 | 1 | 0 | Low-quality | Genome-fragment | 13.3 | AAI-based (high-confidence) | 0 |
| S4-final-viral-combined_k141_76185\|\|full | 4284 | No | NA | 5 | 3 | 0 | Low-quality | Genome-fragment | 8.89 | AAI-based (high-confidence) | 0 |
| S11-final-viral-combined_k141_139681\|\|full | 4285 | No | NA | 3 | 0 | 2 | Low-quality | Genome-fragment | 0.69 | AAI-based (high-confidence) | 0 |
| S2-final-viral-combined_k141_30654\|\|full | 1835 | No | NA | 4 | 0 | 0 | Not-determined | Genome-fragment | NA | NA | 0 |
| S2-final-viral-combined_k141_8780\|\|full | 1588 | No | NA | 3 | 0 | 2 | Not-determined | Genome-fragment | NA | NA | 0 |
| S2-final-viral-combined_k141_134610\|\|full | 2118 | No | NA | 3 | 0 | 1 | Not-determined | Genome-fragment | NA | NA | 0 |
| S11-final-viral-combined_k141_91705\|\|full | 4285 | No | NA | 3 | 0 | 2 | Low-quality | Genome-fragment | 2.46 | AAI-based (medium-confidence) | 0 |
| S6-final-viral-combined_k141_106758\|\|full | 4285 | No | NA | 5 | 0 | 2 | Low-quality | Genome-fragment | 5.06 | AAI-based (medium-confidence) | 0 |
| S1-final-viral-combined_k141_44101\|\|full | 4286 | No | NA | 3 | 0 | 2 | Low-quality | Genome-fragment | 4.42 | AAI-based (medium-confidence) | 0 |
| S2-final-viral-combined_k141_30750\|\|full | 75512 | No | NA | 66 | 1 | 20 | Medium-quality | Genome-fragment | 50.76 | HMM-based (lower-bound) | 0 |
| S17-final-viral-combined_k141_170168\|\|full | 4286 | No | NA | 4 | 0 | 0 | Low-quality | Genome-fragment | 10.64 | AAI-based (high-confidence) | 0 |
| S17-k141_137981 | 4286 | No | NA | 9 | 0 | 2 | Low-quality | Genome-fragment | 6.63 | AAI-based (medium-confidence) | 0 |
| S2-final-viral-combined_k141_8962\|\|full | 1581 | No | NA | 4 | 0 | 1 | Not-determined | Genome-fragment | NA | NA | 0 |
| S2-final-viral-combined_k141_247268\|\|full | 2787 | No | NA | 4 | 0 | 2 | Not-determined | Genome-fragment | NA | NA | 0 |
| S2-final-viral-combined_k141_30796\|\|full | 2425 | No | NA | 3 | 0 | 1 | Not-determined | Genome-fragment | NA | NA | 0 |
| S12-final-viral-combined_k141_114901\|\|full | 4286 | No | NA | 5 | 0 | 4 | Low-quality | Genome-fragment | 1.63 | AAI-based (medium-confidence) | 0 |
| S2-final-viral-combined_k141_247318\|\|full | 1387 | No | NA | 2 | 0 | 0 | Not-determined | Genome-fragment | NA | NA | 0 |
| S7-final-viral-combined_k141_218726\|\|full | 4287 | No | NA | 4 | 0 | 3 | Low-quality | Genome-fragment | 9.73 | AAI-based (medium-confidence) | 0 |
| S2-final-viral-combined_k141_247383\|\|full | 1592 | No | NA | 2 | 0 | 2 | Not-determined | Genome-fragment | NA | NA | 0 |
| S12-final-viral-combined_k141_118186\|\|full | 4287 | No | NA | 5 | 1 | 0 | Low-quality | Genome-fragment | 9 | AAI-based (medium-confidence) | 0 |
| S2-final-viral-combined_k141_247404\|\|full | 2040 | No | NA | 2 | 0 | 1 | Not-determined | Genome-fragment | NA | NA | 0 |
| S6_k141_112731 | 4287 | No | NA | 6 | 1 | 0 | Low-quality | Genome-fragment | 9.84 | AAI-based (high-confidence) | 0 |
| S8_k141_25881 | 4287 | No | NA | 8 | 1 | 0 | Low-quality | Genome-fragment | 11.87 | AAI-based (high-confidence) | 0 |
| S2-final-viral-combined_k141_247494\|\|full | 2502 | No | NA | 2 | 0 | 1 | Not-determined | Genome-fragment | NA | NA | 0 |
| S2-final-viral-combined_k141_9160\|\|full | 3106 | No | NA | 3 | 0 | 3 | Not-determined | Genome-fragment | NA | NA | 0 |
| S2-final-viral-combined_k141_206839\|\|full | 1291 | No | NA | 3 | 0 | 0 | Not-determined | Genome-fragment | NA | NA | 0 |
| S2-final-viral-combined_k141_206841\|\|full | 4159 | No | NA | 6 | 0 | 2 | Not-determined | Genome-fragment | NA | NA | 0 |
| S2-final-viral-combined_k141_206864\|\|full | 1445 | No | NA | 2 | 0 | 1 | Not-determined | Genome-fragment | NA | NA | 0 |
| S2-final-viral-combined_k141_247589\|\|full | 1310 | No | NA | 5 | 0 | 0 | Not-determined | Genome-fragment | NA | NA | 0 |
| S2-final-viral-combined_k141_247601\|\|full | 2451 | No | NA | 2 | 0 | 1 | Not-determined | Genome-fragment | NA | NA | 0 |
| S11_k141_72021 | 4288 | No | NA | 9 | 2 | 0 | Low-quality | Genome-fragment | 10.58 | AAI-based (high-confidence) | 0 |
| S2-final-viral-combined_k141_9320\|\|full | 4656 | No | NA | 4 | 0 | 3 | Not-determined | Genome-fragment | NA | NA | 0 |
| S2-final-viral-combined_k141_247697\|\|full | 2210 | No | NA | 2 | 0 | 1 | Not-determined | Genome-fragment | NA | NA | 0 |
| S2-final-viral-combined_k141_247699\|\|full | 1709 | No | NA | 3 | 0 | 2 | Not-determined | Genome-fragment | NA | NA | 0 |
| S5_k141_4201 | 4288 | No | NA | 5 | 5 | 0 | Low-quality | Genome-fragment | 10.35 | AAI-based (high-confidence) | 0 |
| S2-final-viral-combined_k141_247791\|\|full | 2630 | No | NA | 3 | 0 | 2 | Not-determined | Genome-fragment | NA | NA | 0 |
| S2-final-viral-combined_k141_9365\|\|full | 2579 | No | NA | 2 | 0 | 2 | Not-determined | Genome-fragment | NA | NA | 0 |
| S2-final-viral-combined_k141_247803\|\|full | 33220 | No | NA | 41 | 0 | 18 | Not-determined | Genome-fragment | NA | NA | 0 |
| S12-final-viral-combined_k141_67091\|\|full | 4288 | No | NA | 4 | 2 | 0 | Low-quality | Genome-fragment | 11.7 | AAI-based (high-confidence) | 0 |
| S2-final-viral-combined_k141_247826\|\|full | 4380 | No | NA | 3 | 0 | 1 | Not-determined | Genome-fragment | NA | NA | 0 |
| S2-final-viral-combined_k141_9462\|\|full | 1813 | No | NA | 2 | 0 | 1 | Not-determined | Genome-fragment | NA | NA | 0 |
| S2-final-viral-combined_k141_247915\|\|full | 1266 | No | NA | 3 | 0 | 1 | Not-determined | Genome-fragment | NA | NA | 0 |
| S2-final-viral-combined_k141_9499\|\|full | 2182 | No | NA | 3 | 0 | 2 | Not-determined | Genome-fragment | NA | NA | 0 |
| S2-final-viral-combined_k141_207036\|\|full | 1814 | No | NA | 2 | 0 | 1 | Not-determined | Genome-fragment | NA | NA | 0 |
| S2-final-viral-combined_k141_247998\|\|full | 2969 | No | NA | 5 | 0 | 2 | Not-determined | Genome-fragment | NA | NA | 0 |
| S2-final-viral-combined_k141_248067\|\|full | 1993 | No | NA | 2 | 0 | 0 | Not-determined | Genome-fragment | NA | NA | 0 |
| S2-final-viral-combined_k141_207090\|\|full | 1281 | No | NA | 3 | 0 | 0 | Not-determined | Genome-fragment | NA | NA | 0 |
| S2-final-viral-combined_k141_207108\|\|full | 4966 | No | NA | 5 | 0 | 0 | Not-determined | Genome-fragment | NA | NA | 0 |
| S11-final-viral-combined_k141_147757\|\|full | 4289 | No | NA | 5 | 3 | 0 | Low-quality | Genome-fragment | 11.18 | AAI-based (high-confidence) | 0 |
| S1-final-viral-combined_k141_99972\|\|full | 4289 | No | NA | 4 | 0 | 4 | Low-quality | Genome-fragment | 3.48 | AAI-based (medium-confidence) | 0 |
| S2-final-viral-combined_k141_9644\|\|full | 1898 | No | NA | 2 | 0 | 2 | Not-determined | Genome-fragment | NA | NA | 0 |
| S2-final-viral-combined_k141_9665\|\|full | 3303 | No | NA | 4 | 0 | 0 | Not-determined | Genome-fragment | NA | NA | 0 |
| S17-final-viral-combined_k141_86412\|\|full | 4289 | No | NA | 4 | 3 | 0 | Low-quality | Genome-fragment | 8.17 | AAI-based (high-confidence) | 0 |
| S2-final-viral-combined_k141_248295\|\|full | 1358 | No | NA | 3 | 0 | 1 | Not-determined | Genome-fragment | NA | NA | 0 |
| S2-final-viral-combined_k141_9726\|\|full | 5090 | No | NA | 9 | 0 | 0 | Not-determined | Genome-fragment | NA | NA | 0 |
| S12-final-viral-combined_k141_163920\|\|full | 4289 | No | NA | 3 | 0 | 3 | Low-quality | Genome-fragment | 2.47 | AAI-based (medium-confidence) | 0 |
| S2-final-viral-combined_k141_9888\|\|full | 3600 | No | NA | 4 | 0 | 1 | Not-determined | Genome-fragment | NA | NA | 0 |
| S2-final-viral-combined_k141_9974\|\|full | 2043 | No | NA | 3 | 0 | 0 | Not-determined | Genome-fragment | NA | NA | 0 |
| S2-final-viral-combined_k141_10000\|\|full | 2222 | No | NA | 3 | 0 | 1 | Not-determined | Genome-fragment | NA | NA | 0 |
| S2-final-viral-combined_k141_10005\|\|full | 1673 | No | NA | 4 | 0 | 0 | Not-determined | Genome-fragment | NA | NA | 0 |
| S2-final-viral-combined_k141_10020\|\|full | 2869 | No | NA | 4 | 0 | 0 | Not-determined | Genome-fragment | NA | NA | 0 |
| S2-final-viral-combined_k141_10041\|\|full | 4119 | No | NA | 3 | 0 | 1 | Not-determined | Genome-fragment | NA | NA | 0 |
| S2-final-viral-combined_k141_10064\|\|full | 1655 | No | NA | 3 | 0 | 0 | Not-determined | Genome-fragment | NA | NA | 0 |
| S2_k141_128151 | 4289 | No | NA | 9 | 1 | 0 | Low-quality | Genome-fragment | 4.11 | AAI-based (high-confidence) | 0 |
| S5-final-viral-combined_k141_118657\|\|full | 4290 | No | NA | 4 | 1 | 3 | Low-quality | Genome-fragment | 2.73 | HMM-based (lower-bound) | 0 |
| S2-final-viral-combined_k141_10324\|\|full | 5709 | No | NA | 8 | 0 | 1 | Not-determined | Genome-fragment | NA | NA | 0 |
| S9-final-viral-combined_k141_48592\|\|full | 4292 | No | NA | 5 | 0 | 2 | Low-quality | Genome-fragment | 9.35 | AAI-based (medium-confidence) | 0 |
| S2-final-viral-combined_k141_88248\|\|0_partial | 11320 | No | NA | 14 | 0 | 0 | Not-determined | Genome-fragment | NA | NA | 0 |
| S12-final-viral-combined_k141_66556\|\|full | 4292 | No | NA | 7 | 0 | 0 | Low-quality | Genome-fragment | 7.79 | AAI-based (high-confidence) | 0 |
| S2-final-viral-combined_k141_52006\|\|0_partial | 2711 | No | NA | 3 | 1 | 0 | Medium-quality | Genome-fragment | 60.54 | AAI-based (high-confidence) | 0 |
| S2-final-viral-combined_k141_368\|\|0_partial | 15247 | No | NA | 11 | 0 | 6 | Not-determined | Genome-fragment | NA | NA | 0 |
| S2-final-viral-combined_k141_196980\|\|0_partial | 11954 | No | NA | 25 | 0 | 2 | Not-determined | Genome-fragment | NA | NA | 0 |
| S17-final-viral-combined_k141_39774\|\|full | 4293 | No | NA | 4 | 4 | 0 | Low-quality | Genome-fragment | 11.52 | AAI-based (high-confidence) | 0 |
| S2-final-viral-combined_k141_104174\|\|0_partial | 18019 | No | NA | 17 | 0 | 8 | Not-determined | Genome-fragment | NA | NA | 0 |
| S2-final-viral-combined_k141_218102\|\|0_partial | 13506 | No | NA | 15 | 0 | 3 | Not-determined | Genome-fragment | NA | NA | 0 |
| S11-final-viral-combined_k141_63128\|\|full | 4294 | No | NA | 5 | 0 | 2 | Low-quality | Genome-fragment | 2.8 | AAI-based (medium-confidence) | 0 |
| S12_k141_17404 | 4294 | No | NA | 5 | 2 | 0 | Low-quality | Genome-fragment | 7.42 | AAI-based (high-confidence) | 0 |
| S8-final-viral-combined_k141_75729\|\|full | 4294 | No | NA | 3 | 0 | 0 | Low-quality | Genome-fragment | 9.49 | AAI-based (high-confidence) | 0 |
| S2-final-viral-combined_k141_32320\|\|0_partial | 11930 | No | NA | 16 | 0 | 2 | Not-determined | Genome-fragment | NA | NA | 0 |

| | | | | | | | | | | | | |
|---|---|---|---|---|---|---|---|---|---|---|---|---|
| S7-final-viral-combined_k141_70231\|\|full | 4295 | No | NA | 6 | 0 | 1 | Low-quality | Genome-fragment | | 2.5 | AAI-based (medium-confidence) | 0 |
| S2-final-viral-combined_k141_157082\|\|0_partial | 36662 | No | NA | 28 | 0 | 13 | Not-determined | Genome-fragment | NA | | NA | 0 |
| S2-final-viral-combined_k141_219136\|\|0_partial | 10969 | No | NA | 11 | 0 | 11 | Not-determined | Genome-fragment | NA | | NA | 0 |
| S8-final-viral-combined_k141_219596\|\|full | 4295 | No | NA | 8 | 1 | 0 | Low-quality | Genome-fragment | | 4.72 | HMM-based (lower-bound) | 0 |
| S2-final-viral-combined_k141_198409\|\|0_partial | 14995 | No | NA | 15 | 0 | 10 | Not-determined | Genome-fragment | NA | | NA | 0 |
| S2-final-viral-combined_k141_95583\|\|0_partial | 11644 | No | NA | 9 | 0 | 5 | Not-determined | Genome-fragment | NA | | NA | 0 |
| S2-final-viral-combined_k141_23092\|\|0_partial | 12544 | No | NA | 13 | 0 | 10 | Not-determined | Genome-fragment | NA | | NA | 0 |
| S2-final-viral-combined_k141_33092\|\|0_partial | 10150 | No | NA | 9 | 0 | 1 | Not-determined | Genome-fragment | NA | | NA | 0 |
| S2-final-viral-combined_k141_74730\|\|0_partial | 13460 | No | NA | 11 | 0 | 5 | Not-determined | Genome-fragment | NA | | NA | 0 |
| S11-final-viral-combined_k141_136137\|\|full | 4298 | No | NA | 4 | 0 | 2 | Low-quality | Genome-fragment | | 6.89 | AAI-based (medium-confidence) | 0 |
| S2-final-viral-combined_k141_116405\|\|0_partial | 10769 | No | NA | 11 | 0 | 8 | Not-determined | Genome-fragment | NA | | NA | 0 |
| S2-final-viral-combined_k141_75144\|\|0_partial | 29286 | No | NA | 46 | 1 | 3 | Medium-quality | Genome-fragment | | 76.77 | AAI-based (medium-confidence) | 0 |
| S17-k141_17299 | 4302 | No | NA | 2 | 0 | 2 | Low-quality | Genome-fragment | | 2.11 | AAI-based (medium-confidence) | 0 |
| S2-final-viral-combined_k141_138182\|\|0_partial | 11841 | No | NA | 8 | 0 | 4 | Not-determined | Genome-fragment | NA | | NA | 0 |
| S2-final-viral-combined_k141_55179\|\|0_partial | 11031 | No | NA | 14 | 0 | 9 | Not-determined | Genome-fragment | NA | | NA | 0 |
| S2-final-viral-combined_k141_210534\|\|0_partial | 18489 | No | NA | 21 | 0 | 4 | Not-determined | Genome-fragment | NA | | NA | 0 |
| S2-final-viral-combined_k141_86805\|\|0_partial | 15090 | No | NA | 14 | 0 | 14 | Not-determined | Genome-fragment | NA | | NA | 0 |
| S2-final-viral-combined_k141_210794\|\|0_partial | 11790 | No | NA | 14 | 0 | 9 | Not-determined | Genome-fragment | NA | | NA | 0 |
| S2-final-viral-combined_k141_170048\|\|0_partial | 16779 | No | NA | 18 | 0 | 6 | Not-determined | Genome-fragment | NA | | NA | 0 |
| S2-final-viral-combined_k141_159290\|\|0_partial | 48650 | No | NA | 56 | 0 | 7 | Not-determined | Genome-fragment | NA | | NA | 0 |
| S2-final-viral-combined_k141_181095\|\|0_partial | 3530 | Yes | 2216 | 5 | 1 | 1 | Medium-quality | Genome-fragment | | 53.25 | HMM-based (lower-bound) | 37.22 |
| S6-final-viral-combined_k141_130826\|\|full | 4302 | Yes | 2123 | 7 | 2 | 1 | Low-quality | Genome-fragment | | 4.37 | AAI-based (high-confidence) | 50.65 |
| S2-final-viral-combined_k141_221524\|\|0_partial | 21964 | No | NA | 26 | 0 | 3 | Not-determined | Genome-fragment | NA | | NA | 0 |
| S2-final-viral-combined_k141_181670\|\|0_partial | 13742 | No | NA | 10 | 0 | 7 | Not-determined | Genome-fragment | NA | | NA | 0 |
| S2-final-viral-combined_k141_12128\|\|full | 4304 | No | NA | 5 | 1 | 0 | Low-quality | Genome-fragment | | 10.72 | AAI-based (high-confidence) | 0 |
| S2-final-viral-combined_k141_4685\|\|0_partial | 14637 | No | NA | 18 | 1 | 2 | High-quality | High-quality | | 94.48 | AAI-based (high-confidence) | 0 |
| S11-final-viral-combined_k141_15036\|\|full | 4306 | No | NA | 3 | 3 | 0 | Low-quality | Genome-fragment | | 9.29 | AAI-based (high-confidence) | 0 |
| S4_k141_88718 | 4306 | No | NA | 8 | 0 | 1 | Low-quality | Genome-fragment | | 9.15 | AAI-based (high-confidence) | 0 |
| S2-final-viral-combined_k141_4805\|\|0_partial | 10931 | No | NA | 14 | 0 | 6 | Not-determined | Genome-fragment | NA | | NA | 0 |
| S3-final-viral-combined_k141_38975\|\|full | 4307 | No | NA | 3 | 0 | 3 | Low-quality | Genome-fragment | | 9.76 | AAI-based (medium-confidence) | 0 |
| S8-final-viral-combined_k141_192125\|\|full | 4307 | No | NA | 7 | 1 | 0 | Low-quality | Genome-fragment | | 2.38 | HMM-based (lower-bound) | 0 |
| S2-final-viral-combined_k141_161578\|\|0_partial | 11450 | No | NA | 11 | 0 | 7 | Not-determined | Genome-fragment | NA | | NA | 0 |
| S2-final-viral-combined_k141_121213\|\|0_partial | 15787 | No | NA | 15 | 0 | 11 | Not-determined | Genome-fragment | NA | | NA | 0 |
| S1-final-viral-combined_k141_158714\|\|full | 4309 | No | NA | 5 | 0 | 2 | Low-quality | Genome-fragment | | 4.2 | AAI-based (medium-confidence) | 0 |
| S7_k141_28026 | 4309 | No | NA | 9 | 0 | 0 | Low-quality | Genome-fragment | | 7.81 | AAI-based (high-confidence) | 0 |
| S7-final-viral-combined_k141_170450\|\|full | 4310 | No | NA | 7 | 0 | 0 | Low-quality | Genome-fragment | | 7.04 | AAI-based (medium-confidence) | 0 |
| S2-final-viral-combined_k141_79982\|\|0_partial | 11871 | No | NA | 8 | 0 | 4 | Not-determined | Genome-fragment | NA | | NA | 0 |
| S2-final-viral-combined_k141_51566\|\|0_partial | 21094 | No | NA | 16 | 0 | 4 | Not-determined | Genome-fragment | NA | | NA | 0 |
| S9-final-viral-combined_k141_1655\|\|full | 4311 | No | NA | 4 | 0 | 2 | Low-quality | Genome-fragment | | 6.49 | AAI-based (high-confidence) | 0 |
| S17-final-viral-combined_k141_85675\|\|full | 4311 | No | NA | 11 | 8 | 0 | Low-quality | Genome-fragment | | 10.24 | AAI-based (high-confidence) | 0 |
| S2-final-viral-combined_k141_143737\|\|0_partial | 52742 | Yes | 38952 | 61 | 3 | 10 | Medium-quality | Genome-fragment | | 64.63 | AAI-based (medium-confidence) | 26.15 |
| S2-final-viral-combined_k141_204225\|\|0_partial | 4195 | No | NA | 5 | 1 | 0 | High-quality | High-quality | | 100 | HMM-based (lower-bound) | 0 |
| S4_k141_147766 | 4311 | No | NA | 5 | 0 | 1 | Low-quality | Genome-fragment | | 5.49 | AAI-based (medium-confidence) | 0 |
| S17-k141_64837 | 4312 | No | NA | 7 | 2 | 0 | Low-quality | Genome-fragment | | 9.53 | AAI-based (medium-confidence) | 0 |
| S2-final-viral-combined_k141_61928\|\|0_partial | 17405 | No | NA | 18 | 0 | 11 | Not-determined | Genome-fragment | NA | | NA | 0 |
| S12-final-viral-combined_k141_293098\|\|full | 4312 | No | NA | 4 | 0 | 1 | Low-quality | Genome-fragment | | 8.7 | AAI-based (medium-confidence) | 0 |
| S2-final-viral-combined_k141_7756\|\|0_partial | 63410 | Yes | 48340 | 53 | 8 | 9 | Medium-quality | Genome-fragment | | 84.03 | HMM-based (lower-bound) | 23.77 |
| S7-final-viral-combined_k141_32777\|\|full | 4313 | No | NA | 3 | 0 | 3 | Low-quality | Genome-fragment | | 2.4 | AAI-based (medium-confidence) | 0 |
| S2-final-viral-combined_k141_207000\|\|0_partial | 47238 | Yes | 42306 | 67 | 29 | 6 | High-quality | High-quality | | 95.69 | AAI-based (high-confidence) | 10.44 |
| S11-final-viral-combined_k141_250295\|\|full | 4315 | No | NA | 7 | 2 | 0 | Low-quality | Genome-fragment | | 7.71 | HMM-based (lower-bound) | 0 |
| S7_k141_153285 | 4315 | No | NA | 5 | 0 | 1 | Low-quality | Genome-fragment | | 4.43 | AAI-based (high-confidence) | 0 |
| S2-final-viral-combined_k141_155476\|\|lt2gene | 1616 | No | NA | 1 | 0 | 1 | Not-determined | Genome-fragment | NA | | NA | 0 |
| S11-final-viral-combined_k141_292971\|\|full | 4316 | No | NA | 5 | 3 | 0 | Low-quality | Genome-fragment | | 5.31 | HMM-based (lower-bound) | 0 |
| S1-final-viral-combined_k141_156909\|\|full | 4316 | No | NA | 5 | 0 | 2 | Low-quality | Genome-fragment | | 5.11 | AAI-based (medium-confidence) | 0 |
| S2-final-viral-combined_k141_41792\|\|lt2gene | 1555 | No | NA | 2 | 0 | 0 | Not-determined | Genome-fragment | NA | | NA | 0 |
| S2-final-viral-combined_k141_334\|\|lt2gene | 1892 | No | NA | 2 | 0 | 0 | Not-determined | Genome-fragment | NA | | NA | 0 |
| S5-final-viral-combined_k141_92338\|\|full | 4316 | No | NA | 5 | 1 | 0 | Low-quality | Genome-fragment | | 7.26 | HMM-based (lower-bound) | 0 |
| S2-final-viral-combined_k141_21070\|\|lt2gene | 1022 | No | NA | 1 | 0 | 1 | Not-determined | Genome-fragment | NA | | NA | 0 |
| S2-final-viral-combined_k141_207467\|\|lt2gene | 1061 | No | NA | 1 | 0 | 1 | Not-determined | Genome-fragment | NA | | NA | 0 |
| S5_k141_206352 | 4316 | No | NA | 11 | 0 | 0 | Low-quality | Genome-fragment | | 2.59 | AAI-based (medium-confidence) | 0 |
| S12-final-viral-combined_k141_108936\|\|full | 4317 | No | NA | 9 | 0 | 0 | Low-quality | Genome-fragment | | 9.65 | AAI-based (high-confidence) | 0 |
| S12_k141_352629 | 4317 | No | NA | 3 | 0 | 3 | Low-quality | Genome-fragment | | 1.35 | AAI-based (medium-confidence) | 0 |
| S2-final-viral-combined_k141_114661\|\|lt2gene | 2635 | No | NA | 1 | 0 | 1 | Not-determined | Genome-fragment | NA | | NA | 0 |
| S12_k141_374154 | 4317 | No | NA | 2 | 0 | 0 | Low-quality | Genome-fragment | | 7.16 | AAI-based (medium-confidence) | 0 |
| S7-final-viral-combined_k141_16555\|\|full | 4319 | No | NA | 7 | 2 | 0 | Low-quality | Genome-fragment | | 9.51 | AAI-based (high-confidence) | 0 |
| S4-final-viral-combined_k141_110715\|\|full | 4319 | No | NA | 6 | 6 | 0 | Low-quality | Genome-fragment | | 12.3 | AAI-based (high-confidence) | 0 |
| S5-final-viral-combined_k141_103989\|\|full | 4321 | No | NA | 5 | 3 | 0 | Low-quality | Genome-fragment | | 11.65 | AAI-based (high-confidence) | 0 |
| S2-final-viral-combined_k141_125521\|\|lt2gene | 1130 | No | NA | 1 | 0 | 0 | Not-determined | Genome-fragment | NA | | NA | 0 |
| S2-final-viral-combined_k141_187542\|\|lt2gene | 1040 | No | NA | 2 | 0 | 1 | Not-determined | Genome-fragment | NA | | NA | 0 |
| S11-final-viral-combined_k141_313610\|\|full | 4322 | No | NA | 5 | 0 | 0 | Low-quality | Genome-fragment | | 10.09 | AAI-based (medium-confidence) | 0 |
| S17-final-viral-combined_k141_119322\|\|full | 4322 | No | NA | 3 | 0 | 1 | Low-quality | Genome-fragment | | 4.22 | AAI-based (medium-confidence) | 0 |
| S11-final-viral-combined_k141_43503\|\|full | 4323 | No | NA | 6 | 5 | 0 | Low-quality | Genome-fragment | | 9.21 | AAI-based (high-confidence) | 0 |
| S8-final-viral-combined_k141_234624\|\|full | 4323 | No | NA | 2 | 0 | 2 | Low-quality | Genome-fragment | | 2.4 | AAI-based (medium-confidence) | 0 |
| S4-final-viral-combined_k141_17073\|\|full | 4324 | No | NA | 8 | 0 | 1 | Low-quality | Genome-fragment | | 7.45 | AAI-based (medium-confidence) | 0 |
| S9_k141_11177 | 4325 | No | NA | 6 | 4 | 0 | Low-quality | Genome-fragment | | 10.69 | AAI-based (high-confidence) | 0 |
| S2-final-viral-combined_k141_43598\|\|lt2gene | 2965 | No | NA | 2 | 0 | 1 | Not-determined | Genome-fragment | NA | | NA | 0 |
| S2-final-viral-combined_k141_135976\|\|lt2gene | 1005 | No | NA | 1 | 0 | 0 | Not-determined | Genome-fragment | NA | | NA | 0 |
| S7-final-viral-combined_k141_192003\|\|full | 4326 | No | NA | 4 | 0 | 3 | Low-quality | Genome-fragment | | 2.41 | AAI-based (medium-confidence) | 0 |
| S4_k141_6144 | 4326 | No | NA | 9 | 1 | 0 | Low-quality | Genome-fragment | | 7.39 | HMM-based (lower-bound) | 0 |
| S8-final-viral-combined_k141_199165\|\|full | 4326 | No | NA | 5 | 0 | 1 | Low-quality | Genome-fragment | | 6.97 | AAI-based (medium-confidence) | 0 |
| S1-final-viral-combined_k141_692\|\|full | 4328 | No | NA | 4 | 0 | 0 | Low-quality | Genome-fragment | | 10.31 | AAI-based (high-confidence) | 0 |
| S2-final-viral-combined_k141_188379\|\|lt2gene | 1051 | No | NA | 1 | 0 | 0 | Not-determined | Genome-fragment | NA | | NA | 0 |
| S12_k141_115650 | 4328 | No | NA | 3 | 1 | 0 | Low-quality | Genome-fragment | | 9.51 | AAI-based (high-confidence) | 0 |
| S11-final-viral-combined_k141_5035\|\|full | 4330 | No | NA | 7 | 2 | 1 | Low-quality | Genome-fragment | | 9.09 | AAI-based (high-confidence) | 0 |
| S7-final-viral-combined_k141_6396\|\|full | 4330 | No | NA | 3 | 0 | 2 | Low-quality | Genome-fragment | | 2.8 | AAI-based (medium-confidence) | 0 |
| S2-final-viral-combined_k141_240311\|\|lt2gene | 1726 | No | NA | 2 | 0 | 2 | Not-determined | Genome-fragment | NA | | NA | 0 |
| S11-final-viral-combined_k141_14262\|\|full | 4331 | No | NA | 8 | 0 | 1 | Low-quality | Genome-fragment | | 24.47 | AAI-based (high-confidence) | 0 |
| S17-final-viral-combined_k141_178959\|\|full | 4331 | No | NA | 6 | 1 | 0 | Low-quality | Genome-fragment | | 5.45 | AAI-based (medium-confidence) | 0 |
| S17-k141_200846 | 4332 | No | NA | 3 | 0 | 1 | Low-quality | Genome-fragment | | 5.29 | AAI-based (medium-confidence) | 0 |
| S12-final-viral-combined_k141_144496\|\|full | 4332 | No | NA | 6 | 2 | 0 | Low-quality | Genome-fragment | | 10.09 | AAI-based (medium-confidence) | 0 |
| S4_k141_143298 | 4332 | No | NA | 6 | 3 | 0 | Low-quality | Genome-fragment | | 2.5 | AAI-based (high-confidence) | 0 |
| S2-final-viral-combined_k141_116391\|\|lt2gene | 1167 | No | NA | 2 | 0 | 0 | Not-determined | Genome-fragment | NA | | NA | 0 |
| S2-final-viral-combined_k141_2658\|\|lt2gene | 1089 | No | NA | 1 | 0 | 0 | Not-determined | Genome-fragment | NA | | NA | 0 |
| S7_k141_209514 | 4333 | No | NA | 3 | 0 | 2 | Low-quality | Genome-fragment | | 4.73 | AAI-based (medium-confidence) | 0 |
| S1_k141_107426 | 4334 | No | NA | 6 | 1 | 0 | Low-quality | Genome-fragment | | 2.94 | AAI-based (medium-confidence) | 0 |
| S17-k141_174475 | 4334 | No | NA | 3 | 0 | 0 | Low-quality | Genome-fragment | | 10.83 | AAI-based (high-confidence) | 0 |
| S2-final-viral-combined_k141_34100\|\|lt2gene | 3134 | No | NA | 1 | 0 | 0 | Not-determined | Genome-fragment | NA | | NA | 0 |
| S12-final-viral-combined_k141_102670\|\|full | 4334 | No | NA | 7 | 2 | 1 | Low-quality | Genome-fragment | | 3.15 | AAI-based (high-confidence) | 0 |
| S2-final-viral-combined_k141_158442\|\|lt2gene | 1502 | No | NA | 2 | 0 | 0 | Not-determined | Genome-fragment | NA | | NA | 0 |
| S2-final-viral-combined_k141_13367\|\|lt2gene | 1033 | No | NA | 2 | 0 | 1 | Not-determined | Genome-fragment | NA | | NA | 0 |
| S2-final-viral-combined_k141_65290\|\|lt2gene | 1243 | No | NA | 1 | 0 | 1 | Not-determined | Genome-fragment | NA | | NA | 0 |
| S1-final-viral-combined_k141_232050\|\|full | 4335 | No | NA | 7 | 2 | 0 | Low-quality | Genome-fragment | | 8.96 | AAI-based (high-confidence) | 0 |
| S1-final-viral-combined_k141_179995\|\|full | 4335 | No | NA | 7 | 3 | 0 | Low-quality | Genome-fragment | | 5.75 | HMM-based (lower-bound) | 0 |
| S4-final-viral-combined_k141_160047\|\|full | 4336 | No | NA | 3 | 0 | 2 | Low-quality | Genome-fragment | | 4.3 | AAI-based (high-confidence) | 0 |
| S11-final-viral-combined_k141_122230\|\|full | 4337 | No | NA | 7 | 0 | 1 | Low-quality | Genome-fragment | | 6.97 | AAI-based (medium-confidence) | 0 |
| S2-final-viral-combined_k141_246024\|\|full | 4337 | No | NA | 4 | 0 | 0 | Low-quality | Genome-fragment | | 9.14 | AAI-based (medium-confidence) | 0 |
| S8-final-viral-combined_k141_7678\|\|full | 4337 | No | NA | 5 | 1 | 0 | Low-quality | Genome-fragment | | 9.95 | AAI-based (high-confidence) | 0 |
| S12-final-viral-combined_k141_85785\|\|full | 4340 | No | NA | 3 | 0 | 0 | Low-quality | Genome-fragment | | 4.87 | AAI-based (medium-confidence) | 0 |
| S12-final-viral-combined_k141_295978\|\|full | 4340 | No | NA | 4 | 0 | 1 | Low-quality | Genome-fragment | | 8.31 | AAI-based (high-confidence) | 0 |
| S4-final-viral-combined_k141_136634\|\|lt2gene | 4340 | No | NA | 2 | 2 | 0 | Low-quality | Genome-fragment | | 7.18 | AAI-based (high-confidence) | 0 |
| S17-k141_138870 | 4341 | No | NA | 5 | 0 | 5 | Low-quality | Genome-fragment | | 1.6 | AAI-based (medium-confidence) | 0 |
| S11_k141_274426 | 4342 | No | NA | 4 | 4 | 0 | Low-quality | Genome-fragment | | 5.77 | AAI-based (high-confidence) | 0 |
| S12_k141_67524 | 4343 | No | NA | 7 | 3 | 0 | Low-quality | Genome-fragment | | 9.17 | AAI-based (high-confidence) | 0 |
| S11-final-viral-combined_k141_211605\|\|full | 4344 | No | NA | 5 | 0 | 5 | Low-quality | Genome-fragment | | 2.54 | AAI-based (medium-confidence) | 0 |
| S2-final-viral-combined_k141_138921\|\|lt2gene | 1024 | No | NA | 1 | 0 | 1 | Not-determined | Genome-fragment | NA | | NA | 0 |
| S11_k141_139173 | 4344 | No | NA | 5 | 0 | 0 | Low-quality | Genome-fragment | | 5.21 | AAI-based (medium-confidence) | 0 |
| S9_k141_3381 | 4345 | No | NA | 3 | 0 | 1 | Low-quality | Genome-fragment | | 3.36 | AAI-based (high-confidence) | 0 |
| S7-final-viral-combined_k141_201362\|\|full | 4345 | No | NA | 4 | 0 | 1 | Low-quality | Genome-fragment | | 3.09 | AAI-based (medium-confidence) | 0 |
| S5-final-viral-combined_k141_197577\|\|full | 4346 | No | NA | 5 | 0 | 0 | Low-quality | Genome-fragment | | 8.85 | AAI-based (medium-confidence) | 0 |
| S2-final-viral-combined_k141_118103\|\|lt2gene | 1256 | No | NA | 2 | 0 | 1 | Not-determined | Genome-fragment | NA | | NA | 0 |
| S2-final-viral-combined_k141_220869\|\|lt2gene | 3006 | No | NA | 1 | 0 | 0 | Not-determined | Genome-fragment | NA | | NA | 0 |
| S2-final-viral-combined_k141_56354\|\|lt2gene | 1552 | No | NA | 1 | 0 | 0 | Not-determined | Genome-fragment | NA | | NA | 0 |
| S1_k141_361639 | 4347 | No | NA | 10 | 1 | 0 | Low-quality | Genome-fragment | | 36.01 | AAI-based (high-confidence) | 0 |
| S2-final-viral-combined_k141_128095\|\|lt2gene | 2694 | No | NA | 1 | 0 | 0 | Not-determined | Genome-fragment | NA | | NA | 0 |
| S11_k141_224472 | 4348 | No | NA | 4 | 0 | 0 | Low-quality | Genome-fragment | | 9.94 | AAI-based (medium-confidence) | 0 |
| S5-final-viral-combined_k141_78667\|\|full | 4349 | No | NA | 4 | 0 | 0 | Low-quality | Genome-fragment | | 6.99 | AAI-based (medium-confidence) | 0 |
| S17-k141_251554 | 4349 | No | NA | 5 | 0 | 5 | Low-quality | Genome-fragment | | 1.6 | AAI-based (medium-confidence) | 0 |
| S5_k141_206388 | 4350 | No | NA | 6 | 0 | 0 | Low-quality | Genome-fragment | | 4.27 | AAI-based (high-confidence) | 0 |
| S6-final-viral-combined_k141_9005\|\|full | 4350 | No | NA | 6 | 1 | 4 | Low-quality | Genome-fragment | | 10.28 | AAI-based (medium-confidence) | 0 |
| S1-final-viral-combined_k141_116087\|\|full | 4351 | No | NA | 6 | 3 | 0 | Low-quality | Genome-fragment | | 12.08 | AAI-based (high-confidence) | 0 |
| S17-k141_25407 | 4351 | Yes | 1899 | 4 | 1 | 1 | Low-quality | Genome-fragment | | 3.52 | AAI-based (medium-confidence) | 56.35 |
| S8-final-viral-combined_k141_14211\|\|full | 4351 | No | NA | 3 | 0 | 2 | Low-quality | Genome-fragment | | 5.69 | AAI-based (medium-confidence) | 0 |
| S2-final-viral-combined_k141_36545\|\|lt2gene | 1019 | No | NA | 1 | 0 | 0 | Not-determined | Genome-fragment | NA | | NA | 0 |
| S12-final-viral-combined_k141_281152\|\|full | 4353 | No | NA | 5 | 0 | 2 | Low-quality | Genome-fragment | | 4.99 | AAI-based (medium-confidence) | 0 |
| S8_k141_258722 | 4354 | No | NA | 5 | 3 | 0 | Low-quality | Genome-fragment | | 10.42 | AAI-based (high-confidence) | 0 |
| S8_k141_203772 | 4355 | No | NA | 3 | 0 | 1 | Low-quality | Genome-fragment | | 7.22 | AAI-based (medium-confidence) | 0 |
| S9_k141_111753 | 4356 | No | NA | 2 | 0 | 2 | Low-quality | Genome-fragment | | 8.04 | AAI-based (high-confidence) | 0 |
| S17-final-viral-combined_k141_44875\|\|full | 4356 | No | NA | 7 | 3 | 0 | Low-quality | Genome-fragment | | 11.86 | AAI-based (high-confidence) | 0 |
| S8_k141_161967 | 4358 | No | NA | 3 | 0 | 1 | Low-quality | Genome-fragment | | 4.72 | AAI-based (medium-confidence) | 0 |
| S3-final-viral-combined_k141_123041\|\|full | 4359 | No | NA | 4 | 0 | 0 | Low-quality | Genome-fragment | | 11.49 | AAI-based (high-confidence) | 0 |
| S12-final-viral-combined_k141_318786\|\|full | 4359 | No | NA | 8 | 2 | 0 | Low-quality | Genome-fragment | | 5.31 | AAI-based (medium-confidence) | 0 |
| S2-final-viral-combined_k141_99193\|\|lt2gene | 1078 | No | NA | 2 | 0 | 0 | Not-determined | Genome-fragment | NA | | NA | 0 |
| S12-final-viral-combined_k141_255913\|\|full | 4359 | No | NA | 3 | 0 | 3 | Low-quality | Genome-fragment | | 6.22 | AAI-based (medium-confidence) | 0 |
| S8-final-viral-combined_k141_6979\|\|full | 4360 | No | NA | 6 | 0 | 3 | Low-quality | Genome-fragment | | 3.28 | AAI-based (medium-confidence) | 0 |
| S11-final-viral-combined_k141_193402\|\|full | 4363 | No | NA | 5 | 3 | 0 | Low-quality | Genome-fragment | | 9.09 | AAI-based (high-confidence) | 0 |
| S2-final-viral-combined_k141_49029\|\|lt2gene | 1378 | No | NA | 2 | 0 | 1 | Not-determined | Genome-fragment | NA | | NA | 0 |
| S2-final-viral-combined_k141_172297\|\|lt2gene | 1003 | No | NA | 2 | 0 | 0 | Not-determined | Genome-fragment | NA | | NA | 0 |
| S5_k141_36244 | 4363 | No | NA | 4 | 4 | 0 | Low-quality | Genome-fragment | | 12.54 | AAI-based (high-confidence) | 0 |
| S12-final-viral-combined_k141_287089\|\|full | 4364 | No | NA | 2 | 0 | 1 | Low-quality | Genome-fragment | | 3.96 | AAI-based (medium-confidence) | 0 |
| S12-final-viral-combined_k141_192844\|\|full | 4364 | No | NA | 5 | 2 | 0 | Low-quality | Genome-fragment | | 10.46 | AAI-based (high-confidence) | 0 |
| S12_k141_221670 | 4364 | No | NA | 2 | 0 | 1 | Low-quality | Genome-fragment | | 7.83 | AAI-based (medium-confidence) | 0 |
| S2-final-viral-combined_k141_16348\|\|lt2gene | 1005 | No | NA | 1 | 0 | 0 | Not-determined | Genome-fragment | NA | | NA | 0 |
| S2-final-viral-combined_k141_160804\|\|lt2gene | 1321 | No | NA | 1 | 0 | 0 | Not-determined | Genome-fragment | NA | | NA | 0 |
| S2-final-viral-combined_k141_37547\|\|lt2gene | 2001 | No | NA | 1 | 0 | 1 | Not-determined | Genome-fragment | NA | | NA | 0 |
| S8_k141_65903 | 4364 | No | NA | 11 | 2 | 0 | Low-quality | Genome-fragment | | 9.66 | HMM-based (lower-bound) | 0 |
| S11_k141_181705 | 4365 | No | NA | 8 | 1 | 0 | Low-quality | Genome-fragment | | 8.18 | HMM-based (lower-bound) | 0 |
| S11-final-viral-combined_k141_134527\|\|full | 4366 | No | NA | 5 | 0 | 1 | Low-quality | Genome-fragment | | 2.86 | AAI-based (high-confidence) | 0 |
| S2-final-viral-combined_k141_78925\|\|lt2gene | 3302 | No | NA | 2 | 0 | 0 | Not-determined | Genome-fragment | NA | | NA | 0 |
| S6-final-viral-combined_k141_41169\|\|full | 4366 | No | NA | 3 | 0 | 3 | Low-quality | Genome-fragment | | 2.45 | AAI-based (medium-confidence) | 0 |
| S6-final-viral-combined_k141_23402\|\|full | 4367 | No | NA | 4 | 2 | 0 | Low-quality | Genome-fragment | | 8.84 | AAI-based (high-confidence) | 0 |
| S11-final-viral-combined_k141_166697\|\|full | 4368 | No | NA | 3 | 0 | 0 | Low-quality | Genome-fragment | | 7.78 | AAI-based (high-confidence) | 0 |
| S2-final-viral-combined_k141_141572\|\|lt2gene | 2423 | No | NA | 2 | 0 | 2 | Not-determined | Genome-fragment | NA | | NA | 0 |
| S7-final-viral-combined_k141_191937\|\|lt2gene | 4368 | No | NA | 2 | 1 | 0 | Low-quality | Genome-fragment | | 11.05 | AAI-based (high-confidence) | 0 |
| S2-final-viral-combined_k141_141649\|\|lt2gene | 1867 | No | NA | 1 | 0 | 1 | Not-determined | Genome-fragment | NA | | NA | 0 |
| S2-final-viral-combined_k141_109911\|\|lt2gene | 1447 | No | NA | 1 | 0 | 0 | Not-determined | Genome-fragment | NA | | NA | 0 |
| S7-final-viral-combined_k141_139371\|\|full | 4369 | No | NA | 6 | 3 | 0 | Low-quality | Genome-fragment | | 7.99 | HMM-based (lower-bound) | 0 |
| S12-final-viral-combined_k141_376861\|\|full | 4371 | No | NA | 9 | 0 | 1 | Low-quality | Genome-fragment | | 8.79 | AAI-based (medium-confidence) | 0 |
| S12_k141_361461 | 4372 | No | NA | 3 | 0 | 1 | Low-quality | Genome-fragment | | 9.41 | AAI-based (high-confidence) | 0 |
| S2-final-viral-combined_k141_147531\|\|full | 4372 | No | NA | 5 | 1 | 0 | Low-quality | Genome-fragment | | 2.37 | AAI-based (medium-confidence) | 0 |
| S4_k141_2599 | 4372 | No | NA | 6 | 1 | 0 | Low-quality | Genome-fragment | | 1.43 | HMM-based (lower-bound) | 0 |
| S1-final-viral-combined_k141_276778\|\|full | 4373 | No | NA | 7 | 0 | 4 | Low-quality | Genome-fragment | | 6.04 | AAI-based (medium-confidence) | 0 |
| S12-final-viral-combined_k141_355794\|\|full | 4373 | No | NA | 7 | 0 | 1 | Low-quality | Genome-fragment | | 3.04 | AAI-based (high-confidence) | 0 |
| S4-final-viral-combined_k141_46015\|\|lt2gene | 4373 | No | NA | 1 | 1 | 0 | Low-quality | Genome-fragment | | 6.99 | AAI-based (high-confidence) | 0 |
| S2-final-viral-combined_k141_110706\|\|lt2gene | 1109 | No | NA | 1 | 0 | 1 | Not-determined | Genome-fragment | NA | | NA | 0 |
| S11-final-viral-combined_k141_140288\|\|full | 4374 | No | NA | 7 | 5 | 0 | Low-quality | Genome-fragment | | 12.2 | AAI-based (medium-confidence) | 0 |
| S5-final-viral-combined_k141_137330\|\|full | 4374 | No | NA | 8 | 1 | 0 | Low-quality | Genome-fragment | | 7.29 | AAI-based (high-confidence) | 0 |
| S4_k141_37132 | 4376 | No | NA | 6 | 0 | 2 | Low-quality | Genome-fragment | | 9.57 | AAI-based (medium-confidence) | 0 |
| S2-final-viral-combined_k141_2533\|\|full | 4378 | No | NA | 3 | 0 | 3 | Low-quality | Genome-fragment | | 5.91 | AAI-based (medium-confidence) | 0 |
| S1-final-viral-combined_k141_232725\|\|full | 4382 | No | NA | 6 | 0 | 4 | Low-quality | Genome-fragment | | 8.33 | AAI-based (medium-confidence) | 0 |
| S1_k141_107069 | 4382 | No | NA | 4 | 0 | 3 | Low-quality | Genome-fragment | | 2.02 | AAI-based (medium-confidence) | 0 |
| S3_k141_87147 | 4382 | No | NA | 9 | 0 | 0 | Low-quality | Genome-fragment | | 2.85 | AAI-based (high-confidence) | 0 |
| S7_k141_60976 | 4382 | No | NA | 3 | 0 | 1 | Low-quality | Genome-fragment | | 1.17 | AAI-based (high-confidence) | 0 |
| S2-final-viral-combined_k141_60677\|\|lt2gene | 1115 | No | NA | 2 | 0 | 0 | Not-determined | Genome-fragment | NA | | NA | 0 |
| S12-final-viral-combined_k141_65807\|\|full | 4382 | Yes | 1334 | 7 | 1 | 3 | Low-quality | Genome-fragment | | 1 | AAI-based (medium-confidence) | 69.56 |
| S11-final-viral-combined_k141_181947\|\|full | 4383 | No | NA | 7 | 3 | 0 | Low-quality | Genome-fragment | | 9.26 | AAI-based (high-confidence) | 0 |
| S1_k141_91492 | 4383 | No | NA | 5 | 2 | 0 | Low-quality | Genome-fragment | | 30.53 | AAI-based (medium-confidence) | 0 |
| S2-final-viral-combined_k141_70930\|\|lt2gene | 2745 | No | NA | 1 | 0 | 1 | Not-determined | Genome-fragment | NA | | NA | 0 |
| S9_k141_107136 | 4384 | No | NA | 14 | 3 | 0 | Low-quality | Genome-fragment | | 11.33 | AAI-based (medium-confidence) | 0 |
| S2-final-viral-combined_k141_203874\|\|lt2gene | 1235 | No | NA | 2 | 0 | 1 | Not-determined | Genome-fragment | NA | | NA | 0 |
| S7_k141_57293 | 4384 | No | NA | 5 | 1 | 0 | Low-quality | Genome-fragment | | 3.98 | HMM-based (lower-bound) | 0 |
| S2-final-viral-combined_k141_195333\|\|lt2gene | 1769 | No | NA | 1 | 0 | 1 | Not-determined | Genome-fragment | NA | | NA | 0 |
| S2-final-viral-combined_k141_111553\|\|lt2gene | 2779 | No | NA | 2 | 0 | 0 | Not-determined | Genome-fragment | NA | | NA | 0 |
| S2-final-viral-combined_k141_102861\|\|lt2gene | 1328 | No | NA | 1 | 0 | 0 | Not-determined | Genome-fragment | NA | | NA | 0 |
| S6-final-viral-combined_k141_93580\|\|full | 4384 | No | NA | 9 | 0 | 0 | Low-quality | Genome-fragment | | 6.7 | AAI-based (high-confidence) | 0 |
| S4-final-viral-combined_k141_124936\|\|full | 4385 | No | NA | 3 | 0 | 2 | Low-quality | Genome-fragment | | 4.74 | AAI-based (medium-confidence) | 0 |
| S2-final-viral-combined_k141_143793\|\|lt2gene | 2070 | No | NA | 2 | 0 | 0 | Not-determined | Genome-fragment | NA | | NA | 0 |
| S9-final-viral-combined_k141_8876\|\|full | 4387 | No | NA | 6 | 0 | 1 | Low-quality | Genome-fragment | | 7.78 | AAI-based (medium-confidence) | 0 |
| S2-final-viral-combined_k141_81537\|\|lt2gene | 2873 | No | NA | 2 | 0 | 0 | Not-determined | Genome-fragment | NA | | NA | 0 |
| S8-final-viral-combined_k141_70890\|\|full | 4387 | No | NA | 12 | 1 | 0 | Low-quality | Genome-fragment | | 4.33 | AAI-based (high-confidence) | 0 |
| S4-final-viral-combined_k141_10366\|\|full | 4389 | No | NA | 4 | 0 | 2 | Low-quality | Genome-fragment | | 1.57 | AAI-based (medium-confidence) | 0 |
| S7-final-viral-combined_k141_171335\|\|full | 4390 | No | NA | 9 | 8 | 0 | Low-quality | Genome-fragment | | 10.18 | AAI-based (high-confidence) | 0 |
| S8-final-viral-combined_k141_183291\|\|full | 4390 | No | NA | 8 | 4 | 0 | Low-quality | Genome-fragment | | 9.81 | AAI-based (high-confidence) | 0 |
| S2-final-viral-combined_k141_144248\|\|lt2gene | 1655 | No | NA | 2 | 0 | 1 | Not-determined | Genome-fragment | NA | | NA | 0 |
| S8_k141_207236 | 4390 | No | NA | 5 | 0 | 3 | Low-quality | Genome-fragment | | 1.19 | AAI-based (medium-confidence) | 0 |
| S5-final-viral-combined_k141_140567\|\|full | 4391 | No | NA | 5 | 2 | 0 | Low-quality | Genome-fragment | | 6.65 | HMM-based (lower-bound) | 0 |
| S12_k141_208922 | 4391 | No | NA | 8 | 3 | 0 | Low-quality | Genome-fragment | | 9.59 | AAI-based (high-confidence) | 0 |
| S17-k141_193058 | 4392 | No | NA | 5 | 0 | 1 | Low-quality | Genome-fragment | | 2.75 | AAI-based (medium-confidence) | 0 |
| S17-final-viral-combined_k141_209490\|\|full | 4393 | No | NA | 3 | 0 | 3 | Low-quality | Genome-fragment | | 2.09 | AAI-based (medium-confidence) | 0 |
| S12-final-viral-combined_k141_78866\|\|full | 4393 | No | NA | 6 | 3 | 0 | Low-quality | Genome-fragment | | 5.51 | HMM-based (lower-bound) | 0 |
| S7-final-viral-combined_k141_240636\|\|full | 4394 | No | NA | 4 | 2 | 0 | Low-quality | Genome-fragment | | 11.54 | AAI-based (high-confidence) | 0 |
| S11-final-viral-combined_k141_127320\|\|full | 4395 | No | NA | 11 | 6 | 0 | Low-quality | Genome-fragment | | 7.25 | AAI-based (high-confidence) | 0 |
| S8-final-viral-combined_k141_153016\|\|full | 4395 | No | NA | 6 | 1 | 0 | Low-quality | Genome-fragment | | 3.29 | AAI-based (high-confidence) | 0 |
| S1_k141_200906 | 4396 | No | NA | 5 | 0 | 1 | Low-quality | Genome-fragment | | 1.54 | AAI-based (medium-confidence) | 0 |
| S8-final-viral-combined_k141_38539\|\|full | 4396 | Yes | 393 | 3 | 1 | 2 | Low-quality | Genome-fragment | | 0.61 | HMM-based (lower-bound) | 91.06 |
| S11_k141_38329 | 4397 | No | NA | 5 | 1 | 0 | Low-quality | Genome-fragment | | 9.64 | AAI-based (high-confidence) | 0 |
| S3-final-viral-combined_k141_113116\|\|full | 4397 | No | NA | 4 | 0 | 0 | Low-quality | Genome-fragment | | 3.99 | AAI-based (medium-confidence) | 0 |
| S12_k141_202521 | 4397 | No | NA | 2 | 0 | 1 | Low-quality | Genome-fragment | | 4.59 | AAI-based (medium-confidence) | 0 |
| S12-final-viral-combined_k141_267908\|\|full | 4398 | No | NA | 7 | 1 | 0 | Low-quality | Genome-fragment | | 13.82 | AAI-based (high-confidence) | 0 |
| S2-final-viral-combined_k141_217396\|\|lt2gene | 1040 | No | NA | 2 | 0 | 0 | Not-determined | Genome-fragment | NA | | NA | 0 |
| S11-final-viral-combined_k141_165745\|\|full | 4399 | No | NA | 10 | 4 | 0 | Low-quality | Genome-fragment | | 9.18 | AAI-based (high-confidence) | 0 |
| S2-final-viral-combined_k141_8336\|\|lt2gene | 1612 | No | NA | 1 | 0 | 0 | Not-determined | Genome-fragment | NA | | NA | 0 |
| S2-final-viral-combined_k141_29996\|\|lt2gene | 1353 | No | NA | 2 | 0 | 0 | Not-determined | Genome-fragment | NA | | NA | 0 |
| S11-final-viral-combined_k141_326048\|\|full | 4399 | No | NA | 9 | 4 | 0 | Low-quality | Genome-fragment | | 10.99 | AAI-based (medium-confidence) | 0 |
| S12_k141_169431 | 4399 | Yes | 2703 | 10 | 1 | 1 | Low-quality | Genome-fragment | | 1.64 | HMM-based (lower-bound) | 38.55 |
| S2-final-viral-combined_k141_134533\|\|lt2gene | 1874 | No | NA | 1 | 0 | 1 | Not-determined | Genome-fragment | NA | | NA | 0 |
| S4-final-viral-combined_k141_115498\|\|full | 4399 | Yes | 3734 | 10 | 1 | 1 | Low-quality | Genome-fragment | | 2.15 | AAI-based (high-confidence) | 15.12 |
| S11_k141_146151 | 4401 | No | NA | 9 | 4 | 0 | Low-quality | Genome-fragment | | 9.92 | AAI-based (high-confidence) | 0 |
| S2_k141_72512 | 1526 | No | NA | 2 | 0 | 2 | Not-determined | Genome-fragment | NA | | NA | 0 |
| S2_k141_217502 | 3655 | No | NA | 3 | 0 | 1 | Not-determined | Genome-fragment | NA | | NA | 0 |
| S2_k141_217512 | 1412 | No | NA | 2 | 0 | 0 | Not-determined | Genome-fragment | NA | | NA | 0 |
| S2_k141_53837 | 4401 | No | NA | 3 | 0 | 0 | Low-quality | Genome-fragment | | 7.3 | AAI-based (medium-confidence) | 0 |
| S9-final-viral-combined_k141_5552\|\|full | 4402 | No | NA | 4 | 2 | 0 | Low-quality | Genome-fragment | | 11.79 | AAI-based (medium-confidence) | 0 |
| S2_k141_103640 | 2972 | No | NA | 2 | 0 | 1 | Not-determined | Genome-fragment | NA | | NA | 0 |
| S2_k141_155396 | 1167 | No | NA | 1 | 0 | 1 | Not-determined | Genome-fragment | NA | | NA | 0 |
| S5-final-viral-combined_k141_147604\|\|full | 4402 | No | NA | 4 | 0 | 2 | Low-quality | Genome-fragment | | 5.55 | AAI-based (medium-confidence) | 0 |
| S7-final-viral-combined_k141_144229\|\|full | 4402 | No | NA | 6 | 1 | 0 | Low-quality | Genome-fragment | | 3.52 | HMM-based (lower-bound) | 0 |
| S2_k141_51836 | 1205 | No | NA | 3 | 0 | 0 | Not-determined | Genome-fragment | NA | | NA | 0 |
| S12_k141_369906 | 4402 | No | NA | 10 | 7 | 0 | Low-quality | Genome-fragment | | 7.26 | AAI-based (high-confidence) | 0 |
| S6-final-viral-combined_k141_34542\|\|full | 4402 | No | NA | 7 | 6 | 0 | Low-quality | Genome-fragment | | 9.18 | AAI-based (high-confidence) | 0 |
| S2_k141_41536 | 1779 | No | NA | 2 | 0 | 1 | Not-determined | Genome-fragment | NA | | NA | 0 |
| S2_k141_165743 | 15380 | No | NA | 29 | 0 | 28 | Not-determined | Genome-fragment | NA | | NA | 0 |
| S7_k141_198012 | 4405 | No | NA | 6 | 0 | 2 | Low-quality | Genome-fragment | | 7.15 | AAI-based (medium-confidence) | 0 |
| S1_k141_319675 | 4406 | No | NA | 6 | 1 | 0 | Low-quality | Genome-fragment | | 2.98 | AAI-based (high-confidence) | 0 |
| S2_k141_62216 | 1288 | No | NA | 2 | 0 | 1 | Not-determined | Genome-fragment | NA | | NA | 0 |
| S2_k141_31134 | 1090 | No | NA | 1 | 0 | 0 | Not-determined | Genome-fragment | NA | | NA | 0 |
| S9_k141_6659 | 4407 | No | NA | 3 | 0 | 1 | Low-quality | Genome-fragment | | 4.08 | AAI-based (medium-confidence) | 0 |
| S7-final-viral-combined_k141_200558\|\|full | 4407 | Yes | 2907 | 6 | 4 | 1 | Low-quality | Genome-fragment | | 6.15 | AAI-based (medium-confidence) | 34.04 |
| S2_k141_134779 | 1314 | No | NA | 1 | 0 | 0 | Not-determined | Genome-fragment | NA | | NA | 0 |
| S2_k141_20770 | 15543 | No | NA | 12 | 0 | 8 | Not-determined | Genome-fragment | NA | | NA | 0 |
| S2_k141_186546 | 1402 | No | NA | 1 | 0 | 1 | Not-determined | Genome-fragment | NA | | NA | 0 |
| S2_k141_20789 | 1010 | No | NA | 1 | 0 | 1 | Not-determined | Genome-fragment | NA | | NA | 0 |
| S2_k141_127 | 3726 | No | NA | 2 | 0 | 1 | Not-determined | Genome-fragment | NA | | NA | 0 |
| S2_k141_165821 | 2894 | No | NA | 3 | 0 | 2 | Not-determined | Genome-fragment | NA | | NA | 0 |
| S9-final-viral-combined_k141_17150\|\|full | 4408 | No | NA | 5 | 0 | 1 | Low-quality | Genome-fragment | | 9.51 | AAI-based (medium-confidence) | 0 |
| S2_k141_103814 | 4499 | No | NA | 9 | 0 | 1 | Not-determined | Genome-fragment | NA | | NA | 0 |
| S2_k141_103816 | 1230 | No | NA | 1 | 0 | 0 | Not-determined | Genome-fragment | NA | | NA | 0 |
| S2_k141_51955 | 14311 | No | NA | 10 | 0 | 10 | Not-determined | Genome-fragment | NA | | NA | 0 |
| S2_k141_176280 | 1122 | No | NA | 1 | 0 | 0 | Not-determined | Genome-fragment | NA | | NA | 0 |
| S2_k141_196899 | 1305 | No | NA | 1 | 0 | 0 | Not-determined | Genome-fragment | NA | | NA | 0 |
| S2_k141_134806 | 1159 | No | NA | 1 | 0 | 1 | Not-determined | Genome-fragment | NA | | NA | 0 |
| S2_k141_93304 | 2145 | No | NA | 2 | 0 | 2 | Not-determined | Genome-fragment | NA | | NA | 0 |
| S2_k141_83009 | 1320 | No | NA | 1 | 0 | 0 | Not-determined | Genome-fragment | NA | | NA | 0 |
| S2_k141_72571 | 6314 | No | NA | 8 | 0 | 4 | Not-determined | Genome-fragment | NA | | NA | 0 |
| S2_k141_72578 | 3761 | No | NA | 4 | 0 | 1 | Not-determined | Genome-fragment | NA | | NA | 0 |
| S8-final-viral-combined_k141_161775\|\|full | 4408 | No | NA | 3 | 0 | 2 | Low-quality | Genome-fragment | | 1.46 | AAI-based (medium-confidence) | 0 |
| S12_k141_300377 | 4411 | No | NA | 4 | 0 | 4 | Low-quality | Genome-fragment | | 2.03 | AAI-based (high-confidence) | 0 |
| S2-final-viral-combined_k141_898\|\|full | 4411 | No | NA | 7 | 0 | 2 | Low-quality | Genome-fragment | | 9.35 | AAI-based (medium-confidence) | 0 |
| S2_k141_155516 | 1028 | No | NA | 2 | 0 | 2 | Not-determined | Genome-fragment | NA | | NA | 0 |
| S2_k141_206 | 1498 | No | NA | 2 | 0 | 2 | Not-determined | Genome-fragment | NA | | NA | 0 |
| S2_k141_217735 | 2893 | No | NA | 4 | 0 | 2 | Not-determined | Genome-fragment | NA | | NA | 0 |
| S2_k141_124416 | 58908 | Yes | 46285 | 88 | 23 | 4 | Medium-quality | Genome-fragment | | 72.49 | AAI-based (high-confidence) | 21.43 |
| S2_k141_155547 | 1117 | No | NA | 1 | 0 | 1 | Not-determined | Genome-fragment | NA | | NA | 0 |
| S2_k141_31278 | 2666 | No | NA | 2 | 0 | 2 | Not-determined | Genome-fragment | NA | | NA | 0 |
| S2_k141_52021 | 2283 | No | NA | 2 | 0 | 0 | Not-determined | Genome-fragment | NA | | NA | 0 |
| S6-final-viral-combined_k141_24834\|\|full | 4412 | Yes | 2583 | 7 | 1 | 2 | Low-quality | Genome-fragment | | 5.95 | AAI-based (high-confidence) | 41.46 |
| S5-final-viral-combined_k141_16310\|\|full | 4413 | No | NA | 8 | 7 | 0 | Low-quality | Genome-fragment | | 9.33 | AAI-based (high-confidence) | 0 |
| S2_k141_228112 | 1237 | No | NA | 1 | 0 | 0 | Not-determined | Genome-fragment | NA | | NA | 0 |
| S2_k141_93443 | 1273 | No | NA | 1 | 0 | 1 | Not-determined | Genome-fragment | NA | | NA | 0 |
| S5-final-viral-combined_k141_154913\|\|full | 4414 | No | NA | 5 | 2 | 0 | Low-quality | Genome-fragment | | 2.01 | HMM-based (lower-bound) | 0 |
| S2_k141_93454 | 1084 | No | NA | 1 | 0 | 1 | Not-determined | Genome-fragment | NA | | NA | 0 |
| S2_k141_289 | 2304 | No | NA | 1 | 0 | 1 | Not-determined | Genome-fragment | NA | | NA | 0 |
| S7-final-viral-combined_k141_105060\|\|full | 4414 | No | NA | 3 | 0 | 1 | Low-quality | Genome-fragment | | 7.12 | AAI-based (medium-confidence) | 0 |
| S2_k141_134927 | 1225 | No | NA | 1 | 0 | 0 | Not-determined | Genome-fragment | NA | | NA | 0 |
| S1-final-viral-combined_k141_95202\|\|full | 4416 | Yes | 4201 | 6 | 1 | 1 | Low-quality | Genome-fragment | | 11.12 | AAI-based (high-confidence) | 4.87 |
| S2_k141_41822 | 3179 | No | NA | 4 | 0 | 3 | Not-determined | Genome-fragment | NA | | NA | 0 |
| S2_k141_228162 | 1632 | No | NA | 1 | 0 | 1 | Not-determined | Genome-fragment | NA | | NA | 0 |
| S2_k141_186720 | 3369 | No | NA | 1 | 0 | 1 | Not-determined | Genome-fragment | NA | | NA | 0 |
| S5-final-viral-combined_k141_107301\|\|full | 4416 | No | NA | 9 | 0 | 1 | Low-quality | Genome-fragment | | 4.84 | AAI-based (medium-confidence) | 0 |
| S2_k141_155638 | 1550 | No | NA | 1 | 0 | 1 | Not-determined | Genome-fragment | NA | | NA | 0 |
| S2_k141_186744 | 1590 | No | NA | 1 | 0 | 1 | Not-determined | Genome-fragment | NA | | NA | 0 |
| S9_k141_44894 | 4417 | No | NA | 5 | 0 | 1 | Low-quality | Genome-fragment | | 4.3 | AAI-based (medium-confidence) | 0 |
| S2_k141_103977 | 1521 | No | NA | 2 | 0 | 1 | Not-determined | Genome-fragment | NA | | NA | 0 |
| S7-final-viral-combined_k141_121836\|\|full | 4417 | No | NA | 5 | 0 | 0 | Low-quality | Genome-fragment | | 10.9 | AAI-based (high-confidence) | 0 |
| S2_k141_351 | 3900 | No | NA | 2 | 0 | 0 | Not-determined | Genome-fragment | NA | | NA | 0 |
| S2_k141_72749 | 1025 | No | NA | 1 | 0 | 0 | Not-determined | Genome-fragment | NA | | NA | 0 |
| S2_k141_228214 | 1871 | No | NA | 1 | 0 | 1 | Not-determined | Genome-fragment | NA | | NA | 0 |
| S5-final-viral-combined_k141_51173\|\|full | 4418 | No | NA | 6 | 0 | 4 | Low-quality | Genome-fragment | | 6.02 | AAI-based (medium-confidence) | 0 |
| S2_k141_238487 | 1258 | No | NA | 2 | 0 | 2 | Not-determined | Genome-fragment | NA | | NA | 0 |
| S12-final-viral-combined_k141_332086\|\|full | 4419 | No | NA | 2 | 0 | 2 | Low-quality | Genome-fragment | | 1.48 | AAI-based (medium-confidence) | 0 |
| S1_k141_116432 | 4420 | No | NA | 2 | 1 | 0 | Low-quality | Genome-fragment | | 2.84 | AAI-based (high-confidence) | 0 |
| S2_k141_207359 | 2742 | No | NA | 1 | 0 | 1 | Not-determined | Genome-fragment | NA | | NA | 0 |
| S2_k141_217870 | 1016 | No | NA | 1 | 0 | 1 | Not-determined | Genome-fragment | NA | | NA | 0 |
| S2_k141_124678 | 1121 | No | NA | 1 | 0 | 0 | Not-determined | Genome-fragment | NA | | NA | 0 |
| S2_k141_124680 | 1152 | No | NA | 1 | 0 | 0 | Not-determined | Genome-fragment | NA | | NA | 0 |
| S17-k141_98853 | 4420 | No | NA | 6 | 0 | 1 | Low-quality | Genome-fragment | | 2.35 | AAI-based (medium-confidence) | 0 |
| S2_k141_207388 | 6500 | No | NA | 8 | 0 | 6 | Not-determined | Genome-fragment | NA | | NA | 0 |
| S2_k141_155730 | 1761 | No | NA | 1 | 0 | 0 | Not-determined | Genome-fragment | NA | | NA | 0 |
| S2_k141_166004 | 2019 | No | NA | 2 | 0 | 0 | Not-determined | Genome-fragment | NA | | NA | 0 |
| S2_k141_166005 | 1233 | No | NA | 1 | 0 | 1 | Not-determined | Genome-fragment | NA | | NA | 0 |
| S6_k141_12485 | 4420 | No | NA | 4 | 0 | 0 | Low-quality | Genome-fragment | | 5.23 | AAI-based (medium-confidence) | 0 |
| S2_k141_83195 | 1468 | No | NA | 1 | 0 | 1 | Not-determined | Genome-fragment | NA | | NA | 0 |
| S2_k141_186807 | 7964 | No | NA | 2 | 0 | 1 | Not-determined | Genome-fragment | NA | | NA | 0 |
| S2_k141_217924 | 1251 | No | NA | 1 | 0 | 0 | Not-determined | Genome-fragment | NA | | NA | 0 |
| S2_k141_217927 | 1032 | No | NA | 2 | 0 | 2 | Not-determined | Genome-fragment | NA | | NA | 0 |
| S2_k141_166013 | 4028 | No | NA | 6 | 0 | 3 | Not-determined | Genome-fragment | NA | | NA | 0 |
| S2_k141_83205 | 2643 | No | NA | 2 | 0 | 2 | Not-determined | Genome-fragment | NA | | NA | 0 |
| S2_k141_238558 | 1914 | No | NA | 2 | 0 | 1 | Not-determined | Genome-fragment | NA | | NA | 0 |
| S2_k141_238572 | 1015 | No | NA | 2 | 0 | 2 | Not-determined | Genome-fragment | NA | | NA | 0 |
| S2_k141_114210 | 1024 | No | NA | 2 | 0 | 1 | Not-determined | Genome-fragment | NA | | NA | 0 |
| S2_k141_217962 | 1199 | No | NA | 1 | 0 | 1 | Not-determined | Genome-fragment | NA | | NA | 0 |
| S2_k141_408 | 1172 | No | NA | 1 | 0 | 0 | Not-determined | Genome-fragment | NA | | NA | 0 |
| S2_k141_83253 | 1032 | No | NA | 1 | 0 | 0 | Not-determined | Genome-fragment | NA | | NA | 0 |
| S11-final-viral-combined_k141_302058\|\|full | 4421 | No | NA | 3 | 0 | 3 | Low-quality | Genome-fragment | | 2.86 | AAI-based (medium-confidence) | 0 |
| S2-final-viral-combined_k141_194558\|\|full | 4421 | No | NA | 11 | 0 | 0 | Low-quality | Genome-fragment | | 36.65 | AAI-based (high-confidence) | 0 |
| S2_k141_238617 | 1396 | No | NA | 2 | 0 | 0 | Not-determined | Genome-fragment | NA | | NA | 0 |
| S2_k141_10661 | 3738 | No | NA | 5 | 0 | 4 | Not-determined | Genome-fragment | NA | | NA | 0 |
| S12-final-viral-combined_k141_178743\|\|full | 4424 | No | NA | 5 | 1 | 0 | Low-quality | Genome-fragment | | 9.82 | AAI-based (high-confidence) | 0 |
| S2_k141_145441 | 4140 | No | NA | 4 | 0 | 2 | Not-determined | Genome-fragment | NA | | NA | 0 |
| S2_k141_42131 | 1087 | No | NA | 1 | 0 | 1 | Not-determined | Genome-fragment | NA | | NA | 0 |
| S2_k141_10668 | 1268 | No | NA | 2 | 0 | 0 | Not-determined | Genome-fragment | NA | | NA | 0 |
| S2_k141_10675 | 1589 | No | NA | 1 | 0 | 0 | Not-determined | Genome-fragment | NA | | NA | 0 |
| S2_k141_176558 | 1140 | No | NA | 2 | 0 | 0 | Not-determined | Genome-fragment | NA | | NA | 0 |
| S2_k141_238642 | 1082 | No | NA | 2 | 0 | 0 | Not-determined | Genome-fragment | NA | | NA | 0 |
| S2_k141_238656 | 1160 | No | NA | 3 | 0 | 1 | Not-determined | Genome-fragment | NA | | NA | 0 |
| S2_k141_228333 | 1525 | No | NA | 1 | 0 | 1 | Not-determined | Genome-fragment | NA | | NA | 0 |
| S2_k141_155806 | 2721 | No | NA | 2 | 0 | 0 | Not-determined | Genome-fragment | NA | | NA | 0 |
| S2_k141_238696 | 1672 | No | NA | 2 | 0 | 1 | Not-determined | Genome-fragment | NA | | NA | 0 |
| S2_k141_10723 | 1423 | No | NA | 4 | 0 | 0 | Not-determined | Genome-fragment | NA | | NA | 0 |
| S2_k141_479 | 3183 | No | NA | 3 | 0 | 2 | Not-determined | Genome-fragment | NA | | NA | 0 |
| S2_k141_496 | 1187 | No | NA | 1 | 0 | 0 | Not-determined | Genome-fragment | NA | | NA | 0 |
| S2_k141_21085 | 3126 | No | NA | 4 | 0 | 2 | Not-determined | Genome-fragment | NA | | NA | 0 |
| S12_k141_206021 | 4424 | No | NA | 6 | 0 | 2 | Low-quality | Genome-fragment | | 7.04 | AAI-based (medium-confidence) | 0 |
| S2_k141_238731 | 4711 | No | NA | 2 | 0 | 0 | Not-determined | Genome-fragment | NA | | NA | 0 |
| S2_k141_42163 | 1020 | No | NA | 1 | 0 | 1 | Not-determined | Genome-fragment | NA | | NA | 0 |
| S2_k141_155873 | 1189 | No | NA | 1 | 0 | 1 | Not-determined | Genome-fragment | NA | | NA | 0 |
| S2_k141_155889 | 1457 | No | NA | 1 | 0 | 1 | Not-determined | Genome-fragment | NA | | NA | 0 |
| S2_k141_187001 | 1152 | No | NA | 2 | 0 | 1 | Not-determined | Genome-fragment | NA | | NA | 0 |
| S2_k141_187005 | 1590 | No | NA | 1 | 0 | 0 | Not-determined | Genome-fragment | NA | | NA | 0 |
| S2_k141_73008 | 3724 | No | NA | 3 | 0 | 0 | Not-determined | Genome-fragment | NA | | NA | 0 |
| S2_k141_187009 | 1359 | No | NA | 1 | 0 | 1 | Not-determined | Genome-fragment | NA | | NA | 0 |
| S2_k141_155914 | 1339 | No | NA | 1 | 0 | 1 | Not-determined | Genome-fragment | NA | | NA | 0 |
| S2_k141_155931 | 1174 | No | NA | 1 | 0 | 0 | Not-determined | Genome-fragment | NA | | NA | 0 |
| S8-final-viral-combined_k141_252252\|\|full | 4424 | No | NA | 9 | 1 | 0 | Low-quality | Genome-fragment | | 4.37 | AAI-based (high-confidence) | 0 |
| S2_k141_73036 | 3712 | No | NA | 1 | 0 | 1 | Not-determined | Genome-fragment | NA | | NA | 0 |
| S2_k141_145565 | 1820 | No | NA | 2 | 0 | 1 | Not-determined | Genome-fragment | NA | | NA | 0 |
| S2_k141_42271 | 3588 | No | NA | 6 | 0 | 1 | Not-determined | Genome-fragment | NA | | NA | 0 |
| S2_k141_207622 | 1026 | No | NA | 1 | 0 | 1 | Not-determined | Genome-fragment | NA | | NA | 0 |
| S2_k141_83352 | 1841 | No | NA | 3 | 0 | 0 | Not-determined | Genome-fragment | NA | | NA | 0 |
| S2_k141_21204 | 4502 | No | NA | 5 | 0 | 5 | Not-determined | Genome-fragment | NA | | NA | 0 |
| S2_k141_238867 | 1214 | No | NA | 2 | 0 | 0 | Not-determined | Genome-fragment | NA | | NA | 0 |
| S2_k141_31680 | 3388 | No | NA | 4 | 0 | 0 | Not-determined | Genome-fragment | NA | | NA | 0 |
| S17-k141_163872 | 4425 | No | NA | 3 | 0 | 2 | Low-quality | Genome-fragment | | 2.28 | AAI-based (medium-confidence) | 0 |
| S2_k141_155993 | 1286 | No | NA | 1 | 0 | 0 | Not-determined | Genome-fragment | NA | | NA | 0 |
| S2_k141_176674 | 3685 | No | NA | 7 | 0 | 5 | Not-determined | Genome-fragment | NA | | NA | 0 |
| S2_k141_175924 | 4425 | No | NA | 6 | 0 | 0 | Low-quality | Genome-fragment | | 3.98 | AAI-based (medium-confidence) | 0 |
| S2_k141_601 | 7322 | No | NA | 7 | 0 | 5 | Not-determined | Genome-fragment | NA | | NA | 0 |
| S2_k141_228426 | 3388 | No | NA | 4 | 0 | 0 | Not-determined | Genome-fragment | NA | | NA | 0 |
| S2_k141_135224 | 4816 | No | NA | 6 | 0 | 4 | Not-determined | Genome-fragment | NA | | NA | 0 |
| S2_k141_207659 | 3298 | No | NA | 5 | 0 | 0 | Not-determined | Genome-fragment | NA | | NA | 0 |
| S2_k141_52340 | 2431 | No | NA | 2 | 0 | 2 | Not-determined | Genome-fragment | NA | | NA | 0 |
| S2_k141_21329 | 1097 | No | NA | 1 | 0 | 1 | Not-determined | Genome-fragment | NA | | NA | 0 |
| S2_k141_176723 | 1087 | No | NA | 2 | 0 | 1 | Not-determined | Genome-fragment | NA | | NA | 0 |
| S2_k141_21336 | 4878 | No | NA | 2 | 0 | 0 | Not-determined | Genome-fragment | NA | | NA | 0 |
| S2_k141_228452 | 1042 | No | NA | 1 | 0 | 1 | Not-determined | Genome-fragment | NA | | NA | 0 |
| S2_k141_176743 | 1532 | No | NA | 1 | 0 | 0 | Not-determined | Genome-fragment | NA | | NA | 0 |
| S2_k141_156080 | 2522 | No | NA | 1 | 0 | 0 | Not-determined | Genome-fragment | NA | | NA | 0 |
| S1-final-viral-combined_k141_74632\|\|full | 4426 | No | NA | 5 | 0 | 4 | Low-quality | Genome-fragment | | 2.4 | AAI-based (medium-confidence) | 0 |
| S2_k141_10886 | 1024 | No | NA | 2 | 0 | 2 | Not-determined | Genome-fragment | NA | | NA | 0 |
| S2_k141_93792 | 1208 | No | NA | 1 | 0 | 0 | Not-determined | Genome-fragment | NA | | NA | 0 |
| S2_k141_21361 | 1319 | No | NA | 1 | 0 | 0 | Not-determined | Genome-fragment | NA | | NA | 0 |
| S2_k141_42430 | 1584 | No | NA | 2 | 0 | 0 | Not-determined | Genome-fragment | NA | | NA | 0 |
| S11-final-viral-combined_k141_303365\|\|full | 4428 | No | NA | 5 | 0 | 2 | Low-quality | Genome-fragment | | 5.21 | AAI-based (medium-confidence) | 0 |
| S2_k141_228501 | 1231 | No | NA | 1 | 0 | 0 | Not-determined | Genome-fragment | NA | | NA | 0 |
| S7_k141_131493 | 4428 | No | NA | 5 | 0 | 4 | Low-quality | Genome-fragment | | 0.65 | AAI-based (medium-confidence) | 0 |
| S2_k141_93816 | 2141 | No | NA | 2 | 0 | 2 | Not-determined | Genome-fragment | NA | | NA | 0 |
| S2_k141_42458 | 2298 | No | NA | 3 | 0 | 2 | Not-determined | Genome-fragment | NA | | NA | 0 |
| S2_k141_10979 | 1386 | No | NA | 1 | 0 | 1 | Not-determined | Genome-fragment | NA | | NA | 0 |
| S2_k141_176790 | 1731 | No | NA | 3 | 0 | 2 | Not-determined | Genome-fragment | NA | | NA | 0 |
| S2_k141_42514 | 1357 | No | NA | 2 | 0 | 0 | Not-determined | Genome-fragment | NA | | NA | 0 |
| S2_k141_104287 | 2100 | No | NA | 1 | 0 | 1 | Not-determined | Genome-fragment | NA | | NA | 0 |
| S2_k141_42588 | 2223 | No | NA | 1 | 0 | 0 | Not-determined | Genome-fragment | NA | | NA | 0 |
| S2_k141_52374 | 1626 | No | NA | 1 | 0 | 1 | Not-determined | Genome-fragment | NA | | NA | 0 |
| S4_k141_139834 | 4428 | Yes | 1212 | 9 | 1 | 2 | Low-quality | Genome-fragment | | 0.49 | HMM-based (lower-bound) | 72.63 |
| S2_k141_52382 | 1274 | No | NA | 1 | 0 | 1 | Not-determined | Genome-fragment | NA | | NA | 0 |
| S17-final-viral-combined_k141_91138\|\|full | 4429 | No | NA | 3 | 0 | 3 | Low-quality | Genome-fragment | | 1.47 | AAI-based (medium-confidence) | 0 |
| S2_k141_114398 | 10830 | No | NA | 15 | 0 | 2 | Not-determined | Genome-fragment | NA | | NA | 0 |
| S2_k141_166408 | 1210 | No | NA | 1 | 0 | 1 | Not-determined | Genome-fragment | NA | | NA | 0 |
| S8-final-viral-combined_k141_225295\|\|full | 4429 | No | NA | 4 | 3 | 0 | Low-quality | Genome-fragment | | 9.83 | AAI-based (high-confidence) | 0 |
| S2_k141_52413 | 1072 | No | NA | 1 | 0 | 1 | Not-determined | Genome-fragment | NA | | NA | 0 |
| S2_k141_21558 | 1099 | No | NA | 1 | 0 | 1 | Not-determined | Genome-fragment | NA | | NA | 0 |
| S2_k141_114424 | 3207 | No | NA | 3 | 0 | 2 | Not-determined | Genome-fragment | NA | | NA | 0 |
| S2_k141_826 | 1250 | No | NA | 1 | 0 | 0 | Not-determined | Genome-fragment | NA | | NA | 0 |
| S2_k141_73239 | 1371 | No | NA | 2 | 0 | 0 | Not-determined | Genome-fragment | NA | | NA | 0 |
| S8_k141_122171 | 4429 | No | NA | 8 | 0 | 0 | Low-quality | Genome-fragment | | 10.53 | AAI-based (high-confidence) | 0 |
| S11-final-viral-combined_k141_216757\|\|full | 4430 | No | NA | 5 | 4 | 0 | Low-quality | Genome-fragment | | 12.06 | AAI-based (high-confidence) | 0 |
| S2_k141_109546 | 4431 | No | NA | 3 | 0 | 2 | Low-quality | Genome-fragment | | 6.1 | AAI-based (medium-confidence) | 0 |
| S2_k141_114493 | 1429 | No | NA | 3 | 0 | 0 | Not-determined | Genome-fragment | NA | | NA | 0 |
| S17-final-viral-combined_k141_163126\|\|full | 4433 | No | NA | 7 | 0 | 0 | Low-quality | Genome-fragment | | 10.4 | AAI-based (high-confidence) | 0 |
| S2_k141_166481 | 16861 | No | NA | 30 | 0 | 29 | Not-determined | Genome-fragment | NA | | NA | 0 |
| S2_k141_93858 | 3172 | No | NA | 1 | 0 | 1 | Not-determined | Genome-fragment | NA | | NA | 0 |
| S12_k141_375066 | 4433 | No | NA | 2 | 0 | 2 | Low-quality | Genome-fragment | | 1.94 | AAI-based (medium-confidence) | 0 |
| S17-k141_205323 | 4434 | No | NA | 3 | 0 | 3 | Low-quality | Genome-fragment | | 14.77 | AAI-based (medium-confidence) | 0 |
| S2_k141_62758 | 1452 | No | NA | 3 | 0 | 3 | Not-determined | Genome-fragment | NA | | NA | 0 |

| | | | | | | | | | | | |
|---|---|---|---|---|---|---|---|---|---|---|---|
| S2_k141_218213 | 1823 | No | NA | 1 | 0 | 1 | Not-determined | Genome-fragment | NA | NA | 0 |
| S2_k141_218214 | 2311 | No | NA | 1 | 0 | 1 | Not-determined | Genome-fragment | NA | NA | 0 |
| S2_k141_93890 | 1028 | No | NA | 0 | 0 | 0 | Not-determined | Genome-fragment | NA | NA | 0 |
| S2_k141_177018 | 1288 | No | NA | 2 | 0 | 0 | Not-determined | Genome-fragment | NA | NA | 0 |
| S2_k141_93893 | 5910 | No | NA | 2 | 0 | 1 | Not-determined | Genome-fragment | NA | NA | 0 |
| S2_k141_207892 | 1029 | No | NA | 1 | 0 | 1 | Not-determined | Genome-fragment | NA | NA | 0 |
| S2_k141_104390 | 1436 | No | NA | 3 | 0 | 3 | Not-determined | Genome-fragment | NA | NA | 0 |
| S4-final-viral-combined_k141_52861\|\|full | 4434 | No | NA | 5 | 4 | 0 | Low-quality | Genome-fragment | 9.04 | AAI-based (high-confidence) | 0 |
| S6_k141_98972 | 4435 | No | NA | 4 | 0 | 0 | Low-quality | Genome-fragment | 5.26 | AAI-based (high-confidence) | 0 |
| S2_k141_177054 | 1160 | No | NA | 1 | 0 | 1 | Not-determined | Genome-fragment | NA | NA | 0 |
| S1_k141_306314 | 4438 | No | NA | 3 | 0 | 2 | Low-quality | Genome-fragment | 9.56 | AAI-based (high-confidence) | 0 |
| S2_k141_207919 | 1447 | No | NA | 1 | 0 | 1 | Not-determined | Genome-fragment | NA | NA | 0 |
| S2_k141_73318 | 4240 | No | NA | 2 | 0 | 0 | Not-determined | Genome-fragment | NA | NA | 0 |
| S2_k141_93944 | 1114 | No | NA | 1 | 0 | 1 | Not-determined | Genome-fragment | NA | NA | 0 |
| S2_k141_228891 | 1675 | No | NA | 1 | 0 | 1 | Not-determined | Genome-fragment | NA | NA | 0 |
| S2_k141_135405 | 1467 | No | NA | 1 | 0 | 1 | Not-determined | Genome-fragment | NA | NA | 0 |
| S2_k141_177119 | 3470 | No | NA | 2 | 0 | 2 | Not-determined | Genome-fragment | NA | NA | 0 |
| S2_k141_207931 | 4553 | No | NA | 5 | 0 | 4 | Not-determined | Genome-fragment | NA | NA | 0 |
| S2_k141_218299 | 1222 | No | NA | 1 | 0 | 0 | Not-determined | Genome-fragment | NA | NA | 0 |
| S5_k141_205761 | 4438 | No | NA | 10 | 7 | 0 | Low-quality | Genome-fragment | 4.87 | AAI-based (high-confidence) | 0 |
| S2_k141_145943 | 1004 | No | NA | 2 | 0 | 0 | Not-determined | Genome-fragment | NA | NA | 0 |
| S2_k141_104475 | 1754 | No | NA | 2 | 0 | 2 | Not-determined | Genome-fragment | NA | NA | 0 |
| S2_k141_197306 | 2841 | No | NA | 6 | 0 | 6 | Not-determined | Genome-fragment | NA | NA | 0 |
| S2_k141_228936 | 6252 | No | NA | 8 | 0 | 1 | Not-determined | Genome-fragment | NA | NA | 0 |
| S2_k141_104487 | 4712 | No | NA | 5 | 0 | 0 | Not-determined | Genome-fragment | NA | NA | 0 |
| S2_k141_145993 | 1075 | No | NA | 2 | 0 | 2 | Not-determined | Genome-fragment | NA | NA | 0 |
| S2_k141_231668 | 4438 | No | NA | 11 | 1 | 0 | Low-quality | Genome-fragment | 7.94 | AAI-based (high-confidence) | 0 |
| S2_k141_146027 | 1060 | No | NA | 1 | 0 | 1 | Not-determined | Genome-fragment | NA | NA | 0 |
| S2_k141_166602 | 1249 | No | NA | 1 | 0 | 0 | Not-determined | Genome-fragment | NA | NA | 0 |
| S11_k141_130366 | 4440 | No | NA | 9 | 1 | 0 | Low-quality | Genome-fragment | 22.69 | AAI-based (medium-confidence) | 0 |
| S2_k141_32021 | 1001 | No | NA | 2 | 0 | 1 | Not-determined | Genome-fragment | NA | NA | 0 |
| S2_k141_32025 | 1180 | No | NA | 1 | 0 | 0 | Not-determined | Genome-fragment | NA | NA | 0 |
| S2_k141_52640 | 1091 | No | NA | 2 | 0 | 0 | Not-determined | Genome-fragment | NA | NA | 0 |
| S1-final-viral-combined_k141_68197\|\|full | 4440 | No | NA | 4 | 0 | 2 | Low-quality | Genome-fragment | 2.26 | AAI-based (medium-confidence) | 0 |
| S2_k141_104537 | 4300 | No | NA | 2 | 0 | 1 | Not-determined | Genome-fragment | NA | NA | 0 |
| S12_k141_87728 | 4440 | No | NA | 5 | 0 | 1 | Low-quality | Genome-fragment | 7.41 | AAI-based (medium-confidence) | 0 |
| S2_k141_21769 | 3967 | No | NA | 4 | 0 | 4 | Not-determined | Genome-fragment | NA | NA | 0 |
| S2_k141_166659 | 3936 | No | NA | 4 | 0 | 0 | Not-determined | Genome-fragment | NA | NA | 0 |
| S2_k141_208044 | 1474 | No | NA | 1 | 0 | 0 | Not-determined | Genome-fragment | NA | NA | 0 |
| S2_k141_83753 | 4541 | No | NA | 4 | 0 | 1 | Not-determined | Genome-fragment | NA | NA | 0 |
| S2_k141_104547 | 11248 | No | NA | 12 | 0 | 2 | Not-determined | Genome-fragment | NA | NA | 0 |
| S11_k141_134140 | 4442 | No | NA | 9 | 0 | 1 | Low-quality | Genome-fragment | 6.21 | AAI-based (medium-confidence) | 0 |
| S12-final-viral-combined_k141_379789\|\|full | 4442 | No | NA | 6 | 0 | 2 | Low-quality | Genome-fragment | 6.77 | AAI-based (medium-confidence) | 0 |
| S12_k141_290869 | 4442 | No | NA | 10 | 5 | 0 | Low-quality | Genome-fragment | 7.39 | AAI-based (high-confidence) | 0 |
| S1_k141_313132 | 4443 | No | NA | 3 | 0 | 2 | Low-quality | Genome-fragment | 0.38 | AAI-based (medium-confidence) | 0 |
| S2_k141_135529 | 3986 | No | NA | 3 | 0 | 3 | Not-determined | Genome-fragment | NA | NA | 0 |
| S5_k141_8717 | 4443 | No | NA | 8 | 2 | 0 | Low-quality | Genome-fragment | 4.98 | AAI-based (high-confidence) | 0 |
| S2_k141_166690 | 3163 | No | NA | 7 | 0 | 7 | Not-determined | Genome-fragment | NA | NA | 0 |
| S8-final-viral-combined_k141_161766\|\|full | 4443 | No | NA | 3 | 0 | 1 | Low-quality | Genome-fragment | 3.89 | AAI-based (medium-confidence) | 0 |
| S2_k141_125387 | 2450 | No | NA | 1 | 0 | 1 | Not-determined | Genome-fragment | NA | NA | 0 |
| S2_k141_94067 | 1338 | No | NA | 1 | 0 | 1 | Not-determined | Genome-fragment | NA | NA | 0 |
| S2_k141_197442 | 2663 | No | NA | 2 | 0 | 2 | Not-determined | Genome-fragment | NA | NA | 0 |
| S1-final-viral-combined_k141_22065\|\|full | 4444 | No | NA | 10 | 5 | 0 | Low-quality | Genome-fragment | 12.15 | AAI-based (high-confidence) | 0 |
| S2_k141_146128 | 1546 | No | NA | 2 | 0 | 0 | Not-determined | Genome-fragment | NA | NA | 0 |
| S2_k141_21832 | 7656 | No | NA | 6 | 0 | 2 | Not-determined | Genome-fragment | NA | NA | 0 |
| S2_k141_197462 | 1305 | No | NA | 2 | 0 | 0 | Not-determined | Genome-fragment | NA | NA | 0 |
| S2_k141_156439 | 31389 | Yes | 21599 | 37 | 10 | 8 | Medium-quality | Genome-fragment | 66.55 | AAI-based (high-confidence) | 31.19 |
| S2_k141_166756 | 2117 | No | NA | 1 | 0 | 1 | Not-determined | Genome-fragment | NA | NA | 0 |
| S2_k141_83959 | 1173 | No | NA | 1 | 0 | 1 | Not-determined | Genome-fragment | NA | NA | 0 |
| S1_k141_118841 | 4444 | No | NA | 10 | 2 | 0 | Low-quality | Genome-fragment | 12.98 | AAI-based (high-confidence) | 0 |
| S2_k141_135583 | 3607 | No | NA | 5 | 0 | 2 | Not-determined | Genome-fragment | NA | NA | 0 |
| S2_k141_1070 | 1854 | No | NA | 3 | 0 | 1 | Not-determined | Genome-fragment | NA | NA | 0 |
| S2_k141_11522 | 3257 | No | NA | 2 | 0 | 1 | Not-determined | Genome-fragment | NA | NA | 0 |
| S2_k141_166783 | 4429 | No | NA | 2 | 0 | 2 | Not-determined | Genome-fragment | NA | NA | 0 |
| S1-final-viral-combined_k141_130456\|\|full | 4445 | No | NA | 4 | 0 | 0 | Low-quality | Genome-fragment | 7.15 | AAI-based (medium-confidence) | 0 |
| S2_k141_1092 | 1163 | No | NA | 2 | 0 | 0 | Not-determined | Genome-fragment | NA | NA | 0 |
| S2_k141_83982 | 3943 | No | NA | 7 | 0 | 0 | Not-determined | Genome-fragment | NA | NA | 0 |
| S2_k141_21889 | 4783 | No | NA | 1 | 0 | 0 | Not-determined | Genome-fragment | NA | NA | 0 |
| S2_k141_114862 | 2840 | No | NA | 5 | 0 | 0 | Not-determined | Genome-fragment | NA | NA | 0 |
| S2_k141_52749 | 2129 | No | NA | 2 | 0 | 0 | Not-determined | Genome-fragment | NA | NA | 0 |
| S2_k141_229127 | 3015 | No | NA | 5 | 0 | 4 | Not-determined | Genome-fragment | NA | NA | 0 |
| S1_k141_333110 | 4446 | No | NA | 9 | 0 | 0 | Low-quality | Genome-fragment | 9.7 | AAI-based (high-confidence) | 0 |
| S2_k141_166836 | 1668 | No | NA | 3 | 0 | 0 | Not-determined | Genome-fragment | NA | NA | 0 |
| S2_k141_197525 | 1311 | No | NA | 1 | 0 | 1 | Not-determined | Genome-fragment | NA | NA | 0 |
| S2_k141_104702 | 1406 | No | NA | 2 | 0 | 0 | Not-determined | Genome-fragment | NA | NA | 0 |
| S2_k141_1130 | 1323 | No | NA | 1 | 0 | 1 | Not-determined | Genome-fragment | NA | NA | 0 |
| S12_k141_61771 | 4447 | No | NA | 4 | 0 | 4 | Low-quality | Genome-fragment | 3.2 | AAI-based (medium-confidence) | 0 |
| S2_k141_104735 | 1099 | No | NA | 2 | 0 | 0 | Not-determined | Genome-fragment | NA | NA | 0 |
| S8-final-viral-combined_k141_109759\|\|full | 4447 | No | NA | 8 | 2 | 0 | Low-quality | Genome-fragment | 7.73 | AAI-based (high-confidence) | 0 |
| S1-final-viral-combined_k141_811\|\|full | 4449 | No | NA | 4 | 0 | 0 | Low-quality | Genome-fragment | 15.63 | AAI-based (high-confidence) | 0 |
| S5-final-viral-combined_k141_155736\|\|full | 4450 | No | NA | 5 | 1 | 0 | Low-quality | Genome-fragment | 7.26 | HMM-based (lower-bound) | 0 |
| S2_k141_208213 | 1183 | No | NA | 1 | 0 | 0 | Not-determined | Genome-fragment | NA | NA | 0 |
| S2_k141_156504 | 1490 | No | NA | 3 | 0 | 1 | Not-determined | Genome-fragment | NA | NA | 0 |
| S17-final-viral-combined_k141_217594\|\|full | 4450 | No | NA | 6 | 0 | 2 | Low-quality | Genome-fragment | 4.18 | AAI-based (medium-confidence) | 0 |
| S2_k141_166900 | 1057 | No | NA | 2 | 0 | 1 | Not-determined | Genome-fragment | NA | NA | 0 |
| S8-final-viral-combined_k141_10480\|\|full | 4450 | No | NA | 3 | 1 | 0 | Low-quality | Genome-fragment | 6.6 | HMM-based (lower-bound) | 0 |
| S2_k141_43138 | 5163 | No | NA | 1 | 0 | 0 | Not-determined | Genome-fragment | NA | NA | 0 |
| S1-final-viral-combined_k141_333342\|\|full | 4451 | No | NA | 5 | 0 | 3 | Low-quality | Genome-fragment | 1.8 | AAI-based (medium-confidence) | 0 |
| S2_k141_21976 | 1121 | No | NA | 2 | 0 | 0 | Not-determined | Genome-fragment | NA | NA | 0 |
| S2_k141_239233 | 3995 | No | NA | 6 | 0 | 1 | Not-determined | Genome-fragment | NA | NA | 0 |
| S12_k141_325649 | 4451 | No | NA | 3 | 0 | 2 | Low-quality | Genome-fragment | 3.05 | AAI-based (medium-confidence) | 0 |
| S11-final-viral-combined_k141_138359\|\|full | 4452 | No | NA | 3 | 0 | 0 | Low-quality | Genome-fragment | 8.69 | AAI-based (high-confidence) | 0 |
| S1-final-viral-combined_k141_157175\|\|full | 4452 | No | NA | 6 | 0 | 1 | Low-quality | Genome-fragment | 3.45 | AAI-based (high-confidence) | 0 |
| S2_k141_187506 | 1532 | No | NA | 2 | 0 | 1 | Not-determined | Genome-fragment | NA | NA | 0 |
| S7-final-viral-combined_k141_215304\|\|full | 4452 | No | NA | 4 | 0 | 3 | Low-quality | Genome-fragment | 0.52 | AAI-based (medium-confidence) | 0 |
| S2_k141_239300 | 1892 | No | NA | 2 | 0 | 1 | Not-determined | Genome-fragment | NA | NA | 0 |
| S2_k141_1228 | 4133 | No | NA | 4 | 0 | 1 | Not-determined | Genome-fragment | NA | NA | 0 |
| S2_k141_197599 | 1007 | No | NA | 1 | 0 | 1 | Not-determined | Genome-fragment | NA | NA | 0 |
| S2_k141_52884 | 3847 | No | NA | 7 | 0 | 1 | Not-determined | Genome-fragment | NA | NA | 0 |
| S2_k141_208258 | 1180 | No | NA | 1 | 0 | 0 | Not-determined | Genome-fragment | NA | NA | 0 |
| S7-final-viral-combined_k141_94602\|\|full | 4452 | No | NA | 7 | 0 | 0 | Low-quality | Genome-fragment | 10.33 | AAI-based (high-confidence) | 0 |
| S2_k141_197619 | 4118 | No | NA | 4 | 0 | 3 | Not-determined | Genome-fragment | NA | NA | 0 |
| S5-final-viral-combined_k141_122653\|\|lt2gene | 4453 | No | NA | 2 | 1 | 0 | Low-quality | Genome-fragment | 3.08 | HMM-based (lower-bound) | 0 |
| S2_k141_146401 | 9684 | No | NA | 7 | 0 | 3 | Not-determined | Genome-fragment | NA | NA | 0 |
| S2_k141_197632 | 1096 | No | NA | 2 | 0 | 1 | Not-determined | Genome-fragment | NA | NA | 0 |
| S12-final-viral-combined_k141_65070\|\|full | 4453 | No | NA | 3 | 0 | 0 | Low-quality | Genome-fragment | 11.04 | AAI-based (high-confidence) | 0 |
| S2_k141_114927 | 2286 | No | NA | 2 | 0 | 0 | Not-determined | Genome-fragment | NA | NA | 0 |
| S2_k141_84191 | 1748 | No | NA | 2 | 0 | 1 | Not-determined | Genome-fragment | NA | NA | 0 |
| S2_k141_146409 | 2307 | No | NA | 2 | 0 | 2 | Not-determined | Genome-fragment | NA | NA | 0 |
| S11_k141_215549 | 4454 | No | NA | 11 | 1 | 0 | Low-quality | Genome-fragment | 8.77 | AAI-based (high-confidence) | 0 |
| S2_k141_43331 | 1103 | No | NA | 2 | 0 | 1 | Not-determined | Genome-fragment | NA | NA | 0 |
| S2_k141_146424 | 2405 | No | NA | 1 | 0 | 0 | Not-determined | Genome-fragment | NA | NA | 0 |
| S2_k141_197656 | 4189 | No | NA | 3 | 0 | 3 | Not-determined | Genome-fragment | NA | NA | 0 |
| S2_k141_239392 | 3696 | No | NA | 4 | 0 | 2 | Not-determined | Genome-fragment | NA | NA | 0 |
| S1_k141_51959 | 4454 | No | NA | 6 | 0 | 0 | Low-quality | Genome-fragment | 9.32 | AAI-based (high-confidence) | 0 |
| S2_k141_167159 | 4010 | No | NA | 4 | 0 | 2 | Not-determined | Genome-fragment | NA | NA | 0 |
| S12_k141_375613 | 4454 | No | NA | 11 | 0 | 0 | Low-quality | Genome-fragment | 7.83 | AAI-based (high-confidence) | 0 |
| S2_k141_187585 | 1008 | No | NA | 1 | 0 | 0 | Not-determined | Genome-fragment | NA | NA | 0 |
| S2_k141_177638 | 3826 | No | NA | 5 | 0 | 1 | Not-determined | Genome-fragment | NA | NA | 0 |
| S2_k141_1307 | 5032 | No | NA | 6 | 0 | 0 | Not-determined | Genome-fragment | NA | NA | 0 |
| S2_k141_1308 | 1128 | No | NA | 2 | 0 | 1 | Not-determined | Genome-fragment | NA | NA | 0 |
| S2_k141_135888 | 1064 | No | NA | 2 | 0 | 1 | Not-determined | Genome-fragment | NA | NA | 0 |
| S2_k141_197725 | 1030 | No | NA | 1 | 0 | 0 | Not-determined | Genome-fragment | NA | NA | 0 |
| S2_k141_52951 | 1339 | No | NA | 1 | 0 | 0 | Not-determined | Genome-fragment | NA | NA | 0 |
| S2_k141_177718 | 1248 | No | NA | 1 | 0 | 0 | Not-determined | Genome-fragment | NA | NA | 0 |
| S2_k141_208441 | 1393 | No | NA | 1 | 0 | 0 | Not-determined | Genome-fragment | NA | NA | 0 |
| S2_k141_22170 | 1022 | No | NA | 1 | 0 | 0 | Not-determined | Genome-fragment | NA | NA | 0 |
| S2_k141_218660 | 1240 | No | NA | 1 | 0 | 1 | Not-determined | Genome-fragment | NA | NA | 0 |
| S2_k141_208467 | 1815 | No | NA | 2 | 0 | 1 | Not-determined | Genome-fragment | NA | NA | 0 |
| S2_k141_84333 | 3150 | No | NA | 3 | 0 | 0 | Not-determined | Genome-fragment | NA | NA | 0 |
| S2_k141_115005 | 2234 | No | NA | 4 | 0 | 1 | Not-determined | Genome-fragment | NA | NA | 0 |
| S2_k141_63227 | 1492 | No | NA | 3 | 0 | 2 | Not-determined | Genome-fragment | NA | NA | 0 |
| S2_k141_115042 | 1507 | No | NA | 6 | 0 | 0 | Not-determined | Genome-fragment | NA | NA | 0 |
| S2_k141_22233 | 5132 | No | NA | 8 | 0 | 5 | Not-determined | Genome-fragment | NA | NA | 0 |
| S8-final-viral-combined_k141_253770\|\|full | 4454 | No | NA | 7 | 1 | 1 | Low-quality | Genome-fragment | 2.8 | HMM-based (lower-bound) | 0 |
| S2_k141_84370 | 1583 | No | NA | 2 | 0 | 1 | Not-determined | Genome-fragment | NA | NA | 0 |
| S2_k141_229399 | 1005 | No | NA | 1 | 0 | 0 | Not-determined | Genome-fragment | NA | NA | 0 |
| S2_k141_1355 | 1559 | No | NA | 2 | 0 | 0 | Not-determined | Genome-fragment | NA | NA | 0 |
| S12_k141_276613 | 4456 | No | NA | 8 | 2 | 0 | Low-quality | Genome-fragment | 10.16 | AAI-based (high-confidence) | 0 |
| S2_k141_22281 | 1094 | No | NA | 2 | 0 | 2 | Not-determined | Genome-fragment | NA | NA | 0 |
| S11-final-viral-combined_k141_30549\|\|full | 4458 | No | NA | 6 | 0 | 1 | Low-quality | Genome-fragment | 8.68 | AAI-based (medium-confidence) | 0 |
| S1_k141_131238 | 4458 | No | NA | 6 | 0 | 0 | Low-quality | Genome-fragment | 6.44 | AAI-based (medium-confidence) | 0 |
| S2_k141_115086 | 3957 | No | NA | 3 | 0 | 1 | Not-determined | Genome-fragment | NA | NA | 0 |
| S5-final-viral-combined_k141_18379\|\|full | 4458 | No | NA | 5 | 4 | 0 | Low-quality | Genome-fragment | 10.63 | AAI-based (high-confidence) | 0 |
| S2_k141_115113 | 1006 | No | NA | 1 | 0 | 1 | Not-determined | Genome-fragment | NA | NA | 0 |
| S11_k141_207067 | 4459 | No | NA | 5 | 1 | 0 | Low-quality | Genome-fragment | 4.79 | AAI-based (high-confidence) | 0 |
| S2_k141_167430 | 2917 | No | NA | 4 | 0 | 4 | Not-determined | Genome-fragment | NA | NA | 0 |
| S1-final-viral-combined_k141_282680\|\|full | 4459 | No | NA | 6 | 0 | 2 | Low-quality | Genome-fragment | 10.26 | AAI-based (high-confidence) | 0 |
| S1_k141_195191 | 4459 | No | NA | 4 | 0 | 1 | Low-quality | Genome-fragment | 11.24 | AAI-based (medium-confidence) | 0 |
| S2_k141_11867 | 1190 | No | NA | 1 | 0 | 1 | Not-determined | Genome-fragment | NA | NA | 0 |
| S2_k141_84478 | 3057 | No | NA | 3 | 0 | 1 | Not-determined | Genome-fragment | NA | NA | 0 |
| S7_k141_77080 | 4459 | No | NA | 1 | 0 | 0 | Low-quality | Genome-fragment | 4.57 | AAI-based (medium-confidence) | 0 |
| S12-final-viral-combined_k141_267688\|\|full | 4459 | No | NA | 7 | 0 | 2 | Low-quality | Genome-fragment | 3.09 | AAI-based (medium-confidence) | 0 |
| S4-final-viral-combined_k141_91219\|\|full | 4459 | No | NA | 6 | 6 | 0 | Low-quality | Genome-fragment | 12.74 | AAI-based (high-confidence) | 0 |
| S2_k141_136036 | 1049 | No | NA | 2 | 0 | 1 | Not-determined | Genome-fragment | NA | NA | 0 |
| S2_k141_115207 | 3012 | No | NA | 3 | 0 | 0 | Not-determined | Genome-fragment | NA | NA | 0 |
| S2_k141_32370 | 3458 | No | NA | 3 | 0 | 2 | Not-determined | Genome-fragment | NA | NA | 0 |
| S2_k141_84536 | 4713 | No | NA | 5 | 0 | 2 | Not-determined | Genome-fragment | NA | NA | 0 |
| S2_k141_94706 | 1031 | No | NA | 1 | 0 | 1 | Not-determined | Genome-fragment | NA | NA | 0 |
| S8-final-viral-combined_k141_194176\|\|full | 4459 | No | NA | 9 | 2 | 0 | Low-quality | Genome-fragment | 9.89 | AAI-based (high-confidence) | 0 |
| S2_k141_11979 | 4833 | No | NA | 6 | 0 | 0 | Not-determined | Genome-fragment | NA | NA | 0 |
| S3_k141_47826 | 4461 | Yes | 2756 | 5 | 1 | 1 | Low-quality | Genome-fragment | 1.27 | AAI-based (high-confidence) | 38.22 |
| S6-final-viral-combined_k141_117275\|\|full | 4461 | No | NA | 10 | 2 | 0 | Low-quality | Genome-fragment | 7.03 | AAI-based (high-confidence) | 0 |
| S6-final-viral-combined_k141_91761\|\|full | 4462 | No | NA | 3 | 0 | 3 | Low-quality | Genome-fragment | 6.03 | AAI-based (medium-confidence) | 0 |
| S2_k141_177874 | 1380 | No | NA | 3 | 0 | 2 | Not-determined | Genome-fragment | NA | NA | 0 |
| S2_k141_156767 | 7449 | No | NA | 11 | 0 | 2 | Not-determined | Genome-fragment | NA | NA | 0 |
| S2_k141_63447 | 1032 | No | NA | 1 | 0 | 0 | Not-determined | Genome-fragment | NA | NA | 0 |
| S2_k141_1488 | 1430 | No | NA | 1 | 0 | 1 | Not-determined | Genome-fragment | NA | NA | 0 |
| S8-final-viral-combined_k141_184372\|\|full | 4463 | No | NA | 4 | 1 | 2 | Low-quality | Genome-fragment | 3.58 | HMM-based (lower-bound) | 0 |
| S2_k141_197915 | 3356 | No | NA | 3 | 0 | 1 | Not-determined | Genome-fragment | NA | NA | 0 |
| S2_k141_125776 | 2291 | No | NA | 2 | 0 | 1 | Not-determined | Genome-fragment | NA | NA | 0 |
| S11-final-viral-combined_k141_279565\|\|full | 4464 | No | NA | 3 | 3 | 0 | Low-quality | Genome-fragment | 9.83 | AAI-based (high-confidence) | 0 |
| S3-final-viral-combined_k141_9970\|\|full | 4464 | No | NA | 4 | 0 | 2 | Low-quality | Genome-fragment | 3.09 | AAI-based (medium-confidence) | 0 |
| S2_k141_136132 | 3117 | No | NA | 6 | 0 | 1 | Not-determined | Genome-fragment | NA | NA | 0 |
| S2_k141_187937 | 1485 | No | NA | 2 | 0 | 2 | Not-determined | Genome-fragment | NA | NA | 0 |
| S2_k141_43807 | 3253 | No | NA | 3 | 0 | 2 | Not-determined | Genome-fragment | NA | NA | 0 |
| S2_k141_177961 | 1145 | No | NA | 1 | 0 | 1 | Not-determined | Genome-fragment | NA | NA | 0 |
| S5-final-viral-combined_k141_175421\|\|full | 4464 | No | NA | 3 | 3 | 0 | Low-quality | Genome-fragment | 6.06 | AAI-based (high-confidence) | 0 |
| S2_k141_12051 | 1649 | No | NA | 1 | 0 | 1 | Not-determined | Genome-fragment | NA | NA | 0 |
| S12-final-viral-combined_k141_236942\|\|full | 4464 | No | NA | 4 | 0 | 1 | Low-quality | Genome-fragment | 3.16 | AAI-based (medium-confidence) | 0 |
| S1-final-viral-combined_k141_72127\|\|full | 4466 | No | NA | 6 | 0 | 0 | Low-quality | Genome-fragment | 7.46 | AAI-based (high-confidence) | 0 |
| S2-final-viral-combined_k141_224603\|\|full | 4467 | Yes | 2710 | 8 | 3 | 2 | Low-quality | Genome-fragment | 3.73 | AAI-based (high-confidence) | 39.33 |
| S2_k141_178012 | 2799 | No | NA | 1 | 0 | 0 | Not-determined | Genome-fragment | NA | NA | 0 |
| S2_k141_229574 | 1024 | No | NA | 1 | 0 | 1 | Not-determined | Genome-fragment | NA | NA | 0 |
| S2_k141_98889 | 4467 | No | NA | 5 | 0 | 2 | Low-quality | Genome-fragment | 7.39 | AAI-based (medium-confidence) | 0 |
| S2_k141_12085 | 1391 | No | NA | 2 | 0 | 2 | Not-determined | Genome-fragment | NA | NA | 0 |
| S2_k141_43907 | 1223 | No | NA | 1 | 0 | 1 | Not-determined | Genome-fragment | NA | NA | 0 |
| S2_k141_188021 | 2105 | No | NA | 4 | 0 | 3 | Not-determined | Genome-fragment | NA | NA | 0 |
| S12_k141_363677 | 4468 | No | NA | 2 | 0 | 1 | Low-quality | Genome-fragment | 2.53 | AAI-based (medium-confidence) | 0 |
| S9-final-viral-combined_k141_58105\|\|full | 4471 | No | NA | 6 | 0 | 3 | Low-quality | Genome-fragment | 2.47 | AAI-based (high-confidence) | 0 |
| S2_k141_218949 | 1985 | No | NA | 2 | 0 | 0 | Not-determined | Genome-fragment | NA | NA | 0 |
| S2_k141_218950 | 1553 | No | NA | 1 | 0 | 1 | Not-determined | Genome-fragment | NA | NA | 0 |
| S1_k141_114231 | 4471 | No | NA | 4 | 0 | 3 | Low-quality | Genome-fragment | 0.89 | AAI-based (medium-confidence) | 0 |
| S2_k141_22509 | 1006 | No | NA | 2 | 0 | 2 | Not-determined | Genome-fragment | NA | NA | 0 |
| S2_k141_105172 | 1049 | No | NA | 2 | 0 | 1 | Not-determined | Genome-fragment | NA | NA | 0 |
| S2_k141_53287 | 1500 | No | NA | 2 | 0 | 2 | Not-determined | Genome-fragment | NA | NA | 0 |
| S5-final-viral-combined_k141_29568\|\|lt2gene | 4471 | No | NA | 2 | 2 | 0 | Low-quality | Genome-fragment | 3.1 | HMM-based (lower-bound) | 0 |
| S2_k141_156933 | 1774 | No | NA | 1 | 0 | 0 | Not-determined | Genome-fragment | NA | NA | 0 |
| S2_k141_94914 | 1018 | No | NA | 1 | 0 | 1 | Not-determined | Genome-fragment | NA | NA | 0 |
| S2_k141_22550 | 6786 | No | NA | 2 | 0 | 1 | Not-determined | Genome-fragment | NA | NA | 0 |
| S2_k141_105232 | 1483 | No | NA | 1 | 0 | 1 | Not-determined | Genome-fragment | NA | NA | 0 |
| S2_k141_44030 | 1230 | No | NA | 1 | 0 | 1 | Not-determined | Genome-fragment | NA | NA | 0 |
| S17-k141_216380 | 4471 | No | NA | 7 | 1 | 0 | Low-quality | Genome-fragment | 28.76 | AAI-based (high-confidence) | 0 |
| S2_k141_136341 | 6647 | No | NA | 6 | 0 | 5 | Not-determined | Genome-fragment | NA | NA | 0 |
| S2_k141_219001 | 4388 | No | NA | 4 | 0 | 4 | Not-determined | Genome-fragment | NA | NA | 0 |
| S2_k141_53393 | 1020 | No | NA | 1 | 0 | 0 | Not-determined | Genome-fragment | NA | NA | 0 |
| S2_k141_178121 | 1276 | No | NA | 2 | 0 | 0 | Not-determined | Genome-fragment | NA | NA | 0 |
| S2_k141_156958 | 1013 | No | NA | 1 | 0 | 1 | Not-determined | Genome-fragment | NA | NA | 0 |
| S2_k141_22617 | 1196 | No | NA | 1 | 0 | 1 | Not-determined | Genome-fragment | NA | NA | 0 |
| S2_k141_63717 | 1228 | No | NA | 1 | 0 | 0 | Not-determined | Genome-fragment | NA | NA | 0 |
| S1_k141_258471 | 4472 | No | NA | 2 | 0 | 2 | Low-quality | Genome-fragment | 2.72 | AAI-based (medium-confidence) | 0 |
| S2_k141_178186 | 1111 | No | NA | 1 | 0 | 1 | Not-determined | Genome-fragment | NA | NA | 0 |
| S2_k141_239815 | 3308 | No | NA | 4 | 0 | 1 | Not-determined | Genome-fragment | NA | NA | 0 |
| S2_k141_157021 | 1283 | No | NA | 1 | 0 | 1 | Not-determined | Genome-fragment | NA | NA | 0 |
| S2_k141_239819 | 1347 | No | NA | 2 | 0 | 1 | Not-determined | Genome-fragment | NA | NA | 0 |
| S2-final-viral-combined_k141_172316\|\|full | 4472 | No | NA | 5 | 0 | 2 | Low-quality | Genome-fragment | 5.47 | AAI-based (medium-confidence) | 0 |
| S2_k141_178227 | 1576 | No | NA | 3 | 0 | 1 | Not-determined | Genome-fragment | NA | NA | 0 |
| S8-final-viral-combined_k141_18598\|\|full | 4473 | No | NA | 3 | 2 | 0 | Low-quality | Genome-fragment | 8.14 | AAI-based (high-confidence) | 0 |
| S2_k141_115536 | 1101 | No | NA | 3 | 0 | 0 | Not-determined | Genome-fragment | NA | NA | 0 |
| S1-final-viral-combined_k141_685\|\|full | 4474 | No | NA | 5 | 2 | 0 | Low-quality | Genome-fragment | 7.22 | AAI-based (high-confidence) | 0 |
| S2_k141_32646 | 1330 | No | NA | 1 | 0 | 0 | Not-determined | Genome-fragment | NA | NA | 0 |
| S7-final-viral-combined_k141_17395\|\|full | 4474 | No | NA | 5 | 0 | 2 | Low-quality | Genome-fragment | 8.76 | AAI-based (medium-confidence) | 0 |
| S8-final-viral-combined_k141_19340\|\|full | 4474 | No | NA | 3 | 2 | 0 | Low-quality | Genome-fragment | 6.54 | HMM-based (lower-bound) | 0 |
| S8-final-viral-combined_k141_275009\|\|full | 4475 | No | NA | 5 | 0 | 1 | Low-quality | Genome-fragment | 10.32 | AAI-based (medium-confidence) | 0 |
| S2_k141_1870 | 1008 | No | NA | 1 | 0 | 1 | Not-determined | Genome-fragment | NA | NA | 0 |
| S17-final-viral-combined_k141_118471\|\|full | 4476 | No | NA | 8 | 0 | 0 | Low-quality | Genome-fragment | 7.91 | AAI-based (medium-confidence) | 0 |
| S7_k141_86899 | 4477 | No | NA | 3 | 0 | 2 | Low-quality | Genome-fragment | 1.18 | AAI-based (high-confidence) | 0 |
| S2_k141_136479 | 1092 | No | NA | 1 | 0 | 1 | Not-determined | Genome-fragment | NA | NA | 0 |
| S2_k141_229829 | 1110 | No | NA | 1 | 0 | 1 | Not-determined | Genome-fragment | NA | NA | 0 |
| S2_k141_1877 | 1129 | No | NA | 1 | 0 | 1 | Not-determined | Genome-fragment | NA | NA | 0 |
| S12-final-viral-combined_k141_60625\|\|full | 4477 | No | NA | 10 | 0 | 0 | Low-quality | Genome-fragment | 8.79 | AAI-based (high-confidence) | 0 |
| S2_k141_219064 | 2093 | No | NA | 3 | 0 | 2 | Not-determined | Genome-fragment | NA | NA | 0 |
| S2_k141_32672 | 1376 | No | NA | 1 | 0 | 0 | Not-determined | Genome-fragment | NA | NA | 0 |
| S2_k141_198120 | 5702 | No | NA | 2 | 0 | 0 | Not-determined | Genome-fragment | NA | NA | 0 |
| S2_k141_136527 | 1496 | No | NA | 2 | 0 | 0 | Not-determined | Genome-fragment | NA | NA | 0 |
| S2_k141_126093 | 1554 | No | NA | 3 | 0 | 0 | Not-determined | Genome-fragment | NA | NA | 0 |
| S2_k141_74217 | 3056 | No | NA | 3 | 0 | 1 | Not-determined | Genome-fragment | NA | NA | 0 |
| S2_k141_198154 | 1467 | No | NA | 2 | 0 | 1 | Not-determined | Genome-fragment | NA | NA | 0 |
| S2_k141_126107 | 1288 | No | NA | 1 | 0 | 1 | Not-determined | Genome-fragment | NA | NA | 0 |
| S2_k141_105487 | 4099 | No | NA | 1 | 0 | 1 | Not-determined | Genome-fragment | NA | NA | 0 |
| S2_k141_84775 | 1668 | No | NA | 3 | 0 | 0 | Not-determined | Genome-fragment | NA | NA | 0 |
| S2_k141_126126 | 1155 | No | NA | 3 | 0 | 0 | Not-determined | Genome-fragment | NA | NA | 0 |
| S4_k141_55952 | 4477 | No | NA | 6 | 0 | 6 | Low-quality | Genome-fragment | 1.66 | AAI-based (medium-confidence) | 0 |
| S2_k141_208885 | 6701 | No | NA | 6 | 0 | 4 | Not-determined | Genome-fragment | NA | NA | 0 |
| S5-final-viral-combined_k141_103856\|\|full | 4478 | No | NA | 9 | 3 | 0 | Low-quality | Genome-fragment | 6 | HMM-based (lower-bound) | 0 |
| S17-final-viral-combined_k141_17599\|\|full | 4480 | No | NA | 4 | 0 | 3 | Low-quality | Genome-fragment | 11.7 | AAI-based (medium-confidence) | 0 |
| S2-final-viral-combined_k141_234420\|\|full | 4480 | Yes | 1440 | 6 | 1 | 1 | Low-quality | Genome-fragment | 1.53 | AAI-based (medium-confidence) | 67.86 |
| S2_k141_84862 | 1634 | No | NA | 3 | 0 | 0 | Not-determined | Genome-fragment | NA | NA | 0 |
| S2_k141_53636 | 3731 | No | NA | 4 | 0 | 4 | Not-determined | Genome-fragment | NA | NA | 0 |
| S2_k141_178404 | 1746 | No | NA | 1 | 0 | 1 | Not-determined | Genome-fragment | NA | NA | 0 |
| S2_k141_22754 | 7915 | No | NA | 10 | 0 | 2 | Not-determined | Genome-fragment | NA | NA | 0 |
| S2_k141_229942 | 1317 | No | NA | 2 | 0 | 0 | Not-determined | Genome-fragment | NA | NA | 0 |
| S2_k141_105588 | 10398 | No | NA | 12 | 0 | 2 | Not-determined | Genome-fragment | NA | NA | 0 |
| S2_k141_157150 | 1817 | No | NA | 1 | 0 | 1 | Not-determined | Genome-fragment | NA | NA | 0 |
| S2_k141_74334 | 4350 | No | NA | 3 | 0 | 2 | Not-determined | Genome-fragment | NA | NA | 0 |
| S2-final-viral-combined_k141_93139\|\|full | 4481 | No | NA | 4 | 0 | 1 | Low-quality | Genome-fragment | 9.86 | AAI-based (medium-confidence) | 0 |
| S9-final-viral-combined_k141_34360\|\|full | 4482 | Yes | 1266 | 7 | 4 | 3 | Low-quality | Genome-fragment | 2.21 | AAI-based (high-confidence) | 71.75 |
| S2_k141_229978 | 1410 | No | NA | 2 | 0 | 0 | Not-determined | Genome-fragment | NA | NA | 0 |
| S2_k141_157166 | 8018 | No | NA | 9 | 0 | 6 | Not-determined | Genome-fragment | NA | NA | 0 |
| S2_k141_53717 | 1413 | No | NA | 1 | 0 | 1 | Not-determined | Genome-fragment | NA | NA | 0 |
| S2_k141_74346 | 1122 | No | NA | 1 | 0 | 0 | Not-determined | Genome-fragment | NA | NA | 0 |
| S2_k141_74351 | 3850 | No | NA | 3 | 0 | 2 | Not-determined | Genome-fragment | NA | NA | 0 |
| S2_k141_44380 | 5534 | No | NA | 6 | 0 | 3 | Not-determined | Genome-fragment | NA | NA | 0 |
| S2_k141_105634 | 6237 | No | NA | 7 | 0 | 2 | Not-determined | Genome-fragment | NA | NA | 0 |
| S2_k141_84949 | 5015 | No | NA | 7 | 0 | 4 | Not-determined | Genome-fragment | NA | NA | 0 |
| S2_k141_22794 | 7266 | No | NA | 6 | 0 | 3 | Not-determined | Genome-fragment | NA | NA | 0 |
| S2_k141_115749 | 1342 | No | NA | 2 | 0 | 1 | Not-determined | Genome-fragment | NA | NA | 0 |
| S2_k141_157232 | 1529 | No | NA | 2 | 0 | 1 | Not-determined | Genome-fragment | NA | NA | 0 |
| S2_k141_95179 | 3205 | No | NA | 1 | 0 | 1 | Not-determined | Genome-fragment | NA | NA | 0 |
| S12_k141_351889 | 4482 | Yes | 2253 | 7 | 3 | 2 | Low-quality | Genome-fragment | 4.75 | AAI-based (high-confidence) | 49.73 |
| S2_k141_157246 | 1234 | No | NA | 2 | 0 | 1 | Not-determined | Genome-fragment | NA | NA | 0 |
| S4_k141_3733 | 4483 | No | NA | 10 | 3 | 0 | Low-quality | Genome-fragment | 12.85 | AAI-based (high-confidence) | 0 |
| S8-final-viral-combined_k141_238061\|\|full | 4483 | No | NA | 4 | 0 | 4 | Low-quality | Genome-fragment | 2.82 | AAI-based (medium-confidence) | 0 |
| S2_k141_32757 | 3424 | No | NA | 3 | 0 | 2 | Not-determined | Genome-fragment | NA | NA | 0 |
| S2_k141_53792 | 3024 | No | NA | 3 | 0 | 0 | Not-determined | Genome-fragment | NA | NA | 0 |
| S2_k141_105683 | 1170 | No | NA | 1 | 0 | 1 | Not-determined | Genome-fragment | NA | NA | 0 |
| S2_k141_74392 | 4414 | No | NA | 6 | 0 | 0 | Not-determined | Genome-fragment | NA | NA | 0 |
| S6-final-viral-combined_k141_76684\|\|full | 4484 | No | NA | 4 | 0 | 1 | Low-quality | Genome-fragment | 3.24 | AAI-based (high-confidence) | 0 |
| S2_k141_198281 | 4313 | No | NA | 4 | 0 | 2 | Not-determined | Genome-fragment | NA | NA | 0 |
| S2_k141_219186 | 2758 | No | NA | 3 | 0 | 1 | Not-determined | Genome-fragment | NA | NA | 0 |
| S2_k141_219201 | 1135 | No | NA | 1 | 0 | 0 | Not-determined | Genome-fragment | NA | NA | 0 |
| S8_k141_64886 | 4484 | No | NA | 3 | 0 | 1 | Low-quality | Genome-fragment | 2.08 | AAI-based (high-confidence) | 0 |
| S2_k141_53816 | 3114 | No | NA | 2 | 0 | 1 | Not-determined | Genome-fragment | NA | NA | 0 |
| S9-final-viral-combined_k141_5808\|\|full | 4485 | No | NA | 4 | 0 | 1 | Low-quality | Genome-fragment | 3.5 | AAI-based (high-confidence) | 0 |
| S2_k141_22856 | 1542 | No | NA | 3 | 0 | 0 | Not-determined | Genome-fragment | NA | NA | 0 |
| S2_k141_105739 | 1035 | No | NA | 2 | 0 | 0 | Not-determined | Genome-fragment | NA | NA | 0 |
| S2_k141_22860 | 1266 | No | NA | 2 | 0 | 0 | Not-determined | Genome-fragment | NA | NA | 0 |
| S1_k141_35170 | 4485 | No | NA | 5 | 0 | 0 | Low-quality | Genome-fragment | 9.09 | AAI-based (medium-confidence) | 0 |
| S12-final-viral-combined_k141_238463\|\|full | 4485 | No | NA | 5 | 0 | 3 | Low-quality | Genome-fragment | 1.32 | AAI-based (medium-confidence) | 0 |
| S2_k141_126279 | 4153 | No | NA | 3 | 0 | 3 | Not-determined | Genome-fragment | NA | NA | 0 |
| S12-final-viral-combined_k141_369530\|\|full | 4486 | No | NA | 6 | 0 | 4 | Low-quality | Genome-fragment | 2.06 | AAI-based (medium-confidence) | 0 |
| S2_k141_167998 | 1387 | No | NA | 2 | 0 | 1 | Not-determined | Genome-fragment | NA | NA | 0 |
| S2_k141_230137 | 1605 | No | NA | 2 | 0 | 0 | Not-determined | Genome-fragment | NA | NA | 0 |
| S2_k141_230144 | 1087 | No | NA | 2 | 0 | 1 | Not-determined | Genome-fragment | NA | NA | 0 |
| S6-final-viral-combined_k141_20737\|\|full | 4486 | No | NA | 3 | 0 | 2 | Low-quality | Genome-fragment | 2.77 | AAI-based (medium-confidence) | 0 |
| S11_k141_313153 | 4487 | No | NA | 4 | 0 | 0 | Low-quality | Genome-fragment | 4.77 | AAI-based (high-confidence) | 0 |
| S2_k141_209072 | 1090 | No | NA | 1 | 0 | 1 | Not-determined | Genome-fragment | NA | NA | 0 |
| S2_k141_53881 | 16535 | No | NA | 20 | 7 | 0 | Medium-quality | Genome-fragment | 71.23 | AAI-based (medium-confidence) | 0 |
| S2_k141_115850 | 8069 | No | NA | 3 | 0 | 1 | Not-determined | Genome-fragment | NA | NA | 0 |
| S2_k141_136895 | 1219 | No | NA | 2 | 0 | 1 | Not-determined | Genome-fragment | NA | NA | 0 |
| S2_k141_85123 | 5442 | No | NA | 6 | 0 | 6 | Not-determined | Genome-fragment | NA | NA | 0 |
| S2_k141_95421 | 3156 | No | NA | 7 | 0 | 3 | Not-determined | Genome-fragment | NA | NA | 0 |
| S2_k141_240157 | 7074 | No | NA | 4 | 0 | 1 | Not-determined | Genome-fragment | NA | NA | 0 |
| S8-final-viral-combined_k141_22198\|\|full | 4487 | No | NA | 3 | 0 | 3 | Low-quality | Genome-fragment | 2.09 | AAI-based (medium-confidence) | 0 |
| S8_k141_207684 | 4488 | No | NA | 10 | 2 | 0 | Low-quality | Genome-fragment | 7.27 | AAI-based (high-confidence) | 0 |
| S2_k141_64223 | 2251 | No | NA | 3 | 0 | 2 | Not-determined | Genome-fragment | NA | NA | 0 |
| S2_k141_240169 | 3945 | No | NA | 6 | 0 | 1 | Not-determined | Genome-fragment | NA | NA | 0 |
| S2_k141_95459 | 2952 | No | NA | 1 | 0 | 0 | Not-determined | Genome-fragment | NA | NA | 0 |
| S2_k141_64246 | 1259 | No | NA | 1 | 0 | 0 | Not-determined | Genome-fragment | NA | NA | 0 |
| S17-final-viral-combined_k141_94678\|\|full | 4489 | No | NA | 5 | 0 | 1 | Low-quality | Genome-fragment | 10.55 | AAI-based (medium-confidence) | 0 |
| S12-final-viral-combined_k141_80526\|\|full | 4489 | No | NA | 4 | 0 | 1 | Low-quality | Genome-fragment | 1.35 | AAI-based (medium-confidence) | 0 |
| S2_k141_219313 | 3886 | No | NA | 3 | 0 | 2 | Not-determined | Genome-fragment | NA | NA | 0 |
| S17-final-viral-combined_k141_216449\|\|full | 4490 | No | NA | 4 | 0 | 3 | Low-quality | Genome-fragment | 3.4 | AAI-based (medium-confidence) | 0 |
| S2_k141_95496 | 5671 | No | NA | 9 | 0 | 1 | Not-determined | Genome-fragment | NA | NA | 0 |
| S2_k141_219321 | 11342 | No | NA | 7 | 0 | 2 | Not-determined | Genome-fragment | NA | NA | 0 |
| S2_k141_219323 | 1235 | No | NA | 1 | 0 | 1 | Not-determined | Genome-fragment | NA | NA | 0 |
| S2_k141_74465 | 10075 | No | NA | 13 | 0 | 0 | Not-determined | Genome-fragment | NA | NA | 0 |
| S2_k141_32878 | 2466 | No | NA | 4 | 0 | 2 | Not-determined | Genome-fragment | NA | NA | 0 |
| S2_k141_188540 | 4171 | No | NA | 5 | 0 | 1 | Not-determined | Genome-fragment | NA | NA | 0 |
| S2_k141_2121 | 4841 | No | NA | 2 | 0 | 1 | Not-determined | Genome-fragment | NA | NA | 0 |
| S2_k141_32930 | 1135 | No | NA | 1 | 0 | 0 | Not-determined | Genome-fragment | NA | NA | 0 |
| S2_k141_105887 | 1959 | No | NA | 1 | 0 | 0 | Not-determined | Genome-fragment | NA | NA | 0 |
| S2_k141_2137 | 1634 | No | NA | 3 | 0 | 0 | Not-determined | Genome-fragment | NA | NA | 0 |
| S2_k141_147062 | 1060 | No | NA | 1 | 0 | 0 | Not-determined | Genome-fragment | NA | NA | 0 |
| S2_k141_2155 | 1091 | No | NA | 2 | 0 | 0 | Not-determined | Genome-fragment | NA | NA | 0 |
| S6-final-viral-combined_k141_127219\|\|full | 4490 | No | NA | 4 | 0 | 4 | Low-quality | Genome-fragment | 7.44 | AAI-based (medium-confidence) | 0 |
| S2_k141_85244 | 1249 | No | NA | 1 | 0 | 1 | Not-determined | Genome-fragment | NA | NA | 0 |
| S2_k141_105956 | 2021 | No | NA | 2 | 0 | 0 | Not-determined | Genome-fragment | NA | NA | 0 |
| S7-final-viral-combined_k141_79412\|\|full | 4491 | No | NA | 2 | 0 | 0 | Low-quality | Genome-fragment | 3.9 | AAI-based (medium-confidence) | 0 |
| S2-final-viral-combined_k141_225108\|\|full | 4491 | No | NA | 11 | 2 | 0 | Low-quality | Genome-fragment | 10.87 | AAI-based (medium-confidence) | 0 |
| S2-final-viral-combined_k141_217286\|\|full | 4491 | No | NA | 3 | 0 | 2 | Low-quality | Genome-fragment | 5.01 | AAI-based (medium-confidence) | 0 |
| S8-final-viral-combined_k141_107374\|\|full | 4491 | Yes | 3233 | 4 | 1 | 1 | Low-quality | Genome-fragment | 5.8 | AAI-based (high-confidence) | 28.01 |
| S2_k141_44734 | 1013 | No | NA | 1 | 0 | 1 | Not-determined | Genome-fragment | NA | NA | 0 |
| S2_k141_209111 | 3215 | No | NA | 4 | 0 | 3 | Not-determined | Genome-fragment | NA | NA | 0 |
| S2_k141_54076 | 1465 | No | NA | 2 | 0 | 1 | Not-determined | Genome-fragment | NA | NA | 0 |
| S2_k141_44781 | 4837 | No | NA | 7 | 0 | 1 | Not-determined | Genome-fragment | NA | NA | 0 |
| S2_k141_85286 | 1649 | No | NA | 2 | 0 | 0 | Not-determined | Genome-fragment | NA | NA | 0 |
| S12_k141_86998 | 4492 | No | NA | 3 | 0 | 1 | Low-quality | Genome-fragment | 0.39 | AAI-based (medium-confidence) | 0 |
| S7-final-viral-combined_k141_216381\|\|full | 4494 | No | NA | 5 | 0 | 3 | Low-quality | Genome-fragment | 4.35 | AAI-based (medium-confidence) | 0 |
| S2_k141_240294 | 1104 | No | NA | 1 | 0 | 1 | Not-determined | Genome-fragment | NA | NA | 0 |
| S2_k141_198457 | 3049 | No | NA | 4 | 0 | 0 | Not-determined | Genome-fragment | NA | NA | 0 |
| S2_k141_12544 | 5599 | No | NA | 6 | 0 | 0 | Not-determined | Genome-fragment | NA | NA | 0 |
| S2_k141_135165 | 4494 | No | NA | 4 | 0 | 1 | Low-quality | Genome-fragment | 9.05 | AAI-based (high-confidence) | 0 |
| S2_k141_12549 | 1592 | No | NA | 1 | 0 | 0 | Not-determined | Genome-fragment | NA | NA | 0 |
| S2_k141_116055 | 1776 | No | NA | 1 | 0 | 1 | Not-determined | Genome-fragment | NA | NA | 0 |
| S7-final-viral-combined_k141_121194\|\|full | 4495 | No | NA | 10 | 5 | 0 | Low-quality | Genome-fragment | 10.63 | AAI-based (medium-confidence) | 0 |
| S2_k141_12562 | 1000 | No | NA | 1 | 0 | 1 | Not-determined | Genome-fragment | NA | NA | 0 |
| S2_k141_2235 | 1057 | No | NA | 1 | 0 | 0 | Not-determined | Genome-fragment | NA | NA | 0 |
| S2_k141_188716 | 1118 | No | NA | 3 | 0 | 2 | Not-determined | Genome-fragment | NA | NA | 0 |
| S2_k141_44853 | 1354 | No | NA | 1 | 0 | 1 | Not-determined | Genome-fragment | NA | NA | 0 |
| S2_k141_230529 | 1068 | No | NA | 4 | 0 | 1 | Not-determined | Genome-fragment | NA | NA | 0 |
| S2_k141_95659 | 2492 | No | NA | 5 | 0 | 2 | Not-determined | Genome-fragment | NA | NA | 0 |
| S2_k141_209208 | 1195 | No | NA | 1 | 0 | 1 | Not-determined | Genome-fragment | NA | NA | 0 |
| S2_k141_12577 | 1289 | No | NA | 2 | 0 | 2 | Not-determined | Genome-fragment | NA | NA | 0 |
| S2_k141_188764 | 1283 | No | NA | 1 | 0 | 0 | Not-determined | Genome-fragment | NA | NA | 0 |
| S2_k141_116074 | 4219 | No | NA | 3 | 0 | 1 | Not-determined | Genome-fragment | NA | NA | 0 |
| S2_k141_12600 | 1681 | No | NA | 1 | 0 | 0 | Not-determined | Genome-fragment | NA | NA | 0 |
| S2_k141_12606 | 3123 | No | NA | 1 | 0 | 0 | Not-determined | Genome-fragment | NA | NA | 0 |
| S2_k141_219526 | 1253 | No | NA | 1 | 0 | 1 | Not-determined | Genome-fragment | NA | NA | 0 |
| S2_k141_137160 | 1115 | No | NA | 1 | 0 | 0 | Not-determined | Genome-fragment | NA | NA | 0 |
| S2_k141_198554 | 2070 | No | NA | 3 | 0 | 1 | Not-determined | Genome-fragment | NA | NA | 0 |
| S2_k141_74755 | 1786 | No | NA | 2 | 0 | 2 | Not-determined | Genome-fragment | NA | NA | 0 |
| S2_k141_33141 | 1102 | No | NA | 1 | 0 | 1 | Not-determined | Genome-fragment | NA | NA | 0 |
| S2_k141_54257 | 1116 | No | NA | 1 | 0 | 1 | Not-determined | Genome-fragment | NA | NA | 0 |
| S2_k141_95810 | 12163 | No | NA | 13 | 0 | 2 | Not-determined | Genome-fragment | NA | NA | 0 |
| S2_k141_230583 | 1049 | No | NA | 3 | 0 | 0 | Not-determined | Genome-fragment | NA | NA | 0 |
| S2_k141_2334 | 1498 | No | NA | 2 | 0 | 2 | Not-determined | Genome-fragment | NA | NA | 0 |
| S1_k141_199157 | 4498 | No | NA | 6 | 0 | 2 | Low-quality | Genome-fragment | 7.19 | AAI-based (medium-confidence) | 0 |
| S2_k141_74794 | 4700 | No | NA | 6 | 0 | 4 | Not-determined | Genome-fragment | NA | NA | 0 |
| S2_k141_70463 | 4498 | No | NA | 4 | 0 | 3 | Low-quality | Genome-fragment | 2.19 | AAI-based (medium-confidence) | 0 |
| S2_k141_168445 | 1768 | No | NA | 3 | 0 | 0 | Not-determined | Genome-fragment | NA | NA | 0 |
| S2_k141_240373 | 4787 | No | NA | 2 | 0 | 0 | Not-determined | Genome-fragment | NA | NA | 0 |
| S2_k141_33177 | 1142 | No | NA | 2 | 0 | 2 | Not-determined | Genome-fragment | NA | NA | 0 |
| S2_k141_230612 | 1264 | No | NA | 1 | 0 | 0 | Not-determined | Genome-fragment | NA | NA | 0 |
| S2_k141_116102 | 1108 | No | NA | 2 | 0 | 0 | Not-determined | Genome-fragment | NA | NA | 0 |
| S2_k141_188829 | 1041 | No | NA | 1 | 0 | 1 | Not-determined | Genome-fragment | NA | NA | 0 |
| S5_k141_55010 | 4499 | No | NA | 7 | 4 | 0 | Low-quality | Genome-fragment | 10.36 | AAI-based (high-confidence) | 0 |
| S2_k141_188841 | 1339 | No | NA | 1 | 0 | 0 | Not-determined | Genome-fragment | NA | NA | 0 |
| S2-final-viral-combined_k141_179459\|\|full | 4499 | No | NA | 3 | 0 | 3 | Low-quality | Genome-fragment | 5.83 | AAI-based (medium-confidence) | 0 |
| S2_k141_106043 | 1714 | No | NA | 3 | 0 | 1 | Not-determined | Genome-fragment | NA | NA | 0 |
| S2_k141_45003 | 1394 | No | NA | 2 | 0 | 2 | Not-determined | Genome-fragment | NA | NA | 0 |
| S2_k141_230650 | 3444 | No | NA | 2 | 0 | 1 | Not-determined | Genome-fragment | NA | NA | 0 |
| S17-final-viral-combined_k141_33348\|\|full | 4501 | Yes | 3558 | 6 | 1 | 1 | Low-quality | Genome-fragment | 8.09 | AAI-based (medium-confidence) | 20.95 |
| S8-final-viral-combined_k141_25129\|\|full | 4502 | No | NA | 5 | 0 | 1 | Low-quality | Genome-fragment | 4.57 | AAI-based (medium-confidence) | 0 |
| S2_k141_240397 | 4174 | No | NA | 2 | 0 | 1 | Not-determined | Genome-fragment | NA | NA | 0 |
| S2_k141_116158 | 1167 | No | NA | 1 | 0 | 1 | Not-determined | Genome-fragment | NA | NA | 0 |
| S2_k141_188889 | 1010 | No | NA | 1 | 0 | 0 | Not-determined | Genome-fragment | NA | NA | 0 |
| S2_k141_23238 | 1692 | No | NA | 1 | 0 | 1 | Not-determined | Genome-fragment | NA | NA | 0 |
| S11_k141_95920 | 4503 | No | NA | 7 | 0 | 1 | Low-quality | Genome-fragment | 5.54 | AAI-based (medium-confidence) | 0 |
| S2_k141_240441 | 1314 | No | NA | 1 | 0 | 1 | Not-determined | Genome-fragment | NA | NA | 0 |
| S2_k141_219709 | 1043 | No | NA | 1 | 0 | 0 | Not-determined | Genome-fragment | NA | NA | 0 |
| S1-final-viral-combined_k141_376463\|\|full | 4503 | Yes | 1020 | 5 | 1 | 3 | Low-quality | Genome-fragment | 19.32 | AAI-based (medium-confidence) | 77.35 |
| S2_k141_64423 | 1372 | No | NA | 1 | 0 | 1 | Not-determined | Genome-fragment | NA | NA | 0 |
| S2_k141_74839 | 9555 | No | NA | 10 | 0 | 0 | Not-determined | Genome-fragment | NA | NA | 0 |
| S2_k141_137241 | 1153 | No | NA | 2 | 0 | 2 | Not-determined | Genome-fragment | NA | NA | 0 |
| S2_k141_240474 | 1047 | No | NA | 2 | 0 | 1 | Not-determined | Genome-fragment | NA | NA | 0 |
| S3-final-viral-combined_k141_68189\|\|full | 4503 | No | NA | 4 | 3 | 0 | Low-quality | Genome-fragment | 11.99 | AAI-based (high-confidence) | 0 |
| S7-final-viral-combined_k141_20751\|\|full | 4503 | No | NA | 5 | 0 | 5 | Low-quality | Genome-fragment | 1.86 | AAI-based (medium-confidence) | 0 |
| S2_k141_157899 | 1782 | No | NA | 2 | 0 | 1 | Not-determined | Genome-fragment | NA | NA | 0 |
| S2_k141_2393 | 2441 | No | NA | 1 | 0 | 0 | Not-determined | Genome-fragment | NA | NA | 0 |
| S2_k141_126740 | 1148 | No | NA | 2 | 0 | 0 | Not-determined | Genome-fragment | NA | NA | 0 |
| S2_k141_45122 | 1151 | No | NA | 2 | 0 | 0 | Not-determined | Genome-fragment | NA | NA | 0 |
| S9-final-viral-combined_k141_85217\|\|full | 4506 | No | NA | 5 | 4 | 0 | Low-quality | Genome-fragment | 10.59 | AAI-based (high-confidence) | 0 |
| S2_k141_219783 | 1288 | No | NA | 1 | 0 | 0 | Not-determined | Genome-fragment | NA | NA | 0 |
| S2_k141_198703 | 1740 | No | NA | 4 | 0 | 3 | Not-determined | Genome-fragment | NA | NA | 0 |
| S2_k141_116225 | 3268 | No | NA | 2 | 0 | 2 | Not-determined | Genome-fragment | NA | NA | 0 |
| S4_k141_57680 | 4506 | No | NA | 8 | 1 | 0 | Low-quality | Genome-fragment | 7.8 | AAI-based (medium-confidence) | 0 |
| S2_k141_95923 | 1316 | No | NA | 1 | 0 | 0 | Not-determined | Genome-fragment | NA | NA | 0 |
| S11_k141_206481 | 4507 | No | NA | 6 | 4 | 0 | Low-quality | Genome-fragment | 9.08 | AAI-based (high-confidence) | 0 |
| S3-final-viral-combined_k141_70798\|\|full | 4510 | No | NA | 5 | 0 | 0 | Low-quality | Genome-fragment | 9.07 | AAI-based (high-confidence) | 0 |
| S2_k141_33423 | 1006 | No | NA | 2 | 0 | 0 | Not-determined | Genome-fragment | NA | NA | 0 |
| S2_k141_2464 | 1703 | No | NA | 2 | 0 | 1 | Not-determined | Genome-fragment | NA | NA | 0 |
| S2_k141_147388 | 1019 | No | NA | 2 | 0 | 0 | Not-determined | Genome-fragment | NA | NA | 0 |

| | | | | | | | | | | | |
|---|---|---|---|---|---|---|---|---|---|---|---|
| S12-final-viral-combined_k141_80920\|\|full | 4510 | No | NA | 13 | 2 | 0 | Low-quality | Genome-fragment | 9.53 | AAI-based (medium-confidence) | 0 |
| S2_k141_64580 | 4882 | No | NA | 4 | 0 | 1 | Not-determined | Genome-fragment | NA | NA | 0 |
| S2_k141_178980 | 5306 | No | NA | 3 | 0 | 2 | Not-determined | Genome-fragment | NA | NA | 0 |
| S8_k141_197004 | 4510 | No | NA | 4 | 0 | 3 | Low-quality | Genome-fragment | 1.16 | AAI-based (high-confidence) | 0 |
| S2-final-viral-combined_k141_213017\|\|full | 4511 | No | NA | 4 | 0 | 1 | Low-quality | Genome-fragment | 7.58 | AAI-based (medium-confidence) | 0 |
| S2-final-viral-combined_k141_158036\|\|full | 4512 | No | NA | 9 | 2 | 0 | Low-quality | Genome-fragment | 9.2 | AAI-based (medium-confidence) | 0 |
| S2_k141_96039 | 1135 | No | NA | 2 | 0 | 0 | Not-determined | Genome-fragment | NA | NA | 0 |
| S8-final-viral-combined_k141_108584\|\|full | 4512 | No | NA | 11 | 2 | 0 | Low-quality | Genome-fragment | 8.51 | AAI-based (high-confidence) | 0 |
| S2_k141_209571 | 3127 | No | NA | 3 | 0 | 2 | Not-determined | Genome-fragment | NA | NA | 0 |
| S17-k141_260032 | 4513 | No | NA | 2 | 0 | 2 | Low-quality | Genome-fragment | 1.97 | AAI-based (medium-confidence) | 0 |
| S2-final-viral-combined_k141_12401\|\|full | 4513 | No | NA | 6 | 1 | 0 | Low-quality | Genome-fragment | 6.83 | HMM-based (lower-bound) | 0 |
| S4_k141_34632 | 4513 | No | NA | 2 | 1 | 0 | Low-quality | Genome-fragment | 4.76 | AAI-based (high-confidence) | 0 |
| S2-final-viral-combined_k141_144542\|\|full | 4514 | No | NA | 5 | 1 | 0 | Low-quality | Genome-fragment | 5.97 | HMM-based (lower-bound) | 0 |
| S2_k141_116324 | 1393 | No | NA | 2 | 0 | 0 | Not-determined | Genome-fragment | NA | NA | 0 |
| S2_k141_12980 | 9478 | No | NA | 7 | 0 | 1 | Not-determined | Genome-fragment | NA | NA | 0 |
| S2_k141_230882 | 2325 | No | NA | 2 | 0 | 2 | Not-determined | Genome-fragment | NA | NA | 0 |
| S2_k141_126930 | 1021 | No | NA | 2 | 0 | 1 | Not-determined | Genome-fragment | NA | NA | 0 |
| S12_k141_1424 | 4515 | No | NA | 5 | 0 | 1 | Low-quality | Genome-fragment | 6.77 | AAI-based (medium-confidence) | 0 |
| S2_k141_137479 | 1894 | No | NA | 5 | 0 | 5 | Not-determined | Genome-fragment | NA | NA | 0 |
| S8-final-viral-combined_k141_23543\|\|full | 4518 | No | NA | 8 | 2 | 0 | Low-quality | Genome-fragment | 10.07 | AAI-based (high-confidence) | 0 |
| S2_k141_33585 | 1359 | No | NA | 2 | 0 | 1 | Not-determined | Genome-fragment | NA | NA | 0 |
| S11-final-viral-combined_k141_263038\|\|full | 4519 | Yes | 2188 | 7 | 1 | 1 | Low-quality | Genome-fragment | 3.64 | AAI-based (medium-confidence) | 51.58 |
| S2_k141_168779 | 1183 | No | NA | 2 | 0 | 0 | Not-determined | Genome-fragment | NA | NA | 0 |
| S2_k141_126959 | 1320 | No | NA | 1 | 0 | 1 | Not-determined | Genome-fragment | NA | NA | 0 |
| S17-final-viral-combined_k141_62415\|\|full | 4519 | No | NA | 5 | 0 | 2 | Low-quality | Genome-fragment | 5.42 | AAI-based (medium-confidence) | 0 |
| S2_k141_137528 | 1473 | No | NA | 2 | 0 | 1 | Not-determined | Genome-fragment | NA | NA | 0 |
| S2_k141_168805 | 1210 | No | NA | 2 | 0 | 1 | Not-determined | Genome-fragment | NA | NA | 0 |
| S2_k141_13043 | 1902 | No | NA | 2 | 0 | 1 | Not-determined | Genome-fragment | NA | NA | 0 |
| S2_k141_127036 | 6356 | No | NA | 6 | 0 | 4 | Not-determined | Genome-fragment | NA | NA | 0 |
| S2_k141_75120 | 1328 | No | NA | 2 | 0 | 2 | Not-determined | Genome-fragment | NA | NA | 0 |
| S2_k141_147611 | 1540 | No | NA | 3 | 0 | 1 | Not-determined | Genome-fragment | NA | NA | 0 |
| S2_k141_54727 | 4657 | No | NA | 4 | 0 | 1 | Not-determined | Genome-fragment | NA | NA | 0 |
| S2_k141_189349 | 1062 | No | NA | 2 | 0 | 0 | Not-determined | Genome-fragment | NA | NA | 0 |
| S1-final-viral-combined_k141_301172\|\|full | 4520 | Yes | 1260 | 8 | 1 | 4 | Low-quality | Genome-fragment | 1.08 | HMM-based (lower-bound) | 72.12 |
| S2_k141_85906 | 3695 | No | NA | 3 | 0 | 2 | Not-determined | Genome-fragment | NA | NA | 0 |
| S2_k141_23562 | 13618 | No | NA | 17 | 3 | 0 | High-quality | High-quality | 99.98 | AAI-based (medium-confidence) | 0 |
| S2_k141_116395 | 1125 | No | NA | 2 | 0 | 0 | Not-determined | Genome-fragment | NA | NA | 0 |
| S2_k141_106366 | 4209 | No | NA | 6 | 0 | 1 | Not-determined | Genome-fragment | NA | NA | 0 |
| S2_k141_45489 | 2487 | No | NA | 3 | 0 | 1 | Not-determined | Genome-fragment | NA | NA | 0 |
| S1-final-viral-combined_k141_123208\|\|full | 4520 | No | NA | 5 | 0 | 0 | Low-quality | Genome-fragment | 2.83 | AAI-based (medium-confidence) | 0 |
| S2_k141_106375 | 1296 | No | NA | 3 | 0 | 2 | Not-determined | Genome-fragment | NA | NA | 0 |
| S2_k141_168955 | 7203 | No | NA | 4 | 0 | 4 | Not-determined | Genome-fragment | NA | NA | 0 |
| S7-final-viral-combined_k141_134848\|\|full | 4520 | No | NA | 5 | 0 | 4 | Low-quality | Genome-fragment | 8.99 | AAI-based (medium-confidence) | 0 |
| S2_k141_2694 | 1091 | No | NA | 2 | 0 | 2 | Not-determined | Genome-fragment | NA | NA | 0 |
| S2_k141_127099 | 1389 | No | NA | 3 | 0 | 2 | Not-determined | Genome-fragment | NA | NA | 0 |
| S2_k141_54836 | 1296 | No | NA | 2 | 0 | 2 | Not-determined | Genome-fragment | NA | NA | 0 |
| S2_k141_209794 | 1696 | No | NA | 1 | 0 | 0 | Not-determined | Genome-fragment | NA | NA | 0 |
| S9-final-viral-combined_k141_41747\|\|full | 4521 | No | NA | 4 | 0 | 3 | Low-quality | Genome-fragment | 2.28 | AAI-based (medium-confidence) | 0 |
| S2_k141_116440 | 1399 | No | NA | 2 | 0 | 1 | Not-determined | Genome-fragment | NA | NA | 0 |
| S2_k141_96417 | 1147 | No | NA | 2 | 0 | 2 | Not-determined | Genome-fragment | NA | NA | 0 |
| S2_k141_23670 | 1302 | No | NA | 2 | 0 | 0 | Not-determined | Genome-fragment | NA | NA | 0 |
| S2_k141_158175 | 1242 | No | NA | 2 | 0 | 0 | Not-determined | Genome-fragment | NA | NA | 0 |
| S8-final-viral-combined_k141_45035\|\|full | 4521 | No | NA | 12 | 2 | 0 | Low-quality | Genome-fragment | 7.84 | AAI-based (high-confidence) | 0 |
| S17-final-viral-combined_k141_62306\|\|full | 4522 | No | NA | 6 | 1 | 2 | Low-quality | Genome-fragment | 2.22 | HMM-based (lower-bound) | 0 |
| S2_k141_147742 | 1199 | No | NA | 2 | 0 | 0 | Not-determined | Genome-fragment | NA | NA | 0 |
| S2_k141_13206 | 1136 | No | NA | 2 | 0 | 0 | Not-determined | Genome-fragment | NA | NA | 0 |
| S2_k141_158199 | 3939 | No | NA | 2 | 0 | 2 | Not-determined | Genome-fragment | NA | NA | 0 |
| S2_k141_147751 | 3420 | No | NA | 1 | 0 | 1 | Not-determined | Genome-fragment | NA | NA | 0 |
| S2_k141_116454 | 3069 | No | NA | 2 | 0 | 2 | Not-determined | Genome-fragment | NA | NA | 0 |
| S4_k141_56276 | 4522 | No | NA | 8 | 1 | 2 | Low-quality | Genome-fragment | 4.12 | HMM-based (lower-bound) | 0 |
| S4-final-viral-combined_k141_23135\|\|full | 4523 | No | NA | 5 | 0 | 3 | Low-quality | Genome-fragment | 5.83 | AAI-based (medium-confidence) | 0 |
| S2_k141_127248 | 1401 | No | NA | 1 | 0 | 1 | Not-determined | Genome-fragment | NA | NA | 0 |
| S2_k141_169009 | 2734 | No | NA | 2 | 0 | 2 | Not-determined | Genome-fragment | NA | NA | 0 |
| S2_k141_33968 | 1436 | No | NA | 2 | 0 | 1 | Not-determined | Genome-fragment | NA | NA | 0 |
| S7-final-viral-combined_k141_127901\|\|full | 4524 | No | NA | 6 | 0 | 0 | Low-quality | Genome-fragment | 7.67 | AAI-based (medium-confidence) | 0 |
| S2_k141_179412 | 1255 | No | NA | 4 | 0 | 0 | Not-determined | Genome-fragment | NA | NA | 0 |
| S2_k141_209879 | 1035 | No | NA | 2 | 0 | 2 | Not-determined | Genome-fragment | NA | NA | 0 |
| S2_k141_147788 | 1535 | No | NA | 2 | 0 | 0 | Not-determined | Genome-fragment | NA | NA | 0 |
| S2_k141_179422 | 1193 | No | NA | 1 | 0 | 0 | Not-determined | Genome-fragment | NA | NA | 0 |
| S2_k141_231058 | 1168 | No | NA | 1 | 0 | 1 | Not-determined | Genome-fragment | NA | NA | 0 |
| S2_k141_158249 | 7187 | No | NA | 7 | 0 | 4 | Not-determined | Genome-fragment | NA | NA | 0 |
| S17-k141_229526 | 4525 | No | NA | 5 | 0 | 0 | Low-quality | Genome-fragment | 5.46 | AAI-based (medium-confidence) | 0 |
| S2_k141_169052 | 1338 | No | NA | 2 | 0 | 2 | Not-determined | Genome-fragment | NA | NA | 0 |
| S5-final-viral-combined_k141_16866\|\|full | 4526 | No | NA | 8 | 4 | 0 | Low-quality | Genome-fragment | 9.94 | AAI-based (high-confidence) | 0 |
| S2_k141_64855 | 3291 | No | NA | 4 | 0 | 2 | Not-determined | Genome-fragment | NA | NA | 0 |
| S2_k141_231080 | 1237 | No | NA | 2 | 0 | 1 | Not-determined | Genome-fragment | NA | NA | 0 |
| S2_k141_137781 | 1479 | No | NA | 4 | 0 | 0 | Not-determined | Genome-fragment | NA | NA | 0 |
| S2_k141_45707 | 2280 | No | NA | 3 | 0 | 2 | Not-determined | Genome-fragment | NA | NA | 0 |
| S2_k141_96533 | 1095 | No | NA | 2 | 0 | 1 | Not-determined | Genome-fragment | NA | NA | 0 |
| S2_k141_96543 | 1148 | No | NA | 1 | 0 | 0 | Not-determined | Genome-fragment | NA | NA | 0 |
| S2_k141_199050 | 1534 | No | NA | 1 | 0 | 1 | Not-determined | Genome-fragment | NA | NA | 0 |
| S2-final-viral-combined_k141_236107\|\|full | 4526 | No | NA | 4 | 0 | 3 | Low-quality | Genome-fragment | 2.59 | AAI-based (medium-confidence) | 0 |
| S2_k141_96547 | 3181 | No | NA | 1 | 0 | 1 | Not-determined | Genome-fragment | NA | NA | 0 |
| S2_k141_64900 | 1048 | No | NA | 1 | 0 | 1 | Not-determined | Genome-fragment | NA | NA | 0 |
| S17-final-viral-combined_k141_188626\|\|full | 4527 | No | NA | 10 | 8 | 0 | Low-quality | Genome-fragment | 10.68 | AAI-based (high-confidence) | 0 |
| S2_k141_96550 | 4387 | No | NA | 1 | 0 | 1 | Not-determined | Genome-fragment | NA | NA | 0 |
| S2_k141_2848 | 1642 | No | NA | 3 | 0 | 1 | Not-determined | Genome-fragment | NA | NA | 0 |
| S2_k141_240907 | 1595 | No | NA | 2 | 0 | 0 | Not-determined | Genome-fragment | NA | NA | 0 |
| S2_k141_189676 | 1118 | No | NA | 1 | 0 | 1 | Not-determined | Genome-fragment | NA | NA | 0 |
| S2_k141_106618 | 1162 | No | NA | 1 | 0 | 1 | Not-determined | Genome-fragment | NA | NA | 0 |
| S2_k141_219959 | 1678 | No | NA | 3 | 0 | 0 | Not-determined | Genome-fragment | NA | NA | 0 |
| S2_k141_106636 | 3279 | No | NA | 3 | 0 | 1 | Not-determined | Genome-fragment | NA | NA | 0 |
| S2_k141_127399 | 1448 | No | NA | 2 | 0 | 1 | Not-determined | Genome-fragment | NA | NA | 0 |
| S2_k141_169236 | 1532 | No | NA | 2 | 0 | 0 | Not-determined | Genome-fragment | NA | NA | 0 |
| S8-final-viral-combined_k141_113701\|\|full | 4527 | No | NA | 4 | 1 | 0 | Low-quality | Genome-fragment | 2.94 | AAI-based (high-confidence) | 0 |
| S2_k141_231146 | 2389 | No | NA | 2 | 0 | 1 | Not-determined | Genome-fragment | NA | NA | 0 |
| S2_k141_220005 | 2722 | No | NA | 2 | 0 | 1 | Not-determined | Genome-fragment | NA | NA | 0 |
| S2_k141_34113 | 1046 | No | NA | 2 | 0 | 2 | Not-determined | Genome-fragment | NA | NA | 0 |
| S2_k141_106669 | 1246 | No | NA | 1 | 0 | 1 | Not-determined | Genome-fragment | NA | NA | 0 |
| S2_k141_96579 | 1331 | No | NA | 2 | 0 | 1 | Not-determined | Genome-fragment | NA | NA | 0 |
| S7-final-viral-combined_k141_219253\|\|full | 4529 | Yes | 1343 | 9 | 1 | 3 | Low-quality | Genome-fragment | 2.59 | AAI-based (high-confidence) | 70.35 |
| S2_k141_65029 | 1082 | No | NA | 2 | 0 | 1 | Not-determined | Genome-fragment | NA | NA | 0 |
| S2_k141_2958 | 1050 | No | NA | 1 | 0 | 0 | Not-determined | Genome-fragment | NA | NA | 0 |
| S2_k141_169326 | 1014 | No | NA | 1 | 0 | 1 | Not-determined | Genome-fragment | NA | NA | 0 |
| S2_k141_147903 | 2448 | No | NA | 1 | 0 | 1 | Not-determined | Genome-fragment | NA | NA | 0 |
| S5_k141_7896 | 4531 | No | NA | 7 | 6 | 0 | Low-quality | Genome-fragment | 11.26 | AAI-based (high-confidence) | 0 |
| S2_k141_231232 | 2641 | No | NA | 4 | 0 | 2 | Not-determined | Genome-fragment | NA | NA | 0 |
| S2_k141_2984 | 10803 | No | NA | 6 | 0 | 2 | Not-determined | Genome-fragment | NA | NA | 0 |
| S2_k141_86227 | 2587 | No | NA | 2 | 0 | 0 | Not-determined | Genome-fragment | NA | NA | 0 |
| S2_k141_34159 | 1125 | No | NA | 1 | 0 | 0 | Not-determined | Genome-fragment | NA | NA | 0 |
| S2_k141_169385 | 3466 | No | NA | 5 | 0 | 1 | Not-determined | Genome-fragment | NA | NA | 0 |
| S2_k141_54980 | 4663 | No | NA | 3 | 0 | 2 | Not-determined | Genome-fragment | NA | NA | 0 |
| S2_k141_210092 | 1583 | No | NA | 1 | 0 | 1 | Not-determined | Genome-fragment | NA | NA | 0 |
| S2_k141_106724 | 1113 | No | NA | 2 | 0 | 1 | Not-determined | Genome-fragment | NA | NA | 0 |
| S2_k141_45786 | 13151 | No | NA | 14 | 0 | 10 | Not-determined | Genome-fragment | NA | NA | 0 |
| S2_k141_127552 | 3215 | No | NA | 11 | 0 | 0 | Not-determined | Genome-fragment | NA | NA | 0 |
| S2_k141_148066 | 1308 | No | NA | 2 | 0 | 0 | Not-determined | Genome-fragment | NA | NA | 0 |
| S2_k141_169424 | 2396 | No | NA | 6 | 0 | 1 | Not-determined | Genome-fragment | NA | NA | 0 |
| S2_k141_231331 | 2643 | No | NA | 2 | 0 | 0 | Not-determined | Genome-fragment | NA | NA | 0 |
| S4-final-viral-combined_k141_147780\|\|full | 4531 | Yes | 4237 | 9 | 1 | 1 | Low-quality | Genome-fragment | 1.72 | HMM-based (lower-bound) | 6.49 |
| S2_k141_106794 | 1567 | No | NA | 1 | 0 | 1 | Not-determined | Genome-fragment | NA | NA | 0 |
| S2_k141_86296 | 1564 | No | NA | 6 | 0 | 0 | Not-determined | Genome-fragment | NA | NA | 0 |
| S2_k141_199229 | 1038 | No | NA | 1 | 0 | 1 | Not-determined | Genome-fragment | NA | NA | 0 |
| S2_k141_179732 | 2288 | No | NA | 3 | 0 | 0 | Not-determined | Genome-fragment | NA | NA | 0 |
| S2_k141_9508 | 4532 | No | NA | 1 | 0 | 1 | Low-quality | Genome-fragment | 4.72 | AAI-based (high-confidence) | 0 |
| S2_k141_179739 | 2486 | No | NA | 2 | 0 | 0 | Not-determined | Genome-fragment | NA | NA | 0 |
| S17-final-viral-combined_k141_20424\|\|full | 4533 | No | NA | 7 | 0 | 2 | Low-quality | Genome-fragment | 5.66 | AAI-based (high-confidence) | 0 |
| S2_k141_158458 | 1428 | No | NA | 1 | 0 | 1 | Not-determined | Genome-fragment | NA | NA | 0 |
| S1-final-viral-combined_k141_351604\|\|full | 4534 | No | NA | 4 | 2 | 0 | Low-quality | Genome-fragment | 8.09 | AAI-based (high-confidence) | 0 |
| S2_k141_210244 | 1186 | No | NA | 4 | 0 | 3 | Not-determined | Genome-fragment | NA | NA | 0 |
| S2_k141_106852 | 4051 | No | NA | 3 | 0 | 3 | Not-determined | Genome-fragment | NA | NA | 0 |
| S1_k141_317224 | 4534 | No | NA | 4 | 0 | 1 | Low-quality | Genome-fragment | 7.3 | AAI-based (medium-confidence) | 0 |
| S2_k141_23900 | 4425 | No | NA | 3 | 0 | 1 | Not-determined | Genome-fragment | NA | NA | 0 |
| S2_k141_45997 | 1155 | No | NA | 1 | 0 | 1 | Not-determined | Genome-fragment | NA | NA | 0 |
| S2_k141_45998 | 1063 | No | NA | 1 | 0 | 1 | Not-determined | Genome-fragment | NA | NA | 0 |
| S2_k141_189911 | 1221 | No | NA | 2 | 0 | 1 | Not-determined | Genome-fragment | NA | NA | 0 |
| S11-final-viral-combined_k141_151072\|\|full | 4535 | No | NA | 7 | 5 | 0 | Low-quality | Genome-fragment | 5.79 | HMM-based (lower-bound) | 0 |
| S2_k141_138176 | 3631 | No | NA | 4 | 0 | 3 | Not-determined | Genome-fragment | NA | NA | 0 |
| S2_k141_3192 | 3688 | No | NA | 3 | 0 | 2 | Not-determined | Genome-fragment | NA | NA | 0 |
| S2_k141_46031 | 1535 | No | NA | 2 | 0 | 0 | Not-determined | Genome-fragment | NA | NA | 0 |
| S2_k141_169569 | 1465 | No | NA | 1 | 0 | 1 | Not-determined | Genome-fragment | NA | NA | 0 |
| S9_k141_64765 | 4536 | No | NA | 10 | 1 | 0 | Low-quality | Genome-fragment | 12.86 | AAI-based (high-confidence) | 0 |
| S2_k141_106917 | 1312 | No | NA | 3 | 0 | 2 | Not-determined | Genome-fragment | NA | NA | 0 |
| S2_k141_34419 | 1138 | No | NA | 1 | 0 | 0 | Not-determined | Genome-fragment | NA | NA | 0 |
| S2_k141_96753 | 1551 | No | NA | 2 | 0 | 1 | Not-determined | Genome-fragment | NA | NA | 0 |
| S2_k141_46066 | 6984 | No | NA | 5 | 0 | 3 | Not-determined | Genome-fragment | NA | NA | 0 |
| S3-final-viral-combined_k141_69823\|\|full | 4536 | No | NA | 3 | 0 | 2 | Low-quality | Genome-fragment | 1.59 | AAI-based (medium-confidence) | 0 |
| S2_k141_210372 | 1751 | No | NA | 1 | 0 | 1 | Not-determined | Genome-fragment | NA | NA | 0 |
| S2_k141_241147 | 1030 | No | NA | 2 | 0 | 1 | Not-determined | Genome-fragment | NA | NA | 0 |
| S4-final-viral-combined_k141_99390\|\|full | 4536 | No | NA | 8 | 2 | 0 | Low-quality | Genome-fragment | 3.76 | HMM-based (lower-bound) | 0 |
| S2_k141_231556 | 1019 | No | NA | 2 | 0 | 2 | Not-determined | Genome-fragment | NA | NA | 0 |
| S2_k141_220027 | 1099 | No | NA | 1 | 0 | 1 | Not-determined | Genome-fragment | NA | NA | 0 |
| S2_k141_231564 | 1179 | No | NA | 1 | 0 | 1 | Not-determined | Genome-fragment | NA | NA | 0 |
| S2_k141_46111 | 1048 | No | NA | 1 | 0 | 1 | Not-determined | Genome-fragment | NA | NA | 0 |
| S2_k141_158588 | 1084 | No | NA | 2 | 0 | 0 | Not-determined | Genome-fragment | NA | NA | 0 |
| S2_k141_158592 | 2598 | No | NA | 5 | 0 | 2 | Not-determined | Genome-fragment | NA | NA | 0 |
| S2_k141_117022 | 2896 | No | NA | 4 | 0 | 2 | Not-determined | Genome-fragment | NA | NA | 0 |
| S11-final-viral-combined_k141_85831\|\|full | 4538 | No | NA | 3 | 0 | 3 | Low-quality | Genome-fragment | 6.5 | AAI-based (medium-confidence) | 0 |
| S2_k141_75566 | 1484 | No | NA | 2 | 0 | 1 | Not-determined | Genome-fragment | NA | NA | 0 |
| S11-final-viral-combined_k141_4872\|\|full | 4538 | No | NA | 5 | 0 | 0 | Low-quality | Genome-fragment | 11.15 | AAI-based (medium-confidence) | 0 |
| S2_k141_117060 | 1141 | No | NA | 1 | 0 | 0 | Not-determined | Genome-fragment | NA | NA | 0 |
| S2_k141_179883 | 4482 | No | NA | 6 | 0 | 5 | Not-determined | Genome-fragment | NA | NA | 0 |
| S2_k141_65487 | 1005 | No | NA | 2 | 0 | 0 | Not-determined | Genome-fragment | NA | NA | 0 |
| S2_k141_220076 | 1503 | No | NA | 2 | 0 | 2 | Not-determined | Genome-fragment | NA | NA | 0 |
| S1-final-viral-combined_k141_117096\|\|full | 4539 | No | NA | 4 | 0 | 3 | Low-quality | Genome-fragment | 4.4 | AAI-based (medium-confidence) | 0 |
| S2_k141_75589 | 5917 | No | NA | 6 | 0 | 1 | Not-determined | Genome-fragment | NA | NA | 0 |
| S2_k141_24005 | 1138 | No | NA | 1 | 0 | 0 | Not-determined | Genome-fragment | NA | NA | 0 |
| S2_k141_13570 | 2332 | No | NA | 3 | 0 | 1 | Not-determined | Genome-fragment | NA | NA | 0 |
| S2_k141_55165 | 3303 | No | NA | 4 | 0 | 3 | Not-determined | Genome-fragment | NA | NA | 0 |
| S2_k141_13583 | 1063 | No | NA | 1 | 0 | 1 | Not-determined | Genome-fragment | NA | NA | 0 |
| S2_k141_96858 | 1503 | No | NA | 1 | 0 | 0 | Not-determined | Genome-fragment | NA | NA | 0 |
| S17-k141_182969 | 4539 | No | NA | 4 | 0 | 0 | Low-quality | Genome-fragment | 6.76 | AAI-based (medium-confidence) | 0 |
| S2_k141_148284 | 1181 | No | NA | 3 | 0 | 2 | Not-determined | Genome-fragment | NA | NA | 0 |
| S2_k141_231667 | 1129 | No | NA | 2 | 0 | 1 | Not-determined | Genome-fragment | NA | NA | 0 |
| S7_k141_91983 | 4541 | No | NA | 3 | 0 | 1 | Low-quality | Genome-fragment | 7.58 | AAI-based (high-confidence) | 0 |
| S17-final-viral-combined_k141_92804\|\|full | 4541 | No | NA | 8 | 8 | 0 | Low-quality | Genome-fragment | 9.95 | AAI-based (medium-confidence) | 0 |
| S2_k141_241226 | 1196 | No | NA | 1 | 0 | 0 | Not-determined | Genome-fragment | NA | NA | 0 |
| S2_k141_148336 | 4898 | No | NA | 4 | 0 | 2 | Not-determined | Genome-fragment | NA | NA | 0 |
| S2_k141_117174 | 2537 | No | NA | 3 | 0 | 1 | Not-determined | Genome-fragment | NA | NA | 0 |
| S2_k141_169753 | 1114 | No | NA | 1 | 0 | 0 | Not-determined | Genome-fragment | NA | NA | 0 |
| S2_k141_3358 | 1138 | No | NA | 3 | 0 | 2 | Not-determined | Genome-fragment | NA | NA | 0 |
| S2_k141_86611 | 1300 | No | NA | 2 | 0 | 2 | Not-determined | Genome-fragment | NA | NA | 0 |
| S2_k141_46238 | 2164 | No | NA | 4 | 0 | 1 | Not-determined | Genome-fragment | NA | NA | 0 |
| S2-final-viral-combined_k141_168357\|\|full | 4541 | No | NA | 5 | 0 | 3 | Low-quality | Genome-fragment | 2.2 | AAI-based (medium-confidence) | 0 |
| S2_k141_158693 | 1836 | No | NA | 1 | 0 | 0 | Not-determined | Genome-fragment | NA | NA | 0 |
| S6_k141_130785 | 4541 | No | NA | 3 | 0 | 3 | Low-quality | Genome-fragment | 4.23 | AAI-based (medium-confidence) | 0 |
| S2_k141_138279 | 4064 | No | NA | 6 | 0 | 2 | Not-determined | Genome-fragment | NA | NA | 0 |
| S2_k141_231735 | 2186 | No | NA | 2 | 0 | 2 | Not-determined | Genome-fragment | NA | NA | 0 |
| S7-final-viral-combined_k141_102795\|\|full | 4542 | No | NA | 4 | 0 | 3 | Low-quality | Genome-fragment | 2.15 | AAI-based (medium-confidence) | 0 |
| S12-final-viral-combined_k141_334019\|\|full | 4542 | Yes | 3292 | 10 | 1 | 1 | Low-quality | Genome-fragment | 4.28 | HMM-based (lower-bound) | 27.52 |
| S2_k141_46293 | 1354 | No | NA | 1 | 0 | 0 | Not-determined | Genome-fragment | NA | NA | 0 |
| S2_k141_46308 | 1086 | No | NA | 1 | 0 | 1 | Not-determined | Genome-fragment | NA | NA | 0 |
| S2_k141_241265 | 1045 | No | NA | 2 | 0 | 1 | Not-determined | Genome-fragment | NA | NA | 0 |
| S2_k141_190216 | 4524 | No | NA | 4 | 0 | 4 | Not-determined | Genome-fragment | NA | NA | 0 |
| S2_k141_148485 | 1200 | No | NA | 3 | 0 | 2 | Not-determined | Genome-fragment | NA | NA | 0 |
| S2_k141_231815 | 1234 | No | NA | 1 | 0 | 1 | Not-determined | Genome-fragment | NA | NA | 0 |
| S3-final-viral-combined_k141_129466\|\|full | 4543 | No | NA | 10 | 2 | 0 | Low-quality | Genome-fragment | 6.74 | HMM-based (lower-bound) | 0 |
| S2_k141_13775 | 4288 | No | NA | 3 | 0 | 3 | Not-determined | Genome-fragment | NA | NA | 0 |
| S2_k141_180130 | 1418 | No | NA | 1 | 0 | 0 | Not-determined | Genome-fragment | NA | NA | 0 |
| S2_k141_138359 | 3480 | No | NA | 5 | 0 | 2 | Not-determined | Genome-fragment | NA | NA | 0 |
| S2-final-viral-combined_k141_36882\|\|full | 4544 | No | NA | 4 | 0 | 3 | Low-quality | Genome-fragment | 2.04 | AAI-based (high-confidence) | 0 |
| S2_k141_117271 | 2635 | No | NA | 9 | 0 | 0 | Not-determined | Genome-fragment | NA | NA | 0 |
| S2_k141_210577 | 1167 | No | NA | 3 | 0 | 2 | Not-determined | Genome-fragment | NA | NA | 0 |
| S1-final-viral-combined_k141_387422\|\|full | 4545 | No | NA | 16 | 1 | 0 | Low-quality | Genome-fragment | 7.76 | AAI-based (high-confidence) | 0 |
| S7-final-viral-combined_k141_226924\|\|full | 4545 | No | NA | 5 | 3 | 0 | Low-quality | Genome-fragment | 9.79 | AAI-based (medium-confidence) | 0 |
| S6-final-viral-combined_k141_60003\|\|full | 4548 | No | NA | 5 | 1 | 0 | Low-quality | Genome-fragment | 10.61 | AAI-based (medium-confidence) | 0 |
| S2_k141_148556 | 7059 | No | NA | 5 | 0 | 1 | Not-determined | Genome-fragment | NA | NA | 0 |
| S2_k141_13838 | 1392 | No | NA | 2 | 0 | 0 | Not-determined | Genome-fragment | NA | NA | 0 |
| S2_k141_180200 | 2006 | No | NA | 1 | 0 | 1 | Not-determined | Genome-fragment | NA | NA | 0 |
| S7_k141_40928 | 4550 | No | NA | 13 | 0 | 0 | Low-quality | Genome-fragment | 12.89 | AAI-based (high-confidence) | 0 |
| S2_k141_169945 | 1161 | No | NA | 1 | 0 | 1 | Not-determined | Genome-fragment | NA | NA | 0 |
| S2_k141_241346 | 2183 | No | NA | 1 | 0 | 1 | Not-determined | Genome-fragment | NA | NA | 0 |
| S8-final-viral-combined_k141_190470\|\|full | 4550 | No | NA | 1 | 0 | 1 | Low-quality | Genome-fragment | 4.03 | AAI-based (high-confidence) | 0 |
| S2_k141_190344 | 1162 | No | NA | 1 | 0 | 1 | Not-determined | Genome-fragment | NA | NA | 0 |
| S9_k141_4400 | 4555 | No | NA | 3 | 0 | 0 | Low-quality | Genome-fragment | 6.94 | AAI-based (medium-confidence) | 0 |
| S2_k141_169971 | 2230 | No | NA | 2 | 0 | 2 | Not-determined | Genome-fragment | NA | NA | 0 |
| S2_k141_46435 | 1929 | No | NA | 1 | 0 | 1 | Not-determined | Genome-fragment | NA | NA | 0 |
| S2_k141_241356 | 1571 | No | NA | 2 | 0 | 1 | Not-determined | Genome-fragment | NA | NA | 0 |
| S2_k141_55434 | 3572 | No | NA | 1 | 0 | 1 | Not-determined | Genome-fragment | NA | NA | 0 |
| S2_k141_55440 | 3149 | No | NA | 4 | 0 | 1 | Not-determined | Genome-fragment | NA | NA | 0 |
| S2_k141_65773 | 1233 | No | NA | 2 | 0 | 0 | Not-determined | Genome-fragment | NA | NA | 0 |
| S2_k141_158797 | 4146 | No | NA | 5 | 0 | 1 | Not-determined | Genome-fragment | NA | NA | 0 |
| S7-final-viral-combined_k141_65976\|\|full | 4555 | No | NA | 6 | 2 | 0 | Low-quality | Genome-fragment | 7.45 | AAI-based (high-confidence) | 0 |
| S2_k141_58749 | 4556 | No | NA | 9 | 0 | 0 | Low-quality | Genome-fragment | 8.73 | AAI-based (high-confidence) | 0 |
| S3-final-viral-combined_k141_6730\|\|full | 4557 | Yes | 2244 | 8 | 1 | 1 | Low-quality | Genome-fragment | 2.78 | HMM-based (lower-bound) | 50.76 |
| S2_k141_138468 | 1208 | No | NA | 2 | 0 | 2 | Not-determined | Genome-fragment | NA | NA | 0 |
| S2_k141_34796 | 1042 | No | NA | 2 | 0 | 0 | Not-determined | Genome-fragment | NA | NA | 0 |
| S1-final-viral-combined_k141_163377\|\|full | 4558 | No | NA | 6 | 2 | 0 | Low-quality | Genome-fragment | 11.57 | AAI-based (high-confidence) | 0 |
| S2_k141_148688 | 3719 | No | NA | 1 | 0 | 0 | Not-determined | Genome-fragment | NA | NA | 0 |
| S11-final-viral-combined_k141_235038\|\|full | 4559 | No | NA | 4 | 0 | 1 | Low-quality | Genome-fragment | 5.48 | AAI-based (medium-confidence) | 0 |
| S2_k141_241422 | 1021 | No | NA | 1 | 0 | 0 | Not-determined | Genome-fragment | NA | NA | 0 |
| S12-final-viral-combined_k141_301618\|\|full | 4559 | No | NA | 8 | 4 | 0 | Low-quality | Genome-fragment | 12.08 | AAI-based (medium-confidence) | 0 |
| S1-final-viral-combined_k141_103512\|\|full | 4561 | No | NA | 6 | 0 | 2 | Low-quality | Genome-fragment | 6.2 | AAI-based (medium-confidence) | 0 |
| S2_k141_180325 | 1291 | No | NA | 2 | 0 | 1 | Not-determined | Genome-fragment | NA | NA | 0 |
| S2_k141_3631 | 2344 | No | NA | 6 | 0 | 0 | Not-determined | Genome-fragment | NA | NA | 0 |
| S2_k141_190421 | 3210 | No | NA | 7 | 0 | 2 | Not-determined | Genome-fragment | NA | NA | 0 |
| S2_k141_232022 | 1099 | No | NA | 2 | 0 | 0 | Not-determined | Genome-fragment | NA | NA | 0 |
| S2_k141_107329 | 52666 | Yes | 46224 | 74 | 24 | 5 | Medium-quality | Genome-fragment | 62.78 | AAI-based (high-confidence) | 12.23 |
| S2_k141_210736 | 6725 | No | NA | 9 | 0 | 0 | Not-determined | Genome-fragment | NA | NA | 0 |
| S2_k141_210744 | 1006 | No | NA | 1 | 0 | 1 | Not-determined | Genome-fragment | NA | NA | 0 |
| S12-final-viral-combined_k141_254840\|\|full | 4562 | No | NA | 5 | 1 | 0 | Low-quality | Genome-fragment | 8.41 | HMM-based (lower-bound) | 0 |
| S2_k141_210774 | 1005 | No | NA | 2 | 0 | 0 | Not-determined | Genome-fragment | NA | NA | 0 |
| S2_k141_46544 | 4799 | No | NA | 5 | 0 | 1 | Not-determined | Genome-fragment | NA | NA | 0 |
| S2_k141_75938 | 1089 | No | NA | 1 | 0 | 0 | Not-determined | Genome-fragment | NA | NA | 0 |
| S2_k141_24336 | 16112 | No | NA | 14 | 0 | 10 | Not-determined | Genome-fragment | NA | NA | 0 |
| S2_k141_117522 | 1473 | No | NA | 3 | 0 | 0 | Not-determined | Genome-fragment | NA | NA | 0 |
| S5_k141_100681 | 4563 | No | NA | 5 | 0 | 0 | Low-quality | Genome-fragment | 5.2 | AAI-based (medium-confidence) | 0 |
| S2_k141_24371 | 2129 | No | NA | 1 | 0 | 1 | Not-determined | Genome-fragment | NA | NA | 0 |
| S2_k141_34884 | 1555 | No | NA | 1 | 0 | 1 | Not-determined | Genome-fragment | NA | NA | 0 |
| S11-final-viral-combined_k141_160638\|\|full | 4564 | No | NA | 9 | 1 | 0 | Low-quality | Genome-fragment | 5.52 | HMM-based (lower-bound) | 0 |
| S2_k141_55678 | 1331 | No | NA | 5 | 0 | 0 | Not-determined | Genome-fragment | NA | NA | 0 |
| S2_k141_14045 | 3127 | No | NA | 2 | 0 | 2 | Not-determined | Genome-fragment | NA | NA | 0 |
| S2_k141_180424 | 1187 | No | NA | 1 | 0 | 0 | Not-determined | Genome-fragment | NA | NA | 0 |
| S2_k141_46605 | 3543 | No | NA | 1 | 0 | 0 | Not-determined | Genome-fragment | NA | NA | 0 |
| S2_k141_46607 | 3726 | No | NA | 2 | 0 | 1 | Not-determined | Genome-fragment | NA | NA | 0 |
| S2_k141_46609 | 3797 | No | NA | 5 | 0 | 2 | Not-determined | Genome-fragment | NA | NA | 0 |
| S2_k141_190515 | 2088 | No | NA | 3 | 0 | 1 | Not-determined | Genome-fragment | NA | NA | 0 |
| S5-final-viral-combined_k141_54964\|\|full | 4564 | No | NA | 12 | 0 | 0 | Low-quality | Genome-fragment | 11.55 | AAI-based (high-confidence) | 0 |
| S2_k141_127736 | 2796 | No | NA | 6 | 0 | 1 | Not-determined | Genome-fragment | NA | NA | 0 |
| S1_k141_240090 | 4565 | No | NA | 6 | 0 | 0 | Low-quality | Genome-fragment | 8.14 | AAI-based (medium-confidence) | 0 |
| S2_k141_148838 | 1535 | No | NA | 2 | 0 | 1 | Not-determined | Genome-fragment | NA | NA | 0 |
| S2_k141_55747 | 3385 | No | NA | 2 | 0 | 2 | Not-determined | Genome-fragment | NA | NA | 0 |
| S2_k141_127787 | 4262 | No | NA | 1 | 0 | 1 | Not-determined | Genome-fragment | NA | NA | 0 |
| S2_k141_107542 | 2419 | No | NA | 3 | 0 | 3 | Not-determined | Genome-fragment | NA | NA | 0 |
| S2_k141_46669 | 1507 | No | NA | 1 | 0 | 1 | Not-determined | Genome-fragment | NA | NA | 0 |
| S4-final-viral-combined_k141_116219\|\|full | 4565 | No | NA | 5 | 0 | 1 | Low-quality | Genome-fragment | 12 | AAI-based (medium-confidence) | 0 |
| S2_k141_190604 | 1411 | No | NA | 4 | 0 | 1 | Not-determined | Genome-fragment | NA | NA | 0 |
| S2_k141_232079 | 1176 | No | NA | 1 | 0 | 0 | Not-determined | Genome-fragment | NA | NA | 0 |
| S2_k141_232088 | 1062 | No | NA | 1 | 0 | 0 | Not-determined | Genome-fragment | NA | NA | 0 |
| S2_k141_232109 | 5487 | No | NA | 10 | 0 | 9 | Not-determined | Genome-fragment | NA | NA | 0 |
| S2_k141_107596 | 3183 | No | NA | 2 | 0 | 1 | Not-determined | Genome-fragment | NA | NA | 0 |
| S2_k141_55811 | 3498 | No | NA | 4 | 0 | 2 | Not-determined | Genome-fragment | NA | NA | 0 |
| S12_k141_1113 | 4567 | No | NA | 8 | 3 | 0 | Low-quality | Genome-fragment | 10.28 | AAI-based (high-confidence) | 0 |
| S17-k141_130547 | 4568 | No | NA | 4 | 0 | 2 | Low-quality | Genome-fragment | 7.79 | AAI-based (medium-confidence) | 0 |
| S2_k141_138774 | 1247 | No | NA | 1 | 0 | 0 | Not-determined | Genome-fragment | NA | NA | 0 |
| S2_k141_127853 | 1045 | No | NA | 2 | 0 | 1 | Not-determined | Genome-fragment | NA | NA | 0 |
| S2_k141_241444 | 4029 | No | NA | 1 | 0 | 0 | Not-determined | Genome-fragment | NA | NA | 0 |
| S2_k141_97335 | 3882 | No | NA | 6 | 0 | 2 | Not-determined | Genome-fragment | NA | NA | 0 |
| S1-final-viral-combined_k141_10203\|\|full | 4569 | No | NA | 4 | 0 | 0 | Low-quality | Genome-fragment | 9.14 | AAI-based (medium-confidence) | 0 |
| S2_k141_117716 | 32302 | No | NA | 32 | 0 | 9 | Not-determined | Genome-fragment | NA | NA | 0 |
| S2_k141_76254 | 1830 | No | NA | 2 | 0 | 0 | Not-determined | Genome-fragment | NA | NA | 0 |
| S2_k141_76270 | 5494 | No | NA | 2 | 0 | 1 | Not-determined | Genome-fragment | NA | NA | 0 |
| S2_k141_138841 | 1549 | No | NA | 1 | 0 | 0 | Not-determined | Genome-fragment | NA | NA | 0 |
| S2_k141_65935 | 17917 | No | NA | 26 | 0 | 11 | Not-determined | Genome-fragment | NA | NA | 0 |
| S2_k141_241458 | 1576 | No | NA | 2 | 0 | 0 | Not-determined | Genome-fragment | NA | NA | 0 |
| S2_k141_148944 | 3828 | No | NA | 2 | 0 | 0 | Not-determined | Genome-fragment | NA | NA | 0 |
| S2_k141_14255 | 1211 | No | NA | 2 | 0 | 1 | Not-determined | Genome-fragment | NA | NA | 0 |
| S2_k141_117788 | 1101 | No | NA | 1 | 0 | 1 | Not-determined | Genome-fragment | NA | NA | 0 |
| S17-final-viral-combined_k141_192703\|\|full | 4569 | No | NA | 10 | 0 | 0 | Low-quality | Genome-fragment | 6.86 | AAI-based (high-confidence) | 0 |
| S2_k141_46844 | 1364 | No | NA | 1 | 0 | 1 | Not-determined | Genome-fragment | NA | NA | 0 |
| S2_k141_127228 | 4569 | No | NA | 4 | 0 | 4 | Low-quality | Genome-fragment | 2.08 | AAI-based (high-confidence) | 0 |
| S1-final-viral-combined_k141_37843\|\|full | 4570 | No | NA | 8 | 2 | 0 | Low-quality | Genome-fragment | 31.22 | AAI-based (high-confidence) | 0 |
| S2_k141_241515 | 1202 | No | NA | 1 | 0 | 1 | Not-determined | Genome-fragment | NA | NA | 0 |
| S2_k141_148961 | 1592 | No | NA | 1 | 0 | 0 | Not-determined | Genome-fragment | NA | NA | 0 |
| S2_k141_65995 | 1100 | No | NA | 3 | 0 | 2 | Not-determined | Genome-fragment | NA | NA | 0 |
| S2_k141_76374 | 1604 | No | NA | 1 | 0 | 1 | Not-determined | Genome-fragment | NA | NA | 0 |
| S2_k141_46867 | 1476 | No | NA | 2 | 0 | 1 | Not-determined | Genome-fragment | NA | NA | 0 |
| S2_k141_190718 | 1314 | No | NA | 2 | 0 | 0 | Not-determined | Genome-fragment | NA | NA | 0 |
| S2_k141_220577 | 4159 | No | NA | 3 | 0 | 3 | Not-determined | Genome-fragment | NA | NA | 0 |
| S2_k141_199633 | 1268 | No | NA | 2 | 0 | 2 | Not-determined | Genome-fragment | NA | NA | 0 |
| S2_k141_97449 | 1960 | No | NA | 1 | 0 | 0 | Not-determined | Genome-fragment | NA | NA | 0 |
| S12-final-viral-combined_k141_306484\|\|full | 4570 | No | NA | 3 | 0 | 2 | Low-quality | Genome-fragment | 2.56 | AAI-based (medium-confidence) | 0 |
| S2_k141_55985 | 1056 | No | NA | 1 | 0 | 1 | Not-determined | Genome-fragment | NA | NA | 0 |
| S9-final-viral-combined_k141_11091\|\|full | 4571 | No | NA | 5 | 1 | 0 | Low-quality | Genome-fragment | 11.26 | AAI-based (high-confidence) | 0 |
| S2_k141_199673 | 3189 | No | NA | 3 | 0 | 1 | Not-determined | Genome-fragment | NA | NA | 0 |
| S2_k141_159165 | 1161 | No | NA | 2 | 0 | 1 | Not-determined | Genome-fragment | NA | NA | 0 |
| S2_k141_35249 | 6908 | No | NA | 8 | 0 | 7 | Not-determined | Genome-fragment | NA | NA | 0 |
| S2_k141_35261 | 3016 | No | NA | 2 | 0 | 1 | Not-determined | Genome-fragment | NA | NA | 0 |
| S2_k141_76448 | 1108 | No | NA | 1 | 0 | 0 | Not-determined | Genome-fragment | NA | NA | 0 |
| S2_k141_14387 | 1513 | No | NA | 2 | 0 | 1 | Not-determined | Genome-fragment | NA | NA | 0 |
| S2_k141_138972 | 1963 | No | NA | 2 | 0 | 2 | Not-determined | Genome-fragment | NA | NA | 0 |
| S2-final-viral-combined_k141_155612\|\|full | 4573 | Yes | 2807 | 6 | 1 | 2 | Low-quality | Genome-fragment | 2.93 | HMM-based (lower-bound) | 38.62 |
| S2_k141_47025 | 2008 | No | NA | 2 | 0 | 0 | Not-determined | Genome-fragment | NA | NA | 0 |
| S2_k141_47027 | 1391 | No | NA | 1 | 0 | 0 | Not-determined | Genome-fragment | NA | NA | 0 |
| S2-final-viral-combined_k141_136617\|\|full | 4573 | No | NA | 8 | 0 | 0 | Low-quality | Genome-fragment | 6.98 | AAI-based (high-confidence) | 0 |
| S2_k141_56047 | 1033 | No | NA | 2 | 0 | 2 | Not-determined | Genome-fragment | NA | NA | 0 |
| S2_k141_24690 | 1714 | No | NA | 3 | 0 | 0 | Not-determined | Genome-fragment | NA | NA | 0 |
| S4-final-viral-combined_k141_125412\|\|full | 4573 | No | NA | 5 | 0 | 5 | Low-quality | Genome-fragment | 3.39 | AAI-based (medium-confidence) | 0 |
| S2_k141_220686 | 39926 | No | NA | 41 | 0 | 31 | Not-determined | Genome-fragment | NA | NA | 0 |
| S8-final-viral-combined_k141_243269\|\|full | 4573 | No | NA | 4 | 0 | 2 | Low-quality | Genome-fragment | 2.26 | AAI-based (medium-confidence) | 0 |
| S2_k141_97589 | 1237 | No | NA | 1 | 0 | 0 | Not-determined | Genome-fragment | NA | NA | 0 |
| S2_k141_76540 | 6688 | No | NA | 7 | 0 | 1 | Not-determined | Genome-fragment | NA | NA | 0 |
| S2_k141_170252 | 1614 | No | NA | 2 | 0 | 1 | Not-determined | Genome-fragment | NA | NA | 0 |
| S2_k141_128008 | 1579 | No | NA | 2 | 0 | 2 | Not-determined | Genome-fragment | NA | NA | 0 |
| S5-final-viral-combined_k141_212603\|\|full | 4574 | No | NA | 8 | 1 | 0 | Low-quality | Genome-fragment | 10.58 | AAI-based (medium-confidence) | 0 |
| S2_k141_97619 | 1837 | No | NA | 2 | 0 | 2 | Not-determined | Genome-fragment | NA | NA | 0 |
| S2_k141_170265 | 1095 | No | NA | 1 | 0 | 0 | Not-determined | Genome-fragment | NA | NA | 0 |
| S2_k141_210924 | 3293 | No | NA | 3 | 0 | 0 | Not-determined | Genome-fragment | NA | NA | 0 |
| S2_k141_118034 | 1029 | No | NA | 1 | 0 | 1 | Not-determined | Genome-fragment | NA | NA | 0 |
| S2_k141_56154 | 1193 | No | NA | 3 | 0 | 1 | Not-determined | Genome-fragment | NA | NA | 0 |
| S2_k141_232477 | 1025 | No | NA | 1 | 0 | 0 | Not-determined | Genome-fragment | NA | NA | 0 |
| S2_k141_87431 | 1121 | No | NA | 1 | 0 | 0 | Not-determined | Genome-fragment | NA | NA | 0 |
| S1_k141_104143 | 4576 | No | NA | 5 | 0 | 3 | Low-quality | Genome-fragment | 2.19 | AAI-based (high-confidence) | 0 |
| S2_k141_180876 | 3333 | No | NA | 3 | 0 | 3 | Not-determined | Genome-fragment | NA | NA | 0 |
| S2_k141_56231 | 1133 | No | NA | 1 | 0 | 0 | Not-determined | Genome-fragment | NA | NA | 0 |
| S2_k141_47159 | 1367 | No | NA | 1 | 0 | 0 | Not-determined | Genome-fragment | NA | NA | 0 |
| S11-final-viral-combined_k141_314223\|\|full | 4577 | No | NA | 3 | 0 | 1 | Low-quality | Genome-fragment | 3.03 | AAI-based (medium-confidence) | 0 |
| S17-final-viral-combined_k141_51389\|\|full | 4577 | No | NA | 7 | 3 | 0 | Low-quality | Genome-fragment | 11.22 | AAI-based (high-confidence) | 0 |
| S6-final-viral-combined_k141_55534\|\|full | 4578 | No | NA | 5 | 1 | 0 | Low-quality | Genome-fragment | 11.46 | AAI-based (high-confidence) | 0 |
| S8-final-viral-combined_k141_24503\|\|full | 4578 | No | NA | 5 | 5 | 0 | Low-quality | Genome-fragment | 9.67 | AAI-based (high-confidence) | 0 |
| S2_k141_139193 | 1047 | No | NA | 2 | 0 | 0 | Not-determined | Genome-fragment | NA | NA | 0 |
| S1_k141_25500 | 4579 | No | NA | 6 | 0 | 1 | Low-quality | Genome-fragment | 7.11 | AAI-based (medium-confidence) | 0 |
| S2_k141_180944 | 1179 | No | NA | 1 | 0 | 1 | Not-determined | Genome-fragment | NA | NA | 0 |
| S2_k141_159299 | 4270 | No | NA | 5 | 0 | 4 | Not-determined | Genome-fragment | NA | NA | 0 |
| S11_k141_75663 | 4583 | No | NA | 6 | 2 | 0 | Low-quality | Genome-fragment | 9.57 | AAI-based (high-confidence) | 0 |
| S2_k141_199943 | 1349 | No | NA | 1 | 0 | 0 | Not-determined | Genome-fragment | NA | NA | 0 |
| S2_k141_190815 | 1011 | No | NA | 1 | 0 | 1 | Not-determined | Genome-fragment | NA | NA | 0 |
| S2_k141_220876 | 1387 | No | NA | 1 | 0 | 1 | Not-determined | Genome-fragment | NA | NA | 0 |
| S2_k141_66363 | 1092 | No | NA | 1 | 0 | 1 | Not-determined | Genome-fragment | NA | NA | 0 |
| S3_k141_40445 | 4583 | No | NA | 6 | 0 | 3 | Low-quality | Genome-fragment | 32.72 | AAI-based (high-confidence) | 0 |
| S2_k141_3928 | 1351 | No | NA | 1 | 0 | 0 | Not-determined | Genome-fragment | NA | NA | 0 |
| S2_k141_220896 | 3959 | No | NA | 4 | 0 | 1 | Not-determined | Genome-fragment | NA | NA | 0 |
| S2_k141_190842 | 4377 | No | NA | 6 | 0 | 2 | Not-determined | Genome-fragment | NA | NA | 0 |
| S2_k141_97764 | 1396 | No | NA | 1 | 0 | 1 | Not-determined | Genome-fragment | NA | NA | 0 |
| S2_k141_200005 | 1395 | No | NA | 1 | 0 | 1 | Not-determined | Genome-fragment | NA | NA | 0 |
| S2_k141_190882 | 1237 | No | NA | 1 | 0 | 1 | Not-determined | Genome-fragment | NA | NA | 0 |
| S2_k141_181052 | 1008 | No | NA | 1 | 0 | 1 | Not-determined | Genome-fragment | NA | NA | 0 |
| S2_k141_76754 | 1909 | No | NA | 1 | 0 | 1 | Not-determined | Genome-fragment | NA | NA | 0 |
| S2_k141_211091 | 1047 | No | NA | 2 | 0 | 2 | Not-determined | Genome-fragment | NA | NA | 0 |
| S2_k141_200039 | 4554 | No | NA | 3 | 0 | 3 | Not-determined | Genome-fragment | NA | NA | 0 |
| S2_k141_56466 | 1030 | No | NA | 1 | 0 | 0 | Not-determined | Genome-fragment | NA | NA | 0 |
| S2_k141_153864 | 4583 | No | NA | 11 | 0 | 1 | Low-quality | Genome-fragment | 7.88 | AAI-based (medium-confidence) | 0 |
| S2_k141_3987 | 7196 | No | NA | 9 | 0 | 5 | Not-determined | Genome-fragment | NA | NA | 0 |
| S1_k141_249560 | 4584 | No | NA | 6 | 3 | 0 | Low-quality | Genome-fragment | 9.7 | AAI-based (high-confidence) | 0 |
| S2_k141_118356 | 1124 | No | NA | 1 | 0 | 0 | Not-determined | Genome-fragment | NA | NA | 0 |
| S2_k141_190978 | 1519 | No | NA | 3 | 0 | 3 | Not-determined | Genome-fragment | NA | NA | 0 |
| S2_k141_159447 | 1009 | No | NA | 2 | 0 | 0 | Not-determined | Genome-fragment | NA | NA | 0 |
| S2_k141_170656 | 1266 | No | NA | 1 | 0 | 1 | Not-determined | Genome-fragment | NA | NA | 0 |
| S5-final-viral-combined_k141_29424\|\|full | 4585 | No | NA | 5 | 1 | 0 | Low-quality | Genome-fragment | 6.5 | HMM-based (lower-bound) | 0 |
| S2_k141_139323 | 1489 | No | NA | 1 | 0 | 1 | Not-determined | Genome-fragment | NA | NA | 0 |
| S2_k141_139335 | 3126 | No | NA | 2 | 0 | 0 | Not-determined | Genome-fragment | NA | NA | 0 |
| S2_k141_14608 | 15496 | No | NA | 17 | 0 | 15 | Not-determined | Genome-fragment | NA | NA | 0 |
| S7-final-viral-combined_k141_195170\|\|full | 4585 | Yes | 3534 | 7 | 4 | 1 | Low-quality | Genome-fragment | 7.51 | AAI-based (high-confidence) | 22.92 |
| S2_k141_220958 | 3193 | No | NA | 3 | 0 | 1 | Not-determined | Genome-fragment | NA | NA | 0 |
| S17-final-viral-combined_k141_257628\|\|full | 4585 | No | NA | 5 | 0 | 1 | Low-quality | Genome-fragment | 8.54 | AAI-based (medium-confidence) | 0 |
| S2-final-viral-combined_k141_178106\|\|full | 4585 | No | NA | 4 | 0 | 3 | Low-quality | Genome-fragment | 5.48 | AAI-based (medium-confidence) | 0 |
| S1_k141_264146 | 4587 | No | NA | 4 | 2 | 0 | Low-quality | Genome-fragment | 16.7 | HMM-based (lower-bound) | 0 |
| S2_k141_4113 | 1063 | No | NA | 1 | 0 | 1 | Not-determined | Genome-fragment | NA | NA | 0 |
| S2_k141_76864 | 1482 | No | NA | 1 | 0 | 0 | Not-determined | Genome-fragment | NA | NA | 0 |
| S17-final-viral-combined_k141_147593\|\|full | 4587 | No | NA | 8 | 0 | 5 | Low-quality | Genome-fragment | 3.26 | AAI-based (medium-confidence) | 0 |
| S2_k141_232602 | 2503 | No | NA | 1 | 0 | 0 | Not-determined | Genome-fragment | NA | NA | 0 |
| S1-final-viral-combined_k141_187855\|\|full | 4589 | No | NA | 3 | 0 | 2 | Low-quality | Genome-fragment | 1.45 | AAI-based (medium-confidence) | 0 |
| S2_k141_98044 | 8717 | No | NA | 6 | 0 | 4 | Not-determined | Genome-fragment | NA | NA | 0 |
| S2_k141_128288 | 1287 | No | NA | 2 | 0 | 0 | Not-determined | Genome-fragment | NA | NA | 0 |
| S2_k141_66612 | 1053 | No | NA | 3 | 0 | 3 | Not-determined | Genome-fragment | NA | NA | 0 |
| S12-final-viral-combined_k141_123889\|\|full | 4589 | No | NA | 3 | 0 | 0 | Low-quality | Genome-fragment | 4.68 | AAI-based (medium-confidence) | 0 |
| S2_k141_191116 | 2170 | No | NA | 1 | 0 | 0 | Not-determined | Genome-fragment | NA | NA | 0 |
| S2_k141_98064 | 1118 | No | NA | 1 | 0 | 0 | Not-determined | Genome-fragment | NA | NA | 0 |
| S2_k141_87806 | 1906 | No | NA | 4 | 0 | 1 | Not-determined | Genome-fragment | NA | NA | 0 |
| S2_k141_14735 | 1510 | No | NA | 1 | 0 | 0 | Not-determined | Genome-fragment | NA | NA | 0 |
| S2_k141_118539 | 1108 | No | NA | 1 | 0 | 1 | Not-determined | Genome-fragment | NA | NA | 0 |
| S2_k141_76956 | 1712 | No | NA | 1 | 0 | 0 | Not-determined | Genome-fragment | NA | NA | 0 |
| S9-final-viral-combined_k141_67791\|\|full | 4590 | No | NA | 4 | 0 | 4 | Low-quality | Genome-fragment | 1.88 | AAI-based (medium-confidence) | 0 |
| S2_k141_191140 | 3351 | No | NA | 2 | 0 | 2 | Not-determined | Genome-fragment | NA | NA | 0 |
| S7-final-viral-combined_k141_238024\|\|full | 4590 | No | NA | 8 | 1 | 0 | Low-quality | Genome-fragment | 9.14 | AAI-based (high-confidence) | 0 |
| S2_k141_35887 | 3428 | No | NA | 2 | 0 | 2 | Not-determined | Genome-fragment | NA | NA | 0 |
| S8-final-viral-combined_k141_161289\|\|full | 4590 | No | NA | 5 | 0 | 3 | Low-quality | Genome-fragment | 5.55 | AAI-based (medium-confidence) | 0 |
| S2_k141_66688 | 1095 | No | NA | 2 | 0 | 2 | Not-determined | Genome-fragment | NA | NA | 0 |
| S2_k141_170860 | 1030 | No | NA | 1 | 0 | 1 | Not-determined | Genome-fragment | NA | NA | 0 |
| S2_k141_87860 | 21628 | No | NA | 20 | 0 | 17 | Not-determined | Genome-fragment | NA | NA | 0 |
| S2_k141_170898 | 2064 | No | NA | 3 | 0 | 1 | Not-determined | Genome-fragment | NA | NA | 0 |
| S2_k141_170901 | 7038 | No | NA | 5 | 0 | 5 | Not-determined | Genome-fragment | NA | NA | 0 |
| S2_k141_98212 | 1007 | No | NA | 1 | 0 | 0 | Not-determined | Genome-fragment | NA | NA | 0 |
| S8_k141_243758 | 4590 | No | NA | 9 | 0 | 0 | Low-quality | Genome-fragment | 11.15 | AAI-based (high-confidence) | 0 |
| S2_k141_98220 | 1012 | No | NA | 2 | 0 | 0 | Not-determined | Genome-fragment | NA | NA | 0 |
| S2_k141_118624 | 1065 | No | NA | 1 | 0 | 0 | Not-determined | Genome-fragment | NA | NA | 0 |
| S1-final-viral-combined_k141_354511\|\|full | 4591 | No | NA | 8 | 1 | 2 | Low-quality | Genome-fragment | 2.91 | HMM-based (lower-bound) | 0 |
| S11-final-viral-combined_k141_188689\|\|full | 4592 | No | NA | 3 | 0 | 2 | Low-quality | Genome-fragment | 2.45 | AAI-based (medium-confidence) | 0 |
| S2_k141_159703 | 1360 | No | NA | 1 | 0 | 1 | Not-determined | Genome-fragment | NA | NA | 0 |
| S2_k141_118633 | 1924 | No | NA | 2 | 0 | 2 | Not-determined | Genome-fragment | NA | NA | 0 |
| S2_k141_221198 | 2427 | No | NA | 1 | 0 | 0 | Not-determined | Genome-fragment | NA | NA | 0 |
| S2_k141_56805 | 2180 | No | NA | 2 | 0 | 1 | Not-determined | Genome-fragment | NA | NA | 0 |
| S2_k141_211380 | 1240 | No | NA | 4 | 0 | 1 | Not-determined | Genome-fragment | NA | NA | 0 |

| | | | | | | | | | | | |
|---|---|---|---|---|---|---|---|---|---|---|---|
| S2_k141_14905 | 3162 | No | NA | 1 | 0 | 1 | Not-determined | Genome-fragment | NA | NA | 0 |
| S2_k141_118666 | 1648 | No | NA | 2 | 0 | 2 | Not-determined | Genome-fragment | NA | NA | 0 |
| S2_k141_47676 | 3288 | No | NA | 1 | 0 | 1 | Not-determined | Genome-fragment | NA | NA | 0 |
| S8_k141_138516 | 4592 | No | NA | 5 | 0 | 0 | Low-quality | Genome-fragment | 10.34 | AAI-based (medium-confidence) | 0 |
| S3-final-viral-combined_k141_106139\|\|full | 4593 | No | NA | 5 | 0 | 3 | Low-quality | Genome-fragment | 5.07 | AAI-based (medium-confidence) | 0 |
| S2_k141_241948 | 7142 | No | NA | 11 | 0 | 0 | Not-determined | Genome-fragment | NA | NA | 0 |
| S2_k141_211433 | 1060 | No | NA | 1 | 0 | 0 | Not-determined | Genome-fragment | NA | NA | 0 |
| S2_k141_159777 | 1107 | No | NA | 2 | 0 | 2 | Not-determined | Genome-fragment | NA | NA | 0 |
| S2_k141_211438 | 1002 | No | NA | 1 | 0 | 1 | Not-determined | Genome-fragment | NA | NA | 0 |
| S2_k141_36070 | 11228 | No | NA | 22 | 0 | 22 | Not-determined | Genome-fragment | NA | NA | 0 |
| S2_k141_77161 | 1756 | No | NA | 2 | 0 | 0 | Not-determined | Genome-fragment | NA | NA | 0 |
| S2_k141_139701 | 6014 | No | NA | 4 | 0 | 2 | Not-determined | Genome-fragment | NA | NA | 0 |
| S5-final-viral-combined_k141_111686\|\|full | 4594 | No | NA | 3 | 3 | 0 | Low-quality | Genome-fragment | 11.05 | AAI-based (high-confidence) | 0 |
| S2_k141_241979 | 1466 | No | NA | 1 | 0 | 0 | Not-determined | Genome-fragment | NA | NA | 0 |
| S2_k141_171037 | 4608 | No | NA | 3 | 0 | 3 | Not-determined | Genome-fragment | NA | NA | 0 |
| S8_k141_168369 | 4594 | No | NA | 6 | 0 | 0 | Low-quality | Genome-fragment | 7.48 | AAI-based (medium-confidence) | 0 |
| S2_k141_211481 | 1976 | No | NA | 1 | 0 | 0 | Not-determined | Genome-fragment | NA | NA | 0 |
| S5-final-viral-combined_k141_78543\|\|full | 4595 | No | NA | 4 | 1 | 0 | Low-quality | Genome-fragment | 5.11 | AAI-based (high-confidence) | 0 |
| S2_k141_171050 | 1225 | No | NA | 2 | 0 | 0 | Not-determined | Genome-fragment | NA | NA | 0 |
| S2_k141_159818 | 1443 | No | NA | 2 | 0 | 0 | Not-determined | Genome-fragment | NA | NA | 0 |
| S2_k141_56876 | 2260 | No | NA | 1 | 0 | 1 | Not-determined | Genome-fragment | NA | NA | 0 |
| S2_k141_211510 | 1046 | No | NA | 1 | 0 | 0 | Not-determined | Genome-fragment | NA | NA | 0 |
| S17-k141_116256 | 4596 | No | NA | 3 | 0 | 2 | Low-quality | Genome-fragment | 2.12 | AAI-based (medium-confidence) | 0 |
| S2_k141_221351 | 3471 | No | NA | 4 | 0 | 3 | Not-determined | Genome-fragment | NA | NA | 0 |
| S2_k141_181454 | 1313 | No | NA | 1 | 0 | 1 | Not-determined | Genome-fragment | NA | NA | 0 |
| S2_k141_211529 | 3122 | No | NA | 2 | 0 | 0 | Not-determined | Genome-fragment | NA | NA | 0 |
| S4-final-viral-combined_k141_95345\|\|full | 4596 | No | NA | 2 | 1 | 0 | Low-quality | Genome-fragment | 8.88 | AAI-based (high-confidence) | 0 |
| S2_k141_98391 | 2712 | No | NA | 1 | 0 | 0 | Not-determined | Genome-fragment | NA | NA | 0 |
| S2_k141_232927 | 3137 | No | NA | 5 | 0 | 4 | Not-determined | Genome-fragment | NA | NA | 0 |
| S2_k141_56934 | 4333 | No | NA | 4 | 0 | 2 | Not-determined | Genome-fragment | NA | NA | 0 |
| S2_k141_15052 | 5180 | No | NA | 2 | 0 | 2 | Not-determined | Genome-fragment | NA | NA | 0 |
| S8-final-viral-combined_k141_157984\|\|full | 4596 | Yes | 2316 | 4 | 1 | 1 | Low-quality | Genome-fragment | 1.42 | AAI-based (medium-confidence) | 49.61 |
| S2_k141_36244 | 1083 | No | NA | 3 | 0 | 1 | Not-determined | Genome-fragment | NA | NA | 0 |
| S2_k141_25365 | 8566 | No | NA | 6 | 0 | 3 | Not-determined | Genome-fragment | NA | NA | 0 |
| S3_k141_114696 | 4597 | No | NA | 4 | 3 | 0 | Low-quality | Genome-fragment | 16.22 | HMM-based (lower-bound) | 0 |
| S2_k141_233034 | 3027 | No | NA | 5 | 0 | 0 | Not-determined | Genome-fragment | NA | NA | 0 |
| S2_k141_47939 | 1148 | No | NA | 1 | 0 | 0 | Not-determined | Genome-fragment | NA | NA | 0 |
| S17-final-viral-combined_k141_187076\|\|full | 4597 | No | NA | 3 | 0 | 3 | Low-quality | Genome-fragment | 0.72 | AAI-based (medium-confidence) | 0 |
| S2_k141_200436 | 1214 | No | NA | 1 | 0 | 0 | Not-determined | Genome-fragment | NA | NA | 0 |
| S2_k141_88002 | 1231 | No | NA | 1 | 0 | 1 | Not-determined | Genome-fragment | NA | NA | 0 |
| S11-final-viral-combined_k141_130878\|\|full | 4598 | No | NA | 4 | 3 | 0 | Low-quality | Genome-fragment | 12.68 | AAI-based (high-confidence) | 0 |
| S2_k141_149840 | 3169 | No | NA | 5 | 0 | 3 | Not-determined | Genome-fragment | NA | NA | 0 |
| S2_k141_118831 | 2221 | No | NA | 1 | 0 | 0 | Not-determined | Genome-fragment | NA | NA | 0 |
| S2_k141_15153 | 1316 | No | NA | 2 | 0 | 0 | Not-determined | Genome-fragment | NA | NA | 0 |
| S2_k141_200471 | 1859 | No | NA | 2 | 0 | 2 | Not-determined | Genome-fragment | NA | NA | 0 |
| S2_k141_98611 | 1171 | No | NA | 1 | 0 | 0 | Not-determined | Genome-fragment | NA | NA | 0 |
| S2_k141_191343 | 1463 | No | NA | 1 | 0 | 0 | Not-determined | Genome-fragment | NA | NA | 0 |
| S2_k141_191349 | 3305 | No | NA | 2 | 0 | 1 | Not-determined | Genome-fragment | NA | NA | 0 |
| S2_k141_242187 | 1556 | No | NA | 1 | 0 | 1 | Not-determined | Genome-fragment | NA | NA | 0 |
| S2_k141_242188 | 1154 | No | NA | 2 | 0 | 0 | Not-determined | Genome-fragment | NA | NA | 0 |
| S2_k141_211697 | 1079 | No | NA | 2 | 0 | 0 | Not-determined | Genome-fragment | NA | NA | 0 |
| S2_k141_48023 | 1001 | No | NA | 2 | 0 | 1 | Not-determined | Genome-fragment | NA | NA | 0 |
| S2_k141_25410 | 1127 | No | NA | 1 | 0 | 0 | Not-determined | Genome-fragment | NA | NA | 0 |
| S2_k141_15251 | 8347 | No | NA | 3 | 0 | 1 | Not-determined | Genome-fragment | NA | NA | 0 |
| S2_k141_159985 | 1218 | No | NA | 1 | 0 | 1 | Not-determined | Genome-fragment | NA | NA | 0 |
| S2_k141_48047 | 1231 | No | NA | 2 | 0 | 1 | Not-determined | Genome-fragment | NA | NA | 0 |
| S2_k141_118869 | 1208 | No | NA | 2 | 0 | 1 | Not-determined | Genome-fragment | NA | NA | 0 |
| S2_k141_118872 | 2436 | No | NA | 1 | 0 | 1 | Not-determined | Genome-fragment | NA | NA | 0 |
| S2_k141_15269 | 1039 | No | NA | 2 | 0 | 0 | Not-determined | Genome-fragment | NA | NA | 0 |
| S2_k141_118884 | 1429 | No | NA | 1 | 0 | 1 | Not-determined | Genome-fragment | NA | NA | 0 |
| S2_k141_15301 | 1953 | No | NA | 1 | 0 | 0 | Not-determined | Genome-fragment | NA | NA | 0 |
| S1_k141_307987 | 4598 | No | NA | 6 | 5 | 0 | Low-quality | Genome-fragment | 11.7 | AAI-based (high-confidence) | 0 |
| S2_k141_57077 | 1447 | No | NA | 1 | 0 | 1 | Not-determined | Genome-fragment | NA | NA | 0 |
| S2_k141_36433 | 6324 | No | NA | 6 | 0 | 3 | Not-determined | Genome-fragment | NA | NA | 0 |
| S2_k141_15334 | 1074 | No | NA | 1 | 0 | 1 | Not-determined | Genome-fragment | NA | NA | 0 |
| S2_k141_233161 | 1456 | No | NA | 3 | 0 | 1 | Not-determined | Genome-fragment | NA | NA | 0 |
| S2_k141_57110 | 1127 | No | NA | 1 | 0 | 0 | Not-determined | Genome-fragment | NA | NA | 0 |
| S2_k141_233182 | 1022 | No | NA | 3 | 0 | 2 | Not-determined | Genome-fragment | NA | NA | 0 |
| S3-final-viral-combined_k141_128165\|\|full | 4598 | No | NA | 4 | 0 | 2 | Low-quality | Genome-fragment | 13.3 | AAI-based (high-confidence) | 0 |
| S12-final-viral-combined_k141_319692\|\|full | 4598 | No | NA | 5 | 0 | 1 | Low-quality | Genome-fragment | 5.11 | AAI-based (medium-confidence) | 0 |
| S2-final-viral-combined_k141_217831\|\|full | 4598 | No | NA | 7 | 0 | 0 | Low-quality | Genome-fragment | 2.16 | AAI-based (high-confidence) | 0 |
| S2_k141_15416 | 1541 | No | NA | 2 | 0 | 0 | Not-determined | Genome-fragment | NA | NA | 0 |
| S1-final-viral-combined_k141_284194\|\|full | 4599 | No | NA | 3 | 0 | 3 | Low-quality | Genome-fragment | 3.8 | AAI-based (high-confidence) | 0 |
| S2_k141_200608 | 1348 | No | NA | 1 | 0 | 1 | Not-determined | Genome-fragment | NA | NA | 0 |
| S2_k141_15433 | 2300 | No | NA | 1 | 0 | 0 | Not-determined | Genome-fragment | NA | NA | 0 |
| S7_k141_30074 | 4599 | Yes | 2119 | 3 | 1 | 1 | Low-quality | Genome-fragment | 1 | AAI-based (high-confidence) | 53.92 |
| S2_k141_88196 | 1176 | No | NA | 2 | 0 | 1 | Not-determined | Genome-fragment | NA | NA | 0 |
| S2_k141_191455 | 1115 | No | NA | 1 | 0 | 1 | Not-determined | Genome-fragment | NA | NA | 0 |
| S2_k141_88226 | 1206 | No | NA | 1 | 0 | 0 | Not-determined | Genome-fragment | NA | NA | 0 |
| S2_k141_211886 | 1396 | No | NA | 1 | 0 | 1 | Not-determined | Genome-fragment | NA | NA | 0 |
| S2_k141_128628 | 1329 | No | NA | 1 | 0 | 0 | Not-determined | Genome-fragment | NA | NA | 0 |
| S2_k141_67142 | 1473 | No | NA | 1 | 0 | 1 | Not-determined | Genome-fragment | NA | NA | 0 |
| S2_k141_128630 | 3120 | No | NA | 3 | 0 | 1 | Not-determined | Genome-fragment | NA | NA | 0 |
| S2_k141_139957 | 2140 | No | NA | 2 | 0 | 1 | Not-determined | Genome-fragment | NA | NA | 0 |
| S2_k141_67183 | 1006 | No | NA | 1 | 0 | 0 | Not-determined | Genome-fragment | NA | NA | 0 |
| S2_k141_242404 | 1126 | No | NA | 1 | 0 | 1 | Not-determined | Genome-fragment | NA | NA | 0 |
| S2_k141_171412 | 1557 | No | NA | 2 | 0 | 0 | Not-determined | Genome-fragment | NA | NA | 0 |
| S2_k141_160137 | 2392 | No | NA | 1 | 0 | 1 | Not-determined | Genome-fragment | NA | NA | 0 |
| S2_k141_119048 | 1415 | No | NA | 1 | 0 | 1 | Not-determined | Genome-fragment | NA | NA | 0 |
| S2_k141_233326 | 1076 | No | NA | 1 | 0 | 0 | Not-determined | Genome-fragment | NA | NA | 0 |
| S2_k141_221661 | 3473 | No | NA | 2 | 0 | 1 | Not-determined | Genome-fragment | NA | NA | 0 |
| S2_k141_48197 | 3065 | No | NA | 3 | 0 | 0 | Not-determined | Genome-fragment | NA | NA | 0 |
| S12-final-viral-combined_k141_296790\|\|full | 4599 | No | NA | 13 | 5 | 0 | Low-quality | Genome-fragment | 9.33 | AAI-based (high-confidence) | 0 |
| S2_k141_77655 | 3754 | No | NA | 5 | 0 | 3 | Not-determined | Genome-fragment | NA | NA | 0 |
| S2_k141_15461 | 6749 | No | NA | 7 | 0 | 7 | Not-determined | Genome-fragment | NA | NA | 0 |
| S2_k141_98782 | 1236 | No | NA | 2 | 0 | 1 | Not-determined | Genome-fragment | NA | NA | 0 |
| S2_k141_57272 | 19676 | No | NA | 18 | 0 | 6 | Not-determined | Genome-fragment | NA | NA | 0 |
| S2_k141_119111 | 1353 | No | NA | 2 | 0 | 0 | Not-determined | Genome-fragment | NA | NA | 0 |
| S2-final-viral-combined_k141_242004\|\|lt2gene | 4600 | No | NA | 2 | 2 | 0 | Low-quality | Genome-fragment | 1.65 | HMM-based (lower-bound) | 0 |
| S2_k141_77669 | 13138 | No | NA | 9 | 0 | 6 | Not-determined | Genome-fragment | NA | NA | 0 |
| S2_k141_67223 | 3835 | No | NA | 7 | 0 | 1 | Not-determined | Genome-fragment | NA | NA | 0 |
| S2_k141_119140 | 1072 | No | NA | 1 | 0 | 0 | Not-determined | Genome-fragment | NA | NA | 0 |
| S4_k141_104460 | 4600 | No | NA | 3 | 0 | 2 | Low-quality | Genome-fragment | 2.89 | AAI-based (high-confidence) | 0 |
| S2_k141_181801 | 1050 | No | NA | 1 | 0 | 1 | Not-determined | Genome-fragment | NA | NA | 0 |
| S2_k141_140059 | 1037 | No | NA | 1 | 0 | 0 | Not-determined | Genome-fragment | NA | NA | 0 |
| S2_k141_15582 | 1209 | No | NA | 1 | 0 | 0 | Not-determined | Genome-fragment | NA | NA | 0 |
| S2_k141_98865 | 1623 | No | NA | 2 | 0 | 1 | Not-determined | Genome-fragment | NA | NA | 0 |
| S11-final-viral-combined_k141_321918\|\|full | 4601 | No | NA | 7 | 0 | 1 | Low-quality | Genome-fragment | 8.67 | AAI-based (medium-confidence) | 0 |
| S2_k141_242535 | 2088 | No | NA | 2 | 0 | 1 | Not-determined | Genome-fragment | NA | NA | 0 |
| S4_k141_47574 | 4602 | No | NA | 8 | 0 | 4 | Low-quality | Genome-fragment | 4.9 | AAI-based (medium-confidence) | 0 |
| S2_k141_140148 | 3485 | No | NA | 2 | 0 | 1 | Not-determined | Genome-fragment | NA | NA | 0 |
| S2_k141_15620 | 1254 | No | NA | 1 | 0 | 1 | Not-determined | Genome-fragment | NA | NA | 0 |
| S2_k141_191706 | 1254 | No | NA | 2 | 0 | 0 | Not-determined | Genome-fragment | NA | NA | 0 |
| S2_k141_150277 | 4284 | No | NA | 6 | 0 | 1 | Not-determined | Genome-fragment | NA | NA | 0 |
| S2_k141_128717 | 6578 | No | NA | 14 | 0 | 14 | Not-determined | Genome-fragment | NA | NA | 0 |
| S2_k141_119285 | 3268 | No | NA | 4 | 0 | 1 | Not-determined | Genome-fragment | NA | NA | 0 |
| S2_k141_88450 | 1008 | No | NA | 1 | 0 | 0 | Not-determined | Genome-fragment | NA | NA | 0 |
| S2_k141_57429 | 5651 | No | NA | 4 | 0 | 2 | Not-determined | Genome-fragment | NA | NA | 0 |
| S2_k141_57431 | 1119 | No | NA | 1 | 0 | 0 | Not-determined | Genome-fragment | NA | NA | 0 |
| S11-final-viral-combined_k141_31777\|\|full | 4603 | No | NA | 2 | 1 | 0 | Low-quality | Genome-fragment | 6.48 | HMM-based (lower-bound) | 0 |
| S2_k141_15654 | 1968 | No | NA | 1 | 0 | 1 | Not-determined | Genome-fragment | NA | NA | 0 |
| S2_k141_242642 | 1091 | No | NA | 1 | 0 | 1 | Not-determined | Genome-fragment | NA | NA | 0 |
| S2_k141_191718 | 3243 | No | NA | 1 | 0 | 0 | Not-determined | Genome-fragment | NA | NA | 0 |
| S1-final-viral-combined_k141_22917\|\|full | 4603 | No | NA | 5 | 0 | 0 | Low-quality | Genome-fragment | 5.59 | AAI-based (medium-confidence) | 0 |
| S2_k141_108526 | 1608 | No | NA | 2 | 0 | 2 | Not-determined | Genome-fragment | NA | NA | 0 |
| S2_k141_98959 | 1178 | No | NA | 1 | 0 | 1 | Not-determined | Genome-fragment | NA | NA | 0 |
| S5-final-viral-combined_k141_37035\|\|full | 4603 | No | NA | 6 | 1 | 0 | Low-quality | Genome-fragment | 12.27 | AAI-based (medium-confidence) | 0 |
| S2_k141_88560 | 1890 | No | NA | 1 | 0 | 1 | Not-determined | Genome-fragment | NA | NA | 0 |
| S5_k141_125250 | 4603 | No | NA | 9 | 5 | 0 | Low-quality | Genome-fragment | 9.86 | AAI-based (high-confidence) | 0 |
| S2_k141_15703 | 3176 | No | NA | 5 | 0 | 2 | Not-determined | Genome-fragment | NA | NA | 0 |
| S2_k141_108564 | 3314 | No | NA | 2 | 0 | 2 | Not-determined | Genome-fragment | NA | NA | 0 |
| S2-final-viral-combined_k141_187162\|\|full | 4603 | No | NA | 7 | 5 | 0 | Low-quality | Genome-fragment | 12.36 | AAI-based (high-confidence) | 0 |
| S2_k141_88587 | 3366 | No | NA | 2 | 0 | 2 | Not-determined | Genome-fragment | NA | NA | 0 |
| S2_k141_171711 | 1298 | No | NA | 1 | 0 | 1 | Not-determined | Genome-fragment | NA | NA | 0 |
| S2_k141_77998 | 3519 | No | NA | 5 | 0 | 1 | Not-determined | Genome-fragment | NA | NA | 0 |
| S3-final-viral-combined_k141_30267\|\|full | 4604 | No | NA | 4 | 0 | 0 | Low-quality | Genome-fragment | 2.81 | AAI-based (medium-confidence) | 0 |
| S2_k141_242706 | 4890 | No | NA | 5 | 0 | 4 | Not-determined | Genome-fragment | NA | NA | 0 |
| S2_k141_221774 | 1229 | No | NA | 2 | 0 | 0 | Not-determined | Genome-fragment | NA | NA | 0 |
| S2_k141_212150 | 1075 | No | NA | 1 | 0 | 1 | Not-determined | Genome-fragment | NA | NA | 0 |
| S2_k141_221782 | 1120 | No | NA | 2 | 0 | 2 | Not-determined | Genome-fragment | NA | NA | 0 |
| S2_k141_221783 | 5886 | No | NA | 14 | 0 | 14 | Not-determined | Genome-fragment | NA | NA | 0 |
| S7-final-viral-combined_k141_15534\|\|full | 4605 | No | NA | 7 | 4 | 0 | Low-quality | Genome-fragment | 12.12 | AAI-based (medium-confidence) | 0 |
| S8-final-viral-combined_k141_131986\|\|full | 4605 | No | NA | 4 | 0 | 3 | Low-quality | Genome-fragment | 2.99 | AAI-based (medium-confidence) | 0 |
| S2_k141_78033 | 3512 | No | NA | 3 | 0 | 2 | Not-determined | Genome-fragment | NA | NA | 0 |
| S11-final-viral-combined_k141_268548\|\|full | 4606 | No | NA | 5 | 0 | 3 | Low-quality | Genome-fragment | 1.28 | AAI-based (medium-confidence) | 0 |
| S2_k141_108646 | 5099 | No | NA | 5 | 0 | 4 | Not-determined | Genome-fragment | NA | NA | 0 |
| S1_k141_75913 | 4606 | No | NA | 3 | 0 | 2 | Low-quality | Genome-fragment | 2.97 | AAI-based (medium-confidence) | 0 |
| S5_k141_185020 | 4608 | No | NA | 10 | 6 | 0 | Low-quality | Genome-fragment | 4.34 | AAI-based (high-confidence) | 0 |
| S2_k141_108651 | 1806 | No | NA | 1 | 0 | 0 | Not-determined | Genome-fragment | NA | NA | 0 |
| S2_k141_119554 | 1157 | No | NA | 1 | 0 | 1 | Not-determined | Genome-fragment | NA | NA | 0 |
| S3_k141_80693 | 4609 | No | NA | 8 | 3 | 0 | Low-quality | Genome-fragment | 8.39 | AAI-based (high-confidence) | 0 |
| S17-k141_59313 | 4609 | No | NA | 4 | 0 | 3 | Low-quality | Genome-fragment | 1.92 | AAI-based (medium-confidence) | 0 |
| S3_k141_71012 | 4610 | No | NA | 4 | 0 | 3 | Low-quality | Genome-fragment | 2.4 | AAI-based (medium-confidence) | 0 |
| S8-final-viral-combined_k141_48704\|\|full | 4610 | No | NA | 4 | 0 | 3 | Low-quality | Genome-fragment | 1.31 | AAI-based (high-confidence) | 0 |
| S2_k141_160334 | 1409 | No | NA | 4 | 0 | 3 | Not-determined | Genome-fragment | NA | NA | 0 |
| S1-final-viral-combined_k141_170041\|\|full | 4611 | No | NA | 8 | 3 | 0 | Low-quality | Genome-fragment | 8.8 | AAI-based (high-confidence) | 0 |
| S12-final-viral-combined_k141_381479\|\|full | 4611 | No | NA | 9 | 7 | 0 | Low-quality | Genome-fragment | 7.59 | AAI-based (high-confidence) | 0 |
| S2_k141_48584 | 1993 | No | NA | 1 | 0 | 1 | Not-determined | Genome-fragment | NA | NA | 0 |
| S2_k141_15860 | 4854 | No | NA | 3 | 0 | 3 | Not-determined | Genome-fragment | NA | NA | 0 |
| S2_k141_191890 | 1623 | No | NA | 3 | 0 | 3 | Not-determined | Genome-fragment | NA | NA | 0 |
| S2_k141_201130 | 1411 | No | NA | 1 | 0 | 0 | Not-determined | Genome-fragment | NA | NA | 0 |
| S11_k141_239625 | 4612 | No | NA | 9 | 0 | 1 | Low-quality | Genome-fragment | 7.86 | AAI-based (medium-confidence) | 0 |
| S2_k141_99080 | 2192 | No | NA | 2 | 0 | 0 | Not-determined | Genome-fragment | NA | NA | 0 |
| S2_k141_99083 | 3162 | No | NA | 3 | 0 | 3 | Not-determined | Genome-fragment | NA | NA | 0 |
| S2_k141_212182 | 2040 | No | NA | 2 | 0 | 0 | Not-determined | Genome-fragment | NA | NA | 0 |
| S1_k141_147460 | 4612 | No | NA | 4 | 0 | 3 | Low-quality | Genome-fragment | 1.22 | AAI-based (medium-confidence) | 0 |
| S2_k141_4703 | 1476 | No | NA | 1 | 0 | 0 | Not-determined | Genome-fragment | NA | NA | 0 |
| S2_k141_108774 | 3221 | No | NA | 4 | 0 | 0 | Not-determined | Genome-fragment | NA | NA | 0 |
| S2_k141_128831 | 86290 | No | NA | 110 | 17 | 2 | Complete | High-quality | 100 | DTR (high-confidence) | 0 |
| S2_k141_171973 | 1296 | No | NA | 4 | 0 | 0 | Not-determined | Genome-fragment | NA | NA | 0 |
| S2_k141_88778 | 3824 | No | NA | 6 | 0 | 4 | Not-determined | Genome-fragment | NA | NA | 0 |
| S2_k141_182053 | 1124 | No | NA | 1 | 0 | 0 | Not-determined | Genome-fragment | NA | NA | 0 |
| S7-final-viral-combined_k141_119601\|\|full | 4612 | No | NA | 9 | 8 | 0 | Low-quality | Genome-fragment | 11.01 | AAI-based (high-confidence) | 0 |
| S2_k141_78296 | 5086 | No | NA | 5 | 0 | 1 | Not-determined | Genome-fragment | NA | NA | 0 |
| S2_k141_15960 | 3389 | No | NA | 1 | 0 | 1 | Not-determined | Genome-fragment | NA | NA | 0 |
| S2_k141_88831 | 1443 | No | NA | 3 | 0 | 2 | Not-determined | Genome-fragment | NA | NA | 0 |
| S2_k141_48737 | 1235 | No | NA | 2 | 0 | 0 | Not-determined | Genome-fragment | NA | NA | 0 |
| S2_k141_182116 | 1223 | No | NA | 2 | 0 | 1 | Not-determined | Genome-fragment | NA | NA | 0 |
| S2_k141_15982 | 1065 | No | NA | 1 | 0 | 0 | Not-determined | Genome-fragment | NA | NA | 0 |
| S2_k141_48750 | 1103 | No | NA | 2 | 0 | 1 | Not-determined | Genome-fragment | NA | NA | 0 |
| S2_k141_73133 | 4612 | No | NA | 2 | 0 | 2 | Low-quality | Genome-fragment | 0.4 | AAI-based (medium-confidence) | 0 |
| S2_k141_150620 | 3495 | No | NA | 4 | 0 | 2 | Not-determined | Genome-fragment | NA | NA | 0 |
| S11-final-viral-combined_k141_281679\|\|full | 4613 | No | NA | 6 | 2 | 0 | Low-quality | Genome-fragment | 8.78 | AAI-based (high-confidence) | 0 |
| S2_k141_88860 | 1710 | No | NA | 2 | 0 | 1 | Not-determined | Genome-fragment | NA | NA | 0 |
| S1_k141_171548 | 4613 | No | NA | 4 | 0 | 3 | Low-quality | Genome-fragment | 2.62 | AAI-based (medium-confidence) | 0 |
| S2_k141_119797 | 1019 | No | NA | 1 | 0 | 1 | Not-determined | Genome-fragment | NA | NA | 0 |
| S2_k141_201247 | 1081 | No | NA | 1 | 0 | 0 | Not-determined | Genome-fragment | NA | NA | 0 |
| S2_k141_140679 | 1986 | No | NA | 2 | 0 | 1 | Not-determined | Genome-fragment | NA | NA | 0 |
| S2_k141_128962 | 97342 | No | NA | 97 | 15 | 2 | Complete | High-quality | 100 | DTR (high-confidence) | 0 |
| S2_k141_88965 | 1283 | No | NA | 1 | 0 | 1 | Not-determined | Genome-fragment | NA | NA | 0 |
| S2_k141_182244 | 1110 | No | NA | 2 | 0 | 0 | Not-determined | Genome-fragment | NA | NA | 0 |
| S5-final-viral-combined_k141_35672\|\|full | 4613 | No | NA | 6 | 1 | 0 | Low-quality | Genome-fragment | 11.8 | AAI-based (medium-confidence) | 0 |
| S2_k141_150765 | 1296 | No | NA | 1 | 0 | 0 | Not-determined | Genome-fragment | NA | NA | 0 |
| S2_k141_172173 | 1159 | No | NA | 1 | 0 | 0 | Not-determined | Genome-fragment | NA | NA | 0 |
| S2_k141_140752 | 1378 | No | NA | 1 | 0 | 1 | Not-determined | Genome-fragment | NA | NA | 0 |
| S2_k141_160520 | 1013 | No | NA | 1 | 0 | 0 | Not-determined | Genome-fragment | NA | NA | 0 |
| S2_k141_108963 | 1114 | No | NA | 2 | 0 | 2 | Not-determined | Genome-fragment | NA | NA | 0 |
| S2_k141_49020 | 1188 | No | NA | 3 | 0 | 2 | Not-determined | Genome-fragment | NA | NA | 0 |
| S2_k141_192052 | 5361 | No | NA | 6 | 0 | 2 | Not-determined | Genome-fragment | NA | NA | 0 |
| S2_k141_212313 | 1801 | No | NA | 4 | 0 | 4 | Not-determined | Genome-fragment | NA | NA | 0 |
| S2_k141_192066 | 1291 | No | NA | 2 | 0 | 1 | Not-determined | Genome-fragment | NA | NA | 0 |
| S2_k141_129055 | 1272 | No | NA | 1 | 0 | 0 | Not-determined | Genome-fragment | NA | NA | 0 |
| S2_k141_212334 | 1674 | No | NA | 2 | 0 | 2 | Not-determined | Genome-fragment | NA | NA | 0 |
| S2_k141_108994 | 1672 | No | NA | 3 | 0 | 3 | Not-determined | Genome-fragment | NA | NA | 0 |
| S2_k141_160570 | 6981 | No | NA | 5 | 0 | 0 | Not-determined | Genome-fragment | NA | NA | 0 |
| S2_k141_172247 | 1400 | No | NA | 1 | 0 | 1 | Not-determined | Genome-fragment | NA | NA | 0 |
| S2_k141_37287 | 1445 | No | NA | 1 | 0 | 0 | Not-determined | Genome-fragment | NA | NA | 0 |
| S2_k141_201324 | 1216 | No | NA | 1 | 0 | 1 | Not-determined | Genome-fragment | NA | NA | 0 |
| S9_k141_53700 | 4614 | No | NA | 3 | 0 | 3 | Low-quality | Genome-fragment | 1.91 | AAI-based (medium-confidence) | 0 |
| S11-final-viral-combined_k141_154415\|\|full | 4615 | No | NA | 3 | 0 | 2 | Low-quality | Genome-fragment | 1.45 | AAI-based (medium-confidence) | 0 |
| S2-final-viral-combined_k141_96727\|\|full | 4615 | No | NA | 10 | 1 | 0 | Low-quality | Genome-fragment | 9.04 | AAI-based (high-confidence) | 0 |
| S9_k141_23855 | 4616 | No | NA | 3 | 0 | 0 | Low-quality | Genome-fragment | 7.71 | AAI-based (high-confidence) | 0 |
| S4-final-viral-combined_k141_14852\|\|full | 4616 | No | NA | 11 | 3 | 0 | Low-quality | Genome-fragment | 4.49 | HMM-based (lower-bound) | 0 |
| S2_k141_78523 | 1723 | No | NA | 1 | 0 | 1 | Not-determined | Genome-fragment | NA | NA | 0 |
| S9-final-viral-combined_k141_47413\|\|full | 4617 | No | NA | 4 | 0 | 2 | Low-quality | Genome-fragment | 1.44 | AAI-based (medium-confidence) | 0 |
| S2_k141_192100 | 1094 | No | NA | 2 | 0 | 0 | Not-determined | Genome-fragment | NA | NA | 0 |
| S2_k141_16166 | 2522 | No | NA | 3 | 0 | 2 | Not-determined | Genome-fragment | NA | NA | 0 |
| S2_k141_57881 | 3258 | No | NA | 2 | 0 | 0 | Not-determined | Genome-fragment | NA | NA | 0 |
| S2_k141_172310 | 1269 | No | NA | 1 | 0 | 1 | Not-determined | Genome-fragment | NA | NA | 0 |
| S1-final-viral-combined_k141_65589\|\|full | 4617 | No | NA | 7 | 0 | 1 | Low-quality | Genome-fragment | 2.13 | AAI-based (medium-confidence) | 0 |
| S12_k141_109967 | 4619 | No | NA | 7 | 6 | 0 | Low-quality | Genome-fragment | 11.07 | AAI-based (high-confidence) | 0 |
| S2_k141_67634 | 3046 | No | NA | 6 | 0 | 2 | Not-determined | Genome-fragment | NA | NA | 0 |
| S2_k141_160645 | 1043 | No | NA | 1 | 0 | 0 | Not-determined | Genome-fragment | NA | NA | 0 |
| S2_k141_234025 | 1623 | No | NA | 1 | 0 | 0 | Not-determined | Genome-fragment | NA | NA | 0 |
| S2_k141_99434 | 1574 | No | NA | 2 | 0 | 2 | Not-determined | Genome-fragment | NA | NA | 0 |
| S4-final-viral-combined_k141_36023\|\|full | 4619 | No | NA | 4 | 0 | 3 | Low-quality | Genome-fragment | 6.04 | AAI-based (medium-confidence) | 0 |
| S2_k141_78579 | 8451 | No | NA | 5 | 0 | 2 | Not-determined | Genome-fragment | NA | NA | 0 |
| S2_k141_172413 | 1766 | No | NA | 3 | 0 | 0 | Not-determined | Genome-fragment | NA | NA | 0 |
| S1-final-viral-combined_k141_368001\|\|full | 4620 | No | NA | 3 | 0 | 3 | Low-quality | Genome-fragment | 2.31 | AAI-based (medium-confidence) | 0 |
| S2_k141_78593 | 1327 | No | NA | 1 | 0 | 1 | Not-determined | Genome-fragment | NA | NA | 0 |
| S2_k141_109188 | 1937 | No | NA | 1 | 0 | 0 | Not-determined | Genome-fragment | NA | NA | 0 |
| S2_k141_25671 | 1724 | No | NA | 3 | 0 | 1 | Not-determined | Genome-fragment | NA | NA | 0 |
| S2_k141_37418 | 1286 | No | NA | 2 | 0 | 0 | Not-determined | Genome-fragment | NA | NA | 0 |
| S2_k141_234068 | 2681 | No | NA | 1 | 0 | 1 | Not-determined | Genome-fragment | NA | NA | 0 |
| S1_k141_4686 | 4620 | No | NA | 3 | 0 | 0 | Low-quality | Genome-fragment | 4.87 | AAI-based (medium-confidence) | 0 |
| S2_k141_172463 | 5339 | No | NA | 10 | 0 | 2 | Not-determined | Genome-fragment | NA | NA | 0 |
| S17-final-viral-combined_k141_117412\|\|full | 4620 | No | NA | 7 | 1 | 2 | Low-quality | Genome-fragment | 9.49 | AAI-based (medium-confidence) | 0 |
| S2_k141_120079 | 3871 | No | NA | 5 | 0 | 0 | Not-determined | Genome-fragment | NA | NA | 0 |
| S2_k141_243028 | 1290 | No | NA | 3 | 0 | 2 | Not-determined | Genome-fragment | NA | NA | 0 |
| S2_k141_182467 | 5785 | No | NA | 5 | 0 | 4 | Not-determined | Genome-fragment | NA | NA | 0 |
| S2_k141_120117 | 3407 | No | NA | 4 | 0 | 3 | Not-determined | Genome-fragment | NA | NA | 0 |
| S2_k141_222062 | 1260 | No | NA | 1 | 0 | 1 | Not-determined | Genome-fragment | NA | NA | 0 |
| S2_k141_140933 | 45800 | Yes | 43762 | 75 | 18 | 2 | High-quality | High-quality | 100 | AAI-based (high-confidence) | 4.45 |
| S2_k141_243078 | 1171 | No | NA | 1 | 0 | 1 | Not-determined | Genome-fragment | NA | NA | 0 |
| S12-final-viral-combined_k141_194848\|\|full | 4620 | No | NA | 9 | 8 | 0 | Low-quality | Genome-fragment | 7.62 | AAI-based (high-confidence) | 0 |
| S2_k141_49281 | 2093 | No | NA | 2 | 0 | 2 | Not-determined | Genome-fragment | NA | NA | 0 |
| S2_k141_99489 | 3031 | No | NA | 8 | 0 | 7 | Not-determined | Genome-fragment | NA | NA | 0 |
| S6-final-viral-combined_k141_104329\|\|full | 4620 | No | NA | 5 | 3 | 0 | Low-quality | Genome-fragment | 19.04 | AAI-based (medium-confidence) | 0 |
| S11-final-viral-combined_k141_220386\|\|full | 4621 | No | NA | 8 | 1 | 0 | Low-quality | Genome-fragment | 4.15 | AAI-based (medium-confidence) | 0 |
| S12-final-viral-combined_k141_357681\|\|full | 4621 | No | NA | 3 | 0 | 0 | Low-quality | Genome-fragment | 4.58 | AAI-based (medium-confidence) | 0 |
| S2_k141_99520 | 2271 | No | NA | 1 | 0 | 1 | Not-determined | Genome-fragment | NA | NA | 0 |
| S2_k141_16269 | 4765 | No | NA | 6 | 0 | 2 | Not-determined | Genome-fragment | NA | NA | 0 |
| S2_k141_212459 | 2541 | No | NA | 3 | 0 | 1 | Not-determined | Genome-fragment | NA | NA | 0 |
| S2_k141_243162 | 3307 | No | NA | 5 | 0 | 0 | Not-determined | Genome-fragment | NA | NA | 0 |
| S2_k141_222112 | 3062 | No | NA | 3 | 0 | 3 | Not-determined | Genome-fragment | NA | NA | 0 |
| S4_k141_75138 | 4621 | No | NA | 2 | 0 | 2 | Low-quality | Genome-fragment | 4.19 | AAI-based (medium-confidence) | 0 |
| S2_k141_243176 | 1952 | No | NA | 1 | 0 | 1 | Not-determined | Genome-fragment | NA | NA | 0 |
| S5-final-viral-combined_k141_147678\|\|full | 4622 | No | NA | 2 | 0 | 2 | Low-quality | Genome-fragment | 2.3 | AAI-based (medium-confidence) | 0 |
| S17-final-viral-combined_k141_235644\|\|full | 4622 | No | NA | 4 | 0 | 1 | Low-quality | Genome-fragment | 2.87 | AAI-based (medium-confidence) | 0 |
| S2_k141_222157 | 1022 | No | NA | 1 | 0 | 1 | Not-determined | Genome-fragment | NA | NA | 0 |
| S4-final-viral-combined_k141_89146\|\|full | 4622 | Yes | 3484 | 9 | 2 | 1 | Low-quality | Genome-fragment | 7.47 | AAI-based (high-confidence) | 24.62 |
| S2_k141_212509 | 1215 | No | NA | 1 | 0 | 1 | Not-determined | Genome-fragment | NA | NA | 0 |
| S2_k141_222170 | 1457 | No | NA | 3 | 0 | 1 | Not-determined | Genome-fragment | NA | NA | 0 |
| S2_k141_25759 | 1113 | No | NA | 2 | 0 | 2 | Not-determined | Genome-fragment | NA | NA | 0 |
| S1-final-viral-combined_k141_297422\|\|full | 4623 | No | NA | 4 | 0 | 1 | Low-quality | Genome-fragment | 5.67 | AAI-based (medium-confidence) | 0 |
| S2_k141_16373 | 1708 | No | NA | 3 | 0 | 1 | Not-determined | Genome-fragment | NA | NA | 0 |
| S2_k141_192318 | 1274 | No | NA | 1 | 0 | 1 | Not-determined | Genome-fragment | NA | NA | 0 |
| S3-final-viral-combined_k141_33449\|\|full | 4623 | No | NA | 3 | 0 | 2 | Low-quality | Genome-fragment | 3.02 | AAI-based (high-confidence) | 0 |
| S2_k141_160833 | 1432 | No | NA | 1 | 0 | 1 | Not-determined | Genome-fragment | NA | NA | 0 |
| S2_k141_58094 | 1581 | No | NA | 2 | 0 | 1 | Not-determined | Genome-fragment | NA | NA | 0 |
| S9_k141_22108 | 4624 | No | NA | 7 | 3 | 0 | Low-quality | Genome-fragment | 7.34 | AAI-based (high-confidence) | 0 |
| S11-final-viral-combined_k141_186698\|\|full | 4624 | No | NA | 6 | 3 | 0 | Low-quality | Genome-fragment | 13.29 | AAI-based (high-confidence) | 0 |
| S8-final-viral-combined_k141_139866\|\|full | 4624 | No | NA | 4 | 0 | 0 | Low-quality | Genome-fragment | 7.66 | AAI-based (medium-confidence) | 0 |
| S2_k141_99692 | 3374 | No | NA | 3 | 0 | 3 | Not-determined | Genome-fragment | NA | NA | 0 |
| S8-final-viral-combined_k141_13444\|\|full | 4624 | No | NA | 5 | 3 | 0 | Low-quality | Genome-fragment | 9.54 | AAI-based (high-confidence) | 0 |
| S12-final-viral-combined_k141_119082\|\|full | 4625 | No | NA | 9 | 1 | 0 | Low-quality | Genome-fragment | 2.99 | HMM-based (lower-bound) | 0 |
| S2_k141_120259 | 1444 | No | NA | 2 | 0 | 2 | Not-determined | Genome-fragment | NA | NA | 0 |
| S2_k141_192375 | 1671 | No | NA | 1 | 0 | 1 | Not-determined | Genome-fragment | NA | NA | 0 |
| S2-final-viral-combined_k141_11701\|\|full | 4626 | No | NA | 6 | 1 | 0 | Low-quality | Genome-fragment | 5.19 | HMM-based (lower-bound) | 0 |
| S2_k141_49478 | 1399 | No | NA | 1 | 0 | 1 | Not-determined | Genome-fragment | NA | NA | 0 |
| S2_k141_109412 | 1338 | No | NA | 2 | 0 | 1 | Not-determined | Genome-fragment | NA | NA | 0 |
| S2_k141_222236 | 1255 | No | NA | 3 | 0 | 0 | Not-determined | Genome-fragment | NA | NA | 0 |
| S2_k141_201703 | 8873 | No | NA | 5 | 0 | 3 | Not-determined | Genome-fragment | NA | NA | 0 |
| S1-final-viral-combined_k141_19953\|\|full | 4627 | No | NA | 5 | 0 | 3 | Low-quality | Genome-fragment | 1.53 | AAI-based (medium-confidence) | 0 |
| S2_k141_37517 | 1078 | No | NA | 2 | 0 | 1 | Not-determined | Genome-fragment | NA | NA | 0 |
| S2_k141_222258 | 2566 | No | NA | 1 | 0 | 1 | Not-determined | Genome-fragment | NA | NA | 0 |
| S1-final-viral-combined_k141_13156\|\|full | 4627 | No | NA | 3 | 2 | 0 | Low-quality | Genome-fragment | 13.24 | AAI-based (high-confidence) | 0 |
| S2_k141_129239 | 1132 | No | NA | 1 | 0 | 0 | Not-determined | Genome-fragment | NA | NA | 0 |
| S2_k141_192414 | 1327 | No | NA | 2 | 0 | 1 | Not-determined | Genome-fragment | NA | NA | 0 |
| S2_k141_49534 | 1294 | No | NA | 2 | 0 | 1 | Not-determined | Genome-fragment | NA | NA | 0 |
| S2_k141_234485 | 1182 | No | NA | 1 | 0 | 1 | Not-determined | Genome-fragment | NA | NA | 0 |
| S17-final-viral-combined_k141_17441\|\|full | 4627 | Yes | 2500 | 6 | 1 | 1 | Low-quality | Genome-fragment | 2.81 | HMM-based (lower-bound) | 45.97 |
| S2_k141_160948 | 1015 | No | NA | 1 | 0 | 1 | Not-determined | Genome-fragment | NA | NA | 0 |
| S1-final-viral-combined_k141_20378\|\|full | 4628 | No | NA | 9 | 8 | 0 | Low-quality | Genome-fragment | 10.57 | AAI-based (high-confidence) | 0 |
| S2_k141_222299 | 1176 | No | NA | 1 | 0 | 1 | Not-determined | Genome-fragment | NA | NA | 0 |
| S2_k141_120322 | 1175 | No | NA | 3 | 0 | 1 | Not-determined | Genome-fragment | NA | NA | 0 |
| S2_k141_4954 | 1370 | No | NA | 2 | 0 | 1 | Not-determined | Genome-fragment | NA | NA | 0 |
| S2_k141_141029 | 1366 | No | NA | 1 | 0 | 1 | Not-determined | Genome-fragment | NA | NA | 0 |
| S2_k141_49615 | 4441 | No | NA | 6 | 0 | 2 | Not-determined | Genome-fragment | NA | NA | 0 |
| S17-k141_50917 | 4628 | No | NA | 3 | 0 | 2 | Low-quality | Genome-fragment | 2.49 | AAI-based (medium-confidence) | 0 |
| S2_k141_234546 | 1649 | No | NA | 2 | 0 | 0 | Not-determined | Genome-fragment | NA | NA | 0 |
| S11-final-viral-combined_k141_30701\|\|full | 4629 | No | NA | 3 | 0 | 0 | Low-quality | Genome-fragment | 3.65 | AAI-based (medium-confidence) | 0 |
| S8-final-viral-combined_k141_223141\|\|full | 4629 | No | NA | 4 | 4 | 0 | Low-quality | Genome-fragment | 11.7 | AAI-based (medium-confidence) | 0 |
| S17-final-viral-combined_k141_190317\|\|lt2gene | 4630 | No | NA | 2 | 0 | 1 | Low-quality | Genome-fragment | 10.39 | AAI-based (medium-confidence) | 0 |
| S6_k141_92307 | 4630 | No | NA | 6 | 0 | 1 | Low-quality | Genome-fragment | 9.84 | AAI-based (medium-confidence) | 0 |
| S1-final-viral-combined_k141_208925\|\|full | 4631 | No | NA | 2 | 0 | 0 | Low-quality | Genome-fragment | 10.79 | AAI-based (medium-confidence) | 0 |
| S2_k141_192532 | 1181 | No | NA | 2 | 0 | 2 | Not-determined | Genome-fragment | NA | NA | 0 |
| S2_k141_78799 | 1202 | No | NA | 1 | 0 | 0 | Not-determined | Genome-fragment | NA | NA | 0 |
| S2_k141_49625 | 1048 | No | NA | 2 | 0 | 1 | Not-determined | Genome-fragment | NA | NA | 0 |
| S2_k141_173110 | 3775 | No | NA | 2 | 0 | 2 | Not-determined | Genome-fragment | NA | NA | 0 |
| S11-final-viral-combined_k141_310165\|\|full | 4632 | No | NA | 4 | 0 | 1 | Low-quality | Genome-fragment | 5.97 | AAI-based (medium-confidence) | 0 |
| S11-final-viral-combined_k141_137623\|\|full | 4632 | No | NA | 5 | 0 | 4 | Low-quality | Genome-fragment | 2.01 | AAI-based (medium-confidence) | 0 |
| S1_k141_152171 | 4633 | No | NA | 6 | 0 | 0 | Low-quality | Genome-fragment | 10.09 | AAI-based (high-confidence) | 0 |
| S2_k141_161089 | 1599 | No | NA | 2 | 0 | 2 | Not-determined | Genome-fragment | NA | NA | 0 |
| S2_k141_49653 | 1174 | No | NA | 2 | 0 | 1 | Not-determined | Genome-fragment | NA | NA | 0 |
| S2_k141_182792 | 1199 | No | NA | 1 | 0 | 1 | Not-determined | Genome-fragment | NA | NA | 0 |
| S2_k141_234687 | 2104 | No | NA | 2 | 0 | 2 | Not-determined | Genome-fragment | NA | NA | 0 |
| S17-final-viral-combined_k141_106761\|\|full | 4633 | No | NA | 12 | 2 | 0 | Low-quality | Genome-fragment | 5.12 | HMM-based (lower-bound) | 0 |
| S2_k141_182846 | 2453 | No | NA | 1 | 0 | 1 | Not-determined | Genome-fragment | NA | NA | 0 |
| S2_k141_5071 | 2933 | No | NA | 3 | 0 | 1 | Not-determined | Genome-fragment | NA | NA | 0 |
| S1-final-viral-combined_k141_370168\|\|full | 4636 | Yes | 3592 | 8 | 4 | 1 | Low-quality | Genome-fragment | 5.47 | HMM-based (lower-bound) | 22.52 |
| S5-final-viral-combined_k141_80536\|\|full | 4637 | No | NA | 6 | 0 | 0 | Low-quality | Genome-fragment | 10.6 | AAI-based (high-confidence) | 0 |
| S2_k141_37736 | 1118 | No | NA | 2 | 0 | 1 | Not-determined | Genome-fragment | NA | NA | 0 |
| S6_k141_62908 | 4637 | No | NA | 6 | 2 | 0 | Low-quality | Genome-fragment | 12.03 | AAI-based (high-confidence) | 0 |
| S2_k141_68237 | 1502 | No | NA | 2 | 0 | 0 | Not-determined | Genome-fragment | NA | NA | 0 |
| S2_k141_151608 | 1662 | No | NA | 1 | 0 | 1 | Not-determined | Genome-fragment | NA | NA | 0 |
| S2_k141_89740 | 1809 | No | NA | 2 | 0 | 1 | Not-determined | Genome-fragment | NA | NA | 0 |
| S4-final-viral-combined_k141_115953\|\|full | 4638 | No | NA | 6 | 4 | 0 | Low-quality | Genome-fragment | 11.27 | AAI-based (medium-confidence) | 0 |
| S2_k141_161273 | 1169 | No | NA | 3 | 0 | 0 | Not-determined | Genome-fragment | NA | NA | 0 |
| S9_k141_8725 | 4639 | No | NA | 4 | 0 | 3 | Low-quality | Genome-fragment | 1.23 | AAI-based (high-confidence) | 0 |
| S2_k141_109603 | 1012 | No | NA | 1 | 0 | 1 | Not-determined | Genome-fragment | NA | NA | 0 |
| S2_k141_16815 | 1000 | No | NA | 1 | 0 | 1 | Not-determined | Genome-fragment | NA | NA | 0 |
| S2_k141_212912 | 3640 | No | NA | 2 | 0 | 2 | Not-determined | Genome-fragment | NA | NA | 0 |
| S2_k141_182951 | 3677 | No | NA | 3 | 0 | 2 | Not-determined | Genome-fragment | NA | NA | 0 |
| S2_k141_26086 | 1025 | No | NA | 2 | 0 | 0 | Not-determined | Genome-fragment | NA | NA | 0 |
| S2_k141_78908 | 1575 | No | NA | 1 | 0 | 0 | Not-determined | Genome-fragment | NA | NA | 0 |
| S11-final-viral-combined_k141_322998\|\|full | 4639 | No | NA | 11 | 1 | 0 | Low-quality | Genome-fragment | 10.03 | AAI-based (high-confidence) | 0 |
| S2-final-viral-combined_k141_28491\|\|full | 4640 | No | NA | 11 | 2 | 0 | Low-quality | Genome-fragment | 11.33 | AAI-based (high-confidence) | 0 |
| S2_k141_26098 | 1520 | No | NA | 2 | 0 | 0 | Not-determined | Genome-fragment | NA | NA | 0 |
| S2_k141_49882 | 3933 | No | NA | 5 | 0 | 3 | Not-determined | Genome-fragment | NA | NA | 0 |
| S9-final-viral-combined_k141_32350\|\|full | 4641 | No | NA | 6 | 0 | 0 | Low-quality | Genome-fragment | 9.08 | AAI-based (high-confidence) | 0 |
| S2_k141_89804 | 3955 | No | NA | 1 | 0 | 0 | Not-determined | Genome-fragment | NA | NA | 0 |
| S2_k141_49927 | 2609 | No | NA | 1 | 0 | 1 | Not-determined | Genome-fragment | NA | NA | 0 |
| S2_k141_212974 | 3050 | No | NA | 2 | 0 | 1 | Not-determined | Genome-fragment | NA | NA | 0 |
| S2_k141_49944 | 1061 | No | NA | 1 | 0 | 0 | Not-determined | Genome-fragment | NA | NA | 0 |
| S2_k141_192667 | 26847 | No | NA | 33 | 16 | 2 | Medium-quality | Genome-fragment | 71.79 | AAI-based (high-confidence) | 0 |
| S2_k141_16881 | 3664 | No | NA | 4 | 0 | 2 | Not-determined | Genome-fragment | NA | NA | 0 |
| S2_k141_222562 | 5273 | No | NA | 9 | 0 | 5 | Not-determined | Genome-fragment | NA | NA | 0 |
| S2_k141_151783 | 1500 | No | NA | 2 | 0 | 1 | Not-determined | Genome-fragment | NA | NA | 0 |
| S2_k141_16908 | 1034 | No | NA | 2 | 0 | 0 | Not-determined | Genome-fragment | NA | NA | 0 |
| S2_k141_192731 | 1903 | No | NA | 5 | 0 | 0 | Not-determined | Genome-fragment | NA | NA | 0 |
| S7-final-viral-combined_k141_241461\|\|full | 4641 | No | NA | 7 | 5 | 0 | Low-quality | Genome-fragment | 9.99 | AAI-based (medium-confidence) | 0 |
| S2_k141_222583 | 5452 | No | NA | 9 | 0 | 4 | Not-determined | Genome-fragment | NA | NA | 0 |
| S2_k141_37829 | 1672 | No | NA | 1 | 0 | 0 | Not-determined | Genome-fragment | NA | NA | 0 |
| S17-final-viral-combined_k141_215578\|\|full | 4641 | No | NA | 5 | 0 | 2 | Low-quality | Genome-fragment | 5.28 | AAI-based (high-confidence) | 0 |
| S7-final-viral-combined_k141_170044\|\|full | 4642 | No | NA | 6 | 4 | 0 | Low-quality | Genome-fragment | 10.18 | AAI-based (high-confidence) | 0 |
| S2_k141_141275 | 4040 | No | NA | 8 | 0 | 8 | Not-determined | Genome-fragment | NA | NA | 0 |
| S2_k141_151836 | 3526 | No | NA | 3 | 0 | 3 | Not-determined | Genome-fragment | NA | NA | 0 |
| S17-k141_21344 | 4642 | No | NA | 2 | 0 | 0 | Low-quality | Genome-fragment | 7.7 | AAI-based (high-confidence) | 0 |
| S8-final-viral-combined_k141_247827\|\|full | 4643 | No | NA | 3 | 0 | 2 | Low-quality | Genome-fragment | 3.28 | AAI-based (medium-confidence) | 0 |
| S7-final-viral-combined_k141_11815\|\|full | 4644 | No | NA | 9 | 3 | 0 | Low-quality | Genome-fragment | 9.73 | AAI-based (high-confidence) | 0 |
| S2_k141_141326 | 1397 | No | NA | 2 | 0 | 1 | Not-determined | Genome-fragment | NA | NA | 0 |
| S2_k141_202079 | 1132 | No | NA | 1 | 0 | 0 | Not-determined | Genome-fragment | NA | NA | 0 |
| S2_k141_89953 | 1073 | No | NA | 1 | 0 | 1 | Not-determined | Genome-fragment | NA | NA | 0 |
| S2_k141_161472 | 6053 | No | NA | 10 | 0 | 0 | Not-determined | Genome-fragment | NA | NA | 0 |
| S11-final-viral-combined_k141_245984\|\|full | 4645 | Yes | 3119 | 10 | 2 | 1 | Low-quality | Genome-fragment | 7.93 | AAI-based (high-confidence) | 32.85 |
| S2_k141_120729 | 4469 | No | NA | 1 | 0 | 1 | Not-determined | Genome-fragment | NA | NA | 0 |
| S2_k141_68385 | 2309 | No | NA | 3 | 0 | 1 | Not-determined | Genome-fragment | NA | NA | 0 |
| S2_k141_100135 | 1006 | No | NA | 1 | 0 | 1 | Not-determined | Genome-fragment | NA | NA | 0 |
| S2_k141_89982 | 2802 | No | NA | 4 | 0 | 2 | Not-determined | Genome-fragment | NA | NA | 0 |
| S1-final-viral-combined_k141_27085\|\|full | 4645 | No | NA | 10 | 4 | 0 | Low-quality | Genome-fragment | 7.69 | AAI-based (high-confidence) | 0 |
| S2_k141_202118 | 1331 | No | NA | 2 | 0 | 1 | Not-determined | Genome-fragment | NA | NA | 0 |
| S2_k141_17027 | 4399 | No | NA | 3 | 0 | 0 | Not-determined | Genome-fragment | NA | NA | 0 |
| S2_k141_17056 | 2012 | No | NA | 4 | 0 | 0 | Not-determined | Genome-fragment | NA | NA | 0 |
| S2_k141_17057 | 1313 | No | NA | 4 | 0 | 2 | Not-determined | Genome-fragment | NA | NA | 0 |
| S2_k141_58536 | 3141 | No | NA | 3 | 0 | 1 | Not-determined | Genome-fragment | NA | NA | 0 |
| S2_k141_192811 | 5158 | No | NA | 2 | 0 | 0 | Not-determined | Genome-fragment | NA | NA | 0 |
| S4-final-viral-combined_k141_58159\|\|full | 4645 | No | NA | 3 | 0 | 3 | Low-quality | Genome-fragment | 2.23 | AAI-based (medium-confidence) | 0 |
| S2_k141_222726 | 5522 | No | NA | 6 | 0 | 6 | Not-determined | Genome-fragment | NA | NA | 0 |
| S2_k141_129723 | 1370 | No | NA | 2 | 0 | 1 | Not-determined | Genome-fragment | NA | NA | 0 |
| S2_k141_173606 | 1772 | No | NA | 1 | 0 | 0 | Not-determined | Genome-fragment | NA | NA | 0 |
| S5_k141_46441 | 4647 | No | NA | 7 | 0 | 1 | Low-quality | Genome-fragment | 5.69 | AAI-based (medium-confidence) | 0 |
| S2_k141_234985 | 1920 | No | NA | 2 | 0 | 0 | Not-determined | Genome-fragment | NA | NA | 0 |
| S2_k141_38034 | 1459 | No | NA | 1 | 0 | 0 | Not-determined | Genome-fragment | NA | NA | 0 |
| S2_k141_5295 | 1114 | No | NA | 3 | 0 | 1 | Not-determined | Genome-fragment | NA | NA | 0 |
| S9-final-viral-combined_k141_31418\|\|full | 4649 | No | NA | 8 | 3 | 0 | Low-quality | Genome-fragment | 12.77 | AAI-based (high-confidence) | 0 |
| S2_k141_50182 | 1186 | No | NA | 1 | 0 | 0 | Not-determined | Genome-fragment | NA | NA | 0 |
| S5-final-viral-combined_k141_149945\|\|full | 4651 | No | NA | 7 | 7 | 0 | Low-quality | Genome-fragment | 6.54 | AAI-based (high-confidence) | 0 |
| S2_k141_243613 | 1289 | No | NA | 2 | 0 | 0 | Not-determined | Genome-fragment | NA | NA | 0 |
| S2_k141_129777 | 6304 | No | NA | 5 | 0 | 5 | Not-determined | Genome-fragment | NA | NA | 0 |
| S2_k141_129795 | 1015 | No | NA | 1 | 0 | 1 | Not-determined | Genome-fragment | NA | NA | 0 |
| S2_k141_90140 | 1314 | No | NA | 2 | 0 | 1 | Not-determined | Genome-fragment | NA | NA | 0 |
| S2_k141_192922 | 5066 | No | NA | 7 | 0 | 0 | Not-determined | Genome-fragment | NA | NA | 0 |
| S2-final-viral-combined_k141_1505\|\|full | 4652 | No | NA | 7 | 0 | 4 | Low-quality | Genome-fragment | 2.59 | AAI-based (medium-confidence) | 0 |
| S2_k141_5363 | 1588 | No | NA | 2 | 0 | 1 | Not-determined | Genome-fragment | NA | NA | 0 |
| S7-final-viral-combined_k141_63328\|\|full | 4653 | No | NA | 6 | 0 | 1 | Low-quality | Genome-fragment | 3.58 | AAI-based (high-confidence) | 0 |
| S7-final-viral-combined_k141_13287\|\|full | 4653 | No | NA | 12 | 0 | 0 | Low-quality | Genome-fragment | 10.01 | AAI-based (high-confidence) | 0 |
| S2_k141_100318 | 1714 | No | NA | 3 | 0 | 1 | Not-determined | Genome-fragment | NA | NA | 0 |
| S2_k141_90206 | 1101 | No | NA | 2 | 0 | 1 | Not-determined | Genome-fragment | NA | NA | 0 |
| S1-final-viral-combined_k141_41123\|\|full | 4655 | No | NA | 5 | 0 | 1 | Low-quality | Genome-fragment | 1.48 | AAI-based (medium-confidence) | 0 |
| S2_k141_26229 | 1125 | No | NA | 1 | 0 | 1 | Not-determined | Genome-fragment | NA | NA | 0 |
| S12_k141_356920 | 4655 | No | NA | 4 | 4 | 0 | Low-quality | Genome-fragment | 11.05 | AAI-based (high-confidence) | 0 |
| S2_k141_235124 | 4097 | No | NA | 1 | 0 | 1 | Not-determined | Genome-fragment | NA | NA | 0 |
| S5_k141_135931 | 4656 | No | NA | 9 | 4 | 0 | Low-quality | Genome-fragment | 11.73 | AAI-based (high-confidence) | 0 |
| S2_k141_38221 | 3002 | No | NA | 2 | 0 | 1 | Not-determined | Genome-fragment | NA | NA | 0 |
| S2_k141_213264 | 1067 | No | NA | 1 | 0 | 1 | Not-determined | Genome-fragment | NA | NA | 0 |
| S2_k141_183207 | 6007 | No | NA | 9 | 0 | 8 | Not-determined | Genome-fragment | NA | NA | 0 |
| S2_k141_79304 | 17157 | No | NA | 24 | 7 | 0 | Medium-quality | Genome-fragment | 88.09 | AAI-based (medium-confidence) | 0 |
| S2_k141_100457 | 8042 | No | NA | 4 | 0 | 2 | Not-determined | Genome-fragment | NA | NA | 0 |
| S2_k141_68763 | 1443 | No | NA | 1 | 0 | 1 | Not-determined | Genome-fragment | NA | NA | 0 |
| S2_k141_213330 | 1173 | No | NA | 1 | 0 | 1 | Not-determined | Genome-fragment | NA | NA | 0 |
| S2_k141_100471 | 1333 | No | NA | 1 | 0 | 1 | Not-determined | Genome-fragment | NA | NA | 0 |
| S2_k141_213336 | 1750 | No | NA | 1 | 0 | 1 | Not-determined | Genome-fragment | NA | NA | 0 |

| | | | | | | | | | | | |
|---|---|---|---|---|---|---|---|---|---|---|---|
| S2_k141_68776 | 2386 | No | NA | 3 | 0 | 0 | Not-determined | Genome-fragment | NA | NA | 0 |
| S2_k141_152120 | 3031 | No | NA | 2 | 0 | 1 | Not-determined | Genome-fragment | NA | NA | 0 |
| S11-final-viral-combined_k141_175039\|\|full | 4657 | Yes | 3636 | 4 | 1 | 2 | Low-quality | Genome-fragment | 6.29 | HMM-based (lower-bound) | 21.92 |
| S2_k141_38301 | 1286 | No | NA | 1 | 0 | 0 | Not-determined | Genome-fragment | NA | NA | 0 |
| S2_k141_121092 | 1044 | No | NA | 1 | 0 | 0 | Not-determined | Genome-fragment | NA | NA | 0 |
| S2_k141_173800 | 1049 | No | NA | 2 | 0 | 1 | Not-determined | Genome-fragment | NA | NA | 0 |
| S1-final-viral-combined_k141_376280\|\|full | 4658 | Yes | 3742 | 9 | 2 | 1 | Low-quality | Genome-fragment | 6.36 | AAI-based (medium-confidence) | 19.67 |
| S2_k141_141626 | 2853 | No | NA | 1 | 0 | 0 | Not-determined | Genome-fragment | NA | NA | 0 |
| S2_k141_38349 | 2469 | No | NA | 2 | 0 | 2 | Not-determined | Genome-fragment | NA | NA | 0 |
| S2_k141_100497 | 2803 | No | NA | 4 | 0 | 0 | Not-determined | Genome-fragment | NA | NA | 0 |
| S3_k141_125360 | 4658 | No | NA | 7 | 3 | 0 | Low-quality | Genome-fragment | 8.41 | AAI-based (high-confidence) | 0 |
| S2_k141_141644 | 1192 | No | NA | 1 | 0 | 0 | Not-determined | Genome-fragment | NA | NA | 0 |
| S2_k141_193147 | 2236 | No | NA | 2 | 0 | 1 | Not-determined | Genome-fragment | NA | NA | 0 |
| S2_k141_90429 | 1963 | No | NA | 3 | 0 | 2 | Not-determined | Genome-fragment | NA | NA | 0 |
| S17-final-viral-combined_k141_263869\|\|full | 4658 | No | NA | 9 | 7 | 0 | Low-quality | Genome-fragment | 11.53 | AAI-based (medium-confidence) | 0 |
| S2_k141_50515 | 1261 | No | NA | 1 | 0 | 1 | Not-determined | Genome-fragment | NA | NA | 0 |
| S6-final-viral-combined_k141_79244\|\|full | 4658 | No | NA | 4 | 0 | 3 | Low-quality | Genome-fragment | 2.15 | AAI-based (medium-confidence) | 0 |
| S2_k141_109893 | 1676 | No | NA | 2 | 0 | 2 | Not-determined | Genome-fragment | NA | NA | 0 |
| S2_k141_38402 | 1318 | No | NA | 2 | 0 | 0 | Not-determined | Genome-fragment | NA | NA | 0 |
| S2_k141_50541 | 4065 | No | NA | 6 | 0 | 1 | Not-determined | Genome-fragment | NA | NA | 0 |
| S2_k141_222935 | 1350 | No | NA | 1 | 0 | 0 | Not-determined | Genome-fragment | NA | NA | 0 |
| S2_k141_222956 | 1368 | No | NA | 2 | 0 | 1 | Not-determined | Genome-fragment | NA | NA | 0 |
| S2_k141_100520 | 12823 | No | NA | 5 | 0 | 1 | Not-determined | Genome-fragment | NA | NA | 0 |
| S2_k141_68959 | 6912 | No | NA | 6 | 0 | 2 | Not-determined | Genome-fragment | NA | NA | 0 |
| S12-final-viral-combined_k141_233139\|\|full | 4659 | No | NA | 7 | 1 | 0 | Low-quality | Genome-fragment | 10.63 | AAI-based (high-confidence) | 0 |
| S2_k141_141710 | 1023 | No | NA | 1 | 0 | 1 | Not-determined | Genome-fragment | NA | NA | 0 |
| S8_k141_124844 | 4659 | No | NA | 2 | 0 | 0 | Low-quality | Genome-fragment | 7.7 | AAI-based (medium-confidence) | 0 |
| S2_k141_193217 | 3206 | No | NA | 5 | 0 | 0 | Not-determined | Genome-fragment | NA | NA | 0 |
| S1-final-viral-combined_k141_327862\|\|full | 4660 | No | NA | 4 | 0 | 4 | Low-quality | Genome-fragment | 3.84 | AAI-based (medium-confidence) | 0 |
| S2_k141_109950 | 8903 | No | NA | 2 | 0 | 2 | Not-determined | Genome-fragment | NA | NA | 0 |
| S2_k141_58803 | 1086 | No | NA | 1 | 0 | 1 | Not-determined | Genome-fragment | NA | NA | 0 |
| S2_k141_223013 | 1437 | No | NA | 2 | 0 | 1 | Not-determined | Genome-fragment | NA | NA | 0 |
| S2_k141_50575 | 1452 | No | NA | 1 | 0 | 1 | Not-determined | Genome-fragment | NA | NA | 0 |
| S3-final-viral-combined_k141_35518\|\|full | 4660 | No | NA | 6 | 0 | 4 | Low-quality | Genome-fragment | 2.08 | AAI-based (medium-confidence) | 0 |
| S2_k141_26453 | 1565 | No | NA | 1 | 0 | 0 | Not-determined | Genome-fragment | NA | NA | 0 |
| S12-final-viral-combined_k141_353061\|\|full | 4660 | No | NA | 5 | 0 | 0 | Low-quality | Genome-fragment | 4.16 | AAI-based (medium-confidence) | 0 |
| S2_k141_223057 | 1569 | No | NA | 1 | 0 | 1 | Not-determined | Genome-fragment | NA | NA | 0 |
| S2_k141_235413 | 1230 | No | NA | 1 | 0 | 0 | Not-determined | Genome-fragment | NA | NA | 0 |
| S2_k141_5561 | 6157 | No | NA | 5 | 0 | 2 | Not-determined | Genome-fragment | NA | NA | 0 |
| S2_k141_17410 | 9246 | No | NA | 10 | 0 | 2 | Not-determined | Genome-fragment | NA | NA | 0 |
| S12-final-viral-combined_k141_240329\|\|full | 4660 | No | NA | 6 | 1 | 0 | Low-quality | Genome-fragment | 7.92 | AAI-based (medium-confidence) | 0 |
| S2_k141_235455 | 1354 | No | NA | 2 | 0 | 1 | Not-determined | Genome-fragment | NA | NA | 0 |
| S2_k141_193262 | 1055 | No | NA | 1 | 0 | 0 | Not-determined | Genome-fragment | NA | NA | 0 |
| S2_k141_90472 | 1415 | No | NA | 4 | 0 | 1 | Not-determined | Genome-fragment | NA | NA | 0 |
| S2_k141_17460 | 1502 | No | NA | 1 | 0 | 0 | Not-determined | Genome-fragment | NA | NA | 0 |
| S2_k141_183288 | 1704 | No | NA | 1 | 0 | 1 | Not-determined | Genome-fragment | NA | NA | 0 |
| S2_k141_58884 | 3473 | No | NA | 5 | 0 | 3 | Not-determined | Genome-fragment | NA | NA | 0 |
| S2_k141_213554 | 1003 | No | NA | 1 | 0 | 1 | Not-determined | Genome-fragment | NA | NA | 0 |
| S2_k141_174062 | 1129 | No | NA | 3 | 0 | 2 | Not-determined | Genome-fragment | NA | NA | 0 |
| S2_k141_243834 | 6952 | No | NA | 2 | 0 | 0 | Not-determined | Genome-fragment | NA | NA | 0 |
| S2_k141_243838 | 1499 | No | NA | 2 | 0 | 0 | Not-determined | Genome-fragment | NA | NA | 0 |
| S2_k141_50736 | 1197 | No | NA | 1 | 0 | 0 | Not-determined | Genome-fragment | NA | NA | 0 |
| S2_k141_130152 | 1984 | No | NA | 4 | 0 | 2 | Not-determined | Genome-fragment | NA | NA | 0 |
| S2_k141_121362 | 1146 | No | NA | 2 | 0 | 1 | Not-determined | Genome-fragment | NA | NA | 0 |
| S11_k141_214526 | 4661 | No | NA | 5 | 4 | 0 | Low-quality | Genome-fragment | 11.75 | AAI-based (high-confidence) | 0 |
| S2_k141_174105 | 4585 | No | NA | 1 | 0 | 0 | Not-determined | Genome-fragment | NA | NA | 0 |
| S2_k141_152420 | 1403 | No | NA | 1 | 0 | 1 | Not-determined | Genome-fragment | NA | NA | 0 |
| S2-final-viral-combined_k141_213186\|\|full | 4661 | No | NA | 4 | 1 | 0 | Low-quality | Genome-fragment | 6.29 | AAI-based (high-confidence) | 0 |
| S2_k141_69077 | 3357 | No | NA | 2 | 0 | 2 | Not-determined | Genome-fragment | NA | NA | 0 |
| S17-final-viral-combined_k141_48924\|\|full | 4662 | No | NA | 6 | 1 | 0 | Low-quality | Genome-fragment | 7.05 | HMM-based (lower-bound) | 0 |
| S2_k141_183369 | 1412 | No | NA | 3 | 0 | 0 | Not-determined | Genome-fragment | NA | NA | 0 |
| S2_k141_5681 | 3057 | No | NA | 4 | 0 | 3 | Not-determined | Genome-fragment | NA | NA | 0 |
| S17-k141_154536 | 4663 | No | NA | 5 | 0 | 0 | Low-quality | Genome-fragment | 3.24 | AAI-based (medium-confidence) | 0 |
| S2_k141_213613 | 1003 | No | NA | 2 | 0 | 0 | Not-determined | Genome-fragment | NA | NA | 0 |
| S2_k141_100844 | 3043 | No | NA | 2 | 0 | 1 | Not-determined | Genome-fragment | NA | NA | 0 |
| S2_k141_202695 | 2856 | No | NA | 4 | 0 | 2 | Not-determined | Genome-fragment | NA | NA | 0 |
| S2-final-viral-combined_k141_116543\|\|full | 4663 | No | NA | 5 | 0 | 0 | Low-quality | Genome-fragment | 5.2 | AAI-based (medium-confidence) | 0 |
| S2_k141_43132 | 4663 | No | NA | 7 | 6 | 0 | Low-quality | Genome-fragment | 6.46 | AAI-based (high-confidence) | 0 |
| S2_k141_223215 | 6115 | No | NA | 2 | 0 | 2 | Not-determined | Genome-fragment | NA | NA | 0 |
| S2_k141_100900 | 1393 | No | NA | 2 | 0 | 2 | Not-determined | Genome-fragment | NA | NA | 0 |
| S2_k141_59009 | 1000 | No | NA | 1 | 0 | 1 | Not-determined | Genome-fragment | NA | NA | 0 |
| S12-final-viral-combined_k141_386381\|\|full | 4664 | No | NA | 8 | 1 | 0 | Low-quality | Genome-fragment | 10.65 | AAI-based (high-confidence) | 0 |
| S2_k141_17599 | 3493 | No | NA | 4 | 0 | 3 | Not-determined | Genome-fragment | NA | NA | 0 |
| S9-final-viral-combined_k141_109503\|\|full | 4665 | No | NA | 7 | 1 | 0 | Low-quality | Genome-fragment | 11.88 | AAI-based (high-confidence) | 0 |
| S2_k141_213712 | 1147 | No | NA | 2 | 0 | 1 | Not-determined | Genome-fragment | NA | NA | 0 |
| S11-final-viral-combined_k141_313768\|\|full | 4665 | No | NA | 5 | 0 | 2 | Low-quality | Genome-fragment | 8.21 | AAI-based (high-confidence) | 0 |
| S2_k141_152588 | 1230 | No | NA | 1 | 0 | 0 | Not-determined | Genome-fragment | NA | NA | 0 |
| S2_k141_121480 | 3097 | No | NA | 3 | 0 | 0 | Not-determined | Genome-fragment | NA | NA | 0 |
| S2_k141_193496 | 1244 | No | NA | 2 | 0 | 0 | Not-determined | Genome-fragment | NA | NA | 0 |
| S2_k141_244014 | 1648 | No | NA | 1 | 0 | 1 | Not-determined | Genome-fragment | NA | NA | 0 |
| S12-final-viral-combined_k141_17551\|\|full | 4665 | No | NA | 3 | 0 | 1 | Low-quality | Genome-fragment | 4.92 | AAI-based (medium-confidence) | 0 |
| S12_k141_293746 | 4665 | No | NA | 4 | 0 | 3 | Low-quality | Genome-fragment | 1.05 | AAI-based (medium-confidence) | 0 |
| S2_k141_59094 | 1162 | No | NA | 2 | 0 | 1 | Not-determined | Genome-fragment | NA | NA | 0 |
| S2_k141_183515 | 2742 | No | NA | 2 | 0 | 2 | Not-determined | Genome-fragment | NA | NA | 0 |
| S2_k141_223342 | 1515 | No | NA | 2 | 0 | 2 | Not-determined | Genome-fragment | NA | NA | 0 |
| S2_k141_244065 | 1589 | No | NA | 1 | 0 | 1 | Not-determined | Genome-fragment | NA | NA | 0 |
| S2_k141_235755 | 1800 | No | NA | 3 | 0 | 1 | Not-determined | Genome-fragment | NA | NA | 0 |
| S2-final-viral-combined_k141_131080\|\|full | 4665 | No | NA | 3 | 0 | 1 | Low-quality | Genome-fragment | 2.94 | AAI-based (medium-confidence) | 0 |
| S2_k141_59135 | 1235 | No | NA | 2 | 0 | 1 | Not-determined | Genome-fragment | NA | NA | 0 |
| S2_k141_244090 | 3043 | No | NA | 3 | 0 | 3 | Not-determined | Genome-fragment | NA | NA | 0 |
| S2_k141_161879 | 1139 | No | NA | 1 | 0 | 1 | Not-determined | Genome-fragment | NA | NA | 0 |
| S2_k141_223364 | 1114 | No | NA | 1 | 0 | 1 | Not-determined | Genome-fragment | NA | NA | 0 |
| S9-final-viral-combined_k141_79116\|\|full | 4666 | No | NA | 4 | 4 | 0 | Low-quality | Genome-fragment | 12.33 | AAI-based (high-confidence) | 0 |
| S8-final-viral-combined_k141_106202\|\|full | 4667 | No | NA | 2 | 0 | 1 | Low-quality | Genome-fragment | 4.35 | AAI-based (medium-confidence) | 0 |
| S1_k141_201522 | 4668 | No | NA | 8 | 3 | 0 | Low-quality | Genome-fragment | 11.32 | AAI-based (high-confidence) | 0 |
| S2_k141_183560 | 1121 | No | NA | 2 | 0 | 0 | Not-determined | Genome-fragment | NA | NA | 0 |
| S2_k141_69318 | 1016 | No | NA | 1 | 0 | 0 | Not-determined | Genome-fragment | NA | NA | 0 |
| S2_k141_121571 | 4220 | No | NA | 8 | 0 | 1 | Not-determined | Genome-fragment | NA | NA | 0 |
| S1-final-viral-combined_k141_348682\|\|full | 4670 | No | NA | 3 | 0 | 1 | Low-quality | Genome-fragment | 1.44 | AAI-based (high-confidence) | 0 |
| S2_k141_235886 | 1177 | No | NA | 1 | 0 | 1 | Not-determined | Genome-fragment | NA | NA | 0 |
| S2_k141_183617 | 2638 | No | NA | 1 | 0 | 1 | Not-determined | Genome-fragment | NA | NA | 0 |
| S2_k141_101091 | 1488 | No | NA | 2 | 0 | 1 | Not-determined | Genome-fragment | NA | NA | 0 |
| S5-final-viral-combined_k141_152757\|\|full | 4670 | No | NA | 9 | 1 | 0 | Low-quality | Genome-fragment | 7.26 | HMM-based (lower-bound) | 0 |
| S2_k141_69373 | 2007 | No | NA | 2 | 0 | 0 | Not-determined | Genome-fragment | NA | NA | 0 |
| S2_k141_101105 | 1072 | No | NA | 1 | 0 | 1 | Not-determined | Genome-fragment | NA | NA | 0 |
| S2_k141_59223 | 1002 | No | NA | 1 | 0 | 0 | Not-determined | Genome-fragment | NA | NA | 0 |
| S2_k141_51101 | 3621 | No | NA | 1 | 0 | 1 | Not-determined | Genome-fragment | NA | NA | 0 |
| S12-final-viral-combined_k141_133764\|\|full | 4671 | No | NA | 4 | 1 | 0 | Low-quality | Genome-fragment | 2.65 | AAI-based (high-confidence) | 0 |
| S2-final-viral-combined_k141_198877\|\|full | 4671 | No | NA | 6 | 0 | 2 | Low-quality | Genome-fragment | 3.35 | AAI-based (high-confidence) | 0 |
| S2_k141_235931 | 3111 | No | NA | 2 | 0 | 1 | Not-determined | Genome-fragment | NA | NA | 0 |
| S2_k141_51103 | 19963 | No | NA | 26 | 0 | 5 | Not-determined | Genome-fragment | NA | NA | 0 |
| S2_k141_235933 | 5961 | No | NA | 8 | 0 | 5 | Not-determined | Genome-fragment | NA | NA | 0 |
| S2_k141_235936 | 1535 | No | NA | 2 | 0 | 1 | Not-determined | Genome-fragment | NA | NA | 0 |
| S2_k141_161946 | 1685 | No | NA | 1 | 0 | 1 | Not-determined | Genome-fragment | NA | NA | 0 |
| S2_k141_38985 | 1020 | No | NA | 1 | 0 | 1 | Not-determined | Genome-fragment | NA | NA | 0 |
| S2_k141_90655 | 1520 | No | NA | 2 | 0 | 1 | Not-determined | Genome-fragment | NA | NA | 0 |
| S2_k141_51127 | 1066 | No | NA | 1 | 0 | 1 | Not-determined | Genome-fragment | NA | NA | 0 |
| S2_k141_152726 | 1263 | No | NA | 1 | 0 | 0 | Not-determined | Genome-fragment | NA | NA | 0 |
| S2_k141_130420 | 1958 | No | NA | 1 | 0 | 0 | Not-determined | Genome-fragment | NA | NA | 0 |
| S2_k141_183698 | 1584 | No | NA | 1 | 0 | 0 | Not-determined | Genome-fragment | NA | NA | 0 |
| S9-final-viral-combined_k141_72319\|\|full | 4672 | No | NA | 5 | 0 | 5 | Low-quality | Genome-fragment | 2.19 | AAI-based (high-confidence) | 0 |
| S11-final-viral-combined_k141_82457\|\|full | 4673 | No | NA | 4 | 0 | 3 | Low-quality | Genome-fragment | 10.44 | AAI-based (medium-confidence) | 0 |
| S2_k141_223576 | 4494 | No | NA | 5 | 0 | 1 | Not-determined | Genome-fragment | NA | NA | 0 |
| S2_k141_161986 | 40463 | No | NA | 61 | 17 | 2 | High-quality | High-quality | 100 | AAI-based (high-confidence) | 0 |
| S2_k141_17880 | 3634 | No | NA | 3 | 0 | 2 | Not-determined | Genome-fragment | NA | NA | 0 |
| S2_k141_223609 | 1616 | No | NA | 2 | 0 | 2 | Not-determined | Genome-fragment | NA | NA | 0 |
| S2_k141_244205 | 2907 | No | NA | 1 | 0 | 1 | Not-determined | Genome-fragment | NA | NA | 0 |
| S2_k141_244208 | 1024 | No | NA | 1 | 0 | 0 | Not-determined | Genome-fragment | NA | NA | 0 |
| S2_k141_130437 | 53868 | No | NA | 56 | 0 | 42 | Not-determined | Genome-fragment | NA | NA | 0 |
| S17-final-viral-combined_k141_211701\|\|full | 4674 | No | NA | 8 | 1 | 0 | Low-quality | Genome-fragment | 11.36 | AAI-based (medium-confidence) | 0 |
| S7_k141_188944 | 4675 | No | NA | 4 | 0 | 1 | Low-quality | Genome-fragment | 4.92 | AAI-based (medium-confidence) | 0 |
| S2_k141_110315 | 2362 | No | NA | 1 | 0 | 1 | Not-determined | Genome-fragment | NA | NA | 0 |
| S2_k141_51209 | 1512 | No | NA | 1 | 0 | 0 | Not-determined | Genome-fragment | NA | NA | 0 |
| S2_k141_101205 | 1100 | No | NA | 3 | 0 | 3 | Not-determined | Genome-fragment | NA | NA | 0 |
| S2_k141_214013 | 2779 | No | NA | 2 | 0 | 1 | Not-determined | Genome-fragment | NA | NA | 0 |
| S2_k141_51237 | 1097 | No | NA | 2 | 0 | 2 | Not-determined | Genome-fragment | NA | NA | 0 |
| S2_k141_130472 | 3424 | No | NA | 1 | 0 | 1 | Not-determined | Genome-fragment | NA | NA | 0 |
| S17-final-viral-combined_k141_208764\|\|full | 4675 | No | NA | 4 | 0 | 3 | Low-quality | Genome-fragment | 2.97 | AAI-based (medium-confidence) | 0 |
| S8-final-viral-combined_k141_257029\|\|full | 4676 | No | NA | 7 | 6 | 0 | Low-quality | Genome-fragment | 10.05 | AAI-based (medium-confidence) | 0 |
| S2_k141_193911 | 1372 | No | NA | 1 | 0 | 0 | Not-determined | Genome-fragment | NA | NA | 0 |
| S2_k141_69659 | 3410 | No | NA | 2 | 0 | 1 | Not-determined | Genome-fragment | NA | NA | 0 |
| S2_k141_214051 | 1723 | No | NA | 2 | 0 | 0 | Not-determined | Genome-fragment | NA | NA | 0 |
| S2_k141_17926 | 5325 | No | NA | 5 | 0 | 3 | Not-determined | Genome-fragment | NA | NA | 0 |
| S17-final-viral-combined_k141_191732\|\|full | 4679 | No | NA | 7 | 0 | 0 | Low-quality | Genome-fragment | 7.39 | AAI-based (high-confidence) | 0 |
| S12-final-viral-combined_k141_359283\|\|full | 4680 | No | NA | 4 | 0 | 1 | Low-quality | Genome-fragment | 2.39 | AAI-based (medium-confidence) | 0 |
| S4-final-viral-combined_k141_94046\|\|full | 4680 | No | NA | 8 | 3 | 0 | Low-quality | Genome-fragment | 5.33 | AAI-based (high-confidence) | 0 |
| S2_k141_79944 | 51033 | No | NA | 74 | 24 | 0 | Medium-quality | Genome-fragment | 79.01 | AAI-based (high-confidence) | 0 |
| S2_k141_130553 | 3274 | No | NA | 3 | 0 | 2 | Not-determined | Genome-fragment | NA | NA | 0 |
| S8_k141_131581 | 4680 | No | NA | 5 | 0 | 0 | Low-quality | Genome-fragment | 7.36 | AAI-based (high-confidence) | 0 |
| S2_k141_193951 | 1671 | No | NA | 2 | 0 | 1 | Not-determined | Genome-fragment | NA | NA | 0 |
| S2_k141_5906 | 1172 | No | NA | 1 | 0 | 1 | Not-determined | Genome-fragment | NA | NA | 0 |
| S2_k141_101261 | 1716 | No | NA | 1 | 0 | 0 | Not-determined | Genome-fragment | NA | NA | 0 |
| S2_k141_130619 | 1806 | No | NA | 3 | 0 | 1 | Not-determined | Genome-fragment | NA | NA | 0 |
| S2_k141_130621 | 1204 | No | NA | 2 | 0 | 2 | Not-determined | Genome-fragment | NA | NA | 0 |
| S2_k141_236113 | 1305 | No | NA | 4 | 0 | 2 | Not-determined | Genome-fragment | NA | NA | 0 |
| S2_k141_223740 | 1045 | No | NA | 2 | 0 | 0 | Not-determined | Genome-fragment | NA | NA | 0 |
| S2_k141_110443 | 6464 | No | NA | 4 | 0 | 3 | Not-determined | Genome-fragment | NA | NA | 0 |
| S2_k141_223745 | 1432 | No | NA | 1 | 0 | 1 | Not-determined | Genome-fragment | NA | NA | 0 |
| S2_k141_174576 | 1022 | No | NA | 1 | 0 | 1 | Not-determined | Genome-fragment | NA | NA | 0 |
| S9-final-viral-combined_k141_76595\|\|full | 4682 | Yes | 3290 | 7 | 1 | 2 | Low-quality | Genome-fragment | 8.91 | AAI-based (high-confidence) | 29.73 |
| S9-final-viral-combined_k141_110082\|\|full | 4682 | Yes | 2790 | 5 | 1 | 1 | Low-quality | Genome-fragment | 4.83 | AAI-based (medium-confidence) | 40.41 |
| S12-final-viral-combined_k141_4237\|\|full | 4685 | No | NA | 5 | 0 | 2 | Low-quality | Genome-fragment | 2.61 | AAI-based (high-confidence) | 0 |
| S2_k141_90849 | 4674 | No | NA | 6 | 0 | 3 | Not-determined | Genome-fragment | NA | NA | 0 |
| S2_k141_203269 | 1058 | No | NA | 1 | 0 | 1 | Not-determined | Genome-fragment | NA | NA | 0 |
| S2_k141_130687 | 70128 | Yes | 40699 | 81 | 16 | 21 | High-quality | High-quality | 100 | AAI-based (high-confidence) | 41.96 |
| S6-final-viral-combined_k141_32485\|\|full | 4685 | Yes | 1671 | 3 | 1 | 2 | Low-quality | Genome-fragment | 3.36 | HMM-based (lower-bound) | 64.33 |
| S2_k141_184084 | 1534 | No | NA | 3 | 0 | 1 | Not-determined | Genome-fragment | NA | NA | 0 |
| S2_k141_6077 | 4031 | No | NA | 2 | 0 | 0 | Not-determined | Genome-fragment | NA | NA | 0 |
| S2_k141_69885 | 1119 | No | NA | 2 | 0 | 0 | Not-determined | Genome-fragment | NA | NA | 0 |
| S2_k141_121894 | 1122 | No | NA | 1 | 0 | 0 | Not-determined | Genome-fragment | NA | NA | 0 |
| S2_k141_162155 | 1052 | No | NA | 3 | 0 | 0 | Not-determined | Genome-fragment | NA | NA | 0 |
| S2_k141_184153 | 1003 | No | NA | 2 | 0 | 0 | Not-determined | Genome-fragment | NA | NA | 0 |
| S2_k141_130705 | 3135 | No | NA | 3 | 0 | 3 | Not-determined | Genome-fragment | NA | NA | 0 |
| S2_k141_6094 | 8297 | No | NA | 8 | 0 | 6 | Not-determined | Genome-fragment | NA | NA | 0 |
| S1-final-viral-combined_k141_188991\|\|full | 4688 | No | NA | 10 | 2 | 0 | Low-quality | Genome-fragment | 10.33 | AAI-based (high-confidence) | 0 |
| S2_k141_101479 | 4204 | No | NA | 8 | 0 | 2 | Not-determined | Genome-fragment | NA | NA | 0 |
| S2_k141_153177 | 9596 | No | NA | 15 | 0 | 0 | Not-determined | Genome-fragment | NA | NA | 0 |
| S2_k141_174787 | 3367 | No | NA | 2 | 0 | 1 | Not-determined | Genome-fragment | NA | NA | 0 |
| S2_k141_214256 | 1396 | No | NA | 1 | 0 | 0 | Not-determined | Genome-fragment | NA | NA | 0 |
| S1-final-viral-combined_k141_9218\|\|full | 4688 | No | NA | 3 | 1 | 0 | Low-quality | Genome-fragment | 10.31 | AAI-based (medium-confidence) | 0 |
| S2_k141_101496 | 1502 | No | NA | 2 | 0 | 0 | Not-determined | Genome-fragment | NA | NA | 0 |
| S2_k141_184223 | 1332 | No | NA | 2 | 0 | 1 | Not-determined | Genome-fragment | NA | NA | 0 |
| S1-final-viral-combined_k141_103819\|\|full | 4689 | Yes | 4061 | 10 | 1 | 1 | Low-quality | Genome-fragment | 10.4 | AAI-based (high-confidence) | 13.39 |
| S3_k141_38912 | 4689 | No | NA | 3 | 0 | 1 | Low-quality | Genome-fragment | 0.4 | AAI-based (medium-confidence) | 0 |
| S7-final-viral-combined_k141_216661\|\|full | 4689 | No | NA | 7 | 0 | 1 | Low-quality | Genome-fragment | 12.02 | AAI-based (high-confidence) | 0 |
| S2_k141_162217 | 1219 | No | NA | 2 | 0 | 0 | Not-determined | Genome-fragment | NA | NA | 0 |
| S2_k141_39466 | 1182 | No | NA | 2 | 0 | 0 | Not-determined | Genome-fragment | NA | NA | 0 |
| S2-final-viral-combined_k141_13895\|\|full | 4690 | No | NA | 5 | 4 | 0 | Low-quality | Genome-fragment | 10.78 | AAI-based (high-confidence) | 0 |
| S2_k141_223892 | 3604 | No | NA | 1 | 0 | 1 | Not-determined | Genome-fragment | NA | NA | 0 |
| S2_k141_59676 | 4258 | No | NA | 1 | 0 | 1 | Not-determined | Genome-fragment | NA | NA | 0 |
| S2_k141_184310 | 1560 | No | NA | 1 | 0 | 1 | Not-determined | Genome-fragment | NA | NA | 0 |
| S2_k141_18265 | 2623 | No | NA | 2 | 0 | 2 | Not-determined | Genome-fragment | NA | NA | 0 |
| S7-final-viral-combined_k141_128227\|\|full | 4691 | No | NA | 6 | 0 | 2 | Low-quality | Genome-fragment | 9.92 | AAI-based (medium-confidence) | 0 |
| S11_k141_75740 | 4692 | No | NA | 4 | 0 | 0 | Low-quality | Genome-fragment | 4.42 | AAI-based (high-confidence) | 0 |
| S2_k141_184376 | 1383 | No | NA | 1 | 0 | 1 | Not-determined | Genome-fragment | NA | NA | 0 |
| S2_k141_223915 | 1100 | No | NA | 1 | 0 | 1 | Not-determined | Genome-fragment | NA | NA | 0 |
| S1-final-viral-combined_k141_241850\|\|full | 4692 | No | NA | 5 | 0 | 0 | Low-quality | Genome-fragment | 13.33 | AAI-based (medium-confidence) | 0 |
| S2_k141_91153 | 4427 | No | NA | 2 | 0 | 1 | Not-determined | Genome-fragment | NA | NA | 0 |
| S2_k141_80147 | 1465 | No | NA | 1 | 0 | 1 | Not-determined | Genome-fragment | NA | NA | 0 |
| S2_k141_174968 | 2228 | No | NA | 2 | 0 | 1 | Not-determined | Genome-fragment | NA | NA | 0 |
| S2_k141_110761 | 7856 | No | NA | 11 | 3 | 0 | Medium-quality | Genome-fragment | 57.34 | AAI-based (medium-confidence) | 0 |
| S1_k141_224859 | 4692 | Yes | 3900 | 8 | 2 | 1 | Low-quality | Genome-fragment | 4.74 | AAI-based (high-confidence) | 16.88 |
| S2_k141_91173 | 1118 | No | NA | 2 | 0 | 1 | Not-determined | Genome-fragment | NA | NA | 0 |
| S2_k141_153351 | 2355 | No | NA | 3 | 0 | 0 | Not-determined | Genome-fragment | NA | NA | 0 |
| S2_k141_153361 | 1610 | No | NA | 3 | 0 | 0 | Not-determined | Genome-fragment | NA | NA | 0 |
| S2_k141_162336 | 1049 | No | NA | 1 | 0 | 0 | Not-determined | Genome-fragment | NA | NA | 0 |
| S2_k141_6305 | 2204 | No | NA | 2 | 0 | 2 | Not-determined | Genome-fragment | NA | NA | 0 |
| S2_k141_223960 | 4224 | No | NA | 2 | 0 | 1 | Not-determined | Genome-fragment | NA | NA | 0 |
| S2_k141_51618 | 1118 | No | NA | 3 | 0 | 0 | Not-determined | Genome-fragment | NA | NA | 0 |
| S2_k141_101705 | 1204 | No | NA | 1 | 0 | 0 | Not-determined | Genome-fragment | NA | NA | 0 |
| S2_k141_39667 | 2815 | No | NA | 3 | 0 | 1 | Not-determined | Genome-fragment | NA | NA | 0 |
| S2_k141_101717 | 1414 | No | NA | 2 | 0 | 1 | Not-determined | Genome-fragment | NA | NA | 0 |
| S2_k141_142682 | 1058 | No | NA | 1 | 0 | 1 | Not-determined | Genome-fragment | NA | NA | 0 |
| S2_k141_51641 | 12563 | No | NA | 18 | 0 | 8 | Not-determined | Genome-fragment | NA | NA | 0 |
| S2_k141_162410 | 1452 | No | NA | 1 | 0 | 1 | Not-determined | Genome-fragment | NA | NA | 0 |
| S8-final-viral-combined_k141_271786\|\|full | 4694 | No | NA | 7 | 2 | 0 | Low-quality | Genome-fragment | 9.02 | AAI-based (high-confidence) | 0 |
| S2_k141_162413 | 3332 | No | NA | 3 | 0 | 3 | Not-determined | Genome-fragment | NA | NA | 0 |
| S2_k141_27086 | 1019 | No | NA | 1 | 0 | 1 | Not-determined | Genome-fragment | NA | NA | 0 |
| S2_k141_131004 | 1134 | No | NA | 3 | 0 | 0 | Not-determined | Genome-fragment | NA | NA | 0 |
| S2_k141_70115 | 1515 | No | NA | 2 | 0 | 0 | Not-determined | Genome-fragment | NA | NA | 0 |
| S2_k141_236528 | 1608 | No | NA | 3 | 0 | 0 | Not-determined | Genome-fragment | NA | NA | 0 |
| S2_k141_162428 | 1128 | No | NA | 2 | 0 | 0 | Not-determined | Genome-fragment | NA | NA | 0 |
| S2_k141_236548 | 1552 | No | NA | 2 | 0 | 0 | Not-determined | Genome-fragment | NA | NA | 0 |
| S17-final-viral-combined_k141_81815\|\|full | 4696 | No | NA | 6 | 2 | 0 | Low-quality | Genome-fragment | 8.7 | AAI-based (high-confidence) | 0 |
| S2-final-viral-combined_k141_20776\|\|full | 4696 | No | NA | 6 | 1 | 0 | Low-quality | Genome-fragment | 5.33 | HMM-based (lower-bound) | 0 |
| S2_k141_175101 | 1017 | No | NA | 1 | 0 | 1 | Not-determined | Genome-fragment | NA | NA | 0 |
| S2_k141_27141 | 1718 | No | NA | 4 | 0 | 0 | Not-determined | Genome-fragment | NA | NA | 0 |
| S2_k141_142758 | 1129 | No | NA | 2 | 0 | 0 | Not-determined | Genome-fragment | NA | NA | 0 |
| S2_k141_224071 | 4463 | No | NA | 2 | 0 | 1 | Not-determined | Genome-fragment | NA | NA | 0 |
| S2_k141_59901 | 1375 | No | NA | 3 | 0 | 0 | Not-determined | Genome-fragment | NA | NA | 0 |
| S2_k141_91357 | 4265 | No | NA | 1 | 0 | 0 | Not-determined | Genome-fragment | NA | NA | 0 |
| S2_k141_59928 | 1167 | No | NA | 1 | 0 | 1 | Not-determined | Genome-fragment | NA | NA | 0 |
| S2_k141_59930 | 1506 | No | NA | 1 | 0 | 1 | Not-determined | Genome-fragment | NA | NA | 0 |
| S2_k141_101832 | 1157 | No | NA | 2 | 0 | 0 | Not-determined | Genome-fragment | NA | NA | 0 |
| S2_k141_214488 | 3502 | No | NA | 3 | 0 | 3 | Not-determined | Genome-fragment | NA | NA | 0 |
| S8-final-viral-combined_k141_152341\|\|full | 4697 | No | NA | 9 | 1 | 0 | Low-quality | Genome-fragment | 9.9 | AAI-based (medium-confidence) | 0 |
| S2_k141_122375 | 4097 | No | NA | 6 | 0 | 5 | Not-determined | Genome-fragment | NA | NA | 0 |
| S2_k141_194578 | 1036 | No | NA | 1 | 0 | 1 | Not-determined | Genome-fragment | NA | NA | 0 |
| S2_k141_122383 | 1223 | No | NA | 2 | 0 | 0 | Not-determined | Genome-fragment | NA | NA | 0 |
| S2_k141_80433 | 1015 | No | NA | 3 | 0 | 3 | Not-determined | Genome-fragment | NA | NA | 0 |
| S11-final-viral-combined_k141_81386\|\|full | 4702 | No | NA | 9 | 7 | 0 | Low-quality | Genome-fragment | 10.32 | AAI-based (high-confidence) | 0 |
| S8-final-viral-combined_k141_260770\|\|full | 4705 | No | NA | 3 | 0 | 2 | Low-quality | Genome-fragment | 4.72 | AAI-based (high-confidence) | 0 |
| S8_k141_59074 | 4705 | No | NA | 4 | 0 | 4 | Low-quality | Genome-fragment | 3.37 | AAI-based (medium-confidence) | 0 |
| S2_k141_60010 | 1386 | No | NA | 1 | 0 | 1 | Not-determined | Genome-fragment | NA | NA | 0 |
| S2_k141_27179 | 3330 | No | NA | 4 | 0 | 1 | Not-determined | Genome-fragment | NA | NA | 0 |
| S2_k141_101879 | 1708 | No | NA | 1 | 0 | 0 | Not-determined | Genome-fragment | NA | NA | 0 |
| S12-final-viral-combined_k141_136977\|\|full | 4706 | No | NA | 8 | 0 | 1 | Low-quality | Genome-fragment | 10.08 | AAI-based (high-confidence) | 0 |
| S2_k141_80470 | 3970 | No | NA | 4 | 0 | 3 | Not-determined | Genome-fragment | NA | NA | 0 |
| S2_k141_80495 | 1857 | No | NA | 1 | 0 | 0 | Not-determined | Genome-fragment | NA | NA | 0 |
| S12_k141_160530 | 4706 | No | NA | 11 | 3 | 0 | Low-quality | Genome-fragment | 8.14 | AAI-based (high-confidence) | 0 |
| S2_k141_60097 | 1526 | No | NA | 2 | 0 | 0 | Not-determined | Genome-fragment | NA | NA | 0 |
| S2_k141_203399 | 3937 | No | NA | 6 | 0 | 1 | Not-determined | Genome-fragment | NA | NA | 0 |
| S2_k141_6424 | 73450 | No | NA | 88 | 15 | 1 | Complete | High-quality | 100 | DTR (high-confidence) | 0 |
| S2_k141_60111 | 1633 | No | NA | 1 | 0 | 0 | Not-determined | Genome-fragment | NA | NA | 0 |
| S11-final-viral-combined_k141_186866\|\|full | 4707 | No | NA | 4 | 2 | 0 | Low-quality | Genome-fragment | 2.28 | HMM-based (lower-bound) | 0 |
| S2_k141_224218 | 5821 | No | NA | 2 | 0 | 2 | Not-determined | Genome-fragment | NA | NA | 0 |
| S2_k141_175362 | 1078 | No | NA | 3 | 0 | 1 | Not-determined | Genome-fragment | NA | NA | 0 |
| S2_k141_143010 | 4585 | No | NA | 3 | 0 | 0 | Not-determined | Genome-fragment | NA | NA | 0 |
| S2_k141_203467 | 3500 | No | NA | 4 | 0 | 3 | Not-determined | Genome-fragment | NA | NA | 0 |
| S2_k141_162589 | 1356 | No | NA | 3 | 0 | 3 | Not-determined | Genome-fragment | NA | NA | 0 |
| S2_k141_91596 | 1509 | No | NA | 2 | 0 | 2 | Not-determined | Genome-fragment | NA | NA | 0 |
| S8-final-viral-combined_k141_233682\|\|full | 4708 | No | NA | 6 | 0 | 0 | Low-quality | Genome-fragment | 9.59 | AAI-based (medium-confidence) | 0 |
| S2_k141_6486 | 1979 | No | NA | 1 | 0 | 1 | Not-determined | Genome-fragment | NA | NA | 0 |
| S2_k141_162613 | 1096 | No | NA | 3 | 0 | 0 | Not-determined | Genome-fragment | NA | NA | 0 |
| S11-final-viral-combined_k141_71073\|\|full | 4710 | No | NA | 3 | 0 | 0 | Low-quality | Genome-fragment | 12.05 | AAI-based (medium-confidence) | 0 |
| S1-final-viral-combined_k141_344859\|\|full | 4710 | No | NA | 6 | 0 | 3 | Low-quality | Genome-fragment | 2.19 | AAI-based (medium-confidence) | 0 |
| S2_k141_224254 | 1606 | No | NA | 1 | 0 | 1 | Not-determined | Genome-fragment | NA | NA | 0 |
| S2_k141_224276 | 6536 | No | NA | 13 | 0 | 1 | Not-determined | Genome-fragment | NA | NA | 0 |
| S2_k141_102125 | 2733 | No | NA | 2 | 0 | 0 | Not-determined | Genome-fragment | NA | NA | 0 |
| S12_k141_320495 | 4710 | No | NA | 5 | 4 | 0 | Low-quality | Genome-fragment | 10.38 | AAI-based (high-confidence) | 0 |
| S2_k141_91800 | 1267 | No | NA | 1 | 0 | 0 | Not-determined | Genome-fragment | NA | NA | 0 |
| S2_k141_236830 | 1718 | No | NA | 2 | 0 | 0 | Not-determined | Genome-fragment | NA | NA | 0 |
| S1_k141_359287 | 4711 | No | NA | 7 | 7 | 0 | Low-quality | Genome-fragment | 7.34 | AAI-based (high-confidence) | 0 |
| S11-final-viral-combined_k141_223445\|\|full | 4713 | No | NA | 5 | 0 | 4 | Low-quality | Genome-fragment | 3.93 | AAI-based (medium-confidence) | 0 |
| S2_k141_102174 | 2281 | No | NA | 5 | 0 | 1 | Not-determined | Genome-fragment | NA | NA | 0 |
| S12_k141_42830 | 4713 | No | NA | 5 | 0 | 2 | Low-quality | Genome-fragment | 7.33 | AAI-based (medium-confidence) | 0 |
| S2_k141_40297 | 3237 | No | NA | 2 | 0 | 2 | Not-determined | Genome-fragment | NA | NA | 0 |
| S2_k141_6566 | 1105 | No | NA | 2 | 0 | 1 | Not-determined | Genome-fragment | NA | NA | 0 |
| S4_k141_13631 | 4713 | No | NA | 2 | 0 | 0 | Low-quality | Genome-fragment | 4.66 | AAI-based (high-confidence) | 0 |
| S2_k141_214867 | 4089 | No | NA | 1 | 0 | 1 | Not-determined | Genome-fragment | NA | NA | 0 |
| S2_k141_224360 | 2230 | No | NA | 2 | 0 | 1 | Not-determined | Genome-fragment | NA | NA | 0 |
| S2_k141_131258 | 1298 | No | NA | 1 | 0 | 1 | Not-determined | Genome-fragment | NA | NA | 0 |
| S2_k141_143178 | 2520 | No | NA | 2 | 0 | 1 | Not-determined | Genome-fragment | NA | NA | 0 |
| S8-final-viral-combined_k141_33370\|\|full | 4713 | No | NA | 2 | 0 | 1 | Low-quality | Genome-fragment | 2.64 | AAI-based (medium-confidence) | 0 |
| S2_k141_70630 | 1219 | No | NA | 2 | 0 | 0 | Not-determined | Genome-fragment | NA | NA | 0 |
| S12_k141_310105 | 4714 | No | NA | 3 | 0 | 1 | Low-quality | Genome-fragment | 2.58 | AAI-based (medium-confidence) | 0 |
| S2_k141_111183 | 2058 | No | NA | 1 | 0 | 1 | Not-determined | Genome-fragment | NA | NA | 0 |
| S2_k141_60498 | 1496 | No | NA | 3 | 0 | 0 | Not-determined | Genome-fragment | NA | NA | 0 |
| S2_k141_214914 | 1138 | No | NA | 1 | 0 | 1 | Not-determined | Genome-fragment | NA | NA | 0 |
| S2_k141_236986 | 1114 | No | NA | 2 | 0 | 0 | Not-determined | Genome-fragment | NA | NA | 0 |
| S2_k141_122626 | 1617 | No | NA | 1 | 0 | 1 | Not-determined | Genome-fragment | NA | NA | 0 |
| S2_k141_60538 | 1455 | No | NA | 1 | 0 | 1 | Not-determined | Genome-fragment | NA | NA | 0 |
| S2_k141_214941 | 3149 | No | NA | 2 | 0 | 1 | Not-determined | Genome-fragment | NA | NA | 0 |
| S2_k141_91964 | 2833 | No | NA | 4 | 0 | 4 | Not-determined | Genome-fragment | NA | NA | 0 |
| S2_k141_162809 | 3091 | No | NA | 6 | 0 | 2 | Not-determined | Genome-fragment | NA | NA | 0 |
| S2_k141_162811 | 2535 | No | NA | 2 | 0 | 2 | Not-determined | Genome-fragment | NA | NA | 0 |
| S2_k141_143252 | 6083 | No | NA | 6 | 0 | 6 | Not-determined | Genome-fragment | NA | NA | 0 |
| S17-final-viral-combined_k141_107057\|\|full | 4715 | No | NA | 8 | 5 | 0 | Low-quality | Genome-fragment | 12.68 | AAI-based (high-confidence) | 0 |
| S9-final-viral-combined_k141_36480\|\|full | 4716 | No | NA | 5 | 0 | 2 | Low-quality | Genome-fragment | 3.61 | AAI-based (medium-confidence) | 0 |
| S2_k141_91995 | 1242 | No | NA | 1 | 0 | 0 | Not-determined | Genome-fragment | NA | NA | 0 |
| S2_k141_92009 | 2219 | No | NA | 3 | 0 | 2 | Not-determined | Genome-fragment | NA | NA | 0 |
| S2_k141_214987 | 1197 | No | NA | 1 | 0 | 1 | Not-determined | Genome-fragment | NA | NA | 0 |
| S2_k141_40386 | 5271 | No | NA | 10 | 0 | 0 | Not-determined | Genome-fragment | NA | NA | 0 |
| S1_k141_366623 | 4717 | No | NA | 8 | 0 | 0 | Low-quality | Genome-fragment | 10.99 | AAI-based (high-confidence) | 0 |
| S2_k141_237114 | 1041 | No | NA | 1 | 0 | 0 | Not-determined | Genome-fragment | NA | NA | 0 |
| S2_k141_244725 | 4625 | No | NA | 2 | 0 | 1 | Not-determined | Genome-fragment | NA | NA | 0 |
| S2_k141_175673 | 1145 | No | NA | 1 | 0 | 1 | Not-determined | Genome-fragment | NA | NA | 0 |
| S2_k141_92098 | 2671 | No | NA | 2 | 0 | 2 | Not-determined | Genome-fragment | NA | NA | 0 |
| S17-k141_103215 | 4718 | No | NA | 5 | 0 | 1 | Low-quality | Genome-fragment | 2.51 | AAI-based (medium-confidence) | 0 |
| S2_k141_175689 | 3877 | No | NA | 4 | 0 | 3 | Not-determined | Genome-fragment | NA | NA | 0 |
| S2_k141_237167 | 1337 | No | NA | 2 | 0 | 1 | Not-determined | Genome-fragment | NA | NA | 0 |
| S2_k141_153620 | 3101 | No | NA | 3 | 0 | 2 | Not-determined | Genome-fragment | NA | NA | 0 |
| S2_k141_203702 | 2650 | No | NA | 1 | 0 | 1 | Not-determined | Genome-fragment | NA | NA | 0 |
| S9_k141_56251 | 4719 | Yes | 2724 | 7 | 2 | 1 | Low-quality | Genome-fragment | 7.44 | AAI-based (high-confidence) | 42.28 |
| S2_k141_92146 | 1019 | No | NA | 1 | 0 | 1 | Not-determined | Genome-fragment | NA | NA | 0 |
| S2_k141_60707 | 1240 | No | NA | 2 | 0 | 1 | Not-determined | Genome-fragment | NA | NA | 0 |
| S2_k141_203713 | 5635 | No | NA | 4 | 0 | 2 | Not-determined | Genome-fragment | NA | NA | 0 |
| S7-final-viral-combined_k141_148966\|\|full | 4721 | No | NA | 11 | 0 | 0 | Low-quality | Genome-fragment | 10.52 | AAI-based (high-confidence) | 0 |
| S2_k141_92160 | 2332 | No | NA | 2 | 0 | 0 | Not-determined | Genome-fragment | NA | NA | 0 |
| S2_k141_6726 | 2292 | No | NA | 3 | 0 | 0 | Not-determined | Genome-fragment | NA | NA | 0 |
| S12_k141_136375 | 4722 | Yes | 2477 | 4 | 1 | 1 | Low-quality | Genome-fragment | 1.34 | HMM-based (lower-bound) | 47.54 |
| S2_k141_80859 | 3775 | No | NA | 4 | 0 | 1 | Not-determined | Genome-fragment | NA | NA | 0 |
| S2_k141_244789 | 1247 | No | NA | 1 | 0 | 1 | Not-determined | Genome-fragment | NA | NA | 0 |
| S2_k141_162936 | 1155 | No | NA | 4 | 0 | 3 | Not-determined | Genome-fragment | NA | NA | 0 |
| S2_k141_111361 | 1874 | No | NA | 3 | 0 | 0 | Not-determined | Genome-fragment | NA | NA | 0 |
| S2_k141_47180 | 4722 | No | NA | 8 | 0 | 2 | Low-quality | Genome-fragment | 5.3 | AAI-based (medium-confidence) | 0 |
| S2_k141_70871 | 1071 | No | NA | 2 | 0 | 1 | Not-determined | Genome-fragment | NA | NA | 0 |
| S2_k141_175778 | 2652 | No | NA | 3 | 0 | 0 | Not-determined | Genome-fragment | NA | NA | 0 |
| S9-final-viral-combined_k141_88171\|\|full | 4723 | No | NA | 8 | 0 | 1 | Low-quality | Genome-fragment | 6.32 | AAI-based (medium-confidence) | 0 |
| S2_k141_80906 | 3209 | No | NA | 5 | 0 | 2 | Not-determined | Genome-fragment | NA | NA | 0 |
| S2_k141_102465 | 1246 | No | NA | 1 | 0 | 1 | Not-determined | Genome-fragment | NA | NA | 0 |
| S2_k141_203802 | 1210 | No | NA | 2 | 0 | 1 | Not-determined | Genome-fragment | NA | NA | 0 |
| S4-final-viral-combined_k141_4396\|\|full | 4725 | No | NA | 6 | 0 | 2 | Low-quality | Genome-fragment | 6.97 | AAI-based (medium-confidence) | 0 |
| S2_k141_80953 | 1080 | No | NA | 1 | 0 | 0 | Not-determined | Genome-fragment | NA | NA | 0 |
| S2_k141_60808 | 1255 | No | NA | 1 | 0 | 0 | Not-determined | Genome-fragment | NA | NA | 0 |
| S2_k141_92256 | 3378 | No | NA | 4 | 0 | 0 | Not-determined | Genome-fragment | NA | NA | 0 |
| S2_k141_195076 | 1860 | No | NA | 3 | 0 | 0 | Not-determined | Genome-fragment | NA | NA | 0 |
| S9-final-viral-combined_k141_112260\|\|full | 4726 | No | NA | 5 | 0 | 2 | Low-quality | Genome-fragment | 1.41 | AAI-based (medium-confidence) | 0 |
| S2_k141_81005 | 1598 | No | NA | 3 | 0 | 1 | Not-determined | Genome-fragment | NA | NA | 0 |
| S2_k141_195143 | 1150 | No | NA | 2 | 0 | 0 | Not-determined | Genome-fragment | NA | NA | 0 |
| S2_k141_224582 | 4163 | No | NA | 3 | 0 | 2 | Not-determined | Genome-fragment | NA | NA | 0 |
| S2_k141_185030 | 1161 | No | NA | 1 | 0 | 1 | Not-determined | Genome-fragment | NA | NA | 0 |
| S2_k141_175863 | 2013 | No | NA | 2 | 0 | 2 | Not-determined | Genome-fragment | NA | NA | 0 |
| S2_k141_175917 | 1222 | No | NA | 1 | 0 | 1 | Not-determined | Genome-fragment | NA | NA | 0 |
| S2-final-viral-combined_k141_68165\|\|full | 4726 | No | NA | 3 | 0 | 2 | Low-quality | Genome-fragment | 2.13 | AAI-based (medium-confidence) | 0 |
| S2_k141_19195 | 6728 | No | NA | 6 | 0 | 1 | Not-determined | Genome-fragment | NA | NA | 0 |
| S9-final-viral-combined_k141_15132\|\|full | 4727 | Yes | 3376 | 5 | 2 | 2 | Low-quality | Genome-fragment | 2.09 | HMM-based (lower-bound) | 28.58 |
| S2_k141_81096 | 1521 | No | NA | 2 | 0 | 1 | Not-determined | Genome-fragment | NA | NA | 0 |
| S2_k141_71024 | 1047 | No | NA | 2 | 0 | 0 | Not-determined | Genome-fragment | NA | NA | 0 |
| S2_k141_215402 | 2197 | No | NA | 3 | 0 | 3 | Not-determined | Genome-fragment | NA | NA | 0 |
| S17-final-viral-combined_k141_216353\|\|full | 4727 | No | NA | 7 | 0 | 1 | Low-quality | Genome-fragment | 5.74 | AAI-based (high-confidence) | 0 |
| S2_k141_92327 | 1227 | No | NA | 3 | 0 | 1 | Not-determined | Genome-fragment | NA | NA | 0 |
| S2_k141_143704 | 1382 | No | NA | 3 | 0 | 2 | Not-determined | Genome-fragment | NA | NA | 0 |
| S12-final-viral-combined_k141_385581\|\|full | 4727 | No | NA | 10 | 0 | 0 | Low-quality | Genome-fragment | 7.92 | AAI-based (high-confidence) | 0 |
| S2_k141_81171 | 3339 | No | NA | 3 | 0 | 1 | Not-determined | Genome-fragment | NA | NA | 0 |
| S2_k141_224720 | 1519 | No | NA | 1 | 0 | 0 | Not-determined | Genome-fragment | NA | NA | 0 |
| S2_k141_111511 | 1175 | No | NA | 1 | 0 | 1 | Not-determined | Genome-fragment | NA | NA | 0 |
| S2_k141_102766 | 3796 | No | NA | 8 | 0 | 0 | Not-determined | Genome-fragment | NA | NA | 0 |
| S2_k141_153960 | 1156 | No | NA | 2 | 0 | 2 | Not-determined | Genome-fragment | NA | NA | 0 |
| S2_k141_237579 | 1126 | No | NA | 2 | 0 | 2 | Not-determined | Genome-fragment | NA | NA | 0 |
| S2_k141_81259 | 1005 | No | NA | 1 | 0 | 1 | Not-determined | Genome-fragment | NA | NA | 0 |
| S2_k141_19327 | 1279 | No | NA | 1 | 0 | 0 | Not-determined | Genome-fragment | NA | NA | 0 |
| S2_k141_111545 | 1380 | No | NA | 1 | 0 | 1 | Not-determined | Genome-fragment | NA | NA | 0 |
| S2_k141_92356 | 1075 | No | NA | 1 | 0 | 1 | Not-determined | Genome-fragment | NA | NA | 0 |
| S2_k141_6970 | 1081 | No | NA | 3 | 0 | 0 | Not-determined | Genome-fragment | NA | NA | 0 |
| S2_k141_154009 | 1099 | No | NA | 1 | 0 | 1 | Not-determined | Genome-fragment | NA | NA | 0 |
| S6_k141_9023 | 4728 | No | NA | 5 | 0 | 3 | Low-quality | Genome-fragment | 9.92 | AAI-based (medium-confidence) | 0 |
| S2_k141_195373 | 1276 | No | NA | 1 | 0 | 0 | Not-determined | Genome-fragment | NA | NA | 0 |
| S2_k141_19368 | 1021 | No | NA | 1 | 0 | 1 | Not-determined | Genome-fragment | NA | NA | 0 |
| S2_k141_195382 | 1323 | No | NA | 1 | 0 | 1 | Not-determined | Genome-fragment | NA | NA | 0 |
| S2_k141_204076 | 1182 | No | NA | 1 | 0 | 1 | Not-determined | Genome-fragment | NA | NA | 0 |
| S2_k141_185272 | 2118 | No | NA | 2 | 0 | 2 | Not-determined | Genome-fragment | NA | NA | 0 |
| S2_k141_195404 | 3250 | No | NA | 3 | 0 | 3 | Not-determined | Genome-fragment | NA | NA | 0 |
| S2-final-viral-combined_k141_214718\|\|full | 4729 | No | NA | 8 | 1 | 0 | Low-quality | Genome-fragment | 34.14 | AAI-based (medium-confidence) | 0 |
| S2_k141_81357 | 6760 | No | NA | 14 | 0 | 14 | Not-determined | Genome-fragment | NA | NA | 0 |
| S2_k141_131684 | 1011 | No | NA | 2 | 0 | 1 | Not-determined | Genome-fragment | NA | NA | 0 |
| S2_k141_111660 | 4702 | No | NA | 5 | 0 | 3 | Not-determined | Genome-fragment | NA | NA | 0 |
| S17-final-viral-combined_k141_47862\|\|full | 4730 | No | NA | 10 | 1 | 0 | Low-quality | Genome-fragment | 12.45 | AAI-based (high-confidence) | 0 |
| S2_k141_131727 | 2360 | No | NA | 2 | 0 | 2 | Not-determined | Genome-fragment | NA | NA | 0 |
| S2_k141_185332 | 2259 | No | NA | 2 | 0 | 2 | Not-determined | Genome-fragment | NA | NA | 0 |
| S2_k141_40848 | 1108 | No | NA | 1 | 0 | 1 | Not-determined | Genome-fragment | NA | NA | 0 |
| S12-final-viral-combined_k141_51867\|\|full | 4731 | No | NA | 7 | 0 | 1 | Low-quality | Genome-fragment | 7.14 | AAI-based (high-confidence) | 0 |
| S2_k141_143775 | 1703 | No | NA | 2 | 0 | 2 | Not-determined | Genome-fragment | NA | NA | 0 |
| S2_k141_224945 | 1108 | No | NA | 1 | 0 | 1 | Not-determined | Genome-fragment | NA | NA | 0 |
| S3-final-viral-combined_k141_67400\|\|full | 4733 | No | NA | 5 | 4 | 0 | Low-quality | Genome-fragment | 12.7 | AAI-based (high-confidence) | 0 |
| S7-final-viral-combined_k141_157362\|\|full | 4733 | No | NA | 5 | 0 | 0 | Low-quality | Genome-fragment | 10.9 | AAI-based (medium-confidence) | 0 |
| S2_k141_61220 | 1921 | No | NA | 1 | 0 | 1 | Not-determined | Genome-fragment | NA | NA | 0 |
| S2_k141_185419 | 1126 | No | NA | 2 | 0 | 0 | Not-determined | Genome-fragment | NA | NA | 0 |
| S2_k141_163289 | 1013 | No | NA | 1 | 0 | 1 | Not-determined | Genome-fragment | NA | NA | 0 |
| S4-final-viral-combined_k141_17865\|\|full | 4733 | No | NA | 6 | 0 | 1 | Low-quality | Genome-fragment | 9.3 | AAI-based (high-confidence) | 0 |
| S2_k141_154123 | 1335 | No | NA | 2 | 0 | 1 | Not-determined | Genome-fragment | NA | NA | 0 |
| S6_k141_86121 | 4733 | No | NA | 4 | 0 | 0 | Low-quality | Genome-fragment | 4.93 | AAI-based (medium-confidence) | 0 |
| S2_k141_215680 | 6409 | No | NA | 1 | 0 | 1 | Not-determined | Genome-fragment | NA | NA | 0 |
| S7-final-viral-combined_k141_135017\|\|full | 4734 | No | NA | 6 | 0 | 5 | Low-quality | Genome-fragment | 4.45 | AAI-based (medium-confidence) | 0 |
| S2_k141_237672 | 3234 | No | NA | 2 | 0 | 1 | Not-determined | Genome-fragment | NA | NA | 0 |
| S17-final-viral-combined_k141_201021\|\|full | 4734 | No | NA | 6 | 0 | 3 | Low-quality | Genome-fragment | 5.64 | AAI-based (high-confidence) | 0 |
| S2_k141_215681 | 7559 | No | NA | 7 | 0 | 6 | Not-determined | Genome-fragment | NA | NA | 0 |
| S12-final-viral-combined_k141_205758\|\|full | 4734 | No | NA | 4 | 0 | 2 | Low-quality | Genome-fragment | 3.84 | AAI-based (high-confidence) | 0 |
| S8-final-viral-combined_k141_129488\|\|full | 4734 | No | NA | 5 | 3 | 0 | Low-quality | Genome-fragment | 11.15 | AAI-based (high-confidence) | 0 |
| S2_k141_185478 | 1439 | No | NA | 1 | 0 | 1 | Not-determined | Genome-fragment | NA | NA | 0 |
| S2_k141_224999 | 1140 | No | NA | 1 | 0 | 0 | Not-determined | Genome-fragment | NA | NA | 0 |
| S11_k141_282748 | 4736 | No | NA | 7 | 0 | 1 | Low-quality | Genome-fragment | 4.46 | AAI-based (high-confidence) | 0 |
| S2_k141_123197 | 2142 | No | NA | 2 | 0 | 2 | Not-determined | Genome-fragment | NA | NA | 0 |
| S2_k141_61314 | 1429 | No | NA | 2 | 0 | 1 | Not-determined | Genome-fragment | NA | NA | 0 |
| S2_k141_225025 | 1461 | No | NA | 2 | 0 | 0 | Not-determined | Genome-fragment | NA | NA | 0 |
| S2_k141_92560 | 1464 | No | NA | 2 | 0 | 1 | Not-determined | Genome-fragment | NA | NA | 0 |
| S2_k141_81533 | 3752 | No | NA | 2 | 0 | 1 | Not-determined | Genome-fragment | NA | NA | 0 |
| S2_k141_143916 | 1063 | No | NA | 1 | 0 | 1 | Not-determined | Genome-fragment | NA | NA | 0 |
| S2_k141_27913 | 1109 | No | NA | 1 | 0 | 1 | Not-determined | Genome-fragment | NA | NA | 0 |
| S2_k141_19540 | 1948 | No | NA | 1 | 0 | 1 | Not-determined | Genome-fragment | NA | NA | 0 |
| S2_k141_143934 | 4435 | No | NA | 2 | 0 | 1 | Not-determined | Genome-fragment | NA | NA | 0 |
| S2_k141_154268 | 1669 | No | NA | 1 | 0 | 1 | Not-determined | Genome-fragment | NA | NA | 0 |
| S2_k141_71490 | 4085 | No | NA | 3 | 0 | 2 | Not-determined | Genome-fragment | NA | NA | 0 |

| | | | | | | | | | | | |
|---|---|---|---|---|---|---|---|---|---|---|---|
| S2_k141_123246 | 1320 | No | NA | 1 | 0 | 1 | Not-determined | Genome-fragment | NA | NA | 0 |
| S2_k141_163387 | 1071 | No | NA | 1 | 0 | 1 | Not-determined | Genome-fragment | NA | NA | 0 |
| S1-final-viral-combined_k141_48374\|\|full | 4736 | Yes | 3533 | 9 | 1 | 1 | Low-quality | Genome-fragment | 3.77 | HMM-based (lower-bound) | 25.4 |
| S2_k141_27981 | 1015 | No | NA | 1 | 0 | 1 | Not-determined | Genome-fragment | NA | NA | 0 |
| S17-final-viral-combined_k141_35394\|\|full | 4737 | No | NA | 13 | 3 | 0 | Low-quality | Genome-fragment | 7.14 | AAI-based (high-confidence) | 0 |
| S9_k141_53164 | 4738 | Yes | 3650 | 10 | 1 | 1 | Low-quality | Genome-fragment | 1.69 | HMM-based (lower-bound) | 22.96 |
| S2_k141_144035 | 1970 | No | NA | 1 | 0 | 1 | Not-determined | Genome-fragment | NA | NA | 0 |
| S2_k141_154359 | 1586 | No | NA | 3 | 0 | 0 | Not-determined | Genome-fragment | NA | NA | 0 |
| S2_k141_131954 | 1229 | No | NA | 3 | 0 | 0 | Not-determined | Genome-fragment | NA | NA | 0 |
| S2_k141_144090 | 1053 | No | NA | 1 | 0 | 1 | Not-determined | Genome-fragment | NA | NA | 0 |
| S2_k141_41082 | 1010 | No | NA | 2 | 0 | 1 | Not-determined | Genome-fragment | NA | NA | 0 |
| S2_k141_144094 | 1227 | No | NA | 2 | 0 | 1 | Not-determined | Genome-fragment | NA | NA | 0 |
| S5-final-viral-combined_k141_7576\|\|full | 4738 | No | NA | 9 | 4 | 0 | Low-quality | Genome-fragment | 10.42 | AAI-based (medium-confidence) | 0 |
| S2_k141_28012 | 1032 | No | NA | 2 | 0 | 2 | Not-determined | Genome-fragment | NA | NA | 0 |
| S5_k141_127912 | 4738 | No | NA | 8 | 5 | 0 | Low-quality | Genome-fragment | 10.45 | AAI-based (high-confidence) | 0 |
| S2_k141_61430 | 1015 | No | NA | 1 | 0 | 1 | Not-determined | Genome-fragment | NA | NA | 0 |
| S2_k141_28027 | 3670 | No | NA | 2 | 0 | 0 | Not-determined | Genome-fragment | NA | NA | 0 |
| S2_k141_195914 | 1048 | No | NA | 1 | 0 | 1 | Not-determined | Genome-fragment | NA | NA | 0 |
| S2_k141_28037 | 1587 | No | NA | 1 | 0 | 0 | Not-determined | Genome-fragment | NA | NA | 0 |
| S2_k141_81762 | 2053 | No | NA | 1 | 0 | 1 | Not-determined | Genome-fragment | NA | NA | 0 |
| S2_k141_144135 | 1132 | No | NA | 2 | 0 | 2 | Not-determined | Genome-fragment | NA | NA | 0 |
| S2_k141_154429 | 1528 | No | NA | 2 | 0 | 0 | Not-determined | Genome-fragment | NA | NA | 0 |
| S17-final-viral-combined_k141_57715\|\|full | 4738 | No | NA | 8 | 0 | 0 | Low-quality | Genome-fragment | 13.1 | AAI-based (high-confidence) | 0 |
| S2_k141_92670 | 11256 | No | NA | 12 | 0 | 2 | Not-determined | Genome-fragment | NA | NA | 0 |
| S2_k141_225237 | 2450 | No | NA | 2 | 0 | 0 | Not-determined | Genome-fragment | NA | NA | 0 |
| S2_k141_195950 | 1218 | No | NA | 1 | 0 | 0 | Not-determined | Genome-fragment | NA | NA | 0 |
| S2_k141_245232 | 40791 | No | NA | 54 | 29 | 0 | High-quality | High-quality | 90.66 | AAI-based (high-confidence) | 0 |
| S2_k141_195975 | 1167 | No | NA | 2 | 0 | 0 | Not-determined | Genome-fragment | NA | NA | 0 |
| S2_k141_225273 | 3569 | No | NA | 4 | 0 | 0 | Not-determined | Genome-fragment | NA | NA | 0 |
| S12-final-viral-combined_k141_115942\|\|full | 4738 | No | NA | 6 | 5 | 0 | Low-quality | Genome-fragment | 14.69 | AAI-based (high-confidence) | 0 |
| S2_k141_245258 | 2579 | No | NA | 2 | 0 | 1 | Not-determined | Genome-fragment | NA | NA | 0 |
| S9-final-viral-combined_k141_12453\|\|full | 4739 | No | NA | 6 | 0 | 0 | Low-quality | Genome-fragment | 11.68 | AAI-based (high-confidence) | 0 |
| S2_k141_111899 | 5545 | No | NA | 7 | 0 | 2 | Not-determined | Genome-fragment | NA | NA | 0 |
| S2_k141_92752 | 1800 | No | NA | 1 | 0 | 1 | Not-determined | Genome-fragment | NA | NA | 0 |
| S2_k141_169693 | 4739 | No | NA | 5 | 0 | 0 | Low-quality | Genome-fragment | 7.37 | AAI-based (high-confidence) | 0 |
| S2_k141_111919 | 1056 | No | NA | 2 | 0 | 0 | Not-determined | Genome-fragment | NA | NA | 0 |
| S1-final-viral-combined_k141_362577\|\|full | 4741 | No | NA | 5 | 0 | 0 | Low-quality | Genome-fragment | 13.13 | AAI-based (medium-confidence) | 0 |
| S2_k141_237929 | 1097 | No | NA | 2 | 0 | 2 | Not-determined | Genome-fragment | NA | NA | 0 |
| S2_k141_196108 | 1089 | No | NA | 1 | 0 | 0 | Not-determined | Genome-fragment | NA | NA | 0 |
| S2_k141_154538 | 2494 | No | NA | 4 | 0 | 3 | Not-determined | Genome-fragment | NA | NA | 0 |
| S2_k141_111963 | 1056 | No | NA | 3 | 0 | 0 | Not-determined | Genome-fragment | NA | NA | 0 |
| S2_k141_81891 | 1435 | No | NA | 1 | 0 | 0 | Not-determined | Genome-fragment | NA | NA | 0 |
| S2_k141_71699 | 6730 | No | NA | 4 | 0 | 1 | Not-determined | Genome-fragment | NA | NA | 0 |
| S2_k141_112021 | 2286 | No | NA | 2 | 0 | 2 | Not-determined | Genome-fragment | NA | NA | 0 |
| S2_k141_132099 | 1261 | No | NA | 2 | 0 | 0 | Not-determined | Genome-fragment | NA | NA | 0 |
| S2_k141_196200 | 1059 | No | NA | 1 | 0 | 1 | Not-determined | Genome-fragment | NA | NA | 0 |
| S2_k141_163678 | 2533 | No | NA | 3 | 0 | 1 | Not-determined | Genome-fragment | NA | NA | 0 |
| S2_k141_112041 | 2034 | No | NA | 1 | 0 | 0 | Not-determined | Genome-fragment | NA | NA | 0 |
| S2_k141_7377 | 1849 | No | NA | 2 | 0 | 0 | Not-determined | Genome-fragment | NA | NA | 0 |
| S2_k141_132128 | 1273 | No | NA | 1 | 0 | 1 | Not-determined | Genome-fragment | NA | NA | 0 |
| S2_k141_225416 | 1127 | No | NA | 3 | 0 | 1 | Not-determined | Genome-fragment | NA | NA | 0 |
| S2_k141_41293 | 1314 | No | NA | 2 | 0 | 0 | Not-determined | Genome-fragment | NA | NA | 0 |
| S2_k141_41301 | 1135 | No | NA | 1 | 0 | 1 | Not-determined | Genome-fragment | NA | NA | 0 |
| S2_k141_103216 | 1838 | No | NA | 1 | 0 | 1 | Not-determined | Genome-fragment | NA | NA | 0 |
| S2_k141_103218 | 2698 | No | NA | 1 | 0 | 1 | Not-determined | Genome-fragment | NA | NA | 0 |
| S17-final-viral-combined_k141_173913\|\|full | 4741 | No | NA | 3 | 0 | 0 | Low-quality | Genome-fragment | 1.42 | AAI-based (medium-confidence) | 0 |
| S2_k141_132155 | 9083 | No | NA | 4 | 0 | 0 | Not-determined | Genome-fragment | NA | NA | 0 |
| S2_k141_245325 | 2550 | No | NA | 1 | 0 | 1 | Not-determined | Genome-fragment | NA | NA | 0 |
| S2_k141_7455 | 1052 | No | NA | 1 | 0 | 0 | Not-determined | Genome-fragment | NA | NA | 0 |
| S12-final-viral-combined_k141_66568\|\|full | 4745 | No | NA | 4 | 0 | 3 | Low-quality | Genome-fragment | 0.77 | AAI-based (high-confidence) | 0 |
| S2_k141_123502 | 1752 | No | NA | 1 | 0 | 1 | Not-determined | Genome-fragment | NA | NA | 0 |
| S2_k141_19935 | 1082 | No | NA | 1 | 0 | 0 | Not-determined | Genome-fragment | NA | NA | 0 |
| S2_k141_7485 | 2647 | No | NA | 2 | 0 | 0 | Not-determined | Genome-fragment | NA | NA | 0 |
| S2_k141_61665 | 3340 | No | NA | 2 | 0 | 0 | Not-determined | Genome-fragment | NA | NA | 0 |
| S12-final-viral-combined_k141_256091\|\|full | 4746 | No | NA | 5 | 1 | 0 | Low-quality | Genome-fragment | 11.37 | HMM-based (lower-bound) | 0 |
| S2_k141_154746 | 1474 | No | NA | 1 | 0 | 1 | Not-determined | Genome-fragment | NA | NA | 0 |
| S7-final-viral-combined_k141_59031\|\|full | 4748 | No | NA | 8 | 2 | 0 | Low-quality | Genome-fragment | 10.95 | AAI-based (medium-confidence) | 0 |
| S2_k141_163750 | 4388 | No | NA | 5 | 0 | 0 | Not-determined | Genome-fragment | NA | NA | 0 |
| S2_k141_196272 | 1922 | No | NA | 0 | 0 | 0 | Not-determined | Genome-fragment | NA | NA | 0 |
| S5-final-viral-combined_k141_143528\|\|full | 4750 | No | NA | 6 | 0 | 1 | Low-quality | Genome-fragment | 10.75 | AAI-based (medium-confidence) | 0 |
| S2_k141_216162 | 3791 | No | NA | 5 | 0 | 1 | Not-determined | Genome-fragment | NA | NA | 0 |
| S2_k141_123589 | 4446 | No | NA | 3 | 0 | 1 | Not-determined | Genome-fragment | NA | NA | 0 |
| S2_k141_103322 | 7315 | No | NA | 8 | 0 | 4 | Not-determined | Genome-fragment | NA | NA | 0 |
| S2_k141_216168 | 5214 | No | NA | 4 | 0 | 1 | Not-determined | Genome-fragment | NA | NA | 0 |
| S2_k141_216214 | 1872 | No | NA | 2 | 0 | 2 | Not-determined | Genome-fragment | NA | NA | 0 |
| S2_k141_216224 | 3536 | No | NA | 4 | 0 | 0 | Not-determined | Genome-fragment | NA | NA | 0 |
| S2_k141_61819 | 1294 | No | NA | 3 | 0 | 0 | Not-determined | Genome-fragment | NA | NA | 0 |
| S2_k141_112184 | 1457 | No | NA | 1 | 0 | 1 | Not-determined | Genome-fragment | NA | NA | 0 |
| S2_k141_225584 | 3290 | No | NA | 5 | 0 | 0 | Not-determined | Genome-fragment | NA | NA | 0 |
| S2_k141_225606 | 1342 | No | NA | 3 | 0 | 0 | Not-determined | Genome-fragment | NA | NA | 0 |
| S2_k141_7664 | 1785 | No | NA | 2 | 0 | 2 | Not-determined | Genome-fragment | NA | NA | 0 |
| S2_k141_204594 | 1963 | No | NA | 2 | 0 | 1 | Not-determined | Genome-fragment | NA | NA | 0 |
| S2_k141_112329 | 14130 | No | NA | 19 | 0 | 3 | Not-determined | Genome-fragment | NA | NA | 0 |
| S2_k141_82314 | 1754 | No | NA | 2 | 0 | 0 | Not-determined | Genome-fragment | NA | NA | 0 |
| S7_k141_26306 | 4751 | No | NA | 8 | 1 | 0 | Low-quality | Genome-fragment | 8.7 | AAI-based (medium-confidence) | 0 |
| S2_k141_72197 | 28678 | No | NA | 30 | 0 | 9 | Not-determined | Genome-fragment | NA | NA | 0 |
| S2_k141_231506 | 4751 | No | NA | 2 | 0 | 2 | Low-quality | Genome-fragment | 0.41 | AAI-based (medium-confidence) | 0 |
| S2_k141_103482 | 1196 | No | NA | 1 | 0 | 1 | Not-determined | Genome-fragment | NA | NA | 0 |
| S2_k141_112413 | 1624 | No | NA | 1 | 0 | 0 | Not-determined | Genome-fragment | NA | NA | 0 |
| S1_k141_285530 | 4753 | No | NA | 5 | 0 | 1 | Low-quality | Genome-fragment | 2.82 | AAI-based (medium-confidence) | 0 |
| S2_k141_245496 | 1090 | No | NA | 1 | 0 | 1 | Not-determined | Genome-fragment | NA | NA | 0 |
| S2_k141_61941 | 1037 | No | NA | 2 | 0 | 2 | Not-determined | Genome-fragment | NA | NA | 0 |
| S1-final-viral-combined_k141_187533\|\|full | 4755 | No | NA | 4 | 0 | 2 | Low-quality | Genome-fragment | 3.27 | AAI-based (medium-confidence) | 0 |
| S2_k141_186135 | 1322 | No | NA | 1 | 0 | 0 | Not-determined | Genome-fragment | NA | NA | 0 |
| S2_k141_186162 | 1201 | No | NA | 1 | 0 | 0 | Not-determined | Genome-fragment | NA | NA | 0 |
| S2_k141_82412 | 1809 | No | NA | 1 | 0 | 0 | Not-determined | Genome-fragment | NA | NA | 0 |
| S2_k141_112429 | 1147 | No | NA | 2 | 0 | 2 | Not-determined | Genome-fragment | NA | NA | 0 |
| S2_k141_196616 | 1361 | No | NA | 1 | 0 | 0 | Not-determined | Genome-fragment | NA | NA | 0 |
| S2_k141_144732 | 14307 | No | NA | 10 | 0 | 3 | Not-determined | Genome-fragment | NA | NA | 0 |
| S2_k141_123856 | 1394 | No | NA | 1 | 0 | 1 | Not-determined | Genome-fragment | NA | NA | 0 |
| S7-final-viral-combined_k141_216129\|\|full | 4755 | No | NA | 3 | 0 | 2 | Low-quality | Genome-fragment | 3.53 | AAI-based (medium-confidence) | 0 |
| S2_k141_123859 | 1601 | No | NA | 2 | 0 | 1 | Not-determined | Genome-fragment | NA | NA | 0 |
| S2_k141_216388 | 2816 | No | NA | 3 | 0 | 2 | Not-determined | Genome-fragment | NA | NA | 0 |
| S2_k141_196631 | 1195 | No | NA | 1 | 0 | 1 | Not-determined | Genome-fragment | NA | NA | 0 |
| S4-final-viral-combined_k141_70849\|\|full | 4756 | No | NA | 9 | 8 | 0 | Low-quality | Genome-fragment | 8.92 | AAI-based (high-confidence) | 0 |
| S2_k141_155087 | 1083 | No | NA | 1 | 0 | 0 | Not-determined | Genome-fragment | NA | NA | 0 |
| S2_k141_62062 | 1224 | No | NA | 3 | 0 | 2 | Not-determined | Genome-fragment | NA | NA | 0 |
| S2_k141_155130 | 1243 | No | NA | 1 | 0 | 0 | Not-determined | Genome-fragment | NA | NA | 0 |
| S11_k141_170909 | 4757 | No | NA | 3 | 0 | 1 | Low-quality | Genome-fragment | 5.06 | AAI-based (medium-confidence) | 0 |
| S2_k141_164131 | 1385 | No | NA | 1 | 0 | 1 | Not-determined | Genome-fragment | NA | NA | 0 |
| S11_k141_309726 | 4758 | No | NA | 7 | 2 | 0 | Low-quality | Genome-fragment | 3.58 | AAI-based (high-confidence) | 0 |
| S2_k141_93161 | 1392 | No | NA | 1 | 0 | 1 | Not-determined | Genome-fragment | NA | NA | 0 |
| S2_k141_93193 | 3148 | No | NA | 2 | 0 | 0 | Not-determined | Genome-fragment | NA | NA | 0 |
| S2_k141_28746 | 1471 | No | NA | 2 | 0 | 1 | Not-determined | Genome-fragment | NA | NA | 0 |
| S2_k141_196723 | 1352 | No | NA | 3 | 0 | 1 | Not-determined | Genome-fragment | NA | NA | 0 |
| S2_k141_123935 | 1124 | No | NA | 1 | 0 | 0 | Not-determined | Genome-fragment | NA | NA | 0 |
| S2_k141_72490 | 1006 | No | NA | 1 | 0 | 1 | Not-determined | Genome-fragment | NA | NA | 0 |
| S12-final-viral-combined_k141_179916\|\|full | 4758 | No | NA | 7 | 0 | 0 | Low-quality | Genome-fragment | 10.97 | AAI-based (medium-confidence) | 0 |
| S2_k141_245654 | 3330 | No | NA | 3 | 0 | 3 | Not-determined | Genome-fragment | NA | NA | 0 |
| S5-final-viral-combined_k141_167072\|\|full | 4759 | No | NA | 12 | 3 | 0 | Low-quality | Genome-fragment | 5.33 | AAI-based (high-confidence) | 0 |
| S2-final-viral-combined_k141_111384\|\|full | 4759 | No | NA | 4 | 1 | 0 | Low-quality | Genome-fragment | 10.2 | AAI-based (medium-confidence) | 0 |
| S2_k141_28795 | 1113 | No | NA | 2 | 0 | 1 | Not-determined | Genome-fragment | NA | NA | 0 |
| S2_k141_204922 | 1298 | No | NA | 1 | 0 | 0 | Not-determined | Genome-fragment | NA | NA | 0 |
| S2_k141_245689 | 1020 | No | NA | 2 | 0 | 1 | Not-determined | Genome-fragment | NA | NA | 0 |
| S8-final-viral-combined_k141_58563\|\|full | 4759 | No | NA | 10 | 8 | 0 | Low-quality | Genome-fragment | 11.97 | AAI-based (high-confidence) | 0 |
| S2_k141_132948 | 3224 | No | NA | 3 | 0 | 0 | Not-determined | Genome-fragment | NA | NA | 0 |
| S2_k141_164213 | 1031 | No | NA | 3 | 0 | 0 | Not-determined | Genome-fragment | NA | NA | 0 |
| S2_k141_204992 | 1133 | No | NA | 2 | 0 | 0 | Not-determined | Genome-fragment | NA | NA | 0 |
| S11-final-viral-combined_k141_287373\|\|full | 4760 | No | NA | 6 | 3 | 0 | Low-quality | Genome-fragment | 7.15 | HMM-based (lower-bound) | 0 |
| S4_k141_158890 | 4761 | No | NA | 5 | 1 | 0 | Low-quality | Genome-fragment | 5.31 | AAI-based (medium-confidence) | 0 |
| S7-final-viral-combined_k141_74289\|\|full | 4762 | No | NA | 5 | 0 | 2 | Low-quality | Genome-fragment | 3.29 | AAI-based (high-confidence) | 0 |
| S2_k141_155319 | 1046 | No | NA | 1 | 0 | 1 | Not-determined | Genome-fragment | NA | NA | 0 |
| S2_k141_245712 | 1258 | No | NA | 1 | 0 | 1 | Not-determined | Genome-fragment | NA | NA | 0 |
| S2_k141_196757 | 1594 | No | NA | 1 | 0 | 0 | Not-determined | Genome-fragment | NA | NA | 0 |
| S2_k141_216678 | 1005 | No | NA | 1 | 0 | 0 | Not-determined | Genome-fragment | NA | NA | 0 |
| S2_k141_144859 | 3615 | No | NA | 4 | 0 | 3 | Not-determined | Genome-fragment | NA | NA | 0 |
| S2_k141_28931 | 1017 | No | NA | 1 | 0 | 0 | Not-determined | Genome-fragment | NA | NA | 0 |
| S2_k141_186366 | 1052 | No | NA | 1 | 0 | 0 | Not-determined | Genome-fragment | NA | NA | 0 |
| S17-k141_195181 | 4762 | No | NA | 3 | 0 | 2 | Low-quality | Genome-fragment | 2.13 | AAI-based (medium-confidence) | 0 |
| S2_k141_245760 | 1313 | No | NA | 1 | 0 | 1 | Not-determined | Genome-fragment | NA | NA | 0 |
| S2_k141_186398 | 3057 | No | NA | 2 | 0 | 1 | Not-determined | Genome-fragment | NA | NA | 0 |
| S11-final-viral-combined_k141_237108\|\|full | 4764 | No | NA | 5 | 1 | 0 | Low-quality | Genome-fragment | 11.72 | AAI-based (high-confidence) | 0 |
| S2_k141_225959 | 3538 | No | NA | 4 | 0 | 2 | Not-determined | Genome-fragment | NA | NA | 0 |
| S3_k141_1433 | 4764 | No | NA | 2 | 0 | 0 | Low-quality | Genome-fragment | 5.95 | AAI-based (medium-confidence) | 0 |
| S2_k141_225993 | 1145 | No | NA | 1 | 0 | 0 | Not-determined | Genome-fragment | NA | NA | 0 |
| S2_k141_82635 | 1007 | No | NA | 1 | 0 | 1 | Not-determined | Genome-fragment | NA | NA | 0 |
| S5-final-viral-combined_k141_208220\|\|full | 4765 | No | NA | 3 | 0 | 3 | Low-quality | Genome-fragment | 5.74 | AAI-based (medium-confidence) | 0 |
| S2_k141_144919 | 1403 | No | NA | 3 | 0 | 0 | Not-determined | Genome-fragment | NA | NA | 0 |
| S2_k141_216898 | 3685 | No | NA | 1 | 0 | 0 | Not-determined | Genome-fragment | NA | NA | 0 |
| S2_k141_205111 | 1016 | No | NA | 1 | 0 | 0 | Not-determined | Genome-fragment | NA | NA | 0 |
| S2_k141_205118 | 2164 | No | NA | 1 | 0 | 0 | Not-determined | Genome-fragment | NA | NA | 0 |
| S2_k141_82685 | 1576 | No | NA | 3 | 0 | 3 | Not-determined | Genome-fragment | NA | NA | 0 |
| S2_k141_164360 | 3805 | No | NA | 5 | 0 | 2 | Not-determined | Genome-fragment | NA | NA | 0 |
| S12_k141_200705 | 4765 | No | NA | 5 | 0 | 1 | Low-quality | Genome-fragment | 12.13 | AAI-based (high-confidence) | 0 |
| S2_k141_82717 | 1042 | No | NA | 1 | 0 | 1 | Not-determined | Genome-fragment | NA | NA | 0 |
| S2_k141_20699 | 4752 | No | NA | 8 | 0 | 2 | Not-determined | Genome-fragment | NA | NA | 0 |
| S2_k141_20705 | 1014 | No | NA | 2 | 0 | 2 | Not-determined | Genome-fragment | NA | NA | 0 |
| S2_k141_245891 | 1287 | No | NA | 2 | 0 | 1 | Not-determined | Genome-fragment | NA | NA | 0 |
| S2_k141_133007 | 1421 | No | NA | 1 | 0 | 0 | Not-determined | Genome-fragment | NA | NA | 0 |
| S2_k141_82795 | 1590 | No | NA | 2 | 0 | 2 | Not-determined | Genome-fragment | NA | NA | 0 |
| S11-final-viral-combined_k141_197439\|\|full | 4766 | No | NA | 3 | 0 | 2 | Low-quality | Genome-fragment | 6.8 | AAI-based (medium-confidence) | 0 |
| S17-final-viral-combined_k141_219979\|\|full | 4766 | No | NA | 9 | 0 | 0 | Low-quality | Genome-fragment | 11.05 | AAI-based (medium-confidence) | 0 |
| S2_k141_226206 | 1007 | No | NA | 1 | 0 | 0 | Not-determined | Genome-fragment | NA | NA | 0 |
| S1_k141_374112 | 4767 | Yes | 3023 | 6 | 1 | 1 | Low-quality | Genome-fragment | 6.56 | AAI-based (high-confidence) | 36.58 |
| S2_k141_205354 | 2135 | No | NA | 2 | 0 | 2 | Not-determined | Genome-fragment | NA | NA | 0 |
| S2_k141_29130 | 1050 | No | NA | 1 | 0 | 1 | Not-determined | Genome-fragment | NA | NA | 0 |
| S8-final-viral-combined_k141_5214\|\|full | 4768 | No | NA | 7 | 0 | 2 | Low-quality | Genome-fragment | 11.03 | AAI-based (medium-confidence) | 0 |
| S2_k141_7842 | 2006 | No | NA | 4 | 0 | 0 | Not-determined | Genome-fragment | NA | NA | 0 |
| S2_k141_217127 | 1122 | No | NA | 2 | 0 | 1 | Not-determined | Genome-fragment | NA | NA | 0 |
| S2_k141_133211 | 1862 | No | NA | 2 | 0 | 0 | Not-determined | Genome-fragment | NA | NA | 0 |
| S2_k141_133218 | 1001 | No | NA | 1 | 0 | 1 | Not-determined | Genome-fragment | NA | NA | 0 |
| S5-final-viral-combined_k141_130729\|\|full | 4769 | No | NA | 5 | 2 | 0 | Low-quality | Genome-fragment | 6.54 | AAI-based (high-confidence) | 0 |
| S2_k141_246052 | 2199 | No | NA | 2 | 0 | 1 | Not-determined | Genome-fragment | NA | NA | 0 |
| S7-final-viral-combined_k141_14729\|\|full | 4770 | No | NA | 9 | 1 | 0 | Low-quality | Genome-fragment | 1.94 | HMM-based (lower-bound) | 0 |
| S2_k141_246059 | 1561 | No | NA | 2 | 0 | 2 | Not-determined | Genome-fragment | NA | NA | 0 |
| S2_k141_7889 | 2166 | No | NA | 1 | 0 | 0 | Not-determined | Genome-fragment | NA | NA | 0 |
| S2_k141_226428 | 1802 | No | NA | 2 | 0 | 1 | Not-determined | Genome-fragment | NA | NA | 0 |
| S2_k141_7910 | 1107 | No | NA | 2 | 0 | 0 | Not-determined | Genome-fragment | NA | NA | 0 |
| S2_k141_112997 | 1277 | No | NA | 2 | 0 | 0 | Not-determined | Genome-fragment | NA | NA | 0 |
| S2_k141_133340 | 5306 | No | NA | 10 | 0 | 10 | Not-determined | Genome-fragment | NA | NA | 0 |
| S2_k141_226501 | 3128 | No | NA | 4 | 0 | 2 | Not-determined | Genome-fragment | NA | NA | 0 |
| S2_k141_133361 | 1470 | No | NA | 1 | 0 | 0 | Not-determined | Genome-fragment | NA | NA | 0 |
| S2_k141_164753 | 1173 | No | NA | 2 | 0 | 1 | Not-determined | Genome-fragment | NA | NA | 0 |
| S2_k141_246108 | 1182 | No | NA | 1 | 0 | 0 | Not-determined | Genome-fragment | NA | NA | 0 |
| S2_k141_29282 | 3520 | No | NA | 3 | 0 | 3 | Not-determined | Genome-fragment | NA | NA | 0 |
| S2_k141_226531 | 2797 | No | NA | 2 | 0 | 2 | Not-determined | Genome-fragment | NA | NA | 0 |
| S2_k141_217264 | 2056 | No | NA | 1 | 0 | 1 | Not-determined | Genome-fragment | NA | NA | 0 |
| S5_k141_137119 | 4771 | No | NA | 9 | 1 | 0 | Low-quality | Genome-fragment | 5.57 | HMM-based (lower-bound) | 0 |
| S2_k141_164797 | 1157 | No | NA | 2 | 0 | 0 | Not-determined | Genome-fragment | NA | NA | 0 |
| S2_k141_113175 | 1245 | No | NA | 1 | 0 | 1 | Not-determined | Genome-fragment | NA | NA | 0 |
| S2_k141_7949 | 1332 | No | NA | 2 | 0 | 0 | Not-determined | Genome-fragment | NA | NA | 0 |
| S11-final-viral-combined_k141_268487\|\|full | 4773 | No | NA | 9 | 0 | 0 | Low-quality | Genome-fragment | 9.35 | AAI-based (high-confidence) | 0 |
| S2_k141_113213 | 1323 | No | NA | 2 | 0 | 1 | Not-determined | Genome-fragment | NA | NA | 0 |
| S2_k141_217356 | 1831 | No | NA | 1 | 0 | 0 | Not-determined | Genome-fragment | NA | NA | 0 |
| S2_k141_29343 | 4772 | No | NA | 8 | 0 | 0 | Not-determined | Genome-fragment | NA | NA | 0 |
| S4_k141_132741 | 4773 | No | NA | 12 | 2 | 0 | Low-quality | Genome-fragment | 16.13 | AAI-based (high-confidence) | 0 |
| S2_k141_29444 | 3310 | No | NA | 7 | 0 | 7 | Not-determined | Genome-fragment | NA | NA | 0 |
| S2_k141_165030 | 1938 | No | NA | 2 | 0 | 0 | Not-determined | Genome-fragment | NA | NA | 0 |
| S2_k141_165069 | 1691 | No | NA | 1 | 0 | 1 | Not-determined | Genome-fragment | NA | NA | 0 |
| S11-final-viral-combined_k141_289503\|\|full | 4774 | No | NA | 4 | 2 | 0 | Low-quality | Genome-fragment | 11.48 | AAI-based (high-confidence) | 0 |
| S2_k141_29514 | 1074 | No | NA | 1 | 0 | 1 | Not-determined | Genome-fragment | NA | NA | 0 |
| S2_k141_246296 | 54120 | No | NA | 88 | 14 | 1 | Medium-quality | Genome-fragment | 67.31 | AAI-based (high-confidence) | 0 |
| S2_k141_8163 | 1358 | No | NA | 4 | 0 | 3 | Not-determined | Genome-fragment | NA | NA | 0 |
| S2_k141_217451 | 1562 | No | NA | 4 | 0 | 0 | Not-determined | Genome-fragment | NA | NA | 0 |
| S11-final-viral-combined_k141_288176\|\|full | 4775 | No | NA | 8 | 4 | 0 | Low-quality | Genome-fragment | 6.37 | HMM-based (lower-bound) | 0 |
| S4-final-viral-combined_k141_36170\|\|full | 4775 | No | NA | 8 | 2 | 0 | Low-quality | Genome-fragment | 8.36 | HMM-based (lower-bound) | 0 |
| S2_k141_8201 | 1542 | No | NA | 3 | 0 | 1 | Not-determined | Genome-fragment | NA | NA | 0 |
| S2_k141_8220 | 1098 | No | NA | 2 | 0 | 0 | Not-determined | Genome-fragment | NA | NA | 0 |
| S2_k141_165230 | 4502 | No | NA | 4 | 0 | 2 | Not-determined | Genome-fragment | NA | NA | 0 |
| S7-final-viral-combined_k141_27986\|\|full | 4780 | No | NA | 6 | 0 | 5 | Low-quality | Genome-fragment | 11.23 | AAI-based (medium-confidence) | 0 |
| S2_k141_226985 | 1869 | No | NA | 1 | 0 | 1 | Not-determined | Genome-fragment | NA | NA | 0 |
| S2_k141_165298 | 1308 | No | NA | 1 | 0 | 0 | Not-determined | Genome-fragment | NA | NA | 0 |
| S2_k141_227050 | 2452 | No | NA | 2 | 0 | 1 | Not-determined | Genome-fragment | NA | NA | 0 |
| S12-final-viral-combined_k141_126943\|\|full | 4780 | No | NA | 7 | 2 | 0 | Low-quality | Genome-fragment | 7.84 | AAI-based (high-confidence) | 0 |
| S2_k141_165372 | 5044 | No | NA | 3 | 0 | 1 | Not-determined | Genome-fragment | NA | NA | 0 |
| S9-final-viral-combined_k141_46927\|\|full | 4781 | No | NA | 3 | 0 | 2 | Low-quality | Genome-fragment | 1.5 | AAI-based (medium-confidence) | 0 |
| S2_k141_29759 | 4100 | No | NA | 6 | 0 | 6 | Not-determined | Genome-fragment | NA | NA | 0 |
| S2_k141_133725 | 1329 | No | NA | 1 | 0 | 1 | Not-determined | Genome-fragment | NA | NA | 0 |
| S2_k141_227206 | 1131 | No | NA | 1 | 0 | 0 | Not-determined | Genome-fragment | NA | NA | 0 |
| S2_k141_227246 | 3744 | No | NA | 4 | 0 | 4 | Not-determined | Genome-fragment | NA | NA | 0 |
| S2_k141_227250 | 3295 | No | NA | 6 | 0 | 6 | Not-determined | Genome-fragment | NA | NA | 0 |
| S2_k141_133750 | 2348 | No | NA | 3 | 0 | 0 | Not-determined | Genome-fragment | NA | NA | 0 |
| S2_k141_165558 | 1118 | No | NA | 2 | 0 | 0 | Not-determined | Genome-fragment | NA | NA | 0 |
| S2_k141_206127 | 8078 | No | NA | 11 | 0 | 3 | Not-determined | Genome-fragment | NA | NA | 0 |
| S2_k141_206175 | 1162 | No | NA | 1 | 0 | 1 | Not-determined | Genome-fragment | NA | NA | 0 |
| S2_k141_246768 | 4515 | No | NA | 5 | 0 | 4 | Not-determined | Genome-fragment | NA | NA | 0 |
| S2_k141_133984 | 2315 | No | NA | 2 | 0 | 1 | Not-determined | Genome-fragment | NA | NA | 0 |
| S3_k141_51664 | 4782 | Yes | 1335 | 6 | 1 | 1 | Low-quality | Genome-fragment | 1.75 | AAI-based (medium-confidence) | 72.08 |
| S2_k141_8542 | 25932 | No | NA | 18 | 0 | 12 | Not-determined | Genome-fragment | NA | NA | 0 |
| S2_k141_165625 | 1106 | No | NA | 1 | 0 | 0 | Not-determined | Genome-fragment | NA | NA | 0 |
| S4-final-viral-combined_k141_5543\|\|full | 4782 | No | NA | 6 | 2 | 0 | Low-quality | Genome-fragment | 10.26 | AAI-based (medium-confidence) | 0 |
| S5-final-viral-combined_k141_168816\|\|full | 4783 | No | NA | 6 | 1 | 0 | Low-quality | Genome-fragment | 13.2 | AAI-based (high-confidence) | 0 |
| S2_k141_246886 | 2194 | No | NA | 2 | 0 | 0 | Not-determined | Genome-fragment | NA | NA | 0 |
| S2_k141_113836 | 1582 | No | NA | 1 | 0 | 1 | Not-determined | Genome-fragment | NA | NA | 0 |
| S12-final-viral-combined_k141_224500\|\|full | 4783 | No | NA | 6 | 0 | 3 | Low-quality | Genome-fragment | 4.82 | AAI-based (medium-confidence) | 0 |
| S11-final-viral-combined_k141_138981\|\|full | 4784 | No | NA | 4 | 4 | 0 | Low-quality | Genome-fragment | 12.02 | AAI-based (high-confidence) | 0 |
| S11_k141_224566 | 4784 | No | NA | 11 | 5 | 0 | Low-quality | Genome-fragment | 9.92 | AAI-based (medium-confidence) | 0 |
| S2_k141_246926 | 1083 | No | NA | 2 | 0 | 0 | Not-determined | Genome-fragment | NA | NA | 0 |
| S2_k141_246944 | 3077 | No | NA | 3 | 0 | 2 | Not-determined | Genome-fragment | NA | NA | 0 |
| S2_k141_246951 | 1071 | No | NA | 1 | 0 | 1 | Not-determined | Genome-fragment | NA | NA | 0 |
| S2_k141_227557 | 1786 | No | NA | 1 | 0 | 1 | Not-determined | Genome-fragment | NA | NA | 0 |
| S2_k141_206494 | 1323 | No | NA | 1 | 0 | 1 | Not-determined | Genome-fragment | NA | NA | 0 |
| S2_k141_246995 | 1119 | No | NA | 2 | 0 | 0 | Not-determined | Genome-fragment | NA | NA | 0 |
| S2_k141_246997 | 1476 | No | NA | 2 | 0 | 0 | Not-determined | Genome-fragment | NA | NA | 0 |
| S2_k141_227593 | 1762 | No | NA | 1 | 0 | 1 | Not-determined | Genome-fragment | NA | NA | 0 |
| S8_k141_118190 | 4785 | No | NA | 4 | 2 | 0 | Low-quality | Genome-fragment | 3.59 | AAI-based (high-confidence) | 0 |
| S2_k141_227722 | 1475 | No | NA | 1 | 0 | 1 | Not-determined | Genome-fragment | NA | NA | 0 |
| S5-final-viral-combined_k141_76500\|\|full | 4786 | No | NA | 6 | 0 | 0 | Low-quality | Genome-fragment | 8.02 | AAI-based (medium-confidence) | 0 |
| S12-final-viral-combined_k141_312775\|\|full | 4787 | No | NA | 4 | 0 | 2 | Low-quality | Genome-fragment | 5.21 | AAI-based (medium-confidence) | 0 |
| S2_k141_30593 | 1201 | No | NA | 1 | 0 | 1 | Not-determined | Genome-fragment | NA | NA | 0 |
| S2_k141_134555 | 1079 | No | NA | 1 | 0 | 1 | Not-determined | Genome-fragment | NA | NA | 0 |
| S6-final-viral-combined_k141_130059\|\|full | 4789 | No | NA | 5 | 0 | 3 | Low-quality | Genome-fragment | 2.96 | AAI-based (medium-confidence) | 0 |
| S4-final-viral-combined_k141_94671\|\|full | 4792 | Yes | 4159 | 6 | 1 | 1 | Low-quality | Genome-fragment | 11.8 | AAI-based (high-confidence) | 13.21 |
| S2_k141_134619 | 1754 | No | NA | 2 | 0 | 0 | Not-determined | Genome-fragment | NA | NA | 0 |
| S2_k141_247145 | 1390 | No | NA | 1 | 0 | 0 | Not-determined | Genome-fragment | NA | NA | 0 |
| S2_k141_206776 | 2055 | No | NA | 1 | 0 | 0 | Not-determined | Genome-fragment | NA | NA | 0 |
| S1-final-viral-combined_k141_284183\|\|full | 4793 | No | NA | 9 | 1 | 0 | Low-quality | Genome-fragment | 12.1 | AAI-based (high-confidence) | 0 |
| S2_k141_206791 | 1074 | No | NA | 1 | 0 | 0 | Not-determined | Genome-fragment | NA | NA | 0 |
| S17-final-viral-combined_k141_71697\|\|full | 4793 | No | NA | 4 | 0 | 4 | Low-quality | Genome-fragment | 1.52 | AAI-based (medium-confidence) | 0 |
| S2_k141_247269 | 1161 | No | NA | 1 | 0 | 0 | Not-determined | Genome-fragment | NA | NA | 0 |
| S2_k141_30807 | 1622 | No | NA | 1 | 0 | 0 | Not-determined | Genome-fragment | NA | NA | 0 |
| S2_k141_247288 | 1093 | No | NA | 1 | 0 | 0 | Not-determined | Genome-fragment | NA | NA | 0 |
| S12-final-viral-combined_k141_129808\|\|full | 4793 | No | NA | 5 | 0 | 0 | Low-quality | Genome-fragment | 4.32 | AAI-based (medium-confidence) | 0 |
| S2_k141_247328 | 1262 | No | NA | 2 | 0 | 0 | Not-determined | Genome-fragment | NA | NA | 0 |
| S2_k141_9099 | 1132 | No | NA | 2 | 0 | 1 | Not-determined | Genome-fragment | NA | NA | 0 |
| S2_k141_247422 | 2213 | No | NA | 4 | 0 | 3 | Not-determined | Genome-fragment | NA | NA | 0 |
| S2-final-viral-combined_k141_144140\|\|full | 4793 | No | NA | 3 | 0 | 2 | Low-quality | Genome-fragment | 2.27 | AAI-based (medium-confidence) | 0 |
| S2_k141_247429 | 1292 | No | NA | 2 | 0 | 2 | Not-determined | Genome-fragment | NA | NA | 0 |
| S8-final-viral-combined_k141_168450\|\|full | 4793 | No | NA | 4 | 2 | 0 | Low-quality | Genome-fragment | 11.87 | AAI-based (high-confidence) | 0 |
| S2_k141_247454 | 1033 | No | NA | 1 | 0 | 1 | Not-determined | Genome-fragment | NA | NA | 0 |
| S2_k141_30976 | 4024 | No | NA | 4 | 0 | 1 | Not-determined | Genome-fragment | NA | NA | 0 |
| S5_k141_40858 | 4795 | No | NA | 3 | 0 | 3 | Low-quality | Genome-fragment | 9.76 | AAI-based (medium-confidence) | 0 |
| S2_k141_206843 | 3792 | No | NA | 7 | 0 | 2 | Not-determined | Genome-fragment | NA | NA | 0 |
| S2_k141_247638 | 3878 | No | NA | 7 | 0 | 6 | Not-determined | Genome-fragment | NA | NA | 0 |
| S2_k141_247704 | 1136 | No | NA | 1 | 0 | 1 | Not-determined | Genome-fragment | NA | NA | 0 |
| S5-final-viral-combined_k141_143717\|\|full | 4798 | No | NA | 6 | 0 | 0 | Low-quality | Genome-fragment | 11.16 | AAI-based (medium-confidence) | 0 |
| S7-final-viral-combined_k141_88611\|\|full | 4799 | No | NA | 4 | 0 | 1 | Low-quality | Genome-fragment | 0.41 | AAI-based (medium-confidence) | 0 |
| S2_k141_9370 | 3038 | No | NA | 3 | 0 | 2 | Not-determined | Genome-fragment | NA | NA | 0 |
| S2_k141_247865 | 1020 | No | NA | 3 | 0 | 0 | Not-determined | Genome-fragment | NA | NA | 0 |
| S17-k141_37014 | 4799 | No | NA | 6 | 0 | 4 | Low-quality | Genome-fragment | 2.77 | AAI-based (medium-confidence) | 0 |
| S2_k141_247918 | 7527 | No | NA | 2 | 0 | 1 | Not-determined | Genome-fragment | NA | NA | 0 |
| S2-final-viral-combined_k141_224854\|\|full | 4799 | No | NA | 4 | 0 | 2 | Low-quality | Genome-fragment | 3.3 | AAI-based (high-confidence) | 0 |
| S2_k141_248008 | 1395 | No | NA | 1 | 0 | 0 | Not-determined | Genome-fragment | NA | NA | 0 |
| S2_k141_207046 | 1325 | No | NA | 4 | 0 | 0 | Not-determined | Genome-fragment | NA | NA | 0 |
| S11-final-viral-combined_k141_87518\|\|full | 4800 | No | NA | 2 | 0 | 0 | Low-quality | Genome-fragment | 5.39 | AAI-based (medium-confidence) | 0 |
| S2_k141_248102 | 1146 | No | NA | 2 | 0 | 1 | Not-determined | Genome-fragment | NA | NA | 0 |
| S1-final-viral-combined_k141_139779\|\|full | 4800 | No | NA | 3 | 2 | 0 | Low-quality | Genome-fragment | 4.21 | HMM-based (lower-bound) | 0 |
| S11-final-viral-combined_k141_77086\|\|full | 4804 | No | NA | 7 | 0 | 0 | Low-quality | Genome-fragment | 3.41 | AAI-based (medium-confidence) | 0 |
| S2_k141_9606 | 2527 | No | NA | 2 | 0 | 1 | Not-determined | Genome-fragment | NA | NA | 0 |
| S2_k141_248232 | 2336 | No | NA | 1 | 0 | 0 | Not-determined | Genome-fragment | NA | NA | 0 |
| S2_k141_248252 | 2859 | No | NA | 6 | 0 | 5 | Not-determined | Genome-fragment | NA | NA | 0 |
| S2_k141_248315 | 64248 | No | NA | 96 | 29 | 0 | Complete | High-quality | 100 | DTR (high-confidence) | 0 |
| S2_k141_9780 | 1000 | No | NA | 3 | 0 | 1 | Not-determined | Genome-fragment | NA | NA | 0 |
| S2_k141_248324 | 48927 | No | NA | 73 | 24 | 1 | Complete | High-quality | 100 | DTR (high-confidence) | 0 |
| S2_k141_248327 | 9825 | No | NA | 14 | 0 | 1 | Medium-quality | Genome-fragment | 76.23 | AAI-based (medium-confidence) | 0 |
| S5_k141_199703 | 4805 | No | NA | 8 | 2 | 0 | Low-quality | Genome-fragment | 9.97 | AAI-based (high-confidence) | 0 |
| S2_k141_9831 | 1444 | No | NA | 1 | 0 | 1 | Not-determined | Genome-fragment | NA | NA | 0 |
| S2_k141_248423 | 1029 | No | NA | 2 | 0 | 0 | Not-determined | Genome-fragment | NA | NA | 0 |
| S2_k141_9935 | 3582 | No | NA | 3 | 0 | 2 | Not-determined | Genome-fragment | NA | NA | 0 |
| S2_k141_10035 | 2008 | No | NA | 6 | 0 | 5 | Not-determined | Genome-fragment | NA | NA | 0 |
| S2_k141_10083 | 1216 | No | NA | 3 | 0 | 0 | Not-determined | Genome-fragment | NA | NA | 0 |
| S2_k141_10202 | 1172 | No | NA | 2 | 0 | 0 | Not-determined | Genome-fragment | NA | NA | 0 |
| S2_k141_10214 | 1862 | No | NA | 1 | 0 | 0 | Not-determined | Genome-fragment | NA | NA | 0 |
| S2_k141_10260 | 1067 | No | NA | 2 | 0 | 0 | Not-determined | Genome-fragment | NA | NA | 0 |
| S4-final-viral-combined_k141_143935\|\|full | 9979 | No | NA | 15 | 0 | 3 | Not-determined | Genome-fragment | NA | NA | 0 |
| S4-final-viral-combined_k141_26994\|\|full | 6762 | No | NA | 7 | 0 | 5 | Not-determined | Genome-fragment | NA | NA | 0 |
| S4-final-viral-combined_k141_139314\|\|full | 2019 | No | NA | 3 | 0 | 1 | Not-determined | Genome-fragment | NA | NA | 0 |
| S8-final-viral-combined_k141_38592\|\|full | 4805 | No | NA | 7 | 1 | 0 | Low-quality | Genome-fragment | 11.29 | AAI-based (high-confidence) | 0 |
| S1-final-viral-combined_k141_233346\|\|full | 4806 | No | NA | 5 | 0 | 2 | Low-quality | Genome-fragment | 2.7 | AAI-based (medium-confidence) | 0 |
| S4-final-viral-combined_k141_146307\|\|full | 1673 | No | NA | 2 | 0 | 2 | Not-determined | Genome-fragment | NA | NA | 0 |
| S17-k141_126337 | 4807 | No | NA | 3 | 0 | 2 | Low-quality | Genome-fragment | 1.54 | AAI-based (high-confidence) | 0 |
| S2-final-viral-combined_k141_185685\|\|full | 4808 | No | NA | 4 | 0 | 2 | Low-quality | Genome-fragment | 5.84 | AAI-based (medium-confidence) | 0 |
| S2_k141_26218 | 4808 | No | NA | 5 | 0 | 1 | Low-quality | Genome-fragment | 4.12 | AAI-based (medium-confidence) | 0 |
| S4-final-viral-combined_k141_90589\|\|full | 2910 | No | NA | 5 | 0 | 1 | Not-determined | Genome-fragment | NA | NA | 0 |
| S4-final-viral-combined_k141_104527\|\|full | 2774 | No | NA | 4 | 0 | 0 | Not-determined | Genome-fragment | NA | NA | 0 |
| S4-final-viral-combined_k141_34883\|\|full | 2645 | No | NA | 3 | 0 | 3 | Not-determined | Genome-fragment | NA | NA | 0 |
| S4-final-viral-combined_k141_20951\|\|full | 2901 | No | NA | 4 | 0 | 0 | Not-determined | Genome-fragment | NA | NA | 0 |
| S4-final-viral-combined_k141_69721\|\|full | 3162 | No | NA | 5 | 0 | 1 | Not-determined | Genome-fragment | NA | NA | 0 |
| S4-final-viral-combined_k141_20958\|\|full | 3664 | No | NA | 7 | 0 | 1 | Not-determined | Genome-fragment | NA | NA | 0 |
| S4-final-viral-combined_k141_91041\|\|full | 4808 | No | NA | 3 | 0 | 2 | Low-quality | Genome-fragment | 2.41 | AAI-based (medium-confidence) | 0 |
| S4-final-viral-combined_k141_41833\|\|full | 2007 | No | NA | 3 | 0 | 1 | Not-determined | Genome-fragment | NA | NA | 0 |
| S4-final-viral-combined_k141_118451\|\|full | 4635 | No | NA | 3 | 0 | 3 | Not-determined | Genome-fragment | NA | NA | 0 |
| S1_k141_362208 | 4810 | Yes | 2662 | 6 | 1 | 1 | Low-quality | Genome-fragment | 1.59 | HMM-based (lower-bound) | 44.66 |
| S8-final-viral-combined_k141_155031\|\|full | 4810 | No | NA | 8 | 1 | 0 | Low-quality | Genome-fragment | 7.69 | AAI-based (medium-confidence) | 0 |
| S4-final-viral-combined_k141_153311\|\|full | 1720 | No | NA | 2 | 0 | 0 | Not-determined | Genome-fragment | NA | NA | 0 |
| S4-final-viral-combined_k141_125428\|\|full | 1921 | No | NA | 2 | 0 | 2 | Not-determined | Genome-fragment | NA | NA | 0 |
| S4-final-viral-combined_k141_139388\|\|full | 1314 | No | NA | 3 | 0 | 1 | Not-determined | Genome-fragment | NA | NA | 0 |
| S17-final-viral-combined_k141_66229\|\|full | 4812 | No | NA | 5 | 0 | 3 | Low-quality | Genome-fragment | 1.89 | AAI-based (medium-confidence) | 0 |
| S4-final-viral-combined_k141_153320\|\|full | 1395 | No | NA | 3 | 0 | 3 | Not-determined | Genome-fragment | NA | NA | 0 |
| S4-final-viral-combined_k141_132459\|\|full | 1657 | No | NA | 2 | 0 | 1 | Not-determined | Genome-fragment | NA | NA | 0 |
| S17-k141_160283 | 4813 | No | NA | 5 | 0 | 2 | Low-quality | Genome-fragment | 7.98 | AAI-based (high-confidence) | 0 |
| S4-final-viral-combined_k141_41872\|\|full | 1900 | No | NA | 3 | 0 | 1 | Not-determined | Genome-fragment | NA | NA | 0 |
| S4-final-viral-combined_k141_62813\|\|full | 2785 | No | NA | 7 | 0 | 2 | Not-determined | Genome-fragment | NA | NA | 0 |
| S4-final-viral-combined_k141_125490\|\|full | 2364 | No | NA | 3 | 0 | 2 | Not-determined | Genome-fragment | NA | NA | 0 |
| S4-final-viral-combined_k141_160297\|\|full | 1902 | No | NA | 3 | 0 | 2 | Not-determined | Genome-fragment | NA | NA | 0 |
| S8_k141_270743 | 4813 | No | NA | 17 | 0 | 0 | Low-quality | Genome-fragment | 3.69 | AAI-based (high-confidence) | 0 |
| S4-final-viral-combined_k141_152\|\|full | 1267 | No | NA | 2 | 0 | 0 | Not-determined | Genome-fragment | NA | NA | 0 |
| S4-final-viral-combined_k141_90704\|\|full | 6176 | No | NA | 3 | 0 | 2 | Not-determined | Genome-fragment | NA | NA | 0 |
| S7-final-viral-combined_k141_12381\|\|full | 4816 | No | NA | 5 | 5 | 0 | Low-quality | Genome-fragment | 11.83 | AAI-based (high-confidence) | 0 |
| S4-final-viral-combined_k141_146354\|\|full | 4469 | No | NA | 3 | 0 | 3 | Not-determined | Genome-fragment | NA | NA | 0 |
| S17-final-viral-combined_k141_99399\|\|full | 4816 | No | NA | 3 | 0 | 3 | Low-quality | Genome-fragment | 10.94 | AAI-based (medium-confidence) | 0 |
| S4-final-viral-combined_k141_175\|\|full | 1202 | No | NA | 2 | 0 | 0 | Not-determined | Genome-fragment | NA | NA | 0 |
| S4-final-viral-combined_k141_21036\|\|full | 2028 | No | NA | 3 | 0 | 1 | Not-determined | Genome-fragment | NA | NA | 0 |
| S4-final-viral-combined_k141_27969\|\|full | 1566 | No | NA | 3 | 0 | 2 | Not-determined | Genome-fragment | NA | NA | 0 |
| S9_k141_56691 | 4819 | No | NA | 9 | 3 | 0 | Low-quality | Genome-fragment | 10.13 | AAI-based (high-confidence) | 0 |
| S4-final-viral-combined_k141_14054\|\|full | 1064 | No | NA | 3 | 0 | 1 | Not-determined | Genome-fragment | NA | NA | 0 |
| S4-final-viral-combined_k141_76787\|\|full | 2190 | No | NA | 2 | 0 | 1 | Not-determined | Genome-fragment | NA | NA | 0 |
| S4-final-viral-combined_k141_62860\|\|full | 14210 | No | NA | 17 | 0 | 12 | Not-determined | Genome-fragment | NA | NA | 0 |
| S4-final-viral-combined_k141_76791\|\|full | 4752 | No | NA | 6 | 0 | 2 | Not-determined | Genome-fragment | NA | NA | 0 |
| S1-final-viral-combined_k141_215676\|\|full | 4819 | No | NA | 8 | 0 | 0 | Low-quality | Genome-fragment | 11.01 | AAI-based (medium-confidence) | 0 |
| S4-final-viral-combined_k141_7062\|\|full | 1293 | No | NA | 2 | 0 | 0 | Not-determined | Genome-fragment | NA | NA | 0 |
| S2_k141_2546 | 4819 | No | NA | 8 | 1 | 0 | Low-quality | Genome-fragment | 5.02 | AAI-based (high-confidence) | 0 |
| S4-final-viral-combined_k141_55806\|\|full | 1577 | No | NA | 3 | 0 | 0 | Not-determined | Genome-fragment | NA | NA | 0 |
| S17-final-viral-combined_k141_249456\|\|full | 4821 | Yes | 1818 | 5 | 1 | 1 | Low-quality | Genome-fragment | 0.96 | AAI-based (medium-confidence) | 62.29 |
| S17-final-viral-combined_k141_106398\|\|full | 4822 | No | NA | 7 | 0 | 1 | Low-quality | Genome-fragment | 3.31 | AAI-based (medium-confidence) | 0 |
| S4-final-viral-combined_k141_160361\|\|full | 5132 | No | NA | 6 | 0 | 2 | Not-determined | Genome-fragment | NA | NA | 0 |
| S4-final-viral-combined_k141_125563\|\|full | 3989 | No | NA | 7 | 0 | 4 | Not-determined | Genome-fragment | NA | NA | 0 |
| S4-final-viral-combined_k141_118531\|\|full | 69321 | Yes | 38112 | 89 | 20 | 25 | Medium-quality | Genome-fragment | 81.86 | AAI-based (high-confidence) | 45.02 |
| S8-final-viral-combined_k141_114046\|\|full | 4822 | No | NA | 3 | 0 | 1 | Low-quality | Genome-fragment | 3.45 | AAI-based (high-confidence) | 0 |
| S4-final-viral-combined_k141_14104\|\|full | 1984 | No | NA | 3 | 0 | 1 | Not-determined | Genome-fragment | NA | NA | 0 |
| S1_k141_377899 | 4823 | No | NA | 3 | 0 | 1 | Low-quality | Genome-fragment | 7.99 | AAI-based (medium-confidence) | 0 |
| S4-final-viral-combined_k141_62926\|\|full | 1880 | No | NA | 2 | 0 | 2 | Not-determined | Genome-fragment | NA | NA | 0 |
| S4-final-viral-combined_k141_7146\|\|full | 3241 | No | NA | 3 | 0 | 2 | Not-determined | Genome-fragment | NA | NA | 0 |
| S4-final-viral-combined_k141_97733\|\|full | 3019 | No | NA | 3 | 0 | 0 | Not-determined | Genome-fragment | NA | NA | 0 |
| S4-final-viral-combined_k141_125578\|\|full | 4726 | No | NA | 2 | 0 | 2 | Not-determined | Genome-fragment | NA | NA | 0 |
| S1-final-viral-combined_k141_25031\|\|full | 4825 | No | NA | 10 | 0 | 1 | Low-quality | Genome-fragment | 3.59 | AAI-based (high-confidence) | 0 |
| S17-final-viral-combined_k141_133544\|\|full | 4825 | No | NA | 9 | 7 | 0 | Low-quality | Genome-fragment | 19.08 | AAI-based (high-confidence) | 0 |
| S1_k141_178959 | 4826 | No | NA | 4 | 0 | 2 | Low-quality | Genome-fragment | 0.95 | AAI-based (medium-confidence) | 0 |
| S4-final-viral-combined_k141_28035\|\|full | 1577 | No | NA | 3 | 0 | 1 | Not-determined | Genome-fragment | NA | NA | 0 |
| S4-final-viral-combined_k141_69991\|\|full | 1440 | No | NA | 3 | 0 | 1 | Not-determined | Genome-fragment | NA | NA | 0 |
| S4-final-viral-combined_k141_338\|\|full | 2312 | No | NA | 3 | 0 | 2 | Not-determined | Genome-fragment | NA | NA | 0 |
| S8_k141_223144 | 4827 | Yes | 3994 | 7 | 2 | 1 | Low-quality | Genome-fragment | 10.58 | AAI-based (high-confidence) | 17.26 |
| S4-final-viral-combined_k141_146449\|\|full | 1122 | No | NA | 3 | 0 | 1 | Not-determined | Genome-fragment | NA | NA | 0 |
| S8_k141_40172 | 4827 | No | NA | 11 | 2 | 1 | Low-quality | Genome-fragment | 3.68 | AAI-based (high-confidence) | 0 |
| S4-final-viral-combined_k141_146468\|\|full | 2561 | No | NA | 5 | 0 | 0 | Not-determined | Genome-fragment | NA | NA | 0 |
| S11-final-viral-combined_k141_310844\|\|full | 4828 | No | NA | 5 | 0 | 0 | Low-quality | Genome-fragment | 8.96 | AAI-based (high-confidence) | 0 |
| S17-final-viral-combined_k141_260590\|\|full | 4829 | No | NA | 3 | 0 | 2 | Low-quality | Genome-fragment | 1.99 | AAI-based (medium-confidence) | 0 |
| S4-final-viral-combined_k141_28101\|\|full | 3563 | No | NA | 6 | 0 | 0 | Not-determined | Genome-fragment | NA | NA | 0 |
| S4-final-viral-combined_k141_70030\|\|full | 1064 | No | NA | 2 | 0 | 2 | Not-determined | Genome-fragment | NA | NA | 0 |
| S2_k141_64119 | 4829 | No | NA | 7 | 0 | 0 | Low-quality | Genome-fragment | 4.49 | AAI-based (high-confidence) | 0 |
| S4-final-viral-combined_k141_48977\|\|full | 3185 | No | NA | 4 | 0 | 1 | Not-determined | Genome-fragment | NA | NA | 0 |
| S7-final-viral-combined_k141_95507\|\|full | 4830 | No | NA | 3 | 0 | 2 | Low-quality | Genome-fragment | 4.69 | AAI-based (medium-confidence) | 0 |
| S8_k141_258493 | 4830 | No | NA | 6 | 0 | 6 | Low-quality | Genome-fragment | 1.79 | AAI-based (high-confidence) | 0 |
| S4-final-viral-combined_k141_118651\|\|full | 5283 | No | NA | 7 | 0 | 4 | Not-determined | Genome-fragment | NA | NA | 0 |
| S4-final-viral-combined_k141_63034\|\|full | 1881 | No | NA | 3 | 0 | 2 | Not-determined | Genome-fragment | NA | NA | 0 |
| S3_k141_32237 | 4832 | No | NA | 14 | 1 | 1 | Low-quality | Genome-fragment | 8.8 | AAI-based (high-confidence) | 0 |
| S11_k141_189794 | 4833 | No | NA | 5 | 0 | 0 | Low-quality | Genome-fragment | 18.06 | AAI-based (medium-confidence) | 0 |
| S12-final-viral-combined_k141_231550\|\|full | 4833 | No | NA | 10 | 0 | 1 | Low-quality | Genome-fragment | 12.21 | AAI-based (high-confidence) | 0 |
| S4-final-viral-combined_k141_76931\|\|full | 2428 | No | NA | 4 | 0 | 3 | Not-determined | Genome-fragment | NA | NA | 0 |
| S12-final-viral-combined_k141_181555\|\|full | 4834 | No | NA | 4 | 0 | 2 | Low-quality | Genome-fragment | 2.74 | AAI-based (medium-confidence) | 0 |
| S4-final-viral-combined_k141_104855\|\|full | 1202 | No | NA | 2 | 0 | 1 | Not-determined | Genome-fragment | NA | NA | 0 |
| S8-final-viral-combined_k141_65607\|\|full | 4836 | No | NA | 11 | 2 | 0 | Low-quality | Genome-fragment | 13.4 | AAI-based (high-confidence) | 0 |
| S4-final-viral-combined_k141_76970\|\|full | 1241 | No | NA | 3 | 0 | 0 | Not-determined | Genome-fragment | NA | NA | 0 |
| S4-final-viral-combined_k141_63065\|\|full | 1755 | No | NA | 2 | 0 | 2 | Not-determined | Genome-fragment | NA | NA | 0 |
| S4-final-viral-combined_k141_111878\|\|full | 1269 | No | NA | 2 | 0 | 1 | Not-determined | Genome-fragment | NA | NA | 0 |
| S4-final-viral-combined_k141_56005\|\|full | 5916 | No | NA | 6 | 0 | 3 | Not-determined | Genome-fragment | NA | NA | 0 |
| S4-final-viral-combined_k141_139695\|\|full | 1072 | No | NA | 2 | 0 | 1 | Not-determined | Genome-fragment | NA | NA | 0 |
| S7-final-viral-combined_k141_4987\|\|full | 4837 | No | NA | 9 | 6 | 1 | Low-quality | Genome-fragment | 7.27 | HMM-based (lower-bound) | 0 |
| S4-final-viral-combined_k141_7310\|\|full | 1937 | No | NA | 3 | 0 | 3 | Not-determined | Genome-fragment | NA | NA | 0 |
| S4-final-viral-combined_k141_42153\|\|full | 1244 | No | NA | 3 | 0 | 0 | Not-determined | Genome-fragment | NA | NA | 0 |
| S4-final-viral-combined_k141_28218\|\|full | 2326 | No | NA | 4 | 0 | 0 | Not-determined | Genome-fragment | NA | NA | 0 |
| S12-final-viral-combined_k141_365963\|\|full | 4839 | No | NA | 3 | 0 | 1 | Low-quality | Genome-fragment | 2.69 | AAI-based (high-confidence) | 0 |
| S12-final-viral-combined_k141_140059\|\|full | 4840 | No | NA | 9 | 2 | 0 | Low-quality | Genome-fragment | 5.95 | HMM-based (lower-bound) | 0 |
| S4-final-viral-combined_k141_34429\|\|full | 4840 | No | NA | 2 | 0 | 2 | Low-quality | Genome-fragment | 0.43 | AAI-based (medium-confidence) | 0 |
| S11-final-viral-combined_k141_246009\|\|full | 4841 | No | NA | 6 | 5 | 0 | Low-quality | Genome-fragment | 7.78 | AAI-based (high-confidence) | 0 |
| S4-final-viral-combined_k141_104885\|\|full | 13457 | No | NA | 14 | 0 | 4 | Not-determined | Genome-fragment | NA | NA | 0 |
| S4-final-viral-combined_k141_104886\|\|full | 3680 | No | NA | 6 | 0 | 3 | Not-determined | Genome-fragment | NA | NA | 0 |
| S4-final-viral-combined_k141_42181\|\|full | 2321 | No | NA | 3 | 0 | 2 | Not-determined | Genome-fragment | NA | NA | 0 |
| S1-final-viral-combined_k141_245894\|\|full | 4843 | No | NA | 6 | 0 | 4 | Low-quality | Genome-fragment | 6.15 | AAI-based (medium-confidence) | 0 |
| S4-final-viral-combined_k141_104892\|\|full | 3086 | No | NA | 2 | 0 | 2 | Not-determined | Genome-fragment | NA | NA | 0 |
| S4-final-viral-combined_k141_104903\|\|full | 1993 | No | NA | 3 | 0 | 2 | Not-determined | Genome-fragment | NA | NA | 0 |
| S4-final-viral-combined_k141_160564\|\|full | 1657 | No | NA | 3 | 0 | 0 | Not-determined | Genome-fragment | NA | NA | 0 |
| S12-final-viral-combined_k141_243759\|\|full | 4843 | No | NA | 4 | 0 | 1 | Low-quality | Genome-fragment | 3.06 | AAI-based (medium-confidence) | 0 |

| | | | | | | | | | | | |
|---|---|---|---|---|---|---|---|---|---|---|---|
| S11-final-viral-combined_k141_180228\|\|full | 4845 | No | NA | 7 | 0 | 0 | Low-quality | Genome-fragment | 11.23 | AAI-based (medium-confidence) | 0 |
| S1_k141_267124 | 4845 | No | NA | 6 | 0 | 2 | Low-quality | Genome-fragment | 3.52 | AAI-based (medium-confidence) | 0 |
| S17-final-viral-combined_k141_77366\|\|full | 4847 | No | NA | 4 | 0 | 2 | Low-quality | Genome-fragment | 1.61 | AAI-based (medium-confidence) | 0 |
| S8-final-viral-combined_k141_38289\|\|full | 4848 | Yes | 2925 | 3 | 1 | 1 | Low-quality | Genome-fragment | 1.01 | HMM-based (lower-bound) | 39.67 |
| S4-final-viral-combined_k141_146645\|\|full | 2554 | No | NA | 3 | 0 | 2 | Not-determined | Genome-fragment | NA | NA | 0 |
| S4-final-viral-combined_k141_70134\|\|full | 1207 | No | NA | 2 | 0 | 1 | Not-determined | Genome-fragment | NA | NA | 0 |
| S4-final-viral-combined_k141_14441\|\|full | 4849 | No | NA | 7 | 0 | 3 | Low-quality | Genome-fragment | 2.46 | AAI-based (medium-confidence) | 0 |
| S4-final-viral-combined_k141_104937\|\|full | 1141 | No | NA | 3 | 0 | 1 | Not-determined | Genome-fragment | NA | NA | 0 |
| S4-final-viral-combined_k141_84028\|\|full | 1469 | No | NA | 4 | 0 | 0 | Not-determined | Genome-fragment | NA | NA | 0 |
| S4-final-viral-combined_k141_70148\|\|full | 2639 | No | NA | 4 | 0 | 0 | Not-determined | Genome-fragment | NA | NA | 0 |
| S3_k141_24306 | 4850 | No | NA | 8 | 0 | 0 | Low-quality | Genome-fragment | 12.28 | AAI-based (high-confidence) | 0 |
| S11-final-viral-combined_k141_225086\|\|full | 4852 | No | NA | 3 | 0 | 3 | Low-quality | Genome-fragment | 2.3 | AAI-based (medium-confidence) | 0 |
| S4-final-viral-combined_k141_97939\|\|full | 4024 | No | NA | 3 | 0 | 2 | Not-determined | Genome-fragment | NA | NA | 0 |
| S4-final-viral-combined_k141_21298\|\|full | 1550 | Yes | 430 | 3 | 1 | 1 | Not-determined | Genome-fragment | NA | NA | 72.26 |
| S17-final-viral-combined_k141_187367\|\|full | 4852 | Yes | 3325 | 6 | 1 | 1 | Low-quality | Genome-fragment | 2.13 | AAI-based (medium-confidence) | 31.47 |
| S4-final-viral-combined_k141_139794\|\|full | 1784 | No | NA | 2 | 0 | 2 | Not-determined | Genome-fragment | NA | NA | 0 |
| S4-final-viral-combined_k141_111985\|\|full | 2751 | No | NA | 5 | 0 | 0 | Not-determined | Genome-fragment | NA | NA | 0 |
| S4-final-viral-combined_k141_91095\|\|full | 3484 | No | NA | 6 | 0 | 0 | Not-determined | Genome-fragment | NA | NA | 0 |
| S12-final-viral-combined_k141_168615\|\|full | 4852 | No | NA | 5 | 3 | 0 | Low-quality | Genome-fragment | 7.72 | AAI-based (high-confidence) | 0 |
| S4-final-viral-combined_k141_42236\|\|full | 2452 | No | NA | 3 | 0 | 0 | Not-determined | Genome-fragment | NA | NA | 0 |
| S17-k141_197682 | 4853 | No | NA | 7 | 0 | 5 | Low-quality | Genome-fragment | 1.14 | AAI-based (medium-confidence) | 0 |
| S4-final-viral-combined_k141_28315\|\|full | 1907 | No | NA | 2 | 0 | 2 | Not-determined | Genome-fragment | NA | NA | 0 |
| S4-final-viral-combined_k141_63174\|\|full | 2277 | No | NA | 5 | 0 | 1 | Not-determined | Genome-fragment | NA | NA | 0 |
| S6-final-viral-combined_k141_4038\|\|full | 4853 | No | NA | 4 | 0 | 1 | Low-quality | Genome-fragment | 13.85 | AAI-based (high-confidence) | 0 |
| S12-final-viral-combined_k141_400593\|\|full | 4857 | No | NA | 14 | 1 | 0 | Low-quality | Genome-fragment | 6.04 | AAI-based (high-confidence) | 0 |
| S8-final-viral-combined_k141_192710\|\|full | 4857 | No | NA | 4 | 3 | 0 | Low-quality | Genome-fragment | 7.49 | HMM-based (lower-bound) | 0 |
| S4-final-viral-combined_k141_56157\|\|full | 1978 | No | NA | 6 | 0 | 1 | Not-determined | Genome-fragment | NA | NA | 0 |
| S4-final-viral-combined_k141_84054\|\|full | 1520 | No | NA | 2 | 0 | 1 | Not-determined | Genome-fragment | NA | NA | 0 |
| S4-final-viral-combined_k141_112010\|\|full | 1237 | No | NA | 3 | 0 | 0 | Not-determined | Genome-fragment | NA | NA | 0 |
| S4-final-viral-combined_k141_21329\|\|full | 1207 | No | NA | 2 | 0 | 1 | Not-determined | Genome-fragment | NA | NA | 0 |
| S4-final-viral-combined_k141_118753\|\|full | 7980 | No | NA | 8 | 0 | 0 | Not-determined | Genome-fragment | NA | NA | 0 |
| S7-final-viral-combined_k141_234918\|\|full | 4859 | No | NA | 5 | 0 | 4 | Low-quality | Genome-fragment | 3.04 | AAI-based (medium-confidence) | 0 |
| S2-final-viral-combined_k141_240941\|\|full | 4859 | No | NA | 8 | 2 | 0 | Low-quality | Genome-fragment | 3.44 | HMM-based (lower-bound) | 0 |
| S4-final-viral-combined_k141_125908\|\|full | 1388 | No | NA | 2 | 0 | 1 | Not-determined | Genome-fragment | NA | NA | 0 |
| S4-final-viral-combined_k141_34676\|\|full | 4859 | No | NA | 8 | 2 | 0 | Low-quality | Genome-fragment | 9.39 | AAI-based (high-confidence) | 0 |
| S11-final-viral-combined_k141_252583\|\|full | 4861 | No | NA | 4 | 4 | 0 | Low-quality | Genome-fragment | 11.09 | AAI-based (high-confidence) | 0 |
| S4-final-viral-combined_k141_98019\|\|full | 6068 | No | NA | 3 | 0 | 3 | Not-determined | Genome-fragment | NA | NA | 0 |
| S4-final-viral-combined_k141_118772\|\|full | 1986 | No | NA | 3 | 0 | 2 | Not-determined | Genome-fragment | NA | NA | 0 |
| S4-final-viral-combined_k141_125961\|\|full | 1577 | No | NA | 4 | 0 | 2 | Not-determined | Genome-fragment | NA | NA | 0 |
| S4-final-viral-combined_k141_105011\|\|full | 1032 | No | NA | 3 | 0 | 0 | Not-determined | Genome-fragment | NA | NA | 0 |
| S4-final-viral-combined_k141_21357\|\|full | 1765 | No | NA | 3 | 0 | 2 | Not-determined | Genome-fragment | NA | NA | 0 |
| S4-final-viral-combined_k141_91182\|\|full | 4880 | No | NA | 5 | 0 | 4 | Not-determined | Genome-fragment | NA | NA | 0 |
| S4-final-viral-combined_k141_21360\|\|full | 2599 | No | NA | 2 | 0 | 2 | Not-determined | Genome-fragment | NA | NA | 0 |
| S11-final-viral-combined_k141_35621\|\|full | 4861 | No | NA | 5 | 2 | 0 | Low-quality | Genome-fragment | 10.44 | HMM-based (lower-bound) | 0 |
| S12-final-viral-combined_k141_181326\|\|full | 4861 | No | NA | 5 | 0 | 0 | Low-quality | Genome-fragment | 11.98 | AAI-based (medium-confidence) | 0 |
| S4-final-viral-combined_k141_77220\|\|full | 10744 | No | NA | 13 | 0 | 6 | Not-determined | Genome-fragment | NA | NA | 0 |
| S4-final-viral-combined_k141_70268\|\|full | 1577 | No | NA | 3 | 0 | 1 | Not-determined | Genome-fragment | NA | NA | 0 |
| S4-final-viral-combined_k141_139878\|\|full | 2881 | No | NA | 5 | 0 | 3 | Not-determined | Genome-fragment | NA | NA | 0 |
| S4-final-viral-combined_k141_146711\|\|full | 1883 | No | NA | 3 | 0 | 0 | Not-determined | Genome-fragment | NA | NA | 0 |
| S1_k141_266340 | 4865 | Yes | 3479 | 10 | 1 | 1 | Low-quality | Genome-fragment | 9.41 | AAI-based (high-confidence) | 28.49 |
| S4-final-viral-combined_k141_84170\|\|full | 2378 | No | NA | 2 | 0 | 1 | Not-determined | Genome-fragment | NA | NA | 0 |
| S7-final-viral-combined_k141_174775\|\|full | 4865 | No | NA | 7 | 0 | 4 | Low-quality | Genome-fragment | 9.21 | AAI-based (medium-confidence) | 0 |
| S12-final-viral-combined_k141_106901\|\|full | 4866 | No | NA | 4 | 0 | 4 | Low-quality | Genome-fragment | 9.49 | AAI-based (medium-confidence) | 0 |
| S4-final-viral-combined_k141_91231\|\|full | 1555 | No | NA | 3 | 0 | 2 | Not-determined | Genome-fragment | NA | NA | 0 |
| S4-final-viral-combined_k141_694\|\|full | 5778 | No | NA | 7 | 0 | 4 | Not-determined | Genome-fragment | NA | NA | 0 |
| S11-final-viral-combined_k141_168629\|\|full | 4867 | No | NA | 7 | 4 | 0 | Low-quality | Genome-fragment | 10.06 | AAI-based (medium-confidence) | 0 |
| S4-final-viral-combined_k141_702\|\|full | 1231 | No | NA | 3 | 0 | 0 | Not-determined | Genome-fragment | NA | NA | 0 |
| S4-final-viral-combined_k141_98112\|\|full | 5332 | No | NA | 4 | 0 | 0 | Not-determined | Genome-fragment | NA | NA | 0 |
| S11-final-viral-combined_k141_278912\|\|full | 4867 | No | NA | 3 | 0 | 3 | Low-quality | Genome-fragment | 1.96 | AAI-based (medium-confidence) | 0 |
| S4-final-viral-combined_k141_112079\|\|full | 1420 | No | NA | 3 | 0 | 1 | Not-determined | Genome-fragment | NA | NA | 0 |
| S4-final-viral-combined_k141_77280\|\|full | 2021 | No | NA | 3 | 0 | 1 | Not-determined | Genome-fragment | NA | NA | 0 |
| S5-final-viral-combined_k141_122397\|\|full | 4867 | No | NA | 8 | 1 | 0 | Low-quality | Genome-fragment | 8.37 | HMM-based (lower-bound) | 0 |
| S11-final-viral-combined_k141_275277\|\|full | 4869 | No | NA | 9 | 2 | 1 | Low-quality | Genome-fragment | 13.47 | AAI-based (medium-confidence) | 0 |
| S4-final-viral-combined_k141_14411\|\|full | 43509 | No | NA | 58 | 10 | 0 | High-quality | High-quality | 96.41 | AAI-based (high-confidence) | 0 |
| S4-final-viral-combined_k141_49209\|\|full | 3392 | No | NA | 3 | 0 | 3 | Not-determined | Genome-fragment | NA | NA | 0 |
| S4-final-viral-combined_k141_84214\|\|full | 1489 | No | NA | 4 | 0 | 2 | Not-determined | Genome-fragment | NA | NA | 0 |
| S1-final-viral-combined_k141_328223\|\|full | 4869 | No | NA | 4 | 0 | 2 | Low-quality | Genome-fragment | 14.74 | AAI-based (high-confidence) | 0 |
| S1-final-viral-combined_k141_146235\|\|full | 4872 | Yes | 3575 | 9 | 4 | 2 | Low-quality | Genome-fragment | 7.19 | AAI-based (high-confidence) | 26.62 |
| S4-final-viral-combined_k141_49221\|\|full | 1482 | No | NA | 2 | 0 | 1 | Not-determined | Genome-fragment | NA | NA | 0 |
| S4-final-viral-combined_k141_77319\|\|full | 1664 | No | NA | 3 | 0 | 1 | Not-determined | Genome-fragment | NA | NA | 0 |
| S4-final-viral-combined_k141_112093\|\|full | 25857 | No | NA | 42 | 15 | 2 | Medium-quality | Genome-fragment | 75.64 | AAI-based (high-confidence) | 0 |
| S4-final-viral-combined_k141_118877\|\|full | 1464 | No | NA | 3 | 0 | 0 | Not-determined | Genome-fragment | NA | NA | 0 |
| S11_k141_255440 | 4873 | No | NA | 10 | 1 | 0 | Low-quality | Genome-fragment | 7.55 | AAI-based (high-confidence) | 0 |
| S4-final-viral-combined_k141_56278\|\|full | 7144 | No | NA | 3 | 0 | 2 | Not-determined | Genome-fragment | NA | NA | 0 |
| S3_k141_63446 | 4873 | No | NA | 8 | 0 | 2 | Low-quality | Genome-fragment | 5.16 | AAI-based (high-confidence) | 0 |
| S5_k141_15818 | 4874 | No | NA | 8 | 1 | 0 | Low-quality | Genome-fragment | 5.41 | AAI-based (high-confidence) | 0 |
| S4-final-viral-combined_k141_21512\|\|full | 4757 | No | NA | 4 | 0 | 2 | Not-determined | Genome-fragment | NA | NA | 0 |
| S4-final-viral-combined_k141_118922\|\|full | 1557 | No | NA | 2 | 0 | 1 | Not-determined | Genome-fragment | NA | NA | 0 |
| S4-final-viral-combined_k141_35282\|\|full | 1723 | No | NA | 4 | 0 | 0 | Not-determined | Genome-fragment | NA | NA | 0 |
| S7-final-viral-combined_k141_103381\|\|lt2gene | 4874 | No | NA | 2 | 0 | 0 | Low-quality | Genome-fragment | 8.73 | AAI-based (medium-confidence) | 0 |
| S4-final-viral-combined_k141_91374\|\|full | 2172 | No | NA | 3 | 0 | 1 | Not-determined | Genome-fragment | NA | NA | 0 |
| S12-final-viral-combined_k141_251285\|\|full | 4874 | No | NA | 11 | 4 | 0 | Low-quality | Genome-fragment | 8.16 | HMM-based (lower-bound) | 0 |
| S4-final-viral-combined_k141_14486\|\|full | 1223 | No | NA | 2 | 0 | 0 | Not-determined | Genome-fragment | NA | NA | 0 |
| S4-final-viral-combined_k141_105180\|\|full | 1092 | No | NA | 2 | 0 | 2 | Not-determined | Genome-fragment | NA | NA | 0 |
| S4-final-viral-combined_k141_781\|\|full | 1234 | No | NA | 3 | 0 | 2 | Not-determined | Genome-fragment | NA | NA | 0 |
| S4-final-viral-combined_k141_84355\|\|full | 1877 | No | NA | 3 | 0 | 0 | Not-determined | Genome-fragment | NA | NA | 0 |
| S4-final-viral-combined_k141_105213\|\|full | 1674 | No | NA | 3 | 0 | 1 | Not-determined | Genome-fragment | NA | NA | 0 |
| S4-final-viral-combined_k141_787\|\|full | 5457 | No | NA | 5 | 0 | 3 | Not-determined | Genome-fragment | NA | NA | 0 |
| S4-final-viral-combined_k141_35296\|\|full | 7495 | No | NA | 7 | 0 | 5 | Not-determined | Genome-fragment | NA | NA | 0 |
| S2-final-viral-combined_k141_27607\|\|full | 4874 | No | NA | 5 | 0 | 3 | Low-quality | Genome-fragment | 11.26 | AAI-based (medium-confidence) | 0 |
| S6_k141_77591 | 4874 | No | NA | 12 | 1 | 0 | Low-quality | Genome-fragment | 2.6 | AAI-based (high-confidence) | 0 |
| S4-final-viral-combined_k141_132918\|\|full | 2661 | No | NA | 4 | 0 | 2 | Not-determined | Genome-fragment | NA | NA | 0 |
| S4-final-viral-combined_k141_63345\|\|full | 3918 | No | NA | 4 | 0 | 2 | Not-determined | Genome-fragment | NA | NA | 0 |
| S1-final-viral-combined_k141_209398\|\|full | 4878 | No | NA | 4 | 0 | 3 | Low-quality | Genome-fragment | 2.61 | AAI-based (medium-confidence) | 0 |
| S4-final-viral-combined_k141_140082\|\|full | 1944 | No | NA | 3 | 0 | 2 | Not-determined | Genome-fragment | NA | NA | 0 |
| S2-final-viral-combined_k141_60181\|\|full | 4878 | No | NA | 5 | 0 | 0 | Low-quality | Genome-fragment | 7.6 | AAI-based (medium-confidence) | 0 |
| S1_k141_259639 | 4879 | No | NA | 9 | 2 | 0 | Low-quality | Genome-fragment | 8.94 | AAI-based (high-confidence) | 0 |
| S4-final-viral-combined_k141_153883\|\|full | 2191 | No | NA | 4 | 0 | 0 | Not-determined | Genome-fragment | NA | NA | 0 |
| S11_k141_322964 | 4880 | Yes | 2559 | 6 | 1 | 1 | Low-quality | Genome-fragment | 4.46 | AAI-based (medium-confidence) | 47.56 |
| S4-final-viral-combined_k141_42548\|\|full | 1008 | No | NA | 3 | 0 | 1 | Not-determined | Genome-fragment | NA | NA | 0 |
| S7_k141_157623 | 4881 | No | NA | 5 | 0 | 2 | Low-quality | Genome-fragment | 6.69 | AAI-based (medium-confidence) | 0 |
| S8-final-viral-combined_k141_266077\|\|full | 4883 | No | NA | 8 | 1 | 1 | Low-quality | Genome-fragment | 5.01 | AAI-based (medium-confidence) | 0 |
| S4-final-viral-combined_k141_867\|\|full | 1958 | No | NA | 4 | 0 | 3 | Not-determined | Genome-fragment | NA | NA | 0 |
| S4-final-viral-combined_k141_49342\|\|full | 4307 | No | NA | 4 | 0 | 3 | Not-determined | Genome-fragment | NA | NA | 0 |
| S4-final-viral-combined_k141_7614\|\|full | 4412 | No | NA | 4 | 0 | 4 | Not-determined | Genome-fragment | NA | NA | 0 |
| S17-final-viral-combined_k141_3305\|\|full | 4884 | No | NA | 7 | 1 | 0 | Low-quality | Genome-fragment | 10.98 | AAI-based (high-confidence) | 0 |
| S4-final-viral-combined_k141_112248\|\|full | 1602 | No | NA | 3 | 0 | 0 | Not-determined | Genome-fragment | NA | NA | 0 |
| S4-final-viral-combined_k141_56417\|\|full | 1455 | No | NA | 3 | 0 | 0 | Not-determined | Genome-fragment | NA | NA | 0 |
| S4-final-viral-combined_k141_91472\|\|full | 1549 | No | NA | 2 | 0 | 1 | Not-determined | Genome-fragment | NA | NA | 0 |
| S11_k141_321948 | 4885 | No | NA | 4 | 0 | 3 | Low-quality | Genome-fragment | 0.44 | AAI-based (medium-confidence) | 0 |
| S4-final-viral-combined_k141_91481\|\|full | 1454 | No | NA | 3 | 0 | 1 | Not-determined | Genome-fragment | NA | NA | 0 |
| S4-final-viral-combined_k141_28625\|\|full | 19675 | No | NA | 15 | 0 | 12 | Not-determined | Genome-fragment | NA | NA | 0 |
| S4-final-viral-combined_k141_63455\|\|full | 1611 | No | NA | 3 | 0 | 0 | Not-determined | Genome-fragment | NA | NA | 0 |
| S1-final-viral-combined_k141_226687\|\|full | 4885 | No | NA | 4 | 0 | 3 | Low-quality | Genome-fragment | 3.03 | AAI-based (medium-confidence) | 0 |
| S4-final-viral-combined_k141_91488\|\|full | 1428 | No | NA | 3 | 0 | 3 | Not-determined | Genome-fragment | NA | NA | 0 |
| S3_k141_43394 | 4885 | Yes | 2907 | 8 | 1 | 1 | Low-quality | Genome-fragment | 3.68 | AAI-based (high-confidence) | 40.49 |
| S4-final-viral-combined_k141_119048\|\|full | 1866 | No | NA | 4 | 0 | 0 | Not-determined | Genome-fragment | NA | NA | 0 |
| S11-final-viral-combined_k141_55983\|\|full | 4886 | No | NA | 4 | 0 | 1 | Low-quality | Genome-fragment | 8.66 | AAI-based (high-confidence) | 0 |
| S1-final-viral-combined_k141_17345\|\|full | 4886 | No | NA | 5 | 0 | 0 | Low-quality | Genome-fragment | 2.93 | AAI-based (medium-confidence) | 0 |
| S4-final-viral-combined_k141_35418\|\|full | 1849 | No | NA | 2 | 0 | 0 | Not-determined | Genome-fragment | NA | NA | 0 |
| S4-final-viral-combined_k141_153992\|\|full | 2660 | No | NA | 3 | 0 | 2 | Not-determined | Genome-fragment | NA | NA | 0 |
| S4-final-viral-combined_k141_56476\|\|full | 1208 | No | NA | 2 | 0 | 2 | Not-determined | Genome-fragment | NA | NA | 0 |
| S4-final-viral-combined_k141_126371\|\|full | 3732 | No | NA | 3 | 0 | 2 | Not-determined | Genome-fragment | NA | NA | 0 |
| S4-final-viral-combined_k141_14592\|\|full | 2625 | No | NA | 2 | 0 | 1 | Not-determined | Genome-fragment | NA | NA | 0 |
| S17-k141_96907 | 4886 | No | NA | 3 | 0 | 2 | Low-quality | Genome-fragment | 11.6 | AAI-based (medium-confidence) | 0 |
| S4-final-viral-combined_k141_42647\|\|full | 38640 | Yes | 31372 | 53 | 21 | 5 | High-quality | High-quality | 90.26 | AAI-based (high-confidence) | 18.81 |
| S4-final-viral-combined_k141_35450\|\|full | 2839 | No | NA | 3 | 0 | 1 | Not-determined | Genome-fragment | NA | NA | 0 |
| S4-final-viral-combined_k141_28694\|\|full | 3120 | No | NA | 5 | 0 | 3 | Not-determined | Genome-fragment | NA | NA | 0 |
| S4-final-viral-combined_k141_77496\|\|full | 1559 | No | NA | 3 | 0 | 0 | Not-determined | Genome-fragment | NA | NA | 0 |
| S11-final-viral-combined_k141_253422\|\|full | 4887 | No | NA | 6 | 0 | 0 | Low-quality | Genome-fragment | 11.5 | AAI-based (medium-confidence) | 0 |
| S4_k141_84043 | 4887 | No | NA | 6 | 1 | 0 | Low-quality | Genome-fragment | 9.54 | AAI-based (high-confidence) | 0 |
| S6-final-viral-combined_k141_11662\|\|full | 4887 | No | NA | 9 | 0 | 0 | Low-quality | Genome-fragment | 6.41 | AAI-based (high-confidence) | 0 |
| S4-final-viral-combined_k141_77530\|\|full | 2054 | No | NA | 4 | 0 | 0 | Not-determined | Genome-fragment | NA | NA | 0 |
| S4-final-viral-combined_k141_147089\|\|full | 1785 | No | NA | 3 | 0 | 3 | Not-determined | Genome-fragment | NA | NA | 0 |
| S12-final-viral-combined_k141_86430\|\|full | 4888 | No | NA | 3 | 0 | 1 | Low-quality | Genome-fragment | 1.61 | AAI-based (medium-confidence) | 0 |
| S12-final-viral-combined_k141_220510\|\|full | 4888 | No | NA | 7 | 3 | 0 | Low-quality | Genome-fragment | 12.62 | AAI-based (high-confidence) | 0 |
| S4-final-viral-combined_k141_961\|\|full | 5430 | No | NA | 6 | 0 | 4 | Not-determined | Genome-fragment | NA | NA | 0 |
| S12_k141_209202 | 4888 | No | NA | 7 | 2 | 0 | Low-quality | Genome-fragment | 7.52 | AAI-based (high-confidence) | 0 |
| S7-final-viral-combined_k141_225510\|\|full | 4889 | No | NA | 7 | 3 | 0 | Low-quality | Genome-fragment | 12.36 | AAI-based (high-confidence) | 0 |
| S4-final-viral-combined_k141_70657\|\|full | 3508 | No | NA | 5 | 0 | 3 | Not-determined | Genome-fragment | NA | NA | 0 |
| S4-final-viral-combined_k141_119125\|\|full | 1243 | No | NA | 2 | 0 | 2 | Not-determined | Genome-fragment | NA | NA | 0 |
| S4-final-viral-combined_k141_160940\|\|full | 5268 | No | NA | 7 | 0 | 1 | Not-determined | Genome-fragment | NA | NA | 0 |
| S4-final-viral-combined_k141_70673\|\|full | 1679 | No | NA | 5 | 0 | 0 | Not-determined | Genome-fragment | NA | NA | 0 |
| S2-final-viral-combined_k141_48037\|\|full | 4889 | No | NA | 5 | 1 | 0 | Low-quality | Genome-fragment | 11.58 | AAI-based (medium-confidence) | 0 |
| S4-final-viral-combined_k141_133103\|\|full | 1290 | No | NA | 2 | 0 | 0 | Not-determined | Genome-fragment | NA | NA | 0 |
| S4-final-viral-combined_k141_49445\|\|full | 6816 | No | NA | 5 | 0 | 3 | Not-determined | Genome-fragment | NA | NA | 0 |
| S4-final-viral-combined_k141_105544\|\|full | 1207 | No | NA | 2 | 0 | 1 | Not-determined | Genome-fragment | NA | NA | 0 |
| S2-final-viral-combined_k141_106702\|\|full | 4891 | No | NA | 5 | 0 | 0 | Low-quality | Genome-fragment | 8.25 | AAI-based (medium-confidence) | 0 |
| S2_k141_58105 | 4891 | No | NA | 4 | 0 | 1 | Low-quality | Genome-fragment | 8.77 | AAI-based (medium-confidence) | 0 |
| S4-final-viral-combined_k141_133124\|\|full | 2337 | No | NA | 3 | 0 | 1 | Not-determined | Genome-fragment | NA | NA | 0 |
| S4-final-viral-combined_k141_112441\|\|full | 5108 | No | NA | 7 | 0 | 0 | Not-determined | Genome-fragment | NA | NA | 0 |
| S5_k141_88536 | 4892 | No | NA | 4 | 3 | 0 | Low-quality | Genome-fragment | 9.6 | AAI-based (high-confidence) | 0 |
| S2-final-viral-combined_k141_35763\|\|full | 4892 | No | NA | 4 | 3 | 0 | Low-quality | Genome-fragment | 14.27 | AAI-based (medium-confidence) | 0 |
| S5-final-viral-combined_k141_46462\|\|full | 4894 | Yes | 860 | 6 | 1 | 3 | Low-quality | Genome-fragment | 0.58 | HMM-based (lower-bound) | 82.43 |
| S17-final-viral-combined_k141_184831\|\|full | 4894 | No | NA | 3 | 0 | 1 | Low-quality | Genome-fragment | 2.2 | AAI-based (medium-confidence) | 0 |
| S2-final-viral-combined_k141_160094\|\|full | 4894 | No | NA | 5 | 0 | 0 | Low-quality | Genome-fragment | 8.27 | AAI-based (medium-confidence) | 0 |
| S4-final-viral-combined_k141_126606\|\|full | 3510 | No | NA | 4 | 0 | 2 | Not-determined | Genome-fragment | NA | NA | 0 |
| S4-final-viral-combined_k141_7738\|\|full | 1737 | No | NA | 3 | 0 | 0 | Not-determined | Genome-fragment | NA | NA | 0 |
| S8-final-viral-combined_k141_86576\|\|full | 4894 | No | NA | 9 | 0 | 0 | Low-quality | Genome-fragment | 10.57 | AAI-based (medium-confidence) | 0 |
| S4-final-viral-combined_k141_35605\|\|full | 1828 | No | NA | 3 | 0 | 1 | Not-determined | Genome-fragment | NA | NA | 0 |
| S1-final-viral-combined_k141_130259\|\|full | 4896 | No | NA | 12 | 1 | 0 | Low-quality | Genome-fragment | 2.2 | HMM-based (lower-bound) | 0 |
| S5-final-viral-combined_k141_29821\|\|full | 4897 | No | NA | 8 | 0 | 1 | Low-quality | Genome-fragment | 12.45 | AAI-based (medium-confidence) | 0 |
| S4-final-viral-combined_k141_42751\|\|full | 3495 | No | NA | 3 | 0 | 2 | Not-determined | Genome-fragment | NA | NA | 0 |
| S4-final-viral-combined_k141_119236\|\|full | 2595 | No | NA | 4 | 0 | 2 | Not-determined | Genome-fragment | NA | NA | 0 |
| S12-final-viral-combined_k141_6705\|\|full | 4898 | Yes | 2736 | 7 | 1 | 1 | Low-quality | Genome-fragment | 3.39 | HMM-based (lower-bound) | 44.14 |
| S4-final-viral-combined_k141_140375\|\|full | 4703 | No | NA | 3 | 0 | 0 | Not-determined | Genome-fragment | NA | NA | 0 |
| S4-final-viral-combined_k141_98469\|\|full | 1192 | No | NA | 3 | 0 | 0 | Not-determined | Genome-fragment | NA | NA | 0 |
| S4-final-viral-combined_k141_77682\|\|full | 1146 | No | NA | 2 | 0 | 2 | Not-determined | Genome-fragment | NA | NA | 0 |
| S4-final-viral-combined_k141_98475\|\|full | 2381 | No | NA | 3 | 0 | 2 | Not-determined | Genome-fragment | NA | NA | 0 |
| S12_k141_372914 | 4898 | No | NA | 7 | 3 | 0 | Low-quality | Genome-fragment | 13.63 | AAI-based (high-confidence) | 0 |
| S4-final-viral-combined_k141_147263\|\|full | 1129 | No | NA | 2 | 0 | 2 | Not-determined | Genome-fragment | NA | NA | 0 |
| S4-final-viral-combined_k141_98495\|\|full | 2457 | No | NA | 4 | 0 | 1 | Not-determined | Genome-fragment | NA | NA | 0 |
| S4-final-viral-combined_k141_63720\|\|full | 5862 | No | NA | 7 | 0 | 0 | Not-determined | Genome-fragment | NA | NA | 0 |
| S11-final-viral-combined_k141_193426\|\|full | 4900 | No | NA | 5 | 0 | 1 | Low-quality | Genome-fragment | 3.74 | AAI-based (medium-confidence) | 0 |
| S3-final-viral-combined_k141_130005\|\|full | 4900 | No | NA | 8 | 3 | 0 | Low-quality | Genome-fragment | 10.51 | AAI-based (medium-confidence) | 0 |
| S4-final-viral-combined_k141_119294\|\|full | 3382 | No | NA | 4 | 0 | 0 | Not-determined | Genome-fragment | NA | NA | 0 |
| S4-final-viral-combined_k141_1069\|\|full | 2170 | No | NA | 2 | 0 | 1 | Not-determined | Genome-fragment | NA | NA | 0 |
| S4-final-viral-combined_k141_28977\|\|full | 1679 | No | NA | 2 | 0 | 2 | Not-determined | Genome-fragment | NA | NA | 0 |
| S4-final-viral-combined_k141_140412\|\|full | 1895 | No | NA | 2 | 0 | 1 | Not-determined | Genome-fragment | NA | NA | 0 |
| S4-final-viral-combined_k141_105681\|\|full | 1595 | No | NA | 2 | 0 | 0 | Not-determined | Genome-fragment | NA | NA | 0 |
| S4-final-viral-combined_k141_1074\|\|full | 5256 | No | NA | 4 | 0 | 0 | Not-determined | Genome-fragment | NA | NA | 0 |
| S4-final-viral-combined_k141_140413\|\|full | 2299 | No | NA | 3 | 0 | 1 | Not-determined | Genome-fragment | NA | NA | 0 |
| S4-final-viral-combined_k141_84713\|\|full | 2681 | No | NA | 3 | 0 | 2 | Not-determined | Genome-fragment | NA | NA | 0 |
| S4-final-viral-combined_k141_112548\|\|full | 1465 | No | NA | 2 | 0 | 2 | Not-determined | Genome-fragment | NA | NA | 0 |
| S4-final-viral-combined_k141_28988\|\|full | 2532 | No | NA | 3 | 0 | 0 | Not-determined | Genome-fragment | NA | NA | 0 |
| S4-final-viral-combined_k141_56663\|\|full | 1361 | No | NA | 2 | 0 | 1 | Not-determined | Genome-fragment | NA | NA | 0 |
| S2_k141_235869 | 4902 | No | NA | 4 | 0 | 4 | Low-quality | Genome-fragment | 0.72 | AAI-based (high-confidence) | 0 |
| S4-final-viral-combined_k141_7823\|\|full | 2046 | No | NA | 2 | 0 | 0 | Not-determined | Genome-fragment | NA | NA | 0 |
| S9-final-viral-combined_k141_106822\|\|lt2gene | 4904 | No | NA | 2 | 2 | 0 | Low-quality | Genome-fragment | 11.88 | AAI-based (medium-confidence) | 0 |
| S12-final-viral-combined_k141_213597\|\|full | 4904 | No | NA | 8 | 0 | 0 | Low-quality | Genome-fragment | 6.08 | AAI-based (medium-confidence) | 0 |
| S4-final-viral-combined_k141_91788\|\|full | 1273 | No | NA | 3 | 0 | 1 | Not-determined | Genome-fragment | NA | NA | 0 |
| S4_k141_28679 | 4904 | No | NA | 8 | 0 | 0 | Low-quality | Genome-fragment | 7.59 | AAI-based (high-confidence) | 0 |
| S17-final-viral-combined_k141_222442\|\|full | 4906 | No | NA | 8 | 0 | 0 | Low-quality | Genome-fragment | 4.31 | AAI-based (medium-confidence) | 0 |
| S4-final-viral-combined_k141_119354\|\|full | 8477 | No | NA | 4 | 0 | 4 | Not-determined | Genome-fragment | NA | NA | 0 |
| S17-final-viral-combined_k141_83981\|\|full | 4907 | Yes | 943 | 5 | 1 | 1 | Low-quality | Genome-fragment | 1.02 | AAI-based (high-confidence) | 80.78 |
| S4-final-viral-combined_k141_140461\|\|full | 4156 | No | NA | 7 | 0 | 4 | Not-determined | Genome-fragment | NA | NA | 0 |
| S4-final-viral-combined_k141_63772\|\|full | 1178 | No | NA | 4 | 0 | 0 | Not-determined | Genome-fragment | NA | NA | 0 |
| S9-final-viral-combined_k141_30353\|\|full | 4908 | No | NA | 7 | 0 | 2 | Low-quality | Genome-fragment | 11.29 | AAI-based (high-confidence) | 0 |
| S4-final-viral-combined_k141_126715\|\|full | 2755 | No | NA | 3 | 0 | 3 | Not-determined | Genome-fragment | NA | NA | 0 |
| S4-final-viral-combined_k141_56720\|\|full | 3922 | No | NA | 3 | 0 | 3 | Not-determined | Genome-fragment | NA | NA | 0 |
| S4-final-viral-combined_k141_91816\|\|full | 1624 | No | NA | 2 | 0 | 1 | Not-determined | Genome-fragment | NA | NA | 0 |
| S4-final-viral-combined_k141_133348\|\|full | 1009 | No | NA | 2 | 0 | 1 | Not-determined | Genome-fragment | NA | NA | 0 |
| S4-final-viral-combined_k141_77810\|\|full | 5672 | No | NA | 9 | 0 | 0 | Not-determined | Genome-fragment | NA | NA | 0 |
| S11-final-viral-combined_k141_97632\|\|full | 4909 | No | NA | 6 | 0 | 1 | Low-quality | Genome-fragment | 11.46 | AAI-based (high-confidence) | 0 |
| S4-final-viral-combined_k141_21837\|\|full | 1293 | No | NA | 2 | 0 | 0 | Not-determined | Genome-fragment | NA | NA | 0 |
| S4-final-viral-combined_k141_42896\|\|full | 1607 | No | NA | 3 | 0 | 1 | Not-determined | Genome-fragment | NA | NA | 0 |
| S4_k141_53101 | 4911 | No | NA | 12 | 0 | 0 | Low-quality | Genome-fragment | 8.2 | AAI-based (high-confidence) | 0 |
| S4-final-viral-combined_k141_133363\|\|full | 11960 | No | NA | 14 | 0 | 6 | Not-determined | Genome-fragment | NA | NA | 0 |
| S4-final-viral-combined_k141_154321\|\|full | 6580 | No | NA | 6 | 0 | 3 | Not-determined | Genome-fragment | NA | NA | 0 |
| S4-final-viral-combined_k141_91866\|\|full | 1024 | No | NA | 2 | 0 | 0 | Not-determined | Genome-fragment | NA | NA | 0 |
| S4-final-viral-combined_k141_91869\|\|full | 2096 | No | NA | 3 | 0 | 2 | Not-determined | Genome-fragment | NA | NA | 0 |
| S4-final-viral-combined_k141_140517\|\|full | 4131 | No | NA | 4 | 0 | 3 | Not-determined | Genome-fragment | NA | NA | 0 |
| S4-final-viral-combined_k141_49510\|\|full | 2847 | No | NA | 4 | 0 | 2 | Not-determined | Genome-fragment | NA | NA | 0 |
| S4-final-viral-combined_k141_63845\|\|full | 6441 | No | NA | 8 | 0 | 5 | Not-determined | Genome-fragment | NA | NA | 0 |
| S4-final-viral-combined_k141_140527\|\|full | 2585 | No | NA | 3 | 0 | 1 | Not-determined | Genome-fragment | NA | NA | 0 |
| S11_k141_174486 | 4912 | No | NA | 4 | 0 | 0 | Low-quality | Genome-fragment | 7.87 | AAI-based (medium-confidence) | 0 |
| S4-final-viral-combined_k141_84810\|\|full | 3173 | No | NA | 5 | 0 | 4 | Not-determined | Genome-fragment | NA | NA | 0 |
| S4-final-viral-combined_k141_126798\|\|full | 2277 | No | NA | 1 | 0 | 0 | Not-determined | Genome-fragment | NA | NA | 0 |
| S1-final-viral-combined_k141_380365\|\|full | 4913 | No | NA | 6 | 4 | 0 | Low-quality | Genome-fragment | 11.2 | AAI-based (medium-confidence) | 0 |
| S4-final-viral-combined_k141_140552\|\|full | 3374 | No | NA | 4 | 0 | 1 | Not-determined | Genome-fragment | NA | NA | 0 |
| S4-final-viral-combined_k141_133437\|\|full | 3520 | No | NA | 7 | 0 | 1 | Not-determined | Genome-fragment | NA | NA | 0 |
| S4-final-viral-combined_k141_140565\|\|full | 1297 | No | NA | 3 | 0 | 2 | Not-determined | Genome-fragment | NA | NA | 0 |
| S12_k141_162358 | 4913 | No | NA | 13 | 2 | 0 | Low-quality | Genome-fragment | 8.76 | AAI-based (high-confidence) | 0 |
| S4-final-viral-combined_k141_119517\|\|full | 10068 | No | NA | 8 | 0 | 1 | Not-determined | Genome-fragment | NA | NA | 0 |
| S4-final-viral-combined_k141_98654\|\|full | 1383 | No | NA | 2 | 0 | 0 | Not-determined | Genome-fragment | NA | NA | 0 |
| S11-final-viral-combined_k141_182114\|\|full | 4914 | No | NA | 6 | 0 | 2 | Low-quality | Genome-fragment | 2.94 | AAI-based (medium-confidence) | 0 |
| S6-final-viral-combined_k141_107657\|\|full | 4916 | No | NA | 6 | 0 | 0 | Low-quality | Genome-fragment | 5.84 | AAI-based (high-confidence) | 0 |
| S4-final-viral-combined_k141_119529\|\|full | 2701 | No | NA | 4 | 0 | 1 | Not-determined | Genome-fragment | NA | NA | 0 |
| S4-final-viral-combined_k141_1246\|\|full | 8591 | No | NA | 14 | 0 | 0 | Medium-quality | Genome-fragment | 56.65 | AAI-based (high-confidence) | 0 |
| S1-final-viral-combined_k141_43468\|\|full | 4917 | No | NA | 5 | 0 | 1 | Low-quality | Genome-fragment | 3.84 | AAI-based (medium-confidence) | 0 |
| S4-final-viral-combined_k141_21981\|\|full | 1877 | No | NA | 2 | 0 | 2 | Not-determined | Genome-fragment | NA | NA | 0 |
| S4-final-viral-combined_k141_56863\|\|full | 2368 | No | NA | 3 | 0 | 3 | Not-determined | Genome-fragment | NA | NA | 0 |
| S4-final-viral-combined_k141_78022\|\|full | 1522 | No | NA | 2 | 0 | 1 | Not-determined | Genome-fragment | NA | NA | 0 |
| S11-final-viral-combined_k141_137937\|\|full | 4918 | No | NA | 4 | 2 | 0 | Low-quality | Genome-fragment | 10.25 | AAI-based (high-confidence) | 0 |
| S4-final-viral-combined_k141_126854\|\|full | 6354 | No | NA | 7 | 0 | 5 | Not-determined | Genome-fragment | NA | NA | 0 |
| S4-final-viral-combined_k141_140625\|\|full | 2116 | No | NA | 2 | 0 | 2 | Not-determined | Genome-fragment | NA | NA | 0 |
| S8_k141_164330 | 4921 | No | NA | 3 | 0 | 0 | Low-quality | Genome-fragment | 8.29 | AAI-based (medium-confidence) | 0 |
| S4-final-viral-combined_k141_126868\|\|full | 3030 | No | NA | 4 | 0 | 0 | Not-determined | Genome-fragment | NA | NA | 0 |
| S4-final-viral-combined_k141_105913\|\|full | 3435 | No | NA | 3 | 0 | 1 | Not-determined | Genome-fragment | NA | NA | 0 |
| S4-final-viral-combined_k141_14895\|\|full | 1159 | No | NA | 2 | 0 | 1 | Not-determined | Genome-fragment | NA | NA | 0 |
| S4-final-viral-combined_k141_8086\|\|full | 2359 | No | NA | 3 | 0 | 1 | Not-determined | Genome-fragment | NA | NA | 0 |
| S4-final-viral-combined_k141_98692\|\|full | 2185 | No | NA | 4 | 0 | 2 | Not-determined | Genome-fragment | NA | NA | 0 |
| S4-final-viral-combined_k141_22012\|\|full | 27205 | Yes | 23641 | 32 | 14 | 3 | Medium-quality | Genome-fragment | 59.98 | AAI-based (high-confidence) | 13.1 |
| S9-final-viral-combined_k141_58167\|\|full | 4923 | No | NA | 9 | 3 | 0 | Low-quality | Genome-fragment | 8.95 | AAI-based (high-confidence) | 0 |
| S12_k141_83101 | 4923 | No | NA | 5 | 0 | 4 | Low-quality | Genome-fragment | 0.71 | AAI-based (medium-confidence) | 0 |
| S11_k141_322448 | 4924 | No | NA | 5 | 0 | 4 | Low-quality | Genome-fragment | 8.97 | AAI-based (medium-confidence) | 0 |
| S4-final-viral-combined_k141_98712\|\|full | 1599 | No | NA | 2 | 0 | 0 | Not-determined | Genome-fragment | NA | NA | 0 |
| S4-final-viral-combined_k141_154461\|\|full | 5501 | No | NA | 4 | 0 | 3 | Not-determined | Genome-fragment | NA | NA | 0 |
| S4-final-viral-combined_k141_147514\|\|full | 1102 | No | NA | 2 | 0 | 0 | Not-determined | Genome-fragment | NA | NA | 0 |
| S4-final-viral-combined_k141_154475\|\|full | 2496 | No | NA | 4 | 0 | 1 | Not-determined | Genome-fragment | NA | NA | 0 |
| S4-final-viral-combined_k141_29223\|\|full | 7245 | No | NA | 6 | 0 | 2 | Not-determined | Genome-fragment | NA | NA | 0 |
| S4-final-viral-combined_k141_8139\|\|full | 4089 | No | NA | 5 | 0 | 4 | Not-determined | Genome-fragment | NA | NA | 0 |
| S11-final-viral-combined_k141_112357\|\|full | 4925 | No | NA | 5 | 0 | 4 | Low-quality | Genome-fragment | 2.26 | AAI-based (medium-confidence) | 0 |
| S2-final-viral-combined_k141_145361\|\|full | 4926 | No | NA | 3 | 0 | 1 | Low-quality | Genome-fragment | 3.85 | AAI-based (medium-confidence) | 0 |
| S11-final-viral-combined_k141_190675\|\|full | 4928 | Yes | 2458 | 5 | 1 | 1 | Low-quality | Genome-fragment | 1.18 | HMM-based (lower-bound) | 50.12 |
| S2_k141_191990 | 4929 | No | NA | 5 | 1 | 3 | Low-quality | Genome-fragment | 3 | HMM-based (lower-bound) | 0 |
| S4_k141_51413 | 4930 | No | NA | 3 | 0 | 0 | Low-quality | Genome-fragment | 5.14 | AAI-based (medium-confidence) | 0 |
| S4-final-viral-combined_k141_92076\|\|full | 1808 | No | NA | 4 | 0 | 1 | Not-determined | Genome-fragment | NA | NA | 0 |
| S4_k141_9854 | 4933 | No | NA | 8 | 0 | 0 | Low-quality | Genome-fragment | 8.24 | AAI-based (high-confidence) | 0 |
| S4-final-viral-combined_k141_29283\|\|full | 1869 | No | NA | 2 | 0 | 0 | Not-determined | Genome-fragment | NA | NA | 0 |
| S4-final-viral-combined_k141_84935\|\|full | 3672 | No | NA | 3 | 0 | 2 | Not-determined | Genome-fragment | NA | NA | 0 |
| S4-final-viral-combined_k141_84940\|\|full | 3640 | No | NA | 7 | 0 | 0 | Not-determined | Genome-fragment | NA | NA | 0 |
| S8_k141_191537 | 4935 | Yes | 2300 | 6 | 1 | 2 | Low-quality | Genome-fragment | 0.63 | HMM-based (lower-bound) | 53.39 |
| S4-final-viral-combined_k141_119632\|\|full | 1518 | No | NA | 3 | 0 | 2 | Not-determined | Genome-fragment | NA | NA | 0 |
| S4-final-viral-combined_k141_147584\|\|full | 2491 | No | NA | 3 | 0 | 2 | Not-determined | Genome-fragment | NA | NA | 0 |
| S5-final-viral-combined_k141_187382\|\|full | 4938 | No | NA | 6 | 3 | 0 | Low-quality | Genome-fragment | 11.22 | AAI-based (high-confidence) | 0 |
| S4-final-viral-combined_k141_29297\|\|full | 1543 | No | NA | 3 | 0 | 2 | Not-determined | Genome-fragment | NA | NA | 0 |
| S4-final-viral-combined_k141_8187\|\|full | 1611 | No | NA | 5 | 0 | 0 | Not-determined | Genome-fragment | NA | NA | 0 |
| S12-final-viral-combined_k141_175447\|\|full | 4939 | No | NA | 8 | 0 | 1 | Low-quality | Genome-fragment | 5.54 | AAI-based (medium-confidence) | 0 |
| S4-final-viral-combined_k141_49709\|\|full | 2896 | No | NA | 3 | 0 | 3 | Not-determined | Genome-fragment | NA | NA | 0 |
| S4-final-viral-combined_k141_98759\|\|full | 6345 | No | NA | 5 | 0 | 4 | Not-determined | Genome-fragment | NA | NA | 0 |
| S2-final-viral-combined_k141_218432\|\|full | 4939 | No | NA | 6 | 0 | 2 | Low-quality | Genome-fragment | 7.95 | AAI-based (medium-confidence) | 0 |
| S4-final-viral-combined_k141_92145\|\|full | 2811 | No | NA | 2 | 0 | 0 | Not-determined | Genome-fragment | NA | NA | 0 |
| S4-final-viral-combined_k141_29315\|\|full | 3662 | No | NA | 4 | 0 | 3 | Not-determined | Genome-fragment | NA | NA | 0 |
| S4-final-viral-combined_k141_85002\|\|full | 2471 | No | NA | 3 | 0 | 3 | Not-determined | Genome-fragment | NA | NA | 0 |
| S9-final-viral-combined_k141_40230\|\|full | 4940 | Yes | 1894 | 4 | 1 | 1 | Low-quality | Genome-fragment | 2.13 | HMM-based (lower-bound) | 61.66 |
| S1-final-viral-combined_k141_363003\|\|full | 4940 | No | NA | 5 | 1 | 0 | Low-quality | Genome-fragment | 4.64 | AAI-based (high-confidence) | 0 |
| S4-final-viral-combined_k141_98777\|\|full | 1567 | No | NA | 3 | 0 | 1 | Not-determined | Genome-fragment | NA | NA | 0 |
| S7-final-viral-combined_k141_66762\|\|full | 4940 | No | NA | 7 | 2 | 0 | Low-quality | Genome-fragment | 12.97 | AAI-based (high-confidence) | 0 |
| S4-final-viral-combined_k141_92156\|\|full | 7671 | No | NA | 7 | 0 | 5 | Not-determined | Genome-fragment | NA | NA | 0 |
| S8-final-viral-combined_k141_65861\|\|full | 4942 | No | NA | 3 | 0 | 3 | Low-quality | Genome-fragment | 4.21 | AAI-based (medium-confidence) | 0 |
| S4-final-viral-combined_k141_105979\|\|full | 5382 | No | NA | 6 | 0 | 3 | Not-determined | Genome-fragment | NA | NA | 0 |
| S4-final-viral-combined_k141_1430\|\|full | 1526 | No | NA | 3 | 0 | 2 | Not-determined | Genome-fragment | NA | NA | 0 |
| S4-final-viral-combined_k141_92157\|\|full | 5163 | No | NA | 4 | 0 | 2 | Not-determined | Genome-fragment | NA | NA | 0 |
| S4-final-viral-combined_k141_8219\|\|full | 1018 | No | NA | 3 | 0 | 0 | Not-determined | Genome-fragment | NA | NA | 0 |
| S4-final-viral-combined_k141_49733\|\|full | 1881 | No | NA | 2 | 0 | 0 | Not-determined | Genome-fragment | NA | NA | 0 |
| S4-final-viral-combined_k141_57025\|\|full | 4430 | No | NA | 3 | 0 | 3 | Not-determined | Genome-fragment | NA | NA | 0 |
| S1-final-viral-combined_k141_77444\|\|full | 4943 | No | NA | 5 | 0 | 2 | Low-quality | Genome-fragment | 2.86 | AAI-based (medium-confidence) | 0 |
| S2_k141_140295 | 4943 | No | NA | 8 | 0 | 2 | Low-quality | Genome-fragment | 8.43 | AAI-based (medium-confidence) | 0 |
| S7-final-viral-combined_k141_47539\|\|full | 4944 | No | NA | 3 | 0 | 3 | Low-quality | Genome-fragment | 6.87 | AAI-based (medium-confidence) | 0 |
| S17-final-viral-combined_k141_165509\|\|full | 4946 | No | NA | 5 | 2 | 0 | Low-quality | Genome-fragment | 7.74 | AAI-based (high-confidence) | 0 |
| S2_k141_160918 | 4946 | No | NA | 12 | 0 | 0 | Low-quality | Genome-fragment | 11.03 | AAI-based (high-confidence) | 0 |
| S4-final-viral-combined_k141_105996\|\|full | 2694 | No | NA | 3 | 0 | 2 | Not-determined | Genome-fragment | NA | NA | 0 |
| S7-final-viral-combined_k141_134703\|\|full | 4947 | Yes | 2611 | 5 | 2 | 1 | Low-quality | Genome-fragment | 7.44 | AAI-based (medium-confidence) | 47.22 |
| S4-final-viral-combined_k141_147666\|\|full | 1460 | No | NA | 3 | 0 | 1 | Not-determined | Genome-fragment | NA | NA | 0 |
| S17-final-viral-combined_k141_45520\|\|full | 4947 | No | NA | 6 | 1 | 2 | Low-quality | Genome-fragment | 3.39 | HMM-based (lower-bound) | 0 |
| S4-final-viral-combined_k141_119739\|\|full | 1766 | No | NA | 1 | 0 | 1 | Not-determined | Genome-fragment | NA | NA | 0 |
| S4-final-viral-combined_k141_154622\|\|full | 1611 | No | NA | 2 | 0 | 1 | Not-determined | Genome-fragment | NA | NA | 0 |
| S4-final-viral-combined_k141_8272\|\|full | 3598 | No | NA | 8 | 0 | 5 | Not-determined | Genome-fragment | NA | NA | 0 |
| S4-final-viral-combined_k141_98827\|\|full | 1091 | No | NA | 2 | 0 | 1 | Not-determined | Genome-fragment | NA | NA | 0 |
| S11-final-viral-combined_k141_167462\|\|full | 4948 | No | NA | 4 | 0 | 1 | Low-quality | Genome-fragment | 8.25 | AAI-based (medium-confidence) | 0 |
| S17-k141_145897 | 4948 | No | NA | 1 | 0 | 0 | Low-quality | Genome-fragment | 11.86 | AAI-based (medium-confidence) | 0 |
| S4-final-viral-combined_k141_29385\|\|full | 2792 | No | NA | 3 | 0 | 2 | Not-determined | Genome-fragment | NA | NA | 0 |
| S4-final-viral-combined_k141_49780\|\|full | 1316 | No | NA | 3 | 0 | 0 | Not-determined | Genome-fragment | NA | NA | 0 |
| S6_k141_62995 | 4948 | No | NA | 2 | 0 | 0 | Low-quality | Genome-fragment | 5.29 | AAI-based (high-confidence) | 0 |
| S3_k141_16485 | 4949 | No | NA | 4 | 0 | 0 | Low-quality | Genome-fragment | 2.28 | AAI-based (high-confidence) | 0 |
| S7_k141_55187 | 4949 | No | NA | 1 | 0 | 0 | Low-quality | Genome-fragment | 5.15 | AAI-based (medium-confidence) | 0 |
| S4-final-viral-combined_k141_106061\|\|full | 2887 | No | NA | 3 | 0 | 3 | Not-determined | Genome-fragment | NA | NA | 0 |
| S4-final-viral-combined_k141_43147\|\|full | 1933 | No | NA | 3 | 0 | 1 | Not-determined | Genome-fragment | NA | NA | 0 |
| S7_k141_184986 | 4951 | Yes | 1184 | 5 | 1 | 1 | Low-quality | Genome-fragment | 1.44 | AAI-based (medium-confidence) | 76.09 |
| S11-final-viral-combined_k141_43960\|\|full | 4954 | No | NA | 7 | 4 | 0 | Low-quality | Genome-fragment | 13.48 | AAI-based (high-confidence) | 0 |
| S4-final-viral-combined_k141_98877\|\|full | 2478 | No | NA | 2 | 0 | 1 | Not-determined | Genome-fragment | NA | NA | 0 |
| S8_k141_152836 | 4954 | No | NA | 10 | 2 | 0 | Low-quality | Genome-fragment | 11.2 | AAI-based (high-confidence) | 0 |
| S4-final-viral-combined_k141_127103\|\|full | 1794 | No | NA | 4 | 0 | 0 | Not-determined | Genome-fragment | NA | NA | 0 |
| S4-final-viral-combined_k141_106080\|\|full | 1547 | No | NA | 2 | 0 | 1 | Not-determined | Genome-fragment | NA | NA | 0 |
| S4_k141_43721 | 4955 | No | NA | 9 | 3 | 0 | Low-quality | Genome-fragment | 8.27 | AAI-based (high-confidence) | 0 |
| S4-final-viral-combined_k141_43172\|\|full | 4329 | No | NA | 4 | 0 | 4 | Not-determined | Genome-fragment | NA | NA | 0 |
| S4-final-viral-combined_k141_15176\|\|full | 1169 | No | NA | 2 | 0 | 1 | Not-determined | Genome-fragment | NA | NA | 0 |
| S4-final-viral-combined_k141_43173\|\|full | 3609 | No | NA | 7 | 0 | 0 | Not-determined | Genome-fragment | NA | NA | 0 |
| S4-final-viral-combined_k141_106090\|\|full | 1132 | No | NA | 3 | 0 | 1 | Not-determined | Genome-fragment | NA | NA | 0 |
| S4-final-viral-combined_k141_22189\|\|full | 2577 | No | NA | 3 | 0 | 2 | Not-determined | Genome-fragment | NA | NA | 0 |
| S4-final-viral-combined_k141_140923\|\|full | 1336 | No | NA | 2 | 0 | 1 | Not-determined | Genome-fragment | NA | NA | 0 |
| S7-final-viral-combined_k141_154349\|\|full | 4957 | No | NA | 9 | 0 | 2 | Low-quality | Genome-fragment | 5.98 | AAI-based (high-confidence) | 0 |
| S4-final-viral-combined_k141_71182\|\|full | 4158 | No | NA | 4 | 0 | 1 | Not-determined | Genome-fragment | NA | NA | 0 |
| S4_k141_122495 | 4958 | No | NA | 4 | 0 | 1 | Low-quality | Genome-fragment | 0.85 | AAI-based (high-confidence) | 0 |
| S4-final-viral-combined_k141_98925\|\|full | 1365 | No | NA | 2 | 0 | 1 | Not-determined | Genome-fragment | NA | NA | 0 |
| S4-final-viral-combined_k141_49866\|\|full | 2573 | No | NA | 4 | 0 | 0 | Not-determined | Genome-fragment | NA | NA | 0 |
| S8-final-viral-combined_k141_25089\|\|full | 4958 | No | NA | 3 | 0 | 1 | Low-quality | Genome-fragment | 8.22 | AAI-based (high-confidence) | 0 |
| S4-final-viral-combined_k141_57123\|\|full | 47984 | No | NA | 55 | 2 | 7 | Medium-quality | Genome-fragment | 67.99 | HMM-based (lower-bound) | 0 |
| S4-final-viral-combined_k141_64211\|\|full | 1293 | No | NA | 2 | 0 | 2 | Not-determined | Genome-fragment | NA | NA | 0 |
| S4-final-viral-combined_k141_29503\|\|full | 2613 | No | NA | 3 | 0 | 3 | Not-determined | Genome-fragment | NA | NA | 0 |
| S2_k141_227490 | 4959 | No | NA | 5 | 4 | 0 | Low-quality | Genome-fragment | 8.95 | AAI-based (high-confidence) | 0 |
| S4-final-viral-combined_k141_71204\|\|full | 1161 | No | NA | 3 | 0 | 0 | Not-determined | Genome-fragment | NA | NA | 0 |
| S4-final-viral-combined_k141_85163\|\|full | 1165 | No | NA | 2 | 0 | 2 | Not-determined | Genome-fragment | NA | NA | 0 |
| S4-final-viral-combined_k141_127132\|\|full | 2322 | No | NA | 3 | 0 | 2 | Not-determined | Genome-fragment | NA | NA | 0 |
| S4-final-viral-combined_k141_98976\|\|full | 12063 | No | NA | 20 | 0 | 5 | Not-determined | Genome-fragment | NA | NA | 0 |
| S4-final-viral-combined_k141_98977\|\|full | 1835 | No | NA | 3 | 0 | 2 | Not-determined | Genome-fragment | NA | NA | 0 |
| S1_k141_335285 | 4960 | No | NA | 8 | 3 | 0 | Low-quality | Genome-fragment | 17.5 | HMM-based (lower-bound) | 0 |
| S4-final-viral-combined_k141_1589\|\|full | 5813 | No | NA | 4 | 0 | 2 | Not-determined | Genome-fragment | NA | NA | 0 |
| S9-final-viral-combined_k141_42761\|\|full | 4961 | No | NA | 9 | 6 | 0 | Low-quality | Genome-fragment | 11.95 | AAI-based (high-confidence) | 0 |
| S4-final-viral-combined_k141_71221\|\|full | 2982 | No | NA | 3 | 0 | 1 | Not-determined | Genome-fragment | NA | NA | 0 |
| S17-final-viral-combined_k141_64418\|\|full | 4961 | No | NA | 12 | 6 | 0 | Low-quality | Genome-fragment | 12.46 | AAI-based (high-confidence) | 0 |
| S2_k141_180228 | 4961 | No | NA | 6 | 0 | 2 | Low-quality | Genome-fragment | 7.84 | AAI-based (medium-confidence) | 0 |
| S4-final-viral-combined_k141_154784\|\|full | 2172 | No | NA | 2 | 0 | 1 | Not-determined | Genome-fragment | NA | NA | 0 |
| S4-final-viral-combined_k141_106188\|\|full | 1645 | No | NA | 2 | 0 | 0 | Not-determined | Genome-fragment | NA | NA | 0 |
| S4-final-viral-combined_k141_133853\|\|full | 1214 | No | NA | 3 | 0 | 1 | Not-determined | Genome-fragment | NA | NA | 0 |
| S4-final-viral-combined_k141_8387\|\|full | 9889 | No | NA | 10 | 0 | 10 | Not-determined | Genome-fragment | NA | NA | 0 |
| S4-final-viral-combined_k141_127167\|\|full | 3207 | No | NA | 3 | 0 | 3 | Not-determined | Genome-fragment | NA | NA | 0 |
| S4-final-viral-combined_k141_78417\|\|full | 1779 | No | NA | 2 | 0 | 1 | Not-determined | Genome-fragment | NA | NA | 0 |
| S4-final-viral-combined_k141_127177\|\|full | 1586 | No | NA | 3 | 0 | 2 | Not-determined | Genome-fragment | NA | NA | 0 |
| S6-final-viral-combined_k141_127027\|\|full | 4961 | No | NA | 13 | 0 | 0 | Low-quality | Genome-fragment | 6.17 | AAI-based (high-confidence) | 0 |
| S1_k141_253188 | 4962 | No | NA | 5 | 3 | 0 | Low-quality | Genome-fragment | 11.87 | AAI-based (high-confidence) | 0 |
| S4-final-viral-combined_k141_85224\|\|full | 2236 | No | NA | 2 | 0 | 1 | Not-determined | Genome-fragment | NA | NA | 0 |
| S4-final-viral-combined_k141_133893\|\|full | 2880 | No | NA | 6 | 0 | 0 | Not-determined | Genome-fragment | NA | NA | 0 |
| S4-final-viral-combined_k141_133897\|\|full | 1880 | No | NA | 3 | 0 | 1 | Not-determined | Genome-fragment | NA | NA | 0 |
| S4-final-viral-combined_k141_64284\|\|full | 1334 | No | NA | 2 | 0 | 2 | Not-determined | Genome-fragment | NA | NA | 0 |
| S4-final-viral-combined_k141_141060\|\|full | 3016 | No | NA | 4 | 0 | 0 | Not-determined | Genome-fragment | NA | NA | 0 |
| S4-final-viral-combined_k141_119857\|\|full | 2477 | No | NA | 4 | 0 | 3 | Not-determined | Genome-fragment | NA | NA | 0 |
| S7-final-viral-combined_k141_41223\|\|full | 4962 | No | NA | 5 | 4 | 1 | Low-quality | Genome-fragment | 11.43 | AAI-based (medium-confidence) | 0 |
| S4-final-viral-combined_k141_119867\|\|full | 1670 | No | NA | 2 | 0 | 2 | Not-determined | Genome-fragment | NA | NA | 0 |
| S4-final-viral-combined_k141_154830\|\|full | 1951 | No | NA | 3 | 0 | 1 | Not-determined | Genome-fragment | NA | NA | 0 |
| S17-k141_82058 | 4963 | No | NA | 5 | 5 | 0 | Low-quality | Genome-fragment | 10.67 | AAI-based (high-confidence) | 0 |
| S4-final-viral-combined_k141_15316\|\|full | 2210 | No | NA | 4 | 0 | 1 | Not-determined | Genome-fragment | NA | NA | 0 |
| S4-final-viral-combined_k141_99070\|\|full | 5903 | No | NA | 5 | 0 | 4 | Not-determined | Genome-fragment | NA | NA | 0 |
| S3-final-viral-combined_k141_75451\|\|full | 4964 | No | NA | 5 | 3 | 0 | Low-quality | Genome-fragment | 10.21 | AAI-based (high-confidence) | 0 |
| S4-final-viral-combined_k141_85244\|\|full | 4017 | No | NA | 10 | 0 | 0 | Not-determined | Genome-fragment | NA | NA | 0 |
| S4-final-viral-combined_k141_15325\|\|full | 3946 | No | NA | 8 | 0 | 0 | Not-determined | Genome-fragment | NA | NA | 0 |
| S5_k141_53763 | 4964 | No | NA | 4 | 3 | 0 | Low-quality | Genome-fragment | 10.56 | AAI-based (high-confidence) | 0 |
| S4-final-viral-combined_k141_154845\|\|full | 3063 | No | NA | 4 | 0 | 1 | Not-determined | Genome-fragment | NA | NA | 0 |
| S9-final-viral-combined_k141_9204\|\|full | 4965 | No | NA | 4 | 0 | 3 | Low-quality | Genome-fragment | 7.28 | AAI-based (medium-confidence) | 0 |
| S4-final-viral-combined_k141_127268\|\|full | 1024 | No | NA | 2 | 0 | 1 | Not-determined | Genome-fragment | NA | NA | 0 |
| S4-final-viral-combined_k141_8493\|\|full | 1743 | No | NA | 3 | 0 | 3 | Not-determined | Genome-fragment | NA | NA | 0 |
| S4-final-viral-combined_k141_119890\|\|full | 2514 | No | NA | 4 | 0 | 3 | Not-determined | Genome-fragment | NA | NA | 0 |
| S4-final-viral-combined_k141_78494\|\|full | 1025 | No | NA | 3 | 0 | 0 | Not-determined | Genome-fragment | NA | NA | 0 |
| S8-final-viral-combined_k141_174532\|\|full | 4965 | No | NA | 5 | 2 | 0 | Low-quality | Genome-fragment | 11.4 | AAI-based (high-confidence) | 0 |
| S4-final-viral-combined_k141_119911\|\|full | 2035 | No | NA | 3 | 0 | 1 | Not-determined | Genome-fragment | NA | NA | 0 |
| S2-final-viral-combined_k141_232509\|\|full | 4966 | No | NA | 11 | 3 | 0 | Low-quality | Genome-fragment | 9.3 | AAI-based (medium-confidence) | 0 |
| S4-final-viral-combined_k141_85356\|\|full | 2017 | No | NA | 4 | 0 | 0 | Not-determined | Genome-fragment | NA | NA | 0 |
| S4-final-viral-combined_k141_15403\|\|full | 1076 | No | NA | 2 | 0 | 2 | Not-determined | Genome-fragment | NA | NA | 0 |
| S4-final-viral-combined_k141_8536\|\|full | 7843 | No | NA | 12 | 0 | 1 | Not-determined | Genome-fragment | NA | NA | 0 |
| S4-final-viral-combined_k141_106350\|\|full | 5860 | No | NA | 6 | 0 | 4 | Not-determined | Genome-fragment | NA | NA | 0 |
| S12-final-viral-combined_k141_307626\|\|full | 4969 | No | NA | 6 | 1 | 0 | Low-quality | Genome-fragment | 2.61 | AAI-based (medium-confidence) | 0 |
| S1-final-viral-combined_k141_76307\|\|full | 4973 | No | NA | 12 | 1 | 0 | Low-quality | Genome-fragment | 11.25 | AAI-based (medium-confidence) | 0 |
| S4-final-viral-combined_k141_161683\|\|full | 1463 | No | NA | 3 | 0 | 1 | Not-determined | Genome-fragment | NA | NA | 0 |
| S4-final-viral-combined_k141_29720\|\|full | 3517 | No | NA | 2 | 0 | 2 | Not-determined | Genome-fragment | NA | NA | 0 |
| S4-final-viral-combined_k141_85378\|\|full | 3383 | No | NA | 5 | 0 | 1 | Not-determined | Genome-fragment | NA | NA | 0 |
| S4-final-viral-combined_k141_71395\|\|full | 2791 | No | NA | 4 | 0 | 3 | Not-determined | Genome-fragment | NA | NA | 0 |
| S2-final-viral-combined_k141_228974\|\|full | 4973 | No | NA | 7 | 4 | 0 | Low-quality | Genome-fragment | 9.23 | AAI-based (high-confidence) | 0 |
| S4-final-viral-combined_k141_113216\|\|full | 13488 | No | NA | 9 | 0 | 7 | Not-determined | Genome-fragment | NA | NA | 0 |
| S4-final-viral-combined_k141_127291\|\|full | 2266 | No | NA | 3 | 0 | 2 | Not-determined | Genome-fragment | NA | NA | 0 |
| S4-final-viral-combined_k141_1774\|\|full | 1661 | No | NA | 2 | 0 | 1 | Not-determined | Genome-fragment | NA | NA | 0 |
| S4-final-viral-combined_k141_29744\|\|full | 2375 | No | NA | 2 | 0 | 2 | Not-determined | Genome-fragment | NA | NA | 0 |
| S4-final-viral-combined_k141_36291\|\|full | 1320 | No | NA | 3 | 0 | 0 | Not-determined | Genome-fragment | NA | NA | 0 |
| S4-final-viral-combined_k141_113224\|\|full | 1660 | No | NA | 3 | 0 | 1 | Not-determined | Genome-fragment | NA | NA | 0 |
| S4-final-viral-combined_k141_147927\|\|full | 2158 | No | NA | 3 | 0 | 0 | Not-determined | Genome-fragment | NA | NA | 0 |
| S4-final-viral-combined_k141_71418\|\|full | 2788 | No | NA | 2 | 0 | 1 | Not-determined | Genome-fragment | NA | NA | 0 |
| S4-final-viral-combined_k141_147942\|\|full | 5824 | No | NA | 6 | 0 | 5 | Not-determined | Genome-fragment | NA | NA | 0 |
| S4-final-viral-combined_k141_64450\|\|full | 4050 | No | NA | 3 | 0 | 2 | Not-determined | Genome-fragment | NA | NA | 0 |
| S4-final-viral-combined_k141_92455\|\|full | 7435 | No | NA | 5 | 0 | 1 | Not-determined | Genome-fragment | NA | NA | 0 |
| S4-final-viral-combined_k141_113259\|\|full | 6898 | No | NA | 5 | 0 | 4 | Not-determined | Genome-fragment | NA | NA | 0 |
| S17-final-viral-combined_k141_212268\|\|full | 4974 | No | NA | 6 | 0 | 2 | Low-quality | Genome-fragment | 3.7 | AAI-based (medium-confidence) | 0 |
| S8-final-viral-combined_k141_2053\|\|full | 4974 | No | NA | 3 | 0 | 2 | Low-quality | Genome-fragment | 11.3 | AAI-based (medium-confidence) | 0 |
| S1-final-viral-combined_k141_194827\|\|full | 4975 | No | NA | 6 | 0 | 0 | Low-quality | Genome-fragment | 11.44 | AAI-based (medium-confidence) | 0 |
| S4-final-viral-combined_k141_43499\|\|full | 2456 | No | NA | 4 | 0 | 1 | Not-determined | Genome-fragment | NA | NA | 0 |
| S4-final-viral-combined_k141_127371\|\|full | 1494 | No | NA | 2 | 0 | 1 | Not-determined | Genome-fragment | NA | NA | 0 |
| S4-final-viral-combined_k141_36365\|\|full | 2532 | No | NA | 3 | 0 | 3 | Not-determined | Genome-fragment | NA | NA | 0 |
| S11-final-viral-combined_k141_256045\|\|full | 4976 | No | NA | 4 | 0 | 4 | Low-quality | Genome-fragment | 2.43 | AAI-based (medium-confidence) | 0 |
| S4_k141_43123 | 4976 | No | NA | 9 | 2 | 0 | Low-quality | Genome-fragment | 13.52 | AAI-based (medium-confidence) | 0 |
| S4-final-viral-combined_k141_161832\|\|full | 2478 | No | NA | 3 | 0 | 2 | Not-determined | Genome-fragment | NA | NA | 0 |
| S1-final-viral-combined_k141_381406\|\|full | 4977 | No | NA | 5 | 1 | 0 | Low-quality | Genome-fragment | 4.83 | HMM-based (lower-bound) | 0 |
| S4_k141_76073 | 4979 | No | NA | 6 | 2 | 0 | Low-quality | Genome-fragment | 9.33 | HMM-based (lower-bound) | 0 |
| S4-final-viral-combined_k141_8656\|\|full | 1074 | No | NA | 3 | 0 | 2 | Not-determined | Genome-fragment | NA | NA | 0 |
| S4-final-viral-combined_k141_71496\|\|full | 7037 | No | NA | 6 | 0 | 1 | Not-determined | Genome-fragment | NA | NA | 0 |

| | | | | | | | | | | | |
|---|---|---|---|---|---|---|---|---|---|---|---|
| S4-final-viral-combined_k141_155051\|\|full | 1244 | No | NA | 2 | 0 | 1 | Not-determined | Genome-fragment | NA | NA | 0 |
| S4-final-viral-combined_k141_22434\|\|full | 2608 | No | NA | 3 | 0 | 2 | Not-determined | Genome-fragment | NA | NA | 0 |
| S17-final-viral-combined_k141_178388\|\|full | 4980 | Yes | 1214 | 7 | 1 | 2 | Low-quality | Genome-fragment | 1.87 | AAI-based (medium-confidence) | 75.62 |
| S4-final-viral-combined_k141_22444\|\|full | 2044 | No | NA | 2 | 0 | 2 | Not-determined | Genome-fragment | NA | NA | 0 |
| S4-final-viral-combined_k141_50139\|\|full | 1155 | No | NA | 3 | 0 | 0 | Not-determined | Genome-fragment | NA | NA | 0 |
| S4-final-viral-combined_k141_29879\|\|full | 3034 | No | NA | 4 | 0 | 3 | Not-determined | Genome-fragment | NA | NA | 0 |
| S4-final-viral-combined_k141_85521\|\|full | 10342 | No | NA | 15 | 0 | 5 | Not-determined | Genome-fragment | NA | NA | 0 |
| S4-final-viral-combined_k141_57503\|\|full | 1274 | No | NA | 2 | 0 | 2 | Not-determined | Genome-fragment | NA | NA | 0 |
| S4-final-viral-combined_k141_113338\|\|full | 1887 | No | NA | 3 | 0 | 2 | Not-determined | Genome-fragment | NA | NA | 0 |
| S4-final-viral-combined_k141_8713\|\|full | 5836 | No | NA | 5 | 0 | 4 | Not-determined | Genome-fragment | NA | NA | 0 |
| S5-final-viral-combined_k141_11742\|\|full | 4981 | No | NA | 7 | 6 | 0 | Low-quality | Genome-fragment | 11.77 | AAI-based (high-confidence) | 0 |
| S5-final-viral-combined_k141_181375\|\|full | 4982 | No | NA | 9 | 0 | 4 | Low-quality | Genome-fragment | 5.94 | AAI-based (medium-confidence) | 0 |
| S4-final-viral-combined_k141_161936\|\|full | 2920 | No | NA | 3 | 0 | 3 | Not-determined | Genome-fragment | NA | NA | 0 |
| S4-final-viral-combined_k141_85541\|\|full | 1874 | No | NA | 4 | 0 | 0 | Not-determined | Genome-fragment | NA | NA | 0 |
| S4-final-viral-combined_k141_134187\|\|full | 3245 | No | NA | 3 | 0 | 1 | Not-determined | Genome-fragment | NA | NA | 0 |
| S4-final-viral-combined_k141_92588\|\|full | 4210 | No | NA | 6 | 0 | 2 | Not-determined | Genome-fragment | NA | NA | 0 |
| S17-final-viral-combined_k141_112476\|\|full | 4982 | No | NA | 5 | 0 | 0 | Low-quality | Genome-fragment | 6.05 | AAI-based (high-confidence) | 0 |
| S17-final-viral-combined_k141_246525\|\|full | 4982 | No | NA | 5 | 0 | 1 | Low-quality | Genome-fragment | 4.19 | AAI-based (medium-confidence) | 0 |
| S17-k141_261666 | 4982 | No | NA | 4 | 0 | 4 | Low-quality | Genome-fragment | 8.51 | AAI-based (high-confidence) | 0 |
| S4-final-viral-combined_k141_135538\|\|lt2gene | 4983 | No | NA | 2 | 0 | 0 | Low-quality | Genome-fragment | 5.1 | AAI-based (high-confidence) | 0 |
| S4-final-viral-combined_k141_92624\|\|full | 6608 | No | NA | 6 | 0 | 3 | Not-determined | Genome-fragment | NA | NA | 0 |
| S4-final-viral-combined_k141_106568\|\|full | 9162 | No | NA | 7 | 0 | 6 | Not-determined | Genome-fragment | NA | NA | 0 |
| S4-final-viral-combined_k141_99415\|\|full | 1351 | No | NA | 3 | 0 | 2 | Not-determined | Genome-fragment | NA | NA | 0 |
| S4-final-viral-combined_k141_1967\|\|full | 3808 | No | NA | 5 | 0 | 0 | Not-determined | Genome-fragment | NA | NA | 0 |
| S4-final-viral-combined_k141_22581\|\|full | 1316 | No | NA | 2 | 0 | 2 | Not-determined | Genome-fragment | NA | NA | 0 |
| S1-final-viral-combined_k141_316660\|\|full | 4984 | No | NA | 4 | 0 | 1 | Low-quality | Genome-fragment | 8.16 | AAI-based (medium-confidence) | 0 |
| S4-final-viral-combined_k141_50248\|\|full | 2278 | No | NA | 3 | 0 | 2 | Not-determined | Genome-fragment | NA | NA | 0 |
| S4-final-viral-combined_k141_36489\|\|full | 2025 | No | NA | 3 | 0 | 2 | Not-determined | Genome-fragment | NA | NA | 0 |
| S4-final-viral-combined_k141_85573\|\|full | 9374 | No | NA | 7 | 0 | 6 | Not-determined | Genome-fragment | NA | NA | 0 |
| S4-final-viral-combined_k141_29981\|\|full | 2676 | No | NA | 3 | 0 | 2 | Not-determined | Genome-fragment | NA | NA | 0 |
| S2-final-viral-combined_k141_3334\|\|full | 4985 | No | NA | 5 | 0 | 1 | Low-quality | Genome-fragment | 3.29 | AAI-based (medium-confidence) | 0 |
| S4-final-viral-combined_k141_134262\|\|full | 1699 | No | NA | 3 | 0 | 1 | Not-determined | Genome-fragment | NA | NA | 0 |
| S4-final-viral-combined_k141_50258\|\|full | 2415 | No | NA | 4 | 0 | 1 | Not-determined | Genome-fragment | NA | NA | 0 |
| S4-final-viral-combined_k141_127538\|\|full | 6760 | No | NA | 5 | 0 | 3 | Not-determined | Genome-fragment | NA | NA | 0 |
| S8-final-viral-combined_k141_227403\|\|full | 4985 | No | NA | 4 | 0 | 1 | Low-quality | Genome-fragment | 2.59 | AAI-based (medium-confidence) | 0 |
| S4_k141_28802 | 4986 | No | NA | 5 | 0 | 1 | Low-quality | Genome-fragment | 2.57 | AAI-based (medium-confidence) | 0 |
| S4-final-viral-combined_k141_162013\|\|full | 2953 | No | NA | 3 | 0 | 3 | Not-determined | Genome-fragment | NA | NA | 0 |
| S4-final-viral-combined_k141_15691\|\|full | 1376 | No | NA | 1 | 0 | 1 | Not-determined | Genome-fragment | NA | NA | 0 |
| S4-final-viral-combined_k141_50277\|\|full | 1829 | No | NA | 3 | 0 | 3 | Not-determined | Genome-fragment | NA | NA | 0 |
| S4-final-viral-combined_k141_71631\|\|full | 4473 | No | NA | 5 | 0 | 3 | Not-determined | Genome-fragment | NA | NA | 0 |
| S7-final-viral-combined_k141_215848\|\|full | 4987 | No | NA | 6 | 4 | 0 | Low-quality | Genome-fragment | 14.05 | AAI-based (high-confidence) | 0 |
| S4-final-viral-combined_k141_134298\|\|full | 4165 | No | NA | 4 | 0 | 4 | Not-determined | Genome-fragment | NA | NA | 0 |
| S4-final-viral-combined_k141_25982\|\|full | 4988 | No | NA | 6 | 0 | 0 | Low-quality | Genome-fragment | 3.12 | AAI-based (medium-confidence) | 0 |
| S4-final-viral-combined_k141_15722\|\|full | 1275 | No | NA | 3 | 0 | 0 | Not-determined | Genome-fragment | NA | NA | 0 |
| S4-final-viral-combined_k141_64632\|\|full | 57119 | Yes | 30433 | 78 | 14 | 17 | Medium-quality | Genome-fragment | 56.85 | AAI-based (high-confidence) | 46.72 |
| S12-final-viral-combined_k141_129801\|\|full | 4989 | No | NA | 4 | 0 | 0 | Low-quality | Genome-fragment | 12.39 | AAI-based (medium-confidence) | 0 |
| S4-final-viral-combined_k141_2042\|\|full | 5466 | No | NA | 5 | 0 | 2 | Not-determined | Genome-fragment | NA | NA | 0 |
| S4-final-viral-combined_k141_85644\|\|full | 7861 | No | NA | 7 | 0 | 3 | Not-determined | Genome-fragment | NA | NA | 0 |
| S4-final-viral-combined_k141_15742\|\|full | 1585 | No | NA | 2 | 0 | 0 | Not-determined | Genome-fragment | NA | NA | 0 |
| S1-final-viral-combined_k141_43109\|\|full | 4991 | No | NA | 10 | 0 | 1 | Low-quality | Genome-fragment | 13.63 | AAI-based (high-confidence) | 0 |
| S12-final-viral-combined_k141_57665\|\|full | 4991 | No | NA | 8 | 1 | 6 | Low-quality | Genome-fragment | 8.42 | HMM-based (lower-bound) | 0 |
| S4-final-viral-combined_k141_99499\|\|full | 2100 | No | NA | 2 | 0 | 2 | Not-determined | Genome-fragment | NA | NA | 0 |
| S4-final-viral-combined_k141_43796\|\|full | 2211 | No | NA | 3 | 0 | 0 | Not-determined | Genome-fragment | NA | NA | 0 |
| S4-final-viral-combined_k141_127604\|\|full | 2803 | No | NA | 4 | 0 | 3 | Not-determined | Genome-fragment | NA | NA | 0 |
| S8-final-viral-combined_k141_201456\|\|full | 4992 | Yes | 3413 | 6 | 1 | 1 | Low-quality | Genome-fragment | 6.96 | AAI-based (medium-confidence) | 31.63 |
| S4-final-viral-combined_k141_50369\|\|full | 1984 | No | NA | 2 | 0 | 2 | Not-determined | Genome-fragment | NA | NA | 0 |
| S7-final-viral-combined_k141_50336\|\|full | 4995 | No | NA | 5 | 0 | 2 | Low-quality | Genome-fragment | 8.32 | AAI-based (medium-confidence) | 0 |
| S12-final-viral-combined_k141_176114\|\|full | 4995 | No | NA | 5 | 1 | 0 | Low-quality | Genome-fragment | 7.07 | AAI-based (medium-confidence) | 0 |
| S1-final-viral-combined_k141_144008\|\|full | 4996 | Yes | 2678 | 8 | 1 | 2 | Low-quality | Genome-fragment | 5.81 | AAI-based (high-confidence) | 46.4 |
| S4-final-viral-combined_k141_162147\|\|full | 3712 | No | NA | 2 | 0 | 1 | Not-determined | Genome-fragment | NA | NA | 0 |
| S8-final-viral-combined_k141_272359\|\|full | 4996 | No | NA | 4 | 0 | 2 | Low-quality | Genome-fragment | 2.28 | AAI-based (medium-confidence) | 0 |
| S4-final-viral-combined_k141_22726\|\|full | 2710 | No | NA | 3 | 0 | 2 | Not-determined | Genome-fragment | NA | NA | 0 |
| S4-final-viral-combined_k141_162162\|\|full | 3122 | No | NA | 3 | 0 | 1 | Not-determined | Genome-fragment | NA | NA | 0 |
| S4-final-viral-combined_k141_148282\|\|full | 3080 | No | NA | 3 | 0 | 3 | Not-determined | Genome-fragment | NA | NA | 0 |
| S2-final-viral-combined_k141_117594\|\|full | 4997 | No | NA | 9 | 6 | 0 | Low-quality | Genome-fragment | 6.62 | AAI-based (high-confidence) | 0 |
| S2-final-viral-combined_k141_152126\|\|full | 4997 | No | NA | 6 | 0 | 1 | Low-quality | Genome-fragment | 12.63 | AAI-based (high-confidence) | 0 |
| S5-final-viral-combined_k141_142708\|\|full | 4998 | Yes | 777 | 7 | 1 | 5 | Low-quality | Genome-fragment | 0.73 | AAI-based (medium-confidence) | 84.45 |
| S2-final-viral-combined_k141_40604\|\|full | 4998 | No | NA | 4 | 3 | 0 | Low-quality | Genome-fragment | 11.09 | AAI-based (high-confidence) | 0 |
| S11-final-viral-combined_k141_176980\|\|full | 4999 | No | NA | 11 | 1 | 1 | Low-quality | Genome-fragment | 8.98 | AAI-based (high-confidence) | 0 |
| S2-final-viral-combined_k141_243874\|\|full | 4999 | No | NA | 5 | 0 | 2 | Low-quality | Genome-fragment | 11.87 | AAI-based (medium-confidence) | 0 |
| S4-final-viral-combined_k141_162201\|\|full | 2890 | No | NA | 5 | 0 | 2 | Not-determined | Genome-fragment | NA | NA | 0 |
| S4-final-viral-combined_k141_155307\|\|full | 8485 | No | NA | 6 | 0 | 3 | Not-determined | Genome-fragment | NA | NA | 0 |
| S7-final-viral-combined_k141_40873\|\|full | 5001 | No | NA | 8 | 4 | 0 | Low-quality | Genome-fragment | 11.27 | AAI-based (high-confidence) | 0 |
| S4-final-viral-combined_k141_113465\|\|full | 3352 | No | NA | 4 | 0 | 1 | Not-determined | Genome-fragment | NA | NA | 0 |
| S4-final-viral-combined_k141_85784\|\|full | 2051 | No | NA | 2 | 0 | 2 | Not-determined | Genome-fragment | NA | NA | 0 |
| S4-final-viral-combined_k141_22764\|\|full | 2553 | No | NA | 2 | 0 | 0 | Not-determined | Genome-fragment | NA | NA | 0 |
| S4-final-viral-combined_k141_134450\|\|full | 1313 | No | NA | 2 | 0 | 1 | Not-determined | Genome-fragment | NA | NA | 0 |
| S5_k141_189207 | 5002 | No | NA | 6 | 0 | 0 | Low-quality | Genome-fragment | 5.98 | AAI-based (high-confidence) | 0 |
| S4-final-viral-combined_k141_79008\|\|full | 1680 | No | NA | 2 | 0 | 2 | Not-determined | Genome-fragment | NA | NA | 0 |
| S4-final-viral-combined_k141_57711\|\|full | 1980 | No | NA | 4 | 0 | 0 | Not-determined | Genome-fragment | NA | NA | 0 |
| S11-final-viral-combined_k141_169752\|\|full | 5004 | No | NA | 3 | 0 | 1 | Low-quality | Genome-fragment | 6.14 | AAI-based (medium-confidence) | 0 |
| S4-final-viral-combined_k141_43868\|\|full | 2624 | No | NA | 3 | 0 | 1 | Not-determined | Genome-fragment | NA | NA | 0 |
| S4-final-viral-combined_k141_2156\|\|full | 1307 | No | NA | 2 | 0 | 1 | Not-determined | Genome-fragment | NA | NA | 0 |
| S6_k141_20616 | 5004 | No | NA | 7 | 0 | 0 | Low-quality | Genome-fragment | 3 | AAI-based (medium-confidence) | 0 |
| S4-final-viral-combined_k141_43893\|\|full | 2946 | No | NA | 3 | 0 | 3 | Not-determined | Genome-fragment | NA | NA | 0 |
| S5-final-viral-combined_k141_117393\|\|full | 5005 | Yes | 4590 | 9 | 1 | 1 | Low-quality | Genome-fragment | 0.37 | HMM-based (lower-bound) | 8.29 |
| S4-final-viral-combined_k141_2195\|\|full | 2672 | No | NA | 4 | 0 | 0 | Not-determined | Genome-fragment | NA | NA | 0 |
| S4-final-viral-combined_k141_36674\|\|full | 2211 | No | NA | 3 | 0 | 3 | Not-determined | Genome-fragment | NA | NA | 0 |
| S5-final-viral-combined_k141_110903\|\|full | 5005 | No | NA | 5 | 4 | 0 | Low-quality | Genome-fragment | 11.79 | AAI-based (high-confidence) | 0 |
| S4-final-viral-combined_k141_22838\|\|full | 1719 | No | NA | 2 | 0 | 1 | Not-determined | Genome-fragment | NA | NA | 0 |
| S7-final-viral-combined_k141_117681\|\|full | 5006 | No | NA | 9 | 3 | 0 | Low-quality | Genome-fragment | 11.71 | AAI-based (high-confidence) | 0 |
| S4-final-viral-combined_k141_22847\|\|full | 1177 | No | NA | 2 | 0 | 0 | Not-determined | Genome-fragment | NA | NA | 0 |
| S17-final-viral-combined_k141_39807\|\|full | 5007 | No | NA | 3 | 0 | 2 | Low-quality | Genome-fragment | 3.45 | AAI-based (high-confidence) | 0 |
| S4-final-viral-combined_k141_9027\|\|full | 4155 | No | NA | 4 | 0 | 4 | Not-determined | Genome-fragment | NA | NA | 0 |
| S4-final-viral-combined_k141_57816\|\|full | 6192 | No | NA | 7 | 0 | 6 | Not-determined | Genome-fragment | NA | NA | 0 |
| S4-final-viral-combined_k141_2218\|\|full | 2911 | No | NA | 4 | 0 | 3 | Not-determined | Genome-fragment | NA | NA | 0 |
| S12_k141_388144 | 5007 | No | NA | 3 | 0 | 2 | Low-quality | Genome-fragment | 4.71 | AAI-based (medium-confidence) | 0 |
| S4-final-viral-combined_k141_106783\|\|full | 2368 | No | NA | 2 | 0 | 2 | Not-determined | Genome-fragment | NA | NA | 0 |
| S2-final-viral-combined_k141_186051\|\|full | 5007 | No | NA | 5 | 0 | 4 | Low-quality | Genome-fragment | 4.55 | AAI-based (medium-confidence) | 0 |
| S1-final-viral-combined_k141_278415\|\|full | 5008 | Yes | 923 | 4 | 1 | 1 | Low-quality | Genome-fragment | 2.01 | AAI-based (high-confidence) | 81.57 |
| S4-final-viral-combined_k141_141541\|\|full | 7567 | No | NA | 11 | 0 | 3 | Not-determined | Genome-fragment | NA | NA | 0 |
| S4-final-viral-combined_k141_57861\|\|full | 1099 | No | NA | 2 | 0 | 1 | Not-determined | Genome-fragment | NA | NA | 0 |
| S4-final-viral-combined_k141_64857\|\|full | 2262 | No | NA | 3 | 0 | 2 | Not-determined | Genome-fragment | NA | NA | 0 |
| S4-final-viral-combined_k141_36754\|\|full | 1748 | No | NA | 2 | 0 | 1 | Not-determined | Genome-fragment | NA | NA | 0 |
| S4-final-viral-combined_k141_148437\|\|full | 4107 | No | NA | 5 | 0 | 1 | Not-determined | Genome-fragment | NA | NA | 0 |
| S4-final-viral-combined_k141_64863\|\|full | 3524 | No | NA | 2 | 0 | 1 | Not-determined | Genome-fragment | NA | NA | 0 |
| S4-final-viral-combined_k141_43979\|\|full | 2070 | No | NA | 3 | 0 | 2 | Not-determined | Genome-fragment | NA | NA | 0 |
| S12-final-viral-combined_k141_168810\|\|full | 5009 | No | NA | 5 | 0 | 0 | Low-quality | Genome-fragment | 9.04 | AAI-based (medium-confidence) | 0 |
| S8_k141_182335 | 5010 | No | NA | 6 | 0 | 2 | Low-quality | Genome-fragment | 5.86 | AAI-based (medium-confidence) | 0 |
| S1-final-viral-combined_k141_163498\|\|full | 5011 | No | NA | 6 | 1 | 0 | Low-quality | Genome-fragment | 11.9 | AAI-based (high-confidence) | 0 |
| S4-final-viral-combined_k141_141571\|\|full | 4383 | No | NA | 4 | 0 | 0 | Not-determined | Genome-fragment | NA | NA | 0 |
| S4_k141_51390 | 5011 | No | NA | 5 | 0 | 5 | Low-quality | Genome-fragment | 3.26 | AAI-based (high-confidence) | 0 |
| S8-final-viral-combined_k141_100173\|\|full | 5012 | No | NA | 5 | 0 | 0 | Low-quality | Genome-fragment | 10.82 | AAI-based (high-confidence) | 0 |
| S1_k141_266628 | 5013 | Yes | 1992 | 5 | 1 | 1 | Low-quality | Genome-fragment | 4.95 | AAI-based (medium-confidence) | 60.26 |
| S5-final-viral-combined_k141_129899\|\|full | 5014 | No | NA | 6 | 0 | 2 | Low-quality | Genome-fragment | 8.48 | AAI-based (medium-confidence) | 0 |
| S4-final-viral-combined_k141_85896\|\|full | 1639 | No | NA | 3 | 0 | 0 | Not-determined | Genome-fragment | NA | NA | 0 |
| S4-final-viral-combined_k141_79175\|\|full | 1342 | No | NA | 2 | 0 | 1 | Not-determined | Genome-fragment | NA | NA | 0 |
| S3-final-viral-combined_k141_31510\|\|full | 5015 | No | NA | 4 | 0 | 3 | Low-quality | Genome-fragment | 5.07 | AAI-based (high-confidence) | 0 |
| S4-final-viral-combined_k141_85906\|\|full | 1586 | No | NA | 3 | 0 | 0 | Not-determined | Genome-fragment | NA | NA | 0 |
| S12_k141_51142 | 5015 | No | NA | 3 | 0 | 0 | Low-quality | Genome-fragment | 5.06 | AAI-based (high-confidence) | 0 |
| S2_k141_199482 | 5016 | No | NA | 5 | 0 | 2 | Low-quality | Genome-fragment | 5.46 | AAI-based (medium-confidence) | 0 |
| S11-final-viral-combined_k141_45598\|\|full | 5017 | No | NA | 5 | 0 | 4 | Low-quality | Genome-fragment | 3.26 | AAI-based (medium-confidence) | 0 |
| S4-final-viral-combined_k141_155487\|\|full | 4804 | No | NA | 5 | 0 | 1 | Not-determined | Genome-fragment | NA | NA | 0 |
| S4-final-viral-combined_k141_9174\|\|full | 2738 | No | NA | 2 | 0 | 2 | Not-determined | Genome-fragment | NA | NA | 0 |
| S12-final-viral-combined_k141_218774\|\|full | 5017 | No | NA | 6 | 3 | 1 | Low-quality | Genome-fragment | 22.12 | AAI-based (high-confidence) | 0 |
| S11-final-viral-combined_k141_46556\|\|full | 5018 | No | NA | 6 | 0 | 3 | Low-quality | Genome-fragment | 2.31 | AAI-based (medium-confidence) | 0 |
| S1-final-viral-combined_k141_166368\|\|full | 5018 | No | NA | 8 | 0 | 0 | Low-quality | Genome-fragment | 11.58 | AAI-based (high-confidence) | 0 |
| S4-final-viral-combined_k141_93092\|\|full | 2612 | No | NA | 4 | 0 | 1 | Not-determined | Genome-fragment | NA | NA | 0 |
| S4-final-viral-combined_k141_71917\|\|full | 3602 | No | NA | 2 | 0 | 0 | Not-determined | Genome-fragment | NA | NA | 0 |
| S4-final-viral-combined_k141_44077\|\|full | 2183 | No | NA | 2 | 0 | 1 | Not-determined | Genome-fragment | NA | NA | 0 |
| S1-final-viral-combined_k141_85571\|\|full | 5019 | No | NA | 4 | 0 | 2 | Low-quality | Genome-fragment | 2.78 | AAI-based (medium-confidence) | 0 |
| S2-final-viral-combined_k141_127402\|\|full | 5019 | No | NA | 5 | 0 | 0 | Low-quality | Genome-fragment | 16.63 | AAI-based (high-confidence) | 0 |
| S4-final-viral-combined_k141_79279\|\|full | 3523 | No | NA | 4 | 0 | 1 | Not-determined | Genome-fragment | NA | NA | 0 |
| S4-final-viral-combined_k141_148548\|\|full | 1689 | No | NA | 4 | 0 | 0 | Not-determined | Genome-fragment | NA | NA | 0 |
| S4-final-viral-combined_k141_2300\|\|full | 2959 | No | NA | 3 | 0 | 3 | Not-determined | Genome-fragment | NA | NA | 0 |
| S4-final-viral-combined_k141_85997\|\|full | 2565 | No | NA | 4 | 0 | 1 | Not-determined | Genome-fragment | NA | NA | 0 |
| S4-final-viral-combined_k141_9273\|\|full | 2286 | No | NA | 4 | 0 | 0 | Not-determined | Genome-fragment | NA | NA | 0 |
| S5-final-viral-combined_k141_82056\|\|full | 5020 | No | NA | 6 | 3 | 0 | Low-quality | Genome-fragment | 10.8 | AAI-based (medium-confidence) | 0 |
| S4-final-viral-combined_k141_113721\|\|full | 29681 | No | NA | 29 | 0 | 11 | Medium-quality | Genome-fragment | 53.64 | AAI-based (high-confidence) | 0 |
| S4-final-viral-combined_k141_23107\|\|full | 5814 | No | NA | 9 | 1 | 0 | Not-determined | Genome-fragment | NA | NA | 0 |
| S7_k141_235504 | 5022 | No | NA | 9 | 3 | 0 | Low-quality | Genome-fragment | 8.7 | HMM-based (lower-bound) | 0 |
| S6_k141_40325 | 5022 | Yes | 4760 | 3 | 1 | 1 | Low-quality | Genome-fragment | 2.58 | HMM-based (lower-bound) | 5.22 |
| S11-final-viral-combined_k141_220861\|\|full | 5023 | No | NA | 5 | 3 | 0 | Low-quality | Genome-fragment | 12.2 | AAI-based (high-confidence) | 0 |
| S11_k141_219059 | 5025 | No | NA | 3 | 2 | 0 | Low-quality | Genome-fragment | 11.07 | AAI-based (high-confidence) | 0 |
| S2-final-viral-combined_k141_23369\|\|full | 5025 | No | NA | 12 | 5 | 0 | Low-quality | Genome-fragment | 10.57 | AAI-based (high-confidence) | 0 |
| S17-final-viral-combined_k141_215492\|\|full | 5026 | No | NA | 6 | 0 | 2 | Low-quality | Genome-fragment | 5.89 | AAI-based (medium-confidence) | 0 |
| S4-final-viral-combined_k141_141725\|\|full | 5101 | No | NA | 4 | 0 | 4 | Not-determined | Genome-fragment | NA | NA | 0 |
| S2_k141_101633 | 5026 | No | NA | 7 | 3 | 0 | Low-quality | Genome-fragment | 3.03 | AAI-based (high-confidence) | 0 |
| S4-final-viral-combined_k141_9358\|\|full | 1306 | No | NA | 2 | 0 | 2 | Not-determined | Genome-fragment | NA | NA | 0 |
| S4-final-viral-combined_k141_9369\|\|full | 1541 | No | NA | 2 | 0 | 1 | Not-determined | Genome-fragment | NA | NA | 0 |
| S7-final-viral-combined_k141_176357\|\|full | 5027 | No | NA | 5 | 2 | 1 | Low-quality | Genome-fragment | 10.66 | AAI-based (high-confidence) | 0 |
| S4-final-viral-combined_k141_93177\|\|full | 4284 | No | NA | 6 | 0 | 1 | Not-determined | Genome-fragment | NA | NA | 0 |
| S17-final-viral-combined_k141_44008\|\|full | 5027 | No | NA | 11 | 1 | 0 | Low-quality | Genome-fragment | 3.04 | HMM-based (lower-bound) | 0 |
| S4-final-viral-combined_k141_2384\|\|full | 1123 | No | NA | 2 | 0 | 0 | Not-determined | Genome-fragment | NA | NA | 0 |
| S12-final-viral-combined_k141_389268\|\|full | 5027 | No | NA | 11 | 2 | 0 | Low-quality | Genome-fragment | 7.78 | AAI-based (high-confidence) | 0 |
| S4-final-viral-combined_k141_99572\|\|full | 1477 | No | NA | 2 | 0 | 0 | Not-determined | Genome-fragment | NA | NA | 0 |
| S4-final-viral-combined_k141_30377\|\|full | 1318 | No | NA | 2 | 0 | 1 | Not-determined | Genome-fragment | NA | NA | 0 |
| S12_k141_120099 | 5029 | No | NA | 2 | 0 | 0 | Low-quality | Genome-fragment | 5.93 | AAI-based (high-confidence) | 0 |
| S4-final-viral-combined_k141_44221\|\|full | 1253 | No | NA | 3 | 0 | 0 | Not-determined | Genome-fragment | NA | NA | 0 |
| S4-final-viral-combined_k141_107057\|\|full | 9435 | No | NA | 12 | 0 | 7 | Not-determined | Genome-fragment | NA | NA | 0 |
| S4-final-viral-combined_k141_50645\|\|full | 6267 | No | NA | 6 | 0 | 1 | Not-determined | Genome-fragment | NA | NA | 0 |
| S4-final-viral-combined_k141_72178\|\|full | 1336 | No | NA | 3 | 0 | 1 | Not-determined | Genome-fragment | NA | NA | 0 |
| S4-final-viral-combined_k141_107073\|\|full | 1344 | No | NA | 3 | 0 | 1 | Not-determined | Genome-fragment | NA | NA | 0 |
| S4-final-viral-combined_k141_155633\|\|full | 2157 | No | NA | 3 | 0 | 2 | Not-determined | Genome-fragment | NA | NA | 0 |
| S1_k141_215255 | 5030 | No | NA | 6 | 1 | 0 | Low-quality | Genome-fragment | 7.61 | HMM-based (lower-bound) | 0 |
| S4-final-viral-combined_k141_148666\|\|full | 2210 | No | NA | 5 | 0 | 2 | Not-determined | Genome-fragment | NA | NA | 0 |
| S4-final-viral-combined_k141_155640\|\|full | 5627 | No | NA | 7 | 0 | 2 | Not-determined | Genome-fragment | NA | NA | 0 |
| S17-final-viral-combined_k141_16042\|\|full | 5032 | No | NA | 5 | 3 | 0 | Low-quality | Genome-fragment | 7.74 | HMM-based (lower-bound) | 0 |
| S4-final-viral-combined_k141_57963\|\|full | 161252 | Yes | 34348 | 174 | 26 | 93 | Medium-quality | Genome-fragment | 51.76 | HMM-based (lower-bound) | 78.7 |
| S4-final-viral-combined_k141_134860\|\|full | 7984 | No | NA | 4 | 0 | 4 | Not-determined | Genome-fragment | NA | NA | 0 |
| S7_k141_145375 | 5038 | Yes | 1694 | 10 | 2 | 3 | Low-quality | Genome-fragment | 3.8 | AAI-based (high-confidence) | 66.38 |
| S4-final-viral-combined_k141_44293\|\|full | 3903 | No | NA | 4 | 0 | 4 | Not-determined | Genome-fragment | NA | NA | 0 |
| S2_k141_103579 | 5039 | No | NA | 6 | 0 | 0 | Low-quality | Genome-fragment | 7.7 | AAI-based (medium-confidence) | 0 |
| S4-final-viral-combined_k141_86203\|\|full | 1968 | No | NA | 3 | 0 | 1 | Not-determined | Genome-fragment | NA | NA | 0 |
| S8-final-viral-combined_k141_11568\|\|full | 5039 | No | NA | 8 | 0 | 2 | Low-quality | Genome-fragment | 10.8 | AAI-based (high-confidence) | 0 |
| S4-final-viral-combined_k141_23217\|\|full | 2567 | No | NA | 6 | 0 | 1 | Not-determined | Genome-fragment | NA | NA | 0 |
| S4-final-viral-combined_k141_37098\|\|full | 4103 | No | NA | 3 | 0 | 0 | Not-determined | Genome-fragment | NA | NA | 0 |
| S6-final-viral-combined_k141_66880\|\|full | 5041 | No | NA | 6 | 0 | 3 | Low-quality | Genome-fragment | 3.69 | AAI-based (high-confidence) | 0 |
| S4-final-viral-combined_k141_38073\|\|full | 5042 | No | NA | 8 | 0 | 4 | Low-quality | Genome-fragment | 15.77 | AAI-based (high-confidence) | 0 |
| S4-final-viral-combined_k141_99665\|\|full | 1052 | No | NA | 2 | 0 | 1 | Not-determined | Genome-fragment | NA | NA | 0 |
| S4-final-viral-combined_k141_148711\|\|full | 8531 | No | NA | 6 | 0 | 3 | Not-determined | Genome-fragment | NA | NA | 0 |
| S4-final-viral-combined_k141_141902\|\|full | 2911 | No | NA | 4 | 0 | 3 | Not-determined | Genome-fragment | NA | NA | 0 |
| S4-final-viral-combined_k141_37121\|\|full | 3513 | No | NA | 3 | 0 | 2 | Not-determined | Genome-fragment | NA | NA | 0 |
| S3-final-viral-combined_k141_137799\|\|full | 5043 | No | NA | 7 | 4 | 0 | Low-quality | Genome-fragment | 12.44 | AAI-based (medium-confidence) | 0 |
| S11_k141_4155 | 5044 | No | NA | 10 | 1 | 0 | Low-quality | Genome-fragment | 13.11 | AAI-based (high-confidence) | 0 |
| S4-final-viral-combined_k141_23260\|\|full | 3264 | No | NA | 3 | 0 | 3 | Not-determined | Genome-fragment | NA | NA | 0 |
| S8-final-viral-combined_k141_177858\|\|full | 5044 | Yes | 1933 | 5 | 1 | 2 | Low-quality | Genome-fragment | 4.12 | AAI-based (high-confidence) | 61.68 |
| S11_k141_66568 | 5045 | No | NA | 10 | 1 | 0 | Low-quality | Genome-fragment | 4.75 | AAI-based (high-confidence) | 0 |
| S4-final-viral-combined_k141_155724\|\|full | 1315 | No | NA | 3 | 0 | 1 | Not-determined | Genome-fragment | NA | NA | 0 |
| S4-final-viral-combined_k141_30499\|\|full | 3690 | No | NA | 6 | 0 | 2 | Not-determined | Genome-fragment | NA | NA | 0 |
| S5-final-viral-combined_k141_77464\|\|full | 5045 | No | NA | 10 | 0 | 0 | Low-quality | Genome-fragment | 11.63 | AAI-based (medium-confidence) | 0 |
| S4-final-viral-combined_k141_37136\|\|full | 3791 | No | NA | 3 | 0 | 2 | Not-determined | Genome-fragment | NA | NA | 0 |
| S2-final-viral-combined_k141_117545\|\|full | 5046 | No | NA | 3 | 0 | 1 | Low-quality | Genome-fragment | 7.31 | AAI-based (medium-confidence) | 0 |
| S11-final-viral-combined_k141_284068\|\|full | 5049 | No | NA | 3 | 0 | 2 | Low-quality | Genome-fragment | 13.55 | AAI-based (medium-confidence) | 0 |
| S4-final-viral-combined_k141_162604\|\|full | 1190 | No | NA | 4 | 0 | 0 | Not-determined | Genome-fragment | NA | NA | 0 |
| S3-final-viral-combined_k141_5647\|\|full | 5049 | No | NA | 4 | 3 | 0 | Low-quality | Genome-fragment | 10.47 | AAI-based (medium-confidence) | 0 |
| S4-final-viral-combined_k141_120484\|\|full | 1141 | No | NA | 2 | 0 | 1 | Not-determined | Genome-fragment | NA | NA | 0 |
| S4-final-viral-combined_k141_141952\|\|full | 1543 | No | NA | 2 | 0 | 1 | Not-determined | Genome-fragment | NA | NA | 0 |
| S4-final-viral-combined_k141_155766\|\|full | 2174 | No | NA | 3 | 0 | 2 | Not-determined | Genome-fragment | NA | NA | 0 |
| S1-final-viral-combined_k141_4265\|\|full | 5050 | No | NA | 3 | 0 | 3 | Low-quality | Genome-fragment | 8.52 | AAI-based (medium-confidence) | 0 |
| S11-final-viral-combined_k141_275358\|\|full | 5052 | No | NA | 9 | 2 | 1 | Low-quality | Genome-fragment | 13.39 | AAI-based (high-confidence) | 0 |
| S4-final-viral-combined_k141_93282\|\|full | 2549 | No | NA | 4 | 0 | 1 | Not-determined | Genome-fragment | NA | NA | 0 |
| S4-final-viral-combined_k141_72335\|\|full | 2282 | No | NA | 6 | 0 | 5 | Not-determined | Genome-fragment | NA | NA | 0 |
| S4-final-viral-combined_k141_155817\|\|full | 4337 | No | NA | 10 | 0 | 2 | Not-determined | Genome-fragment | NA | NA | 0 |
| S4-final-viral-combined_k141_120537\|\|full | 6609 | No | NA | 3 | 0 | 2 | Not-determined | Genome-fragment | NA | NA | 0 |
| S4-final-viral-combined_k141_37190\|\|full | 1875 | No | NA | 2 | 0 | 1 | Not-determined | Genome-fragment | NA | NA | 0 |
| S4-final-viral-combined_k141_79625\|\|full | 1483 | No | NA | 3 | 0 | 1 | Not-determined | Genome-fragment | NA | NA | 0 |
| S4-final-viral-combined_k141_99748\|\|full | 1200 | No | NA | 3 | 0 | 2 | Not-determined | Genome-fragment | NA | NA | 0 |
| S4-final-viral-combined_k141_16397\|\|full | 2331 | No | NA | 2 | 0 | 1 | Not-determined | Genome-fragment | NA | NA | 0 |
| S12-final-viral-combined_k141_167764\|\|full | 5053 | No | NA | 9 | 2 | 0 | Low-quality | Genome-fragment | 11.64 | AAI-based (medium-confidence) | 0 |
| S4-final-viral-combined_k141_155859\|\|full | 2779 | No | NA | 5 | 0 | 0 | Not-determined | Genome-fragment | NA | NA | 0 |
| S4-final-viral-combined_k141_44419\|\|full | 1414 | No | NA | 3 | 0 | 1 | Not-determined | Genome-fragment | NA | NA | 0 |
| S1-final-viral-combined_k141_312305\|\|full | 5055 | No | NA | 4 | 0 | 2 | Low-quality | Genome-fragment | 3.55 | AAI-based (medium-confidence) | 0 |
| S8_k141_220180 | 5055 | No | NA | 6 | 3 | 0 | Low-quality | Genome-fragment | 10.79 | AAI-based (high-confidence) | 0 |
| S17-final-viral-combined_k141_183963\|\|full | 5056 | No | NA | 4 | 0 | 3 | Low-quality | Genome-fragment | 4.19 | AAI-based (medium-confidence) | 0 |
| S4-final-viral-combined_k141_86408\|\|full | 3711 | No | NA | 3 | 0 | 2 | Not-determined | Genome-fragment | NA | NA | 0 |
| S4-final-viral-combined_k141_148860\|\|full | 5074 | No | NA | 8 | 0 | 4 | Not-determined | Genome-fragment | NA | NA | 0 |
| S1-final-viral-combined_k141_103482\|\|full | 5058 | No | NA | 7 | 0 | 1 | Low-quality | Genome-fragment | 6.38 | AAI-based (medium-confidence) | 0 |
| S7-final-viral-combined_k141_9309\|\|full | 5058 | No | NA | 5 | 0 | 3 | Low-quality | Genome-fragment | 13.2 | AAI-based (medium-confidence) | 0 |
| S4-final-viral-combined_k141_113984\|\|full | 1602 | No | NA | 3 | 0 | 1 | Not-determined | Genome-fragment | NA | NA | 0 |
| S4-final-viral-combined_k141_2748\|\|full | 2239 | No | NA | 2 | 0 | 1 | Not-determined | Genome-fragment | NA | NA | 0 |
| S17-final-viral-combined_k141_186265\|\|full | 5059 | No | NA | 9 | 1 | 0 | Low-quality | Genome-fragment | 7.63 | HMM-based (lower-bound) | 0 |
| S2-final-viral-combined_k141_108280\|\|full | 5059 | No | NA | 4 | 0 | 3 | Low-quality | Genome-fragment | 0.58 | AAI-based (medium-confidence) | 0 |
| S4-final-viral-combined_k141_148888\|\|full | 1506 | No | NA | 3 | 0 | 3 | Not-determined | Genome-fragment | NA | NA | 0 |
| S1-final-viral-combined_k141_279629\|\|full | 5060 | No | NA | 4 | 0 | 3 | Low-quality | Genome-fragment | 5.8 | AAI-based (medium-confidence) | 0 |
| S17-k141_85486 | 5060 | No | NA | 6 | 0 | 3 | Low-quality | Genome-fragment | 6.05 | AAI-based (high-confidence) | 0 |
| S4-final-viral-combined_k141_30707\|\|full | 1198 | No | NA | 3 | 0 | 1 | Not-determined | Genome-fragment | NA | NA | 0 |
| S4-final-viral-combined_k141_30719\|\|full | 1372 | No | NA | 4 | 0 | 0 | Not-determined | Genome-fragment | NA | NA | 0 |
| S4-final-viral-combined_k141_72466\|\|full | 1212 | No | NA | 2 | 0 | 2 | Not-determined | Genome-fragment | NA | NA | 0 |
| S4-final-viral-combined_k141_37268\|\|full | 4152 | No | NA | 5 | 0 | 2 | Not-determined | Genome-fragment | NA | NA | 0 |
| S4-final-viral-combined_k141_120657\|\|full | 6377 | No | NA | 8 | 0 | 6 | Not-determined | Genome-fragment | NA | NA | 0 |
| S4-final-viral-combined_k141_107258\|\|full | 2165 | No | NA | 3 | 0 | 1 | Not-determined | Genome-fragment | NA | NA | 0 |
| S4-final-viral-combined_k141_37278\|\|full | 1250 | No | NA | 1 | 0 | 1 | Not-determined | Genome-fragment | NA | NA | 0 |
| S7-final-viral-combined_k141_88246\|\|full | 5062 | No | NA | 8 | 0 | 0 | Low-quality | Genome-fragment | 6 | AAI-based (medium-confidence) | 0 |
| S17-final-viral-combined_k141_83579\|\|full | 5062 | No | NA | 6 | 0 | 3 | Low-quality | Genome-fragment | 2.86 | AAI-based (medium-confidence) | 0 |
| S4-final-viral-combined_k141_120669\|\|full | 4907 | No | NA | 5 | 0 | 0 | Not-determined | Genome-fragment | NA | NA | 0 |
| S8-final-viral-combined_k141_241323\|\|lt2gene | 5064 | No | NA | 2 | 1 | 0 | Low-quality | Genome-fragment | 3.49 | HMM-based (lower-bound) | 0 |
| S5-final-viral-combined_k141_71446\|\|full | 5065 | No | NA | 8 | 3 | 0 | Low-quality | Genome-fragment | 5.68 | AAI-based (high-confidence) | 0 |
| S4-final-viral-combined_k141_155987\|\|full | 1224 | No | NA | 3 | 0 | 0 | Not-determined | Genome-fragment | NA | NA | 0 |
| S11_k141_314171 | 5066 | No | NA | 5 | 0 | 2 | Low-quality | Genome-fragment | 6.1 | AAI-based (medium-confidence) | 0 |
| S4-final-viral-combined_k141_135099\|\|full | 1864 | No | NA | 2 | 0 | 2 | Not-determined | Genome-fragment | NA | NA | 0 |
| S4-final-viral-combined_k141_93450\|\|full | 1374 | No | NA | 3 | 0 | 0 | Not-determined | Genome-fragment | NA | NA | 0 |
| S17-final-viral-combined_k141_189075\|\|full | 5067 | No | NA | 5 | 0 | 4 | Low-quality | Genome-fragment | 1.19 | AAI-based (medium-confidence) | 0 |
| S8_k141_34925 | 5067 | No | NA | 7 | 2 | 0 | Low-quality | Genome-fragment | 8.21 | AAI-based (high-confidence) | 0 |
| S1-final-viral-combined_k141_174622\|\|full | 5068 | No | NA | 5 | 1 | 0 | Low-quality | Genome-fragment | 10.14 | AAI-based (medium-confidence) | 0 |
| S4-final-viral-combined_k141_156049\|\|full | 1333 | No | NA | 3 | 0 | 3 | Not-determined | Genome-fragment | NA | NA | 0 |
| S4-final-viral-combined_k141_51009\|\|full | 4974 | No | NA | 5 | 0 | 3 | Not-determined | Genome-fragment | NA | NA | 0 |
| S4-final-viral-combined_k141_99943\|\|full | 4821 | No | NA | 5 | 0 | 2 | Not-determined | Genome-fragment | NA | NA | 0 |
| S8-final-viral-combined_k141_256135\|\|full | 5068 | No | NA | 5 | 4 | 0 | Low-quality | Genome-fragment | 12.3 | AAI-based (high-confidence) | 0 |
| S4-final-viral-combined_k141_99950\|\|full | 2853 | No | NA | 5 | 0 | 0 | Not-determined | Genome-fragment | NA | NA | 0 |
| S4-final-viral-combined_k141_142220\|\|full | 1865 | No | NA | 2 | 0 | 1 | Not-determined | Genome-fragment | NA | NA | 0 |
| S8_k141_166057 | 5068 | No | NA | 10 | 3 | 0 | Low-quality | Genome-fragment | 11.14 | AAI-based (high-confidence) | 0 |
| S2-final-viral-combined_k141_7979\|\|full | 5069 | No | NA | 7 | 0 | 0 | Low-quality | Genome-fragment | 5.9 | AAI-based (medium-confidence) | 0 |
| S7-final-viral-combined_k141_178495\|\|full | 5071 | No | NA | 5 | 0 | 2 | Low-quality | Genome-fragment | 4.61 | AAI-based (medium-confidence) | 0 |
| S2_k141_30792 | 5071 | No | NA | 5 | 0 | 4 | Low-quality | Genome-fragment | 1.89 | AAI-based (medium-confidence) | 0 |
| S8-final-viral-combined_k141_31631\|\|full | 5073 | No | NA | 8 | 6 | 0 | Low-quality | Genome-fragment | 20.06 | AAI-based (high-confidence) | 0 |
| S12-final-viral-combined_k141_225589\|\|full | 5074 | No | NA | 8 | 2 | 0 | Low-quality | Genome-fragment | 12.08 | AAI-based (medium-confidence) | 0 |
| S2-final-viral-combined_k141_221062\|\|full | 5074 | No | NA | 8 | 0 | 3 | Low-quality | Genome-fragment | 5.19 | AAI-based (high-confidence) | 0 |
| S2_k141_35076 | 5076 | No | NA | 3 | 0 | 2 | Low-quality | Genome-fragment | 4.6 | AAI-based (medium-confidence) | 0 |
| S1-final-viral-combined_k141_366518\|\|full | 5077 | No | NA | 15 | 1 | 0 | Low-quality | Genome-fragment | 7.96 | AAI-based (medium-confidence) | 0 |
| S7-final-viral-combined_k141_46949\|\|full | 5077 | No | NA | 5 | 0 | 1 | Low-quality | Genome-fragment | 9.63 | AAI-based (medium-confidence) | 0 |
| S4-final-viral-combined_k141_142293\|\|full | 1661 | No | NA | 3 | 0 | 1 | Not-determined | Genome-fragment | NA | NA | 0 |
| S4-final-viral-combined_k141_79805\|\|full | 2301 | No | NA | 3 | 0 | 1 | Not-determined | Genome-fragment | NA | NA | 0 |
| S8_k141_41524 | 5077 | No | NA | 3 | 0 | 1 | Low-quality | Genome-fragment | 2.46 | AAI-based (medium-confidence) | 0 |
| S4-final-viral-combined_k141_100023\|\|full | 1670 | No | NA | 3 | 0 | 1 | Not-determined | Genome-fragment | NA | NA | 0 |
| S4-final-viral-combined_k141_86578\|\|full | 1576 | No | NA | 2 | 0 | 1 | Not-determined | Genome-fragment | NA | NA | 0 |
| S4-final-viral-combined_k141_107416\|\|full | 2074 | No | NA | 3 | 0 | 3 | Not-determined | Genome-fragment | NA | NA | 0 |
| S4-final-viral-combined_k141_79827\|\|full | 6210 | No | NA | 5 | 0 | 4 | Not-determined | Genome-fragment | NA | NA | 0 |
| S4-final-viral-combined_k141_58387\|\|full | 2400 | No | NA | 3 | 0 | 0 | Not-determined | Genome-fragment | NA | NA | 0 |
| S4-final-viral-combined_k141_107422\|\|full | 3039 | No | NA | 3 | 0 | 1 | Not-determined | Genome-fragment | NA | NA | 0 |
| S4-final-viral-combined_k141_37417\|\|full | 3018 | No | NA | 6 | 0 | 2 | Not-determined | Genome-fragment | NA | NA | 0 |
| S4-final-viral-combined_k141_58408\|\|full | 1964 | No | NA | 2 | 0 | 0 | Not-determined | Genome-fragment | NA | NA | 0 |
| S4-final-viral-combined_k141_128108\|\|full | 2058 | No | NA | 3 | 0 | 1 | Not-determined | Genome-fragment | NA | NA | 0 |
| S7-final-viral-combined_k141_192956\|\|full | 5078 | No | NA | 10 | 1 | 0 | Low-quality | Genome-fragment | 5.55 | HMM-based (lower-bound) | 0 |
| S4-final-viral-combined_k141_128131\|\|full | 1771 | No | NA | 2 | 0 | 1 | Not-determined | Genome-fragment | NA | NA | 0 |
| S17-k141_215918 | 5079 | No | NA | 8 | 0 | 0 | Low-quality | Genome-fragment | 7.91 | AAI-based (medium-confidence) | 0 |
| S1-final-viral-combined_k141_26996\|\|full | 5080 | No | NA | 5 | 0 | 3 | Low-quality | Genome-fragment | 2.38 | AAI-based (medium-confidence) | 0 |
| S4-final-viral-combined_k141_9842\|\|full | 1959 | No | NA | 2 | 0 | 2 | Not-determined | Genome-fragment | NA | NA | 0 |
| S12-final-viral-combined_k141_61389\|\|full | 5080 | No | NA | 6 | 1 | 0 | Low-quality | Genome-fragment | 11.08 | AAI-based (medium-confidence) | 0 |
| S2_k141_57511 | 5080 | No | NA | 11 | 0 | 0 | Low-quality | Genome-fragment | 12.01 | AAI-based (medium-confidence) | 0 |
| S4-final-viral-combined_k141_31015\|\|full | 12692 | No | NA | 11 | 0 | 8 | Not-determined | Genome-fragment | NA | NA | 0 |
| S4-final-viral-combined_k141_156252\|\|full | 2431 | No | NA | 2 | 0 | 0 | Not-determined | Genome-fragment | NA | NA | 0 |
| S4-final-viral-combined_k141_65629\|\|full | 1121 | No | NA | 3 | 0 | 0 | Not-determined | Genome-fragment | NA | NA | 0 |
| S5-final-viral-combined_k141_157443\|\|full | 5081 | No | NA | 11 | 9 | 0 | Low-quality | Genome-fragment | 12.01 | AAI-based (high-confidence) | 0 |
| S4-final-viral-combined_k141_156300\|\|full | 2356 | No | NA | 4 | 0 | 0 | Not-determined | Genome-fragment | NA | NA | 0 |
| S4-final-viral-combined_k141_163002\|\|full | 7206 | No | NA | 6 | 0 | 5 | Not-determined | Genome-fragment | NA | NA | 0 |
| S4-final-viral-combined_k141_114194\|\|full | 43825 | No | NA | 44 | 0 | 5 | Not-determined | Genome-fragment | NA | NA | 0 |
| S4-final-viral-combined_k141_163010\|\|full | 3026 | No | NA | 5 | 0 | 0 | Not-determined | Genome-fragment | NA | NA | 0 |
| S4-final-viral-combined_k141_156322\|\|full | 1476 | No | NA | 3 | 0 | 1 | Not-determined | Genome-fragment | NA | NA | 0 |
| S4-final-viral-combined_k141_44768\|\|full | 2296 | No | NA | 4 | 0 | 2 | Not-determined | Genome-fragment | NA | NA | 0 |
| S17-final-viral-combined_k141_198411\|\|full | 5087 | No | NA | 6 | 1 | 0 | Low-quality | Genome-fragment | 21.96 | AAI-based (high-confidence) | 0 |
| S4_k141_137764 | 5087 | No | NA | 5 | 0 | 4 | Low-quality | Genome-fragment | 4.25 | AAI-based (high-confidence) | 0 |
| S4-final-viral-combined_k141_107573\|\|full | 1136 | No | NA | 2 | 0 | 1 | Not-determined | Genome-fragment | NA | NA | 0 |
| S4-final-viral-combined_k141_163038\|\|full | 2431 | No | NA | 2 | 0 | 1 | Not-determined | Genome-fragment | NA | NA | 0 |
| S8-final-viral-combined_k141_129368\|\|full | 5089 | No | NA | 3 | 3 | 0 | Low-quality | Genome-fragment | 8.75 | AAI-based (medium-confidence) | 0 |
| S4-final-viral-combined_k141_3092\|\|full | 2636 | No | NA | 3 | 0 | 2 | Not-determined | Genome-fragment | NA | NA | 0 |
| S4-final-viral-combined_k141_120958\|\|full | 1884 | No | NA | 5 | 0 | 2 | Not-determined | Genome-fragment | NA | NA | 0 |
| S4-final-viral-combined_k141_156371\|\|full | 1670 | No | NA | 3 | 0 | 1 | Not-determined | Genome-fragment | NA | NA | 0 |
| S4-final-viral-combined_k141_44816\|\|full | 3470 | No | NA | 6 | 0 | 0 | Not-determined | Genome-fragment | NA | NA | 0 |
| S4-final-viral-combined_k141_156381\|\|full | 2609 | No | NA | 3 | 0 | 1 | Not-determined | Genome-fragment | NA | NA | 0 |
| S12-final-viral-combined_k141_203161\|\|full | 5095 | No | NA | 5 | 0 | 1 | Low-quality | Genome-fragment | 1.78 | AAI-based (medium-confidence) | 0 |
| S12-final-viral-combined_k141_174808\|\|full | 5095 | No | NA | 5 | 0 | 4 | Low-quality | Genome-fragment | 3.21 | AAI-based (medium-confidence) | 0 |
| S4-final-viral-combined_k141_163089\|\|full | 1794 | No | NA | 2 | 0 | 2 | Not-determined | Genome-fragment | NA | NA | 0 |
| S2-final-viral-combined_k141_181171\|\|full | 5096 | No | NA | 4 | 0 | 3 | Low-quality | Genome-fragment | 4.72 | AAI-based (medium-confidence) | 0 |
| S4-final-viral-combined_k141_80019\|\|full | 2711 | No | NA | 5 | 0 | 2 | Not-determined | Genome-fragment | NA | NA | 0 |
| S9-final-viral-combined_k141_55146\|\|full | 5098 | No | NA | 9 | 0 | 2 | Low-quality | Genome-fragment | 6.6 | AAI-based (medium-confidence) | 0 |
| S12_k141_250493 | 5100 | No | NA | 6 | 3 | 0 | Low-quality | Genome-fragment | 10.64 | AAI-based (high-confidence) | 0 |
| S1-final-viral-combined_k141_105989\|\|full | 5101 | Yes | 3545 | 10 | 1 | 1 | Low-quality | Genome-fragment | 2.54 | HMM-based (lower-bound) | 30.5 |
| S4-final-viral-combined_k141_31156\|\|full | 7021 | No | NA | 6 | 0 | 0 | Not-determined | Genome-fragment | NA | NA | 0 |
| S1_k141_52604 | 5101 | No | NA | 8 | 3 | 0 | Low-quality | Genome-fragment | 11.56 | AAI-based (high-confidence) | 0 |
| S4-final-viral-combined_k141_128353\|\|full | 3190 | No | NA | 4 | 0 | 4 | Not-determined | Genome-fragment | NA | NA | 0 |
| S12-final-viral-combined_k141_251062\|\|full | 5102 | No | NA | 6 | 0 | 1 | Low-quality | Genome-fragment | 6.3 | AAI-based (high-confidence) | 0 |
| S4-final-viral-combined_k141_23845\|\|full | 1299 | No | NA | 3 | 0 | 1 | Not-determined | Genome-fragment | NA | NA | 0 |
| S4-final-viral-combined_k141_142612\|\|full | 5406 | No | NA | 5 | 0 | 3 | Not-determined | Genome-fragment | NA | NA | 0 |
| S4-final-viral-combined_k141_10046\|\|full | 2080 | No | NA | 3 | 0 | 3 | Not-determined | Genome-fragment | NA | NA | 0 |
| S4-final-viral-combined_k141_149308\|\|full | 5215 | No | NA | 2 | 0 | 0 | Not-determined | Genome-fragment | NA | NA | 0 |
| S4-final-viral-combined_k141_156443\|\|full | 3448 | No | NA | 4 | 0 | 3 | Not-determined | Genome-fragment | NA | NA | 0 |
| S2_k141_202978 | 5102 | No | NA | 2 | 0 | 2 | Low-quality | Genome-fragment | 3.03 | AAI-based (medium-confidence) | 0 |
| S7_k141_241654 | 5103 | No | NA | 5 | 0 | 2 | Low-quality | Genome-fragment | 2.56 | AAI-based (high-confidence) | 0 |
| S4-final-viral-combined_k141_114360\|\|full | 2377 | No | NA | 2 | 0 | 2 | Not-determined | Genome-fragment | NA | NA | 0 |
| S4-final-viral-combined_k141_37693\|\|full | 2281 | No | NA | 3 | 0 | 1 | Not-determined | Genome-fragment | NA | NA | 0 |
| S4-final-viral-combined_k141_107770\|\|full | 1043 | No | NA | 2 | 0 | 0 | Not-determined | Genome-fragment | NA | NA | 0 |
| S4-final-viral-combined_k141_72798\|\|full | 2483 | No | NA | 2 | 0 | 0 | Not-determined | Genome-fragment | NA | NA | 0 |
| S4-final-viral-combined_k141_120999\|\|full | 2771 | No | NA | 3 | 0 | 3 | Not-determined | Genome-fragment | NA | NA | 0 |
| S2_k141_65800 | 5103 | No | NA | 10 | 2 | 1 | Low-quality | Genome-fragment | 8.4 | AAI-based (high-confidence) | 0 |
| S12_k141_106888 | 5104 | No | NA | 7 | 1 | 0 | Low-quality | Genome-fragment | 6.8 | AAI-based (high-confidence) | 0 |
| S9-final-viral-combined_k141_88019\|\|full | 5105 | No | NA | 10 | 2 | 0 | Low-quality | Genome-fragment | 8.32 | AAI-based (high-confidence) | 0 |
| S7-final-viral-combined_k141_80085\|\|full | 5105 | No | NA | 12 | 8 | 0 | Low-quality | Genome-fragment | 12.33 | HMM-based (lower-bound) | 0 |
| S17-final-viral-combined_k141_172365\|\|full | 5106 | No | NA | 7 | 1 | 1 | Low-quality | Genome-fragment | 1.92 | HMM-based (lower-bound) | 0 |
| S4-final-viral-combined_k141_149379\|\|full | 1100 | No | NA | 3 | 0 | 2 | Not-determined | Genome-fragment | NA | NA | 0 |
| S4-final-viral-combined_k141_156491\|\|full | 6740 | No | NA | 5 | 0 | 4 | Not-determined | Genome-fragment | NA | NA | 0 |
| S4-final-viral-combined_k141_86889\|\|full | 2990 | No | NA | 3 | 0 | 0 | Not-determined | Genome-fragment | NA | NA | 0 |
| S4-final-viral-combined_k141_51313\|\|full | 7741 | No | NA | 2 | 0 | 2 | Not-determined | Genome-fragment | NA | NA | 0 |
| S5-final-viral-combined_k141_187042\|\|full | 5108 | No | NA | 4 | 0 | 2 | Low-quality | Genome-fragment | 1.63 | AAI-based (medium-confidence) | 0 |
| S12-final-viral-combined_k141_325559\|\|full | 5108 | No | NA | 8 | 2 | 0 | Low-quality | Genome-fragment | 11.6 | AAI-based (high-confidence) | 0 |
| S4-final-viral-combined_k141_86900\|\|full | 2614 | No | NA | 4 | 0 | 2 | Not-determined | Genome-fragment | NA | NA | 0 |
| S8-final-viral-combined_k141_124626\|\|full | 5108 | No | NA | 3 | 1 | 0 | Low-quality | Genome-fragment | 10.07 | AAI-based (medium-confidence) | 0 |
| S4-final-viral-combined_k141_58565\|\|full | 1852 | No | NA | 5 | 0 | 3 | Not-determined | Genome-fragment | NA | NA | 0 |
| S4-final-viral-combined_k141_51328\|\|full | 1408 | No | NA | 3 | 0 | 2 | Not-determined | Genome-fragment | NA | NA | 0 |
| S4-final-viral-combined_k141_114409\|\|full | 4722 | No | NA | 4 | 0 | 4 | Not-determined | Genome-fragment | NA | NA | 0 |
| S4-final-viral-combined_k141_156523\|\|full | 2214 | No | NA | 3 | 0 | 2 | Not-determined | Genome-fragment | NA | NA | 0 |
| S4-final-viral-combined_k141_128475\|\|full | 7559 | No | NA | 6 | 0 | 5 | Not-determined | Genome-fragment | NA | NA | 0 |
| S4-final-viral-combined_k141_10152\|\|full | 3422 | No | NA | 4 | 0 | 2 | Not-determined | Genome-fragment | NA | NA | 0 |
| S11-final-viral-combined_k141_319130\|\|full | 5113 | No | NA | 5 | 0 | 1 | Low-quality | Genome-fragment | 8.45 | AAI-based (medium-confidence) | 0 |
| S4-final-viral-combined_k141_10165\|\|full | 3615 | No | NA | 4 | 0 | 2 | Not-determined | Genome-fragment | NA | NA | 0 |
| S4-final-viral-combined_k141_135508\|\|full | 1498 | No | NA | 3 | 0 | 0 | Not-determined | Genome-fragment | NA | NA | 0 |
| S4-final-viral-combined_k141_45073\|\|full | 1330 | No | NA | 2 | 0 | 2 | Not-determined | Genome-fragment | NA | NA | 0 |
| S4-final-viral-combined_k141_10175\|\|full | 2876 | No | NA | 2 | 0 | 1 | Not-determined | Genome-fragment | NA | NA | 0 |
| S4-final-viral-combined_k141_149420\|\|full | 1708 | No | NA | 3 | 0 | 2 | Not-determined | Genome-fragment | NA | NA | 0 |
| S12-final-viral-combined_k141_325470\|\|full | 5113 | No | NA | 8 | 2 | 0 | Low-quality | Genome-fragment | 11.56 | AAI-based (high-confidence) | 0 |
| S9-final-viral-combined_k141_90325\|\|full | 5114 | No | NA | 4 | 0 | 2 | Low-quality | Genome-fragment | 1.6 | AAI-based (medium-confidence) | 0 |
| S4-final-viral-combined_k141_121108\|\|full | 2863 | No | NA | 2 | 0 | 1 | Not-determined | Genome-fragment | NA | NA | 0 |
| S4-final-viral-combined_k141_23997\|\|full | 1400 | No | NA | 3 | 0 | 2 | Not-determined | Genome-fragment | NA | NA | 0 |
| S11-final-viral-combined_k141_51073\|\|full | 5114 | No | NA | 5 | 0 | 4 | Low-quality | Genome-fragment | 2.53 | AAI-based (medium-confidence) | 0 |
| S11-final-viral-combined_k141_234017\|\|full | 5117 | No | NA | 6 | 3 | 0 | Low-quality | Genome-fragment | 10.78 | AAI-based (high-confidence) | 0 |
| S4-final-viral-combined_k141_156566\|\|full | 5145 | No | NA | 4 | 0 | 0 | Not-determined | Genome-fragment | NA | NA | 0 |
| S4-final-viral-combined_k141_45126\|\|full | 1201 | No | NA | 3 | 0 | 1 | Not-determined | Genome-fragment | NA | NA | 0 |
| S1-final-viral-combined_k141_327301\|\|full | 5117 | No | NA | 10 | 0 | 0 | Low-quality | Genome-fragment | 27.69 | AAI-based (high-confidence) | 0 |
| S4-final-viral-combined_k141_86948\|\|full | 2152 | No | NA | 5 | 0 | 0 | Not-determined | Genome-fragment | NA | NA | 0 |
| S4-final-viral-combined_k141_86961\|\|full | 1885 | No | NA | 3 | 0 | 2 | Not-determined | Genome-fragment | NA | NA | 0 |
| S4-final-viral-combined_k141_128571\|\|full | 6951 | No | NA | 8 | 0 | 4 | Not-determined | Genome-fragment | NA | NA | 0 |
| S4_k141_88819 | 5117 | No | NA | 3 | 0 | 2 | Low-quality | Genome-fragment | 2.75 | AAI-based (medium-confidence) | 0 |
| S4-final-viral-combined_k141_26946\|\|full | 5118 | No | NA | 10 | 1 | 4 | Low-quality | Genome-fragment | 7.59 | AAI-based (high-confidence) | 0 |
| S12-final-viral-combined_k141_379886\|\|full | 5119 | No | NA | 6 | 0 | 3 | Low-quality | Genome-fragment | 9.09 | AAI-based (medium-confidence) | 0 |
| S12_k141_18970 | 5119 | No | NA | 10 | 2 | 0 | Low-quality | Genome-fragment | 15.14 | HMM-based (lower-bound) | 0 |
| S4-final-viral-combined_k141_10264\|\|full | 3090 | No | NA | 4 | 0 | 3 | Not-determined | Genome-fragment | NA | NA | 0 |
| S2-final-viral-combined_k141_200952\|\|full | 5121 | No | NA | 5 | 0 | 0 | Low-quality | Genome-fragment | 3.51 | AAI-based (medium-confidence) | 0 |
| S4-final-viral-combined_k141_163352\|\|full | 3006 | No | NA | 4 | 0 | 3 | Not-determined | Genome-fragment | NA | NA | 0 |
| S4-final-viral-combined_k141_45136\|\|full | 13488 | No | NA | 7 | 0 | 4 | Not-determined | Genome-fragment | NA | NA | 0 |
| S4-final-viral-combined_k141_73034\|\|full | 2439 | No | NA | 2 | 0 | 0 | Not-determined | Genome-fragment | NA | NA | 0 |
| S4-final-viral-combined_k141_114534\|\|full | 2903 | No | NA | 4 | 0 | 2 | Not-determined | Genome-fragment | NA | NA | 0 |
| S8-final-viral-combined_k141_2821\|\|full | 5121 | Yes | 864 | 2 | 1 | 1 | Low-quality | Genome-fragment | 1.12 | HMM-based (lower-bound) | 83.13 |
| S4-final-viral-combined_k141_73062\|\|full | 1893 | No | NA | 3 | 0 | 3 | Not-determined | Genome-fragment | NA | NA | 0 |
| S4-final-viral-combined_k141_58771\|\|full | 1740 | No | NA | 2 | 0 | 1 | Not-determined | Genome-fragment | NA | NA | 0 |
| S9_k141_58465 | 5122 | No | NA | 5 | 0 | 1 | Low-quality | Genome-fragment | 3.83 | AAI-based (medium-confidence) | 0 |
| S4-final-viral-combined_k141_149565\|\|full | 2638 | No | NA | 4 | 0 | 3 | Not-determined | Genome-fragment | NA | NA | 0 |
| S4-final-viral-combined_k141_100566\|\|full | 4520 | No | NA | 4 | 0 | 3 | Not-determined | Genome-fragment | NA | NA | 0 |
| S4-final-viral-combined_k141_37874\|\|full | 1160 | No | NA | 3 | 0 | 0 | Not-determined | Genome-fragment | NA | NA | 0 |
| S4-final-viral-combined_k141_45195\|\|full | 1073 | No | NA | 2 | 0 | 2 | Not-determined | Genome-fragment | NA | NA | 0 |
| S4-final-viral-combined_k141_58798\|\|full | 9029 | No | NA | 12 | 0 | 2 | Not-determined | Genome-fragment | NA | NA | 0 |
| S4-final-viral-combined_k141_121187\|\|full | 3413 | No | NA | 3 | 0 | 2 | Not-determined | Genome-fragment | NA | NA | 0 |
| S1-final-viral-combined_k141_247161\|\|full | 5123 | No | NA | 3 | 0 | 2 | Low-quality | Genome-fragment | 4.64 | AAI-based (medium-confidence) | 0 |
| S3-final-viral-combined_k141_18305\|\|full | 5123 | No | NA | 6 | 0 | 0 | Low-quality | Genome-fragment | 8.09 | AAI-based (medium-confidence) | 0 |
| S12-final-viral-combined_k141_240194\|\|full | 5126 | No | NA | 6 | 4 | 0 | Low-quality | Genome-fragment | 14.04 | AAI-based (high-confidence) | 0 |
| S4-final-viral-combined_k141_65998\|\|full | 2420 | No | NA | 3 | 0 | 3 | Not-determined | Genome-fragment | NA | NA | 0 |
| S4-final-viral-combined_k141_135673\|\|full | 1858 | No | NA | 3 | 0 | 0 | Not-determined | Genome-fragment | NA | NA | 0 |
| S4-final-viral-combined_k141_66000\|\|full | 2306 | No | NA | 2 | 0 | 2 | Not-determined | Genome-fragment | NA | NA | 0 |
| S4-final-viral-combined_k141_45222\|\|full | 4111 | No | NA | 8 | 0 | 0 | Not-determined | Genome-fragment | NA | NA | 0 |
| S4-final-viral-combined_k141_73110\|\|full | 9174 | No | NA | 6 | 0 | 6 | Not-determined | Genome-fragment | NA | NA | 0 |
| S2_k141_7787 | 5126 | No | NA | 4 | 0 | 0 | Low-quality | Genome-fragment | 3.12 | AAI-based (high-confidence) | 0 |
| S17-final-viral-combined_k141_6181\|\|full | 5128 | No | NA | 7 | 6 | 0 | Low-quality | Genome-fragment | 14.06 | AAI-based (high-confidence) | 0 |
| S4-final-viral-combined_k141_135686\|\|full | 11322 | No | NA | 10 | 0 | 8 | Not-determined | Genome-fragment | NA | NA | 0 |
| S4-final-viral-combined_k141_10414\|\|full | 2411 | No | NA | 2 | 0 | 1 | Not-determined | Genome-fragment | NA | NA | 0 |
| S4-final-viral-combined_k141_108063\|\|full | 1032 | No | NA | 2 | 0 | 2 | Not-determined | Genome-fragment | NA | NA | 0 |
| S7-final-viral-combined_k141_155388\|\|full | 5129 | No | NA | 6 | 0 | 1 | Low-quality | Genome-fragment | 13.74 | AAI-based (high-confidence) | 0 |
| S17-k141_139594 | 5129 | Yes | 1806 | 8 | 1 | 3 | Low-quality | Genome-fragment | 3 | HMM-based (lower-bound) | 64.79 |
| S17-final-viral-combined_k141_248732\|\|full | 5131 | No | NA | 8 | 0 | 1 | Low-quality | Genome-fragment | 11.95 | AAI-based (medium-confidence) | 0 |
| S4-final-viral-combined_k141_108092\|\|full | 1913 | No | NA | 3 | 0 | 1 | Not-determined | Genome-fragment | NA | NA | 0 |
| S4-final-viral-combined_k141_80368\|\|full | 1799 | No | NA | 3 | 0 | 2 | Not-determined | Genome-fragment | NA | NA | 0 |
| S4-final-viral-combined_k141_163450\|\|full | 1130 | No | NA | 2 | 0 | 0 | Not-determined | Genome-fragment | NA | NA | 0 |
| S4-final-viral-combined_k141_16953\|\|full | 1385 | No | NA | 3 | 0 | 2 | Not-determined | Genome-fragment | NA | NA | 0 |
| S4-final-viral-combined_k141_73226\|\|full | 2221 | No | NA | 2 | 0 | 1 | Not-determined | Genome-fragment | NA | NA | 0 |
| S4-final-viral-combined_k141_142924\|\|full | 3194 | No | NA | 2 | 0 | 0 | Not-determined | Genome-fragment | NA | NA | 0 |
| S12-final-viral-combined_k141_186081\|\|full | 5132 | No | NA | 7 | 2 | 0 | Low-quality | Genome-fragment | 14.01 | AAI-based (high-confidence) | 0 |
| S4-final-viral-combined_k141_128668\|\|full | 2651 | No | NA | 2 | 0 | 1 | Not-determined | Genome-fragment | NA | NA | 0 |
| S4-final-viral-combined_k141_162190\|\|full | 5132 | Yes | 3355 | 9 | 1 | 2 | Low-quality | Genome-fragment | 3.01 | AAI-based (medium-confidence) | 34.63 |
| S2_k141_38630 | 5134 | No | NA | 6 | 0 | 4 | Low-quality | Genome-fragment | 2.87 | AAI-based (high-confidence) | 0 |
| S3_k141_84560 | 5136 | No | NA | 12 | 1 | 0 | Low-quality | Genome-fragment | 6.66 | AAI-based (medium-confidence) | 0 |
| S7-final-viral-combined_k141_144671\|\|full | 5136 | No | NA | 5 | 0 | 3 | Low-quality | Genome-fragment | 1.24 | AAI-based (medium-confidence) | 0 |
| S12_k141_285564 | 5137 | No | NA | 3 | 0 | 0 | Low-quality | Genome-fragment | 8.51 | AAI-based (medium-confidence) | 0 |
| S4-final-viral-combined_k141_10461\|\|full | 2114 | No | NA | 4 | 0 | 3 | Not-determined | Genome-fragment | NA | NA | 0 |
| S4-final-viral-combined_k141_149706\|\|full | 2710 | No | NA | 3 | 0 | 1 | Not-determined | Genome-fragment | NA | NA | 0 |
| S4-final-viral-combined_k141_51569\|\|full | 1852 | No | NA | 2 | 0 | 0 | Not-determined | Genome-fragment | NA | NA | 0 |

| | | | | | | | | | | | |
|---|---|---|---|---|---|---|---|---|---|---|---|
| S17-k141_224040 | 5138 | No | NA | 4 | 0 | 3 | Low-quality | Genome-fragment | 2.77 | AAI-based (medium-confidence) | 0 |
| S4-final-viral-combined_k141_58949\|\|full | 1205 | No | NA | 2 | 0 | 1 | Not-determined | Genome-fragment | NA | NA | 0 |
| S12_k141_159329 | 5138 | No | NA | 9 | 3 | 0 | Low-quality | Genome-fragment | 9.1 | AAI-based (high-confidence) | 0 |
| S4-final-viral-combined_k141_58973\|\|full | 6201 | No | NA | 7 | 0 | 1 | Not-determined | Genome-fragment | NA | NA | 0 |
| S17-final-viral-combined_k141_9722\|\|full | 5139 | No | NA | 4 | 3 | 0 | Low-quality | Genome-fragment | 11.47 | AAI-based (high-confidence) | 0 |
| S17-k141_94180 | 5139 | No | NA | 3 | 0 | 1 | Low-quality | Genome-fragment | 7.91 | AAI-based (high-confidence) | 0 |
| S4_k141_2247 | 5142 | No | NA | 6 | 4 | 0 | Low-quality | Genome-fragment | 9.01 | AAI-based (high-confidence) | 0 |
| S4-final-viral-combined_k141_58981\|\|full | 1129 | No | NA | 2 | 0 | 0 | Not-determined | Genome-fragment | NA | NA | 0 |
| S7-final-viral-combined_k141_64783\|\|full | 5143 | No | NA | 5 | 2 | 0 | Low-quality | Genome-fragment | 10.94 | AAI-based (medium-confidence) | 0 |
| S6_k141_101634 | 5143 | No | NA | 5 | 0 | 4 | Low-quality | Genome-fragment | 1.77 | AAI-based (medium-confidence) | 0 |
| S7-final-viral-combined_k141_78864\|\|full | 5144 | No | NA | 6 | 0 | 2 | Low-quality | Genome-fragment | 10.47 | AAI-based (medium-confidence) | 0 |
| S6-final-viral-combined_k141_79367\|\|full | 5144 | No | NA | 4 | 0 | 3 | Low-quality | Genome-fragment | 4 | AAI-based (high-confidence) | 0 |
| S4-final-viral-combined_k141_3548\|\|full | 1461 | No | NA | 3 | 0 | 1 | Not-determined | Genome-fragment | NA | NA | 0 |
| S4-final-viral-combined_k141_31559\|\|full | 2950 | No | NA | 2 | 0 | 1 | Not-determined | Genome-fragment | NA | NA | 0 |
| S11-final-viral-combined_k141_126914\|\|full | 5147 | No | NA | 8 | 4 | 0 | Low-quality | Genome-fragment | 8.89 | AAI-based (high-confidence) | 0 |
| S17-k141_139946 | 5147 | No | NA | 11 | 8 | 0 | Low-quality | Genome-fragment | 10.92 | AAI-based (high-confidence) | 0 |
| S2_k141_136099 | 5147 | No | NA | 5 | 0 | 3 | Low-quality | Genome-fragment | 1.38 | AAI-based (high-confidence) | 0 |
| S4-final-viral-combined_k141_94101\|\|full | 3786 | No | NA | 7 | 0 | 0 | Not-determined | Genome-fragment | NA | NA | 0 |
| S4-final-viral-combined_k141_51578\|\|full | 7887 | No | NA | 7 | 0 | 5 | Not-determined | Genome-fragment | NA | NA | 0 |
| S8-final-viral-combined_k141_156015\|\|full | 5147 | No | NA | 9 | 0 | 1 | Low-quality | Genome-fragment | 12.02 | AAI-based (high-confidence) | 0 |
| S4-final-viral-combined_k141_94104\|\|full | 1374 | No | NA | 2 | 0 | 0 | Not-determined | Genome-fragment | NA | NA | 0 |
| S4-final-viral-combined_k141_108251\|\|full | 1307 | No | NA | 2 | 0 | 0 | Not-determined | Genome-fragment | NA | NA | 0 |
| S4-final-viral-combined_k141_24315\|\|full | 1585 | No | NA | 3 | 0 | 2 | Not-determined | Genome-fragment | NA | NA | 0 |
| S17-k141_132044 | 5148 | No | NA | 7 | 0 | 3 | Low-quality | Genome-fragment | 6.21 | AAI-based (medium-confidence) | 0 |
| S4-final-viral-combined_k141_80454\|\|full | 3540 | No | NA | 3 | 0 | 1 | Not-determined | Genome-fragment | NA | NA | 0 |
| S4-final-viral-combined_k141_80462\|\|full | 2912 | No | NA | 3 | 0 | 2 | Not-determined | Genome-fragment | NA | NA | 0 |
| S4-final-viral-combined_k141_73346\|\|full | 1037 | No | NA | 3 | 0 | 1 | Not-determined | Genome-fragment | NA | NA | 0 |
| S8_k141_1211 | 5148 | No | NA | 6 | 0 | 5 | Low-quality | Genome-fragment | 1.65 | AAI-based (medium-confidence) | 0 |
| S7-final-viral-combined_k141_2744\|\|full | 5149 | No | NA | 4 | 0 | 2 | Low-quality | Genome-fragment | 1.6 | AAI-based (medium-confidence) | 0 |
| S7_k141_91537 | 5149 | No | NA | 2 | 0 | 2 | Low-quality | Genome-fragment | 4.66 | AAI-based (medium-confidence) | 0 |
| S4-final-viral-combined_k141_80480\|\|full | 2692 | No | NA | 3 | 0 | 3 | Not-determined | Genome-fragment | NA | NA | 0 |
| S4-final-viral-combined_k141_149816\|\|full | 2288 | No | NA | 6 | 0 | 0 | Not-determined | Genome-fragment | NA | NA | 0 |
| S7_k141_127084 | 5151 | No | NA | 2 | 0 | 1 | Low-quality | Genome-fragment | 7.71 | AAI-based (medium-confidence) | 0 |
| S4-final-viral-combined_k141_87206\|\|full | 1948 | No | NA | 3 | 0 | 2 | Not-determined | Genome-fragment | NA | NA | 0 |
| S4-final-viral-combined_k141_108311\|\|full | 3718 | No | NA | 7 | 0 | 1 | Not-determined | Genome-fragment | NA | NA | 0 |
| S12_k141_162235 | 5151 | No | NA | 9 | 3 | 0 | Low-quality | Genome-fragment | 11.37 | AAI-based (high-confidence) | 0 |
| S4-final-viral-combined_k141_45487\|\|full | 2776 | No | NA | 3 | 0 | 3 | Not-determined | Genome-fragment | NA | NA | 0 |
| S4-final-viral-combined_k141_10619\|\|full | 17050 | No | NA | 11 | 0 | 10 | Not-determined | Genome-fragment | NA | NA | 0 |
| S4-final-viral-combined_k141_80525\|\|full | 2623 | No | NA | 2 | 0 | 1 | Not-determined | Genome-fragment | NA | NA | 0 |
| S7-final-viral-combined_k141_2452\|\|full | 5152 | Yes | 4795 | 8 | 2 | 2 | Low-quality | Genome-fragment | 8.1 | AAI-based (medium-confidence) | 6.93 |
| S17-k141_173334 | 5155 | Yes | 2483 | 7 | 1 | 1 | Low-quality | Genome-fragment | 1.83 | HMM-based (lower-bound) | 51.83 |
| S6_k141_88773 | 5156 | No | NA | 5 | 0 | 3 | Low-quality | Genome-fragment | 2.14 | AAI-based (medium-confidence) | 0 |
| S4-final-viral-combined_k141_80541\|\|full | 3774 | No | NA | 4 | 0 | 2 | Not-determined | Genome-fragment | NA | NA | 0 |
| S2-final-viral-combined_k141_11046\|\|full | 5157 | No | NA | 9 | 0 | 0 | Low-quality | Genome-fragment | 10.82 | AAI-based (high-confidence) | 0 |
| S4-final-viral-combined_k141_108322\|\|full | 2134 | No | NA | 4 | 0 | 0 | Not-determined | Genome-fragment | NA | NA | 0 |
| S4-final-viral-combined_k141_143116\|\|full | 3803 | No | NA | 5 | 0 | 3 | Not-determined | Genome-fragment | NA | NA | 0 |
| S6-final-viral-combined_k141_78377\|\|full | 5157 | No | NA | 6 | 0 | 2 | Low-quality | Genome-fragment | 5.17 | AAI-based (medium-confidence) | 0 |
| S4-final-viral-combined_k141_143127\|\|full | 1600 | No | NA | 2 | 0 | 2 | Not-determined | Genome-fragment | NA | NA | 0 |
| S4-final-viral-combined_k141_100904\|\|full | 2105 | No | NA | 3 | 0 | 0 | Not-determined | Genome-fragment | NA | NA | 0 |
| S4-final-viral-combined_k141_149900\|\|full | 3475 | No | NA | 4 | 0 | 4 | Not-determined | Genome-fragment | NA | NA | 0 |
| S17-k141_207989 | 5158 | No | NA | 8 | 0 | 0 | Low-quality | Genome-fragment | 6.87 | AAI-based (high-confidence) | 0 |
| S6-final-viral-combined_k141_98092\|\|full | 5158 | No | NA | 3 | 0 | 2 | Low-quality | Genome-fragment | 1.73 | AAI-based (medium-confidence) | 0 |
| S4-final-viral-combined_k141_87262\|\|full | 1898 | No | NA | 3 | 0 | 1 | Not-determined | Genome-fragment | NA | NA | 0 |
| S4-final-viral-combined_k141_100926\|\|full | 2638 | No | NA | 3 | 0 | 1 | Not-determined | Genome-fragment | NA | NA | 0 |
| S4-final-viral-combined_k141_59152\|\|full | 1621 | No | NA | 4 | 0 | 2 | Not-determined | Genome-fragment | NA | NA | 0 |
| S4-final-viral-combined_k141_66344\|\|full | 1891 | No | NA | 3 | 0 | 1 | Not-determined | Genome-fragment | NA | NA | 0 |
| S4-final-viral-combined_k141_114887\|\|full | 4905 | No | NA | 4 | 0 | 1 | Not-determined | Genome-fragment | NA | NA | 0 |
| S9-final-viral-combined_k141_81418\|\|full | 5159 | No | NA | 4 | 0 | 1 | Low-quality | Genome-fragment | 9.95 | AAI-based (medium-confidence) | 0 |
| S17-final-viral-combined_k141_122398\|\|full | 5160 | No | NA | 7 | 0 | 4 | Low-quality | Genome-fragment | 3.41 | AAI-based (medium-confidence) | 0 |
| S1_k141_235037 | 5162 | No | NA | 6 | 0 | 2 | Low-quality | Genome-fragment | 3.54 | AAI-based (medium-confidence) | 0 |
| S4-final-viral-combined_k141_108399\|\|full | 1362 | No | NA | 3 | 0 | 1 | Not-determined | Genome-fragment | NA | NA | 0 |
| S4-final-viral-combined_k141_31695\|\|full | 2689 | No | NA | 3 | 0 | 3 | Not-determined | Genome-fragment | NA | NA | 0 |
| S5-final-viral-combined_k141_141462\|\|full | 5162 | No | NA | 6 | 2 | 0 | Low-quality | Genome-fragment | 13.67 | AAI-based (medium-confidence) | 0 |
| S7_k141_209798 | 5163 | No | NA | 13 | 2 | 0 | Low-quality | Genome-fragment | 11.39 | AAI-based (high-confidence) | 0 |
| S5-final-viral-combined_k141_61367\|\|full | 5166 | No | NA | 5 | 3 | 0 | Low-quality | Genome-fragment | 10.18 | AAI-based (high-confidence) | 0 |
| S7-final-viral-combined_k141_85991\|\|full | 5167 | No | NA | 4 | 0 | 3 | Low-quality | Genome-fragment | 3.87 | AAI-based (medium-confidence) | 0 |
| S7-final-viral-combined_k141_203503\|\|full | 5169 | No | NA | 9 | 1 | 0 | Low-quality | Genome-fragment | 2.33 | HMM-based (lower-bound) | 0 |
| S12-final-viral-combined_k141_125255\|\|full | 5169 | No | NA | 4 | 0 | 0 | Low-quality | Genome-fragment | 13.54 | AAI-based (medium-confidence) | 0 |
| S4-final-viral-combined_k141_121549\|\|full | 1885 | No | NA | 2 | 0 | 1 | Not-determined | Genome-fragment | NA | NA | 0 |
| S4-final-viral-combined_k141_73521\|\|full | 2050 | No | NA | 4 | 0 | 1 | Not-determined | Genome-fragment | NA | NA | 0 |
| S2-final-viral-combined_k141_32453\|\|full | 5169 | No | NA | 10 | 2 | 0 | Low-quality | Genome-fragment | 9.43 | AAI-based (high-confidence) | 0 |
| S9-final-viral-combined_k141_18835\|\|full | 5171 | No | NA | 5 | 0 | 3 | Low-quality | Genome-fragment | 3.56 | AAI-based (high-confidence) | 0 |
| S4-final-viral-combined_k141_143247\|\|full | 2662 | No | NA | 2 | 0 | 1 | Not-determined | Genome-fragment | NA | NA | 0 |
| S4-final-viral-combined_k141_45573\|\|full | 2142 | No | NA | 2 | 0 | 1 | Not-determined | Genome-fragment | NA | NA | 0 |
| S17-final-viral-combined_k141_84337\|\|full | 5171 | No | NA | 5 | 0 | 0 | Low-quality | Genome-fragment | 11.65 | AAI-based (medium-confidence) | 0 |
| S4-final-viral-combined_k141_121580\|\|full | 2637 | No | NA | 4 | 0 | 3 | Not-determined | Genome-fragment | NA | NA | 0 |
| S4-final-viral-combined_k141_114930\|\|full | 13297 | No | NA | 9 | 0 | 6 | Not-determined | Genome-fragment | NA | NA | 0 |
| S4-final-viral-combined_k141_94319\|\|full | 1909 | No | NA | 2 | 0 | 1 | Not-determined | Genome-fragment | NA | NA | 0 |
| S17-k141_203610 | 5171 | No | NA | 11 | 1 | 0 | Low-quality | Genome-fragment | 10.58 | AAI-based (high-confidence) | 0 |
| S1_k141_305143 | 5172 | No | NA | 8 | 1 | 0 | Low-quality | Genome-fragment | 8.91 | AAI-based (high-confidence) | 0 |
| S4-final-viral-combined_k141_157148\|\|full | 10241 | No | NA | 7 | 0 | 6 | Not-determined | Genome-fragment | NA | NA | 0 |
| S4-final-viral-combined_k141_59218\|\|full | 1903 | No | NA | 2 | 0 | 2 | Not-determined | Genome-fragment | NA | NA | 0 |
| S4-final-viral-combined_k141_31782\|\|full | 1267 | No | NA | 3 | 0 | 1 | Not-determined | Genome-fragment | NA | NA | 0 |
| S8_k141_46023 | 5172 | No | NA | 10 | 0 | 0 | Low-quality | Genome-fragment | 6.54 | AAI-based (medium-confidence) | 0 |
| S4-final-viral-combined_k141_87349\|\|full | 1914 | No | NA | 3 | 0 | 1 | Not-determined | Genome-fragment | NA | NA | 0 |
| S2-final-viral-combined_k141_4052\|\|full | 5174 | No | NA | 9 | 4 | 0 | Low-quality | Genome-fragment | 10.98 | AAI-based (high-confidence) | 0 |
| S7-final-viral-combined_k141_75383\|\|full | 5175 | Yes | 3190 | 6 | 1 | 1 | Low-quality | Genome-fragment | 4.98 | AAI-based (medium-confidence) | 38.36 |
| S4-final-viral-combined_k141_150034\|\|full | 1214 | No | NA | 3 | 0 | 2 | Not-determined | Genome-fragment | NA | NA | 0 |
| S4-final-viral-combined_k141_87371\|\|full | 1888 | No | NA | 3 | 0 | 0 | Not-determined | Genome-fragment | NA | NA | 0 |
| S2-final-viral-combined_k141_228435\|\|full | 5176 | No | NA | 4 | 0 | 2 | Low-quality | Genome-fragment | 0.73 | AAI-based (medium-confidence) | 0 |
| S4-final-viral-combined_k141_128952\|\|full | 2979 | No | NA | 3 | 0 | 3 | Not-determined | Genome-fragment | NA | NA | 0 |
| S12_k141_74423 | 5177 | Yes | 378 | 2 | 1 | 1 | Low-quality | Genome-fragment | 0.65 | HMM-based (lower-bound) | 92.7 |
| S4-final-viral-combined_k141_136140\|\|full | 3714 | No | NA | 4 | 0 | 1 | Not-determined | Genome-fragment | NA | NA | 0 |
| S4-final-viral-combined_k141_10758\|\|full | 83535 | No | NA | 76 | 13 | 2 | Medium-quality | Genome-fragment | 55.35 | AAI-based (high-confidence) | 0 |
| S4-final-viral-combined_k141_45697\|\|full | 2932 | No | NA | 4 | 0 | 3 | Not-determined | Genome-fragment | NA | NA | 0 |
| S4-final-viral-combined_k141_51805\|\|full | 3613 | No | NA | 2 | 0 | 2 | Not-determined | Genome-fragment | NA | NA | 0 |
| S4-final-viral-combined_k141_94428\|\|full | 4334 | No | NA | 5 | 0 | 2 | Not-determined | Genome-fragment | NA | NA | 0 |
| S4-final-viral-combined_k141_129003\|\|full | 3563 | No | NA | 5 | 0 | 4 | Not-determined | Genome-fragment | NA | NA | 0 |
| S4-final-viral-combined_k141_51832\|\|full | 1202 | No | NA | 3 | 0 | 1 | Not-determined | Genome-fragment | NA | NA | 0 |
| S4-final-viral-combined_k141_51843\|\|full | 1901 | No | NA | 2 | 0 | 0 | Not-determined | Genome-fragment | NA | NA | 0 |
| S4-final-viral-combined_k141_38394\|\|full | 2158 | No | NA | 3 | 0 | 0 | Not-determined | Genome-fragment | NA | NA | 0 |
| S6-final-viral-combined_k141_47490\|\|full | 5177 | No | NA | 7 | 0 | 1 | Low-quality | Genome-fragment | 3.33 | AAI-based (medium-confidence) | 0 |
| S4-final-viral-combined_k141_45757\|\|full | 4138 | No | NA | 4 | 0 | 0 | Not-determined | Genome-fragment | NA | NA | 0 |
| S12_k141_283848 | 5179 | No | NA | 5 | 4 | 0 | Low-quality | Genome-fragment | 12.86 | AAI-based (medium-confidence) | 0 |
| S2-final-viral-combined_k141_83017\|\|full | 5183 | No | NA | 4 | 1 | 0 | Low-quality | Genome-fragment | 5.7 | HMM-based (lower-bound) | 0 |
| S1-final-viral-combined_k141_237518\|\|full | 5184 | No | NA | 7 | 1 | 0 | Low-quality | Genome-fragment | 13.41 | AAI-based (medium-confidence) | 0 |
| S4-final-viral-combined_k141_51888\|\|full | 3041 | No | NA | 4 | 0 | 1 | Not-determined | Genome-fragment | NA | NA | 0 |
| S4-final-viral-combined_k141_136195\|\|full | 6111 | No | NA | 7 | 0 | 7 | Not-determined | Genome-fragment | NA | NA | 0 |
| S4-final-viral-combined_k141_17284\|\|full | 73022 | Yes | 36004 | 85 | 19 | 17 | Complete | High-quality | 100 | Provirus (medium-confidence) | 50.69 |
| S4-final-viral-combined_k141_150097\|\|full | 3692 | No | NA | 5 | 0 | 4 | Not-determined | Genome-fragment | NA | NA | 0 |
| S2-final-viral-combined_k141_53322\|\|full | 5185 | No | NA | 6 | 0 | 0 | Low-quality | Genome-fragment | 11.74 | AAI-based (high-confidence) | 0 |
| S4-final-viral-combined_k141_163860\|\|full | 3130 | No | NA | 5 | 0 | 3 | Not-determined | Genome-fragment | NA | NA | 0 |
| S4_k141_644 | 5188 | Yes | 1985 | 5 | 1 | 3 | Low-quality | Genome-fragment | 1.28 | HMM-based (lower-bound) | 61.74 |
| S4-final-viral-combined_k141_51904\|\|full | 3235 | No | NA | 3 | 0 | 0 | Not-determined | Genome-fragment | NA | NA | 0 |
| S17-final-viral-combined_k141_84605\|\|full | 5192 | No | NA | 5 | 0 | 2 | Low-quality | Genome-fragment | 1.74 | AAI-based (medium-confidence) | 0 |
| S4-final-viral-combined_k141_108577\|\|full | 2558 | No | NA | 2 | 0 | 1 | Not-determined | Genome-fragment | NA | NA | 0 |
| S4-final-viral-combined_k141_80780\|\|full | 2489 | No | NA | 4 | 0 | 1 | Not-determined | Genome-fragment | NA | NA | 0 |
| S4-final-viral-combined_k141_24659\|\|full | 9268 | No | NA | 11 | 1 | 0 | Medium-quality | Genome-fragment | 55.68 | AAI-based (high-confidence) | 0 |
| S4-final-viral-combined_k141_31922\|\|full | 1207 | No | NA | 2 | 0 | 2 | Not-determined | Genome-fragment | NA | NA | 0 |
| S12-final-viral-combined_k141_398449\|\|full | 5192 | No | NA | 6 | 1 | 0 | Low-quality | Genome-fragment | 13.95 | AAI-based (medium-confidence) | 0 |
| S2_k141_52976 | 5192 | No | NA | 8 | 0 | 1 | Low-quality | Genome-fragment | 13.82 | AAI-based (high-confidence) | 0 |
| S4-final-viral-combined_k141_87470\|\|full | 1161 | No | NA | 2 | 0 | 2 | Not-determined | Genome-fragment | NA | NA | 0 |
| S3-final-viral-combined_k141_134750\|\|full | 5194 | Yes | 4044 | 8 | 1 | 1 | Low-quality | Genome-fragment | 2.58 | HMM-based (lower-bound) | 22.14 |
| S4-final-viral-combined_k141_101181\|\|full | 3502 | No | NA | 3 | 0 | 1 | Not-determined | Genome-fragment | NA | NA | 0 |
| S11-final-viral-combined_k141_94158\|\|full | 5196 | Yes | 3687 | 8 | 1 | 1 | Low-quality | Genome-fragment | 5.94 | AAI-based (high-confidence) | 29.04 |
| S4-final-viral-combined_k141_31946\|\|full | 3432 | No | NA | 5 | 0 | 3 | Not-determined | Genome-fragment | NA | NA | 0 |
| S4-final-viral-combined_k141_143474\|\|full | 8021 | No | NA | 8 | 0 | 7 | Not-determined | Genome-fragment | NA | NA | 0 |
| S11-final-viral-combined_k141_130456\|\|full | 5199 | No | NA | 8 | 1 | 0 | Low-quality | Genome-fragment | 9.42 | HMM-based (lower-bound) | 0 |
| S12-final-viral-combined_k141_206757\|\|full | 5199 | No | NA | 9 | 1 | 0 | Low-quality | Genome-fragment | 7.4 | HMM-based (lower-bound) | 0 |
| S4-final-viral-combined_k141_3980\|\|full | 2777 | No | NA | 2 | 0 | 2 | Not-determined | Genome-fragment | NA | NA | 0 |
| S4-final-viral-combined_k141_59429\|\|full | 1831 | No | NA | 3 | 0 | 3 | Not-determined | Genome-fragment | NA | NA | 0 |
| S1-final-viral-combined_k141_123338\|\|full | 5202 | No | NA | 6 | 0 | 4 | Low-quality | Genome-fragment | 8.92 | AAI-based (high-confidence) | 0 |
| S4-final-viral-combined_k141_66718\|\|full | 4142 | No | NA | 7 | 0 | 0 | Not-determined | Genome-fragment | NA | NA | 0 |
| S12-final-viral-combined_k141_349489\|\|full | 5202 | No | NA | 7 | 0 | 2 | Low-quality | Genome-fragment | 3.2 | AAI-based (medium-confidence) | 0 |
| S12-final-viral-combined_k141_312915\|\|full | 5203 | No | NA | 4 | 1 | 0 | Low-quality | Genome-fragment | 12.47 | AAI-based (medium-confidence) | 0 |
| S4-final-viral-combined_k141_51995\|\|full | 4244 | No | NA | 5 | 0 | 5 | Not-determined | Genome-fragment | NA | NA | 0 |
| S17-k141_13418 | 5206 | No | NA | 3 | 1 | 0 | Low-quality | Genome-fragment | 8.63 | AAI-based (medium-confidence) | 0 |
| S12-final-viral-combined_k141_121853\|\|full | 5209 | No | NA | 4 | 0 | 2 | Low-quality | Genome-fragment | 2.89 | AAI-based (medium-confidence) | 0 |
| S4-final-viral-combined_k141_38581\|\|full | 1195 | No | NA | 2 | 0 | 1 | Not-determined | Genome-fragment | NA | NA | 0 |
| S4-final-viral-combined_k141_73831\|\|full | 1719 | No | NA | 2 | 0 | 1 | Not-determined | Genome-fragment | NA | NA | 0 |
| S4-final-viral-combined_k141_143554\|\|full | 5339 | No | NA | 4 | 0 | 2 | Not-determined | Genome-fragment | NA | NA | 0 |
| S12_k141_269802 | 5210 | No | NA | 7 | 1 | 0 | Low-quality | Genome-fragment | 5.99 | HMM-based (lower-bound) | 0 |
| S4-final-viral-combined_k141_143555\|\|full | 2796 | No | NA | 3 | 0 | 0 | Not-determined | Genome-fragment | NA | NA | 0 |
| S4-final-viral-combined_k141_129236\|\|full | 2402 | No | NA | 4 | 0 | 0 | Not-determined | Genome-fragment | NA | NA | 0 |
| S4-final-viral-combined_k141_52002\|\|full | 2778 | No | NA | 2 | 0 | 2 | Not-determined | Genome-fragment | NA | NA | 0 |
| S4-final-viral-combined_k141_10967\|\|full | 2402 | No | NA | 4 | 0 | 2 | Not-determined | Genome-fragment | NA | NA | 0 |
| S4-final-viral-combined_k141_129251\|\|full | 4971 | No | NA | 5 | 0 | 3 | Not-determined | Genome-fragment | NA | NA | 0 |
| S4-final-viral-combined_k141_52032\|\|full | 2609 | No | NA | 4 | 0 | 2 | Not-determined | Genome-fragment | NA | NA | 0 |
| S4-final-viral-combined_k141_121821\|\|full | 3204 | No | NA | 3 | 0 | 2 | Not-determined | Genome-fragment | NA | NA | 0 |
| S8-final-viral-combined_k141_188886\|\|full | 5212 | No | NA | 5 | 1 | 0 | Low-quality | Genome-fragment | 12.18 | AAI-based (high-confidence) | 0 |
| S1-final-viral-combined_k141_370912\|\|full | 5213 | No | NA | 6 | 0 | 1 | Low-quality | Genome-fragment | 3.69 | AAI-based (high-confidence) | 0 |
| S4-final-viral-combined_k141_81006\|\|full | 1376 | No | NA | 4 | 0 | 1 | Not-determined | Genome-fragment | NA | NA | 0 |
| S2-final-viral-combined_k141_137489\|\|full | 5213 | No | NA | 4 | 0 | 0 | Low-quality | Genome-fragment | 2.66 | AAI-based (medium-confidence) | 0 |
| S4-final-viral-combined_k141_115277\|\|full | 8060 | No | NA | 8 | 0 | 0 | Not-determined | Genome-fragment | NA | NA | 0 |
| S4-final-viral-combined_k141_24858\|\|full | 1269 | No | NA | 2 | 0 | 1 | Not-determined | Genome-fragment | NA | NA | 0 |
| S1_k141_282827 | 5214 | No | NA | 4 | 0 | 4 | Low-quality | Genome-fragment | 0.88 | AAI-based (medium-confidence) | 0 |
| S4-final-viral-combined_k141_52055\|\|full | 4861 | No | NA | 6 | 0 | 1 | Not-determined | Genome-fragment | NA | NA | 0 |
| S3_k141_78767 | 5215 | No | NA | 11 | 1 | 1 | Low-quality | Genome-fragment | 13.43 | AAI-based (high-confidence) | 0 |
| S7_k141_157175 | 5219 | No | NA | 4 | 0 | 1 | Low-quality | Genome-fragment | 3.12 | AAI-based (medium-confidence) | 0 |
| S2_k141_29550 | 5219 | No | NA | 8 | 5 | 0 | Low-quality | Genome-fragment | 10.84 | HMM-based (lower-bound) | 0 |
| S4-final-viral-combined_k141_164127\|\|full | 1323 | No | NA | 4 | 0 | 0 | Not-determined | Genome-fragment | NA | NA | 0 |
| S6_k141_74360 | 5220 | No | NA | 5 | 0 | 2 | Low-quality | Genome-fragment | 12.42 | AAI-based (high-confidence) | 0 |
| S7_k141_192546 | 5221 | No | NA | 5 | 3 | 0 | Low-quality | Genome-fragment | 10.24 | AAI-based (medium-confidence) | 0 |
| S2-final-viral-combined_k141_174800\|\|full | 5221 | No | NA | 10 | 7 | 0 | Low-quality | Genome-fragment | 12.02 | AAI-based (high-confidence) | 0 |
| S4-final-viral-combined_k141_164140\|\|full | 1523 | No | NA | 2 | 0 | 0 | Not-determined | Genome-fragment | NA | NA | 0 |
| S4-final-viral-combined_k141_38658\|\|full | 4701 | No | NA | 5 | 0 | 5 | Not-determined | Genome-fragment | NA | NA | 0 |
| S2_k141_235375 | 5221 | No | NA | 12 | 3 | 0 | Low-quality | Genome-fragment | 34.82 | AAI-based (high-confidence) | 0 |
| S5-final-viral-combined_k141_161149\|\|full | 5224 | No | NA | 10 | 3 | 0 | Low-quality | Genome-fragment | 9.54 | HMM-based (lower-bound) | 0 |
| S4-final-viral-combined_k141_59567\|\|full | 1160 | No | NA | 2 | 0 | 2 | Not-determined | Genome-fragment | NA | NA | 0 |
| S4-final-viral-combined_k141_66929\|\|full | 1737 | No | NA | 3 | 0 | 0 | Not-determined | Genome-fragment | NA | NA | 0 |
| S4-final-viral-combined_k141_24970\|\|full | 1415 | No | NA | 3 | 0 | 2 | Not-determined | Genome-fragment | NA | NA | 0 |
| S9_k141_48364 | 5225 | No | NA | 6 | 2 | 0 | Low-quality | Genome-fragment | 9.09 | AAI-based (medium-confidence) | 0 |
| S4-final-viral-combined_k141_87641\|\|full | 3251 | No | NA | 3 | 0 | 1 | Not-determined | Genome-fragment | NA | NA | 0 |
| S4-final-viral-combined_k141_143683\|\|full | 2303 | No | NA | 5 | 0 | 2 | Not-determined | Genome-fragment | NA | NA | 0 |
| S4-final-viral-combined_k141_4171\|\|full | 4500 | No | NA | 5 | 0 | 4 | Not-determined | Genome-fragment | NA | NA | 0 |
| S4-final-viral-combined_k141_101485\|\|full | 1521 | No | NA | 4 | 0 | 1 | Not-determined | Genome-fragment | NA | NA | 0 |
| S4-final-viral-combined_k141_66954\|\|full | 10155 | No | NA | 5 | 0 | 5 | Not-determined | Genome-fragment | NA | NA | 0 |
| S4-final-viral-combined_k141_150396\|\|full | 1797 | No | NA | 3 | 0 | 2 | Not-determined | Genome-fragment | NA | NA | 0 |
| S11_k141_8585 | 5227 | No | NA | 10 | 8 | 0 | Low-quality | Genome-fragment | 13.36 | AAI-based (high-confidence) | 0 |
| S4-final-viral-combined_k141_59628\|\|full | 2353 | No | NA | 3 | 0 | 0 | Not-determined | Genome-fragment | NA | NA | 0 |
| S4-final-viral-combined_k141_11241\|\|full | 1125 | No | NA | 3 | 0 | 1 | Not-determined | Genome-fragment | NA | NA | 0 |
| S4-final-viral-combined_k141_32175\|\|full | 2498 | No | NA | 3 | 0 | 2 | Not-determined | Genome-fragment | NA | NA | 0 |
| S4-final-viral-combined_k141_157451\|\|full | 10852 | No | NA | 19 | 0 | 1 | Not-determined | Genome-fragment | NA | NA | 0 |
| S6-final-viral-combined_k141_102876\|\|full | 5227 | No | NA | 5 | 0 | 2 | Low-quality | Genome-fragment | 1.65 | AAI-based (medium-confidence) | 0 |
| S4-final-viral-combined_k141_25049\|\|full | 1801 | No | NA | 3 | 0 | 1 | Not-determined | Genome-fragment | NA | NA | 0 |
| S4-final-viral-combined_k141_81187\|\|full | 1394 | No | NA | 3 | 0 | 1 | Not-determined | Genome-fragment | NA | NA | 0 |
| S4-final-viral-combined_k141_46192\|\|full | 1423 | No | NA | 3 | 0 | 2 | Not-determined | Genome-fragment | NA | NA | 0 |
| S2-final-viral-combined_k141_24636\|\|full | 5230 | No | NA | 8 | 0 | 1 | Low-quality | Genome-fragment | 5.57 | AAI-based (medium-confidence) | 0 |
| S4-final-viral-combined_k141_38825\|\|full | 6821 | No | NA | 6 | 0 | 6 | Not-determined | Genome-fragment | NA | NA | 0 |
| S17-final-viral-combined_k141_38250\|\|full | 5232 | No | NA | 7 | 2 | 0 | Low-quality | Genome-fragment | 7.52 | AAI-based (high-confidence) | 0 |
| S4-final-viral-combined_k141_59693\|\|full | 3212 | No | NA | 4 | 0 | 3 | Not-determined | Genome-fragment | NA | NA | 0 |
| S4-final-viral-combined_k141_116863\|\|full | 5234 | No | NA | 4 | 0 | 0 | Low-quality | Genome-fragment | 1.7 | AAI-based (medium-confidence) | 0 |
| S4-final-viral-combined_k141_94878\|\|full | 2755 | No | NA | 3 | 0 | 2 | Not-determined | Genome-fragment | NA | NA | 0 |
| S1_k141_159141 | 5235 | Yes | 3956 | 8 | 1 | 1 | Low-quality | Genome-fragment | 5.68 | HMM-based (lower-bound) | 24.43 |
| S4-final-viral-combined_k141_101570\|\|full | 2011 | No | NA | 3 | 0 | 0 | Not-determined | Genome-fragment | NA | NA | 0 |
| S4-final-viral-combined_k141_136592\|\|full | 2044 | No | NA | 3 | 0 | 0 | Not-determined | Genome-fragment | NA | NA | 0 |
| S7_k141_11341 | 5235 | No | NA | 4 | 0 | 3 | Low-quality | Genome-fragment | 6.34 | AAI-based (medium-confidence) | 0 |
| S4-final-viral-combined_k141_121978\|\|full | 129425 | Yes | 45051 | 137 | 24 | 53 | Complete | High-quality | 100 | Provirus (high-confidence) | 65.19 |
| S9_k141_44463 | 5237 | No | NA | 2 | 0 | 2 | Low-quality | Genome-fragment | 3.41 | AAI-based (high-confidence) | 0 |
| S4-final-viral-combined_k141_4291\|\|full | 1021 | No | NA | 2 | 0 | 0 | Not-determined | Genome-fragment | NA | NA | 0 |
| S4-final-viral-combined_k141_32288\|\|full | 2495 | No | NA | 3 | 0 | 2 | Not-determined | Genome-fragment | NA | NA | 0 |
| S11-final-viral-combined_k141_80893\|\|full | 5240 | Yes | 4389 | 4 | 2 | 1 | Low-quality | Genome-fragment | 6.36 | AAI-based (high-confidence) | 16.24 |
| S12_k141_191313 | 5241 | No | NA | 1 | 0 | 0 | Low-quality | Genome-fragment | 5.61 | AAI-based (medium-confidence) | 0 |
| S4-final-viral-combined_k141_101599\|\|full | 1739 | No | NA | 3 | 0 | 3 | Not-determined | Genome-fragment | NA | NA | 0 |
| S2-final-viral-combined_k141_187928\|\|full | 5242 | No | NA | 5 | 0 | 0 | Low-quality | Genome-fragment | 9.03 | AAI-based (medium-confidence) | 0 |
| S2-final-viral-combined_k141_98947\|\|full | 5242 | No | NA | 10 | 0 | 0 | Low-quality | Genome-fragment | 3.07 | AAI-based (medium-confidence) | 0 |
| S4-final-viral-combined_k141_164405\|\|full | 1222 | No | NA | 2 | 0 | 1 | Not-determined | Genome-fragment | NA | NA | 0 |
| S17-final-viral-combined_k141_68583\|\|full | 5243 | No | NA | 8 | 0 | 0 | Low-quality | Genome-fragment | 12.12 | AAI-based (medium-confidence) | 0 |
| S4-final-viral-combined_k141_143826\|\|full | 2813 | No | NA | 3 | 0 | 2 | Not-determined | Genome-fragment | NA | NA | 0 |
| S4-final-viral-combined_k141_32302\|\|full | 3220 | No | NA | 4 | 0 | 2 | Not-determined | Genome-fragment | NA | NA | 0 |
| S4-final-viral-combined_k141_121996\|\|full | 2818 | No | NA | 3 | 0 | 1 | Not-determined | Genome-fragment | NA | NA | 0 |
| S4-final-viral-combined_k141_25139\|\|full | 1608 | No | NA | 2 | 0 | 0 | Not-determined | Genome-fragment | NA | NA | 0 |
| S11-final-viral-combined_k141_180522\|\|full | 5244 | No | NA | 7 | 3 | 0 | Low-quality | Genome-fragment | 13.48 | AAI-based (high-confidence) | 0 |
| S4-final-viral-combined_k141_101622\|\|full | 1083 | No | NA | 3 | 0 | 2 | Not-determined | Genome-fragment | NA | NA | 0 |
| S2-final-viral-combined_k141_20663\|\|full | 5244 | No | NA | 7 | 0 | 3 | Low-quality | Genome-fragment | 3.59 | AAI-based (medium-confidence) | 0 |
| S2-final-viral-combined_k141_82703\|\|full | 5244 | No | NA | 7 | 0 | 1 | Low-quality | Genome-fragment | 9.51 | AAI-based (medium-confidence) | 0 |
| S2_k141_185531 | 5244 | No | NA | 8 | 0 | 1 | Low-quality | Genome-fragment | 10.88 | AAI-based (high-confidence) | 0 |
| S7_k141_51856 | 5247 | Yes | 3914 | 4 | 1 | 2 | Low-quality | Genome-fragment | 6.3 | AAI-based (high-confidence) | 25.4 |
| S4-final-viral-combined_k141_115463\|\|full | 1595 | No | NA | 2 | 0 | 1 | Not-determined | Genome-fragment | NA | NA | 0 |
| S12_k141_169861 | 5247 | No | NA | 3 | 0 | 0 | Low-quality | Genome-fragment | 2.87 | AAI-based (medium-confidence) | 0 |
| S6-final-viral-combined_k141_100776\|\|full | 5247 | No | NA | 3 | 0 | 0 | Low-quality | Genome-fragment | 6.52 | AAI-based (medium-confidence) | 0 |
| S4-final-viral-combined_k141_1562\|\|full | 5249 | No | NA | 8 | 1 | 0 | Low-quality | Genome-fragment | 3.53 | HMM-based (lower-bound) | 0 |
| S4-final-viral-combined_k141_11347\|\|full | 1888 | No | NA | 2 | 0 | 2 | Not-determined | Genome-fragment | NA | NA | 0 |
| S4-final-viral-combined_k141_59787\|\|full | 2533 | No | NA | 4 | 0 | 0 | Not-determined | Genome-fragment | NA | NA | 0 |
| S4-final-viral-combined_k141_38945\|\|full | 1661 | No | NA | 3 | 0 | 0 | Not-determined | Genome-fragment | NA | NA | 0 |
| S6-final-viral-combined_k141_91243\|\|full | 5255 | No | NA | 3 | 0 | 0 | Low-quality | Genome-fragment | 5.45 | AAI-based (medium-confidence) | 0 |
| S11_k141_323586 | 5256 | No | NA | 5 | 0 | 3 | Low-quality | Genome-fragment | 8.67 | AAI-based (medium-confidence) | 0 |
| S17-final-viral-combined_k141_212437\|\|full | 5256 | No | NA | 9 | 1 | 0 | Low-quality | Genome-fragment | 12.51 | AAI-based (medium-confidence) | 0 |
| S4_k141_40441 | 5259 | No | NA | 10 | 3 | 0 | Low-quality | Genome-fragment | 7.55 | AAI-based (high-confidence) | 0 |
| S17-final-viral-combined_k141_70561\|\|full | 5262 | No | NA | 5 | 0 | 4 | Low-quality | Genome-fragment | 4.3 | AAI-based (medium-confidence) | 0 |
| S4-final-viral-combined_k141_109173\|\|full | 3523 | No | NA | 5 | 0 | 4 | Not-determined | Genome-fragment | NA | NA | 0 |
| S4-final-viral-combined_k141_122099\|\|full | 4015 | No | NA | 4 | 0 | 3 | Not-determined | Genome-fragment | NA | NA | 0 |
| S4-final-viral-combined_k141_4344\|\|full | 1684 | No | NA | 2 | 0 | 1 | Not-determined | Genome-fragment | NA | NA | 0 |
| S1_k141_115603 | 5264 | No | NA | 8 | 4 | 0 | Low-quality | Genome-fragment | 9.38 | AAI-based (high-confidence) | 0 |
| S4-final-viral-combined_k141_67185\|\|full | 1335 | No | NA | 2 | 0 | 0 | Not-determined | Genome-fragment | NA | NA | 0 |
| S4-final-viral-combined_k141_136700\|\|full | 1172 | No | NA | 2 | 0 | 0 | Not-determined | Genome-fragment | NA | NA | 0 |
| S4-final-viral-combined_k141_38995\|\|full | 4258 | No | NA | 5 | 0 | 2 | Not-determined | Genome-fragment | NA | NA | 0 |
| S2_k141_68048 | 5264 | No | NA | 4 | 0 | 4 | Low-quality | Genome-fragment | 4.24 | AAI-based (high-confidence) | 0 |
| S4-final-viral-combined_k141_67197\|\|full | 3177 | No | NA | 3 | 0 | 1 | Not-determined | Genome-fragment | NA | NA | 0 |
| S4-final-viral-combined_k141_122112\|\|full | 3797 | No | NA | 2 | 0 | 0 | Not-determined | Genome-fragment | NA | NA | 0 |
| S6_k141_39916 | 5264 | No | NA | 8 | 2 | 0 | Low-quality | Genome-fragment | 5.26 | AAI-based (high-confidence) | 0 |
| S4-final-viral-combined_k141_25201\|\|full | 44355 | Yes | 26360 | 57 | 17 | 11 | Medium-quality | Genome-fragment | 76.6 | AAI-based (high-confidence) | 40.57 |
| S8_k141_136692 | 5264 | No | NA | 9 | 3 | 0 | Low-quality | Genome-fragment | 3.96 | AAI-based (high-confidence) | 0 |
| S4-final-viral-combined_k141_17608\|\|full | 1615 | No | NA | 2 | 0 | 1 | Not-determined | Genome-fragment | NA | NA | 0 |
| S4-final-viral-combined_k141_52508\|\|full | 3005 | No | NA | 7 | 0 | 1 | Not-determined | Genome-fragment | NA | NA | 0 |
| S4-final-viral-combined_k141_150632\|\|full | 2228 | No | NA | 3 | 0 | 0 | Not-determined | Genome-fragment | NA | NA | 0 |
| S4-final-viral-combined_k141_129664\|\|full | 5036 | No | NA | 4 | 0 | 0 | Not-determined | Genome-fragment | NA | NA | 0 |
| S4-final-viral-combined_k141_164551\|\|full | 4063 | No | NA | 4 | 0 | 1 | Not-determined | Genome-fragment | NA | NA | 0 |
| S4-final-viral-combined_k141_25242\|\|full | 6693 | No | NA | 5 | 0 | 4 | Not-determined | Genome-fragment | NA | NA | 0 |
| S4-final-viral-combined_k141_32458\|\|full | 2924 | No | NA | 3 | 0 | 3 | Not-determined | Genome-fragment | NA | NA | 0 |
| S4-final-viral-combined_k141_95060\|\|full | 1252 | No | NA | 3 | 0 | 1 | Not-determined | Genome-fragment | NA | NA | 0 |
| S11_k141_219485 | 5265 | No | NA | 5 | 1 | 0 | Low-quality | Genome-fragment | 7.64 | AAI-based (high-confidence) | 0 |
| S4-final-viral-combined_k141_95067\|\|full | 5119 | No | NA | 3 | 0 | 3 | Not-determined | Genome-fragment | NA | NA | 0 |
| S6-final-viral-combined_k141_43386\|\|full | 5265 | No | NA | 8 | 5 | 0 | Low-quality | Genome-fragment | 7.13 | AAI-based (high-confidence) | 0 |
| S4-final-viral-combined_k141_95072\|\|full | 3804 | No | NA | 2 | 0 | 2 | Not-determined | Genome-fragment | NA | NA | 0 |
| S11-final-viral-combined_k141_59814\|\|full | 5266 | No | NA | 5 | 5 | 0 | Low-quality | Genome-fragment | 11.65 | AAI-based (high-confidence) | 0 |
| S4-final-viral-combined_k141_25282\|\|full | 1715 | No | NA | 4 | 0 | 0 | Not-determined | Genome-fragment | NA | NA | 0 |
| S1-final-viral-combined_k141_247550\|\|full | 5266 | No | NA | 6 | 1 | 0 | Low-quality | Genome-fragment | 8.58 | AAI-based (medium-confidence) | 0 |
| S4-final-viral-combined_k141_157811\|\|full | 1554 | No | NA | 3 | 0 | 0 | Not-determined | Genome-fragment | NA | NA | 0 |
| S4-final-viral-combined_k141_122206\|\|full | 14436 | No | NA | 17 | 0 | 4 | Not-determined | Genome-fragment | NA | NA | 0 |
| S12_k141_127403 | 5269 | Yes | 1884 | 10 | 1 | 2 | Low-quality | Genome-fragment | 3.27 | AAI-based (high-confidence) | 64.24 |
| S4-final-viral-combined_k141_81516\|\|full | 1888 | No | NA | 3 | 0 | 0 | Not-determined | Genome-fragment | NA | NA | 0 |
| S4-final-viral-combined_k141_74383\|\|full | 1669 | No | NA | 3 | 0 | 1 | Not-determined | Genome-fragment | NA | NA | 0 |
| S4-final-viral-combined_k141_32497\|\|full | 1730 | No | NA | 3 | 0 | 2 | Not-determined | Genome-fragment | NA | NA | 0 |
| S4-final-viral-combined_k141_52587\|\|full | 4254 | No | NA | 5 | 0 | 2 | Not-determined | Genome-fragment | NA | NA | 0 |
| S17-final-viral-combined_k141_20178\|\|full | 5270 | No | NA | 10 | 1 | 0 | Low-quality | Genome-fragment | 15.23 | AAI-based (high-confidence) | 0 |
| S4-final-viral-combined_k141_87997\|\|full | 1814 | No | NA | 3 | 0 | 0 | Not-determined | Genome-fragment | NA | NA | 0 |
| S4-final-viral-combined_k141_46556\|\|full | 4125 | No | NA | 3 | 0 | 1 | Not-determined | Genome-fragment | NA | NA | 0 |
| S4-final-viral-combined_k141_81533\|\|full | 2873 | No | NA | 3 | 0 | 3 | Not-determined | Genome-fragment | NA | NA | 0 |
| S4-final-viral-combined_k141_115646\|\|full | 2029 | No | NA | 3 | 0 | 0 | Not-determined | Genome-fragment | NA | NA | 0 |
| S4_k141_104765 | 5270 | No | NA | 7 | 0 | 5 | Low-quality | Genome-fragment | 2.35 | AAI-based (medium-confidence) | 0 |
| S4-final-viral-combined_k141_59948\|\|full | 35193 | No | NA | 26 | 0 | 18 | Not-determined | Genome-fragment | NA | NA | 0 |
| S12-final-viral-combined_k141_169455\|\|full | 5271 | No | NA | 8 | 4 | 0 | Low-quality | Genome-fragment | 11.15 | AAI-based (high-confidence) | 0 |
| S4-final-viral-combined_k141_39139\|\|full | 2123 | No | NA | 2 | 0 | 1 | Not-determined | Genome-fragment | NA | NA | 0 |
| S4-final-viral-combined_k141_164636\|\|full | 2745 | No | NA | 3 | 0 | 1 | Not-determined | Genome-fragment | NA | NA | 0 |
| S4-final-viral-combined_k141_115676\|\|full | 1188 | No | NA | 2 | 0 | 1 | Not-determined | Genome-fragment | NA | NA | 0 |
| S4-final-viral-combined_k141_32549\|\|full | 2728 | No | NA | 2 | 0 | 0 | Not-determined | Genome-fragment | NA | NA | 0 |
| S4-final-viral-combined_k141_52635\|\|full | 5000 | No | NA | 6 | 0 | 4 | Not-determined | Genome-fragment | NA | NA | 0 |
| S2_k141_20375 | 5273 | No | NA | 5 | 4 | 0 | Low-quality | Genome-fragment | 3.68 | AAI-based (high-confidence) | 0 |
| S4-final-viral-combined_k141_59972\|\|full | 2540 | No | NA | 2 | 0 | 1 | Not-determined | Genome-fragment | NA | NA | 0 |
| S4-final-viral-combined_k141_25356\|\|full | 1397 | No | NA | 2 | 0 | 0 | Not-determined | Genome-fragment | NA | NA | 0 |
| S11_k141_34666 | 5274 | No | NA | 10 | 0 | 0 | Low-quality | Genome-fragment | 11.51 | AAI-based (high-confidence) | 0 |
| S1-final-viral-combined_k141_211508\|\|full | 5274 | No | NA | 3 | 0 | 2 | Low-quality | Genome-fragment | 1.74 | AAI-based (medium-confidence) | 0 |
| S4-final-viral-combined_k141_129730\|\|full | 2795 | No | NA | 5 | 0 | 0 | Not-determined | Genome-fragment | NA | NA | 0 |
| S4-final-viral-combined_k141_39175\|\|full | 1142 | No | NA | 2 | 0 | 1 | Not-determined | Genome-fragment | NA | NA | 0 |
| S4-final-viral-combined_k141_115717\|\|full | 1161 | No | NA | 3 | 0 | 0 | Not-determined | Genome-fragment | NA | NA | 0 |
| S7-final-viral-combined_k141_187919\|\|full | 5274 | No | NA | 7 | 0 | 0 | Low-quality | Genome-fragment | 12.3 | AAI-based (medium-confidence) | 0 |
| S12-final-viral-combined_k141_386572\|\|full | 5274 | No | NA | 5 | 0 | 4 | Low-quality | Genome-fragment | 12.19 | AAI-based (medium-confidence) | 0 |
| S1-final-viral-combined_k141_367301\|\|full | 5275 | Yes | 4308 | 9 | 1 | 1 | Low-quality | Genome-fragment | 3.3 | HMM-based (lower-bound) | 18.33 |
| S4-final-viral-combined_k141_17790\|\|full | 12637 | No | NA | 10 | 0 | 7 | Not-determined | Genome-fragment | NA | NA | 0 |
| S4-final-viral-combined_k141_109403\|\|full | 1544 | No | NA | 3 | 0 | 0 | Not-determined | Genome-fragment | NA | NA | 0 |
| S4-final-viral-combined_k141_4491\|\|full | 2214 | No | NA | 3 | 0 | 1 | Not-determined | Genome-fragment | NA | NA | 0 |
| S4-final-viral-combined_k141_144115\|\|full | 3517 | No | NA | 6 | 0 | 3 | Not-determined | Genome-fragment | NA | NA | 0 |
| S12-final-viral-combined_k141_140039\|\|full | 5275 | No | NA | 10 | 2 | 1 | Low-quality | Genome-fragment | 11.22 | AAI-based (medium-confidence) | 0 |
| S4-final-viral-combined_k141_101945\|\|full | 2653 | No | NA | 3 | 0 | 2 | Not-determined | Genome-fragment | NA | NA | 0 |
| S4-final-viral-combined_k141_46634\|\|full | 1165 | No | NA | 3 | 0 | 0 | Not-determined | Genome-fragment | NA | NA | 0 |
| S4-final-viral-combined_k141_129778\|\|full | 4979 | No | NA | 2 | 0 | 2 | Not-determined | Genome-fragment | NA | NA | 0 |
| S11_k141_87185 | 5276 | Yes | 3661 | 7 | 1 | 1 | Low-quality | Genome-fragment | 9.08 | AAI-based (medium-confidence) | 30.61 |
| S4-final-viral-combined_k141_52731\|\|full | 2755 | No | NA | 2 | 0 | 2 | Not-determined | Genome-fragment | NA | NA | 0 |
| S4-final-viral-combined_k141_115766\|\|full | 1332 | No | NA | 3 | 0 | 1 | Not-determined | Genome-fragment | NA | NA | 0 |
| S4-final-viral-combined_k141_39236\|\|full | 2389 | No | NA | 3 | 0 | 0 | Not-determined | Genome-fragment | NA | NA | 0 |
| S4-final-viral-combined_k141_95173\|\|full | 3283 | No | NA | 3 | 0 | 2 | Not-determined | Genome-fragment | NA | NA | 0 |
| S4-final-viral-combined_k141_136943\|\|full | 2758 | No | NA | 3 | 0 | 1 | Not-determined | Genome-fragment | NA | NA | 0 |
| S4-final-viral-combined_k141_115787\|\|full | 2027 | No | NA | 2 | 0 | 1 | Not-determined | Genome-fragment | NA | NA | 0 |
| S4-final-viral-combined_k141_150777\|\|full | 2261 | No | NA | 3 | 0 | 1 | Not-determined | Genome-fragment | NA | NA | 0 |
| S4-final-viral-combined_k141_32628\|\|full | 6351 | No | NA | 6 | 0 | 5 | Not-determined | Genome-fragment | NA | NA | 0 |
| S4-final-viral-combined_k141_88132\|\|full | 2526 | No | NA | 3 | 0 | 1 | Not-determined | Genome-fragment | NA | NA | 0 |
| S8_k141_47877 | 5276 | No | NA | 6 | 0 | 1 | Low-quality | Genome-fragment | 3.84 | AAI-based (high-confidence) | 0 |
| S5-final-viral-combined_k141_207484\|\|full | 5279 | Yes | 3906 | 10 | 1 | 1 | Low-quality | Genome-fragment | 9.71 | AAI-based (medium-confidence) | 26.01 |
| S5-final-viral-combined_k141_156754\|\|full | 5279 | No | NA | 12 | 1 | 0 | Low-quality | Genome-fragment | 10.24 | AAI-based (medium-confidence) | 0 |
| S11-final-viral-combined_k141_302875\|\|full | 5281 | No | NA | 16 | 1 | 0 | Low-quality | Genome-fragment | 8.77 | AAI-based (high-confidence) | 0 |
| S4-final-viral-combined_k141_102007\|\|full | 7038 | No | NA | 11 | 0 | 3 | Not-determined | Genome-fragment | NA | NA | 0 |
| S4-final-viral-combined_k141_115830\|\|full | 2019 | No | NA | 2 | 0 | 1 | Not-determined | Genome-fragment | NA | NA | 0 |
| S7-final-viral-combined_k141_177401\|\|full | 5284 | No | NA | 6 | 3 | 0 | Low-quality | Genome-fragment | 12.04 | AAI-based (medium-confidence) | 0 |
| S4-final-viral-combined_k141_109474\|\|full | 1292 | No | NA | 3 | 0 | 0 | Not-determined | Genome-fragment | NA | NA | 0 |
| S4-final-viral-combined_k141_115839\|\|full | 1713 | No | NA | 4 | 0 | 2 | Not-determined | Genome-fragment | NA | NA | 0 |
| S4-final-viral-combined_k141_164391\|\|full | 5284 | No | NA | 10 | 1 | 0 | Low-quality | Genome-fragment | 5.24 | HMM-based (lower-bound) | 0 |
| S6-final-viral-combined_k141_40727\|\|full | 5284 | Yes | 5030 | 9 | 4 | 1 | Low-quality | Genome-fragment | 8.35 | AAI-based (high-confidence) | 4.81 |
| S4-final-viral-combined_k141_88180\|\|full | 2259 | No | NA | 2 | 0 | 2 | Not-determined | Genome-fragment | NA | NA | 0 |
| S4-final-viral-combined_k141_60148\|\|full | 4800 | No | NA | 5 | 0 | 5 | Not-determined | Genome-fragment | NA | NA | 0 |
| S11-final-viral-combined_k141_10435\|\|full | 5285 | No | NA | 6 | 0 | 1 | Low-quality | Genome-fragment | 8.29 | AAI-based (medium-confidence) | 0 |
| S12-final-viral-combined_k141_351914\|\|full | 5285 | No | NA | 4 | 0 | 3 | Low-quality | Genome-fragment | 2.55 | AAI-based (medium-confidence) | 0 |
| S8-final-viral-combined_k141_38007\|\|full | 5285 | No | NA | 5 | 3 | 0 | Low-quality | Genome-fragment | 6.69 | AAI-based (medium-confidence) | 0 |
| S11-final-viral-combined_k141_45866\|\|full | 5286 | No | NA | 6 | 0 | 3 | Low-quality | Genome-fragment | 8.12 | AAI-based (medium-confidence) | 0 |
| S4-final-viral-combined_k141_158084\|\|full | 2170 | No | NA | 3 | 0 | 0 | Not-determined | Genome-fragment | NA | NA | 0 |
| S4-final-viral-combined_k141_81774\|\|full | 1185 | No | NA | 2 | 0 | 1 | Not-determined | Genome-fragment | NA | NA | 0 |
| S4-final-viral-combined_k141_60160\|\|full | 2219 | No | NA | 4 | 0 | 1 | Not-determined | Genome-fragment | NA | NA | 0 |
| S11-final-viral-combined_k141_87721\|\|full | 5286 | No | NA | 6 | 3 | 0 | Low-quality | Genome-fragment | 11.45 | AAI-based (high-confidence) | 0 |
| S17-final-viral-combined_k141_151748\|\|full | 5286 | No | NA | 5 | 0 | 2 | Low-quality | Genome-fragment | 8.86 | AAI-based (high-confidence) | 0 |
| S4-final-viral-combined_k141_150907\|\|full | 2258 | No | NA | 3 | 0 | 1 | Not-determined | Genome-fragment | NA | NA | 0 |
| S4-final-viral-combined_k141_74630\|\|full | 1848 | No | NA | 5 | 0 | 0 | Not-determined | Genome-fragment | NA | NA | 0 |
| S4-final-viral-combined_k141_88262\|\|full | 2072 | No | NA | 2 | 0 | 2 | Not-determined | Genome-fragment | NA | NA | 0 |
| S12-final-viral-combined_k141_48466\|\|full | 5288 | No | NA | 5 | 0 | 5 | Low-quality | Genome-fragment | 1.56 | AAI-based (medium-confidence) | 0 |
| S4-final-viral-combined_k141_11854\|\|full | 2255 | No | NA | 3 | 0 | 1 | Not-determined | Genome-fragment | NA | NA | 0 |
| S4-final-viral-combined_k141_144304\|\|full | 1073 | No | NA | 2 | 0 | 0 | Not-determined | Genome-fragment | NA | NA | 0 |
| S4-final-viral-combined_k141_102069\|\|full | 2938 | No | NA | 5 | 0 | 2 | Not-determined | Genome-fragment | NA | NA | 0 |
| S9-final-viral-combined_k141_104792\|\|full | 5290 | No | NA | 12 | 3 | 0 | Low-quality | Genome-fragment | 12.47 | AAI-based (high-confidence) | 0 |
| S7-final-viral-combined_k141_217731\|\|full | 5290 | No | NA | 8 | 3 | 0 | Low-quality | Genome-fragment | 14.81 | AAI-based (high-confidence) | 0 |
| S4-final-viral-combined_k141_109638\|\|full | 1977 | No | NA | 3 | 0 | 3 | Not-determined | Genome-fragment | NA | NA | 0 |
| S8-final-viral-combined_k141_157283\|\|full | 5290 | No | NA | 6 | 0 | 5 | Low-quality | Genome-fragment | 4.32 | AAI-based (medium-confidence) | 0 |
| S4-final-viral-combined_k141_81863\|\|full | 2816 | No | NA | 6 | 0 | 1 | Not-determined | Genome-fragment | NA | NA | 0 |
| S11_k141_9258 | 5291 | No | NA | 7 | 0 | 2 | Low-quality | Genome-fragment | 2.75 | AAI-based (medium-confidence) | 0 |
| S4-final-viral-combined_k141_25627\|\|full | 2406 | No | NA | 2 | 0 | 1 | Not-determined | Genome-fragment | NA | NA | 0 |
| S4-final-viral-combined_k141_164909\|\|full | 1958 | No | NA | 2 | 0 | 1 | Not-determined | Genome-fragment | NA | NA | 0 |
| S6_k141_119373 | 5292 | No | NA | 4 | 0 | 4 | Low-quality | Genome-fragment | 6.81 | AAI-based (medium-confidence) | 0 |
| S4-final-viral-combined_k141_116015\|\|full | 1012 | No | NA | 2 | 0 | 0 | Not-determined | Genome-fragment | NA | NA | 0 |
| S4-final-viral-combined_k141_122525\|\|full | 1900 | No | NA | 5 | 0 | 0 | Not-determined | Genome-fragment | NA | NA | 0 |
| S4-final-viral-combined_k141_151007\|\|full | 2204 | No | NA | 3 | 0 | 3 | Not-determined | Genome-fragment | NA | NA | 0 |
| S4-final-viral-combined_k141_144342\|\|full | 9681 | No | NA | 5 | 0 | 5 | Not-determined | Genome-fragment | NA | NA | 0 |
| S4-final-viral-combined_k141_109677\|\|full | 10634 | No | NA | 12 | 0 | 1 | Not-determined | Genome-fragment | NA | NA | 0 |
| S4-final-viral-combined_k141_11895\|\|full | 1238 | No | NA | 3 | 0 | 2 | Not-determined | Genome-fragment | NA | NA | 0 |
| S4-final-viral-combined_k141_46783\|\|full | 4920 | No | NA | 6 | 0 | 3 | Not-determined | Genome-fragment | NA | NA | 0 |
| S7-final-viral-combined_k141_200001\|\|full | 5297 | No | NA | 9 | 2 | 0 | Low-quality | Genome-fragment | 6.01 | AAI-based (high-confidence) | 0 |
| S11-final-viral-combined_k141_295242\|\|full | 5298 | No | NA | 8 | 0 | 0 | Low-quality | Genome-fragment | 11.28 | AAI-based (medium-confidence) | 0 |
| S4-final-viral-combined_k141_158205\|\|full | 1511 | No | NA | 2 | 0 | 2 | Not-determined | Genome-fragment | NA | NA | 0 |
| S4-final-viral-combined_k141_122555\|\|full | 2031 | No | NA | 3 | 0 | 2 | Not-determined | Genome-fragment | NA | NA | 0 |
| S12_k141_394185 | 5298 | No | NA | 7 | 3 | 0 | Low-quality | Genome-fragment | 18.47 | HMM-based (lower-bound) | 0 |
| S2-final-viral-combined_k141_225239\|\|full | 5298 | No | NA | 6 | 0 | 0 | Low-quality | Genome-fragment | 10.42 | AAI-based (medium-confidence) | 0 |
| S4-final-viral-combined_k141_46803\|\|full | 1317 | No | NA | 4 | 0 | 4 | Not-determined | Genome-fragment | NA | NA | 0 |
| S17-final-viral-combined_k141_238722\|\|full | 5299 | No | NA | 5 | 0 | 1 | Low-quality | Genome-fragment | 2.88 | AAI-based (medium-confidence) | 0 |
| S4_k141_122773 | 5300 | No | NA | 5 | 0 | 1 | Low-quality | Genome-fragment | 7.95 | AAI-based (medium-confidence) | 0 |
| S12_k141_203551 | 5302 | No | NA | 6 | 1 | 0 | Low-quality | Genome-fragment | 12.44 | AAI-based (high-confidence) | 0 |
| S4-final-viral-combined_k141_116057\|\|full | 1868 | No | NA | 2 | 0 | 1 | Not-determined | Genome-fragment | NA | NA | 0 |
| S3_k141_65241 | 5303 | Yes | 1008 | 6 | 1 | 2 | Low-quality | Genome-fragment | 0.64 | HMM-based (lower-bound) | 80.99 |
| S4-final-viral-combined_k141_144395\|\|full | 1612 | No | NA | 3 | 0 | 1 | Not-determined | Genome-fragment | NA | NA | 0 |
| S12-final-viral-combined_k141_374432\|\|full | 5303 | No | NA | 4 | 0 | 2 | Low-quality | Genome-fragment | 13.84 | AAI-based (medium-confidence) | 0 |
| S1-final-viral-combined_k141_169647\|\|full | 5310 | No | NA | 8 | 0 | 0 | Low-quality | Genome-fragment | 6.2 | AAI-based (medium-confidence) | 0 |
| S12-final-viral-combined_k141_85358\|\|full | 5310 | Yes | 4404 | 7 | 2 | 1 | Low-quality | Genome-fragment | 11.19 | AAI-based (high-confidence) | 17.06 |
| S4-final-viral-combined_k141_95450\|\|full | 1990 | No | NA | 3 | 0 | 0 | Not-determined | Genome-fragment | NA | NA | 0 |
| S4-final-viral-combined_k141_130066\|\|full | 2292 | No | NA | 4 | 0 | 1 | Not-determined | Genome-fragment | NA | NA | 0 |
| S4-final-viral-combined_k141_46817\|\|full | 2704 | No | NA | 3 | 0 | 0 | Not-determined | Genome-fragment | NA | NA | 0 |
| S7_k141_58244 | 5312 | No | NA | 7 | 0 | 6 | Low-quality | Genome-fragment | 1.96 | AAI-based (medium-confidence) | 0 |
| S17-final-viral-combined_k141_254754\|\|full | 5312 | No | NA | 12 | 2 | 1 | Low-quality | Genome-fragment | 13.48 | AAI-based (high-confidence) | 0 |
| S11-final-viral-combined_k141_891\|\|full | 5313 | No | NA | 8 | 8 | 0 | Low-quality | Genome-fragment | 8.4 | AAI-based (high-confidence) | 0 |
| S4-final-viral-combined_k141_46849\|\|full | 1099 | No | NA | 3 | 0 | 1 | Not-determined | Genome-fragment | NA | NA | 0 |
| S4-final-viral-combined_k141_158296\|\|full | 2661 | No | NA | 2 | 0 | 2 | Not-determined | Genome-fragment | NA | NA | 0 |
| S5-final-viral-combined_k141_200334\|\|full | 5313 | No | NA | 6 | 2 | 0 | Low-quality | Genome-fragment | 12.75 | AAI-based (high-confidence) | 0 |
| S4-final-viral-combined_k141_130109\|\|full | 2524 | No | NA | 3 | 0 | 1 | Not-determined | Genome-fragment | NA | NA | 0 |
| S4-final-viral-combined_k141_25751\|\|full | 1450 | No | NA | 3 | 0 | 0 | Not-determined | Genome-fragment | NA | NA | 0 |
| S4-final-viral-combined_k141_53008\|\|full | 1552 | No | NA | 3 | 0 | 1 | Not-determined | Genome-fragment | NA | NA | 0 |
| S4-final-viral-combined_k141_18100\|\|full | 1151 | No | NA | 2 | 0 | 0 | Not-determined | Genome-fragment | NA | NA | 0 |
| S4-final-viral-combined_k141_39521\|\|full | 1469 | No | NA | 2 | 0 | 1 | Not-determined | Genome-fragment | NA | NA | 0 |
| S4-final-viral-combined_k141_39525\|\|full | 2656 | No | NA | 3 | 0 | 2 | Not-determined | Genome-fragment | NA | NA | 0 |
| S4-final-viral-combined_k141_95493\|\|full | 5825 | No | NA | 5 | 0 | 5 | Not-determined | Genome-fragment | NA | NA | 0 |
| S11_k141_149922 | 5314 | No | NA | 1 | 1 | 0 | Low-quality | Genome-fragment | 1.3 | HMM-based (lower-bound) | 0 |
| S17-final-viral-combined_k141_246054\|\|full | 5317 | No | NA | 7 | 4 | 0 | Low-quality | Genome-fragment | 13.61 | AAI-based (high-confidence) | 0 |
| S11_k141_137228 | 5318 | No | NA | 13 | 0 | 0 | Low-quality | Genome-fragment | 11.85 | AAI-based (high-confidence) | 0 |
| S4-final-viral-combined_k141_53033\|\|full | 2280 | No | NA | 2 | 0 | 1 | Not-determined | Genome-fragment | NA | NA | 0 |
| S4-final-viral-combined_k141_95505\|\|full | 1040 | No | NA | 2 | 0 | 1 | Not-determined | Genome-fragment | NA | NA | 0 |
| S4-final-viral-combined_k141_81978\|\|full | 3218 | No | NA | 4 | 0 | 3 | Not-determined | Genome-fragment | NA | NA | 0 |
| S4-final-viral-combined_k141_95512\|\|full | 2683 | No | NA | 4 | 0 | 1 | Not-determined | Genome-fragment | NA | NA | 0 |
| S4-final-viral-combined_k141_109797\|\|full | 1787 | No | NA | 2 | 0 | 1 | Not-determined | Genome-fragment | NA | NA | 0 |
| S4-final-viral-combined_k141_60361\|\|full | 1610 | No | NA | 3 | 0 | 1 | Not-determined | Genome-fragment | NA | NA | 0 |
| S4-final-viral-combined_k141_102271\|\|full | 2193 | No | NA | 4 | 0 | 0 | Not-determined | Genome-fragment | NA | NA | 0 |
| S4-final-viral-combined_k141_4875\|\|full | 2181 | No | NA | 3 | 0 | 0 | Not-determined | Genome-fragment | NA | NA | 0 |
| S1_k141_195504 | 5321 | No | NA | 5 | 4 | 0 | Low-quality | Genome-fragment | 13.21 | AAI-based (high-confidence) | 0 |
| S6_k141_41787 | 5321 | No | NA | 6 | 6 | 0 | Low-quality | Genome-fragment | 11.54 | AAI-based (high-confidence) | 0 |
| S4-final-viral-combined_k141_116159\|\|full | 1191 | No | NA | 2 | 0 | 0 | Not-determined | Genome-fragment | NA | NA | 0 |
| S4-final-viral-combined_k141_46916\|\|full | 1839 | No | NA | 3 | 0 | 2 | Not-determined | Genome-fragment | NA | NA | 0 |
| S4-final-viral-combined_k141_60384\|\|full | 1908 | No | NA | 2 | 0 | 2 | Not-determined | Genome-fragment | NA | NA | 0 |
| S11-final-viral-combined_k141_253539\|\|full | 5322 | No | NA | 6 | 4 | 0 | Low-quality | Genome-fragment | 14.42 | AAI-based (high-confidence) | 0 |
| S11-final-viral-combined_k141_174246\|\|full | 5322 | No | NA | 10 | 0 | 2 | Low-quality | Genome-fragment | 11.9 | AAI-based (medium-confidence) | 0 |
| S4-final-viral-combined_k141_122718\|\|full | 3451 | No | NA | 3 | 0 | 0 | Not-determined | Genome-fragment | NA | NA | 0 |
| S4-final-viral-combined_k141_4918\|\|full | 1319 | No | NA | 3 | 0 | 2 | Not-determined | Genome-fragment | NA | NA | 0 |
| S4-final-viral-combined_k141_116212\|\|full | 10089 | No | NA | 7 | 0 | 6 | Not-determined | Genome-fragment | NA | NA | 0 |
| S11-final-viral-combined_k141_134535\|\|full | 5323 | No | NA | 6 | 1 | 0 | Low-quality | Genome-fragment | 4.96 | AAI-based (medium-confidence) | 0 |
| S4-final-viral-combined_k141_109841\|\|full | 1625 | No | NA | 2 | 0 | 1 | Not-determined | Genome-fragment | NA | NA | 0 |
| S7_k141_30838 | 5323 | No | NA | 7 | 1 | 0 | Low-quality | Genome-fragment | 4.78 | HMM-based (lower-bound) | 0 |
| S4-final-viral-combined_k141_67828\|\|full | 2520 | No | NA | 4 | 0 | 1 | Not-determined | Genome-fragment | NA | NA | 0 |
| S4-final-viral-combined_k141_46974\|\|full | 2491 | No | NA | 2 | 0 | 0 | Not-determined | Genome-fragment | NA | NA | 0 |
| S3-final-viral-combined_k141_118445\|\|full | 5324 | No | NA | 11 | 10 | 0 | Low-quality | Genome-fragment | 14.35 | AAI-based (high-confidence) | 0 |
| S4-final-viral-combined_k141_4959\|\|full | 2062 | No | NA | 3 | 0 | 2 | Not-determined | Genome-fragment | NA | NA | 0 |
| S4-final-viral-combined_k141_39594\|\|full | 1990 | No | NA | 2 | 0 | 1 | Not-determined | Genome-fragment | NA | NA | 0 |

| | | | | | | | | | | | |
|---|---|---|---|---|---|---|---|---|---|---|---|
| S4-final-viral-combined_k141_53112\|\|full | 1886 | No | NA | 2 | 0 | 2 | Not-determined | Genome-fragment | NA | NA | 0 |
| S4-final-viral-combined_k141_151176\|\|full | 4033 | No | NA | 8 | 0 | 0 | Not-determined | Genome-fragment | NA | NA | 0 |
| S4-final-viral-combined_k141_137341\|\|full | 2802 | No | NA | 4 | 0 | 2 | Not-determined | Genome-fragment | NA | NA | 0 |
| S4-final-viral-combined_k141_74958\|\|full | 1907 | No | NA | 2 | 0 | 2 | Not-determined | Genome-fragment | NA | NA | 0 |
| S17-final-viral-combined_k141_27262\|\|full | 5324 | No | NA | 5 | 0 | 2 | Low-quality | Genome-fragment | 1.66 | AAI-based (medium-confidence) | 0 |
| S4-final-viral-combined_k141_109889\|\|full | 1262 | No | NA | 2 | 0 | 0 | Not-determined | Genome-fragment | NA | NA | 0 |
| S4-final-viral-combined_k141_33001\|\|full | 3491 | No | NA | 2 | 0 | 0 | Not-determined | Genome-fragment | NA | NA | 0 |
| S4-final-viral-combined_k141_18266\|\|full | 1228 | No | NA | 2 | 0 | 1 | Not-determined | Genome-fragment | NA | NA | 0 |
| S4-final-viral-combined_k141_67853\|\|full | 1412 | No | NA | 2 | 0 | 1 | Not-determined | Genome-fragment | NA | NA | 0 |
| S12-final-viral-combined_k141_401325\|\|full | 5324 | No | NA | 9 | 1 | 0 | Low-quality | Genome-fragment | 9.32 | AAI-based (high-confidence) | 0 |
| S4-final-viral-combined_k141_116281\|\|full | 13945 | No | NA | 10 | 0 | 3 | Not-determined | Genome-fragment | NA | NA | 0 |
| S4-final-viral-combined_k141_25906\|\|full | 8522 | No | NA | 8 | 0 | 0 | Not-determined | Genome-fragment | NA | NA | 0 |
| S4-final-viral-combined_k141_12085\|\|full | 1130 | No | NA | 3 | 0 | 0 | Not-determined | Genome-fragment | NA | NA | 0 |
| S7-final-viral-combined_k141_51992\|\|full | 5325 | No | NA | 12 | 1 | 0 | Low-quality | Genome-fragment | 15.12 | AAI-based (high-confidence) | 0 |
| S4-final-viral-combined_k141_39629\|\|full | 4391 | No | NA | 9 | 0 | 1 | Not-determined | Genome-fragment | NA | NA | 0 |
| S4-final-viral-combined_k141_116290\|\|full | 2385 | No | NA | 3 | 0 | 3 | Not-determined | Genome-fragment | NA | NA | 0 |
| S4-final-viral-combined_k141_82096\|\|full | 1319 | No | NA | 3 | 0 | 0 | Not-determined | Genome-fragment | NA | NA | 0 |
| S4-final-viral-combined_k141_165258\|\|full | 1417 | No | NA | 2 | 0 | 1 | Not-determined | Genome-fragment | NA | NA | 0 |
| S4-final-viral-combined_k141_95659\|\|full | 1349 | No | NA | 2 | 0 | 0 | Not-determined | Genome-fragment | NA | NA | 0 |
| S4-final-viral-combined_k141_95660\|\|full | 2153 | No | NA | 2 | 0 | 0 | Not-determined | Genome-fragment | NA | NA | 0 |
| S11-final-viral-combined_k141_59920\|\|full | 5329 | No | NA | 6 | 3 | 0 | Low-quality | Genome-fragment | 6.47 | AAI-based (high-confidence) | 0 |
| S4-final-viral-combined_k141_151264\|\|full | 1603 | No | NA | 3 | 0 | 0 | Not-determined | Genome-fragment | NA | NA | 0 |
| S11-final-viral-combined_k141_161588\|\|full | 5330 | No | NA | 8 | 2 | 0 | Low-quality | Genome-fragment | 11.09 | AAI-based (high-confidence) | 0 |
| S5_k141_64075 | 5331 | No | NA | 7 | 2 | 0 | Low-quality | Genome-fragment | 11.85 | AAI-based (high-confidence) | 0 |
| S4-final-viral-combined_k141_67937\|\|full | 1875 | No | NA | 4 | 0 | 3 | Not-determined | Genome-fragment | NA | NA | 0 |
| S4_k141_139357 | 5331 | No | NA | 7 | 2 | 0 | Low-quality | Genome-fragment | 5.28 | AAI-based (high-confidence) | 0 |
| S7-final-viral-combined_k141_187358\|\|full | 5338 | No | NA | 4 | 1 | 0 | Low-quality | Genome-fragment | 2.95 | HMM-based (lower-bound) | 0 |
| S12-final-viral-combined_k141_180396\|\|full | 5338 | No | NA | 9 | 2 | 0 | Low-quality | Genome-fragment | 13.32 | AAI-based (high-confidence) | 0 |
| S2-final-viral-combined_k141_190982\|\|full | 5338 | No | NA | 6 | 4 | 0 | Low-quality | Genome-fragment | 15.85 | AAI-based (high-confidence) | 0 |
| S4-final-viral-combined_k141_39697\|\|full | 3026 | No | NA | 6 | 0 | 1 | Not-determined | Genome-fragment | NA | NA | 0 |
| S4-final-viral-combined_k141_39699\|\|full | 1340 | No | NA | 3 | 0 | 1 | Not-determined | Genome-fragment | NA | NA | 0 |
| S8_k141_92737 | 5338 | No | NA | 6 | 0 | 5 | Low-quality | Genome-fragment | 2.51 | AAI-based (medium-confidence) | 0 |
| S4-final-viral-combined_k141_53252\|\|full | 5411 | No | NA | 7 | 0 | 4 | Not-determined | Genome-fragment | NA | NA | 0 |
| S2_k141_189350 | 5340 | No | NA | 8 | 0 | 4 | Low-quality | Genome-fragment | 10.54 | AAI-based (medium-confidence) | 0 |
| S1-final-viral-combined_k141_89851\|\|full | 5342 | No | NA | 3 | 0 | 0 | Low-quality | Genome-fragment | 12.95 | AAI-based (high-confidence) | 0 |
| S4-final-viral-combined_k141_122866\|\|full | 2827 | No | NA | 3 | 0 | 2 | Not-determined | Genome-fragment | NA | NA | 0 |
| S4-final-viral-combined_k141_165320\|\|full | 8660 | No | NA | 7 | 0 | 6 | Not-determined | Genome-fragment | NA | NA | 0 |
| S4-final-viral-combined_k141_158663\|\|full | 5607 | No | NA | 7 | 0 | 1 | Not-determined | Genome-fragment | NA | NA | 0 |
| S4-final-viral-combined_k141_88641\|\|full | 1731 | No | NA | 5 | 0 | 0 | Not-determined | Genome-fragment | NA | NA | 0 |
| S4-final-viral-combined_k141_39721\|\|full | 2311 | No | NA | 3 | 0 | 2 | Not-determined | Genome-fragment | NA | NA | 0 |
| S17-final-viral-combined_k141_229541\|\|full | 5343 | No | NA | 6 | 0 | 6 | Low-quality | Genome-fragment | 10.1 | AAI-based (high-confidence) | 0 |
| S1-final-viral-combined_k141_275459\|\|full | 5344 | No | NA | 4 | 0 | 1 | Low-quality | Genome-fragment | 3.86 | AAI-based (high-confidence) | 0 |
| S17-final-viral-combined_k141_265045\|\|full | 5344 | No | NA | 6 | 1 | 0 | Low-quality | Genome-fragment | 8.09 | HMM-based (lower-bound) | 0 |
| S12-final-viral-combined_k141_255883\|\|full | 5349 | No | NA | 6 | 1 | 0 | Low-quality | Genome-fragment | 9.64 | AAI-based (medium-confidence) | 0 |
| S4-final-viral-combined_k141_18439\|\|full | 3569 | No | NA | 3 | 0 | 1 | Not-determined | Genome-fragment | NA | NA | 0 |
| S4-final-viral-combined_k141_12202\|\|full | 3262 | No | NA | 3 | 0 | 3 | Not-determined | Genome-fragment | NA | NA | 0 |
| S12-final-viral-combined_k141_344653\|\|full | 5349 | No | NA | 4 | 0 | 2 | Low-quality | Genome-fragment | 1.67 | AAI-based (medium-confidence) | 0 |
| S4-final-viral-combined_k141_110083\|\|full | 1887 | No | NA | 3 | 0 | 1 | Not-determined | Genome-fragment | NA | NA | 0 |
| S4-final-viral-combined_k141_26048\|\|full | 7625 | No | NA | 12 | 0 | 2 | Not-determined | Genome-fragment | NA | NA | 0 |
| S2-final-viral-combined_k141_206749\|\|full | 5350 | No | NA | 7 | 1 | 0 | Low-quality | Genome-fragment | 9.76 | AAI-based (high-confidence) | 0 |
| S4-final-viral-combined_k141_144801\|\|full | 2027 | No | NA | 2 | 0 | 0 | Not-determined | Genome-fragment | NA | NA | 0 |
| S4-final-viral-combined_k141_110097\|\|full | 1677 | No | NA | 3 | 0 | 1 | Not-determined | Genome-fragment | NA | NA | 0 |
| S7-final-viral-combined_k141_78168\|\|full | 5353 | No | NA | 7 | 3 | 0 | Low-quality | Genome-fragment | 13.28 | AAI-based (high-confidence) | 0 |
| S7_k141_178829 | 5353 | No | NA | 7 | 2 | 0 | Low-quality | Genome-fragment | 13.36 | AAI-based (high-confidence) | 0 |
| S4-final-viral-combined_k141_53319\|\|full | 1848 | No | NA | 5 | 0 | 1 | Not-determined | Genome-fragment | NA | NA | 0 |
| S4-final-viral-combined_k141_33150\|\|full | 1171 | No | NA | 3 | 0 | 0 | Not-determined | Genome-fragment | NA | NA | 0 |
| S4-final-viral-combined_k141_75096\|\|full | 2999 | No | NA | 3 | 0 | 1 | Not-determined | Genome-fragment | NA | NA | 0 |
| S4-final-viral-combined_k141_18480\|\|full | 2043 | No | NA | 3 | 0 | 2 | Not-determined | Genome-fragment | NA | NA | 0 |
| S1-final-viral-combined_k141_87910\|\|full | 5354 | No | NA | 3 | 0 | 3 | Low-quality | Genome-fragment | 3.68 | AAI-based (medium-confidence) | 0 |
| S4-final-viral-combined_k141_151398\|\|full | 1278 | No | NA | 2 | 0 | 0 | Not-determined | Genome-fragment | NA | NA | 0 |
| S4-final-viral-combined_k141_82224\|\|full | 1516 | No | NA | 3 | 0 | 0 | Not-determined | Genome-fragment | NA | NA | 0 |
| S12-final-viral-combined_k141_140471\|\|full | 5354 | No | NA | 4 | 0 | 0 | Low-quality | Genome-fragment | 4.31 | AAI-based (medium-confidence) | 0 |
| S8-final-viral-combined_k141_115710\|\|full | 5354 | Yes | 700 | 8 | 1 | 2 | Low-quality | Genome-fragment | 0.47 | HMM-based (lower-bound) | 86.93 |
| S4-final-viral-combined_k141_110143\|\|full | 2271 | No | NA | 3 | 0 | 3 | Not-determined | Genome-fragment | NA | NA | 0 |
| S4-final-viral-combined_k141_39856\|\|full | 1966 | No | NA | 3 | 0 | 1 | Not-determined | Genome-fragment | NA | NA | 0 |
| S4-final-viral-combined_k141_18506\|\|full | 5505 | No | NA | 2 | 0 | 2 | Not-determined | Genome-fragment | NA | NA | 0 |
| S2-final-viral-combined_k141_241591\|\|full | 5355 | No | NA | 12 | 2 | 0 | Low-quality | Genome-fragment | 15.03 | AAI-based (high-confidence) | 0 |
| S1-final-viral-combined_k141_100721\|\|full | 5359 | No | NA | 7 | 0 | 5 | Low-quality | Genome-fragment | 3.8 | AAI-based (medium-confidence) | 0 |
| S4-final-viral-combined_k141_82241\|\|full | 1046 | No | NA | 2 | 0 | 1 | Not-determined | Genome-fragment | NA | NA | 0 |
| S4-final-viral-combined_k141_151432\|\|full | 2543 | No | NA | 2 | 0 | 1 | Not-determined | Genome-fragment | NA | NA | 0 |
| S2_k141_119474 | 5359 | No | NA | 2 | 0 | 2 | Low-quality | Genome-fragment | 5.73 | AAI-based (medium-confidence) | 0 |
| S4-final-viral-combined_k141_144863\|\|full | 1600 | No | NA | 3 | 0 | 2 | Not-determined | Genome-fragment | NA | NA | 0 |
| S4-final-viral-combined_k141_151446\|\|full | 1111 | No | NA | 3 | 0 | 1 | Not-determined | Genome-fragment | NA | NA | 0 |
| S7-final-viral-combined_k141_14215\|\|full | 5361 | Yes | 2074 | 7 | 1 | 1 | Low-quality | Genome-fragment | 2.99 | HMM-based (lower-bound) | 61.31 |
| S4-final-viral-combined_k141_26140\|\|full | 4585 | No | NA | 3 | 0 | 3 | Not-determined | Genome-fragment | NA | NA | 0 |
| S4-final-viral-combined_k141_5096\|\|full | 1735 | No | NA | 3 | 0 | 2 | Not-determined | Genome-fragment | NA | NA | 0 |
| S4-final-viral-combined_k141_75154\|\|full | 2748 | No | NA | 2 | 0 | 0 | Not-determined | Genome-fragment | NA | NA | 0 |
| S4-final-viral-combined_k141_82279\|\|full | 1545 | No | NA | 3 | 0 | 0 | Not-determined | Genome-fragment | NA | NA | 0 |
| S4-final-viral-combined_k141_47213\|\|full | 4074 | No | NA | 3 | 0 | 2 | Not-determined | Genome-fragment | NA | NA | 0 |
| S4-final-viral-combined_k141_33246\|\|full | 2939 | No | NA | 2 | 0 | 0 | Not-determined | Genome-fragment | NA | NA | 0 |
| S4-final-viral-combined_k141_151465\|\|full | 2148 | No | NA | 3 | 0 | 3 | Not-determined | Genome-fragment | NA | NA | 0 |
| S4-final-viral-combined_k141_5102\|\|full | 1596 | No | NA | 3 | 0 | 0 | Not-determined | Genome-fragment | NA | NA | 0 |
| S4-final-viral-combined_k141_82284\|\|full | 5729 | No | NA | 7 | 0 | 6 | Not-determined | Genome-fragment | NA | NA | 0 |
| S7-final-viral-combined_k141_218757\|\|full | 5362 | Yes | 2104 | 4 | 1 | 1 | Low-quality | Genome-fragment | 0.97 | AAI-based (high-confidence) | 60.76 |
| S4-final-viral-combined_k141_88822\|\|full | 2799 | No | NA | 5 | 0 | 0 | Not-determined | Genome-fragment | NA | NA | 0 |
| S4-final-viral-combined_k141_82299\|\|full | 1328 | No | NA | 2 | 0 | 2 | Not-determined | Genome-fragment | NA | NA | 0 |
| S4-final-viral-combined_k141_53447\|\|full | 2749 | No | NA | 3 | 0 | 1 | Not-determined | Genome-fragment | NA | NA | 0 |
| S4-final-viral-combined_k141_95964\|\|full | 1301 | No | NA | 3 | 0 | 1 | Not-determined | Genome-fragment | NA | NA | 0 |
| S17-final-viral-combined_k141_122469\|\|full | 5364 | No | NA | 10 | 9 | 0 | Low-quality | Genome-fragment | 16.7 | AAI-based (high-confidence) | 0 |
| S4-final-viral-combined_k141_137534\|\|full | 1364 | No | NA | 2 | 0 | 1 | Not-determined | Genome-fragment | NA | NA | 0 |
| S4-final-viral-combined_k141_68138\|\|full | 1407 | No | NA | 2 | 0 | 0 | Not-determined | Genome-fragment | NA | NA | 0 |
| S4-final-viral-combined_k141_110268\|\|full | 3163 | No | NA | 4 | 0 | 3 | Not-determined | Genome-fragment | NA | NA | 0 |
| S4-final-viral-combined_k141_47260\|\|full | 1492 | No | NA | 3 | 0 | 2 | Not-determined | Genome-fragment | NA | NA | 0 |
| S2-final-viral-combined_k141_96287\|\|full | 5364 | No | NA | 9 | 1 | 0 | Low-quality | Genome-fragment | 8.48 | HMM-based (lower-bound) | 0 |
| S2-final-viral-combined_k141_212466\|\|full | 5365 | No | NA | 5 | 0 | 2 | Low-quality | Genome-fragment | 8.88 | AAI-based (high-confidence) | 0 |
| S4-final-viral-combined_k141_75237\|\|full | 4661 | No | NA | 4 | 0 | 4 | Not-determined | Genome-fragment | NA | NA | 0 |
| S2-final-viral-combined_k141_217239\|\|full | 5365 | No | NA | 5 | 0 | 3 | Low-quality | Genome-fragment | 1.68 | AAI-based (medium-confidence) | 0 |
| S1-final-viral-combined_k141_288519\|\|full | 5366 | No | NA | 5 | 0 | 4 | Low-quality | Genome-fragment | 6.95 | AAI-based (medium-confidence) | 0 |
| S4-final-viral-combined_k141_110295\|\|full | 2390 | No | NA | 3 | 0 | 1 | Not-determined | Genome-fragment | NA | NA | 0 |
| S4-final-viral-combined_k141_88855\|\|full | 4034 | No | NA | 6 | 0 | 1 | Not-determined | Genome-fragment | NA | NA | 0 |
| S1_k141_298881 | 5366 | No | NA | 5 | 0 | 2 | Low-quality | Genome-fragment | 7.18 | AAI-based (medium-confidence) | 0 |
| S7-final-viral-combined_k141_100585\|\|full | 5367 | No | NA | 6 | 0 | 3 | Low-quality | Genome-fragment | 3.78 | AAI-based (medium-confidence) | 0 |
| S4-final-viral-combined_k141_137570\|\|full | 5683 | No | NA | 8 | 0 | 4 | Not-determined | Genome-fragment | NA | NA | 0 |
| S8-final-viral-combined_k141_100404\|\|full | 5368 | No | NA | 7 | 7 | 0 | Low-quality | Genome-fragment | 11.78 | AAI-based (high-confidence) | 0 |
| S11-final-viral-combined_k141_281622\|\|full | 5369 | No | NA | 9 | 2 | 0 | Low-quality | Genome-fragment | 12.08 | AAI-based (high-confidence) | 0 |
| S4-final-viral-combined_k141_33350\|\|full | 2411 | No | NA | 3 | 0 | 2 | Not-determined | Genome-fragment | NA | NA | 0 |
| S4-final-viral-combined_k141_144981\|\|full | 2358 | No | NA | 2 | 0 | 1 | Not-determined | Genome-fragment | NA | NA | 0 |
| S4-final-viral-combined_k141_12426\|\|full | 2464 | No | NA | 2 | 0 | 0 | Not-determined | Genome-fragment | NA | NA | 0 |
| S11-final-viral-combined_k141_127282\|\|full | 5370 | No | NA | 3 | 0 | 3 | Low-quality | Genome-fragment | 4.8 | AAI-based (medium-confidence) | 0 |
| S4-final-viral-combined_k141_18696\|\|full | 6151 | No | NA | 7 | 0 | 3 | Not-determined | Genome-fragment | NA | NA | 0 |
| S8-final-viral-combined_k141_54310\|\|full | 5371 | No | NA | 5 | 0 | 3 | Low-quality | Genome-fragment | 8.2 | AAI-based (high-confidence) | 0 |
| S12-final-viral-combined_k141_142527\|\|full | 5376 | Yes | 3032 | 8 | 2 | 2 | Low-quality | Genome-fragment | 2.17 | AAI-based (medium-confidence) | 43.6 |
| S4-final-viral-combined_k141_130632\|\|full | 4415 | No | NA | 3 | 0 | 3 | Not-determined | Genome-fragment | NA | NA | 0 |
| S1-final-viral-combined_k141_354705\|\|full | 5380 | No | NA | 8 | 0 | 0 | Low-quality | Genome-fragment | 11.33 | AAI-based (high-confidence) | 0 |
| S4-final-viral-combined_k141_82367\|\|full | 2074 | No | NA | 2 | 0 | 1 | Not-determined | Genome-fragment | NA | NA | 0 |
| S4-final-viral-combined_k141_137650\|\|full | 1208 | No | NA | 2 | 0 | 2 | Not-determined | Genome-fragment | NA | NA | 0 |
| S1-final-viral-combined_k141_363924\|\|full | 5382 | No | NA | 3 | 0 | 2 | Low-quality | Genome-fragment | 6.88 | AAI-based (medium-confidence) | 0 |
| S4-final-viral-combined_k141_68277\|\|full | 1029 | No | NA | 2 | 0 | 1 | Not-determined | Genome-fragment | NA | NA | 0 |
| S12_k141_333860 | 5383 | No | NA | 5 | 0 | 3 | Low-quality | Genome-fragment | 1.43 | AAI-based (medium-confidence) | 0 |
| S5-final-viral-combined_k141_109547\|\|full | 5385 | No | NA | 8 | 2 | 0 | Low-quality | Genome-fragment | 3.94 | HMM-based (lower-bound) | 0 |
| S4-final-viral-combined_k141_53582\|\|full | 1148 | No | NA | 2 | 0 | 1 | Not-determined | Genome-fragment | NA | NA | 0 |
| S12-final-viral-combined_k141_40637\|\|full | 5385 | Yes | 2169 | 10 | 4 | 4 | Low-quality | Genome-fragment | 5.53 | AAI-based (medium-confidence) | 59.72 |
| S6-final-viral-combined_k141_57301\|\|full | 5385 | No | NA | 15 | 2 | 0 | Low-quality | Genome-fragment | 12.32 | AAI-based (high-confidence) | 0 |
| S4-final-viral-combined_k141_26308\|\|full | 2049 | No | NA | 3 | 0 | 2 | Not-determined | Genome-fragment | NA | NA | 0 |
| S4-final-viral-combined_k141_33424\|\|full | 5217 | No | NA | 5 | 0 | 4 | Not-determined | Genome-fragment | NA | NA | 0 |
| S4-final-viral-combined_k141_110429\|\|full | 3755 | No | NA | 4 | 0 | 3 | Not-determined | Genome-fragment | NA | NA | 0 |
| S12-final-viral-combined_k141_140676\|\|full | 5389 | No | NA | 7 | 0 | 1 | Low-quality | Genome-fragment | 3.74 | AAI-based (high-confidence) | 0 |
| S4-final-viral-combined_k141_21761\|\|full | 5390 | No | NA | 3 | 0 | 3 | Low-quality | Genome-fragment | 3.12 | AAI-based (medium-confidence) | 0 |
| S11-final-viral-combined_k141_180682\|\|full | 5391 | No | NA | 6 | 2 | 0 | Low-quality | Genome-fragment | 8.98 | AAI-based (high-confidence) | 0 |
| S4-final-viral-combined_k141_130698\|\|full | 2641 | No | NA | 3 | 0 | 0 | Not-determined | Genome-fragment | NA | NA | 0 |
| S2_k141_151850 | 5392 | No | NA | 5 | 0 | 2 | Low-quality | Genome-fragment | 2.43 | AAI-based (medium-confidence) | 0 |
| S4-final-viral-combined_k141_68324\|\|full | 2388 | No | NA | 5 | 0 | 2 | Not-determined | Genome-fragment | NA | NA | 0 |
| S4-final-viral-combined_k141_116745\|\|full | 1286 | No | NA | 2 | 0 | 0 | Not-determined | Genome-fragment | NA | NA | 0 |
| S4-final-viral-combined_k141_53616\|\|full | 1615 | No | NA | 3 | 0 | 2 | Not-determined | Genome-fragment | NA | NA | 0 |
| S12-final-viral-combined_k141_117595\|\|full | 5393 | Yes | 1891 | 6 | 1 | 1 | Low-quality | Genome-fragment | 0.72 | HMM-based (lower-bound) | 64.94 |
| S4-final-viral-combined_k141_137737\|\|full | 1805 | No | NA | 2 | 0 | 1 | Not-determined | Genome-fragment | NA | NA | 0 |
| S4-final-viral-combined_k141_123285\|\|full | 1445 | No | NA | 3 | 0 | 1 | Not-determined | Genome-fragment | NA | NA | 0 |
| S4-final-viral-combined_k141_53630\|\|full | 1984 | No | NA | 2 | 0 | 1 | Not-determined | Genome-fragment | NA | NA | 0 |
| S4-final-viral-combined_k141_33512\|\|full | 2287 | No | NA | 4 | 0 | 1 | Not-determined | Genome-fragment | NA | NA | 0 |
| S4-final-viral-combined_k141_130742\|\|full | 2070 | No | NA | 5 | 0 | 0 | Not-determined | Genome-fragment | NA | NA | 0 |
| S12-final-viral-combined_k141_336341\|\|full | 5395 | No | NA | 5 | 0 | 2 | Low-quality | Genome-fragment | 3.74 | AAI-based (medium-confidence) | 0 |
| S8-final-viral-combined_k141_230948\|\|full | 5396 | No | NA | 10 | 0 | 3 | Low-quality | Genome-fragment | 30.74 | AAI-based (medium-confidence) | 0 |
| S9-final-viral-combined_k141_79414\|\|full | 5399 | No | NA | 8 | 0 | 0 | Low-quality | Genome-fragment | 13.08 | AAI-based (high-confidence) | 0 |
| S7-final-viral-combined_k141_76287\|\|full | 5404 | No | NA | 13 | 0 | 0 | Low-quality | Genome-fragment | 9.75 | AAI-based (high-confidence) | 0 |
| S2_k141_144682 | 5404 | No | NA | 4 | 0 | 2 | Low-quality | Genome-fragment | 1.46 | AAI-based (high-confidence) | 0 |
| S4-final-viral-combined_k141_47479\|\|full | 1600 | No | NA | 3 | 0 | 1 | Not-determined | Genome-fragment | NA | NA | 0 |
| S9-final-viral-combined_k141_73968\|\|full | 5405 | No | NA | 15 | 8 | 0 | Low-quality | Genome-fragment | 13.55 | AAI-based (high-confidence) | 0 |
| S4-final-viral-combined_k141_60984\|\|full | 1349 | No | NA | 3 | 0 | 0 | Not-determined | Genome-fragment | NA | NA | 0 |
| S4-final-viral-combined_k141_26463\|\|full | 1582 | No | NA | 2 | 0 | 2 | Not-determined | Genome-fragment | NA | NA | 0 |
| S5-final-viral-combined_k141_167983\|\|full | 5405 | No | NA | 5 | 0 | 2 | Low-quality | Genome-fragment | 10.54 | AAI-based (medium-confidence) | 0 |
| S4-final-viral-combined_k141_5522\|\|full | 1389 | No | NA | 3 | 0 | 0 | Not-determined | Genome-fragment | NA | NA | 0 |
| S8_k141_81399 | 5405 | No | NA | 4 | 0 | 0 | Low-quality | Genome-fragment | 5.72 | AAI-based (high-confidence) | 0 |
| S4-final-viral-combined_k141_33601\|\|full | 3007 | No | NA | 2 | 0 | 1 | Not-determined | Genome-fragment | NA | NA | 0 |
| S4-final-viral-combined_k141_47494\|\|full | 2860 | No | NA | 3 | 0 | 2 | Not-determined | Genome-fragment | NA | NA | 0 |
| S7-final-viral-combined_k141_192086\|\|full | 5410 | No | NA | 7 | 0 | 0 | Low-quality | Genome-fragment | 8.2 | AAI-based (high-confidence) | 0 |
| S7_k141_130292 | 5411 | No | NA | 3 | 2 | 0 | Low-quality | Genome-fragment | 8.73 | HMM-based (lower-bound) | 0 |
| S1-final-viral-combined_k141_322319\|\|full | 5412 | No | NA | 3 | 0 | 1 | Low-quality | Genome-fragment | 4.34 | AAI-based (high-confidence) | 0 |
| S4-final-viral-combined_k141_33611\|\|full | 1960 | No | NA | 2 | 0 | 2 | Not-determined | Genome-fragment | NA | NA | 0 |
| S4-final-viral-combined_k141_12684\|\|full | 1284 | No | NA | 3 | 0 | 0 | Not-determined | Genome-fragment | NA | NA | 0 |
| S2-final-viral-combined_k141_227635\|\|full | 5415 | No | NA | 11 | 1 | 0 | Low-quality | Genome-fragment | 11 | AAI-based (high-confidence) | 0 |
| S4-final-viral-combined_k141_159162\|\|full | 2391 | No | NA | 4 | 0 | 2 | Not-determined | Genome-fragment | NA | NA | 0 |
| S4-final-viral-combined_k141_61011\|\|full | 8321 | No | NA | 6 | 0 | 2 | Not-determined | Genome-fragment | NA | NA | 0 |
| S4-final-viral-combined_k141_18958\|\|full | 1333 | No | NA | 3 | 0 | 1 | Not-determined | Genome-fragment | NA | NA | 0 |
| S4-final-viral-combined_k141_145244\|\|full | 1493 | No | NA | 3 | 0 | 1 | Not-determined | Genome-fragment | NA | NA | 0 |
| S8-final-viral-combined_k141_22140\|\|full | 5416 | No | NA | 7 | 2 | 0 | Low-quality | Genome-fragment | 14.02 | AAI-based (high-confidence) | 0 |
| S17-k141_45045 | 5417 | No | NA | 7 | 0 | 5 | Low-quality | Genome-fragment | 1.54 | AAI-based (medium-confidence) | 0 |
| S4-final-viral-combined_k141_89191\|\|full | 2536 | No | NA | 3 | 0 | 0 | Not-determined | Genome-fragment | NA | NA | 0 |
| S4-final-viral-combined_k141_159187\|\|full | 3045 | No | NA | 3 | 0 | 3 | Not-determined | Genome-fragment | NA | NA | 0 |
| S4-final-viral-combined_k141_47548\|\|full | 2694 | No | NA | 3 | 0 | 2 | Not-determined | Genome-fragment | NA | NA | 0 |
| S12-final-viral-combined_k141_388990\|\|full | 5417 | No | NA | 10 | 2 | 0 | Low-quality | Genome-fragment | 24.59 | AAI-based (high-confidence) | 0 |
| S12_k141_14844 | 5417 | Yes | 2988 | 5 | 1 | 1 | Low-quality | Genome-fragment | 2.75 | AAI-based (high-confidence) | 44.84 |
| S11-final-viral-combined_k141_122963\|\|full | 5418 | No | NA | 4 | 2 | 0 | Low-quality | Genome-fragment | 9.81 | AAI-based (high-confidence) | 0 |
| S12_k141_118674 | 5419 | No | NA | 4 | 4 | 0 | Low-quality | Genome-fragment | 13.89 | AAI-based (high-confidence) | 0 |
| S8_k141_133694 | 5419 | No | NA | 7 | 3 | 0 | Low-quality | Genome-fragment | 8.52 | HMM-based (lower-bound) | 0 |
| S4-final-viral-combined_k141_130821\|\|full | 1661 | No | NA | 2 | 0 | 2 | Not-determined | Genome-fragment | NA | NA | 0 |
| S2-final-viral-combined_k141_17670\|\|full | 5420 | No | NA | 11 | 7 | 0 | Low-quality | Genome-fragment | 12.44 | AAI-based (high-confidence) | 0 |
| S17-final-viral-combined_k141_211542\|\|full | 5422 | No | NA | 6 | 0 | 2 | Low-quality | Genome-fragment | 3.45 | AAI-based (medium-confidence) | 0 |
| S2-final-viral-combined_k141_101211\|\|full | 5423 | No | NA | 7 | 0 | 1 | Low-quality | Genome-fragment | 8.93 | AAI-based (medium-confidence) | 0 |
| S4-final-viral-combined_k141_47565\|\|full | 27062 | No | NA | 33 | 0 | 2 | Medium-quality | Genome-fragment | 61.48 | AAI-based (medium-confidence) | 0 |
| S4-final-viral-combined_k141_123490\|\|full | 1455 | No | NA | 3 | 0 | 1 | Not-determined | Genome-fragment | NA | NA | 0 |
| S1_k141_326139 | 5426 | No | NA | 9 | 5 | 0 | Low-quality | Genome-fragment | 12.77 | AAI-based (high-confidence) | 0 |
| S4-final-viral-combined_k141_47594\|\|full | 1546 | No | NA | 2 | 0 | 1 | Not-determined | Genome-fragment | NA | NA | 0 |
| S2_k141_93834 | 5426 | No | NA | 10 | 0 | 1 | Low-quality | Genome-fragment | 5.2 | AAI-based (medium-confidence) | 0 |
| S4-final-viral-combined_k141_96406\|\|full | 2252 | No | NA | 3 | 0 | 1 | Not-determined | Genome-fragment | NA | NA | 0 |
| S4-final-viral-combined_k141_110691\|\|full | 8656 | No | NA | 13 | 0 | 6 | Not-determined | Genome-fragment | NA | NA | 0 |
| S4-final-viral-combined_k141_33711\|\|full | 4565 | No | NA | 4 | 0 | 1 | Not-determined | Genome-fragment | NA | NA | 0 |
| S1-final-viral-combined_k141_126510\|\|full | 5427 | No | NA | 10 | 10 | 0 | Low-quality | Genome-fragment | 12.13 | AAI-based (high-confidence) | 0 |
| S4-final-viral-combined_k141_89292\|\|full | 1442 | No | NA | 2 | 0 | 1 | Not-determined | Genome-fragment | NA | NA | 0 |
| S12-final-viral-combined_k141_266992\|\|full | 5428 | No | NA | 3 | 0 | 1 | Low-quality | Genome-fragment | 2.74 | AAI-based (medium-confidence) | 0 |
| S4-final-viral-combined_k141_26574\|\|full | 4483 | No | NA | 5 | 0 | 3 | Not-determined | Genome-fragment | NA | NA | 0 |
| S4-final-viral-combined_k141_116996\|\|full | 1583 | No | NA | 3 | 0 | 2 | Not-determined | Genome-fragment | NA | NA | 0 |
| S4-final-viral-combined_k141_82796\|\|full | 1026 | No | NA | 3 | 0 | 0 | Not-determined | Genome-fragment | NA | NA | 0 |
| S4-final-viral-combined_k141_40443\|\|full | 1817 | No | NA | 2 | 0 | 0 | Not-determined | Genome-fragment | NA | NA | 0 |
| S4-final-viral-combined_k141_159263\|\|full | 3230 | No | NA | 2 | 0 | 2 | Not-determined | Genome-fragment | NA | NA | 0 |
| S4_k141_146506 | 5431 | No | NA | 8 | 4 | 0 | Low-quality | Genome-fragment | 13.91 | AAI-based (high-confidence) | 0 |
| S4-final-viral-combined_k141_5646\|\|full | 1319 | No | NA | 2 | 0 | 0 | Not-determined | Genome-fragment | NA | NA | 0 |
| S4-final-viral-combined_k141_165657\|\|full | 1626 | No | NA | 3 | 0 | 0 | Not-determined | Genome-fragment | NA | NA | 0 |
| S5-final-viral-combined_k141_108267\|\|full | 5432 | No | NA | 3 | 3 | 0 | Low-quality | Genome-fragment | 9.18 | AAI-based (high-confidence) | 0 |
| S4-final-viral-combined_k141_47662\|\|full | 1492 | No | NA | 2 | 0 | 0 | Not-determined | Genome-fragment | NA | NA | 0 |
| S1-final-viral-combined_k141_335950\|\|full | 5436 | No | NA | 9 | 5 | 0 | Low-quality | Genome-fragment | 9.51 | AAI-based (high-confidence) | 0 |
| S7-final-viral-combined_k141_97378\|\|full | 5436 | No | NA | 6 | 0 | 3 | Low-quality | Genome-fragment | 7.95 | AAI-based (high-confidence) | 0 |
| S8-final-viral-combined_k141_59849\|\|full | 5436 | No | NA | 6 | 0 | 0 | Low-quality | Genome-fragment | 3.94 | AAI-based (medium-confidence) | 0 |
| S4-final-viral-combined_k141_40476\|\|full | 1382 | No | NA | 2 | 0 | 2 | Not-determined | Genome-fragment | NA | NA | 0 |
| S4-final-viral-combined_k141_110725\|\|full | 1431 | No | NA | 3 | 0 | 0 | Not-determined | Genome-fragment | NA | NA | 0 |
| S4-final-viral-combined_k141_82843\|\|full | 1856 | No | NA | 2 | 0 | 0 | Not-determined | Genome-fragment | NA | NA | 0 |
| S11-final-viral-combined_k141_74017\|\|full | 5437 | No | NA | 8 | 1 | 0 | Low-quality | Genome-fragment | 11.4 | AAI-based (high-confidence) | 0 |
| S4-final-viral-combined_k141_123563\|\|full | 2455 | No | NA | 3 | 0 | 3 | Not-determined | Genome-fragment | NA | NA | 0 |
| S6_k141_48173 | 5442 | No | NA | 3 | 1 | 0 | Low-quality | Genome-fragment | 9.01 | AAI-based (high-confidence) | 0 |
| S4-final-viral-combined_k141_33764\|\|full | 9966 | No | NA | 18 | 0 | 0 | Not-determined | Genome-fragment | NA | NA | 0 |
| S8_k141_86727 | 5443 | No | NA | 10 | 3 | 0 | Low-quality | Genome-fragment | 12.27 | AAI-based (high-confidence) | 0 |
| S4-final-viral-combined_k141_159325\|\|full | 1345 | No | NA | 3 | 0 | 1 | Not-determined | Genome-fragment | NA | NA | 0 |
| S4-final-viral-combined_k141_47732\|\|full | 1152 | No | NA | 3 | 0 | 0 | Not-determined | Genome-fragment | NA | NA | 0 |
| S3_k141_61289 | 5444 | No | NA | 4 | 0 | 4 | Low-quality | Genome-fragment | 3.24 | AAI-based (medium-confidence) | 0 |
| S9_k141_122359 | 5446 | No | NA | 5 | 3 | 0 | Low-quality | Genome-fragment | 13.11 | AAI-based (high-confidence) | 0 |
| S4-final-viral-combined_k141_145500\|\|full | 4793 | No | NA | 7 | 0 | 2 | Not-determined | Genome-fragment | NA | NA | 0 |
| S4-final-viral-combined_k141_165742\|\|full | 1279 | No | NA | 2 | 0 | 1 | Not-determined | Genome-fragment | NA | NA | 0 |
| S4-final-viral-combined_k141_151985\|\|full | 1674 | No | NA | 2 | 0 | 0 | Not-determined | Genome-fragment | NA | NA | 0 |
| S4-final-viral-combined_k141_12879\|\|full | 1803 | No | NA | 3 | 0 | 1 | Not-determined | Genome-fragment | NA | NA | 0 |
| S4-final-viral-combined_k141_152002\|\|full | 1547 | No | NA | 2 | 0 | 0 | Not-determined | Genome-fragment | NA | NA | 0 |
| S4-final-viral-combined_k141_89457\|\|full | 1217 | No | NA | 3 | 0 | 0 | Not-determined | Genome-fragment | NA | NA | 0 |
| S4-final-viral-combined_k141_12881\|\|full | 2953 | No | NA | 3 | 0 | 2 | Not-determined | Genome-fragment | NA | NA | 0 |
| S4-final-viral-combined_k141_145545\|\|full | 2047 | No | NA | 3 | 0 | 0 | Not-determined | Genome-fragment | NA | NA | 0 |
| S4-final-viral-combined_k141_165768\|\|full | 1208 | No | NA | 2 | 0 | 1 | Not-determined | Genome-fragment | NA | NA | 0 |
| S4-final-viral-combined_k141_53938\|\|full | 2643 | No | NA | 3 | 0 | 3 | Not-determined | Genome-fragment | NA | NA | 0 |
| S17-final-viral-combined_k141_199621\|\|full | 5446 | No | NA | 9 | 0 | 2 | Low-quality | Genome-fragment | 14.64 | AAI-based (medium-confidence) | 0 |
| S3-final-viral-combined_k141_133056\|\|full | 5449 | No | NA | 7 | 0 | 1 | Low-quality | Genome-fragment | 9.79 | AAI-based (medium-confidence) | 0 |
| S4-final-viral-combined_k141_47792\|\|full | 2227 | No | NA | 4 | 0 | 0 | Not-determined | Genome-fragment | NA | NA | 0 |
| S4-final-viral-combined_k141_82991\|\|full | 1514 | No | NA | 3 | 0 | 1 | Not-determined | Genome-fragment | NA | NA | 0 |
| S2-final-viral-combined_k141_66794\|\|full | 5449 | No | NA | 5 | 0 | 0 | Low-quality | Genome-fragment | 9.07 | AAI-based (high-confidence) | 0 |
| S4-final-viral-combined_k141_138056\|\|full | 1831 | No | NA | 3 | 0 | 1 | Not-determined | Genome-fragment | NA | NA | 0 |
| S5_k141_86609 | 5452 | No | NA | 5 | 0 | 0 | Low-quality | Genome-fragment | 6.06 | AAI-based (medium-confidence) | 0 |
| S4-final-viral-combined_k141_110887\|\|full | 1655 | No | NA | 3 | 0 | 2 | Not-determined | Genome-fragment | NA | NA | 0 |
| S4-final-viral-combined_k141_103219\|\|full | 8948 | No | NA | 6 | 0 | 5 | Not-determined | Genome-fragment | NA | NA | 0 |
| S4-final-viral-combined_k141_145609\|\|full | 3463 | No | NA | 3 | 0 | 2 | Not-determined | Genome-fragment | NA | NA | 0 |
| S12-final-viral-combined_k141_192589\|\|full | 5452 | Yes | 2482 | 8 | 2 | 4 | Low-quality | Genome-fragment | 5.54 | HMM-based (lower-bound) | 54.48 |
| S4-final-viral-combined_k141_75783\|\|full | 4450 | No | NA | 6 | 0 | 2 | Not-determined | Genome-fragment | NA | NA | 0 |
| S2-final-viral-combined_k141_200575\|\|full | 5452 | Yes | 3194 | 7 | 1 | 1 | Low-quality | Genome-fragment | 2.92 | HMM-based (lower-bound) | 41.42 |
| S4-final-viral-combined_k141_68792\|\|full | 1337 | No | NA | 2 | 0 | 1 | Not-determined | Genome-fragment | NA | NA | 0 |
| S4-final-viral-combined_k141_83037\|\|full | 1024 | No | NA | 2 | 0 | 0 | Not-determined | Genome-fragment | NA | NA | 0 |
| S4-final-viral-combined_k141_165814\|\|full | 3010 | No | NA | 4 | 0 | 2 | Not-determined | Genome-fragment | NA | NA | 0 |
| S8-final-viral-combined_k141_199148\|\|full | 5452 | No | NA | 3 | 2 | 0 | Low-quality | Genome-fragment | 5.67 | AAI-based (high-confidence) | 0 |
| S4-final-viral-combined_k141_40606\|\|full | 3443 | No | NA | 3 | 0 | 3 | Not-determined | Genome-fragment | NA | NA | 0 |
| S4-final-viral-combined_k141_103244\|\|full | 3480 | No | NA | 6 | 0 | 1 | Not-determined | Genome-fragment | NA | NA | 0 |
| S4-final-viral-combined_k141_5797\|\|full | 2823 | No | NA | 3 | 0 | 2 | Not-determined | Genome-fragment | NA | NA | 0 |
| S4-final-viral-combined_k141_159484\|\|full | 1731 | No | NA | 2 | 0 | 1 | Not-determined | Genome-fragment | NA | NA | 0 |
| S4-final-viral-combined_k141_138114\|\|full | 2351 | No | NA | 4 | 0 | 1 | Not-determined | Genome-fragment | NA | NA | 0 |
| S7-final-viral-combined_k141_214389\|\|full | 5453 | No | NA | 8 | 4 | 0 | Low-quality | Genome-fragment | 8.7 | HMM-based (lower-bound) | 0 |
| S4-final-viral-combined_k141_19262\|\|full | 1166 | No | NA | 3 | 0 | 0 | Not-determined | Genome-fragment | NA | NA | 0 |
| S8-final-viral-combined_k141_243907\|\|full | 5454 | No | NA | 4 | 0 | 2 | Low-quality | Genome-fragment | 1.33 | AAI-based (high-confidence) | 0 |
| S4-final-viral-combined_k141_159494\|\|full | 1682 | No | NA | 3 | 0 | 2 | Not-determined | Genome-fragment | NA | NA | 0 |
| S4-final-viral-combined_k141_165838\|\|full | 1113 | No | NA | 2 | 0 | 1 | Not-determined | Genome-fragment | NA | NA | 0 |
| S17-final-viral-combined_k141_211122\|\|full | 5455 | No | NA | 8 | 1 | 1 | Low-quality | Genome-fragment | 3.06 | HMM-based (lower-bound) | 0 |
| S4-final-viral-combined_k141_83079\|\|full | 4173 | No | NA | 5 | 0 | 3 | Not-determined | Genome-fragment | NA | NA | 0 |
| S4-final-viral-combined_k141_138146\|\|full | 2355 | No | NA | 3 | 0 | 1 | Not-determined | Genome-fragment | NA | NA | 0 |
| S4-final-viral-combined_k141_138153\|\|full | 1834 | No | NA | 2 | 0 | 1 | Not-determined | Genome-fragment | NA | NA | 0 |
| S4-final-viral-combined_k141_40668\|\|full | 4328 | No | NA | 5 | 0 | 1 | Not-determined | Genome-fragment | NA | NA | 0 |
| S4-final-viral-combined_k141_19287\|\|full | 1974 | No | NA | 2 | 0 | 2 | Not-determined | Genome-fragment | NA | NA | 0 |
| S4-final-viral-combined_k141_96656\|\|full | 1512 | No | NA | 2 | 0 | 1 | Not-determined | Genome-fragment | NA | NA | 0 |
| S7-final-viral-combined_k141_117531\|\|full | 5456 | Yes | 2901 | 6 | 1 | 1 | Low-quality | Genome-fragment | 4.39 | HMM-based (lower-bound) | 46.83 |
| S4-final-viral-combined_k141_103308\|\|full | 2834 | No | NA | 2 | 0 | 1 | Not-determined | Genome-fragment | NA | NA | 0 |
| S8-final-viral-combined_k141_168476\|\|full | 5456 | No | NA | 6 | 0 | 5 | Low-quality | Genome-fragment | 7.41 | AAI-based (medium-confidence) | 0 |
| S4-final-viral-combined_k141_117289\|\|full | 1520 | No | NA | 2 | 0 | 0 | Not-determined | Genome-fragment | NA | NA | 0 |
| S7_k141_199272 | 5458 | No | NA | 6 | 0 | 5 | Low-quality | Genome-fragment | 2.39 | AAI-based (high-confidence) | 0 |
| S4-final-viral-combined_k141_117294\|\|full | 2314 | No | NA | 4 | 0 | 2 | Not-determined | Genome-fragment | NA | NA | 0 |
| S4-final-viral-combined_k141_5893\|\|full | 1703 | No | NA | 3 | 0 | 1 | Not-determined | Genome-fragment | NA | NA | 0 |
| S2_k141_119951 | 5458 | No | NA | 12 | 3 | 0 | Low-quality | Genome-fragment | 13.3 | AAI-based (medium-confidence) | 0 |
| S1-final-viral-combined_k141_73310\|\|full | 5459 | No | NA | 7 | 0 | 4 | Low-quality | Genome-fragment | 1.49 | AAI-based (medium-confidence) | 0 |
| S4-final-viral-combined_k141_83166\|\|full | 1940 | No | NA | 3 | 0 | 0 | Not-determined | Genome-fragment | NA | NA | 0 |
| S6-final-viral-combined_k141_69074\|\|full | 5466 | Yes | 2374 | 5 | 1 | 2 | Low-quality | Genome-fragment | 1.31 | HMM-based (lower-bound) | 56.57 |
| S4-final-viral-combined_k141_83180\|\|full | 1793 | No | NA | 2 | 0 | 2 | Not-determined | Genome-fragment | NA | NA | 0 |
| S6-final-viral-combined_k141_99376\|\|full | 5466 | No | NA | 13 | 1 | 0 | Low-quality | Genome-fragment | 8.46 | AAI-based (high-confidence) | 0 |
| S4-final-viral-combined_k141_61427\|\|full | 2607 | No | NA | 2 | 0 | 0 | Not-determined | Genome-fragment | NA | NA | 0 |
| S4-final-viral-combined_k141_117331\|\|full | 1325 | No | NA | 1 | 0 | 1 | Not-determined | Genome-fragment | NA | NA | 0 |
| S4-final-viral-combined_k141_75963\|\|full | 4173 | No | NA | 3 | 0 | 2 | Not-determined | Genome-fragment | NA | NA | 0 |
| S4-final-viral-combined_k141_89583\|\|full | 1617 | No | NA | 3 | 0 | 0 | Not-determined | Genome-fragment | NA | NA | 0 |
| S4-final-viral-combined_k141_152218\|\|full | 8058 | No | NA | 8 | 0 | 2 | Not-determined | Genome-fragment | NA | NA | 0 |
| S4-final-viral-combined_k141_145834\|\|full | 2078 | No | NA | 3 | 0 | 2 | Not-determined | Genome-fragment | NA | NA | 0 |
| S4-final-viral-combined_k141_61454\|\|full | 2760 | No | NA | 3 | 0 | 0 | Not-determined | Genome-fragment | NA | NA | 0 |
| S4-final-viral-combined_k141_165974\|\|full | 1334 | No | NA | 2 | 0 | 0 | Not-determined | Genome-fragment | NA | NA | 0 |
| S4-final-viral-combined_k141_40824\|\|full | 2674 | No | NA | 3 | 0 | 1 | Not-determined | Genome-fragment | NA | NA | 0 |
| S11_k141_117286 | 5467 | No | NA | 12 | 3 | 0 | Low-quality | Genome-fragment | 16.96 | AAI-based (high-confidence) | 0 |
| S4-final-viral-combined_k141_61466\|\|full | 6362 | No | NA | 7 | 0 | 7 | Not-determined | Genome-fragment | NA | NA | 0 |
| S4-final-viral-combined_k141_5977\|\|full | 4316 | No | NA | 1 | 0 | 1 | Not-determined | Genome-fragment | NA | NA | 0 |
| S4-final-viral-combined_k141_61472\|\|full | 1537 | No | NA | 3 | 0 | 0 | Not-determined | Genome-fragment | NA | NA | 0 |
| S4-final-viral-combined_k141_54202\|\|full | 1093 | No | NA | 2 | 0 | 2 | Not-determined | Genome-fragment | NA | NA | 0 |
| S4-final-viral-combined_k141_40865\|\|full | 1968 | No | NA | 2 | 0 | 0 | Not-determined | Genome-fragment | NA | NA | 0 |
| S17-k141_257683 | 5467 | No | NA | 3 | 0 | 1 | Low-quality | Genome-fragment | 2.96 | AAI-based (high-confidence) | 0 |
| S4-final-viral-combined_k141_89626\|\|full | 1140 | No | NA | 3 | 0 | 1 | Not-determined | Genome-fragment | NA | NA | 0 |
| S4-final-viral-combined_k141_61506\|\|full | 1159 | No | NA | 3 | 0 | 0 | Not-determined | Genome-fragment | NA | NA | 0 |
| S4-final-viral-combined_k141_76034\|\|full | 2712 | No | NA | 6 | 0 | 0 | Not-determined | Genome-fragment | NA | NA | 0 |
| S4-final-viral-combined_k141_117407\|\|full | 1514 | No | NA | 2 | 0 | 2 | Not-determined | Genome-fragment | NA | NA | 0 |
| S12-final-viral-combined_k141_117779\|\|full | 5468 | No | NA | 12 | 1 | 0 | Low-quality | Genome-fragment | 10.24 | AAI-based (high-confidence) | 0 |
| S12-final-viral-combined_k141_167298\|\|full | 5470 | No | NA | 5 | 2 | 0 | Low-quality | Genome-fragment | 16.27 | AAI-based (medium-confidence) | 0 |
| S1_k141_343034 | 5471 | No | NA | 7 | 0 | 0 | Low-quality | Genome-fragment | 11.29 | AAI-based (medium-confidence) | 0 |
| S4-final-viral-combined_k141_40903\|\|full | 1958 | No | NA | 4 | 0 | 0 | Not-determined | Genome-fragment | NA | NA | 0 |
| S5-final-viral-combined_k141_103958\|\|full | 5471 | Yes | 2900 | 6 | 1 | 3 | Low-quality | Genome-fragment | 6.54 | AAI-based (medium-confidence) | 46.99 |
| S6-final-viral-combined_k141_66514\|\|full | 5471 | No | NA | 9 | 0 | 2 | Low-quality | Genome-fragment | 4.28 | AAI-based (high-confidence) | 0 |
| S4-final-viral-combined_k141_13177\|\|full | 2571 | No | NA | 2 | 0 | 1 | Not-determined | Genome-fragment | NA | NA | 0 |
| S8-final-viral-combined_k141_69538\|\|full | 5471 | No | NA | 7 | 1 | 0 | Low-quality | Genome-fragment | 14.05 | AAI-based (medium-confidence) | 0 |
| S9_k141_48325 | 5472 | No | NA | 5 | 0 | 0 | Low-quality | Genome-fragment | 4.62 | AAI-based (medium-confidence) | 0 |
| S12-final-viral-combined_k141_261677\|\|full | 5472 | No | NA | 15 | 2 | 0 | Low-quality | Genome-fragment | 13.76 | AAI-based (high-confidence) | 0 |
| S4-final-viral-combined_k141_76061\|\|full | 2028 | No | NA | 4 | 0 | 1 | Not-determined | Genome-fragment | NA | NA | 0 |
| S7-final-viral-combined_k141_124833\|\|full | 5474 | No | NA | 11 | 1 | 2 | Low-quality | Genome-fragment | 7.43 | AAI-based (medium-confidence) | 0 |
| S4-final-viral-combined_k141_145859\|\|full | 1486 | No | NA | 2 | 0 | 0 | Not-determined | Genome-fragment | NA | NA | 0 |
| S4-final-viral-combined_k141_103353\|\|full | 4811 | No | NA | 6 | 0 | 1 | Not-determined | Genome-fragment | NA | NA | 0 |
| S4-final-viral-combined_k141_111180\|\|full | 3522 | No | NA | 5 | 0 | 3 | Not-determined | Genome-fragment | NA | NA | 0 |
| S4-final-viral-combined_k141_48146\|\|full | 2045 | No | NA | 3 | 0 | 1 | Not-determined | Genome-fragment | NA | NA | 0 |
| S4-final-viral-combined_k141_123982\|\|full | 3132 | No | NA | 4 | 0 | 0 | Not-determined | Genome-fragment | NA | NA | 0 |
| S5-final-viral-combined_k141_27431\|\|full | 5476 | No | NA | 8 | 4 | 0 | Low-quality | Genome-fragment | 14.81 | AAI-based (high-confidence) | 0 |
| S4-final-viral-combined_k141_26901\|\|full | 1707 | No | NA | 2 | 0 | 1 | Not-determined | Genome-fragment | NA | NA | 0 |
| S4-final-viral-combined_k141_34188\|\|full | 1213 | No | NA | 2 | 0 | 1 | Not-determined | Genome-fragment | NA | NA | 0 |
| S4-final-viral-combined_k141_96815\|\|full | 2648 | No | NA | 3 | 0 | 0 | Not-determined | Genome-fragment | NA | NA | 0 |
| S4-final-viral-combined_k141_103413\|\|full | 1408 | No | NA | 2 | 0 | 2 | Not-determined | Genome-fragment | NA | NA | 0 |
| S4-final-viral-combined_k141_54309\|\|full | 1884 | No | NA | 3 | 0 | 1 | Not-determined | Genome-fragment | NA | NA | 0 |
| S4-final-viral-combined_k141_124007\|\|full | 1523 | No | NA | 2 | 0 | 0 | Not-determined | Genome-fragment | NA | NA | 0 |
| S4-final-viral-combined_k141_111204\|\|full | 2866 | No | NA | 4 | 0 | 0 | Not-determined | Genome-fragment | NA | NA | 0 |
| S4-final-viral-combined_k141_69046\|\|full | 1804 | No | NA | 2 | 0 | 1 | Not-determined | Genome-fragment | NA | NA | 0 |
| S4-final-viral-combined_k141_138407\|\|full | 9016 | No | NA | 10 | 0 | 9 | Not-determined | Genome-fragment | NA | NA | 0 |
| S4-final-viral-combined_k141_19514\|\|full | 3566 | No | NA | 6 | 0 | 1 | Not-determined | Genome-fragment | NA | NA | 0 |
| S6_k141_14827 | 5476 | No | NA | 3 | 0 | 2 | Low-quality | Genome-fragment | 3.41 | AAI-based (high-confidence) | 0 |
| S4-final-viral-combined_k141_48186\|\|full | 1171 | No | NA | 2 | 0 | 1 | Not-determined | Genome-fragment | NA | NA | 0 |
| S1-final-viral-combined_k141_45195\|\|full | 5477 | No | NA | 13 | 1 | 0 | Low-quality | Genome-fragment | 4.08 | AAI-based (medium-confidence) | 0 |
| S4-final-viral-combined_k141_19533\|\|full | 4155 | No | NA | 3 | 0 | 3 | Not-determined | Genome-fragment | NA | NA | 0 |
| S7_k141_11305 | 5477 | No | NA | 9 | 1 | 0 | Low-quality | Genome-fragment | 2.2 | HMM-based (lower-bound) | 0 |
| S4-final-viral-combined_k141_103471\|\|full | 1773 | No | NA | 3 | 0 | 1 | Not-determined | Genome-fragment | NA | NA | 0 |
| S4-final-viral-combined_k141_89765\|\|full | 1102 | No | NA | 2 | 0 | 1 | Not-determined | Genome-fragment | NA | NA | 0 |
| S2-final-viral-combined_k141_156272\|\|full | 5477 | No | NA | 8 | 1 | 0 | Low-quality | Genome-fragment | 7.86 | HMM-based (lower-bound) | 0 |
| S4-final-viral-combined_k141_61645\|\|full | 3127 | No | NA | 7 | 0 | 1 | Not-determined | Genome-fragment | NA | NA | 0 |
| S4-final-viral-combined_k141_152335\|\|full | 1180 | No | NA | 2 | 0 | 1 | Not-determined | Genome-fragment | NA | NA | 0 |
| S4-final-viral-combined_k141_117557\|\|full | 1019 | No | NA | 2 | 0 | 1 | Not-determined | Genome-fragment | NA | NA | 0 |
| S4-final-viral-combined_k141_19558\|\|full | 4946 | No | NA | 5 | 0 | 3 | Not-determined | Genome-fragment | NA | NA | 0 |
| S4-final-viral-combined_k141_166146\|\|full | 2968 | No | NA | 3 | 0 | 3 | Not-determined | Genome-fragment | NA | NA | 0 |
| S17-final-viral-combined_k141_120117\|\|full | 5479 | No | NA | 7 | 6 | 0 | Low-quality | Genome-fragment | 13.81 | AAI-based (high-confidence) | 0 |
| S4-final-viral-combined_k141_152349\|\|full | 8208 | No | NA | 9 | 0 | 2 | Not-determined | Genome-fragment | NA | NA | 0 |
| S8-final-viral-combined_k141_68852\|\|full | 5480 | No | NA | 5 | 0 | 3 | Low-quality | Genome-fragment | 5.15 | AAI-based (medium-confidence) | 0 |
| S2-final-viral-combined_k141_30443\|\|full | 5482 | No | NA | 12 | 1 | 1 | Low-quality | Genome-fragment | 10.17 | AAI-based (medium-confidence) | 0 |
| S17-final-viral-combined_k141_104478\|\|full | 5484 | Yes | 4184 | 7 | 5 | 2 | Low-quality | Genome-fragment | 9.06 | AAI-based (medium-confidence) | 23.71 |
| S4-final-viral-combined_k141_41048\|\|full | 1251 | No | NA | 2 | 0 | 2 | Not-determined | Genome-fragment | NA | NA | 0 |
| S4-final-viral-combined_k141_96883\|\|full | 2515 | No | NA | 4 | 0 | 2 | Not-determined | Genome-fragment | NA | NA | 0 |
| S4-final-viral-combined_k141_159804\|\|full | 1862 | No | NA | 4 | 0 | 1 | Not-determined | Genome-fragment | NA | NA | 0 |
| S4-final-viral-combined_k141_166199\|\|full | 2240 | No | NA | 2 | 0 | 1 | Not-determined | Genome-fragment | NA | NA | 0 |
| S17-final-viral-combined_k141_181782\|\|full | 5485 | No | NA | 5 | 0 | 2 | Low-quality | Genome-fragment | 14.98 | AAI-based (high-confidence) | 0 |
| S4-final-viral-combined_k141_145984\|\|full | 25693 | No | NA | 40 | 0 | 2 | Not-determined | Genome-fragment | NA | NA | 0 |
| S12-final-viral-combined_k141_53346\|\|full | 5485 | No | NA | 4 | 0 | 3 | Low-quality | Genome-fragment | 2.58 | AAI-based (medium-confidence) | 0 |
| S4-final-viral-combined_k141_166208\|\|full | 1464 | No | NA | 4 | 0 | 2 | Not-determined | Genome-fragment | NA | NA | 0 |
| S4-final-viral-combined_k141_48288\|\|full | 2300 | No | NA | 3 | 0 | 3 | Not-determined | Genome-fragment | NA | NA | 0 |
| S4-final-viral-combined_k141_41073\|\|full | 3264 | No | NA | 4 | 0 | 3 | Not-determined | Genome-fragment | NA | NA | 0 |
| S4-final-viral-combined_k141_41074\|\|full | 1019 | No | NA | 2 | 0 | 0 | Not-determined | Genome-fragment | NA | NA | 0 |
| S2_k141_247739 | 5485 | No | NA | 5 | 0 | 5 | Low-quality | Genome-fragment | 2.52 | AAI-based (high-confidence) | 0 |
| S4-final-viral-combined_k141_41078\|\|full | 4233 | No | NA | 3 | 0 | 3 | Not-determined | Genome-fragment | NA | NA | 0 |
| S6_k141_140272 | 5487 | No | NA | 7 | 1 | 0 | Low-quality | Genome-fragment | 10.81 | AAI-based (medium-confidence) | 0 |
| S4-final-viral-combined_k141_61715\|\|full | 2596 | No | NA | 3 | 0 | 2 | Not-determined | Genome-fragment | NA | NA | 0 |
| S4-final-viral-combined_k141_41086\|\|full | 2284 | No | NA | 2 | 0 | 1 | Not-determined | Genome-fragment | NA | NA | 0 |
| S4-final-viral-combined_k141_103540\|\|full | 1738 | No | NA | 3 | 0 | 0 | Not-determined | Genome-fragment | NA | NA | 0 |
| S4-final-viral-combined_k141_34294\|\|full | 6117 | No | NA | 8 | 0 | 4 | Not-determined | Genome-fragment | NA | NA | 0 |
| S2_k141_215541 | 5488 | No | NA | 5 | 0 | 4 | Low-quality | Genome-fragment | 0.79 | AAI-based (medium-confidence) | 0 |
| S17-final-viral-combined_k141_219349\|\|full | 5490 | No | NA | 9 | 6 | 0 | Low-quality | Genome-fragment | 16.22 | AAI-based (high-confidence) | 0 |
| S4-final-viral-combined_k141_153178\|\|full | 5491 | No | NA | 11 | 1 | 0 | Low-quality | Genome-fragment | 4.64 | HMM-based (lower-bound) | 0 |
| S9-final-viral-combined_k141_115394\|\|full | 5493 | No | NA | 6 | 3 | 0 | Low-quality | Genome-fragment | 8.66 | AAI-based (high-confidence) | 0 |
| S4-final-viral-combined_k141_117648\|\|full | 1649 | No | NA | 2 | 0 | 2 | Not-determined | Genome-fragment | NA | NA | 0 |
| S2-final-viral-combined_k141_184812\|\|full | 5498 | No | NA | 7 | 5 | 0 | Low-quality | Genome-fragment | 12.74 | AAI-based (medium-confidence) | 0 |
| S1-final-viral-combined_k141_36751\|\|full | 5499 | No | NA | 5 | 1 | 0 | Low-quality | Genome-fragment | 14.22 | AAI-based (medium-confidence) | 0 |
| S4-final-viral-combined_k141_6262\|\|full | 7045 | No | NA | 7 | 0 | 5 | Not-determined | Genome-fragment | NA | NA | 0 |
| S4-final-viral-combined_k141_166281\|\|full | 2109 | No | NA | 3 | 0 | 1 | Not-determined | Genome-fragment | NA | NA | 0 |
| S4-final-viral-combined_k141_19652\|\|full | 4996 | No | NA | 5 | 0 | 4 | Not-determined | Genome-fragment | NA | NA | 0 |
| S4-final-viral-combined_k141_83476\|\|full | 2361 | No | NA | 3 | 0 | 1 | Not-determined | Genome-fragment | NA | NA | 0 |
| S7-final-viral-combined_k141_148923\|\|full | 5499 | No | NA | 10 | 4 | 0 | Low-quality | Genome-fragment | 14.94 | AAI-based (high-confidence) | 0 |
| S4-final-viral-combined_k141_83485\|\|full | 4616 | No | NA | 5 | 0 | 1 | Not-determined | Genome-fragment | NA | NA | 0 |
| S7_k141_99609 | 5499 | No | NA | 5 | 5 | 0 | Low-quality | Genome-fragment | 13.48 | AAI-based (high-confidence) | 0 |
| S4-final-viral-combined_k141_98935\|\|full | 5502 | No | NA | 2 | 0 | 1 | Low-quality | Genome-fragment | 19.18 | AAI-based (medium-confidence) | 0 |
| S7-final-viral-combined_k141_202722\|\|full | 5503 | No | NA | 10 | 0 | 0 | Low-quality | Genome-fragment | 10.66 | AAI-based (medium-confidence) | 0 |
| S7_k141_103832 | 5503 | No | NA | 7 | 0 | 1 | Low-quality | Genome-fragment | 6.69 | AAI-based (medium-confidence) | 0 |
| S7_k141_157831 | 5506 | No | NA | 3 | 0 | 2 | Low-quality | Genome-fragment | 11.84 | AAI-based (high-confidence) | 0 |
| S12-final-viral-combined_k141_300052\|\|full | 5506 | No | NA | 17 | 2 | 0 | Low-quality | Genome-fragment | 11.55 | AAI-based (medium-confidence) | 0 |
| S4-final-viral-combined_k141_83492\|\|full | 38154 | No | NA | 52 | 9 | 2 | Medium-quality | Genome-fragment | 75.07 | HMM-based (lower-bound) | 0 |
| S4-final-viral-combined_k141_61860\|\|full | 2279 | No | NA | 5 | 0 | 0 | Not-determined | Genome-fragment | NA | NA | 0 |
| S4-final-viral-combined_k141_117754\|\|full | 1325 | No | NA | 3 | 0 | 0 | Not-determined | Genome-fragment | NA | NA | 0 |
| S2_k141_7221 | 5506 | No | NA | 7 | 0 | 6 | Low-quality | Genome-fragment | 2.31 | AAI-based (medium-confidence) | 0 |
| S4-final-viral-combined_k141_152452\|\|full | 1275 | No | NA | 3 | 0 | 0 | Not-determined | Genome-fragment | NA | NA | 0 |
| S5-final-viral-combined_k141_190577\|\|full | 5509 | No | NA | 7 | 5 | 0 | Low-quality | Genome-fragment | 9.09 | AAI-based (medium-confidence) | 0 |
| S7-final-viral-combined_k141_30140\|\|full | 5509 | No | NA | 7 | 1 | 2 | Low-quality | Genome-fragment | 8.34 | HMM-based (lower-bound) | 0 |
| S12_k141_266372 | 5510 | No | NA | 2 | 0 | 2 | Low-quality | Genome-fragment | 2.97 | AAI-based (medium-confidence) | 0 |
| S7-final-viral-combined_k141_98073\|\|full | 5511 | Yes | 2636 | 4 | 1 | 1 | Low-quality | Genome-fragment | 1.22 | HMM-based (lower-bound) | 52.17 |
| S9-final-viral-combined_k141_26972\|\|full | 5512 | No | NA | 8 | 0 | 2 | Low-quality | Genome-fragment | 6.97 | AAI-based (medium-confidence) | 0 |
| S1_k141_391726 | 5512 | No | NA | 3 | 0 | 1 | Low-quality | Genome-fragment | 6.23 | AAI-based (medium-confidence) | 0 |
| S4-final-viral-combined_k141_89950\|\|full | 3762 | No | NA | 3 | 0 | 2 | Not-determined | Genome-fragment | NA | NA | 0 |
| S4-final-viral-combined_k141_13535\|\|full | 1129 | No | NA | 2 | 0 | 1 | Not-determined | Genome-fragment | NA | NA | 0 |
| S4-final-viral-combined_k141_41214\|\|full | 2901 | No | NA | 3 | 0 | 1 | Not-determined | Genome-fragment | NA | NA | 0 |
| S4-final-viral-combined_k141_152488\|\|full | 1810 | No | NA | 2 | 0 | 0 | Not-determined | Genome-fragment | NA | NA | 0 |
| S2-final-viral-combined_k141_138006\|\|full | 5512 | No | NA | 5 | 0 | 4 | Low-quality | Genome-fragment | 9.39 | AAI-based (medium-confidence) | 0 |
| S17-final-viral-combined_k141_240045\|\|full | 5513 | No | NA | 7 | 3 | 0 | Low-quality | Genome-fragment | 7.46 | AAI-based (high-confidence) | 0 |
| S4-final-viral-combined_k141_111418\|\|full | 2567 | No | NA | 5 | 0 | 0 | Not-determined | Genome-fragment | NA | NA | 0 |
| S4-final-viral-combined_k141_160034\|\|full | 5369 | No | NA | 4 | 0 | 2 | Not-determined | Genome-fragment | NA | NA | 0 |
| S4-final-viral-combined_k141_146228\|\|full | 1087 | No | NA | 2 | 0 | 0 | Not-determined | Genome-fragment | NA | NA | 0 |
| S4-final-viral-combined_k141_131533\|\|full | 1519 | No | NA | 2 | 0 | 1 | Not-determined | Genome-fragment | NA | NA | 0 |
| S1_k141_214115 | 5517 | Yes | 1707 | 8 | 1 | 3 | Low-quality | Genome-fragment | 2.58 | HMM-based (lower-bound) | 69.06 |
| S4-final-viral-combined_k141_160043\|\|full | 1124 | No | NA | 3 | 0 | 1 | Not-determined | Genome-fragment | NA | NA | 0 |
| S11-final-viral-combined_k141_213591\|\|full | 5518 | No | NA | 8 | 4 | 0 | Low-quality | Genome-fragment | 11.83 | AAI-based (high-confidence) | 0 |
| S1-final-viral-combined_k141_109305\|\|full | 5521 | No | NA | 6 | 0 | 0 | Low-quality | Genome-fragment | 9.4 | AAI-based (medium-confidence) | 0 |
| S1_k141_330138 | 5522 | No | NA | 4 | 0 | 2 | Low-quality | Genome-fragment | 0.95 | AAI-based (medium-confidence) | 0 |
| S4-final-viral-combined_k141_138751\|\|full | 2209 | No | NA | 3 | 0 | 2 | Not-determined | Genome-fragment | NA | NA | 0 |
| S4-final-viral-combined_k141_124376\|\|full | 1202 | No | NA | 2 | 0 | 2 | Not-determined | Genome-fragment | NA | NA | 0 |
| S4-final-viral-combined_k141_27264\|\|full | 8646 | No | NA | 5 | 0 | 4 | Not-determined | Genome-fragment | NA | NA | 0 |
| S12-final-viral-combined_k141_199062\|\|full | 5523 | No | NA | 5 | 2 | 0 | Low-quality | Genome-fragment | 13.31 | AAI-based (high-confidence) | 0 |
| S4-final-viral-combined_k141_6469\|\|full | 1831 | No | NA | 2 | 0 | 1 | Not-determined | Genome-fragment | NA | NA | 0 |
| S4-final-viral-combined_k141_97111\|\|full | 4718 | No | NA | 8 | 0 | 2 | Not-determined | Genome-fragment | NA | NA | 0 |
| S4-final-viral-combined_k141_160086\|\|full | 1066 | No | NA | 2 | 0 | 0 | Not-determined | Genome-fragment | NA | NA | 0 |
| S4-final-viral-combined_k141_166447\|\|full | 3300 | No | NA | 5 | 0 | 1 | Not-determined | Genome-fragment | NA | NA | 0 |
| S4-final-viral-combined_k141_69435\|\|full | 3101 | No | NA | 4 | 0 | 4 | Not-determined | Genome-fragment | NA | NA | 0 |
| S11-final-viral-combined_k141_157151\|\|full | 5524 | No | NA | 4 | 0 | 2 | Low-quality | Genome-fragment | 7.2 | AAI-based (medium-confidence) | 0 |
| S12-final-viral-combined_k141_140499\|\|full | 5525 | Yes | 3465 | 6 | 2 | 1 | Low-quality | Genome-fragment | 5.22 | HMM-based (lower-bound) | 37.29 |
| S4-final-viral-combined_k141_97117\|\|full | 1082 | No | NA | 2 | 0 | 1 | Not-determined | Genome-fragment | NA | NA | 0 |
| S4-final-viral-combined_k141_69450\|\|full | 5825 | No | NA | 3 | 0 | 2 | Not-determined | Genome-fragment | NA | NA | 0 |

| | | | | | | | | | | | |
|---|---|---|---|---|---|---|---|---|---|---|---|
| S4-final-viral-combined_k141_90036\|\|full | 2058 | No | NA | 2 | 0 | 2 | Not-determined | Genome-fragment | NA | NA | 0 |
| S4-final-viral-combined_k141_97171\|\|full | 2734 | No | NA | 2 | 0 | 2 | Not-determined | Genome-fragment | NA | NA | 0 |
| S4-final-viral-combined_k141_54808\|\|full | 1444 | No | NA | 2 | 0 | 2 | Not-determined | Genome-fragment | NA | NA | 0 |
| S4-final-viral-combined_k141_62071\|\|full | 2231 | No | NA | 3 | 0 | 3 | Not-determined | Genome-fragment | NA | NA | 0 |
| S4-final-viral-combined_k141_166488\|\|full | 4039 | No | NA | 3 | 0 | 3 | Not-determined | Genome-fragment | NA | NA | 0 |
| S4-final-viral-combined_k141_97198\|\|full | 1634 | No | NA | 2 | 0 | 1 | Not-determined | Genome-fragment | NA | NA | 0 |
| S6-final-viral-combined_k141_92028\|\|full | 5525 | No | NA | 9 | 4 | 0 | Low-quality | Genome-fragment | 10.3 | AAI-based (medium-confidence) | 0 |
| S4-final-viral-combined_k141_19920\|\|full | 3246 | No | NA | 4 | 0 | 4 | Not-determined | Genome-fragment | NA | NA | 0 |
| S4-final-viral-combined_k141_117869\|\|full | 2304 | No | NA | 3 | 0 | 1 | Not-determined | Genome-fragment | NA | NA | 0 |
| S4-final-viral-combined_k141_4306\|\|full | 5526 | No | NA | 11 | 2 | 0 | Low-quality | Genome-fragment | 16.17 | AAI-based (high-confidence) | 0 |
| S3_k141_20915 | 5529 | No | NA | 8 | 5 | 0 | Low-quality | Genome-fragment | 10.27 | AAI-based (high-confidence) | 0 |
| S4-final-viral-combined_k141_166539\|\|full | 2054 | No | NA | 2 | 0 | 1 | Not-determined | Genome-fragment | NA | NA | 0 |
| S4-final-viral-combined_k141_54893\|\|full | 3132 | No | NA | 3 | 0 | 0 | Not-determined | Genome-fragment | NA | NA | 0 |
| S4-final-viral-combined_k141_138929\|\|full | 4490 | No | NA | 5 | 0 | 1 | Not-determined | Genome-fragment | NA | NA | 0 |
| S1-final-viral-combined_k141_116595\|\|full | 5532 | No | NA | 7 | 0 | 0 | Low-quality | Genome-fragment | 7.86 | AAI-based (medium-confidence) | 0 |
| S12-final-viral-combined_k141_94018\|\|full | 5532 | No | NA | 5 | 0 | 2 | Low-quality | Genome-fragment | 1.93 | AAI-based (medium-confidence) | 0 |
| S9_k141_66042 | 5533 | No | NA | 2 | 0 | 1 | Low-quality | Genome-fragment | 0.48 | AAI-based (medium-confidence) | 0 |
| S4-final-viral-combined_k141_6627\|\|full | 1202 | No | NA | 3 | 0 | 2 | Not-determined | Genome-fragment | NA | NA | 0 |
| S1_k141_172548 | 5534 | No | NA | 10 | 1 | 0 | Low-quality | Genome-fragment | 43.55 | AAI-based (high-confidence) | 0 |
| S4-final-viral-combined_k141_34481\|\|full | 1589 | No | NA | 2 | 0 | 1 | Not-determined | Genome-fragment | NA | NA | 0 |
| S4-final-viral-combined_k141_19955\|\|full | 3336 | No | NA | 3 | 0 | 3 | Not-determined | Genome-fragment | NA | NA | 0 |
| S4-final-viral-combined_k141_62182\|\|full | 1345 | No | NA | 3 | 0 | 3 | Not-determined | Genome-fragment | NA | NA | 0 |
| S4-final-viral-combined_k141_69575\|\|full | 4837 | No | NA | 6 | 0 | 6 | Not-determined | Genome-fragment | NA | NA | 0 |
| S8-final-viral-combined_k141_161057\|\|full | 5534 | No | NA | 8 | 0 | 0 | Low-quality | Genome-fragment | 14.34 | AAI-based (high-confidence) | 0 |
| S17-final-viral-combined_k141_35944\|\|full | 5535 | No | NA | 13 | 2 | 0 | Low-quality | Genome-fragment | 9.09 | AAI-based (high-confidence) | 0 |
| S11-final-viral-combined_k141_254271\|\|full | 5536 | No | NA | 4 | 0 | 2 | Low-quality | Genome-fragment | 3.2 | AAI-based (medium-confidence) | 0 |
| S4-final-viral-combined_k141_48625\|\|full | 1537 | No | NA | 2 | 0 | 0 | Not-determined | Genome-fragment | NA | NA | 0 |
| S7-final-viral-combined_k141_110472\|\|full | 5536 | No | NA | 6 | 0 | 5 | Low-quality | Genome-fragment | 4.83 | AAI-based (medium-confidence) | 0 |
| S4-final-viral-combined_k141_13723\|\|full | 3061 | No | NA | 4 | 0 | 0 | Not-determined | Genome-fragment | NA | NA | 0 |
| S12_k141_71977 | 5536 | No | NA | 6 | 0 | 0 | Low-quality | Genome-fragment | 8.86 | AAI-based (high-confidence) | 0 |
| S9_k141_97274 | 5537 | No | NA | 2 | 0 | 2 | Low-quality | Genome-fragment | 2.17 | AAI-based (medium-confidence) | 0 |
| S4-final-viral-combined_k141_90218\|\|full | 2861 | No | NA | 2 | 0 | 2 | Not-determined | Genome-fragment | NA | NA | 0 |
| S4-final-viral-combined_k141_48654\|\|full | 2183 | No | NA | 6 | 0 | 0 | Not-determined | Genome-fragment | NA | NA | 0 |
| S4-final-viral-combined_k141_166660\|\|full | 2042 | No | NA | 3 | 0 | 1 | Not-determined | Genome-fragment | NA | NA | 0 |
| S4-final-viral-combined_k141_34582\|\|full | 2124 | No | NA | 3 | 0 | 1 | Not-determined | Genome-fragment | NA | NA | 0 |
| S4-final-viral-combined_k141_139068\|\|full | 1363 | No | NA | 2 | 0 | 2 | Not-determined | Genome-fragment | NA | NA | 0 |
| S1_k141_27081 | 5539 | No | NA | 6 | 1 | 0 | Low-quality | Genome-fragment | 5.95 | HMM-based (lower-bound) | 0 |
| S5_k141_76337 | 5540 | No | NA | 9 | 2 | 0 | Low-quality | Genome-fragment | 16.75 | AAI-based (medium-confidence) | 0 |
| S4-final-viral-combined_k141_13770\|\|full | 3398 | No | NA | 3 | 0 | 1 | Not-determined | Genome-fragment | NA | NA | 0 |
| S4-final-viral-combined_k141_118000\|\|full | 1115 | No | NA | 3 | 0 | 1 | Not-determined | Genome-fragment | NA | NA | 0 |
| S4-final-viral-combined_k141_41468\|\|full | 2738 | No | NA | 4 | 0 | 1 | Not-determined | Genome-fragment | NA | NA | 0 |
| S2-final-viral-combined_k141_62712\|\|full | 5542 | Yes | 2007 | 4 | 1 | 1 | Low-quality | Genome-fragment | 1.54 | AAI-based (medium-confidence) | 63.79 |
| S12-final-viral-combined_k141_162078\|\|full | 5543 | No | NA | 10 | 0 | 0 | Low-quality | Genome-fragment | 12.92 | AAI-based (medium-confidence) | 0 |
| S5-final-viral-combined_k141_84219\|\|full | 5544 | No | NA | 4 | 0 | 0 | Low-quality | Genome-fragment | 3.26 | AAI-based (high-confidence) | 0 |
| S4-final-viral-combined_k141_139092\|\|full | 1141 | No | NA | 2 | 0 | 0 | Not-determined | Genome-fragment | NA | NA | 0 |
| S4-final-viral-combined_k141_103842\|\|full | 1135 | No | NA | 2 | 0 | 0 | Not-determined | Genome-fragment | NA | NA | 0 |
| S4-final-viral-combined_k141_62283\|\|full | 7929 | No | NA | 6 | 0 | 1 | Not-determined | Genome-fragment | NA | NA | 0 |
| S4-final-viral-combined_k141_118013\|\|full | 2266 | No | NA | 7 | 0 | 0 | Not-determined | Genome-fragment | NA | NA | 0 |
| S4-final-viral-combined_k141_166701\|\|full | 5669 | No | NA | 5 | 0 | 4 | Not-determined | Genome-fragment | NA | NA | 0 |
| S4-final-viral-combined_k141_90316\|\|full | 3351 | No | NA | 3 | 0 | 2 | Not-determined | Genome-fragment | NA | NA | 0 |
| S4-final-viral-combined_k141_48718\|\|full | 1055 | No | NA | 3 | 0 | 0 | Not-determined | Genome-fragment | NA | NA | 0 |
| S4_k141_55504 | 5544 | No | NA | 5 | 0 | 1 | Low-quality | Genome-fragment | 12.71 | AAI-based (medium-confidence) | 0 |
| S2-final-viral-combined_k141_114669\|\|full | 5546 | No | NA | 4 | 0 | 3 | Low-quality | Genome-fragment | 12.39 | AAI-based (medium-confidence) | 0 |
| S4-final-viral-combined_k141_103881\|\|full | 1627 | No | NA | 2 | 0 | 0 | Not-determined | Genome-fragment | NA | NA | 0 |
| S4-final-viral-combined_k141_145005\|\|full | 5546 | No | NA | 7 | 2 | 0 | Low-quality | Genome-fragment | 3.93 | HMM-based (lower-bound) | 0 |
| S17-final-viral-combined_k141_102956\|\|full | 5549 | No | NA | 5 | 0 | 1 | Low-quality | Genome-fragment | 4.31 | AAI-based (medium-confidence) | 0 |
| S17-k141_115367 | 5549 | No | NA | 7 | 1 | 2 | Low-quality | Genome-fragment | 9.64 | AAI-based (medium-confidence) | 0 |
| S4-final-viral-combined_k141_55020\|\|full | 23595 | No | NA | 10 | 0 | 8 | Not-determined | Genome-fragment | NA | NA | 0 |
| S4-final-viral-combined_k141_131844\|\|full | 1843 | No | NA | 3 | 0 | 0 | Not-determined | Genome-fragment | NA | NA | 0 |
| S1-final-viral-combined_k141_134515\|\|full | 5550 | No | NA | 5 | 0 | 2 | Low-quality | Genome-fragment | 6.19 | AAI-based (medium-confidence) | 0 |
| S4-final-viral-combined_k141_6764\|\|full | 3778 | No | NA | 4 | 0 | 2 | Not-determined | Genome-fragment | NA | NA | 0 |
| S4-final-viral-combined_k141_34715\|\|full | 2122 | No | NA | 2 | 0 | 0 | Not-determined | Genome-fragment | NA | NA | 0 |
| S4-final-viral-combined_k141_139192\|\|full | 1286 | No | NA | 4 | 0 | 3 | Not-determined | Genome-fragment | NA | NA | 0 |
| S4-final-viral-combined_k141_13842\|\|full | 12975 | No | NA | 10 | 0 | 5 | Not-determined | Genome-fragment | NA | NA | 0 |
| S4-final-viral-combined_k141_41490\|\|full | 87229 | Yes | 33472 | 89 | 16 | 33 | Medium-quality | Genome-fragment | 55.23 | HMM-based (lower-bound) | 61.63 |
| S4-final-viral-combined_k141_62378\|\|full | 2895 | No | NA | 3 | 0 | 1 | Not-determined | Genome-fragment | NA | NA | 0 |
| S4-final-viral-combined_k141_34740\|\|full | 2602 | No | NA | 3 | 0 | 2 | Not-determined | Genome-fragment | NA | NA | 0 |
| [illegible] | [illegible] | [illegible] | [illegible] | [illegible] | [illegible] | [illegible] | [illegible] | [illegible] | [illegible] | [illegible] | [illegible] |

| | | | | | | | | | | | |
|---|---|---|---|---|---|---|---|---|---|---|---|
| S2-final-viral-combined_k141_49196\|\|full | 5732 | No | NA | 7 | 2 | 0 | Low-quality | Genome-fragment | 7.06 | AAI-based (medium-confidence) | 0 |
| S2-final-viral-combined_k141_47327\|\|full | 5734 | Yes | 3365 | 9 | 1 | 2 | Low-quality | Genome-fragment | 1.43 | HMM-based (lower-bound) | 41.31 |
| S4_k141_84257 | 2687 | No | NA | 4 | 0 | 2 | Not-determined | Genome-fragment | NA | NA | 0 |
| S4_k141_84260 | 1042 | No | NA | 1 | 0 | 1 | Not-determined | Genome-fragment | NA | NA | 0 |
| S7-final-viral-combined_k141_218664\|\|full | 5736 | No | NA | 5 | 1 | 0 | Low-quality | Genome-fragment | 10.32 | AAI-based (high-confidence) | 0 |
| S1_k141_137680 | 5738 | Yes | 3392 | 6 | 1 | 2 | Low-quality | Genome-fragment | 4.2 | HMM-based (lower-bound) | 40.89 |
| S4_k141_91343 | 1351 | No | NA | 1 | 0 | 1 | Not-determined | Genome-fragment | NA | NA | 0 |
| S4_k141_28520 | 7453 | No | NA | 5 | 0 | 3 | Not-determined | Genome-fragment | NA | NA | 0 |
| S4_k141_7476 | 1535 | No | NA | 3 | 0 | 2 | Not-determined | Genome-fragment | NA | NA | 0 |
| S4_k141_77399 | 1263 | No | NA | 2 | 0 | 1 | Not-determined | Genome-fragment | NA | NA | 0 |
| S4_k141_153774 | 1582 | No | NA | 1 | 0 | 0 | Not-determined | Genome-fragment | NA | NA | 0 |
| S17-final-viral-combined_k141_246097\|\|full | 5739 | No | NA | 5 | 0 | 2 | Low-quality | Genome-fragment | 1.78 | AAI-based (medium-confidence) | 0 |
| S4_k141_105169 | 1350 | No | NA | 3 | 0 | 1 | Not-determined | Genome-fragment | NA | NA | 0 |
| S4_k141_772 | 1901 | No | NA | 4 | 0 | 3 | Not-determined | Genome-fragment | NA | NA | 0 |
| S4_k141_146886 | 1119 | No | NA | 1 | 0 | 0 | Not-determined | Genome-fragment | NA | NA | 0 |
| S4_k141_91382 | 1303 | No | NA | 3 | 0 | 2 | Not-determined | Genome-fragment | NA | NA | 0 |
| S6_k141_68687 | 5740 | No | NA | 9 | 0 | 0 | Low-quality | Genome-fragment | 3.07 | AAI-based (high-confidence) | 0 |
| S4_k141_777 | 1453 | No | NA | 1 | 0 | 0 | Not-determined | Genome-fragment | NA | NA | 0 |
| S4_k141_126199 | 1407 | No | NA | 2 | 0 | 2 | Not-determined | Genome-fragment | NA | NA | 0 |
| S4_k141_132874 | 1476 | No | NA | 1 | 0 | 1 | Not-determined | Genome-fragment | NA | NA | 0 |
| S4_k141_49308 | 4174 | No | NA | 6 | 0 | 1 | Not-determined | Genome-fragment | NA | NA | 0 |
| S4_k141_112161 | 1790 | No | NA | 2 | 0 | 1 | Not-determined | Genome-fragment | NA | NA | 0 |
| S4_k141_126205 | 5430 | No | NA | 6 | 0 | 1 | Not-determined | Genome-fragment | NA | NA | 0 |
| S7-final-viral-combined_k141_158695\|\|full | 5741 | No | NA | 3 | 0 | 0 | Low-quality | Genome-fragment | 5.96 | AAI-based (medium-confidence) | 0 |
| S4_k141_91401 | 1227 | No | NA | 3 | 0 | 1 | Not-determined | Genome-fragment | NA | NA | 0 |
| S4_k141_105224 | 1198 | No | NA | 2 | 0 | 1 | Not-determined | Genome-fragment | NA | NA | 0 |
| S4_k141_28550 | 1252 | No | NA | 2 | 0 | 1 | Not-determined | Genome-fragment | NA | NA | 0 |
| S4_k141_14521 | 2751 | No | NA | 2 | 0 | 1 | Not-determined | Genome-fragment | NA | NA | 0 |
| S4_k141_14522 | 1080 | No | NA | 1 | 0 | 1 | Not-determined | Genome-fragment | NA | NA | 0 |
| S4_k141_146916 | 1201 | No | NA | 1 | 0 | 1 | Not-determined | Genome-fragment | NA | NA | 0 |
| S4_k141_153833 | 3291 | No | NA | 2 | 0 | 1 | Not-determined | Genome-fragment | NA | NA | 0 |
| S4_k141_153838 | 1404 | No | NA | 1 | 0 | 1 | Not-determined | Genome-fragment | NA | NA | 0 |
| S4_k141_795 | 1834 | No | NA | 4 | 0 | 3 | Not-determined | Genome-fragment | NA | NA | 0 |
| S4_k141_21575 | 3196 | No | NA | 8 | 0 | 3 | Not-determined | Genome-fragment | NA | NA | 0 |
| S4_k141_21583 | 1142 | No | NA | 1 | 0 | 1 | Not-determined | Genome-fragment | NA | NA | 0 |
| S4_k141_84391 | 1124 | No | NA | 1 | 0 | 1 | Not-determined | Genome-fragment | NA | NA | 0 |
| S17-k141_250663 | 5742 | No | NA | 6 | 0 | 0 | Low-quality | Genome-fragment | 3.51 | AAI-based (medium-confidence) | 0 |
| S4_k141_42519 | 1187 | No | NA | 3 | 0 | 2 | Not-determined | Genome-fragment | NA | NA | 0 |
| S4_k141_821 | 5378 | No | NA | 2 | 0 | 0 | Not-determined | Genome-fragment | NA | NA | 0 |
| S6_k141_114894 | 5746 | No | NA | 6 | 0 | 3 | Low-quality | Genome-fragment | 19.92 | AAI-based (medium-confidence) | 0 |
| S4_k141_98264 | 1233 | No | NA | 1 | 0 | 1 | Not-determined | Genome-fragment | NA | NA | 0 |
| S11-final-viral-combined_k141_277850\|\|full | 5747 | No | NA | 8 | 0 | 0 | Low-quality | Genome-fragment | 13.47 | AAI-based (high-confidence) | 0 |
| S4_k141_21601 | 1208 | No | NA | 1 | 0 | 0 | Not-determined | Genome-fragment | NA | NA | 0 |
| S11-final-viral-combined_k141_61406\|\|full | 5747 | No | NA | 6 | 0 | 3 | Low-quality | Genome-fragment | 6.12 | AAI-based (medium-confidence) | 0 |
| S5-final-viral-combined_k141_3188\|\|full | 5748 | No | NA | 12 | 1 | 0 | Low-quality | Genome-fragment | 8.95 | HMM-based (lower-bound) | 0 |
| S12-final-viral-combined_k141_200867\|\|full | 5748 | No | NA | 5 | 0 | 2 | Low-quality | Genome-fragment | 9.78 | AAI-based (high-confidence) | 0 |
| S4_k141_21615 | 1275 | No | NA | 2 | 0 | 1 | Not-determined | Genome-fragment | NA | NA | 0 |
| S1-final-viral-combined_k141_358535\|\|full | 5749 | Yes | 2958 | 7 | 1 | 2 | Low-quality | Genome-fragment | 6.74 | AAI-based (medium-confidence) | 48.55 |
| S4_k141_140144 | 4486 | No | NA | 4 | 0 | 3 | Not-determined | Genome-fragment | NA | NA | 0 |
| S4_k141_140148 | 1309 | No | NA | 3 | 0 | 0 | Not-determined | Genome-fragment | NA | NA | 0 |
| S4_k141_49373 | 1449 | No | NA | 1 | 0 | 0 | Not-determined | Genome-fragment | NA | NA | 0 |
| S9-final-viral-combined_k141_126647\|\|full | 5750 | No | NA | 5 | 0 | 3 | Low-quality | Genome-fragment | 3.84 | AAI-based (high-confidence) | 0 |
| S4_k141_42606 | 16960 | No | NA | 10 | 0 | 8 | Not-determined | Genome-fragment | NA | NA | 0 |
| S7_k141_145875 | 5755 | No | NA | 12 | 1 | 0 | Low-quality | Genome-fragment | 5.41 | AAI-based (high-confidence) | 0 |
| S4_k141_70563 | 1031 | No | NA | 1 | 0 | 0 | Not-determined | Genome-fragment | NA | NA | 0 |
| S4_k141_21633 | 1015 | No | NA | 1 | 0 | 0 | Not-determined | Genome-fragment | NA | NA | 0 |
| S4_k141_14551 | 1404 | No | NA | 2 | 0 | 2 | Not-determined | Genome-fragment | NA | NA | 0 |
| S4_k141_28636 | 3673 | No | NA | 4 | 0 | 3 | Not-determined | Genome-fragment | NA | NA | 0 |
| S4_k141_42643 | 1624 | No | NA | 1 | 0 | 1 | Not-determined | Genome-fragment | NA | NA | 0 |
| S4_k141_119051 | 5930 | No | NA | 3 | 0 | 3 | Not-determined | Genome-fragment | NA | NA | 0 |
| S4_k141_112293 | 1071 | No | NA | 1 | 0 | 1 | Not-determined | Genome-fragment | NA | NA | 0 |
| S4_k141_119061 | 4200 | No | NA | 4 | 0 | 1 | Not-determined | Genome-fragment | NA | NA | 0 |
| S4_k141_70616 | 3140 | No | NA | 7 | 0 | 3 | Not-determined | Genome-fragment | NA | NA | 0 |
| S4_k141_133003 | 1747 | No | NA | 1 | 0 | 1 | Not-determined | Genome-fragment | NA | NA | 0 |
| S4_k141_153988 | 1040 | No | NA | 1 | 0 | 1 | Not-determined | Genome-fragment | NA | NA | 0 |
| S4_k141_21671 | 1238 | No | NA | 1 | 0 | 1 | Not-determined | Genome-fragment | NA | NA | 0 |
| S4_k141_133015 | 1423 | No | NA | 2 | 0 | 1 | Not-determined | Genome-fragment | NA | NA | 0 |
| S4_k141_56471 | 4103 | No | NA | 5 | 0 | 3 | Not-determined | Genome-fragment | NA | NA | 0 |
| S4_k141_70639 | 1185 | No | NA | 1 | 0 | 0 | Not-determined | Genome-fragment | NA | NA | 0 |
| S4_k141_898 | 1244 | No | NA | 2 | 0 | 2 | Not-determined | Genome-fragment | NA | NA | 0 |
| S2-final-viral-combined_k141_195361\|\|full | 5756 | Yes | 3918 | 6 | 1 | 2 | Low-quality | Genome-fragment | 1.85 | AAI-based (high-confidence) | 31.93 |
| S3-final-viral-combined_k141_128342\|\|full | 5757 | No | NA | 9 | 0 | 1 | Low-quality | Genome-fragment | 13.44 | AAI-based (medium-confidence) | 0 |
| S8-final-viral-combined_k141_270503\|\|full | 5757 | No | NA | 9 | 1 | 1 | Low-quality | Genome-fragment | 3.21 | HMM-based (lower-bound) | 0 |
| S4_k141_910 | 2162 | No | NA | 2 | 0 | 1 | Not-determined | Genome-fragment | NA | NA | 0 |
| S4_k141_63485 | 1537 | No | NA | 2 | 0 | 2 | Not-determined | Genome-fragment | NA | NA | 0 |
| S4_k141_140192 | 1853 | No | NA | 2 | 0 | 0 | Not-determined | Genome-fragment | NA | NA | 0 |
| S4_k141_133026 | 5317 | No | NA | 3 | 0 | 2 | Not-determined | Genome-fragment | NA | NA | 0 |
| S4_k141_119077 | 1466 | No | NA | 1 | 0 | 1 | Not-determined | Genome-fragment | NA | NA | 0 |
| S4_k141_21684 | 1115 | No | NA | 1 | 0 | 1 | Not-determined | Genome-fragment | NA | NA | 0 |
| S4_k141_14615 | 12982 | No | NA | 6 | 0 | 4 | Not-determined | Genome-fragment | NA | NA | 0 |
| S1-final-viral-combined_k141_302985\|\|full | 5758 | No | NA | 4 | 0 | 3 | Low-quality | Genome-fragment | 5.22 | AAI-based (medium-confidence) | 0 |
| S4_k141_133055 | 6623 | No | NA | 2 | 0 | 1 | Not-determined | Genome-fragment | NA | NA | 0 |
| S4_k141_133070 | 1164 | No | NA | 1 | 0 | 0 | Not-determined | Genome-fragment | NA | NA | 0 |
| S4_k141_119111 | 1088 | No | NA | 2 | 0 | 1 | Not-determined | Genome-fragment | NA | NA | 0 |
| S4_k141_77540 | 1031 | No | NA | 2 | 0 | 2 | Not-determined | Genome-fragment | NA | NA | 0 |
| S4_k141_42676 | 5407 | No | NA | 3 | 0 | 3 | Not-determined | Genome-fragment | NA | NA | 0 |
| S7-final-viral-combined_k141_139764\|\|full | 5758 | No | NA | 8 | 0 | 1 | Low-quality | Genome-fragment | 5.6 | AAI-based (high-confidence) | 0 |
| S7-final-viral-combined_k141_213614\|\|full | 5758 | No | NA | 6 | 3 | 0 | Low-quality | Genome-fragment | 16.2 | AAI-based (high-confidence) | 0 |
| S4_k141_7682 | 1253 | No | NA | 2 | 0 | 0 | Not-determined | Genome-fragment | NA | NA | 0 |
| S4_k141_70655 | 3353 | No | NA | 3 | 0 | 1 | Not-determined | Genome-fragment | NA | NA | 0 |
| S4_k141_105509 | 1109 | No | NA | 1 | 0 | 1 | Not-determined | Genome-fragment | NA | NA | 0 |
| S4_k141_98352 | 1092 | No | NA | 1 | 0 | 1 | Not-determined | Genome-fragment | NA | NA | 0 |
| S4_k141_126483 | 1666 | No | NA | 1 | 0 | 0 | Not-determined | Genome-fragment | NA | NA | 0 |
| S4_k141_140245 | 1390 | No | NA | 2 | 0 | 1 | Not-determined | Genome-fragment | NA | NA | 0 |
| S12-final-viral-combined_k141_109590\|\|full | 5758 | No | NA | 3 | 0 | 3 | Low-quality | Genome-fragment | 2.3 | AAI-based (medium-confidence) | 0 |
| S4_k141_147133 | 8361 | No | NA | 5 | 0 | 3 | Not-determined | Genome-fragment | NA | NA | 0 |
| S2_k141_95554 | 5759 | No | NA | 10 | 4 | 0 | Low-quality | Genome-fragment | 5.58 | AAI-based (high-confidence) | 0 |
| S8-final-viral-combined_k141_244196\|\|full | 5761 | No | NA | 6 | 0 | 3 | Low-quality | Genome-fragment | 7.02 | AAI-based (medium-confidence) | 0 |
| S4_k141_21701 | 1177 | No | NA | 2 | 0 | 0 | Not-determined | Genome-fragment | NA | NA | 0 |
| S4_k141_49448 | 7461 | No | NA | 7 | 0 | 5 | Not-determined | Genome-fragment | NA | NA | 0 |
| S17-k141_5812 | 5763 | No | NA | 3 | 0 | 2 | Low-quality | Genome-fragment | 2.39 | AAI-based (high-confidence) | 0 |
| S4_k141_119184 | 3144 | No | NA | 4 | 0 | 3 | Not-determined | Genome-fragment | NA | NA | 0 |
| S4_k141_126561 | 1052 | No | NA | 1 | 0 | 1 | Not-determined | Genome-fragment | NA | NA | 0 |
| S4_k141_21723 | 1671 | No | NA | 2 | 0 | 0 | Not-determined | Genome-fragment | NA | NA | 0 |
| S4_k141_28834 | 5049 | No | NA | 5 | 0 | 3 | Not-determined | Genome-fragment | NA | NA | 0 |
| S4_k141_21741 | 1292 | No | NA | 2 | 0 | 2 | Not-determined | Genome-fragment | NA | NA | 0 |
| S4_k141_140303 | 3319 | No | NA | 4 | 0 | 3 | Not-determined | Genome-fragment | NA | NA | 0 |
| S2-final-viral-combined_k141_198481\|\|full | 5764 | No | NA | 12 | 1 | 0 | Low-quality | Genome-fragment | 14.26 | AAI-based (high-confidence) | 0 |
| S9-final-viral-combined_k141_110452\|\|full | 5765 | No | NA | 4 | 0 | 0 | Low-quality | Genome-fragment | 6.1 | AAI-based (medium-confidence) | 0 |
| S4_k141_21779 | 1413 | No | NA | 3 | 0 | 0 | Not-determined | Genome-fragment | NA | NA | 0 |
| S1-final-viral-combined_k141_194245\|\|full | 5767 | No | NA | 8 | 5 | 0 | Low-quality | Genome-fragment | 13.39 | AAI-based (high-confidence) | 0 |
| S9-final-viral-combined_k141_106800\|\|full | 5771 | No | NA | 10 | 7 | 0 | Low-quality | Genome-fragment | 14.41 | AAI-based (medium-confidence) | 0 |
| S4_k141_140351 | 1337 | No | NA | 1 | 0 | 1 | Not-determined | Genome-fragment | NA | NA | 0 |
| S4-final-viral-combined_k141_27156\|\|full | 5771 | Yes | 835 | 6 | 1 | 4 | Low-quality | Genome-fragment | 1.37 | HMM-based (lower-bound) | 85.53 |
| S4_k141_140360 | 1247 | No | NA | 1 | 0 | 1 | Not-determined | Genome-fragment | NA | NA | 0 |
| S4_k141_98460 | 1721 | No | NA | 1 | 0 | 1 | Not-determined | Genome-fragment | NA | NA | 0 |
| S4_k141_7763 | 6477 | No | NA | 9 | 0 | 5 | Not-determined | Genome-fragment | NA | NA | 0 |
| S4_k141_1057 | 1421 | No | NA | 1 | 0 | 0 | Not-determined | Genome-fragment | NA | NA | 0 |
| S4_k141_126638 | 1417 | No | NA | 1 | 0 | 1 | Not-determined | Genome-fragment | NA | NA | 0 |
| S4_k141_84677 | 1383 | No | NA | 1 | 0 | 0 | Not-determined | Genome-fragment | NA | NA | 0 |
| S11_k141_297511 | 5772 | No | NA | 2 | 0 | 1 | Low-quality | Genome-fragment | 1.99 | AAI-based (medium-confidence) | 0 |
| S11_k141_53788 | 5772 | No | NA | 3 | 0 | 3 | Low-quality | Genome-fragment | 18.43 | AAI-based (medium-confidence) | 0 |
| S9_k141_96609 | 5773 | No | NA | 7 | 3 | 0 | Low-quality | Genome-fragment | 14.04 | AAI-based (high-confidence) | 0 |
| S4_k141_14752 | 5485 | No | NA | 8 | 0 | 2 | Not-determined | Genome-fragment | NA | NA | 0 |
| S4_k141_84701 | 7052 | No | NA | 7 | 0 | 3 | Not-determined | Genome-fragment | NA | NA | 0 |
| S4_k141_7801 | 1004 | No | NA | 1 | 0 | 1 | Not-determined | Genome-fragment | NA | NA | 0 |
| S4_k141_105676 | 1456 | No | NA | 1 | 0 | 0 | Not-determined | Genome-fragment | NA | NA | 0 |
| S4_k141_63744 | 1605 | No | NA | 1 | 0 | 0 | Not-determined | Genome-fragment | NA | NA | 0 |
| S5-final-viral-combined_k141_3108\|\|full | 5774 | No | NA | 12 | 3 | 0 | Low-quality | Genome-fragment | 14.83 | AAI-based (medium-confidence) | 0 |
| S4_k141_147299 | 2207 | No | NA | 3 | 0 | 1 | Not-determined | Genome-fragment | NA | NA | 0 |
| S4_k141_1083 | 1114 | No | NA | 1 | 0 | 0 | Not-determined | Genome-fragment | NA | NA | 0 |
| S4_k141_154264 | 1221 | No | NA | 2 | 0 | 1 | Not-determined | Genome-fragment | NA | NA | 0 |
| S4_k141_63749 | 1741 | No | NA | 1 | 0 | 0 | Not-determined | Genome-fragment | NA | NA | 0 |
| S4_k141_1090 | 1267 | No | NA | 2 | 0 | 2 | Not-determined | Genome-fragment | NA | NA | 0 |
| S4_k141_49489 | 4720 | No | NA | 8 | 0 | 2 | Not-determined | Genome-fragment | NA | NA | 0 |
| S4_k141_42840 | 1122 | No | NA | 1 | 0 | 1 | Not-determined | Genome-fragment | NA | NA | 0 |
| S4_k141_77769 | 3261 | No | NA | 5 | 0 | 5 | Not-determined | Genome-fragment | NA | NA | 0 |
| S4_k141_35684 | 2126 | No | NA | 2 | 0 | 2 | Not-determined | Genome-fragment | NA | NA | 0 |
| S4_k141_29010 | 3694 | No | NA | 7 | 0 | 1 | Not-determined | Genome-fragment | NA | NA | 0 |
| S4_k141_119358 | 1020 | No | NA | 2 | 0 | 1 | Not-determined | Genome-fragment | NA | NA | 0 |
| S4_k141_1127 | 2385 | No | NA | 1 | 0 | 1 | Not-determined | Genome-fragment | NA | NA | 0 |
| S1_k141_109044 | 5776 | Yes | 4745 | 8 | 1 | 1 | Low-quality | Genome-fragment | 34.62 | AAI-based (medium-confidence) | 17.85 |
| S4_k141_56698 | 1133 | No | NA | 2 | 0 | 1 | Not-determined | Genome-fragment | NA | NA | 0 |
| S4_k141_35701 | 1971 | No | NA | 2 | 0 | 2 | Not-determined | Genome-fragment | NA | NA | 0 |
| S4_k141_35714 | 1322 | No | NA | 2 | 0 | 2 | Not-determined | Genome-fragment | NA | NA | 0 |
| S8-final-viral-combined_k141_131782\|\|full | 5786 | No | NA | 10 | 7 | 0 | Low-quality | Genome-fragment | 17.31 | AAI-based (medium-confidence) | 0 |
| S4_k141_112636 | 1693 | No | NA | 2 | 0 | 0 | Not-determined | Genome-fragment | NA | NA | 0 |
| S4_k141_39988 | 5790 | No | NA | 5 | 0 | 4 | Low-quality | Genome-fragment | 1.51 | AAI-based (medium-confidence) | 0 |
| S4_k141_14754 | 2060 | No | NA | 2 | 0 | 2 | Not-determined | Genome-fragment | NA | NA | 0 |
| S17-k141_260606 | 5791 | No | NA | 6 | 2 | 0 | Low-quality | Genome-fragment | 14.66 | AAI-based (high-confidence) | 0 |
| S5-final-viral-combined_k141_69458\|\|full | 5792 | No | NA | 6 | 2 | 0 | Low-quality | Genome-fragment | 6.18 | AAI-based (high-confidence) | 0 |
| S4_k141_112671 | 2204 | No | NA | 5 | 0 | 2 | Not-determined | Genome-fragment | NA | NA | 0 |
| S4_k141_29043 | 1565 | No | NA | 3 | 0 | 2 | Not-determined | Genome-fragment | NA | NA | 0 |
| S9_k141_333 | 5793 | No | NA | 9 | 1 | 0 | Low-quality | Genome-fragment | 12.79 | HMM-based (lower-bound) | 0 |
| S5-final-viral-combined_k141_139118\|\|full | 5794 | No | NA | 7 | 0 | 1 | Low-quality | Genome-fragment | 11.25 | AAI-based (medium-confidence) | 0 |
| S4_k141_84794 | 1030 | No | NA | 2 | 0 | 0 | Not-determined | Genome-fragment | NA | NA | 0 |
| S4_k141_126768 | 1542 | No | NA | 1 | 0 | 0 | Not-determined | Genome-fragment | NA | NA | 0 |
| S4_k141_98610 | 10039 | No | NA | 10 | 0 | 4 | Not-determined | Genome-fragment | NA | NA | 0 |
| S4_k141_42946 | 1646 | No | NA | 4 | 0 | 1 | Not-determined | Genome-fragment | NA | NA | 0 |
| S4_k141_29071 | 3848 | No | NA | 6 | 0 | 5 | Not-determined | Genome-fragment | NA | NA | 0 |
| S4_k141_126776 | 3989 | No | NA | 2 | 0 | 2 | Not-determined | Genome-fragment | NA | NA | 0 |
| S4_k141_140528 | 1864 | No | NA | 1 | 0 | 1 | Not-determined | Genome-fragment | NA | NA | 0 |
| S1-final-viral-combined_k141_119405\|\|full | 5796 | No | NA | 9 | 0 | 2 | Low-quality | Genome-fragment | 4.66 | AAI-based (medium-confidence) | 0 |
| S4_k141_119476 | 3900 | No | NA | 2 | 0 | 1 | Not-determined | Genome-fragment | NA | NA | 0 |
| S4_k141_42992 | 1396 | No | NA | 1 | 0 | 0 | Not-determined | Genome-fragment | NA | NA | 0 |
| S4_k141_91946 | 1781 | No | NA | 2 | 0 | 1 | Not-determined | Genome-fragment | NA | NA | 0 |
| S4_k141_147411 | 5525 | No | NA | 2 | 0 | 1 | Not-determined | Genome-fragment | NA | NA | 0 |
| S4_k141_56819 | 1084 | No | NA | 2 | 0 | 1 | Not-determined | Genome-fragment | NA | NA | 0 |
| S4_k141_56820 | 1055 | No | NA | 1 | 0 | 1 | Not-determined | Genome-fragment | NA | NA | 0 |
| S5_k141_198438 | 5797 | No | NA | 10 | 8 | 0 | Low-quality | Genome-fragment | 18.11 | AAI-based (high-confidence) | 0 |
| S4_k141_35821 | 1101 | No | NA | 1 | 0 | 1 | Not-determined | Genome-fragment | NA | NA | 0 |
| S4_k141_77969 | 1098 | No | NA | 1 | 0 | 0 | Not-determined | Genome-fragment | NA | NA | 0 |
| S4_k141_63897 | 1020 | No | NA | 1 | 0 | 1 | Not-determined | Genome-fragment | NA | NA | 0 |
| S4_k141_14845 | 1020 | No | NA | 1 | 0 | 0 | Not-determined | Genome-fragment | NA | NA | 0 |
| S4_k141_133458 | 2892 | No | NA | 2 | 0 | 2 | Not-determined | Genome-fragment | NA | NA | 0 |
| S4_k141_154361 | 1055 | No | NA | 2 | 0 | 1 | Not-determined | Genome-fragment | NA | NA | 0 |
| S9_k141_89276 | 5798 | No | NA | 3 | 0 | 2 | Low-quality | Genome-fragment | 5.37 | AAI-based (medium-confidence) | 0 |
| S4_k141_154371 | 1025 | No | NA | 2 | 0 | 0 | Not-determined | Genome-fragment | NA | NA | 0 |
| S4_k141_78011 | 1846 | No | NA | 2 | 0 | 0 | Not-determined | Genome-fragment | NA | NA | 0 |
| S4_k141_119559 | 1004 | No | NA | 1 | 0 | 0 | Not-determined | Genome-fragment | NA | NA | 0 |
| S4_k141_35839 | 1133 | No | NA | 1 | 0 | 0 | Not-determined | Genome-fragment | NA | NA | 0 |
| S1_k141_212478 | 5798 | No | NA | 6 | 0 | 5 | Low-quality | Genome-fragment | 3.27 | AAI-based (medium-confidence) | 0 |
| S4_k141_78051 | 1858 | No | NA | 2 | 0 | 0 | Not-determined | Genome-fragment | NA | NA | 0 |
| S4_k141_49566 | 3530 | No | NA | 4 | 0 | 0 | Not-determined | Genome-fragment | NA | NA | 0 |
| S12_k141_309577 | 5799 | No | NA | 9 | 0 | 4 | Low-quality | Genome-fragment | 3.16 | AAI-based (high-confidence) | 0 |
| S6-final-viral-combined_k141_25350\|\|full | 5799 | No | NA | 8 | 0 | 0 | Low-quality | Genome-fragment | 8.98 | AAI-based (high-confidence) | 0 |
| S4_k141_63962 | 1612 | No | NA | 3 | 0 | 1 | Not-determined | Genome-fragment | NA | NA | 0 |
| S8-final-viral-combined_k141_66841\|\|full | 5799 | No | NA | 11 | 2 | 0 | Low-quality | Genome-fragment | 6.82 | HMM-based (lower-bound) | 0 |
| S4_k141_29193 | 3166 | No | NA | 3 | 0 | 3 | Not-determined | Genome-fragment | NA | NA | 0 |
| S4_k141_1315 | 5371 | No | NA | 3 | 0 | 2 | Not-determined | Genome-fragment | NA | NA | 0 |
| S2-final-viral-combined_k141_62294\|\|full | 5800 | No | NA | 4 | 0 | 4 | Low-quality | Genome-fragment | 2.61 | AAI-based (medium-confidence) | 0 |
| S4_k141_63994 | 1047 | No | NA | 1 | 0 | 1 | Not-determined | Genome-fragment | NA | NA | 0 |
| S4_k141_35870 | 1061 | No | NA | 3 | 0 | 0 | Not-determined | Genome-fragment | NA | NA | 0 |
| S4_k141_22053 | 3988 | No | NA | 6 | 0 | 2 | Not-determined | Genome-fragment | NA | NA | 0 |
| S3_k141_52355 | 5801 | No | NA | 15 | 1 | 0 | Low-quality | Genome-fragment | 10.01 | AAI-based (high-confidence) | 0 |
| S4_k141_8144 | 1022 | No | NA | 1 | 0 | 1 | Not-determined | Genome-fragment | NA | NA | 0 |
| S4_k141_35914 | 1179 | No | NA | 2 | 0 | 2 | Not-determined | Genome-fragment | NA | NA | 0 |
| S4_k141_119590 | 5982 | No | NA | 3 | 0 | 2 | Not-determined | Genome-fragment | NA | NA | 0 |
| S4_k141_70958 | 4004 | No | NA | 5 | 0 | 0 | Not-determined | Genome-fragment | NA | NA | 0 |
| S4_k141_35923 | 1789 | No | NA | 2 | 0 | 1 | Not-determined | Genome-fragment | NA | NA | 0 |
| S4_k141_161286 | 1291 | No | NA | 2 | 0 | 2 | Not-determined | Genome-fragment | NA | NA | 0 |
| S4-final-viral-combined_k141_21592\|\|full | 5801 | No | NA | 7 | 3 | 0 | Low-quality | Genome-fragment | 9.52 | AAI-based (high-confidence) | 0 |
| S4_k141_92067 | 4972 | No | NA | 5 | 0 | 0 | Not-determined | Genome-fragment | NA | NA | 0 |
| S4_k141_147559 | 9868 | No | NA | 8 | 0 | 5 | Not-determined | Genome-fragment | NA | NA | 0 |
| S8-final-viral-combined_k141_94121\|\|full | 5801 | No | NA | 11 | 1 | 2 | Low-quality | Genome-fragment | 14.68 | AAI-based (medium-confidence) | 0 |
| S1_k141_370963 | 5803 | No | NA | 4 | 0 | 3 | Low-quality | Genome-fragment | 4.01 | AAI-based (medium-confidence) | 0 |
| S4_k141_49678 | 1150 | No | NA | 2 | 0 | 1 | Not-determined | Genome-fragment | NA | NA | 0 |
| S12-final-viral-combined_k141_212946\|\|full | 5803 | No | NA | 11 | 2 | 0 | Low-quality | Genome-fragment | 3.38 | HMM-based (lower-bound) | 0 |
| S4_k141_147570 | 7681 | No | NA | 7 | 0 | 6 | Not-determined | Genome-fragment | NA | NA | 0 |
| S4_k141_161322 | 1123 | No | NA | 1 | 0 | 1 | Not-determined | Genome-fragment | NA | NA | 0 |
| S4_k141_1396 | 1477 | No | NA | 1 | 0 | 1 | Not-determined | Genome-fragment | NA | NA | 0 |
| S11-final-viral-combined_k141_37477\|\|full | 5805 | No | NA | 5 | 0 | 4 | Low-quality | Genome-fragment | 5.03 | AAI-based (medium-confidence) | 0 |
| S4_k141_43036 | 1016 | No | NA | 2 | 0 | 0 | Not-determined | Genome-fragment | NA | NA | 0 |
| S4_k141_64059 | 1744 | No | NA | 3 | 0 | 0 | Not-determined | Genome-fragment | NA | NA | 0 |
| S5_k141_28473 | 5807 | No | NA | 4 | 0 | 4 | Low-quality | Genome-fragment | 2.91 | AAI-based (medium-confidence) | 0 |
| S4_k141_140815 | 1102 | No | NA | 1 | 0 | 1 | Not-determined | Genome-fragment | NA | NA | 0 |
| S4_k141_119655 | 1136 | No | NA | 1 | 0 | 1 | Not-determined | Genome-fragment | NA | NA | 0 |
| S4_k141_71042 | 4835 | No | NA | 6 | 0 | 1 | Not-determined | Genome-fragment | NA | NA | 0 |
| S4_k141_84995 | 1414 | No | NA | 1 | 0 | 1 | Not-determined | Genome-fragment | NA | NA | 0 |
| S4_k141_85010 | 1404 | No | NA | 2 | 0 | 1 | Not-determined | Genome-fragment | NA | NA | 0 |
| S4_k141_22137 | 2612 | No | NA | 2 | 0 | 2 | Not-determined | Genome-fragment | NA | NA | 0 |
| S4_k141_29336 | 1115 | No | NA | 1 | 0 | 0 | Not-determined | Genome-fragment | NA | NA | 0 |
| S4_k141_105993 | 1001 | No | NA | 2 | 0 | 0 | Not-determined | Genome-fragment | NA | NA | 0 |
| S4_k141_78222 | 3014 | No | NA | 4 | 0 | 3 | Not-determined | Genome-fragment | NA | NA | 0 |
| S4_k141_126999 | 1100 | No | NA | 2 | 0 | 2 | Not-determined | Genome-fragment | NA | NA | 0 |
| S4_k141_49742 | 1074 | No | NA | 1 | 0 | 0 | Not-determined | Genome-fragment | NA | NA | 0 |
| S4_k141_78226 | 3411 | No | NA | 4 | 0 | 1 | Not-determined | Genome-fragment | NA | NA | 0 |
| S17-final-viral-combined_k141_50982\|\|full | 5808 | No | NA | 6 | 0 | 5 | Low-quality | Genome-fragment | 4.36 | AAI-based (medium-confidence) | 0 |
| S4_k141_8250 | 1053 | No | NA | 1 | 0 | 1 | Not-determined | Genome-fragment | NA | NA | 0 |
| S11-final-viral-combined_k141_58031\|\|full | 5809 | No | NA | 6 | 0 | 2 | Low-quality | Genome-fragment | 2.73 | AAI-based (medium-confidence) | 0 |
| S17-k141_187340 | 5816 | No | NA | 5 | 0 | 4 | Low-quality | Genome-fragment | 0.64 | AAI-based (medium-confidence) | 0 |
| S11-final-viral-combined_k141_70432\|\|full | 5818 | No | NA | 11 | 0 | 0 | Low-quality | Genome-fragment | 13.19 | AAI-based (high-confidence) | 0 |
| S4_k141_127045 | 1428 | No | NA | 5 | 0 | 1 | Not-determined | Genome-fragment | NA | NA | 0 |
| S4_k141_57080 | 3751 | No | NA | 5 | 0 | 0 | Not-determined | Genome-fragment | NA | NA | 0 |
| S4_k141_64106 | 2571 | No | NA | 3 | 0 | 3 | Not-determined | Genome-fragment | NA | NA | 0 |
| S4_k141_36085 | 5631 | No | NA | 2 | 0 | 0 | Not-determined | Genome-fragment | NA | NA | 0 |
| S4_k141_98857 | 4613 | No | NA | 5 | 0 | 0 | Not-determined | Genome-fragment | NA | NA | 0 |
| S4_k141_119767 | 8716 | No | NA | 5 | 0 | 3 | Not-determined | Genome-fragment | NA | NA | 0 |
| S4_k141_43152 | 1090 | No | NA | 1 | 0 | 1 | Not-determined | Genome-fragment | NA | NA | 0 |
| S4_k141_161443 | 15678 | No | NA | 18 | 4 | 0 | Medium-quality | Genome-fragment | 52.58 | HMM-based (lower-bound) | 0 |
| S4_k141_1471 | 17951 | No | NA | 10 | 0 | 7 | Not-determined | Genome-fragment | NA | NA | 0 |
| S4_k141_140895 | 4346 | No | NA | 2 | 0 | 0 | Not-determined | Genome-fragment | NA | NA | 0 |
| S4_k141_8313 | 1742 | No | NA | 1 | 0 | 1 | Not-determined | Genome-fragment | NA | NA | 0 |
| S1-final-viral-combined_k141_345468\|\|full | 5819 | No | NA | 8 | 0 | 1 | Low-quality | Genome-fragment | 16.65 | AAI-based (high-confidence) | 0 |
| S4_k141_112903 | 1295 | No | NA | 3 | 0 | 1 | Not-determined | Genome-fragment | NA | NA | 0 |
| S4_k141_85122 | 4051 | No | NA | 1 | 0 | 0 | Not-determined | Genome-fragment | NA | NA | 0 |
| S2-final-viral-combined_k141_49536\|\|full | 5820 | No | NA | 3 | 0 | 3 | Low-quality | Genome-fragment | 3.06 | AAI-based (medium-confidence) | 0 |
| S4_k141_112912 | 2247 | No | NA | 4 | 0 | 1 | Not-determined | Genome-fragment | NA | NA | 0 |
| S4_k141_147709 | 1705 | No | NA | 2 | 0 | 2 | Not-determined | Genome-fragment | NA | NA | 0 |
| S4_k141_154737 | 2071 | No | NA | 2 | 0 | 1 | Not-determined | Genome-fragment | NA | NA | 0 |
| S2-final-viral-combined_k141_162748\|\|full | 5822 | No | NA | 5 | 1 | 0 | Low-quality | Genome-fragment | 9.91 | AAI-based (medium-confidence) | 0 |
| S4_k141_133804 | 1138 | No | NA | 1 | 0 | 0 | Not-determined | Genome-fragment | NA | NA | 0 |
| S4_k141_43220 | 10678 | No | NA | 6 | 0 | 6 | Not-determined | Genome-fragment | NA | NA | 0 |
| S4_k141_161540 | 1036 | No | NA | 1 | 0 | 0 | Not-determined | Genome-fragment | NA | NA | 0 |
| S17-final-viral-combined_k141_70037\|\|full | 5824 | No | NA | 8 | 1 | 0 | Low-quality | Genome-fragment | 8.38 | AAI-based (high-confidence) | 0 |
| S4_k141_22222 | 1145 | No | NA | 4 | 0 | 2 | Not-determined | Genome-fragment | NA | NA | 0 |
| S4_k141_1585 | 5263 | No | NA | 6 | 0 | 2 | Not-determined | Genome-fragment | NA | NA | 0 |
| S4_k141_161550 | 5346 | No | NA | 3 | 0 | 2 | Not-determined | Genome-fragment | NA | NA | 0 |
| S4_k141_1590 | 1041 | No | NA | 1 | 0 | 0 | Not-determined | Genome-fragment | NA | NA | 0 |
| S4_k141_64257 | 1536 | No | NA | 2 | 0 | 0 | Not-determined | Genome-fragment | NA | NA | 0 |
| S4_k141_106182 | 1624 | No | NA | 2 | 0 | 1 | Not-determined | Genome-fragment | NA | NA | 0 |
| S4_k141_133848 | 1729 | No | NA | 2 | 0 | 2 | Not-determined | Genome-fragment | NA | NA | 0 |
| S4_k141_29548 | 1117 | No | NA | 1 | 0 | 0 | Not-determined | Genome-fragment | NA | NA | 0 |
| S4_k141_49940 | 1400 | No | NA | 1 | 0 | 0 | Not-determined | Genome-fragment | NA | NA | 0 |
| S4_k141_22271 | 4615 | No | NA | 7 | 0 | 4 | Not-determined | Genome-fragment | NA | NA | 0 |
| S4_k141_85208 | 1892 | No | NA | 2 | 0 | 2 | Not-determined | Genome-fragment | NA | NA | 0 |
| S4_k141_99008 | 4226 | No | NA | 2 | 0 | 1 | Not-determined | Genome-fragment | NA | NA | 0 |
| S4_k141_161584 | 1155 | No | NA | 1 | 0 | 1 | Not-determined | Genome-fragment | NA | NA | 0 |
| S4_k141_99012 | 1744 | No | NA | 2 | 0 | 1 | Not-determined | Genome-fragment | NA | NA | 0 |
| S4_k141_1622 | 3461 | No | NA | 6 | 0 | 0 | Not-determined | Genome-fragment | NA | NA | 0 |
| S8-final-viral-combined_k141_60351\|\|full | 5825 | No | NA | 10 | 5 | 0 | Low-quality | Genome-fragment | 16.11 | AAI-based (medium-confidence) | 0 |
| S4_k141_1625 | 1239 | No | NA | 3 | 0 | 1 | Not-determined | Genome-fragment | NA | NA | 0 |
| S1-final-viral-combined_k141_357211\|\|full | 5827 | No | NA | 6 | 0 | 1 | Low-quality | Genome-fragment | 37.72 | AAI-based (medium-confidence) | 0 |
| S4_k141_57230 | 1149 | No | NA | 2 | 0 | 0 | Not-determined | Genome-fragment | NA | NA | 0 |
| S4_k141_57231 | 1013 | No | NA | 2 | 0 | 0 | Not-determined | Genome-fragment | NA | NA | 0 |
| S4_k141_147829 | 1501 | No | NA | 1 | 0 | 0 | Not-determined | Genome-fragment | NA | NA | 0 |
| S8_k141_239141 | 5827 | No | NA | 4 | 0 | 4 | Low-quality | Genome-fragment | 9.65 | AAI-based (medium-confidence) | 0 |
| S4_k141_119863 | 1111 | No | NA | 2 | 0 | 0 | Not-determined | Genome-fragment | NA | NA | 0 |
| S4_k141_43311 | 1001 | No | NA | 1 | 0 | 0 | Not-determined | Genome-fragment | NA | NA | 0 |
| S4_k141_29620 | 4694 | No | NA | 1 | 0 | 0 | Not-determined | Genome-fragment | NA | NA | 0 |
| S4_k141_127217 | 1602 | No | NA | 1 | 0 | 1 | Not-determined | Genome-fragment | NA | NA | 0 |
| S4_k141_99068 | 3286 | No | NA | 7 | 0 | 0 | Not-determined | Genome-fragment | NA | NA | 0 |
| S17-final-viral-combined_k141_34627\|\|full | 5829 | No | NA | 6 | 4 | 0 | Low-quality | Genome-fragment | 14.26 | AAI-based (high-confidence) | 0 |
| S4_k141_1649 | 4036 | No | NA | 3 | 0 | 1 | Not-determined | Genome-fragment | NA | NA | 0 |
| S11-final-viral-combined_k141_186370\|\|full | 5831 | No | NA | 8 | 3 | 0 | Low-quality | Genome-fragment | 12.13 | AAI-based (high-confidence) | 0 |
| S11_k141_141828 | 5831 | No | NA | 2 | 0 | 0 | Low-quality | Genome-fragment | 6.06 | AAI-based (medium-confidence) | 0 |
| S4_k141_78472 | 1245 | No | NA | 1 | 0 | 1 | Not-determined | Genome-fragment | NA | NA | 0 |
| S4_k141_99078 | 1639 | No | NA | 2 | 0 | 2 | Not-determined | Genome-fragment | NA | NA | 0 |
| S4_k141_1670 | 10554 | No | NA | 5 | 0 | 4 | Not-determined | Genome-fragment | NA | NA | 0 |
| S4_k141_106292 | 3327 | No | NA | 4 | 0 | 3 | Not-determined | Genome-fragment | NA | NA | 0 |
| S4_k141_127259 | 5810 | No | NA | 7 | 0 | 1 | Not-determined | Genome-fragment | NA | NA | 0 |
| S4_k141_1672 | 11563 | No | NA | 15 | 0 | 4 | Not-determined | Genome-fragment | NA | NA | 0 |
| S4_k141_113109 | 1740 | No | NA | 2 | 0 | 2 | Not-determined | Genome-fragment | NA | NA | 0 |
| S1_k141_165343 | 5831 | No | NA | 7 | 4 | 0 | Low-quality | Genome-fragment | 18.53 | HMM-based (lower-bound) | 0 |
| S4_k141_1685 | 5107 | No | NA | 4 | 0 | 1 | Not-determined | Genome-fragment | NA | NA | 0 |
| S4_k141_119882 | 13252 | No | NA | 9 | 0 | 3 | Not-determined | Genome-fragment | NA | NA | 0 |
| S4_k141_71334 | 1126 | No | NA | 1 | 0 | 1 | Not-determined | Genome-fragment | NA | NA | 0 |
| S4_k141_113138 | 1119 | No | NA | 1 | 0 | 0 | Not-determined | Genome-fragment | NA | NA | 0 |
| S4_k141_64313 | 9719 | No | NA | 6 | 0 | 6 | Not-determined | Genome-fragment | NA | NA | 0 |
| S4_k141_1701 | 1539 | No | NA | 2 | 0 | 1 | Not-determined | Genome-fragment | NA | NA | 0 |
| S4_k141_154880 | 1074 | No | NA | 2 | 0 | 1 | Not-determined | Genome-fragment | NA | NA | 0 |
| S4_k141_15379 | 3927 | No | NA | 2 | 0 | 1 | Not-determined | Genome-fragment | NA | NA | 0 |
| S4_k141_78524 | 1060 | No | NA | 2 | 0 | 1 | Not-determined | Genome-fragment | NA | NA | 0 |
| S4_k141_1717 | 1264 | No | NA | 1 | 0 | 0 | Not-determined | Genome-fragment | NA | NA | 0 |
| S17-final-viral-combined_k141_89480\|\|full | 5831 | No | NA | 9 | 0 | 0 | Low-quality | Genome-fragment | 11.5 | AAI-based (high-confidence) | 0 |
| S12_k141_252781 | 5832 | No | NA | 9 | 0 | 3 | Low-quality | Genome-fragment | 4.13 | AAI-based (high-confidence) | 0 |
| S4_k141_154889 | 4455 | No | NA | 5 | 0 | 4 | Not-determined | Genome-fragment | NA | NA | 0 |
| S4_k141_71365 | 1757 | No | NA | 1 | 0 | 0 | Not-determined | Genome-fragment | NA | NA | 0 |
| S8_k141_127532 | 5832 | No | NA | 1 | 0 | 0 | Low-quality | Genome-fragment | 11.02 | AAI-based (high-confidence) | 0 |
| S7-final-viral-combined_k141_26873\|\|full | 5833 | No | NA | 5 | 3 | 0 | Low-quality | Genome-fragment | 14.87 | AAI-based (medium-confidence) | 0 |
| S4_k141_64340 | 1535 | No | NA | 3 | 0 | 3 | Not-determined | Genome-fragment | NA | NA | 0 |
| S4_k141_141132 | 5980 | No | NA | 6 | 0 | 6 | Not-determined | Genome-fragment | NA | NA | 0 |
| S4_k141_147902 | 1581 | No | NA | 1 | 0 | 1 | Not-determined | Genome-fragment | NA | NA | 0 |
| S4_k141_161698 | 2029 | No | NA | 1 | 0 | 1 | Not-determined | Genome-fragment | NA | NA | 0 |
| S4_k141_99212 | 1281 | No | NA | 3 | 0 | 0 | Not-determined | Genome-fragment | NA | NA | 0 |
| S7-final-viral-combined_k141_205914\|\|full | 5834 | No | NA | 9 | 1 | 0 | Low-quality | Genome-fragment | 37.71 | AAI-based (high-confidence) | 0 |
| S4_k141_161727 | 1010 | No | NA | 1 | 0 | 1 | Not-determined | Genome-fragment | NA | NA | 0 |
| S8-final-viral-combined_k141_21844\|\|full | 5838 | No | NA | 7 | 0 | 1 | Low-quality | Genome-fragment | 8.95 | AAI-based (medium-confidence) | 0 |
| S4_k141_85430 | 3167 | No | NA | 3 | 0 | 2 | Not-determined | Genome-fragment | NA | NA | 0 |
| S4_k141_106387 | 2304 | No | NA | 4 | 0 | 1 | Not-determined | Genome-fragment | NA | NA | 0 |
| S11-final-viral-combined_k141_34596\|\|full | 5840 | Yes | 4544 | 9 | 1 | 2 | Low-quality | Genome-fragment | 8.25 | AAI-based (high-confidence) | 22.19 |
| S8-final-viral-combined_k141_217127\|\|full | 5842 | No | NA | 7 | 1 | 0 | Low-quality | Genome-fragment | 12.73 | AAI-based (high-confidence) | 0 |
| S2-final-viral-combined_k141_115240\|\|full | 5843 | No | NA | 6 | 4 | 0 | Low-quality | Genome-fragment | 13.93 | AAI-based (high-confidence) | 0 |
| S4_k141_29784 | 1140 | No | NA | 1 | 0 | 0 | Not-determined | Genome-fragment | NA | NA | 0 |
| S4_k141_29802 | 1023 | No | NA | 3 | 0 | 0 | Not-determined | Genome-fragment | NA | NA | 0 |
| S17-final-viral-combined_k141_122833\|\|full | 5844 | No | NA | 9 | 2 | 0 | Low-quality | Genome-fragment | 12.52 | AAI-based (high-confidence) | 0 |
| S8_k141_171482 | 5845 | No | NA | 10 | 0 | 1 | Low-quality | Genome-fragment | 3.36 | AAI-based (high-confidence) | 0 |
| S4_k141_134046 | 1103 | No | NA | 2 | 0 | 1 | Not-determined | Genome-fragment | NA | NA | 0 |
| S4_k141_85454 | 1099 | No | NA | 2 | 0 | 0 | Not-determined | Genome-fragment | NA | NA | 0 |
| S6-final-viral-combined_k141_103750\|\|full | 5846 | No | NA | 7 | 0 | 5 | Low-quality | Genome-fragment | 15.02 | AAI-based (medium-confidence) | 0 |
| S4_k141_106447 | 2848 | No | NA | 4 | 0 | 0 | Not-determined | Genome-fragment | NA | NA | 0 |
| S4_k141_155009 | 1857 | No | NA | 3 | 0 | 0 | Not-determined | Genome-fragment | NA | NA | 0 |
| S4_k141_64493 | 1040 | No | NA | 2 | 0 | 0 | Not-determined | Genome-fragment | NA | NA | 0 |
| S4_k141_99286 | 1295 | No | NA | 2 | 0 | 0 | Not-determined | Genome-fragment | NA | NA | 0 |
| S1_k141_393055 | 5848 | No | NA | 14 | 1 | 0 | Low-quality | Genome-fragment | 4.35 | AAI-based (high-confidence) | 0 |
| S4_k141_1861 | 1638 | No | NA | 3 | 0 | 1 | Not-determined | Genome-fragment | NA | NA | 0 |
| S4_k141_147981 | 2726 | No | NA | 4 | 0 | 3 | Not-determined | Genome-fragment | NA | NA | 0 |
| S4_k141_155038 | 1320 | No | NA | 1 | 0 | 1 | Not-determined | Genome-fragment | NA | NA | 0 |
| S4_k141_15516 | 1039 | No | NA | 1 | 0 | 1 | Not-determined | Genome-fragment | NA | NA | 0 |
| S4_k141_64508 | 11116 | No | NA | 7 | 0 | 5 | Not-determined | Genome-fragment | NA | NA | 0 |
| S4_k141_106474 | 4734 | No | NA | 3 | 0 | 3 | Not-determined | Genome-fragment | NA | NA | 0 |
| S4_k141_1878 | 1331 | No | NA | 2 | 0 | 0 | Not-determined | Genome-fragment | NA | NA | 0 |
| S4_k141_64531 | 6256 | No | NA | 4 | 0 | 1 | Not-determined | Genome-fragment | NA | NA | 0 |
| S4_k141_64533 | 1142 | No | NA | 2 | 0 | 0 | Not-determined | Genome-fragment | NA | NA | 0 |
| S6-final-viral-combined_k141_128260\|\|full | 5848 | No | NA | 10 | 1 | 0 | Low-quality | Genome-fragment | 16.18 | AAI-based (medium-confidence) | 0 |
| S4_k141_64534 | 1291 | No | NA | 1 | 0 | 1 | Not-determined | Genome-fragment | NA | NA | 0 |
| S8-final-viral-combined_k141_10019\|\|full | 5848 | No | NA | 6 | 2 | 0 | Low-quality | Genome-fragment | 42.11 | AAI-based (medium-confidence) | 0 |
| S4_k141_127427 | 1256 | No | NA | 1 | 0 | 1 | Not-determined | Genome-fragment | NA | NA | 0 |
| S4_k141_155085 | 9541 | No | NA | 8 | 0 | 7 | Not-determined | Genome-fragment | NA | NA | 0 |
| S4_k141_36412 | 4111 | No | NA | 5 | 0 | 4 | Not-determined | Genome-fragment | NA | NA | 0 |
| S4_k141_1904 | 1629 | No | NA | 1 | 0 | 1 | Not-determined | Genome-fragment | NA | NA | 0 |
| S4_k141_71505 | 4946 | No | NA | 8 | 0 | 1 | Not-determined | Genome-fragment | NA | NA | 0 |
| S4_k141_113326 | 2319 | No | NA | 4 | 0 | 3 | Not-determined | Genome-fragment | NA | NA | 0 |
| S4_k141_43587 | 1513 | No | NA | 2 | 0 | 2 | Not-determined | Genome-fragment | NA | NA | 0 |
| S4_k141_71514 | 1066 | No | NA | 2 | 0 | 0 | Not-determined | Genome-fragment | NA | NA | 0 |
| S4_k141_36432 | 1345 | No | NA | 2 | 0 | 2 | Not-determined | Genome-fragment | NA | NA | 0 |
| S4_k141_29922 | 1301 | No | NA | 1 | 0 | 1 | Not-determined | Genome-fragment | NA | NA | 0 |
| S4_k141_113340 | 2660 | No | NA | 4 | 0 | 1 | Not-determined | Genome-fragment | NA | NA | 0 |
| S4_k141_43606 | 1086 | No | NA | 1 | 0 | 1 | Not-determined | Genome-fragment | NA | NA | 0 |
| S4_k141_99357 | 3488 | No | NA | 2 | 0 | 1 | Not-determined | Genome-fragment | NA | NA | 0 |
| S4_k141_78719 | 3951 | No | NA | 4 | 0 | 4 | Not-determined | Genome-fragment | NA | NA | 0 |
| S4_k141_127467 | 12669 | No | NA | 11 | 0 | 9 | Not-determined | Genome-fragment | NA | NA | 0 |
| S4_k141_155108 | 12166 | No | NA | 8 | 0 | 2 | Not-determined | Genome-fragment | NA | NA | 0 |
| S4_k141_92580 | 1787 | No | NA | 3 | 0 | 2 | Not-determined | Genome-fragment | NA | NA | 0 |
| S4_k141_148063 | 5527 | No | NA | 7 | 0 | 0 | Not-determined | Genome-fragment | NA | NA | 0 |
| S4_k141_8729 | 2476 | No | NA | 3 | 0 | 0 | Not-determined | Genome-fragment | NA | NA | 0 |
| S4_k141_113346 | 2459 | No | NA | 4 | 0 | 0 | Not-determined | Genome-fragment | NA | NA | 0 |
| S4_k141_78729 | 3943 | No | NA | 4 | 0 | 1 | Not-determined | Genome-fragment | NA | NA | 0 |
| S4_k141_22507 | 5723 | No | NA | 4 | 0 | 3 | Not-determined | Genome-fragment | NA | NA | 0 |
| S4_k141_43646 | 2024 | No | NA | 1 | 0 | 1 | Not-determined | Genome-fragment | NA | NA | 0 |
| S4_k141_22521 | 1005 | No | NA | 2 | 0 | 0 | Not-determined | Genome-fragment | NA | NA | 0 |
| S4_k141_92602 | 2458 | No | NA | 4 | 0 | 3 | Not-determined | Genome-fragment | NA | NA | 0 |
| S4_k141_99394 | 2177 | No | NA | 2 | 0 | 1 | Not-determined | Genome-fragment | NA | NA | 0 |
| S4_k141_15636 | 1156 | No | NA | 1 | 0 | 1 | Not-determined | Genome-fragment | NA | NA | 0 |
| S9-final-viral-combined_k141_58912\|\|full | 5849 | No | NA | 7 | 5 | 0 | Low-quality | Genome-fragment | 8.09 | AAI-based (medium-confidence) | 0 |
| S4_k141_78767 | 1671 | No | NA | 2 | 0 | 1 | Not-determined | Genome-fragment | NA | NA | 0 |
| S4_k141_8766 | 9884 | No | NA | 6 | 0 | 3 | Not-determined | Genome-fragment | NA | NA | 0 |
| S4_k141_148127 | 1750 | No | NA | 3 | 0 | 2 | Not-determined | Genome-fragment | NA | NA | 0 |
| S4_k141_8776 | 2148 | No | NA | 1 | 0 | 0 | Not-determined | Genome-fragment | NA | NA | 0 |
| S4_k141_161990 | 1909 | No | NA | 5 | 0 | 0 | Not-determined | Genome-fragment | NA | NA | 0 |
| S4_k141_127525 | 1230 | No | NA | 1 | 0 | 1 | Not-determined | Genome-fragment | NA | NA | 0 |
| S4_k141_155168 | 1132 | No | NA | 1 | 0 | 0 | Not-determined | Genome-fragment | NA | NA | 0 |
| S8-final-viral-combined_k141_122179\|\|full | 5851 | No | NA | 5 | 0 | 5 | Low-quality | Genome-fragment | 5.2 | AAI-based (medium-confidence) | 0 |
| S12-final-viral-combined_k141_73592\|\|full | 5852 | No | NA | 5 | 4 | 0 | Low-quality | Genome-fragment | 18.17 | AAI-based (high-confidence) | 0 |
| S12-final-viral-combined_k141_203344\|\|full | 5853 | Yes | 3151 | 9 | 1 | 1 | Low-quality | Genome-fragment | 8.33 | AAI-based (high-confidence) | 46.16 |
| S4-final-viral-combined_k141_150686\|\|full | 5854 | No | NA | 3 | 0 | 2 | Low-quality | Genome-fragment | 2.37 | AAI-based (medium-confidence) | 0 |
| S1-final-viral-combined_k141_59940\|\|full | 5856 | No | NA | 7 | 0 | 3 | Low-quality | Genome-fragment | 0.68 | AAI-based (medium-confidence) | 0 |
| S4_k141_141348 | 1555 | No | NA | 2 | 0 | 1 | Not-determined | Genome-fragment | NA | NA | 0 |
| S4_k141_141354 | 1077 | No | NA | 1 | 0 | 1 | Not-determined | Genome-fragment | NA | NA | 0 |
| S4_k141_30053 | 3910 | No | NA | 3 | 0 | 2 | Not-determined | Genome-fragment | NA | NA | 0 |
| S4_k141_127578 | 2680 | No | NA | 1 | 0 | 1 | Not-determined | Genome-fragment | NA | NA | 0 |
| S4_k141_134318 | 1041 | No | NA | 2 | 0 | 2 | Not-determined | Genome-fragment | NA | NA | 0 |
| S4_k141_43785 | 1424 | No | NA | 2 | 0 | 2 | Not-determined | Genome-fragment | NA | NA | 0 |
| S4_k141_127612 | 1426 | No | NA | 2 | 0 | 0 | Not-determined | Genome-fragment | NA | NA | 0 |
| S8_k141_72791 | 5857 | No | NA | 12 | 1 | 0 | Low-quality | Genome-fragment | 6.76 | HMM-based (lower-bound) | 0 |
| S4_k141_127613 | 1155 | No | NA | 1 | 0 | 1 | Not-determined | Genome-fragment | NA | NA | 0 |
| S6_k141_65888 | 5858 | Yes | 3204 | 9 | 1 | 1 | Low-quality | Genome-fragment | 2.15 | HMM-based (lower-bound) | 45.31 |
| S4_k141_106649 | 2096 | No | NA | 4 | 0 | 1 | Not-determined | Genome-fragment | NA | NA | 0 |
| S1_k141_19883 | 5859 | No | NA | 7 | 0 | 0 | Low-quality | Genome-fragment | 9.79 | AAI-based (high-confidence) | 0 |
| S4_k141_134355 | 1005 | No | NA | 2 | 0 | 2 | Not-determined | Genome-fragment | NA | NA | 0 |
| S4_k141_2086 | 18384 | No | NA | 9 | 0 | 4 | Not-determined | Genome-fragment | NA | NA | 0 |
| S4_k141_30078 | 1280 | No | NA | 1 | 0 | 0 | Not-determined | Genome-fragment | NA | NA | 0 |
| S4_k141_50405 | 1316 | No | NA | 1 | 0 | 1 | Not-determined | Genome-fragment | NA | NA | 0 |
| S4_k141_71719 | 1850 | No | NA | 3 | 0 | 0 | Not-determined | Genome-fragment | NA | NA | 0 |
| S4_k141_127659 | 4999 | No | NA | 3 | 0 | 1 | Not-determined | Genome-fragment | NA | NA | 0 |
| S4_k141_162158 | 2228 | No | NA | 2 | 0 | 0 | Not-determined | Genome-fragment | NA | NA | 0 |
| S7-final-viral-combined_k141_177500\|\|full | 5861 | No | NA | 15 | 3 | 0 | Low-quality | Genome-fragment | 13.39 | AAI-based (high-confidence) | 0 |
| S4_k141_64710 | 1808 | No | NA | 1 | 0 | 1 | Not-determined | Genome-fragment | NA | NA | 0 |
| S4_k141_71737 | 1913 | No | NA | 6 | 0 | 6 | Not-determined | Genome-fragment | NA | NA | 0 |
| S4_k141_155275 | 3743 | No | NA | 3 | 0 | 1 | Not-determined | Genome-fragment | NA | NA | 0 |
| S4_k141_92824 | 1205 | No | NA | 2 | 0 | 2 | Not-determined | Genome-fragment | NA | NA | 0 |
| S1-final-viral-combined_k141_165273\|\|full | 5862 | No | NA | 7 | 6 | 0 | Low-quality | Genome-fragment | 13.78 | AAI-based (high-confidence) | 0 |
| S4_k141_119970 | 1812 | No | NA | 2 | 0 | 0 | Not-determined | Genome-fragment | NA | NA | 0 |

| | | | | | | | | | | | |
|---|---|---|---|---|---|---|---|---|---|---|---|
| S7_k141_158751 | 5863 | No | NA | 5 | 2 | 0 | Low-quality | Genome-fragment | 9.08 | AAI-based (high-confidence) | 0 |
| S4_k141_92841 | 2149 | No | NA | 2 | 0 | 1 | Not-determined | Genome-fragment | NA | NA | 0 |
| S4_k141_92847 | 1295 | No | NA | 2 | 0 | 1 | Not-determined | Genome-fragment | NA | NA | 0 |
| S4_k141_162211 | 1664 | No | NA | 1 | 0 | 1 | Not-determined | Genome-fragment | NA | NA | 0 |
| S4_k141_162214 | 3130 | No | NA | 2 | 0 | 0 | Not-determined | Genome-fragment | NA | NA | 0 |
| S4_k141_141454 | 3776 | No | NA | 1 | 0 | 0 | Not-determined | Genome-fragment | NA | NA | 0 |
| S4_k141_43852 | 1050 | No | NA | 2 | 0 | 2 | Not-determined | Genome-fragment | NA | NA | 0 |
| S9-final-viral-combined_k141_62513\|\|full | 5865 | No | NA | 8 | 4 | 0 | Low-quality | Genome-fragment | 14.13 | AAI-based (high-confidence) | 0 |
| S11_k141_112259 | 5868 | No | NA | 9 | 3 | 0 | Low-quality | Genome-fragment | 12.48 | AAI-based (high-confidence) | 0 |
| S4_k141_162239 | 8305 | No | NA | 6 | 0 | 5 | Not-determined | Genome-fragment | NA | NA | 0 |
| S4_k141_22801 | 3044 | No | NA | 3 | 0 | 1 | Not-determined | Genome-fragment | NA | NA | 0 |
| S4_k141_148330 | 1158 | No | NA | 2 | 0 | 1 | Not-determined | Genome-fragment | NA | NA | 0 |
| S4_k141_134491 | 1903 | No | NA | 1 | 0 | 1 | Not-determined | Genome-fragment | NA | NA | 0 |
| S4_k141_79042 | 5287 | No | NA | 4 | 0 | 4 | Not-determined | Genome-fragment | NA | NA | 0 |
| S4_k141_8984 | 1583 | No | NA | 2 | 0 | 2 | Not-determined | Genome-fragment | NA | NA | 0 |
| S8_k141_14145 | 5869 | No | NA | 8 | 2 | 0 | Low-quality | Genome-fragment | 9.51 | AAI-based (high-confidence) | 0 |
| S4_k141_162278 | 2079 | No | NA | 3 | 0 | 0 | Not-determined | Genome-fragment | NA | NA | 0 |
| S4_k141_9012 | 1111 | No | NA | 1 | 0 | 0 | Not-determined | Genome-fragment | NA | NA | 0 |
| S4_k141_148364 | 1391 | No | NA | 2 | 0 | 1 | Not-determined | Genome-fragment | NA | NA | 0 |
| S4_k141_57795 | 1152 | No | NA | 1 | 0 | 1 | Not-determined | Genome-fragment | NA | NA | 0 |
| S4_k141_71784 | 3990 | No | NA | 6 | 0 | 1 | Not-determined | Genome-fragment | NA | NA | 0 |
| S4_k141_15961 | 1479 | No | NA | 2 | 0 | 1 | Not-determined | Genome-fragment | NA | NA | 0 |
| S17-final-viral-combined_k141_170607\|\|full | 5870 | No | NA | 7 | 0 | 0 | Low-quality | Genome-fragment | 13.82 | AAI-based (medium-confidence) | 0 |
| S4_k141_36727 | 6306 | No | NA | 2 | 0 | 1 | Not-determined | Genome-fragment | NA | NA | 0 |
| S4_k141_113516 | 2321 | No | NA | 3 | 0 | 0 | Not-determined | Genome-fragment | NA | NA | 0 |
| S4_k141_79109 | 2889 | No | NA | 3 | 0 | 2 | Not-determined | Genome-fragment | NA | NA | 0 |
| S4_k141_127752 | 1730 | No | NA | 1 | 0 | 1 | Not-determined | Genome-fragment | NA | NA | 0 |
| S4_k141_57836 | 1275 | No | NA | 3 | 0 | 0 | Not-determined | Genome-fragment | NA | NA | 0 |
| S4_k141_15999 | 3257 | No | NA | 2 | 0 | 1 | Not-determined | Genome-fragment | NA | NA | 0 |
| S4_k141_127761 | 1122 | No | NA | 1 | 0 | 1 | Not-determined | Genome-fragment | NA | NA | 0 |
| S4_k141_141536 | 1816 | No | NA | 1 | 0 | 1 | Not-determined | Genome-fragment | NA | NA | 0 |
| S12-final-viral-combined_k141_139430\|\|full | 5870 | No | NA | 11 | 1 | 0 | Low-quality | Genome-fragment | 9.5 | AAI-based (high-confidence) | 0 |
| S2-final-viral-combined_k141_25989\|\|full | 5870 | No | NA | 15 | 2 | 0 | Low-quality | Genome-fragment | 14.63 | AAI-based (high-confidence) | 0 |
| S4_k141_50475 | 99598 | Yes | 68255 | 140 | 17 | 20 | Complete | High-quality | 100 | Provirus (medium-confidence) | 31.47 |
| S4_k141_134588 | 1250 | No | NA | 1 | 0 | 1 | Not-determined | Genome-fragment | NA | NA | 0 |
| S4_k141_50476 | 2219 | No | NA | 2 | 0 | 0 | Not-determined | Genome-fragment | NA | NA | 0 |
| S4_k141_9087 | 2220 | No | NA | 3 | 0 | 1 | Not-determined | Genome-fragment | NA | NA | 0 |
| S12_k141_48000 | 5871 | No | NA | 13 | 2 | 0 | Low-quality | Genome-fragment | 12.82 | AAI-based (high-confidence) | 0 |
| S7_k141_191636 | 5872 | No | NA | 3 | 0 | 2 | Low-quality | Genome-fragment | 6.12 | AAI-based (medium-confidence) | 0 |
| S1-final-viral-combined_k141_392539\|\|full | 5873 | No | NA | 16 | 1 | 0 | Low-quality | Genome-fragment | 9.67 | AAI-based (high-confidence) | 0 |
| S4_k141_127802 | 1452 | No | NA | 1 | 0 | 1 | Not-determined | Genome-fragment | NA | NA | 0 |
| S4_k141_16050 | 2801 | No | NA | 5 | 0 | 0 | Not-determined | Genome-fragment | NA | NA | 0 |
| S4_k141_71862 | 2006 | No | NA | 1 | 0 | 1 | Not-determined | Genome-fragment | NA | NA | 0 |
| S4_k141_30180 | 1000 | No | NA | 1 | 0 | 1 | Not-determined | Genome-fragment | NA | NA | 0 |
| S4_k141_9123 | 1001 | No | NA | 2 | 0 | 1 | Not-determined | Genome-fragment | NA | NA | 0 |
| S4_k141_93035 | 1304 | No | NA | 1 | 0 | 1 | Not-determined | Genome-fragment | NA | NA | 0 |
| S4_k141_64902 | 2099 | No | NA | 1 | 0 | 1 | Not-determined | Genome-fragment | NA | NA | 0 |
| S5-final-viral-combined_k141_64169\|\|full | 5873 | No | NA | 12 | 3 | 0 | Low-quality | Genome-fragment | 13.42 | AAI-based (high-confidence) | 0 |
| S4_k141_134646 | 1355 | No | NA | 1 | 0 | 1 | Not-determined | Genome-fragment | NA | NA | 0 |
| S4_k141_23004 | 1524 | No | NA | 3 | 0 | 2 | Not-determined | Genome-fragment | NA | NA | 0 |
| S4_k141_50523 | 1714 | No | NA | 2 | 0 | 2 | Not-determined | Genome-fragment | NA | NA | 0 |
| S4_k141_93077 | 1177 | No | NA | 1 | 0 | 1 | Not-determined | Genome-fragment | NA | NA | 0 |
| S4_k141_120167 | 3628 | No | NA | 1 | 0 | 1 | Not-determined | Genome-fragment | NA | NA | 0 |
| S4_k141_50537 | 3162 | No | NA | 2 | 0 | 2 | Not-determined | Genome-fragment | NA | NA | 0 |
| S4_k141_85930 | 20880 | No | NA | 9 | 0 | 8 | Not-determined | Genome-fragment | NA | NA | 0 |
| S4_k141_141607 | 35335 | No | NA | 37 | 14 | 3 | Medium-quality | Genome-fragment | 86.12 | AAI-based (high-confidence) | 0 |
| S4_k141_44058 | 1096 | No | NA | 1 | 0 | 0 | Not-determined | Genome-fragment | NA | NA | 0 |
| S4_k141_71913 | 1505 | No | NA | 2 | 0 | 0 | Not-determined | Genome-fragment | NA | NA | 0 |
| S2-final-viral-combined_k141_18895\|\|full | 5873 | No | NA | 4 | 0 | 1 | Low-quality | Genome-fragment | 6.93 | AAI-based (medium-confidence) | 0 |
| S4_k141_106881 | 1746 | No | NA | 2 | 0 | 1 | Not-determined | Genome-fragment | NA | NA | 0 |
| S4_k141_93102 | 1320 | No | NA | 1 | 0 | 0 | Not-determined | Genome-fragment | NA | NA | 0 |
| S2_k141_126003 | 5873 | No | NA | 11 | 2 | 0 | Low-quality | Genome-fragment | 10.62 | AAI-based (high-confidence) | 0 |
| S3-final-viral-combined_k141_65152\|\|full | 5875 | No | NA | 4 | 0 | 3 | Low-quality | Genome-fragment | 6.2 | AAI-based (high-confidence) | 0 |
| S5-final-viral-combined_k141_64016\|\|full | 5875 | No | NA | 4 | 0 | 2 | Low-quality | Genome-fragment | 13.74 | AAI-based (medium-confidence) | 0 |
| S4_k141_36880 | 1196 | No | NA | 1 | 0 | 1 | Not-determined | Genome-fragment | NA | NA | 0 |
| S4_k141_148538 | 1128 | No | NA | 1 | 0 | 1 | Not-determined | Genome-fragment | NA | NA | 0 |
| S4_k141_36898 | 3117 | No | NA | 3 | 0 | 2 | Not-determined | Genome-fragment | NA | NA | 0 |
| S3-final-viral-combined_k141_125893\|\|full | 5878 | No | NA | 3 | 0 | 2 | Low-quality | Genome-fragment | 1.83 | AAI-based (medium-confidence) | 0 |
| S4_k141_71951 | 1466 | No | NA | 2 | 0 | 0 | Not-determined | Genome-fragment | NA | NA | 0 |
| S4_k141_36905 | 4853 | No | NA | 1 | 0 | 0 | Not-determined | Genome-fragment | NA | NA | 0 |
| S4_k141_30245 | 1670 | No | NA | 2 | 0 | 0 | Not-determined | Genome-fragment | NA | NA | 0 |
| S4_k141_155548 | 1022 | No | NA | 2 | 0 | 0 | Not-determined | Genome-fragment | NA | NA | 0 |
| S4_k141_16119 | 1079 | No | NA | 1 | 0 | 1 | Not-determined | Genome-fragment | NA | NA | 0 |
| S4_k141_71979 | 1908 | No | NA | 3 | 0 | 1 | Not-determined | Genome-fragment | NA | NA | 0 |
| S4_k141_155557 | 1036 | No | NA | 3 | 0 | 1 | Not-determined | Genome-fragment | NA | NA | 0 |
| S4_k141_65001 | 1467 | No | NA | 1 | 0 | 1 | Not-determined | Genome-fragment | NA | NA | 0 |
| S4_k141_2329 | 1252 | No | NA | 1 | 0 | 1 | Not-determined | Genome-fragment | NA | NA | 0 |
| S4_k141_23103 | 1072 | No | NA | 3 | 0 | 0 | Not-determined | Genome-fragment | NA | NA | 0 |
| S4_k141_86032 | 1270 | No | NA | 1 | 0 | 1 | Not-determined | Genome-fragment | NA | NA | 0 |
| S4_k141_141683 | 4561 | No | NA | 3 | 0 | 2 | Not-determined | Genome-fragment | NA | NA | 0 |
| S4_k141_2343 | 1363 | No | NA | 2 | 0 | 2 | Not-determined | Genome-fragment | NA | NA | 0 |
| S12-final-viral-combined_k141_395235\|\|full | 5878 | No | NA | 11 | 2 | 0 | Low-quality | Genome-fragment | 15.57 | AAI-based (high-confidence) | 0 |
| S4_k141_127862 | 3261 | No | NA | 4 | 0 | 3 | Not-determined | Genome-fragment | NA | NA | 0 |
| S4_k141_36976 | 3212 | No | NA | 1 | 0 | 1 | Not-determined | Genome-fragment | NA | NA | 0 |
| S4_k141_30202 | 5878 | No | NA | 11 | 1 | 0 | Low-quality | Genome-fragment | 12.81 | AAI-based (high-confidence) | 0 |
| S4_k141_50588 | 50250 | No | NA | 56 | 0 | 52 | Not-determined | Genome-fragment | NA | NA | 0 |
| S4_k141_127872 | 6351 | No | NA | 6 | 0 | 6 | Not-determined | Genome-fragment | NA | NA | 0 |
| S4_k141_113738 | 3716 | No | NA | 5 | 0 | 1 | Not-determined | Genome-fragment | NA | NA | 0 |
| S4_k141_86059 | 1384 | No | NA | 2 | 0 | 0 | Not-determined | Genome-fragment | NA | NA | 0 |
| S4_k141_141716 | 4615 | No | NA | 5 | 0 | 1 | Not-determined | Genome-fragment | NA | NA | 0 |
| S4_k141_134766 | 1263 | No | NA | 1 | 0 | 1 | Not-determined | Genome-fragment | NA | NA | 0 |
| S4_k141_16185 | 1565 | No | NA | 1 | 0 | 0 | Not-determined | Genome-fragment | NA | NA | 0 |
| S4_k141_16189 | 3524 | No | NA | 4 | 0 | 3 | Not-determined | Genome-fragment | NA | NA | 0 |
| S4_k141_106994 | 1254 | No | NA | 2 | 0 | 0 | Not-determined | Genome-fragment | NA | NA | 0 |
| S4_k141_79366 | 3593 | No | NA | 4 | 0 | 1 | Not-determined | Genome-fragment | NA | NA | 0 |
| S6-final-viral-combined_k141_9175\|\|full | 5878 | No | NA | 3 | 0 | 2 | Low-quality | Genome-fragment | 2.41 | AAI-based (medium-confidence) | 0 |
| S4_k141_79380 | 1125 | No | NA | 2 | 0 | 1 | Not-determined | Genome-fragment | NA | NA | 0 |
| S4_k141_86097 | 1030 | No | NA | 1 | 0 | 1 | Not-determined | Genome-fragment | NA | NA | 0 |
| S4_k141_30336 | 5362 | No | NA | 1 | 0 | 1 | Not-determined | Genome-fragment | NA | NA | 0 |
| S4_k141_72093 | 1171 | No | NA | 2 | 0 | 2 | Not-determined | Genome-fragment | NA | NA | 0 |
| S8-final-viral-combined_k141_224814\|\|full | 5879 | No | NA | 9 | 0 | 1 | Low-quality | Genome-fragment | 4.59 | AAI-based (medium-confidence) | 0 |
| S4_k141_162464 | 3645 | No | NA | 4 | 0 | 4 | Not-determined | Genome-fragment | NA | NA | 0 |
| S6_k141_10489 | 5880 | No | NA | 16 | 4 | 1 | Low-quality | Genome-fragment | 18.23 | AAI-based (high-confidence) | 0 |
| S4_k141_23136 | 1791 | No | NA | 1 | 0 | 0 | Not-determined | Genome-fragment | NA | NA | 0 |
| S12-final-viral-combined_k141_358314\|\|full | 5885 | No | NA | 5 | 0 | 2 | Low-quality | Genome-fragment | 4.98 | AAI-based (medium-confidence) | 0 |
| S4_k141_141769 | 1068 | No | NA | 2 | 0 | 0 | Not-determined | Genome-fragment | NA | NA | 0 |
| S9-final-viral-combined_k141_18268\|\|full | 5886 | No | NA | 9 | 1 | 0 | Low-quality | Genome-fragment | 4.33 | HMM-based (lower-bound) | 0 |
| S4_k141_44205 | 1167 | No | NA | 1 | 0 | 1 | Not-determined | Genome-fragment | NA | NA | 0 |
| S4_k141_127926 | 1303 | No | NA | 1 | 0 | 0 | Not-determined | Genome-fragment | NA | NA | 0 |
| S4_k141_30371 | 1623 | No | NA | 1 | 0 | 0 | Not-determined | Genome-fragment | NA | NA | 0 |
| S2-final-viral-combined_k141_199126\|\|full | 5889 | No | NA | 12 | 0 | 0 | Low-quality | Genome-fragment | 7.58 | AAI-based (high-confidence) | 0 |
| S4_k141_99579 | 1348 | No | NA | 2 | 0 | 0 | Not-determined | Genome-fragment | NA | NA | 0 |
| S6_k141_9987 | 5889 | No | NA | 9 | 0 | 0 | Low-quality | Genome-fragment | 5.84 | AAI-based (high-confidence) | 0 |
| S4_k141_9421 | 1498 | No | NA | 1 | 0 | 1 | Not-determined | Genome-fragment | NA | NA | 0 |
| S9-final-viral-combined_k141_66831\|\|full | 5892 | No | NA | 7 | 0 | 1 | Low-quality | Genome-fragment | 8.43 | AAI-based (medium-confidence) | 0 |
| S4_k141_2433 | 1512 | No | NA | 1 | 0 | 1 | Not-determined | Genome-fragment | NA | NA | 0 |
| S4_k141_134838 | 3344 | No | NA | 4 | 0 | 4 | Not-determined | Genome-fragment | NA | NA | 0 |
| S7-final-viral-combined_k141_134450\|\|full | 5892 | No | NA | 10 | 4 | 0 | Low-quality | Genome-fragment | 10.54 | AAI-based (high-confidence) | 0 |
| S4_k141_155627 | 4081 | No | NA | 5 | 0 | 0 | Not-determined | Genome-fragment | NA | NA | 0 |
| S4_k141_113840 | 1661 | No | NA | 1 | 0 | 1 | Not-determined | Genome-fragment | NA | NA | 0 |
| S4_k141_120397 | 1162 | No | NA | 1 | 0 | 1 | Not-determined | Genome-fragment | NA | NA | 0 |
| S4_k141_72183 | 2818 | No | NA | 2 | 0 | 2 | Not-determined | Genome-fragment | NA | NA | 0 |
| S4_k141_44264 | 1699 | No | NA | 1 | 0 | 0 | Not-determined | Genome-fragment | NA | NA | 0 |
| S17_k141_148946 | 5892 | No | NA | 8 | 1 | 0 | Low-quality | Genome-fragment | 7.2 | HMM-based (lower-bound) | 0 |
| S4_k141_134856 | 1772 | No | NA | 2 | 0 | 1 | Not-determined | Genome-fragment | NA | NA | 0 |
| S11-final-viral-combined_k141_51182\|\|full | 5896 | No | NA | 8 | 5 | 0 | Low-quality | Genome-fragment | 12.81 | AAI-based (high-confidence) | 0 |
| S4_k141_86178 | 1111 | No | NA | 1 | 0 | 0 | Not-determined | Genome-fragment | NA | NA | 0 |
| S4_k141_57966 | 8172 | No | NA | 5 | 0 | 1 | Not-determined | Genome-fragment | NA | NA | 0 |
| S4_k141_57968 | 1256 | No | NA | 1 | 0 | 1 | Not-determined | Genome-fragment | NA | NA | 0 |
| S4_k141_107107 | 1389 | No | NA | 4 | 0 | 1 | Not-determined | Genome-fragment | NA | NA | 0 |
| S4_k141_9469 | 1261 | No | NA | 1 | 0 | 1 | Not-determined | Genome-fragment | NA | NA | 0 |
| S4_k141_57977 | 1009 | No | NA | 1 | 0 | 1 | Not-determined | Genome-fragment | NA | NA | 0 |
| S11-final-viral-combined_k141_78599\|\|full | 5899 | No | NA | 11 | 1 | 0 | Low-quality | Genome-fragment | 10.12 | HMM-based (lower-bound) | 0 |
| S4_k141_79510 | 1091 | No | NA | 1 | 0 | 1 | Not-determined | Genome-fragment | NA | NA | 0 |
| S4_k141_99656 | 1449 | No | NA | 1 | 0 | 1 | Not-determined | Genome-fragment | NA | NA | 0 |
| S4_k141_86206 | 1462 | No | NA | 2 | 0 | 1 | Not-determined | Genome-fragment | NA | NA | 0 |
| S4_k141_79532 | 1672 | No | NA | 3 | 0 | 1 | Not-determined | Genome-fragment | NA | NA | 0 |
| S6_k141_110387 | 5900 | No | NA | 5 | 0 | 4 | Low-quality | Genome-fragment | 3.49 | AAI-based (medium-confidence) | 0 |
| S8-final-viral-combined_k141_70389\|\|full | 5900 | No | NA | 5 | 4 | 0 | Low-quality | Genome-fragment | 13.82 | AAI-based (high-confidence) | 0 |
| S4_k141_141904 | 2438 | No | NA | 4 | 0 | 0 | Not-determined | Genome-fragment | NA | NA | 0 |
| S4_k141_107136 | 6056 | No | NA | 3 | 0 | 0 | Not-determined | Genome-fragment | NA | NA | 0 |
| S4_k141_162576 | 1150 | No | NA | 1 | 0 | 1 | Not-determined | Genome-fragment | NA | NA | 0 |
| S4_k141_65253 | 1116 | No | NA | 1 | 0 | 0 | Not-determined | Genome-fragment | NA | NA | 0 |
| S4_k141_2553 | 7210 | No | NA | 5 | 0 | 4 | Not-determined | Genome-fragment | NA | NA | 0 |
| S4_k141_107143 | 2904 | No | NA | 3 | 0 | 3 | Not-determined | Genome-fragment | NA | NA | 0 |
| S4_k141_120455 | 1360 | No | NA | 2 | 0 | 2 | Not-determined | Genome-fragment | NA | NA | 0 |
| S3-final-viral-combined_k141_59652\|\|full | 5901 | Yes | 1915 | 7 | 1 | 1 | Low-quality | Genome-fragment | 1.35 | AAI-based (medium-confidence) | 67.55 |
| S2-final-viral-combined_k141_64503\|\|full | 5901 | Yes | 1041 | 6 | 1 | 3 | Low-quality | Genome-fragment | 1.48 | HMM-based (lower-bound) | 82.36 |
| S4_k141_58038 | 1483 | No | NA | 1 | 0 | 1 | Not-determined | Genome-fragment | NA | NA | 0 |
| S4_k141_9508 | 1459 | No | NA | 3 | 0 | 0 | Not-determined | Genome-fragment | NA | NA | 0 |
| S4_k141_162609 | 1038 | No | NA | 2 | 0 | 0 | Not-determined | Genome-fragment | NA | NA | 0 |
| S2_k141_123962 | 5901 | No | NA | 6 | 0 | 5 | Low-quality | Genome-fragment | 19.84 | AAI-based (medium-confidence) | 0 |
| S4_k141_155752 | 1041 | No | NA | 1 | 0 | 1 | Not-determined | Genome-fragment | NA | NA | 0 |
| S4_k141_99692 | 1859 | No | NA | 2 | 0 | 1 | Not-determined | Genome-fragment | NA | NA | 0 |
| S4_k141_23289 | 1098 | No | NA | 1 | 0 | 1 | Not-determined | Genome-fragment | NA | NA | 0 |
| S5-final-viral-combined_k141_170829\|\|full | 5902 | No | NA | 6 | 0 | 2 | Low-quality | Genome-fragment | 15.67 | AAI-based (high-confidence) | 0 |
| S4_k141_30531 | 1014 | No | NA | 2 | 0 | 2 | Not-determined | Genome-fragment | NA | NA | 0 |
| S4_k141_44367 | 1118 | No | NA | 1 | 0 | 0 | Not-determined | Genome-fragment | NA | NA | 0 |
| S4_k141_99708 | 2612 | No | NA | 1 | 0 | 0 | Not-determined | Genome-fragment | NA | NA | 0 |
| S4_k141_58079 | 1270 | No | NA | 1 | 0 | 0 | Not-determined | Genome-fragment | NA | NA | 0 |
| S4_k141_79588 | 5086 | No | NA | 7 | 0 | 4 | Not-determined | Genome-fragment | NA | NA | 0 |
| S4_k141_162632 | 1312 | No | NA | 2 | 0 | 1 | Not-determined | Genome-fragment | NA | NA | 0 |
| S7_k141_162706 | 5908 | No | NA | 2 | 0 | 2 | Low-quality | Genome-fragment | 2.07 | AAI-based (medium-confidence) | 0 |
| S4_k141_120513 | 1148 | No | NA | 3 | 0 | 0 | Not-determined | Genome-fragment | NA | NA | 0 |
| S4_k141_9537 | 2241 | No | NA | 2 | 0 | 2 | Not-determined | Genome-fragment | NA | NA | 0 |
| S4_k141_30561 | 1463 | No | NA | 4 | 0 | 0 | Not-determined | Genome-fragment | NA | NA | 0 |
| S4_k141_65314 | 1105 | No | NA | 1 | 0 | 0 | Not-determined | Genome-fragment | NA | NA | 0 |
| S4_k141_9549 | 2817 | No | NA | 2 | 0 | 0 | Not-determined | Genome-fragment | NA | NA | 0 |
| S4_k141_135013 | 1584 | No | NA | 2 | 0 | 1 | Not-determined | Genome-fragment | NA | NA | 0 |
| S4_k141_65324 | 1481 | No | NA | 2 | 0 | 2 | Not-determined | Genome-fragment | NA | NA | 0 |
| S4_k141_30574 | 1252 | No | NA | 3 | 0 | 2 | Not-determined | Genome-fragment | NA | NA | 0 |
| S12_k141_342054 | 5908 | No | NA | 1 | 0 | 0 | Low-quality | Genome-fragment | 7.85 | AAI-based (high-confidence) | 0 |
| S4_k141_50803 | 5379 | No | NA | 2 | 0 | 2 | Not-determined | Genome-fragment | NA | NA | 0 |
| S4_k141_79617 | 1098 | No | NA | 2 | 0 | 1 | Not-determined | Genome-fragment | NA | NA | 0 |
| S11-final-viral-combined_k141_300700\|\|full | 5911 | No | NA | 10 | 4 | 0 | Low-quality | Genome-fragment | 9.99 | AAI-based (medium-confidence) | 0 |
| S4_k141_79628 | 1892 | No | NA | 2 | 0 | 2 | Not-determined | Genome-fragment | NA | NA | 0 |
| S4_k141_107182 | 17322 | Yes | 15593 | 25 | 7 | 3 | Medium-quality | Genome-fragment | 52.71 | AAI-based (high-confidence) | 9.98 |
| S4_k141_30596 | 1452 | No | NA | 1 | 0 | 0 | Not-determined | Genome-fragment | NA | NA | 0 |
| S4_k141_30598 | 1282 | No | NA | 1 | 0 | 1 | Not-determined | Genome-fragment | NA | NA | 0 |
| S4_k141_107190 | 3554 | No | NA | 2 | 0 | 1 | Not-determined | Genome-fragment | NA | NA | 0 |
| S4_k141_2686 | 1080 | No | NA | 1 | 0 | 1 | Not-determined | Genome-fragment | NA | NA | 0 |
| S4_k141_148815 | 1350 | No | NA | 1 | 0 | 1 | Not-determined | Genome-fragment | NA | NA | 0 |
| S4_k141_120554 | 5447 | No | NA | 6 | 0 | 5 | Not-determined | Genome-fragment | NA | NA | 0 |
| S4_k141_120567 | 1137 | No | NA | 3 | 0 | 0 | Not-determined | Genome-fragment | NA | NA | 0 |
| S4_k141_135031 | 1249 | No | NA | 1 | 0 | 1 | Not-determined | Genome-fragment | NA | NA | 0 |
| S9-final-viral-combined_k141_33471\|\|full | 5913 | No | NA | 6 | 0 | 0 | Low-quality | Genome-fragment | 9.09 | AAI-based (medium-confidence) | 0 |
| S4_k141_30637 | 1971 | No | NA | 1 | 0 | 1 | Not-determined | Genome-fragment | NA | NA | 0 |
| S4_k141_93362 | 1007 | No | NA | 1 | 0 | 0 | Not-determined | Genome-fragment | NA | NA | 0 |
| S4_k141_142060 | 1065 | No | NA | 1 | 0 | 1 | Not-determined | Genome-fragment | NA | NA | 0 |
| S4_k141_148843 | 1924 | No | NA | 3 | 0 | 2 | Not-determined | Genome-fragment | NA | NA | 0 |
| S4_k141_44449 | 1256 | No | NA | 1 | 0 | 1 | Not-determined | Genome-fragment | NA | NA | 0 |
| S4_k141_93382 | 1011 | No | NA | 1 | 0 | 0 | Not-determined | Genome-fragment | NA | NA | 0 |
| S1-final-viral-combined_k141_103224\|\|full | 5914 | No | NA | 7 | 0 | 1 | Low-quality | Genome-fragment | 9.43 | AAI-based (medium-confidence) | 0 |
| S4_k141_148854 | 3126 | No | NA | 3 | 0 | 3 | Not-determined | Genome-fragment | NA | NA | 0 |
| S4_k141_99794 | 1123 | No | NA | 2 | 0 | 1 | Not-determined | Genome-fragment | NA | NA | 0 |
| S4_k141_30677 | 1184 | No | NA | 2 | 0 | 1 | Not-determined | Genome-fragment | NA | NA | 0 |
| S4_k141_135076 | 1573 | No | NA | 2 | 0 | 2 | Not-determined | Genome-fragment | NA | NA | 0 |
| S4_k141_2734 | 7424 | No | NA | 5 | 0 | 4 | Not-determined | Genome-fragment | NA | NA | 0 |
| S1-final-viral-combined_k141_372498\|\|full | 5914 | Yes | 5076 | 2 | 1 | 1 | Low-quality | Genome-fragment | 1.64 | HMM-based (lower-bound) | 14.17 |
| S4_k141_50877 | 3499 | No | NA | 4 | 0 | 0 | Not-determined | Genome-fragment | NA | NA | 0 |
| S6_k141_115701 | 5919 | No | NA | 8 | 0 | 0 | Low-quality | Genome-fragment | 7.03 | AAI-based (high-confidence) | 0 |
| S4_k141_37230 | 3087 | No | NA | 4 | 0 | 2 | Not-determined | Genome-fragment | NA | NA | 0 |
| S4_k141_2771 | 1215 | No | NA | 1 | 0 | 1 | Not-determined | Genome-fragment | NA | NA | 0 |
| S4_k141_114001 | 4141 | No | NA | 2 | 0 | 2 | Not-determined | Genome-fragment | NA | NA | 0 |
| S12-final-viral-combined_k141_106722\|\|full | 5920 | No | NA | 4 | 1 | 0 | Low-quality | Genome-fragment | 4.09 | HMM-based (lower-bound) | 0 |
| S4_k141_142115 | 5016 | No | NA | 5 | 0 | 4 | Not-determined | Genome-fragment | NA | NA | 0 |
| S4_k141_30724 | 3890 | No | NA | 6 | 0 | 5 | Not-determined | Genome-fragment | NA | NA | 0 |
| S4_k141_72489 | 1065 | No | NA | 1 | 0 | 0 | Not-determined | Genome-fragment | NA | NA | 0 |
| S4_k141_99872 | 3035 | No | NA | 3 | 0 | 1 | Not-determined | Genome-fragment | NA | NA | 0 |
| S4_k141_107263 | 1035 | No | NA | 1 | 0 | 1 | Not-determined | Genome-fragment | NA | NA | 0 |
| S4_k141_148931 | 1208 | No | NA | 1 | 0 | 0 | Not-determined | Genome-fragment | NA | NA | 0 |
| S7_k141_226865 | 5923 | No | NA | 3 | 1 | 0 | Low-quality | Genome-fragment | 9.82 | AAI-based (medium-confidence) | 0 |
| S4_k141_142142 | 2096 | No | NA | 4 | 0 | 3 | Not-determined | Genome-fragment | NA | NA | 0 |
| S4_k141_16484 | 1032 | No | NA | 1 | 0 | 1 | Not-determined | Genome-fragment | NA | NA | 0 |
| S4_k141_148952 | 2868 | No | NA | 3 | 0 | 1 | Not-determined | Genome-fragment | NA | NA | 0 |
| S2_k141_53968 | 5923 | No | NA | 7 | 2 | 0 | Low-quality | Genome-fragment | 8.73 | HMM-based (lower-bound) | 0 |
| S4_k141_148962 | 1091 | No | NA | 1 | 0 | 1 | Not-determined | Genome-fragment | NA | NA | 0 |
| S4_k141_72532 | 2100 | No | NA | 2 | 0 | 0 | Not-determined | Genome-fragment | NA | NA | 0 |
| S4_k141_2823 | 1161 | No | NA | 1 | 0 | 1 | Not-determined | Genome-fragment | NA | NA | 0 |
| S4_k141_16500 | 1187 | No | NA | 2 | 0 | 1 | Not-determined | Genome-fragment | NA | NA | 0 |
| S4_k141_148966 | 1548 | No | NA | 2 | 0 | 2 | Not-determined | Genome-fragment | NA | NA | 0 |
| S1-final-viral-combined_k141_96416\|\|full | 5933 | No | NA | 5 | 0 | 4 | Low-quality | Genome-fragment | 12.74 | AAI-based (medium-confidence) | 0 |
| S4_k141_40840 | 5941 | No | NA | 13 | 4 | 0 | Low-quality | Genome-fragment | 12.62 | AAI-based (high-confidence) | 0 |
| S4_k141_2836 | 2852 | No | NA | 1 | 0 | 1 | Not-determined | Genome-fragment | NA | NA | 0 |
| S4-final-viral-combined_k141_29306\|\|full | 5943 | No | NA | 11 | 0 | 2 | Low-quality | Genome-fragment | 8.55 | AAI-based (medium-confidence) | 0 |
| S4_k141_79679 | 1592 | No | NA | 2 | 0 | 1 | Not-determined | Genome-fragment | NA | NA | 0 |
| S2-final-viral-combined_k141_134682\|\|full | 5944 | Yes | 857 | 7 | 1 | 5 | Low-quality | Genome-fragment | 1.87 | HMM-based (lower-bound) | 85.58 |
| S4_k141_156038 | 1314 | No | NA | 1 | 0 | 1 | Not-determined | Genome-fragment | NA | NA | 0 |
| S4_k141_142191 | 2529 | No | NA | 4 | 0 | 2 | Not-determined | Genome-fragment | NA | NA | 0 |
| S4_k141_9652 | 3971 | No | NA | 2 | 0 | 2 | Not-determined | Genome-fragment | NA | NA | 0 |
| S4_k141_114067 | 1608 | No | NA | 3 | 0 | 2 | Not-determined | Genome-fragment | NA | NA | 0 |
| S4_k141_135128 | 1637 | No | NA | 3 | 0 | 3 | Not-determined | Genome-fragment | NA | NA | 0 |
| S1-final-viral-combined_k141_175299\|\|full | 5947 | No | NA | 5 | 0 | 4 | Low-quality | Genome-fragment | 1.86 | AAI-based (medium-confidence) | 0 |
| S1_k141_245583 | 5950 | No | NA | 3 | 1 | 0 | Low-quality | Genome-fragment | 6.8 | AAI-based (high-confidence) | 0 |
| S8-final-viral-combined_k141_136694\|\|full | 5951 | No | NA | 12 | 5 | 0 | Low-quality | Genome-fragment | 12.95 | AAI-based (medium-confidence) | 0 |
| S4_k141_9669 | 1285 | No | NA | 1 | 0 | 0 | Not-determined | Genome-fragment | NA | NA | 0 |
| S8_k141_47229 | 5954 | No | NA | 10 | 4 | 0 | Low-quality | Genome-fragment | 12.97 | AAI-based (high-confidence) | 0 |
| S4_k141_65492 | 1280 | No | NA | 1 | 0 | 0 | Not-determined | Genome-fragment | NA | NA | 0 |
| S4_k141_79754 | 1252 | No | NA | 1 | 0 | 1 | Not-determined | Genome-fragment | NA | NA | 0 |
| S4_k141_37373 | 1100 | No | NA | 3 | 0 | 2 | Not-determined | Genome-fragment | NA | NA | 0 |
| S4_k141_107383 | 1613 | No | NA | 3 | 0 | 0 | Not-determined | Genome-fragment | NA | NA | 0 |
| S4_k141_9714 | 1698 | No | NA | 1 | 0 | 1 | Not-determined | Genome-fragment | NA | NA | 0 |
| S4_k141_44623 | 2715 | No | NA | 5 | 0 | 0 | Not-determined | Genome-fragment | NA | NA | 0 |
| S4_k141_128045 | 1356 | No | NA | 2 | 0 | 0 | Not-determined | Genome-fragment | NA | NA | 0 |
| S4_k141_37392 | 1536 | No | NA | 1 | 0 | 1 | Not-determined | Genome-fragment | NA | NA | 0 |
| S4_k141_72618 | 1134 | No | NA | 1 | 0 | 0 | Not-determined | Genome-fragment | NA | NA | 0 |
| S4_k141_156136 | 1190 | No | NA | 1 | 0 | 0 | Not-determined | Genome-fragment | NA | NA | 0 |
| S4_k141_23636 | 2256 | No | NA | 1 | 0 | 1 | Not-determined | Genome-fragment | NA | NA | 0 |
| S4_k141_44645 | 1262 | No | NA | 2 | 0 | 1 | Not-determined | Genome-fragment | NA | NA | 0 |
| S4_k141_128066 | 1049 | No | NA | 1 | 0 | 1 | Not-determined | Genome-fragment | NA | NA | 0 |
| S11-final-viral-combined_k141_174067\|\|full | 5955 | Yes | 2718 | 4 | 1 | 1 | Low-quality | Genome-fragment | 1.36 | HMM-based (lower-bound) | 54.36 |
| S4_k141_37414 | 1157 | No | NA | 2 | 0 | 0 | Not-determined | Genome-fragment | NA | NA | 0 |
| S11-final-viral-combined_k141_203843\|\|full | 5956 | No | NA | 4 | 0 | 3 | Low-quality | Genome-fragment | 16.57 | AAI-based (medium-confidence) | 0 |
| S4_k141_23656 | 1045 | No | NA | 1 | 0 | 1 | Not-determined | Genome-fragment | NA | NA | 0 |
| S4_k141_114120 | 11434 | No | NA | 13 | 5 | 0 | High-quality | High-quality | 99.64 | AAI-based (medium-confidence) | 0 |
| S4_k141_2985 | 1290 | No | NA | 2 | 0 | 0 | Not-determined | Genome-fragment | NA | NA | 0 |
| S4_k141_135224 | 1138 | No | NA | 2 | 0 | 1 | Not-determined | Genome-fragment | NA | NA | 0 |
| S4_k141_142329 | 4436 | No | NA | 3 | 0 | 3 | Not-determined | Genome-fragment | NA | NA | 0 |
| S4_k141_72639 | 1170 | No | NA | 3 | 0 | 0 | Not-determined | Genome-fragment | NA | NA | 0 |
| S4_k141_107454 | 3684 | No | NA | 4 | 0 | 3 | Not-determined | Genome-fragment | NA | NA | 0 |
| S4_k141_162923 | 14544 | No | NA | 7 | 0 | 6 | Not-determined | Genome-fragment | NA | NA | 0 |
| S4_k141_30986 | 1392 | No | NA | 1 | 0 | 1 | Not-determined | Genome-fragment | NA | NA | 0 |
| S4_k141_23693 | 2104 | No | NA | 1 | 0 | 1 | Not-determined | Genome-fragment | NA | NA | 0 |
| S4_k141_128133 | 1211 | No | NA | 2 | 0 | 1 | Not-determined | Genome-fragment | NA | NA | 0 |
| S4_k141_51102 | 1587 | No | NA | 1 | 0 | 1 | Not-determined | Genome-fragment | NA | NA | 0 |
| S2_k141_117905 | 5956 | No | NA | 6 | 4 | 0 | Low-quality | Genome-fragment | 19.58 | AAI-based (high-confidence) | 0 |
| S4_k141_65556 | 45143 | No | NA | 31 | 0 | 25 | Not-determined | Genome-fragment | NA | NA | 0 |
| S4_k141_86617 | 1761 | No | NA | 1 | 0 | 1 | Not-determined | Genome-fragment | NA | NA | 0 |
| S4_k141_57954 | 5958 | No | NA | 9 | 0 | 0 | Low-quality | Genome-fragment | 13.27 | AAI-based (high-confidence) | 0 |
| S8-final-viral-combined_k141_119637\|\|full | 5962 | No | NA | 4 | 4 | 0 | Low-quality | Genome-fragment | 8.85 | HMM-based (lower-bound) | 0 |
| S4_k141_16656 | 1391 | No | NA | 1 | 0 | 1 | Not-determined | Genome-fragment | NA | NA | 0 |
| S4_k141_128155 | 1105 | No | NA | 1 | 0 | 1 | Not-determined | Genome-fragment | NA | NA | 0 |
| S4_k141_162946 | 5471 | No | NA | 12 | 0 | 11 | Not-determined | Genome-fragment | NA | NA | 0 |
| S4_k141_162950 | 5183 | No | NA | 3 | 0 | 0 | Not-determined | Genome-fragment | NA | NA | 0 |
| S4_k141_156225 | 3720 | No | NA | 1 | 0 | 1 | Not-determined | Genome-fragment | NA | NA | 0 |
| S4_k141_100115 | 1507 | No | NA | 1 | 0 | 1 | Not-determined | Genome-fragment | NA | NA | 0 |
| S4_k141_94950 | 5966 | No | NA | 6 | 0 | 4 | Low-quality | Genome-fragment | 3.1 | AAI-based (high-confidence) | 0 |
| S9-final-viral-combined_k141_109220\|\|full | 5967 | No | NA | 7 | 2 | 1 | Low-quality | Genome-fragment | 4.7 | HMM-based (lower-bound) | 0 |
| S4_k141_100135 | 1424 | No | NA | 2 | 0 | 1 | Not-determined | Genome-fragment | NA | NA | 0 |
| S4_k141_114192 | 1262 | No | NA | 1 | 0 | 1 | Not-determined | Genome-fragment | NA | NA | 0 |
| S9-final-viral-combined_k141_51407\|\|full | 5969 | No | NA | 8 | 7 | 0 | Low-quality | Genome-fragment | 13.13 | AAI-based (high-confidence) | 0 |
| S4_k141_86706 | 1334 | No | NA | 1 | 0 | 0 | Not-determined | Genome-fragment | NA | NA | 0 |
| S4_k141_51180 | 1357 | No | NA | 1 | 0 | 0 | Not-determined | Genome-fragment | NA | NA | 0 |
| S4_k141_51181 | 1173 | No | NA | 2 | 0 | 1 | Not-determined | Genome-fragment | NA | NA | 0 |
| S4_k141_163007 | 1303 | No | NA | 1 | 0 | 0 | Not-determined | Genome-fragment | NA | NA | 0 |
| S17-final-viral-combined_k141_18755\|\|full | 5969 | No | NA | 6 | 1 | 0 | Low-quality | Genome-fragment | 8.6 | AAI-based (high-confidence) | 0 |
| S1-final-viral-combined_k141_241653\|\|full | 5970 | No | NA | 11 | 5 | 0 | Low-quality | Genome-fragment | 9.78 | AAI-based (high-confidence) | 0 |
| S1-final-viral-combined_k141_199170\|\|full | 5980 | No | NA | 12 | 1 | 0 | Low-quality | Genome-fragment | 3.85 | HMM-based (lower-bound) | 0 |
| S4_k141_100167 | 6079 | No | NA | 13 | 0 | 13 | Not-determined | Genome-fragment | NA | NA | 0 |
| S4_k141_135293 | 1196 | No | NA | 2 | 0 | 0 | Not-determined | Genome-fragment | NA | NA | 0 |
| S4_k141_114251 | 1219 | No | NA | 2 | 0 | 0 | Not-determined | Genome-fragment | NA | NA | 0 |
| S4_k141_9927 | 2613 | No | NA | 2 | 0 | 1 | Not-determined | Genome-fragment | NA | NA | 0 |
| S4_k141_163040 | 5218 | No | NA | 7 | 0 | 0 | Not-determined | Genome-fragment | NA | NA | 0 |
| S4_k141_3078 | 1033 | No | NA | 1 | 0 | 1 | Not-determined | Genome-fragment | NA | NA | 0 |
| S3-final-viral-combined_k141_32690\|\|full | 5980 | No | NA | 3 | 0 | 0 | Low-quality | Genome-fragment | 2.75 | AAI-based (high-confidence) | 0 |
| S4_k141_114266 | 1507 | No | NA | 2 | 0 | 1 | Not-determined | Genome-fragment | NA | NA | 0 |
| S4_k141_58535 | 3473 | No | NA | 2 | 0 | 1 | Not-determined | Genome-fragment | NA | NA | 0 |
| S4_k141_107587 | 3442 | No | NA | 4 | 0 | 1 | Not-determined | Genome-fragment | NA | NA | 0 |
| S4_k141_149230 | 1068 | No | NA | 2 | 0 | 0 | Not-determined | Genome-fragment | NA | NA | 0 |
| S4_k141_142555 | 3670 | No | NA | 3 | 0 | 2 | Not-determined | Genome-fragment | NA | NA | 0 |
| S4_k141_72731 | 1545 | No | NA | 1 | 0 | 1 | Not-determined | Genome-fragment | NA | NA | 0 |
| S11-final-viral-combined_k141_134599\|\|full | 5982 | No | NA | 6 | 0 | 0 | Low-quality | Genome-fragment | 13.27 | AAI-based (medium-confidence) | 0 |
| S4_k141_128329 | 1368 | No | NA | 4 | 0 | 3 | Not-determined | Genome-fragment | NA | NA | 0 |
| S7-final-viral-combined_k141_153406\|\|full | 5983 | No | NA | 11 | 4 | 0 | Low-quality | Genome-fragment | 11.1 | AAI-based (medium-confidence) | 0 |
| S4_k141_120990 | 2004 | No | NA | 3 | 0 | 2 | Not-determined | Genome-fragment | NA | NA | 0 |
| S4_k141_31169 | 6499 | No | NA | 7 | 0 | 6 | Not-determined | Genome-fragment | NA | NA | 0 |
| S4_k141_100251 | 1528 | No | NA | 3 | 0 | 0 | Not-determined | Genome-fragment | NA | NA | 0 |
| S4_k141_107688 | 1054 | No | NA | 2 | 0 | 1 | Not-determined | Genome-fragment | NA | NA | 0 |
| S4_k141_37596 | 1054 | No | NA | 1 | 0 | 1 | Not-determined | Genome-fragment | NA | NA | 0 |
| S4_k141_135422 | 1754 | No | NA | 3 | 0 | 3 | Not-determined | Genome-fragment | NA | NA | 0 |
| S4_k141_44909 | 1641 | No | NA | 2 | 0 | 0 | Not-determined | Genome-fragment | NA | NA | 0 |
| S4_k141_10062 | 1068 | No | NA | 1 | 0 | 1 | Not-determined | Genome-fragment | NA | NA | 0 |
| S11-final-viral-combined_k141_27630\|\|full | 5984 | No | NA | 10 | 1 | 0 | Low-quality | Genome-fragment | 3.87 | HMM-based (lower-bound) | 0 |
| S4_k141_23859 | 1238 | No | NA | 1 | 0 | 1 | Not-determined | Genome-fragment | NA | NA | 0 |
| S4_k141_93772 | 3645 | No | NA | 1 | 0 | 0 | Not-determined | Genome-fragment | NA | NA | 0 |
| S4_k141_107754 | 1258 | No | NA | 1 | 0 | 1 | Not-determined | Genome-fragment | NA | NA | 0 |
| S4_k141_86856 | 4431 | No | NA | 2 | 0 | 2 | Not-determined | Genome-fragment | NA | NA | 0 |
| S4_k141_149338 | 2661 | No | NA | 3 | 0 | 0 | Not-determined | Genome-fragment | NA | NA | 0 |
| S4_k141_100312 | 1913 | No | NA | 1 | 0 | 1 | Not-determined | Genome-fragment | NA | NA | 0 |
| S11_k141_182219 | 5985 | No | NA | 8 | 0 | 6 | Low-quality | Genome-fragment | 2.42 | AAI-based (medium-confidence) | 0 |
| S4_k141_114363 | 10802 | No | NA | 1 | 0 | 0 | Not-determined | Genome-fragment | NA | NA | 0 |
| S4_k141_3232 | 8772 | No | NA | 4 | 0 | 3 | Not-determined | Genome-fragment | NA | NA | 0 |
| S4-final-viral-combined_k141_6242\|\|full | 5986 | No | NA | 6 | 0 | 3 | Low-quality | Genome-fragment | 11.69 | AAI-based (medium-confidence) | 0 |
| S4_k141_149366 | 1675 | No | NA | 3 | 0 | 3 | Not-determined | Genome-fragment | NA | NA | 0 |
| S8_k141_17650 | 5986 | No | NA | 9 | 3 | 0 | Low-quality | Genome-fragment | 13.06 | AAI-based (high-confidence) | 0 |
| S4_k141_107812 | 5496 | No | NA | 3 | 0 | 3 | Not-determined | Genome-fragment | NA | NA | 0 |
| S2-final-viral-combined_k141_220011\|\|full | 5988 | No | NA | 4 | 0 | 1 | Low-quality | Genome-fragment | 16.47 | AAI-based (medium-confidence) | 0 |
| S4_k141_65919 | 1352 | No | NA | 1 | 0 | 1 | Not-determined | Genome-fragment | NA | NA | 0 |
| S4_k141_3251 | 1076 | No | NA | 1 | 0 | 1 | Not-determined | Genome-fragment | NA | NA | 0 |
| S4_k141_121060 | 2131 | No | NA | 2 | 0 | 1 | Not-determined | Genome-fragment | NA | NA | 0 |
| S6-final-viral-combined_k141_17428\|\|full | 5992 | No | NA | 7 | 0 | 2 | Low-quality | Genome-fragment | 11.2 | AAI-based (high-confidence) | 0 |
| S4_k141_121072 | 1424 | No | NA | 2 | 0 | 1 | Not-determined | Genome-fragment | NA | NA | 0 |
| S11-final-viral-combined_k141_292678\|\|full | 5993 | No | NA | 8 | 2 | 0 | Low-quality | Genome-fragment | 13 | AAI-based (high-confidence) | 0 |
| S4_k141_114422 | 1101 | No | NA | 1 | 0 | 0 | Not-determined | Genome-fragment | NA | NA | 0 |
| S4_k141_80187 | 1007 | No | NA | 1 | 0 | 1 | Not-determined | Genome-fragment | NA | NA | 0 |
| S4-final-viral-combined_k141_2882\|\|full | 5996 | Yes | 4157 | 5 | 1 | 1 | Low-quality | Genome-fragment | 3.41 | AAI-based (high-confidence) | 30.67 |
| S4_k141_51342 | 7038 | No | NA | 7 | 0 | 5 | Not-determined | Genome-fragment | NA | NA | 0 |
| S9-final-viral-combined_k141_45168\|\|full | 5997 | Yes | 3996 | 5 | 1 | 1 | Low-quality | Genome-fragment | 10.06 | AAI-based (high-confidence) | 33.37 |
| S4_k141_114449 | 7321 | No | NA | 6 | 0 | 3 | Not-determined | Genome-fragment | NA | NA | 0 |
| S9-final-viral-combined_k141_72428\|\|full | 5999 | No | NA | 5 | 0 | 4 | Low-quality | Genome-fragment | 13.55 | AAI-based (medium-confidence) | 0 |
| S4-final-viral-combined_k141_104298\|\|full | 6000 | No | NA | 11 | 2 | 1 | Low-quality | Genome-fragment | 10.02 | AAI-based (high-confidence) | 0 |
| S4_k141_58672 | 3583 | No | NA | 6 | 0 | 1 | Not-determined | Genome-fragment | NA | NA | 0 |
| S4_k141_163322 | 7080 | No | NA | 3 | 0 | 1 | Not-determined | Genome-fragment | NA | NA | 0 |
| S4_k141_72967 | 1773 | No | NA | 2 | 0 | 2 | Not-determined | Genome-fragment | NA | NA | 0 |
| S8_k141_63523 | 6000 | No | NA | 8 | 3 | 0 | Low-quality | Genome-fragment | 9.17 | HMM-based (lower-bound) | 0 |
| S4_k141_51418 | 1914 | No | NA | 2 | 0 | 0 | Not-determined | Genome-fragment | NA | NA | 0 |
| S4_k141_128565 | 4244 | No | NA | 5 | 0 | 5 | Not-determined | Genome-fragment | NA | NA | 0 |
| S4_k141_31367 | 2078 | No | NA | 5 | 0 | 4 | Not-determined | Genome-fragment | NA | NA | 0 |
| S4_k141_135570 | 1049 | No | NA | 1 | 0 | 1 | Not-determined | Genome-fragment | NA | NA | 0 |
| S4_k141_58695 | 2895 | No | NA | 3 | 0 | 2 | Not-determined | Genome-fragment | NA | NA | 0 |
| S4_k141_24050 | 1721 | No | NA | 2 | 0 | 1 | Not-determined | Genome-fragment | NA | NA | 0 |
| S4_k141_51427 | 3294 | No | NA | 2 | 0 | 1 | Not-determined | Genome-fragment | NA | NA | 0 |
| S11-final-viral-combined_k141_210426\|\|full | 6002 | No | NA | 9 | 4 | 0 | Low-quality | Genome-fragment | 12.5 | AAI-based (high-confidence) | 0 |
| S4_k141_37817 | 1083 | No | NA | 1 | 0 | 1 | Not-determined | Genome-fragment | NA | NA | 0 |
| S1_k141_37300 | 6005 | No | NA | 3 | 0 | 3 | Low-quality | Genome-fragment | 10.94 | AAI-based (medium-confidence) | 0 |
| S4_k141_45128 | 41161 | No | NA | 67 | 10 | 2 | Medium-quality | Genome-fragment | 61.22 | AAI-based (high-confidence) | 0 |
| S4_k141_163332 | 1090 | No | NA | 2 | 0 | 2 | Not-determined | Genome-fragment | NA | NA | 0 |
| S4_k141_135610 | 4313 | No | NA | 5 | 0 | 4 | Not-determined | Genome-fragment | NA | NA | 0 |
| S4_k141_37839 | 1438 | No | NA | 1 | 0 | 0 | Not-determined | Genome-fragment | NA | NA | 0 |
| S4_k141_135618 | 1056 | No | NA | 2 | 0 | 2 | Not-determined | Genome-fragment | NA | NA | 0 |
| S4_k141_24087 | 1420 | No | NA | 1 | 0 | 0 | Not-determined | Genome-fragment | NA | NA | 0 |
| S4_k141_128609 | 1392 | No | NA | 2 | 0 | 1 | Not-determined | Genome-fragment | NA | NA | 0 |
| S4_k141_114526 | 1225 | No | NA | 1 | 0 | 1 | Not-determined | Genome-fragment | NA | NA | 0 |
| S4_k141_135630 | 3924 | No | NA | 7 | 0 | 2 | Not-determined | Genome-fragment | NA | NA | 0 |
| S12_k141_89929 | 6008 | No | NA | 6 | 1 | 0 | Low-quality | Genome-fragment | 14.24 | AAI-based (medium-confidence) | 0 |
| S4_k141_107957 | 1056 | No | NA | 2 | 0 | 1 | Not-determined | Genome-fragment | NA | NA | 0 |
| S4_k141_121157 | 1823 | No | NA | 2 | 0 | 2 | Not-determined | Genome-fragment | NA | NA | 0 |
| S4_k141_45149 | 1497 | No | NA | 2 | 0 | 2 | Not-determined | Genome-fragment | NA | NA | 0 |
| S8-final-viral-combined_k141_81153\|\|full | 6008 | No | NA | 5 | 0 | 3 | Low-quality | Genome-fragment | 5.47 | AAI-based (medium-confidence) | 0 |
| S4_k141_100565 | 1042 | No | NA | 3 | 0 | 0 | Not-determined | Genome-fragment | NA | NA | 0 |
| S4_k141_114566 | 1920 | No | NA | 3 | 0 | 0 | Not-determined | Genome-fragment | NA | NA | 0 |
| S4_k141_114570 | 1083 | No | NA | 1 | 0 | 1 | Not-determined | Genome-fragment | NA | NA | 0 |
| S4_k141_58778 | 5703 | No | NA | 2 | 0 | 2 | Not-determined | Genome-fragment | NA | NA | 0 |
| S4_k141_100572 | 1229 | No | NA | 2 | 0 | 2 | Not-determined | Genome-fragment | NA | NA | 0 |
| S4_k141_10320 | 1998 | No | NA | 3 | 0 | 0 | Not-determined | Genome-fragment | NA | NA | 0 |
| S4_k141_3349 | 7304 | No | NA | 6 | 0 | 4 | Not-determined | Genome-fragment | NA | NA | 0 |
| S4_k141_107994 | 3710 | No | NA | 4 | 0 | 3 | Not-determined | Genome-fragment | NA | NA | 0 |
| S4_k141_37869 | 1428 | No | NA | 1 | 0 | 0 | Not-determined | Genome-fragment | NA | NA | 0 |
| S4_k141_142876 | 2099 | No | NA | 4 | 0 | 1 | Not-determined | Genome-fragment | NA | NA | 0 |
| S12-final-viral-combined_k141_172668\|\|full | 6009 | No | NA | 9 | 2 | 1 | Low-quality | Genome-fragment | 4.48 | AAI-based (high-confidence) | 0 |
| S4_k141_149590 | 4873 | No | NA | 3 | 0 | 0 | Not-determined | Genome-fragment | NA | NA | 0 |
| S4_k141_51450 | 1406 | No | NA | 2 | 0 | 1 | Not-determined | Genome-fragment | NA | NA | 0 |
| S17-final-viral-combined_k141_100733\|\|full | 6010 | No | NA | 4 | 0 | 4 | Low-quality | Genome-fragment | 5.33 | AAI-based (medium-confidence) | 0 |
| S4_k141_135665 | 1031 | No | NA | 1 | 0 | 1 | Not-determined | Genome-fragment | NA | NA | 0 |
| S4_k141_51457 | 3275 | No | NA | 4 | 0 | 2 | Not-determined | Genome-fragment | NA | NA | 0 |
| S4_k141_93961 | 1923 | No | NA | 2 | 0 | 0 | Not-determined | Genome-fragment | NA | NA | 0 |
| S4_k141_31496 | 1512 | No | NA | 2 | 0 | 1 | Not-determined | Genome-fragment | NA | NA | 0 |
| S4_k141_163416 | 3528 | No | NA | 6 | 0 | 1 | Not-determined | Genome-fragment | NA | NA | 0 |
| S4_k141_87052 | 1098 | No | NA | 2 | 0 | 2 | Not-determined | Genome-fragment | NA | NA | 0 |
| S4_k141_93976 | 2210 | No | NA | 1 | 0 | 0 | Not-determined | Genome-fragment | NA | NA | 0 |
| S8-final-viral-combined_k141_105244\|\|full | 6011 | Yes | 1060 | 6 | 1 | 2 | Low-quality | Genome-fragment | 0.67 | HMM-based (lower-bound) | 82.37 |
| S4_k141_24152 | 1554 | No | NA | 2 | 0 | 1 | Not-determined | Genome-fragment | NA | NA | 0 |
| S4_k141_51482 | 3105 | No | NA | 4 | 0 | 2 | Not-determined | Genome-fragment | NA | NA | 0 |
| S4_k141_114613 | 1168 | No | NA | 2 | 0 | 1 | Not-determined | Genome-fragment | NA | NA | 0 |
| S4_k141_87078 | 1015 | No | NA | 3 | 0 | 0 | Not-determined | Genome-fragment | NA | NA | 0 |
| S4_k141_31518 | 1054 | No | NA | 1 | 0 | 1 | Not-determined | Genome-fragment | NA | NA | 0 |
| S4_k141_3416 | 1103 | No | NA | 1 | 0 | 1 | Not-determined | Genome-fragment | NA | NA | 0 |
| S4_k141_16913 | 1165 | No | NA | 1 | 0 | 0 | Not-determined | Genome-fragment | NA | NA | 0 |
| S4_k141_51489 | 1583 | No | NA | 1 | 0 | 0 | Not-determined | Genome-fragment | NA | NA | 0 |
| S4_k141_156701 | 1012 | No | NA | 1 | 0 | 1 | Not-determined | Genome-fragment | NA | NA | 0 |
| S4_k141_156704 | 3108 | No | NA | 5 | 0 | 2 | Not-determined | Genome-fragment | NA | NA | 0 |
| S2-final-viral-combined_k141_237374\|\|full | 6013 | No | NA | 8 | 0 | 0 | Low-quality | Genome-fragment | 10.53 | AAI-based (high-confidence) | 0 |
| S11-final-viral-combined_k141_234538\|\|full | 6014 | No | NA | 11 | 4 | 0 | Low-quality | Genome-fragment | 8.1 | HMM-based (lower-bound) | 0 |
| S1_k141_2418 | 6014 | No | NA | 10 | 1 | 0 | Low-quality | Genome-fragment | 9.44 | AAI-based (high-confidence) | 0 |
| S4_k141_58879 | 3157 | No | NA | 3 | 0 | 0 | Not-determined | Genome-fragment | NA | NA | 0 |
| S8-final-viral-combined_k141_59906\|\|full | 6014 | No | NA | 15 | 1 | 0 | Low-quality | Genome-fragment | 14.05 | AAI-based (high-confidence) | 0 |
| S7_k141_50495 | 6015 | No | NA | 6 | 0 | 0 | Low-quality | Genome-fragment | 15.41 | AAI-based (medium-confidence) | 0 |
| S4_k141_58905 | 3793 | No | NA | 3 | 0 | 3 | Not-determined | Genome-fragment | NA | NA | 0 |
| S4_k141_114661 | 1240 | No | NA | 1 | 0 | 0 | Not-determined | Genome-fragment | NA | NA | 0 |
| S4_k141_149688 | 1721 | No | NA | 1 | 0 | 1 | Not-determined | Genome-fragment | NA | NA | 0 |
| S4_k141_149701 | 1245 | No | NA | 1 | 0 | 1 | Not-determined | Genome-fragment | NA | NA | 0 |
| S4_k141_37978 | 3178 | No | NA | 3 | 0 | 3 | Not-determined | Genome-fragment | NA | NA | 0 |
| S4_k141_24243 | 1614 | No | NA | 2 | 0 | 1 | Not-determined | Genome-fragment | NA | NA | 0 |

| | | | | | | | | | | | |
|---|---|---|---|---|---|---|---|---|---|---|---|
| S4_k141_108179 | 1859 | No | NA | 2 | 0 | 1 | Not-determined | Genome-fragment | NA | NA | 0 |
| S4_k141_108183 | 1110 | No | NA | 2 | 0 | 1 | Not-determined | Genome-fragment | NA | NA | 0 |
| S4_k141_135809 | 1452 | No | NA | 1 | 0 | 1 | Not-determined | Genome-fragment | NA | NA | 0 |
| S4_k141_114723 | 4152 | No | NA | 4 | 0 | 4 | Not-determined | Genome-fragment | NA | NA | 0 |
| S4_k141_121356 | 3717 | No | NA | 3 | 0 | 2 | Not-determined | Genome-fragment | NA | NA | 0 |
| S4_k141_10503 | 1439 | No | NA | 1 | 0 | 0 | Not-determined | Genome-fragment | NA | NA | 0 |
| S4_k141_10521 | 1952 | No | NA | 2 | 0 | 0 | Not-determined | Genome-fragment | NA | NA | 0 |
| S4_k141_73303 | 1968 | No | NA | 1 | 0 | 0 | Not-determined | Genome-fragment | NA | NA | 0 |
| S9-final-viral-combined_k141_37634\|\|full | 6018 | No | NA | 6 | 0 | 2 | Low-quality | Genome-fragment | 12.82 | AAI-based (medium-confidence) | 0 |
| S4_k141_114760 | 1212 | No | NA | 3 | 0 | 3 | Not-determined | Genome-fragment | NA | NA | 0 |
| S4_k141_66192 | 1446 | No | NA | 2 | 0 | 1 | Not-determined | Genome-fragment | NA | NA | 0 |
| S4_k141_3549 | 1241 | No | NA | 1 | 0 | 0 | Not-determined | Genome-fragment | NA | NA | 0 |
| S4_k141_73313 | 1330 | No | NA | 1 | 0 | 0 | Not-determined | Genome-fragment | NA | NA | 0 |
| S4_k141_163544 | 1039 | No | NA | 1 | 0 | 1 | Not-determined | Genome-fragment | NA | NA | 0 |
| S4_k141_24290 | 2477 | No | NA | 2 | 0 | 1 | Not-determined | Genome-fragment | NA | NA | 0 |
| S4-final-viral-combined_k141_83960\|\|full | 6020 | No | NA | 13 | 3 | 0 | Low-quality | Genome-fragment | 5.22 | HMM-based (lower-bound) | 0 |
| S6_k141_49254 | 6022 | No | NA | 6 | 0 | 6 | Low-quality | Genome-fragment | 4.13 | AAI-based (high-confidence) | 0 |
| S4_k141_156858 | 1400 | No | NA | 1 | 0 | 1 | Not-determined | Genome-fragment | NA | NA | 0 |
| S17-k141_176999 | 6023 | No | NA | 7 | 0 | 1 | Low-quality | Genome-fragment | 6.23 | AAI-based (medium-confidence) | 0 |
| S4_k141_114772 | 1016 | No | NA | 2 | 0 | 2 | Not-determined | Genome-fragment | NA | NA | 0 |
| S9-final-viral-combined_k141_21658\|\|full | 6025 | Yes | 1706 | 6 | 1 | 1 | Low-quality | Genome-fragment | 1.1 | HMM-based (lower-bound) | 71.68 |
| S4_k141_24317 | 1164 | No | NA | 1 | 0 | 0 | Not-determined | Genome-fragment | NA | NA | 0 |
| S4_k141_100790 | 1424 | No | NA | 1 | 0 | 1 | Not-determined | Genome-fragment | NA | NA | 0 |
| S4_k141_156887 | 1046 | No | NA | 1 | 0 | 1 | Not-determined | Genome-fragment | NA | NA | 0 |
| S8_k141_4206 | 6026 | No | NA | 5 | 0 | 1 | Low-quality | Genome-fragment | 10.07 | AAI-based (high-confidence) | 0 |
| S4_k141_94118 | 2778 | No | NA | 7 | 0 | 6 | Not-determined | Genome-fragment | NA | NA | 0 |
| S7_k141_94629 | 6030 | No | NA | 11 | 1 | 2 | Low-quality | Genome-fragment | 3.11 | HMM-based (lower-bound) | 0 |
| S4_k141_94129 | 1152 | No | NA | 2 | 0 | 1 | Not-determined | Genome-fragment | NA | NA | 0 |
| S4_k141_80479 | 1267 | No | NA | 1 | 0 | 0 | Not-determined | Genome-fragment | NA | NA | 0 |
| S2_k141_248115 | 6031 | No | NA | 10 | 0 | 0 | Low-quality | Genome-fragment | 10.07 | AAI-based (high-confidence) | 0 |
| S4_k141_80487 | 4592 | No | NA | 8 | 0 | 2 | Not-determined | Genome-fragment | NA | NA | 0 |
| S4_k141_17100 | 8051 | No | NA | 5 | 0 | 5 | Not-determined | Genome-fragment | NA | NA | 0 |
| S4_k141_166214 | 6032 | No | NA | 4 | 0 | 4 | Low-quality | Genome-fragment | 2.81 | AAI-based (medium-confidence) | 0 |
| S4_k141_156954 | 1075 | No | NA | 2 | 0 | 2 | Not-determined | Genome-fragment | NA | NA | 0 |
| S8-final-viral-combined_k141_110416\|\|full | 6032 | No | NA | 5 | 0 | 3 | Low-quality | Genome-fragment | 6.32 | AAI-based (medium-confidence) | 0 |
| S4_k141_17119 | 2413 | No | NA | 4 | 0 | 3 | Not-determined | Genome-fragment | NA | NA | 0 |
| S4_k141_59073 | 11739 | No | NA | 7 | 0 | 7 | Not-determined | Genome-fragment | NA | NA | 0 |
| S4_k141_17125 | 1730 | No | NA | 1 | 0 | 1 | Not-determined | Genome-fragment | NA | NA | 0 |
| S4_k141_163609 | 1539 | No | NA | 1 | 0 | 0 | Not-determined | Genome-fragment | NA | NA | 0 |
| S4_k141_163612 | 1007 | No | NA | 2 | 0 | 2 | Not-determined | Genome-fragment | NA | NA | 0 |
| S12-final-viral-combined_k141_3634\|\|full | 6033 | No | NA | 14 | 4 | 1 | Low-quality | Genome-fragment | 13.94 | AAI-based (medium-confidence) | 0 |
| S4_k141_59077 | 10082 | No | NA | 6 | 0 | 2 | Not-determined | Genome-fragment | NA | NA | 0 |
| S4_k141_108318 | 1003 | No | NA | 1 | 0 | 1 | Not-determined | Genome-fragment | NA | NA | 0 |
| S4_k141_80539 | 1758 | No | NA | 2 | 0 | 2 | Not-determined | Genome-fragment | NA | NA | 0 |
| S4_k141_10643 | 1067 | No | NA | 2 | 0 | 1 | Not-determined | Genome-fragment | NA | NA | 0 |
| S4_k141_114834 | 1144 | No | NA | 1 | 0 | 0 | Not-determined | Genome-fragment | NA | NA | 0 |
| S4_k141_17155 | 4977 | No | NA | 5 | 0 | 4 | Not-determined | Genome-fragment | NA | NA | 0 |
| S4_k141_108323 | 1287 | No | NA | 1 | 0 | 1 | Not-determined | Genome-fragment | NA | NA | 0 |
| S4_k141_45531 | 1074 | No | NA | 1 | 0 | 0 | Not-determined | Genome-fragment | NA | NA | 0 |
| S4_k141_45533 | 2047 | No | NA | 3 | 0 | 3 | Not-determined | Genome-fragment | NA | NA | 0 |
| S4_k141_108329 | 2432 | No | NA | 2 | 0 | 1 | Not-determined | Genome-fragment | NA | NA | 0 |
| S4_k141_80568 | 1453 | No | NA | 1 | 0 | 0 | Not-determined | Genome-fragment | NA | NA | 0 |
| S4_k141_59119 | 1056 | No | NA | 3 | 0 | 3 | Not-determined | Genome-fragment | NA | NA | 0 |
| S4_k141_108342 | 1100 | No | NA | 3 | 0 | 0 | Not-determined | Genome-fragment | NA | NA | 0 |
| S11-final-viral-combined_k141_279152\|\|full | 6034 | No | NA | 10 | 4 | 0 | Low-quality | Genome-fragment | 10.35 | HMM-based (lower-bound) | 0 |
| S4_k141_31667 | 1587 | No | NA | 1 | 0 | 1 | Not-determined | Genome-fragment | NA | NA | 0 |
| S4_k141_157056 | 1119 | No | NA | 2 | 0 | 0 | Not-determined | Genome-fragment | NA | NA | 0 |
| S4_k141_143141 | 1189 | No | NA | 2 | 0 | 0 | Not-determined | Genome-fragment | NA | NA | 0 |
| S4_k141_24458 | 1048 | No | NA | 1 | 0 | 1 | Not-determined | Genome-fragment | NA | NA | 0 |
| S4_k141_136008 | 8933 | No | NA | 8 | 0 | 3 | Not-determined | Genome-fragment | NA | NA | 0 |
| S4_k141_24465 | 2480 | No | NA | 4 | 0 | 4 | Not-determined | Genome-fragment | NA | NA | 0 |
| S1_k141_170112 | 6034 | No | NA | 7 | 0 | 0 | Low-quality | Genome-fragment | 13.61 | AAI-based (medium-confidence) | 0 |
| S4_k141_87266 | 5452 | No | NA | 7 | 0 | 0 | Not-determined | Genome-fragment | NA | NA | 0 |
| S4_k141_108391 | 1145 | No | NA | 2 | 0 | 0 | Not-determined | Genome-fragment | NA | NA | 0 |
| S4_k141_100941 | 4766 | No | NA | 4 | 0 | 4 | Not-determined | Genome-fragment | NA | NA | 0 |
| S4_k141_73505 | 2250 | No | NA | 2 | 0 | 1 | Not-determined | Genome-fragment | NA | NA | 0 |
| S4_k141_149966 | 1528 | No | NA | 3 | 0 | 0 | Not-determined | Genome-fragment | NA | NA | 0 |
| S4_k141_51709 | 1470 | No | NA | 3 | 0 | 1 | Not-determined | Genome-fragment | NA | NA | 0 |
| S4_k141_38234 | 1028 | No | NA | 1 | 0 | 1 | Not-determined | Genome-fragment | NA | NA | 0 |
| S4_k141_94283 | 4002 | No | NA | 3 | 0 | 2 | Not-determined | Genome-fragment | NA | NA | 0 |
| S4_k141_3686 | 2746 | No | NA | 1 | 0 | 0 | Not-determined | Genome-fragment | NA | NA | 0 |
| S4_k141_45568 | 1007 | No | NA | 3 | 0 | 0 | Not-determined | Genome-fragment | NA | NA | 0 |
| S4_k141_121582 | 2758 | No | NA | 2 | 0 | 1 | Not-determined | Genome-fragment | NA | NA | 0 |
| S4_k141_157136 | 2261 | No | NA | 3 | 0 | 0 | Not-determined | Genome-fragment | NA | NA | 0 |
| S2-final-viral-combined_k141_34133\|\|full | 6034 | No | NA | 5 | 0 | 0 | Low-quality | Genome-fragment | 4.4 | AAI-based (medium-confidence) | 0 |
| S4_k141_66433 | 3049 | No | NA | 5 | 0 | 0 | Not-determined | Genome-fragment | NA | NA | 0 |
| S4_k141_94325 | 1754 | No | NA | 1 | 0 | 1 | Not-determined | Genome-fragment | NA | NA | 0 |
| S4_k141_128940 | 2007 | No | NA | 3 | 0 | 0 | Not-determined | Genome-fragment | NA | NA | 0 |
| S4_k141_45607 | 1127 | No | NA | 2 | 0 | 0 | Not-determined | Genome-fragment | NA | NA | 0 |
| S4_k141_121611 | 1555 | No | NA | 3 | 0 | 1 | Not-determined | Genome-fragment | NA | NA | 0 |
| S4_k141_45616 | 1024 | No | NA | 1 | 0 | 0 | Not-determined | Genome-fragment | NA | NA | 0 |
| S8-final-viral-combined_k141_186939\|\|full | 6036 | No | NA | 6 | 0 | 2 | Low-quality | Genome-fragment | 10.98 | AAI-based (medium-confidence) | 0 |
| S4_k141_24552 | 4399 | No | NA | 2 | 0 | 2 | Not-determined | Genome-fragment | NA | NA | 0 |
| S4_k141_136124 | 1291 | No | NA | 2 | 0 | 0 | Not-determined | Genome-fragment | NA | NA | 0 |
| S1-final-viral-combined_k141_316933\|\|full | 6039 | No | NA | 5 | 0 | 5 | Low-quality | Genome-fragment | 5.58 | AAI-based (medium-confidence) | 0 |
| S1-final-viral-combined_k141_121379\|\|full | 6039 | No | NA | 7 | 2 | 0 | Low-quality | Genome-fragment | 4.78 | HMM-based (lower-bound) | 0 |
| S4_k141_150020 | 1670 | No | NA | 1 | 0 | 1 | Not-determined | Genome-fragment | NA | NA | 0 |
| S4_k141_143312 | 2513 | No | NA | 2 | 0 | 0 | Not-determined | Genome-fragment | NA | NA | 0 |
| S17-k141_230048 | 6043 | No | NA | 7 | 1 | 5 | Low-quality | Genome-fragment | 1.79 | HMM-based (lower-bound) | 0 |
| S4_k141_163738 | 6535 | No | NA | 8 | 0 | 5 | Not-determined | Genome-fragment | NA | NA | 0 |
| S11-final-viral-combined_k141_309011\|\|full | 6045 | No | NA | 3 | 0 | 3 | Low-quality | Genome-fragment | 2 | AAI-based (medium-confidence) | 0 |
| S4_k141_163754 | 1282 | No | NA | 1 | 0 | 1 | Not-determined | Genome-fragment | NA | NA | 0 |
| S4_k141_66500 | 2632 | No | NA | 7 | 0 | 7 | Not-determined | Genome-fragment | NA | NA | 0 |
| S12-final-viral-combined_k141_224151\|\|full | 6046 | No | NA | 9 | 5 | 0 | Low-quality | Genome-fragment | 13.02 | AAI-based (high-confidence) | 0 |
| S4_k141_38340 | 1540 | No | NA | 2 | 0 | 0 | Not-determined | Genome-fragment | NA | NA | 0 |
| S4_k141_66513 | 1018 | No | NA | 1 | 0 | 1 | Not-determined | Genome-fragment | NA | NA | 0 |
| S4_k141_121653 | 1108 | No | NA | 1 | 0 | 1 | Not-determined | Genome-fragment | NA | NA | 0 |
| S4_k141_3764 | 4661 | No | NA | 5 | 0 | 1 | Not-determined | Genome-fragment | NA | NA | 0 |
| S4_k141_163766 | 1699 | No | NA | 1 | 0 | 0 | Not-determined | Genome-fragment | NA | NA | 0 |
| S4_k141_143331 | 1933 | No | NA | 4 | 0 | 1 | Not-determined | Genome-fragment | NA | NA | 0 |
| S4_k141_45698 | 1093 | No | NA | 1 | 0 | 0 | Not-determined | Genome-fragment | NA | NA | 0 |
| S4_k141_143342 | 2644 | No | NA | 1 | 0 | 1 | Not-determined | Genome-fragment | NA | NA | 0 |
| S4_k141_143343 | 1084 | No | NA | 2 | 0 | 1 | Not-determined | Genome-fragment | NA | NA | 0 |
| S4_k141_38367 | 5456 | No | NA | 2 | 0 | 2 | Not-determined | Genome-fragment | NA | NA | 0 |
| S4_k141_115056 | 7303 | No | NA | 8 | 0 | 7 | Not-determined | Genome-fragment | NA | NA | 0 |
| S4_k141_129018 | 2098 | No | NA | 2 | 0 | 1 | Not-determined | Genome-fragment | NA | NA | 0 |
| S4_k141_150055 | 2040 | No | NA | 2 | 0 | 0 | Not-determined | Genome-fragment | NA | NA | 0 |
| S4_k141_45730 | 1159 | No | NA | 2 | 0 | 1 | Not-determined | Genome-fragment | NA | NA | 0 |
| S1_k141_208229 | 6049 | No | NA | 6 | 0 | 4 | Low-quality | Genome-fragment | 1.92 | AAI-based (medium-confidence) | 0 |
| S11-final-viral-combined_k141_76736\|\|full | 6051 | No | NA | 8 | 7 | 0 | Low-quality | Genome-fragment | 15.04 | AAI-based (high-confidence) | 0 |
| S4_k141_150061 | 1428 | No | NA | 2 | 0 | 1 | Not-determined | Genome-fragment | NA | NA | 0 |
| S4_k141_94459 | 1866 | No | NA | 3 | 0 | 0 | Not-determined | Genome-fragment | NA | NA | 0 |
| S4_k141_163816 | 7411 | No | NA | 3 | 0 | 2 | Not-determined | Genome-fragment | NA | NA | 0 |
| S4_k141_115095 | 2546 | No | NA | 1 | 0 | 1 | Not-determined | Genome-fragment | NA | NA | 0 |
| S4_k141_163839 | 1048 | No | NA | 1 | 0 | 1 | Not-determined | Genome-fragment | NA | NA | 0 |
| S4_k141_136197 | 1285 | No | NA | 1 | 0 | 1 | Not-determined | Genome-fragment | NA | NA | 0 |
| S1-final-viral-combined_k141_54040\|\|full | 6051 | No | NA | 12 | 2 | 0 | Low-quality | Genome-fragment | 41.25 | AAI-based (high-confidence) | 0 |
| S4_k141_51898 | 5288 | No | NA | 4 | 0 | 3 | Not-determined | Genome-fragment | NA | NA | 0 |
| S4_k141_136207 | 1024 | No | NA | 2 | 0 | 0 | Not-determined | Genome-fragment | NA | NA | 0 |
| S4_k141_59355 | 1151 | No | NA | 1 | 0 | 1 | Not-determined | Genome-fragment | NA | NA | 0 |
| S4_k141_115107 | 7888 | No | NA | 1 | 0 | 0 | Not-determined | Genome-fragment | NA | NA | 0 |
| S2-final-viral-combined_k141_24432\|\|full | 6059 | Yes | 4066 | 8 | 1 | 1 | Low-quality | Genome-fragment | 12.17 | HMM-based (lower-bound) | 32.89 |
| S4_k141_3896 | 1102 | No | NA | 1 | 0 | 1 | Not-determined | Genome-fragment | NA | NA | 0 |
| S2-final-viral-combined_k141_113900\|\|full | 6059 | No | NA | 7 | 1 | 0 | Low-quality | Genome-fragment | 4.64 | HMM-based (lower-bound) | 0 |
| S4_k141_73706 | 1121 | No | NA | 2 | 0 | 1 | Not-determined | Genome-fragment | NA | NA | 0 |
| S1-final-viral-combined_k141_357126\|\|full | 6062 | No | NA | 7 | 0 | 1 | Low-quality | Genome-fragment | 39.21 | AAI-based (medium-confidence) | 0 |
| S4_k141_101135 | 6398 | No | NA | 8 | 0 | 0 | Not-determined | Genome-fragment | NA | NA | 0 |
| S4_k141_121705 | 1461 | No | NA | 2 | 0 | 0 | Not-determined | Genome-fragment | NA | NA | 0 |
| S4_k141_66647 | 1121 | No | NA | 2 | 0 | 1 | Not-determined | Genome-fragment | NA | NA | 0 |
| S4_k141_108583 | 3029 | No | NA | 4 | 0 | 2 | Not-determined | Genome-fragment | NA | NA | 0 |
| S4_k141_94510 | 3142 | No | NA | 4 | 0 | 4 | Not-determined | Genome-fragment | NA | NA | 0 |
| S11-final-viral-combined_k141_246962\|\|full | 6063 | No | NA | 6 | 0 | 4 | Low-quality | Genome-fragment | 6.44 | AAI-based (medium-confidence) | 0 |
| S2-final-viral-combined_k141_128980\|\|full | 6063 | No | NA | 10 | 2 | 0 | Low-quality | Genome-fragment | 14.6 | AAI-based (medium-confidence) | 0 |
| S4_k141_3957 | 1306 | No | NA | 3 | 0 | 2 | Not-determined | Genome-fragment | NA | NA | 0 |
| S4_k141_80827 | 2038 | No | NA | 1 | 0 | 0 | Not-determined | Genome-fragment | NA | NA | 0 |
| S4_k141_108609 | 1403 | No | NA | 1 | 0 | 1 | Not-determined | Genome-fragment | NA | NA | 0 |
| S4_k141_3968 | 2700 | No | NA | 4 | 0 | 1 | Not-determined | Genome-fragment | NA | NA | 0 |
| S4_k141_163918 | 13542 | No | NA | 9 | 0 | 6 | Not-determined | Genome-fragment | NA | NA | 0 |
| S4_k141_157295 | 1540 | No | NA | 2 | 0 | 2 | Not-determined | Genome-fragment | NA | NA | 0 |
| S4_k141_38487 | 1096 | No | NA | 1 | 0 | 0 | Not-determined | Genome-fragment | NA | NA | 0 |
| S4_k141_59409 | 5779 | No | NA | 6 | 0 | 5 | Not-determined | Genome-fragment | NA | NA | 0 |
| S4_k141_94561 | 1054 | No | NA | 1 | 0 | 0 | Not-determined | Genome-fragment | NA | NA | 0 |
| S4_k141_73768 | 4798 | No | NA | 3 | 0 | 2 | Not-determined | Genome-fragment | NA | NA | 0 |
| S4_k141_129157 | 1200 | No | NA | 1 | 0 | 1 | Not-determined | Genome-fragment | NA | NA | 0 |
| S4_k141_163983 | 1319 | No | NA | 4 | 0 | 3 | Not-determined | Genome-fragment | NA | NA | 0 |
| S4_k141_143503 | 1789 | No | NA | 2 | 0 | 1 | Not-determined | Genome-fragment | NA | NA | 0 |
| S4_k141_94575 | 1081 | No | NA | 1 | 0 | 1 | Not-determined | Genome-fragment | NA | NA | 0 |
| S4_k141_80899 | 1774 | No | NA | 2 | 0 | 1 | Not-determined | Genome-fragment | NA | NA | 0 |
| S4_k141_38516 | 1467 | No | NA | 3 | 0 | 3 | Not-determined | Genome-fragment | NA | NA | 0 |
| S2_k141_130789 | 6064 | No | NA | 13 | 0 | 1 | Low-quality | Genome-fragment | 7.52 | AAI-based (medium-confidence) | 0 |
| S4_k141_143515 | 4884 | No | NA | 4 | 0 | 3 | Not-determined | Genome-fragment | NA | NA | 0 |
| S4_k141_80903 | 2034 | No | NA | 1 | 0 | 0 | Not-determined | Genome-fragment | NA | NA | 0 |
| S4_k141_4002 | 1224 | No | NA | 1 | 0 | 0 | Not-determined | Genome-fragment | NA | NA | 0 |
| S4_k141_121774 | 1030 | No | NA | 1 | 0 | 1 | Not-determined | Genome-fragment | NA | NA | 0 |
| S4_k141_136338 | 1205 | No | NA | 2 | 0 | 1 | Not-determined | Genome-fragment | NA | NA | 0 |
| S4_k141_17338 | 1104 | No | NA | 1 | 0 | 1 | Not-determined | Genome-fragment | NA | NA | 0 |
| S1-final-viral-combined_k141_297766\|\|full | 6065 | No | NA | 9 | 5 | 0 | Low-quality | Genome-fragment | 13.84 | AAI-based (medium-confidence) | 0 |
| S4_k141_80939 | 1436 | No | NA | 1 | 0 | 0 | Not-determined | Genome-fragment | NA | NA | 0 |
| S4_k141_24795 | 3018 | No | NA | 3 | 0 | 3 | Not-determined | Genome-fragment | NA | NA | 0 |
| S4-final-viral-combined_k141_33890\|\|full | 6068 | No | NA | 5 | 0 | 4 | Low-quality | Genome-fragment | 2.54 | AAI-based (medium-confidence) | 0 |
| S4_k141_87482 | 1495 | No | NA | 2 | 0 | 2 | Not-determined | Genome-fragment | NA | NA | 0 |
| S4_k141_80956 | 1236 | No | NA | 1 | 0 | 0 | Not-determined | Genome-fragment | NA | NA | 0 |
| S4_k141_94633 | 6435 | No | NA | 5 | 0 | 3 | Not-determined | Genome-fragment | NA | NA | 0 |
| S4_k141_31996 | 1050 | No | NA | 2 | 0 | 0 | Not-determined | Genome-fragment | NA | NA | 0 |
| S4_k141_4051 | 1055 | No | NA | 2 | 0 | 1 | Not-determined | Genome-fragment | NA | NA | 0 |
| S4_k141_66754 | 4601 | No | NA | 4 | 0 | 4 | Not-determined | Genome-fragment | NA | NA | 0 |
| S4_k141_121805 | 1225 | No | NA | 2 | 0 | 1 | Not-determined | Genome-fragment | NA | NA | 0 |
| S4_k141_94654 | 3125 | No | NA | 5 | 0 | 2 | Not-determined | Genome-fragment | NA | NA | 0 |
| S4_k141_101293 | 3261 | No | NA | 2 | 0 | 1 | Not-determined | Genome-fragment | NA | NA | 0 |
| S11-final-viral-combined_k141_225302\|\|full | 6069 | No | NA | 10 | 1 | 0 | Low-quality | Genome-fragment | 12.56 | AAI-based (medium-confidence) | 0 |
| S4_k141_32027 | 12765 | No | NA | 10 | 0 | 7 | Not-determined | Genome-fragment | NA | NA | 0 |
| S4_k141_150218 | 1166 | No | NA | 2 | 0 | 1 | Not-determined | Genome-fragment | NA | NA | 0 |
| S2-final-viral-combined_k141_232748\|\|full | 6069 | No | NA | 5 | 2 | 0 | Low-quality | Genome-fragment | 12.68 | AAI-based (high-confidence) | 0 |
| S4_k141_38605 | 1040 | No | NA | 1 | 0 | 0 | Not-determined | Genome-fragment | NA | NA | 0 |
| S4_k141_32040 | 1254 | No | NA | 1 | 0 | 1 | Not-determined | Genome-fragment | NA | NA | 0 |
| S4_k141_59485 | 1564 | No | NA | 2 | 0 | 1 | Not-determined | Genome-fragment | NA | NA | 0 |
| S4_k141_11036 | 1080 | No | NA | 1 | 0 | 1 | Not-determined | Genome-fragment | NA | NA | 0 |
| S4_k141_46000 | 4224 | No | NA | 2 | 0 | 2 | Not-determined | Genome-fragment | NA | NA | 0 |
| S3-final-viral-combined_k141_135181\|\|full | 6070 | No | NA | 9 | 1 | 0 | Low-quality | Genome-fragment | 9.74 | HMM-based (lower-bound) | 0 |
| S4_k141_157345 | 2025 | No | NA | 4 | 0 | 4 | Not-determined | Genome-fragment | NA | NA | 0 |
| S4_k141_46006 | 1356 | No | NA | 3 | 0 | 1 | Not-determined | Genome-fragment | NA | NA | 0 |
| S4_k141_81033 | 1403 | No | NA | 1 | 0 | 0 | Not-determined | Genome-fragment | NA | NA | 0 |
| S11_k141_330503 | 6071 | No | NA | 7 | 0 | 7 | Low-quality | Genome-fragment | 5.33 | AAI-based (medium-confidence) | 0 |
| S4_k141_101394 | 4152 | No | NA | 5 | 0 | 3 | Not-determined | Genome-fragment | NA | NA | 0 |
| S4_k141_157375 | 8419 | No | NA | 6 | 0 | 4 | Not-determined | Genome-fragment | NA | NA | 0 |
| S4_k141_157383 | 1573 | No | NA | 2 | 0 | 2 | Not-determined | Genome-fragment | NA | NA | 0 |
| S4_k141_81051 | 3783 | No | NA | 5 | 0 | 3 | Not-determined | Genome-fragment | NA | NA | 0 |
| S4_k141_129341 | 1096 | No | NA | 2 | 0 | 0 | Not-determined | Genome-fragment | NA | NA | 0 |
| S4_k141_136360 | 1095 | No | NA | 2 | 0 | 1 | Not-determined | Genome-fragment | NA | NA | 0 |
| S4_k141_11149 | 1335 | No | NA | 1 | 0 | 1 | Not-determined | Genome-fragment | NA | NA | 0 |
| S4_k141_38664 | 3461 | No | NA | 1 | 0 | 1 | Not-determined | Genome-fragment | NA | NA | 0 |
| S4_k141_87621 | 1814 | No | NA | 2 | 0 | 0 | Not-determined | Genome-fragment | NA | NA | 0 |
| S4_k141_59551 | 1208 | No | NA | 1 | 0 | 1 | Not-determined | Genome-fragment | NA | NA | 0 |
| S4_k141_11170 | 8012 | No | NA | 8 | 0 | 1 | Not-determined | Genome-fragment | NA | NA | 0 |
| S8_k141_63278 | 6073 | No | NA | 5 | 0 | 2 | Low-quality | Genome-fragment | 4.35 | AAI-based (high-confidence) | 0 |
| S4_k141_38695 | 5528 | No | NA | 4 | 0 | 3 | Not-determined | Genome-fragment | NA | NA | 0 |
| S4_k141_66926 | 1072 | No | NA | 2 | 0 | 0 | Not-determined | Genome-fragment | NA | NA | 0 |
| S4-final-viral-combined_k141_93204\|\|full | 6074 | No | NA | 6 | 0 | 3 | Low-quality | Genome-fragment | 6.83 | AAI-based (high-confidence) | 0 |
| S4_k141_11195 | 1300 | No | NA | 1 | 0 | 1 | Not-determined | Genome-fragment | NA | NA | 0 |
| S4_k141_101463 | 8554 | No | NA | 8 | 0 | 4 | Not-determined | Genome-fragment | NA | NA | 0 |
| S4_k141_121971 | 1363 | No | NA | 3 | 0 | 2 | Not-determined | Genome-fragment | NA | NA | 0 |
| S1-final-viral-combined_k141_341489\|\|full | 6076 | No | NA | 11 | 1 | 0 | Low-quality | Genome-fragment | 38.67 | AAI-based (high-confidence) | 0 |
| S4_k141_17503 | 1370 | No | NA | 2 | 0 | 0 | Not-determined | Genome-fragment | NA | NA | 0 |
| S3_k141_56708 | 6076 | No | NA | 2 | 0 | 0 | Low-quality | Genome-fragment | 2.8 | AAI-based (high-confidence) | 0 |
| S4_k141_32126 | 1192 | No | NA | 1 | 0 | 0 | Not-determined | Genome-fragment | NA | NA | 0 |
| S2-final-viral-combined_k141_115833\|\|lt2gene | 6078 | No | NA | 1 | 1 | 0 | Low-quality | Genome-fragment | 14.37 | AAI-based (high-confidence) | 0 |
| S4_k141_139882 | 6078 | No | NA | 4 | 0 | 3 | Low-quality | Genome-fragment | 3.41 | AAI-based (medium-confidence) | 0 |
| S4_k141_32155 | 1215 | No | NA | 1 | 0 | 1 | Not-determined | Genome-fragment | NA | NA | 0 |
| S4_k141_164272 | 1753 | No | NA | 2 | 0 | 0 | Not-determined | Genome-fragment | NA | NA | 0 |
| S4_k141_81135 | 11723 | No | NA | 10 | 0 | 7 | Not-determined | Genome-fragment | NA | NA | 0 |
| S4_k141_11230 | 1171 | No | NA | 2 | 0 | 1 | Not-determined | Genome-fragment | NA | NA | 0 |
| S4_k141_73990 | 67580 | No | NA | 45 | 7 | 0 | Medium-quality | Genome-fragment | 67.53 | AAI-based (high-confidence) | 0 |
| S4_k141_25026 | 1273 | No | NA | 2 | 0 | 1 | Not-determined | Genome-fragment | NA | NA | 0 |
| S4_k141_38755 | 1114 | No | NA | 1 | 0 | 0 | Not-determined | Genome-fragment | NA | NA | 0 |
| S4_k141_143719 | 4022 | No | NA | 6 | 0 | 2 | Not-determined | Genome-fragment | NA | NA | 0 |
| S4_k141_81152 | 1408 | No | NA | 2 | 0 | 1 | Not-determined | Genome-fragment | NA | NA | 0 |
| S4_k141_87694 | 1092 | No | NA | 1 | 0 | 1 | Not-determined | Genome-fragment | NA | NA | 0 |
| S4_k141_32188 | 2819 | No | NA | 3 | 0 | 1 | Not-determined | Genome-fragment | NA | NA | 0 |
| S1-final-viral-combined_k141_41039\|\|full | 6079 | No | NA | 5 | 4 | 0 | Low-quality | Genome-fragment | 12.84 | AAI-based (high-confidence) | 0 |
| S4_k141_143756 | 1553 | No | NA | 3 | 0 | 0 | Not-determined | Genome-fragment | NA | NA | 0 |
| S4_k141_136503 | 1227 | No | NA | 1 | 0 | 0 | Not-determined | Genome-fragment | NA | NA | 0 |
| S4_k141_164312 | 5453 | No | NA | 4 | 0 | 2 | Not-determined | Genome-fragment | NA | NA | 0 |
| S1-final-viral-combined_k141_32406\|\|full | 6083 | No | NA | 19 | 3 | 0 | Low-quality | Genome-fragment | 15.58 | AAI-based (high-confidence) | 0 |
| S4_k141_32218 | 1477 | No | NA | 1 | 0 | 0 | Not-determined | Genome-fragment | NA | NA | 0 |
| S4_k141_109018 | 1046 | No | NA | 2 | 0 | 1 | Not-determined | Genome-fragment | NA | NA | 0 |
| S4_k141_4235 | 1055 | No | NA | 3 | 0 | 1 | Not-determined | Genome-fragment | NA | NA | 0 |
| S4_k141_94837 | 3221 | No | NA | 3 | 0 | 2 | Not-determined | Genome-fragment | NA | NA | 0 |
| S4_k141_136527 | 1484 | No | NA | 1 | 0 | 0 | Not-determined | Genome-fragment | NA | NA | 0 |
| S4_k141_109033 | 1405 | No | NA | 1 | 0 | 1 | Not-determined | Genome-fragment | NA | NA | 0 |
| S4_k141_32230 | 4113 | No | NA | 4 | 0 | 3 | Not-determined | Genome-fragment | NA | NA | 0 |
| S4_k141_109039 | 3612 | No | NA | 2 | 0 | 1 | Not-determined | Genome-fragment | NA | NA | 0 |
| S4_k141_32262 | 4662 | No | NA | 3 | 0 | 3 | Not-determined | Genome-fragment | NA | NA | 0 |
| S17-final-viral-combined_k141_197927\|\|full | 6084 | No | NA | 5 | 0 | 1 | Low-quality | Genome-fragment | 1.17 | AAI-based (medium-confidence) | 0 |
| S7-final-viral-combined_k141_84022\|\|full | 6085 | No | NA | 10 | 4 | 0 | Low-quality | Genome-fragment | 17.11 | AAI-based (high-confidence) | 0 |
| S4_k141_67063 | 1369 | No | NA | 1 | 0 | 1 | Not-determined | Genome-fragment | NA | NA | 0 |
| S4_k141_52337 | 3177 | No | NA | 3 | 0 | 2 | Not-determined | Genome-fragment | NA | NA | 0 |
| S8-final-viral-combined_k141_131375\|\|full | 6088 | No | NA | 6 | 3 | 0 | Low-quality | Genome-fragment | 8.57 | AAI-based (high-confidence) | 0 |
| S4_k141_38861 | 3718 | No | NA | 5 | 0 | 4 | Not-determined | Genome-fragment | NA | NA | 0 |
| S4_k141_164380 | 3973 | No | NA | 2 | 0 | 0 | Not-determined | Genome-fragment | NA | NA | 0 |
| S4_k141_11319 | 3950 | No | NA | 2 | 0 | 2 | Not-determined | Genome-fragment | NA | NA | 0 |
| S4_k141_87790 | 2274 | No | NA | 4 | 0 | 0 | Not-determined | Genome-fragment | NA | NA | 0 |
| S4_k141_129568 | 1708 | No | NA | 6 | 0 | 5 | Not-determined | Genome-fragment | NA | NA | 0 |
| S4_k141_164397 | 1352 | No | NA | 1 | 0 | 1 | Not-determined | Genome-fragment | NA | NA | 0 |
| S4_k141_4305 | 1048 | No | NA | 3 | 0 | 0 | Not-determined | Genome-fragment | NA | NA | 0 |
| S4_k141_94925 | 1217 | No | NA | 1 | 0 | 1 | Not-determined | Genome-fragment | NA | NA | 0 |
| S4_k141_46290 | 1052 | No | NA | 3 | 0 | 2 | Not-determined | Genome-fragment | NA | NA | 0 |
| S4_k141_25144 | 1561 | No | NA | 1 | 0 | 1 | Not-determined | Genome-fragment | NA | NA | 0 |
| S4_k141_109121 | 1330 | No | NA | 3 | 0 | 1 | Not-determined | Genome-fragment | NA | NA | 0 |
| S4_k141_17540 | 1008 | No | NA | 3 | 0 | 0 | Not-determined | Genome-fragment | NA | NA | 0 |
| S4_k141_122021 | 3402 | No | NA | 3 | 0 | 2 | Not-determined | Genome-fragment | NA | NA | 0 |
| S4_k141_67111 | 1784 | No | NA | 2 | 0 | 1 | Not-determined | Genome-fragment | NA | NA | 0 |
| S2-final-viral-combined_k141_151204\|\|full | 6094 | No | NA | 7 | 1 | 0 | Low-quality | Genome-fragment | 11.2 | AAI-based (high-confidence) | 0 |
| S2-final-viral-combined_k141_165218\|\|full | 6096 | No | NA | 5 | 1 | 0 | Low-quality | Genome-fragment | 12.49 | HMM-based (lower-bound) | 0 |
| S4_k141_74146 | 1128 | No | NA | 2 | 0 | 1 | Not-determined | Genome-fragment | NA | NA | 0 |
| S12_k141_172207 | 6099 | No | NA | 4 | 0 | 3 | Low-quality | Genome-fragment | 2.28 | AAI-based (medium-confidence) | 0 |
| S4_k141_46328 | 1001 | No | NA | 2 | 0 | 1 | Not-determined | Genome-fragment | NA | NA | 0 |
| S8_k141_116822 | 6099 | No | NA | 3 | 0 | 1 | Low-quality | Genome-fragment | 6.05 | AAI-based (medium-confidence) | 0 |
| S4_k141_150537 | 1645 | No | NA | 3 | 0 | 2 | Not-determined | Genome-fragment | NA | NA | 0 |
| S12-final-viral-combined_k141_304113\|\|full | 6100 | No | NA | 9 | 1 | 2 | Low-quality | Genome-fragment | 8.55 | HMM-based (lower-bound) | 0 |
| S4_k141_164424 | 1708 | No | NA | 2 | 0 | 0 | Not-determined | Genome-fragment | NA | NA | 0 |
| S4_k141_129629 | 3219 | No | NA | 2 | 0 | 2 | Not-determined | Genome-fragment | NA | NA | 0 |
| S4_k141_74179 | 1153 | No | NA | 2 | 0 | 1 | Not-determined | Genome-fragment | NA | NA | 0 |
| S1-final-viral-combined_k141_3157\|\|full | 6102 | No | NA | 11 | 1 | 2 | Low-quality | Genome-fragment | 14.25 | AAI-based (medium-confidence) | 0 |
| S8_k141_92218 | 6103 | No | NA | 11 | 1 | 2 | Low-quality | Genome-fragment | 7.78 | HMM-based (lower-bound) | 0 |
| S4_k141_115509 | 3089 | No | NA | 2 | 0 | 1 | Not-determined | Genome-fragment | NA | NA | 0 |
| S4_k141_11405 | 1019 | No | NA | 2 | 0 | 1 | Not-determined | Genome-fragment | NA | NA | 0 |
| S4_k141_81374 | 1023 | No | NA | 2 | 0 | 0 | Not-determined | Genome-fragment | NA | NA | 0 |
| S4_k141_115516 | 1145 | No | NA | 3 | 0 | 0 | Not-determined | Genome-fragment | NA | NA | 0 |
| S4_k141_46384 | 1240 | No | NA | 1 | 0 | 1 | Not-determined | Genome-fragment | NA | NA | 0 |
| S4_k141_52427 | 6118 | No | NA | 4 | 0 | 4 | Not-determined | Genome-fragment | NA | NA | 0 |
| S4_k141_101669 | 4353 | No | NA | 4 | 0 | 4 | Not-determined | Genome-fragment | NA | NA | 0 |
| S4_k141_46389 | 2273 | No | NA | 1 | 0 | 1 | Not-determined | Genome-fragment | NA | NA | 0 |
| S2_k141_108079 | 6104 | No | NA | 9 | 1 | 0 | Low-quality | Genome-fragment | 12.5 | AAI-based (medium-confidence) | 0 |
| S4_k141_115536 | 6150 | No | NA | 4 | 0 | 1 | Not-determined | Genome-fragment | NA | NA | 0 |
| S4_k141_67219 | 1857 | No | NA | 4 | 0 | 2 | Not-determined | Genome-fragment | NA | NA | 0 |
| S4_k141_74254 | 1543 | No | NA | 2 | 0 | 0 | Not-determined | Genome-fragment | NA | NA | 0 |
| S4_k141_11438 | 7165 | No | NA | 5 | 0 | 2 | Not-determined | Genome-fragment | NA | NA | 0 |
| S4_k141_136734 | 1328 | No | NA | 2 | 0 | 1 | Not-determined | Genome-fragment | NA | NA | 0 |
| S4_k141_143917 | 1016 | No | NA | 2 | 0 | 2 | Not-determined | Genome-fragment | NA | NA | 0 |
| S4_k141_74263 | 1657 | No | NA | 1 | 0 | 1 | Not-determined | Genome-fragment | NA | NA | 0 |
| S4_k141_109220 | 1730 | No | NA | 2 | 0 | 2 | Not-determined | Genome-fragment | NA | NA | 0 |
| S4_k141_74267 | 1119 | No | NA | 1 | 0 | 1 | Not-determined | Genome-fragment | NA | NA | 0 |
| S3-final-viral-combined_k141_134100\|\|full | 6107 | No | NA | 8 | 0 | 0 | Low-quality | Genome-fragment | 12.42 | AAI-based (high-confidence) | 0 |
| S4_k141_143932 | 1898 | No | NA | 3 | 0 | 3 | Not-determined | Genome-fragment | NA | NA | 0 |
| S11-final-viral-combined_k141_293485\|\|full | 6108 | No | NA | 8 | 0 | 3 | Low-quality | Genome-fragment | 13.87 | AAI-based (medium-confidence) | 0 |
| S4_k141_101718 | 1043 | No | NA | 2 | 0 | 2 | Not-determined | Genome-fragment | NA | NA | 0 |
| S4_k141_46418 | 1039 | No | NA | 1 | 0 | 0 | Not-determined | Genome-fragment | NA | NA | 0 |
| S8_k141_87489 | 6109 | No | NA | 5 | 0 | 1 | Low-quality | Genome-fragment | 10.94 | AAI-based (medium-confidence) | 0 |
| S4_k141_17613 | 8245 | No | NA | 4 | 0 | 3 | Not-determined | Genome-fragment | NA | NA | 0 |
| S4_k141_17614 | 1803 | No | NA | 2 | 0 | 0 | Not-determined | Genome-fragment | NA | NA | 0 |
| S4_k141_74289 | 1008 | No | NA | 1 | 0 | 1 | Not-determined | Genome-fragment | NA | NA | 0 |
| S4_k141_136749 | 2735 | No | NA | 2 | 0 | 2 | Not-determined | Genome-fragment | NA | NA | 0 |
| S4_k141_11451 | 1108 | No | NA | 2 | 0 | 2 | Not-determined | Genome-fragment | NA | NA | 0 |
| S4_k141_74301 | 1123 | No | NA | 2 | 0 | 0 | Not-determined | Genome-fragment | NA | NA | 0 |
| S4_k141_11453 | 1185 | No | NA | 1 | 0 | 0 | Not-determined | Genome-fragment | NA | NA | 0 |
| S4_k141_136755 | 1276 | No | NA | 1 | 0 | 1 | Not-determined | Genome-fragment | NA | NA | 0 |
| S4_k141_52518 | 4414 | No | NA | 2 | 0 | 2 | Not-determined | Genome-fragment | NA | NA | 0 |
| S4_k141_157747 | 4559 | No | NA | 4 | 0 | 3 | Not-determined | Genome-fragment | NA | NA | 0 |
| S17-k141_20876 | 6111 | No | NA | 3 | 0 | 2 | Low-quality | Genome-fragment | 3.72 | AAI-based (high-confidence) | 0 |
| S4_k141_129678 | 1351 | No | NA | 1 | 0 | 1 | Not-determined | Genome-fragment | NA | NA | 0 |
| S4_k141_129679 | 1189 | No | NA | 2 | 0 | 1 | Not-determined | Genome-fragment | NA | NA | 0 |
| S11-final-viral-combined_k141_292248\|\|full | 6112 | No | NA | 5 | 0 | 3 | Low-quality | Genome-fragment | 2.95 | AAI-based (medium-confidence) | 0 |
| S4_k141_143967 | 1253 | No | NA | 2 | 0 | 1 | Not-determined | Genome-fragment | NA | NA | 0 |
| S4_k141_39062 | 1231 | No | NA | 2 | 0 | 1 | Not-determined | Genome-fragment | NA | NA | 0 |
| S4_k141_157789 | 1120 | No | NA | 3 | 0 | 0 | Not-determined | Genome-fragment | NA | NA | 0 |
| S4_k141_59914 | 2073 | No | NA | 2 | 0 | 1 | Not-determined | Genome-fragment | NA | NA | 0 |
| S4_k141_25255 | 1829 | No | NA | 1 | 0 | 1 | Not-determined | Genome-fragment | NA | NA | 0 |
| S4_k141_164575 | 1138 | No | NA | 2 | 0 | 0 | Not-determined | Genome-fragment | NA | NA | 0 |
| S7_k141_111365 | 6112 | No | NA | 3 | 0 | 1 | Low-quality | Genome-fragment | 11.14 | AAI-based (medium-confidence) | 0 |
| S4_k141_74343 | 1723 | No | NA | 2 | 0 | 2 | Not-determined | Genome-fragment | NA | NA | 0 |
| S1-final-viral-combined_k141_52457\|\|full | 6114 | No | NA | 9 | 0 | 1 | Low-quality | Genome-fragment | 15.19 | AAI-based (medium-confidence) | 0 |
| S6-final-viral-combined_k141_129984\|\|full | 6114 | No | NA | 2 | 0 | 0 | Low-quality | Genome-fragment | 15.07 | AAI-based (medium-confidence) | 0 |
| S4_k141_17690 | 1962 | No | NA | 4 | 0 | 1 | Not-determined | Genome-fragment | NA | NA | 0 |
| S4_k141_52549 | 1199 | No | NA | 1 | 0 | 0 | Not-determined | Genome-fragment | NA | NA | 0 |
| S4_k141_52551 | 1114 | No | NA | 2 | 0 | 0 | Not-determined | Genome-fragment | NA | NA | 0 |
| S4_k141_150664 | 1327 | No | NA | 1 | 0 | 1 | Not-determined | Genome-fragment | NA | NA | 0 |
| S11_k141_170635 | 6120 | No | NA | 13 | 1 | 0 | Low-quality | Genome-fragment | 12.93 | AAI-based (high-confidence) | 0 |
| S4_k141_122212 | 2769 | No | NA | 4 | 0 | 2 | Not-determined | Genome-fragment | NA | NA | 0 |
| S4_k141_115629 | 1015 | No | NA | 2 | 0 | 1 | Not-determined | Genome-fragment | NA | NA | 0 |
| S4_k141_136820 | 1618 | No | NA | 1 | 0 | 1 | Not-determined | Genome-fragment | NA | NA | 0 |
| S4_k141_157843 | 1123 | No | NA | 2 | 0 | 0 | Not-determined | Genome-fragment | NA | NA | 0 |
| S4_k141_52604 | 1123 | No | NA | 2 | 0 | 1 | Not-determined | Genome-fragment | NA | NA | 0 |
| S4_k141_144044 | 1309 | No | NA | 2 | 0 | 1 | Not-determined | Genome-fragment | NA | NA | 0 |
| S4_k141_52610 | 4295 | No | NA | 3 | 0 | 3 | Not-determined | Genome-fragment | NA | NA | 0 |
| S12_k141_260841 | 6123 | No | NA | 7 | 3 | 0 | Low-quality | Genome-fragment | 10.73 | AAI-based (high-confidence) | 0 |
| S4_k141_32517 | 4255 | No | NA | 7 | 0 | 3 | Not-determined | Genome-fragment | NA | NA | 0 |
| S4_k141_88019 | 1281 | No | NA | 2 | 0 | 0 | Not-determined | Genome-fragment | NA | NA | 0 |
| S4_k141_25334 | 1894 | No | NA | 3 | 0 | 0 | Not-determined | Genome-fragment | NA | NA | 0 |
| S4_k141_32534 | 1334 | No | NA | 1 | 0 | 1 | Not-determined | Genome-fragment | NA | NA | 0 |
| S4_k141_129696 | 6064 | No | NA | 5 | 0 | 4 | Not-determined | Genome-fragment | NA | NA | 0 |
| S4_k141_101866 | 1928 | No | NA | 3 | 0 | 0 | Not-determined | Genome-fragment | NA | NA | 0 |
| S1-final-viral-combined_k141_340440\|\|full | 6133 | No | NA | 7 | 1 | 0 | Low-quality | Genome-fragment | 39.55 | AAI-based (high-confidence) | 0 |
| S6_k141_45688 | 6133 | No | NA | 5 | 0 | 3 | Low-quality | Genome-fragment | 1.02 | AAI-based (high-confidence) | 0 |
| S4_k141_164646 | 3381 | No | NA | 4 | 0 | 0 | Not-determined | Genome-fragment | NA | NA | 0 |
| S17-final-viral-combined_k141_157167\|\|full | 6140 | Yes | 4472 | 8 | 3 | 1 | Low-quality | Genome-fragment | 5.43 | AAI-based (high-confidence) | 27.17 |
| S6-final-viral-combined_k141_11983\|\|full | 6143 | No | NA | 12 | 3 | 0 | Low-quality | Genome-fragment | 5.83 | HMM-based (lower-bound) | 0 |
| S4_k141_32563 | 1003 | No | NA | 2 | 0 | 1 | Not-determined | Genome-fragment | NA | NA | 0 |
| S5-final-viral-combined_k141_153728\|\|full | 6144 | No | NA | 6 | 0 | 5 | Low-quality | Genome-fragment | 4.11 | AAI-based (medium-confidence) | 0 |
| S4_k141_81599 | 1092 | No | NA | 1 | 0 | 0 | Not-determined | Genome-fragment | NA | NA | 0 |
| S4_k141_150716 | 1579 | No | NA | 1 | 0 | 1 | Not-determined | Genome-fragment | NA | NA | 0 |
| S1-final-viral-combined_k141_332984\|\|full | 6146 | No | NA | 9 | 5 | 0 | Low-quality | Genome-fragment | 14.66 | AAI-based (high-confidence) | 0 |
| S2-final-viral-combined_k141_4741\|\|full | 6147 | No | NA | 14 | 7 | 0 | Low-quality | Genome-fragment | 11.19 | AAI-based (high-confidence) | 0 |
| S4_k141_164662 | 1735 | No | NA | 2 | 0 | 0 | Not-determined | Genome-fragment | NA | NA | 0 |
| S4_k141_157893 | 12743 | No | NA | 7 | 0 | 7 | Not-determined | Genome-fragment | NA | NA | 0 |
| S4_k141_59995 | 1642 | No | NA | 2 | 0 | 0 | Not-determined | Genome-fragment | NA | NA | 0 |
| S4_k141_25383 | 1156 | No | NA | 2 | 0 | 1 | Not-determined | Genome-fragment | NA | NA | 0 |
| S4_k141_52679 | 1078 | No | NA | 2 | 0 | 0 | Not-determined | Genome-fragment | NA | NA | 0 |
| S4_k141_157911 | 4086 | No | NA | 5 | 0 | 3 | Not-determined | Genome-fragment | NA | NA | 0 |
| S4_k141_67386 | 2827 | No | NA | 4 | 0 | 0 | Not-determined | Genome-fragment | NA | NA | 0 |
| S4_k141_74451 | 1168 | No | NA | 2 | 0 | 0 | Not-determined | Genome-fragment | NA | NA | 0 |
| S4_k141_74455 | 1081 | No | NA | 2 | 0 | 2 | Not-determined | Genome-fragment | NA | NA | 0 |
| S4_k141_11654 | 2401 | No | NA | 2 | 0 | 2 | Not-determined | Genome-fragment | NA | NA | 0 |
| S8-final-viral-combined_k141_207503\|\|full | 6147 | No | NA | 7 | 1 | 1 | Low-quality | Genome-fragment | 10.6 | AAI-based (medium-confidence) | 0 |
| S6-final-viral-combined_k141_140230\|\|full | 6148 | No | NA | 7 | 0 | 2 | Low-quality | Genome-fragment | 6.27 | AAI-based (medium-confidence) | 0 |
| S1-final-viral-combined_k141_159320\|\|full | 6150 | Yes | 3148 | 8 | 1 | 1 | Low-quality | Genome-fragment | 2.34 | AAI-based (high-confidence) | 48.81 |
| S4_k141_4509 | 1952 | No | NA | 2 | 0 | 2 | Not-determined | Genome-fragment | NA | NA | 0 |
| S4_k141_101963 | 1802 | No | NA | 3 | 0 | 2 | Not-determined | Genome-fragment | NA | NA | 0 |
| S4_k141_17819 | 1286 | No | NA | 2 | 0 | 1 | Not-determined | Genome-fragment | NA | NA | 0 |
| S4_k141_81668 | 1091 | No | NA | 2 | 0 | 0 | Not-determined | Genome-fragment | NA | NA | 0 |
| S4_k141_52736 | 1967 | No | NA | 6 | 0 | 2 | Not-determined | Genome-fragment | NA | NA | 0 |
| S4_k141_39228 | 1138 | No | NA | 1 | 0 | 1 | Not-determined | Genome-fragment | NA | NA | 0 |
| S4_k141_81687 | 1750 | No | NA | 2 | 0 | 2 | Not-determined | Genome-fragment | NA | NA | 0 |
| S4_k141_11694 | 1369 | No | NA | 1 | 0 | 0 | Not-determined | Genome-fragment | NA | NA | 0 |
| S4_k141_164748 | 2180 | No | NA | 2 | 0 | 1 | Not-determined | Genome-fragment | NA | NA | 0 |
| S17-final-viral-combined_k141_104931\|\|full | 6152 | No | NA | 7 | 3 | 0 | Low-quality | Genome-fragment | 13.95 | AAI-based (high-confidence) | 0 |
| S4_k141_109461 | 1021 | No | NA | 2 | 0 | 1 | Not-determined | Genome-fragment | NA | NA | 0 |
| S2-final-viral-combined_k141_196024\|\|full | 6152 | No | NA | 5 | 0 | 1 | Low-quality | Genome-fragment | 15.11 | AAI-based (medium-confidence) | 0 |
| S4_k141_60085 | 1592 | No | NA | 3 | 0 | 2 | Not-determined | Genome-fragment | NA | NA | 0 |
| S4_k141_122391 | 1422 | No | NA | 2 | 0 | 0 | Not-determined | Genome-fragment | NA | NA | 0 |
| S4_k141_4578 | 1170 | No | NA | 1 | 0 | 0 | Not-determined | Genome-fragment | NA | NA | 0 |
| S4_k141_60115 | 1636 | No | NA | 1 | 0 | 1 | Not-determined | Genome-fragment | NA | NA | 0 |
| S4_k141_150828 | 1212 | No | NA | 1 | 0 | 0 | Not-determined | Genome-fragment | NA | NA | 0 |
| S4_k141_158045 | 1034 | No | NA | 2 | 0 | 0 | Not-determined | Genome-fragment | NA | NA | 0 |
| S4_k141_67528 | 3174 | No | NA | 4 | 0 | 3 | Not-determined | Genome-fragment | NA | NA | 0 |
| S4_k141_95242 | 1101 | No | NA | 2 | 0 | 1 | Not-determined | Genome-fragment | NA | NA | 0 |
| S4_k141_17890 | 3127 | No | NA | 3 | 0 | 3 | Not-determined | Genome-fragment | NA | NA | 0 |
| S4_k141_46680 | 3511 | No | NA | 5 | 0 | 5 | Not-determined | Genome-fragment | NA | NA | 0 |
| S4_k141_11803 | 1772 | No | NA | 2 | 0 | 2 | Not-determined | Genome-fragment | NA | NA | 0 |
| S4_k141_88216 | 2394 | No | NA | 2 | 0 | 0 | Not-determined | Genome-fragment | NA | NA | 0 |
| S4_k141_4655 | 1033 | No | NA | 1 | 0 | 1 | Not-determined | Genome-fragment | NA | NA | 0 |
| S4_k141_60164 | 3140 | No | NA | 4 | 0 | 0 | Not-determined | Genome-fragment | NA | NA | 0 |
| S4_k141_67575 | 1405 | No | NA | 1 | 0 | 1 | Not-determined | Genome-fragment | NA | NA | 0 |
| S4_k141_109556 | 3258 | No | NA | 4 | 0 | 0 | Not-determined | Genome-fragment | NA | NA | 0 |
| S4_k141_74621 | 3337 | No | NA | 6 | 0 | 0 | Not-determined | Genome-fragment | NA | NA | 0 |
| S4_k141_32714 | 1108 | No | NA | 2 | 0 | 1 | Not-determined | Genome-fragment | NA | NA | 0 |
| S4_k141_4689 | 1347 | No | NA | 1 | 0 | 0 | Not-determined | Genome-fragment | NA | NA | 0 |
| S4_k141_39378 | 1025 | No | NA | 1 | 0 | 1 | Not-determined | Genome-fragment | NA | NA | 0 |
| S2_k141_44971 | 6152 | No | NA | 6 | 0 | 5 | Low-quality | Genome-fragment | 0.72 | AAI-based (medium-confidence) | 0 |
| S12_k141_4758 | 6154 | No | NA | 9 | 5 | 0 | Low-quality | Genome-fragment | 13.85 | AAI-based (high-confidence) | 0 |
| S4_k141_39399 | 1063 | No | NA | 1 | 0 | 0 | Not-determined | Genome-fragment | NA | NA | 0 |
| S12_k141_94115 | 6154 | Yes | 4250 | 8 | 2 | 1 | Low-quality | Genome-fragment | 9.83 | AAI-based (high-confidence) | 30.94 |
| S4_k141_95343 | 4031 | No | NA | 6 | 0 | 5 | Not-determined | Genome-fragment | NA | NA | 0 |
| S4_k141_32749 | 1455 | No | NA | 1 | 0 | 1 | Not-determined | Genome-fragment | NA | NA | 0 |
| S4_k141_88280 | 1018 | No | NA | 1 | 0 | 0 | Not-determined | Genome-fragment | NA | NA | 0 |
| S4_k141_74662 | 10207 | No | NA | 9 | 0 | 2 | Not-determined | Genome-fragment | NA | NA | 0 |
| S12-final-viral-combined_k141_102344\|\|full | 6160 | No | NA | 6 | 3 | 0 | Low-quality | Genome-fragment | 10.05 | HMM-based (lower-bound) | 0 |
| S4_k141_102075 | 1291 | No | NA | 3 | 0 | 0 | Not-determined | Genome-fragment | NA | NA | 0 |
| S8_k141_139595 | 6160 | No | NA | 6 | 0 | 1 | Low-quality | Genome-fragment | 19 | AAI-based (high-confidence) | 0 |
| S4_k141_88315 | 10755 | No | NA | 12 | 0 | 7 | Not-determined | Genome-fragment | NA | NA | 0 |
| S17-final-viral-combined_k141_100598\|\|full | 6166 | No | NA | 13 | 1 | 0 | Low-quality | Genome-fragment | 7.11 | HMM-based (lower-bound) | 0 |
| S4_k141_102113 | 2305 | No | NA | 2 | 0 | 1 | Not-determined | Genome-fragment | NA | NA | 0 |
| S4_k141_52916 | 1306 | No | NA | 3 | 0 | 3 | Not-determined | Genome-fragment | NA | NA | 0 |
| S4_k141_137129 | 5316 | No | NA | 5 | 0 | 5 | Not-determined | Genome-fragment | NA | NA | 0 |
| S4_k141_116027 | 4525 | No | NA | 2 | 0 | 0 | Not-determined | Genome-fragment | NA | NA | 0 |
| S4_k141_109688 | 1126 | No | NA | 1 | 0 | 0 | Not-determined | Genome-fragment | NA | NA | 0 |
| S4_k141_164944 | 1392 | No | NA | 3 | 0 | 2 | Not-determined | Genome-fragment | NA | NA | 0 |
| S4_k141_46793 | 3372 | No | NA | 5 | 0 | 4 | Not-determined | Genome-fragment | NA | NA | 0 |
| S4_k141_109689 | 7669 | No | NA | 5 | 0 | 3 | Not-determined | Genome-fragment | NA | NA | 0 |
| S7-final-viral-combined_k141_173995\|\|full | 6168 | No | NA | 8 | 4 | 0 | Low-quality | Genome-fragment | 13.44 | AAI-based (high-confidence) | 0 |
| S4_k141_46798 | 2695 | No | NA | 3 | 0 | 0 | Not-determined | Genome-fragment | NA | NA | 0 |
| S4_k141_4805 | 1078 | No | NA | 1 | 0 | 1 | Not-determined | Genome-fragment | NA | NA | 0 |
| S4_k141_102175 | 1630 | No | NA | 3 | 0 | 0 | Not-determined | Genome-fragment | NA | NA | 0 |
| S4_k141_11936 | 1075 | No | NA | 1 | 0 | 0 | Not-determined | Genome-fragment | NA | NA | 0 |

| | | | | | | | | | | | |
|---|---|---|---|---|---|---|---|---|---|---|---|
| S4_k141_109706 | 1961 | No | NA | 3 | 0 | 2 | Not-determined | Genome-fragment | NA | NA | 0 |
| S4_k141_164975 | 1416 | No | NA | 3 | 0 | 3 | Not-determined | Genome-fragment | NA | NA | 0 |
| S4_k141_164979 | 1131 | No | NA | 2 | 0 | 1 | Not-determined | Genome-fragment | NA | NA | 0 |
| S17-final-viral-combined_k141_224633\|\|full | 6172 | No | NA | 7 | 1 | 2 | Low-quality | Genome-fragment | 5.62 | HMM-based (lower-bound) | 0 |
| S4_k141_52992 | 1669 | No | NA | 3 | 0 | 2 | Not-determined | Genome-fragment | NA | NA | 0 |
| S4_k141_151011 | 1059 | No | NA | 2 | 0 | 1 | Not-determined | Genome-fragment | NA | NA | 0 |
| S4_k141_39492 | 1212 | No | NA | 2 | 0 | 0 | Not-determined | Genome-fragment | NA | NA | 0 |
| S7-final-viral-combined_k141_166416\|\|full | 6174 | No | NA | 14 | 6 | 1 | Low-quality | Genome-fragment | 9.68 | HMM-based (lower-bound) | 0 |
| S4_k141_144433 | 1071 | No | NA | 1 | 0 | 0 | Not-determined | Genome-fragment | NA | NA | 0 |
| S4_k141_74791 | 1325 | No | NA | 1 | 0 | 0 | Not-determined | Genome-fragment | NA | NA | 0 |
| S4_k141_18108 | 1046 | No | NA | 2 | 0 | 1 | Not-determined | Genome-fragment | NA | NA | 0 |
| S4_k141_95497 | 1274 | No | NA | 2 | 0 | 0 | Not-determined | Genome-fragment | NA | NA | 0 |
| S4_k141_158313 | 3665 | No | NA | 3 | 0 | 3 | Not-determined | Genome-fragment | NA | NA | 0 |
| S4_k141_11967 | 3724 | No | NA | 2 | 0 | 2 | Not-determined | Genome-fragment | NA | NA | 0 |
| S4_k141_18125 | 3083 | No | NA | 3 | 0 | 2 | Not-determined | Genome-fragment | NA | NA | 0 |
| S4_k141_46881 | 1818 | No | NA | 2 | 0 | 1 | Not-determined | Genome-fragment | NA | NA | 0 |
| S4_k141_144488 | 3755 | No | NA | 6 | 0 | 1 | Not-determined | Genome-fragment | NA | NA | 0 |
| S4_k141_60363 | 1611 | No | NA | 2 | 0 | 0 | Not-determined | Genome-fragment | NA | NA | 0 |
| S4_k141_158362 | 1107 | No | NA | 1 | 0 | 1 | Not-determined | Genome-fragment | NA | NA | 0 |
| S8-final-viral-combined_k141_269147\|\|full | 6175 | No | NA | 5 | 5 | 0 | Low-quality | Genome-fragment | 16.98 | AAI-based (high-confidence) | 0 |
| S4_k141_32854 | 1537 | No | NA | 2 | 0 | 2 | Not-determined | Genome-fragment | NA | NA | 0 |
| S11-final-viral-combined_k141_192109\|\|full | 6178 | No | NA | 7 | 0 | 0 | Low-quality | Genome-fragment | 13.88 | AAI-based (high-confidence) | 0 |
| S4_k141_82010 | 1404 | No | NA | 2 | 0 | 2 | Not-determined | Genome-fragment | NA | NA | 0 |
| S4_k141_60380 | 4375 | No | NA | 1 | 0 | 0 | Not-determined | Genome-fragment | NA | NA | 0 |
| S4_k141_102281 | 1818 | No | NA | 2 | 0 | 1 | Not-determined | Genome-fragment | NA | NA | 0 |
| S4_k141_109810 | 1659 | No | NA | 2 | 0 | 1 | Not-determined | Genome-fragment | NA | NA | 0 |
| S1_k141_50204 | 6179 | No | NA | 8 | 0 | 0 | Low-quality | Genome-fragment | 14.26 | AAI-based (medium-confidence) | 0 |
| S4_k141_74868 | 1252 | No | NA | 1 | 0 | 0 | Not-determined | Genome-fragment | NA | NA | 0 |
| S4_k141_102295 | 1199 | No | NA | 1 | 0 | 0 | Not-determined | Genome-fragment | NA | NA | 0 |
| S4_k141_18176 | 1581 | No | NA | 2 | 0 | 0 | Not-determined | Genome-fragment | NA | NA | 0 |
| S4_k141_158394 | 1576 | No | NA | 2 | 0 | 1 | Not-determined | Genome-fragment | NA | NA | 0 |
| S4_k141_95534 | 3781 | No | NA | 4 | 0 | 2 | Not-determined | Genome-fragment | NA | NA | 0 |
| S4_k141_109821 | 1565 | No | NA | 2 | 0 | 2 | Not-determined | Genome-fragment | NA | NA | 0 |
| S4_k141_74881 | 1134 | No | NA | 2 | 0 | 2 | Not-determined | Genome-fragment | NA | NA | 0 |
| S4_k141_95540 | 2599 | No | NA | 3 | 0 | 0 | Not-determined | Genome-fragment | NA | NA | 0 |
| S4_k141_67800 | 1365 | No | NA | 1 | 0 | 0 | Not-determined | Genome-fragment | NA | NA | 0 |
| S4_k141_95550 | 1211 | No | NA | 1 | 0 | 1 | Not-determined | Genome-fragment | NA | NA | 0 |
| S4_k141_18197 | 2028 | No | NA | 2 | 0 | 0 | Not-determined | Genome-fragment | NA | NA | 0 |
| S4_k141_25821 | 1214 | No | NA | 2 | 0 | 2 | Not-determined | Genome-fragment | NA | NA | 0 |
| S17-final-viral-combined_k141_157809\|\|full | 6179 | No | NA | 7 | 0 | 2 | Low-quality | Genome-fragment | 13.57 | AAI-based (medium-confidence) | 0 |
| S4_k141_137294 | 1401 | No | NA | 1 | 0 | 1 | Not-determined | Genome-fragment | NA | NA | 0 |
| S4_k141_67813 | 3722 | No | NA | 1 | 0 | 1 | Not-determined | Genome-fragment | NA | NA | 0 |
| S4_k141_102315 | 3933 | No | NA | 1 | 0 | 1 | Not-determined | Genome-fragment | NA | NA | 0 |
| S12-final-viral-combined_k141_377699\|\|full | 6180 | Yes | 525 | 4 | 1 | 3 | Low-quality | Genome-fragment | 0.21 | HMM-based (lower-bound) | 91.5 |
| S4_k141_46957 | 3460 | No | NA | 6 | 0 | 0 | Not-determined | Genome-fragment | NA | NA | 0 |
| S4_k141_122744 | 1056 | No | NA | 2 | 0 | 1 | Not-determined | Genome-fragment | NA | NA | 0 |
| S4_k141_109849 | 6083 | No | NA | 4 | 0 | 4 | Not-determined | Genome-fragment | NA | NA | 0 |
| S2-final-viral-combined_k141_83063\|\|full | 6181 | No | NA | 8 | 2 | 0 | Low-quality | Genome-fragment | 4.38 | HMM-based (lower-bound) | 0 |
| S4_k141_32974 | 1421 | No | NA | 3 | 0 | 3 | Not-determined | Genome-fragment | NA | NA | 0 |
| S4_k141_144599 | 1335 | No | NA | 1 | 0 | 1 | Not-determined | Genome-fragment | NA | NA | 0 |
| S4_k141_116256 | 8583 | No | NA | 2 | 0 | 2 | Not-determined | Genome-fragment | NA | NA | 0 |
| S2-final-viral-combined_k141_175906\|\|full | 6182 | No | NA | 15 | 3 | 0 | Low-quality | Genome-fragment | 6.9 | HMM-based (lower-bound) | 0 |
| S4_k141_165218 | 1010 | No | NA | 1 | 0 | 0 | Not-determined | Genome-fragment | NA | NA | 0 |
| S4_k141_116271 | 1112 | No | NA | 1 | 0 | 1 | Not-determined | Genome-fragment | NA | NA | 0 |
| S4_k141_158514 | 1032 | No | NA | 3 | 0 | 0 | Not-determined | Genome-fragment | NA | NA | 0 |
| S4_k141_144631 | 2841 | No | NA | 3 | 0 | 0 | Not-determined | Genome-fragment | NA | NA | 0 |
| S4_k141_122763 | 1324 | No | NA | 5 | 0 | 4 | Not-determined | Genome-fragment | NA | NA | 0 |
| S4_k141_144632 | 1185 | No | NA | 2 | 0 | 1 | Not-determined | Genome-fragment | NA | NA | 0 |
| S4_k141_33026 | 1575 | No | NA | 2 | 0 | 1 | Not-determined | Genome-fragment | NA | NA | 0 |
| S4_k141_67883 | 1214 | No | NA | 0 | 0 | 0 | Not-determined | Genome-fragment | NA | NA | 0 |
| S8-final-viral-combined_k141_69377\|\|full | 6182 | No | NA | 6 | 5 | 0 | Low-quality | Genome-fragment | 14.57 | AAI-based (high-confidence) | 0 |
| S4_k141_158580 | 1000 | No | NA | 2 | 0 | 1 | Not-determined | Genome-fragment | NA | NA | 0 |
| S8-final-viral-combined_k141_22007\|\|full | 6183 | No | NA | 13 | 1 | 0 | Low-quality | Genome-fragment | 18.51 | HMM-based (lower-bound) | 0 |
| S4-final-viral-combined_k141_124930\|\|full | 6184 | No | NA | 7 | 1 | 0 | Low-quality | Genome-fragment | 15.39 | AAI-based (high-confidence) | 0 |
| S4_k141_67935 | 1305 | No | NA | 1 | 0 | 1 | Not-determined | Genome-fragment | NA | NA | 0 |
| S4_k141_25941 | 1308 | No | NA | 1 | 0 | 1 | Not-determined | Genome-fragment | NA | NA | 0 |
| S4_k141_39665 | 1078 | No | NA | 1 | 0 | 0 | Not-determined | Genome-fragment | NA | NA | 0 |
| S4_k141_144691 | 1840 | No | NA | 1 | 0 | 1 | Not-determined | Genome-fragment | NA | NA | 0 |
| S4_k141_102481 | 1084 | No | NA | 2 | 0 | 2 | Not-determined | Genome-fragment | NA | NA | 0 |
| S11-final-viral-combined_k141_212492\|\|full | 6192 | No | NA | 8 | 3 | 0 | Low-quality | Genome-fragment | 10.94 | AAI-based (high-confidence) | 0 |
| S4_k141_18370 | 3299 | No | NA | 3 | 0 | 3 | Not-determined | Genome-fragment | NA | NA | 0 |
| S5-final-viral-combined_k141_46240\|\|full | 6192 | No | NA | 4 | 0 | 2 | Low-quality | Genome-fragment | 2.53 | AAI-based (medium-confidence) | 0 |
| S4_k141_82157 | 1520 | No | NA | 2 | 0 | 1 | Not-determined | Genome-fragment | NA | NA | 0 |
| S4_k141_74983 | 4300 | No | NA | 6 | 0 | 4 | Not-determined | Genome-fragment | NA | NA | 0 |
| S4_k141_5006 | 1244 | No | NA | 1 | 0 | 0 | Not-determined | Genome-fragment | NA | NA | 0 |
| S4_k141_12170 | 1025 | No | NA | 1 | 0 | 0 | Not-determined | Genome-fragment | NA | NA | 0 |
| S7-final-viral-combined_k141_21342\|\|full | 6195 | No | NA | 7 | 0 | 5 | Low-quality | Genome-fragment | 2.84 | AAI-based (medium-confidence) | 0 |
| S4_k141_18375 | 1159 | No | NA | 2 | 0 | 2 | Not-determined | Genome-fragment | NA | NA | 0 |
| S4_k141_5018 | 1176 | No | NA | 1 | 0 | 1 | Not-determined | Genome-fragment | NA | NA | 0 |
| S4_k141_122859 | 1192 | No | NA | 1 | 0 | 0 | Not-determined | Genome-fragment | NA | NA | 0 |
| S4_k141_18387 | 1327 | No | NA | 1 | 0 | 1 | Not-determined | Genome-fragment | NA | NA | 0 |
| S4_k141_122873 | 1578 | No | NA | 2 | 0 | 1 | Not-determined | Genome-fragment | NA | NA | 0 |
| S4_k141_60487 | 3143 | No | NA | 1 | 0 | 1 | Not-determined | Genome-fragment | NA | NA | 0 |
| S4_k141_144732 | 1017 | No | NA | 2 | 0 | 1 | Not-determined | Genome-fragment | NA | NA | 0 |
| S1-final-viral-combined_k141_169159\|\|full | 6196 | No | NA | 6 | 0 | 0 | Low-quality | Genome-fragment | 8.41 | AAI-based (medium-confidence) | 0 |
| S4_k141_18421 | 1820 | No | NA | 2 | 0 | 2 | Not-determined | Genome-fragment | NA | NA | 0 |
| S8_k141_84139 | 6197 | No | NA | 2 | 0 | 0 | Low-quality | Genome-fragment | 10.27 | AAI-based (medium-confidence) | 0 |
| S1-final-viral-combined_k141_84831\|\|full | 6199 | No | NA | 14 | 8 | 1 | Low-quality | Genome-fragment | 11.33 | AAI-based (high-confidence) | 0 |
| S4_k141_88654 | 1175 | No | NA | 1 | 0 | 1 | Not-determined | Genome-fragment | NA | NA | 0 |
| S4_k141_67968 | 1138 | No | NA | 2 | 0 | 1 | Not-determined | Genome-fragment | NA | NA | 0 |
| S4_k141_47074 | 1320 | No | NA | 4 | 0 | 1 | Not-determined | Genome-fragment | NA | NA | 0 |
| S4_k141_67972 | 1563 | No | NA | 2 | 0 | 2 | Not-determined | Genome-fragment | NA | NA | 0 |
| S4-final-viral-combined_k141_138318\|\|full | 6199 | No | NA | 6 | 0 | 5 | Low-quality | Genome-fragment | 3.39 | AAI-based (medium-confidence) | 0 |
| S4_k141_47089 | 2851 | No | NA | 5 | 0 | 5 | Not-determined | Genome-fragment | NA | NA | 0 |
| S4_k141_60535 | 1326 | No | NA | 1 | 0 | 1 | Not-determined | Genome-fragment | NA | NA | 0 |
| S1-final-viral-combined_k141_265395\|\|full | 6203 | Yes | 3807 | 10 | 2 | 2 | Low-quality | Genome-fragment | 8.61 | AAI-based (medium-confidence) | 38.63 |
| S1-final-viral-combined_k141_305629\|\|full | 6203 | No | NA | 7 | 0 | 1 | Low-quality | Genome-fragment | 2.51 | AAI-based (medium-confidence) | 0 |
| S7_k141_11245 | 6204 | No | NA | 12 | 1 | 0 | Low-quality | Genome-fragment | 7.52 | AAI-based (medium-confidence) | 0 |
| S2-final-viral-combined_k141_142361\|\|full | 6204 | No | NA | 11 | 1 | 0 | Low-quality | Genome-fragment | 1.6 | HMM-based (lower-bound) | 0 |
| S4_k141_68028 | 1008 | No | NA | 2 | 0 | 2 | Not-determined | Genome-fragment | NA | NA | 0 |
| S4_k141_12234 | 2515 | No | NA | 2 | 0 | 2 | Not-determined | Genome-fragment | NA | NA | 0 |
| S8-final-viral-combined_k141_168430\|\|full | 6206 | No | NA | 10 | 0 | 1 | Low-quality | Genome-fragment | 15.89 | AAI-based (medium-confidence) | 0 |
| S4_k141_116477 | 1072 | No | NA | 2 | 0 | 2 | Not-determined | Genome-fragment | NA | NA | 0 |
| S4_k141_60589 | 1976 | No | NA | 3 | 0 | 2 | Not-determined | Genome-fragment | NA | NA | 0 |
| S4_k141_39803 | 1044 | No | NA | 2 | 0 | 1 | Not-determined | Genome-fragment | NA | NA | 0 |
| S4_k141_33151 | 1101 | No | NA | 1 | 0 | 1 | Not-determined | Genome-fragment | NA | NA | 0 |
| S4_k141_33153 | 1195 | No | NA | 2 | 0 | 1 | Not-determined | Genome-fragment | NA | NA | 0 |
| S1-final-viral-combined_k141_23765\|\|full | 6207 | No | NA | 9 | 0 | 0 | Low-quality | Genome-fragment | 10.12 | AAI-based (medium-confidence) | 0 |
| S4_k141_116508 | 3128 | No | NA | 1 | 0 | 1 | Not-determined | Genome-fragment | NA | NA | 0 |
| S7-final-viral-combined_k141_111650\|\|full | 6208 | No | NA | 7 | 1 | 0 | Low-quality | Genome-fragment | 12.69 | AAI-based (medium-confidence) | 0 |
| S4_k141_53361 | 1495 | No | NA | 2 | 0 | 0 | Not-determined | Genome-fragment | NA | NA | 0 |
| S4_k141_47130 | 1837 | No | NA | 2 | 0 | 1 | Not-determined | Genome-fragment | NA | NA | 0 |
| S4_k141_158799 | 3012 | No | NA | 2 | 0 | 2 | Not-determined | Genome-fragment | NA | NA | 0 |
| S4_k141_12309 | 3594 | No | NA | 4 | 0 | 4 | Not-determined | Genome-fragment | NA | NA | 0 |
| S12_k141_214414 | 6210 | No | NA | 3 | 0 | 1 | Low-quality | Genome-fragment | 14.71 | AAI-based (medium-confidence) | 0 |
| S4_k141_47179 | 1166 | No | NA | 1 | 0 | 0 | Not-determined | Genome-fragment | NA | NA | 0 |
| S8-final-viral-combined_k141_254969\|\|full | 6213 | No | NA | 3 | 0 | 1 | Low-quality | Genome-fragment | 21.66 | AAI-based (medium-confidence) | 0 |
| S12-final-viral-combined_k141_352251\|\|full | 6216 | No | NA | 3 | 1 | 0 | Low-quality | Genome-fragment | 4.26 | HMM-based (lower-bound) | 0 |
| S8_k141_217079 | 6219 | No | NA | 11 | 0 | 0 | Low-quality | Genome-fragment | 6.5 | AAI-based (high-confidence) | 0 |
| S4_k141_18536 | 1498 | No | NA | 2 | 0 | 0 | Not-determined | Genome-fragment | NA | NA | 0 |
| S4_k141_158832 | 2162 | No | NA | 1 | 0 | 1 | Not-determined | Genome-fragment | NA | NA | 0 |
| S4_k141_82266 | 1008 | No | NA | 1 | 0 | 0 | Not-determined | Genome-fragment | NA | NA | 0 |
| S4_k141_110212 | 1956 | No | NA | 1 | 0 | 1 | Not-determined | Genome-fragment | NA | NA | 0 |
| S4_k141_5101 | 1823 | No | NA | 1 | 0 | 1 | Not-determined | Genome-fragment | NA | NA | 0 |
| S4_k141_33251 | 1045 | No | NA | 2 | 0 | 2 | Not-determined | Genome-fragment | NA | NA | 0 |
| S4_k141_12360 | 1393 | No | NA | 1 | 0 | 1 | Not-determined | Genome-fragment | NA | NA | 0 |
| S4_k141_82285 | 1000 | No | NA | 1 | 0 | 0 | Not-determined | Genome-fragment | NA | NA | 0 |
| S2_k141_106041 | 6220 | No | NA | 18 | 3 | 0 | Low-quality | Genome-fragment | 14.63 | AAI-based (high-confidence) | 0 |
| S4_k141_53432 | 1447 | No | NA | 2 | 0 | 1 | Not-determined | Genome-fragment | NA | NA | 0 |
| S12-final-viral-combined_k141_135434\|\|full | 6224 | Yes | 3215 | 7 | 1 | 3 | Low-quality | Genome-fragment | 2.16 | HMM-based (lower-bound) | 48.35 |
| S4_k141_18590 | 3105 | No | NA | 3 | 0 | 3 | Not-determined | Genome-fragment | NA | NA | 0 |
| S4_k141_75207 | 1131 | No | NA | 2 | 0 | 2 | Not-determined | Genome-fragment | NA | NA | 0 |
| S2-final-viral-combined_k141_8893\|\|full | 6227 | No | NA | 6 | 3 | 0 | Low-quality | Genome-fragment | 9.7 | AAI-based (high-confidence) | 0 |
| S4_k141_18596 | 1325 | No | NA | 2 | 0 | 1 | Not-determined | Genome-fragment | NA | NA | 0 |
| S4_k141_75222 | 1144 | No | NA | 1 | 0 | 0 | Not-determined | Genome-fragment | NA | NA | 0 |
| S4_k141_123068 | 2224 | No | NA | 2 | 0 | 2 | Not-determined | Genome-fragment | NA | NA | 0 |
| S4_k141_47252 | 1249 | No | NA | 1 | 0 | 1 | Not-determined | Genome-fragment | NA | NA | 0 |
| S4_k141_39961 | 1212 | No | NA | 1 | 0 | 0 | Not-determined | Genome-fragment | NA | NA | 0 |
| S4_k141_123079 | 1516 | No | NA | 1 | 0 | 1 | Not-determined | Genome-fragment | NA | NA | 0 |
| S4_k141_137537 | 1027 | No | NA | 2 | 0 | 0 | Not-determined | Genome-fragment | NA | NA | 0 |
| S4_k141_110271 | 1540 | No | NA | 1 | 0 | 1 | Not-determined | Genome-fragment | NA | NA | 0 |
| S2-final-viral-combined_k141_145713\|\|full | 6228 | No | NA | 15 | 4 | 0 | Low-quality | Genome-fragment | 10.25 | AAI-based (high-confidence) | 0 |
| S4_k141_26194 | 3842 | No | NA | 4 | 0 | 2 | Not-determined | Genome-fragment | NA | NA | 0 |
| S2-final-viral-combined_k141_29534\|\|full | 6228 | No | NA | 9 | 6 | 0 | Low-quality | Genome-fragment | 11.7 | AAI-based (high-confidence) | 0 |
| S4_k141_39982 | 16769 | No | NA | 8 | 0 | 4 | Not-determined | Genome-fragment | NA | NA | 0 |
| S4_k141_60707 | 1477 | No | NA | 1 | 0 | 1 | Not-determined | Genome-fragment | NA | NA | 0 |
| S12-final-viral-combined_k141_65836\|\|full | 6229 | Yes | 3213 | 6 | 1 | 1 | Low-quality | Genome-fragment | 3.51 | HMM-based (lower-bound) | 48.42 |
| S4_k141_75278 | 1557 | No | NA | 1 | 0 | 1 | Not-determined | Genome-fragment | NA | NA | 0 |
| S17-k141_211653 | 6231 | No | NA | 11 | 2 | 0 | Low-quality | Genome-fragment | 13.04 | AAI-based (high-confidence) | 0 |
| S3-final-viral-combined_k141_68979\|\|full | 6232 | No | NA | 9 | 1 | 0 | Low-quality | Genome-fragment | 6.84 | AAI-based (medium-confidence) | 0 |
| S4_k141_123128 | 1026 | No | NA | 1 | 0 | 1 | Not-determined | Genome-fragment | NA | NA | 0 |
| S4_k141_47300 | 5067 | No | NA | 3 | 0 | 2 | Not-determined | Genome-fragment | NA | NA | 0 |
| S4_k141_75284 | 1438 | No | NA | 1 | 0 | 1 | Not-determined | Genome-fragment | NA | NA | 0 |
| S4_k141_116625 | 1363 | No | NA | 2 | 0 | 2 | Not-determined | Genome-fragment | NA | NA | 0 |
| S4_k141_82321 | 7062 | No | NA | 10 | 0 | 0 | Not-determined | Genome-fragment | NA | NA | 0 |
| S4_k141_116632 | 1353 | No | NA | 4 | 0 | 0 | Not-determined | Genome-fragment | NA | NA | 0 |
| S4_k141_5217 | 1380 | No | NA | 2 | 0 | 2 | Not-determined | Genome-fragment | NA | NA | 0 |
| S4_k141_26248 | 1138 | No | NA | 1 | 0 | 1 | Not-determined | Genome-fragment | NA | NA | 0 |
| S4_k141_82347 | 1663 | No | NA | 1 | 0 | 0 | Not-determined | Genome-fragment | NA | NA | 0 |
| S4_k141_137620 | 1374 | No | NA | 1 | 0 | 1 | Not-determined | Genome-fragment | NA | NA | 0 |
| S4_k141_158988 | 1026 | No | NA | 3 | 0 | 2 | Not-determined | Genome-fragment | NA | NA | 0 |
| S4_k141_110348 | 3786 | No | NA | 4 | 0 | 4 | Not-determined | Genome-fragment | NA | NA | 0 |
| S5-final-viral-combined_k141_2351\|\|full | 6232 | No | NA | 9 | 0 | 4 | Low-quality | Genome-fragment | 12.09 | AAI-based (high-confidence) | 0 |
| S11-final-viral-combined_k141_323889\|\|full | 6235 | No | NA | 5 | 0 | 4 | Low-quality | Genome-fragment | 2.96 | AAI-based (medium-confidence) | 0 |
| S12-final-viral-combined_k141_360499\|\|full | 6235 | No | NA | 8 | 2 | 0 | Low-quality | Genome-fragment | 9.83 | AAI-based (high-confidence) | 0 |
| S4_k141_137635 | 1339 | No | NA | 2 | 0 | 2 | Not-determined | Genome-fragment | NA | NA | 0 |
| S4_k141_47358 | 4914 | No | NA | 5 | 0 | 1 | Not-determined | Genome-fragment | NA | NA | 0 |
| S4_k141_75297 | 1312 | No | NA | 1 | 0 | 1 | Not-determined | Genome-fragment | NA | NA | 0 |
| S8-final-viral-combined_k141_110899\|\|full | 6235 | Yes | 5197 | 4 | 1 | 1 | Low-quality | Genome-fragment | 11.09 | AAI-based (high-confidence) | 16.65 |
| S1-final-viral-combined_k141_90822\|\|full | 6237 | No | NA | 7 | 0 | 4 | Low-quality | Genome-fragment | 3.23 | AAI-based (medium-confidence) | 0 |
| S5-final-viral-combined_k141_192433\|\|full | 6240 | No | NA | 10 | 2 | 1 | Low-quality | Genome-fragment | 4.15 | HMM-based (lower-bound) | 0 |
| S4_k141_137663 | 2437 | No | NA | 2 | 0 | 2 | Not-determined | Genome-fragment | NA | NA | 0 |
| S4_k141_145059 | 3656 | No | NA | 4 | 0 | 1 | Not-determined | Genome-fragment | NA | NA | 0 |
| S4_k141_53584 | 1083 | No | NA | 1 | 0 | 0 | Not-determined | Genome-fragment | NA | NA | 0 |
| S4_k141_145062 | 3524 | No | NA | 3 | 0 | 3 | Not-determined | Genome-fragment | NA | NA | 0 |
| S8_k141_86532 | 6243 | No | NA | 5 | 0 | 0 | Low-quality | Genome-fragment | 7.2 | AAI-based (high-confidence) | 0 |
| S11_k141_169652 | 6245 | No | NA | 17 | 3 | 0 | Low-quality | Genome-fragment | 7.54 | AAI-based (high-confidence) | 0 |
| S4_k141_110422 | 3314 | No | NA | 3 | 0 | 2 | Not-determined | Genome-fragment | NA | NA | 0 |
| S2-final-viral-combined_k141_421\|\|full | 6245 | No | NA | 6 | 0 | 5 | Low-quality | Genome-fragment | 2.95 | AAI-based (medium-confidence) | 0 |
| S4_k141_82435 | 1556 | No | NA | 1 | 0 | 0 | Not-determined | Genome-fragment | NA | NA | 0 |
| S4_k141_53603 | 3962 | No | NA | 1 | 0 | 1 | Not-determined | Genome-fragment | NA | NA | 0 |
| S2-final-viral-combined_k141_18352\|\|full | 6245 | No | NA | 8 | 0 | 0 | Low-quality | Genome-fragment | 11 | AAI-based (medium-confidence) | 0 |
| S4_k141_53606 | 1000 | No | NA | 2 | 0 | 0 | Not-determined | Genome-fragment | NA | NA | 0 |
| S4_k141_60863 | 1034 | No | NA | 1 | 0 | 1 | Not-determined | Genome-fragment | NA | NA | 0 |
| S4_k141_40144 | 1014 | No | NA | 1 | 0 | 1 | Not-determined | Genome-fragment | NA | NA | 0 |
| S11-final-viral-combined_k141_270129\|\|full | 6249 | Yes | 1950 | 4 | 1 | 3 | Low-quality | Genome-fragment | 2.98 | AAI-based (medium-confidence) | 68.8 |
| S4_k141_18792 | 1978 | No | NA | 1 | 0 | 1 | Not-determined | Genome-fragment | NA | NA | 0 |
| S4_k141_18793 | 1319 | No | NA | 2 | 0 | 0 | Not-determined | Genome-fragment | NA | NA | 0 |
| S4_k141_75346 | 4195 | No | NA | 2 | 0 | 1 | Not-determined | Genome-fragment | NA | NA | 0 |
| S4_k141_89010 | 6222 | No | NA | 6 | 0 | 5 | Not-determined | Genome-fragment | NA | NA | 0 |
| S4_k141_33456 | 1321 | No | NA | 1 | 0 | 1 | Not-determined | Genome-fragment | NA | NA | 0 |
| S1-final-viral-combined_k141_145309\|\|full | 6249 | No | NA | 10 | 6 | 0 | Low-quality | Genome-fragment | 10.21 | AAI-based (high-confidence) | 0 |
| S4_k141_130704 | 1339 | No | NA | 2 | 0 | 0 | Not-determined | Genome-fragment | NA | NA | 0 |
| S4_k141_123270 | 1388 | No | NA | 1 | 0 | 0 | Not-determined | Genome-fragment | NA | NA | 0 |
| S4_k141_68351 | 3511 | No | NA | 4 | 0 | 4 | Not-determined | Genome-fragment | NA | NA | 0 |
| S1-final-viral-combined_k141_257934\|\|full | 6251 | No | NA | 6 | 3 | 0 | Low-quality | Genome-fragment | 9.61 | AAI-based (high-confidence) | 0 |
| S4_k141_130741 | 4035 | No | NA | 4 | 0 | 4 | Not-determined | Genome-fragment | NA | NA | 0 |
| S4_k141_89064 | 3585 | No | NA | 2 | 0 | 0 | Not-determined | Genome-fragment | NA | NA | 0 |
| S4_k141_123314 | 1072 | No | NA | 1 | 0 | 1 | Not-determined | Genome-fragment | NA | NA | 0 |
| S4_k141_26403 | 1234 | No | NA | 2 | 0 | 2 | Not-determined | Genome-fragment | NA | NA | 0 |
| S1-final-viral-combined_k141_63831\|\|full | 6252 | No | NA | 15 | 0 | 1 | Low-quality | Genome-fragment | 11.42 | AAI-based (high-confidence) | 0 |
| S4_k141_82560 | 1761 | No | NA | 1 | 0 | 1 | Not-determined | Genome-fragment | NA | NA | 0 |
| S4_k141_12605 | 1277 | No | NA | 1 | 0 | 0 | Not-determined | Genome-fragment | NA | NA | 0 |
| S4_k141_33559 | 6295 | No | NA | 4 | 0 | 2 | Not-determined | Genome-fragment | NA | NA | 0 |
| S4_k141_110507 | 4866 | No | NA | 3 | 0 | 3 | Not-determined | Genome-fragment | NA | NA | 0 |
| S4_k141_137783 | 1064 | No | NA | 1 | 0 | 0 | Not-determined | Genome-fragment | NA | NA | 0 |
| S4_k141_5461 | 1229 | No | NA | 1 | 0 | 1 | Not-determined | Genome-fragment | NA | NA | 0 |
| S4_k141_89112 | 4804 | No | NA | 1 | 0 | 1 | Not-determined | Genome-fragment | NA | NA | 0 |
| S11-final-viral-combined_k141_224105\|\|full | 6253 | No | NA | 6 | 4 | 0 | Low-quality | Genome-fragment | 15.43 | AAI-based (medium-confidence) | 0 |
| S12_k141_334972 | 6253 | No | NA | 7 | 0 | 1 | Low-quality | Genome-fragment | 10 | AAI-based (medium-confidence) | 0 |
| S7-final-viral-combined_k141_146466\|\|full | 6254 | No | NA | 4 | 0 | 1 | Low-quality | Genome-fragment | 5.47 | AAI-based (medium-confidence) | 0 |
| S4_k141_75439 | 2434 | No | NA | 3 | 0 | 2 | Not-determined | Genome-fragment | NA | NA | 0 |
| S4_k141_18920 | 1586 | No | NA | 4 | 0 | 0 | Not-determined | Genome-fragment | NA | NA | 0 |
| S4_k141_33600 | 1174 | No | NA | 1 | 0 | 1 | Not-determined | Genome-fragment | NA | NA | 0 |
| S4_k141_18930 | 1138 | No | NA | 1 | 0 | 1 | Not-determined | Genome-fragment | NA | NA | 0 |
| S6_k141_5412 | 6256 | No | NA | 12 | 3 | 0 | Low-quality | Genome-fragment | 3.34 | AAI-based (high-confidence) | 0 |
| S4_k141_165534 | 12967 | No | NA | 9 | 0 | 5 | Not-determined | Genome-fragment | NA | NA | 0 |
| S4_k141_116850 | 1189 | No | NA | 2 | 0 | 1 | Not-determined | Genome-fragment | NA | NA | 0 |
| S4_k141_40313 | 2651 | No | NA | 1 | 0 | 0 | Not-determined | Genome-fragment | NA | NA | 0 |
| S4_k141_40315 | 1256 | No | NA | 2 | 0 | 1 | Not-determined | Genome-fragment | NA | NA | 0 |
| S4_k141_102905 | 1086 | No | NA | 1 | 0 | 0 | Not-determined | Genome-fragment | NA | NA | 0 |
| S4_k141_18950 | 1046 | No | NA | 1 | 0 | 1 | Not-determined | Genome-fragment | NA | NA | 0 |
| S4_k141_130781 | 1358 | No | NA | 1 | 0 | 0 | Not-determined | Genome-fragment | NA | NA | 0 |
| S17-final-viral-combined_k141_132062\|\|full | 6258 | No | NA | 13 | 1 | 0 | Low-quality | Genome-fragment | 7.02 | HMM-based (lower-bound) | 0 |
| S4_k141_40326 | 3307 | No | NA | 3 | 0 | 0 | Not-determined | Genome-fragment | NA | NA | 0 |
| S4_k141_82674 | 3635 | No | NA | 6 | 0 | 3 | Not-determined | Genome-fragment | NA | NA | 0 |
| S4_k141_33621 | 1632 | No | NA | 4 | 0 | 0 | Not-determined | Genome-fragment | NA | NA | 0 |
| S4_k141_145249 | 3361 | No | NA | 3 | 0 | 3 | Not-determined | Genome-fragment | NA | NA | 0 |
| S4_k141_33626 | 1654 | No | NA | 2 | 0 | 1 | Not-determined | Genome-fragment | NA | NA | 0 |
| S4_k141_75496 | 1490 | No | NA | 2 | 0 | 2 | Not-determined | Genome-fragment | NA | NA | 0 |
| S4_k141_26484 | 3352 | No | NA | 2 | 0 | 1 | Not-determined | Genome-fragment | NA | NA | 0 |
| S4_k141_110610 | 3112 | No | NA | 2 | 0 | 2 | Not-determined | Genome-fragment | NA | NA | 0 |
| S4_k141_82695 | 1062 | No | NA | 1 | 0 | 0 | Not-determined | Genome-fragment | NA | NA | 0 |
| S4_k141_75502 | 2594 | No | NA | 4 | 0 | 3 | Not-determined | Genome-fragment | NA | NA | 0 |
| S4_k141_137854 | 2268 | No | NA | 3 | 0 | 1 | Not-determined | Genome-fragment | NA | NA | 0 |
| S4_k141_75515 | 1363 | No | NA | 2 | 0 | 0 | Not-determined | Genome-fragment | NA | NA | 0 |
| S11_k141_26726 | 6265 | No | NA | 4 | 0 | 1 | Low-quality | Genome-fragment | 10.46 | AAI-based (high-confidence) | 0 |
| S4_k141_102935 | 1185 | No | NA | 2 | 0 | 2 | Not-determined | Genome-fragment | NA | NA | 0 |
| S4_k141_61055 | 5062 | No | NA | 6 | 0 | 3 | Not-determined | Genome-fragment | NA | NA | 0 |
| S4_k141_123471 | 1888 | No | NA | 4 | 0 | 0 | Not-determined | Genome-fragment | NA | NA | 0 |
| S4_k141_123479 | 2626 | No | NA | 3 | 0 | 3 | Not-determined | Genome-fragment | NA | NA | 0 |
| S4_k141_40371 | 3455 | No | NA | 3 | 0 | 2 | Not-determined | Genome-fragment | NA | NA | 0 |
| S4_k141_40385 | 1678 | No | NA | 2 | 0 | 2 | Not-determined | Genome-fragment | NA | NA | 0 |
| S8_k141_109599 | 6268 | No | NA | 13 | 1 | 0 | Low-quality | Genome-fragment | 2.93 | HMM-based (lower-bound) | 0 |
| S12-final-viral-combined_k141_52857\|\|full | 6270 | No | NA | 11 | 3 | 0 | Low-quality | Genome-fragment | 14.19 | AAI-based (high-confidence) | 0 |
| S4_k141_137910 | 3376 | No | NA | 2 | 0 | 2 | Not-determined | Genome-fragment | NA | NA | 0 |
| S4_k141_130868 | 1010 | No | NA | 1 | 0 | 0 | Not-determined | Genome-fragment | NA | NA | 0 |
| S4_k141_137915 | 2123 | No | NA | 4 | 0 | 3 | Not-determined | Genome-fragment | NA | NA | 0 |
| S4_k141_103005 | 1198 | No | NA | 1 | 0 | 0 | Not-determined | Genome-fragment | NA | NA | 0 |
| S4_k141_89283 | 1137 | No | NA | 2 | 0 | 1 | Not-determined | Genome-fragment | NA | NA | 0 |
| S4_k141_19049 | 1169 | No | NA | 3 | 0 | 2 | Not-determined | Genome-fragment | NA | NA | 0 |
| S4_k141_130904 | 1093 | No | NA | 1 | 0 | 1 | Not-determined | Genome-fragment | NA | NA | 0 |
| S4_k141_145348 | 1213 | No | NA | 3 | 0 | 1 | Not-determined | Genome-fragment | NA | NA | 0 |
| S4_k141_53796 | 1287 | No | NA | 1 | 0 | 0 | Not-determined | Genome-fragment | NA | NA | 0 |
| S4_k141_68521 | 1659 | No | NA | 1 | 0 | 0 | Not-determined | Genome-fragment | NA | NA | 0 |
| S7_k141_138092 | 6273 | No | NA | 12 | 6 | 0 | Low-quality | Genome-fragment | 11.4 | AAI-based (high-confidence) | 0 |
| S4_k141_116997 | 2130 | No | NA | 1 | 0 | 1 | Not-determined | Genome-fragment | NA | NA | 0 |
| S12-final-viral-combined_k141_194649\|\|full | 6274 | No | NA | 5 | 0 | 3 | Low-quality | Genome-fragment | 1.79 | AAI-based (high-confidence) | 0 |
| S4_k141_53816 | 1802 | No | NA | 1 | 0 | 1 | Not-determined | Genome-fragment | NA | NA | 0 |
| S4_k141_53817 | 1130 | No | NA | 2 | 0 | 1 | Not-determined | Genome-fragment | NA | NA | 0 |
| S4_k141_5638 | 1162 | No | NA | 1 | 0 | 1 | Not-determined | Genome-fragment | NA | NA | 0 |
| S4_k141_5642 | 1104 | No | NA | 1 | 0 | 0 | Not-determined | Genome-fragment | NA | NA | 0 |
| S4_k141_159270 | 1076 | No | NA | 2 | 0 | 2 | Not-determined | Genome-fragment | NA | NA | 0 |
| S4_k141_40464 | 2000 | No | NA | 2 | 0 | 1 | Not-determined | Genome-fragment | NA | NA | 0 |
| S4_k141_48124 | 6278 | No | NA | 8 | 4 | 0 | Low-quality | Genome-fragment | 13.44 | AAI-based (high-confidence) | 0 |
| S4_k141_68577 | 2576 | No | NA | 1 | 0 | 1 | Not-determined | Genome-fragment | NA | NA | 0 |
| S4_k141_40473 | 1958 | No | NA | 1 | 0 | 1 | Not-determined | Genome-fragment | NA | NA | 0 |
| S4_k141_12741 | 1687 | No | NA | 3 | 0 | 0 | Not-determined | Genome-fragment | NA | NA | 0 |
| S4_k141_68589 | 1308 | No | NA | 1 | 0 | 1 | Not-determined | Genome-fragment | NA | NA | 0 |
| S4_k141_151902 | 14445 | No | NA | 11 | 0 | 6 | Not-determined | Genome-fragment | NA | NA | 0 |
| S4_k141_53831 | 1482 | No | NA | 1 | 0 | 1 | Not-determined | Genome-fragment | NA | NA | 0 |
| S4_k141_68609 | 1298 | No | NA | 2 | 0 | 1 | Not-determined | Genome-fragment | NA | NA | 0 |
| S4_k141_89372 | 1902 | No | NA | 3 | 0 | 1 | Not-determined | Genome-fragment | NA | NA | 0 |
| S4_k141_19150 | 3039 | No | NA | 1 | 0 | 1 | Not-determined | Genome-fragment | NA | NA | 0 |
| S4_k141_47700 | 5015 | No | NA | 3 | 0 | 1 | Not-determined | Genome-fragment | NA | NA | 0 |
| S6_k141_36247 | 6285 | No | NA | 11 | 1 | 0 | Low-quality | Genome-fragment | 3.36 | AAI-based (high-confidence) | 0 |
| S4_k141_137995 | 1269 | No | NA | 1 | 0 | 1 | Not-determined | Genome-fragment | NA | NA | 0 |
| S4_k141_47743 | 5524 | No | NA | 3 | 0 | 3 | Not-determined | Genome-fragment | NA | NA | 0 |
| S4_k141_19211 | 3511 | No | NA | 4 | 0 | 3 | Not-determined | Genome-fragment | NA | NA | 0 |
| S4_k141_165741 | 1233 | No | NA | 1 | 0 | 1 | Not-determined | Genome-fragment | NA | NA | 0 |
| S4_k141_117125 | 1031 | No | NA | 1 | 0 | 1 | Not-determined | Genome-fragment | NA | NA | 0 |
| S4_k141_19225 | 1139 | No | NA | 5 | 0 | 0 | Not-determined | Genome-fragment | NA | NA | 0 |
| S4_k141_12882 | 1023 | No | NA | 2 | 0 | 1 | Not-determined | Genome-fragment | NA | NA | 0 |
| S4_k141_131041 | 1411 | No | NA | 1 | 0 | 1 | Not-determined | Genome-fragment | NA | NA | 0 |
| S4_k141_53931 | 1161 | No | NA | 1 | 0 | 1 | Not-determined | Genome-fragment | NA | NA | 0 |
| S4_k141_165770 | 1122 | No | NA | 1 | 0 | 1 | Not-determined | Genome-fragment | NA | NA | 0 |
| S8-final-viral-combined_k141_50732\|\|full | 6286 | No | NA | 4 | 0 | 3 | Low-quality | Genome-fragment | 1.02 | AAI-based (high-confidence) | 0 |
| S4_k141_5749 | 1542 | No | NA | 3 | 0 | 0 | Not-determined | Genome-fragment | NA | NA | 0 |
| S4_k141_145571 | 1935 | No | NA | 2 | 0 | 0 | Not-determined | Genome-fragment | NA | NA | 0 |
| S4_k141_33886 | 3370 | No | NA | 4 | 0 | 4 | Not-determined | Genome-fragment | NA | NA | 0 |
| S4_k141_68769 | 3201 | No | NA | 3 | 0 | 3 | Not-determined | Genome-fragment | NA | NA | 0 |
| S4_k141_159474 | 1258 | No | NA | 1 | 0 | 1 | Not-determined | Genome-fragment | NA | NA | 0 |
| S4_k141_61303 | 1398 | No | NA | 1 | 0 | 1 | Not-determined | Genome-fragment | NA | NA | 0 |
| S4_k141_40611 | 4148 | No | NA | 3 | 0 | 1 | Not-determined | Genome-fragment | NA | NA | 0 |
| S4_k141_61311 | 1147 | No | NA | 1 | 0 | 1 | Not-determined | Genome-fragment | NA | NA | 0 |
| S4_k141_165831 | 1134 | No | NA | 2 | 0 | 0 | Not-determined | Genome-fragment | NA | NA | 0 |
| S11-final-viral-combined_k141_166522\|\|full | 6287 | No | NA | 2 | 1 | 0 | Low-quality | Genome-fragment | 10.9 | HMM-based (lower-bound) | 0 |
| S4_k141_83071 | 3952 | No | NA | 6 | 0 | 1 | Not-determined | Genome-fragment | NA | NA | 0 |
| S4_k141_12959 | 1187 | No | NA | 1 | 0 | 0 | Not-determined | Genome-fragment | NA | NA | 0 |
| S4_k141_159510 | 1257 | No | NA | 1 | 0 | 1 | Not-determined | Genome-fragment | NA | NA | 0 |
| S4_k141_75834 | 1014 | No | NA | 1 | 0 | 1 | Not-determined | Genome-fragment | NA | NA | 0 |
| S4_k141_165848 | 1034 | No | NA | 2 | 0 | 1 | Not-determined | Genome-fragment | NA | NA | 0 |
| S4_k141_12974 | 3461 | No | NA | 2 | 0 | 1 | Not-determined | Genome-fragment | NA | NA | 0 |
| S4_k141_103259 | 1096 | No | NA | 1 | 0 | 0 | Not-determined | Genome-fragment | NA | NA | 0 |
| S4_k141_96651 | 1213 | No | NA | 2 | 0 | 1 | Not-determined | Genome-fragment | NA | NA | 0 |
| S4_k141_159537 | 4333 | No | NA | 5 | 0 | 0 | Not-determined | Genome-fragment | NA | NA | 0 |
| S11_k141_149316 | 6287 | No | NA | 3 | 0 | 2 | Low-quality | Genome-fragment | 1.99 | AAI-based (high-confidence) | 0 |
| S8_k141_779 | 6288 | No | NA | 6 | 0 | 2 | Low-quality | Genome-fragment | 12.1 | AAI-based (medium-confidence) | 0 |
| S4_k141_138179 | 4021 | No | NA | 3 | 0 | 2 | Not-determined | Genome-fragment | NA | NA | 0 |
| S1_k141_251166 | 6290 | No | NA | 8 | 6 | 0 | Low-quality | Genome-fragment | 15.61 | AAI-based (high-confidence) | 0 |
| S4_k141_75870 | 1789 | No | NA | 1 | 0 | 0 | Not-determined | Genome-fragment | NA | NA | 0 |
| S7_k141_39180 | 6292 | No | NA | 12 | 2 | 0 | Low-quality | Genome-fragment | 15.45 | AAI-based (high-confidence) | 0 |
| S4_k141_96664 | 5358 | No | NA | 3 | 0 | 3 | Not-determined | Genome-fragment | NA | NA | 0 |
| S4_k141_152117 | 2335 | No | NA | 2 | 0 | 2 | Not-determined | Genome-fragment | NA | NA | 0 |
| S4_k141_75880 | 1089 | No | NA | 2 | 0 | 2 | Not-determined | Genome-fragment | NA | NA | 0 |
| S4_k141_165893 | 3617 | No | NA | 2 | 0 | 2 | Not-determined | Genome-fragment | NA | NA | 0 |
| S4_k141_131156 | 4654 | No | NA | 3 | 0 | 1 | Not-determined | Genome-fragment | NA | NA | 0 |
| S4_k141_96681 | 2135 | No | NA | 3 | 0 | 3 | Not-determined | Genome-fragment | NA | NA | 0 |
| S4_k141_5887 | 1207 | No | NA | 3 | 0 | 0 | Not-determined | Genome-fragment | NA | NA | 0 |
| S7-final-viral-combined_k141_113695\|\|full | 6294 | No | NA | 7 | 1 | 0 | Low-quality | Genome-fragment | 4.53 | HMM-based (lower-bound) | 0 |
| S4_k141_138231 | 1168 | No | NA | 1 | 0 | 0 | Not-determined | Genome-fragment | NA | NA | 0 |
| S4_k141_138239 | 1020 | No | NA | 2 | 0 | 1 | Not-determined | Genome-fragment | NA | NA | 0 |
| S4_k141_165914 | 1090 | No | NA | 1 | 0 | 1 | Not-determined | Genome-fragment | NA | NA | 0 |
| S4_k141_111034 | 1757 | No | NA | 2 | 0 | 2 | Not-determined | Genome-fragment | NA | NA | 0 |
| S11-final-viral-combined_k141_196664\|\|full | 6297 | No | NA | 10 | 5 | 0 | Low-quality | Genome-fragment | 14.03 | AAI-based (high-confidence) | 0 |
| S4_k141_152183 | 2596 | No | NA | 1 | 0 | 0 | Not-determined | Genome-fragment | NA | NA | 0 |
| S4_k141_152191 | 1575 | No | NA | 1 | 0 | 0 | Not-determined | Genome-fragment | NA | NA | 0 |
| S4_k141_13050 | 2055 | No | NA | 3 | 0 | 0 | Not-determined | Genome-fragment | NA | NA | 0 |
| S4_k141_26763 | 1219 | No | NA | 1 | 0 | 1 | Not-determined | Genome-fragment | NA | NA | 0 |
| S5-final-viral-combined_k141_55606\|\|full | 6302 | No | NA | 8 | 5 | 0 | Low-quality | Genome-fragment | 14.99 | AAI-based (high-confidence) | 0 |
| S12-final-viral-combined_k141_297025\|\|full | 6302 | No | NA | 8 | 0 | 7 | Low-quality | Genome-fragment | 2.91 | AAI-based (medium-confidence) | 0 |
| S4_k141_165949 | 1082 | No | NA | 2 | 0 | 2 | Not-determined | Genome-fragment | NA | NA | 0 |
| S4_k141_5934 | 1134 | No | NA | 1 | 0 | 0 | Not-determined | Genome-fragment | NA | NA | 0 |
| S6-final-viral-combined_k141_86986\|\|full | 6307 | No | NA | 9 | 3 | 0 | Low-quality | Genome-fragment | 16.2 | AAI-based (medium-confidence) | 0 |
| S4_k141_145830 | 1424 | No | NA | 1 | 0 | 1 | Not-determined | Genome-fragment | NA | NA | 0 |
| S4_k141_68951 | 1025 | No | NA | 3 | 0 | 1 | Not-determined | Genome-fragment | NA | NA | 0 |
| S4_k141_68955 | 1715 | No | NA | 2 | 0 | 2 | Not-determined | Genome-fragment | NA | NA | 0 |
| S4_k141_48006 | 1120 | No | NA | 2 | 0 | 0 | Not-determined | Genome-fragment | NA | NA | 0 |
| S4_k141_54174 | 1012 | No | NA | 2 | 0 | 0 | Not-determined | Genome-fragment | NA | NA | 0 |
| S11-final-viral-combined_k141_214104\|\|full | 6308 | No | NA | 10 | 0 | 0 | Low-quality | Genome-fragment | 7.66 | AAI-based (high-confidence) | 0 |
| S4_k141_34077 | 1179 | No | NA | 1 | 0 | 1 | Not-determined | Genome-fragment | NA | NA | 0 |
| S11-final-viral-combined_k141_284700\|\|full | 6309 | No | NA | 9 | 4 | 0 | Low-quality | Genome-fragment | 7 | HMM-based (lower-bound) | 0 |
| S4_k141_48045 | 1051 | No | NA | 2 | 0 | 0 | Not-determined | Genome-fragment | NA | NA | 0 |
| S4_k141_131194 | 1463 | No | NA | 1 | 0 | 1 | Not-determined | Genome-fragment | NA | NA | 0 |
| S4_k141_26834 | 1088 | No | NA | 2 | 0 | 2 | Not-determined | Genome-fragment | NA | NA | 0 |
| S4_k141_166029 | 1844 | No | NA | 2 | 0 | 0 | Not-determined | Genome-fragment | NA | NA | 0 |
| S4_k141_89624 | 1272 | No | NA | 2 | 0 | 2 | Not-determined | Genome-fragment | NA | NA | 0 |
| S4_k141_89639 | 1317 | No | NA | 2 | 0 | 1 | Not-determined | Genome-fragment | NA | NA | 0 |
| S11-final-viral-combined_k141_214754\|\|full | 6310 | No | NA | 6 | 5 | 0 | Low-quality | Genome-fragment | 15.42 | AAI-based (high-confidence) | 0 |
| S4_k141_145857 | 1301 | No | NA | 3 | 0 | 1 | Not-determined | Genome-fragment | NA | NA | 0 |
| S4_k141_96766 | 1019 | No | NA | 1 | 0 | 1 | Not-determined | Genome-fragment | NA | NA | 0 |
| S4_k141_68980 | 1178 | No | NA | 2 | 0 | 0 | Not-determined | Genome-fragment | NA | NA | 0 |
| S4_k141_61524 | 1105 | No | NA | 1 | 0 | 0 | Not-determined | Genome-fragment | NA | NA | 0 |
| S4_k141_40940 | 2455 | No | NA | 3 | 0 | 1 | Not-determined | Genome-fragment | NA | NA | 0 |
| S1-final-viral-combined_k141_120356\|\|full | 6311 | No | NA | 6 | 0 | 5 | Low-quality | Genome-fragment | 4.01 | AAI-based (medium-confidence) | 0 |
| S11-final-viral-combined_k141_146057\|\|full | 6317 | No | NA | 10 | 7 | 0 | Low-quality | Genome-fragment | 10.15 | AAI-based (high-confidence) | 0 |
| S4_k141_103364 | 1052 | No | NA | 1 | 0 | 0 | Not-determined | Genome-fragment | NA | NA | 0 |
| S1-final-viral-combined_k141_185240\|\|full | 6318 | No | NA | 10 | 7 | 0 | Low-quality | Genome-fragment | 18.09 | AAI-based (high-confidence) | 0 |
| S7_k141_189382 | 6319 | No | NA | 8 | 3 | 0 | Low-quality | Genome-fragment | 14.71 | AAI-based (high-confidence) | 0 |
| S4_k141_83342 | 1056 | No | NA | 2 | 0 | 1 | Not-determined | Genome-fragment | NA | NA | 0 |
| S4_k141_13257 | 1143 | No | NA | 3 | 0 | 0 | Not-determined | Genome-fragment | NA | NA | 0 |
| S4_k141_96824 | 1019 | No | NA | 2 | 0 | 1 | Not-determined | Genome-fragment | NA | NA | 0 |
| S17-final-viral-combined_k141_93802\|\|full | 6325 | No | NA | 5 | 0 | 3 | Low-quality | Genome-fragment | 4.83 | AAI-based (high-confidence) | 0 |
| S4_k141_76104 | 1446 | No | NA | 2 | 0 | 1 | Not-determined | Genome-fragment | NA | NA | 0 |
| S4_k141_34203 | 8511 | No | NA | 6 | 0 | 3 | Not-determined | Genome-fragment | NA | NA | 0 |
| S7-final-viral-combined_k141_194783\|\|full | 6328 | No | NA | 11 | 0 | 1 | Low-quality | Genome-fragment | 11.27 | AAI-based (medium-confidence) | 0 |
| S4_k141_13287 | 1329 | No | NA | 2 | 0 | 2 | Not-determined | Genome-fragment | NA | NA | 0 |
| S1_k141_256411 | 6331 | No | NA | 13 | 8 | 0 | Low-quality | Genome-fragment | 15.88 | AAI-based (high-confidence) | 0 |
| S2-final-viral-combined_k141_206249\|\|full | 6332 | Yes | 5176 | 8 | 1 | 1 | Low-quality | Genome-fragment | 3.95 | AAI-based (high-confidence) | 18.26 |
| S4_k141_159720 | 1513 | No | NA | 3 | 0 | 1 | Not-determined | Genome-fragment | NA | NA | 0 |
| S4_k141_34214 | 1077 | No | NA | 1 | 0 | 1 | Not-determined | Genome-fragment | NA | NA | 0 |
| S4_k141_76150 | 1075 | No | NA | 2 | 0 | 0 | Not-determined | Genome-fragment | NA | NA | 0 |
| S1_k141_101303 | 6335 | No | NA | 5 | 0 | 0 | Low-quality | Genome-fragment | 15 | AAI-based (high-confidence) | 0 |
| S4_k141_152319 | 1247 | No | NA | 3 | 0 | 1 | Not-determined | Genome-fragment | NA | NA | 0 |
| S17-final-viral-combined_k141_168764\|\|full | 6336 | No | NA | 5 | 0 | 5 | Low-quality | Genome-fragment | 2.14 | AAI-based (medium-confidence) | 0 |
| S4_k141_13322 | 1088 | No | NA | 3 | 0 | 1 | Not-determined | Genome-fragment | NA | NA | 0 |
| S4_k141_48244 | 1299 | No | NA | 1 | 0 | 1 | Not-determined | Genome-fragment | NA | NA | 0 |
| S4_k141_89800 | 3034 | No | NA | 2 | 0 | 1 | Not-determined | Genome-fragment | NA | NA | 0 |
| S4_k141_34253 | 1433 | No | NA | 1 | 0 | 1 | Not-determined | Genome-fragment | NA | NA | 0 |
| S2-final-viral-combined_k141_51432\|\|full | 6336 | No | NA | 9 | 1 | 1 | Low-quality | Genome-fragment | 4.28 | AAI-based (high-confidence) | 0 |
| S11-final-viral-combined_k141_248753\|\|full | 6338 | No | NA | 13 | 3 | 0 | Low-quality | Genome-fragment | 10.37 | AAI-based (high-confidence) | 0 |
| S4_k141_76226 | 1040 | No | NA | 1 | 0 | 1 | Not-determined | Genome-fragment | NA | NA | 0 |
| S4_k141_96896 | 1852 | No | NA | 2 | 0 | 2 | Not-determined | Genome-fragment | NA | NA | 0 |
| S4_k141_34277 | 1132 | No | NA | 1 | 0 | 0 | Not-determined | Genome-fragment | NA | NA | 0 |
| S4_k141_13372 | 1154 | No | NA | 3 | 0 | 1 | Not-determined | Genome-fragment | NA | NA | 0 |
| S4_k141_145999 | 1471 | No | NA | 1 | 0 | 0 | Not-determined | Genome-fragment | NA | NA | 0 |
| S12-final-viral-combined_k141_87515\|\|full | 6338 | No | NA | 8 | 1 | 3 | Low-quality | Genome-fragment | 13.82 | AAI-based (medium-confidence) | 0 |
| S4_k141_146003 | 1050 | No | NA | 2 | 0 | 0 | Not-determined | Genome-fragment | NA | NA | 0 |
| S8-final-viral-combined_k141_57362\|\|full | 6344 | Yes | 1759 | 8 | 1 | 3 | Low-quality | Genome-fragment | 3.59 | AAI-based (medium-confidence) | 72.27 |
| S4_k141_111274 | 1119 | No | NA | 1 | 0 | 1 | Not-determined | Genome-fragment | NA | NA | 0 |
| S4_k141_89836 | 1069 | No | NA | 2 | 0 | 2 | Not-determined | Genome-fragment | NA | NA | 0 |
| S4_k141_103543 | 2818 | No | NA | 2 | 0 | 0 | Not-determined | Genome-fragment | NA | NA | 0 |
| S4_k141_138552 | 2471 | No | NA | 2 | 0 | 1 | Not-determined | Genome-fragment | NA | NA | 0 |
| S4_k141_124196 | 1088 | No | NA | 1 | 0 | 0 | Not-determined | Genome-fragment | NA | NA | 0 |
| S4_k141_6235 | 1006 | No | NA | 1 | 0 | 1 | Not-determined | Genome-fragment | NA | NA | 0 |
| S4_k141_34303 | 1617 | No | NA | 2 | 0 | 0 | Not-determined | Genome-fragment | NA | NA | 0 |
| S4_k141_13419 | 5239 | No | NA | 4 | 0 | 4 | Not-determined | Genome-fragment | NA | NA | 0 |
| S7-final-viral-combined_k141_217480\|\|full | 6345 | No | NA | 9 | 0 | 0 | Low-quality | Genome-fragment | 17.48 | AAI-based (high-confidence) | 0 |
| S4_k141_83454 | 1128 | No | NA | 1 | 0 | 1 | Not-determined | Genome-fragment | NA | NA | 0 |
| S2-final-viral-combined_k141_134976\|\|full | 6346 | No | NA | 11 | 0 | 2 | Low-quality | Genome-fragment | 16.85 | AAI-based (high-confidence) | 0 |
| S4_k141_159874 | 1554 | No | NA | 1 | 0 | 1 | Not-determined | Genome-fragment | NA | NA | 0 |
| S4_k141_117652 | 4568 | No | NA | 5 | 0 | 4 | Not-determined | Genome-fragment | NA | NA | 0 |
| S11-final-viral-combined_k141_13159\|\|full | 6360 | No | NA | 9 | 2 | 0 | Low-quality | Genome-fragment | 8.46 | HMM-based (lower-bound) | 0 |
| S4_k141_61783 | 1411 | No | NA | 1 | 0 | 1 | Not-determined | Genome-fragment | NA | NA | 0 |
| S4_k141_89886 | 1189 | No | NA | 1 | 0 | 0 | Not-determined | Genome-fragment | NA | NA | 0 |
| S4_k141_103564 | 1009 | No | NA | 1 | 0 | 1 | Not-determined | Genome-fragment | NA | NA | 0 |
| S4_k141_138594 | 3856 | No | NA | 2 | 0 | 2 | Not-determined | Genome-fragment | NA | NA | 0 |
| S4_k141_117692 | 3140 | No | NA | 2 | 0 | 1 | Not-determined | Genome-fragment | NA | NA | 0 |
| S4_k141_146093 | 4344 | No | NA | 4 | 0 | 4 | Not-determined | Genome-fragment | NA | NA | 0 |
| S4_k141_76314 | 5425 | No | NA | 5 | 0 | 4 | Not-determined | Genome-fragment | NA | NA | 0 |
| S4_k141_138599 | 4849 | No | NA | 5 | 0 | 4 | Not-determined | Genome-fragment | NA | NA | 0 |
| S4_k141_97012 | 1011 | No | NA | 1 | 0 | 0 | Not-determined | Genome-fragment | NA | NA | 0 |
| S4_k141_13461 | 1068 | No | NA | 1 | 0 | 0 | Not-determined | Genome-fragment | NA | NA | 0 |

| | | | | | | | | | | | |
|---|---|---|---|---|---|---|---|---|---|---|---|
| S4_k141_138601 | 2003 | No | NA | 3 | 0 | 1 | Not-determined | Genome-fragment | NA | NA | 0 |
| S4_k141_117707 | 1034 | No | NA | 1 | 0 | 0 | Not-determined | Genome-fragment | NA | NA | 0 |
| S4_k141_138612 | 7674 | No | NA | 4 | 0 | 2 | Not-determined | Genome-fragment | NA | NA | 0 |
| S4_k141_61830 | 1404 | No | NA | 1 | 0 | 0 | Not-determined | Genome-fragment | NA | NA | 0 |
| S4_k141_146124 | 6608 | No | NA | 4 | 0 | 4 | Not-determined | Genome-fragment | NA | NA | 0 |
| S1-final-viral-combined_k141_122914\|\|full | 6360 | No | NA | 7 | 0 | 4 | Low-quality | Genome-fragment | 2.11 | AAI-based (medium-confidence) | 0 |
| S4-final-viral-combined_k141_100742\|\|full | 6360 | Yes | 5384 | 10 | 3 | 1 | Low-quality | Genome-fragment | 12.57 | AAI-based (high-confidence) | 15.35 |
| S4_k141_41165 | 1414 | No | NA | 2 | 0 | 0 | Not-determined | Genome-fragment | NA | NA | 0 |
| S4_k141_54635 | 1492 | No | NA | 1 | 0 | 0 | Not-determined | Genome-fragment | NA | NA | 0 |
| S4_k141_19743 | 1838 | No | NA | 3 | 0 | 0 | Not-determined | Genome-fragment | NA | NA | 0 |
| S4_k141_6359 | 1141 | No | NA | 2 | 0 | 0 | Not-determined | Genome-fragment | NA | NA | 0 |
| S4_k141_69286 | 2631 | No | NA | 2 | 0 | 1 | Not-determined | Genome-fragment | NA | NA | 0 |
| S4_k141_166330 | 1382 | No | NA | 2 | 0 | 2 | Not-determined | Genome-fragment | NA | NA | 0 |
| S4_k141_159993 | 1160 | No | NA | 2 | 0 | 0 | Not-determined | Genome-fragment | NA | NA | 0 |
| S4_k141_103659 | 1345 | No | NA | 1 | 0 | 1 | Not-determined | Genome-fragment | NA | NA | 0 |
| S4_k141_13534 | 6110 | No | NA | 4 | 0 | 3 | Not-determined | Genome-fragment | NA | NA | 0 |
| S4_k141_152477 | 2103 | No | NA | 2 | 0 | 2 | Not-determined | Genome-fragment | NA | NA | 0 |
| S4_k141_48420 | 1031 | No | NA | 2 | 0 | 2 | Not-determined | Genome-fragment | NA | NA | 0 |
| S4_k141_83575 | 1037 | No | NA | 1 | 0 | 1 | Not-determined | Genome-fragment | NA | NA | 0 |
| S8-final-viral-combined_k141_95200\|\|full | 6362 | No | NA | 5 | 0 | 1 | Low-quality | Genome-fragment | 3.85 | AAI-based (high-confidence) | 0 |
| S4_k141_166363 | 1107 | No | NA | 1 | 0 | 1 | Not-determined | Genome-fragment | NA | NA | 0 |
| S4_k141_111416 | 3279 | No | NA | 2 | 0 | 2 | Not-determined | Genome-fragment | NA | NA | 0 |
| S4_k141_76391 | 1228 | No | NA | 1 | 0 | 0 | Not-determined | Genome-fragment | NA | NA | 0 |
| S7-final-viral-combined_k141_135182\|\|full | 6365 | No | NA | 8 | 4 | 0 | Low-quality | Genome-fragment | 13.22 | AAI-based (high-confidence) | 0 |
| S4_k141_103720 | 2461 | No | NA | 2 | 0 | 1 | Not-determined | Genome-fragment | NA | NA | 0 |
| S4_k141_34313 | 2427 | No | NA | 2 | 0 | 2 | Not-determined | Genome-fragment | NA | NA | 0 |
| S4_k141_19795 | 1082 | No | NA | 1 | 0 | 0 | Not-determined | Genome-fragment | NA | NA | 0 |
| S2_k141_91497 | 6367 | No | NA | 11 | 4 | 0 | Low-quality | Genome-fragment | 13.06 | AAI-based (high-confidence) | 0 |
| S4_k141_138759 | 1500 | No | NA | 3 | 0 | 2 | Not-determined | Genome-fragment | NA | NA | 0 |
| S4_k141_124387 | 1264 | No | NA | 4 | 0 | 3 | Not-determined | Genome-fragment | NA | NA | 0 |
| S4_k141_138786 | 1585 | No | NA | 2 | 0 | 0 | Not-determined | Genome-fragment | NA | NA | 0 |
| S4_k141_27301 | 1263 | No | NA | 2 | 0 | 1 | Not-determined | Genome-fragment | NA | NA | 0 |
| S4_k141_54767 | 2823 | No | NA | 1 | 0 | 1 | Not-determined | Genome-fragment | NA | NA | 0 |
| S4_k141_76464 | 1535 | No | NA | 1 | 0 | 0 | Not-determined | Genome-fragment | NA | NA | 0 |
| S4_k141_19867 | 1100 | No | NA | 1 | 0 | 1 | Not-determined | Genome-fragment | NA | NA | 0 |
| S4_k141_34356 | 1278 | No | NA | 2 | 0 | 0 | Not-determined | Genome-fragment | NA | NA | 0 |
| S4_k141_6500 | 1265 | No | NA | 1 | 0 | 1 | Not-determined | Genome-fragment | NA | NA | 0 |
| S4_k141_90039 | 1104 | No | NA | 1 | 0 | 1 | Not-determined | Genome-fragment | NA | NA | 0 |
| S4_k141_138825 | 1123 | No | NA | 1 | 0 | 1 | Not-determined | Genome-fragment | NA | NA | 0 |
| S4_k141_62050 | 1898 | No | NA | 1 | 0 | 0 | Not-determined | Genome-fragment | NA | NA | 0 |
| S5-final-viral-combined_k141_108478\|\|full | 6368 | No | NA | 9 | 0 | 1 | Low-quality | Genome-fragment | 6.62 | AAI-based (medium-confidence) | 0 |
| S2-final-viral-combined_k141_63228\|\|full | 6369 | Yes | 2556 | 5 | 2 | 1 | Low-quality | Genome-fragment | 3.98 | HMM-based (lower-bound) | 59.87 |
| S11_k141_239018 | 6372 | No | NA | 14 | 0 | 2 | Low-quality | Genome-fragment | 11 | AAI-based (high-confidence) | 0 |
| S4_k141_13651 | 3854 | No | NA | 6 | 0 | 2 | Not-determined | Genome-fragment | NA | NA | 0 |
| S4_k141_6560 | 3012 | No | NA | 3 | 0 | 0 | Not-determined | Genome-fragment | NA | NA | 0 |
| S4_k141_166513 | 2248 | No | NA | 1 | 0 | 1 | Not-determined | Genome-fragment | NA | NA | 0 |
| S4_k141_131656 | 3299 | No | NA | 4 | 0 | 4 | Not-determined | Genome-fragment | NA | NA | 0 |
| S4_k141_48585 | 11027 | No | NA | 7 | 0 | 6 | Not-determined | Genome-fragment | NA | NA | 0 |
| S4_k141_76525 | 9307 | No | NA | 5 | 0 | 4 | Not-determined | Genome-fragment | NA | NA | 0 |
| S4_k141_90141 | 1131 | No | NA | 4 | 0 | 0 | Not-determined | Genome-fragment | NA | NA | 0 |
| S4_k141_117894 | 1208 | No | NA | 2 | 0 | 1 | Not-determined | Genome-fragment | NA | NA | 0 |
| S4_k141_62147 | 3407 | No | NA | 8 | 0 | 1 | Not-determined | Genome-fragment | NA | NA | 0 |
| S4_k141_103724 | 1126 | No | NA | 2 | 0 | 1 | Not-determined | Genome-fragment | NA | NA | 0 |
| S11_k141_213274 | 6381 | No | NA | 4 | 0 | 3 | Low-quality | Genome-fragment | 5.79 | AAI-based (medium-confidence) | 0 |
| S4_k141_166579 | 1613 | No | NA | 1 | 0 | 1 | Not-determined | Genome-fragment | NA | NA | 0 |
| S4_k141_13693 | 1157 | No | NA | 2 | 0 | 1 | Not-determined | Genome-fragment | NA | NA | 0 |
| S4_k141_19961 | 1273 | No | NA | 1 | 0 | 1 | Not-determined | Genome-fragment | NA | NA | 0 |
| S17-final-viral-combined_k141_208533\|\|full | 6382 | No | NA | 7 | 0 | 5 | Low-quality | Genome-fragment | 5.13 | AAI-based (medium-confidence) | 0 |
| S4_k141_54931 | 1201 | No | NA | 1 | 0 | 0 | Not-determined | Genome-fragment | NA | NA | 0 |
| S4_k141_41401 | 1263 | No | NA | 2 | 0 | 0 | Not-determined | Genome-fragment | NA | NA | 0 |
| S4_k141_166595 | 1256 | No | NA | 2 | 0 | 0 | Not-determined | Genome-fragment | NA | NA | 0 |
| S4_k141_152614 | 1085 | No | NA | 1 | 0 | 1 | Not-determined | Genome-fragment | NA | NA | 0 |
| S4_k141_97249 | 1106 | No | NA | 2 | 0 | 0 | Not-determined | Genome-fragment | NA | NA | 0 |
| S4_k141_62199 | 1347 | No | NA | 2 | 0 | 2 | Not-determined | Genome-fragment | NA | NA | 0 |
| S8-final-viral-combined_k141_132026\|\|full | 6384 | No | NA | 7 | 0 | 6 | Low-quality | Genome-fragment | 2.7 | AAI-based (medium-confidence) | 0 |
| S4_k141_19981 | 3108 | No | NA | 4 | 0 | 4 | Not-determined | Genome-fragment | NA | NA | 0 |
| S4_k141_90201 | 1879 | No | NA | 2 | 0 | 1 | Not-determined | Genome-fragment | NA | NA | 0 |
| S4_k141_54948 | 4781 | No | NA | 5 | 0 | 2 | Not-determined | Genome-fragment | NA | NA | 0 |
| S8-final-viral-combined_k141_232498\|\|full | 6385 | No | NA | 5 | 0 | 3 | Low-quality | Genome-fragment | 2 | AAI-based (medium-confidence) | 0 |
| S4_k141_48652 | 1013 | No | NA | 2 | 0 | 1 | Not-determined | Genome-fragment | NA | NA | 0 |
| S4_k141_97293 | 4273 | No | NA | 6 | 0 | 5 | Not-determined | Genome-fragment | NA | NA | 0 |
| S7-final-viral-combined_k141_93578\|\|full | 6392 | No | NA | 6 | 0 | 5 | Low-quality | Genome-fragment | 3.44 | AAI-based (medium-confidence) | 0 |
| S4_k141_41441 | 1785 | No | NA | 3 | 0 | 0 | Not-determined | Genome-fragment | NA | NA | 0 |
| S4_k141_103814 | 1018 | No | NA | 1 | 0 | 0 | Not-determined | Genome-fragment | NA | NA | 0 |
| S1-final-viral-combined_k141_279137\|\|full | 6401 | No | NA | 8 | 6 | 0 | Low-quality | Genome-fragment | 14.74 | AAI-based (high-confidence) | 0 |
| S4_k141_41461 | 1694 | No | NA | 4 | 0 | 4 | Not-determined | Genome-fragment | NA | NA | 0 |
| S4_k141_166677 | 2441 | No | NA | 1 | 0 | 1 | Not-determined | Genome-fragment | NA | NA | 0 |
| S1-final-viral-combined_k141_264254\|\|full | 6403 | No | NA | 7 | 0 | 1 | Low-quality | Genome-fragment | 12.62 | AAI-based (medium-confidence) | 0 |
| S12-final-viral-combined_k141_358036\|\|full | 6406 | No | NA | 9 | 1 | 0 | Low-quality | Genome-fragment | 11.94 | AAI-based (high-confidence) | 0 |
| S7-final-viral-combined_k141_19917\|\|full | 6408 | No | NA | 6 | 0 | 0 | Low-quality | Genome-fragment | 5.16 | AAI-based (medium-confidence) | 0 |
| S4_k141_20048 | 3570 | No | NA | 5 | 0 | 2 | Not-determined | Genome-fragment | NA | NA | 0 |
| S4_k141_27497 | 1715 | No | NA | 1 | 0 | 1 | Not-determined | Genome-fragment | NA | NA | 0 |
| S4_k141_139090 | 1001 | No | NA | 1 | 0 | 1 | Not-determined | Genome-fragment | NA | NA | 0 |
| S4_k141_103844 | 1160 | No | NA | 2 | 0 | 0 | Not-determined | Genome-fragment | NA | NA | 0 |
| S4_k141_90279 | 1870 | No | NA | 2 | 0 | 1 | Not-determined | Genome-fragment | NA | NA | 0 |
| S4_k141_20064 | 1161 | No | NA | 1 | 0 | 1 | Not-determined | Genome-fragment | NA | NA | 0 |
| S17-final-viral-combined_k141_170786\|\|full | 6410 | No | NA | 7 | 0 | 2 | Low-quality | Genome-fragment | 3.69 | AAI-based (high-confidence) | 0 |
| S4_k141_166721 | 1436 | No | NA | 1 | 0 | 1 | Not-determined | Genome-fragment | NA | NA | 0 |
| S4_k141_90326 | 1208 | No | NA | 1 | 0 | 0 | Not-determined | Genome-fragment | NA | NA | 0 |
| S4_k141_124687 | 1471 | No | NA | 1 | 0 | 1 | Not-determined | Genome-fragment | NA | NA | 0 |
| S4_k141_69656 | 1050 | No | NA | 1 | 0 | 1 | Not-determined | Genome-fragment | NA | NA | 0 |
| S11-final-viral-combined_k141_193173\|\|full | 6414 | No | NA | 5 | 2 | 0 | Low-quality | Genome-fragment | 10.47 | HMM-based (lower-bound) | 0 |
| S4_k141_97387 | 1003 | No | NA | 1 | 0 | 1 | Not-determined | Genome-fragment | NA | NA | 0 |
| S4_k141_131838 | 1167 | No | NA | 1 | 0 | 1 | Not-determined | Genome-fragment | NA | NA | 0 |
| S4_k141_166776 | 1107 | No | NA | 1 | 0 | 0 | Not-determined | Genome-fragment | NA | NA | 0 |
| S4_k141_55028 | 1805 | No | NA | 2 | 0 | 0 | Not-determined | Genome-fragment | NA | NA | 0 |
| S4_k141_131861 | 1108 | No | NA | 2 | 0 | 1 | Not-determined | Genome-fragment | NA | NA | 0 |
| S4_k141_48759 | 4642 | No | NA | 6 | 0 | 5 | Not-determined | Genome-fragment | NA | NA | 0 |
| S4_k141_124724 | 1808 | No | NA | 1 | 0 | 1 | Not-determined | Genome-fragment | NA | NA | 0 |
| S2-final-viral-combined_k141_813\|\|full | 6414 | No | NA | 12 | 2 | 1 | Low-quality | Genome-fragment | 6.38 | HMM-based (lower-bound) | 0 |
| S4_k141_34742 | 1021 | No | NA | 2 | 0 | 1 | Not-determined | Genome-fragment | NA | NA | 0 |
| S4_k141_62389 | 1265 | No | NA | 1 | 0 | 1 | Not-determined | Genome-fragment | NA | NA | 0 |
| S4_k141_27626 | 2687 | No | NA | 3 | 0 | 1 | Not-determined | Genome-fragment | NA | NA | 0 |
| S4_k141_152895 | 1500 | No | NA | 1 | 0 | 0 | Not-determined | Genome-fragment | NA | NA | 0 |
| S4_k141_166797 | 3505 | No | NA | 3 | 0 | 3 | Not-determined | Genome-fragment | NA | NA | 0 |
| S4_k141_152902 | 1151 | No | NA | 2 | 0 | 1 | Not-determined | Genome-fragment | NA | NA | 0 |
| S4_k141_124778 | 2292 | No | NA | 1 | 0 | 1 | Not-determined | Genome-fragment | NA | NA | 0 |
| S4_k141_124780 | 1707 | No | NA | 2 | 0 | 0 | Not-determined | Genome-fragment | NA | NA | 0 |
| S4_k141_152906 | 2560 | No | NA | 3 | 0 | 3 | Not-determined | Genome-fragment | NA | NA | 0 |
| S8-final-viral-combined_k141_155622\|\|full | 6419 | Yes | 4108 | 5 | 1 | 1 | Low-quality | Genome-fragment | 4.61 | HMM-based (lower-bound) | 36 |
| S4_k141_20165 | 10553 | No | NA | 10 | 0 | 3 | Not-determined | Genome-fragment | NA | NA | 0 |
| S4_k141_90432 | 1042 | No | NA | 2 | 0 | 1 | Not-determined | Genome-fragment | NA | NA | 0 |
| S4_k141_13912 | 1676 | No | NA | 4 | 0 | 3 | Not-determined | Genome-fragment | NA | NA | 0 |
| S4_k141_6829 | 1071 | No | NA | 1 | 0 | 1 | Not-determined | Genome-fragment | NA | NA | 0 |
| S4_k141_131908 | 4526 | No | NA | 6 | 0 | 4 | Not-determined | Genome-fragment | NA | NA | 0 |
| S4_k141_27653 | 7982 | No | NA | 6 | 0 | 2 | Not-determined | Genome-fragment | NA | NA | 0 |
| S4_k141_139259 | 1157 | No | NA | 2 | 0 | 1 | Not-determined | Genome-fragment | NA | NA | 0 |
| S7-final-viral-combined_k141_24233\|\|full | 6421 | No | NA | 11 | 0 | 0 | Low-quality | Genome-fragment | 13.46 | AAI-based (high-confidence) | 0 |
| S4_k141_20182 | 2793 | No | NA | 1 | 0 | 1 | Not-determined | Genome-fragment | NA | NA | 0 |
| S12-final-viral-combined_k141_112023\|\|full | 6422 | No | NA | 4 | 0 | 0 | Low-quality | Genome-fragment | 10.64 | AAI-based (medium-confidence) | 0 |
| S11-final-viral-combined_k141_242686\|\|full | 6424 | No | NA | 9 | 3 | 0 | Low-quality | Genome-fragment | 10.57 | HMM-based (lower-bound) | 0 |
| S4_k141_41572 | 1929 | No | NA | 2 | 0 | 2 | Not-determined | Genome-fragment | NA | NA | 0 |
| S4_k141_152969 | 1058 | No | NA | 2 | 0 | 1 | Not-determined | Genome-fragment | NA | NA | 0 |
| S4_k141_139282 | 3272 | No | NA | 4 | 0 | 3 | Not-determined | Genome-fragment | NA | NA | 0 |
| S4_k141_34819 | 1249 | No | NA | 1 | 0 | 1 | Not-determined | Genome-fragment | NA | NA | 0 |
| S4_k141_166837 | 1176 | No | NA | 2 | 0 | 0 | Not-determined | Genome-fragment | NA | NA | 0 |
| S4_k141_104004 | 3989 | No | NA | 2 | 0 | 2 | Not-determined | Genome-fragment | NA | NA | 0 |
| S4_k141_27683 | 1793 | No | NA | 2 | 0 | 2 | Not-determined | Genome-fragment | NA | NA | 0 |
| S4_k141_139293 | 1611 | No | NA | 2 | 0 | 0 | Not-determined | Genome-fragment | NA | NA | 0 |
| S6_k141_17841 | 6425 | No | NA | 8 | 1 | 0 | Low-quality | Genome-fragment | 17.95 | AAI-based (medium-confidence) | 0 |
| S3-final-viral-combined_k141_59461\|\|full | 6427 | No | NA | 8 | 0 | 4 | Low-quality | Genome-fragment | 4.23 | AAI-based (high-confidence) | 0 |
| S4_k141_104050 | 1761 | No | NA | 1 | 0 | 1 | Not-determined | Genome-fragment | NA | NA | 0 |
| S4_k141_118242 | 2360 | No | NA | 2 | 0 | 1 | Not-determined | Genome-fragment | NA | NA | 0 |
| S12_k141_199703 | 6427 | No | NA | 5 | 0 | 0 | Low-quality | Genome-fragment | 10.93 | AAI-based (medium-confidence) | 0 |
| S4_k141_27727 | 1037 | No | NA | 1 | 0 | 1 | Not-determined | Genome-fragment | NA | NA | 0 |
| S4_k141_21165 | 6430 | No | NA | 3 | 0 | 1 | Low-quality | Genome-fragment | 6.63 | AAI-based (medium-confidence) | 0 |
| S11_k141_145072 | 6432 | No | NA | 9 | 2 | 0 | Low-quality | Genome-fragment | 4.84 | AAI-based (high-confidence) | 0 |
| S4_k141_118298 | 1611 | No | NA | 2 | 0 | 1 | Not-determined | Genome-fragment | NA | NA | 0 |
| S4_k141_62512 | 1697 | No | NA | 1 | 0 | 0 | Not-determined | Genome-fragment | NA | NA | 0 |
| S4_k141_41660 | 1463 | No | NA | 1 | 0 | 1 | Not-determined | Genome-fragment | NA | NA | 0 |
| S4_k141_41665 | 3262 | No | NA | 5 | 0 | 5 | Not-determined | Genome-fragment | NA | NA | 0 |
| S4_k141_90539 | 1552 | No | NA | 1 | 0 | 1 | Not-determined | Genome-fragment | NA | NA | 0 |
| S4_k141_132090 | 1303 | No | NA | 2 | 0 | 0 | Not-determined | Genome-fragment | NA | NA | 0 |
| S4_k141_166945 | 1371 | No | NA | 1 | 0 | 1 | Not-determined | Genome-fragment | NA | NA | 0 |
| S17-final-viral-combined_k141_18855\|\|full | 6434 | No | NA | 7 | 5 | 0 | Low-quality | Genome-fragment | 18.16 | AAI-based (medium-confidence) | 0 |
| S4_k141_118313 | 25309 | No | NA | 15 | 0 | 9 | Not-determined | Genome-fragment | NA | NA | 0 |
| S4_k141_132117 | 1325 | No | NA | 2 | 0 | 2 | Not-determined | Genome-fragment | NA | NA | 0 |
| S11-final-viral-combined_k141_105573\|\|full | 6435 | No | NA | 10 | 0 | 1 | Low-quality | Genome-fragment | 8.37 | AAI-based (medium-confidence) | 0 |
| S4_k141_132137 | 1637 | No | NA | 2 | 0 | 1 | Not-determined | Genome-fragment | NA | NA | 0 |
| S4_k141_132140 | 1205 | No | NA | 2 | 0 | 2 | Not-determined | Genome-fragment | NA | NA | 0 |
| S17-final-viral-combined_k141_224251\|\|full | 6437 | No | NA | 3 | 0 | 2 | Low-quality | Genome-fragment | 3.02 | AAI-based (medium-confidence) | 0 |
| S4_k141_153096 | 1013 | No | NA | 1 | 0 | 0 | Not-determined | Genome-fragment | NA | NA | 0 |
| S4_k141_118375 | 3049 | No | NA | 4 | 0 | 3 | Not-determined | Genome-fragment | NA | NA | 0 |
| S4_k141_41732 | 2296 | No | NA | 5 | 0 | 1 | Not-determined | Genome-fragment | NA | NA | 0 |
| S4_k141_41749 | 1790 | No | NA | 2 | 0 | 1 | Not-determined | Genome-fragment | NA | NA | 0 |
| S4_k141_55197 | 1167 | No | NA | 2 | 1 | 0 | Not-determined | Genome-fragment | NA | NA | 0 |
| S4_k141_153165 | 3371 | No | NA | 4 | 0 | 1 | Not-determined | Genome-fragment | NA | NA | 0 |
| S4_k141_55210 | 1828 | No | NA | 1 | 0 | 1 | Not-determined | Genome-fragment | NA | NA | 0 |
| S12_k141_235276 | 6440 | No | NA | 5 | 0 | 2 | Low-quality | Genome-fragment | 10.41 | AAI-based (high-confidence) | 0 |
| S4_k141_153173 | 1093 | No | NA | 1 | 0 | 0 | Not-determined | Genome-fragment | NA | NA | 0 |
| S4_k141_104261 | 1327 | No | NA | 2 | 0 | 2 | Not-determined | Genome-fragment | NA | NA | 0 |
| S4_k141_62635 | 44295 | No | NA | 69 | 20 | 0 | High-quality | High-quality | 98.17 | AAI-based (high-confidence) | 0 |
| S4_k141_55273 | 1392 | No | NA | 1 | 0 | 1 | Not-determined | Genome-fragment | NA | NA | 0 |
| S4_k141_55280 | 1745 | No | NA | 2 | 0 | 0 | Not-determined | Genome-fragment | NA | NA | 0 |
| S5-final-viral-combined_k141_47368\|\|full | 6444 | No | NA | 13 | 6 | 0 | Low-quality | Genome-fragment | 17.05 | AAI-based (medium-confidence) | 0 |
| S4_k141_125154 | 1101 | No | NA | 1 | 0 | 0 | Not-determined | Genome-fragment | NA | NA | 0 |
| S4_k141_20472 | 3072 | No | NA | 2 | 0 | 1 | Not-determined | Genome-fragment | NA | NA | 0 |
| S4_k141_125165 | 1158 | No | NA | 2 | 0 | 1 | Not-determined | Genome-fragment | NA | NA | 0 |
| S4_k141_132336 | 1208 | No | NA | 1 | 0 | 1 | Not-determined | Genome-fragment | NA | NA | 0 |
| S17-final-viral-combined_k141_100117\|\|full | 6444 | No | NA | 12 | 1 | 0 | Low-quality | Genome-fragment | 17.77 | AAI-based (medium-confidence) | 0 |
| S4_k141_125232 | 2809 | No | NA | 3 | 0 | 3 | Not-determined | Genome-fragment | NA | NA | 0 |
| S12-final-viral-combined_k141_112289\|\|full | 6444 | No | NA | 9 | 4 | 0 | Low-quality | Genome-fragment | 19.43 | AAI-based (high-confidence) | 0 |
| S4_k141_20571 | 1188 | No | NA | 1 | 0 | 0 | Not-determined | Genome-fragment | NA | NA | 0 |
| S4_k141_132342 | 1333 | No | NA | 2 | 0 | 2 | Not-determined | Genome-fragment | NA | NA | 0 |
| S12-final-viral-combined_k141_358759\|\|full | 6448 | No | NA | 6 | 0 | 5 | Low-quality | Genome-fragment | 4.7 | AAI-based (medium-confidence) | 0 |
| S4_k141_125277 | 1085 | No | NA | 3 | 0 | 0 | Not-determined | Genome-fragment | NA | NA | 0 |
| S3-final-viral-combined_k141_75824\|\|full | 6449 | No | NA | 7 | 0 | 5 | Low-quality | Genome-fragment | 4.72 | AAI-based (medium-confidence) | 0 |
| S4_k141_55604 | 1686 | No | NA | 2 | 0 | 2 | Not-determined | Genome-fragment | NA | NA | 0 |
| S4_k141_125348 | 7345 | No | NA | 9 | 0 | 3 | Not-determined | Genome-fragment | NA | NA | 0 |
| S4_k141_55685 | 1154 | No | NA | 1 | 0 | 0 | Not-determined | Genome-fragment | NA | NA | 0 |
| S4_k141_20848 | 1334 | No | NA | 2 | 0 | 1 | Not-determined | Genome-fragment | NA | NA | 0 |
| S6-final-viral-combined_k141_117756\|\|full | 51257 | No | NA | 76 | 20 | 0 | High-quality | High-quality | 100 | AAI-based (high-confidence) | 0 |
| S6-final-viral-combined_k141_6450\|\|full | 3539 | No | NA | 2 | 0 | 2 | Not-determined | Genome-fragment | NA | NA | 0 |
| S6-final-viral-combined_k141_136044\|\|full | 3740 | No | NA | 3 | 0 | 3 | Not-determined | Genome-fragment | NA | NA | 0 |
| S6-final-viral-combined_k141_14387\|\|full | 33637 | No | NA | 37 | 0 | 13 | Not-determined | Genome-fragment | NA | NA | 0 |
| S1-final-viral-combined_k141_56221\|\|full | 6451 | No | NA | 3 | 0 | 1 | Low-quality | Genome-fragment | 18.6 | AAI-based (medium-confidence) | 0 |
| S6-final-viral-combined_k141_123301\|\|full | 1650 | No | NA | 2 | 0 | 1 | Not-determined | Genome-fragment | NA | NA | 0 |
| S6-final-viral-combined_k141_64598\|\|full | 2207 | No | NA | 2 | 0 | 2 | Not-determined | Genome-fragment | NA | NA | 0 |
| S6-final-viral-combined_k141_11753\|\|full | 4173 | No | NA | 3 | 0 | 2 | Not-determined | Genome-fragment | NA | NA | 0 |
| S6-final-viral-combined_k141_123304\|\|full | 1016 | No | NA | 3 | 0 | 0 | Not-determined | Genome-fragment | NA | NA | 0 |
| S3_k141_74835 | 6451 | No | NA | 11 | 1 | 0 | Low-quality | Genome-fragment | 8.17 | AAI-based (high-confidence) | 0 |
| S11_k141_234216 | 6454 | No | NA | 6 | 0 | 5 | Low-quality | Genome-fragment | 3.36 | AAI-based (medium-confidence) | 0 |
| S6-final-viral-combined_k141_23502\|\|full | 1638 | No | NA | 3 | 0 | 0 | Not-determined | Genome-fragment | NA | NA | 0 |
| S6-final-viral-combined_k141_93968\|\|full | 2132 | No | NA | 3 | 0 | 3 | Not-determined | Genome-fragment | NA | NA | 0 |
| S2-final-viral-combined_k141_76888\|\|full | 6454 | No | NA | 11 | 4 | 0 | Low-quality | Genome-fragment | 7.31 | HMM-based (lower-bound) | 0 |
| S6-final-viral-combined_k141_82240\|\|full | 2670 | No | NA | 3 | 0 | 2 | Not-determined | Genome-fragment | NA | NA | 0 |
| S6-final-viral-combined_k141_135061\|\|full | 1128 | No | NA | 3 | 0 | 2 | Not-determined | Genome-fragment | NA | NA | 0 |
| S2_k141_25678 | 6456 | No | NA | 4 | 0 | 0 | Low-quality | Genome-fragment | 35.19 | AAI-based (high-confidence) | 0 |
| S6-final-viral-combined_k141_82256\|\|full | 1467 | No | NA | 4 | 0 | 0 | Not-determined | Genome-fragment | NA | NA | 0 |
| S6-final-viral-combined_k141_60\|\|full | 2202 | No | NA | 4 | 0 | 2 | Not-determined | Genome-fragment | NA | NA | 0 |
| S6-final-viral-combined_k141_23525\|\|full | 1844 | No | NA | 4 | 0 | 2 | Not-determined | Genome-fragment | NA | NA | 0 |
| S6-final-viral-combined_k141_11789\|\|full | 2833 | No | NA | 4 | 0 | 0 | Not-determined | Genome-fragment | NA | NA | 0 |
| S6-final-viral-combined_k141_76381\|\|full | 1156 | No | NA | 3 | 0 | 1 | Not-determined | Genome-fragment | NA | NA | 0 |
| S11-final-viral-combined_k141_84762\|\|full | 6457 | No | NA | 8 | 6 | 0 | Low-quality | Genome-fragment | 14.45 | AAI-based (high-confidence) | 0 |
| S6-final-viral-combined_k141_88\|\|full | 2183 | No | NA | 3 | 0 | 1 | Not-determined | Genome-fragment | NA | NA | 0 |
| S6-final-viral-combined_k141_17670\|\|full | 1510 | No | NA | 3 | 0 | 0 | Not-determined | Genome-fragment | NA | NA | 0 |
| S11_k141_35698 | 6458 | No | NA | 3 | 0 | 0 | Low-quality | Genome-fragment | 7.13 | AAI-based (medium-confidence) | 0 |
| S11-final-viral-combined_k141_308832\|\|full | 6459 | No | NA | 12 | 4 | 0 | Low-quality | Genome-fragment | 9.71 | HMM-based (lower-bound) | 0 |
| S6-final-viral-combined_k141_117447\|\|full | 3149 | No | NA | 4 | 0 | 1 | Not-determined | Genome-fragment | NA | NA | 0 |
| S2-final-viral-combined_k141_228280\|\|full | 6460 | No | NA | 4 | 0 | 3 | Low-quality | Genome-fragment | 3.27 | AAI-based (medium-confidence) | 0 |
| S6-final-viral-combined_k141_46990\|\|full | 5429 | No | NA | 8 | 0 | 3 | Not-determined | Genome-fragment | NA | NA | 0 |
| S6-final-viral-combined_k141_17690\|\|full | 3078 | No | NA | 5 | 0 | 0 | Not-determined | Genome-fragment | NA | NA | 0 |
| S6-final-viral-combined_k141_117461\|\|full | 1683 | No | NA | 4 | 0 | 0 | Not-determined | Genome-fragment | NA | NA | 0 |
| S11-final-viral-combined_k141_183090\|\|full | 6462 | No | NA | 11 | 6 | 0 | Low-quality | Genome-fragment | 14.39 | AAI-based (high-confidence) | 0 |
| S6-final-viral-combined_k141_88170\|\|full | 1445 | No | NA | 3 | 0 | 2 | Not-determined | Genome-fragment | NA | NA | 0 |
| S6-final-viral-combined_k141_35319\|\|full | 18923 | No | NA | 18 | 0 | 10 | Not-determined | Genome-fragment | NA | NA | 0 |
| S1-final-viral-combined_k141_221393\|\|full | 6464 | No | NA | 5 | 0 | 5 | Low-quality | Genome-fragment | 4.82 | AAI-based (high-confidence) | 0 |
| S6-final-viral-combined_k141_40097\|\|full | 6464 | No | NA | 8 | 0 | 0 | Low-quality | Genome-fragment | 12.71 | AAI-based (medium-confidence) | 0 |
| S6-final-viral-combined_k141_111656\|\|full | 1631 | No | NA | 2 | 0 | 1 | Not-determined | Genome-fragment | NA | NA | 0 |
| S6_k141_32435 | 6466 | No | NA | 9 | 2 | 0 | Low-quality | Genome-fragment | 15.35 | AAI-based (high-confidence) | 0 |
| S6-final-viral-combined_k141_129263\|\|full | 4049 | No | NA | 4 | 0 | 2 | Not-determined | Genome-fragment | NA | NA | 0 |
| S6-final-viral-combined_k141_111657\|\|full | 2691 | No | NA | 3 | 0 | 2 | Not-determined | Genome-fragment | NA | NA | 0 |
| S6-final-viral-combined_k141_94058\|\|full | 1485 | No | NA | 2 | 0 | 1 | Not-determined | Genome-fragment | NA | NA | 0 |
| S6-final-viral-combined_k141_111661\|\|full | 3978 | No | NA | 5 | 0 | 3 | Not-determined | Genome-fragment | NA | NA | 0 |
| S6-final-viral-combined_k141_135156\|\|full | 4211 | No | NA | 3 | 0 | 1 | Not-determined | Genome-fragment | NA | NA | 0 |
| S8-final-viral-combined_k141_31797\|\|full | 6468 | No | NA | 8 | 0 | 0 | Low-quality | Genome-fragment | 11.89 | AAI-based (medium-confidence) | 0 |
| S6-final-viral-combined_k141_64669\|\|full | 3736 | No | NA | 3 | 0 | 1 | Not-determined | Genome-fragment | NA | NA | 0 |
| S6-final-viral-combined_k141_117500\|\|full | 3712 | No | NA | 5 | 0 | 3 | Not-determined | Genome-fragment | NA | NA | 0 |
| S11_k141_275970 | 6469 | No | NA | 5 | 0 | 1 | Low-quality | Genome-fragment | 11.33 | AAI-based (medium-confidence) | 0 |
| S6-final-viral-combined_k141_88242\|\|full | 1237 | No | NA | 2 | 0 | 1 | Not-determined | Genome-fragment | NA | NA | 0 |
| S1-final-viral-combined_k141_218397\|\|full | 6470 | No | NA | 5 | 0 | 1 | Low-quality | Genome-fragment | 19.59 | AAI-based (medium-confidence) | 0 |
| S6-final-viral-combined_k141_41237\|\|full | 3004 | No | NA | 4 | 0 | 3 | Not-determined | Genome-fragment | NA | NA | 0 |
| S6-final-viral-combined_k141_123420\|\|full | 1757 | No | NA | 2 | 0 | 2 | Not-determined | Genome-fragment | NA | NA | 0 |
| S1_k141_71183 | 6471 | No | NA | 13 | 1 | 1 | Low-quality | Genome-fragment | 3.45 | HMM-based (lower-bound) | 0 |
| S6-final-viral-combined_k141_64713\|\|full | 1162 | No | NA | 3 | 0 | 1 | Not-determined | Genome-fragment | NA | NA | 0 |
| S5-final-viral-combined_k141_176632\|\|full | 6472 | Yes | 3012 | 8 | 1 | 1 | Low-quality | Genome-fragment | 2.86 | AAI-based (medium-confidence) | 53.46 |
| S6-final-viral-combined_k141_105845\|\|full | 6754 | No | NA | 6 | 0 | 5 | Not-determined | Genome-fragment | NA | NA | 0 |
| S6-final-viral-combined_k141_23580\|\|full | 2395 | No | NA | 3 | 0 | 1 | Not-determined | Genome-fragment | NA | NA | 0 |
| S6-final-viral-combined_k141_58906\|\|full | 1611 | No | NA | 4 | 0 | 1 | Not-determined | Genome-fragment | NA | NA | 0 |
| S6-final-viral-combined_k141_255\|\|full | 1995 | No | NA | 2 | 0 | 1 | Not-determined | Genome-fragment | NA | NA | 0 |
| S6-final-viral-combined_k141_129383\|\|full | 3209 | No | NA | 3 | 0 | 2 | Not-determined | Genome-fragment | NA | NA | 0 |
| S6-final-viral-combined_k141_41246\|\|full | 40602 | No | NA | 46 | 1 | 17 | High-quality | High-quality | 100 | AAI-based (medium-confidence) | 0 |
| S9-final-viral-combined_k141_49306\|\|full | 6473 | No | NA | 9 | 0 | 0 | Low-quality | Genome-fragment | 7.86 | AAI-based (medium-confidence) | 0 |
| S6-final-viral-combined_k141_76480\|\|full | 1256 | No | NA | 3 | 0 | 0 | Not-determined | Genome-fragment | NA | NA | 0 |
| S6-final-viral-combined_k141_88297\|\|full | 2349 | No | NA | 4 | 0 | 3 | Not-determined | Genome-fragment | NA | NA | 0 |
| S2-final-viral-combined_k141_41215\|\|full | 6473 | No | NA | 11 | 7 | 0 | Low-quality | Genome-fragment | 14.87 | AAI-based (high-confidence) | 0 |
| S6-final-viral-combined_k141_35394\|\|full | 3112 | No | NA | 3 | 0 | 1 | Not-determined | Genome-fragment | NA | NA | 0 |
| S6-final-viral-combined_k141_88339\|\|full | 2209 | No | NA | 4 | 0 | 0 | Not-determined | Genome-fragment | NA | NA | 0 |
| S1-final-viral-combined_k141_222156\|\|full | 6477 | No | NA | 4 | 0 | 2 | Low-quality | Genome-fragment | 1.9 | AAI-based (medium-confidence) | 0 |
| S9-final-viral-combined_k141_63062\|\|full | 6478 | No | NA | 8 | 1 | 5 | Low-quality | Genome-fragment | 0.72 | HMM-based (lower-bound) | 0 |
| S6-final-viral-combined_k141_111823\|\|full | 2729 | No | NA | 3 | 0 | 0 | Not-determined | Genome-fragment | NA | NA | 0 |
| S6-final-viral-combined_k141_129415\|\|full | 1379 | No | NA | 2 | 0 | 1 | Not-determined | Genome-fragment | NA | NA | 0 |
| S12-final-viral-combined_k141_197856\|\|full | 6478 | No | NA | 10 | 4 | 0 | Low-quality | Genome-fragment | 12.5 | AAI-based (high-confidence) | 0 |
| S6-final-viral-combined_k141_123490\|\|full | 3397 | No | NA | 3 | 0 | 1 | Not-determined | Genome-fragment | NA | NA | 0 |
| S8-final-viral-combined_k141_13520\|\|full | 6480 | No | NA | 9 | 4 | 0 | Low-quality | Genome-fragment | 14.28 | AAI-based (medium-confidence) | 0 |
| S6-final-viral-combined_k141_94215\|\|full | 1038 | No | NA | 3 | 0 | 1 | Not-determined | Genome-fragment | NA | NA | 0 |
| S6-final-viral-combined_k141_123521\|\|full | 1826 | No | NA | 2 | 0 | 2 | Not-determined | Genome-fragment | NA | NA | 0 |
| S6-final-viral-combined_k141_58998\|\|full | 9717 | No | NA | 9 | 0 | 9 | Not-determined | Genome-fragment | NA | NA | 0 |
| S6-final-viral-combined_k141_76592\|\|full | 1907 | No | NA | 3 | 0 | 1 | Not-determined | Genome-fragment | NA | NA | 0 |
| S6-final-viral-combined_k141_12028\|\|full | 2945 | No | NA | 5 | 0 | 0 | Not-determined | Genome-fragment | NA | NA | 0 |
| S6-final-viral-combined_k141_106003\|\|full | 1548 | No | NA | 3 | 0 | 2 | Not-determined | Genome-fragment | NA | NA | 0 |
| S6-final-viral-combined_k141_12059\|\|full | 3319 | No | NA | 3 | 0 | 1 | Not-determined | Genome-fragment | NA | NA | 0 |
| S6-final-viral-combined_k141_106030\|\|full | 1822 | No | NA | 5 | 0 | 4 | Not-determined | Genome-fragment | NA | NA | 0 |
| S6-final-viral-combined_k141_6044\|\|full | 2159 | No | NA | 2 | 0 | 1 | Not-determined | Genome-fragment | NA | NA | 0 |
| S6-final-viral-combined_k141_64885\|\|full | 1169 | No | NA | 4 | 0 | 0 | Not-determined | Genome-fragment | NA | NA | 0 |
| S12-final-viral-combined_k141_45356\|\|full | 6483 | No | NA | 15 | 1 | 0 | Low-quality | Genome-fragment | 17.55 | AAI-based (high-confidence) | 0 |
| S6-final-viral-combined_k141_59044\|\|full | 5578 | No | NA | 7 | 0 | 0 | Not-determined | Genome-fragment | NA | NA | 0 |
| S9_k141_102224 | 6490 | No | NA | 7 | 4 | 0 | Low-quality | Genome-fragment | 17.26 | AAI-based (high-confidence) | 0 |
| S6-final-viral-combined_k141_35522\|\|full | 3247 | No | NA | 4 | 0 | 4 | Not-determined | Genome-fragment | NA | NA | 0 |
| S17_k141_256456 | 6492 | No | NA | 9 | 0 | 2 | Low-quality | Genome-fragment | 8.77 | AAI-based (high-confidence) | 0 |
| S6-final-viral-combined_k141_41365\|\|full | 2808 | No | NA | 5 | 0 | 0 | Not-determined | Genome-fragment | NA | NA | 0 |
| S6-final-viral-combined_k141_12127\|\|full | 1098 | No | NA | 2 | 0 | 0 | Not-determined | Genome-fragment | NA | NA | 0 |
| S6-final-viral-combined_k141_135223\|\|full | 147913 | No | NA | 138 | 2 | 29 | Medium-quality | Genome-fragment | 79.46 | HMM-based (lower-bound) | 0 |
| S6-final-viral-combined_k141_59082\|\|full | 2065 | No | NA | 4 | 0 | 2 | Not-determined | Genome-fragment | NA | NA | 0 |
| S12-final-viral-combined_k141_203299\|\|full | 6494 | No | NA | 6 | 0 | 1 | Low-quality | Genome-fragment | 5.29 | AAI-based (medium-confidence) | 0 |
| S11_k141_22328 | 6500 | Yes | 2885 | 9 | 1 | 2 | Low-quality | Genome-fragment | 6.2 | AAI-based (high-confidence) | 55.62 |
| S1-final-viral-combined_k141_309353\|\|full | 6500 | No | NA | 3 | 0 | 0 | Low-quality | Genome-fragment | 7.32 | AAI-based (medium-confidence) | 0 |
| S6-final-viral-combined_k141_59093\|\|full | 1366 | No | NA | 3 | 0 | 1 | Not-determined | Genome-fragment | NA | NA | 0 |
| S6-final-viral-combined_k141_94344\|\|full | 2901 | No | NA | 4 | 0 | 0 | Not-determined | Genome-fragment | NA | NA | 0 |
| S6_k141_110309 | 6502 | No | NA | 11 | 4 | 1 | Low-quality | Genome-fragment | 18.21 | AAI-based (high-confidence) | 0 |
| S6-final-viral-combined_k141_88414\|\|full | 101171 | Yes | 30132 | 115 | 14 | 48 | Complete | High-quality | 100 | Provirus (medium-confidence) | 70.22 |
| S6-final-viral-combined_k141_88422\|\|full | 1610 | No | NA | 2 | 0 | 1 | Not-determined | Genome-fragment | NA | NA | 0 |
| S6-final-viral-combined_k141_6096\|\|full | 1239 | No | NA | 3 | 0 | 0 | Not-determined | Genome-fragment | NA | NA | 0 |
| S6-final-viral-combined_k141_82444\|\|full | 5380 | No | NA | 5 | 0 | 4 | Not-determined | Genome-fragment | NA | NA | 0 |
| S6-final-viral-combined_k141_59157\|\|full | 3952 | No | NA | 3 | 0 | 3 | Not-determined | Genome-fragment | NA | NA | 0 |
| S6-final-viral-combined_k141_23773\|\|full | 2158 | No | NA | 2 | 0 | 1 | Not-determined | Genome-fragment | NA | NA | 0 |
| S2-final-viral-combined_k141_62159\|\|full | 6504 | No | NA | 15 | 1 | 4 | Low-quality | Genome-fragment | 16.27 | AAI-based (high-confidence) | 0 |
| S2_k141_134607 | 6504 | No | NA | 14 | 2 | 0 | Low-quality | Genome-fragment | 13.56 | HMM-based (lower-bound) | 0 |
| S11-final-viral-combined_k141_140493\|\|full | 6506 | No | NA | 6 | 1 | 0 | Low-quality | Genome-fragment | 4.02 | AAI-based (medium-confidence) | 0 |
| S6-final-viral-combined_k141_6122\|\|full | 1611 | No | NA | 3 | 0 | 1 | Not-determined | Genome-fragment | NA | NA | 0 |
| S5_k141_128692 | 6507 | No | NA | 11 | 3 | 1 | Low-quality | Genome-fragment | 18.39 | AAI-based (high-confidence) | 0 |
| S6-final-viral-combined_k141_94427\|\|full | 1364 | No | NA | 2 | 0 | 1 | Not-determined | Genome-fragment | NA | NA | 0 |
| S7-final-viral-combined_k141_115568\|\|full | 6508 | No | NA | 5 | 0 | 0 | Low-quality | Genome-fragment | 7.49 | AAI-based (medium-confidence) | 0 |
| S9-final-viral-combined_k141_110381\|\|full | 6509 | No | NA | 13 | 4 | 0 | Low-quality | Genome-fragment | 16 | AAI-based (high-confidence) | 0 |
| S1-final-viral-combined_k141_63940\|\|full | 6509 | No | NA | 9 | 6 | 0 | Low-quality | Genome-fragment | 12.41 | AAI-based (high-confidence) | 0 |
| S6-final-viral-combined_k141_41549\|\|full | 2138 | No | NA | 4 | 0 | 2 | Not-determined | Genome-fragment | NA | NA | 0 |
| S6-final-viral-combined_k141_76715\|\|full | 1905 | No | NA | 3 | 0 | 0 | Not-determined | Genome-fragment | NA | NA | 0 |
| S6-final-viral-combined_k141_12272\|\|full | 3829 | No | NA | 4 | 0 | 2 | Not-determined | Genome-fragment | NA | NA | 0 |
| S6-final-viral-combined_k141_82513\|\|full | 2255 | No | NA | 2 | 0 | 1 | Not-determined | Genome-fragment | NA | NA | 0 |
| S6-final-viral-combined_k141_123743\|\|full | 4185 | No | NA | 5 | 0 | 0 | Not-determined | Genome-fragment | NA | NA | 0 |
| S1-final-viral-combined_k141_382608\|\|full | 6512 | No | NA | 5 | 0 | 1 | Low-quality | Genome-fragment | 8.1 | AAI-based (medium-confidence) | 0 |
| S2-final-viral-combined_k141_160731\|\|full | 6512 | No | NA | 4 | 1 | 0 | Low-quality | Genome-fragment | 4.5 | HMM-based (lower-bound) | 0 |
| S12-final-viral-combined_k141_51084\|\|full | 6513 | Yes | 4351 | 7 | 1 | 2 | Low-quality | Genome-fragment | 9.37 | AAI-based (medium-confidence) | 33.2 |
| S17-final-viral-combined_k141_209140\|\|full | 6515 | No | NA | 6 | 1 | 4 | Low-quality | Genome-fragment | 0.54 | HMM-based (lower-bound) | 0 |
| S6-final-viral-combined_k141_52935\|\|full | 5970 | No | NA | 8 | 0 | 3 | Not-determined | Genome-fragment | NA | NA | 0 |
| S2_k141_228760 | 6515 | No | NA | 7 | 0 | 4 | Low-quality | Genome-fragment | 15.5 | AAI-based (high-confidence) | 0 |
| S6-final-viral-combined_k141_88628\|\|full | 2668 | No | NA | 4 | 0 | 1 | Not-determined | Genome-fragment | NA | NA | 0 |
| S6-final-viral-combined_k141_601\|\|full | 1674 | No | NA | 3 | 0 | 1 | Not-determined | Genome-fragment | NA | NA | 0 |
| S6-final-viral-combined_k141_123834\|\|full | 3079 | No | NA | 1 | 0 | 1 | Not-determined | Genome-fragment | NA | NA | 0 |
| S3_k141_49064 | 6518 | No | NA | 13 | 2 | 0 | Low-quality | Genome-fragment | 17.69 | AAI-based (high-confidence) | 0 |
| S6-final-viral-combined_k141_123859\|\|full | 2425 | No | NA | 3 | 0 | 3 | Not-determined | Genome-fragment | NA | NA | 0 |
| S12-final-viral-combined_k141_290588\|\|full | 6522 | No | NA | 17 | 1 | 0 | Low-quality | Genome-fragment | 11.63 | AAI-based (high-confidence) | 0 |
| S6-final-viral-combined_k141_6274\|\|full | 3216 | No | NA | 3 | 0 | 2 | Not-determined | Genome-fragment | NA | NA | 0 |
| S6-final-viral-combined_k141_10821\|\|full | 6522 | No | NA | 3 | 1 | 0 | Low-quality | Genome-fragment | 14.46 | AAI-based (high-confidence) | 0 |
| S6-final-viral-combined_k141_117852\|\|full | 3607 | No | NA | 2 | 0 | 2 | Not-determined | Genome-fragment | NA | NA | 0 |
| S6-final-viral-combined_k141_117873\|\|full | 2528 | No | NA | 3 | 0 | 3 | Not-determined | Genome-fragment | NA | NA | 0 |
| S6-final-viral-combined_k141_117881\|\|full | 1350 | No | NA | 2 | 0 | 1 | Not-determined | Genome-fragment | NA | NA | 0 |
| S6-final-viral-combined_k141_112081\|\|full | 2259 | No | NA | 3 | 0 | 1 | Not-determined | Genome-fragment | NA | NA | 0 |
| S6-final-viral-combined_k141_676\|\|full | 1225 | No | NA | 3 | 0 | 0 | Not-determined | Genome-fragment | NA | NA | 0 |
| S2_k141_192332 | 6523 | No | NA | 9 | 2 | 0 | Low-quality | Genome-fragment | 35.88 | AAI-based (high-confidence) | 0 |
| S6-final-viral-combined_k141_88696\|\|full | 1303 | No | NA | 4 | 0 | 0 | Not-determined | Genome-fragment | NA | NA | 0 |
| S4_k141_128270 | 6524 | No | NA | 8 | 0 | 0 | Low-quality | Genome-fragment | 11.68 | AAI-based (medium-confidence) | 0 |
| S6-final-viral-combined_k141_71023\|\|full | 5341 | No | NA | 6 | 0 | 1 | Not-determined | Genome-fragment | NA | NA | 0 |
| S1-final-viral-combined_k141_338985\|\|full | 6526 | No | NA | 8 | 0 | 3 | Low-quality | Genome-fragment | 4.26 | AAI-based (high-confidence) | 0 |
| S6-final-viral-combined_k141_59333\|\|full | 2349 | No | NA | 5 | 0 | 1 | Not-determined | Genome-fragment | NA | NA | 0 |
| S6-final-viral-combined_k141_66184\|\|full | 6527 | No | NA | 5 | 0 | 3 | Low-quality | Genome-fragment | 4.6 | AAI-based (medium-confidence) | 0 |
| S6-final-viral-combined_k141_47467\|\|full | 1732 | No | NA | 2 | 0 | 2 | Not-determined | Genome-fragment | NA | NA | 0 |
| S4_k141_112351 | 6529 | No | NA | 12 | 1 | 1 | Low-quality | Genome-fragment | 3.54 | HMM-based (lower-bound) | 0 |
| S12-final-viral-combined_k141_184510\|\|full | 6537 | No | NA | 6 | 0 | 5 | Low-quality | Genome-fragment | 1.93 | AAI-based (medium-confidence) | 0 |
| S12_k141_36593 | 6541 | No | NA | 4 | 0 | 3 | Low-quality | Genome-fragment | 5.28 | AAI-based (medium-confidence) | 0 |
| S11_k141_210386 | 6543 | No | NA | 7 | 1 | 0 | Low-quality | Genome-fragment | 15.06 | AAI-based (high-confidence) | 0 |
| S6_k141_129745 | 6545 | No | NA | 9 | 0 | 1 | Low-quality | Genome-fragment | 10.93 | AAI-based (high-confidence) | 0 |
| S6-final-viral-combined_k141_123985\|\|full | 9605 | No | NA | 5 | 0 | 4 | Not-determined | Genome-fragment | NA | NA | 0 |
| S6-final-viral-combined_k141_94664\|\|full | 1724 | No | NA | 4 | 0 | 0 | Not-determined | Genome-fragment | NA | NA | 0 |
| S6-final-viral-combined_k141_124003\|\|full | 1621 | No | NA | 2 | 0 | 1 | Not-determined | Genome-fragment | NA | NA | 0 |
| S9-final-viral-combined_k141_96377\|\|full | 6546 | No | NA | 5 | 0 | 1 | Low-quality | Genome-fragment | 6.75 | AAI-based (medium-confidence) | 0 |
| S11_k141_146857 | 6548 | No | NA | 4 | 0 | 2 | Low-quality | Genome-fragment | 3.52 | AAI-based (medium-confidence) | 0 |
| S6-final-viral-combined_k141_71067\|\|full | 1845 | No | NA | 4 | 0 | 0 | Not-determined | Genome-fragment | NA | NA | 0 |
| S6-final-viral-combined_k141_94701\|\|full | 6800 | No | NA | 5 | 0 | 3 | Not-determined | Genome-fragment | NA | NA | 0 |
| S6-final-viral-combined_k141_129788\|\|full | 2297 | No | NA | 3 | 0 | 2 | Not-determined | Genome-fragment | NA | NA | 0 |
| S6-final-viral-combined_k141_745\|\|full | 36814 | No | NA | 52 | 11 | 1 | High-quality | High-quality | 91.45 | AAI-based (high-confidence) | 0 |
| S9_k141_115414 | 6550 | No | NA | 6 | 0 | 2 | Low-quality | Genome-fragment | 6.15 | AAI-based (medium-confidence) | 0 |
| S6-final-viral-combined_k141_6479\|\|full | 1545 | No | NA | 3 | 0 | 0 | Not-determined | Genome-fragment | NA | NA | 0 |
| S6-final-viral-combined_k141_59372\|\|full | 1064 | No | NA | 2 | 0 | 0 | Not-determined | Genome-fragment | NA | NA | 0 |
| S6-final-viral-combined_k141_88823\|\|full | 8036 | No | NA | 7 | 0 | 7 | Not-determined | Genome-fragment | NA | NA | 0 |
| S6-final-viral-combined_k141_88836\|\|full | 3138 | No | NA | 5 | 0 | 1 | Not-determined | Genome-fragment | NA | NA | 0 |
| S6-final-viral-combined_k141_94772\|\|full | 1213 | No | NA | 3 | 0 | 2 | Not-determined | Genome-fragment | NA | NA | 0 |
| S6-final-viral-combined_k141_65208\|\|full | 2061 | No | NA | 2 | 0 | 2 | Not-determined | Genome-fragment | NA | NA | 0 |
| S6-final-viral-combined_k141_100353\|\|full | 10784 | No | NA | 8 | 0 | 6 | Not-determined | Genome-fragment | NA | NA | 0 |
| S6-final-viral-combined_k141_88854\|\|full | 1504 | No | NA | 3 | 0 | 1 | Not-determined | Genome-fragment | NA | NA | 0 |
| S6-final-viral-combined_k141_59403\|\|full | 3611 | No | NA | 5 | 0 | 0 | Not-determined | Genome-fragment | NA | NA | 0 |
| S6-final-viral-combined_k141_808\|\|full | 1701 | No | NA | 3 | 0 | 0 | Not-determined | Genome-fragment | NA | NA | 0 |
| S6-final-viral-combined_k141_82597\|\|full | 215227 | Yes | 38568 | 191 | 13 | 101 | Complete | High-quality | 100 | Provirus (high-confidence) | 82.08 |
| S6-final-viral-combined_k141_65225\|\|full | 23171 | No | NA | 36 | 18 | 0 | Medium-quality | Genome-fragment | 62.64 | AAI-based (high-confidence) | 0 |
| S12_k141_216804 | 6553 | No | NA | 12 | 8 | 1 | Low-quality | Genome-fragment | 12.24 | AAI-based (high-confidence) | 0 |
| S6-final-viral-combined_k141_47611\|\|full | 5270 | No | NA | 6 | 0 | 4 | Not-determined | Genome-fragment | NA | NA | 0 |
| S6-final-viral-combined_k141_100378\|\|full | 6749 | No | NA | 5 | 0 | 3 | Not-determined | Genome-fragment | NA | NA | 0 |
| S4-final-viral-combined_k141_97391\|\|full | 6553 | No | NA | 11 | 1 | 0 | Low-quality | Genome-fragment | 13.27 | AAI-based (high-confidence) | 0 |
| S6-final-viral-combined_k141_100388\|\|full | 3730 | No | NA | 8 | 0 | 0 | Not-determined | Genome-fragment | NA | NA | 0 |
| S6-final-viral-combined_k141_129848\|\|full | 1991 | No | NA | 4 | 0 | 0 | Not-determined | Genome-fragment | NA | NA | 0 |
| S6-final-viral-combined_k141_53259\|\|full | 2785 | No | NA | 4 | 0 | 3 | Not-determined | Genome-fragment | NA | NA | 0 |
| S6-final-viral-combined_k141_65322\|\|full | 1842 | No | NA | 2 | 0 | 1 | Not-determined | Genome-fragment | NA | NA | 0 |
| S6-final-viral-combined_k141_6606\|\|full | 6880 | No | NA | 5 | 0 | 1 | Not-determined | Genome-fragment | NA | NA | 0 |
| S7-final-viral-combined_k141_104120\|\|full | 6556 | No | NA | 6 | 0 | 1 | Low-quality | Genome-fragment | 5.37 | AAI-based (medium-confidence) | 0 |
| S6-final-viral-combined_k141_82734\|\|full | 1373 | No | NA | 2 | 0 | 1 | Not-determined | Genome-fragment | NA | NA | 0 |
| S12-final-viral-combined_k141_201969\|\|full | 6559 | No | NA | 8 | 3 | 0 | Low-quality | Genome-fragment | 15.56 | AAI-based (high-confidence) | 0 |
| S2-final-viral-combined_k141_184870\|\|full | 6559 | No | NA | 7 | 5 | 0 | Low-quality | Genome-fragment | 15.37 | AAI-based (high-confidence) | 0 |
| S6-final-viral-combined_k141_76955\|\|full | 1699 | No | NA | 4 | 0 | 0 | Not-determined | Genome-fragment | NA | NA | 0 |
| S6-final-viral-combined_k141_17983\|\|full | 1042 | No | NA | 1 | 0 | 1 | Not-determined | Genome-fragment | NA | NA | 0 |
| S6-final-viral-combined_k141_53309\|\|full | 1240 | No | NA | 2 | 0 | 1 | Not-determined | Genome-fragment | NA | NA | 0 |
| S6-final-viral-combined_k141_65385\|\|full | 1122 | No | NA | 2 | 0 | 2 | Not-determined | Genome-fragment | NA | NA | 0 |
| S1-final-viral-combined_k141_189547\|\|full | 6561 | No | NA | 7 | 0 | 1 | Low-quality | Genome-fragment | 6.98 | AAI-based (medium-confidence) | 0 |
| S7_k141_129898 | 6563 | No | NA | 7 | 3 | 0 | Low-quality | Genome-fragment | 18.57 | AAI-based (high-confidence) | 0 |
| S8-final-viral-combined_k141_249675\|\|full | 6566 | No | NA | 6 | 0 | 4 | Low-quality | Genome-fragment | 3.96 | AAI-based (medium-confidence) | 0 |
| S6-final-viral-combined_k141_6705\|\|full | 2110 | No | NA | 6 | 0 | 0 | Not-determined | Genome-fragment | NA | NA | 0 |
| S1-final-viral-combined_k141_345521\|\|full | 6569 | No | NA | 9 | 0 | 0 | Low-quality | Genome-fragment | 6.84 | AAI-based (medium-confidence) | 0 |
| S6-final-viral-combined_k141_30162\|\|full | 2421 | No | NA | 4 | 0 | 3 | Not-determined | Genome-fragment | NA | NA | 0 |
| S6-final-viral-combined_k141_77001\|\|full | 1530 | No | NA | 3 | 0 | 0 | Not-determined | Genome-fragment | NA | NA | 0 |
| S6-final-viral-combined_k141_47715\|\|full | 2167 | No | NA | 2 | 0 | 2 | Not-determined | Genome-fragment | NA | NA | 0 |
| S6-final-viral-combined_k141_135593\|\|full | 4990 | No | NA | 7 | 0 | 1 | Not-determined | Genome-fragment | NA | NA | 0 |
| S12_k141_161020 | 6569 | No | NA | 5 | 0 | 5 | Low-quality | Genome-fragment | 3.08 | AAI-based (medium-confidence) | 0 |
| S1-final-viral-combined_k141_274459\|\|full | 6574 | No | NA | 10 | 4 | 0 | Low-quality | Genome-fragment | 10.9 | AAI-based (high-confidence) | 0 |
| S6-final-viral-combined_k141_65516\|\|full | 1182 | No | NA | 3 | 0 | 0 | Not-determined | Genome-fragment | NA | NA | 0 |
| S6-final-viral-combined_k141_71292\|\|full | 2582 | No | NA | 2 | 0 | 2 | Not-determined | Genome-fragment | NA | NA | 0 |
| S6-final-viral-combined_k141_18072\|\|full | 1063 | No | NA | 4 | 0 | 0 | Not-determined | Genome-fragment | NA | NA | 0 |
| S6-final-viral-combined_k141_89197\|\|full | 6282 | No | NA | 9 | 0 | 5 | Not-determined | Genome-fragment | NA | NA | 0 |
| S6-final-viral-combined_k141_95083\|\|full | 1104 | No | NA | 2 | 0 | 1 | Not-determined | Genome-fragment | NA | NA | 0 |
| S5-final-viral-combined_k141_33534\|\|full | 6574 | No | NA | 9 | 4 | 0 | Low-quality | Genome-fragment | 14.08 | AAI-based (high-confidence) | 0 |
| S6-final-viral-combined_k141_12613\|\|full | 13128 | No | NA | 11 | 0 | 7 | Not-determined | Genome-fragment | NA | NA | 0 |
| S6-final-viral-combined_k141_24230\|\|full | 1928 | No | NA | 4 | 0 | 0 | Not-determined | Genome-fragment | NA | NA | 0 |
| S6-final-viral-combined_k141_89233\|\|full | 1503 | No | NA | 3 | 0 | 0 | Not-determined | Genome-fragment | NA | NA | 0 |
| S6_k141_45921 | 6576 | No | NA | 1 | 0 | 0 | Low-quality | Genome-fragment | 6.88 | AAI-based (medium-confidence) | 0 |
| S6-final-viral-combined_k141_18118\|\|full | 1747 | No | NA | 3 | 0 | 1 | Not-determined | Genome-fragment | NA | NA | 0 |
| S6-final-viral-combined_k141_89236\|\|full | 1470 | No | NA | 2 | 0 | 2 | Not-determined | Genome-fragment | NA | NA | 0 |
| S6-final-viral-combined_k141_89241\|\|full | 2176 | No | NA | 3 | 0 | 2 | Not-determined | Genome-fragment | NA | NA | 0 |
| S9_k141_98221 | 6578 | No | NA | 7 | 0 | 1 | Low-quality | Genome-fragment | 13.02 | AAI-based (high-confidence) | 0 |
| S6-final-viral-combined_k141_30297\|\|full | 8521 | No | NA | 10 | 0 | 2 | Not-determined | Genome-fragment | NA | NA | 0 |
| S6-final-viral-combined_k141_35840\|\|full | 6373 | No | NA | 8 | 0 | 6 | Not-determined | Genome-fragment | NA | NA | 0 |
| S6-final-viral-combined_k141_6857\|\|full | 2028 | No | NA | 3 | 0 | 1 | Not-determined | Genome-fragment | NA | NA | 0 |
| S6-final-viral-combined_k141_82926\|\|full | 3363 | No | NA | 3 | 0 | 2 | Not-determined | Genome-fragment | NA | NA | 0 |
| S7-final-viral-combined_k141_106310\|\|full | 6580 | No | NA | 7 | 3 | 0 | Low-quality | Genome-fragment | 15.72 | AAI-based (high-confidence) | 0 |
| S6-final-viral-combined_k141_124203\|\|full | 2512 | No | NA | 2 | 0 | 2 | Not-determined | Genome-fragment | NA | NA | 0 |
| S9_k141_57555 | 6581 | No | NA | 12 | 5 | 0 | Low-quality | Genome-fragment | 16.33 | AAI-based (high-confidence) | 0 |
| S1-final-viral-combined_k141_247356\|\|full | 6581 | Yes | 6044 | 10 | 1 | 1 | Low-quality | Genome-fragment | 9.77 | AAI-based (high-confidence) | 8.16 |
| S5-final-viral-combined_k141_53700\|\|full | 6581 | No | NA | 10 | 8 | 0 | Low-quality | Genome-fragment | 14.47 | AAI-based (medium-confidence) | 0 |
| S6-final-viral-combined_k141_135649\|\|full | 1346 | No | NA | 3 | 0 | 0 | Not-determined | Genome-fragment | NA | NA | 0 |
| S6-final-viral-combined_k141_42141\|\|full | 7428 | No | NA | 5 | 0 | 2 | Not-determined | Genome-fragment | NA | NA | 0 |
| S6-final-viral-combined_k141_18172\|\|full | 6776 | No | NA | 6 | 0 | 4 | Not-determined | Genome-fragment | NA | NA | 0 |
| S6-final-viral-combined_k141_100516\|\|full | 1060 | No | NA | 3 | 0 | 1 | Not-determined | Genome-fragment | NA | NA | 0 |
| S6-final-viral-combined_k141_82977\|\|full | 3970 | No | NA | 5 | 0 | 2 | Not-determined | Genome-fragment | NA | NA | 0 |
| S6-final-viral-combined_k141_18178\|\|full | 2160 | No | NA | 4 | 0 | 0 | Not-determined | Genome-fragment | NA | NA | 0 |
| S6-final-viral-combined_k141_59743\|\|full | 5874 | No | NA | 8 | 0 | 0 | Not-determined | Genome-fragment | NA | NA | 0 |
| S2-final-viral-combined_k141_74946\|\|full | 6583 | No | NA | 13 | 2 | 0 | Low-quality | Genome-fragment | 10.82 | AAI-based (high-confidence) | 0 |

| | | | | | | | | | | | |
|---|---|---|---|---|---|---|---|---|---|---|---|
| S6-final-viral-combined_k141_71444\|\|full | 2379 | No | NA | 4 | 0 | 0 | Not-determined | Genome-fragment | NA | NA | 0 |
| S4-final-viral-combined_k141_65441\|\|full | 6587 | No | NA | 10 | 4 | 0 | Low-quality | Genome-fragment | 11.45 | AAI-based (high-confidence) | 0 |
| S6-final-viral-combined_k141_82987\|\|full | 19110 | No | NA | 12 | 0 | 9 | Not-determined | Genome-fragment | NA | NA | 0 |
| S17-final-viral-combined_k141_71049\|\|full | 6591 | Yes | 4810 | 10 | 2 | 2 | Low-quality | Genome-fragment | 7.03 | AAI-based (high-confidence) | 27.02 |
| S1-final-viral-combined_k141_193839\|\|full | 6592 | Yes | 5410 | 9 | 1 | 1 | Low-quality | Genome-fragment | 6.63 | AAI-based (medium-confidence) | 17.93 |
| S6-final-viral-combined_k141_6944\|\|full | 1308 | No | NA | 2 | 0 | 0 | Not-determined | Genome-fragment | NA | NA | 0 |
| S11-final-viral-combined_k141_294629\|\|full | 6595 | No | NA | 10 | 6 | 0 | Low-quality | Genome-fragment | 9.87 | AAI-based (high-confidence) | 0 |
| S6-final-viral-combined_k141_12678\|\|full | 10086 | No | NA | 13 | 0 | 6 | Not-determined | Genome-fragment | NA | NA | 0 |
| S12-final-viral-combined_k141_206513\|\|full | 6595 | No | NA | 12 | 2 | 0 | Low-quality | Genome-fragment | 20.46 | AAI-based (high-confidence) | 0 |
| S6-final-viral-combined_k141_112337\|\|full | 1047 | No | NA | 2 | 0 | 1 | Not-determined | Genome-fragment | NA | NA | 0 |
| S8_k141_35910 | 6599 | No | NA | 10 | 3 | 0 | Low-quality | Genome-fragment | 12.46 | AAI-based (high-confidence) | 0 |
| S6-final-viral-combined_k141_83007\|\|full | 3206 | No | NA | 3 | 0 | 2 | Not-determined | Genome-fragment | NA | NA | 0 |
| S6-final-viral-combined_k141_112339\|\|full | 6797 | No | NA | 6 | 0 | 4 | Not-determined | Genome-fragment | NA | NA | 0 |
| S6-final-viral-combined_k141_106691\|\|full | 1789 | No | NA | 3 | 0 | 2 | Not-determined | Genome-fragment | NA | NA | 0 |
| S5-final-viral-combined_k141_137342\|\|full | 6605 | No | NA | 5 | 5 | 0 | Low-quality | Genome-fragment | 14.29 | AAI-based (high-confidence) | 0 |
| S6-final-viral-combined_k141_6985\|\|full | 1726 | No | NA | 3 | 0 | 0 | Not-determined | Genome-fragment | NA | NA | 0 |
| S17-final-viral-combined_k141_238908\|\|full | 6605 | No | NA | 11 | 0 | 2 | Low-quality | Genome-fragment | 11.76 | AAI-based (medium-confidence) | 0 |
| S6-final-viral-combined_k141_53430\|\|full | 2993 | No | NA | 4 | 0 | 3 | Not-determined | Genome-fragment | NA | NA | 0 |
| S12_k141_315695 | 6608 | No | NA | 3 | 0 | 0 | Low-quality | Genome-fragment | 11.53 | AAI-based (high-confidence) | 0 |
| S6-final-viral-combined_k141_36000\|\|full | 5303 | No | NA | 6 | 0 | 5 | Not-determined | Genome-fragment | NA | NA | 0 |
| S6-final-viral-combined_k141_106727\|\|full | 2860 | No | NA | 5 | 0 | 2 | Not-determined | Genome-fragment | NA | NA | 0 |
| S6-final-viral-combined_k141_36021\|\|full | 1769 | No | NA | 4 | 0 | 1 | Not-determined | Genome-fragment | NA | NA | 0 |
| S17-final-viral-combined_k141_94524\|\|full | 6613 | No | NA | 8 | 6 | 0 | Low-quality | Genome-fragment | 15.74 | AAI-based (high-confidence) | 0 |
| S8_k141_120922 | 6619 | No | NA | 7 | 0 | 5 | Low-quality | Genome-fragment | 5.39 | AAI-based (medium-confidence) | 0 |
| S1_k141_148844 | 6624 | No | NA | 11 | 6 | 0 | Low-quality | Genome-fragment | 13.96 | AAI-based (high-confidence) | 0 |
| S6-final-viral-combined_k141_124326\|\|full | 1446 | No | NA | 4 | 0 | 0 | Not-determined | Genome-fragment | NA | NA | 0 |
| S8-final-viral-combined_k141_138596\|\|full | 6626 | No | NA | 3 | 0 | 0 | Low-quality | Genome-fragment | 16.16 | AAI-based (high-confidence) | 0 |
| S6-final-viral-combined_k141_71541\|\|full | 11121 | No | NA | 10 | 0 | 6 | Not-determined | Genome-fragment | NA | NA | 0 |
| S6-final-viral-combined_k141_36070\|\|full | 3707 | No | NA | 7 | 0 | 1 | Not-determined | Genome-fragment | NA | NA | 0 |
| S6-final-viral-combined_k141_71546\|\|full | 1232 | No | NA | 2 | 0 | 1 | Not-determined | Genome-fragment | NA | NA | 0 |
| S6-final-viral-combined_k141_77222\|\|full | 2034 | No | NA | 4 | 0 | 1 | Not-determined | Genome-fragment | NA | NA | 0 |
| S6_k141_30410 | 6627 | No | NA | 12 | 5 | 0 | Low-quality | Genome-fragment | 11.1 | AAI-based (high-confidence) | 0 |
| S6-final-viral-combined_k141_118159\|\|full | 1491 | No | NA | 4 | 0 | 1 | Not-determined | Genome-fragment | NA | NA | 0 |
| S6-final-viral-combined_k141_18333\|\|full | 3270 | No | NA | 4 | 0 | 2 | Not-determined | Genome-fragment | NA | NA | 0 |
| S6-final-viral-combined_k141_77244\|\|full | 1380 | No | NA | 3 | 0 | 2 | Not-determined | Genome-fragment | NA | NA | 0 |
| S6-final-viral-combined_k141_112448\|\|full | 1010 | No | NA | 3 | 0 | 2 | Not-determined | Genome-fragment | NA | NA | 0 |
| S6-final-viral-combined_k141_124365\|\|full | 9976 | No | NA | 9 | 0 | 7 | Not-determined | Genome-fragment | NA | NA | 0 |
| S8-final-viral-combined_k141_114524\|\|full | 6627 | No | NA | 19 | 0 | 0 | Low-quality | Genome-fragment | 19.21 | AAI-based (high-confidence) | 0 |
| S1-final-viral-combined_k141_325559\|\|full | 6628 | No | NA | 8 | 7 | 0 | Low-quality | Genome-fragment | 12.67 | AAI-based (high-confidence) | 0 |
| S9_k141_48380 | 6629 | No | NA | 6 | 0 | 3 | Low-quality | Genome-fragment | 5.83 | AAI-based (medium-confidence) | 0 |
| S6-final-viral-combined_k141_12738\|\|full | 1980 | No | NA | 4 | 0 | 1 | Not-determined | Genome-fragment | NA | NA | 0 |
| S6-final-viral-combined_k141_112475\|\|full | 3949 | No | NA | 3 | 0 | 0 | Not-determined | Genome-fragment | NA | NA | 0 |
| S6-final-viral-combined_k141_1516\|\|full | 1910 | No | NA | 2 | 0 | 2 | Not-determined | Genome-fragment | NA | NA | 0 |
| S6-final-viral-combined_k141_12742\|\|full | 1166 | No | NA | 2 | 0 | 1 | Not-determined | Genome-fragment | NA | NA | 0 |
| S6-final-viral-combined_k141_30510\|\|full | 3753 | No | NA | 4 | 0 | 4 | Not-determined | Genome-fragment | NA | NA | 0 |
| S6-final-viral-combined_k141_59888\|\|full | 1112 | No | NA | 3 | 0 | 0 | Not-determined | Genome-fragment | NA | NA | 0 |
| S6-final-viral-combined_k141_18384\|\|full | 1037 | No | NA | 2 | 0 | 0 | Not-determined | Genome-fragment | NA | NA | 0 |
| S6-final-viral-combined_k141_95237\|\|full | 3213 | No | NA | 3 | 0 | 2 | Not-determined | Genome-fragment | NA | NA | 0 |
| S2-final-viral-combined_k141_36395\|\|full | 6630 | No | NA | 19 | 1 | 1 | Low-quality | Genome-fragment | 12.62 | HMM-based (lower-bound) | 0 |
| S5_k141_116840 | 6632 | No | NA | 8 | 0 | 0 | Low-quality | Genome-fragment | 8.54 | AAI-based (medium-confidence) | 0 |
| S6-final-viral-combined_k141_30553\|\|full | 1012 | No | NA | 2 | 0 | 0 | Not-determined | Genome-fragment | NA | NA | 0 |
| S6-final-viral-combined_k141_77296\|\|full | 2852 | No | NA | 3 | 0 | 1 | Not-determined | Genome-fragment | NA | NA | 0 |
| S6-final-viral-combined_k141_112521\|\|full | 1131 | No | NA | 2 | 0 | 2 | Not-determined | Genome-fragment | NA | NA | 0 |
| S9-final-viral-combined_k141_28041\|\|full | 6633 | No | NA | 7 | 0 | 1 | Low-quality | Genome-fragment | 7.42 | AAI-based (medium-confidence) | 0 |
| S6-final-viral-combined_k141_30577\|\|full | 1303 | No | NA | 2 | 0 | 2 | Not-determined | Genome-fragment | NA | NA | 0 |
| S6-final-viral-combined_k141_48188\|\|full | 2171 | No | NA | 3 | 0 | 2 | Not-determined | Genome-fragment | NA | NA | 0 |
| S6-final-viral-combined_k141_53504\|\|full | 6290 | No | NA | 8 | 0 | 5 | Not-determined | Genome-fragment | NA | NA | 0 |
| S6-final-viral-combined_k141_124477\|\|full | 1856 | No | NA | 3 | 0 | 2 | Not-determined | Genome-fragment | NA | NA | 0 |
| S6-final-viral-combined_k141_24426\|\|full | 2662 | No | NA | 4 | 0 | 2 | Not-determined | Genome-fragment | NA | NA | 0 |
| S5_k141_49454 | 6639 | No | NA | 8 | 5 | 0 | Low-quality | Genome-fragment | 14.77 | AAI-based (high-confidence) | 0 |
| S6-final-viral-combined_k141_59955\|\|full | 2566 | No | NA | 3 | 0 | 2 | Not-determined | Genome-fragment | NA | NA | 0 |
| S6-final-viral-combined_k141_36230\|\|full | 2870 | No | NA | 4 | 0 | 1 | Not-determined | Genome-fragment | NA | NA | 0 |
| S6-final-viral-combined_k141_59967\|\|full | 3594 | No | NA | 4 | 0 | 3 | Not-determined | Genome-fragment | NA | NA | 0 |
| S17-final-viral-combined_k141_163623\|\|full | 6639 | No | NA | 5 | 1 | 1 | Low-quality | Genome-fragment | 4.87 | HMM-based (lower-bound) | 0 |
| S6-final-viral-combined_k141_42413\|\|full | 1125 | No | NA | 3 | 0 | 1 | Not-determined | Genome-fragment | NA | NA | 0 |
| S1-final-viral-combined_k141_123547\|\|full | 6643 | No | NA | 6 | 0 | 4 | Low-quality | Genome-fragment | 21.21 | AAI-based (medium-confidence) | 0 |
| S12-final-viral-combined_k141_208077\|\|full | 6648 | No | NA | 9 | 7 | 0 | Low-quality | Genome-fragment | 11.16 | AAI-based (high-confidence) | 0 |
| S6-final-viral-combined_k141_48253\|\|full | 1866 | No | NA | 4 | 0 | 2 | Not-determined | Genome-fragment | NA | NA | 0 |
| S12_k141_327346 | 6650 | No | NA | 7 | 0 | 0 | Low-quality | Genome-fragment | 6.69 | AAI-based (high-confidence) | 0 |
| S6-final-viral-combined_k141_95325\|\|full | 1312 | No | NA | 2 | 0 | 1 | Not-determined | Genome-fragment | NA | NA | 0 |
| S8-final-viral-combined_k141_196057\|\|full | 6652 | No | NA | 4 | 0 | 2 | Low-quality | Genome-fragment | 8.54 | AAI-based (medium-confidence) | 0 |
| S12_k141_253488 | 6653 | No | NA | 14 | 3 | 0 | Low-quality | Genome-fragment | 5.37 | HMM-based (lower-bound) | 0 |
| S2-final-viral-combined_k141_145344\|\|full | 6653 | No | NA | 7 | 3 | 0 | Low-quality | Genome-fragment | 17.05 | AAI-based (high-confidence) | 0 |
| S2_k141_134076 | 6654 | No | NA | 5 | 3 | 0 | Low-quality | Genome-fragment | 15.74 | AAI-based (high-confidence) | 0 |
| S8-final-viral-combined_k141_159754\|\|full | 6658 | No | NA | 5 | 0 | 2 | Low-quality | Genome-fragment | 3.95 | AAI-based (medium-confidence) | 0 |
| S6-final-viral-combined_k141_106955\|\|full | 5809 | No | NA | 4 | 0 | 3 | Not-determined | Genome-fragment | NA | NA | 0 |
| S1-final-viral-combined_k141_211412\|\|full | 6660 | No | NA | 9 | 1 | 0 | Low-quality | Genome-fragment | 16.64 | AAI-based (high-confidence) | 0 |
| S6-final-viral-combined_k141_83297\|\|full | 2712 | No | NA | 2 | 0 | 2 | Not-determined | Genome-fragment | NA | NA | 0 |
| S4-final-viral-combined_k141_2750\|\|full | 6660 | No | NA | 10 | 0 | 4 | Low-quality | Genome-fragment | 6.25 | AAI-based (medium-confidence) | 0 |
| S6-final-viral-combined_k141_1682\|\|full | 3510 | No | NA | 3 | 0 | 1 | Not-determined | Genome-fragment | NA | NA | 0 |
| S6-final-viral-combined_k141_60077\|\|full | 2127 | No | NA | 3 | 0 | 1 | Not-determined | Genome-fragment | NA | NA | 0 |
| S6-final-viral-combined_k141_48323\|\|full | 2713 | No | NA | 3 | 0 | 1 | Not-determined | Genome-fragment | NA | NA | 0 |
| S6-final-viral-combined_k141_36349\|\|full | 1127 | No | NA | 2 | 0 | 1 | Not-determined | Genome-fragment | NA | NA | 0 |
| S6-final-viral-combined_k141_130310\|\|full | 1467 | No | NA | 3 | 0 | 1 | Not-determined | Genome-fragment | NA | NA | 0 |
| S6-final-viral-combined_k141_65938\|\|full | 1189 | No | NA | 3 | 0 | 0 | Not-determined | Genome-fragment | NA | NA | 0 |
| S17-final-viral-combined_k141_77111\|\|full | 6661 | Yes | 3235 | 8 | 1 | 1 | Low-quality | Genome-fragment | 4.84 | HMM-based (lower-bound) | 51.43 |
| S6-final-viral-combined_k141_135866\|\|full | 1428 | No | NA | 3 | 0 | 1 | Not-determined | Genome-fragment | NA | NA | 0 |
| S2-final-viral-combined_k141_222222\|\|full | 6663 | No | NA | 7 | 0 | 1 | Low-quality | Genome-fragment | 14.34 | AAI-based (medium-confidence) | 0 |
| S6-final-viral-combined_k141_60099\|\|full | 2154 | No | NA | 3 | 0 | 2 | Not-determined | Genome-fragment | NA | NA | 0 |
| S6-final-viral-combined_k141_24496\|\|full | 2602 | No | NA | 3 | 0 | 2 | Not-determined | Genome-fragment | NA | NA | 0 |
| S6-final-viral-combined_k141_83344\|\|full | 1839 | No | NA | 2 | 0 | 2 | Not-determined | Genome-fragment | NA | NA | 0 |
| S1-final-viral-combined_k141_180292\|\|full | 6669 | No | NA | 13 | 4 | 0 | Low-quality | Genome-fragment | 16.39 | AAI-based (high-confidence) | 0 |
| S6-final-viral-combined_k141_60112\|\|full | 1916 | No | NA | 3 | 0 | 2 | Not-determined | Genome-fragment | NA | NA | 0 |
| S6-final-viral-combined_k141_95422\|\|full | 3677 | No | NA | 4 | 0 | 0 | Not-determined | Genome-fragment | NA | NA | 0 |
| S6-final-viral-combined_k141_83395\|\|full | 1001 | No | NA | 3 | 0 | 0 | Not-determined | Genome-fragment | NA | NA | 0 |
| S6-final-viral-combined_k141_89679\|\|full | 2720 | No | NA | 3 | 0 | 1 | Not-determined | Genome-fragment | NA | NA | 0 |
| S6-final-viral-combined_k141_77485\|\|full | 1016 | No | NA | 2 | 0 | 1 | Not-determined | Genome-fragment | NA | NA | 0 |
| S6-final-viral-combined_k141_18620\|\|full | 1558 | No | NA | 3 | 0 | 1 | Not-determined | Genome-fragment | NA | NA | 0 |
| S6-final-viral-combined_k141_77492\|\|full | 3363 | No | NA | 3 | 0 | 2 | Not-determined | Genome-fragment | NA | NA | 0 |
| S6-final-viral-combined_k141_18645\|\|full | 1288 | No | NA | 3 | 0 | 0 | Not-determined | Genome-fragment | NA | NA | 0 |
| S9_k141_53735 | 6671 | No | NA | 6 | 4 | 0 | Low-quality | Genome-fragment | 14.67 | AAI-based (high-confidence) | 0 |
| S4-final-viral-combined_k141_153097\|\|full | 6674 | No | NA | 6 | 0 | 1 | Low-quality | Genome-fragment | 11.03 | AAI-based (medium-confidence) | 0 |
| S6-final-viral-combined_k141_100905\|\|full | 2044 | No | NA | 5 | 0 | 0 | Not-determined | Genome-fragment | NA | NA | 0 |
| S5-final-viral-combined_k141_91703\|\|full | 6679 | No | NA | 8 | 5 | 0 | Low-quality | Genome-fragment | 18.27 | AAI-based (high-confidence) | 0 |
| S6-final-viral-combined_k141_42644\|\|full | 1398 | No | NA | 2 | 0 | 1 | Not-determined | Genome-fragment | NA | NA | 0 |
| S5_k141_35838 | 6683 | No | NA | 6 | 4 | 0 | Low-quality | Genome-fragment | 10.51 | AAI-based (high-confidence) | 0 |
| S12_k141_214950 | 6684 | No | NA | 8 | 7 | 0 | Low-quality | Genome-fragment | 19.51 | AAI-based (high-confidence) | 0 |
| S6-final-viral-combined_k141_30797\|\|full | 1474 | No | NA | 3 | 0 | 2 | Not-determined | Genome-fragment | NA | NA | 0 |
| S6-final-viral-combined_k141_30800\|\|full | 1163 | No | NA | 2 | 0 | 2 | Not-determined | Genome-fragment | NA | NA | 0 |
| S6-final-viral-combined_k141_130412\|\|full | 2873 | No | NA | 4 | 0 | 3 | Not-determined | Genome-fragment | NA | NA | 0 |
| S6-final-viral-combined_k141_53694\|\|full | 7373 | No | NA | 3 | 0 | 2 | Not-determined | Genome-fragment | NA | NA | 0 |
| S12-final-viral-combined_k141_379629\|\|full | 6686 | Yes | 3849 | 7 | 1 | 1 | Low-quality | Genome-fragment | 6.17 | HMM-based (lower-bound) | 42.43 |
| S6-final-viral-combined_k141_100943\|\|full | 1375 | No | NA | 2 | 0 | 0 | Not-determined | Genome-fragment | NA | NA | 0 |
| S6-final-viral-combined_k141_71717\|\|full | 1035 | No | NA | 2 | 0 | 0 | Not-determined | Genome-fragment | NA | NA | 0 |
| S6-final-viral-combined_k141_30821\|\|full | 3270 | No | NA | 5 | 0 | 4 | Not-determined | Genome-fragment | NA | NA | 0 |
| S17-final-viral-combined_k141_38158\|\|full | 6688 | No | NA | 8 | 1 | 0 | Low-quality | Genome-fragment | 15.2 | AAI-based (medium-confidence) | 0 |
| S6-final-viral-combined_k141_1825\|\|full | 1247 | No | NA | 3 | 0 | 1 | Not-determined | Genome-fragment | NA | NA | 0 |
| S6-final-viral-combined_k141_83526\|\|full | 4154 | No | NA | 5 | 0 | 3 | Not-determined | Genome-fragment | NA | NA | 0 |
| S6-final-viral-combined_k141_18771\|\|full | 2047 | No | NA | 3 | 0 | 3 | Not-determined | Genome-fragment | NA | NA | 0 |
| S6-final-viral-combined_k141_18775\|\|full | 1813 | No | NA | 2 | 0 | 2 | Not-determined | Genome-fragment | NA | NA | 0 |
| S12-final-viral-combined_k141_5564\|\|full | 6691 | No | NA | 15 | 1 | 0 | Low-quality | Genome-fragment | 17.39 | AAI-based (high-confidence) | 0 |
| S3-final-viral-combined_k141_88465\|\|full | 6692 | No | NA | 7 | 0 | 6 | Low-quality | Genome-fragment | 6.28 | AAI-based (medium-confidence) | 0 |
| S6-final-viral-combined_k141_36376\|\|full | 2177 | No | NA | 3 | 0 | 1 | Not-determined | Genome-fragment | NA | NA | 0 |
| S6-final-viral-combined_k141_53747\|\|full | 1395 | No | NA | 2 | 0 | 1 | Not-determined | Genome-fragment | NA | NA | 0 |
| S6-final-viral-combined_k141_118427\|\|full | 176709 | Yes | 49596 | 205 | 23 | 93 | Complete | High-quality | 100 | Provirus (high-confidence) | 71.93 |
| S6-final-viral-combined_k141_112725\|\|full | 1784 | No | NA | 3 | 0 | 1 | Not-determined | Genome-fragment | NA | NA | 0 |
| S6-final-viral-combined_k141_1923\|\|full | 1115 | No | NA | 3 | 0 | 1 | Not-determined | Genome-fragment | NA | NA | 0 |
| S5-final-viral-combined_k141_105612\|\|full | 6697 | No | NA | 8 | 5 | 0 | Low-quality | Genome-fragment | 7.65 | HMM-based (lower-bound) | 0 |
| S6-final-viral-combined_k141_89825\|\|full | 1481 | No | NA | 2 | 0 | 1 | Not-determined | Genome-fragment | NA | NA | 0 |
| S6_k141_24742 | 6697 | No | NA | 14 | 1 | 1 | Low-quality | Genome-fragment | 3.63 | HMM-based (lower-bound) | 0 |
| S6-final-viral-combined_k141_53771\|\|full | 2988 | No | NA | 4 | 0 | 3 | Not-determined | Genome-fragment | NA | NA | 0 |
| S17-k141_67818 | 6698 | No | NA | 5 | 0 | 5 | Low-quality | Genome-fragment | 3.6 | AAI-based (medium-confidence) | 0 |
| S6-final-viral-combined_k141_66146\|\|full | 1874 | No | NA | 2 | 0 | 1 | Not-determined | Genome-fragment | NA | NA | 0 |
| S6-final-viral-combined_k141_13071\|\|full | 1106 | No | NA | 3 | 0 | 0 | Not-determined | Genome-fragment | NA | NA | 0 |
| S6-final-viral-combined_k141_118484\|\|full | 18804 | No | NA | 26 | 14 | 1 | Medium-quality | Genome-fragment | 55.01 | AAI-based (medium-confidence) | 0 |
| S6-final-viral-combined_k141_13091\|\|full | 3460 | No | NA | 2 | 0 | 2 | Not-determined | Genome-fragment | NA | NA | 0 |
| S12-final-viral-combined_k141_113126\|\|full | 6699 | No | NA | 9 | 0 | 1 | Low-quality | Genome-fragment | 10.54 | AAI-based (medium-confidence) | 0 |
| S6-final-viral-combined_k141_30988\|\|full | 1919 | No | NA | 3 | 0 | 2 | Not-determined | Genome-fragment | NA | NA | 0 |
| S6-final-viral-combined_k141_130587\|\|full | 1757 | No | NA | 3 | 0 | 3 | Not-determined | Genome-fragment | NA | NA | 0 |
| S8-final-viral-combined_k141_190591\|\|full | 6699 | No | NA | 10 | 5 | 0 | Low-quality | Genome-fragment | 16.66 | AAI-based (high-confidence) | 0 |
| S12-final-viral-combined_k141_293913\|\|full | 6704 | Yes | 3596 | 7 | 1 | 1 | Low-quality | Genome-fragment | 2.46 | HMM-based (lower-bound) | 46.36 |
| S3_k141_135776 | 6708 | No | NA | 6 | 1 | 2 | Low-quality | Genome-fragment | 10.15 | HMM-based (lower-bound) | 0 |
| S6-final-viral-combined_k141_130612\|\|full | 1235 | No | NA | 2 | 0 | 1 | Not-determined | Genome-fragment | NA | NA | 0 |
| S6-final-viral-combined_k141_124685\|\|full | 1040 | No | NA | 2 | 0 | 1 | Not-determined | Genome-fragment | NA | NA | 0 |
| S6-final-viral-combined_k141_130615\|\|full | 5341 | No | NA | 6 | 0 | 1 | Not-determined | Genome-fragment | NA | NA | 0 |
| S6-final-viral-combined_k141_2061\|\|full | 1328 | No | NA | 2 | 0 | 2 | Not-determined | Genome-fragment | NA | NA | 0 |
| S6-final-viral-combined_k141_112781\|\|full | 2111 | No | NA | 2 | 0 | 1 | Not-determined | Genome-fragment | NA | NA | 0 |
| S6-final-viral-combined_k141_24723\|\|full | 1032 | No | NA | 2 | 0 | 0 | Not-determined | Genome-fragment | NA | NA | 0 |
| S6-final-viral-combined_k141_2064\|\|full | 1639 | No | NA | 3 | 0 | 2 | Not-determined | Genome-fragment | NA | NA | 0 |
| S6-final-viral-combined_k141_130653\|\|full | 2366 | No | NA | 4 | 0 | 1 | Not-determined | Genome-fragment | NA | NA | 0 |
| S6-final-viral-combined_k141_2076\|\|full | 1775 | No | NA | 3 | 0 | 0 | Not-determined | Genome-fragment | NA | NA | 0 |
| S6-final-viral-combined_k141_130658\|\|full | 2978 | No | NA | 2 | 0 | 2 | Not-determined | Genome-fragment | NA | NA | 0 |
| S6-final-viral-combined_k141_71850\|\|full | 1313 | No | NA | 3 | 0 | 0 | Not-determined | Genome-fragment | NA | NA | 0 |
| S6-final-viral-combined_k141_31051\|\|full | 2520 | No | NA | 4 | 0 | 2 | Not-determined | Genome-fragment | NA | NA | 0 |
| S6-final-viral-combined_k141_95739\|\|full | 5414 | No | NA | 3 | 0 | 2 | Not-determined | Genome-fragment | NA | NA | 0 |
| S8-final-viral-combined_k141_206425\|\|full | 6708 | Yes | 5494 | 10 | 2 | 1 | Low-quality | Genome-fragment | 12.84 | AAI-based (medium-confidence) | 18.1 |
| S8_k141_2873 | 6708 | No | NA | 13 | 2 | 0 | Low-quality | Genome-fragment | 15.7 | AAI-based (high-confidence) | 0 |
| S6-final-viral-combined_k141_89939\|\|full | 3913 | No | NA | 4 | 0 | 1 | Not-determined | Genome-fragment | NA | NA | 0 |
| S2-final-viral-combined_k141_2465\|\|full | 6709 | Yes | 1865 | 12 | 2 | 4 | Low-quality | Genome-fragment | 2.73 | HMM-based (lower-bound) | 72.2 |
| S6-final-viral-combined_k141_13275\|\|full | 2675 | No | NA | 3 | 0 | 3 | Not-determined | Genome-fragment | NA | NA | 0 |
| S6-final-viral-combined_k141_71875\|\|full | 11226 | No | NA | 12 | 0 | 8 | Not-determined | Genome-fragment | NA | NA | 0 |
| S6-final-viral-combined_k141_31093\|\|full | 13254 | No | NA | 7 | 0 | 5 | Not-determined | Genome-fragment | NA | NA | 0 |
| S6-final-viral-combined_k141_77729\|\|full | 7527 | No | NA | 8 | 0 | 7 | Not-determined | Genome-fragment | NA | NA | 0 |
| S8-final-viral-combined_k141_259326\|\|full | 6710 | No | NA | 10 | 0 | 3 | Low-quality | Genome-fragment | 14.08 | AAI-based (high-confidence) | 0 |
| S6-final-viral-combined_k141_71880\|\|full | 1599 | No | NA | 3 | 0 | 1 | Not-determined | Genome-fragment | NA | NA | 0 |
| S5_k141_111346 | 6711 | No | NA | 6 | 6 | 0 | Low-quality | Genome-fragment | 16.58 | AAI-based (high-confidence) | 0 |
| S6-final-viral-combined_k141_60561\|\|full | 2421 | No | NA | 4 | 0 | 0 | Not-determined | Genome-fragment | NA | NA | 0 |
| S6-final-viral-combined_k141_118600\|\|full | 1620 | No | NA | 4 | 0 | 1 | Not-determined | Genome-fragment | NA | NA | 0 |
| S6-final-viral-combined_k141_13286\|\|full | 1615 | No | NA | 2 | 0 | 0 | Not-determined | Genome-fragment | NA | NA | 0 |
| S6-final-viral-combined_k141_60587\|\|full | 1522 | No | NA | 3 | 0 | 2 | Not-determined | Genome-fragment | NA | NA | 0 |
| S6-final-viral-combined_k141_77772\|\|full | 4857 | No | NA | 6 | 0 | 4 | Not-determined | Genome-fragment | NA | NA | 0 |
| S9_k141_71736 | 6716 | No | NA | 1 | 0 | 0 | Low-quality | Genome-fragment | 2.5 | AAI-based (high-confidence) | 0 |
| S6-final-viral-combined_k141_130736\|\|full | 12831 | No | NA | 14 | 0 | 6 | Not-determined | Genome-fragment | NA | NA | 0 |
| S6-final-viral-combined_k141_107440\|\|full | 3096 | No | NA | 4 | 0 | 3 | Not-determined | Genome-fragment | NA | NA | 0 |
| S6-final-viral-combined_k141_136237\|\|full | 2025 | No | NA | 3 | 0 | 1 | Not-determined | Genome-fragment | NA | NA | 0 |
| S6-final-viral-combined_k141_36699\|\|full | 2317 | No | NA | 1 | 0 | 0 | Not-determined | Genome-fragment | NA | NA | 0 |
| S4-final-viral-combined_k141_48096\|\|full | 6719 | No | NA | 9 | 6 | 0 | Low-quality | Genome-fragment | 14.2 | AAI-based (medium-confidence) | 0 |
| S6-final-viral-combined_k141_136245\|\|full | 1364 | No | NA | 3 | 0 | 1 | Not-determined | Genome-fragment | NA | NA | 0 |
| S6-final-viral-combined_k141_13308\|\|full | 9762 | No | NA | 13 | 0 | 3 | Not-determined | Genome-fragment | NA | NA | 0 |
| S12-final-viral-combined_k141_188407\|\|full | 6723 | No | NA | 7 | 4 | 0 | Low-quality | Genome-fragment | 16.43 | AAI-based (high-confidence) | 0 |
| S12-final-viral-combined_k141_242298\|\|full | 6724 | No | NA | 10 | 0 | 1 | Low-quality | Genome-fragment | 15.72 | AAI-based (medium-confidence) | 0 |
| S11-final-viral-combined_k141_300283\|\|full | 6725 | No | NA | 9 | 2 | 0 | Low-quality | Genome-fragment | 14.35 | AAI-based (high-confidence) | 0 |
| S6-final-viral-combined_k141_134863\|\|full | 6729 | No | NA | 7 | 1 | 4 | Low-quality | Genome-fragment | 3.72 | HMM-based (lower-bound) | 0 |
| S6-final-viral-combined_k141_71940\|\|full | 2391 | No | NA | 4 | 0 | 1 | Not-determined | Genome-fragment | NA | NA | 0 |
| S6-final-viral-combined_k141_130792\|\|full | 2203 | No | NA | 3 | 0 | 2 | Not-determined | Genome-fragment | NA | NA | 0 |
| S6-final-viral-combined_k141_31193\|\|full | 1097 | No | NA | 2 | 0 | 0 | Not-determined | Genome-fragment | NA | NA | 0 |
| S5-final-viral-combined_k141_2856\|\|full | 6733 | No | NA | 8 | 4 | 0 | Low-quality | Genome-fragment | 14.46 | AAI-based (high-confidence) | 0 |
| S6-final-viral-combined_k141_95846\|\|full | 1964 | No | NA | 3 | 0 | 0 | Not-determined | Genome-fragment | NA | NA | 0 |
| S6-final-viral-combined_k141_71987\|\|full | 2236 | No | NA | 3 | 0 | 1 | Not-determined | Genome-fragment | NA | NA | 0 |
| S11_k141_320767 | 6734 | No | NA | 11 | 1 | 0 | Low-quality | Genome-fragment | 14.71 | AAI-based (high-confidence) | 0 |
| S7-final-viral-combined_k141_95205\|\|full | 6737 | No | NA | 5 | 0 | 1 | Low-quality | Genome-fragment | 4.96 | AAI-based (medium-confidence) | 0 |
| S6-final-viral-combined_k141_43031\|\|full | 2003 | No | NA | 4 | 0 | 1 | Not-determined | Genome-fragment | NA | NA | 0 |
| S2-final-viral-combined_k141_157887\|\|full | 6739 | No | NA | 7 | 4 | 0 | Low-quality | Genome-fragment | 13.53 | AAI-based (high-confidence) | 0 |
| S6-final-viral-combined_k141_83850\|\|full | 1333 | No | NA | 3 | 0 | 1 | Not-determined | Genome-fragment | NA | NA | 0 |
| S9-final-viral-combined_k141_117244\|\|full | 6744 | No | NA | 8 | 2 | 0 | Low-quality | Genome-fragment | 7.51 | HMM-based (lower-bound) | 0 |
| S6-final-viral-combined_k141_13423\|\|full | 1564 | No | NA | 2 | 0 | 0 | Not-determined | Genome-fragment | NA | NA | 0 |
| S6-final-viral-combined_k141_77813\|\|full | 6351 | No | NA | 3 | 0 | 3 | Not-determined | Genome-fragment | NA | NA | 0 |
| S9-final-viral-combined_k141_34916\|\|full | 6746 | No | NA | 16 | 0 | 0 | Low-quality | Genome-fragment | 16.28 | AAI-based (high-confidence) | 0 |
| S7-final-viral-combined_k141_63945\|\|full | 6746 | No | NA | 11 | 0 | 3 | Low-quality | Genome-fragment | 12.53 | AAI-based (medium-confidence) | 0 |
| S8-final-viral-combined_k141_255214\|\|full | 6748 | No | NA | 9 | 0 | 1 | Low-quality | Genome-fragment | 15.42 | AAI-based (medium-confidence) | 0 |
| S6-final-viral-combined_k141_113057\|\|full | 3062 | No | NA | 3 | 0 | 1 | Not-determined | Genome-fragment | NA | NA | 0 |
| S6-final-viral-combined_k141_13494\|\|full | 5282 | No | NA | 3 | 0 | 2 | Not-determined | Genome-fragment | NA | NA | 0 |
| S6-final-viral-combined_k141_24927\|\|full | 1742 | No | NA | 2 | 0 | 1 | Not-determined | Genome-fragment | NA | NA | 0 |
| S6-final-viral-combined_k141_72082\|\|full | 1863 | No | NA | 3 | 0 | 0 | Not-determined | Genome-fragment | NA | NA | 0 |
| S6-final-viral-combined_k141_60749\|\|full | 2928 | No | NA | 2 | 0 | 2 | Not-determined | Genome-fragment | NA | NA | 0 |
| S6-final-viral-combined_k141_43139\|\|full | 1510 | No | NA | 3 | 0 | 1 | Not-determined | Genome-fragment | NA | NA | 0 |
| S6-final-viral-combined_k141_136430\|\|full | 1367 | No | NA | 2 | 0 | 1 | Not-determined | Genome-fragment | NA | NA | 0 |
| S6-final-viral-combined_k141_2327\|\|full | 1936 | No | NA | 3 | 0 | 1 | Not-determined | Genome-fragment | NA | NA | 0 |
| S6-final-viral-combined_k141_136434\|\|full | 1622 | No | NA | 3 | 0 | 0 | Not-determined | Genome-fragment | NA | NA | 0 |
| S6-final-viral-combined_k141_24945\|\|full | 1438 | No | NA | 2 | 0 | 0 | Not-determined | Genome-fragment | NA | NA | 0 |
| S3-final-viral-combined_k141_28899\|\|full | 6750 | No | NA | 4 | 0 | 1 | Low-quality | Genome-fragment | 6.49 | AAI-based (medium-confidence) | 0 |
| S6-final-viral-combined_k141_95884\|\|full | 4505 | No | NA | 5 | 0 | 4 | Not-determined | Genome-fragment | NA | NA | 0 |
| S6-final-viral-combined_k141_136465\|\|full | 1516 | No | NA | 2 | 0 | 0 | Not-determined | Genome-fragment | NA | NA | 0 |
| S6-final-viral-combined_k141_95908\|\|full | 2149 | No | NA | 3 | 0 | 0 | Not-determined | Genome-fragment | NA | NA | 0 |
| S17-final-viral-combined_k141_25498\|\|full | 6750 | No | NA | 11 | 6 | 0 | Low-quality | Genome-fragment | 15.58 | AAI-based (high-confidence) | 0 |
| S11_k141_14066 | 6751 | No | NA | 8 | 0 | 5 | Low-quality | Genome-fragment | 22.37 | AAI-based (high-confidence) | 0 |
| S6-final-viral-combined_k141_95919\|\|full | 5589 | No | NA | 6 | 0 | 4 | Not-determined | Genome-fragment | NA | NA | 0 |
| S6-final-viral-combined_k141_124973\|\|full | 1712 | No | NA | 4 | 0 | 0 | Not-determined | Genome-fragment | NA | NA | 0 |
| S6-final-viral-combined_k141_7596\|\|full | 2489 | No | NA | 3 | 0 | 1 | Not-determined | Genome-fragment | NA | NA | 0 |
| S6-final-viral-combined_k141_31371\|\|full | 6853 | No | NA | 8 | 0 | 5 | Not-determined | Genome-fragment | NA | NA | 0 |
| S6_k141_28193 | 6751 | No | NA | 10 | 2 | 0 | Low-quality | Genome-fragment | 15.71 | AAI-based (high-confidence) | 0 |
| S6-final-viral-combined_k141_25009\|\|full | 2973 | No | NA | 3 | 0 | 3 | Not-determined | Genome-fragment | NA | NA | 0 |
| S6-final-viral-combined_k141_136551\|\|full | 1586 | No | NA | 3 | 0 | 2 | Not-determined | Genome-fragment | NA | NA | 0 |
| S6-final-viral-combined_k141_36964\|\|full | 3580 | No | NA | 3 | 0 | 2 | Not-determined | Genome-fragment | NA | NA | 0 |
| S6-final-viral-combined_k141_113167\|\|full | 2818 | No | NA | 3 | 0 | 1 | Not-determined | Genome-fragment | NA | NA | 0 |
| S6-final-viral-combined_k141_77888\|\|full | 4196 | No | NA | 6 | 0 | 1 | Not-determined | Genome-fragment | NA | NA | 0 |
| S6-final-viral-combined_k141_72190\|\|full | 1699 | No | NA | 3 | 0 | 0 | Not-determined | Genome-fragment | NA | NA | 0 |
| S6-final-viral-combined_k141_31418\|\|full | 3856 | No | NA | 4 | 0 | 3 | Not-determined | Genome-fragment | NA | NA | 0 |
| S6-final-viral-combined_k141_113203\|\|full | 1407 | No | NA | 4 | 0 | 1 | Not-determined | Genome-fragment | NA | NA | 0 |
| S6-final-viral-combined_k141_2525\|\|full | 4005 | No | NA | 6 | 0 | 4 | Not-determined | Genome-fragment | NA | NA | 0 |
| S6-final-viral-combined_k141_77932\|\|full | 4636 | No | NA | 5 | 0 | 0 | Not-determined | Genome-fragment | NA | NA | 0 |
| S6-final-viral-combined_k141_136621\|\|full | 3237 | No | NA | 4 | 0 | 0 | Not-determined | Genome-fragment | NA | NA | 0 |
| S6-final-viral-combined_k141_2537\|\|full | 8321 | No | NA | 7 | 0 | 3 | Not-determined | Genome-fragment | NA | NA | 0 |
| S6-final-viral-combined_k141_101045\|\|full | 5953 | No | NA | 5 | 0 | 3 | Not-determined | Genome-fragment | NA | NA | 0 |
| S6-final-viral-combined_k141_130980\|\|full | 7022 | No | NA | 5 | 0 | 4 | Not-determined | Genome-fragment | NA | NA | 0 |
| S6-final-viral-combined_k141_136635\|\|full | 1541 | No | NA | 2 | 0 | 1 | Not-determined | Genome-fragment | NA | NA | 0 |
| S6-final-viral-combined_k141_136636\|\|full | 4302 | No | NA | 2 | 0 | 2 | Not-determined | Genome-fragment | NA | NA | 0 |
| S6-final-viral-combined_k141_101058\|\|full | 2810 | No | NA | 2 | 0 | 1 | Not-determined | Genome-fragment | NA | NA | 0 |
| S6-final-viral-combined_k141_77954\|\|full | 1097 | No | NA | 3 | 0 | 1 | Not-determined | Genome-fragment | NA | NA | 0 |
| S6-final-viral-combined_k141_95972\|\|full | 16470 | No | NA | 17 | 0 | 15 | Not-determined | Genome-fragment | NA | NA | 0 |
| S6-final-viral-combined_k141_118858\|\|full | 2290 | No | NA | 4 | 0 | 0 | Not-determined | Genome-fragment | NA | NA | 0 |
| S2-final-viral-combined_k141_59505\|\|full | 6753 | No | NA | 15 | 2 | 0 | Low-quality | Genome-fragment | 15.31 | AAI-based (high-confidence) | 0 |
| S6-final-viral-combined_k141_54138\|\|full | 1884 | No | NA | 2 | 0 | 0 | Not-determined | Genome-fragment | NA | NA | 0 |
| S11-final-viral-combined_k141_36437\|\|full | 6756 | No | NA | 9 | 4 | 0 | Low-quality | Genome-fragment | 12.86 | AAI-based (high-confidence) | 0 |
| S6-final-viral-combined_k141_83909\|\|full | 4410 | No | NA | 4 | 0 | 1 | Not-determined | Genome-fragment | NA | NA | 0 |
| S7_k141_59851 | 6763 | No | NA | 7 | 1 | 0 | Low-quality | Genome-fragment | 6.92 | AAI-based (high-confidence) | 0 |
| S6-final-viral-combined_k141_113301\|\|full | 1209 | No | NA | 2 | 0 | 2 | Not-determined | Genome-fragment | NA | NA | 0 |
| S5-final-viral-combined_k141_118036\|\|full | 6770 | No | NA | 11 | 2 | 0 | Low-quality | Genome-fragment | 5.35 | HMM-based (lower-bound) | 0 |
| S5-final-viral-combined_k141_151429\|\|full | 6770 | No | NA | 7 | 1 | 0 | Low-quality | Genome-fragment | 13.76 | AAI-based (medium-confidence) | 0 |
| S6-final-viral-combined_k141_54178\|\|full | 1334 | No | NA | 2 | 0 | 1 | Not-determined | Genome-fragment | NA | NA | 0 |
| S3-final-viral-combined_k141_7115\|\|full | 6772 | Yes | 4978 | 5 | 1 | 1 | Low-quality | Genome-fragment | 2.92 | HMM-based (lower-bound) | 26.49 |
| S6-final-viral-combined_k141_72228\|\|full | 91672 | Yes | 44908 | 86 | 12 | 29 | Medium-quality | Genome-fragment | 70.39 | HMM-based (lower-bound) | 51.01 |
| S6-final-viral-combined_k141_83923\|\|full | 20495 | No | NA | 11 | 0 | 9 | Not-determined | Genome-fragment | NA | NA | 0 |
| S6-final-viral-combined_k141_96063\|\|full | 1730 | No | NA | 2 | 0 | 1 | Not-determined | Genome-fragment | NA | NA | 0 |
| S6-final-viral-combined_k141_101102\|\|full | 1282 | No | NA | 2 | 0 | 0 | Not-determined | Genome-fragment | NA | NA | 0 |
| S6-final-viral-combined_k141_72231\|\|full | 4668 | No | NA | 6 | 0 | 2 | Not-determined | Genome-fragment | NA | NA | 0 |
| S6-final-viral-combined_k141_2636\|\|full | 1463 | No | NA | 3 | 0 | 0 | Not-determined | Genome-fragment | NA | NA | 0 |
| S6-final-viral-combined_k141_107671\|\|full | 2236 | No | NA | 3 | 0 | 2 | Not-determined | Genome-fragment | NA | NA | 0 |
| S6-final-viral-combined_k141_96075\|\|full | 3040 | No | NA | 2 | 0 | 1 | Not-determined | Genome-fragment | NA | NA | 0 |
| S4-final-viral-combined_k141_46371\|\|full | 6773 | No | NA | 11 | 1 | 0 | Low-quality | Genome-fragment | 19.77 | AAI-based (high-confidence) | 0 |
| S17-final-viral-combined_k141_147410\|\|full | 6777 | No | NA | 9 | 1 | 0 | Low-quality | Genome-fragment | 4.14 | HMM-based (lower-bound) | 0 |
| S4-final-viral-combined_k141_100887\|\|full | 6780 | No | NA | 11 | 3 | 1 | Low-quality | Genome-fragment | 15.83 | AAI-based (high-confidence) | 0 |
| S2_k141_34659 | 6782 | No | NA | 16 | 1 | 0 | Low-quality | Genome-fragment | 3.47 | HMM-based (lower-bound) | 0 |
| S6-final-viral-combined_k141_60952\|\|full | 2455 | No | NA | 3 | 0 | 0 | Not-determined | Genome-fragment | NA | NA | 0 |
| S6-final-viral-combined_k141_43443\|\|full | 1449 | No | NA | 3 | 0 | 2 | Not-determined | Genome-fragment | NA | NA | 0 |
| S17-final-viral-combined_k141_93712\|\|full | 6785 | No | NA | 8 | 6 | 0 | Low-quality | Genome-fragment | 16.56 | AAI-based (high-confidence) | 0 |
| S6-final-viral-combined_k141_54248\|\|full | 1505 | No | NA | 3 | 0 | 1 | Not-determined | Genome-fragment | NA | NA | 0 |
| S12_k141_234871 | 6789 | No | NA | 8 | 6 | 0 | Low-quality | Genome-fragment | 15.49 | AAI-based (high-confidence) | 0 |
| S6-final-viral-combined_k141_101171\|\|full | 1687 | No | NA | 4 | 0 | 0 | Not-determined | Genome-fragment | NA | NA | 0 |
| S6-final-viral-combined_k141_125299\|\|full | 2873 | No | NA | 3 | 0 | 2 | Not-determined | Genome-fragment | NA | NA | 0 |
| S12_k141_252041 | 6789 | No | NA | 4 | 0 | 2 | Low-quality | Genome-fragment | 6.67 | AAI-based (high-confidence) | 0 |
| S6-final-viral-combined_k141_101175\|\|full | 1829 | No | NA | 6 | 0 | 0 | Not-determined | Genome-fragment | NA | NA | 0 |
| S6-final-viral-combined_k141_131064\|\|full | 2453 | No | NA | 2 | 0 | 2 | Not-determined | Genome-fragment | NA | NA | 0 |
| S6-final-viral-combined_k141_60987\|\|full | 6200 | No | NA | 5 | 0 | 4 | Not-determined | Genome-fragment | NA | NA | 0 |
| S7-final-viral-combined_k141_83974\|\|full | 6791 | Yes | 3100 | 11 | 2 | 4 | Low-quality | Genome-fragment | 2.71 | HMM-based (lower-bound) | 54.35 |
| S6-final-viral-combined_k141_2725\|\|full | 2527 | No | NA | 3 | 0 | 2 | Not-determined | Genome-fragment | NA | NA | 0 |
| S6-final-viral-combined_k141_19053\|\|full | 2048 | No | NA | 3 | 0 | 1 | Not-determined | Genome-fragment | NA | NA | 0 |
| S6-final-viral-combined_k141_84033\|\|full | 1882 | No | NA | 3 | 0 | 1 | Not-determined | Genome-fragment | NA | NA | 0 |
| S1_k141_161854 | 6795 | No | NA | 11 | 8 | 0 | Low-quality | Genome-fragment | 14.36 | AAI-based (high-confidence) | 0 |
| S6-final-viral-combined_k141_31609\|\|full | 2674 | No | NA | 2 | 0 | 1 | Not-determined | Genome-fragment | NA | NA | 0 |
| S6-final-viral-combined_k141_54294\|\|full | 3597 | No | NA | 5 | 0 | 2 | Not-determined | Genome-fragment | NA | NA | 0 |
| S6-final-viral-combined_k141_37283\|\|full | 1694 | No | NA | 2 | 0 | 2 | Not-determined | Genome-fragment | NA | NA | 0 |
| S6-final-viral-combined_k141_31636\|\|full | 1633 | No | NA | 2 | 0 | 2 | Not-determined | Genome-fragment | NA | NA | 0 |
| S12_k141_250015 | 6796 | No | NA | 6 | 0 | 5 | Low-quality | Genome-fragment | 15.18 | AAI-based (medium-confidence) | 0 |
| S6-final-viral-combined_k141_119017\|\|full | 25955 | No | NA | 37 | 18 | 0 | Medium-quality | Genome-fragment | 71.5 | AAI-based (high-confidence) | 0 |
| S6-final-viral-combined_k141_61041\|\|full | 4914 | No | NA | 5 | 0 | 0 | Not-determined | Genome-fragment | NA | NA | 0 |
| S4-final-viral-combined_k141_161277\|\|full | 6796 | No | NA | 5 | 1 | 0 | Low-quality | Genome-fragment | 44.13 | AAI-based (high-confidence) | 0 |
| S4_k141_79869 | 6797 | No | NA | 6 | 0 | 1 | Low-quality | Genome-fragment | 7.17 | AAI-based (medium-confidence) | 0 |
| S11-final-viral-combined_k141_279164\|\|full | 6798 | No | NA | 6 | 1 | 0 | Low-quality | Genome-fragment | 4.98 | AAI-based (high-confidence) | 0 |
| S9-final-viral-combined_k141_97394\|\|full | 6799 | No | NA | 8 | 7 | 0 | Low-quality | Genome-fragment | 13.88 | AAI-based (high-confidence) | 0 |
| S6-final-viral-combined_k141_31700\|\|full | 1328 | No | NA | 1 | 0 | 1 | Not-determined | Genome-fragment | NA | NA | 0 |
| S8-final-viral-combined_k141_81804\|\|full | 6799 | No | NA | 9 | 1 | 0 | Low-quality | Genome-fragment | 10.66 | AAI-based (medium-confidence) | 0 |
| S6-final-viral-combined_k141_66939\|\|full | 1857 | No | NA | 3 | 0 | 0 | Not-determined | Genome-fragment | NA | NA | 0 |
| S6-final-viral-combined_k141_25253\|\|full | 1366 | No | NA | 2 | 0 | 2 | Not-determined | Genome-fragment | NA | NA | 0 |
| S6-final-viral-combined_k141_43571\|\|full | 1699 | No | NA | 3 | 0 | 2 | Not-determined | Genome-fragment | NA | NA | 0 |
| S6-final-viral-combined_k141_78135\|\|full | 4855 | No | NA | 5 | 0 | 3 | Not-determined | Genome-fragment | NA | NA | 0 |
| S6-final-viral-combined_k141_78144\|\|full | 2336 | No | NA | 4 | 0 | 1 | Not-determined | Genome-fragment | NA | NA | 0 |
| S11-final-viral-combined_k141_235827\|\|full | 6801 | No | NA | 7 | 0 | 5 | Low-quality | Genome-fragment | 5.53 | AAI-based (medium-confidence) | 0 |
| S6-final-viral-combined_k141_7958\|\|full | 1404 | No | NA | 2 | 0 | 2 | Not-determined | Genome-fragment | NA | NA | 0 |
| S11-final-viral-combined_k141_214464\|\|full | 6802 | No | NA | 10 | 4 | 0 | Low-quality | Genome-fragment | 13.05 | AAI-based (high-confidence) | 0 |
| S12-final-viral-combined_k141_4061\|\|full | 6809 | No | NA | 9 | 4 | 0 | Low-quality | Genome-fragment | 19.02 | AAI-based (high-confidence) | 0 |
| S6-final-viral-combined_k141_78159\|\|full | 7710 | No | NA | 6 | 0 | 3 | Not-determined | Genome-fragment | NA | NA | 0 |
| S6-final-viral-combined_k141_19149\|\|full | 4332 | No | NA | 5 | 0 | 4 | Not-determined | Genome-fragment | NA | NA | 0 |
| S1_k141_131012 | 6811 | No | NA | 9 | 1 | 0 | Low-quality | Genome-fragment | 5.47 | HMM-based (lower-bound) | 0 |
| S12-final-viral-combined_k141_87967\|\|full | 6813 | No | NA | 8 | 6 | 0 | Low-quality | Genome-fragment | 15.28 | AAI-based (medium-confidence) | 0 |
| S6-final-viral-combined_k141_101340\|\|full | 3411 | No | NA | 4 | 0 | 2 | Not-determined | Genome-fragment | NA | NA | 0 |
| S2_k141_86684 | 6813 | No | NA | 10 | 2 | 1 | Low-quality | Genome-fragment | 5.73 | AAI-based (high-confidence) | 0 |
| S6-final-viral-combined_k141_2914\|\|full | 1515 | No | NA | 2 | 0 | 1 | Not-determined | Genome-fragment | NA | NA | 0 |
| S6-final-viral-combined_k141_72460\|\|full | 53620 | No | NA | 44 | 0 | 13 | Not-determined | Genome-fragment | NA | NA | 0 |
| S6-final-viral-combined_k141_72462\|\|full | 8159 | No | NA | 14 | 0 | 0 | Not-determined | Genome-fragment | NA | NA | 0 |
| S2-final-viral-combined_k141_35435\|\|full | 6824 | No | NA | 10 | 4 | 0 | Low-quality | Genome-fragment | 18.69 | AAI-based (high-confidence) | 0 |
| S6-final-viral-combined_k141_25389\|\|full | 1629 | No | NA | 3 | 0 | 2 | Not-determined | Genome-fragment | NA | NA | 0 |
| S6-final-viral-combined_k141_2968\|\|full | 2541 | No | NA | 3 | 0 | 1 | Not-determined | Genome-fragment | NA | NA | 0 |
| S6-final-viral-combined_k141_72468\|\|full | 1590 | No | NA | 2 | 0 | 1 | Not-determined | Genome-fragment | NA | NA | 0 |
| S6-final-viral-combined_k141_96320\|\|full | 1544 | No | NA | 3 | 0 | 1 | Not-determined | Genome-fragment | NA | NA | 0 |
| S6-final-viral-combined_k141_90377\|\|full | 1131 | No | NA | 2 | 0 | 1 | Not-determined | Genome-fragment | NA | NA | 0 |
| S6-final-viral-combined_k141_2999\|\|full | 3079 | No | NA | 4 | 0 | 2 | Not-determined | Genome-fragment | NA | NA | 0 |
| S6-final-viral-combined_k141_90388\|\|full | 4420 | No | NA | 5 | 0 | 1 | Not-determined | Genome-fragment | NA | NA | 0 |
| S9_k141_120591 | 6826 | No | NA | 6 | 0 | 4 | Low-quality | Genome-fragment | 5.53 | AAI-based (high-confidence) | 0 |
| S3_k141_100237 | 6832 | No | NA | 4 | 0 | 0 | Low-quality | Genome-fragment | 3.15 | AAI-based (high-confidence) | 0 |
| S12-final-viral-combined_k141_297984\|\|full | 6833 | No | NA | 2 | 0 | 0 | Low-quality | Genome-fragment | 7.12 | AAI-based (medium-confidence) | 0 |
| S6-final-viral-combined_k141_113502\|\|full | 1863 | No | NA | 3 | 0 | 2 | Not-determined | Genome-fragment | NA | NA | 0 |
| S12_k141_174342 | 6833 | No | NA | 5 | 0 | 4 | Low-quality | Genome-fragment | 3.68 | AAI-based (medium-confidence) | 0 |
| S3-final-viral-combined_k141_122102\|\|full | 6841 | No | NA | 5 | 0 | 1 | Low-quality | Genome-fragment | 7.89 | AAI-based (medium-confidence) | 0 |
| S12-final-viral-combined_k141_95302\|\|full | 6842 | No | NA | 11 | 9 | 0 | Low-quality | Genome-fragment | 17.07 | AAI-based (medium-confidence) | 0 |
| S6-final-viral-combined_k141_72558\|\|full | 1343 | No | NA | 3 | 0 | 1 | Not-determined | Genome-fragment | NA | NA | 0 |
| S6-final-viral-combined_k141_37583\|\|full | 2921 | No | NA | 4 | 0 | 3 | Not-determined | Genome-fragment | NA | NA | 0 |
| S6-final-viral-combined_k141_90408\|\|full | 28400 | No | NA | 24 | 0 | 14 | Not-determined | Genome-fragment | NA | NA | 0 |
| S6-final-viral-combined_k141_119138\|\|full | 61583 | Yes | 32460 | 81 | 20 | 19 | Medium-quality | Genome-fragment | 54.9 | HMM-based (lower-bound) | 47.29 |
| S1-final-viral-combined_k141_203367\|\|full | 6846 | No | NA | 7 | 3 | 0 | Low-quality | Genome-fragment | 4.53 | HMM-based (lower-bound) | 0 |
| S6-final-viral-combined_k141_3071\|\|full | 4290 | No | NA | 6 | 0 | 1 | Not-determined | Genome-fragment | NA | NA | 0 |
| S6-final-viral-combined_k141_96396\|\|full | 3597 | No | NA | 3 | 0 | 1 | Not-determined | Genome-fragment | NA | NA | 0 |
| S3-final-viral-combined_k141_54496\|\|full | 6847 | No | NA | 6 | 0 | 6 | Low-quality | Genome-fragment | 3.83 | AAI-based (medium-confidence) | 0 |
| S6-final-viral-combined_k141_113571\|\|full | 1948 | No | NA | 2 | 0 | 2 | Not-determined | Genome-fragment | NA | NA | 0 |
| S6-final-viral-combined_k141_108059\|\|full | 3736 | No | NA | 3 | 0 | 3 | Not-determined | Genome-fragment | NA | NA | 0 |
| S6-final-viral-combined_k141_78340\|\|full | 4351 | No | NA | 4 | 0 | 3 | Not-determined | Genome-fragment | NA | NA | 0 |
| S6-final-viral-combined_k141_108064\|\|full | 6289 | No | NA | 9 | 0 | 4 | Not-determined | Genome-fragment | NA | NA | 0 |
| S11_k141_151732 | 6848 | No | NA | 6 | 0 | 2 | Low-quality | Genome-fragment | 6.33 | AAI-based (medium-confidence) | 0 |
| S6-final-viral-combined_k141_84278\|\|full | 1145 | No | NA | 2 | 0 | 2 | Not-determined | Genome-fragment | NA | NA | 0 |
| S6-final-viral-combined_k141_19364\|\|full | 13300 | No | NA | 8 | 0 | 1 | Not-determined | Genome-fragment | NA | NA | 0 |
| S17-final-viral-combined_k141_227640\|\|full | 6858 | No | NA | 6 | 0 | 2 | Low-quality | Genome-fragment | 3.69 | AAI-based (medium-confidence) | 0 |
| S6-final-viral-combined_k141_96437\|\|full | 3950 | No | NA | 3 | 0 | 1 | Not-determined | Genome-fragment | NA | NA | 0 |
| S4-final-viral-combined_k141_44054\|\|full | 6860 | No | NA | 11 | 0 | 1 | Low-quality | Genome-fragment | 12.67 | AAI-based (high-confidence) | 0 |
| S6-final-viral-combined_k141_90524\|\|full | 6227 | No | NA | 3 | 0 | 2 | Not-determined | Genome-fragment | NA | NA | 0 |
| S11-final-viral-combined_k141_231084\|\|full | 6861 | No | NA | 7 | 0 | 5 | Low-quality | Genome-fragment | 3.46 | AAI-based (medium-confidence) | 0 |
| S6-final-viral-combined_k141_108096\|\|full | 1739 | No | NA | 2 | 0 | 1 | Not-determined | Genome-fragment | NA | NA | 0 |
| S6-final-viral-combined_k141_31854\|\|full | 1338 | No | NA | 2 | 0 | 0 | Not-determined | Genome-fragment | NA | NA | 0 |
| S8-final-viral-combined_k141_210709\|\|full | 6861 | No | NA | 2 | 0 | 0 | Low-quality | Genome-fragment | 6.94 | AAI-based (high-confidence) | 0 |
| S6-final-viral-combined_k141_37754\|\|full | 2184 | No | NA | 4 | 0 | 2 | Not-determined | Genome-fragment | NA | NA | 0 |
| S8_k141_237380 | 6861 | No | NA | 13 | 3 | 0 | Low-quality | Genome-fragment | 13.95 | HMM-based (lower-bound) | 0 |
| S6-final-viral-combined_k141_49582\|\|full | 2552 | No | NA | 3 | 0 | 2 | Not-determined | Genome-fragment | NA | NA | 0 |
| S6-final-viral-combined_k141_19466\|\|full | 3002 | No | NA | 3 | 0 | 0 | Not-determined | Genome-fragment | NA | NA | 0 |
| S6-final-viral-combined_k141_119330\|\|full | 3675 | No | NA | 4 | 0 | 2 | Not-determined | Genome-fragment | NA | NA | 0 |
| S12_k141_49556 | 6862 | No | NA | 5 | 0 | 1 | Low-quality | Genome-fragment | 2.17 | AAI-based (medium-confidence) | 0 |
| S6-final-viral-combined_k141_90657\|\|full | 1877 | No | NA | 2 | 0 | 2 | Not-determined | Genome-fragment | NA | NA | 0 |
| S6-final-viral-combined_k141_25708\|\|full | 19414 | No | NA | 15 | 0 | 7 | Not-determined | Genome-fragment | NA | NA | 0 |
| S6-final-viral-combined_k141_119382\|\|full | 1511 | No | NA | 3 | 0 | 0 | Not-determined | Genome-fragment | NA | NA | 0 |
| S6-final-viral-combined_k141_90686\|\|full | 1815 | No | NA | 3 | 0 | 3 | Not-determined | Genome-fragment | NA | NA | 0 |
| S8_k141_231410 | 6867 | No | NA | 11 | 3 | 0 | Low-quality | Genome-fragment | 5.24 | AAI-based (high-confidence) | 0 |
| S6-final-viral-combined_k141_101641\|\|full | 3057 | No | NA | 3 | 0 | 2 | Not-determined | Genome-fragment | NA | NA | 0 |
| S6-final-viral-combined_k141_131393\|\|full | 2395 | No | NA | 3 | 0 | 1 | Not-determined | Genome-fragment | NA | NA | 0 |
| S6-final-viral-combined_k141_8217\|\|full | 21125 | No | NA | 20 | 0 | 15 | Not-determined | Genome-fragment | NA | NA | 0 |
| S12-final-viral-combined_k141_138238\|\|full | 6868 | No | NA | 4 | 0 | 3 | Low-quality | Genome-fragment | 3.7 | AAI-based (medium-confidence) | 0 |
| S6-final-viral-combined_k141_137031\|\|full | 1813 | No | NA | 2 | 0 | 2 | Not-determined | Genome-fragment | NA | NA | 0 |
| S12-final-viral-combined_k141_390771\|\|full | 6872 | No | NA | 3 | 0 | 0 | Low-quality | Genome-fragment | 6.74 | AAI-based (medium-confidence) | 0 |
| S6-final-viral-combined_k141_67371\|\|full | 1595 | No | NA | 3 | 0 | 2 | Not-determined | Genome-fragment | NA | NA | 0 |
| S6-final-viral-combined_k141_101691\|\|full | 2007 | No | NA | 2 | 0 | 1 | Not-determined | Genome-fragment | NA | NA | 0 |
| S6-final-viral-combined_k141_78591\|\|full | 1080 | No | NA | 2 | 0 | 1 | Not-determined | Genome-fragment | NA | NA | 0 |
| S8_k141_169159 | 6872 | No | NA | 10 | 2 | 0 | Low-quality | Genome-fragment | 9.55 | HMM-based (lower-bound) | 0 |
| S6-final-viral-combined_k141_54693\|\|full | 5613 | No | NA | 6 | 0 | 3 | Not-determined | Genome-fragment | NA | NA | 0 |
| S6-final-viral-combined_k141_25833\|\|full | 1112 | No | NA | 3 | 0 | 0 | Not-determined | Genome-fragment | NA | NA | 0 |
| S6-final-viral-combined_k141_37960\|\|full | 16326 | No | NA | 26 | 0 | 5 | Not-determined | Genome-fragment | NA | NA | 0 |
| S6-final-viral-combined_k141_54708\|\|full | 5827 | No | NA | 4 | 0 | 3 | Not-determined | Genome-fragment | NA | NA | 0 |
| S1_k141_203650 | 6878 | Yes | 4035 | 17 | 3 | 2 | Low-quality | Genome-fragment | 7.72 | AAI-based (medium-confidence) | 41.33 |
| S11-final-viral-combined_k141_190288\|\|full | 6883 | No | NA | 11 | 4 | 0 | Low-quality | Genome-fragment | 12.49 | AAI-based (high-confidence) | 0 |
| S6-final-viral-combined_k141_14174\|\|full | 17997 | No | NA | 13 | 0 | 7 | Not-determined | Genome-fragment | NA | NA | 0 |
| S6-final-viral-combined_k141_131507\|\|full | 5446 | No | NA | 7 | 0 | 6 | Not-determined | Genome-fragment | NA | NA | 0 |
| S17-final-viral-combined_k141_244480\|\|full | 6886 | Yes | 2769 | 10 | 1 | 2 | Low-quality | Genome-fragment | 5.62 | AAI-based (medium-confidence) | 59.79 |
| S2-final-viral-combined_k141_231375\|\|full | 6889 | No | NA | 11 | 2 | 0 | Low-quality | Genome-fragment | 11.46 | AAI-based (high-confidence) | 0 |
| S6-final-viral-combined_k141_90791\|\|full | 4494 | No | NA | 5 | 0 | 5 | Not-determined | Genome-fragment | NA | NA | 0 |
| S12-final-viral-combined_k141_38043\|\|full | 6890 | No | NA | 12 | 4 | 0 | Low-quality | Genome-fragment | 11.42 | AAI-based (high-confidence) | 0 |
| S11-final-viral-combined_k141_229283\|\|full | 6898 | No | NA | 10 | 0 | 1 | Low-quality | Genome-fragment | 16.08 | AAI-based (medium-confidence) | 0 |
| S6-final-viral-combined_k141_19629\|\|full | 2199 | No | NA | 3 | 0 | 1 | Not-determined | Genome-fragment | NA | NA | 0 |
| S6-final-viral-combined_k141_32071\|\|full | 3526 | No | NA | 4 | 0 | 1 | Not-determined | Genome-fragment | NA | NA | 0 |
| S17-final-viral-combined_k141_213444\|\|full | 6904 | No | NA | 5 | 0 | 4 | Low-quality | Genome-fragment | 3.72 | AAI-based (medium-confidence) | 0 |
| S6-final-viral-combined_k141_61496\|\|full | 1446 | No | NA | 3 | 0 | 2 | Not-determined | Genome-fragment | NA | NA | 0 |
| S6-final-viral-combined_k141_38046\|\|full | 1288 | No | NA | 3 | 0 | 2 | Not-determined | Genome-fragment | NA | NA | 0 |
| S6-final-viral-combined_k141_32094\|\|full | 1356 | No | NA | 3 | 0 | 2 | Not-determined | Genome-fragment | NA | NA | 0 |
| S6-final-viral-combined_k141_72969\|\|full | 1178 | No | NA | 2 | 0 | 2 | Not-determined | Genome-fragment | NA | NA | 0 |
| S8-final-viral-combined_k141_268594\|\|full | 6904 | Yes | 2223 | 5 | 1 | 2 | Low-quality | Genome-fragment | 2.69 | HMM-based (lower-bound) | 67.8 |
| S6-final-viral-combined_k141_49909\|\|full | 28186 | No | NA | 48 | 18 | 1 | Medium-quality | Genome-fragment | 82.47 | AAI-based (high-confidence) | 0 |
| S6-final-viral-combined_k141_25846\|\|full | 3453 | No | NA | 4 | 0 | 1 | Not-determined | Genome-fragment | NA | NA | 0 |
| S6-final-viral-combined_k141_84584\|\|full | 1041 | No | NA | 2 | 0 | 0 | Not-determined | Genome-fragment | NA | NA | 0 |
| S6-final-viral-combined_k141_78658\|\|full | 2263 | No | NA | 5 | 0 | 1 | Not-determined | Genome-fragment | NA | NA | 0 |
| S2-final-viral-combined_k141_131640\|\|full | 6909 | No | NA | 8 | 5 | 0 | Low-quality | Genome-fragment | 29.17 | AAI-based (medium-confidence) | 0 |
| S6-final-viral-combined_k141_119572\|\|full | 1472 | No | NA | 3 | 0 | 1 | Not-determined | Genome-fragment | NA | NA | 0 |
| S3_k141_46401 | 6911 | No | NA | 16 | 1 | 1 | Low-quality | Genome-fragment | 3.4 | HMM-based (lower-bound) | 0 |
| S6-final-viral-combined_k141_137271\|\|full | 1639 | No | NA | 4 | 0 | 0 | Not-determined | Genome-fragment | NA | NA | 0 |
| S6-final-viral-combined_k141_101827\|\|full | 2522 | No | NA | 4 | 0 | 0 | Not-determined | Genome-fragment | NA | NA | 0 |
| S1-final-viral-combined_k141_11429\|\|full | 6912 | No | NA | 8 | 1 | 0 | Low-quality | Genome-fragment | 15.34 | HMM-based (lower-bound) | 0 |
| S8-final-viral-combined_k141_74267\|\|full | 6913 | No | NA | 4 | 0 | 0 | Low-quality | Genome-fragment | 7.26 | AAI-based (medium-confidence) | 0 |
| S6-final-viral-combined_k141_19719\|\|full | 5340 | No | NA | 6 | 0 | 4 | Not-determined | Genome-fragment | NA | NA | 0 |
| S6-final-viral-combined_k141_38123\|\|full | 1337 | No | NA | 2 | 0 | 1 | Not-determined | Genome-fragment | NA | NA | 0 |
| S6-final-viral-combined_k141_61556\|\|full | 2279 | No | NA | 4 | 0 | 3 | Not-determined | Genome-fragment | NA | NA | 0 |
| S6-final-viral-combined_k141_96895\|\|full | 5544 | No | NA | 9 | 0 | 4 | Not-determined | Genome-fragment | NA | NA | 0 |
| S1-final-viral-combined_k141_267686\|\|full | 6920 | No | NA | 16 | 8 | 1 | Low-quality | Genome-fragment | 14.65 | AAI-based (medium-confidence) | 0 |
| S12_k141_129910 | 6920 | No | NA | 6 | 0 | 1 | Low-quality | Genome-fragment | 14.7 | AAI-based (high-confidence) | 0 |
| S12-final-viral-combined_k141_244823\|\|full | 6922 | No | NA | 9 | 2 | 0 | Low-quality | Genome-fragment | 9.01 | HMM-based (lower-bound) | 0 |
| S6-final-viral-combined_k141_44205\|\|full | 3425 | No | NA | 6 | 0 | 1 | Not-determined | Genome-fragment | NA | NA | 0 |
| S6-final-viral-combined_k141_14359\|\|full | 6154 | No | NA | 5 | 0 | 3 | Not-determined | Genome-fragment | NA | NA | 0 |
| S6-final-viral-combined_k141_14367\|\|full | 1129 | No | NA | 3 | 0 | 1 | Not-determined | Genome-fragment | NA | NA | 0 |
| S6-final-viral-combined_k141_108363\|\|full | 1769 | No | NA | 2 | 0 | 1 | Not-determined | Genome-fragment | NA | NA | 0 |
| S6-final-viral-combined_k141_113986\|\|full | 34147 | No | NA | 55 | 14 | 0 | Medium-quality | Genome-fragment | 62.23 | AAI-based (high-confidence) | 0 |

| | | | | | | | | | | | |
|---|---|---|---|---|---|---|---|---|---|---|---|
| S2-final-viral-combined_k141_230588\|\|full | 6926 | No | NA | 13 | 1 | 1 | Low-quality | Genome-fragment | 3.72 | HMM-based (lower-bound) | 0 |
| S6-final-viral-combined_k141_44235\|\|full | 1347 | No | NA | 4 | 0 | 1 | Not-determined | Genome-fragment | NA | NA | 0 |
| S2-final-viral-combined_k141_192346\|\|full | 6926 | No | NA | 7 | 1 | 1 | Low-quality | Genome-fragment | 9.59 | AAI-based (medium-confidence) | 0 |
| S6-final-viral-combined_k141_3584\|\|full | 2684 | No | NA | 3 | 0 | 1 | Not-determined | Genome-fragment | NA | NA | 0 |
| S6-final-viral-combined_k141_73150\|\|full | 1847 | No | NA | 2 | 0 | 1 | Not-determined | Genome-fragment | NA | NA | 0 |
| S6-final-viral-combined_k141_67663\|\|full | 4454 | No | NA | 5 | 0 | 4 | Not-determined | Genome-fragment | NA | NA | 0 |
| S6-final-viral-combined_k141_44276\|\|full | 1952 | No | NA | 3 | 0 | 1 | Not-determined | Genome-fragment | NA | NA | 0 |
| S6-final-viral-combined_k141_38224\|\|full | 1947 | No | NA | 3 | 0 | 1 | Not-determined | Genome-fragment | NA | NA | 0 |
| S6-final-viral-combined_k141_32221\|\|full | 2661 | No | NA | 2 | 0 | 1 | Not-determined | Genome-fragment | NA | NA | 0 |
| S17-final-viral-combined_k141_212390\|\|full | 6930 | No | NA | 12 | 1 | 0 | Low-quality | Genome-fragment | 4.19 | HMM-based (lower-bound) | 0 |
| S6-final-viral-combined_k141_96982\|\|full | 1657 | No | NA | 4 | 0 | 0 | Not-determined | Genome-fragment | NA | NA | 0 |
| S6-final-viral-combined_k141_32236\|\|full | 2685 | No | NA | 3 | 0 | 2 | Not-determined | Genome-fragment | NA | NA | 0 |
| S6-final-viral-combined_k141_50048\|\|full | 1022 | No | NA | 2 | 0 | 0 | Not-determined | Genome-fragment | NA | NA | 0 |
| S6-final-viral-combined_k141_3634\|\|full | 1341 | No | NA | 3 | 0 | 1 | Not-determined | Genome-fragment | NA | NA | 0 |
| S12-final-viral-combined_k141_382004\|\|full | 6931 | No | NA | 13 | 4 | 1 | Low-quality | Genome-fragment | 8.9 | AAI-based (high-confidence) | 0 |
| S6-final-viral-combined_k141_102018\|\|full | 1384 | No | NA | 2 | 0 | 1 | Not-determined | Genome-fragment | NA | NA | 0 |
| S6-final-viral-combined_k141_14441\|\|full | 1780 | No | NA | 1 | 0 | 1 | Not-determined | Genome-fragment | NA | NA | 0 |
| S2-final-viral-combined_k141_122813\|\|full | 6931 | No | NA | 7 | 1 | 0 | Low-quality | Genome-fragment | 16.23 | HMM-based (lower-bound) | 0 |
| S6-final-viral-combined_k141_67746\|\|full | 1736 | No | NA | 4 | 0 | 4 | Not-determined | Genome-fragment | NA | NA | 0 |
| S12-final-viral-combined_k141_53993\|\|full | 6936 | No | NA | 11 | 1 | 4 | Low-quality | Genome-fragment | 12.64 | AAI-based (high-confidence) | 0 |
| S6-final-viral-combined_k141_32284\|\|full | 2298 | No | NA | 4 | 0 | 1 | Not-determined | Genome-fragment | NA | NA | 0 |
| S8-final-viral-combined_k141_275616\|\|full | 6936 | No | NA | 14 | 1 | 0 | Low-quality | Genome-fragment | 10.81 | HMM-based (lower-bound) | 0 |
| S6-final-viral-combined_k141_137438\|\|full | 1391 | No | NA | 2 | 0 | 1 | Not-determined | Genome-fragment | NA | NA | 0 |
| S6-final-viral-combined_k141_61699\|\|full | 7934 | No | NA | 10 | 0 | 3 | Not-determined | Genome-fragment | NA | NA | 0 |
| S3-final-viral-combined_k141_24948\|\|full | 6937 | No | NA | 5 | 0 | 2 | Low-quality | Genome-fragment | 4.66 | AAI-based (medium-confidence) | 0 |
| S6-final-viral-combined_k141_131860\|\|full | 7039 | No | NA | 6 | 0 | 3 | Not-determined | Genome-fragment | NA | NA | 0 |
| S2-final-viral-combined_k141_242259\|\|full | 6938 | No | NA | 9 | 8 | 0 | Low-quality | Genome-fragment | 11.39 | AAI-based (high-confidence) | 0 |
| S6-final-viral-combined_k141_10472\|\|full | 6939 | No | NA | 13 | 4 | 1 | Low-quality | Genome-fragment | 16.29 | AAI-based (high-confidence) | 0 |
| S6-final-viral-combined_k141_102116\|\|full | 3173 | No | NA | 5 | 0 | 0 | Not-determined | Genome-fragment | NA | NA | 0 |
| S6-final-viral-combined_k141_73260\|\|full | 1709 | No | NA | 2 | 0 | 2 | Not-determined | Genome-fragment | NA | NA | 0 |
| S6-final-viral-combined_k141_119713\|\|full | 1476 | No | NA | 2 | 0 | 0 | Not-determined | Genome-fragment | NA | NA | 0 |
| S12-final-viral-combined_k141_270561\|\|full | 6945 | No | NA | 7 | 0 | 0 | Low-quality | Genome-fragment | 11.86 | AAI-based (medium-confidence) | 0 |
| S6-final-viral-combined_k141_55090\|\|full | 1890 | No | NA | 6 | 0 | 5 | Not-determined | Genome-fragment | NA | NA | 0 |
| S6-final-viral-combined_k141_8452\|\|full | 1901 | No | NA | 2 | 0 | 1 | Not-determined | Genome-fragment | NA | NA | 0 |
| S11-final-viral-combined_k141_322692\|\|full | 6952 | Yes | 3320 | 9 | 3 | 3 | Low-quality | Genome-fragment | 8.53 | AAI-based (high-confidence) | 52.24 |
| S6-final-viral-combined_k141_97147\|\|full | 1485 | No | NA | 3 | 0 | 0 | Not-determined | Genome-fragment | NA | NA | 0 |
| S12_k141_122504 | 6953 | No | NA | 12 | 9 | 1 | Low-quality | Genome-fragment | 8.65 | AAI-based (high-confidence) | 0 |
| S2-final-viral-combined_k141_40628\|\|full | 6956 | No | NA | 8 | 0 | 5 | Low-quality | Genome-fragment | 1.74 | AAI-based (medium-confidence) | 0 |
| S5-final-viral-combined_k141_7254\|\|full | 6963 | No | NA | 12 | 0 | 4 | Low-quality | Genome-fragment | 10.42 | AAI-based (high-confidence) | 0 |
| S6-final-viral-combined_k141_32441\|\|full | 1425 | No | NA | 2 | 0 | 0 | Not-determined | Genome-fragment | NA | NA | 0 |
| S6-final-viral-combined_k141_84887\|\|full | 1612 | No | NA | 2 | 0 | 0 | Not-determined | Genome-fragment | NA | NA | 0 |
| S6-final-viral-combined_k141_97256\|\|full | 2854 | No | NA | 3 | 0 | 2 | Not-determined | Genome-fragment | NA | NA | 0 |
| S6-final-viral-combined_k141_26037\|\|full | 1947 | No | NA | 4 | 0 | 0 | Not-determined | Genome-fragment | NA | NA | 0 |
| S6-final-viral-combined_k141_55210\|\|full | 1992 | No | NA | 3 | 0 | 0 | Not-determined | Genome-fragment | NA | NA | 0 |
| S6-final-viral-combined_k141_8508\|\|full | 87084 | Yes | 32441 | 109 | 16 | 36 | Medium-quality | Genome-fragment | 72.28 | AAI-based (high-confidence) | 62.75 |
| S2-final-viral-combined_k141_150293\|\|full | 6964 | No | NA | 15 | 3 | 0 | Low-quality | Genome-fragment | 17.77 | AAI-based (medium-confidence) | 0 |
| S12-final-viral-combined_k141_249540\|\|full | 6966 | No | NA | 15 | 2 | 1 | Low-quality | Genome-fragment | 11.39 | AAI-based (high-confidence) | 0 |
| S6-final-viral-combined_k141_97296\|\|full | 1133 | No | NA | 3 | 0 | 1 | Not-determined | Genome-fragment | NA | NA | 0 |
| S6-final-viral-combined_k141_32506\|\|full | 1308 | No | NA | 4 | 0 | 0 | Not-determined | Genome-fragment | NA | NA | 0 |
| S12-final-viral-combined_k141_200841\|\|full | 6966 | No | NA | 13 | 0 | 0 | Low-quality | Genome-fragment | 19.2 | AAI-based (high-confidence) | 0 |
| S7-final-viral-combined_k141_70365\|\|full | 6968 | No | NA | 11 | 3 | 0 | Low-quality | Genome-fragment | 14.83 | AAI-based (high-confidence) | 0 |
| S6-final-viral-combined_k141_3949\|\|full | 7982 | No | NA | 6 | 0 | 5 | Not-determined | Genome-fragment | NA | NA | 0 |
| S6-final-viral-combined_k141_102269\|\|full | 6962 | No | NA | 6 | 0 | 4 | Not-determined | Genome-fragment | NA | NA | 0 |
| S6-final-viral-combined_k141_3967\|\|full | 2055 | No | NA | 2 | 0 | 1 | Not-determined | Genome-fragment | NA | NA | 0 |
| S12-final-viral-combined_k141_38315\|\|full | 6970 | Yes | 3939 | 9 | 1 | 4 | Low-quality | Genome-fragment | 3.53 | HMM-based (lower-bound) | 43.49 |
| S6-final-viral-combined_k141_44531\|\|full | 1929 | No | NA | 3 | 0 | 0 | Not-determined | Genome-fragment | NA | NA | 0 |
| S6-final-viral-combined_k141_108787\|\|full | 2989 | No | NA | 5 | 0 | 1 | Not-determined | Genome-fragment | NA | NA | 0 |
| S6-final-viral-combined_k141_38499\|\|full | 1646 | No | NA | 3 | 0 | 1 | Not-determined | Genome-fragment | NA | NA | 0 |
| S6-final-viral-combined_k141_55298\|\|full | 5220 | No | NA | 8 | 1 | 0 | Not-determined | Genome-fragment | NA | NA | 0 |
| S9-final-viral-combined_k141_81091\|\|full | 6971 | No | NA | 8 | 7 | 0 | Low-quality | Genome-fragment | 14.97 | AAI-based (high-confidence) | 0 |
| S6-final-viral-combined_k141_32530\|\|full | 46060 | Yes | 33901 | 64 | 17 | 4 | Medium-quality | Genome-fragment | 59.41 | HMM-based (lower-bound) | 26.4 |
| S6-final-viral-combined_k141_26151\|\|full | 2380 | No | NA | 3 | 0 | 2 | Not-determined | Genome-fragment | NA | NA | 0 |
| S6-final-viral-combined_k141_126236\|\|full | 69368 | Yes | 38859 | 79 | 18 | 27 | Medium-quality | Genome-fragment | 74.68 | AAI-based (high-confidence) | 43.98 |
| S9-final-viral-combined_k141_48156\|\|full | 6974 | No | NA | 5 | 0 | 5 | Low-quality | Genome-fragment | 18.21 | AAI-based (medium-confidence) | 0 |
| S6-final-viral-combined_k141_102339\|\|full | 1926 | No | NA | 6 | 0 | 0 | Not-determined | Genome-fragment | NA | NA | 0 |
| S6-final-viral-combined_k141_20089\|\|full | 1491 | No | NA | 2 | 0 | 2 | Not-determined | Genome-fragment | NA | NA | 0 |
| S1-final-viral-combined_k141_280621\|\|full | 6975 | Yes | 5424 | 9 | 1 | 1 | Low-quality | Genome-fragment | 4.35 | HMM-based (lower-bound) | 22.24 |
| S2_k141_145306 | 6982 | No | NA | 6 | 0 | 2 | Low-quality | Genome-fragment | 6.49 | AAI-based (high-confidence) | 0 |
| S6_k141_57955 | 7001 | No | NA | 3 | 1 | 0 | Low-quality | Genome-fragment | 5.03 | AAI-based (medium-confidence) | 0 |
| S9-final-viral-combined_k141_43275\|\|full | 7002 | No | NA | 10 | 5 | 0 | Low-quality | Genome-fragment | 14.95 | AAI-based (high-confidence) | 0 |
| S6-final-viral-combined_k141_132139\|\|full | 3067 | No | NA | 5 | 0 | 0 | Not-determined | Genome-fragment | NA | NA | 0 |
| S6-final-viral-combined_k141_85070\|\|full | 4529 | No | NA | 5 | 0 | 3 | Not-determined | Genome-fragment | NA | NA | 0 |
| S7-final-viral-combined_k141_227400\|\|full | 7003 | No | NA | 15 | 2 | 0 | Low-quality | Genome-fragment | 14.17 | AAI-based (medium-confidence) | 0 |
| S6-final-viral-combined_k141_4097\|\|full | 7426 | No | NA | 6 | 0 | 6 | Not-determined | Genome-fragment | NA | NA | 0 |
| S8_k141_56441 | 7006 | No | NA | 12 | 3 | 0 | Low-quality | Genome-fragment | 7.42 | AAI-based (high-confidence) | 0 |
| S11_k141_314598 | 7007 | No | NA | 10 | 3 | 0 | Low-quality | Genome-fragment | 14.51 | AAI-based (high-confidence) | 0 |
| S6-final-viral-combined_k141_108928\|\|full | 2103 | No | NA | 4 | 0 | 0 | Not-determined | Genome-fragment | NA | NA | 0 |
| S12_k141_174158 | 7014 | No | NA | 17 | 0 | 0 | Low-quality | Genome-fragment | 11.63 | AAI-based (high-confidence) | 0 |
| S6-final-viral-combined_k141_97502\|\|full | 3487 | No | NA | 4 | 0 | 0 | Not-determined | Genome-fragment | NA | NA | 0 |
| S11-final-viral-combined_k141_266557\|\|full | 7016 | No | NA | 10 | 7 | 0 | Low-quality | Genome-fragment | 15.92 | AAI-based (high-confidence) | 0 |
| S6-final-viral-combined_k141_120043\|\|full | 5603 | No | NA | 6 | 0 | 5 | Not-determined | Genome-fragment | NA | NA | 0 |
| S6-final-viral-combined_k141_97515\|\|full | 2516 | No | NA | 5 | 0 | 2 | Not-determined | Genome-fragment | NA | NA | 0 |
| S11-final-viral-combined_k141_187585\|\|full | 7016 | No | NA | 5 | 0 | 2 | Low-quality | Genome-fragment | 2.88 | AAI-based (medium-confidence) | 0 |
| S3-final-viral-combined_k141_92639\|\|full | 7016 | No | NA | 6 | 0 | 4 | Low-quality | Genome-fragment | 6.09 | AAI-based (medium-confidence) | 0 |
| S6-final-viral-combined_k141_26237\|\|full | 2250 | No | NA | 3 | 0 | 1 | Not-determined | Genome-fragment | NA | NA | 0 |
| S6-final-viral-combined_k141_32632\|\|full | 2655 | No | NA | 3 | 0 | 3 | Not-determined | Genome-fragment | NA | NA | 0 |
| S6-final-viral-combined_k141_20199\|\|full | 3617 | No | NA | 4 | 0 | 4 | Not-determined | Genome-fragment | NA | NA | 0 |
| S6-final-viral-combined_k141_97544\|\|full | 4509 | No | NA | 4 | 0 | 0 | Not-determined | Genome-fragment | NA | NA | 0 |
| S6-final-viral-combined_k141_132260\|\|full | 1391 | No | NA | 2 | 0 | 2 | Not-determined | Genome-fragment | NA | NA | 0 |
| S6-final-viral-combined_k141_120066\|\|full | 1151 | No | NA | 2 | 0 | 1 | Not-determined | Genome-fragment | NA | NA | 0 |
| S6-final-viral-combined_k141_73621\|\|full | 1567 | No | NA | 2 | 0 | 2 | Not-determined | Genome-fragment | NA | NA | 0 |
| S6-final-viral-combined_k141_68229\|\|full | 2790 | No | NA | 3 | 0 | 1 | Not-determined | Genome-fragment | NA | NA | 0 |
| S6-final-viral-combined_k141_62139\|\|full | 1102 | No | NA | 2 | 0 | 0 | Not-determined | Genome-fragment | NA | NA | 0 |
| S1-final-viral-combined_k141_354384\|\|full | 7018 | No | NA | 11 | 0 | 0 | Low-quality | Genome-fragment | 6.94 | AAI-based (high-confidence) | 0 |
| S6-final-viral-combined_k141_126482\|\|full | 7501 | No | NA | 7 | 0 | 6 | Not-determined | Genome-fragment | NA | NA | 0 |
| S12_k141_74627 | 7019 | No | NA | 8 | 1 | 0 | Low-quality | Genome-fragment | 10.62 | HMM-based (lower-bound) | 0 |
| S6-final-viral-combined_k141_62217\|\|full | 2701 | No | NA | 3 | 0 | 0 | Not-determined | Genome-fragment | NA | NA | 0 |
| S6-final-viral-combined_k141_44679\|\|full | 1405 | No | NA | 2 | 0 | 1 | Not-determined | Genome-fragment | NA | NA | 0 |
| S6-final-viral-combined_k141_137806\|\|full | 22096 | No | NA | 22 | 0 | 15 | Not-determined | Genome-fragment | NA | NA | 0 |
| S4-final-viral-combined_k141_98644\|\|full | 7019 | No | NA | 9 | 5 | 0 | Low-quality | Genome-fragment | 20.29 | AAI-based (high-confidence) | 0 |
| S4-final-viral-combined_k141_154655\|\|full | 7022 | No | NA | 5 | 0 | 3 | Low-quality | Genome-fragment | 7.73 | AAI-based (high-confidence) | 0 |
| S4_k141_138226 | 7025 | No | NA | 8 | 6 | 0 | Low-quality | Genome-fragment | 19.28 | AAI-based (high-confidence) | 0 |
| S6-final-viral-combined_k141_4275\|\|full | 2166 | No | NA | 3 | 0 | 3 | Not-determined | Genome-fragment | NA | NA | 0 |
| S6-final-viral-combined_k141_38778\|\|full | 1860 | No | NA | 2 | 0 | 0 | Not-determined | Genome-fragment | NA | NA | 0 |
| S6-final-viral-combined_k141_137846\|\|full | 2065 | No | NA | 2 | 0 | 2 | Not-determined | Genome-fragment | NA | NA | 0 |
| S17-final-viral-combined_k141_180981\|\|full | 7027 | No | NA | 7 | 6 | 0 | Low-quality | Genome-fragment | 21.42 | AAI-based (high-confidence) | 0 |
| S2_k141_156864 | 7027 | No | NA | 10 | 1 | 1 | Low-quality | Genome-fragment | 5.42 | AAI-based (high-confidence) | 0 |
| S6-final-viral-combined_k141_102644\|\|full | 1240 | No | NA | 3 | 0 | 0 | Not-determined | Genome-fragment | NA | NA | 0 |
| S6-final-viral-combined_k141_73674\|\|full | 3262 | No | NA | 3 | 0 | 3 | Not-determined | Genome-fragment | NA | NA | 0 |
| S4_k141_118842 | 7027 | No | NA | 9 | 3 | 0 | Low-quality | Genome-fragment | 4.05 | AAI-based (high-confidence) | 0 |
| S2-final-viral-combined_k141_202253\|\|full | 7028 | No | NA | 6 | 0 | 1 | Low-quality | Genome-fragment | 4.26 | AAI-based (high-confidence) | 0 |
| S6-final-viral-combined_k141_97616\|\|full | 4992 | No | NA | 7 | 0 | 5 | Not-determined | Genome-fragment | NA | NA | 0 |
| S6-final-viral-combined_k141_20369\|\|full | 6514 | No | NA | 5 | 0 | 2 | Not-determined | Genome-fragment | NA | NA | 0 |
| S12-final-viral-combined_k141_254754\|\|full | 7033 | No | NA | 8 | 1 | 4 | Low-quality | Genome-fragment | 3.74 | HMM-based (lower-bound) | 0 |
| S6-final-viral-combined_k141_126584\|\|full | 2854 | No | NA | 3 | 0 | 1 | Not-determined | Genome-fragment | NA | NA | 0 |
| S5-final-viral-combined_k141_104100\|\|full | 7034 | No | NA | 12 | 1 | 0 | Low-quality | Genome-fragment | 4.35 | HMM-based (lower-bound) | 0 |
| S6-final-viral-combined_k141_32743\|\|full | 1979 | No | NA | 3 | 0 | 2 | Not-determined | Genome-fragment | NA | NA | 0 |
| S6-final-viral-combined_k141_8845\|\|full | 1411 | No | NA | 2 | 0 | 0 | Not-determined | Genome-fragment | NA | NA | 0 |
| S6-final-viral-combined_k141_132360\|\|full | 3264 | No | NA | 3 | 0 | 1 | Not-determined | Genome-fragment | NA | NA | 0 |
| S6-final-viral-combined_k141_79276\|\|full | 1710 | No | NA | 3 | 0 | 2 | Not-determined | Genome-fragment | NA | NA | 0 |
| S5-final-viral-combined_k141_194204\|\|full | 7034 | No | NA | 9 | 0 | 0 | Low-quality | Genome-fragment | 12.08 | AAI-based (medium-confidence) | 0 |
| S1_k141_375254 | 7035 | No | NA | 11 | 5 | 0 | Low-quality | Genome-fragment | 18.85 | AAI-based (high-confidence) | 0 |
| S2-final-viral-combined_k141_158916\|\|full | 7038 | No | NA | 7 | 1 | 0 | Low-quality | Genome-fragment | 11.72 | AAI-based (high-confidence) | 0 |
| S6-final-viral-combined_k141_4408\|\|full | 2312 | No | NA | 3 | 0 | 2 | Not-determined | Genome-fragment | NA | NA | 0 |
| S6-final-viral-combined_k141_44770\|\|full | 2874 | No | NA | 3 | 0 | 2 | Not-determined | Genome-fragment | NA | NA | 0 |
| S2-final-viral-combined_k141_155751\|\|full | 7040 | No | NA | 14 | 1 | 1 | Low-quality | Genome-fragment | 1.85 | HMM-based (lower-bound) | 0 |
| S1-final-viral-combined_k141_225670\|\|full | 7041 | No | NA | 7 | 0 | 0 | Low-quality | Genome-fragment | 16.75 | AAI-based (high-confidence) | 0 |
| S6-final-viral-combined_k141_26481\|\|full | 1346 | No | NA | 3 | 0 | 1 | Not-determined | Genome-fragment | NA | NA | 0 |
| S6-final-viral-combined_k141_73732\|\|full | 6108 | No | NA | 7 | 0 | 5 | Not-determined | Genome-fragment | NA | NA | 0 |
| S17-final-viral-combined_k141_253340\|\|full | 7041 | No | NA | 5 | 3 | 0 | Low-quality | Genome-fragment | 8.55 | AAI-based (high-confidence) | 0 |
| S4-final-viral-combined_k141_105037\|\|full | 7044 | No | NA | 8 | 0 | 5 | Low-quality | Genome-fragment | 4.22 | AAI-based (medium-confidence) | 0 |
| S6-final-viral-combined_k141_137951\|\|full | 9331 | No | NA | 9 | 0 | 6 | Not-determined | Genome-fragment | NA | NA | 0 |
| S6-final-viral-combined_k141_62410\|\|full | 5522 | No | NA | 5 | 0 | 4 | Not-determined | Genome-fragment | NA | NA | 0 |
| S6-final-viral-combined_k141_132432\|\|full | 1877 | No | NA | 2 | 0 | 1 | Not-determined | Genome-fragment | NA | NA | 0 |
| S6-final-viral-combined_k141_91348\|\|full | 1516 | No | NA | 3 | 0 | 0 | Not-determined | Genome-fragment | NA | NA | 0 |
| S6-final-viral-combined_k141_50479\|\|full | 2004 | No | NA | 4 | 0 | 0 | Not-determined | Genome-fragment | NA | NA | 0 |
| S11-final-viral-combined_k141_243184\|\|full | 7047 | No | NA | 13 | 3 | 0 | Low-quality | Genome-fragment | 18.08 | AAI-based (high-confidence) | 0 |
| S6-final-viral-combined_k141_8931\|\|full | 1877 | No | NA | 3 | 0 | 2 | Not-determined | Genome-fragment | NA | NA | 0 |
| S6-final-viral-combined_k141_97686\|\|full | 1132 | No | NA | 3 | 0 | 1 | Not-determined | Genome-fragment | NA | NA | 0 |
| S1-final-viral-combined_k141_123711\|\|full | 7048 | No | NA | 11 | 1 | 1 | Low-quality | Genome-fragment | 3.83 | AAI-based (medium-confidence) | 0 |
| S6-final-viral-combined_k141_26523\|\|full | 1813 | No | NA | 3 | 0 | 0 | Not-determined | Genome-fragment | NA | NA | 0 |
| S6-final-viral-combined_k141_79354\|\|full | 6674 | No | NA | 5 | 0 | 4 | Not-determined | Genome-fragment | NA | NA | 0 |
| S6-final-viral-combined_k141_114754\|\|full | 4570 | No | NA | 6 | 0 | 1 | Not-determined | Genome-fragment | NA | NA | 0 |
| S6-final-viral-combined_k141_50513\|\|full | 2570 | No | NA | 2 | 0 | 1 | Not-determined | Genome-fragment | NA | NA | 0 |
| S12_k141_210447 | 7048 | No | NA | 8 | 0 | 2 | Low-quality | Genome-fragment | 6.22 | AAI-based (medium-confidence) | 0 |
| S5-final-viral-combined_k141_151552\|\|full | 7051 | No | NA | 10 | 0 | 2 | Low-quality | Genome-fragment | 46.23 | AAI-based (medium-confidence) | 0 |
| S8-final-viral-combined_k141_73280\|\|full | 7052 | No | NA | 13 | 1 | 0 | Low-quality | Genome-fragment | 12.25 | AAI-based (high-confidence) | 0 |
| S11-final-viral-combined_k141_121054\|\|full | 7061 | No | NA | 5 | 1 | 0 | Low-quality | Genome-fragment | 4.29 | AAI-based (high-confidence) | 0 |
| S6-final-viral-combined_k141_39003\|\|full | 3435 | No | NA | 3 | 0 | 2 | Not-determined | Genome-fragment | NA | NA | 0 |
| S6-final-viral-combined_k141_126668\|\|full | 1579 | No | NA | 4 | 0 | 0 | Not-determined | Genome-fragment | NA | NA | 0 |
| S6-final-viral-combined_k141_55639\|\|full | 1032 | No | NA | 2 | 0 | 0 | Not-determined | Genome-fragment | NA | NA | 0 |
| S6-final-viral-combined_k141_114792\|\|full | 1036 | No | NA | 2 | 0 | 0 | Not-determined | Genome-fragment | NA | NA | 0 |
| S6-final-viral-combined_k141_73805\|\|full | 10643 | No | NA | 6 | 0 | 6 | Not-determined | Genome-fragment | NA | NA | 0 |
| S11_k141_174584 | 7063 | No | NA | 1 | 0 | 0 | Low-quality | Genome-fragment | 7.51 | AAI-based (high-confidence) | 0 |
| S6-final-viral-combined_k141_73808\|\|full | 1268 | No | NA | 2 | 0 | 1 | Not-determined | Genome-fragment | NA | NA | 0 |
| S8-final-viral-combined_k141_210045\|\|full | 7063 | No | NA | 12 | 8 | 0 | Low-quality | Genome-fragment | 20.26 | AAI-based (high-confidence) | 0 |
| S11_k141_174285 | 7066 | No | NA | 15 | 5 | 0 | Low-quality | Genome-fragment | 19.53 | AAI-based (high-confidence) | 0 |
| S6-final-viral-combined_k141_55675\|\|full | 3054 | No | NA | 3 | 0 | 2 | Not-determined | Genome-fragment | NA | NA | 0 |
| S6-final-viral-combined_k141_9002\|\|full | 1614 | No | NA | 3 | 0 | 2 | Not-determined | Genome-fragment | NA | NA | 0 |
| S12-final-viral-combined_k141_363300\|\|full | 7073 | No | NA | 7 | 0 | 0 | Low-quality | Genome-fragment | 11.53 | AAI-based (medium-confidence) | 0 |
| S11-final-viral-combined_k141_328345\|\|full | 7074 | No | NA | 14 | 3 | 0 | Low-quality | Genome-fragment | 13.87 | AAI-based (high-confidence) | 0 |
| S5-final-viral-combined_k141_153553\|\|full | 7076 | No | NA | 7 | 0 | 1 | Low-quality | Genome-fragment | 14.01 | AAI-based (medium-confidence) | 0 |
| S6-final-viral-combined_k141_79421\|\|full | 6508 | No | NA | 4 | 0 | 2 | Not-determined | Genome-fragment | NA | NA | 0 |
| S6-final-viral-combined_k141_97800\|\|full | 2347 | No | NA | 3 | 0 | 1 | Not-determined | Genome-fragment | NA | NA | 0 |
| S6-final-viral-combined_k141_73845\|\|full | 1485 | No | NA | 3 | 0 | 3 | Not-determined | Genome-fragment | NA | NA | 0 |
| S6-final-viral-combined_k141_102837\|\|full | 1357 | No | NA | 3 | 0 | 1 | Not-determined | Genome-fragment | NA | NA | 0 |
| S6-final-viral-combined_k141_97810\|\|full | 2703 | No | NA | 3 | 0 | 0 | Not-determined | Genome-fragment | NA | NA | 0 |
| S12-final-viral-combined_k141_18041\|\|full | 7076 | No | NA | 11 | 2 | 0 | Low-quality | Genome-fragment | 18.03 | AAI-based (medium-confidence) | 0 |
| S6-final-viral-combined_k141_102854\|\|full | 2618 | No | NA | 4 | 0 | 0 | Not-determined | Genome-fragment | NA | NA | 0 |
| S5-final-viral-combined_k141_44631\|\|full | 7079 | No | NA | 10 | 1 | 0 | Low-quality | Genome-fragment | 9.68 | HMM-based (lower-bound) | 0 |
| S6-final-viral-combined_k141_85523\|\|full | 2204 | No | NA | 3 | 0 | 1 | Not-determined | Genome-fragment | NA | NA | 0 |
| S11-final-viral-combined_k141_27829\|\|full | 7080 | No | NA | 13 | 4 | 0 | Low-quality | Genome-fragment | 8.02 | HMM-based (lower-bound) | 0 |
| S6-final-viral-combined_k141_102872\|\|full | 1607 | No | NA | 3 | 0 | 0 | Not-determined | Genome-fragment | NA | NA | 0 |
| S6-final-viral-combined_k141_85550\|\|full | 1730 | No | NA | 2 | 0 | 2 | Not-determined | Genome-fragment | NA | NA | 0 |
| S6-final-viral-combined_k141_79432\|\|full | 3068 | No | NA | 3 | 0 | 2 | Not-determined | Genome-fragment | NA | NA | 0 |
| S6-final-viral-combined_k141_62504\|\|full | 2251 | No | NA | 4 | 0 | 1 | Not-determined | Genome-fragment | NA | NA | 0 |
| S17-final-viral-combined_k141_223404\|\|full | 7082 | No | NA | 4 | 0 | 2 | Low-quality | Genome-fragment | 3.81 | AAI-based (medium-confidence) | 0 |
| S12_k141_158830 | 7086 | No | NA | 16 | 0 | 0 | Low-quality | Genome-fragment | 11.67 | AAI-based (high-confidence) | 0 |
| S11-final-viral-combined_k141_135601\|\|full | 7090 | No | NA | 16 | 5 | 0 | Low-quality | Genome-fragment | 20.87 | AAI-based (high-confidence) | 0 |
| S6-final-viral-combined_k141_132608\|\|full | 1153 | No | NA | 2 | 0 | 1 | Not-determined | Genome-fragment | NA | NA | 0 |
| S12-final-viral-combined_k141_162562\|\|full | 7090 | No | NA | 5 | 0 | 4 | Low-quality | Genome-fragment | 1.61 | AAI-based (medium-confidence) | 0 |
| S6-final-viral-combined_k141_62576\|\|full | 6144 | No | NA | 7 | 0 | 2 | Not-determined | Genome-fragment | NA | NA | 0 |
| S9_k141_79042 | 7093 | No | NA | 11 | 0 | 0 | Low-quality | Genome-fragment | 7.49 | AAI-based (medium-confidence) | 0 |
| S6-final-viral-combined_k141_79497\|\|full | 2460 | No | NA | 5 | 0 | 0 | Not-determined | Genome-fragment | NA | NA | 0 |
| S6-final-viral-combined_k141_120461\|\|full | 1782 | No | NA | 3 | 0 | 1 | Not-determined | Genome-fragment | NA | NA | 0 |
| S1-final-viral-combined_k141_33448\|\|full | 7095 | No | NA | 9 | 4 | 0 | Low-quality | Genome-fragment | 14.61 | AAI-based (high-confidence) | 0 |
| S7-final-viral-combined_k141_242340\|\|full | 7095 | No | NA | 6 | 3 | 0 | Low-quality | Genome-fragment | 17.07 | AAI-based (high-confidence) | 0 |
| S6-final-viral-combined_k141_120472\|\|full | 2936 | No | NA | 4 | 0 | 3 | Not-determined | Genome-fragment | NA | NA | 0 |
| S6-final-viral-combined_k141_45088\|\|full | 3276 | No | NA | 4 | 0 | 3 | Not-determined | Genome-fragment | NA | NA | 0 |
| S6-final-viral-combined_k141_79544\|\|full | 1519 | No | NA | 2 | 0 | 0 | Not-determined | Genome-fragment | NA | NA | 0 |
| S7-final-viral-combined_k141_3979\|\|full | 7096 | No | NA | 10 | 0 | 7 | Low-quality | Genome-fragment | 5.7 | AAI-based (medium-confidence) | 0 |
| S1_k141_22097 | 7101 | No | NA | 6 | 0 | 6 | Low-quality | Genome-fragment | 4.21 | AAI-based (medium-confidence) | 0 |
| S6-final-viral-combined_k141_45111\|\|full | 2570 | No | NA | 2 | 0 | 1 | Not-determined | Genome-fragment | NA | NA | 0 |
| S1_k141_387868 | 7101 | No | NA | 10 | 0 | 3 | Low-quality | Genome-fragment | 18.8 | AAI-based (medium-confidence) | 0 |
| S6-final-viral-combined_k141_62660\|\|full | 1198 | No | NA | 2 | 0 | 0 | Not-determined | Genome-fragment | NA | NA | 0 |
| S6-final-viral-combined_k141_91631\|\|full | 1137 | No | NA | 2 | 0 | 2 | Not-determined | Genome-fragment | NA | NA | 0 |
| S6-final-viral-combined_k141_91632\|\|full | 3391 | No | NA | 4 | 0 | 1 | Not-determined | Genome-fragment | NA | NA | 0 |
| S6-final-viral-combined_k141_68693\|\|full | 1272 | No | NA | 2 | 0 | 0 | Not-determined | Genome-fragment | NA | NA | 0 |
| S11_k141_263316 | 7111 | No | NA | 6 | 4 | 0 | Low-quality | Genome-fragment | 19.79 | AAI-based (high-confidence) | 0 |
| S6-final-viral-combined_k141_4709\|\|full | 1883 | No | NA | 2 | 0 | 2 | Not-determined | Genome-fragment | NA | NA | 0 |
| S6-final-viral-combined_k141_26770\|\|full | 1350 | No | NA | 2 | 0 | 2 | Not-determined | Genome-fragment | NA | NA | 0 |
| S6-final-viral-combined_k141_85702\|\|full | 5737 | No | NA | 5 | 0 | 4 | Not-determined | Genome-fragment | NA | NA | 0 |
| S6-final-viral-combined_k141_55890\|\|full | 2357 | No | NA | 2 | 0 | 0 | Not-determined | Genome-fragment | NA | NA | 0 |
| S6-final-viral-combined_k141_114971\|\|full | 2427 | No | NA | 6 | 0 | 0 | Not-determined | Genome-fragment | NA | NA | 0 |
| S1-final-viral-combined_k141_120673\|\|full | 7112 | No | NA | 13 | 1 | 1 | Low-quality | Genome-fragment | 11.14 | HMM-based (lower-bound) | 0 |
| S6-final-viral-combined_k141_26829\|\|full | 2056 | No | NA | 4 | 0 | 1 | Not-determined | Genome-fragment | NA | NA | 0 |
| S6-final-viral-combined_k141_91708\|\|full | 2223 | No | NA | 2 | 0 | 1 | Not-determined | Genome-fragment | NA | NA | 0 |
| S7-final-viral-combined_k141_187556\|\|full | 7115 | No | NA | 10 | 7 | 0 | Low-quality | Genome-fragment | 17.57 | AAI-based (high-confidence) | 0 |
| S6-final-viral-combined_k141_50721\|\|full | 132102 | Yes | 37019 | 132 | 15 | 61 | Medium-quality | Genome-fragment | 63.84 | HMM-based (lower-bound) | 71.98 |
| S6-final-viral-combined_k141_91711\|\|full | 1089 | No | NA | 3 | 0 | 1 | Not-determined | Genome-fragment | NA | NA | 0 |
| S8-final-viral-combined_k141_77045\|\|full | 7115 | No | NA | 8 | 2 | 0 | Low-quality | Genome-fragment | 18.4 | AAI-based (high-confidence) | 0 |
| S11_k141_131843 | 7121 | No | NA | 18 | 3 | 0 | Low-quality | Genome-fragment | 11.03 | AAI-based (high-confidence) | 0 |
| S12-final-viral-combined_k141_58028\|\|full | 7121 | No | NA | 7 | 1 | 0 | Low-quality | Genome-fragment | 16.72 | AAI-based (high-confidence) | 0 |
| S5_k141_87863 | 7123 | No | NA | 19 | 0 | 0 | Low-quality | Genome-fragment | 4.62 | AAI-based (high-confidence) | 0 |
| S1-final-viral-combined_k141_225074\|\|full | 7130 | No | NA | 9 | 4 | 0 | Low-quality | Genome-fragment | 15.57 | AAI-based (high-confidence) | 0 |
| S6-final-viral-combined_k141_91768\|\|full | 3878 | No | NA | 4 | 0 | 2 | Not-determined | Genome-fragment | NA | NA | 0 |
| S11-final-viral-combined_k141_132442\|\|full | 7133 | No | NA | 10 | 2 | 0 | Low-quality | Genome-fragment | 16.8 | AAI-based (medium-confidence) | 0 |
| S6-final-viral-combined_k141_115058\|\|full | 1278 | No | NA | 3 | 0 | 2 | Not-determined | Genome-fragment | NA | NA | 0 |
| S6-final-viral-combined_k141_79664\|\|full | 1027 | No | NA | 2 | 0 | 2 | Not-determined | Genome-fragment | NA | NA | 0 |
| S12-final-viral-combined_k141_357360\|\|full | 7134 | No | NA | 7 | 0 | 0 | Low-quality | Genome-fragment | 12.81 | AAI-based (high-confidence) | 0 |
| S7_k141_108466 | 7139 | No | NA | 7 | 0 | 6 | Low-quality | Genome-fragment | 2.11 | AAI-based (medium-confidence) | 0 |
| S6-final-viral-combined_k141_85854\|\|full | 2102 | No | NA | 2 | 0 | 1 | Not-determined | Genome-fragment | NA | NA | 0 |
| S6-final-viral-combined_k141_74023\|\|full | 1354 | No | NA | 2 | 0 | 2 | Not-determined | Genome-fragment | NA | NA | 0 |
| S6-final-viral-combined_k141_26952\|\|full | 3077 | No | NA | 4 | 0 | 3 | Not-determined | Genome-fragment | NA | NA | 0 |
| S6-final-viral-combined_k141_74027\|\|full | 1411 | No | NA | 2 | 0 | 2 | Not-determined | Genome-fragment | NA | NA | 0 |
| S6-final-viral-combined_k141_103108\|\|full | 2226 | No | NA | 4 | 0 | 1 | Not-determined | Genome-fragment | NA | NA | 0 |
| S6-final-viral-combined_k141_126936\|\|full | 1712 | No | NA | 3 | 0 | 0 | Not-determined | Genome-fragment | NA | NA | 0 |
| S12-final-viral-combined_k141_394230\|\|full | 7143 | No | NA | 10 | 1 | 0 | Low-quality | Genome-fragment | 17.31 | HMM-based (lower-bound) | 0 |
| S6-final-viral-combined_k141_85883\|\|full | 4360 | No | NA | 7 | 0 | 4 | Not-determined | Genome-fragment | NA | NA | 0 |
| S6_k141_25809 | 7143 | No | NA | 3 | 0 | 0 | Low-quality | Genome-fragment | 11.84 | AAI-based (medium-confidence) | 0 |
| S4-final-viral-combined_k141_31990\|\|full | 7144 | No | NA | 10 | 3 | 0 | Low-quality | Genome-fragment | 12.02 | AAI-based (high-confidence) | 0 |
| S1-final-viral-combined_k141_148159\|\|full | 7148 | No | NA | 8 | 3 | 0 | Low-quality | Genome-fragment | 12.59 | AAI-based (high-confidence) | 0 |
| S7-final-viral-combined_k141_191294\|\|full | 7149 | No | NA | 4 | 0 | 3 | Low-quality | Genome-fragment | 2.69 | AAI-based (medium-confidence) | 0 |
| S6-final-viral-combined_k141_26973\|\|full | 1352 | No | NA | 2 | 0 | 1 | Not-determined | Genome-fragment | NA | NA | 0 |
| S11-final-viral-combined_k141_210884\|\|full | 7151 | No | NA | 7 | 4 | 0 | Low-quality | Genome-fragment | 4.17 | AAI-based (medium-confidence) | 0 |
| S6-final-viral-combined_k141_115138\|\|full | 3599 | No | NA | 8 | 0 | 0 | Not-determined | Genome-fragment | NA | NA | 0 |
| S6-final-viral-combined_k141_62924\|\|full | 3424 | No | NA | 4 | 0 | 2 | Not-determined | Genome-fragment | NA | NA | 0 |
| S2_k141_5702 | 7151 | No | NA | 10 | 2 | 0 | Low-quality | Genome-fragment | 8.78 | AAI-based (high-confidence) | 0 |
| S6-final-viral-combined_k141_45284\|\|full | 1756 | No | NA | 3 | 0 | 0 | Not-determined | Genome-fragment | NA | NA | 0 |
| S6-final-viral-combined_k141_27001\|\|full | 3708 | No | NA | 4 | 0 | 3 | Not-determined | Genome-fragment | NA | NA | 0 |
| S11-final-viral-combined_k141_215631\|\|full | 7159 | No | NA | 8 | 1 | 0 | Low-quality | Genome-fragment | 3.23 | HMM-based (lower-bound) | 0 |
| S8-final-viral-combined_k141_252857\|\|full | 7163 | No | NA | 14 | 3 | 0 | Low-quality | Genome-fragment | 16.32 | AAI-based (medium-confidence) | 0 |
| S6-final-viral-combined_k141_103188\|\|full | 1685 | No | NA | 3 | 0 | 0 | Not-determined | Genome-fragment | NA | NA | 0 |
| S7-final-viral-combined_k141_235803\|\|full | 7167 | No | NA | 11 | 3 | 1 | Low-quality | Genome-fragment | 21.38 | AAI-based (high-confidence) | 0 |
| S6-final-viral-combined_k141_45287\|\|full | 12529 | No | NA | 10 | 0 | 8 | Not-determined | Genome-fragment | NA | NA | 0 |
| S6-final-viral-combined_k141_45289\|\|full | 2213 | No | NA | 2 | 0 | 0 | Not-determined | Genome-fragment | NA | NA | 0 |
| S6-final-viral-combined_k141_33068\|\|full | 1583 | No | NA | 2 | 0 | 2 | Not-determined | Genome-fragment | NA | NA | 0 |
| S6-final-viral-combined_k141_74117\|\|full | 1739 | No | NA | 3 | 0 | 1 | Not-determined | Genome-fragment | NA | NA | 0 |
| S6-final-viral-combined_k141_79776\|\|full | 3440 | No | NA | 4 | 0 | 2 | Not-determined | Genome-fragment | NA | NA | 0 |
| S6-final-viral-combined_k141_39320\|\|full | 1307 | No | NA | 2 | 0 | 0 | Not-determined | Genome-fragment | NA | NA | 0 |
| S6-final-viral-combined_k141_33080\|\|full | 8183 | No | NA | 6 | 0 | 4 | Not-determined | Genome-fragment | NA | NA | 0 |
| S6-final-viral-combined_k141_98201\|\|full | 17095 | No | NA | 19 | 0 | 11 | Not-determined | Genome-fragment | NA | NA | 0 |
| S5-final-viral-combined_k141_170073\|\|full | 7168 | No | NA | 9 | 5 | 0 | Low-quality | Genome-fragment | 17.58 | AAI-based (high-confidence) | 0 |
| S6_k141_94059 | 7170 | No | NA | 4 | 0 | 4 | Low-quality | Genome-fragment | 3.86 | AAI-based (medium-confidence) | 0 |
| S6-final-viral-combined_k141_109578\|\|full | 1274 | No | NA | 2 | 0 | 2 | Not-determined | Genome-fragment | NA | NA | 0 |
| S6-final-viral-combined_k141_79812\|\|full | 1445 | No | NA | 2 | 0 | 1 | Not-determined | Genome-fragment | NA | NA | 0 |
| S12_k141_229570 | 7172 | No | NA | 10 | 0 | 3 | Low-quality | Genome-fragment | 7.35 | AAI-based (medium-confidence) | 0 |
| S6-final-viral-combined_k141_103232\|\|full | 1461 | No | NA | 2 | 0 | 0 | Not-determined | Genome-fragment | NA | NA | 0 |
| S8-final-viral-combined_k141_149891\|\|full | 7177 | No | NA | 11 | 6 | 0 | Low-quality | Genome-fragment | 18.02 | AAI-based (medium-confidence) | 0 |
| S6-final-viral-combined_k141_33132\|\|full | 7192 | No | NA | 6 | 0 | 4 | Not-determined | Genome-fragment | NA | NA | 0 |
| S6-final-viral-combined_k141_15520\|\|full | 3038 | No | NA | 3 | 0 | 2 | Not-determined | Genome-fragment | NA | NA | 0 |
| S17-final-viral-combined_k141_44329\|\|full | 7178 | No | NA | 16 | 1 | 0 | Low-quality | Genome-fragment | 38.37 | HMM-based (lower-bound) | 0 |
| S11-final-viral-combined_k141_169750\|\|full | 7180 | No | NA | 5 | 2 | 0 | Low-quality | Genome-fragment | 15.72 | HMM-based (lower-bound) | 0 |
| S6-final-viral-combined_k141_103277\|\|full | 4316 | No | NA | 3 | 0 | 2 | Not-determined | Genome-fragment | NA | NA | 0 |
| S6_k141_118357 | 7180 | No | NA | 4 | 0 | 0 | Low-quality | Genome-fragment | 8.52 | AAI-based (high-confidence) | 0 |
| S7-final-viral-combined_k141_116042\|\|full | 7181 | No | NA | 7 | 0 | 6 | Low-quality | Genome-fragment | 5.75 | AAI-based (medium-confidence) | 0 |
| S6-final-viral-combined_k141_27114\|\|full | 1025 | No | NA | 1 | 0 | 0 | Not-determined | Genome-fragment | NA | NA | 0 |
| S6-final-viral-combined_k141_127060\|\|full | 2580 | No | NA | 4 | 0 | 1 | Not-determined | Genome-fragment | NA | NA | 0 |
| S6-final-viral-combined_k141_74204\|\|full | 1456 | No | NA | 2 | 0 | 1 | Not-determined | Genome-fragment | NA | NA | 0 |
| S6-final-viral-combined_k141_39459\|\|full | 2439 | No | NA | 3 | 0 | 2 | Not-determined | Genome-fragment | NA | NA | 0 |
| S17-final-viral-combined_k141_157676\|\|full | 7183 | No | NA | 13 | 3 | 0 | Low-quality | Genome-fragment | 15.88 | AAI-based (high-confidence) | 0 |
| S11-final-viral-combined_k141_157333\|\|full | 7185 | No | NA | 9 | 7 | 0 | Low-quality | Genome-fragment | 16.5 | AAI-based (high-confidence) | 0 |
| S6-final-viral-combined_k141_39484\|\|full | 2337 | No | NA | 2 | 0 | 0 | Not-determined | Genome-fragment | NA | NA | 0 |
| S6-final-viral-combined_k141_103335\|\|full | 3466 | No | NA | 3 | 0 | 1 | Not-determined | Genome-fragment | NA | NA | 0 |
| S6-final-viral-combined_k141_74245\|\|full | 7348 | No | NA | 5 | 0 | 4 | Not-determined | Genome-fragment | NA | NA | 0 |
| S6-final-viral-combined_k141_127118\|\|full | 1711 | No | NA | 3 | 0 | 0 | Not-determined | Genome-fragment | NA | NA | 0 |
| S6-final-viral-combined_k141_63180\|\|full | 1453 | No | NA | 3 | 0 | 1 | Not-determined | Genome-fragment | NA | NA | 0 |
| S6-final-viral-combined_k141_9462\|\|full | 1095 | No | NA | 3 | 0 | 1 | Not-determined | Genome-fragment | NA | NA | 0 |
| S6-final-viral-combined_k141_20943\|\|full | 14047 | No | NA | 11 | 0 | 5 | Not-determined | Genome-fragment | NA | NA | 0 |
| S6-final-viral-combined_k141_63201\|\|full | 4118 | No | NA | 3 | 0 | 2 | Not-determined | Genome-fragment | NA | NA | 0 |
| S6-final-viral-combined_k141_138523\|\|full | 1413 | No | NA | 2 | 0 | 1 | Not-determined | Genome-fragment | NA | NA | 0 |
| S1-final-viral-combined_k141_67298\|\|full | 7198 | No | NA | 5 | 0 | 0 | Low-quality | Genome-fragment | 12.99 | AAI-based (high-confidence) | 0 |
| S6-final-viral-combined_k141_103411\|\|full | 2540 | No | NA | 3 | 0 | 1 | Not-determined | Genome-fragment | NA | NA | 0 |
| S6-final-viral-combined_k141_9507\|\|full | 4557 | No | NA | 3 | 0 | 2 | Not-determined | Genome-fragment | NA | NA | 0 |
| S6-final-viral-combined_k141_33359\|\|full | 2153 | No | NA | 3 | 0 | 1 | Not-determined | Genome-fragment | NA | NA | 0 |
| S6-final-viral-combined_k141_63235\|\|full | 2258 | No | NA | 3 | 0 | 1 | Not-determined | Genome-fragment | NA | NA | 0 |
| S8-final-viral-combined_k141_225189\|\|full | 7199 | No | NA | 10 | 0 | 1 | Low-quality | Genome-fragment | 4.85 | AAI-based (high-confidence) | 0 |
| S1-final-viral-combined_k141_359862\|\|full | 7208 | No | NA | 7 | 0 | 0 | Low-quality | Genome-fragment | 17.33 | AAI-based (medium-confidence) | 0 |
| S2-final-viral-combined_k141_228233\|\|full | 7208 | No | NA | 6 | 0 | 5 | Low-quality | Genome-fragment | 3.58 | AAI-based (medium-confidence) | 0 |
| S6-final-viral-combined_k141_33406\|\|full | 1547 | No | NA | 3 | 0 | 2 | Not-determined | Genome-fragment | NA | NA | 0 |
| S17-final-viral-combined_k141_49931\|\|full | 7220 | No | NA | 12 | 0 | 0 | Low-quality | Genome-fragment | 17.9 | AAI-based (medium-confidence) | 0 |
| S12_k141_124338 | 7221 | No | NA | 17 | 2 | 1 | Low-quality | Genome-fragment | 22.24 | AAI-based (high-confidence) | 0 |
| S12_k141_393348 | 7221 | No | NA | 7 | 0 | 1 | Low-quality | Genome-fragment | 8.09 | AAI-based (medium-confidence) | 0 |
| S6-final-viral-combined_k141_33415\|\|full | 1076 | No | NA | 2 | 0 | 0 | Not-determined | Genome-fragment | NA | NA | 0 |
| S6-final-viral-combined_k141_120901\|\|full | 3294 | No | NA | 4 | 0 | 3 | Not-determined | Genome-fragment | NA | NA | 0 |
| S12_k141_216781 | 7221 | No | NA | 20 | 1 | 0 | Low-quality | Genome-fragment | 4.84 | HMM-based (lower-bound) | 0 |
| S6-final-viral-combined_k141_33427\|\|full | 4950 | No | NA | 5 | 0 | 3 | Not-determined | Genome-fragment | NA | NA | 0 |
| S12_k141_179926 | 7224 | No | NA | 12 | 2 | 0 | Low-quality | Genome-fragment | 4.49 | HMM-based (lower-bound) | 0 |
| S4-final-viral-combined_k141_33086\|\|full | 7225 | Yes | 4760 | 9 | 2 | 1 | Low-quality | Genome-fragment | 5.4 | HMM-based (lower-bound) | 34.12 |
| S6-final-viral-combined_k141_9595\|\|full | 1290 | No | NA | 2 | 0 | 0 | Not-determined | Genome-fragment | NA | NA | 0 |
| S1_k141_378559 | 7228 | Yes | 957 | 6 | 1 | 5 | Low-quality | Genome-fragment | 0.58 | HMM-based (lower-bound) | 86.76 |
| S1_k141_55245 | 7231 | No | NA | 9 | 1 | 0 | Low-quality | Genome-fragment | 5.34 | AAI-based (high-confidence) | 0 |
| S6-final-viral-combined_k141_109709\|\|full | 1055 | No | NA | 3 | 0 | 0 | Not-determined | Genome-fragment | NA | NA | 0 |
| S6-final-viral-combined_k141_79976\|\|full | 2203 | No | NA | 4 | 0 | 1 | Not-determined | Genome-fragment | NA | NA | 0 |
| S11-final-viral-combined_k141_179909\|\|full | 7233 | No | NA | 9 | 1 | 0 | Low-quality | Genome-fragment | 10.94 | HMM-based (lower-bound) | 0 |
| S6-final-viral-combined_k141_33479\|\|full | 1496 | No | NA | 3 | 0 | 1 | Not-determined | Genome-fragment | NA | NA | 0 |
| S1_k141_277486 | 7237 | No | NA | 4 | 1 | 3 | Low-quality | Genome-fragment | 4.85 | HMM-based (lower-bound) | 0 |
| S5-final-viral-combined_k141_128924\|\|full | 7240 | No | NA | 6 | 2 | 0 | Low-quality | Genome-fragment | 8.12 | AAI-based (high-confidence) | 0 |
| S6-final-viral-combined_k141_69103\|\|full | 9972 | No | NA | 10 | 0 | 6 | Not-determined | Genome-fragment | NA | NA | 0 |
| S7-final-viral-combined_k141_223650\|\|full | 7242 | Yes | 4918 | 6 | 2 | 1 | Low-quality | Genome-fragment | 3.26 | AAI-based (high-confidence) | 32.09 |
| S8-final-viral-combined_k141_94508\|\|full | 7250 | No | NA | 11 | 9 | 0 | Low-quality | Genome-fragment | 18.14 | AAI-based (medium-confidence) | 0 |
| S6-final-viral-combined_k141_39668\|\|full | 1924 | No | NA | 2 | 0 | 1 | Not-determined | Genome-fragment | NA | NA | 0 |
| S6-final-viral-combined_k141_98488\|\|full | 2240 | No | NA | 3 | 0 | 2 | Not-determined | Genome-fragment | NA | NA | 0 |
| S6-final-viral-combined_k141_109778\|\|full | 1625 | No | NA | 2 | 0 | 0 | Not-determined | Genome-fragment | NA | NA | 0 |
| S5_k141_204367 | 7253 | No | NA | 9 | 8 | 0 | Low-quality | Genome-fragment | 17.25 | AAI-based (high-confidence) | 0 |
| S6-final-viral-combined_k141_21011\|\|full | 2973 | No | NA | 3 | 0 | 2 | Not-determined | Genome-fragment | NA | NA | 0 |
| S6-final-viral-combined_k141_5419\|\|full | 4724 | No | NA | 9 | 0 | 0 | Not-determined | Genome-fragment | NA | NA | 0 |
| S6-final-viral-combined_k141_121049\|\|full | 1485 | No | NA | 3 | 0 | 1 | Not-determined | Genome-fragment | NA | NA | 0 |
| S6-final-viral-combined_k141_45666\|\|full | 1223 | No | NA | 3 | 0 | 1 | Not-determined | Genome-fragment | NA | NA | 0 |
| S6-final-viral-combined_k141_5465\|\|full | 6805 | No | NA | 6 | 0 | 5 | Not-determined | Genome-fragment | NA | NA | 0 |
| S6-final-viral-combined_k141_80100\|\|full | 3009 | No | NA | 3 | 0 | 2 | Not-determined | Genome-fragment | NA | NA | 0 |
| S6-final-viral-combined_k141_27429\|\|full | 1283 | No | NA | 3 | 0 | 3 | Not-determined | Genome-fragment | NA | NA | 0 |
| S6-final-viral-combined_k141_80124\|\|full | 2932 | No | NA | 6 | 0 | 0 | Not-determined | Genome-fragment | NA | NA | 0 |
| S6-final-viral-combined_k141_15922\|\|full | 3095 | No | NA | 5 | 0 | 1 | Not-determined | Genome-fragment | NA | NA | 0 |
| S12-final-viral-combined_k141_14090\|\|full | 7259 | Yes | 5942 | 9 | 1 | 1 | Low-quality | Genome-fragment | 6.19 | AAI-based (high-confidence) | 18.14 |
| S9-final-viral-combined_k141_29300\|\|full | 7260 | No | NA | 7 | 0 | 1 | Low-quality | Genome-fragment | 8.96 | AAI-based (medium-confidence) | 0 |
| S6-final-viral-combined_k141_27438\|\|full | 3245 | No | NA | 3 | 0 | 2 | Not-determined | Genome-fragment | NA | NA | 0 |
| S6-final-viral-combined_k141_9718\|\|full | 1284 | No | NA | 3 | 0 | 1 | Not-determined | Genome-fragment | NA | NA | 0 |
| S6-final-viral-combined_k141_56206\|\|full | 7608 | No | NA | 5 | 0 | 2 | Not-determined | Genome-fragment | NA | NA | 0 |
| S6-final-viral-combined_k141_15941\|\|full | 1917 | No | NA | 3 | 0 | 1 | Not-determined | Genome-fragment | NA | NA | 0 |
| S6-final-viral-combined_k141_45707\|\|full | 2688 | No | NA | 3 | 0 | 1 | Not-determined | Genome-fragment | NA | NA | 0 |
| S12_k141_191454 | 7263 | No | NA | 8 | 0 | 1 | Low-quality | Genome-fragment | 14.22 | AAI-based (medium-confidence) | 0 |
| S6-final-viral-combined_k141_69201\|\|full | 3016 | No | NA | 3 | 0 | 1 | Not-determined | Genome-fragment | NA | NA | 0 |
| S6-final-viral-combined_k141_27146\|\|full | 7266 | No | NA | 4 | 0 | 0 | Low-quality | Genome-fragment | 8.62 | AAI-based (high-confidence) | 0 |
| S12-final-viral-combined_k141_136153\|\|full | 7269 | No | NA | 8 | 2 | 0 | Low-quality | Genome-fragment | 21.24 | AAI-based (high-confidence) | 0 |
| S11-final-viral-combined_k141_221159\|\|full | 7274 | No | NA | 12 | 1 | 0 | Low-quality | Genome-fragment | 13.13 | AAI-based (medium-confidence) | 0 |
| S6-final-viral-combined_k141_5473\|\|full | 1224 | No | NA | 2 | 0 | 0 | Not-determined | Genome-fragment | NA | NA | 0 |
| S8-final-viral-combined_k141_36101\|\|full | 7274 | No | NA | 7 | 0 | 4 | Low-quality | Genome-fragment | 3.52 | AAI-based (medium-confidence) | 0 |
| S6-final-viral-combined_k141_74514\|\|full | 1682 | No | NA | 2 | 0 | 2 | Not-determined | Genome-fragment | NA | NA | 0 |
| S6-final-viral-combined_k141_80177\|\|full | 2687 | No | NA | 4 | 0 | 2 | Not-determined | Genome-fragment | NA | NA | 0 |
| S11-final-viral-combined_k141_142281\|\|full | 7275 | Yes | 1731 | 5 | 1 | 3 | Low-quality | Genome-fragment | 0.27 | AAI-based (medium-confidence) | 76.21 |
| S6-final-viral-combined_k141_39815\|\|full | 1960 | No | NA | 3 | 0 | 3 | Not-determined | Genome-fragment | NA | NA | 0 |
| S8-final-viral-combined_k141_70542\|\|full | 7277 | No | NA | 14 | 1 | 0 | Low-quality | Genome-fragment | 12.63 | HMM-based (lower-bound) | 0 |
| S6-final-viral-combined_k141_16010\|\|full | 3054 | No | NA | 2 | 0 | 1 | Not-determined | Genome-fragment | NA | NA | 0 |
| S12_k141_280034 | 7279 | No | NA | 6 | 0 | 3 | Low-quality | Genome-fragment | 6.73 | AAI-based (medium-confidence) | 0 |
| S6-final-viral-combined_k141_56322\|\|full | 1762 | No | NA | 2 | 0 | 1 | Not-determined | Genome-fragment | NA | NA | 0 |
| S6-final-viral-combined_k141_51431\|\|full | 1785 | No | NA | 3 | 0 | 0 | Not-determined | Genome-fragment | NA | NA | 0 |
| S17-k141_143834 | 7281 | No | NA | 4 | 0 | 3 | Low-quality | Genome-fragment | 7.09 | AAI-based (medium-confidence) | 0 |
| S6-final-viral-combined_k141_51439\|\|full | 1731 | No | NA | 3 | 0 | 1 | Not-determined | Genome-fragment | NA | NA | 0 |
| S6-final-viral-combined_k141_56336\|\|full | 1892 | No | NA | 3 | 0 | 2 | Not-determined | Genome-fragment | NA | NA | 0 |
| S6-final-viral-combined_k141_133048\|\|full | 3998 | No | NA | 5 | 0 | 2 | Not-determined | Genome-fragment | NA | NA | 0 |
| S6-final-viral-combined_k141_9790\|\|full | 6911 | No | NA | 2 | 0 | 1 | Not-determined | Genome-fragment | NA | NA | 0 |
| S6-final-viral-combined_k141_138710\|\|full | 39594 | No | NA | 35 | 3 | 16 | High-quality | High-quality | 96.85 | AAI-based (high-confidence) | 0 |
| S6-final-viral-combined_k141_92264\|\|full | 2215 | No | NA | 3 | 0 | 1 | Not-determined | Genome-fragment | NA | NA | 0 |
| S6-final-viral-combined_k141_115722\|\|full | 4684 | No | NA | 5 | 0 | 1 | Not-determined | Genome-fragment | NA | NA | 0 |
| S6-final-viral-combined_k141_69294\|\|full | 1274 | No | NA | 2 | 0 | 0 | Not-determined | Genome-fragment | NA | NA | 0 |
| S6-final-viral-combined_k141_110001\|\|full | 1196 | No | NA | 3 | 0 | 1 | Not-determined | Genome-fragment | NA | NA | 0 |
| S6_k141_46001 | 7283 | No | NA | 14 | 0 | 0 | Low-quality | Genome-fragment | 16.35 | AAI-based (medium-confidence) | 0 |
| S6-final-viral-combined_k141_21228\|\|full | 1061 | No | NA | 3 | 0 | 1 | Not-determined | Genome-fragment | NA | NA | 0 |
| S6-final-viral-combined_k141_74601\|\|full | 5064 | No | NA | 6 | 0 | 3 | Not-determined | Genome-fragment | NA | NA | 0 |
| S6-final-viral-combined_k141_133126\|\|full | 2018 | No | NA | 3 | 0 | 2 | Not-determined | Genome-fragment | NA | NA | 0 |
| S6-final-viral-combined_k141_69337\|\|full | 14216 | No | NA | 10 | 0 | 7 | Not-determined | Genome-fragment | NA | NA | 0 |
| S6-final-viral-combined_k141_133133\|\|full | 1150 | No | NA | 3 | 0 | 1 | Not-determined | Genome-fragment | NA | NA | 0 |
| S11-final-viral-combined_k141_181591\|\|full | 7287 | No | NA | 9 | 1 | 0 | Low-quality | Genome-fragment | 17.01 | AAI-based (high-confidence) | 0 |
| S12-final-viral-combined_k141_400707\|\|full | 7287 | No | NA | 7 | 0 | 0 | Low-quality | Genome-fragment | 11.98 | AAI-based (medium-confidence) | 0 |
| S2_k141_101542 | 7288 | No | NA | 9 | 2 | 0 | Low-quality | Genome-fragment | 17.27 | AAI-based (high-confidence) | 0 |
| S8-final-viral-combined_k141_12691\|\|full | 7288 | No | NA | 11 | 2 | 0 | Low-quality | Genome-fragment | 4.5 | HMM-based (lower-bound) | 0 |
| S6-final-viral-combined_k141_51529\|\|full | 5037 | No | NA | 6 | 0 | 5 | Not-determined | Genome-fragment | NA | NA | 0 |
| S12_k141_171883 | 7289 | No | NA | 7 | 0 | 7 | Low-quality | Genome-fragment | 5.84 | AAI-based (high-confidence) | 0 |
| S1-final-viral-combined_k141_170398\|\|full | 7291 | Yes | 4732 | 9 | 1 | 1 | Low-quality | Genome-fragment | 11.11 | AAI-based (medium-confidence) | 35.1 |
| S6-final-viral-combined_k141_63644\|\|full | 2270 | No | NA | 3 | 0 | 0 | Not-determined | Genome-fragment | NA | NA | 0 |
| S12_k141_271947 | 7291 | No | NA | 7 | 0 | 3 | Low-quality | Genome-fragment | 13.72 | AAI-based (medium-confidence) | 0 |
| S6-final-viral-combined_k141_45970\|\|full | 2054 | No | NA | 3 | 0 | 3 | Not-determined | Genome-fragment | NA | NA | 0 |
| S2-final-viral-combined_k141_129016\|\|full | 7291 | No | NA | 12 | 1 | 3 | Low-quality | Genome-fragment | 17.71 | HMM-based (lower-bound) | 0 |
| S17-final-viral-combined_k141_13804\|\|full | 7294 | No | NA | 11 | 4 | 0 | Low-quality | Genome-fragment | 19.6 | AAI-based (high-confidence) | 0 |
| S6-final-viral-combined_k141_98832\|\|full | 3147 | No | NA | 4 | 0 | 3 | Not-determined | Genome-fragment | NA | NA | 0 |
| S6-final-viral-combined_k141_133189\|\|full | 2234 | No | NA | 2 | 0 | 2 | Not-determined | Genome-fragment | NA | NA | 0 |
| S6-final-viral-combined_k141_86472\|\|full | 3477 | No | NA | 5 | 0 | 5 | Not-determined | Genome-fragment | NA | NA | 0 |
| S6-final-viral-combined_k141_133191\|\|full | 5174 | No | NA | 6 | 0 | 3 | Not-determined | Genome-fragment | NA | NA | 0 |
| S6-final-viral-combined_k141_21305\|\|full | 5252 | No | NA | 8 | 0 | 0 | Not-determined | Genome-fragment | NA | NA | 0 |
| S1_k141_77019 | 7297 | No | NA | 9 | 1 | 1 | Low-quality | Genome-fragment | 8.17 | HMM-based (lower-bound) | 0 |
| S2-final-viral-combined_k141_191300\|\|full | 7306 | No | NA | 14 | 2 | 0 | Low-quality | Genome-fragment | 16.04 | AAI-based (high-confidence) | 0 |
| S11-final-viral-combined_k141_88348\|\|full | 7308 | No | NA | 6 | 0 | 4 | Low-quality | Genome-fragment | 5.6 | AAI-based (medium-confidence) | 0 |
| S12_k141_166905 | 7308 | No | NA | 11 | 2 | 0 | Low-quality | Genome-fragment | 3.36 | AAI-based (high-confidence) | 0 |
| S6-final-viral-combined_k141_5739\|\|full | 6101 | No | NA | 5 | 0 | 3 | Not-determined | Genome-fragment | NA | NA | 0 |
| S8-final-viral-combined_k141_885\|\|full | 7311 | No | NA | 6 | 0 | 5 | Low-quality | Genome-fragment | 11.21 | AAI-based (medium-confidence) | 0 |
| S2-final-viral-combined_k141_92277\|\|full | 7316 | No | NA | 10 | 1 | 0 | Low-quality | Genome-fragment | 11.07 | HMM-based (lower-bound) | 0 |
| S6-final-viral-combined_k141_27785\|\|full | 5587 | No | NA | 4 | 0 | 4 | Not-determined | Genome-fragment | NA | NA | 0 |
| S6-final-viral-combined_k141_63726\|\|full | 2006 | No | NA | 4 | 0 | 3 | Not-determined | Genome-fragment | NA | NA | 0 |
| S6-final-viral-combined_k141_74847\|\|full | 2349 | No | NA | 3 | 0 | 2 | Not-determined | Genome-fragment | NA | NA | 0 |
| S6-final-viral-combined_k141_80126\|\|full | 7317 | No | NA | 6 | 0 | 4 | Low-quality | Genome-fragment | 14.13 | AAI-based (medium-confidence) | 0 |
| S4_k141_136411 | 7319 | No | NA | 10 | 1 | 1 | Low-quality | Genome-fragment | 10.51 | HMM-based (lower-bound) | 0 |
| S2-final-viral-combined_k141_178335\|\|full | 7322 | No | NA | 9 | 5 | 0 | Low-quality | Genome-fragment | 10.11 | HMM-based (lower-bound) | 0 |
| S6-final-viral-combined_k141_110243\|\|full | 3077 | No | NA | 6 | 0 | 0 | Not-determined | Genome-fragment | NA | NA | 0 |
| S12-final-viral-combined_k141_198239\|\|full | 7326 | Yes | 5135 | 10 | 1 | 1 | Low-quality | Genome-fragment | 3.45 | HMM-based (lower-bound) | 29.91 |
| S6-final-viral-combined_k141_56464\|\|full | 35272 | Yes | 24442 | 42 | 12 | 8 | Medium-quality | Genome-fragment | 69.66 | AAI-based (high-confidence) | 30.7 |
| S6-final-viral-combined_k141_92577\|\|full | 3664 | No | NA | 4 | 0 | 0 | Not-determined | Genome-fragment | NA | NA | 0 |
| S2-final-viral-combined_k141_10740\|\|full | 7332 | No | NA | 10 | 0 | 1 | Low-quality | Genome-fragment | 15.05 | AAI-based (medium-confidence) | 0 |
| S17-k141_115619 | 7333 | No | NA | 12 | 1 | 0 | Low-quality | Genome-fragment | 21.96 | HMM-based (lower-bound) | 0 |
| S5-final-viral-combined_k141_6801\|\|full | 7335 | No | NA | 11 | 2 | 1 | Low-quality | Genome-fragment | 8.16 | AAI-based (high-confidence) | 0 |
| S11-final-viral-combined_k141_144607\|\|full | 7338 | No | NA | 6 | 0 | 1 | Low-quality | Genome-fragment | 7.71 | AAI-based (medium-confidence) | 0 |
| S6-final-viral-combined_k141_92637\|\|full | 1790 | No | NA | 2 | 0 | 1 | Not-determined | Genome-fragment | NA | NA | 0 |
| S6-final-viral-combined_k141_133382\|\|full | 1571 | No | NA | 3 | 0 | 2 | Not-determined | Genome-fragment | NA | NA | 0 |
| S8-final-viral-combined_k141_110362\|\|full | 7338 | No | NA | 18 | 4 | 0 | Low-quality | Genome-fragment | 17.37 | AAI-based (high-confidence) | 0 |
| S6-final-viral-combined_k141_51695\|\|full | 7905 | No | NA | 7 | 0 | 6 | Not-determined | Genome-fragment | NA | NA | 0 |
| S7_k141_87267 | 7348 | No | NA | 19 | 3 | 0 | Low-quality | Genome-fragment | 6.19 | HMM-based (lower-bound) | 0 |
| S6-final-viral-combined_k141_133394\|\|full | 1315 | No | NA | 2 | 0 | 1 | Not-determined | Genome-fragment | NA | NA | 0 |
| S2-final-viral-combined_k141_188090\|\|full | 7348 | No | NA | 16 | 2 | 0 | Low-quality | Genome-fragment | 17.1 | AAI-based (high-confidence) | 0 |
| S9-final-viral-combined_k141_115069\|\|full | 7349 | No | NA | 11 | 5 | 0 | Low-quality | Genome-fragment | 17.36 | HMM-based (lower-bound) | 0 |
| S1-final-viral-combined_k141_214205\|\|full | 7368 | Yes | 4835 | 9 | 1 | 3 | Low-quality | Genome-fragment | 11.22 | AAI-based (medium-confidence) | 34.38 |
| S6-final-viral-combined_k141_9984\|\|full | 2324 | No | NA | 3 | 0 | 0 | Not-determined | Genome-fragment | NA | NA | 0 |
| S6-final-viral-combined_k141_85936\|\|full | 7368 | No | NA | 7 | 0 | 7 | Low-quality | Genome-fragment | 6.15 | AAI-based (medium-confidence) | 0 |
| S6-final-viral-combined_k141_99091\|\|full | 2043 | No | NA | 2 | 0 | 1 | Not-determined | Genome-fragment | NA | NA | 0 |

| | | | | | | | | | | | |
|---|---|---|---|---|---|---|---|---|---|---|---|
| S6-final-viral-combined_k141_46228\|\|full | 2152 | No | NA | 2 | 0 | 2 | Not-determined | Genome-fragment | NA | NA | 0 |
| S6-final-viral-combined_k141_63914\|\|full | 1151 | No | NA | 2 | 0 | 0 | Not-determined | Genome-fragment | NA | NA | 0 |
| S12-final-viral-combined_k141_401160\|\|full | 7369 | No | NA | 12 | 1 | 1 | Low-quality | Genome-fragment | 17.2 | AAI-based (medium-confidence) | 0 |
| S2_k141_125322 | 7371 | No | NA | 12 | 4 | 0 | Low-quality | Genome-fragment | 5.04 | AAI-based (high-confidence) | 0 |
| S6_k141_119695 | 7372 | No | NA | 11 | 2 | 0 | Low-quality | Genome-fragment | 7.3 | AAI-based (high-confidence) | 0 |
| S6-final-viral-combined_k141_10027\|\|full | 1199 | No | NA | 2 | 0 | 1 | Not-determined | Genome-fragment | NA | NA | 0 |
| S6-final-viral-combined_k141_74934\|\|full | 2512 | No | NA | 3 | 0 | 2 | Not-determined | Genome-fragment | NA | NA | 0 |
| S6-final-viral-combined_k141_21478\|\|full | 1933 | No | NA | 3 | 0 | 2 | Not-determined | Genome-fragment | NA | NA | 0 |
| S6-final-viral-combined_k141_51798\|\|full | 1721 | No | NA | 3 | 0 | 1 | Not-determined | Genome-fragment | NA | NA | 0 |
| S6-final-viral-combined_k141_86740\|\|full | 3452 | No | NA | 2 | 0 | 1 | Not-determined | Genome-fragment | NA | NA | 0 |
| S9-final-viral-combined_k141_111982\|\|full | 7374 | No | NA | 14 | 6 | 0 | Low-quality | Genome-fragment | 8.08 | HMM-based (lower-bound) | 0 |
| S17-final-viral-combined_k141_14571\|\|full | 7374 | No | NA | 12 | 9 | 0 | Low-quality | Genome-fragment | 8.96 | AAI-based (high-confidence) | 0 |
| S11-final-viral-combined_k141_141316\|\|full | 7377 | No | NA | 10 | 3 | 0 | Low-quality | Genome-fragment | 25.71 | AAI-based (high-confidence) | 0 |
| S6-final-viral-combined_k141_116108\|\|full | 7382 | No | NA | 8 | 0 | 6 | Not-determined | Genome-fragment | NA | NA | 0 |
| S12-final-viral-combined_k141_281902\|\|full | 7383 | No | NA | 14 | 2 | 0 | Low-quality | Genome-fragment | 12.55 | AAI-based (high-confidence) | 0 |
| S6-final-viral-combined_k141_127741\|\|full | 10963 | No | NA | 12 | 0 | 8 | Not-determined | Genome-fragment | NA | NA | 0 |
| S6-final-viral-combined_k141_27996\|\|full | 2514 | No | NA | 5 | 0 | 1 | Not-determined | Genome-fragment | NA | NA | 0 |
| S6-final-viral-combined_k141_99177\|\|full | 1376 | No | NA | 2 | 0 | 1 | Not-determined | Genome-fragment | NA | NA | 0 |
| S6-final-viral-combined_k141_10094\|\|full | 4634 | No | NA | 4 | 0 | 2 | Not-determined | Genome-fragment | NA | NA | 0 |
| S1-final-viral-combined_k141_3510\|\|full | 7392 | Yes | 3136 | 11 | 3 | 3 | Low-quality | Genome-fragment | 6.66 | AAI-based (high-confidence) | 57.58 |
| S6-final-viral-combined_k141_46325\|\|full | 1791 | No | NA | 3 | 0 | 0 | Not-determined | Genome-fragment | NA | NA | 0 |
| S7-final-viral-combined_k141_64740\|\|full | 7399 | No | NA | 18 | 3 | 0 | Low-quality | Genome-fragment | 21 | AAI-based (high-confidence) | 0 |
| S6-final-viral-combined_k141_139168\|\|full | 2024 | No | NA | 3 | 0 | 2 | Not-determined | Genome-fragment | NA | NA | 0 |
| S6-final-viral-combined_k141_16518\|\|full | 2527 | No | NA | 5 | 0 | 1 | Not-determined | Genome-fragment | NA | NA | 0 |
| S6-final-viral-combined_k141_21578\|\|full | 3124 | No | NA | 3 | 0 | 2 | Not-determined | Genome-fragment | NA | NA | 0 |
| S6-final-viral-combined_k141_21583\|\|full | 1624 | No | NA | 3 | 0 | 0 | Not-determined | Genome-fragment | NA | NA | 0 |
| S6-final-viral-combined_k141_51871\|\|full | 43498 | No | NA | 42 | 0 | 14 | Not-determined | Genome-fragment | NA | NA | 0 |
| S6-final-viral-combined_k141_133540\|\|full | 2172 | No | NA | 3 | 0 | 2 | Not-determined | Genome-fragment | NA | NA | 0 |
| S9-final-viral-combined_k141_74179\|\|full | 7404 | No | NA | 6 | 0 | 1 | Low-quality | Genome-fragment | 7.38 | AAI-based (medium-confidence) | 0 |
| S11_k141_94935 | 7412 | No | NA | 3 | 0 | 0 | Low-quality | Genome-fragment | 7.99 | AAI-based (medium-confidence) | 0 |
| S11-final-viral-combined_k141_157235\|\|full | 7413 | No | NA | 10 | 4 | 0 | Low-quality | Genome-fragment | 10.37 | HMM-based (lower-bound) | 0 |
| S11-final-viral-combined_k141_239416\|\|full | 7414 | No | NA | 13 | 6 | 0 | Low-quality | Genome-fragment | 19.95 | AAI-based (high-confidence) | 0 |
| S6-final-viral-combined_k141_121484\|\|full | 2124 | No | NA | 4 | 0 | 1 | Not-determined | Genome-fragment | NA | NA | 0 |
| S7_k141_156336 | 7415 | No | NA | 6 | 3 | 1 | Low-quality | Genome-fragment | 11.72 | AAI-based (high-confidence) | 0 |
| S8-final-viral-combined_k141_261330\|\|full | 7417 | No | NA | 10 | 4 | 0 | Low-quality | Genome-fragment | 17.63 | AAI-based (high-confidence) | 0 |
| S6-final-viral-combined_k141_99266\|\|full | 1450 | No | NA | 3 | 0 | 1 | Not-determined | Genome-fragment | NA | NA | 0 |
| S2-final-viral-combined_k141_31161\|\|full | 7418 | Yes | 6182 | 10 | 1 | 2 | Low-quality | Genome-fragment | 5.65 | AAI-based (medium-confidence) | 16.66 |
| S6-final-viral-combined_k141_99273\|\|full | 1752 | No | NA | 4 | 0 | 0 | Not-determined | Genome-fragment | NA | NA | 0 |
| S6-final-viral-combined_k141_121493\|\|full | 1480 | No | NA | 2 | 0 | 0 | Not-determined | Genome-fragment | NA | NA | 0 |
| S6-final-viral-combined_k141_99281\|\|full | 5650 | No | NA | 5 | 0 | 2 | Not-determined | Genome-fragment | NA | NA | 0 |
| S11-final-viral-combined_k141_140792\|\|full | 7419 | No | NA | 14 | 2 | 0 | Low-quality | Genome-fragment | 11.96 | AAI-based (medium-confidence) | 0 |
| S6-final-viral-combined_k141_139254\|\|full | 8754 | No | NA | 7 | 0 | 5 | Not-determined | Genome-fragment | NA | NA | 0 |
| S6-final-viral-combined_k141_16644\|\|full | 3834 | No | NA | 5 | 0 | 2 | Not-determined | Genome-fragment | NA | NA | 0 |
| S11_k141_95476 | 7424 | No | NA | 6 | 1 | 0 | Low-quality | Genome-fragment | 14.66 | AAI-based (high-confidence) | 0 |
| S6-final-viral-combined_k141_133626\|\|full | 3170 | No | NA | 3 | 0 | 0 | Not-determined | Genome-fragment | NA | NA | 0 |
| S12-final-viral-combined_k141_120350\|\|full | 7425 | No | NA | 11 | 1 | 0 | Low-quality | Genome-fragment | 8.11 | HMM-based (lower-bound) | 0 |
| S6-final-viral-combined_k141_121571\|\|full | 3348 | No | NA | 3 | 0 | 2 | Not-determined | Genome-fragment | NA | NA | 0 |
| S6-final-viral-combined_k141_10230\|\|full | 6101 | No | NA | 7 | 0 | 7 | Not-determined | Genome-fragment | NA | NA | 0 |
| S6-final-viral-combined_k141_121578\|\|full | 2247 | No | NA | 3 | 0 | 1 | Not-determined | Genome-fragment | NA | NA | 0 |
| S6-final-viral-combined_k141_64168\|\|full | 2457 | No | NA | 4 | 0 | 2 | Not-determined | Genome-fragment | NA | NA | 0 |
| S6-final-viral-combined_k141_40478\|\|full | 1041 | No | NA | 1 | 0 | 0 | Not-determined | Genome-fragment | NA | NA | 0 |
| S3_k141_61573 | 7434 | No | NA | 11 | 1 | 0 | Low-quality | Genome-fragment | 5.42 | HMM-based (lower-bound) | 0 |
| S2-final-viral-combined_k141_168665\|\|full | 7434 | No | NA | 11 | 9 | 1 | Low-quality | Genome-fragment | 15.58 | AAI-based (high-confidence) | 0 |
| S6-final-viral-combined_k141_99327\|\|full | 2308 | No | NA | 3 | 0 | 1 | Not-determined | Genome-fragment | NA | NA | 0 |
| S12_k141_146816 | 7438 | No | NA | 12 | 4 | 0 | Low-quality | Genome-fragment | 12.38 | AAI-based (high-confidence) | 0 |
| S6-final-viral-combined_k141_69919\|\|full | 4624 | No | NA | 7 | 0 | 2 | Not-determined | Genome-fragment | NA | NA | 0 |
| S6-final-viral-combined_k141_121619\|\|full | 1956 | No | NA | 3 | 0 | 3 | Not-determined | Genome-fragment | NA | NA | 0 |
| S6-final-viral-combined_k141_139364\|\|full | 6792 | No | NA | 5 | 0 | 5 | Not-determined | Genome-fragment | NA | NA | 0 |
| S12-final-viral-combined_k141_165279\|\|full | 7441 | No | NA | 11 | 0 | 0 | Low-quality | Genome-fragment | 13.5 | AAI-based (medium-confidence) | 0 |
| S6-final-viral-combined_k141_103961\|\|full | 1622 | No | NA | 3 | 0 | 2 | Not-determined | Genome-fragment | NA | NA | 0 |
| S6-final-viral-combined_k141_128000\|\|full | 1187 | No | NA | 3 | 0 | 0 | Not-determined | Genome-fragment | NA | NA | 0 |
| S11-final-viral-combined_k141_170495\|\|full | 7445 | No | NA | 14 | 1 | 0 | Low-quality | Genome-fragment | 12.18 | AAI-based (high-confidence) | 0 |
| S4-final-viral-combined_k141_130575\|\|full | 7446 | No | NA | 13 | 0 | 0 | Low-quality | Genome-fragment | 17.46 | AAI-based (high-confidence) | 0 |
| S6-final-viral-combined_k141_69972\|\|full | 1755 | No | NA | 3 | 0 | 0 | Not-determined | Genome-fragment | NA | NA | 0 |
| S6-final-viral-combined_k141_110648\|\|full | 1642 | No | NA | 3 | 0 | 3 | Not-determined | Genome-fragment | NA | NA | 0 |
| S6-final-viral-combined_k141_10343\|\|full | 3037 | No | NA | 3 | 0 | 0 | Not-determined | Genome-fragment | NA | NA | 0 |
| S12-final-viral-combined_k141_57851\|\|full | 7447 | No | NA | 9 | 3 | 0 | Low-quality | Genome-fragment | 25.95 | AAI-based (high-confidence) | 0 |
| S11-final-viral-combined_k141_271215\|\|full | 7453 | No | NA | 8 | 0 | 1 | Low-quality | Genome-fragment | 11.93 | AAI-based (high-confidence) | 0 |
| S2-final-viral-combined_k141_71556\|\|full | 7457 | No | NA | 4 | 0 | 1 | Low-quality | Genome-fragment | 5.34 | AAI-based (medium-confidence) | 0 |
| S6-final-viral-combined_k141_133804\|\|full | 2547 | No | NA | 4 | 0 | 3 | Not-determined | Genome-fragment | NA | NA | 0 |
| S6-final-viral-combined_k141_99471\|\|full | 1189 | No | NA | 3 | 0 | 0 | Not-determined | Genome-fragment | NA | NA | 0 |
| S6-final-viral-combined_k141_116450\|\|full | 2115 | No | NA | 3 | 0 | 0 | Not-determined | Genome-fragment | NA | NA | 0 |
| S6-final-viral-combined_k141_46451\|\|full | 2595 | No | NA | 4 | 0 | 2 | Not-determined | Genome-fragment | NA | NA | 0 |
| S6-final-viral-combined_k141_70073\|\|full | 1668 | No | NA | 4 | 0 | 0 | Not-determined | Genome-fragment | NA | NA | 0 |
| S1_k141_297882 | 7464 | No | NA | 14 | 2 | 0 | Low-quality | Genome-fragment | 13.74 | AAI-based (high-confidence) | 0 |
| S1_k141_158541 | 7464 | No | NA | 8 | 1 | 0 | Low-quality | Genome-fragment | 4.45 | AAI-based (high-confidence) | 0 |
| S6-final-viral-combined_k141_110710\|\|full | 1990 | No | NA | 2 | 0 | 2 | Not-determined | Genome-fragment | NA | NA | 0 |
| S6-final-viral-combined_k141_133869\|\|full | 2221 | No | NA | 3 | 0 | 1 | Not-determined | Genome-fragment | NA | NA | 0 |
| S7-final-viral-combined_k141_27469\|\|full | 7464 | No | NA | 6 | 0 | 3 | Low-quality | Genome-fragment | 3.44 | AAI-based (medium-confidence) | 0 |
| S6-final-viral-combined_k141_133870\|\|full | 1472 | No | NA | 3 | 0 | 0 | Not-determined | Genome-fragment | NA | NA | 0 |
| S6-final-viral-combined_k141_64404\|\|full | 1789 | No | NA | 2 | 0 | 0 | Not-determined | Genome-fragment | NA | NA | 0 |
| S6-final-viral-combined_k141_40688\|\|full | 1851 | No | NA | 3 | 0 | 1 | Not-determined | Genome-fragment | NA | NA | 0 |
| S12-final-viral-combined_k141_200505\|\|full | 7465 | No | NA | 10 | 0 | 1 | Low-quality | Genome-fragment | 13.27 | AAI-based (medium-confidence) | 0 |
| S6-final-viral-combined_k141_92980\|\|full | 3467 | No | NA | 5 | 0 | 4 | Not-determined | Genome-fragment | NA | NA | 0 |
| S12_k141_27135 | 7466 | No | NA | 4 | 0 | 2 | Low-quality | Genome-fragment | 4.13 | AAI-based (high-confidence) | 0 |
| S6-final-viral-combined_k141_121776\|\|full | 3410 | No | NA | 3 | 0 | 1 | Not-determined | Genome-fragment | NA | NA | 0 |
| S6-final-viral-combined_k141_10466\|\|full | 1040 | No | NA | 2 | 0 | 1 | Not-determined | Genome-fragment | NA | NA | 0 |
| S6-final-viral-combined_k141_133918\|\|full | 1644 | No | NA | 2 | 0 | 2 | Not-determined | Genome-fragment | NA | NA | 0 |
| S6-final-viral-combined_k141_75261\|\|full | 10416 | No | NA | 7 | 0 | 4 | Not-determined | Genome-fragment | NA | NA | 0 |
| S6-final-viral-combined_k141_56893\|\|full | 4766 | No | NA | 8 | 0 | 3 | Not-determined | Genome-fragment | NA | NA | 0 |
| S8-final-viral-combined_k141_194073\|\|full | 7471 | No | NA | 7 | 3 | 0 | Low-quality | Genome-fragment | 12.19 | HMM-based (lower-bound) | 0 |
| S4_k141_24096 | 7473 | No | NA | 6 | 0 | 0 | Low-quality | Genome-fragment | 7.7 | AAI-based (medium-confidence) | 0 |
| S1-final-viral-combined_k141_65913\|\|full | 7475 | Yes | 5300 | 8 | 1 | 1 | Low-quality | Genome-fragment | 5.95 | HMM-based (lower-bound) | 29.1 |
| S6-final-viral-combined_k141_93014\|\|full | 1089 | No | NA | 2 | 0 | 0 | Not-determined | Genome-fragment | NA | NA | 0 |
| S1-final-viral-combined_k141_263106\|\|full | 7475 | No | NA | 5 | 0 | 2 | Low-quality | Genome-fragment | 4.02 | AAI-based (medium-confidence) | 0 |
| S17-final-viral-combined_k141_202024\|\|full | 7482 | No | NA | 11 | 3 | 0 | Low-quality | Genome-fragment | 13.57 | AAI-based (high-confidence) | 0 |
| S2-final-viral-combined_k141_24455\|\|full | 7483 | No | NA | 11 | 1 | 3 | Low-quality | Genome-fragment | 15.34 | HMM-based (lower-bound) | 0 |
| S12_k141_116869 | 7485 | No | NA | 12 | 1 | 1 | Low-quality | Genome-fragment | 4.06 | HMM-based (lower-bound) | 0 |
| S11-final-viral-combined_k141_187620\|\|full | 7487 | No | NA | 11 | 1 | 2 | Low-quality | Genome-fragment | 5.03 | HMM-based (lower-bound) | 0 |
| S6-final-viral-combined_k141_40792\|\|full | 8801 | No | NA | 7 | 0 | 6 | Not-determined | Genome-fragment | NA | NA | 0 |
| S8-final-viral-combined_k141_178001\|\|full | 7488 | No | NA | 11 | 8 | 0 | Low-quality | Genome-fragment | 23.09 | AAI-based (high-confidence) | 0 |
| S6_k141_15800 | 7489 | No | NA | 5 | 0 | 4 | Low-quality | Genome-fragment | 4.03 | AAI-based (medium-confidence) | 0 |
| S17-k141_163172 | 7491 | No | NA | 6 | 0 | 1 | Low-quality | Genome-fragment | 12.51 | AAI-based (medium-confidence) | 0 |
| S6-final-viral-combined_k141_46628\|\|full | 1863 | No | NA | 4 | 0 | 3 | Not-determined | Genome-fragment | NA | NA | 0 |
| S6-final-viral-combined_k141_121863\|\|full | 1017 | No | NA | 3 | 0 | 0 | Not-determined | Genome-fragment | NA | NA | 0 |
| S6-final-viral-combined_k141_99604\|\|full | 1477 | No | NA | 3 | 0 | 3 | Not-determined | Genome-fragment | NA | NA | 0 |
| S6-final-viral-combined_k141_104252\|\|full | 1053 | No | NA | 2 | 0 | 0 | Not-determined | Genome-fragment | NA | NA | 0 |
| S5_k141_18280 | 7494 | No | NA | 6 | 0 | 6 | Low-quality | Genome-fragment | 6.01 | AAI-based (high-confidence) | 0 |
| S12-final-viral-combined_k141_282590\|\|full | 7501 | No | NA | 8 | 0 | 6 | Low-quality | Genome-fragment | 22.56 | AAI-based (high-confidence) | 0 |
| S6-final-viral-combined_k141_139652\|\|full | 1354 | No | NA | 3 | 0 | 0 | Not-determined | Genome-fragment | NA | NA | 0 |
| S5_k141_180920 | 7506 | No | NA | 6 | 5 | 0 | Low-quality | Genome-fragment | 12.58 | AAI-based (high-confidence) | 0 |
| S6-final-viral-combined_k141_46672\|\|full | 4673 | No | NA | 6 | 0 | 4 | Not-determined | Genome-fragment | NA | NA | 0 |
| S2-final-viral-combined_k141_171058\|\|full | 7508 | No | NA | 13 | 7 | 0 | Low-quality | Genome-fragment | 15.63 | AAI-based (high-confidence) | 0 |
| S6-final-viral-combined_k141_99657\|\|full | 3444 | No | NA | 4 | 0 | 2 | Not-determined | Genome-fragment | NA | NA | 0 |
| S6-final-viral-combined_k141_52327\|\|full | 1583 | No | NA | 3 | 0 | 0 | Not-determined | Genome-fragment | NA | NA | 0 |
| S6-final-viral-combined_k141_57089\|\|full | 4030 | No | NA | 5 | 0 | 2 | Not-determined | Genome-fragment | NA | NA | 0 |
| S6-final-viral-combined_k141_52355\|\|full | 4425 | No | NA | 4 | 0 | 1 | Not-determined | Genome-fragment | NA | NA | 0 |
| S2_k141_126577 | 7510 | No | NA | 9 | 8 | 0 | Low-quality | Genome-fragment | 12.47 | AAI-based (high-confidence) | 0 |
| S12-final-viral-combined_k141_61245\|\|full | 7511 | Yes | 7259 | 8 | 1 | 1 | Low-quality | Genome-fragment | 14.89 | AAI-based (medium-confidence) | 3.36 |
| S6-final-viral-combined_k141_57140\|\|full | 5079 | No | NA | 4 | 0 | 4 | Not-determined | Genome-fragment | NA | NA | 0 |
| S6-final-viral-combined_k141_10638\|\|full | 4982 | No | NA | 3 | 0 | 0 | Not-determined | Genome-fragment | NA | NA | 0 |
| S17-k141_158973 | 7514 | No | NA | 9 | 0 | 1 | Low-quality | Genome-fragment | 14 | AAI-based (medium-confidence) | 0 |
| S6-final-viral-combined_k141_121919\|\|full | 1195 | No | NA | 4 | 0 | 0 | Not-determined | Genome-fragment | NA | NA | 0 |
| S12_k141_174106 | 7515 | No | NA | 10 | 1 | 0 | Low-quality | Genome-fragment | 15.64 | AAI-based (high-confidence) | 0 |
| S6-final-viral-combined_k141_87449\|\|full | 2123 | No | NA | 3 | 0 | 2 | Not-determined | Genome-fragment | NA | NA | 0 |
| S1-final-viral-combined_k141_237198\|\|full | 7519 | No | NA | 11 | 1 | 2 | Low-quality | Genome-fragment | 18 | AAI-based (high-confidence) | 0 |
| S6-final-viral-combined_k141_110950\|\|full | 33443 | No | NA | 31 | 4 | 0 | Medium-quality | Genome-fragment | 86.94 | AAI-based (high-confidence) | 0 |
| S6-final-viral-combined_k141_34327\|\|full | 5242 | No | NA | 7 | 0 | 4 | Not-determined | Genome-fragment | NA | NA | 0 |
| S5-final-viral-combined_k141_193500\|\|full | 7519 | No | NA | 6 | 0 | 5 | Low-quality | Genome-fragment | 4.13 | AAI-based (medium-confidence) | 0 |
| S6-final-viral-combined_k141_22260\|\|full | 1211 | No | NA | 2 | 0 | 1 | Not-determined | Genome-fragment | NA | NA | 0 |
| S6-final-viral-combined_k141_121985\|\|full | 3169 | No | NA | 5 | 0 | 1 | Not-determined | Genome-fragment | NA | NA | 0 |
| S6-final-viral-combined_k141_93280\|\|full | 3028 | No | NA | 4 | 0 | 3 | Not-determined | Genome-fragment | NA | NA | 0 |
| S6-final-viral-combined_k141_116799\|\|full | 2910 | No | NA | 3 | 0 | 3 | Not-determined | Genome-fragment | NA | NA | 0 |
| S6-final-viral-combined_k141_104460\|\|full | 2755 | No | NA | 3 | 0 | 1 | Not-determined | Genome-fragment | NA | NA | 0 |
| S2-final-viral-combined_k141_81136\|\|full | 7520 | No | NA | 15 | 0 | 0 | Low-quality | Genome-fragment | 13.98 | AAI-based (high-confidence) | 0 |
| S5-final-viral-combined_k141_82886\|\|full | 7527 | No | NA | 10 | 0 | 1 | Low-quality | Genome-fragment | 11.49 | AAI-based (medium-confidence) | 0 |
| S5_k141_121290 | 7528 | No | NA | 5 | 2 | 0 | Low-quality | Genome-fragment | 8.38 | AAI-based (high-confidence) | 0 |
| S6-final-viral-combined_k141_22306\|\|full | 1068 | No | NA | 3 | 0 | 1 | Not-determined | Genome-fragment | NA | NA | 0 |
| S4-final-viral-combined_k141_64864\|\|full | 7530 | No | NA | 7 | 0 | 5 | Low-quality | Genome-fragment | 3.56 | AAI-based (medium-confidence) | 0 |
| S6-final-viral-combined_k141_81122\|\|full | 6834 | No | NA | 12 | 0 | 1 | Not-determined | Genome-fragment | NA | NA | 0 |
| S6-final-viral-combined_k141_134134\|\|full | 1761 | No | NA | 3 | 0 | 1 | Not-determined | Genome-fragment | NA | NA | 0 |
| S6-final-viral-combined_k141_93340\|\|full | 1721 | No | NA | 2 | 0 | 1 | Not-determined | Genome-fragment | NA | NA | 0 |
| S6-final-viral-combined_k141_52550\|\|full | 1319 | No | NA | 3 | 0 | 1 | Not-determined | Genome-fragment | NA | NA | 0 |
| S6-final-viral-combined_k141_75586\|\|full | 7054 | No | NA | 6 | 0 | 6 | Not-determined | Genome-fragment | NA | NA | 0 |
| S8-final-viral-combined_k141_223240\|\|full | 7535 | No | NA | 11 | 3 | 0 | Low-quality | Genome-fragment | 15.72 | AAI-based (high-confidence) | 0 |
| S6-final-viral-combined_k141_70451\|\|full | 1121 | No | NA | 3 | 0 | 0 | Not-determined | Genome-fragment | NA | NA | 0 |
| S6-final-viral-combined_k141_116874\|\|full | 2011 | No | NA | 4 | 0 | 1 | Not-determined | Genome-fragment | NA | NA | 0 |
| S6-final-viral-combined_k141_134190\|\|full | 1872 | No | NA | 1 | 0 | 1 | Not-determined | Genome-fragment | NA | NA | 0 |
| S6-final-viral-combined_k141_93380\|\|full | 6009 | No | NA | 13 | 0 | 1 | Not-determined | Genome-fragment | NA | NA | 0 |
| S6-final-viral-combined_k141_52602\|\|full | 1476 | No | NA | 2 | 0 | 1 | Not-determined | Genome-fragment | NA | NA | 0 |
| S6-final-viral-combined_k141_87597\|\|full | 1900 | No | NA | 2 | 0 | 0 | Not-determined | Genome-fragment | NA | NA | 0 |
| S12-final-viral-combined_k141_238685\|\|full | 7538 | No | NA | 9 | 3 | 0 | Low-quality | Genome-fragment | 17.98 | AAI-based (medium-confidence) | 0 |
| S17-final-viral-combined_k141_56461\|\|full | 7539 | No | NA | 4 | 0 | 3 | Low-quality | Genome-fragment | 7.47 | AAI-based (medium-confidence) | 0 |
| S2_k141_244585 | 7540 | Yes | 4496 | 8 | 1 | 1 | Low-quality | Genome-fragment | 2.65 | AAI-based (medium-confidence) | 40.37 |
| S2_k141_82606 | 7542 | No | NA | 12 | 6 | 0 | Low-quality | Genome-fragment | 20.06 | AAI-based (high-confidence) | 0 |
| S6-final-viral-combined_k141_10722\|\|full | 2273 | No | NA | 2 | 0 | 2 | Not-determined | Genome-fragment | NA | NA | 0 |
| S6-final-viral-combined_k141_17227\|\|full | 2300 | No | NA | 3 | 0 | 3 | Not-determined | Genome-fragment | NA | NA | 0 |
| S6-final-viral-combined_k141_104558\|\|full | 2105 | No | NA | 2 | 0 | 1 | Not-determined | Genome-fragment | NA | NA | 0 |
| S7-final-viral-combined_k141_208542\|\|full | 7547 | No | NA | 5 | 0 | 2 | Low-quality | Genome-fragment | 0.65 | AAI-based (medium-confidence) | 0 |
| S6-final-viral-combined_k141_104565\|\|full | 2416 | No | NA | 3 | 0 | 1 | Not-determined | Genome-fragment | NA | NA | 0 |
| S6-final-viral-combined_k141_34515\|\|full | 4451 | No | NA | 5 | 0 | 0 | Not-determined | Genome-fragment | NA | NA | 0 |
| S6-final-viral-combined_k141_52711\|\|full | 1235 | No | NA | 3 | 0 | 0 | Not-determined | Genome-fragment | NA | NA | 0 |
| S17-final-viral-combined_k141_265594\|\|full | 7552 | No | NA | 9 | 0 | 3 | Low-quality | Genome-fragment | 12.2 | AAI-based (medium-confidence) | 0 |
| S6-final-viral-combined_k141_22347\|\|full | 21617 | No | NA | 19 | 0 | 14 | Not-determined | Genome-fragment | NA | NA | 0 |
| S11-final-viral-combined_k141_157870\|\|full | 7556 | No | NA | 5 | 0 | 3 | Low-quality | Genome-fragment | 4.09 | AAI-based (medium-confidence) | 0 |
| S12-final-viral-combined_k141_386785\|\|full | 7559 | No | NA | 5 | 0 | 5 | Low-quality | Genome-fragment | 3.28 | AAI-based (medium-confidence) | 0 |
| S6-final-viral-combined_k141_87628\|\|full | 1163 | No | NA | 2 | 0 | 2 | Not-determined | Genome-fragment | NA | NA | 0 |
| S4-final-viral-combined_k141_27687\|\|full | 7562 | No | NA | 11 | 3 | 0 | Low-quality | Genome-fragment | 17.64 | AAI-based (medium-confidence) | 0 |
| S5-final-viral-combined_k141_87262\|\|full | 7564 | No | NA | 10 | 0 | 1 | Low-quality | Genome-fragment | 18.36 | AAI-based (high-confidence) | 0 |
| S6-final-viral-combined_k141_128604\|\|full | 20343 | No | NA | 21 | 0 | 11 | Not-determined | Genome-fragment | NA | NA | 0 |
| S6-final-viral-combined_k141_104616\|\|full | 1083 | No | NA | 3 | 0 | 1 | Not-determined | Genome-fragment | NA | NA | 0 |
| S4_k141_136635 | 7568 | No | NA | 15 | 1 | 0 | Low-quality | Genome-fragment | 12.61 | HMM-based (lower-bound) | 0 |
| S6-final-viral-combined_k141_117017\|\|full | 2205 | No | NA | 5 | 0 | 2 | Not-determined | Genome-fragment | NA | NA | 0 |
| S6-final-viral-combined_k141_34577\|\|full | 1912 | No | NA | 3 | 0 | 0 | Not-determined | Genome-fragment | NA | NA | 0 |
| S17-final-viral-combined_k141_207580\|\|full | 7570 | No | NA | 7 | 2 | 0 | Low-quality | Genome-fragment | 13.28 | AAI-based (medium-confidence) | 0 |
| S7_k141_102462 | 7571 | Yes | 5167 | 9 | 2 | 2 | Low-quality | Genome-fragment | 8.64 | AAI-based (high-confidence) | 31.75 |
| S6-final-viral-combined_k141_52736\|\|full | 1652 | No | NA | 3 | 0 | 1 | Not-determined | Genome-fragment | NA | NA | 0 |
| S11-final-viral-combined_k141_32566\|\|full | 7573 | No | NA | 7 | 0 | 0 | Low-quality | Genome-fragment | 21.65 | AAI-based (medium-confidence) | 0 |
| S2-final-viral-combined_k141_207345\|\|full | 7573 | No | NA | 7 | 0 | 4 | Low-quality | Genome-fragment | 6.09 | AAI-based (high-confidence) | 0 |
| S12-final-viral-combined_k141_43296\|\|full | 7575 | No | NA | 7 | 0 | 5 | Low-quality | Genome-fragment | 4.52 | AAI-based (medium-confidence) | 0 |
| S8-final-viral-combined_k141_81836\|\|full | 7575 | Yes | 1503 | 9 | 1 | 4 | Low-quality | Genome-fragment | 1.08 | HMM-based (lower-bound) | 80.16 |
| S6-final-viral-combined_k141_57416\|\|full | 4091 | No | NA | 3 | 0 | 2 | Not-determined | Genome-fragment | NA | NA | 0 |
| S1-final-viral-combined_k141_180037\|\|full | 7576 | Yes | 243 | 9 | 1 | 5 | Low-quality | Genome-fragment | 0.14 | HMM-based (lower-bound) | 96.79 |
| S11-final-viral-combined_k141_193356\|\|full | 7580 | No | NA | 9 | 0 | 3 | Low-quality | Genome-fragment | 12.8 | AAI-based (medium-confidence) | 0 |
| S6-final-viral-combined_k141_104691\|\|full | 2377 | No | NA | 4 | 0 | 3 | Not-determined | Genome-fragment | NA | NA | 0 |
| S6-final-viral-combined_k141_10867\|\|full | 5851 | No | NA | 6 | 0 | 4 | Not-determined | Genome-fragment | NA | NA | 0 |
| S2_k141_99079 | 7581 | No | NA | 1 | 1 | 0 | Low-quality | Genome-fragment | 8.06 | HMM-based (lower-bound) | 0 |
| S7-final-viral-combined_k141_218351\|\|full | 7583 | No | NA | 11 | 0 | 4 | Low-quality | Genome-fragment | 12.67 | AAI-based (high-confidence) | 0 |
| S6-final-viral-combined_k141_17399\|\|full | 1481 | No | NA | 3 | 0 | 0 | Not-determined | Genome-fragment | NA | NA | 0 |
| S2-final-viral-combined_k141_226688\|\|full | 7585 | No | NA | 8 | 4 | 1 | Low-quality | Genome-fragment | 18.83 | AAI-based (high-confidence) | 0 |
| S6-final-viral-combined_k141_28819\|\|full | 3020 | No | NA | 2 | 0 | 1 | Not-determined | Genome-fragment | NA | NA | 0 |
| S8-final-viral-combined_k141_11018\|\|full | 7587 | No | NA | 5 | 3 | 0 | Low-quality | Genome-fragment | 17.58 | AAI-based (medium-confidence) | 0 |
| S6-final-viral-combined_k141_104813\|\|full | 2263 | No | NA | 3 | 0 | 1 | Not-determined | Genome-fragment | NA | NA | 0 |
| S11-final-viral-combined_k141_84859\|\|full | 7590 | No | NA | 11 | 0 | 3 | Low-quality | Genome-fragment | 3.69 | AAI-based (medium-confidence) | 0 |
| S6-final-viral-combined_k141_28835\|\|full | 12194 | No | NA | 10 | 0 | 8 | Not-determined | Genome-fragment | NA | NA | 0 |
| S6-final-viral-combined_k141_17452\|\|full | 1432 | No | NA | 3 | 0 | 1 | Not-determined | Genome-fragment | NA | NA | 0 |
| S6-final-viral-combined_k141_28846\|\|full | 1468 | No | NA | 2 | 0 | 1 | Not-determined | Genome-fragment | NA | NA | 0 |
| S6-final-viral-combined_k141_111313\|\|full | 3487 | No | NA | 3 | 0 | 3 | Not-determined | Genome-fragment | NA | NA | 0 |
| S6-final-viral-combined_k141_10981\|\|full | 2879 | No | NA | 3 | 0 | 1 | Not-determined | Genome-fragment | NA | NA | 0 |
| S6-final-viral-combined_k141_87822\|\|full | 2300 | No | NA | 2 | 0 | 1 | Not-determined | Genome-fragment | NA | NA | 0 |
| S6-final-viral-combined_k141_22573\|\|full | 2002 | No | NA | 4 | 0 | 0 | Not-determined | Genome-fragment | NA | NA | 0 |
| S6-final-viral-combined_k141_75834\|\|full | 2199 | No | NA | 3 | 0 | 1 | Not-determined | Genome-fragment | NA | NA | 0 |
| S6-final-viral-combined_k141_81527\|\|full | 3986 | No | NA | 3 | 0 | 2 | Not-determined | Genome-fragment | NA | NA | 0 |
| S6-final-viral-combined_k141_10988\|\|full | 7544 | No | NA | 8 | 0 | 3 | Not-determined | Genome-fragment | NA | NA | 0 |
| S5-final-viral-combined_k141_113629\|\|full | 7590 | No | NA | 13 | 2 | 0 | Low-quality | Genome-fragment | 21.67 | AAI-based (high-confidence) | 0 |
| S12-final-viral-combined_k141_260417\|\|full | 7590 | No | NA | 11 | 0 | 1 | Low-quality | Genome-fragment | 12.86 | AAI-based (medium-confidence) | 0 |
| S17-final-viral-combined_k141_123460\|\|full | 7592 | No | NA | 5 | 0 | 2 | Low-quality | Genome-fragment | 2.43 | AAI-based (medium-confidence) | 0 |
| S6-final-viral-combined_k141_34649\|\|full | 1940 | No | NA | 3 | 0 | 2 | Not-determined | Genome-fragment | NA | NA | 0 |
| S17-final-viral-combined_k141_76985\|\|full | 7594 | Yes | 943 | 9 | 1 | 3 | Low-quality | Genome-fragment | 2.22 | AAI-based (medium-confidence) | 87.58 |
| S5-final-viral-combined_k141_17152\|\|full | 7595 | No | NA | 9 | 0 | 7 | Low-quality | Genome-fragment | 7.13 | AAI-based (medium-confidence) | 0 |
| S12_k141_109397 | 7596 | No | NA | 9 | 0 | 2 | Low-quality | Genome-fragment | 6.42 | AAI-based (medium-confidence) | 0 |
| S8-final-viral-combined_k141_168300\|\|full | 7598 | No | NA | 5 | 0 | 4 | Low-quality | Genome-fragment | 3.62 | AAI-based (high-confidence) | 0 |
| S6-final-viral-combined_k141_87862\|\|full | 1373 | No | NA | 2 | 0 | 0 | Not-determined | Genome-fragment | NA | NA | 0 |
| S6-final-viral-combined_k141_104928\|\|full | 1171 | No | NA | 4 | 0 | 1 | Not-determined | Genome-fragment | NA | NA | 0 |
| S6-final-viral-combined_k141_57587\|\|full | 1237 | No | NA | 2 | 0 | 0 | Not-determined | Genome-fragment | NA | NA | 0 |
| S6-final-viral-combined_k141_111358\|\|full | 1594 | No | NA | 3 | 0 | 0 | Not-determined | Genome-fragment | NA | NA | 0 |
| S5-final-viral-combined_k141_82022\|\|full | 7599 | No | NA | 12 | 6 | 0 | Low-quality | Genome-fragment | 12.53 | AAI-based (medium-confidence) | 0 |
| S6-final-viral-combined_k141_128922\|\|full | 1663 | No | NA | 4 | 0 | 2 | Not-determined | Genome-fragment | NA | NA | 0 |
| S6-final-viral-combined_k141_34715\|\|full | 1484 | No | NA | 2 | 0 | 2 | Not-determined | Genome-fragment | NA | NA | 0 |
| S6-final-viral-combined_k141_140203\|\|full | 14013 | No | NA | 9 | 0 | 4 | Not-determined | Genome-fragment | NA | NA | 0 |
| S6-final-viral-combined_k141_81655\|\|full | 4045 | No | NA | 2 | 0 | 1 | Not-determined | Genome-fragment | NA | NA | 0 |
| S12-final-viral-combined_k141_99807\|\|full | 7600 | No | NA | 5 | 5 | 0 | Low-quality | Genome-fragment | 18.55 | AAI-based (high-confidence) | 0 |
| S3_k141_62971 | 7603 | No | NA | 14 | 3 | 0 | Low-quality | Genome-fragment | 21.71 | AAI-based (high-confidence) | 0 |
| S12_k141_261783 | 7606 | No | NA | 7 | 0 | 4 | Low-quality | Genome-fragment | 6.17 | AAI-based (high-confidence) | 0 |
| S6-final-viral-combined_k141_93777\|\|full | 3310 | No | NA | 2 | 0 | 2 | Not-determined | Genome-fragment | NA | NA | 0 |
| S6-final-viral-combined_k141_22704\|\|full | 2096 | No | NA | 4 | 0 | 3 | Not-determined | Genome-fragment | NA | NA | 0 |
| S6-final-viral-combined_k141_22708\|\|full | 5155 | No | NA | 6 | 0 | 4 | Not-determined | Genome-fragment | NA | NA | 0 |
| S12-final-viral-combined_k141_351217\|\|full | 7608 | No | NA | 2 | 0 | 0 | Low-quality | Genome-fragment | 17.45 | AAI-based (medium-confidence) | 0 |
| S6-final-viral-combined_k141_128972\|\|full | 22130 | No | NA | 35 | 11 | 0 | Medium-quality | Genome-fragment | 57.05 | AAI-based (medium-confidence) | 0 |
| S6-final-viral-combined_k141_34780\|\|full | 2891 | No | NA | 5 | 0 | 0 | Not-determined | Genome-fragment | NA | NA | 0 |
| S11_k141_42028 | 7614 | No | NA | 16 | 1 | 2 | Low-quality | Genome-fragment | 4.13 | HMM-based (lower-bound) | 0 |
| S6-final-viral-combined_k141_28980\|\|full | 1069 | No | NA | 3 | 0 | 2 | Not-determined | Genome-fragment | NA | NA | 0 |
| S2-final-viral-combined_k141_235836\|\|full | 7615 | No | NA | 10 | 2 | 0 | Low-quality | Genome-fragment | 17.93 | AAI-based (high-confidence) | 0 |
| S5_k141_47963 | 7628 | No | NA | 14 | 1 | 1 | Low-quality | Genome-fragment | 12.89 | AAI-based (medium-confidence) | 0 |
| S3-final-viral-combined_k141_60444\|\|full | 7631 | No | NA | 8 | 4 | 0 | Low-quality | Genome-fragment | 20.33 | AAI-based (medium-confidence) | 0 |
| S6-final-viral-combined_k141_117347\|\|full | 4370 | No | NA | 3 | 0 | 0 | Not-determined | Genome-fragment | NA | NA | 0 |
| S11-final-viral-combined_k141_91725\|\|full | 7637 | No | NA | 12 | 7 | 0 | Low-quality | Genome-fragment | 18.91 | AAI-based (medium-confidence) | 0 |
| S6-final-viral-combined_k141_57772\|\|full | 1276 | No | NA | 2 | 0 | 0 | Not-determined | Genome-fragment | NA | NA | 0 |
| S6-final-viral-combined_k141_57797\|\|full | 1341 | No | NA | 1 | 0 | 0 | Not-determined | Genome-fragment | NA | NA | 0 |
| S6-final-viral-combined_k141_122647\|\|full | 4806 | No | NA | 6 | 0 | 3 | Not-determined | Genome-fragment | NA | NA | 0 |
| S6-final-viral-combined_k141_34865\|\|full | 2063 | No | NA | 3 | 0 | 1 | Not-determined | Genome-fragment | NA | NA | 0 |
| S5-final-viral-combined_k141_6576\|\|full | 7645 | Yes | 2628 | 9 | 1 | 3 | Low-quality | Genome-fragment | 5.22 | AAI-based (medium-confidence) | 65.62 |
| S6-final-viral-combined_k141_140460\|\|full | 4759 | No | NA | 7 | 0 | 1 | Not-determined | Genome-fragment | NA | NA | 0 |
| S6-final-viral-combined_k141_105086\|\|full | 5791 | No | NA | 6 | 0 | 6 | Not-determined | Genome-fragment | NA | NA | 0 |
| S17-final-viral-combined_k141_180330\|\|full | 7646 | No | NA | 11 | 5 | 0 | Low-quality | Genome-fragment | 19.87 | AAI-based (medium-confidence) | 0 |
| S6-final-viral-combined_k141_11212\|\|full | 1909 | No | NA | 2 | 0 | 0 | Not-determined | Genome-fragment | NA | NA | 0 |
| S6_k141_77919 | 7661 | No | NA | 14 | 1 | 1 | Low-quality | Genome-fragment | 13.01 | AAI-based (medium-confidence) | 0 |
| S6-final-viral-combined_k141_11223\|\|full | 4203 | No | NA | 4 | 0 | 2 | Not-determined | Genome-fragment | NA | NA | 0 |
| S11-final-viral-combined_k141_225740\|\|full | 7668 | No | NA | 11 | 1 | 1 | Low-quality | Genome-fragment | 18 | AAI-based (medium-confidence) | 0 |
| S6-final-viral-combined_k141_111470\|\|full | 1140 | No | NA | 2 | 0 | 0 | Not-determined | Genome-fragment | NA | NA | 0 |
| S6-final-viral-combined_k141_134869\|\|full | 4525 | No | NA | 3 | 0 | 2 | Not-determined | Genome-fragment | NA | NA | 0 |
| S6-final-viral-combined_k141_11280\|\|full | 2337 | No | NA | 3 | 0 | 3 | Not-determined | Genome-fragment | NA | NA | 0 |
| S6-final-viral-combined_k141_34987\|\|full | 5411 | No | NA | 5 | 0 | 4 | Not-determined | Genome-fragment | NA | NA | 0 |
| S1-final-viral-combined_k141_301406\|\|full | 7668 | No | NA | 6 | 0 | 5 | Low-quality | Genome-fragment | 17.39 | AAI-based (medium-confidence) | 0 |
| S6-final-viral-combined_k141_81864\|\|full | 5381 | No | NA | 8 | 0 | 4 | Not-determined | Genome-fragment | NA | NA | 0 |
| S6-final-viral-combined_k141_81872\|\|full | 3517 | No | NA | 3 | 0 | 1 | Not-determined | Genome-fragment | NA | NA | 0 |
| S6-final-viral-combined_k141_29291\|\|full | 1038 | No | NA | 2 | 0 | 0 | Not-determined | Genome-fragment | NA | NA | 0 |
| S6-final-viral-combined_k141_81879\|\|full | 5455 | No | NA | 6 | 0 | 1 | Not-determined | Genome-fragment | NA | NA | 0 |
| S4_k141_157792 | 7670 | No | NA | 15 | 1 | 1 | Low-quality | Genome-fragment | 12.94 | AAI-based (medium-confidence) | 0 |
| S6-final-viral-combined_k141_81899\|\|full | 2331 | No | NA | 3 | 0 | 0 | Not-determined | Genome-fragment | NA | NA | 0 |
| S1-final-viral-combined_k141_61643\|\|full | 7676 | No | NA | 14 | 2 | 1 | Low-quality | Genome-fragment | 12.79 | AAI-based (medium-confidence) | 0 |
| S12_k141_161678 | 7678 | No | NA | 9 | 2 | 0 | Low-quality | Genome-fragment | 5.68 | AAI-based (medium-confidence) | 0 |
| S6-final-viral-combined_k141_105241\|\|full | 7371 | No | NA | 5 | 0 | 5 | Not-determined | Genome-fragment | NA | NA | 0 |
| S6-final-viral-combined_k141_105252\|\|full | 1787 | No | NA | 2 | 0 | 0 | Not-determined | Genome-fragment | NA | NA | 0 |
| S6-final-viral-combined_k141_81971\|\|full | 3702 | No | NA | 4 | 0 | 2 | Not-determined | Genome-fragment | NA | NA | 0 |
| S6-final-viral-combined_k141_122976\|\|full | 1601 | No | NA | 3 | 0 | 2 | Not-determined | Genome-fragment | NA | NA | 0 |
| S6-final-viral-combined_k141_81986\|\|full | 3731 | No | NA | 3 | 0 | 2 | Not-determined | Genome-fragment | NA | NA | 0 |
| S6-final-viral-combined_k141_105350\|\|full | 2287 | No | NA | 3 | 0 | 2 | Not-determined | Genome-fragment | NA | NA | 0 |
| S6-final-viral-combined_k141_76238\|\|full | 2770 | No | NA | 6 | 0 | 0 | Not-determined | Genome-fragment | NA | NA | 0 |
| S6-final-viral-combined_k141_82057\|\|full | 2213 | No | NA | 3 | 0 | 1 | Not-determined | Genome-fragment | NA | NA | 0 |
| S6-final-viral-combined_k141_140731\|\|full | 33579 | No | NA | 55 | 13 | 1 | Medium-quality | Genome-fragment | 55.31 | AAI-based (high-confidence) | 0 |
| S7_k141_82777 | 7679 | No | NA | 16 | 1 | 0 | Low-quality | Genome-fragment | 3.08 | HMM-based (lower-bound) | 0 |
| S6-final-viral-combined_k141_123116\|\|full | 1764 | No | NA | 3 | 0 | 1 | Not-determined | Genome-fragment | NA | NA | 0 |
| S6-final-viral-combined_k141_123121\|\|full | 5597 | No | NA | 5 | 0 | 5 | Not-determined | Genome-fragment | NA | NA | 0 |
| S6-final-viral-combined_k141_123140\|\|full | 2682 | No | NA | 2 | 0 | 2 | Not-determined | Genome-fragment | NA | NA | 0 |
| S6-final-viral-combined_k141_23352\|\|full | 1231 | No | NA | 3 | 0 | 2 | Not-determined | Genome-fragment | NA | NA | 0 |
| S6-final-viral-combined_k141_105483\|\|full | 1387 | No | NA | 3 | 0 | 0 | Not-determined | Genome-fragment | NA | NA | 0 |
| S6-final-viral-combined_k141_58243\|\|full | 4577 | No | NA | 4 | 0 | 3 | Not-determined | Genome-fragment | NA | NA | 0 |
| S1-final-viral-combined_k141_386492\|\|full | 7683 | No | NA | 6 | 0 | 4 | Low-quality | Genome-fragment | 4.51 | AAI-based (medium-confidence) | 0 |
| S3-final-viral-combined_k141_121034\|\|full | 7683 | No | NA | 3 | 0 | 0 | Low-quality | Genome-fragment | 8.93 | AAI-based (medium-confidence) | 0 |
| S6-final-viral-combined_k141_23465\|\|full | 1036 | No | NA | 2 | 0 | 1 | Not-determined | Genome-fragment | NA | NA | 0 |
| S6-final-viral-combined_k141_105649\|\|full | 1263 | No | NA | 2 | 0 | 1 | Not-determined | Genome-fragment | NA | NA | 0 |
| S1-final-viral-combined_k141_34894\|\|full | 7684 | No | NA | 10 | 0 | 4 | Low-quality | Genome-fragment | 5.07 | AAI-based (medium-confidence) | 0 |
| S7-final-viral-combined_k141_10585\|\|full | 7685 | No | NA | 3 | 0 | 3 | Low-quality | Genome-fragment | 0.9 | AAI-based (medium-confidence) | 0 |
| S17-final-viral-combined_k141_38993\|\|full | 7695 | No | NA | 6 | 0 | 6 | Low-quality | Genome-fragment | 18.31 | AAI-based (high-confidence) | 0 |
| S1-final-viral-combined_k141_131576\|\|full | 7700 | Yes | 1332 | 8 | 1 | 5 | Low-quality | Genome-fragment | 2.07 | HMM-based (lower-bound) | 82.7 |
| S6-final-viral-combined_k141_11645\|\|full | 5878 | No | NA | 4 | 0 | 4 | Not-determined | Genome-fragment | NA | NA | 0 |
| S17-final-viral-combined_k141_37767\|\|full | 7702 | No | NA | 9 | 2 | 0 | Low-quality | Genome-fragment | 11.07 | AAI-based (high-confidence) | 0 |
| S7-final-viral-combined_k141_151756\|\|full | 7707 | No | NA | 5 | 2 | 0 | Low-quality | Genome-fragment | 9.77 | AAI-based (medium-confidence) | 0 |
| S6-final-viral-combined_k141_11714\|\|full | 4057 | No | NA | 5 | 0 | 2 | Not-determined | Genome-fragment | NA | NA | 0 |
| S1-final-viral-combined_k141_247437\|\|full | 7711 | Yes | 6769 | 10 | 2 | 1 | Low-quality | Genome-fragment | 5.04 | AAI-based (high-confidence) | 12.22 |
| S7_k141_4042 | 7712 | No | NA | 7 | 0 | 3 | Low-quality | Genome-fragment | 12.78 | AAI-based (medium-confidence) | 0 |
| S2-final-viral-combined_k141_82735\|\|full | 7716 | No | NA | 12 | 5 | 0 | Low-quality | Genome-fragment | 20.52 | AAI-based (high-confidence) | 0 |
| S1_k141_35650 | 7718 | No | NA | 2 | 0 | 0 | Low-quality | Genome-fragment | 9.97 | AAI-based (medium-confidence) | 0 |
| S6-final-viral-combined_k141_118037\|\|0_partial | 22598 | No | NA | 17 | 0 | 4 | Not-determined | Genome-fragment | NA | NA | 0 |
| S6-final-viral-combined_k141_12636\|\|0_partial | 13080 | No | NA | 7 | 0 | 6 | Not-determined | Genome-fragment | NA | NA | 0 |
| S6-final-viral-combined_k141_12728\|\|0_partial | 11859 | No | NA | 12 | 0 | 9 | Not-determined | Genome-fragment | NA | NA | 0 |
| S6-final-viral-combined_k141_107641\|\|0_partial | 57623 | No | NA | 72 | 0 | 14 | Not-determined | Genome-fragment | NA | NA | 0 |
| S8-final-viral-combined_k141_253463\|\|full | 7725 | No | NA | 16 | 5 | 0 | Low-quality | Genome-fragment | 18.31 | AAI-based (medium-confidence) | 0 |
| S6-final-viral-combined_k141_78615\|\|0_partial | 22584 | No | NA | 17 | 0 | 11 | Not-determined | Genome-fragment | NA | NA | 0 |
| S6-final-viral-combined_k141_125904\|\|0_partial | 24438 | No | NA | 15 | 0 | 12 | Not-determined | Genome-fragment | NA | NA | 0 |
| S6-final-viral-combined_k141_91039\|\|0_partial | 92936 | Yes | 35633 | 87 | 7 | 30 | Complete | High-quality | 100 | Provirus (medium-confidence) | 61.66 |
| S6-final-viral-combined_k141_109628\|\|0_partial | 18107 | No | NA | 21 | 0 | 3 | Not-determined | Genome-fragment | NA | NA | 0 |
| S6-final-viral-combined_k141_138695\|\|full | 7729 | No | NA | 5 | 0 | 1 | Low-quality | Genome-fragment | 2.41 | AAI-based (medium-confidence) | 0 |
| S6-final-viral-combined_k141_69378\|\|0_partial | 10444 | No | NA | 10 | 0 | 7 | Not-determined | Genome-fragment | NA | NA | 0 |
| S7-final-viral-combined_k141_176071\|\|full | 7734 | No | NA | 10 | 0 | 4 | Low-quality | Genome-fragment | 13.66 | AAI-based (medium-confidence) | 0 |
| S5-final-viral-combined_k141_96478\|\|full | 7735 | No | NA | 14 | 0 | 1 | Low-quality | Genome-fragment | 17.23 | AAI-based (high-confidence) | 0 |
| S6-final-viral-combined_k141_100173\|\|lt2gene | 1381 | No | NA | 2 | 0 | 0 | Not-determined | Genome-fragment | NA | NA | 0 |
| S11-final-viral-combined_k141_292947\|\|full | 7738 | No | NA | 4 | 0 | 2 | Low-quality | Genome-fragment | 9.33 | AAI-based (medium-confidence) | 0 |
| S5-final-viral-combined_k141_131999\|\|full | 7740 | No | NA | 16 | 3 | 0 | Low-quality | Genome-fragment | 19.89 | AAI-based (medium-confidence) | 0 |
| S2-final-viral-combined_k141_109623\|\|full | 7740 | No | NA | 10 | 0 | 6 | Low-quality | Genome-fragment | 3.69 | AAI-based (medium-confidence) | 0 |
| S1-final-viral-combined_k141_201535\|\|full | 7745 | No | NA | 14 | 2 | 0 | Low-quality | Genome-fragment | 18.18 | AAI-based (medium-confidence) | 0 |
| S3-final-viral-combined_k141_133504\|\|full | 7746 | No | NA | 8 | 0 | 2 | Low-quality | Genome-fragment | 6.48 | AAI-based (medium-confidence) | 0 |
| S7-final-viral-combined_k141_82701\|\|full | 7747 | No | NA | 9 | 6 | 0 | Low-quality | Genome-fragment | 12.46 | AAI-based (high-confidence) | 0 |
| S9_k141_42987 | 7748 | No | NA | 8 | 0 | 4 | Low-quality | Genome-fragment | 5.33 | AAI-based (high-confidence) | 0 |
| S11-final-viral-combined_k141_304586\|\|full | 7753 | Yes | 6408 | 8 | 2 | 1 | Low-quality | Genome-fragment | 16.36 | AAI-based (medium-confidence) | 17.35 |
| S1-final-viral-combined_k141_75767\|\|full | 7757 | No | NA | 5 | 0 | 4 | Low-quality | Genome-fragment | 3.83 | AAI-based (medium-confidence) | 0 |
| S6-final-viral-combined_k141_7682\|\|lt2gene | 1102 | No | NA | 1 | 0 | 1 | Not-determined | Genome-fragment | NA | NA | 0 |
| S2_k141_215826 | 7758 | No | NA | 2 | 0 | 2 | Low-quality | Genome-fragment | 3.15 | AAI-based (high-confidence) | 0 |
| S8-final-viral-combined_k141_58647\|\|full | 7760 | No | NA | 5 | 0 | 3 | Low-quality | Genome-fragment | 1.21 | AAI-based (medium-confidence) | 0 |
| S6-final-viral-combined_k141_78212\|\|lt2gene | 1849 | No | NA | 2 | 0 | 0 | Not-determined | Genome-fragment | NA | NA | 0 |
| S6-final-viral-combined_k141_37531\|\|lt2gene | 1404 | No | NA | 1 | 0 | 0 | Not-determined | Genome-fragment | NA | NA | 0 |
| S17-final-viral-combined_k141_441\|\|full | 7762 | No | NA | 3 | 0 | 2 | Low-quality | Genome-fragment | 0.9 | AAI-based (medium-confidence) | 0 |
| S6-final-viral-combined_k141_58662\|\|full | 7765 | No | NA | 8 | 2 | 0 | Low-quality | Genome-fragment | 9.87 | AAI-based (high-confidence) | 0 |
| S6-final-viral-combined_k141_44045\|\|lt2gene | 1146 | No | NA | 1 | 0 | 0 | Not-determined | Genome-fragment | NA | NA | 0 |
| S6-final-viral-combined_k141_96871\|\|lt2gene | 1761 | No | NA | 1 | 0 | 0 | Not-determined | Genome-fragment | NA | NA | 0 |
| S3_k141_83252 | 7767 | No | NA | 3 | 0 | 1 | Low-quality | Genome-fragment | 7.9 | AAI-based (medium-confidence) | 0 |
| S6-final-viral-combined_k141_19820\|\|lt2gene | 1100 | No | NA | 1 | 0 | 0 | Not-determined | Genome-fragment | NA | NA | 0 |
| S11-final-viral-combined_k141_222276\|\|full | 7778 | No | NA | 14 | 9 | 0 | Low-quality | Genome-fragment | 15.85 | AAI-based (high-confidence) | 0 |
| S1-final-viral-combined_k141_254386\|\|full | 7783 | No | NA | 13 | 6 | 0 | Low-quality | Genome-fragment | 20.04 | AAI-based (high-confidence) | 0 |
| S8-final-viral-combined_k141_43976\|\|full | 7783 | Yes | 1631 | 8 | 1 | 6 | Low-quality | Genome-fragment | 0.67 | AAI-based (medium-confidence) | 79.04 |
| S6-final-viral-combined_k141_126217\|\|lt2gene | 2285 | No | NA | 1 | 0 | 0 | Not-determined | Genome-fragment | NA | NA | 0 |
| S3-final-viral-combined_k141_40344\|\|full | 7784 | No | NA | 6 | 0 | 3 | Low-quality | Genome-fragment | 25.97 | AAI-based (medium-confidence) | 0 |
| S11-final-viral-combined_k141_152916\|\|full | 7786 | No | NA | 11 | 0 | 1 | Low-quality | Genome-fragment | 14.79 | AAI-based (medium-confidence) | 0 |
| S6-final-viral-combined_k141_109023\|\|lt2gene | 1206 | No | NA | 2 | 0 | 0 | Not-determined | Genome-fragment | NA | NA | 0 |
| S2-final-viral-combined_k141_112101\|\|full | 7786 | Yes | 4925 | 7 | 1 | 1 | Low-quality | Genome-fragment | 8.57 | AAI-based (medium-confidence) | 36.75 |
| S6-final-viral-combined_k141_91822\|\|lt2gene | 1462 | No | NA | 2 | 0 | 0 | Not-determined | Genome-fragment | NA | NA | 0 |
| S6-final-viral-combined_k141_15667\|\|lt2gene | 1206 | No | NA | 1 | 0 | 0 | Not-determined | Genome-fragment | NA | NA | 0 |
| S11-final-viral-combined_k141_14500\|\|full | 7794 | No | NA | 11 | 5 | 0 | Low-quality | Genome-fragment | 19.05 | AAI-based (high-confidence) | 0 |
| S6-final-viral-combined_k141_86345\|\|lt2gene | 1325 | No | NA | 2 | 0 | 0 | Not-determined | Genome-fragment | NA | NA | 0 |
| S12_k141_351719 | 7794 | No | NA | 7 | 0 | 3 | Low-quality | Genome-fragment | 2.24 | AAI-based (medium-confidence) | 0 |
| S17-final-viral-combined_k141_194993\|\|full | 7795 | No | NA | 5 | 0 | 0 | Low-quality | Genome-fragment | 6.9 | AAI-based (medium-confidence) | 0 |
| S4-final-viral-combined_k141_86034\|\|full | 7796 | No | NA | 14 | 8 | 2 | Low-quality | Genome-fragment | 17.5 | AAI-based (medium-confidence) | 0 |
| S6-final-viral-combined_k141_121082\|\|lt2gene | 1128 | No | NA | 1 | 0 | 0 | Not-determined | Genome-fragment | NA | NA | 0 |
| S6-final-viral-combined_k141_80170\|\|lt2gene | 1150 | No | NA | 2 | 0 | 2 | Not-determined | Genome-fragment | NA | NA | 0 |
| S11_k141_155691 | 7805 | No | NA | 5 | 0 | 2 | Low-quality | Genome-fragment | 3.15 | AAI-based (medium-confidence) | 0 |
| S6-final-viral-combined_k141_5704\|\|lt2gene | 1067 | No | NA | 1 | 0 | 0 | Not-determined | Genome-fragment | NA | NA | 0 |
| S17-k141_129986 | 7806 | No | NA | 13 | 0 | 3 | Low-quality | Genome-fragment | 17.38 | AAI-based (medium-confidence) | 0 |
| S8_k141_110493 | 7806 | No | NA | 10 | 7 | 0 | Low-quality | Genome-fragment | 16.77 | AAI-based (high-confidence) | 0 |
| S6-final-viral-combined_k141_69854\|\|lt2gene | 1012 | No | NA | 2 | 0 | 0 | Not-determined | Genome-fragment | NA | NA | 0 |
| S8_k141_274052 | 7806 | No | NA | 12 | 1 | 0 | Low-quality | Genome-fragment | 17.21 | HMM-based (lower-bound) | 0 |
| S11_k141_184386 | 7811 | No | NA | 15 | 1 | 1 | Low-quality | Genome-fragment | 4.24 | HMM-based (lower-bound) | 0 |
| S2-final-viral-combined_k141_240629\|\|full | 7811 | No | NA | 7 | 0 | 4 | Low-quality | Genome-fragment | 3.98 | AAI-based (medium-confidence) | 0 |
| S9-final-viral-combined_k141_127271\|\|full | 7814 | No | NA | 11 | 2 | 1 | Low-quality | Genome-fragment | 2.88 | HMM-based (lower-bound) | 0 |
| S6-final-viral-combined_k141_93372\|\|lt2gene | 2096 | No | NA | 2 | 0 | 0 | Not-determined | Genome-fragment | NA | NA | 0 |
| S8-final-viral-combined_k141_255638\|\|full | 7816 | No | NA | 7 | 0 | 4 | Low-quality | Genome-fragment | 2.43 | AAI-based (medium-confidence) | 0 |
| S6-final-viral-combined_k141_122136\|\|lt2gene | 1491 | No | NA | 1 | 0 | 0 | Not-determined | Genome-fragment | NA | NA | 0 |
| S6-final-viral-combined_k141_128607\|\|lt2gene | 1084 | No | NA | 2 | 0 | 0 | Not-determined | Genome-fragment | NA | NA | 0 |
| S6-final-viral-combined_k141_81398\|\|lt2gene | 1094 | No | NA | 1 | 0 | 0 | Not-determined | Genome-fragment | NA | NA | 0 |
| S6-final-viral-combined_k141_134451\|\|lt2gene | 1839 | No | NA | 1 | 0 | 0 | Not-determined | Genome-fragment | NA | NA | 0 |
| S6-final-viral-combined_k141_117211\|\|lt2gene | 1674 | No | NA | 1 | 0 | 1 | Not-determined | Genome-fragment | NA | NA | 0 |
| S1_k141_166724 | 7823 | No | NA | 7 | 0 | 4 | Low-quality | Genome-fragment | 18.61 | AAI-based (high-confidence) | 0 |
| S6_k141_41111 | 1855 | No | NA | 1 | 0 | 1 | Not-determined | Genome-fragment | NA | NA | 0 |
| S6_k141_82215 | 1131 | No | NA | 1 | 0 | 1 | Not-determined | Genome-fragment | NA | NA | 0 |
| S6_k141_129185 | 3650 | No | NA | 6 | 0 | 6 | Not-determined | Genome-fragment | NA | NA | 0 |
| S6_k141_58742 | 1197 | No | NA | 1 | 0 | 0 | Not-determined | Genome-fragment | NA | NA | 0 |
| S1_k141_340842 | 7826 | No | NA | 5 | 0 | 3 | Low-quality | Genome-fragment | 7.06 | AAI-based (medium-confidence) | 0 |
| S6_k141_129193 | 1656 | No | NA | 1 | 0 | 0 | Not-determined | Genome-fragment | NA | NA | 0 |
| S6_k141_123324 | 2863 | No | NA | 2 | 0 | 2 | Not-determined | Genome-fragment | NA | NA | 0 |
| S6_k141_129221 | 1014 | No | NA | 3 | 0 | 1 | Not-determined | Genome-fragment | NA | NA | 0 |
| S4_k141_144978 | 7829 | No | NA | 14 | 3 | 0 | Low-quality | Genome-fragment | 16.61 | AAI-based (high-confidence) | 0 |
| S6_k141_135115 | 6814 | No | NA | 7 | 0 | 5 | Not-determined | Genome-fragment | NA | NA | 0 |
| S6_k141_114 | 2050 | No | NA | 1 | 0 | 0 | Not-determined | Genome-fragment | NA | NA | 0 |
| S6_k141_41203 | 4234 | No | NA | 3 | 0 | 3 | Not-determined | Genome-fragment | NA | NA | 0 |
| S6_k141_111635 | 1027 | No | NA | 3 | 0 | 0 | Not-determined | Genome-fragment | NA | NA | 0 |
| S1-final-viral-combined_k141_148079\|\|full | 7830 | No | NA | 10 | 4 | 0 | Low-quality | Genome-fragment | 16.74 | AAI-based (medium-confidence) | 0 |
| S6_k141_11847 | 7361 | No | NA | 7 | 0 | 0 | Not-determined | Genome-fragment | NA | NA | 0 |
| S6_k141_105785 | 1061 | No | NA | 1 | 0 | 0 | Not-determined | Genome-fragment | NA | NA | 0 |
| S6_k141_117475 | 1451 | No | NA | 1 | 0 | 1 | Not-determined | Genome-fragment | NA | NA | 0 |
| S6_k141_17743 | 7744 | No | NA | 7 | 0 | 6 | Not-determined | Genome-fragment | NA | NA | 0 |
| S6_k141_123402 | 1387 | No | NA | 2 | 0 | 1 | Not-determined | Genome-fragment | NA | NA | 0 |
| S8_k141_14279 | 7831 | No | NA | 12 | 7 | 0 | Low-quality | Genome-fragment | 12.41 | HMM-based (lower-bound) | 0 |
| S12-final-viral-combined_k141_176128\|\|full | 7835 | No | NA | 9 | 2 | 0 | Low-quality | Genome-fragment | 11.55 | HMM-based (lower-bound) | 0 |
| S6_k141_239 | 3558 | No | NA | 4 | 0 | 2 | Not-determined | Genome-fragment | NA | NA | 0 |
| S12_k141_368517 | 7835 | No | NA | 11 | 1 | 3 | Low-quality | Genome-fragment | 4.04 | HMM-based (lower-bound) | 0 |
| S6_k141_94118 | 1367 | No | NA | 2 | 0 | 1 | Not-determined | Genome-fragment | NA | NA | 0 |
| S6_k141_94134 | 1378 | No | NA | 1 | 0 | 0 | Not-determined | Genome-fragment | NA | NA | 0 |
| S6_k141_58920 | 3129 | No | NA | 3 | 0 | 3 | Not-determined | Genome-fragment | NA | NA | 0 |
| S6_k141_105887 | 1450 | No | NA | 2 | 0 | 0 | Not-determined | Genome-fragment | NA | NA | 0 |
| S6_k141_35385 | 1349 | No | NA | 2 | 0 | 0 | Not-determined | Genome-fragment | NA | NA | 0 |
| S6_k141_41263 | 3534 | No | NA | 8 | 0 | 0 | Not-determined | Genome-fragment | NA | NA | 0 |
| S6_k141_70623 | 2717 | No | NA | 2 | 0 | 1 | Not-determined | Genome-fragment | NA | NA | 0 |
| S6_k141_76538 | 1988 | No | NA | 1 | 0 | 0 | Not-determined | Genome-fragment | NA | NA | 0 |
| S6_k141_70637 | 5239 | No | NA | 6 | 0 | 0 | Not-determined | Genome-fragment | NA | NA | 0 |
| S6_k141_117615 | 1080 | No | NA | 2 | 0 | 2 | Not-determined | Genome-fragment | NA | NA | 0 |
| S6_k141_64799 | 5362 | No | NA | 4 | 0 | 1 | Not-determined | Genome-fragment | NA | NA | 0 |
| S6_k141_47204 | 3407 | No | NA | 4 | 0 | 0 | Not-determined | Genome-fragment | NA | NA | 0 |
| S6_k141_64812 | 2544 | No | NA | 1 | 0 | 0 | Not-determined | Genome-fragment | NA | NA | 0 |
| S6_k141_12004 | 3686 | No | NA | 4 | 0 | 1 | Not-determined | Genome-fragment | NA | NA | 0 |
| S6_k141_76571 | 3412 | No | NA | 4 | 0 | 1 | Not-determined | Genome-fragment | NA | NA | 0 |
| S6_k141_111852 | 4683 | No | NA | 3 | 0 | 2 | Not-determined | Genome-fragment | NA | NA | 0 |
| S1_k141_74706 | 7836 | No | NA | 4 | 0 | 1 | Low-quality | Genome-fragment | 7.78 | AAI-based (medium-confidence) | 0 |
| S6_k141_64859 | 1318 | No | NA | 1 | 0 | 0 | Not-determined | Genome-fragment | NA | NA | 0 |
| S6_k141_105995 | 1792 | No | NA | 2 | 0 | 0 | Not-determined | Genome-fragment | NA | NA | 0 |
| S6_k141_105996 | 1117 | No | NA | 1 | 0 | 0 | Not-determined | Genome-fragment | NA | NA | 0 |
| S6_k141_106008 | 1282 | No | NA | 1 | 0 | 0 | Not-determined | Genome-fragment | NA | NA | 0 |
| S6_k141_70699 | 1650 | No | NA | 1 | 0 | 0 | Not-determined | Genome-fragment | NA | NA | 0 |
| S6_k141_70706 | 3344 | No | NA | 4 | 0 | 2 | Not-determined | Genome-fragment | NA | NA | 0 |
| S17-final-viral-combined_k141_49153\|\|full | 7840 | No | NA | 6 | 0 | 5 | Low-quality | Genome-fragment | 4.22 | AAI-based (medium-confidence) | 0 |
| S6_k141_59062 | 1561 | No | NA | 2 | 0 | 2 | Not-determined | Genome-fragment | NA | NA | 0 |
| S6_k141_70745 | 1653 | No | NA | 1 | 0 | 1 | Not-determined | Genome-fragment | NA | NA | 0 |
| S6_k141_35541 | 5300 | No | NA | 5 | 0 | 2 | Not-determined | Genome-fragment | NA | NA | 0 |
| S6_k141_12142 | 3226 | No | NA | 3 | 0 | 3 | Not-determined | Genome-fragment | NA | NA | 0 |

| | | | | | | | | | | | |
|---|---|---|---|---|---|---|---|---|---|---|---|
| S6_k141_29549 | 2449 | No | NA | 3 | 0 | 0 | Not-determined | Genome-fragment | NA | NA | 0 |
| S6_k141_450 | 1086 | No | NA | 1 | 0 | 1 | Not-determined | Genome-fragment | NA | NA | 0 |
| S11_k141_126114 | 7841 | No | NA | 9 | 1 | 0 | Low-quality | Genome-fragment | 16.06 | AAI-based (medium-confidence) | 0 |
| S6_k141_70812 | 5425 | No | NA | 4 | 0 | 2 | Not-determined | Genome-fragment | NA | NA | 0 |
| S6_k141_59098 | 2100 | No | NA | 1 | 0 | 1 | Not-determined | Genome-fragment | NA | NA | 0 |
| S6_k141_6087 | 1708 | No | NA | 1 | 0 | 1 | Not-determined | Genome-fragment | NA | NA | 0 |
| S6_k141_12184 | 3708 | No | NA | 5 | 0 | 3 | Not-determined | Genome-fragment | NA | NA | 0 |
| S6_k141_64906 | 1046 | No | NA | 1 | 0 | 1 | Not-determined | Genome-fragment | NA | NA | 0 |
| S7_k141_47264 | 7845 | Yes | 5117 | 2 | 1 | 1 | Low-quality | Genome-fragment | 0.41 | HMM-based (lower-bound) | 34.77 |
| S6_k141_29712 | 3250 | No | NA | 1 | 0 | 1 | Not-determined | Genome-fragment | NA | NA | 0 |
| S6_k141_117763 | 1023 | No | NA | 1 | 0 | 0 | Not-determined | Genome-fragment | NA | NA | 0 |
| S6_k141_542 | 3497 | No | NA | 4 | 0 | 2 | Not-determined | Genome-fragment | NA | NA | 0 |
| S6_k141_29722 | 3050 | No | NA | 4 | 0 | 0 | Not-determined | Genome-fragment | NA | NA | 0 |
| S6_k141_88508 | 1778 | No | NA | 2 | 0 | 2 | Not-determined | Genome-fragment | NA | NA | 0 |
| S6_k141_123710 | 1094 | No | NA | 1 | 0 | 1 | Not-determined | Genome-fragment | NA | NA | 0 |
| S6_k141_94446 | 4668 | No | NA | 5 | 0 | 3 | Not-determined | Genome-fragment | NA | NA | 0 |
| S5-final-viral-combined_k141_102211\|\|full | 7850 | No | NA | 6 | 3 | 0 | Low-quality | Genome-fragment | 21.1 | AAI-based (high-confidence) | 0 |
| S6_k141_76712 | 1460 | No | NA | 2 | 0 | 1 | Not-determined | Genome-fragment | NA | NA | 0 |
| S2-final-viral-combined_k141_34081\|\|full | 7850 | No | NA | 11 | 3 | 0 | Low-quality | Genome-fragment | 20.78 | AAI-based (high-confidence) | 0 |
| S6_k141_12278 | 3377 | No | NA | 4 | 0 | 0 | Not-determined | Genome-fragment | NA | NA | 0 |
| S5-final-viral-combined_k141_77012\|\|full | 7852 | No | NA | 14 | 3 | 0 | Low-quality | Genome-fragment | 13.24 | AAI-based (high-confidence) | 0 |
| S8-final-viral-combined_k141_146184\|\|full | 7855 | No | NA | 5 | 2 | 0 | Low-quality | Genome-fragment | 17.69 | AAI-based (high-confidence) | 0 |
| S1-final-viral-combined_k141_167448\|\|full | 7856 | No | NA | 7 | 0 | 2 | Low-quality | Genome-fragment | 10.2 | AAI-based (medium-confidence) | 0 |
| S6_k141_71001 | 4426 | No | NA | 4 | 0 | 3 | Not-determined | Genome-fragment | NA | NA | 0 |
| S6_k141_12328 | 4099 | No | NA | 6 | 0 | 1 | Not-determined | Genome-fragment | NA | NA | 0 |
| S9_k141_59480 | 7857 | No | NA | 14 | 6 | 0 | Low-quality | Genome-fragment | 13.52 | AAI-based (medium-confidence) | 0 |
| S6_k141_82588 | 3275 | No | NA | 2 | 0 | 1 | Not-determined | Genome-fragment | NA | NA | 0 |
| S11-final-viral-combined_k141_105940\|\|full | 7857 | No | NA | 17 | 4 | 0 | Low-quality | Genome-fragment | 14.44 | AAI-based (high-confidence) | 0 |
| S6_k141_82590 | 3335 | No | NA | 3 | 0 | 3 | Not-determined | Genome-fragment | NA | NA | 0 |
| S6_k141_135293 | 3037 | No | NA | 4 | 0 | 3 | Not-determined | Genome-fragment | NA | NA | 0 |
| S6_k141_82592 | 2689 | No | NA | 3 | 0 | 2 | Not-determined | Genome-fragment | NA | NA | 0 |
| S6_k141_643 | 8283 | No | NA | 10 | 0 | 8 | Not-determined | Genome-fragment | NA | NA | 0 |
| S6-final-viral-combined_k141_454\|\|full | 7857 | No | NA | 8 | 0 | 5 | Low-quality | Genome-fragment | 2.99 | AAI-based (medium-confidence) | 0 |
| S4_k141_127765 | 7858 | No | NA | 5 | 1 | 0 | Low-quality | Genome-fragment | 14.43 | AAI-based (high-confidence) | 0 |
| S7-final-viral-combined_k141_233435\|\|full | 7859 | No | NA | 13 | 9 | 1 | Low-quality | Genome-fragment | 19.33 | AAI-based (medium-confidence) | 0 |
| S6_k141_117906 | 1590 | No | NA | 2 | 0 | 0 | Not-determined | Genome-fragment | NA | NA | 0 |
| S1-final-viral-combined_k141_137853\|\|full | 7860 | No | NA | 15 | 2 | 0 | Low-quality | Genome-fragment | 23.87 | AAI-based (high-confidence) | 0 |
| S6_k141_88726 | 3446 | No | NA | 5 | 0 | 3 | Not-determined | Genome-fragment | NA | NA | 0 |
| S6_k141_100306 | 1022 | No | NA | 2 | 0 | 1 | Not-determined | Genome-fragment | NA | NA | 0 |
| S11-final-viral-combined_k141_279098\|\|full | 7862 | No | NA | 7 | 0 | 4 | Low-quality | Genome-fragment | 4.14 | AAI-based (medium-confidence) | 0 |
| S11_k141_115256 | 7862 | No | NA | 14 | 1 | 0 | Low-quality | Genome-fragment | 4.26 | HMM-based (lower-bound) | 0 |
| S9-final-viral-combined_k141_73816\|\|full | 7870 | No | NA | 7 | 0 | 4 | Low-quality | Genome-fragment | 3.88 | AAI-based (medium-confidence) | 0 |
| S6_k141_23954 | 2436 | No | NA | 1 | 0 | 1 | Not-determined | Genome-fragment | NA | NA | 0 |
| S17-final-viral-combined_k141_253679\|\|full | 7870 | No | NA | 11 | 2 | 0 | Low-quality | Genome-fragment | 8.3 | HMM-based (lower-bound) | 0 |
| S6_k141_65170 | 1336 | No | NA | 1 | 0 | 1 | Not-determined | Genome-fragment | NA | NA | 0 |
| S6_k141_94745 | 5060 | No | NA | 1 | 0 | 0 | Not-determined | Genome-fragment | NA | NA | 0 |
| S4-final-viral-combined_k141_102460\|\|full | 7870 | No | NA | 6 | 0 | 1 | Low-quality | Genome-fragment | 10.19 | AAI-based (medium-confidence) | 0 |
| S6_k141_88844 | 1007 | No | NA | 1 | 0 | 1 | Not-determined | Genome-fragment | NA | NA | 0 |
| S6_k141_71136 | 1497 | No | NA | 3 | 0 | 3 | Not-determined | Genome-fragment | NA | NA | 0 |
| S6_k141_786 | 1232 | No | NA | 2 | 0 | 1 | Not-determined | Genome-fragment | NA | NA | 0 |
| S6_k141_65218 | 1100 | No | NA | 2 | 0 | 2 | Not-determined | Genome-fragment | NA | NA | 0 |
| S11-final-viral-combined_k141_151871\|\|full | 7873 | Yes | 6259 | 10 | 5 | 1 | Low-quality | Genome-fragment | 13.95 | AAI-based (high-confidence) | 20.5 |
| S6_k141_135512 | 4607 | No | NA | 4 | 0 | 1 | Not-determined | Genome-fragment | NA | NA | 0 |
| S6_k141_799 | 1035 | No | NA | 2 | 0 | 0 | Not-determined | Genome-fragment | NA | NA | 0 |
| S6_k141_71181 | 1103 | No | NA | 2 | 0 | 2 | Not-determined | Genome-fragment | NA | NA | 0 |
| S6_k141_41812 | 6264 | No | NA | 4 | 0 | 3 | Not-determined | Genome-fragment | NA | NA | 0 |
| S6_k141_82612 | 4783 | No | NA | 6 | 0 | 0 | Not-determined | Genome-fragment | NA | NA | 0 |
| S6_k141_76896 | 1921 | No | NA | 3 | 0 | 2 | Not-determined | Genome-fragment | NA | NA | 0 |
| S6_k141_59432 | 1001 | No | NA | 1 | 0 | 0 | Not-determined | Genome-fragment | NA | NA | 0 |
| S1_k141_381812 | 7874 | No | NA | 22 | 7 | 1 | Low-quality | Genome-fragment | 20.18 | AAI-based (high-confidence) | 0 |
| S6_k141_53226 | 3438 | No | NA | 6 | 0 | 5 | Not-determined | Genome-fragment | NA | NA | 0 |
| S6_k141_24032 | 2789 | No | NA | 4 | 0 | 0 | Not-determined | Genome-fragment | NA | NA | 0 |
| S6_k141_53236 | 2640 | No | NA | 2 | 0 | 2 | Not-determined | Genome-fragment | NA | NA | 0 |
| S6_k141_65281 | 3110 | No | NA | 8 | 0 | 7 | Not-determined | Genome-fragment | NA | NA | 0 |
| S6_k141_82658 | 1866 | No | NA | 2 | 0 | 2 | Not-determined | Genome-fragment | NA | NA | 0 |
| S6_k141_6565 | 1895 | No | NA | 1 | 0 | 1 | Not-determined | Genome-fragment | NA | NA | 0 |
| S6_k141_30045 | 1209 | No | NA | 3 | 0 | 1 | Not-determined | Genome-fragment | NA | NA | 0 |
| S12-final-viral-combined_k141_295161\|\|full | 7878 | No | NA | 8 | 0 | 4 | Low-quality | Genome-fragment | 8.2 | AAI-based (medium-confidence) | 0 |
| S1-final-viral-combined_k141_31849\|\|full | 7884 | No | NA | 10 | 4 | 0 | Low-quality | Genome-fragment | 23.05 | AAI-based (high-confidence) | 0 |
| S6_k141_82701 | 1286 | No | NA | 1 | 0 | 1 | Not-determined | Genome-fragment | NA | NA | 0 |
| S6_k141_94874 | 1985 | No | NA | 3 | 0 | 0 | Not-determined | Genome-fragment | NA | NA | 0 |
| S6_k141_12571 | 1047 | No | NA | 1 | 0 | 1 | Not-determined | Genome-fragment | NA | NA | 0 |
| S2_k141_126147 | 7884 | No | NA | 13 | 1 | 2 | Low-quality | Genome-fragment | 18.35 | AAI-based (medium-confidence) | 0 |
| S1-final-viral-combined_k141_138114\|\|full | 7887 | No | NA | 7 | 0 | 4 | Low-quality | Genome-fragment | 11.92 | AAI-based (medium-confidence) | 0 |
| S6_k141_129928 | 1921 | No | NA | 1 | 0 | 1 | Not-determined | Genome-fragment | NA | NA | 0 |
| S6_k141_53342 | 1230 | No | NA | 1 | 0 | 1 | Not-determined | Genome-fragment | NA | NA | 0 |
| S6_k141_6669 | 1277 | No | NA | 3 | 0 | 1 | Not-determined | Genome-fragment | NA | NA | 0 |
| S6_k141_30133 | 3326 | No | NA | 5 | 0 | 1 | Not-determined | Genome-fragment | NA | NA | 0 |
| S6_k141_24140 | 1836 | No | NA | 1 | 0 | 1 | Not-determined | Genome-fragment | NA | NA | 0 |
| S6_k141_65479 | 2139 | No | NA | 2 | 0 | 2 | Not-determined | Genome-fragment | NA | NA | 0 |
| S6_k141_89182 | 2941 | No | NA | 2 | 0 | 2 | Not-determined | Genome-fragment | NA | NA | 0 |
| S6_k141_77043 | 5198 | No | NA | 4 | 0 | 1 | Not-determined | Genome-fragment | NA | NA | 0 |
| S6_k141_6792 | 1005 | No | NA | 2 | 0 | 1 | Not-determined | Genome-fragment | NA | NA | 0 |
| S4-final-viral-combined_k141_31986\|\|full | 7887 | No | NA | 16 | 7 | 0 | Low-quality | Genome-fragment | 21.67 | AAI-based (high-confidence) | 0 |
| S6_k141_95046 | 4010 | No | NA | 4 | 0 | 2 | Not-determined | Genome-fragment | NA | NA | 0 |
| S6_k141_82907 | 1214 | No | NA | 2 | 0 | 1 | Not-determined | Genome-fragment | NA | NA | 0 |
| S6_k141_12616 | 7485 | No | NA | 7 | 0 | 6 | Not-determined | Genome-fragment | NA | NA | 0 |
| S6_k141_12618 | 3686 | No | NA | 3 | 0 | 0 | Not-determined | Genome-fragment | NA | NA | 0 |
| S6_k141_124191 | 3333 | No | NA | 3 | 0 | 0 | Not-determined | Genome-fragment | NA | NA | 0 |
| S5_k141_204523 | 7895 | No | NA | 4 | 0 | 4 | Low-quality | Genome-fragment | 6.99 | AAI-based (medium-confidence) | 0 |
| S6_k141_130016 | 1220 | No | NA | 2 | 0 | 0 | Not-determined | Genome-fragment | NA | NA | 0 |
| S7-final-viral-combined_k141_37304\|\|full | 7898 | No | NA | 14 | 3 | 0 | Low-quality | Genome-fragment | 17.1 | AAI-based (high-confidence) | 0 |
| S7-final-viral-combined_k141_196091\|\|full | 7899 | No | NA | 13 | 1 | 0 | Low-quality | Genome-fragment | 18.55 | AAI-based (high-confidence) | 0 |
| S6_k141_135637 | 5683 | No | NA | 11 | 0 | 3 | Not-determined | Genome-fragment | NA | NA | 0 |
| S6_k141_30330 | 1112 | No | NA | 3 | 0 | 2 | Not-determined | Genome-fragment | NA | NA | 0 |
| S1_k141_295557 | 7901 | No | NA | 8 | 0 | 2 | Low-quality | Genome-fragment | 7.42 | AAI-based (medium-confidence) | 0 |
| S6_k141_65657 | 3261 | No | NA | 1 | 0 | 1 | Not-determined | Genome-fragment | NA | NA | 0 |
| S17-final-viral-combined_k141_32208\|\|full | 7904 | No | NA | 8 | 0 | 5 | Low-quality | Genome-fragment | 3.94 | AAI-based (medium-confidence) | 0 |
| S6_k141_30349 | 1278 | No | NA | 1 | 0 | 0 | Not-determined | Genome-fragment | NA | NA | 0 |
| S6_k141_124254 | 1381 | No | NA | 2 | 0 | 1 | Not-determined | Genome-fragment | NA | NA | 0 |
| S1_k141_17777 | 7905 | No | NA | 13 | 1 | 0 | Low-quality | Genome-fragment | 9.78 | HMM-based (lower-bound) | 0 |
| S11-final-viral-combined_k141_216831\|\|full | 7906 | No | NA | 6 | 0 | 5 | Low-quality | Genome-fragment | 13.04 | AAI-based (medium-confidence) | 0 |
| S6_k141_6914 | 1870 | No | NA | 1 | 0 | 1 | Not-determined | Genome-fragment | NA | NA | 0 |
| S6_k141_71445 | 7138 | No | NA | 7 | 0 | 7 | Not-determined | Genome-fragment | NA | NA | 0 |
| S6_k141_65671 | 1202 | No | NA | 2 | 0 | 2 | Not-determined | Genome-fragment | NA | NA | 0 |
| S6_k141_65673 | 2705 | No | NA | 2 | 0 | 1 | Not-determined | Genome-fragment | NA | NA | 0 |
| S6_k141_124282 | 1324 | No | NA | 1 | 0 | 0 | Not-determined | Genome-fragment | NA | NA | 0 |
| S6_k141_118084 | 6462 | No | NA | 6 | 0 | 1 | Not-determined | Genome-fragment | NA | NA | 0 |
| S6_k141_18214 | 4069 | No | NA | 8 | 0 | 1 | Not-determined | Genome-fragment | NA | NA | 0 |
| S6_k141_112320 | 1225 | No | NA | 2 | 0 | 2 | Not-determined | Genome-fragment | NA | NA | 0 |
| S6_k141_65689 | 5965 | No | NA | 3 | 0 | 3 | Not-determined | Genome-fragment | NA | NA | 0 |
| S8-final-viral-combined_k141_127137\|\|full | 7906 | No | NA | 12 | 0 | 0 | Low-quality | Genome-fragment | 15.69 | AAI-based (high-confidence) | 0 |
| S9-final-viral-combined_k141_117641\|\|full | 7910 | No | NA | 16 | 1 | 1 | Low-quality | Genome-fragment | 18.72 | AAI-based (medium-confidence) | 0 |
| S6_k141_118107 | 1023 | No | NA | 2 | 0 | 1 | Not-determined | Genome-fragment | NA | NA | 0 |
| S6_k141_83046 | 1206 | No | NA | 2 | 0 | 0 | Not-determined | Genome-fragment | NA | NA | 0 |
| S6_k141_1390 | 2276 | No | NA | 4 | 0 | 1 | Not-determined | Genome-fragment | NA | NA | 0 |
| S6_k141_106733 | 1637 | No | NA | 2 | 0 | 0 | Not-determined | Genome-fragment | NA | NA | 0 |
| S6_k141_77199 | 10430 | No | NA | 8 | 0 | 1 | Not-determined | Genome-fragment | NA | NA | 0 |
| S6_k141_59845 | 3344 | No | NA | 4 | 0 | 0 | Not-determined | Genome-fragment | NA | NA | 0 |
| S6_k141_36044 | 1707 | No | NA | 4 | 0 | 4 | Not-determined | Genome-fragment | NA | NA | 0 |
| S2_k141_175509 | 7917 | No | NA | 11 | 0 | 1 | Low-quality | Genome-fragment | 17.72 | AAI-based (medium-confidence) | 0 |
| S17-final-viral-combined_k141_88375\|\|full | 7920 | No | NA | 11 | 5 | 0 | Low-quality | Genome-fragment | 23.3 | AAI-based (medium-confidence) | 0 |
| S6_k141_135704 | 5821 | No | NA | 7 | 0 | 4 | Not-determined | Genome-fragment | NA | NA | 0 |
| S6_k141_118176 | 1010 | No | NA | 1 | 0 | 0 | Not-determined | Genome-fragment | NA | NA | 0 |
| S8-final-viral-combined_k141_245402\|\|full | 7921 | No | NA | 9 | 2 | 0 | Low-quality | Genome-fragment | 17.25 | AAI-based (high-confidence) | 0 |
| S6_k141_124364 | 1021 | No | NA | 1 | 0 | 1 | Not-determined | Genome-fragment | NA | NA | 0 |
| S6_k141_83124 | 5810 | No | NA | 4 | 0 | 3 | Not-determined | Genome-fragment | NA | NA | 0 |
| S6_k141_30508 | 1037 | No | NA | 1 | 0 | 1 | Not-determined | Genome-fragment | NA | NA | 0 |
| S9_k141_96110 | 7923 | No | NA | 2 | 0 | 2 | Low-quality | Genome-fragment | 2.95 | AAI-based (high-confidence) | 0 |
| S6_k141_59905 | 1126 | No | NA | 2 | 0 | 0 | Not-determined | Genome-fragment | NA | NA | 0 |
| S6_k141_12776 | 4233 | No | NA | 1 | 0 | 1 | Not-determined | Genome-fragment | NA | NA | 0 |
| S11-final-viral-combined_k141_297297\|\|full | 7924 | No | NA | 14 | 1 | 1 | Low-quality | Genome-fragment | 12.97 | AAI-based (high-confidence) | 0 |
| S6_k141_12781 | 6165 | No | NA | 9 | 0 | 1 | Not-determined | Genome-fragment | NA | NA | 0 |
| S6_k141_12789 | 10820 | No | NA | 8 | 0 | 4 | Not-determined | Genome-fragment | NA | NA | 0 |
| S6_k141_36201 | 1531 | No | NA | 3 | 0 | 1 | Not-determined | Genome-fragment | NA | NA | 0 |
| S6_k141_42393 | 1273 | No | NA | 3 | 0 | 0 | Not-determined | Genome-fragment | NA | NA | 0 |
| S1_k141_129700 | 7924 | No | NA | 12 | 1 | 1 | Low-quality | Genome-fragment | 9.92 | AAI-based (medium-confidence) | 0 |
| S6_k141_100775 | 3091 | No | NA | 2 | 0 | 2 | Not-determined | Genome-fragment | NA | NA | 0 |
| S6_k141_94551 | 7924 | No | NA | 14 | 3 | 0 | Low-quality | Genome-fragment | 18.86 | AAI-based (medium-confidence) | 0 |
| S6_k141_135812 | 1459 | No | NA | 1 | 0 | 1 | Not-determined | Genome-fragment | NA | NA | 0 |
| S6_k141_48241 | 1075 | No | NA | 1 | 0 | 1 | Not-determined | Genome-fragment | NA | NA | 0 |
| S6_k141_42480 | 2477 | No | NA | 3 | 0 | 1 | Not-determined | Genome-fragment | NA | NA | 0 |
| S6_k141_95327 | 1814 | No | NA | 1 | 0 | 1 | Not-determined | Genome-fragment | NA | NA | 0 |
| S6-final-viral-combined_k141_31647\|\|full | 7927 | No | NA | 8 | 0 | 3 | Low-quality | Genome-fragment | 3.09 | AAI-based (medium-confidence) | 0 |
| S6_k141_18488 | 1028 | No | NA | 1 | 0 | 0 | Not-determined | Genome-fragment | NA | NA | 0 |
| S6_k141_48279 | 1014 | No | NA | 3 | 0 | 2 | Not-determined | Genome-fragment | NA | NA | 0 |
| S6_k141_42502 | 3722 | No | NA | 4 | 0 | 1 | Not-determined | Genome-fragment | NA | NA | 0 |
| S1-final-viral-combined_k141_457\|\|full | 7928 | No | NA | 7 | 0 | 4 | Low-quality | Genome-fragment | 3.99 | AAI-based (medium-confidence) | 0 |
| S6-final-viral-combined_k141_39441\|\|full | 7932 | No | NA | 18 | 1 | 1 | Low-quality | Genome-fragment | 13.86 | AAI-based (medium-confidence) | 0 |
| S6_k141_12874 | 1053 | No | NA | 2 | 0 | 1 | Not-determined | Genome-fragment | NA | NA | 0 |
| S6_k141_89576 | 1044 | No | NA | 2 | 0 | 1 | Not-determined | Genome-fragment | NA | NA | 0 |
| S6_k141_95367 | 7603 | No | NA | 7 | 0 | 7 | Not-determined | Genome-fragment | NA | NA | 0 |
| S6_k141_77461 | 1879 | No | NA | 3 | 0 | 3 | Not-determined | Genome-fragment | NA | NA | 0 |
| S8_k141_108395 | 7934 | No | NA | 5 | 0 | 3 | Low-quality | Genome-fragment | 13.16 | AAI-based (high-confidence) | 0 |
| S1-final-viral-combined_k141_85248\|\|full | 7935 | No | NA | 10 | 0 | 0 | Low-quality | Genome-fragment | 18.82 | AAI-based (medium-confidence) | 0 |
| S6_k141_112637 | 2923 | No | NA | 2 | 0 | 1 | Not-determined | Genome-fragment | NA | NA | 0 |
| S12-final-viral-combined_k141_329184\|\|full | 7940 | No | NA | 11 | 9 | 0 | Low-quality | Genome-fragment | 19.73 | AAI-based (medium-confidence) | 0 |
| S6_k141_53584 | 1646 | No | NA | 3 | 0 | 1 | Not-determined | Genome-fragment | NA | NA | 0 |
| S6_k141_60137 | 1048 | No | NA | 2 | 0 | 1 | Not-determined | Genome-fragment | NA | NA | 0 |
| S6_k141_12925 | 3560 | No | NA | 2 | 0 | 2 | Not-determined | Genome-fragment | NA | NA | 0 |
| S6_k141_18598 | 1698 | No | NA | 2 | 0 | 2 | Not-determined | Genome-fragment | NA | NA | 0 |
| S6_k141_24522 | 1499 | No | NA | 1 | 0 | 1 | Not-determined | Genome-fragment | NA | NA | 0 |
| S6_k141_24533 | 3395 | No | NA | 3 | 0 | 1 | Not-determined | Genome-fragment | NA | NA | 0 |
| S7-final-viral-combined_k141_135377\|\|full | 7945 | No | NA | 5 | 1 | 0 | Low-quality | Genome-fragment | 18.36 | AAI-based (high-confidence) | 0 |
| S6_k141_7284 | 2229 | No | NA | 1 | 0 | 1 | Not-determined | Genome-fragment | NA | NA | 0 |
| S6_k141_83437 | 6148 | No | NA | 5 | 0 | 3 | Not-determined | Genome-fragment | NA | NA | 0 |
| S6_k141_1762 | 3975 | No | NA | 5 | 0 | 4 | Not-determined | Genome-fragment | NA | NA | 0 |
| S6_k141_42635 | 5929 | No | NA | 8 | 0 | 0 | Not-determined | Genome-fragment | NA | NA | 0 |
| S6_k141_77517 | 2123 | No | NA | 2 | 0 | 2 | Not-determined | Genome-fragment | NA | NA | 0 |
| S17-final-viral-combined_k141_72422\|\|full | 7946 | No | NA | 12 | 5 | 0 | Low-quality | Genome-fragment | 20.99 | AAI-based (high-confidence) | 0 |
| S1-final-viral-combined_k141_231087\|\|full | 7951 | No | NA | 7 | 0 | 6 | Low-quality | Genome-fragment | 13.99 | AAI-based (medium-confidence) | 0 |
| S3-final-viral-combined_k141_60447\|\|full | 7956 | No | NA | 6 | 3 | 0 | Low-quality | Genome-fragment | 5.24 | AAI-based (high-confidence) | 0 |
| S6_k141_95491 | 4281 | No | NA | 5 | 0 | 0 | Not-determined | Genome-fragment | NA | NA | 0 |
| S17-final-viral-combined_k141_123600\|\|full | 7957 | No | NA | 6 | 0 | 3 | Low-quality | Genome-fragment | 4.88 | AAI-based (medium-confidence) | 0 |
| S6_k141_42678 | 1161 | No | NA | 1 | 0 | 0 | Not-determined | Genome-fragment | NA | NA | 0 |
| S6_k141_107173 | 1065 | No | NA | 2 | 0 | 0 | Not-determined | Genome-fragment | NA | NA | 0 |
| S12-final-viral-combined_k141_5646\|\|full | 7960 | No | NA | 10 | 0 | 1 | Low-quality | Genome-fragment | 12.22 | AAI-based (medium-confidence) | 0 |
| S6_k141_71704 | 1553 | No | NA | 1 | 0 | 1 | Not-determined | Genome-fragment | NA | NA | 0 |
| S1_k141_389486 | 7963 | No | NA | 15 | 2 | 0 | Low-quality | Genome-fragment | 21.99 | AAI-based (high-confidence) | 0 |
| S2-final-viral-combined_k141_100234\|\|full | 7965 | No | NA | 9 | 3 | 0 | Low-quality | Genome-fragment | 16.51 | AAI-based (medium-confidence) | 0 |
| S12_k141_297325 | 7968 | No | NA | 13 | 7 | 1 | Low-quality | Genome-fragment | 18.66 | AAI-based (high-confidence) | 0 |
| S6_k141_71741 | 9193 | No | NA | 3 | 0 | 3 | Not-determined | Genome-fragment | NA | NA | 0 |
| S6_k141_53724 | 1264 | No | NA | 1 | 0 | 0 | Not-determined | Genome-fragment | NA | NA | 0 |
| S6_k141_42726 | 1182 | No | NA | 1 | 0 | 0 | Not-determined | Genome-fragment | NA | NA | 0 |
| S6_k141_71744 | 4010 | No | NA | 3 | 0 | 1 | Not-determined | Genome-fragment | NA | NA | 0 |
| S9-final-viral-combined_k141_127211\|\|full | 7971 | No | NA | 13 | 10 | 0 | Low-quality | Genome-fragment | 23.25 | AAI-based (high-confidence) | 0 |
| S6_k141_42738 | 1440 | No | NA | 3 | 0 | 1 | Not-determined | Genome-fragment | NA | NA | 0 |
| S6_k141_7358 | 1183 | No | NA | 2 | 0 | 1 | Not-determined | Genome-fragment | NA | NA | 0 |
| S6_k141_66301 | 7980 | No | NA | 6 | 0 | 3 | Low-quality | Genome-fragment | 8.29 | AAI-based (medium-confidence) | 0 |
| S6_k141_30883 | 3439 | No | NA | 4 | 0 | 1 | Not-determined | Genome-fragment | NA | NA | 0 |
| S6_k141_7381 | 1603 | No | NA | 2 | 0 | 0 | Not-determined | Genome-fragment | NA | NA | 0 |
| S2_k141_192737 | 7982 | No | NA | 11 | 2 | 1 | Low-quality | Genome-fragment | 13.15 | AAI-based (high-confidence) | 0 |
| S8-final-viral-combined_k141_252243\|\|full | 7984 | No | NA | 7 | 0 | 1 | Low-quality | Genome-fragment | 15.19 | AAI-based (medium-confidence) | 0 |
| S6_k141_95596 | 1006 | No | NA | 2 | 0 | 1 | Not-determined | Genome-fragment | NA | NA | 0 |
| S6_k141_60400 | 1804 | No | NA | 1 | 0 | 0 | Not-determined | Genome-fragment | NA | NA | 0 |
| S12-final-viral-combined_k141_243408\|\|full | 7985 | No | NA | 8 | 2 | 0 | Low-quality | Genome-fragment | 13.56 | AAI-based (high-confidence) | 0 |
| S6_k141_13061 | 1545 | No | NA | 1 | 0 | 1 | Not-determined | Genome-fragment | NA | NA | 0 |
| S6_k141_89866 | 3526 | No | NA | 4 | 0 | 2 | Not-determined | Genome-fragment | NA | NA | 0 |
| S6_k141_136106 | 3609 | No | NA | 3 | 0 | 2 | Not-determined | Genome-fragment | NA | NA | 0 |
| S6_k141_24616 | 1008 | No | NA | 3 | 0 | 2 | Not-determined | Genome-fragment | NA | NA | 0 |
| S6_k141_36476 | 1067 | No | NA | 2 | 0 | 1 | Not-determined | Genome-fragment | NA | NA | 0 |
| S12_k141_339999 | 7986 | No | NA | 15 | 1 | 0 | Low-quality | Genome-fragment | 3.67 | AAI-based (high-confidence) | 0 |
| S2_k141_44033 | 7986 | No | NA | 2 | 0 | 2 | Low-quality | Genome-fragment | 2.8 | AAI-based (high-confidence) | 0 |
| S6_k141_124650 | 1029 | No | NA | 1 | 0 | 0 | Not-determined | Genome-fragment | NA | NA | 0 |
| S6_k141_42809 | 3486 | No | NA | 4 | 0 | 0 | Not-determined | Genome-fragment | NA | NA | 0 |
| S6_k141_42833 | 1060 | No | NA | 2 | 0 | 1 | Not-determined | Genome-fragment | NA | NA | 0 |
| S6_k141_18813 | 1047 | No | NA | 2 | 0 | 0 | Not-determined | Genome-fragment | NA | NA | 0 |
| S7-final-viral-combined_k141_22167\|\|full | 7987 | No | NA | 15 | 1 | 1 | Low-quality | Genome-fragment | 17.05 | AAI-based (medium-confidence) | 0 |
| S6_k141_107400 | 1183 | No | NA | 4 | 0 | 1 | Not-determined | Genome-fragment | NA | NA | 0 |
| S6_k141_130642 | 1829 | No | NA | 1 | 0 | 1 | Not-determined | Genome-fragment | NA | NA | 0 |
| S5-final-viral-combined_k141_176302\|\|full | 7990 | No | NA | 9 | 3 | 0 | Low-quality | Genome-fragment | 17.32 | AAI-based (medium-confidence) | 0 |
| S6_k141_60503 | 1003 | No | NA | 1 | 0 | 1 | Not-determined | Genome-fragment | NA | NA | 0 |
| S6_k141_7464 | 1310 | No | NA | 2 | 0 | 0 | Not-determined | Genome-fragment | NA | NA | 0 |
| S6_k141_77688 | 1690 | No | NA | 3 | 0 | 2 | Not-determined | Genome-fragment | NA | NA | 0 |
| S6_k141_112826 | 1373 | No | NA | 2 | 0 | 0 | Not-determined | Genome-fragment | NA | NA | 0 |
| S5-final-viral-combined_k141_132999\|\|full | 7991 | No | NA | 14 | 1 | 1 | Low-quality | Genome-fragment | 5.2 | AAI-based (high-confidence) | 0 |
| S6_k141_60527 | 1451 | No | NA | 1 | 0 | 0 | Not-determined | Genome-fragment | NA | NA | 0 |
| S11-final-viral-combined_k141_327936\|\|full | 7999 | No | NA | 12 | 0 | 3 | Low-quality | Genome-fragment | 20.53 | AAI-based (high-confidence) | 0 |
| S6_k141_77728 | 4335 | No | NA | 6 | 0 | 1 | Not-determined | Genome-fragment | NA | NA | 0 |
| S6_k141_112850 | 3168 | No | NA | 3 | 0 | 0 | Not-determined | Genome-fragment | NA | NA | 0 |
| S6_k141_130711 | 1204 | No | NA | 1 | 0 | 1 | Not-determined | Genome-fragment | NA | NA | 0 |
| S12_k141_301769 | 7999 | No | NA | 11 | 9 | 0 | Low-quality | Genome-fragment | 13.28 | AAI-based (high-confidence) | 0 |
| S6_k141_13288 | 1670 | No | NA | 3 | 0 | 1 | Not-determined | Genome-fragment | NA | NA | 0 |
| S6_k141_36672 | 4369 | No | NA | 6 | 0 | 2 | Not-determined | Genome-fragment | NA | NA | 0 |
| S11_k141_200693 | 8000 | No | NA | 9 | 2 | 0 | Low-quality | Genome-fragment | 16.5 | AAI-based (medium-confidence) | 0 |
| S1-final-viral-combined_k141_18767\|\|full | 8007 | No | NA | 8 | 2 | 0 | Low-quality | Genome-fragment | 5.28 | HMM-based (lower-bound) | 0 |
| S6_k141_107437 | 1356 | No | NA | 3 | 0 | 1 | Not-determined | Genome-fragment | NA | NA | 0 |
| S2_k141_142330 | 8009 | No | NA | 14 | 2 | 1 | Low-quality | Genome-fragment | 18.78 | AAI-based (medium-confidence) | 0 |
| S6_k141_90010 | 1083 | No | NA | 1 | 0 | 0 | Not-determined | Genome-fragment | NA | NA | 0 |
| S6_k141_118620 | 1174 | No | NA | 2 | 0 | 1 | Not-determined | Genome-fragment | NA | NA | 0 |
| S6_k141_66287 | 1081 | No | NA | 1 | 0 | 1 | Not-determined | Genome-fragment | NA | NA | 0 |
| S6_k141_136271 | 1211 | No | NA | 1 | 0 | 1 | Not-determined | Genome-fragment | NA | NA | 0 |
| S6_k141_107476 | 1276 | No | NA | 3 | 0 | 1 | Not-determined | Genome-fragment | NA | NA | 0 |
| S11-final-viral-combined_k141_167797\|\|full | 8010 | No | NA | 8 | 0 | 7 | Low-quality | Genome-fragment | 5.04 | AAI-based (medium-confidence) | 0 |
| S6_k141_90048 | 6648 | No | NA | 5 | 0 | 4 | Not-determined | Genome-fragment | NA | NA | 0 |
| S12-final-viral-combined_k141_240009\|\|full | 8011 | No | NA | 7 | 0 | 2 | Low-quality | Genome-fragment | 12.86 | AAI-based (medium-confidence) | 0 |
| S6_k141_18827 | 3630 | No | NA | 7 | 0 | 0 | Not-determined | Genome-fragment | NA | NA | 0 |
| S6_k141_77779 | 4640 | No | NA | 4 | 0 | 2 | Not-determined | Genome-fragment | NA | NA | 0 |
| S6_k141_112964 | 1261 | No | NA | 2 | 0 | 0 | Not-determined | Genome-fragment | NA | NA | 0 |
| S6_k141_2240 | 3896 | No | NA | 7 | 0 | 0 | Not-determined | Genome-fragment | NA | NA | 0 |
| S6_k141_66395 | 3521 | No | NA | 3 | 0 | 3 | Not-determined | Genome-fragment | NA | NA | 0 |
| S2-final-viral-combined_k141_84640\|\|full | 8012 | No | NA | 14 | 1 | 1 | Low-quality | Genome-fragment | 3.71 | HMM-based (lower-bound) | 0 |
| S1-final-viral-combined_k141_108980\|\|full | 8015 | No | NA | 18 | 5 | 1 | Low-quality | Genome-fragment | 9.97 | AAI-based (high-confidence) | 0 |
| S6_k141_107557 | 1054 | No | NA | 2 | 0 | 0 | Not-determined | Genome-fragment | NA | NA | 0 |
| S6_k141_95856 | 1691 | No | NA | 2 | 0 | 1 | Not-determined | Genome-fragment | NA | NA | 0 |
| S6_k141_60708 | 8546 | No | NA | 15 | 0 | 13 | Not-determined | Genome-fragment | NA | NA | 0 |
| S6_k141_36911 | 1162 | No | NA | 2 | 0 | 2 | Not-determined | Genome-fragment | NA | NA | 0 |
| S6_k141_36937 | 1524 | No | NA | 2 | 0 | 1 | Not-determined | Genome-fragment | NA | NA | 0 |
| S6_k141_77835 | 3228 | No | NA | 3 | 0 | 3 | Not-determined | Genome-fragment | NA | NA | 0 |
| S6_k141_36948 | 5047 | No | NA | 2 | 0 | 1 | Not-determined | Genome-fragment | NA | NA | 0 |
| S5-final-viral-combined_k141_197118\|\|full | 8018 | No | NA | 18 | 5 | 0 | Low-quality | Genome-fragment | 18.96 | AAI-based (medium-confidence) | 0 |
| S6_k141_77841 | 13228 | No | NA | 8 | 0 | 6 | Not-determined | Genome-fragment | NA | NA | 0 |
| S6_k141_95869 | 1315 | No | NA | 3 | 0 | 3 | Not-determined | Genome-fragment | NA | NA | 0 |
| S6_k141_31331 | 1811 | No | NA | 2 | 0 | 1 | Not-determined | Genome-fragment | NA | NA | 0 |
| S6_k141_24944 | 1947 | No | NA | 3 | 0 | 0 | Not-determined | Genome-fragment | NA | NA | 0 |
| S6_k141_95883 | 5626 | No | NA | 6 | 0 | 3 | Not-determined | Genome-fragment | NA | NA | 0 |
| S17-final-viral-combined_k141_65800\|\|full | 8019 | No | NA | 8 | 3 | 0 | Low-quality | Genome-fragment | 19.27 | AAI-based (high-confidence) | 0 |
| S6_k141_7562 | 1175 | No | NA | 3 | 0 | 1 | Not-determined | Genome-fragment | NA | NA | 0 |
| S6_k141_18977 | 1072 | No | NA | 2 | 0 | 0 | Not-determined | Genome-fragment | NA | NA | 0 |
| S6_k141_2376 | 3004 | No | NA | 3 | 0 | 2 | Not-determined | Genome-fragment | NA | NA | 0 |
| S6_k141_24970 | 3166 | No | NA | 4 | 0 | 2 | Not-determined | Genome-fragment | NA | NA | 0 |
| S1_k141_238907 | 8020 | No | NA | 5 | 0 | 0 | Low-quality | Genome-fragment | 19.88 | AAI-based (high-confidence) | 0 |
| S4_k141_116045 | 8026 | No | NA | 7 | 0 | 1 | Low-quality | Genome-fragment | 13.41 | AAI-based (high-confidence) | 0 |
| S6_k141_31370 | 1072 | No | NA | 1 | 0 | 0 | Not-determined | Genome-fragment | NA | NA | 0 |
| S6_k141_43247 | 7317 | No | NA | 7 | 0 | 6 | Not-determined | Genome-fragment | NA | NA | 0 |
| S6_k141_60834 | 2100 | No | NA | 3 | 0 | 3 | Not-determined | Genome-fragment | NA | NA | 0 |
| S6_k141_7631 | 3476 | No | NA | 6 | 0 | 3 | Not-determined | Genome-fragment | NA | NA | 0 |
| S6_k141_118786 | 3521 | No | NA | 5 | 0 | 1 | Not-determined | Genome-fragment | NA | NA | 0 |
| S6_k141_66554 | 1219 | No | NA | 2 | 0 | 2 | Not-determined | Genome-fragment | NA | NA | 0 |
| S6_k141_43303 | 3673 | No | NA | 4 | 0 | 3 | Not-determined | Genome-fragment | NA | NA | 0 |
| S12_k141_348840 | 8035 | No | NA | 16 | 3 | 2 | Low-quality | Genome-fragment | 20.4 | AAI-based (high-confidence) | 0 |
| S6_k141_136600 | 6407 | No | NA | 10 | 0 | 2 | Not-determined | Genome-fragment | NA | NA | 0 |
| S6_k141_113194 | 4228 | No | NA | 5 | 0 | 2 | Not-determined | Genome-fragment | NA | NA | 0 |
| S6_k141_77927 | 2325 | No | NA | 6 | 0 | 0 | Not-determined | Genome-fragment | NA | NA | 0 |
| S6_k141_77941 | 1059 | No | NA | 1 | 0 | 1 | Not-determined | Genome-fragment | NA | NA | 0 |
| S6_k141_77967 | 1337 | No | NA | 1 | 0 | 0 | Not-determined | Genome-fragment | NA | NA | 0 |
| S6_k141_113262 | 3784 | No | NA | 4 | 0 | 3 | Not-determined | Genome-fragment | NA | NA | 0 |
| S2_k141_165691 | 8038 | No | NA | 7 | 4 | 0 | Low-quality | Genome-fragment | 7.83 | AAI-based (high-confidence) | 0 |
| S6_k141_66699 | 7413 | No | NA | 7 | 0 | 5 | Not-determined | Genome-fragment | NA | NA | 0 |
| S6_k141_125199 | 1423 | No | NA | 1 | 0 | 0 | Not-determined | Genome-fragment | NA | NA | 0 |
| S6_k141_43406 | 2338 | No | NA | 3 | 0 | 1 | Not-determined | Genome-fragment | NA | NA | 0 |
| S6_k141_25100 | 4822 | No | NA | 2 | 0 | 2 | Not-determined | Genome-fragment | NA | NA | 0 |
| S6_k141_2649 | 3185 | No | NA | 4 | 0 | 4 | Not-determined | Genome-fragment | NA | NA | 0 |
| S6_k141_37160 | 1220 | No | NA | 2 | 0 | 2 | Not-determined | Genome-fragment | NA | NA | 0 |
| S6_k141_83941 | 3718 | No | NA | 3 | 0 | 1 | Not-determined | Genome-fragment | NA | NA | 0 |
| S4_k141_91986 | 8038 | No | NA | 6 | 0 | 2 | Low-quality | Genome-fragment | 17.29 | AAI-based (high-confidence) | 0 |
| S6_k141_83942 | 1815 | No | NA | 2 | 0 | 2 | Not-determined | Genome-fragment | NA | NA | 0 |
| S6_k141_78068 | 10262 | No | NA | 6 | 0 | 3 | Not-determined | Genome-fragment | NA | NA | 0 |
| S6_k141_83959 | 1456 | No | NA | 2 | 0 | 1 | Not-determined | Genome-fragment | NA | NA | 0 |
| S6_k141_66795 | 1052 | No | NA | 1 | 0 | 0 | Not-determined | Genome-fragment | NA | NA | 0 |
| S6_k141_25161 | 1052 | No | NA | 2 | 0 | 2 | Not-determined | Genome-fragment | NA | NA | 0 |
| S6_k141_37224 | 1502 | No | NA | 1 | 0 | 0 | Not-determined | Genome-fragment | NA | NA | 0 |
| S6_k141_84000 | 1259 | No | NA | 2 | 0 | 1 | Not-determined | Genome-fragment | NA | NA | 0 |
| S6_k141_60993 | 3550 | No | NA | 7 | 0 | 7 | Not-determined | Genome-fragment | NA | NA | 0 |
| S6_k141_107754 | 1282 | No | NA | 1 | 0 | 1 | Not-determined | Genome-fragment | NA | NA | 0 |
| S12_k141_62305 | 8050 | No | NA | 16 | 1 | 0 | Low-quality | Genome-fragment | 25.06 | AAI-based (high-confidence) | 0 |
| S6_k141_118996 | 1170 | No | NA | 3 | 0 | 0 | Not-determined | Genome-fragment | NA | NA | 0 |
| S6_k141_101199 | 1947 | No | NA | 4 | 0 | 2 | Not-determined | Genome-fragment | NA | NA | 0 |
| S6_k141_72335 | 2078 | No | NA | 2 | 0 | 1 | Not-determined | Genome-fragment | NA | NA | 0 |
| S8_k141_19531 | 8055 | No | NA | 9 | 6 | 2 | Low-quality | Genome-fragment | 18.16 | AAI-based (medium-confidence) | 0 |
| S6_k141_61023 | 21758 | No | NA | 19 | 0 | 14 | Not-determined | Genome-fragment | NA | NA | 0 |
| S6_k141_101254 | 1324 | No | NA | 3 | 0 | 3 | Not-determined | Genome-fragment | NA | NA | 0 |
| S6_k141_131111 | 8500 | No | NA | 14 | 0 | 13 | Not-determined | Genome-fragment | NA | NA | 0 |
| S17-k141_222848 | 8062 | No | NA | 9 | 0 | 2 | Low-quality | Genome-fragment | 7.98 | AAI-based (medium-confidence) | 0 |
| S12-final-viral-combined_k141_2501\|\|full | 8063 | No | NA | 15 | 2 | 0 | Low-quality | Genome-fragment | 16.5 | AAI-based (high-confidence) | 0 |
| S1-final-viral-combined_k141_394456\|\|full | 8070 | No | NA | 8 | 0 | 2 | Low-quality | Genome-fragment | 10.61 | AAI-based (medium-confidence) | 0 |
| S8-final-viral-combined_k141_219336\|\|full | 8075 | No | NA | 12 | 7 | 0 | Low-quality | Genome-fragment | 22.9 | AAI-based (medium-confidence) | 0 |
| S6_k141_49264 | 6203 | No | NA | 7 | 0 | 3 | Not-determined | Genome-fragment | NA | NA | 0 |
| S6_k141_31727 | 1121 | No | NA | 1 | 0 | 1 | Not-determined | Genome-fragment | NA | NA | 0 |
| S7-final-viral-combined_k141_109254\|\|full | 8078 | No | NA | 8 | 0 | 4 | Low-quality | Genome-fragment | 1.56 | AAI-based (medium-confidence) | 0 |
| S6_k141_7948 | 1136 | No | NA | 1 | 0 | 0 | Not-determined | Genome-fragment | NA | NA | 0 |
| S5-final-viral-combined_k141_66610\|\|full | 8082 | Yes | 5704 | 10 | 1 | 1 | Low-quality | Genome-fragment | 12.67 | AAI-based (medium-confidence) | 29.42 |
| S6_k141_132783 | 8082 | No | NA | 8 | 0 | 6 | Low-quality | Genome-fragment | 4.95 | AAI-based (medium-confidence) | 0 |
| S6_k141_78164 | 1118 | No | NA | 2 | 0 | 1 | Not-determined | Genome-fragment | NA | NA | 0 |
| S6_k141_49347 | 1209 | No | NA | 1 | 0 | 1 | Not-determined | Genome-fragment | NA | NA | 0 |
| S6_k141_61078 | 1540 | No | NA | 2 | 0 | 0 | Not-determined | Genome-fragment | NA | NA | 0 |
| S6_k141_107908 | 1115 | No | NA | 1 | 0 | 0 | Not-determined | Genome-fragment | NA | NA | 0 |
| S6_k141_84099 | 3893 | No | NA | 6 | 0 | 0 | Not-determined | Genome-fragment | NA | NA | 0 |
| S6_k141_90340 | 1850 | No | NA | 3 | 0 | 1 | Not-determined | Genome-fragment | NA | NA | 0 |
| S6_k141_37497 | 1148 | No | NA | 2 | 0 | 2 | Not-determined | Genome-fragment | NA | NA | 0 |
| S6_k141_84113 | 1848 | No | NA | 1 | 0 | 1 | Not-determined | Genome-fragment | NA | NA | 0 |
| S11-final-viral-combined_k141_152609\|\|full | 8085 | No | NA | 22 | 2 | 0 | Low-quality | Genome-fragment | 21.09 | AAI-based (medium-confidence) | 0 |
| S6_k141_101382 | 1340 | No | NA | 3 | 0 | 3 | Not-determined | Genome-fragment | NA | NA | 0 |
| S11-final-viral-combined_k141_67873\|\|full | 8099 | No | NA | 6 | 0 | 4 | Low-quality | Genome-fragment | 15.78 | AAI-based (medium-confidence) | 0 |
| S6_k141_25441 | 8300 | No | NA | 7 | 0 | 5 | Not-determined | Genome-fragment | NA | NA | 0 |
| S1-final-viral-combined_k141_381004\|\|full | 8107 | No | NA | 12 | 9 | 0 | Low-quality | Genome-fragment | 17.53 | AAI-based (medium-confidence) | 0 |
| S6_k141_84162 | 1627 | No | NA | 3 | 0 | 1 | Not-determined | Genome-fragment | NA | NA | 0 |
| S8_k141_274705 | 8109 | No | NA | 9 | 1 | 0 | Low-quality | Genome-fragment | 8.71 | AAI-based (high-confidence) | 0 |
| S11-final-viral-combined_k141_218830\|\|full | 8110 | No | NA | 3 | 0 | 2 | Low-quality | Genome-fragment | 23.39 | AAI-based (medium-confidence) | 0 |
| S6_k141_19291 | 1145 | No | NA | 2 | 0 | 0 | Not-determined | Genome-fragment | NA | NA | 0 |
| S6_k141_19302 | 2386 | No | NA | 3 | 0 | 1 | Not-determined | Genome-fragment | NA | NA | 0 |
| S6_k141_84211 | 1820 | No | NA | 1 | 0 | 1 | Not-determined | Genome-fragment | NA | NA | 0 |
| S6_k141_43734 | 1274 | No | NA | 1 | 0 | 1 | Not-determined | Genome-fragment | NA | NA | 0 |
| S6_k141_25486 | 3730 | No | NA | 1 | 0 | 0 | Not-determined | Genome-fragment | NA | NA | 0 |
| S6_k141_72584 | 1215 | No | NA | 2 | 0 | 1 | Not-determined | Genome-fragment | NA | NA | 0 |
| S11-final-viral-combined_k141_58598\|\|full | 8119 | No | NA | 6 | 0 | 4 | Low-quality | Genome-fragment | 1.27 | AAI-based (medium-confidence) | 0 |
| S11-final-viral-combined_k141_185767\|\|full | 8121 | No | NA | 10 | 7 | 0 | Low-quality | Genome-fragment | 18.69 | AAI-based (high-confidence) | 0 |
| S6_k141_119163 | 3494 | No | NA | 2 | 0 | 1 | Not-determined | Genome-fragment | NA | NA | 0 |
| S6_k141_67113 | 1019 | No | NA | 1 | 0 | 0 | Not-determined | Genome-fragment | NA | NA | 0 |
| S6_k141_43795 | 1003 | No | NA | 1 | 0 | 1 | Not-determined | Genome-fragment | NA | NA | 0 |
| S11_k141_34142 | 8123 | Yes | 406 | 6 | 1 | 4 | Low-quality | Genome-fragment | 0.17 | HMM-based (lower-bound) | 95 |
| S6_k141_108081 | 3854 | No | NA | 6 | 0 | 3 | Not-determined | Genome-fragment | NA | NA | 0 |
| S6_k141_14074 | 4197 | No | NA | 4 | 0 | 2 | Not-determined | Genome-fragment | NA | NA | 0 |
| S6_k141_113624 | 3461 | No | NA | 2 | 0 | 1 | Not-determined | Genome-fragment | NA | NA | 0 |
| S6_k141_14088 | 1326 | No | NA | 1 | 0 | 0 | Not-determined | Genome-fragment | NA | NA | 0 |
| S6_k141_78413 | 1033 | No | NA | 2 | 0 | 2 | Not-determined | Genome-fragment | NA | NA | 0 |
| S6_k141_54534 | 2356 | No | NA | 1 | 0 | 1 | Not-determined | Genome-fragment | NA | NA | 0 |
| S11_k141_190543 | 8124 | No | NA | 3 | 0 | 1 | Low-quality | Genome-fragment | 13.07 | AAI-based (medium-confidence) | 0 |
| S6_k141_61240 | 1556 | No | NA | 2 | 0 | 2 | Not-determined | Genome-fragment | NA | NA | 0 |
| S9-final-viral-combined_k141_122632\|\|full | 8126 | No | NA | 13 | 3 | 1 | Low-quality | Genome-fragment | 13.36 | HMM-based (lower-bound) | 0 |
| S6_k141_113667 | 1143 | No | NA | 2 | 0 | 0 | Not-determined | Genome-fragment | NA | NA | 0 |
| S6_k141_119291 | 1205 | No | NA | 1 | 0 | 1 | Not-determined | Genome-fragment | NA | NA | 0 |
| S6_k141_90589 | 1182 | No | NA | 2 | 0 | 1 | Not-determined | Genome-fragment | NA | NA | 0 |
| S6_k141_113695 | 1533 | No | NA | 2 | 0 | 0 | Not-determined | Genome-fragment | NA | NA | 0 |
| S6_k141_54578 | 34993 | No | NA | 55 | 11 | 1 | Medium-quality | Genome-fragment | 77.49 | AAI-based (high-confidence) | 0 |
| S6_k141_67285 | 1099 | No | NA | 2 | 0 | 0 | Not-determined | Genome-fragment | NA | NA | 0 |
| S6_k141_78471 | 1453 | No | NA | 1 | 0 | 1 | Not-determined | Genome-fragment | NA | NA | 0 |
| S6_k141_67305 | 1197 | No | NA | 1 | 0 | 1 | Not-determined | Genome-fragment | NA | NA | 0 |
| S11-final-viral-combined_k141_1162\|\|full | 8132 | No | NA | 12 | 2 | 1 | Low-quality | Genome-fragment | 14.11 | AAI-based (high-confidence) | 0 |
| S3-final-viral-combined_k141_114975\|\|full | 8137 | No | NA | 8 | 0 | 5 | Low-quality | Genome-fragment | 4.14 | AAI-based (medium-confidence) | 0 |
| S6_k141_101635 | 1054 | No | NA | 1 | 0 | 1 | Not-determined | Genome-fragment | NA | NA | 0 |
| S6_k141_37901 | 2570 | No | NA | 1 | 0 | 1 | Not-determined | Genome-fragment | NA | NA | 0 |
| S6_k141_78546 | 5225 | No | NA | 4 | 0 | 2 | Not-determined | Genome-fragment | NA | NA | 0 |
| S6_k141_72757 | 3477 | No | NA | 2 | 0 | 2 | Not-determined | Genome-fragment | NA | NA | 0 |
| S6_k141_108231 | 1029 | No | NA | 1 | 0 | 1 | Not-determined | Genome-fragment | NA | NA | 0 |
| S6_k141_108242 | 1528 | No | NA | 1 | 0 | 0 | Not-determined | Genome-fragment | NA | NA | 0 |
| S6_k141_25808 | 1012 | No | NA | 2 | 0 | 0 | Not-determined | Genome-fragment | NA | NA | 0 |
| S8-final-viral-combined_k141_172184\|\|full | 8140 | No | NA | 7 | 2 | 0 | Low-quality | Genome-fragment | 18.61 | AAI-based (medium-confidence) | 0 |
| S6_k141_25811 | 3503 | No | NA | 4 | 0 | 2 | Not-determined | Genome-fragment | NA | NA | 0 |
| S6_k141_14164 | 4071 | No | NA | 3 | 0 | 1 | Not-determined | Genome-fragment | NA | NA | 0 |
| S9_k141_42662 | 8142 | No | NA | 9 | 0 | 3 | Low-quality | Genome-fragment | 7.65 | AAI-based (medium-confidence) | 0 |
| S6_k141_101722 | 2311 | No | NA | 3 | 0 | 2 | Not-determined | Genome-fragment | NA | NA | 0 |
| S6_k141_67403 | 1419 | No | NA | 1 | 0 | 1 | Not-determined | Genome-fragment | NA | NA | 0 |
| S6_k141_44053 | 5061 | No | NA | 5 | 0 | 2 | Not-determined | Genome-fragment | NA | NA | 0 |
| S6_k141_72853 | 2013 | No | NA | 1 | 0 | 1 | Not-determined | Genome-fragment | NA | NA | 0 |
| S6_k141_137123 | 4277 | No | NA | 6 | 0 | 3 | Not-determined | Genome-fragment | NA | NA | 0 |
| S6_k141_49821 | 1296 | No | NA | 1 | 0 | 0 | Not-determined | Genome-fragment | NA | NA | 0 |
| S6_k141_137128 | 2633 | No | NA | 3 | 0 | 1 | Not-determined | Genome-fragment | NA | NA | 0 |
| S6_k141_3449 | 3754 | No | NA | 4 | 0 | 2 | Not-determined | Genome-fragment | NA | NA | 0 |
| S6_k141_3452 | 1635 | No | NA | 1 | 0 | 1 | Not-determined | Genome-fragment | NA | NA | 0 |
| S6_k141_72918 | 1080 | No | NA | 2 | 0 | 0 | Not-determined | Genome-fragment | NA | NA | 0 |
| S6_k141_14235 | 3290 | No | NA | 2 | 0 | 2 | Not-determined | Genome-fragment | NA | NA | 0 |
| S6_k141_101748 | 5408 | No | NA | 7 | 0 | 1 | Not-determined | Genome-fragment | NA | NA | 0 |
| S6_k141_101754 | 1094 | No | NA | 1 | 0 | 1 | Not-determined | Genome-fragment | NA | NA | 0 |
| S2-final-viral-combined_k141_214609\|\|full | 8143 | No | NA | 8 | 0 | 0 | Low-quality | Genome-fragment | 9.89 | AAI-based (medium-confidence) | 0 |
| S6_k141_67487 | 2161 | No | NA | 2 | 0 | 1 | Not-determined | Genome-fragment | NA | NA | 0 |
| S6_k141_72985 | 1156 | No | NA | 1 | 0 | 1 | Not-determined | Genome-fragment | NA | NA | 0 |
| S3-final-viral-combined_k141_107254\|\|full | 8145 | No | NA | 6 | 0 | 5 | Low-quality | Genome-fragment | 1.81 | AAI-based (medium-confidence) | 0 |
| S6_k141_113899 | 4727 | No | NA | 4 | 0 | 3 | Not-determined | Genome-fragment | NA | NA | 0 |
| S6_k141_3512 | 1196 | No | NA | 2 | 0 | 2 | Not-determined | Genome-fragment | NA | NA | 0 |
| S6_k141_8309 | 1027 | No | NA | 2 | 0 | 2 | Not-determined | Genome-fragment | NA | NA | 0 |
| S17-k141_250404 | 8150 | No | NA | 12 | 2 | 0 | Low-quality | Genome-fragment | 12.21 | AAI-based (high-confidence) | 0 |
| S6_k141_78684 | 2439 | No | NA | 4 | 0 | 2 | Not-determined | Genome-fragment | NA | NA | 0 |
| S6_k141_73059 | 1554 | No | NA | 1 | 0 | 1 | Not-determined | Genome-fragment | NA | NA | 0 |
| S6_k141_131601 | 1533 | No | NA | 2 | 0 | 1 | Not-determined | Genome-fragment | NA | NA | 0 |
| S6_k141_67584 | 3002 | No | NA | 6 | 0 | 3 | Not-determined | Genome-fragment | NA | NA | 0 |
| S6_k141_84639 | 1405 | No | NA | 2 | 0 | 2 | Not-determined | Genome-fragment | NA | NA | 0 |
| S6_k141_38144 | 3694 | No | NA | 2 | 0 | 0 | Not-determined | Genome-fragment | NA | NA | 0 |
| S6_k141_44212 | 1121 | No | NA | 1 | 0 | 1 | Not-determined | Genome-fragment | NA | NA | 0 |
| S6_k141_101887 | 4253 | No | NA | 4 | 0 | 1 | Not-determined | Genome-fragment | NA | NA | 0 |
| S6_k141_73107 | 3764 | No | NA | 4 | 0 | 1 | Not-determined | Genome-fragment | NA | NA | 0 |
| S8_k141_149604 | 8150 | No | NA | 9 | 7 | 0 | Low-quality | Genome-fragment | 18.24 | AAI-based (high-confidence) | 0 |
| S1_k141_390170 | 8151 | No | NA | 8 | 0 | 6 | Low-quality | Genome-fragment | 2.35 | AAI-based (high-confidence) | 0 |
| S12-final-viral-combined_k141_241092\|\|full | 8152 | No | NA | 10 | 0 | 8 | Low-quality | Genome-fragment | 1.61 | AAI-based (medium-confidence) | 0 |
| S6_k141_14392 | 1202 | No | NA | 2 | 0 | 1 | Not-determined | Genome-fragment | NA | NA | 0 |
| S11-final-viral-combined_k141_320814\|\|full | 8158 | No | NA | 6 | 0 | 4 | Low-quality | Genome-fragment | 2.55 | AAI-based (medium-confidence) | 0 |
| S6_k141_32245 | 7818 | No | NA | 11 | 0 | 8 | Not-determined | Genome-fragment | NA | NA | 0 |
| S6_k141_32261 | 3639 | No | NA | 7 | 0 | 0 | Not-determined | Genome-fragment | NA | NA | 0 |
| S6_k141_3655 | 3531 | No | NA | 3 | 0 | 3 | Not-determined | Genome-fragment | NA | NA | 0 |
| S6_k141_119646 | 8390 | No | NA | 9 | 0 | 1 | Not-determined | Genome-fragment | NA | NA | 0 |
| S11-final-viral-combined_k141_24265\|\|full | 8171 | No | NA | 11 | 5 | 0 | Low-quality | Genome-fragment | 15.8 | AAI-based (medium-confidence) | 0 |
| S6_k141_114036 | 1396 | No | NA | 2 | 0 | 1 | Not-determined | Genome-fragment | NA | NA | 0 |
| S6_k141_114037 | 1160 | No | NA | 1 | 0 | 0 | Not-determined | Genome-fragment | NA | NA | 0 |

| | | | | | | | | | | | |
|---|---|---|---|---|---|---|---|---|---|---|---|
| S6_k141_125970 | 1484 | No | NA | 2 | 0 | 1 | Not-determined | Genome-fragment | NA | NA | 0 |
| S6_k141_32275 | 2136 | No | NA | 1 | 0 | 1 | Not-determined | Genome-fragment | NA | NA | 0 |
| S11-final-viral-combined_k141_270097\|\|full | 8175 | No | NA | 11 | 1 | 3 | Low-quality | Genome-fragment | 2.66 | HMM-based (lower-bound) | 0 |
| S6_k141_84782 | 1659 | No | NA | 1 | 0 | 1 | Not-determined | Genome-fragment | NA | NA | 0 |
| S3_k141_117638 | 8175 | No | NA | 7 | 1 | 0 | Low-quality | Genome-fragment | 13.88 | AAI-based (medium-confidence) | 0 |
| S12-final-viral-combined_k141_357114\|\|full | 8177 | No | NA | 9 | 0 | 0 | Low-quality | Genome-fragment | 4.98 | AAI-based (medium-confidence) | 0 |
| S6_k141_38321 | 1266 | No | NA | 2 | 0 | 2 | Not-determined | Genome-fragment | NA | NA | 0 |
| S6_k141_3704 | 1070 | No | NA | 2 | 0 | 0 | Not-determined | Genome-fragment | NA | NA | 0 |
| S2_k141_187786 | 8177 | No | NA | 12 | 7 | 0 | Low-quality | Genome-fragment | 25.08 | HMM-based (lower-bound) | 0 |
| S1-final-viral-combined_k141_97767\|\|full | 8178 | No | NA | 13 | 0 | 3 | Low-quality | Genome-fragment | 24.42 | AAI-based (medium-confidence) | 0 |
| S6_k141_126018 | 4962 | No | NA | 6 | 0 | 5 | Not-determined | Genome-fragment | NA | NA | 0 |
| S6_k141_119703 | 1452 | No | NA | 2 | 0 | 2 | Not-determined | Genome-fragment | NA | NA | 0 |
| S6_k141_32342 | 1049 | No | NA | 1 | 0 | 1 | Not-determined | Genome-fragment | NA | NA | 0 |
| S6_k141_126066 | 1163 | No | NA | 1 | 0 | 1 | Not-determined | Genome-fragment | NA | NA | 0 |
| S6_k141_8500 | 6157 | No | NA | 7 | 0 | 2 | Not-determined | Genome-fragment | NA | NA | 0 |
| S6_k141_97167 | 3860 | No | NA | 2 | 0 | 2 | Not-determined | Genome-fragment | NA | NA | 0 |
| S6_k141_3824 | 15632 | No | NA | 19 | 0 | 7 | Not-determined | Genome-fragment | NA | NA | 0 |
| S6_k141_61790 | 1278 | No | NA | 2 | 0 | 1 | Not-determined | Genome-fragment | NA | NA | 0 |
| S6_k141_97173 | 5926 | No | NA | 6 | 0 | 6 | Not-determined | Genome-fragment | NA | NA | 0 |
| S5-final-viral-combined_k141_203702\|\|full | 8184 | No | NA | 7 | 0 | 5 | Low-quality | Genome-fragment | 14.45 | AAI-based (medium-confidence) | 0 |
| S6_k141_102173 | 2753 | No | NA | 1 | 0 | 1 | Not-determined | Genome-fragment | NA | NA | 0 |
| S6_k141_3856 | 1149 | No | NA | 2 | 0 | 1 | Not-determined | Genome-fragment | NA | NA | 0 |
| S6_k141_32443 | 1279 | No | NA | 5 | 0 | 4 | Not-determined | Genome-fragment | NA | NA | 0 |
| S6_k141_97201 | 1150 | No | NA | 2 | 0 | 0 | Not-determined | Genome-fragment | NA | NA | 0 |
| S6_k141_55197 | 1878 | No | NA | 2 | 0 | 2 | Not-determined | Genome-fragment | NA | NA | 0 |
| S6_k141_108668 | 3513 | No | NA | 4 | 0 | 3 | Not-determined | Genome-fragment | NA | NA | 0 |
| S6_k141_14577 | 1294 | No | NA | 2 | 0 | 2 | Not-determined | Genome-fragment | NA | NA | 0 |
| S17-final-viral-combined_k141_90143\|\|full | 8184 | No | NA | 12 | 10 | 0 | Low-quality | Genome-fragment | 19.15 | AAI-based (high-confidence) | 0 |
| S6_k141_32474 | 2294 | No | NA | 2 | 0 | 1 | Not-determined | Genome-fragment | NA | NA | 0 |
| S11-final-viral-combined_k141_171699\|\|full | 8185 | No | NA | 16 | 10 | 0 | Low-quality | Genome-fragment | 18.72 | AAI-based (high-confidence) | 0 |
| S6_k141_20040 | 1515 | No | NA | 4 | 0 | 3 | Not-determined | Genome-fragment | NA | NA | 0 |
| S6_k141_137561 | 1056 | No | NA | 1 | 0 | 0 | Not-determined | Genome-fragment | NA | NA | 0 |
| S6_k141_44501 | 1579 | No | NA | 2 | 0 | 0 | Not-determined | Genome-fragment | NA | NA | 0 |
| S6_k141_44515 | 1715 | No | NA | 2 | 0 | 2 | Not-determined | Genome-fragment | NA | NA | 0 |
| S6_k141_84966 | 4099 | No | NA | 12 | 0 | 0 | Not-determined | Genome-fragment | NA | NA | 0 |
| S6_k141_102278 | 3845 | No | NA | 6 | 0 | 6 | Not-determined | Genome-fragment | NA | NA | 0 |
| S6_k141_137591 | 13884 | No | NA | 15 | 6 | 0 | Complete | High-quality | 100 | ITR (high-confidence) | 0 |
| S6_k141_73419 | 1152 | No | NA | 2 | 0 | 2 | Not-determined | Genome-fragment | NA | NA | 0 |
| S6_k141_102331 | 1631 | No | NA | 2 | 0 | 1 | Not-determined | Genome-fragment | NA | NA | 0 |
| S6_k141_14653 | 2527 | No | NA | 4 | 0 | 2 | Not-determined | Genome-fragment | NA | NA | 0 |
| S6_k141_97349 | 1562 | No | NA | 2 | 0 | 1 | Not-determined | Genome-fragment | NA | NA | 0 |
| S6_k141_126257 | 1031 | No | NA | 1 | 0 | 1 | Not-determined | Genome-fragment | NA | NA | 0 |
| S6_k141_114391 | 1071 | No | NA | 2 | 0 | 1 | Not-determined | Genome-fragment | NA | NA | 0 |
| S6_k141_8603 | 1023 | No | NA | 3 | 0 | 2 | Not-determined | Genome-fragment | NA | NA | 0 |
| S6_k141_4064 | 2012 | No | NA | 1 | 0 | 0 | Not-determined | Genome-fragment | NA | NA | 0 |
| S12-final-viral-combined_k141_103343\|\|full | 8186 | No | NA | 13 | 7 | 0 | Low-quality | Genome-fragment | 22.34 | AAI-based (high-confidence) | 0 |
| S6_k141_68042 | 1566 | No | NA | 2 | 0 | 2 | Not-determined | Genome-fragment | NA | NA | 0 |
| S6_k141_14683 | 2855 | No | NA | 4 | 0 | 1 | Not-determined | Genome-fragment | NA | NA | 0 |
| S6_k141_102400 | 2555 | No | NA | 4 | 0 | 1 | Not-determined | Genome-fragment | NA | NA | 0 |
| S6_k141_14725 | 1093 | No | NA | 1 | 0 | 0 | Not-determined | Genome-fragment | NA | NA | 0 |
| S6_k141_14740 | 1264 | No | NA | 1 | 0 | 1 | Not-determined | Genome-fragment | NA | NA | 0 |
| S6_k141_4115 | 1601 | No | NA | 2 | 0 | 0 | Not-determined | Genome-fragment | NA | NA | 0 |
| S6_k141_79041 | 4030 | No | NA | 5 | 0 | 2 | Not-determined | Genome-fragment | NA | NA | 0 |
| S6_k141_108940 | 1324 | No | NA | 4 | 0 | 3 | Not-determined | Genome-fragment | NA | NA | 0 |
| S6_k141_120051 | 1329 | No | NA | 1 | 0 | 1 | Not-determined | Genome-fragment | NA | NA | 0 |
| S6_k141_120071 | 1389 | No | NA | 2 | 0 | 0 | Not-determined | Genome-fragment | NA | NA | 0 |
| S6_k141_62122 | 3275 | No | NA | 1 | 0 | 1 | Not-determined | Genome-fragment | NA | NA | 0 |
| S6_k141_20239 | 1284 | No | NA | 2 | 0 | 1 | Not-determined | Genome-fragment | NA | NA | 0 |
| S6_k141_120107 | 2218 | No | NA | 2 | 0 | 2 | Not-determined | Genome-fragment | NA | NA | 0 |
| S8-final-viral-combined_k141_158208\|\|full | 8187 | No | NA | 8 | 0 | 1 | Low-quality | Genome-fragment | 12.4 | AAI-based (medium-confidence) | 0 |
| S6_k141_14800 | 1790 | No | NA | 2 | 0 | 2 | Not-determined | Genome-fragment | NA | NA | 0 |
| S6_k141_8641 | 1103 | No | NA | 1 | 0 | 1 | Not-determined | Genome-fragment | NA | NA | 0 |
| S6_k141_91133 | 1387 | No | NA | 2 | 0 | 2 | Not-determined | Genome-fragment | NA | NA | 0 |
| S6_k141_14818 | 1321 | No | NA | 3 | 0 | 0 | Not-determined | Genome-fragment | NA | NA | 0 |
| S8_k141_74066 | 8188 | No | NA | 5 | 0 | 3 | Low-quality | Genome-fragment | 14.68 | AAI-based (medium-confidence) | 0 |
| S6_k141_91168 | 1234 | No | NA | 2 | 0 | 0 | Not-determined | Genome-fragment | NA | NA | 0 |
| S6_k141_14874 | 1123 | No | NA | 2 | 0 | 2 | Not-determined | Genome-fragment | NA | NA | 0 |
| S5-final-viral-combined_k141_117630\|\|full | 8193 | No | NA | 6 | 1 | 0 | Low-quality | Genome-fragment | 20.77 | AAI-based (medium-confidence) | 0 |
| S6_k141_79218 | 3472 | No | NA | 4 | 0 | 3 | Not-determined | Genome-fragment | NA | NA | 0 |
| S6_k141_26390 | 1243 | No | NA | 2 | 0 | 0 | Not-determined | Genome-fragment | NA | NA | 0 |
| S5-final-viral-combined_k141_28796\|\|full | 8194 | No | NA | 17 | 3 | 0 | Low-quality | Genome-fragment | 7.85 | HMM-based (lower-bound) | 0 |
| S6_k141_8786 | 1103 | No | NA | 1 | 0 | 1 | Not-determined | Genome-fragment | NA | NA | 0 |
| S6_k141_4315 | 3072 | No | NA | 4 | 0 | 4 | Not-determined | Genome-fragment | NA | NA | 0 |
| S6_k141_8796 | 16963 | No | NA | 17 | 0 | 6 | Not-determined | Genome-fragment | NA | NA | 0 |
| S6_k141_38880 | 1278 | No | NA | 2 | 0 | 1 | Not-determined | Genome-fragment | NA | NA | 0 |
| S6_k141_91257 | 1133 | No | NA | 1 | 0 | 0 | Not-determined | Genome-fragment | NA | NA | 0 |
| S6_k141_8826 | 1677 | No | NA | 1 | 0 | 1 | Not-determined | Genome-fragment | NA | NA | 0 |
| S6_k141_55509 | 5672 | No | NA | 4 | 0 | 0 | Not-determined | Genome-fragment | NA | NA | 0 |
| S6_k141_126588 | 4622 | No | NA | 4 | 0 | 0 | Not-determined | Genome-fragment | NA | NA | 0 |
| S6_k141_137917 | 9417 | No | NA | 9 | 0 | 8 | Not-determined | Genome-fragment | NA | NA | 0 |
| S6_k141_137925 | 3281 | No | NA | 2 | 0 | 1 | Not-determined | Genome-fragment | NA | NA | 0 |
| S6_k141_73730 | 4192 | No | NA | 1 | 0 | 1 | Not-determined | Genome-fragment | NA | NA | 0 |
| S6_k141_102667 | 1093 | No | NA | 1 | 0 | 1 | Not-determined | Genome-fragment | NA | NA | 0 |
| S9-final-viral-combined_k141_82096\|\|full | 8195 | No | NA | 12 | 6 | 0 | Low-quality | Genome-fragment | 18.11 | AAI-based (high-confidence) | 0 |
| S6_k141_114737 | 5312 | No | NA | 6 | 0 | 3 | Not-determined | Genome-fragment | NA | NA | 0 |
| S6_k141_55564 | 3844 | No | NA | 4 | 0 | 3 | Not-determined | Genome-fragment | NA | NA | 0 |
| S6_k141_132443 | 1388 | No | NA | 1 | 0 | 1 | Not-determined | Genome-fragment | NA | NA | 0 |
| S6_k141_102704 | 1890 | No | NA | 2 | 0 | 2 | Not-determined | Genome-fragment | NA | NA | 0 |
| S6_k141_79336 | 1412 | No | NA | 2 | 0 | 1 | Not-determined | Genome-fragment | NA | NA | 0 |
| S6_k141_4462 | 4312 | No | NA | 1 | 0 | 0 | Not-determined | Genome-fragment | NA | NA | 0 |
| S6_k141_62450 | 1634 | No | NA | 1 | 0 | 0 | Not-determined | Genome-fragment | NA | NA | 0 |
| S6_k141_20412 | 1250 | No | NA | 1 | 0 | 1 | Not-determined | Genome-fragment | NA | NA | 0 |
| S6_k141_114773 | 1904 | No | NA | 3 | 0 | 3 | Not-determined | Genome-fragment | NA | NA | 0 |
| S6_k141_15029 | 1285 | No | NA | 2 | 0 | 2 | Not-determined | Genome-fragment | NA | NA | 0 |
| S6_k141_50538 | 1071 | No | NA | 2 | 0 | 1 | Not-determined | Genome-fragment | NA | NA | 0 |
| S6_k141_44901 | 1147 | No | NA | 1 | 0 | 0 | Not-determined | Genome-fragment | NA | NA | 0 |
| S6_k141_85481 | 1375 | No | NA | 1 | 0 | 1 | Not-determined | Genome-fragment | NA | NA | 0 |
| S6_k141_138029 | 6087 | No | NA | 9 | 0 | 1 | Not-determined | Genome-fragment | NA | NA | 0 |
| S12-final-viral-combined_k141_305153\|\|full | 8196 | Yes | 1433 | 8 | 1 | 2 | Low-quality | Genome-fragment | 0.45 | AAI-based (medium-confidence) | 82.52 |
| S17-final-viral-combined_k141_26353\|\|full | 8200 | No | NA | 7 | 1 | 2 | Low-quality | Genome-fragment | 6.02 | HMM-based (lower-bound) | 0 |
| S6_k141_9037 | 4620 | No | NA | 3 | 0 | 1 | Not-determined | Genome-fragment | NA | NA | 0 |
| S6_k141_138082 | 1129 | No | NA | 2 | 0 | 0 | Not-determined | Genome-fragment | NA | NA | 0 |
| S6_k141_132588 | 52822 | No | NA | 69 | 22 | 0 | Medium-quality | Genome-fragment | 87.02 | AAI-based (high-confidence) | 0 |
| S6_k141_32935 | 3073 | No | NA | 5 | 0 | 4 | Not-determined | Genome-fragment | NA | NA | 0 |
| S6_k141_50662 | 8017 | No | NA | 6 | 0 | 1 | Not-determined | Genome-fragment | NA | NA | 0 |
| S6_k141_4602 | 1133 | No | NA | 2 | 0 | 0 | Not-determined | Genome-fragment | NA | NA | 0 |
| S6_k141_91543 | 1533 | No | NA | 2 | 0 | 0 | Not-determined | Genome-fragment | NA | NA | 0 |
| S6_k141_4625 | 1241 | No | NA | 3 | 0 | 0 | Not-determined | Genome-fragment | NA | NA | 0 |
| S11-final-viral-combined_k141_116261\|\|full | 8201 | No | NA | 13 | 4 | 0 | Low-quality | Genome-fragment | 18.13 | AAI-based (high-confidence) | 0 |
| S6_k141_85657 | 1208 | No | NA | 3 | 0 | 3 | Not-determined | Genome-fragment | NA | NA | 0 |
| S6_k141_85664 | 1188 | No | NA | 1 | 0 | 0 | Not-determined | Genome-fragment | NA | NA | 0 |
| S6_k141_120476 | 3087 | No | NA | 2 | 0 | 1 | Not-determined | Genome-fragment | NA | NA | 0 |
| S6_k141_9089 | 1190 | No | NA | 0 | 0 | 0 | Not-determined | Genome-fragment | NA | NA | 0 |
| S6_k141_138194 | 1189 | No | NA | 1 | 0 | 1 | Not-determined | Genome-fragment | NA | NA | 0 |
| S6_k141_9100 | 4914 | No | NA | 3 | 0 | 3 | Not-determined | Genome-fragment | NA | NA | 0 |
| S7-final-viral-combined_k141_134467\|\|full | 8203 | No | NA | 13 | 0 | 0 | Low-quality | Genome-fragment | 19.51 | AAI-based (medium-confidence) | 0 |
| S6_k141_138214 | 1258 | No | NA | 1 | 0 | 1 | Not-determined | Genome-fragment | NA | NA | 0 |
| S2_k141_166892 | 8203 | No | NA | 12 | 0 | 4 | Low-quality | Genome-fragment | 9.82 | AAI-based (medium-confidence) | 0 |
| S6_k141_102965 | 1982 | No | NA | 3 | 0 | 2 | Not-determined | Genome-fragment | NA | NA | 0 |
| S6_k141_4720 | 2646 | No | NA | 2 | 0 | 2 | Not-determined | Genome-fragment | NA | NA | 0 |
| S6_k141_114967 | 1040 | No | NA | 1 | 0 | 0 | Not-determined | Genome-fragment | NA | NA | 0 |
| S11-final-viral-combined_k141_266156\|\|full | 8204 | No | NA | 19 | 6 | 0 | Low-quality | Genome-fragment | 10.93 | AAI-based (high-confidence) | 0 |
| S6_k141_4739 | 3436 | No | NA | 3 | 0 | 1 | Not-determined | Genome-fragment | NA | NA | 0 |
| S6_k141_26815 | 3948 | No | NA | 6 | 0 | 2 | Not-determined | Genome-fragment | NA | NA | 0 |
| S6_k141_85741 | 1168 | No | NA | 2 | 0 | 2 | Not-determined | Genome-fragment | NA | NA | 0 |
| S6_k141_62746 | 1217 | No | NA | 2 | 0 | 0 | Not-determined | Genome-fragment | NA | NA | 0 |
| S6_k141_109320 | 2591 | No | NA | 3 | 0 | 2 | Not-determined | Genome-fragment | NA | NA | 0 |
| S6_k141_20671 | 1063 | No | NA | 1 | 0 | 1 | Not-determined | Genome-fragment | NA | NA | 0 |
| S6_k141_68762 | 1047 | No | NA | 3 | 0 | 2 | Not-determined | Genome-fragment | NA | NA | 0 |
| S3_k141_128569 | 8205 | No | NA | 7 | 0 | 0 | Low-quality | Genome-fragment | 7.78 | AAI-based (high-confidence) | 0 |
| S6_k141_15274 | 3171 | No | NA | 2 | 0 | 1 | Not-determined | Genome-fragment | NA | NA | 0 |
| S6_k141_91733 | 2110 | No | NA | 4 | 0 | 0 | Not-determined | Genome-fragment | NA | NA | 0 |
| S12-final-viral-combined_k141_244553\|\|full | 8207 | No | NA | 13 | 0 | 1 | Low-quality | Genome-fragment | 22.82 | AAI-based (medium-confidence) | 0 |
| S6_k141_62801 | 4918 | No | NA | 1 | 0 | 0 | Not-determined | Genome-fragment | NA | NA | 0 |
| S6_k141_39195 | 5102 | No | NA | 9 | 0 | 2 | Not-determined | Genome-fragment | NA | NA | 0 |
| S6_k141_26887 | 1892 | No | NA | 4 | 0 | 4 | Not-determined | Genome-fragment | NA | NA | 0 |
| S6_k141_62828 | 3409 | No | NA | 3 | 0 | 1 | Not-determined | Genome-fragment | NA | NA | 0 |
| S6_k141_39215 | 1045 | No | NA | 2 | 0 | 2 | Not-determined | Genome-fragment | NA | NA | 0 |
| S7-final-viral-combined_k141_216761\|\|full | 8208 | No | NA | 6 | 0 | 6 | Low-quality | Genome-fragment | 2.26 | AAI-based (medium-confidence) | 0 |
| S6_k141_68819 | 1683 | No | NA | 1 | 0 | 1 | Not-determined | Genome-fragment | NA | NA | 0 |
| S6_k141_109427 | 1488 | No | NA | 2 | 0 | 1 | Not-determined | Genome-fragment | NA | NA | 0 |
| S6_k141_138331 | 1188 | No | NA | 1 | 0 | 0 | Not-determined | Genome-fragment | NA | NA | 0 |
| S6_k141_138332 | 1407 | No | NA | 1 | 0 | 1 | Not-determined | Genome-fragment | NA | NA | 0 |
| S6_k141_4911 | 1184 | No | NA | 3 | 0 | 2 | Not-determined | Genome-fragment | NA | NA | 0 |
| S6_k141_9201 | 2824 | No | NA | 2 | 0 | 2 | Not-determined | Genome-fragment | NA | NA | 0 |
| S6_k141_120637 | 2854 | No | NA | 3 | 0 | 2 | Not-determined | Genome-fragment | NA | NA | 0 |
| S6_k141_9210 | 5927 | No | NA | 9 | 0 | 7 | Not-determined | Genome-fragment | NA | NA | 0 |
| S6_k141_62903 | 1142 | No | NA | 2 | 0 | 2 | Not-determined | Genome-fragment | NA | NA | 0 |
| S6_k141_50819 | 1582 | No | NA | 3 | 0 | 2 | Not-determined | Genome-fragment | NA | NA | 0 |
| S2-final-viral-combined_k141_145788\|\|full | 8211 | No | NA | 12 | 1 | 1 | Low-quality | Genome-fragment | 10.87 | HMM-based (lower-bound) | 0 |
| S1-final-viral-combined_k141_315918\|\|full | 8214 | No | NA | 14 | 3 | 0 | Low-quality | Genome-fragment | 22.9 | AAI-based (high-confidence) | 0 |
| S6_k141_115201 | 1281 | No | NA | 4 | 0 | 1 | Not-determined | Genome-fragment | NA | NA | 0 |
| S6_k141_15470 | 1054 | No | NA | 2 | 0 | 2 | Not-determined | Genome-fragment | NA | NA | 0 |
| S8_k141_2595 | 8214 | No | NA | 19 | 0 | 1 | Low-quality | Genome-fragment | 20.81 | AAI-based (high-confidence) | 0 |
| S6_k141_85943 | 3304 | No | NA | 4 | 0 | 0 | Not-determined | Genome-fragment | NA | NA | 0 |
| S6_k141_85944 | 1517 | No | NA | 3 | 0 | 3 | Not-determined | Genome-fragment | NA | NA | 0 |
| S6_k141_138342 | 99625 | No | NA | 102 | 15 | 2 | Complete | High-quality | 100 | DTR (high-confidence) | 0 |
| S6_k141_126996 | 1060 | No | NA | 1 | 0 | 1 | Not-determined | Genome-fragment | NA | NA | 0 |
| S1-final-viral-combined_k141_35591\|\|full | 8217 | No | NA | 18 | 4 | 0 | Low-quality | Genome-fragment | 19.09 | AAI-based (high-confidence) | 0 |
| S6-final-viral-combined_k141_25004\|\|full | 8217 | No | NA | 5 | 0 | 1 | Low-quality | Genome-fragment | 7.46 | AAI-based (medium-confidence) | 0 |
| S6_k141_9298 | 1215 | No | NA | 1 | 0 | 0 | Not-determined | Genome-fragment | NA | NA | 0 |
| S6_k141_120763 | 1446 | No | NA | 1 | 0 | 0 | Not-determined | Genome-fragment | NA | NA | 0 |
| S7_k141_152605 | 8219 | Yes | 247 | 4 | 1 | 1 | Low-quality | Genome-fragment | 0.24 | AAI-based (high-confidence) | 96.99 |
| S6_k141_109594 | 4953 | No | NA | 7 | 0 | 1 | Not-determined | Genome-fragment | NA | NA | 0 |
| S6_k141_91938 | 1430 | No | NA | 2 | 0 | 1 | Not-determined | Genome-fragment | NA | NA | 0 |
| S1-final-viral-combined_k141_244190\|\|full | 8220 | No | NA | 7 | 0 | 2 | Low-quality | Genome-fragment | 5.36 | AAI-based (medium-confidence) | 0 |
| S6_k141_20850 | 1524 | No | NA | 2 | 0 | 1 | Not-determined | Genome-fragment | NA | NA | 0 |
| S6_k141_50963 | 1063 | No | NA | 1 | 0 | 1 | Not-determined | Genome-fragment | NA | NA | 0 |
| S6_k141_5110 | 1549 | No | NA | 1 | 0 | 1 | Not-determined | Genome-fragment | NA | NA | 0 |
| S6_k141_69010 | 1162 | No | NA | 1 | 0 | 0 | Not-determined | Genome-fragment | NA | NA | 0 |
| S6_k141_27115 | 1378 | No | NA | 1 | 0 | 1 | Not-determined | Genome-fragment | NA | NA | 0 |
| S6_k141_20898 | 1313 | No | NA | 2 | 0 | 1 | Not-determined | Genome-fragment | NA | NA | 0 |
| S6_k141_120813 | 6146 | No | NA | 7 | 0 | 6 | Not-determined | Genome-fragment | NA | NA | 0 |
| S6_k141_63105 | 1975 | No | NA | 1 | 0 | 1 | Not-determined | Genome-fragment | NA | NA | 0 |
| S6_k141_9428 | 2073 | No | NA | 2 | 0 | 2 | Not-determined | Genome-fragment | NA | NA | 0 |
| S6_k141_127079 | 4365 | No | NA | 2 | 0 | 2 | Not-determined | Genome-fragment | NA | NA | 0 |
| S6_k141_127081 | 2044 | No | NA | 2 | 0 | 2 | Not-determined | Genome-fragment | NA | NA | 0 |
| S1_k141_321793 | 8224 | No | NA | 13 | 1 | 2 | Low-quality | Genome-fragment | 5.01 | HMM-based (lower-bound) | 0 |
| S6_k141_74247 | 1690 | No | NA | 2 | 0 | 0 | Not-determined | Genome-fragment | NA | NA | 0 |
| S6_k141_39539 | 8134 | No | NA | 8 | 0 | 7 | Not-determined | Genome-fragment | NA | NA | 0 |
| S6_k141_115407 | 3525 | No | NA | 1 | 0 | 1 | Not-determined | Genome-fragment | NA | NA | 0 |
| S6_k141_5244 | 1229 | No | NA | 1 | 0 | 1 | Not-determined | Genome-fragment | NA | NA | 0 |
| S6_k141_15674 | 3899 | No | NA | 7 | 0 | 6 | Not-determined | Genome-fragment | NA | NA | 0 |
| S6_k141_127204 | 1208 | No | NA | 2 | 0 | 1 | Not-determined | Genome-fragment | NA | NA | 0 |
| S6_k141_98387 | 1149 | No | NA | 2 | 0 | 0 | Not-determined | Genome-fragment | NA | NA | 0 |
| S1-final-viral-combined_k141_389755\|\|full | 8231 | No | NA | 7 | 1 | 2 | Low-quality | Genome-fragment | 9.24 | HMM-based (lower-bound) | 0 |
| S6_k141_63265 | 2322 | No | NA | 3 | 0 | 1 | Not-determined | Genome-fragment | NA | NA | 0 |
| S6_k141_63285 | 3040 | No | NA | 3 | 0 | 1 | Not-determined | Genome-fragment | NA | NA | 0 |
| S2-final-viral-combined_k141_74248\|\|full | 8233 | No | NA | 11 | 3 | 2 | Low-quality | Genome-fragment | 19.8 | AAI-based (high-confidence) | 0 |
| S8-final-viral-combined_k141_36444\|\|full | 8234 | No | NA | 6 | 0 | 6 | Low-quality | Genome-fragment | 11.27 | AAI-based (medium-confidence) | 0 |
| S6_k141_51154 | 2211 | No | NA | 6 | 0 | 6 | Not-determined | Genome-fragment | NA | NA | 0 |
| S6_k141_109708 | 1112 | No | NA | 3 | 0 | 0 | Not-determined | Genome-fragment | NA | NA | 0 |
| S6_k141_15779 | 1059 | No | NA | 2 | 0 | 0 | Not-determined | Genome-fragment | NA | NA | 0 |
| S6_k141_63321 | 2497 | No | NA | 3 | 0 | 1 | Not-determined | Genome-fragment | NA | NA | 0 |
| S6_k141_15783 | 1081 | No | NA | 1 | 0 | 1 | Not-determined | Genome-fragment | NA | NA | 0 |
| S6_k141_98449 | 3058 | No | NA | 6 | 0 | 2 | Not-determined | Genome-fragment | NA | NA | 0 |
| S6_k141_74389 | 1007 | No | NA | 1 | 0 | 0 | Not-determined | Genome-fragment | NA | NA | 0 |
| S12-final-viral-combined_k141_158555\|\|full | 8242 | No | NA | 12 | 0 | 0 | Low-quality | Genome-fragment | 15.45 | AAI-based (high-confidence) | 0 |
| S9-final-viral-combined_k141_110068\|\|full | 8244 | No | NA | 9 | 4 | 0 | Low-quality | Genome-fragment | 13.15 | AAI-based (high-confidence) | 0 |
| S6_k141_56151 | 1185 | No | NA | 1 | 0 | 0 | Not-determined | Genome-fragment | NA | NA | 0 |
| S6_k141_27334 | 7987 | No | NA | 13 | 0 | 2 | Not-determined | Genome-fragment | NA | NA | 0 |
| S6_k141_5402 | 1385 | No | NA | 2 | 0 | 2 | Not-determined | Genome-fragment | NA | NA | 0 |
| S6_k141_132885 | 4498 | No | NA | 3 | 0 | 1 | Not-determined | Genome-fragment | NA | NA | 0 |
| S6_k141_103534 | 5058 | No | NA | 4 | 0 | 3 | Not-determined | Genome-fragment | NA | NA | 0 |
| S7_k141_79865 | 8251 | No | NA | 12 | 3 | 0 | Low-quality | Genome-fragment | 17.01 | AAI-based (high-confidence) | 0 |
| S17-final-viral-combined_k141_161995\|\|full | 8252 | No | NA | 18 | 1 | 0 | Low-quality | Genome-fragment | 19.44 | AAI-based (high-confidence) | 0 |
| S8_k141_192222 | 8254 | No | NA | 6 | 0 | 4 | Low-quality | Genome-fragment | 4.27 | AAI-based (medium-confidence) | 0 |
| S6_k141_27372 | 2402 | No | NA | 3 | 0 | 3 | Not-determined | Genome-fragment | NA | NA | 0 |
| S6_k141_5431 | 1171 | No | NA | 2 | 0 | 0 | Not-determined | Genome-fragment | NA | NA | 0 |
| S6_k141_86321 | 1362 | No | NA | 3 | 0 | 2 | Not-determined | Genome-fragment | NA | NA | 0 |
| S6_k141_63367 | 1211 | No | NA | 1 | 0 | 0 | Not-determined | Genome-fragment | NA | NA | 0 |
| S6_k141_121057 | 3687 | No | NA | 3 | 0 | 1 | Not-determined | Genome-fragment | NA | NA | 0 |
| S12_k141_241604 | 8267 | No | NA | 5 | 0 | 1 | Low-quality | Genome-fragment | 7.42 | AAI-based (medium-confidence) | 0 |
| S3-final-viral-combined_k141_37545\|\|full | 8268 | Yes | 4750 | 11 | 4 | 2 | Low-quality | Genome-fragment | 7.87 | AAI-based (high-confidence) | 42.55 |
| S6_k141_21084 | 1021 | No | NA | 2 | 0 | 0 | Not-determined | Genome-fragment | NA | NA | 0 |
| S8-final-viral-combined_k141_51431\|\|full | 8268 | No | NA | 7 | 5 | 0 | Low-quality | Genome-fragment | 16.25 | AAI-based (medium-confidence) | 0 |
| S6_k141_86409 | 4121 | No | NA | 4 | 0 | 3 | Not-determined | Genome-fragment | NA | NA | 0 |
| S17-final-viral-combined_k141_26240\|\|full | 8270 | No | NA | 10 | 1 | 0 | Low-quality | Genome-fragment | 21.4 | AAI-based (medium-confidence) | 0 |
| S17-final-viral-combined_k141_148074\|\|full | 8271 | No | NA | 12 | 7 | 0 | Low-quality | Genome-fragment | 20.34 | AAI-based (high-confidence) | 0 |
| S6_k141_63498 | 1188 | No | NA | 1 | 0 | 0 | Not-determined | Genome-fragment | NA | NA | 0 |
| S6_k141_63516 | 1000 | No | NA | 1 | 0 | 1 | Not-determined | Genome-fragment | NA | NA | 0 |
| S6_k141_109985 | 1848 | No | NA | 2 | 0 | 2 | Not-determined | Genome-fragment | NA | NA | 0 |
| S6_k141_63542 | 1166 | No | NA | 3 | 0 | 0 | Not-determined | Genome-fragment | NA | NA | 0 |
| S17-final-viral-combined_k141_226354\|\|full | 8271 | No | NA | 8 | 4 | 0 | Low-quality | Genome-fragment | 20.43 | AAI-based (medium-confidence) | 0 |
| S6_k141_45864 | 1398 | No | NA | 1 | 0 | 1 | Not-determined | Genome-fragment | NA | NA | 0 |
| S6_k141_45877 | 1261 | No | NA | 1 | 0 | 1 | Not-determined | Genome-fragment | NA | NA | 0 |
| S6_k141_110020 | 1088 | No | NA | 2 | 0 | 1 | Not-determined | Genome-fragment | NA | NA | 0 |
| S3_k141_41295 | 8274 | No | NA | 5 | 0 | 4 | Low-quality | Genome-fragment | 0.71 | AAI-based (medium-confidence) | 0 |
| S7_k141_138826 | 8281 | No | NA | 6 | 1 | 4 | Low-quality | Genome-fragment | 2.2 | AAI-based (high-confidence) | 0 |
| S6_k141_98738 | 1292 | No | NA | 2 | 0 | 2 | Not-determined | Genome-fragment | NA | NA | 0 |
| S6_k141_21243 | 3612 | No | NA | 4 | 0 | 4 | Not-determined | Genome-fragment | NA | NA | 0 |
| S8-final-viral-combined_k141_257197\|\|full | 8282 | No | NA | 13 | 1 | 0 | Low-quality | Genome-fragment | 5.01 | HMM-based (lower-bound) | 0 |
| S5-final-viral-combined_k141_29462\|\|full | 8285 | No | NA | 14 | 6 | 1 | Low-quality | Genome-fragment | 17.44 | AAI-based (high-confidence) | 0 |
| S8_k141_127423 | 8287 | No | NA | 12 | 7 | 0 | Low-quality | Genome-fragment | 24.15 | AAI-based (high-confidence) | 0 |
| S6_k141_51542 | 8157 | No | NA | 13 | 0 | 2 | Not-determined | Genome-fragment | NA | NA | 0 |
| S6_k141_16163 | 4488 | No | NA | 6 | 0 | 1 | Not-determined | Genome-fragment | NA | NA | 0 |
| S6_k141_138950 | 5155 | No | NA | 7 | 0 | 2 | Not-determined | Genome-fragment | NA | NA | 0 |
| S6_k141_103710 | 4787 | No | NA | 5 | 0 | 5 | Not-determined | Genome-fragment | NA | NA | 0 |
| S6_k141_80393 | 2093 | No | NA | 3 | 0 | 2 | Not-determined | Genome-fragment | NA | NA | 0 |
| S11_k141_205397 | 8289 | No | NA | 11 | 4 | 0 | Low-quality | Genome-fragment | 37.08 | AAI-based (medium-confidence) | 0 |
| S6_k141_74772 | 1517 | No | NA | 3 | 0 | 3 | Not-determined | Genome-fragment | NA | NA | 0 |
| S6_k141_21323 | 5192 | No | NA | 3 | 0 | 2 | Not-determined | Genome-fragment | NA | NA | 0 |
| S6_k141_5738 | 4396 | No | NA | 3 | 0 | 3 | Not-determined | Genome-fragment | NA | NA | 0 |
| S6_k141_74785 | 1088 | No | NA | 2 | 0 | 1 | Not-determined | Genome-fragment | NA | NA | 0 |
| S1-final-viral-combined_k141_219989\|\|full | 8289 | No | NA | 15 | 1 | 1 | Low-quality | Genome-fragment | 8.29 | AAI-based (medium-confidence) | 0 |
| S6_k141_45998 | 6629 | No | NA | 6 | 0 | 4 | Not-determined | Genome-fragment | NA | NA | 0 |
| S6_k141_74809 | 2362 | No | NA | 1 | 0 | 0 | Not-determined | Genome-fragment | NA | NA | 0 |
| S1-final-viral-combined_k141_217110\|\|full | 8291 | No | NA | 11 | 1 | 1 | Low-quality | Genome-fragment | 5.21 | HMM-based (lower-bound) | 0 |
| S6_k141_133245 | 1561 | No | NA | 2 | 0 | 2 | Not-determined | Genome-fragment | NA | NA | 0 |
| S6_k141_139010 | 1387 | No | NA | 1 | 0 | 1 | Not-determined | Genome-fragment | NA | NA | 0 |
| S6_k141_46025 | 1347 | No | NA | 1 | 0 | 1 | Not-determined | Genome-fragment | NA | NA | 0 |
| S6_k141_98916 | 1927 | No | NA | 2 | 0 | 1 | Not-determined | Genome-fragment | NA | NA | 0 |
| S6_k141_98925 | 1213 | No | NA | 2 | 0 | 1 | Not-determined | Genome-fragment | NA | NA | 0 |
| S6_k141_40086 | 4500 | No | NA | 2 | 0 | 2 | Not-determined | Genome-fragment | NA | NA | 0 |
| S6_k141_139049 | 1032 | No | NA | 2 | 0 | 2 | Not-determined | Genome-fragment | NA | NA | 0 |
| S1-final-viral-combined_k141_347664\|\|full | 8294 | No | NA | 13 | 1 | 2 | Low-quality | Genome-fragment | 16.75 | AAI-based (high-confidence) | 0 |
| S6_k141_133325 | 1875 | No | NA | 3 | 0 | 0 | Not-determined | Genome-fragment | NA | NA | 0 |
| S12-final-viral-combined_k141_245693\|\|full | 8298 | No | NA | 9 | 0 | 6 | Low-quality | Genome-fragment | 1.29 | AAI-based (medium-confidence) | 0 |
| S6_k141_121383 | 1105 | No | NA | 3 | 0 | 2 | Not-determined | Genome-fragment | NA | NA | 0 |
| S6_k141_46155 | 1874 | No | NA | 2 | 0 | 1 | Not-determined | Genome-fragment | NA | NA | 0 |
| S6_k141_121387 | 4186 | No | NA | 3 | 0 | 3 | Not-determined | Genome-fragment | NA | NA | 0 |
| S6_k141_63814 | 1402 | No | NA | 2 | 0 | 2 | Not-determined | Genome-fragment | NA | NA | 0 |
| S6_k141_133371 | 1125 | No | NA | 1 | 0 | 1 | Not-determined | Genome-fragment | NA | NA | 0 |
| S6_k141_127581 | 3964 | No | NA | 2 | 0 | 0 | Not-determined | Genome-fragment | NA | NA | 0 |
| S7_k141_74002 | 8301 | No | NA | 17 | 3 | 0 | Low-quality | Genome-fragment | 17.17 | AAI-based (high-confidence) | 0 |
| S6_k141_115984 | 1463 | No | NA | 1 | 0 | 1 | Not-determined | Genome-fragment | NA | NA | 0 |
| S6_k141_27863 | 7083 | No | NA | 7 | 0 | 1 | Not-determined | Genome-fragment | NA | NA | 0 |
| S6_k141_86651 | 6555 | No | NA | 8 | 0 | 4 | Not-determined | Genome-fragment | NA | NA | 0 |
| S6_k141_99037 | 1509 | No | NA | 3 | 0 | 0 | Not-determined | Genome-fragment | NA | NA | 0 |
| S6_k141_9983 | 2580 | No | NA | 1 | 0 | 1 | Not-determined | Genome-fragment | NA | NA | 0 |
| S8-final-viral-combined_k141_44160\|\|full | 8301 | No | NA | 5 | 0 | 2 | Low-quality | Genome-fragment | 4.24 | AAI-based (medium-confidence) | 0 |
| S6_k141_99074 | 4692 | No | NA | 6 | 0 | 3 | Not-determined | Genome-fragment | NA | NA | 0 |
| S6_k141_86680 | 3587 | No | NA | 3 | 0 | 3 | Not-determined | Genome-fragment | NA | NA | 0 |
| S6_k141_9994 | 2665 | No | NA | 1 | 0 | 1 | Not-determined | Genome-fragment | NA | NA | 0 |
| S2_k141_420 | 8302 | No | NA | 11 | 1 | 1 | Low-quality | Genome-fragment | 11.08 | AAI-based (high-confidence) | 0 |
| S6_k141_127685 | 3326 | No | NA | 4 | 0 | 0 | Not-determined | Genome-fragment | NA | NA | 0 |
| S6_k141_139078 | 5308 | No | NA | 7 | 0 | 3 | Not-determined | Genome-fragment | NA | NA | 0 |
| S6_k141_10013 | 4472 | No | NA | 5 | 0 | 2 | Not-determined | Genome-fragment | NA | NA | 0 |
| S6_k141_69532 | 1319 | No | NA | 1 | 0 | 0 | Not-determined | Genome-fragment | NA | NA | 0 |
| S7-final-viral-combined_k141_109212\|\|full | 8308 | No | NA | 8 | 0 | 3 | Low-quality | Genome-fragment | 20.93 | AAI-based (medium-confidence) | 0 |
| S2-final-viral-combined_k141_171965\|\|full | 8308 | Yes | 5658 | 10 | 1 | 2 | Low-quality | Genome-fragment | 8.46 | HMM-based (lower-bound) | 31.9 |
| S2_k141_164187 | 8309 | No | NA | 7 | 0 | 6 | Low-quality | Genome-fragment | 3.41 | AAI-based (high-confidence) | 0 |
| S6_k141_117717 | 8310 | Yes | 2826 | 6 | 1 | 4 | Low-quality | Genome-fragment | 3.95 | AAI-based (medium-confidence) | 65.99 |
| S12-final-viral-combined_k141_170251\|\|full | 8312 | No | NA | 3 | 0 | 0 | Low-quality | Genome-fragment | 21.81 | AAI-based (high-confidence) | 0 |
| S6_k141_27986 | 1108 | No | NA | 1 | 0 | 1 | Not-determined | Genome-fragment | NA | NA | 0 |
| S6_k141_40283 | 1438 | No | NA | 1 | 0 | 1 | Not-determined | Genome-fragment | NA | NA | 0 |
| S6_k141_92698 | 1164 | No | NA | 2 | 0 | 1 | Not-determined | Genome-fragment | NA | NA | 0 |
| S6_k141_21519 | 1755 | No | NA | 2 | 0 | 1 | Not-determined | Genome-fragment | NA | NA | 0 |
| S12-final-viral-combined_k141_297646\|\|full | 8319 | No | NA | 7 | 0 | 0 | Low-quality | Genome-fragment | 7.56 | AAI-based (medium-confidence) | 0 |
| S9-final-viral-combined_k141_108225\|\|full | 8321 | Yes | 2412 | 5 | 1 | 3 | Low-quality | Genome-fragment | 1.44 | HMM-based (lower-bound) | 71.01 |
| S6_k141_46316 | 3299 | No | NA | 1 | 0 | 0 | Not-determined | Genome-fragment | NA | NA | 0 |
| S6_k141_40344 | 7668 | No | NA | 5 | 0 | 3 | Not-determined | Genome-fragment | NA | NA | 0 |
| S6_k141_116182 | 4632 | No | NA | 4 | 0 | 3 | Not-determined | Genome-fragment | NA | NA | 0 |
| S6_k141_116214 | 3099 | No | NA | 3 | 0 | 1 | Not-determined | Genome-fragment | NA | NA | 0 |
| S6_k141_69731 | 1010 | No | NA | 2 | 0 | 2 | Not-determined | Genome-fragment | NA | NA | 0 |
| S6_k141_33856 | 3252 | No | NA | 1 | 0 | 1 | Not-determined | Genome-fragment | NA | NA | 0 |
| S6_k141_28115 | 8352 | No | NA | 5 | 0 | 3 | Not-determined | Genome-fragment | NA | NA | 0 |
| S2-final-viral-combined_k141_245454\|\|full | 8321 | No | NA | 5 | 0 | 4 | Low-quality | Genome-fragment | 3.5 | AAI-based (high-confidence) | 0 |
| S6_k141_64114 | 1121 | No | NA | 1 | 0 | 1 | Not-determined | Genome-fragment | NA | NA | 0 |
| S6_k141_56698 | 3290 | No | NA | 3 | 0 | 3 | Not-determined | Genome-fragment | NA | NA | 0 |
| S6_k141_86911 | 6083 | No | NA | 4 | 0 | 3 | Not-determined | Genome-fragment | NA | NA | 0 |
| S6_k141_40422 | 1235 | No | NA | 1 | 0 | 1 | Not-determined | Genome-fragment | NA | NA | 0 |
| S5-final-viral-combined_k141_167222\|\|full | 8323 | No | NA | 10 | 7 | 0 | Low-quality | Genome-fragment | 19.3 | AAI-based (medium-confidence) | 0 |
| S7-final-viral-combined_k141_88654\|\|full | 8332 | No | NA | 9 | 7 | 0 | Low-quality | Genome-fragment | 16.48 | AAI-based (high-confidence) | 0 |
| S6_k141_33982 | 3362 | No | NA | 2 | 0 | 2 | Not-determined | Genome-fragment | NA | NA | 0 |
| S6_k141_64162 | 1304 | No | NA | 1 | 0 | 1 | Not-determined | Genome-fragment | NA | NA | 0 |
| S6_k141_133653 | 4319 | No | NA | 3 | 0 | 3 | Not-determined | Genome-fragment | NA | NA | 0 |
| S6_k141_21719 | 1947 | No | NA | 3 | 0 | 0 | Not-determined | Genome-fragment | NA | NA | 0 |
| S6_k141_86969 | 3270 | No | NA | 1 | 0 | 0 | Not-determined | Genome-fragment | NA | NA | 0 |
| S6_k141_64183 | 1005 | No | NA | 1 | 0 | 1 | Not-determined | Genome-fragment | NA | NA | 0 |
| S6_k141_40496 | 3971 | No | NA | 3 | 0 | 1 | Not-determined | Genome-fragment | NA | NA | 0 |
| S2_k141_109446 | 8336 | No | NA | 2 | 0 | 0 | Low-quality | Genome-fragment | 22.06 | AAI-based (medium-confidence) | 0 |
| S6_k141_121606 | 1118 | No | NA | 2 | 0 | 0 | Not-determined | Genome-fragment | NA | NA | 0 |
| S6_k141_133715 | 1207 | No | NA | 5 | 0 | 0 | Not-determined | Genome-fragment | NA | NA | 0 |
| S11-final-viral-combined_k141_278488\|\|full | 8337 | No | NA | 12 | 1 | 3 | Low-quality | Genome-fragment | 12.47 | HMM-based (lower-bound) | 0 |
| S6_k141_128002 | 2212 | No | NA | 4 | 0 | 0 | Not-determined | Genome-fragment | NA | NA | 0 |
| S6_k141_40572 | 2225 | No | NA | 3 | 0 | 3 | Not-determined | Genome-fragment | NA | NA | 0 |
| S6_k141_110617 | 4153 | No | NA | 3 | 0 | 3 | Not-determined | Genome-fragment | NA | NA | 0 |
| S6_k141_110623 | 2985 | No | NA | 3 | 0 | 1 | Not-determined | Genome-fragment | NA | NA | 0 |
| S6_k141_116408 | 1329 | No | NA | 1 | 0 | 0 | Not-determined | Genome-fragment | NA | NA | 0 |
| S9-final-viral-combined_k141_27648\|\|full | 8340 | No | NA | 11 | 10 | 0 | Low-quality | Genome-fragment | 17.79 | AAI-based (high-confidence) | 0 |
| S17-final-viral-combined_k141_115883\|\|full | 8344 | No | NA | 5 | 0 | 2 | Low-quality | Genome-fragment | 6.77 | AAI-based (medium-confidence) | 0 |
| S6_k141_121697 | 6254 | No | NA | 5 | 0 | 2 | Not-determined | Genome-fragment | NA | NA | 0 |
| S6_k141_87046 | 1011 | No | NA | 1 | 0 | 1 | Not-determined | Genome-fragment | NA | NA | 0 |
| S6_k141_70014 | 1062 | No | NA | 1 | 0 | 0 | Not-determined | Genome-fragment | NA | NA | 0 |
| S6_k141_128040 | 2045 | No | NA | 1 | 0 | 1 | Not-determined | Genome-fragment | NA | NA | 0 |
| S6_k141_139446 | 1601 | No | NA | 2 | 0 | 1 | Not-determined | Genome-fragment | NA | NA | 0 |
| S6_k141_99451 | 3435 | No | NA | 1 | 0 | 1 | Not-determined | Genome-fragment | NA | NA | 0 |
| S11-final-viral-combined_k141_74766\|\|full | 8345 | No | NA | 10 | 5 | 0 | Low-quality | Genome-fragment | 17.51 | AAI-based (high-confidence) | 0 |
| S6_k141_87066 | 4884 | No | NA | 3 | 0 | 2 | Not-determined | Genome-fragment | NA | NA | 0 |
| S6_k141_128059 | 1396 | No | NA | 1 | 0 | 1 | Not-determined | Genome-fragment | NA | NA | 0 |
| S6_k141_28294 | 1600 | No | NA | 1 | 0 | 1 | Not-determined | Genome-fragment | NA | NA | 0 |
| S6_k141_21842 | 1078 | No | NA | 2 | 0 | 1 | Not-determined | Genome-fragment | NA | NA | 0 |
| S3-final-viral-combined_k141_112937\|\|full | 8345 | No | NA | 5 | 0 | 5 | Low-quality | Genome-fragment | 10.8 | AAI-based (high-confidence) | 0 |
| S6_k141_70075 | 1043 | No | NA | 2 | 0 | 1 | Not-determined | Genome-fragment | NA | NA | 0 |
| S6_k141_116464 | 1327 | No | NA | 1 | 0 | 0 | Not-determined | Genome-fragment | NA | NA | 0 |
| S6_k141_40675 | 3173 | No | NA | 2 | 0 | 2 | Not-determined | Genome-fragment | NA | NA | 0 |
| S17-k141_93224 | 8360 | No | NA | 8 | 0 | 6 | Low-quality | Genome-fragment | 2.62 | AAI-based (medium-confidence) | 0 |
| S6_k141_116483 | 1223 | No | NA | 1 | 0 | 1 | Not-determined | Genome-fragment | NA | NA | 0 |
| S6_k141_104110 | 5305 | No | NA | 2 | 0 | 2 | Not-determined | Genome-fragment | NA | NA | 0 |
| S6_k141_34197 | 1122 | No | NA | 1 | 0 | 1 | Not-determined | Genome-fragment | NA | NA | 0 |
| S6_k141_56877 | 4833 | No | NA | 3 | 0 | 2 | Not-determined | Genome-fragment | NA | NA | 0 |
| S6_k141_75246 | 1169 | No | NA | 1 | 0 | 1 | Not-determined | Genome-fragment | NA | NA | 0 |
| S6_k141_21909 | 1189 | No | NA | 1 | 0 | 1 | Not-determined | Genome-fragment | NA | NA | 0 |
| S6_k141_40740 | 4179 | No | NA | 6 | 0 | 3 | Not-determined | Genome-fragment | NA | NA | 0 |
| S11-final-viral-combined_k141_2567\|\|full | 8362 | No | NA | 21 | 6 | 0 | Low-quality | Genome-fragment | 27.06 | AAI-based (medium-confidence) | 0 |
| S6_k141_139516 | 1529 | No | NA | 2 | 0 | 1 | Not-determined | Genome-fragment | NA | NA | 0 |
| S6_k141_139517 | 3311 | No | NA | 4 | 0 | 0 | Not-determined | Genome-fragment | NA | NA | 0 |
| S6_k141_139518 | 3599 | No | NA | 4 | 0 | 0 | Not-determined | Genome-fragment | NA | NA | 0 |
| S6_k141_16950 | 2438 | No | NA | 2 | 0 | 1 | Not-determined | Genome-fragment | NA | NA | 0 |
| S6_k141_21965 | 1088 | No | NA | 1 | 0 | 1 | Not-determined | Genome-fragment | NA | NA | 0 |
| S8-final-viral-combined_k141_231058\|\|full | 8366 | No | NA | 8 | 0 | 3 | Low-quality | Genome-fragment | 18.2 | AAI-based (high-confidence) | 0 |
| S6_k141_87248 | 1729 | No | NA | 1 | 0 | 0 | Not-determined | Genome-fragment | NA | NA | 0 |
| S6-final-viral-combined_k141_102250\|\|full | 8375 | No | NA | 10 | 4 | 0 | Low-quality | Genome-fragment | 17.1 | AAI-based (high-confidence) | 0 |
| S6_k141_40801 | 1083 | No | NA | 1 | 0 | 1 | Not-determined | Genome-fragment | NA | NA | 0 |
| S6_k141_80933 | 1431 | No | NA | 1 | 0 | 0 | Not-determined | Genome-fragment | NA | NA | 0 |
| S6_k141_56991 | 4596 | No | NA | 6 | 0 | 3 | Not-determined | Genome-fragment | NA | NA | 0 |
| S6_k141_110871 | 1316 | No | NA | 2 | 0 | 1 | Not-determined | Genome-fragment | NA | NA | 0 |
| S6_k141_116606 | 1543 | No | NA | 1 | 0 | 1 | Not-determined | Genome-fragment | NA | NA | 0 |
| S6_k141_22040 | 1091 | No | NA | 2 | 0 | 2 | Not-determined | Genome-fragment | NA | NA | 0 |
| S6_k141_99600 | 1367 | No | NA | 5 | 0 | 0 | Not-determined | Genome-fragment | NA | NA | 0 |
| S6_k141_110889 | 3507 | No | NA | 3 | 0 | 2 | Not-determined | Genome-fragment | NA | NA | 0 |
| S6_k141_128284 | 6173 | No | NA | 6 | 0 | 5 | Not-determined | Genome-fragment | NA | NA | 0 |
| S2-final-viral-combined_k141_187089\|\|full | 8376 | No | NA | 10 | 0 | 1 | Low-quality | Genome-fragment | 18.53 | AAI-based (medium-confidence) | 0 |
| S6_k141_64524 | 1431 | No | NA | 2 | 0 | 0 | Not-determined | Genome-fragment | NA | NA | 0 |
| S6_k141_139636 | 1499 | No | NA | 2 | 0 | 1 | Not-determined | Genome-fragment | NA | NA | 0 |
| S6_k141_40851 | 1593 | No | NA | 3 | 0 | 3 | Not-determined | Genome-fragment | NA | NA | 0 |
| S6_k141_41320 | 8376 | No | NA | 5 | 1 | 0 | Low-quality | Genome-fragment | 8.85 | AAI-based (high-confidence) | 0 |
| S6_k141_80991 | 2006 | No | NA | 2 | 0 | 0 | Not-determined | Genome-fragment | NA | NA | 0 |
| S6_k141_116655 | 7740 | No | NA | 8 | 0 | 1 | Not-determined | Genome-fragment | NA | NA | 0 |
| S1-final-viral-combined_k141_298996\|\|full | 8378 | No | NA | 8 | 0 | 4 | Low-quality | Genome-fragment | 12.76 | AAI-based (medium-confidence) | 0 |
| S12-final-viral-combined_k141_192293\|\|full | 8386 | No | NA | 22 | 1 | 1 | Low-quality | Genome-fragment | 26.11 | AAI-based (high-confidence) | 0 |
| S12-final-viral-combined_k141_39486\|\|full | 8395 | No | NA | 14 | 1 | 2 | Low-quality | Genome-fragment | 7.52 | HMM-based (lower-bound) | 0 |
| S6_k141_139701 | 1004 | No | NA | 2 | 0 | 0 | Not-determined | Genome-fragment | NA | NA | 0 |
| S6_k141_104319 | 1410 | No | NA | 2 | 0 | 1 | Not-determined | Genome-fragment | NA | NA | 0 |
| S6-final-viral-combined_k141_76640\|\|full | 8396 | No | NA | 7 | 0 | 2 | Low-quality | Genome-fragment | 7.31 | AAI-based (medium-confidence) | 0 |
| S6_k141_70318 | 1521 | No | NA | 1 | 0 | 0 | Not-determined | Genome-fragment | NA | NA | 0 |
| S6_k141_17063 | 3708 | No | NA | 4 | 0 | 2 | Not-determined | Genome-fragment | NA | NA | 0 |
| S6_k141_99674 | 1231 | No | NA | 1 | 0 | 0 | Not-determined | Genome-fragment | NA | NA | 0 |
| S8-final-viral-combined_k141_81782\|\|full | 8399 | No | NA | 7 | 0 | 6 | Low-quality | Genome-fragment | 3.79 | AAI-based (medium-confidence) | 0 |
| S6_k141_52383 | 3022 | No | NA | 4 | 0 | 2 | Not-determined | Genome-fragment | NA | NA | 0 |
| S6_k141_116746 | 1051 | No | NA | 1 | 0 | 0 | Not-determined | Genome-fragment | NA | NA | 0 |
| S6_k141_46759 | 3267 | No | NA | 4 | 0 | 4 | Not-determined | Genome-fragment | NA | NA | 0 |
| S6_k141_93172 | 3474 | No | NA | 2 | 0 | 2 | Not-determined | Genome-fragment | NA | NA | 0 |
| S2-final-viral-combined_k141_241558\|\|full | 8400 | No | NA | 9 | 0 | 2 | Low-quality | Genome-fragment | 13.62 | AAI-based (medium-confidence) | 0 |
| S6_k141_87434 | 2713 | No | NA | 3 | 0 | 2 | Not-determined | Genome-fragment | NA | NA | 0 |
| S3-final-viral-combined_k141_27154\|\|full | 8403 | No | NA | 11 | 3 | 0 | Low-quality | Genome-fragment | 20.17 | AAI-based (high-confidence) | 0 |
| S6_k141_22212 | 1058 | No | NA | 2 | 0 | 1 | Not-determined | Genome-fragment | NA | NA | 0 |
| S2-final-viral-combined_k141_81697\|\|full | 8403 | No | NA | 15 | 6 | 0 | Low-quality | Genome-fragment | 22.42 | AAI-based (high-confidence) | 0 |
| S8-final-viral-combined_k141_244096\|\|full | 8408 | No | NA | 13 | 0 | 4 | Low-quality | Genome-fragment | 17.73 | AAI-based (high-confidence) | 0 |
| S6_k141_10666 | 3812 | No | NA | 2 | 0 | 2 | Not-determined | Genome-fragment | NA | NA | 0 |
| S6_k141_104423 | 1114 | No | NA | 1 | 0 | 0 | Not-determined | Genome-fragment | NA | NA | 0 |
| S6_k141_22237 | 1180 | No | NA | 1 | 0 | 1 | Not-determined | Genome-fragment | NA | NA | 0 |
| S6_k141_10678 | 1792 | No | NA | 2 | 0 | 0 | Not-determined | Genome-fragment | NA | NA | 0 |
| S7-final-viral-combined_k141_230524\|\|full | 8413 | No | NA | 15 | 3 | 0 | Low-quality | Genome-fragment | 17.21 | AAI-based (medium-confidence) | 0 |
| S6_k141_87460 | 3621 | No | NA | 2 | 0 | 1 | Not-determined | Genome-fragment | NA | NA | 0 |
| S7-final-viral-combined_k141_73251\|\|full | 8414 | No | NA | 3 | 0 | 1 | Low-quality | Genome-fragment | 12.75 | AAI-based (medium-confidence) | 0 |
| S6_k141_139848 | 1154 | No | NA | 2 | 0 | 1 | Not-determined | Genome-fragment | NA | NA | 0 |
| S1_k141_84760 | 8422 | No | NA | 7 | 0 | 5 | Low-quality | Genome-fragment | 4.76 | AAI-based (medium-confidence) | 0 |
| S6_k141_52523 | 4752 | No | NA | 3 | 0 | 3 | Not-determined | Genome-fragment | NA | NA | 0 |
| S2_k141_87476 | 8425 | No | NA | 5 | 0 | 3 | Low-quality | Genome-fragment | 5.15 | AAI-based (high-confidence) | 0 |
| S6_k141_122031 | 1559 | No | NA | 1 | 0 | 0 | Not-determined | Genome-fragment | NA | NA | 0 |
| S6_k141_104475 | 2904 | No | NA | 5 | 0 | 4 | Not-determined | Genome-fragment | NA | NA | 0 |
| S17-final-viral-combined_k141_67475\|\|full | 8426 | No | NA | 6 | 0 | 4 | Low-quality | Genome-fragment | 6.32 | AAI-based (medium-confidence) | 0 |
| S6_k141_52561 | 1180 | No | NA | 1 | 0 | 1 | Not-determined | Genome-fragment | NA | NA | 0 |
| S6_k141_111007 | 3111 | No | NA | 3 | 0 | 3 | Not-determined | Genome-fragment | NA | NA | 0 |
| S6_k141_57305 | 2160 | No | NA | 2 | 0 | 1 | Not-determined | Genome-fragment | NA | NA | 0 |
| S6_k141_34465 | 1301 | No | NA | 1 | 0 | 1 | Not-determined | Genome-fragment | NA | NA | 0 |
| S6_k141_57317 | 1584 | No | NA | 1 | 0 | 1 | Not-determined | Genome-fragment | NA | NA | 0 |
| S8-final-viral-combined_k141_232141\|\|full | 8427 | Yes | 2484 | 9 | 2 | 3 | Low-quality | Genome-fragment | 4.57 | HMM-based (lower-bound) | 70.52 |
| S6_k141_81205 | 1117 | No | NA | 2 | 0 | 0 | Not-determined | Genome-fragment | NA | NA | 0 |
| S17-final-viral-combined_k141_56937\|\|full | 8431 | No | NA | 14 | 1 | 0 | Low-quality | Genome-fragment | 2.31 | HMM-based (lower-bound) | 0 |
| S17-final-viral-combined_k141_76219\|\|full | 8434 | No | NA | 10 | 6 | 0 | Low-quality | Genome-fragment | 24.4 | AAI-based (high-confidence) | 0 |

| | | | | | | | | | | | |
|---|---|---|---|---|---|---|---|---|---|---|---|
| S6_k141_75644 | 3672 | No | NA | 1 | 0 | 1 | Not-determined | Genome-fragment | NA | NA | 0 |
| S6_k141_10772 | 2914 | No | NA | 3 | 0 | 2 | Not-determined | Genome-fragment | NA | NA | 0 |
| S6_k141_41039 | 12158 | No | NA | 13 | 0 | 11 | Not-determined | Genome-fragment | NA | NA | 0 |
| S6_k141_116938 | 3524 | No | NA | 10 | 0 | 8 | Not-determined | Genome-fragment | NA | NA | 0 |
| S17-final-viral-combined_k141_142770\|\|full | 8436 | No | NA | 12 | 8 | 1 | Low-quality | Genome-fragment | 18.77 | AAI-based (medium-confidence) | 0 |
| S6_k141_81300 | 1581 | No | NA | 2 | 0 | 1 | Not-determined | Genome-fragment | NA | NA | 0 |
| S7_k141_240950 | 8437 | No | NA | 15 | 0 | 0 | Low-quality | Genome-fragment | 14.09 | AAI-based (high-confidence) | 0 |
| S6_k141_140004 | 1396 | No | NA | 1 | 0 | 1 | Not-determined | Genome-fragment | NA | NA | 0 |
| S12-final-viral-combined_k141_12710\|\|full | 8439 | No | NA | 14 | 3 | 1 | Low-quality | Genome-fragment | 9.47 | HMM-based (lower-bound) | 0 |
| S6_k141_93468 | 7054 | No | NA | 5 | 0 | 5 | Not-determined | Genome-fragment | NA | NA | 0 |
| S11-final-viral-combined_k141_79383\|\|full | 8442 | Yes | 5450 | 8 | 1 | 3 | Low-quality | Genome-fragment | 4.05 | HMM-based (lower-bound) | 35.44 |
| S6_k141_128716 | 1714 | No | NA | 2 | 0 | 0 | Not-determined | Genome-fragment | NA | NA | 0 |
| S5-final-viral-combined_k141_115985\|\|full | 8450 | Yes | 740 | 8 | 1 | 5 | Low-quality | Genome-fragment | 17.78 | HMM-based (lower-bound) | 91.24 |
| S12-final-viral-combined_k141_153394\|\|full | 8457 | No | NA | 11 | 1 | 0 | Low-quality | Genome-fragment | 7.71 | AAI-based (medium-confidence) | 0 |
| S6_k141_104713 | 1173 | No | NA | 1 | 0 | 1 | Not-determined | Genome-fragment | NA | NA | 0 |
| S6_k141_52786 | 1587 | No | NA | 2 | 0 | 1 | Not-determined | Genome-fragment | NA | NA | 0 |
| S6_k141_10893 | 1167 | No | NA | 2 | 0 | 0 | Not-determined | Genome-fragment | NA | NA | 0 |
| S6_k141_22475 | 3859 | No | NA | 5 | 0 | 4 | Not-determined | Genome-fragment | NA | NA | 0 |
| S12_k141_13942 | 8465 | Yes | 2190 | 10 | 4 | 2 | Low-quality | Genome-fragment | 3.39 | AAI-based (high-confidence) | 74.13 |
| S6_k141_111261 | 1501 | No | NA | 2 | 0 | 1 | Not-determined | Genome-fragment | NA | NA | 0 |
| S6_k141_10954 | 4833 | No | NA | 4 | 0 | 4 | Not-determined | Genome-fragment | NA | NA | 0 |
| S6_k141_22507 | 1049 | No | NA | 2 | 0 | 1 | Not-determined | Genome-fragment | NA | NA | 0 |
| S6_k141_134470 | 1075 | No | NA | 1 | 0 | 1 | Not-determined | Genome-fragment | NA | NA | 0 |
| S6_k141_75806 | 1955 | No | NA | 2 | 0 | 0 | Not-determined | Genome-fragment | NA | NA | 0 |
| S9_k141_76730 | 8467 | No | NA | 9 | 8 | 0 | Low-quality | Genome-fragment | 21 | AAI-based (high-confidence) | 0 |
| S7-final-viral-combined_k141_75194\|\|full | 8467 | No | NA | 15 | 3 | 1 | Low-quality | Genome-fragment | 11.75 | HMM-based (lower-bound) | 0 |
| S6_k141_104865 | 1176 | No | NA | 1 | 0 | 0 | Not-determined | Genome-fragment | NA | NA | 0 |
| S4-final-viral-combined_k141_67837\|\|full | 8467 | Yes | 4867 | 10 | 1 | 1 | Low-quality | Genome-fragment | 11.39 | AAI-based (high-confidence) | 42.52 |
| S6_k141_81534 | 1123 | No | NA | 2 | 0 | 1 | Not-determined | Genome-fragment | NA | NA | 0 |
| S1-final-viral-combined_k141_255309\|\|full | 8468 | No | NA | 8 | 0 | 1 | Low-quality | Genome-fragment | 12.97 | AAI-based (medium-confidence) | 0 |
| S6_k141_75863 | 3011 | No | NA | 4 | 0 | 1 | Not-determined | Genome-fragment | NA | NA | 0 |
| S1-final-viral-combined_k141_126852\|\|full | 8471 | No | NA | 15 | 2 | 0 | Low-quality | Genome-fragment | 24.17 | AAI-based (medium-confidence) | 0 |
| S6_k141_104921 | 1188 | No | NA | 1 | 0 | 1 | Not-determined | Genome-fragment | NA | NA | 0 |
| S6_k141_104924 | 1262 | No | NA | 2 | 0 | 1 | Not-determined | Genome-fragment | NA | NA | 0 |
| S6_k141_134594 | 1560 | No | NA | 2 | 0 | 1 | Not-determined | Genome-fragment | NA | NA | 0 |
| S6_k141_117197 | 3531 | No | NA | 5 | 0 | 5 | Not-determined | Genome-fragment | NA | NA | 0 |
| S6_k141_122436 | 3065 | No | NA | 2 | 0 | 1 | Not-determined | Genome-fragment | NA | NA | 0 |
| S6_k141_128897 | 1010 | No | NA | 1 | 0 | 0 | Not-determined | Genome-fragment | NA | NA | 0 |
| S5-final-viral-combined_k141_114605\|\|full | 8476 | No | NA | 9 | 0 | 5 | Low-quality | Genome-fragment | 2.18 | AAI-based (high-confidence) | 0 |
| S6_k141_111384 | 1336 | No | NA | 2 | 0 | 1 | Not-determined | Genome-fragment | NA | NA | 0 |
| S6_k141_34714 | 1141 | No | NA | 2 | 0 | 1 | Not-determined | Genome-fragment | NA | NA | 0 |
| S6_k141_122474 | 3257 | No | NA | 5 | 0 | 3 | Not-determined | Genome-fragment | NA | NA | 0 |
| S6_k141_87893 | 1418 | No | NA | 2 | 0 | 1 | Not-determined | Genome-fragment | NA | NA | 0 |
| S1_k141_140344 | 8482 | No | NA | 10 | 1 | 3 | Low-quality | Genome-fragment | 9.82 | AAI-based (medium-confidence) | 0 |
| S6_k141_93761 | 5924 | No | NA | 8 | 0 | 2 | Not-determined | Genome-fragment | NA | NA | 0 |
| S6_k141_87914 | 5579 | No | NA | 4 | 0 | 3 | Not-determined | Genome-fragment | NA | NA | 0 |
| S12-final-viral-combined_k141_326327\|\|full | 8483 | No | NA | 5 | 0 | 5 | Low-quality | Genome-fragment | 9.04 | AAI-based (medium-confidence) | 0 |
| S6_k141_87947 | 2436 | No | NA | 2 | 0 | 2 | Not-determined | Genome-fragment | NA | NA | 0 |
| S6_k141_129032 | 3971 | No | NA | 4 | 0 | 4 | Not-determined | Genome-fragment | NA | NA | 0 |
| S6_k141_104996 | 1034 | No | NA | 2 | 0 | 2 | Not-determined | Genome-fragment | NA | NA | 0 |
| S6_k141_11119 | 1087 | No | NA | 2 | 0 | 1 | Not-determined | Genome-fragment | NA | NA | 0 |
| S6_k141_140389 | 1431 | No | NA | 2 | 0 | 2 | Not-determined | Genome-fragment | NA | NA | 0 |
| S6_k141_75966 | 4890 | No | NA | 6 | 0 | 2 | Not-determined | Genome-fragment | NA | NA | 0 |
| S8-final-viral-combined_k141_132807\|\|full | 8486 | No | NA | 12 | 9 | 0 | Low-quality | Genome-fragment | 21.84 | AAI-based (medium-confidence) | 0 |
| S6_k141_34923 | 1960 | No | NA | 2 | 0 | 1 | Not-determined | Genome-fragment | NA | NA | 0 |
| S6_k141_105093 | 5421 | No | NA | 3 | 0 | 1 | Not-determined | Genome-fragment | NA | NA | 0 |
| S6_k141_140496 | 1503 | No | NA | 4 | 0 | 1 | Not-determined | Genome-fragment | NA | NA | 0 |
| S6_k141_57861 | 4647 | No | NA | 5 | 0 | 0 | Not-determined | Genome-fragment | NA | NA | 0 |
| S2_k141_46484 | 8488 | No | NA | 13 | 1 | 1 | Low-quality | Genome-fragment | 12.84 | HMM-based (lower-bound) | 0 |
| S4-final-viral-combined_k141_29007\|\|full | 8492 | Yes | 5426 | 11 | 2 | 2 | Low-quality | Genome-fragment | 6.59 | AAI-based (medium-confidence) | 36.1 |
| S6_k141_105110 | 1179 | No | NA | 3 | 0 | 3 | Not-determined | Genome-fragment | NA | NA | 0 |
| S6_k141_76091 | 4415 | No | NA | 4 | 0 | 2 | Not-determined | Genome-fragment | NA | NA | 0 |
| S17-final-viral-combined_k141_17066\|\|full | 8493 | No | NA | 13 | 0 | 0 | Low-quality | Genome-fragment | 28.2 | AAI-based (medium-confidence) | 0 |
| S1_k141_192699 | 8494 | No | NA | 10 | 0 | 6 | Low-quality | Genome-fragment | 2.49 | AAI-based (medium-confidence) | 0 |
| S6_k141_134898 | 1534 | No | NA | 3 | 0 | 1 | Not-determined | Genome-fragment | NA | NA | 0 |
| S5-final-viral-combined_k141_169632\|\|full | 8496 | No | NA | 8 | 8 | 0 | Low-quality | Genome-fragment | 21.61 | AAI-based (high-confidence) | 0 |
| S6_k141_29319 | 1045 | No | NA | 2 | 0 | 0 | Not-determined | Genome-fragment | NA | NA | 0 |
| S6_k141_134925 | 1603 | No | NA | 2 | 0 | 1 | Not-determined | Genome-fragment | NA | NA | 0 |
| S6_k141_35109 | 2217 | No | NA | 2 | 0 | 2 | Not-determined | Genome-fragment | NA | NA | 0 |
| S6_k141_35127 | 19224 | No | NA | 5 | 0 | 3 | Not-determined | Genome-fragment | NA | NA | 0 |
| S6_k141_122981 | 1386 | No | NA | 2 | 0 | 2 | Not-determined | Genome-fragment | NA | NA | 0 |
| S6_k141_81987 | 1153 | No | NA | 2 | 0 | 0 | Not-determined | Genome-fragment | NA | NA | 0 |
| S6_k141_35148 | 1119 | No | NA | 1 | 0 | 1 | Not-determined | Genome-fragment | NA | NA | 0 |
| S6_k141_35158 | 3768 | No | NA | 4 | 0 | 4 | Not-determined | Genome-fragment | NA | NA | 0 |
| S6_k141_23240 | 5060 | No | NA | 1 | 0 | 1 | Not-determined | Genome-fragment | NA | NA | 0 |
| S6_k141_11421 | 1642 | No | NA | 1 | 0 | 1 | Not-determined | Genome-fragment | NA | NA | 0 |
| S6_k141_135024 | 1563 | No | NA | 3 | 0 | 3 | Not-determined | Genome-fragment | NA | NA | 0 |
| S6_k141_58068 | 4236 | No | NA | 3 | 0 | 0 | Not-determined | Genome-fragment | NA | NA | 0 |
| S6_k141_123071 | 1026 | No | NA | 2 | 0 | 1 | Not-determined | Genome-fragment | NA | NA | 0 |
| S6_k141_76257 | 4603 | No | NA | 3 | 0 | 1 | Not-determined | Genome-fragment | NA | NA | 0 |
| S12-final-viral-combined_k141_268888\|\|full | 8504 | No | NA | 10 | 6 | 0 | Low-quality | Genome-fragment | 20.44 | AAI-based (high-confidence) | 0 |
| S6_k141_23407 | 3637 | No | NA | 1 | 0 | 1 | Not-determined | Genome-fragment | NA | NA | 0 |
| S6_k141_140743 | 65184 | No | NA | 93 | 14 | 0 | Complete | High-quality | 100 | DTR (high-confidence) | 0 |
| S8-final-viral-combined_k141_246389\|\|full | 41205 | No | NA | 78 | 17 | 0 | High-quality | High-quality | 100 | AAI-based (high-confidence) | 0 |
| S8-final-viral-combined_k141_86965\|\|full | 4784 | No | NA | 8 | 0 | 2 | Not-determined | Genome-fragment | NA | NA | 0 |
| S7-final-viral-combined_k141_132132\|\|full | 8513 | No | NA | 12 | 2 | 0 | Low-quality | Genome-fragment | 9.61 | HMM-based (lower-bound) | 0 |
| S8-final-viral-combined_k141_215102\|\|full | 66504 | Yes | 37446 | 66 | 15 | 16 | High-quality | High-quality | 100 | AAI-based (high-confidence) | 43.69 |
| S8-final-viral-combined_k141_192515\|\|full | 12161 | No | NA | 9 | 0 | 4 | Not-determined | Genome-fragment | NA | NA | 0 |
| S8-final-viral-combined_k141_276204\|\|full | 40180 | No | NA | 67 | 7 | 1 | High-quality | High-quality | 100 | AAI-based (high-confidence) | 0 |
| S8-final-viral-combined_k141_276211\|\|full | 42456 | No | NA | 75 | 16 | 0 | High-quality | High-quality | 100 | AAI-based (high-confidence) | 0 |
| S9-final-viral-combined_k141_71074\|\|full | 8515 | Yes | 5272 | 10 | 1 | 1 | Low-quality | Genome-fragment | 12.2 | AAI-based (medium-confidence) | 38.09 |
| S8-final-viral-combined_k141_172791\|\|full | 1312 | No | NA | 2 | 0 | 1 | Not-determined | Genome-fragment | NA | NA | 0 |
| S1-final-viral-combined_k141_213346\|\|full | 8517 | No | NA | 5 | 0 | 5 | Low-quality | Genome-fragment | 18.78 | AAI-based (medium-confidence) | 0 |
| S2-final-viral-combined_k141_185482\|\|full | 8517 | No | NA | 9 | 2 | 0 | Low-quality | Genome-fragment | 7.99 | HMM-based (lower-bound) | 0 |
| S8-final-viral-combined_k141_218886\|\|full | 1161 | No | NA | 3 | 0 | 1 | Not-determined | Genome-fragment | NA | NA | 0 |
| S7-final-viral-combined_k141_55368\|\|full | 8519 | No | NA | 9 | 1 | 1 | Low-quality | Genome-fragment | 15.41 | AAI-based (medium-confidence) | 0 |
| S8-final-viral-combined_k141_11540\|\|full | 1953 | No | NA | 2 | 0 | 1 | Not-determined | Genome-fragment | NA | NA | 0 |
| S8-final-viral-combined_k141_149780\|\|full | 3923 | No | NA | 3 | 0 | 0 | Not-determined | Genome-fragment | NA | NA | 0 |
| S3-final-viral-combined_k141_33355\|\|full | 8522 | No | NA | 10 | 1 | 0 | Low-quality | Genome-fragment | 12.89 | HMM-based (lower-bound) | 0 |
| S8-final-viral-combined_k141_230439\|\|full | 3062 | No | NA | 4 | 0 | 2 | Not-determined | Genome-fragment | NA | NA | 0 |
| S8-final-viral-combined_k141_34589\|\|full | 6620 | No | NA | 4 | 0 | 4 | Not-determined | Genome-fragment | NA | NA | 0 |
| S2-final-viral-combined_k141_93988\|\|full | 8522 | No | NA | 7 | 1 | 1 | Low-quality | Genome-fragment | 9.54 | HMM-based (lower-bound) | 0 |
| S2-final-viral-combined_k141_229220\|\|full | 8524 | No | NA | 17 | 3 | 1 | Low-quality | Genome-fragment | 18.29 | AAI-based (medium-confidence) | 0 |
| S8-final-viral-combined_k141_253456\|\|full | 2223 | No | NA | 3 | 0 | 0 | Not-determined | Genome-fragment | NA | NA | 0 |
| S8-final-viral-combined_k141_138323\|\|full | 1298 | No | NA | 3 | 0 | 0 | Not-determined | Genome-fragment | NA | NA | 0 |
| S9-final-viral-combined_k141_46536\|\|full | 8528 | No | NA | 10 | 5 | 0 | Low-quality | Genome-fragment | 17.79 | AAI-based (high-confidence) | 0 |
| S1-final-viral-combined_k141_361731\|\|full | 8529 | No | NA | 16 | 4 | 0 | Low-quality | Genome-fragment | 11.37 | HMM-based (lower-bound) | 0 |
| S8-final-viral-combined_k141_29343\|\|full | 8532 | No | NA | 8 | 1 | 4 | Low-quality | Genome-fragment | 4.71 | HMM-based (lower-bound) | 0 |
| S8-final-viral-combined_k141_213539\|\|full | 8535 | Yes | 1233 | 10 | 1 | 3 | Low-quality | Genome-fragment | 2.28 | AAI-based (high-confidence) | 85.55 |
| S8-final-viral-combined_k141_253464\|\|full | 1343 | No | NA | 2 | 0 | 2 | Not-determined | Genome-fragment | NA | NA | 0 |
| S8-final-viral-combined_k141_23053\|\|full | 18177 | No | NA | 11 | 0 | 9 | Not-determined | Genome-fragment | NA | NA | 0 |
| S12-final-viral-combined_k141_76936\|\|full | 8539 | No | NA | 13 | 4 | 0 | Low-quality | Genome-fragment | 14.46 | AAI-based (medium-confidence) | 0 |
| S4-final-viral-combined_k141_165832\|\|full | 8558 | No | NA | 12 | 7 | 0 | Low-quality | Genome-fragment | 15.53 | HMM-based (lower-bound) | 0 |
| S8-final-viral-combined_k141_265058\|\|full | 1652 | No | NA | 2 | 0 | 0 | Not-determined | Genome-fragment | NA | NA | 0 |
| S8-final-viral-combined_k141_115288\|\|full | 2704 | No | NA | 5 | 0 | 5 | Not-determined | Genome-fragment | NA | NA | 0 |
| S8-final-viral-combined_k141_103711\|\|full | 1409 | No | NA | 2 | 0 | 0 | Not-determined | Genome-fragment | NA | NA | 0 |
| S8-final-viral-combined_k141_138381\|\|full | 1064 | No | NA | 3 | 0 | 1 | Not-determined | Genome-fragment | NA | NA | 0 |
| S3_k141_123518 | 8561 | No | NA | 4 | 2 | 0 | Low-quality | Genome-fragment | 15.93 | AAI-based (high-confidence) | 0 |
| S8-final-viral-combined_k141_103718\|\|full | 1710 | No | NA | 3 | 0 | 2 | Not-determined | Genome-fragment | NA | NA | 0 |
| S8-final-viral-combined_k141_218941\|\|full | 3417 | No | NA | 8 | 0 | 0 | Not-determined | Genome-fragment | NA | NA | 0 |
| S8-final-viral-combined_k141_207414\|\|full | 1400 | No | NA | 2 | 0 | 1 | Not-determined | Genome-fragment | NA | NA | 0 |
| S17-final-viral-combined_k141_92287\|\|full | 8565 | No | NA | 15 | 0 | 8 | Low-quality | Genome-fragment | 18.82 | AAI-based (medium-confidence) | 0 |
| S8-final-viral-combined_k141_113\|\|full | 1441 | No | NA | 2 | 0 | 1 | Not-determined | Genome-fragment | NA | NA | 0 |
| S8-final-viral-combined_k141_149844\|\|full | 1116 | No | NA | 3 | 0 | 0 | Not-determined | Genome-fragment | NA | NA | 0 |
| S1-final-viral-combined_k141_231600\|\|full | 8566 | No | NA | 18 | 7 | 0 | Low-quality | Genome-fragment | 14.07 | AAI-based (high-confidence) | 0 |
| S8-final-viral-combined_k141_218953\|\|full | 9236 | No | NA | 8 | 0 | 5 | Not-determined | Genome-fragment | NA | NA | 0 |
| S1-final-viral-combined_k141_88045\|\|full | 8569 | No | NA | 11 | 1 | 2 | Low-quality | Genome-fragment | 5.76 | HMM-based (lower-bound) | 0 |
| S11-final-viral-combined_k141_323976\|\|full | 8574 | No | NA | 7 | 0 | 6 | Low-quality | Genome-fragment | 3.71 | AAI-based (medium-confidence) | 0 |
| S8-final-viral-combined_k141_23119\|\|full | 3137 | No | NA | 7 | 0 | 0 | Not-determined | Genome-fragment | NA | NA | 0 |
| S12-final-viral-combined_k141_320793\|\|full | 8574 | No | NA | 9 | 0 | 6 | Low-quality | Genome-fragment | 2.83 | AAI-based (medium-confidence) | 0 |
| S8-final-viral-combined_k141_135\|\|full | 4046 | No | NA | 3 | 0 | 1 | Not-determined | Genome-fragment | NA | NA | 0 |
| S8-final-viral-combined_k141_211140\|\|full | 8582 | No | NA | 9 | 0 | 5 | Low-quality | Genome-fragment | 5.3 | AAI-based (medium-confidence) | 0 |
| S8-final-viral-combined_k141_184415\|\|full | 5150 | No | NA | 4 | 0 | 3 | Not-determined | Genome-fragment | NA | NA | 0 |
| S8-final-viral-combined_k141_172899\|\|full | 1342 | No | NA | 3 | 0 | 1 | Not-determined | Genome-fragment | NA | NA | 0 |
| S8-final-viral-combined_k141_242079\|\|full | 1424 | No | NA | 2 | 0 | 0 | Not-determined | Genome-fragment | NA | NA | 0 |
| S1-final-viral-combined_k141_255336\|\|full | 8585 | No | NA | 7 | 0 | 4 | Low-quality | Genome-fragment | 5.54 | AAI-based (medium-confidence) | 0 |
| S6-final-viral-combined_k141_36114\|\|full | 8589 | No | NA | 3 | 0 | 1 | Low-quality | Genome-fragment | 8.47 | AAI-based (medium-confidence) | 0 |
| S11-final-viral-combined_k141_168518\|\|full | 8593 | No | NA | 10 | 1 | 0 | Low-quality | Genome-fragment | 10.23 | HMM-based (lower-bound) | 0 |
| S8-final-viral-combined_k141_172941\|\|full | 2722 | No | NA | 3 | 0 | 2 | Not-determined | Genome-fragment | NA | NA | 0 |
| S8-final-viral-combined_k141_23224\|\|full | 3646 | No | NA | 6 | 0 | 0 | Not-determined | Genome-fragment | NA | NA | 0 |
| S8-final-viral-combined_k141_230656\|\|full | 8515 | No | NA | 4 | 0 | 3 | Not-determined | Genome-fragment | NA | NA | 0 |
| S17-final-viral-combined_k141_17313\|\|full | 8596 | No | NA | 9 | 0 | 4 | Low-quality | Genome-fragment | 4.14 | AAI-based (medium-confidence) | 0 |
| S8-final-viral-combined_k141_138485\|\|full | 4038 | No | NA | 3 | 0 | 2 | Not-determined | Genome-fragment | NA | NA | 0 |
| S8-final-viral-combined_k141_11578\|\|full | 2075 | No | NA | 2 | 0 | 1 | Not-determined | Genome-fragment | NA | NA | 0 |
| S8_k141_111834 | 8597 | No | NA | 14 | 4 | 0 | Low-quality | Genome-fragment | 14.29 | AAI-based (high-confidence) | 0 |
| S2-final-viral-combined_k141_45082\|\|full | 8598 | Yes | 4485 | 10 | 3 | 3 | Low-quality | Genome-fragment | 2.64 | AAI-based (medium-confidence) | 47.84 |
| S2_k141_87857 | 8598 | No | NA | 16 | 3 | 0 | Low-quality | Genome-fragment | 22.16 | HMM-based (lower-bound) | 0 |
| S8-final-viral-combined_k141_230695\|\|full | 3233 | No | NA | 3 | 0 | 2 | Not-determined | Genome-fragment | NA | NA | 0 |
| S7-final-viral-combined_k141_1081\|\|full | 8602 | No | NA | 7 | 0 | 3 | Low-quality | Genome-fragment | 8.25 | AAI-based (medium-confidence) | 0 |
| S8-final-viral-combined_k141_138520\|\|full | 1777 | No | NA | 5 | 0 | 1 | Not-determined | Genome-fragment | NA | NA | 0 |
| S8-final-viral-combined_k141_80913\|\|full | 1789 | No | NA | 3 | 0 | 2 | Not-determined | Genome-fragment | NA | NA | 0 |
| S8-final-viral-combined_k141_253554\|\|full | 2223 | No | NA | 3 | 0 | 2 | Not-determined | Genome-fragment | NA | NA | 0 |
| S17-final-viral-combined_k141_25588\|\|full | 8604 | No | NA | 8 | 0 | 3 | Low-quality | Genome-fragment | 9.88 | AAI-based (medium-confidence) | 0 |
| S8-final-viral-combined_k141_23271\|\|full | 3597 | No | NA | 4 | 0 | 3 | Not-determined | Genome-fragment | NA | NA | 0 |
| S8-final-viral-combined_k141_213\|\|full | 2382 | No | NA | 4 | 0 | 0 | Not-determined | Genome-fragment | NA | NA | 0 |
| S1-final-viral-combined_k141_186887\|\|full | 8605 | No | NA | 10 | 2 | 0 | Low-quality | Genome-fragment | 15.47 | AAI-based (medium-confidence) | 0 |
| S8-final-viral-combined_k141_126914\|\|full | 4326 | No | NA | 4 | 0 | 4 | Not-determined | Genome-fragment | NA | NA | 0 |
| S8-final-viral-combined_k141_196025\|\|full | 2560 | No | NA | 5 | 0 | 1 | Not-determined | Genome-fragment | NA | NA | 0 |
| S8-final-viral-combined_k141_243281\|\|full | 8610 | No | NA | 6 | 0 | 6 | Low-quality | Genome-fragment | 7.26 | AAI-based (medium-confidence) | 0 |
| S2-final-viral-combined_k141_37218\|\|full | 8614 | No | NA | 5 | 0 | 1 | Low-quality | Genome-fragment | 8.77 | AAI-based (medium-confidence) | 0 |
| S3_k141_88873 | 8617 | No | NA | 20 | 4 | 0 | Low-quality | Genome-fragment | 21.88 | AAI-based (high-confidence) | 0 |
| S8-final-viral-combined_k141_80959\|\|full | 1645 | No | NA | 3 | 0 | 2 | Not-determined | Genome-fragment | NA | NA | 0 |
| S8-final-viral-combined_k141_23302\|\|full | 2503 | No | NA | 2 | 0 | 1 | Not-determined | Genome-fragment | NA | NA | 0 |
| S8-final-viral-combined_k141_230731\|\|full | 3325 | No | NA | 6 | 0 | 1 | Not-determined | Genome-fragment | NA | NA | 0 |
| S8-final-viral-combined_k141_257\|\|full | 4120 | No | NA | 6 | 0 | 1 | Not-determined | Genome-fragment | NA | NA | 0 |
| S8-final-viral-combined_k141_242171\|\|full | 2362 | No | NA | 4 | 0 | 2 | Not-determined | Genome-fragment | NA | NA | 0 |
| S11-final-viral-combined_k141_310877\|\|full | 8619 | No | NA | 11 | 5 | 0 | Low-quality | Genome-fragment | 17.94 | AAI-based (high-confidence) | 0 |
| S8-final-viral-combined_k141_103785\|\|full | 2329 | No | NA | 3 | 0 | 2 | Not-determined | Genome-fragment | NA | NA | 0 |
| S8-final-viral-combined_k141_57858\|\|full | 2553 | No | NA | 2 | 0 | 2 | Not-determined | Genome-fragment | NA | NA | 0 |
| S8-final-viral-combined_k141_207589\|\|full | 1662 | No | NA | 4 | 0 | 0 | Not-determined | Genome-fragment | NA | NA | 0 |
| S8-final-viral-combined_k141_103788\|\|full | 3607 | No | NA | 5 | 0 | 4 | Not-determined | Genome-fragment | NA | NA | 0 |
| S8-final-viral-combined_k141_161357\|\|full | 2906 | No | NA | 2 | 0 | 2 | Not-determined | Genome-fragment | NA | NA | 0 |
| S2-final-viral-combined_k141_229077\|\|full | 8619 | No | NA | 10 | 0 | 5 | Low-quality | Genome-fragment | 5.47 | AAI-based (high-confidence) | 0 |
| S8-final-viral-combined_k141_196078\|\|full | 3027 | No | NA | 5 | 0 | 2 | Not-determined | Genome-fragment | NA | NA | 0 |
| S8-final-viral-combined_k141_11660\|\|full | 8170 | No | NA | 9 | 0 | 6 | Not-determined | Genome-fragment | NA | NA | 0 |
| S8-final-viral-combined_k141_184559\|\|full | 1120 | No | NA | 3 | 0 | 0 | Not-determined | Genome-fragment | NA | NA | 0 |
| S8-final-viral-combined_k141_11667\|\|full | 7172 | No | NA | 9 | 0 | 7 | Not-determined | Genome-fragment | NA | NA | 0 |
| S12-final-viral-combined_k141_19556\|\|full | 8623 | Yes | 5739 | 11 | 2 | 2 | Low-quality | Genome-fragment | 10.42 | AAI-based (medium-confidence) | 33.45 |
| S7-final-viral-combined_k141_215362\|\|full | 8627 | No | NA | 16 | 3 | 1 | Low-quality | Genome-fragment | 24.3 | AAI-based (high-confidence) | 0 |
| S8-final-viral-combined_k141_230823\|\|full | 1439 | No | NA | 3 | 0 | 2 | Not-determined | Genome-fragment | NA | NA | 0 |
| S8-final-viral-combined_k141_46382\|\|full | 6415 | No | NA | 3 | 0 | 2 | Not-determined | Genome-fragment | NA | NA | 0 |
| S8-final-viral-combined_k141_230837\|\|full | 1117 | No | NA | 2 | 0 | 1 | Not-determined | Genome-fragment | NA | NA | 0 |
| S8-final-viral-combined_k141_23370\|\|full | 6667 | No | NA | 4 | 0 | 1 | Not-determined | Genome-fragment | NA | NA | 0 |
| S17-final-viral-combined_k141_106310\|\|full | 8631 | No | NA | 13 | 1 | 0 | Low-quality | Genome-fragment | 11.67 | AAI-based (high-confidence) | 0 |
| S8-final-viral-combined_k141_11700\|\|full | 1112 | No | NA | 3 | 0 | 0 | Not-determined | Genome-fragment | NA | NA | 0 |
| S11-final-viral-combined_k141_100692\|\|full | 8634 | No | NA | 8 | 2 | 0 | Low-quality | Genome-fragment | 15.14 | AAI-based (medium-confidence) | 0 |
| S8-final-viral-combined_k141_138666\|\|full | 7560 | No | NA | 7 | 0 | 7 | Not-determined | Genome-fragment | NA | NA | 0 |
| S8-final-viral-combined_k141_57952\|\|full | 1622 | No | NA | 3 | 0 | 1 | Not-determined | Genome-fragment | NA | NA | 0 |
| S8-final-viral-combined_k141_207644\|\|full | 3551 | No | NA | 3 | 0 | 2 | Not-determined | Genome-fragment | NA | NA | 0 |
| S8-final-viral-combined_k141_69477\|\|full | 5079 | No | NA | 4 | 0 | 2 | Not-determined | Genome-fragment | NA | NA | 0 |
| S3_k141_134600 | 8637 | No | NA | 13 | 7 | 1 | Low-quality | Genome-fragment | 15.95 | AAI-based (high-confidence) | 0 |
| S8-final-viral-combined_k141_103855\|\|full | 6586 | No | NA | 6 | 0 | 5 | Not-determined | Genome-fragment | NA | NA | 0 |
| S8-final-viral-combined_k141_242285\|\|full | 8653 | No | NA | 8 | 0 | 3 | Not-determined | Genome-fragment | NA | NA | 0 |
| S8-final-viral-combined_k141_219132\|\|full | 7431 | No | NA | 8 | 0 | 7 | Not-determined | Genome-fragment | NA | NA | 0 |
| S8-final-viral-combined_k141_161405\|\|full | 1104 | No | NA | 3 | 0 | 0 | Not-determined | Genome-fragment | NA | NA | 0 |
| S1_k141_154304 | 8640 | No | NA | 13 | 4 | 0 | Low-quality | Genome-fragment | 14.56 | AAI-based (high-confidence) | 0 |
| S8-final-viral-combined_k141_138711\|\|full | 1158 | No | NA | 2 | 0 | 0 | Not-determined | Genome-fragment | NA | NA | 0 |
| S8-final-viral-combined_k141_196155\|\|full | 2203 | No | NA | 3 | 0 | 2 | Not-determined | Genome-fragment | NA | NA | 0 |
| S8-final-viral-combined_k141_127042\|\|full | 2817 | No | NA | 3 | 0 | 1 | Not-determined | Genome-fragment | NA | NA | 0 |
| S12-final-viral-combined_k141_156504\|\|full | 8642 | No | NA | 11 | 4 | 0 | Low-quality | Genome-fragment | 19.59 | AAI-based (medium-confidence) | 0 |
| S8-final-viral-combined_k141_196165\|\|full | 1076 | No | NA | 3 | 0 | 1 | Not-determined | Genome-fragment | NA | NA | 0 |
| S8-final-viral-combined_k141_23405\|\|full | 4065 | No | NA | 6 | 0 | 3 | Not-determined | Genome-fragment | NA | NA | 0 |
| S8-final-viral-combined_k141_265263\|\|full | 2665 | No | NA | 3 | 0 | 0 | Not-determined | Genome-fragment | NA | NA | 0 |
| S9-final-viral-combined_k141_113640\|\|full | 8643 | Yes | 7350 | 12 | 3 | 3 | Low-quality | Genome-fragment | 18.2 | AAI-based (medium-confidence) | 14.96 |
| S3_k141_80234 | 8644 | No | NA | 13 | 6 | 0 | Low-quality | Genome-fragment | 13.57 | AAI-based (high-confidence) | 0 |
| S11-final-viral-combined_k141_317588\|\|full | 8646 | No | NA | 9 | 0 | 1 | Low-quality | Genome-fragment | 20.06 | AAI-based (medium-confidence) | 0 |
| S3_k141_119166 | 8646 | No | NA | 19 | 0 | 0 | Low-quality | Genome-fragment | 3.98 | AAI-based (high-confidence) | 0 |
| S17-final-viral-combined_k141_147673\|\|full | 8646 | No | NA | 12 | 1 | 1 | Low-quality | Genome-fragment | 13.08 | HMM-based (lower-bound) | 0 |
| S8-final-viral-combined_k141_92546\|\|full | 2443 | No | NA | 3 | 0 | 1 | Not-determined | Genome-fragment | NA | NA | 0 |
| S8-final-viral-combined_k141_242362\|\|full | 1930 | No | NA | 3 | 0 | 0 | Not-determined | Genome-fragment | NA | NA | 0 |
| S8-final-viral-combined_k141_173183\|\|full | 4757 | No | NA | 6 | 0 | 1 | Not-determined | Genome-fragment | NA | NA | 0 |
| S8-final-viral-combined_k141_127103\|\|full | 2052 | No | NA | 6 | 0 | 0 | Not-determined | Genome-fragment | NA | NA | 0 |
| S8-final-viral-combined_k141_265301\|\|full | 4064 | No | NA | 4 | 0 | 3 | Not-determined | Genome-fragment | NA | NA | 0 |
| S17-final-viral-combined_k141_192391\|\|full | 8651 | Yes | 1916 | 9 | 1 | 5 | Low-quality | Genome-fragment | 1.56 | HMM-based (lower-bound) | 77.85 |
| S8-final-viral-combined_k141_219149\|\|full | 1644 | No | NA | 3 | 0 | 1 | Not-determined | Genome-fragment | NA | NA | 0 |
| S8-final-viral-combined_k141_92602\|\|full | 2114 | No | NA | 2 | 0 | 1 | Not-determined | Genome-fragment | NA | NA | 0 |
| S8-final-viral-combined_k141_115565\|\|full | 1685 | No | NA | 3 | 0 | 0 | Not-determined | Genome-fragment | NA | NA | 0 |
| S8-final-viral-combined_k141_173250\|\|full | 3020 | No | NA | 3 | 0 | 1 | Not-determined | Genome-fragment | NA | NA | 0 |
| S11-final-viral-combined_k141_7321\|\|full | 8653 | No | NA | 13 | 4 | 0 | Low-quality | Genome-fragment | 19.57 | HMM-based (lower-bound) | 0 |
| S8-final-viral-combined_k141_103941\|\|full | 4540 | No | NA | 3 | 0 | 3 | Not-determined | Genome-fragment | NA | NA | 0 |
| S6-final-viral-combined_k141_140565\|\|full | 8654 | No | NA | 10 | 1 | 1 | Low-quality | Genome-fragment | 9.71 | HMM-based (lower-bound) | 0 |
| S8_k141_201720 | 8655 | No | NA | 12 | 0 | 4 | Low-quality | Genome-fragment | 8.43 | AAI-based (medium-confidence) | 0 |
| S6-final-viral-combined_k141_140087\|\|full | 8656 | No | NA | 14 | 2 | 0 | Low-quality | Genome-fragment | 23.93 | AAI-based (medium-confidence) | 0 |
| S8-final-viral-combined_k141_439\|\|full | 2684 | No | NA | 3 | 0 | 2 | Not-determined | Genome-fragment | NA | NA | 0 |
| S8-final-viral-combined_k141_115631\|\|full | 4597 | No | NA | 5 | 0 | 0 | Not-determined | Genome-fragment | NA | NA | 0 |
| S8-final-viral-combined_k141_447\|\|full | 2614 | No | NA | 3 | 0 | 0 | Not-determined | Genome-fragment | NA | NA | 0 |
| S8-final-viral-combined_k141_69667\|\|full | 1558 | No | NA | 4 | 0 | 0 | Not-determined | Genome-fragment | NA | NA | 0 |
| S5-final-viral-combined_k141_106116\|\|full | 8659 | Yes | 7248 | 4 | 1 | 1 | Low-quality | Genome-fragment | 10.71 | AAI-based (high-confidence) | 16.3 |
| S8-final-viral-combined_k141_81209\|\|full | 1238 | No | NA | 2 | 0 | 1 | Not-determined | Genome-fragment | NA | NA | 0 |
| S8-final-viral-combined_k141_92659\|\|full | 3495 | No | NA | 8 | 0 | 0 | Not-determined | Genome-fragment | NA | NA | 0 |
| S8-final-viral-combined_k141_219255\|\|full | 4734 | No | NA | 5 | 0 | 1 | Not-determined | Genome-fragment | NA | NA | 0 |
| S8-final-viral-combined_k141_81224\|\|full | 6295 | No | NA | 7 | 0 | 6 | Not-determined | Genome-fragment | NA | NA | 0 |
| S9-final-viral-combined_k141_28591\|\|full | 8672 | No | NA | 11 | 4 | 1 | Low-quality | Genome-fragment | 17.69 | AAI-based (high-confidence) | 0 |
| S5-final-viral-combined_k141_138120\|\|full | 8685 | No | NA | 17 | 2 | 0 | Low-quality | Genome-fragment | 17.94 | AAI-based (medium-confidence) | 0 |
| S1-final-viral-combined_k141_10521\|\|full | 8686 | Yes | 3874 | 10 | 2 | 3 | Low-quality | Genome-fragment | 4.19 | HMM-based (lower-bound) | 55.4 |
| S5-final-viral-combined_k141_65262\|\|full | 8686 | No | NA | 14 | 0 | 1 | Low-quality | Genome-fragment | 26.86 | AAI-based (medium-confidence) | 0 |
| S9-final-viral-combined_k141_86868\|\|full | 8690 | Yes | 5348 | 8 | 1 | 2 | Low-quality | Genome-fragment | 0.85 | HMM-based (lower-bound) | 38.46 |
| S1_k141_96145 | 8693 | No | NA | 10 | 0 | 1 | Low-quality | Genome-fragment | 19.31 | AAI-based (high-confidence) | 0 |
| S8-final-viral-combined_k141_508\|\|full | 4719 | No | NA | 4 | 0 | 1 | Not-determined | Genome-fragment | NA | NA | 0 |
| S8-final-viral-combined_k141_196329\|\|full | 3331 | No | NA | 5 | 0 | 2 | Not-determined | Genome-fragment | NA | NA | 0 |
| S6-final-viral-combined_k141_30707\|\|full | 8698 | Yes | 1760 | 5 | 1 | 3 | Low-quality | Genome-fragment | 3.75 | AAI-based (medium-confidence) | 79.77 |
| S8-final-viral-combined_k141_141434\|\|full | 8699 | No | NA | 13 | 7 | 0 | Low-quality | Genome-fragment | 24.58 | AAI-based (medium-confidence) | 0 |
| S12-final-viral-combined_k141_312831\|\|full | 8700 | Yes | 1341 | 12 | 2 | 4 | Low-quality | Genome-fragment | 2.97 | HMM-based (lower-bound) | 84.59 |
| S8-final-viral-combined_k141_196362\|\|full | 1764 | No | NA | 3 | 0 | 1 | Not-determined | Genome-fragment | NA | NA | 0 |
| S8-final-viral-combined_k141_69706\|\|full | 1294 | No | NA | 2 | 0 | 1 | Not-determined | Genome-fragment | NA | NA | 0 |
| S8-final-viral-combined_k141_196366\|\|full | 2697 | No | NA | 4 | 0 | 4 | Not-determined | Genome-fragment | NA | NA | 0 |
| S8-final-viral-combined_k141_127176\|\|full | 1938 | No | NA | 1 | 0 | 0 | Not-determined | Genome-fragment | NA | NA | 0 |
| S8-final-viral-combined_k141_242591\|\|full | 19053 | No | NA | 13 | 0 | 7 | Not-determined | Genome-fragment | NA | NA | 0 |
| S8-final-viral-combined_k141_46621\|\|full | 2264 | No | NA | 5 | 0 | 3 | Not-determined | Genome-fragment | NA | NA | 0 |
| S5-final-viral-combined_k141_173014\|\|full | 8701 | No | NA | 9 | 3 | 0 | Low-quality | Genome-fragment | 13.78 | HMM-based (lower-bound) | 0 |
| S8-final-viral-combined_k141_265523\|\|full | 2414 | No | NA | 3 | 0 | 1 | Not-determined | Genome-fragment | NA | NA | 0 |
| S8-final-viral-combined_k141_242624\|\|full | 2118 | No | NA | 4 | 0 | 2 | Not-determined | Genome-fragment | NA | NA | 0 |
| S8-final-viral-combined_k141_173408\|\|full | 2192 | No | NA | 3 | 0 | 1 | Not-determined | Genome-fragment | NA | NA | 0 |
| S8-final-viral-combined_k141_184808\|\|full | 2386 | No | NA | 4 | 0 | 2 | Not-determined | Genome-fragment | NA | NA | 0 |
| S8-final-viral-combined_k141_34807\|\|full | 2263 | No | NA | 4 | 0 | 1 | Not-determined | Genome-fragment | NA | NA | 0 |
| S8-final-viral-combined_k141_92834\|\|full | 2190 | No | NA | 2 | 0 | 1 | Not-determined | Genome-fragment | NA | NA | 0 |
| S6-final-viral-combined_k141_127719\|\|full | 8704 | No | NA | 11 | 0 | 10 | Low-quality | Genome-fragment | 3.61 | AAI-based (medium-confidence) | 0 |
| S8-final-viral-combined_k141_23583\|\|full | 105620 | Yes | 36199 | 117 | 9 | 48 | Complete | High-quality | 100 | Provirus (medium-confidence) | 65.73 |
| S1-final-viral-combined_k141_114627\|\|full | 8706 | Yes | 4563 | 10 | 3 | 4 | Low-quality | Genome-fragment | 15.9 | AAI-based (high-confidence) | 47.59 |
| S8-final-viral-combined_k141_196455\|\|full | 1715 | No | NA | 2 | 0 | 1 | Not-determined | Genome-fragment | NA | NA | 0 |
| S8-final-viral-combined_k141_173456\|\|full | 4236 | No | NA | 5 | 0 | 2 | Not-determined | Genome-fragment | NA | NA | 0 |
| S8-final-viral-combined_k141_12027\|\|full | 6950 | No | NA | 6 | 0 | 5 | Not-determined | Genome-fragment | NA | NA | 0 |
| S8-final-viral-combined_k141_242661\|\|full | 1088 | No | NA | 3 | 0 | 0 | Not-determined | Genome-fragment | NA | NA | 0 |
| S4-final-viral-combined_k141_14616\|\|full | 8707 | No | NA | 10 | 0 | 4 | Low-quality | Genome-fragment | 5.82 | AAI-based (high-confidence) | 0 |
| S8-final-viral-combined_k141_674\|\|full | 2050 | No | NA | 3 | 0 | 0 | Not-determined | Genome-fragment | NA | NA | 0 |
| S8-final-viral-combined_k141_231055\|\|full | 2002 | No | NA | 2 | 0 | 1 | Not-determined | Genome-fragment | NA | NA | 0 |
| S12_k141_258637 | 8712 | No | NA | 6 | 0 | 5 | Low-quality | Genome-fragment | 7.73 | AAI-based (medium-confidence) | 0 |
| S8-final-viral-combined_k141_150257\|\|full | 2183 | No | NA | 3 | 0 | 1 | Not-determined | Genome-fragment | NA | NA | 0 |
| S8-final-viral-combined_k141_92892\|\|full | 1176 | No | NA | 3 | 0 | 1 | Not-determined | Genome-fragment | NA | NA | 0 |
| S12-final-viral-combined_k141_103484\|\|full | 8722 | No | NA | 11 | 4 | 1 | Low-quality | Genome-fragment | 17.8 | AAI-based (medium-confidence) | 0 |
| S7-final-viral-combined_k141_78914\|\|full | 8726 | No | NA | 14 | 4 | 0 | Low-quality | Genome-fragment | 9.58 | AAI-based (medium-confidence) | 0 |
| S8-final-viral-combined_k141_34877\|\|full | 3855 | No | NA | 2 | 0 | 0 | Not-determined | Genome-fragment | NA | NA | 0 |
| S8-final-viral-combined_k141_58352\|\|full | 1357 | No | NA | 3 | 0 | 2 | Not-determined | Genome-fragment | NA | NA | 0 |
| S8-final-viral-combined_k141_150283\|\|full | 1487 | No | NA | 2 | 0 | 0 | Not-determined | Genome-fragment | NA | NA | 0 |
| S2-final-viral-combined_k141_89284\|\|full | 8730 | No | NA | 15 | 0 | 0 | Low-quality | Genome-fragment | 24.17 | AAI-based (medium-confidence) | 0 |
| S17-k141_67305 | 8731 | No | NA | 10 | 4 | 0 | Low-quality | Genome-fragment | 11.04 | AAI-based (medium-confidence) | 0 |
| S8-final-viral-combined_k141_23653\|\|full | 1792 | No | NA | 4 | 0 | 1 | Not-determined | Genome-fragment | NA | NA | 0 |
| S8-final-viral-combined_k141_688\|\|full | 1501 | No | NA | 2 | 0 | 0 | Not-determined | Genome-fragment | NA | NA | 0 |
| S12_k141_243542 | 8731 | No | NA | 12 | 3 | 1 | Low-quality | Genome-fragment | 7.19 | AAI-based (high-confidence) | 0 |
| S11-final-viral-combined_k141_53919\|\|full | 8734 | No | NA | 12 | 4 | 0 | Low-quality | Genome-fragment | 23.33 | AAI-based (high-confidence) | 0 |
| S8-final-viral-combined_k141_199113\|\|full | 8734 | No | NA | 9 | 1 | 4 | Low-quality | Genome-fragment | 4.7 | HMM-based (lower-bound) | 0 |
| S8-final-viral-combined_k141_12063\|\|full | 4638 | No | NA | 4 | 0 | 2 | Not-determined | Genome-fragment | NA | NA | 0 |
| S2-final-viral-combined_k141_189883\|\|full | 8736 | No | NA | 12 | 1 | 3 | Low-quality | Genome-fragment | 21.14 | AAI-based (medium-confidence) | 0 |
| S8-final-viral-combined_k141_161796\|\|full | 1105 | No | NA | 2 | 0 | 0 | Not-determined | Genome-fragment | NA | NA | 0 |
| S8-final-viral-combined_k141_104223\|\|full | 3827 | No | NA | 3 | 0 | 1 | Not-determined | Genome-fragment | NA | NA | 0 |
| S8-final-viral-combined_k141_23718\|\|full | 1852 | No | NA | 2 | 0 | 0 | Not-determined | Genome-fragment | NA | NA | 0 |
| S8-final-viral-combined_k141_92971\|\|full | 7073 | No | NA | 7 | 0 | 3 | Not-determined | Genome-fragment | NA | NA | 0 |
| S12-final-viral-combined_k141_292510\|\|full | 8746 | No | NA | 11 | 0 | 1 | Low-quality | Genome-fragment | 14.13 | AAI-based (medium-confidence) | 0 |
| S7-final-viral-combined_k141_36494\|\|full | 8753 | No | NA | 14 | 9 | 0 | Low-quality | Genome-fragment | 23.75 | AAI-based (high-confidence) | 0 |
| S1-final-viral-combined_k141_214966\|\|full | 8756 | Yes | 7636 | 9 | 2 | 1 | Low-quality | Genome-fragment | 19.79 | AAI-based (high-confidence) | 12.79 |
| S2-final-viral-combined_k141_198596\|\|full | 8760 | No | NA | 13 | 0 | 0 | Low-quality | Genome-fragment | 21.66 | AAI-based (medium-confidence) | 0 |
| S8-final-viral-combined_k141_92997\|\|full | 3183 | No | NA | 2 | 0 | 2 | Not-determined | Genome-fragment | NA | NA | 0 |
| S8-final-viral-combined_k141_115815\|\|full | 5030 | No | NA | 4 | 0 | 1 | Not-determined | Genome-fragment | NA | NA | 0 |
| S8-final-viral-combined_k141_69829\|\|full | 1831 | No | NA | 3 | 0 | 2 | Not-determined | Genome-fragment | NA | NA | 0 |
| S8-final-viral-combined_k141_46677\|\|full | 1258 | No | NA | 3 | 0 | 0 | Not-determined | Genome-fragment | NA | NA | 0 |
| S2-final-viral-combined_k141_80803\|\|full | 8760 | No | NA | 11 | 0 | 2 | Low-quality | Genome-fragment | 27.22 | AAI-based (medium-confidence) | 0 |
| S1-final-viral-combined_k141_21721\|\|full | 8771 | No | NA | 13 | 11 | 0 | Low-quality | Genome-fragment | 19.02 | AAI-based (high-confidence) | 0 |
| S8-final-viral-combined_k141_127331\|\|full | 1665 | No | NA | 4 | 0 | 1 | Not-determined | Genome-fragment | NA | NA | 0 |
| S8-final-viral-combined_k141_752\|\|full | 2118 | No | NA | 1 | 0 | 0 | Not-determined | Genome-fragment | NA | NA | 0 |
| S2-final-viral-combined_k141_179334\|\|full | 8775 | Yes | 6775 | 7 | 1 | 2 | Low-quality | Genome-fragment | 10.37 | AAI-based (medium-confidence) | 22.79 |
| S2_k141_31426 | 8776 | No | NA | 3 | 0 | 0 | Low-quality | Genome-fragment | 9 | AAI-based (medium-confidence) | 0 |
| S8-final-viral-combined_k141_69899\|\|full | 1849 | No | NA | 2 | 0 | 2 | Not-determined | Genome-fragment | NA | NA | 0 |
| S11-final-viral-combined_k141_232643\|\|full | 8791 | No | NA | 16 | 1 | 0 | Low-quality | Genome-fragment | 8.54 | HMM-based (lower-bound) | 0 |
| S4_k141_75002 | 8796 | No | NA | 3 | 0 | 3 | Low-quality | Genome-fragment | 3.65 | AAI-based (high-confidence) | 0 |
| S8-final-viral-combined_k141_253985\|\|full | 2196 | No | NA | 3 | 0 | 0 | Not-determined | Genome-fragment | NA | NA | 0 |
| S11-final-viral-combined_k141_239628\|\|full | 8799 | No | NA | 6 | 0 | 3 | Low-quality | Genome-fragment | 11.54 | AAI-based (medium-confidence) | 0 |
| S6-final-viral-combined_k141_64607\|\|full | 8812 | No | NA | 12 | 1 | 1 | Low-quality | Genome-fragment | 16.5 | AAI-based (medium-confidence) | 0 |
| S8-final-viral-combined_k141_46747\|\|full | 2067 | No | NA | 2 | 0 | 1 | Not-determined | Genome-fragment | NA | NA | 0 |
| S1-final-viral-combined_k141_124564\|\|full | 8816 | No | NA | 11 | 0 | 5 | Low-quality | Genome-fragment | 28.15 | AAI-based (medium-confidence) | 0 |
| S8-final-viral-combined_k141_58404\|\|full | 1428 | No | NA | 3 | 0 | 0 | Not-determined | Genome-fragment | NA | NA | 0 |
| S8-final-viral-combined_k141_68533\|\|full | 8817 | No | NA | 13 | 4 | 0 | Low-quality | Genome-fragment | 17.94 | AAI-based (medium-confidence) | 0 |
| S12-final-viral-combined_k141_7270\|\|full | 8822 | No | NA | 15 | 5 | 0 | Low-quality | Genome-fragment | 27.38 | AAI-based (high-confidence) | 0 |
| S5_k141_119010 | 8824 | No | NA | 8 | 5 | 0 | Low-quality | Genome-fragment | 21.87 | AAI-based (high-confidence) | 0 |
| S2_k141_212515 | 8824 | No | NA | 4 | 0 | 1 | Low-quality | Genome-fragment | 13.84 | AAI-based (medium-confidence) | 0 |
| S8-final-viral-combined_k141_127426\|\|full | 6049 | No | NA | 6 | 0 | 1 | Not-determined | Genome-fragment | NA | NA | 0 |
| S8-final-viral-combined_k141_115884\|\|full | 2460 | No | NA | 3 | 0 | 1 | Not-determined | Genome-fragment | NA | NA | 0 |
| S8-final-viral-combined_k141_20238\|\|full | 8827 | No | NA | 9 | 0 | 0 | Low-quality | Genome-fragment | 13.36 | AAI-based (medium-confidence) | 0 |
| S8-final-viral-combined_k141_139116\|\|full | 2169 | No | NA | 3 | 0 | 3 | Not-determined | Genome-fragment | NA | NA | 0 |
| S8-final-viral-combined_k141_93127\|\|full | 2731 | No | NA | 6 | 0 | 1 | Not-determined | Genome-fragment | NA | NA | 0 |
| S11-final-viral-combined_k141_192075\|\|full | 8837 | No | NA | 16 | 2 | 0 | Low-quality | Genome-fragment | 8.8 | HMM-based (lower-bound) | 0 |
| S8-final-viral-combined_k141_127464\|\|full | 2208 | No | NA | 2 | 0 | 0 | Not-determined | Genome-fragment | NA | NA | 0 |
| S8-final-viral-combined_k141_46812\|\|full | 1436 | No | NA | 3 | 0 | 1 | Not-determined | Genome-fragment | NA | NA | 0 |
| S11_k141_76291 | 8844 | No | NA | 9 | 3 | 0 | Low-quality | Genome-fragment | 18.7 | AAI-based (high-confidence) | 0 |
| S1-final-viral-combined_k141_122127\|\|full | 8852 | No | NA | 13 | 1 | 0 | Low-quality | Genome-fragment | 42.43 | AAI-based (high-confidence) | 0 |
| S8-final-viral-combined_k141_185085\|\|full | 2326 | No | NA | 2 | 0 | 1 | Not-determined | Genome-fragment | NA | NA | 0 |
| S2_k141_154092 | 8855 | No | NA | 9 | 1 | 1 | Low-quality | Genome-fragment | 17.91 | AAI-based (medium-confidence) | 0 |
| S4-final-viral-combined_k141_88801\|\|full | 8856 | Yes | 3121 | 6 | 1 | 2 | Low-quality | Genome-fragment | 1.08 | HMM-based (lower-bound) | 64.76 |
| S8-final-viral-combined_k141_231227\|\|full | 1652 | No | NA | 3 | 0 | 1 | Not-determined | Genome-fragment | NA | NA | 0 |
| S8-final-viral-combined_k141_826\|\|full | 4330 | No | NA | 5 | 0 | 4 | Not-determined | Genome-fragment | NA | NA | 0 |
| S8-final-viral-combined_k141_161971\|\|full | 2650 | No | NA | 3 | 0 | 0 | Not-determined | Genome-fragment | NA | NA | 0 |
| S11-final-viral-combined_k141_68804\|\|full | 8858 | No | NA | 16 | 5 | 0 | Low-quality | Genome-fragment | 22.9 | AAI-based (high-confidence) | 0 |
| S8-final-viral-combined_k141_219628\|\|full | 2079 | No | NA | 3 | 0 | 0 | Not-determined | Genome-fragment | NA | NA | 0 |
| S8-final-viral-combined_k141_833\|\|full | 1532 | No | NA | 2 | 0 | 1 | Not-determined | Genome-fragment | NA | NA | 0 |
| S8-final-viral-combined_k141_139170\|\|full | 1835 | No | NA | 2 | 0 | 1 | Not-determined | Genome-fragment | NA | NA | 0 |
| S12-final-viral-combined_k141_154145\|\|full | 8859 | No | NA | 19 | 1 | 1 | Low-quality | Genome-fragment | 20.6 | HMM-based (lower-bound) | 0 |
| S8-final-viral-combined_k141_173732\|\|full | 2029 | No | NA | 2 | 0 | 2 | Not-determined | Genome-fragment | NA | NA | 0 |
| S8-final-viral-combined_k141_115995\|\|full | 1316 | No | NA | 3 | 0 | 1 | Not-determined | Genome-fragment | NA | NA | 0 |
| S8-final-viral-combined_k141_127521\|\|full | 2123 | No | NA | 4 | 0 | 1 | Not-determined | Genome-fragment | NA | NA | 0 |
| S12-final-viral-combined_k141_311448\|\|full | 8862 | No | NA | 10 | 5 | 0 | Low-quality | Genome-fragment | 26.07 | AAI-based (high-confidence) | 0 |
| S8-final-viral-combined_k141_175198\|\|full | 8862 | No | NA | 9 | 3 | 0 | Low-quality | Genome-fragment | 14.62 | AAI-based (medium-confidence) | 0 |
| S1-final-viral-combined_k141_212913\|\|full | 8869 | No | NA | 9 | 2 | 0 | Low-quality | Genome-fragment | 7.21 | HMM-based (lower-bound) | 0 |
| S8-final-viral-combined_k141_857\|\|full | 2204 | No | NA | 2 | 0 | 0 | Not-determined | Genome-fragment | NA | NA | 0 |
| S9_k141_11239 | 8870 | No | NA | 13 | 7 | 0 | Low-quality | Genome-fragment | 20.34 | AAI-based (high-confidence) | 0 |
| S8-final-viral-combined_k141_81597\|\|full | 24279 | No | NA | 19 | 3 | 1 | Medium-quality | Genome-fragment | 56.69 | AAI-based (medium-confidence) | 0 |
| S8-final-viral-combined_k141_70067\|\|full | 1123 | No | NA | 2 | 0 | 0 | Not-determined | Genome-fragment | NA | NA | 0 |
| S12-final-viral-combined_k141_167650\|\|full | 8871 | No | NA | 16 | 1 | 0 | Low-quality | Genome-fragment | 11.92 | HMM-based (lower-bound) | 0 |
| S8-final-viral-combined_k141_70070\|\|full | 2835 | No | NA | 3 | 0 | 0 | Not-determined | Genome-fragment | NA | NA | 0 |
| S8-final-viral-combined_k141_35208\|\|full | 1197 | No | NA | 3 | 0 | 1 | Not-determined | Genome-fragment | NA | NA | 0 |
| S2-final-viral-combined_k141_188530\|\|full | 8873 | No | NA | 9 | 0 | 1 | Low-quality | Genome-fragment | 21.75 | AAI-based (medium-confidence) | 0 |
| S4-final-viral-combined_k141_64074\|\|full | 8874 | No | NA | 9 | 4 | 0 | Low-quality | Genome-fragment | 13.11 | AAI-based (high-confidence) | 0 |
| S11-final-viral-combined_k141_16121\|\|full | 8878 | No | NA | 11 | 4 | 0 | Low-quality | Genome-fragment | 14.31 | AAI-based (medium-confidence) | 0 |
| S8-final-viral-combined_k141_254110\|\|full | 2766 | No | NA | 3 | 0 | 3 | Not-determined | Genome-fragment | NA | NA | 0 |
| S8-final-viral-combined_k141_116015\|\|full | 6035 | No | NA | 7 | 0 | 7 | Not-determined | Genome-fragment | NA | NA | 0 |
| S8-final-viral-combined_k141_104445\|\|full | 4369 | No | NA | 2 | 0 | 1 | Not-determined | Genome-fragment | NA | NA | 0 |
| S3-final-viral-combined_k141_65772\|\|full | 8883 | No | NA | 12 | 0 | 1 | Low-quality | Genome-fragment | 21.5 | AAI-based (high-confidence) | 0 |
| S8-final-viral-combined_k141_162036\|\|full | 2305 | No | NA | 3 | 0 | 2 | Not-determined | Genome-fragment | NA | NA | 0 |
| S12-final-viral-combined_k141_107016\|\|full | 8885 | No | NA | 12 | 1 | 5 | Low-quality | Genome-fragment | 11 | HMM-based (lower-bound) | 0 |
| S8-final-viral-combined_k141_150630\|\|full | 1218 | No | NA | 3 | 0 | 0 | Not-determined | Genome-fragment | NA | NA | 0 |
| S8-final-viral-combined_k141_35250\|\|full | 2073 | No | NA | 3 | 0 | 2 | Not-determined | Genome-fragment | NA | NA | 0 |
| S12-final-viral-combined_k141_37350\|\|full | 8898 | No | NA | 14 | 2 | 1 | Low-quality | Genome-fragment | 9.39 | HMM-based (lower-bound) | 0 |
| S1-final-viral-combined_k141_33687\|\|full | 8900 | No | NA | 11 | 3 | 1 | Low-quality | Genome-fragment | 18.1 | AAI-based (high-confidence) | 0 |
| S12-final-viral-combined_k141_309214\|\|full | 8903 | No | NA | 9 | 1 | 0 | Low-quality | Genome-fragment | 23.51 | AAI-based (high-confidence) | 0 |
| S8-final-viral-combined_k141_58565\|\|full | 1450 | No | NA | 3 | 0 | 1 | Not-determined | Genome-fragment | NA | NA | 0 |
| S11-final-viral-combined_k141_128335\|\|full | 8904 | No | NA | 16 | 3 | 0 | Low-quality | Genome-fragment | 22.76 | AAI-based (medium-confidence) | 0 |
| S8-final-viral-combined_k141_127559\|\|full | 7779 | No | NA | 4 | 0 | 4 | Not-determined | Genome-fragment | NA | NA | 0 |
| S8-final-viral-combined_k141_70160\|\|full | 2200 | No | NA | 3 | 0 | 2 | Not-determined | Genome-fragment | NA | NA | 0 |
| S8-final-viral-combined_k141_231426\|\|full | 1687 | No | NA | 3 | 0 | 0 | Not-determined | Genome-fragment | NA | NA | 0 |
| S11_k141_286267 | 8908 | No | NA | 14 | 11 | 0 | Low-quality | Genome-fragment | 20.13 | AAI-based (high-confidence) | 0 |
| S8-final-viral-combined_k141_150706\|\|full | 5095 | No | NA | 7 | 0 | 0 | Not-determined | Genome-fragment | NA | NA | 0 |
| S8-final-viral-combined_k141_173926\|\|full | 1702 | No | NA | 3 | 0 | 1 | Not-determined | Genome-fragment | NA | NA | 0 |
| S1-final-viral-combined_k141_369073\|\|full | 8908 | No | NA | 5 | 0 | 0 | Low-quality | Genome-fragment | 11.46 | AAI-based (medium-confidence) | 0 |
| S3-final-viral-combined_k141_70401\|\|full | 8909 | No | NA | 14 | 0 | 0 | Low-quality | Genome-fragment | 30.83 | AAI-based (medium-confidence) | 0 |
| S8-final-viral-combined_k141_93191\|\|full | 1269 | No | NA | 3 | 0 | 0 | Not-determined | Genome-fragment | NA | NA | 0 |
| S8-final-viral-combined_k141_231471\|\|full | 4976 | No | NA | 5 | 0 | 3 | Not-determined | Genome-fragment | NA | NA | 0 |
| S1-final-viral-combined_k141_97642\|\|full | 8911 | No | NA | 9 | 0 | 8 | Low-quality | Genome-fragment | 3.95 | AAI-based (medium-confidence) | 0 |
| S11-final-viral-combined_k141_75233\|\|full | 8920 | No | NA | 15 | 6 | 0 | Low-quality | Genome-fragment | 21.51 | AAI-based (high-confidence) | 0 |
| S8-final-viral-combined_k141_46948\|\|full | 4117 | No | NA | 7 | 0 | 3 | Not-determined | Genome-fragment | NA | NA | 0 |
| S8-final-viral-combined_k141_231491\|\|full | 1616 | No | NA | 3 | 0 | 1 | Not-determined | Genome-fragment | NA | NA | 0 |
| S5_k141_115446 | 8923 | No | NA | 17 | 5 | 0 | Low-quality | Genome-fragment | 19.92 | AAI-based (high-confidence) | 0 |
| S8-final-viral-combined_k141_162152\|\|full | 1400 | No | NA | 3 | 0 | 2 | Not-determined | Genome-fragment | NA | NA | 0 |
| S1-final-viral-combined_k141_271332\|\|full | 8929 | No | NA | 7 | 0 | 5 | Low-quality | Genome-fragment | 5.32 | AAI-based (high-confidence) | 0 |
| S1-final-viral-combined_k141_76465\|\|full | 8932 | Yes | 4552 | 13 | 3 | 4 | Low-quality | Genome-fragment | 6.03 | HMM-based (lower-bound) | 49.04 |
| S4-final-viral-combined_k141_88335\|\|full | 8942 | Yes | 2378 | 7 | 1 | 3 | Low-quality | Genome-fragment | 5.34 | AAI-based (high-confidence) | 73.41 |
| S8-final-viral-combined_k141_231512\|\|full | 2589 | No | NA | 3 | 0 | 3 | Not-determined | Genome-fragment | NA | NA | 0 |
| S8-final-viral-combined_k141_981\|\|full | 2895 | No | NA | 4 | 0 | 3 | Not-determined | Genome-fragment | NA | NA | 0 |
| S11-final-viral-combined_k141_325427\|\|full | 8944 | No | NA | 20 | 4 | 0 | Low-quality | Genome-fragment | 19.5 | AAI-based (high-confidence) | 0 |
| S8-final-viral-combined_k141_208421\|\|full | 1250 | No | NA | 2 | 0 | 0 | Not-determined | Genome-fragment | NA | NA | 0 |
| S8-final-viral-combined_k141_70248\|\|full | 1291 | No | NA | 3 | 0 | 1 | Not-determined | Genome-fragment | NA | NA | 0 |
| S8-final-viral-combined_k141_196865\|\|full | 15459 | No | NA | 19 | 0 | 16 | Not-determined | Genome-fragment | NA | NA | 0 |
| S8-final-viral-combined_k141_116131\|\|full | 3208 | No | NA | 2 | 0 | 2 | Not-determined | Genome-fragment | NA | NA | 0 |
| S8-final-viral-combined_k141_244898\|\|full | 8946 | No | NA | 11 | 2 | 0 | Low-quality | Genome-fragment | 18.37 | AAI-based (high-confidence) | 0 |
| S3-final-viral-combined_k141_102138\|\|full | 8949 | No | NA | 16 | 3 | 0 | Low-quality | Genome-fragment | 15.14 | AAI-based (high-confidence) | 0 |
| S9-final-viral-combined_k141_75346\|\|full | 8950 | No | NA | 12 | 5 | 0 | Low-quality | Genome-fragment | 20.52 | AAI-based (medium-confidence) | 0 |
| S11-final-viral-combined_k141_53794\|\|full | 8968 | No | NA | 13 | 4 | 0 | Low-quality | Genome-fragment | 24.35 | AAI-based (medium-confidence) | 0 |
| S1-final-viral-combined_k141_184231\|\|full | 8976 | No | NA | 14 | 2 | 1 | Low-quality | Genome-fragment | 20.04 | HMM-based (lower-bound) | 0 |
| S8-final-viral-combined_k141_139476\|\|full | 3263 | No | NA | 4 | 0 | 3 | Not-determined | Genome-fragment | NA | NA | 0 |
| S8-final-viral-combined_k141_70295\|\|full | 2107 | No | NA | 4 | 0 | 0 | Not-determined | Genome-fragment | NA | NA | 0 |
| S8-final-viral-combined_k141_213255\|\|full | 8985 | No | NA | 10 | 1 | 5 | Low-quality | Genome-fragment | 7.37 | HMM-based (lower-bound) | 0 |
| S9-final-viral-combined_k141_64838\|\|full | 8987 | No | NA | 3 | 0 | 0 | Low-quality | Genome-fragment | 10.14 | AAI-based (medium-confidence) | 0 |
| S2_k141_134497 | 8988 | No | NA | 7 | 0 | 2 | Low-quality | Genome-fragment | 11.2 | AAI-based (medium-confidence) | 0 |
| S8-final-viral-combined_k141_127725\|\|full | 1946 | No | NA | 3 | 0 | 1 | Not-determined | Genome-fragment | NA | NA | 0 |
| S9-final-viral-combined_k141_35469\|\|full | 8994 | No | NA | 11 | 4 | 0 | Low-quality | Genome-fragment | 22.06 | AAI-based (medium-confidence) | 0 |
| S2-final-viral-combined_k141_219799\|\|full | 8994 | No | NA | 14 | 5 | 0 | Low-quality | Genome-fragment | 17.88 | HMM-based (lower-bound) | 0 |
| S8-final-viral-combined_k141_231619\|\|full | 2719 | No | NA | 5 | 0 | 1 | Not-determined | Genome-fragment | NA | NA | 0 |
| S8-final-viral-combined_k141_104645\|\|full | 1488 | No | NA | 2 | 0 | 2 | Not-determined | Genome-fragment | NA | NA | 0 |
| S8-final-viral-combined_k141_127729\|\|full | 2386 | No | NA | 3 | 0 | 1 | Not-determined | Genome-fragment | NA | NA | 0 |
| S7-final-viral-combined_k141_65398\|\|full | 8997 | No | NA | 8 | 0 | 1 | Low-quality | Genome-fragment | 8.45 | AAI-based (medium-confidence) | 0 |
| S17-k141_260959 | 8997 | No | NA | 7 | 0 | 0 | Low-quality | Genome-fragment | 16.59 | AAI-based (medium-confidence) | 0 |
| S8-final-viral-combined_k141_24140\|\|full | 1246 | No | NA | 3 | 0 | 0 | Not-determined | Genome-fragment | NA | NA | 0 |
| S8-final-viral-combined_k141_104703\|\|full | 1972 | No | NA | 5 | 0 | 0 | Not-determined | Genome-fragment | NA | NA | 0 |
| S2-final-viral-combined_k141_125414\|\|full | 9000 | No | NA | 9 | 0 | 2 | Low-quality | Genome-fragment | 6.35 | AAI-based (medium-confidence) | 0 |
| S8-final-viral-combined_k141_162260\|\|full | 5815 | No | NA | 8 | 0 | 6 | Not-determined | Genome-fragment | NA | NA | 0 |
| S8-final-viral-combined_k141_162261\|\|full | 1972 | No | NA | 3 | 0 | 2 | Not-determined | Genome-fragment | NA | NA | 0 |

| | | | | | | | | | | | |
|---|---|---|---|---|---|---|---|---|---|---|---|
| S8-final-viral-combined_k141_162263\|\|full | 2458 | No | NA | 2 | 0 | 0 | Not-determined | Genome-fragment | NA | NA | 0 |
| S8-final-viral-combined_k141_185495\|\|full | 1438 | No | NA | 3 | 0 | 2 | Not-determined | Genome-fragment | NA | NA | 0 |
| S6-final-viral-combined_k141_70036\|\|full | 9000 | No | NA | 7 | 1 | 0 | Low-quality | Genome-fragment | 10.13 | AAI-based (high-confidence) | 0 |
| S5-final-viral-combined_k141_54278\|\|full | 9013 | No | NA | 7 | 6 | 0 | Low-quality | Genome-fragment | 28.11 | AAI-based (high-confidence) | 0 |
| S8-final-viral-combined_k141_47125\|\|full | 4188 | No | NA | 3 | 0 | 0 | Not-determined | Genome-fragment | NA | NA | 0 |
| S8-final-viral-combined_k141_265933\|\|full | 6692 | No | NA | 8 | 0 | 4 | Not-determined | Genome-fragment | NA | NA | 0 |
| S17-k141_63588 | 9047 | No | NA | 12 | 0 | 2 | Low-quality | Genome-fragment | 11.4 | AAI-based (medium-confidence) | 0 |
| S8_k141_82485 | 9052 | No | NA | 14 | 7 | 0 | Low-quality | Genome-fragment | 22.38 | AAI-based (high-confidence) | 0 |
| S8-final-viral-combined_k141_47134\|\|full | 3510 | No | NA | 5 | 0 | 4 | Not-determined | Genome-fragment | NA | NA | 0 |
| S5-final-viral-combined_k141_116216\|\|full | 9054 | No | NA | 12 | 1 | 1 | Low-quality | Genome-fragment | 22.72 | AAI-based (medium-confidence) | 0 |
| S11-final-viral-combined_k141_13267\|\|full | 9057 | No | NA | 10 | 0 | 6 | Low-quality | Genome-fragment | 2.21 | AAI-based (medium-confidence) | 0 |
| S8-final-viral-combined_k141_162276\|\|full | 2737 | No | NA | 4 | 0 | 0 | Not-determined | Genome-fragment | NA | NA | 0 |
| S8-final-viral-combined_k141_58855\|\|full | 3162 | No | NA | 3 | 0 | 1 | Not-determined | Genome-fragment | NA | NA | 0 |
| S8-final-viral-combined_k141_58860\|\|full | 1554 | No | NA | 2 | 0 | 0 | Not-determined | Genome-fragment | NA | NA | 0 |
| S8-final-viral-combined_k141_24217\|\|full | 5424 | No | NA | 6 | 0 | 2 | Not-determined | Genome-fragment | NA | NA | 0 |
| S8-final-viral-combined_k141_197001\|\|full | 1410 | No | NA | 2 | 0 | 0 | Not-determined | Genome-fragment | NA | NA | 0 |
| S8-final-viral-combined_k141_104804\|\|full | 1725 | No | NA | 3 | 0 | 2 | Not-determined | Genome-fragment | NA | NA | 0 |
| S8-final-viral-combined_k141_58871\|\|full | 2001 | No | NA | 3 | 0 | 0 | Not-determined | Genome-fragment | NA | NA | 0 |
| S8-final-viral-combined_k141_70435\|\|full | 4623 | No | NA | 2 | 0 | 1 | Not-determined | Genome-fragment | NA | NA | 0 |
| S9-final-viral-combined_k141_45544\|\|full | 9058 | No | NA | 9 | 0 | 7 | Low-quality | Genome-fragment | 2.83 | AAI-based (medium-confidence) | 0 |
| S8-final-viral-combined_k141_58885\|\|full | 1429 | No | NA | 3 | 0 | 1 | Not-determined | Genome-fragment | NA | NA | 0 |
| S8-final-viral-combined_k141_185595\|\|full | 1477 | No | NA | 3 | 0 | 2 | Not-determined | Genome-fragment | NA | NA | 0 |
| S5-final-viral-combined_k141_63657\|\|full | 9059 | No | NA | 12 | 1 | 0 | Low-quality | Genome-fragment | 5.86 | HMM-based (lower-bound) | 0 |
| S8-final-viral-combined_k141_197019\|\|full | 3248 | No | NA | 4 | 0 | 2 | Not-determined | Genome-fragment | NA | NA | 0 |
| S1-final-viral-combined_k141_84686\|\|full | 9061 | No | NA | 9 | 0 | 0 | Low-quality | Genome-fragment | 7.91 | AAI-based (medium-confidence) | 0 |
| S1_k141_378913 | 9063 | No | NA | 7 | 1 | 2 | Low-quality | Genome-fragment | 5.74 | HMM-based (lower-bound) | 0 |
| S8-final-viral-combined_k141_231689\|\|full | 2122 | No | NA | 4 | 0 | 1 | Not-determined | Genome-fragment | NA | NA | 0 |
| S2_k141_52842 | 9063 | No | NA | 10 | 0 | 2 | Low-quality | Genome-fragment | 8.84 | AAI-based (medium-confidence) | 0 |
| S8-final-viral-combined_k141_219796\|\|full | 26572 | No | NA | 39 | 21 | 1 | Medium-quality | Genome-fragment | 57.29 | AAI-based (medium-confidence) | 0 |
| S8-final-viral-combined_k141_152116\|\|full | 9065 | No | NA | 12 | 1 | 0 | Low-quality | Genome-fragment | 17.22 | AAI-based (high-confidence) | 0 |
| S1-final-viral-combined_k141_380763\|\|full | 9067 | No | NA | 15 | 13 | 0 | Low-quality | Genome-fragment | 26.07 | AAI-based (high-confidence) | 0 |
| S8-final-viral-combined_k141_58939\|\|full | 3978 | No | NA | 5 | 0 | 3 | Not-determined | Genome-fragment | NA | NA | 0 |
| S8-final-viral-combined_k141_1122\|\|full | 1324 | No | NA | 2 | 0 | 0 | Not-determined | Genome-fragment | NA | NA | 0 |
| S8-final-viral-combined_k141_242003\|\|full | 9072 | No | NA | 12 | 2 | 0 | Low-quality | Genome-fragment | 6.8 | AAI-based (high-confidence) | 0 |
| S11-final-viral-combined_k141_96949\|\|full | 9076 | No | NA | 16 | 3 | 0 | Low-quality | Genome-fragment | 21.36 | AAI-based (high-confidence) | 0 |
| S8-final-viral-combined_k141_100278\|\|full | 9084 | No | NA | 23 | 2 | 2 | Low-quality | Genome-fragment | 29.18 | AAI-based (medium-confidence) | 0 |
| S8-final-viral-combined_k141_185611\|\|full | 36010 | No | NA | 31 | 5 | 2 | Medium-quality | Genome-fragment | 62.09 | HMM-based (lower-bound) | 0 |
| S8-final-viral-combined_k141_165601\|\|full | 9089 | No | NA | 7 | 0 | 3 | Low-quality | Genome-fragment | 3.99 | AAI-based (medium-confidence) | 0 |
| S8-final-viral-combined_k141_24299\|\|full | 3375 | No | NA | 5 | 0 | 2 | Not-determined | Genome-fragment | NA | NA | 0 |
| S1_k141_260402 | 9091 | No | NA | 9 | 0 | 5 | Low-quality | Genome-fragment | 4.28 | AAI-based (medium-confidence) | 0 |
| S8-final-viral-combined_k141_47207\|\|full | 2061 | No | NA | 3 | 0 | 0 | Not-determined | Genome-fragment | NA | NA | 0 |
| S17-final-viral-combined_k141_1962\|\|full | 9091 | No | NA | 11 | 0 | 0 | Low-quality | Genome-fragment | 8.67 | AAI-based (medium-confidence) | 0 |
| S8-final-viral-combined_k141_70527\|\|full | 1897 | No | NA | 2 | 0 | 0 | Not-determined | Genome-fragment | NA | NA | 0 |
| S8-final-viral-combined_k141_58967\|\|full | 3558 | No | NA | 4 | 0 | 0 | Not-determined | Genome-fragment | NA | NA | 0 |
| S8-final-viral-combined_k141_231786\|\|full | 1366 | No | NA | 2 | 0 | 2 | Not-determined | Genome-fragment | NA | NA | 0 |
| S8-final-viral-combined_k141_60591\|\|full | 9091 | No | NA | 9 | 1 | 0 | Low-quality | Genome-fragment | 8.96 | AAI-based (high-confidence) | 0 |
| S17-k141_68635 | 9095 | No | NA | 17 | 0 | 3 | Low-quality | Genome-fragment | 13.41 | AAI-based (high-confidence) | 0 |
| S8-final-viral-combined_k141_127877\|\|full | 1148 | No | NA | 2 | 0 | 1 | Not-determined | Genome-fragment | NA | NA | 0 |
| S12_k141_194721 | 9096 | No | NA | 13 | 3 | 0 | Low-quality | Genome-fragment | 17.68 | AAI-based (high-confidence) | 0 |
| S8-final-viral-combined_k141_185652\|\|full | 1981 | No | NA | 3 | 0 | 2 | Not-determined | Genome-fragment | NA | NA | 0 |
| S8-final-viral-combined_k141_82043\|\|full | 3319 | No | NA | 2 | 0 | 2 | Not-determined | Genome-fragment | NA | NA | 0 |
| S8-final-viral-combined_k141_93624\|\|full | 1013 | No | NA | 2 | 0 | 1 | Not-determined | Genome-fragment | NA | NA | 0 |
| S11-final-viral-combined_k141_289579\|\|full | 9097 | No | NA | 14 | 5 | 0 | Low-quality | Genome-fragment | 19.29 | AAI-based (high-confidence) | 0 |
| S8-final-viral-combined_k141_219846\|\|full | 6438 | No | NA | 8 | 0 | 1 | Not-determined | Genome-fragment | NA | NA | 0 |
| S8-final-viral-combined_k141_243360\|\|full | 1014 | No | NA | 3 | 0 | 1 | Not-determined | Genome-fragment | NA | NA | 0 |
| S8-final-viral-combined_k141_70572\|\|full | 7156 | No | NA | 7 | 0 | 6 | Not-determined | Genome-fragment | NA | NA | 0 |
| S4_k141_69815 | 9102 | No | NA | 16 | 4 | 1 | Low-quality | Genome-fragment | 6.03 | AAI-based (high-confidence) | 0 |
| S7-final-viral-combined_k141_100522\|\|full | 9118 | Yes | 3528 | 10 | 1 | 1 | Low-quality | Genome-fragment | 1.01 | HMM-based (lower-bound) | 61.31 |
| S8-final-viral-combined_k141_254463\|\|full | 2096 | No | NA | 3 | 0 | 1 | Not-determined | Genome-fragment | NA | NA | 0 |
| S2_k141_160025 | 9128 | No | NA | 19 | 2 | 1 | Low-quality | Genome-fragment | 28.1 | AAI-based (high-confidence) | 0 |
| S8-final-viral-combined_k141_185683\|\|full | 1262 | No | NA | 3 | 0 | 1 | Not-determined | Genome-fragment | NA | NA | 0 |
| S8-final-viral-combined_k141_186819\|\|full | 9128 | No | NA | 14 | 8 | 0 | Low-quality | Genome-fragment | 21.29 | AAI-based (high-confidence) | 0 |
| S8-final-viral-combined_k141_139737\|\|full | 2391 | No | NA | 4 | 0 | 1 | Not-determined | Genome-fragment | NA | NA | 0 |
| S8-final-viral-combined_k141_12455\|\|full | 1727 | No | NA | 3 | 0 | 3 | Not-determined | Genome-fragment | NA | NA | 0 |
| S17-final-viral-combined_k141_254248\|\|full | 9129 | No | NA | 9 | 0 | 3 | Low-quality | Genome-fragment | 6.56 | AAI-based (medium-confidence) | 0 |
| S8-final-viral-combined_k141_35695\|\|full | 2469 | No | NA | 4 | 0 | 2 | Not-determined | Genome-fragment | NA | NA | 0 |
| S17-final-viral-combined_k141_159480\|\|full | 9132 | No | NA | 13 | 7 | 0 | Low-quality | Genome-fragment | 23.46 | AAI-based (high-confidence) | 0 |
| S2_k141_130858 | 9133 | No | NA | 12 | 3 | 1 | Low-quality | Genome-fragment | 5.67 | AAI-based (medium-confidence) | 0 |
| S17-final-viral-combined_k141_149074\|\|full | 9134 | No | NA | 7 | 0 | 4 | Low-quality | Genome-fragment | 4.79 | AAI-based (high-confidence) | 0 |
| S8-final-viral-combined_k141_104938\|\|full | 8833 | No | NA | 11 | 0 | 5 | Not-determined | Genome-fragment | NA | NA | 0 |
| S4-final-viral-combined_k141_52994\|\|full | 9137 | No | NA | 17 | 2 | 1 | Low-quality | Genome-fragment | 12.11 | AAI-based (high-confidence) | 0 |
| S8-final-viral-combined_k141_231895\|\|full | 3012 | No | NA | 2 | 0 | 1 | Not-determined | Genome-fragment | NA | NA | 0 |
| S12_k141_4487 | 9139 | Yes | 6509 | 13 | 5 | 2 | Low-quality | Genome-fragment | 14.55 | AAI-based (high-confidence) | 28.78 |
| S8-final-viral-combined_k141_12497\|\|full | 1969 | No | NA | 3 | 0 | 0 | Not-determined | Genome-fragment | NA | NA | 0 |
| S8-final-viral-combined_k141_24438\|\|full | 21855 | No | NA | 23 | 0 | 19 | Not-determined | Genome-fragment | NA | NA | 0 |
| S8-final-viral-combined_k141_151117\|\|full | 1250 | No | NA | 3 | 0 | 1 | Not-determined | Genome-fragment | NA | NA | 0 |
| S11-final-viral-combined_k141_177049\|\|full | 9141 | No | NA | 11 | 2 | 0 | Low-quality | Genome-fragment | 11.9 | HMM-based (lower-bound) | 0 |
| S8-final-viral-combined_k141_243433\|\|full | 1196 | No | NA | 2 | 0 | 1 | Not-determined | Genome-fragment | NA | NA | 0 |
| S8-final-viral-combined_k141_35758\|\|full | 4477 | No | NA | 5 | 0 | 3 | Not-determined | Genome-fragment | NA | NA | 0 |
| S8-final-viral-combined_k141_60069\|\|full | 9160 | No | NA | 20 | 2 | 0 | Low-quality | Genome-fragment | 21.27 | AAI-based (high-confidence) | 0 |
| S8-final-viral-combined_k141_162581\|\|full | 1248 | No | NA | 3 | 0 | 2 | Not-determined | Genome-fragment | NA | NA | 0 |
| S8-final-viral-combined_k141_254495\|\|full | 5867 | No | NA | 3 | 0 | 0 | Not-determined | Genome-fragment | NA | NA | 0 |
| S11-final-viral-combined_k141_87215\|\|full | 9164 | No | NA | 15 | 0 | 0 | Low-quality | Genome-fragment | 19.99 | AAI-based (high-confidence) | 0 |
| S8-final-viral-combined_k141_266073\|\|full | 1204 | No | NA | 2 | 0 | 0 | Not-determined | Genome-fragment | NA | NA | 0 |
| S8-final-viral-combined_k141_82161\|\|full | 1839 | No | NA | 4 | 0 | 1 | Not-determined | Genome-fragment | NA | NA | 0 |
| S17-final-viral-combined_k141_109199\|\|full | 9165 | No | NA | 12 | 8 | 0 | Low-quality | Genome-fragment | 24.71 | AAI-based (high-confidence) | 0 |
| S8-final-viral-combined_k141_151138\|\|full | 2753 | No | NA | 2 | 0 | 2 | Not-determined | Genome-fragment | NA | NA | 0 |
| S7-final-viral-combined_k141_106934\|\|full | 9168 | No | NA | 12 | 1 | 0 | Low-quality | Genome-fragment | 20.64 | AAI-based (high-confidence) | 0 |
| S17-final-viral-combined_k141_234067\|\|full | 9168 | No | NA | 9 | 2 | 0 | Low-quality | Genome-fragment | 11.14 | AAI-based (high-confidence) | 0 |
| S8-final-viral-combined_k141_219945\|\|full | 1068 | No | NA | 2 | 0 | 1 | Not-determined | Genome-fragment | NA | NA | 0 |
| S17-k141_191299 | 9168 | No | NA | 14 | 2 | 0 | Low-quality | Genome-fragment | 16.81 | AAI-based (high-confidence) | 0 |
| S1-final-viral-combined_k141_155936\|\|full | 9169 | No | NA | 5 | 0 | 4 | Low-quality | Genome-fragment | 5.84 | AAI-based (high-confidence) | 0 |
| S8-final-viral-combined_k141_59247\|\|full | 4209 | No | NA | 4 | 0 | 1 | Not-determined | Genome-fragment | NA | NA | 0 |
| S8-final-viral-combined_k141_254525\|\|full | 1804 | No | NA | 2 | 0 | 1 | Not-determined | Genome-fragment | NA | NA | 0 |
| S12-final-viral-combined_k141_49332\|\|full | 9171 | No | NA | 13 | 0 | 5 | Low-quality | Genome-fragment | 16.95 | AAI-based (medium-confidence) | 0 |
| S8-final-viral-combined_k141_105025\|\|full | 1279 | No | NA | 2 | 0 | 2 | Not-determined | Genome-fragment | NA | NA | 0 |
| S8-final-viral-combined_k141_208837\|\|full | 1618 | No | NA | 2 | 0 | 2 | Not-determined | Genome-fragment | NA | NA | 0 |
| S8-final-viral-combined_k141_59274\|\|full | 8168 | No | NA | 8 | 0 | 5 | Not-determined | Genome-fragment | NA | NA | 0 |
| S8-final-viral-combined_k141_174270\|\|full | 2180 | No | NA | 2 | 0 | 1 | Not-determined | Genome-fragment | NA | NA | 0 |
| S8-final-viral-combined_k141_197047\|\|full | 2354 | No | NA | 4 | 0 | 1 | Not-determined | Genome-fragment | NA | NA | 0 |
| S1-final-viral-combined_k141_1470\|\|full | 9178 | Yes | 1864 | 10 | 1 | 5 | Low-quality | Genome-fragment | 2.82 | HMM-based (lower-bound) | 79.69 |
| S8-final-viral-combined_k141_162632\|\|full | 2351 | No | NA | 4 | 0 | 1 | Not-determined | Genome-fragment | NA | NA | 0 |
| S8-final-viral-combined_k141_139937\|\|full | 1871 | No | NA | 2 | 0 | 1 | Not-determined | Genome-fragment | NA | NA | 0 |
| S8-final-viral-combined_k141_197059\|\|full | 2194 | No | NA | 5 | 0 | 0 | Not-determined | Genome-fragment | NA | NA | 0 |
| S9-final-viral-combined_k141_64967\|\|full | 9182 | No | NA | 11 | 0 | 5 | Low-quality | Genome-fragment | 18.37 | AAI-based (medium-confidence) | 0 |
| S8-final-viral-combined_k141_266156\|\|full | 1437 | No | NA | 2 | 0 | 1 | Not-determined | Genome-fragment | NA | NA | 0 |
| S2-final-viral-combined_k141_211355\|\|full | 9193 | No | NA | 12 | 1 | 0 | Low-quality | Genome-fragment | 5.82 | HMM-based (lower-bound) | 0 |
| S8-final-viral-combined_k141_185890\|\|full | 4186 | No | NA | 3 | 0 | 3 | Not-determined | Genome-fragment | NA | NA | 0 |
| S11_k141_173181 | 9195 | No | NA | 12 | 0 | 3 | Low-quality | Genome-fragment | 13.98 | AAI-based (medium-confidence) | 0 |
| S8-final-viral-combined_k141_24596\|\|full | 1277 | No | NA | 2 | 0 | 1 | Not-determined | Genome-fragment | NA | NA | 0 |
| S1_k141_172749 | 9198 | No | NA | 6 | 3 | 0 | Low-quality | Genome-fragment | 14.4 | AAI-based (high-confidence) | 0 |
| S17-final-viral-combined_k141_84316\|\|full | 9203 | No | NA | 4 | 0 | 2 | Low-quality | Genome-fragment | 7.56 | AAI-based (medium-confidence) | 0 |
| S8-final-viral-combined_k141_59346\|\|full | 3964 | No | NA | 3 | 0 | 1 | Not-determined | Genome-fragment | NA | NA | 0 |
| S8-final-viral-combined_k141_35908\|\|full | 13532 | No | NA | 4 | 0 | 2 | Not-determined | Genome-fragment | NA | NA | 0 |
| S8-final-viral-combined_k141_266214\|\|full | 1780 | No | NA | 3 | 0 | 1 | Not-determined | Genome-fragment | NA | NA | 0 |
| S8-final-viral-combined_k141_139965\|\|full | 2582 | No | NA | 4 | 0 | 4 | Not-determined | Genome-fragment | NA | NA | 0 |
| S1_k141_266931 | 9208 | No | NA | 14 | 3 | 0 | Low-quality | Genome-fragment | 23.86 | AAI-based (high-confidence) | 0 |
| S8-final-viral-combined_k141_139968\|\|full | 2215 | No | NA | 4 | 0 | 0 | Not-determined | Genome-fragment | NA | NA | 0 |
| S5-final-viral-combined_k141_73626\|\|full | 9212 | No | NA | 9 | 0 | 0 | Low-quality | Genome-fragment | 10.27 | AAI-based (medium-confidence) | 0 |
| S8-final-viral-combined_k141_266250\|\|full | 1285 | No | NA | 2 | 0 | 1 | Not-determined | Genome-fragment | NA | NA | 0 |
| S17-final-viral-combined_k141_23584\|\|full | 9213 | No | NA | 9 | 1 | 0 | Low-quality | Genome-fragment | 11.19 | AAI-based (high-confidence) | 0 |
| S8-final-viral-combined_k141_59395\|\|full | 1377 | No | NA | 2 | 0 | 0 | Not-determined | Genome-fragment | NA | NA | 0 |
| S12_k141_23919 | 9219 | No | NA | 17 | 1 | 0 | Low-quality | Genome-fragment | 1.47 | HMM-based (lower-bound) | 0 |
| S8-final-viral-combined_k141_35928\|\|full | 2389 | No | NA | 5 | 0 | 3 | Not-determined | Genome-fragment | NA | NA | 0 |
| S2-final-viral-combined_k141_163450\|\|full | 9219 | No | NA | 12 | 5 | 0 | Low-quality | Genome-fragment | 13.86 | AAI-based (high-confidence) | 0 |
| S2-final-viral-combined_k141_89495\|\|full | 9222 | No | NA | 6 | 0 | 1 | Low-quality | Genome-fragment | 8.51 | AAI-based (medium-confidence) | 0 |
| S1_k141_83798 | 9227 | No | NA | 7 | 0 | 1 | Low-quality | Genome-fragment | 12.98 | AAI-based (medium-confidence) | 0 |
| S8-final-viral-combined_k141_70878\|\|full | 2142 | No | NA | 2 | 0 | 1 | Not-determined | Genome-fragment | NA | NA | 0 |
| S1_k141_20026 | 9232 | No | NA | 12 | 8 | 0 | Low-quality | Genome-fragment | 15.01 | AAI-based (high-confidence) | 0 |
| S17-final-viral-combined_k141_163902\|\|full | 9233 | No | NA | 16 | 5 | 0 | Low-quality | Genome-fragment | 21.73 | AAI-based (high-confidence) | 0 |
| S7-final-viral-combined_k141_92661\|\|full | 9245 | No | NA | 16 | 1 | 4 | Low-quality | Genome-fragment | 18.91 | AAI-based (medium-confidence) | 0 |
| S8-final-viral-combined_k141_266285\|\|full | 1768 | No | NA | 4 | 0 | 1 | Not-determined | Genome-fragment | NA | NA | 0 |
| S8-final-viral-combined_k141_197226\|\|full | 2985 | No | NA | 4 | 0 | 3 | Not-determined | Genome-fragment | NA | NA | 0 |
| S8-final-viral-combined_k141_59469\|\|full | 1251 | No | NA | 3 | 0 | 3 | Not-determined | Genome-fragment | NA | NA | 0 |
| S8-final-viral-combined_k141_82266\|\|full | 1385 | No | NA | 3 | 0 | 1 | Not-determined | Genome-fragment | NA | NA | 0 |
| S3_k141_48311 | 9247 | No | NA | 8 | 0 | 7 | Low-quality | Genome-fragment | 8.67 | AAI-based (medium-confidence) | 0 |
| S8-final-viral-combined_k141_1485\|\|full | 5132 | No | NA | 8 | 0 | 1 | Not-determined | Genome-fragment | NA | NA | 0 |
| S8-final-viral-combined_k141_116647\|\|full | 2140 | No | NA | 3 | 0 | 1 | Not-determined | Genome-fragment | NA | NA | 0 |
| S8-final-viral-combined_k141_12603\|\|full | 1632 | No | NA | 4 | 0 | 2 | Not-determined | Genome-fragment | NA | NA | 0 |
| S5-final-viral-combined_k141_88696\|\|full | 9254 | No | NA | 11 | 0 | 8 | Low-quality | Genome-fragment | 4.1 | AAI-based (medium-confidence) | 0 |
| S8-final-viral-combined_k141_186014\|\|full | 1296 | No | NA | 2 | 0 | 0 | Not-determined | Genome-fragment | NA | NA | 0 |
| S8-final-viral-combined_k141_140059\|\|full | 31825 | No | NA | 45 | 11 | 1 | High-quality | High-quality | 100 | AAI-based (high-confidence) | 0 |
| S8-final-viral-combined_k141_174450\|\|full | 3494 | No | NA | 2 | 0 | 2 | Not-determined | Genome-fragment | NA | NA | 0 |
| S2-final-viral-combined_k141_136758\|\|full | 9257 | No | NA | 12 | 1 | 0 | Low-quality | Genome-fragment | 20.19 | AAI-based (medium-confidence) | 0 |
| S7-final-viral-combined_k141_69630\|\|full | 9258 | No | NA | 9 | 0 | 7 | Low-quality | Genome-fragment | 7.46 | AAI-based (medium-confidence) | 0 |
| S8-final-viral-combined_k141_105184\|\|full | 1831 | No | NA | 2 | 0 | 0 | Not-determined | Genome-fragment | NA | NA | 0 |
| S8-final-viral-combined_k141_140097\|\|full | 2722 | No | NA | 4 | 0 | 1 | Not-determined | Genome-fragment | NA | NA | 0 |
| S8-final-viral-combined_k141_128240\|\|full | 2246 | No | NA | 3 | 0 | 0 | Not-determined | Genome-fragment | NA | NA | 0 |
| S8-final-viral-combined_k141_220189\|\|full | 1832 | No | NA | 2 | 0 | 0 | Not-determined | Genome-fragment | NA | NA | 0 |
| S8-final-viral-combined_k141_59499\|\|full | 3241 | No | NA | 4 | 0 | 0 | Not-determined | Genome-fragment | NA | NA | 0 |
| S8-final-viral-combined_k141_59501\|\|full | 1612 | No | NA | 4 | 0 | 1 | Not-determined | Genome-fragment | NA | NA | 0 |
| S8-final-viral-combined_k141_105232\|\|full | 1089 | No | NA | 2 | 0 | 1 | Not-determined | Genome-fragment | NA | NA | 0 |
| S9-final-viral-combined_k141_23284\|\|full | 9259 | No | NA | 9 | 1 | 0 | Low-quality | Genome-fragment | 5.66 | HMM-based (lower-bound) | 0 |
| S8-final-viral-combined_k141_174518\|\|full | 3382 | No | NA | 5 | 0 | 1 | Not-determined | Genome-fragment | NA | NA | 0 |
| S2-final-viral-combined_k141_206609\|\|full | 9259 | No | NA | 7 | 0 | 2 | Low-quality | Genome-fragment | 11.25 | AAI-based (medium-confidence) | 0 |
| S8-final-viral-combined_k141_93920\|\|full | 4198 | No | NA | 9 | 0 | 0 | Not-determined | Genome-fragment | NA | NA | 0 |
| S6-final-viral-combined_k141_93006\|\|full | 9260 | No | NA | 11 | 1 | 0 | Low-quality | Genome-fragment | 3.94 | HMM-based (lower-bound) | 0 |
| S8-final-viral-combined_k141_253850\|\|full | 9261 | No | NA | 20 | 1 | 0 | Low-quality | Genome-fragment | 14.5 | HMM-based (lower-bound) | 0 |
| S8-final-viral-combined_k141_186054\|\|full | 53551 | Yes | 52057 | 67 | 25 | 4 | High-quality | High-quality | 100 | AAI-based (high-confidence) | 2.79 |
| S8-final-viral-combined_k141_254757\|\|full | 2923 | No | NA | 4 | 0 | 3 | Not-determined | Genome-fragment | NA | NA | 0 |
| S11-final-viral-combined_k141_110341\|\|full | 9265 | No | NA | 11 | 0 | 7 | Low-quality | Genome-fragment | 9.66 | AAI-based (medium-confidence) | 0 |
| S8-final-viral-combined_k141_174574\|\|full | 1478 | No | NA | 2 | 0 | 2 | Not-determined | Genome-fragment | NA | NA | 0 |
| S8_k141_176363 | 9265 | No | NA | 14 | 1 | 0 | Low-quality | Genome-fragment | 4.54 | HMM-based (lower-bound) | 0 |
| S5_k141_127132 | 9268 | No | NA | 13 | 9 | 0 | Low-quality | Genome-fragment | 22.87 | AAI-based (high-confidence) | 0 |
| S8-final-viral-combined_k141_151547\|\|full | 2179 | No | NA | 3 | 0 | 2 | Not-determined | Genome-fragment | NA | NA | 0 |
| S8-final-viral-combined_k141_1594\|\|full | 1734 | No | NA | 2 | 0 | 1 | Not-determined | Genome-fragment | NA | NA | 0 |
| S8-final-viral-combined_k141_1596\|\|full | 2228 | No | NA | 2 | 0 | 1 | Not-determined | Genome-fragment | NA | NA | 0 |
| S8-final-viral-combined_k141_140224\|\|full | 4515 | No | NA | 5 | 0 | 2 | Not-determined | Genome-fragment | NA | NA | 0 |
| S8-final-viral-combined_k141_243767\|\|full | 2431 | No | NA | 4 | 0 | 0 | Not-determined | Genome-fragment | NA | NA | 0 |
| S8-final-viral-combined_k141_151577\|\|full | 3248 | No | NA | 3 | 0 | 2 | Not-determined | Genome-fragment | NA | NA | 0 |
| S8-final-viral-combined_k141_59569\|\|full | 4848 | No | NA | 7 | 0 | 0 | Not-determined | Genome-fragment | NA | NA | 0 |
| S7-final-viral-combined_k141_3512\|\|full | 9275 | Yes | 1401 | 8 | 1 | 3 | Low-quality | Genome-fragment | 33.67 | HMM-based (lower-bound) | 84.89 |
| S4-final-viral-combined_k141_161822\|\|full | 9275 | No | NA | 6 | 0 | 5 | Low-quality | Genome-fragment | 1.38 | AAI-based (medium-confidence) | 0 |
| S8-final-viral-combined_k141_23561\|\|full | 9275 | No | NA | 5 | 0 | 3 | Low-quality | Genome-fragment | 4.04 | AAI-based (medium-confidence) | 0 |
| S8-final-viral-combined_k141_91888\|\|full | 9275 | No | NA | 16 | 2 | 0 | Low-quality | Genome-fragment | 22.29 | AAI-based (high-confidence) | 0 |
| S8-final-viral-combined_k141_93973\|\|full | 8423 | No | NA | 8 | 0 | 5 | Not-determined | Genome-fragment | NA | NA | 0 |
| S8-final-viral-combined_k141_151633\|\|full | 2312 | No | NA | 4 | 0 | 1 | Not-determined | Genome-fragment | NA | NA | 0 |
| S12-final-viral-combined_k141_95853\|\|full | 9277 | No | NA | 8 | 0 | 5 | Low-quality | Genome-fragment | 3.1 | AAI-based (medium-confidence) | 0 |
| S11-final-viral-combined_k141_240647\|\|full | 9279 | No | NA | 11 | 5 | 1 | Low-quality | Genome-fragment | 15.17 | AAI-based (medium-confidence) | 0 |
| S11-final-viral-combined_k141_116240\|\|full | 9283 | No | NA | 6 | 0 | 6 | Low-quality | Genome-fragment | 5.78 | AAI-based (medium-confidence) | 0 |
| S1-final-viral-combined_k141_135162\|\|full | 9285 | No | NA | 10 | 0 | 3 | Low-quality | Genome-fragment | 7.94 | AAI-based (medium-confidence) | 0 |
| S8-final-viral-combined_k141_154739\|\|full | 9285 | No | NA | 9 | 0 | 1 | Low-quality | Genome-fragment | 15.46 | AAI-based (medium-confidence) | 0 |
| S8-final-viral-combined_k141_232297\|\|full | 2002 | No | NA | 4 | 0 | 1 | Not-determined | Genome-fragment | NA | NA | 0 |
| S11-final-viral-combined_k141_142244\|\|full | 9286 | Yes | 3104 | 8 | 2 | 3 | Low-quality | Genome-fragment | 4.78 | HMM-based (lower-bound) | 66.57 |
| S1-final-viral-combined_k141_306814\|\|full | 9286 | No | NA | 10 | 1 | 0 | Low-quality | Genome-fragment | 14.21 | AAI-based (medium-confidence) | 0 |
| S8-final-viral-combined_k141_36104\|\|full | 6073 | No | NA | 5 | 0 | 3 | Not-determined | Genome-fragment | NA | NA | 0 |
| S8-final-viral-combined_k141_151674\|\|full | 1428 | No | NA | 3 | 0 | 1 | Not-determined | Genome-fragment | NA | NA | 0 |
| S8-final-viral-combined_k141_47539\|\|full | 1412 | No | NA | 3 | 0 | 2 | Not-determined | Genome-fragment | NA | NA | 0 |
| S8-final-viral-combined_k141_209250\|\|full | 1446 | No | NA | 3 | 0 | 1 | Not-determined | Genome-fragment | NA | NA | 0 |
| S17-final-viral-combined_k141_175464\|\|full | 9289 | No | NA | 12 | 2 | 0 | Low-quality | Genome-fragment | 13.72 | AAI-based (high-confidence) | 0 |
| S8-final-viral-combined_k141_241294\|\|full | 9295 | Yes | 570 | 8 | 1 | 4 | Low-quality | Genome-fragment | 0.93 | HMM-based (lower-bound) | 93.87 |
| S8-final-viral-combined_k141_186211\|\|full | 3908 | No | NA | 3 | 0 | 2 | Not-determined | Genome-fragment | NA | NA | 0 |
| S8-final-viral-combined_k141_149833\|\|full | 9297 | No | NA | 23 | 0 | 0 | Low-quality | Genome-fragment | 9.18 | AAI-based (high-confidence) | 0 |
| S7-final-viral-combined_k141_61501\|\|full | 9299 | No | NA | 11 | 6 | 0 | Low-quality | Genome-fragment | 19.14 | AAI-based (medium-confidence) | 0 |
| S8-final-viral-combined_k141_186236\|\|full | 1349 | No | NA | 3 | 0 | 1 | Not-determined | Genome-fragment | NA | NA | 0 |
| S5_k141_142911 | 9300 | No | NA | 9 | 1 | 2 | Low-quality | Genome-fragment | 12.72 | HMM-based (lower-bound) | 0 |
| S8-final-viral-combined_k141_140361\|\|full | 2767 | No | NA | 3 | 0 | 2 | Not-determined | Genome-fragment | NA | NA | 0 |
| S11-final-viral-combined_k141_55233\|\|full | 9306 | No | NA | 10 | 7 | 0 | Low-quality | Genome-fragment | 20.45 | AAI-based (high-confidence) | 0 |
| S8-final-viral-combined_k141_215996\|\|full | 9314 | No | NA | 19 | 1 | 1 | Low-quality | Genome-fragment | 26.74 | AAI-based (high-confidence) | 0 |
| S8-final-viral-combined_k141_94027\|\|full | 16865 | No | NA | 21 | 0 | 3 | Not-determined | Genome-fragment | NA | NA | 0 |
| S9_k141_71289 | 9317 | No | NA | 14 | 3 | 0 | Low-quality | Genome-fragment | 22.3 | AAI-based (high-confidence) | 0 |
| S8-final-viral-combined_k141_186269\|\|full | 2711 | No | NA | 4 | 0 | 2 | Not-determined | Genome-fragment | NA | NA | 0 |
| S8-final-viral-combined_k141_162989\|\|full | 4423 | No | NA | 6 | 0 | 1 | Not-determined | Genome-fragment | NA | NA | 0 |
| S8_k141_34117 | 9324 | No | NA | 7 | 0 | 4 | Low-quality | Genome-fragment | 5.67 | AAI-based (medium-confidence) | 0 |
| S8-final-viral-combined_k141_163000\|\|full | 1335 | No | NA | 3 | 0 | 2 | Not-determined | Genome-fragment | NA | NA | 0 |
| S8-final-viral-combined_k141_151787\|\|full | 1688 | No | NA | 2 | 0 | 1 | Not-determined | Genome-fragment | NA | NA | 0 |
| S8-final-viral-combined_k141_140423\|\|full | 1043 | No | NA | 2 | 0 | 0 | Not-determined | Genome-fragment | NA | NA | 0 |
| S6_k141_60172 | 9332 | No | NA | 4 | 0 | 2 | Low-quality | Genome-fragment | 2.83 | AAI-based (medium-confidence) | 0 |
| S12-final-viral-combined_k141_16811\|\|full | 9338 | No | NA | 16 | 3 | 1 | Low-quality | Genome-fragment | 21.11 | AAI-based (medium-confidence) | 0 |
| S5-final-viral-combined_k141_177862\|\|full | 9341 | No | NA | 12 | 1 | 0 | Low-quality | Genome-fragment | 6.73 | HMM-based (lower-bound) | 0 |
| S8-final-viral-combined_k141_266543\|\|full | 1763 | No | NA | 3 | 0 | 2 | Not-determined | Genome-fragment | NA | NA | 0 |
| S1-final-viral-combined_k141_11702\|\|full | 9345 | Yes | 6241 | 14 | 2 | 2 | Low-quality | Genome-fragment | 7.19 | HMM-based (lower-bound) | 33.22 |
| S5-final-viral-combined_k141_197591\|\|full | 9349 | No | NA | 16 | 9 | 0 | Low-quality | Genome-fragment | 37.82 | AAI-based (high-confidence) | 0 |
| S8-final-viral-combined_k141_220408\|\|full | 4326 | No | NA | 3 | 0 | 3 | Not-determined | Genome-fragment | NA | NA | 0 |
| S8_k141_47800 | 9361 | No | NA | 13 | 1 | 1 | Low-quality | Genome-fragment | 20.56 | AAI-based (medium-confidence) | 0 |
| S8-final-viral-combined_k141_151812\|\|full | 2770 | No | NA | 3 | 0 | 2 | Not-determined | Genome-fragment | NA | NA | 0 |
| S8-final-viral-combined_k141_117002\|\|full | 1249 | No | NA | 2 | 0 | 1 | Not-determined | Genome-fragment | NA | NA | 0 |
| S17-final-viral-combined_k141_182347\|\|full | 9369 | No | NA | 9 | 1 | 0 | Low-quality | Genome-fragment | 6.9 | AAI-based (medium-confidence) | 0 |
| S17-final-viral-combined_k141_177817\|\|full | 9371 | No | NA | 13 | 7 | 0 | Low-quality | Genome-fragment | 22.89 | AAI-based (high-confidence) | 0 |
| S8-final-viral-combined_k141_254918\|\|full | 16272 | No | NA | 16 | 0 | 4 | Not-determined | Genome-fragment | NA | NA | 0 |
| S8-final-viral-combined_k141_243965\|\|full | 1345 | No | NA | 3 | 0 | 1 | Not-determined | Genome-fragment | NA | NA | 0 |
| S8-final-viral-combined_k141_160357\|\|full | 9375 | No | NA | 13 | 0 | 5 | Low-quality | Genome-fragment | 16.26 | AAI-based (medium-confidence) | 0 |
| S8-final-viral-combined_k141_94130\|\|full | 5213 | No | NA | 4 | 0 | 4 | Not-determined | Genome-fragment | NA | NA | 0 |
| S8-final-viral-combined_k141_117031\|\|full | 7806 | No | NA | 5 | 0 | 4 | Not-determined | Genome-fragment | NA | NA | 0 |
| S8-final-viral-combined_k141_232430\|\|full | 2108 | No | NA | 2 | 0 | 1 | Not-determined | Genome-fragment | NA | NA | 0 |
| S8-final-viral-combined_k141_163066\|\|full | 3068 | No | NA | 4 | 0 | 0 | Not-determined | Genome-fragment | NA | NA | 0 |
| S8-final-viral-combined_k141_209343\|\|full | 1685 | No | NA | 3 | 0 | 0 | Not-determined | Genome-fragment | NA | NA | 0 |
| S8-final-viral-combined_k141_82543\|\|full | 1816 | No | NA | 2 | 0 | 2 | Not-determined | Genome-fragment | NA | NA | 0 |
| S6-final-viral-combined_k141_718\|\|full | 9378 | No | NA | 11 | 0 | 3 | Low-quality | Genome-fragment | 6.18 | AAI-based (medium-confidence) | 0 |
| S11_k141_309285 | 9379 | No | NA | 7 | 0 | 3 | Low-quality | Genome-fragment | 8.68 | AAI-based (medium-confidence) | 0 |
| S8-final-viral-combined_k141_163104\|\|full | 2999 | No | NA | 2 | 0 | 2 | Not-determined | Genome-fragment | NA | NA | 0 |
| S1-final-viral-combined_k141_159695\|\|full | 9379 | No | NA | 7 | 2 | 0 | Low-quality | Genome-fragment | 15.15 | HMM-based (lower-bound) | 0 |
| S2-final-viral-combined_k141_206256\|\|full | 9381 | Yes | 3481 | 14 | 2 | 3 | Low-quality | Genome-fragment | 7.27 | AAI-based (high-confidence) | 62.89 |
| S12_k141_360070 | 9385 | No | NA | 24 | 2 | 0 | Low-quality | Genome-fragment | 6.99 | AAI-based (high-confidence) | 0 |
| S8-final-viral-combined_k141_254940\|\|full | 2234 | No | NA | 4 | 0 | 2 | Not-determined | Genome-fragment | NA | NA | 0 |
| S8-final-viral-combined_k141_244043\|\|full | 1333 | No | NA | 2 | 0 | 0 | Not-determined | Genome-fragment | NA | NA | 0 |
| S6-final-viral-combined_k141_139263\|\|full | 9386 | No | NA | 12 | 1 | 1 | Low-quality | Genome-fragment | 11.07 | AAI-based (high-confidence) | 0 |
| S8-final-viral-combined_k141_36335\|\|full | 2787 | No | NA | 3 | 0 | 0 | Not-determined | Genome-fragment | NA | NA | 0 |
| S6_k141_7842 | 9386 | No | NA | 15 | 1 | 2 | Low-quality | Genome-fragment | 5.09 | HMM-based (lower-bound) | 0 |
| S8-final-viral-combined_k141_162899\|\|full | 9389 | No | NA | 19 | 8 | 0 | Low-quality | Genome-fragment | 28.95 | AAI-based (high-confidence) | 0 |
| S8-final-viral-combined_k141_128584\|\|full | 1778 | No | NA | 2 | 0 | 2 | Not-determined | Genome-fragment | NA | NA | 0 |
| S1_k141_345605 | 9390 | No | NA | 10 | 3 | 0 | Low-quality | Genome-fragment | 23.28 | AAI-based (high-confidence) | 0 |
| S8-final-viral-combined_k141_209441\|\|full | 3211 | No | NA | 3 | 0 | 0 | Not-determined | Genome-fragment | NA | NA | 0 |
| S5-final-viral-combined_k141_201657\|\|full | 9396 | No | NA | 11 | 0 | 2 | Low-quality | Genome-fragment | 12.34 | AAI-based (medium-confidence) | 0 |
| S1-final-viral-combined_k141_202188\|\|full | 9403 | No | NA | 16 | 3 | 0 | Low-quality | Genome-fragment | 16.32 | AAI-based (high-confidence) | 0 |
| S11-final-viral-combined_k141_66244\|\|full | 9410 | No | NA | 9 | 0 | 4 | Low-quality | Genome-fragment | 6.4 | AAI-based (high-confidence) | 0 |
| S8-final-viral-combined_k141_151863\|\|full | 6409 | No | NA | 7 | 0 | 2 | Not-determined | Genome-fragment | NA | NA | 0 |
| S12_k141_257209 | 9414 | No | NA | 4 | 0 | 4 | Low-quality | Genome-fragment | 3.57 | AAI-based (high-confidence) | 0 |
| S8-final-viral-combined_k141_209458\|\|full | 1811 | No | NA | 3 | 0 | 0 | Not-determined | Genome-fragment | NA | NA | 0 |
| S17-final-viral-combined_k141_143135\|\|full | 9431 | No | NA | 13 | 2 | 0 | Low-quality | Genome-fragment | 23.84 | AAI-based (medium-confidence) | 0 |
| S7-final-viral-combined_k141_118634\|\|full | 9436 | No | NA | 12 | 6 | 0 | Low-quality | Genome-fragment | 25.99 | AAI-based (high-confidence) | 0 |
| S6-final-viral-combined_k141_20462\|\|full | 9438 | No | NA | 15 | 3 | 0 | Low-quality | Genome-fragment | 13.49 | HMM-based (lower-bound) | 0 |
| S8-final-viral-combined_k141_59803\|\|full | 2112 | No | NA | 2 | 0 | 1 | Not-determined | Genome-fragment | NA | NA | 0 |
| S8-final-viral-combined_k141_105614\|\|full | 1646 | No | NA | 2 | 0 | 0 | Not-determined | Genome-fragment | NA | NA | 0 |
| S8-final-viral-combined_k141_266704\|\|full | 6348 | No | NA | 8 | 0 | 2 | Not-determined | Genome-fragment | NA | NA | 0 |
| S8-final-viral-combined_k141_174941\|\|full | 1322 | No | NA | 3 | 0 | 0 | Not-determined | Genome-fragment | NA | NA | 0 |
| S11-final-viral-combined_k141_208545\|\|full | 9442 | No | NA | 14 | 2 | 1 | Low-quality | Genome-fragment | 5.44 | HMM-based (lower-bound) | 0 |
| S8-final-viral-combined_k141_174948\|\|full | 1067 | No | NA | 4 | 0 | 1 | Not-determined | Genome-fragment | NA | NA | 0 |
| S8-final-viral-combined_k141_255001\|\|full | 3599 | No | NA | 2 | 0 | 2 | Not-determined | Genome-fragment | NA | NA | 0 |
| S5_k141_37972 | 9446 | No | NA | 26 | 4 | 0 | Low-quality | Genome-fragment | 10.44 | HMM-based (lower-bound) | 0 |
| S8-final-viral-combined_k141_59817\|\|full | 2029 | No | NA | 2 | 0 | 2 | Not-determined | Genome-fragment | NA | NA | 0 |
| S8-final-viral-combined_k141_25303\|\|full | 1035 | No | NA | 3 | 0 | 0 | Not-determined | Genome-fragment | NA | NA | 0 |
| S8-final-viral-combined_k141_59835\|\|full | 7368 | No | NA | 9 | 0 | 1 | Not-determined | Genome-fragment | NA | NA | 0 |
| S8-final-viral-combined_k141_117092\|\|full | 7259 | No | NA | 9 | 0 | 0 | Not-determined | Genome-fragment | NA | NA | 0 |
| S12-final-viral-combined_k141_323473\|\|full | 9449 | No | NA | 12 | 0 | 1 | Low-quality | Genome-fragment | 22.75 | AAI-based (medium-confidence) | 0 |
| S8-final-viral-combined_k141_47807\|\|full | 2177 | No | NA | 3 | 0 | 1 | Not-determined | Genome-fragment | NA | NA | 0 |
| S8-final-viral-combined_k141_151927\|\|full | 2176 | No | NA | 4 | 0 | 3 | Not-determined | Genome-fragment | NA | NA | 0 |
| S8-final-viral-combined_k141_175016\|\|full | 2013 | No | NA | 3 | 0 | 1 | Not-determined | Genome-fragment | NA | NA | 0 |
| S8-final-viral-combined_k141_59862\|\|full | 4269 | No | NA | 7 | 0 | 5 | Not-determined | Genome-fragment | NA | NA | 0 |
| S8-final-viral-combined_k141_197697\|\|full | 3035 | No | NA | 3 | 0 | 0 | Not-determined | Genome-fragment | NA | NA | 0 |
| S8-final-viral-combined_k141_232589\|\|full | 1996 | No | NA | 3 | 0 | 1 | Not-determined | Genome-fragment | NA | NA | 0 |
| S8-final-viral-combined_k141_163249\|\|full | 3612 | No | NA | 5 | 0 | 3 | Not-determined | Genome-fragment | NA | NA | 0 |
| S2-final-viral-combined_k141_171550\|\|full | 9458 | No | NA | 9 | 0 | 6 | Low-quality | Genome-fragment | 2.29 | AAI-based (medium-confidence) | 0 |
| S4_k141_155873 | 9459 | Yes | 5218 | 10 | 2 | 1 | Low-quality | Genome-fragment | 9.58 | AAI-based (high-confidence) | 44.84 |
| S8-final-viral-combined_k141_94305\|\|full | 1081 | No | NA | 3 | 0 | 1 | Not-determined | Genome-fragment | NA | NA | 0 |
| S8-final-viral-combined_k141_47847\|\|full | 2697 | No | NA | 7 | 0 | 5 | Not-determined | Genome-fragment | NA | NA | 0 |
| S4-final-viral-combined_k141_120754\|\|full | 9474 | No | NA | 12 | 1 | 0 | Low-quality | Genome-fragment | 14.34 | HMM-based (lower-bound) | 0 |
| S7-final-viral-combined_k141_168524\|\|full | 9478 | No | NA | 9 | 0 | 1 | Low-quality | Genome-fragment | 12.15 | AAI-based (medium-confidence) | 0 |
| S12_k141_94852 | 9483 | No | NA | 6 | 0 | 6 | Low-quality | Genome-fragment | 2.52 | AAI-based (medium-confidence) | 0 |
| S8-final-viral-combined_k141_220932\|\|full | 9492 | No | NA | 8 | 0 | 5 | Low-quality | Genome-fragment | 5.82 | AAI-based (medium-confidence) | 0 |
| S3-final-viral-combined_k141_137869\|\|full | 9497 | No | NA | 11 | 1 | 0 | Low-quality | Genome-fragment | 6.29 | AAI-based (high-confidence) | 0 |
| S8-final-viral-combined_k141_240545\|\|full | 9507 | No | NA | 10 | 2 | 0 | Low-quality | Genome-fragment | 6.89 | HMM-based (lower-bound) | 0 |
| S8-final-viral-combined_k141_25414\|\|full | 2433 | No | NA | 3 | 0 | 2 | Not-determined | Genome-fragment | NA | NA | 0 |
| S12-final-viral-combined_k141_181185\|\|full | 9514 | Yes | 4332 | 13 | 2 | 3 | Low-quality | Genome-fragment | 10.9 | AAI-based (medium-confidence) | 54.47 |
| S7-final-viral-combined_k141_51529\|\|full | 9519 | No | NA | 6 | 0 | 2 | Low-quality | Genome-fragment | 13.67 | AAI-based (medium-confidence) | 0 |
| S8-final-viral-combined_k141_197763\|\|full | 1478 | No | NA | 2 | 0 | 2 | Not-determined | Genome-fragment | NA | NA | 0 |
| S8-final-viral-combined_k141_220595\|\|full | 4061 | No | NA | 4 | 0 | 2 | Not-determined | Genome-fragment | NA | NA | 0 |
| S12-final-viral-combined_k141_84659\|\|full | 9519 | No | NA | 18 | 1 | 0 | Low-quality | Genome-fragment | 7.09 | AAI-based (high-confidence) | 0 |
| S7-final-viral-combined_k141_219255\|\|full | 9521 | No | NA | 13 | 0 | 5 | Low-quality | Genome-fragment | 10.22 | AAI-based (medium-confidence) | 0 |
| S4_k141_42890 | 9522 | No | NA | 10 | 0 | 6 | Low-quality | Genome-fragment | 9.33 | AAI-based (medium-confidence) | 0 |
| S12_k141_221321 | 9529 | No | NA | 22 | 3 | 0 | Low-quality | Genome-fragment | 14.24 | AAI-based (high-confidence) | 0 |
| S2-final-viral-combined_k141_155615\|\|full | 9546 | No | NA | 16 | 2 | 2 | Low-quality | Genome-fragment | 9.5 | HMM-based (lower-bound) | 0 |
| S9-final-viral-combined_k141_72458\|\|full | 9548 | No | NA | 14 | 8 | 0 | Low-quality | Genome-fragment | 23.55 | AAI-based (medium-confidence) | 0 |
| S8_k141_249323 | 9554 | No | NA | 10 | 1 | 1 | Low-quality | Genome-fragment | 14.09 | HMM-based (lower-bound) | 0 |
| S8-final-viral-combined_k141_94367\|\|full | 2260 | No | NA | 3 | 0 | 1 | Not-determined | Genome-fragment | NA | NA | 0 |
| S12_k141_225972 | 9557 | No | NA | 8 | 2 | 0 | Low-quality | Genome-fragment | 9.47 | AAI-based (high-confidence) | 0 |
| S8-final-viral-combined_k141_186540\|\|full | 1313 | No | NA | 3 | 0 | 1 | Not-determined | Genome-fragment | NA | NA | 0 |
| S8-final-viral-combined_k141_266856\|\|full | 6420 | No | NA | 8 | 0 | 0 | Not-determined | Genome-fragment | NA | NA | 0 |
| S12-final-viral-combined_k141_38581\|\|full | 9558 | No | NA | 11 | 1 | 2 | Low-quality | Genome-fragment | 16.92 | AAI-based (high-confidence) | 0 |
| S8-final-viral-combined_k141_105731\|\|full | 3248 | No | NA | 3 | 0 | 3 | Not-determined | Genome-fragment | NA | NA | 0 |
| S5-final-viral-combined_k141_52560\|\|full | 9561 | No | NA | 12 | 4 | 0 | Low-quality | Genome-fragment | 25.06 | AAI-based (medium-confidence) | 0 |
| S8-final-viral-combined_k141_255164\|\|full | 2443 | No | NA | 3 | 0 | 1 | Not-determined | Genome-fragment | NA | NA | 0 |
| S8-final-viral-combined_k141_197804\|\|full | 2141 | No | NA | 4 | 0 | 0 | Not-determined | Genome-fragment | NA | NA | 0 |
| S8-final-viral-combined_k141_209555\|\|full | 6309 | No | NA | 7 | 0 | 2 | Not-determined | Genome-fragment | NA | NA | 0 |
| S2_k141_176190 | 9578 | No | NA | 14 | 6 | 0 | Low-quality | Genome-fragment | 21.22 | AAI-based (high-confidence) | 0 |
| S8-final-viral-combined_k141_244375\|\|full | 1483 | No | NA | 2 | 0 | 0 | Not-determined | Genome-fragment | NA | NA | 0 |
| S8-final-viral-combined_k141_59946\|\|full | 7592 | No | NA | 10 | 0 | 7 | Not-determined | Genome-fragment | NA | NA | 0 |
| S8-final-viral-combined_k141_220705\|\|full | 4181 | No | NA | 7 | 0 | 6 | Not-determined | Genome-fragment | NA | NA | 0 |
| S12-final-viral-combined_k141_343440\|\|full | 9587 | No | NA | 12 | 9 | 0 | Low-quality | Genome-fragment | 15.79 | AAI-based (high-confidence) | 0 |
| S12_k141_387291 | 9597 | No | NA | 11 | 1 | 1 | Low-quality | Genome-fragment | 4.13 | HMM-based (lower-bound) | 0 |
| S1-final-viral-combined_k141_235947\|\|full | 9599 | No | NA | 5 | 0 | 3 | Low-quality | Genome-fragment | 3.67 | AAI-based (medium-confidence) | 0 |
| S12-final-viral-combined_k141_256457\|\|full | 9604 | No | NA | 15 | 1 | 0 | Low-quality | Genome-fragment | 5.38 | HMM-based (lower-bound) | 0 |
| S12-final-viral-combined_k141_88289\|\|full | 9606 | Yes | 6764 | 10 | 5 | 4 | Low-quality | Genome-fragment | 9.85 | AAI-based (high-confidence) | 29.59 |
| S8-final-viral-combined_k141_71674\|\|full | 3676 | No | NA | 4 | 0 | 0 | Not-determined | Genome-fragment | NA | NA | 0 |
| S8-final-viral-combined_k141_186614\|\|full | 1928 | No | NA | 2 | 0 | 0 | Not-determined | Genome-fragment | NA | NA | 0 |
| S12_k141_226861 | 9611 | No | NA | 17 | 3 | 1 | Low-quality | Genome-fragment | 15.71 | HMM-based (lower-bound) | 0 |
| S8-final-viral-combined_k141_209612\|\|full | 3309 | No | NA | 2 | 0 | 1 | Not-determined | Genome-fragment | NA | NA | 0 |
| S17-k141_101205 | 9612 | No | NA | 1 | 0 | 0 | Low-quality | Genome-fragment | 17.76 | AAI-based (medium-confidence) | 0 |
| S8-final-viral-combined_k141_71686\|\|full | 1906 | No | NA | 5 | 0 | 0 | Not-determined | Genome-fragment | NA | NA | 0 |
| S8-final-viral-combined_k141_82781\|\|full | 5332 | No | NA | 9 | 0 | 6 | Not-determined | Genome-fragment | NA | NA | 0 |
| S2_k141_109942 | 9612 | No | NA | 8 | 0 | 3 | Low-quality | Genome-fragment | 13.01 | AAI-based (high-confidence) | 0 |
| S8-final-viral-combined_k141_209630\|\|full | 2258 | No | NA | 3 | 0 | 1 | Not-determined | Genome-fragment | NA | NA | 0 |
| S8-final-viral-combined_k141_232796\|\|full | 1941 | No | NA | 2 | 0 | 1 | Not-determined | Genome-fragment | NA | NA | 0 |
| S8-final-viral-combined_k141_209631\|\|full | 4046 | No | NA | 4 | 0 | 1 | Not-determined | Genome-fragment | NA | NA | 0 |
| S8_k141_177518 | 9612 | No | NA | 12 | 2 | 0 | Low-quality | Genome-fragment | 28.08 | AAI-based (medium-confidence) | 0 |
| S8-final-viral-combined_k141_232809\|\|full | 1914 | No | NA | 3 | 0 | 0 | Not-determined | Genome-fragment | NA | NA | 0 |
| S2-final-viral-combined_k141_181852\|\|full | 9613 | Yes | 1801 | 8 | 1 | 4 | Low-quality | Genome-fragment | 0.53 | HMM-based (lower-bound) | 81.26 |
| S5_k141_51234 | 9615 | No | NA | 9 | 0 | 7 | Low-quality | Genome-fragment | 4.74 | AAI-based (medium-confidence) | 0 |
| S8-final-viral-combined_k141_255215\|\|full | 2042 | No | NA | 5 | 0 | 2 | Not-determined | Genome-fragment | NA | NA | 0 |
| S17-final-viral-combined_k141_213329\|\|full | 9619 | No | NA | 11 | 0 | 1 | Low-quality | Genome-fragment | 15.7 | AAI-based (medium-confidence) | 0 |
| S8-final-viral-combined_k141_1980\|\|full | 1251 | No | NA | 3 | 0 | 1 | Not-determined | Genome-fragment | NA | NA | 0 |
| S8-final-viral-combined_k141_36583\|\|full | 2200 | No | NA | 4 | 0 | 2 | Not-determined | Genome-fragment | NA | NA | 0 |
| S4_k141_125871 | 9625 | No | NA | 12 | 3 | 1 | Low-quality | Genome-fragment | 17.03 | AAI-based (medium-confidence) | 0 |
| S7-final-viral-combined_k141_234311\|\|full | 9628 | No | NA | 13 | 4 | 0 | Low-quality | Genome-fragment | 20.76 | AAI-based (medium-confidence) | 0 |
| S5-final-viral-combined_k141_172978\|\|full | 9633 | No | NA | 16 | 1 | 0 | Low-quality | Genome-fragment | 5.73 | HMM-based (lower-bound) | 0 |
| S12-final-viral-combined_k141_218079\|\|full | 9639 | No | NA | 15 | 4 | 0 | Low-quality | Genome-fragment | 21.16 | AAI-based (high-confidence) | 0 |
| S17-final-viral-combined_k141_74446\|\|full | 9640 | No | NA | 12 | 5 | 0 | Low-quality | Genome-fragment | 23.14 | AAI-based (high-confidence) | 0 |
| S8-final-viral-combined_k141_92542\|\|full | 9649 | No | NA | 14 | 1 | 4 | Low-quality | Genome-fragment | 21.76 | AAI-based (medium-confidence) | 0 |
| S8-final-viral-combined_k141_94534\|\|full | 2027 | No | NA | 3 | 0 | 0 | Not-determined | Genome-fragment | NA | NA | 0 |
| S8-final-viral-combined_k141_60071\|\|full | 1675 | No | NA | 3 | 0 | 1 | Not-determined | Genome-fragment | NA | NA | 0 |
| S8-final-viral-combined_k141_117281\|\|full | 27542 | No | NA | 37 | 13 | 0 | Medium-quality | Genome-fragment | 57.51 | AAI-based (medium-confidence) | 0 |
| S8-final-viral-combined_k141_244465\|\|full | 2756 | No | NA | 3 | 0 | 2 | Not-determined | Genome-fragment | NA | NA | 0 |
| S7-final-viral-combined_k141_64054\|\|full | 9656 | No | NA | 6 | 0 | 6 | Low-quality | Genome-fragment | 7.83 | AAI-based (medium-confidence) | 0 |
| S8-final-viral-combined_k141_220802\|\|full | 1565 | No | NA | 2 | 0 | 1 | Not-determined | Genome-fragment | NA | NA | 0 |
| S8-final-viral-combined_k141_152142\|\|full | 1044 | No | NA | 2 | 0 | 0 | Not-determined | Genome-fragment | NA | NA | 0 |
| S8-final-viral-combined_k141_82852\|\|full | 1478 | No | NA | 2 | 0 | 0 | Not-determined | Genome-fragment | NA | NA | 0 |
| S8_k141_155816 | 9660 | No | NA | 16 | 3 | 0 | Low-quality | Genome-fragment | 17.28 | AAI-based (high-confidence) | 0 |
| S11-final-viral-combined_k141_22783\|\|full | 9665 | No | NA | 12 | 6 | 0 | Low-quality | Genome-fragment | 20.15 | AAI-based (high-confidence) | 0 |
| S8-final-viral-combined_k141_94566\|\|full | 6032 | No | NA | 6 | 0 | 4 | Not-determined | Genome-fragment | NA | NA | 0 |
| S7_k141_196033 | 9669 | No | NA | 8 | 0 | 5 | Low-quality | Genome-fragment | 4.95 | AAI-based (medium-confidence) | 0 |
| S8-final-viral-combined_k141_105838\|\|full | 2405 | No | NA | 2 | 0 | 1 | Not-determined | Genome-fragment | NA | NA | 0 |
| S8-final-viral-combined_k141_25607\|\|full | 42754 | No | NA | 46 | 7 | 0 | High-quality | High-quality | 96.36 | AAI-based (high-confidence) | 0 |
| S2-final-viral-combined_k141_12955\|\|full | 9676 | No | NA | 11 | 0 | 2 | Low-quality | Genome-fragment | 20.2 | AAI-based (medium-confidence) | 0 |
| S2-final-viral-combined_k141_239811\|\|full | 9678 | No | NA | 12 | 0 | 2 | Low-quality | Genome-fragment | 12.48 | AAI-based (high-confidence) | 0 |
| S8-final-viral-combined_k141_140825\|\|full | 2221 | No | NA | 4 | 0 | 2 | Not-determined | Genome-fragment | NA | NA | 0 |
| S8-final-viral-combined_k141_60129\|\|full | 3369 | No | NA | 3 | 0 | 2 | Not-determined | Genome-fragment | NA | NA | 0 |
| S8-final-viral-combined_k141_685\|\|full | 9680 | No | NA | 13 | 2 | 0 | Low-quality | Genome-fragment | 11.32 | HMM-based (lower-bound) | 0 |
| S8-final-viral-combined_k141_48046\|\|full | 66596 | Yes | 24470 | 67 | 10 | 32 | Medium-quality | Genome-fragment | 51.77 | AAI-based (high-confidence) | 63.26 |
| S8-final-viral-combined_k141_117348\|\|full | 3277 | No | NA | 5 | 0 | 1 | Not-determined | Genome-fragment | NA | NA | 0 |
| S7_k141_46272 | 9682 | No | NA | 9 | 0 | 4 | Low-quality | Genome-fragment | 21.74 | AAI-based (medium-confidence) | 0 |
| S8-final-viral-combined_k141_220840\|\|full | 1267 | No | NA | 2 | 0 | 1 | Not-determined | Genome-fragment | NA | NA | 0 |
| S8-final-viral-combined_k141_36667\|\|full | 1235 | No | NA | 3 | 0 | 2 | Not-determined | Genome-fragment | NA | NA | 0 |
| S8-final-viral-combined_k141_2076\|\|full | 3934 | No | NA | 4 | 0 | 3 | Not-determined | Genome-fragment | NA | NA | 0 |
| S8-final-viral-combined_k141_220853\|\|full | 4433 | No | NA | 6 | 0 | 1 | Not-determined | Genome-fragment | NA | NA | 0 |
| S8-final-viral-combined_k141_2098\|\|full | 2969 | No | NA | 4 | 0 | 1 | Not-determined | Genome-fragment | NA | NA | 0 |
| S8-final-viral-combined_k141_209744\|\|full | 2215 | No | NA | 3 | 0 | 2 | Not-determined | Genome-fragment | NA | NA | 0 |
| S11-final-viral-combined_k141_166308\|\|full | 9685 | No | NA | 17 | 2 | 0 | Low-quality | Genome-fragment | 19.49 | HMM-based (lower-bound) | 0 |
| S1-final-viral-combined_k141_358306\|\|full | 9688 | No | NA | 7 | 2 | 0 | Low-quality | Genome-fragment | 8.72 | AAI-based (medium-confidence) | 0 |
| S8-final-viral-combined_k141_197985\|\|full | 1136 | No | NA | 3 | 0 | 0 | Not-determined | Genome-fragment | NA | NA | 0 |
| S8-final-viral-combined_k141_62162\|\|full | 9701 | No | NA | 14 | 4 | 0 | Low-quality | Genome-fragment | 33.81 | AAI-based (high-confidence) | 0 |
| S9-final-viral-combined_k141_81286\|\|full | 9714 | No | NA | 12 | 11 | 0 | Low-quality | Genome-fragment | 21.21 | AAI-based (medium-confidence) | 0 |
| S8-final-viral-combined_k141_186703\|\|full | 3281 | No | NA | 4 | 0 | 0 | Not-determined | Genome-fragment | NA | NA | 0 |
| S7-final-viral-combined_k141_11494\|\|full | 9716 | No | NA | 13 | 6 | 0 | Low-quality | Genome-fragment | 15.49 | HMM-based (lower-bound) | 0 |
| S11-final-viral-combined_k141_297475\|\|full | 9718 | No | NA | 19 | 1 | 0 | Low-quality | Genome-fragment | 18.19 | AAI-based (high-confidence) | 0 |
| S12_k141_253353 | 9729 | No | NA | 19 | 1 | 0 | Low-quality | Genome-fragment | 17.68 | AAI-based (high-confidence) | 0 |
| S8-final-viral-combined_k141_244530\|\|full | 3259 | No | NA | 2 | 0 | 2 | Not-determined | Genome-fragment | NA | NA | 0 |
| S8-final-viral-combined_k141_71845\|\|full | 2077 | No | NA | 3 | 0 | 1 | Not-determined | Genome-fragment | NA | NA | 0 |
| S8-final-viral-combined_k141_24694\|\|full | 9732 | Yes | 7308 | 10 | 3 | 2 | Low-quality | Genome-fragment | 17.56 | AAI-based (medium-confidence) | 24.91 |
| S12-final-viral-combined_k141_51930\|\|full | 9733 | No | NA | 4 | 0 | 0 | Low-quality | Genome-fragment | 12.57 | AAI-based (medium-confidence) | 0 |
| S8-final-viral-combined_k141_13155\|\|full | 5109 | No | NA | 5 | 0 | 4 | Not-determined | Genome-fragment | NA | NA | 0 |
| S8-final-viral-combined_k141_140932\|\|full | 2312 | No | NA | 4 | 0 | 0 | Not-determined | Genome-fragment | NA | NA | 0 |
| S2-final-viral-combined_k141_185718\|\|full | 9744 | No | NA | 14 | 6 | 0 | Low-quality | Genome-fragment | 22.89 | AAI-based (medium-confidence) | 0 |
| S3-final-viral-combined_k141_30561\|\|full | 9746 | No | NA | 11 | 3 | 0 | Low-quality | Genome-fragment | 13.5 | HMM-based (lower-bound) | 0 |
| S17-k141_244862 | 9751 | Yes | 8649 | 9 | 2 | 2 | Low-quality | Genome-fragment | 3.45 | HMM-based (lower-bound) | 11.3 |
| S12_k141_55369 | 9767 | No | NA | 18 | 1 | 0 | Low-quality | Genome-fragment | 4.5 | AAI-based (high-confidence) | 0 |
| S3-final-viral-combined_k141_117313\|\|full | 9787 | No | NA | 9 | 0 | 0 | Low-quality | Genome-fragment | 17.11 | AAI-based (high-confidence) | 0 |
| S1-final-viral-combined_k141_257788\|\|full | 9788 | No | NA | 10 | 3 | 0 | Low-quality | Genome-fragment | 7.94 | AAI-based (high-confidence) | 0 |
| S8-final-viral-combined_k141_48219\|\|full | 1308 | No | NA | 4 | 0 | 3 | Not-determined | Genome-fragment | NA | NA | 0 |
| S8_k141_265910 | 9789 | No | NA | 4 | 0 | 4 | Low-quality | Genome-fragment | 5.96 | AAI-based (high-confidence) | 0 |
| S8-final-viral-combined_k141_220971\|\|full | 5394 | No | NA | 4 | 0 | 2 | Not-determined | Genome-fragment | NA | NA | 0 |
| S1-final-viral-combined_k141_163378\|\|full | 9797 | Yes | 8783 | 10 | 2 | 1 | Low-quality | Genome-fragment | 6.37 | HMM-based (lower-bound) | 10.35 |
| S8-final-viral-combined_k141_94719\|\|full | 1213 | No | NA | 2 | 0 | 1 | Not-determined | Genome-fragment | NA | NA | 0 |
| S8-final-viral-combined_k141_198138\|\|full | 1634 | No | NA | 2 | 0 | 1 | Not-determined | Genome-fragment | NA | NA | 0 |
| S8-final-viral-combined_k141_25706\|\|full | 7206 | No | NA | 6 | 0 | 2 | Not-determined | Genome-fragment | NA | NA | 0 |
| S2-final-viral-combined_k141_4724\|\|full | 9799 | No | NA | 21 | 5 | 0 | Low-quality | Genome-fragment | 23.65 | AAI-based (high-confidence) | 0 |
| S7-final-viral-combined_k141_86003\|\|full | 9807 | Yes | 2846 | 13 | 3 | 6 | Low-quality | Genome-fragment | 5.34 | AAI-based (high-confidence) | 70.98 |
| S8-final-viral-combined_k141_221006\|\|full | 1100 | No | NA | 2 | 0 | 1 | Not-determined | Genome-fragment | NA | NA | 0 |
| S8-final-viral-combined_k141_105867\|\|full | 2923 | No | NA | 4 | 0 | 0 | Not-determined | Genome-fragment | NA | NA | 0 |
| S8-final-viral-combined_k141_146892\|\|full | 9818 | No | NA | 15 | 8 | 0 | Low-quality | Genome-fragment | 21.23 | AAI-based (medium-confidence) | 0 |
| S12-final-viral-combined_k141_238428\|\|full | 9823 | No | NA | 12 | 7 | 0 | Low-quality | Genome-fragment | 24.58 | AAI-based (medium-confidence) | 0 |
| S7-final-viral-combined_k141_37574\|\|full | 9828 | No | NA | 17 | 4 | 0 | Low-quality | Genome-fragment | 21.33 | AAI-based (high-confidence) | 0 |
| S8-final-viral-combined_k141_221016\|\|full | 1491 | No | NA | 2 | 0 | 1 | Not-determined | Genome-fragment | NA | NA | 0 |
| S6-final-viral-combined_k141_120988\|\|full | 9831 | No | NA | 13 | 1 | 4 | Low-quality | Genome-fragment | 4.53 | HMM-based (lower-bound) | 0 |
| S1-final-viral-combined_k141_337775\|\|full | 9842 | No | NA | 10 | 3 | 0 | Low-quality | Genome-fragment | 30.93 | AAI-based (high-confidence) | 0 |
| S8-final-viral-combined_k141_209925\|\|full | 4162 | No | NA | 4 | 0 | 1 | Not-determined | Genome-fragment | NA | NA | 0 |
| S1-final-viral-combined_k141_380771\|\|full | 9843 | No | NA | 14 | 5 | 0 | Low-quality | Genome-fragment | 13.61 | HMM-based (lower-bound) | 0 |
| S9_k141_50113 | 9869 | No | NA | 9 | 1 | 4 | Low-quality | Genome-fragment | 22.84 | HMM-based (lower-bound) | 0 |
| S8-final-viral-combined_k141_186851\|\|full | 3073 | No | NA | 2 | 0 | 2 | Not-determined | Genome-fragment | NA | NA | 0 |
| S8-final-viral-combined_k141_36912\|\|full | 2497 | No | NA | 2 | 0 | 2 | Not-determined | Genome-fragment | NA | NA | 0 |
| S11_k141_210341 | 9869 | No | NA | 12 | 1 | 1 | Low-quality | Genome-fragment | 5.24 | HMM-based (lower-bound) | 0 |
| S8-final-viral-combined_k141_71938\|\|full | 2810 | No | NA | 3 | 0 | 2 | Not-determined | Genome-fragment | NA | NA | 0 |
| S2-final-viral-combined_k141_45767\|\|full | 9872 | No | NA | 12 | 1 | 0 | Low-quality | Genome-fragment | 19.16 | AAI-based (medium-confidence) | 0 |
| S8-final-viral-combined_k141_48314\|\|full | 1441 | No | NA | 4 | 0 | 1 | Not-determined | Genome-fragment | NA | NA | 0 |

| | | | | | | | | | | | | | |
|---|---|---|---|---|---|---|---|---|---|---|---|---|---|
| S8-final-viral-combined_k141_152425\|\|full | 6784 | No | NA | | 5 | 0 | 5 | Not-determined | Genome-fragment | NA | | NA | 0 |
| S12-final-viral-combined_k141_351811\|\|full | 9875 | Yes | | 3637 | 9 | 1 | 3 | Low-quality | Genome-fragment | | 5.18 | HMM-based (lower-bound) | 63.17 |
| S11-final-viral-combined_k141_4908\|\|full | 9876 | No | NA | | 7 | 0 | 3 | Low-quality | Genome-fragment | | 4.27 | AAI-based (medium-confidence) | 0 |
| S11_k141_256624 | 9882 | No | NA | | 13 | 1 | 8 | Low-quality | Genome-fragment | | 6.65 | HMM-based (lower-bound) | 0 |
| S8-final-viral-combined_k141_163717\|\|full | 1654 | No | NA | | 3 | 0 | 0 | Not-determined | Genome-fragment | NA | | NA | 0 |
| S8-final-viral-combined_k141_2290\|\|full | 1265 | No | NA | | 3 | 0 | 2 | Not-determined | Genome-fragment | NA | | NA | 0 |
| S8-final-viral-combined_k141_94823\|\|full | 4761 | No | NA | | 3 | 0 | 3 | Not-determined | Genome-fragment | NA | | NA | 0 |
| S2-final-viral-combined_k141_2078\|\|full | 9891 | No | NA | | 13 | 6 | 0 | Low-quality | Genome-fragment | | 24.26 | AAI-based (high-confidence) | 0 |
| S8-final-viral-combined_k141_244768\|\|full | 1264 | No | NA | | 3 | 0 | 2 | Not-determined | Genome-fragment | NA | | NA | 0 |
| S1_k141_356369 | 9897 | No | NA | | 18 | 2 | 1 | Low-quality | Genome-fragment | | 25.52 | AAI-based (high-confidence) | 0 |
| S8-final-viral-combined_k141_221097\|\|full | 3842 | No | NA | | 4 | 0 | 4 | Not-determined | Genome-fragment | NA | | NA | 0 |
| S7_k141_58293 | 9904 | No | NA | | 8 | 0 | 6 | Low-quality | Genome-fragment | | 1.49 | AAI-based (medium-confidence) | 0 |
| S1-final-viral-combined_k141_199643\|\|full | 9908 | No | NA | | 22 | 4 | 0 | Low-quality | Genome-fragment | | 22.34 | AAI-based (high-confidence) | 0 |
| S8-final-viral-combined_k141_94868\|\|full | 3390 | No | NA | | 3 | 0 | 3 | Not-determined | Genome-fragment | NA | | NA | 0 |
| S8-final-viral-combined_k141_48385\|\|full | 1580 | No | NA | | 2 | 0 | 2 | Not-determined | Genome-fragment | NA | | NA | 0 |
| S8-final-viral-combined_k141_129143\|\|full | 1653 | No | NA | | 2 | 0 | 1 | Not-determined | Genome-fragment | NA | | NA | 0 |
| S8-final-viral-combined_k141_163769\|\|full | 2560 | No | NA | | 4 | 0 | 1 | Not-determined | Genome-fragment | NA | | NA | 0 |
| S8-final-viral-combined_k141_186952\|\|full | 6505 | No | NA | | 7 | 0 | 2 | Not-determined | Genome-fragment | NA | | NA | 0 |
| S1_k141_102320 | 9908 | No | NA | | 12 | 5 | 0 | Low-quality | Genome-fragment | | 24.16 | AAI-based (high-confidence) | 0 |
| S17-final-viral-combined_k141_144277\|\|full | 9908 | No | NA | | 16 | 3 | 0 | Low-quality | Genome-fragment | | 15.1 | AAI-based (high-confidence) | 0 |
| S8-final-viral-combined_k141_233135\|\|full | 1466 | No | NA | | 3 | 0 | 0 | Not-determined | Genome-fragment | NA | | NA | 0 |
| S8-final-viral-combined_k141_175617\|\|full | 4323 | No | NA | | 4 | 0 | 0 | Not-determined | Genome-fragment | NA | | NA | 0 |
| S12-final-viral-combined_k141_334721\|\|full | 9908 | No | NA | | 14 | 5 | 1 | Low-quality | Genome-fragment | | 27.46 | AAI-based (high-confidence) | 0 |
| S8-final-viral-combined_k141_13282\|\|full | 1431 | No | NA | | 4 | 0 | 1 | Not-determined | Genome-fragment | NA | | NA | 0 |
| S7-final-viral-combined_k141_3090\|\|full | 9915 | No | NA | | 16 | 0 | 1 | Low-quality | Genome-fragment | | 22.09 | AAI-based (high-confidence) | 0 |
| S7_k141_11012 | 9915 | Yes | | 6554 | 5 | 1 | 1 | Low-quality | Genome-fragment | | 11 | HMM-based (lower-bound) | 33.9 |
| S8-final-viral-combined_k141_244832\|\|full | 1013 | No | NA | | 2 | 0 | 0 | Not-determined | Genome-fragment | NA | | NA | 0 |
| S8-final-viral-combined_k141_152549\|\|full | 1148 | No | NA | | 3 | 0 | 1 | Not-determined | Genome-fragment | NA | | NA | 0 |
| S8-final-viral-combined_k141_105953\|\|full | 2891 | No | NA | | 3 | 0 | 2 | Not-determined | Genome-fragment | NA | | NA | 0 |
| S12-final-viral-combined_k141_79062\|\|full | 9922 | No | NA | | 9 | 1 | 0 | Low-quality | Genome-fragment | | 11.14 | HMM-based (lower-bound) | 0 |
| S2-final-viral-combined_k141_237107\|\|full | 9926 | No | NA | | 6 | 1 | 0 | Low-quality | Genome-fragment | | 23.32 | AAI-based (high-confidence) | 0 |
| S5-final-viral-combined_k141_112046\|\|full | 9933 | No | NA | | 12 | 2 | 0 | Low-quality | Genome-fragment | | 25.74 | AAI-based (medium-confidence) | 0 |
| S8-final-viral-combined_k141_198327\|\|full | 4319 | No | NA | | 5 | 0 | 0 | Not-determined | Genome-fragment | NA | | NA | 0 |
| S8-final-viral-combined_k141_267314\|\|full | 2420 | No | NA | | 2 | 0 | 1 | Not-determined | Genome-fragment | NA | | NA | 0 |
| S2_k141_189443 | 9933 | No | NA | | 9 | 0 | 5 | Low-quality | Genome-fragment | | 6.73 | AAI-based (high-confidence) | 0 |
| S8-final-viral-combined_k141_141241\|\|full | 2069 | No | NA | | 4 | 0 | 1 | Not-determined | Genome-fragment | NA | | NA | 0 |
| S8-final-viral-combined_k141_267339\|\|full | 1061 | No | NA | | 2 | 0 | 0 | Not-determined | Genome-fragment | NA | | NA | 0 |
| S7-final-viral-combined_k141_232639\|\|full | 9938 | No | NA | | 13 | 1 | 3 | Low-quality | Genome-fragment | | 4.04 | HMM-based (lower-bound) | 0 |
| S8-final-viral-combined_k141_267360\|\|full | 4532 | No | NA | | 6 | 0 | 3 | Not-determined | Genome-fragment | NA | | NA | 0 |
| S8-final-viral-combined_k141_152619\|\|full | 2641 | No | NA | | 3 | 0 | 3 | Not-determined | Genome-fragment | NA | | NA | 0 |
| S8-final-viral-combined_k141_129264\|\|full | 1035 | No | NA | | 2 | 0 | 0 | Not-determined | Genome-fragment | NA | | NA | 0 |
| S8-final-viral-combined_k141_72197\|\|full | 1737 | No | NA | | 2 | 0 | 1 | Not-determined | Genome-fragment | NA | | NA | 0 |
| S8-final-viral-combined_k141_13381\|\|full | 5864 | No | NA | | 7 | 0 | 2 | Not-determined | Genome-fragment | NA | | NA | 0 |
| S12-final-viral-combined_k141_274051\|\|full | 9943 | No | NA | | 25 | 2 | 0 | Low-quality | Genome-fragment | | 11.9 | AAI-based (high-confidence) | 0 |
| S17-final-viral-combined_k141_76772\|\|full | 9949 | No | NA | | 4 | 0 | 2 | Low-quality | Genome-fragment | | 22.51 | AAI-based (medium-confidence) | 0 |
| S8-final-viral-combined_k141_198381\|\|full | 1466 | No | NA | | 2 | 0 | 1 | Not-determined | Genome-fragment | NA | | NA | 0 |
| S17-final-viral-combined_k141_133816\|\|full | 9951 | No | NA | | 9 | 0 | 5 | Low-quality | Genome-fragment | | 7.42 | AAI-based (high-confidence) | 0 |
| S8-final-viral-combined_k141_72219\|\|full | 1593 | No | NA | | 3 | 0 | 1 | Not-determined | Genome-fragment | NA | | NA | 0 |
| S8-final-viral-combined_k141_129285\|\|full | 9953 | No | NA | | 12 | 0 | 12 | Not-determined | Genome-fragment | NA | | NA | 0 |
| S8-final-viral-combined_k141_175764\|\|full | 4203 | No | NA | | 4 | 0 | 2 | Not-determined | Genome-fragment | NA | | NA | 0 |
| S8-final-viral-combined_k141_233257\|\|full | 9000 | No | NA | | 8 | 0 | 5 | Not-determined | Genome-fragment | NA | | NA | 0 |
| S8-final-viral-combined_k141_175786\|\|full | 3467 | No | NA | | 4 | 0 | 1 | Not-determined | Genome-fragment | NA | | NA | 0 |
| S8-final-viral-combined_k141_141331\|\|full | 1339 | No | NA | | 2 | 0 | 1 | Not-determined | Genome-fragment | NA | | NA | 0 |
| S11-final-viral-combined_k141_189365\|\|full | 9956 | No | NA | | 11 | 2 | 0 | Low-quality | Genome-fragment | | 7.51 | AAI-based (high-confidence) | 0 |
| S12-final-viral-combined_k141_327196\|\|full | 9968 | No | NA | | 15 | 4 | 0 | Low-quality | Genome-fragment | | 20.92 | AAI-based (medium-confidence) | 0 |
| S8-final-viral-combined_k141_26019\|\|full | 1344 | No | NA | | 3 | 0 | 1 | Not-determined | Genome-fragment | NA | | NA | 0 |
| S7-final-viral-combined_k141_147109\|\|full | 9976 | No | NA | | 13 | 8 | 0 | Low-quality | Genome-fragment | | 28.48 | AAI-based (high-confidence) | 0 |
| S8-final-viral-combined_k141_26024\|\|full | 1066 | No | NA | | 3 | 0 | 1 | Not-determined | Genome-fragment | NA | | NA | 0 |
| S5-final-viral-combined_k141_85321\|\|full | 9979 | Yes | | 1877 | 15 | 3 | 6 | Low-quality | Genome-fragment | | 3.77 | HMM-based (lower-bound) | 81.19 |
| S4-final-viral-combined_k141_14721\|\|full | 9984 | No | NA | | 9 | 0 | 6 | Low-quality | Genome-fragment | | 1.56 | AAI-based (medium-confidence) | 0 |
| S5-final-viral-combined_k141_156180\|\|full | 9990 | No | NA | | 17 | 2 | 0 | Low-quality | Genome-fragment | | 26.7 | AAI-based (high-confidence) | 0 |
| S11-final-viral-combined_k141_28286\|\|full | 9994 | No | NA | | 13 | 4 | 1 | Low-quality | Genome-fragment | | 25.68 | AAI-based (medium-confidence) | 0 |
| S8-final-viral-combined_k141_244992\|\|full | 1694 | No | NA | | 3 | 0 | 1 | Not-determined | Genome-fragment | NA | | NA | 0 |
| S8-final-viral-combined_k141_141357\|\|full | 5086 | No | NA | | 6 | 0 | 0 | Not-determined | Genome-fragment | NA | | NA | 0 |
| S8-final-viral-combined_k141_175868\|\|full | 1580 | No | NA | | 3 | 0 | 1 | Not-determined | Genome-fragment | NA | | NA | 0 |
| S8-final-viral-combined_k141_55140\|\|full | 9995 | No | NA | | 16 | 12 | 0 | Low-quality | Genome-fragment | | 16.2 | AAI-based (high-confidence) | 0 |
| S8-final-viral-combined_k141_141364\|\|full | 2587 | No | NA | | 3 | 0 | 3 | Not-determined | Genome-fragment | NA | | NA | 0 |
| S8-final-viral-combined_k141_37208\|\|full | 2487 | No | NA | | 3 | 0 | 2 | Not-determined | Genome-fragment | NA | | NA | 0 |
| S1_k141_195193 | 9996 | No | NA | | 4 | 0 | 4 | Low-quality | Genome-fragment | | 3.72 | AAI-based (high-confidence) | 0 |
| S8-final-viral-combined_k141_50744\|\|full | 10008 | Yes | | 6461 | 13 | 2 | 2 | Low-quality | Genome-fragment | | 10.65 | HMM-based (lower-bound) | 35.44 |
| S8-final-viral-combined_k141_221201\|\|full | 4421 | No | NA | | 4 | 0 | 2 | Not-determined | Genome-fragment | NA | | NA | 0 |
| S8-final-viral-combined_k141_255744\|\|full | 1622 | No | NA | | 2 | 0 | 1 | Not-determined | Genome-fragment | NA | | NA | 0 |
| S8-final-viral-combined_k141_37221\|\|full | 3253 | No | NA | | 4 | 0 | 2 | Not-determined | Genome-fragment | NA | | NA | 0 |
| S8-final-viral-combined_k141_210160\|\|full | 2644 | No | NA | | 2 | 0 | 2 | Not-determined | Genome-fragment | NA | | NA | 0 |
| S8-final-viral-combined_k141_267504\|\|full | 1074 | No | NA | | 3 | 0 | 0 | Not-determined | Genome-fragment | NA | | NA | 0 |
| S8-final-viral-combined_k141_106104\|\|full | 2967 | No | NA | | 3 | 0 | 1 | Not-determined | Genome-fragment | NA | | NA | 0 |
| S17-k141_182681 | 10011 | No | NA | | 16 | 4 | 1 | Low-quality | Genome-fragment | | 15.49 | AAI-based (high-confidence) | 0 |
| S8-final-viral-combined_k141_13522\|\|full | 3315 | No | NA | | 3 | 0 | 2 | Not-determined | Genome-fragment | NA | | NA | 0 |
| S8-final-viral-combined_k141_106119\|\|full | 4623 | No | NA | | 5 | 0 | 5 | Not-determined | Genome-fragment | NA | | NA | 0 |
| S8-final-viral-combined_k141_216894\|\|0_partial | 10015 | No | NA | | 11 | 1 | 2 | Low-quality | Genome-fragment | | 16.06 | HMM-based (lower-bound) | 0 |
| S5-final-viral-combined_k141_57709\|\|full | 10021 | No | NA | | 9 | 1 | 4 | Low-quality | Genome-fragment | | 3.58 | HMM-based (lower-bound) | 0 |
| S8-final-viral-combined_k141_117707\|\|full | 1897 | No | NA | | 3 | 0 | 0 | Not-determined | Genome-fragment | NA | | NA | 0 |
| S8-final-viral-combined_k141_117709\|\|full | 2438 | No | NA | | 2 | 0 | 2 | Not-determined | Genome-fragment | NA | | NA | 0 |
| S12-final-viral-combined_k141_34243\|\|0_partial | 10033 | Yes | | 1625 | 10 | 1 | 4 | Low-quality | Genome-fragment | | 1.65 | HMM-based (lower-bound) | 83.8 |
| S8-final-viral-combined_k141_152775\|\|full | 1181 | No | NA | | 4 | 0 | 1 | Not-determined | Genome-fragment | NA | | NA | 0 |
| S8-final-viral-combined_k141_13568\|\|full | 2631 | No | NA | | 2 | 0 | 2 | Not-determined | Genome-fragment | NA | | NA | 0 |
| S8-final-viral-combined_k141_175974\|\|full | 3106 | No | NA | | 5 | 0 | 0 | Not-determined | Genome-fragment | NA | | NA | 0 |
| S8-final-viral-combined_k141_233432\|\|full | 1158 | No | NA | | 3 | 0 | 0 | Not-determined | Genome-fragment | NA | | NA | 0 |
| S8-final-viral-combined_k141_255774\|\|full | 3360 | No | NA | | 4 | 0 | 2 | Not-determined | Genome-fragment | NA | | NA | 0 |
| S2-final-viral-combined_k141_89508\|\|full | 10035 | No | NA | | 18 | 4 | 1 | Low-quality | Genome-fragment | | 24.06 | AAI-based (high-confidence) | 0 |
| S9-final-viral-combined_k141_109959\|\|full | 10037 | No | NA | | 10 | 5 | 0 | Low-quality | Genome-fragment | | 24.58 | AAI-based (high-confidence) | 0 |
| S8-final-viral-combined_k141_106161\|\|full | 2690 | No | NA | | 3 | 0 | 2 | Not-determined | Genome-fragment | NA | | NA | 0 |
| S5-final-viral-combined_k141_80044\|\|full | 10038 | No | NA | | 6 | 0 | 1 | Low-quality | Genome-fragment | | 12.16 | AAI-based (high-confidence) | 0 |
| S8-final-viral-combined_k141_221271\|\|full | 2677 | No | NA | | 3 | 0 | 2 | Not-determined | Genome-fragment | NA | | NA | 0 |
| S8-final-viral-combined_k141_210200\|\|full | 2536 | No | NA | | 5 | 0 | 0 | Not-determined | Genome-fragment | NA | | NA | 0 |
| S1-final-viral-combined_k141_59807\|\|full | 10040 | Yes | | 6659 | 12 | 6 | 3 | Low-quality | Genome-fragment | | 17.87 | AAI-based (medium-confidence) | 33.68 |
| S8-final-viral-combined_k141_176026\|\|full | 3375 | No | NA | | 4 | 0 | 2 | Not-determined | Genome-fragment | NA | | NA | 0 |
| S2_k141_82804 | 10051 | No | NA | | 13 | 0 | 1 | Low-quality | Genome-fragment | | 13.69 | AAI-based (medium-confidence) | 0 |
| S17-final-viral-combined_k141_248634\|\|full | 10057 | No | NA | | 10 | 3 | 0 | Low-quality | Genome-fragment | | 24.62 | AAI-based (high-confidence) | 0 |
| S8-final-viral-combined_k141_184511\|\|full | 10068 | No | NA | | 16 | 4 | 1 | Low-quality | Genome-fragment | | 21.51 | HMM-based (lower-bound) | 0 |
| S8-final-viral-combined_k141_26087\|\|full | 73390 | Yes | | 47543 | 82 | 17 | 15 | High-quality | High-quality | | 100 | AAI-based (high-confidence) | 35.22 |
| S4-final-viral-combined_k141_49713\|\|full | 10070 | No | NA | | 3 | 0 | 1 | Low-quality | Genome-fragment | | 13.42 | AAI-based (medium-confidence) | 0 |
| S11_k141_153539 | 10076 | No | NA | | 20 | 7 | 0 | Low-quality | Genome-fragment | | 22.36 | AAI-based (high-confidence) | 0 |
| S8-final-viral-combined_k141_233529\|\|full | 3179 | No | NA | | 3 | 0 | 1 | Not-determined | Genome-fragment | NA | | NA | 0 |
| S8-final-viral-combined_k141_141535\|\|full | 2245 | No | NA | | 3 | 0 | 0 | Not-determined | Genome-fragment | NA | | NA | 0 |
| S8-final-viral-combined_k141_221310\|\|full | 2913 | No | NA | | 5 | 0 | 0 | Not-determined | Genome-fragment | NA | | NA | 0 |
| S11-final-viral-combined_k141_140846\|\|full | 10086 | No | NA | | 13 | 5 | 0 | Low-quality | Genome-fragment | | 21.23 | AAI-based (high-confidence) | 0 |
| S8-final-viral-combined_k141_129565\|\|full | 2647 | No | NA | | 3 | 0 | 1 | Not-determined | Genome-fragment | NA | | NA | 0 |
| S8-final-viral-combined_k141_267673\|\|full | 1030 | No | NA | | 3 | 0 | 1 | Not-determined | Genome-fragment | NA | | NA | 0 |
| S7-final-viral-combined_k141_73453\|\|full | 10087 | No | NA | | 19 | 5 | 0 | Low-quality | Genome-fragment | | 13.11 | HMM-based (lower-bound) | 0 |
| S1-final-viral-combined_k141_389300\|\|full | 10091 | No | NA | | 14 | 1 | 1 | Low-quality | Genome-fragment | | 19.55 | AAI-based (medium-confidence) | 0 |
| S17-final-viral-combined_k141_136786\|\|full | 10094 | No | NA | | 11 | 7 | 0 | Low-quality | Genome-fragment | | 25.44 | AAI-based (high-confidence) | 0 |
| S8-final-viral-combined_k141_163928\|\|full | 2424 | No | NA | | 4 | 0 | 3 | Not-determined | Genome-fragment | NA | | NA | 0 |
| S2-final-viral-combined_k141_241319\|\|full | 10121 | No | NA | | 4 | 0 | 1 | Low-quality | Genome-fragment | | 10.17 | AAI-based (high-confidence) | 0 |
| S8-final-viral-combined_k141_26189\|\|full | 6545 | No | NA | | 5 | 0 | 4 | Not-determined | Genome-fragment | NA | | NA | 0 |
| S8-final-viral-combined_k141_221376\|\|full | 5099 | No | NA | | 8 | 0 | 2 | Not-determined | Genome-fragment | NA | | NA | 0 |
| S8-final-viral-combined_k141_255956\|\|full | 2712 | No | NA | | 3 | 0 | 3 | Not-determined | Genome-fragment | NA | | NA | 0 |
| S11-final-viral-combined_k141_101947\|\|full | 10123 | No | NA | | 14 | 0 | 1 | Low-quality | Genome-fragment | | 23.22 | AAI-based (high-confidence) | 0 |
| S17-k141_12496 | 10124 | No | NA | | 4 | 0 | 0 | Low-quality | Genome-fragment | | 17.06 | AAI-based (medium-confidence) | 0 |
| S8-final-viral-combined_k141_129632\|\|full | 3480 | No | NA | | 3 | 0 | 3 | Not-determined | Genome-fragment | NA | | NA | 0 |
| S8-final-viral-combined_k141_26234\|\|full | 4646 | No | NA | | 5 | 0 | 4 | Not-determined | Genome-fragment | NA | | NA | 0 |
| S8-final-viral-combined_k141_267733\|\|full | 2671 | No | NA | | 2 | 0 | 0 | Not-determined | Genome-fragment | NA | | NA | 0 |
| S8-final-viral-combined_k141_95435\|\|full | 2693 | No | NA | | 3 | 0 | 3 | Not-determined | Genome-fragment | NA | | NA | 0 |
| S8-final-viral-combined_k141_95454\|\|full | 2297 | No | NA | | 2 | 0 | 1 | Not-determined | Genome-fragment | NA | | NA | 0 |
| S1-final-viral-combined_k141_168803\|\|full | 10132 | No | NA | | 13 | 8 | 0 | Low-quality | Genome-fragment | | 16.32 | AAI-based (high-confidence) | 0 |
| S8_k141_161458 | 10139 | No | NA | | 11 | 0 | 3 | Low-quality | Genome-fragment | | 12.33 | AAI-based (medium-confidence) | 0 |
| S8-final-viral-combined_k141_48682\|\|full | 5747 | No | NA | | 6 | 0 | 3 | Not-determined | Genome-fragment | NA | | NA | 0 |
| S4-final-viral-combined_k141_74442\|\|full | 10143 | No | NA | | 10 | 0 | 2 | Low-quality | Genome-fragment | | 23.68 | AAI-based (high-confidence) | 0 |
| S8-final-viral-combined_k141_164078\|\|full | 3510 | No | NA | | 4 | 0 | 4 | Not-determined | Genome-fragment | NA | | NA | 0 |
| S11-final-viral-combined_k141_160007\|\|full | 10145 | No | NA | | 13 | 9 | 0 | Low-quality | Genome-fragment | | 14.19 | AAI-based (high-confidence) | 0 |
| S8-final-viral-combined_k141_245162\|\|full | 10230 | No | NA | | 9 | 0 | 0 | Not-determined | Genome-fragment | NA | | NA | 0 |
| S8-final-viral-combined_k141_255987\|\|full | 33715 | No | NA | | 28 | 0 | 18 | Not-determined | Genome-fragment | NA | | NA | 0 |
| S8-final-viral-combined_k141_210374\|\|full | 2538 | No | NA | | 3 | 0 | 2 | Not-determined | Genome-fragment | NA | | NA | 0 |
| S8-final-viral-combined_k141_245180\|\|full | 2685 | No | NA | | 3 | 0 | 0 | Not-determined | Genome-fragment | NA | | NA | 0 |
| S8-final-viral-combined_k141_164116\|\|full | 4328 | No | NA | | 6 | 0 | 3 | Not-determined | Genome-fragment | NA | | NA | 0 |
| S8-final-viral-combined_k141_2771\|\|full | 1446 | No | NA | | 2 | 0 | 1 | Not-determined | Genome-fragment | NA | | NA | 0 |
| S8-final-viral-combined_k141_37431\|\|full | 10856 | No | NA | | 8 | 0 | 7 | Not-determined | Genome-fragment | NA | | NA | 0 |
| S8-final-viral-combined_k141_245191\|\|full | 7389 | No | NA | | 7 | 0 | 5 | Not-determined | Genome-fragment | NA | | NA | 0 |
| S17-final-viral-combined_k141_145409\|\|full | 10157 | No | NA | | 13 | 1 | 6 | Low-quality | Genome-fragment | | 8.33 | HMM-based (lower-bound) | 0 |
| S8-final-viral-combined_k141_48739\|\|full | 1223 | No | NA | | 2 | 0 | 0 | Not-determined | Genome-fragment | NA | | NA | 0 |
| S8-final-viral-combined_k141_256013\|\|full | 4755 | No | NA | | 4 | 0 | 4 | Not-determined | Genome-fragment | NA | | NA | 0 |
| S6-final-viral-combined_k141_110836\|\|full | 10159 | No | NA | | 4 | 0 | 0 | Low-quality | Genome-fragment | | 4.13 | AAI-based (medium-confidence) | 0 |
| S8-final-viral-combined_k141_117932\|\|full | 3717 | No | NA | | 5 | 0 | 4 | Not-determined | Genome-fragment | NA | | NA | 0 |
| S8-final-viral-combined_k141_129714\|\|full | 8694 | No | NA | | 7 | 0 | 5 | Not-determined | Genome-fragment | NA | | NA | 0 |
| S1-final-viral-combined_k141_49960\|\|0_partial | 10172 | No | NA | | 11 | 1 | 7 | Low-quality | Genome-fragment | | 4.52 | HMM-based (lower-bound) | 0 |
| S5-final-viral-combined_k141_201845\|\|full | 10177 | No | NA | | 15 | 0 | 1 | Low-quality | Genome-fragment | | 9.24 | AAI-based (medium-confidence) | 0 |
| S5-final-viral-combined_k141_38863\|\|full | 10179 | No | NA | | 8 | 0 | 8 | Low-quality | Genome-fragment | | 4.61 | AAI-based (medium-confidence) | 0 |
| S8-final-viral-combined_k141_187349\|\|full | 1114 | No | NA | | 1 | 0 | 0 | Not-determined | Genome-fragment | NA | | NA | 0 |
| S8-final-viral-combined_k141_48770\|\|full | 2507 | No | NA | | 6 | 0 | 1 | Not-determined | Genome-fragment | NA | | NA | 0 |
| S6_k141_101758 | 10179 | No | NA | | 18 | 1 | 0 | Low-quality | Genome-fragment | | 5.44 | AAI-based (high-confidence) | 0 |
| S11-final-viral-combined_k141_290231\|\|full | 10190 | No | NA | | 11 | 1 | 2 | Low-quality | Genome-fragment | | 11.44 | HMM-based (lower-bound) | 0 |
| S8-final-viral-combined_k141_72676\|\|full | 2947 | No | NA | | 3 | 0 | 2 | Not-determined | Genome-fragment | NA | | NA | 0 |
| S12_k141_316914 | 10191 | No | NA | | 19 | 4 | 1 | Low-quality | Genome-fragment | | 21.77 | AAI-based (high-confidence) | 0 |
| S8-final-viral-combined_k141_245278\|\|full | 1863 | No | NA | | 2 | 0 | 2 | Not-determined | Genome-fragment | NA | | NA | 0 |
| S8-final-viral-combined_k141_106250\|\|full | 2782 | No | NA | | 4 | 0 | 1 | Not-determined | Genome-fragment | NA | | NA | 0 |
| S8-final-viral-combined_k141_37499\|\|full | 1752 | No | NA | | 4 | 0 | 1 | Not-determined | Genome-fragment | NA | | NA | 0 |
| S12_k141_219874 | 10207 | No | NA | | 10 | 0 | 2 | Low-quality | Genome-fragment | | 21.96 | AAI-based (medium-confidence) | 0 |
| S4-final-viral-combined_k141_38640\|\|full | 10214 | No | NA | | 15 | 0 | 0 | Low-quality | Genome-fragment | | 9.6 | AAI-based (medium-confidence) | 0 |
| S7-final-viral-combined_k141_112674\|\|full | 10217 | Yes | | 2679 | 10 | 2 | 4 | Low-quality | Genome-fragment | | 3.14 | HMM-based (lower-bound) | 73.78 |
| S8-final-viral-combined_k141_245293\|\|full | 1911 | No | NA | | 5 | 0 | 0 | Not-determined | Genome-fragment | NA | | NA | 0 |
| S8-final-viral-combined_k141_229346\|\|full | 10221 | No | NA | | 20 | 3 | 1 | Low-quality | Genome-fragment | | 25.8 | AAI-based (high-confidence) | 0 |
| S8-final-viral-combined_k141_72722\|\|full | 3440 | No | NA | | 5 | 0 | 1 | Not-determined | Genome-fragment | NA | | NA | 0 |
| S2_k141_3137 | 10223 | No | NA | | 18 | 3 | 0 | Low-quality | Genome-fragment | | 20.38 | AAI-based (high-confidence) | 0 |
| S1-final-viral-combined_k141_210843\|\|full | 10230 | No | NA | | 16 | 7 | 0 | Low-quality | Genome-fragment | | 29.27 | AAI-based (high-confidence) | 0 |
| S8-final-viral-combined_k141_13752\|\|full | 2126 | No | NA | | 3 | 0 | 0 | Not-determined | Genome-fragment | NA | | NA | 0 |
| S8-final-viral-combined_k141_221601\|\|full | 2361 | No | NA | | 2 | 0 | 1 | Not-determined | Genome-fragment | NA | | NA | 0 |
| S11-final-viral-combined_k141_100009\|\|full | 10234 | No | NA | | 9 | 0 | 5 | Low-quality | Genome-fragment | | 9.36 | AAI-based (medium-confidence) | 0 |
| S17-k141_40213 | 10235 | No | NA | | 14 | 5 | 0 | Low-quality | Genome-fragment | | 14.1 | HMM-based (lower-bound) | 0 |
| S8-final-viral-combined_k141_118063\|\|full | 2226 | No | NA | | 3 | 0 | 1 | Not-determined | Genome-fragment | NA | | NA | 0 |
| S2_k141_146888 | 10238 | No | NA | | 13 | 4 | 0 | Low-quality | Genome-fragment | | 25.84 | AAI-based (high-confidence) | 0 |
| S8-final-viral-combined_k141_129845\|\|full | 2394 | No | NA | | 3 | 0 | 0 | Not-determined | Genome-fragment | NA | | NA | 0 |
| S8-final-viral-combined_k141_233844\|\|full | 1804 | No | NA | | 3 | 0 | 1 | Not-determined | Genome-fragment | NA | | NA | 0 |
| S8-final-viral-combined_k141_95595\|\|full | 3294 | No | NA | | 6 | 0 | 2 | Not-determined | Genome-fragment | NA | | NA | 0 |
| S8-final-viral-combined_k141_83235\|\|full | 2425 | No | NA | | 4 | 0 | 1 | Not-determined | Genome-fragment | NA | | NA | 0 |
| S8-final-viral-combined_k141_256120\|\|full | 1322 | No | NA | | 3 | 0 | 0 | Not-determined | Genome-fragment | NA | | NA | 0 |
| S8-final-viral-combined_k141_141905\|\|full | 13640 | No | NA | | 10 | 0 | 7 | Not-determined | Genome-fragment | NA | | NA | 0 |
| S12-final-viral-combined_k141_314092\|\|full | 10245 | No | NA | | 4 | 1 | 0 | Low-quality | Genome-fragment | | 14.62 | HMM-based (lower-bound) | 0 |
| S8-final-viral-combined_k141_48886\|\|full | 3984 | No | NA | | 3 | 0 | 2 | Not-determined | Genome-fragment | NA | | NA | 0 |
| S9-final-viral-combined_k141_40304\|\|full | 10247 | No | NA | | 15 | 0 | 1 | Low-quality | Genome-fragment | | 23.59 | AAI-based (high-confidence) | 0 |
| S2_k141_236273 | 10251 | No | NA | | 9 | 0 | 3 | Low-quality | Genome-fragment | | 3.08 | AAI-based (medium-confidence) | 0 |
| S8-final-viral-combined_k141_245392\|\|full | 1211 | No | NA | | 2 | 0 | 1 | Not-determined | Genome-fragment | NA | | NA | 0 |
| S8-final-viral-combined_k141_129904\|\|full | 10191 | No | NA | | 11 | 0 | 7 | Not-determined | Genome-fragment | NA | | NA | 0 |
| S8-final-viral-combined_k141_245400\|\|full | 2055 | No | NA | | 2 | 0 | 0 | Not-determined | Genome-fragment | NA | | NA | 0 |
| S8-final-viral-combined_k141_153078\|\|full | 2088 | No | NA | | 3 | 0 | 1 | Not-determined | Genome-fragment | NA | | NA | 0 |
| S8-final-viral-combined_k141_129908\|\|full | 2230 | No | NA | | 2 | 0 | 1 | Not-determined | Genome-fragment | NA | | NA | 0 |
| S8-final-viral-combined_k141_164289\|\|full | 33007 | No | NA | | 55 | 18 | 0 | High-quality | High-quality | | 92.4 | AAI-based (high-confidence) | 0 |
| S8-final-viral-combined_k141_106361\|\|full | 3048 | No | NA | | 2 | 0 | 2 | Not-determined | Genome-fragment | NA | | NA | 0 |
| S1-final-viral-combined_k141_28568\|\|full | 10266 | No | NA | | 12 | 2 | 0 | Low-quality | Genome-fragment | | 25.19 | AAI-based (high-confidence) | 0 |
| S8-final-viral-combined_k141_153089\|\|full | 2396 | No | NA | | 3 | 0 | 2 | Not-determined | Genome-fragment | NA | | NA | 0 |
| S8-final-viral-combined_k141_72840\|\|full | 3106 | No | NA | | 5 | 0 | 0 | Not-determined | Genome-fragment | NA | | NA | 0 |
| S8-final-viral-combined_k141_198803\|\|full | 9588 | No | NA | | 10 | 0 | 8 | Not-determined | Genome-fragment | NA | | NA | 0 |
| S3_k141_123109 | 10276 | No | NA | | 17 | 6 | 0 | Low-quality | Genome-fragment | | 28.41 | AAI-based (high-confidence) | 0 |
| S8-final-viral-combined_k141_268054\|\|full | 9596 | No | NA | | 11 | 0 | 0 | Not-determined | Genome-fragment | NA | | NA | 0 |
| S8-final-viral-combined_k141_26436\|\|full | 1703 | No | NA | | 2 | 0 | 2 | Not-determined | Genome-fragment | NA | | NA | 0 |
| S8-final-viral-combined_k141_118136\|\|full | 5759 | No | NA | | 5 | 0 | 4 | Not-determined | Genome-fragment | NA | | NA | 0 |
| S8-final-viral-combined_k141_221690\|\|full | 2757 | No | NA | | 5 | 0 | 2 | Not-determined | Genome-fragment | NA | | NA | 0 |
| S8-final-viral-combined_k141_95615\|\|full | 1392 | No | NA | | 3 | 0 | 0 | Not-determined | Genome-fragment | NA | | NA | 0 |
| S8-final-viral-combined_k141_176504\|\|full | 3642 | No | NA | | 3 | 0 | 2 | Not-determined | Genome-fragment | NA | | NA | 0 |
| S8-final-viral-combined_k141_164347\|\|full | 1897 | No | NA | | 2 | 0 | 2 | Not-determined | Genome-fragment | NA | | NA | 0 |
| S8-final-viral-combined_k141_2996\|\|full | 1846 | No | NA | | 3 | 0 | 0 | Not-determined | Genome-fragment | NA | | NA | 0 |
| S8-final-viral-combined_k141_95628\|\|full | 1509 | No | NA | | 2 | 0 | 1 | Not-determined | Genome-fragment | NA | | NA | 0 |
| S8-final-viral-combined_k141_268107\|\|full | 1703 | No | NA | | 3 | 0 | 0 | Not-determined | Genome-fragment | NA | | NA | 0 |
| S8-final-viral-combined_k141_95637\|\|full | 5890 | No | NA | | 5 | 0 | 4 | Not-determined | Genome-fragment | NA | | NA | 0 |
| S8-final-viral-combined_k141_72891\|\|full | 5785 | No | NA | | 5 | 0 | 2 | Not-determined | Genome-fragment | NA | | NA | 0 |
| S8-final-viral-combined_k141_187622\|\|full | 1288 | No | NA | | 2 | 0 | 1 | Not-determined | Genome-fragment | NA | | NA | 0 |
| S8-final-viral-combined_k141_95641\|\|full | 3095 | No | NA | | 3 | 0 | 3 | Not-determined | Genome-fragment | NA | | NA | 0 |
| S8-final-viral-combined_k141_72896\|\|full | 1125 | No | NA | | 2 | 0 | 1 | Not-determined | Genome-fragment | NA | | NA | 0 |
| S5-final-viral-combined_k141_45678\|\|full | 10288 | No | NA | | 10 | 0 | 9 | Low-quality | Genome-fragment | | 2.5 | AAI-based (medium-confidence) | 0 |
| S8-final-viral-combined_k141_83362\|\|full | 1449 | No | NA | | 2 | 0 | 0 | Not-determined | Genome-fragment | NA | | NA | 0 |
| S7-final-viral-combined_k141_8392\|\|full | 10295 | No | NA | | 19 | 3 | 0 | Low-quality | Genome-fragment | | 22.4 | AAI-based (high-confidence) | 0 |
| S8-final-viral-combined_k141_256265\|\|full | 1105 | No | NA | | 3 | 0 | 0 | Not-determined | Genome-fragment | NA | | NA | 0 |
| S8-final-viral-combined_k141_48939\|\|full | 6541 | No | NA | | 7 | 0 | 3 | Not-determined | Genome-fragment | NA | | NA | 0 |
| S8-final-viral-combined_k141_268118\|\|full | 2856 | No | NA | | 3 | 0 | 2 | Not-determined | Genome-fragment | NA | | NA | 0 |
| S8-final-viral-combined_k141_268119\|\|full | 1128 | No | NA | | 2 | 0 | 1 | Not-determined | Genome-fragment | NA | | NA | 0 |
| S11_k141_142891 | 10298 | No | NA | | 13 | 2 | 0 | Low-quality | Genome-fragment | | 19.12 | AAI-based (high-confidence) | 0 |
| S8-final-viral-combined_k141_95675\|\|full | 4612 | No | NA | | 3 | 0 | 1 | Not-determined | Genome-fragment | NA | | NA | 0 |
| S8-final-viral-combined_k141_210614\|\|full | 1173 | No | NA | | 3 | 0 | 0 | Not-determined | Genome-fragment | NA | | NA | 0 |
| S8-final-viral-combined_k141_48968\|\|full | 1494 | No | NA | | 3 | 0 | 0 | Not-determined | Genome-fragment | NA | | NA | 0 |
| S12_k141_127626 | 10301 | No | NA | | 4 | 0 | 3 | Low-quality | Genome-fragment | | 3.71 | AAI-based (high-confidence) | 0 |
| S8-final-viral-combined_k141_153167\|\|full | 1418 | No | NA | | 2 | 0 | 1 | Not-determined | Genome-fragment | NA | | NA | 0 |
| S12-final-viral-combined_k141_218837\|\|full | 10307 | No | NA | | 11 | 0 | 5 | Low-quality | Genome-fragment | | 7.66 | AAI-based (medium-confidence) | 0 |
| S2_k141_143075 | 10307 | No | NA | | 15 | 6 | 0 | Low-quality | Genome-fragment | | 23.91 | AAI-based (high-confidence) | 0 |
| S8-final-viral-combined_k141_26595\|\|full | 3039 | No | NA | | 3 | 0 | 0 | Not-determined | Genome-fragment | NA | | NA | 0 |
| S12-final-viral-combined_k141_169609\|\|full | 10316 | No | NA | | 15 | 3 | 0 | Low-quality | Genome-fragment | | 27.02 | AAI-based (high-confidence) | 0 |
| S8-final-viral-combined_k141_114736\|\|full | 10318 | No | NA | | 17 | 7 | 0 | Low-quality | Genome-fragment | | 28.23 | AAI-based (medium-confidence) | 0 |
| S5_k141_131282 | 10322 | No | NA | | 7 | 0 | 7 | Low-quality | Genome-fragment | | 21.67 | AAI-based (medium-confidence) | 0 |
| S7-final-viral-combined_k141_27877\|\|full | 10322 | No | NA | | 9 | 2 | 0 | Low-quality | Genome-fragment | | 24.64 | AAI-based (high-confidence) | 0 |
| S8-final-viral-combined_k141_153198\|\|full | 1548 | No | NA | | 3 | 0 | 0 | Not-determined | Genome-fragment | NA | | NA | 0 |
| S8-final-viral-combined_k141_48995\|\|full | 2967 | No | NA | | 7 | 0 | 0 | Not-determined | Genome-fragment | NA | | NA | 0 |
| S7-final-viral-combined_k141_167452\|\|full | 10324 | No | NA | | 11 | 1 | 2 | Low-quality | Genome-fragment | | 7.44 | HMM-based (lower-bound) | 0 |
| S8-final-viral-combined_k141_3085\|\|full | 2429 | No | NA | | 3 | 0 | 0 | Not-determined | Genome-fragment | NA | | NA | 0 |
| S8-final-viral-combined_k141_49009\|\|full | 2202 | No | NA | | 3 | 0 | 2 | Not-determined | Genome-fragment | NA | | NA | 0 |
| S8-final-viral-combined_k141_72911\|\|full | 60264 | Yes | | 44670 | 74 | 20 | 8 | Medium-quality | Genome-fragment | | 75.82 | AAI-based (high-confidence) | 25.88 |
| S12-final-viral-combined_k141_85582\|\|full | 10328 | No | NA | | 11 | 0 | 0 | Low-quality | Genome-fragment | | 18.8 | AAI-based (medium-confidence) | 0 |
| S8-final-viral-combined_k141_49027\|\|full | 2012 | No | NA | | 4 | 0 | 1 | Not-determined | Genome-fragment | NA | | NA | 0 |
| S9-final-viral-combined_k141_13758\|\|full | 10339 | No | NA | | 14 | 11 | 0 | Low-quality | Genome-fragment | | 25.7 | AAI-based (medium-confidence) | 0 |
| S8-final-viral-combined_k141_37817\|\|full | 1458 | No | NA | | 3 | 0 | 1 | Not-determined | Genome-fragment | NA | | NA | 0 |
| S12-final-viral-combined_k141_116208\|\|full | 10341 | No | NA | | 14 | 1 | 4 | Low-quality | Genome-fragment | | 7.7 | AAI-based (high-confidence) | 0 |
| S8-final-viral-combined_k141_106563\|\|full | 4970 | No | NA | | 7 | 0 | 1 | Not-determined | Genome-fragment | NA | | NA | 0 |
| S8-final-viral-combined_k141_245504\|\|full | 3633 | No | NA | | 4 | 0 | 1 | Not-determined | Genome-fragment | NA | | NA | 0 |
| S8-final-viral-combined_k141_130116\|\|full | 1207 | No | NA | | 2 | 0 | 1 | Not-determined | Genome-fragment | NA | | NA | 0 |
| S7_k141_93176 | 10369 | No | NA | | 10 | 0 | 4 | Low-quality | Genome-fragment | | 20.61 | AAI-based (medium-confidence) | 0 |
| S8-final-viral-combined_k141_221840\|\|full | 1004 | No | NA | | 2 | 0 | 1 | Not-determined | Genome-fragment | NA | | NA | 0 |
| S8-final-viral-combined_k141_72977\|\|full | 3017 | No | NA | | 6 | 0 | 5 | Not-determined | Genome-fragment | NA | | NA | 0 |
| S3_k141_65729 | 10371 | No | NA | | 20 | 6 | 0 | Low-quality | Genome-fragment | | 19.24 | AAI-based (high-confidence) | 0 |
| S2-final-viral-combined_k141_171832\|\|0_partial | 10374 | No | NA | | 8 | 0 | 8 | Low-quality | Genome-fragment | | 1.27 | AAI-based (medium-confidence) | 0 |
| S8-final-viral-combined_k141_13906\|\|full | 2157 | No | NA | | 3 | 0 | 1 | Not-determined | Genome-fragment | NA | | NA | 0 |
| S8-final-viral-combined_k141_221848\|\|full | 3799 | No | NA | | 3 | 0 | 2 | Not-determined | Genome-fragment | NA | | NA | 0 |
| S8-final-viral-combined_k141_210674\|\|full | 1012 | No | NA | | 4 | 0 | 0 | Not-determined | Genome-fragment | NA | | NA | 0 |
| S8-final-viral-combined_k141_49107\|\|full | 5402 | No | NA | | 3 | 0 | 0 | Not-determined | Genome-fragment | NA | | NA | 0 |
| S11-final-viral-combined_k141_297347\|\|full | 10379 | No | NA | | 15 | 3 | 1 | Low-quality | Genome-fragment | | 22.76 | AAI-based (high-confidence) | 0 |
| S4_k141_154470 | 10381 | No | NA | | 9 | 0 | 9 | Low-quality | Genome-fragment | | 1.51 | AAI-based (medium-confidence) | 0 |
| S8-final-viral-combined_k141_130212\|\|full | 1104 | No | NA | | 3 | 0 | 1 | Not-determined | Genome-fragment | NA | | NA | 0 |
| S8-final-viral-combined_k141_164515\|\|full | 1429 | No | NA | | 3 | 0 | 0 | Not-determined | Genome-fragment | NA | | NA | 0 |
| S11-final-viral-combined_k141_24936\|\|full | 10384 | No | NA | | 17 | 7 | 1 | Low-quality | Genome-fragment | | 26.37 | AAI-based (medium-confidence) | 0 |
| S1_k141_56696 | 10385 | No | NA | | 6 | 0 | 5 | Low-quality | Genome-fragment | | 9.26 | AAI-based (medium-confidence) | 0 |
| S8-final-viral-combined_k141_210720\|\|full | 1726 | No | NA | | 2 | 0 | 1 | Not-determined | Genome-fragment | NA | | NA | 0 |
| S8-final-viral-combined_k141_199013\|\|full | 1078 | No | NA | | 4 | 0 | 1 | Not-determined | Genome-fragment | NA | | NA | 0 |
| S8-final-viral-combined_k141_268325\|\|full | 2445 | No | NA | | 5 | 0 | 1 | Not-determined | Genome-fragment | NA | | NA | 0 |
| S4-final-viral-combined_k141_24944\|\|full | 10389 | No | NA | | 17 | 8 | 0 | Low-quality | Genome-fragment | | 29.22 | AAI-based (high-confidence) | 0 |
| S17-final-viral-combined_k141_185264\|\|full | 10390 | Yes | | 3994 | 8 | 2 | 4 | Low-quality | Genome-fragment | | 7.53 | AAI-based (medium-confidence) | 61.56 |
| S7-final-viral-combined_k141_42391\|\|full | 10398 | No | NA | | 20 | 4 | 1 | Low-quality | Genome-fragment | | 17.88 | AAI-based (high-confidence) | 0 |
| S8-final-viral-combined_k141_176817\|\|full | 2436 | No | NA | | 7 | 0 | 2 | Not-determined | Genome-fragment | NA | | NA | 0 |
| S8-final-viral-combined_k141_187903\|\|full | 1493 | No | NA | | 3 | 0 | 0 | Not-determined | Genome-fragment | NA | | NA | 0 |
| S8-final-viral-combined_k141_210796\|\|full | 3380 | No | NA | | 3 | 0 | 1 | Not-determined | Genome-fragment | NA | | NA | 0 |
| S8-final-viral-combined_k141_37970\|\|full | 14080 | No | NA | | 8 | 0 | 6 | Not-determined | Genome-fragment | NA | | NA | 0 |
| S12-final-viral-combined_k141_124618\|\|full | 10410 | No | NA | | 15 | 3 | 1 | Low-quality | Genome-fragment | | 22.88 | HMM-based (lower-bound) | 0 |
| S8-final-viral-combined_k141_49211\|\|full | 4159 | No | NA | | 5 | 0 | 3 | Not-determined | Genome-fragment | NA | | NA | 0 |
| S8-final-viral-combined_k141_268400\|\|full | 1187 | No | NA | | 2 | 0 | 1 | Not-determined | Genome-fragment | NA | | NA | 0 |
| S8-final-viral-combined_k141_210830\|\|full | 4512 | No | NA | | 2 | 0 | 2 | Not-determined | Genome-fragment | NA | | NA | 0 |
| S8-final-viral-combined_k141_118462\|\|full | 1212 | No | NA | | 3 | 0 | 2 | Not-determined | Genome-fragment | NA | | NA | 0 |
| S8-final-viral-combined_k141_210832\|\|full | 6871 | No | NA | | 4 | 0 | 1 | Not-determined | Genome-fragment | NA | | NA | 0 |
| S11_k141_235215 | 10419 | No | NA | | 16 | 10 | 0 | Low-quality | Genome-fragment | | 28.83 | AAI-based (high-confidence) | 0 |
| S12-final-viral-combined_k141_382222\|\|full | 10419 | No | NA | | 23 | 3 | 0 | Low-quality | Genome-fragment | | 16.44 | AAI-based (high-confidence) | 0 |
| S8-final-viral-combined_k141_73160\|\|full | 3051 | No | NA | | 3 | 0 | 1 | Not-determined | Genome-fragment | NA | | NA | 0 |
| S8-final-viral-combined_k141_176896\|\|full | 1673 | No | NA | | 2 | 0 | 2 | Not-determined | Genome-fragment | NA | | NA | 0 |
| S6-final-viral-combined_k141_84322\|\|full | 10424 | No | NA | | 13 | 0 | 7 | Low-quality | Genome-fragment | | 7.72 | AAI-based (medium-confidence) | 0 |
| S2-final-viral-combined_k141_151921\|\|full | 10427 | No | NA | | 20 | 1 | 0 | Low-quality | Genome-fragment | | 28.66 | AAI-based (high-confidence) | 0 |
| S12_k141_388449 | 10428 | No | NA | | 9 | 0 | 5 | Low-quality | Genome-fragment | | 9.64 | AAI-based (medium-confidence) | 0 |
| S8-final-viral-combined_k141_153417\|\|full | 2224 | No | NA | | 3 | 0 | 0 | Not-determined | Genome-fragment | NA | | NA | 0 |
| S8-final-viral-combined_k141_106774\|\|full | 6637 | No | NA | | 8 | 0 | 8 | Not-determined | Genome-fragment | NA | | NA | 0 |
| S1-final-viral-combined_k141_210164\|\|full | 10433 | No | NA | | 13 | 8 | 0 | Low-quality | Genome-fragment | | 26.31 | AAI-based (medium-confidence) | 0 |
| S8_k141_26766 | 10438 | No | NA | | 10 | 1 | 2 | Low-quality | Genome-fragment | | 9.51 | HMM-based (lower-bound) | 0 |
| S8-final-viral-combined_k141_210858\|\|full | 2153 | No | NA | | 5 | 0 | 0 | Not-determined | Genome-fragment | NA | | NA | 0 |
| S5-final-viral-combined_k141_30713\|\|full | 10439 | No | NA | | 11 | 0 | 4 | Low-quality | Genome-fragment | | 16.45 | AAI-based (medium-confidence) | 0 |
| S12_k141_131498 | 10446 | No | NA | | 10 | 1 | 0 | Low-quality | Genome-fragment | | 3.72 | HMM-based (lower-bound) | 0 |
| S8_k141_148203 | 10447 | No | NA | | 14 | 2 | 0 | Low-quality | Genome-fragment | | 19.77 | AAI-based (high-confidence) | 0 |
| S8-final-viral-combined_k141_118538\|\|full | 1592 | No | NA | | 3 | 0 | 0 | Not-determined | Genome-fragment | NA | | NA | 0 |
| S8-final-viral-combined_k141_106790\|\|full | 3989 | No | NA | | 5 | 0 | 0 | Not-determined | Genome-fragment | NA | | NA | 0 |
| S1-final-viral-combined_k141_85258\|\|0_partial | 10449 | Yes | | 2359 | 13 | 2 | 7 | Low-quality | Genome-fragment | | 2.64 | HMM-based (lower-bound) | 77.42 |
| S4_k141_63561 | 10450 | No | NA | | 14 | 0 | 0 | Low-quality | Genome-fragment | | 17.45 | AAI-based (high-confidence) | 0 |
| S8-final-viral-combined_k141_26879\|\|full | 2890 | No | NA | | 5 | 0 | 4 | Not-determined | Genome-fragment | NA | | NA | 0 |
| S8-final-viral-combined_k141_61112\|\|full | 4348 | No | NA | | 4 | 0 | 2 | Not-determined | Genome-fragment | NA | | NA | 0 |
| S7-final-viral-combined_k141_132808\|\|full | 10452 | No | NA | | 4 | 1 | 0 | Low-quality | Genome-fragment | | 5.04 | AAI-based (high-confidence) | 0 |
| S8-final-viral-combined_k141_118586\|\|full | 1051 | No | NA | | 2 | 0 | 1 | Not-determined | Genome-fragment | NA | | NA | 0 |
| S4_k141_72293 | 10469 | No | NA | | 13 | 0 | 7 | Low-quality | Genome-fragment | | 19.38 | AAI-based (medium-confidence) | 0 |
| S17-final-viral-combined_k141_127160\|\|full | 10470 | No | NA | | 19 | 2 | 0 | Low-quality | Genome-fragment | | 15.62 | AAI-based (medium-confidence) | 0 |
| S6_k141_45908 | 10470 | No | NA | | 11 | 2 | 0 | Low-quality | Genome-fragment | | 13.3 | AAI-based (high-confidence) | 0 |
| S8_k141_186183 | 10473 | Yes | | 6439 | 20 | 3 | 2 | Low-quality | Genome-fragment | | 4.54 | AAI-based (high-confidence) | 38.52 |
| S8-final-viral-combined_k141_245777\|\|full | 6530 | No | NA | | 7 | 0 | 4 | Not-determined | Genome-fragment | NA | | NA | 0 |
| S11-final-viral-combined_k141_79315\|\|full | 10483 | No | NA | | 13 | 0 | 0 | Low-quality | Genome-fragment | | 11.15 | AAI-based (medium-confidence) | 0 |
| S17-final-viral-combined_k141_109840\|\|0_partial | 10493 | Yes | | 5986 | 13 | 3 | 2 | Low-quality | Genome-fragment | | 7.94 | HMM-based (lower-bound) | 42.95 |
| S8-final-viral-combined_k141_153564\|\|full | 3174 | No | NA | | 3 | 0 | 3 | Not-determined | Genome-fragment | NA | | NA | 0 |
| S8-final-viral-combined_k141_245820\|\|full | 4263 | No | NA | | 3 | 0 | 2 | Not-determined | Genome-fragment | NA | | NA | 0 |
| S7-final-viral-combined_k141_174634\|\|full | 10494 | No | NA | | 21 | 7 | 0 | Low-quality | Genome-fragment | | 26.54 | AAI-based (high-confidence) | 0 |
| S1-final-viral-combined_k141_296974\|\|full | 10499 | No | NA | | 18 | 3 | 1 | Low-quality | Genome-fragment | | 29.52 | AAI-based (high-confidence) | 0 |
| S7-final-viral-combined_k141_199406\|\|full | 10506 | No | NA | | 15 | 1 | 0 | Low-quality | Genome-fragment | | 27.54 | AAI-based (medium-confidence) | 0 |
| S8-final-viral-combined_k141_83694\|\|full | 1410 | No | NA | | 3 | 0 | 2 | Not-determined | Genome-fragment | NA | | NA | 0 |
| S8-final-viral-combined_k141_207428\|\|full | 10511 | No | NA | | 17 | 0 | 5 | Low-quality | Genome-fragment | | 32.81 | AAI-based (high-confidence) | 0 |
| S5_k141_104787 | 10514 | Yes | | 8591 | 9 | 1 | 1 | Low-quality | Genome-fragment | | 5.2 | HMM-based (lower-bound) | 18.29 |
| S2_k141_97899 | 10517 | No | NA | | 25 | 2 | 0 | Low-quality | Genome-fragment | | 10.07 | AAI-based (medium-confidence) | 0 |
| S8_k141_152574 | 10523 | No | NA | | 17 | 2 | 0 | Low-quality | Genome-fragment | | 20.73 | AAI-based (high-confidence) | 0 |
| S11-final-viral-combined_k141_159229\|\|full | 10550 | No | NA | | 10 | 1 | 0 | Low-quality | Genome-fragment | | 13.96 | HMM-based (lower-bound) | 0 |
| S8-final-viral-combined_k141_164765\|\|full | 1605 | No | NA | | 3 | 0 | 2 | Not-determined | Genome-fragment | NA | | NA | 0 |
| S8-final-viral-combined_k141_268601\|\|full | 2410 | No | NA | | 2 | 0 | 2 | Not-determined | Genome-fragment | NA | | NA | 0 |
| S7-final-viral-combined_k141_201379\|\|full | 10560 | No | NA | | 14 | 1 | 1 | Low-quality | Genome-fragment | | 7.1 | HMM-based (lower-bound) | 0 |
| S8-final-viral-combined_k141_164769\|\|full | 1643 | No | NA | | 3 | 0 | 0 | Not-determined | Genome-fragment | NA | | NA | 0 |
| S9-final-viral-combined_k141_42136\|\|0_partial | 10562 | No | NA | | 11 | 0 | 7 | Low-quality | Genome-fragment | | 25.13 | AAI-based (high-confidence) | 0 |
| S8-final-viral-combined_k141_49369\|\|full | 57772 | Yes | | 41920 | 57 | 4 | 13 | High-quality | High-quality | | 100 | AAI-based (medium-confidence) | 27.44 |
| S8-final-viral-combined_k141_61281\|\|full | 3080 | No | NA | | 3 | 0 | 1 | Not-determined | Genome-fragment | NA | | NA | 0 |
| S8-final-viral-combined_k141_234391\|\|full | 2493 | No | NA | | 4 | 0 | 1 | Not-determined | Genome-fragment | NA | | NA | 0 |
| S8-final-viral-combined_k141_96055\|\|full | 20847 | No | NA | | 17 | 0 | 9 | Not-determined | Genome-fragment | NA | | NA | 0 |
| S8-final-viral-combined_k141_210993\|\|full | 2057 | No | NA | | 5 | 0 | 3 | Not-determined | Genome-fragment | NA | | NA | 0 |
| S8-final-viral-combined_k141_164821\|\|full | 3093 | No | NA | | 4 | 0 | 2 | Not-determined | Genome-fragment | NA | | NA | 0 |
| S8-final-viral-combined_k141_83756\|\|full | 3265 | No | NA | | 5 | 0 | 4 | Not-determined | Genome-fragment | NA | | NA | 0 |
| S4_k141_140185 | 10568 | No | NA | | 6 | 2 | 0 | Low-quality | Genome-fragment | | 17.05 | AAI-based (medium-confidence) | 0 |
| S4-final-viral-combined_k141_132852\|\|full | 10570 | No | NA | | 7 | 0 | 6 | Low-quality | Genome-fragment | | 6.18 | AAI-based (medium-confidence) | 0 |
| S12-final-viral-combined_k141_181135\|\|0_partial | 10574 | No | NA | | 17 | 1 | 6 | Low-quality | Genome-fragment | | 27.2 | AAI-based (medium-confidence) | 0 |
| S8-final-viral-combined_k141_164839\|\|full | 2365 | No | NA | | 4 | 0 | 2 | Not-determined | Genome-fragment | NA | | NA | 0 |
| S8-final-viral-combined_k141_27115\|\|full | 1394 | No | NA | | 3 | 0 | 1 | Not-determined | Genome-fragment | NA | | NA | 0 |
| S8-final-viral-combined_k141_142497\|\|full | 7133 | No | NA | | 8 | 0 | 1 | Not-determined | Genome-fragment | NA | | NA | 0 |
| S8-final-viral-combined_k141_27131\|\|full | 1835 | No | NA | | 3 | 0 | 1 | Not-determined | Genome-fragment | NA | | NA | 0 |
| S4-final-viral-combined_k141_86501\|\|full | 10588 | No | NA | | 19 | 1 | 0 | Low-quality | Genome-fragment | | 23.59 | AAI-based (high-confidence) | 0 |
| S12_k141_228849 | 10591 | No | NA | | 12 | 2 | 1 | Low-quality | Genome-fragment | | 17.3 | AAI-based (medium-confidence) | 0 |
| S8-final-viral-combined_k141_234448\|\|full | 2710 | No | NA | | 4 | 0 | 3 | Not-determined | Genome-fragment | NA | | NA | 0 |
| S8-final-viral-combined_k141_7420\|\|full | 10595 | No | NA | | 18 | 3 | 1 | Low-quality | Genome-fragment | | 8.13 | AAI-based (high-confidence) | 0 |
| S8-final-viral-combined_k141_246007\|\|full | 3825 | No | NA | | 4 | 0 | 2 | Not-determined | Genome-fragment | NA | | NA | 0 |
| S8-final-viral-combined_k141_61328\|\|full | 2194 | No | NA | | 4 | 0 | 1 | Not-determined | Genome-fragment | NA | | NA | 0 |
| S5-final-viral-combined_k141_18807\|\|full | 10597 | No | NA | | 18 | 1 | 1 | Low-quality | Genome-fragment | | 6.84 | HMM-based (lower-bound) | 0 |
| S8-final-viral-combined_k141_222264\|\|full | 2342 | No | NA | | 3 | 0 | 3 | Not-determined | Genome-fragment | NA | | NA | 0 |
| S8-final-viral-combined_k141_256401\|\|full | 2240 | No | NA | | 3 | 0 | 0 | Not-determined | Genome-fragment | NA | | NA | 0 |
| S6_k141_121334 | 10597 | No | NA | | 15 | 1 | 2 | Low-quality | Genome-fragment | | 5.75 | HMM-based (lower-bound) | 0 |
| S8-final-viral-combined_k141_61341\|\|full | 2253 | No | NA | | 3 | 0 | 1 | Not-determined | Genome-fragment | NA | | NA | 0 |
| S6-final-viral-combined_k141_96082\|\|full | 10607 | No | NA | | 14 | 5 | 0 | Low-quality | Genome-fragment | | 26.41 | AAI-based (high-confidence) | 0 |
| S8-final-viral-combined_k141_61342\|\|full | 2403 | No | NA | | 3 | 0 | 2 | Not-determined | Genome-fragment | NA | | NA | 0 |
| S8-final-viral-combined_k141_107094\|\|full | 5458 | No | NA | | 2 | 0 | 2 | Not-determined | Genome-fragment | NA | | NA | 0 |
| S8-final-viral-combined_k141_234473\|\|full | 3037 | No | NA | | 4 | 1 | 0 | Medium-quality | Genome-fragment | | 55.04 | AAI-based (medium-confidence) | 0 |
| S1-final-viral-combined_k141_3623\|\|full | 10609 | No | NA | | 21 | 4 | 1 | Low-quality | Genome-fragment | | 24.85 | AAI-based (high-confidence) | 0 |
| S1-final-viral-combined_k141_109127\|\|full | 10610 | No | NA | | 10 | 5 | 0 | Low-quality | Genome-fragment | | 18.11 | AAI-based (high-confidence) | 0 |
| S8-final-viral-combined_k141_27189\|\|full | 1265 | No | NA | | 3 | 0 | 3 | Not-determined | Genome-fragment | NA | | NA | 0 |
| S17-final-viral-combined_k141_183098\|\|full | 10610 | No | NA | | 21 | 1 | 0 | Low-quality | Genome-fragment | | 29.02 | AAI-based (high-confidence) | 0 |
| S8-final-viral-combined_k141_3483\|\|full | 11175 | No | NA | | 10 | 0 | 7 | Not-determined | Genome-fragment | NA | | NA | 0 |
| S8-final-viral-combined_k141_234507\|\|full | 2466 | No | NA | | 3 | 0 | 0 | Not-determined | Genome-fragment | NA | | NA | 0 |
| S8-final-viral-combined_k141_130623\|\|full | 1997 | No | NA | | 3 | 0 | 1 | Not-determined | Genome-fragment | NA | | NA | 0 |
| S8-final-viral-combined_k141_153730\|\|full | 1646 | No | NA | | 2 | 0 | 0 | Not-determined | Genome-fragment | NA | | NA | 0 |
| S8-final-viral-combined_k141_199267\|\|full | 1928 | No | NA | | 3 | 0 | 2 | Not-determined | Genome-fragment | NA | | NA | 0 |
| S6_k141_57436 | 10613 | No | NA | | 15 | 13 | 0 | Low-quality | Genome-fragment | | 23.29 | AAI-based (high-confidence) | 0 |
| S8-final-viral-combined_k141_162592\|\|0_partial | 10621 | No | NA | | 16 | 1 | 6 | Low-quality | Genome-fragment | | 7.14 | HMM-based (lower-bound) | 0 |
| S11-final-viral-combined_k141_96729\|\|full | 10626 | No | NA | | 17 | 7 | 0 | Low-quality | Genome-fragment | | 26.01 | AAI-based (high-confidence) | 0 |
| S8-final-viral-combined_k141_3511\|\|full | 3953 | No | NA | | 8 | 0 | 1 | Not-determined | Genome-fragment | NA | | NA | 0 |
| S12_k141_375093 | 10628 | No | NA | | 8 | 0 | 7 | Low-quality | Genome-fragment | | 9.64 | AAI-based (medium-confidence) | 0 |
| S8-final-viral-combined_k141_234548\|\|full | 2597 | No | NA | | 3 | 0 | 2 | Not-determined | Genome-fragment | NA | | NA | 0 |
| S1-final-viral-combined_k141_290285\|\|full | 10636 | No | NA | | 14 | 11 | 0 | Low-quality | Genome-fragment | | 14.93 | AAI-based (medium-confidence) | 0 |
| S8-final-viral-combined_k141_177251\|\|full | 3729 | No | NA | | 4 | 0 | 2 | Not-determined | Genome-fragment | NA | | NA | 0 |
| S8-final-viral-combined_k141_211119\|\|full | 25630 | No | NA | | 33 | 7 | 0 | Medium-quality | Genome-fragment | | 61.51 | AAI-based (high-confidence) | 0 |
| S17-final-viral-combined_k141_89914\|\|full | 10652 | No | NA | | 10 | 0 | 6 | Low-quality | Genome-fragment | | 5.73 | AAI-based (medium-confidence) | 0 |
| S8-final-viral-combined_k141_142597\|\|full | 3737 | No | NA | | 6 | 0 | 1 | Not-determined | Genome-fragment | NA | | NA | 0 |
| S11-final-viral-combined_k141_315528\|\|0_partial | 10662 | Yes | | 4172 | 10 | 2 | 6 | Low-quality | Genome-fragment | | 5.05 | HMM-based (lower-bound) | 60.87 |
| S8-final-viral-combined_k141_130665\|\|full | 1676 | No | NA | | 2 | 0 | 1 | Not-determined | Genome-fragment | NA | | NA | 0 |
| S8-final-viral-combined_k141_222362\|\|full | 1414 | No | NA | | 2 | 0 | 0 | Not-determined | Genome-fragment | NA | | NA | 0 |
| S8-final-viral-combined_k141_73378\|\|full | 1634 | No | NA | | 2 | 0 | 1 | Not-determined | Genome-fragment | NA | | NA | 0 |
| S2_k141_105039 | 10666 | No | NA | | 8 | 0 | 1 | Low-quality | Genome-fragment | | 13.34 | AAI-based (medium-confidence) | 0 |
| S8-final-viral-combined_k141_177294\|\|full | 1272 | No | NA | | 4 | 0 | 2 | Not-determined | Genome-fragment | NA | | NA | 0 |
| S2-final-viral-combined_k141_230278\|\|full | 10673 | No | NA | | 12 | 1 | 4 | Low-quality | Genome-fragment | | 6.76 | HMM-based (lower-bound) | 0 |
| S8-final-viral-combined_k141_177309\|\|full | 2997 | No | NA | | 4 | 0 | 1 | Not-determined | Genome-fragment | NA | | NA | 0 |
| S7-final-viral-combined_k141_154606\|\|full | 10679 | No | NA | | 9 | 0 | 1 | Low-quality | Genome-fragment | | 24.56 | AAI-based (medium-confidence) | 0 |
| S8-final-viral-combined_k141_177347\|\|full | 1995 | No | NA | | 2 | 0 | 0 | Not-determined | Genome-fragment | NA | | NA | 0 |
| S8-final-viral-combined_k141_107235\|\|full | 1218 | No | NA | | 3 | 0 | 0 | Not-determined | Genome-fragment | NA | | NA | 0 |
| S8-final-viral-combined_k141_96197\|\|full | 1672 | No | NA | | 2 | 0 | 0 | Not-determined | Genome-fragment | NA | | NA | 0 |
| S8-final-viral-combined_k141_153767\|\|full | 1866 | No | NA | | 4 | 0 | 0 | Not-determined | Genome-fragment | NA | | NA | 0 |
| S17-final-viral-combined_k141_257936\|\|full | 10690 | No | NA | | 16 | 1 | 4 | Low-quality | Genome-fragment | | 9.71 | HMM-based (lower-bound) | 0 |
| S8-final-viral-combined_k141_83929\|\|full | 2003 | No | NA | | 5 | 0 | 0 | Not-determined | Genome-fragment | NA | | NA | 0 |
| S8-final-viral-combined_k141_268766\|\|full | 1033 | No | NA | | 3 | 0 | 1 | Not-determined | Genome-fragment | NA | | NA | 0 |
| S8-final-viral-combined_k141_144269\|\|full | 10698 | No | NA | | 12 | 3 | 1 | Low-quality | Genome-fragment | | 26 | AAI-based (high-confidence) | 0 |
| S1-final-viral-combined_k141_145295\|\|full | 10711 | No | NA | | 17 | 5 | 0 | Low-quality | Genome-fragment | | 22.08 | AAI-based (high-confidence) | 0 |
| S8-final-viral-combined_k141_38370\|\|full | 4459 | No | NA | | 5 | 0 | 5 | Not-determined | Genome-fragment | NA | | NA | 0 |
| S11-final-viral-combined_k141_198187\|\|full | 10713 | No | NA | | 17 | 10 | 0 | Low-quality | Genome-fragment | | 32.79 | AAI-based (medium-confidence) | 0 |
| S1-final-viral-combined_k141_321604\|\|full | 10718 | No | NA | | 22 | 4 | 0 | Low-quality | Genome-fragment | | 23.4 | AAI-based (high-confidence) | 0 |
| S11_k141_318371 | 10719 | No | NA | | 11 | 6 | 1 | Low-quality | Genome-fragment | | 18.02 | HMM-based (lower-bound) | 0 |
| S8-final-viral-combined_k141_61493\|\|full | 1137 | No | NA | | 3 | 0 | 1 | Not-determined | Genome-fragment | NA | | NA | 0 |
| S8-final-viral-combined_k141_49659\|\|full | 4375 | No | NA | | 3 | 0 | 1 | Not-determined | Genome-fragment | NA | | NA | 0 |
| S4-final-viral-combined_k141_65995\|\|0_partial | 10739 | No | NA | | 12 | 1 | 5 | Low-quality | Genome-fragment | | 19.66 | HMM-based (lower-bound) | 0 |
| S8-final-viral-combined_k141_61496\|\|full | 4364 | No | NA | | 5 | 0 | 3 | Not-determined | Genome-fragment | NA | | NA | 0 |
| S8-final-viral-combined_k141_83998\|\|full | 8540 | No | NA | | 5 | 0 | 5 | Not-determined | Genome-fragment | NA | | NA | 0 |
| S9_k141_3036 | 10744 | No | NA | | 16 | 1 | 4 | Low-quality | Genome-fragment | | 16.26 | HMM-based (lower-bound) | 0 |
| S11-final-viral-combined_k141_45081\|\|full | 10754 | No | NA | | 12 | 1 | 7 | Low-quality | Genome-fragment | | 5.83 | HMM-based (lower-bound) | 0 |
| S8_k141_266646 | 10757 | No | NA | | 15 | 8 | 0 | Low-quality | Genome-fragment | | 33.1 | AAI-based (high-confidence) | 0 |
| S8-final-viral-combined_k141_165130\|\|full | 4473 | No | NA | | 5 | 0 | 3 | Not-determined | Genome-fragment | NA | | NA | 0 |
| S8-final-viral-combined_k141_84009\|\|full | 3807 | No | NA | | 3 | 0 | 2 | Not-determined | Genome-fragment | NA | | NA | 0 |
| S2-final-viral-combined_k141_209184\|\|full | 10761 | No | NA | | 15 | 0 | 0 | Low-quality | Genome-fragment | | 24.73 | AAI-based (high-confidence) | 0 |
| S4_k141_27422 | 10761 | Yes | | 9655 | 10 | 1 | 1 | Low-quality | Genome-fragment | | 11.73 | AAI-based (high-confidence) | 10.28 |
| S1-final-viral-combined_k141_183133\|\|full | 10768 | No | NA | | 19 | 2 | 3 | Low-quality | Genome-fragment | | 13.03 | AAI-based (medium-confidence) | 0 |
| S8-final-viral-combined_k141_246152\|\|full | 4321 | No | NA | | 3 | 0 | 3 | Not-determined | Genome-fragment | NA | | NA | 0 |
| S8-final-viral-combined_k141_38501\|\|full | 1631 | No | NA | | 3 | 0 | 2 | Not-determined | Genome-fragment | NA | | NA | 0 |
| S8-final-viral-combined_k141_153864\|\|full | 2691 | No | NA | | 5 | 0 | 2 | Not-determined | Genome-fragment | NA | | NA | 0 |
| S8-final-viral-combined_k141_84046\|\|full | 2188 | No | NA | | 3 | 0 | 3 | Not-determined | Genome-fragment | NA | | NA | 0 |
| S6-final-viral-combined_k141_20271\|\|full | 10773 | No | NA | | 14 | 4 | 1 | Low-quality | Genome-fragment | | 31.43 | AAI-based (high-confidence) | 0 |
| S8-final-viral-combined_k141_256576\|\|full | 1863 | No | NA | | 4 | 0 | 1 | Not-determined | Genome-fragment | NA | | NA | 0 |
| S8-final-viral-combined_k141_268887\|\|full | 1397 | No | NA | | 3 | 0 | 1 | Not-determined | Genome-fragment | NA | | NA | 0 |
| S8-final-viral-combined_k141_65204\|\|full | 10774 | No | NA | | 11 | 4 | 0 | Low-quality | Genome-fragment | | 25.95 | AAI-based (medium-confidence) | 0 |
| S8-final-viral-combined_k141_61567\|\|full | 1770 | No | NA | | 2 | 0 | 1 | Not-determined | Genome-fragment | NA | | NA | 0 |
| S9-final-viral-combined_k141_63218\|\|full | 10777 | No | NA | | 14 | 0 | 4 | Low-quality | Genome-fragment | | 21.19 | AAI-based (medium-confidence) | 0 |
| S2_k141_30457 | 10817 | No | NA | | 5 | 0 | 5 | Low-quality | Genome-fragment | | 4.08 | AAI-based (high-confidence) | 0 |

| | | | | | | | | | | | | | |
|---|---|---|---|---|---|---|---|---|---|---|---|---|---|
| S8-final-viral-combined_k141_73539\|\|full | 3863 | No | NA | | 4 | 0 | 3 | Not-determined | Genome-fragment | NA | | NA | 0 |
| S8-final-viral-combined_k141_49726\|\|full | 3145 | No | NA | | 5 | 0 | 0 | Not-determined | Genome-fragment | NA | | NA | 0 |
| S4-final-viral-combined_k141_36148\|\|0_partial | 10820 | No | NA | | 15 | 1 | 6 | Low-quality | Genome-fragment | | 7.27 | HMM-based (lower-bound) | 0 |
| S8-final-viral-combined_k141_177505\|\|full | 1641 | No | NA | | 3 | 0 | 1 | Not-determined | Genome-fragment | NA | | NA | 0 |
| S8-final-viral-combined_k141_177507\|\|full | 1332 | No | NA | | 2 | 0 | 1 | Not-determined | Genome-fragment | NA | | NA | 0 |
| S8-final-viral-combined_k141_130847\|\|full | 3571 | No | NA | | 3 | 0 | 2 | Not-determined | Genome-fragment | NA | | NA | 0 |
| S7-final-viral-combined_k141_189339\|\|full | 10829 | No | NA | | 14 | 0 | 0 | Low-quality | Genome-fragment | | 21.93 | AAI-based (medium-confidence) | 0 |
| S8-final-viral-combined_k141_153912\|\|full | 4708 | No | NA | | 5 | 0 | 2 | Not-determined | Genome-fragment | NA | | NA | 0 |
| S5-final-viral-combined_k141_121501\|\|full | 10838 | Yes | | 7397 | 9 | 1 | 2 | Low-quality | Genome-fragment | | 13.24 | AAI-based (high-confidence) | 31.75 |
| S17-k141_122708 | 10839 | No | NA | | 22 | 4 | 0 | Low-quality | Genome-fragment | | 16.75 | AAI-based (high-confidence) | 0 |
| S8-final-viral-combined_k141_38572\|\|full | 4784 | No | NA | | 7 | 0 | 2 | Not-determined | Genome-fragment | NA | | NA | 0 |
| S8-final-viral-combined_k141_119094\|\|full | 2411 | No | NA | | 3 | 0 | 0 | Not-determined | Genome-fragment | NA | | NA | 0 |
| S8-final-viral-combined_k141_177526\|\|full | 5339 | No | NA | | 7 | 0 | 1 | Not-determined | Genome-fragment | NA | | NA | 0 |
| S8-final-viral-combined_k141_268958\|\|full | 3091 | No | NA | | 3 | 0 | 2 | Not-determined | Genome-fragment | NA | | NA | 0 |
| S12-final-viral-combined_k141_38330\|\|full | 10844 | No | NA | | 12 | 1 | 1 | Low-quality | Genome-fragment | | 15.44 | HMM-based (lower-bound) | 0 |
| S1-final-viral-combined_k141_22082\|\|full | 10849 | Yes | | 5930 | 14 | 3 | 4 | Low-quality | Genome-fragment | | 4.47 | HMM-based (lower-bound) | 45.34 |
| S8-final-viral-combined_k141_153930\|\|full | 2296 | No | NA | | 5 | 0 | 0 | Not-determined | Genome-fragment | NA | | NA | 0 |
| S8-final-viral-combined_k141_130872\|\|full | 1778 | No | NA | | 4 | 0 | 4 | Not-determined | Genome-fragment | NA | | NA | 0 |
| S17-final-viral-combined_k141_183065\|\|full | 10866 | No | NA | | 8 | 4 | 0 | Low-quality | Genome-fragment | | 26.99 | AAI-based (high-confidence) | 0 |
| S8-final-viral-combined_k141_73579\|\|full | 2758 | No | NA | | 3 | 0 | 0 | Not-determined | Genome-fragment | NA | | NA | 0 |
| S8-final-viral-combined_k141_268980\|\|full | 1082 | No | NA | | 2 | 0 | 1 | Not-determined | Genome-fragment | NA | | NA | 0 |
| S8-final-viral-combined_k141_142745\|\|full | 1717 | No | NA | | 3 | 0 | 2 | Not-determined | Genome-fragment | NA | | NA | 0 |
| S8-final-viral-combined_k141_49824\|\|full | 1307 | No | NA | | 2 | 0 | 0 | Not-determined | Genome-fragment | NA | | NA | 0 |
| S8-final-viral-combined_k141_107424\|\|full | 1199 | No | NA | | 2 | 0 | 0 | Not-determined | Genome-fragment | NA | | NA | 0 |
| S8-final-viral-combined_k141_38602\|\|full | 4651 | No | NA | | 3 | 0 | 2 | Not-determined | Genome-fragment | NA | | NA | 0 |
| S7-final-viral-combined_k141_193445\|\|full | 10868 | No | NA | | 9 | 0 | 3 | Low-quality | Genome-fragment | | 11.72 | AAI-based (medium-confidence) | 0 |
| S8-final-viral-combined_k141_61694\|\|full | 1694 | No | NA | | 3 | 0 | 0 | Not-determined | Genome-fragment | NA | | NA | 0 |
| S8-final-viral-combined_k141_14433\|\|full | 1306 | No | NA | | 3 | 0 | 2 | Not-determined | Genome-fragment | NA | | NA | 0 |
| S2-final-viral-combined_k141_154612\|\|0_partial | 10887 | Yes | | 7001 | 15 | 2 | 5 | Low-quality | Genome-fragment | | 4.96 | HMM-based (lower-bound) | 35.69 |
| S1-final-viral-combined_k141_340270\|\|full | 10891 | No | NA | | 17 | 0 | 7 | Low-quality | Genome-fragment | | 22.53 | AAI-based (high-confidence) | 0 |
| S8-final-viral-combined_k141_256653\|\|full | 3148 | No | NA | | 3 | 0 | 2 | Not-determined | Genome-fragment | NA | | NA | 0 |
| S8-final-viral-combined_k141_107453\|\|full | 5222 | No | NA | | 5 | 0 | 4 | Not-determined | Genome-fragment | NA | | NA | 0 |
| S8-final-viral-combined_k141_61699\|\|full | 9412 | No | NA | | 10 | 0 | 2 | Not-determined | Genome-fragment | NA | | NA | 0 |
| S5-final-viral-combined_k141_40893\|\|full | 10891 | No | NA | | 22 | 9 | 0 | Low-quality | Genome-fragment | | 29.36 | AAI-based (high-confidence) | 0 |
| S7_k141_31477 | 10895 | No | NA | | 10 | 4 | 0 | Low-quality | Genome-fragment | | 29.15 | AAI-based (high-confidence) | 0 |
| S8-final-viral-combined_k141_84207\|\|full | 21394 | No | NA | | 13 | 0 | 10 | Not-determined | Genome-fragment | NA | | NA | 0 |
| S8-final-viral-combined_k141_107489\|\|full | 2406 | No | NA | | 3 | 0 | 3 | Not-determined | Genome-fragment | NA | | NA | 0 |
| S8-final-viral-combined_k141_188566\|\|full | 9986 | No | NA | | 5 | 0 | 0 | Not-determined | Genome-fragment | NA | | NA | 0 |
| S8-final-viral-combined_k141_96480\|\|full | 1703 | No | NA | | 2 | 0 | 0 | Not-determined | Genome-fragment | NA | | NA | 0 |
| S8-final-viral-combined_k141_107498\|\|full | 3830 | No | NA | | 3 | 0 | 1 | Not-determined | Genome-fragment | NA | | NA | 0 |
| S2-final-viral-combined_k141_232295\|\|full | 10896 | No | NA | | 15 | 1 | 0 | Low-quality | Genome-fragment | | 17.89 | AAI-based (medium-confidence) | 0 |
| S8-final-viral-combined_k141_269082\|\|full | 2170 | No | NA | | 2 | 0 | 1 | Not-determined | Genome-fragment | NA | | NA | 0 |
| S3-final-viral-combined_k141_132729\|\|full | 10905 | No | NA | | 16 | 10 | 0 | Low-quality | Genome-fragment | | 31.13 | AAI-based (high-confidence) | 0 |
| S1-final-viral-combined_k141_127794\|\|full | 10906 | No | NA | | 9 | 1 | 4 | Low-quality | Genome-fragment | | 12.36 | HMM-based (lower-bound) | 0 |
| S8-final-viral-combined_k141_211451\|\|full | 2063 | No | NA | | 3 | 0 | 2 | Not-determined | Genome-fragment | NA | | NA | 0 |
| S5-final-viral-combined_k141_16811\|\|full | 10908 | No | NA | | 16 | 8 | 0 | Low-quality | Genome-fragment | | 26.74 | AAI-based (high-confidence) | 0 |
| S9_k141_8129 | 10915 | No | NA | | 13 | 12 | 0 | Low-quality | Genome-fragment | | 27.06 | AAI-based (high-confidence) | 0 |
| S8-final-viral-combined_k141_234873\|\|full | 4391 | No | NA | | 9 | 0 | 1 | Not-determined | Genome-fragment | NA | | NA | 0 |
| S8-final-viral-combined_k141_96493\|\|full | 3591 | No | NA | | 3 | 0 | 2 | Not-determined | Genome-fragment | NA | | NA | 0 |
| S8-final-viral-combined_k141_246288\|\|full | 1800 | No | NA | | 3 | 0 | 1 | Not-determined | Genome-fragment | NA | | NA | 0 |
| S12-final-viral-combined_k141_202288\|\|full | 10920 | No | NA | | 22 | 3 | 1 | Low-quality | Genome-fragment | | 30.51 | AAI-based (high-confidence) | 0 |
| S8-final-viral-combined_k141_234885\|\|full | 6934 | No | NA | | 5 | 0 | 1 | Not-determined | Genome-fragment | NA | | NA | 0 |
| S8-final-viral-combined_k141_199584\|\|full | 1428 | No | NA | | 2 | 0 | 2 | Not-determined | Genome-fragment | NA | | NA | 0 |
| S2-final-viral-combined_k141_197284\|\|full | 10926 | No | NA | | 9 | 1 | 5 | Low-quality | Genome-fragment | | 6.89 | HMM-based (lower-bound) | 0 |
| S8-final-viral-combined_k141_154065\|\|full | 2395 | No | NA | | 5 | 0 | 2 | Not-determined | Genome-fragment | NA | | NA | 0 |
| S8-final-viral-combined_k141_78629\|\|full | 10931 | No | NA | | 15 | 6 | 1 | Low-quality | Genome-fragment | | 23.79 | AAI-based (high-confidence) | 0 |
| S8-final-viral-combined_k141_154087\|\|full | 1382 | No | NA | | 2 | 0 | 1 | Not-determined | Genome-fragment | NA | | NA | 0 |
| S8-final-viral-combined_k141_84247\|\|full | 1950 | No | NA | | 2 | 0 | 1 | Not-determined | Genome-fragment | NA | | NA | 0 |
| S8-final-viral-combined_k141_84260\|\|full | 1056 | No | NA | | 3 | 0 | 0 | Not-determined | Genome-fragment | NA | | NA | 0 |
| S8-final-viral-combined_k141_222848\|\|full | 2095 | No | NA | | 3 | 0 | 2 | Not-determined | Genome-fragment | NA | | NA | 0 |
| S5-final-viral-combined_k141_157993\|\|full | 10932 | No | NA | | 9 | 0 | 3 | Low-quality | Genome-fragment | | 8.3 | AAI-based (medium-confidence) | 0 |
| S8-final-viral-combined_k141_199623\|\|full | 4258 | No | NA | | 8 | 0 | 1 | Not-determined | Genome-fragment | NA | | NA | 0 |
| S8-final-viral-combined_k141_177673\|\|full | 3492 | No | NA | | 5 | 0 | 3 | Not-determined | Genome-fragment | NA | | NA | 0 |
| S5_k141_103467 | 10935 | No | NA | | 19 | 12 | 1 | Low-quality | Genome-fragment | | 27.32 | AAI-based (high-confidence) | 0 |
| S2-final-viral-combined_k141_239431\|\|0_partial | 10946 | No | NA | | 12 | 1 | 6 | Low-quality | Genome-fragment | | 7.44 | AAI-based (medium-confidence) | 0 |
| S8-final-viral-combined_k141_27614\|\|full | 1698 | No | NA | | 3 | 0 | 0 | Not-determined | Genome-fragment | NA | | NA | 0 |
| S8-final-viral-combined_k141_14592\|\|full | 1967 | No | NA | | 2 | 0 | 0 | Not-determined | Genome-fragment | NA | | NA | 0 |
| S8-final-viral-combined_k141_27627\|\|full | 2962 | No | NA | | 4 | 0 | 2 | Not-determined | Genome-fragment | NA | | NA | 0 |
| S2-final-viral-combined_k141_214152\|\|full | 10958 | No | NA | | 13 | 1 | 2 | Low-quality | Genome-fragment | | 6.11 | HMM-based (lower-bound) | 0 |
| S8-final-viral-combined_k141_107537\|\|full | 1573 | No | NA | | 4 | 0 | 0 | Not-determined | Genome-fragment | NA | | NA | 0 |
| S8-final-viral-combined_k141_131104\|\|full | 17805 | No | NA | | 18 | 0 | 7 | Not-determined | Genome-fragment | NA | | NA | 0 |
| S8-final-viral-combined_k141_177766\|\|full | 3163 | No | NA | | 4 | 0 | 4 | Not-determined | Genome-fragment | NA | | NA | 0 |
| S8-final-viral-combined_k141_107581\|\|full | 1898 | No | NA | | 5 | 0 | 1 | Not-determined | Genome-fragment | NA | | NA | 0 |
| S6-final-viral-combined_k141_20586\|\|full | 10958 | No | NA | | 5 | 0 | 0 | Low-quality | Genome-fragment | | 27.56 | AAI-based (high-confidence) | 0 |
| S8-final-viral-combined_k141_107590\|\|full | 1770 | No | NA | | 2 | 0 | 0 | Not-determined | Genome-fragment | NA | | NA | 0 |
| S8-final-viral-combined_k141_256786\|\|full | 1376 | No | NA | | 2 | 0 | 1 | Not-determined | Genome-fragment | NA | | NA | 0 |
| S3_k141_99318 | 10962 | No | NA | | 17 | 10 | 0 | Low-quality | Genome-fragment | | 10.49 | AAI-based (high-confidence) | 0 |
| S8-final-viral-combined_k141_188760\|\|full | 2367 | No | NA | | 3 | 0 | 1 | Not-determined | Genome-fragment | NA | | NA | 0 |
| S12_k141_116916 | 10962 | No | NA | | 15 | 1 | 3 | Low-quality | Genome-fragment | | 5.89 | HMM-based (lower-bound) | 0 |
| S8-final-viral-combined_k141_235045\|\|full | 1523 | No | NA | | 2 | 0 | 2 | Not-determined | Genome-fragment | NA | | NA | 0 |
| S12_k141_112191 | 10978 | No | NA | | 9 | 2 | 1 | Low-quality | Genome-fragment | | 14.85 | AAI-based (high-confidence) | 0 |
| S8-final-viral-combined_k141_61982\|\|full | 3202 | No | NA | | 3 | 0 | 2 | Not-determined | Genome-fragment | NA | | NA | 0 |
| S7-final-viral-combined_k141_212263\|\|full | 10985 | No | NA | | 16 | 4 | 0 | Low-quality | Genome-fragment | | 8.83 | HMM-based (lower-bound) | 0 |
| S2_k141_197876 | 10985 | No | NA | | 5 | 0 | 3 | Low-quality | Genome-fragment | | 1.87 | AAI-based (medium-confidence) | 0 |
| S8-final-viral-combined_k141_119317\|\|full | 3336 | No | NA | | 3 | 0 | 2 | Not-determined | Genome-fragment | NA | | NA | 0 |
| S8-final-viral-combined_k141_96652\|\|full | 1284 | No | NA | | 3 | 0 | 1 | Not-determined | Genome-fragment | NA | | NA | 0 |
| S8-final-viral-combined_k141_50020\|\|full | 5364 | No | NA | | 5 | 0 | 4 | Not-determined | Genome-fragment | NA | | NA | 0 |
| S2_k141_62776 | 10992 | No | NA | | 14 | 5 | 0 | Low-quality | Genome-fragment | | 38.37 | HMM-based (lower-bound) | 0 |
| S9-final-viral-combined_k141_85775\|\|full | 11004 | No | NA | | 9 | 5 | 0 | Low-quality | Genome-fragment | | 28.56 | AAI-based (medium-confidence) | 0 |
| S5-final-viral-combined_k141_143458\|\|full | 11005 | No | NA | | 14 | 7 | 0 | Low-quality | Genome-fragment | | 24.49 | AAI-based (high-confidence) | 0 |
| S8-final-viral-combined_k141_188883\|\|full | 2676 | No | NA | | 2 | 0 | 2 | Not-determined | Genome-fragment | NA | | NA | 0 |
| S12-final-viral-combined_k141_106299\|\|full | 11016 | No | NA | | 13 | 6 | 0 | Low-quality | Genome-fragment | | 6.28 | AAI-based (medium-confidence) | 0 |
| S12-final-viral-combined_k141_268660\|\|full | 11046 | No | NA | | 16 | 5 | 0 | Low-quality | Genome-fragment | | 19.45 | HMM-based (lower-bound) | 0 |
| S8-final-viral-combined_k141_4027\|\|full | 1900 | No | NA | | 3 | 0 | 0 | Not-determined | Genome-fragment | NA | | NA | 0 |
| S8-final-viral-combined_k141_131224\|\|full | 2558 | No | NA | | 4 | 0 | 0 | Not-determined | Genome-fragment | NA | | NA | 0 |
| S8-final-viral-combined_k141_73767\|\|full | 1014 | No | NA | | 3 | 0 | 1 | Not-determined | Genome-fragment | NA | | NA | 0 |
| S8-final-viral-combined_k141_62124\|\|full | 3235 | No | NA | | 4 | 0 | 2 | Not-determined | Genome-fragment | NA | | NA | 0 |
| S2-final-viral-combined_k141_145250\|\|full | 11046 | No | NA | | 12 | 4 | 0 | Low-quality | Genome-fragment | | 26.34 | AAI-based (high-confidence) | 0 |
| S9-final-viral-combined_k141_109742\|\|0_partial | 11055 | No | NA | | 12 | 1 | 7 | Low-quality | Genome-fragment | | 9.92 | HMM-based (lower-bound) | 0 |
| S8-final-viral-combined_k141_269329\|\|full | 1181 | No | NA | | 4 | 0 | 0 | Not-determined | Genome-fragment | NA | | NA | 0 |
| S1-final-viral-combined_k141_118100\|\|full | 11056 | No | NA | | 9 | 0 | 5 | Low-quality | Genome-fragment | | 5.66 | AAI-based (medium-confidence) | 0 |
| S8_k141_246316 | 11057 | No | NA | | 8 | 0 | 0 | Low-quality | Genome-fragment | | 7.63 | AAI-based (high-confidence) | 0 |
| S8-final-viral-combined_k141_165662\|\|full | 2399 | No | NA | | 2 | 0 | 1 | Not-determined | Genome-fragment | NA | | NA | 0 |
| S8-final-viral-combined_k141_14794\|\|full | 1435 | No | NA | | 2 | 0 | 2 | Not-determined | Genome-fragment | NA | | NA | 0 |
| S12-final-viral-combined_k141_301396\|\|full | 11059 | No | NA | | 13 | 1 | 3 | Low-quality | Genome-fragment | | 18.89 | AAI-based (medium-confidence) | 0 |
| S11-final-viral-combined_k141_26341\|\|full | 11067 | No | NA | | 15 | 1 | 2 | Low-quality | Genome-fragment | | 7.15 | HMM-based (lower-bound) | 0 |
| S8-final-viral-combined_k141_131283\|\|full | 1200 | No | NA | | 2 | 0 | 0 | Not-determined | Genome-fragment | NA | | NA | 0 |
| S8-final-viral-combined_k141_165673\|\|full | 1091 | No | NA | | 2 | 0 | 1 | Not-determined | Genome-fragment | NA | | NA | 0 |
| S1-final-viral-combined_k141_156653\|\|full | 11068 | No | NA | | 13 | 10 | 0 | Low-quality | Genome-fragment | | 26.19 | AAI-based (medium-confidence) | 0 |
| S8-final-viral-combined_k141_177882\|\|full | 1066 | No | NA | | 2 | 0 | 0 | Not-determined | Genome-fragment | NA | | NA | 0 |
| S8-final-viral-combined_k141_154342\|\|full | 1528 | No | NA | | 2 | 0 | 1 | Not-determined | Genome-fragment | NA | | NA | 0 |
| S9_k141_50916 | 11072 | No | NA | | 15 | 0 | 4 | Low-quality | Genome-fragment | | 8.95 | AAI-based (high-confidence) | 0 |
| S8-final-viral-combined_k141_177897\|\|full | 2600 | No | NA | | 2 | 0 | 1 | Not-determined | Genome-fragment | NA | | NA | 0 |
| S2-final-viral-combined_k141_65567\|\|full | 11073 | No | NA | | 15 | 1 | 0 | Low-quality | Genome-fragment | | 16.5 | AAI-based (high-confidence) | 0 |
| S12_k141_371328 | 11075 | No | NA | | 15 | 0 | 4 | Low-quality | Genome-fragment | | 7.77 | AAI-based (medium-confidence) | 0 |
| S8-final-viral-combined_k141_143135\|\|full | 7488 | No | NA | | 6 | 0 | 5 | Not-determined | Genome-fragment | NA | | NA | 0 |
| S4-final-viral-combined_k141_89288\|\|full | 11078 | No | NA | | 10 | 0 | 6 | Low-quality | Genome-fragment | | 3.61 | AAI-based (medium-confidence) | 0 |
| S8-final-viral-combined_k141_14840\|\|full | 1193 | No | NA | | 3 | 0 | 2 | Not-determined | Genome-fragment | NA | | NA | 0 |
| S8-final-viral-combined_k141_246447\|\|full | 1443 | No | NA | | 2 | 0 | 2 | Not-determined | Genome-fragment | NA | | NA | 0 |
| S11-final-viral-combined_k141_215562\|\|full | 11080 | No | NA | | 9 | 3 | 0 | Low-quality | Genome-fragment | | 8.36 | AAI-based (high-confidence) | 0 |
| S8-final-viral-combined_k141_14872\|\|full | 2370 | No | NA | | 3 | 0 | 0 | Not-determined | Genome-fragment | NA | | NA | 0 |
| S8-final-viral-combined_k141_62226\|\|full | 1777 | No | NA | | 4 | 0 | 1 | Not-determined | Genome-fragment | NA | | NA | 0 |
| S8-final-viral-combined_k141_188950\|\|full | 1092 | No | NA | | 2 | 0 | 1 | Not-determined | Genome-fragment | NA | | NA | 0 |
| S8-final-viral-combined_k141_165736\|\|full | 1869 | No | NA | | 2 | 0 | 1 | Not-determined | Genome-fragment | NA | | NA | 0 |
| S3-final-viral-combined_k141_2481\|\|full | 11084 | No | NA | | 9 | 0 | 6 | Low-quality | Genome-fragment | | 3.46 | AAI-based (medium-confidence) | 0 |
| S3-final-viral-combined_k141_115283\|\|full | 11085 | Yes | | 10548 | 9 | 2 | 1 | Low-quality | Genome-fragment | | 6.9 | AAI-based (medium-confidence) | 4.84 |
| S11-final-viral-combined_k141_47214\|\|full | 11094 | No | NA | | 18 | 2 | 1 | Low-quality | Genome-fragment | | 24.15 | AAI-based (high-confidence) | 0 |
| S9-final-viral-combined_k141_39747\|\|full | 11098 | Yes | | 6992 | 12 | 3 | 2 | Low-quality | Genome-fragment | | 11.6 | AAI-based (medium-confidence) | 37 |
| S8-final-viral-combined_k141_39136\|\|full | 2148 | No | NA | | 2 | 0 | 1 | Not-determined | Genome-fragment | NA | | NA | 0 |
| S8-final-viral-combined_k141_165748\|\|full | 17947 | No | NA | | 17 | 0 | 9 | Not-determined | Genome-fragment | NA | | NA | 0 |
| S8-final-viral-combined_k141_207717\|\|0_partial | 11103 | Yes | | 2633 | 11 | 2 | 7 | Low-quality | Genome-fragment | | 4.55 | HMM-based (lower-bound) | 76.29 |
| S12-final-viral-combined_k141_64423\|\|full | 11111 | No | NA | | 10 | 0 | 6 | Low-quality | Genome-fragment | | 6.59 | AAI-based (medium-confidence) | 0 |
| S8-final-viral-combined_k141_39145\|\|full | 3250 | No | NA | | 3 | 0 | 1 | Not-determined | Genome-fragment | NA | | NA | 0 |
| S12-final-viral-combined_k141_273721\|\|0_partial | 11120 | No | NA | | 14 | 1 | 2 | Low-quality | Genome-fragment | | 17.36 | HMM-based (lower-bound) | 0 |
| S8-final-viral-combined_k141_235236\|\|full | 5165 | No | NA | | 4 | 0 | 3 | Not-determined | Genome-fragment | NA | | NA | 0 |
| S8_k141_253777 | 11146 | No | NA | | 14 | 2 | 1 | Low-quality | Genome-fragment | | 21.78 | AAI-based (high-confidence) | 0 |
| S7-final-viral-combined_k141_95028\|\|0_partial | 11147 | No | NA | | 12 | 1 | 4 | Low-quality | Genome-fragment | | 16.68 | HMM-based (lower-bound) | 0 |
| S12-final-viral-combined_k141_186337\|\|full | 11156 | No | NA | | 9 | 0 | 1 | Low-quality | Genome-fragment | | 13.58 | AAI-based (medium-confidence) | 0 |
| S12-final-viral-combined_k141_171971\|\|0_partial | 11165 | No | NA | | 12 | 1 | 1 | Low-quality | Genome-fragment | | 15.9 | HMM-based (lower-bound) | 0 |
| S8-final-viral-combined_k141_269525\|\|full | 3080 | No | NA | | 3 | 0 | 2 | Not-determined | Genome-fragment | NA | | NA | 0 |
| S8-final-viral-combined_k141_14975\|\|full | 2874 | No | NA | | 4 | 0 | 2 | Not-determined | Genome-fragment | NA | | NA | 0 |
| S8-final-viral-combined_k141_269537\|\|full | 2192 | No | NA | | 2 | 0 | 2 | Not-determined | Genome-fragment | NA | | NA | 0 |
| S2_k141_199244 | 11170 | Yes | | 8741 | 15 | 5 | 2 | Low-quality | Genome-fragment | | 5.16 | AAI-based (medium-confidence) | 21.75 |
| S8-final-viral-combined_k141_15009\|\|full | 1613 | No | NA | | 2 | 0 | 2 | Not-determined | Genome-fragment | NA | | NA | 0 |
| S8-final-viral-combined_k141_143239\|\|full | 1037 | No | NA | | 2 | 0 | 0 | Not-determined | Genome-fragment | NA | | NA | 0 |
| S8-final-viral-combined_k141_15011\|\|full | 1245 | No | NA | | 2 | 0 | 0 | Not-determined | Genome-fragment | NA | | NA | 0 |
| S8-final-viral-combined_k141_96982\|\|full | 1660 | No | NA | | 2 | 0 | 1 | Not-determined | Genome-fragment | NA | | NA | 0 |
| S8-final-viral-combined_k141_39228\|\|full | 1994 | No | NA | | 2 | 0 | 2 | Not-determined | Genome-fragment | NA | | NA | 0 |
| S12-final-viral-combined_k141_50426\|\|0_partial | 11177 | No | NA | | 11 | 0 | 9 | Low-quality | Genome-fragment | | 3.15 | AAI-based (medium-confidence) | 0 |
| S4-final-viral-combined_k141_125813\|\|full | 11178 | No | NA | | 10 | 2 | 1 | Low-quality | Genome-fragment | | 15.91 | HMM-based (lower-bound) | 0 |
| S11-final-viral-combined_k141_212078\|\|0_partial | 11182 | No | NA | | 15 | 0 | 4 | Low-quality | Genome-fragment | | 10.38 | AAI-based (medium-confidence) | 0 |
| S8-final-viral-combined_k141_178108\|\|full | 2671 | No | NA | | 3 | 0 | 1 | Not-determined | Genome-fragment | NA | | NA | 0 |
| S8-final-viral-combined_k141_235381\|\|full | 1960 | No | NA | | 3 | 0 | 2 | Not-determined | Genome-fragment | NA | | NA | 0 |
| S8-final-viral-combined_k141_62417\|\|full | 4345 | No | NA | | 4 | 0 | 2 | Not-determined | Genome-fragment | NA | | NA | 0 |
| S8-final-viral-combined_k141_27829\|\|full | 3183 | No | NA | | 3 | 0 | 2 | Not-determined | Genome-fragment | NA | | NA | 0 |
| S8-final-viral-combined_k141_223263\|\|full | 4558 | No | NA | | 4 | 0 | 3 | Not-determined | Genome-fragment | NA | | NA | 0 |
| S8-final-viral-combined_k141_246593\|\|full | 2930 | No | NA | | 3 | 0 | 2 | Not-determined | Genome-fragment | NA | | NA | 0 |
| S8-final-viral-combined_k141_15044\|\|full | 5480 | No | NA | | 5 | 0 | 1 | Not-determined | Genome-fragment | NA | | NA | 0 |
| S8-final-viral-combined_k141_235394\|\|full | 3389 | No | NA | | 4 | 0 | 3 | Not-determined | Genome-fragment | NA | | NA | 0 |
| S8-final-viral-combined_k141_154550\|\|full | 2502 | No | NA | | 3 | 0 | 3 | Not-determined | Genome-fragment | NA | | NA | 0 |
| S8-final-viral-combined_k141_50396\|\|full | 2574 | No | NA | | 3 | 0 | 0 | Not-determined | Genome-fragment | NA | | NA | 0 |
| S11_k141_29840 | 11190 | No | NA | | 13 | 8 | 0 | Low-quality | Genome-fragment | | 21.18 | AAI-based (high-confidence) | 0 |
| S8-final-viral-combined_k141_212056\|\|full | 1877 | No | NA | | 4 | 0 | 0 | Not-determined | Genome-fragment | NA | | NA | 0 |
| S11_k141_249433 | 11199 | No | NA | | 5 | 0 | 5 | Low-quality | Genome-fragment | | 4.92 | AAI-based (high-confidence) | 0 |
| S8-final-viral-combined_k141_235439\|\|full | 1297 | No | NA | | 3 | 0 | 0 | Not-determined | Genome-fragment | NA | | NA | 0 |
| S8-final-viral-combined_k141_246664\|\|full | 2501 | No | NA | | 2 | 0 | 0 | Not-determined | Genome-fragment | NA | | NA | 0 |
| S8-final-viral-combined_k141_74205\|\|full | 2469 | No | NA | | 4 | 0 | 2 | Not-determined | Genome-fragment | NA | | NA | 0 |
| S9-final-viral-combined_k141_40313\|\|full | 11201 | No | NA | | 23 | 5 | 0 | Low-quality | Genome-fragment | | 27.68 | AAI-based (high-confidence) | 0 |
| S8-final-viral-combined_k141_235459\|\|full | 1687 | No | NA | | 2 | 0 | 0 | Not-determined | Genome-fragment | NA | | NA | 0 |
| S8-final-viral-combined_k141_244219\|\|full | 11206 | No | NA | | 12 | 6 | 0 | Low-quality | Genome-fragment | | 11.07 | AAI-based (high-confidence) | 0 |
| S8-final-viral-combined_k141_189197\|\|full | 2255 | No | NA | | 2 | 0 | 1 | Not-determined | Genome-fragment | NA | | NA | 0 |
| S8-final-viral-combined_k141_133746\|\|full | 11226 | No | NA | | 16 | 1 | 1 | Low-quality | Genome-fragment | | 24.81 | AAI-based (high-confidence) | 0 |
| S8-final-viral-combined_k141_88389\|\|0_partial | 11232 | Yes | | 4464 | 8 | 1 | 3 | Low-quality | Genome-fragment | | 2.89 | HMM-based (lower-bound) | 60.26 |
| S8-final-viral-combined_k141_199815\|\|full | 7879 | No | NA | | 8 | 0 | 5 | Not-determined | Genome-fragment | NA | | NA | 0 |
| S8-final-viral-combined_k141_235514\|\|full | 1576 | No | NA | | 2 | 0 | 2 | Not-determined | Genome-fragment | NA | | NA | 0 |
| S2-final-viral-combined_k141_135149\|\|full | 11261 | No | NA | | 18 | 6 | 1 | Low-quality | Genome-fragment | | 18.83 | AAI-based (high-confidence) | 0 |
| S8-final-viral-combined_k141_212170\|\|full | 1003 | No | NA | | 2 | 0 | 0 | Not-determined | Genome-fragment | NA | | NA | 0 |
| S8-final-viral-combined_k141_212171\|\|full | 2201 | No | NA | | 3 | 0 | 3 | Not-determined | Genome-fragment | NA | | NA | 0 |
| S1-final-viral-combined_k141_99434\|\|full | 11266 | No | NA | | 13 | 1 | 1 | Low-quality | Genome-fragment | | 17.05 | HMM-based (lower-bound) | 0 |
| S8-final-viral-combined_k141_199835\|\|full | 1070 | No | NA | | 2 | 0 | 0 | Not-determined | Genome-fragment | NA | | NA | 0 |
| S3-final-viral-combined_k141_41523\|\|full | 11267 | No | NA | | 13 | 9 | 0 | Low-quality | Genome-fragment | | 30.97 | AAI-based (high-confidence) | 0 |
| S8-final-viral-combined_k141_257058\|\|full | 3697 | No | NA | | 5 | 0 | 3 | Not-determined | Genome-fragment | NA | | NA | 0 |
| S8-final-viral-combined_k141_74661\|\|full | 11270 | No | NA | | 8 | 0 | 6 | Low-quality | Genome-fragment | | 6.14 | AAI-based (medium-confidence) | 0 |
| S8-final-viral-combined_k141_97112\|\|full | 4909 | No | NA | | 4 | 0 | 4 | Not-determined | Genome-fragment | NA | | NA | 0 |
| S8-final-viral-combined_k141_212193\|\|full | 1970 | No | NA | | 3 | 0 | 0 | Not-determined | Genome-fragment | NA | | NA | 0 |
| S8-final-viral-combined_k141_165914\|\|full | 4737 | No | NA | | 4 | 0 | 3 | Not-determined | Genome-fragment | NA | | NA | 0 |
| S8-final-viral-combined_k141_143445\|\|full | 1875 | No | NA | | 2 | 0 | 1 | Not-determined | Genome-fragment | NA | | NA | 0 |
| S8-final-viral-combined_k141_4374\|\|full | 1796 | No | NA | | 2 | 0 | 2 | Not-determined | Genome-fragment | NA | | NA | 0 |
| S1_k141_35582 | 11281 | No | NA | | 23 | 4 | 1 | Low-quality | Genome-fragment | | 26.93 | AAI-based (high-confidence) | 0 |
| S8-final-viral-combined_k141_119752\|\|full | 3798 | No | NA | | 7 | 0 | 2 | Not-determined | Genome-fragment | NA | | NA | 0 |
| S8-final-viral-combined_k141_235587\|\|full | 1672 | No | NA | | 2 | 0 | 0 | Not-determined | Genome-fragment | NA | | NA | 0 |
| S8-final-viral-combined_k141_269790\|\|full | 2229 | No | NA | | 3 | 0 | 1 | Not-determined | Genome-fragment | NA | | NA | 0 |
| S12-final-viral-combined_k141_54497\|\|full | 11285 | No | NA | | 17 | 4 | 0 | Low-quality | Genome-fragment | | 26.42 | AAI-based (high-confidence) | 0 |
| S4-final-viral-combined_k141_85444\|\|0_partial | 11294 | No | NA | | 11 | 1 | 5 | Low-quality | Genome-fragment | | 7.31 | HMM-based (lower-bound) | 0 |
| S5-final-viral-combined_k141_136478\|\|full | 11301 | No | NA | | 6 | 0 | 5 | Low-quality | Genome-fragment | | 4.46 | AAI-based (medium-confidence) | 0 |
| S8-final-viral-combined_k141_269806\|\|full | 1213 | No | NA | | 2 | 0 | 1 | Not-determined | Genome-fragment | NA | | NA | 0 |
| S2-final-viral-combined_k141_43699\|\|0_partial | 11304 | No | NA | | 14 | 1 | 11 | Low-quality | Genome-fragment | | 7.66 | HMM-based (lower-bound) | 0 |
| S8-final-viral-combined_k141_97179\|\|full | 2298 | No | NA | | 2 | 0 | 1 | Not-determined | Genome-fragment | NA | | NA | 0 |
| S8-final-viral-combined_k141_15235\|\|full | 1573 | No | NA | | 2 | 0 | 1 | Not-determined | Genome-fragment | NA | | NA | 0 |
| S8-final-viral-combined_k141_74275\|\|full | 39935 | Yes | | 22987 | 36 | 2 | 6 | Medium-quality | Genome-fragment | | 50.81 | AAI-based (medium-confidence) | 42.44 |
| S8-final-viral-combined_k141_62705\|\|full | 1179 | No | NA | | 2 | 0 | 2 | Not-determined | Genome-fragment | NA | | NA | 0 |
| S8-final-viral-combined_k141_97205\|\|full | 2884 | No | NA | | 2 | 0 | 2 | Not-determined | Genome-fragment | NA | | NA | 0 |
| S8-final-viral-combined_k141_223445\|\|full | 1559 | No | NA | | 3 | 0 | 0 | Not-determined | Genome-fragment | NA | | NA | 0 |
| S8-final-viral-combined_k141_212267\|\|full | 1332 | No | NA | | 3 | 0 | 0 | Not-determined | Genome-fragment | NA | | NA | 0 |
| S8-final-viral-combined_k141_74309\|\|full | 2524 | No | NA | | 2 | 0 | 2 | Not-determined | Genome-fragment | NA | | NA | 0 |
| S6_k141_120175 | 11308 | No | NA | | 14 | 1 | 4 | Low-quality | Genome-fragment | | 4.87 | HMM-based (lower-bound) | 0 |
| S8-final-viral-combined_k141_15286\|\|full | 2559 | No | NA | | 3 | 0 | 2 | Not-determined | Genome-fragment | NA | | NA | 0 |
| S7-final-viral-combined_k141_40737\|\|full | 11309 | No | NA | | 15 | 12 | 0 | Low-quality | Genome-fragment | | 23.6 | AAI-based (high-confidence) | 0 |
| S9-final-viral-combined_k141_11616\|\|full | 11311 | No | NA | | 7 | 0 | 6 | Low-quality | Genome-fragment | | 4.89 | AAI-based (medium-confidence) | 0 |
| S8-final-viral-combined_k141_246861\|\|full | 2473 | No | NA | | 4 | 0 | 1 | Not-determined | Genome-fragment | NA | | NA | 0 |
| S5-final-viral-combined_k141_187294\|\|full | 11313 | No | NA | | 23 | 6 | 0 | Low-quality | Genome-fragment | | 28.03 | AAI-based (medium-confidence) | 0 |
| S8-final-viral-combined_k141_15316\|\|full | 1255 | No | NA | | 2 | 0 | 0 | Not-determined | Genome-fragment | NA | | NA | 0 |
| S7-final-viral-combined_k141_6280\|\|full | 11313 | No | NA | | 11 | 5 | 0 | Low-quality | Genome-fragment | | 8.35 | AAI-based (medium-confidence) | 0 |
| S8-final-viral-combined_k141_50685\|\|full | 2149 | No | NA | | 3 | 0 | 1 | Not-determined | Genome-fragment | NA | | NA | 0 |
| S17-final-viral-combined_k141_233852\|\|full | 11318 | No | NA | | 10 | 0 | 6 | Low-quality | Genome-fragment | | 6.3 | AAI-based (medium-confidence) | 0 |
| S8-final-viral-combined_k141_269906\|\|full | 1644 | No | NA | | 3 | 0 | 2 | Not-determined | Genome-fragment | NA | | NA | 0 |
| S8-final-viral-combined_k141_131735\|\|full | 2118 | No | NA | | 3 | 0 | 2 | Not-determined | Genome-fragment | NA | | NA | 0 |
| S8-final-viral-combined_k141_50700\|\|full | 3371 | No | NA | | 5 | 0 | 2 | Not-determined | Genome-fragment | NA | | NA | 0 |
| S2-final-viral-combined_k141_171237\|\|full | 11323 | No | NA | | 10 | 1 | 0 | Low-quality | Genome-fragment | | 20.85 | HMM-based (lower-bound) | 0 |
| S8-final-viral-combined_k141_223495\|\|full | 3134 | No | NA | | 2 | 0 | 1 | Not-determined | Genome-fragment | NA | | NA | 0 |
| S7_k141_82868 | 11324 | No | NA | | 15 | 0 | 1 | Low-quality | Genome-fragment | | 18.91 | AAI-based (high-confidence) | 0 |
| S8-final-viral-combined_k141_223503\|\|full | 4074 | No | NA | | 3 | 0 | 0 | Not-determined | Genome-fragment | NA | | NA | 0 |
| S8-final-viral-combined_k141_189401\|\|full | 1513 | No | NA | | 3 | 0 | 0 | Not-determined | Genome-fragment | NA | | NA | 0 |
| S1-final-viral-combined_k141_172194\|\|0_partial | 11326 | No | NA | | 11 | 0 | 10 | Low-quality | Genome-fragment | | 5.13 | AAI-based (high-confidence) | 0 |
| S8-final-viral-combined_k141_178503\|\|full | 2631 | No | NA | | 3 | 0 | 3 | Not-determined | Genome-fragment | NA | | NA | 0 |
| S12_k141_218683 | 11327 | No | NA | | 12 | 1 | 3 | Low-quality | Genome-fragment | | 16.09 | AAI-based (medium-confidence) | 0 |
| S8-final-viral-combined_k141_189427\|\|full | 2060 | No | NA | | 2 | 0 | 2 | Not-determined | Genome-fragment | NA | | NA | 0 |
| S8-final-viral-combined_k141_28147\|\|full | 8572 | No | NA | | 8 | 0 | 5 | Not-determined | Genome-fragment | NA | | NA | 0 |
| S8-final-viral-combined_k141_108248\|\|full | 24739 | No | NA | | 25 | 0 | 18 | Not-determined | Genome-fragment | NA | | NA | 0 |
| S1_k141_237750 | 11328 | No | NA | | 18 | 3 | 0 | Low-quality | Genome-fragment | | 12.48 | HMM-based (lower-bound) | 0 |
| S11-final-viral-combined_k141_269687\|\|full | 11329 | No | NA | | 16 | 5 | 1 | Low-quality | Genome-fragment | | 26.45 | AAI-based (medium-confidence) | 0 |
| S8-final-viral-combined_k141_178515\|\|full | 2390 | No | NA | | 4 | 0 | 0 | Not-determined | Genome-fragment | NA | | NA | 0 |
| S8-final-viral-combined_k141_74369\|\|full | 2512 | No | NA | | 3 | 0 | 3 | Not-determined | Genome-fragment | NA | | NA | 0 |
| S7-final-viral-combined_k141_141459\|\|full | 11330 | No | NA | | 11 | 0 | 4 | Low-quality | Genome-fragment | | 18.72 | AAI-based (medium-confidence) | 0 |
| S2-final-viral-combined_k141_133823\|\|full | 11330 | No | NA | | 7 | 0 | 5 | Low-quality | Genome-fragment | | 7.26 | AAI-based (medium-confidence) | 0 |
| S11-final-viral-combined_k141_95918\|\|full | 11347 | No | NA | | 10 | 4 | 0 | Low-quality | Genome-fragment | | 16.51 | HMM-based (lower-bound) | 0 |
| S8-final-viral-combined_k141_62907\|\|full | 2028 | No | NA | | 3 | 0 | 1 | Not-determined | Genome-fragment | NA | | NA | 0 |
| S8-final-viral-combined_k141_108344\|\|full | 1640 | No | NA | | 2 | 0 | 2 | Not-determined | Genome-fragment | NA | | NA | 0 |
| S8-final-viral-combined_k141_235844\|\|full | 7578 | No | NA | | 7 | 0 | 2 | Not-determined | Genome-fragment | NA | | NA | 0 |
| S11-final-viral-combined_k141_275698\|\|full | 11350 | No | NA | | 12 | 7 | 0 | Low-quality | Genome-fragment | | 24.48 | AAI-based (high-confidence) | 0 |
| S12_k141_4192 | 11350 | No | NA | | 10 | 3 | 0 | Low-quality | Genome-fragment | | 21.58 | AAI-based (medium-confidence) | 0 |
| S8-final-viral-combined_k141_199888\|\|full | 15830 | No | NA | | 13 | 0 | 7 | Not-determined | Genome-fragment | NA | | NA | 0 |
| S8-final-viral-combined_k141_15444\|\|full | 2081 | No | NA | | 2 | 0 | 2 | Not-determined | Genome-fragment | NA | | NA | 0 |
| S8-final-viral-combined_k141_108382\|\|full | 2835 | No | NA | | 3 | 0 | 1 | Not-determined | Genome-fragment | NA | | NA | 0 |
| S11-final-viral-combined_k141_102922\|\|full | 11355 | No | NA | | 14 | 2 | 0 | Low-quality | Genome-fragment | | 26.53 | AAI-based (medium-confidence) | 0 |
| S8-final-viral-combined_k141_199900\|\|full | 3791 | No | NA | | 5 | 0 | 1 | Not-determined | Genome-fragment | NA | | NA | 0 |
| S8-final-viral-combined_k141_108393\|\|full | 1923 | No | NA | | 2 | 0 | 0 | Not-determined | Genome-fragment | NA | | NA | 0 |
| S8-final-viral-combined_k141_178632\|\|full | 1162 | No | NA | | 3 | 0 | 0 | Not-determined | Genome-fragment | NA | | NA | 0 |
| S8-final-viral-combined_k141_97267\|\|full | 2890 | No | NA | | 4 | 0 | 3 | Not-determined | Genome-fragment | NA | | NA | 0 |
| S8-final-viral-combined_k141_22358\|\|full | 11359 | No | NA | | 20 | 5 | 0 | Low-quality | Genome-fragment | | 28.74 | AAI-based (high-confidence) | 0 |
| S8-final-viral-combined_k141_189547\|\|full | 2012 | No | NA | | 3 | 0 | 0 | Not-determined | Genome-fragment | NA | | NA | 0 |
| S8-final-viral-combined_k141_247115\|\|full | 3040 | No | NA | | 3 | 0 | 1 | Not-determined | Genome-fragment | NA | | NA | 0 |
| S8-final-viral-combined_k141_199955\|\|full | 5867 | No | NA | | 6 | 0 | 2 | Not-determined | Genome-fragment | NA | | NA | 0 |
| S11-final-viral-combined_k141_320815\|\|full | 11360 | No | NA | | 19 | 1 | 2 | Low-quality | Genome-fragment | | 30.22 | AAI-based (medium-confidence) | 0 |
| S8-final-viral-combined_k141_97336\|\|full | 1557 | No | NA | | 2 | 0 | 0 | Not-determined | Genome-fragment | NA | | NA | 0 |
| S8-final-viral-combined_k141_50856\|\|full | 1544 | No | NA | | 2 | 0 | 1 | Not-determined | Genome-fragment | NA | | NA | 0 |
| S2-final-viral-combined_k141_197733\|\|full | 11365 | No | NA | | 12 | 1 | 2 | Low-quality | Genome-fragment | | 8.34 | HMM-based (lower-bound) | 0 |
| S8-final-viral-combined_k141_50870\|\|full | 1604 | No | NA | | 3 | 0 | 0 | Not-determined | Genome-fragment | NA | | NA | 0 |
| S1-final-viral-combined_k141_233894\|\|full | 11369 | No | NA | | 18 | 1 | 1 | Low-quality | Genome-fragment | | 6.95 | HMM-based (lower-bound) | 0 |
| S8-final-viral-combined_k141_247181\|\|full | 2706 | No | NA | | 3 | 0 | 2 | Not-determined | Genome-fragment | NA | | NA | 0 |
| S8-final-viral-combined_k141_97367\|\|full | 2576 | No | NA | | 4 | 0 | 2 | Not-determined | Genome-fragment | NA | | NA | 0 |
| S8-final-viral-combined_k141_235979\|\|full | 1526 | No | NA | | 2 | 0 | 2 | Not-determined | Genome-fragment | NA | | NA | 0 |
| S8-final-viral-combined_k141_108464\|\|full | 1625 | No | NA | | 3 | 0 | 3 | Not-determined | Genome-fragment | NA | | NA | 0 |
| S8-final-viral-combined_k141_212490\|\|full | 2328 | No | NA | | 5 | 0 | 2 | Not-determined | Genome-fragment | NA | | NA | 0 |
| S1-final-viral-combined_k141_4987\|\|full | 11381 | No | NA | | 14 | 8 | 0 | Low-quality | Genome-fragment | | 24.58 | AAI-based (high-confidence) | 0 |
| S8-final-viral-combined_k141_257335\|\|full | 1485 | No | NA | | 2 | 0 | 0 | Not-determined | Genome-fragment | NA | | NA | 0 |
| S8-final-viral-combined_k141_97388\|\|full | 1541 | No | NA | | 4 | 0 | 0 | Not-determined | Genome-fragment | NA | | NA | 0 |
| S8-final-viral-combined_k141_189595\|\|full | 3819 | No | NA | | 5 | 0 | 4 | Not-determined | Genome-fragment | NA | | NA | 0 |
| S8-final-viral-combined_k141_257345\|\|full | 1804 | No | NA | | 1 | 0 | 0 | Not-determined | Genome-fragment | NA | | NA | 0 |
| S1-final-viral-combined_k141_309919\|\|full | 11382 | No | NA | | 10 | 0 | 8 | Low-quality | Genome-fragment | | 1.77 | AAI-based (medium-confidence) | 0 |
| S8-final-viral-combined_k141_28390\|\|full | 1235 | No | NA | | 3 | 0 | 1 | Not-determined | Genome-fragment | NA | | NA | 0 |
| S8-final-viral-combined_k141_4555\|\|full | 1864 | No | NA | | 3 | 0 | 0 | Not-determined | Genome-fragment | NA | | NA | 0 |
| S8-final-viral-combined_k141_74508\|\|full | 1800 | No | NA | | 2 | 0 | 2 | Not-determined | Genome-fragment | NA | | NA | 0 |
| S11-final-viral-combined_k141_231594\|\|full | 11391 | No | NA | | 9 | 2 | 0 | Low-quality | Genome-fragment | | 26.24 | AAI-based (medium-confidence) | 0 |
| S8-final-viral-combined_k141_200019\|\|full | 1506 | No | NA | | 2 | 0 | 2 | Not-determined | Genome-fragment | NA | | NA | 0 |
| S8-final-viral-combined_k141_257361\|\|full | 1671 | No | NA | | 3 | 0 | 2 | Not-determined | Genome-fragment | NA | | NA | 0 |
| S4-final-viral-combined_k141_107856\|\|full | 11396 | No | NA | | 16 | 1 | 1 | Low-quality | Genome-fragment | | 13.57 | HMM-based (lower-bound) | 0 |
| S8-final-viral-combined_k141_223674\|\|full | 1614 | No | NA | | 2 | 0 | 2 | Not-determined | Genome-fragment | NA | | NA | 0 |
| S8-final-viral-combined_k141_131945\|\|full | 2120 | No | NA | | 3 | 0 | 2 | Not-determined | Genome-fragment | NA | | NA | 0 |
| S8-final-viral-combined_k141_189651\|\|full | 2358 | No | NA | | 3 | 0 | 0 | Not-determined | Genome-fragment | NA | | NA | 0 |
| S2_k141_227428 | 11403 | No | NA | | 22 | 10 | 0 | Low-quality | Genome-fragment | | 37.64 | AAI-based (high-confidence) | 0 |
| S8-final-viral-combined_k141_131953\|\|full | 4231 | No | NA | | 6 | 0 | 0 | Not-determined | Genome-fragment | NA | | NA | 0 |
| S8-final-viral-combined_k141_97437\|\|full | 10770 | No | NA | | 7 | 0 | 5 | Not-determined | Genome-fragment | NA | | NA | 0 |
| S8-final-viral-combined_k141_270192\|\|full | 4376 | No | NA | | 7 | 0 | 4 | Not-determined | Genome-fragment | NA | | NA | 0 |
| S1-final-viral-combined_k141_52605\|\|0_partial | 11405 | No | NA | | 13 | 1 | 7 | Low-quality | Genome-fragment | | 19.77 | AAI-based (medium-confidence) | 0 |
| S8-final-viral-combined_k141_97440\|\|full | 2995 | No | NA | | 2 | 0 | 1 | Not-determined | Genome-fragment | NA | | NA | 0 |
| S11-final-viral-combined_k141_102842\|\|0_partial | 11406 | No | NA | | 8 | 1 | 0 | Low-quality | Genome-fragment | | 21.31 | HMM-based (lower-bound) | 0 |
| S8-final-viral-combined_k141_236065\|\|full | 4320 | No | NA | | 6 | 0 | 5 | Not-determined | Genome-fragment | NA | | NA | 0 |
| S8-final-viral-combined_k141_39868\|\|full | 1209 | No | NA | | 2 | 0 | 0 | Not-determined | Genome-fragment | NA | | NA | 0 |
| S17-final-viral-combined_k141_235527\|\|full | 11418 | No | NA | | 17 | 5 | 0 | Low-quality | Genome-fragment | | 14.33 | AAI-based (high-confidence) | 0 |
| S8-final-viral-combined_k141_212537\|\|full | 5247 | No | NA | | 4 | 0 | 1 | Not-determined | Genome-fragment | NA | | NA | 0 |
| S7_k141_199397 | 11427 | No | NA | | 10 | 7 | 0 | Low-quality | Genome-fragment | | 29.3 | AAI-based (high-confidence) | 0 |
| S6_k141_48799 | 11429 | No | NA | | 8 | 0 | 1 | Low-quality | Genome-fragment | | 11.51 | AAI-based (medium-confidence) | 0 |
| S5-final-viral-combined_k141_471\|\|full | 11449 | No | NA | | 16 | 6 | 0 | Low-quality | Genome-fragment | | 30.55 | AAI-based (high-confidence) | 0 |
| S11-final-viral-combined_k141_119641\|\|full | 11454 | No | NA | | 14 | 6 | 0 | Low-quality | Genome-fragment | | 31.08 | AAI-based (high-confidence) | 0 |
| S8-final-viral-combined_k141_257421\|\|full | 2065 | No | NA | | 2 | 0 | 2 | Not-determined | Genome-fragment | NA | | NA | 0 |
| S1-final-viral-combined_k141_302539\|\|full | 11455 | Yes | | 7834 | 9 | 1 | 1 | Low-quality | Genome-fragment | | 4.04 | HMM-based (lower-bound) | 31.61 |
| S8-final-viral-combined_k141_39911\|\|full | 2567 | No | NA | | 4 | 0 | 1 | Not-determined | Genome-fragment | NA | | NA | 0 |
| S9_k141_70965 | 11456 | Yes | | 367 | 10 | 1 | 7 | Low-quality | Genome-fragment | | 0.24 | AAI-based (high-confidence) | 96.8 |
| S3-final-viral-combined_k141_58440\|\|full | 11462 | No | NA | | 7 | 0 | 7 | Low-quality | Genome-fragment | | 7.09 | AAI-based (medium-confidence) | 0 |
| S2-final-viral-combined_k141_129815\|\|full | 11466 | Yes | | 4152 | 10 | 1 | 7 | Low-quality | Genome-fragment | | 1.49 | HMM-based (lower-bound) | 63.79 |
| S8-final-viral-combined_k141_189701\|\|full | 2286 | No | NA | | 3 | 0 | 2 | Not-determined | Genome-fragment | NA | | NA | 0 |
| S8-final-viral-combined_k141_155075\|\|full | 3019 | No | NA | | 2 | 0 | 1 | Not-determined | Genome-fragment | NA | | NA | 0 |
| S8-final-viral-combined_k141_236164\|\|full | 3910 | No | NA | | 3 | 0 | 3 | Not-determined | Genome-fragment | NA | | NA | 0 |
| S8-final-viral-combined_k141_247410\|\|full | 1109 | No | NA | | 2 | 0 | 0 | Not-determined | Genome-fragment | NA | | NA | 0 |
| S8-final-viral-combined_k141_143923\|\|full | 2149 | No | NA | | 2 | 0 | 0 | Not-determined | Genome-fragment | NA | | NA | 0 |
| S8-final-viral-combined_k141_28613\|\|full | 1303 | No | NA | | 3 | 0 | 2 | Not-determined | Genome-fragment | NA | | NA | 0 |
| S9-final-viral-combined_k141_123844\|\|0_partial | 11477 | Yes | | 1741 | 10 | 1 | 5 | Low-quality | Genome-fragment | | 0.66 | HMM-based (lower-bound) | 84.83 |
| S8-final-viral-combined_k141_143942\|\|full | 1682 | No | NA | | 3 | 0 | 0 | Not-determined | Genome-fragment | NA | | NA | 0 |
| S8-final-viral-combined_k141_28620\|\|full | 6412 | No | NA | | 12 | 0 | 1 | Not-determined | Genome-fragment | NA | | NA | 0 |
| S8-final-viral-combined_k141_143958\|\|full | 2194 | No | NA | | 3 | 0 | 1 | Not-determined | Genome-fragment | NA | | NA | 0 |
| S8-final-viral-combined_k141_178934\|\|full | 3116 | No | NA | | 5 | 0 | 2 | Not-determined | Genome-fragment | NA | | NA | 0 |
| S1-final-viral-combined_k141_148194\|\|full | 11479 | No | NA | | 17 | 3 | 1 | Low-quality | Genome-fragment | | 22.95 | AAI-based (high-confidence) | 0 |
| S8-final-viral-combined_k141_247447\|\|full | 9665 | No | NA | | 9 | 0 | 0 | Not-determined | Genome-fragment | NA | | NA | 0 |
| S8-final-viral-combined_k141_63247\|\|full | 2286 | No | NA | | 6 | 0 | 0 | Not-determined | Genome-fragment | NA | | NA | 0 |
| S8-final-viral-combined_k141_51135\|\|full | 3820 | No | NA | | 4 | 0 | 3 | Not-determined | Genome-fragment | NA | | NA | 0 |
| S8-final-viral-combined_k141_257494\|\|full | 1786 | No | NA | | 3 | 0 | 1 | Not-determined | Genome-fragment | NA | | NA | 0 |
| S1-final-viral-combined_k141_282383\|\|full | 11492 | No | NA | | 16 | 0 | 6 | Low-quality | Genome-fragment | | 27.51 | AAI-based (high-confidence) | 0 |
| S8-final-viral-combined_k141_247463\|\|full | 3103 | No | NA | | 3 | 0 | 1 | Not-determined | Genome-fragment | NA | | NA | 0 |
| S8-final-viral-combined_k141_85409\|\|full | 2126 | No | NA | | 4 | 0 | 2 | Not-determined | Genome-fragment | NA | | NA | 0 |
| S8-final-viral-combined_k141_166375\|\|full | 2233 | No | NA | | 3 | 0 | 2 | Not-determined | Genome-fragment | NA | | NA | 0 |
| S7-final-viral-combined_k141_73358\|\|full | 11499 | No | NA | | 18 | 1 | 5 | Low-quality | Genome-fragment | | 19.81 | HMM-based (lower-bound) | 0 |
| S4-final-viral-combined_k141_114312\|\|full | 11511 | No | NA | | 15 | 2 | 0 | Low-quality | Genome-fragment | | 19.22 | AAI-based (high-confidence) | 0 |
| S8-final-viral-combined_k141_166415\|\|full | 1518 | No | NA | | 2 | 0 | 1 | Not-determined | Genome-fragment | NA | | NA | 0 |
| S6_k141_61139 | 11515 | No | NA | | 19 | 3 | 0 | Low-quality | Genome-fragment | | 11.44 | AAI-based (high-confidence) | 0 |
| S12-final-viral-combined_k141_305384\|\|full | 11517 | No | NA | | 13 | 3 | 2 | Low-quality | Genome-fragment | | 21.92 | HMM-based (lower-bound) | 0 |
| S11_k141_170816 | 11528 | No | NA | | 21 | 1 | 0 | Low-quality | Genome-fragment | | 12.25 | AAI-based (high-confidence) | 0 |
| S2-final-viral-combined_k141_55238\|\|full | 11543 | No | NA | | 5 | 3 | 0 | Low-quality | Genome-fragment | | 30.44 | AAI-based (medium-confidence) | 0 |
| S8-final-viral-combined_k141_97656\|\|full | 1544 | No | NA | | 2 | 0 | 1 | Not-determined | Genome-fragment | NA | | NA | 0 |
| S8-final-viral-combined_k141_4710\|\|full | 7061 | No | NA | | 8 | 0 | 6 | Not-determined | Genome-fragment | NA | | NA | 0 |
| S8-final-viral-combined_k141_85485\|\|full | 1677 | No | NA | | 3 | 0 | 1 | Not-determined | Genome-fragment | NA | | NA | 0 |
| S8-final-viral-combined_k141_144052\|\|full | 7035 | No | NA | | 5 | 0 | 3 | Not-determined | Genome-fragment | NA | | NA | 0 |
| S11_k141_247103 | 11551 | No | NA | | 12 | 1 | 3 | Low-quality | Genome-fragment | | 18.04 | HMM-based (lower-bound) | 0 |
| S8-final-viral-combined_k141_4720\|\|full | 2067 | No | NA | | 2 | 0 | 1 | Not-determined | Genome-fragment | NA | | NA | 0 |
| S2-final-viral-combined_k141_119820\|\|full | 11553 | No | NA | | 26 | 6 | 0 | Low-quality | Genome-fragment | | 17.89 | AAI-based (high-confidence) | 0 |
| S8-final-viral-combined_k141_144080\|\|full | 1564 | No | NA | | 3 | 0 | 0 | Not-determined | Genome-fragment | NA | | NA | 0 |
| S1-final-viral-combined_k141_180675\|\|0_partial | 11570 | No | NA | | 11 | 1 | 4 | Low-quality | Genome-fragment | | 22.77 | AAI-based (medium-confidence) | 0 |
| S8-final-viral-combined_k141_257546\|\|full | 5740 | No | NA | | 8 | 0 | 3 | Not-determined | Genome-fragment | NA | | NA | 0 |
| S8-final-viral-combined_k141_74809\|\|full | 4365 | No | NA | | 5 | 0 | 5 | Not-determined | Genome-fragment | NA | | NA | 0 |
| S17-final-viral-combined_k141_91489\|\|full | 11574 | No | NA | | 13 | 2 | 1 | Low-quality | Genome-fragment | | 25.93 | AAI-based (medium-confidence) | 0 |
| S8-final-viral-combined_k141_132212\|\|full | 8339 | No | NA | | 9 | 0 | 3 | Not-determined | Genome-fragment | NA | | NA | 0 |
| S8-final-viral-combined_k141_85501\|\|full | 14522 | No | NA | | 11 | 0 | 8 | Not-determined | Genome-fragment | NA | | NA | 0 |
| S11-final-viral-combined_k141_187496\|\|full | 11579 | No | NA | | 21 | 2 | 0 | Low-quality | Genome-fragment | | 31.71 | AAI-based (medium-confidence) | 0 |
| S8-final-viral-combined_k141_97668\|\|full | 1061 | No | NA | | 3 | 0 | 1 | Not-determined | Genome-fragment | NA | | NA | 0 |
| S12-final-viral-combined_k141_166684\|\|0_partial | 11602 | No | NA | | 11 | 1 | 4 | Low-quality | Genome-fragment | | 5.48 | AAI-based (medium-confidence) | 0 |
| S8-final-viral-combined_k141_155239\|\|full | 3175 | No | NA | | 4 | 0 | 3 | Not-determined | Genome-fragment | NA | | NA | 0 |
| S8-final-viral-combined_k141_236275\|\|full | 1484 | No | NA | | 2 | 0 | 2 | Not-determined | Genome-fragment | NA | | NA | 0 |
| S8-final-viral-combined_k141_257581\|\|full | 2680 | No | NA | | 2 | 0 | 1 | Not-determined | Genome-fragment | NA | | NA | 0 |
| S8-final-viral-combined_k141_166485\|\|full | 3066 | No | NA | | 2 | 0 | 0 | Not-determined | Genome-fragment | NA | | NA | 0 |
| S8-final-viral-combined_k141_28758\|\|full | 2978 | No | NA | | 6 | 0 | 1 | Not-determined | Genome-fragment | NA | | NA | 0 |
| S17-final-viral-combined_k141_174905\|\|full | 11608 | No | NA | | 14 | 5 | 0 | Low-quality | Genome-fragment | | 23.15 | HMM-based (lower-bound) | 0 |
| S8-final-viral-combined_k141_166488\|\|full | 7109 | No | NA | | 5 | 0 | 2 | Not-determined | Genome-fragment | NA | | NA | 0 |
| S8-final-viral-combined_k141_270362\|\|full | 69096 | Yes | | 57344 | 93 | 12 | 12 | High-quality | High-quality | | 98.49 | AAI-based (high-confidence) | 17.01 |
| S8-final-viral-combined_k141_97714\|\|full | 1420 | No | NA | | 2 | 0 | 0 | Not-determined | Genome-fragment | NA | | NA | 0 |
| S12-final-viral-combined_k141_328463\|\|0_partial | 11612 | No | NA | | 16 | 4 | 0 | Low-quality | Genome-fragment | | 40.48 | AAI-based (high-confidence) | 0 |
| S7_k141_152900 | 11625 | No | NA | | 13 | 5 | 0 | Low-quality | Genome-fragment | | 11.13 | AAI-based (high-confidence) | 0 |
| S5-final-viral-combined_k141_192025\|\|full | 11648 | No | NA | | 18 | 7 | 0 | Low-quality | Genome-fragment | | 32.42 | AAI-based (medium-confidence) | 0 |
| S8-final-viral-combined_k141_4777\|\|full | 1130 | No | NA | | 1 | 0 | 0 | Not-determined | Genome-fragment | NA | | NA | 0 |
| S1_k141_389114 | 11649 | No | NA | | 12 | 1 | 3 | Low-quality | Genome-fragment | | 10.61 | HMM-based (lower-bound) | 0 |
| S8-final-viral-combined_k141_108746\|\|full | 1172 | No | NA | | 3 | 0 | 1 | Not-determined | Genome-fragment | NA | | NA | 0 |
| S2-final-viral-combined_k141_54488\|\|full | 11649 | No | NA | | 11 | 1 | 5 | Low-quality | Genome-fragment | | 6.76 | AAI-based (medium-confidence) | 0 |
| S12_k141_315563 | 11651 | No | NA | | 14 | 5 | 0 | Low-quality | Genome-fragment | | 38.78 | HMM-based (lower-bound) | 0 |
| S8-final-viral-combined_k141_85603\|\|full | 2528 | No | NA | | 3 | 0 | 1 | Not-determined | Genome-fragment | NA | | NA | 0 |
| S8-final-viral-combined_k141_155288\|\|full | 3833 | No | NA | | 3 | 0 | 2 | Not-determined | Genome-fragment | NA | | NA | 0 |
| S8-final-viral-combined_k141_166515\|\|full | 4241 | No | NA | | 4 | 0 | 2 | Not-determined | Genome-fragment | NA | | NA | 0 |
| S8-final-viral-combined_k141_85606\|\|full | 2079 | No | NA | | 3 | 0 | 2 | Not-determined | Genome-fragment | NA | | NA | 0 |
| S8-final-viral-combined_k141_15816\|\|full | 1937 | No | NA | | 3 | 0 | 1 | Not-determined | Genome-fragment | NA | | NA | 0 |
| S8-final-viral-combined_k141_200168\|\|full | 78244 | Yes | | 60907 | 102 | 15 | 15 | High-quality | High-quality | | 100 | AAI-based (high-confidence) | 22.16 |
| S17-final-viral-combined_k141_79582\|\|full | 11653 | No | NA | | 14 | 0 | 1 | Low-quality | Genome-fragment | | 18.09 | AAI-based (high-confidence) | 0 |
| S8-final-viral-combined_k141_236370\|\|full | 1728 | No | NA | | 4 | 0 | 1 | Not-determined | Genome-fragment | NA | | NA | 0 |
| S11-final-viral-combined_k141_207561\|\|0_partial | 11657 | Yes | | 6453 | 13 | 4 | 4 | Low-quality | Genome-fragment | | 11.73 | AAI-based (medium-confidence) | 44.64 |
| S8-final-viral-combined_k141_51264\|\|full | 2454 | No | NA | | 3 | 0 | 2 | Not-determined | Genome-fragment | NA | | NA | 0 |
| S8-final-viral-combined_k141_240188\|\|full | 11657 | No | NA | | 10 | 6 | 0 | Low-quality | Genome-fragment | | 49.86 | AAI-based (medium-confidence) | 0 |
| S17-final-viral-combined_k141_73126\|\|0_partial | 11673 | No | NA | | 14 | 1 | 3 | Low-quality | Genome-fragment | | 13.9 | HMM-based (lower-bound) | 0 |
| S12_k141_183036 | 11673 | No | NA | | 10 | 1 | 0 | Low-quality | Genome-fragment | | 12.21 | AAI-based (high-confidence) | 0 |
| S8-final-viral-combined_k141_189982\|\|full | 1110 | No | NA | | 2 | 0 | 0 | Not-determined | Genome-fragment | NA | | NA | 0 |
| S8-final-viral-combined_k141_63462\|\|full | 2070 | No | NA | | 3 | 0 | 0 | Not-determined | Genome-fragment | NA | | NA | 0 |
| S8-final-viral-combined_k141_51302\|\|full | 1164 | No | NA | | 3 | 0 | 1 | Not-determined | Genome-fragment | NA | | NA | 0 |
| S1-final-viral-combined_k141_764\|\|full | 11687 | No | NA | | 31 | 5 | 0 | Low-quality | Genome-fragment | | 28.43 | AAI-based (high-confidence) | 0 |
| S8-final-viral-combined_k141_270448\|\|full | 9983 | No | NA | | 7 | 0 | 6 | Not-determined | Genome-fragment | NA | | NA | 0 |
| S8-final-viral-combined_k141_179179\|\|full | 3528 | No | NA | | 5 | 0 | 2 | Not-determined | Genome-fragment | NA | | NA | 0 |
| S8-final-viral-combined_k141_270455\|\|full | 1512 | No | NA | | 3 | 0 | 0 | Not-determined | Genome-fragment | NA | | NA | 0 |
| S1-final-viral-combined_k141_34823\|\|full | 11692 | Yes | | 8275 | 8 | 2 | 2 | Low-quality | Genome-fragment | | 22.04 | AAI-based (medium-confidence) | 29.23 |
| S8-final-viral-combined_k141_166603\|\|full | 2121 | No | NA | | 4 | 0 | 3 | Not-determined | Genome-fragment | NA | | NA | 0 |
| S8-final-viral-combined_k141_120402\|\|full | 2254 | No | NA | | 3 | 0 | 3 | Not-determined | Genome-fragment | NA | | NA | 0 |
| S1_k141_340458 | 11692 | No | NA | | 16 | 4 | 0 | Low-quality | Genome-fragment | | 14.54 | AAI-based (high-confidence) | 0 |
| S8-final-viral-combined_k141_4887\|\|full | 1144 | No | NA | | 1 | 0 | 0 | Not-determined | Genome-fragment | NA | | NA | 0 |
| S1-final-viral-combined_k141_33421\|\|full | 11694 | No | NA | | 18 | 1 | 1 | Low-quality | Genome-fragment | | 24.25 | AAI-based (high-confidence) | 0 |
| S12-final-viral-combined_k141_77521\|\|full | 11697 | No | NA | | 18 | 1 | 5 | Low-quality | Genome-fragment | | 12.63 | HMM-based (lower-bound) | 0 |
| S12_k141_372961 | 11704 | No | NA | | 16 | 1 | 5 | Low-quality | Genome-fragment | | 17.71 | HMM-based (lower-bound) | 0 |
| S8-final-viral-combined_k141_74985\|\|full | 1128 | No | NA | | 3 | 0 | 1 | Not-determined | Genome-fragment | NA | | NA | 0 |
| S8-final-viral-combined_k141_270470\|\|full | 3380 | No | NA | | 4 | 0 | 0 | Not-determined | Genome-fragment | NA | | NA | 0 |
| S8-final-viral-combined_k141_257726\|\|full | 7180 | No | NA | | 7 | 0 | 6 | Not-determined | Genome-fragment | NA | | NA | 0 |
| S8-final-viral-combined_k141_166651\|\|full | 1081 | No | NA | | 2 | 0 | 1 | Not-determined | Genome-fragment | NA | | NA | 0 |
| S8-final-viral-combined_k141_270482\|\|full | 1116 | No | NA | | 2 | 0 | 0 | Not-determined | Genome-fragment | NA | | NA | 0 |
| S12-final-viral-combined_k141_345009\|\|full | 11714 | No | NA | | 29 | 0 | 1 | Low-quality | Genome-fragment | | 26.05 | AAI-based (high-confidence) | 0 |
| S12-final-viral-combined_k141_288525\|\|full | 11720 | Yes | | 8754 | 10 | 2 | 2 | Low-quality | Genome-fragment | | 23.81 | AAI-based (high-confidence) | 25.31 |
| S8-final-viral-combined_k141_257731\|\|full | 4995 | No | NA | | 7 | 0 | 0 | Not-determined | Genome-fragment | NA | | NA | 0 |
| S8-final-viral-combined_k141_108824\|\|full | 1736 | No | NA | | 3 | 0 | 0 | Not-determined | Genome-fragment | NA | | NA | 0 |
| S8-final-viral-combined_k141_40398\|\|full | 11720 | No | NA | | 24 | 4 | 0 | Low-quality | Genome-fragment | | 36.47 | AAI-based (high-confidence) | 0 |
| S1-final-viral-combined_k141_335375\|\|full | 11723 | No | NA | | 21 | 5 | 0 | Low-quality | Genome-fragment | | 16.81 | HMM-based (lower-bound) | 0 |
| S12_k141_55446 | 11723 | No | NA | | 16 | 0 | 6 | Low-quality | Genome-fragment | | 8.31 | AAI-based (medium-confidence) | 0 |
| S8-final-viral-combined_k141_224114\|\|full | 7175 | No | NA | | 6 | 0 | 3 | Not-determined | Genome-fragment | NA | | NA | 0 |
| S8-final-viral-combined_k141_63568\|\|full | 1391 | No | NA | | 3 | 0 | 2 | Not-determined | Genome-fragment | NA | | NA | 0 |
| S5-final-viral-combined_k141_26849\|\|0_partial | 11724 | No | NA | | 14 | 1 | 6 | Low-quality | Genome-fragment | | 0.94 | HMM-based (lower-bound) | 0 |
| S17-final-viral-combined_k141_212188\|\|full | 11731 | No | NA | | 5 | 0 | 0 | Low-quality | Genome-fragment | | 26.69 | AAI-based (high-confidence) | 0 |
| S2-final-viral-combined_k141_100489\|\|full | 11731 | No | NA | | 14 | 1 | 1 | Low-quality | Genome-fragment | | 13.17 | HMM-based (lower-bound) | 0 |
| S8-final-viral-combined_k141_179266\|\|full | 1825 | No | NA | | 4 | 0 | 0 | Not-determined | Genome-fragment | NA | | NA | 0 |
| S11_k141_322046 | 11732 | No | NA | | 14 | 9 | 0 | Low-quality | Genome-fragment | | 30.52 | AAI-based (high-confidence) | 0 |
| S8-final-viral-combined_k141_40312\|\|full | 12877 | No | NA | | 8 | 0 | 8 | Not-determined | Genome-fragment | NA | | NA | 0 |
| S8-final-viral-combined_k141_179289\|\|full | 3126 | No | NA | | 3 | 0 | 1 | Not-determined | Genome-fragment | NA | | NA | 0 |
| S8-final-viral-combined_k141_166764\|\|full | 2536 | No | NA | | 3 | 0 | 3 | Not-determined | Genome-fragment | NA | | NA | 0 |
| S11-final-viral-combined_k141_188903\|\|full | 11739 | No | NA | | 9 | 0 | 6 | Low-quality | Genome-fragment | | 3.67 | AAI-based (medium-confidence) | 0 |

| | | | | | | | | | | | |
|---|---|---|---|---|---|---|---|---|---|---|---|
| S8-final-viral-combined_k141_97848\|\|full | 40473 | Yes | 27882 | 45 | 8 | 3 | Medium-quality | Genome-fragment | 65.1 | AAI-based (high-confidence) | 31.11 |
| S8-final-viral-combined_k141_29118\|\|full | 1058 | No | NA | 2 | 0 | 0 | Not-determined | Genome-fragment | NA | NA | 0 |
| S8-final-viral-combined_k141_236576\|\|full | 2358 | No | NA | 3 | 0 | 0 | Not-determined | Genome-fragment | NA | NA | 0 |
| S8-final-viral-combined_k141_63626\|\|full | 1175 | No | NA | 2 | 0 | 0 | Not-determined | Genome-fragment | NA | NA | 0 |
| S8-final-viral-combined_k141_212931\|\|full | 2610 | No | NA | 3 | 0 | 2 | Not-determined | Genome-fragment | NA | NA | 0 |
| S8-final-viral-combined_k141_247778\|\|full | 2907 | No | NA | 3 | 0 | 3 | Not-determined | Genome-fragment | NA | NA | 0 |
| S11-final-viral-combined_k141_326268\|\|full | 11759 | Yes | 4297 | 11 | 3 | 6 | Low-quality | Genome-fragment | 10.23 | AAI-based (medium-confidence) | 63.46 |
| S8-final-viral-combined_k141_15983\|\|full | 1988 | No | NA | 4 | 0 | 2 | Not-determined | Genome-fragment | NA | NA | 0 |
| S12-final-viral-combined_k141_331312\|\|0_partial | 11774 | No | NA | 7 | 0 | 7 | Low-quality | Genome-fragment | 11.08 | AAI-based (medium-confidence) | 0 |
| S1-final-viral-combined_k141_37233\|\|full | 11775 | No | NA | 21 | 1 | 4 | Low-quality | Genome-fragment | 20.28 | HMM-based (lower-bound) | 0 |
| S5-final-viral-combined_k141_108991\|\|full | 11781 | No | NA | 21 | 8 | 1 | Low-quality | Genome-fragment | 32.15 | AAI-based (high-confidence) | 0 |
| S8-final-viral-combined_k141_190182\|\|full | 2330 | No | NA | 4 | 0 | 1 | Not-determined | Genome-fragment | NA | NA | 0 |
| S8-final-viral-combined_k141_51530\|\|full | 3337 | No | NA | 3 | 0 | 1 | Not-determined | Genome-fragment | NA | NA | 0 |
| S2-final-viral-combined_k141_239862\|\|full | 11795 | No | NA | 15 | 1 | 1 | Low-quality | Genome-fragment | 27.46 | AAI-based (high-confidence) | 0 |
| S9_k141_59229 | 11805 | No | NA | 6 | 0 | 6 | Low-quality | Genome-fragment | 6.66 | AAI-based (high-confidence) | 0 |
| S8-final-viral-combined_k141_155513\|\|full | 1269 | No | NA | 2 | 0 | 2 | Not-determined | Genome-fragment | NA | NA | 0 |
| S8-final-viral-combined_k141_212969\|\|full | 1802 | No | NA | 3 | 0 | 1 | Not-determined | Genome-fragment | NA | NA | 0 |
| S8-final-viral-combined_k141_273181\|\|0_partial | 11815 | No | NA | 12 | 1 | 5 | Low-quality | Genome-fragment | 6.4 | HMM-based (lower-bound) | 0 |
| S8-final-viral-combined_k141_270659\|\|full | 2340 | No | NA | 3 | 0 | 2 | Not-determined | Genome-fragment | NA | NA | 0 |
| S17-final-viral-combined_k141_211076\|\|full | 11824 | No | NA | 6 | 0 | 0 | Low-quality | Genome-fragment | 27.61 | AAI-based (medium-confidence) | 0 |
| S8-final-viral-combined_k141_236635\|\|full | 3887 | No | NA | 8 | 0 | 2 | Not-determined | Genome-fragment | NA | NA | 0 |
| S8-final-viral-combined_k141_120574\|\|full | 6228 | No | NA | 7 | 0 | 7 | Not-determined | Genome-fragment | NA | NA | 0 |
| S11-final-viral-combined_k141_130862\|\|full | 11829 | Yes | 4169 | 13 | 2 | 2 | Low-quality | Genome-fragment | 2.65 | HMM-based (lower-bound) | 64.76 |
| S8-final-viral-combined_k141_190208\|\|full | 2517 | No | NA | 2 | 0 | 2 | Not-determined | Genome-fragment | NA | NA | 0 |
| S8-final-viral-combined_k141_16045\|\|full | 2198 | No | NA | 3 | 0 | 0 | Not-determined | Genome-fragment | NA | NA | 0 |
| S7-final-viral-combined_k141_57787\|\|full | 11833 | No | NA | 11 | 0 | 2 | Low-quality | Genome-fragment | 9.7 | AAI-based (medium-confidence) | 0 |
| S8-final-viral-combined_k141_51582\|\|full | 10762 | No | NA | 10 | 0 | 8 | Not-determined | Genome-fragment | NA | NA | 0 |
| S17-final-viral-combined_k141_191558\|\|full | 11841 | No | NA | 6 | 0 | 0 | Low-quality | Genome-fragment | 28.01 | AAI-based (medium-confidence) | 0 |
| S8-final-viral-combined_k141_29262\|\|full | 1384 | No | NA | 3 | 0 | 2 | Not-determined | Genome-fragment | NA | NA | 0 |
| S1-final-viral-combined_k141_264734\|\|full | 11845 | No | NA | 25 | 2 | 1 | Low-quality | Genome-fragment | 22.64 | AAI-based (high-confidence) | 0 |
| S8-final-viral-combined_k141_120611\|\|full | 2462 | No | NA | 3 | 0 | 1 | Not-determined | Genome-fragment | NA | NA | 0 |
| S8-final-viral-combined_k141_108917\|\|full | 70457 | No | NA | 86 | 27 | 1 | High-quality | High-quality | 100 | AAI-based (high-confidence) | 0 |
| S8-final-viral-combined_k141_247840\|\|full | 4352 | No | NA | 3 | 0 | 3 | Not-determined | Genome-fragment | NA | NA | 0 |
| S11-final-viral-combined_k141_87230\|\|full | 11865 | No | NA | 8 | 0 | 7 | Low-quality | Genome-fragment | 10.98 | AAI-based (medium-confidence) | 0 |
| S8-final-viral-combined_k141_120636\|\|full | 2628 | No | NA | 4 | 0 | 1 | Not-determined | Genome-fragment | NA | NA | 0 |
| S11-final-viral-combined_k141_219612\|\|full | 11865 | No | NA | 18 | 2 | 0 | Low-quality | Genome-fragment | 13.55 | HMM-based (lower-bound) | 0 |
| S8-final-viral-combined_k141_200478\|\|full | 2844 | No | NA | 4 | 0 | 1 | Not-determined | Genome-fragment | NA | NA | 0 |
| S11-final-viral-combined_k141_144214\|\|full | 11873 | No | NA | 20 | 13 | 0 | Low-quality | Genome-fragment | 29.4 | AAI-based (high-confidence) | 0 |
| S8-final-viral-combined_k141_40538\|\|full | 1871 | No | NA | 2 | 0 | 2 | Not-determined | Genome-fragment | NA | NA | 0 |
| S2-final-viral-combined_k141_115553\|\|full | 11874 | Yes | 8512 | 18 | 2 | 3 | Low-quality | Genome-fragment | 18.43 | HMM-based (lower-bound) | 28.31 |
| S2-final-viral-combined_k141_180380\|\|full | 11875 | No | NA | 17 | 9 | 0 | Low-quality | Genome-fragment | 30.33 | AAI-based (high-confidence) | 0 |
| S8-final-viral-combined_k141_190284\|\|full | 3164 | No | NA | 6 | 0 | 1 | Not-determined | Genome-fragment | NA | NA | 0 |
| S5-final-viral-combined_k141_103844\|\|full | 11879 | No | NA | 16 | 10 | 0 | Low-quality | Genome-fragment | 32.49 | AAI-based (high-confidence) | 0 |
| S8-final-viral-combined_k141_155656\|\|full | 1747 | No | NA | 2 | 0 | 1 | Not-determined | Genome-fragment | NA | NA | 0 |
| S12-final-viral-combined_k141_142123\|\|full | 11883 | Yes | 4812 | 10 | 1 | 4 | Low-quality | Genome-fragment | 23.07 | AAI-based (high-confidence) | 59.51 |
| S8-final-viral-combined_k141_75098\|\|full | 2047 | No | NA | 5 | 0 | 1 | Not-determined | Genome-fragment | NA | NA | 0 |
| S8-final-viral-combined_k141_179484\|\|full | 2147 | No | NA | 2 | 0 | 2 | Not-determined | Genome-fragment | NA | NA | 0 |
| S8-final-viral-combined_k141_155691\|\|full | 1476 | No | NA | 3 | 0 | 1 | Not-determined | Genome-fragment | NA | NA | 0 |
| S12-final-viral-combined_k141_169418\|\|full | 11887 | No | NA | 16 | 6 | 0 | Low-quality | Genome-fragment | 20.21 | AAI-based (medium-confidence) | 0 |
| S8-final-viral-combined_k141_236715\|\|full | 1799 | No | NA | 2 | 0 | 2 | Not-determined | Genome-fragment | NA | NA | 0 |
| S8-final-viral-combined_k141_75139\|\|full | 2931 | No | NA | 3 | 0 | 2 | Not-determined | Genome-fragment | NA | NA | 0 |
| S8-final-viral-combined_k141_224390\|\|full | 1719 | No | NA | 2 | 0 | 2 | Not-determined | Genome-fragment | NA | NA | 0 |
| S8-final-viral-combined_k141_270881\|\|full | 10307 | No | NA | 14 | 0 | 0 | Not-determined | Genome-fragment | NA | NA | 0 |
| S8-final-viral-combined_k141_155722\|\|full | 5869 | No | NA | 7 | 0 | 0 | Not-determined | Genome-fragment | NA | NA | 0 |
| S8-final-viral-combined_k141_247976\|\|full | 1066 | No | NA | 3 | 0 | 2 | Not-determined | Genome-fragment | NA | NA | 0 |
| S8-final-viral-combined_k141_109016\|\|full | 1084 | No | NA | 2 | 0 | 0 | Not-determined | Genome-fragment | NA | NA | 0 |
| S8-final-viral-combined_k141_5261\|\|full | 1106 | No | NA | 3 | 0 | 0 | Not-determined | Genome-fragment | NA | NA | 0 |
| S8-final-viral-combined_k141_224419\|\|full | 2275 | No | NA | 2 | 0 | 0 | Not-determined | Genome-fragment | NA | NA | 0 |
| S1-final-viral-combined_k141_164281\|\|full | 11888 | No | NA | 17 | 1 | 2 | Low-quality | Genome-fragment | 7.69 | HMM-based (lower-bound) | 0 |
| S12-final-viral-combined_k141_6507\|\|full | 11901 | No | NA | 16 | 0 | 3 | Low-quality | Genome-fragment | 7.65 | AAI-based (medium-confidence) | 0 |
| S8-final-viral-combined_k141_155748\|\|full | 3198 | No | NA | 3 | 0 | 1 | Not-determined | Genome-fragment | NA | NA | 0 |
| S5-final-viral-combined_k141_201140\|\|full | 11904 | No | NA | 11 | 1 | 0 | Low-quality | Genome-fragment | 4.6 | HMM-based (lower-bound) | 0 |
| S8-final-viral-combined_k141_75205\|\|full | 8404 | No | NA | 9 | 0 | 8 | Not-determined | Genome-fragment | NA | NA | 0 |
| S9_k141_90206 | 11907 | No | NA | 12 | 0 | 6 | Low-quality | Genome-fragment | 3.14 | AAI-based (medium-confidence) | 0 |
| S4-final-viral-combined_k141_67066\|\|full | 11913 | No | NA | 28 | 5 | 0 | Low-quality | Genome-fragment | 23.51 | AAI-based (high-confidence) | 0 |
| S8-final-viral-combined_k141_63775\|\|full | 1848 | No | NA | 3 | 0 | 2 | Not-determined | Genome-fragment | NA | NA | 0 |
| S8-final-viral-combined_k141_270947\|\|full | 2969 | No | NA | 4 | 0 | 2 | Not-determined | Genome-fragment | NA | NA | 0 |
| S8-final-viral-combined_k141_270949\|\|full | 5148 | No | NA | 7 | 0 | 6 | Not-determined | Genome-fragment | NA | NA | 0 |
| S8-final-viral-combined_k141_155802\|\|full | 2888 | No | NA | 5 | 0 | 0 | Not-determined | Genome-fragment | NA | NA | 0 |
| S8-final-viral-combined_k141_29533\|\|full | 1195 | No | NA | 3 | 0 | 1 | Not-determined | Genome-fragment | NA | NA | 0 |
| S11-final-viral-combined_k141_228673\|\|full | 11926 | No | NA | 10 | 0 | 9 | Low-quality | Genome-fragment | 2.88 | AAI-based (medium-confidence) | 0 |
| S8-final-viral-combined_k141_179612\|\|full | 1363 | No | NA | 3 | 0 | 0 | Not-determined | Genome-fragment | NA | NA | 0 |
| S2_k141_67277 | 11942 | No | NA | 8 | 1 | 0 | Low-quality | Genome-fragment | 9.24 | AAI-based (medium-confidence) | 0 |
| S8-final-viral-combined_k141_179622\|\|full | 1806 | No | NA | 3 | 0 | 1 | Not-determined | Genome-fragment | NA | NA | 0 |
| S12-final-viral-combined_k141_24247\|\|full | 11945 | No | NA | 14 | 7 | 0 | Low-quality | Genome-fragment | 19.98 | HMM-based (lower-bound) | 0 |
| S8-final-viral-combined_k141_248107\|\|full | 2084 | No | NA | 2 | 0 | 1 | Not-determined | Genome-fragment | NA | NA | 0 |
| S11_k141_265514 | 11949 | No | NA | 5 | 0 | 0 | Low-quality | Genome-fragment | 12.7 | AAI-based (high-confidence) | 0 |
| S11-final-viral-combined_k141_162851\|\|full | 11953 | No | NA | 18 | 3 | 0 | Low-quality | Genome-fragment | 18.66 | AAI-based (medium-confidence) | 0 |
| S8-final-viral-combined_k141_179655\|\|full | 2097 | No | NA | 3 | 0 | 2 | Not-determined | Genome-fragment | NA | NA | 0 |
| S8-final-viral-combined_k141_155871\|\|full | 1347 | No | NA | 4 | 0 | 0 | Not-determined | Genome-fragment | NA | NA | 0 |
| S12-final-viral-combined_k141_390363\|\|full | 11953 | No | NA | 9 | 1 | 2 | Low-quality | Genome-fragment | 8.61 | HMM-based (lower-bound) | 0 |
| S8-final-viral-combined_k141_51895\|\|full | 1096 | No | NA | 3 | 0 | 3 | Not-determined | Genome-fragment | NA | NA | 0 |
| S8-final-viral-combined_k141_40666\|\|full | 71582 | Yes | 23651 | 79 | 5 | 21 | Medium-quality | Genome-fragment | 60.72 | AAI-based (medium-confidence) | 66.96 |
| S12_k141_103755 | 11955 | No | NA | 10 | 0 | 9 | Low-quality | Genome-fragment | 5.13 | AAI-based (high-confidence) | 0 |
| S2-final-viral-combined_k141_12309\|\|full | 11955 | No | NA | 18 | 0 | 0 | Low-quality | Genome-fragment | 32.64 | AAI-based (medium-confidence) | 0 |
| S8-final-viral-combined_k141_109169\|\|full | 3246 | No | NA | 3 | 0 | 2 | Not-determined | Genome-fragment | NA | NA | 0 |
| S5-final-viral-combined_k141_133589\|\|full | 11963 | No | NA | 26 | 3 | 0 | Low-quality | Genome-fragment | 7.78 | AAI-based (high-confidence) | 0 |
| S17-final-viral-combined_k141_125171\|\|full | 11963 | No | NA | 16 | 7 | 0 | Low-quality | Genome-fragment | 32.82 | AAI-based (high-confidence) | 0 |
| S1-final-viral-combined_k141_191223\|\|full | 11971 | No | NA | 20 | 5 | 0 | Low-quality | Genome-fragment | 26.79 | AAI-based (medium-confidence) | 0 |
| S8-final-viral-combined_k141_75325\|\|full | 1587 | No | NA | 2 | 0 | 1 | Not-determined | Genome-fragment | NA | NA | 0 |
| S12-final-viral-combined_k141_51696\|\|full | 11983 | No | NA | 16 | 3 | 0 | Low-quality | Genome-fragment | 27.11 | AAI-based (medium-confidence) | 0 |
| S8-final-viral-combined_k141_167153\|\|full | 4997 | No | NA | 5 | 0 | 2 | Not-determined | Genome-fragment | NA | NA | 0 |
| S4_k141_135748 | 11984 | No | NA | 11 | 0 | 9 | Low-quality | Genome-fragment | 7.62 | AAI-based (high-confidence) | 0 |
| S8-final-viral-combined_k141_144352\|\|full | 2280 | No | NA | 4 | 0 | 0 | Not-determined | Genome-fragment | NA | NA | 0 |
| S8-final-viral-combined_k141_120920\|\|full | 2246 | No | NA | 2 | 0 | 2 | Not-determined | Genome-fragment | NA | NA | 0 |
| S8-final-viral-combined_k141_63876\|\|full | 3998 | No | NA | 5 | 0 | 0 | Not-determined | Genome-fragment | NA | NA | 0 |
| S8-final-viral-combined_k141_155955\|\|full | 1964 | No | NA | 5 | 0 | 3 | Not-determined | Genome-fragment | NA | NA | 0 |
| S8-final-viral-combined_k141_86109\|\|full | 1921 | No | NA | 3 | 0 | 1 | Not-determined | Genome-fragment | NA | NA | 0 |
| S8-final-viral-combined_k141_236915\|\|full | 4270 | No | NA | 7 | 0 | 1 | Not-determined | Genome-fragment | NA | NA | 0 |
| S8-final-viral-combined_k141_16385\|\|full | 2678 | No | NA | 4 | 0 | 3 | Not-determined | Genome-fragment | NA | NA | 0 |
| S8-final-viral-combined_k141_271016\|\|full | 1153 | No | NA | 2 | 0 | 0 | Not-determined | Genome-fragment | NA | NA | 0 |
| S2-final-viral-combined_k141_240746\|\|full | 11985 | No | NA | 7 | 0 | 6 | Low-quality | Genome-fragment | 3.8 | AAI-based (medium-confidence) | 0 |
| S8-final-viral-combined_k141_248226\|\|full | 2153 | No | NA | 2 | 0 | 2 | Not-determined | Genome-fragment | NA | NA | 0 |
| S12_k141_253656 | 11994 | No | NA | 9 | 0 | 7 | Low-quality | Genome-fragment | 11.24 | AAI-based (medium-confidence) | 0 |
| S3-final-viral-combined_k141_121752\|\|full | 11999 | No | NA | 11 | 0 | 1 | Low-quality | Genome-fragment | 13.87 | AAI-based (medium-confidence) | 0 |
| S8-final-viral-combined_k141_271034\|\|full | 10200 | No | NA | 12 | 0 | 7 | Not-determined | Genome-fragment | NA | NA | 0 |
| S17-final-viral-combined_k141_31510\|\|full | 12006 | No | NA | 16 | 2 | 0 | Low-quality | Genome-fragment | 8.32 | AAI-based (medium-confidence) | 0 |
| S8-final-viral-combined_k141_132855\|\|full | 1660 | No | NA | 2 | 0 | 0 | Not-determined | Genome-fragment | NA | NA | 0 |
| S4-final-viral-combined_k141_126313\|\|full | 12006 | No | NA | 15 | 9 | 0 | Low-quality | Genome-fragment | 25.11 | AAI-based (medium-confidence) | 0 |
| S8-final-viral-combined_k141_144418\|\|full | 1742 | No | NA | 2 | 0 | 1 | Not-determined | Genome-fragment | NA | NA | 0 |
| S4_k141_76784 | 12016 | No | NA | 17 | 6 | 0 | Low-quality | Genome-fragment | 40.62 | AAI-based (high-confidence) | 0 |
| S6-final-viral-combined_k141_117996\|\|0_partial | 12025 | No | NA | 8 | 0 | 7 | Low-quality | Genome-fragment | 15.88 | AAI-based (medium-confidence) | 0 |
| S2-final-viral-combined_k141_64909\|\|full | 12026 | No | NA | 14 | 6 | 0 | Low-quality | Genome-fragment | 34.2 | AAI-based (high-confidence) | 0 |
| S8-final-viral-combined_k141_16440\|\|full | 2457 | No | NA | 3 | 0 | 0 | Not-determined | Genome-fragment | NA | NA | 0 |
| S8-final-viral-combined_k141_132904\|\|full | 2595 | No | NA | 5 | 0 | 2 | Not-determined | Genome-fragment | NA | NA | 0 |
| S7-final-viral-combined_k141_142495\|\|full | 12030 | No | NA | 10 | 3 | 0 | Low-quality | Genome-fragment | 26.68 | AAI-based (medium-confidence) | 0 |
| S8-final-viral-combined_k141_29729\|\|full | 2750 | No | NA | 6 | 0 | 0 | Not-determined | Genome-fragment | NA | NA | 0 |
| S8-final-viral-combined_k141_144457\|\|full | 1122 | No | NA | 3 | 0 | 1 | Not-determined | Genome-fragment | NA | NA | 0 |
| S8-final-viral-combined_k141_179815\|\|full | 2121 | No | NA | 3 | 0 | 1 | Not-determined | Genome-fragment | NA | NA | 0 |
| S8-final-viral-combined_k141_16455\|\|full | 13670 | No | NA | 13 | 0 | 5 | Not-determined | Genome-fragment | NA | NA | 0 |
| S8-final-viral-combined_k141_224776\|\|full | 2585 | No | NA | 2 | 0 | 1 | Not-determined | Genome-fragment | NA | NA | 0 |
| S8-final-viral-combined_k141_86237\|\|full | 1573 | No | NA | 3 | 0 | 3 | Not-determined | Genome-fragment | NA | NA | 0 |
| S8-final-viral-combined_k141_190587\|\|full | 1894 | No | NA | 4 | 0 | 0 | Not-determined | Genome-fragment | NA | NA | 0 |
| S8-final-viral-combined_k141_271099\|\|full | 2487 | No | NA | 4 | 0 | 3 | Not-determined | Genome-fragment | NA | NA | 0 |
| S12-final-viral-combined_k141_91479\|\|full | 12031 | Yes | 5514 | 15 | 2 | 4 | Low-quality | Genome-fragment | 12.33 | AAI-based (medium-confidence) | 54.17 |
| S11-final-viral-combined_k141_27722\|\|full | 12036 | No | NA | 11 | 0 | 8 | Low-quality | Genome-fragment | 5.9 | AAI-based (medium-confidence) | 0 |
| S8-final-viral-combined_k141_121041\|\|full | 2082 | No | NA | 2 | 0 | 2 | Not-determined | Genome-fragment | NA | NA | 0 |
| S8-final-viral-combined_k141_224798\|\|full | 2736 | No | NA | 2 | 0 | 1 | Not-determined | Genome-fragment | NA | NA | 0 |
| S9-final-viral-combined_k141_95436\|\|full | 12063 | No | NA | 10 | 5 | 0 | Low-quality | Genome-fragment | 26.11 | AAI-based (high-confidence) | 0 |
| S12_k141_286243 | 12063 | No | NA | 14 | 1 | 0 | Low-quality | Genome-fragment | 26.07 | AAI-based (high-confidence) | 0 |
| S4_k141_50956 | 12064 | No | NA | 6 | 0 | 6 | Low-quality | Genome-fragment | 7.81 | AAI-based (high-confidence) | 0 |
| S8-final-viral-combined_k141_5580\|\|full | 6017 | No | NA | 8 | 0 | 1 | Not-determined | Genome-fragment | NA | NA | 0 |
| S8-final-viral-combined_k141_75433\|\|full | 1274 | No | NA | 3 | 0 | 1 | Not-determined | Genome-fragment | NA | NA | 0 |
| S8-final-viral-combined_k141_237129\|\|full | 4857 | No | NA | 4 | 0 | 2 | Not-determined | Genome-fragment | NA | NA | 0 |
| S4_k141_132725 | 12084 | No | NA | 17 | 0 | 5 | Low-quality | Genome-fragment | 8.36 | AAI-based (medium-confidence) | 0 |
| S2_k141_179922 | 12088 | Yes | 3217 | 10 | 1 | 3 | Low-quality | Genome-fragment | 5.73 | HMM-based (lower-bound) | 73.39 |
| S8-final-viral-combined_k141_190638\|\|full | 1135 | No | NA | 2 | 0 | 0 | Not-determined | Genome-fragment | NA | NA | 0 |
| S8-final-viral-combined_k141_156027\|\|full | 3683 | No | NA | 3 | 0 | 3 | Not-determined | Genome-fragment | NA | NA | 0 |
| S8-final-viral-combined_k141_109423\|\|full | 8236 | No | NA | 12 | 0 | 8 | Not-determined | Genome-fragment | NA | NA | 0 |
| S8-final-viral-combined_k141_167439\|\|full | 1387 | No | NA | 3 | 0 | 0 | Not-determined | Genome-fragment | NA | NA | 0 |
| S12_k141_241630 | 12092 | No | NA | 13 | 1 | 4 | Low-quality | Genome-fragment | 11.35 | AAI-based (medium-confidence) | 0 |
| S5_k141_115698 | 12097 | No | NA | 14 | 7 | 0 | Low-quality | Genome-fragment | 19.72 | AAI-based (high-confidence) | 0 |
| S8-final-viral-combined_k141_167449\|\|full | 2838 | No | NA | 3 | 0 | 2 | Not-determined | Genome-fragment | NA | NA | 0 |
| S8-final-viral-combined_k141_224898\|\|full | 1092 | No | NA | 2 | 0 | 0 | Not-determined | Genome-fragment | NA | NA | 0 |
| S8-final-viral-combined_k141_156050\|\|full | 8746 | No | NA | 6 | 0 | 4 | Not-determined | Genome-fragment | NA | NA | 0 |
| S5_k141_135457 | 12109 | No | NA | 22 | 1 | 1 | Low-quality | Genome-fragment | 26.98 | AAI-based (high-confidence) | 0 |
| S8-final-viral-combined_k141_156096\|\|full | 1266 | No | NA | 3 | 0 | 1 | Not-determined | Genome-fragment | NA | NA | 0 |
| S8-final-viral-combined_k141_167487\|\|full | 3421 | No | NA | 3 | 0 | 2 | Not-determined | Genome-fragment | NA | NA | 0 |
| S9-final-viral-combined_k141_57695\|\|0_partial | 12111 | No | NA | 16 | 1 | 2 | Low-quality | Genome-fragment | 8.14 | HMM-based (lower-bound) | 0 |
| S8-final-viral-combined_k141_200920\|\|full | 2929 | No | NA | 2 | 0 | 2 | Not-determined | Genome-fragment | NA | NA | 0 |
| S12-final-viral-combined_k141_352665\|\|full | 12115 | No | NA | 11 | 1 | 9 | Low-quality | Genome-fragment | 14 | HMM-based (lower-bound) | 0 |
| S1_k141_17290 | 12120 | No | NA | 8 | 0 | 8 | Low-quality | Genome-fragment | 8.34 | AAI-based (medium-confidence) | 0 |
| S8-final-viral-combined_k141_258257\|\|full | 2149 | No | NA | 3 | 0 | 2 | Not-determined | Genome-fragment | NA | NA | 0 |
| S2-final-viral-combined_k141_114092\|\|full | 12133 | No | NA | 16 | 6 | 0 | Low-quality | Genome-fragment | 23.57 | AAI-based (high-confidence) | 0 |
| S17-k141_112295 | 12141 | No | NA | 7 | 0 | 2 | Low-quality | Genome-fragment | 10.59 | AAI-based (medium-confidence) | 0 |
| S8_k141_108880 | 12147 | No | NA | 6 | 3 | 0 | Low-quality | Genome-fragment | 9.18 | AAI-based (high-confidence) | 0 |
| S8-final-viral-combined_k141_41027\|\|full | 7216 | No | NA | 9 | 0 | 0 | Not-determined | Genome-fragment | NA | NA | 0 |
| S17-k141_156129 | 12156 | No | NA | 16 | 0 | 2 | Low-quality | Genome-fragment | 23.4 | AAI-based (medium-confidence) | 0 |
| S8-final-viral-combined_k141_52213\|\|full | 9795 | No | NA | 12 | 0 | 2 | Not-determined | Genome-fragment | NA | NA | 0 |
| S8-final-viral-combined_k141_271249\|\|full | 4601 | No | NA | 5 | 0 | 1 | Not-determined | Genome-fragment | NA | NA | 0 |
| S11-final-viral-combined_k141_186153\|\|0_partial | 12159 | No | NA | 9 | 0 | 7 | Low-quality | Genome-fragment | 11.31 | AAI-based (medium-confidence) | 0 |
| S8-final-viral-combined_k141_208252\|\|full | 12168 | No | NA | 12 | 1 | 5 | Low-quality | Genome-fragment | 18.41 | HMM-based (lower-bound) | 0 |
| S8-final-viral-combined_k141_29833\|\|full | 4984 | No | NA | 6 | 0 | 5 | Not-determined | Genome-fragment | NA | NA | 0 |
| S8-final-viral-combined_k141_5646\|\|full | 3756 | No | NA | 6 | 0 | 1 | Not-determined | Genome-fragment | NA | NA | 0 |
| S2-final-viral-combined_k141_125493\|\|full | 12170 | No | NA | 19 | 7 | 0 | Low-quality | Genome-fragment | 19.47 | AAI-based (high-confidence) | 0 |
| S8-final-viral-combined_k141_86414\|\|full | 2031 | No | NA | 3 | 0 | 2 | Not-determined | Genome-fragment | NA | NA | 0 |
| S8-final-viral-combined_k141_41071\|\|full | 1404 | No | NA | 4 | 0 | 0 | Not-determined | Genome-fragment | NA | NA | 0 |
| S8-final-viral-combined_k141_190785\|\|full | 1404 | No | NA | 3 | 0 | 0 | Not-determined | Genome-fragment | NA | NA | 0 |
| S11-final-viral-combined_k141_160329\|\|full | 12174 | No | NA | 20 | 6 | 0 | Low-quality | Genome-fragment | 19.44 | AAI-based (medium-confidence) | 0 |
| S8-final-viral-combined_k141_52291\|\|full | 1556 | No | NA | 2 | 0 | 0 | Not-determined | Genome-fragment | NA | NA | 0 |
| S2_k141_122280 | 12185 | No | NA | 11 | 2 | 0 | Low-quality | Genome-fragment | 12.7 | AAI-based (high-confidence) | 0 |
| S8-final-viral-combined_k141_258330\|\|full | 8362 | No | NA | 7 | 0 | 6 | Not-determined | Genome-fragment | NA | NA | 0 |
| S8-final-viral-combined_k141_190805\|\|full | 2783 | No | NA | 3 | 0 | 2 | Not-determined | Genome-fragment | NA | NA | 0 |
| S8-final-viral-combined_k141_41121\|\|full | 3673 | No | NA | 3 | 0 | 1 | Not-determined | Genome-fragment | NA | NA | 0 |
| S8-final-viral-combined_k141_190810\|\|full | 1364 | No | NA | 3 | 0 | 1 | Not-determined | Genome-fragment | NA | NA | 0 |
| S1-final-viral-combined_k141_350335\|\|full | 12186 | Yes | 2109 | 10 | 1 | 6 | Low-quality | Genome-fragment | 2.36 | HMM-based (lower-bound) | 82.69 |
| S4_k141_112547 | 12196 | No | NA | 17 | 9 | 0 | Low-quality | Genome-fragment | 32.67 | AAI-based (high-confidence) | 0 |
| S8-final-viral-combined_k141_201063\|\|full | 6515 | No | NA | 7 | 0 | 5 | Not-determined | Genome-fragment | NA | NA | 0 |
| S8-final-viral-combined_k141_180065\|\|full | 2810 | No | NA | 5 | 0 | 4 | Not-determined | Genome-fragment | NA | NA | 0 |
| S3-final-viral-combined_k141_35554\|\|full | 12197 | Yes | 8247 | 18 | 2 | 3 | Low-quality | Genome-fragment | 8.21 | HMM-based (lower-bound) | 32.39 |
| S8-final-viral-combined_k141_29974\|\|full | 1590 | No | NA | 1 | 0 | 1 | Not-determined | Genome-fragment | NA | NA | 0 |
| S8-final-viral-combined_k141_64387\|\|full | 2034 | No | NA | 2 | 0 | 1 | Not-determined | Genome-fragment | NA | NA | 0 |
| S8-final-viral-combined_k141_52389\|\|full | 5248 | No | NA | 8 | 0 | 1 | Not-determined | Genome-fragment | NA | NA | 0 |
| S8-final-viral-combined_k141_5726\|\|full | 3647 | No | NA | 7 | 0 | 1 | Not-determined | Genome-fragment | NA | NA | 0 |
| S8-final-viral-combined_k141_213617\|\|full | 1059 | No | NA | 2 | 0 | 0 | Not-determined | Genome-fragment | NA | NA | 0 |
| S8-final-viral-combined_k141_237249\|\|full | 2434 | No | NA | 4 | 0 | 1 | Not-determined | Genome-fragment | NA | NA | 0 |
| S5-final-viral-combined_k141_197664\|\|0_partial | 12204 | Yes | 5948 | 14 | 2 | 4 | Low-quality | Genome-fragment | 4.32 | HMM-based (lower-bound) | 51.26 |
| S8-final-viral-combined_k141_201104\|\|full | 4405 | No | NA | 5 | 0 | 2 | Not-determined | Genome-fragment | NA | NA | 0 |
| S8-final-viral-combined_k141_225109\|\|full | 2172 | No | NA | 2 | 0 | 0 | Not-determined | Genome-fragment | NA | NA | 0 |
| S1-final-viral-combined_k141_213332\|\|full | 12217 | No | NA | 14 | 0 | 1 | Low-quality | Genome-fragment | 9.17 | AAI-based (medium-confidence) | 0 |
| S8-final-viral-combined_k141_98639\|\|full | 1048 | No | NA | 2 | 0 | 1 | Not-determined | Genome-fragment | NA | NA | 0 |
| S8-final-viral-combined_k141_98648\|\|full | 5936 | No | NA | 6 | 0 | 5 | Not-determined | Genome-fragment | NA | NA | 0 |
| S8-final-viral-combined_k141_41246\|\|full | 3345 | No | NA | 2 | 0 | 1 | Not-determined | Genome-fragment | NA | NA | 0 |
| S1-final-viral-combined_k141_106034\|\|full | 12224 | No | NA | 18 | 4 | 0 | Low-quality | Genome-fragment | 13.38 | HMM-based (lower-bound) | 0 |
| S8-final-viral-combined_k141_144857\|\|full | 9058 | No | NA | 6 | 0 | 4 | Not-determined | Genome-fragment | NA | NA | 0 |
| S8-final-viral-combined_k141_248566\|\|full | 4125 | No | NA | 6 | 0 | 1 | Not-determined | Genome-fragment | NA | NA | 0 |
| S5-final-viral-combined_k141_35379\|\|0_partial | 12227 | Yes | 4314 | 14 | 4 | 4 | Low-quality | Genome-fragment | 6.22 | HMM-based (lower-bound) | 64.72 |
| S8-final-viral-combined_k141_248568\|\|full | 1234 | No | NA | 4 | 0 | 0 | Not-determined | Genome-fragment | NA | NA | 0 |
| S8-final-viral-combined_k141_258423\|\|full | 1125 | No | NA | 2 | 0 | 1 | Not-determined | Genome-fragment | NA | NA | 0 |
| S11-final-viral-combined_k141_180726\|\|full | 12228 | Yes | 1786 | 13 | 4 | 8 | Low-quality | Genome-fragment | 2.27 | HMM-based (lower-bound) | 85.39 |
| S12-final-viral-combined_k141_74622\|\|0_partial | 12228 | No | NA | 14 | 1 | 10 | Low-quality | Genome-fragment | 23.53 | HMM-based (lower-bound) | 0 |
| S1_k141_368077 | 12235 | No | NA | 8 | 0 | 8 | Low-quality | Genome-fragment | 5.34 | AAI-based (medium-confidence) | 0 |
| S8-final-viral-combined_k141_225114\|\|full | 51279 | No | NA | 77 | 28 | 0 | High-quality | High-quality | 96.08 | AAI-based (high-confidence) | 0 |
| S7-final-viral-combined_k141_54196\|\|full | 12235 | No | NA | 7 | 0 | 6 | Low-quality | Genome-fragment | 6.52 | AAI-based (medium-confidence) | 0 |
| S11_k141_302522 | 12245 | No | NA | 14 | 3 | 0 | Low-quality | Genome-fragment | 26.64 | AAI-based (high-confidence) | 0 |
| S9_k141_37713 | 12248 | No | NA | 18 | 7 | 0 | Low-quality | Genome-fragment | 29.13 | AAI-based (high-confidence) | 0 |
| S12_k141_353288 | 12255 | No | NA | 17 | 2 | 1 | Low-quality | Genome-fragment | 10.88 | HMM-based (lower-bound) | 0 |
| S8-final-viral-combined_k141_271422\|\|full | 2120 | No | NA | 2 | 0 | 2 | Not-determined | Genome-fragment | NA | NA | 0 |
| S8-final-viral-combined_k141_64539\|\|full | 3229 | No | NA | 4 | 0 | 2 | Not-determined | Genome-fragment | NA | NA | 0 |
| S8-final-viral-combined_k141_5829\|\|full | 1331 | No | NA | 2 | 0 | 1 | Not-determined | Genome-fragment | NA | NA | 0 |
| S12-final-viral-combined_k141_836\|\|full | 12263 | No | NA | 11 | 1 | 1 | Low-quality | Genome-fragment | 18.56 | HMM-based (lower-bound) | 0 |
| S12-final-viral-combined_k141_319588\|\|full | 12264 | No | NA | 19 | 8 | 0 | Low-quality | Genome-fragment | 20.44 | AAI-based (high-confidence) | 0 |
| S1-final-viral-combined_k141_22044\|\|full | 12270 | No | NA | 22 | 4 | 0 | Low-quality | Genome-fragment | 31.63 | AAI-based (high-confidence) | 0 |
| S8-final-viral-combined_k141_258477\|\|full | 1503 | No | NA | 3 | 0 | 1 | Not-determined | Genome-fragment | NA | NA | 0 |
| S8-final-viral-combined_k141_121444\|\|full | 1785 | No | NA | 3 | 0 | 0 | Not-determined | Genome-fragment | NA | NA | 0 |
| S8-final-viral-combined_k141_248637\|\|full | 2014 | No | NA | 3 | 0 | 0 | Not-determined | Genome-fragment | NA | NA | 0 |
| S8-final-viral-combined_k141_75850\|\|full | 3300 | No | NA | 4 | 0 | 2 | Not-determined | Genome-fragment | NA | NA | 0 |
| S11-final-viral-combined_k141_110390\|\|full | 12276 | No | NA | 14 | 1 | 5 | Low-quality | Genome-fragment | 8.25 | HMM-based (lower-bound) | 0 |
| S8-final-viral-combined_k141_180279\|\|full | 4448 | No | NA | 4 | 0 | 3 | Not-determined | Genome-fragment | NA | NA | 0 |
| S8-final-viral-combined_k141_156301\|\|full | 2529 | No | NA | 4 | 0 | 3 | Not-determined | Genome-fragment | NA | NA | 0 |
| S5-final-viral-combined_k141_113189\|\|full | 12281 | No | NA | 16 | 1 | 8 | Low-quality | Genome-fragment | 20.36 | AAI-based (medium-confidence) | 0 |
| S8-final-viral-combined_k141_86622\|\|full | 1725 | No | NA | 3 | 0 | 1 | Not-determined | Genome-fragment | NA | NA | 0 |
| S8-final-viral-combined_k141_109765\|\|full | 2675 | No | NA | 2 | 0 | 0 | Not-determined | Genome-fragment | NA | NA | 0 |
| S8-final-viral-combined_k141_52477\|\|full | 2302 | No | NA | 3 | 0 | 0 | Not-determined | Genome-fragment | NA | NA | 0 |
| S8-final-viral-combined_k141_167766\|\|full | 3242 | No | NA | 5 | 0 | 0 | Not-determined | Genome-fragment | NA | NA | 0 |
| S8-final-viral-combined_k141_144975\|\|full | 1128 | No | NA | 3 | 0 | 0 | Not-determined | Genome-fragment | NA | NA | 0 |
| S17-final-viral-combined_k141_133121\|\|full | 12281 | No | NA | 17 | 7 | 1 | Low-quality | Genome-fragment | 18.8 | AAI-based (high-confidence) | 0 |
| S8-final-viral-combined_k141_86661\|\|full | 1844 | No | NA | 2 | 0 | 1 | Not-determined | Genome-fragment | NA | NA | 0 |
| S1-final-viral-combined_k141_105813\|\|full | 12283 | No | NA | 13 | 3 | 0 | Low-quality | Genome-fragment | 32.25 | AAI-based (high-confidence) | 0 |
| S5-final-viral-combined_k141_178725\|\|full | 12290 | No | NA | 16 | 7 | 0 | Low-quality | Genome-fragment | 26.09 | AAI-based (high-confidence) | 0 |
| S8_k141_67145 | 12293 | No | NA | 16 | 3 | 0 | Low-quality | Genome-fragment | 12.87 | AAI-based (high-confidence) | 0 |
| S3-final-viral-combined_k141_10860\|\|full | 12295 | No | NA | 16 | 6 | 0 | Low-quality | Genome-fragment | 21.36 | AAI-based (high-confidence) | 0 |
| S8-final-viral-combined_k141_271528\|\|full | 5994 | No | NA | 6 | 0 | 2 | Not-determined | Genome-fragment | NA | NA | 0 |
| S8-final-viral-combined_k141_213694\|\|full | 10065 | No | NA | 5 | 0 | 4 | Not-determined | Genome-fragment | NA | NA | 0 |
| S8-final-viral-combined_k141_237494\|\|full | 15991 | No | NA | 15 | 0 | 10 | Not-determined | Genome-fragment | NA | NA | 0 |
| S8-final-viral-combined_k141_41388\|\|full | 3787 | No | NA | 5 | 0 | 5 | Not-determined | Genome-fragment | NA | NA | 0 |
| S8-final-viral-combined_k141_201327\|\|full | 1105 | No | NA | 3 | 0 | 1 | Not-determined | Genome-fragment | NA | NA | 0 |
| S8-final-viral-combined_k141_180382\|\|full | 1064 | No | NA | 2 | 0 | 1 | Not-determined | Genome-fragment | NA | NA | 0 |
| S8-final-viral-combined_k141_30131\|\|full | 2551 | No | NA | 6 | 0 | 0 | Not-determined | Genome-fragment | NA | NA | 0 |
| S8-final-viral-combined_k141_248719\|\|full | 1718 | No | NA | 2 | 0 | 2 | Not-determined | Genome-fragment | NA | NA | 0 |
| S8-final-viral-combined_k141_225268\|\|full | 1556 | No | NA | 2 | 0 | 1 | Not-determined | Genome-fragment | NA | NA | 0 |
| S5-final-viral-combined_k141_124904\|\|0_partial | 12298 | Yes | 6755 | 14 | 2 | 3 | Low-quality | Genome-fragment | 9.85 | HMM-based (lower-bound) | 45.07 |
| S8-final-viral-combined_k141_201366\|\|full | 2110 | No | NA | 2 | 0 | 0 | Not-determined | Genome-fragment | NA | NA | 0 |
| S8-final-viral-combined_k141_167869\|\|full | 2843 | No | NA | 2 | 0 | 1 | Not-determined | Genome-fragment | NA | NA | 0 |
| S8-final-viral-combined_k141_201367\|\|full | 1873 | No | NA | 2 | 0 | 1 | Not-determined | Genome-fragment | NA | NA | 0 |
| S8-final-viral-combined_k141_258578\|\|full | 1359 | No | NA | 2 | 0 | 1 | Not-determined | Genome-fragment | NA | NA | 0 |
| S8-final-viral-combined_k141_133466\|\|full | 12489 | No | NA | 12 | 0 | 9 | Not-determined | Genome-fragment | NA | NA | 0 |
| S8-final-viral-combined_k141_75963\|\|full | 1208 | No | NA | 2 | 0 | 1 | Not-determined | Genome-fragment | NA | NA | 0 |
| S1-final-viral-combined_k141_128375\|\|full | 12300 | No | NA | 28 | 8 | 0 | Low-quality | Genome-fragment | 25.87 | AAI-based (high-confidence) | 0 |
| S8-final-viral-combined_k141_225299\|\|full | 1614 | No | NA | 2 | 0 | 0 | Not-determined | Genome-fragment | NA | NA | 0 |
| S8-final-viral-combined_k141_145084\|\|full | 6301 | No | NA | 7 | 0 | 5 | Not-determined | Genome-fragment | NA | NA | 0 |
| S5-final-viral-combined_k141_155902\|\|0_partial | 12307 | No | NA | 14 | 1 | 3 | Low-quality | Genome-fragment | 31.28 | AAI-based (high-confidence) | 0 |
| S8-final-viral-combined_k141_6000\|\|full | 4066 | No | NA | 4 | 0 | 1 | Not-determined | Genome-fragment | NA | NA | 0 |
| S8-final-viral-combined_k141_30224\|\|full | 3730 | No | NA | 8 | 0 | 1 | Not-determined | Genome-fragment | NA | NA | 0 |
| S1_k141_311018 | 12312 | No | NA | 17 | 13 | 0 | Low-quality | Genome-fragment | 27.12 | AAI-based (high-confidence) | 0 |
| S8-final-viral-combined_k141_145138\|\|full | 1008 | No | NA | 2 | 0 | 1 | Not-determined | Genome-fragment | NA | NA | 0 |
| S8-final-viral-combined_k141_191048\|\|full | 2003 | No | NA | 3 | 0 | 0 | Not-determined | Genome-fragment | NA | NA | 0 |
| S8-final-viral-combined_k141_30234\|\|full | 1208 | No | NA | 3 | 0 | 2 | Not-determined | Genome-fragment | NA | NA | 0 |
| S5-final-viral-combined_k141_73929\|\|0_partial | 12328 | Yes | 9210 | 15 | 2 | 4 | Low-quality | Genome-fragment | 6.23 | AAI-based (medium-confidence) | 25.29 |
| S8-final-viral-combined_k141_237584\|\|full | 1489 | No | NA | 3 | 0 | 2 | Not-determined | Genome-fragment | NA | NA | 0 |
| S8-final-viral-combined_k141_213858\|\|full | 2037 | No | NA | 4 | 0 | 0 | Not-determined | Genome-fragment | NA | NA | 0 |
| S11-final-viral-combined_k141_148271\|\|full | 12341 | No | NA | 11 | 5 | 0 | Low-quality | Genome-fragment | 20.36 | AAI-based (medium-confidence) | 0 |
| S8-final-viral-combined_k141_237585\|\|full | 1795 | No | NA | 3 | 0 | 0 | Not-determined | Genome-fragment | NA | NA | 0 |
| S17-final-viral-combined_k141_42139\|\|full | 12341 | No | NA | 26 | 3 | 0 | Low-quality | Genome-fragment | 31.31 | AAI-based (high-confidence) | 0 |
| S8-final-viral-combined_k141_225410\|\|full | 2471 | No | NA | 3 | 0 | 3 | Not-determined | Genome-fragment | NA | NA | 0 |
| S8-final-viral-combined_k141_213876\|\|full | 2803 | No | NA | 5 | 0 | 4 | Not-determined | Genome-fragment | NA | NA | 0 |
| S8-final-viral-combined_k141_271684\|\|full | 2918 | No | NA | 4 | 0 | 1 | Not-determined | Genome-fragment | NA | NA | 0 |
| S8-final-viral-combined_k141_41535\|\|full | 6041 | No | NA | 6 | 0 | 3 | Not-determined | Genome-fragment | NA | NA | 0 |
| S8-final-viral-combined_k141_191110\|\|full | 1297 | No | NA | 3 | 0 | 2 | Not-determined | Genome-fragment | NA | NA | 0 |
| S8-final-viral-combined_k141_98929\|\|full | 5762 | No | NA | 6 | 0 | 3 | Not-determined | Genome-fragment | NA | NA | 0 |
| S8-final-viral-combined_k141_110007\|\|full | 3163 | No | NA | 5 | 0 | 0 | Not-determined | Genome-fragment | NA | NA | 0 |
| S8-final-viral-combined_k141_110009\|\|full | 1278 | No | NA | 2 | 0 | 0 | Not-determined | Genome-fragment | NA | NA | 0 |
| S12-final-viral-combined_k141_393497\|\|full | 12359 | No | NA | 15 | 1 | 7 | Low-quality | Genome-fragment | 20.85 | HMM-based (lower-bound) | 0 |
| S11_k141_208877 | 12368 | No | NA | 13 | 3 | 0 | Low-quality | Genome-fragment | 13.18 | AAI-based (high-confidence) | 0 |
| S8-final-viral-combined_k141_133599\|\|full | 1264 | No | NA | 2 | 0 | 0 | Not-determined | Genome-fragment | NA | NA | 0 |
| S8-final-viral-combined_k141_237616\|\|full | 2202 | No | NA | 3 | 0 | 2 | Not-determined | Genome-fragment | NA | NA | 0 |
| S11-final-viral-combined_k141_97207\|\|full | 12379 | No | NA | 16 | 1 | 5 | Low-quality | Genome-fragment | 7.84 | HMM-based (lower-bound) | 0 |
| S8-final-viral-combined_k141_156563\|\|full | 4466 | No | NA | 4 | 0 | 4 | Not-determined | Genome-fragment | NA | NA | 0 |
| S8-final-viral-combined_k141_23909\|\|full | 12386 | No | NA | 8 | 0 | 0 | Low-quality | Genome-fragment | 17.44 | AAI-based (medium-confidence) | 0 |
| S8-final-viral-combined_k141_168032\|\|full | 3106 | No | NA | 3 | 0 | 2 | Not-determined | Genome-fragment | NA | NA | 0 |
| S5-final-viral-combined_k141_191791\|\|0_partial | 12415 | No | NA | 16 | 0 | 5 | Low-quality | Genome-fragment | 30.57 | AAI-based (medium-confidence) | 0 |
| S7-final-viral-combined_k141_4932\|\|full | 12415 | No | NA | 6 | 0 | 0 | Low-quality | Genome-fragment | 32.81 | AAI-based (high-confidence) | 0 |
| S11-final-viral-combined_k141_273104\|\|full | 12430 | No | NA | 25 | 5 | 0 | Low-quality | Genome-fragment | 27.1 | AAI-based (high-confidence) | 0 |
| S8-final-viral-combined_k141_201513\|\|full | 1521 | No | NA | 3 | 0 | 1 | Not-determined | Genome-fragment | NA | NA | 0 |
| S8-final-viral-combined_k141_86869\|\|full | 4440 | No | NA | 3 | 0 | 3 | Not-determined | Genome-fragment | NA | NA | 0 |
| S8-final-viral-combined_k141_76073\|\|full | 6507 | No | NA | 6 | 0 | 2 | Not-determined | Genome-fragment | NA | NA | 0 |
| S8-final-viral-combined_k141_6145\|\|full | 1644 | No | NA | 3 | 0 | 1 | Not-determined | Genome-fragment | NA | NA | 0 |
| S8-final-viral-combined_k141_213967\|\|full | 2051 | No | NA | 4 | 0 | 1 | Not-determined | Genome-fragment | NA | NA | 0 |
| S2-final-viral-combined_k141_142899\|\|full | 12452 | No | NA | 12 | 1 | 2 | Low-quality | Genome-fragment | 8.97 | HMM-based (lower-bound) | 0 |
| S8-final-viral-combined_k141_41580\|\|full | 9068 | No | NA | 14 | 0 | 1 | Not-determined | Genome-fragment | NA | NA | 0 |
| S1-final-viral-combined_k141_38210\|\|full | 12464 | No | NA | 23 | 3 | 1 | Low-quality | Genome-fragment | 20.14 | AAI-based (high-confidence) | 0 |
| S8-final-viral-combined_k141_226044\|\|0_partial | 12467 | No | NA | 12 | 1 | 3 | Low-quality | Genome-fragment | 32.65 | AAI-based (medium-confidence) | 0 |
| S8-final-viral-combined_k141_258738\|\|full | 1683 | No | NA | 3 | 0 | 2 | Not-determined | Genome-fragment | NA | NA | 0 |
| S8-final-viral-combined_k141_76107\|\|full | 3095 | No | NA | 4 | 0 | 4 | Not-determined | Genome-fragment | NA | NA | 0 |
| S1-final-viral-combined_k141_303217\|\|full | 12482 | No | NA | 18 | 10 | 0 | Low-quality | Genome-fragment | 22.04 | HMM-based (lower-bound) | 0 |
| S12_k141_313973 | 12496 | Yes | 6504 | 10 | 1 | 1 | Low-quality | Genome-fragment | 7.23 | AAI-based (medium-confidence) | 47.95 |
| S8-final-viral-combined_k141_110084\|\|full | 3036 | No | NA | 3 | 0 | 2 | Not-determined | Genome-fragment | NA | NA | 0 |
| S2-final-viral-combined_k141_20388\|\|full | 12497 | No | NA | 16 | 9 | 0 | Low-quality | Genome-fragment | 32.54 | AAI-based (medium-confidence) | 0 |
| S1-final-viral-combined_k141_86054\|\|full | 12511 | No | NA | 21 | 3 | 1 | Low-quality | Genome-fragment | 33.46 | AAI-based (high-confidence) | 0 |
| S8-final-viral-combined_k141_110102\|\|full | 3411 | No | NA | 3 | 0 | 2 | Not-determined | Genome-fragment | NA | NA | 0 |
| S8_k141_131219 | 12534 | No | NA | 19 | 1 | 0 | Low-quality | Genome-fragment | 23.68 | AAI-based (high-confidence) | 0 |
| S8-final-viral-combined_k141_225586\|\|full | 1962 | No | NA | 3 | 0 | 1 | Not-determined | Genome-fragment | NA | NA | 0 |
| S2-final-viral-combined_k141_94265\|\|full | 12535 | Yes | 9041 | 16 | 10 | 3 | Low-quality | Genome-fragment | 19.22 | AAI-based (high-confidence) | 27.87 |
| S8-final-viral-combined_k141_225590\|\|full | 4183 | No | NA | 6 | 0 | 2 | Not-determined | Genome-fragment | NA | NA | 0 |
| S8-final-viral-combined_k141_41661\|\|full | 4733 | No | NA | 4 | 0 | 4 | Not-determined | Genome-fragment | NA | NA | 0 |
| S8-final-viral-combined_k141_156736\|\|full | 18096 | No | NA | 15 | 0 | 11 | Not-determined | Genome-fragment | NA | NA | 0 |
| S8-final-viral-combined_k141_237730\|\|full | 5143 | No | NA | 3 | 0 | 3 | Not-determined | Genome-fragment | NA | NA | 0 |
| S12-final-viral-combined_k141_66124\|\|full | 12538 | No | NA | 15 | 1 | 0 | Low-quality | Genome-fragment | 20.26 | AAI-based (high-confidence) | 0 |
| S17-final-viral-combined_k141_241675\|\|full | 12540 | No | NA | 12 | 0 | 1 | Low-quality | Genome-fragment | 43.47 | AAI-based (high-confidence) | 0 |
| S3-final-viral-combined_k141_60628\|\|full | 12541 | No | NA | 17 | 12 | 0 | Low-quality | Genome-fragment | 35.13 | AAI-based (high-confidence) | 0 |
| S8-final-viral-combined_k141_168194\|\|full | 1696 | No | NA | 3 | 0 | 0 | Not-determined | Genome-fragment | NA | NA | 0 |
| S6-final-viral-combined_k141_30386\|\|full | 12541 | No | NA | 20 | 7 | 1 | Low-quality | Genome-fragment | 19.87 | HMM-based (lower-bound) | 0 |
| S8-final-viral-combined_k141_6242\|\|full | 2455 | No | NA | 3 | 0 | 0 | Not-determined | Genome-fragment | NA | NA | 0 |
| S8-final-viral-combined_k141_99151\|\|full | 2666 | No | NA | 3 | 0 | 0 | Not-determined | Genome-fragment | NA | NA | 0 |
| S8-final-viral-combined_k141_52779\|\|full | 4450 | No | NA | 4 | 0 | 2 | Not-determined | Genome-fragment | NA | NA | 0 |
| S1_k141_254826 | 12542 | No | NA | 14 | 0 | 3 | Low-quality | Genome-fragment | 20.08 | AAI-based (medium-confidence) | 0 |
| S8-final-viral-combined_k141_99156\|\|full | 6508 | No | NA | 4 | 0 | 4 | Not-determined | Genome-fragment | NA | NA | 0 |
| S8-final-viral-combined_k141_133779\|\|full | 1054 | No | NA | 2 | 0 | 0 | Not-determined | Genome-fragment | NA | NA | 0 |
| S8-final-viral-combined_k141_52806\|\|full | 1459 | No | NA | 1 | 0 | 0 | Not-determined | Genome-fragment | NA | NA | 0 |
| S8-final-viral-combined_k141_258826\|\|full | 4642 | No | NA | 5 | 0 | 0 | Not-determined | Genome-fragment | NA | NA | 0 |
| S8-final-viral-combined_k141_52815\|\|full | 1100 | No | NA | 3 | 0 | 1 | Not-determined | Genome-fragment | NA | NA | 0 |
| S8-final-viral-combined_k141_110137\|\|full | 3732 | No | NA | 3 | 0 | 1 | Not-determined | Genome-fragment | NA | NA | 0 |
| S8-final-viral-combined_k141_99192\|\|full | 1732 | No | NA | 2 | 0 | 1 | Not-determined | Genome-fragment | NA | NA | 0 |
| S8-final-viral-combined_k141_86981\|\|full | 1418 | No | NA | 3 | 0 | 0 | Not-determined | Genome-fragment | NA | NA | 0 |
| S8-final-viral-combined_k141_214092\|\|full | 1543 | No | NA | 2 | 0 | 0 | Not-determined | Genome-fragment | NA | NA | 0 |
| S8-final-viral-combined_k141_145369\|\|full | 5965 | No | NA | 4 | 0 | 3 | Not-determined | Genome-fragment | NA | NA | 0 |
| S8-final-viral-combined_k141_145375\|\|full | 1031 | No | NA | 3 | 0 | 1 | Not-determined | Genome-fragment | NA | NA | 0 |
| S8-final-viral-combined_k141_258849\|\|full | 1984 | No | NA | 2 | 0 | 1 | Not-determined | Genome-fragment | NA | NA | 0 |
| S5-final-viral-combined_k141_20957\|\|0_partial | 12547 | No | NA | 17 | 1 | 8 | Low-quality | Genome-fragment | 14.08 | HMM-based (lower-bound) | 0 |
| S8-final-viral-combined_k141_191341\|\|full | 1838 | No | NA | 4 | 0 | 2 | Not-determined | Genome-fragment | NA | NA | 0 |
| S8-final-viral-combined_k141_145381\|\|full | 3097 | No | NA | 3 | 0 | 1 | Not-determined | Genome-fragment | NA | NA | 0 |
| S8-final-viral-combined_k141_30540\|\|full | 4547 | No | NA | 3 | 0 | 3 | Not-determined | Genome-fragment | NA | NA | 0 |
| S5-final-viral-combined_k141_171975\|\|full | 12548 | No | NA | 13 | 1 | 1 | Low-quality | Genome-fragment | 14.09 | HMM-based (lower-bound) | 0 |
| S8-final-viral-combined_k141_133844\|\|full | 2811 | No | NA | 4 | 0 | 0 | Not-determined | Genome-fragment | NA | NA | 0 |
| S5-final-viral-combined_k141_147938\|\|0_partial | 12549 | No | NA | 13 | 3 | 0 | Low-quality | Genome-fragment | 21.77 | AAI-based (medium-confidence) | 0 |
| S8-final-viral-combined_k141_225671\|\|full | 1246 | No | NA | 2 | 0 | 0 | Not-determined | Genome-fragment | NA | NA | 0 |
| S8-final-viral-combined_k141_258886\|\|full | 2060 | No | NA | 3 | 0 | 1 | Not-determined | Genome-fragment | NA | NA | 0 |
| S8-final-viral-combined_k141_30585\|\|full | 1539 | No | NA | 3 | 0 | 0 | Not-determined | Genome-fragment | NA | NA | 0 |
| S11_k141_325353 | 12562 | No | NA | 10 | 9 | 0 | Low-quality | Genome-fragment | 31.15 | AAI-based (high-confidence) | 0 |
| S8-final-viral-combined_k141_145424\|\|full | 1770 | No | NA | 3 | 0 | 0 | Not-determined | Genome-fragment | NA | NA | 0 |
| S8-final-viral-combined_k141_99216\|\|full | 4578 | No | NA | 5 | 0 | 2 | Not-determined | Genome-fragment | NA | NA | 0 |
| S8-final-viral-combined_k141_271929\|\|full | 3648 | No | NA | 4 | 0 | 4 | Not-determined | Genome-fragment | NA | NA | 0 |
| S8-final-viral-combined_k141_249008\|\|full | 1198 | No | NA | 2 | 0 | 1 | Not-determined | Genome-fragment | NA | NA | 0 |
| S17-final-viral-combined_k141_80637\|\|full | 12563 | Yes | 7875 | 18 | 4 | 3 | Low-quality | Genome-fragment | 9.38 | HMM-based (lower-bound) | 37.32 |
| S8-final-viral-combined_k141_145445\|\|full | 1222 | No | NA | 3 | 0 | 0 | Not-determined | Genome-fragment | NA | NA | 0 |
| S8-final-viral-combined_k141_168336\|\|full | 1543 | No | NA | 2 | 0 | 1 | Not-determined | Genome-fragment | NA | NA | 0 |
| S8-final-viral-combined_k141_191417\|\|full | 1290 | No | NA | 2 | 0 | 1 | Not-determined | Genome-fragment | NA | NA | 0 |
| S7-final-viral-combined_k141_243236\|\|full | 12572 | Yes | 5995 | 11 | 3 | 4 | Low-quality | Genome-fragment | 12.9 | AAI-based (high-confidence) | 52.31 |
| S8-final-viral-combined_k141_30669\|\|full | 1473 | No | NA | 3 | 0 | 0 | Not-determined | Genome-fragment | NA | NA | 0 |
| S8-final-viral-combined_k141_214183\|\|full | 3196 | No | NA | 4 | 0 | 3 | Not-determined | Genome-fragment | NA | NA | 0 |
| S8-final-viral-combined_k141_133892\|\|full | 28115 | No | NA | 24 | 0 | 16 | Not-determined | Genome-fragment | NA | NA | 0 |
| S1-final-viral-combined_k141_126717\|\|full | 12593 | No | NA | 8 | 0 | 8 | Low-quality | Genome-fragment | 4.78 | AAI-based (medium-confidence) | 0 |
| S2-final-viral-combined_k141_156323\|\|full | 12593 | No | NA | 16 | 1 | 1 | Low-quality | Genome-fragment | 7.21 | HMM-based (lower-bound) | 0 |
| S8-final-viral-combined_k141_237823\|\|full | 3640 | No | NA | 3 | 0 | 3 | Not-determined | Genome-fragment | NA | NA | 0 |
| S8-final-viral-combined_k141_237831\|\|full | 1088 | No | NA | 2 | 0 | 1 | Not-determined | Genome-fragment | NA | NA | 0 |
| S8-final-viral-combined_k141_156936\|\|full | 1868 | No | NA | 3 | 0 | 0 | Not-determined | Genome-fragment | NA | NA | 0 |
| S8-final-viral-combined_k141_87156\|\|full | 1865 | No | NA | 3 | 0 | 0 | Not-determined | Genome-fragment | NA | NA | 0 |
| S8-final-viral-combined_k141_214200\|\|full | 2558 | No | NA | 4 | 0 | 2 | Not-determined | Genome-fragment | NA | NA | 0 |
| S12-final-viral-combined_k141_376584\|\|full | 12597 | No | NA | 16 | 6 | 0 | Low-quality | Genome-fragment | 19.08 | HMM-based (lower-bound) | 0 |
| S8-final-viral-combined_k141_156983\|\|full | 3374 | No | NA | 4 | 0 | 1 | Not-determined | Genome-fragment | NA | NA | 0 |
| S12-final-viral-combined_k141_383230\|\|full | 12608 | No | NA | 10 | 0 | 4 | Low-quality | Genome-fragment | 10.56 | AAI-based (medium-confidence) | 0 |
| S1_k141_158392 | 12616 | No | NA | 24 | 2 | 0 | Low-quality | Genome-fragment | 34.52 | AAI-based (high-confidence) | 0 |
| S7-final-viral-combined_k141_56372\|\|0_partial | 12618 | Yes | 2935 | 13 | 2 | 5 | Low-quality | Genome-fragment | 7.32 | AAI-based (medium-confidence) | 76.74 |
| S8-final-viral-combined_k141_70390\|\|full | 12636 | No | NA | 19 | 7 | 0 | Low-quality | Genome-fragment | 28.76 | AAI-based (high-confidence) | 0 |
| S8-final-viral-combined_k141_65021\|\|full | 1485 | No | NA | 3 | 0 | 0 | Not-determined | Genome-fragment | NA | NA | 0 |
| S8-final-viral-combined_k141_87256\|\|full | 2377 | No | NA | 4 | 0 | 2 | Not-determined | Genome-fragment | NA | NA | 0 |
| S8-final-viral-combined_k141_3252\|\|full | 12656 | No | NA | 13 | 0 | 7 | Low-quality | Genome-fragment | 9.84 | AAI-based (medium-confidence) | 0 |
| S12-final-viral-combined_k141_236737\|\|0_partial | 12683 | No | NA | 18 | 1 | 7 | Low-quality | Genome-fragment | 8.53 | HMM-based (lower-bound) | 0 |
| S3-final-viral-combined_k141_64496\|\|full | 12699 | No | NA | 23 | 2 | 1 | Low-quality | Genome-fragment | 8.07 | HMM-based (lower-bound) | 0 |
| S8-final-viral-combined_k141_30785\|\|full | 1366 | No | NA | 2 | 0 | 1 | Not-determined | Genome-fragment | NA | NA | 0 |
| S17-final-viral-combined_k141_198228\|\|full | 12708 | No | NA | 21 | 4 | 0 | Low-quality | Genome-fragment | 28.93 | AAI-based (high-confidence) | 0 |
| S8-final-viral-combined_k141_157105\|\|full | 1875 | No | NA | 3 | 0 | 2 | Not-determined | Genome-fragment | NA | NA | 0 |
| S8-final-viral-combined_k141_30804\|\|full | 7487 | No | NA | 7 | 0 | 2 | Not-determined | Genome-fragment | NA | NA | 0 |
| S12-final-viral-combined_k141_366345\|\|full | 12710 | No | NA | 15 | 1 | 0 | Low-quality | Genome-fragment | 28.43 | AAI-based (medium-confidence) | 0 |
| S8-final-viral-combined_k141_17247\|\|full | 2206 | No | NA | 2 | 0 | 0 | Not-determined | Genome-fragment | NA | NA | 0 |
| S8-final-viral-combined_k141_272120\|\|full | 1279 | No | NA | 2 | 0 | 0 | Not-determined | Genome-fragment | NA | NA | 0 |
| S8-final-viral-combined_k141_65100\|\|full | 2306 | No | NA | 2 | 0 | 0 | Not-determined | Genome-fragment | NA | NA | 0 |
| S8-final-viral-combined_k141_225836\|\|full | 6141 | No | NA | 4 | 0 | 3 | Not-determined | Genome-fragment | NA | NA | 0 |
| S1-final-viral-combined_k141_228010\|\|full | 12717 | No | NA | 19 | 1 | 3 | Low-quality | Genome-fragment | 9.77 | AAI-based (high-confidence) | 0 |
| S8-final-viral-combined_k141_17259\|\|full | 2158 | No | NA | 3 | 0 | 3 | Not-determined | Genome-fragment | NA | NA | 0 |
| S8-final-viral-combined_k141_272127\|\|full | 1078 | No | NA | 3 | 0 | 0 | Not-determined | Genome-fragment | NA | NA | 0 |
| S11-final-viral-combined_k141_36324\|\|full | 12719 | Yes | 7947 | 22 | 2 | 2 | Low-quality | Genome-fragment | 22.4 | AAI-based (high-confidence) | 37.52 |
| S8-final-viral-combined_k141_180942\|\|full | 1032 | No | NA | 2 | 0 | 1 | Not-determined | Genome-fragment | NA | NA | 0 |
| S8-final-viral-combined_k141_134098\|\|full | 23377 | No | NA | 38 | 7 | 0 | Medium-quality | Genome-fragment | 54.32 | AAI-based (high-confidence) | 0 |
| S11-final-viral-combined_k141_183174\|\|full | 12726 | No | NA | 17 | 9 | 0 | Low-quality | Genome-fragment | 21.79 | AAI-based (high-confidence) | 0 |
| S8-final-viral-combined_k141_65127\|\|full | 11260 | No | NA | 10 | 0 | 6 | Not-determined | Genome-fragment | NA | NA | 0 |
| S8-final-viral-combined_k141_30868\|\|full | 1082 | No | NA | 3 | 0 | 0 | Not-determined | Genome-fragment | NA | NA | 0 |
| S17-final-viral-combined_k141_97582\|\|full | 12731 | No | NA | 19 | 6 | 0 | Low-quality | Genome-fragment | 31.26 | AAI-based (high-confidence) | 0 |
| S8-final-viral-combined_k141_238031\|\|full | 2322 | No | NA | 3 | 0 | 1 | Not-determined | Genome-fragment | NA | NA | 0 |
| S8-final-viral-combined_k141_180957\|\|full | 20853 | No | NA | 22 | 4 | 0 | Medium-quality | Genome-fragment | 51.7 | AAI-based (medium-confidence) | 0 |
| S11-final-viral-combined_k141_187733\|\|full | 12764 | No | NA | 16 | 6 | 0 | Low-quality | Genome-fragment | 35.05 | AAI-based (medium-confidence) | 0 |
| S8-final-viral-combined_k141_41803\|\|full | 1580 | No | NA | 3 | 0 | 2 | Not-determined | Genome-fragment | NA | NA | 0 |
| S6-final-viral-combined_k141_106449\|\|full | 12776 | No | NA | 16 | 5 | 0 | Low-quality | Genome-fragment | 37.22 | AAI-based (high-confidence) | 0 |
| S8-final-viral-combined_k141_201844\|\|full | 1211 | No | NA | 3 | 0 | 3 | Not-determined | Genome-fragment | NA | NA | 0 |
| S11_k141_244323 | 12780 | No | NA | 15 | 10 | 1 | Low-quality | Genome-fragment | 22.07 | AAI-based (high-confidence) | 0 |
| S12-final-viral-combined_k141_123991\|\|0_partial | 12783 | No | NA | 13 | 2 | 1 | Low-quality | Genome-fragment | 23.58 | AAI-based (medium-confidence) | 0 |
| S11_k141_326002 | 12794 | No | NA | 16 | 1 | 3 | Low-quality | Genome-fragment | 6.94 | HMM-based (lower-bound) | 0 |
| S7_k141_108203 | 12808 | No | NA | 18 | 1 | 2 | Low-quality | Genome-fragment | 5.51 | HMM-based (lower-bound) | 0 |
| S8-final-viral-combined_k141_201849\|\|full | 1172 | No | NA | 2 | 0 | 2 | Not-determined | Genome-fragment | NA | NA | 0 |
| S8-final-viral-combined_k141_87465\|\|full | 4092 | No | NA | 4 | 0 | 0 | Not-determined | Genome-fragment | NA | NA | 0 |
| S8-final-viral-combined_k141_238067\|\|full | 2478 | No | NA | 3 | 0 | 3 | Not-determined | Genome-fragment | NA | NA | 0 |
| S1-final-viral-combined_k141_239427\|\|full | 12811 | No | NA | 26 | 1 | 0 | Low-quality | Genome-fragment | 21.5 | AAI-based (high-confidence) | 0 |
| S12-final-viral-combined_k141_218262\|\|0_partial | 12811 | Yes | 10613 | 10 | 1 | 1 | Low-quality | Genome-fragment | 24.8 | AAI-based (medium-confidence) | 17.16 |
| S8-final-viral-combined_k141_115875\|\|full | 12821 | No | NA | 18 | 8 | 0 | Low-quality | Genome-fragment | 22.02 | AAI-based (high-confidence) | 0 |
| S4-final-viral-combined_k141_154378\|\|full | 12828 | No | NA | 22 | 4 | 2 | Low-quality | Genome-fragment | 35.91 | AAI-based (high-confidence) | 0 |
| S4-final-viral-combined_k141_6948\|\|0_partial | 12828 | No | NA | 16 | 3 | 1 | Low-quality | Genome-fragment | 44.72 | AAI-based (high-confidence) | 0 |
| S8-final-viral-combined_k141_6522\|\|full | 4167 | No | NA | 2 | 0 | 2 | Not-determined | Genome-fragment | NA | NA | 0 |
| S12_k141_240870 | 12833 | No | NA | 13 | 0 | 0 | Low-quality | Genome-fragment | 30.6 | AAI-based (high-confidence) | 0 |
| S8-final-viral-combined_k141_76613\|\|full | 3666 | No | NA | 4 | 0 | 2 | Not-determined | Genome-fragment | NA | NA | 0 |
| S12-final-viral-combined_k141_35106\|\|0_partial | 12834 | No | NA | 13 | 1 | 2 | Low-quality | Genome-fragment | 5.11 | HMM-based (lower-bound) | 0 |
| S2-final-viral-combined_k141_90860\|\|full | 12840 | Yes | 7795 | 25 | 3 | 3 | Low-quality | Genome-fragment | 18.5 | AAI-based (high-confidence) | 39.29 |
| S8-final-viral-combined_k141_214398\|\|full | 1608 | No | NA | 4 | 0 | 0 | Not-determined | Genome-fragment | NA | NA | 0 |
| S8-final-viral-combined_k141_65222\|\|full | 1250 | No | NA | 2 | 0 | 2 | Not-determined | Genome-fragment | NA | NA | 0 |

| | | | | | | | | | | | |
|---|---|---|---|---|---|---|---|---|---|---|---|
| S1-final-viral-combined_k141_184295\|\|full | 12848 | Yes | 9761 | 10 | 1 | 1 | Low-quality | Genome-fragment | 3.5 | HMM-based (lower-bound) | 24.03 |
| S8-final-viral-combined_k141_87570\|\|full | 1896 | No | NA | 4 | 0 | 0 | Not-determined | Genome-fragment | NA | NA | 0 |
| S1-final-viral-combined_k141_219614\|\|full | 12857 | No | NA | 12 | 1 | 0 | Low-quality | Genome-fragment | 31.33 | AAI-based (high-confidence) | 0 |
| S1-final-viral-combined_k141_85217\|\|full | 12864 | No | NA | 19 | 0 | 5 | Low-quality | Genome-fragment | 20.59 | AAI-based (medium-confidence) | 0 |
| S8-final-viral-combined_k141_238154\|\|full | 3106 | No | NA | 4 | 0 | 2 | Not-determined | Genome-fragment | NA | NA | 0 |
| S8-final-viral-combined_k141_122200\|\|full | 1726 | No | NA | 2 | 0 | 0 | Not-determined | Genome-fragment | NA | NA | 0 |
| S8-final-viral-combined_k141_181120\|\|full | 4025 | No | NA | 3 | 0 | 2 | Not-determined | Genome-fragment | NA | NA | 0 |
| S7-final-viral-combined_k141_187480\|\|full | 12865 | No | NA | 13 | 1 | 7 | Low-quality | Genome-fragment | 17.29 | HMM-based (lower-bound) | 0 |
| S8-final-viral-combined_k141_168637\|\|full | 1081 | No | NA | 3 | 0 | 1 | Not-determined | Genome-fragment | NA | NA | 0 |
| S8-final-viral-combined_k141_225941\|\|full | 1983 | No | NA | 3 | 0 | 0 | Not-determined | Genome-fragment | NA | NA | 0 |
| S8-final-viral-combined_k141_65269\|\|full | 1664 | No | NA | 5 | 0 | 4 | Not-determined | Genome-fragment | NA | NA | 0 |
| S8-final-viral-combined_k141_214453\|\|full | 1614 | No | NA | 3 | 0 | 2 | Not-determined | Genome-fragment | NA | NA | 0 |
| S8-final-viral-combined_k141_155052\|\|full | 12866 | No | NA | 18 | 6 | 1 | Low-quality | Genome-fragment | 35.97 | AAI-based (high-confidence) | 0 |
| S3-final-viral-combined_k141_129583\|\|full | 12869 | No | NA | 17 | 4 | 1 | Low-quality | Genome-fragment | 30.7 | AAI-based (high-confidence) | 0 |
| S8-final-viral-combined_k141_272268\|\|full | 3387 | No | NA | 3 | 0 | 0 | Not-determined | Genome-fragment | NA | NA | 0 |
| S1-final-viral-combined_k141_347934\|\|full | 12874 | No | NA | 3 | 0 | 0 | Low-quality | Genome-fragment | 33.72 | AAI-based (high-confidence) | 0 |
| S7-final-viral-combined_k141_11408\|\|full | 12878 | No | NA | 14 | 1 | 0 | Low-quality | Genome-fragment | 7.94 | HMM-based (lower-bound) | 0 |
| S8-final-viral-combined_k141_122252\|\|full | 5047 | No | NA | 5 | 0 | 1 | Not-determined | Genome-fragment | NA | NA | 0 |
| S8-final-viral-combined_k141_145864\|\|full | 1225 | No | NA | 2 | 0 | 0 | Not-determined | Genome-fragment | NA | NA | 0 |
| S9-final-viral-combined_k141_103478\|\|0_partial | 12881 | Yes | 9657 | 20 | 3 | 5 | Low-quality | Genome-fragment | 18.92 | HMM-based (lower-bound) | 25.03 |
| S8-final-viral-combined_k141_168694\|\|full | 1676 | No | NA | 2 | 0 | 2 | Not-determined | Genome-fragment | NA | NA | 0 |
| S8-final-viral-combined_k141_134284\|\|full | 26978 | No | NA | 21 | 5 | 0 | Medium-quality | Genome-fragment | 65.74 | AAI-based (high-confidence) | 0 |
| S7-final-viral-combined_k141_163781\|\|0_partial | 12903 | Yes | 4910 | 14 | 2 | 4 | Low-quality | Genome-fragment | 1.62 | HMM-based (lower-bound) | 61.95 |
| S8-final-viral-combined_k141_249367\|\|full | 2483 | No | NA | 4 | 0 | 2 | Not-determined | Genome-fragment | NA | NA | 0 |
| S8-final-viral-combined_k141_76704\|\|full | 1243 | No | NA | 2 | 0 | 0 | Not-determined | Genome-fragment | NA | NA | 0 |
| S8-final-viral-combined_k141_201961\|\|full | 3245 | No | NA | 5 | 0 | 2 | Not-determined | Genome-fragment | NA | NA | 0 |
| S4_k141_501 | 12903 | No | NA | 31 | 6 | 0 | Low-quality | Genome-fragment | 30.52 | AAI-based (high-confidence) | 0 |
| S8-final-viral-combined_k141_214510\|\|full | 1812 | No | NA | 3 | 0 | 1 | Not-determined | Genome-fragment | NA | NA | 0 |
| S8-final-viral-combined_k141_214541\|\|full | 1739 | No | NA | 2 | 0 | 2 | Not-determined | Genome-fragment | NA | NA | 0 |
| S8-final-viral-combined_k141_17401\|\|full | 6393 | No | NA | 3 | 0 | 3 | Not-determined | Genome-fragment | NA | NA | 0 |
| S8-final-viral-combined_k141_168757\|\|full | 3507 | No | NA | 4 | 0 | 1 | Not-determined | Genome-fragment | NA | NA | 0 |
| S8-final-viral-combined_k141_53307\|\|full | 3345 | No | NA | 5 | 0 | 0 | Not-determined | Genome-fragment | NA | NA | 0 |
| S8_k141_11757 | 12904 | No | NA | 18 | 9 | 0 | Low-quality | Genome-fragment | 30.34 | AAI-based (high-confidence) | 0 |
| S7-final-viral-combined_k141_132590\|\|full | 12920 | No | NA | 14 | 1 | 1 | Low-quality | Genome-fragment | 19.55 | HMM-based (lower-bound) | 0 |
| S8-final-viral-combined_k141_6630\|\|full | 1594 | No | NA | 3 | 0 | 2 | Not-determined | Genome-fragment | NA | NA | 0 |
| S8-final-viral-combined_k141_202032\|\|full | 2499 | No | NA | 3 | 0 | 1 | Not-determined | Genome-fragment | NA | NA | 0 |
| S8-final-viral-combined_k141_238391\|\|full | 1706 | No | NA | 3 | 0 | 2 | Not-determined | Genome-fragment | NA | NA | 0 |
| S5_k141_170727 | 12922 | No | NA | 12 | 0 | 3 | Low-quality | Genome-fragment | 23.58 | AAI-based (medium-confidence) | 0 |
| S8-final-viral-combined_k141_42075\|\|full | 6190 | No | NA | 6 | 0 | 4 | Not-determined | Genome-fragment | NA | NA | 0 |
| S12-final-viral-combined_k141_106251\|\|full | 12924 | No | NA | 18 | 3 | 0 | Low-quality | Genome-fragment | 10.77 | AAI-based (medium-confidence) | 0 |
| S2_k141_119739 | 12927 | No | NA | 12 | 2 | 0 | Low-quality | Genome-fragment | 7.38 | AAI-based (high-confidence) | 0 |
| S17-k141_50561 | 12929 | No | NA | 15 | 1 | 3 | Low-quality | Genome-fragment | 19.56 | HMM-based (lower-bound) | 0 |
| S6-final-viral-combined_k141_76046\|\|full | 12940 | No | NA | 13 | 1 | 2 | Low-quality | Genome-fragment | 11.69 | AAI-based (medium-confidence) | 0 |
| S8-final-viral-combined_k141_146005\|\|full | 1100 | No | NA | 2 | 0 | 0 | Not-determined | Genome-fragment | NA | NA | 0 |
| S11-final-viral-combined_k141_182056\|\|full | 12945 | No | NA | 10 | 0 | 7 | Low-quality | Genome-fragment | 6.47 | AAI-based (medium-confidence) | 0 |
| S8-final-viral-combined_k141_99718\|\|full | 1115 | No | NA | 2 | 0 | 1 | Not-determined | Genome-fragment | NA | NA | 0 |
| S8-final-viral-combined_k141_87689\|\|full | 3465 | No | NA | 3 | 0 | 2 | Not-determined | Genome-fragment | NA | NA | 0 |
| S8-final-viral-combined_k141_226076\|\|full | 52058 | No | NA | 60 | 7 | 0 | High-quality | High-quality | 99.81 | AAI-based (high-confidence) | 0 |
| S8-final-viral-combined_k141_214626\|\|full | 16833 | No | NA | 12 | 0 | 6 | Not-determined | Genome-fragment | NA | NA | 0 |
| S2-final-viral-combined_k141_212690\|\|full | 12945 | No | NA | 13 | 3 | 0 | Low-quality | Genome-fragment | 7.83 | HMM-based (lower-bound) | 0 |
| S8-final-viral-combined_k141_2267\|\|full | 12957 | No | NA | 15 | 1 | 0 | Low-quality | Genome-fragment | 29.74 | AAI-based (high-confidence) | 0 |
| S8-final-viral-combined_k141_259324\|\|full | 2675 | No | NA | 4 | 0 | 1 | Not-determined | Genome-fragment | NA | NA | 0 |
| S8-final-viral-combined_k141_134493\|\|full | 4524 | No | NA | 9 | 0 | 2 | Not-determined | Genome-fragment | NA | NA | 0 |
| S2-final-viral-combined_k141_110728\|\|full | 12958 | Yes | 5467 | 20 | 2 | 5 | Low-quality | Genome-fragment | 9.76 | HMM-based (lower-bound) | 57.81 |
| S9-final-viral-combined_k141_71960\|\|full | 12965 | No | NA | 17 | 1 | 5 | Low-quality | Genome-fragment | 8.72 | HMM-based (lower-bound) | 0 |
| S11-final-viral-combined_k141_113055\|\|0_partial | 12976 | No | NA | 11 | 1 | 1 | Low-quality | Genome-fragment | 18.08 | HMM-based (lower-bound) | 0 |
| S8-final-viral-combined_k141_238450\|\|full | 2159 | No | NA | 4 | 0 | 3 | Not-determined | Genome-fragment | NA | NA | 0 |
| S8-final-viral-combined_k141_53450\|\|full | 2272 | No | NA | 2 | 0 | 1 | Not-determined | Genome-fragment | NA | NA | 0 |
| S8-final-viral-combined_k141_76858\|\|full | 2811 | No | NA | 3 | 0 | 1 | Not-determined | Genome-fragment | NA | NA | 0 |
| S8-final-viral-combined_k141_168930\|\|full | 3061 | No | NA | 3 | 0 | 3 | Not-determined | Genome-fragment | NA | NA | 0 |
| S1_k141_184140 | 12978 | No | NA | 16 | 9 | 0 | Low-quality | Genome-fragment | 20.86 | AAI-based (high-confidence) | 0 |
| S8-final-viral-combined_k141_99755\|\|full | 1871 | No | NA | 2 | 0 | 1 | Not-determined | Genome-fragment | NA | NA | 0 |
| S8-final-viral-combined_k141_259363\|\|full | 1269 | No | NA | 3 | 0 | 0 | Not-determined | Genome-fragment | NA | NA | 0 |
| S8-final-viral-combined_k141_110641\|\|full | 4268 | No | NA | 4 | 0 | 2 | Not-determined | Genome-fragment | NA | NA | 0 |
| S8-final-viral-combined_k141_31323\|\|full | 1423 | No | NA | 2 | 0 | 0 | Not-determined | Genome-fragment | NA | NA | 0 |
| S8-final-viral-combined_k141_214667\|\|full | 2902 | No | NA | 4 | 0 | 4 | Not-determined | Genome-fragment | NA | NA | 0 |
| S8-final-viral-combined_k141_226162\|\|full | 2051 | No | NA | 2 | 0 | 2 | Not-determined | Genome-fragment | NA | NA | 0 |
| S5-final-viral-combined_k141_206359\|\|0_partial | 12999 | Yes | 2868 | 16 | 3 | 6 | Low-quality | Genome-fragment | 3.61 | HMM-based (lower-bound) | 77.94 |
| S8-final-viral-combined_k141_6783\|\|full | 6855 | No | NA | 5 | 0 | 2 | Not-determined | Genome-fragment | NA | NA | 0 |
| S8-final-viral-combined_k141_249573\|\|full | 2103 | No | NA | 2 | 0 | 0 | Not-determined | Genome-fragment | NA | NA | 0 |
| S8-final-viral-combined_k141_76945\|\|full | 1604 | No | NA | 3 | 0 | 1 | Not-determined | Genome-fragment | NA | NA | 0 |
| S8-final-viral-combined_k141_99792\|\|full | 3789 | No | NA | 4 | 0 | 1 | Not-determined | Genome-fragment | NA | NA | 0 |
| S8-final-viral-combined_k141_53555\|\|full | 2925 | No | NA | 3 | 0 | 3 | Not-determined | Genome-fragment | NA | NA | 0 |
| S12-final-viral-combined_k141_363126\|\|0_partial | 12999 | No | NA | 16 | 1 | 5 | Low-quality | Genome-fragment | 14.59 | HMM-based (lower-bound) | 0 |
| S8-final-viral-combined_k141_249608\|\|full | 1094 | No | NA | 2 | 0 | 1 | Not-determined | Genome-fragment | NA | NA | 0 |
| S7-final-viral-combined_k141_119732\|\|0_partial | 13006 | No | NA | 9 | 0 | 4 | Low-quality | Genome-fragment | 13.97 | AAI-based (medium-confidence) | 0 |
| S8-final-viral-combined_k141_259437\|\|full | 1062 | No | NA | 3 | 0 | 3 | Not-determined | Genome-fragment | NA | NA | 0 |
| S8-final-viral-combined_k141_134587\|\|full | 3518 | No | NA | 4 | 0 | 2 | Not-determined | Genome-fragment | NA | NA | 0 |
| S8-final-viral-combined_k141_259438\|\|full | 12246 | No | NA | 12 | 0 | 6 | Not-determined | Genome-fragment | NA | NA | 0 |
| S8-final-viral-combined_k141_110801\|\|full | 1519 | No | NA | 2 | 0 | 0 | Not-determined | Genome-fragment | NA | NA | 0 |
| S8-final-viral-combined_k141_157607\|\|full | 2046 | No | NA | 4 | 0 | 1 | Not-determined | Genome-fragment | NA | NA | 0 |
| S8-final-viral-combined_k141_146233\|\|full | 1522 | No | NA | 2 | 0 | 1 | Not-determined | Genome-fragment | NA | NA | 0 |
| S1-final-viral-combined_k141_156458\|\|full | 13009 | No | NA | 15 | 7 | 0 | Low-quality | Genome-fragment | 34.03 | AAI-based (high-confidence) | 0 |
| S1_k141_197349 | 13021 | No | NA | 18 | 1 | 1 | Low-quality | Genome-fragment | 16.2 | AAI-based (high-confidence) | 0 |
| S3_k141_25214 | 13023 | No | NA | 22 | 5 | 0 | Low-quality | Genome-fragment | 12.47 | AAI-based (high-confidence) | 0 |
| S1_k141_68105 | 13049 | No | NA | 11 | 0 | 4 | Low-quality | Genome-fragment | 12.94 | AAI-based (medium-confidence) | 0 |
| S8_k141_187987 | 13049 | No | NA | 9 | 2 | 0 | Low-quality | Genome-fragment | 9.18 | AAI-based (high-confidence) | 0 |
| S8-final-viral-combined_k141_65689\|\|full | 1042 | No | NA | 2 | 0 | 1 | Not-determined | Genome-fragment | NA | NA | 0 |
| S7-final-viral-combined_k141_53754\|\|0_partial | 13050 | Yes | 8671 | 13 | 3 | 4 | Low-quality | Genome-fragment | 12.86 | HMM-based (lower-bound) | 33.56 |
| S8-final-viral-combined_k141_146274\|\|full | 1219 | No | NA | 4 | 0 | 0 | Not-determined | Genome-fragment | NA | NA | 0 |
| S8-final-viral-combined_k141_92329\|\|0_partial | 13055 | Yes | 7431 | 16 | 4 | 7 | Low-quality | Genome-fragment | 14.63 | AAI-based (medium-confidence) | 43.08 |
| S8-final-viral-combined_k141_6955\|\|full | 1253 | No | NA | 2 | 0 | 2 | Not-determined | Genome-fragment | NA | NA | 0 |
| S8-final-viral-combined_k141_157703\|\|full | 2043 | No | NA | 5 | 0 | 1 | Not-determined | Genome-fragment | NA | NA | 0 |
| S8-final-viral-combined_k141_272563\|\|full | 6384 | No | NA | 4 | 0 | 3 | Not-determined | Genome-fragment | NA | NA | 0 |
| S8-final-viral-combined_k141_214864\|\|full | 2661 | No | NA | 4 | 0 | 1 | Not-determined | Genome-fragment | NA | NA | 0 |
| S8-final-viral-combined_k141_214866\|\|full | 2607 | No | NA | 3 | 0 | 3 | Not-determined | Genome-fragment | NA | NA | 0 |
| S8-final-viral-combined_k141_17713\|\|full | 6868 | No | NA | 6 | 0 | 4 | Not-determined | Genome-fragment | NA | NA | 0 |
| S1-final-viral-combined_k141_192780\|\|full | 13063 | No | NA | 20 | 7 | 0 | Low-quality | Genome-fragment | 29.94 | HMM-based (lower-bound) | 0 |
| S8-final-viral-combined_k141_226313\|\|full | 6671 | No | NA | 5 | 0 | 4 | Not-determined | Genome-fragment | NA | NA | 0 |
| S5-final-viral-combined_k141_19526\|\|full | 13071 | No | NA | 18 | 7 | 0 | Low-quality | Genome-fragment | 31.27 | AAI-based (medium-confidence) | 0 |
| S8-final-viral-combined_k141_214872\|\|full | 7186 | No | NA | 12 | 0 | 2 | Not-determined | Genome-fragment | NA | NA | 0 |
| S8-final-viral-combined_k141_99988\|\|full | 1552 | No | NA | 4 | 0 | 0 | Not-determined | Genome-fragment | NA | NA | 0 |
| S8-final-viral-combined_k141_259507\|\|full | 1408 | No | NA | 1 | 0 | 0 | Not-determined | Genome-fragment | NA | NA | 0 |
| S12-final-viral-combined_k141_6407\|\|full | 13075 | No | NA | 17 | 2 | 0 | Low-quality | Genome-fragment | 25.31 | AAI-based (medium-confidence) | 0 |
| S8-final-viral-combined_k141_191981\|\|full | 2051 | No | NA | 4 | 0 | 0 | Not-determined | Genome-fragment | NA | NA | 0 |
| S2-final-viral-combined_k141_30395\|\|full | 13081 | No | NA | 15 | 2 | 1 | Low-quality | Genome-fragment | 9.49 | HMM-based (lower-bound) | 0 |
| S8-final-viral-combined_k141_214925\|\|full | 1826 | No | NA | 2 | 0 | 1 | Not-determined | Genome-fragment | NA | NA | 0 |
| S17-final-viral-combined_k141_56878\|\|full | 13091 | No | NA | 9 | 0 | 1 | Low-quality | Genome-fragment | 15.9 | AAI-based (high-confidence) | 0 |
| S11-final-viral-combined_k141_53204\|\|0_partial | 13105 | No | NA | 18 | 3 | 0 | Low-quality | Genome-fragment | 7.95 | AAI-based (medium-confidence) | 0 |
| S11-final-viral-combined_k141_98981\|\|0_partial | 13121 | No | NA | 13 | 1 | 6 | Low-quality | Genome-fragment | 31.41 | AAI-based (high-confidence) | 0 |
| S8-final-viral-combined_k141_202428\|\|full | 6439 | No | NA | 7 | 0 | 4 | Not-determined | Genome-fragment | NA | NA | 0 |
| S8-final-viral-combined_k141_157794\|\|full | 2782 | No | NA | 4 | 0 | 1 | Not-determined | Genome-fragment | NA | NA | 0 |
| S12-final-viral-combined_k141_108456\|\|full | 13122 | No | NA | 18 | 13 | 0 | Low-quality | Genome-fragment | 32.37 | AAI-based (medium-confidence) | 0 |
| S8-final-viral-combined_k141_7099\|\|full | 2489 | No | NA | 3 | 0 | 0 | Not-determined | Genome-fragment | NA | NA | 0 |
| S8-final-viral-combined_k141_238810\|\|full | 11496 | No | NA | 11 | 0 | 10 | Not-determined | Genome-fragment | NA | NA | 0 |
| S8-final-viral-combined_k141_272663\|\|full | 3293 | No | NA | 5 | 0 | 1 | Not-determined | Genome-fragment | NA | NA | 0 |
| S1-final-viral-combined_k141_216985\|\|0_partial | 13139 | Yes | 7239 | 17 | 4 | 3 | Low-quality | Genome-fragment | 21.68 | AAI-based (high-confidence) | 44.9 |
| S8-final-viral-combined_k141_7111\|\|full | 5068 | No | NA | 3 | 0 | 2 | Not-determined | Genome-fragment | NA | NA | 0 |
| S8-final-viral-combined_k141_226435\|\|full | 1034 | No | NA | 2 | 0 | 1 | Not-determined | Genome-fragment | NA | NA | 0 |
| S7_k141_88997 | 13141 | No | NA | 20 | 12 | 0 | Low-quality | Genome-fragment | 36.2 | AAI-based (high-confidence) | 0 |
| S8-final-viral-combined_k141_31689\|\|full | 2423 | No | NA | 3 | 0 | 1 | Not-determined | Genome-fragment | NA | NA | 0 |
| S8-final-viral-combined_k141_146490\|\|full | 12467 | No | NA | 5 | 0 | 5 | Not-determined | Genome-fragment | NA | NA | 0 |
| S8-final-viral-combined_k141_122795\|\|full | 1314 | No | NA | 4 | 0 | 1 | Not-determined | Genome-fragment | NA | NA | 0 |
| S7-final-viral-combined_k141_69467\|\|full | 13144 | No | NA | 13 | 1 | 2 | Low-quality | Genome-fragment | 9.09 | AAI-based (high-confidence) | 0 |
| S8-final-viral-combined_k141_238884\|\|full | 2906 | No | NA | 6 | 0 | 0 | Not-determined | Genome-fragment | NA | NA | 0 |
| S8-final-viral-combined_k141_259653\|\|full | 3595 | No | NA | 4 | 0 | 2 | Not-determined | Genome-fragment | NA | NA | 0 |
| S8-final-viral-combined_k141_122828\|\|full | 5215 | No | NA | 7 | 0 | 5 | Not-determined | Genome-fragment | NA | NA | 0 |
| S8-final-viral-combined_k141_134781\|\|full | 4713 | No | NA | 4 | 0 | 3 | Not-determined | Genome-fragment | NA | NA | 0 |
| S8-final-viral-combined_k141_215025\|\|full | 3119 | No | NA | 3 | 0 | 2 | Not-determined | Genome-fragment | NA | NA | 0 |
| S8-final-viral-combined_k141_226476\|\|full | 2826 | No | NA | 3 | 0 | 2 | Not-determined | Genome-fragment | NA | NA | 0 |
| S1_k141_80302 | 13147 | No | NA | 17 | 9 | 0 | Low-quality | Genome-fragment | 33.98 | AAI-based (high-confidence) | 0 |
| S8-final-viral-combined_k141_53845\|\|full | 7738 | No | NA | 5 | 0 | 5 | Not-determined | Genome-fragment | NA | NA | 0 |
| S8-final-viral-combined_k141_7145\|\|full | 1529 | No | NA | 3 | 0 | 0 | Not-determined | Genome-fragment | NA | NA | 0 |
| S8-final-viral-combined_k141_42451\|\|full | 1663 | No | NA | 3 | 0 | 0 | Not-determined | Genome-fragment | NA | NA | 0 |
| S7-final-viral-combined_k141_63256\|\|0_partial | 13155 | No | NA | 12 | 1 | 3 | Low-quality | Genome-fragment | 1.09 | HMM-based (lower-bound) | 0 |
| S8-final-viral-combined_k141_31728\|\|full | 2273 | No | NA | 2 | 0 | 1 | Not-determined | Genome-fragment | NA | NA | 0 |
| S1-final-viral-combined_k141_5759\|\|0_partial | 13164 | No | NA | 11 | 1 | 9 | Low-quality | Genome-fragment | 5.52 | AAI-based (medium-confidence) | 0 |
| S4-final-viral-combined_k141_93625\|\|0_partial | 13171 | No | NA | 12 | 3 | 1 | Low-quality | Genome-fragment | 8.98 | HMM-based (lower-bound) | 0 |
| S8-final-viral-combined_k141_215056\|\|full | 11597 | No | NA | 14 | 0 | 2 | Not-determined | Genome-fragment | NA | NA | 0 |
| S8-final-viral-combined_k141_100090\|\|full | 1190 | No | NA | 2 | 0 | 1 | Not-determined | Genome-fragment | NA | NA | 0 |
| S8-final-viral-combined_k141_106223\|\|0_partial | 13184 | No | NA | 10 | 1 | 8 | Low-quality | Genome-fragment | 7.09 | HMM-based (lower-bound) | 0 |
| S4-final-viral-combined_k141_151281\|\|full | 13189 | No | NA | 17 | 3 | 2 | Low-quality | Genome-fragment | 46.84 | HMM-based (lower-bound) | 0 |
| S8-final-viral-combined_k141_226502\|\|full | 9884 | No | NA | 9 | 0 | 3 | Not-determined | Genome-fragment | NA | NA | 0 |
| S5-final-viral-combined_k141_39517\|\|full | 13193 | No | NA | 15 | 1 | 4 | Low-quality | Genome-fragment | 10.36 | HMM-based (lower-bound) | 0 |
| S11-final-viral-combined_k141_311238\|\|full | 13202 | No | NA | 19 | 7 | 0 | Low-quality | Genome-fragment | 27.51 | AAI-based (high-confidence) | 0 |
| S8-final-viral-combined_k141_65877\|\|full | 1423 | No | NA | 2 | 0 | 1 | Not-determined | Genome-fragment | NA | NA | 0 |
| S8-final-viral-combined_k141_100117\|\|full | 4874 | No | NA | 8 | 0 | 7 | Not-determined | Genome-fragment | NA | NA | 0 |
| S8-final-viral-combined_k141_7195\|\|full | 2598 | No | NA | 8 | 0 | 0 | Not-determined | Genome-fragment | NA | NA | 0 |
| S8-final-viral-combined_k141_146562\|\|full | 27866 | No | NA | 39 | 19 | 2 | Medium-quality | Genome-fragment | 53.28 | AAI-based (medium-confidence) | 0 |
| S7_k141_107523 | 13218 | No | NA | 23 | 5 | 0 | Low-quality | Genome-fragment | 26.23 | AAI-based (high-confidence) | 0 |
| S8-final-viral-combined_k141_202482\|\|full | 2024 | No | NA | 3 | 0 | 0 | Not-determined | Genome-fragment | NA | NA | 0 |
| S3-final-viral-combined_k141_55397\|\|full | 13245 | No | NA | 20 | 7 | 0 | Low-quality | Genome-fragment | 40.44 | AAI-based (high-confidence) | 0 |
| S8-final-viral-combined_k141_157988\|\|full | 2382 | No | NA | 4 | 0 | 2 | Not-determined | Genome-fragment | NA | NA | 0 |
| S17-k141_229317 | 13261 | No | NA | 19 | 1 | 0 | Low-quality | Genome-fragment | 48.78 | HMM-based (lower-bound) | 0 |
| S8-final-viral-combined_k141_53973\|\|full | 7641 | No | NA | 5 | 0 | 5 | Not-determined | Genome-fragment | NA | NA | 0 |
| S8-final-viral-combined_k141_192183\|\|full | 2837 | No | NA | 2 | 0 | 2 | Not-determined | Genome-fragment | NA | NA | 0 |
| S9-final-viral-combined_k141_39807\|\|0_partial | 13266 | No | NA | 10 | 0 | 7 | Low-quality | Genome-fragment | 1.99 | AAI-based (medium-confidence) | 0 |
| S8-final-viral-combined_k141_17971\|\|full | 2704 | No | NA | 3 | 0 | 2 | Not-determined | Genome-fragment | NA | NA | 0 |
| S12_k141_158887 | 13275 | No | NA | 12 | 9 | 0 | Low-quality | Genome-fragment | 32.9 | AAI-based (medium-confidence) | 0 |
| S8-final-viral-combined_k141_31813\|\|full | 9338 | No | NA | 7 | 0 | 5 | Not-determined | Genome-fragment | NA | NA | 0 |
| S2-final-viral-combined_k141_26313\|\|full | 13281 | No | NA | 17 | 7 | 0 | Low-quality | Genome-fragment | 41.69 | HMM-based (lower-bound) | 0 |
| S8-final-viral-combined_k141_202521\|\|full | 2334 | No | NA | 5 | 0 | 2 | Not-determined | Genome-fragment | NA | NA | 0 |
| S8-final-viral-combined_k141_54003\|\|full | 2362 | No | NA | 2 | 0 | 2 | Not-determined | Genome-fragment | NA | NA | 0 |
| S8-final-viral-combined_k141_202529\|\|full | 2722 | No | NA | 3 | 0 | 3 | Not-determined | Genome-fragment | NA | NA | 0 |
| S8-final-viral-combined_k141_272859\|\|full | 1226 | No | NA | 2 | 0 | 1 | Not-determined | Genome-fragment | NA | NA | 0 |
| S1-final-viral-combined_k141_24555\|\|full | 13287 | No | NA | 15 | 1 | 1 | Low-quality | Genome-fragment | 31.45 | AAI-based (high-confidence) | 0 |
| S8-final-viral-combined_k141_54011\|\|full | 2326 | No | NA | 3 | 0 | 3 | Not-determined | Genome-fragment | NA | NA | 0 |
| S8-final-viral-combined_k141_239065\|\|full | 5350 | No | NA | 5 | 0 | 2 | Not-determined | Genome-fragment | NA | NA | 0 |
| S8-final-viral-combined_k141_122979\|\|full | 1553 | No | NA | 3 | 0 | 1 | Not-determined | Genome-fragment | NA | NA | 0 |
| S12-final-viral-combined_k141_46526\|\|full | 13292 | Yes | 5753 | 7 | 2 | 2 | Low-quality | Genome-fragment | 2.42 | HMM-based (lower-bound) | 56.72 |
| S8-final-viral-combined_k141_239070\|\|full | 4463 | No | NA | 4 | 0 | 4 | Not-determined | Genome-fragment | NA | NA | 0 |
| S8-final-viral-combined_k141_31829\|\|full | 1031 | No | NA | 2 | 0 | 0 | Not-determined | Genome-fragment | NA | NA | 0 |
| S8-final-viral-combined_k141_181844\|\|full | 2063 | No | NA | 2 | 0 | 2 | Not-determined | Genome-fragment | NA | NA | 0 |
| S12-final-viral-combined_k141_361390\|\|0_partial | 13293 | Yes | 460 | 7 | 1 | 4 | Low-quality | Genome-fragment | 0.41 | HMM-based (lower-bound) | 96.54 |
| S8-final-viral-combined_k141_202560\|\|full | 8439 | No | NA | 6 | 0 | 4 | Not-determined | Genome-fragment | NA | NA | 0 |
| S17-final-viral-combined_k141_71030\|\|full | 13295 | No | NA | 15 | 1 | 5 | Low-quality | Genome-fragment | 8.56 | HMM-based (lower-bound) | 0 |
| S8-final-viral-combined_k141_165765\|\|full | 13297 | No | NA | 19 | 5 | 1 | Low-quality | Genome-fragment | 16 | HMM-based (lower-bound) | 0 |
| S8-final-viral-combined_k141_42579\|\|full | 2709 | No | NA | 3 | 0 | 2 | Not-determined | Genome-fragment | NA | NA | 0 |
| S8-final-viral-combined_k141_192227\|\|full | 1758 | No | NA | 2 | 0 | 0 | Not-determined | Genome-fragment | NA | NA | 0 |
| S8-final-viral-combined_k141_31864\|\|full | 2130 | No | NA | 3 | 0 | 0 | Not-determined | Genome-fragment | NA | NA | 0 |
| S11-final-viral-combined_k141_328403\|\|full | 13299 | No | NA | 17 | 1 | 1 | Low-quality | Genome-fragment | 5.1 | HMM-based (lower-bound) | 0 |
| S5_k141_38576 | 13320 | No | NA | 15 | 13 | 0 | Low-quality | Genome-fragment | 33.06 | AAI-based (high-confidence) | 0 |
| S8-final-viral-combined_k141_181882\|\|full | 5884 | No | NA | 5 | 0 | 3 | Not-determined | Genome-fragment | NA | NA | 0 |
| S8-final-viral-combined_k141_7278\|\|full | 2160 | No | NA | 5 | 0 | 1 | Not-determined | Genome-fragment | NA | NA | 0 |
| S8-final-viral-combined_k141_239129\|\|full | 3270 | No | NA | 4 | 0 | 3 | Not-determined | Genome-fragment | NA | NA | 0 |
| S8-final-viral-combined_k141_100214\|\|full | 4060 | No | NA | 4 | 0 | 3 | Not-determined | Genome-fragment | NA | NA | 0 |
| S8-final-viral-combined_k141_77446\|\|full | 2313 | No | NA | 4 | 0 | 3 | Not-determined | Genome-fragment | NA | NA | 0 |
| S5-final-viral-combined_k141_172722\|\|0_partial | 13336 | Yes | 8048 | 18 | 2 | 2 | Low-quality | Genome-fragment | 3.43 | HMM-based (lower-bound) | 39.65 |
| S11-final-viral-combined_k141_157987\|\|full | 13345 | No | NA | 20 | 5 | 1 | Low-quality | Genome-fragment | 16.55 | HMM-based (lower-bound) | 0 |
| S12-final-viral-combined_k141_28763\|\|full | 13345 | No | NA | 19 | 1 | 6 | Low-quality | Genome-fragment | 19.16 | HMM-based (lower-bound) | 0 |
| S11-final-viral-combined_k141_15522\|\|full | 13348 | No | NA | 22 | 2 | 1 | Low-quality | Genome-fragment | 8.48 | HMM-based (lower-bound) | 0 |
| S5-final-viral-combined_k141_213553\|\|0_partial | 13360 | Yes | 10450 | 14 | 2 | 2 | Low-quality | Genome-fragment | 5.87 | AAI-based (medium-confidence) | 21.78 |
| S8-final-viral-combined_k141_226645\|\|full | 3870 | No | NA | 4 | 0 | 1 | Not-determined | Genome-fragment | NA | NA | 0 |
| S8-final-viral-combined_k141_178559\|\|0_partial | 13363 | No | NA | 11 | 1 | 6 | Low-quality | Genome-fragment | 19.02 | HMM-based (lower-bound) | 0 |
| S8-final-viral-combined_k141_42714\|\|full | 1708 | No | NA | 2 | 0 | 1 | Not-determined | Genome-fragment | NA | NA | 0 |
| S8-final-viral-combined_k141_146759\|\|full | 1478 | No | NA | 3 | 0 | 1 | Not-determined | Genome-fragment | NA | NA | 0 |
| S12_k141_5449 | 13366 | No | NA | 20 | 9 | 0 | Low-quality | Genome-fragment | 46.33 | AAI-based (high-confidence) | 0 |
| S8-final-viral-combined_k141_135088\|\|full | 2599 | No | NA | 2 | 0 | 2 | Not-determined | Genome-fragment | NA | NA | 0 |
| S8-final-viral-combined_k141_31954\|\|full | 1121 | No | NA | 3 | 0 | 1 | Not-determined | Genome-fragment | NA | NA | 0 |
| S7_k141_154409 | 13385 | No | NA | 11 | 2 | 0 | Low-quality | Genome-fragment | 22.94 | AAI-based (high-confidence) | 0 |
| S8-final-viral-combined_k141_273063\|\|full | 2621 | No | NA | 3 | 0 | 1 | Not-determined | Genome-fragment | NA | NA | 0 |
| S8-final-viral-combined_k141_42760\|\|full | 5087 | No | NA | 7 | 0 | 2 | Not-determined | Genome-fragment | NA | NA | 0 |
| S9-final-viral-combined_k141_30377\|\|full | 13389 | No | NA | 14 | 2 | 0 | Low-quality | Genome-fragment | 11.7 | HMM-based (lower-bound) | 0 |
| S1_k141_386742 | 13396 | No | NA | 16 | 9 | 1 | Low-quality | Genome-fragment | 27.88 | AAI-based (high-confidence) | 0 |
| S5-final-viral-combined_k141_161907\|\|full | 13398 | No | NA | 17 | 2 | 0 | Low-quality | Genome-fragment | 42.33 | AAI-based (high-confidence) | 0 |
| S4_k141_8591 | 13410 | No | NA | 26 | 8 | 1 | Low-quality | Genome-fragment | 34.02 | AAI-based (high-confidence) | 0 |
| S1_k141_275179 | 13417 | No | NA | 12 | 4 | 0 | Low-quality | Genome-fragment | 22.46 | AAI-based (high-confidence) | 0 |
| S2_k141_208802 | 13417 | No | NA | 11 | 0 | 9 | Low-quality | Genome-fragment | 7.7 | AAI-based (medium-confidence) | 0 |
| S8-final-viral-combined_k141_54183\|\|full | 2047 | No | NA | 2 | 0 | 1 | Not-determined | Genome-fragment | NA | NA | 0 |
| S8-final-viral-combined_k141_239337\|\|full | 1868 | No | NA | 4 | 0 | 1 | Not-determined | Genome-fragment | NA | NA | 0 |
| S8-final-viral-combined_k141_241916\|\|full | 13421 | No | NA | 16 | 4 | 0 | Low-quality | Genome-fragment | 10.13 | AAI-based (high-confidence) | 0 |
| S8-final-viral-combined_k141_239359\|\|full | 1404 | No | NA | 3 | 0 | 2 | Not-determined | Genome-fragment | NA | NA | 0 |
| S5-final-viral-combined_k141_12116\|\|full | 13423 | No | NA | 13 | 3 | 0 | Low-quality | Genome-fragment | 7.97 | HMM-based (lower-bound) | 0 |
| S8-final-viral-combined_k141_135188\|\|full | 6504 | No | NA | 7 | 0 | 0 | Not-determined | Genome-fragment | NA | NA | 0 |
| S8-final-viral-combined_k141_192339\|\|full | 1433 | No | NA | 3 | 0 | 0 | Not-determined | Genome-fragment | NA | NA | 0 |
| S17-final-viral-combined_k141_78587\|\|full | 13434 | No | NA | 16 | 1 | 4 | Low-quality | Genome-fragment | 16.19 | AAI-based (medium-confidence) | 0 |
| S2_k141_25353 | 13436 | No | NA | 29 | 8 | 0 | Low-quality | Genome-fragment | 29.57 | AAI-based (high-confidence) | 0 |
| S8-final-viral-combined_k141_111335\|\|full | 2514 | No | NA | 3 | 0 | 2 | Not-determined | Genome-fragment | NA | NA | 0 |
| S8-final-viral-combined_k141_243625\|\|full | 13439 | Yes | 10453 | 14 | 4 | 3 | Low-quality | Genome-fragment | 26.08 | AAI-based (high-confidence) | 22.22 |
| S11-final-viral-combined_k141_75879\|\|full | 13443 | No | NA | 26 | 3 | 0 | Low-quality | Genome-fragment | 29.95 | AAI-based (high-confidence) | 0 |
| S8-final-viral-combined_k141_77712\|\|full | 3479 | No | NA | 2 | 0 | 2 | Not-determined | Genome-fragment | NA | NA | 0 |
| S8-final-viral-combined_k141_42869\|\|full | 1693 | No | NA | 3 | 0 | 0 | Not-determined | Genome-fragment | NA | NA | 0 |
| S8-final-viral-combined_k141_202688\|\|full | 8292 | No | NA | 8 | 0 | 8 | Not-determined | Genome-fragment | NA | NA | 0 |
| S8-final-viral-combined_k141_202693\|\|full | 1985 | No | NA | 4 | 0 | 1 | Not-determined | Genome-fragment | NA | NA | 0 |
| S1-final-viral-combined_k141_51436\|\|full | 13448 | No | NA | 19 | 4 | 1 | Low-quality | Genome-fragment | 32.29 | AAI-based (medium-confidence) | 0 |
| S8-final-viral-combined_k141_111386\|\|full | 1789 | No | NA | 3 | 0 | 1 | Not-determined | Genome-fragment | NA | NA | 0 |
| S8-final-viral-combined_k141_192410\|\|full | 1972 | No | NA | 3 | 0 | 1 | Not-determined | Genome-fragment | NA | NA | 0 |
| S17-final-viral-combined_k141_132430\|\|full | 13452 | No | NA | 19 | 8 | 1 | Low-quality | Genome-fragment | 33.37 | AAI-based (medium-confidence) | 0 |
| S9-final-viral-combined_k141_118456\|\|full | 13455 | No | NA | 22 | 9 | 0 | Low-quality | Genome-fragment | 31.05 | AAI-based (high-confidence) | 0 |
| S8-final-viral-combined_k141_18185\|\|full | 3517 | No | NA | 5 | 0 | 0 | Not-determined | Genome-fragment | NA | NA | 0 |
| S8-final-viral-combined_k141_4659\|\|full | 13476 | No | NA | 18 | 9 | 0 | Low-quality | Genome-fragment | 34.44 | AAI-based (high-confidence) | 0 |
| S8-final-viral-combined_k141_111410\|\|full | 1772 | No | NA | 2 | 0 | 1 | Not-determined | Genome-fragment | NA | NA | 0 |
| S7-final-viral-combined_k141_27553\|\|0_partial | 13488 | No | NA | 11 | 1 | 2 | Low-quality | Genome-fragment | 20.69 | HMM-based (lower-bound) | 0 |
| S8-final-viral-combined_k141_169688\|\|full | 1066 | No | NA | 2 | 0 | 1 | Not-determined | Genome-fragment | NA | NA | 0 |
| S4-final-viral-combined_k141_116046\|\|full | 13504 | No | NA | 25 | 1 | 0 | Low-quality | Genome-fragment | 8.87 | AAI-based (high-confidence) | 0 |
| S8-final-viral-combined_k141_18217\|\|full | 1745 | No | NA | 2 | 0 | 1 | Not-determined | Genome-fragment | NA | NA | 0 |
| S8-final-viral-combined_k141_239436\|\|full | 2909 | No | NA | 4 | 0 | 4 | Not-determined | Genome-fragment | NA | NA | 0 |
| S2_k141_188028 | 13518 | No | NA | 12 | 1 | 0 | Low-quality | Genome-fragment | 30.44 | AAI-based (medium-confidence) | 0 |
| S8-final-viral-combined_k141_192468\|\|full | 1094 | No | NA | 3 | 0 | 1 | Not-determined | Genome-fragment | NA | NA | 0 |
| S8-final-viral-combined_k141_32196\|\|full | 1764 | No | NA | 2 | 0 | 1 | Not-determined | Genome-fragment | NA | NA | 0 |
| S8-final-viral-combined_k141_42984\|\|full | 5004 | No | NA | 7 | 0 | 1 | Not-determined | Genome-fragment | NA | NA | 0 |
| S8-final-viral-combined_k141_226847\|\|full | 2444 | No | NA | 2 | 0 | 1 | Not-determined | Genome-fragment | NA | NA | 0 |
| S8-final-viral-combined_k141_169741\|\|full | 3618 | No | NA | 4 | 0 | 1 | Not-determined | Genome-fragment | NA | NA | 0 |
| S7-final-viral-combined_k141_144094\|\|full | 13527 | No | NA | 13 | 1 | 5 | Low-quality | Genome-fragment | 8.71 | HMM-based (lower-bound) | 0 |
| S8-final-viral-combined_k141_169745\|\|full | 1374 | No | NA | 3 | 0 | 1 | Not-determined | Genome-fragment | NA | NA | 0 |
| S8-final-viral-combined_k141_43022\|\|full | 3594 | No | NA | 5 | 0 | 2 | Not-determined | Genome-fragment | NA | NA | 0 |
| S2-final-viral-combined_k141_7057\|\|full | 13533 | No | NA | 26 | 5 | 3 | Low-quality | Genome-fragment | 22.73 | AAI-based (high-confidence) | 0 |
| S8-final-viral-combined_k141_32235\|\|full | 1683 | No | NA | 4 | 0 | 1 | Not-determined | Genome-fragment | NA | NA | 0 |
| S17-k141_264801 | 13534 | No | NA | 14 | 0 | 0 | Low-quality | Genome-fragment | 37.03 | AAI-based (medium-confidence) | 0 |
| S8-final-viral-combined_k141_192538\|\|full | 1452 | No | NA | 2 | 0 | 0 | Not-determined | Genome-fragment | NA | NA | 0 |
| S8-final-viral-combined_k141_273107\|\|full | 1175 | No | NA | 2 | 0 | 0 | Not-determined | Genome-fragment | NA | NA | 0 |
| S8-final-viral-combined_k141_169768\|\|full | 3290 | No | NA | 3 | 0 | 3 | Not-determined | Genome-fragment | NA | NA | 0 |
| S11_k141_308145 | 13550 | No | NA | 18 | 0 | 2 | Low-quality | Genome-fragment | 28.66 | AAI-based (medium-confidence) | 0 |
| S5-final-viral-combined_k141_110550\|\|0_partial | 13552 | No | NA | 19 | 1 | 5 | Low-quality | Genome-fragment | 23.35 | HMM-based (lower-bound) | 0 |
| S11_k141_236731 | 13572 | No | NA | 16 | 10 | 0 | Low-quality | Genome-fragment | 26.35 | AAI-based (high-confidence) | 0 |
| S8-final-viral-combined_k141_111512\|\|full | 1589 | No | NA | 3 | 0 | 1 | Not-determined | Genome-fragment | NA | NA | 0 |
| S2_k141_131484 | 13577 | No | NA | 21 | 8 | 0 | Low-quality | Genome-fragment | 27.16 | AAI-based (high-confidence) | 0 |
| S2-final-viral-combined_k141_239885\|\|full | 13580 | No | NA | 19 | 9 | 0 | Low-quality | Genome-fragment | 21.8 | AAI-based (high-confidence) | 0 |
| S2-final-viral-combined_k141_166678\|\|0_partial | 13595 | No | NA | 15 | 1 | 6 | Low-quality | Genome-fragment | 11.14 | HMM-based (lower-bound) | 0 |
| S8-final-viral-combined_k141_100573\|\|full | 1099 | No | NA | 3 | 0 | 0 | Not-determined | Genome-fragment | NA | NA | 0 |
| S1-final-viral-combined_k141_47461\|\|full | 13599 | No | NA | 7 | 1 | 0 | Low-quality | Genome-fragment | 17.83 | AAI-based (high-confidence) | 0 |
| S8_k141_221515 | 13607 | No | NA | 27 | 0 | 2 | Low-quality | Genome-fragment | 20.57 | AAI-based (medium-confidence) | 0 |
| S8-final-viral-combined_k141_123439\|\|full | 1279 | No | NA | 4 | 0 | 1 | Not-determined | Genome-fragment | NA | NA | 0 |
| S8-final-viral-combined_k141_192626\|\|full | 1035 | No | NA | 3 | 0 | 0 | Not-determined | Genome-fragment | NA | NA | 0 |
| S11_k141_292449 | 13620 | No | NA | 8 | 1 | 3 | Low-quality | Genome-fragment | 18.63 | HMM-based (lower-bound) | 0 |
| S8-final-viral-combined_k141_111570\|\|full | 2690 | No | NA | 3 | 0 | 0 | Not-determined | Genome-fragment | NA | NA | 0 |
| S8-final-viral-combined_k141_147204\|\|full | 1888 | No | NA | 3 | 0 | 1 | Not-determined | Genome-fragment | NA | NA | 0 |
| S8-final-viral-combined_k141_7418\|\|full | 3821 | No | NA | 3 | 0 | 2 | Not-determined | Genome-fragment | NA | NA | 0 |
| S8-final-viral-combined_k141_123493\|\|full | 2387 | No | NA | 4 | 0 | 0 | Not-determined | Genome-fragment | NA | NA | 0 |
| S8-final-viral-combined_k141_147213\|\|full | 5675 | No | NA | 8 | 0 | 1 | Not-determined | Genome-fragment | NA | NA | 0 |
| S4-final-viral-combined_k141_132283\|\|full | 13624 | No | NA | 16 | 2 | 2 | Low-quality | Genome-fragment | 12.19 | HMM-based (lower-bound) | 0 |
| S2-final-viral-combined_k141_111661\|\|full | 13640 | No | NA | 21 | 6 | 1 | Low-quality | Genome-fragment | 36.28 | AAI-based (high-confidence) | 0 |
| S3_k141_61814 | 13645 | No | NA | 18 | 1 | 2 | Low-quality | Genome-fragment | 9.04 | AAI-based (high-confidence) | 0 |
| S7-final-viral-combined_k141_224688\|\|0_partial | 13654 | No | NA | 14 | 1 | 6 | Low-quality | Genome-fragment | 19.19 | HMM-based (lower-bound) | 0 |
| S8-final-viral-combined_k141_250331\|\|full | 1586 | No | NA | 3 | 0 | 0 | Not-determined | Genome-fragment | NA | NA | 0 |
| S12-final-viral-combined_k141_125760\|\|full | 13656 | No | NA | 15 | 5 | 0 | Low-quality | Genome-fragment | 21.28 | AAI-based (high-confidence) | 0 |
| S8-final-viral-combined_k141_169835\|\|full | 52956 | Yes | 19532 | 59 | 12 | 16 | Medium-quality | Genome-fragment | 60.76 | AAI-based (high-confidence) | 63.12 |
| S1-final-viral-combined_k141_310670\|\|full | 13667 | Yes | 7931 | 11 | 2 | 4 | Low-quality | Genome-fragment | 18.52 | AAI-based (high-confidence) | 41.97 |
| S9-final-viral-combined_k141_5150\|\|full | 13671 | No | NA | 15 | 1 | 3 | Low-quality | Genome-fragment | 15.35 | HMM-based (lower-bound) | 0 |
| S8-final-viral-combined_k141_100674\|\|full | 7452 | No | NA | 4 | 0 | 3 | Not-determined | Genome-fragment | NA | NA | 0 |
| S8-final-viral-combined_k141_111631\|\|full | 1993 | No | NA | 3 | 0 | 2 | Not-determined | Genome-fragment | NA | NA | 0 |
| S12-final-viral-combined_k141_322930\|\|full | 13684 | No | NA | 29 | 4 | 0 | Low-quality | Genome-fragment | 33.2 | AAI-based (high-confidence) | 0 |
| S8-final-viral-combined_k141_259930\|\|full | 1749 | No | NA | 3 | 0 | 1 | Not-determined | Genome-fragment | NA | NA | 0 |
| S2-final-viral-combined_k141_220063\|\|full | 13687 | No | NA | 17 | 1 | 0 | Low-quality | Genome-fragment | 31.45 | AAI-based (high-confidence) | 0 |
| S1-final-viral-combined_k141_23998\|\|full | 13689 | No | NA | 23 | 0 | 0 | Low-quality | Genome-fragment | 31.55 | AAI-based (medium-confidence) | 0 |
| S12-final-viral-combined_k141_317446\|\|full | 13712 | No | NA | 10 | 0 | 7 | Low-quality | Genome-fragment | 4.42 | AAI-based (medium-confidence) | 0 |
| S8-final-viral-combined_k141_135505\|\|full | 2226 | No | NA | 3 | 0 | 1 | Not-determined | Genome-fragment | NA | NA | 0 |
| S6_k141_46770 | 13727 | No | NA | 14 | 6 | 0 | Low-quality | Genome-fragment | 19.89 | AAI-based (high-confidence) | 0 |
| S12-final-viral-combined_k141_230358\|\|full | 13731 | No | NA | 19 | 8 | 1 | Low-quality | Genome-fragment | 40.75 | AAI-based (high-confidence) | 0 |
| S8-final-viral-combined_k141_63752\|\|full | 13737 | No | NA | 21 | 5 | 0 | Low-quality | Genome-fragment | 16.54 | HMM-based (lower-bound) | 0 |
| S8-final-viral-combined_k141_135528\|\|full | 2548 | No | NA | 2 | 0 | 1 | Not-determined | Genome-fragment | NA | NA | 0 |
| S8-final-viral-combined_k141_32403\|\|full | 1663 | No | NA | 3 | 0 | 1 | Not-determined | Genome-fragment | NA | NA | 0 |
| S8-final-viral-combined_k141_54559\|\|full | 1650 | No | NA | 4 | 0 | 0 | Not-determined | Genome-fragment | NA | NA | 0 |
| S8-final-viral-combined_k141_32411\|\|full | 4206 | No | NA | 6 | 0 | 3 | Not-determined | Genome-fragment | NA | NA | 0 |
| S8-final-viral-combined_k141_78011\|\|full | 1780 | No | NA | 2 | 0 | 2 | Not-determined | Genome-fragment | NA | NA | 0 |
| S8-final-viral-combined_k141_18490\|\|full | 2494 | No | NA | 3 | 0 | 1 | Not-determined | Genome-fragment | NA | NA | 0 |
| S17-final-viral-combined_k141_167117\|\|0_partial | 13756 | No | NA | 13 | 2 | 1 | Low-quality | Genome-fragment | 34.3 | AAI-based (medium-confidence) | 0 |
| S8-final-viral-combined_k141_147366\|\|full | 2241 | No | NA | 4 | 0 | 3 | Not-determined | Genome-fragment | NA | NA | 0 |
| S9_k141_122453 | 13784 | No | NA | 20 | 1 | 2 | Low-quality | Genome-fragment | 28.25 | HMM-based (lower-bound) | 0 |
| S8-final-viral-combined_k141_123652\|\|full | 2993 | No | NA | 3 | 0 | 1 | Not-determined | Genome-fragment | NA | NA | 0 |
| S8-final-viral-combined_k141_100787\|\|full | 1472 | No | NA | 3 | 0 | 2 | Not-determined | Genome-fragment | NA | NA | 0 |
| S8-final-viral-combined_k141_182290\|\|full | 2431 | No | NA | 3 | 0 | 1 | Not-determined | Genome-fragment | NA | NA | 0 |
| S8-final-viral-combined_k141_43298\|\|full | 4067 | No | NA | 3 | 0 | 3 | Not-determined | Genome-fragment | NA | NA | 0 |
| S8-final-viral-combined_k141_54625\|\|full | 1991 | No | NA | 2 | 0 | 1 | Not-determined | Genome-fragment | NA | NA | 0 |
| S8-final-viral-combined_k141_260029\|\|full | 4893 | No | NA | 3 | 0 | 2 | Not-determined | Genome-fragment | NA | NA | 0 |
| S8-final-viral-combined_k141_203063\|\|full | 10933 | No | NA | 9 | 0 | 6 | Not-determined | Genome-fragment | NA | NA | 0 |
| S1_k141_208752 | 13795 | No | NA | 19 | 17 | 0 | Low-quality | Genome-fragment | 35.74 | AAI-based (high-confidence) | 0 |
| S8-final-viral-combined_k141_227151\|\|full | 6544 | No | NA | 7 | 0 | 5 | Not-determined | Genome-fragment | NA | NA | 0 |
| S8-final-viral-combined_k141_203070\|\|full | 1358 | No | NA | 2 | 0 | 0 | Not-determined | Genome-fragment | NA | NA | 0 |
| S8-final-viral-combined_k141_135665\|\|full | 2469 | No | NA | 3 | 0 | 0 | Not-determined | Genome-fragment | NA | NA | 0 |
| S8-final-viral-combined_k141_147458\|\|full | 1592 | No | NA | 2 | 0 | 0 | Not-determined | Genome-fragment | NA | NA | 0 |
| S8-final-viral-combined_k141_158523\|\|full | 3225 | No | NA | 4 | 0 | 2 | Not-determined | Genome-fragment | NA | NA | 0 |
| S4-final-viral-combined_k141_935\|\|full | 13795 | No | NA | 19 | 1 | 6 | Low-quality | Genome-fragment | 19.34 | AAI-based (high-confidence) | 0 |
| S8-final-viral-combined_k141_273353\|\|full | 1233 | No | NA | 2 | 0 | 0 | Not-determined | Genome-fragment | NA | NA | 0 |
| S8-final-viral-combined_k141_169963\|\|full | 2339 | No | NA | 2 | 0 | 1 | Not-determined | Genome-fragment | NA | NA | 0 |
| S8-final-viral-combined_k141_43333\|\|full | 1796 | No | NA | 2 | 0 | 0 | Not-determined | Genome-fragment | NA | NA | 0 |
| S8-final-viral-combined_k141_182332\|\|full | 1910 | No | NA | 3 | 0 | 0 | Not-determined | Genome-fragment | NA | NA | 0 |
| S8-final-viral-combined_k141_43349\|\|full | 1173 | No | NA | 2 | 0 | 0 | Not-determined | Genome-fragment | NA | NA | 0 |
| S3-final-viral-combined_k141_29087\|\|full | 13834 | No | NA | 10 | 0 | 1 | Low-quality | Genome-fragment | 28.13 | AAI-based (high-confidence) | 0 |
| S8-final-viral-combined_k141_18577\|\|full | 6896 | No | NA | 9 | 0 | 3 | Not-determined | Genome-fragment | NA | NA | 0 |
| S7_k141_67056 | 13836 | No | NA | 17 | 10 | 0 | Low-quality | Genome-fragment | 39.76 | AAI-based (high-confidence) | 0 |
| S8-final-viral-combined_k141_239791\|\|full | 33258 | No | NA | 48 | 13 | 0 | High-quality | High-quality | 97.67 | AAI-based (high-confidence) | 0 |
| S11-final-viral-combined_k141_253889\|\|full | 13841 | No | NA | 30 | 3 | 0 | Low-quality | Genome-fragment | 31.06 | AAI-based (high-confidence) | 0 |
| S7-final-viral-combined_k141_222689\|\|full | 13855 | No | NA | 22 | 1 | 2 | Low-quality | Genome-fragment | 3.93 | HMM-based (lower-bound) | 0 |
| S8-final-viral-combined_k141_182355\|\|full | 5023 | No | NA | 5 | 0 | 4 | Not-determined | Genome-fragment | NA | NA | 0 |
| S8-final-viral-combined_k141_123782\|\|full | 1219 | No | NA | 2 | 0 | 0 | Not-determined | Genome-fragment | NA | NA | 0 |
| S11-final-viral-combined_k141_70339\|\|0_partial | 13856 | Yes | 10298 | 13 | 3 | 3 | Low-quality | Genome-fragment | 19.6 | HMM-based (lower-bound) | 25.68 |
| S8-final-viral-combined_k141_89414\|\|full | 13861 | Yes | 11509 | 17 | 2 | 2 | Low-quality | Genome-fragment | 25.94 | AAI-based (high-confidence) | 16.97 |
| S8-final-viral-combined_k141_169996\|\|full | 2109 | No | NA | 4 | 0 | 3 | Not-determined | Genome-fragment | NA | NA | 0 |
| S8-final-viral-combined_k141_224663\|\|full | 13863 | No | NA | 26 | 6 | 1 | Low-quality | Genome-fragment | 26.72 | AAI-based (high-confidence) | 0 |
| S8-final-viral-combined_k141_135768\|\|full | 2005 | No | NA | 3 | 0 | 0 | Not-determined | Genome-fragment | NA | NA | 0 |
| S9-final-viral-combined_k141_46865\|\|full | 13864 | No | NA | 16 | 1 | 7 | Low-quality | Genome-fragment | 9.32 | HMM-based (lower-bound) | 0 |
| S11-final-viral-combined_k141_192004\|\|full | 13874 | No | NA | 13 | 1 | 0 | Low-quality | Genome-fragment | 12.5 | AAI-based (medium-confidence) | 0 |
| S8-final-viral-combined_k141_54754\|\|full | 22989 | No | NA | 23 | 0 | 19 | Not-determined | Genome-fragment | NA | NA | 0 |
| S8-final-viral-combined_k141_239802\|\|full | 4245 | No | NA | 2 | 0 | 1 | Not-determined | Genome-fragment | NA | NA | 0 |
| S11-final-viral-combined_k141_254703\|\|full | 13884 | Yes | 11592 | 14 | 3 | 2 | Low-quality | Genome-fragment | 30.39 | AAI-based (high-confidence) | 16.51 |
| S8-final-viral-combined_k141_88620\|\|full | 1699 | No | NA | 3 | 0 | 0 | Not-determined | Genome-fragment | NA | NA | 0 |
| S8-final-viral-combined_k141_170051\|\|full | 3007 | No | NA | 3 | 0 | 0 | Not-determined | Genome-fragment | NA | NA | 0 |
| S7-final-viral-combined_k141_37846\|\|full | 13900 | No | NA | 17 | 10 | 0 | Low-quality | Genome-fragment | 28.33 | AAI-based (high-confidence) | 0 |
| S8-final-viral-combined_k141_78166\|\|full | 1313 | No | NA | 3 | 0 | 2 | Not-determined | Genome-fragment | NA | NA | 0 |
| S8-final-viral-combined_k141_66432\|\|full | 2117 | No | NA | 3 | 0 | 0 | Not-determined | Genome-fragment | NA | NA | 0 |
| S8-final-viral-combined_k141_54783\|\|full | 1344 | No | NA | 5 | 0 | 0 | Not-determined | Genome-fragment | NA | NA | 0 |
| S2_k141_243335 | 13901 | No | NA | 22 | 4 | 0 | Low-quality | Genome-fragment | 8.92 | AAI-based (high-confidence) | 0 |
| S8-final-viral-combined_k141_100913\|\|full | 2790 | No | NA | 4 | 0 | 1 | Not-determined | Genome-fragment | NA | NA | 0 |
| S8-final-viral-combined_k141_123855\|\|full | 1593 | No | NA | 3 | 0 | 0 | Not-determined | Genome-fragment | NA | NA | 0 |
| S8-final-viral-combined_k141_158638\|\|full | 1945 | No | NA | 4 | 0 | 0 | Not-determined | Genome-fragment | NA | NA | 0 |
| S8-final-viral-combined_k141_260174\|\|full | 3118 | No | NA | 4 | 0 | 3 | Not-determined | Genome-fragment | NA | NA | 0 |
| S17-k141_253220 | 13911 | No | NA | 17 | 3 | 1 | Low-quality | Genome-fragment | 12.44 | AAI-based (high-confidence) | 0 |
| S8-final-viral-combined_k141_273503\|\|full | 3000 | No | NA | 3 | 0 | 1 | Not-determined | Genome-fragment | NA | NA | 0 |
| S8-final-viral-combined_k141_54823\|\|full | 2925 | No | NA | 3 | 0 | 0 | Not-determined | Genome-fragment | NA | NA | 0 |
| S8-final-viral-combined_k141_54832\|\|full | 5419 | No | NA | 7 | 0 | 5 | Not-determined | Genome-fragment | NA | NA | 0 |
| S8-final-viral-combined_k141_273521\|\|full | 1304 | No | NA | 2 | 0 | 0 | Not-determined | Genome-fragment | NA | NA | 0 |
| S8-final-viral-combined_k141_192963\|\|full | 1602 | No | NA | 4 | 0 | 0 | Not-determined | Genome-fragment | NA | NA | 0 |
| S8-final-viral-combined_k141_7740\|\|full | 2914 | No | NA | 5 | 0 | 4 | Not-determined | Genome-fragment | NA | NA | 0 |
| S9-final-viral-combined_k141_42795\|\|full | 13947 | No | NA | 19 | 2 | 1 | Low-quality | Genome-fragment | 14.02 | HMM-based (lower-bound) | 0 |
| S8-final-viral-combined_k141_260196\|\|full | 2385 | No | NA | 4 | 0 | 0 | Not-determined | Genome-fragment | NA | NA | 0 |
| S3-final-viral-combined_k141_40315\|\|full | 13949 | No | NA | 13 | 3 | 1 | Low-quality | Genome-fragment | 9.51 | HMM-based (lower-bound) | 0 |
| S8-final-viral-combined_k141_88668\|\|full | 1608 | No | NA | 3 | 0 | 1 | Not-determined | Genome-fragment | NA | NA | 0 |
| S8-final-viral-combined_k141_192969\|\|full | 4624 | No | NA | 9 | 0 | 0 | Not-determined | Genome-fragment | NA | NA | 0 |
| S8-final-viral-combined_k141_135901\|\|full | 10537 | No | NA | 11 | 0 | 10 | Not-determined | Genome-fragment | NA | NA | 0 |
| S8-final-viral-combined_k141_147624\|\|full | 1741 | No | NA | 3 | 0 | 0 | Not-determined | Genome-fragment | NA | NA | 0 |
| S8-final-viral-combined_k141_43506\|\|full | 3749 | No | NA | 5 | 0 | 1 | Not-determined | Genome-fragment | NA | NA | 0 |
| S8-final-viral-combined_k141_66488\|\|full | 3525 | No | NA | 3 | 0 | 2 | Not-determined | Genome-fragment | NA | NA | 0 |
| S8-final-viral-combined_k141_7787\|\|full | 1063 | No | NA | 2 | 0 | 1 | Not-determined | Genome-fragment | NA | NA | 0 |
| S8-final-viral-combined_k141_147642\|\|full | 2968 | No | NA | 3 | 0 | 0 | Not-determined | Genome-fragment | NA | NA | 0 |
| S8-final-viral-combined_k141_182538\|\|full | 2178 | No | NA | 3 | 0 | 0 | Not-determined | Genome-fragment | NA | NA | 0 |
| S8-final-viral-combined_k141_170057\|\|full | 1107 | No | NA | 3 | 0 | 0 | Not-determined | Genome-fragment | NA | NA | 0 |
| S8-final-viral-combined_k141_260239\|\|full | 2008 | No | NA | 2 | 0 | 0 | Not-determined | Genome-fragment | NA | NA | 0 |
| S11-final-viral-combined_k141_293182\|\|0_partial | 13950 | Yes | 9277 | 14 | 3 | 2 | Low-quality | Genome-fragment | 10.81 | HMM-based (lower-bound) | 33.5 |
| S7-final-viral-combined_k141_168841\|\|full | 13952 | No | NA | 17 | 1 | 2 | Low-quality | Genome-fragment | 20.03 | HMM-based (lower-bound) | 0 |
| S8-final-viral-combined_k141_18740\|\|full | 2784 | No | NA | 6 | 0 | 1 | Not-determined | Genome-fragment | NA | NA | 0 |
| S17-final-viral-combined_k141_70437\|\|full | 13957 | No | NA | 24 | 8 | 0 | Low-quality | Genome-fragment | 35.73 | AAI-based (high-confidence) | 0 |
| S5-final-viral-combined_k141_2952\|\|full | 13959 | No | NA | 15 | 10 | 0 | Low-quality | Genome-fragment | 34.36 | AAI-based (high-confidence) | 0 |
| S8-final-viral-combined_k141_18752\|\|full | 2511 | No | NA | 5 | 0 | 2 | Not-determined | Genome-fragment | NA | NA | 0 |
| S11-final-viral-combined_k141_76187\|\|full | 13970 | No | NA | 22 | 6 | 0 | Low-quality | Genome-fragment | 17.64 | HMM-based (lower-bound) | 0 |
| S1-final-viral-combined_k141_248139\|\|full | 13970 | No | NA | 14 | 1 | 9 | Low-quality | Genome-fragment | 8.99 | HMM-based (lower-bound) | 0 |
| S8-final-viral-combined_k141_18769\|\|full | 2468 | No | NA | 3 | 0 | 1 | Not-determined | Genome-fragment | NA | NA | 0 |
| S8-final-viral-combined_k141_66566\|\|full | 1144 | No | NA | 2 | 0 | 1 | Not-determined | Genome-fragment | NA | NA | 0 |
| S11-final-viral-combined_k141_36015\|\|full | 13976 | No | NA | 22 | 6 | 0 | Low-quality | Genome-fragment | 23.95 | HMM-based (lower-bound) | 0 |
| S5-final-viral-combined_k141_179168\|\|0_partial | 13977 | No | NA | 15 | 0 | 6 | Low-quality | Genome-fragment | 14.88 | AAI-based (medium-confidence) | 0 |
| S8-final-viral-combined_k141_239915\|\|full | 1262 | No | NA | 4 | 0 | 0 | Not-determined | Genome-fragment | NA | NA | 0 |
| S5-final-viral-combined_k141_27009\|\|0_partial | 13978 | Yes | 9341 | 19 | 3 | 3 | Low-quality | Genome-fragment | 10.67 | HMM-based (lower-bound) | 33.17 |
| S12-final-viral-combined_k141_378713\|\|0_partial | 13981 | No | NA | 15 | 1 | 4 | Low-quality | Genome-fragment | 35.89 | AAI-based (high-confidence) | 0 |
| S8-final-viral-combined_k141_88800\|\|full | 2379 | No | NA | 4 | 0 | 2 | Not-determined | Genome-fragment | NA | NA | 0 |
| S8-final-viral-combined_k141_66607\|\|full | 1394 | No | NA | 3 | 0 | 1 | Not-determined | Genome-fragment | NA | NA | 0 |
| S8-final-viral-combined_k141_112034\|\|full | 1729 | No | NA | 3 | 0 | 2 | Not-determined | Genome-fragment | NA | NA | 0 |
| S8-final-viral-combined_k141_170181\|\|full | 1938 | No | NA | 2 | 0 | 0 | Not-determined | Genome-fragment | NA | NA | 0 |
| S1-final-viral-combined_k141_330703\|\|full | 13992 | Yes | 12001 | 22 | 5 | 2 | Low-quality | Genome-fragment | 14.44 | HMM-based (lower-bound) | 14.23 |
| S17-final-viral-combined_k141_50505\|\|0_partial | 13998 | No | NA | 12 | 1 | 3 | Low-quality | Genome-fragment | 8.78 | HMM-based (lower-bound) | 0 |
| S8-final-viral-combined_k141_32769\|\|full | 1637 | No | NA | 4 | 0 | 2 | Not-determined | Genome-fragment | NA | NA | 0 |
| S8-final-viral-combined_k141_66624\|\|full | 24713 | No | NA | 20 | 0 | 12 | Not-determined | Genome-fragment | NA | NA | 0 |
| S8-final-viral-combined_k141_182740\|\|full | 1977 | No | NA | 3 | 0 | 1 | Not-determined | Genome-fragment | NA | NA | 0 |
| S8-final-viral-combined_k141_182741\|\|full | 3969 | No | NA | 5 | 0 | 4 | Not-determined | Genome-fragment | NA | NA | 0 |
| S8-final-viral-combined_k141_250737\|\|full | 1290 | No | NA | 3 | 0 | 0 | Not-determined | Genome-fragment | NA | NA | 0 |
| S8-final-viral-combined_k141_250740\|\|full | 2484 | No | NA | 2 | 0 | 1 | Not-determined | Genome-fragment | NA | NA | 0 |
| S17-final-viral-combined_k141_213306\|\|full | 14002 | No | NA | 22 | 6 | 0 | Low-quality | Genome-fragment | 30.57 | AAI-based (high-confidence) | 0 |
| S8-final-viral-combined_k141_203376\|\|full | 2748 | No | NA | 3 | 0 | 1 | Not-determined | Genome-fragment | NA | NA | 0 |
| S8-final-viral-combined_k141_260318\|\|full | 1831 | No | NA | 4 | 0 | 0 | Not-determined | Genome-fragment | NA | NA | 0 |
| S8-final-viral-combined_k141_227501\|\|full | 1120 | No | NA | 2 | 0 | 1 | Not-determined | Genome-fragment | NA | NA | 0 |
| S8-final-viral-combined_k141_18890\|\|full | 2008 | No | NA | 2 | 0 | 2 | Not-determined | Genome-fragment | NA | NA | 0 |
| S8-final-viral-combined_k141_66669\|\|full | 2315 | No | NA | 5 | 0 | 0 | Not-determined | Genome-fragment | NA | NA | 0 |
| S6-final-viral-combined_k141_7554\|\|full | 14004 | No | NA | 22 | 6 | 1 | Low-quality | Genome-fragment | 18.84 | HMM-based (lower-bound) | 0 |
| S7-final-viral-combined_k141_214665\|\|full | 14006 | No | NA | 10 | 0 | 1 | Low-quality | Genome-fragment | 23.32 | AAI-based (medium-confidence) | 0 |
| S2-final-viral-combined_k141_33298\|\|full | 14016 | No | NA | 19 | 7 | 0 | Low-quality | Genome-fragment | 34.38 | AAI-based (high-confidence) | 0 |
| S6_k141_83556 | 14033 | No | NA | 12 | 2 | 0 | Low-quality | Genome-fragment | 7.48 | AAI-based (medium-confidence) | 0 |
| S8-final-viral-combined_k141_147840\|\|full | 1177 | No | NA | 2 | 0 | 0 | Not-determined | Genome-fragment | NA | NA | 0 |
| S7-final-viral-combined_k141_60173\|\|full | 14043 | No | NA | 15 | 1 | 12 | Low-quality | Genome-fragment | 8.4 | HMM-based (lower-bound) | 0 |
| S1-final-viral-combined_k141_167803\|\|full | 14057 | Yes | 5216 | 16 | 2 | 7 | Low-quality | Genome-fragment | 7.64 | HMM-based (lower-bound) | 62.89 |
| S8-final-viral-combined_k141_273792\|\|full | 2201 | No | NA | 2 | 0 | 0 | Not-determined | Genome-fragment | NA | NA | 0 |
| S8-final-viral-combined_k141_170311\|\|full | 4876 | No | NA | 3 | 0 | 2 | Not-determined | Genome-fragment | NA | NA | 0 |
| S12_k141_320351 | 14061 | No | NA | 14 | 1 | 2 | Low-quality | Genome-fragment | 27.3 | AAI-based (medium-confidence) | 0 |
| S2-final-viral-combined_k141_97494\|\|full | 14061 | No | NA | 19 | 8 | 0 | Low-quality | Genome-fragment | 38.53 | AAI-based (medium-confidence) | 0 |
| S8-final-viral-combined_k141_216265\|\|full | 7211 | No | NA | 7 | 0 | 5 | Not-determined | Genome-fragment | NA | NA | 0 |

| | | | | | | | | | | | |
|---|---|---|---|---|---|---|---|---|---|---|---|
| S8-final-viral-combined_k141_250814\|\|full | 2298 | No | NA | 3 | 0 | 2 | Not-determined | Genome-fragment | NA | NA | 0 |
| S9-final-viral-combined_k141_78363\|\|full | 14065 | No | NA | 8 | 0 | 0 | Low-quality | Genome-fragment | 35.99 | AAI-based (high-confidence) | 0 |
| S8-final-viral-combined_k141_18970\|\|full | 5564 | No | NA | 4 | 0 | 4 | Not-determined | Genome-fragment | NA | NA | 0 |
| S8-final-viral-combined_k141_240151\|\|full | 1713 | No | NA | 2 | 0 | 1 | Not-determined | Genome-fragment | NA | NA | 0 |
| S2-final-viral-combined_k141_180391\|\|full | 14065 | No | NA | 17 | 1 | 0 | Low-quality | Genome-fragment | 28.37 | AAI-based (medium-confidence) | 0 |
| S8-final-viral-combined_k141_101336\|\|full | 1817 | No | NA | 6 | 0 | 0 | Not-determined | Genome-fragment | NA | NA | 0 |
| S8-final-viral-combined_k141_112212\|\|full | 4263 | No | NA | 7 | 0 | 4 | Not-determined | Genome-fragment | NA | NA | 0 |
| S1-final-viral-combined_k141_175144\|\|full | 14079 | Yes | 8387 | 8 | 2 | 4 | Low-quality | Genome-fragment | 12.18 | HMM-based (lower-bound) | 40.43 |
| S8-final-viral-combined_k141_273845\|\|full | 1457 | No | NA | 3 | 0 | 1 | Not-determined | Genome-fragment | NA | NA | 0 |
| S7-final-viral-combined_k141_177981\|\|full | 14095 | No | NA | 16 | 1 | 6 | Low-quality | Genome-fragment | 21.33 | HMM-based (lower-bound) | 0 |
| S1-final-viral-combined_k141_386941\|\|full | 14100 | No | NA | 14 | 3 | 2 | Low-quality | Genome-fragment | 34.51 | AAI-based (high-confidence) | 0 |
| S8-final-viral-combined_k141_43890\|\|full | 3059 | No | NA | 4 | 0 | 3 | Not-determined | Genome-fragment | NA | NA | 0 |
| S8-final-viral-combined_k141_112240\|\|full | 2241 | No | NA | 3 | 0 | 0 | Not-determined | Genome-fragment | NA | NA | 0 |
| S1-final-viral-combined_k141_28946\|\|full | 14105 | Yes | 9900 | 16 | 2 | 2 | Low-quality | Genome-fragment | 25.06 | AAI-based (high-confidence) | 29.81 |
| S8-final-viral-combined_k141_273855\|\|full | 1039 | No | NA | 2 | 0 | 1 | Not-determined | Genome-fragment | NA | NA | 0 |
| S1-final-viral-combined_k141_264065\|\|0_partial | 14121 | No | NA | 10 | 0 | 6 | Low-quality | Genome-fragment | 13.08 | AAI-based (medium-confidence) | 0 |
| S8-final-viral-combined_k141_112252\|\|full | 2513 | No | NA | 3 | 0 | 2 | Not-determined | Genome-fragment | NA | NA | 0 |
| S12-final-viral-combined_k141_324947\|\|0_partial | 14124 | No | NA | 11 | 1 | 2 | Low-quality | Genome-fragment | 8.86 | HMM-based (lower-bound) | 0 |
| S9-final-viral-combined_k141_16127\|\|full | 14125 | Yes | 3595 | 17 | 3 | 6 | Low-quality | Genome-fragment | 8.91 | AAI-based (high-confidence) | 74.55 |
| S8-final-viral-combined_k141_19071\|\|full | 5556 | Yes | 3812 | 4 | 1 | 1 | Not-determined | Genome-fragment | NA | NA | 31.39 |
| S11_k141_303288 | 14126 | No | NA | 28 | 7 | 1 | Low-quality | Genome-fragment | 27.78 | AAI-based (high-confidence) | 0 |
| S8-final-viral-combined_k141_124176\|\|full | 2713 | No | NA | 3 | 0 | 2 | Not-determined | Genome-fragment | NA | NA | 0 |
| S2-final-viral-combined_k141_115451\|\|full | 14144 | No | NA | 12 | 1 | 6 | Low-quality | Genome-fragment | 1.14 | HMM-based (lower-bound) | 0 |
| S8-final-viral-combined_k141_170435\|\|full | 6080 | No | NA | 4 | 0 | 4 | Not-determined | Genome-fragment | NA | NA | 0 |
| S8-final-viral-combined_k141_170436\|\|full | 1288 | No | NA | 3 | 0 | 0 | Not-determined | Genome-fragment | NA | NA | 0 |
| S8-final-viral-combined_k141_33061\|\|full | 1170 | No | NA | 4 | 0 | 0 | Not-determined | Genome-fragment | NA | NA | 0 |
| S9-final-viral-combined_k141_56989\|\|full | 14155 | Yes | 5688 | 21 | 4 | 7 | Low-quality | Genome-fragment | 6.52 | HMM-based (lower-bound) | 59.82 |
| S8-final-viral-combined_k141_159024\|\|full | 1688 | No | NA | 3 | 0 | 1 | Not-determined | Genome-fragment | NA | NA | 0 |
| S8-final-viral-combined_k141_33077\|\|full | 2792 | No | NA | 5 | 0 | 0 | Not-determined | Genome-fragment | NA | NA | 0 |
| S8-final-viral-combined_k141_89196\|\|full | 2450 | No | NA | 2 | 0 | 2 | Not-determined | Genome-fragment | NA | NA | 0 |
| S8-final-viral-combined_k141_33094\|\|full | 6179 | No | NA | 2 | 0 | 2 | Not-determined | Genome-fragment | NA | NA | 0 |
| S8-final-viral-combined_k141_136400\|\|full | 4142 | No | NA | 4 | 0 | 1 | Not-determined | Genome-fragment | NA | NA | 0 |
| S8-final-viral-combined_k141_89227\|\|full | 1531 | No | NA | 4 | 0 | 0 | Not-determined | Genome-fragment | NA | NA | 0 |
| S8-final-viral-combined_k141_101433\|\|full | 7553 | No | NA | 7 | 0 | 3 | Not-determined | Genome-fragment | NA | NA | 0 |
| S8-final-viral-combined_k141_148064\|\|full | 2502 | No | NA | 4 | 0 | 4 | Not-determined | Genome-fragment | NA | NA | 0 |
| S5-final-viral-combined_k141_180393\|\|0_partial | 14156 | No | NA | 19 | 5 | 1 | Low-quality | Genome-fragment | 20.19 | HMM-based (lower-bound) | 0 |
| S17-final-viral-combined_k141_243257\|\|full | 14156 | No | NA | 19 | 6 | 0 | Low-quality | Genome-fragment | 26.13 | AAI-based (high-confidence) | 0 |
| S1-final-viral-combined_k141_170104\|\|full | 14183 | Yes | 7704 | 20 | 3 | 5 | Low-quality | Genome-fragment | 12.61 | AAI-based (high-confidence) | 45.68 |
| S17-final-viral-combined_k141_262590\|\|full | 14188 | No | NA | 14 | 0 | 3 | Low-quality | Genome-fragment | 19.6 | AAI-based (medium-confidence) | 0 |
| S5_k141_163550 | 14196 | No | NA | 14 | 4 | 0 | Low-quality | Genome-fragment | 31.49 | AAI-based (high-confidence) | 0 |
| S2-final-viral-combined_k141_99753\|\|0_partial | 14201 | No | NA | 16 | 1 | 5 | Low-quality | Genome-fragment | 16.91 | HMM-based (lower-bound) | 0 |
| S8-final-viral-combined_k141_136473\|\|full | 1008 | No | NA | 3 | 0 | 1 | Not-determined | Genome-fragment | NA | NA | 0 |
| S8-final-viral-combined_k141_33161\|\|full | 1813 | No | NA | 2 | 0 | 1 | Not-determined | Genome-fragment | NA | NA | 0 |
| S8-final-viral-combined_k141_240410\|\|full | 10937 | No | NA | 11 | 0 | 7 | Not-determined | Genome-fragment | NA | NA | 0 |
| S8-final-viral-combined_k141_183088\|\|full | 2684 | No | NA | 3 | 0 | 2 | Not-determined | Genome-fragment | NA | NA | 0 |
| S8_k141_203842 | 14202 | No | NA | 13 | 3 | 0 | Low-quality | Genome-fragment | 28.56 | AAI-based (high-confidence) | 0 |
| S8-final-viral-combined_k141_124335\|\|full | 1498 | No | NA | 2 | 0 | 0 | Not-determined | Genome-fragment | NA | NA | 0 |
| S11_k141_21871 | 14204 | No | NA | 11 | 0 | 9 | Low-quality | Genome-fragment | 7.64 | AAI-based (medium-confidence) | 0 |
| S8-final-viral-combined_k141_55166\|\|full | 2395 | No | NA | 3 | 0 | 0 | Not-determined | Genome-fragment | NA | NA | 0 |
| S11-final-viral-combined_k141_19223\|\|full | 14216 | No | NA | 22 | 4 | 1 | Low-quality | Genome-fragment | 33.19 | AAI-based (high-confidence) | 0 |
| S8-final-viral-combined_k141_19257\|\|full | 1957 | No | NA | 3 | 0 | 1 | Not-determined | Genome-fragment | NA | NA | 0 |
| S11-final-viral-combined_k141_95621\|\|full | 14217 | No | NA | 16 | 1 | 8 | Low-quality | Genome-fragment | 15.63 | HMM-based (lower-bound) | 0 |
| S8-final-viral-combined_k141_89320\|\|full | 1285 | No | NA | 3 | 0 | 0 | Not-determined | Genome-fragment | NA | NA | 0 |
| S8-final-viral-combined_k141_251029\|\|full | 1392 | No | NA | 3 | 0 | 0 | Not-determined | Genome-fragment | NA | NA | 0 |
| S8-final-viral-combined_k141_101578\|\|full | 4121 | No | NA | 4 | 0 | 4 | Not-determined | Genome-fragment | NA | NA | 0 |
| S8-final-viral-combined_k141_159166\|\|full | 5990 | No | NA | 7 | 0 | 1 | Not-determined | Genome-fragment | NA | NA | 0 |
| S8-final-viral-combined_k141_148178\|\|full | 4223 | No | NA | 2 | 0 | 2 | Not-determined | Genome-fragment | NA | NA | 0 |
| S8-final-viral-combined_k141_183160\|\|full | 1292 | No | NA | 2 | 0 | 2 | Not-determined | Genome-fragment | NA | NA | 0 |
| S8-final-viral-combined_k141_112431\|\|full | 2219 | No | NA | 3 | 0 | 1 | Not-determined | Genome-fragment | NA | NA | 0 |
| S1-final-viral-combined_k141_116925\|\|full | 14231 | No | NA | 9 | 0 | 2 | Low-quality | Genome-fragment | 30.97 | AAI-based (high-confidence) | 0 |
| S8-final-viral-combined_k141_170630\|\|full | 2503 | No | NA | 1 | 0 | 1 | Not-determined | Genome-fragment | NA | NA | 0 |
| S11-final-viral-combined_k141_301313\|\|full | 14234 | No | NA | 17 | 14 | 0 | Low-quality | Genome-fragment | 33.28 | AAI-based (high-confidence) | 0 |
| S12-final-viral-combined_k141_303580\|\|0_partial | 14234 | No | NA | 15 | 1 | 8 | Low-quality | Genome-fragment | 29.27 | AAI-based (medium-confidence) | 0 |
| S8-final-viral-combined_k141_89386\|\|full | 3477 | No | NA | 4 | 0 | 2 | Not-determined | Genome-fragment | NA | NA | 0 |
| S8-final-viral-combined_k141_159212\|\|full | 3136 | No | NA | 3 | 0 | 3 | Not-determined | Genome-fragment | NA | NA | 0 |
| S12_k141_213320 | 14238 | No | NA | 16 | 1 | 2 | Low-quality | Genome-fragment | 7.72 | HMM-based (lower-bound) | 0 |
| S8-final-viral-combined_k141_89387\|\|full | 4002 | No | NA | 2 | 0 | 2 | Not-determined | Genome-fragment | NA | NA | 0 |
| S8-final-viral-combined_k141_67081\|\|full | 1185 | No | NA | 3 | 0 | 1 | Not-determined | Genome-fragment | NA | NA | 0 |
| S8-final-viral-combined_k141_8310\|\|full | 1483 | No | NA | 3 | 0 | 0 | Not-determined | Genome-fragment | NA | NA | 0 |
| S4_k141_56920 | 14239 | No | NA | 11 | 1 | 6 | Low-quality | Genome-fragment | 5.07 | HMM-based (lower-bound) | 0 |
| S8-final-viral-combined_k141_148240\|\|full | 1007 | No | NA | 2 | 0 | 0 | Not-determined | Genome-fragment | NA | NA | 0 |
| S8-final-viral-combined_k141_8312\|\|full | 6933 | No | NA | 6 | 0 | 5 | Not-determined | Genome-fragment | NA | NA | 0 |
| S8-final-viral-combined_k141_96917\|\|full | 14239 | No | NA | 17 | 3 | 0 | Low-quality | Genome-fragment | 26.66 | AAI-based (high-confidence) | 0 |
| S11_k141_116497 | 14251 | No | NA | 9 | 1 | 2 | Low-quality | Genome-fragment | 21.72 | AAI-based (medium-confidence) | 0 |
| S8-final-viral-combined_k141_227930\|\|full | 1221 | No | NA | 2 | 0 | 2 | Not-determined | Genome-fragment | NA | NA | 0 |
| S2-final-viral-combined_k141_213750\|\|full | 14262 | No | NA | 25 | 9 | 0 | Low-quality | Genome-fragment | 38.34 | AAI-based (high-confidence) | 0 |
| S7-final-viral-combined_k141_10161\|\|full | 14280 | No | NA | 8 | 1 | 0 | Low-quality | Genome-fragment | 32.56 | AAI-based (high-confidence) | 0 |
| S8-final-viral-combined_k141_136661\|\|full | 1040 | No | NA | 2 | 0 | 1 | Not-determined | Genome-fragment | NA | NA | 0 |
| S8_k141_121654 | 14280 | No | NA | 17 | 10 | 0 | Low-quality | Genome-fragment | 27.77 | AAI-based (medium-confidence) | 0 |
| S8-final-viral-combined_k141_203875\|\|full | 2548 | No | NA | 3 | 0 | 0 | Not-determined | Genome-fragment | NA | NA | 0 |
| S8-final-viral-combined_k141_67115\|\|full | 9630 | No | NA | 11 | 0 | 2 | Not-determined | Genome-fragment | NA | NA | 0 |
| S8-final-viral-combined_k141_274119\|\|full | 5319 | No | NA | 3 | 0 | 2 | Not-determined | Genome-fragment | NA | NA | 0 |
| S11-final-viral-combined_k141_180333\|\|full | 14292 | No | NA | 10 | 0 | 4 | Low-quality | Genome-fragment | 6.31 | AAI-based (high-confidence) | 0 |
| S12-final-viral-combined_k141_219943\|\|full | 14306 | No | NA | 21 | 4 | 0 | Low-quality | Genome-fragment | 27.28 | AAI-based (medium-confidence) | 0 |
| S8-final-viral-combined_k141_183294\|\|full | 1624 | No | NA | 2 | 0 | 2 | Not-determined | Genome-fragment | NA | NA | 0 |
| S5-final-viral-combined_k141_179362\|\|full | 14320 | No | NA | 21 | 5 | 1 | Low-quality | Genome-fragment | 17.59 | HMM-based (lower-bound) | 0 |
| S8-final-viral-combined_k141_148332\|\|full | 1236 | No | NA | 3 | 0 | 2 | Not-determined | Genome-fragment | NA | NA | 0 |
| S8_k141_78538 | 14334 | Yes | 10402 | 19 | 2 | 2 | Low-quality | Genome-fragment | 25.66 | AAI-based (high-confidence) | 27.43 |
| S8-final-viral-combined_k141_216647\|\|full | 3890 | No | NA | 3 | 0 | 1 | Not-determined | Genome-fragment | NA | NA | 0 |
| S8-final-viral-combined_k141_170727\|\|full | 6003 | No | NA | 7 | 0 | 6 | Not-determined | Genome-fragment | NA | NA | 0 |
| S8-final-viral-combined_k141_228010\|\|full | 5167 | No | NA | 5 | 0 | 4 | Not-determined | Genome-fragment | NA | NA | 0 |
| S8-final-viral-combined_k141_8429\|\|full | 1134 | No | NA | 3 | 0 | 1 | Not-determined | Genome-fragment | NA | NA | 0 |
| S7-final-viral-combined_k141_200677\|\|full | 14341 | No | NA | 9 | 0 | 6 | Low-quality | Genome-fragment | 33.83 | AAI-based (medium-confidence) | 0 |
| S8-final-viral-combined_k141_183351\|\|full | 1531 | No | NA | 2 | 0 | 1 | Not-determined | Genome-fragment | NA | NA | 0 |
| S11-final-viral-combined_k141_21419\|\|full | 14358 | No | NA | 17 | 10 | 0 | Low-quality | Genome-fragment | 30.56 | AAI-based (high-confidence) | 0 |
| S11-final-viral-combined_k141_165692\|\|0_partial | 14360 | Yes | 9727 | 16 | 2 | 3 | Low-quality | Genome-fragment | 13.51 | HMM-based (lower-bound) | 32.26 |
| S8-final-viral-combined_k141_55316\|\|full | 1166 | No | NA | 3 | 0 | 1 | Not-determined | Genome-fragment | NA | NA | 0 |
| S8-final-viral-combined_k141_79032\|\|full | 3692 | No | NA | 4 | 0 | 2 | Not-determined | Genome-fragment | NA | NA | 0 |
| S9-final-viral-combined_k141_22573\|\|full | 14381 | Yes | 8310 | 23 | 2 | 4 | Low-quality | Genome-fragment | 14.27 | AAI-based (medium-confidence) | 42.22 |
| S3_k141_71709 | 14382 | No | NA | 17 | 3 | 2 | Low-quality | Genome-fragment | 13.77 | AAI-based (high-confidence) | 0 |
| S8-final-viral-combined_k141_8462\|\|full | 1024 | No | NA | 2 | 0 | 1 | Not-determined | Genome-fragment | NA | NA | 0 |
| S8-final-viral-combined_k141_33391\|\|full | 1672 | No | NA | 2 | 0 | 2 | Not-determined | Genome-fragment | NA | NA | 0 |
| S1-final-viral-combined_k141_385431\|\|full | 14392 | No | NA | 18 | 6 | 0 | Low-quality | Genome-fragment | 20.61 | HMM-based (lower-bound) | 0 |
| S8-final-viral-combined_k141_260643\|\|full | 2091 | No | NA | 2 | 0 | 2 | Not-determined | Genome-fragment | NA | NA | 0 |
| S8-final-viral-combined_k141_251207\|\|full | 4827 | No | NA | 5 | 0 | 2 | Not-determined | Genome-fragment | NA | NA | 0 |
| S8-final-viral-combined_k141_55338\|\|full | 1099 | No | NA | 2 | 0 | 0 | Not-determined | Genome-fragment | NA | NA | 0 |
| S8-final-viral-combined_k141_148408\|\|full | 2935 | No | NA | 5 | 0 | 1 | Not-determined | Genome-fragment | NA | NA | 0 |
| S2-final-viral-combined_k141_105666\|\|full | 14398 | No | NA | 14 | 1 | 3 | Low-quality | Genome-fragment | 5.49 | HMM-based (lower-bound) | 0 |
| S8-final-viral-combined_k141_228080\|\|full | 1731 | No | NA | 3 | 0 | 0 | Not-determined | Genome-fragment | NA | NA | 0 |
| S2-final-viral-combined_k141_167133\|\|full | 14400 | No | NA | 16 | 1 | 5 | Low-quality | Genome-fragment | 21.79 | HMM-based (lower-bound) | 0 |
| S8-final-viral-combined_k141_228091\|\|full | 1244 | No | NA | 2 | 0 | 1 | Not-determined | Genome-fragment | NA | NA | 0 |
| S2_k141_2569 | 14400 | Yes | 5968 | 20 | 4 | 5 | Low-quality | Genome-fragment | 9.4 | HMM-based (lower-bound) | 58.56 |
| S7-final-viral-combined_k141_177541\|\|full | 14409 | No | NA | 15 | 0 | 2 | Low-quality | Genome-fragment | 25.02 | AAI-based (medium-confidence) | 0 |
| S8-final-viral-combined_k141_67166\|\|full | 2638 | No | NA | 3 | 0 | 1 | Not-determined | Genome-fragment | NA | NA | 0 |
| S8-final-viral-combined_k141_8516\|\|full | 2545 | No | NA | 2 | 0 | 1 | Not-determined | Genome-fragment | NA | NA | 0 |
| S2-final-viral-combined_k141_96816\|\|full | 14410 | No | NA | 28 | 5 | 0 | Low-quality | Genome-fragment | 23.96 | AAI-based (high-confidence) | 0 |
| S17-k141_156650 | 14426 | No | NA | 18 | 0 | 0 | Low-quality | Genome-fragment | 18.52 | AAI-based (medium-confidence) | 0 |
| S11-final-viral-combined_k141_170745\|\|0_partial | 14434 | Yes | 11196 | 15 | 2 | 2 | Low-quality | Genome-fragment | 7.13 | HMM-based (lower-bound) | 22.43 |
| S1-final-viral-combined_k141_109362\|\|full | 14437 | No | NA | 7 | 0 | 0 | Low-quality | Genome-fragment | 19.55 | AAI-based (medium-confidence) | 0 |
| S8-final-viral-combined_k141_136791\|\|full | 2531 | No | NA | 4 | 0 | 2 | Not-determined | Genome-fragment | NA | NA | 0 |
| S8-final-viral-combined_k141_251274\|\|full | 1650 | No | NA | 3 | 0 | 2 | Not-determined | Genome-fragment | NA | NA | 0 |
| S8-final-viral-combined_k141_260701\|\|full | 4486 | No | NA | 5 | 0 | 2 | Not-determined | Genome-fragment | NA | NA | 0 |
| S8-final-viral-combined_k141_159466\|\|full | 2469 | No | NA | 3 | 0 | 2 | Not-determined | Genome-fragment | NA | NA | 0 |
| S8-final-viral-combined_k141_148528\|\|full | 1971 | No | NA | 3 | 0 | 2 | Not-determined | Genome-fragment | NA | NA | 0 |
| S9_k141_91332 | 14443 | No | NA | 15 | 0 | 3 | Low-quality | Genome-fragment | 24.12 | AAI-based (high-confidence) | 0 |
| S8-final-viral-combined_k141_67224\|\|full | 3433 | No | NA | 4 | 0 | 2 | Not-determined | Genome-fragment | NA | NA | 0 |
| S8-final-viral-combined_k141_216867\|\|full | 2802 | No | NA | 4 | 0 | 2 | Not-determined | Genome-fragment | NA | NA | 0 |
| S8-final-viral-combined_k141_55457\|\|full | 2978 | No | NA | 4 | 0 | 1 | Not-determined | Genome-fragment | NA | NA | 0 |
| S8-final-viral-combined_k141_55462\|\|full | 1548 | No | NA | 4 | 0 | 1 | Not-determined | Genome-fragment | NA | NA | 0 |
| S8-final-viral-combined_k141_204063\|\|full | 2543 | No | NA | 3 | 0 | 1 | Not-determined | Genome-fragment | NA | NA | 0 |
| S7-final-viral-combined_k141_22609\|\|full | 14448 | No | NA | 18 | 10 | 1 | Low-quality | Genome-fragment | 29.12 | AAI-based (high-confidence) | 0 |
| S8-final-viral-combined_k141_240796\|\|full | 1586 | No | NA | 3 | 0 | 0 | Not-determined | Genome-fragment | NA | NA | 0 |
| S8-final-viral-combined_k141_170864\|\|full | 2439 | No | NA | 4 | 0 | 2 | Not-determined | Genome-fragment | NA | NA | 0 |
| S8-final-viral-combined_k141_67292\|\|full | 1212 | No | NA | 3 | 0 | 0 | Not-determined | Genome-fragment | NA | NA | 0 |
| S8-final-viral-combined_k141_159576\|\|full | 4353 | No | NA | 3 | 0 | 2 | Not-determined | Genome-fragment | NA | NA | 0 |
| S8-final-viral-combined_k141_148599\|\|full | 2362 | No | NA | 3 | 0 | 2 | Not-determined | Genome-fragment | NA | NA | 0 |
| S12-final-viral-combined_k141_373131\|\|full | 14468 | Yes | 8059 | 15 | 4 | 4 | Low-quality | Genome-fragment | 18.31 | AAI-based (medium-confidence) | 44.3 |
| S8-final-viral-combined_k141_19632\|\|full | 2906 | No | NA | 4 | 0 | 0 | Not-determined | Genome-fragment | NA | NA | 0 |
| S8-final-viral-combined_k141_240848\|\|full | 7093 | No | NA | 7 | 0 | 3 | Not-determined | Genome-fragment | NA | NA | 0 |
| S8-final-viral-combined_k141_251332\|\|full | 6794 | No | NA | 13 | 0 | 1 | Not-determined | Genome-fragment | NA | NA | 0 |
| S8-final-viral-combined_k141_101824\|\|full | 2968 | No | NA | 3 | 0 | 3 | Not-determined | Genome-fragment | NA | NA | 0 |
| S4_k141_68963 | 14481 | No | NA | 18 | 5 | 1 | Low-quality | Genome-fragment | 26.52 | AAI-based (high-confidence) | 0 |
| S8-final-viral-combined_k141_19657\|\|full | 2358 | No | NA | 4 | 0 | 2 | Not-determined | Genome-fragment | NA | NA | 0 |
| S8-final-viral-combined_k141_193866\|\|full | 3244 | No | NA | 2 | 0 | 2 | Not-determined | Genome-fragment | NA | NA | 0 |
| S8-final-viral-combined_k141_55559\|\|full | 3372 | No | NA | 3 | 0 | 3 | Not-determined | Genome-fragment | NA | NA | 0 |
| S8-final-viral-combined_k141_101869\|\|full | 8730 | No | NA | 10 | 0 | 2 | Not-determined | Genome-fragment | NA | NA | 0 |
| S8-final-viral-combined_k141_79337\|\|full | 1532 | No | NA | 3 | 0 | 0 | Not-determined | Genome-fragment | NA | NA | 0 |
| S1_k141_150429 | 14491 | No | NA | 9 | 0 | 7 | Low-quality | Genome-fragment | 5.42 | AAI-based (medium-confidence) | 0 |
| S1-final-viral-combined_k141_46244\|\|0_partial | 14510 | Yes | 11168 | 19 | 2 | 3 | Low-quality | Genome-fragment | 14.87 | HMM-based (lower-bound) | 23.03 |
| S8-final-viral-combined_k141_63641\|\|full | 14515 | No | NA | 19 | 2 | 1 | Low-quality | Genome-fragment | 40.55 | AAI-based (medium-confidence) | 0 |
| S8-final-viral-combined_k141_148736\|\|full | 2878 | No | NA | 4 | 0 | 2 | Not-determined | Genome-fragment | NA | NA | 0 |
| S8-final-viral-combined_k141_112930\|\|full | 1765 | No | NA | 2 | 0 | 2 | Not-determined | Genome-fragment | NA | NA | 0 |
| S8-final-viral-combined_k141_193922\|\|full | 2379 | No | NA | 4 | 0 | 2 | Not-determined | Genome-fragment | NA | NA | 0 |
| S8-final-viral-combined_k141_159705\|\|full | 1705 | No | NA | 3 | 0 | 1 | Not-determined | Genome-fragment | NA | NA | 0 |
| S8-final-viral-combined_k141_251394\|\|full | 8461 | No | NA | 9 | 0 | 7 | Not-determined | Genome-fragment | NA | NA | 0 |
| S11-final-viral-combined_k141_119802\|\|0_partial | 14518 | No | NA | 16 | 1 | 5 | Low-quality | Genome-fragment | 8.97 | HMM-based (lower-bound) | 0 |
| S1_k141_238579 | 14523 | No | NA | 23 | 3 | 0 | Low-quality | Genome-fragment | 31.91 | AAI-based (high-confidence) | 0 |
| S2-final-viral-combined_k141_119517\|\|full | 14547 | No | NA | 21 | 5 | 1 | Low-quality | Genome-fragment | 23.04 | AAI-based (high-confidence) | 0 |
| S12_k141_373608 | 14548 | No | NA | 19 | 10 | 1 | Low-quality | Genome-fragment | 27.3 | AAI-based (high-confidence) | 0 |
| S8-final-viral-combined_k141_183713\|\|full | 1279 | No | NA | 2 | 0 | 1 | Not-determined | Genome-fragment | NA | NA | 0 |
| S7-final-viral-combined_k141_111548\|\|full | 14565 | No | NA | 10 | 0 | 1 | Low-quality | Genome-fragment | 18.46 | AAI-based (medium-confidence) | 0 |
| S17-final-viral-combined_k141_190763\|\|full | 14569 | Yes | 10601 | 14 | 3 | 4 | Low-quality | Genome-fragment | 26.04 | AAI-based (high-confidence) | 27.24 |
| S2_k141_130220 | 14572 | No | NA | 19 | 7 | 0 | Low-quality | Genome-fragment | 26.37 | AAI-based (high-confidence) | 0 |
| S2-final-viral-combined_k141_119169\|\|full | 14579 | No | NA | 16 | 0 | 12 | Low-quality | Genome-fragment | 6.26 | AAI-based (medium-confidence) | 0 |
| S11-final-viral-combined_k141_209542\|\|full | 14591 | No | NA | 18 | 7 | 0 | Low-quality | Genome-fragment | 24.16 | HMM-based (lower-bound) | 0 |
| S2-final-viral-combined_k141_185186\|\|full | 14624 | No | NA | 16 | 1 | 0 | Low-quality | Genome-fragment | 33.85 | AAI-based (medium-confidence) | 0 |
| S8-final-viral-combined_k141_79453\|\|full | 6164 | No | NA | 7 | 0 | 2 | Not-determined | Genome-fragment | NA | NA | 0 |
| S8-final-viral-combined_k141_124919\|\|full | 2150 | No | NA | 3 | 0 | 3 | Not-determined | Genome-fragment | NA | NA | 0 |
| S8-final-viral-combined_k141_194031\|\|full | 1876 | No | NA | 3 | 0 | 2 | Not-determined | Genome-fragment | NA | NA | 0 |
| S8-final-viral-combined_k141_148851\|\|full | 1510 | No | NA | 2 | 0 | 1 | Not-determined | Genome-fragment | NA | NA | 0 |
| S8-final-viral-combined_k141_148854\|\|full | 2120 | No | NA | 2 | 0 | 1 | Not-determined | Genome-fragment | NA | NA | 0 |
| S12_k141_157923 | 14625 | Yes | 3657 | 18 | 2 | 5 | Low-quality | Genome-fragment | 1.12 | HMM-based (lower-bound) | 74.99 |
| S11_k141_314239 | 14632 | No | NA | 20 | 9 | 0 | Low-quality | Genome-fragment | 32.06 | AAI-based (high-confidence) | 0 |
| S11-final-viral-combined_k141_322838\|\|full | 14636 | No | NA | 25 | 4 | 1 | Low-quality | Genome-fragment | 17.43 | HMM-based (lower-bound) | 0 |
| S8-final-viral-combined_k141_44758\|\|full | 4193 | No | NA | 8 | 0 | 2 | Not-determined | Genome-fragment | NA | NA | 0 |
| S8-final-viral-combined_k141_33824\|\|full | 1728 | No | NA | 4 | 0 | 1 | Not-determined | Genome-fragment | NA | NA | 0 |
| S8-final-viral-combined_k141_124995\|\|full | 2745 | No | NA | 4 | 0 | 1 | Not-determined | Genome-fragment | NA | NA | 0 |
| S11-final-viral-combined_k141_28478\|\|full | 14641 | No | NA | 22 | 1 | 2 | Low-quality | Genome-fragment | 8.66 | HMM-based (lower-bound) | 0 |
| S2-final-viral-combined_k141_96601\|\|full | 14672 | No | NA | 27 | 4 | 0 | Low-quality | Genome-fragment | 24.9 | AAI-based (high-confidence) | 0 |
| S9-final-viral-combined_k141_61803\|\|full | 14691 | No | NA | 16 | 4 | 0 | Low-quality | Genome-fragment | 41.48 | AAI-based (medium-confidence) | 0 |
| S8-final-viral-combined_k141_89952\|\|full | 2956 | No | NA | 6 | 0 | 2 | Not-determined | Genome-fragment | NA | NA | 0 |
| S8-final-viral-combined_k141_19924\|\|full | 4771 | No | NA | 3 | 0 | 2 | Not-determined | Genome-fragment | NA | NA | 0 |
| S8-final-viral-combined_k141_274284\|\|full | 2837 | No | NA | 2 | 0 | 1 | Not-determined | Genome-fragment | NA | NA | 0 |
| S8-final-viral-combined_k141_241168\|\|full | 1178 | No | NA | 3 | 0 | 0 | Not-determined | Genome-fragment | NA | NA | 0 |
| S7_k141_56098 | 14691 | No | NA | 7 | 0 | 6 | Low-quality | Genome-fragment | 5.6 | AAI-based (medium-confidence) | 0 |
| S2-final-viral-combined_k141_166548\|\|full | 14721 | No | NA | 22 | 4 | 0 | Low-quality | Genome-fragment | 37.16 | AAI-based (high-confidence) | 0 |
| S8-final-viral-combined_k141_274314\|\|full | 2763 | No | NA | 3 | 0 | 1 | Not-determined | Genome-fragment | NA | NA | 0 |
| S8-final-viral-combined_k141_8768\|\|full | 1104 | No | NA | 3 | 0 | 0 | Not-determined | Genome-fragment | NA | NA | 0 |
| S9_k141_25087 | 14745 | No | NA | 7 | 0 | 7 | Low-quality | Genome-fragment | 9.92 | AAI-based (medium-confidence) | 0 |
| S8-final-viral-combined_k141_216978\|\|full | 1586 | No | NA | 5 | 0 | 1 | Not-determined | Genome-fragment | NA | NA | 0 |
| S8-final-viral-combined_k141_14080\|\|full | 14753 | No | NA | 20 | 1 | 2 | Low-quality | Genome-fragment | 11.18 | AAI-based (high-confidence) | 0 |
| S12_k141_352472 | 14758 | No | NA | 21 | 8 | 2 | Low-quality | Genome-fragment | 25.25 | AAI-based (high-confidence) | 0 |
| S8-final-viral-combined_k141_55789\|\|full | 1712 | No | NA | 3 | 0 | 2 | Not-determined | Genome-fragment | NA | NA | 0 |
| S5-final-viral-combined_k141_175353\|\|full | 14760 | No | NA | 22 | 1 | 6 | Low-quality | Genome-fragment | 16.57 | HMM-based (lower-bound) | 0 |
| S11-final-viral-combined_k141_247486\|\|full | 14763 | No | NA | 19 | 6 | 0 | Low-quality | Genome-fragment | 42.54 | AAI-based (medium-confidence) | 0 |
| S8-final-viral-combined_k141_228512\|\|full | 2518 | No | NA | 2 | 0 | 2 | Not-determined | Genome-fragment | NA | NA | 0 |
| S8-final-viral-combined_k141_20010\|\|full | 1833 | No | NA | 2 | 0 | 1 | Not-determined | Genome-fragment | NA | NA | 0 |
| S1-final-viral-combined_k141_154009\|\|0_partial | 14766 | Yes | 9652 | 17 | 2 | 3 | Low-quality | Genome-fragment | 13.62 | HMM-based (lower-bound) | 34.63 |
| S8-final-viral-combined_k141_175448\|\|full | 14777 | No | NA | 8 | 0 | 7 | Low-quality | Genome-fragment | 4.18 | AAI-based (medium-confidence) | 0 |
| S12-final-viral-combined_k141_259497\|\|0_partial | 14781 | No | NA | 15 | 2 | 8 | Low-quality | Genome-fragment | 27.23 | HMM-based (lower-bound) | 0 |
| S8-final-viral-combined_k141_194186\|\|full | 2657 | No | NA | 5 | 0 | 2 | Not-determined | Genome-fragment | NA | NA | 0 |
| S8-final-viral-combined_k141_113216\|\|full | 1717 | No | NA | 2 | 0 | 1 | Not-determined | Genome-fragment | NA | NA | 0 |
| S8_k141_162823 | 14787 | No | NA | 17 | 1 | 5 | Low-quality | Genome-fragment | 22.37 | HMM-based (lower-bound) | 0 |
| S8-final-viral-combined_k141_274374\|\|full | 1591 | No | NA | 3 | 0 | 2 | Not-determined | Genome-fragment | NA | NA | 0 |
| S8-final-viral-combined_k141_183906\|\|full | 1807 | No | NA | 3 | 0 | 0 | Not-determined | Genome-fragment | NA | NA | 0 |
| S11-final-viral-combined_k141_299741\|\|full | 14790 | No | NA | 20 | 5 | 1 | Low-quality | Genome-fragment | 8.32 | HMM-based (lower-bound) | 0 |
| S17-final-viral-combined_k141_113412\|\|full | 14804 | No | NA | 16 | 11 | 0 | Low-quality | Genome-fragment | 42.04 | AAI-based (medium-confidence) | 0 |
| S8-final-viral-combined_k141_204367\|\|full | 1068 | No | NA | 3 | 0 | 0 | Not-determined | Genome-fragment | NA | NA | 0 |
| S1-final-viral-combined_k141_351087\|\|full | 14818 | No | NA | 16 | 3 | 0 | Low-quality | Genome-fragment | 39.06 | AAI-based (high-confidence) | 0 |
| S17-final-viral-combined_k141_99077\|\|full | 14824 | No | NA | 18 | 9 | 0 | Low-quality | Genome-fragment | 38.04 | AAI-based (medium-confidence) | 0 |
| S8-final-viral-combined_k141_194199\|\|full | 1030 | No | NA | 3 | 0 | 0 | Not-determined | Genome-fragment | NA | NA | 0 |
| S12-final-viral-combined_k141_174292\|\|0_partial | 14861 | No | NA | 12 | 1 | 3 | Low-quality | Genome-fragment | 23.23 | AAI-based (medium-confidence) | 0 |
| S2-final-viral-combined_k141_135100\|\|full | 14891 | No | NA | 16 | 1 | 6 | Low-quality | Genome-fragment | 9.17 | HMM-based (lower-bound) | 0 |
| S8-final-viral-combined_k141_20056\|\|full | 2327 | No | NA | 3 | 0 | 0 | Not-determined | Genome-fragment | NA | NA | 0 |
| S8-final-viral-combined_k141_44955\|\|full | 4336 | No | NA | 11 | 0 | 1 | Not-determined | Genome-fragment | NA | NA | 0 |
| S8-final-viral-combined_k141_79718\|\|full | 7382 | No | NA | 6 | 0 | 4 | Not-determined | Genome-fragment | NA | NA | 0 |
| S8-final-viral-combined_k141_102202\|\|full | 1725 | No | NA | 3 | 0 | 1 | Not-determined | Genome-fragment | NA | NA | 0 |
| S8-final-viral-combined_k141_159934\|\|full | 1337 | No | NA | 2 | 0 | 0 | Not-determined | Genome-fragment | NA | NA | 0 |
| S1-final-viral-combined_k141_232694\|\|full | 14903 | No | NA | 31 | 5 | 0 | Low-quality | Genome-fragment | 34.64 | AAI-based (high-confidence) | 0 |
| S1-final-viral-combined_k141_187505\|\|0_partial | 14930 | Yes | 2240 | 10 | 1 | 5 | Low-quality | Genome-fragment | 3.19 | HMM-based (lower-bound) | 85 |
| S11-final-viral-combined_k141_57682\|\|full | 14937 | No | NA | 17 | 1 | 14 | Low-quality | Genome-fragment | 8.1 | HMM-based (lower-bound) | 0 |
| S8-final-viral-combined_k141_241320\|\|full | 1386 | No | NA | 2 | 0 | 1 | Not-determined | Genome-fragment | NA | NA | 0 |
| S8-final-viral-combined_k141_34054\|\|full | 1346 | No | NA | 1 | 0 | 0 | Not-determined | Genome-fragment | NA | NA | 0 |
| S17-final-viral-combined_k141_168815\|\|full | 14940 | No | NA | 18 | 6 | 1 | Low-quality | Genome-fragment | 18.15 | AAI-based (high-confidence) | 0 |
| S8-final-viral-combined_k141_79768\|\|full | 5348 | No | NA | 6 | 0 | 3 | Not-determined | Genome-fragment | NA | NA | 0 |
| S8-final-viral-combined_k141_137406\|\|full | 9665 | No | NA | 8 | 0 | 5 | Not-determined | Genome-fragment | NA | NA | 0 |
| S8-final-viral-combined_k141_241329\|\|full | 1065 | No | NA | 3 | 0 | 0 | Not-determined | Genome-fragment | NA | NA | 0 |
| S12_k141_47756 | 14946 | No | NA | 19 | 7 | 0 | Low-quality | Genome-fragment | 28.13 | AAI-based (high-confidence) | 0 |
| S3-final-viral-combined_k141_46465\|\|full | 14952 | No | NA | 11 | 0 | 3 | Low-quality | Genome-fragment | 15.58 | AAI-based (medium-confidence) | 0 |
| S8-final-viral-combined_k141_102323\|\|full | 5227 | No | NA | 5 | 0 | 2 | Not-determined | Genome-fragment | NA | NA | 0 |
| S8-final-viral-combined_k141_204473\|\|full | 10800 | No | NA | 11 | 0 | 9 | Not-determined | Genome-fragment | NA | NA | 0 |
| S8-final-viral-combined_k141_125132\|\|full | 1136 | No | NA | 2 | 0 | 1 | Not-determined | Genome-fragment | NA | NA | 0 |
| S12-final-viral-combined_k141_216293\|\|full | 14960 | No | NA | 23 | 6 | 0 | Low-quality | Genome-fragment | 32.89 | AAI-based (high-confidence) | 0 |
| S1-final-viral-combined_k141_220842\|\|full | 14961 | Yes | 9314 | 20 | 2 | 3 | Low-quality | Genome-fragment | 27.42 | AAI-based (medium-confidence) | 37.74 |
| S2-final-viral-combined_k141_45479\|\|full | 14964 | No | NA | 20 | 10 | 0 | Low-quality | Genome-fragment | 41.66 | AAI-based (high-confidence) | 0 |
| S8-final-viral-combined_k141_79838\|\|full | 1343 | No | NA | 3 | 0 | 1 | Not-determined | Genome-fragment | NA | NA | 0 |
| S8-final-viral-combined_k141_90291\|\|full | 3023 | No | NA | 4 | 0 | 2 | Not-determined | Genome-fragment | NA | NA | 0 |
| S8-final-viral-combined_k141_184015\|\|full | 1732 | No | NA | 3 | 0 | 1 | Not-determined | Genome-fragment | NA | NA | 0 |
| S5-final-viral-combined_k141_32173\|\|full | 14965 | Yes | 7667 | 19 | 3 | 4 | Low-quality | Genome-fragment | 8.46 | HMM-based (lower-bound) | 48.77 |
| S8-final-viral-combined_k141_137490\|\|full | 1177 | No | NA | 3 | 0 | 1 | Not-determined | Genome-fragment | NA | NA | 0 |
| S1-final-viral-combined_k141_393219\|\|full | 14970 | No | NA | 20 | 10 | 0 | Low-quality | Genome-fragment | 31.02 | AAI-based (high-confidence) | 0 |
| S8-final-viral-combined_k141_56032\|\|full | 2086 | No | NA | 2 | 0 | 1 | Not-determined | Genome-fragment | NA | NA | 0 |
| S8-final-viral-combined_k141_125176\|\|full | 3345 | No | NA | 2 | 0 | 2 | Not-determined | Genome-fragment | NA | NA | 0 |
| S1-final-viral-combined_k141_143857\|\|full | 14978 | No | NA | 15 | 1 | 10 | Low-quality | Genome-fragment | 27.72 | HMM-based (lower-bound) | 0 |
| S8-final-viral-combined_k141_194365\|\|full | 4100 | No | NA | 7 | 0 | 1 | Not-determined | Genome-fragment | NA | NA | 0 |
| S8-final-viral-combined_k141_56044\|\|full | 1132 | No | NA | 3 | 0 | 2 | Not-determined | Genome-fragment | NA | NA | 0 |
| S1-final-viral-combined_k141_359134\|\|full | 14979 | No | NA | 8 | 1 | 0 | Low-quality | Genome-fragment | 25.47 | AAI-based (medium-confidence) | 0 |
| S8-final-viral-combined_k141_67990\|\|full | 1140 | No | NA | 3 | 0 | 0 | Not-determined | Genome-fragment | NA | NA | 0 |
| S5-final-viral-combined_k141_204940\|\|0_partial | 14982 | Yes | 8290 | 15 | 3 | 3 | Low-quality | Genome-fragment | 7.39 | AAI-based (high-confidence) | 44.67 |
| S1-final-viral-combined_k141_104344\|\|full | 14989 | No | NA | 16 | 12 | 1 | Low-quality | Genome-fragment | 42.88 | AAI-based (high-confidence) | 0 |
| S8-final-viral-combined_k141_274642\|\|full | 4131 | No | NA | 5 | 0 | 4 | Not-determined | Genome-fragment | NA | NA | 0 |
| S8-final-viral-combined_k141_113451\|\|full | 1160 | No | NA | 3 | 0 | 1 | Not-determined | Genome-fragment | NA | NA | 0 |
| S2-final-viral-combined_k141_200797\|\|full | 15007 | No | NA | 12 | 1 | 9 | Low-quality | Genome-fragment | 10.14 | AAI-based (medium-confidence) | 0 |
| S4-final-viral-combined_k141_146670\|\|full | 15025 | No | NA | 19 | 0 | 2 | Low-quality | Genome-fragment | 22.73 | AAI-based (medium-confidence) | 0 |
| S8-final-viral-combined_k141_45254\|\|full | 1400 | No | NA | 2 | 0 | 0 | Not-determined | Genome-fragment | NA | NA | 0 |
| S8-final-viral-combined_k141_34165\|\|full | 1534 | No | NA | 3 | 0 | 2 | Not-determined | Genome-fragment | NA | NA | 0 |
| S8-final-viral-combined_k141_194408\|\|full | 3563 | No | NA | 3 | 0 | 3 | Not-determined | Genome-fragment | NA | NA | 0 |
| S8-final-viral-combined_k141_45281\|\|full | 1809 | No | NA | 2 | 0 | 2 | Not-determined | Genome-fragment | NA | NA | 0 |
| S8-final-viral-combined_k141_56094\|\|full | 3254 | No | NA | 3 | 0 | 2 | Not-determined | Genome-fragment | NA | NA | 0 |
| S11-final-viral-combined_k141_294946\|\|full | 15026 | No | NA | 25 | 8 | 0 | Low-quality | Genome-fragment | 30.65 | AAI-based (medium-confidence) | 0 |
| S11-final-viral-combined_k141_244786\|\|0_partial | 15027 | No | NA | 13 | 2 | 1 | Low-quality | Genome-fragment | 6.77 | HMM-based (lower-bound) | 0 |
| S8-final-viral-combined_k141_160105\|\|full | 1966 | No | NA | 3 | 0 | 1 | Not-determined | Genome-fragment | NA | NA | 0 |
| S8-final-viral-combined_k141_56117\|\|full | 6120 | No | NA | 7 | 0 | 4 | Not-determined | Genome-fragment | NA | NA | 0 |
| S8-final-viral-combined_k141_102407\|\|full | 1569 | No | NA | 2 | 0 | 2 | Not-determined | Genome-fragment | NA | NA | 0 |
| S8-final-viral-combined_k141_137618\|\|full | 2061 | No | NA | 3 | 0 | 2 | Not-determined | Genome-fragment | NA | NA | 0 |
| S8-final-viral-combined_k141_274675\|\|full | 4104 | No | NA | 4 | 0 | 3 | Not-determined | Genome-fragment | NA | NA | 0 |
| S11_k141_278866 | 15032 | No | NA | 19 | 7 | 0 | Low-quality | Genome-fragment | 36.71 | AAI-based (medium-confidence) | 0 |
| S8-final-viral-combined_k141_45336\|\|full | 1074 | No | NA | 1 | 0 | 0 | Not-determined | Genome-fragment | NA | NA | 0 |
| S2-final-viral-combined_k141_125151\|\|full | 15034 | No | NA | 16 | 9 | 0 | Low-quality | Genome-fragment | 37.01 | AAI-based (high-confidence) | 0 |
| S8-final-viral-combined_k141_68078\|\|full | 3499 | No | NA | 3 | 0 | 2 | Not-determined | Genome-fragment | NA | NA | 0 |
| S8-final-viral-combined_k141_137659\|\|full | 1199 | No | NA | 2 | 0 | 0 | Not-determined | Genome-fragment | NA | NA | 0 |
| S8-final-viral-combined_k141_90466\|\|full | 2278 | No | NA | 4 | 0 | 3 | Not-determined | Genome-fragment | NA | NA | 0 |
| S8-final-viral-combined_k141_184194\|\|full | 2278 | No | NA | 4 | 0 | 3 | Not-determined | Genome-fragment | NA | NA | 0 |
| S8-final-viral-combined_k141_160163\|\|full | 2360 | No | NA | 2 | 0 | 2 | Not-determined | Genome-fragment | NA | NA | 0 |
| S7-final-viral-combined_k141_233162\|\|full | 15055 | No | NA | 15 | 1 | 5 | Low-quality | Genome-fragment | 22.78 | HMM-based (lower-bound) | 0 |
| S5-final-viral-combined_k141_131113\|\|full | 15062 | Yes | 7339 | 20 | 5 | 7 | Low-quality | Genome-fragment | 15.79 | AAI-based (medium-confidence) | 51.27 |
| S8-final-viral-combined_k141_194523\|\|full | 3223 | No | NA | 3 | 0 | 1 | Not-determined | Genome-fragment | NA | NA | 0 |
| S8-final-viral-combined_k141_45387\|\|full | 1392 | No | NA | 3 | 0 | 2 | Not-determined | Genome-fragment | NA | NA | 0 |
| S8-final-viral-combined_k141_149454\|\|full | 2924 | No | NA | 4 | 0 | 1 | Not-determined | Genome-fragment | NA | NA | 0 |
| S8_k141_172748 | 15084 | No | NA | 10 | 0 | 2 | Low-quality | Genome-fragment | 19.27 | AAI-based (medium-confidence) | 0 |
| S8-final-viral-combined_k141_68155\|\|full | 1066 | No | NA | 2 | 0 | 0 | Not-determined | Genome-fragment | NA | NA | 0 |
| S8-final-viral-combined_k141_194553\|\|full | 2806 | No | NA | 3 | 0 | 1 | Not-determined | Genome-fragment | NA | NA | 0 |
| S8-final-viral-combined_k141_274747\|\|full | 1406 | No | NA | 2 | 0 | 1 | Not-determined | Genome-fragment | NA | NA | 0 |
| S8-final-viral-combined_k141_90502\|\|full | 27579 | No | NA | 22 | 0 | 21 | Not-determined | Genome-fragment | NA | NA | 0 |
| S9_k141_2318 | 15131 | No | NA | 27 | 11 | 0 | Low-quality | Genome-fragment | 33.66 | AAI-based (medium-confidence) | 0 |
| S8-final-viral-combined_k141_184270\|\|full | 2281 | No | NA | 3 | 0 | 1 | Not-determined | Genome-fragment | NA | NA | 0 |
| S8-final-viral-combined_k141_9157\|\|full | 1901 | No | NA | 5 | 0 | 0 | Not-determined | Genome-fragment | NA | NA | 0 |
| S7-final-viral-combined_k141_231311\|\|full | 15133 | No | NA | 24 | 7 | 0 | Low-quality | Genome-fragment | 30.45 | AAI-based (medium-confidence) | 0 |
| S8-final-viral-combined_k141_204705\|\|full | 3663 | No | NA | 5 | 0 | 1 | Not-determined | Genome-fragment | NA | NA | 0 |
| S8-final-viral-combined_k141_68191\|\|full | 1267 | No | NA | 2 | 0 | 0 | Not-determined | Genome-fragment | NA | NA | 0 |
| S1_k141_368784 | 15135 | No | NA | 24 | 1 | 3 | Low-quality | Genome-fragment | 29.57 | AAI-based (high-confidence) | 0 |
| S8-final-viral-combined_k141_252017\|\|full | 6164 | No | NA | 8 | 0 | 1 | Not-determined | Genome-fragment | NA | NA | 0 |
| S8-final-viral-combined_k141_90521\|\|full | 7102 | No | NA | 15 | 0 | 0 | Not-determined | Genome-fragment | NA | NA | 0 |
| S8-final-viral-combined_k141_171585\|\|full | 1859 | No | NA | 2 | 0 | 2 | Not-determined | Genome-fragment | NA | NA | 0 |
| S17-k141_87167 | 15141 | No | NA | 15 | 0 | 9 | Low-quality | Genome-fragment | 6.11 | AAI-based (medium-confidence) | 0 |
| S1-final-viral-combined_k141_37915\|\|0_partial | 15161 | Yes | 3077 | 12 | 2 | 4 | Low-quality | Genome-fragment | 2.07 | HMM-based (lower-bound) | 79.7 |
| S7-final-viral-combined_k141_121474\|\|full | 15166 | No | NA | 21 | 11 | 0 | Low-quality | Genome-fragment | 31.65 | AAI-based (high-confidence) | 0 |
| S1_k141_259291 | 15174 | No | NA | 17 | 10 | 0 | Low-quality | Genome-fragment | 25.4 | AAI-based (high-confidence) | 0 |
| S8-final-viral-combined_k141_217369\|\|full | 1352 | No | NA | 2 | 0 | 2 | Not-determined | Genome-fragment | NA | NA | 0 |
| S8-final-viral-combined_k141_274815\|\|full | 1598 | No | NA | 2 | 0 | 0 | Not-determined | Genome-fragment | NA | NA | 0 |
| S8-final-viral-combined_k141_9193\|\|full | 4662 | No | NA | 4 | 0 | 3 | Not-determined | Genome-fragment | NA | NA | 0 |
| S7-final-viral-combined_k141_32346\|\|0_partial | 15208 | Yes | 4615 | 17 | 2 | 5 | Low-quality | Genome-fragment | 12.58 | AAI-based (medium-confidence) | 69.65 |
| S8-final-viral-combined_k141_34370\|\|full | 6568 | No | NA | 6 | 0 | 3 | Not-determined | Genome-fragment | NA | NA | 0 |
| S7_k141_227845 | 15216 | No | NA | 15 | 1 | 4 | Low-quality | Genome-fragment | 18.75 | HMM-based (lower-bound) | 0 |
| S9_k141_72652 | 15239 | No | NA | 23 | 12 | 0 | Low-quality | Genome-fragment | 36.64 | AAI-based (high-confidence) | 0 |
| S8-final-viral-combined_k141_217390\|\|full | 1178 | No | NA | 3 | 0 | 1 | Not-determined | Genome-fragment | NA | NA | 0 |
| S8-final-viral-combined_k141_20305\|\|full | 1872 | No | NA | 4 | 0 | 0 | Not-determined | Genome-fragment | NA | NA | 0 |
| S8-final-viral-combined_k141_68750\|\|full | 15248 | No | NA | 14 | 1 | 8 | Low-quality | Genome-fragment | 25.76 | HMM-based (lower-bound) | 0 |
| S8-final-viral-combined_k141_204807\|\|full | 4000 | No | NA | 4 | 0 | 3 | Not-determined | Genome-fragment | NA | NA | 0 |
| S8-final-viral-combined_k141_100540\|\|full | 15303 | No | NA | 17 | 2 | 0 | Low-quality | Genome-fragment | 15.12 | AAI-based (high-confidence) | 0 |
| S2_k141_120870 | 15319 | No | NA | 29 | 10 | 2 | Low-quality | Genome-fragment | 19.06 | AAI-based (high-confidence) | 0 |
| S8-final-viral-combined_k141_125462\|\|full | 2112 | No | NA | 3 | 0 | 0 | Not-determined | Genome-fragment | NA | NA | 0 |
| S8-final-viral-combined_k141_274912\|\|full | 1748 | No | NA | 2 | 0 | 1 | Not-determined | Genome-fragment | NA | NA | 0 |
| S8-final-viral-combined_k141_194696\|\|full | 8192 | No | NA | 3 | 0 | 1 | Not-determined | Genome-fragment | NA | NA | 0 |
| S1-final-viral-combined_k141_221925\|\|0_partial | 15321 | No | NA | 14 | 0 | 9 | Low-quality | Genome-fragment | 20.74 | AAI-based (high-confidence) | 0 |
| S8-final-viral-combined_k141_68354\|\|full | 6008 | No | NA | 4 | 0 | 3 | Not-determined | Genome-fragment | NA | NA | 0 |
| S8-final-viral-combined_k141_90674\|\|full | 21396 | No | NA | 16 | 0 | 7 | Not-determined | Genome-fragment | NA | NA | 0 |
| S8-final-viral-combined_k141_217468\|\|full | 1553 | No | NA | 4 | 0 | 0 | Not-determined | Genome-fragment | NA | NA | 0 |
| S8-final-viral-combined_k141_90677\|\|full | 3492 | No | NA | 4 | 0 | 3 | Not-determined | Genome-fragment | NA | NA | 0 |
| S4-final-viral-combined_k141_26399\|\|full | 15324 | No | NA | 7 | 0 | 0 | Low-quality | Genome-fragment | 35.78 | AAI-based (high-confidence) | 0 |
| S8-final-viral-combined_k141_204902\|\|full | 4870 | No | NA | 4 | 0 | 3 | Not-determined | Genome-fragment | NA | NA | 0 |
| S8-final-viral-combined_k141_252184\|\|full | 2016 | No | NA | 2 | 0 | 0 | Not-determined | Genome-fragment | NA | NA | 0 |
| S8-final-viral-combined_k141_171708\|\|full | 2745 | No | NA | 4 | 0 | 3 | Not-determined | Genome-fragment | NA | NA | 0 |
| S3_k141_45917 | 15325 | No | NA | 31 | 1 | 0 | Low-quality | Genome-fragment | 8.98 | HMM-based (lower-bound) | 0 |
| S8-final-viral-combined_k141_9324\|\|full | 1174 | No | NA | 2 | 0 | 0 | Not-determined | Genome-fragment | NA | NA | 0 |
| S8-final-viral-combined_k141_274974\|\|full | 2532 | No | NA | 3 | 0 | 3 | Not-determined | Genome-fragment | NA | NA | 0 |
| S8-final-viral-combined_k141_20404\|\|full | 3776 | No | NA | 4 | 0 | 3 | Not-determined | Genome-fragment | NA | NA | 0 |
| S8-final-viral-combined_k141_102656\|\|full | 3516 | No | NA | 4 | 0 | 1 | Not-determined | Genome-fragment | NA | NA | 0 |
| S8-final-viral-combined_k141_160466\|\|full | 4428 | No | NA | 5 | 0 | 1 | Not-determined | Genome-fragment | NA | NA | 0 |
| S8-final-viral-combined_k141_241632\|\|full | 1221 | No | NA | 3 | 0 | 0 | Not-determined | Genome-fragment | NA | NA | 0 |
| S1-final-viral-combined_k141_287155\|\|full | 15330 | No | NA | 14 | 0 | 0 | Low-quality | Genome-fragment | 25.97 | AAI-based (medium-confidence) | 0 |
| S8-final-viral-combined_k141_252230\|\|full | 4443 | No | NA | 3 | 0 | 2 | Not-determined | Genome-fragment | NA | NA | 0 |
| S8-final-viral-combined_k141_194761\|\|full | 1482 | No | NA | 2 | 0 | 1 | Not-determined | Genome-fragment | NA | NA | 0 |
| S5-final-viral-combined_k141_126730\|\|full | 15341 | No | NA | 10 | 0 | 1 | Low-quality | Genome-fragment | 17.61 | AAI-based (medium-confidence) | 0 |
| S12_k141_135169 | 15344 | Yes | 9544 | 14 | 2 | 2 | Low-quality | Genome-fragment | 14.79 | HMM-based (lower-bound) | 37.8 |
| S8-final-viral-combined_k141_275001\|\|full | 2898 | No | NA | 3 | 0 | 3 | Not-determined | Genome-fragment | NA | NA | 0 |
| S8-final-viral-combined_k141_90781\|\|full | 2191 | No | NA | 3 | 0 | 2 | Not-determined | Genome-fragment | NA | NA | 0 |
| S8-final-viral-combined_k141_45688\|\|full | 1643 | No | NA | 2 | 0 | 2 | Not-determined | Genome-fragment | NA | NA | 0 |
| S6_k141_3027 | 15363 | No | NA | 15 | 2 | 0 | Low-quality | Genome-fragment | 8.36 | AAI-based (medium-confidence) | 0 |
| S5-final-viral-combined_k141_165363\|\|full | 15364 | Yes | 8023 | 17 | 2 | 6 | Low-quality | Genome-fragment | 8.51 | HMM-based (lower-bound) | 47.78 |
| S8-final-viral-combined_k141_275013\|\|full | 4221 | No | NA | 2 | 0 | 2 | Not-determined | Genome-fragment | NA | NA | 0 |
| S8-final-viral-combined_k141_68474\|\|full | 2048 | No | NA | 2 | 0 | 1 | Not-determined | Genome-fragment | NA | NA | 0 |
| S8-final-viral-combined_k141_113787\|\|full | 3960 | No | NA | 5 | 0 | 3 | Not-determined | Genome-fragment | NA | NA | 0 |
| S12_k141_253276 | 15367 | No | NA | 16 | 0 | 5 | Low-quality | Genome-fragment | 15.02 | AAI-based (medium-confidence) | 0 |
| S8-final-viral-combined_k141_229048\|\|full | 2324 | No | NA | 2 | 0 | 1 | Not-determined | Genome-fragment | NA | NA | 0 |
| S11_k141_176128 | 15405 | No | NA | 17 | 9 | 0 | Low-quality | Genome-fragment | 25.44 | AAI-based (medium-confidence) | 0 |
| S11-final-viral-combined_k141_291057\|\|full | 15406 | No | NA | 21 | 12 | 0 | Low-quality | Genome-fragment | 40.33 | AAI-based (medium-confidence) | 0 |
| S1-final-viral-combined_k141_28652\|\|full | 15408 | No | NA | 19 | 0 | 0 | Low-quality | Genome-fragment | 24.91 | AAI-based (high-confidence) | 0 |
| S12_k141_273191 | 15413 | No | NA | 31 | 4 | 0 | Low-quality | Genome-fragment | 20.49 | AAI-based (high-confidence) | 0 |
| S2-final-viral-combined_k141_91443\|\|full | 15418 | Yes | 9073 | 14 | 3 | 4 | Low-quality | Genome-fragment | 23.01 | AAI-based (medium-confidence) | 41.15 |
| S8-final-viral-combined_k141_252272\|\|full | 2116 | No | NA | 3 | 0 | 2 | Not-determined | Genome-fragment | NA | NA | 0 |
| S8-final-viral-combined_k141_90881\|\|full | 2521 | No | NA | 2 | 0 | 0 | Not-determined | Genome-fragment | NA | NA | 0 |
| S1-final-viral-combined_k141_54654\|\|0_partial | 15425 | No | NA | 19 | 1 | 4 | Low-quality | Genome-fragment | 18.74 | AAI-based (medium-confidence) | 0 |
| S8-final-viral-combined_k141_229099\|\|full | 1409 | No | NA | 2 | 0 | 1 | Not-determined | Genome-fragment | NA | NA | 0 |
| S5-final-viral-combined_k141_192791\|\|full | 15467 | No | NA | 13 | 2 | 1 | Low-quality | Genome-fragment | 23.33 | HMM-based (lower-bound) | 0 |
| S8-final-viral-combined_k141_229104\|\|full | 1465 | No | NA | 2 | 0 | 2 | Not-determined | Genome-fragment | NA | NA | 0 |
| S8-final-viral-combined_k141_275068\|\|full | 2819 | No | NA | 4 | 0 | 1 | Not-determined | Genome-fragment | NA | NA | 0 |
| S8-final-viral-combined_k141_241778\|\|full | 3163 | No | NA | 5 | 0 | 0 | Not-determined | Genome-fragment | NA | NA | 0 |
| S1-final-viral-combined_k141_364547\|\|full | 15471 | No | NA | 21 | 1 | 1 | Low-quality | Genome-fragment | 9.86 | HMM-based (lower-bound) | 0 |
| S8-final-viral-combined_k141_8521\|\|0_partial | 15475 | No | NA | 16 | 1 | 6 | Low-quality | Genome-fragment | 6.83 | HMM-based (lower-bound) | 0 |
| S8-final-viral-combined_k141_125691\|\|full | 1337 | No | NA | 2 | 0 | 1 | Not-determined | Genome-fragment | NA | NA | 0 |
| S5-final-viral-combined_k141_119787\|\|full | 15481 | No | NA | 13 | 1 | 6 | Low-quality | Genome-fragment | 10.6 | HMM-based (lower-bound) | 0 |
| S8-final-viral-combined_k141_88768\|\|full | 15481 | No | NA | 21 | 15 | 0 | Low-quality | Genome-fragment | 34.79 | AAI-based (medium-confidence) | 0 |
| S17-final-viral-combined_k141_91500\|\|full | 15491 | No | NA | 25 | 7 | 0 | Low-quality | Genome-fragment | 40.6 | AAI-based (high-confidence) | 0 |
| S8-final-viral-combined_k141_179662\|\|full | 15504 | No | NA | 15 | 0 | 1 | Low-quality | Genome-fragment | 36.43 | AAI-based (high-confidence) | 0 |
| S8-final-viral-combined_k141_125721\|\|full | 2488 | No | NA | 2 | 0 | 1 | Not-determined | Genome-fragment | NA | NA | 0 |
| S8-final-viral-combined_k141_149670\|\|full | 2081 | No | NA | 2 | 0 | 2 | Not-determined | Genome-fragment | NA | NA | 0 |
| S12-final-viral-combined_k141_72985\|\|full | 15509 | No | NA | 32 | 2 | 2 | Low-quality | Genome-fragment | 28.24 | AAI-based (high-confidence) | 0 |
| S7-final-viral-combined_k141_20704\|\|full | 15515 | No | NA | 18 | 1 | 2 | Low-quality | Genome-fragment | 13.45 | AAI-based (medium-confidence) | 0 |
| S4-final-viral-combined_k141_104993\|\|full | 15535 | No | NA | 9 | 0 | 8 | Low-quality | Genome-fragment | 5.99 | AAI-based (high-confidence) | 0 |
| S8-final-viral-combined_k141_9507\|\|full | 2149 | No | NA | 4 | 0 | 1 | Not-determined | Genome-fragment | NA | NA | 0 |
| S11-final-viral-combined_k141_255796\|\|full | 15548 | No | NA | 16 | 0 | 5 | Low-quality | Genome-fragment | 37.34 | AAI-based (medium-confidence) | 0 |
| S8-final-viral-combined_k141_275163\|\|full | 2593 | No | NA | 3 | 0 | 2 | Not-determined | Genome-fragment | NA | NA | 0 |
| S8-final-viral-combined_k141_229197\|\|full | 1472 | No | NA | 3 | 0 | 0 | Not-determined | Genome-fragment | NA | NA | 0 |
| S8-final-viral-combined_k141_241823\|\|full | 1226 | No | NA | 2 | 0 | 0 | Not-determined | Genome-fragment | NA | NA | 0 |
| S8-final-viral-combined_k141_138118\|\|full | 22100 | No | NA | 19 | 0 | 5 | Not-determined | Genome-fragment | NA | NA | 0 |
| S12-final-viral-combined_k141_248567\|\|0_partial | 15558 | No | NA | 13 | 0 | 3 | Low-quality | Genome-fragment | 18.33 | AAI-based (medium-confidence) | 0 |
| S2_k141_232833 | 15564 | No | NA | 30 | 11 | 0 | Low-quality | Genome-fragment | 44.42 | AAI-based (high-confidence) | 0 |
| S8-final-viral-combined_k141_91097\|\|full | 4176 | No | NA | 3 | 0 | 3 | Not-determined | Genome-fragment | NA | NA | 0 |
| S8-final-viral-combined_k141_45848\|\|full | 1571 | No | NA | 3 | 0 | 2 | Not-determined | Genome-fragment | NA | NA | 0 |
| S12-final-viral-combined_k141_133322\|\|full | 15568 | No | NA | 12 | 10 | 0 | Low-quality | Genome-fragment | 26.31 | AAI-based (high-confidence) | 0 |
| S12-final-viral-combined_k141_109895\|\|full | 15574 | No | NA | 18 | 14 | 0 | Low-quality | Genome-fragment | 48.45 | AAI-based (high-confidence) | 0 |
| S11-final-viral-combined_k141_17108\|\|full | 15575 | No | NA | 31 | 4 | 0 | Low-quality | Genome-fragment | 34.7 | AAI-based (high-confidence) | 0 |
| S8-final-viral-combined_k141_217841\|\|full | 1867 | No | NA | 3 | 0 | 1 | Not-determined | Genome-fragment | NA | NA | 0 |
| S8-final-viral-combined_k141_20634\|\|full | 2318 | No | NA | 2 | 0 | 2 | Not-determined | Genome-fragment | NA | NA | 0 |
| S8-final-viral-combined_k141_252446\|\|full | 3246 | No | NA | 3 | 0 | 2 | Not-determined | Genome-fragment | NA | NA | 0 |
| S8-final-viral-combined_k141_229308\|\|full | 2331 | No | NA | 3 | 0 | 2 | Not-determined | Genome-fragment | NA | NA | 0 |
| S12-final-viral-combined_k141_385599\|\|0_partial | 15588 | Yes | 8837 | 23 | 3 | 4 | Low-quality | Genome-fragment | 21.93 | AAI-based (medium-confidence) | 43.31 |
| S1-final-viral-combined_k141_103012\|\|full | 15620 | No | NA | 15 | 0 | 10 | Low-quality | Genome-fragment | 13.12 | AAI-based (medium-confidence) | 0 |
| S8-final-viral-combined_k141_125948\|\|full | 2950 | No | NA | 2 | 0 | 1 | Not-determined | Genome-fragment | NA | NA | 0 |
| S8-final-viral-combined_k141_160712\|\|full | 4332 | No | NA | 5 | 0 | 2 | Not-determined | Genome-fragment | NA | NA | 0 |
| S8-final-viral-combined_k141_261809\|\|full | 2663 | No | NA | 3 | 0 | 1 | Not-determined | Genome-fragment | NA | NA | 0 |
| S5-final-viral-combined_k141_145151\|\|full | 15624 | No | NA | 22 | 18 | 0 | Low-quality | Genome-fragment | 33.55 | AAI-based (high-confidence) | 0 |
| S12-final-viral-combined_k141_351322\|\|full | 15632 | No | NA | 20 | 5 | 0 | Low-quality | Genome-fragment | 32.31 | AAI-based (high-confidence) | 0 |
| S12-final-viral-combined_k141_330633\|\|0_partial | 15641 | Yes | 10125 | 17 | 3 | 2 | Low-quality | Genome-fragment | 17.46 | AAI-based (medium-confidence) | 35.27 |
| S11-final-viral-combined_k141_197700\|\|0_partial | 15668 | Yes | 11978 | 18 | 3 | 3 | Low-quality | Genome-fragment | 14.2 | HMM-based (lower-bound) | 23.55 |
| S8-final-viral-combined_k141_125971\|\|full | 6950 | No | NA | 6 | 0 | 5 | Not-determined | Genome-fragment | NA | NA | 0 |
| S2-final-viral-combined_k141_109161\|\|full | 15701 | No | NA | 21 | 1 | 3 | Low-quality | Genome-fragment | 6.31 | HMM-based (lower-bound) | 0 |
| S8-final-viral-combined_k141_217866\|\|full | 50303 | No | NA | 69 | 19 | 1 | High-quality | High-quality | 97.21 | AAI-based (high-confidence) | 0 |
| S8-final-viral-combined_k141_126004\|\|full | 2370 | No | NA | 3 | 0 | 1 | Not-determined | Genome-fragment | NA | NA | 0 |
| S4-final-viral-combined_k141_104233\|\|full | 15721 | No | NA | 25 | 5 | 1 | Low-quality | Genome-fragment | 26.62 | AAI-based (medium-confidence) | 0 |
| S11-final-viral-combined_k141_143125\|\|0_partial | 15728 | No | NA | 14 | 2 | 7 | Low-quality | Genome-fragment | 23.96 | AAI-based (medium-confidence) | 0 |
| S8-final-viral-combined_k141_68634\|\|full | 1936 | No | NA | 3 | 0 | 0 | Not-determined | Genome-fragment | NA | NA | 0 |
| S8-final-viral-combined_k141_91221\|\|full | 1050 | No | NA | 2 | 0 | 0 | Not-determined | Genome-fragment | NA | NA | 0 |
| S8-final-viral-combined_k141_9658\|\|full | 1247 | No | NA | 2 | 0 | 1 | Not-determined | Genome-fragment | NA | NA | 0 |
| S8-final-viral-combined_k141_275391\|\|full | 2704 | No | NA | 6 | 0 | 2 | Not-determined | Genome-fragment | NA | NA | 0 |
| S8-final-viral-combined_k141_229406\|\|full | 3464 | No | NA | 4 | 0 | 3 | Not-determined | Genome-fragment | NA | NA | 0 |
| S8-final-viral-combined_k141_80657\|\|full | 15728 | No | NA | 10 | 0 | 1 | Low-quality | Genome-fragment | 16.95 | AAI-based (medium-confidence) | 0 |
| S8-final-viral-combined_k141_20800\|\|full | 2092 | No | NA | 4 | 0 | 0 | Not-determined | Genome-fragment | NA | NA | 0 |
| S8-final-viral-combined_k141_252638\|\|full | 1199 | No | NA | 3 | 0 | 0 | Not-determined | Genome-fragment | NA | NA | 0 |
| S8-final-viral-combined_k141_45985\|\|full | 5701 | No | NA | 7 | 0 | 0 | Not-determined | Genome-fragment | NA | NA | 0 |
| S8-final-viral-combined_k141_217911\|\|full | 2114 | No | NA | 2 | 0 | 1 | Not-determined | Genome-fragment | NA | NA | 0 |
| S9_k141_99266 | 15737 | No | NA | 29 | 8 | 0 | Low-quality | Genome-fragment | 33.46 | AAI-based (high-confidence) | 0 |

| | | | | | | | | | | | |
|---|---|---|---|---|---|---|---|---|---|---|---|
| S5_k141_100029 | 15739 | Yes | 5274 | 14 | 2 | 4 | Low-quality | Genome-fragment | 2.9 | HMM-based (lower-bound) | 66.49 |
| S2-final-viral-combined_k141_72534\|\|0_partial | 15743 | No | NA | 8 | 0 | 6 | Low-quality | Genome-fragment | 17.49 | AAI-based (medium-confidence) | 0 |
| S8-final-viral-combined_k141_185132\|\|0_partial | 15744 | Yes | 8373 | 15 | 3 | 6 | Low-quality | Genome-fragment | 8.06 | HMM-based (lower-bound) | 46.82 |
| S11-final-viral-combined_k141_322993\|\|full | 15749 | No | NA | 6 | 0 | 2 | Low-quality | Genome-fragment | 28.41 | AAI-based (medium-confidence) | 0 |
| S8-final-viral-combined_k141_275448\|\|full | 1768 | No | NA | 3 | 0 | 1 | Not-determined | Genome-fragment | NA | NA | 0 |
| S8-final-viral-combined_k141_229477\|\|full | 1943 | No | NA | 3 | 0 | 0 | Not-determined | Genome-fragment | NA | NA | 0 |
| S8-final-viral-combined_k141_252688\|\|full | 1649 | No | NA | 3 | 0 | 2 | Not-determined | Genome-fragment | NA | NA | 0 |
| S8-final-viral-combined_k141_259282\|\|0_partial | 15749 | No | NA | 16 | 1 | 8 | Low-quality | Genome-fragment | 36.67 | AAI-based (medium-confidence) | 0 |
| S8-final-viral-combined_k141_91303\|\|full | 5701 | No | NA | 7 | 0 | 2 | Not-determined | Genome-fragment | NA | NA | 0 |
| S8-final-viral-combined_k141_114135\|\|full | 5300 | No | NA | 5 | 0 | 4 | Not-determined | Genome-fragment | NA | NA | 0 |
| S8-final-viral-combined_k141_80580\|\|full | 3536 | No | NA | 5 | 0 | 4 | Not-determined | Genome-fragment | NA | NA | 0 |
| S8-final-viral-combined_k141_68714\|\|full | 1184 | No | NA | 3 | 0 | 2 | Not-determined | Genome-fragment | NA | NA | 0 |
| S8-final-viral-combined_k141_114150\|\|full | 24125 | No | NA | 16 | 0 | 14 | Not-determined | Genome-fragment | NA | NA | 0 |
| S6-final-viral-combined_k141_76252\|\|full | 15751 | No | NA | 24 | 18 | 0 | Low-quality | Genome-fragment | 32.68 | AAI-based (high-confidence) | 0 |
| S8-final-viral-combined_k141_56874\|\|full | 1381 | No | NA | 2 | 0 | 2 | Not-determined | Genome-fragment | NA | NA | 0 |
| S12-final-viral-combined_k141_160344\|\|full | 15764 | No | NA | 26 | 7 | 1 | Low-quality | Genome-fragment | 24.42 | AAI-based (high-confidence) | 0 |
| S8-final-viral-combined_k141_91399\|\|full | 1915 | No | NA | 3 | 0 | 1 | Not-determined | Genome-fragment | NA | NA | 0 |
| S12_k141_156500 | 15781 | No | NA | 20 | 1 | 3 | Low-quality | Genome-fragment | 11.12 | HMM-based (lower-bound) | 0 |
| S8-final-viral-combined_k141_275535\|\|full | 4002 | No | NA | 6 | 0 | 5 | Not-determined | Genome-fragment | NA | NA | 0 |
| S2_k141_202014 | 15798 | No | NA | 20 | 5 | 0 | Low-quality | Genome-fragment | 40.68 | AAI-based (medium-confidence) | 0 |
| S8-final-viral-combined_k141_160946\|\|full | 4412 | No | NA | 5 | 0 | 3 | Not-determined | Genome-fragment | NA | NA | 0 |
| S2-final-viral-combined_k141_83469\|\|0_partial | 15810 | Yes | 450 | 10 | 1 | 5 | Low-quality | Genome-fragment | 0.83 | HMM-based (lower-bound) | 97.15 |
| S5-final-viral-combined_k141_211161\|\|full | 15811 | No | NA | 20 | 12 | 0 | Low-quality | Genome-fragment | 33.53 | AAI-based (high-confidence) | 0 |
| S4_k141_27636 | 15824 | No | NA | 23 | 4 | 1 | Low-quality | Genome-fragment | 47.37 | HMM-based (lower-bound) | 0 |
| S8-final-viral-combined_k141_103107\|\|full | 1691 | No | NA | 3 | 0 | 2 | Not-determined | Genome-fragment | NA | NA | 0 |
| S7-final-viral-combined_k141_21711\|\|full | 15829 | No | NA | 18 | 1 | 1 | Low-quality | Genome-fragment | 11.4 | HMM-based (lower-bound) | 0 |
| S7-final-viral-combined_k141_87022\|\|full | 15829 | No | NA | 27 | 11 | 0 | Low-quality | Genome-fragment | 25.7 | AAI-based (medium-confidence) | 0 |
| S8-final-viral-combined_k141_103130\|\|full | 6617 | No | NA | 8 | 0 | 0 | Not-determined | Genome-fragment | NA | NA | 0 |
| S3-final-viral-combined_k141_35740\|\|full | 15835 | No | NA | 10 | 0 | 8 | Low-quality | Genome-fragment | 1.66 | AAI-based (medium-confidence) | 0 |
| S8-final-viral-combined_k141_91518\|\|full | 1486 | No | NA | 2 | 0 | 1 | Not-determined | Genome-fragment | NA | NA | 0 |
| S8-final-viral-combined_k141_126236\|\|full | 3157 | No | NA | 3 | 0 | 2 | Not-determined | Genome-fragment | NA | NA | 0 |
| S8-final-viral-combined_k141_161004\|\|full | 1314 | No | NA | 4 | 0 | 0 | Not-determined | Genome-fragment | NA | NA | 0 |
| S8-final-viral-combined_k141_218183\|\|full | 2095 | No | NA | 3 | 0 | 0 | Not-determined | Genome-fragment | NA | NA | 0 |
| S12-final-viral-combined_k141_31488\|\|full | 15845 | No | NA | 25 | 7 | 0 | Low-quality | Genome-fragment | 36.99 | AAI-based (medium-confidence) | 0 |
| S8-final-viral-combined_k141_20999\|\|full | 1322 | No | NA | 3 | 0 | 0 | Not-determined | Genome-fragment | NA | NA | 0 |
| S5-final-viral-combined_k141_139224\|\|full | 15872 | No | NA | 13 | 1 | 7 | Low-quality | Genome-fragment | 9.82 | HMM-based (lower-bound) | 0 |
| S8-final-viral-combined_k141_126278\|\|full | 1196 | No | NA | 3 | 0 | 0 | Not-determined | Genome-fragment | NA | NA | 0 |
| S8-final-viral-combined_k141_261975\|\|full | 1787 | No | NA | 3 | 0 | 1 | Not-determined | Genome-fragment | NA | NA | 0 |
| S8-final-viral-combined_k141_9897\|\|full | 1311 | No | NA | 2 | 0 | 0 | Not-determined | Genome-fragment | NA | NA | 0 |
| S4-final-viral-combined_k141_105849\|\|full | 15888 | No | NA | 19 | 1 | 7 | Low-quality | Genome-fragment | 9.39 | HMM-based (lower-bound) | 0 |
| S8-final-viral-combined_k141_68780\|\|full | 73585 | Yes | 51556 | 95 | 21 | 18 | High-quality | High-quality | 100 | AAI-based (high-confidence) | 29.94 |
| S8-final-viral-combined_k141_218237\|\|full | 3100 | No | NA | 2 | 0 | 0 | Not-determined | Genome-fragment | NA | NA | 0 |
| S12-final-viral-combined_k141_338509\|\|full | 15889 | No | NA | 13 | 1 | 0 | Low-quality | Genome-fragment | 33.52 | AAI-based (medium-confidence) | 0 |
| S9-final-viral-combined_k141_35014\|\|full | 15910 | No | NA | 24 | 13 | 0 | Low-quality | Genome-fragment | 33.97 | AAI-based (high-confidence) | 0 |
| S8-final-viral-combined_k141_9954\|\|full | 2002 | No | NA | 3 | 0 | 2 | Not-determined | Genome-fragment | NA | NA | 0 |
| S8-final-viral-combined_k141_114361\|\|full | 2009 | No | NA | 3 | 0 | 0 | Not-determined | Genome-fragment | NA | NA | 0 |
| S8-final-viral-combined_k141_68811\|\|full | 3916 | No | NA | 3 | 0 | 1 | Not-determined | Genome-fragment | NA | NA | 0 |
| S12-final-viral-combined_k141_121851\|\|full | 15910 | No | NA | 14 | 4 | 0 | Low-quality | Genome-fragment | 38.19 | AAI-based (medium-confidence) | 0 |
| S8-final-viral-combined_k141_88253\|\|0_partial | 15920 | Yes | 9376 | 17 | 3 | 4 | Low-quality | Genome-fragment | 22.54 | AAI-based (high-confidence) | 41.11 |
| S8-final-viral-combined_k141_195288\|\|full | 4701 | No | NA | 4 | 0 | 2 | Not-determined | Genome-fragment | NA | NA | 0 |
| S12_k141_50919 | 15926 | No | NA | 31 | 1 | 0 | Low-quality | Genome-fragment | 22.07 | AAI-based (high-confidence) | 0 |
| S1_k141_50450 | 15942 | No | NA | 16 | 10 | 0 | Low-quality | Genome-fragment | 23.26 | AAI-based (high-confidence) | 0 |
| S8-final-viral-combined_k141_126429\|\|full | 1886 | No | NA | 2 | 0 | 2 | Not-determined | Genome-fragment | NA | NA | 0 |
| S8-final-viral-combined_k141_91658\|\|full | 3557 | No | NA | 4 | 0 | 3 | Not-determined | Genome-fragment | NA | NA | 0 |
| S8-final-viral-combined_k141_218309\|\|full | 1450 | No | NA | 2 | 0 | 1 | Not-determined | Genome-fragment | NA | NA | 0 |
| S2_k141_197833 | 15944 | No | NA | 33 | 3 | 2 | Low-quality | Genome-fragment | 10.69 | AAI-based (high-confidence) | 0 |
| S8-final-viral-combined_k141_91700\|\|full | 1055 | No | NA | 4 | 0 | 0 | Not-determined | Genome-fragment | NA | NA | 0 |
| S17-k141_237656 | 15951 | No | NA | 19 | 1 | 4 | Low-quality | Genome-fragment | 28.18 | AAI-based (medium-confidence) | 0 |
| S8-final-viral-combined_k141_91759\|\|full | 1028 | No | NA | 2 | 0 | 2 | Not-determined | Genome-fragment | NA | NA | 0 |
| S8-final-viral-combined_k141_252880\|\|full | 2253 | No | NA | 4 | 0 | 3 | Not-determined | Genome-fragment | NA | NA | 0 |
| S8-final-viral-combined_k141_10070\|\|full | 1185 | No | NA | 3 | 0 | 1 | Not-determined | Genome-fragment | NA | NA | 0 |
| S8-final-viral-combined_k141_103321\|\|full | 1819 | No | NA | 4 | 0 | 0 | Not-determined | Genome-fragment | NA | NA | 0 |
| S8-final-viral-combined_k141_252927\|\|full | 3442 | No | NA | 3 | 0 | 1 | Not-determined | Genome-fragment | NA | NA | 0 |
| S8-final-viral-combined_k141_114468\|\|full | 1856 | No | NA | 3 | 0 | 2 | Not-determined | Genome-fragment | NA | NA | 0 |
| S8-final-viral-combined_k141_218406\|\|full | 2589 | No | NA | 3 | 0 | 2 | Not-determined | Genome-fragment | NA | NA | 0 |
| S2_k141_26369 | 15951 | No | NA | 14 | 4 | 0 | Low-quality | Genome-fragment | 9.1 | AAI-based (high-confidence) | 0 |
| S8-final-viral-combined_k141_252977\|\|full | 2487 | No | NA | 3 | 0 | 0 | Not-determined | Genome-fragment | NA | NA | 0 |
| S8-final-viral-combined_k141_57173\|\|full | 3523 | No | NA | 4 | 0 | 2 | Not-determined | Genome-fragment | NA | NA | 0 |
| S8-final-viral-combined_k141_21256\|\|full | 8652 | No | NA | 9 | 0 | 3 | Not-determined | Genome-fragment | NA | NA | 0 |
| S5-final-viral-combined_k141_26983\|\|0_partial | 15955 | Yes | 2997 | 15 | 3 | 9 | Low-quality | Genome-fragment | 3.54 | HMM-based (lower-bound) | 81.22 |
| S8-final-viral-combined_k141_161181\|\|full | 51085 | Yes | 33165 | 64 | 19 | 7 | Medium-quality | Genome-fragment | 57.63 | AAI-based (high-confidence) | 35.08 |
| S17-final-viral-combined_k141_120194\|\|0_partial | 15955 | No | NA | 9 | 0 | 3 | Low-quality | Genome-fragment | 15.8 | AAI-based (medium-confidence) | 0 |
| S8-final-viral-combined_k141_103440\|\|full | 2080 | No | NA | 2 | 0 | 2 | Not-determined | Genome-fragment | NA | NA | 0 |
| S8-final-viral-combined_k141_275884\|\|full | 9990 | No | NA | 11 | 0 | 1 | Not-determined | Genome-fragment | NA | NA | 0 |
| S8-final-viral-combined_k141_21280\|\|full | 1817 | No | NA | 2 | 0 | 2 | Not-determined | Genome-fragment | NA | NA | 0 |
| S8-final-viral-combined_k141_91917\|\|full | 1925 | No | NA | 1 | 0 | 1 | Not-determined | Genome-fragment | NA | NA | 0 |
| S8-final-viral-combined_k141_126565\|\|full | 3061 | No | NA | 6 | 0 | 4 | Not-determined | Genome-fragment | NA | NA | 0 |
| S8-final-viral-combined_k141_57207\|\|full | 19034 | No | NA | 22 | 0 | 19 | Not-determined | Genome-fragment | NA | NA | 0 |
| S6-final-viral-combined_k141_112635\|\|full | 15964 | Yes | 12035 | 16 | 6 | 3 | Low-quality | Genome-fragment | 32.95 | AAI-based (medium-confidence) | 24.61 |
| S5-final-viral-combined_k141_203080\|\|0_partial | 15972 | No | NA | 15 | 1 | 9 | Low-quality | Genome-fragment | 10.7 | HMM-based (lower-bound) | 0 |
| S11-final-viral-combined_k141_199280\|\|full | 15992 | Yes | 10966 | 29 | 2 | 4 | Low-quality | Genome-fragment | 16.72 | HMM-based (lower-bound) | 31.43 |
| S8-final-viral-combined_k141_253051\|\|full | 4856 | No | NA | 6 | 0 | 0 | Not-determined | Genome-fragment | NA | NA | 0 |
| S5-final-viral-combined_k141_178287\|\|full | 16006 | No | NA | 21 | 14 | 0 | Low-quality | Genome-fragment | 42.93 | AAI-based (high-confidence) | 0 |
| S4_k141_125980 | 16047 | No | NA | 17 | 10 | 0 | Low-quality | Genome-fragment | 29.25 | AAI-based (high-confidence) | 0 |
| S12-final-viral-combined_k141_313233\|\|full | 16058 | No | NA | 32 | 3 | 1 | Low-quality | Genome-fragment | 31.3 | AAI-based (medium-confidence) | 0 |
| S8-final-viral-combined_k141_21376\|\|full | 1696 | No | NA | 4 | 0 | 0 | Not-determined | Genome-fragment | NA | NA | 0 |
| S11-final-viral-combined_k141_266918\|\|0_partial | 16063 | No | NA | 18 | 3 | 7 | Low-quality | Genome-fragment | 19.21 | HMM-based (lower-bound) | 0 |
| S8-final-viral-combined_k141_103563\|\|full | 1552 | No | NA | 1 | 0 | 1 | Not-determined | Genome-fragment | NA | NA | 0 |
| S8-final-viral-combined_k141_57346\|\|full | 1361 | No | NA | 3 | 0 | 1 | Not-determined | Genome-fragment | NA | NA | 0 |
| S11-final-viral-combined_k141_299568\|\|0_partial | 16069 | Yes | 9968 | 18 | 3 | 2 | Low-quality | Genome-fragment | 6.13 | AAI-based (medium-confidence) | 37.97 |
| S9-final-viral-combined_k141_118243\|\|full | 16073 | No | NA | 10 | 0 | 3 | Low-quality | Genome-fragment | 17.02 | AAI-based (medium-confidence) | 0 |
| S8_k141_148039 | 16075 | No | NA | 13 | 0 | 11 | Low-quality | Genome-fragment | 29.29 | AAI-based (medium-confidence) | 0 |
| S12_k141_57989 | 16133 | No | NA | 18 | 9 | 0 | Low-quality | Genome-fragment | 34.47 | AAI-based (high-confidence) | 0 |
| S2-final-viral-combined_k141_65444\|\|full | 16137 | No | NA | 20 | 1 | 3 | Low-quality | Genome-fragment | 4.84 | HMM-based (lower-bound) | 0 |
| S8-final-viral-combined_k141_92007\|\|full | 4611 | No | NA | 5 | 0 | 5 | Not-determined | Genome-fragment | NA | NA | 0 |
| S8-final-viral-combined_k141_256295\|\|0_partial | 16145 | No | NA | 19 | 0 | 2 | Low-quality | Genome-fragment | 48.42 | AAI-based (medium-confidence) | 0 |
| S12-final-viral-combined_k141_52961\|\|full | 16152 | Yes | 10858 | 12 | 2 | 3 | Low-quality | Genome-fragment | 6.97 | HMM-based (lower-bound) | 32.78 |
| S8-final-viral-combined_k141_262405\|\|full | 3812 | No | NA | 5 | 0 | 1 | Not-determined | Genome-fragment | NA | NA | 0 |
| S4-final-viral-combined_k141_39193\|\|full | 16179 | No | NA | 23 | 9 | 0 | Low-quality | Genome-fragment | 33.14 | AAI-based (high-confidence) | 0 |
| S8-final-viral-combined_k141_195693\|\|full | 1216 | No | NA | 2 | 0 | 1 | Not-determined | Genome-fragment | NA | NA | 0 |
| S8_k141_116278 | 16192 | No | NA | 13 | 0 | 2 | Low-quality | Genome-fragment | 16.15 | AAI-based (medium-confidence) | 0 |
| S8-final-viral-combined_k141_195702\|\|full | 6283 | No | NA | 5 | 0 | 4 | Not-determined | Genome-fragment | NA | NA | 0 |
| S11-final-viral-combined_k141_244799\|\|full | 16200 | Yes | 13886 | 15 | 6 | 2 | Low-quality | Genome-fragment | 35.84 | AAI-based (medium-confidence) | 14.28 |
| S8-final-viral-combined_k141_10320\|\|full | 1166 | No | NA | 2 | 0 | 1 | Not-determined | Genome-fragment | NA | NA | 0 |
| S9-final-viral-combined_k141_24172\|\|0_partial | 16220 | Yes | 14846 | 6 | 3 | 1 | Low-quality | Genome-fragment | 30.26 | AAI-based (high-confidence) | 8.47 |
| S5-final-viral-combined_k141_90785\|\|full | 16221 | Yes | 10355 | 13 | 2 | 3 | Low-quality | Genome-fragment | 32.71 | AAI-based (high-confidence) | 36.16 |
| S2-final-viral-combined_k141_128045\|\|full | 16240 | Yes | 8601 | 23 | 3 | 5 | Low-quality | Genome-fragment | 10.82 | HMM-based (lower-bound) | 47.04 |
| S8-final-viral-combined_k141_206195\|\|full | 2483 | No | NA | 2 | 0 | 0 | Not-determined | Genome-fragment | NA | NA | 0 |
| S1-final-viral-combined_k141_252402\|\|full | 16259 | No | NA | 19 | 4 | 1 | Low-quality | Genome-fragment | 36.46 | AAI-based (medium-confidence) | 0 |
| S8-final-viral-combined_k141_114714\|\|full | 4476 | No | NA | 3 | 0 | 1 | Not-determined | Genome-fragment | NA | NA | 0 |
| S8-final-viral-combined_k141_21620\|\|full | 2315 | No | NA | 2 | 0 | 1 | Not-determined | Genome-fragment | NA | NA | 0 |
| S8-final-viral-combined_k141_21621\|\|full | 2004 | No | NA | 3 | 0 | 1 | Not-determined | Genome-fragment | NA | NA | 0 |
| S8-final-viral-combined_k141_21624\|\|full | 1276 | No | NA | 4 | 0 | 0 | Not-determined | Genome-fragment | NA | NA | 0 |
| S8-final-viral-combined_k141_230359\|\|full | 2470 | No | NA | 4 | 0 | 2 | Not-determined | Genome-fragment | NA | NA | 0 |
| S8-final-viral-combined_k141_57558\|\|full | 5948 | No | NA | 7 | 0 | 2 | Not-determined | Genome-fragment | NA | NA | 0 |
| S5-final-viral-combined_k141_202237\|\|0_partial | 16266 | No | NA | 21 | 0 | 5 | Low-quality | Genome-fragment | 23.41 | AAI-based (medium-confidence) | 0 |
| S8-final-viral-combined_k141_161889\|\|full | 16277 | No | NA | 15 | 5 | 0 | Low-quality | Genome-fragment | 22.76 | AAI-based (medium-confidence) | 0 |
| S12-final-viral-combined_k141_18256\|\|full | 16284 | No | NA | 19 | 1 | 2 | Low-quality | Genome-fragment | 9.95 | AAI-based (medium-confidence) | 0 |
| S8-final-viral-combined_k141_114780\|\|full | 2141 | No | NA | 3 | 0 | 0 | Not-determined | Genome-fragment | NA | NA | 0 |
| S8-final-viral-combined_k141_92148\|\|full | 1919 | No | NA | 2 | 0 | 1 | Not-determined | Genome-fragment | NA | NA | 0 |
| S8-final-viral-combined_k141_21708\|\|full | 1345 | No | NA | 2 | 0 | 1 | Not-determined | Genome-fragment | NA | NA | 0 |
| S1-final-viral-combined_k141_389133\|\|full | 16294 | No | NA | 16 | 1 | 0 | Low-quality | Genome-fragment | 14.67 | AAI-based (medium-confidence) | 0 |
| S2-final-viral-combined_k141_103941\|\|full | 16300 | No | NA | 33 | 5 | 0 | Low-quality | Genome-fragment | 16.48 | HMM-based (lower-bound) | 0 |
| S8-final-viral-combined_k141_194197\|\|0_partial | 16320 | No | NA | 15 | 1 | 2 | Low-quality | Genome-fragment | 32.48 | AAI-based (medium-confidence) | 0 |
| S8-final-viral-combined_k141_206338\|\|full | 2645 | No | NA | 3 | 0 | 1 | Not-determined | Genome-fragment | NA | NA | 0 |
| S12-final-viral-combined_k141_301117\|\|full | 16346 | No | NA | 22 | 1 | 7 | Low-quality | Genome-fragment | 10.99 | HMM-based (lower-bound) | 0 |
| S12_k141_191626 | 16353 | No | NA | 19 | 4 | 1 | Low-quality | Genome-fragment | 20.34 | AAI-based (high-confidence) | 0 |
| S12-final-viral-combined_k141_84980\|\|full | 16409 | No | NA | 16 | 4 | 0 | Low-quality | Genome-fragment | 24.11 | AAI-based (medium-confidence) | 0 |
| S8-final-viral-combined_k141_10754\|\|full | 2586 | No | NA | 3 | 0 | 1 | Not-determined | Genome-fragment | NA | NA | 0 |
| S1-final-viral-combined_k141_222691\|\|full | 16415 | No | NA | 19 | 0 | 16 | Low-quality | Genome-fragment | 8.55 | AAI-based (medium-confidence) | 0 |
| S8-final-viral-combined_k141_262828\|\|full | 2608 | No | NA | 3 | 0 | 1 | Not-determined | Genome-fragment | NA | NA | 0 |
| S8-final-viral-combined_k141_21875\|\|full | 2097 | No | NA | 2 | 0 | 1 | Not-determined | Genome-fragment | NA | NA | 0 |
| S8-final-viral-combined_k141_206667\|\|full | 6294 | No | NA | 7 | 0 | 4 | Not-determined | Genome-fragment | NA | NA | 0 |
| S11-final-viral-combined_k141_324356\|\|0_partial | 16416 | No | NA | 17 | 0 | 13 | Low-quality | Genome-fragment | 27.21 | AAI-based (high-confidence) | 0 |
| S5-final-viral-combined_k141_193235\|\|full | 16420 | No | NA | 22 | 12 | 0 | Low-quality | Genome-fragment | 34.81 | AAI-based (high-confidence) | 0 |
| S8-final-viral-combined_k141_263002\|\|full | 1385 | No | NA | 3 | 0 | 1 | Not-determined | Genome-fragment | NA | NA | 0 |
| S5-final-viral-combined_k141_93249\|\|full | 16423 | No | NA | 25 | 1 | 0 | Low-quality | Genome-fragment | 11.23 | HMM-based (lower-bound) | 0 |
| S12-final-viral-combined_k141_148608\|\|full | 16446 | No | NA | 21 | 18 | 0 | Low-quality | Genome-fragment | 36.58 | AAI-based (high-confidence) | 0 |
| S2_k141_70158 | 16446 | No | NA | 25 | 2 | 0 | Low-quality | Genome-fragment | 27.09 | AAI-based (high-confidence) | 0 |
| S8-final-viral-combined_k141_212554\|\|0_partial | 16465 | Yes | 5512 | 16 | 2 | 8 | Low-quality | Genome-fragment | 13.27 | AAI-based (medium-confidence) | 66.52 |
| S8-final-viral-combined_k141_10960\|\|full | 7807 | No | NA | 7 | 0 | 7 | Not-determined | Genome-fragment | NA | NA | 0 |
| S8-final-viral-combined_k141_10975\|\|full | 1455 | No | NA | 2 | 0 | 1 | Not-determined | Genome-fragment | NA | NA | 0 |
| S5-final-viral-combined_k141_198368\|\|full | 16472 | No | NA | 20 | 10 | 0 | Low-quality | Genome-fragment | 40.71 | AAI-based (high-confidence) | 0 |
| S8-final-viral-combined_k141_263126\|\|full | 2776 | No | NA | 3 | 0 | 1 | Not-determined | Genome-fragment | NA | NA | 0 |
| S8-final-viral-combined_k141_223339\|\|0_partial | 16474 | Yes | 9686 | 17 | 3 | 7 | Low-quality | Genome-fragment | 18.43 | HMM-based (lower-bound) | 41.2 |
| S8-final-viral-combined_k141_263142\|\|full | 5061 | No | NA | 4 | 0 | 4 | Not-determined | Genome-fragment | NA | NA | 0 |
| S7-final-viral-combined_k141_226173\|\|full | 16488 | No | NA | 25 | 2 | 4 | Low-quality | Genome-fragment | 34.86 | AAI-based (medium-confidence) | 0 |
| S7-final-viral-combined_k141_81262\|\|0_partial | 16489 | No | NA | 30 | 7 | 1 | Low-quality | Genome-fragment | 41.99 | AAI-based (high-confidence) | 0 |
| S5_k141_15129 | 16494 | No | NA | 24 | 0 | 4 | Low-quality | Genome-fragment | 15.49 | AAI-based (medium-confidence) | 0 |
| S12_k141_201691 | 16499 | No | NA | 27 | 9 | 0 | Low-quality | Genome-fragment | 45.13 | AAI-based (high-confidence) | 0 |
| S7-final-viral-combined_k141_126514\|\|full | 16508 | Yes | 8251 | 24 | 2 | 3 | Low-quality | Genome-fragment | 16.9 | AAI-based (high-confidence) | 50.02 |
| S8-final-viral-combined_k141_206861\|\|full | 4365 | No | NA | 3 | 0 | 3 | Not-determined | Genome-fragment | NA | NA | 0 |
| S11-final-viral-combined_k141_88102\|\|full | 16512 | No | NA | 26 | 14 | 0 | Low-quality | Genome-fragment | 37.55 | AAI-based (high-confidence) | 0 |
| S8-final-viral-combined_k141_263419\|\|full | 5579 | No | NA | 3 | 0 | 3 | Not-determined | Genome-fragment | NA | NA | 0 |
| S8-final-viral-combined_k141_22476\|\|full | 2994 | No | NA | 4 | 0 | 3 | Not-determined | Genome-fragment | NA | NA | 0 |
| S8-final-viral-combined_k141_206987\|\|full | 1955 | No | NA | 4 | 0 | 2 | Not-determined | Genome-fragment | NA | NA | 0 |
| S8-final-viral-combined_k141_263507\|\|full | 3157 | No | NA | 4 | 0 | 2 | Not-determined | Genome-fragment | NA | NA | 0 |
| S2-final-viral-combined_k141_133733\|\|0_partial | 16518 | No | NA | 15 | 1 | 3 | Low-quality | Genome-fragment | 7.62 | HMM-based (lower-bound) | 0 |
| S8-final-viral-combined_k141_263572\|\|full | 2819 | No | NA | 3 | 0 | 2 | Not-determined | Genome-fragment | NA | NA | 0 |
| S8-final-viral-combined_k141_11130\|\|full | 6990 | No | NA | 9 | 0 | 7 | Not-determined | Genome-fragment | NA | NA | 0 |
| S7-final-viral-combined_k141_173587\|\|0_partial | 16523 | No | NA | 13 | 1 | 3 | Low-quality | Genome-fragment | 15.49 | HMM-based (lower-bound) | 0 |
| S8-final-viral-combined_k141_11165\|\|full | 1751 | No | NA | 2 | 0 | 1 | Not-determined | Genome-fragment | NA | NA | 0 |
| S8-final-viral-combined_k141_11173\|\|full | 6252 | No | NA | 7 | 0 | 6 | Not-determined | Genome-fragment | NA | NA | 0 |
| S11-final-viral-combined_k141_291525\|\|0_partial | 16532 | No | NA | 12 | 1 | 1 | Low-quality | Genome-fragment | 13.23 | HMM-based (lower-bound) | 0 |
| S2-final-viral-combined_k141_108203\|\|0_partial | 16539 | Yes | 10314 | 16 | 3 | 7 | Low-quality | Genome-fragment | 19.63 | HMM-based (lower-bound) | 37.64 |
| S8-final-viral-combined_k141_263752\|\|full | 3004 | No | NA | 4 | 0 | 0 | Not-determined | Genome-fragment | NA | NA | 0 |
| S8-final-viral-combined_k141_22659\|\|full | 2488 | No | NA | 6 | 0 | 0 | Not-determined | Genome-fragment | NA | NA | 0 |
| S8-final-viral-combined_k141_263758\|\|full | 1059 | No | NA | 2 | 0 | 1 | Not-determined | Genome-fragment | NA | NA | 0 |
| S11_k141_257474 | 16551 | No | NA | 22 | 18 | 0 | Low-quality | Genome-fragment | 36.73 | AAI-based (high-confidence) | 0 |
| S8-final-viral-combined_k141_140822\|\|full | 16571 | Yes | 8053 | 23 | 7 | 8 | Low-quality | Genome-fragment | 20.41 | AAI-based (medium-confidence) | 51.4 |
| S3-final-viral-combined_k141_24572\|\|full | 16584 | Yes | 10218 | 17 | 3 | 4 | Low-quality | Genome-fragment | 25.06 | AAI-based (high-confidence) | 38.39 |
| S8-final-viral-combined_k141_22728\|\|full | 2671 | No | NA | 3 | 0 | 1 | Not-determined | Genome-fragment | NA | NA | 0 |
| S8-final-viral-combined_k141_11334\|\|full | 1367 | No | NA | 2 | 0 | 1 | Not-determined | Genome-fragment | NA | NA | 0 |
| S8-final-viral-combined_k141_207302\|\|full | 1101 | No | NA | 2 | 0 | 2 | Not-determined | Genome-fragment | NA | NA | 0 |
| S8-final-viral-combined_k141_207308\|\|full | 1613 | No | NA | 3 | 0 | 0 | Not-determined | Genome-fragment | NA | NA | 0 |
| S8-final-viral-combined_k141_207339\|\|full | 1094 | No | NA | 2 | 0 | 2 | Not-determined | Genome-fragment | NA | NA | 0 |
| S8-final-viral-combined_k141_157464\|\|full | 16584 | No | NA | 21 | 1 | 1 | Low-quality | Genome-fragment | 36.66 | AAI-based (high-confidence) | 0 |
| S8-final-viral-combined_k141_22948\|\|full | 2454 | No | NA | 5 | 0 | 0 | Not-determined | Genome-fragment | NA | NA | 0 |
| S8-final-viral-combined_k141_264158\|\|full | 3148 | No | NA | 7 | 0 | 0 | Not-determined | Genome-fragment | NA | NA | 0 |
| S1-final-viral-combined_k141_282151\|\|0_partial | 16608 | Yes | 8324 | 17 | 2 | 6 | Low-quality | Genome-fragment | 21.1 | AAI-based (high-confidence) | 49.88 |
| S8-final-viral-combined_k141_264319\|\|full | 1745 | No | NA | 3 | 0 | 1 | Not-determined | Genome-fragment | NA | NA | 0 |
| S12-final-viral-combined_k141_153141\|\|0_partial | 16615 | No | NA | 16 | 2 | 6 | Low-quality | Genome-fragment | 25.31 | AAI-based (medium-confidence) | 0 |
| S7-final-viral-combined_k141_232064\|\|full | 16617 | No | NA | 24 | 5 | 0 | Low-quality | Genome-fragment | 44.33 | AAI-based (high-confidence) | 0 |
| S12-final-viral-combined_k141_267354\|\|full | 16659 | No | NA | 29 | 10 | 0 | Low-quality | Genome-fragment | 27.08 | AAI-based (high-confidence) | 0 |
| S8-final-viral-combined_k141_264685\|\|full | 1132 | No | NA | 2 | 0 | 2 | Not-determined | Genome-fragment | NA | NA | 0 |
| S12_k141_98495 | 16682 | No | NA | 19 | 16 | 0 | Low-quality | Genome-fragment | 36.97 | AAI-based (high-confidence) | 0 |
| S8-final-viral-combined_k141_264711\|\|full | 3537 | No | NA | 6 | 1 | 1 | Medium-quality | Genome-fragment | 85 | HMM-based (lower-bound) | 0 |
| S12-final-viral-combined_k141_338158\|\|0_partial | 16684 | No | NA | 17 | 1 | 8 | Low-quality | Genome-fragment | 40.08 | AAI-based (medium-confidence) | 0 |
| S8-final-viral-combined_k141_264865\|\|full | 3653 | No | NA | 5 | 0 | 4 | Not-determined | Genome-fragment | NA | NA | 0 |
| S3-final-viral-combined_k141_14046\|\|full | 16697 | No | NA | 13 | 0 | 11 | Low-quality | Genome-fragment | 13.5 | AAI-based (medium-confidence) | 0 |
| S17-final-viral-combined_k141_100459\|\|full | 16700 | No | NA | 17 | 2 | 1 | Low-quality | Genome-fragment | 38.99 | AAI-based (high-confidence) | 0 |
| S12-final-viral-combined_k141_73729\|\|full | 16704 | Yes | 8692 | 19 | 2 | 5 | Low-quality | Genome-fragment | 19.85 | AAI-based (medium-confidence) | 47.96 |
| S5-final-viral-combined_k141_202147\|\|0_partial | 16705 | No | NA | 12 | 1 | 1 | Low-quality | Genome-fragment | 12.8 | HMM-based (lower-bound) | 0 |
| S12-final-viral-combined_k141_256799\|\|full | 16714 | No | NA | 23 | 5 | 0 | Low-quality | Genome-fragment | 46.3 | AAI-based (medium-confidence) | 0 |
| S2_k141_199451 | 16741 | No | NA | 20 | 9 | 1 | Low-quality | Genome-fragment | 11.36 | AAI-based (high-confidence) | 0 |
| S8-final-viral-combined_k141_219482\|\|0_partial | 12441 | No | NA | 15 | 0 | 9 | Not-determined | Genome-fragment | NA | NA | 0 |
| S12-final-viral-combined_k141_294260\|\|0_partial | 16798 | Yes | 9342 | 17 | 2 | 4 | Low-quality | Genome-fragment | 13.2 | HMM-based (lower-bound) | 44.39 |
| S8-final-viral-combined_k141_151303\|\|full | 16820 | No | NA | 12 | 0 | 11 | Low-quality | Genome-fragment | 6.71 | AAI-based (medium-confidence) | 0 |
| S12-final-viral-combined_k141_236407\|\|0_partial | 16821 | No | NA | 18 | 1 | 6 | Low-quality | Genome-fragment | 33.2 | AAI-based (medium-confidence) | 0 |
| S8-final-viral-combined_k141_12543\|\|0_partial | 14505 | No | NA | 8 | 0 | 6 | Not-determined | Genome-fragment | NA | NA | 0 |
| S7-final-viral-combined_k141_157284\|\|full | 16824 | No | NA | 21 | 6 | 0 | Low-quality | Genome-fragment | 31.19 | AAI-based (medium-confidence) | 0 |
| S5-final-viral-combined_k141_120070\|\|0_partial | 16841 | Yes | 12873 | 20 | 7 | 3 | Low-quality | Genome-fragment | 31.42 | AAI-based (medium-confidence) | 23.56 |
| S8-final-viral-combined_k141_105853\|\|0_partial | 11265 | No | NA | 11 | 0 | 4 | Not-determined | Genome-fragment | NA | NA | 0 |
| S17-final-viral-combined_k141_91575\|\|full | 16850 | No | NA | 24 | 14 | 0 | Low-quality | Genome-fragment | 48.85 | AAI-based (high-confidence) | 0 |
| S8-final-viral-combined_k141_100775\|\|full | 16868 | Yes | 10327 | 18 | 3 | 3 | Low-quality | Genome-fragment | 21.97 | AAI-based (high-confidence) | 38.78 |
| S17-final-viral-combined_k141_162054\|\|0_partial | 16882 | Yes | 4489 | 14 | 2 | 10 | Low-quality | Genome-fragment | 6.74 | HMM-based (lower-bound) | 73.41 |
| S1-final-viral-combined_k141_73122\|\|full | 16892 | No | NA | 26 | 0 | 8 | Low-quality | Genome-fragment | 19.39 | AAI-based (medium-confidence) | 0 |
| S11-final-viral-combined_k141_318155\|\|full | 16916 | No | NA | 22 | 12 | 0 | Low-quality | Genome-fragment | 49.79 | AAI-based (medium-confidence) | 0 |
| S7-final-viral-combined_k141_22291\|\|0_partial | 16941 | No | NA | 21 | 5 | 1 | Low-quality | Genome-fragment | 23.02 | HMM-based (lower-bound) | 0 |
| S8-final-viral-combined_k141_220187\|\|0_partial | 16956 | No | NA | 16 | 2 | 6 | Low-quality | Genome-fragment | 38.55 | AAI-based (high-confidence) | 0 |
| S6_k141_134328 | 16994 | Yes | 13661 | 23 | 10 | 2 | Low-quality | Genome-fragment | 22.67 | AAI-based (high-confidence) | 19.61 |
| S8-final-viral-combined_k141_246185\|\|0_partial | 43201 | Yes | 35887 | 39 | 6 | 7 | High-quality | High-quality | 93.99 | AAI-based (high-confidence) | 16.93 |
| S9-final-viral-combined_k141_72213\|\|0_partial | 17007 | No | NA | 12 | 0 | 9 | Low-quality | Genome-fragment | 1.58 | AAI-based (medium-confidence) | 0 |
| S2-final-viral-combined_k141_188796\|\|0_partial | 17020 | No | NA | 26 | 5 | 1 | Low-quality | Genome-fragment | 21.1 | HMM-based (lower-bound) | 0 |
| S11-final-viral-combined_k141_241798\|\|0_partial | 17044 | No | NA | 18 | 1 | 9 | Low-quality | Genome-fragment | 7.32 | HMM-based (lower-bound) | 0 |
| S7-final-viral-combined_k141_128401\|\|full | 17064 | No | NA | 17 | 1 | 6 | Low-quality | Genome-fragment | 19.15 | HMM-based (lower-bound) | 0 |
| S7-final-viral-combined_k141_3928\|\|full | 17075 | No | NA | 21 | 15 | 0 | Low-quality | Genome-fragment | 48.77 | AAI-based (high-confidence) | 0 |
| S12-final-viral-combined_k141_155528\|\|0_partial | 17078 | No | NA | 12 | 1 | 4 | Low-quality | Genome-fragment | 7.18 | HMM-based (lower-bound) | 0 |
| S2-final-viral-combined_k141_156287\|\|full | 17085 | No | NA | 18 | 5 | 0 | Low-quality | Genome-fragment | 30.55 | AAI-based (high-confidence) | 0 |
| S7-final-viral-combined_k141_102796\|\|full | 17093 | No | NA | 12 | 0 | 2 | Low-quality | Genome-fragment | 24.11 | AAI-based (medium-confidence) | 0 |
| S17-final-viral-combined_k141_22549\|\|full | 17107 | No | NA | 29 | 8 | 1 | Low-quality | Genome-fragment | 46.48 | AAI-based (medium-confidence) | 0 |
| S8-final-viral-combined_k141_78097\|\|0_partial | 23269 | No | NA | 19 | 0 | 13 | Not-determined | Genome-fragment | NA | NA | 0 |
| S1-final-viral-combined_k141_219519\|\|full | 17112 | No | NA | 20 | 1 | 1 | Low-quality | Genome-fragment | 12.25 | HMM-based (lower-bound) | 0 |
| S6_k141_119537 | 17118 | No | NA | 24 | 7 | 0 | Low-quality | Genome-fragment | 38.72 | AAI-based (medium-confidence) | 0 |
| S8_k141_31107 | 17124 | No | NA | 32 | 0 | 0 | Low-quality | Genome-fragment | 32.47 | AAI-based (high-confidence) | 0 |
| S7-final-viral-combined_k141_243145\|\|0_partial | 17158 | No | NA | 19 | 1 | 2 | Low-quality | Genome-fragment | 35.17 | HMM-based (lower-bound) | 0 |
| S8-final-viral-combined_k141_228797\|\|0_partial | 10539 | No | NA | 12 | 0 | 10 | Not-determined | Genome-fragment | NA | NA | 0 |
| S8-final-viral-combined_k141_20235\|\|0_partial | 10078 | No | NA | 8 | 0 | 3 | Not-determined | Genome-fragment | NA | NA | 0 |
| S8-final-viral-combined_k141_229038\|\|0_partial | 10968 | No | NA | 11 | 0 | 7 | Not-determined | Genome-fragment | NA | NA | 0 |
| S6-final-viral-combined_k141_602\|\|full | 17202 | Yes | 12367 | 15 | 4 | 6 | Low-quality | Genome-fragment | 18.36 | HMM-based (lower-bound) | 28.11 |
| S8-final-viral-combined_k141_92109\|\|0_partial | 10825 | No | NA | 8 | 0 | 7 | Not-determined | Genome-fragment | NA | NA | 0 |
| S2-final-viral-combined_k141_62655\|\|full | 17224 | No | NA | 34 | 5 | 0 | Low-quality | Genome-fragment | 44.46 | AAI-based (high-confidence) | 0 |
| S8-final-viral-combined_k141_126965\|\|lt2gene | 1099 | No | NA | 2 | 0 | 1 | Not-determined | Genome-fragment | NA | NA | 0 |
| S8-final-viral-combined_k141_88408\|\|full | 17225 | No | NA | 19 | 11 | 1 | Low-quality | Genome-fragment | 22.76 | AAI-based (medium-confidence) | 0 |
| S8-final-viral-combined_k141_131922\|\|full | 17231 | No | NA | 27 | 5 | 0 | Low-quality | Genome-fragment | 37.61 | AAI-based (high-confidence) | 0 |
| S11-final-viral-combined_k141_174593\|\|0_partial | 17235 | Yes | 8109 | 17 | 2 | 4 | Low-quality | Genome-fragment | 5.11 | HMM-based (lower-bound) | 52.95 |
| S8-final-viral-combined_k141_173251\|\|lt2gene | 3190 | No | NA | 1 | 0 | 0 | Not-determined | Genome-fragment | NA | NA | 0 |
| S8-final-viral-combined_k141_220747\|\|full | 17236 | No | NA | 27 | 14 | 0 | Low-quality | Genome-fragment | 36.74 | AAI-based (high-confidence) | 0 |
| S8_k141_149733 | 17241 | No | NA | 19 | 9 | 0 | Low-quality | Genome-fragment | 31.12 | AAI-based (high-confidence) | 0 |
| S8-final-viral-combined_k141_34765\|\|lt2gene | 2238 | No | NA | 1 | 0 | 0 | Not-determined | Genome-fragment | NA | NA | 0 |
| S7-final-viral-combined_k141_207936\|\|full | 17245 | No | NA | 26 | 7 | 0 | Low-quality | Genome-fragment | 29.44 | AAI-based (medium-confidence) | 0 |
| S8-final-viral-combined_k141_184907\|\|lt2gene | 1157 | No | NA | 2 | 0 | 0 | Not-determined | Genome-fragment | NA | NA | 0 |
| S7-final-viral-combined_k141_199567\|\|full | 17266 | No | NA | 27 | 0 | 1 | Low-quality | Genome-fragment | 38.39 | AAI-based (high-confidence) | 0 |
| S8-final-viral-combined_k141_46792\|\|lt2gene | 1188 | No | NA | 2 | 0 | 0 | Not-determined | Genome-fragment | NA | NA | 0 |
| S1-final-viral-combined_k141_295022\|\|full | 17283 | No | NA | 22 | 16 | 0 | Low-quality | Genome-fragment | 49.15 | AAI-based (high-confidence) | 0 |
| S8-final-viral-combined_k141_231186\|\|lt2gene | 1285 | No | NA | 1 | 0 | 0 | Not-determined | Genome-fragment | NA | NA | 0 |
| S8-final-viral-combined_k141_242853\|\|lt2gene | 1520 | No | NA | 2 | 0 | 0 | Not-determined | Genome-fragment | NA | NA | 0 |
| S8-final-viral-combined_k141_139398\|\|lt2gene | 2158 | No | NA | 1 | 0 | 0 | Not-determined | Genome-fragment | NA | NA | 0 |
| S5_k141_155507 | 17291 | No | NA | 20 | 11 | 0 | Low-quality | Genome-fragment | 43.8 | AAI-based (medium-confidence) | 0 |
| S4_k141_48184 | 17303 | No | NA | 15 | 0 | 7 | Low-quality | Genome-fragment | 5.25 | AAI-based (medium-confidence) | 0 |
| S8-final-viral-combined_k141_24056\|\|lt2gene | 1615 | No | NA | 1 | 0 | 0 | Not-determined | Genome-fragment | NA | NA | 0 |
| S11-final-viral-combined_k141_319509\|\|0_partial | 17304 | Yes | 11832 | 17 | 3 | 7 | Low-quality | Genome-fragment | 22.52 | HMM-based (lower-bound) | 31.62 |
| S17-final-viral-combined_k141_44707\|\|full | 17341 | No | NA | 9 | 3 | 0 | Low-quality | Genome-fragment | 21.06 | AAI-based (high-confidence) | 0 |
| S9-final-viral-combined_k141_88664\|\|full | 17345 | No | NA | 31 | 5 | 0 | Low-quality | Genome-fragment | 44.09 | AAI-based (high-confidence) | 0 |
| S8-final-viral-combined_k141_197085\|\|lt2gene | 1081 | No | NA | 2 | 0 | 0 | Not-determined | Genome-fragment | NA | NA | 0 |
| S17-final-viral-combined_k141_128756\|\|0_partial | 17345 | No | NA | 17 | 1 | 13 | Low-quality | Genome-fragment | 9.57 | HMM-based (lower-bound) | 0 |
| S17-final-viral-combined_k141_124065\|\|full | 17361 | No | NA | 24 | 3 | 0 | Low-quality | Genome-fragment | 21.09 | AAI-based (high-confidence) | 0 |
| S2-final-viral-combined_k141_231102\|\|full | 17373 | No | NA | 20 | 10 | 0 | Low-quality | Genome-fragment | 46.97 | AAI-based (high-confidence) | 0 |
| S8-final-viral-combined_k141_162894\|\|lt2gene | 2211 | No | NA | 1 | 0 | 0 | Not-determined | Genome-fragment | NA | NA | 0 |
| S8-final-viral-combined_k141_204683\|\|full | 17375 | No | NA | 14 | 2 | 1 | Low-quality | Genome-fragment | 32.04 | AAI-based (medium-confidence) | 0 |
| S5-final-viral-combined_k141_197371\|\|full | 17393 | No | NA | 12 | 0 | 3 | Low-quality | Genome-fragment | 13.08 | AAI-based (medium-confidence) | 0 |
| S5-final-viral-combined_k141_41485\|\|0_partial | 17393 | No | NA | 12 | 2 | 5 | Low-quality | Genome-fragment | 28.39 | AAI-based (high-confidence) | 0 |
| S8-final-viral-combined_k141_105601\|\|lt2gene | 1199 | No | NA | 2 | 0 | 1 | Not-determined | Genome-fragment | NA | NA | 0 |
| S5-final-viral-combined_k141_48525\|\|0_partial | 17395 | No | NA | 19 | 2 | 9 | Low-quality | Genome-fragment | 9.71 | HMM-based (lower-bound) | 0 |
| S7_k141_17917 | 17434 | No | NA | 33 | 6 | 1 | Low-quality | Genome-fragment | 31.13 | AAI-based (high-confidence) | 0 |
| S8-final-viral-combined_k141_36373\|\|lt2gene | 1059 | No | NA | 1 | 0 | 0 | Not-determined | Genome-fragment | NA | NA | 0 |
| S7-final-viral-combined_k141_212403\|\|0_partial | 17447 | No | NA | 21 | 1 | 1 | Low-quality | Genome-fragment | 38.04 | AAI-based (high-confidence) | 0 |
| S8-final-viral-combined_k141_189681\|\|full | 17458 | No | NA | 27 | 11 | 0 | Low-quality | Genome-fragment | 43.1 | AAI-based (high-confidence) | 0 |
| S1-final-viral-combined_k141_114540\|\|full | 17481 | No | NA | 13 | 0 | 11 | Low-quality | Genome-fragment | 4.82 | AAI-based (medium-confidence) | 0 |
| S8-final-viral-combined_k141_158562\|\|full | 17489 | No | NA | 14 | 2 | 1 | Low-quality | Genome-fragment | 40.55 | AAI-based (high-confidence) | 0 |
| S17-final-viral-combined_k141_31713\|\|full | 17518 | Yes | 14875 | 24 | 4 | 4 | Low-quality | Genome-fragment | 16.93 | HMM-based (lower-bound) | 15.09 |
| S8-final-viral-combined_k141_198260\|\|lt2gene | 1176 | No | NA | 2 | 0 | 0 | Not-determined | Genome-fragment | NA | NA | 0 |
| S12-final-viral-combined_k141_348927\|\|full | 17531 | No | NA | 23 | 16 | 0 | Low-quality | Genome-fragment | 22.44 | AAI-based (high-confidence) | 0 |
| S7-final-viral-combined_k141_28721\|\|full | 17564 | Yes | 10648 | 27 | 3 | 7 | Low-quality | Genome-fragment | 3.79 | AAI-based (medium-confidence) | 39.38 |
| S1-final-viral-combined_k141_363977\|\|full | 17586 | No | NA | 12 | 0 | 2 | Low-quality | Genome-fragment | 18.15 | AAI-based (medium-confidence) | 0 |
| S7-final-viral-combined_k141_185773\|\|full | 17589 | No | NA | 19 | 14 | 1 | Low-quality | Genome-fragment | 42.51 | AAI-based (high-confidence) | 0 |
| S8-final-viral-combined_k141_118022\|\|lt2gene | 1415 | No | NA | 1 | 0 | 0 | Not-determined | Genome-fragment | NA | NA | 0 |
| S2-final-viral-combined_k141_72911\|\|full | 17596 | No | NA | 23 | 11 | 0 | Low-quality | Genome-fragment | 36.18 | AAI-based (high-confidence) | 0 |
| S11-final-viral-combined_k141_266464\|\|full | 17598 | No | NA | 19 | 10 | 0 | Low-quality | Genome-fragment | 48.49 | AAI-based (high-confidence) | 0 |
| S8-final-viral-combined_k141_60946\|\|lt2gene | 1118 | No | NA | 2 | 0 | 0 | Not-determined | Genome-fragment | NA | NA | 0 |
| S8-final-viral-combined_k141_63699\|\|0_partial | 17613 | Yes | 11556 | 23 | 3 | 2 | Low-quality | Genome-fragment | 13.32 | HMM-based (lower-bound) | 34.39 |
| S11-final-viral-combined_k141_53705\|\|full | 17616 | No | NA | 17 | 2 | 1 | Low-quality | Genome-fragment | 39.97 | AAI-based (high-confidence) | 0 |
| S2-final-viral-combined_k141_156069\|\|full | 17623 | No | NA | 20 | 8 | 0 | Low-quality | Genome-fragment | 27.57 | AAI-based (high-confidence) | 0 |
| S9-final-viral-combined_k141_48407\|\|full | 17648 | No | NA | 23 | 14 | 0 | Low-quality | Genome-fragment | 38.43 | AAI-based (high-confidence) | 0 |
| S12_k141_23928 | 17680 | No | NA | 17 | 1 | 3 | Low-quality | Genome-fragment | 27.48 | AAI-based (medium-confidence) | 0 |
| S8-final-viral-combined_k141_34871\|\|full | 17681 | No | NA | 15 | 0 | 3 | Low-quality | Genome-fragment | 18.15 | AAI-based (medium-confidence) | 0 |
| S8-final-viral-combined_k141_130531\|\|lt2gene | 1761 | No | NA | 1 | 0 | 0 | Not-determined | Genome-fragment | NA | NA | 0 |
| S8-final-viral-combined_k141_49421\|\|lt2gene | 1474 | No | NA | 2 | 0 | 0 | Not-determined | Genome-fragment | NA | NA | 0 |
| S8-final-viral-combined_k141_3463\|\|lt2gene | 1468 | No | NA | 1 | 0 | 0 | Not-determined | Genome-fragment | NA | NA | 0 |
| S8-final-viral-combined_k141_83841\|\|lt2gene | 1275 | No | NA | 2 | 0 | 0 | Not-determined | Genome-fragment | NA | NA | 0 |
| S8-final-viral-combined_k141_130696\|\|lt2gene | 1218 | No | NA | 1 | 0 | 0 | Not-determined | Genome-fragment | NA | NA | 0 |
| S1-final-viral-combined_k141_205178\|\|full | 17687 | No | NA | 25 | 17 | 1 | Low-quality | Genome-fragment | 38 | AAI-based (high-confidence) | 0 |
| S8-final-viral-combined_k141_14803\|\|lt2gene | 1281 | No | NA | 1 | 0 | 0 | Not-determined | Genome-fragment | NA | NA | 0 |
| S2-final-viral-combined_k141_132212\|\|full | 17697 | No | NA | 15 | 1 | 3 | Low-quality | Genome-fragment | 19.82 | HMM-based (lower-bound) | 0 |
| S7-final-viral-combined_k141_150174\|\|full | 17704 | No | NA | 26 | 1 | 2 | Low-quality | Genome-fragment | 12.75 | HMM-based (lower-bound) | 0 |
| S8-final-viral-combined_k141_62253\|\|lt2gene | 1603 | No | NA | 1 | 0 | 0 | Not-determined | Genome-fragment | NA | NA | 0 |
| S12_k141_326806 | 17720 | No | NA | 16 | 1 | 3 | Low-quality | Genome-fragment | 29.75 | HMM-based (lower-bound) | 0 |
| S8-final-viral-combined_k141_27797\|\|lt2gene | 2313 | No | NA | 2 | 0 | 2 | Not-determined | Genome-fragment | NA | NA | 0 |
| S8-final-viral-combined_k141_154500\|\|lt2gene | 1106 | No | NA | 1 | 0 | 0 | Not-determined | Genome-fragment | NA | NA | 0 |
| S7-final-viral-combined_k141_74742\|\|full | 17723 | No | NA | 17 | 6 | 0 | Low-quality | Genome-fragment | 25.67 | AAI-based (medium-confidence) | 0 |
| S8-final-viral-combined_k141_15236\|\|lt2gene | 1039 | No | NA | 1 | 0 | 0 | Not-determined | Genome-fragment | NA | NA | 0 |
| S2_k141_138761 | 17746 | No | NA | 19 | 17 | 0 | Low-quality | Genome-fragment | 42.37 | AAI-based (high-confidence) | 0 |
| S11-final-viral-combined_k141_121561\|\|full | 17751 | Yes | 9532 | 19 | 3 | 9 | Low-quality | Genome-fragment | 18.14 | HMM-based (lower-bound) | 46.3 |
| S17-final-viral-combined_k141_241082\|\|full | 17771 | No | NA | 15 | 0 | 3 | Low-quality | Genome-fragment | 23.58 | AAI-based (medium-confidence) | 0 |
| S1-final-viral-combined_k141_92192\|\|full | 17827 | No | NA | 13 | 1 | 9 | Low-quality | Genome-fragment | 10.98 | HMM-based (lower-bound) | 0 |
| S1_k141_216462 | 17865 | No | NA | 21 | 10 | 0 | Low-quality | Genome-fragment | 27.82 | AAI-based (high-confidence) | 0 |
| S17-final-viral-combined_k141_201345\|\|full | 17876 | No | NA | 17 | 0 | 4 | Low-quality | Genome-fragment | 23.16 | AAI-based (medium-confidence) | 0 |
| S8-final-viral-combined_k141_16105\|\|lt2gene | 1643 | No | NA | 1 | 0 | 0 | Not-determined | Genome-fragment | NA | NA | 0 |
| S7-final-viral-combined_k141_134974\|\|1_partial | 17880 | No | NA | 25 | 1 | 2 | Low-quality | Genome-fragment | 7.52 | HMM-based (lower-bound) | 0 |
| S2_k141_162889 | 17885 | No | NA | 26 | 5 | 1 | Low-quality | Genome-fragment | 22.81 | AAI-based (high-confidence) | 0 |
| S5-final-viral-combined_k141_104194\|\|0_partial | 17901 | Yes | 7175 | 21 | 2 | 5 | Low-quality | Genome-fragment | 5.11 | HMM-based (lower-bound) | 59.92 |
| S2-final-viral-combined_k141_167195\|\|full | 17908 | No | NA | 20 | 11 | 0 | Low-quality | Genome-fragment | 40.88 | AAI-based (high-confidence) | 0 |
| S2-final-viral-combined_k141_6365\|\|full | 17933 | No | NA | 11 | 0 | 8 | Low-quality | Genome-fragment | 9.65 | AAI-based (medium-confidence) | 0 |
| S8-final-viral-combined_k141_75322\|\|lt2gene | 1817 | No | NA | 1 | 0 | 0 | Not-determined | Genome-fragment | NA | NA | 0 |
| S7-final-viral-combined_k141_72443\|\|full | 17951 | No | NA | 26 | 9 | 0 | Low-quality | Genome-fragment | 49.79 | AAI-based (high-confidence) | 0 |
| S8-final-viral-combined_k141_121074\|\|lt2gene | 1746 | No | NA | 2 | 0 | 1 | Not-determined | Genome-fragment | NA | NA | 0 |
| S8-final-viral-combined_k141_40885\|\|lt2gene | 1461 | No | NA | 1 | 0 | 1 | Not-determined | Genome-fragment | NA | NA | 0 |
| S8-final-viral-combined_k141_98593\|\|lt2gene | 1711 | No | NA | 1 | 0 | 1 | Not-determined | Genome-fragment | NA | NA | 0 |
| S12-final-viral-combined_k141_119843\|\|full | 17988 | No | NA | 20 | 1 | 1 | Low-quality | Genome-fragment | 40.75 | AAI-based (medium-confidence) | 0 |
| S12_k141_343028 | 17994 | No | NA | 14 | 2 | 1 | Low-quality | Genome-fragment | 41.16 | AAI-based (medium-confidence) | 0 |
| S17-final-viral-combined_k141_134075\|\|full | 18008 | No | NA | 21 | 11 | 0 | Low-quality | Genome-fragment | 42.5 | AAI-based (high-confidence) | 0 |
| S12-final-viral-combined_k141_129555\|\|full | 18011 | No | NA | 12 | 0 | 8 | Low-quality | Genome-fragment | 6.92 | AAI-based (medium-confidence) | 0 |
| S8-final-viral-combined_k141_86546\|\|lt2gene | 1120 | No | NA | 2 | 0 | 1 | Not-determined | Genome-fragment | NA | NA | 0 |
| S8-final-viral-combined_k141_27195\|\|full | 18021 | No | NA | 25 | 6 | 0 | Low-quality | Genome-fragment | 38.8 | AAI-based (medium-confidence) | 0 |
| S8-final-viral-combined_k141_258492\|\|lt2gene | 1262 | No | NA | 1 | 0 | 0 | Not-determined | Genome-fragment | NA | NA | 0 |
| S1-final-viral-combined_k141_201898\|\|full | 18036 | Yes | 13777 | 18 | 4 | 2 | Low-quality | Genome-fragment | 21.84 | AAI-based (medium-confidence) | 23.61 |
| S7-final-viral-combined_k141_13200\|\|0_partial | 18039 | No | NA | 15 | 0 | 11 | Low-quality | Genome-fragment | 19.46 | AAI-based (medium-confidence) | 0 |
| S8-final-viral-combined_k141_214653\|\|full | 18066 | No | NA | 28 | 18 | 2 | Low-quality | Genome-fragment | 48.74 | AAI-based (high-confidence) | 0 |
| S8-final-viral-combined_k141_19380\|\|0_partial | 18104 | No | NA | 11 | 1 | 10 | Low-quality | Genome-fragment | 9.73 | HMM-based (lower-bound) | 0 |
| S3-final-viral-combined_k141_134912\|\|0_partial | 18106 | Yes | 4436 | 17 | 3 | 7 | Low-quality | Genome-fragment | 3.35 | HMM-based (lower-bound) | 75.5 |
| S1-final-viral-combined_k141_15423\|\|full | 18117 | No | NA | 11 | 0 | 9 | Low-quality | Genome-fragment | 6.88 | AAI-based (medium-confidence) | 0 |
| S4_k141_114651 | 18132 | No | NA | 29 | 11 | 1 | Low-quality | Genome-fragment | 47.64 | AAI-based (high-confidence) | 0 |
| S4-final-viral-combined_k141_66328\|\|full | 18134 | No | NA | 17 | 1 | 1 | Low-quality | Genome-fragment | 42.66 | AAI-based (medium-confidence) | 0 |
| S8-final-viral-combined_k141_258903\|\|lt2gene | 1571 | No | NA | 1 | 0 | 0 | Not-determined | Genome-fragment | NA | NA | 0 |
| S8-final-viral-combined_k141_164484\|\|0_partial | 18142 | No | NA | 19 | 3 | 4 | Low-quality | Genome-fragment | 34.87 | HMM-based (lower-bound) | 0 |
| S4-final-viral-combined_k141_19767\|\|0_partial | 18155 | Yes | 2647 | 19 | 2 | 4 | Low-quality | Genome-fragment | 7.85 | AAI-based (medium-confidence) | 85.42 |
| S7-final-viral-combined_k141_164983\|\|full | 18168 | No | NA | 25 | 10 | 0 | Low-quality | Genome-fragment | 30.69 | AAI-based (high-confidence) | 0 |
| S2-final-viral-combined_k141_145631\|\|full | 18195 | No | NA | 14 | 2 | 0 | Low-quality | Genome-fragment | 10.39 | AAI-based (high-confidence) | 0 |
| S1-final-viral-combined_k141_313762\|\|full | 18201 | No | NA | 22 | 1 | 6 | Low-quality | Genome-fragment | 40.78 | AAI-based (medium-confidence) | 0 |
| S8-final-viral-combined_k141_146017\|\|lt2gene | 1403 | No | NA | 2 | 0 | 1 | Not-determined | Genome-fragment | NA | NA | 0 |
| S1-final-viral-combined_k141_313149\|\|0_partial | 18203 | Yes | 12262 | 19 | 3 | 8 | Low-quality | Genome-fragment | 23.33 | HMM-based (lower-bound) | 32.64 |
| S9-final-viral-combined_k141_30805\|\|0_partial | 18205 | No | NA | 22 | 1 | 11 | Low-quality | Genome-fragment | 11.78 | HMM-based (lower-bound) | 0 |
| S8-final-viral-combined_k141_100283\|\|lt2gene | 1076 | No | NA | 1 | 0 | 1 | Not-determined | Genome-fragment | NA | NA | 0 |
| S5-final-viral-combined_k141_137171\|\|0_partial | 18209 | No | NA | 19 | 1 | 5 | Low-quality | Genome-fragment | 11.52 | HMM-based (lower-bound) | 0 |
| S9_k141_77933 | 18226 | No | NA | 17 | 1 | 2 | Low-quality | Genome-fragment | 6.95 | HMM-based (lower-bound) | 0 |
| S8-final-viral-combined_k141_100482\|\|lt2gene | 2358 | No | NA | 1 | 0 | 0 | Not-determined | Genome-fragment | NA | NA | 0 |
| S17-final-viral-combined_k141_225711\|\|full | 18232 | Yes | 14749 | 24 | 8 | 7 | Low-quality | Genome-fragment | 24.45 | AAI-based (medium-confidence) | 19.1 |
| S8-final-viral-combined_k141_66192\|\|lt2gene | 1046 | No | NA | 1 | 0 | 0 | Not-determined | Genome-fragment | NA | NA | 0 |
| S2-final-viral-combined_k141_207251\|\|full | 18247 | No | NA | 37 | 6 | 2 | Low-quality | Genome-fragment | 30.28 | AAI-based (high-confidence) | 0 |
| S17-k141_88603 | 18266 | No | NA | 14 | 0 | 7 | Low-quality | Genome-fragment | 25.38 | AAI-based (medium-confidence) | 0 |
| S8-final-viral-combined_k141_202994\|\|lt2gene | 1163 | No | NA | 2 | 0 | 0 | Not-determined | Genome-fragment | NA | NA | 0 |
| S12-final-viral-combined_k141_357120\|\|0_partial | 18268 | No | NA | 16 | 1 | 13 | Low-quality | Genome-fragment | 10.43 | HMM-based (lower-bound) | 0 |
| S17-final-viral-combined_k141_16146\|\|0_partial | 18304 | No | NA | 16 | 1 | 9 | Low-quality | Genome-fragment | 16.37 | AAI-based (high-confidence) | 0 |
| S1-final-viral-combined_k141_13091\|\|full | 18344 | Yes | 12705 | 25 | 6 | 4 | Low-quality | Genome-fragment | 17.09 | HMM-based (lower-bound) | 30.74 |
| S4-final-viral-combined_k141_27848\|\|full | 18387 | No | NA | 31 | 5 | 0 | Low-quality | Genome-fragment | 43.55 | AAI-based (high-confidence) | 0 |
| S8-final-viral-combined_k141_216081\|\|lt2gene | 1602 | No | NA | 1 | 0 | 0 | Not-determined | Genome-fragment | NA | NA | 0 |
| S7-final-viral-combined_k141_242241\|\|0_partial | 18392 | No | NA | 23 | 1 | 2 | Low-quality | Genome-fragment | 12.36 | HMM-based (lower-bound) | 0 |
| S12-final-viral-combined_k141_99590\|\|full | 18396 | Yes | 12861 | 18 | 2 | 3 | Low-quality | Genome-fragment | 31.93 | AAI-based (medium-confidence) | 30.09 |
| S8-final-viral-combined_k141_112147\|\|lt2gene | 6883 | No | NA | 1 | 0 | 0 | Not-determined | Genome-fragment | NA | NA | 0 |
| S2-final-viral-combined_k141_53022\|\|full | 18404 | No | NA | 20 | 0 | 10 | Low-quality | Genome-fragment | 20.28 | AAI-based (medium-confidence) | 0 |
| S2-final-viral-combined_k141_142224\|\|0_partial | 18419 | No | NA | 26 | 1 | 2 | Low-quality | Genome-fragment | 7.75 | HMM-based (lower-bound) | 0 |
| S8-final-viral-combined_k141_216484\|\|lt2gene | 1069 | No | NA | 1 | 0 | 1 | Not-determined | Genome-fragment | NA | NA | 0 |
| S8-final-viral-combined_k141_8450\|\|lt2gene | 1667 | No | NA | 1 | 0 | 1 | Not-determined | Genome-fragment | NA | NA | 0 |
| S2-final-viral-combined_k141_103506\|\|0_partial | 18463 | Yes | 10130 | 28 | 2 | 6 | Low-quality | Genome-fragment | 4.54 | HMM-based (lower-bound) | 45.13 |
| S17-final-viral-combined_k141_79391\|\|full | 18503 | No | NA | 26 | 12 | 0 | Low-quality | Genome-fragment | 49.74 | AAI-based (high-confidence) | 0 |
| S4_k141_104468 | 18507 | No | NA | 18 | 0 | 6 | Low-quality | Genome-fragment | 24.32 | AAI-based (medium-confidence) | 0 |
| S12-final-viral-combined_k141_295365\|\|full | 18510 | No | NA | 25 | 1 | 0 | Low-quality | Genome-fragment | 11.73 | HMM-based (lower-bound) | 0 |
| S12-final-viral-combined_k141_98930\|\|0_partial | 18517 | Yes | 4031 | 22 | 2 | 12 | Low-quality | Genome-fragment | 6.61 | HMM-based (lower-bound) | 78.23 |
| S8-final-viral-combined_k141_175582\|\|full | 18517 | No | NA | 24 | 12 | 1 | Low-quality | Genome-fragment | 47.79 | AAI-based (high-confidence) | 0 |
| S7_k141_235684 | 18545 | No | NA | 19 | 0 | 2 | Low-quality | Genome-fragment | 19.48 | AAI-based (medium-confidence) | 0 |
| S5-final-viral-combined_k141_38656\|\|full | 18578 | No | NA | 12 | 0 | 8 | Low-quality | Genome-fragment | 7.19 | AAI-based (medium-confidence) | 0 |
| S17-final-viral-combined_k141_83747\|\|full | 18612 | No | NA | 16 | 0 | 4 | Low-quality | Genome-fragment | 22.53 | AAI-based (medium-confidence) | 0 |
| S8-final-viral-combined_k141_8801\|\|lt2gene | 1572 | No | NA | 1 | 0 | 0 | Not-determined | Genome-fragment | NA | NA | 0 |
| S11-final-viral-combined_k141_110117\|\|full | 18620 | No | NA | 17 | 4 | 1 | Low-quality | Genome-fragment | 24.08 | AAI-based (high-confidence) | 0 |
| S5-final-viral-combined_k141_109582\|\|full | 18625 | No | NA | 26 | 10 | 1 | Low-quality | Genome-fragment | 45.31 | AAI-based (high-confidence) | 0 |
| S11-final-viral-combined_k141_49444\|\|full | 18660 | No | NA | 18 | 5 | 1 | Low-quality | Genome-fragment | 44.49 | AAI-based (medium-confidence) | 0 |
| S8-final-viral-combined_k141_149129\|\|lt2gene | 1986 | No | NA | 1 | 0 | 0 | Not-determined | Genome-fragment | NA | NA | 0 |
| S1-final-viral-combined_k141_73202\|\|full | 18660 | No | NA | 16 | 12 | 0 | Low-quality | Genome-fragment | 30.89 | AAI-based (high-confidence) | 0 |
| S8-final-viral-combined_k141_204550\|\|lt2gene | 3052 | No | NA | 1 | 0 | 0 | Not-determined | Genome-fragment | NA | NA | 0 |
| S8-final-viral-combined_k141_9149\|\|lt2gene | 2013 | No | NA | 1 | 0 | 0 | Not-determined | Genome-fragment | NA | NA | 0 |
| S3-final-viral-combined_k141_7588\|\|0_partial | 18664 | Yes | 13984 | 20 | 3 | 4 | Low-quality | Genome-fragment | 33.62 | AAI-based (medium-confidence) | 25.08 |
| S5-final-viral-combined_k141_168988\|\|full | 18667 | No | NA | 24 | 12 | 0 | Low-quality | Genome-fragment | 23.61 | AAI-based (medium-confidence) | 0 |
| S5-final-viral-combined_k141_7742\|\|0_partial | 18676 | No | NA | 12 | 0 | 3 | Low-quality | Genome-fragment | 18.5 | AAI-based (medium-confidence) | 0 |
| S8-final-viral-combined_k141_172192\|\|lt2gene | 1078 | No | NA | 2 | 0 | 1 | Not-determined | Genome-fragment | NA | NA | 0 |
| S8-final-viral-combined_k141_217951\|\|lt2gene | 1039 | No | NA | 2 | 0 | 0 | Not-determined | Genome-fragment | NA | NA | 0 |
| S1_k141_104373 | 18717 | No | NA | 20 | 6 | 0 | Low-quality | Genome-fragment | 41.69 | AAI-based (high-confidence) | 0 |
| S5-final-viral-combined_k141_163801\|\|0_partial | 18730 | Yes | 5608 | 16 | 2 | 3 | Low-quality | Genome-fragment | 2.98 | HMM-based (lower-bound) | 70.06 |
| S8-final-viral-combined_k141_205925\|\|lt2gene | 1551 | No | NA | 2 | 0 | 0 | Not-determined | Genome-fragment | NA | NA | 0 |
| S8-final-viral-combined_k141_253261\|\|lt2gene | 1019 | No | NA | 1 | 0 | 1 | Not-determined | Genome-fragment | NA | NA | 0 |
| S12-final-viral-combined_k141_337033\|\|full | 18746 | Yes | 13017 | 30 | 6 | 3 | Low-quality | Genome-fragment | 16.43 | HMM-based (lower-bound) | 30.56 |
| S8_k141_245079 | 18784 | No | NA | 23 | 1 | 7 | Low-quality | Genome-fragment | 11.5 | AAI-based (medium-confidence) | 0 |
| S5-final-viral-combined_k141_208807\|\|full | 18788 | No | NA | 18 | 1 | 0 | Low-quality | Genome-fragment | 12.63 | HMM-based (lower-bound) | 0 |
| S8_k141_92163 | 5700 | No | NA | 5 | 0 | 0 | Not-determined | Genome-fragment | NA | NA | 0 |
| S8_k141_138257 | 2755 | No | NA | 3 | 0 | 2 | Not-determined | Genome-fragment | NA | NA | 0 |
| S5-final-viral-combined_k141_189786\|\|0_partial | 18800 | Yes | 8259 | 16 | 2 | 5 | Low-quality | Genome-fragment | 5.36 | HMM-based (lower-bound) | 56.07 |
| S8_k141_253427 | 4078 | No | NA | 4 | 0 | 4 | Not-determined | Genome-fragment | NA | NA | 0 |
| S8_k141_46109 | 1044 | No | NA | 2 | 0 | 1 | Not-determined | Genome-fragment | NA | NA | 0 |
| S8_k141_138303 | 1074 | No | NA | 1 | 0 | 1 | Not-determined | Genome-fragment | NA | NA | 0 |
| S8_k141_218920 | 1641 | No | NA | 4 | 0 | 0 | Not-determined | Genome-fragment | NA | NA | 0 |
| S1_k141_19414 | 18813 | No | NA | 32 | 1 | 0 | Low-quality | Genome-fragment | 27.97 | AAI-based (high-confidence) | 0 |
| S8_k141_80650 | 1294 | No | NA | 3 | 0 | 2 | Not-determined | Genome-fragment | NA | NA | 0 |
| S8_k141_265005 | 1016 | No | NA | 2 | 0 | 0 | Not-determined | Genome-fragment | NA | NA | 0 |
| S8_k141_11557 | 1765 | No | NA | 3 | 0 | 2 | Not-determined | Genome-fragment | NA | NA | 0 |
| S8_k141_138377 | 2302 | No | NA | 2 | 0 | 2 | Not-determined | Genome-fragment | NA | NA | 0 |
| S8_k141_195915 | 3266 | No | NA | 2 | 0 | 2 | Not-determined | Genome-fragment | NA | NA | 0 |
| S8_k141_195923 | 1095 | No | NA | 1 | 0 | 1 | Not-determined | Genome-fragment | NA | NA | 0 |
| S4_k141_32115 | 18819 | No | NA | 21 | 6 | 0 | Low-quality | Genome-fragment | 29.04 | AAI-based (high-confidence) | 0 |
| S8_k141_80720 | 2032 | No | NA | 1 | 0 | 1 | Not-determined | Genome-fragment | NA | NA | 0 |
| S12_k141_74112 | 18828 | No | NA | 23 | 1 | 3 | Low-quality | Genome-fragment | 10.21 | HMM-based (lower-bound) | 0 |
| S8_k141_230590 | 5350 | No | NA | 3 | 0 | 2 | Not-determined | Genome-fragment | NA | NA | 0 |
| S8_k141_253509 | 1428 | No | NA | 2 | 0 | 0 | Not-determined | Genome-fragment | NA | NA | 0 |
| S8_k141_46243 | 1019 | No | NA | 1 | 0 | 1 | Not-determined | Genome-fragment | NA | NA | 0 |
| S8_k141_184434 | 1854 | No | NA | 2 | 0 | 1 | Not-determined | Genome-fragment | NA | NA | 0 |

| | | | | | | | | | | | |
|---|---|---|---|---|---|---|---|---|---|---|---|
| S8_k141_242099 | 1230 | No | NA | 3 | 0 | 3 | Not-determined | Genome-fragment | NA | NA | 0 |
| S12-final-viral-combined_k141_111628||full | 18856 | No | NA | 26 | 10 | 0 | Low-quality | Genome-fragment | 49.73 | AAI-based (high-confidence) | 0 |
| S8_k141_80862 | 1036 | No | NA | 2 | 0 | 0 | Not-determined | Genome-fragment | NA | NA | 0 |
| S8_k141_184459 | 5936 | No | NA | 7 | 0 | 3 | Not-determined | Genome-fragment | NA | NA | 0 |
| S8_k141_230690 | 1528 | No | NA | 2 | 0 | 0 | Not-determined | Genome-fragment | NA | NA | 0 |
| S8-final-viral-combined_k141_52198||full | 18856 | No | NA | 28 | 11 | 0 | Low-quality | Genome-fragment | 40.92 | AAI-based (high-confidence) | 0 |
| S8_k141_23277 | 1013 | No | NA | 1 | 0 | 0 | Not-determined | Genome-fragment | NA | NA | 0 |
| S8_k141_173007 | 1307 | No | NA | 2 | 0 | 2 | Not-determined | Genome-fragment | NA | NA | 0 |
| S8_k141_222 | 1316 | No | NA | 1 | 0 | 1 | Not-determined | Genome-fragment | NA | NA | 0 |
| S8_k141_92403 | 3293 | No | NA | 3 | 0 | 2 | Not-determined | Genome-fragment | NA | NA | 0 |
| S8_k141_229 | 1632 | No | NA | 2 | 0 | 1 | Not-determined | Genome-fragment | NA | NA | 0 |
| S8_k141_138552 | 3115 | No | NA | 1 | 0 | 0 | Not-determined | Genome-fragment | NA | NA | 0 |
| S8_k141_161339 | 1371 | No | NA | 2 | 0 | 0 | Not-determined | Genome-fragment | NA | NA | 0 |
| S8_k141_138615 | 1200 | No | NA | 1 | 0 | 0 | Not-determined | Genome-fragment | NA | NA | 0 |
| S8_k141_270 | 5400 | No | NA | 7 | 2 | 0 | High-quality | High-quality | 97.67 | AAI-based (high-confidence) | 0 |
| S5-final-viral-combined_k141_63378||0_partial | 18880 | Yes | 11688 | 20 | 3 | 7 | Low-quality | Genome-fragment | 22.24 | HMM-based (lower-bound) | 38.09 |
| S8_k141_81005 | 4305 | No | NA | 3 | 0 | 1 | Not-determined | Genome-fragment | NA | NA | 0 |
| S8_k141_184560 | 1678 | No | NA | 1 | 0 | 0 | Not-determined | Genome-fragment | NA | NA | 0 |
| S7_k141_152458 | 18889 | No | NA | 30 | 8 | 0 | Low-quality | Genome-fragment | 30.36 | AAI-based (high-confidence) | 0 |
| S8_k141_23350 | 1023 | No | NA | 2 | 0 | 1 | Not-determined | Genome-fragment | NA | NA | 0 |
| S5-final-viral-combined_k141_9528||0_partial | 18892 | No | NA | 20 | 2 | 8 | Low-quality | Genome-fragment | 9.54 | HMM-based (lower-bound) | 0 |
| S8_k141_242246 | 1227 | No | NA | 2 | 0 | 2 | Not-determined | Genome-fragment | NA | NA | 0 |
| S8_k141_219089 | 1846 | No | NA | 2 | 0 | 1 | Not-determined | Genome-fragment | NA | NA | 0 |
| S11-final-viral-combined_k141_48716||full | 18904 | No | NA | 23 | 12 | 1 | Low-quality | Genome-fragment | 39.11 | AAI-based (high-confidence) | 0 |
| S6-final-viral-combined_k141_134039||full | 18926 | No | NA | 23 | 8 | 0 | Low-quality | Genome-fragment | 46.02 | AAI-based (medium-confidence) | 0 |
| S8_k141_57935 | 1277 | No | NA | 2 | 0 | 1 | Not-determined | Genome-fragment | NA | NA | 0 |
| S11-final-viral-combined_k141_236944||full | 18951 | No | NA | 21 | 13 | 0 | Low-quality | Genome-fragment | 39.93 | AAI-based (high-confidence) | 0 |
| S8_k141_173163 | 4682 | No | NA | 2 | 0 | 2 | Not-determined | Genome-fragment | NA | NA | 0 |
| S8_k141_69507 | 1252 | No | NA | 1 | 0 | 1 | Not-determined | Genome-fragment | NA | NA | 0 |
| S8_k141_81045 | 1095 | No | NA | 1 | 0 | 0 | Not-determined | Genome-fragment | NA | NA | 0 |
| S8_k141_253667 | 1048 | No | NA | 2 | 0 | 0 | Not-determined | Genome-fragment | NA | NA | 0 |
| S8_k141_115516 | 1639 | No | NA | 2 | 0 | 2 | Not-determined | Genome-fragment | NA | NA | 0 |
| S8_k141_230914 | 2118 | No | NA | 1 | 0 | 0 | Not-determined | Genome-fragment | NA | NA | 0 |
| S7-final-viral-combined_k141_172366||full | 18956 | No | NA | 14 | 2 | 0 | Low-quality | Genome-fragment | 32.12 | AAI-based (medium-confidence) | 0 |
| S8_k141_58021 | 1997 | No | NA | 1 | 0 | 1 | Not-determined | Genome-fragment | NA | NA | 0 |
| S9-final-viral-combined_k141_88706||full | 18963 | No | NA | 23 | 1 | 6 | Low-quality | Genome-fragment | 21.29 | HMM-based (lower-bound) | 0 |
| S12_k141_202330 | 18969 | No | NA | 30 | 7 | 1 | Low-quality | Genome-fragment | 45.03 | AAI-based (high-confidence) | 0 |
| S8_k141_127075 | 3035 | No | NA | 5 | 0 | 1 | Not-determined | Genome-fragment | NA | NA | 0 |
| S8_k141_161455 | 1939 | No | NA | 3 | 0 | 1 | Not-determined | Genome-fragment | NA | NA | 0 |
| S2-final-viral-combined_k141_9600||0_partial | 18974 | No | NA | 22 | 1 | 8 | Low-quality | Genome-fragment | 42.63 | AAI-based (high-confidence) | 0 |
| S2-final-viral-combined_k141_116441||full | 18975 | Yes | 12161 | 21 | 3 | 3 | Low-quality | Genome-fragment | 11.88 | HMM-based (lower-bound) | 35.91 |
| S8_k141_103929 | 9270 | No | NA | 17 | 0 | 4 | Not-determined | Genome-fragment | NA | NA | 0 |
| S8_k141_196198 | 4718 | No | NA | 9 | 0 | 0 | Not-determined | Genome-fragment | NA | NA | 0 |
| S8_k141_11823 | 4619 | No | NA | 5 | 0 | 3 | Not-determined | Genome-fragment | NA | NA | 0 |
| S8_k141_161463 | 1668 | No | NA | 1 | 0 | 1 | Not-determined | Genome-fragment | NA | NA | 0 |
| S8_k141_173228 | 3077 | No | NA | 4 | 0 | 2 | Not-determined | Genome-fragment | NA | NA | 0 |
| S8_k141_219177 | 1365 | No | NA | 2 | 0 | 2 | Not-determined | Genome-fragment | NA | NA | 0 |
| S8_k141_115584 | 1746 | No | NA | 1 | 0 | 1 | Not-determined | Genome-fragment | NA | NA | 0 |
| S8_k141_103948 | 3380 | No | NA | 3 | 0 | 3 | Not-determined | Genome-fragment | NA | NA | 0 |
| S8_k141_23519 | 1407 | No | NA | 1 | 0 | 0 | Not-determined | Genome-fragment | NA | NA | 0 |
| S5-final-viral-combined_k141_42369||full | 18980 | No | NA | 20 | 5 | 0 | Low-quality | Genome-fragment | 45.87 | AAI-based (high-confidence) | 0 |
| S8_k141_81191 | 2859 | No | NA | 3 | 0 | 1 | Not-determined | Genome-fragment | NA | NA | 0 |
| S8_k141_81205 | 3788 | No | NA | 3 | 0 | 1 | Not-determined | Genome-fragment | NA | NA | 0 |
| S8_k141_173323 | 1043 | No | NA | 1 | 0 | 1 | Not-determined | Genome-fragment | NA | NA | 0 |
| S8_k141_219282 | 1183 | No | NA | 3 | 0 | 0 | Not-determined | Genome-fragment | NA | NA | 0 |
| S12-final-viral-combined_k141_389739||full | 18987 | No | NA | 25 | 11 | 0 | Low-quality | Genome-fragment | 45.73 | AAI-based (high-confidence) | 0 |
| S2_k141_112556 | 19005 | No | NA | 37 | 1 | 2 | Low-quality | Genome-fragment | 17.36 | AAI-based (high-confidence) | 0 |
| S8_k141_219299 | 1063 | No | NA | 1 | 0 | 1 | Not-determined | Genome-fragment | NA | NA | 0 |
| S8_k141_242570 | 1376 | No | NA | 2 | 0 | 0 | Not-determined | Genome-fragment | NA | NA | 0 |
| S17-final-viral-combined_k141_112180||0_partial | 19006 | Yes | 13186 | 26 | 5 | 2 | Low-quality | Genome-fragment | 16.35 | HMM-based (lower-bound) | 30.62 |
| S8_k141_127149 | 1106 | No | NA | 2 | 0 | 1 | Not-determined | Genome-fragment | NA | NA | 0 |
| S5-final-viral-combined_k141_107308||full | 19018 | No | NA | 28 | 7 | 0 | Low-quality | Genome-fragment | 48.53 | AAI-based (high-confidence) | 0 |
| S2-final-viral-combined_k141_132917||full | 19029 | No | NA | 19 | 0 | 10 | Low-quality | Genome-fragment | 10.24 | AAI-based (medium-confidence) | 0 |
| S8_k141_253780 | 3976 | No | NA | 6 | 0 | 0 | Not-determined | Genome-fragment | NA | NA | 0 |
| S8_k141_11962 | 4833 | No | NA | 3 | 0 | 1 | Not-determined | Genome-fragment | NA | NA | 0 |
| S5-final-viral-combined_k141_109133||0_partial | 19032 | No | NA | 17 | 1 | 7 | Low-quality | Genome-fragment | 10.5 | HMM-based (lower-bound) | 0 |
| S2-final-viral-combined_k141_155788||full | 19081 | No | NA | 16 | 0 | 12 | Low-quality | Genome-fragment | 16.79 | AAI-based (medium-confidence) | 0 |
| S8_k141_219376 | 3265 | No | NA | 2 | 0 | 1 | Not-determined | Genome-fragment | NA | NA | 0 |
| S9-final-viral-combined_k141_45779||0_partial | 19122 | Yes | 6372 | 14 | 5 | 7 | Low-quality | Genome-fragment | 13.58 | AAI-based (high-confidence) | 66.68 |
| S8_k141_81318 | 1149 | No | NA | 2 | 0 | 0 | Not-determined | Genome-fragment | NA | NA | 0 |
| S8_k141_265530 | 2606 | No | NA | 2 | 0 | 2 | Not-determined | Genome-fragment | NA | NA | 0 |
| S8_k141_173421 | 1198 | No | NA | 1 | 0 | 0 | Not-determined | Genome-fragment | NA | NA | 0 |
| S8_k141_12006 | 5363 | No | NA | 7 | 0 | 5 | Not-determined | Genome-fragment | NA | NA | 0 |
| S8_k141_161685 | 5210 | No | NA | 3 | 0 | 2 | Not-determined | Genome-fragment | NA | NA | 0 |
| S8_k141_173430 | 3107 | No | NA | 3 | 0 | 0 | Not-determined | Genome-fragment | NA | NA | 0 |
| S8_k141_219410 | 5829 | No | NA | 3 | 0 | 3 | Not-determined | Genome-fragment | NA | NA | 0 |
| S8_k141_92847 | 1908 | No | NA | 2 | 0 | 1 | Not-determined | Genome-fragment | NA | NA | 0 |
| S8_k141_127191 | 59987 | No | NA | 93 | 31 | 0 | High-quality | High-quality | 97.01 | AAI-based (high-confidence) | 0 |
| S8-final-viral-combined_k141_31732||full | 19133 | No | NA | 23 | 2 | 0 | Low-quality | Genome-fragment | 42.55 | AAI-based (medium-confidence) | 0 |
| S8_k141_231044 | 4349 | No | NA | 7 | 0 | 3 | Not-determined | Genome-fragment | NA | NA | 0 |
| S5-final-viral-combined_k141_188623||full | 19188 | Yes | 9680 | 22 | 3 | 4 | Low-quality | Genome-fragment | 8.94 | HMM-based (lower-bound) | 49.55 |
| S8_k141_265574 | 7953 | No | NA | 3 | 0 | 0 | Not-determined | Genome-fragment | NA | NA | 0 |
| S8_k141_23645 | 1206 | No | NA | 2 | 0 | 0 | Not-determined | Genome-fragment | NA | NA | 0 |
| S8_k141_161763 | 5106 | No | NA | 3 | 0 | 2 | Not-determined | Genome-fragment | NA | NA | 0 |
| S2_k141_109866 | 19228 | No | NA | 43 | 1 | 0 | Low-quality | Genome-fragment | 12.9 | AAI-based (medium-confidence) | 0 |
| S5-final-viral-combined_k141_29589||full | 19233 | No | NA | 20 | 0 | 6 | Low-quality | Genome-fragment | 41.37 | AAI-based (high-confidence) | 0 |
| S8_k141_58371 | 1580 | No | NA | 1 | 0 | 1 | Not-determined | Genome-fragment | NA | NA | 0 |
| S8_k141_265620 | 1088 | No | NA | 2 | 0 | 1 | Not-determined | Genome-fragment | NA | NA | 0 |
| S8_k141_150291 | 4130 | No | NA | 4 | 0 | 2 | Not-determined | Genome-fragment | NA | NA | 0 |
| S1-final-viral-combined_k141_36686||full | 19246 | Yes | 13597 | 18 | 4 | 6 | Low-quality | Genome-fragment | 32.68 | AAI-based (medium-confidence) | 29.35 |
| S5-final-viral-combined_k141_40688||0_partial | 19292 | No | NA | 11 | 0 | 9 | Low-quality | Genome-fragment | 9.27 | AAI-based (medium-confidence) | 0 |
| S8_k141_150316 | 2094 | No | NA | 2 | 0 | 0 | Not-determined | Genome-fragment | NA | NA | 0 |
| S8-final-viral-combined_k141_60264||0_partial | 19302 | Yes | 11409 | 18 | 5 | 6 | Low-quality | Genome-fragment | 28.31 | AAI-based (medium-confidence) | 40.89 |
| S7-final-viral-combined_k141_180474||full | 19306 | Yes | 12153 | 31 | 10 | 6 | Low-quality | Genome-fragment | 22.77 | AAI-based (high-confidence) | 37.05 |
| S12-final-viral-combined_k141_272503||full | 19344 | No | NA | 25 | 5 | 2 | Low-quality | Genome-fragment | 46.03 | AAI-based (medium-confidence) | 0 |
| S11-final-viral-combined_k141_105008||full | 19356 | No | NA | 28 | 9 | 1 | Low-quality | Genome-fragment | 43.13 | AAI-based (high-confidence) | 0 |
| S6-final-viral-combined_k141_3454||full | 19378 | No | NA | 22 | 4 | 1 | Low-quality | Genome-fragment | 17.15 | HMM-based (lower-bound) | 0 |
| S5_k141_37388 | 19385 | No | NA | 14 | 0 | 5 | Low-quality | Genome-fragment | 32.16 | AAI-based (medium-confidence) | 0 |
| S8_k141_23748 | 1146 | No | NA | 2 | 0 | 1 | Not-determined | Genome-fragment | NA | NA | 0 |
| S8_k141_139075 | 1453 | No | NA | 1 | 0 | 0 | Not-determined | Genome-fragment | NA | NA | 0 |
| S8_k141_219523 | 1762 | No | NA | 2 | 0 | 1 | Not-determined | Genome-fragment | NA | NA | 0 |
| S8_k141_150395 | 1478 | No | NA | 1 | 0 | 1 | Not-determined | Genome-fragment | NA | NA | 0 |
| S8_k141_150396 | 1338 | No | NA | 2 | 0 | 1 | Not-determined | Genome-fragment | NA | NA | 0 |
| S8_k141_173645 | 1610 | No | NA | 2 | 0 | 1 | Not-determined | Genome-fragment | NA | NA | 0 |
| S8_k141_242745 | 1526 | No | NA | 3 | 0 | 1 | Not-determined | Genome-fragment | NA | NA | 0 |
| S8_k141_208135 | 3166 | No | NA | 4 | 0 | 1 | Not-determined | Genome-fragment | NA | NA | 0 |
| S8_k141_253995 | 4153 | No | NA | 4 | 0 | 1 | Not-determined | Genome-fragment | NA | NA | 0 |
| S8_k141_208154 | 1521 | No | NA | 1 | 0 | 1 | Not-determined | Genome-fragment | NA | NA | 0 |
| S8_k141_196625 | 1042 | No | NA | 2 | 0 | 2 | Not-determined | Genome-fragment | NA | NA | 0 |
| S7-final-viral-combined_k141_123861||0_partial | 19392 | No | NA | 14 | 1 | 1 | Low-quality | Genome-fragment | 15.52 | HMM-based (lower-bound) | 0 |
| S8_k141_231164 | 1202 | No | NA | 1 | 0 | 1 | Not-determined | Genome-fragment | NA | NA | 0 |
| S7-final-viral-combined_k141_53197||full | 19399 | No | NA | 15 | 1 | 4 | Low-quality | Genome-fragment | 12.97 | AAI-based (medium-confidence) | 0 |
| S7-final-viral-combined_k141_10237||full | 19425 | No | NA | 26 | 2 | 0 | Low-quality | Genome-fragment | 42.9 | AAI-based (high-confidence) | 0 |
| S8_k141_242799 | 1221 | No | NA | 1 | 0 | 0 | Not-determined | Genome-fragment | NA | NA | 0 |
| S8_k141_35099 | 2304 | No | NA | 1 | 0 | 1 | Not-determined | Genome-fragment | NA | NA | 0 |
| S8_k141_115921 | 1234 | No | NA | 2 | 0 | 0 | Not-determined | Genome-fragment | NA | NA | 0 |
| S8_k141_208200 | 1208 | No | NA | 1 | 0 | 0 | Not-determined | Genome-fragment | NA | NA | 0 |
| S8_k141_69998 | 4738 | No | NA | 3 | 0 | 0 | Not-determined | Genome-fragment | NA | NA | 0 |
| S8-final-viral-combined_k141_51225||full | 19431 | No | NA | 21 | 3 | 1 | Low-quality | Genome-fragment | 19.19 | AAI-based (high-confidence) | 0 |
| S2_k141_104975 | 19508 | Yes | 16620 | 28 | 5 | 2 | Low-quality | Genome-fragment | 26.64 | AAI-based (high-confidence) | 14.8 |
| S17-final-viral-combined_k141_242634||0_partial | 19534 | Yes | 13713 | 25 | 6 | 5 | Low-quality | Genome-fragment | 18.45 | HMM-based (lower-bound) | 29.8 |
| S8_k141_254043 | 5204 | No | NA | 7 | 0 | 4 | Not-determined | Genome-fragment | NA | NA | 0 |
| S8_k141_254045 | 1099 | No | NA | 1 | 0 | 1 | Not-determined | Genome-fragment | NA | NA | 0 |
| S8_k141_23891 | 1702 | No | NA | 1 | 0 | 1 | Not-determined | Genome-fragment | NA | NA | 0 |
| S8_k141_81584 | 1905 | No | NA | 2 | 0 | 2 | Not-determined | Genome-fragment | NA | NA | 0 |
| S8_k141_219643 | 1243 | No | NA | 1 | 0 | 1 | Not-determined | Genome-fragment | NA | NA | 0 |
| S8_k141_162003 | 1904 | No | NA | 3 | 0 | 2 | Not-determined | Genome-fragment | NA | NA | 0 |
| S8_k141_208240 | 3044 | No | NA | 1 | 0 | 1 | Not-determined | Genome-fragment | NA | NA | 0 |
| S8_k141_265774 | 3029 | No | NA | 3 | 0 | 3 | Not-determined | Genome-fragment | NA | NA | 0 |
| S8_k141_58515 | 22508 | No | NA | 19 | 0 | 7 | Medium-quality | Genome-fragment | 55.52 | AAI-based (high-confidence) | 0 |
| S8_k141_265789 | 1012 | No | NA | 1 | 0 | 0 | Not-determined | Genome-fragment | NA | NA | 0 |
| S5-final-viral-combined_k141_108718||0_partial | 19536 | No | NA | 33 | 11 | 0 | Low-quality | Genome-fragment | 47.99 | AAI-based (medium-confidence) | 0 |
| S8_k141_35216 | 1235 | No | NA | 1 | 0 | 0 | Not-determined | Genome-fragment | NA | NA | 0 |
| S8_k141_116020 | 1006 | No | NA | 1 | 0 | 1 | Not-determined | Genome-fragment | NA | NA | 0 |
| S8_k141_139289 | 2255 | No | NA | 3 | 0 | 1 | Not-determined | Genome-fragment | NA | NA | 0 |
| S8_k141_242952 | 3131 | No | NA | 2 | 0 | 1 | Not-determined | Genome-fragment | NA | NA | 0 |
| S9_k141_51486 | 19550 | No | NA | 29 | 1 | 10 | Low-quality | Genome-fragment | 5.53 | HMM-based (lower-bound) | 0 |
| S8_k141_937 | 1687 | No | NA | 2 | 0 | 2 | Not-determined | Genome-fragment | NA | NA | 0 |
| S8_k141_150663 | 1007 | No | NA | 1 | 0 | 0 | Not-determined | Genome-fragment | NA | NA | 0 |
| S8_k141_173847 | 3624 | No | NA | 1 | 0 | 0 | Not-determined | Genome-fragment | NA | NA | 0 |
| S8_k141_173849 | 5631 | No | NA | 3 | 0 | 2 | Not-determined | Genome-fragment | NA | NA | 0 |
| S2-final-viral-combined_k141_120238||full | 19574 | No | NA | 27 | 13 | 1 | Low-quality | Genome-fragment | 36.58 | AAI-based (high-confidence) | 0 |
| S8_k141_254160 | 2217 | No | NA | 1 | 0 | 0 | Not-determined | Genome-fragment | NA | NA | 0 |
| S12-final-viral-combined_k141_110869||full | 19603 | No | NA | 21 | 3 | 1 | Low-quality | Genome-fragment | 29.33 | AAI-based (high-confidence) | 0 |
| S8_k141_81657 | 1117 | No | NA | 1 | 0 | 1 | Not-determined | Genome-fragment | NA | NA | 0 |
| S8_k141_242989 | 1068 | No | NA | 2 | 0 | 0 | Not-determined | Genome-fragment | NA | NA | 0 |
| S8_k141_196788 | 2199 | No | NA | 3 | 0 | 1 | Not-determined | Genome-fragment | NA | NA | 0 |
| S8_k141_150705 | 3019 | No | NA | 3 | 0 | 3 | Not-determined | Genome-fragment | NA | NA | 0 |
| S8_k141_58600 | 1157 | No | NA | 1 | 0 | 0 | Not-determined | Genome-fragment | NA | NA | 0 |
| S8_k141_127587 | 2053 | No | NA | 2 | 0 | 1 | Not-determined | Genome-fragment | NA | NA | 0 |
| S4-final-viral-combined_k141_163530||full | 19625 | No | NA | 24 | 18 | 0 | Low-quality | Genome-fragment | 47.81 | AAI-based (high-confidence) | 0 |
| S2-final-viral-combined_k141_22422||full | 19643 | No | NA | 17 | 1 | 4 | Low-quality | Genome-fragment | 30.14 | AAI-based (medium-confidence) | 0 |
| S8_k141_162176 | 1731 | No | NA | 2 | 0 | 0 | Not-determined | Genome-fragment | NA | NA | 0 |
| S8_k141_243079 | 1081 | No | NA | 1 | 0 | 0 | Not-determined | Genome-fragment | NA | NA | 0 |
| S8_k141_185329 | 1232 | No | NA | 2 | 0 | 2 | Not-determined | Genome-fragment | NA | NA | 0 |
| S11-final-viral-combined_k141_318161||full | 19686 | No | NA | 28 | 9 | 1 | Low-quality | Genome-fragment | 35.58 | AAI-based (high-confidence) | 0 |
| S2-final-viral-combined_k141_63018||full | 19713 | No | NA | 24 | 2 | 6 | Low-quality | Genome-fragment | 12.53 | HMM-based (lower-bound) | 0 |
| S8_k141_231580 | 1372 | No | NA | 2 | 0 | 1 | Not-determined | Genome-fragment | NA | NA | 0 |
| S8_k141_254310 | 1929 | No | NA | 2 | 0 | 2 | Not-determined | Genome-fragment | NA | NA | 0 |
| S11-final-viral-combined_k141_34424||full | 19809 | No | NA | 29 | 18 | 2 | Low-quality | Genome-fragment | 45.4 | AAI-based (high-confidence) | 0 |
| S8_k141_58755 | 1711 | No | NA | 1 | 0 | 1 | Not-determined | Genome-fragment | NA | NA | 0 |
| S7-final-viral-combined_k141_80611||full | 19831 | No | NA | 25 | 10 | 0 | Low-quality | Genome-fragment | 40.96 | AAI-based (high-confidence) | 0 |
| S8_k141_261150 | 19845 | No | NA | 13 | 0 | 11 | Low-quality | Genome-fragment | 7.23 | AAI-based (medium-confidence) | 0 |
| S11-final-viral-combined_k141_306975||0_partial | 19848 | No | NA | 17 | 1 | 8 | Low-quality | Genome-fragment | 42.69 | AAI-based (high-confidence) | 0 |
| S12-final-viral-combined_k141_222408||full | 19857 | Yes | 9760 | 22 | 3 | 11 | Low-quality | Genome-fragment | 23.5 | AAI-based (medium-confidence) | 50.85 |
| S8_k141_116229 | 9095 | No | NA | 4 | 0 | 3 | Not-determined | Genome-fragment | NA | NA | 0 |
| S8_k141_139536 | 1360 | No | NA | 2 | 0 | 2 | Not-determined | Genome-fragment | NA | NA | 0 |
| S2-final-viral-combined_k141_163855||full | 19885 | No | NA | 43 | 5 | 0 | Low-quality | Genome-fragment | 37.39 | AAI-based (high-confidence) | 0 |
| S5-final-viral-combined_k141_71542||full | 19893 | No | NA | 15 | 1 | 10 | Low-quality | Genome-fragment | 10.79 | HMM-based (lower-bound) | 0 |
| S8_k141_127774 | 1470 | No | NA | 1 | 0 | 1 | Not-determined | Genome-fragment | NA | NA | 0 |
| S8_k141_208682 | 1014 | No | NA | 3 | 0 | 3 | Not-determined | Genome-fragment | NA | NA | 0 |
| S7_k141_385 | 19914 | No | NA | 32 | 3 | 0 | Low-quality | Genome-fragment | 21.37 | AAI-based (high-confidence) | 0 |
| S7-final-viral-combined_k141_8554||full | 19919 | Yes | 11474 | 26 | 5 | 4 | Low-quality | Genome-fragment | 19.64 | AAI-based (high-confidence) | 42.4 |
| S8_k141_127805 | 3009 | No | NA | 3 | 0 | 1 | Not-determined | Genome-fragment | NA | NA | 0 |
| S8_k141_162312 | 1154 | No | NA | 2 | 0 | 0 | Not-determined | Genome-fragment | NA | NA | 0 |
| S8_k141_24234 | 3080 | No | NA | 5 | 0 | 2 | Not-determined | Genome-fragment | NA | NA | 0 |
| S9_k141_108429 | 19922 | No | NA | 23 | 16 | 0 | Low-quality | Genome-fragment | 42.88 | AAI-based (medium-confidence) | 0 |
| S8_k141_127810 | 1787 | No | NA | 2 | 0 | 0 | Not-determined | Genome-fragment | NA | NA | 0 |
| S8_k141_24241 | 1054 | No | NA | 1 | 0 | 0 | Not-determined | Genome-fragment | NA | NA | 0 |
| S8_k141_243270 | 1529 | No | NA | 1 | 0 | 1 | Not-determined | Genome-fragment | NA | NA | 0 |
| S5-final-viral-combined_k141_11978||full | 19928 | No | NA | 20 | 2 | 1 | Low-quality | Genome-fragment | 12.37 | HMM-based (lower-bound) | 0 |
| S7-final-viral-combined_k141_198152||0_partial | 19931 | Yes | 9260 | 29 | 5 | 7 | Low-quality | Genome-fragment | 23.74 | AAI-based (high-confidence) | 53.54 |
| S9-final-viral-combined_k141_14106||0_partial | 19935 | Yes | 10212 | 26 | 2 | 6 | Low-quality | Genome-fragment | 10.45 | AAI-based (medium-confidence) | 48.77 |
| S8_k141_116347 | 1155 | No | NA | 1 | 0 | 1 | Not-determined | Genome-fragment | NA | NA | 0 |
| S8_k141_93552 | 1413 | No | NA | 1 | 0 | 1 | Not-determined | Genome-fragment | NA | NA | 0 |
| S8_k141_243310 | 1684 | No | NA | 1 | 0 | 1 | Not-determined | Genome-fragment | NA | NA | 0 |
| S8_k141_70495 | 1905 | No | NA | 3 | 0 | 1 | Not-determined | Genome-fragment | NA | NA | 0 |
| S8_k141_93558 | 4868 | No | NA | 7 | 0 | 1 | Not-determined | Genome-fragment | NA | NA | 0 |
| S8_k141_70503 | 1051 | No | NA | 1 | 0 | 1 | Not-determined | Genome-fragment | NA | NA | 0 |
| S8_k141_116370 | 10016 | No | NA | 10 | 0 | 2 | Not-determined | Genome-fragment | NA | NA | 0 |
| S8_k141_116393 | 1040 | No | NA | 2 | 0 | 0 | Not-determined | Genome-fragment | NA | NA | 0 |
| S17-final-viral-combined_k141_122180||0_partial | 19939 | No | NA | 21 | 1 | 8 | Low-quality | Genome-fragment | 8.1 | HMM-based (lower-bound) | 0 |
| S7-final-viral-combined_k141_157107||full | 19957 | Yes | 9754 | 23 | 3 | 8 | Low-quality | Genome-fragment | 21.84 | AAI-based (medium-confidence) | 51.12 |
| S8_k141_70567 | 1002 | No | NA | 1 | 0 | 1 | Not-determined | Genome-fragment | NA | NA | 0 |
| S8_k141_162433 | 1178 | No | NA | 1 | 0 | 0 | Not-determined | Genome-fragment | NA | NA | 0 |
| S8_k141_59027 | 4331 | No | NA | 6 | 0 | 2 | Not-determined | Genome-fragment | NA | NA | 0 |
| S8_k141_243382 | 4541 | No | NA | 2 | 0 | 1 | Not-determined | Genome-fragment | NA | NA | 0 |
| S8_k141_139732 | 4560 | No | NA | 4 | 0 | 1 | Not-determined | Genome-fragment | NA | NA | 0 |
| S5-final-viral-combined_k141_183414||full | 19974 | No | NA | 24 | 9 | 0 | Low-quality | Genome-fragment | 43.32 | AAI-based (medium-confidence) | 0 |
| S7_k141_241878 | 19993 | No | NA | 22 | 0 | 6 | Low-quality | Genome-fragment | 18.77 | AAI-based (medium-confidence) | 0 |
| S12-final-viral-combined_k141_282695||full | 19997 | Yes | 8609 | 24 | 5 | 7 | Low-quality | Genome-fragment | 16.94 | HMM-based (lower-bound) | 56.95 |
| S8_k141_35554 | 20024 | No | NA | 46 | 6 | 1 | Low-quality | Genome-fragment | 46.6 | AAI-based (high-confidence) | 0 |
| S8_k141_162492 | 1095 | No | NA | 1 | 0 | 1 | Not-determined | Genome-fragment | NA | NA | 0 |
| S8_k141_12475 | 3663 | No | NA | 1 | 0 | 0 | Not-determined | Genome-fragment | NA | NA | 0 |
| S8_k141_139778 | 4507 | No | NA | 5 | 0 | 4 | Not-determined | Genome-fragment | NA | NA | 0 |
| S7-final-viral-combined_k141_227291||full | 20070 | No | NA | 24 | 6 | 2 | Low-quality | Genome-fragment | 29.1 | HMM-based (lower-bound) | 0 |
| S8_k141_162524 | 1624 | No | NA | 2 | 0 | 1 | Not-determined | Genome-fragment | NA | NA | 0 |
| S8_k141_1230 | 1472 | No | NA | 1 | 0 | 1 | Not-determined | Genome-fragment | NA | NA | 0 |
| S8_k141_82127 | 1627 | No | NA | 1 | 0 | 0 | Not-determined | Genome-fragment | NA | NA | 0 |
| S8_k141_82145 | 1248 | No | NA | 1 | 0 | 1 | Not-determined | Genome-fragment | NA | NA | 0 |
| S8_k141_24471 | 3228 | No | NA | 2 | 0 | 2 | Not-determined | Genome-fragment | NA | NA | 0 |
| S8_k141_82165 | 1250 | No | NA | 2 | 0 | 1 | Not-determined | Genome-fragment | NA | NA | 0 |
| S8_k141_266078 | 1258 | No | NA | 1 | 0 | 1 | Not-determined | Genome-fragment | NA | NA | 0 |
| S8_k141_151152 | 1323 | No | NA | 2 | 0 | 1 | Not-determined | Genome-fragment | NA | NA | 0 |
| S8_k141_1292 | 1304 | No | NA | 2 | 0 | 1 | Not-determined | Genome-fragment | NA | NA | 0 |
| S1-final-viral-combined_k141_73553||full | 20073 | Yes | 9701 | 31 | 4 | 8 | Low-quality | Genome-fragment | 11.55 | HMM-based (lower-bound) | 51.67 |
| S8_k141_24513 | 1011 | No | NA | 2 | 0 | 0 | Not-determined | Genome-fragment | NA | NA | 0 |
| S8_k141_162631 | 4544 | No | NA | 3 | 0 | 2 | Not-determined | Genome-fragment | NA | NA | 0 |
| S8_k141_47350 | 1027 | No | NA | 2 | 0 | 1 | Not-determined | Genome-fragment | NA | NA | 0 |
| S8_k141_185867 | 1137 | No | NA | 1 | 0 | 1 | Not-determined | Genome-fragment | NA | NA | 0 |
| S12-final-viral-combined_k141_83483||full | 20074 | No | NA | 23 | 2 | 0 | Low-quality | Genome-fragment | 25.9 | HMM-based (lower-bound) | 0 |
| S1-final-viral-combined_k141_356732||full | 20118 | Yes | 6901 | 20 | 2 | 9 | Low-quality | Genome-fragment | 11.44 | AAI-based (high-confidence) | 65.7 |
| S12-final-viral-combined_k141_228954||full | 20121 | Yes | 13681 | 40 | 3 | 8 | Low-quality | Genome-fragment | 27.25 | AAI-based (high-confidence) | 32.01 |
| S8_k141_12520 | 4142 | No | NA | 5 | 0 | 4 | Not-determined | Genome-fragment | NA | NA | 0 |
| S8_k141_24581 | 1300 | No | NA | 2 | 0 | 0 | Not-determined | Genome-fragment | NA | NA | 0 |
| S8_k141_151277 | 1014 | No | NA | 2 | 0 | 2 | Not-determined | Genome-fragment | NA | NA | 0 |
| S8_k141_243565 | 1579 | No | NA | 1 | 0 | 1 | Not-determined | Genome-fragment | NA | NA | 0 |
| S8_k141_232029 | 1007 | No | NA | 3 | 0 | 2 | Not-determined | Genome-fragment | NA | NA | 0 |
| S4-final-viral-combined_k141_30070||0_partial | 20134 | No | NA | 14 | 0 | 7 | Low-quality | Genome-fragment | 6.15 | AAI-based (medium-confidence) | 0 |
| S5-final-viral-combined_k141_141803||0_partial | 20141 | Yes | 12949 | 24 | 3 | 7 | Low-quality | Genome-fragment | 28.04 | AAI-based (medium-confidence) | 35.71 |
| S8_k141_232046 | 1224 | No | NA | 1 | 0 | 1 | Not-determined | Genome-fragment | NA | NA | 0 |
| S8_k141_174356 | 1018 | No | NA | 1 | 0 | 0 | Not-determined | Genome-fragment | NA | NA | 0 |
| S8_k141_120720 | 20153 | No | NA | 21 | 3 | 0 | Low-quality | Genome-fragment | 39.73 | AAI-based (high-confidence) | 0 |
| S8_k141_59387 | 1225 | No | NA | 1 | 0 | 0 | Not-determined | Genome-fragment | NA | NA | 0 |
| S8_k141_139981 | 1402 | No | NA | 2 | 0 | 0 | Not-determined | Genome-fragment | NA | NA | 0 |
| S8-final-viral-combined_k141_219682||0_partial | 20163 | No | NA | 19 | 2 | 8 | Low-quality | Genome-fragment | 14.8 | HMM-based (lower-bound) | 0 |
| S5-final-viral-combined_k141_174931||full | 20174 | No | NA | 10 | 0 | 9 | Low-quality | Genome-fragment | 1.8 | AAI-based (medium-confidence) | 0 |
| S8_k141_105141 | 3850 | No | NA | 5 | 0 | 2 | Not-determined | Genome-fragment | NA | NA | 0 |
| S8_k141_93834 | 1460 | No | NA | 3 | 0 | 2 | Not-determined | Genome-fragment | NA | NA | 0 |
| S8_k141_185934 | 2302 | No | NA | 3 | 0 | 1 | Not-determined | Genome-fragment | NA | NA | 0 |
| S8_k141_266274 | 1036 | No | NA | 1 | 0 | 1 | Not-determined | Genome-fragment | NA | NA | 0 |
| S9-final-viral-combined_k141_71523||0_partial | 20195 | No | NA | 21 | 0 | 11 | Low-quality | Genome-fragment | 13.04 | AAI-based (high-confidence) | 0 |
| S8_k141_243651 | 1515 | No | NA | 2 | 0 | 0 | Not-determined | Genome-fragment | NA | NA | 0 |
| S8_k141_116600 | 1190 | No | NA | 1 | 0 | 1 | Not-determined | Genome-fragment | NA | NA | 0 |
| S8_k141_151342 | 1211 | No | NA | 1 | 0 | 0 | Not-determined | Genome-fragment | NA | NA | 0 |
| S8_k141_266282 | 1289 | No | NA | 2 | 0 | 2 | Not-determined | Genome-fragment | NA | NA | 0 |
| S8_k141_174394 | 1729 | No | NA | 2 | 0 | 2 | Not-determined | Genome-fragment | NA | NA | 0 |
| S8_k141_70891 | 1227 | No | NA | 1 | 0 | 0 | Not-determined | Genome-fragment | NA | NA | 0 |
| S5-final-viral-combined_k141_41953||0_partial | 20203 | Yes | 10977 | 24 | 2 | 7 | Low-quality | Genome-fragment | 28.42 | AAI-based (medium-confidence) | 45.67 |
| S8_k141_116634 | 2237 | No | NA | 4 | 0 | 2 | Not-determined | Genome-fragment | NA | NA | 0 |
| S8_k141_105169 | 1084 | No | NA | 2 | 0 | 1 | Not-determined | Genome-fragment | NA | NA | 0 |
| S8_k141_254663 | 1808 | No | NA | 2 | 0 | 2 | Not-determined | Genome-fragment | NA | NA | 0 |
| S8_k141_70926 | 1071 | No | NA | 1 | 0 | 0 | Not-determined | Genome-fragment | NA | NA | 0 |
| S8-final-viral-combined_k141_186488||full | 20245 | No | NA | 23 | 11 | 0 | Low-quality | Genome-fragment | 45.14 | AAI-based (medium-confidence) | 0 |
| S8_k141_151434 | 4545 | No | NA | 1 | 0 | 1 | Not-determined | Genome-fragment | NA | NA | 0 |
| S8_k141_186040 | 1380 | No | NA | 1 | 0 | 1 | Not-determined | Genome-fragment | NA | NA | 0 |
| S8_k141_151438 | 1360 | No | NA | 1 | 0 | 0 | Not-determined | Genome-fragment | NA | NA | 0 |
| S8_k141_47395 | 5054 | No | NA | 4 | 0 | 3 | Not-determined | Genome-fragment | NA | NA | 0 |
| S8_k141_1513 | 4219 | No | NA | 1 | 0 | 1 | Not-determined | Genome-fragment | NA | NA | 0 |
| S11-final-viral-combined_k141_325982||full | 20262 | Yes | 13222 | 26 | 4 | 3 | Low-quality | Genome-fragment | 15.24 | HMM-based (lower-bound) | 34.74 |
| S8_k141_59522 | 1029 | No | NA | 1 | 0 | 0 | Not-determined | Genome-fragment | NA | NA | 0 |
| S8_k141_105262 | 1481 | No | NA | 1 | 0 | 0 | Not-determined | Genome-fragment | NA | NA | 0 |
| S8_k141_140190 | 1133 | No | NA | 1 | 0 | 1 | Not-determined | Genome-fragment | NA | NA | 0 |
| S8_k141_24893 | 4460 | No | NA | 6 | 0 | 0 | Not-determined | Genome-fragment | NA | NA | 0 |
| S12-final-viral-combined_k141_230453||full | 20293 | No | NA | 22 | 9 | 0 | Low-quality | Genome-fragment | 29.4 | AAI-based (medium-confidence) | 0 |
| S8_k141_105281 | 1086 | No | NA | 2 | 0 | 0 | Not-determined | Genome-fragment | NA | NA | 0 |
| S9-final-viral-combined_k141_88193||full | 20318 | No | NA | 35 | 5 | 1 | Low-quality | Genome-fragment | 24.19 | HMM-based (lower-bound) | 0 |
| S8_k141_24913 | 3230 | No | NA | 1 | 0 | 1 | Not-determined | Genome-fragment | NA | NA | 0 |
| S8_k141_197381 | 1220 | No | NA | 2 | 0 | 1 | Not-determined | Genome-fragment | NA | NA | 0 |
| S8_k141_59572 | 5486 | No | NA | 5 | 0 | 5 | Not-determined | Genome-fragment | NA | NA | 0 |
| S8_k141_243787 | 4151 | No | NA | 3 | 0 | 2 | Not-determined | Genome-fragment | NA | NA | 0 |
| S8_k141_209196 | 25946 | No | NA | 17 | 0 | 13 | Not-determined | Genome-fragment | NA | NA | 0 |
| S8_k141_232265 | 2439 | No | NA | 3 | 0 | 1 | Not-determined | Genome-fragment | NA | NA | 0 |
| S1-final-viral-combined_k141_18465||0_partial | 20359 | Yes | 12149 | 25 | 5 | 8 | Low-quality | Genome-fragment | 23.91 | HMM-based (lower-bound) | 40.33 |
| S12-final-viral-combined_k141_274850||0_partial | 20393 | Yes | 12361 | 24 | 5 | 6 | Low-quality | Genome-fragment | 16.37 | HMM-based (lower-bound) | 39.39 |
| S8_k141_59610 | 3673 | No | NA | 4 | 0 | 3 | Not-determined | Genome-fragment | NA | NA | 0 |
| S8_k141_59630 | 1570 | No | NA | 2 | 0 | 2 | Not-determined | Genome-fragment | NA | NA | 0 |
| S1-final-viral-combined_k141_78972||full | 20435 | No | NA | 23 | 15 | 0 | Low-quality | Genome-fragment | 33.36 | AAI-based (high-confidence) | 0 |
| S12-final-viral-combined_k141_242718||full | 20440 | No | NA | 20 | 1 | 9 | Low-quality | Genome-fragment | 30.58 | HMM-based (lower-bound) | 0 |
| S8_k141_116840 | 2059 | No | NA | 2 | 0 | 2 | Not-determined | Genome-fragment | NA | NA | 0 |
| S4_k141_121837 | 20454 | No | NA | 26 | 16 | 0 | Low-quality | Genome-fragment | 11.78 | AAI-based (high-confidence) | 0 |
| S8_k141_12773 | 1211 | No | NA | 1 | 0 | 1 | Not-determined | Genome-fragment | NA | NA | 0 |
| S8_k141_36133 | 1009 | No | NA | 2 | 0 | 1 | Not-determined | Genome-fragment | NA | NA | 0 |
| S4-final-viral-combined_k141_152228||0_partial | 20513 | No | NA | 18 | 0 | 10 | Low-quality | Genome-fragment | 11.63 | AAI-based (medium-confidence) | 0 |
| S8_k141_162937 | 2955 | No | NA | 4 | 0 | 0 | Not-determined | Genome-fragment | NA | NA | 0 |
| S8_k141_1645 | 1418 | No | NA | 1 | 0 | 1 | Not-determined | Genome-fragment | NA | NA | 0 |
| S8_k141_82430 | 1267 | No | NA | 1 | 0 | 1 | Not-determined | Genome-fragment | NA | NA | 0 |
| S8_k141_162961 | 1189 | No | NA | 2 | 0 | 1 | Not-determined | Genome-fragment | NA | NA | 0 |
| S8_k141_25001 | 1520 | No | NA | 2 | 0 | 1 | Not-determined | Genome-fragment | NA | NA | 0 |
| S8_k141_197445 | 3327 | No | NA | 6 | 0 | 1 | Not-determined | Genome-fragment | NA | NA | 0 |
| S8_k141_151749 | 1175 | No | NA | 3 | 0 | 0 | Not-determined | Genome-fragment | NA | NA | 0 |
| S8_k141_116922 | 1202 | No | NA | 1 | 0 | 1 | Not-determined | Genome-fragment | NA | NA | 0 |
| S8_k141_25039 | 1163 | No | NA | 1 | 0 | 1 | Not-determined | Genome-fragment | NA | NA | 0 |
| S12-final-viral-combined_k141_27677||0_partial | 20540 | No | NA | 22 | 1 | 4 | Low-quality | Genome-fragment | 23.06 | HMM-based (lower-bound) | 0 |
| S8_k141_174702 | 1435 | No | NA | 2 | 0 | 2 | Not-determined | Genome-fragment | NA | NA | 0 |
| S9-final-viral-combined_k141_51419||full | 20547 | No | NA | 37 | 9 | 0 | Low-quality | Genome-fragment | 49.66 | AAI-based (high-confidence) | 0 |
| S8_k141_220396 | 10913 | No | NA | 11 | 0 | 10 | Not-determined | Genome-fragment | NA | NA | 0 |
| S8_k141_232377 | 3996 | No | NA | 7 | 0 | 2 | Not-determined | Genome-fragment | NA | NA | 0 |
| S8_k141_128493 | 2370 | No | NA | 2 | 0 | 2 | Not-determined | Genome-fragment | NA | NA | 0 |
| S5-final-viral-combined_k141_168456||full | 20562 | No | NA | 26 | 1 | 0 | Low-quality | Genome-fragment | 13.18 | HMM-based (lower-bound) | 0 |
| S8_k141_140462 | 1719 | No | NA | 2 | 0 | 1 | Not-determined | Genome-fragment | NA | NA | 0 |
| S8_k141_140475 | 1336 | No | NA | 1 | 0 | 1 | Not-determined | Genome-fragment | NA | NA | 0 |
| S8_k141_186349 | 1110 | No | NA | 2 | 0 | 2 | Not-determined | Genome-fragment | NA | NA | 0 |
| S8_k141_232449 | 2112 | No | NA | 1 | 0 | 0 | Not-determined | Genome-fragment | NA | NA | 0 |
| S8_k141_71260 | 1464 | No | NA | 1 | 0 | 0 | Not-determined | Genome-fragment | NA | NA | 0 |
| S1-final-viral-combined_k141_16499||full | 20581 | Yes | 14685 | 21 | 4 | 7 | Low-quality | Genome-fragment | 35.43 | AAI-based (medium-confidence) | 28.65 |
| S8_k141_105499 | 2675 | No | NA | 3 | 0 | 2 | Not-determined | Genome-fragment | NA | NA | 0 |
| S8_k141_209400 | 3255 | No | NA | 1 | 0 | 1 | Not-determined | Genome-fragment | NA | NA | 0 |
| S8_k141_209416 | 3496 | No | NA | 1 | 0 | 1 | Not-determined | Genome-fragment | NA | NA | 0 |
| S5-final-viral-combined_k141_138631||full | 20629 | No | NA | 43 | 1 | 1 | Low-quality | Genome-fragment | 13.41 | AAI-based (high-confidence) | 0 |
| S8_k141_244076 | 1317 | No | NA | 2 | 0 | 1 | Not-determined | Genome-fragment | NA | NA | 0 |
| S7-final-viral-combined_k141_27659||full | 20720 | Yes | 7485 | 31 | 7 | 8 | Low-quality | Genome-fragment | 21.19 | AAI-based (high-confidence) | 63.88 |
| S8_k141_82664 | 7647 | No | NA | 5 | 0 | 0 | Not-determined | Genome-fragment | NA | NA | 0 |
| S5-final-viral-combined_k141_73800||full | 20831 | Yes | 18193 | 26 | 14 | 2 | Low-quality | Genome-fragment | 47.47 | AAI-based (medium-confidence) | 12.66 |
| S8_k141_209449 | 1165 | No | NA | 1 | 0 | 1 | Not-determined | Genome-fragment | NA | NA | 0 |
| S8_k141_266667 | 1307 | No | NA | 1 | 0 | 1 | Not-determined | Genome-fragment | NA | NA | 0 |
| S8_k141_47717 | 3207 | No | NA | 2 | 0 | 2 | Not-determined | Genome-fragment | NA | NA | 0 |
| S12-final-viral-combined_k141_305612||full | 20840 | No | NA | 20 | 14 | 0 | Low-quality | Genome-fragment | 35.15 | AAI-based (high-confidence) | 0 |
| S8_k141_25254 | 2180 | No | NA | 2 | 0 | 1 | Not-determined | Genome-fragment | NA | NA | 0 |
| S5-final-viral-combined_k141_163215||full | 20846 | Yes | 8653 | 19 | 2 | 9 | Low-quality | Genome-fragment | 6.36 | HMM-based (lower-bound) | 58.49 |
| S8_k141_244155 | 9417 | No | NA | 8 | 0 | 7 | Not-determined | Genome-fragment | NA | NA | 0 |
| S8_k141_105631 | 1050 | No | NA | 1 | 0 | 1 | Not-determined | Genome-fragment | NA | NA | 0 |
| S1-final-viral-combined_k141_135020||full | 20862 | No | NA | 22 | 8 | 2 | Low-quality | Genome-fragment | 42.98 | AAI-based (medium-confidence) | 0 |
| S8_k141_197635 | 6079 | No | NA | 9 | 0 | 2 | Not-determined | Genome-fragment | NA | NA | 0 |
| S8_k141_266727 | 1177 | No | NA | 1 | 0 | 0 | Not-determined | Genome-fragment | NA | NA | 0 |
| S17-final-viral-combined_k141_233855||full | 20873 | Yes | 14086 | 21 | 2 | 6 | Low-quality | Genome-fragment | 17.5 | HMM-based (lower-bound) | 32.52 |
| S8_k141_244214 | 2155 | No | NA | 3 | 0 | 2 | Not-determined | Genome-fragment | NA | NA | 0 |
| S8_k141_232610 | 2333 | No | NA | 2 | 0 | 2 | Not-determined | Genome-fragment | NA | NA | 0 |
| S5-final-viral-combined_k141_200471||full | 20889 | Yes | 12756 | 29 | 5 | 11 | Low-quality | Genome-fragment | 17.91 | HMM-based (lower-bound) | 38.93 |
| S8_k141_59915 | 3174 | No | NA | 3 | 0 | 1 | Not-determined | Genome-fragment | NA | NA | 0 |
| S17-final-viral-combined_k141_223200||full | 20922 | No | NA | 23 | 6 | 2 | Low-quality | Genome-fragment | 28.14 | AAI-based (high-confidence) | 0 |
| S17-final-viral-combined_k141_147336||full | 20924 | No | NA | 27 | 1 | 6 | Low-quality | Genome-fragment | 18.92 | HMM-based (lower-bound) | 0 |
| S8_k141_232664 | 1001 | No | NA | 2 | 0 | 1 | Not-determined | Genome-fragment | NA | NA | 0 |
| S8_k141_12920 | 1083 | No | NA | 2 | 0 | 1 | Not-determined | Genome-fragment | NA | NA | 0 |
| S8_k141_266863 | 1548 | No | NA | 1 | 0 | 1 | Not-determined | Genome-fragment | NA | NA | 0 |
| S11-final-viral-combined_k141_14713||full | 20936 | No | NA | 32 | 6 | 1 | Low-quality | Genome-fragment | 48.88 | AAI-based (medium-confidence) | 0 |
| S5_k141_29349 | 21041 | No | NA | 20 | 0 | 5 | Low-quality | Genome-fragment | 13.7 | AAI-based (medium-confidence) | 0 |
| S17-final-viral-combined_k141_41932||full | 21052 | No | NA | 33 | 1 | 4 | Low-quality | Genome-fragment | 15.17 | HMM-based (lower-bound) | 0 |
| S8_k141_255135 | 2006 | No | NA | 1 | 0 | 0 | Not-determined | Genome-fragment | NA | NA | 0 |
| S8_k141_266891 | 1541 | No | NA | 2 | 0 | 1 | Not-determined | Genome-fragment | NA | NA | 0 |
| S2-final-viral-combined_k141_188890||full | 21068 | Yes | 11889 | 30 | 6 | 7 | Low-quality | Genome-fragment | 30.47 | AAI-based (high-confidence) | 43.57 |
| S8_k141_117177 | 1086 | No | NA | 1 | 0 | 0 | Not-determined | Genome-fragment | NA | NA | 0 |
| S8_k141_71619 | 1052 | No | NA | 2 | 0 | 1 | Not-determined | Genome-fragment | NA | NA | 0 |
| S8_k141_94421 | 2295 | No | NA | 1 | 0 | 1 | Not-determined | Genome-fragment | NA | NA | 0 |
| S8_k141_197829 | 1278 | No | NA | 2 | 0 | 2 | Not-determined | Genome-fragment | NA | NA | 0 |
| S2_k141_42313 | 21151 | No | NA | 11 | 2 | 0 | Low-quality | Genome-fragment | 17.02 | AAI-based (medium-confidence) | 0 |
| S2-final-viral-combined_k141_44523||full | 21152 | No | NA | 26 | 4 | 0 | Low-quality | Genome-fragment | 37.86 | AAI-based (high-confidence) | 0 |
| S8_k141_232744 | 1139 | No | NA | 1 | 0 | 1 | Not-determined | Genome-fragment | NA | NA | 0 |
| S8_k141_186589 | 2853 | No | NA | 1 | 0 | 1 | Not-determined | Genome-fragment | NA | NA | 0 |
| S8_k141_36530 | 1524 | No | NA | 2 | 0 | 0 | Not-determined | Genome-fragment | NA | NA | 0 |
| S8_k141_232758 | 1030 | No | NA | 1 | 0 | 0 | Not-determined | Genome-fragment | NA | NA | 0 |
| S8_k141_255195 | 1140 | No | NA | 3 | 0 | 2 | Not-determined | Genome-fragment | NA | NA | 0 |
| S17-final-viral-combined_k141_224415||full | 21157 | Yes | 8125 | 25 | 2 | 7 | Low-quality | Genome-fragment | 21.49 | AAI-based (medium-confidence) | 61.6 |
| S7-final-viral-combined_k141_52860||full | 21160 | No | NA | 22 | 2 | 3 | Low-quality | Genome-fragment | 14.44 | HMM-based (lower-bound) | 0 |
| S8_k141_232774 | 1149 | No | NA | 2 | 0 | 2 | Not-determined | Genome-fragment | NA | NA | 0 |
| S8_k141_25548 | 1172 | No | NA | 1 | 0 | 0 | Not-determined | Genome-fragment | NA | NA | 0 |
| S8-final-viral-combined_k141_242759||full | 21174 | No | NA | 32 | 12 | 0 | Low-quality | Genome-fragment | 44.38 | AAI-based (medium-confidence) | 0 |
| S2-final-viral-combined_k141_198899||full | 21202 | No | NA | 14 | 0 | 3 | Low-quality | Genome-fragment | 28.38 | AAI-based (medium-confidence) | 0 |
| S8_k141_197844 | 7676 | No | NA | 8 | 0 | 3 | Not-determined | Genome-fragment | NA | NA | 0 |
| S8_k141_71721 | 1002 | No | NA | 1 | 0 | 0 | Not-determined | Genome-fragment | NA | NA | 0 |
| S8_k141_175277 | 1389 | No | NA | 1 | 0 | 0 | Not-determined | Genome-fragment | NA | NA | 0 |
| S11-final-viral-combined_k141_241698||full | 21276 | No | NA | 17 | 2 | 2 | Low-quality | Genome-fragment | 14.78 | AAI-based (medium-confidence) | 0 |
| S8_k141_1978 | 1000 | No | NA | 1 | 0 | 0 | Not-determined | Genome-fragment | NA | NA | 0 |
| S8_k141_197873 | 3836 | No | NA | 4 | 0 | 4 | Not-determined | Genome-fragment | NA | NA | 0 |
| S5-final-viral-combined_k141_69272||0_partial | 21316 | No | NA | 19 | 1 | 4 | Low-quality | Genome-fragment | 17.06 | HMM-based (lower-bound) | 0 |
| S8_k141_244460 | 3439 | No | NA | 1 | 0 | 1 | Not-determined | Genome-fragment | NA | NA | 0 |
| S8_k141_186669 | 1484 | No | NA | 2 | 0 | 2 | Not-determined | Genome-fragment | NA | NA | 0 |
| S8-final-viral-combined_k141_270960||full | 21344 | Yes | 8611 | 22 | 3 | 8 | Low-quality | Genome-fragment | 11.36 | HMM-based (lower-bound) | 59.66 |
| S8_k141_2010 | 1169 | No | NA | 2 | 0 | 2 | Not-determined | Genome-fragment | NA | NA | 0 |
| S8_k141_232885 | 3028 | No | NA | 3 | 0 | 0 | Not-determined | Genome-fragment | NA | NA | 0 |
| S8_k141_117293 | 4063 | No | NA | 4 | 0 | 4 | Not-determined | Genome-fragment | NA | NA | 0 |
| S8_k141_163167 | 21354 | No | NA | 33 | 2 | 3 | Low-quality | Genome-fragment | 9.79 | HMM-based (lower-bound) | 0 |
| S8_k141_255279 | 1200 | No | NA | 3 | 0 | 0 | Not-determined | Genome-fragment | NA | NA | 0 |
| S8_k141_267060 | 1017 | No | NA | 1 | 0 | 0 | Not-determined | Genome-fragment | NA | NA | 0 |
| S1_k141_113935 | 21357 | No | NA | 27 | 4 | 4 | Low-quality | Genome-fragment | 32.5 | AAI-based (high-confidence) | 0 |
| S8_k141_232898 | 8770 | No | NA | 6 | 0 | 5 | Not-determined | Genome-fragment | NA | NA | 0 |
| S8_k141_25630 | 4162 | No | NA | 5 | 0 | 1 | Not-determined | Genome-fragment | NA | NA | 0 |
| S8_k141_267090 | 2044 | No | NA | 3 | 0 | 0 | Not-determined | Genome-fragment | NA | NA | 0 |
| S8_k141_163562 | 3621 | No | NA | 6 | 0 | 0 | Not-determined | Genome-fragment | NA | NA | 0 |
| S8_k141_267109 | 1420 | No | NA | 1 | 0 | 1 | Not-determined | Genome-fragment | NA | NA | 0 |
| S8_k141_82880 | 47863 | No | NA | 74 | 27 | 0 | Medium-quality | Genome-fragment | 63.31 | AAI-based (medium-confidence) | 0 |
| S8_k141_36697 | 1029 | No | NA | 1 | 0 | 1 | Not-determined | Genome-fragment | NA | NA | 0 |
| S8_k141_60174 | 1360 | No | NA | 1 | 0 | 0 | Not-determined | Genome-fragment | NA | NA | 0 |
| S8_k141_2101 | 19563 | No | NA | 13 | 0 | 11 | Not-determined | Genome-fragment | NA | NA | 0 |
| S8_k141_82901 | 1219 | No | NA | 1 | 0 | 1 | Not-determined | Genome-fragment | NA | NA | 0 |
| S8_k141_140918 | 1092 | No | NA | 2 | 0 | 1 | Not-determined | Genome-fragment | NA | NA | 0 |
| S8_k141_94642 | 1113 | No | NA | 1 | 0 | 1 | Not-determined | Genome-fragment | NA | NA | 0 |
| S8_k141_244525 | 17376 | No | NA | 22 | 7 | 0 | Medium-quality | Genome-fragment | 81.36 | AAI-based (medium-confidence) | 0 |
| S8_k141_36753 | 1225 | No | NA | 1 | 0 | 1 | Not-determined | Genome-fragment | NA | NA | 0 |
| S8_k141_186716 | 2535 | No | NA | 1 | 0 | 1 | Not-determined | Genome-fragment | NA | NA | 0 |
| S8_k141_48164 | 1009 | No | NA | 2 | 0 | 1 | Not-determined | Genome-fragment | NA | NA | 0 |
| S8_k141_198048 | 3744 | No | NA | 5 | 0 | 3 | Not-determined | Genome-fragment | NA | NA | 0 |
| S8_k141_13150 | 1217 | No | NA | 1 | 0 | 1 | Not-determined | Genome-fragment | NA | NA | 0 |
| S8_k141_128929 | 1439 | No | NA | 1 | 0 | 0 | Not-determined | Genome-fragment | NA | NA | 0 |
| S8_k141_36773 | 1391 | No | NA | 1 | 0 | 1 | Not-determined | Genome-fragment | NA | NA | 0 |
| S8_k141_232952 | 3480 | No | NA | 1 | 0 | 0 | Not-determined | Genome-fragment | NA | NA | 0 |
| S8_k141_186668 | 21361 | No | NA | 33 | 9 | 0 | Low-quality | Genome-fragment | 48.36 | AAI-based (high-confidence) | 0 |
| S8_k141_152288 | 1330 | No | NA | 2 | 0 | 2 | Not-determined | Genome-fragment | NA | NA | 0 |
| S8_k141_232966 | 1559 | No | NA | 1 | 0 | 1 | Not-determined | Genome-fragment | NA | NA | 0 |
| S17-final-viral-combined_k141_248661||full | 21371 | No | NA | 23 | 2 | 0 | Low-quality | Genome-fragment | 49.9 | AAI-based (medium-confidence) | 0 |
| S1_k141_197296 | 21386 | No | NA | 37 | 2 | 2 | Low-quality | Genome-fragment | 36.8 | AAI-based (high-confidence) | 0 |
| S8_k141_2230 | 3217 | No | NA | 4 | 0 | 0 | Not-determined | Genome-fragment | NA | NA | 0 |
| S12_k141_398986 | 21417 | No | NA | 17 | 0 | 16 | Low-quality | Genome-fragment | 7.37 | AAI-based (high-confidence) | 0 |
| S8_k141_141071 | 1210 | No | NA | 1 | 0 | 1 | Not-determined | Genome-fragment | NA | NA | 0 |
| S8_k141_255400 | 1157 | No | NA | 1 | 0 | 0 | Not-determined | Genome-fragment | NA | NA | 0 |
| S8_k141_267227 | 2072 | No | NA | 2 | 0 | 1 | Not-determined | Genome-fragment | NA | NA | 0 |
| S8_k141_2253 | 3180 | No | NA | 4 | 0 | 1 | Not-determined | Genome-fragment | NA | NA | 0 |
| S8_k141_36900 | 1317 | No | NA | 1 | 0 | 1 | Not-determined | Genome-fragment | NA | NA | 0 |
| S8_k141_2259 | 1044 | No | NA | 2 | 0 | 1 | Not-determined | Genome-fragment | NA | NA | 0 |
| S8_k141_267234 | 2330 | No | NA | 1 | 0 | 1 | Not-determined | Genome-fragment | NA | NA | 0 |
| S1-final-viral-combined_k141_218685||full | 21429 | Yes | 3689 | 20 | 2 | 7 | Low-quality | Genome-fragment | 8.01 | AAI-based (medium-confidence) | 82.79 |
| S8_k141_255420 | 2138 | No | NA | 2 | 0 | 0 | Not-determined | Genome-fragment | NA | NA | 0 |
| S8_k141_129062 | 2215 | No | NA | 3 | 0 | 2 | Not-determined | Genome-fragment | NA | NA | 0 |

| | | | | | | | | | | | |
|---|---|---|---|---|---|---|---|---|---|---|---|
| S8_k141_2281 | 1009 | No | NA | 1 | 0 | 0 | Not-determined | Genome-fragment | NA | NA | 0 |
| S8_k141_175569 | 1711 | No | NA | 2 | 0 | 2 | Not-determined | Genome-fragment | NA | NA | 0 |
| S8_k141_198221 | 1214 | No | NA | 4 | 0 | 2 | Not-determined | Genome-fragment | NA | NA | 0 |
| S8_k141_13246 | 10943 | No | NA | 17 | 0 | 6 | Not-determined | Genome-fragment | NA | NA | 0 |
| S8_k141_2299 | 1062 | No | NA | 1 | 0 | 0 | Not-determined | Genome-fragment | NA | NA | 0 |
| S8_k141_221081 | 2355 | No | NA | 1 | 0 | 1 | Not-determined | Genome-fragment | NA | NA | 0 |
| S8_k141_186941 | 1224 | No | NA | 2 | 0 | 2 | Not-determined | Genome-fragment | NA | NA | 0 |
| S8_k141_36987 | 1368 | No | NA | 2 | 0 | 2 | Not-determined | Genome-fragment | NA | NA | 0 |
| S8_k141_152483 | 1504 | No | NA | 3 | 0 | 3 | Not-determined | Genome-fragment | NA | NA | 0 |
| S8_k141_152512 | 1010 | No | NA | 1 | 0 | 1 | Not-determined | Genome-fragment | NA | NA | 0 |
| S8_k141_72062 | 2768 | No | NA | 1 | 0 | 1 | Not-determined | Genome-fragment | NA | NA | 0 |
| S7-final-viral-combined_k141_209817\|\|full | 21436 | No | NA | 30 | 1 | 4 | Low-quality | Genome-fragment | 12.67 | HMM-based (lower-bound) | 0 |
| S8_k141_141204 | 1855 | No | NA | 3 | 0 | 3 | Not-determined | Genome-fragment | NA | NA | 0 |
| S8_k141_117573 | 10366 | No | NA | 8 | 0 | 8 | Not-determined | Genome-fragment | NA | NA | 0 |
| S5-final-viral-combined_k141_1481\|\|full | 21590 | Yes | 5045 | 24 | 4 | 13 | Low-quality | Genome-fragment | 10.72 | HMM-based (lower-bound) | 76.63 |
| S8_k141_152545 | 1427 | No | NA | 1 | 0 | 0 | Not-determined | Genome-fragment | NA | NA | 0 |
| S8_k141_25888 | 2364 | No | NA | 2 | 0 | 0 | Not-determined | Genome-fragment | NA | NA | 0 |
| S8_k141_163842 | 3195 | No | NA | 3 | 0 | 2 | Not-determined | Genome-fragment | NA | NA | 0 |
| S8_k141_132046 | 21608 | No | NA | 23 | 8 | 0 | Low-quality | Genome-fragment | 16.31 | AAI-based (high-confidence) | 0 |
| S8_k141_255570 | 1173 | No | NA | 1 | 0 | 1 | Not-determined | Genome-fragment | NA | NA | 0 |
| S8_k141_210100 | 1364 | No | NA | 1 | 0 | 0 | Not-determined | Genome-fragment | NA | NA | 0 |
| S7-final-viral-combined_k141_139306\|\|0_partial | 21609 | Yes | 12280 | 31 | 5 | 9 | Low-quality | Genome-fragment | 14.79 | HMM-based (lower-bound) | 43.17 |
| S8-final-viral-combined_k141_83144\|\|0_partial | 21617 | Yes | 16195 | 21 | 3 | 3 | Low-quality | Genome-fragment | 30.96 | HMM-based (lower-bound) | 25.08 |
| S8_k141_37066 | 1016 | No | NA | 1 | 0 | 1 | Not-determined | Genome-fragment | NA | NA | 0 |
| S8_k141_72131 | 5189 | No | NA | 1 | 0 | 1 | Not-determined | Genome-fragment | NA | NA | 0 |
| S8_k141_186995 | 3666 | No | NA | 4 | 0 | 0 | Not-determined | Genome-fragment | NA | NA | 0 |
| S8_k141_83049 | 1732 | No | NA | 2 | 0 | 2 | Not-determined | Genome-fragment | NA | NA | 0 |
| S8_k141_175689 | 1142 | No | NA | 1 | 0 | 1 | Not-determined | Genome-fragment | NA | NA | 0 |
| S7-final-viral-combined_k141_119188\|\|full | 21703 | No | NA | 38 | 13 | 0 | Low-quality | Genome-fragment | 34.2 | AAI-based (medium-confidence) | 0 |
| S7-final-viral-combined_k141_165331\|\|1_partial | 21711 | No | NA | 15 | 1 | 7 | Low-quality | Genome-fragment | 32.19 | HMM-based (lower-bound) | 0 |
| S8_k141_117637 | 1424 | No | NA | 2 | 0 | 0 | Not-determined | Genome-fragment | NA | NA | 0 |
| S8_k141_37080 | 1245 | No | NA | 1 | 0 | 0 | Not-determined | Genome-fragment | NA | NA | 0 |
| S8_k141_105977 | 1241 | No | NA | 2 | 0 | 1 | Not-determined | Genome-fragment | NA | NA | 0 |
| S8_k141_141284 | 1264 | No | NA | 1 | 0 | 1 | Not-determined | Genome-fragment | NA | NA | 0 |
| S8_k141_152629 | 1310 | No | NA | 2 | 0 | 1 | Not-determined | Genome-fragment | NA | NA | 0 |
| S1-final-viral-combined_k141_334456\|\|full | 21798 | Yes | 13642 | 25 | 4 | 7 | Low-quality | Genome-fragment | 32.76 | AAI-based (medium-confidence) | 37.42 |
| S8_k141_25973 | 3003 | No | NA | 4 | 0 | 0 | Not-determined | Genome-fragment | NA | NA | 0 |
| S8_k141_117664 | 3307 | No | NA | 1 | 0 | 1 | Not-determined | Genome-fragment | NA | NA | 0 |
| S8_k141_13395 | 1156 | No | NA | 2 | 0 | 1 | Not-determined | Genome-fragment | NA | NA | 0 |
| S17-final-viral-combined_k141_132603\|\|full | 21808 | No | NA | 14 | 5 | 1 | Low-quality | Genome-fragment | 27.04 | AAI-based (high-confidence) | 0 |
| S8_k141_163869 | 1304 | No | NA | 4 | 0 | 0 | Not-determined | Genome-fragment | NA | NA | 0 |
| S8_k141_25996 | 1227 | No | NA | 1 | 0 | 1 | Not-determined | Genome-fragment | NA | NA | 0 |
| S8_k141_267401 | 1646 | No | NA | 2 | 0 | 2 | Not-determined | Genome-fragment | NA | NA | 0 |
| S17-final-viral-combined_k141_259683\|\|full | 21818 | No | NA | 24 | 2 | 1 | Low-quality | Genome-fragment | 10.7 | HMM-based (lower-bound) | 0 |
| S8_k141_267426 | 1225 | No | NA | 3 | 0 | 1 | Not-determined | Genome-fragment | NA | NA | 0 |
| S8_k141_187038 | 1112 | No | NA | 2 | 0 | 2 | Not-determined | Genome-fragment | NA | NA | 0 |
| S8_k141_60337 | 1324 | No | NA | 1 | 0 | 0 | Not-determined | Genome-fragment | NA | NA | 0 |
| S8_k141_175825 | 1230 | No | NA | 1 | 0 | 1 | Not-determined | Genome-fragment | NA | NA | 0 |
| S6_k141_106244 | 21933 | No | NA | 27 | 4 | 0 | Low-quality | Genome-fragment | 32.52 | AAI-based (high-confidence) | 0 |
| S8_k141_37171 | 1063 | No | NA | 1 | 0 | 1 | Not-determined | Genome-fragment | NA | NA | 0 |
| S8_k141_267480 | 1276 | No | NA | 2 | 0 | 0 | Not-determined | Genome-fragment | NA | NA | 0 |
| S8_k141_13507 | 1137 | No | NA | 1 | 0 | 1 | Not-determined | Genome-fragment | NA | NA | 0 |
| S8_k141_60417 | 1480 | No | NA | 3 | 0 | 0 | Not-determined | Genome-fragment | NA | NA | 0 |
| S17-final-viral-combined_k141_203274\|\|full | 21957 | No | NA | 36 | 13 | 1 | Low-quality | Genome-fragment | 46.64 | AAI-based (high-confidence) | 0 |
| S8_k141_255759 | 1366 | No | NA | 2 | 0 | 2 | Not-determined | Genome-fragment | NA | NA | 0 |
| S8_k141_72391 | 1027 | No | NA | 1 | 0 | 0 | Not-determined | Genome-fragment | NA | NA | 0 |
| S8_k141_13591 | 1370 | No | NA | 1 | 0 | 1 | Not-determined | Genome-fragment | NA | NA | 0 |
| S9-final-viral-combined_k141_102994\|\|0_partial | 22007 | Yes | 10735 | 27 | 4 | 6 | Low-quality | Genome-fragment | 13.66 | HMM-based (lower-bound) | 51.22 |
| S8_k141_187205 | 3441 | No | NA | 3 | 0 | 3 | Not-determined | Genome-fragment | NA | NA | 0 |
| S9-final-viral-combined_k141_28998\|\|full | 22063 | Yes | 13623 | 24 | 6 | 3 | Low-quality | Genome-fragment | 24.64 | HMM-based (lower-bound) | 38.25 |
| S1-final-viral-combined_k141_31949\|\|0_partial | 22074 | No | NA | 17 | 1 | 4 | Low-quality | Genome-fragment | 17.66 | HMM-based (lower-bound) | 0 |
| S8_k141_152825 | 1809 | No | NA | 2 | 0 | 0 | Not-determined | Genome-fragment | NA | NA | 0 |
| S12-final-viral-combined_k141_203191\|\|full | 22108 | No | NA | 19 | 4 | 1 | Low-quality | Genome-fragment | 28.18 | AAI-based (high-confidence) | 0 |
| S8_k141_26095 | 3868 | No | NA | 3 | 0 | 1 | Not-determined | Genome-fragment | NA | NA | 0 |
| S8_k141_117762 | 2400 | No | NA | 1 | 0 | 0 | Not-determined | Genome-fragment | NA | NA | 0 |
| S8_k141_198502 | 1150 | No | NA | 1 | 0 | 0 | Not-determined | Genome-fragment | NA | NA | 0 |
| S6-final-viral-combined_k141_35594\|\|0_partial | 22136 | Yes | 8301 | 19 | 3 | 8 | Low-quality | Genome-fragment | 15.8 | HMM-based (lower-bound) | 62.5 |
| S8_k141_198514 | 1004 | No | NA | 1 | 0 | 0 | Not-determined | Genome-fragment | NA | NA | 0 |
| S8_k141_152831 | 28619 | No | NA | 33 | 16 | 0 | Medium-quality | Genome-fragment | 52.11 | AAI-based (high-confidence) | 0 |
| S8-final-viral-combined_k141_4771\|\|full | 22146 | Yes | 13063 | 25 | 5 | 5 | Low-quality | Genome-fragment | 29.13 | AAI-based (high-confidence) | 41.01 |
| S8_k141_95287 | 1013 | No | NA | 1 | 0 | 1 | Not-determined | Genome-fragment | NA | NA | 0 |
| S8_k141_267665 | 1041 | No | NA | 1 | 0 | 1 | Not-determined | Genome-fragment | NA | NA | 0 |
| S1-final-viral-combined_k141_149201\|\|full | 22148 | No | NA | 25 | 8 | 0 | Low-quality | Genome-fragment | 45.69 | AAI-based (high-confidence) | 0 |
| S8_k141_95328 | 1048 | No | NA | 2 | 0 | 2 | Not-determined | Genome-fragment | NA | NA | 0 |
| S12_k141_327700 | 22191 | No | NA | 28 | 1 | 0 | Low-quality | Genome-fragment | 18.59 | AAI-based (high-confidence) | 0 |
| S5-final-viral-combined_k141_99254\|\|0_partial | 22325 | No | NA | 23 | 2 | 10 | Low-quality | Genome-fragment | 41.12 | HMM-based (lower-bound) | 0 |
| S8_k141_129573 | 1457 | No | NA | 2 | 0 | 1 | Not-determined | Genome-fragment | NA | NA | 0 |
| S8_k141_129577 | 1229 | No | NA | 1 | 0 | 0 | Not-determined | Genome-fragment | NA | NA | 0 |
| S8_k141_72445 | 1572 | No | NA | 2 | 0 | 2 | Not-determined | Genome-fragment | NA | NA | 0 |
| S8_k141_255909 | 1359 | No | NA | 3 | 0 | 2 | Not-determined | Genome-fragment | NA | NA | 0 |
| S8_k141_221337 | 5766 | No | NA | 10 | 0 | 4 | Not-determined | Genome-fragment | NA | NA | 0 |
| S8_k141_163972 | 2171 | No | NA | 1 | 0 | 1 | Not-determined | Genome-fragment | NA | NA | 0 |
| S5-final-viral-combined_k141_205305\|\|full | 22331 | No | NA | 21 | 1 | 4 | Low-quality | Genome-fragment | 49.51 | AAI-based (medium-confidence) | 0 |
| S11-final-viral-combined_k141_178671\|\|full | 22337 | No | NA | 27 | 17 | 0 | Low-quality | Genome-fragment | 47.23 | AAI-based (medium-confidence) | 0 |
| S8_k141_141657 | 7648 | No | NA | 6 | 0 | 5 | Not-determined | Genome-fragment | NA | NA | 0 |
| S8_k141_198609 | 4264 | No | NA | 4 | 0 | 3 | Not-determined | Genome-fragment | NA | NA | 0 |
| S8_k141_164050 | 1597 | No | NA | 1 | 0 | 1 | Not-determined | Genome-fragment | NA | NA | 0 |
| S8-final-viral-combined_k141_164670\|\|full | 22428 | No | NA | 24 | 1 | 7 | Low-quality | Genome-fragment | 25.18 | HMM-based (lower-bound) | 0 |
| S8_k141_233677 | 2009 | No | NA | 3 | 0 | 0 | Not-determined | Genome-fragment | NA | NA | 0 |
| S7-final-viral-combined_k141_102731\|\|0_partial | 22476 | Yes | 15049 | 24 | 3 | 8 | Low-quality | Genome-fragment | 28.64 | HMM-based (lower-bound) | 33.04 |
| S8_k141_129645 | 1005 | No | NA | 2 | 0 | 1 | Not-determined | Genome-fragment | NA | NA | 0 |
| S2_k141_5030 | 22479 | No | NA | 14 | 0 | 10 | Low-quality | Genome-fragment | 16.48 | AAI-based (medium-confidence) | 0 |
| S8_k141_141746 | 1662 | No | NA | 2 | 0 | 1 | Not-determined | Genome-fragment | NA | NA | 0 |
| S8_k141_48747 | 1117 | No | NA | 2 | 0 | 1 | Not-determined | Genome-fragment | NA | NA | 0 |
| S8_k141_106205 | 3130 | No | NA | 2 | 0 | 2 | Not-determined | Genome-fragment | NA | NA | 0 |
| S8_k141_256033 | 1455 | No | NA | 1 | 0 | 0 | Not-determined | Genome-fragment | NA | NA | 0 |
| S8_k141_26324 | 3142 | No | NA | 2 | 0 | 1 | Not-determined | Genome-fragment | NA | NA | 0 |
| S2-final-viral-combined_k141_213786\|\|0_partial | 22506 | No | NA | 15 | 1 | 5 | Low-quality | Genome-fragment | 9.46 | HMM-based (lower-bound) | 0 |
| S8_k141_210448 | 1022 | No | NA | 1 | 0 | 1 | Not-determined | Genome-fragment | NA | NA | 0 |
| S8_k141_198735 | 1273 | No | NA | 3 | 0 | 0 | Not-determined | Genome-fragment | NA | NA | 0 |
| S7-final-viral-combined_k141_21929\|\|full | 22538 | Yes | 10170 | 27 | 5 | 6 | Low-quality | Genome-fragment | 27.71 | AAI-based (high-confidence) | 54.88 |
| S9-final-viral-combined_k141_97514\|\|full | 22551 | Yes | 12485 | 15 | 3 | 3 | Low-quality | Genome-fragment | 18.35 | AAI-based (medium-confidence) | 44.64 |
| S5-final-viral-combined_k141_91926\|\|0_partial | 22574 | No | NA | 16 | 2 | 8 | Low-quality | Genome-fragment | 35.6 | HMM-based (lower-bound) | 0 |
| S8_k141_267929 | 3256 | No | NA | 1 | 0 | 0 | Not-determined | Genome-fragment | NA | NA | 0 |
| S8_k141_37501 | 1609 | No | NA | 4 | 0 | 2 | Not-determined | Genome-fragment | NA | NA | 0 |
| S12_k141_262219 | 22582 | Yes | 18730 | 24 | 2 | 3 | Low-quality | Genome-fragment | 12.79 | HMM-based (lower-bound) | 17.06 |
| S8_k141_72721 | 1702 | No | NA | 1 | 0 | 1 | Not-determined | Genome-fragment | NA | NA | 0 |
| S8_k141_129794 | 2317 | No | NA | 2 | 0 | 0 | Not-determined | Genome-fragment | NA | NA | 0 |
| S8_k141_164211 | 3556 | No | NA | 3 | 0 | 2 | Not-determined | Genome-fragment | NA | NA | 0 |
| S8_k141_118052 | 3467 | No | NA | 2 | 0 | 1 | Not-determined | Genome-fragment | NA | NA | 0 |
| S17-final-viral-combined_k141_127505\|\|full | 22583 | Yes | 5975 | 26 | 3 | 10 | Low-quality | Genome-fragment | 13.82 | AAI-based (medium-confidence) | 73.54 |
| S8_k141_245320 | 5568 | No | NA | 9 | 0 | 1 | Not-determined | Genome-fragment | NA | NA | 0 |
| S11-final-viral-combined_k141_108043\|\|full | 22602 | No | NA | 24 | 4 | 0 | Low-quality | Genome-fragment | 32.68 | AAI-based (medium-confidence) | 0 |
| S1-final-viral-combined_k141_28120\|\|full | 22620 | No | NA | 20 | 1 | 5 | Low-quality | Genome-fragment | 26.65 | AAI-based (medium-confidence) | 0 |
| S8_k141_48868 | 9355 | No | NA | 5 | 0 | 4 | Not-determined | Genome-fragment | NA | NA | 0 |
| S5-final-viral-combined_k141_180367\|\|0_partial | 22680 | Yes | 14516 | 27 | 6 | 6 | Low-quality | Genome-fragment | 19.53 | HMM-based (lower-bound) | 36 |
| S8_k141_245360 | 1198 | No | NA | 1 | 0 | 1 | Not-determined | Genome-fragment | NA | NA | 0 |
| S8_k141_83275 | 1978 | No | NA | 2 | 0 | 2 | Not-determined | Genome-fragment | NA | NA | 0 |
| S8_k141_106339 | 2728 | No | NA | 3 | 0 | 3 | Not-determined | Genome-fragment | NA | NA | 0 |
| S8_k141_141950 | 1375 | No | NA | 1 | 0 | 1 | Not-determined | Genome-fragment | NA | NA | 0 |
| S8_k141_26403 | 3265 | No | NA | 6 | 0 | 2 | Not-determined | Genome-fragment | NA | NA | 0 |
| S8_k141_37648 | 3999 | No | NA | 3 | 0 | 1 | Not-determined | Genome-fragment | NA | NA | 0 |
| S17-final-viral-combined_k141_202941\|\|full | 22685 | Yes | 16772 | 27 | 4 | 5 | Low-quality | Genome-fragment | 38 | AAI-based (high-confidence) | 26.07 |
| S8_k141_129931 | 1076 | No | NA | 2 | 0 | 2 | Not-determined | Genome-fragment | NA | NA | 0 |
| S12-final-viral-combined_k141_34886\|\|0_partial | 22706 | Yes | 12794 | 23 | 5 | 7 | Low-quality | Genome-fragment | 16.95 | HMM-based (lower-bound) | 43.65 |
| S8_k141_72850 | 3388 | No | NA | 5 | 0 | 2 | Not-determined | Genome-fragment | NA | NA | 0 |
| S8_k141_245409 | 7401 | No | NA | 3 | 0 | 2 | Not-determined | Genome-fragment | NA | NA | 0 |
| S2-final-viral-combined_k141_59390\|\|full | 22712 | No | NA | 18 | 0 | 8 | Low-quality | Genome-fragment | 11.04 | AAI-based (medium-confidence) | 0 |
| S1-final-viral-combined_k141_127310\|\|0_partial | 22732 | No | NA | 15 | 1 | 9 | Low-quality | Genome-fragment | 20.7 | HMM-based (lower-bound) | 0 |
| S8_k141_268091 | 1200 | No | NA | 1 | 0 | 0 | Not-determined | Genome-fragment | NA | NA | 0 |
| S8_k141_187596 | 2400 | No | NA | 3 | 0 | 1 | Not-determined | Genome-fragment | NA | NA | 0 |
| S8_k141_72879 | 3698 | No | NA | 3 | 0 | 0 | Not-determined | Genome-fragment | NA | NA | 0 |
| S8_k141_48935 | 58944 | Yes | 50535 | 81 | 19 | 4 | High-quality | High-quality | 100 | AAI-based (high-confidence) | 14.27 |
| S8_k141_164381 | 1195 | No | NA | 2 | 0 | 1 | Not-determined | Genome-fragment | NA | NA | 0 |
| S8_k141_268114 | 9594 | No | NA | 8 | 0 | 5 | Not-determined | Genome-fragment | NA | NA | 0 |
| S8-final-viral-combined_k141_163586\|\|0_partial | 22732 | Yes | 13507 | 26 | 3 | 7 | Low-quality | Genome-fragment | 13.91 | HMM-based (lower-bound) | 40.58 |
| S8_k141_95661 | 1332 | No | NA | 2 | 0 | 1 | Not-determined | Genome-fragment | NA | NA | 0 |
| S8_k141_210591 | 1574 | No | NA | 1 | 0 | 1 | Not-determined | Genome-fragment | NA | NA | 0 |
| S11-final-viral-combined_k141_295134\|\|0_partial | 22733 | Yes | 14917 | 21 | 3 | 8 | Low-quality | Genome-fragment | 35.95 | AAI-based (medium-confidence) | 34.38 |
| S8_k141_176563 | 1168 | No | NA | 1 | 0 | 1 | Not-determined | Genome-fragment | NA | NA | 0 |
| S8_k141_268139 | 3301 | No | NA | 5 | 0 | 1 | Not-determined | Genome-fragment | NA | NA | 0 |
| S8_k141_26584 | 1074 | No | NA | 1 | 0 | 0 | Not-determined | Genome-fragment | NA | NA | 0 |
| S8_k141_48987 | 1231 | No | NA | 1 | 0 | 0 | Not-determined | Genome-fragment | NA | NA | 0 |
| S8_k141_187688 | 3141 | No | NA | 4 | 0 | 3 | Not-determined | Genome-fragment | NA | NA | 0 |
| S1-final-viral-combined_k141_198056\|\|full | 22733 | No | NA | 20 | 1 | 15 | Low-quality | Genome-fragment | 42.59 | HMM-based (lower-bound) | 0 |
| S8_k141_118288 | 1558 | No | NA | 2 | 0 | 0 | Not-determined | Genome-fragment | NA | NA | 0 |
| S8_k141_106526 | 3290 | No | NA | 3 | 0 | 1 | Not-determined | Genome-fragment | NA | NA | 0 |
| S8_k141_245461 | 1873 | No | NA | 1 | 0 | 1 | Not-determined | Genome-fragment | NA | NA | 0 |
| S8_k141_153214 | 1107 | No | NA | 1 | 0 | 1 | Not-determined | Genome-fragment | NA | NA | 0 |
| S6-final-viral-combined_k141_34613\|\|0_partial | 22747 | No | NA | 16 | 2 | 10 | Low-quality | Genome-fragment | 9.68 | HMM-based (lower-bound) | 0 |
| S8_k141_106557 | 1106 | No | NA | 1 | 0 | 0 | Not-determined | Genome-fragment | NA | NA | 0 |
| S8_k141_49036 | 1739 | No | NA | 2 | 0 | 1 | Not-determined | Genome-fragment | NA | NA | 0 |
| S1-final-viral-combined_k141_379439\|\|full | 22753 | No | NA | 29 | 10 | 0 | Low-quality | Genome-fragment | 39.03 | AAI-based (medium-confidence) | 0 |
| S8_k141_187721 | 4140 | No | NA | 1 | 0 | 1 | Not-determined | Genome-fragment | NA | NA | 0 |
| S8_k141_176687 | 7659 | No | NA | 4 | 0 | 1 | Not-determined | Genome-fragment | NA | NA | 0 |
| S8_k141_245524 | 2756 | No | NA | 3 | 0 | 2 | Not-determined | Genome-fragment | NA | NA | 0 |
| S8_k141_210650 | 1967 | No | NA | 3 | 0 | 3 | Not-determined | Genome-fragment | NA | NA | 0 |
| S8_k141_142124 | 7600 | No | NA | 10 | 0 | 2 | Not-determined | Genome-fragment | NA | NA | 0 |
| S8_k141_130171 | 1368 | No | NA | 1 | 0 | 1 | Not-determined | Genome-fragment | NA | NA | 0 |
| S8_k141_234115 | 3069 | No | NA | 2 | 0 | 1 | Not-determined | Genome-fragment | NA | NA | 0 |
| S8_k141_73082 | 1097 | No | NA | 1 | 0 | 0 | Not-determined | Genome-fragment | NA | NA | 0 |
| S8_k141_187837 | 1236 | No | NA | 1 | 0 | 0 | Not-determined | Genome-fragment | NA | NA | 0 |
| S8_k141_130243 | 1076 | No | NA | 2 | 0 | 2 | Not-determined | Genome-fragment | NA | NA | 0 |
| S8_k141_164549 | 1177 | No | NA | 1 | 0 | 1 | Not-determined | Genome-fragment | NA | NA | 0 |
| S8_k141_106681 | 1483 | No | NA | 2 | 0 | 2 | Not-determined | Genome-fragment | NA | NA | 0 |
| S8_k141_210729 | 2540 | No | NA | 7 | 0 | 0 | Not-determined | Genome-fragment | NA | NA | 0 |
| S8_k141_245599 | 9351 | No | NA | 8 | 0 | 6 | Not-determined | Genome-fragment | NA | NA | 0 |
| S8_k141_199042 | 1055 | No | NA | 2 | 0 | 1 | Not-determined | Genome-fragment | NA | NA | 0 |
| S8_k141_73100 | 1486 | No | NA | 2 | 0 | 1 | Not-determined | Genome-fragment | NA | NA | 0 |
| S2-final-viral-combined_k141_168785\|\|full | 22764 | Yes | 18068 | 22 | 2 | 3 | Low-quality | Genome-fragment | 35.52 | AAI-based (medium-confidence) | 20.63 |
| S8_k141_106704 | 1024 | No | NA | 1 | 0 | 1 | Not-determined | Genome-fragment | NA | NA | 0 |
| S8_k141_83525 | 2140 | No | NA | 1 | 0 | 0 | Not-determined | Genome-fragment | NA | NA | 0 |
| S8_k141_83532 | 1546 | No | NA | 2 | 0 | 1 | Not-determined | Genome-fragment | NA | NA | 0 |
| S8_k141_187935 | 3202 | No | NA | 5 | 0 | 2 | Not-determined | Genome-fragment | NA | NA | 0 |
| S4-final-viral-combined_k141_162311\|\|full | 22767 | No | NA | 22 | 1 | 2 | Low-quality | Genome-fragment | 12.35 | HMM-based (lower-bound) | 0 |
| S8_k141_26780 | 2054 | No | NA | 1 | 0 | 1 | Not-determined | Genome-fragment | NA | NA | 0 |
| S8_k141_3250 | 1107 | No | NA | 1 | 0 | 1 | Not-determined | Genome-fragment | NA | NA | 0 |
| S8_k141_142221 | 3152 | No | NA | 5 | 0 | 1 | Not-determined | Genome-fragment | NA | NA | 0 |
| S12-final-viral-combined_k141_310910\|\|0_partial | 22812 | Yes | 16009 | 22 | 5 | 3 | Low-quality | Genome-fragment | 38.49 | AAI-based (medium-confidence) | 29.82 |
| S8_k141_245712 | 3339 | No | NA | 2 | 0 | 1 | Not-determined | Genome-fragment | NA | NA | 0 |
| S6-final-viral-combined_k141_3948\|\|full | 22831 | No | NA | 35 | 8 | 0 | Low-quality | Genome-fragment | 37.61 | AAI-based (high-confidence) | 0 |
| S8_k141_38014 | 3411 | No | NA | 2 | 0 | 2 | Not-determined | Genome-fragment | NA | NA | 0 |
| S8_k141_118528 | 1541 | No | NA | 3 | 0 | 1 | Not-determined | Genome-fragment | NA | NA | 0 |
| S8_k141_199126 | 1126 | No | NA | 1 | 0 | 1 | Not-determined | Genome-fragment | NA | NA | 0 |
| S8_k141_222056 | 2987 | No | NA | 1 | 0 | 0 | Not-determined | Genome-fragment | NA | NA | 0 |
| S8_k141_187978 | 1605 | No | NA | 2 | 0 | 2 | Not-determined | Genome-fragment | NA | NA | 0 |
| S8_k141_164645 | 1028 | No | NA | 1 | 0 | 1 | Not-determined | Genome-fragment | NA | NA | 0 |
| S8_k141_222076 | 1120 | No | NA | 2 | 0 | 1 | Not-determined | Genome-fragment | NA | NA | 0 |
| S8_k141_142296 | 1011 | No | NA | 1 | 0 | 1 | Not-determined | Genome-fragment | NA | NA | 0 |
| S3-final-viral-combined_k141_112743\|\|full | 22841 | Yes | 5452 | 24 | 2 | 11 | Low-quality | Genome-fragment | 12.24 | AAI-based (medium-confidence) | 76.13 |
| S8_k141_83618 | 8285 | No | NA | 3 | 0 | 1 | Not-determined | Genome-fragment | NA | NA | 0 |
| S8_k141_153518 | 1048 | No | NA | 1 | 0 | 1 | Not-determined | Genome-fragment | NA | NA | 0 |
| S8_k141_234276 | 1079 | No | NA | 1 | 0 | 1 | Not-determined | Genome-fragment | NA | NA | 0 |
| S8_k141_95939 | 3549 | No | NA | 2 | 0 | 2 | Not-determined | Genome-fragment | NA | NA | 0 |
| S8_k141_14087 | 1286 | No | NA | 1 | 0 | 1 | Not-determined | Genome-fragment | NA | NA | 0 |
| S8_k141_188037 | 1041 | No | NA | 2 | 0 | 1 | Not-determined | Genome-fragment | NA | NA | 0 |
| S8_k141_188044 | 1289 | No | NA | 1 | 0 | 1 | Not-determined | Genome-fragment | NA | NA | 0 |
| S8_k141_177032 | 1440 | No | NA | 1 | 0 | 0 | Not-determined | Genome-fragment | NA | NA | 0 |
| S8_k141_199190 | 1537 | No | NA | 1 | 0 | 1 | Not-determined | Genome-fragment | NA | NA | 0 |
| S8_k141_245781 | 1035 | No | NA | 2 | 0 | 0 | Not-determined | Genome-fragment | NA | NA | 0 |
| S8_k141_26964 | 4706 | No | NA | 5 | 0 | 0 | Not-determined | Genome-fragment | NA | NA | 0 |
| S8_k141_142376 | 1653 | No | NA | 3 | 0 | 1 | Not-determined | Genome-fragment | NA | NA | 0 |
| S8_k141_201884 | 22895 | No | NA | 17 | 3 | 0 | Low-quality | Genome-fragment | 22.6 | AAI-based (high-confidence) | 0 |
| S8_k141_142385 | 1184 | No | NA | 2 | 0 | 0 | Not-determined | Genome-fragment | NA | NA | 0 |
| S8_k141_268530 | 1304 | No | NA | 2 | 0 | 1 | Not-determined | Genome-fragment | NA | NA | 0 |
| S8_k141_188085 | 1013 | No | NA | 2 | 0 | 0 | Not-determined | Genome-fragment | NA | NA | 0 |
| S8_k141_118665 | 1042 | No | NA | 1 | 0 | 1 | Not-determined | Genome-fragment | NA | NA | 0 |
| S11-final-viral-combined_k141_247298\|\|full | 22906 | No | NA | 18 | 6 | 2 | Low-quality | Genome-fragment | 30.99 | HMM-based (lower-bound) | 0 |
| S8_k141_96044 | 1559 | No | NA | 1 | 0 | 1 | Not-determined | Genome-fragment | NA | NA | 0 |
| S8_k141_83735 | 1462 | No | NA | 2 | 0 | 2 | Not-determined | Genome-fragment | NA | NA | 0 |
| S8_k141_234394 | 1521 | No | NA | 1 | 0 | 1 | Not-determined | Genome-fragment | NA | NA | 0 |
| S8_k141_27073 | 1049 | No | NA | 1 | 0 | 0 | Not-determined | Genome-fragment | NA | NA | 0 |
| S8_k141_245948 | 1433 | No | NA | 1 | 0 | 0 | Not-determined | Genome-fragment | NA | NA | 0 |
| S8_k141_38196 | 3425 | No | NA | 3 | 0 | 1 | Not-determined | Genome-fragment | NA | NA | 0 |
| S7-final-viral-combined_k141_119356\|\|0_partial | 22912 | No | NA | 17 | 1 | 3 | Low-quality | Genome-fragment | 38.91 | AAI-based (medium-confidence) | 0 |
| S8_k141_211035 | 1228 | No | NA | 1 | 0 | 1 | Not-determined | Genome-fragment | NA | NA | 0 |
| S8_k141_96071 | 1432 | No | NA | 1 | 0 | 1 | Not-determined | Genome-fragment | NA | NA | 0 |
| S9-final-viral-combined_k141_24310\|\|0_partial | 22940 | No | NA | 28 | 2 | 4 | Low-quality | Genome-fragment | 35.5 | HMM-based (lower-bound) | 0 |
| S8_k141_153681 | 1488 | No | NA | 2 | 0 | 2 | Not-determined | Genome-fragment | NA | NA | 0 |
| S8_k141_96099 | 1082 | No | NA | 2 | 0 | 2 | Not-determined | Genome-fragment | NA | NA | 0 |
| S8_k141_246011 | 1018 | No | NA | 2 | 0 | 0 | Not-determined | Genome-fragment | NA | NA | 0 |
| S7-final-viral-combined_k141_128307\|\|full | 23052 | No | NA | 19 | 0 | 4 | Low-quality | Genome-fragment | 41.34 | AAI-based (medium-confidence) | 0 |
| S8_k141_256404 | 4475 | No | NA | 5 | 0 | 1 | Not-determined | Genome-fragment | NA | NA | 0 |
| S8_k141_38272 | 3312 | No | NA | 3 | 0 | 2 | Not-determined | Genome-fragment | NA | NA | 0 |
| S8_k141_27180 | 1033 | No | NA | 1 | 0 | 1 | Not-determined | Genome-fragment | NA | NA | 0 |
| S8_k141_107102 | 1327 | No | NA | 2 | 0 | 0 | Not-determined | Genome-fragment | NA | NA | 0 |
| S1-final-viral-combined_k141_2595\|\|0_partial | 23058 | Yes | 18254 | 30 | 4 | 3 | Low-quality | Genome-fragment | 40.43 | AAI-based (high-confidence) | 20.83 |
| S17-final-viral-combined_k141_51741\|\|full | 23060 | No | NA | 25 | 0 | 5 | Low-quality | Genome-fragment | 35.53 | AAI-based (high-confidence) | 0 |
| S8_k141_256416 | 3746 | No | NA | 4 | 0 | 3 | Not-determined | Genome-fragment | NA | NA | 0 |
| S12_k141_154504 | 23060 | No | NA | 21 | 3 | 3 | Low-quality | Genome-fragment | 27.59 | AAI-based (medium-confidence) | 0 |
| S8_k141_142570 | 1112 | No | NA | 1 | 0 | 1 | Not-determined | Genome-fragment | NA | NA | 0 |
| S8_k141_3503 | 1176 | No | NA | 1 | 0 | 1 | Not-determined | Genome-fragment | NA | NA | 0 |
| S8_k141_177225 | 1062 | No | NA | 2 | 0 | 2 | Not-determined | Genome-fragment | NA | NA | 0 |
| S8_k141_246060 | 1202 | No | NA | 2 | 0 | 1 | Not-determined | Genome-fragment | NA | NA | 0 |
| S6_k141_74427 | 23142 | No | NA | 42 | 5 | 1 | Low-quality | Genome-fragment | 12.37 | AAI-based (high-confidence) | 0 |
| S8_k141_83816 | 3097 | No | NA | 3 | 0 | 2 | Not-determined | Genome-fragment | NA | NA | 0 |
| S8_k141_142586 | 1407 | No | NA | 1 | 0 | 1 | Not-determined | Genome-fragment | NA | NA | 0 |
| S8_k141_142593 | 8302 | No | NA | 4 | 0 | 2 | Not-determined | Genome-fragment | NA | NA | 0 |
| S12-final-viral-combined_k141_71200\|\|full | 23199 | No | NA | 27 | 1 | 6 | Low-quality | Genome-fragment | 13.71 | HMM-based (lower-bound) | 0 |
| S2-final-viral-combined_k141_118728\|\|full | 23266 | No | NA | 35 | 16 | 1 | Low-quality | Genome-fragment | 49.3 | AAI-based (medium-confidence) | 0 |
| S8_k141_142618 | 1377 | No | NA | 1 | 0 | 1 | Not-determined | Genome-fragment | NA | NA | 0 |
| S8_k141_199318 | 2234 | No | NA | 1 | 0 | 0 | Not-determined | Genome-fragment | NA | NA | 0 |
| S8_k141_118897 | 1166 | No | NA | 2 | 0 | 1 | Not-determined | Genome-fragment | NA | NA | 0 |
| S8_k141_199344 | 1102 | No | NA | 2 | 0 | 0 | Not-determined | Genome-fragment | NA | NA | 0 |
| S8_k141_61403 | 9828 | No | NA | 7 | 0 | 6 | Not-determined | Genome-fragment | NA | NA | 0 |
| S8_k141_165044 | 1300 | No | NA | 1 | 0 | 0 | Not-determined | Genome-fragment | NA | NA | 0 |
| S8_k141_188278 | 6243 | No | NA | 5 | 0 | 3 | Not-determined | Genome-fragment | NA | NA | 0 |
| S8_k141_177367 | 1922 | No | NA | 4 | 0 | 1 | Not-determined | Genome-fragment | NA | NA | 0 |
| S8_k141_234625 | 4761 | No | NA | 3 | 0 | 3 | Not-determined | Genome-fragment | NA | NA | 0 |
| S8_k141_38359 | 3646 | No | NA | 7 | 0 | 0 | Not-determined | Genome-fragment | NA | NA | 0 |
| S8_k141_222474 | 1813 | No | NA | 2 | 0 | 0 | Not-determined | Genome-fragment | NA | NA | 0 |
| S8_k141_188301 | 1246 | No | NA | 1 | 0 | 1 | Not-determined | Genome-fragment | NA | NA | 0 |
| S8_k141_199396 | 2754 | No | NA | 2 | 0 | 1 | Not-determined | Genome-fragment | NA | NA | 0 |
| S8_k141_268790 | 5991 | No | NA | 4 | 0 | 3 | Not-determined | Genome-fragment | NA | NA | 0 |
| S5-final-viral-combined_k141_15798\|\|0_partial | 23273 | Yes | 12459 | 26 | 5 | 6 | Low-quality | Genome-fragment | 16.93 | HMM-based (lower-bound) | 46.47 |
| S8_k141_119001 | 4198 | No | NA | 2 | 0 | 2 | Not-determined | Genome-fragment | NA | NA | 0 |
| S8_k141_119008 | 1736 | No | NA | 2 | 0 | 2 | Not-determined | Genome-fragment | NA | NA | 0 |
| S8_k141_268821 | 1060 | No | NA | 2 | 0 | 0 | Not-determined | Genome-fragment | NA | NA | 0 |
| S12-final-viral-combined_k141_270292\|\|full | 23358 | Yes | 19063 | 23 | 2 | 4 | Low-quality | Genome-fragment | 37.17 | AAI-based (medium-confidence) | 18.39 |
| S8_k141_199449 | 1193 | No | NA | 2 | 0 | 2 | Not-determined | Genome-fragment | NA | NA | 0 |
| S8_k141_38447 | 3186 | No | NA | 3 | 0 | 1 | Not-determined | Genome-fragment | NA | NA | 0 |
| S8_k141_199467 | 1086 | No | NA | 1 | 0 | 1 | Not-determined | Genome-fragment | NA | NA | 0 |
| S8_k141_222581 | 1281 | No | NA | 3 | 0 | 0 | Not-determined | Genome-fragment | NA | NA | 0 |
| S8_k141_38486 | 1323 | No | NA | 2 | 0 | 0 | Not-determined | Genome-fragment | NA | NA | 0 |
| S8_k141_38493 | 2718 | No | NA | 2 | 0 | 1 | Not-determined | Genome-fragment | NA | NA | 0 |
| S8_k141_256565 | 3252 | No | NA | 1 | 0 | 1 | Not-determined | Genome-fragment | NA | NA | 0 |
| S8_k141_199503 | 1847 | No | NA | 4 | 0 | 2 | Not-determined | Genome-fragment | NA | NA | 0 |
| S8_k141_222642 | 1059 | No | NA | 2 | 0 | 2 | Not-determined | Genome-fragment | NA | NA | 0 |
| S8_k141_130819 | 3089 | No | NA | 5 | 0 | 0 | Not-determined | Genome-fragment | NA | NA | 0 |
| S8_k141_153892 | 1416 | No | NA | 3 | 0 | 0 | Not-determined | Genome-fragment | NA | NA | 0 |
| S8_k141_211345 | 1269 | No | NA | 1 | 0 | 0 | Not-determined | Genome-fragment | NA | NA | 0 |
| S8_k141_73559 | 1302 | No | NA | 2 | 0 | 0 | Not-determined | Genome-fragment | NA | NA | 0 |
| S8_k141_211362 | 2022 | No | NA | 3 | 0 | 2 | Not-determined | Genome-fragment | NA | NA | 0 |
| S11-final-viral-combined_k141_206634\|\|full | 23375 | Yes | 12283 | 33 | 4 | 9 | Low-quality | Genome-fragment | 14.15 | HMM-based (lower-bound) | 47.45 |
| S8_k141_27438 | 1079 | No | NA | 1 | 0 | 1 | Not-determined | Genome-fragment | NA | NA | 0 |
| S8_k141_268951 | 3359 | No | NA | 3 | 0 | 1 | Not-determined | Genome-fragment | NA | NA | 0 |
| S8_k141_27443 | 1572 | No | NA | 1 | 0 | 1 | Not-determined | Genome-fragment | NA | NA | 0 |
| S7-final-viral-combined_k141_157811\|\|full | 23381 | Yes | 14932 | 23 | 9 | 2 | Low-quality | Genome-fragment | 31.16 | AAI-based (high-confidence) | 36.14 |
| S8_k141_130885 | 1047 | No | NA | 2 | 0 | 1 | Not-determined | Genome-fragment | NA | NA | 0 |
| S8_k141_222720 | 2079 | No | NA | 2 | 0 | 1 | Not-determined | Genome-fragment | NA | NA | 0 |
| S8-final-viral-combined_k141_114651\|\|0_partial | 23411 | Yes | 13327 | 23 | 4 | 8 | Low-quality | Genome-fragment | 33.83 | AAI-based (medium-confidence) | 43.07 |
| S12_k141_399744 | 23441 | No | NA | 18 | 0 | 14 | Low-quality | Genome-fragment | 10.78 | AAI-based (medium-confidence) | 0 |
| S1-final-viral-combined_k141_106283\|\|full | 23459 | Yes | 16096 | 40 | 7 | 8 | Low-quality | Genome-fragment | 29.44 | AAI-based (high-confidence) | 31.39 |
| S8_k141_165358 | 1089 | No | NA | 1 | 0 | 1 | Not-determined | Genome-fragment | NA | NA | 0 |
| S8_k141_165363 | 1157 | No | NA | 1 | 0 | 1 | Not-determined | Genome-fragment | NA | NA | 0 |
| S5-final-viral-combined_k141_98307\|\|0_partial | 23489 | Yes | 12656 | 32 | 4 | 10 | Low-quality | Genome-fragment | 12.5 | HMM-based (lower-bound) | 46.12 |
| S8_k141_234868 | 1049 | No | NA | 1 | 0 | 1 | Not-determined | Genome-fragment | NA | NA | 0 |
| S8_k141_165383 | 1136 | No | NA | 2 | 0 | 0 | Not-determined | Genome-fragment | NA | NA | 0 |
| S8_k141_199557 | 1377 | No | NA | 2 | 0 | 2 | Not-determined | Genome-fragment | NA | NA | 0 |
| S8_k141_49887 | 1649 | No | NA | 3 | 0 | 0 | Not-determined | Genome-fragment | NA | NA | 0 |
| S8_k141_199568 | 3898 | No | NA | 2 | 0 | 1 | Not-determined | Genome-fragment | NA | NA | 0 |
| S8_k141_256703 | 1704 | No | NA | 1 | 0 | 1 | Not-determined | Genome-fragment | NA | NA | 0 |
| S8_k141_84238 | 3469 | No | NA | 3 | 0 | 0 | Not-determined | Genome-fragment | NA | NA | 0 |
| S8_k141_269120 | 1198 | No | NA | 1 | 0 | 1 | Not-determined | Genome-fragment | NA | NA | 0 |
| S8_k141_61757 | 2935 | No | NA | 2 | 0 | 1 | Not-determined | Genome-fragment | NA | NA | 0 |
| S8_k141_269132 | 1574 | No | NA | 1 | 0 | 1 | Not-determined | Genome-fragment | NA | NA | 0 |
| S8_k141_177638 | 1712 | No | NA | 2 | 0 | 1 | Not-determined | Genome-fragment | NA | NA | 0 |
| S8_k141_61771 | 1012 | No | NA | 1 | 0 | 1 | Not-determined | Genome-fragment | NA | NA | 0 |
| S8-final-viral-combined_k141_161901\|\|full | 23490 | No | NA | 34 | 7 | 1 | Low-quality | Genome-fragment | 49.63 | AAI-based (high-confidence) | 0 |
| S8_k141_188641 | 1370 | No | NA | 1 | 0 | 0 | Not-determined | Genome-fragment | NA | NA | 0 |
| S7-final-viral-combined_k141_152511\|\|full | 23519 | No | NA | 26 | 1 | 10 | Low-quality | Genome-fragment | 26.4 | HMM-based (lower-bound) | 0 |
| S8_k141_177681 | 1075 | No | NA | 1 | 0 | 0 | Not-determined | Genome-fragment | NA | NA | 0 |
| S8_k141_222919 | 1231 | No | NA | 1 | 0 | 0 | Not-determined | Genome-fragment | NA | NA | 0 |
| S8_k141_96591 | 1454 | No | NA | 1 | 0 | 1 | Not-determined | Genome-fragment | NA | NA | 0 |
| S8_k141_84317 | 3040 | No | NA | 2 | 0 | 2 | Not-determined | Genome-fragment | NA | NA | 0 |
| S8_k141_38814 | 1103 | No | NA | 1 | 0 | 0 | Not-determined | Genome-fragment | NA | NA | 0 |
| S17-final-viral-combined_k141_150874\|\|full | 23534 | Yes | 10890 | 30 | 7 | 8 | Low-quality | Genome-fragment | 20.68 | HMM-based (lower-bound) | 53.73 |
| S8_k141_107538 | 1283 | No | NA | 1 | 0 | 1 | Not-determined | Genome-fragment | NA | NA | 0 |
| S8_k141_14629 | 9683 | No | NA | 5 | 0 | 5 | Not-determined | Genome-fragment | NA | NA | 0 |
| S8_k141_119273 | 3087 | No | NA | 1 | 0 | 0 | Not-determined | Genome-fragment | NA | NA | 0 |
| S12-final-viral-combined_k141_153404\|\|full | 23559 | Yes | 14541 | 33 | 15 | 4 | Low-quality | Genome-fragment | 41.72 | AAI-based (high-confidence) | 38.28 |
| S8_k141_119304 | 2047 | No | NA | 2 | 0 | 0 | Not-determined | Genome-fragment | NA | NA | 0 |
| S8_k141_234978 | 1409 | No | NA | 1 | 0 | 1 | Not-determined | Genome-fragment | NA | NA | 0 |
| S8_k141_211645 | 1049 | No | NA | 1 | 0 | 0 | Not-determined | Genome-fragment | NA | NA | 0 |
| S8_k141_131148 | 1280 | No | NA | 1 | 0 | 1 | Not-determined | Genome-fragment | NA | NA | 0 |
| S8_k141_38900 | 1054 | No | NA | 1 | 0 | 0 | Not-determined | Genome-fragment | NA | NA | 0 |
| S8_k141_235055 | 1429 | No | NA | 2 | 0 | 2 | Not-determined | Genome-fragment | NA | NA | 0 |
| S8_k141_188820 | 1185 | No | NA | 1 | 0 | 0 | Not-determined | Genome-fragment | NA | NA | 0 |
| S8_k141_246348 | 1768 | No | NA | 1 | 0 | 1 | Not-determined | Genome-fragment | NA | NA | 0 |
| S8_k141_269243 | 5047 | No | NA | 7 | 0 | 0 | Not-determined | Genome-fragment | NA | NA | 0 |
| S7-final-viral-combined_k141_138418\|\|full | 23668 | No | NA | 24 | 11 | 0 | Low-quality | Genome-fragment | 36.73 | AAI-based (medium-confidence) | 0 |
| S8_k141_3983 | 1132 | No | NA | 1 | 0 | 1 | Not-determined | Genome-fragment | NA | NA | 0 |
| S8_k141_256880 | 2569 | No | NA | 2 | 0 | 1 | Not-determined | Genome-fragment | NA | NA | 0 |
| S8_k141_131204 | 1216 | No | NA | 2 | 0 | 1 | Not-determined | Genome-fragment | NA | NA | 0 |
| S8_k141_269303 | 2063 | No | NA | 1 | 0 | 1 | Not-determined | Genome-fragment | NA | NA | 0 |
| S5-final-viral-combined_k141_144698\|\|0_partial | 23714 | No | NA | 22 | 1 | 8 | Low-quality | Genome-fragment | 40.74 | HMM-based (lower-bound) | 0 |
| S1-final-viral-combined_k141_95394\|\|0_partial | 23717 | Yes | 14989 | 21 | 3 | 9 | Low-quality | Genome-fragment | 36.76 | AAI-based (high-confidence) | 36.8 |
| S4-final-viral-combined_k141_102196\|\|full | 23888 | No | NA | 19 | 0 | 12 | Low-quality | Genome-fragment | 9.29 | AAI-based (medium-confidence) | 0 |
| S8_k141_269320 | 3006 | No | NA | 5 | 0 | 3 | Not-determined | Genome-fragment | NA | NA | 0 |
| S8_k141_143075 | 1561 | No | NA | 1 | 0 | 1 | Not-determined | Genome-fragment | NA | NA | 0 |
| S8_k141_131240 | 1863 | No | NA | 1 | 0 | 1 | Not-determined | Genome-fragment | NA | NA | 0 |
| S8_k141_131242 | 1232 | No | NA | 1 | 0 | 1 | Not-determined | Genome-fragment | NA | NA | 0 |
| S9-final-viral-combined_k141_57441\|\|0_partial | 23889 | No | NA | 25 | 0 | 17 | Low-quality | Genome-fragment | 22.13 | AAI-based (medium-confidence) | 0 |
| S8_k141_269347 | 1705 | No | NA | 1 | 0 | 1 | Not-determined | Genome-fragment | NA | NA | 0 |
| S8_k141_39075 | 1212 | No | NA | 1 | 0 | 1 | Not-determined | Genome-fragment | NA | NA | 0 |
| S8_k141_246405 | 2477 | No | NA | 3 | 0 | 1 | Not-determined | Genome-fragment | NA | NA | 0 |
| S8_k141_269377 | 1812 | No | NA | 1 | 0 | 0 | Not-determined | Genome-fragment | NA | NA | 0 |
| S8_k141_256953 | 1437 | No | NA | 2 | 0 | 2 | Not-determined | Genome-fragment | NA | NA | 0 |
| S8_k141_269400 | 1719 | No | NA | 1 | 0 | 1 | Not-determined | Genome-fragment | NA | NA | 0 |
| S8_k141_62190 | 1524 | No | NA | 1 | 0 | 1 | Not-determined | Genome-fragment | NA | NA | 0 |
| S8_k141_131332 | 1524 | No | NA | 2 | 0 | 0 | Not-determined | Genome-fragment | NA | NA | 0 |
| S8_k141_119412 | 1329 | No | NA | 1 | 0 | 1 | Not-determined | Genome-fragment | NA | NA | 0 |
| S8_k141_119424 | 1189 | No | NA | 2 | 0 | 0 | Not-determined | Genome-fragment | NA | NA | 0 |
| S8_k141_223083 | 1108 | No | NA | 2 | 0 | 2 | Not-determined | Genome-fragment | NA | NA | 0 |
| S8_k141_177943 | 1080 | No | NA | 1 | 0 | 0 | Not-determined | Genome-fragment | NA | NA | 0 |
| S17-final-viral-combined_k141_178208\|\|0_partial | 23942 | No | NA | 21 | 2 | 3 | Low-quality | Genome-fragment | 14.56 | AAI-based (medium-confidence) | 0 |
| S6-final-viral-combined_k141_107641\|\|1_partial | 24099 | Yes | 21450 | 20 | 2 | 3 | Low-quality | Genome-fragment | 27.64 | AAI-based (medium-confidence) | 10.99 |
| S8_k141_96811 | 1348 | No | NA | 1 | 0 | 0 | Not-determined | Genome-fragment | NA | NA | 0 |
| S8_k141_188964 | 1060 | No | NA | 1 | 0 | 1 | Not-determined | Genome-fragment | NA | NA | 0 |
| S8_k141_235210 | 1009 | No | NA | 1 | 0 | 0 | Not-determined | Genome-fragment | NA | NA | 0 |
| S8_k141_73895 | 1951 | No | NA | 1 | 0 | 0 | Not-determined | Genome-fragment | NA | NA | 0 |
| S4-final-viral-combined_k141_132997\|\|full | 24125 | No | NA | 10 | 0 | 2 | Low-quality | Genome-fragment | 47.52 | AAI-based (medium-confidence) | 0 |
| S8_k141_154399 | 1754 | No | NA | 1 | 0 | 1 | Not-determined | Genome-fragment | NA | NA | 0 |
| S8_k141_96886 | 2105 | No | NA | 1 | 0 | 1 | Not-determined | Genome-fragment | NA | NA | 0 |
| S8-final-viral-combined_k141_91209\|\|full | 24143 | Yes | 12416 | 27 | 5 | 4 | Low-quality | Genome-fragment | 16.66 | HMM-based (lower-bound) | 48.57 |
| S7-final-viral-combined_k141_92909\|\|0_partial | 24164 | No | NA | 15 | 0 | 12 | Low-quality | Genome-fragment | 32.92 | AAI-based (medium-confidence) | 0 |
| S8_k141_269526 | 1181 | No | NA | 2 | 0 | 2 | Not-determined | Genome-fragment | NA | NA | 0 |
| S8_k141_131428 | 1111 | No | NA | 1 | 0 | 0 | Not-determined | Genome-fragment | NA | NA | 0 |
| S8_k141_154452 | 4807 | No | NA | 4 | 0 | 4 | Not-determined | Genome-fragment | NA | NA | 0 |
| S8_k141_96968 | 1038 | No | NA | 1 | 0 | 0 | Not-determined | Genome-fragment | NA | NA | 0 |
| S8_k141_84683 | 1026 | No | NA | 1 | 0 | 1 | Not-determined | Genome-fragment | NA | NA | 0 |
| S8_k141_269572 | 1090 | No | NA | 1 | 0 | 0 | Not-determined | Genome-fragment | NA | NA | 0 |
| S4_k141_44821 | 24205 | No | NA | 44 | 3 | 1 | Low-quality | Genome-fragment | 24.16 | AAI-based (high-confidence) | 0 |
| S8_k141_235375 | 1165 | No | NA | 1 | 0 | 1 | Not-determined | Genome-fragment | NA | NA | 0 |
| S8_k141_235380 | 1594 | No | NA | 1 | 0 | 1 | Not-determined | Genome-fragment | NA | NA | 0 |
| S8_k141_211976 | 3286 | No | NA | 4 | 0 | 3 | Not-determined | Genome-fragment | NA | NA | 0 |
| S8_k141_97025 | 3256 | No | NA | 3 | 0 | 3 | Not-determined | Genome-fragment | NA | NA | 0 |
| S8_k141_131547 | 1176 | No | NA | 2 | 0 | 2 | Not-determined | Genome-fragment | NA | NA | 0 |
| S8_k141_131553 | 1119 | No | NA | 2 | 0 | 1 | Not-determined | Genome-fragment | NA | NA | 0 |
| S8_k141_62462 | 1308 | No | NA | 3 | 0 | 1 | Not-determined | Genome-fragment | NA | NA | 0 |
| S8_k141_50398 | 1013 | No | NA | 1 | 0 | 0 | Not-determined | Genome-fragment | NA | NA | 0 |
| S11-final-viral-combined_k141_4014\|\|full | 24247 | Yes | 11713 | 18 | 2 | 6 | Low-quality | Genome-fragment | 25.6 | HMM-based (lower-bound) | 51.69 |
| S2-final-viral-combined_k141_114586\|\|full | 24253 | No | NA | 32 | 1 | 3 | Low-quality | Genome-fragment | 38.7 | AAI-based (high-confidence) | 0 |
| S8_k141_84757 | 4359 | No | NA | 4 | 0 | 3 | Not-determined | Genome-fragment | NA | NA | 0 |
| S2-final-viral-combined_k141_1184\|\|full | 24374 | No | NA | 22 | 0 | 15 | Low-quality | Genome-fragment | 14.05 | AAI-based (medium-confidence) | 0 |
| S17-k141_227198 | 24410 | No | NA | 24 | 0 | 8 | Low-quality | Genome-fragment | 40.77 | AAI-based (medium-confidence) | 0 |
| S8_k141_235456 | 5738 | No | NA | 8 | 0 | 3 | Not-determined | Genome-fragment | NA | NA | 0 |
| S11_k141_129008 | 24466 | No | NA | 40 | 10 | 0 | Low-quality | Genome-fragment | 42.25 | AAI-based (high-confidence) | 0 |
| S8_k141_15123 | 1081 | No | NA | 2 | 0 | 2 | Not-determined | Genome-fragment | NA | NA | 0 |
| S8_k141_143388 | 3997 | No | NA | 2 | 0 | 1 | Not-determined | Genome-fragment | NA | NA | 0 |
| S7_k141_76641 | 24550 | No | NA | 31 | 5 | 1 | Low-quality | Genome-fragment | 34.72 | AAI-based (high-confidence) | 0 |
| S8_k141_27918 | 1551 | No | NA | 2 | 0 | 2 | Not-determined | Genome-fragment | NA | NA | 0 |
| S8_k141_27919 | 4007 | No | NA | 5 | 0 | 0 | Not-determined | Genome-fragment | NA | NA | 0 |
| S8_k141_84796 | 7980 | No | NA | 6 | 0 | 3 | Not-determined | Genome-fragment | NA | NA | 0 |
| S8_k141_269756 | 1537 | No | NA | 3 | 0 | 2 | Not-determined | Genome-fragment | NA | NA | 0 |
| S8_k141_212177 | 7867 | No | NA | 2 | 0 | 1 | Not-determined | Genome-fragment | NA | NA | 0 |
| S8_k141_257059 | 1759 | No | NA | 2 | 0 | 1 | Not-determined | Genome-fragment | NA | NA | 0 |
| S7-final-viral-combined_k141_217026\|\|full | 24557 | Yes | 11219 | 30 | 4 | 6 | Low-quality | Genome-fragment | 29.12 | AAI-based (medium-confidence) | 54.31 |
| S8_k141_74248 | 1051 | No | NA | 2 | 0 | 0 | Not-determined | Genome-fragment | NA | NA | 0 |
| S8_k141_235567 | 1407 | No | NA | 2 | 0 | 0 | Not-determined | Genome-fragment | NA | NA | 0 |
| S8_k141_212194 | 1159 | No | NA | 1 | 0 | 1 | Not-determined | Genome-fragment | NA | NA | 0 |
| S8_k141_27997 | 1571 | No | NA | 1 | 0 | 1 | Not-determined | Genome-fragment | NA | NA | 0 |
| S2-final-viral-combined_k141_4315\|\|full | 24568 | No | NA | 14 | 1 | 8 | Low-quality | Genome-fragment | 36.75 | HMM-based (lower-bound) | 0 |
| S8_k141_269799 | 2139 | No | NA | 1 | 0 | 1 | Not-determined | Genome-fragment | NA | NA | 0 |
| S8_k141_143487 | 1373 | No | NA | 1 | 0 | 1 | Not-determined | Genome-fragment | NA | NA | 0 |
| S8_k141_97166 | 1920 | No | NA | 3 | 0 | 1 | Not-determined | Genome-fragment | NA | NA | 0 |
| S8_k141_269813 | 2741 | No | NA | 2 | 0 | 2 | Not-determined | Genome-fragment | NA | NA | 0 |
| S8_k141_50607 | 4343 | No | NA | 6 | 0 | 3 | Not-determined | Genome-fragment | NA | NA | 0 |
| S8_k141_257148 | 2017 | No | NA | 3 | 0 | 2 | Not-determined | Genome-fragment | NA | NA | 0 |
| S8_k141_212228 | 1006 | No | NA | 1 | 0 | 0 | Not-determined | Genome-fragment | NA | NA | 0 |
| S8_k141_74289 | 1174 | No | NA | 1 | 0 | 1 | Not-determined | Genome-fragment | NA | NA | 0 |
| S8_k141_119786 | 3553 | No | NA | 2 | 0 | 1 | Not-determined | Genome-fragment | NA | NA | 0 |
| S8_k141_189358 | 3058 | No | NA | 5 | 0 | 0 | Not-determined | Genome-fragment | NA | NA | 0 |
| S9-final-viral-combined_k141_66252\|\|full | 24622 | Yes | 18083 | 39 | 15 | 5 | Low-quality | Genome-fragment | 48.41 | AAI-based (high-confidence) | 26.56 |
| S8_k141_235680 | 1039 | No | NA | 3 | 0 | 1 | Not-determined | Genome-fragment | NA | NA | 0 |
| S7-final-viral-combined_k141_228257\|\|full | 24639 | Yes | 8114 | 23 | 2 | 9 | Low-quality | Genome-fragment | 18.83 | HMM-based (lower-bound) | 67.07 |
| S8_k141_246879 | 1041 | No | NA | 1 | 0 | 1 | Not-determined | Genome-fragment | NA | NA | 0 |
| S8_k141_28078 | 4979 | No | NA | 4 | 0 | 1 | Not-determined | Genome-fragment | NA | NA | 0 |
| S8_k141_166082 | 1521 | No | NA | 2 | 0 | 1 | Not-determined | Genome-fragment | NA | NA | 0 |
| S8_k141_269917 | 4185 | No | NA | 3 | 0 | 1 | Not-determined | Genome-fragment | NA | NA | 0 |
| S17-final-viral-combined_k141_158852\|\|0_partial | 24690 | Yes | 10924 | 28 | 4 | 5 | Low-quality | Genome-fragment | 13.24 | HMM-based (lower-bound) | 55.76 |
| S8_k141_246891 | 18213 | No | NA | 20 | 0 | 14 | Not-determined | Genome-fragment | NA | NA | 0 |
| S8_k141_246896 | 1351 | No | NA | 1 | 0 | 1 | Not-determined | Genome-fragment | NA | NA | 0 |

| | | | | | | | | | | | |
|---|---|---|---|---|---|---|---|---|---|---|---|
| S8_k141_143555 | 3096 | No | NA | 3 | 0 | 1 | Not-determined | Genome-fragment | NA | NA | 0 |
| S8_k141_178526 | 1247 | No | NA | 2 | 0 | 0 | Not-determined | Genome-fragment | NA | NA | 0 |
| S8_k141_74379 | 1317 | No | NA | 1 | 0 | 0 | Not-determined | Genome-fragment | NA | NA | 0 |
| S8_k141_212334 | 1995 | No | NA | 2 | 0 | 1 | Not-determined | Genome-fragment | NA | NA | 0 |
| S8_k141_74395 | 2196 | No | NA | 5 | 0 | 0 | Not-determined | Genome-fragment | NA | NA | 0 |
| S8_k141_108303 | 1059 | No | NA | 1 | 0 | 0 | Not-determined | Genome-fragment | NA | NA | 0 |
| S8_k141_50746 | 1111 | No | NA | 2 | 0 | 1 | Not-determined | Genome-fragment | NA | NA | 0 |
| S8_k141_166182 | 1068 | No | NA | 1 | 0 | 1 | Not-determined | Genome-fragment | NA | NA | 0 |
| S8_k141_189495 | 5359 | No | NA | 6 | 0 | 0 | Not-determined | Genome-fragment | NA | NA | 0 |
| S8_k141_62892 | 1337 | No | NA | 1 | 0 | 1 | Not-determined | Genome-fragment | NA | NA | 0 |
| S8-final-viral-combined_k141_188165\|\|full | 24794 | No | NA | 28 | 1 | 19 | Low-quality | Genome-fragment | 8.88 | HMM-based (lower-bound) | 0 |
| S8_k141_62935 | 1837 | No | NA | 6 | 0 | 0 | Not-determined | Genome-fragment | NA | NA | 0 |
| S8_k141_131859 | 1243 | No | NA | 1 | 0 | 0 | Not-determined | Genome-fragment | NA | NA | 0 |
| S8_k141_257272 | 1048 | No | NA | 4 | 0 | 0 | Not-determined | Genome-fragment | NA | NA | 0 |
| S8_k141_97265 | 1826 | No | NA | 3 | 0 | 3 | Not-determined | Genome-fragment | NA | NA | 0 |
| S8_k141_62981 | 1696 | No | NA | 1 | 0 | 1 | Not-determined | Genome-fragment | NA | NA | 0 |
| S9_k141_60841 | 24835 | Yes | 12842 | 32 | 4 | 4 | Low-quality | Genome-fragment | 25.65 | HMM-based (lower-bound) | 48.29 |
| S8_k141_119960 | 5296 | No | NA | 3 | 0 | 2 | Not-determined | Genome-fragment | NA | NA | 0 |
| S4-final-viral-combined_k141_80415\|\|full | 24876 | No | NA | 31 | 10 | 0 | Low-quality | Genome-fragment | 41.52 | AAI-based (high-confidence) | 0 |
| S12-final-viral-combined_k141_394734\|\|full | 24926 | Yes | 10859 | 23 | 3 | 10 | Low-quality | Genome-fragment | 26.69 | AAI-based (high-confidence) | 56.44 |
| S8_k141_247114 | 1493 | No | NA | 2 | 0 | 1 | Not-determined | Genome-fragment | NA | NA | 0 |
| S2-final-viral-combined_k141_187023\|\|0_partial | 24939 | Yes | 20789 | 18 | 3 | 2 | Low-quality | Genome-fragment | 48.01 | AAI-based (high-confidence) | 16.64 |
| S4_k141_160656 | 24955 | No | NA | 16 | 3 | 0 | Low-quality | Genome-fragment | 37.1 | AAI-based (high-confidence) | 0 |
| S8_k141_143637 | 2526 | No | NA | 1 | 0 | 0 | Not-determined | Genome-fragment | NA | NA | 0 |
| S8_k141_178684 | 1120 | No | NA | 1 | 0 | 1 | Not-determined | Genome-fragment | NA | NA | 0 |
| S8_k141_247140 | 5988 | No | NA | 3 | 0 | 1 | Not-determined | Genome-fragment | NA | NA | 0 |
| S8_k141_15514 | 1283 | No | NA | 2 | 0 | 1 | Not-determined | Genome-fragment | NA | NA | 0 |
| S1-final-viral-combined_k141_191141\|\|full | 25112 | No | NA | 32 | 3 | 0 | Low-quality | Genome-fragment | 39.78 | HMM-based (lower-bound) | 0 |
| S8_k141_120016 | 1160 | No | NA | 1 | 0 | 0 | Not-determined | Genome-fragment | NA | NA | 0 |
| S4-final-viral-combined_k141_511\|\|full | 25127 | Yes | 7096 | 31 | 3 | 14 | Low-quality | Genome-fragment | 11.57 | HMM-based (lower-bound) | 71.76 |
| S2-final-viral-combined_k141_196261\|\|0_partial | 25211 | Yes | 9919 | 29 | 3 | 14 | Low-quality | Genome-fragment | 15.35 | HMM-based (lower-bound) | 60.66 |
| S11-final-viral-combined_k141_248571\|\|0_partial | 25336 | Yes | 13668 | 22 | 3 | 5 | Low-quality | Genome-fragment | 32.83 | AAI-based (medium-confidence) | 46.05 |
| S8_k141_212495 | 1099 | No | NA | 1 | 0 | 0 | Not-determined | Genome-fragment | NA | NA | 0 |
| S8_k141_200059 | 1098 | No | NA | 2 | 0 | 1 | Not-determined | Genome-fragment | NA | NA | 0 |
| S11-final-viral-combined_k141_229554\|\|full | 25355 | Yes | 9071 | 29 | 3 | 11 | Low-quality | Genome-fragment | 8.17 | AAI-based (medium-confidence) | 64.22 |
| S9-final-viral-combined_k141_32355\|\|0_partial | 25360 | No | NA | 20 | 3 | 8 | Low-quality | Genome-fragment | 48.74 | HMM-based (lower-bound) | 0 |
| S8_k141_132013 | 1020 | No | NA | 1 | 0 | 1 | Not-determined | Genome-fragment | NA | NA | 0 |
| S8_k141_155041 | 1540 | No | NA | 2 | 0 | 2 | Not-determined | Genome-fragment | NA | NA | 0 |
| S8_k141_74618 | 1249 | No | NA | 1 | 0 | 0 | Not-determined | Genome-fragment | NA | NA | 0 |
| S11_k141_186395 | 25407 | No | NA | 31 | 12 | 0 | Low-quality | Genome-fragment | 39.89 | AAI-based (high-confidence) | 0 |
| S8_k141_257458 | 1183 | No | NA | 1 | 0 | 0 | Not-determined | Genome-fragment | NA | NA | 0 |
| S5-final-viral-combined_k141_42162\|\|full | 25450 | Yes | 10599 | 32 | 2 | 8 | Low-quality | Genome-fragment | 8.23 | HMM-based (lower-bound) | 58.35 |
| S8_k141_51041 | 1178 | No | NA | 3 | 0 | 0 | Not-determined | Genome-fragment | NA | NA | 0 |
| S8_k141_74682 | 7254 | No | NA | 6 | 0 | 3 | Not-determined | Genome-fragment | NA | NA | 0 |
| S2-final-viral-combined_k141_104574\|\|full | 25468 | No | NA | 26 | 0 | 1 | Low-quality | Genome-fragment | 24.35 | AAI-based (medium-confidence) | 0 |
| S8_k141_189722 | 1469 | No | NA | 2 | 0 | 2 | Not-determined | Genome-fragment | NA | NA | 0 |
| S11-final-viral-combined_k141_23572\|\|0_partial | 25474 | Yes | 13158 | 32 | 6 | 11 | Low-quality | Genome-fragment | 17.83 | HMM-based (lower-bound) | 48.35 |
| S8_k141_143929 | 1163 | No | NA | 2 | 0 | 0 | Not-determined | Genome-fragment | NA | NA | 0 |
| S17-final-viral-combined_k141_122280\|\|0_partial | 25488 | Yes | 13572 | 30 | 9 | 5 | Low-quality | Genome-fragment | 35.15 | AAI-based (medium-confidence) | 46.75 |
| S8_k141_143961 | 1431 | No | NA | 3 | 0 | 3 | Not-determined | Genome-fragment | NA | NA | 0 |
| S7-final-viral-combined_k141_104726\|\|full | 25535 | Yes | 9282 | 21 | 4 | 9 | Low-quality | Genome-fragment | 21.11 | AAI-based (medium-confidence) | 63.65 |
| S8_k141_247477 | 2993 | No | NA | 5 | 0 | 1 | Not-determined | Genome-fragment | NA | NA | 0 |
| S8_k141_74739 | 1236 | No | NA | 2 | 0 | 2 | Not-determined | Genome-fragment | NA | NA | 0 |
| S8_k141_108571 | 1175 | No | NA | 2 | 0 | 2 | Not-determined | Genome-fragment | NA | NA | 0 |
| S3_k141_40590 | 25640 | No | NA | 57 | 3 | 0 | Low-quality | Genome-fragment | 24.54 | AAI-based (high-confidence) | 0 |
| S8_k141_189780 | 2318 | No | NA | 2 | 0 | 0 | Not-determined | Genome-fragment | NA | NA | 0 |
| S8_k141_63281 | 3428 | No | NA | 2 | 0 | 2 | Not-determined | Genome-fragment | NA | NA | 0 |
| S8_k141_108613 | 1107 | No | NA | 1 | 0 | 1 | Not-determined | Genome-fragment | NA | NA | 0 |
| S8_k141_51148 | 4073 | No | NA | 3 | 0 | 2 | Not-determined | Genome-fragment | NA | NA | 0 |
| S11-final-viral-combined_k141_48227\|\|full | 25658 | Yes | 16039 | 30 | 8 | 7 | Low-quality | Genome-fragment | 32.07 | AAI-based (medium-confidence) | 37.49 |
| S8_k141_28693 | 1459 | No | NA | 1 | 0 | 0 | Not-determined | Genome-fragment | NA | NA | 0 |
| S8_k141_63337 | 1439 | No | NA | 2 | 0 | 2 | Not-determined | Genome-fragment | NA | NA | 0 |
| S2-final-viral-combined_k141_209819\|\|full | 25672 | No | NA | 36 | 9 | 0 | Low-quality | Genome-fragment | 49.81 | AAI-based (high-confidence) | 0 |
| S8_k141_179012 | 1311 | No | NA | 1 | 0 | 1 | Not-determined | Genome-fragment | NA | NA | 0 |
| S8_k141_132216 | 1400 | No | NA | 2 | 0 | 2 | Not-determined | Genome-fragment | NA | NA | 0 |
| S8_k141_144100 | 1477 | No | NA | 1 | 0 | 1 | Not-determined | Genome-fragment | NA | NA | 0 |
| S8_k141_63398 | 1231 | No | NA | 1 | 0 | 1 | Not-determined | Genome-fragment | NA | NA | 0 |
| S11-final-viral-combined_k141_59049\|\|full | 25737 | No | NA | 37 | 3 | 6 | Low-quality | Genome-fragment | 46.92 | AAI-based (high-confidence) | 0 |
| S8_k141_15762 | 3326 | No | NA | 2 | 0 | 1 | Not-determined | Genome-fragment | NA | NA | 0 |
| S8_k141_236289 | 5463 | No | NA | 2 | 0 | 2 | Not-determined | Genome-fragment | NA | NA | 0 |
| S8_k141_15777 | 1070 | No | NA | 1 | 0 | 1 | Not-determined | Genome-fragment | NA | NA | 0 |
| S8_k141_212707 | 1381 | No | NA | 4 | 0 | 0 | Not-determined | Genome-fragment | NA | NA | 0 |
| S8_k141_132246 | 3316 | No | NA | 2 | 0 | 1 | Not-determined | Genome-fragment | NA | NA | 0 |
| S8_k141_223983 | 1226 | No | NA | 1 | 0 | 0 | Not-determined | Genome-fragment | NA | NA | 0 |
| S8_k141_144173 | 1109 | No | NA | 1 | 0 | 1 | Not-determined | Genome-fragment | NA | NA | 0 |
| S8_k141_155273 | 1092 | No | NA | 1 | 0 | 0 | Not-determined | Genome-fragment | NA | NA | 0 |
| S8_k141_155283 | 1137 | No | NA | 2 | 0 | 0 | Not-determined | Genome-fragment | NA | NA | 0 |
| S4-final-viral-combined_k141_156635\|\|full | 25753 | Yes | 12471 | 29 | 3 | 9 | Low-quality | Genome-fragment | 5.24 | AAI-based (medium-confidence) | 51.57 |
| S8_k141_108769 | 5758 | No | NA | 4 | 0 | 2 | Not-determined | Genome-fragment | NA | NA | 0 |
| S8_k141_28814 | 2493 | No | NA | 1 | 0 | 1 | Not-determined | Genome-fragment | NA | NA | 0 |
| S8_k141_132299 | 1194 | No | NA | 1 | 0 | 0 | Not-determined | Genome-fragment | NA | NA | 0 |
| S8_k141_4806 | 1119 | No | NA | 2 | 0 | 1 | Not-determined | Genome-fragment | NA | NA | 0 |
| S8_k141_179134 | 2030 | No | NA | 1 | 0 | 1 | Not-determined | Genome-fragment | NA | NA | 0 |
| S8_k141_51263 | 1148 | No | NA | 1 | 0 | 0 | Not-determined | Genome-fragment | NA | NA | 0 |
| S8_k141_132312 | 4913 | No | NA | 1 | 0 | 1 | Not-determined | Genome-fragment | NA | NA | 0 |
| S12-final-viral-combined_k141_141246\|\|full | 25775 | No | NA | 28 | 1 | 20 | Low-quality | Genome-fragment | 39 | HMM-based (lower-bound) | 0 |
| S8_k141_63444 | 3692 | No | NA | 3 | 0 | 2 | Not-determined | Genome-fragment | NA | NA | 0 |
| S8_k141_120349 | 1060 | No | NA | 2 | 0 | 2 | Not-determined | Genome-fragment | NA | NA | 0 |
| S8_k141_85682 | 5328 | No | NA | 8 | 0 | 1 | Not-determined | Genome-fragment | NA | NA | 0 |
| S8_k141_4836 | 3473 | No | NA | 3 | 0 | 2 | Not-determined | Genome-fragment | NA | NA | 0 |
| S11_k141_41490 | 25852 | No | NA | 23 | 0 | 8 | Low-quality | Genome-fragment | 31.93 | AAI-based (medium-confidence) | 0 |
| S8_k141_200220 | 1219 | No | NA | 1 | 0 | 1 | Not-determined | Genome-fragment | NA | NA | 0 |
| S8_k141_247613 | 1532 | No | NA | 1 | 0 | 1 | Not-determined | Genome-fragment | NA | NA | 0 |
| S8_k141_120374 | 1012 | No | NA | 1 | 0 | 0 | Not-determined | Genome-fragment | NA | NA | 0 |
| S8_k141_224024 | 9132 | No | NA | 6 | 0 | 1 | Not-determined | Genome-fragment | NA | NA | 0 |
| S8_k141_236433 | 2224 | No | NA | 5 | 0 | 0 | Not-determined | Genome-fragment | NA | NA | 0 |
| S8_k141_120393 | 1427 | No | NA | 1 | 0 | 1 | Not-determined | Genome-fragment | NA | NA | 0 |
| S8_k141_190019 | 1541 | No | NA | 2 | 0 | 1 | Not-determined | Genome-fragment | NA | NA | 0 |
| S8_k141_85733 | 1097 | No | NA | 1 | 0 | 1 | Not-determined | Genome-fragment | NA | NA | 0 |
| S8_k141_4877 | 1187 | No | NA | 2 | 0 | 2 | Not-determined | Genome-fragment | NA | NA | 0 |
| S8_k141_190055 | 1256 | No | NA | 3 | 0 | 0 | Not-determined | Genome-fragment | NA | NA | 0 |
| S8_k141_74977 | 1575 | No | NA | 1 | 0 | 1 | Not-determined | Genome-fragment | NA | NA | 0 |
| S8_k141_179211 | 1191 | No | NA | 2 | 0 | 1 | Not-determined | Genome-fragment | NA | NA | 0 |
| S8_k141_212847 | 1945 | No | NA | 4 | 0 | 2 | Not-determined | Genome-fragment | NA | NA | 0 |
| S3-final-viral-combined_k141_120607\|\|0_partial | 25868 | Yes | 13870 | 31 | 5 | 6 | Low-quality | Genome-fragment | 22.3 | HMM-based (lower-bound) | 46.38 |
| S6-final-viral-combined_k141_139992\|\|full | 25964 | Yes | 8855 | 28 | 3 | 14 | Low-quality | Genome-fragment | 16.85 | HMM-based (lower-bound) | 65.9 |
| S2-final-viral-combined_k141_105844\|\|full | 25991 | Yes | 15638 | 36 | 9 | 4 | Low-quality | Genome-fragment | 41.4 | AAI-based (high-confidence) | 39.83 |
| S7-final-viral-combined_k141_83431\|\|full | 26021 | No | NA | 20 | 1 | 2 | Low-quality | Genome-fragment | 34.45 | HMM-based (lower-bound) | 0 |
| S6-final-viral-combined_k141_17119\|\|full | 26042 | No | NA | 22 | 13 | 0 | Low-quality | Genome-fragment | 32.26 | AAI-based (high-confidence) | 0 |
| S8_k141_270538 | 2215 | No | NA | 2 | 0 | 1 | Not-determined | Genome-fragment | NA | NA | 0 |
| S8_k141_166725 | 11468 | No | NA | 6 | 0 | 2 | Not-determined | Genome-fragment | NA | NA | 0 |
| S12-final-viral-combined_k141_8659\|\|full | 26073 | No | NA | 50 | 13 | 0 | Low-quality | Genome-fragment | 31.21 | AAI-based (high-confidence) | 0 |
| S12-final-viral-combined_k141_240489\|\|full | 26099 | Yes | 13614 | 24 | 4 | 3 | Low-quality | Genome-fragment | 24.93 | HMM-based (lower-bound) | 47.84 |
| S8_k141_236566 | 1307 | No | NA | 1 | 0 | 0 | Not-determined | Genome-fragment | NA | NA | 0 |
| S2-final-viral-combined_k141_202570\|\|0_partial | 26152 | Yes | 14189 | 35 | 7 | 9 | Low-quality | Genome-fragment | 17.38 | HMM-based (lower-bound) | 45.74 |
| S5_k141_819 | 26171 | No | NA | 38 | 7 | 2 | Low-quality | Genome-fragment | 44.34 | AAI-based (high-confidence) | 0 |
| S8_k141_166779 | 5626 | No | NA | 3 | 0 | 3 | Not-determined | Genome-fragment | NA | NA | 0 |
| S4-final-viral-combined_k141_131178\|\|0_partial | 26228 | Yes | 12944 | 34 | 6 | 8 | Low-quality | Genome-fragment | 16.34 | HMM-based (lower-bound) | 50.65 |
| S8_k141_190168 | 3552 | No | NA | 3 | 0 | 3 | Not-determined | Genome-fragment | NA | NA | 0 |
| S8_k141_97881 | 1017 | No | NA | 2 | 0 | 0 | Not-determined | Genome-fragment | NA | NA | 0 |
| S3-final-viral-combined_k141_16291\|\|full | 26366 | No | NA | 28 | 16 | 0 | Low-quality | Genome-fragment | 43.51 | AAI-based (high-confidence) | 0 |
| S8_k141_108901 | 1344 | No | NA | 1 | 0 | 1 | Not-determined | Genome-fragment | NA | NA | 0 |
| S8_k141_212957 | 1040 | No | NA | 1 | 0 | 1 | Not-determined | Genome-fragment | NA | NA | 0 |
| S8_k141_200429 | 4534 | No | NA | 4 | 0 | 1 | Not-determined | Genome-fragment | NA | NA | 0 |
| S8_k141_166822 | 1541 | No | NA | 1 | 0 | 0 | Not-determined | Genome-fragment | NA | NA | 0 |
| S8_k141_120562 | 4054 | No | NA | 3 | 0 | 1 | Not-determined | Genome-fragment | NA | NA | 0 |
| S8_k141_5091 | 1347 | No | NA | 1 | 0 | 1 | Not-determined | Genome-fragment | NA | NA | 0 |
| S3-final-viral-combined_k141_3474\|\|full | 26555 | Yes | 15048 | 36 | 6 | 6 | Low-quality | Genome-fragment | 19 | HMM-based (lower-bound) | 43.33 |
| S7-final-viral-combined_k141_127766\|\|full | 26560 | Yes | 17929 | 40 | 5 | 4 | Low-quality | Genome-fragment | 30.61 | AAI-based (high-confidence) | 32.5 |
| S8_k141_155590 | 1276 | No | NA | 2 | 0 | 2 | Not-determined | Genome-fragment | NA | NA | 0 |
| S8_k141_213010 | 3452 | No | NA | 4 | 0 | 3 | Not-determined | Genome-fragment | NA | NA | 0 |
| S8_k141_236662 | 3196 | No | NA | 2 | 0 | 2 | Not-determined | Genome-fragment | NA | NA | 0 |
| S8_k141_270733 | 1243 | No | NA | 3 | 0 | 0 | Not-determined | Genome-fragment | NA | NA | 0 |
| S8_k141_40452 | 4326 | No | NA | 2 | 0 | 2 | Not-determined | Genome-fragment | NA | NA | 0 |
| S8_k141_224253 | 1274 | No | NA | 2 | 0 | 2 | Not-determined | Genome-fragment | NA | NA | 0 |
| S5-final-viral-combined_k141_10686\|\|full | 26680 | Yes | 9384 | 29 | 3 | 12 | Low-quality | Genome-fragment | 24.99 | AAI-based (medium-confidence) | 64.83 |
| S8_k141_257883 | 5379 | No | NA | 4 | 0 | 2 | Not-determined | Genome-fragment | NA | NA | 0 |
| S8_k141_257887 | 4716 | No | NA | 6 | 0 | 1 | Not-determined | Genome-fragment | NA | NA | 0 |
| S3_k141_59235 | 26715 | No | NA | 25 | 3 | 0 | Low-quality | Genome-fragment | 17.7 | AAI-based (high-confidence) | 0 |
| S8_k141_98050 | 1164 | No | NA | 1 | 0 | 0 | Not-determined | Genome-fragment | NA | NA | 0 |
| S8_k141_132494 | 3293 | No | NA | 3 | 0 | 3 | Not-determined | Genome-fragment | NA | NA | 0 |
| S2-final-viral-combined_k141_87977\|\|full | 26727 | Yes | 15147 | 28 | 9 | 6 | Low-quality | Genome-fragment | 40.07 | AAI-based (high-confidence) | 43.33 |
| S8_k141_40545 | 1320 | No | NA | 2 | 0 | 1 | Not-determined | Genome-fragment | NA | NA | 0 |
| S8_k141_224339 | 2496 | No | NA | 1 | 0 | 0 | Not-determined | Genome-fragment | NA | NA | 0 |
| S8_k141_224341 | 2862 | No | NA | 3 | 0 | 2 | Not-determined | Genome-fragment | NA | NA | 0 |
| S8_k141_132549 | 1485 | No | NA | 2 | 0 | 1 | Not-determined | Genome-fragment | NA | NA | 0 |
| S8_k141_29366 | 1000 | No | NA | 2 | 0 | 1 | Not-determined | Genome-fragment | NA | NA | 0 |
| S7_k141_108011 | 26865 | No | NA | 22 | 18 | 0 | Low-quality | Genome-fragment | 44.79 | AAI-based (medium-confidence) | 0 |
| S8_k141_213066 | 1355 | No | NA | 1 | 0 | 1 | Not-determined | Genome-fragment | NA | NA | 0 |
| S9-final-viral-combined_k141_103062\|\|full | 26951 | Yes | 13645 | 37 | 7 | 12 | Low-quality | Genome-fragment | 30.74 | AAI-based (high-confidence) | 49.37 |
| S8_k141_98155 | 1084 | No | NA | 1 | 0 | 0 | Not-determined | Genome-fragment | NA | NA | 0 |
| S8_k141_108983 | 1051 | No | NA | 1 | 0 | 1 | Not-determined | Genome-fragment | NA | NA | 0 |
| S8_k141_190339 | 1206 | No | NA | 1 | 0 | 1 | Not-determined | Genome-fragment | NA | NA | 0 |
| S8_k141_224389 | 1434 | No | NA | 1 | 0 | 0 | Not-determined | Genome-fragment | NA | NA | 0 |
| S17-final-viral-combined_k141_77807\|\|full | 26977 | No | NA | 32 | 9 | 0 | Low-quality | Genome-fragment | 41.23 | AAI-based (high-confidence) | 0 |
| S8_k141_98177 | 1392 | No | NA | 1 | 0 | 1 | Not-determined | Genome-fragment | NA | NA | 0 |
| S8_k141_190361 | 1452 | No | NA | 1 | 0 | 1 | Not-determined | Genome-fragment | NA | NA | 0 |
| S8_k141_200582 | 2556 | No | NA | 2 | 0 | 2 | Not-determined | Genome-fragment | NA | NA | 0 |
| S8_k141_51752 | 2545 | No | NA | 3 | 0 | 0 | Not-determined | Genome-fragment | NA | NA | 0 |
| S11_k141_181614 | 27093 | No | NA | 32 | 4 | 1 | Low-quality | Genome-fragment | 25.5 | AAI-based (high-confidence) | 0 |
| S8_k141_29405 | 1287 | No | NA | 1 | 0 | 1 | Not-determined | Genome-fragment | NA | NA | 0 |
| S8_k141_120748 | 3569 | No | NA | 4 | 0 | 3 | Not-determined | Genome-fragment | NA | NA | 0 |
| S8_k141_236757 | 1246 | No | NA | 1 | 0 | 1 | Not-determined | Genome-fragment | NA | NA | 0 |
| S8_k141_75210 | 1080 | No | NA | 2 | 0 | 1 | Not-determined | Genome-fragment | NA | NA | 0 |
| S8_k141_29468 | 4206 | No | NA | 8 | 0 | 8 | Not-determined | Genome-fragment | NA | NA | 0 |
| S8_k141_224474 | 1448 | No | NA | 1 | 0 | 1 | Not-determined | Genome-fragment | NA | NA | 0 |
| S8_k141_248028 | 1562 | No | NA | 1 | 0 | 0 | Not-determined | Genome-fragment | NA | NA | 0 |
| S8_k141_5323 | 2114 | No | NA | 1 | 0 | 0 | Not-determined | Genome-fragment | NA | NA | 0 |
| S8_k141_29513 | 1194 | No | NA | 2 | 0 | 1 | Not-determined | Genome-fragment | NA | NA | 0 |
| S8_k141_29524 | 1261 | No | NA | 1 | 0 | 1 | Not-determined | Genome-fragment | NA | NA | 0 |
| S8_k141_5339 | 1447 | No | NA | 1 | 0 | 1 | Not-determined | Genome-fragment | NA | NA | 0 |
| S8_k141_144256 | 1325 | No | NA | 1 | 0 | 1 | Not-determined | Genome-fragment | NA | NA | 0 |
| S5-final-viral-combined_k141_18702\|\|full | 27098 | Yes | 9819 | 31 | 3 | 7 | Low-quality | Genome-fragment | 11.43 | HMM-based (lower-bound) | 63.76 |
| S8_k141_5348 | 1202 | No | NA | 2 | 0 | 0 | Not-determined | Genome-fragment | NA | NA | 0 |
| S2-final-viral-combined_k141_103450\|\|full | 27098 | Yes | 14356 | 30 | 6 | 9 | Low-quality | Genome-fragment | 18.13 | HMM-based (lower-bound) | 47.02 |
| S8_k141_224530 | 1782 | No | NA | 1 | 0 | 1 | Not-determined | Genome-fragment | NA | NA | 0 |
| S8_k141_167075 | 1708 | No | NA | 1 | 0 | 0 | Not-determined | Genome-fragment | NA | NA | 0 |
| S8_k141_248123 | 1195 | No | NA | 2 | 0 | 1 | Not-determined | Genome-fragment | NA | NA | 0 |
| S8_k141_51886 | 4285 | No | NA | 4 | 0 | 2 | Not-determined | Genome-fragment | NA | NA | 0 |
| S8_k141_98284 | 2182 | No | NA | 1 | 0 | 1 | Not-determined | Genome-fragment | NA | NA | 0 |
| S8_k141_155902 | 1252 | No | NA | 1 | 0 | 0 | Not-determined | Genome-fragment | NA | NA | 0 |
| S8_k141_109170 | 1023 | No | NA | 1 | 0 | 1 | Not-determined | Genome-fragment | NA | NA | 0 |
| S8_k141_248191 | 1210 | No | NA | 2 | 0 | 2 | Not-determined | Genome-fragment | NA | NA | 0 |
| S11-final-viral-combined_k141_106106\|\|full | 27181 | Yes | 13924 | 32 | 3 | 8 | Low-quality | Genome-fragment | 32.97 | AAI-based (medium-confidence) | 48.77 |
| S8_k141_29650 | 1426 | No | NA | 1 | 0 | 1 | Not-determined | Genome-fragment | NA | NA | 0 |
| S8_k141_120942 | 1066 | No | NA | 1 | 0 | 1 | Not-determined | Genome-fragment | NA | NA | 0 |
| S8_k141_63894 | 4883 | No | NA | 2 | 0 | 1 | Not-determined | Genome-fragment | NA | NA | 0 |
| S8_k141_63897 | 2053 | No | NA | 2 | 0 | 1 | Not-determined | Genome-fragment | NA | NA | 0 |
| S8_k141_40720 | 1125 | No | NA | 2 | 0 | 1 | Not-determined | Genome-fragment | NA | NA | 0 |
| S8_k141_132837 | 1091 | No | NA | 2 | 0 | 1 | Not-determined | Genome-fragment | NA | NA | 0 |
| S8_k141_86148 | 1033 | No | NA | 2 | 0 | 0 | Not-determined | Genome-fragment | NA | NA | 0 |
| S8_k141_179736 | 1071 | No | NA | 1 | 0 | 0 | Not-determined | Genome-fragment | NA | NA | 0 |
| S8_k141_224682 | 1100 | No | NA | 1 | 0 | 1 | Not-determined | Genome-fragment | NA | NA | 0 |
| S8_k141_258202 | 1018 | No | NA | 1 | 0 | 0 | Not-determined | Genome-fragment | NA | NA | 0 |
| S8_k141_5515 | 1235 | No | NA | 1 | 0 | 0 | Not-determined | Genome-fragment | NA | NA | 0 |
| S5-final-viral-combined_k141_182127\|\|0_partial | 27398 | Yes | 20784 | 30 | 3 | 6 | Low-quality | Genome-fragment | 48.72 | AAI-based (medium-confidence) | 24.14 |
| S8_k141_109311 | 1040 | No | NA | 1 | 0 | 1 | Not-determined | Genome-fragment | NA | NA | 0 |
| S8_k141_105856 | 27400 | Yes | 23601 | 44 | 3 | 3 | Low-quality | Genome-fragment | 43.03 | AAI-based (high-confidence) | 13.86 |
| S8_k141_155998 | 1222 | No | NA | 2 | 0 | 1 | Not-determined | Genome-fragment | NA | NA | 0 |
| S8_k141_224757 | 1148 | No | NA | 3 | 0 | 0 | Not-determined | Genome-fragment | NA | NA | 0 |
| S8_k141_144496 | 1025 | No | NA | 1 | 0 | 1 | Not-determined | Genome-fragment | NA | NA | 0 |
| S9-final-viral-combined_k141_93806\|\|0_partial | 27451 | Yes | 9264 | 26 | 3 | 12 | Low-quality | Genome-fragment | 17.63 | HMM-based (lower-bound) | 66.25 |
| S8_k141_248317 | 1126 | No | NA | 1 | 0 | 1 | Not-determined | Genome-fragment | NA | NA | 0 |
| S11-final-viral-combined_k141_148610\|\|full | 27598 | No | NA | 24 | 3 | 0 | Low-quality | Genome-fragment | 24.92 | AAI-based (medium-confidence) | 0 |
| S8_k141_40865 | 6015 | No | NA | 8 | 0 | 1 | Not-determined | Genome-fragment | NA | NA | 0 |
| S1_k141_187812 | 27618 | No | NA | 37 | 7 | 0 | Low-quality | Genome-fragment | 40.87 | AAI-based (high-confidence) | 0 |
| S8_k141_179859 | 1017 | No | NA | 1 | 0 | 0 | Not-determined | Genome-fragment | NA | NA | 0 |
| S8_k141_98414 | 1192 | No | NA | 2 | 0 | 1 | Not-determined | Genome-fragment | NA | NA | 0 |
| S8_k141_109393 | 1327 | No | NA | 2 | 0 | 1 | Not-determined | Genome-fragment | NA | NA | 0 |
| S8_k141_121090 | 1014 | No | NA | 1 | 0 | 1 | Not-determined | Genome-fragment | NA | NA | 0 |
| S8_k141_52117 | 4888 | No | NA | 5 | 0 | 0 | Not-determined | Genome-fragment | NA | NA | 0 |
| S8_k141_190666 | 1483 | No | NA | 2 | 0 | 1 | Not-determined | Genome-fragment | NA | NA | 0 |
| S8_k141_5594 | 1088 | No | NA | 2 | 0 | 2 | Not-determined | Genome-fragment | NA | NA | 0 |
| S8_k141_156051 | 1128 | No | NA | 1 | 0 | 0 | Not-determined | Genome-fragment | NA | NA | 0 |
| S8_k141_190691 | 1660 | No | NA | 1 | 0 | 1 | Not-determined | Genome-fragment | NA | NA | 0 |
| S7-final-viral-combined_k141_173684\|\|full | 27626 | Yes | 11330 | 42 | 7 | 11 | Low-quality | Genome-fragment | 30.68 | AAI-based (medium-confidence) | 58.99 |
| S11-final-viral-combined_k141_233266\|\|full | 27649 | Yes | 16140 | 35 | 17 | 8 | Low-quality | Genome-fragment | 43.6 | AAI-based (high-confidence) | 41.63 |
| S8_k141_41025 | 1164 | No | NA | 2 | 0 | 2 | Not-determined | Genome-fragment | NA | NA | 0 |
| S8_k141_144647 | 1631 | No | NA | 1 | 0 | 1 | Not-determined | Genome-fragment | NA | NA | 0 |
| S8_k141_200949 | 1084 | No | NA | 2 | 0 | 2 | Not-determined | Genome-fragment | NA | NA | 0 |
| S11-final-viral-combined_k141_181500\|\|0_partial | 27696 | No | NA | 21 | 2 | 6 | Low-quality | Genome-fragment | 20.33 | HMM-based (lower-bound) | 0 |
| S8_k141_224958 | 2087 | No | NA | 2 | 0 | 2 | Not-determined | Genome-fragment | NA | NA | 0 |
| S8_k141_190763 | 2051 | No | NA | 2 | 0 | 0 | Not-determined | Genome-fragment | NA | NA | 0 |
| S8_k141_16523 | 1037 | No | NA | 2 | 0 | 2 | Not-determined | Genome-fragment | NA | NA | 0 |
| S8_k141_271263 | 1597 | No | NA | 2 | 0 | 0 | Not-determined | Genome-fragment | NA | NA | 0 |
| S12_k141_355194 | 27763 | No | NA | 39 | 1 | 2 | Low-quality | Genome-fragment | 15.06 | HMM-based (lower-bound) | 0 |
| S8_k141_41059 | 2793 | No | NA | 1 | 0 | 1 | Not-determined | Genome-fragment | NA | NA | 0 |
| S8_k141_29872 | 1580 | No | NA | 2 | 0 | 1 | Not-determined | Genome-fragment | NA | NA | 0 |
| S8_k141_98559 | 1954 | No | NA | 2 | 0 | 1 | Not-determined | Genome-fragment | NA | NA | 0 |
| S2-final-viral-combined_k141_74070\|\|full | 27837 | Yes | 18216 | 33 | 5 | 5 | Low-quality | Genome-fragment | 23.69 | HMM-based (lower-bound) | 34.56 |
| S8_k141_190781 | 31564 | No | NA | 45 | 23 | 0 | Medium-quality | Genome-fragment | 69.89 | AAI-based (high-confidence) | 0 |
| S8_k141_156182 | 1448 | No | NA | 1 | 0 | 1 | Not-determined | Genome-fragment | NA | NA | 0 |
| S2-final-viral-combined_k141_224593\|\|full | 27891 | No | NA | 38 | 2 | 1 | Low-quality | Genome-fragment | 16.04 | HMM-based (lower-bound) | 0 |
| S8_k141_201049 | 1065 | No | NA | 1 | 0 | 1 | Not-determined | Genome-fragment | NA | NA | 0 |
| S11_k141_125234 | 28039 | No | NA | 39 | 3 | 0 | Low-quality | Genome-fragment | 26.39 | AAI-based (high-confidence) | 0 |
| S8_k141_180044 | 2003 | No | NA | 1 | 0 | 1 | Not-determined | Genome-fragment | NA | NA | 0 |
| S8_k141_201062 | 2011 | No | NA | 3 | 0 | 1 | Not-determined | Genome-fragment | NA | NA | 0 |
| S8_k141_64364 | 2498 | No | NA | 2 | 0 | 0 | Not-determined | Genome-fragment | NA | NA | 0 |
| S8_k141_248503 | 8670 | No | NA | 4 | 0 | 3 | Not-determined | Genome-fragment | NA | NA | 0 |
| S8_k141_121287 | 1093 | No | NA | 2 | 0 | 1 | Not-determined | Genome-fragment | NA | NA | 0 |
| S8_k141_41205 | 3691 | No | NA | 1 | 0 | 0 | Not-determined | Genome-fragment | NA | NA | 0 |
| S8_k141_5725 | 1516 | No | NA | 2 | 0 | 0 | Not-determined | Genome-fragment | NA | NA | 0 |
| S8_k141_271341 | 1392 | No | NA | 2 | 0 | 1 | Not-determined | Genome-fragment | NA | NA | 0 |
| S8_k141_167609 | 1275 | No | NA | 2 | 0 | 2 | Not-determined | Genome-fragment | NA | NA | 0 |
| S8_k141_213612 | 1376 | No | NA | 2 | 0 | 1 | Not-determined | Genome-fragment | NA | NA | 0 |
| S8_k141_133250 | 1049 | No | NA | 1 | 0 | 0 | Not-determined | Genome-fragment | NA | NA | 0 |
| S2-final-viral-combined_k141_130098\|\|full | 28097 | No | NA | 31 | 1 | 15 | Low-quality | Genome-fragment | 31.54 | HMM-based (lower-bound) | 0 |
| S8_k141_144835 | 1428 | No | NA | 3 | 0 | 0 | Not-determined | Genome-fragment | NA | NA | 0 |
| S8_k141_64471 | 2052 | No | NA | 1 | 0 | 1 | Not-determined | Genome-fragment | NA | NA | 0 |
| S8_k141_180176 | 1693 | No | NA | 1 | 0 | 1 | Not-determined | Genome-fragment | NA | NA | 0 |
| S5-final-viral-combined_k141_98955\|\|full | 28123 | No | NA | 26 | 2 | 11 | Low-quality | Genome-fragment | 49.32 | HMM-based (lower-bound) | 0 |
| S9-final-viral-combined_k141_88140\|\|0_partial | 28132 | Yes | 9645 | 26 | 3 | 12 | Low-quality | Genome-fragment | 18.36 | HMM-based (lower-bound) | 65.72 |
| S8_k141_52398 | 14079 | No | NA | 8 | 0 | 5 | Not-determined | Genome-fragment | NA | NA | 0 |
| S17-final-viral-combined_k141_115233\|\|0_partial | 28148 | No | NA | 21 | 2 | 1 | Low-quality | Genome-fragment | 44.31 | AAI-based (high-confidence) | 0 |
| S8_k141_225150 | 1184 | No | NA | 1 | 0 | 0 | Not-determined | Genome-fragment | NA | NA | 0 |
| S8_k141_156283 | 3190 | No | NA | 1 | 0 | 0 | Not-determined | Genome-fragment | NA | NA | 0 |
| S8_k141_237400 | 3620 | No | NA | 7 | 0 | 2 | Not-determined | Genome-fragment | NA | NA | 0 |
| S8_k141_109732 | 3009 | No | NA | 3 | 0 | 1 | Not-determined | Genome-fragment | NA | NA | 0 |
| S8_k141_144935 | 1740 | No | NA | 5 | 0 | 4 | Not-determined | Genome-fragment | NA | NA | 0 |
| S8_k141_52462 | 1110 | No | NA | 1 | 0 | 1 | Not-determined | Genome-fragment | NA | NA | 0 |
| S8-final-viral-combined_k141_71889\|\|full | 28151 | Yes | 12471 | 32 | 8 | 14 | Low-quality | Genome-fragment | 19.71 | HMM-based (lower-bound) | 55.7 |
| S9-final-viral-combined_k141_86623\|\|full | 28154 | Yes | 13307 | 31 | 19 | 10 | Low-quality | Genome-fragment | 28.22 | AAI-based (medium-confidence) | 52.73 |
| S7-final-viral-combined_k141_179127\|\|full | 28156 | Yes | 7400 | 27 | 4 | 13 | Low-quality | Genome-fragment | 8.18 | HMM-based (lower-bound) | 73.72 |
| S8_k141_180300 | 1007 | No | NA | 1 | 0 | 1 | Not-determined | Genome-fragment | NA | NA | 0 |
| S5-final-viral-combined_k141_91275\|\|0_partial | 28179 | No | NA | 15 | 1 | 10 | Low-quality | Genome-fragment | 15.28 | HMM-based (lower-bound) | 0 |
| S11-final-viral-combined_k141_41630\|\|full | 28311 | No | NA | 21 | 0 | 17 | Low-quality | Genome-fragment | 6.95 | AAI-based (medium-confidence) | 0 |
| S8_k141_248656 | 1405 | No | NA | 1 | 0 | 0 | Not-determined | Genome-fragment | NA | NA | 0 |
| S8_k141_75907 | 1240 | No | NA | 1 | 0 | 1 | Not-determined | Genome-fragment | NA | NA | 0 |
| S8_k141_98755 | 5879 | No | NA | 4 | 0 | 2 | Not-determined | Genome-fragment | NA | NA | 0 |
| S8_k141_156395 | 1034 | No | NA | 1 | 0 | 0 | Not-determined | Genome-fragment | NA | NA | 0 |
| S8_k141_30102 | 1792 | No | NA | 1 | 0 | 1 | Not-determined | Genome-fragment | NA | NA | 0 |
| S8_k141_213713 | 2347 | No | NA | 3 | 0 | 0 | Not-determined | Genome-fragment | NA | NA | 0 |
| S8_k141_133445 | 1597 | No | NA | 1 | 0 | 1 | Not-determined | Genome-fragment | NA | NA | 0 |
| S8_k141_180393 | 3057 | No | NA | 1 | 0 | 1 | Not-determined | Genome-fragment | NA | NA | 0 |
| S2-final-viral-combined_k141_206808\|\|full | 28331 | Yes | 6187 | 31 | 3 | 9 | Low-quality | Genome-fragment | 5.01 | AAI-based (high-confidence) | 78.16 |
| S8_k141_52559 | 5034 | No | NA | 5 | 0 | 2 | Not-determined | Genome-fragment | NA | NA | 0 |
| S8_k141_258577 | 1513 | No | NA | 1 | 0 | 0 | Not-determined | Genome-fragment | NA | NA | 0 |
| S8_k141_5980 | 1013 | No | NA | 1 | 0 | 1 | Not-determined | Genome-fragment | NA | NA | 0 |
| S8_k141_30206 | 1280 | No | NA | 1 | 0 | 0 | Not-determined | Genome-fragment | NA | NA | 0 |
| S1-final-viral-combined_k141_100645\|\|full | 28346 | No | NA | 31 | 2 | 16 | Low-quality | Genome-fragment | 44.89 | HMM-based (lower-bound) | 0 |
| S8_k141_64635 | 1163 | No | NA | 2 | 0 | 2 | Not-determined | Genome-fragment | NA | NA | 0 |
| S2-final-viral-combined_k141_76645\|\|full | 28413 | No | NA | 53 | 6 | 1 | Low-quality | Genome-fragment | 30.81 | AAI-based (high-confidence) | 0 |
| S8_k141_30227 | 3582 | No | NA | 2 | 0 | 0 | Not-determined | Genome-fragment | NA | NA | 0 |
| S1_k141_218094 | 28437 | No | NA | 25 | 1 | 3 | Low-quality | Genome-fragment | 8.04 | HMM-based (lower-bound) | 0 |
| S8_k141_248815 | 1010 | No | NA | 1 | 0 | 1 | Not-determined | Genome-fragment | NA | NA | 0 |
| S2-final-viral-combined_k141_178485\|\|full | 28492 | Yes | 13785 | 39 | 6 | 7 | Low-quality | Genome-fragment | 17.4 | HMM-based (lower-bound) | 51.62 |
| S8_k141_109959 | 1497 | No | NA | 1 | 0 | 1 | Not-determined | Genome-fragment | NA | NA | 0 |
| S8_k141_201452 | 1120 | No | NA | 1 | 0 | 0 | Not-determined | Genome-fragment | NA | NA | 0 |
| S8-final-viral-combined_k141_34759\|\|1_partial | 28603 | Yes | 12963 | 31 | 4 | 5 | Low-quality | Genome-fragment | 17.62 | HMM-based (lower-bound) | 54.68 |
| S12-final-viral-combined_k141_57174\|\|full | 28677 | Yes | 10572 | 26 | 7 | 8 | Low-quality | Genome-fragment | 15.02 | HMM-based (lower-bound) | 63.13 |
| S5_k141_101569 | 28783 | No | NA | 27 | 1 | 6 | Low-quality | Genome-fragment | 15.61 | HMM-based (lower-bound) | 0 |
| S8_k141_201479 | 1218 | No | NA | 1 | 0 | 1 | Not-determined | Genome-fragment | NA | NA | 0 |
| S8_k141_41541 | 3062 | No | NA | 2 | 0 | 1 | Not-determined | Genome-fragment | NA | NA | 0 |
| S2-final-viral-combined_k141_240006\|\|full | 29031 | No | NA | 29 | 1 | 8 | Low-quality | Genome-fragment | 32.59 | HMM-based (lower-bound) | 0 |
| S8_k141_213901 | 1049 | No | NA | 2 | 0 | 2 | Not-determined | Genome-fragment | NA | NA | 0 |
| S8_k141_213916 | 1839 | No | NA | 2 | 0 | 0 | Not-determined | Genome-fragment | NA | NA | 0 |
| S9-final-viral-combined_k141_65073\|\|0_partial | 29054 | Yes | 17512 | 29 | 2 | 7 | Low-quality | Genome-fragment | 38.27 | HMM-based (lower-bound) | 39.73 |
| S8_k141_225446 | 1260 | No | NA | 1 | 0 | 0 | Not-determined | Genome-fragment | NA | NA | 0 |
| S8_k141_237621 | 3310 | No | NA | 2 | 0 | 2 | Not-determined | Genome-fragment | NA | NA | 0 |
| S8_k141_156586 | 1840 | No | NA | 1 | 0 | 0 | Not-determined | Genome-fragment | NA | NA | 0 |
| S8_k141_168042 | 1062 | No | NA | 2 | 0 | 2 | Not-determined | Genome-fragment | NA | NA | 0 |
| S2_k141_131222 | 29149 | No | NA | 35 | 8 | 1 | Low-quality | Genome-fragment | 29.4 | AAI-based (high-confidence) | 0 |
| S8_k141_201508 | 3864 | No | NA | 3 | 0 | 0 | Not-determined | Genome-fragment | NA | NA | 0 |
| S8_k141_213953 | 1017 | No | NA | 1 | 0 | 1 | Not-determined | Genome-fragment | NA | NA | 0 |
| S9-final-viral-combined_k141_83924\|\|0_partial | 29298 | Yes | 18823 | 28 | 2 | 4 | Low-quality | Genome-fragment | 8.19 | AAI-based (medium-confidence) | 35.75 |
| S8_k141_86882 | 1210 | No | NA | 1 | 0 | 0 | Not-determined | Genome-fragment | NA | NA | 0 |
| S8_k141_237640 | 1484 | No | NA | 2 | 0 | 0 | Not-determined | Genome-fragment | NA | NA | 0 |
| S8_k141_271753 | 1018 | No | NA | 2 | 0 | 2 | Not-determined | Genome-fragment | NA | NA | 0 |
| S17-final-viral-combined_k141_156069\|\|full | 29358 | No | NA | 40 | 6 | 3 | Low-quality | Genome-fragment | 42.89 | HMM-based (lower-bound) | 0 |
| S8_k141_156639 | 1606 | No | NA | 1 | 0 | 1 | Not-determined | Genome-fragment | NA | NA | 0 |
| S8_k141_76104 | 1919 | No | NA | 2 | 0 | 0 | Not-determined | Genome-fragment | NA | NA | 0 |
| S11-final-viral-combined_k141_320503\|\|full | 29459 | Yes | 12828 | 36 | 6 | 12 | Low-quality | Genome-fragment | 16.2 | HMM-based (lower-bound) | 56.45 |
| S8_k141_237666 | 1157 | No | NA | 1 | 0 | 0 | Not-determined | Genome-fragment | NA | NA | 0 |
| S8_k141_191219 | 2338 | No | NA | 3 | 0 | 2 | Not-determined | Genome-fragment | NA | NA | 0 |
| S2_k141_133661 | 29631 | No | NA | 15 | 1 | 0 | Low-quality | Genome-fragment | 29.64 | AAI-based (high-confidence) | 0 |
| S5-final-viral-combined_k141_173239\|\|full | 29634 | Yes | 12361 | 28 | 2 | 10 | Low-quality | Genome-fragment | 13.75 | HMM-based (lower-bound) | 58.29 |
| S8_k141_180583 | 2054 | No | NA | 1 | 0 | 0 | Not-determined | Genome-fragment | NA | NA | 0 |
| S17-final-viral-combined_k141_9123\|\|full | 29706 | Yes | 19581 | 35 | 7 | 5 | Low-quality | Genome-fragment | 38.15 | HMM-based (lower-bound) | 34.08 |
| S8_k141_271803 | 1087 | No | NA | 2 | 0 | 1 | Not-determined | Genome-fragment | NA | NA | 0 |
| S8_k141_180607 | 4904 | No | NA | 5 | 0 | 1 | Not-determined | Genome-fragment | NA | NA | 0 |
| S7-final-viral-combined_k141_8773\|\|0_partial | 29726 | Yes | 16919 | 27 | 3 | 9 | Low-quality | Genome-fragment | 32.2 | HMM-based (lower-bound) | 43.08 |
| S8_k141_76156 | 1612 | No | NA | 2 | 0 | 2 | Not-determined | Genome-fragment | NA | NA | 0 |
| S8_k141_6219 | 1111 | No | NA | 2 | 0 | 0 | Not-determined | Genome-fragment | NA | NA | 0 |
| S8_k141_145293 | 1802 | No | NA | 1 | 0 | 1 | Not-determined | Genome-fragment | NA | NA | 0 |
| S4-final-viral-combined_k141_152020\|\|full | 29773 | No | NA | 31 | 1 | 6 | Low-quality | Genome-fragment | 20.01 | HMM-based (lower-bound) | 0 |
| S8_k141_168199 | 1007 | No | NA | 1 | 0 | 1 | Not-determined | Genome-fragment | NA | NA | 0 |
| S8_k141_145320 | 1275 | No | NA | 1 | 0 | 1 | Not-determined | Genome-fragment | NA | NA | 0 |
| S8_k141_30498 | 31308 | No | NA | 40 | 13 | 2 | Medium-quality | Genome-fragment | 87.47 | AAI-based (medium-confidence) | 0 |
| S8_k141_52795 | 1211 | No | NA | 2 | 0 | 1 | Not-determined | Genome-fragment | NA | NA | 0 |
| S17-final-viral-combined_k141_91111\|\|0_partial | 29941 | Yes | 13690 | 33 | 5 | 7 | Low-quality | Genome-fragment | 16.97 | HMM-based (lower-bound) | 54.28 |
| S8-final-viral-combined_k141_249145\|\|full | 30006 | No | NA | 37 | 1 | 5 | Low-quality | Genome-fragment | 40.32 | HMM-based (lower-bound) | 0 |
| S8_k141_121805 | 12945 | No | NA | 12 | 0 | 9 | Not-determined | Genome-fragment | NA | NA | 0 |
| S8_k141_52819 | 1171 | No | NA | 1 | 0 | 1 | Not-determined | Genome-fragment | NA | NA | 0 |
| S12-final-viral-combined_k141_340844\|\|full | 30009 | No | NA | 30 | 1 | 22 | Low-quality | Genome-fragment | 34.01 | HMM-based (lower-bound) | 0 |
| S8_k141_271874 | 3175 | No | NA | 1 | 0 | 1 | Not-determined | Genome-fragment | NA | NA | 0 |
| S8_k141_41750 | 1129 | No | NA | 1 | 0 | 1 | Not-determined | Genome-fragment | NA | NA | 0 |
| S8_k141_156833 | 1925 | No | NA | 1 | 0 | 1 | Not-determined | Genome-fragment | NA | NA | 0 |
| S8_k141_271894 | 3847 | No | NA | 5 | 0 | 0 | Not-determined | Genome-fragment | NA | NA | 0 |
| S8_k141_271898 | 1332 | No | NA | 2 | 0 | 1 | Not-determined | Genome-fragment | NA | NA | 0 |
| S8_k141_168296 | 1067 | No | NA | 1 | 0 | 1 | Not-determined | Genome-fragment | NA | NA | 0 |
| S8_k141_17130 | 1595 | No | NA | 2 | 0 | 0 | Not-determined | Genome-fragment | NA | NA | 0 |
| S8_k141_76297 | 1035 | No | NA | 1 | 0 | 1 | Not-determined | Genome-fragment | NA | NA | 0 |
| S8_k141_99212 | 7177 | No | NA | 6 | 0 | 2 | Not-determined | Genome-fragment | NA | NA | 0 |
| S8_k141_237774 | 1313 | No | NA | 1 | 0 | 1 | Not-determined | Genome-fragment | NA | NA | 0 |
| S3_k141_36297 | 30156 | No | NA | 50 | 13 | 0 | Low-quality | Genome-fragment | 28.87 | AAI-based (high-confidence) | 0 |
| S8_k141_168339 | 1907 | No | NA | 2 | 0 | 1 | Not-determined | Genome-fragment | NA | NA | 0 |
| S8_k141_30654 | 1009 | No | NA | 3 | 0 | 3 | Not-determined | Genome-fragment | NA | NA | 0 |
| S8_k141_258960 | 3493 | No | NA | 3 | 0 | 3 | Not-determined | Genome-fragment | NA | NA | 0 |
| S17-final-viral-combined_k141_251356\|\|full | 30298 | No | NA | 20 | 4 | 0 | Low-quality | Genome-fragment | 36.8 | AAI-based (high-confidence) | 0 |
| S8_k141_76324 | 3532 | No | NA | 3 | 0 | 3 | Not-determined | Genome-fragment | NA | NA | 0 |
| S8_k141_76326 | 1184 | No | NA | 2 | 0 | 2 | Not-determined | Genome-fragment | NA | NA | 0 |
| S2-final-viral-combined_k141_26158\|\|0_partial | 30424 | No | NA | 36 | 2 | 8 | Low-quality | Genome-fragment | 19.2 | HMM-based (lower-bound) | 0 |
| S8_k141_133929 | 1090 | No | NA | 1 | 0 | 1 | Not-determined | Genome-fragment | NA | NA | 0 |
| S8_k141_99298 | 1163 | No | NA | 2 | 0 | 2 | Not-determined | Genome-fragment | NA | NA | 0 |
| S1-final-viral-combined_k141_194818\|\|full | 30487 | Yes | 10120 | 30 | 4 | 14 | Low-quality | Genome-fragment | 7.21 | HMM-based (lower-bound) | 66.81 |
| S8_k141_30736 | 1020 | No | NA | 1 | 0 | 1 | Not-determined | Genome-fragment | NA | NA | 0 |
| S8_k141_201683 | 1033 | No | NA | 1 | 0 | 1 | Not-determined | Genome-fragment | NA | NA | 0 |
| S17-final-viral-combined_k141_127300\|\|full | 30549 | Yes | 13047 | 33 | 5 | 13 | Low-quality | Genome-fragment | 16.18 | HMM-based (lower-bound) | 57.29 |
| S8_k141_53027 | 1346 | No | NA | 2 | 0 | 1 | Not-determined | Genome-fragment | NA | NA | 0 |
| S8_k141_122026 | 1192 | No | NA | 2 | 0 | 1 | Not-determined | Genome-fragment | NA | NA | 0 |
| S8_k141_225819 | 2043 | No | NA | 3 | 0 | 1 | Not-determined | Genome-fragment | NA | NA | 0 |
| S11_k141_258059 | 30608 | No | NA | 35 | 8 | 2 | Low-quality | Genome-fragment | 28.8 | AAI-based (high-confidence) | 0 |
| S8_k141_134029 | 1217 | No | NA | 2 | 0 | 2 | Not-determined | Genome-fragment | NA | NA | 0 |
| S4_k141_17373 | 30725 | Yes | 25244 | 30 | 4 | 5 | Low-quality | Genome-fragment | 43.56 | AAI-based (high-confidence) | 17.84 |
| S17-final-viral-combined_k141_138075\|\|full | 31014 | No | NA | 41 | 7 | 0 | Low-quality | Genome-fragment | 49.43 | HMM-based (lower-bound) | 0 |
| S8_k141_99376 | 3517 | No | NA | 2 | 0 | 1 | Not-determined | Genome-fragment | NA | NA | 0 |
| S2-final-viral-combined_k141_75144\|\|1_partial | 31028 | Yes | 21077 | 24 | 2 | 9 | Low-quality | Genome-fragment | 34.07 | AAI-based (high-confidence) | 32.07 |
| S8_k141_134049 | 1576 | No | NA | 2 | 0 | 2 | Not-determined | Genome-fragment | NA | NA | 0 |
| S8_k141_65065 | 1227 | No | NA | 1 | 0 | 1 | Not-determined | Genome-fragment | NA | NA | 0 |
| S8_k141_237960 | 2027 | No | NA | 2 | 0 | 2 | Not-determined | Genome-fragment | NA | NA | 0 |
| S8_k141_214281 | 4232 | No | NA | 4 | 0 | 3 | Not-determined | Genome-fragment | NA | NA | 0 |
| S8-final-viral-combined_k141_115862\|\|full | 31472 | No | NA | 37 | 1 | 3 | Low-quality | Genome-fragment | 25.51 | HMM-based (lower-bound) | 0 |
| S8_k141_30842 | 1132 | No | NA | 1 | 0 | 1 | Not-determined | Genome-fragment | NA | NA | 0 |
| S8_k141_87370 | 1260 | No | NA | 2 | 0 | 1 | Not-determined | Genome-fragment | NA | NA | 0 |
| S8_k141_53090 | 1048 | No | NA | 1 | 0 | 0 | Not-determined | Genome-fragment | NA | NA | 0 |
| S8_k141_110386 | 1000 | No | NA | 1 | 0 | 1 | Not-determined | Genome-fragment | NA | NA | 0 |
| S11-final-viral-combined_k141_129364\|\|0_partial | 31684 | Yes | 20699 | 24 | 2 | 4 | Low-quality | Genome-fragment | 15.32 | HMM-based (lower-bound) | 34.67 |
| S8_k141_180967 | 1508 | No | NA | 2 | 0 | 2 | Not-determined | Genome-fragment | NA | NA | 0 |
| S8_k141_65147 | 1745 | No | NA | 3 | 0 | 1 | Not-determined | Genome-fragment | NA | NA | 0 |
| S8_k141_201839 | 1252 | No | NA | 1 | 0 | 0 | Not-determined | Genome-fragment | NA | NA | 0 |
| S8_k141_145708 | 1066 | No | NA | 1 | 0 | 1 | Not-determined | Genome-fragment | NA | NA | 0 |
| S8_k141_134165 | 1333 | No | NA | 1 | 0 | 0 | Not-determined | Genome-fragment | NA | NA | 0 |
| S1-final-viral-combined_k141_217597\|\|full | 32007 | No | NA | 35 | 1 | 7 | Low-quality | Genome-fragment | 37.01 | AAI-based (medium-confidence) | 0 |
| S8_k141_134195 | 1391 | No | NA | 1 | 0 | 0 | Not-determined | Genome-fragment | NA | NA | 0 |
| S8_k141_110491 | 1285 | No | NA | 2 | 0 | 1 | Not-determined | Genome-fragment | NA | NA | 0 |
| S8_k141_191702 | 1144 | No | NA | 1 | 0 | 0 | Not-determined | Genome-fragment | NA | NA | 0 |
| S7-final-viral-combined_k141_84489\|\|full | 32148 | No | NA | 33 | 3 | 4 | Low-quality | Genome-fragment | 17.13 | AAI-based (medium-confidence) | 0 |
| S2-final-viral-combined_k141_128426\|\|full | 32301 | No | NA | 14 | 1 | 0 | Low-quality | Genome-fragment | 18.57 | AAI-based (high-confidence) | 0 |
| S1-final-viral-combined_k141_53913\|\|full | 32611 | Yes | 11846 | 35 | 2 | 10 | Low-quality | Genome-fragment | 10.67 | AAI-based (medium-confidence) | 63.67 |
| S8_k141_249296 | 1455 | No | NA | 2 | 0 | 1 | Not-determined | Genome-fragment | NA | NA | 0 |
| S8_k141_238142 | 1801 | No | NA | 1 | 0 | 1 | Not-determined | Genome-fragment | NA | NA | 0 |
| S8_k141_238169 | 1546 | No | NA | 1 | 0 | 0 | Not-determined | Genome-fragment | NA | NA | 0 |
| S8_k141_249310 | 2204 | No | NA | 3 | 0 | 1 | Not-determined | Genome-fragment | NA | NA | 0 |
| S8_k141_65258 | 3890 | No | NA | 4 | 0 | 1 | Not-determined | Genome-fragment | NA | NA | 0 |
| S8_k141_201926 | 1607 | No | NA | 1 | 0 | 1 | Not-determined | Genome-fragment | NA | NA | 0 |
| S8_k141_214445 | 1685 | No | NA | 1 | 0 | 0 | Not-determined | Genome-fragment | NA | NA | 0 |
| S1-final-viral-combined_k141_237598\|\|full | 32658 | Yes | 18453 | 41 | 15 | 7 | Low-quality | Genome-fragment | 42.89 | AAI-based (medium-confidence) | 43.5 |
| S17-final-viral-combined_k141_213914\|\|full | 32760 | Yes | 12861 | 42 | 5 | 14 | Low-quality | Genome-fragment | 15.94 | HMM-based (lower-bound) | 60.74 |
| S8_k141_214462 | 1258 | No | NA | 3 | 0 | 0 | Not-determined | Genome-fragment | NA | NA | 0 |
| S8_k141_225981 | 2076 | No | NA | 1 | 0 | 1 | Not-determined | Genome-fragment | NA | NA | 0 |
| S8_k141_76674 | 1125 | No | NA | 1 | 0 | 1 | Not-determined | Genome-fragment | NA | NA | 0 |
| S8_k141_41893 | 2728 | No | NA | 2 | 0 | 0 | Not-determined | Genome-fragment | NA | NA | 0 |
| S8_k141_238275 | 1509 | No | NA | 1 | 0 | 0 | Not-determined | Genome-fragment | NA | NA | 0 |
| S5-final-viral-combined_k141_126862\|\|full | 32860 | No | NA | 33 | 1 | 16 | Low-quality | Genome-fragment | 22.09 | HMM-based (lower-bound) | 0 |
| S8_k141_122308 | 4104 | No | NA | 3 | 0 | 0 | Not-determined | Genome-fragment | NA | NA | 0 |
| S1-final-viral-combined_k141_218385\|\|full | 32950 | Yes | 17134 | 41 | 5 | 10 | Low-quality | Genome-fragment | 43.56 | AAI-based (high-confidence) | 48 |
| S8_k141_145899 | 2737 | No | NA | 2 | 0 | 2 | Not-determined | Genome-fragment | NA | NA | 0 |
| S8_k141_122324 | 1255 | No | NA | 3 | 0 | 0 | Not-determined | Genome-fragment | NA | NA | 0 |
| S8_k141_53272 | 1056 | No | NA | 1 | 0 | 1 | Not-determined | Genome-fragment | NA | NA | 0 |
| S8_k141_31140 | 1438 | No | NA | 2 | 0 | 0 | Not-determined | Genome-fragment | NA | NA | 0 |
| S8_k141_76745 | 1475 | No | NA | 1 | 0 | 1 | Not-determined | Genome-fragment | NA | NA | 0 |
| S17-final-viral-combined_k141_48445\|\|full | 33049 | No | NA | 28 | 17 | 1 | Low-quality | Genome-fragment | 40.98 | AAI-based (high-confidence) | 0 |
| S8_k141_201996 | 1188 | No | NA | 1 | 0 | 0 | Not-determined | Genome-fragment | NA | NA | 0 |

| | | | | | | | | | | | |
|---|---|---|---|---|---|---|---|---|---|---|---|
| S8_k141_202001 | 1118 | No | NA | 1 | 0 | 1 | Not-determined | Genome-fragment | NA | NA | 0 |
| S8_k141_259268 | 1022 | No | NA | 1 | 0 | 0 | Not-determined | Genome-fragment | NA | NA | 0 |
| S8_k141_122350 | 2084 | No | NA | 2 | 0 | 2 | Not-determined | Genome-fragment | NA | NA | 0 |
| S7-final-viral-combined_k141_74802\|\|0_partial | 33124 | No | NA | 42 | 1 | 2 | Low-quality | Genome-fragment | 37.18 | HMM-based (lower-bound) | 0 |
| S8_k141_65393 | 1102 | No | NA | 1 | 0 | 1 | Not-determined | Genome-fragment | NA | NA | 0 |
| S8_k141_238321 | 1675 | No | NA | 3 | 0 | 2 | Not-determined | Genome-fragment | NA | NA | 0 |
| S2-final-viral-combined_k141_150929\|\|0_partial | 33303 | Yes | 18560 | 41 | 4 | 7 | Low-quality | Genome-fragment | 41.1 | AAI-based (medium-confidence) | 44.27 |
| S8_k141_272320 | 1247 | No | NA | 1 | 0 | 1 | Not-determined | Genome-fragment | NA | NA | 0 |
| S8_k141_42026 | 4505 | No | NA | 2 | 0 | 0 | Not-determined | Genome-fragment | NA | NA | 0 |
| S8_k141_134413 | 1375 | No | NA | 2 | 0 | 1 | Not-determined | Genome-fragment | NA | NA | 0 |
| S8_k141_202049 | 1277 | No | NA | 1 | 0 | 1 | Not-determined | Genome-fragment | NA | NA | 0 |
| S8_k141_202054 | 1791 | No | NA | 3 | 0 | 2 | Not-determined | Genome-fragment | NA | NA | 0 |
| S8_k141_238380 | 1053 | No | NA | 1 | 0 | 1 | Not-determined | Genome-fragment | NA | NA | 0 |
| S8_k141_99691 | 1390 | No | NA | 2 | 0 | 1 | Not-determined | Genome-fragment | NA | NA | 0 |
| S8_k141_145985 | 1987 | No | NA | 2 | 0 | 1 | Not-determined | Genome-fragment | NA | NA | 0 |
| S8_k141_31228 | 1452 | No | NA | 2 | 0 | 0 | Not-determined | Genome-fragment | NA | NA | 0 |
| S8_k141_122410 | 1111 | No | NA | 1 | 0 | 0 | Not-determined | Genome-fragment | NA | NA | 0 |
| S8_k141_146002 | 4204 | No | NA | 3 | 0 | 2 | Not-determined | Genome-fragment | NA | NA | 0 |
| S2-final-viral-combined_k141_16054\|\|0_partial | 33318 | No | NA | 25 | 1 | 5 | Low-quality | Genome-fragment | 48.41 | AAI-based (medium-confidence) | 0 |
| S8_k141_53394 | 1734 | No | NA | 3 | 0 | 1 | Not-determined | Genome-fragment | NA | NA | 0 |
| S8_k141_146024 | 1107 | No | NA | 2 | 0 | 2 | Not-determined | Genome-fragment | NA | NA | 0 |
| S8_k141_181243 | 1152 | No | NA | 1 | 0 | 0 | Not-determined | Genome-fragment | NA | NA | 0 |
| S8_k141_168862 | 1156 | No | NA | 1 | 0 | 0 | Not-determined | Genome-fragment | NA | NA | 0 |
| S8_k141_259318 | 1263 | No | NA | 1 | 0 | 1 | Not-determined | Genome-fragment | NA | NA | 0 |
| S8_k141_110578 | 1151 | No | NA | 1 | 0 | 1 | Not-determined | Genome-fragment | NA | NA | 0 |
| S1_k141_146020 | 33329 | No | NA | 47 | 1 | 2 | Low-quality | Genome-fragment | 23.86 | HMM-based (lower-bound) | 0 |
| S8_k141_31286 | 2616 | No | NA | 3 | 0 | 2 | Not-determined | Genome-fragment | NA | NA | 0 |
| S8_k141_249494 | 4564 | No | NA | 3 | 0 | 0 | Not-determined | Genome-fragment | NA | NA | 0 |
| S12_k141_335092 | 33424 | No | NA | 37 | 2 | 4 | Low-quality | Genome-fragment | 14.97 | HMM-based (lower-bound) | 0 |
| S5-final-viral-combined_k141_164620\|\|0_partial | 33704 | No | NA | 39 | 1 | 4 | Low-quality | Genome-fragment | 37.83 | HMM-based (lower-bound) | 0 |
| S8_k141_146063 | 1170 | No | NA | 2 | 0 | 0 | Not-determined | Genome-fragment | NA | NA | 0 |
| S8_k141_146088 | 1263 | No | NA | 4 | 0 | 2 | Not-determined | Genome-fragment | NA | NA | 0 |
| S8_k141_146090 | 1718 | No | NA | 1 | 0 | 1 | Not-determined | Genome-fragment | NA | NA | 0 |
| S8_k141_259354 | 1110 | No | NA | 1 | 0 | 1 | Not-determined | Genome-fragment | NA | NA | 0 |
| S8_k141_238489 | 1198 | No | NA | 1 | 0 | 1 | Not-determined | Genome-fragment | NA | NA | 0 |
| S8_k141_202194 | 1295 | No | NA | 1 | 0 | 1 | Not-determined | Genome-fragment | NA | NA | 0 |
| S8_k141_226143 | 3614 | No | NA | 2 | 0 | 2 | Not-determined | Genome-fragment | NA | NA | 0 |
| S7-final-viral-combined_k141_103212\|\|0_partial | 33716 | Yes | 17224 | 50 | 7 | 10 | Low-quality | Genome-fragment | 22.32 | HMM-based (lower-bound) | 48.91 |
| S8_k141_202213 | 1552 | No | NA | 1 | 0 | 1 | Not-determined | Genome-fragment | NA | NA | 0 |
| S8_k141_87751 | 1775 | No | NA | 2 | 0 | 0 | Not-determined | Genome-fragment | NA | NA | 0 |
| S8_k141_181307 | 3153 | No | NA | 2 | 0 | 0 | Not-determined | Genome-fragment | NA | NA | 0 |
| S1-final-viral-combined_k141_22297\|\|full | 33798 | Yes | 16187 | 29 | 5 | 15 | Low-quality | Genome-fragment | 40.93 | AAI-based (medium-confidence) | 52.11 |
| S8_k141_31338 | 1260 | No | NA | 1 | 0 | 0 | Not-determined | Genome-fragment | NA | NA | 0 |
| S8_k141_6772 | 1090 | No | NA | 1 | 0 | 1 | Not-determined | Genome-fragment | NA | NA | 0 |
| S11-final-viral-combined_k141_21942\|\|0_partial | 33800 | Yes | 14488 | 34 | 6 | 13 | Low-quality | Genome-fragment | 23.25 | HMM-based (lower-bound) | 57.14 |
| S8_k141_146161 | 1806 | No | NA | 1 | 0 | 0 | Not-determined | Genome-fragment | NA | NA | 0 |
| S8_k141_42217 | 3210 | No | NA | 6 | 0 | 4 | Not-determined | Genome-fragment | NA | NA | 0 |
| S11-final-viral-combined_k141_141301\|\|full | 33879 | No | NA | 48 | 4 | 1 | Low-quality | Genome-fragment | 34.71 | AAI-based (medium-confidence) | 0 |
| S8_k141_42237 | 1106 | No | NA | 2 | 0 | 0 | Not-determined | Genome-fragment | NA | NA | 0 |
| S8_k141_42242 | 1063 | No | NA | 1 | 0 | 0 | Not-determined | Genome-fragment | NA | NA | 0 |
| S8_k141_122485 | 1077 | No | NA | 1 | 0 | 1 | Not-determined | Genome-fragment | NA | NA | 0 |
| S8_k141_53562 | 1140 | No | NA | 1 | 0 | 0 | Not-determined | Genome-fragment | NA | NA | 0 |
| S8_k141_6824 | 1292 | No | NA | 1 | 0 | 1 | Not-determined | Genome-fragment | NA | NA | 0 |
| S8_k141_87812 | 1085 | No | NA | 2 | 0 | 0 | Not-determined | Genome-fragment | NA | NA | 0 |
| S2-final-viral-combined_k141_53444\|\|full | 33902 | Yes | 12823 | 38 | 5 | 13 | Low-quality | Genome-fragment | 15.9 | HMM-based (lower-bound) | 62.18 |
| S8_k141_181413 | 1269 | No | NA | 1 | 0 | 0 | Not-determined | Genome-fragment | NA | NA | 0 |
| S8_k141_77014 | 2237 | No | NA | 1 | 0 | 0 | Not-determined | Genome-fragment | NA | NA | 0 |
| S8_k141_87827 | 1360 | No | NA | 2 | 0 | 1 | Not-determined | Genome-fragment | NA | NA | 0 |
| S8_k141_134612 | 1329 | No | NA | 5 | 0 | 0 | Not-determined | Genome-fragment | NA | NA | 0 |
| S8_k141_169053 | 3576 | No | NA | 5 | 0 | 1 | Not-determined | Genome-fragment | NA | NA | 0 |
| S8_k141_202299 | 1173 | No | NA | 1 | 0 | 1 | Not-determined | Genome-fragment | NA | NA | 0 |
| S7-final-viral-combined_k141_80334\|\|full | 33913 | Yes | 22223 | 36 | 12 | 3 | Low-quality | Genome-fragment | 46.78 | AAI-based (high-confidence) | 34.47 |
| S17-final-viral-combined_k141_140294\|\|0_partial | 33987 | Yes | 24521 | 29 | 2 | 4 | Low-quality | Genome-fragment | 16.38 | HMM-based (lower-bound) | 27.85 |
| S8_k141_122608 | 1121 | No | NA | 2 | 0 | 2 | Not-determined | Genome-fragment | NA | NA | 0 |
| S8_k141_146286 | 1037 | No | NA | 1 | 0 | 0 | Not-determined | Genome-fragment | NA | NA | 0 |
| S11-final-viral-combined_k141_313284\|\|full | 34171 | Yes | 11779 | 30 | 3 | 13 | Low-quality | Genome-fragment | 41.05 | AAI-based (medium-confidence) | 65.53 |
| S8-final-viral-combined_k141_34759\|\|0_partial | 34226 | No | NA | 43 | 2 | 2 | Low-quality | Genome-fragment | 36.98 | HMM-based (lower-bound) | 0 |
| S8_k141_53641 | 2853 | No | NA | 4 | 0 | 3 | Not-determined | Genome-fragment | NA | NA | 0 |
| S8_k141_122627 | 1085 | No | NA | 1 | 0 | 0 | Not-determined | Genome-fragment | NA | NA | 0 |
| S7-final-viral-combined_k141_149187\|\|full | 34315 | Yes | 13671 | 40 | 6 | 14 | Low-quality | Genome-fragment | 25.99 | HMM-based (lower-bound) | 60.16 |
| S8_k141_249683 | 1006 | No | NA | 1 | 0 | 1 | Not-determined | Genome-fragment | NA | NA | 0 |
| S8_k141_87902 | 2304 | No | NA | 2 | 0 | 1 | Not-determined | Genome-fragment | NA | NA | 0 |
| S7-final-viral-combined_k141_165331\|\|0_partial | 34540 | No | NA | 34 | 1 | 11 | Low-quality | Genome-fragment | 19.84 | HMM-based (lower-bound) | 0 |
| S8_k141_191877 | 1205 | No | NA | 1 | 0 | 0 | Not-determined | Genome-fragment | NA | NA | 0 |
| S8_k141_87934 | 1259 | No | NA | 1 | 0 | 0 | Not-determined | Genome-fragment | NA | NA | 0 |
| S8_k141_272564 | 8011 | No | NA | 10 | 0 | 5 | Not-determined | Genome-fragment | NA | NA | 0 |
| S17-final-viral-combined_k141_61420\|\|full | 34597 | Yes | 14585 | 41 | 5 | 16 | Low-quality | Genome-fragment | 18.08 | HMM-based (lower-bound) | 57.84 |
| S11-final-viral-combined_k141_247023\|\|full | 34912 | Yes | 13299 | 38 | 6 | 15 | Low-quality | Genome-fragment | 14.15 | HMM-based (lower-bound) | 61.91 |
| S8_k141_110924 | 1168 | No | NA | 1 | 0 | 1 | Not-determined | Genome-fragment | NA | NA | 0 |
| S8_k141_53721 | 2411 | No | NA | 1 | 0 | 1 | Not-determined | Genome-fragment | NA | NA | 0 |
| S8_k141_249733 | 3772 | No | NA | 1 | 0 | 1 | Not-determined | Genome-fragment | NA | NA | 0 |
| S8_k141_192005 | 1518 | No | NA | 1 | 0 | 0 | Not-determined | Genome-fragment | NA | NA | 0 |
| S8_k141_110955 | 1326 | No | NA | 2 | 0 | 0 | Not-determined | Genome-fragment | NA | NA | 0 |
| S8_k141_249779 | 3122 | No | NA | 2 | 0 | 1 | Not-determined | Genome-fragment | NA | NA | 0 |
| S8_k141_122736 | 1014 | No | NA | 1 | 0 | 1 | Not-determined | Genome-fragment | NA | NA | 0 |
| S7-final-viral-combined_k141_14783\|\|full | 35015 | Yes | 16316 | 48 | 6 | 14 | Low-quality | Genome-fragment | 32.04 | AAI-based (high-confidence) | 53.4 |
| S8_k141_88043 | 1054 | No | NA | 1 | 0 | 0 | Not-determined | Genome-fragment | NA | NA | 0 |
| S8_k141_157837 | 4320 | No | NA | 3 | 0 | 1 | Not-determined | Genome-fragment | NA | NA | 0 |
| S8_k141_88061 | 5515 | No | NA | 4 | 0 | 4 | Not-determined | Genome-fragment | NA | NA | 0 |
| S8_k141_134765 | 1033 | No | NA | 1 | 0 | 0 | Not-determined | Genome-fragment | NA | NA | 0 |
| S8_k141_77164 | 1154 | No | NA | 2 | 0 | 1 | Not-determined | Genome-fragment | NA | NA | 0 |
| S8_k141_134773 | 1712 | No | NA | 2 | 0 | 0 | Not-determined | Genome-fragment | NA | NA | 0 |
| S8_k141_122806 | 1361 | No | NA | 2 | 0 | 1 | Not-determined | Genome-fragment | NA | NA | 0 |
| S8_k141_42420 | 7688 | No | NA | 7 | 0 | 6 | Not-determined | Genome-fragment | NA | NA | 0 |
| S1-final-viral-combined_k141_250445\|\|0_partial | 35087 | Yes | 21320 | 44 | 2 | 4 | Low-quality | Genome-fragment | 23.03 | HMM-based (lower-bound) | 39.24 |
| S8_k141_42430 | 1780 | No | NA | 3 | 0 | 0 | Not-determined | Genome-fragment | NA | NA | 0 |
| S8_k141_121758 | 35170 | No | NA | 25 | 3 | 0 | Low-quality | Genome-fragment | 36.91 | AAI-based (high-confidence) | 0 |
| S6-final-viral-combined_k141_20158\|\|full | 35204 | Yes | 18255 | 44 | 9 | 9 | Low-quality | Genome-fragment | 33.27 | AAI-based (high-confidence) | 48.15 |
| S8_k141_42487 | 1115 | No | NA | 1 | 0 | 1 | Not-determined | Genome-fragment | NA | NA | 0 |
| S8_k141_215083 | 1002 | No | NA | 1 | 0 | 0 | Not-determined | Genome-fragment | NA | NA | 0 |
| S8-final-viral-combined_k141_196892\|\|full | 35467 | Yes | 8105 | 38 | 5 | 22 | Low-quality | Genome-fragment | 11.74 | HMM-based (lower-bound) | 77.15 |
| S8_k141_215087 | 2891 | No | NA | 1 | 0 | 0 | Not-determined | Genome-fragment | NA | NA | 0 |
| S9-final-viral-combined_k141_47\|\|full | 35555 | No | NA | 32 | 0 | 23 | Low-quality | Genome-fragment | 30.54 | AAI-based (medium-confidence) | 0 |
| S8_k141_111056 | 54171 | No | NA | 48 | 4 | 0 | Medium-quality | Genome-fragment | 56.87 | AAI-based (high-confidence) | 0 |
| S8_k141_157975 | 2607 | No | NA | 4 | 0 | 2 | Not-determined | Genome-fragment | NA | NA | 0 |
| S8_k141_53944 | 1781 | No | NA | 3 | 0 | 0 | Not-determined | Genome-fragment | NA | NA | 0 |
| S8_k141_192177 | 1061 | No | NA | 1 | 0 | 0 | Not-determined | Genome-fragment | NA | NA | 0 |
| S8_k141_259721 | 1068 | No | NA | 2 | 0 | 1 | Not-determined | Genome-fragment | NA | NA | 0 |
| S8_k141_17960 | 1474 | No | NA | 1 | 0 | 0 | Not-determined | Genome-fragment | NA | NA | 0 |
| S8_k141_202507 | 1252 | No | NA | 2 | 0 | 2 | Not-determined | Genome-fragment | NA | NA | 0 |
| S8_k141_259742 | 1812 | No | NA | 6 | 0 | 5 | Not-determined | Genome-fragment | NA | NA | 0 |
| S8_k141_192193 | 7389 | No | NA | 6 | 0 | 3 | Not-determined | Genome-fragment | NA | NA | 0 |
| S8_k141_42553 | 1708 | No | NA | 1 | 0 | 1 | Not-determined | Genome-fragment | NA | NA | 0 |
| S12-final-viral-combined_k141_183146\|\|full | 35615 | No | NA | 50 | 1 | 4 | Low-quality | Genome-fragment | 26.35 | AAI-based (medium-confidence) | 0 |
| S12-final-viral-combined_k141_287341\|\|full | 35881 | Yes | 15801 | 38 | 4 | 13 | Low-quality | Genome-fragment | 31.72 | AAI-based (high-confidence) | 55.96 |
| S17-final-viral-combined_k141_36601\|\|full | 35897 | Yes | 12572 | 49 | 5 | 18 | Low-quality | Genome-fragment | 33.72 | AAI-based (medium-confidence) | 64.98 |
| S8_k141_123035 | 1116 | No | NA | 2 | 0 | 0 | Not-determined | Genome-fragment | NA | NA | 0 |
| S8_k141_215126 | 5530 | No | NA | 2 | 0 | 2 | Not-determined | Genome-fragment | NA | NA | 0 |
| S8_k141_134994 | 1755 | No | NA | 1 | 0 | 0 | Not-determined | Genome-fragment | NA | NA | 0 |
| S8_k141_7291 | 1115 | No | NA | 2 | 0 | 1 | Not-determined | Genome-fragment | NA | NA | 0 |
| S8_k141_259786 | 1000 | No | NA | 1 | 0 | 1 | Not-determined | Genome-fragment | NA | NA | 0 |
| S12-final-viral-combined_k141_275038\|\|full | 36003 | Yes | 8418 | 42 | 6 | 24 | Low-quality | Genome-fragment | 13.62 | HMM-based (lower-bound) | 76.62 |
| S8_k141_146704 | 1003 | No | NA | 1 | 0 | 0 | Not-determined | Genome-fragment | NA | NA | 0 |
| S8_k141_54091 | 1427 | No | NA | 1 | 0 | 1 | Not-determined | Genome-fragment | NA | NA | 0 |
| S8_k141_31913 | 1521 | No | NA | 3 | 0 | 1 | Not-determined | Genome-fragment | NA | NA | 0 |
| S8_k141_135058 | 1494 | No | NA | 1 | 0 | 1 | Not-determined | Genome-fragment | NA | NA | 0 |
| S8_k141_100254 | 1052 | No | NA | 1 | 0 | 1 | Not-determined | Genome-fragment | NA | NA | 0 |
| S8_k141_123100 | 1167 | No | NA | 1 | 0 | 1 | Not-determined | Genome-fragment | NA | NA | 0 |
| S8_k141_192295 | 1127 | No | NA | 2 | 0 | 0 | Not-determined | Genome-fragment | NA | NA | 0 |
| S8_k141_181970 | 9784 | No | NA | 3 | 0 | 1 | Not-determined | Genome-fragment | NA | NA | 0 |
| S8_k141_54153 | 1093 | No | NA | 1 | 0 | 1 | Not-determined | Genome-fragment | NA | NA | 0 |
| S8_k141_158126 | 1669 | No | NA | 2 | 0 | 0 | Not-determined | Genome-fragment | NA | NA | 0 |
| S17-final-viral-combined_k141_234491\|\|0_partial | 36153 | Yes | 19781 | 47 | 7 | 13 | Low-quality | Genome-fragment | 45.97 | AAI-based (high-confidence) | 45.29 |
| S8_k141_239313 | 3822 | No | NA | 4 | 0 | 1 | Not-determined | Genome-fragment | NA | NA | 0 |
| S8_k141_66041 | 1212 | No | NA | 1 | 0 | 1 | Not-determined | Genome-fragment | NA | NA | 0 |
| S4-final-viral-combined_k141_114177\|\|full | 36170 | No | NA | 39 | 1 | 0 | Low-quality | Genome-fragment | 23.97 | AAI-based (high-confidence) | 0 |
| S5-final-viral-combined_k141_169295\|\|full | 36199 | Yes | 9153 | 39 | 2 | 17 | Low-quality | Genome-fragment | 28.21 | AAI-based (medium-confidence) | 74.71 |
| S8_k141_100348 | 5221 | No | NA | 7 | 0 | 2 | Not-determined | Genome-fragment | NA | NA | 0 |
| S8_k141_135179 | 7048 | No | NA | 4 | 0 | 2 | Not-determined | Genome-fragment | NA | NA | 0 |
| S8_k141_226726 | 1082 | No | NA | 1 | 0 | 0 | Not-determined | Genome-fragment | NA | NA | 0 |
| S8_k141_239371 | 4205 | No | NA | 3 | 0 | 2 | Not-determined | Genome-fragment | NA | NA | 0 |
| S8_k141_100381 | 1729 | No | NA | 4 | 0 | 0 | Not-determined | Genome-fragment | NA | NA | 0 |
| S8_k141_66112 | 3258 | No | NA | 4 | 0 | 1 | Not-determined | Genome-fragment | NA | NA | 0 |
| S8_k141_202684 | 1031 | No | NA | 1 | 0 | 1 | Not-determined | Genome-fragment | NA | NA | 0 |
| S9-final-viral-combined_k141_10161\|\|full | 36618 | Yes | 13096 | 47 | 10 | 21 | Low-quality | Genome-fragment | 34.87 | AAI-based (high-confidence) | 64.24 |
| S8_k141_77716 | 2947 | No | NA | 2 | 0 | 2 | Not-determined | Genome-fragment | NA | NA | 0 |
| S8_k141_259850 | 1004 | No | NA | 1 | 0 | 0 | Not-determined | Genome-fragment | NA | NA | 0 |
| S8_k141_135208 | 1080 | No | NA | 2 | 0 | 0 | Not-determined | Genome-fragment | NA | NA | 0 |
| S8_k141_226765 | 3101 | No | NA | 2 | 0 | 2 | Not-determined | Genome-fragment | NA | NA | 0 |
| S8_k141_42944 | 1319 | No | NA | 1 | 0 | 1 | Not-determined | Genome-fragment | NA | NA | 0 |
| S11-final-viral-combined_k141_116141\|\|full | 36819 | No | NA | 38 | 1 | 28 | Low-quality | Genome-fragment | 17.86 | HMM-based (lower-bound) | 0 |
| S8_k141_135240 | 1824 | No | NA | 3 | 0 | 0 | Not-determined | Genome-fragment | NA | NA | 0 |
| S8_k141_146987 | 1250 | No | NA | 1 | 0 | 1 | Not-determined | Genome-fragment | NA | NA | 0 |
| S8_k141_77787 | 1197 | No | NA | 2 | 0 | 1 | Not-determined | Genome-fragment | NA | NA | 0 |
| S8_k141_215398 | 1254 | No | NA | 1 | 0 | 0 | Not-determined | Genome-fragment | NA | NA | 0 |
| S8_k141_158281 | 1236 | No | NA | 3 | 0 | 1 | Not-determined | Genome-fragment | NA | NA | 0 |
| S17-final-viral-combined_k141_189865\|\|0_partial | 36875 | Yes | 14420 | 39 | 6 | 12 | Low-quality | Genome-fragment | 19.4 | HMM-based (lower-bound) | 60.89 |
| S8_k141_169697 | 3148 | No | NA | 6 | 0 | 0 | Not-determined | Genome-fragment | NA | NA | 0 |
| S8_k141_169711 | 1302 | No | NA | 1 | 0 | 1 | Not-determined | Genome-fragment | NA | NA | 0 |
| S8_k141_202828 | 1063 | No | NA | 3 | 0 | 0 | Not-determined | Genome-fragment | NA | NA | 0 |
| S8_k141_239464 | 2051 | No | NA | 1 | 0 | 1 | Not-determined | Genome-fragment | NA | NA | 0 |
| S8_k141_100496 | 1054 | No | NA | 3 | 0 | 0 | Not-determined | Genome-fragment | NA | NA | 0 |
| S8_k141_32246 | 1030 | No | NA | 2 | 0 | 1 | Not-determined | Genome-fragment | NA | NA | 0 |
| S8_k141_32259 | 1604 | No | NA | 1 | 0 | 1 | Not-determined | Genome-fragment | NA | NA | 0 |
| S7-final-viral-combined_k141_1795\|\|full | 36997 | No | NA | 48 | 1 | 9 | Low-quality | Genome-fragment | 41.53 | HMM-based (lower-bound) | 0 |
| S8_k141_273131 | 1183 | No | NA | 2 | 0 | 1 | Not-determined | Genome-fragment | NA | NA | 0 |
| S8_k141_147170 | 1349 | No | NA | 1 | 0 | 1 | Not-determined | Genome-fragment | NA | NA | 0 |
| S8_k141_7403 | 5776 | No | NA | 6 | 0 | 2 | Not-determined | Genome-fragment | NA | NA | 0 |
| S8_k141_88394 | 1453 | No | NA | 1 | 0 | 1 | Not-determined | Genome-fragment | NA | NA | 0 |
| S8_k141_169828 | 6096 | No | NA | 2 | 0 | 2 | Not-determined | Genome-fragment | NA | NA | 0 |
| S8_k141_135399 | 1332 | No | NA | 1 | 0 | 1 | Not-determined | Genome-fragment | NA | NA | 0 |
| S6-final-viral-combined_k141_103573\|\|0_partial | 37129 | No | NA | 21 | 1 | 9 | Low-quality | Genome-fragment | 47.45 | AAI-based (medium-confidence) | 0 |
| S5-final-viral-combined_k141_102913\|\|full | 37260 | No | NA | 36 | 3 | 1 | Low-quality | Genome-fragment | 24.23 | AAI-based (high-confidence) | 0 |
| S8_k141_250288 | 1228 | No | NA | 1 | 0 | 0 | Not-determined | Genome-fragment | NA | NA | 0 |
| S8_k141_239554 | 1205 | No | NA | 1 | 0 | 0 | Not-determined | Genome-fragment | NA | NA | 0 |
| S2-final-viral-combined_k141_160149\|\|full | 37277 | No | NA | 41 | 1 | 12 | Low-quality | Genome-fragment | 41.84 | HMM-based (lower-bound) | 0 |
| S8_k141_126168 | 37399 | No | NA | 29 | 0 | 27 | Low-quality | Genome-fragment | 13.17 | AAI-based (medium-confidence) | 0 |
| S8_k141_250300 | 2547 | No | NA | 2 | 0 | 2 | Not-determined | Genome-fragment | NA | NA | 0 |
| S8_k141_88426 | 1156 | No | NA | 1 | 0 | 1 | Not-determined | Genome-fragment | NA | NA | 0 |
| S8_k141_192653 | 1553 | No | NA | 4 | 0 | 3 | Not-determined | Genome-fragment | NA | NA | 0 |
| S8_k141_111577 | 3531 | No | NA | 3 | 0 | 3 | Not-determined | Genome-fragment | NA | NA | 0 |
| S8_k141_100635 | 1145 | No | NA | 1 | 0 | 1 | Not-determined | Genome-fragment | NA | NA | 0 |
| S8_k141_88436 | 1319 | No | NA | 2 | 0 | 1 | Not-determined | Genome-fragment | NA | NA | 0 |
| S8_k141_226989 | 3439 | No | NA | 3 | 0 | 2 | Not-determined | Genome-fragment | NA | NA | 0 |
| S11-final-viral-combined_k141_75058\|\|full | 37739 | Yes | 10764 | 40 | 5 | 19 | Low-quality | Genome-fragment | 30.97 | AAI-based (medium-confidence) | 71.48 |
| S8_k141_135428 | 9838 | No | NA | 10 | 0 | 6 | Not-determined | Genome-fragment | NA | NA | 0 |
| S11-final-viral-combined_k141_306671\|\|full | 37835 | Yes | 10781 | 39 | 4 | 17 | Low-quality | Genome-fragment | 14.24 | HMM-based (lower-bound) | 71.51 |
| S8_k141_147221 | 1040 | No | NA | 1 | 0 | 0 | Not-determined | Genome-fragment | NA | NA | 0 |
| S8_k141_192687 | 2857 | No | NA | 6 | 0 | 6 | Not-determined | Genome-fragment | NA | NA | 0 |
| S8_k141_100666 | 1520 | No | NA | 2 | 0 | 1 | Not-determined | Genome-fragment | NA | NA | 0 |
| S8_k141_123515 | 1013 | No | NA | 1 | 0 | 1 | Not-determined | Genome-fragment | NA | NA | 0 |
| S17-final-viral-combined_k141_191266\|\|full | 37902 | No | NA | 37 | 1 | 23 | Low-quality | Genome-fragment | 25.95 | HMM-based (lower-bound) | 0 |
| S8_k141_227034 | 1532 | No | NA | 1 | 0 | 0 | Not-determined | Genome-fragment | NA | NA | 0 |
| S8_k141_18444 | 5537 | No | NA | 4 | 0 | 1 | Not-determined | Genome-fragment | NA | NA | 0 |
| S8_k141_215642 | 1119 | No | NA | 2 | 0 | 1 | Not-determined | Genome-fragment | NA | NA | 0 |
| S8_k141_135536 | 1450 | No | NA | 1 | 0 | 0 | Not-determined | Genome-fragment | NA | NA | 0 |
| S8-final-viral-combined_k141_139022\|\|0_partial | 37967 | Yes | 18263 | 41 | 2 | 15 | Low-quality | Genome-fragment | 11.36 | HMM-based (lower-bound) | 51.9 |
| S8_k141_227088 | 1096 | No | NA | 1 | 0 | 1 | Not-determined | Genome-fragment | NA | NA | 0 |
| S8_k141_273280 | 1324 | No | NA | 2 | 0 | 0 | Not-determined | Genome-fragment | NA | NA | 0 |
| S8_k141_32430 | 1115 | No | NA | 1 | 0 | 1 | Not-determined | Genome-fragment | NA | NA | 0 |
| S1-final-viral-combined_k141_355800\|\|full | 38229 | Yes | 19875 | 48 | 6 | 10 | Low-quality | Genome-fragment | 39.83 | AAI-based (high-confidence) | 48.01 |
| S8_k141_54623 | 1609 | No | NA | 2 | 0 | 2 | Not-determined | Genome-fragment | NA | NA | 0 |
| S8_k141_18504 | 1042 | No | NA | 1 | 0 | 1 | Not-determined | Genome-fragment | NA | NA | 0 |
| S9-final-viral-combined_k141_53997\|\|full | 38272 | Yes | 19050 | 29 | 3 | 7 | Low-quality | Genome-fragment | 9.83 | HMM-based (lower-bound) | 50.22 |
| S8_k141_147444 | 2164 | No | NA | 2 | 0 | 2 | Not-determined | Genome-fragment | NA | NA | 0 |
| S1-final-viral-combined_k141_334278\|\|full | 38307 | Yes | 27385 | 46 | 7 | 3 | Low-quality | Genome-fragment | 38.87 | HMM-based (lower-bound) | 28.51 |
| S1_k141_231066 | 38309 | No | NA | 36 | 2 | 5 | Low-quality | Genome-fragment | 21.36 | HMM-based (lower-bound) | 0 |
| S5-final-viral-combined_k141_132583\|\|full | 38513 | Yes | 16604 | 33 | 2 | 12 | Low-quality | Genome-fragment | 34.4 | AAI-based (medium-confidence) | 56.89 |
| S8_k141_258737 | 39402 | No | NA | 23 | 2 | 0 | Low-quality | Genome-fragment | 41.02 | AAI-based (high-confidence) | 0 |
| S7-final-viral-combined_k141_34924\|\|full | 39528 | Yes | 22736 | 38 | 13 | 5 | Low-quality | Genome-fragment | 47.25 | AAI-based (high-confidence) | 42.48 |
| S8_k141_111817 | 1679 | No | NA | 1 | 0 | 0 | Not-determined | Genome-fragment | NA | NA | 0 |
| S7-final-viral-combined_k141_45014\|\|full | 39761 | No | NA | 35 | 1 | 5 | Low-quality | Genome-fragment | 25.93 | AAI-based (medium-confidence) | 0 |
| S8_k141_111832 | 1139 | No | NA | 1 | 0 | 1 | Not-determined | Genome-fragment | NA | NA | 0 |
| S8_k141_259900 | 39845 | No | NA | 68 | 9 | 1 | Low-quality | Genome-fragment | 29.92 | AAI-based (high-confidence) | 0 |
| S8_k141_192891 | 1379 | No | NA | 1 | 0 | 0 | Not-determined | Genome-fragment | NA | NA | 0 |
| S8_k141_7629 | 1278 | No | NA | 2 | 0 | 1 | Not-determined | Genome-fragment | NA | NA | 0 |
| S8_k141_250429 | 1616 | No | NA | 3 | 0 | 1 | Not-determined | Genome-fragment | NA | NA | 0 |
| S8_k141_123801 | 4784 | No | NA | 5 | 0 | 2 | Not-determined | Genome-fragment | NA | NA | 0 |
| S8_k141_273426 | 1338 | No | NA | 2 | 0 | 0 | Not-determined | Genome-fragment | NA | NA | 0 |
| S8_k141_54764 | 1578 | No | NA | 1 | 0 | 0 | Not-determined | Genome-fragment | NA | NA | 0 |
| S8_k141_239811 | 1107 | No | NA | 2 | 0 | 0 | Not-determined | Genome-fragment | NA | NA | 0 |
| S2_k141_239668 | 39863 | No | NA | 47 | 0 | 3 | Low-quality | Genome-fragment | 25.86 | AAI-based (medium-confidence) | 0 |
| S8_k141_7696 | 1068 | No | NA | 1 | 0 | 0 | Not-determined | Genome-fragment | NA | NA | 0 |
| S8_k141_182454 | 1740 | No | NA | 1 | 0 | 0 | Not-determined | Genome-fragment | NA | NA | 0 |
| S8_k141_18652 | 3961 | No | NA | 7 | 0 | 2 | Not-determined | Genome-fragment | NA | NA | 0 |
| S8_k141_7731 | 1135 | No | NA | 2 | 0 | 1 | Not-determined | Genome-fragment | NA | NA | 0 |
| S8_k141_54835 | 2679 | No | NA | 2 | 0 | 1 | Not-determined | Genome-fragment | NA | NA | 0 |
| S8_k141_88666 | 1049 | No | NA | 1 | 0 | 0 | Not-determined | Genome-fragment | NA | NA | 0 |
| S8_k141_18667 | 1777 | No | NA | 3 | 0 | 1 | Not-determined | Genome-fragment | NA | NA | 0 |
| S8_k141_123904 | 1408 | No | NA | 1 | 0 | 0 | Not-determined | Genome-fragment | NA | NA | 0 |
| S8_k141_78233 | 2024 | No | NA | 2 | 0 | 2 | Not-determined | Genome-fragment | NA | NA | 0 |
| S8_k141_18678 | 1335 | No | NA | 2 | 0 | 1 | Not-determined | Genome-fragment | NA | NA | 0 |
| S8_k141_182488 | 2128 | No | NA | 2 | 0 | 2 | Not-determined | Genome-fragment | NA | NA | 0 |
| S8_k141_18698 | 1413 | No | NA | 1 | 0 | 1 | Not-determined | Genome-fragment | NA | NA | 0 |
| S11_k141_232412 | 40225 | Yes | 7182 | 44 | 6 | 22 | Low-quality | Genome-fragment | 11.47 | AAI-based (medium-confidence) | 82.15 |
| S8_k141_66496 | 1013 | No | NA | 1 | 0 | 1 | Not-determined | Genome-fragment | NA | NA | 0 |
| S2_k141_67467 | 40371 | No | NA | 42 | 11 | 2 | Low-quality | Genome-fragment | 37.27 | AAI-based (high-confidence) | 0 |
| S8_k141_182569 | 1276 | No | NA | 2 | 0 | 1 | Not-determined | Genome-fragment | NA | NA | 0 |
| S8_k141_215984 | 1412 | No | NA | 1 | 0 | 1 | Not-determined | Genome-fragment | NA | NA | 0 |
| S8_k141_135964 | 1239 | No | NA | 1 | 0 | 0 | Not-determined | Genome-fragment | NA | NA | 0 |
| S8_k141_112002 | 1053 | No | NA | 1 | 0 | 0 | Not-determined | Genome-fragment | NA | NA | 0 |
| S8_k141_170114 | 5089 | No | NA | 4 | 0 | 3 | Not-determined | Genome-fragment | NA | NA | 0 |
| S8_k141_66572 | 1441 | No | NA | 2 | 0 | 2 | Not-determined | Genome-fragment | NA | NA | 0 |
| S8_k141_32686 | 1295 | No | NA | 1 | 0 | 1 | Not-determined | Genome-fragment | NA | NA | 0 |
| S8_k141_250654 | 1663 | No | NA | 2 | 0 | 1 | Not-determined | Genome-fragment | NA | NA | 0 |
| S8_k141_250660 | 4456 | No | NA | 3 | 0 | 3 | Not-determined | Genome-fragment | NA | NA | 0 |
| S8_k141_101096 | 1540 | No | NA | 2 | 0 | 0 | Not-determined | Genome-fragment | NA | NA | 0 |
| S8_k141_78324 | 1497 | No | NA | 2 | 0 | 1 | Not-determined | Genome-fragment | NA | NA | 0 |
| S8_k141_193021 | 2847 | No | NA | 7 | 0 | 6 | Not-determined | Genome-fragment | NA | NA | 0 |
| S8_k141_273635 | 1178 | No | NA | 1 | 0 | 0 | Not-determined | Genome-fragment | NA | NA | 0 |
| S8_k141_32731 | 5593 | No | NA | 1 | 0 | 1 | Not-determined | Genome-fragment | NA | NA | 0 |
| S8_k141_54960 | 3796 | No | NA | 5 | 0 | 0 | Not-determined | Genome-fragment | NA | NA | 0 |
| S8_k141_43668 | 1817 | No | NA | 2 | 0 | 0 | Not-determined | Genome-fragment | NA | NA | 0 |
| S8_k141_182705 | 1217 | No | NA | 1 | 0 | 1 | Not-determined | Genome-fragment | NA | NA | 0 |
| S8-final-viral-combined_k141_120497\|\|full | 40514 | Yes | 12423 | 41 | 4 | 16 | Low-quality | Genome-fragment | 30.57 | AAI-based (high-confidence) | 69.34 |
| S8_k141_227472 | 1683 | No | NA | 5 | 0 | 4 | Not-determined | Genome-fragment | NA | NA | 0 |
| S8_k141_216105 | 1930 | No | NA | 1 | 0 | 0 | Not-determined | Genome-fragment | NA | NA | 0 |
| S8_k141_136083 | 1490 | No | NA | 1 | 0 | 1 | Not-determined | Genome-fragment | NA | NA | 0 |
| S8_k141_66631 | 1412 | No | NA | 1 | 0 | 1 | Not-determined | Genome-fragment | NA | NA | 0 |
| S8_k141_54988 | 10404 | No | NA | 9 | 0 | 8 | Not-determined | Genome-fragment | NA | NA | 0 |
| S8_k141_18884 | 1703 | No | NA | 1 | 0 | 0 | Not-determined | Genome-fragment | NA | NA | 0 |
| S11-final-viral-combined_k141_262567\|\|full | 40624 | Yes | 9083 | 39 | 3 | 19 | Low-quality | Genome-fragment | 22.94 | AAI-based (high-confidence) | 77.64 |
| S8_k141_216136 | 5350 | No | NA | 2 | 0 | 2 | Not-determined | Genome-fragment | NA | NA | 0 |
| S8_k141_136144 | 4646 | No | NA | 6 | 0 | 3 | Not-determined | Genome-fragment | NA | NA | 0 |
| S8_k141_170237 | 1010 | No | NA | 1 | 0 | 1 | Not-determined | Genome-fragment | NA | NA | 0 |
| S5_k141_205181 | 40783 | No | NA | 47 | 1 | 12 | Low-quality | Genome-fragment | 20 | HMM-based (lower-bound) | 0 |
| S8_k141_124008 | 1396 | No | NA | 1 | 0 | 1 | Not-determined | Genome-fragment | NA | NA | 0 |
| S17-final-viral-combined_k141_68781\|\|full | 41038 | Yes | 14257 | 49 | 7 | 23 | Low-quality | Genome-fragment | 15.78 | HMM-based (lower-bound) | 65.26 |
| S8_k141_203418 | 2357 | No | NA | 3 | 0 | 1 | Not-determined | Genome-fragment | NA | NA | 0 |
| S2_k141_178405 | 41112 | No | NA | 52 | 8 | 0 | Low-quality | Genome-fragment | 40.42 | AAI-based (high-confidence) | 0 |
| S8_k141_124031 | 1096 | No | NA | 1 | 0 | 1 | Not-determined | Genome-fragment | NA | NA | 0 |
| S8_k141_147867 | 4246 | No | NA | 2 | 0 | 2 | Not-determined | Genome-fragment | NA | NA | 0 |
| S8_k141_18950 | 3017 | No | NA | 3 | 0 | 2 | Not-determined | Genome-fragment | NA | NA | 0 |
| S8_k141_240100 | 1142 | No | NA | 1 | 0 | 1 | Not-determined | Genome-fragment | NA | NA | 0 |
| S8_k141_101274 | 1384 | No | NA | 1 | 0 | 1 | Not-determined | Genome-fragment | NA | NA | 0 |
| S8_k141_147894 | 8831 | No | NA | 14 | 0 | 1 | Not-determined | Genome-fragment | NA | NA | 0 |
| S8_k141_240131 | 1024 | No | NA | 1 | 0 | 0 | Not-determined | Genome-fragment | NA | NA | 0 |
| S8_k141_216290 | 3415 | No | NA | 3 | 0 | 1 | Not-determined | Genome-fragment | NA | NA | 0 |
| S8_k141_8028 | 4790 | No | NA | 2 | 0 | 2 | Not-determined | Genome-fragment | NA | NA | 0 |
| S17-final-viral-combined_k141_70838\|\|full | 41252 | Yes | 13001 | 45 | 6 | 17 | Low-quality | Genome-fragment | 16.41 | HMM-based (lower-bound) | 68.48 |
| S8_k141_216316 | 1327 | No | NA | 1 | 0 | 1 | Not-determined | Genome-fragment | NA | NA | 0 |
| S6-final-viral-combined_k141_55357\|\|full | 41474 | Yes | 14429 | 39 | 10 | 18 | Low-quality | Genome-fragment | 16.84 | HMM-based (lower-bound) | 65.21 |
| S8_k141_193257 | 1066 | No | NA | 1 | 0 | 1 | Not-determined | Genome-fragment | NA | NA | 0 |
| S8_k141_260444 | 1127 | No | NA | 1 | 0 | 1 | Not-determined | Genome-fragment | NA | NA | 0 |
| S8_k141_112262 | 3125 | No | NA | 1 | 0 | 0 | Not-determined | Genome-fragment | NA | NA | 0 |
| S5-final-viral-combined_k141_39649\|\|full | 41529 | No | NA | 44 | 2 | 8 | Low-quality | Genome-fragment | 49.04 | HMM-based (lower-bound) | 0 |
| S8_k141_148006 | 1595 | No | NA | 1 | 0 | 1 | Not-determined | Genome-fragment | NA | NA | 0 |
| S8_k141_78607 | 1171 | No | NA | 2 | 0 | 1 | Not-determined | Genome-fragment | NA | NA | 0 |
| S8_k141_124182 | 1348 | No | NA | 1 | 0 | 1 | Not-determined | Genome-fragment | NA | NA | 0 |
| S8_k141_89152 | 1410 | No | NA | 1 | 0 | 1 | Not-determined | Genome-fragment | NA | NA | 0 |
| S8_k141_136363 | 1687 | No | NA | 1 | 0 | 0 | Not-determined | Genome-fragment | NA | NA | 0 |
| S8_k141_227743 | 1386 | No | NA | 1 | 0 | 0 | Not-determined | Genome-fragment | NA | NA | 0 |
| S8_k141_19114 | 1065 | No | NA | 1 | 0 | 1 | Not-determined | Genome-fragment | NA | NA | 0 |
| S8_k141_193354 | 1472 | No | NA | 3 | 0 | 1 | Not-determined | Genome-fragment | NA | NA | 0 |
| S8_k141_136378 | 1865 | No | NA | 2 | 0 | 1 | Not-determined | Genome-fragment | NA | NA | 0 |
| S8_k141_250918 | 4478 | No | NA | 9 | 0 | 0 | Not-determined | Genome-fragment | NA | NA | 0 |
| S8_k141_159039 | 1166 | No | NA | 2 | 0 | 2 | Not-determined | Genome-fragment | NA | NA | 0 |
| S8_k141_44012 | 1572 | No | NA | 2 | 0 | 0 | Not-determined | Genome-fragment | NA | NA | 0 |
| S8_k141_182987 | 3301 | No | NA | 3 | 0 | 1 | Not-determined | Genome-fragment | NA | NA | 0 |
| S8_k141_170473 | 3349 | No | NA | 3 | 0 | 2 | Not-determined | Genome-fragment | NA | NA | 0 |
| S8-final-viral-combined_k141_183742\|\|full | 41665 | Yes | 12280 | 47 | 6 | 21 | Low-quality | Genome-fragment | 14.24 | HMM-based (lower-bound) | 70.53 |
| S8_k141_148056 | 1041 | No | NA | 2 | 0 | 0 | Not-determined | Genome-fragment | NA | NA | 0 |
| S8_k141_203664 | 2073 | No | NA | 2 | 0 | 0 | Not-determined | Genome-fragment | NA | NA | 0 |
| S5-final-viral-combined_k141_52731\|\|full | 41772 | No | NA | 50 | 1 | 7 | Low-quality | Genome-fragment | 16.98 | HMM-based (lower-bound) | 0 |
| S8_k141_193467 | 3092 | No | NA | 2 | 0 | 1 | Not-determined | Genome-fragment | NA | NA | 0 |
| S8_k141_170549 | 3619 | No | NA | 2 | 0 | 2 | Not-determined | Genome-fragment | NA | NA | 0 |
| S2-final-viral-combined_k141_84659\|\|full | 41944 | No | NA | 37 | 3 | 2 | Low-quality | Genome-fragment | 37.88 | AAI-based (high-confidence) | 0 |
| S8_k141_227849 | 1175 | No | NA | 4 | 0 | 0 | Not-determined | Genome-fragment | NA | NA | 0 |
| S1-final-viral-combined_k141_262801\|\|full | 42016 | Yes | 20335 | 54 | 10 | 9 | Low-quality | Genome-fragment | 45.31 | AAI-based (high-confidence) | 51.6 |
| S8_k141_55153 | 1065 | No | NA | 1 | 0 | 0 | Not-determined | Genome-fragment | NA | NA | 0 |
| S8_k141_227861 | 1031 | No | NA | 1 | 0 | 1 | Not-determined | Genome-fragment | NA | NA | 0 |
| S8-final-viral-combined_k141_80639\|\|full | 42028 | Yes | 11768 | 40 | 6 | 23 | Low-quality | Genome-fragment | 19.41 | AAI-based (high-confidence) | 72 |
| S8_k141_159162 | 1185 | No | NA | 2 | 0 | 1 | Not-determined | Genome-fragment | NA | NA | 0 |
| S8_k141_8231 | 1325 | No | NA | 1 | 0 | 0 | Not-determined | Genome-fragment | NA | NA | 0 |
| S8_k141_170581 | 7107 | No | NA | 5 | 0 | 5 | Not-determined | Genome-fragment | NA | NA | 0 |
| S8_k141_193528 | 1594 | No | NA | 2 | 0 | 1 | Not-determined | Genome-fragment | NA | NA | 0 |
| S8_k141_136572 | 1525 | No | NA | 2 | 0 | 2 | Not-determined | Genome-fragment | NA | NA | 0 |
| S8_k141_216506 | 11021 | No | NA | 9 | 0 | 7 | Not-determined | Genome-fragment | NA | NA | 0 |
| S8_k141_67014 | 1532 | No | NA | 2 | 0 | 1 | Not-determined | Genome-fragment | NA | NA | 0 |
| S5-final-viral-combined_k141_179447\|\|full | 42059 | Yes | 20303 | 35 | 2 | 19 | Low-quality | Genome-fragment | 31.33 | AAI-based (medium-confidence) | 51.73 |
| S8-final-viral-combined_k141_132363\|\|full | 42292 | No | NA | 36 | 0 | 27 | Low-quality | Genome-fragment | 12.78 | AAI-based (medium-confidence) | 0 |
| S1-final-viral-combined_k141_130851\|\|full | 42384 | Yes | 20276 | 51 | 15 | 15 | Low-quality | Genome-fragment | 35.57 | AAI-based (medium-confidence) | 52.16 |
| S8_k141_78887 | 1360 | No | NA | 1 | 0 | 0 | Not-determined | Genome-fragment | NA | NA | 0 |
| S17-final-viral-combined_k141_47162\|\|0_partial | 43510 | Yes | 21909 | 63 | 13 | 15 | Low-quality | Genome-fragment | 44.66 | AAI-based (high-confidence) | 49.65 |
| S8_k141_148207 | 1294 | No | NA | 2 | 0 | 0 | Not-determined | Genome-fragment | NA | NA | 0 |
| S5-final-viral-combined_k141_13568\|\|full | 44532 | Yes | 11078 | 44 | 7 | 10 | Low-quality | Genome-fragment | 18.37 | HMM-based (lower-bound) | 75.12 |
| S8_k141_148233 | 1070 | No | NA | 2 | 0 | 1 | Not-determined | Genome-fragment | NA | NA | 0 |
| S8_k141_251082 | 1232 | No | NA | 2 | 0 | 0 | Not-determined | Genome-fragment | NA | NA | 0 |
| S8_k141_55221 | 5239 | No | NA | 10 | 0 | 10 | Not-determined | Genome-fragment | NA | NA | 0 |
| S8_k141_78917 | 1646 | No | NA | 1 | 0 | 1 | Not-determined | Genome-fragment | NA | NA | 0 |
| S2-final-viral-combined_k141_239032\|\|full | 44560 | No | NA | 35 | 1 | 23 | Low-quality | Genome-fragment | 17 | HMM-based (lower-bound) | 0 |
| S1-final-viral-combined_k141_227515\|\|full | 44610 | Yes | 8596 | 49 | 5 | 20 | Low-quality | Genome-fragment | 13.21 | HMM-based (lower-bound) | 80.73 |
| S8_k141_240520 | 4543 | No | NA | 6 | 0 | 1 | Not-determined | Genome-fragment | NA | NA | 0 |
| S8_k141_78947 | 1722 | No | NA | 2 | 0 | 1 | Not-determined | Genome-fragment | NA | NA | 0 |
| S7-final-viral-combined_k141_10758\|\|full | 44860 | No | NA | 44 | 1 | 4 | Low-quality | Genome-fragment | 12.68 | HMM-based (lower-bound) | 0 |
| S8_k141_19353 | 1405 | No | NA | 1 | 0 | 0 | Not-determined | Genome-fragment | NA | NA | 0 |
| S3-final-viral-combined_k141_15467\|\|full | 45105 | No | NA | 49 | 6 | 1 | Low-quality | Genome-fragment | 29.88 | AAI-based (high-confidence) | 0 |
| S8_k141_274105 | 7214 | No | NA | 4 | 0 | 3 | Not-determined | Genome-fragment | NA | NA | 0 |
| S8_k141_8358 | 1821 | No | NA | 1 | 0 | 1 | Not-determined | Genome-fragment | NA | NA | 0 |
| S8_k141_78971 | 1987 | No | NA | 3 | 0 | 2 | Not-determined | Genome-fragment | NA | NA | 0 |
| S8_k141_136687 | 1229 | No | NA | 1 | 0 | 1 | Not-determined | Genome-fragment | NA | NA | 0 |
| S2-final-viral-combined_k141_125615\|\|0_partial | 46036 | Yes | 16016 | 61 | 5 | 17 | Low-quality | Genome-fragment | 15.54 | AAI-based (medium-confidence) | 65.21 |
| S8_k141_33360 | 1105 | No | NA | 1 | 0 | 0 | Not-determined | Genome-fragment | NA | NA | 0 |
| S6-final-viral-combined_k141_134905\|\|full | 46301 | Yes | 12108 | 48 | 5 | 19 | Low-quality | Genome-fragment | 18.62 | HMM-based (lower-bound) | 73.85 |
| S8_k141_79021 | 4536 | No | NA | 4 | 0 | 1 | Not-determined | Genome-fragment | NA | NA | 0 |
| S8-final-viral-combined_k141_257014\|\|0_partial | 46422 | Yes | 27747 | 48 | 12 | 13 | Low-quality | Genome-fragment | 37.17 | HMM-based (lower-bound) | 40.23 |
| S12-final-viral-combined_k141_154703\|\|full | 46507 | No | NA | 52 | 3 | 11 | Low-quality | Genome-fragment | 23.61 | HMM-based (lower-bound) | 0 |
| S8_k141_170757 | 2382 | No | NA | 3 | 0 | 2 | Not-determined | Genome-fragment | NA | NA | 0 |
| S8_k141_136725 | 1830 | No | NA | 2 | 0 | 0 | Not-determined | Genome-fragment | NA | NA | 0 |
| S8_k141_203905 | 6887 | No | NA | 3 | 0 | 3 | Not-determined | Genome-fragment | NA | NA | 0 |
| S9-final-viral-combined_k141_5120\|\|full | 46706 | No | NA | 56 | 2 | 7 | Low-quality | Genome-fragment | 28.98 | HMM-based (lower-bound) | 0 |
| S8_k141_33399 | 1157 | No | NA | 1 | 0 | 1 | Not-determined | Genome-fragment | NA | NA | 0 |
| S8_k141_203921 | 2606 | No | NA | 4 | 0 | 0 | Not-determined | Genome-fragment | NA | NA | 0 |
| S8_k141_79063 | 1907 | No | NA | 1 | 0 | 1 | Not-determined | Genome-fragment | NA | NA | 0 |
| S1_k141_99716 | 46798 | No | NA | 49 | 12 | 0 | Low-quality | Genome-fragment | 34.19 | AAI-based (high-confidence) | 0 |
| S8_k141_240673 | 5184 | No | NA | 5 | 0 | 3 | Not-determined | Genome-fragment | NA | NA | 0 |
| S8_k141_240686 | 1110 | No | NA | 1 | 0 | 0 | Not-determined | Genome-fragment | NA | NA | 0 |
| S8_k141_216774 | 1126 | No | NA | 3 | 0 | 2 | Not-determined | Genome-fragment | NA | NA | 0 |
| S8_k141_251244 | 1000 | No | NA | 2 | 0 | 0 | Not-determined | Genome-fragment | NA | NA | 0 |
| S8_k141_136781 | 3342 | No | NA | 1 | 0 | 1 | Not-determined | Genome-fragment | NA | NA | 0 |
| S2_k141_230976 | 47181 | No | NA | 55 | 12 | 1 | Low-quality | Genome-fragment | 43.56 | AAI-based (high-confidence) | 0 |
| S8_k141_33472 | 1087 | No | NA | 1 | 0 | 0 | Not-determined | Genome-fragment | NA | NA | 0 |
| S8_k141_79156 | 4941 | No | NA | 2 | 0 | 1 | Not-determined | Genome-fragment | NA | NA | 0 |
| S8-final-viral-combined_k141_243662\|\|full | 47613 | Yes | 15607 | 51 | 5 | 19 | Low-quality | Genome-fragment | 23.66 | HMM-based (lower-bound) | 67.22 |
| S8_k141_240725 | 1720 | No | NA | 1 | 0 | 1 | Not-determined | Genome-fragment | NA | NA | 0 |
| S8_k141_112748 | 1890 | No | NA | 1 | 0 | 1 | Not-determined | Genome-fragment | NA | NA | 0 |
| S2-final-viral-combined_k141_131116\|\|0_partial | 47724 | Yes | 15946 | 58 | 4 | 19 | Low-quality | Genome-fragment | 34.81 | AAI-based (high-confidence) | 66.59 |
| S8_k141_136856 | 4764 | No | NA | 1 | 0 | 1 | Not-determined | Genome-fragment | NA | NA | 0 |
| S8_k141_44453 | 1097 | No | NA | 2 | 0 | 0 | Not-determined | Genome-fragment | NA | NA | 0 |
| S6_k141_101258 | 48465 | No | NA | 39 | 7 | 0 | Low-quality | Genome-fragment | 48.3 | AAI-based (high-confidence) | 0 |
| S1-final-viral-combined_k141_371708\|\|full | 48526 | Yes | 11650 | 48 | 9 | 27 | Low-quality | Genome-fragment | 25.43 | AAI-based (high-confidence) | 75.99 |
| S8_k141_251326 | 1365 | No | NA | 3 | 0 | 3 | Not-determined | Genome-fragment | NA | NA | 0 |
| S8_k141_33601 | 1009 | No | NA | 1 | 0 | 0 | Not-determined | Genome-fragment | NA | NA | 0 |
| S8_k141_101846 | 4842 | No | NA | 5 | 0 | 2 | Not-determined | Genome-fragment | NA | NA | 0 |
| S8_k141_44582 | 1204 | No | NA | 1 | 0 | 1 | Not-determined | Genome-fragment | NA | NA | 0 |
| S8_k141_79303 | 1249 | No | NA | 2 | 0 | 0 | Not-determined | Genome-fragment | NA | NA | 0 |
| S8_k141_204136 | 1253 | No | NA | 1 | 0 | 1 | Not-determined | Genome-fragment | NA | NA | 0 |
| S8_k141_260856 | 1229 | No | NA | 2 | 0 | 1 | Not-determined | Genome-fragment | NA | NA | 0 |
| S8_k141_89817 | 1560 | No | NA | 2 | 0 | 2 | Not-determined | Genome-fragment | NA | NA | 0 |
| S2-final-viral-combined_k141_125370\|\|full | 48761 | Yes | 13802 | 52 | 6 | 21 | Low-quality | Genome-fragment | 28.23 | HMM-based (lower-bound) | 71.69 |
| S8_k141_112918 | 1213 | No | NA | 2 | 0 | 1 | Not-determined | Genome-fragment | NA | NA | 0 |
| S8_k141_204168 | 3053 | No | NA | 1 | 0 | 1 | Not-determined | Genome-fragment | NA | NA | 0 |
| S8_k141_204174 | 3000 | No | NA | 2 | 0 | 2 | Not-determined | Genome-fragment | NA | NA | 0 |
| S17-final-viral-combined_k141_186805\|\|full | 48935 | No | NA | 45 | 1 | 18 | Low-quality | Genome-fragment | 33.82 | HMM-based (lower-bound) | 0 |
| S8-final-viral-combined_k141_189169\|\|full | 49202 | Yes | 24143 | 53 | 11 | 13 | Low-quality | Genome-fragment | 40.02 | AAI-based (high-confidence) | 50.93 |
| S8_k141_193942 | 1336 | No | NA | 1 | 0 | 1 | Not-determined | Genome-fragment | NA | NA | 0 |
| S8_k141_148782 | 5608 | No | NA | 7 | 0 | 1 | Not-determined | Genome-fragment | NA | NA | 0 |
| S12-final-viral-combined_k141_88126\|\|full | 49396 | Yes | 12299 | 52 | 6 | 25 | Low-quality | Genome-fragment | 15.53 | HMM-based (lower-bound) | 75.1 |
| S8_k141_183721 | 1686 | No | NA | 1 | 0 | 0 | Not-determined | Genome-fragment | NA | NA | 0 |
| S8_k141_137058 | 1035 | No | NA | 1 | 0 | 0 | Not-determined | Genome-fragment | NA | NA | 0 |
| S8_k141_193981 | 1739 | No | NA | 1 | 0 | 1 | Not-determined | Genome-fragment | NA | NA | 0 |
| S8_k141_228285 | 3216 | No | NA | 1 | 0 | 0 | Not-determined | Genome-fragment | NA | NA | 0 |
| S8_k141_8619 | 1069 | No | NA | 2 | 0 | 0 | Not-determined | Genome-fragment | NA | NA | 0 |
| S8_k141_8620 | 1262 | No | NA | 1 | 0 | 1 | Not-determined | Genome-fragment | NA | NA | 0 |
| S8_k141_274271 | 1150 | No | NA | 2 | 0 | 0 | Not-determined | Genome-fragment | NA | NA | 0 |
| S8_k141_241130 | 1741 | No | NA | 1 | 0 | 0 | Not-determined | Genome-fragment | NA | NA | 0 |
| S8_k141_55753 | 1133 | No | NA | 1 | 0 | 1 | Not-determined | Genome-fragment | NA | NA | 0 |
| S8_k141_79555 | 1117 | No | NA | 1 | 0 | 0 | Not-determined | Genome-fragment | NA | NA | 0 |
| S8_k141_44765 | 1167 | No | NA | 2 | 0 | 0 | Not-determined | Genome-fragment | NA | NA | 0 |
| S8_k141_67612 | 1048 | No | NA | 2 | 0 | 1 | Not-determined | Genome-fragment | NA | NA | 0 |
| S8_k141_89970 | 1482 | No | NA | 2 | 0 | 1 | Not-determined | Genome-fragment | NA | NA | 0 |
| S2_k141_141503 | 49431 | No | NA | 84 | 11 | 4 | Low-quality | Genome-fragment | 28.23 | AAI-based (high-confidence) | 0 |
| S11_k141_267198 | 49437 | No | NA | 43 | 0 | 34 | Low-quality | Genome-fragment | 16.08 | AAI-based (medium-confidence) | 0 |
| S8_k141_102134 | 1061 | No | NA | 1 | 0 | 1 | Not-determined | Genome-fragment | NA | NA | 0 |
| S8_k141_148936 | 1072 | No | NA | 1 | 0 | 1 | Not-determined | Genome-fragment | NA | NA | 0 |
| S8_k141_137248 | 1734 | No | NA | 3 | 0 | 3 | Not-determined | Genome-fragment | NA | NA | 0 |
| S8_k141_171162 | 1899 | No | NA | 2 | 0 | 2 | Not-determined | Genome-fragment | NA | NA | 0 |
| S8_k141_19985 | 1227 | No | NA | 2 | 0 | 0 | Not-determined | Genome-fragment | NA | NA | 0 |
| S12-final-viral-combined_k141_221051\|\|full | 49651 | Yes | 23025 | 52 | 7 | 17 | Low-quality | Genome-fragment | 25.62 | HMM-based (lower-bound) | 53.63 |
| S8_k141_137277 | 1819 | No | NA | 1 | 0 | 1 | Not-determined | Genome-fragment | NA | NA | 0 |
| S8_k141_125008 | 1066 | No | NA | 2 | 0 | 1 | Not-determined | Genome-fragment | NA | NA | 0 |
| S8_k141_274352 | 1746 | No | NA | 2 | 0 | 1 | Not-determined | Genome-fragment | NA | NA | 0 |
| S8_k141_44886 | 1447 | No | NA | 1 | 0 | 1 | Not-determined | Genome-fragment | NA | NA | 0 |

| | | | | | | | | | | | |
|---|---|---|---|---|---|---|---|---|---|---|---|
| S8_k141_241227 | 2236 | No | NA | 4 | 0 | 2 | Not-determined | Genome-fragment | NA | NA | 0 |
| S8_k141_55874 | 1889 | No | NA | 2 | 0 | 1 | Not-determined | Genome-fragment | NA | NA | 0 |
| S8_k141_90131 | 1114 | No | NA | 1 | 0 | 1 | Not-determined | Genome-fragment | NA | NA | 0 |
| S8_k141_217047 | 1062 | No | NA | 2 | 0 | 2 | Not-determined | Genome-fragment | NA | NA | 0 |
| S8_k141_102220 | 1338 | No | NA | 1 | 0 | 1 | Not-determined | Genome-fragment | NA | NA | 0 |
| S8_k141_171300 | 4409 | No | NA | 2 | 0 | 2 | Not-determined | Genome-fragment | NA | NA | 0 |
| S17-final-viral-combined_k141_211644\|\|full | 49961 | Yes | 17882 | 55 | 12 | 21 | Low-quality | Genome-fragment | 33.6 | HMM-based (lower-bound) | 64.21 |
| S2_k141_223642 | 50340 | No | NA | 73 | 4 | 0 | Low-quality | Genome-fragment | 28.74 | AAI-based (high-confidence) | 0 |
| S6-final-viral-combined_k141_33563\|\|full | 50540 | Yes | 13572 | 51 | 10 | 21 | Low-quality | Genome-fragment | 26.96 | AAI-based (medium-confidence) | 73.15 |
| S17-k141_256069 | 50697 | No | NA | 60 | 4 | 4 | Low-quality | Genome-fragment | 38.01 | HMM-based (lower-bound) | 0 |
| S8_k141_149139 | 1021 | No | NA | 2 | 0 | 2 | Not-determined | Genome-fragment | NA | NA | 0 |
| S8_k141_228647 | 1194 | No | NA | 1 | 0 | 1 | Not-determined | Genome-fragment | NA | NA | 0 |
| S8_k141_55981 | 1744 | No | NA | 1 | 0 | 1 | Not-determined | Genome-fragment | NA | NA | 0 |
| S8_k141_261201 | 1230 | No | NA | 1 | 0 | 1 | Not-determined | Genome-fragment | NA | NA | 0 |
| S8_k141_8926 | 1223 | No | NA | 1 | 0 | 1 | Not-determined | Genome-fragment | NA | NA | 0 |
| S8_k141_55998 | 1034 | No | NA | 1 | 0 | 0 | Not-determined | Genome-fragment | NA | NA | 0 |
| S8_k141_56005 | 3681 | No | NA | 1 | 0 | 1 | Not-determined | Genome-fragment | NA | NA | 0 |
| S4-final-viral-combined_k141_21132\|\|full | 51590 | Yes | 15289 | 48 | 8 | 22 | Low-quality | Genome-fragment | 30.45 | HMM-based (lower-bound) | 70.36 |
| S8-final-viral-combined_k141_253766\|\|full | 52823 | Yes | 18479 | 48 | 6 | 19 | Low-quality | Genome-fragment | 44.11 | AAI-based (high-confidence) | 65.02 |
| S8_k141_274593 | 1445 | No | NA | 1 | 0 | 0 | Not-determined | Genome-fragment | NA | NA | 0 |
| S7-final-viral-combined_k141_65892\|\|full | 52939 | Yes | 27019 | 57 | 8 | 18 | Low-quality | Genome-fragment | 46.25 | AAI-based (high-confidence) | 48.96 |
| S8_k141_67980 | 1148 | No | NA | 2 | 0 | 2 | Not-determined | Genome-fragment | NA | NA | 0 |
| S8_k141_56060 | 1456 | No | NA | 1 | 0 | 1 | Not-determined | Genome-fragment | NA | NA | 0 |
| S1-final-viral-combined_k141_128221\|\|full | 53298 | Yes | 16600 | 44 | 7 | 13 | Low-quality | Genome-fragment | 26.65 | HMM-based (lower-bound) | 68.85 |
| S8_k141_194399 | 1415 | No | NA | 1 | 0 | 0 | Not-determined | Genome-fragment | NA | NA | 0 |
| S8_k141_56077 | 3212 | No | NA | 4 | 0 | 4 | Not-determined | Genome-fragment | NA | NA | 0 |
| S8_k141_228728 | 1203 | No | NA | 1 | 0 | 1 | Not-determined | Genome-fragment | NA | NA | 0 |
| S8_k141_34176 | 2225 | No | NA | 1 | 0 | 1 | Not-determined | Genome-fragment | NA | NA | 0 |
| S12-final-viral-combined_k141_2715\|\|full | 53351 | Yes | 12512 | 61 | 6 | 25 | Low-quality | Genome-fragment | 25.43 | HMM-based (lower-bound) | 76.55 |
| S8_k141_149341 | 1091 | No | NA | 1 | 0 | 0 | Not-determined | Genome-fragment | NA | NA | 0 |
| S17-final-viral-combined_k141_12762\|\|full | 53645 | Yes | 16547 | 48 | 6 | 13 | Low-quality | Genome-fragment | 26.55 | HMM-based (lower-bound) | 69.15 |
| S12-final-viral-combined_k141_119829\|\|full | 54392 | No | NA | 59 | 1 | 30 | Low-quality | Genome-fragment | 30.01 | HMM-based (lower-bound) | 0 |
| S8_k141_251966 | 1209 | No | NA | 1 | 0 | 1 | Not-determined | Genome-fragment | NA | NA | 0 |
| S8_k141_217235 | 3258 | No | NA | 5 | 0 | 2 | Not-determined | Genome-fragment | NA | NA | 0 |
| S8_k141_160136 | 4291 | No | NA | 7 | 0 | 2 | Not-determined | Genome-fragment | NA | NA | 0 |
| S8_k141_204638 | 1270 | No | NA | 1 | 0 | 1 | Not-determined | Genome-fragment | NA | NA | 0 |
| S8_k141_160148 | 1015 | No | NA | 1 | 0 | 0 | Not-determined | Genome-fragment | NA | NA | 0 |
| S8_k141_9092 | 1201 | No | NA | 1 | 0 | 1 | Not-determined | Genome-fragment | NA | NA | 0 |
| S5_k141_82331 | 54701 | No | NA | 43 | 1 | 28 | Low-quality | Genome-fragment | 31.53 | HMM-based (lower-bound) | 0 |
| S8_k141_160173 | 1397 | No | NA | 2 | 0 | 0 | Not-determined | Genome-fragment | NA | NA | 0 |
| S8_k141_228829 | 3063 | No | NA | 5 | 0 | 2 | Not-determined | Genome-fragment | NA | NA | 0 |
| S8_k141_204675 | 1200 | No | NA | 1 | 0 | 1 | Not-determined | Genome-fragment | NA | NA | 0 |
| S17-final-viral-combined_k141_175742\|\|full | 54877 | Yes | 7128 | 62 | 6 | 28 | Low-quality | Genome-fragment | 18.33 | AAI-based (medium-confidence) | 87.01 |
| S8_k141_113590 | 2116 | No | NA | 2 | 0 | 2 | Not-determined | Genome-fragment | NA | NA | 0 |
| S8_k141_211385 | 55168 | No | NA | 58 | 1 | 9 | Low-quality | Genome-fragment | 22.41 | HMM-based (lower-bound) | 0 |
| S8_k141_9134 | 2069 | No | NA | 1 | 0 | 1 | Not-determined | Genome-fragment | NA | NA | 0 |
| S8_k141_45414 | 1194 | No | NA | 1 | 0 | 1 | Not-determined | Genome-fragment | NA | NA | 0 |
| S8_k141_137721 | 2063 | No | NA | 1 | 0 | 1 | Not-determined | Genome-fragment | NA | NA | 0 |
| S2_k141_30628 | 55427 | No | NA | 55 | 7 | 2 | Low-quality | Genome-fragment | 49.56 | AAI-based (high-confidence) | 0 |
| S8_k141_252015 | 2420 | No | NA | 2 | 0 | 2 | Not-determined | Genome-fragment | NA | NA | 0 |
| S8_k141_184280 | 1948 | No | NA | 2 | 0 | 0 | Not-determined | Genome-fragment | NA | NA | 0 |
| S8_k141_9161 | 2797 | No | NA | 3 | 0 | 2 | Not-determined | Genome-fragment | NA | NA | 0 |
| S8_k141_68211 | 1479 | No | NA | 1 | 0 | 1 | Not-determined | Genome-fragment | NA | NA | 0 |
| S5-final-viral-combined_k141_71959\|\|full | 55574 | No | NA | 56 | 2 | 28 | Low-quality | Genome-fragment | 32.75 | AAI-based (medium-confidence) | 0 |
| S8_k141_90540 | 1030 | No | NA | 2 | 0 | 0 | Not-determined | Genome-fragment | NA | NA | 0 |
| S1-final-viral-combined_k141_318869\|\|full | 56055 | Yes | 25620 | 69 | 3 | 18 | Low-quality | Genome-fragment | 13 | HMM-based (lower-bound) | 54.29 |
| S8_k141_274787 | 1247 | No | NA | 2 | 0 | 0 | Not-determined | Genome-fragment | NA | NA | 0 |
| S8_k141_90546 | 1068 | No | NA | 1 | 0 | 1 | Not-determined | Genome-fragment | NA | NA | 0 |
| S8_k141_68231 | 1033 | No | NA | 1 | 0 | 0 | Not-determined | Genome-fragment | NA | NA | 0 |
| S8_k141_20283 | 1136 | No | NA | 1 | 0 | 1 | Not-determined | Genome-fragment | NA | NA | 0 |
| S11-final-viral-combined_k141_205111\|\|full | 56280 | Yes | 11289 | 56 | 11 | 24 | Low-quality | Genome-fragment | 18.71 | AAI-based (high-confidence) | 79.94 |
| S2-final-viral-combined_k141_158151\|\|full | 56365 | Yes | 21866 | 49 | 2 | 17 | Low-quality | Genome-fragment | 25.86 | HMM-based (lower-bound) | 61.21 |
| S8_k141_252132 | 2085 | No | NA | 3 | 0 | 2 | Not-determined | Genome-fragment | NA | NA | 0 |
| S1-final-viral-combined_k141_179384\|\|full | 56428 | Yes | 24624 | 62 | 4 | 17 | Low-quality | Genome-fragment | 46.1 | AAI-based (high-confidence) | 56.36 |
| S8_k141_113693 | 19665 | No | NA | 23 | 10 | 1 | Medium-quality | Genome-fragment | 57.35 | AAI-based (medium-confidence) | 0 |
| S8_k141_228999 | 1932 | No | NA | 3 | 0 | 0 | Not-determined | Genome-fragment | NA | NA | 0 |
| S8_k141_204826 | 1089 | No | NA | 1 | 0 | 1 | Not-determined | Genome-fragment | NA | NA | 0 |
| S8_k141_261474 | 1896 | No | NA | 2 | 0 | 2 | Not-determined | Genome-fragment | NA | NA | 0 |
| S8_k141_160365 | 1193 | No | NA | 1 | 0 | 1 | Not-determined | Genome-fragment | NA | NA | 0 |
| S8_k141_241503 | 2933 | No | NA | 2 | 0 | 1 | Not-determined | Genome-fragment | NA | NA | 0 |
| S8_k141_90668 | 1267 | No | NA | 1 | 0 | 0 | Not-determined | Genome-fragment | NA | NA | 0 |
| S7_k141_40754 | 56723 | No | NA | 52 | 1 | 12 | Low-quality | Genome-fragment | 30.77 | HMM-based (lower-bound) | 0 |
| S8_k141_241535 | 1167 | No | NA | 1 | 0 | 1 | Not-determined | Genome-fragment | NA | NA | 0 |
| S8-final-viral-combined_k141_217031\|\|full | 57516 | Yes | 14334 | 55 | 3 | 36 | Low-quality | Genome-fragment | 34.58 | AAI-based (medium-confidence) | 75.08 |
| S9-final-viral-combined_k141_103057\|\|full | 57518 | Yes | 10772 | 60 | 6 | 29 | Low-quality | Genome-fragment | 16.72 | HMM-based (lower-bound) | 81.27 |
| S8_k141_204901 | 1040 | No | NA | 1 | 0 | 1 | Not-determined | Genome-fragment | NA | NA | 0 |
| S8_k141_9296 | 1375 | No | NA | 1 | 0 | 1 | Not-determined | Genome-fragment | NA | NA | 0 |
| S8_k141_241583 | 1447 | No | NA | 1 | 0 | 1 | Not-determined | Genome-fragment | NA | NA | 0 |
| S8-final-viral-combined_k141_105882\|\|full | 57691 | Yes | 17957 | 57 | 4 | 34 | Low-quality | Genome-fragment | 26.05 | HMM-based (lower-bound) | 68.87 |
| S8_k141_171739 | 1705 | No | NA | 1 | 0 | 1 | Not-determined | Genome-fragment | NA | NA | 0 |
| S5_k141_33219 | 58260 | No | NA | 47 | 5 | 0 | Low-quality | Genome-fragment | 37.88 | AAI-based (high-confidence) | 0 |
| S1-final-viral-combined_k141_23655\|\|full | 58420 | Yes | 19786 | 54 | 11 | 18 | Low-quality | Genome-fragment | 32.8 | AAI-based (high-confidence) | 66.13 |
| S8_k141_160473 | 1164 | No | NA | 2 | 0 | 1 | Not-determined | Genome-fragment | NA | NA | 0 |
| S9-final-viral-combined_k141_108121\|\|full | 58563 | Yes | 13906 | 72 | 25 | 37 | Low-quality | Genome-fragment | 19.26 | HMM-based (lower-bound) | 76.25 |
| S7-final-viral-combined_k141_41216\|\|full | 58752 | Yes | 17890 | 52 | 2 | 24 | Low-quality | Genome-fragment | 39 | AAI-based (high-confidence) | 69.55 |
| S8_k141_171757 | 1052 | No | NA | 3 | 0 | 1 | Not-determined | Genome-fragment | NA | NA | 0 |
| S17-final-viral-combined_k141_141841\|\|full | 58804 | Yes | 16182 | 59 | 14 | 24 | Low-quality | Genome-fragment | 44.41 | AAI-based (medium-confidence) | 72.48 |
| S8_k141_68457 | 1032 | No | NA | 2 | 0 | 1 | Not-determined | Genome-fragment | NA | NA | 0 |
| S8_k141_217552 | 1357 | No | NA | 2 | 0 | 1 | Not-determined | Genome-fragment | NA | NA | 0 |
| S8_k141_160503 | 1662 | No | NA | 2 | 0 | 1 | Not-determined | Genome-fragment | NA | NA | 0 |
| S8_k141_137983 | 4256 | No | NA | 2 | 0 | 2 | Not-determined | Genome-fragment | NA | NA | 0 |
| S8_k141_217564 | 1095 | No | NA | 2 | 0 | 2 | Not-determined | Genome-fragment | NA | NA | 0 |
| S9-final-viral-combined_k141_113207\|\|full | 59358 | No | NA | 58 | 2 | 30 | Low-quality | Genome-fragment | 46.34 | AAI-based (medium-confidence) | 0 |
| S8_k141_217574 | 1502 | No | NA | 2 | 0 | 0 | Not-determined | Genome-fragment | NA | NA | 0 |
| S8_k141_204975 | 1182 | No | NA | 2 | 0 | 1 | Not-determined | Genome-fragment | NA | NA | 0 |
| S8_k141_102752 | 1574 | No | NA | 2 | 0 | 0 | Not-determined | Genome-fragment | NA | NA | 0 |
| S8_k141_125621 | 1559 | No | NA | 2 | 0 | 2 | Not-determined | Genome-fragment | NA | NA | 0 |
| S8_k141_275028 | 1320 | No | NA | 3 | 0 | 1 | Not-determined | Genome-fragment | NA | NA | 0 |
| S5-final-viral-combined_k141_18783\|\|full | 59609 | Yes | 16696 | 60 | 6 | 21 | Low-quality | Genome-fragment | 19.24 | HMM-based (lower-bound) | 71.99 |
| S8_k141_80267 | 1337 | No | NA | 2 | 0 | 1 | Not-determined | Genome-fragment | NA | NA | 0 |
| S12-final-viral-combined_k141_170610\|\|full | 60149 | Yes | 20318 | 70 | 9 | 33 | Low-quality | Genome-fragment | 48.48 | AAI-based (high-confidence) | 66.22 |
| S8_k141_194926 | 1280 | No | NA | 2 | 0 | 1 | Not-determined | Genome-fragment | NA | NA | 0 |
| S9-final-viral-combined_k141_27514\|\|full | 60397 | Yes | 31680 | 75 | 14 | 23 | Low-quality | Genome-fragment | 46.93 | HMM-based (lower-bound) | 47.55 |
| S5-final-viral-combined_k141_68979\|\|full | 60791 | No | NA | 61 | 3 | 15 | Low-quality | Genome-fragment | 29.4 | AAI-based (medium-confidence) | 0 |
| S8_k141_160572 | 4347 | No | NA | 1 | 0 | 0 | Not-determined | Genome-fragment | NA | NA | 0 |
| S8_k141_68515 | 1116 | No | NA | 1 | 0 | 1 | Not-determined | Genome-fragment | NA | NA | 0 |
| S8_k141_20528 | 43375 | No | NA | 57 | 31 | 0 | High-quality | High-quality | 98.6 | AAI-based (high-confidence) | 0 |
| S8_k141_56481 | 1557 | No | NA | 1 | 0 | 0 | Not-determined | Genome-fragment | NA | NA | 0 |
| S8_k141_229154 | 1968 | No | NA | 1 | 0 | 0 | Not-determined | Genome-fragment | NA | NA | 0 |
| S12_k141_108569 | 60967 | Yes | 56966 | 86 | 15 | 9 | Low-quality | Genome-fragment | 47.27 | AAI-based (high-confidence) | 6.56 |
| S8_k141_80376 | 1131 | No | NA | 1 | 0 | 0 | Not-determined | Genome-fragment | NA | NA | 0 |
| S8_k141_9533 | 2463 | No | NA | 2 | 0 | 1 | Not-determined | Genome-fragment | NA | NA | 0 |
| S8_k141_261706 | 1226 | No | NA | 2 | 0 | 2 | Not-determined | Genome-fragment | NA | NA | 0 |
| S1-final-viral-combined_k141_313409\|\|full | 61049 | Yes | 10610 | 65 | 2 | 33 | Low-quality | Genome-fragment | 7.79 | HMM-based (lower-bound) | 82.62 |
| S8_k141_45836 | 1242 | No | NA | 2 | 0 | 1 | Not-determined | Genome-fragment | NA | NA | 0 |
| S8_k141_195016 | 2269 | No | NA | 2 | 0 | 1 | Not-determined | Genome-fragment | NA | NA | 0 |
| S8_k141_252411 | 1067 | No | NA | 1 | 0 | 1 | Not-determined | Genome-fragment | NA | NA | 0 |
| S8_k141_125852 | 1525 | No | NA | 3 | 0 | 3 | Not-determined | Genome-fragment | NA | NA | 0 |
| S8_k141_20605 | 1176 | No | NA | 1 | 0 | 1 | Not-determined | Genome-fragment | NA | NA | 0 |
| S8_k141_171961 | 1234 | No | NA | 1 | 0 | 0 | Not-determined | Genome-fragment | NA | NA | 0 |
| S6_k141_12993 | 62126 | No | NA | 36 | 4 | 0 | Low-quality | Genome-fragment | 34.98 | AAI-based (high-confidence) | 0 |
| S6-final-viral-combined_k141_124845\|\|full | 63201 | No | NA | 75 | 3 | 17 | Low-quality | Genome-fragment | 49.45 | HMM-based (lower-bound) | 0 |
| S2-final-viral-combined_k141_218586\|\|full | 63434 | Yes | 20703 | 64 | 3 | 21 | Low-quality | Genome-fragment | 34.79 | AAI-based (medium-confidence) | 67.36 |
| S8_k141_138168 | 1537 | No | NA | 1 | 0 | 1 | Not-determined | Genome-fragment | NA | NA | 0 |
| S8_k141_56554 | 8120 | No | NA | 6 | 0 | 2 | Not-determined | Genome-fragment | NA | NA | 0 |
| S8_k141_45863 | 1016 | No | NA | 2 | 0 | 1 | Not-determined | Genome-fragment | NA | NA | 0 |
| S8_k141_205162 | 1007 | No | NA | 1 | 0 | 1 | Not-determined | Genome-fragment | NA | NA | 0 |
| S8_k141_275232 | 1316 | No | NA | 1 | 0 | 0 | Not-determined | Genome-fragment | NA | NA | 0 |
| S8_k141_275234 | 1183 | No | NA | 1 | 0 | 1 | Not-determined | Genome-fragment | NA | NA | 0 |
| S8_k141_68549 | 5977 | No | NA | 7 | 0 | 0 | Not-determined | Genome-fragment | NA | NA | 0 |
| S8_k141_252463 | 1139 | No | NA | 2 | 0 | 1 | Not-determined | Genome-fragment | NA | NA | 0 |
| S8_k141_20669 | 1077 | No | NA | 4 | 0 | 0 | Not-determined | Genome-fragment | NA | NA | 0 |
| S11-final-viral-combined_k141_28934\|\|full | 64287 | Yes | 20121 | 75 | 12 | 32 | Low-quality | Genome-fragment | 26.06 | HMM-based (lower-bound) | 68.7 |
| S8_k141_113997 | 1790 | No | NA | 3 | 0 | 2 | Not-determined | Genome-fragment | NA | NA | 0 |
| S8_k141_160716 | 1199 | No | NA | 1 | 0 | 0 | Not-determined | Genome-fragment | NA | NA | 0 |
| S8_k141_275273 | 6219 | No | NA | 4 | 0 | 2 | Not-determined | Genome-fragment | NA | NA | 0 |
| S8_k141_68601 | 1041 | No | NA | 1 | 0 | 1 | Not-determined | Genome-fragment | NA | NA | 0 |
| S8_k141_45961 | 1364 | No | NA | 1 | 0 | 0 | Not-determined | Genome-fragment | NA | NA | 0 |
| S8_k141_172156 | 1156 | No | NA | 1 | 0 | 1 | Not-determined | Genome-fragment | NA | NA | 0 |
| S8_k141_114080 | 1272 | No | NA | 2 | 0 | 0 | Not-determined | Genome-fragment | NA | NA | 0 |
| S8_k141_172164 | 1509 | No | NA | 1 | 0 | 1 | Not-determined | Genome-fragment | NA | NA | 0 |
| S8_k141_195101 | 1257 | No | NA | 1 | 0 | 1 | Not-determined | Genome-fragment | NA | NA | 0 |
| S8_k141_20816 | 1166 | No | NA | 3 | 0 | 0 | Not-determined | Genome-fragment | NA | NA | 0 |
| S8_k141_275434 | 1009 | No | NA | 2 | 0 | 2 | Not-determined | Genome-fragment | NA | NA | 0 |
| S8_k141_205321 | 2004 | No | NA | 1 | 0 | 1 | Not-determined | Genome-fragment | NA | NA | 0 |
| S8_k141_217944 | 1621 | No | NA | 2 | 0 | 1 | Not-determined | Genome-fragment | NA | NA | 0 |
| S12-final-viral-combined_k141_3824\|\|full | 64336 | Yes | 13997 | 54 | 6 | 24 | Low-quality | Genome-fragment | 22.46 | HMM-based (lower-bound) | 78.24 |
| S17-final-viral-combined_k141_184906\|\|full | 66539 | Yes | 15238 | 85 | 10 | 19 | Low-quality | Genome-fragment | 30.13 | HMM-based (lower-bound) | 77.1 |
| S8_k141_217954 | 1235 | No | NA | 2 | 0 | 2 | Not-determined | Genome-fragment | NA | NA | 0 |
| S12-final-viral-combined_k141_295794\|\|full | 67638 | Yes | 18946 | 63 | 4 | 33 | Low-quality | Genome-fragment | 14.11 | AAI-based (medium-confidence) | 71.99 |
| S8_k141_172239 | 1117 | No | NA | 1 | 0 | 1 | Not-determined | Genome-fragment | NA | NA | 0 |
| S8_k141_205349 | 1168 | No | NA | 1 | 0 | 1 | Not-determined | Genome-fragment | NA | NA | 0 |
| S8_k141_261868 | 1160 | No | NA | 2 | 0 | 1 | Not-determined | Genome-fragment | NA | NA | 0 |
| S8_k141_91318 | 1645 | No | NA | 1 | 0 | 1 | Not-determined | Genome-fragment | NA | NA | 0 |
| S1-final-viral-combined_k141_200256\|\|full | 69601 | Yes | 8549 | 57 | 2 | 39 | Low-quality | Genome-fragment | 18.68 | HMM-based (lower-bound) | 87.72 |
| S8_k141_205379 | 1157 | No | NA | 1 | 0 | 0 | Not-determined | Genome-fragment | NA | NA | 0 |
| S11-final-viral-combined_k141_138533\|\|full | 71410 | Yes | 11703 | 78 | 4 | 36 | Low-quality | Genome-fragment | 22.29 | HMM-based (lower-bound) | 83.61 |
| S8_k141_46058 | 1690 | No | NA | 2 | 0 | 0 | Not-determined | Genome-fragment | NA | NA | 0 |
| S8_k141_91341 | 1205 | No | NA | 2 | 0 | 1 | Not-determined | Genome-fragment | NA | NA | 0 |
| S8_k141_229511 | 5190 | No | NA | 6 | 0 | 3 | Not-determined | Genome-fragment | NA | NA | 0 |
| S7-final-viral-combined_k141_27337\|\|full | 72311 | Yes | 55695 | 88 | 5 | 9 | Low-quality | Genome-fragment | 49.83 | HMM-based (lower-bound) | 22.98 |
| S4_k141_55031 | 74293 | No | NA | 164 | 51 | 0 | Low-quality | Genome-fragment | 42.8 | AAI-based (high-confidence) | 0 |
| S8_k141_275511 | 1190 | No | NA | 3 | 0 | 2 | Not-determined | Genome-fragment | NA | NA | 0 |
| S5-final-viral-combined_k141_54548\|\|full | 74676 | No | NA | 88 | 2 | 16 | Low-quality | Genome-fragment | 46.3 | HMM-based (lower-bound) | 0 |
| S8_k141_218112 | 1722 | No | NA | 1 | 0 | 1 | Not-determined | Genome-fragment | NA | NA | 0 |
| S6-final-viral-combined_k141_89744\|\|full | 75785 | Yes | 14866 | 93 | 11 | 31 | Low-quality | Genome-fragment | 42.49 | AAI-based (high-confidence) | 80.38 |
| S17-final-viral-combined_k141_66285\|\|full | 78479 | Yes | 22300 | 82 | 7 | 48 | Low-quality | Genome-fragment | 41.36 | HMM-based (lower-bound) | 71.58 |
| S8_k141_172451 | 1181 | No | NA | 2 | 0 | 1 | Not-determined | Genome-fragment | NA | NA | 0 |
| S8_k141_9880 | 4305 | No | NA | 2 | 0 | 2 | Not-determined | Genome-fragment | NA | NA | 0 |
| S8_k141_126265 | 1677 | No | NA | 1 | 0 | 1 | Not-determined | Genome-fragment | NA | NA | 0 |
| S11-final-viral-combined_k141_307157\|\|full | 78492 | Yes | 17704 | 84 | 9 | 44 | Low-quality | Genome-fragment | 39.66 | HMM-based (lower-bound) | 77.44 |
| S8_k141_20989 | 1872 | No | NA | 1 | 0 | 1 | Not-determined | Genome-fragment | NA | NA | 0 |
| S8_k141_114259 | 4431 | No | NA | 3 | 0 | 1 | Not-determined | Genome-fragment | NA | NA | 0 |
| S8_k141_161016 | 1684 | No | NA | 1 | 0 | 0 | Not-determined | Genome-fragment | NA | NA | 0 |
| S8_k141_103165 | 6589 | No | NA | 9 | 0 | 1 | Not-determined | Genome-fragment | NA | NA | 0 |
| S8_k141_229697 | 1696 | No | NA | 1 | 0 | 1 | Not-determined | Genome-fragment | NA | NA | 0 |
| S8_k141_161035 | 2235 | No | NA | 6 | 0 | 0 | Not-determined | Genome-fragment | NA | NA | 0 |
| S8_k141_103201 | 2295 | No | NA | 3 | 0 | 1 | Not-determined | Genome-fragment | NA | NA | 0 |
| S8_k141_275648 | 1153 | No | NA | 2 | 0 | 0 | Not-determined | Genome-fragment | NA | NA | 0 |
| S8_k141_229747 | 1005 | No | NA | 1 | 0 | 1 | Not-determined | Genome-fragment | NA | NA | 0 |
| S8_k141_9990 | 1042 | No | NA | 1 | 0 | 1 | Not-determined | Genome-fragment | NA | NA | 0 |
| S2-final-viral-combined_k141_80784\|\|full | 79666 | Yes | 24009 | 81 | 2 | 37 | Low-quality | Genome-fragment | 21.65 | AAI-based (high-confidence) | 69.86 |
| S8_k141_195322 | 1233 | No | NA | 1 | 0 | 1 | Not-determined | Genome-fragment | NA | NA | 0 |
| S8_k141_229789 | 1046 | No | NA | 1 | 0 | 0 | Not-determined | Genome-fragment | NA | NA | 0 |
| S8_k141_275710 | 3356 | No | NA | 3 | 0 | 1 | Not-determined | Genome-fragment | NA | NA | 0 |
| S8_k141_195329 | 1078 | No | NA | 1 | 0 | 1 | Not-determined | Genome-fragment | NA | NA | 0 |
| S8_k141_229847 | 13929 | No | NA | 16 | 0 | 3 | Not-determined | Genome-fragment | NA | NA | 0 |
| S8_k141_68907 | 2452 | No | NA | 1 | 0 | 0 | Not-determined | Genome-fragment | NA | NA | 0 |
| S8_k141_205791 | 1157 | No | NA | 1 | 0 | 0 | Not-determined | Genome-fragment | NA | NA | 0 |
| S8_k141_218380 | 1100 | No | NA | 1 | 0 | 1 | Not-determined | Genome-fragment | NA | NA | 0 |
| S8_k141_68950 | 2056 | No | NA | 1 | 0 | 0 | Not-determined | Genome-fragment | NA | NA | 0 |
| S8_k141_252933 | 1579 | No | NA | 2 | 0 | 2 | Not-determined | Genome-fragment | NA | NA | 0 |
| S8_k141_218404 | 4421 | No | NA | 6 | 0 | 1 | Not-determined | Genome-fragment | NA | NA | 0 |
| S8_k141_57121 | 35030 | No | NA | 51 | 11 | 0 | High-quality | High-quality | 95.35 | AAI-based (high-confidence) | 0 |
| S8_k141_91838 | 1087 | No | NA | 2 | 0 | 1 | Not-determined | Genome-fragment | NA | NA | 0 |
| S8_k141_57124 | 1018 | No | NA | 2 | 0 | 1 | Not-determined | Genome-fragment | NA | NA | 0 |
| S8_k141_205835 | 1056 | No | NA | 2 | 0 | 0 | Not-determined | Genome-fragment | NA | NA | 0 |
| S8_k141_252967 | 1028 | No | NA | 2 | 0 | 0 | Not-determined | Genome-fragment | NA | NA | 0 |
| S8_k141_68996 | 1100 | No | NA | 2 | 0 | 0 | Not-determined | Genome-fragment | NA | NA | 0 |
| S7-final-viral-combined_k141_225455\|\|full | 79825 | Yes | 23509 | 76 | 13 | 37 | Low-quality | Genome-fragment | 36.54 | AAI-based (high-confidence) | 70.55 |
| S8_k141_103393 | 1100 | No | NA | 1 | 0 | 1 | Not-determined | Genome-fragment | NA | NA | 0 |
| S8_k141_205863 | 3427 | No | NA | 3 | 0 | 2 | Not-determined | Genome-fragment | NA | NA | 0 |
| S8_k141_103402 | 2855 | No | NA | 4 | 0 | 2 | Not-determined | Genome-fragment | NA | NA | 0 |
| S8-final-viral-combined_k141_249907\|\|full | 81356 | No | NA | 87 | 1 | 37 | Low-quality | Genome-fragment | 48.09 | HMM-based (lower-bound) | 0 |
| S8_k141_230000 | 1841 | No | NA | 1 | 0 | 0 | Not-determined | Genome-fragment | NA | NA | 0 |
| S8_k141_195448 | 2324 | No | NA | 1 | 0 | 0 | Not-determined | Genome-fragment | NA | NA | 0 |
| S8_k141_230014 | 3439 | No | NA | 3 | 0 | 3 | Not-determined | Genome-fragment | NA | NA | 0 |
| S8_k141_275929 | 1118 | No | NA | 1 | 0 | 0 | Not-determined | Genome-fragment | NA | NA | 0 |
| S8_k141_57219 | 3770 | No | NA | 3 | 0 | 1 | Not-determined | Genome-fragment | NA | NA | 0 |
| S2-final-viral-combined_k141_106420\|\|full | 84026 | Yes | 17717 | 93 | 7 | 43 | Low-quality | Genome-fragment | 39.47 | AAI-based (high-confidence) | 78.91 |
| S17-final-viral-combined_k141_45110\|\|full | 84635 | Yes | 8510 | 81 | 2 | 51 | Low-quality | Genome-fragment | 8.98 | HMM-based (lower-bound) | 89.95 |
| S8_k141_230058 | 1215 | No | NA | 2 | 0 | 1 | Not-determined | Genome-fragment | NA | NA | 0 |
| S11-final-viral-combined_k141_195758\|\|full | 87232 | Yes | 27458 | 82 | 11 | 43 | Low-quality | Genome-fragment | 42.39 | HMM-based (lower-bound) | 68.52 |
| S8_k141_195504 | 5505 | No | NA | 6 | 0 | 1 | Not-determined | Genome-fragment | NA | NA | 0 |
| S8_k141_230081 | 1307 | No | NA | 4 | 0 | 4 | Not-determined | Genome-fragment | NA | NA | 0 |
| S5-final-viral-combined_k141_163318\|\|full | 88243 | No | NA | 93 | 1 | 15 | Low-quality | Genome-fragment | 47.86 | HMM-based (lower-bound) | 0 |
| S8_k141_262319 | 1187 | No | NA | 2 | 0 | 1 | Not-determined | Genome-fragment | NA | NA | 0 |
| S8_k141_205942 | 2000 | No | NA | 1 | 0 | 0 | Not-determined | Genome-fragment | NA | NA | 0 |
| S8_k141_57316 | 3680 | No | NA | 5 | 0 | 0 | Not-determined | Genome-fragment | NA | NA | 0 |
| S17-final-viral-combined_k141_249003\|\|full | 90402 | Yes | 19532 | 81 | 7 | 34 | Low-quality | Genome-fragment | 27.74 | HMM-based (lower-bound) | 78.39 |
| S2-final-viral-combined_k141_87140\|\|full | 90928 | Yes | 33522 | 107 | 12 | 39 | Low-quality | Genome-fragment | 33.47 | HMM-based (lower-bound) | 63.13 |
| S8_k141_230141 | 3180 | No | NA | 3 | 0 | 2 | Not-determined | Genome-fragment | NA | NA | 0 |
| S8_k141_92005 | 1006 | No | NA | 1 | 0 | 1 | Not-determined | Genome-fragment | NA | NA | 0 |
| S8_k141_262408 | 3331 | No | NA | 3 | 0 | 3 | Not-determined | Genome-fragment | NA | NA | 0 |
| S5-final-viral-combined_k141_27904\|\|full | 91336 | Yes | 18540 | 83 | 7 | 43 | Low-quality | Genome-fragment | 33.1 | HMM-based (lower-bound) | 79.7 |
| S5-final-viral-combined_k141_94195\|\|full | 92378 | Yes | 18130 | 86 | 12 | 52 | Low-quality | Genome-fragment | 25.21 | HMM-based (lower-bound) | 80.37 |
| S8_k141_230227 | 3000 | No | NA | 1 | 0 | 1 | Not-determined | Genome-fragment | NA | NA | 0 |
| S8_k141_57441 | 2330 | No | NA | 2 | 0 | 2 | Not-determined | Genome-fragment | NA | NA | 0 |
| S8_k141_253270 | 1319 | No | NA | 2 | 0 | 2 | Not-determined | Genome-fragment | NA | NA | 0 |
| S8_k141_262439 | 1298 | No | NA | 1 | 0 | 1 | Not-determined | Genome-fragment | NA | NA | 0 |
| S7-final-viral-combined_k141_41336\|\|full | 95933 | Yes | 22680 | 110 | 10 | 29 | Low-quality | Genome-fragment | 41.15 | AAI-based (high-confidence) | 76.36 |
| S8_k141_276090 | 1110 | No | NA | 1 | 0 | 0 | Not-determined | Genome-fragment | NA | NA | 0 |
| S8_k141_230299 | 1059 | No | NA | 1 | 0 | 0 | Not-determined | Genome-fragment | NA | NA | 0 |
| S8_k141_218786 | 3380 | No | NA | 3 | 0 | 2 | Not-determined | Genome-fragment | NA | NA | 0 |
| S12-final-viral-combined_k141_259195\|\|full | 97360 | Yes | 51756 | 101 | 22 | 35 | Low-quality | Genome-fragment | 43.3 | AAI-based (high-confidence) | 46.84 |
| S8_k141_276202 | 11517 | No | NA | 15 | 0 | 1 | Not-determined | Genome-fragment | NA | NA | 0 |
| S7-final-viral-combined_k141_195842\|\|full | 98191 | Yes | 21525 | 100 | 7 | 51 | Low-quality | Genome-fragment | 38.51 | HMM-based (lower-bound) | 78.08 |
| S8_k141_10514 | 1444 | No | NA | 3 | 0 | 2 | Not-determined | Genome-fragment | NA | NA | 0 |
| S8_k141_276231 | 3709 | No | NA | 5 | 0 | 0 | Not-determined | Genome-fragment | NA | NA | 0 |
| S8_k141_262643 | 1905 | No | NA | 1 | 0 | 1 | Not-determined | Genome-fragment | NA | NA | 0 |
| S8_k141_10622 | 1060 | No | NA | 1 | 0 | 1 | Not-determined | Genome-fragment | NA | NA | 0 |
| S5-final-viral-combined_k141_32467\|\|full | 102578 | Yes | 64498 | 104 | 10 | 26 | Low-quality | Genome-fragment | 42.85 | HMM-based (lower-bound) | 37.12 |
| S8_k141_206419 | 1066 | No | NA | 2 | 0 | 1 | Not-determined | Genome-fragment | NA | NA | 0 |
| S8_k141_114979 | 1009 | No | NA | 2 | 0 | 1 | Not-determined | Genome-fragment | NA | NA | 0 |
| S6-final-viral-combined_k141_73644\|\|full | 105447 | Yes | 27309 | 101 | 5 | 47 | Low-quality | Genome-fragment | 43.4 | HMM-based (lower-bound) | 74.1 |
| S6-final-viral-combined_k141_10694\|\|full | 106838 | Yes | 20725 | 112 | 12 | 58 | Low-quality | Genome-fragment | 24.89 | HMM-based (lower-bound) | 80.6 |
| S2-final-viral-combined_k141_217413\|\|full | 115084 | Yes | 20266 | 118 | 19 | 68 | Low-quality | Genome-fragment | 30.03 | HMM-based (lower-bound) | 82.39 |
| S8_k141_262856 | 1479 | No | NA | 2 | 0 | 0 | Not-determined | Genome-fragment | NA | NA | 0 |
| S8_k141_262897 | 2800 | No | NA | 4 | 0 | 0 | Not-determined | Genome-fragment | NA | NA | 0 |
| S8_k141_21959 | 2211 | No | NA | 3 | 0 | 2 | Not-determined | Genome-fragment | NA | NA | 0 |
| S8_k141_115184 | 3838 | No | NA | 1 | 0 | 1 | Not-determined | Genome-fragment | NA | NA | 0 |
| S8_k141_21989 | 3119 | No | NA | 5 | 0 | 2 | Not-determined | Genome-fragment | NA | NA | 0 |
| S2-final-viral-combined_k141_16239\|\|full | 116261 | Yes | 19844 | 107 | 3 | 49 | Low-quality | Genome-fragment | 46.33 | AAI-based (high-confidence) | 82.93 |
| S8_k141_206724 | 5111 | No | NA | 5 | 0 | 5 | Not-determined | Genome-fragment | NA | NA | 0 |
| S8_k141_22079 | 3243 | No | NA | 1 | 0 | 0 | Not-determined | Genome-fragment | NA | NA | 0 |
| S2-final-viral-combined_k141_83135\|\|full | 119786 | Yes | 20368 | 111 | 6 | 60 | Low-quality | Genome-fragment | 14.58 | HMM-based (lower-bound) | 83 |
| S5-final-viral-combined_k141_174100\|\|full | 127560 | Yes | 24661 | 128 | 11 | 58 | Low-quality | Genome-fragment | 36.86 | HMM-based (lower-bound) | 80.67 |
| S5-final-viral-combined_k141_41618\|\|full | 134913 | Yes | 17541 | 145 | 18 | 71 | Low-quality | Genome-fragment | 25.94 | HMM-based (lower-bound) | 87 |
| S8_k141_22331 | 1206 | No | NA | 5 | 0 | 0 | Not-determined | Genome-fragment | NA | NA | 0 |
| S8_k141_263340 | 2320 | No | NA | 2 | 0 | 0 | Not-determined | Genome-fragment | NA | NA | 0 |
| S2-final-viral-combined_k141_156116\|\|full | 142752 | Yes | 33447 | 163 | 17 | 78 | Low-quality | Genome-fragment | 48.88 | HMM-based (lower-bound) | 76.57 |
| S8_k141_22478 | 2270 | No | NA | 2 | 0 | 1 | Not-determined | Genome-fragment | NA | NA | 0 |
| S8_k141_263535 | 3069 | No | NA | 4 | 0 | 4 | Not-determined | Genome-fragment | NA | NA | 0 |
| S8_k141_207135 | 1492 | No | NA | 2 | 0 | 1 | Not-determined | Genome-fragment | NA | NA | 0 |
| S12-final-viral-combined_k141_120161\|\|full | 143327 | Yes | 23850 | 148 | 13 | 77 | Low-quality | Genome-fragment | 37.9 | HMM-based (lower-bound) | 83.36 |
| S8_k141_22622 | 1958 | No | NA | 2 | 0 | 2 | Not-determined | Genome-fragment | NA | NA | 0 |
| S1-final-viral-combined_k141_278285\|\|full | 158368 | Yes | 40349 | 154 | 8 | 78 | Low-quality | Genome-fragment | 44.07 | HMM-based (lower-bound) | 74.52 |
| S8_k141_22657 | 1233 | No | NA | 1 | 0 | 1 | Not-determined | Genome-fragment | NA | NA | 0 |
| S8_k141_263769 | 1040 | No | NA | 1 | 0 | 0 | Not-determined | Genome-fragment | NA | NA | 0 |
| S8_k141_11313 | 4631 | No | NA | 8 | 0 | 3 | Not-determined | Genome-fragment | NA | NA | 0 |
| S8_k141_11327 | 3103 | No | NA | 5 | 0 | 2 | Not-determined | Genome-fragment | NA | NA | 0 |
| S8_k141_11358 | 1038 | No | NA | 2 | 0 | 1 | Not-determined | Genome-fragment | NA | NA | 0 |
| S1-final-viral-combined_k141_230950\|\|full | 161096 | Yes | 36081 | 153 | 9 | 73 | Low-quality | Genome-fragment | 41.22 | HMM-based (lower-bound) | 77.6 |
| S8_k141_264050 | 1138 | No | NA | 1 | 0 | 0 | Not-determined | Genome-fragment | NA | NA | 0 |
| S8_k141_264100 | 1271 | No | NA | 1 | 0 | 0 | Not-determined | Genome-fragment | NA | NA | 0 |
| S8_k141_264116 | 2239 | No | NA | 1 | 0 | 1 | Not-determined | Genome-fragment | NA | NA | 0 |
| S8_k141_264225 | 1570 | No | NA | 2 | 0 | 2 | Not-determined | Genome-fragment | NA | NA | 0 |
| S8_k141_264431 | 1113 | No | NA | 2 | 0 | 1 | Not-determined | Genome-fragment | NA | NA | 0 |
| S8_k141_264449 | 1075 | No | NA | 2 | 0 | 2 | Not-determined | Genome-fragment | NA | NA | 0 |
| S5-final-viral-combined_k141_63625\|\|full | 224706 | Yes | 32735 | 240 | 18 | 129 | Low-quality | Genome-fragment | 47.75 | HMM-based (lower-bound) | 85.43 |
| S8_k141_264841 | 1116 | No | NA | 1 | 0 | 1 | Not-determined | Genome-fragment | NA | NA | 0 |
| S8_k141_264900 | 1031 | No | NA | 1 | 0 | 0 | Not-determined | Genome-fragment | NA | NA | 0 |

| | Count | Mean | Median | SD |
|---|---|---|---|---|
| Complete (100% completeness) | 63 | 90560.9 | 83257 | 63003.9 |
| High quality (90-100% completeness) | 201 | 49058.3 | 44314 | 30073.6 |
| Medium quality (50-90% completeness) | 424 | 43571.4 | 31658 | 39059 |
| Low quality (< 50% completeness) | 21576 | 3939 | 2116.5 | 6663 |
| Undetermined | 37489 | 2877.2 | 1899 | 3250.7 |
| Total vOTUs >= 1kb | 59753 | 38001.4 | 31658 | 24590.8 |
| Sum of 50-100% genome completeness | 688 | 61063.5 | 44314 | 17022.05 |
