## Supplementary material for "Comprehensive profiling of the human intestinal DNA virome and prediction of disease-associated bacterial hosts in severe Myalgic Encephalomyelitis/Chronic Fatigue Syndrome (ME/CFS)": S Table 6 proviruses and plasmids

| vlp_seq_name | n_genes_with_taxonomy | agreement | taxid | lineage |
|---|---|---|---|---|
| S17_vs_110666\|provirus_6805_70348 | 58 | 1 | 2561 | Viruses;Duplodnaviria;Heunggongvirae;Uroviricota;Caudoviricetes |
| S9_vf_13507\|provirus_13823_120230 | 88 | 1 | 2561 | Viruses;Duplodnaviria;Heunggongvirae;Uroviricota;Caudoviricetes |
| S9_vs_46498\|provirus_1704_20897 | 20 | 1 | 2561 | Viruses;Duplodnaviria;Heunggongvirae;Uroviricota;Caudoviricetes |
| S9_vs_67792\|provirus_23562_67089 | 41 | 1 | 2561 | Viruses;Duplodnaviria;Heunggongvirae;Uroviricota;Caudoviricetes |
| S9_vs_90727\|provirus_109214_150749 | 38 | 1 | 2561 | Viruses;Duplodnaviria;Heunggongvirae;Uroviricota;Caudoviricetes |
| S11_vs_45658\|provirus_1_50991 | 48 | 1 | 2561 | Viruses;Duplodnaviria;Heunggongvirae;Uroviricota;Caudoviricetes |
| S11_vs_48109\|provirus_19503_33025 | 11 | 1 | 2561 | Viruses;Duplodnaviria;Heunggongvirae;Uroviricota;Caudoviricetes |
| S11_vs_48109\|provirus_37443_52410 | 17 | 1 | 2561 | Viruses;Duplodnaviria;Heunggongvirae;Uroviricota;Caudoviricetes |
| S11_vs_82331\|provirus_1_39593 | 24 | 1 | 2561 | Viruses;Duplodnaviria;Heunggongvirae;Uroviricota;Caudoviricetes |
| S13_vs_9438\|provirus_1_26027 | 15 | 1 | 2561 | Viruses;Duplodnaviria;Heunggongvirae;Uroviricota;Caudoviricetes |
| S13_vs_23314\|provirus_1_43630 | 42 | 1 | 2561 | Viruses;Duplodnaviria;Heunggongvirae;Uroviricota;Caudoviricetes |
| S13_vs_26756\|provirus_1_36490 | 34 | 1 | 2561 | Viruses;Duplodnaviria;Heunggongvirae;Uroviricota;Caudoviricetes |
| S13_vs_30101\|provirus_4484_51177 | 33 | 0.9912 | 2561 | Viruses;Duplodnaviria;Heunggongvirae;Uroviricota;Caudoviricetes |
| S13_vs_30167\|provirus_1_53839 | 46 | 1 | 2561 | Viruses;Duplodnaviria;Heunggongvirae;Uroviricota;Caudoviricetes |
| S13_vs_33947\|provirus_1_26796 | 14 | 1 | 2561 | Viruses;Duplodnaviria;Heunggongvirae;Uroviricota;Caudoviricetes |
| S13_vs_35816\|provirus_9638_57224 | 59 | 1 | 2561 | Viruses;Duplodnaviria;Heunggongvirae;Uroviricota;Caudoviricetes |
| S13_vs_45488\|provirus_49188_88826 | 27 | 1 | 2561 | Viruses;Duplodnaviria;Heunggongvirae;Uroviricota;Caudoviricetes |
| S13_vs_67429\|provirus_51626_91295 | 36 | 1 | 2561 | Viruses;Duplodnaviria;Heunggongvirae;Uroviricota;Caudoviricetes |
| S13_vs_114213\|provirus_1_28952 | 23 | 1 | 2561 | Viruses;Duplodnaviria;Heunggongvirae;Uroviricota;Caudoviricetes |
| S14_vs_66139\|provirus_119631_178690 | 40 | 1 | 2561 | Viruses;Duplodnaviria;Heunggongvirae;Uroviricota;Caudoviricetes |
| S14_vs_66208\|provirus_10049_67174 | 49 | 1 | 2561 | Viruses;Duplodnaviria;Heunggongvirae;Uroviricota;Caudoviricetes |
| S14_vs_96111\|provirus_1_24143 | 26 | 1 | 2561 | Viruses;Duplodnaviria;Heunggongvirae;Uroviricota;Caudoviricetes |
| S14_vs_74067\|provirus_149062_181135 | 27 | 1 | 2561 | Viruses;Duplodnaviria;Heunggongvirae;Uroviricota;Caudoviricetes |
| S14_vs_74067\|provirus_193206_252822 | 56 | 1 | 2561 | Viruses;Duplodnaviria;Heunggongvirae;Uroviricota;Caudoviricetes |
| S14_vs_9162\|provirus_77661_102634 | 28 | 1 | 2561 | Viruses;Duplodnaviria;Heunggongvirae;Uroviricota;Caudoviricetes |
| S14_vs_112345\|provirus_25548_38263 | 8 | 1 | 2561 | Viruses;Duplodnaviria;Heunggongvirae;Uroviricota;Caudoviricetes |
| S14_vs_39266\|provirus_8204_37426 | 25 | 1 | 2561 | Viruses;Duplodnaviria;Heunggongvirae;Uroviricota;Caudoviricetes |
| S14_vs_2870\|provirus_1_47896 | 42 | 1 | 2561 | Viruses;Duplodnaviria;Heunggongvirae;Uroviricota;Caudoviricetes |
| S14_vs_32499\|provirus_16787_41581 | 28 | 1 | 2561 | Viruses;Duplodnaviria;Heunggongvirae;Uroviricota;Caudoviricetes |
| S14_vs_25852\|provirus_1_35257 | 9 | 1 | 2561 | Viruses;Duplodnaviria;Heunggongvirae;Uroviricota;Caudoviricetes |
| S14_vs_25852\|provirus_71427_107963 | 13 | 1 | 2561 | Viruses;Duplodnaviria;Heunggongvirae;Uroviricota;Caudoviricetes |
| S14_vs_11510\|provirus_1_37329 | 10 | 1 | 2561 | Viruses;Duplodnaviria;Heunggongvirae;Uroviricota;Caudoviricetes |
| S14_vs_78164\|provirus_1_33442 | 35 | 1 | 2561 | Viruses;Duplodnaviria;Heunggongvirae;Uroviricota;Caudoviricetes |
| S14_vs_70985\|provirus_15261_57392 | 12 | 1 | 2561 | Viruses;Duplodnaviria;Heunggongvirae;Uroviricota;Caudoviricetes |
| S14_vs_70985\|provirus_96748_158000 | 14 | 1 | 2561 | Viruses;Duplodnaviria;Heunggongvirae;Uroviricota;Caudoviricetes |
| S1_vs_117317\|provirus_1_31247 | 49 | 1 | 2561 | Viruses;Duplodnaviria;Heunggongvirae;Uroviricota;Caudoviricetes |
| S3_vf_106401\|provirus_6294_39808 | 21 | 1 | 2561 | Viruses;Duplodnaviria;Heunggongvirae;Uroviricota;Caudoviricetes |
| S3_vs_119605\|provirus_10462_56346 | 45 | 1 | 2561 | Viruses;Duplodnaviria;Heunggongvirae;Uroviricota;Caudoviricetes |
| S3_vs_9158\|provirus_1_28511 | 24 | 0.9779 | 2561 | Viruses;Duplodnaviria;Heunggongvirae;Uroviricota;Caudoviricetes |
| S3_vs_64424\|provirus_1_35095 | 28 | 1 | 2561 | Viruses;Duplodnaviria;Heunggongvirae;Uroviricota;Caudoviricetes |
| S3_vs_17168\|provirus_62162_103365 | 30 | 1 | 2561 | Viruses;Duplodnaviria;Heunggongvirae;Uroviricota;Caudoviricetes |
| S3_vs_42569\|provirus_1_55470 | 64 | 1 | 2561 | Viruses;Duplodnaviria;Heunggongvirae;Uroviricota;Caudoviricetes |
| S3_vs_18858\|provirus_7640_40921 | 21 | 1 | 2561 | Viruses;Duplodnaviria;Heunggongvirae;Uroviricota;Caudoviricetes |
| S3_vs_68031\|provirus_1_31195 | 35 | 1 | 2561 | Viruses;Duplodnaviria;Heunggongvirae;Uroviricota;Caudoviricetes |
| S3_vs_6604\|provirus_44093_78643 | 21 | 1 | 2561 | Viruses;Duplodnaviria;Heunggongvirae;Uroviricota;Caudoviricetes |
| S3_vs_70508\|provirus_10985_57171 | 30 | 1 | 2561 | Viruses;Duplodnaviria;Heunggongvirae;Uroviricota;Caudoviricetes |
| S3_vs_117894\|provirus_1_42562 | 23 | 1 | 2561 | Viruses;Duplodnaviria;Heunggongvirae;Uroviricota;Caudoviricetes |
| S5_vs_30490\|provirus_19631_56501 | 24 | 1 | 2561 | Viruses;Duplodnaviria;Heunggongvirae;Uroviricota;Caudoviricetes |
| S5_vs_109527\|provirus_86293_134147 | 16 | 1 | 2561 | Viruses;Duplodnaviria;Heunggongvirae;Uroviricota;Caudoviricetes |
| S5_vs_123256\|provirus_35835_71330 | 29 | 1 | 2561 | Viruses;Duplodnaviria;Heunggongvirae;Uroviricota;Caudoviricetes |
| S5_vs_117319\|provirus_37947_108449 | 65 | 1 | 2561 | Viruses;Duplodnaviria;Heunggongvirae;Uroviricota;Caudoviricetes |
| S5_vs_79992\|provirus_1_37142 | 42 | 1 | 2561 | Viruses;Duplodnaviria;Heunggongvirae;Uroviricota;Caudoviricetes |
| S5_vs_20500\|provirus_26729_71704 | 33 | 1 | 2561 | Viruses;Duplodnaviria;Heunggongvirae;Uroviricota;Caudoviricetes |
| S5_vs_3501\|provirus_235_23805 | 21 | 1 | 2561 | Viruses;Duplodnaviria;Heunggongvirae;Uroviricota;Caudoviricetes |

| | | | | |
|---|---|---|---|---|
| S5_vs_124677\|provirus_1_41828 | 41 | 1 | 2561 | Viruses;Duplodnaviria;Heunggongvirae;Uroviricota;Caudoviricetes |
| S5_vs_130294\|provirus_21565_31442 | 9 | 1 | 2561 | Viruses;Duplodnaviria;Heunggongvirae;Uroviricota;Caudoviricetes |
| S5_vs_80496\|provirus_58106_119911 | 51 | 1 | 2561 | Viruses;Duplodnaviria;Heunggongvirae;Uroviricota;Caudoviricetes |
| S5_vs_142195\|provirus_1_52517 | 17 | 1 | 2561 | Viruses;Duplodnaviria;Heunggongvirae;Uroviricota;Caudoviricetes |
| S5_vs_9747\|provirus_14158_64437 | 20 | 1 | 2561 | Viruses;Duplodnaviria;Heunggongvirae;Uroviricota;Caudoviricetes |
| S5_vs_21778\|provirus_68731_157098 | 14 | 1 | 2561 | Viruses;Duplodnaviria;Heunggongvirae;Uroviricota;Caudoviricetes |
| S5_vs_131510\|provirus_92040_133423 | 42 | 1 | 2561 | Viruses;Duplodnaviria;Heunggongvirae;Uroviricota;Caudoviricetes |
| S5_vs_46394\|provirus_33205_42387 | 9 | 1 | 2561 | Viruses;Duplodnaviria;Heunggongvirae;Uroviricota;Caudoviricetes |
| S5_vs_40848\|provirus_22909_78371 | 40 | 1 | 2561 | Viruses;Duplodnaviria;Heunggongvirae;Uroviricota;Caudoviricetes |
| S5_vs_72532\|provirus_8356_50725 | 36 | 1 | 2561 | Viruses;Duplodnaviria;Heunggongvirae;Uroviricota;Caudoviricetes |
| S5_vs_77593\|provirus_49321_99338 | 54 | 1 | 2561 | Viruses;Duplodnaviria;Heunggongvirae;Uroviricota;Caudoviricetes |
| S5_vs_138980\|provirus_1_47926 | 40 | 1 | 2561 | Viruses;Duplodnaviria;Heunggongvirae;Uroviricota;Caudoviricetes |
| S5_vs_74199\|provirus_14632_30526 | 9 | 1 | 2561 | Viruses;Duplodnaviria;Heunggongvirae;Uroviricota;Caudoviricetes |
| S5_vs_76500\|provirus_1_36552 | 29 | 1 | 2561 | Viruses;Duplodnaviria;Heunggongvirae;Uroviricota;Caudoviricetes |
| S7_vf_36627\|provirus_69_11085 | 11 | 1 | 2561 | Viruses;Duplodnaviria;Heunggongvirae;Uroviricota;Caudoviricetes |
| S7_vs_11081\|provirus_1_46169 | 41 | 1 | 2561 | Viruses;Duplodnaviria;Heunggongvirae;Uroviricota;Caudoviricetes |
| S7_vs_16344\|provirus_1_34983 | 32 | 1 | 2561 | Viruses;Duplodnaviria;Heunggongvirae;Uroviricota;Caudoviricetes |
| S7_vs_47050\|provirus_1_34528 | 29 | 1 | 2561 | Viruses;Duplodnaviria;Heunggongvirae;Uroviricota;Caudoviricetes |
| S7_vs_49426\|provirus_19260_57556 | 30 | 1 | 2561 | Viruses;Duplodnaviria;Heunggongvirae;Uroviricota;Caudoviricetes |
| S7_vs_51332\|provirus_1_12316 | 11 | 1 | 2561 | Viruses;Duplodnaviria;Heunggongvirae;Uroviricota;Caudoviricetes |
| S7_vs_97737\|provirus_53372_63197 | 9 | 1 | 2561 | Viruses;Duplodnaviria;Heunggongvirae;Uroviricota;Caudoviricetes |
| S7_vs_119679\|provirus_14434_51206 | 39 | 1 | 2561 | Viruses;Duplodnaviria;Heunggongvirae;Uroviricota;Caudoviricetes |
| S16_vs_19968\|provirus_18399_54417 | 27 | 1 | 2561 | Viruses;Duplodnaviria;Heunggongvirae;Uroviricota;Caudoviricetes |
| S16_vs_32764\|provirus_43014_55386 | 7 | 1 | 2561 | Viruses;Duplodnaviria;Heunggongvirae;Uroviricota;Caudoviricetes |
| S16_vs_34021\|provirus_1_29929 | 29 | 1 | 2561 | Viruses;Duplodnaviria;Heunggongvirae;Uroviricota;Caudoviricetes |
| S16_vs_48714\|provirus_9547_23424 | 5 | 0.9272 | 8016 | Viruses;Monodnaviria;Loebvirae;Hofneiviricota;Faserviricetes;Tubulavirales;Inoviridae |
| S16_vs_82116\|provirus_1_40122 | 45 | 0.9942 | 2561 | Viruses;Duplodnaviria;Heunggongvirae;Uroviricota;Caudoviricetes |
| S16_vs_89511\|provirus_1_61265 | 42 | 1 | 2561 | Viruses;Duplodnaviria;Heunggongvirae;Uroviricota;Caudoviricetes |
| S16_vs_101072\|provirus_6845_31451 | 27 | 1 | 2561 | Viruses;Duplodnaviria;Heunggongvirae;Uroviricota;Caudoviricetes |
| S16_vs_107235\|provirus_1_37912 | 35 | 1 | 2561 | Viruses;Duplodnaviria;Heunggongvirae;Uroviricota;Caudoviricetes |
| S16_vs_132979\|provirus_1_22166 | 19 | 0.9814 | 2561 | Viruses;Duplodnaviria;Heunggongvirae;Uroviricota;Caudoviricetes |
| S16_vs_140468\|provirus_7020_20506 | 12 | 1 | 2561 | Viruses;Duplodnaviria;Heunggongvirae;Uroviricota;Caudoviricetes |
| S16_vs_141218\|provirus_1_41081 | 30 | 0.9878 | 2561 | Viruses;Duplodnaviria;Heunggongvirae;Uroviricota;Caudoviricetes |
| S16_vs_142561\|provirus_46226_82728 | 33 | 1 | 2561 | Viruses;Duplodnaviria;Heunggongvirae;Uroviricota;Caudoviricetes |
| S10_vs_14557\|provirus_24735_75557 | 31 | 1 | 2561 | Viruses;Duplodnaviria;Heunggongvirae;Uroviricota;Caudoviricetes |
| S12_vf_60224\|provirus_3687_54526 | 45 | 1 | 2561 | Viruses;Duplodnaviria;Heunggongvirae;Uroviricota;Caudoviricetes |
| S12_vs_71525\|provirus_5502_31779 | 16 | 1 | 2561 | Viruses;Duplodnaviria;Heunggongvirae;Uroviricota;Caudoviricetes |
| S12_vs_142625\|provirus_29520_70930 | 33 | 1 | 2561 | Viruses;Duplodnaviria;Heunggongvirae;Uroviricota;Caudoviricetes |
| S12_vs_26336\|provirus_16576_67092 | 16 | 1 | 2561 | Viruses;Duplodnaviria;Heunggongvirae;Uroviricota;Caudoviricetes |
| S12_vs_121913\|provirus_20147_123530 | 81 | 1 | 2561 | Viruses;Duplodnaviria;Heunggongvirae;Uroviricota;Caudoviricetes |
| S12_vs_153780\|provirus_11265_61281 | 31 | 1 | 2561 | Viruses;Duplodnaviria;Heunggongvirae;Uroviricota;Caudoviricetes |
| S12_vs_123620\|provirus_40480_78363 | 29 | 1 | 2561 | Viruses;Duplodnaviria;Heunggongvirae;Uroviricota;Caudoviricetes |
| S12_vs_133070\|provirus_55125_132222 | 44 | 1 | 2561 | Viruses;Duplodnaviria;Heunggongvirae;Uroviricota;Caudoviricetes |
| S12_vs_172379\|provirus_92122_132266 | 25 | 0.9882 | 2561 | Viruses;Duplodnaviria;Heunggongvirae;Uroviricota;Caudoviricetes |
| S12_vs_23796\|provirus_1_38428 | 33 | 1 | 2561 | Viruses;Duplodnaviria;Heunggongvirae;Uroviricota;Caudoviricetes |
| S15_vs_42043\|provirus_134410_193486 | 49 | 1 | 2561 | Viruses;Duplodnaviria;Heunggongvirae;Uroviricota;Caudoviricetes |
| S15_vs_43913\|provirus_11358_44889 | 34 | 1 | 2561 | Viruses;Duplodnaviria;Heunggongvirae;Uroviricota;Caudoviricetes |
| S15_vs_71968\|provirus_18124_39396 | 17 | 1 | 2561 | Viruses;Duplodnaviria;Heunggongvirae;Uroviricota;Caudoviricetes |
| S2_vs_10289\|provirus_7396_46594 | 37 | 1 | 2561 | Viruses;Duplodnaviria;Heunggongvirae;Uroviricota;Caudoviricetes |
| S6_vs_9784\|provirus_9084_47596 | 38 | 1 | 2561 | Viruses;Duplodnaviria;Heunggongvirae;Uroviricota;Caudoviricetes |
| S6_vs_54606\|provirus_1_21709 | 30 | 1 | 2561 | Viruses;Duplodnaviria;Heunggongvirae;Uroviricota;Caudoviricetes |
| S6_vs_29152\|provirus_10612_64693 | 57 | 1 | 2561 | Viruses;Duplodnaviria;Heunggongvirae;Uroviricota;Caudoviricetes |
| S6_vs_10314\|provirus_44149_83752 | 42 | 1 | 2561 | Viruses;Duplodnaviria;Heunggongvirae;Uroviricota;Caudoviricetes |
| S6_vs_1146\|provirus_1_36227 | 36 | 1 | 2561 | Viruses;Duplodnaviria;Heunggongvirae;Uroviricota;Caudoviricetes |
| S6_vs_46039\|provirus_1_38172 | 12 | 1 | 2561 | Viruses;Duplodnaviria;Heunggongvirae;Uroviricota;Caudoviricetes |

| | | | | |
|---|---|---|---|---|
| S6_vs_46039\|provirus_69634_111911 | 14 | 1 | 2561 | Viruses;Duplodnaviria;Heunggongvirae;Uroviricota;Caudoviricetes |
| S6_vs_34453\|provirus_4536_65412 | 58 | 1 | 2561 | Viruses;Duplodnaviria;Heunggongvirae;Uroviricota;Caudoviricetes |
| S6_vs_39025\|provirus_18844_51286 | 38 | 1 | 2561 | Viruses;Duplodnaviria;Heunggongvirae;Uroviricota;Caudoviricetes |
| S6_vs_39065\|provirus_14912_41773 | 9 | 1 | 2561 | Viruses;Duplodnaviria;Heunggongvirae;Uroviricota;Caudoviricetes |
| S8_vf_28956\|provirus_172_25419 | 22 | 1 | 2561 | Viruses;Duplodnaviria;Heunggongvirae;Uroviricota;Caudoviricetes |
| S8_vf_12212\|provirus_3_31196 | 31 | 1 | 2561 | Viruses;Duplodnaviria;Heunggongvirae;Uroviricota;Caudoviricetes |
| S8_vf_170878\|provirus_6035_59917 | 54 | 1 | 2561 | Viruses;Duplodnaviria;Heunggongvirae;Uroviricota;Caudoviricetes |
| S8_vs_149700\|provirus_19406_69356 | 42 | 1 | 2561 | Viruses;Duplodnaviria;Heunggongvirae;Uroviricota;Caudoviricetes |
| S8_vs_115958\|provirus_68174_127950 | 45 | 1 | 2561 | Viruses;Duplodnaviria;Heunggongvirae;Uroviricota;Caudoviricetes |
| S8_vs_125717\|provirus_34678_52956 | 23 | 1 | 2561 | Viruses;Duplodnaviria;Heunggongvirae;Uroviricota;Caudoviricetes |
| S8_vs_4153\|provirus_1_12586 | 10 | 1 | 2561 | Viruses;Duplodnaviria;Heunggongvirae;Uroviricota;Caudoviricetes |
| S8_vs_47265\|provirus_19038_69505 | 46 | 1 | 2561 | Viruses;Duplodnaviria;Heunggongvirae;Uroviricota;Caudoviricetes |
| S8_vs_179208\|provirus_1_52017 | 20 | 1 | 2561 | Viruses;Duplodnaviria;Heunggongvirae;Uroviricota;Caudoviricetes |
| S8_vs_205257\|provirus_1_42712 | 28 | 1 | 2561 | Viruses;Duplodnaviria;Heunggongvirae;Uroviricota;Caudoviricetes |

| wms_seq_name | n_genes_with_taxonomy | agreement | taxid | lineage |
|---|---|---|---|---|
| S9-wms-combined_64277\|provirus_1_50273 | 40 | 1 | 2561 | Viruses;Duplodnaviria;Heunggongvirae;Uroviricota;Caudoviricetes |
| S9-wms-combined_53991\|provirus_1_38377 | 39 | 1 | 2561 | Viruses;Duplodnaviria;Heunggongvirae;Uroviricota;Caudoviricetes |
| S9-wms-combined_97755\|provirus_1_31801 | 35 | 1 | 2561 | Viruses;Duplodnaviria;Heunggongvirae;Uroviricota;Caudoviricetes |
| S9-wms-combined_33230\|provirus_1_29168 | 30 | 1 | 2561 | Viruses;Duplodnaviria;Heunggongvirae;Uroviricota;Caudoviricetes |
| S9-wms-combined_124857\|provirus_24439_80893 | 56 | 1 | 2561 | Viruses;Duplodnaviria;Heunggongvirae;Uroviricota;Caudoviricetes |
| S9-wms-combined_28805\|provirus_23826_59739 | 38 | 1 | 2561 | Viruses;Duplodnaviria;Heunggongvirae;Uroviricota;Caudoviricetes |
| S9-wms-combined_93232\|provirus_5433_50669 | 48 | 1 | 2561 | Viruses;Duplodnaviria;Heunggongvirae;Uroviricota;Caudoviricetes |
| S9-wms-combined_109322\|provirus_1_43439 | 37 | 1 | 2561 | Viruses;Duplodnaviria;Heunggongvirae;Uroviricota;Caudoviricetes |
| S9-wms-combined_29611\|provirus_307713_413955 | 93 | 1 | 2561 | Viruses;Duplodnaviria;Heunggongvirae;Uroviricota;Caudoviricetes |
| S9-wms-combined_20485\|provirus_49228_90295 | 46 | 1 | 2561 | Viruses;Duplodnaviria;Heunggongvirae;Uroviricota;Caudoviricetes |
| S9-wms-combined_9931\|provirus_43028_90630 | 39 | 1 | 2561 | Viruses;Duplodnaviria;Heunggongvirae;Uroviricota;Caudoviricetes |
| S9-wms-combined_37587\|provirus_1_47703 | 32 | 1 | 2561 | Viruses;Duplodnaviria;Heunggongvirae;Uroviricota;Caudoviricetes |
| S9-wms-combined_5526\|provirus_1_38027 | 32 | 1 | 2561 | Viruses;Duplodnaviria;Heunggongvirae;Uroviricota;Caudoviricetes |
| S9-wms-combined_76274\|provirus_3258_36781 | 35 | 1 | 2561 | Viruses;Duplodnaviria;Heunggongvirae;Uroviricota;Caudoviricetes |
| S11-wms-combined_193148\|provirus_8658_39231 | 9 | 1 | 2561 | Viruses;Duplodnaviria;Heunggongvirae;Uroviricota;Caudoviricetes |
| S11-wms-combined_179605\|provirus_18936_43615 | 24 | 1 | 2561 | Viruses;Duplodnaviria;Heunggongvirae;Uroviricota;Caudoviricetes |
| S11-wms-combined_83571\|provirus_9777_40621 | 36 | 1 | 2561 | Viruses;Duplodnaviria;Heunggongvirae;Uroviricota;Caudoviricetes |
| S11-wms-combined_125176\|provirus_1_23922 | 22 | 1 | 2561 | Viruses;Duplodnaviria;Heunggongvirae;Uroviricota;Caudoviricetes |
| S11-wms-combined_236214\|provirus_60975_93358 | 29 | 1 | 2561 | Viruses;Duplodnaviria;Heunggongvirae;Uroviricota;Caudoviricetes |
| S11-wms-combined_264050\|provirus_28059_77647 | 49 | 1 | 2561 | Viruses;Duplodnaviria;Heunggongvirae;Uroviricota;Caudoviricetes |
| S11-wms-combined_140009\|provirus_19523_54887 | 37 | 1 | 2561 | Viruses;Duplodnaviria;Heunggongvirae;Uroviricota;Caudoviricetes |
| S11-wms-combined_168048\|provirus_1_41217 | 31 | 1 | 2561 | Viruses;Duplodnaviria;Heunggongvirae;Uroviricota;Caudoviricetes |
| S11-wms-combined_3507\|provirus_29062_70964 | 52 | 1 | 2561 | Viruses;Duplodnaviria;Heunggongvirae;Uroviricota;Caudoviricetes |
| S11-wms-combined_101337\|provirus_1_25287 | 19 | 1 | 2561 | Viruses;Duplodnaviria;Heunggongvirae;Uroviricota;Caudoviricetes |
| S11-wms-combined_73204\|provirus_23691_55764 | 27 | 1 | 2561 | Viruses;Duplodnaviria;Heunggongvirae;Uroviricota;Caudoviricetes |
| S11-wms-combined_198512\|provirus_3772_41546 | 31 | 1 | 2561 | Viruses;Duplodnaviria;Heunggongvirae;Uroviricota;Caudoviricetes |
| S11-wms-combined_267993\|provirus_1_65458 | 54 | 1 | 2561 | Viruses;Duplodnaviria;Heunggongvirae;Uroviricota;Caudoviricetes |
| S11-wms-combined_116589\|provirus_12509_25615 | 10 | 1 | 2561 | Viruses;Duplodnaviria;Heunggongvirae;Uroviricota;Caudoviricetes |
| S11-wms-combined_48246\|provirus_1_44551 | 43 | 1 | 2561 | Viruses;Duplodnaviria;Heunggongvirae;Uroviricota;Caudoviricetes |
| S11-wms-combined_63818\|provirus_8325_62965 | 45 | 1 | 2561 | Viruses;Duplodnaviria;Heunggongvirae;Uroviricota;Caudoviricetes |
| S11-wms-combined_215723\|provirus_1_42332 | 30 | 1 | 2561 | Viruses;Duplodnaviria;Heunggongvirae;Uroviricota;Caudoviricetes |
| S11-wms-combined_92114\|provirus_1_26924 | 20 | 1 | 2561 | Viruses;Duplodnaviria;Heunggongvirae;Uroviricota;Caudoviricetes |
| S11-wms-combined_120257\|provirus_8529_35507 | 33 | 1 | 2561 | Viruses;Duplodnaviria;Heunggongvirae;Uroviricota;Caudoviricetes |
| S11-wms-combined_217790\|provirus_1_43023 | 13 | 1 | 2561 | Viruses;Duplodnaviria;Heunggongvirae;Uroviricota;Caudoviricetes |
| S11-wms_32691\|provirus_3_142121 | 9 | 1 | 2561 | Viruses;Duplodnaviria;Heunggongvirae;Uroviricota;Caudoviricetes |
| S1-wms-combined_52708\|provirus_6059_16025 | 9 | 1 | 2561 | Viruses;Duplodnaviria;Heunggongvirae;Uroviricota;Caudoviricetes |
| S1-wms-combined_68998\|provirus_3499_56973 | 63 | 1 | 2561 | Viruses;Duplodnaviria;Heunggongvirae;Uroviricota;Caudoviricetes |
| S1-wms-combined_102801\|provirus_3620_28850 | 7 | 1 | 2561 | Viruses;Duplodnaviria;Heunggongvirae;Uroviricota;Caudoviricetes |
| S1-wms-combined_302166\|provirus_11917_75458 | 42 | 1 | 2561 | Viruses;Duplodnaviria;Heunggongvirae;Uroviricota;Caudoviricetes |
| S1-wms-combined_285012\|provirus_1_46216 | 53 | 1 | 2561 | Viruses;Duplodnaviria;Heunggongvirae;Uroviricota;Caudoviricetes |
| S1-wms-combined_220022\|provirus_1_85675 | 4 | 1 | 2561 | Viruses;Duplodnaviria;Heunggongvirae;Uroviricota;Caudoviricetes |
| S1-wms-combined_220022\|provirus_128734_155946 | 4 | 1 | 2561 | Viruses;Duplodnaviria;Heunggongvirae;Uroviricota;Caudoviricetes |
| S1-wms-combined_23248\|provirus_27603_62426 | 36 | 1 | 2561 | Viruses;Duplodnaviria;Heunggongvirae;Uroviricota;Caudoviricetes |
| S1-wms-combined_304377\|provirus_1_66513 | 60 | 1 | 2561 | Viruses;Duplodnaviria;Heunggongvirae;Uroviricota;Caudoviricetes |
| S1-wms-combined_57370\|provirus_1_21189 | 19 | 1 | 2561 | Viruses;Duplodnaviria;Heunggongvirae;Uroviricota;Caudoviricetes |
| S1-wms-combined_107766\|provirus_1_36184 | 27 | 1 | 2561 | Viruses;Duplodnaviria;Heunggongvirae;Uroviricota;Caudoviricetes |
| S1-wms-combined_92523\|provirus_13796_31793 | 13 | 1 | 2561 | Viruses;Duplodnaviria;Heunggongvirae;Uroviricota;Caudoviricetes |
| S1-wms-combined_331544\|provirus_1_23231 | 21 | 1 | 2561 | Viruses;Duplodnaviria;Heunggongvirae;Uroviricota;Caudoviricetes |
| S1-wms-combined_85225\|provirus_1_39579 | 48 | 1 | 2561 | Viruses;Duplodnaviria;Heunggongvirae;Uroviricota;Caudoviricetes |
| S1-wms-combined_136097\|provirus_8755_19804 | 10 | 1 | 2561 | Viruses;Duplodnaviria;Heunggongvirae;Uroviricota;Caudoviricetes |
| S1-wms-combined_307323\|provirus_1_42035 | 40 | 1 | 2561 | Viruses;Duplodnaviria;Heunggongvirae;Uroviricota;Caudoviricetes |
| S1-wms-combined_94988\|provirus_10302_22992 | 10 | 1 | 2561 | Viruses;Duplodnaviria;Heunggongvirae;Uroviricota;Caudoviricetes |
| S5-wms-combined_18356\|provirus_1_22757 | 23 | 1 | 2561 | Viruses;Duplodnaviria;Heunggongvirae;Uroviricota;Caudoviricetes |
| S5-wms-combined_161191\|provirus_134452_175715 | 44 | 1 | 2561 | Viruses;Duplodnaviria;Heunggongvirae;Uroviricota;Caudoviricetes |

| | | | | |
|---|---|---|---|---|
| S5-wms-combined_134381\|provirus_32274_74956 | 57 | 1 | 2561 | Viruses;Duplodnaviria;Heunggongvirae;Uroviricota;Caudoviricetes |
| S5-wms-combined_90315\|provirus_11740_73154 | 59 | 1 | 2561 | Viruses;Duplodnaviria;Heunggongvirae;Uroviricota;Caudoviricetes |
| S5-wms-combined_134743\|provirus_30511_101552 | 44 | 0.9919 | 2561 | Viruses;Duplodnaviria;Heunggongvirae;Uroviricota;Caudoviricetes |
| S5-wms-combined_72410\|provirus_71571_129551 | 66 | 1 | 2561 | Viruses;Duplodnaviria;Heunggongvirae;Uroviricota;Caudoviricetes |
| S5-wms-combined_206050\|provirus_15780_57403 | 41 | 1 | 2561 | Viruses;Duplodnaviria;Heunggongvirae;Uroviricota;Caudoviricetes |
| S5-wms-combined_108794\|provirus_19443_74905 | 40 | 1 | 2561 | Viruses;Duplodnaviria;Heunggongvirae;Uroviricota;Caudoviricetes |
| S5-wms-combined_207291\|provirus_13547_57653 | 31 | 1 | 2561 | Viruses;Duplodnaviria;Heunggongvirae;Uroviricota;Caudoviricetes |
| S5-wms-combined_29447\|provirus_1_32335 | 37 | 1 | 2561 | Viruses;Duplodnaviria;Heunggongvirae;Uroviricota;Caudoviricetes |
| S5-wms-combined_83370\|provirus_1_42289 | 36 | 1 | 2561 | Viruses;Duplodnaviria;Heunggongvirae;Uroviricota;Caudoviricetes |
| S5-wms-combined_66662\|provirus_52916_98874 | 33 | 1 | 2561 | Viruses;Duplodnaviria;Heunggongvirae;Uroviricota;Caudoviricetes |
| S5-wms-combined_59064\|provirus_35710_77093 | 42 | 1 | 2561 | Viruses;Duplodnaviria;Heunggongvirae;Uroviricota;Caudoviricetes |
| S5-wms-combined_94482\|provirus_9030_59881 | 34 | 1 | 2561 | Viruses;Duplodnaviria;Heunggongvirae;Uroviricota;Caudoviricetes |
| S5-wms-combined_67650\|provirus_42189_77684 | 29 | 1 | 2561 | Viruses;Duplodnaviria;Heunggongvirae;Uroviricota;Caudoviricetes |
| S5-wms-combined_94786\|provirus_4677_40532 | 25 | 1 | 2561 | Viruses;Duplodnaviria;Heunggongvirae;Uroviricota;Caudoviricetes |
| S5-wms-combined_210959\|provirus_126240_159070 | 27 | 1 | 2561 | Viruses;Duplodnaviria;Heunggongvirae;Uroviricota;Caudoviricetes |
| S5-wms-combined_78434\|provirus_1_90452 | 15 | 1 | 2561 | Viruses;Duplodnaviria;Heunggongvirae;Uroviricota;Caudoviricetes |
| S5-wms-combined_98202\|provirus_1_61764 | 20 | 0.9818 | 2561 | Viruses;Duplodnaviria;Heunggongvirae;Uroviricota;Caudoviricetes |
| S5-wms-combined_26971\|provirus_4471_34173 | 35 | 1 | 2561 | Viruses;Duplodnaviria;Heunggongvirae;Uroviricota;Caudoviricetes |
| S5-wms-combined_178432\|provirus_1_33423 | 25 | 1 | 2561 | Viruses;Duplodnaviria;Heunggongvirae;Uroviricota;Caudoviricetes |
| S5-wms-combined_179241\|provirus_5356_19361 | 9 | 1 | 2561 | Viruses;Duplodnaviria;Heunggongvirae;Uroviricota;Caudoviricetes |
| S5-wms-combined_117213\|provirus_1_35094 | 35 | 1 | 2561 | Viruses;Duplodnaviria;Heunggongvirae;Uroviricota;Caudoviricetes |
| S5-wms_94965\|provirus_4329_41461 | 34 | 1 | 2561 | Viruses;Duplodnaviria;Heunggongvirae;Uroviricota;Caudoviricetes |
| S7-wms-combined_71592\|provirus_9021_38751 | 32 | 1 | 2561 | Viruses;Duplodnaviria;Heunggongvirae;Uroviricota;Caudoviricetes |
| S7-wms-combined_20921\|provirus_1_139822 | 92 | 1 | 2561 | Viruses;Duplodnaviria;Heunggongvirae;Uroviricota;Caudoviricetes |
| S7-wms-combined_62119\|provirus_55121_129634 | 73 | 1 | 2561 | Viruses;Duplodnaviria;Heunggongvirae;Uroviricota;Caudoviricetes |
| S7-wms-combined_185016\|provirus_8013_41583 | 35 | 1 | 2561 | Viruses;Duplodnaviria;Heunggongvirae;Uroviricota;Caudoviricetes |
| S7-wms-combined_174935\|provirus_1_23182 | 22 | 1 | 2561 | Viruses;Duplodnaviria;Heunggongvirae;Uroviricota;Caudoviricetes |
| S7-wms-combined_134458\|provirus_1_46614 | 49 | 1 | 2561 | Viruses;Duplodnaviria;Heunggongvirae;Uroviricota;Caudoviricetes |
| S7-wms-combined_2122\|provirus_17514_50765 | 28 | 1 | 2561 | Viruses;Duplodnaviria;Heunggongvirae;Uroviricota;Caudoviricetes |
| S7-wms-combined_135154\|provirus_1_28157 | 17 | 1 | 2561 | Viruses;Duplodnaviria;Heunggongvirae;Uroviricota;Caudoviricetes |
| S7-wms-combined_238231\|provirus_41225_71249 | 26 | 1 | 2561 | Viruses;Duplodnaviria;Heunggongvirae;Uroviricota;Caudoviricetes |
| S7-wms-combined_43950\|provirus_8198_57129 | 41 | 1 | 2561 | Viruses;Duplodnaviria;Heunggongvirae;Uroviricota;Caudoviricetes |
| S7-wms-combined_3697\|provirus_66701_124033 | 46 | 1 | 2561 | Viruses;Duplodnaviria;Heunggongvirae;Uroviricota;Caudoviricetes |
| S7-wms-combined_157260\|provirus_19304_75227 | 58 | 1 | 2561 | Viruses;Duplodnaviria;Heunggongvirae;Uroviricota;Caudoviricetes |
| S7-wms-combined_74829\|provirus_2475_38322 | 29 | 1 | 2561 | Viruses;Duplodnaviria;Heunggongvirae;Uroviricota;Caudoviricetes |
| S7-wms-combined_14267\|provirus_25528_51357 | 23 | 1 | 2561 | Viruses;Duplodnaviria;Heunggongvirae;Uroviricota;Caudoviricetes |
| S7-wms-combined_167867\|provirus_1_81146 | 11 | 1 | 2561 | Viruses;Duplodnaviria;Heunggongvirae;Uroviricota;Caudoviricetes |
| S7-wms-combined_106253\|provirus_1_36567 | 28 | 0.9688 | 2561 | Viruses;Duplodnaviria;Heunggongvirae;Uroviricota;Caudoviricetes |
| S7-wms-combined_14529\|provirus_20870_62786 | 27 | 1 | 2561 | Viruses;Duplodnaviria;Heunggongvirae;Uroviricota;Caudoviricetes |
| S7-wms-combined_137858\|provirus_28744_38569 | 9 | 1 | 2561 | Viruses;Duplodnaviria;Heunggongvirae;Uroviricota;Caudoviricetes |
| S7-wms-combined_5625\|provirus_26522_67320 | 41 | 1 | 2561 | Viruses;Duplodnaviria;Heunggongvirae;Uroviricota;Caudoviricetes |
| S7-wms-combined_190281\|provirus_28520_63064 | 33 | 1 | 2561 | Viruses;Duplodnaviria;Heunggongvirae;Uroviricota;Caudoviricetes |
| S7-wms-combined_26217\|provirus_37948_88555 | 44 | 1 | 2561 | Viruses;Duplodnaviria;Heunggongvirae;Uroviricota;Caudoviricetes |
| S7-wms-combined_7205\|provirus_1_62239 | 58 | 1 | 2561 | Viruses;Duplodnaviria;Heunggongvirae;Uroviricota;Caudoviricetes |
| S7-wms-combined_72070\|provirus_3771_55473 | 32 | 1 | 2561 | Viruses;Duplodnaviria;Heunggongvirae;Uroviricota;Caudoviricetes |
| S7-wms-combined_136642\|provirus_1_17488 | 21 | 1 | 2561 | Viruses;Duplodnaviria;Heunggongvirae;Uroviricota;Caudoviricetes |
| S7-wms-combined_147158\|provirus_1_43742 | 42 | 1 | 2561 | Viruses;Duplodnaviria;Heunggongvirae;Uroviricota;Caudoviricetes |
| S7-wms-combined_218379\|provirus_3331_36800 | 28 | 1 | 2561 | Viruses;Duplodnaviria;Heunggongvirae;Uroviricota;Caudoviricetes |
| S7-wms-combined_107679_0\|provirus_1_37921 | 39 | 1 | 2561 | Viruses;Duplodnaviria;Heunggongvirae;Uroviricota;Caudoviricetes |
| S7-wms-combined_107679_1\|provirus_4391_36183 | 18 | 1 | 2561 | Viruses;Duplodnaviria;Heunggongvirae;Uroviricota;Caudoviricetes |
| S17-wms-combined_233761\|provirus_1_38860 | 25 | 1 | 2561 | Viruses;Duplodnaviria;Heunggongvirae;Uroviricota;Caudoviricetes |
| S17-wms-combined_56006\|provirus_18810_96019 | 60 | 1 | 2561 | Viruses;Duplodnaviria;Heunggongvirae;Uroviricota;Caudoviricetes |
| S17-wms-combined_56061\|provirus_96601_107131 | 9 | 1 | 2561 | Viruses;Duplodnaviria;Heunggongvirae;Uroviricota;Caudoviricetes |
| S17-wms-combined_2234\|provirus_16885_70511 | 61 | 1 | 2561 | Viruses;Duplodnaviria;Heunggongvirae;Uroviricota;Caudoviricetes |
| S17-wms-combined_203701\|provirus_1_59819 | 21 | 1 | 2561 | Viruses;Duplodnaviria;Heunggongvirae;Uroviricota;Caudoviricetes |

| | | | | |
|---|---|---|---|---|
| S17-wms-combined_247494\|provirus_1_40245 | 36 | 1 | 2561 | Viruses;Duplodnaviria;Heunggongvirae;Uroviricota;Caudoviricetes |
| S17-wms-combined_126774\|provirus_33403_66657 | 6 | 1 | 2561 | Viruses;Duplodnaviria;Heunggongvirae;Uroviricota;Caudoviricetes |
| S17-wms-combined_248321\|provirus_1_64257 | 60 | 1 | 2561 | Viruses;Duplodnaviria;Heunggongvirae;Uroviricota;Caudoviricetes |
| S17-wms-combined_248321\|provirus_78214_127137 | 61 | 1 | 2561 | Viruses;Duplodnaviria;Heunggongvirae;Uroviricota;Caudoviricetes |
| S17-wms-combined_193958\|provirus_41974_101082 | 50 | 1 | 2561 | Viruses;Duplodnaviria;Heunggongvirae;Uroviricota;Caudoviricetes |
| S17-wms-combined_259811\|provirus_1_48861 | 60 | 1 | 2561 | Viruses;Duplodnaviria;Heunggongvirae;Uroviricota;Caudoviricetes |
| S17-wms-combined_185239\|provirus_13026_23450 | 4 | 1 | 2561 | Viruses;Duplodnaviria;Heunggongvirae;Uroviricota;Caudoviricetes |
| S17-wms-combined_186314\|provirus_28354_68895 | 54 | 1 | 2561 | Viruses;Duplodnaviria;Heunggongvirae;Uroviricota;Caudoviricetes |
| S17-wms-combined_162786\|provirus_1_130535 | 29 | 1 | 2561 | Viruses;Duplodnaviria;Heunggongvirae;Uroviricota;Caudoviricetes |
| S17-wms-combined_95360\|provirus_1_43743 | 30 | 1 | 2561 | Viruses;Duplodnaviria;Heunggongvirae;Uroviricota;Caudoviricetes |
| S17-wms-combined_199746\|provirus_311_40236 | 41 | 1 | 2561 | Viruses;Duplodnaviria;Heunggongvirae;Uroviricota;Caudoviricetes |
| S12-wms-combined_334806\|provirus_1_39697 | 21 | 1 | 2561 | Viruses;Duplodnaviria;Heunggongvirae;Uroviricota;Caudoviricetes |
| S12-wms-combined_184338\|provirus_7702_50077 | 45 | 1 | 2561 | Viruses;Duplodnaviria;Heunggongvirae;Uroviricota;Caudoviricetes |
| S12-wms-combined_368462\|provirus_3828_55485 | 40 | 1 | 2561 | Viruses;Duplodnaviria;Heunggongvirae;Uroviricota;Caudoviricetes |
| S12-wms-combined_201125\|provirus_169478_224193 | 46 | 1 | 2561 | Viruses;Duplodnaviria;Heunggongvirae;Uroviricota;Caudoviricetes |
| S12-wms-combined_368562\|provirus_5352_23237 | 11 | 1 | 2561 | Viruses;Duplodnaviria;Heunggongvirae;Uroviricota;Caudoviricetes |
| S12-wms-combined_117628\|provirus_37358_58691 | 22 | 1 | 2561 | Viruses;Duplodnaviria;Heunggongvirae;Uroviricota;Caudoviricetes |
| S12-wms-combined_17617\|provirus_14012_54837 | 45 | 1 | 2561 | Viruses;Duplodnaviria;Heunggongvirae;Uroviricota;Caudoviricetes |
| S12-wms-combined_51076\|provirus_1_52990 | 7 | 1 | 2561 | Viruses;Duplodnaviria;Heunggongvirae;Uroviricota;Caudoviricetes |
| S12-wms-combined_51076\|provirus_63671_110930 | 11 | 0.9503 | 2561 | Viruses;Duplodnaviria;Heunggongvirae;Uroviricota;Caudoviricetes |
| S12-wms-combined_151843\|provirus_1_54070 | 53 | 1 | 2561 | Viruses;Duplodnaviria;Heunggongvirae;Uroviricota;Caudoviricetes |
| S12-wms-combined_18287\|provirus_31039_78190 | 48 | 1 | 2561 | Viruses;Duplodnaviria;Heunggongvirae;Uroviricota;Caudoviricetes |
| S12-wms-combined_2450\|provirus_49617_81938 | 36 | 1 | 2561 | Viruses;Duplodnaviria;Heunggongvirae;Uroviricota;Caudoviricetes |
| S12-wms-combined_320727\|provirus_43373_74615 | 26 | 1 | 2561 | Viruses;Duplodnaviria;Heunggongvirae;Uroviricota;Caudoviricetes |
| S12-wms-combined_320727\|provirus_86494_158985 | 72 | 1 | 2561 | Viruses;Duplodnaviria;Heunggongvirae;Uroviricota;Caudoviricetes |
| S12-wms-combined_371541\|provirus_27093_40221 | 11 | 1 | 2561 | Viruses;Duplodnaviria;Heunggongvirae;Uroviricota;Caudoviricetes |
| S12-wms-combined_120133\|provirus_1_28638 | 29 | 1 | 2561 | Viruses;Duplodnaviria;Heunggongvirae;Uroviricota;Caudoviricetes |
| S12-wms-combined_189323\|provirus_1_38428 | 33 | 1 | 2561 | Viruses;Duplodnaviria;Heunggongvirae;Uroviricota;Caudoviricetes |
| S12-wms-combined_256299\|provirus_6861_53411 | 38 | 1 | 2561 | Viruses;Duplodnaviria;Heunggongvirae;Uroviricota;Caudoviricetes |
| S12-wms-combined_156234\|provirus_1_28774 | 30 | 1 | 2561 | Viruses;Duplodnaviria;Heunggongvirae;Uroviricota;Caudoviricetes |
| S12-wms-combined_23517\|provirus_1_43179 | 61 | 1 | 2561 | Viruses;Duplodnaviria;Heunggongvirae;Uroviricota;Caudoviricetes |
| S12-wms-combined_375343\|provirus_1_29654 | 26 | 1 | 2561 | Viruses;Duplodnaviria;Heunggongvirae;Uroviricota;Caudoviricetes |
| S12-wms-combined_358010\|provirus_6040_57083 | 36 | 1 | 2561 | Viruses;Duplodnaviria;Heunggongvirae;Uroviricota;Caudoviricetes |
| S12-wms-combined_358797\|provirus_1_26270 | 22 | 1 | 2561 | Viruses;Duplodnaviria;Heunggongvirae;Uroviricota;Caudoviricetes |
| S12-wms-combined_359190\|provirus_24482_64793 | 34 | 1 | 2561 | Viruses;Duplodnaviria;Heunggongvirae;Uroviricota;Caudoviricetes |
| S12-wms-combined_328554\|provirus_9763_43415 | 32 | 1 | 2561 | Viruses;Duplodnaviria;Heunggongvirae;Uroviricota;Caudoviricetes |
| S12-wms-combined_329242\|provirus_1_27761 | 6 | 1 | 2561 | Viruses;Duplodnaviria;Heunggongvirae;Uroviricota;Caudoviricetes |
| S12-wms-combined_313953\|provirus_1_52155 | 31 | 1 | 2561 | Viruses;Duplodnaviria;Heunggongvirae;Uroviricota;Caudoviricetes |
| S12-wms-combined_111524\|provirus_1_48545 | 17 | 1 | 2561 | Viruses;Duplodnaviria;Heunggongvirae;Uroviricota;Caudoviricetes |
| S12-wms-combined_297545\|provirus_1_54974 | 66 | 1 | 2561 | Viruses;Duplodnaviria;Heunggongvirae;Uroviricota;Caudoviricetes |
| S12-wms-combined_129799\|provirus_25526_72680 | 52 | 1 | 2561 | Viruses;Duplodnaviria;Heunggongvirae;Uroviricota;Caudoviricetes |
| S12-wms-combined_282594\|provirus_1_43487 | 25 | 1 | 2561 | Viruses;Duplodnaviria;Heunggongvirae;Uroviricota;Caudoviricetes |
| S12-wms-combined_147628\|provirus_1_66934 | 47 | 1 | 2561 | Viruses;Duplodnaviria;Heunggongvirae;Uroviricota;Caudoviricetes |
| S12-wms-combined_264845\|provirus_13087_19738 | 5 | 1 | 2561 | Viruses;Duplodnaviria;Heunggongvirae;Uroviricota;Caudoviricetes |
| S12-wms-combined_299293\|provirus_20365_74597 | 49 | 1 | 2561 | Viruses;Duplodnaviria;Heunggongvirae;Uroviricota;Caudoviricetes |
| S12-wms-combined_98046\|provirus_43964_89205 | 38 | 1 | 2561 | Viruses;Duplodnaviria;Heunggongvirae;Uroviricota;Caudoviricetes |
| S12-wms-combined_31761\|provirus_4908_40932 | 30 | 1 | 2561 | Viruses;Duplodnaviria;Heunggongvirae;Uroviricota;Caudoviricetes |
| S12-wms-combined_368190\|provirus_4987_62142 | 68 | 1 | 2561 | Viruses;Duplodnaviria;Heunggongvirae;Uroviricota;Caudoviricetes |
| S12-wms-combined_168259\|provirus_1_29820 | 12 | 1 | 2561 | Viruses;Duplodnaviria;Heunggongvirae;Uroviricota;Caudoviricetes |
| S12-wms-combined_270839\|provirus_3921_42253 | 29 | 1 | 2561 | Viruses;Duplodnaviria;Heunggongvirae;Uroviricota;Caudoviricetes |
| S12-wms_161239\|provirus_5587_25370 | 24 | 1 | 2561 | Viruses;Duplodnaviria;Heunggongvirae;Uroviricota;Caudoviricetes |
| S2-wms-combined_196783\|provirus_1_43247 | 38 | 1 | 2561 | Viruses;Duplodnaviria;Heunggongvirae;Uroviricota;Caudoviricetes |
| S2-wms-combined_62519\|provirus_6893_41704 | 41 | 1 | 2561 | Viruses;Duplodnaviria;Heunggongvirae;Uroviricota;Caudoviricetes |
| S2-wms-combined_135139\|provirus_1_34436 | 9 | 1 | 2561 | Viruses;Duplodnaviria;Heunggongvirae;Uroviricota;Caudoviricetes |
| S2-wms-combined_218002\|provirus_41237_76226 | 35 | 1 | 2561 | Viruses;Duplodnaviria;Heunggongvirae;Uroviricota;Caudoviricetes |

| | | | | |
|---|---|---|---|---|
| S2-wms-combined_135154\|provirus_11137_34738 | 13 | 1 | 2561 | Viruses;Duplodnaviria;Heunggongvirae;Uroviricota;Caudoviricetes |
| S2-wms-combined_93845\|provirus_1_41258 | 39 | 1 | 2561 | Viruses;Duplodnaviria;Heunggongvirae;Uroviricota;Caudoviricetes |
| S2-wms-combined_52568\|provirus_1_46833 | 39 | 1 | 2561 | Viruses;Duplodnaviria;Heunggongvirae;Uroviricota;Caudoviricetes |
| S2-wms-combined_166665\|provirus_1_31971 | 32 | 1 | 2561 | Viruses;Duplodnaviria;Heunggongvirae;Uroviricota;Caudoviricetes |
| S2-wms-combined_229529\|provirus_28533_87711 | 49 | 1 | 2561 | Viruses;Duplodnaviria;Heunggongvirae;Uroviricota;Caudoviricetes |
| S2-wms-combined_84608\|provirus_18423_96376 | 30 | 1 | 2561 | Viruses;Duplodnaviria;Heunggongvirae;Uroviricota;Caudoviricetes |
| S2-wms-combined_74387\|provirus_8639_47478 | 38 | 1 | 2561 | Viruses;Duplodnaviria;Heunggongvirae;Uroviricota;Caudoviricetes |
| S2-wms-combined_147065\|provirus_99157_133534 | 36 | 1 | 2561 | Viruses;Duplodnaviria;Heunggongvirae;Uroviricota;Caudoviricetes |
| S2-wms-combined_137119\|provirus_23866_96227 | 57 | 1 | 2561 | Viruses;Duplodnaviria;Heunggongvirae;Uroviricota;Caudoviricetes |
| S2-wms-combined_85381\|provirus_1_35531 | 30 | 1 | 2561 | Viruses;Duplodnaviria;Heunggongvirae;Uroviricota;Caudoviricetes |
| S2-wms-combined_116221\|provirus_11792_43877 | 36 | 1 | 2561 | Viruses;Duplodnaviria;Heunggongvirae;Uroviricota;Caudoviricetes |
| S2-wms-combined_116221\|provirus_59302_90374 | 30 | 1 | 2561 | Viruses;Duplodnaviria;Heunggongvirae;Uroviricota;Caudoviricetes |
| S2-wms-combined_157934\|provirus_4946_38145 | 38 | 1 | 2561 | Viruses;Duplodnaviria;Heunggongvirae;Uroviricota;Caudoviricetes |
| S2-wms-combined_13246\|provirus_142843_195068 | 54 | 1 | 2561 | Viruses;Duplodnaviria;Heunggongvirae;Uroviricota;Caudoviricetes |
| S2-wms-combined_210822\|provirus_1_50976 | 18 | 1 | 2561 | Viruses;Duplodnaviria;Heunggongvirae;Uroviricota;Caudoviricetes |
| S2-wms-combined_24549\|provirus_1_19009 | 20 | 1 | 2561 | Viruses;Duplodnaviria;Heunggongvirae;Uroviricota;Caudoviricetes |
| S2-wms-combined_24549\|provirus_52727_65681 | 13 | 1 | 2561 | Viruses;Duplodnaviria;Heunggongvirae;Uroviricota;Caudoviricetes |
| S2-wms-combined_170845\|provirus_1_34623 | 30 | 1 | 2561 | Viruses;Duplodnaviria;Heunggongvirae;Uroviricota;Caudoviricetes |
| S2-wms-combined_159670\|provirus_4693_36742 | 33 | 1 | 2561 | Viruses;Duplodnaviria;Heunggongvirae;Uroviricota;Caudoviricetes |
| S2-wms-combined_181391\|provirus_93678_123498 | 27 | 1 | 2561 | Viruses;Duplodnaviria;Heunggongvirae;Uroviricota;Caudoviricetes |
| S2-wms-combined_200390\|provirus_1_42588 | 37 | 1 | 2561 | Viruses;Duplodnaviria;Heunggongvirae;Uroviricota;Caudoviricetes |
| S2-wms-combined_57293\|provirus_12687_28064 | 9 | 1 | 2561 | Viruses;Duplodnaviria;Heunggongvirae;Uroviricota;Caudoviricetes |
| S2-wms-combined_191613\|provirus_1_40741 | 24 | 1 | 2561 | Viruses;Duplodnaviria;Heunggongvirae;Uroviricota;Caudoviricetes |
| S2-wms-combined_98888\|provirus_46109_107937 | 81 | 1 | 2561 | Viruses;Duplodnaviria;Heunggongvirae;Uroviricota;Caudoviricetes |
| S2-wms-combined_181849\|provirus_4199_47914 | 28 | 1 | 2561 | Viruses;Duplodnaviria;Heunggongvirae;Uroviricota;Caudoviricetes |
| S2-wms-combined_99446\|provirus_17627_43104 | 25 | 1 | 2561 | Viruses;Duplodnaviria;Heunggongvirae;Uroviricota;Caudoviricetes |
| S2-wms-combined_25727\|provirus_4849_37113 | 22 | 1 | 2561 | Viruses;Duplodnaviria;Heunggongvirae;Uroviricota;Caudoviricetes |
| S2-wms-combined_58121\|provirus_18687_27276 | 5 | 1 | 2561 | Viruses;Duplodnaviria;Heunggongvirae;Uroviricota;Caudoviricetes |
| S2-wms-combined_140996\|provirus_89093_146281 | 65 | 1 | 2561 | Viruses;Duplodnaviria;Heunggongvirae;Uroviricota;Caudoviricetes |
| S2-wms-combined_140996\|provirus_204609_222314 | 14 | 1 | 2561 | Viruses;Duplodnaviria;Heunggongvirae;Uroviricota;Caudoviricetes |
| S2-wms-combined_222535\|provirus_7864_38886 | 5 | 1 | 2561 | Viruses;Duplodnaviria;Heunggongvirae;Uroviricota;Caudoviricetes |
| S2-wms-combined_58563\|provirus_21097_53508 | 35 | 1 | 2561 | Viruses;Duplodnaviria;Heunggongvirae;Uroviricota;Caudoviricetes |
| S2-wms-combined_110215\|provirus_1_66879 | 60 | 1 | 2561 | Viruses;Duplodnaviria;Heunggongvirae;Uroviricota;Caudoviricetes |
| S2-wms-combined_5791\|provirus_106816_143252 | 34 | 1 | 2561 | Viruses;Duplodnaviria;Heunggongvirae;Uroviricota;Caudoviricetes |
| S2-wms-combined_101826\|provirus_12326_53091 | 34 | 1 | 2561 | Viruses;Duplodnaviria;Heunggongvirae;Uroviricota;Caudoviricetes |
| S2-wms-combined_122457\|provirus_1_24741 | 30 | 1 | 2561 | Viruses;Duplodnaviria;Heunggongvirae;Uroviricota;Caudoviricetes |
| S2-wms-combined_185494\|provirus_1_41920 | 29 | 1 | 2561 | Viruses;Duplodnaviria;Heunggongvirae;Uroviricota;Caudoviricetes |
| S2-wms-combined_225908\|provirus_81930_115597 | 26 | 1 | 2561 | Viruses;Duplodnaviria;Heunggongvirae;Uroviricota;Caudoviricetes |
| S2-wms-combined_225908\|provirus_180828_195686 | 7 | 1 | 2561 | Viruses;Duplodnaviria;Heunggongvirae;Uroviricota;Caudoviricetes |
| S2-wms-combined_227800\|provirus_1_24245 | 25 | 1 | 2561 | Viruses;Duplodnaviria;Heunggongvirae;Uroviricota;Caudoviricetes |
| S2-wms-combined_143737\|provirus_1_42150 | 35 | 1 | 2561 | Viruses;Duplodnaviria;Heunggongvirae;Uroviricota;Caudoviricetes |
| S2-wms-combined_7756\|provirus_1_48340 | 17 | 1 | 2561 | Viruses;Duplodnaviria;Heunggongvirae;Uroviricota;Caudoviricetes |
| S2-wms_124416\|provirus_5437_58906 | 48 | 1 | 2561 | Viruses;Duplodnaviria;Heunggongvirae;Uroviricota;Caudoviricetes |
| S2-wms_156439\|provirus_1_21599 | 19 | 1 | 2561 | Viruses;Duplodnaviria;Heunggongvirae;Uroviricota;Caudoviricetes |
| S2-wms_107329\|provirus_5393_52664 | 55 | 1 | 2561 | Viruses;Duplodnaviria;Heunggongvirae;Uroviricota;Caudoviricetes |
| S2-wms_130687\|provirus_27517_69355 | 42 | 0.994 | 2561 | Viruses;Duplodnaviria;Heunggongvirae;Uroviricota;Caudoviricetes |
| S4-wms-combined_118531\|provirus_26963_69321 | 33 | 1 | 2561 | Viruses;Duplodnaviria;Heunggongvirae;Uroviricota;Caudoviricetes |
| S4-wms-combined_22012\|provirus_3895_27205 | 24 | 1 | 2561 | Viruses;Duplodnaviria;Heunggongvirae;Uroviricota;Caudoviricetes |
| S4-wms-combined_57123\|provirus_1_36340 | 18 | 1 | 2561 | Viruses;Duplodnaviria;Heunggongvirae;Uroviricota;Caudoviricetes |
| S4-wms-combined_64632\|provirus_24726_57119 | 38 | 1 | 2561 | Viruses;Duplodnaviria;Heunggongvirae;Uroviricota;Caudoviricetes |
| S4-wms-combined_113721\|provirus_1_14776 | 10 | 1 | 2561 | Viruses;Duplodnaviria;Heunggongvirae;Uroviricota;Caudoviricetes |
| S4-wms-combined_57963\|provirus_66556_101297 | 39 | 1 | 2561 | Viruses;Duplodnaviria;Heunggongvirae;Uroviricota;Caudoviricetes |
| S4-wms-combined_17284\|provirus_1_54754 | 44 | 1 | 2561 | Viruses;Duplodnaviria;Heunggongvirae;Uroviricota;Caudoviricetes |
| S4-wms-combined_121978\|provirus_55570_110001 | 44 | 1 | 2561 | Viruses;Duplodnaviria;Heunggongvirae;Uroviricota;Caudoviricetes |
| S4-wms-combined_25201\|provirus_1_27684 | 32 | 1 | 2561 | Viruses;Duplodnaviria;Heunggongvirae;Uroviricota;Caudoviricetes |

| | | | | |
|---|---|---|---|---|
| S4-wms-combined_41490\|provirus_33118_73219 | 31 | 1 | 2561 | Viruses;Duplodnaviria;Heunggongvirae;Uroviricota;Caudoviricetes |
| S4-wms-combined_51079\|provirus_1_19968 | 22 | 1 | 2561 | Viruses;Duplodnaviria;Heunggongvirae;Uroviricota;Caudoviricetes |
| S4-wms_50475\|provirus_24295_99597 | 77 | 0.9974 | 2561 | Viruses;Duplodnaviria;Heunggongvirae;Uroviricota;Caudoviricetes |
| S6-wms-combined_41246\|provirus_15790_40602 | 5 | 0.8534 | 2561 | Viruses;Duplodnaviria;Heunggongvirae;Uroviricota;Caudoviricetes |
| S6-wms-combined_135223\|provirus_91822_139676 | 16 | 1 | 2561 | Viruses;Duplodnaviria;Heunggongvirae;Uroviricota;Caudoviricetes |
| S6-wms-combined_88414\|provirus_44149_78550 | 41 | 1 | 2561 | Viruses;Duplodnaviria;Heunggongvirae;Uroviricota;Caudoviricetes |
| S6-wms-combined_82597\|provirus_164408_202920 | 38 | 1 | 2561 | Viruses;Duplodnaviria;Heunggongvirae;Uroviricota;Caudoviricetes |
| S6-wms-combined_118427\|provirus_96034_109457 | 12 | 1 | 2561 | Viruses;Duplodnaviria;Heunggongvirae;Uroviricota;Caudoviricetes |
| S6-wms-combined_118427\|provirus_117014_176709 | 53 | 1 | 2561 | Viruses;Duplodnaviria;Heunggongvirae;Uroviricota;Caudoviricetes |
| S6-wms-combined_72228\|provirus_24529_52555 | 15 | 1 | 2561 | Viruses;Duplodnaviria;Heunggongvirae;Uroviricota;Caudoviricetes |
| S6-wms-combined_119138\|provirus_23040_55482 | 38 | 1 | 2561 | Viruses;Duplodnaviria;Heunggongvirae;Uroviricota;Caudoviricetes |
| S6-wms-combined_8508\|provirus_1_42230 | 49 | 1 | 2561 | Viruses;Duplodnaviria;Heunggongvirae;Uroviricota;Caudoviricetes |
| S6-wms-combined_32530\|provirus_8460_46060 | 35 | 1 | 2561 | Viruses;Duplodnaviria;Heunggongvirae;Uroviricota;Caudoviricetes |
| S6-wms-combined_126236\|provirus_31179_69368 | 37 | 1 | 2561 | Viruses;Duplodnaviria;Heunggongvirae;Uroviricota;Caudoviricetes |
| S6-wms-combined_50721\|provirus_71105_115106 | 43 | 1 | 2561 | Viruses;Duplodnaviria;Heunggongvirae;Uroviricota;Caudoviricetes |
| S6-wms-combined_138710\|provirus_7149_17026 | 9 | 1 | 2561 | Viruses;Duplodnaviria;Heunggongvirae;Uroviricota;Caudoviricetes |
| S6-wms-combined_56464\|provirus_1_23497 | 25 | 0.9875 | 2561 | Viruses;Duplodnaviria;Heunggongvirae;Uroviricota;Caudoviricetes |
| S6-wms-combined_91039\|provirus_1_44817 | 38 | 1 | 2561 | Viruses;Duplodnaviria;Heunggongvirae;Uroviricota;Caudoviricetes |
| S8-wms-combined_215102\|provirus_23618_66504 | 26 | 1 | 2561 | Viruses;Duplodnaviria;Heunggongvirae;Uroviricota;Caudoviricetes |
| S8-wms-combined_23583\|provirus_43975_85815 | 41 | 1 | 2561 | Viruses;Duplodnaviria;Heunggongvirae;Uroviricota;Caudoviricetes |
| S8-wms-combined_48046\|provirus_1_29855 | 22 | 1 | 2561 | Viruses;Duplodnaviria;Heunggongvirae;Uroviricota;Caudoviricetes |
| S8-wms-combined_26087\|provirus_1_49451 | 33 | 1 | 2561 | Viruses;Duplodnaviria;Heunggongvirae;Uroviricota;Caudoviricetes |
| S8-wms-combined_72911\|provirus_1_47687 | 33 | 1 | 2561 | Viruses;Duplodnaviria;Heunggongvirae;Uroviricota;Caudoviricetes |
| S8-wms-combined_49369\|provirus_1_39890 | 24 | 1 | 2561 | Viruses;Duplodnaviria;Heunggongvirae;Uroviricota;Caudoviricetes |
| S8-wms-combined_74275\|provirus_1_26939 | 25 | 1 | 2561 | Viruses;Duplodnaviria;Heunggongvirae;Uroviricota;Caudoviricetes |
| S8-wms-combined_270362\|provirus_10571_69096 | 55 | 1 | 2561 | Viruses;Duplodnaviria;Heunggongvirae;Uroviricota;Caudoviricetes |
| S8-wms-combined_200168\|provirus_1_66777 | 45 | 1 | 2561 | Viruses;Duplodnaviria;Heunggongvirae;Uroviricota;Caudoviricetes |
| S8-wms-combined_108917\|provirus_9767_70457 | 48 | 1 | 2561 | Viruses;Duplodnaviria;Heunggongvirae;Uroviricota;Caudoviricetes |
| S8-wms-combined_40666\|provirus_44493_71582 | 21 | 1 | 2561 | Viruses;Duplodnaviria;Heunggongvirae;Uroviricota;Caudoviricetes |
| S8-wms-combined_169835\|provirus_34678_52956 | 23 | 1 | 2561 | Viruses;Duplodnaviria;Heunggongvirae;Uroviricota;Caudoviricetes |
| S8-wms-combined_68780\|provirus_22210_73585 | 54 | 1 | 2561 | Viruses;Duplodnaviria;Heunggongvirae;Uroviricota;Caudoviricetes |
| S8-wms-combined_161181\|provirus_9043_44282 | 39 | 1 | 2561 | Viruses;Duplodnaviria;Heunggongvirae;Uroviricota;Caudoviricetes |
| S8-wms-combined_246185\|provirus_1_35887 | 11 | 1 | 2561 | Viruses;Duplodnaviria;Heunggongvirae;Uroviricota;Caudoviricetes |
| S8-wms_58515\|provirus_15402_22507 | 6 | 1 | 2561 | Viruses;Duplodnaviria;Heunggongvirae;Uroviricota;Caudoviricetes |
| S8-wms_48935\|provirus_1_52721 | 54 | 1 | 2561 | Viruses;Duplodnaviria;Heunggongvirae;Uroviricota;Caudoviricetes |

**VLP-HQ-vOTUs:**

| seq_name | length | topology | n_genes | genetic_code | plasmid_score | fdr | n_hallmarks | marker_enrichment | conjugation_genes | amr_genes |
|---|---|---|---|---|---|---|---|---|---|---|
| S16_vs_173141 | 3774 | No terminal repeats | 3 | 11 | 0.9899 | NA | 1 | 0.9293 | NA | NA |
| S5_vs_109527 | 142384 | No terminal repeats | 133 | 11 | 0.9873 | NA | 0 | 9.1116 | NA | NA |
| S17_vs_52890 | 60210 | No terminal repeats | 53 | 11 | 0.9832 | NA | 1 | 1.8946 | NA | NA |
| S6_vs_46039 | 111911 | No terminal repeats | 121 | 11 | 0.9823 | NA | 2 | 0.0396 | MOBV | NF000153 |
| S14_vs_25852 | 107963 | No terminal repeats | 113 | 11 | 0.9815 | NA | 3 | 1.395 | MOBV | NA |
| S12_vs_164056 | 6664 | No terminal repeats | 9 | 11 | 0.9612 | NA | 0 | 0.5113 | NA | NA |

**MWS-HQ-vOTUs:**

| seq_name | length | topology | n_genes | genetic_code | plasmid_score | fdr | n_hallmarks | marker_enrichment | conjugation_genes | amr_genes |
|---|---|---|---|---|---|---|---|---|---|---|
| S6-wms-combined_135223 | 147913 | No terminal repeats | 138 | 11 | 0.9866 | NA | 0 | 9.7754 | B_traF | NA |
| S1-wms-combined_394628 | 10807 | No terminal repeats | 13 | 11 | 0.9673 | NA | 0 | 1.1593 | NA | NA |
| S2-wms-combined_233107 | 16186 | No terminal repeats | 16 | 11 | 0.9548 | NA | 1 | 0.4814 | NA | NA |
| S2-wms-combined_115352 | 3640 | No terminal repeats | 6 | 11 | 0.9387 | NA | 0 | 1.196 | NA | NA |
| S2-wms-combined_132966 | 4461 | No terminal repeats | 11 | 11 | 0.9346 | NA | 0 | 0.8148 | NA | NA |
| S1-wms-combined_169788 | 4712 | No terminal repeats | 8 | 11 | 0.8482 | NA | 0 | 0.876 | NA | NA |

Note:

**Provirus detected**
