## Supplementary material for "Comprehensive profiling of the human intestinal DNA virome and prediction of disease-associated bacterial hosts in severe Myalgic Encephalomyelitis/Chronic Fatigue Syndrome (ME/CFS)": S Table 7 Contaminant monitor

| Sample_IDs | # Raw_reads (PE) | # cleaned_reads (PE) | # Total_raw_contigs | N50 (bp) | Longest_contig_length (bp) |
|---|---|---|---|---|---|
| NC1 | 306630 | 360240 | 3597 | 433 | 13109 |
| NC2 | 16623 | 18592 | 19 | 446 | 910 |
| NC3 | 72714 | 111664 | 2128 | 495 | 8397 |
| Sum | 395967 | 490496 | 5744 | 1374 | 22416 |
| Mean | 131989.00 | 163498.67 | 1914.67 | 458.00 | 7472.00 |
| SD | 153821.84 | 176623.79 | 1798.51 | 32.70 | 6151.88 |

| | | | NC1-3 | | |
|---|---|---|---|---|---|
| | Merged_UViGs | LQ_undetermined_NC-UViGs_checkv | nr-vOTUs_size_filtration_1kb_cut_off | LQ_undetermined_nr-vOTUs_checkv | Notes |
| # Viral genomes | 5953 | 5953 | 103 | 103 | All NC viral genomes are low quality and undetermined |
| # Viral genomes >= 1 kb | 255 | 255 | 103 | 103 | |
| # Viral genomes >= 5 kb | 13 | 13 | 3 | 3 | |
| # Viral genomes >= 10 kb | 4 | 4 | 0 | 0 | |
| N50 (bp) | 468 | 468 | NA | NA | |
| Longest representative (bp) | 13109 | 13109 | 8080 | 8080 | Not-determined |
| Shortest representative (bp) | 239 | 239 | 1040 | 1040 | Not-determined |

NC1

| | | | | | |
|---|---|---|---|---|---|
| 25.04 | 45105 | 45105 | U | 0 | unclassified |
| 74.96 | 135015 | 859 | R | 1 | root |
| 74 | 133297 | 185 | R1 | 131567 | cellular organisms |
| 72.32 | 130261 | 475 | D | 2 | Bacteria |
| 58.69 | 105718 | 1452 | P | 1224 | Proteobacteria |
| 27.47 | 49478 | 53 | C | 28216 | Betaproteobacteria |
| 27.4 | 49360 | 1152 | O | 80840 | Burkholderiales |
| 25.49 | 45914 | 1107 | F | 80864 | Comamonadaceae |
| 24.53 | 44185 | 28928 | G | 80865 | Delftia |
| 6.82 | 12277 | 12277 | S | 558537 | Delftia lacustris |
| 1.01 | 1814 | 1814 | S | 180282 | Delftia tsuruhatensis |
| 0.47 | 855 | 854 | S | 80866 | Delftia acidovorans |
| 0 | 1 | 1 | S1 | 398578 | Delftia acidovorans SPH-1 |
| 0.17 | 311 | 67 | G1 | 2613839 | unclassified Delftia |
| 0.08 | 148 | 148 | S | 742013 | Delftia sp. Cs1-4 |
| 0.05 | 96 | 96 | S | 1920191 | Delftia sp. HK171 |
| 0.07 | 134 | 9 | G | 12916 | Acidovorax |
| 0.06 | 108 | 0 | G1 | 2684926 | unclassified Acidovorax |
| 0.05 | 82 | 82 | S | 358220 | Acidovorax sp. KKS102 |
| 0.01 | 11 | 11 | S | 232721 | Acidovorax sp. JS42 |
| 0 | 7 | 7 | S | 2478662 | Acidovorax sp. 1608163 |
| 0 | 4 | 4 | S | 1842533 | Acidovorax sp. RAC01 |
| 0 | 4 | 4 | S | 2518343 | Acidovorax sp. JMULE5 |
| 0.01 | 14 | 14 | S | 553814 | Acidovorax carolinensis |
| 0 | 3 | 3 | S | 2743470 | Acidovorax antarcticus |
| 0.07 | 133 | 0 | G | 201096 | Alicycliphilus |
| 0.07 | 133 | 0 | S | 179636 | Alicycliphilus denitrificans |
| 0.07 | 133 | 133 | S1 | 596154 | Alicycliphilus denitrificans K601 |
| 0.07 | 125 | 22 | G | 34072 | Variovorax |
| 0.05 | 95 | 10 | G1 | 663243 | unclassified Variovorax |
| 0.03 | 63 | 63 | S | 1034889 | Variovorax sp. HW608 |
| 0.01 | 14 | 14 | S | 207745 | Variovorax sp. WDL1 |

| | | | | | |
|---|---|---|---|---|---|
| 0 | 5 | 5 | S | 662548 | Variovorax sp. RA8 |
| 0 | 3 | 3 | S | 2762322 | Variovorax sp. PAMC26660 |
| 0 | 5 | 5 | S | 34073 | Variovorax paradoxus |
| 0 | 3 | 3 | S | 436515 | Variovorax boronicumulans |
| 0.06 | 105 | 14 | G | 283 | Comamonas |
| 0.01 | 22 | 22 | S | 160825 | Comamonas koreensis |
| 0.01 | 21 | 10 | S | 285 | Comamonas testosteroni |
| 0.01 | 11 | 11 | S1 | 1392005 | Comamonas testosteroni TK102 |
| 0.01 | 20 | 20 | S | 1562974 | Comamonas piscis |
| 0.01 | 18 | 18 | S | 225991 | Comamonas aquatica |
| 0 | 9 | 0 | G1 | 2638500 | unclassified Comamonas |
| 0 | 9 | 9 | S | 1232667 | Comamonas sp. 7D-2 |
| 0 | 1 | 1 | S | 1082851 | Comamonas serinivorans |
| 0.05 | 93 | 0 | G | 238749 | Diaphorobacter |
| 0.04 | 80 | 1 | G1 | 2649760 | unclassified Diaphorobacter |
| 0.03 | 46 | 46 | S | 2714924 | Diaphorobacter sp. HDW4A |
| 0.01 | 26 | 26 | S | 2792224 | Diaphorobacter sp. JS3051 |
| 0 | 6 | 6 | S | 2735554 | Diaphorobacter sp. JS3050 |
| 0 | 1 | 1 | S | 2714925 | Diaphorobacter sp. HDW4B |
| 0 | 8 | 8 | S | 1288495 | Diaphorobacter aerolatus |
| 0 | 3 | 3 | S | 1715720 | Diaphorobacter ruginosibacter |
| 0 | 2 | 2 | S | 1546149 | Diaphorobacter polyhydroxybutyrativorans |
| 0.01 | 13 | 1 | G | 47420 | Hydrogenophaga |
| 0.01 | 12 | 3 | G1 | 2610897 | unclassified Hydrogenophaga |
| 0 | 8 | 8 | S | 795665 | Hydrogenophaga sp. PBC |
| 0 | 1 | 1 | S | 434010 | Hydrogenophaga sp. PBL-H3 |
| 0 | 9 | 0 | G | 28065 | Rhodoferax |
| 0 | 5 | 0 | G1 | 2627954 | unclassified Rhodoferax |
| 0 | 5 | 5 | S | 2741720 | Rhodoferax sp. BAB1 |
| 0 | 4 | 4 | S | 1484693 | Rhodoferax saidenbachensis |
| 0 | 6 | 0 | G | 665874 | Limnohabitans |
| 0 | 6 | 0 | G1 | 2626134 | unclassified Limnohabitans |
| 0 | 5 | 5 | S | 1678129 | Limnohabitans sp. 103DPR2 |

| | | | | | |
|---|---|---|---|---|---|
| 0 | 1 | 1 | S | 1678128 | Limnohabitans sp. 63ED37-2 |
| 0 | 1 | 0 | G | 215579 | Schlegelella |
| 0 | 1 | 1 | S | 215580 | Schlegelella thermodepolymerans |
| 0 | 1 | 0 | G | 364316 | Verminephrobacter |
| 0 | 1 | 0 | S | 364317 | Verminephrobacter eiseniae |
| 0 | 1 | 1 | S1 | 391735 | Verminephrobacter eiseniae EF01-2 |
| 0 | 1 | 0 | G | 683756 | Kinneretia |
| 0 | 1 | 0 | G1 | 2632542 | unclassified Kinneretia |
| 0 | 1 | 1 | S | 2714952 | Kinneretia sp. DAIF2 |
| 0 | 1 | 0 | G | 1649468 | Melaminivora |
| 0 | 1 | 0 | G1 | 2641902 | unclassified Melaminivora |
| 0 | 1 | 1 | S | 2109913 | Melaminivora sp. SC2-9 |
| 0.63 | 1138 | 200 | F | 119060 | Burkholderiaceae |
| 0.38 | 691 | 327 | G | 106589 | Cupriavidus |
| 0.03 | 57 | 57 | S | 82633 | Cupriavidus pauculus |
| 0.03 | 54 | 54 | S | 164546 | Cupriavidus taiwanensis |
| 0.03 | 52 | 48 | S | 82541 | Cupriavidus gilardii |
| 0 | 4 | 4 | S1 | 1267562 | Cupriavidus gilardii CR3 |
| 0.02 | 34 | 34 | S | 1796606 | Cupriavidus nantongensis |
| 0.02 | 31 | 30 | S | 106590 | Cupriavidus necator |
| 0 | 1 | 1 | S1 | 1042878 | Cupriavidus necator N-1 |
| 0.02 | 29 | 29 | S | 96344 | Cupriavidus oxalaticus |
| 0.01 | 26 | 26 | S | 119219 | Cupriavidus metallidurans |
| 0.01 | 25 | 25 | S | 151783 | Cupriavidus campinensis |
| 0.01 | 20 | 20 | S | 68895 | Cupriavidus basilensis |
| 0.01 | 13 | 13 | S | 1040979 | Cupriavidus neocaledonicus |
| 0.01 | 12 | 0 | G1 | 2640874 | unclassified Cupriavidus |
| 0 | 6 | 6 | S | 876364 | Cupriavidus sp. USMAA2-4 |
| 0 | 6 | 6 | S | 2771360 | Cupriavidus sp. ISTL7 |
| 0.01 | 11 | 0 | S | 248026 | Cupriavidus pinatubonensis |
| 0.01 | 11 | 11 | S1 | 264198 | Cupriavidus pinatubonensis JMP134 |
| 0.06 | 104 | 19 | G | 48736 | Ralstonia |
| 0.03 | 47 | 41 | S | 329 | Ralstonia pickettii |

| | | | | | |
|---|---|---|---|---|---|
| 0 | 5 | 5 | S1 | 428406 | Ralstonia pickettii 12D |
| 0 | 1 | 1 | S1 | 402626 | Ralstonia pickettii 12J |
| 0.01 | 19 | 19 | S | 305 | Ralstonia solanacearum |
| 0.01 | 13 | 13 | S | 105219 | Ralstonia mannitolilytica |
| 0 | 6 | 6 | S | 190721 | Ralstonia insidiosa |
| 0.05 | 86 | 3 | G | 32008 | Burkholderia |
| 0.03 | 59 | 4 | G1 | 87882 | Burkholderia cepacia complex |
| 0.02 | 35 | 35 | S | 95485 | Burkholderia stabilis |
| 0.01 | 14 | 14 | S | 95486 | Burkholderia cenocepacia |
| 0 | 4 | 4 | S | 488731 | Burkholderia seminalis |
| 0 | 1 | 0 | S | 292 | Burkholderia cepacia |
| 0 | 1 | 1 | S1 | 1395570 | Burkholderia cepacia JBK9 |
| 0 | 1 | 1 | S | 488732 | Burkholderia diffusa |
| 0.01 | 20 | 1 | G1 | 2613784 | unclassified Burkholderia |
| 0 | 7 | 7 | S | 1097668 | Burkholderia sp. YI23 |
| 0 | 5 | 5 | S | 1855726 | Burkholderia sp. KK1 |
| 0 | 4 | 4 | S | 1705310 | Burkholderia sp. IDO3 |
| 0 | 2 | 2 | S | 1740163 | Burkholderia sp. Bp7605 |
| 0 | 1 | 1 | S | 758782 | Burkholderia sp. THE68 |
| 0 | 2 | 2 | S | 28095 | Burkholderia gladioli |
| 0 | 1 | 1 | S | 337 | Burkholderia glumae |
| 0 | 1 | 0 | G1 | 111527 | pseudomallei group |
| 0 | 1 | 1 | S | 28450 | Burkholderia pseudomallei |
| 0.02 | 40 | 7 | G | 1822464 | Paraburkholderia |
| 0 | 9 | 0 | G1 | 2615204 | unclassified Paraburkholderia |
| 0 | 7 | 7 | S | 1926494 | Paraburkholderia sp. SOS3 |
| 0 | 2 | 2 | S | 2604047 | Paraburkholderia sp. Msb3 |
| 0 | 8 | 8 | S | 311230 | Paraburkholderia terrae |
| 0 | 5 | 5 | S | 92647 | Paraburkholderia tropica |
| 0 | 4 | 4 | S | 311231 | Paraburkholderia ginsengisoli |
| 0 | 3 | 3 | S | 2654982 | Paraburkholderia atlantica |
| 0 | 2 | 0 | S | 36873 | Paraburkholderia xenovorans |
| 0 | 2 | 2 | S1 | 266265 | Paraburkholderia xenovorans LB400 |

| | | | | | |
|---|---|---|---|---|---|
| 0 | 2 | 0 | S | 261302 | Paraburkholderia phytofirmans |
| 0 | 2 | 2 | S1 | 398527 | Paraburkholderia phytofirmans PsJN |
| 0.01 | 14 | 0 | G | 93217 | Pandoraea |
| 0 | 6 | 6 | S | 93220 | Pandoraea pnomenusa |
| 0 | 5 | 5 | S | 1891094 | Pandoraea fibrosis |
| 0 | 1 | 1 | S | 445709 | Pandoraea thiooxydans |
| 0 | 1 | 1 | S | 656179 | Pandoraea faecigallinarum |
| 0 | 1 | 0 | G1 | 2624094 | unclassified Pandoraea |
| 0 | 1 | 1 | S | 2518599 | Pandoraea sp. XY-2 |
| 0 | 2 | 0 | G | 1827195 | Caballeronia |
| 0 | 2 | 0 | G1 | 2646786 | unclassified Caballeronia |
| 0 | 2 | 2 | S | 2814653 | Caballeronia sp. M1242 |
| 0 | 1 | 0 | G | 2571159 | Mycetohabitans |
| 0 | 1 | 0 | S | 412963 | Mycetohabitans rhizoxinica |
| 0 | 1 | 1 | S1 | 882378 | Mycetohabitans rhizoxinica HKI 454 |
| 0.55 | 982 | 114 | F | 75682 | Oxalobacteraceae |
| 0.38 | 680 | 115 | G | 149698 | Massilia |
| 0.15 | 272 | 40 | G1 | 2609279 | unclassified Massilia |
| 0.06 | 110 | 110 | S | 1707785 | Massilia sp. WG5 |
| 0.04 | 72 | 72 | S | 2769491 | Massilia sp. LPB0304 |
| 0.01 | 19 | 19 | S | 2765360 | Massilia sp. CCM 8941 |
| 0.01 | 15 | 15 | S | 1678028 | Massilia sp. NR 4-1 |
| 0 | 9 | 9 | S | 1593482 | Massilia sp. YMA4 |
| 0 | 7 | 7 | S | 2709302 | Massilia sp. Dwa41.01b |
| 0.1 | 177 | 177 | S | 945844 | Massilia oculi |
| 0.02 | 42 | 42 | S | 1141883 | Massilia putida |
| 0.01 | 21 | 21 | S | 864828 | Massilia umbonata |
| 0.01 | 20 | 20 | S | 2072590 | Massilia armeniaca |
| 0.01 | 19 | 19 | S | 871742 | Massilia flava |
| 0.01 | 11 | 11 | S | 321984 | Massilia plicata |
| 0 | 3 | 3 | S | 2045208 | Massilia violaceinigra |
| 0.05 | 99 | 0 | G | 75654 | Duganella |
| 0.05 | 99 | 1 | G1 | 2636909 | unclassified Duganella |

| | | | | | |
|---|---|---|---|---|---|
| 0.04 | 72 | 72 | S | 2728020 | Duganella sp. GN2-R2 |
| 0.01 | 26 | 26 | S | 2728021 | Duganella sp. AF9R3 |
| 0.03 | 46 | 34 | G | 29580 | Janthinobacterium |
| 0 | 4 | 0 | G1 | 2610881 | unclassified Janthinobacterium |
| 0 | 2 | 2 | S | 1644131 | Janthinobacterium sp. 1_2014MBL_MicDiv |
| 0 | 2 | 2 | S | 2497863 | Janthinobacterium sp. 17J80-10 |
| 0 | 3 | 0 | S | 55508 | Janthinobacterium agaricidamnosum |
| 0 | 3 | 3 | S1 | 1349767 | Janthinobacterium agaricidamnosum NBRC 102515 = DSM 9628 |
| 0 | 3 | 3 | S | 368607 | Janthinobacterium svalbardensis |
| 0 | 1 | 1 | S | 29581 | Janthinobacterium lividum |
| 0 | 1 | 1 | S | 2590869 | Janthinobacterium tructae |
| 0.01 | 16 | 0 | G | 963 | Herbaspirillum |
| 0.01 | 11 | 11 | S | 863372 | Herbaspirillum huttiense |
| 0 | 3 | 3 | S | 964 | Herbaspirillum seropedicae |
| 0 | 1 | 0 | S | 80842 | Herbaspirillum rubrisubalbicans |
| 0 | 1 | 1 | S1 | 1235827 | Herbaspirillum rubrisubalbicans Os34 |
| 0 | 1 | 0 | S | 341045 | Herbaspirillum hiltneri |
| 0 | 1 | 1 | S1 | 1262470 | Herbaspirillum hiltneri N3 |
| 0.01 | 13 | 0 | G | 401469 | Undibacterium |
| 0.01 | 13 | 5 | G1 | 2630295 | unclassified Undibacterium |
| 0 | 6 | 6 | S | 2058625 | Undibacterium sp. YM2 |
| 0 | 2 | 2 | S | 2058624 | Undibacterium sp. KW1 |
| 0.01 | 11 | 0 | G | 1344552 | Noviherbaspirillum |
| 0.01 | 11 | 0 | G1 | 2617509 | unclassified Noviherbaspirillum |
| 0.01 | 11 | 11 | S | 2601898 | Noviherbaspirillum sp. UKPF54 |
| 0 | 3 | 0 | G | 202907 | Collimonas |
| 0 | 2 | 2 | S | 279058 | Collimonas arenae |
| 0 | 1 | 1 | S | 158899 | Collimonas fungivorans |
| 0.05 | 93 | 0 | F | 506 | Alcaligenaceae |
| 0.02 | 39 | 0 | G | 507 | Alcaligenes |
| 0.02 | 39 | 39 | S | 511 | Alcaligenes faecalis |
| 0.01 | 20 | 10 | G | 222 | Achromobacter |
| 0 | 7 | 6 | S | 85698 | Achromobacter xylosoxidans |

| | | | | | |
|---|---|---|---|---|---|
| 0 | 1 | 1 | S1 | 762376 | Achromobacter xylosoxidans A8 |
| 0 | 1 | 1 | S | 217203 | Achromobacter spanius |
| 0 | 1 | 1 | S | 1353891 | Achromobacter deleyi |
| 0 | 1 | 0 | G1 | 2626865 | unclassified Achromobacter |
| 0 | 1 | 1 | S | 1758194 | Achromobacter sp. AONIH1 |
| 0.01 | 17 | 0 | G | 517 | Bordetella |
| 0 | 8 | 8 | S | 103855 | Bordetella hinzii |
| 0 | 5 | 5 | S | 94624 | Bordetella petrii |
| 0 | 2 | 2 | S | 1331258 | Bordetella pseudohinzii |
| 0 | 1 | 1 | S | 1416803 | Bordetella genomosp. 9 |
| 0 | 1 | 0 | G1 | 2630031 | unclassified Bordetella |
| 0 | 1 | 1 | S | 1746199 | Bordetella sp. N |
| 0.01 | 12 | 0 | G | 305976 | Pusillimonas |
| 0 | 7 | 7 | S | 2028345 | Pusillimonas thiosulfatoxidans |
| 0 | 5 | 0 | G1 | 2640016 | unclassified Pusillimonas |
| 0 | 5 | 5 | S | 1007105 | Pusillimonas sp. T7-7 |
| 0 | 5 | 0 | G | 152267 | Pigmentiphaga |
| 0 | 5 | 0 | G1 | 2626614 | unclassified Pigmentiphaga |
| 0 | 5 | 5 | S | 2488560 | Pigmentiphaga sp. H8 |
| 0.03 | 51 | 0 | F | 995019 | Sutterellaceae |
| 0.03 | 48 | 0 | G | 40544 | Sutterella |
| 0.02 | 43 | 43 | S | 40545 | Sutterella wadsworthensis |
| 0 | 5 | 5 | S | 2494234 | Sutterella megalosphaeroides |
| 0 | 3 | 0 | G | 1918598 | Turicimonas |
| 0 | 3 | 3 | S | 1796652 | Turicimonas muris |
| 0.02 | 30 | 0 | O1 | 224471 | Burkholderiales genera incertae sedis |
| 0.01 | 11 | 0 | G | 88 | Leptothrix |
| 0.01 | 11 | 0 | S | 34029 | Leptothrix cholodnii |
| 0.01 | 11 | 11 | S1 | 395495 | Leptothrix cholodnii SP-6 |
| 0 | 6 | 3 | G | 316612 | Methylibium |
| 0 | 3 | 0 | G1 | 2633235 | unclassified Methylibium |
| 0 | 3 | 3 | S | 2082386 | Methylibium sp. Pch-M |
| 0 | 5 | 0 | G | 54066 | Xylophilus |

| | | | | | |
|---|---|---|---|---|---|
| 0 | 5 | 5 | S | 2697032 | Xylophilus rhododendri |
| 0 | 4 | 0 | G | 318147 | Paucibacter |
| 0 | 4 | 0 | G1 | 2642959 | unclassified Paucibacter |
| 0 | 4 | 4 | S | 1768242 | Paucibacter sp. KCTC 42545 |
| 0 | 3 | 0 | G | 212743 | Rhizobacter |
| 0 | 2 | 2 | S | 946333 | Rhizobacter gummiphilus |
| 0 | 1 | 0 | G1 | 2640088 | unclassified Rhizobacter |
| 0 | 1 | 1 | S | 2753607 | Rhizobacter sp. AJA081-3 |
| 0 | 1 | 0 | G | 92793 | Aquabacterium |
| 0 | 1 | 1 | S | 1296669 | Aquabacterium olei |
| 0.03 | 50 | 0 | O | 206351 | Neisseriales |
| 0.02 | 34 | 0 | F | 481 | Neisseriaceae |
| 0.01 | 18 | 0 | G | 32257 | Kingella |
| 0.01 | 18 | 18 | S | 502 | Kingella denitrificans |
| 0.01 | 15 | 0 | G | 482 | Neisseria |
| 0 | 7 | 7 | S | 490 | Neisseria sicca |
| 0 | 5 | 5 | S | 28449 | Neisseria subflava |
| 0 | 2 | 2 | S | 495 | Neisseria elongata |
| 0 | 1 | 1 | S | 488 | Neisseria mucosa |
| 0 | 1 | 0 | G | 538 | Eikenella |
| 0 | 1 | 1 | S | 539 | Eikenella corrodens |
| 0.01 | 16 | 0 | F | 1499392 | Chromobacteriaceae |
| 0.01 | 10 | 0 | G | 568394 | Pseudogulbenkiania |
| 0.01 | 10 | 0 | G1 | 2642558 | unclassified Pseudogulbenkiania |
| 0.01 | 10 | 10 | S | 748280 | Pseudogulbenkiania sp. NH8B |
| 0 | 5 | 0 | F1 | 90153 | Chromobacterium group |
| 0 | 5 | 0 | G | 535 | Chromobacterium |
| 0 | 2 | 2 | S | 536 | Chromobacterium violaceum |
| 0 | 2 | 2 | S | 2202141 | Chromobacterium phragmitis |
| 0 | 1 | 1 | S | 1108595 | Chromobacterium vaccinii |
| 0 | 1 | 0 | G | 407217 | Aquitalea |
| 0 | 1 | 1 | S | 1537400 | Aquitalea aquatilis |
| 0.01 | 11 | 2 | O | 206389 | Rhodocyclales |

| | | | | | |
|---|---|---|---|---|---|
| 0 | 7 | 0 | F | 2008794 | Zoogloeaceae |
| 0 | 4 | 0 | G | 33057 | Thauera |
| 0 | 2 | 0 | S | 59405 | Thauera aromatica |
| 0 | 2 | 2 | S1 | 44139 | Thauera aromatica K172 |
| 0 | 1 | 1 | S | 1134435 | Thauera humireducens |
| 0 | 1 | 1 | G1 | 2609274 | unclassified Thauera |
| 0 | 3 | 0 | G | 12960 | Azoarcus |
| 0 | 3 | 0 | G1 | 2629479 | unclassified Azoarcus |
| 0 | 3 | 3 | S | 2016596 | Azoarcus sp. M9-3-2 |
| 0 | 2 | 0 | F | 75787 | Rhodocyclaceae |
| 0 | 2 | 1 | G | 146937 | Azospira |
| 0 | 1 | 0 | S | 146939 | Azospira oryzae |
| 0 | 1 | 1 | S1 | 640081 | Azospira oryzae PS |
| 0 | 4 | 0 | O | 32003 | Nitrosomonadales |
| 0 | 2 | 0 | F | 2772226 | Sulfuricellaceae |
| 0 | 1 | 0 | G | 1778653 | Sulfuriferula |
| 0 | 1 | 1 | S | 171865 | Sulfuriferula plumbiphila |
| 0 | 1 | 0 | G | 2772198 | Sulfurimicrobium |
| 0 | 1 | 1 | S | 2715678 | Sulfurimicrobium lacus |
| 0 | 1 | 0 | F | 206379 | Nitrosomonadaceae |
| 0 | 1 | 0 | G | 914 | Nitrosomonas |
| 0 | 1 | 0 | S | 916 | Nitrosomonas eutropha |
| 0 | 1 | 1 | S1 | 335283 | Nitrosomonas eutropha C91 |
| 0 | 1 | 0 | F | 2008790 | Thiobacillaceae |
| 0 | 1 | 0 | G | 1938335 | Sulfuritortus |
| 0 | 1 | 1 | S | 1914471 | Sulfuritortus calidifontis |
| 22.39 | 40331 | 353 | C | 28211 | Alphaproteobacteria |
| 19.55 | 35209 | 79 | O | 204457 | Sphingomonadales |
| 19.47 | 35061 | 1294 | F | 41297 | Sphingomonadaceae |
| 17.77 | 32003 | 626 | G | 13687 | Sphingomonas |
| 16.89 | 30418 | 30418 | S | 13689 | Sphingomonas paucimobilis |
| 0.15 | 277 | 277 | S | 93064 | Sphingomonas koreensis |
| 0.13 | 236 | 5 | G1 | 196159 | unclassified Sphingomonas |

| | | | | | |
|---|---|---|---|---|---|
| 0.03 | 56 | 56 | S | 1961362 | Sphingomonas sp. NIC1 |
| 0.02 | 44 | 44 | S | 2219696 | Sphingomonas sp. FARSPH |
| 0.02 | 34 | 34 | S | 2596913 | Sphingomonas sp. NBWT7 |
| 0.01 | 17 | 17 | S | 2759526 | Sphingomonas sp. DH-S5 |
| 0.01 | 16 | 16 | S | 304378 | Sphingomonas sp. IC081 |
| 0.01 | 13 | 13 | S | 2735134 | Sphingomonas sp. AP4-R1 |
| 0.01 | 13 | 13 | S | 1523415 | Sphingomonas sp. AAP5 |
| 0.01 | 10 | 10 | S | 1938607 | Sphingomonas sp. LM7 |
| 0 | 8 | 8 | S | 1517554 | Sphingomonas sp. HMP9 |
| 0 | 7 | 7 | S | 2565555 | Sphingomonas sp. PAMC26645 |
| 0 | 7 | 7 | S | 1390395 | Sphingomonas sp. LK11 |
| 0 | 4 | 4 | S | 28214 | Sphingomonas sp. |
| 0 | 1 | 1 | S | 849864 | Sphingomonas sp. SH |
| 0 | 1 | 1 | S | 2653203 | Sphingomonas sp. CL5.1 |
| 0.12 | 211 | 0 | S | 397260 | Sphingomonas sanxanigenens |
| 0.12 | 211 | 211 | S1 | 1123269 | Sphingomonas sanxanigenens DSM 19645 = NX02 |
| 0.09 | 155 | 13 | S | 152682 | Sphingomonas melonis |
| 0.08 | 142 | 142 | S1 | 621456 | Sphingomonas melonis TY |
| 0.02 | 32 | 32 | S | 424800 | Sphingomonas insulae |
| 0.01 | 20 | 0 | S | 160791 | Sphingomonas wittichii |
| 0.01 | 20 | 20 | S1 | 392499 | Sphingomonas wittichii RW1 |
| 0.01 | 17 | 17 | S | 1609977 | Sphingomonas hengshuiensis |
| 0 | 5 | 5 | S | 653931 | Sphingomonas alpina |
| 0 | 4 | 4 | S | 1549858 | Sphingomonas taxi |
| 0 | 2 | 2 | S | 1813879 | Sphingomonas panacisoli |
| 0.69 | 1248 | 130 | G | 165695 | Sphingobium |
| 0.17 | 302 | 302 | S | 1673076 | Sphingobium hydrophobicum |
| 0.12 | 222 | 222 | S | 13690 | Sphingobium yanoikuyae |
| 0.09 | 169 | 47 | G1 | 2611147 | unclassified Sphingobium |
| 0.02 | 40 | 40 | S | 520734 | Sphingobium sp. RSMS |
| 0.02 | 32 | 32 | S | 1315974 | Sphingobium sp. TKS |
| 0.02 | 28 | 28 | S | 484429 | Sphingobium sp. YBL2 |
| 0.01 | 19 | 19 | S | 2082188 | Sphingobium sp. YG1 |

| | | | | | |
|---|---|---|---|---|---|
| 0 | 2 | 2 | S | 2565554 | Sphingobium sp. PAMC28499 |
| 0 | 1 | 1 | S | 1843368 | Sphingobium sp. RAC03 |
| 0.08 | 143 | 143 | S | 120107 | Sphingobium cloacae |
| 0.07 | 126 | 126 | S | 76947 | Sphingobium herbicidovorans |
| 0.05 | 82 | 82 | S | 135719 | Sphingobium amiense |
| 0.04 | 65 | 65 | S | 1332080 | Sphingobium baderi |
| 0 | 5 | 0 | S | 336203 | Sphingobium fuliginis |
| 0 | 5 | 5 | S1 | 1208342 | Sphingobium fuliginis ATCC 27551 |
| 0 | 2 | 0 | S | 332056 | Sphingobium japonicum |
| 0 | 2 | 2 | S1 | 452662 | Sphingobium japonicum UT26S |
| 0 | 2 | 2 | S | 1054037 | Sphingobium barthaii |
| 0.11 | 204 | 0 | G | 165696 | Novosphingobium |
| 0.1 | 177 | 45 | G1 | 2644732 | unclassified Novosphingobium |
| 0.05 | 98 | 98 | S | 164608 | Novosphingobium sp. KA1 |
| 0.01 | 16 | 16 | S | 702113 | Novosphingobium sp. PP1Y |
| 0.01 | 10 | 10 | S | 2780074 | Novosphingobium sp. ES2-1 |
| 0 | 7 | 7 | S | 2571749 | Novosphingobium sp. ABRDHK2 |
| 0 | 1 | 1 | S | 1609758 | Novosphingobium sp. P6W |
| 0.01 | 22 | 0 | S | 205844 | Novosphingobium pentaromativorans |
| 0.01 | 22 | 22 | S1 | 1088721 | Novosphingobium pentaromativorans US6-1 |
| 0 | 5 | 0 | S | 48935 | Novosphingobium aromaticivorans |
| 0 | 5 | 5 | S1 | 279238 | Novosphingobium aromaticivorans DSM 12444 |
| 0.1 | 186 | 16 | G | 165697 | Sphingopyxis |
| 0.07 | 122 | 1 | G1 | 2614943 | unclassified Sphingopyxis |
| 0.05 | 89 | 89 | S | 292913 | Sphingopyxis sp. 113P3 |
| 0.01 | 10 | 10 | S | 1914525 | Sphingopyxis sp. FD7 |
| 0 | 9 | 9 | S | 1874061 | Sphingopyxis sp. EG6 |
| 0 | 6 | 6 | S | 1357916 | Sphingopyxis sp. QXT-31 |
| 0 | 4 | 4 | S | 2486273 | Sphingopyxis sp. OPL5 |
| 0 | 2 | 2 | S | 1866325 | Sphingopyxis sp. MG |
| 0 | 1 | 1 | S | 2565556 | Sphingopyxis sp. PAMC25046 |
| 0.02 | 32 | 32 | S | 33050 | Sphingopyxis macrogoltabida |
| 0 | 9 | 0 | S | 117207 | Sphingopyxis alaskensis |

| | | | | | |
|---|---|---|---|---|---|
| 0 | 9 | 9 | S1 | 317655 | Sphingopyxis alaskensis RB2256 |
| 0 | 5 | 5 | S | 1515612 | Sphingopyxis fribergensis |
| 0 | 2 | 2 | S | 267128 | Sphingopyxis granuli |
| 0.07 | 126 | 0 | G | 1649486 | Rhizorhabdus |
| 0.07 | 126 | 126 | S | 1850238 | Rhizorhabdus dicambivorans |
| 0.04 | 68 | 4 | F | 335929 | Erythrobacteraceae |
| 0.02 | 36 | 0 | F1 | 2800788 | Erythrobacter/Porphyrobacter group |
| 0.02 | 32 | 0 | G | 1041 | Erythrobacter |
| 0.02 | 28 | 0 | G1 | 2633097 | unclassified Erythrobacter |
| 0.01 | 26 | 26 | S | 2785907 | Erythrobacter sp. A30-3 |
| 0 | 2 | 2 | S | 1798193 | Erythrobacter sp. HL-111 |
| 0 | 3 | 0 | S | 39960 | Erythrobacter litoralis |
| 0 | 3 | 3 | S1 | 314225 | Erythrobacter litoralis HTCC2594 |
| 0 | 1 | 1 | S | 1112 | Erythrobacter neustonensis |
| 0 | 4 | 0 | G | 1111 | Porphyrobacter |
| 0 | 4 | 0 | G1 | 2683265 | unclassified Porphyrobacter |
| 0 | 2 | 2 | S | 2003315 | Porphyrobacter sp. CACIAM 03H1 |
| 0 | 2 | 2 | S | 2023229 | Porphyrobacter sp. HT-58-2 |
| 0.01 | 13 | 0 | G | 1295327 | Croceicoccus |
| 0 | 9 | 9 | S | 1348774 | Croceicoccus naphthovorans |
| 0 | 4 | 0 | G1 | 2629967 | unclassified Croceicoccus |
| 0 | 4 | 4 | S | 2798724 | Croceicoccus sp. YJ47 |
| 0.01 | 10 | 0 | G | 361177 | Altererythrobacter |
| 0 | 9 | 9 | S | 1267766 | Altererythrobacter atlanticus |
| 0 | 1 | 0 | G1 | 2614945 | unclassified Altererythrobacter |
| 0 | 1 | 1 | S | 2060312 | Altererythrobacter sp. B11 |
| 0 | 5 | 0 | G | 2800681 | Aurantiacibacter |
| 0 | 3 | 3 | S | 1648404 | Aurantiacibacter atlanticus |
| 0 | 2 | 2 | S | 502682 | Aurantiacibacter gangjinensis |
| 0 | 1 | 0 | F | 2820280 | Sphingosinicellaceae |
| 0 | 1 | 0 | G | 335405 | Sphingosinicella |
| 0 | 1 | 0 | G1 | 2644549 | unclassified Sphingosinicella |
| 0 | 1 | 1 | S | 1892855 | Sphingosinicella sp. BN140058 |

| | | | | | |
|---|---|---|---|---|---|
| 2.25 | 4051 | 75 | O | 356 | Hyphomicrobiales |
| 1.87 | 3371 | 419 | F | 119045 | Methylobacteriaceae |
| 1.25 | 2247 | 176 | G | 2282523 | Methylorubrum |
| 0.85 | 1523 | 1433 | S | 223967 | Methylorubrum populi |
| 0.05 | 90 | 90 | S1 | 441620 | Methylorubrum populi BJ001 |
| 0.3 | 535 | 55 | S | 408 | Methylorubrum extorquens |
| 0.21 | 386 | 386 | S1 | 272630 | Methylorubrum extorquens AM1 |
| 0.05 | 83 | 83 | S1 | 440085 | Methylorubrum extorquens CM4 |
| 0.01 | 11 | 11 | S1 | 419610 | Methylorubrum extorquens PA1 |
| 0.01 | 13 | 13 | S | 29429 | Methylorubrum zatmanii |
| 0.39 | 697 | 107 | G | 407 | Methylobacterium |
| 0.27 | 478 | 232 | G1 | 2615210 | unclassified Methylobacterium |
| 0.06 | 111 | 111 | S | 2067957 | Methylobacterium sp. DM1 |
| 0.04 | 67 | 67 | S | 925818 | Methylobacterium sp. AMS5 |
| 0.02 | 28 | 28 | S | 2202828 | Methylobacterium sp. 17Sr1-43 |
| 0.01 | 23 | 23 | S | 739141 | Methylobacterium sp. XJLW |
| 0 | 9 | 9 | S | 426117 | Methylobacterium sp. 4-46 |
| 0 | 7 | 7 | S | 2603276 | Methylobacterium sp. WL1 |
| 0 | 1 | 1 | S | 2202826 | Methylobacterium sp. 17Sr1-1 |
| 0.02 | 39 | 39 | S | 2202825 | Methylobacterium durans |
| 0.01 | 25 | 25 | S | 269660 | Methylobacterium brachiatum |
| 0.01 | 18 | 0 | S | 39956 | Methylobacterium mesophilicum |
| 0.01 | 18 | 18 | S1 | 908290 | Methylobacterium mesophilicum SR1.6/6 |
| 0.01 | 12 | 12 | S | 270351 | Methylobacterium aquaticum |
| 0 | 7 | 0 | S | 31998 | Methylobacterium radiotolerans |
| 0 | 7 | 7 | S1 | 426355 | Methylobacterium radiotolerans JCM 2831 |
| 0 | 5 | 5 | S | 418223 | Methylobacterium phyllosphaerae |
| 0 | 3 | 3 | S | 2051553 | Methylobacterium currus |
| 0 | 2 | 0 | S | 114616 | Methylobacterium nodulans |
| 0 | 2 | 2 | S1 | 460265 | Methylobacterium nodulans ORS 2060 |
| 0 | 1 | 0 | S | 334852 | Methylobacterium oryzae |
| 0 | 1 | 1 | S1 | 693986 | Methylobacterium oryzae CBMB20 |
| 0 | 8 | 0 | G | 186650 | Microvirga |

| | | | | | |
|---|---|---|---|---|---|
| 0 | 5 | 5 | S | 1882682 | Microvirga ossetica |
| 0 | 3 | 0 | G1 | 2617746 | unclassified Microvirga |
| 0 | 3 | 3 | S | 2807101 | Microvirga sp. VF16 |
| 0.13 | 226 | 3 | F | 41294 | Bradyrhizobiaceae |
| 0.06 | 117 | 4 | G | 85413 | Bosea |
| 0.06 | 106 | 13 | G1 | 2653178 | unclassified Bosea |
| 0.02 | 45 | 45 | S | 1842539 | Bosea sp. RAC05 |
| 0.01 | 13 | 13 | S | 1867715 | Bosea sp. Tri-49 |
| 0.01 | 10 | 10 | S | 2020412 | Bosea sp. ANAM02 |
| 0 | 9 | 9 | S | 2015316 | Bosea sp. AS-1 |
| 0 | 8 | 8 | S | 1792307 | Bosea sp. PAMC 26642 |
| 0 | 8 | 8 | S | 2599640 | Bosea sp. F3-2 |
| 0 | 7 | 7 | S | 1526658 | Bosea vaviloviae |
| 0.05 | 94 | 39 | G | 374 | Bradyrhizobium |
| 0.02 | 33 | 9 | G1 | 2631580 | unclassified Bradyrhizobium |
| 0 | 9 | 9 | S | 2057741 | Bradyrhizobium sp. SK17 |
| 0 | 6 | 6 | S | 2715960 | Bradyrhizobium sp. PSBB068 |
| 0 | 3 | 3 | S | 1325102 | Bradyrhizobium sp. CCBAU 51765 |
| 0 | 3 | 3 | S | 2493093 | Bradyrhizobium sp. LCT2 |
| 0 | 2 | 2 | S | 1325114 | Bradyrhizobium sp. CCBAU 53351 |
| 0 | 1 | 1 | S | 858422 | Bradyrhizobium sp. CCBAU 051011 |
| 0 | 5 | 5 | S | 190148 | Bradyrhizobium paxllaeri |
| 0 | 5 | 5 | S | 1437360 | Bradyrhizobium erythrophlei |
| 0 | 3 | 0 | S | 44255 | Bradyrhizobium oligotrophicum |
| 0 | 3 | 3 | S1 | 1245469 | Bradyrhizobium oligotrophicum S58 |
| 0 | 3 | 3 | S | 244734 | Bradyrhizobium betae |
| 0 | 3 | 3 | S | 858423 | Bradyrhizobium arachidis |
| 0 | 2 | 2 | S | 83637 | Bradyrhizobium genosp. L |
| 0 | 1 | 1 | S | 1404864 | Bradyrhizobium cosmicum |
| 0 | 6 | 0 | G | 1073 | Rhodopseudomonas |
| 0 | 6 | 0 | S | 1076 | Rhodopseudomonas palustris |
| 0 | 6 | 6 | S1 | 316056 | Rhodopseudomonas palustris BisB18 |
| 0 | 3 | 0 | G | 1033 | Afipia |

| | | | | | |
|---|---|---|---|---|---|
| 0 | 3 | 3 | S | 40137 | Afipia carboxidovorans |
| 0 | 3 | 0 | G | 1395974 | Tardiphaga |
| 0 | 2 | 2 | S | 943830 | Tardiphaga robiniae |
| 0 | 1 | 1 | G1 | 2631404 | unclassified Tardiphaga |
| 0.12 | 223 | 23 | F | 82115 | Rhizobiaceae |
| 0.1 | 172 | 23 | F1 | 227290 | Rhizobium/Agrobacterium group |
| 0.04 | 80 | 9 | G | 357 | Agrobacterium |
| 0.03 | 63 | 5 | G1 | 1183400 | Agrobacterium tumefaciens complex |
| 0.03 | 58 | 55 | S | 358 | Agrobacterium tumefaciens |
| 0 | 3 | 3 | S1 | 311403 | Agrobacterium radiobacter K84 |
| 0 | 5 | 5 | S | 359 | Agrobacterium rhizogenes |
| 0 | 3 | 3 | S | 160699 | Agrobacterium larrymoorei |
| 0.04 | 68 | 30 | G | 379 | Rhizobium |
| 0.01 | 16 | 10 | G1 | 2613769 | unclassified Rhizobium |
| 0 | 3 | 3 | S | 1869170 | Rhizobium sp. S41 |
| 0 | 2 | 2 | S | 1571470 | Rhizobium sp. ACO-34A |
| 0 | 1 | 1 | S | 1981173 | Rhizobium sp. NXC14 |
| 0 | 7 | 7 | S | 648995 | Rhizobium pusense |
| 0 | 6 | 6 | S | 2267833 | Rhizobium oryzihabitans |
| 0 | 4 | 4 | S | 384 | Rhizobium leguminosarum |
| 0 | 2 | 2 | S | 396 | Rhizobium phaseoli |
| 0 | 2 | 2 | S | 1138190 | Rhizobium binae |
| 0 | 1 | 1 | S | 56730 | Rhizobium gallicum |
| 0 | 1 | 0 | G | 1525371 | Neorhizobium |
| 0 | 1 | 0 | S | 399 | Neorhizobium galegae |
| 0 | 1 | 0 | S1 | 323655 | Neorhizobium galegae bv. orientalis |
| 0 | 1 | 1 | S2 | 1028800 | Neorhizobium galegae bv. orientalis str. HAMBI 540 |
| 0.01 | 16 | 0 | G | 323620 | Shinella |
| 0.01 | 16 | 0 | G1 | 2643062 | unclassified Shinella |
| 0.01 | 12 | 12 | S | 2715959 | Shinella sp. PSBB067 |
| 0 | 4 | 4 | S | 879274 | Shinella sp. HZN7 |
| 0.01 | 12 | 3 | F1 | 227292 | Sinorhizobium/Ensifer group |
| 0 | 8 | 0 | G | 28105 | Sinorhizobium |

| | | | | | |
|---|---|---|---|---|---|
| 0 | 3 | 3 | S | 194963 | Sinorhizobium americanum |
| 0 | 2 | 2 | S | 382 | Sinorhizobium meliloti |
| 0 | 2 | 0 | G1 | 2613772 | unclassified Sinorhizobium |
| 0 | 2 | 2 | S | 1449788 | Sinorhizobium sp. LM21 |
| 0 | 1 | 0 | S | 110321 | Sinorhizobium medicae |
| 0 | 1 | 1 | S1 | 366394 | Sinorhizobium medicae WSM419 |
| 0 | 1 | 1 | G | 106591 | Ensifer |
| 0.05 | 88 | 0 | F | 69277 | Phyllobacteriaceae |
| 0.03 | 48 | 0 | G | 31988 | Aminobacter |
| 0.03 | 48 | 0 | G1 | 2644704 | unclassified Aminobacter |
| 0.03 | 46 | 46 | S | 2774562 | Aminobacter sp. SR38 |
| 0 | 2 | 2 | S | 374606 | Aminobacter sp. MSH1 |
| 0.02 | 39 | 7 | G | 68287 | Mesorhizobium |
| 0.01 | 16 | 5 | G1 | 325217 | unclassified Mesorhizobium |
| 0 | 6 | 6 | S | 2744521 | Mesorhizobium sp. L-2-11 |
| 0 | 3 | 3 | S | 2744518 | Mesorhizobium sp. 131-2-1 |
| 0 | 1 | 1 | S | 1854057 | Mesorhizobium sp. AA22 |
| 0 | 1 | 1 | S | 2493673 | Mesorhizobium sp. M1B.F.Ca.ET.045.04.1.1 |
| 0 | 5 | 0 | S | 39645 | Mesorhizobium ciceri |
| 0 | 5 | 5 | S1 | 682633 | Mesorhizobium ciceri ca181 |
| 0 | 4 | 4 | S | 1670800 | Mesorhizobium oceanicum |
| 0 | 3 | 0 | S | 381 | Mesorhizobium loti |
| 0 | 3 | 3 | S1 | 935548 | Mesorhizobium loti R88b |
| 0 | 2 | 2 | S | 28104 | Mesorhizobium huakuii |
| 0 | 2 | 0 | S | 593909 | Mesorhizobium opportunistum |
| 0 | 2 | 2 | S1 | 536019 | Mesorhizobium opportunistum WSM2075 |
| 0 | 1 | 0 | G | 449972 | Chelativorans |
| 0 | 1 | 0 | G1 | 2643807 | unclassified Chelativorans |
| 0 | 1 | 1 | S | 266779 | Chelativorans sp. BNC1 |
| 0.01 | 23 | 0 | F | 118882 | Brucellaceae |
| 0.01 | 23 | 9 | F1 | 2826938 | Brucella/Ochrobactrum group |
| 0 | 8 | 0 | G | 234 | Brucella |
| 0 | 7 | 7 | S | 271865 | [Ochrobactrum] quorumnocens |

| | | | | | |
|---|---|---|---|---|---|
| 0 | 1 | 1 | S | 529 | Brucella anthropi |
| 0 | 6 | 0 | G | 528 | Ochrobactrum |
| 0 | 6 | 0 | G1 | 239106 | unclassified Ochrobactrum |
| 0 | 5 | 5 | S | 1449781 | Ochrobactrum sp. LM19 |
| 0 | 1 | 1 | S | 2726427 | Ochrobactrum sp. MT180101 |
| 0.01 | 14 | 0 | F | 255475 | Aurantimonadaceae |
| 0.01 | 12 | 2 | G | 293088 | Martelella |
| 0.01 | 10 | 0 | S | 293089 | Martelella mediterranea |
| 0.01 | 10 | 10 | S1 | 1122214 | Martelella mediterranea DSM 17316 |
| 0 | 2 | 0 | G | 414371 | Aureimonas |
| 0 | 1 | 1 | S | 1701758 | Aureimonas populi |
| 0 | 1 | 0 | G1 | 2615206 | unclassified Aureimonas |
| 0 | 1 | 1 | S | 2816454 | Aureimonas sp. OT7 |
| 0.01 | 10 | 5 | F | 335928 | Xanthobacteraceae |
| 0 | 2 | 0 | G | 6 | Azorhizobium |
| 0 | 2 | 0 | S | 7 | Azorhizobium caulinodans |
| 0 | 2 | 2 | S1 | 438753 | Azorhizobium caulinodans ORS 571 |
| 0 | 2 | 0 | G | 556257 | Pseudolabrys |
| 0 | 2 | 2 | S | 331696 | Pseudolabrys taiwanensis |
| 0 | 1 | 0 | G | 99 | Ancylobacter |
| 0 | 1 | 0 | G1 | 2626613 | unclassified Ancylobacter |
| 0 | 1 | 1 | S | 1850374 | Ancylobacter sp. TS-1 |
| 0.01 | 10 | 0 | F | 2821832 | Stappiaceae |
| 0 | 7 | 0 | G | 227873 | Pannonibacter |
| 0 | 7 | 2 | S | 121719 | Pannonibacter phragmitetus |
| 0 | 5 | 5 | S1 | 1402210 | Pannonibacter phragmitetus BB |
| 0 | 3 | 0 | G | 478070 | Labrenzia |
| 0 | 3 | 3 | G1 | 2648686 | unclassified Labrenzia |
| 0 | 4 | 0 | F | 45401 | Hyphomicrobiaceae |
| 0 | 3 | 0 | G | 46913 | Devosia |
| 0 | 3 | 1 | G1 | 196773 | unclassified Devosia |
| 0 | 2 | 2 | S | 2499144 | Devosia sp. 1566 |
| 0 | 1 | 0 | G | 45402 | Aquabacter |

| | | | | | |
|---|---|---|---|---|---|
| 0 | 1 | 0 | G1 | 2663263 | unclassified Aquabacter |
| 0 | 1 | 1 | S | 2820278 | Aquabacter sp. L1I39 |
| 0 | 4 | 0 | O1 | 119042 | Rhizobiales incertae sedis |
| 0 | 4 | 0 | G | 1734920 | Pseudorhodoplanes |
| 0 | 4 | 4 | S | 1235591 | Pseudorhodoplanes sinuspersici |
| 0 | 2 | 0 | F | 45404 | Beijerinckiaceae |
| 0 | 2 | 0 | G | 120652 | Methylocella |
| 0 | 2 | 0 | S | 199596 | Methylocella silvestris |
| 0 | 2 | 2 | S1 | 395965 | Methylocella silvestris BL2 |
| 0 | 1 | 0 | F | 655351 | Cohaesibacteraceae |
| 0 | 1 | 0 | G | 1406135 | Breoghania |
| 0 | 1 | 0 | G1 | 2624130 | unclassified Breoghania |
| 0 | 1 | 1 | S | 2304600 | Breoghania sp. L-A4 |
| 0.24 | 424 | 0 | O | 204458 | Caulobacterales |
| 0.24 | 424 | 18 | F | 76892 | Caulobacteraceae |
| 0.21 | 377 | 112 | G | 41275 | Brevundimonas |
| 0.09 | 165 | 38 | G1 | 2622653 | unclassified Brevundimonas |
| 0.01 | 21 | 21 | S | 2774190 | Brevundimonas sp. LVF2 |
| 0.01 | 21 | 21 | S | 2813778 | Brevundimonas sp. CS1 |
| 0.01 | 20 | 20 | S | 2591463 | Brevundimonas sp. M20 |
| 0.01 | 16 | 16 | S | 2579977 | Brevundimonas sp. SGAir0440 |
| 0.01 | 15 | 15 | S | 2752515 | Brevundimonas sp. AJA228-03 |
| 0.01 | 12 | 12 | S | 2774189 | Brevundimonas sp. LVF1 |
| 0 | 7 | 7 | S | 2800818 | Brevundimonas sp. GRTSA-9 |
| 0 | 6 | 6 | S | 1938605 | Brevundimonas sp. LM2 |
| 0 | 4 | 4 | S | 2561924 | Brevundimonas sp. MF30-B |
| 0 | 4 | 4 | S | 2562582 | Brevundimonas sp. 'scallop' |
| 0 | 1 | 1 | S | 1827469 | Brevundimonas sp. GW460-12-10-14-LB2 |
| 0.02 | 44 | 44 | S | 588932 | Brevundimonas naejangsanensis |
| 0.02 | 32 | 32 | S | 293 | Brevundimonas diminuta |
| 0.01 | 11 | 0 | S | 74313 | Brevundimonas subvibrioides |
| 0.01 | 11 | 11 | S1 | 633149 | Brevundimonas subvibrioides ATCC 15264 |
| 0.01 | 11 | 11 | S | 74329 | Brevundimonas mediterranea |

| | | | | | |
|---|---|---|---|---|---|
| 0 | 1 | 1 | S | 41276 | Brevundimonas vesicularis |
| 0 | 1 | 1 | S | 1325724 | Brevundimonas vancanneytii |
| 0.01 | 25 | 9 | G | 75 | Caulobacter |
| 0 | 8 | 8 | S | 155892 | Caulobacter vibrioides |
| 0 | 4 | 0 | G1 | 2648921 | unclassified Caulobacter |
| 0 | 4 | 4 | S | 69665 | Caulobacter sp. FWC26 |
| 0 | 3 | 3 | S | 2708539 | Caulobacter soli |
| 0 | 1 | 1 | S | 1679497 | Caulobacter flavus |
| 0 | 4 | 0 | G | 76890 | Asticcacaulis |
| 0 | 4 | 1 | S | 78587 | Asticcacaulis excentricus |
| 0 | 3 | 3 | S1 | 573065 | Asticcacaulis excentricus CB 48 |
| 0.11 | 206 | 0 | O | 204455 | Rhodobacterales |
| 0.11 | 206 | 13 | F | 31989 | Rhodobacteraceae |
| 0.07 | 125 | 14 | G | 265 | Paracoccus |
| 0.03 | 54 | 0 | G1 | 246570 | unclassified Paracoccus |
| 0.03 | 49 | 49 | S | 2500532 | Paracoccus sp. Arc7-R13 |
| 0 | 3 | 3 | S | 2589076 | Paracoccus sp. AK26 |
| 0 | 2 | 2 | S | 2812658 | Paracoccus sp. H4-D09 |
| 0.02 | 34 | 34 | S | 147645 | Paracoccus yeei |
| 0 | 9 | 9 | S | 266 | Paracoccus denitrificans |
| 0 | 5 | 0 | S | 34003 | Paracoccus aminophilus |
| 0 | 5 | 5 | S1 | 1367847 | Paracoccus aminophilus JCM 7686 |
| 0 | 3 | 3 | S | 453842 | Paracoccus aestuarii |
| 0 | 3 | 3 | S | 2560053 | Paracoccus liaowanqingii |
| 0 | 1 | 1 | S | 82367 | Paracoccus pantotrophus |
| 0 | 1 | 1 | S | 1545044 | Paracoccus sanguinis |
| 0 | 1 | 1 | S | 1945662 | Paracoccus contaminans |
| 0.01 | 18 | 0 | G | 366614 | Haematobacter |
| 0.01 | 18 | 18 | S | 195105 | Haematobacter massiliensis |
| 0.01 | 12 | 0 | G | 1097466 | Defluviimonas |
| 0.01 | 12 | 12 | S | 1335048 | Defluviimonas alba |
| 0 | 9 | 0 | G | 92944 | Ketogulonicigenium |
| 0 | 9 | 9 | S | 92945 | Ketogulonicigenium vulgare |

| | | | | | |
|---|---|---|---|---|---|
| 0 | 5 | 0 | G | 2816884 | Qingshengfaniella |
| 0 | 5 | 5 | S | 2599296 | Qingshengfaniella alkalisoli |
| 0 | 5 | 0 | G | 1060 | Rhodobacter |
| 0 | 5 | 5 | S | 1075 | Rhodobacter blasticus |
| 0 | 5 | 5 | G | 302485 | Phaeobacter |
| 0 | 3 | 0 | G | 991903 | Polymorphum |
| 0 | 3 | 0 | S | 991904 | Polymorphum gilvum |
| 0 | 3 | 3 | S1 | 991905 | Polymorphum gilvum SL003B-26A1 |
| 0 | 2 | 0 | G | 2613960 | Pseudopuniceibacterium |
| 0 | 2 | 2 | S | 2613965 | Pseudopuniceibacterium antarcticum |
| 0 | 2 | 0 | G | 1855413 | Brevirhabdus |
| 0 | 2 | 2 | S | 1267768 | Brevirhabdus pacifica |
| 0 | 2 | 0 | G | 875170 | Celeribacter |
| 0 | 2 | 2 | S | 1208324 | Celeribacter indicus |
| 0 | 1 | 0 | G | 74030 | Roseovarius |
| 0 | 1 | 0 | G1 | 2614913 | unclassified Roseovarius |
| 0 | 1 | 0 | S | 391613 | Roseovarius sp. TM1035 |
| 0 | 1 | 1 | S1 | 2203213 | Roseovarius sp. AK1035 |
| 0 | 1 | 0 | G | 74032 | Antarctobacter |
| 0 | 1 | 1 | S | 74033 | Antarctobacter heliothermus |
| 0 | 1 | 0 | G | 1653176 | Cereibacter |
| 0 | 1 | 1 | S | 1063 | Cereibacter sphaeroides |
| 0 | 1 | 0 | G | 159345 | Roseibacterium |
| 0 | 1 | 0 | S | 159346 | Roseibacterium elongatum |
| 0 | 1 | 1 | S1 | 1294273 | Roseibacterium elongatum DSM 19469 |
| 0 | 1 | 0 | G | 335927 | Thalassobius |
| 0 | 1 | 1 | S | 53501 | Thalassobius gelatinovorus |
| 0.04 | 75 | 2 | O | 204441 | Rhodospirillales |
| 0.03 | 57 | 0 | F | 41295 | Rhodospirillaceae |
| 0.02 | 31 | 0 | G | 1543704 | Niveispirillum |
| 0.02 | 31 | 31 | S | 1612173 | Niveispirillum cyanobacteriorum |
| 0 | 8 | 0 | G | 1612157 | Pararhodospirillum |
| 0 | 8 | 0 | S | 1084 | Pararhodospirillum photometricum |

| | | | | | |
|---|---|---|---|---|---|
| 0 | 8 | 8 | S1 | 1150469 | Pararhodospirillum photometricum DSM 122 |
| 0 | 7 | 0 | G | 2705399 | Hypericibacter |
| 0 | 7 | 7 | S | 2602015 | Hypericibacter terrae |
| 0 | 6 | 1 | G | 191 | Azospirillum |
| 0 | 4 | 4 | S | 192 | Azospirillum brasilense |
| 0 | 1 | 0 | G1 | 2630922 | unclassified Azospirillum |
| 0 | 1 | 1 | S | 652764 | Azospirillum sp. TSH100 |
| 0 | 4 | 0 | G | 2478349 | Indioceanicola |
| 0 | 4 | 4 | S | 2220096 | Indioceanicola profundi |
| 0 | 1 | 0 | G | 1543705 | Nitrospirillum |
| 0 | 1 | 0 | S | 28077 | Nitrospirillum amazonense |
| 0 | 1 | 1 | S1 | 1441467 | Nitrospirillum amazonense CBAmc |
| 0.01 | 16 | 3 | F | 433 | Acetobacteraceae |
| 0 | 4 | 0 | G | 2603324 | Acidibrevibacterium |
| 0 | 4 | 4 | S | 1969806 | Acidibrevibacterium fodinaquatile |
| 0 | 3 | 0 | G | 1434011 | Komagataeibacter |
| 0 | 2 | 2 | S | 28448 | Komagataeibacter xylinus |
| 0 | 1 | 1 | S | 265960 | Komagataeibacter nataicola |
| 0 | 2 | 0 | G | 125216 | Roseomonas |
| 0 | 1 | 1 | S | 207340 | Roseomonas mucosa |
| 0 | 1 | 0 | G1 | 2617492 | unclassified Roseomonas |
| 0 | 1 | 1 | S | 2768161 | Roseomonas sp. 1318 |
| 0 | 2 | 0 | G | 2804525 | Lichenicola |
| 0 | 2 | 2 | S | 1484109 | Lichenicola cladoniae |
| 0 | 1 | 1 | G | 441 | Gluconobacter |
| 0 | 1 | 0 | G | 35812 | Roseococcus |
| 0 | 1 | 0 | G1 | 2639982 | unclassified Roseococcus |
| 0 | 1 | 1 | S | 2771361 | Roseococcus sp. NIBR12 |
| 0 | 9 | 0 | C1 | 82117 | Alphaproteobacteria incertae sedis |
| 0 | 9 | 0 | G | 1632780 | Phreatobacter |
| 0 | 9 | 9 | S | 1940610 | Phreatobacter stygius |
| 0 | 2 | 0 | O | 766 | Rickettsiales |
| 0 | 1 | 0 | F | 942 | Anaplasmataceae |

| | | | | | |
|---|---|---|---|---|---|
| 0 | 1 | 0 | F1 | 952 | Wolbachieae |
| 0 | 1 | 0 | G | 953 | Wolbachia |
| 0 | 1 | 0 | G1 | 2640676 | unclassified Wolbachia |
| 0 | 1 | 1 | S | 169402 | Wolbachia endosymbiont of Folsomia candida |
| 0 | 1 | 0 | F | 1328881 | Candidatus Midichloriaceae |
| 0 | 1 | 0 | G | 1802983 | Candidatus Fokinia |
| 0 | 1 | 1 | S | 1802984 | Candidatus Fokinia solitaria |
| 0 | 2 | 0 | O | 2800059 | Maricaulales |
| 0 | 2 | 0 | F | 2800061 | Maricaulaceae |
| 0 | 2 | 0 | G | 1649466 | Marinicauda |
| 0 | 2 | 2 | S | 2029849 | Marinicauda algicola |
| 8 | 14411 | 119 | C | 1236 | Gammaproteobacteria |
| 6.02 | 10837 | 12 | O | 135614 | Xanthomonadales |
| 6.01 | 10820 | 247 | F | 32033 | Xanthomonadaceae |
| 5.83 | 10504 | 1358 | G | 40323 | Stenotrophomonas |
| 5 | 9008 | 162 | G1 | 995085 | Stenotrophomonas maltophilia group |
| 4.9 | 8832 | 7952 | S | 40324 | Stenotrophomonas maltophilia |
| 0.44 | 784 | 784 | S1 | 391008 | Stenotrophomonas maltophilia R551-3 |
| 0.05 | 86 | 86 | S1 | 522373 | Stenotrophomonas maltophilia K279a |
| 0 | 8 | 8 | S1 | 868597 | Stenotrophomonas maltophilia JV3 |
| 0 | 2 | 2 | S1 | 1163399 | Stenotrophomonas maltophilia D457 |
| 0 | 8 | 8 | S | 2072413 | Stenotrophomonas sp. SAU14A_NAIMI4_5 |
| 0 | 3 | 3 | S | 2072408 | Stenotrophomonas sp. YAU14A_MKIMI4_1 |
| 0 | 1 | 1 | S | 2072407 | Stenotrophomonas sp. YAU14D1_LEIMI4_1 |
| 0 | 1 | 1 | S | 2072412 | Stenotrophomonas sp. ZAC14A_NAIMI4_1 |
| 0 | 1 | 1 | S | 2072414 | Stenotrophomonas sp. ESTM1D_MKCIP4_1 |
| 0.05 | 85 | 4 | G1 | 196198 | unclassified Stenotrophomonas |
| 0.01 | 24 | 24 | S | 1904944 | Stenotrophomonas sp. LM091 |
| 0.01 | 18 | 18 | S | 2005046 | Stenotrophomonas sp. WZN-1 |
| 0.01 | 15 | 15 | S | 2682487 | Stenotrophomonas sp. SXG-1 |
| 0 | 9 | 9 | S | 2282124 | Stenotrophomonas sp. ASS1 |
| 0 | 7 | 7 | S | 2691571 | Stenotrophomonas sp. 364 |
| 0 | 6 | 6 | S | 2742129 | Stenotrophomonas sp. NA06056 |

| | | | | | |
|---|---|---|---|---|---|
| 0 | 1 | 1 | S | 1827305 | Stenotrophomonas sp. MYb57 |
| 0 | 1 | 1 | S | 2303750 | Stenotrophomonas sp. G4 |
| 0.02 | 37 | 37 | S | 216778 | Stenotrophomonas rhizophila |
| 0.01 | 16 | 16 | S | 128780 | Stenotrophomonas acidaminiphila |
| 0.02 | 28 | 10 | G | 338 | Xanthomonas |
| 0 | 9 | 0 | G1 | 643453 | Xanthomonas citri group |
| 0 | 9 | 6 | S | 346 | Xanthomonas citri |
| 0 | 2 | 2 | S1 | 473421 | Xanthomonas citri pv. glycines |
| 0 | 1 | 1 | S1 | 76802 | Xanthomonas citri pv. aurantifolii |
| 0 | 6 | 6 | S | 442694 | Xanthomonas perforans |
| 0 | 3 | 3 | S | 347 | Xanthomonas oryzae |
| 0.01 | 23 | 0 | G | 83618 | Pseudoxanthomonas |
| 0.01 | 23 | 23 | S | 128785 | Pseudoxanthomonas mexicana |
| 0.01 | 11 | 5 | G | 68 | Lysobacter |
| 0 | 2 | 2 | S | 69 | Lysobacter enzymogenes |
| 0 | 2 | 0 | G1 | 2635362 | unclassified Lysobacter |
| 0 | 2 | 2 | S | 2290922 | Lysobacter sp. TY2-98 |
| 0 | 1 | 1 | S | 2591633 | Lysobacter alkalisoli |
| 0 | 1 | 1 | S | 2763317 | Lysobacter solisilvae |
| 0 | 4 | 0 | G | 141948 | Thermomonas |
| 0 | 2 | 2 | S | 1463158 | Thermomonas carbonis |
| 0 | 2 | 0 | G1 | 2633315 | unclassified Thermomonas |
| 0 | 2 | 2 | S | 2771436 | Thermomonas sp. XSG |
| 0 | 3 | 0 | G | 83614 | Luteimonas |
| 0 | 3 | 0 | G1 | 2629088 | unclassified Luteimonas |
| 0 | 3 | 3 | S | 1896164 | Luteimonas sp. JM171 |
| 0 | 5 | 0 | F | 1775411 | Rhodanobacteraceae |
| 0 | 3 | 0 | G | 242605 | Luteibacter |
| 0 | 2 | 0 | S | 242606 | Luteibacter rhizovicinus |
| 0 | 2 | 2 | S1 | 1440763 | Luteibacter rhizovicinus DSM 16549 |
| 0 | 1 | 1 | S | 2589080 | Luteibacter pinisoli |
| 0 | 2 | 0 | G | 231454 | Dyella |
| 0 | 1 | 1 | S | 445710 | Dyella thiooxydans |

| | | | | | |
|---|---|---|---|---|---|
| 0 | 1 | 1 | S | 1849581 | Dyella caseinilytica |
| 1.62 | 2919 | 4 | O | 72274 | Pseudomonadales |
| 1.09 | 1962 | 12 | F | 468 | Moraxellaceae |
| 1.04 | 1866 | 665 | G | 469 | Acinetobacter |
| 0.43 | 779 | 766 | S | 40214 | Acinetobacter johnsonii |
| 0.01 | 13 | 13 | S1 | 1242245 | Acinetobacter johnsonii XBB1 |
| 0.05 | 87 | 87 | S | 40215 | Acinetobacter junii |
| 0.04 | 76 | 2 | G1 | 196816 | unclassified Acinetobacter |
| 0.01 | 20 | 20 | S | 2743575 | Acinetobacter sp. NEB 394 |
| 0.01 | 17 | 17 | S | 2004646 | Acinetobacter sp. WCHA55 |
| 0.01 | 17 | 17 | S | 2004644 | Acinetobacter sp. WCHA45 |
| 0 | 8 | 8 | S | 2601122 | Acinetobacter sp. YH12138 |
| 0 | 5 | 5 | S | 1636603 | Acinetobacter sp. ACNIH1 |
| 0 | 2 | 2 | S | 2545797 | Acinetobacter sp. FDAARGOS_724 |
| 0 | 2 | 2 | S | 1646498 | Acinetobacter sp. TTH0-4 |
| 0 | 1 | 1 | S | 1280052 | Acinetobacter sp. M131 |
| 0 | 1 | 1 | S | 1879049 | Acinetobacter sp. WCHAc010034 |
| 0 | 1 | 1 | S | 1758189 | Acinetobacter sp. ACNIH2 |
| 0.03 | 62 | 15 | G1 | 909768 | Acinetobacter calcoaceticus/baumannii complex |
| 0.02 | 40 | 40 | S | 470 | Acinetobacter baumannii |
| 0 | 4 | 4 | S | 1530123 | Acinetobacter seifertii |
| 0 | 3 | 1 | S | 48296 | Acinetobacter pittii |
| 0 | 2 | 2 | S1 | 871585 | Acinetobacter pittii PHEA-2 |
| 0.02 | 30 | 30 | S | 202956 | Acinetobacter towneri |
| 0.01 | 23 | 23 | S | 756892 | Acinetobacter indicus |
| 0.01 | 21 | 19 | S | 28090 | Acinetobacter lwoffii |
| 0 | 2 | 2 | S1 | 1046625 | Acinetobacter lwoffii WJ10621 |
| 0.01 | 21 | 21 | S | 29430 | Acinetobacter haemolyticus |
| 0.01 | 19 | 19 | S | 108980 | Acinetobacter ursingii |
| 0.01 | 16 | 16 | S | 108981 | Acinetobacter schindleri |
| 0.01 | 16 | 16 | S | 134534 | Acinetobacter gyllenbergii |
| 0.01 | 14 | 14 | S | 106648 | Acinetobacter bereziniae |
| 0.01 | 13 | 13 | S | 1879050 | Acinetobacter wuhouensis |

| | | | | | |
|---|---|---|---|---|---|
| 0 | 9 | 0 | S | 52133 | Acinetobacter venetianus |
| 0 | 9 | 9 | S1 | 1197884 | Acinetobacter venetianus VE-C3 |
| 0 | 6 | 6 | S | 2715164 | Acinetobacter shaoyimingii |
| 0 | 5 | 5 | S | 40216 | Acinetobacter radioresistens |
| 0 | 2 | 2 | S | 70348 | Acinetobacter dispersus |
| 0 | 1 | 1 | S | 487316 | Acinetobacter soli |
| 0 | 1 | 1 | S | 2006115 | Acinetobacter piscicola |
| 0.04 | 69 | 0 | G | 475 | Moraxella |
| 0.04 | 69 | 69 | S | 34062 | Moraxella osloensis |
| 0.01 | 14 | 0 | G | 497 | Psychrobacter |
| 0 | 9 | 0 | G1 | 196806 | unclassified Psychrobacter |
| 0 | 7 | 7 | S | 2733866 | Psychrobacter sp. KCTC 72983 |
| 0 | 2 | 2 | S | 1699621 | Psychrobacter sp. P11F6 |
| 0 | 5 | 0 | S | 334543 | Psychrobacter arcticus |
| 0 | 5 | 5 | S1 | 259536 | Psychrobacter arcticus 273-4 |
| 0 | 1 | 0 | G | 2824158 | Aquirhabdus |
| 0 | 1 | 1 | S | 2283318 | Aquirhabdus parva |
| 0.53 | 953 | 0 | F | 135621 | Pseudomonadaceae |
| 0.53 | 953 | 219 | G | 286 | Pseudomonas |
| 0.22 | 402 | 0 | G1 | 136841 | Pseudomonas aeruginosa group |
| 0.18 | 320 | 320 | S | 287 | Pseudomonas aeruginosa |
| 0.02 | 40 | 28 | S | 300 | Pseudomonas mendocina |
| 0.01 | 11 | 11 | S1 | 1225174 | Pseudomonas mendocina S5.2 |
| 0 | 1 | 1 | S1 | 399739 | Pseudomonas mendocina ymp |
| 0.02 | 40 | 0 | G2 | 1232139 | Pseudomonas oleovorans/pseudoalcaligenes group |
| 0.02 | 40 | 1 | S | 301 | Pseudomonas oleovorans |
| 0.02 | 39 | 39 | S1 | 1182590 | Pseudomonas pseudoalcaligenes CECT 5344 |
| 0 | 2 | 0 | G2 | 627141 | Pseudomonas nitroreducens/multiresinivorans group |
| 0 | 2 | 2 | S | 46680 | Pseudomonas nitroreducens |
| 0.08 | 152 | 6 | G1 | 136843 | Pseudomonas fluorescens group |
| 0.07 | 121 | 120 | S | 294 | Pseudomonas fluorescens |
| 0 | 1 | 1 | S1 | 463794 | Pseudomonas fluorescens BBc6R8 |
| 0 | 9 | 9 | S | 78543 | Pseudomonas migulae |

| | | | | | |
|---|---|---|---|---|---|
| 0 | 9 | 9 | S | 380021 | Pseudomonas protegens |
| 0 | 7 | 7 | S | 29442 | Pseudomonas tolaasii |
| 0.03 | 61 | 21 | G1 | 196821 | unclassified Pseudomonas |
| 0 | 5 | 5 | S | 253237 | Pseudomonas sp. phDV1 |
| 0 | 4 | 4 | S | 2735906 | Pseudomonas sp. B11D7D |
| 0 | 3 | 3 | S | 2774873 | Pseudomonas sp. ADPe |
| 0 | 3 | 3 | S | 2762896 | Pseudomonas sp. MPDS |
| 0 | 3 | 3 | S | 2731681 | Pseudomonas sp. S1-A32-2 |
| 0 | 3 | 3 | S | 2614442 | Pseudomonas sp. LPB0260 |
| 0 | 3 | 3 | S | 2201356 | Pseudomonas sp. 31-12 |
| 0 | 3 | 3 | S | 1294143 | Pseudomonas sp. ATCC 13867 |
| 0 | 2 | 2 | S | 2812000 | Pseudomonas sp. SDM007 |
| 0 | 2 | 2 | S | 2811422 | Pseudomonas sp. PDNC002 |
| 0 | 1 | 1 | S | 2049589 | Pseudomonas sp. HLS-6 |
| 0 | 1 | 1 | S | 76885 | Pseudomonas sp. K-62 |
| 0 | 1 | 1 | S | 2725477 | Pseudomonas sp. TUM18999 |
| 0 | 1 | 1 | S | 2605424 | Pseudomonas sp. J380 |
| 0 | 1 | 1 | S | 2587845 | Pseudomonas sp. THAF7b |
| 0 | 1 | 1 | S | 2054919 | Pseudomonas sp. S09G 359 |
| 0 | 1 | 1 | S | 2025658 | Pseudomonas sp. NS1(2017) |
| 0 | 1 | 1 | S | 1931241 | Pseudomonas sp. S-6-2 |
| 0 | 1 | 1 | S | 1898684 | Pseudomonas sp. LPH1 |
| 0.02 | 35 | 0 | G1 | 136845 | Pseudomonas putida group |
| 0.02 | 33 | 25 | S | 303 | Pseudomonas putida |
| 0 | 6 | 6 | S1 | 1211579 | Pseudomonas putida NBRC 14164 |
| 0 | 2 | 2 | S1 | 1384061 | Pseudomonas putida S13.1.2 |
| 0 | 2 | 2 | S | 47885 | Pseudomonas oryzihabitans |
| 0.01 | 19 | 0 | G1 | 136846 | Pseudomonas stutzeri group |
| 0.01 | 11 | 0 | G2 | 578833 | Pseudomonas stutzeri subgroup |
| 0.01 | 11 | 9 | S | 316 | Pseudomonas stutzeri |
| 0 | 2 | 2 | S1 | 1123519 | Pseudomonas stutzeri DSM 10701 |
| 0 | 8 | 8 | S | 74829 | Pseudomonas balearica |
| 0.01 | 11 | 0 | S | 101564 | Pseudomonas alcaliphila |

| | | | | | |
|---|---|---|---|---|---|
| 0.01 | 11 | 11 | S1 | 741155 | Pseudomonas alcaliphila JAB1 |
| 0.01 | 10 | 10 | S | 359110 | Pseudomonas extremaustralis |
| 0 | 8 | 8 | S | 237610 | Pseudomonas psychrotolerans |
| 0 | 6 | 6 | S | 65741 | Pseudomonas knackmussii |
| 0 | 6 | 6 | S | 216142 | Pseudomonas rhizosphaerae |
| 0 | 4 | 4 | S | 104087 | Pseudomonas frederiksbergensis |
| 0 | 3 | 3 | S | 2502979 | Pseudomonas khazarica |
| 0 | 3 | 3 | S | 1615674 | Pseudomonas lactis |
| 0 | 3 | 3 | S | 1499686 | Pseudomonas saudiphocaensis |
| 0 | 3 | 3 | S | 237609 | Pseudomonas alkylphenolica |
| 0 | 3 | 3 | S | 157782 | Pseudomonas parafulva |
| 0 | 2 | 2 | S | 219572 | Pseudomonas antarctica |
| 0 | 1 | 1 | S | 1691904 | Pseudomonas sediminis |
| 0 | 1 | 1 | S | 395598 | Pseudomonas reinekei |
| 0 | 1 | 0 | G1 | 136844 | Pseudomonas pertucinogena group |
| 0 | 1 | 1 | S | 43306 | Pseudomonas denitrificans (nom. rej.) |
| 0.17 | 302 | 2 | O | 91347 | Enterobacterales |
| 0.09 | 168 | 51 | F | 543 | Enterobacteriaceae |
| 0.04 | 68 | 4 | G | 561 | Escherichia |
| 0.03 | 63 | 61 | S | 562 | Escherichia coli |
| 0 | 1 | 1 | S1 | 2778655 | Escherichia coli O167:H26 |
| 0 | 1 | 1 | S1 | 2763104 | Escherichia coli O150:H6 |
| 0 | 1 | 1 | S | 208962 | Escherichia albertii |
| 0.02 | 33 | 22 | G | 570 | Klebsiella |
| 0.01 | 10 | 10 | S | 548 | Klebsiella aerogenes |
| 0 | 1 | 1 | S | 571 | Klebsiella oxytoca |
| 0 | 6 | 0 | G | 83654 | Leclercia |
| 0 | 5 | 5 | S | 83655 | Leclercia adecarboxylata |
| 0 | 1 | 0 | G1 | 2627398 | unclassified Leclercia |
| 0 | 1 | 1 | S | 1920114 | Leclercia sp. LSNIH1 |
| 0 | 4 | 1 | G | 590 | Salmonella |
| 0 | 3 | 1 | S | 28901 | Salmonella enterica |
| 0 | 2 | 0 | S1 | 59201 | Salmonella enterica subsp. enterica |

| | | | | | |
|---|---|---|---|---|---|
| 0 | 1 | 1 | S2 | 189201 | Salmonella enterica subsp. enterica serovar Cubana |
| 0 | 1 | 0 | S2 | 598 | Salmonella enterica subsp. enterica serovar Rubislaw |
| 0 | 1 | 1 | S3 | 938143 | Salmonella enterica subsp. enterica serovar Rubislaw str. ATCC 10717 |
| 0 | 3 | 0 | G | 544 | Citrobacter |
| 0 | 3 | 0 | G1 | 1344959 | Citrobacter freundii complex |
| 0 | 3 | 3 | S | 546 | Citrobacter freundii |
| 0 | 2 | 1 | G | 547 | Enterobacter |
| 0 | 1 | 1 | G1 | 354276 | Enterobacter cloacae complex |
| 0 | 1 | 1 | G | 160674 | Raoultella |
| 0.07 | 124 | 0 | F | 1903411 | Yersiniaceae |
| 0.07 | 120 | 56 | G | 613 | Serratia |
| 0.03 | 53 | 50 | S | 614 | Serratia liquefaciens |
| 0 | 3 | 3 | S1 | 1346614 | Serratia liquefaciens ATCC 27592 |
| 0 | 4 | 4 | S | 615 | Serratia marcescens |
| 0 | 4 | 0 | G1 | 2647522 | unclassified Serratia |
| 0 | 4 | 4 | S | 488142 | Serratia sp. SCBI |
| 0 | 3 | 3 | S | 2741499 | Serratia surfactantfaciens |
| 0 | 4 | 0 | G | 629 | Yersinia |
| 0 | 4 | 4 | S | 2607663 | Yersinia canariae |
| 0 | 4 | 0 | F | 1903414 | Morganellaceae |
| 0 | 4 | 0 | G | 586 | Providencia |
| 0 | 4 | 4 | S | 333965 | Providencia vermicola |
| 0 | 2 | 0 | F | 1903410 | Pectobacteriaceae |
| 0 | 2 | 0 | G | 204037 | Dickeya |
| 0 | 2 | 2 | S | 204042 | Dickeya zeae |
| 0 | 1 | 0 | O1 | 451511 | Enterobacterales incertae sedis |
| 0 | 1 | 0 | G | 702 | Plesiomonas |
| 0 | 1 | 1 | S | 703 | Plesiomonas shigelloides |
| 0 | 1 | 0 | F | 1903409 | Erwiniaceae |
| 0 | 1 | 0 | G | 53335 | Pantoea |
| 0 | 1 | 1 | S | 66269 | Pantoea stewartii |
| 0.04 | 66 | 0 | O | 135624 | Aeromonadales |
| 0.04 | 66 | 0 | F | 84642 | Aeromonadaceae |

| | | | | | |
|---|---|---|---|---|---|
| 0.04 | 66 | 30 | G | 642 | Aeromonas |
| 0.01 | 13 | 13 | S | 644 | Aeromonas hydrophila |
| 0 | 8 | 8 | S | 651 | Aeromonas media |
| 0 | 7 | 7 | S | 648 | Aeromonas caviae |
| 0 | 4 | 4 | S | 218936 | Aeromonas simiae |
| 0 | 2 | 2 | S | 73010 | Aeromonas encheleia |
| 0 | 1 | 1 | S | 654 | Aeromonas veronii |
| 0 | 1 | 0 | G1 | 257493 | unclassified Aeromonas |
| 0 | 1 | 1 | S | 2675707 | Aeromonas sp. WP2-W18-CRE-05 |
| 0.03 | 59 | 0 | O | 135625 | Pasteurellales |
| 0.03 | 59 | 0 | F | 712 | Pasteurellaceae |
| 0.03 | 58 | 0 | G | 724 | Haemophilus |
| 0.03 | 48 | 41 | S | 729 | Haemophilus parainfluenzae |
| 0 | 7 | 7 | S1 | 862965 | Haemophilus parainfluenzae T3T1 |
| 0 | 7 | 7 | S | 726 | Haemophilus haemolyticus |
| 0 | 3 | 3 | S | 730 | [Haemophilus] ducreyi |
| 0 | 1 | 0 | G | 416916 | Aggregatibacter |
| 0 | 1 | 1 | S | 739 | Aggregatibacter segnis |
| 0.02 | 35 | 0 | O | 135619 | Oceanospirillales |
| 0.01 | 19 | 0 | F | 224372 | Alcanivoracaceae |
| 0.01 | 19 | 0 | G | 59753 | Alcanivorax |
| 0.01 | 10 | 10 | S | 1094342 | Alcanivorax xenomutans |
| 0 | 6 | 0 | S | 285091 | Alcanivorax dieselolei |
| 0 | 6 | 6 | S1 | 930169 | Alcanivorax dieselolei B5 |
| 0 | 3 | 0 | S | 1306787 | Alcanivorax pacificus |
| 0 | 3 | 3 | S1 | 391936 | Alcanivorax pacificus W11-5 |
| 0.01 | 16 | 5 | F | 28256 | Halomonadaceae |
| 0 | 9 | 0 | G | 2745 | Halomonas |
| 0 | 9 | 5 | G1 | 2609666 | unclassified Halomonas |
| 0 | 3 | 3 | S | 2733487 | Halomonas sp. MCCC 1A13316 |
| 0 | 1 | 1 | S | 2733488 | Halomonas sp. MCCC 1A13718 |
| 0 | 1 | 0 | G | 42054 | Chromohalobacter |
| 0 | 1 | 0 | S | 158080 | Chromohalobacter salexigens |

| | | | | | |
|---|---|---|---|---|---|
| 0 | 1 | 1 | S1 | 290398 | Chromohalobacter salexigens DSM 3043 |
| 0 | 1 | 0 | G | 204286 | Cobetia |
| 0 | 1 | 0 | G1 | 2609414 | unclassified Cobetia |
| 0 | 1 | 1 | S | 2686360 | Cobetia sp. L2A1 |
| 0.01 | 24 | 0 | O | 135613 | Chromatiales |
| 0.01 | 13 | 0 | F | 1046 | Chromatiaceae |
| 0.01 | 13 | 0 | G | 67575 | Rheinheimera |
| 0.01 | 13 | 0 | G1 | 115860 | unclassified Rheinheimera |
| 0.01 | 10 | 10 | S | 2498451 | Rheinheimera sp. LHK132 |
| 0 | 3 | 3 | S | 1763998 | Rheinheimera sp. F8 |
| 0 | 9 | 0 | F | 1096778 | Thioalkalispiraceae |
| 0 | 9 | 0 | G | 2034504 | Sulfurivermis |
| 0 | 9 | 9 | S | 1972068 | Sulfurivermis fontis |
| 0 | 2 | 0 | F | 72276 | Ectothiorhodospiraceae |
| 0 | 2 | 0 | G | 106633 | Thioalkalivibrio |
| 0 | 2 | 0 | S | 1033854 | Thioalkalivibrio sulfidiphilus |
| 0 | 2 | 2 | S1 | 396588 | Thioalkalivibrio sulfidiphilus HL-EbGr7 |
| 0.01 | 17 | 0 | O | 135622 | Alteromonadales |
| 0 | 9 | 0 | F | 267888 | Pseudoalteromonadaceae |
| 0 | 6 | 3 | G | 53246 | Pseudoalteromonas |
| 0 | 2 | 2 | S | 166935 | Pseudoalteromonas translucida |
| 0 | 1 | 1 | S | 874454 | Pseudoalteromonas arabiensis |
| 0 | 3 | 0 | G | 907197 | Psychrosphaera |
| 0 | 3 | 3 | S | 1266052 | Psychrosphaera aestuarii |
| 0 | 6 | 0 | F | 267890 | Shewanellaceae |
| 0 | 6 | 0 | G | 22 | Shewanella |
| 0 | 4 | 0 | G1 | 196818 | unclassified Shewanella |
| 0 | 4 | 4 | S | 94122 | Shewanella sp. ANA-3 |
| 0 | 2 | 0 | S | 271098 | Shewanella halifaxensis |
| 0 | 2 | 2 | S1 | 458817 | Shewanella halifaxensis HAW-EB4 |
| 0 | 2 | 0 | F | 72275 | Alteromonadaceae |
| 0 | 2 | 0 | G | 2742 | Marinobacter |
| 0 | 2 | 2 | S | 418719 | Marinobacter salsuginis |

| | | | | | |
|---|---|---|---|---|---|
| 0 | 9 | 0 | O | 72273 | Thiotrichales |
| 0 | 7 | 0 | F | 135616 | Piscirickettsiaceae |
| 0 | 7 | 0 | G | 28884 | Hydrogenovibrio |
| 0 | 7 | 7 | S | 28885 | Hydrogenovibrio marinus |
| 0 | 2 | 0 | F | 135617 | Thiotrichaceae |
| 0 | 2 | 0 | G | 1030 | Thiothrix |
| 0 | 2 | 0 | G1 | 2636184 | unclassified Thiothrix |
| 0 | 2 | 2 | S | 1032 | Thiothrix sp. |
| 0 | 9 | 0 | O | 135615 | Cardiobacteriales |
| 0 | 9 | 0 | F | 868 | Cardiobacteriaceae |
| 0 | 9 | 0 | G | 2717 | Cardiobacterium |
| 0 | 9 | 9 | S | 2718 | Cardiobacterium hominis |
| 0 | 6 | 0 | O | 1706369 | Cellvibrionales |
| 0 | 3 | 0 | F | 1706371 | Cellvibrionaceae |
| 0 | 3 | 0 | G | 10 | Cellvibrio |
| 0 | 3 | 0 | G1 | 2624793 | unclassified Cellvibrio |
| 0 | 3 | 3 | S | 454662 | Cellvibrio sp. KY-YJ-3 |
| 0 | 3 | 0 | F | 1706373 | Microbulbiferaceae |
| 0 | 3 | 0 | G | 48073 | Microbulbifer |
| 0 | 3 | 0 | G1 | 2619833 | unclassified Microbulbifer |
| 0 | 3 | 3 | S | 2745199 | Microbulbifer sp. YPW1 |
| 0 | 4 | 0 | O | 1934945 | Immundisolibacterales |
| 0 | 4 | 0 | F | 1934946 | Immundisolibacteraceae |
| 0 | 4 | 0 | G | 1934947 | Immundisolibacter |
| 0 | 4 | 4 | S | 1810504 | Immundisolibacter cernigliae |
| 0 | 2 | 0 | O | 135618 | Methylococcales |
| 0 | 2 | 0 | F | 403 | Methylococcaceae |
| 0 | 2 | 0 | G | 416 | Methylomonas |
| 0 | 2 | 0 | S | 421 | Methylomonas methanica |
| 0 | 2 | 2 | S1 | 857087 | Methylomonas methanica MC09 |
| 0 | 2 | 0 | O | 135623 | Vibrionales |
| 0 | 2 | 0 | F | 641 | Vibrionaceae |
| 0 | 2 | 0 | G | 662 | Vibrio |

| | | | | | |
|---|---|---|---|---|---|
| 0 | 1 | 1 | S | 666 | Vibrio cholerae |
| 0 | 1 | 0 | G1 | 717610 | Vibrio harveyi group |
| 0 | 1 | 1 | S | 663 | Vibrio alginolyticus |
| 0 | 1 | 0 | O | 1775403 | Nevskiales |
| 0 | 1 | 0 | F | 568386 | Sinobacteraceae |
| 0 | 1 | 0 | G | 1861863 | Sinimarinibacterium |
| 0 | 1 | 0 | G1 | 2621506 | unclassified Sinimarinibacterium |
| 0 | 1 | 1 | S | 2698684 | Sinimarinibacterium sp. NLF-5-8 |
| 0.02 | 28 | 0 | P1 | 68525 | delta/epsilon subdivisions |
| 0.01 | 15 | 0 | C | 28221 | Deltaproteobacteria |
| 0.01 | 14 | 0 | O | 29 | Myxococcales |
| 0.01 | 10 | 0 | O1 | 80812 | Sorangiineae |
| 0 | 8 | 0 | F | 1524216 | Labilitrichaceae |
| 0 | 8 | 0 | G | 1524217 | Labilithrix |
| 0 | 8 | 8 | S | 1391654 | Labilithrix luteola |
| 0 | 2 | 0 | F | 49 | Polyangiaceae |
| 0 | 2 | 0 | G | 39643 | Sorangium |
| 0 | 2 | 1 | S | 56 | Sorangium cellulosum |
| 0 | 1 | 1 | S1 | 1254432 | Sorangium cellulosum So0157-2 |
| 0 | 2 | 0 | O1 | 80811 | Cystobacterineae |
| 0 | 2 | 0 | F | 31 | Myxococcaceae |
| 0 | 2 | 0 | G | 32 | Myxococcus |
| 0 | 2 | 2 | S | 34 | Myxococcus xanthus |
| 0 | 2 | 0 | O1 | 224462 | Nannocystineae |
| 0 | 2 | 0 | F | 224464 | Kofleriaceae |
| 0 | 2 | 0 | G | 162027 | Haliangium |
| 0 | 2 | 0 | S | 80816 | Haliangium ochraceum |
| 0 | 2 | 2 | S1 | 502025 | Haliangium ochraceum DSM 14365 |
| 0 | 1 | 0 | O | 213115 | Desulfovibrionales |
| 0 | 1 | 0 | F | 194924 | Desulfovibrionaceae |
| 0 | 1 | 0 | G | 2773294 | Desulfolutivibrio |
| 0 | 1 | 1 | S | 2773299 | Desulfolutivibrio sulfoxidireducens |
| 0.01 | 13 | 0 | C | 29547 | Epsilonproteobacteria |

| | | | | | |
|---|---|---|---|---|---|
| 0.01 | 13 | 0 | O | 213849 | Campylobacterales |
| 0.01 | 13 | 0 | F | 72294 | Campylobacteraceae |
| 0.01 | 13 | 2 | G | 194 | Campylobacter |
| 0.01 | 11 | 10 | S | 195 | Campylobacter coli |
| 0 | 1 | 1 | S1 | 1367491 | Campylobacter coli 76339 |
| 0.01 | 11 | 0 | C | 2008785 | Hydrogenophilalia |
| 0.01 | 11 | 0 | O | 119069 | Hydrogenophilales |
| 0.01 | 11 | 0 | F | 206349 | Hydrogenophilaceae |
| 0.01 | 11 | 0 | G | 70774 | Hydrogenophilus |
| 0.01 | 11 | 11 | S | 297 | Hydrogenophilus thermoluteolus |
| 0 | 7 | 0 | C | 1553900 | Oligoflexia |
| 0 | 7 | 0 | O | 213481 | Bdellovibrionales |
| 0 | 7 | 0 | F | 213483 | Bdellovibrionaceae |
| 0 | 7 | 0 | G | 958 | Bdellovibrio |
| 0 | 5 | 0 | S | 959 | Bdellovibrio bacteriovorus |
| 0 | 5 | 5 | S1 | 765869 | Bdellovibrio bacteriovorus W |
| 0 | 2 | 0 | G1 | 2633795 | unclassified Bdellovibrio |
| 0 | 2 | 2 | S | 2231053 | Bdellovibrio sp. ZAP7 |
| 9.57 | 17233 | 29 | D1 | 1783272 | Terrabacteria group |
| 7.72 | 13900 | 2 | P | 201174 | Actinobacteria |
| 7.69 | 13850 | 191 | C | 1760 | Actinomycetia |
| 5.72 | 10299 | 5 | O | 85009 | Propionibacteriales |
| 5.69 | 10254 | 105 | F | 31957 | Propionibacteriaceae |
| 5.56 | 10008 | 72 | G | 1912216 | Cutibacterium |
| 5.44 | 9805 | 9781 | S | 1747 | Cutibacterium acnes |
| 0.01 | 14 | 1 | S1 | 1734925 | Cutibacterium acnes subsp. acnes |
| 0.01 | 11 | 11 | S2 | 1114967 | Cutibacterium acnes TypeIA2 P.acn17 |
| 0 | 2 | 2 | S2 | 1114966 | Cutibacterium acnes TypeIA2 P.acn33 |
| 0 | 7 | 7 | S1 | 1134454 | Cutibacterium acnes HL096PA1 |
| 0 | 2 | 2 | S1 | 1234380 | Cutibacterium acnes C1 |
| 0 | 1 | 0 | S1 | 1905725 | Cutibacterium acnes subsp. defendens |
| 0 | 1 | 1 | S2 | 1091045 | Cutibacterium acnes subsp. defendens ATCC 11828 |
| 0.07 | 121 | 121 | S | 33011 | Cutibacterium granulosum |

| | | | | | |
|---|---|---|---|---|---|
| 0.01 | 10 | 9 | S | 33010 | Cutibacterium avidum |
| 0 | 1 | 1 | S1 | 1170318 | Cutibacterium avidum 44067 |
| 0.05 | 96 | 0 | G | 1743 | Propionibacterium |
| 0.03 | 49 | 0 | G1 | 2608905 | unclassified Propionibacterium |
| 0.03 | 49 | 49 | S | 671223 | Propionibacterium sp. oral taxon 193 |
| 0.02 | 43 | 17 | S | 1744 | Propionibacterium freudenreichii |
| 0.01 | 26 | 26 | S1 | 1752 | Propionibacterium freudenreichii subsp. shermanii |
| 0 | 4 | 4 | S | 556499 | Propionibacterium acidifaciens |
| 0.01 | 18 | 0 | G | 72763 | Tessaracoccus |
| 0.01 | 14 | 14 | S | 399497 | Tessaracoccus flavescens |
| 0 | 3 | 3 | S | 1285901 | Tessaracoccus defluvii |
| 0 | 1 | 1 | S | 1332264 | Tessaracoccus aquimaris |
| 0.01 | 14 | 0 | G | 1912215 | Acidipropionibacterium |
| 0.01 | 13 | 13 | S | 1749 | Acidipropionibacterium jensenii |
| 0 | 1 | 1 | S | 2057246 | Acidipropionibacterium virtanenii |
| 0.01 | 10 | 0 | G | 29404 | Microlunatus |
| 0 | 5 | 0 | S | 29405 | Microlunatus phosphovorus |
| 0 | 5 | 5 | S1 | 1032480 | Microlunatus phosphovorus NM-1 |
| 0 | 5 | 5 | S | 546874 | Microlunatus sagamiharensis |
| 0 | 3 | 0 | G | 1085622 | Propioniciclava |
| 0 | 3 | 0 | G1 | 2642922 | unclassified Propioniciclava |
| 0 | 3 | 3 | S | 2714937 | Propioniciclava sp. HDW11 |
| 0.02 | 40 | 1 | F | 85015 | Nocardioidaceae |
| 0.01 | 22 | 7 | G | 1839 | Nocardioides |
| 0 | 7 | 0 | G1 | 2615069 | unclassified Nocardioides |
| 0 | 4 | 4 | S | 2736757 | Nocardioides sp. MC1495 |
| 0 | 1 | 1 | S | 2017486 | Nocardioides sp. S5 |
| 0 | 1 | 1 | S | 2663857 | Nocardioides sp. zg-579 |
| 0 | 1 | 1 | S | 2760089 | Nocardioides sp. S-713 |
| 0 | 4 | 4 | S | 200618 | Nocardioides aromaticivorans |
| 0 | 2 | 2 | S | 402297 | Nocardioides daphniae |
| 0 | 1 | 1 | S | 449461 | Nocardioides humi |
| 0 | 1 | 1 | S | 2712223 | Nocardioides anomalus |

| | | | | | |
|---|---|---|---|---|---|
| 0.01 | 10 | 1 | G | 2040 | Aeromicrobium |
| 0 | 3 | 0 | S | 219314 | Aeromicrobium marinum |
| 0 | 3 | 3 | S1 | 585531 | Aeromicrobium marinum DSM 15272 |
| 0 | 3 | 3 | S | 1736691 | Aeromicrobium choanae |
| 0 | 2 | 0 | G1 | 2633570 | unclassified Aeromicrobium |
| 0 | 2 | 2 | S | 2663859 | Aeromicrobium sp. zg-629 |
| 0 | 1 | 1 | S | 2041 | Aeromicrobium erythreum |
| 0 | 7 | 0 | G | 116071 | Micropruina |
| 0 | 7 | 7 | S | 75385 | Micropruina glycogenica |
| 1.33 | 2389 | 68 | O | 85006 | Micrococcales |
| 1.1 | 1989 | 151 | F | 85023 | Microbacteriaceae |
| 0.85 | 1540 | 438 | G | 33882 | Microbacterium |
| 0.24 | 428 | 59 | G1 | 2609290 | unclassified Microbacterium |
| 0.02 | 40 | 40 | S | 2483401 | Microbacterium sp. 10M-3C3 |
| 0.02 | 37 | 37 | S | 2014534 | Microbacterium sp. PM5 |
| 0.01 | 27 | 27 | S | 2567934 | Microbacterium sp. 4R-513 |
| 0.01 | 25 | 25 | S | 367477 | Microbacterium sp. XT11 |
| 0.01 | 25 | 25 | S | 2603598 | Microbacterium sp. CBA3102 |
| 0.01 | 25 | 25 | S | 2782167 | Microbacterium sp. A18JL200 |
| 0.01 | 24 | 24 | S | 1714373 | Microbacterium sp. No. 7 |
| 0.01 | 24 | 24 | S | 2268461 | Microbacterium sp. ABRD_28 |
| 0.01 | 22 | 22 | S | 2782169 | Microbacterium sp. NY27 |
| 0.01 | 20 | 20 | S | 2782166 | Microbacterium sp. A18JL241 |
| 0.01 | 14 | 14 | S | 1795053 | Microbacterium sp. PAMC 28756 |
| 0.01 | 14 | 14 | S | 2763257 | Microbacterium sp. YJN-G |
| 0.01 | 12 | 12 | S | 1906742 | Microbacterium sp. BH-3-3-3 |
| 0.01 | 11 | 11 | S | 2782168 | Microbacterium sp. WY121 |
| 0.01 | 10 | 10 | S | 2489212 | Microbacterium sp. RG1 |
| 0 | 8 | 8 | S | 1906274 | Microbacterium sp. JZ31 |
| 0 | 7 | 7 | S | 2103230 | Microbacterium sp. str. 'China' |
| 0 | 6 | 6 | S | 1938334 | Microbacterium sp. TPU 3598 |
| 0 | 5 | 5 | S | 912630 | Microbacterium sp. LKL04 |
| 0 | 5 | 5 | S | 2766784 | Microbacterium sp. Nx66 |

| | | | | | |
|---|---|---|---|---|---|
| 0 | 4 | 4 | S | 2048898 | Microbacterium sp. Y-01 |
| 0 | 2 | 2 | S | 1916917 | Microbacterium sp. 1.5R |
| 0 | 1 | 1 | S | 2606451 | Microbacterium sp. 1S1 |
| 0 | 1 | 1 | S | 2709304 | Microbacterium sp. Se63.02b |
| 0.13 | 240 | 240 | S | 36805 | Microbacterium aurum |
| 0.08 | 138 | 138 | S | 162426 | Microbacterium hominis |
| 0.03 | 63 | 63 | S | 199592 | Microbacterium paraoxydans |
| 0.02 | 42 | 42 | S | 1072463 | Microbacterium lemovicicum |
| 0.02 | 33 | 33 | S | 104336 | Microbacterium foliorum |
| 0.02 | 30 | 30 | S | 273677 | Microbacterium oleivorans |
| 0.01 | 21 | 21 | S | 82380 | Microbacterium oxydans |
| 0.01 | 21 | 21 | S | 2541726 | Microbacterium wangchenii |
| 0.01 | 18 | 18 | S | 84292 | Microbacterium chocolatum |
| 0.01 | 16 | 16 | S | 370764 | Microbacterium pygmaeum |
| 0.01 | 11 | 11 | S | 2614638 | Microbacterium caowuchunii |
| 0.01 | 10 | 10 | S | 904291 | Microbacterium sediminis |
| 0 | 7 | 7 | S | 69362 | Microbacterium schleiferi |
| 0 | 7 | 7 | S | 743009 | Microbacterium oryzae |
| 0 | 5 | 5 | S | 2614639 | Microbacterium lushaniae |
| 0 | 4 | 4 | S | 273678 | Microbacterium hydrocarbonoxydans |
| 0 | 3 | 3 | S | 2509458 | Microbacterium protaetiae |
| 0 | 2 | 2 | S | 57043 | Microbacterium esteraromaticum |
| 0 | 2 | 2 | S | 1526412 | Microbacterium endophyticum |
| 0 | 1 | 0 | S | 2033 | Microbacterium testaceum |
| 0 | 1 | 1 | S1 | 979556 | Microbacterium testaceum StLB037 |
| 0.05 | 98 | 2 | G | 2034 | Curtobacterium |
| 0.03 | 60 | 0 | S | 2035 | Curtobacterium flaccumfaciens |
| 0.03 | 60 | 60 | S1 | 138532 | Curtobacterium flaccumfaciens pv. flaccumfaciens |
| 0.02 | 36 | 3 | G1 | 257496 | unclassified Curtobacterium |
| 0.01 | 16 | 16 | S | 1561023 | Curtobacterium sp. MR_MD2014 |
| 0.01 | 10 | 10 | S | 2070337 | Curtobacterium sp. SGAir0471 |
| 0 | 3 | 3 | S | 2495430 | Curtobacterium sp. Csp2 |
| 0 | 2 | 2 | S | 1905847 | Curtobacterium sp. BH-2-1-1 |

| | | | | | |
|---|---|---|---|---|---|
| 0 | 1 | 1 | S | 2795488 | Curtobacterium sp. YC1 |
| 0 | 1 | 1 | S | 2804758 | Curtobacterium sp. 24E2 |
| 0.02 | 40 | 0 | G | 33877 | Agromyces |
| 0.02 | 34 | 0 | G1 | 2639701 | unclassified Agromyces |
| 0.01 | 16 | 16 | S | 2592652 | Agromyces sp. KACC 19306 |
| 0 | 9 | 9 | S | 2498704 | Agromyces sp. LHK192 |
| 0 | 5 | 5 | S | 2781962 | Agromyces sp. G127AT |
| 0 | 4 | 4 | S | 2585717 | Agromyces sp. HY052 |
| 0 | 3 | 3 | S | 589382 | Agromyces flavus |
| 0 | 2 | 2 | S | 2509455 | Agromyces protaetiae |
| 0 | 1 | 1 | S | 2080742 | Agromyces badenianii |
| 0.02 | 30 | 0 | G | 33886 | Rathayibacter |
| 0.01 | 21 | 2 | G1 | 2609250 | unclassified Rathayibacter |
| 0.01 | 16 | 16 | S | 2609253 | Rathayibacter sp. VKM Ac-2760 |
| 0 | 2 | 2 | S | 2609257 | Rathayibacter sp. VKM Ac-2804 |
| 0 | 1 | 1 | S | 2609254 | Rathayibacter sp. VKM Ac-2762 |
| 0 | 8 | 8 | S | 59737 | Rathayibacter iranicus |
| 0 | 1 | 1 | S | 1671680 | Rathayibacter tanaceti |
| 0.01 | 27 | 0 | G | 55968 | Leucobacter |
| 0.01 | 16 | 16 | S | 1784719 | Leucobacter triazinivorans |
| 0.01 | 11 | 11 | S | 1935379 | Leucobacter muris |
| 0.01 | 20 | 0 | G | 110932 | Leifsonia |
| 0.01 | 12 | 4 | G1 | 2663824 | unclassified Leifsonia |
| 0 | 4 | 4 | S | 1798223 | Leifsonia sp. 21MFCrub1.1 |
| 0 | 4 | 4 | S | 2724914 | Leifsonia sp. PS1209 |
| 0 | 8 | 8 | S | 150026 | Leifsonia shinshuensis |
| 0.01 | 14 | 0 | G | 1573 | Clavibacter |
| 0.01 | 11 | 0 | S | 28447 | Clavibacter michiganensis |
| 0 | 6 | 6 | S1 | 31963 | Clavibacter michiganensis subsp. nebraskensis |
| 0 | 3 | 3 | S1 | 33013 | Clavibacter michiganensis subsp. michiganensis |
| 0 | 2 | 2 | S1 | 1874630 | Clavibacter michiganensis subsp. capsici |
| 0 | 3 | 0 | G1 | 2626594 | unclassified Clavibacter |
| 0 | 3 | 3 | S | 2768071 | Clavibacter sp. DM1 |

| | | | | | |
|---|---|---|---|---|---|
| 0.01 | 12 | 0 | G | 1433997 | Diaminobutyricimonas |
| 0.01 | 12 | 0 | G1 | 2643261 | unclassified Diaminobutyricimonas |
| 0.01 | 12 | 12 | S | 2683590 | Diaminobutyricimonas sp. LJ205 |
| 0 | 9 | 0 | G | 235888 | Salinibacterium |
| 0 | 9 | 0 | G1 | 2632331 | unclassified Salinibacterium |
| 0 | 4 | 4 | S | 2708338 | Salinibacterium sp. ZJ450 |
| 0 | 3 | 3 | S | 2603292 | Salinibacterium sp. dk2585 |
| 0 | 2 | 2 | S | 2708084 | Salinibacterium sp. ZJ70 |
| 0 | 7 | 0 | G | 76634 | Mycetocola |
| 0 | 6 | 0 | G1 | 2685235 | unclassified Mycetocola |
| 0 | 6 | 6 | S | 2116510 | Mycetocola sp. JXN-3 |
| 0 | 1 | 1 | S | 2079792 | Mycetocola zhujimingii |
| 0 | 7 | 0 | G | 190323 | Plantibacter |
| 0 | 6 | 6 | S | 150123 | Plantibacter flavus |
| 0 | 1 | 1 | G1 | 2624265 | unclassified Plantibacter |
| 0 | 6 | 0 | G | 1195526 | Gryllotalpicola |
| 0 | 6 | 6 | S | 2419771 | Gryllotalpicola protaetiae |
| 0 | 5 | 0 | G | 69578 | Cryobacterium |
| 0 | 3 | 0 | G1 | 2649013 | unclassified Cryobacterium |
| 0 | 3 | 3 | S | 1978566 | Cryobacterium sp. LW097 |
| 0 | 2 | 2 | S | 670052 | Cryobacterium arcticum |
| 0 | 5 | 0 | G | 881616 | Herbiconiux |
| 0 | 5 | 0 | G1 | 2618217 | unclassified Herbiconiux |
| 0 | 5 | 5 | S | 2735133 | Herbiconiux sp. SALV-R1 |
| 0 | 4 | 0 | G | 337004 | Microcella |
| 0 | 4 | 4 | S | 279828 | Microcella alkaliphila |
| 0 | 3 | 0 | G | 46352 | Agrococcus |
| 0 | 2 | 2 | S | 399736 | Agrococcus jejuensis |
| 0 | 1 | 0 | G1 | 2615065 | unclassified Agrococcus |
| 0 | 1 | 1 | S | 2070347 | Agrococcus sp. SGAir0287 |
| 0 | 3 | 0 | G | 96492 | Frigoribacterium |
| 0 | 3 | 0 | G1 | 2627005 | unclassified Frigoribacterium |
| 0 | 3 | 3 | S | 2596916 | Frigoribacterium sp. NBH87 |

| | | | | | |
|---|---|---|---|---|---|
| 0 | 3 | 0 | G | 447237 | Frondihabitans |
| 0 | 3 | 0 | G1 | 2626248 | unclassified Frondihabitans |
| 0 | 3 | 3 | S | 1795630 | Frondihabitans sp. PAMC 28766 |
| 0 | 3 | 0 | G | 1331730 | Chryseoglobus |
| 0 | 3 | 3 | S | 2750620 | Chryseoglobus indicus |
| 0 | 2 | 0 | G | 110934 | Agreia |
| 0 | 2 | 0 | G1 | 2641148 | unclassified Agreia |
| 0 | 2 | 2 | S | 2773266 | Agreia sp. COWG |
| 0.09 | 171 | 38 | F | 1268 | Micrococcaceae |
| 0.02 | 44 | 9 | G | 1269 | Micrococcus |
| 0.02 | 34 | 34 | S | 1270 | Micrococcus luteus |
| 0 | 1 | 0 | G1 | 2620948 | unclassified Micrococcus |
| 0 | 1 | 1 | S | 376418 | Micrococcus sp. A7 |
| 0.02 | 28 | 0 | G | 32207 | Rothia |
| 0.01 | 15 | 1 | S | 2047 | Rothia dentocariosa |
| 0.01 | 14 | 14 | S1 | 762948 | Rothia dentocariosa ATCC 17931 |
| 0 | 8 | 8 | S | 37923 | Rothia kristinae |
| 0 | 4 | 4 | S | 43675 | Rothia mucilaginosa |
| 0 | 1 | 1 | S | 169480 | Rothia amarae |
| 0.01 | 25 | 1 | G | 57493 | Kocuria |
| 0 | 9 | 9 | S | 71999 | Kocuria palustris |
| 0 | 8 | 8 | S | 1272 | Kocuria varians |
| 0 | 7 | 7 | S | 72000 | Kocuria rhizophila |
| 0.01 | 12 | 0 | G | 1742989 | Glutamicibacter |
| 0.01 | 12 | 12 | S | 37929 | Glutamicibacter nicotianae |
| 0.01 | 11 | 0 | G | 1663 | Arthrobacter |
| 0 | 9 | 0 | G1 | 235627 | unclassified Arthrobacter |
| 0 | 6 | 6 | S | 1357915 | Arthrobacter sp. QXT-31 |
| 0 | 3 | 3 | S | 904039 | Arthrobacter sp. NEB 688 |
| 0 | 2 | 2 | S | 37921 | Arthrobacter agilis |
| 0 | 8 | 0 | G | 370735 | Zhihengliuella |
| 0 | 8 | 0 | G1 | 2626589 | unclassified Zhihengliuella |
| 0 | 8 | 8 | S | 2058657 | Zhihengliuella sp. ISTPL4 |

| | | | | | |
|---|---|---|---|---|---|
| 0 | 4 | 0 | G | 1645 | Renibacterium |
| 0 | 4 | 4 | S | 1646 | Renibacterium salmoninarum |
| 0 | 1 | 0 | G | 1742993 | Pseudarthrobacter |
| 0 | 1 | 1 | S | 121292 | Pseudarthrobacter sulfonivorans |
| 0.03 | 58 | 0 | F | 85019 | Brevibacteriaceae |
| 0.03 | 58 | 1 | G | 1696 | Brevibacterium |
| 0.02 | 30 | 0 | G1 | 2614124 | unclassified Brevibacterium |
| 0.01 | 12 | 12 | S | 1496080 | Brevibacterium sp. YB235 |
| 0.01 | 12 | 12 | S | 2575923 | Brevibacterium sp. CS2 |
| 0 | 6 | 6 | S | 1406197 | Brevibacterium sp. Ap13 |
| 0.01 | 14 | 14 | S | 273384 | Brevibacterium aurantiacum |
| 0 | 8 | 8 | S | 1703 | Brevibacterium linens |
| 0 | 5 | 5 | S | 629680 | Brevibacterium sandarakinum |
| 0.02 | 38 | 0 | F | 85021 | Intrasporangiaceae |
| 0.02 | 37 | 10 | G | 53457 | Janibacter |
| 0.01 | 13 | 13 | S | 857417 | Janibacter indicus |
| 0.01 | 10 | 10 | S | 53458 | Janibacter limosus |
| 0 | 3 | 3 | S | 262209 | Janibacter melonis |
| 0 | 1 | 0 | G1 | 2649294 | unclassified Janibacter |
| 0 | 1 | 1 | S | 2761047 | Janibacter sp. YB324 |
| 0 | 1 | 0 | G | 367298 | Phycicoccus |
| 0 | 1 | 0 | G1 | 2637926 | unclassified Phycicoccus |
| 0 | 1 | 1 | S | 2714941 | Phycicoccus sp. HDW14 |
| 0.01 | 21 | 0 | F | 85016 | Cellulomonadaceae |
| 0.01 | 20 | 0 | G | 1707 | Cellulomonas |
| 0 | 9 | 0 | S | 1711 | Cellulomonas flavigena |
| 0 | 9 | 9 | S1 | 446466 | Cellulomonas flavigena DSM 20109 |
| 0 | 7 | 0 | G1 | 2620175 | unclassified Cellulomonas |
| 0 | 5 | 5 | S | 2654191 | Cellulomonas sp. JZ18 |
| 0 | 2 | 2 | S | 2704467 | Cellulomonas sp. H30R-01 |
| 0 | 4 | 4 | S | 2729175 | Cellulomonas taurus |
| 0 | 1 | 0 | G | 162491 | Oerskovia |
| 0 | 1 | 0 | G1 | 2619021 | unclassified Oerskovia |

| | | | | | |
|---|---|---|---|---|---|
| 0 | 1 | 1 | S | 1179673 | Oerskovia sp. KBS0722 |
| 0.01 | 17 | 0 | F | 2805426 | Kytococcaceae |
| 0.01 | 17 | 0 | G | 57499 | Kytococcus |
| 0.01 | 17 | 17 | S | 1276 | Kytococcus sedentarius |
| 0.01 | 10 | 0 | F | 85017 | Promicromonosporaceae |
| 0 | 7 | 0 | G | 157920 | Cellulosimicrobium |
| 0 | 5 | 5 | S | 1710 | Cellulosimicrobium cellulans |
| 0 | 2 | 0 | G1 | 2624466 | unclassified Cellulosimicrobium |
| 0 | 1 | 1 | S | 1906273 | Cellulosimicrobium sp. JZ28 |
| 0 | 1 | 1 | S | 2731680 | Cellulosimicrobium sp. 72-3 |
| 0 | 3 | 0 | G | 186188 | Xylanimonas |
| 0 | 3 | 3 | S | 2509459 | Xylanimonas allomyrinae |
| 0 | 9 | 0 | F | 85020 | Dermabacteraceae |
| 0 | 8 | 0 | G | 43668 | Brachybacterium |
| 0 | 4 | 0 | S | 43669 | Brachybacterium faecium |
| 0 | 4 | 4 | S1 | 446465 | Brachybacterium faecium DSM 4810 |
| 0 | 4 | 4 | S | 1331682 | Brachybacterium ginsengisoli |
| 0 | 1 | 0 | G | 36739 | Dermabacter |
| 0 | 1 | 1 | S | 1630135 | Dermabacter vaginalis |
| 0 | 3 | 0 | F | 145357 | Dermacoccaceae |
| 0 | 3 | 0 | G | 57495 | Dermacoccus |
| 0 | 3 | 3 | S | 1274 | Dermacoccus nishinomiyaensis |
| 0 | 2 | 0 | F | 145358 | Bogoriellaceae |
| 0 | 2 | 0 | G | 154116 | Georgenia |
| 0 | 1 | 1 | S | 2483799 | Georgenia faecalis |
| 0 | 1 | 0 | G1 | 2626815 | unclassified Georgenia |
| 0 | 1 | 1 | S | 2589797 | Georgenia sp. Z443 |
| 0 | 2 | 0 | F | 145360 | Sanguibacteraceae |
| 0 | 2 | 0 | G | 60919 | Sanguibacter |
| 0 | 2 | 0 | S | 60920 | Sanguibacter keddieii |
| 0 | 2 | 2 | S1 | 446469 | Sanguibacter keddieii DSM 10542 |
| 0 | 1 | 0 | F | 2805590 | Ornithinimicrobiaceae |
| 0 | 1 | 0 | G | 265976 | Serinicoccus |

| | | | | | |
|---|---|---|---|---|---|
| 0 | 1 | 1 | S | 1078471 | Serinicoccus profundi |
| 0.43 | 775 | 11 | O | 85007 | Corynebacteriales |
| 0.29 | 517 | 0 | F | 1653 | Corynebacteriaceae |
| 0.29 | 517 | 33 | G | 1716 | Corynebacterium |
| 0.11 | 194 | 194 | S | 1979527 | Corynebacterium kefirresidentii |
| 0.06 | 117 | 117 | S | 38304 | Corynebacterium tuberculostearicum |
| 0.03 | 46 | 46 | S | 43990 | Corynebacterium segmentosum |
| 0.01 | 24 | 24 | S | 161879 | Corynebacterium kroppenstedtii |
| 0.01 | 20 | 20 | S | 2594913 | Corynebacterium sanguinis |
| 0.01 | 18 | 18 | S | 38290 | Corynebacterium macginleyi |
| 0.01 | 10 | 0 | S | 1404244 | Corynebacterium glyciniphilum |
| 0.01 | 10 | 10 | S1 | 1404245 | Corynebacterium glyciniphilum AJ 3170 |
| 0.01 | 10 | 10 | S | 401472 | Corynebacterium ureicelerivorans |
| 0 | 6 | 0 | G1 | 2624378 | unclassified Corynebacterium |
| 0 | 3 | 3 | S | 2675216 | Corynebacterium sp. 1959 |
| 0 | 2 | 2 | S | 2754725 | Corynebacterium sp. Marseille-Q3630 |
| 0 | 1 | 1 | S | 702967 | Corynebacterium sp. NML98-0116 |
| 0 | 6 | 6 | S | 169292 | Corynebacterium aurimucosum |
| 0 | 5 | 5 | S | 35757 | Corynebacterium cystitidis |
| 0 | 5 | 5 | S | 43770 | Corynebacterium striatum |
| 0 | 4 | 4 | S | 43768 | Corynebacterium matruchotii |
| 0 | 3 | 0 | S | 1230998 | Corynebacterium frankenforstense |
| 0 | 3 | 3 | S1 | 1437875 | Corynebacterium frankenforstense DSM 45800 |
| 0 | 3 | 0 | S | 225326 | Corynebacterium halotolerans |
| 0 | 3 | 3 | S1 | 1121362 | Corynebacterium halotolerans YIM 70093 = DSM 44683 |
| 0 | 3 | 3 | S | 161899 | Corynebacterium singulare |
| 0 | 2 | 0 | S | 575200 | Corynebacterium maris |
| 0 | 2 | 2 | S1 | 1224163 | Corynebacterium maris DSM 45190 |
| 0 | 2 | 2 | S | 156976 | Corynebacterium riegelii |
| 0 | 2 | 2 | S | 146827 | Corynebacterium simulans |
| 0 | 1 | 1 | S | 1471400 | Corynebacterium pelargi |
| 0 | 1 | 1 | S | 35755 | Corynebacterium kutscheri |
| 0 | 1 | 1 | S | 38289 | Corynebacterium jeikeium |

| | | | | | |
|---|---|---|---|---|---|
| 0 | 1 | 0 | S | 38305 | Corynebacterium vitaeruminis |
| 0 | 1 | 1 | S1 | 1224164 | Corynebacterium vitaeruminis DSM 20294 |
| 0.05 | 86 | 0 | F | 85025 | Nocardiaceae |
| 0.05 | 85 | 15 | G | 1827 | Rhodococcus |
| 0.03 | 51 | 0 | G1 | 192944 | unclassified Rhodococcus |
| 0.02 | 41 | 41 | S | 2795031 | Rhodococcus sp. P-2 |
| 0 | 5 | 5 | S | 2507582 | Rhodococcus sp. ABRD24 |
| 0 | 4 | 4 | S | 1045808 | Rhodococcus sp. YL-1 |
| 0 | 1 | 1 | S | 2499145 | Rhodococcus sp. X156 |
| 0.01 | 13 | 13 | S | 334542 | Rhodococcus qingshengii |
| 0 | 5 | 5 | S | 1833 | Rhodococcus erythropolis |
| 0 | 1 | 1 | S | 1830 | Rhodococcus ruber |
| 0 | 1 | 0 | G | 1817 | Nocardia |
| 0 | 1 | 1 | S | 37329 | Nocardia farcinica |
| 0.04 | 80 | 3 | F | 1762 | Mycobacteriaceae |
| 0.03 | 52 | 0 | G | 1763 | Mycobacterium |
| 0.02 | 36 | 36 | S | 1789 | Mycobacterium xenopi |
| 0 | 9 | 9 | S | 1389713 | Mycobacterium paragordonae |
| 0 | 5 | 5 | S | 1768 | Mycobacterium kansasii |
| 0 | 1 | 1 | S | 2492438 | Mycobacterium novum |
| 0 | 1 | 0 | G1 | 77643 | Mycobacterium tuberculosis complex |
| 0 | 1 | 1 | S | 78331 | Mycobacterium canettii |
| 0.01 | 25 | 2 | G | 1866885 | Mycolicibacterium |
| 0.01 | 11 | 11 | S | 67081 | Mycolicibacterium alvei |
| 0 | 5 | 5 | S | 1794 | Mycolicibacterium gadium |
| 0 | 1 | 1 | S | 1431246 | Mycolicibacterium anyangense |
| 0 | 1 | 1 | S | 1286181 | Mycolicibacterium arabiense |
| 0 | 1 | 1 | S | 1286180 | Mycolicibacterium sediminis |
| 0 | 1 | 1 | S | 1791 | Mycolicibacterium aurum |
| 0 | 1 | 1 | S | 319706 | Mycolicibacterium phocaicum |
| 0 | 1 | 1 | S | 53462 | Mycolicibacterium mageritense |
| 0 | 1 | 0 | S | 1804 | Mycolicibacterium gilvum |
| 0 | 1 | 1 | S1 | 350054 | Mycolicibacterium gilvum PYR-GCK |

| | | | | | |
|---|---|---|---|---|---|
| 0.02 | 32 | 0 | F | 85026 | Gordoniaceae |
| 0.02 | 32 | 10 | G | 2053 | Gordonia |
| 0.01 | 12 | 12 | S | 2055 | Gordonia terrae |
| 0 | 7 | 7 | S | 2054 | Gordonia bronchialis |
| 0 | 2 | 2 | S | 36822 | Gordonia rubripertincta |
| 0 | 1 | 0 | S | 84595 | Gordonia polyisoprenivorans |
| 0 | 1 | 1 | S1 | 1112204 | Gordonia polyisoprenivorans VH2 |
| 0.01 | 21 | 0 | F | 85028 | Tsukamurellaceae |
| 0.01 | 21 | 1 | G | 2060 | Tsukamurella |
| 0.01 | 12 | 12 | S | 57704 | Tsukamurella tyrosinosolvens |
| 0 | 8 | 8 | S | 2061 | Tsukamurella paurometabola |
| 0.01 | 18 | 0 | F | 2805586 | Lawsonellaceae |
| 0.01 | 18 | 0 | G | 1847725 | Lawsonella |
| 0.01 | 18 | 18 | S | 1528099 | Lawsonella clevelandensis |
| 0.01 | 10 | 0 | F | 85029 | Dietziaceae |
| 0.01 | 10 | 1 | G | 37914 | Dietzia |
| 0 | 5 | 0 | G1 | 2617939 | unclassified Dietzia |
| 0 | 5 | 5 | S | 712270 | Dietzia sp. oral taxon 368 |
| 0 | 2 | 2 | S | 499555 | Dietzia timorensis |
| 0 | 2 | 2 | S | 546160 | Dietzia lutea |
| 0.04 | 65 | 0 | O | 85011 | Streptomycetales |
| 0.04 | 65 | 3 | F | 2062 | Streptomycetaceae |
| 0.03 | 62 | 34 | G | 1883 | Streptomyces |
| 0.01 | 13 | 2 | G1 | 2593676 | unclassified Streptomyces |
| 0 | 3 | 3 | S | 2721173 | Streptomyces sp. DSM 40868 |
| 0 | 3 | 3 | S | 444103 | Streptomyces sp. CNQ-509 |
| 0 | 2 | 2 | S | 2203205 | Streptomyces sp. WAC 01529 |
| 0 | 1 | 1 | S | 2768069 | Streptomyces sp. CRXT-Y-14 |
| 0 | 1 | 1 | S | 2695266 | Streptomyces sp. HM190 |
| 0 | 1 | 1 | S | 2059884 | Streptomyces sp. CMB-StM0423 |
| 0 | 4 | 0 | S | 379067 | Streptomyces bingchenggensis |
| 0 | 4 | 4 | S1 | 749414 | Streptomyces bingchenggensis BCW-1 |
| 0 | 4 | 4 | S | 67267 | Streptomyces alboflavus |

| | | | | | |
|---|---|---|---|---|---|
| 0 | 3 | 3 | S | 51201 | Streptomyces griseocarneus |
| 0 | 2 | 2 | S | 80860 | Streptomyces cyanogenus |
| 0 | 1 | 1 | S | 40318 | Streptomyces nodosus |
| 0 | 1 | 0 | S | 33903 | Streptomyces avermitilis |
| 0 | 1 | 1 | S1 | 227882 | Streptomyces avermitilis MA-4680 = NBRC 14893 |
| 0.02 | 41 | 0 | O | 85004 | Bifidobacteriales |
| 0.02 | 41 | 0 | F | 31953 | Bifidobacteriaceae |
| 0.02 | 41 | 6 | G | 1678 | Bifidobacterium |
| 0.01 | 13 | 13 | S | 1681 | Bifidobacterium bifidum |
| 0.01 | 11 | 11 | S | 216816 | Bifidobacterium longum |
| 0 | 6 | 0 | S | 78448 | Bifidobacterium pullorum |
| 0 | 6 | 6 | S1 | 78344 | Bifidobacterium pullorum subsp. gallinarum |
| 0 | 3 | 3 | S | 1680 | Bifidobacterium adolescentis |
| 0 | 2 | 2 | S | 28026 | Bifidobacterium pseudocatenulatum |
| 0.02 | 36 | 0 | O | 85010 | Pseudonocardiales |
| 0.02 | 36 | 1 | F | 2070 | Pseudonocardiaceae |
| 0.01 | 15 | 0 | G | 1835 | Saccharopolyspora |
| 0 | 9 | 9 | S | 1836 | Saccharopolyspora erythraea |
| 0 | 4 | 4 | S | 2665642 | Saccharopolyspora coralli |
| 0 | 1 | 1 | S | 60894 | Saccharopolyspora spinosa |
| 0 | 1 | 1 | S | 333966 | Saccharopolyspora pogona |
| 0 | 8 | 4 | G | 40566 | Actinosynnema |
| 0 | 4 | 4 | S | 42197 | Actinosynnema pretiosum |
| 0 | 3 | 3 | G | 1851 | Saccharomonospora |
| 0 | 2 | 0 | G | 1813 | Amycolatopsis |
| 0 | 1 | 1 | S | 33910 | Amycolatopsis mediterranei |
| 0 | 1 | 0 | G1 | 2618356 | unclassified Amycolatopsis |
| 0 | 1 | 1 | S | 2742131 | Amycolatopsis sp. Hca4 |
| 0 | 2 | 0 | G | 1847 | Pseudonocardia |
| 0 | 1 | 0 | S | 240495 | Pseudonocardia dioxanivorans |
| 0 | 1 | 1 | S1 | 675635 | Pseudonocardia dioxanivorans CB1190 |
| 0 | 1 | 0 | G1 | 2619320 | unclassified Pseudonocardia |
| 0 | 1 | 1 | S | 1641402 | Pseudonocardia sp. HH130629-09 |

| | | | | | |
|---|---|---|---|---|---|
| 0 | 2 | 0 | G | 2071 | Saccharothrix |
| 0 | 1 | 0 | S | 103731 | Saccharothrix espanaensis |
| 0 | 1 | 1 | S1 | 1179773 | Saccharothrix espanaensis DSM 44229 |
| 0 | 1 | 0 | G1 | 2593673 | unclassified Saccharothrix |
| 0 | 1 | 1 | S | 2781735 | Saccharothrix sp. 6-C |
| 0 | 2 | 0 | G | 1137960 | Allokutzneria |
| 0 | 2 | 2 | S | 211114 | Allokutzneria albata |
| 0 | 1 | 1 | G | 65496 | Actinoalloteichus |
| 0.01 | 20 | 0 | O | 85012 | Streptosporangiales |
| 0.01 | 10 | 0 | F | 2004 | Streptosporangiaceae |
| 0 | 6 | 3 | G | 2000 | Streptosporangium |
| 0 | 2 | 0 | G1 | 2632669 | unclassified Streptosporangium |
| 0 | 2 | 2 | S | 2202249 | Streptosporangium sp. 'caverna' |
| 0 | 1 | 0 | S | 2001 | Streptosporangium roseum |
| 0 | 1 | 1 | S1 | 479432 | Streptosporangium roseum DSM 43021 |
| 0 | 4 | 2 | G | 83681 | Nonomuraea |
| 0 | 1 | 0 | G1 | 2593643 | unclassified Nonomuraea |
| 0 | 1 | 1 | S | 1909395 | Nonomuraea sp. ATCC 55076 |
| 0 | 1 | 1 | S | 2656914 | Nonomuraea nitratireducens |
| 0.01 | 10 | 0 | F | 2012 | Thermomonosporaceae |
| 0.01 | 10 | 0 | G | 1988 | Actinomadura |
| 0.01 | 10 | 5 | G1 | 2626254 | unclassified Actinomadura |
| 0 | 3 | 3 | S | 2742128 | Actinomadura sp. NAK00032 |
| 0 | 1 | 1 | S | 1219491 | Actinomadura sp. WMMB 499 |
| 0 | 1 | 1 | S | 2591108 | Actinomadura sp. WMMA1423 |
| 0.01 | 15 | 0 | O | 2037 | Actinomycetales |
| 0.01 | 15 | 1 | F | 2049 | Actinomycetaceae |
| 0.01 | 12 | 6 | G | 1654 | Actinomyces |
| 0 | 4 | 4 | S | 1655 | Actinomyces naeslundii |
| 0 | 2 | 2 | S | 2057800 | Actinomyces qiguomingii |
| 0 | 1 | 0 | G | 1522056 | Flaviflexus |
| 0 | 1 | 1 | S | 1282737 | Flaviflexus salsibiostraticola |
| 0 | 1 | 0 | G | 2529408 | Schaalia |

| | | | | | |
|---|---|---|---|---|---|
| 0 | 1 | 0 | G1 | 2691889 | unclassified Schaalia |
| 0 | 1 | 1 | S | 2758572 | Schaalia sp. JY-X169 |
| 0.01 | 10 | 0 | O | 85008 | Micromonosporales |
| 0.01 | 10 | 3 | F | 28056 | Micromonosporaceae |
| 0 | 4 | 0 | G | 1865 | Actinoplanes |
| 0 | 4 | 0 | G1 | 2626549 | unclassified Actinoplanes |
| 0 | 4 | 4 | S | 649831 | Actinoplanes sp. N902-109 |
| 0 | 3 | 0 | G | 1873 | Micromonospora |
| 0 | 2 | 2 | S | 47875 | Micromonospora sagamiensis |
| 0 | 1 | 1 | S | 1881 | Micromonospora viridifaciens |
| 0 | 5 | 0 | O | 1643682 | Geodermatophilales |
| 0 | 5 | 0 | F | 85030 | Geodermatophilaceae |
| 0 | 5 | 0 | G | 88138 | Modestobacter |
| 0 | 5 | 5 | S | 477641 | Modestobacter marinus |
| 0 | 4 | 0 | O | 85013 | Frankiales |
| 0 | 4 | 0 | F | 74712 | Frankiaceae |
| 0 | 4 | 0 | G | 1854 | Frankia |
| 0 | 3 | 0 | S | 1859 | Frankia alni |
| 0 | 3 | 3 | S1 | 326424 | Frankia alni ACN14a |
| 0 | 1 | 1 | S | 106370 | Frankia casuarinae |
| 0.02 | 29 | 0 | C | 84995 | Rubrobacteria |
| 0.02 | 29 | 0 | O | 84996 | Rubrobacterales |
| 0.02 | 29 | 0 | F | 84997 | Rubrobacteraceae |
| 0.02 | 29 | 0 | G | 42255 | Rubrobacter |
| 0.02 | 29 | 14 | S | 49319 | Rubrobacter xylanophilus |
| 0.01 | 15 | 15 | S1 | 266117 | Rubrobacter xylanophilus DSM 9941 |
| 0.01 | 10 | 0 | C | 1497346 | Thermoleophilia |
| 0.01 | 10 | 0 | O | 588673 | Solirubrobacterales |
| 0.01 | 10 | 0 | F | 320583 | Conexibacteraceae |
| 0.01 | 10 | 0 | G | 191494 | Conexibacter |
| 0.01 | 10 | 0 | S | 191495 | Conexibacter woesei |
| 0.01 | 10 | 10 | S1 | 469383 | Conexibacter woesei DSM 14684 |
| 0 | 9 | 0 | C | 84998 | Coriobacteriia |

| | | | | | |
|---|---|---|---|---|---|
| 0 | 6 | 0 | O | 84999 | Coriobacteriales |
| 0 | 6 | 0 | F | 84107 | Coriobacteriaceae |
| 0 | 6 | 0 | G | 102106 | Collinsella |
| 0 | 6 | 6 | S | 74426 | Collinsella aerofaciens |
| 0 | 3 | 0 | O | 1643822 | Eggerthellales |
| 0 | 3 | 0 | F | 1643826 | Eggerthellaceae |
| 0 | 2 | 0 | G | 84111 | Eggerthella |
| 0 | 2 | 2 | S | 84112 | Eggerthella lenta |
| 0 | 1 | 1 | G | 447020 | Adlercreutzia |
| 1.79 | 3219 | 5 | P | 1239 | Firmicutes |
| 1.4 | 2522 | 0 | C | 91061 | Bacilli |
| 0.79 | 1417 | 3 | O | 186826 | Lactobacillales |
| 0.68 | 1221 | 0 | F | 33958 | Lactobacillaceae |
| 0.66 | 1195 | 0 | G | 1578 | Lactobacillus |
| 0.65 | 1171 | 282 | S | 1584 | Lactobacillus delbrueckii |
| 0.49 | 888 | 888 | S1 | 29397 | Lactobacillus delbrueckii subsp. lactis |
| 0 | 1 | 0 | S1 | 1537158 | Lactobacillus delbrueckii subsp. jakobsenii |
| 0 | 1 | 1 | S2 | 1217420 | Lactobacillus delbrueckii subsp. jakobsenii ZN7a-9 = DSM 26046 |
| 0.01 | 16 | 16 | S | 47770 | Lactobacillus crispatus |
| 0 | 6 | 6 | S | 147802 | Lactobacillus iners |
| 0 | 1 | 0 | S | 33959 | Lactobacillus johnsonii |
| 0 | 1 | 1 | S1 | 633699 | Lactobacillus johnsonii FI9785 |
| 0 | 1 | 1 | S | 109790 | Lactobacillus jensenii |
| 0.01 | 17 | 0 | G | 2742598 | Limosilactobacillus |
| 0.01 | 15 | 15 | S | 1598 | Limosilactobacillus reuteri |
| 0 | 2 | 2 | S | 1633 | Limosilactobacillus vaginalis |
| 0 | 9 | 1 | G | 1243 | Leuconostoc |
| 0 | 7 | 7 | S | 33964 | Leuconostoc citreum |
| 0 | 1 | 1 | S | 1245 | Leuconostoc mesenteroides |
| 0.09 | 165 | 0 | F | 1300 | Streptococcaceae |
| 0.09 | 162 | 38 | G | 1301 | Streptococcus |
| 0.02 | 41 | 41 | S | 28037 | Streptococcus mitis |
| 0.01 | 21 | 6 | G1 | 2608887 | unclassified Streptococcus |

| | | | | | |
|---|---|---|---|---|---|
| 0.01 | 14 | 14 | S | 2610896 | Streptococcus sp. LPB0220 |
| 0 | 1 | 1 | S | 2598453 | Streptococcus sp. 116-D4 |
| 0.01 | 16 | 16 | S | 1313 | Streptococcus pneumoniae |
| 0.01 | 12 | 12 | S | 1308 | Streptococcus thermophilus |
| 0.01 | 11 | 11 | S | 1305 | Streptococcus sanguinis |
| 0.01 | 10 | 10 | S | 1304 | Streptococcus salivarius |
| 0 | 7 | 7 | S | 1302 | Streptococcus gordonii |
| 0 | 4 | 0 | S | 1318 | Streptococcus parasanguinis |
| 0 | 2 | 2 | S1 | 760570 | Streptococcus parasanguinis ATCC 15912 |
| 0 | 2 | 2 | S1 | 1114965 | Streptococcus parasanguinis FW213 |
| 0 | 2 | 2 | S | 1303 | Streptococcus oralis |
| 0 | 3 | 0 | G | 1357 | Lactococcus |
| 0 | 2 | 2 | S | 1358 | Lactococcus lactis |
| 0 | 1 | 1 | S | 1359 | Lactococcus cremoris |
| 0.01 | 17 | 0 | F | 81852 | Enterococcaceae |
| 0.01 | 17 | 0 | G | 1350 | Enterococcus |
| 0.01 | 15 | 15 | S | 44008 | Enterococcus cecorum |
| 0 | 2 | 2 | S | 1351 | Enterococcus faecalis |
| 0 | 6 | 0 | F | 186827 | Aerococcaceae |
| 0 | 5 | 0 | G | 46123 | Abiotrophia |
| 0 | 5 | 5 | S | 46125 | Abiotrophia defectiva |
| 0 | 1 | 0 | G | 1375 | Aerococcus |
| 0 | 1 | 1 | S | 1377 | Aerococcus viridans |
| 0 | 5 | 0 | F | 186828 | Carnobacteriaceae |
| 0 | 5 | 0 | G | 29393 | Dolosigranulum |
| 0 | 5 | 5 | S | 29394 | Dolosigranulum pigrum |
| 0.61 | 1105 | 6 | O | 1385 | Bacillales |
| 0.56 | 1015 | 3 | F | 90964 | Staphylococcaceae |
| 0.55 | 991 | 73 | G | 1279 | Staphylococcus |
| 0.2 | 366 | 366 | S | 29388 | Staphylococcus capitis |
| 0.18 | 332 | 316 | S | 1282 | Staphylococcus epidermidis |
| 0.01 | 16 | 16 | S1 | 1449752 | Staphylococcus epidermidis PM221 |
| 0.06 | 113 | 113 | S | 33028 | Staphylococcus saccharolyticus |

| | | | | | |
|---|---|---|---|---|---|
| 0.04 | 67 | 67 | S | 1292 | Staphylococcus warneri |
| 0.01 | 13 | 13 | S | 1280 | Staphylococcus aureus |
| 0.01 | 10 | 10 | S | 1283 | Staphylococcus haemolyticus |
| 0 | 5 | 5 | S | 29385 | Staphylococcus saprophyticus |
| 0 | 5 | 5 | S | 1290 | Staphylococcus hominis |
| 0 | 3 | 3 | S | 45972 | Staphylococcus pasteuri |
| 0 | 1 | 1 | S | 170573 | Staphylococcus pettenkoferi |
| 0 | 1 | 0 | G1 | 91994 | unclassified Staphylococcus |
| 0 | 1 | 1 | S | 2799680 | Staphylococcus sp. 11-B-312 |
| 0 | 1 | 1 | S | 28035 | Staphylococcus lugdunensis |
| 0 | 1 | 0 | S | 1286 | Staphylococcus simulans |
| 0 | 1 | 1 | S1 | 1287 | Staphylococcus simulans bv. staphylolyticus |
| 0.01 | 13 | 0 | G | 2803850 | Mammaliicoccus |
| 0.01 | 13 | 13 | S | 42858 | Mammaliicoccus lentus |
| 0 | 8 | 0 | G | 227979 | Jeotgalicoccus |
| 0 | 8 | 8 | S | 1461582 | Jeotgalicoccus saudimassiliensis |
| 0.03 | 50 | 6 | F | 186817 | Bacillaceae |
| 0.01 | 21 | 0 | G | 1276290 | Caldibacillus |
| 0.01 | 21 | 21 | S | 35841 | Caldibacillus thermoamylovorans |
| 0.01 | 11 | 0 | G | 2817139 | Weizmannia |
| 0.01 | 11 | 11 | S | 1398 | Weizmannia coagulans |
| 0 | 9 | 0 | G | 1386 | Bacillus |
| 0 | 5 | 5 | S | 38875 | Bacillus oleronius |
| 0 | 4 | 0 | G1 | 86661 | Bacillus cereus group |
| 0 | 4 | 4 | S | 1392 | Bacillus anthracis |
| 0 | 1 | 0 | G | 2675232 | Neobacillus |
| 0 | 1 | 1 | S | 1215031 | Neobacillus thermocopriae |
| 0 | 1 | 0 | G | 1329200 | Fictibacillus |
| 0 | 1 | 1 | S | 255247 | Fictibacillus arsenicus |
| 0 | 1 | 0 | G | 150247 | Anoxybacillus |
| 0 | 1 | 1 | S | 33934 | Anoxybacillus flavithermus |
| 0.01 | 16 | 0 | O1 | 539002 | Bacillales incertae sedis |
| 0.01 | 16 | 0 | O2 | 539738 | Bacillales Family XI. Incertae Sedis |

| | | | | | |
|---|---|---|---|---|---|
| 0.01 | 16 | 3 | G | 1378 | Gemella |
| 0.01 | 12 | 12 | S | 29391 | Gemella morbillorum |
| 0 | 1 | 1 | S | 1379 | Gemella haemolysans |
| 0 | 7 | 1 | F | 186822 | Paenibacillaceae |
| 0 | 6 | 2 | G | 44249 | Paenibacillus |
| 0 | 4 | 0 | G1 | 185978 | unclassified Paenibacillus |
| 0 | 2 | 2 | S | 2660554 | Paenibacillus sp. B01 |
| 0 | 2 | 2 | S | 2023772 | Paenibacillus sp. RUD330 |
| 0 | 7 | 0 | F | 186823 | Alicyclobacillaceae |
| 0 | 7 | 0 | G | 29330 | Alicyclobacillus |
| 0 | 7 | 0 | G1 | 2622400 | unclassified Alicyclobacillus |
| 0 | 7 | 7 | S | 2665646 | Alicyclobacillus sp. SO9 |
| 0 | 4 | 0 | F | 186818 | Planococcaceae |
| 0 | 4 | 0 | G | 1372 | Planococcus |
| 0 | 4 | 4 | S | 1215089 | Planococcus halocryophilus |
| 0.23 | 408 | 0 | C | 186801 | Clostridia |
| 0.23 | 408 | 32 | O | 186802 | Eubacteriales |
| 0.12 | 209 | 1 | F | 216572 | Oscillospiraceae |
| 0.09 | 167 | 0 | G | 216851 | Faecalibacterium |
| 0.09 | 167 | 129 | S | 853 | Faecalibacterium prausnitzii |
| 0.01 | 26 | 26 | S1 | 718252 | Faecalibacterium prausnitzii L2-6 |
| 0.01 | 12 | 12 | S1 | 657322 | Faecalibacterium prausnitzii SL3/3 |
| 0.02 | 34 | 0 | G | 1263 | Ruminococcus |
| 0.02 | 33 | 33 | S | 1160721 | Ruminococcus bicirculans |
| 0 | 1 | 0 | S | 1264 | Ruminococcus albus |
| 0 | 1 | 1 | S1 | 697329 | Ruminococcus albus 7 = DSM 20455 |
| 0 | 3 | 0 | G | 459786 | Oscillibacter |
| 0 | 3 | 0 | G1 | 2629304 | unclassified Oscillibacter |
| 0 | 2 | 2 | S | 2763056 | Oscillibacter sp. NSJ-62 |
| 0 | 1 | 1 | S | 2109687 | Oscillibacter sp. PEA192 |
| 0 | 2 | 0 | G | 2591381 | Dysosmobacter |
| 0 | 2 | 2 | S | 2093857 | Dysosmobacter welbionis |
| 0 | 1 | 0 | G | 1905344 | Ruthenibacterium |

| | | | | | |
|---|---|---|---|---|---|
| 0 | 1 | 1 | S | 1550024 | Ruthenibacterium lactatiformans |
| 0 | 1 | 0 | G | 2304691 | Thermoclostridium |
| 0 | 1 | 1 | S | 1510 | Thermoclostridium stercorarium |
| 0.07 | 134 | 12 | F | 186803 | Lachnospiraceae |
| 0.03 | 50 | 0 | G | 841 | Roseburia |
| 0.03 | 47 | 19 | S | 166486 | Roseburia intestinalis |
| 0.01 | 19 | 19 | S1 | 657315 | Roseburia intestinalis M50/1 |
| 0 | 8 | 8 | S1 | 536231 | Roseburia intestinalis L1-82 |
| 0 | 1 | 1 | S1 | 718255 | Roseburia intestinalis XB6B4 |
| 0 | 3 | 3 | S | 301301 | Roseburia hominis |
| 0.01 | 17 | 0 | G | 572511 | Blautia |
| 0.01 | 13 | 0 | G1 | 2648079 | unclassified Blautia |
| 0.01 | 13 | 13 | S | 2479767 | Blautia sp. SC05B48 |
| 0 | 3 | 0 | S | 40520 | Blautia obeum |
| 0 | 3 | 3 | S1 | 657314 | Blautia obeum A2-162 |
| 0 | 1 | 1 | S | 33035 | Blautia producta |
| 0.01 | 16 | 0 | G | 28050 | Lachnospira |
| 0.01 | 16 | 0 | S | 39485 | Lachnospira eligens |
| 0.01 | 16 | 16 | S1 | 515620 | [Eubacterium] eligens ATCC 27750 |
| 0.01 | 11 | 0 | G | 2316020 | Mediterraneibacter |
| 0.01 | 10 | 0 | S | 33039 | [Ruminococcus] torques |
| 0.01 | 10 | 10 | S1 | 657313 | [Ruminococcus] torques L2-14 |
| 0 | 1 | 1 | S | 33038 | [Ruminococcus] gnavus |
| 0 | 8 | 0 | G | 1506553 | Lachnoclostridium |
| 0 | 5 | 5 | S | 29347 | [Clostridium] scindens |
| 0 | 3 | 3 | S | 1871021 | Lachnoclostridium phocaeense |
| 0 | 7 | 0 | G | 207244 | Anaerostipes |
| 0 | 5 | 5 | S | 649756 | Anaerostipes hadrus |
| 0 | 2 | 2 | S | 105841 | Anaerostipes caccae |
| 0 | 6 | 0 | G | 33042 | Coprococcus |
| 0 | 4 | 0 | G1 | 2684943 | unclassified Coprococcus |
| 0 | 4 | 4 | S | 751585 | Coprococcus sp. ART55/1 |
| 0 | 2 | 2 | S | 116085 | Coprococcus catus |

| | | | | | |
|---|---|---|---|---|---|
| 0 | 3 | 0 | G | 830 | Butyrivibrio |
| 0 | 3 | 3 | S | 831 | Butyrivibrio fibrisolvens |
| 0 | 2 | 0 | G | 2719313 | Enterocloster |
| 0 | 2 | 2 | S | 208479 | Enterocloster bolteae |
| 0 | 1 | 0 | G | 2569097 | Anaerobutyricum |
| 0 | 1 | 1 | S | 39488 | Anaerobutyricum hallii |
| 0 | 1 | 0 | G | 2719231 | Lacrimispora |
| 0 | 1 | 0 | S | 84030 | Lacrimispora saccharolytica |
| 0 | 1 | 1 | S1 | 717608 | [Clostridium] cf. saccharolyticum K10 |
| 0.01 | 17 | 0 | O1 | 538999 | Eubacteriales incertae sedis |
| 0 | 8 | 0 | G | 2039302 | Monoglobus |
| 0 | 8 | 8 | S | 1981510 | Monoglobus pectinilyticus |
| 0 | 7 | 0 | G | 1918454 | Flintibacter |
| 0 | 7 | 0 | G1 | 2610894 | unclassified Flintibacter |
| 0 | 7 | 7 | S | 2610895 | Flintibacter sp. KGMB00164 |
| 0 | 2 | 0 | G | 1392389 | Intestinimonas |
| 0 | 2 | 2 | S | 1297617 | Intestinimonas butyriciproducens |
| 0.01 | 11 | 0 | F | 31979 | Clostridiaceae |
| 0.01 | 11 | 4 | G | 1485 | Clostridium |
| 0 | 3 | 3 | S | 1504 | Clostridium septicum |
| 0 | 2 | 2 | S | 1491 | Clostridium botulinum |
| 0 | 2 | 2 | S | 1502 | Clostridium perfringens |
| 0 | 2 | 1 | F | 186804 | Peptostreptococcaceae |
| 0 | 1 | 0 | G | 1870884 | Clostridioides |
| 0 | 1 | 1 | S | 1496 | Clostridioides difficile |
| 0 | 2 | 0 | F | 990719 | Christensenellaceae |
| 0 | 2 | 0 | G | 990721 | Christensenella |
| 0 | 2 | 2 | S | 626937 | Christensenella minuta |
| 0 | 1 | 0 | F | 186806 | Eubacteriaceae |
| 0 | 1 | 0 | G | 1730 | Eubacterium |
| 0 | 1 | 1 | S | 2041044 | Eubacterium maltosivorans |
| 0.14 | 260 | 0 | C | 526524 | Erysipelotrichia |
| 0.14 | 260 | 0 | O | 526525 | Erysipelotrichales |

| | | | | | |
|---|---|---|---|---|---|
| 0.14 | 258 | 0 | F | 2810281 | Turicibacteraceae |
| 0.14 | 258 | 0 | G | 191303 | Turicibacter |
| 0.09 | 168 | 168 | S | 154288 | Turicibacter sanguinis |
| 0.05 | 90 | 0 | G1 | 2638206 | unclassified Turicibacter |
| 0.05 | 90 | 90 | S | 1712675 | Turicibacter sp. H121 |
| 0 | 2 | 0 | F | 128827 | Erysipelotrichaceae |
| 0 | 1 | 0 | G | 1647 | Erysipelothrix |
| 0 | 1 | 1 | S | 1514105 | Erysipelothrix larvae |
| 0 | 1 | 0 | G | 1505663 | Erysipelatoclostridium |
| 0 | 1 | 1 | S | 1522 | [Clostridium] innocuum |
| 0.01 | 17 | 0 | C | 909932 | Negativicutes |
| 0.01 | 10 | 0 | O | 1843489 | Veillonellales |
| 0.01 | 10 | 0 | F | 31977 | Veillonellaceae |
| 0.01 | 10 | 0 | G | 29465 | Veillonella |
| 0 | 5 | 5 | S | 39778 | Veillonella dispar |
| 0 | 4 | 4 | S | 29466 | Veillonella parvula |
| 0 | 1 | 1 | S | 2682456 | Veillonella nakazawae |
| 0 | 5 | 0 | O | 909929 | Selenomonadales |
| 0 | 5 | 0 | F | 1843491 | Selenomonadaceae |
| 0 | 5 | 0 | G | 158846 | Megamonas |
| 0 | 5 | 5 | S | 437897 | Megamonas funiformis |
| 0 | 2 | 0 | O | 1843488 | Acidaminococcales |
| 0 | 2 | 0 | F | 909930 | Acidaminococcaceae |
| 0 | 2 | 0 | G | 33024 | Phascolarctobacterium |
| 0 | 2 | 0 | G1 | 2639160 | unclassified Phascolarctobacterium |
| 0 | 2 | 2 | S | 2823317 | Phascolarctobacterium sp. Marseille-Q4147 |
| 0 | 7 | 3 | C | 1737404 | Tissierellia |
| 0 | 4 | 0 | O | 1737405 | Tissierellales |
| 0 | 4 | 0 | F | 1570339 | Peptoniphilaceae |
| 0 | 4 | 0 | G | 150022 | Finegoldia |
| 0 | 4 | 4 | S | 1260 | Finegoldia magna |
| 0.03 | 56 | 0 | P | 1297 | Deinococcus-Thermus |
| 0.03 | 56 | 0 | C | 188787 | Deinococci |

| | | | | | |
|---|---|---|---|---|---|
| 0.03 | 53 | 0 | O | 118964 | Deinococcales |
| 0.03 | 53 | 0 | F | 183710 | Deinococcaceae |
| 0.03 | 53 | 12 | G | 1298 | Deinococcus |
| 0.01 | 11 | 11 | S | 1211322 | Deinococcus metallilatus |
| 0 | 8 | 8 | S | 1768108 | Deinococcus actinosclerus |
| 0 | 6 | 5 | S | 980427 | Deinococcus wulumuqiensis |
| 0 | 1 | 1 | S1 | 1288484 | Deinococcus wulumuqiensis R12 |
| 0 | 5 | 0 | S | 309887 | Deinococcus maricopensis |
| 0 | 5 | 5 | S1 | 709986 | Deinococcus maricopensis DSM 21211 |
| 0 | 3 | 0 | S | 55148 | Deinococcus proteolyticus |
| 0 | 3 | 3 | S1 | 693977 | Deinococcus proteolyticus MRP |
| 0 | 2 | 0 | S | 310783 | Deinococcus deserti |
| 0 | 2 | 2 | S1 | 546414 | Deinococcus deserti VCD115 |
| 0 | 2 | 2 | S | 317577 | Deinococcus ficus |
| 0 | 2 | 0 | S | 432329 | Deinococcus peraridilitoris |
| 0 | 2 | 2 | S1 | 937777 | Deinococcus peraridilitoris DSM 19664 |
| 0 | 2 | 0 | G1 | 2623546 | unclassified Deinococcus |
| 0 | 2 | 2 | S | 2652443 | Deinococcus sp. AJ005 |
| 0 | 3 | 0 | O | 68933 | Thermales |
| 0 | 3 | 0 | F | 188786 | Thermaceae |
| 0 | 3 | 0 | G | 270 | Thermus |
| 0 | 2 | 0 | S | 37636 | Thermus scotoductus |
| 0 | 2 | 2 | S1 | 743525 | Thermus scotoductus SA-01 |
| 0 | 1 | 0 | S | 56957 | Thermus oshimai |
| 0 | 1 | 1 | S1 | 751945 | Thermus oshimai JL-2 |
| 0.01 | 26 | 0 | D2 | 1798711 | Cyanobacteria/Melainabacteria group |
| 0.01 | 26 | 0 | P | 1117 | Cyanobacteria |
| 0.01 | 14 | 0 | O | 1890424 | Synechococcales |
| 0.01 | 13 | 0 | F | 1890426 | Synechococcaceae |
| 0.01 | 13 | 0 | G | 1129 | Synechococcus |
| 0.01 | 13 | 6 | G1 | 2626047 | unclassified Synechococcus |
| 0 | 7 | 7 | S | 1916956 | Synechococcus sp. SynAce01 |
| 0 | 1 | 0 | F | 2303730 | Prochlorotrichaceae |

| | | | | | |
|---|---|---|---|---|---|
| 0 | 1 | 0 | G | 170610 | Halomicronema |
| 0 | 1 | 0 | S | 1209493 | Halomicronema hongdechloris |
| 0 | 1 | 1 | S1 | 1641165 | Halomicronema hongdechloris C2206 |
| 0.01 | 10 | 0 | P1 | 1301283 | Oscillatoriophycideae |
| 0.01 | 10 | 0 | O | 1150 | Oscillatoriales |
| 0 | 9 | 0 | F | 1892252 | Microcoleaceae |
| 0 | 9 | 0 | G | 44471 | Microcoleus |
| 0 | 9 | 0 | G1 | 2642155 | unclassified Microcoleus |
| 0 | 9 | 9 | S | 1173027 | Microcoleus sp. PCC 7113 |
| 0 | 1 | 0 | F | 1892254 | Oscillatoriaceae |
| 0 | 1 | 0 | G | 1155738 | Moorea |
| 0 | 1 | 0 | S | 1155739 | Moorea producens |
| 0 | 1 | 1 | S1 | 1454205 | Moorea producens JHB |
| 0 | 1 | 0 | O | 1161 | Nostocales |
| 0 | 1 | 0 | F | 1162 | Nostocaceae |
| 0 | 1 | 0 | G | 1177 | Nostoc |
| 0 | 1 | 0 | G1 | 2593658 | unclassified Nostoc |
| 0 | 1 | 1 | S | 1618022 | Nostoc sp. 'Lobaria pulmonaria (5183) cyanobiont' |
| 0 | 1 | 0 | C | 307596 | Gloeobacteria |
| 0 | 1 | 0 | O | 307595 | Gloeobacterales |
| 0 | 1 | 0 | F | 1890422 | Gloeobacteraceae |
| 0 | 1 | 0 | G | 33071 | Gloeobacter |
| 0 | 1 | 0 | S | 33072 | Gloeobacter violaceus |
| 0 | 1 | 1 | S1 | 251221 | Gloeobacter violaceus PCC 7421 |
| 0 | 2 | 0 | P | 200795 | Chloroflexi |
| 0 | 2 | 0 | C | 189775 | Thermomicrobia |
| 0 | 2 | 0 | C1 | 85000 | Sphaerobacteridae |
| 0 | 2 | 0 | O | 85001 | Sphaerobacterales |
| 0 | 2 | 0 | O1 | 255728 | Sphaerobacterineae |
| 0 | 2 | 0 | F | 85002 | Sphaerobacteraceae |
| 0 | 2 | 0 | G | 2056 | Sphaerobacter |
| 0 | 2 | 0 | S | 2057 | Sphaerobacter thermophilus |
| 0 | 2 | 2 | S1 | 479434 | Sphaerobacter thermophilus DSM 20745 |

| | | | | | |
|---|---|---|---|---|---|
| 0 | 1 | 0 | P | 544448 | Tenericutes |
| 0 | 1 | 0 | C | 31969 | Mollicutes |
| 0 | 1 | 0 | O | 186329 | Acholeplasmatales |
| 0 | 1 | 0 | F | 2146 | Acholeplasmataceae |
| 0 | 1 | 0 | G | 2147 | Acholeplasma |
| 0 | 1 | 1 | S | 2148 | Acholeplasma laidlawii |
| 3.77 | 6791 | 0 | D1 | 1783270 | FCB group |
| 3.77 | 6791 | 0 | D2 | 68336 | Bacteroidetes/Chlorobi group |
| 3.77 | 6791 | 17 | P | 976 | Bacteroidetes |
| 2.46 | 4437 | 0 | C | 200643 | Bacteroidia |
| 2.46 | 4437 | 280 | O | 171549 | Bacteroidales |
| 1.88 | 3381 | 0 | F | 815 | Bacteroidaceae |
| 1.88 | 3381 | 341 | G | 816 | Bacteroides |
| 1.24 | 2228 | 2228 | S | 818 | Bacteroides thetaiotaomicron |
| 0.22 | 401 | 399 | S | 817 | Bacteroides fragilis |
| 0 | 1 | 1 | S1 | 295405 | Bacteroides fragilis YCH46 |
| 0 | 1 | 1 | S1 | 862962 | Bacteroides fragilis 638R |
| 0.08 | 138 | 138 | S | 820 | Bacteroides uniformis |
| 0.07 | 127 | 14 | G1 | 2646097 | unclassified Bacteroides |
| 0.02 | 44 | 44 | S | 2785531 | Bacteroides sp. HF-162 |
| 0.01 | 26 | 26 | S | 2650157 | Bacteroides sp. HF-5287 |
| 0.01 | 18 | 18 | S | 2763022 | Bacteroides sp. M10 |
| 0.01 | 13 | 13 | S | 2528203 | Bacteroides sp. A1C1 |
| 0 | 8 | 8 | S | 2650158 | Bacteroides sp. HF-5141 |
| 0 | 4 | 4 | S | 2755405 | Bacteroides sp. CACC 737 |
| 0.04 | 69 | 69 | S | 28116 | Bacteroides ovatus |
| 0.02 | 29 | 29 | S | 28111 | Bacteroides eggerthii |
| 0.02 | 29 | 27 | S | 371601 | Bacteroides xylanisolvens |
| 0 | 2 | 2 | S1 | 657309 | Bacteroides xylanisolvens XB1A |
| 0.01 | 15 | 15 | S | 246787 | Bacteroides cellulosilyticus |
| 0 | 2 | 2 | S | 1796613 | Bacteroides caecimuris |
| 0 | 1 | 1 | S | 47678 | Bacteroides caccae |
| 0 | 1 | 1 | S | 329854 | Bacteroides intestinalis |

| | | | | | |
|---|---|---|---|---|---|
| 0.34 | 619 | 0 | O1 | 333046 | Bacteroidales incertae sedis |
| 0.34 | 619 | 71 | G | 909656 | Phocaeicola |
| 0.21 | 373 | 373 | S | 821 | Phocaeicola vulgatus |
| 0.09 | 167 | 80 | S | 357276 | Phocaeicola dorei |
| 0.05 | 87 | 87 | S1 | 997877 | Bacteroides dorei CL03T12C01 |
| 0 | 7 | 7 | S | 387090 | Phocaeicola coprophilus |
| 0 | 1 | 0 | S | 376805 | Phocaeicola salanitronis |
| 0 | 1 | 1 | S1 | 667015 | Phocaeicola salanitronis DSM 18170 |
| 0.03 | 57 | 0 | F | 171550 | Rikenellaceae |
| 0.03 | 57 | 4 | G | 239759 | Alistipes |
| 0.01 | 25 | 0 | S | 328813 | Alistipes onderdonkii |
| 0.01 | 25 | 25 | S1 | 2585117 | Alistipes onderdonkii subsp. vulgaris |
| 0.01 | 14 | 14 | S | 2585118 | Alistipes communis |
| 0 | 8 | 0 | S | 214856 | Alistipes finegoldii |
| 0 | 8 | 8 | S1 | 679935 | Alistipes finegoldii DSM 17242 |
| 0 | 2 | 0 | S | 328814 | Alistipes shahii |
| 0 | 2 | 2 | S1 | 717959 | Alistipes shahii WAL 8301 |
| 0 | 2 | 2 | S | 2364787 | Alistipes megaguti |
| 0 | 1 | 1 | S | 626932 | Alistipes indistinctus |
| 0 | 1 | 1 | S | 2585119 | Alistipes dispar |
| 0.03 | 57 | 0 | F | 1853231 | Odoribacteraceae |
| 0.02 | 34 | 0 | G | 283168 | Odoribacter |
| 0.02 | 34 | 34 | S | 28118 | Odoribacter splanchnicus |
| 0.01 | 23 | 9 | G | 574697 | Butyricimonas |
| 0.01 | 14 | 14 | S | 544645 | Butyricimonas virosa |
| 0.01 | 21 | 0 | F | 171552 | Prevotellaceae |
| 0.01 | 18 | 0 | G | 838 | Prevotella |
| 0 | 7 | 7 | S | 165179 | Prevotella copri |
| 0 | 6 | 6 | S | 28132 | Prevotella melaninogenica |
| 0 | 4 | 4 | S | 28135 | Prevotella oris |
| 0 | 1 | 0 | S | 589437 | Prevotella scopos |
| 0 | 1 | 1 | S1 | 1236518 | Prevotella scopos JCM 17725 |
| 0 | 3 | 0 | G | 577309 | Paraprevotella |

| | | | | | |
|---|---|---|---|---|---|
| 0 | 3 | 0 | S | 454155 | Paraprevotella xylaniphila |
| 0 | 3 | 3 | S1 | 762982 | Paraprevotella xylaniphila YIT 11841 |
| 0.01 | 19 | 0 | F | 2005525 | Tannerellaceae |
| 0.01 | 19 | 4 | G | 375288 | Parabacteroides |
| 0.01 | 14 | 14 | S | 823 | Parabacteroides distasonis |
| 0 | 1 | 1 | S | 328812 | Parabacteroides goldsteinii |
| 0 | 2 | 0 | F | 2005473 | Muribaculaceae |
| 0 | 2 | 0 | G | 2518495 | Duncaniella |
| 0 | 2 | 2 | S | 2518971 | Duncaniella dubosii |
| 0 | 1 | 0 | F | 171551 | Porphyromonadaceae |
| 0 | 1 | 0 | G | 836 | Porphyromonas |
| 0 | 1 | 1 | S | 837 | Porphyromonas gingivalis |
| 0.74 | 1334 | 0 | C | 117743 | Flavobacteriia |
| 0.74 | 1334 | 7 | O | 200644 | Flavobacteriales |
| 0.72 | 1295 | 11 | F | 2762318 | Weeksellaceae |
| 0.38 | 690 | 39 | F1 | 2782232 | Chryseobacterium group |
| 0.29 | 529 | 92 | G | 59732 | Chryseobacterium |
| 0.09 | 156 | 156 | S | 246 | Chryseobacterium balustinum |
| 0.08 | 143 | 143 | S | 254 | Chryseobacterium indoltheticum |
| 0.04 | 67 | 0 | G1 | 2593645 | unclassified Chryseobacterium |
| 0.01 | 11 | 11 | S | 2547600 | Chryseobacterium sp. NBC 122 |
| 0.01 | 10 | 10 | S | 1721091 | Chryseobacterium sp. IHB B 17019 |
| 0.01 | 10 | 10 | S | 2015076 | Chryseobacterium sp. T16E-39 |
| 0.01 | 10 | 10 | S | 2478663 | Chryseobacterium sp. 3008163 |
| 0 | 8 | 8 | S | 2487065 | Chryseobacterium sp. G0201 |
| 0 | 7 | 7 | S | 2724619 | Chryseobacterium sp. NEB161 |
| 0 | 6 | 6 | S | 2758037 | Chryseobacterium sp. cx-624 |
| 0 | 3 | 3 | S | 1932669 | Chryseobacterium sp. JV274 |
| 0 | 2 | 2 | S | 2039166 | Chryseobacterium sp. 6424 |
| 0.01 | 24 | 24 | S | 1493872 | Chryseobacterium shandongense |
| 0.01 | 21 | 21 | S | 1685010 | Chryseobacterium glaciei |
| 0.01 | 10 | 10 | S | 1241982 | Chryseobacterium nakagawai |
| 0 | 7 | 7 | S | 2754694 | Chryseobacterium manosquense |

| | | | | | |
|---|---|---|---|---|---|
| 0 | 4 | 4 | S | 250 | Chryseobacterium gleum |
| 0 | 3 | 3 | S | 1324352 | Chryseobacterium gallinarum |
| 0 | 1 | 1 | S | 253 | Chryseobacterium indologenes |
| 0 | 1 | 1 | S | 1124835 | Chryseobacterium carnipullorum |
| 0.05 | 95 | 0 | G | 2782229 | Epilithonimonas |
| 0.05 | 95 | 95 | S | 2487072 | Epilithonimonas vandammei |
| 0.01 | 26 | 0 | G | 2782231 | Kaistella |
| 0.01 | 23 | 23 | S | 421525 | Chryseobacterium haifense |
| 0 | 3 | 3 | S | 266748 | Kaistella antarctica |
| 0 | 1 | 0 | G | 2782228 | Planobacterium |
| 0 | 1 | 1 | S | 536441 | Chryseobacterium taklimakanense |
| 0.31 | 553 | 39 | G | 59734 | Empedobacter |
| 0.27 | 480 | 480 | S | 343874 | Empedobacter falsenii |
| 0.02 | 28 | 28 | S | 1628248 | Empedobacter stercoris |
| 0 | 6 | 6 | S | 247 | Empedobacter brevis |
| 0.02 | 29 | 0 | G | 501783 | Cloacibacterium |
| 0.02 | 29 | 29 | S | 237258 | Cloacibacterium normanense |
| 0 | 6 | 0 | G | 308865 | Elizabethkingia |
| 0 | 5 | 5 | S | 1756150 | Elizabethkingia ursingii |
| 0 | 1 | 1 | S | 1117645 | Elizabethkingia anophelis |
| 0 | 6 | 0 | G | 1433995 | Cruoricaptor |
| 0 | 6 | 6 | S | 1118202 | Cruoricaptor ignavus |
| 0.02 | 32 | 0 | F | 49546 | Flavobacteriaceae |
| 0.01 | 14 | 1 | G | 1016 | Capnocytophaga |
| 0 | 7 | 7 | S | 2708117 | Capnocytophaga endodontalis |
| 0 | 5 | 0 | G1 | 2640652 | unclassified Capnocytophaga |
| 0 | 5 | 5 | S | 2748316 | Capnocytophaga sp. oral taxon 902 |
| 0 | 1 | 1 | S | 1019 | Capnocytophaga sputigena |
| 0 | 9 | 0 | G | 237 | Flavobacterium |
| 0 | 5 | 5 | S | 2175091 | Flavobacterium album |
| 0 | 2 | 2 | S | 2172098 | Flavobacterium pallidum |
| 0 | 1 | 0 | G1 | 196869 | unclassified Flavobacterium |
| 0 | 1 | 1 | S | 2294119 | Flavobacterium sp. CJ74 |

| | | | | | |
|---|---|---|---|---|---|
| 0 | 1 | 1 | S | 1306519 | Flavobacterium commune |
| 0 | 4 | 0 | G | 363408 | Nonlabens |
| 0 | 4 | 4 | S | 331648 | Nonlabens spongiae |
| 0 | 3 | 0 | G | 76831 | Myroides |
| 0 | 3 | 3 | S | 76832 | Myroides odoratimimus |
| 0 | 2 | 0 | G | 104267 | Tenacibaculum |
| 0 | 2 | 0 | G1 | 2635139 | unclassified Tenacibaculum |
| 0 | 2 | 2 | S | 2358479 | Tenacibaculum sp. DSM 106434 |
| 0.36 | 643 | 0 | C | 768503 | Cytophagia |
| 0.36 | 643 | 1 | O | 768507 | Cytophagales |
| 0.35 | 629 | 2 | F | 1853232 | Hymenobacteraceae |
| 0.35 | 625 | 11 | G | 89966 | Hymenobacter |
| 0.33 | 587 | 25 | G1 | 2615202 | unclassified Hymenobacter |
| 0.28 | 506 | 506 | S | 2584940 | Hymenobacter sp. DG01 |
| 0.01 | 19 | 19 | S | 2735321 | Hymenobacter sp. TS19 |
| 0.01 | 14 | 14 | S | 2596915 | Hymenobacter sp. NBH84 |
| 0 | 7 | 7 | S | 1356852 | Hymenobacter sp. APR13 |
| 0 | 5 | 5 | S | 1385664 | Hymenobacter sp. DG25B |
| 0 | 4 | 4 | S | 2675877 | Hymenobacter sp. BRD67 |
| 0 | 4 | 4 | S | 2675878 | Hymenobacter sp. BRD128 |
| 0 | 1 | 1 | S | 1484116 | Hymenobacter sp. PAMC 26554 |
| 0 | 1 | 1 | S | 1484118 | Hymenobacter sp. PAMC 26628 |
| 0 | 1 | 1 | S | 2761579 | Hymenobacter sp. S2-20-2 |
| 0.01 | 10 | 10 | S | 2724192 | Hymenobacter russus |
| 0 | 8 | 0 | S | 1446467 | Hymenobacter swuensis |
| 0 | 8 | 8 | S1 | 1227739 | Hymenobacter swuensis DY53 |
| 0 | 6 | 6 | S | 2319843 | Hymenobacter oligotrophus |
| 0 | 3 | 3 | S | 1385715 | Hymenobacter qilianensis |
| 0 | 2 | 0 | G | 323449 | Pontibacter |
| 0 | 2 | 2 | S | 2694929 | Pontibacter russatus |
| 0 | 9 | 0 | F | 89373 | Cytophagaceae |
| 0 | 9 | 0 | G | 107 | Spirosoma |
| 0 | 8 | 8 | S | 2666025 | Spirosoma endbachense |

| | | | | | |
|---|---|---|---|---|---|
| 0 | 1 | 0 | G1 | 2621999 | unclassified Spirosoma |
| 0 | 1 | 1 | S | 2710596 | Spirosoma sp. PL0136 |
| 0 | 4 | 0 | F | 563798 | Cyclobacteriaceae |
| 0 | 4 | 0 | G | 390846 | Echinicola |
| 0 | 4 | 0 | S | 390884 | Echinicola vietnamensis |
| 0 | 4 | 4 | S1 | 926556 | Echinicola vietnamensis DSM 17526 |
| 0.19 | 350 | 0 | C | 117747 | Sphingobacteriia |
| 0.19 | 350 | 0 | O | 200666 | Sphingobacteriales |
| 0.19 | 350 | 0 | F | 84566 | Sphingobacteriaceae |
| 0.19 | 343 | 13 | G | 28453 | Sphingobacterium |
| 0.11 | 190 | 190 | S | 28454 | Sphingobacterium multivorum |
| 0.08 | 139 | 13 | G1 | 2609468 | unclassified Sphingobacterium |
| 0.06 | 112 | 112 | S | 2795738 | Sphingobacterium sp. UDSM-2020 |
| 0 | 7 | 7 | S | 2003121 | Sphingobacterium sp. G1-14 |
| 0 | 6 | 6 | S | 1933220 | Sphingobacterium sp. B29 |
| 0 | 1 | 1 | S | 1538644 | Sphingobacterium sp. ML3W |
| 0 | 1 | 1 | S | 371142 | Sphingobacterium daejeonense |
| 0 | 5 | 0 | G | 84567 | Pedobacter |
| 0 | 5 | 5 | S | 363852 | Pedobacter ginsengisoli |
| 0 | 2 | 0 | G | 1649482 | Pseudopedobacter |
| 0 | 2 | 0 | S | 151895 | Pseudopedobacter saltans |
| 0 | 2 | 2 | S1 | 762903 | Pseudopedobacter saltans DSM 12145 |
| 0.01 | 10 | 0 | C | 1853228 | Chitinophagia |
| 0.01 | 10 | 0 | O | 1853229 | Chitinophagales |
| 0.01 | 10 | 1 | F | 563835 | Chitinophagaceae |
| 0 | 6 | 0 | G | 79328 | Chitinophaga |
| 0 | 4 | 4 | S | 2029983 | Chitinophaga caeni |
| 0 | 2 | 2 | S | 2203219 | Chitinophaga alhagiae |
| 0 | 2 | 0 | G | 379899 | Niabella |
| 0 | 2 | 0 | S | 446683 | Niabella soli |
| 0 | 2 | 2 | S1 | 929713 | Niabella soli DSM 19437 |
| 0 | 1 | 0 | G | 1860196 | Pseudobacter |
| 0 | 1 | 1 | S | 661488 | Pseudobacter ginsenosidimutans |

| | | | | | |
|---|---|---|---|---|---|
| 0.01 | 20 | 0 | D1 | 1783257 | PVC group |
| 0.01 | 12 | 0 | P | 204428 | Chlamydiae |
| 0.01 | 12 | 1 | C | 204429 | Chlamydiia |
| 0.01 | 11 | 0 | O | 1963360 | Parachlamydiales |
| 0.01 | 11 | 11 | F | 92713 | Parachlamydiaceae |
| 0 | 4 | 0 | P | 74201 | Verrucomicrobia |
| 0 | 2 | 0 | C | 203494 | Verrucomicrobiae |
| 0 | 2 | 0 | O | 48461 | Verrucomicrobiales |
| 0 | 2 | 0 | F | 1647988 | Akkermansiaceae |
| 0 | 2 | 0 | G | 239934 | Akkermansia |
| 0 | 2 | 2 | S | 239935 | Akkermansia muciniphila |
| 0 | 1 | 0 | P1 | 326457 | Verrucomicrobia incertae sedis |
| 0 | 1 | 0 | G | 1541670 | Methylacidimicrobium |
| 0 | 1 | 0 | G1 | 2730358 | unclassified Methylacidimicrobium |
| 0 | 1 | 1 | S | 2730359 | Methylacidimicrobium sp. AP8 |
| 0 | 1 | 0 | C | 414999 | Opitutae |
| 0 | 1 | 0 | O | 415000 | Opitutales |
| 0 | 1 | 0 | F | 134623 | Opitutaceae |
| 0 | 1 | 0 | G | 178440 | Opitutus |
| 0 | 1 | 0 | G1 | 2649488 | unclassified Opitutus |
| 0 | 1 | 1 | S | 1882749 | Opitutus sp. GAS368 |
| 0 | 4 | 0 | P | 203682 | Planctomycetes |
| 0 | 4 | 0 | C | 203683 | Planctomycetia |
| 0 | 4 | 0 | O | 2691355 | Gemmatales |
| 0 | 4 | 0 | F | 1914233 | Gemmataceae |
| 0 | 4 | 0 | G | 113 | Gemmata |
| 0 | 4 | 4 | S | 114 | Gemmata obscuriglobus |
| 0.01 | 11 | 0 | P | 203691 | Spirochaetes |
| 0.01 | 11 | 0 | C | 203692 | Spirochaetia |
| 0 | 8 | 0 | O | 1643686 | Brachyspirales |
| 0 | 8 | 0 | F | 143786 | Brachyspiraceae |
| 0 | 8 | 0 | G | 29521 | Brachyspira |
| 0 | 8 | 8 | S | 52584 | Brachyspira pilosicoli |

| | | | | | |
|---|---|---|---|---|---|
| 0 | 3 | 0 | O | 136 | Spirochaetales |
| 0 | 3 | 0 | F | 137 | Spirochaetaceae |
| 0 | 2 | 2 | G | 157 | Treponema |
| 0 | 1 | 0 | G | 146 | Spirochaeta |
| 0 | 1 | 0 | S | 46355 | Spirochaeta africana |
| 0 | 1 | 1 | S1 | 889378 | Spirochaeta africana DSM 8902 |
| 0 | 8 | 0 | P | 32066 | Fusobacteria |
| 0 | 8 | 0 | C | 203490 | Fusobacteriia |
| 0 | 8 | 0 | O | 203491 | Fusobacteriales |
| 0 | 8 | 0 | F | 203492 | Fusobacteriaceae |
| 0 | 8 | 0 | G | 848 | Fusobacterium |
| 0 | 8 | 0 | S | 851 | Fusobacterium nucleatum |
| 0 | 8 | 8 | S1 | 155615 | Fusobacterium nucleatum subsp. vincentii |
| 0 | 3 | 0 | P | 40117 | Nitrospirae |
| 0 | 3 | 0 | C | 203693 | Nitrospira |
| 0 | 3 | 0 | O | 189778 | Nitrospirales |
| 0 | 3 | 0 | F | 189779 | Nitrospiraceae |
| 0 | 3 | 0 | G | 1234 | Nitrospira |
| 0 | 3 | 3 | S | 42253 | Nitrospira moscoviensis |
| 0 | 2 | 0 | D1 | 2323 | Bacteria incertae sedis |
| 0 | 2 | 0 | D2 | 1783234 | Bacteria candidate phyla |
| 0 | 2 | 0 | D3 | 95901 | Candidatus Dependentiae |
| 0 | 2 | 0 | C | 2497643 | Candidatus Babeliae |
| 0 | 2 | 0 | O | 2497644 | Candidatus Babeliales |
| 0 | 2 | 0 | F | 2497645 | Candidatus Babeliaceae |
| 0 | 2 | 0 | G | 1551504 | Candidatus Babela |
| 0 | 2 | 2 | S | 673862 | Candidatus Babela massiliensis |
| 1.58 | 2844 | 21 | D | 2759 | Eukaryota |
| 1.35 | 2424 | 0 | D1 | 33154 | Opisthokonta |
| 0.86 | 1553 | 0 | K | 4751 | Fungi |
| 0.86 | 1553 | 0 | K1 | 451864 | Dikarya |
| 0.86 | 1541 | 0 | P | 5204 | Basidiomycota |
| 0.86 | 1541 | 1 | P1 | 452284 | Ustilaginomycotina |

| | | | | | |
|---|---|---|---|---|---|
| 0.84 | 1519 | 0 | C | 1538075 | Malasseziomycetes |
| 0.84 | 1519 | 0 | O | 162474 | Malasseziales |
| 0.84 | 1519 | 0 | F | 742845 | Malasseziaceae |
| 0.84 | 1519 | 0 | G | 55193 | Malassezia |
| 0.84 | 1519 | 1519 | S | 76775 | Malassezia restricta |
| 0.01 | 21 | 0 | C | 5257 | Ustilaginomycetes |
| 0.01 | 21 | 0 | O | 5267 | Ustilaginales |
| 0.01 | 21 | 0 | F | 5268 | Ustilaginaceae |
| 0.01 | 17 | 0 | G | 63265 | Sporisorium |
| 0.01 | 17 | 17 | S | 280036 | Sporisorium graminicola |
| 0 | 4 | 0 | G | 5269 | Ustilago |
| 0 | 4 | 0 | S | 5270 | Ustilago maydis |
| 0 | 4 | 4 | S1 | 237631 | Ustilago maydis 521 |
| 0.01 | 12 | 0 | P | 4890 | Ascomycota |
| 0.01 | 12 | 0 | P1 | 716545 | saccharomyceta |
| 0 | 9 | 0 | P2 | 147537 | Saccharomycotina |
| 0 | 9 | 0 | C | 4891 | Saccharomycetes |
| 0 | 9 | 0 | O | 4892 | Saccharomycetales |
| 0 | 7 | 0 | F | 766764 | Debaryomycetaceae |
| 0 | 7 | 0 | G | 4958 | Debaryomyces |
| 0 | 7 | 0 | S | 4959 | Debaryomyces hansenii |
| 0 | 7 | 7 | S1 | 284592 | Debaryomyces hansenii CBS767 |
| 0 | 2 | 0 | F | 4893 | Saccharomycetaceae |
| 0 | 2 | 0 | G | 4930 | Saccharomyces |
| 0 | 2 | 0 | S | 4932 | Saccharomyces cerevisiae |
| 0 | 2 | 2 | S1 | 559292 | Saccharomyces cerevisiae S288C |
| 0 | 3 | 0 | P2 | 147538 | Pezizomycotina |
| 0 | 3 | 0 | P3 | 716546 | leotiomyceta |
| 0 | 2 | 0 | P4 | 715989 | sordariomyceta |
| 0 | 1 | 0 | C | 147548 | Leotiomycetes |
| 0 | 1 | 0 | O | 5178 | Helotiales |
| 0 | 1 | 0 | F | 28983 | Sclerotiniaceae |
| 0 | 1 | 0 | G | 33196 | Botrytis |

| | | | | | |
|---|---|---|---|---|---|
| 0 | 1 | 0 | S | 40559 | Botrytis cinerea |
| 0 | 1 | 1 | S1 | 332648 | Botrytis cinerea B05.10 |
| 0 | 1 | 0 | C | 147550 | Sordariomycetes |
| 0 | 1 | 0 | C1 | 222543 | Hypocreomycetidae |
| 0 | 1 | 0 | O | 5125 | Hypocreales |
| 0 | 1 | 0 | F | 474942 | Ophiocordycipitaceae |
| 0 | 1 | 0 | G | 98402 | Drechmeria |
| 0 | 1 | 1 | S | 98403 | Drechmeria coniospora |
| 0 | 1 | 0 | P4 | 715962 | dothideomyceta |
| 0 | 1 | 0 | C | 147541 | Dothideomycetes |
| 0 | 1 | 0 | C1 | 451867 | Dothideomycetidae |
| 0 | 1 | 0 | O | 2726947 | Mycosphaerellales |
| 0 | 1 | 0 | F | 93133 | Mycosphaerellaceae |
| 0 | 1 | 0 | G | 29002 | Cercospora |
| 0 | 1 | 1 | S | 122368 | Cercospora beticola |
| 0.48 | 871 | 0 | K | 33208 | Metazoa |
| 0.48 | 871 | 0 | K1 | 6072 | Eumetazoa |
| 0.48 | 871 | 0 | K2 | 33213 | Bilateria |
| 0.48 | 871 | 0 | K3 | 33511 | Deuterostomia |
| 0.48 | 871 | 0 | P | 7711 | Chordata |
| 0.48 | 871 | 0 | P1 | 89593 | Craniata |
| 0.48 | 871 | 0 | P2 | 7742 | Vertebrata |
| 0.48 | 871 | 0 | P3 | 7776 | Gnathostomata |
| 0.48 | 871 | 0 | P4 | 117570 | Teleostomi |
| 0.48 | 871 | 0 | P5 | 117571 | Euteleostomi |
| 0.48 | 871 | 0 | P6 | 8287 | Sarcopterygii |
| 0.48 | 871 | 0 | C | 1338369 | Dipnotetrapodomorpha |
| 0.48 | 871 | 0 | C1 | 32523 | Tetrapoda |
| 0.48 | 871 | 0 | C2 | 32524 | Amniota |
| 0.48 | 871 | 0 | C | 40674 | Mammalia |
| 0.48 | 871 | 0 | C1 | 32525 | Theria |
| 0.48 | 871 | 0 | C2 | 9347 | Eutheria |
| 0.48 | 871 | 0 | C3 | 1437010 | Boreoeutheria |

| | | | | | |
|---|---|---|---|---|---|
| 0.48 | 871 | 0 | C4 | 314146 | Euarchontoglires |
| 0.48 | 871 | 0 | O | 9443 | Primates |
| 0.48 | 871 | 0 | O1 | 376913 | Haplorrhini |
| 0.48 | 871 | 0 | O2 | 314293 | Simiiformes |
| 0.48 | 871 | 0 | O3 | 9526 | Catarrhini |
| 0.48 | 871 | 0 | O4 | 314295 | Hominoidea |
| 0.48 | 871 | 0 | F | 9604 | Hominidae |
| 0.48 | 871 | 0 | F1 | 207598 | Homininae |
| 0.48 | 871 | 0 | G | 9605 | Homo |
| 0.48 | 871 | 871 | S | 9606 | Homo sapiens |
| 0.2 | 359 | 0 | K | 33090 | Viridiplantae |
| 0.2 | 359 | 0 | P | 35493 | Streptophyta |
| 0.2 | 359 | 0 | P1 | 131221 | Streptophytina |
| 0.2 | 359 | 0 | P2 | 3193 | Embryophyta |
| 0.2 | 355 | 0 | P3 | 58023 | Tracheophyta |
| 0.2 | 355 | 0 | P4 | 78536 | Euphyllophyta |
| 0.2 | 355 | 0 | P5 | 58024 | Spermatophyta |
| 0.2 | 355 | 0 | C | 3398 | Magnoliopsida |
| 0.19 | 350 | 1 | C1 | 1437183 | Mesangiospermae |
| 0.1 | 185 | 0 | C2 | 71240 | eudicotyledons |
| 0.1 | 185 | 0 | C3 | 91827 | Gunneridae |
| 0.1 | 185 | 4 | C4 | 1437201 | Pentapetalae |
| 0.07 | 124 | 0 | C5 | 71275 | rosids |
| 0.06 | 110 | 3 | C6 | 91835 | fabids |
| 0.05 | 84 | 0 | O | 72025 | Fabales |
| 0.05 | 84 | 0 | F | 3803 | Fabaceae |
| 0.05 | 84 | 0 | F1 | 3814 | Papilionoideae |
| 0.05 | 84 | 0 | F2 | 2231393 | 50 kb inversion clade |
| 0.04 | 79 | 0 | F3 | 2231382 | NPAAA clade |
| 0.04 | 78 | 0 | F4 | 2233855 | indigoferoid/millettioid clade |
| 0.04 | 78 | 0 | F5 | 163735 | Phaseoleae |
| 0.04 | 69 | 0 | G | 3846 | Glycine |
| 0.04 | 69 | 0 | G1 | 1462606 | Glycine subgen. Soja |

| | | | | | |
|---|---|---|---|---|---|
| 0.04 | 69 | 69 | S | 3848 | Glycine soja |
| 0 | 8 | 0 | G | 3820 | Cajanus |
| 0 | 8 | 8 | S | 3821 | Cajanus cajan |
| 0 | 1 | 0 | G | 3883 | Phaseolus |
| 0 | 1 | 1 | S | 3885 | Phaseolus vulgaris |
| 0 | 1 | 0 | F4 | 2233838 | Hologalegina |
| 0 | 1 | 0 | F5 | 2233839 | IRL clade |
| 0 | 1 | 0 | F6 | 163742 | Trifolieae |
| 0 | 1 | 0 | G | 3877 | Medicago |
| 0 | 1 | 1 | S | 3880 | Medicago truncatula |
| 0 | 5 | 0 | F3 | 2231387 | dalbergioids sensu lato |
| 0 | 5 | 0 | F4 | 163725 | Dalbergieae |
| 0 | 5 | 0 | F5 | 2231390 | Pterocarpus clade |
| 0 | 5 | 5 | G | 3817 | Arachis |
| 0.01 | 13 | 0 | O | 71239 | Cucurbitales |
| 0.01 | 13 | 3 | F | 3650 | Cucurbitaceae |
| 0 | 7 | 0 | F1 | 1003878 | Cucurbiteae |
| 0 | 7 | 0 | G | 3660 | Cucurbita |
| 0 | 7 | 0 | S | 3663 | Cucurbita pepo |
| 0 | 7 | 7 | S1 | 3664 | Cucurbita pepo subsp. pepo |
| 0 | 3 | 0 | F1 | 1003877 | Benincaseae |
| 0 | 3 | 0 | G | 102210 | Benincasa |
| 0 | 3 | 3 | S | 102211 | Benincasa hispida |
| 0 | 5 | 0 | O | 3502 | Fagales |
| 0 | 5 | 0 | F | 3503 | Fagaceae |
| 0 | 5 | 0 | G | 3511 | Quercus |
| 0 | 5 | 5 | S | 97700 | Quercus lobata |
| 0 | 5 | 0 | O | 3744 | Rosales |
| 0 | 5 | 0 | F | 3745 | Rosaceae |
| 0 | 5 | 0 | F1 | 171638 | Rosoideae |
| 0 | 5 | 0 | F2 | 721789 | Potentilleae |
| 0 | 5 | 0 | F3 | 1184124 | Fragariinae |
| 0 | 5 | 0 | G | 3746 | Fragaria |

| | | | | | |
|---|---|---|---|---|---|
| 0 | 5 | 0 | S | 57918 | Fragaria vesca |
| 0 | 5 | 5 | S1 | 101020 | Fragaria vesca subsp. vesca |
| 0.01 | 14 | 0 | C6 | 91836 | malvids |
| 0 | 5 | 0 | O | 41937 | Sapindales |
| 0 | 5 | 0 | F | 23513 | Rutaceae |
| 0 | 5 | 0 | F1 | 1728959 | Aurantioideae |
| 0 | 5 | 0 | G | 2706 | Citrus |
| 0 | 5 | 5 | S | 2711 | Citrus sinensis |
| 0 | 3 | 0 | O | 3699 | Brassicales |
| 0 | 3 | 0 | F | 3700 | Brassicaceae |
| 0 | 2 | 0 | F1 | 981071 | Brassiceae |
| 0 | 2 | 2 | G | 3705 | Brassica |
| 0 | 1 | 0 | F1 | 980083 | Camelineae |
| 0 | 1 | 0 | G | 71323 | Camelina |
| 0 | 1 | 1 | S | 90675 | Camelina sativa |
| 0 | 3 | 0 | O | 41938 | Malvales |
| 0 | 3 | 0 | F | 3629 | Malvaceae |
| 0 | 2 | 0 | F1 | 214907 | Malvoideae |
| 0 | 2 | 1 | G | 3633 | Gossypium |
| 0 | 1 | 1 | S | 29730 | Gossypium raimondii |
| 0 | 1 | 0 | F1 | 214909 | Byttnerioideae |
| 0 | 1 | 0 | G | 3640 | Theobroma |
| 0 | 1 | 1 | S | 3641 | Theobroma cacao |
| 0 | 3 | 0 | O | 41944 | Myrtales |
| 0 | 2 | 0 | F | 3931 | Myrtaceae |
| 0 | 2 | 0 | F1 | 1699513 | Myrtoideae |
| 0 | 2 | 0 | F2 | 1699524 | Eucalypteae |
| 0 | 2 | 0 | G | 3932 | Eucalyptus |
| 0 | 2 | 2 | S | 71139 | Eucalyptus grandis |
| 0 | 1 | 0 | F | 3928 | Lythraceae |
| 0 | 1 | 0 | G | 22662 | Punica |
| 0 | 1 | 1 | S | 22663 | Punica granatum |
| 0.03 | 57 | 0 | C5 | 71274 | asterids |

| | | | | | |
|---|---|---|---|---|---|
| 0.03 | 50 | 0 | C6 | 91888 | lamiids |
| 0.03 | 50 | 0 | O | 4069 | Solanales |
| 0.03 | 50 | 0 | F | 4070 | Solanaceae |
| 0.03 | 50 | 6 | F1 | 424551 | Solanoideae |
| 0.02 | 41 | 0 | F2 | 424564 | Capsiceae |
| 0.02 | 41 | 0 | G | 4071 | Capsicum |
| 0.02 | 41 | 41 | S | 4072 | Capsicum annuum |
| 0 | 3 | 0 | F2 | 424574 | Solaneae |
| 0 | 3 | 0 | G | 4107 | Solanum |
| 0 | 3 | 3 | G1 | 49274 | Solanum subgen. Lycopersicon |
| 0 | 7 | 0 | C6 | 91882 | campanulids |
| 0 | 4 | 0 | O | 4209 | Asterales |
| 0 | 4 | 0 | F | 4210 | Asteraceae |
| 0 | 2 | 0 | F1 | 102804 | Asteroideae |
| 0 | 2 | 0 | F2 | 911341 | Heliantheae alliance |
| 0 | 2 | 0 | F3 | 102814 | Heliantheae |
| 0 | 2 | 0 | G | 4231 | Helianthus |
| 0 | 2 | 2 | S | 4232 | Helianthus annuus |
| 0 | 2 | 0 | F1 | 219103 | Carduoideae |
| 0 | 2 | 0 | F2 | 102818 | Cardueae |
| 0 | 2 | 0 | F3 | 742010 | Carduinae |
| 0 | 2 | 0 | G | 4264 | Cynara |
| 0 | 2 | 0 | S | 4265 | Cynara cardunculus |
| 0 | 2 | 0 | S1 | 309979 | Cynara cardunculus subsp. cardunculus |
| 0 | 2 | 2 | S2 | 59895 | Cynara cardunculus var. scolymus |
| 0 | 3 | 0 | O | 4036 | Apiales |
| 0 | 3 | 0 | O1 | 364270 | Apiineae |
| 0 | 3 | 0 | F | 4037 | Apiaceae |
| 0 | 3 | 0 | F1 | 241778 | Apioideae |
| 0 | 3 | 0 | F2 | 241789 | Scandiceae |
| 0 | 3 | 0 | F3 | 241799 | Daucinae |
| 0 | 3 | 0 | G | 4038 | Daucus |
| 0 | 3 | 0 | G1 | 1873447 | Daucus sect. Daucus |

| | | | | | |
|---|---|---|---|---|---|
| 0 | 3 | 0 | S | 4039 | Daucus carota |
| 0 | 3 | 3 | S1 | 79200 | Daucus carota subsp. sativus |
| 0.07 | 126 | 0 | C2 | 4447 | Liliopsida |
| 0.07 | 126 | 0 | C3 | 1437197 | Petrosaviidae |
| 0.07 | 126 | 2 | C4 | 4734 | commelinids |
| 0.03 | 57 | 0 | O | 4618 | Zingiberales |
| 0.03 | 57 | 0 | F | 4637 | Musaceae |
| 0.03 | 57 | 0 | G | 4640 | Musa |
| 0.03 | 57 | 0 | S | 4641 | Musa acuminata |
| 0.03 | 57 | 57 | S1 | 214687 | Musa acuminata subsp. malaccensis |
| 0.03 | 51 | 0 | O | 38820 | Poales |
| 0.03 | 51 | 0 | F | 4479 | Poaceae |
| 0.03 | 47 | 0 | F1 | 359160 | BOP clade |
| 0.02 | 36 | 0 | F2 | 147368 | Pooideae |
| 0.02 | 34 | 0 | F3 | 1648038 | Triticodae |
| 0.02 | 34 | 0 | F4 | 147389 | Triticeae |
| 0.02 | 34 | 9 | F5 | 1648030 | Triticinae |
| 0.01 | 25 | 0 | G | 4564 | Triticum |
| 0.01 | 25 | 25 | S | 85692 | Triticum dicoccoides |
| 0 | 2 | 0 | F3 | 2822797 | Stipodae |
| 0 | 2 | 0 | F4 | 147385 | Brachypodieae |
| 0 | 2 | 0 | G | 15367 | Brachypodium |
| 0 | 2 | 2 | S | 15368 | Brachypodium distachyon |
| 0.01 | 11 | 0 | F2 | 147367 | Oryzoideae |
| 0.01 | 11 | 0 | F3 | 147380 | Oryzeae |
| 0.01 | 11 | 0 | F4 | 1648021 | Oryzinae |
| 0.01 | 11 | 0 | G | 4527 | Oryza |
| 0.01 | 11 | 0 | S | 4530 | Oryza sativa |
| 0.01 | 11 | 11 | S1 | 39947 | Oryza sativa Japonica Group |
| 0 | 4 | 0 | F1 | 147370 | PACMAD clade |
| 0 | 4 | 0 | F2 | 147369 | Panicoideae |
| 0 | 3 | 0 | F3 | 1648033 | Andropogonodae |
| 0 | 3 | 0 | F4 | 147429 | Andropogoneae |

| | | | | | |
|---|---|---|---|---|---|
| 0 | 3 | 0 | F5 | 1648029 | Tripsacinae |
| 0 | 3 | 0 | G | 4575 | Zea |
| 0 | 3 | 3 | S | 4577 | Zea mays |
| 0 | 1 | 0 | F3 | 1648036 | Panicodae |
| 0 | 1 | 0 | F4 | 147428 | Paniceae |
| 0 | 1 | 0 | F5 | 1293365 | Panicinae |
| 0 | 1 | 0 | G | 4539 | Panicum |
| 0 | 1 | 0 | G1 | 2100772 | Panicum sect. Panicum |
| 0 | 1 | 1 | S | 206008 | Panicum hallii |
| 0.01 | 16 | 0 | O | 40551 | Arecales |
| 0.01 | 16 | 0 | F | 4710 | Arecaceae |
| 0.01 | 16 | 0 | F1 | 169697 | Arecoideae |
| 0.01 | 16 | 0 | F2 | 169705 | Cocoseae |
| 0.01 | 16 | 0 | F3 | 169729 | Elaeidinae |
| 0.01 | 16 | 0 | G | 51952 | Elaeis |
| 0.01 | 16 | 16 | S | 51953 | Elaeis guineensis |
| 0.02 | 38 | 0 | O | 41768 | Ranunculales |
| 0.02 | 38 | 0 | F | 3465 | Papaveraceae |
| 0.02 | 38 | 0 | F1 | 1462614 | Papaveroideae |
| 0.02 | 38 | 0 | G | 3468 | Papaver |
| 0.02 | 38 | 38 | S | 3469 | Papaver somniferum |
| 0 | 5 | 0 | O | 261007 | Nymphaeales |
| 0 | 5 | 0 | F | 4410 | Nymphaeaceae |
| 0 | 5 | 0 | G | 4418 | Nymphaea |
| 0 | 5 | 5 | S | 210225 | Nymphaea colorata |
| 0 | 4 | 0 | P3 | 3208 | Bryophyta |
| 0 | 4 | 0 | P4 | 404260 | Bryophytina |
| 0 | 4 | 0 | C | 3214 | Bryopsida |
| 0 | 4 | 0 | C1 | 114656 | Funariidae |
| 0 | 4 | 0 | O | 3215 | Funariales |
| 0 | 4 | 0 | F | 3216 | Funariaceae |
| 0 | 4 | 0 | G | 37414 | Physcomitrium |
| 0 | 4 | 4 | S | 3218 | Physcomitrium patens |

| | | | | | |
|---|---|---|---|---|---|
| 0.02 | 35 | 0 | D1 | 2698737 | Sar |
| 0.02 | 35 | 0 | D2 | 33630 | Alveolata |
| 0.02 | 35 | 0 | P | 5794 | Apicomplexa |
| 0.02 | 33 | 0 | C | 1280412 | Conoidasida |
| 0.02 | 33 | 0 | C1 | 5796 | Coccidia |
| 0.02 | 33 | 0 | O | 75739 | Eucoccidiorida |
| 0.02 | 33 | 0 | O1 | 423054 | Eimeriorina |
| 0.02 | 33 | 0 | F | 5809 | Sarcocystidae |
| 0.02 | 33 | 0 | G | 5810 | Toxoplasma |
| 0.02 | 33 | 0 | S | 5811 | Toxoplasma gondii |
| 0.02 | 33 | 33 | S1 | 508771 | Toxoplasma gondii ME49 |
| 0 | 2 | 0 | C | 422676 | Aconoidasida |
| 0 | 2 | 0 | O | 5863 | Piroplasmida |
| 0 | 2 | 0 | F | 32594 | Babesiidae |
| 0 | 2 | 0 | G | 5864 | Babesia |
| 0 | 2 | 2 | S | 5866 | Babesia bigemina |
| 0 | 4 | 0 | D1 | 2611352 | Discoba |
| 0 | 4 | 0 | P | 33682 | Euglenozoa |
| 0 | 4 | 0 | C | 5653 | Kinetoplastea |
| 0 | 4 | 0 | C1 | 2704647 | Metakinetoplastina |
| 0 | 4 | 0 | O | 2704949 | Trypanosomatida |
| 0 | 4 | 0 | F | 5654 | Trypanosomatidae |
| 0 | 4 | 0 | F1 | 1286322 | Leishmaniinae |
| 0 | 4 | 0 | G | 5658 | Leishmania |
| 0 | 4 | 0 | G1 | 38568 | Leishmania |
| 0 | 4 | 0 | G2 | 38582 | Leishmania mexicana species complex |
| 0 | 4 | 0 | S | 5665 | Leishmania mexicana |
| 0 | 4 | 4 | S1 | 929439 | Leishmania mexicana MHOM/GT/2001/U1103 |
| 0 | 1 | 0 | P | 2763 | Rhodophyta |
| 0 | 1 | 0 | C | 2797 | Bangiophyceae |
| 0 | 1 | 0 | O | 265318 | Cyanidiales |
| 0 | 1 | 0 | F | 265316 | Cyanidiaceae |
| 0 | 1 | 0 | G | 45156 | Cyanidioschyzon |

| | | | | | |
|---|---|---|---|---|---|
| 0 | 1 | 0 | S | 45157 | Cyanidioschyzon merolae |
| 0 | 1 | 1 | S1 | 280699 | Cyanidioschyzon merolae strain 10D |
| 0 | 7 | 0 | D | 2157 | Archaea |
| 0 | 7 | 0 | P | 28890 | Euryarchaeota |
| 0 | 6 | 0 | P1 | 2290931 | Stenosarchaea group |
| 0 | 3 | 0 | C | 183963 | Halobacteria |
| 0 | 3 | 0 | O | 1644055 | Haloferacales |
| 0 | 3 | 0 | F | 1644056 | Haloferacaceae |
| 0 | 2 | 0 | G | 1073986 | Halobellus |
| 0 | 2 | 2 | S | 699433 | Halobellus limi |
| 0 | 1 | 0 | G | 376170 | Haloplanus |
| 0 | 1 | 1 | S | 660522 | Haloplanus aerogenes |
| 0 | 3 | 0 | C | 224756 | Methanomicrobia |
| 0 | 3 | 0 | O | 94695 | Methanosarcinales |
| 0 | 3 | 0 | F | 143067 | Methanotrichaceae |
| 0 | 3 | 0 | G | 2222 | Methanothrix |
| 0 | 3 | 0 | S | 2223 | Methanothrix soehngenii |
| 0 | 3 | 3 | S1 | 990316 | Methanothrix soehngenii GP6 |
| 0 | 1 | 0 | P1 | 2283794 | Methanomada group |
| 0 | 1 | 0 | C | 183925 | Methanobacteria |
| 0 | 1 | 0 | O | 2158 | Methanobacteriales |
| 0 | 1 | 0 | F | 2159 | Methanobacteriaceae |
| 0 | 1 | 0 | G | 2172 | Methanobrevibacter |
| 0 | 1 | 1 | S | 2173 | Methanobrevibacter smithii |
| 0.48 | 859 | 0 | D | 10239 | Viruses |
| 0.47 | 854 | 0 | D1 | 2731341 | Duplodnaviria |
| 0.47 | 854 | 0 | D2 | 2731360 | Heunggongvirae |
| 0.47 | 854 | 0 | P | 2731618 | Uroviricota |
| 0.47 | 854 | 0 | C | 2731619 | Caudoviricetes |
| 0.47 | 854 | 0 | O | 28883 | Caudovirales |
| 0.37 | 665 | 0 | F | 10744 | Podoviridae |
| 0.37 | 665 | 0 | F1 | 196895 | unclassified Podoviridae |
| 0.37 | 665 | 0 | F2 | 1978007 | crAss-like viruses |

| | | | | | |
|---|---|---|---|---|---|
| 0.37 | 665 | 0 | F3 | 2315857 | environmental samples |
| 0.37 | 665 | 665 | S | 1211417 | uncultured crAssphage |
| 0.1 | 176 | 0 | F | 10699 | Siphoviridae |
| 0.08 | 141 | 20 | G | 1623305 | Skunavirus |
| 0.06 | 117 | 56 | G1 | 2050979 | unclassified Skunavirus |
| 0 | 6 | 6 | S | 2029658 | Lactococcus phage 05601 |
| 0 | 4 | 4 | S | 2675246 | Lactococcus phage CHPC129 |
| 0 | 4 | 4 | S | 2029674 | Lactococcus phage 96401 |
| 0 | 4 | 4 | S | 2029664 | Lactococcus phage 51701 |
| 0 | 3 | 3 | S | 2029662 | Lactococcus phage 38503 |
| 0 | 3 | 3 | S | 1636570 | Lactococcus phage 936 group phage Phi5.12 |
| 0 | 3 | 3 | S | 1636566 | Lactococcus phage 936 group phage Phi4.2 |
| 0 | 2 | 2 | S | 2675257 | Lactococcus phage CHPC965 |
| 0 | 2 | 2 | S | 2029672 | Lactococcus phage 79201 |
| 0 | 2 | 2 | S | 2029671 | Lactococcus phage 66901 |
| 0 | 2 | 2 | S | 2029668 | Lactococcus phage 62601 |
| 0 | 2 | 2 | S | 2029666 | Lactococcus phage 56301 |
| 0 | 2 | 2 | S | 2029661 | Lactococcus phage 37201 |
| 0 | 2 | 2 | S | 2029659 | Lactococcus phage 16802 |
| 0 | 2 | 2 | S | 1874593 | Lactococcus phage i0139 |
| 0 | 2 | 2 | S | 1636573 | Lactococcus phage 936 group phage PhiB1127 |
| 0 | 1 | 1 | S | 39838 | Lactococcus phage 936 |
| 0 | 1 | 1 | S | 2675256 | Lactococcus phage CHPC964 |
| 0 | 1 | 1 | S | 2675251 | Lactococcus phage CHPC52 |
| 0 | 1 | 1 | S | 1636560 | Lactococcus phage 936 group phage Phi155 |
| 0 | 1 | 1 | S | 1636576 | Lactococcus phage 936 group phage PhiE1127 |
| 0 | 1 | 1 | S | 2675250 | Lactococcus phage CHPC362 |
| 0 | 1 | 1 | S | 2662298 | Lactococcus phage P656 |
| 0 | 1 | 1 | S | 2029675 | Lactococcus phage 96403 |
| 0 | 1 | 1 | S | 213777 | Lactococcus lactis phage p272 |
| 0 | 1 | 1 | S | 1636583 | Lactococcus phage 936 group phage PhiLj |
| 0 | 1 | 1 | S | 213782 | Lactococcus phage fd13 |
| 0 | 1 | 1 | S | 1262537 | Lactococcus phage P680 |

| | | | | | |
|---|---|---|---|---|---|
| 0 | 1 | 1 | S | 1636724 | Lactococcus phage 936 group phage PhiA.16 |
| 0 | 1 | 1 | S | 1289467 | Lactococcus phage CaseusJM1 |
| 0 | 1 | 1 | S | 2029660 | Lactococcus phage 30804 |
| 0 | 1 | 1 | S | 2027272 | Lactococcus phage LP0903 |
| 0 | 2 | 2 | S | 287412 | Lactococcus virus Sl4 |
| 0 | 2 | 2 | S | 665883 | Lactococcus virus CB13 |
| 0.02 | 28 | 22 | G | 1982251 | Pahexavirus |
| 0 | 3 | 0 | S | 1982287 | Propionibacterium virus PHL114L00 |
| 0 | 3 | 3 | S1 | 1235656 | Propionibacterium phage PHL114L00 |
| 0 | 2 | 0 | S | 1982297 | Propionibacterium virus PHL171M01 |
| 0 | 2 | 2 | S1 | 1500827 | Propionibacterium phage PHL171M01 |
| 0 | 1 | 0 | S | 1982282 | Propionibacterium virus PHL092M00 |
| 0 | 1 | 1 | S1 | 1500813 | Propionibacterium phage PHL092M00 |
| 0 | 7 | 3 | F1 | 196894 | unclassified Siphoviridae |
| 0 | 3 | 3 | S | 1105171 | Bacteroides phage B124-14 |
| 0 | 1 | 1 | S | 881953 | Lactococcus phage 949 |
| 0.01 | 13 | 0 | F | 10662 | Myoviridae |
| 0.01 | 12 | 0 | F1 | 196896 | unclassified Myoviridae |
| 0.01 | 12 | 12 | S | 12402 | Streptococcus phage EJ-1 |
| 0 | 1 | 0 | G | 2733134 | Taranisvirus |
| 0 | 1 | 0 | S | 2734146 | Faecalibacterium virus Taranis |
| 0 | 1 | 1 | S1 | 2070186 | Faecalibacterium phage FP_Taranis |
| 0 | 5 | 0 | D1 | 2731342 | Monodnaviria |
| 0 | 5 | 0 | K | 2732092 | Shotokuvirae |
| 0 | 5 | 0 | P | 2732415 | Cossaviricota |
| 0 | 5 | 0 | C | 2732421 | Papovaviricetes |
| 0 | 5 | 0 | O | 2732532 | Sepolyvirales |
| 0 | 5 | 0 | F | 151341 | Polyomaviridae |
| 0 | 5 | 0 | G | 1891713 | Alphapolyomavirus |
| 0 | 5 | 0 | S | 1891727 | Human polyomavirus 8 |
| 0 | 5 | 5 | S1 | 862909 | Trichodysplasia spinulosa-associated polyomavirus |

NC2

| | | | | | |
|---|---|---|---|---|---|
| 54.1 | 5029 | 5029 | U | 0 | unclassified |
| 45.9 | 4267 | 99 | R | 1 | root |
| 44.83 | 4167 | 5 | R1 | 131567 | cellular organisms |
| 42.99 | 3996 | 45 | D | 2 | Bacteria |
| 19.95 | 1855 | 7 | P | 1224 | Proteobacteria |
| 8.85 | 823 | 0 | C | 28216 | Betaproteobacteria |
| 8.84 | 822 | 22 | O | 80840 | Burkholderiales |
| 7.99 | 743 | 6 | F | 80864 | Comamonadaceae |
| 7.85 | 730 | 451 | G | 80865 | Delftia |
| 2.09 | 194 | 194 | S | 558537 | Delftia lacustris |
| 0.77 | 72 | 72 | S | 180282 | Delftia tsuruhatensis |
| 0.12 | 11 | 11 | S | 80866 | Delftia acidovorans |
| 0.02 | 2 | 0 | G1 | 2613839 | unclassified Delftia |
| 0.01 | 1 | 1 | S | 742013 | Delftia sp. Cs1-4 |
| 0.01 | 1 | 1 | S | 1920191 | Delftia sp. HK171 |
| 0.03 | 3 | 0 | G | 201096 | Alicycliphilus |
| 0.03 | 3 | 0 | S | 179636 | Alicycliphilus denitrificans |
| 0.03 | 3 | 3 | S1 | 596154 | Alicycliphilus denitrificans K601 |
| 0.02 | 2 | 0 | G | 215579 | Schlegelella |
| 0.02 | 2 | 2 | S | 215580 | Schlegelella thermodepolymerans |
| 0.01 | 1 | 0 | G | 34072 | Variovorax |
| 0.01 | 1 | 1 | S | 34073 | Variovorax paradoxus |
| 0.01 | 1 | 0 | G | 47420 | Hydrogenophaga |
| 0.01 | 1 | 1 | S | 1763535 | Hydrogenophaga crassostreae |
| 0.26 | 24 | 6 | F | 75682 | Oxalobacteraceae |
| 0.19 | 18 | 0 | G | 149698 | Massilia |
| 0.12 | 11 | 0 | G1 | 2609279 | unclassified Massilia |
| 0.09 | 8 | 8 | S | 2769491 | Massilia sp. LPB0304 |
| 0.02 | 2 | 2 | S | 2765360 | Massilia sp. CCM 8941 |
| 0.01 | 1 | 1 | S | 1707785 | Massilia sp. WG5 |
| 0.05 | 5 | 5 | S | 871742 | Massilia flava |
| 0.01 | 1 | 1 | S | 321985 | Massilia lutea |

| | | | | | |
|---|---|---|---|---|---|
| 0.01 | 1 | 1 | S | 1141883 | Massilia putida |
| 0.19 | 18 | 11 | F | 119060 | Burkholderiaceae |
| 0.08 | 7 | 5 | G | 106589 | Cupriavidus |
| 0.02 | 2 | 2 | S | 82633 | Cupriavidus pauculus |
| 0.12 | 11 | 0 | F | 995019 | Sutterellaceae |
| 0.12 | 11 | 0 | G | 40544 | Sutterella |
| 0.12 | 11 | 11 | S | 40545 | Sutterella wadsworthensis |
| 0.03 | 3 | 0 | F | 506 | Alcaligenaceae |
| 0.03 | 3 | 1 | G | 517 | Bordetella |
| 0.02 | 2 | 2 | S | 123899 | Bordetella trematum |
| 0.01 | 1 | 0 | O1 | 224471 | Burkholderiales genera incertae sedis |
| 0.01 | 1 | 1 | G | 32012 | Thiomonas |
| 0.01 | 1 | 0 | O | 206389 | Rhodocyclales |
| 0.01 | 1 | 0 | F | 2008794 | Zoogloeaceae |
| 0.01 | 1 | 0 | G | 33057 | Thauera |
| 0.01 | 1 | 1 | S | 96773 | Thauera chlorobenzoica |
| 7.87 | 732 | 18 | C | 28211 | Alphaproteobacteria |
| 5.02 | 467 | 0 | O | 204457 | Sphingomonadales |
| 4.99 | 464 | 15 | F | 41297 | Sphingomonadaceae |
| 4.59 | 427 | 19 | G | 13687 | Sphingomonas |
| 4.21 | 391 | 391 | S | 13689 | Sphingomonas paucimobilis |
| 0.05 | 5 | 0 | G1 | 196159 | unclassified Sphingomonas |
| 0.05 | 5 | 5 | S | 2698679 | Sphingomonas sp. C33 |
| 0.04 | 4 | 4 | S | 93064 | Sphingomonas koreensis |
| 0.03 | 3 | 3 | S | 152682 | Sphingomonas melonis |
| 0.03 | 3 | 0 | S | 397260 | Sphingomonas sanxanigenens |
| 0.03 | 3 | 3 | S1 | 1123269 | Sphingomonas sanxanigenens DSM 19645 = NX02 |
| 0.02 | 2 | 2 | S | 2698828 | Sphingomonas lacunae |
| 0.13 | 12 | 3 | G | 165695 | Sphingobium |
| 0.06 | 6 | 6 | S | 13690 | Sphingobium yanoikuyae |
| 0.03 | 3 | 3 | S | 135719 | Sphingobium amiense |
| 0.08 | 7 | 3 | G | 165696 | Novosphingobium |
| 0.02 | 2 | 2 | S | 158500 | Novosphingobium resinovorum |

| | | | | | |
|---|---|---|---|---|---|
| 0.02 | 2 | 0 | G1 | 2644732 | unclassified Novosphingobium |
| 0.02 | 2 | 2 | S | 164608 | Novosphingobium sp. KA1 |
| 0.02 | 2 | 0 | G | 1649486 | Rhizorhabdus |
| 0.02 | 2 | 2 | S | 1850238 | Rhizorhabdus dicambivorans |
| 0.01 | 1 | 0 | G | 165697 | Sphingopyxis |
| 0.01 | 1 | 1 | S | 33050 | Sphingopyxis macrogoltabida |
| 0.03 | 3 | 0 | F | 335929 | Erythrobacteraceae |
| 0.03 | 3 | 0 | F1 | 2800788 | Erythrobacter/Porphyrobacter group |
| 0.03 | 3 | 0 | G | 1041 | Erythrobacter |
| 0.03 | 3 | 0 | G1 | 2633097 | unclassified Erythrobacter |
| 0.03 | 3 | 3 | S | 2785907 | Erythrobacter sp. A30-3 |
| 1.91 | 178 | 1 | O | 356 | Hyphomicrobiales |
| 1.61 | 150 | 27 | F | 119045 | Methylobacteriaceae |
| 1.17 | 109 | 14 | G | 2282523 | Methylorubrum |
| 0.55 | 51 | 42 | S | 223967 | Methylorubrum populi |
| 0.1 | 9 | 9 | S1 | 441620 | Methylorubrum populi BJ001 |
| 0.47 | 44 | 7 | S | 408 | Methylorubrum extorquens |
| 0.29 | 27 | 27 | S1 | 272630 | Methylorubrum extorquens AM1 |
| 0.11 | 10 | 10 | S1 | 440085 | Methylorubrum extorquens CM4 |
| 0.15 | 14 | 1 | G | 407 | Methylobacterium |
| 0.11 | 10 | 4 | G1 | 2615210 | unclassified Methylobacterium |
| 0.06 | 6 | 6 | S | 2067957 | Methylobacterium sp. DM1 |
| 0.02 | 2 | 2 | S | 270351 | Methylobacterium aquaticum |
| 0.01 | 1 | 1 | S | 2202825 | Methylobacterium durans |
| 0.11 | 10 | 0 | F | 82115 | Rhizobiaceae |
| 0.06 | 6 | 0 | F1 | 227292 | Sinorhizobium/Ensifer group |
| 0.06 | 6 | 0 | G | 28105 | Sinorhizobium |
| 0.06 | 6 | 0 | G1 | 663276 | Sinorhizobium fredii group |
| 0.06 | 6 | 6 | S | 380 | Sinorhizobium fredii |
| 0.04 | 4 | 0 | F1 | 227290 | Rhizobium/Agrobacterium group |
| 0.02 | 2 | 1 | G | 357 | Agrobacterium |
| 0.01 | 1 | 1 | S | 359 | Agrobacterium rhizogenes |
| 0.02 | 2 | 0 | G | 1525371 | Neorhizobium |

| | | | | | |
|---|---|---|---|---|---|
| 0.02 | 2 | 0 | G1 | 2629175 | unclassified Neorhizobium |
| 0.02 | 2 | 2 | S | 1825976 | Neorhizobium sp. NCHU2750 |
| 0.06 | 6 | 0 | F | 41294 | Bradyrhizobiaceae |
| 0.04 | 4 | 2 | G | 374 | Bradyrhizobium |
| 0.02 | 2 | 0 | G1 | 2631580 | unclassified Bradyrhizobium |
| 0.02 | 2 | 2 | S | 319017 | Bradyrhizobium sp. WSM471 |
| 0.02 | 2 | 0 | G | 85413 | Bosea |
| 0.02 | 2 | 0 | G1 | 2653178 | unclassified Bosea |
| 0.02 | 2 | 2 | S | 1867715 | Bosea sp. Tri-49 |
| 0.06 | 6 | 0 | F | 69277 | Phyllobacteriaceae |
| 0.06 | 6 | 6 | G | 68287 | Mesorhizobium |
| 0.03 | 3 | 0 | F | 118882 | Brucellaceae |
| 0.03 | 3 | 0 | F1 | 2826938 | Brucella/Ochrobactrum group |
| 0.03 | 3 | 0 | G | 234 | Brucella |
| 0.03 | 3 | 3 | S | 271865 | [Ochrobactrum] quorumnocens |
| 0.01 | 1 | 0 | F | 31993 | Methylocystaceae |
| 0.01 | 1 | 0 | G | 133 | Methylocystis |
| 0.01 | 1 | 1 | S | 391905 | Methylocystis heyeri |
| 0.01 | 1 | 0 | F | 45401 | Hyphomicrobiaceae |
| 0.01 | 1 | 0 | G | 1484898 | Methyloceanibacter |
| 0.01 | 1 | 0 | G1 | 2617503 | unclassified Methyloceanibacter |
| 0.01 | 1 | 1 | S | 2170729 | Methyloceanibacter sp. wino2 |
| 0.48 | 45 | 0 | O | 204455 | Rhodobacterales |
| 0.48 | 45 | 19 | F | 31989 | Rhodobacteraceae |
| 0.12 | 11 | 0 | G | 1060 | Rhodobacter |
| 0.12 | 11 | 0 | G1 | 196779 | unclassified Rhodobacter |
| 0.12 | 11 | 11 | S | 1850250 | Rhodobacter sp. LPB0142 |
| 0.05 | 5 | 3 | G | 265 | Paracoccus |
| 0.01 | 1 | 1 | S | 135740 | Paracoccus kondratievae |
| 0.01 | 1 | 0 | G1 | 246570 | unclassified Paracoccus |
| 0.01 | 1 | 1 | S | 2500532 | Paracoccus sp. Arc7-R13 |
| 0.04 | 4 | 0 | G | 366614 | Haematobacter |
| 0.04 | 4 | 4 | S | 195105 | Haematobacter massiliensis |

| | | | | | |
|---|---|---|---|---|---|
| 0.02 | 2 | 0 | G | 1653176 | Cereibacter |
| 0.02 | 2 | 2 | S | 1063 | Cereibacter sphaeroides |
| 0.02 | 2 | 0 | G | 74030 | Roseovarius |
| 0.02 | 2 | 2 | G1 | 2614913 | unclassified Roseovarius |
| 0.01 | 1 | 0 | G | 53945 | Octadecabacter |
| 0.01 | 1 | 0 | S | 53946 | Octadecabacter arcticus |
| 0.01 | 1 | 1 | S1 | 391616 | Octadecabacter arcticus 238 |
| 0.01 | 1 | 0 | G | 204456 | Gemmobacter |
| 0.01 | 1 | 0 | G1 | 2636157 | unclassified Gemmobacter |
| 0.01 | 1 | 1 | S | 2169400 | Gemmobacter sp. HYN0069 |
| 0.24 | 22 | 0 | O | 204458 | Caulobacterales |
| 0.24 | 22 | 1 | F | 76892 | Caulobacteraceae |
| 0.19 | 18 | 1 | G | 41275 | Brevundimonas |
| 0.16 | 15 | 0 | G1 | 2622653 | unclassified Brevundimonas |
| 0.06 | 6 | 6 | S | 1532555 | Brevundimonas sp. DS20 |
| 0.05 | 5 | 5 | S | 2752515 | Brevundimonas sp. AJA228-03 |
| 0.02 | 2 | 2 | S | 2579977 | Brevundimonas sp. SGAir0440 |
| 0.02 | 2 | 2 | S | 2774189 | Brevundimonas sp. LVF1 |
| 0.02 | 2 | 2 | S | 293 | Brevundimonas diminuta |
| 0.03 | 3 | 1 | G | 75 | Caulobacter |
| 0.02 | 2 | 0 | G1 | 2648921 | unclassified Caulobacter |
| 0.02 | 2 | 2 | S | 69665 | Caulobacter sp. FWC26 |
| 0.01 | 1 | 0 | O | 204441 | Rhodospirillales |
| 0.01 | 1 | 0 | F | 433 | Acetobacteraceae |
| 0.01 | 1 | 0 | G | 1654741 | Bombella |
| 0.01 | 1 | 0 | G1 | 2644098 | unclassified Bombella |
| 0.01 | 1 | 1 | S | 2697033 | Bombella sp. KACC 21507 |
| 0.01 | 1 | 0 | O | 255473 | Parvularculales |
| 0.01 | 1 | 0 | F | 255474 | Parvularculaceae |
| 0.01 | 1 | 0 | G | 208215 | Parvularcula |
| 0.01 | 1 | 0 | S | 208216 | Parvularcula bermudensis |
| 0.01 | 1 | 1 | S1 | 314260 | Parvularcula bermudensis HTCC2503 |
| 3 | 279 | 0 | C | 1236 | Gammaproteobacteria |

| | | | | | |
|---|---|---|---|---|---|
| 1.96 | 182 | 2 | O | 135614 | Xanthomonadales |
| 1.94 | 180 | 0 | F | 32033 | Xanthomonadaceae |
| 1.93 | 179 | 31 | G | 40323 | Stenotrophomonas |
| 1.55 | 144 | 0 | G1 | 995085 | Stenotrophomonas maltophilia group |
| 1.55 | 144 | 137 | S | 40324 | Stenotrophomonas maltophilia |
| 0.08 | 7 | 7 | S1 | 391008 | Stenotrophomonas maltophilia R551-3 |
| 0.03 | 3 | 0 | G1 | 196198 | unclassified Stenotrophomonas |
| 0.03 | 3 | 3 | S | 2282124 | Stenotrophomonas sp. ASS1 |
| 0.01 | 1 | 1 | S | 128780 | Stenotrophomonas acidaminiphila |
| 0.01 | 1 | 0 | G | 68 | Lysobacter |
| 0.01 | 1 | 1 | S | 1324796 | Lysobacter lycopersici |
| 0.9 | 84 | 0 | O | 72274 | Pseudomonadales |
| 0.55 | 51 | 0 | F | 468 | Moraxellaceae |
| 0.55 | 51 | 12 | G | 469 | Acinetobacter |
| 0.16 | 15 | 15 | S | 28090 | Acinetobacter lwoffii |
| 0.12 | 11 | 0 | S | 52133 | Acinetobacter venetianus |
| 0.12 | 11 | 11 | S1 | 1197884 | Acinetobacter venetianus VE-C3 |
| 0.08 | 7 | 7 | S | 40214 | Acinetobacter johnsonii |
| 0.04 | 4 | 0 | G1 | 196816 | unclassified Acinetobacter |
| 0.03 | 3 | 3 | S | 2725684 | Acinetobacter sp. NEB149 |
| 0.01 | 1 | 1 | S | 2004644 | Acinetobacter sp. WCHA45 |
| 0.02 | 2 | 0 | G1 | 909768 | Acinetobacter calcoaceticus/baumannii complex |
| 0.02 | 2 | 2 | S | 470 | Acinetobacter baumannii |
| 0.35 | 33 | 0 | F | 135621 | Pseudomonadaceae |
| 0.35 | 33 | 2 | G | 286 | Pseudomonas |
| 0.22 | 20 | 0 | G1 | 136841 | Pseudomonas aeruginosa group |
| 0.22 | 20 | 20 | S | 287 | Pseudomonas aeruginosa |
| 0.05 | 5 | 0 | G1 | 136843 | Pseudomonas fluorescens group |
| 0.05 | 5 | 5 | S | 294 | Pseudomonas fluorescens |
| 0.03 | 3 | 3 | S | 157782 | Pseudomonas parafulva |
| 0.02 | 2 | 0 | G1 | 136845 | Pseudomonas putida group |
| 0.01 | 1 | 1 | S | 303 | Pseudomonas putida |
| 0.01 | 1 | 0 | S | 47880 | Pseudomonas fulva |

| | | | | | |
|---|---|---|---|---|---|
| 0.01 | 1 | 1 | S1 | 743720 | Pseudomonas fulva 12-X |
| 0.01 | 1 | 0 | G1 | 196821 | unclassified Pseudomonas |
| 0.01 | 1 | 1 | S | 2083054 | Pseudomonas sp. LG1D9 |
| 0.09 | 8 | 1 | O | 91347 | Enterobacterales |
| 0.08 | 7 | 3 | F | 543 | Enterobacteriaceae |
| 0.04 | 4 | 0 | G | 561 | Escherichia |
| 0.04 | 4 | 4 | S | 562 | Escherichia coli |
| 0.04 | 4 | 0 | O | 135624 | Aeromonadales |
| 0.04 | 4 | 0 | F | 84642 | Aeromonadaceae |
| 0.04 | 4 | 0 | G | 642 | Aeromonas |
| 0.04 | 4 | 0 | G1 | 257493 | unclassified Aeromonas |
| 0.04 | 4 | 4 | S | 1636607 | Aeromonas sp. ASNIH2 |
| 0.01 | 1 | 0 | O | 135625 | Pasteurellales |
| 0.01 | 1 | 0 | F | 712 | Pasteurellaceae |
| 0.01 | 1 | 0 | G | 724 | Haemophilus |
| 0.01 | 1 | 1 | S | 729 | Haemophilus parainfluenzae |
| 0.15 | 14 | 0 | P1 | 68525 | delta/epsilon subdivisions |
| 0.11 | 10 | 0 | C | 28221 | Deltaproteobacteria |
| 0.08 | 7 | 0 | O | 29 | Myxococcales |
| 0.08 | 7 | 0 | O1 | 80811 | Cystobacterineae |
| 0.06 | 6 | 0 | F | 31 | Myxococcaceae |
| 0.06 | 6 | 0 | G | 83461 | Corallococcus |
| 0.06 | 6 | 6 | S | 35 | Corallococcus macrosporus |
| 0.01 | 1 | 0 | F | 39 | Archangiaceae |
| 0.01 | 1 | 0 | G | 40 | Stigmatella |
| 0.01 | 1 | 0 | S | 41 | Stigmatella aurantiaca |
| 0.01 | 1 | 1 | S1 | 378806 | Stigmatella aurantiaca DW4/3-1 |
| 0.03 | 3 | 0 | O | 213115 | Desulfovibrionales |
| 0.03 | 3 | 0 | F | 194924 | Desulfovibrionaceae |
| 0.03 | 3 | 0 | G | 872 | Desulfovibrio |
| 0.03 | 3 | 0 | S | 345370 | Desulfovibrio carbinoliphilus |
| 0.03 | 3 | 3 | S1 | 694327 | Desulfovibrio carbinoliphilus subsp. oakridgensis |
| 0.04 | 4 | 0 | C | 29547 | Epsilonproteobacteria |

| | | | | | |
|---|---|---|---|---|---|
| 0.04 | 4 | 0 | O | 213849 | Campylobacterales |
| 0.04 | 4 | 0 | F | 72294 | Campylobacteraceae |
| 0.04 | 4 | 0 | G | 194 | Campylobacter |
| 0.04 | 4 | 4 | S | 195 | Campylobacter coli |
| 14.68 | 1365 | 0 | D1 | 1783270 | FCB group |
| 14.68 | 1365 | 0 | D2 | 68336 | Bacteroidetes/Chlorobi group |
| 14.68 | 1365 | 1 | P | 976 | Bacteroidetes |
| 14.09 | 1310 | 0 | C | 200643 | Bacteroidia |
| 14.09 | 1310 | 110 | O | 171549 | Bacteroidales |
| 10.83 | 1007 | 0 | F | 815 | Bacteroidaceae |
| 10.83 | 1007 | 93 | G | 816 | Bacteroides |
| 6.81 | 633 | 633 | S | 818 | Bacteroides thetaiotaomicron |
| 0.75 | 70 | 70 | S | 246787 | Bacteroides cellulosilyticus |
| 0.65 | 60 | 60 | S | 820 | Bacteroides uniformis |
| 0.43 | 40 | 40 | S | 28111 | Bacteroides eggerthii |
| 0.4 | 37 | 9 | G1 | 2646097 | unclassified Bacteroides |
| 0.11 | 10 | 10 | S | 2650157 | Bacteroides sp. HF-5287 |
| 0.11 | 10 | 10 | S | 2785531 | Bacteroides sp. HF-162 |
| 0.05 | 5 | 5 | S | 2528203 | Bacteroides sp. A1C1 |
| 0.02 | 2 | 2 | S | 2755405 | Bacteroides sp. CACC 737 |
| 0.01 | 1 | 1 | S | 2763022 | Bacteroides sp. M10 |
| 0.38 | 35 | 35 | S | 817 | Bacteroides fragilis |
| 0.24 | 22 | 22 | S | 28116 | Bacteroides ovatus |
| 0.08 | 7 | 7 | S | 329854 | Bacteroides intestinalis |
| 0.05 | 5 | 5 | S | 47678 | Bacteroides caccae |
| 0.04 | 4 | 3 | S | 371601 | Bacteroides xylanisolvens |
| 0.01 | 1 | 1 | S1 | 657309 | Bacteroides xylanisolvens XB1A |
| 0.01 | 1 | 1 | S | 1796613 | Bacteroides caecimuris |
| 1.38 | 128 | 0 | O1 | 333046 | Bacteroidales incertae sedis |
| 1.38 | 128 | 15 | G | 909656 | Phocaeicola |
| 0.73 | 68 | 63 | S | 357276 | Phocaeicola dorei |
| 0.05 | 5 | 5 | S1 | 997877 | Bacteroides dorei CL03T12C01 |
| 0.46 | 43 | 43 | S | 821 | Phocaeicola vulgatus |

| | | | | | |
|---|---|---|---|---|---|
| 0.01 | 1 | 0 | S | 376805 | Phocaeicola salanitronis |
| 0.01 | 1 | 1 | S1 | 667015 | Phocaeicola salanitronis DSM 18170 |
| 0.01 | 1 | 1 | S | 387090 | Phocaeicola coprophilus |
| 0.47 | 44 | 0 | F | 171550 | Rikenellaceae |
| 0.47 | 44 | 0 | G | 239759 | Alistipes |
| 0.32 | 30 | 0 | S | 328813 | Alistipes onderdonkii |
| 0.32 | 30 | 30 | S1 | 2585117 | Alistipes onderdonkii subsp. vulgaris |
| 0.06 | 6 | 6 | S | 2585118 | Alistipes communis |
| 0.04 | 4 | 0 | S | 214856 | Alistipes finegoldii |
| 0.04 | 4 | 4 | S1 | 679935 | Alistipes finegoldii DSM 17242 |
| 0.02 | 2 | 0 | S | 328814 | Alistipes shahii |
| 0.02 | 2 | 2 | S1 | 717959 | Alistipes shahii WAL 8301 |
| 0.01 | 1 | 1 | S | 2585119 | Alistipes dispar |
| 0.01 | 1 | 0 | G1 | 2608932 | unclassified Alistipes |
| 0.01 | 1 | 1 | S | 2662363 | Alistipes sp. dk3624 |
| 0.11 | 10 | 0 | F | 2005525 | Tannerellaceae |
| 0.11 | 10 | 3 | G | 375288 | Parabacteroides |
| 0.06 | 6 | 6 | S | 823 | Parabacteroides distasonis |
| 0.01 | 1 | 1 | S | 328812 | Parabacteroides goldsteinii |
| 0.1 | 9 | 0 | F | 1853231 | Odoribacteraceae |
| 0.1 | 9 | 0 | G | 574697 | Butyricimonas |
| 0.08 | 7 | 7 | S | 544645 | Butyricimonas virosa |
| 0.02 | 2 | 2 | S | 2093856 | Butyricimonas faecalis |
| 0.01 | 1 | 0 | F | 171552 | Prevotellaceae |
| 0.01 | 1 | 0 | G | 838 | Prevotella |
| 0.01 | 1 | 1 | S | 1177574 | Prevotella jejuni |
| 0.01 | 1 | 0 | F | 2005473 | Muribaculaceae |
| 0.01 | 1 | 0 | G | 1918540 | Muribaculum |
| 0.01 | 1 | 1 | S | 2530390 | Muribaculum gordoncarteri |
| 0.34 | 32 | 0 | C | 768503 | Cytophagia |
| 0.34 | 32 | 0 | O | 768507 | Cytophagales |
| 0.34 | 32 | 0 | F | 1853232 | Hymenobacteraceae |
| 0.34 | 32 | 12 | G | 89966 | Hymenobacter |

| | | | | | |
|---|---|---|---|---|---|
| 0.15 | 14 | 0 | G1 | 2615202 | unclassified Hymenobacter |
| 0.12 | 11 | 11 | S | 2675878 | Hymenobacter sp. BRD128 |
| 0.02 | 2 | 2 | S | 1356852 | Hymenobacter sp. APR13 |
| 0.01 | 1 | 1 | S | 2584940 | Hymenobacter sp. DG01 |
| 0.05 | 5 | 5 | S | 2607656 | Hymenobacter busanensis |
| 0.01 | 1 | 1 | S | 1385715 | Hymenobacter qilianensis |
| 0.24 | 22 | 0 | C | 117743 | Flavobacteriia |
| 0.24 | 22 | 0 | O | 200644 | Flavobacteriales |
| 0.24 | 22 | 0 | F | 2762318 | Weeksellaceae |
| 0.19 | 18 | 0 | F1 | 2782232 | Chryseobacterium group |
| 0.19 | 18 | 1 | G | 59732 | Chryseobacterium |
| 0.08 | 7 | 7 | S | 246 | Chryseobacterium balustinum |
| 0.08 | 7 | 7 | S | 558152 | Chryseobacterium piperi |
| 0.03 | 3 | 0 | G1 | 2593645 | unclassified Chryseobacterium |
| 0.03 | 3 | 3 | S | 2487065 | Chryseobacterium sp. G0201 |
| 0.04 | 4 | 4 | G | 59734 | Empedobacter |
| 7.86 | 731 | 3 | D1 | 1783272 | Terrabacteria group |
| 5.78 | 537 | 0 | P | 201174 | Actinobacteria |
| 5.66 | 526 | 33 | C | 1760 | Actinomycetia |
| 2.8 | 260 | 0 | O | 85009 | Propionibacteriales |
| 2.47 | 230 | 4 | F | 31957 | Propionibacteriaceae |
| 2.38 | 221 | 0 | G | 1912216 | Cutibacterium |
| 2.37 | 220 | 220 | S | 1747 | Cutibacterium acnes |
| 0.01 | 1 | 1 | S | 33011 | Cutibacterium granulosum |
| 0.05 | 5 | 0 | G | 1743 | Propionibacterium |
| 0.04 | 4 | 4 | S | 1744 | Propionibacterium freudenreichii |
| 0.01 | 1 | 0 | G1 | 2608905 | unclassified Propionibacterium |
| 0.01 | 1 | 1 | S | 671223 | Propionibacterium sp. oral taxon 193 |
| 0.32 | 30 | 3 | F | 85015 | Nocardioidaceae |
| 0.27 | 25 | 0 | G | 1839 | Nocardioides |
| 0.12 | 11 | 11 | S | 2518370 | Nocardioides euryhalodurans |
| 0.08 | 7 | 7 | S | 433659 | Nocardioides mesophilus |
| 0.08 | 7 | 0 | G1 | 2615069 | unclassified Nocardioides |

| | | | | | |
|---|---|---|---|---|---|
| 0.04 | 4 | 4 | S | 2763008 | Nocardioides sp. zg-1228 |
| 0.02 | 2 | 2 | S | 110319 | Nocardioides sp. CF8 |
| 0.01 | 1 | 1 | S | 2714939 | Nocardioides sp. HDW12B |
| 0.02 | 2 | 0 | G | 86795 | Marmoricola |
| 0.02 | 2 | 2 | S | 642780 | Marmoricola scoriae |
| 1.42 | 132 | 5 | O | 85006 | Micrococcales |
| 0.71 | 66 | 0 | F | 85023 | Microbacteriaceae |
| 0.66 | 61 | 16 | G | 33882 | Microbacterium |
| 0.17 | 16 | 16 | S | 57043 | Microbacterium esteraromaticum |
| 0.11 | 10 | 10 | S | 162426 | Microbacterium hominis |
| 0.09 | 8 | 0 | G1 | 2609290 | unclassified Microbacterium |
| 0.05 | 5 | 5 | S | 2782166 | Microbacterium sp. A18JL241 |
| 0.02 | 2 | 2 | S | 2048898 | Microbacterium sp. Y-01 |
| 0.01 | 1 | 1 | S | 2603598 | Microbacterium sp. CBA3102 |
| 0.05 | 5 | 5 | S | 104336 | Microbacterium foliorum |
| 0.03 | 3 | 3 | S | 273677 | Microbacterium oleivorans |
| 0.02 | 2 | 2 | S | 69362 | Microbacterium schleiferi |
| 0.01 | 1 | 1 | S | 936337 | Microbacterium amylolyticum |
| 0.02 | 2 | 0 | G | 110932 | Leifsonia |
| 0.02 | 2 | 1 | G1 | 2663824 | unclassified Leifsonia |
| 0.01 | 1 | 1 | S | 1798223 | Leifsonia sp. 21MFCrub1.1 |
| 0.01 | 1 | 0 | G | 2034 | Curtobacterium |
| 0.01 | 1 | 0 | S | 2035 | Curtobacterium flaccumfaciens |
| 0.01 | 1 | 1 | S1 | 138532 | Curtobacterium flaccumfaciens pv. flaccumfaciens |
| 0.01 | 1 | 0 | G | 33877 | Agromyces |
| 0.01 | 1 | 0 | G1 | 2639701 | unclassified Agromyces |
| 0.01 | 1 | 1 | S | 2498704 | Agromyces sp. LHK192 |
| 0.01 | 1 | 0 | G | 235888 | Salinibacterium |
| 0.01 | 1 | 0 | G1 | 2632331 | unclassified Salinibacterium |
| 0.01 | 1 | 1 | S | 2708084 | Salinibacterium sp. ZJ70 |
| 0.3 | 28 | 0 | F | 2805590 | Ornithinimicrobiaceae |
| 0.2 | 19 | 0 | G | 265976 | Serinicoccus |
| 0.2 | 19 | 19 | S | 1078471 | Serinicoccus profundi |

| | | | | | |
|---|---|---|---|---|---|
| 0.1 | 9 | 0 | G | 125287 | Ornithinimicrobium |
| 0.06 | 6 | 6 | S | 1288636 | Ornithinimicrobium flavum |
| 0.03 | 3 | 0 | G1 | 2615080 | unclassified Ornithinimicrobium |
| 0.02 | 2 | 2 | S | 2594265 | Ornithinimicrobium sp. H23M54 |
| 0.01 | 1 | 1 | S | 2508882 | Ornithinimicrobium sp. HY006 |
| 0.28 | 26 | 0 | F | 1268 | Micrococcaceae |
| 0.19 | 18 | 10 | G | 1269 | Micrococcus |
| 0.09 | 8 | 8 | S | 1270 | Micrococcus luteus |
| 0.08 | 7 | 0 | G | 1663 | Arthrobacter |
| 0.04 | 4 | 4 | S | 37921 | Arthrobacter agilis |
| 0.03 | 3 | 0 | G1 | 235627 | unclassified Arthrobacter |
| 0.03 | 3 | 3 | S | 2575374 | Arthrobacter sp. 24S4-2 |
| 0.01 | 1 | 0 | G | 57493 | Kocuria |
| 0.01 | 1 | 1 | S | 72000 | Kocuria rhizophila |
| 0.04 | 4 | 0 | F | 85019 | Brevibacteriaceae |
| 0.04 | 4 | 1 | G | 1696 | Brevibacterium |
| 0.03 | 3 | 3 | S | 629680 | Brevibacterium sandarakinum |
| 0.03 | 3 | 0 | F | 85021 | Intrasporangiaceae |
| 0.03 | 3 | 0 | G | 53457 | Janibacter |
| 0.03 | 3 | 3 | S | 53458 | Janibacter limosus |
| 0.58 | 54 | 0 | O | 85007 | Corynebacteriales |
| 0.33 | 31 | 0 | F | 1653 | Corynebacteriaceae |
| 0.33 | 31 | 0 | G | 1716 | Corynebacterium |
| 0.13 | 12 | 12 | S | 43990 | Corynebacterium segmentosum |
| 0.11 | 10 | 0 | S | 1404244 | Corynebacterium glyciniphilum |
| 0.11 | 10 | 10 | S1 | 1404245 | Corynebacterium glyciniphilum AJ 3170 |
| 0.06 | 6 | 6 | S | 1979527 | Corynebacterium kefirresidentii |
| 0.03 | 3 | 3 | S | 38304 | Corynebacterium tuberculostearicum |
| 0.13 | 12 | 0 | F | 1762 | Mycobacteriaceae |
| 0.08 | 7 | 5 | G | 1866885 | Mycolicibacterium |
| 0.01 | 1 | 1 | S | 2487344 | Mycolicibacterium nivoides |
| 0.01 | 1 | 1 | S | 1794 | Mycolicibacterium gadium |
| 0.03 | 3 | 3 | G | 1763 | Mycobacterium |

| | | | | | |
|---|---|---|---|---|---|
| 0.02 | 2 | 0 | G | 670516 | Mycobacteroides |
| 0.02 | 2 | 2 | S | 1774 | Mycobacteroides chelonae |
| 0.05 | 5 | 0 | F | 2805586 | Lawsonellaceae |
| 0.05 | 5 | 0 | G | 1847725 | Lawsonella |
| 0.05 | 5 | 5 | S | 1528099 | Lawsonella clevelandensis |
| 0.04 | 4 | 0 | F | 85025 | Nocardiaceae |
| 0.04 | 4 | 4 | G | 1827 | Rhodococcus |
| 0.02 | 2 | 0 | F | 85029 | Dietziaceae |
| 0.02 | 2 | 0 | G | 37914 | Dietzia |
| 0.02 | 2 | 0 | G1 | 2617939 | unclassified Dietzia |
| 0.02 | 2 | 2 | S | 712270 | Dietzia sp. oral taxon 368 |
| 0.17 | 16 | 0 | O | 85011 | Streptomycetales |
| 0.17 | 16 | 0 | F | 2062 | Streptomycetaceae |
| 0.17 | 16 | 8 | G | 1883 | Streptomyces |
| 0.02 | 2 | 0 | G1 | 2593676 | unclassified Streptomyces |
| 0.01 | 1 | 1 | S | 2695266 | Streptomyces sp. HM190 |
| 0.01 | 1 | 1 | S | 646637 | Streptomyces sp. M2 |
| 0.02 | 2 | 2 | S | 1169025 | Streptomyces pratensis |
| 0.02 | 2 | 2 | S | 1927 | Streptomyces rimosus |
| 0.01 | 1 | 1 | S | 68270 | Streptomyces spectabilis |
| 0.01 | 1 | 0 | S | 1912 | Streptomyces hygroscopicus |
| 0.01 | 1 | 1 | S1 | 264445 | Streptomyces hygroscopicus subsp. limoneus |
| 0.13 | 12 | 0 | O | 85008 | Micromonosporales |
| 0.13 | 12 | 6 | F | 28056 | Micromonosporaceae |
| 0.06 | 6 | 0 | G | 1873 | Micromonospora |
| 0.04 | 4 | 0 | G1 | 2617518 | unclassified Micromonospora |
| 0.04 | 4 | 4 | S | 2583243 | Micromonospora sp. HM134 |
| 0.02 | 2 | 2 | S | 261654 | Micromonospora auratinigra |
| 0.11 | 10 | 0 | O | 85010 | Pseudonocardiales |
| 0.11 | 10 | 0 | F | 2070 | Pseudonocardiaceae |
| 0.06 | 6 | 0 | G | 1813 | Amycolatopsis |
| 0.06 | 6 | 6 | S | 31958 | Amycolatopsis orientalis |
| 0.04 | 4 | 0 | G | 1847 | Pseudonocardia |

| | | | | | |
|---|---|---|---|---|---|
| 0.04 | 4 | 3 | G1 | 2619320 | unclassified Pseudonocardia |
| 0.01 | 1 | 1 | S | 2761535 | Pseudonocardia sp. CGMCC 4.1532 |
| 0.02 | 2 | 0 | O | 2037 | Actinomycetales |
| 0.02 | 2 | 0 | F | 2049 | Actinomycetaceae |
| 0.02 | 2 | 0 | G | 1654 | Actinomyces |
| 0.02 | 2 | 2 | S | 675090 | Actinomyces weissii |
| 0.02 | 2 | 0 | O | 85004 | Bifidobacteriales |
| 0.02 | 2 | 0 | F | 31953 | Bifidobacteriaceae |
| 0.02 | 2 | 0 | G | 1678 | Bifidobacterium |
| 0.01 | 1 | 1 | S | 1680 | Bifidobacterium adolescentis |
| 0.01 | 1 | 1 | S | 28025 | Bifidobacterium animalis |
| 0.02 | 2 | 0 | O | 85013 | Frankiales |
| 0.02 | 2 | 0 | F | 74712 | Frankiaceae |
| 0.02 | 2 | 0 | G | 1854 | Frankia |
| 0.02 | 2 | 0 | G1 | 2632575 | unclassified Frankia |
| 0.02 | 2 | 2 | S | 710111 | Frankia sp. QA3 |
| 0.02 | 2 | 0 | O | 1217098 | Jiangellales |
| 0.02 | 2 | 0 | F | 1217100 | Jiangellaceae |
| 0.02 | 2 | 0 | G | 281472 | Jiangella |
| 0.02 | 2 | 0 | G1 | 2624397 | unclassified Jiangella |
| 0.02 | 2 | 2 | S | 1798224 | Jiangella sp. DSM 45060 |
| 0.01 | 1 | 0 | O | 85014 | Glycomycetales |
| 0.01 | 1 | 0 | F | 85034 | Glycomycetaceae |
| 0.01 | 1 | 0 | G | 283810 | Stackebrandtia |
| 0.01 | 1 | 0 | S | 283811 | Stackebrandtia nassauensis |
| 0.01 | 1 | 1 | S1 | 446470 | Stackebrandtia nassauensis DSM 44728 |
| 0.12 | 11 | 0 | C | 84995 | Rubrobacteria |
| 0.12 | 11 | 0 | O | 84996 | Rubrobacterales |
| 0.11 | 10 | 0 | F | 84997 | Rubrobacteraceae |
| 0.11 | 10 | 0 | G | 42255 | Rubrobacter |
| 0.11 | 10 | 0 | G1 | 2637691 | unclassified Rubrobacter |
| 0.11 | 10 | 10 | S | 2653851 | Rubrobacter sp. SCSIO 52909 |
| 0.01 | 1 | 0 | F | 2600303 | Baekduiaceae |

| | | | | | |
|---|---|---|---|---|---|
| 0.01 | 1 | 0 | G | 2600304 | Baekduia |
| 0.01 | 1 | 1 | S | 496014 | Baekduia soli |
| 1.85 | 172 | 3 | P | 1239 | Firmicutes |
| 0.79 | 73 | 2 | C | 91061 | Bacilli |
| 0.53 | 49 | 0 | O | 1385 | Bacillales |
| 0.53 | 49 | 0 | F | 90964 | Staphylococcaceae |
| 0.53 | 49 | 8 | G | 1279 | Staphylococcus |
| 0.25 | 23 | 23 | S | 1282 | Staphylococcus epidermidis |
| 0.14 | 13 | 13 | S | 29388 | Staphylococcus capitis |
| 0.05 | 5 | 5 | S | 1292 | Staphylococcus warneri |
| 0.24 | 22 | 3 | O | 186826 | Lactobacillales |
| 0.11 | 10 | 0 | F | 1300 | Streptococcaceae |
| 0.11 | 10 | 3 | G | 1301 | Streptococcus |
| 0.02 | 2 | 2 | S | 28037 | Streptococcus mitis |
| 0.02 | 2 | 2 | S | 1308 | Streptococcus thermophilus |
| 0.02 | 2 | 2 | S | 1304 | Streptococcus salivarius |
| 0.01 | 1 | 1 | S | 1305 | Streptococcus sanguinis |
| 0.1 | 9 | 0 | F | 33958 | Lactobacillaceae |
| 0.08 | 7 | 0 | G | 1578 | Lactobacillus |
| 0.08 | 7 | 0 | S | 1584 | Lactobacillus delbrueckii |
| 0.08 | 7 | 7 | S1 | 29397 | Lactobacillus delbrueckii subsp. lactis |
| 0.01 | 1 | 0 | G | 2767842 | Lactiplantibacillus |
| 0.01 | 1 | 1 | S | 1590 | Lactiplantibacillus plantarum |
| 0.01 | 1 | 0 | G | 2767885 | Latilactobacillus |
| 0.01 | 1 | 1 | S | 1599 | Latilactobacillus sakei |
| 0.72 | 67 | 0 | C | 186801 | Clostridia |
| 0.72 | 67 | 3 | O | 186802 | Eubacteriales |
| 0.37 | 34 | 2 | F | 186803 | Lachnospiraceae |
| 0.18 | 17 | 1 | G | 841 | Roseburia |
| 0.15 | 14 | 7 | S | 166486 | Roseburia intestinalis |
| 0.03 | 3 | 3 | S1 | 536231 | Roseburia intestinalis L1-82 |
| 0.03 | 3 | 3 | S1 | 657315 | Roseburia intestinalis M50/1 |
| 0.01 | 1 | 1 | S1 | 718255 | Roseburia intestinalis XB6B4 |

| | | | | | |
|---|---|---|---|---|---|
| 0.02 | 2 | 2 | S | 301301 | Roseburia hominis |
| 0.04 | 4 | 0 | G | 28050 | Lachnospira |
| 0.04 | 4 | 0 | S | 39485 | Lachnospira eligens |
| 0.04 | 4 | 4 | S1 | 515620 | [Eubacterium] eligens ATCC 27750 |
| 0.03 | 3 | 0 | G | 572511 | Blautia |
| 0.02 | 2 | 0 | G1 | 2648079 | unclassified Blautia |
| 0.02 | 2 | 2 | S | 2479767 | Blautia sp. SC05B48 |
| 0.01 | 1 | 0 | S | 40520 | Blautia obeum |
| 0.01 | 1 | 1 | S1 | 657314 | Blautia obeum A2-162 |
| 0.02 | 2 | 0 | G | 2569097 | Anaerobutyricum |
| 0.02 | 2 | 2 | S | 39488 | Anaerobutyricum hallii |
| 0.02 | 2 | 1 | G | 2719313 | Enterocloster |
| 0.01 | 1 | 1 | S | 1531 | Enterocloster clostridioformis |
| 0.01 | 1 | 0 | G | 33042 | Coprococcus |
| 0.01 | 1 | 1 | S | 410072 | Coprococcus comes |
| 0.01 | 1 | 1 | G | 1506553 | Lachnoclostridium |
| 0.01 | 1 | 0 | G | 2316020 | Mediterraneibacter |
| 0.01 | 1 | 0 | S | 33039 | [Ruminococcus] torques |
| 0.01 | 1 | 1 | S1 | 657313 | [Ruminococcus] torques L2-14 |
| 0.01 | 1 | 0 | G | 2719231 | Lacrimispora |
| 0.01 | 1 | 1 | S | 84030 | Lacrimispora saccharolytica |
| 0.25 | 23 | 0 | F | 216572 | Oscillospiraceae |
| 0.18 | 17 | 0 | G | 216851 | Faecalibacterium |
| 0.18 | 17 | 11 | S | 853 | Faecalibacterium prausnitzii |
| 0.04 | 4 | 4 | S1 | 718252 | Faecalibacterium prausnitzii L2-6 |
| 0.02 | 2 | 2 | S1 | 657322 | Faecalibacterium prausnitzii SL3/3 |
| 0.02 | 2 | 0 | G | 1263 | Ruminococcus |
| 0.02 | 2 | 2 | S | 1160721 | Ruminococcus bicirculans |
| 0.02 | 2 | 0 | G | 459786 | Oscillibacter |
| 0.02 | 2 | 0 | G1 | 2629304 | unclassified Oscillibacter |
| 0.01 | 1 | 1 | S | 2109687 | Oscillibacter sp. PEA192 |
| 0.01 | 1 | 1 | S | 2763056 | Oscillibacter sp. NSJ-62 |
| 0.01 | 1 | 0 | G | 1905344 | Ruthenibacterium |

| | | | | | |
|---|---|---|---|---|---|
| 0.01 | 1 | 1 | S | 1550024 | Ruthenibacterium lactatiformans |
| 0.01 | 1 | 0 | G | 2591381 | Dysosmobacter |
| 0.01 | 1 | 1 | S | 2093857 | Dysosmobacter welbionis |
| 0.03 | 3 | 0 | F | 186804 | Peptostreptococcaceae |
| 0.02 | 2 | 0 | G | 1870884 | Clostridioides |
| 0.02 | 2 | 2 | S | 1496 | Clostridioides difficile |
| 0.01 | 1 | 1 | G | 1501226 | Romboutsia |
| 0.02 | 2 | 0 | O1 | 538999 | Eubacteriales incertae sedis |
| 0.01 | 1 | 0 | G | 1392389 | Intestinimonas |
| 0.01 | 1 | 1 | S | 1297617 | Intestinimonas butyriciproducens |
| 0.01 | 1 | 0 | G | 2039302 | Monoglobus |
| 0.01 | 1 | 1 | S | 1981510 | Monoglobus pectinilyticus |
| 0.01 | 1 | 0 | F | 31979 | Clostridiaceae |
| 0.01 | 1 | 0 | G | 1485 | Clostridium |
| 0.01 | 1 | 1 | S | 1561 | Clostridium baratii |
| 0.01 | 1 | 0 | F | 186806 | Eubacteriaceae |
| 0.01 | 1 | 0 | G | 33951 | Acetobacterium |
| 0.01 | 1 | 0 | S | 33952 | Acetobacterium woodii |
| 0.01 | 1 | 1 | S1 | 931626 | Acetobacterium woodii DSM 1030 |
| 0.16 | 15 | 0 | C | 909932 | Negativicutes |
| 0.13 | 12 | 0 | O | 1843489 | Veillonellales |
| 0.13 | 12 | 0 | F | 31977 | Veillonellaceae |
| 0.13 | 12 | 0 | G | 29465 | Veillonella |
| 0.13 | 12 | 7 | S | 29466 | Veillonella parvula |
| 0.05 | 5 | 5 | S1 | 1316254 | Veillonella parvula HSIVP1 |
| 0.03 | 3 | 0 | O | 1843488 | Acidaminococcales |
| 0.03 | 3 | 0 | F | 909930 | Acidaminococcaceae |
| 0.03 | 3 | 0 | G | 33024 | Phascolarctobacterium |
| 0.03 | 3 | 3 | S | 33025 | Phascolarctobacterium faecium |
| 0.15 | 14 | 0 | C | 526524 | Erysipelotrichia |
| 0.15 | 14 | 0 | O | 526525 | Erysipelotrichales |
| 0.15 | 14 | 0 | F | 2810281 | Turicibacteraceae |
| 0.15 | 14 | 0 | G | 191303 | Turicibacter |

| | | | | | |
|---|---|---|---|---|---|
| 0.13 | 12 | 12 | S | 154288 | Turicibacter sanguinis |
| 0.02 | 2 | 0 | G1 | 2638206 | unclassified Turicibacter |
| 0.02 | 2 | 2 | S | 1712675 | Turicibacter sp. H121 |
| 0.2 | 19 | 0 | D2 | 1798711 | Cyanobacteria/Melainabacteria group |
| 0.2 | 19 | 0 | P | 1117 | Cyanobacteria |
| 0.12 | 11 | 1 | O | 1161 | Nostocales |
| 0.05 | 5 | 0 | F | 1162 | Nostocaceae |
| 0.05 | 5 | 5 | G | 1177 | Nostoc |
| 0.05 | 5 | 0 | F | 1182 | Scytonemataceae |
| 0.05 | 5 | 0 | G | 1203 | Scytonema |
| 0.05 | 5 | 0 | G1 | 2618749 | unclassified Scytonema |
| 0.05 | 5 | 5 | S | 1137095 | Scytonema sp. HK-05 |
| 0.08 | 7 | 0 | O | 52604 | Pleurocapsales |
| 0.08 | 7 | 0 | F | 1890500 | Hyellaceae |
| 0.08 | 7 | 0 | G | 44474 | Pleurocapsa |
| 0.08 | 7 | 0 | S | 54308 | Pleurocapsa minor |
| 0.08 | 7 | 7 | S1 | 118163 | Pleurocapsa sp. PCC 7327 |
| 0.01 | 1 | 0 | O | 1890424 | Synechococcales |
| 0.01 | 1 | 0 | F | 1890438 | Leptolyngbyaceae |
| 0.01 | 1 | 0 | G | 47251 | Leptolyngbya |
| 0.01 | 1 | 0 | G1 | 2650499 | unclassified Leptolyngbya |
| 0.01 | 1 | 1 | S | 1752064 | Leptolyngbya sp. NIES-3755 |
| 1.79 | 166 | 0 | D | 2759 | Eukaryota |
| 1.2 | 112 | 0 | D1 | 33154 | Opisthokonta |
| 1.09 | 101 | 0 | K | 4751 | Fungi |
| 1.09 | 101 | 0 | K1 | 451864 | Dikarya |
| 1.09 | 101 | 0 | P | 5204 | Basidiomycota |
| 1.09 | 101 | 0 | P1 | 452284 | Ustilaginomycotina |
| 1.09 | 101 | 0 | C | 1538075 | Malasseziomycetes |
| 1.09 | 101 | 0 | O | 162474 | Malasseziales |
| 1.09 | 101 | 0 | F | 742845 | Malasseziaceae |
| 1.09 | 101 | 0 | G | 55193 | Malassezia |
| 1.09 | 101 | 101 | S | 76775 | Malassezia restricta |

| | | | | | |
|---|---|---|---|---|---|
| 0.12 | 11 | 0 | K | 33208 | Metazoa |
| 0.12 | 11 | 0 | K1 | 6072 | Eumetazoa |
| 0.12 | 11 | 0 | K2 | 33213 | Bilateria |
| 0.12 | 11 | 0 | K3 | 33511 | Deuterostomia |
| 0.12 | 11 | 0 | P | 7711 | Chordata |
| 0.12 | 11 | 0 | P1 | 89593 | Craniata |
| 0.12 | 11 | 0 | P2 | 7742 | Vertebrata |
| 0.12 | 11 | 0 | P3 | 7776 | Gnathostomata |
| 0.12 | 11 | 0 | P4 | 117570 | Teleostomi |
| 0.12 | 11 | 0 | P5 | 117571 | Euteleostomi |
| 0.12 | 11 | 0 | P6 | 8287 | Sarcopterygii |
| 0.12 | 11 | 0 | C | 1338369 | Dipnotetrapodomorpha |
| 0.12 | 11 | 0 | C1 | 32523 | Tetrapoda |
| 0.12 | 11 | 0 | C2 | 32524 | Amniota |
| 0.12 | 11 | 0 | C | 40674 | Mammalia |
| 0.12 | 11 | 0 | C1 | 32525 | Theria |
| 0.12 | 11 | 0 | C2 | 9347 | Eutheria |
| 0.12 | 11 | 0 | C3 | 1437010 | Boreoeutheria |
| 0.12 | 11 | 0 | C4 | 314146 | Euarchontoglires |
| 0.12 | 11 | 0 | O | 9443 | Primates |
| 0.12 | 11 | 0 | O1 | 376913 | Haplorrhini |
| 0.12 | 11 | 0 | O2 | 314293 | Simiiformes |
| 0.12 | 11 | 0 | O3 | 9526 | Catarrhini |
| 0.12 | 11 | 0 | O4 | 314295 | Hominoidea |
| 0.12 | 11 | 0 | F | 9604 | Hominidae |
| 0.12 | 11 | 0 | F1 | 207598 | Homininae |
| 0.12 | 11 | 0 | G | 9605 | Homo |
| 0.12 | 11 | 11 | S | 9606 | Homo sapiens |
| 0.58 | 54 | 0 | K | 33090 | Viridiplantae |
| 0.58 | 54 | 0 | P | 35493 | Streptophyta |
| 0.58 | 54 | 0 | P1 | 131221 | Streptophytina |
| 0.58 | 54 | 0 | P2 | 3193 | Embryophyta |
| 0.58 | 54 | 0 | P3 | 58023 | Tracheophyta |

| | | | | | |
|---|---|---|---|---|---|
| 0.58 | 54 | 0 | P4 | 78536 | Euphyllophyta |
| 0.58 | 54 | 0 | P5 | 58024 | Spermatophyta |
| 0.58 | 54 | 0 | C | 3398 | Magnoliopsida |
| 0.58 | 54 | 0 | C1 | 1437183 | Mesangiospermae |
| 0.42 | 39 | 0 | C2 | 4447 | Liliopsida |
| 0.42 | 39 | 0 | C3 | 1437197 | Petrosaviidae |
| 0.42 | 39 | 0 | C4 | 4734 | commelinids |
| 0.42 | 39 | 0 | O | 38820 | Poales |
| 0.42 | 39 | 0 | F | 4479 | Poaceae |
| 0.42 | 39 | 0 | F1 | 359160 | BOP clade |
| 0.42 | 39 | 0 | F2 | 147368 | Pooideae |
| 0.42 | 39 | 0 | F3 | 1648038 | Triticodae |
| 0.42 | 39 | 0 | F4 | 147389 | Triticeae |
| 0.42 | 39 | 12 | F5 | 1648030 | Triticinae |
| 0.29 | 27 | 0 | G | 4564 | Triticum |
| 0.29 | 27 | 27 | S | 85692 | Triticum dicoccoides |
| 0.16 | 15 | 0 | C2 | 71240 | eudicotyledons |
| 0.16 | 15 | 0 | C3 | 91827 | Gunneridae |
| 0.16 | 15 | 0 | C4 | 1437201 | Pentapetalae |
| 0.15 | 14 | 0 | C5 | 71275 | rosids |
| 0.14 | 13 | 1 | C6 | 91836 | malvids |
| 0.13 | 12 | 0 | O | 41937 | Sapindales |
| 0.13 | 12 | 0 | F | 23513 | Rutaceae |
| 0.13 | 12 | 0 | F1 | 1728959 | Aurantioideae |
| 0.13 | 12 | 0 | G | 2706 | Citrus |
| 0.13 | 12 | 12 | S | 2711 | Citrus sinensis |
| 0.01 | 1 | 0 | C6 | 91835 | fabids |
| 0.01 | 1 | 0 | O | 72025 | Fabales |
| 0.01 | 1 | 0 | F | 3803 | Fabaceae |
| 0.01 | 1 | 0 | F1 | 3814 | Papilionoideae |
| 0.01 | 1 | 0 | F2 | 2231393 | 50 kb inversion clade |
| 0.01 | 1 | 0 | F3 | 2231387 | dalbergioids sensu lato |
| 0.01 | 1 | 0 | F4 | 163725 | Dalbergieae |

| | | | | | |
|---|---|---|---|---|---|
| 0.01 | 1 | 0 | F5 | 2231390 | Pterocarpus clade |
| 0.01 | 1 | 1 | G | 3817 | Arachis |
| 0.01 | 1 | 0 | C5 | 71274 | asterids |
| 0.01 | 1 | 0 | C6 | 91882 | campanulids |
| 0.01 | 1 | 0 | O | 4209 | Asterales |
| 0.01 | 1 | 0 | F | 4210 | Asteraceae |
| 0.01 | 1 | 0 | F1 | 102804 | Asteroideae |
| 0.01 | 1 | 0 | F2 | 911341 | Heliantheae alliance |
| 0.01 | 1 | 0 | F3 | 102814 | Heliantheae |
| 0.01 | 1 | 0 | G | 4231 | Helianthus |
| 0.01 | 1 | 1 | S | 4232 | Helianthus annuus |
| 0.01 | 1 | 0 | D | 10239 | Viruses |
| 0.01 | 1 | 0 | D1 | 2731341 | Duplodnaviria |
| 0.01 | 1 | 0 | D2 | 2731360 | Heunggongvirae |
| 0.01 | 1 | 0 | P | 2731618 | Uroviricota |
| 0.01 | 1 | 0 | C | 2731619 | Caudoviricetes |
| 0.01 | 1 | 0 | O | 28883 | Caudovirales |
| 0.01 | 1 | 0 | F | 10699 | Siphoviridae |
| 0.01 | 1 | 0 | G | 1982251 | Pahexavirus |
| 0.01 | 1 | 0 | S | 1982277 | Propionibacterium virus PHL060L00 |
| 0.01 | 1 | 1 | S1 | 1235647 | Propionibacterium phage PHL060L00 |

NC3

| | | | | | |
|---|---|---|---|---|---|
| 16.8 | 9381 | 9381 | U | 0 | unclassified |
| 83.2 | 46451 | 0 | R | 1 | root |
| 82.04 | 45804 | 13 | R1 | 131567 | cellular organisms |
| 75.39 | 42092 | 273 | D | 2 | Bacteria |
| 71.67 | 40013 | 830 | P | 1224 | Proteobacteria |
| 68.6 | 38302 | 8 | C | 28216 | Betaproteobacteria |
| 68.58 | 38290 | 1000 | O | 80840 | Burkholderiales |
| 66.65 | 37212 | 927 | F | 80864 | Comamonadaceae |
| 64.2 | 35844 | 23321 | G | 80865 | Delftia |
| 19.75 | 11025 | 11025 | S | 558537 | Delftia lacustris |
| 1.33 | 742 | 742 | S | 80866 | Delftia acidovorans |
| 1.01 | 564 | 564 | S | 180282 | Delftia tsuruhatensis |
| 0.34 | 192 | 25 | G1 | 2613839 | unclassified Delftia |
| 0.22 | 123 | 123 | S | 742013 | Delftia sp. Cs1-4 |
| 0.08 | 44 | 44 | S | 1920191 | Delftia sp. HK171 |
| 0.28 | 154 | 24 | G | 12916 | Acidovorax |
| 0.22 | 122 | 5 | G1 | 2684926 | unclassified Acidovorax |
| 0.18 | 102 | 102 | S | 358220 | Acidovorax sp. KKS102 |
| 0.02 | 9 | 9 | S | 232721 | Acidovorax sp. JS42 |
| 0.01 | 5 | 5 | S | 2518343 | Acidovorax sp. JMULE5 |
| 0 | 1 | 1 | S | 1842533 | Acidovorax sp. RAC01 |
| 0.01 | 8 | 8 | S | 553814 | Acidovorax carolinensis |
| 0.22 | 125 | 0 | G | 201096 | Alicycliphilus |
| 0.22 | 125 | 1 | S | 179636 | Alicycliphilus denitrificans |
| 0.22 | 124 | 124 | S1 | 596154 | Alicycliphilus denitrificans K601 |
| 0.09 | 48 | 2 | G | 34072 | Variovorax |
| 0.08 | 46 | 1 | G1 | 663243 | unclassified Variovorax |
| 0.05 | 27 | 27 | S | 1034889 | Variovorax sp. HW608 |
| 0.02 | 10 | 10 | S | 662548 | Variovorax sp. RA8 |
| 0.01 | 8 | 8 | S | 207745 | Variovorax sp. WDL1 |
| 0.09 | 48 | 0 | G | 238749 | Diaphorobacter |
| 0.09 | 48 | 0 | G1 | 2649760 | unclassified Diaphorobacter |

| | | | | | |
|---|---|---|---|---|---|
| 0.06 | 34 | 34 | S | 2714924 | Diaphorobacter sp. HDW4A |
| 0.03 | 14 | 14 | S | 2792224 | Diaphorobacter sp. JS3051 |
| 0.08 | 45 | 0 | G | 283 | Comamonas |
| 0.05 | 26 | 26 | S | 160825 | Comamonas koreensis |
| 0.01 | 6 | 0 | S | 285 | Comamonas testosteroni |
| 0.01 | 6 | 6 | S1 | 1392005 | Comamonas testosteroni TK102 |
| 0.01 | 4 | 4 | S | 225992 | Comamonas kerstersii |
| 0.01 | 4 | 0 | G1 | 2638500 | unclassified Comamonas |
| 0.01 | 4 | 4 | S | 1232667 | Comamonas sp. 7D-2 |
| 0.01 | 3 | 3 | S | 1082851 | Comamonas serinivorans |
| 0 | 1 | 1 | S | 225991 | Comamonas aquatica |
| 0 | 1 | 1 | S | 363952 | Comamonas thiooxydans |
| 0.02 | 12 | 0 | G | 47420 | Hydrogenophaga |
| 0.02 | 12 | 0 | G1 | 2610897 | unclassified Hydrogenophaga |
| 0.02 | 12 | 12 | S | 795665 | Hydrogenophaga sp. PBC |
| 0 | 2 | 0 | G | 28065 | Rhodoferax |
| 0 | 1 | 0 | S | 192843 | Rhodoferax ferrireducens |
| 0 | 1 | 1 | S1 | 338969 | Rhodoferax ferrireducens T118 |
| 0 | 1 | 0 | G1 | 2627954 | unclassified Rhodoferax |
| 0 | 1 | 1 | S | 2741720 | Rhodoferax sp. BAB1 |
| 0 | 2 | 0 | G | 52972 | Polaromonas |
| 0 | 2 | 2 | G1 | 2638319 | unclassified Polaromonas |
| 0 | 2 | 0 | G | 215579 | Schlegelella |
| 0 | 2 | 2 | S | 215580 | Schlegelella thermodepolymerans |
| 0 | 2 | 0 | G | 1649468 | Melaminivora |
| 0 | 2 | 0 | G1 | 2641902 | unclassified Melaminivora |
| 0 | 2 | 2 | S | 2109913 | Melaminivora sp. SC2-9 |
| 0 | 1 | 0 | G | 2678886 | Pulveribacter |
| 0 | 1 | 1 | S | 2116657 | Pulveribacter suum |
| 0.09 | 50 | 0 | F | 119060 | Burkholderiaceae |
| 0.06 | 31 | 0 | G | 106589 | Cupriavidus |
| 0.03 | 17 | 17 | S | 119219 | Cupriavidus metallidurans |
| 0.02 | 13 | 13 | S | 1796606 | Cupriavidus nantongensis |

| | | | | | |
|---|---|---|---|---|---|
| 0 | 1 | 1 | S | 68895 | Cupriavidus basilensis |
| 0.02 | 12 | 0 | G | 48736 | Ralstonia |
| 0.02 | 11 | 11 | S | 305 | Ralstonia solanacearum |
| 0 | 1 | 0 | S | 329 | Ralstonia pickettii |
| 0 | 1 | 1 | S1 | 428406 | Ralstonia pickettii 12D |
| 0.01 | 5 | 0 | G | 32008 | Burkholderia |
| 0.01 | 5 | 5 | G1 | 87882 | Burkholderia cepacia complex |
| 0 | 2 | 0 | G | 93217 | Pandoraea |
| 0 | 1 | 1 | S | 93220 | Pandoraea pnomenusa |
| 0 | 1 | 1 | S | 573737 | Pandoraea oxalativorans |
| 0.02 | 13 | 0 | F | 75682 | Oxalobacteraceae |
| 0.02 | 12 | 0 | G | 75654 | Duganella |
| 0.02 | 12 | 0 | G1 | 2636909 | unclassified Duganella |
| 0.02 | 12 | 12 | S | 2728020 | Duganella sp. GN2-R2 |
| 0 | 1 | 0 | G | 1344552 | Noviherbaspirillum |
| 0 | 1 | 0 | G1 | 2617509 | unclassified Noviherbaspirillum |
| 0 | 1 | 1 | S | 2601898 | Noviherbaspirillum sp. UKPF54 |
| 0.02 | 9 | 0 | F | 506 | Alcaligenaceae |
| 0.01 | 5 | 0 | G | 517 | Bordetella |
| 0.01 | 5 | 5 | S | 94624 | Bordetella petrii |
| 0 | 2 | 1 | G | 222 | Achromobacter |
| 0 | 1 | 1 | S | 85698 | Achromobacter xylosoxidans |
| 0 | 2 | 0 | G | 507 | Alcaligenes |
| 0 | 2 | 2 | S | 511 | Alcaligenes faecalis |
| 0.01 | 4 | 0 | O1 | 224471 | Burkholderiales genera incertae sedis |
| 0.01 | 4 | 0 | G | 88 | Leptothrix |
| 0.01 | 4 | 0 | S | 34029 | Leptothrix cholodnii |
| 0.01 | 4 | 4 | S1 | 395495 | Leptothrix cholodnii SP-6 |
| 0 | 2 | 0 | F | 995019 | Sutterellaceae |
| 0 | 2 | 0 | G | 40544 | Sutterella |
| 0 | 2 | 2 | S | 40545 | Sutterella wadsworthensis |
| 0 | 2 | 0 | O | 32003 | Nitrosomonadales |
| 0 | 2 | 0 | F | 2008793 | Sterolibacteriaceae |

| | | | | | |
|---|---|---|---|---|---|
| 0 | 2 | 0 | G | 311181 | Denitratisoma |
| 0 | 2 | 2 | S | 311182 | Denitratisoma oestradiolicum |
| 0 | 2 | 0 | O | 206389 | Rhodocyclales |
| 0 | 2 | 0 | F | 2008794 | Zoogloeaceae |
| 0 | 2 | 0 | G | 12960 | Azoarcus |
| 0 | 2 | 2 | G1 | 2629479 | unclassified Azoarcus |
| 1.39 | 775 | 16 | C | 1236 | Gammaproteobacteria |
| 0.81 | 453 | 0 | O | 135614 | Xanthomonadales |
| 0.81 | 453 | 0 | F | 32033 | Xanthomonadaceae |
| 0.8 | 449 | 71 | G | 40323 | Stenotrophomonas |
| 0.68 | 377 | 0 | G1 | 995085 | Stenotrophomonas maltophilia group |
| 0.68 | 377 | 374 | S | 40324 | Stenotrophomonas maltophilia |
| 0 | 2 | 2 | S1 | 868597 | Stenotrophomonas maltophilia JV3 |
| 0 | 1 | 1 | S1 | 391008 | Stenotrophomonas maltophilia R551-3 |
| 0 | 1 | 0 | G1 | 196198 | unclassified Stenotrophomonas |
| 0 | 1 | 1 | S | 2282124 | Stenotrophomonas sp. ASS1 |
| 0.01 | 4 | 0 | G | 338 | Xanthomonas |
| 0.01 | 3 | 3 | S | 339 | Xanthomonas campestris |
| 0 | 1 | 0 | G1 | 643453 | Xanthomonas citri group |
| 0 | 1 | 0 | S | 346 | Xanthomonas citri |
| 0 | 1 | 1 | S1 | 473421 | Xanthomonas citri pv. glycines |
| 0.53 | 297 | 0 | O | 72274 | Pseudomonadales |
| 0.52 | 290 | 0 | F | 135621 | Pseudomonadaceae |
| 0.52 | 290 | 2 | G | 286 | Pseudomonas |
| 0.51 | 284 | 0 | G1 | 136841 | Pseudomonas aeruginosa group |
| 0.51 | 283 | 283 | S | 287 | Pseudomonas aeruginosa |
| 0 | 1 | 0 | G2 | 1232139 | Pseudomonas oleovorans/pseudoalcaligenes group |
| 0 | 1 | 1 | S | 301 | Pseudomonas oleovorans |
| 0 | 2 | 0 | G1 | 136842 | Pseudomonas chlororaphis group |
| 0 | 2 | 2 | S | 587753 | Pseudomonas chlororaphis |
| 0 | 1 | 0 | G1 | 136849 | Pseudomonas syringae group |
| 0 | 1 | 1 | G2 | 251698 | Pseudomonas syringae group genomosp. 2 |
| 0 | 1 | 0 | G1 | 136846 | Pseudomonas stutzeri group |

| | | | | | |
|---|---|---|---|---|---|
| 0 | 1 | 0 | G2 | 578833 | Pseudomonas stutzeri subgroup |
| 0 | 1 | 1 | S | 316 | Pseudomonas stutzeri |
| 0.01 | 7 | 0 | F | 468 | Moraxellaceae |
| 0.01 | 7 | 0 | G | 469 | Acinetobacter |
| 0.01 | 7 | 7 | S | 40214 | Acinetobacter johnsonii |
| 0.01 | 5 | 0 | O | 91347 | Enterobacterales |
| 0.01 | 5 | 2 | F | 543 | Enterobacteriaceae |
| 0 | 1 | 0 | G | 547 | Enterobacter |
| 0 | 1 | 0 | G1 | 354276 | Enterobacter cloacae complex |
| 0 | 1 | 1 | S | 208224 | Enterobacter kobei |
| 0 | 1 | 0 | G | 561 | Escherichia |
| 0 | 1 | 1 | S | 562 | Escherichia coli |
| 0 | 1 | 1 | G | 570 | Klebsiella |
| 0.01 | 4 | 0 | O | 135625 | Pasteurellales |
| 0.01 | 4 | 0 | F | 712 | Pasteurellaceae |
| 0.01 | 4 | 0 | G | 2094023 | Glaesserella |
| 0.01 | 4 | 4 | S | 738 | Glaesserella parasuis |
| 0.15 | 85 | 0 | C | 28211 | Alphaproteobacteria |
| 0.13 | 70 | 0 | O | 356 | Hyphomicrobiales |
| 0.09 | 53 | 0 | F | 69277 | Phyllobacteriaceae |
| 0.08 | 46 | 0 | G | 31988 | Aminobacter |
| 0.08 | 46 | 0 | G1 | 2644704 | unclassified Aminobacter |
| 0.08 | 46 | 46 | S | 2774562 | Aminobacter sp. SR38 |
| 0.01 | 7 | 0 | G | 28100 | Phyllobacterium |
| 0.01 | 5 | 5 | S | 1867719 | Phyllobacterium zundukense |
| 0 | 2 | 0 | G1 | 2638441 | unclassified Phyllobacterium |
| 0 | 2 | 2 | S | 2718938 | Phyllobacterium sp. 628 |
| 0.02 | 11 | 0 | F | 82115 | Rhizobiaceae |
| 0.02 | 11 | 0 | F1 | 227290 | Rhizobium/Agrobacterium group |
| 0.01 | 7 | 0 | G | 1525371 | Neorhizobium |
| 0.01 | 7 | 0 | S | 399 | Neorhizobium galegae |
| 0.01 | 7 | 0 | S1 | 323655 | Neorhizobium galegae bv. orientalis |
| 0.01 | 7 | 7 | S2 | 1028800 | Neorhizobium galegae bv. orientalis str. HAMBI 540 |

| | | | | | |
|---|---|---|---|---|---|
| 0.01 | 4 | 0 | G | 357 | Agrobacterium |
| 0 | 2 | 2 | S | 28099 | Agrobacterium rubi |
| 0 | 2 | 0 | G1 | 1183400 | Agrobacterium tumefaciens complex |
| 0 | 2 | 2 | S | 358 | Agrobacterium tumefaciens |
| 0.01 | 5 | 2 | F | 119045 | Methylobacteriaceae |
| 0 | 2 | 0 | G | 2282523 | Methylorubrum |
| 0 | 1 | 0 | S | 408 | Methylorubrum extorquens |
| 0 | 1 | 1 | S1 | 272630 | Methylorubrum extorquens AM1 |
| 0 | 1 | 1 | S | 223967 | Methylorubrum populi |
| 0 | 1 | 1 | G | 407 | Methylobacterium |
| 0 | 1 | 0 | F | 41294 | Bradyrhizobiaceae |
| 0 | 1 | 0 | G | 374 | Bradyrhizobium |
| 0 | 1 | 0 | G1 | 2631580 | unclassified Bradyrhizobium |
| 0 | 1 | 1 | S | 2493093 | Bradyrhizobium sp. LCT2 |
| 0.03 | 14 | 0 | O | 204457 | Sphingomonadales |
| 0.03 | 14 | 0 | F | 41297 | Sphingomonadaceae |
| 0.01 | 7 | 0 | G | 13687 | Sphingomonas |
| 0.01 | 5 | 5 | S | 13689 | Sphingomonas paucimobilis |
| 0 | 2 | 0 | G1 | 196159 | unclassified Sphingomonas |
| 0 | 1 | 1 | S | 2698679 | Sphingomonas sp. C33 |
| 0 | 1 | 1 | S | 304378 | Sphingomonas sp. IC081 |
| 0.01 | 4 | 3 | G | 165695 | Sphingobium |
| 0 | 1 | 0 | G1 | 2611147 | unclassified Sphingobium |
| 0 | 1 | 1 | S | 2082188 | Sphingobium sp. YG1 |
| 0.01 | 3 | 2 | G | 165697 | Sphingopyxis |
| 0 | 1 | 0 | S | 117207 | Sphingopyxis alaskensis |
| 0 | 1 | 1 | S1 | 317655 | Sphingopyxis alaskensis RB2256 |
| 0 | 1 | 0 | O | 204458 | Caulobacterales |
| 0 | 1 | 0 | F | 76892 | Caulobacteraceae |
| 0 | 1 | 1 | G | 41275 | Brevundimonas |
| 0.04 | 21 | 0 | P1 | 68525 | delta/epsilon subdivisions |
| 0.03 | 18 | 0 | C | 29547 | Epsilonproteobacteria |
| 0.03 | 18 | 0 | O | 213849 | Campylobacterales |

| | | | | | |
|---|---|---|---|---|---|
| 0.03 | 18 | 0 | F | 72294 | Campylobacteraceae |
| 0.03 | 18 | 7 | G | 194 | Campylobacter |
| 0.02 | 11 | 11 | S | 195 | Campylobacter coli |
| 0.01 | 3 | 0 | C | 28221 | Deltaproteobacteria |
| 0 | 2 | 0 | O | 69541 | Desulfuromonadales |
| 0 | 2 | 0 | F | 2812024 | Syntrophotaleaceae |
| 0 | 2 | 0 | G | 2812025 | Syntrophotalea |
| 0 | 2 | 2 | S | 1842532 | Syntrophotalea acetylenivorans |
| 0 | 1 | 0 | O | 29 | Myxococcales |
| 0 | 1 | 0 | O1 | 80811 | Cystobacterineae |
| 0 | 1 | 0 | F | 31 | Myxococcaceae |
| 0 | 1 | 0 | G | 83461 | Corallococcus |
| 0 | 1 | 1 | S | 184914 | Corallococcus coralloides |
| 2.34 | 1304 | 0 | D1 | 1783270 | FCB group |
| 2.34 | 1304 | 0 | D2 | 68336 | Bacteroidetes/Chlorobi group |
| 2.34 | 1304 | 3 | P | 976 | Bacteroidetes |
| 2.31 | 1288 | 0 | C | 200643 | Bacteroidia |
| 2.31 | 1288 | 318 | O | 171549 | Bacteroidales |
| 0.77 | 429 | 0 | F | 815 | Bacteroidaceae |
| 0.77 | 429 | 80 | G | 816 | Bacteroides |
| 0.19 | 107 | 107 | S | 820 | Bacteroides uniformis |
| 0.11 | 64 | 20 | G1 | 2646097 | unclassified Bacteroides |
| 0.04 | 24 | 24 | S | 2785531 | Bacteroides sp. HF-162 |
| 0.01 | 7 | 7 | S | 2528203 | Bacteroides sp. A1C1 |
| 0.01 | 5 | 5 | S | 2650157 | Bacteroides sp. HF-5287 |
| 0.01 | 5 | 5 | S | 2755405 | Bacteroides sp. CACC 737 |
| 0 | 1 | 1 | S | 2650158 | Bacteroides sp. HF-5141 |
| 0 | 1 | 1 | S | 2715212 | Bacteroides sp. CBA7301 |
| 0 | 1 | 1 | S | 2763022 | Bacteroides sp. M10 |
| 0.07 | 40 | 40 | S | 28116 | Bacteroides ovatus |
| 0.07 | 38 | 38 | S | 28111 | Bacteroides eggerthii |
| 0.06 | 35 | 35 | S | 246787 | Bacteroides cellulosilyticus |
| 0.05 | 27 | 27 | S | 818 | Bacteroides thetaiotaomicron |

| | | | | | |
|---|---|---|---|---|---|
| 0.02 | 10 | 8 | S | 371601 | Bacteroides xylanisolvens |
| 0 | 2 | 2 | S1 | 657309 | Bacteroides xylanisolvens XB1A |
| 0.02 | 9 | 9 | S | 47678 | Bacteroides caccae |
| 0.01 | 7 | 6 | S | 817 | Bacteroides fragilis |
| 0 | 1 | 1 | S1 | 295405 | Bacteroides fragilis YCH46 |
| 0.01 | 7 | 7 | S | 329854 | Bacteroides intestinalis |
| 0 | 2 | 0 | S | 290053 | Bacteroides helcogenes |
| 0 | 2 | 2 | S1 | 693979 | Bacteroides helcogenes P 36-108 |
| 0 | 2 | 2 | S | 1796613 | Bacteroides caecimuris |
| 0 | 1 | 1 | S | 28113 | Bacteroides heparinolyticus |
| 0.59 | 328 | 0 | O1 | 333046 | Bacteroidales incertae sedis |
| 0.59 | 328 | 35 | G | 909656 | Phocaeicola |
| 0.28 | 154 | 25 | S | 357276 | Phocaeicola dorei |
| 0.23 | 129 | 129 | S1 | 997877 | Bacteroides dorei CL03T12C01 |
| 0.25 | 139 | 139 | S | 821 | Phocaeicola vulgatus |
| 0.2 | 111 | 1 | F | 1853231 | Odoribacteraceae |
| 0.14 | 79 | 2 | G | 574697 | Butyricimonas |
| 0.13 | 74 | 74 | S | 544645 | Butyricimonas virosa |
| 0.01 | 3 | 3 | S | 2093856 | Butyricimonas faecalis |
| 0.06 | 31 | 0 | G | 283168 | Odoribacter |
| 0.06 | 31 | 31 | S | 28118 | Odoribacter splanchnicus |
| 0.09 | 53 | 0 | F | 171550 | Rikenellaceae |
| 0.09 | 53 | 1 | G | 239759 | Alistipes |
| 0.03 | 17 | 0 | S | 328813 | Alistipes onderdonkii |
| 0.03 | 17 | 17 | S1 | 2585117 | Alistipes onderdonkii subsp. vulgaris |
| 0.03 | 16 | 0 | S | 328814 | Alistipes shahii |
| 0.03 | 16 | 16 | S1 | 717959 | Alistipes shahii WAL 8301 |
| 0.02 | 9 | 0 | S | 214856 | Alistipes finegoldii |
| 0.02 | 9 | 9 | S1 | 679935 | Alistipes finegoldii DSM 17242 |
| 0.01 | 8 | 8 | S | 2585118 | Alistipes communis |
| 0 | 2 | 2 | S | 2585119 | Alistipes dispar |
| 0.07 | 39 | 0 | F | 171552 | Prevotellaceae |
| 0.07 | 39 | 0 | G | 838 | Prevotella |

| | | | | | |
|---|---|---|---|---|---|
| 0.07 | 38 | 38 | S | 165179 | Prevotella copri |
| 0 | 1 | 1 | S | 28132 | Prevotella melaninogenica |
| 0.01 | 7 | 0 | F | 2005525 | Tannerellaceae |
| 0.01 | 7 | 3 | G | 375288 | Parabacteroides |
| 0 | 2 | 2 | S | 823 | Parabacteroides distasonis |
| 0 | 2 | 2 | S | 328812 | Parabacteroides goldsteinii |
| 0 | 2 | 0 | F | 2005519 | Barnesiellaceae |
| 0 | 2 | 0 | G | 397864 | Barnesiella |
| 0 | 2 | 0 | S | 397865 | Barnesiella viscericola |
| 0 | 2 | 2 | S1 | 880074 | Barnesiella viscericola DSM 18177 |
| 0 | 1 | 0 | F | 171551 | Porphyromonadaceae |
| 0 | 1 | 0 | G | 836 | Porphyromonas |
| 0 | 1 | 1 | S | 837 | Porphyromonas gingivalis |
| 0.01 | 8 | 0 | C | 768503 | Cytophagia |
| 0.01 | 8 | 5 | O | 768507 | Cytophagales |
| 0.01 | 3 | 0 | F | 1853232 | Hymenobacteraceae |
| 0.01 | 3 | 0 | G | 89966 | Hymenobacter |
| 0.01 | 3 | 0 | G1 | 2615202 | unclassified Hymenobacter |
| 0.01 | 3 | 3 | S | 2584940 | Hymenobacter sp. DG01 |
| 0.01 | 5 | 0 | C | 117743 | Flavobacteriia |
| 0.01 | 5 | 0 | O | 200644 | Flavobacteriales |
| 0.01 | 4 | 0 | F | 2762318 | Weeksellaceae |
| 0.01 | 4 | 0 | F1 | 2782232 | Chryseobacterium group |
| 0.01 | 4 | 0 | G | 59732 | Chryseobacterium |
| 0.01 | 4 | 0 | G1 | 2593645 | unclassified Chryseobacterium |
| 0.01 | 4 | 4 | S | 2487065 | Chryseobacterium sp. G0201 |
| 0 | 1 | 0 | F | 49546 | Flavobacteriaceae |
| 0 | 1 | 1 | G | 237 | Flavobacterium |
| 0.9 | 502 | 1 | D1 | 1783272 | Terrabacteria group |
| 0.59 | 329 | 3 | P | 1239 | Firmicutes |
| 0.29 | 162 | 0 | C | 186801 | Clostridia |
| 0.29 | 161 | 12 | O | 186802 | Eubacteriales |
| 0.18 | 101 | 3 | F | 216572 | Oscillospiraceae |

| | | | | | |
|---|---|---|---|---|---|
| 0.16 | 90 | 0 | G | 216851 | Faecalibacterium |
| 0.16 | 90 | 51 | S | 853 | Faecalibacterium prausnitzii |
| 0.04 | 21 | 21 | S1 | 657322 | Faecalibacterium prausnitzii SL3/3 |
| 0.03 | 18 | 18 | S1 | 718252 | Faecalibacterium prausnitzii L2-6 |
| 0.01 | 6 | 1 | G | 1263 | Ruminococcus |
| 0.01 | 5 | 5 | S | 1160721 | Ruminococcus bicirculans |
| 0 | 1 | 0 | G | 459786 | Oscillibacter |
| 0 | 1 | 0 | G1 | 2629304 | unclassified Oscillibacter |
| 0 | 1 | 1 | S | 2109687 | Oscillibacter sp. PEA192 |
| 0 | 1 | 0 | G | 1905344 | Ruthenibacterium |
| 0 | 1 | 1 | S | 1550024 | Ruthenibacterium lactatiformans |
| 0.04 | 21 | 3 | F | 186803 | Lachnospiraceae |
| 0.01 | 7 | 0 | G | 841 | Roseburia |
| 0.01 | 5 | 5 | S | 166486 | Roseburia intestinalis |
| 0 | 2 | 2 | S | 301301 | Roseburia hominis |
| 0 | 2 | 0 | G | 33042 | Coprococcus |
| 0 | 1 | 1 | S | 116085 | Coprococcus catus |
| 0 | 1 | 0 | G1 | 2684943 | unclassified Coprococcus |
| 0 | 1 | 1 | S | 751585 | Coprococcus sp. ART55/1 |
| 0 | 2 | 0 | G | 207244 | Anaerostipes |
| 0 | 2 | 2 | S | 649756 | Anaerostipes hadrus |
| 0 | 2 | 0 | G | 2719231 | Lacrimispora |
| 0 | 2 | 0 | S | 84030 | Lacrimispora saccharolytica |
| 0 | 2 | 2 | S1 | 717608 | [Clostridium] cf. saccharolyticum K10 |
| 0 | 1 | 0 | G | 830 | Butyrivibrio |
| 0 | 1 | 1 | S | 831 | Butyrivibrio fibrisolvens |
| 0 | 1 | 0 | G | 28050 | Lachnospira |
| 0 | 1 | 0 | S | 39485 | Lachnospira eligens |
| 0 | 1 | 1 | S1 | 515620 | [Eubacterium] eligens ATCC 27750 |
| 0 | 1 | 0 | G | 1506553 | Lachnoclostridium |
| 0 | 1 | 1 | S | 29347 | [Clostridium] scindens |
| 0 | 1 | 0 | G | 1843210 | Anaerocolumna |
| 0 | 1 | 1 | S | 433286 | Anaerocolumna cellulosilytica |

| | | | | | |
|---|---|---|---|---|---|
| 0 | 1 | 0 | G | 2316020 | Mediterraneibacter |
| 0 | 1 | 1 | S | 33038 | [Ruminococcus] gnavus |
| 0.03 | 18 | 3 | F | 186804 | Peptostreptococcaceae |
| 0.03 | 14 | 1 | G | 1501226 | Romboutsia |
| 0.02 | 12 | 12 | S | 1115758 | Romboutsia ilealis |
| 0 | 1 | 1 | S | 1507512 | Romboutsia hominis |
| 0 | 1 | 0 | G | 2743582 | Peptacetobacter |
| 0 | 1 | 1 | S | 89152 | Peptacetobacter hiranonis |
| 0.01 | 5 | 0 | F | 31979 | Clostridiaceae |
| 0.01 | 4 | 1 | G | 1485 | Clostridium |
| 0 | 1 | 0 | G1 | 2614128 | unclassified Clostridium |
| 0 | 1 | 1 | S | 641107 | Clostridium sp. DL-VIII |
| 0 | 1 | 1 | S | 1504 | Clostridium septicum |
| 0 | 1 | 1 | S | 1502 | Clostridium perfringens |
| 0 | 1 | 0 | G | 1649459 | Hungatella |
| 0 | 1 | 0 | S | 154046 | Hungatella hathewayi |
| 0 | 1 | 1 | S1 | 742737 | Hungatella hathewayi WAL-18680 |
| 0 | 2 | 0 | O1 | 538999 | Eubacteriales incertae sedis |
| 0 | 2 | 0 | G | 2039302 | Monoglobus |
| 0 | 2 | 2 | S | 1981510 | Monoglobus pectinilyticus |
| 0 | 2 | 0 | F | 990719 | Christensenellaceae |
| 0 | 2 | 0 | G | 990721 | Christensenella |
| 0 | 1 | 1 | S | 626937 | Christensenella minuta |
| 0 | 1 | 0 | G1 | 2649046 | unclassified Christensenella |
| 0 | 1 | 1 | S | 2086585 | Christensenella sp. Marseille-P3954 |
| 0 | 1 | 0 | O | 485256 | Natranaerobiales |
| 0 | 1 | 0 | F | 485255 | Natranaerobiaceae |
| 0 | 1 | 0 | G | 375928 | Natranaerobius |
| 0 | 1 | 0 | S | 375929 | Natranaerobius thermophilus |
| 0 | 1 | 1 | S1 | 457570 | Natranaerobius thermophilus JW/NM-WN-LF |
| 0.17 | 95 | 0 | C | 526524 | Erysipelotrichia |
| 0.17 | 95 | 0 | O | 526525 | Erysipelotrichales |
| 0.17 | 93 | 0 | F | 2810281 | Turicibacteraceae |

| | | | | | |
|---|---|---|---|---|---|
| 0.17 | 93 | 0 | G | 191303 | Turicibacter |
| 0.12 | 65 | 65 | S | 154288 | Turicibacter sanguinis |
| 0.05 | 28 | 0 | G1 | 2638206 | unclassified Turicibacter |
| 0.05 | 28 | 28 | S | 1712675 | Turicibacter sp. H121 |
| 0 | 2 | 0 | F | 128827 | Erysipelotrichaceae |
| 0 | 1 | 0 | G | 1918538 | Longibaculum |
| 0 | 1 | 0 | G1 | 2636055 | unclassified Longibaculum |
| 0 | 1 | 1 | S | 2584943 | Longibaculum sp. KGMB06250 |
| 0 | 1 | 0 | G | 2678885 | Faecalibacillus |
| 0 | 1 | 1 | S | 1982626 | Faecalibacillus intestinalis |
| 0.11 | 63 | 0 | C | 91061 | Bacilli |
| 0.09 | 53 | 0 | O | 1385 | Bacillales |
| 0.09 | 50 | 0 | F | 90964 | Staphylococcaceae |
| 0.09 | 50 | 3 | G | 1279 | Staphylococcus |
| 0.07 | 40 | 40 | S | 1282 | Staphylococcus epidermidis |
| 0.01 | 3 | 3 | S | 1290 | Staphylococcus hominis |
| 0 | 2 | 2 | S | 29388 | Staphylococcus capitis |
| 0 | 2 | 2 | S | 1283 | Staphylococcus haemolyticus |
| 0 | 2 | 0 | F | 186817 | Bacillaceae |
| 0 | 2 | 0 | G | 1386 | Bacillus |
| 0 | 2 | 0 | G1 | 86661 | Bacillus cereus group |
| 0 | 2 | 2 | S | 64104 | Bacillus pseudomycoides |
| 0 | 1 | 0 | O1 | 539002 | Bacillales incertae sedis |
| 0 | 1 | 0 | O2 | 539738 | Bacillales Family XI. Incertae Sedis |
| 0 | 1 | 0 | G | 1378 | Gemella |
| 0 | 1 | 1 | S | 1379 | Gemella haemolysans |
| 0.02 | 10 | 1 | O | 186826 | Lactobacillales |
| 0.01 | 7 | 0 | F | 1300 | Streptococcaceae |
| 0.01 | 7 | 2 | G | 1301 | Streptococcus |
| 0 | 2 | 0 | G1 | 2608887 | unclassified Streptococcus |
| 0 | 2 | 2 | S | 1759399 | Streptococcus sp. A12 |
| 0 | 1 | 0 | S | 257758 | Streptococcus pseudopneumoniae |
| 0 | 1 | 1 | S1 | 1054460 | Streptococcus pseudopneumoniae IS7493 |

| | | | | | |
|---|---|---|---|---|---|
| 0 | 1 | 1 | S | 1304 | Streptococcus salivarius |
| 0 | 1 | 1 | S | 1343 | Streptococcus vestibularis |
| 0 | 2 | 0 | F | 81852 | Enterococcaceae |
| 0 | 2 | 0 | G | 1350 | Enterococcus |
| 0 | 2 | 2 | S | 1351 | Enterococcus faecalis |
| 0.01 | 6 | 0 | C | 1737404 | Tissierellia |
| 0.01 | 6 | 0 | O | 1737405 | Tissierellales |
| 0.01 | 6 | 0 | F | 1570339 | Peptoniphilaceae |
| 0.01 | 6 | 0 | G | 165779 | Anaerococcus |
| 0.01 | 6 | 6 | S | 1870984 | Anaerococcus mediterraneensis |
| 0.31 | 171 | 0 | P | 201174 | Actinobacteria |
| 0.29 | 164 | 0 | C | 1760 | Actinomycetia |
| 0.24 | 136 | 0 | O | 85009 | Propionibacteriales |
| 0.24 | 133 | 3 | F | 31957 | Propionibacteriaceae |
| 0.23 | 130 | 3 | G | 1912216 | Cutibacterium |
| 0.22 | 121 | 121 | S | 1747 | Cutibacterium acnes |
| 0.01 | 5 | 5 | S | 33011 | Cutibacterium granulosum |
| 0 | 1 | 1 | S | 33010 | Cutibacterium avidum |
| 0.01 | 3 | 0 | F | 85015 | Nocardioidaceae |
| 0.01 | 3 | 0 | G | 1839 | Nocardioides |
| 0.01 | 3 | 0 | G1 | 2615069 | unclassified Nocardioides |
| 0 | 2 | 2 | S | 2763008 | Nocardioides sp. zg-1228 |
| 0 | 1 | 1 | S | 2017486 | Nocardioides sp. S5 |
| 0.03 | 14 | 0 | O | 85006 | Micrococcales |
| 0.01 | 6 | 0 | F | 1268 | Micrococcaceae |
| 0.01 | 6 | 0 | G | 32207 | Rothia |
| 0.01 | 5 | 5 | S | 172042 | Rothia aeria |
| 0 | 1 | 1 | S | 2047 | Rothia dentocariosa |
| 0.01 | 5 | 0 | F | 85020 | Dermabacteraceae |
| 0.01 | 5 | 0 | G | 36739 | Dermabacter |
| 0.01 | 5 | 5 | S | 1630135 | Dermabacter vaginalis |
| 0.01 | 3 | 2 | F | 85023 | Microbacteriaceae |
| 0 | 1 | 0 | G | 33882 | Microbacterium |

| | | | | | |
|---|---|---|---|---|---|
| 0 | 1 | 1 | S | 104336 | Microbacterium foliorum |
| 0.02 | 9 | 0 | O | 85007 | Corynebacteriales |
| 0.02 | 9 | 0 | F | 1653 | Corynebacteriaceae |
| 0.02 | 9 | 0 | G | 1716 | Corynebacterium |
| 0.01 | 5 | 5 | S | 38304 | Corynebacterium tuberculostearicum |
| 0 | 2 | 2 | S | 161879 | Corynebacterium kroppenstedtii |
| 0 | 1 | 0 | G1 | 2624378 | unclassified Corynebacterium |
| 0 | 1 | 1 | S | 2778078 | Corynebacterium sp. FDAARGOS 1242 |
| 0 | 1 | 1 | S | 161899 | Corynebacterium singulare |
| 0.01 | 5 | 0 | O | 85004 | Bifidobacteriales |
| 0.01 | 5 | 0 | F | 31953 | Bifidobacteriaceae |
| 0.01 | 4 | 0 | G | 1678 | Bifidobacterium |
| 0 | 2 | 2 | S | 1681 | Bifidobacterium bifidum |
| 0 | 1 | 1 | S | 1680 | Bifidobacterium adolescentis |
| 0 | 1 | 1 | S | 1685 | Bifidobacterium breve |
| 0 | 1 | 0 | G | 2701 | Gardnerella |
| 0 | 1 | 1 | S | 2702 | Gardnerella vaginalis |
| 0.01 | 7 | 0 | C | 84998 | Coriobacteriia |
| 0.01 | 6 | 0 | O | 84999 | Coriobacteriales |
| 0.01 | 6 | 0 | F | 84107 | Coriobacteriaceae |
| 0.01 | 6 | 0 | G | 102106 | Collinsella |
| 0.01 | 6 | 6 | S | 74426 | Collinsella aerofaciens |
| 0 | 1 | 0 | O | 1643822 | Eggerthellales |
| 0 | 1 | 0 | F | 1643826 | Eggerthellaceae |
| 0 | 1 | 0 | G | 644652 | Gordonibacter |
| 0 | 1 | 0 | S | 471189 | Gordonibacter pamelaeae |
| 0 | 1 | 1 | S1 | 657308 | Gordonibacter pamelaeae 7-10-1-b |
| 0 | 1 | 0 | P | 544448 | Tenericutes |
| 0 | 1 | 0 | C | 31969 | Mollicutes |
| 0 | 1 | 0 | O | 186328 | Entomoplasmatales |
| 0 | 1 | 0 | F | 2131 | Spiroplasmataceae |
| 0 | 1 | 1 | G | 2132 | Spiroplasma |
| 6.63 | 3699 | 2 | D | 2759 | Eukaryota |

| | | | | | |
|---|---|---|---|---|---|
| 6.59 | 3680 | 0 | D1 | 33154 | Opisthokonta |
| 6.55 | 3655 | 0 | K | 4751 | Fungi |
| 6.55 | 3655 | 0 | K1 | 451864 | Dikarya |
| 6.54 | 3654 | 0 | P | 5204 | Basidiomycota |
| 6.54 | 3654 | 0 | P1 | 452284 | Ustilaginomycotina |
| 6.54 | 3654 | 0 | C | 1538075 | Malasseziomycetes |
| 6.54 | 3654 | 0 | O | 162474 | Malasseziales |
| 6.54 | 3654 | 0 | F | 742845 | Malasseziaceae |
| 6.54 | 3654 | 0 | G | 55193 | Malassezia |
| 6.54 | 3654 | 3654 | S | 76775 | Malassezia restricta |
| 0 | 1 | 0 | P | 4890 | Ascomycota |
| 0 | 1 | 0 | P1 | 716545 | saccharomyceta |
| 0 | 1 | 0 | P2 | 147538 | Pezizomycotina |
| 0 | 1 | 0 | P3 | 716546 | leotiomyceta |
| 0 | 1 | 0 | P4 | 715962 | dothideomyceta |
| 0 | 1 | 0 | C | 147541 | Dothideomycetes |
| 0 | 1 | 0 | C1 | 451867 | Dothideomycetidae |
| 0 | 1 | 0 | O | 2726947 | Mycosphaerellales |
| 0 | 1 | 0 | F | 93133 | Mycosphaerellaceae |
| 0 | 1 | 0 | G | 1047167 | Zymoseptoria |
| 0 | 1 | 0 | S | 1047171 | Zymoseptoria tritici |
| 0 | 1 | 1 | S1 | 336722 | Zymoseptoria tritici IPO323 |
| 0.04 | 25 | 0 | K | 33208 | Metazoa |
| 0.04 | 25 | 0 | K1 | 6072 | Eumetazoa |
| 0.04 | 25 | 0 | K2 | 33213 | Bilateria |
| 0.04 | 25 | 0 | K3 | 33511 | Deuterostomia |
| 0.04 | 25 | 0 | P | 7711 | Chordata |
| 0.04 | 25 | 0 | P1 | 89593 | Craniata |
| 0.04 | 25 | 0 | P2 | 7742 | Vertebrata |
| 0.04 | 25 | 0 | P3 | 7776 | Gnathostomata |
| 0.04 | 25 | 0 | P4 | 117570 | Teleostomi |
| 0.04 | 25 | 0 | P5 | 117571 | Euteleostomi |
| 0.04 | 25 | 0 | P6 | 8287 | Sarcopterygii |

| | | | | | |
|---|---|---|---|---|---|
| 0.04 | 25 | 0 | C | 1338369 | Dipnotetrapodomorpha |
| 0.04 | 25 | 0 | C1 | 32523 | Tetrapoda |
| 0.04 | 25 | 0 | C2 | 32524 | Amniota |
| 0.04 | 25 | 0 | C | 40674 | Mammalia |
| 0.04 | 25 | 0 | C1 | 32525 | Theria |
| 0.04 | 25 | 0 | C2 | 9347 | Eutheria |
| 0.04 | 25 | 0 | C3 | 1437010 | Boreoeutheria |
| 0.04 | 25 | 0 | C4 | 314146 | Euarchontoglires |
| 0.04 | 25 | 0 | O | 9443 | Primates |
| 0.04 | 25 | 0 | O1 | 376913 | Haplorrhini |
| 0.04 | 25 | 0 | O2 | 314293 | Simiiformes |
| 0.04 | 25 | 0 | O3 | 9526 | Catarrhini |
| 0.04 | 25 | 0 | O4 | 314295 | Hominoidea |
| 0.04 | 25 | 0 | F | 9604 | Hominidae |
| 0.04 | 25 | 0 | F1 | 207598 | Homininae |
| 0.04 | 25 | 0 | G | 9605 | Homo |
| 0.04 | 25 | 25 | S | 9606 | Homo sapiens |
| 0.03 | 17 | 0 | K | 33090 | Viridiplantae |
| 0.03 | 17 | 0 | P | 35493 | Streptophyta |
| 0.03 | 17 | 0 | P1 | 131221 | Streptophytina |
| 0.03 | 17 | 0 | P2 | 3193 | Embryophyta |
| 0.03 | 17 | 0 | P3 | 58023 | Tracheophyta |
| 0.03 | 17 | 0 | P4 | 78536 | Euphyllophyta |
| 0.03 | 17 | 0 | P5 | 58024 | Spermatophyta |
| 0.03 | 17 | 0 | C | 3398 | Magnoliopsida |
| 0.03 | 17 | 0 | C1 | 1437183 | Mesangiospermae |
| 0.01 | 8 | 0 | C2 | 71240 | eudicotyledons |
| 0.01 | 8 | 0 | C3 | 91827 | Gunneridae |
| 0.01 | 8 | 0 | C4 | 1437201 | Pentapetalae |
| 0.01 | 5 | 0 | C5 | 71275 | rosids |
| 0.01 | 4 | 0 | C6 | 91835 | fabids |
| 0 | 1 | 0 | O | 3502 | Fagales |
| 0 | 1 | 0 | F | 3503 | Fagaceae |

| | | | | | |
|---|---|---|---|---|---|
| 0 | 1 | 0 | G | 3511 | Quercus |
| 0 | 1 | 1 | S | 97700 | Quercus lobata |
| 0 | 1 | 0 | O | 3744 | Rosales |
| 0 | 1 | 0 | F | 3745 | Rosaceae |
| 0 | 1 | 0 | F1 | 171638 | Rosoideae |
| 0 | 1 | 0 | F2 | 1176516 | Rosoideae incertae sedis |
| 0 | 1 | 0 | G | 3764 | Rosa |
| 0 | 1 | 1 | S | 74649 | Rosa chinensis |
| 0 | 1 | 0 | O | 72025 | Fabales |
| 0 | 1 | 0 | F | 3803 | Fabaceae |
| 0 | 1 | 0 | F1 | 3814 | Papilionoideae |
| 0 | 1 | 0 | F2 | 2231393 | 50 kb inversion clade |
| 0 | 1 | 0 | F3 | 2231382 | NPAAA clade |
| 0 | 1 | 0 | F4 | 2233855 | indigoferoid/millettioid clade |
| 0 | 1 | 0 | F5 | 163735 | Phaseoleae |
| 0 | 1 | 0 | G | 3820 | Cajanus |
| 0 | 1 | 1 | S | 3821 | Cajanus cajan |
| 0 | 1 | 0 | O | 233875 | Celastrales |
| 0 | 1 | 0 | F | 4305 | Celastraceae |
| 0 | 1 | 0 | G | 123484 | Tripterygium |
| 0 | 1 | 1 | S | 458696 | Tripterygium wilfordii |
| 0 | 1 | 0 | C6 | 91834 | rosids incertae sedis |
| 0 | 1 | 0 | O | 403667 | Vitales |
| 0 | 1 | 0 | F | 3602 | Vitaceae |
| 0 | 1 | 0 | F1 | 2304100 | Viteae |
| 0 | 1 | 1 | G | 3603 | Vitis |
| 0.01 | 3 | 0 | C5 | 71274 | asterids |
| 0.01 | 3 | 0 | C6 | 91888 | lamiids |
| 0.01 | 3 | 0 | O | 4069 | Solanales |
| 0.01 | 3 | 0 | F | 4070 | Solanaceae |
| 0.01 | 3 | 0 | F1 | 424551 | Solanoideae |
| 0.01 | 3 | 0 | F2 | 424564 | Capsiceae |
| 0.01 | 3 | 0 | G | 4071 | Capsicum |

| | | | | | |
|---|---|---|---|---|---|
| 0.01 | 3 | 3 | S | 4072 | Capsicum annuum |
| 0.01 | 5 | 0 | C2 | 4447 | Liliopsida |
| 0.01 | 5 | 0 | C3 | 1437197 | Petrosaviidae |
| 0.01 | 5 | 0 | C4 | 4734 | commelinids |
| 0.01 | 4 | 0 | O | 38820 | Poales |
| 0.01 | 4 | 0 | F | 4479 | Poaceae |
| 0.01 | 3 | 0 | F1 | 359160 | BOP clade |
| 0.01 | 3 | 0 | F2 | 147368 | Pooideae |
| 0.01 | 3 | 0 | F3 | 1648038 | Triticodae |
| 0.01 | 3 | 0 | F4 | 147389 | Triticeae |
| 0.01 | 3 | 0 | F5 | 1648030 | Triticinae |
| 0 | 2 | 0 | G | 4564 | Triticum |
| 0 | 2 | 2 | S | 85692 | Triticum dicoccoides |
| 0 | 1 | 0 | G | 4480 | Aegilops |
| 0 | 1 | 0 | S | 37682 | Aegilops tauschii |
| 0 | 1 | 1 | S1 | 200361 | Aegilops tauschii subsp. strangulata |
| 0 | 1 | 0 | F1 | 147370 | PACMAD clade |
| 0 | 1 | 0 | F2 | 147369 | Panicoideae |
| 0 | 1 | 0 | F3 | 1648033 | Andropogonodae |
| 0 | 1 | 0 | F4 | 147429 | Andropogoneae |
| 0 | 1 | 0 | F5 | 1648029 | Tripsacinae |
| 0 | 1 | 0 | G | 4575 | Zea |
| 0 | 1 | 1 | S | 4577 | Zea mays |
| 0 | 1 | 0 | O | 4618 | Zingiberales |
| 0 | 1 | 0 | F | 4637 | Musaceae |
| 0 | 1 | 0 | G | 4640 | Musa |
| 0 | 1 | 0 | S | 4641 | Musa acuminata |
| 0 | 1 | 1 | S1 | 214687 | Musa acuminata subsp. malaccensis |
| 0.01 | 4 | 0 | O | 41768 | Ranunculales |
| 0.01 | 4 | 0 | F | 3465 | Papaveraceae |
| 0.01 | 4 | 0 | F1 | 1462614 | Papaveroideae |
| 0.01 | 4 | 0 | G | 3468 | Papaver |
| 0.01 | 4 | 4 | S | 3469 | Papaver somniferum |

| | | | | | |
|---|---|---|---|---|---|
| 1.16 | 647 | 0 | D | 10239 | Viruses |
| 1.16 | 647 | 0 | D1 | 2731341 | Duplodnaviria |
| 1.16 | 647 | 0 | D2 | 2731360 | Heunggongvirae |
| 1.16 | 647 | 0 | P | 2731618 | Uroviricota |
| 1.16 | 647 | 0 | C | 2731619 | Caudoviricetes |
| 1.16 | 647 | 0 | O | 28883 | Caudovirales |
| 1.12 | 626 | 0 | F | 10744 | Podoviridae |
| 1.12 | 626 | 0 | F1 | 196895 | unclassified Podoviridae |
| 1.12 | 626 | 0 | F2 | 1978007 | crAss-like viruses |
| 1.12 | 626 | 0 | F3 | 2315857 | environmental samples |
| 1.12 | 626 | 626 | S | 1211417 | uncultured crAssphage |
| 0.04 | 21 | 0 | F | 10699 | Siphoviridae |
| 0.02 | 10 | 1 | G | 186532 | Ceduovirus |
| 0.01 | 7 | 4 | G1 | 329156 | unclassified Ceduovirus |
| 0 | 2 | 2 | S | 1862960 | Lactococcus phage M5938 |
| 0 | 1 | 1 | S | 1862959 | Lactococcus phage D4412 |
| 0 | 2 | 2 | S | 36343 | Lactococcus virus bIL67 |
| 0.02 | 9 | 2 | G | 1623305 | Skunavirus |
| 0.01 | 7 | 3 | G1 | 2050979 | unclassified Skunavirus |
| 0 | 2 | 2 | S | 2029660 | Lactococcus phage 30804 |
| 0 | 1 | 1 | S | 39838 | Lactococcus phage 936 |
| 0 | 1 | 1 | S | 2029671 | Lactococcus phage 66901 |
| 0 | 1 | 1 | G | 1982251 | Pahexavirus |
| 0 | 1 | 1 | G | 1623304 | Moineauvirus |
