## Supplementary material for "Comprehensive profiling of the human intestinal DNA virome and prediction of disease-associated bacterial hosts in severe Myalgic Encephalomyelitis/Chronic Fatigue Syndrome (ME/CFS)": S Table 8 Virla taxonomy

| VLP_Scaffold | Closer | Accession | Status | VC | Level | Weight | Host | BaltimoreGroup | Realm | Kingdom | Phylum | Class | Order | Family | Subfamily | Genus |
|---|---|---|---|---|---|---|---|---|---|---|---|---|---|---|---|---|
| S10_vf_18671 | Parabacteroides phage PDS1 | MN929097 | Clustered | VC_855_0 | C1 | 169 | Parabacteroides | Group I | Duplodnaviria | Heunggongvirae | Uroviricota | Caudoviricetes | n.a. | Unclassified | Unclassified | Unclassified |
| S10_vf_24791 | Faecalibacterium phage FP_Epona | MG711462 | Clustered | VC_723_0 | C2 | 95 | Faecalibacterium | Group I | Duplodnaviria | Heunggongvirae | Uroviricota | Caudoviricetes | n.a. | Unclassified | Unclassified | Eponavirus |
| S10_vf_25241 | n.a. | n.a. | Clustered | VC_1558_0 | F | n.a. | n.a. | n.a. | n.a. | n.a. | n.a. | n.a. | n.a. | n.a. | n.a. | n.a. |
| S10_vf_3644 | uncultured phage cr109_1 | MT774399 | Clustered | VC_519_7 | N2 | 277 | Unspecified | Group I | Duplodnaviria | Heunggongvirae | Uroviricota | Caudoviricetes | Crassvirales | Suoliviridae | O | O |
| S10_vf_36880 | uncultured phage cr127_1 | MT774393 | Clustered | VC_693_0 | C1 | 105 | Unspecified | Group I | Duplodnaviria | Heunggongvirae | Uroviricota | Caudoviricetes | Crassvirales | Crevaviridae | Doltivirinae | Kahucivirus |
| S10_vf_64371 | n.a. | n.a. | Clustered | VC_1564_0 | F | n.a. | n.a. | n.a. | n.a. | n.a. | n.a. | n.a. | n.a. | n.a. | n.a. | n.a. |
| S10_vf_80712 | n.a. | n.a. | Clustered | VC_219_0 | A | n.a. | n.a. | n.a. | n.a. | n.a. | n.a. | n.a. | n.a. | n.a. | n.a. | n.a. |
| S10_vs_120489 | n.a. | n.a. | Clustered | VC_1566_0 | A | n.a. | n.a. | n.a. | n.a. | n.a. | n.a. | n.a. | n.a. | n.a. | n.a. | n.a. |
| S10_vs_120541 | n.a. | n.a. | Clustered | VC_1567_0 | A | n.a. | n.a. | n.a. | n.a. | n.a. | n.a. | n.a. | n.a. | n.a. | n.a. | n.a. |
| S10_vs_14557 | Flavobacterium phage vB_FspS_laban6-1 | MN812211 | Clustered | VC_1062_0 | N5 | 6 | Flavobacterium | Group I | Duplodnaviria | Heunggongvirae | Uroviricota | Caudoviricetes | n.a. | Duneviridae | O | O |
| S10_vs_18901 | n.a. | n.a. | Clustered | VC_1291_0 | A | n.a. | n.a. | n.a. | n.a. | n.a. | n.a. | n.a. | n.a. | n.a. | n.a. | n.a. |
| S10_vs_2538 | Lactobacillus phage c5 | EU340421 | Clustered | VC_658_0 | N10 | 6 | Lactobacillus | Group I | Duplodnaviria | Heunggongvirae | Uroviricota | Caudoviricetes | n.a. | Unclassified | O | O |
| S10_vs_26365 | Faecalibacterium phage FP_oengus | MG711463 | Clustered | VC_1085_1 | N20 | 67 | Faecalibacterium | Group I | Duplodnaviria | Heunggongvirae | Uroviricota | Caudoviricetes | n.a. | Unclassified | O | O |
| S10_vs_35854 | Bacteroides phage LoVEphage | MW660583 | Clustered | VC_1527_0 | N4 | 24 | Bacteroides | Group I | Duplodnaviria | Heunggongvirae | Uroviricota | Caudoviricetes | n.a. | Unclassified | O | O |
| S10_vs_36407 | n.a. | n.a. | Clustered | VC_1578_0 | A | n.a. | n.a. | n.a. | n.a. | n.a. | n.a. | n.a. | n.a. | n.a. | n.a. | n.a. |
| S10_vs_36598 | n.a. | n.a. | Clustered | VC_1579_0 | A | n.a. | n.a. | n.a. | n.a. | n.a. | n.a. | n.a. | n.a. | n.a. | n.a. | n.a. |
| S10_vs_43473 | Microviridae sp. | MG945488 | Clustered | VC_1334_0 | C1 | 19 | Unspecified | Group II | Monodnaviria | Sangervirae | Phixviricota | Malgrandaviricetes | Petitvirales | Microviridae | Unclassified | Unclassified |
| S10_vs_58014 | Bacteroides phage p00 | BK010646 | Clustered | VC_695_0 | C2 | 93 | Bacteroides | Group I | Duplodnaviria | Heunggongvirae | Uroviricota | Caudoviricetes | n.a. | Unclassified | Unclassified | Unclassified |
| S10_vs_67471 | n.a. | n.a. | Clustered | VC_950_0 | A | n.a. | n.a. | n.a. | n.a. | n.a. | n.a. | n.a. | n.a. | n.a. | n.a. | n.a. |
| S10_vs_71900 | n.a. | n.a. | Clustered | VC_1576_0 | A | n.a. | n.a. | n.a. | n.a. | n.a. | n.a. | n.a. | n.a. | n.a. | n.a. | n.a. |
| S11_vf_104035 | uncultured phage cr53_1 | MT774396 | Clustered | VC_519_0 | N2 | 233 | Unspecified | Group I | Duplodnaviria | Heunggongvirae | Uroviricota | Caudoviricetes | Crassvirales | Suoliviridae | O | O |
| S11_vf_104044 | uncultured phage cr85_1 | MT774390 | Clustered | VC_1054_0 | C1 | 300 | Unspecified | Group I | Duplodnaviria | Heunggongvirae | Uroviricota | Caudoviricetes | Crassvirales | Steigviridae | Asinivirinae | Kahnovirus |
| S11_vf_104048 | Akkermansia phage DTMo-2021a | CP084202 | Clustered | VC_750_0 | N2 | 1 | Akkermansia | Group I | Duplodnaviria | Heunggongvirae | Uroviricota | Caudoviricetes | n.a. | Unclassified | O | O |
| S11_vf_104052 | Bacillus phage vB_BtS_BMBtp15 | KX190835 | Clustered | VC_759_0 | N6 | 9 | Bacillus | Group I | Duplodnaviria | Heunggongvirae | Uroviricota | Caudoviricetes | n.a. | Unclassified | O | O |
| S11_vf_104059 | uncultured phage cr110_1 | MT774386 | Clustered | VC_691_0 | C1 | 193 | Unspecified | Group I | Duplodnaviria | Heunggongvirae | Uroviricota | Caudoviricetes | Crassvirales | Intestiviridae | Crudevirinae | Delmidovirus |
| S11_vf_104070 | Clostridium phage phiMMP01 | LN681541 | Clustered | VC_150_0 | N17 | 13 | Clostridium | Group I | Duplodnaviria | Heunggongvirae | Uroviricota | Caudoviricetes | n.a. | Unclassified | O | O |
| S11_vf_104071 | Croceibacter phage P2559Y | KC688701 | Clustered | VC_1045_0 | N2 | 16 | Croceibacter | Group I | Duplodnaviria | Heunggongvirae | Uroviricota | Caudoviricetes | n.a. | Unclassified | O | O |
| S11_vf_10907 | Staphylococcus phage PI-Sepi-HH2 | MT880871 | Clustered | VC_759_0 | N11 | 3 | Staphylococcus | Group I | Duplodnaviria | Heunggongvirae | Uroviricota | Caudoviricetes | n.a. | Unclassified | O | O |
| S11_vf_13033 | Faecalibacterium phage FP_oengus | MG711463 | Clustered | VC_1085_1 | N20 | 65 | Faecalibacterium | Group I | Duplodnaviria | Heunggongvirae | Uroviricota | Caudoviricetes | n.a. | Unclassified | O | O |
| S11_vf_27845 | n.a. | n.a. | Clustered | VC_1564_0 | F | n.a. | n.a. | n.a. | n.a. | n.a. | n.a. | n.a. | n.a. | n.a. | n.a. | n.a. |
| S11_vf_28515 | Bacillus phage BUCT082 | MZ969646 | Clustered | VC_219_0 | N9 | 5 | Bacillus | Group I | Duplodnaviria | Heunggongvirae | Uroviricota | Caudoviricetes | n.a. | Unclassified | O | O |
| S11_vf_28645 | n.a. | n.a. | Clustered | VC_1558_0 | F | n.a. | n.a. | n.a. | n.a. | n.a. | n.a. | n.a. | n.a. | n.a. | n.a. | n.a. |
| S11_vf_34224 | Clostridium phage phiMMP01 | LN681541 | Clustered | VC_150_0 | N17 | 13 | Clostridium | Group I | Duplodnaviria | Heunggongvirae | Uroviricota | Caudoviricetes | n.a. | Unclassified | O | O |
| S11_vf_43997 | CrAssphage sp. O-152 | MK069403 | Clustered | VC_691_0 | C1 | 131 | Unspecified | Group I | Duplodnaviria | Heunggongvirae | Uroviricota | Caudoviricetes | Crassvirales | Unclassified | Unclassified | Unclassified |
| S11_vf_47632 | Butyrivibrio phage Idris | MN882554 | Clustered | VC_1289_0 | N7 | 31 | Butyrivibrio | Group I | Duplodnaviria | Heunggongvirae | Uroviricota | Caudoviricetes | n.a. | Unclassified | O | O |
| S11_vf_66160 | Roseburia phage Shimadzu | LR596903 | Clustered | VC_1256_0 | N5 | 4 | Roseburia | Group I | Duplodnaviria | Heunggongvirae | Uroviricota | Caudoviricetes | n.a. | Unclassified | O | O |
| S11_vf_66522 | Actinomyces phage LC001 | ON920540 | Clustered | VC_1435_0 | C1 | 131 | Actinomyces | Group I | Duplodnaviria | Heunggongvirae | Uroviricota | Caudoviricetes | n.a. | Unclassified | Unclassified | Unclassified |
| S11_vf_68693 | n.a. | n.a. | Clustered | VC_1559_0 | A | n.a. | n.a. | n.a. | n.a. | n.a. | n.a. | n.a. | n.a. | n.a. | n.a. | n.a. |
| S11_vf_84303 | n.a. | n.a. | Clustered | VC_1552_0 | A | n.a. | n.a. | n.a. | n.a. | n.a. | n.a. | n.a. | n.a. | n.a. | n.a. | n.a. |
| S11_vf_92391 | n.a. | n.a. | Clustered | VC_1585_0 | A | n.a. | n.a. | n.a. | n.a. | n.a. | n.a. | n.a. | n.a. | n.a. | n.a. | n.a. |
| S11_vf_95067 | n.a. | n.a. | Clustered | VC_1552_0 | A | n.a. | n.a. | n.a. | n.a. | n.a. | n.a. | n.a. | n.a. | n.a. | n.a. | n.a. |
| S11_vs_10172 | Faecalibacterium phage FP_Epona | MG711462 | Clustered | VC_723_0 | C13 | 55 | Faecalibacterium | Group I | Duplodnaviria | Heunggongvirae | Uroviricota | Caudoviricetes | n.a. | Unclassified | Unclassified | Eponavirus |
| S11_vs_104032 | Bdellovibrio phage phi1422 | JQ177062 | Clustered | VC_940_0 | C1 | 26 | Bdellovibrio | Group I | Duplodnaviria | Heunggongvirae | Uroviricota | Caudoviricetes | n.a. | Unclassified | Unclassified | Unclassified |
| S11_vs_104038 | Bacillus phage vB_BtS_BMBtp13 | KX190832 | Clustered | VC_667_0 | C4 | 10 | Bacillus | Group I | Duplodnaviria | Heunggongvirae | Uroviricota | Caudoviricetes | n.a. | Unclassified | Unclassified | Unclassified |
| S11_vs_104041 | Clostridioides phage CD1801 | MW512570 | Clustered | VC_669_0 | N4 | 5 | Clostridioides | Group I | Duplodnaviria | Heunggongvirae | Uroviricota | Caudoviricetes | n.a. | Unclassified | O | O |
| S11_vs_104047 | Streptococcus phage D4446 | MN938931 | Clustered | VC_643_0 | N8 | 9 | Streptococcus | Group I | Duplodnaviria | Heunggongvirae | Uroviricota | Caudoviricetes | n.a. | Unclassified | O | O |
| S11_vs_104053 | Paenibacillus phage PG1 | HQ332138 | Clustered | VC_946_0 | N10 | 2 | Paenibacillus | Group I | Duplodnaviria | Heunggongvirae | Uroviricota | Caudoviricetes | n.a. | Unclassified | O | O |
| S11_vs_104054 | n.a. | n.a. | Clustered | VC_1567_0 | A | n.a. | n.a. | n.a. | n.a. | n.a. | n.a. | n.a. | n.a. | n.a. | n.a. | n.a. |
| S11_vs_104064 | Faecalibacterium phage FP_oengus | MG711463 | Clustered | VC_1085_1 | N19 | 69 | Faecalibacterium | Group I | Duplodnaviria | Heunggongvirae | Uroviricota | Caudoviricetes | n.a. | Unclassified | O | O |
| S11_vs_11267 | Geobacillus phage GBSV1 | DQ340064 | Clustered | VC_778_0 | N24 | 8 | Geobacillus | Group I | Duplodnaviria | Heunggongvirae | Uroviricota | Caudoviricetes | n.a. | Unclassified | O | O |
| S11_vs_11994 | Faecalibacterium phage FP_Taranis | MG711467 | Clustered | VC_732_0 | N3 | 96 | Faecalibacterium | Group I | Duplodnaviria | Heunggongvirae | Uroviricota | Caudoviricetes | n.a. | Unclassified | O | O |
| S11_vs_14882 | n.a. | n.a. | Clustered | VC_1570_0 | A | n.a. | n.a. | n.a. | n.a. | n.a. | n.a. | n.a. | n.a. | n.a. | n.a. | n.a. |
| S11_vs_1937 | Phage vB_RanS_PJN03 | ON245412 | Clustered | VC_951_0 | N13 | 6 | Unspecified | Group I | Duplodnaviria | Heunggongvirae | Uroviricota | Caudoviricetes | n.a. | Unclassified | O | O |
| S11_vs_19892 | Faecalibacterium phage FP_Epona | MG711462 | Clustered | VC_723_0 | C13 | 50 | Faecalibacterium | Group I | Duplodnaviria | Heunggongvirae | Uroviricota | Caudoviricetes | n.a. | Unclassified | Unclassified | Eponavirus |
| S11_vs_23188 | n.a. | n.a. | Clustered | VC_1559_0 | A | n.a. | n.a. | n.a. | n.a. | n.a. | n.a. | n.a. | n.a. | n.a. | n.a. | n.a. |
| S11_vs_23357 | Faecalibacterium phage FP_Epona | MG711462 | Clustered | VC_723_0 | C13 | 52 | Faecalibacterium | Group I | Duplodnaviria | Heunggongvirae | Uroviricota | Caudoviricetes | n.a. | Unclassified | Unclassified | Eponavirus |
| S11_vs_26767 | Streptococcus phage MismyG | OL774867 | Clustered | VC_152_0 | N13 | 9 | Streptococcus | Group I | Duplodnaviria | Heunggongvirae | Uroviricota | Caudoviricetes | n.a. | Unclassified | O | O |
| S11_vs_30739 | Faecalibacterium phage FP_Lagaffe | MG711461 | Clustered | VC_1289_0 | N24 | 3 | Faecalibacterium | Group I | Duplodnaviria | Heunggongvirae | Uroviricota | Caudoviricetes | n.a. | Unclassified | O | O |
| S11_vs_3277 | Bacteroides phage p00 | BK010646 | Clustered | VC_695_0 | C4 | 43 | Bacteroides | Group I | Duplodnaviria | Heunggongvirae | Uroviricota | Caudoviricetes | n.a. | Unclassified | Unclassified | Unclassified |
| S11_vs_34137 | Geobacillus phage GBSV1 | DQ340064 | Clustered | VC_779_0 | N25 | 6 | Geobacillus | Group I | Duplodnaviria | Heunggongvirae | Uroviricota | Caudoviricetes | n.a. | Unclassified | O | O |
| S11_vs_37666 | Clostridium phage HM2 | LT600745 | Clustered | VC_1258_0 | N48 | 4 | Clostridium | Group I | Duplodnaviria | Heunggongvirae | Uroviricota | Caudoviricetes | n.a. | Salasmaviridae | O | O |
| S11_vs_40945 | Faecalibacterium phage FP_Epona | MG711462 | Clustered | VC_723_0 | C13 | 55 | Faecalibacterium | Group I | Duplodnaviria | Heunggongvirae | Uroviricota | Caudoviricetes | n.a. | Unclassified | Unclassified | Eponavirus |
| S11_vs_41107 | Clostridium phage phiMMP01 | LN681541 | Clustered | VC_151_0 | N20 | 16 | Clostridium | Group I | Duplodnaviria | Heunggongvirae | Uroviricota | Caudoviricetes | n.a. | Unclassified | O | O |
| S11_vs_42927 | Streptococcus phage Javan630 | MK448997 | Clustered | VC_649_1 | C1 | 104 | Streptococcus | Group I | Duplodnaviria | Heunggongvirae | Uroviricota | Caudoviricetes | n.a. | Unclassified | Unclassified | Unclassified |
| S11_vs_5077 | Escherichia phage ESSI2_ev015 | LR597649 | Clustered | VC_5_0 | C1 | 115 | Escherichia | Group I | Duplodnaviria | Heunggongvirae | Uroviricota | Caudoviricetes | n.a. | Peduoviridae | Unclassified | Quadragintavirus |
| S11_vs_52577 | Faecalibacterium phage FP_Mushu | MG711460 | Clustered | VC_1288_0 | C1 | 145 | Faecalibacterium | Group I | Duplodnaviria | Heunggongvirae | Uroviricota | Caudoviricetes | n.a. | Unclassified | Unclassified | Mushuvirus |
| S11_vs_58702 | Clostridium phage phiMMP01 | LN681541 | Clustered | VC_151_0 | N17 | 32 | Clostridium | Group I | Duplodnaviria | Heunggongvirae | Uroviricota | Caudoviricetes | n.a. | Unclassified | O | O |
| S11_vs_58930 | Bacteroides phage p00 | BK010646 | Clustered | VC_695_0 | C3 | 46 | Bacteroides | Group I | Duplodnaviria | Heunggongvirae | Uroviricota | Caudoviricetes | n.a. | Unclassified | Unclassified | Unclassified |
| S11_vs_62573 | Roseburia phage Shimadzu | LR596903 | Clustered | VC_1256_0 | N5 | 2 | Roseburia | Group I | Duplodnaviria | Heunggongvirae | Uroviricota | Caudoviricetes | n.a. | Unclassified | O | O |
| S11_vs_6458 | n.a. | n.a. | Clustered | VC_1595_0 | A | n.a. | n.a. | n.a. | n.a. | n.a. | n.a. | n.a. | n.a. | n.a. | n.a. | n.a. |
| S11_vs_68151 | Faecalibacterium phage FP_oengus | MG711463 | Clustered | VC_1085_1 | N19 | 65 | Faecalibacterium | Group I | Duplodnaviria | Heunggongvirae | Uroviricota | Caudoviricetes | n.a. | Unclassified | O | O |
| S11_vs_76784 | Streptococcus phage Javan580 | MK448989 | Clustered | VC_558_0 | N12 | 8 | Streptococcus | Group I | Duplodnaviria | Heunggongvirae | Uroviricota | Caudoviricetes | n.a. | Unclassified | O | O |
| S11_vs_79275 | n.a. | n.a. | Clustered | VC_1570_0 | A | n.a. | n.a. | n.a. | n.a. | n.a. | n.a. | n.a. | n.a. | n.a. | n.a. | n.a. |
| S11_vs_81775 | Bacillus phage vB_BtS_BMBtp13 | KX190832 | Clustered | VC_667_0 | C4 | 10 | Bacillus | Group I | Duplodnaviria | Heunggongvirae | Uroviricota | Caudoviricetes | n.a. | Unclassified | Unclassified | Unclassified |
| S11_vs_8456 | Caldibacillus phage CBP1 | MF595878 | Clustered | VC_349_0 | C2 | 5 | Caldibacillus | Group I | Duplodnaviria | Heunggongvirae | Uroviricota | Caudoviricetes | n.a. | Unclassified | Unclassified | Unclassified |
| S11_vs_84627 | n.a. | n.a. | Clustered | VC_1596_0 | A | n.a. | n.a. | n.a. | n.a. | n.a. | n.a. | n.a. | n.a. | n.a. | n.a. | n.a. |
| S11_vs_84848 | Bacteroides phage p00 | BK010646 | Clustered | VC_695_0 | C1 | 73 | Bacteroides | Group I | Duplodnaviria | Heunggongvirae | Uroviricota | Caudoviricetes | n.a. | Unclassified | Unclassified | Unclassified |
| S11_vs_93101 | Bdellovibrio phage phi1402 | JF344709 | Clustered | VC_967_0 | C4 | 37 | Bdellovibrio | Group I | Duplodnaviria | Heunggongvirae | Uroviricota | Caudoviricetes | n.a. | Unclassified | Unclassified | Unclassified |
| S11_vs_98193 | Bacteroides phage p00 | BK010646 | Clustered | VC_695_0 | C6 | 22 | Bacteroides | Group I | Duplodnaviria | Heunggongvirae | Uroviricota | Caudoviricetes | n.a. | Unclassified | Unclassified | Unclassified |
| S12_vf_10887 | n.a. | n.a. | Clustered | VC_1597_0 | F | n.a. | n.a. | n.a. | n.a. | n.a. | n.a. | n.a. | n.a. | n.a. | n.a. | n.a. |
| S12_vf_128794 | n.a. | n.a. | Clustered | VC_1552_0 | A | n.a. | n.a. | n.a. | n.a. | n.a. | n.a. | n.a. | n.a. | n.a. | n.a. | n.a. |
| S12_vf_169796 | Bacillus phage vB_BcoS-136 | MH884508 | Clustered | VC_491_0 | N22 | 2 | Bacillus | Group I | Duplodnaviria | Heunggongvirae | Uroviricota | Caudoviricetes | n.a. | Unclassified | O | O |
| S12_vf_173208 | n.a. | n.a. | Clustered | VC_1274_0 | A | n.a. | n.a. | n.a. | n.a. | n.a. | n.a. | n.a. | n.a. | n.a. | n.a. | n.a. |
| S12_vf_174374 | n.a. | n.a. | Clustered | VC_1576_0 | A | n.a. | n.a. | n.a. | n.a. | n.a. | n.a. | n.a. | n.a. | n.a. | n.a. | n.a. |
| S12_vf_176437 | Clostridium phage HM2 | LT600745 | Clustered | VC_1261_0 | N43 | 4 | Clostridium | Group I | Duplodnaviria | Heunggongvirae | Uroviricota | Caudoviricetes | n.a. | Salasmaviridae | O | O |
| S12_vf_181024 | Bacillus phage BUCT082 | MZ969646 | Clustered | VC_1040_0 | N19 | 1 | Bacillus | Group I | Duplodnaviria | Heunggongvirae | Uroviricota | Caudoviricetes | n.a. | Unclassified | O | O |

| | | | | | | | | | | | | | | | | |
|---|---|---|---|---|---|---|---|---|---|---|---|---|---|---|---|---|
| S12_vf_188822 | n.a. | n.a. | Clustered | VC_1597_0 | F | n.a. | n.a. | n.a. | n.a. | n.a. | n.a. | n.a. | n.a. | n.a. | n.a. | n.a. |
| S12_vf_188824 | Phage DP SC_6_H4_2017 | MT121964 | Clustered | VC_1490_0 | N7 | 121 | Unspecified | Group I | Duplodnaviria | Heunggongvirae | Uroviricota | Caudoviricetes | n.a. | Unclassified | O | O |
| S12_vf_188830 | n.a. | n.a. | Clustered | VC_1598_0 | F | n.a. | n.a. | n.a. | n.a. | n.a. | n.a. | n.a. | n.a. | n.a. | n.a. | n.a. |
| S12_vf_188838 | Inovirus sp. | MT135293 | Clustered | VC_1495_0 | C1 | 12 | Unspecified | Group II | Monodnaviria | Loebvirae | Hofneiviricota | Faserviricetes | Tubulavirales | Inoviridae | Unclassified | Inovirus |
| S12_vf_48278 | Myoviridae sp. | MW202455 | Clustered | VC_439_0 | N3 | 3 | Unspecified | Group I | Duplodnaviria | Heunggongvirae | Uroviricota | Caudoviricetes | n.a. | Unclassified | O | O |
| S12_vf_55175 | Faecalibacterium phage FP_Lagaffe | MG711461 | Clustered | VC_1290_0 | N30 | 3 | Faecalibacterium | Group I | Duplodnaviria | Heunggongvirae | Uroviricota | Caudoviricetes | n.a. | Unclassified | O | O |
| S12_vf_60224 | Phage vB_RanS_PJN03 | ON245412 | Clustered | VC_950_0 | N14 | 7 | Unspecified | Group I | Duplodnaviria | Heunggongvirae | Uroviricota | Caudoviricetes | n.a. | Unclassified | O | O |
| S12_vf_69136 | Phage vB_RanS_PJN03 | ON245412 | Clustered | VC_950_0 | N14 | 4 | Unspecified | Group I | Duplodnaviria | Heunggongvirae | Uroviricota | Caudoviricetes | n.a. | Unclassified | O | O |
| S12_vf_79666 | n.a. | n.a. | Clustered | VC_1552_0 | A | n.a. | n.a. | n.a. | n.a. | n.a. | n.a. | n.a. | n.a. | n.a. | n.a. | n.a. |
| S12_vs_106329 | Geobacillus phage GBSV1 | DQ340064 | Clustered | VC_779_0 | N44 | 1 | Geobacillus | Group I | Duplodnaviria | Heunggongvirae | Uroviricota | Caudoviricetes | n.a. | Unclassified | O | O |
| S12_vs_111815 | n.a. | n.a. | Clustered | VC_1553_0 | A | n.a. | n.a. | n.a. | n.a. | n.a. | n.a. | n.a. | n.a. | n.a. | n.a. | n.a. |
| S12_vs_121913 | n.a. | n.a. | Clustered | VC_1582_0 | A | n.a. | n.a. | n.a. | n.a. | n.a. | n.a. | n.a. | n.a. | n.a. | n.a. | n.a. |
| S12_vs_137069 | n.a. | n.a. | Clustered | VC_1601_0 | F | n.a. | n.a. | n.a. | n.a. | n.a. | n.a. | n.a. | n.a. | n.a. | n.a. | n.a. |
| S12_vs_138261 | n.a. | n.a. | Clustered | VC_1583_0 | A | n.a. | n.a. | n.a. | n.a. | n.a. | n.a. | n.a. | n.a. | n.a. | n.a. | n.a. |
| S12_vs_141337 | Bacteroides phage p00 | BK010646 | Clustered | VC_695_0 | C1 | 127 | Bacteroides | Group I | Duplodnaviria | Heunggongvirae | Uroviricota | Caudoviricetes | n.a. | Unclassified | Unclassified | Unclassified |
| S12_vs_14927 | uncultured phage cr3_1 | MT774381 | Clustered | VC_691_0 | C1 | 165 | Unspecified | Group I | Duplodnaviria | Heunggongvirae | Uroviricota | Caudoviricetes | Crassvirales | Intestiviridae | Crudevirinae | Diorhovirus |
| S12_vs_150812 | Clostridium phage phiMMP01 | LN681541 | Clustered | VC_151_0 | N18 | 25 | Clostridium | Group I | Duplodnaviria | Heunggongvirae | Uroviricota | Caudoviricetes | n.a. | Unclassified | O | O |
| S12_vs_151292 | Bacteroides phage LoVEphage | MW660583 | Clustered | VC_1526_0 | C2 | 199 | Bacteroides | Group I | Duplodnaviria | Heunggongvirae | Uroviricota | Caudoviricetes | n.a. | Unclassified | Unclassified | Unclassified |
| S12_vs_163990 | Faecalibacterium phage FP_oengus | MG711463 | Clustered | VC_1085_1 | N18 | 66 | Faecalibacterium | Group I | Duplodnaviria | Heunggongvirae | Uroviricota | Caudoviricetes | n.a. | Unclassified | O | O |
| S12_vs_183752 | Clostridium phage HM2 | LT600745 | Clustered | VC_1261_0 | N43 | 4 | Clostridium | Group I | Duplodnaviria | Heunggongvirae | Uroviricota | Caudoviricetes | n.a. | Salasmaviridae | O | O |
| S12_vs_183891 | n.a. | n.a. | Clustered | VC_1603_0 | F | n.a. | n.a. | n.a. | n.a. | n.a. | n.a. | n.a. | n.a. | n.a. | n.a. | n.a. |
| S12_vs_188820 | n.a. | n.a. | Clustered | VC_1553_0 | A | n.a. | n.a. | n.a. | n.a. | n.a. | n.a. | n.a. | n.a. | n.a. | n.a. | n.a. |
| S12_vs_188821 | Faecalibacterium phage FP_oengus | MG711463 | Clustered | VC_1085_1 | N15 | 79 | Faecalibacterium | Group I | Duplodnaviria | Heunggongvirae | Uroviricota | Caudoviricetes | n.a. | Unclassified | O | O |
| S12_vs_188823 | n.a. | n.a. | Clustered | VC_1566_0 | A | n.a. | n.a. | n.a. | n.a. | n.a. | n.a. | n.a. | n.a. | n.a. | n.a. | n.a. |
| S12_vs_188828 | n.a. | n.a. | Clustered | VC_1568_0 | A | n.a. | n.a. | n.a. | n.a. | n.a. | n.a. | n.a. | n.a. | n.a. | n.a. | n.a. |
| S12_vs_188832 | Tortoise microvirus 9 | MK765559 | Clustered | VC_1161_0 | C1 | 9 | Unspecified | Group II | Monodnaviria | Sangervirae | Phixviricota | Malgrandaviricetes | Petitvirales | Microviridae | Unclassified | Unclassified |
| S12_vs_188833 | n.a. | n.a. | Clustered | VC_1274_0 | A | n.a. | n.a. | n.a. | n.a. | n.a. | n.a. | n.a. | n.a. | n.a. | n.a. | n.a. |
| S12_vs_23796 | Bacteroides phage p00 | BK010646 | Clustered | VC_695_0 | C3 | 48 | Bacteroides | Group I | Duplodnaviria | Heunggongvirae | Uroviricota | Caudoviricetes | n.a. | Unclassified | Unclassified | Unclassified |
| S12_vs_26336 | n.a. | n.a. | Clustered | VC_1605_0 | A | n.a. | n.a. | n.a. | n.a. | n.a. | n.a. | n.a. | n.a. | n.a. | n.a. | n.a. |
| S12_vs_48045 | Exiguobacterium phage vB_EauM-23 | MH844558 | Clustered | VC_145_0 | C2 | 24 | Exiguobacterium | Group I | Duplodnaviria | Heunggongvirae | Uroviricota | Caudoviricetes | n.a. | Unclassified | Unclassified | Unclassified |
| S12_vs_58560 | Faecalibacterium phage FP_oengus | MG711463 | Clustered | VC_1085_1 | N20 | 40 | Faecalibacterium | Group I | Duplodnaviria | Heunggongvirae | Uroviricota | Caudoviricetes | n.a. | Unclassified | O | O |
| S12_vs_5857 | Bacillus phage vB_BtS_BMBtp13 | KX190832 | Clustered | VC_1230_0 | N12 | 5 | Bacillus | Group I | Duplodnaviria | Heunggongvirae | Uroviricota | Caudoviricetes | n.a. | Unclassified | O | O |
| S12_vs_6836 | Clostridium phage phiMMP01 | LN681541 | Clustered | VC_151_0 | N10 | 26 | Clostridium | Group I | Duplodnaviria | Heunggongvirae | Uroviricota | Caudoviricetes | n.a. | Unclassified | O | O |
| S12_vs_71525 | n.a. | n.a. | Clustered | VC_1606_0 | A | n.a. | n.a. | n.a. | n.a. | n.a. | n.a. | n.a. | n.a. | n.a. | n.a. | n.a. |
| S12_vs_79785 | Clostridium phage phiMMP01 | LN681541 | Clustered | VC_151_0 | N21 | 23 | Clostridium | Group I | Duplodnaviria | Heunggongvirae | Uroviricota | Caudoviricetes | n.a. | Unclassified | O | O |
| S12_vs_86928 | Akkermansia phage DTMo-2021a | CP084202 | Clustered | VC_752_0 | N2 | 7 | Akkermansia | Group I | Duplodnaviria | Heunggongvirae | Uroviricota | Caudoviricetes | n.a. | Unclassified | O | O |
| S12_vs_92868 | Faecalibacterium phage FP_oengus | MG711463 | Clustered | VC_1085_0 | N18 | 72 | Faecalibacterium | Group I | Duplodnaviria | Heunggongvirae | Uroviricota | Caudoviricetes | n.a. | Unclassified | O | O |
| S12_vs_93357 | Acetobacter phage phiAO1 | LC644971 | Clustered | VC_383_0 | N4 | 18 | Acetobacter | Group I | Duplodnaviria | Heunggongvirae | Uroviricota | Caudoviricetes | n.a. | Unclassified | O | O |
| S12_vs_99092 | Geobacillus phage GBSV1 | DQ340064 | Clustered | VC_778_0 | N35 | 6 | Geobacillus | Group I | Duplodnaviria | Heunggongvirae | Uroviricota | Caudoviricetes | n.a. | Unclassified | O | O |
| S13_vf_109235 | n.a. | n.a. | Clustered | VC_1552_0 | A | n.a. | n.a. | n.a. | n.a. | n.a. | n.a. | n.a. | n.a. | n.a. | n.a. | n.a. |
| S13_vf_11246 | uncultured phage cr53_1 | MT774396 | Clustered | VC_519_0 | N2 | 225 | Unspecified | Group I | Duplodnaviria | Heunggongvirae | Uroviricota | Caudoviricetes | Crassvirales | Suoliviridae | O | O |
| S13_vf_131208 | Clostridium phage HM2 | LT600745 | Clustered | VC_1261_0 | N40 | 4 | Clostridium | Group I | Duplodnaviria | Heunggongvirae | Uroviricota | Caudoviricetes | n.a. | Salasmaviridae | O | O |
| S13_vf_143422 | uncultured phage cr10_1 | MT774382 | Clustered | VC_519_11 | C3 | 112 | Unspecified | Group I | Duplodnaviria | Heunggongvirae | Uroviricota | Caudoviricetes | Crassvirales | Suoliviridae | Boorivirinae | Canhaevirus |
| S13_vf_150407 | Inovirus sp. | MT135292 | Clustered | VC_1495_0 | C1 | 23 | Unspecified | Group II | Monodnaviria | Loebvirae | Hofneiviricota | Faserviricetes | Tubulavirales | Inoviridae | Unclassified | Inovirus |
| S13_vf_151045 | Phage DP SC_6_H4_2017 | MT121964 | Clustered | VC_1490_0 | N6 | 122 | Unspecified | Group I | Duplodnaviria | Heunggongvirae | Uroviricota | Caudoviricetes | n.a. | Unclassified | O | O |
| S13_vf_151755 | Delftia phage IME-DE1 | KR153873 | Clustered | VC_86_0 | N3 | 6 | Delftia | Group I | Duplodnaviria | Heunggongvirae | Uroviricota | Caudoviricetes | n.a. | Autographiviridae | O | O |
| S13_vf_22175 | Exiguobacterium phage vB_EauS-123 | KU160495 | Clustered | VC_779_0 | N38 | 3 | Exiguobacterium | Group I | Duplodnaviria | Heunggongvirae | Uroviricota | Caudoviricetes | n.a. | Unclassified | O | O |
| S13_vf_28309 | Clostridium phage HM2 | LT600745 | Clustered | VC_1258_0 | N39 | 4 | Clostridium | Group I | Duplodnaviria | Heunggongvirae | Uroviricota | Caudoviricetes | n.a. | Salasmaviridae | O | O |
| S13_vf_56074 | Phage DP SC_6_H4_2017 | MT121964 | Clustered | VC_1490_1 | C3 | 269 | Unspecified | Group I | Duplodnaviria | Heunggongvirae | Uroviricota | Caudoviricetes | n.a. | Unclassified | Unclassified | Unclassified |
| S13_vf_7670 | n.a. | n.a. | Clustered | VC_1598_0 | F | n.a. | n.a. | n.a. | n.a. | n.a. | n.a. | n.a. | n.a. | n.a. | n.a. | n.a. |
| S13_vf_87873 | Parabacteroides phage PDS1 | MN929097 | Clustered | VC_855_0 | C4 | 115 | Parabacteroides | Group I | Duplodnaviria | Heunggongvirae | Uroviricota | Caudoviricetes | n.a. | Unclassified | Unclassified | Unclassified |
| S13_vs_113593 | Bifidobacterium phage BitterVaud1 | MT006236 | Clustered | VC_826_0 | C1 | 52 | Bifidobacterium | Group I | Duplodnaviria | Heunggongvirae | Uroviricota | Caudoviricetes | n.a. | Unclassified | Unclassified | Unclassified |
| S13_vs_121307 | n.a. | n.a. | Clustered | VC_905_0 | A | n.a. | n.a. | n.a. | n.a. | n.a. | n.a. | n.a. | n.a. | n.a. | n.a. | n.a. |
| S13_vs_12957 | Bacteriophage sp. | OP549880 | Clustered | VC_1549_0 | N5 | 13 | Unspecified | Unclassified | Unclassified | n.a. | n.a. | n.a. | n.a. | Unclassified | O | O |
| S13_vs_148294 | Streptococcus phage Javan336 | MK448910 | Clustered | VC_152_0 | N13 | 7 | Streptococcus | Group I | Duplodnaviria | Heunggongvirae | Uroviricota | Caudoviricetes | n.a. | Unclassified | O | O |
| S13_vs_151510 | uncultured phage cr128_1 | MT774392 | Clustered | VC_1054_0 | C1 | 175 | Unspecified | Group I | Duplodnaviria | Heunggongvirae | Uroviricota | Caudoviricetes | Crassvirales | Steigviridae | Asinivirinae | Mahlunavirus |
| S13_vs_152223 | Bifidobacterium phage BitterVaud1 | MT006236 | Clustered | VC_826_0 | C1 | 30 | Bifidobacterium | Group I | Duplodnaviria | Heunggongvirae | Uroviricota | Caudoviricetes | n.a. | Unclassified | Unclassified | Unclassified |
| S13_vs_15645 | Cellulophaga phage phi17:2 | KC821609 | Clustered | VC_1055_0 | N8 | 6 | Cellulophaga | Group I | Duplodnaviria | Heunggongvirae | Uroviricota | Caudoviricetes | n.a. | Unclassified | O | O |
| S13_vs_23314 | Streptococcus phage Javan191 | MK448700 | Clustered | VC_649_1 | C2 | 72 | Streptococcus | Group I | Duplodnaviria | Heunggongvirae | Uroviricota | Caudoviricetes | n.a. | Unclassified | Unclassified | Unclassified |
| S13_vs_30101 | Streptococcus phage Javan630 | MK448997 | Clustered | VC_649_3 | N6 | 37 | Streptococcus | Group I | Duplodnaviria | Heunggongvirae | Uroviricota | Caudoviricetes | n.a. | Unclassified | O | O |
| S13_vs_30167 | Streptococcus phage Javan630 | MK448997 | Clustered | VC_649_2 | N3 | 71 | Streptococcus | Group I | Duplodnaviria | Heunggongvirae | Uroviricota | Caudoviricetes | n.a. | Unclassified | O | O |
| S13_vs_33947 | n.a. | n.a. | Clustered | VC_1291_0 | A | n.a. | n.a. | n.a. | n.a. | n.a. | n.a. | n.a. | n.a. | n.a. | n.a. | n.a. |
| S13_vs_35816 | Faecalibacterium phage FP_Lagaffe | MG711461 | Clustered | VC_727_0 | C4 | 107 | Faecalibacterium | Group I | Duplodnaviria | Heunggongvirae | Uroviricota | Caudoviricetes | n.a. | Unclassified | Unclassified | Lagaffevirus |
| S13_vs_67429 | n.a. | n.a. | Clustered | VC_153_0 | A | n.a. | n.a. | n.a. | n.a. | n.a. | n.a. | n.a. | n.a. | n.a. | n.a. | n.a. |
| S13_vs_68102 | Bacillus phage vB_BsuP-Goe1 | KU831549 | Clustered | VC_847_0 | N5 | 7 | Bacillus | Group I | Duplodnaviria | Heunggongvirae | Uroviricota | Caudoviricetes | n.a. | Salasmaviridae | O | O |
| S13_vs_83447 | n.a. | n.a. | Clustered | VC_1565_0 | A | n.a. | n.a. | n.a. | n.a. | n.a. | n.a. | n.a. | n.a. | n.a. | n.a. | n.a. |
| S13_vs_87108 | n.a. | n.a. | Clustered | VC_1607_0 | F | n.a. | n.a. | n.a. | n.a. | n.a. | n.a. | n.a. | n.a. | n.a. | n.a. | n.a. |
| S13_vs_90967 | Tortoise microvirus 19 | MK765569 | Clustered | VC_1300_0 | N4 | 5 | Unspecified | Group II | Monodnaviria | Sangervirae | Phixviricota | Malgrandaviricetes | Petitvirales | Microviridae | O | O |
| S13_vs_9438 | n.a. | n.a. | Clustered | VC_1596_0 | A | n.a. | n.a. | n.a. | n.a. | n.a. | n.a. | n.a. | n.a. | n.a. | n.a. | n.a. |
| S13_vs_98591 | Myoviridae sp. | MW202455 | Clustered | VC_439_0 | N3 | 5 | Unspecified | Group I | Duplodnaviria | Heunggongvirae | Uroviricota | Caudoviricetes | n.a. | Unclassified | O | O |
| S14_vf_106156 | Lactobacillus phage JNU_P11 | MN830257 | Clustered | VC_275_0 | C1 | 88 | Lactobacillus | Group I | Duplodnaviria | Heunggongvirae | Uroviricota | Caudoviricetes | n.a. | Unclassified | Unclassified | Unclassified |
| S14_vf_109545 | n.a. | n.a. | Clustered | VC_1421_0 | A | n.a. | n.a. | n.a. | n.a. | n.a. | n.a. | n.a. | n.a. | n.a. | n.a. | n.a. |
| S14_vf_114491 | n.a. | n.a. | Clustered | VC_1258_0 | A | n.a. | n.a. | n.a. | n.a. | n.a. | n.a. | n.a. | n.a. | n.a. | n.a. | n.a. |
| S14_vf_117150 | Clostridium phage phiCp-B | MW478290 | Clustered | VC_21_0 | N4 | 11 | Clostridium | Group I | Duplodnaviria | Heunggongvirae | Uroviricota | Caudoviricetes | n.a. | Unclassified | O | O |
| S14_vf_117157 | n.a. | n.a. | Clustered | VC_1564_0 | F | n.a. | n.a. | n.a. | n.a. | n.a. | n.a. | n.a. | n.a. | n.a. | n.a. | n.a. |
| S14_vf_117164 | n.a. | n.a. | Clustered | VC_946_0 | A | n.a. | n.a. | n.a. | n.a. | n.a. | n.a. | n.a. | n.a. | n.a. | n.a. | n.a. |
| S14_vf_37500 | Microviridae sp. | MG945488 | Clustered | VC_1334_0 | C1 | 19 | Unspecified | Group II | Monodnaviria | Sangervirae | Phixviricota | Malgrandaviricetes | Petitvirales | Microviridae | Unclassified | Unclassified |
| S14_vf_44452 | n.a. | n.a. | Clustered | VC_1511_0 | A | n.a. | n.a. | n.a. | n.a. | n.a. | n.a. | n.a. | n.a. | n.a. | n.a. | n.a. |
| S14_vf_48775 | n.a. | n.a. | Clustered | VC_1597_0 | F | n.a. | n.a. | n.a. | n.a. | n.a. | n.a. | n.a. | n.a. | n.a. | n.a. | n.a. |
| S14_vf_56130 | Clostridium phage phiMMP01 | LN681541 | Clustered | VC_151_0 | N22 | 20 | Clostridium | Group I | Duplodnaviria | Heunggongvirae | Uroviricota | Caudoviricetes | n.a. | Unclassified | O | O |
| S14_vf_58919 | Faecalibacterium phage FP_Mushu | MG711460 | Clustered | VC_1288_0 | C1 | 76 | Faecalibacterium | Group I | Duplodnaviria | Heunggongvirae | Uroviricota | Caudoviricetes | n.a. | Unclassified | Unclassified | Mushuvirus |
| S14_vf_72335 | Butyrivibrio virus Arian | MN882551 | Clustered | VC_720_0 | N6 | 14 | Butyrivibrio | Group I | Duplodnaviria | Heunggongvirae | Uroviricota | Caudoviricetes | n.a. | Unclassified | O | O |
| S14_vf_72953 | Lactobacillus phage phiEF-1.1 | MF041990 | Clustered | VC_315_0 | C1 | 23 | Lactobacillus | Group I | Duplodnaviria | Heunggongvirae | Uroviricota | Caudoviricetes | n.a. | Unclassified | Unclassified | Unclassified |
| S14_vf_78139 | Lactobacillus phage JCL1032 | EU409559 | Clustered | VC_390_0 | C1 | 155 | Lactobacillus | Group I | Duplodnaviria | Heunggongvirae | Uroviricota | Caudoviricetes | n.a. | Unclassified | Unclassified | Unclassified |
| S14_vf_92240 | Butyrivibrio phage Idris | MN882554 | Clustered | VC_1289_0 | N4 | 48 | Butyrivibrio | Group I | Duplodnaviria | Heunggongvirae | Uroviricota | Caudoviricetes | n.a. | Unclassified | O | O |
| S14_vf_99344 | n.a. | n.a. | Clustered | VC_1258_0 | A | n.a. | n.a. | n.a. | n.a. | n.a. | n.a. | n.a. | n.a. | n.a. | n.a. | n.a. |
| S14_vs_102698 | Bacillus phage 250 | GU229986 | Clustered | VC_782_0 | N8 | 3 | Bacillus | Group I | Duplodnaviria | Heunggongvirae | Uroviricota | Caudoviricetes | n.a. | Unclassified | O | O |

| | | | | | | | | | | | | | | | | |
|---|---|---|---|---|---|---|---|---|---|---|---|---|---|---|---|---|
| S14_vs_110290 | n.a. | n.a. | Clustered | VC_782_0 | A | n.a. | n.a. | n.a. | n.a. | n.a. | n.a. | n.a. | n.a. | n.a. | n.a. | n.a. |
| S14_vs_11510 | n.a. | n.a. | Clustered | VC_1612_1 | A | n.a. | n.a. | n.a. | n.a. | n.a. | n.a. | n.a. | n.a. | n.a. | n.a. | n.a. |
| S14_vs_117158 | Streptococcus phage MismyG | OL774867 | Clustered | VC_152_0 | N11 | 7 | Streptococcus | Group I | Duplodnaviria | Heunggongvirae | Uroviricota | Caudoviricetes | n.a. | Unclassified | O | O |
| S14_vs_117160 | n.a. | n.a. | Clustered | VC_658_0 | A | n.a. | n.a. | n.a. | n.a. | n.a. | n.a. | n.a. | n.a. | n.a. | n.a. | n.a. |
| S14_vs_117161 | Butyrivibrio phage Idris | MN882554 | Clustered | VC_1289_0 | N8 | 34 | Butyrivibrio | Group I | Duplodnaviria | Heunggongvirae | Uroviricota | Caudoviricetes | n.a. | Unclassified | O | O |
| S14_vs_117165 | Butyrivibrio virus Ceridwen | MN882553 | Clustered | VC_1479_0 | N39 | 2 | Butyrivibrio | Group I | Duplodnaviria | Heunggongvirae | Uroviricota | Caudoviricetes | n.a. | Unclassified | O | O |
| S14_vs_117166 | n.a. | n.a. | Clustered | VC_779_0 | A | n.a. | n.a. | n.a. | n.a. | n.a. | n.a. | n.a. | n.a. | n.a. | n.a. | n.a. |
| S14_vs_14737 | Faecalibacterium phage FP_Lugh | MG711464 | Clustered | VC_923_0 | N4 | 12 | Faecalibacterium | Group I | Duplodnaviria | Heunggongvirae | Uroviricota | Caudoviricetes | n.a. | Unclassified | O | O |
| S14_vs_15228 | Cellulophaga phage phi17:2 | KC821609 | Clustered | VC_1055_1 | N8 | 6 | Cellulophaga | Group I | Duplodnaviria | Heunggongvirae | Uroviricota | Caudoviricetes | n.a. | Unclassified | O | O |
| S14_vs_25852 | n.a. | n.a. | Clustered | VC_1612_0 | A | n.a. | n.a. | n.a. | n.a. | n.a. | n.a. | n.a. | n.a. | n.a. | n.a. | n.a. |
| S14_vs_2870 | Acetobacter phage phiAO1 | LC644971 | Clustered | VC_383_0 | N4 | 16 | Acetobacter | Group I | Duplodnaviria | Heunggongvirae | Uroviricota | Caudoviricetes | n.a. | Unclassified | O | O |
| S14_vs_32499 | Bacteroides phage F2 | MT806187 | Clustered | VC_1505_0 | N11 | 5 | Bacteroides | Group I | Duplodnaviria | Heunggongvirae | Uroviricota | Caudoviricetes | n.a. | Unclassified | O | O |
| S14_vs_3251 | n.a. | n.a. | Clustered | VC_1294_0 | A | n.a. | n.a. | n.a. | n.a. | n.a. | n.a. | n.a. | n.a. | n.a. | n.a. | n.a. |
| S14_vs_39797 | Clostridium phage phiCD111 | LN681535 | Clustered | VC_923_0 | N5 | 11 | Clostridium | Group I | Duplodnaviria | Heunggongvirae | Uroviricota | Caudoviricetes | n.a. | Unclassified | O | O |
| S14_vs_59347 | Butyrivibrio virus Arian | MN882551 | Clustered | VC_720_0 | N5 | 17 | Butyrivibrio | Group I | Duplodnaviria | Heunggongvirae | Uroviricota | Caudoviricetes | n.a. | Unclassified | O | O |
| S14_vs_61530 | n.a. | n.a. | Clustered | VC_779_0 | A | n.a. | n.a. | n.a. | n.a. | n.a. | n.a. | n.a. | n.a. | n.a. | n.a. | n.a. |
| S14_vs_62178 | Geobacillus phage GBSV1 | DQ340064 | Clustered | VC_778_0 | N43 | 6 | Geobacillus | Group I | Duplodnaviria | Heunggongvirae | Uroviricota | Caudoviricetes | n.a. | Unclassified | O | O |
| S14_vs_632 | Clostridium phage phiMMP01 | LN681541 | Clustered | VC_151_0 | N18 | 24 | Clostridium | Group I | Duplodnaviria | Heunggongvirae | Uroviricota | Caudoviricetes | n.a. | Unclassified | O | O |
| S14_vs_66139 | Streptococcus phage Javan630 | MK448997 | Clustered | VC_649_0 | N6 | 52 | Streptococcus | Group I | Duplodnaviria | Heunggongvirae | Uroviricota | Caudoviricetes | n.a. | Unclassified | O | O |
| S14_vs_66208 | Phage vB_RanS_PJN03 | ON245412 | Clustered | VC_950_0 | N13 | 7 | Unspecified | Group I | Duplodnaviria | Heunggongvirae | Uroviricota | Caudoviricetes | n.a. | Unclassified | O | O |
| S14_vs_66841 | Erysipelothrix phage SE-1 | KR816341 | Clustered | VC_1149_0 | N3 | 15 | Erysipelothrix | Group I | Duplodnaviria | Heunggongvirae | Uroviricota | Caudoviricetes | n.a. | Unclassified | O | O |
| S14_vs_67094 | n.a. | n.a. | Clustered | VC_1614_0 | F | n.a. | n.a. | n.a. | n.a. | n.a. | n.a. | n.a. | n.a. | n.a. | n.a. | n.a. |
| S14_vs_73301 | Clostridium phage phiCTP1 | HM159959 | Clustered | VC_920_0 | N5 | 7 | Clostridium | Group I | Duplodnaviria | Heunggongvirae | Uroviricota | Caudoviricetes | n.a. | Unclassified | O | O |
| S14_vs_78164 | Winogradskyella phage Peternella_1 | MT732475 | Clustered | VC_694_0 | N5 | 17 | Winogradskyella | Group I | Duplodnaviria | Heunggongvirae | Uroviricota | Caudoviricetes | n.a. | Winoviridae | O | O |
| S14_vs_88666 | Clostridium phage phiCp-A | MW478289 | Clustered | VC_1035_0 | N2 | 14 | Clostridium | Group I | Duplodnaviria | Heunggongvirae | Uroviricota | Caudoviricetes | n.a. | Unclassified | O | O |
| S14_vs_96111 | Streptococcus phage Javan105 | MK448667 | Clustered | VC_649_1 | C1 | 58 | Streptococcus | Group I | Duplodnaviria | Heunggongvirae | Uroviricota | Caudoviricetes | n.a. | Unclassified | Unclassified | Unclassified |
| S14_vs_9676 | Faecalibacterium phage FP_Lagaffe | MG711461 | Clustered | VC_1291_0 | N43 | 3 | Faecalibacterium | Group I | Duplodnaviria | Heunggongvirae | Uroviricota | Caudoviricetes | n.a. | Unclassified | O | O |
| S15_vf_2032 | Bacteroides phage LoVEphage | MW660583 | Clustered | VC_1527_0 | N3 | 36 | Bacteroides | Group I | Duplodnaviria | Heunggongvirae | Uroviricota | Caudoviricetes | n.a. | Unclassified | O | O |
| S15_vf_20644 | Phage DP SC_6_H4_2017 | MT121964 | Clustered | VC_1490_1 | C3 | 268 | Unspecified | Group I | Duplodnaviria | Heunggongvirae | Uroviricota | Caudoviricetes | n.a. | Unclassified | Unclassified | Unclassified |
| S15_vf_2513 | n.a. | n.a. | Clustered | VC_1254_2 | A | n.a. | n.a. | n.a. | n.a. | n.a. | n.a. | n.a. | n.a. | n.a. | n.a. | n.a. |
| S15_vf_55639 | n.a. | n.a. | Clustered | VC_946_0 | A | n.a. | n.a. | n.a. | n.a. | n.a. | n.a. | n.a. | n.a. | n.a. | n.a. | n.a. |
| S15_vf_74586 | Phage DP SC_6_H4_2017 | MT121964 | Clustered | VC_1490_0 | N8 | 118 | Unspecified | Group I | Duplodnaviria | Heunggongvirae | Uroviricota | Caudoviricetes | n.a. | Unclassified | O | O |
| S15_vs_1407 | n.a. | n.a. | Clustered | VC_946_0 | A | n.a. | n.a. | n.a. | n.a. | n.a. | n.a. | n.a. | n.a. | n.a. | n.a. | n.a. |
| S15_vs_18274 | n.a. | n.a. | Clustered | VC_1602_0 | A | n.a. | n.a. | n.a. | n.a. | n.a. | n.a. | n.a. | n.a. | n.a. | n.a. | n.a. |
| S15_vs_27510 | n.a. | n.a. | Clustered | VC_1559_1 | A | n.a. | n.a. | n.a. | n.a. | n.a. | n.a. | n.a. | n.a. | n.a. | n.a. | n.a. |
| S15_vs_35445 | n.a. | n.a. | Clustered | VC_1599_0 | A | n.a. | n.a. | n.a. | n.a. | n.a. | n.a. | n.a. | n.a. | n.a. | n.a. | n.a. |
| S15_vs_7103 | Eggerthella phage PMBT5 | MH626557 | Clustered | VC_1421_0 | N35 | 3 | Eggerthella | Group I | Duplodnaviria | Heunggongvirae | Uroviricota | Caudoviricetes | n.a. | Unclassified | O | O |
| S15_vs_71968 | Pasteurella phage AFS-2018a | MH238466 | Clustered | VC_630_0 | N4 | 6 | Pasteurella | Group I | Duplodnaviria | Heunggongvirae | Uroviricota | Caudoviricetes | n.a. | Unclassified | O | O |
| S15_vs_76390 | Bdellovibrio phage phi1402 | JF344709 | Clustered | VC_967_0 | C4 | 30 | Bdellovibrio | Group I | Duplodnaviria | Heunggongvirae | Uroviricota | Caudoviricetes | n.a. | Unclassified | Unclassified | Unclassified |
| S16_vf_100235 | n.a. | n.a. | Clustered | VC_1258_0 | A | n.a. | n.a. | n.a. | n.a. | n.a. | n.a. | n.a. | n.a. | n.a. | n.a. | n.a. |
| S16_vf_100378 | Bifidobacterium phage BitterVaud1 | MT006236 | Clustered | VC_826_0 | C3 | 22 | Bifidobacterium | Group I | Duplodnaviria | Heunggongvirae | Uroviricota | Caudoviricetes | n.a. | Unclassified | Unclassified | Unclassified |
| S16_vf_1037 | n.a. | n.a. | Clustered | VC_1254_1 | A | n.a. | n.a. | n.a. | n.a. | n.a. | n.a. | n.a. | n.a. | n.a. | n.a. | n.a. |
| S16_vf_119834 | Clostridium phage HM2 | LT600745 | Clustered | VC_1261_0 | N34 | 4 | Clostridium | Group I | Duplodnaviria | Heunggongvirae | Uroviricota | Caudoviricetes | n.a. | Salasmaviridae | O | O |
| S16_vf_151450 | uncultured phage cr127_1 | MT774393 | Clustered | VC_693_0 | C2 | 64 | Unspecified | Group I | Duplodnaviria | Heunggongvirae | Uroviricota | Caudoviricetes | Crassvirales | Crevaviridae | Doltivirinae | Kahucivirus |
| S16_vf_164913 | n.a. | n.a. | Clustered | VC_1607_0 | F | n.a. | n.a. | n.a. | n.a. | n.a. | n.a. | n.a. | n.a. | n.a. | n.a. | n.a. |
| S16_vf_169895 | Clostridium phage HM2 | LT600745 | Clustered | VC_1261_0 | N41 | 3 | Clostridium | Group I | Duplodnaviria | Heunggongvirae | Uroviricota | Caudoviricetes | n.a. | Salasmaviridae | O | O |
| S16_vf_169985 | Phage DP SC_6_H4_2017 | MT121964 | Clustered | VC_1490_0 | N8 | 118 | Unspecified | Group I | Duplodnaviria | Heunggongvirae | Uroviricota | Caudoviricetes | n.a. | Unclassified | O | O |
| S16_vf_173109 | Bifidobacterium phage BlindBasel1 | MT006235 | Clustered | VC_1485_0 | C1 | 48 | Bifidobacterium | Group I | Duplodnaviria | Heunggongvirae | Uroviricota | Caudoviricetes | n.a. | Unclassified | Unclassified | Unclassified |
| S16_vf_36146 | Clostridium phage phiMMP01 | LN681541 | Clustered | VC_151_0 | N18 | 18 | Clostridium | Group I | Duplodnaviria | Heunggongvirae | Uroviricota | Caudoviricetes | n.a. | Unclassified | O | O |
| S16_vf_5270 | Faecalibacterium phage FP_oengus | MG711463 | Clustered | VC_1085_1 | N19 | 46 | Faecalibacterium | Group I | Duplodnaviria | Heunggongvirae | Uroviricota | Caudoviricetes | n.a. | Unclassified | O | O |
| S16_vf_6102 | Faecalibacterium phage FP_Toutatis | MG711466 | Clustered | VC_728_0 | C4 | 65 | Faecalibacterium | Group I | Duplodnaviria | Heunggongvirae | Uroviricota | Caudoviricetes | n.a. | Unclassified | Unclassified | Toutatisvirus |
| S16_vf_87190 | Cellulophaga phage phi13:1 | KC821625 | Clustered | VC_963_0 | N5 | 17 | Cellulophaga | Group I | Duplodnaviria | Heunggongvirae | Uroviricota | Caudoviricetes | n.a. | Unclassified | O | O |
| S16_vf_90328 | Butyrivibrio phage Idris | MN882554 | Clustered | VC_1477_0 | N16 | 1 | Butyrivibrio | Group I | Duplodnaviria | Heunggongvirae | Uroviricota | Caudoviricetes | n.a. | Unclassified | O | O |
| S16_vf_97279 | Faecalibacterium phage FP_Epona | MG711462 | Clustered | VC_723_0 | C1 | 113 | Faecalibacterium | Group I | Duplodnaviria | Heunggongvirae | Uroviricota | Caudoviricetes | n.a. | Unclassified | Unclassified | Eponavirus |
| S16_vs_103726 | n.a. | n.a. | Clustered | VC_779_0 | A | n.a. | n.a. | n.a. | n.a. | n.a. | n.a. | n.a. | n.a. | n.a. | n.a. | n.a. |
| S16_vs_116956 | n.a. | n.a. | Clustered | VC_442_0 | A | n.a. | n.a. | n.a. | n.a. | n.a. | n.a. | n.a. | n.a. | n.a. | n.a. | n.a. |
| S16_vs_120622 | Acetobacter phage phiAO1 | LC644971 | Clustered | VC_383_0 | N4 | 14 | Acetobacter | Group I | Duplodnaviria | Heunggongvirae | Uroviricota | Caudoviricetes | n.a. | Unclassified | O | O |
| S16_vs_124703 | Bacillus phage vB_BtS_BMBtp13 | KX190832 | Clustered | VC_667_0 | C9 | 8 | Bacillus | Group I | Duplodnaviria | Heunggongvirae | Uroviricota | Caudoviricetes | n.a. | Unclassified | Unclassified | Unclassified |
| S16_vs_127669 | Bacteroides phage LoVEphage | MW660583 | Clustered | VC_1526_0 | C1 | 297 | Bacteroides | Group I | Duplodnaviria | Heunggongvirae | Uroviricota | Caudoviricetes | n.a. | Unclassified | Unclassified | Unclassified |
| S16_vs_127905 | n.a. | n.a. | Clustered | VC_1559_1 | A | n.a. | n.a. | n.a. | n.a. | n.a. | n.a. | n.a. | n.a. | n.a. | n.a. | n.a. |
| S16_vs_136935 | n.a. | n.a. | Clustered | VC_1063_0 | A | n.a. | n.a. | n.a. | n.a. | n.a. | n.a. | n.a. | n.a. | n.a. | n.a. | n.a. |
| S16_vs_142561 | Pasteurella phage AFS-2018a | MH238466 | Clustered | VC_630_0 | N5 | 4 | Pasteurella | Group I | Duplodnaviria | Heunggongvirae | Uroviricota | Caudoviricetes | n.a. | Unclassified | O | O |
| S16_vs_173079 | Faecalibacterium phage FP_Brigit | MG711465 | Clustered | VC_442_0 | N4 | 34 | Faecalibacterium | Group I | Duplodnaviria | Heunggongvirae | Uroviricota | Caudoviricetes | n.a. | Unclassified | O | O |
| S16_vs_173096 | Akkermansia phage DTMo-2021a | CP084202 | Clustered | VC_751_0 | C1 | 90 | Akkermansia | Group I | Duplodnaviria | Heunggongvirae | Uroviricota | Caudoviricetes | n.a. | Unclassified | Unclassified | Unclassified |
| S16_vs_173102 | Parabacteroides phage PDS1 | MN929097 | Clustered | VC_855_0 | C4 | 114 | Parabacteroides | Group I | Duplodnaviria | Heunggongvirae | Uroviricota | Caudoviricetes | n.a. | Unclassified | Unclassified | Unclassified |
| S16_vs_18440 | Faecalibacterium phage FP_Toutatis | MG711466 | Clustered | VC_728_0 | C1 | 121 | Faecalibacterium | Group I | Duplodnaviria | Heunggongvirae | Uroviricota | Caudoviricetes | n.a. | Unclassified | Unclassified | Toutatisvirus |
| S16_vs_19320 | Bacillus phage vB_BtS_BMBtp13 | KX190832 | Clustered | VC_1230_0 | N9 | 5 | Bacillus | Group I | Duplodnaviria | Heunggongvirae | Uroviricota | Caudoviricetes | n.a. | Unclassified | O | O |
| S16_vs_19968 | n.a. | n.a. | Clustered | VC_1591_0 | A | n.a. | n.a. | n.a. | n.a. | n.a. | n.a. | n.a. | n.a. | n.a. | n.a. | n.a. |
| S16_vs_26078 | Bacillus phage 1 | DQ840344 | Clustered | VC_778_0 | N28 | 8 | Bacillus | Group I | Duplodnaviria | Heunggongvirae | Uroviricota | Caudoviricetes | n.a. | Unclassified | O | O |
| S16_vs_32764 | n.a. | n.a. | Clustered | VC_1599_0 | A | n.a. | n.a. | n.a. | n.a. | n.a. | n.a. | n.a. | n.a. | n.a. | n.a. | n.a. |
| S16_vs_33281 | n.a. | n.a. | Clustered | VC_1591_0 | A | n.a. | n.a. | n.a. | n.a. | n.a. | n.a. | n.a. | n.a. | n.a. | n.a. | n.a. |
| S16_vs_4662 | Bacillus phage vB_BtS_BMBtp13 | KX190832 | Clustered | VC_667_0 | C7 | 5 | Bacillus | Group I | Duplodnaviria | Heunggongvirae | Uroviricota | Caudoviricetes | n.a. | Unclassified | Unclassified | Unclassified |
| S16_vs_48714 | n.a. | n.a. | Clustered | VC_1616_0 | F | n.a. | n.a. | n.a. | n.a. | n.a. | n.a. | n.a. | n.a. | n.a. | n.a. | n.a. |
| S16_vs_51901 | n.a. | n.a. | Clustered | VC_1602_0 | A | n.a. | n.a. | n.a. | n.a. | n.a. | n.a. | n.a. | n.a. | n.a. | n.a. | n.a. |
| S16_vs_64584 | n.a. | n.a. | Clustered | VC_1611_0 | A | n.a. | n.a. | n.a. | n.a. | n.a. | n.a. | n.a. | n.a. | n.a. | n.a. | n.a. |
| S16_vs_75108 | n.a. | n.a. | Clustered | VC_1290_0 | A | n.a. | n.a. | n.a. | n.a. | n.a. | n.a. | n.a. | n.a. | n.a. | n.a. | n.a. |
| S16_vs_78361 | Bifidobacterium phage PMBT6 | MH444512 | Clustered | VC_878_0 | N2 | 52 | Bifidobacterium | Group I | Duplodnaviria | Heunggongvirae | Uroviricota | Caudoviricetes | n.a. | Unclassified | O | O |
| S16_vs_80244 | n.a. | n.a. | Clustered | VC_1579_0 | A | n.a. | n.a. | n.a. | n.a. | n.a. | n.a. | n.a. | n.a. | n.a. | n.a. | n.a. |
| S16_vs_86116 | Faecalibacterium phage FP_Lagaffe | MG711461 | Clustered | VC_727_0 | C1 | 169 | Faecalibacterium | Group I | Duplodnaviria | Heunggongvirae | Uroviricota | Caudoviricetes | n.a. | Unclassified | Unclassified | Lagaffevirus |
| S16_vs_89511 | Clostridioides phage CD1801 | MW512570 | Clustered | VC_670_0 | N12 | 5 | Clostridioides | Group I | Duplodnaviria | Heunggongvirae | Uroviricota | Caudoviricetes | n.a. | Unclassified | O | O |
| S16_vs_92150 | Faecalibacterium phage FP_Lugh | MG711464 | Clustered | VC_779_0 | N27 | 7 | Faecalibacterium | Group I | Duplodnaviria | Heunggongvirae | Uroviricota | Caudoviricetes | n.a. | Unclassified | O | O |
| S17_vf_148303 | n.a. | n.a. | Clustered | VC_1258_0 | A | n.a. | n.a. | n.a. | n.a. | n.a. | n.a. | n.a. | n.a. | n.a. | n.a. | n.a. |
| S17_vf_172132 | n.a. | n.a. | Clustered | VC_1607_0 | F | n.a. | n.a. | n.a. | n.a. | n.a. | n.a. | n.a. | n.a. | n.a. | n.a. | n.a. |
| S17_vf_174033 | Bacillus phage SPR | OM236515 | Clustered | VC_1040_0 | N10 | 3 | Bacillus | Group I | Duplodnaviria | Heunggongvirae | Uroviricota | Caudoviricetes | n.a. | Unclassified | O | O |
| S17_vf_174203 | Clostridium phage HM2 | LT600745 | Clustered | VC_1258_0 | N48 | 4 | Clostridium | Group I | Duplodnaviria | Heunggongvirae | Uroviricota | Caudoviricetes | n.a. | Salasmaviridae | O | O |
| S17_vf_177197 | n.a. | n.a. | Clustered | VC_1617_0 | F | n.a. | n.a. | n.a. | n.a. | n.a. | n.a. | n.a. | n.a. | n.a. | n.a. | n.a. |
| S17_vf_177199 | Enterobacter phage phiEap-2 | KT287080 | Clustered | VC_560_0 | C1 | 93 | Enterobacter | Group I | Duplodnaviria | Heunggongvirae | Uroviricota | Caudoviricetes | n.a. | Unclassified | Guernseyvirinae | Cornellvirus |

| | | | | | | | | | | | | | | | | |
|---|---|---|---|---|---|---|---|---|---|---|---|---|---|---|---|---|
| S17_vf_35138 | Phage vB_RanS_PJN03 | ON245412 | Clustered | VC_949_0 | N12 | 2 | Unspecified | Group I | Duplodnaviria | Heunggongvirae | Uroviricota | Caudoviricetes | n.a. | Unclassified | O | O |
| S17_vf_67042 | Clostridium phage HM2 | LT600745 | Clustered | VC_1261_0 | N27 | 8 | Clostridium | Group I | Duplodnaviria | Heunggongvirae | Uroviricota | Caudoviricetes | n.a. | Salasmaviridae | O | O |
| S17_vf_77057 | n.a. | n.a. | Clustered | VC_1585_0 | A | n.a. | n.a. | n.a. | n.a. | n.a. | n.a. | n.a. | n.a. | n.a. | n.a. | n.a. |
| S17_vf_79151 | n.a. | n.a. | Clustered | VC_1258_0 | A | n.a. | n.a. | n.a. | n.a. | n.a. | n.a. | n.a. | n.a. | n.a. | n.a. | n.a. |
| S17_vf_81232 | uncultured phage cr10_1 | MT774382 | Clustered | VC_519_11 | C1 | 292 | Unspecified | Group I | Duplodnaviria | Heunggongvirae | Uroviricota | Caudoviricetes | Crassvirales | Suoliviridae | Boorivirinae | Canhaevirus |
| S17_vf_97671 | Faecalibacterium phage FP_Mushu | MG711460 | Clustered | VC_1288_0 | C3 | 78 | Faecalibacterium | Group I | Duplodnaviria | Heunggongvirae | Uroviricota | Caudoviricetes | n.a. | Unclassified | Unclassified | Mushuvirus |
| S17_vs_103009 | n.a. | n.a. | Clustered | VC_1291_0 | A | n.a. | n.a. | n.a. | n.a. | n.a. | n.a. | n.a. | n.a. | n.a. | n.a. | n.a. |
| S17_vs_116640 | Bacteroides phage F4 | MT806185 | Clustered | VC_1505_0 | N12 | 2 | Bacteroides | Group I | Duplodnaviria | Heunggongvirae | Uroviricota | Caudoviricetes | n.a. | Unclassified | O | O |
| S17_vs_121041 | Geobacillus phage GBSV1 | DQ340064 | Clustered | VC_778_0 | N52 | 6 | Geobacillus | Group I | Duplodnaviria | Heunggongvirae | Uroviricota | Caudoviricetes | n.a. | Unclassified | O | O |
| S17_vs_127519 | n.a. | n.a. | Clustered | VC_1290_0 | A | n.a. | n.a. | n.a. | n.a. | n.a. | n.a. | n.a. | n.a. | n.a. | n.a. | n.a. |
| S17_vs_138036 | Bdellovibrio phage phi1402 | JF344709 | Clustered | VC_967_0 | C4 | 31 | Bdellovibrio | Group I | Duplodnaviria | Heunggongvirae | Uroviricota | Caudoviricetes | n.a. | Unclassified | Unclassified | Unclassified |
| S17_vs_160502 | uncultured phage cr116_1 | MT774389 | Clustered | VC_948_0 | N13 | 4 | Unspecified | Group I | Duplodnaviria | Heunggongvirae | Uroviricota | Caudoviricetes | Crassvirales | Steigviridae | O | O |
| S17_vs_177196 | n.a. | n.a. | Clustered | VC_1602_0 | A | n.a. | n.a. | n.a. | n.a. | n.a. | n.a. | n.a. | n.a. | n.a. | n.a. | n.a. |
| S17_vs_2079 | Bacteroides phage p00 | BK010646 | Clustered | VC_695_0 | C1 | 38 | Bacteroides | Group I | Duplodnaviria | Heunggongvirae | Uroviricota | Caudoviricetes | n.a. | Unclassified | Unclassified | Unclassified |
| S17_vs_39515 | uncultured phage cr271_1 | MT774394 | Clustered | VC_948_0 | N12 | 2 | Unspecified | Group I | Duplodnaviria | Heunggongvirae | Uroviricota | Caudoviricetes | Crassvirales | Intestiviridae | O | O |
| S17_vs_41845 | Ruminococcus phage phiRM10 | MT980841 | Clustered | VC_779_0 | N55 | 3 | Ruminococcus | Group I | Duplodnaviria | Heunggongvirae | Uroviricota | Caudoviricetes | n.a. | Unclassified | O | O |
| S17_vs_43307 | n.a. | n.a. | Clustered | VC_1583_0 | A | n.a. | n.a. | n.a. | n.a. | n.a. | n.a. | n.a. | n.a. | n.a. | n.a. | n.a. |
| S17_vs_58025 | n.a. | n.a. | Clustered | VC_1607_0 | F | n.a. | n.a. | n.a. | n.a. | n.a. | n.a. | n.a. | n.a. | n.a. | n.a. | n.a. |
| S17_vs_73830 | n.a. | n.a. | Clustered | VC_779_0 | A | n.a. | n.a. | n.a. | n.a. | n.a. | n.a. | n.a. | n.a. | n.a. | n.a. | n.a. |
| S1_vf_125650 | n.a. | n.a. | Clustered | VC_1607_0 | F | n.a. | n.a. | n.a. | n.a. | n.a. | n.a. | n.a. | n.a. | n.a. | n.a. | n.a. |
| S1_vf_158961 | Faecalibacterium phage FP_Lagaffe | MG711461 | Clustered | VC_1256_0 | N4 | 8 | Faecalibacterium | Group I | Duplodnaviria | Heunggongvirae | Uroviricota | Caudoviricetes | n.a. | Unclassified | O | O |
| S1_vf_159951 | Bacteroides phage SJC03 | MT074146 | Clustered | VC_1489_0 | N6 | 15 | Bacteroides | Group I | Duplodnaviria | Heunggongvirae | Uroviricota | Caudoviricetes | n.a. | Unclassified | O | O |
| S1_vf_22149 | n.a. | n.a. | Clustered | VC_1552_0 | A | n.a. | n.a. | n.a. | n.a. | n.a. | n.a. | n.a. | n.a. | n.a. | n.a. | n.a. |
| S1_vf_50591 | Streptococcus phage Javan261 | MK448720 | Clustered | VC_905_0 | N4 | 10 | Streptococcus | Group I | Duplodnaviria | Heunggongvirae | Uroviricota | Caudoviricetes | n.a. | Unclassified | O | O |
| S1_vs_100019 | Tortoise microvirus 19 | MK765569 | Clustered | VC_1300_0 | N4 | 4 | Unspecified | Group II | Monodnaviria | Sangervirae | Phixviricota | Malgrandaviricetes | Petitvirales | Microviridae | O | O |
| S1_vs_143929 | n.a. | n.a. | Clustered | VC_1554_0 | A | n.a. | n.a. | n.a. | n.a. | n.a. | n.a. | n.a. | n.a. | n.a. | n.a. | n.a. |
| S1_vs_3435 | Microviridae sp. | MG945598 | Clustered | VC_1385_0 | C1 | 4 | Unspecified | Group II | Monodnaviria | Sangervirae | Phixviricota | Malgrandaviricetes | Petitvirales | Microviridae | Unclassified | Unclassified |
| S1_vs_62409 | Geobacillus phage GBSV1 | DQ340064 | Clustered | VC_778_0 | N20 | 9 | Geobacillus | Group I | Duplodnaviria | Heunggongvirae | Uroviricota | Caudoviricetes | n.a. | Unclassified | O | O |
| S1_vs_91270 | n.a. | n.a. | Clustered | VC_782_0 | A | n.a. | n.a. | n.a. | n.a. | n.a. | n.a. | n.a. | n.a. | n.a. | n.a. | n.a. |
| S2_vf_20910 | n.a. | n.a. | Clustered | VC_1585_0 | A | n.a. | n.a. | n.a. | n.a. | n.a. | n.a. | n.a. | n.a. | n.a. | n.a. | n.a. |
| S2_vf_21511 | Faecalibacterium phage FP_oengus | MG711463 | Clustered | VC_1085_1 | N21 | 70 | Faecalibacterium | Group I | Duplodnaviria | Heunggongvirae | Uroviricota | Caudoviricetes | n.a. | Unclassified | O | O |
| S2_vf_21630 | Clostridium phage phiCTP1 | HM159959 | Clustered | VC_921_0 | N5 | 6 | Clostridium | Group I | Duplodnaviria | Heunggongvirae | Uroviricota | Caudoviricetes | n.a. | Unclassified | O | O |
| S2_vf_3356 | n.a. | n.a. | Clustered | VC_1607_0 | F | n.a. | n.a. | n.a. | n.a. | n.a. | n.a. | n.a. | n.a. | n.a. | n.a. | n.a. |
| S2_vf_4910 | Clostridium phage HM2 | LT600745 | Clustered | VC_1261_0 | N27 | 7 | Clostridium | Group I | Duplodnaviria | Heunggongvirae | Uroviricota | Caudoviricetes | n.a. | Salasmaviridae | O | O |
| S2_vs_10289 | Bacteroides phage p00 | BK010646 | Clustered | VC_695_0 | C1 | 47 | Bacteroides | Group I | Duplodnaviria | Heunggongvirae | Uroviricota | Caudoviricetes | n.a. | Unclassified | Unclassified | Unclassified |
| S2_vs_7306 | Parabacteroides phage PDS1 | MN929097 | Clustered | VC_855_0 | C2 | 160 | Parabacteroides | Group I | Duplodnaviria | Heunggongvirae | Uroviricota | Caudoviricetes | n.a. | Unclassified | Unclassified | Unclassified |
| S3_vf_100440 | Clostridium phage phiMMP01 | LN681541 | Clustered | VC_151_0 | N20 | 17 | Clostridium | Group I | Duplodnaviria | Heunggongvirae | Uroviricota | Caudoviricetes | n.a. | Unclassified | O | O |
| S3_vf_106401 | Clostridium phage phiCD6356 | GU949551 | Clustered | VC_430_0 | N3 | 17 | Clostridium | Group I | Duplodnaviria | Heunggongvirae | Uroviricota | Caudoviricetes | n.a. | Unclassified | O | O |
| S3_vf_109277 | n.a. | n.a. | Clustered | VC_1558_0 | F | n.a. | n.a. | n.a. | n.a. | n.a. | n.a. | n.a. | n.a. | n.a. | n.a. | n.a. |
| S3_vf_111776 | Clostridium phage HM2 | LT600745 | Clustered | VC_1261_0 | N36 | 4 | Clostridium | Group I | Duplodnaviria | Heunggongvirae | Uroviricota | Caudoviricetes | n.a. | Salasmaviridae | O | O |
| S3_vf_115304 | uncultured phage cr125_1 | MT774407 | Clustered | VC_519_2 | C1 | 239 | Unspecified | Group I | Duplodnaviria | Heunggongvirae | Uroviricota | Caudoviricetes | Crassvirales | Suoliviridae | Uncouvirinae | Aurodevirus |
| S3_vf_127168 | n.a. | n.a. | Clustered | VC_1607_0 | F | n.a. | n.a. | n.a. | n.a. | n.a. | n.a. | n.a. | n.a. | n.a. | n.a. | n.a. |
| S3_vf_127399 | Bacteroides phage SJC03 | MT074146 | Clustered | VC_1489_0 | N6 | 13 | Bacteroides | Group I | Duplodnaviria | Heunggongvirae | Uroviricota | Caudoviricetes | n.a. | Unclassified | O | O |
| S3_vf_127405 | Clostridium phage phiSM101 | CP000315 | Clustered | VC_21_0 | N4 | 11 | Clostridium | Group I | Duplodnaviria | Heunggongvirae | Uroviricota | Caudoviricetes | n.a. | Unclassified | O | O |
| S3_vf_127406 | Phage DP SC_6_H4_2017 | MT121964 | Clustered | VC_1490_1 | C3 | 269 | Unspecified | Group I | Duplodnaviria | Heunggongvirae | Uroviricota | Caudoviricetes | n.a. | Unclassified | Unclassified | Unclassified |
| S3_vf_127409 | Clostridium phage phiMMP01 | LN681541 | Clustered | VC_151_0 | N19 | 21 | Clostridium | Group I | Duplodnaviria | Heunggongvirae | Uroviricota | Caudoviricetes | n.a. | Unclassified | O | O |
| S3_vf_127423 | n.a. | n.a. | Clustered | VC_1618_0 | F | n.a. | n.a. | n.a. | n.a. | n.a. | n.a. | n.a. | n.a. | n.a. | n.a. | n.a. |
| S3_vf_127426 | Gokushovirus WZ-2015a | KT264756 | Clustered | VC_1145_0 | C1 | 14 | Unspecified | Group II | Monodnaviria | Sangervirae | Phixviricota | Malgrandaviricetes | Petitvirales | Microviridae | Gokushovirinae | Gokushovirus |
| S3_vf_127431 | Inovirus sp. | MT135308 | Clustered | VC_1492_0 | C1 | 36 | Unspecified | Group II | Monodnaviria | Loebvirae | Hofneiviricota | Faserviricetes | Tubulavirales | Inoviridae | Unclassified | Inovirus |
| S3_vf_127447 | uncultured phage cr126_1 | MT774391 | Clustered | VC_1054_1 | C1 | 233 | Unspecified | Group I | Duplodnaviria | Heunggongvirae | Uroviricota | Caudoviricetes | Crassvirales | Steigviridae | Asinivirinae | Kolpuevirus |
| S3_vf_17748 | Carjivirus communis | BK010471 | Clustered | VC_691_0 | C1 | 284 | Unspecified | Group I | Duplodnaviria | Heunggongvirae | Uroviricota | Caudoviricetes | Crassvirales | Intestiviridae | Crudevirinae | Carjivirus |
| S3_vf_18080 | n.a. | n.a. | Clustered | VC_1576_0 | A | n.a. | n.a. | n.a. | n.a. | n.a. | n.a. | n.a. | n.a. | n.a. | n.a. | n.a. |
| S3_vf_18829 | Phage DP SC_6_H4_2017 | MT121964 | Clustered | VC_1490_0 | N6 | 121 | Unspecified | Group I | Duplodnaviria | Heunggongvirae | Uroviricota | Caudoviricetes | n.a. | Unclassified | O | O |
| S3_vf_25163 | uncultured phage cr109_1 | MT774399 | Clustered | VC_519_7 | N1 | 300 | Unspecified | Group I | Duplodnaviria | Heunggongvirae | Uroviricota | Caudoviricetes | Crassvirales | Suoliviridae | O | O |
| S3_vf_35073 | Faecalibacterium phage FP_oengus | MG711463 | Clustered | VC_1085_1 | N20 | 53 | Faecalibacterium | Group I | Duplodnaviria | Heunggongvirae | Uroviricota | Caudoviricetes | n.a. | Unclassified | O | O |
| S3_vf_44405 | Siphovirus 29632 | KY053532 | Clustered | VC_826_0 | C2 | 16 | Unspecified | Group I | Duplodnaviria | Heunggongvirae | Uroviricota | Caudoviricetes | n.a. | Unclassified | Unclassified | Unclassified |
| S3_vf_54353 | Clavibacter phage CMP1 | GQ241246 | Clustered | VC_880_0 | C2 | 14 | Clavibacter | Group I | Duplodnaviria | Heunggongvirae | Uroviricota | Caudoviricetes | n.a. | Unclassified | Unclassified | Cimpunavirus |
| S3_vf_54498 | Microviridae sp. | MG945511 | Clustered | VC_1299_0 | C1 | 17 | Unspecified | Group II | Monodnaviria | Sangervirae | Phixviricota | Malgrandaviricetes | Petitvirales | Microviridae | Unclassified | Unclassified |
| S3_vf_57422 | n.a. | n.a. | Clustered | VC_1258_0 | A | n.a. | n.a. | n.a. | n.a. | n.a. | n.a. | n.a. | n.a. | n.a. | n.a. | n.a. |
| S3_vf_67333 | Roseburia phage Jekyll | LR596902 | Clustered | VC_1254_0 | N20 | 6 | Roseburia | Group I | Duplodnaviria | Heunggongvirae | Uroviricota | Caudoviricetes | n.a. | Unclassified | O | O |
| S3_vf_67828 | Tortoise microvirus 13 | MK765563 | Clustered | VC_1299_0 | C1 | 21 | Unspecified | Group II | Monodnaviria | Sangervirae | Phixviricota | Malgrandaviricetes | Petitvirales | Microviridae | Unclassified | Unclassified |
| S3_vf_70746 | Bifidobacterium phage BigBern1 | MT006237 | Clustered | VC_879_0 | C1 | 17 | Bifidobacterium | Group I | Duplodnaviria | Heunggongvirae | Uroviricota | Caudoviricetes | n.a. | Unclassified | Unclassified | Unclassified |
| S3_vf_73597 | Phage vB_RanS_PJN03 | ON245412 | Clustered | VC_948_0 | N21 | 2 | Unspecified | Group I | Duplodnaviria | Heunggongvirae | Uroviricota | Caudoviricetes | n.a. | Unclassified | O | O |
| S3_vf_80287 | Clostridium phage phiCT453A | KM983327 | Clustered | VC_151_0 | N23 | 22 | Clostridium | Group I | Duplodnaviria | Heunggongvirae | Uroviricota | Caudoviricetes | n.a. | Unclassified | O | O |
| S3_vf_8058 | Faecalibacterium phage FP_Epona | MG711462 | Clustered | VC_723_0 | C5 | 91 | Faecalibacterium | Group I | Duplodnaviria | Heunggongvirae | Uroviricota | Caudoviricetes | n.a. | Unclassified | Unclassified | Eponavirus |
| S3_vf_82571 | Caldibacillus phage CBP1 | MF595878 | Clustered | VC_1274_0 | N18 | 4 | Caldibacillus | Group I | Duplodnaviria | Heunggongvirae | Uroviricota | Caudoviricetes | n.a. | Unclassified | O | O |
| S3_vf_83536 | n.a. | n.a. | Clustered | VC_1258_0 | A | n.a. | n.a. | n.a. | n.a. | n.a. | n.a. | n.a. | n.a. | n.a. | n.a. | n.a. |
| S3_vf_84832 | n.a. | n.a. | Clustered | VC_1258_0 | A | n.a. | n.a. | n.a. | n.a. | n.a. | n.a. | n.a. | n.a. | n.a. | n.a. | n.a. |
| S3_vf_96222 | Lactococcus phage CaseusJM1 | KC522412 | Clustered | VC_210_0 | C1 | 46 | Lactococcus | Group I | Duplodnaviria | Heunggongvirae | Uroviricota | Caudoviricetes | n.a. | Unclassified | Unclassified | Skunavirus |
| S3_vf_99734 | Faecalibacterium phage FP_Lagaffe | MG711461 | Clustered | VC_727_0 | C5 | 112 | Faecalibacterium | Group I | Duplodnaviria | Heunggongvirae | Uroviricota | Caudoviricetes | n.a. | Unclassified | Unclassified | Lagaffevirus |
| S3_vs_100785 | Bacillus phage vB_BtS_BMBtp13 | KX190832 | Clustered | VC_905_0 | N21 | 4 | Bacillus | Group I | Duplodnaviria | Heunggongvirae | Uroviricota | Caudoviricetes | n.a. | Unclassified | O | O |
| S3_vs_108076 | Streptococcus phage MismyG | OL774867 | Clustered | VC_152_0 | N12 | 5 | Streptococcus | Group I | Duplodnaviria | Heunggongvirae | Uroviricota | Caudoviricetes | n.a. | Unclassified | O | O |
| S3_vs_114271 | n.a. | n.a. | Clustered | VC_1274_0 | A | n.a. | n.a. | n.a. | n.a. | n.a. | n.a. | n.a. | n.a. | n.a. | n.a. | n.a. |
| S3_vs_1143 | Faecalibacterium phage FP_Epona | MG711462 | Clustered | VC_723_0 | C5 | 92 | Faecalibacterium | Group I | Duplodnaviria | Heunggongvirae | Uroviricota | Caudoviricetes | n.a. | Unclassified | Unclassified | Eponavirus |
| S3_vs_117894 | n.a. | n.a. | Clustered | VC_1606_0 | A | n.a. | n.a. | n.a. | n.a. | n.a. | n.a. | n.a. | n.a. | n.a. | n.a. | n.a. |
| S3_vs_119605 | Streptococcus phage Javan630 | MK448997 | Clustered | VC_649_2 | N10 | 41 | Streptococcus | Group I | Duplodnaviria | Heunggongvirae | Uroviricota | Caudoviricetes | n.a. | Unclassified | O | O |
| S3_vs_119660 | Bacteroides phage p00 | BK010646 | Clustered | VC_695_0 | C3 | 62 | Bacteroides | Group I | Duplodnaviria | Heunggongvirae | Uroviricota | Caudoviricetes | n.a. | Unclassified | Unclassified | Unclassified |
| S3_vs_123834 | Bifidobacterium phage PMBT6 | MH444512 | Clustered | VC_878_0 | N2 | 52 | Bifidobacterium | Group I | Duplodnaviria | Heunggongvirae | Uroviricota | Caudoviricetes | n.a. | Unclassified | O | O |
| S3_vs_127387 | Butyrivibrio virus Arian | MN882551 | Clustered | VC_720_0 | N6 | 16 | Butyrivibrio | Group I | Duplodnaviria | Heunggongvirae | Uroviricota | Caudoviricetes | n.a. | Unclassified | O | O |
| S3_vs_127391 | n.a. | n.a. | Clustered | VC_641_0 | A | n.a. | n.a. | n.a. | n.a. | n.a. | n.a. | n.a. | n.a. | n.a. | n.a. | n.a. |
| S3_vs_127404 | Parabacteroides phage PDS1 | MN929097 | Clustered | VC_855_0 | C1 | 84 | Parabacteroides | Group I | Duplodnaviria | Heunggongvirae | Uroviricota | Caudoviricetes | n.a. | Unclassified | Unclassified | Unclassified |
| S3_vs_127413 | Croceibacter phage P2559Y | KC688701 | Clustered | VC_1045_0 | N2 | 13 | Croceibacter | Group I | Duplodnaviria | Heunggongvirae | Uroviricota | Caudoviricetes | n.a. | Unclassified | O | O |
| S3_vs_127442 | Poophage MBI-2016a | LT986678 | Clustered | VC_1267_0 | C1 | 24 | Unspecified | Group II | Monodnaviria | Sangervirae | Phixviricota | Malgrandaviricetes | Petitvirales | Microviridae | Unclassified | Unclassified |
| S3_vs_127445 | Phage DP SC_6_H4_2017 | MT121964 | Clustered | VC_1490_1 | C2 | 172 | Unspecified | Group I | Duplodnaviria | Heunggongvirae | Uroviricota | Caudoviricetes | n.a. | Unclassified | Unclassified | Unclassified |
| S3_vs_12973 | n.a. | n.a. | Clustered | VC_1611_0 | A | n.a. | n.a. | n.a. | n.a. | n.a. | n.a. | n.a. | n.a. | n.a. | n.a. | n.a. |
| S3_vs_12985 | Butyrivibrio phage Idris | MN882554 | Clustered | VC_1289_0 | N5 | 30 | Butyrivibrio | Group I | Duplodnaviria | Heunggongvirae | Uroviricota | Caudoviricetes | n.a. | Unclassified | O | O |
| S3_vs_17168 | Pasteurella phage AFS-2018a | MH238466 | Clustered | VC_630_0 | N4 | 4 | Pasteurella | Group I | Duplodnaviria | Heunggongvirae | Uroviricota | Caudoviricetes | n.a. | Unclassified | O | O |

| | | | | | | | | | | | | | | | | |
|---|---|---|---|---|---|---|---|---|---|---|---|---|---|---|---|---|
| S3_vs_18858 | n.a. | n.a. | Clustered | VC_1291_0 | A | n.a. | n.a. | n.a. | n.a. | n.a. | n.a. | n.a. | n.a. | n.a. | n.a. | n.a. |
| S3_vs_28402 | n.a. | n.a. | Clustered | VC_779_0 | A | n.a. | n.a. | n.a. | n.a. | n.a. | n.a. | n.a. | n.a. | n.a. | n.a. | n.a. |
| S3_vs_29077 | Streptococcus phage Javan90 | MK449010 | Clustered | VC_643_0 | N5 | 15 | Streptococcus | Group I | Duplodnaviria | Heunggongvirae | Uroviricota | Caudoviricetes | n.a. | Unclassified | O | O |
| S3_vs_42569 | Faecalibacterium phage FP_Toutatis | MG711466 | Clustered | VC_728_0 | C1 | 146 | Faecalibacterium | Group I | Duplodnaviria | Heunggongvirae | Uroviricota | Caudoviricetes | n.a. | Unclassified | Unclassified | Toutatisvirus |
| S3_vs_45147 | n.a. | n.a. | Clustered | VC_1560_0 | A | n.a. | n.a. | n.a. | n.a. | n.a. | n.a. | n.a. | n.a. | n.a. | n.a. | n.a. |
| S3_vs_49835 | n.a. | n.a. | Clustered | VC_1291_0 | A | n.a. | n.a. | n.a. | n.a. | n.a. | n.a. | n.a. | n.a. | n.a. | n.a. | n.a. |
| S3_vs_50924 | n.a. | n.a. | Clustered | VC_1613_0 | A | n.a. | n.a. | n.a. | n.a. | n.a. | n.a. | n.a. | n.a. | n.a. | n.a. | n.a. |
| S3_vs_54303 | Faecalibacterium phage FP_Epona | MG711462 | Clustered | VC_723_0 | C13 | 56 | Faecalibacterium | Group I | Duplodnaviria | Heunggongvirae | Uroviricota | Caudoviricetes | n.a. | Unclassified | Unclassified | Eponavirus |
| S3_vs_64424 | n.a. | n.a. | Clustered | VC_1063_0 | A | n.a. | n.a. | n.a. | n.a. | n.a. | n.a. | n.a. | n.a. | n.a. | n.a. | n.a. |
| S3_vs_70508 | n.a. | n.a. | Clustered | VC_1572_0 | A | n.a. | n.a. | n.a. | n.a. | n.a. | n.a. | n.a. | n.a. | n.a. | n.a. | n.a. |
| S3_vs_72118 | n.a. | n.a. | Clustered | VC_1570_0 | A | n.a. | n.a. | n.a. | n.a. | n.a. | n.a. | n.a. | n.a. | n.a. | n.a. | n.a. |
| S3_vs_73361 | n.a. | n.a. | Clustered | VC_1231_0 | A | n.a. | n.a. | n.a. | n.a. | n.a. | n.a. | n.a. | n.a. | n.a. | n.a. | n.a. |
| S3_vs_77156 | Microviridae sp. | MG945455 | Clustered | VC_1267_0 | C1 | 23 | Unspecified | Group II | Monodnaviria | Sangervirae | Phixviricota | Malgrandaviricetes | Petitvirales | Microviridae | Unclassified | Unclassified |
| S3_vs_80151 | Cellulophaga phage phi17:2 | KC821609 | Clustered | VC_1056_0 | N8 | 4 | Cellulophaga | Group I | Duplodnaviria | Heunggongvirae | Uroviricota | Caudoviricetes | n.a. | Unclassified | O | O |
| S3_vs_82895 | Cellulophaga phage phi17:2 | KC821609 | Clustered | VC_1055_1 | N8 | 6 | Cellulophaga | Group I | Duplodnaviria | Heunggongvirae | Uroviricota | Caudoviricetes | n.a. | Unclassified | O | O |
| S3_vs_89898 | Parabacteroides phage PDS1 | MN929097 | Clustered | VC_855_0 | C5 | 106 | Parabacteroides | Group I | Duplodnaviria | Heunggongvirae | Uroviricota | Caudoviricetes | n.a. | Unclassified | Unclassified | Unclassified |
| S3_vs_90341 | n.a. | n.a. | Clustered | VC_1620_0 | F | n.a. | n.a. | n.a. | n.a. | n.a. | n.a. | n.a. | n.a. | n.a. | n.a. | n.a. |
| S3_vs_95574 | n.a. | n.a. | Clustered | VC_1291_0 | A | n.a. | n.a. | n.a. | n.a. | n.a. | n.a. | n.a. | n.a. | n.a. | n.a. | n.a. |
| S3_vs_98675 | Clostridium phage CWou-2020a | CP063964 | Clustered | VC_747_0 | N2 | 5 | Clostridium | Group I | Duplodnaviria | Heunggongvirae | Uroviricota | Caudoviricetes | n.a. | Unclassified | O | O |
| S4_vf_10506 | n.a. | n.a. | Clustered | VC_1617_0 | F | n.a. | n.a. | n.a. | n.a. | n.a. | n.a. | n.a. | n.a. | n.a. | n.a. | n.a. |
| S4_vf_12614 | n.a. | n.a. | Clustered | VC_949_0 | A | n.a. | n.a. | n.a. | n.a. | n.a. | n.a. | n.a. | n.a. | n.a. | n.a. | n.a. |
| S4_vf_12624 | Curtobacterium phage Penoan | MZ333133 | Clustered | VC_1195_0 | N3 | 4 | Curtobacterium | Group I | Duplodnaviria | Heunggongvirae | Uroviricota | Caudoviricetes | n.a. | Unclassified | O | O |
| S4_vf_13627 | n.a. | n.a. | Clustered | VC_1258_0 | A | n.a. | n.a. | n.a. | n.a. | n.a. | n.a. | n.a. | n.a. | n.a. | n.a. | n.a. |
| S4_vf_13759 | Bacillus phage vB_BtS_BMBtp13 | KX190832 | Clustered | VC_667_0 | C1 | 13 | Bacillus | Group I | Duplodnaviria | Heunggongvirae | Uroviricota | Caudoviricetes | n.a. | Unclassified | Unclassified | Unclassified |
| S4_vf_18803 | Bacteroides phage F4 | MT806185 | Clustered | VC_1044_0 | C2 | 59 | Bacteroides | Group I | Duplodnaviria | Heunggongvirae | Uroviricota | Caudoviricetes | n.a. | Unclassified | Unclassified | Unclassified |
| S4_vf_22219 | Bacillus phage BCD7 | JN712910 | Clustered | VC_995_0 | C1 | 49 | Bacillus | Group I | Duplodnaviria | Heunggongvirae | Uroviricota | Caudoviricetes | n.a. | Unclassified | Unclassified | Becedseptimavirus |
| S4_vf_2524 | n.a. | n.a. | Clustered | VC_946_0 | A | n.a. | n.a. | n.a. | n.a. | n.a. | n.a. | n.a. | n.a. | n.a. | n.a. | n.a. |
| S4_vf_26417 | uncultured phage cr127_1 | MT774393 | Clustered | VC_693_0 | C1 | 264 | Unspecified | Group I | Duplodnaviria | Heunggongvirae | Uroviricota | Caudoviricetes | Crassvirales | Crevaviridae | Doltivirinae | Kahucivirus |
| S4_vf_27861 | Phage DP SC_6_H4_2017 | MT121964 | Clustered | VC_1490_0 | N7 | 120 | Unspecified | Group I | Duplodnaviria | Heunggongvirae | Uroviricota | Caudoviricetes | n.a. | Unclassified | O | O |
| S4_vf_28601 | Bifidobacterium phage BigBern1 | MT006237 | Clustered | VC_879_0 | C4 | 6 | Bifidobacterium | Group I | Duplodnaviria | Heunggongvirae | Uroviricota | Caudoviricetes | n.a. | Unclassified | Unclassified | Unclassified |
| S4_vf_31978 | n.a. | n.a. | Clustered | VC_1254_1 | A | n.a. | n.a. | n.a. | n.a. | n.a. | n.a. | n.a. | n.a. | n.a. | n.a. | n.a. |
| S4_vf_31981 | uncultured phage cr118_1 | MT774379 | Clustered | VC_519_10 | C1 | 201 | Unspecified | Group I | Duplodnaviria | Heunggongvirae | Uroviricota | Caudoviricetes | Crassvirales | Suoliviridae | Uncouvirinae | Besingivirus |
| S4_vf_4838 | uncultured phage cr50_1 | MT774375 | Clustered | VC_519_11 | C1 | 272 | Unspecified | Group I | Duplodnaviria | Heunggongvirae | Uroviricota | Caudoviricetes | Crassvirales | Suoliviridae | Boorivirinae | Cohcovirus |
| S4_vf_4891 | n.a. | n.a. | Clustered | VC_1561_0 | A | n.a. | n.a. | n.a. | n.a. | n.a. | n.a. | n.a. | n.a. | n.a. | n.a. | n.a. |
| S4_vf_5021 | Clostridium phage HM2 | LT600745 | Clustered | VC_1258_0 | N38 | 3 | Clostridium | Group I | Duplodnaviria | Heunggongvirae | Uroviricota | Caudoviricetes | n.a. | Salasmaviridae | O | O |
| S4_vs_VIRSorter_1135-cat_2 | Clostridium phage phiMMP03 | LN681542 | Clustered | VC_151_0 | N19 | 22 | Clostridium | Group I | Duplodnaviria | Heunggongvirae | Uroviricota | Caudoviricetes | n.a. | Unclassified | O | O |
| S4_vs_VIRSorter_12831-circular-cat_2 | Ruminococcus phage phiRgPS_6 | MT980839 | Clustered | VC_779_0 | N31 | 5 | Ruminococcus | Group I | Duplodnaviria | Heunggongvirae | Uroviricota | Caudoviricetes | n.a. | Unclassified | O | O |
| S4_vs_VIRSorter_13190-cat_2 | Cellulophaga phage phi17:2 | KC821609 | Clustered | VC_1056_0 | N8 | 3 | Cellulophaga | Group I | Duplodnaviria | Heunggongvirae | Uroviricota | Caudoviricetes | n.a. | Unclassified | O | O |
| S4_vs_VIRSorter_13488-cat_2 | n.a. | n.a. | Clustered | VC_1611_0 | A | n.a. | n.a. | n.a. | n.a. | n.a. | n.a. | n.a. | n.a. | n.a. | n.a. | n.a. |
| S4_vs_VIRSorter_15112-cat_2 | n.a. | n.a. | Clustered | VC_1604_0 | A | n.a. | n.a. | n.a. | n.a. | n.a. | n.a. | n.a. | n.a. | n.a. | n.a. | n.a. |
| S4_vs_VIRSorter_15837-cat_2 | Clostridium phage phiCT453A | KM983327 | Clustered | VC_152_0 | N19 | 8 | Clostridium | Group I | Duplodnaviria | Heunggongvirae | Uroviricota | Caudoviricetes | n.a. | Unclassified | O | O |
| S4_vs_VIRSorter_16493-cat_2 | Bifidobacterium phage Bbif-1 | GQ141189 | Clustered | VC_876_0 | C1 | 150 | Bifidobacterium | Group I | Duplodnaviria | Heunggongvirae | Uroviricota | Caudoviricetes | n.a. | Unclassified | Unclassified | Unclassified |
| S4_vs_VIRSorter_19324-cat_2 | Erysipelothrix phage SE-1 | KR816341 | Clustered | VC_1149_0 | N3 | 18 | Erysipelothrix | Group I | Duplodnaviria | Heunggongvirae | Uroviricota | Caudoviricetes | n.a. | Unclassified | O | O |
| S4_vs_VIRSorter_3006-cat_1 | Faecalibacterium phage FP_oengus | MG711463 | Clustered | VC_1085_1 | N21 | 64 | Faecalibacterium | Group I | Duplodnaviria | Heunggongvirae | Uroviricota | Caudoviricetes | n.a. | Unclassified | O | O |
| S4_vs_VIRSorter_30222-cat_2 | Faecalibacterium phage FP_oengus | MG711463 | Clustered | VC_1085_0 | N17 | 71 | Faecalibacterium | Group I | Duplodnaviria | Heunggongvirae | Uroviricota | Caudoviricetes | n.a. | Unclassified | O | O |
| S4_vs_VIRSorter_31171-circular-cat_2 | Faecalibacterium phage FP_Lugh | MG711464 | Clustered | VC_923_0 | N5 | 13 | Faecalibacterium | Group I | Duplodnaviria | Heunggongvirae | Uroviricota | Caudoviricetes | n.a. | Unclassified | O | O |
| S4_vs_VIRSorter_31196-circular-cat_2 | n.a. | n.a. | Clustered | VC_1481_0 | A | n.a. | n.a. | n.a. | n.a. | n.a. | n.a. | n.a. | n.a. | n.a. | n.a. | n.a. |
| S4_vs_VIRSorter_31560-circular-cat_2 | Butyrivibrio virus Ceridwen | MN882553 | Clustered | VC_1481_1 | N49 | 1 | Butyrivibrio | Group I | Duplodnaviria | Heunggongvirae | Uroviricota | Caudoviricetes | n.a. | Unclassified | O | O |
| S4_vs_VIRSorter_31976-circular-cat_2 | n.a. | n.a. | Clustered | VC_641_0 | A | n.a. | n.a. | n.a. | n.a. | n.a. | n.a. | n.a. | n.a. | n.a. | n.a. | n.a. |
| S4_vs_VIRSorter_31977-circular-cat_2 | Faecalibacterium phage FP_Lugh | MG711464 | Clustered | VC_923_0 | N23 | 3 | Faecalibacterium | Group I | Duplodnaviria | Heunggongvirae | Uroviricota | Caudoviricetes | n.a. | Unclassified | O | O |
| S4_vs_VIRSorter_31980-circular-cat_2 | Phage WC36-2 | OL791266 | Clustered | VC_249_1 | C1 | 37 | Unspecified | Group I | Duplodnaviria | Heunggongvirae | Uroviricota | Caudoviricetes | n.a. | Unclassified | Unclassified | Unclassified |
| S4_vs_VIRSorter_4746-cat_2 | Clostridium phage HM2 | LT600745 | Clustered | VC_1265_0 | N19 | 4 | Clostridium | Group I | Duplodnaviria | Heunggongvirae | Uroviricota | Caudoviricetes | n.a. | Salasmaviridae | O | O |
| S4_vs_VIRSorter_4839-cat_2 | Faecalibacterium phage FP_oengus | MG711463 | Clustered | VC_1085_1 | N20 | 65 | Faecalibacterium | Group I | Duplodnaviria | Heunggongvirae | Uroviricota | Caudoviricetes | n.a. | Unclassified | O | O |
| S4_vs_VIRSorter_4959-cat_2 | Clostridioides phage CD1801 | MW512570 | Clustered | VC_670_0 | N7 | 13 | Clostridioides | Group I | Duplodnaviria | Heunggongvirae | Uroviricota | Caudoviricetes | n.a. | Unclassified | O | O |
| S4_vs_VIRSorter_6022-circular-cat_1 | Parabacteroides phage PDS1 | MN929097 | Clustered | VC_855_0 | C3 | 147 | Parabacteroides | Group I | Duplodnaviria | Heunggongvirae | Uroviricota | Caudoviricetes | n.a. | Unclassified | Unclassified | Unclassified |
| S4_vs_VIRSorter_7386-circular-cat_2 | Faecalibacterium phage FP_Epona | MG711462 | Clustered | VC_723_0 | C14 | 48 | Faecalibacterium | Group I | Duplodnaviria | Heunggongvirae | Uroviricota | Caudoviricetes | n.a. | Unclassified | Unclassified | Eponavirus |
| S4_vs_VIRSorter_8622-cat_2 | Faecalibacterium phage FP_Lagaffe | MG711461 | Clustered | VC_727_0 | C5 | 99 | Faecalibacterium | Group I | Duplodnaviria | Heunggongvirae | Uroviricota | Caudoviricetes | n.a. | Unclassified | Unclassified | Lagaffevirus |
| S4_vs_VIRSorter_9913-cat_1 | Parabacteroides phage PDS1 | MN929097 | Clustered | VC_855_0 | C5 | 83 | Parabacteroides | Group I | Duplodnaviria | Heunggongvirae | Uroviricota | Caudoviricetes | n.a. | Unclassified | Unclassified | Unclassified |
| S5_vf_110171 | n.a. | n.a. | Clustered | VC_1552_0 | A | n.a. | n.a. | n.a. | n.a. | n.a. | n.a. | n.a. | n.a. | n.a. | n.a. | n.a. |
| S5_vf_11450 | Geobacillus phage GBSV1 | DQ340064 | Clustered | VC_778_0 | N37 | 6 | Geobacillus | Group I | Duplodnaviria | Heunggongvirae | Uroviricota | Caudoviricetes | n.a. | Unclassified | O | O |
| S5_vf_122568 | Faecalibacterium phage FP_Taranis | MG711467 | Clustered | VC_732_0 | N3 | 98 | Faecalibacterium | Group I | Duplodnaviria | Heunggongvirae | Uroviricota | Caudoviricetes | n.a. | Unclassified | O | O |
| S5_vf_123990 | n.a. | n.a. | Clustered | VC_1258_0 | A | n.a. | n.a. | n.a. | n.a. | n.a. | n.a. | n.a. | n.a. | n.a. | n.a. | n.a. |
| S5_vf_134040 | Faecalibacterium phage FP_Mushu | MG711460 | Clustered | VC_1288_0 | C3 | 100 | Faecalibacterium | Group I | Duplodnaviria | Heunggongvirae | Uroviricota | Caudoviricetes | n.a. | Unclassified | Unclassified | Mushuvirus |
| S5_vf_136076 | n.a. | n.a. | Clustered | VC_1552_0 | A | n.a. | n.a. | n.a. | n.a. | n.a. | n.a. | n.a. | n.a. | n.a. | n.a. | n.a. |
| S5_vf_137260 | Bacteriophage sp. | OP549880 | Clustered | VC_1549_0 | N5 | 16 | Unspecified | Unclassified | Unclassified | n.a. | n.a. | n.a. | n.a. | Unclassified | O | O |
| S5_vf_145585 | Bacillus phage vB_BpsS-140 | MH884512 | Clustered | VC_638_0 | C1 | 6 | Bacillus | Group I | Duplodnaviria | Heunggongvirae | Uroviricota | Caudoviricetes | n.a. | Unclassified | Unclassified | Unclassified |
| S5_vf_145588 | Clostridium phage phiCTP1 | HM159959 | Clustered | VC_921_0 | N4 | 8 | Clostridium | Group I | Duplodnaviria | Heunggongvirae | Uroviricota | Caudoviricetes | n.a. | Unclassified | O | O |
| S5_vf_18461 | Phage vB_RanS_PJN03 | ON245412 | Clustered | VC_951_1 | N15 | 4 | Unspecified | Group I | Duplodnaviria | Heunggongvirae | Uroviricota | Caudoviricetes | n.a. | Unclassified | O | O |
| S5_vf_25423 | Lactococcus phage 936 group phage Phi4.2 | KP793123 | Clustered | VC_210_0 | C1 | 76 | Lactococcus | Group I | Duplodnaviria | Heunggongvirae | Uroviricota | Caudoviricetes | n.a. | Unclassified | Unclassified | Skunavirus |
| S5_vf_28061 | n.a. | n.a. | Clustered | VC_430_0 | A | n.a. | n.a. | n.a. | n.a. | n.a. | n.a. | n.a. | n.a. | n.a. | n.a. | n.a. |
| S5_vf_55415 | Bacteroides phage crAss001 | MH675552 | Clustered | VC_1054_1 | C1 | 261 | Bacteroides | Group I | Duplodnaviria | Heunggongvirae | Uroviricota | Caudoviricetes | Crassvirales | Steigviridae | Asinivirinae | Kehishuvirus |
| S5_vf_6032 | n.a. | n.a. | Clustered | VC_1511_0 | A | n.a. | n.a. | n.a. | n.a. | n.a. | n.a. | n.a. | n.a. | n.a. | n.a. | n.a. |
| S5_vf_6506 | Clostridium phage vB_CpeS-1181 | MW478292 | Clustered | VC_923_0 | N1 | 28 | Clostridium | Group I | Duplodnaviria | Heunggongvirae | Uroviricota | Caudoviricetes | n.a. | Unclassified | O | O |
| S5_vf_74661 | Escherichia phage TL-2011b | JQ011317 | Clustered | VC_492_0 | C1 | 112 | Escherichia | Group I | Duplodnaviria | Heunggongvirae | Uroviricota | Caudoviricetes | n.a. | Unclassified | Unclassified | Unclassified |
| S5_vf_77701 | Bifidobacterium phage BitterVaud1 | MT006236 | Clustered | VC_826_0 | C2 | 11 | Bifidobacterium | Group I | Duplodnaviria | Heunggongvirae | Uroviricota | Caudoviricetes | n.a. | Unclassified | Unclassified | Unclassified |
| S5_vf_8965 | Bacteroides phage F4 | MT806185 | Clustered | VC_1505_0 | N9 | 11 | Bacteroides | Group I | Duplodnaviria | Heunggongvirae | Uroviricota | Caudoviricetes | n.a. | Unclassified | O | O |
| S5_vs_107105 | Butyrivibrio phage Idris | MN882554 | Clustered | VC_724_0 | N15 | 4 | Butyrivibrio | Group I | Duplodnaviria | Heunggongvirae | Uroviricota | Caudoviricetes | n.a. | Unclassified | O | O |
| S5_vs_116235 | Streptococcus phage Javan261 | MK448720 | Clustered | VC_905_0 | N10 | 7 | Streptococcus | Group I | Duplodnaviria | Heunggongvirae | Uroviricota | Caudoviricetes | n.a. | Unclassified | O | O |
| S5_vs_120079 | Roseburia phage Jekyll | LR596902 | Clustered | VC_1252_0 | C1 | 68 | Roseburia | Group I | Duplodnaviria | Heunggongvirae | Uroviricota | Caudoviricetes | n.a. | Unclassified | Unclassified | Unclassified |
| S5_vs_121419 | n.a. | n.a. | Clustered | VC_1570_0 | A | n.a. | n.a. | n.a. | n.a. | n.a. | n.a. | n.a. | n.a. | n.a. | n.a. | n.a. |
| S5_vs_123256 | Bacteroides phage p00 | BK010646 | Clustered | VC_695_0 | C5 | 15 | Bacteroides | Group I | Duplodnaviria | Heunggongvirae | Uroviricota | Caudoviricetes | n.a. | Unclassified | Unclassified | Unclassified |
| S5_vs_124035 | n.a. | n.a. | Clustered | VC_1614_0 | F | n.a. | n.a. | n.a. | n.a. | n.a. | n.a. | n.a. | n.a. | n.a. | n.a. | n.a. |
| S5_vs_129970 | n.a. | n.a. | Clustered | VC_1572_0 | A | n.a. | n.a. | n.a. | n.a. | n.a. | n.a. | n.a. | n.a. | n.a. | n.a. | n.a. |
| S5_vs_13090 | n.a. | n.a. | Clustered | VC_1587_0 | A | n.a. | n.a. | n.a. | n.a. | n.a. | n.a. | n.a. | n.a. | n.a. | n.a. | n.a. |
| S5_vs_130958 | Faecalibacterium phage FP_Taranis | MG711467 | Clustered | VC_732_0 | N3 | 75 | Faecalibacterium | Group I | Duplodnaviria | Heunggongvirae | Uroviricota | Caudoviricetes | n.a. | Unclassified | O | O |
| S5_vs_1338 | n.a. | n.a. | Clustered | VC_1582_0 | A | n.a. | n.a. | n.a. | n.a. | n.a. | n.a. | n.a. | n.a. | n.a. | n.a. | n.a. |
| S5_vs_136679 | n.a. | n.a. | Clustered | VC_1291_0 | A | n.a. | n.a. | n.a. | n.a. | n.a. | n.a. | n.a. | n.a. | n.a. | n.a. | n.a. |

| | | | | | | | | | | | | | | | | |
|---|---|---|---|---|---|---|---|---|---|---|---|---|---|---|---|---|
| S5_vs_137154 | Clostridium phage PhiS63 | JQ660954 | Clustered | VC_747_0 | N2 | 5 | Clostridium | Group I | Duplodnaviria | Heunggongvirae | Uroviricota | Caudoviricetes | n.a. | Unclassified | O | O |
| S5_vs_138980 | Clostridium phage phiCT19406B | KM983331 | Clustered | VC_660_0 | N9 | 9 | Clostridium | Group I | Duplodnaviria | Heunggongvirae | Uroviricota | Caudoviricetes | n.a. | Unclassified | O | O |
| S5_vs_139401 | n.a. | n.a. | Clustered | VC_1587_0 | A | n.a. | n.a. | n.a. | n.a. | n.a. | n.a. | n.a. | n.a. | n.a. | n.a. | n.a. |
| S5_vs_140946 | n.a. | n.a. | Clustered | VC_1613_0 | A | n.a. | n.a. | n.a. | n.a. | n.a. | n.a. | n.a. | n.a. | n.a. | n.a. | n.a. |
| S5_vs_142195 | n.a. | n.a. | Clustered | VC_1605_0 | A | n.a. | n.a. | n.a. | n.a. | n.a. | n.a. | n.a. | n.a. | n.a. | n.a. | n.a. |
| S5_vs_142450 | Clostridium phage HM2 | LT600745 | Clustered | VC_1261_0 | N36 | 4 | Clostridium | Group I | Duplodnaviria | Heunggongvirae | Uroviricota | Caudoviricetes | n.a. | Salasmaviridae | O | O |
| S5_vs_145026 | n.a. | n.a. | Clustered | VC_1291_0 | A | n.a. | n.a. | n.a. | n.a. | n.a. | n.a. | n.a. | n.a. | n.a. | n.a. | n.a. |
| S5_vs_145581 | n.a. | n.a. | Clustered | VC_658_0 | A | n.a. | n.a. | n.a. | n.a. | n.a. | n.a. | n.a. | n.a. | n.a. | n.a. | n.a. |
| S5_vs_145590 | Enterococcus phage EFC-1 | KJ608188 | Clustered | VC_278_0 | N12 | 14 | Enterococcus | Group I | Duplodnaviria | Heunggongvirae | Uroviricota | Caudoviricetes | n.a. | Unclassified | O | O |
| S5_vs_145592 | n.a. | n.a. | Clustered | VC_1554_0 | A | n.a. | n.a. | n.a. | n.a. | n.a. | n.a. | n.a. | n.a. | n.a. | n.a. | n.a. |
| S5_vs_145593 | n.a. | n.a. | Clustered | VC_1615_0 | F | n.a. | n.a. | n.a. | n.a. | n.a. | n.a. | n.a. | n.a. | n.a. | n.a. | n.a. |
| S5_vs_145597 | Thermoanaerobacterium phage THSA-485A | CP003186 | Clustered | VC_724_0 | N12 | 3 | Thermoanaerobacterium | Group I | Duplodnaviria | Heunggongvirae | Uroviricota | Caudoviricetes | n.a. | Unclassified | O | O |
| S5_vs_145598 | Clostridium phage CDMH1 | HG531805 | Clustered | VC_151_0 | N11 | 25 | Clostridium | Group I | Duplodnaviria | Heunggongvirae | Uroviricota | Caudoviricetes | n.a. | Unclassified | O | O |
| S5_vs_145599 | Bacteriophage sp. | LT996078 | Clustered | VC_278_0 | N26 | 4 | Unspecified | Unclassified | Unclassified | n.a. | n.a. | n.a. | n.a. | Unclassified | O | O |
| S5_vs_145601 | Streptococcus phage MismyG | OL774867 | Clustered | VC_152_0 | N11 | 7 | Streptococcus | Group I | Duplodnaviria | Heunggongvirae | Uroviricota | Caudoviricetes | n.a. | Unclassified | O | O |
| S5_vs_145602 | Enterococcus phage EFC-1 | KJ608188 | Clustered | VC_278_0 | N11 | 13 | Enterococcus | Group I | Duplodnaviria | Heunggongvirae | Uroviricota | Caudoviricetes | n.a. | Unclassified | O | O |
| S5_vs_145605 | Bacillus phage vB_BanS_Athena | OK500002 | Clustered | VC_946_0 | N8 | 2 | Bacillus | Group I | Duplodnaviria | Heunggongvirae | Uroviricota | Caudoviricetes | n.a. | Unclassified | O | O |
| S5_vs_1572 | Parabacteroides phage PDS1 | MN929097 | Clustered | VC_855_0 | C4 | 120 | Parabacteroides | Group I | Duplodnaviria | Heunggongvirae | Uroviricota | Caudoviricetes | n.a. | Unclassified | Unclassified | Unclassified |
| S5_vs_17520 | n.a. | n.a. | Clustered | VC_1587_0 | A | n.a. | n.a. | n.a. | n.a. | n.a. | n.a. | n.a. | n.a. | n.a. | n.a. | n.a. |
| S5_vs_20268 | Paenibacillus phage PG1 | HQ332138 | Clustered | VC_349_0 | C1 | 11 | Paenibacillus | Group I | Duplodnaviria | Heunggongvirae | Uroviricota | Caudoviricetes | n.a. | Unclassified | Unclassified | Unclassified |
| S5_vs_20500 | Pasteurella phage AFS-2018a | MH238466 | Clustered | VC_630_0 | N4 | 5 | Pasteurella | Group I | Duplodnaviria | Heunggongvirae | Uroviricota | Caudoviricetes | n.a. | Unclassified | O | O |
| S5_vs_28263 | n.a. | n.a. | Clustered | VC_1587_0 | A | n.a. | n.a. | n.a. | n.a. | n.a. | n.a. | n.a. | n.a. | n.a. | n.a. | n.a. |
| S5_vs_2876 | Lactococcus phage CAP | MZ308445 | Clustered | VC_278_0 | N13 | 5 | Lactococcus | Group I | Duplodnaviria | Heunggongvirae | Uroviricota | Caudoviricetes | n.a. | Unclassified | O | O |
| S5_vs_3183 | Bacillus phage vB_BpsS-140 | MH884512 | Clustered | VC_782_0 | N11 | 1 | Bacillus | Group I | Duplodnaviria | Heunggongvirae | Uroviricota | Caudoviricetes | n.a. | Unclassified | O | O |
| S5_vs_3501 | Clostridium phage vB_CpeS-CP51 | KC237729 | Clustered | VC_1035_0 | N2 | 11 | Clostridium | Group I | Duplodnaviria | Heunggongvirae | Uroviricota | Caudoviricetes | n.a. | Unclassified | O | O |
| S5_vs_38949 | Faecalibacterium phage FP_Epona | MG711462 | Clustered | VC_723_0 | C13 | 52 | Faecalibacterium | Group I | Duplodnaviria | Heunggongvirae | Uroviricota | Caudoviricetes | n.a. | Unclassified | Unclassified | Eponavirus |
| S5_vs_40848 | Streptococcus phage Javan630 | MK448997 | Clustered | VC_649_0 | N6 | 56 | Streptococcus | Group I | Duplodnaviria | Heunggongvirae | Uroviricota | Caudoviricetes | n.a. | Unclassified | O | O |
| S5_vs_49480 | Geobacillus phage GBSV1 | DQ340064 | Clustered | VC_778_0 | N37 | 6 | Geobacillus | Group I | Duplodnaviria | Heunggongvirae | Uroviricota | Caudoviricetes | n.a. | Unclassified | O | O |
| S5_vs_56083 | n.a. | n.a. | Clustered | VC_1274_0 | A | n.a. | n.a. | n.a. | n.a. | n.a. | n.a. | n.a. | n.a. | n.a. | n.a. | n.a. |
| S5_vs_58766 | Clostridium phage phiCD38-2 | HM568888 | Clustered | VC_923_0 | N14 | 8 | Clostridium | Group I | Duplodnaviria | Heunggongvirae | Uroviricota | Caudoviricetes | n.a. | Unclassified | O | O |
| S5_vs_58859 | Geobacillus phage GBSV1 | DQ340064 | Clustered | VC_779_0 | N41 | 2 | Geobacillus | Group I | Duplodnaviria | Heunggongvirae | Uroviricota | Caudoviricetes | n.a. | Unclassified | O | O |
| S5_vs_63924 | Bifidobacterium phage BigBern1 | MT006237 | Clustered | VC_879_0 | C2 | 32 | Bifidobacterium | Group I | Duplodnaviria | Heunggongvirae | Uroviricota | Caudoviricetes | n.a. | Unclassified | Unclassified | Unclassified |
| S5_vs_67958 | n.a. | n.a. | Clustered | VC_1554_0 | A | n.a. | n.a. | n.a. | n.a. | n.a. | n.a. | n.a. | n.a. | n.a. | n.a. | n.a. |
| S5_vs_69083 | Winogradskyella phage Peternella_1 | MT732475 | Clustered | VC_696_0 | N3 | 27 | Winogradskyella | Group I | Duplodnaviria | Heunggongvirae | Uroviricota | Caudoviricetes | n.a. | Winoviridae | O | O |
| S5_vs_74199 | n.a. | n.a. | Clustered | VC_1291_0 | A | n.a. | n.a. | n.a. | n.a. | n.a. | n.a. | n.a. | n.a. | n.a. | n.a. | n.a. |
| S5_vs_7444 | n.a. | n.a. | Clustered | VC_1614_0 | F | n.a. | n.a. | n.a. | n.a. | n.a. | n.a. | n.a. | n.a. | n.a. | n.a. | n.a. |
| S5_vs_76500 | Bacteroides phage p00 | BK010646 | Clustered | VC_695_0 | C6 | 20 | Bacteroides | Group I | Duplodnaviria | Heunggongvirae | Uroviricota | Caudoviricetes | n.a. | Unclassified | Unclassified | Unclassified |
| S5_vs_79992 | Winogradskyella phage Peternella_1 | MT732475 | Clustered | VC_696_0 | N3 | 24 | Winogradskyella | Group I | Duplodnaviria | Heunggongvirae | Uroviricota | Caudoviricetes | n.a. | Winoviridae | O | O |
| S5_vs_80498 | Lactococcus phage Tuc2009 | AF109874 | Clustered | VC_278_0 | N9 | 11 | Lactococcus | Group I | Duplodnaviria | Heunggongvirae | Uroviricota | Caudoviricetes | n.a. | Unclassified | O | O |
| S5_vs_83886 | n.a. | n.a. | Clustered | VC_920_0 | A | n.a. | n.a. | n.a. | n.a. | n.a. | n.a. | n.a. | n.a. | n.a. | n.a. | n.a. |
| S5_vs_84946 | Streptococcus phage Javan290 | MK448900 | Clustered | VC_332_0 | N27 | 5 | Streptococcus | Group I | Duplodnaviria | Heunggongvirae | Uroviricota | Caudoviricetes | n.a. | Unclassified | O | O |
| S5_vs_85065 | Faecalibacterium phage FP_oengus | MG711463 | Clustered | VC_1085_1 | N17 | 70 | Faecalibacterium | Group I | Duplodnaviria | Heunggongvirae | Uroviricota | Caudoviricetes | n.a. | Unclassified | O | O |
| S5_vs_86562 | Akkermansia phage DTMo-2021a | CP084202 | Clustered | VC_752_0 | N2 | 15 | Akkermansia | Group I | Duplodnaviria | Heunggongvirae | Uroviricota | Caudoviricetes | n.a. | Unclassified | O | O |
| S5_vs_8847 | Streptococcus phage Javan191 | MK448700 | Clustered | VC_649_1 | C2 | 63 | Streptococcus | Group I | Duplodnaviria | Heunggongvirae | Uroviricota | Caudoviricetes | n.a. | Unclassified | Unclassified | Unclassified |
| S5_vs_93839 | Flavobacterium phage fF4 | MN850656 | Clustered | VC_694_0 | N2 | 23 | Flavobacterium | Group I | Duplodnaviria | Heunggongvirae | Uroviricota | Caudoviricetes | n.a. | Unclassified | O | O |
| S5_vs_9747 | n.a. | n.a. | Clustered | VC_1595_0 | A | n.a. | n.a. | n.a. | n.a. | n.a. | n.a. | n.a. | n.a. | n.a. | n.a. | n.a. |
| S6_vf_25710 | n.a. | n.a. | Clustered | VC_750_0 | A | n.a. | n.a. | n.a. | n.a. | n.a. | n.a. | n.a. | n.a. | n.a. | n.a. | n.a. |
| S6_vf_3435 | Lactococcus phage 62403 | MF443120 | Clustered | VC_706_0 | C1 | 75 | Lactococcus | Group I | Duplodnaviria | Heunggongvirae | Uroviricota | Caudoviricetes | n.a. | Unclassified | Unclassified | Ceduovirus |
| S6_vf_45228 | Clostridium phage HM2 | LT600745 | Clustered | VC_1261_0 | N33 | 4 | Clostridium | Group I | Duplodnaviria | Heunggongvirae | Uroviricota | Caudoviricetes | n.a. | Salasmaviridae | O | O |
| S6_vf_45618 | Eggerthella phage PMBT5 | MH626557 | Clustered | VC_1421_0 | N42 | 2 | Eggerthella | Group I | Duplodnaviria | Heunggongvirae | Uroviricota | Caudoviricetes | n.a. | Unclassified | O | O |
| S6_vf_56486 | Bacteroides phage SJC03 | MT074146 | Clustered | VC_963_0 | N5 | 49 | Bacteroides | Group I | Duplodnaviria | Heunggongvirae | Uroviricota | Caudoviricetes | n.a. | Unclassified | O | O |
| S6_vf_6314 | n.a. | n.a. | Clustered | VC_1607_0 | F | n.a. | n.a. | n.a. | n.a. | n.a. | n.a. | n.a. | n.a. | n.a. | n.a. | n.a. |
| S6_vs_10314 | Butyrivibrio phage Idris | MN882554 | Clustered | VC_1289_0 | N4 | 38 | Butyrivibrio | Group I | Duplodnaviria | Heunggongvirae | Uroviricota | Caudoviricetes | n.a. | Unclassified | O | O |
| S6_vs_12945 | Clostridioides phage CD1801 | MW512570 | Clustered | VC_670_0 | N10 | 7 | Clostridioides | Group I | Duplodnaviria | Heunggongvirae | Uroviricota | Caudoviricetes | n.a. | Unclassified | O | O |
| S6_vs_15813 | n.a. | n.a. | Clustered | VC_1481_0 | A | n.a. | n.a. | n.a. | n.a. | n.a. | n.a. | n.a. | n.a. | n.a. | n.a. | n.a. |
| S6_vs_32886 | n.a. | n.a. | Clustered | VC_1590_0 | A | n.a. | n.a. | n.a. | n.a. | n.a. | n.a. | n.a. | n.a. | n.a. | n.a. | n.a. |
| S6_vs_33711 | Bacillus phage vB_BtS_BMBtp13 | KX190832 | Clustered | VC_667_0 | C7 | 7 | Bacillus | Group I | Duplodnaviria | Heunggongvirae | Uroviricota | Caudoviricetes | n.a. | Unclassified | Unclassified | Unclassified |
| S6_vs_38250 | Bacillus phage vB_BtS_BMBtp13 | KX190832 | Clustered | VC_667_0 | C8 | 6 | Bacillus | Group I | Duplodnaviria | Heunggongvirae | Uroviricota | Caudoviricetes | n.a. | Unclassified | Unclassified | Unclassified |
| S6_vs_39025 | Clostridioides phage CD1801 | MW512570 | Clustered | VC_670_0 | N11 | 6 | Clostridioides | Group I | Duplodnaviria | Heunggongvirae | Uroviricota | Caudoviricetes | n.a. | Unclassified | O | O |
| S6_vs_46039 | n.a. | n.a. | Clustered | VC_1612_0 | A | n.a. | n.a. | n.a. | n.a. | n.a. | n.a. | n.a. | n.a. | n.a. | n.a. | n.a. |
| S6_vs_54606 | Butyrivibrio phage Idris | MN882554 | Clustered | VC_1289_0 | N4 | 33 | Butyrivibrio | Group I | Duplodnaviria | Heunggongvirae | Uroviricota | Caudoviricetes | n.a. | Unclassified | O | O |
| S6_vs_56490 | Geobacillus phage GBSV1 | DQ340064 | Clustered | VC_778_0 | N39 | 6 | Geobacillus | Group I | Duplodnaviria | Heunggongvirae | Uroviricota | Caudoviricetes | n.a. | Unclassified | O | O |
| S6_vs_56491 | Lactococcus phage CAP | MZ308445 | Clustered | VC_278_0 | N22 | 2 | Lactococcus | Group I | Duplodnaviria | Heunggongvirae | Uroviricota | Caudoviricetes | n.a. | Unclassified | O | O |
| S6_vs_56493 | n.a. | n.a. | Clustered | VC_1601_0 | F | n.a. | n.a. | n.a. | n.a. | n.a. | n.a. | n.a. | n.a. | n.a. | n.a. | n.a. |
| S6_vs_56496 | Faecalibacterium phage FP_Epona | MG711462 | Clustered | VC_723_0 | C2 | 104 | Faecalibacterium | Group I | Duplodnaviria | Heunggongvirae | Uroviricota | Caudoviricetes | n.a. | Unclassified | Unclassified | Eponavirus |
| S6_vs_56498 | Butyrivibrio phage Idris | MN882554 | Clustered | VC_1289_0 | N5 | 41 | Butyrivibrio | Group I | Duplodnaviria | Heunggongvirae | Uroviricota | Caudoviricetes | n.a. | Unclassified | O | O |
| S6_vs_9784 | n.a. | n.a. | Clustered | VC_1505_0 | A | n.a. | n.a. | n.a. | n.a. | n.a. | n.a. | n.a. | n.a. | n.a. | n.a. | n.a. |
| S7_vf_114476 | Butyrivibrio virus Arian | MN882551 | Clustered | VC_720_0 | N6 | 14 | Butyrivibrio | Group I | Duplodnaviria | Heunggongvirae | Uroviricota | Caudoviricetes | n.a. | Unclassified | O | O |
| S7_vf_117697 | n.a. | n.a. | Clustered | VC_1576_0 | A | n.a. | n.a. | n.a. | n.a. | n.a. | n.a. | n.a. | n.a. | n.a. | n.a. | n.a. |
| S7_vf_134023 | n.a. | n.a. | Clustered | VC_1590_0 | A | n.a. | n.a. | n.a. | n.a. | n.a. | n.a. | n.a. | n.a. | n.a. | n.a. | n.a. |
| S7_vf_135778 | Bacillus phage vB_BanS_Athena | OK500002 | Clustered | VC_946_0 | N8 | 2 | Bacillus | Group I | Duplodnaviria | Heunggongvirae | Uroviricota | Caudoviricetes | n.a. | Unclassified | O | O |
| S7_vf_138019 | n.a. | n.a. | Clustered | VC_1615_0 | F | n.a. | n.a. | n.a. | n.a. | n.a. | n.a. | n.a. | n.a. | n.a. | n.a. | n.a. |
| S7_vf_138069 | n.a. | n.a. | Clustered | VC_1564_0 | F | n.a. | n.a. | n.a. | n.a. | n.a. | n.a. | n.a. | n.a. | n.a. | n.a. | n.a. |
| S7_vf_19866 | Clostridium phage HM2 | LT600745 | Clustered | VC_1258_0 | N44 | 3 | Clostridium | Group I | Duplodnaviria | Heunggongvirae | Uroviricota | Caudoviricetes | n.a. | Salasmaviridae | O | O |
| S7_vf_20129 | Bacteriophage sp. | OP549880 | Clustered | VC_1549_0 | N5 | 16 | Unspecified | Unclassified | Unclassified | n.a. | n.a. | n.a. | n.a. | Unclassified | O | O |
| S7_vf_27437 | n.a. | n.a. | Clustered | VC_1607_0 | F | n.a. | n.a. | n.a. | n.a. | n.a. | n.a. | n.a. | n.a. | n.a. | n.a. | n.a. |
| S7_vf_28429 | Cellulophaga phage phi17:2 | KC821609 | Clustered | VC_1055_0 | N8 | 4 | Cellulophaga | Group I | Duplodnaviria | Heunggongvirae | Uroviricota | Caudoviricetes | n.a. | Unclassified | O | O |
| S7_vf_29196 | n.a. | n.a. | Clustered | VC_1554_0 | A | n.a. | n.a. | n.a. | n.a. | n.a. | n.a. | n.a. | n.a. | n.a. | n.a. | n.a. |
| S7_vf_32559 | n.a. | n.a. | Clustered | VC_1258_0 | A | n.a. | n.a. | n.a. | n.a. | n.a. | n.a. | n.a. | n.a. | n.a. | n.a. | n.a. |
| S7_vf_36627 | n.a. | n.a. | Clustered | VC_1258_0 | A | n.a. | n.a. | n.a. | n.a. | n.a. | n.a. | n.a. | n.a. | n.a. | n.a. | n.a. |
| S7_vf_38697 | Faecalibacterium phage FP_oengus | MG711463 | Clustered | VC_1085_1 | N18 | 59 | Faecalibacterium | Group I | Duplodnaviria | Heunggongvirae | Uroviricota | Caudoviricetes | n.a. | Unclassified | O | O |
| S7_vf_39724 | Faecalibacterium phage FP_Mushu | MG711460 | Clustered | VC_1288_0 | C1 | 171 | Faecalibacterium | Group I | Duplodnaviria | Heunggongvirae | Uroviricota | Caudoviricetes | n.a. | Unclassified | Unclassified | Mushuvirus |
| S7_vf_5439 | Faecalibacterium phage FP_Epona | MG711462 | Clustered | VC_723_0 | C1 | 118 | Faecalibacterium | Group I | Duplodnaviria | Heunggongvirae | Uroviricota | Caudoviricetes | n.a. | Unclassified | Unclassified | Eponavirus |
| S7_vf_5843 | Curtobacterium phage Penoan | MZ333133 | Clustered | VC_1195_0 | N3 | 4 | Curtobacterium | Group I | Duplodnaviria | Heunggongvirae | Uroviricota | Caudoviricetes | n.a. | Unclassified | O | O |
| S7_vf_60968 | Ruminococcus phage phiRgPS_6 | MT980839 | Clustered | VC_278_0 | N1 | 47 | Ruminococcus | Group I | Duplodnaviria | Heunggongvirae | Uroviricota | Caudoviricetes | n.a. | Unclassified | O | O |
| S7_vf_63894 | Leuconostoc phage 1-A4 | GQ451696 | Clustered | VC_889_0 | C2 | 116 | Leuconostoc | Group I | Duplodnaviria | Heunggongvirae | Uroviricota | Caudoviricetes | n.a. | Unclassified | Mccleskeyvirinae | Unaquatrovirus |
| S7_vf_68546 | Clostridium phage phiCp-D | MW478291 | Clustered | VC_921_0 | N4 | 12 | Clostridium | Group I | Duplodnaviria | Heunggongvirae | Uroviricota | Caudoviricetes | n.a. | Unclassified | O | O |

| | | | | | | | | | | | | | | | | |
|---|---|---|---|---|---|---|---|---|---|---|---|---|---|---|---|---|
| S7_vf_69348 | n.a. | n.a. | Clustered | VC_1258_0 | A | n.a. | n.a. | n.a. | n.a. | n.a. | n.a. | n.a. | n.a. | n.a. | n.a. | n.a. |
| S7_vf_71527 | n.a. | n.a. | Clustered | VC_1616_0 | F | n.a. | n.a. | n.a. | n.a. | n.a. | n.a. | n.a. | n.a. | n.a. | n.a. | n.a. |
| S7_vf_76382 | Clostridium phage HM2 | LT600745 | Clustered | VC_1261_0 | N20 | 7 | Clostridium | Group I | Duplodnaviria | Heunggongvirae | Uroviricota | Caudoviricetes | n.a. | Salasmaviridae | O | O |
| S7_vf_76463 | n.a. | n.a. | Clustered | VC_1552_0 | A | n.a. | n.a. | n.a. | n.a. | n.a. | n.a. | n.a. | n.a. | n.a. | n.a. | n.a. |
| S7_vf_84974 | Clostridioides phage ES-S-0173-01 | CP067352 | Clustered | VC_748_0 | N12 | 1 | Clostridioides | Group I | Duplodnaviria | Heunggongvirae | Uroviricota | Caudoviricetes | n.a. | Unclassified | O | O |
| S7_vf_89197 | Lactococcus phage vB_LLc_bIBB14 | MH779519 | Clustered | VC_706_0 | C1 | 113 | Lactococcus | Group I | Duplodnaviria | Heunggongvirae | Uroviricota | Caudoviricetes | n.a. | Unclassified | Unclassified | Ceduovirus |
| S7_vs_100640 | n.a. | n.a. | Clustered | VC_1587_0 | A | n.a. | n.a. | n.a. | n.a. | n.a. | n.a. | n.a. | n.a. | n.a. | n.a. | n.a. |
| S7_vs_11081 | Bacteroides phage p00 | BK010646 | Clustered | VC_695_0 | C5 | 43 | Bacteroides | Group I | Duplodnaviria | Heunggongvirae | Uroviricota | Caudoviricetes | n.a. | Unclassified | Unclassified | Unclassified |
| S7_vs_111831 | Streptococcus phage Javan580 | MK448989 | Clustered | VC_558_0 | N10 | 8 | Streptococcus | Group I | Duplodnaviria | Heunggongvirae | Uroviricota | Caudoviricetes | n.a. | Unclassified | O | O |
| S7_vs_118295 | n.a. | n.a. | Clustered | VC_1481_0 | A | n.a. | n.a. | n.a. | n.a. | n.a. | n.a. | n.a. | n.a. | n.a. | n.a. | n.a. |
| S7_vs_119492 | Clostridium phage HM2 | LT600745 | Clustered | VC_1261_0 | N36 | 4 | Clostridium | Group I | Duplodnaviria | Heunggongvirae | Uroviricota | Caudoviricetes | n.a. | Salasmaviridae | O | O |
| S7_vs_119679 | Winogradskyella phage Peternella_1 | MT732475 | Clustered | VC_696_0 | N4 | 22 | Winogradskyella | Group I | Duplodnaviria | Heunggongvirae | Uroviricota | Caudoviricetes | n.a. | Winoviridae | O | O |
| S7_vs_122151 | n.a. | n.a. | Clustered | VC_779_0 | A | n.a. | n.a. | n.a. | n.a. | n.a. | n.a. | n.a. | n.a. | n.a. | n.a. | n.a. |
| S7_vs_129271 | n.a. | n.a. | Clustered | VC_1578_0 | A | n.a. | n.a. | n.a. | n.a. | n.a. | n.a. | n.a. | n.a. | n.a. | n.a. | n.a. |
| S7_vs_129858 | n.a. | n.a. | Clustered | VC_1620_0 | F | n.a. | n.a. | n.a. | n.a. | n.a. | n.a. | n.a. | n.a. | n.a. | n.a. | n.a. |
| S7_vs_13041 | Butyrivibrio virus Arian | MN882551 | Clustered | VC_720_0 | N6 | 14 | Butyrivibrio | Group I | Duplodnaviria | Heunggongvirae | Uroviricota | Caudoviricetes | n.a. | Unclassified | O | O |
| S7_vs_134391 | n.a. | n.a. | Clustered | VC_1568_0 | A | n.a. | n.a. | n.a. | n.a. | n.a. | n.a. | n.a. | n.a. | n.a. | n.a. | n.a. |
| S7_vs_134583 | Faecalibacterium phage FP_Toutatis | MG711466 | Clustered | VC_728_0 | C4 | 56 | Faecalibacterium | Group I | Duplodnaviria | Heunggongvirae | Uroviricota | Caudoviricetes | n.a. | Unclassified | Unclassified | Toutatisvirus |
| S7_vs_135828 | Microviridae sp. | MG945563 | Clustered | VC_1335_0 | C1 | 9 | Unspecified | Group II | Monodnaviria | Sangervirae | Phixviricota | Malgrandaviricetes | Petitvirales | Microviridae | Unclassified | Unclassified |
| S7_vs_136164 | n.a. | n.a. | Clustered | VC_1570_0 | A | n.a. | n.a. | n.a. | n.a. | n.a. | n.a. | n.a. | n.a. | n.a. | n.a. | n.a. |
| S7_vs_137915 | Bacteroides phage DAC16 | MT074137 | Clustered | VC_963_0 | N5 | 42 | Bacteroides | Group I | Duplodnaviria | Heunggongvirae | Uroviricota | Caudoviricetes | n.a. | Unclassified | O | O |
| S7_vs_13799 | Roseburia phage Jekyll | LR596902 | Clustered | VC_278_0 | N14 | 5 | Roseburia | Group I | Duplodnaviria | Heunggongvirae | Uroviricota | Caudoviricetes | n.a. | Unclassified | O | O |
| S7_vs_138096 | n.a. | n.a. | Clustered | VC_1570_0 | A | n.a. | n.a. | n.a. | n.a. | n.a. | n.a. | n.a. | n.a. | n.a. | n.a. | n.a. |
| S7_vs_138109 | Lactococcus phage CAP | MZ308445 | Clustered | VC_278_0 | N13 | 8 | Lactococcus | Group I | Duplodnaviria | Heunggongvirae | Uroviricota | Caudoviricetes | n.a. | Unclassified | O | O |
| S7_vs_138173 | Phage vB_RanS_PJN03 | ON245412 | Clustered | VC_950_0 | N13 | 5 | Unspecified | Group I | Duplodnaviria | Heunggongvirae | Uroviricota | Caudoviricetes | n.a. | Unclassified | O | O |
| S7_vs_20200 | n.a. | n.a. | Clustered | VC_1291_0 | A | n.a. | n.a. | n.a. | n.a. | n.a. | n.a. | n.a. | n.a. | n.a. | n.a. | n.a. |
| S7_vs_2599 | n.a. | n.a. | Clustered | VC_1572_0 | A | n.a. | n.a. | n.a. | n.a. | n.a. | n.a. | n.a. | n.a. | n.a. | n.a. | n.a. |
| S7_vs_27530 | n.a. | n.a. | Clustered | VC_1567_0 | A | n.a. | n.a. | n.a. | n.a. | n.a. | n.a. | n.a. | n.a. | n.a. | n.a. | n.a. |
| S7_vs_28733 | Faecalibacterium phage FP_Lugh | MG711464 | Clustered | VC_923_0 | N19 | 4 | Faecalibacterium | Group I | Duplodnaviria | Heunggongvirae | Uroviricota | Caudoviricetes | n.a. | Unclassified | O | O |
| S7_vs_38113 | Geobacillus phage GBSV1 | DQ340064 | Clustered | VC_779_0 | N42 | 2 | Geobacillus | Group I | Duplodnaviria | Heunggongvirae | Uroviricota | Caudoviricetes | n.a. | Unclassified | O | O |
| S7_vs_38834 | Exiguobacterium phage vB_EauS-123 | KU160495 | Clustered | VC_779_0 | N30 | 5 | Exiguobacterium | Group I | Duplodnaviria | Heunggongvirae | Uroviricota | Caudoviricetes | n.a. | Unclassified | O | O |
| S7_vs_41098 | Faecalibacterium phage FP_oengus | MG711463 | Clustered | VC_1085_1 | N20 | 65 | Faecalibacterium | Group I | Duplodnaviria | Heunggongvirae | Uroviricota | Caudoviricetes | n.a. | Unclassified | O | O |
| S7_vs_41534 | Clostridium phage HM2 | LT600745 | Clustered | VC_1265_0 | N19 | 4 | Clostridium | Group I | Duplodnaviria | Heunggongvirae | Uroviricota | Caudoviricetes | n.a. | Salasmaviridae | O | O |
| S7_vs_42563 | n.a. | n.a. | Clustered | VC_1291_0 | A | n.a. | n.a. | n.a. | n.a. | n.a. | n.a. | n.a. | n.a. | n.a. | n.a. | n.a. |
| S7_vs_47050 | Butyrivibrio virus Arian | MN882551 | Clustered | VC_720_0 | N1 | 32 | Butyrivibrio | Group I | Duplodnaviria | Heunggongvirae | Uroviricota | Caudoviricetes | n.a. | Unclassified | O | O |
| S7_vs_48523 | Streptococcus phage MismyG | OL774867 | Clustered | VC_152_0 | N34 | 2 | Streptococcus | Group I | Duplodnaviria | Heunggongvirae | Uroviricota | Caudoviricetes | n.a. | Unclassified | O | O |
| S7_vs_48708 | Clostridium phage Clo-PEP-1 | KY206887 | Clustered | VC_442_0 | N3 | 25 | Clostridium | Group I | Duplodnaviria | Heunggongvirae | Uroviricota | Caudoviricetes | n.a. | Unclassified | O | O |
| S7_vs_48832 | Bacteriophage sp. | LT996078 | Clustered | VC_278_0 | N14 | 5 | Unspecified | Unclassified | Unclassified | n.a. | n.a. | n.a. | n.a. | Unclassified | O | O |
| S7_vs_49426 | Erysipelothrix phage SE-1 | KR816341 | Clustered | VC_1149_0 | N3 | 11 | Erysipelothrix | Group I | Duplodnaviria | Heunggongvirae | Uroviricota | Caudoviricetes | n.a. | Unclassified | O | O |
| S7_vs_4956 | Faecalibacterium phage FP_Epona | MG711462 | Clustered | VC_723_0 | C13 | 52 | Faecalibacterium | Group I | Duplodnaviria | Heunggongvirae | Uroviricota | Caudoviricetes | n.a. | Unclassified | Unclassified | Eponavirus |
| S7_vs_50194 | n.a. | n.a. | Clustered | VC_1602_0 | A | n.a. | n.a. | n.a. | n.a. | n.a. | n.a. | n.a. | n.a. | n.a. | n.a. | n.a. |
| S7_vs_5236 | Butyrivibrio phage Idris | MN882554 | Clustered | VC_1289_0 | N7 | 31 | Butyrivibrio | Group I | Duplodnaviria | Heunggongvirae | Uroviricota | Caudoviricetes | n.a. | Unclassified | O | O |
| S7_vs_55409 | Bacillus phage 1 | DQ840344 | Clustered | VC_778_0 | N28 | 7 | Bacillus | Group I | Duplodnaviria | Heunggongvirae | Uroviricota | Caudoviricetes | n.a. | Unclassified | O | O |
| S7_vs_557 | Bacteroides phage F2 | MT806187 | Clustered | VC_1505_0 | N12 | 9 | Bacteroides | Group I | Duplodnaviria | Heunggongvirae | Uroviricota | Caudoviricetes | n.a. | Unclassified | O | O |
| S7_vs_56529 | Bacillus phage vB_BtS_BMBtp13 | KX190832 | Clustered | VC_667_0 | C9 | 5 | Bacillus | Group I | Duplodnaviria | Heunggongvirae | Uroviricota | Caudoviricetes | n.a. | Unclassified | Unclassified | Unclassified |
| S7_vs_59465 | Clostridium phage phiMMP01 | LN681541 | Clustered | VC_151_0 | N16 | 27 | Clostridium | Group I | Duplodnaviria | Heunggongvirae | Uroviricota | Caudoviricetes | n.a. | Unclassified | O | O |
| S7_vs_6626 | Faecalibacterium phage FP_Epona | MG711462 | Clustered | VC_723_0 | C13 | 49 | Faecalibacterium | Group I | Duplodnaviria | Heunggongvirae | Uroviricota | Caudoviricetes | n.a. | Unclassified | Unclassified | Eponavirus |
| S7_vs_69630 | Clostridium phage phiCT19406B | KM983331 | Clustered | VC_660_0 | N8 | 10 | Clostridium | Group I | Duplodnaviria | Heunggongvirae | Uroviricota | Caudoviricetes | n.a. | Unclassified | O | O |
| S7_vs_73280 | Microbacterium phage Min1 | EF579802 | Clustered | VC_642_0 | C1 | 13 | Microbacterium | Group I | Duplodnaviria | Heunggongvirae | Uroviricota | Caudoviricetes | n.a. | Unclassified | Unclassified | Minunavirus |
| S7_vs_82140 | Geobacillus phage GBSV1 | DQ340064 | Clustered | VC_779_0 | N41 | 2 | Geobacillus | Group I | Duplodnaviria | Heunggongvirae | Uroviricota | Caudoviricetes | n.a. | Unclassified | O | O |
| S7_vs_83100 | Streptococcus phage MismyG | OL774867 | Clustered | VC_152_0 | N17 | 5 | Streptococcus | Group I | Duplodnaviria | Heunggongvirae | Uroviricota | Caudoviricetes | n.a. | Unclassified | O | O |
| S7_vs_86377 | Streptococcus phage Javan290 | MK448900 | Clustered | VC_332_0 | N27 | 2 | Streptococcus | Group I | Duplodnaviria | Heunggongvirae | Uroviricota | Caudoviricetes | n.a. | Unclassified | O | O |
| S7_vs_87439 | n.a. | n.a. | Clustered | VC_1479_0 | A | n.a. | n.a. | n.a. | n.a. | n.a. | n.a. | n.a. | n.a. | n.a. | n.a. | n.a. |
| S7_vs_90915 | Streptococcus phage MismyG | OL774867 | Clustered | VC_152_0 | N15 | 6 | Streptococcus | Group I | Duplodnaviria | Heunggongvirae | Uroviricota | Caudoviricetes | n.a. | Unclassified | O | O |
| S7_vs_944 | n.a. | n.a. | Clustered | VC_1505_0 | A | n.a. | n.a. | n.a. | n.a. | n.a. | n.a. | n.a. | n.a. | n.a. | n.a. | n.a. |
| S8_vf_101283 | Faecalibacterium phage FP_Toutatis | MG711466 | Clustered | VC_728_0 | C1 | 101 | Faecalibacterium | Group I | Duplodnaviria | Heunggongvirae | Uroviricota | Caudoviricetes | n.a. | Unclassified | Unclassified | Toutatisvirus |
| S8_vf_106946 | Ralstonia phage RPSC1 | MF893341 | Clustered | VC_86_0 | N3 | 7 | Ralstonia | Group I | Duplodnaviria | Heunggongvirae | Uroviricota | Caudoviricetes | n.a. | Autographiviridae | O | O |
| S8_vf_111790 | Eggerthella phage PMBT5 | MH626557 | Clustered | VC_1421_0 | N20 | 4 | Eggerthella | Group I | Duplodnaviria | Heunggongvirae | Uroviricota | Caudoviricetes | n.a. | Unclassified | O | O |
| S8_vf_117796 | Bacteroides phage p00 | BK010646 | Clustered | VC_695_0 | C5 | 43 | Bacteroides | Group I | Duplodnaviria | Heunggongvirae | Uroviricota | Caudoviricetes | n.a. | Unclassified | Unclassified | Unclassified |
| S8_vf_12212 | n.a. | n.a. | Clustered | VC_153_0 | A | n.a. | n.a. | n.a. | n.a. | n.a. | n.a. | n.a. | n.a. | n.a. | n.a. | n.a. |
| S8_vf_129423 | Leuconostoc phage 1-A4 | GQ451696 | Clustered | VC_889_0 | C1 | 127 | Leuconostoc | Group I | Duplodnaviria | Heunggongvirae | Uroviricota | Caudoviricetes | n.a. | Unclassified | Mccleskeyvirinae | Unaquatrovirus |
| S8_vf_131009 | Roseburia phage Jekyll | LR596902 | Clustered | VC_1254_2 | N26 | 1 | Roseburia | Group I | Duplodnaviria | Heunggongvirae | Uroviricota | Caudoviricetes | n.a. | Unclassified | O | O |
| S8_vf_131802 | uncultured phage cr56_1 | MT774397 | Clustered | VC_519_5 | C1 | 300 | Unspecified | Group I | Duplodnaviria | Heunggongvirae | Uroviricota | Caudoviricetes | Crassvirales | Suoliviridae | Loutivirinae | Buchavirus |
| S8_vf_132697 | Roseburia phage Jekyll | LR596902 | Clustered | VC_1254_0 | N19 | 10 | Roseburia | Group I | Duplodnaviria | Heunggongvirae | Uroviricota | Caudoviricetes | n.a. | Unclassified | O | O |
| S8_vf_134943 | n.a. | n.a. | Clustered | VC_1564_0 | F | n.a. | n.a. | n.a. | n.a. | n.a. | n.a. | n.a. | n.a. | n.a. | n.a. | n.a. |
| S8_vf_136235 | Bacteriophage sp. | OP549880 | Clustered | VC_1549_0 | N5 | 16 | Unspecified | Unclassified | Unclassified | n.a. | n.a. | n.a. | n.a. | Unclassified | O | O |
| S8_vf_13758 | Butyrivibrio phage Idris | MN882554 | Clustered | VC_1477_0 | N13 | 2 | Butyrivibrio | Group I | Duplodnaviria | Heunggongvirae | Uroviricota | Caudoviricetes | n.a. | Unclassified | O | O |
| S8_vf_142700 | Bacteriophage sp. | OP549880 | Clustered | VC_1549_0 | N5 | 17 | Unspecified | Unclassified | Unclassified | n.a. | n.a. | n.a. | n.a. | Unclassified | O | O |
| S8_vf_147128 | Phage DP SC_6_H4_2017 | MT121964 | Clustered | VC_1490_1 | C2 | 174 | Unspecified | Group I | Duplodnaviria | Heunggongvirae | Uroviricota | Caudoviricetes | n.a. | Unclassified | Unclassified | Unclassified |
| S8_vf_151384 | Faecalibacterium phage FP_Epona | MG711462 | Clustered | VC_723_0 | C2 | 83 | Faecalibacterium | Group I | Duplodnaviria | Heunggongvirae | Uroviricota | Caudoviricetes | n.a. | Unclassified | Unclassified | Eponavirus |
| S8_vf_158225 | uncultured phage cr3_1 | MT774381 | Clustered | VC_691_0 | C1 | 148 | Unspecified | Group I | Duplodnaviria | Heunggongvirae | Uroviricota | Caudoviricetes | Crassvirales | Intestiviridae | Crudevirinae | Diorhovirus |
| S8_vf_16172 | n.a. | n.a. | Clustered | VC_1583_0 | A | n.a. | n.a. | n.a. | n.a. | n.a. | n.a. | n.a. | n.a. | n.a. | n.a. | n.a. |
| S8_vf_163760 | Faecalibacterium phage FP_oengus | MG711463 | Clustered | VC_1085_1 | N15 | 73 | Faecalibacterium | Group I | Duplodnaviria | Heunggongvirae | Uroviricota | Caudoviricetes | n.a. | Unclassified | O | O |
| S8_vf_166403 | n.a. | n.a. | Clustered | VC_1294_0 | A | n.a. | n.a. | n.a. | n.a. | n.a. | n.a. | n.a. | n.a. | n.a. | n.a. | n.a. |
| S8_vf_166677 | uncultured phage cr8_1 | MT774384 | Clustered | VC_691_1 | C1 | 191 | Unspecified | Group I | Duplodnaviria | Heunggongvirae | Uroviricota | Caudoviricetes | Crassvirales | Intestiviridae | Obtuvirinae | Fohxhuevirus |
| S8_vf_170430 | Clavibacter phage CMP1 | GQ241246 | Clustered | VC_880_0 | C2 | 14 | Clavibacter | Group I | Duplodnaviria | Heunggongvirae | Uroviricota | Caudoviricetes | n.a. | Unclassified | Unclassified | Cimpunavirus |
| S8_vf_170878 | Faecalibacterium phage FP_Toutatis | MG711466 | Clustered | VC_728_0 | C4 | 72 | Faecalibacterium | Group I | Duplodnaviria | Heunggongvirae | Uroviricota | Caudoviricetes | n.a. | Unclassified | Unclassified | Toutatisvirus |
| S8_vf_174559 | Clostridium phage HM2 | LT600745 | Clustered | VC_1261_0 | N32 | 4 | Clostridium | Group I | Duplodnaviria | Heunggongvirae | Uroviricota | Caudoviricetes | n.a. | Salasmaviridae | O | O |
| S8_vf_183045 | Bacteroides phage F4 | MT806185 | Clustered | VC_1044_0 | C1 | 94 | Bacteroides | Group I | Duplodnaviria | Heunggongvirae | Uroviricota | Caudoviricetes | n.a. | Unclassified | Unclassified | Unclassified |
| S8_vf_187742 | Geobacillus virus E3 | KP144388 | Clustered | VC_748_0 | N17 | 2 | Geobacillus | Group I | Duplodnaviria | Heunggongvirae | Uroviricota | Caudoviricetes | n.a. | Unclassified | O | O |
| S8_vf_196670 | n.a. | n.a. | Clustered | VC_1258_0 | A | n.a. | n.a. | n.a. | n.a. | n.a. | n.a. | n.a. | n.a. | n.a. | n.a. | n.a. |
| S8_vf_197061 | Bifidobacterium phage BigBern1 | MT006237 | Clustered | VC_879_0 | C2 | 34 | Bifidobacterium | Group I | Duplodnaviria | Heunggongvirae | Uroviricota | Caudoviricetes | n.a. | Unclassified | Unclassified | Unclassified |
| S8_vf_203044 | Bacillus phage vB_BsuP-Goe1 | KU831549 | Clustered | VC_847_0 | N20 | 3 | Bacillus | Group I | Duplodnaviria | Heunggongvirae | Uroviricota | Caudoviricetes | n.a. | Salasmaviridae | O | O |
| S8_vf_208102 | n.a. | n.a. | Clustered | VC_1619_0 | F | n.a. | n.a. | n.a. | n.a. | n.a. | n.a. | n.a. | n.a. | n.a. | n.a. | n.a. |
| S8_vf_208107 | n.a. | n.a. | Clustered | VC_1619_0 | F | n.a. | n.a. | n.a. | n.a. | n.a. | n.a. | n.a. | n.a. | n.a. | n.a. | n.a. |
| S8_vf_208110 | Bacillus phage vB_BcoS-136 | MH884508 | Clustered | VC_491_0 | N19 | 3 | Bacillus | Group I | Duplodnaviria | Heunggongvirae | Uroviricota | Caudoviricetes | n.a. | Unclassified | O | O |
| S8_vf_208119 | Gokushovirus WZ-2015a | KT264793 | Clustered | VC_1170_0 | C1 | 27 | Unspecified | Group II | Monodnaviria | Sangervirae | Phixviricota | Malgrandaviricetes | Petitvirales | Microviridae | Gokushovirinae | Gokushovirus |

| | | | | | | | | | | | | | | | | |
|---|---|---|---|---|---|---|---|---|---|---|---|---|---|---|---|---|
| S8_vf_208120 | Phage vB_RanS_PJN03 | ON245412 | Clustered | VC_951_1 | N14 | 12 | Unspecified | Group I | Duplodnaviria | Heunggongvirae | Uroviricota | Caudoviricetes | n.a. | Unclassified | O | O |
| S8_vf_208126 | Clostridium phage phiMMP01 | LN681541 | Clustered | VC_151_0 | N19 | 27 | Clostridium | Group I | Duplodnaviria | Heunggongvirae | Uroviricota | Caudoviricetes | n.a. | Unclassified | O | O |
| S8_vf_208146 | Human gut gokushovirus | KX513867 | Clustered | VC_1184_0 | C1 | 28 | Unspecified | Group II | Monodnaviria | Sangervirae | Phixviricota | Malgrandaviricetes | Petitvirales | Microviridae | Gokushovirinae | Unclassified |
| S8_vf_41376 | Faecalibacterium phage FP_oengus | MG711463 | Clustered | VC_1085_1 | N21 | 59 | Faecalibacterium | Group I | Duplodnaviria | Heunggongvirae | Uroviricota | Caudoviricetes | n.a. | Unclassified | O | O |
| S8_vf_43483 | n.a. | n.a. | Clustered | VC_1607_0 | F | n.a. | n.a. | n.a. | n.a. | n.a. | n.a. | n.a. | n.a. | n.a. | n.a. | n.a. |
| S8_vf_45776 | Lactococcus phage 62605 | MF448565 | Clustered | VC_210_0 | C1 | 141 | Lactococcus | Group I | Duplodnaviria | Heunggongvirae | Uroviricota | Caudoviricetes | n.a. | Unclassified | Unclassified | Skunavirus |
| S8_vf_52050 | Clostridium phage HM2 | LT600745 | Clustered | VC_1258_0 | N38 | 4 | Clostridium | Group I | Duplodnaviria | Heunggongvirae | Uroviricota | Caudoviricetes | n.a. | Salasmaviridae | O | O |
| S8_vf_55476 | n.a. | n.a. | Clustered | VC_1481_1 | A | n.a. | n.a. | n.a. | n.a. | n.a. | n.a. | n.a. | n.a. | n.a. | n.a. | n.a. |
| S8_vf_71214 | Arthrobacter phage Orcanus | MT952853 | Clustered | VC_1511_0 | N3 | 1 | Arthrobacter | Group I | Duplodnaviria | Heunggongvirae | Uroviricota | Caudoviricetes | n.a. | Unclassified | O | O |
| S8_vf_72773 | Faecalibacterium phage FP_Lagaffe | MG711461 | Clustered | VC_1291_0 | N40 | 3 | Faecalibacterium | Group I | Duplodnaviria | Heunggongvirae | Uroviricota | Caudoviricetes | n.a. | Unclassified | O | O |
| S8_vf_79769 | n.a. | n.a. | Clustered | VC_1561_0 | A | n.a. | n.a. | n.a. | n.a. | n.a. | n.a. | n.a. | n.a. | n.a. | n.a. | n.a. |
| S8_vf_87755 | CrAssphage sp. C0521BD4 | MW067000 | Clustered | VC_691_0 | C2 | 168 | Unspecified | Group I | Duplodnaviria | Heunggongvirae | Uroviricota | Caudoviricetes | Crassvirales | Unclassified | Unclassified | Unclassified |
| S8_vf_93976 | n.a. | n.a. | Clustered | VC_1570_0 | A | n.a. | n.a. | n.a. | n.a. | n.a. | n.a. | n.a. | n.a. | n.a. | n.a. | n.a. |
| S8_vf_94361 | Microviridae sp. | MG945227 | Clustered | VC_1145_0 | C1 | 20 | Unspecified | Group II | Monodnaviria | Sangervirae | Phixviricota | Malgrandaviricetes | Petitvirales | Microviridae | Unclassified | Unclassified |
| S8_vs_125717 | n.a. | n.a. | Clustered | VC_1274_0 | A | n.a. | n.a. | n.a. | n.a. | n.a. | n.a. | n.a. | n.a. | n.a. | n.a. | n.a. |
| S8_vs_132359 | n.a. | n.a. | Clustered | VC_1618_0 | F | n.a. | n.a. | n.a. | n.a. | n.a. | n.a. | n.a. | n.a. | n.a. | n.a. | n.a. |
| S8_vs_134805 | Faecalibacterium phage FP_Mushu | MG711460 | Clustered | VC_1288_0 | C1 | 147 | Faecalibacterium | Group I | Duplodnaviria | Heunggongvirae | Uroviricota | Caudoviricetes | n.a. | Unclassified | Unclassified | Mushuvirus |
| S8_vs_136922 | Clostridium phage phiCTP1 | HM159959 | Clustered | VC_921_0 | N7 | 4 | Clostridium | Group I | Duplodnaviria | Heunggongvirae | Uroviricota | Caudoviricetes | n.a. | Unclassified | O | O |
| S8_vs_142097 | n.a. | n.a. | Clustered | VC_1258_0 | A | n.a. | n.a. | n.a. | n.a. | n.a. | n.a. | n.a. | n.a. | n.a. | n.a. | n.a. |
| S8_vs_143608 | Streptococcus phage Javan261 | MK448720 | Clustered | VC_778_0 | N20 | 6 | Streptococcus | Group I | Duplodnaviria | Heunggongvirae | Uroviricota | Caudoviricetes | n.a. | Unclassified | O | O |
| S8_vs_145173 | Faecalibacterium phage FP_Taranis | MG711467 | Clustered | VC_1290_0 | N36 | 2 | Faecalibacterium | Group I | Duplodnaviria | Heunggongvirae | Uroviricota | Caudoviricetes | n.a. | Unclassified | O | O |
| S8_vs_148389 | Bacteroides phage F2 | MT806186 | Clustered | VC_1507_0 | N4 | 4 | Bacteroides | Group I | Duplodnaviria | Heunggongvirae | Uroviricota | Caudoviricetes | n.a. | Unclassified | O | O |
| S8_vs_149700 | Streptococcus phage Javan630 | MK448997 | Clustered | VC_649_3 | N2 | 71 | Streptococcus | Group I | Duplodnaviria | Heunggongvirae | Uroviricota | Caudoviricetes | n.a. | Unclassified | O | O |
| S8_vs_154231 | n.a. | n.a. | Clustered | VC_779_0 | A | n.a. | n.a. | n.a. | n.a. | n.a. | n.a. | n.a. | n.a. | n.a. | n.a. | n.a. |
| S8_vs_162053 | Clostridium phage HM2 | LT600745 | Clustered | VC_1258_0 | N38 | 4 | Clostridium | Group I | Duplodnaviria | Heunggongvirae | Uroviricota | Caudoviricetes | n.a. | Salasmaviridae | O | O |
| S8_vs_175235 | Bacillus phage vB_BtS_BMBtp13 | KX190832 | Clustered | VC_667_0 | C10 | 7 | Bacillus | Group I | Duplodnaviria | Heunggongvirae | Uroviricota | Caudoviricetes | n.a. | Unclassified | Unclassified | Unclassified |
| S8_vs_176604 | n.a. | n.a. | Clustered | VC_1231_0 | A | n.a. | n.a. | n.a. | n.a. | n.a. | n.a. | n.a. | n.a. | n.a. | n.a. | n.a. |
| S8_vs_176720 | Bacillus phage vB_BtS_BMBtp13 | KX190832 | Clustered | VC_667_0 | C7 | 9 | Bacillus | Group I | Duplodnaviria | Heunggongvirae | Uroviricota | Caudoviricetes | n.a. | Unclassified | Unclassified | Unclassified |
| S8_vs_179208 | n.a. | n.a. | Clustered | VC_1612_1 | A | n.a. | n.a. | n.a. | n.a. | n.a. | n.a. | n.a. | n.a. | n.a. | n.a. | n.a. |
| S8_vs_182288 | Cellulophaga phage phi17:2 | KC821609 | Clustered | VC_1055_1 | N8 | 4 | Cellulophaga | Group I | Duplodnaviria | Heunggongvirae | Uroviricota | Caudoviricetes | n.a. | Unclassified | O | O |
| S8_vs_182331 | Clostridium phage phiMMP04 | JX145342 | Clustered | VC_669_0 | N2 | 4 | Clostridium | Group I | Duplodnaviria | Heunggongvirae | Uroviricota | Caudoviricetes | n.a. | Unclassified | O | O |
| S8_vs_193843 | n.a. | n.a. | Clustered | VC_1566_0 | A | n.a. | n.a. | n.a. | n.a. | n.a. | n.a. | n.a. | n.a. | n.a. | n.a. | n.a. |
| S8_vs_204260 | n.a. | n.a. | Clustered | VC_1291_0 | A | n.a. | n.a. | n.a. | n.a. | n.a. | n.a. | n.a. | n.a. | n.a. | n.a. | n.a. |
| S8_vs_205257 | n.a. | n.a. | Clustered | VC_1291_0 | A | n.a. | n.a. | n.a. | n.a. | n.a. | n.a. | n.a. | n.a. | n.a. | n.a. | n.a. |
| S8_vs_206526 | Roseburia phage Shimadzu | LR596903 | Clustered | VC_727_0 | C1 | 214 | Roseburia | Group I | Duplodnaviria | Heunggongvirae | Uroviricota | Caudoviricetes | n.a. | Unclassified | Unclassified | Unclassified |
| S8_vs_208103 | Bacteroides phage SJC03 | MT074146 | Clustered | VC_963_0 | N5 | 35 | Bacteroides | Group I | Duplodnaviria | Heunggongvirae | Uroviricota | Caudoviricetes | n.a. | Unclassified | O | O |
| S8_vs_208113 | n.a. | n.a. | Clustered | VC_1291_0 | A | n.a. | n.a. | n.a. | n.a. | n.a. | n.a. | n.a. | n.a. | n.a. | n.a. | n.a. |
| S8_vs_208114 | Psychrobacter phage pOW20-A | JF974295 | Clustered | VC_439_0 | N3 | 2 | Psychrobacter | Group I | Duplodnaviria | Heunggongvirae | Uroviricota | Caudoviricetes | n.a. | Unclassified | O | O |
| S8_vs_208118 | Streptococcus phage MismyG | OL774867 | Clustered | VC_152_0 | N15 | 4 | Streptococcus | Group I | Duplodnaviria | Heunggongvirae | Uroviricota | Caudoviricetes | n.a. | Unclassified | O | O |
| S8_vs_208129 | Bdellovibrio phage phi1402 | JF344709 | Clustered | VC_967_0 | C4 | 37 | Bdellovibrio | Group I | Duplodnaviria | Heunggongvirae | Uroviricota | Caudoviricetes | n.a. | Unclassified | Unclassified | Unclassified |
| S8_vs_28311 | Flavobacterium phage vB_FspS_laban6-1 | MN812211 | Clustered | VC_1062_0 | N5 | 8 | Flavobacterium | Group I | Duplodnaviria | Heunggongvirae | Uroviricota | Caudoviricetes | n.a. | Duneviridae | O | O |
| S8_vs_4153 | n.a. | n.a. | Clustered | VC_1507_0 | A | n.a. | n.a. | n.a. | n.a. | n.a. | n.a. | n.a. | n.a. | n.a. | n.a. | n.a. |
| S8_vs_47265 | Faecalibacterium phage FP_Epona | MG711462 | Clustered | VC_723_0 | C5 | 81 | Faecalibacterium | Group I | Duplodnaviria | Heunggongvirae | Uroviricota | Caudoviricetes | n.a. | Unclassified | Unclassified | Eponavirus |
| S8_vs_4883 | n.a. | n.a. | Clustered | VC_1554_0 | A | n.a. | n.a. | n.a. | n.a. | n.a. | n.a. | n.a. | n.a. | n.a. | n.a. | n.a. |
| S8_vs_50431 | n.a. | n.a. | Clustered | VC_1567_0 | A | n.a. | n.a. | n.a. | n.a. | n.a. | n.a. | n.a. | n.a. | n.a. | n.a. | n.a. |
| S8_vs_5211 | n.a. | n.a. | Clustered | VC_1611_0 | A | n.a. | n.a. | n.a. | n.a. | n.a. | n.a. | n.a. | n.a. | n.a. | n.a. | n.a. |
| S8_vs_71212 | n.a. | n.a. | Clustered | VC_1479_0 | A | n.a. | n.a. | n.a. | n.a. | n.a. | n.a. | n.a. | n.a. | n.a. | n.a. | n.a. |
| S8_vs_76852 | Streptococcus phage Javan261 | MK448720 | Clustered | VC_724_0 | N30 | 2 | Streptococcus | Group I | Duplodnaviria | Heunggongvirae | Uroviricota | Caudoviricetes | n.a. | Unclassified | O | O |
| S8_vs_9114 | n.a. | n.a. | Clustered | VC_1576_0 | A | n.a. | n.a. | n.a. | n.a. | n.a. | n.a. | n.a. | n.a. | n.a. | n.a. | n.a. |
| S8_vs_99362 | Bacteroides phage p00 | BK010646 | Clustered | VC_695_0 | C5 | 45 | Bacteroides | Group I | Duplodnaviria | Heunggongvirae | Uroviricota | Caudoviricetes | n.a. | Unclassified | Unclassified | Unclassified |
| S8_vs_9987 | Phage vB_RanS_PJN03 | ON245412 | Clustered | VC_951_0 | N14 | 7 | Unspecified | Group I | Duplodnaviria | Heunggongvirae | Uroviricota | Caudoviricetes | n.a. | Unclassified | O | O |
| S9_vf_15012 | Streptococcus phage Javan372 | MK448920 | Clustered | VC_344_0 | C1 | 124 | Streptococcus | Group I | Duplodnaviria | Heunggongvirae | Uroviricota | Caudoviricetes | n.a. | Unclassified | Unclassified | Unclassified |
| S9_vf_23901 | Tortoise microvirus 13 | MK765563 | Clustered | VC_1299_0 | C1 | 26 | Unspecified | Group II | Monodnaviria | Sangervirae | Phixviricota | Malgrandaviricetes | Petitvirales | Microviridae | Unclassified | Unclassified |
| S9_vf_26805 | Microviridae sp. | MG945488 | Clustered | VC_1334_0 | C1 | 20 | Unspecified | Group II | Monodnaviria | Sangervirae | Phixviricota | Malgrandaviricetes | Petitvirales | Microviridae | Unclassified | Unclassified |
| S9_vf_34736 | Clostridium phage HM2 | LT600745 | Clustered | VC_1261_0 | N28 | 8 | Clostridium | Group I | Duplodnaviria | Heunggongvirae | Uroviricota | Caudoviricetes | n.a. | Salasmaviridae | O | O |
| S9_vf_4115 | Streptococcus phage Javan318 | MK448905 | Clustered | VC_425_0 | C1 | 98 | Streptococcus | Group I | Duplodnaviria | Heunggongvirae | Uroviricota | Caudoviricetes | n.a. | Unclassified | Unclassified | Unclassified |
| S9_vf_51656 | uncultured phage cr8_1 | MT774384 | Clustered | VC_691_1 | C1 | 246 | Unspecified | Group I | Duplodnaviria | Heunggongvirae | Uroviricota | Caudoviricetes | Crassvirales | Intestiviridae | Obtuvirinae | Fohxhuevirus |
| S9_vf_64844 | Faecalibacterium phage FP_Lagaffe | MG711461 | Clustered | VC_727_0 | C4 | 104 | Faecalibacterium | Group I | Duplodnaviria | Heunggongvirae | Uroviricota | Caudoviricetes | n.a. | Unclassified | Unclassified | Lagaffevirus |
| S9_vf_70236 | n.a. | n.a. | Clustered | VC_1258_0 | A | n.a. | n.a. | n.a. | n.a. | n.a. | n.a. | n.a. | n.a. | n.a. | n.a. | n.a. |
| S9_vf_73311 | Exiguobacterium phage vB_EauM-23 | MH844558 | Clustered | VC_145_0 | C2 | 22 | Exiguobacterium | Group I | Duplodnaviria | Heunggongvirae | Uroviricota | Caudoviricetes | n.a. | Unclassified | Unclassified | Unclassified |
| S9_vf_75363 | n.a. | n.a. | Clustered | VC_1258_0 | A | n.a. | n.a. | n.a. | n.a. | n.a. | n.a. | n.a. | n.a. | n.a. | n.a. | n.a. |
| S9_vf_76769 | uncultured phage cr52_1 | MT774395 | Clustered | VC_519_6 | C1 | 199 | Unspecified | Group I | Duplodnaviria | Heunggongvirae | Uroviricota | Caudoviricetes | Crassvirales | Suoliviridae | Loutivirinae | Buchavirus |
| S9_vf_79919 | n.a. | n.a. | Clustered | VC_779_0 | A | n.a. | n.a. | n.a. | n.a. | n.a. | n.a. | n.a. | n.a. | n.a. | n.a. | n.a. |
| S9_vf_90852 | Inovirus sp. | MT135289 | Clustered | VC_1492_0 | C1 | 31 | Unspecified | Group II | Monodnaviria | Loebvirae | Hofneiviricota | Faserviricetes | Tubulavirales | Inoviridae | Unclassified | Inovirus |
| S9_vf_90866 | Streptococcus phage phiZJ20091101-2 | KX077893 | Clustered | VC_1229_0 | C1 | 16 | Streptococcus | Unclassified | Unclassified | n.a. | n.a. | n.a. | n.a. | Unclassified | Unclassified | Unclassified |
| S9_vf_90871 | Clostridium phage phiMMP01 | LN681541 | Clustered | VC_150_0 | N16 | 10 | Clostridium | Group I | Duplodnaviria | Heunggongvirae | Uroviricota | Caudoviricetes | n.a. | Unclassified | O | O |
| S9_vs_1966 | Streptococcus phage Javan290 | MK448900 | Clustered | VC_332_0 | N19 | 3 | Streptococcus | Group I | Duplodnaviria | Heunggongvirae | Uroviricota | Caudoviricetes | n.a. | Unclassified | O | O |
| S9_vs_21850 | Bacteroides phage p00 | BK010646 | Clustered | VC_695_0 | C4 | 42 | Bacteroides | Group I | Duplodnaviria | Heunggongvirae | Uroviricota | Caudoviricetes | n.a. | Unclassified | Unclassified | Unclassified |
| S9_vs_26179 | n.a. | n.a. | Clustered | VC_782_0 | A | n.a. | n.a. | n.a. | n.a. | n.a. | n.a. | n.a. | n.a. | n.a. | n.a. | n.a. |
| S9_vs_27813 | Faecalibacterium phage FP_Epona | MG711462 | Clustered | VC_723_0 | C13 | 52 | Faecalibacterium | Group I | Duplodnaviria | Heunggongvirae | Uroviricota | Caudoviricetes | n.a. | Unclassified | Unclassified | Eponavirus |
| S9_vs_28197 | Bacteroides phage F4 | MT806185 | Clustered | VC_1505_0 | N7 | 7 | Bacteroides | Group I | Duplodnaviria | Heunggongvirae | Uroviricota | Caudoviricetes | n.a. | Unclassified | O | O |
| S9_vs_32588 | Faecalibacterium phage FP_oengus | MG711463 | Clustered | VC_1085_1 | N16 | 70 | Faecalibacterium | Group I | Duplodnaviria | Heunggongvirae | Uroviricota | Caudoviricetes | n.a. | Unclassified | O | O |
| S9_vs_34898 | Lactococcus phage CAP | MZ308445 | Clustered | VC_278_0 | N12 | 9 | Lactococcus | Group I | Duplodnaviria | Heunggongvirae | Uroviricota | Caudoviricetes | n.a. | Unclassified | O | O |
| S9_vs_4343 | n.a. | n.a. | Clustered | VC_1291_0 | A | n.a. | n.a. | n.a. | n.a. | n.a. | n.a. | n.a. | n.a. | n.a. | n.a. | n.a. |
| S9_vs_4558 | Faecalibacterium phage FP_oengus | MG711463 | Clustered | VC_1085_1 | N18 | 68 | Faecalibacterium | Group I | Duplodnaviria | Heunggongvirae | Uroviricota | Caudoviricetes | n.a. | Unclassified | O | O |
| S9_vs_45812 | Bacillus phage vB_BtS_BMBtp13 | KX190832 | Clustered | VC_667_0 | C7 | 7 | Bacillus | Group I | Duplodnaviria | Heunggongvirae | Uroviricota | Caudoviricetes | n.a. | Unclassified | Unclassified | Unclassified |
| S9_vs_46498 | Geobacillus phage GBSV1 | DQ340064 | Clustered | VC_779_0 | N29 | 4 | Geobacillus | Group I | Duplodnaviria | Heunggongvirae | Uroviricota | Caudoviricetes | n.a. | Unclassified | O | O |
| S9_vs_48424 | n.a. | n.a. | Clustered | VC_1565_0 | A | n.a. | n.a. | n.a. | n.a. | n.a. | n.a. | n.a. | n.a. | n.a. | n.a. | n.a. |
| S9_vs_56354 | Faecalibacterium phage FP_Epona | MG711462 | Clustered | VC_723_0 | C13 | 56 | Faecalibacterium | Group I | Duplodnaviria | Heunggongvirae | Uroviricota | Caudoviricetes | n.a. | Unclassified | Unclassified | Eponavirus |
| S9_vs_5676 | Streptococcus phage Javan261 | MK448720 | Clustered | VC_724_0 | N45 | 1 | Streptococcus | Group I | Duplodnaviria | Heunggongvirae | Uroviricota | Caudoviricetes | n.a. | Unclassified | O | O |
| S9_vs_58402 | n.a. | n.a. | Clustered | VC_1613_0 | A | n.a. | n.a. | n.a. | n.a. | n.a. | n.a. | n.a. | n.a. | n.a. | n.a. | n.a. |
| S9_vs_63485 | n.a. | n.a. | Clustered | VC_1604_0 | A | n.a. | n.a. | n.a. | n.a. | n.a. | n.a. | n.a. | n.a. | n.a. | n.a. | n.a. |
| S9_vs_73764 | Geobacillus phage GBSV1 | DQ340064 | Clustered | VC_778_0 | N36 | 7 | Geobacillus | Group I | Duplodnaviria | Heunggongvirae | Uroviricota | Caudoviricetes | n.a. | Unclassified | O | O |
| S9_vs_75050 | Geobacillus phage GBSV1 | DQ340064 | Clustered | VC_778_0 | N20 | 9 | Geobacillus | Group I | Duplodnaviria | Heunggongvirae | Uroviricota | Caudoviricetes | n.a. | Unclassified | O | O |
| S9_vs_75290 | n.a. | n.a. | Clustered | VC_1607_0 | F | n.a. | n.a. | n.a. | n.a. | n.a. | n.a. | n.a. | n.a. | n.a. | n.a. | n.a. |
| S9_vs_7881 | Clostridium phage phiMMP01 | LN681541 | Clustered | VC_151_0 | N12 | 27 | Clostridium | Group I | Duplodnaviria | Heunggongvirae | Uroviricota | Caudoviricetes | n.a. | Unclassified | O | O |
| S9_vs_83108 | n.a. | n.a. | Clustered | VC_1560_0 | A | n.a. | n.a. | n.a. | n.a. | n.a. | n.a. | n.a. | n.a. | n.a. | n.a. | n.a. |

| | | | | | | | | | | | | | | | | |
|---|---|---|---|---|---|---|---|---|---|---|---|---|---|---|---|---|
| S9_vs_84046 | Butyrivibrio phage Idris | MN882554 | Clustered | VC_724_0 | N27 | 4 | Butyrivibrio | Group I | Duplodnaviria | Heunggongvirae | Uroviricota | Caudoviricetes | n.a. | Unclassified | O | O |
| S9_vs_90849 | n.a. | n.a. | Clustered | VC_1291_0 | A | n.a. | n.a. | n.a. | n.a. | n.a. | n.a. | n.a. | n.a. | n.a. | n.a. | n.a. |
| S9_vs_90851 | Clostridium phage phiMMP01 | LN681541 | Clustered | VC_151_0 | N21 | 23 | Clostridium | Group I | Duplodnaviria | Heunggongvirae | Uroviricota | Caudoviricetes | n.a. | Unclassified | O | O |
| S9_vs_90857 | n.a. | n.a. | Clustered | VC_1479_0 | A | n.a. | n.a. | n.a. | n.a. | n.a. | n.a. | n.a. | n.a. | n.a. | n.a. | n.a. |
| S9_vs_90860 | Butyrivibrio phage Idris | MN882554 | Clustered | VC_724_0 | N36 | 4 | Butyrivibrio | Group I | Duplodnaviria | Heunggongvirae | Uroviricota | Caudoviricetes | n.a. | Unclassified | O | O |
| S9_vs_90863 | n.a. | n.a. | Clustered | VC_1603_0 | F | n.a. | n.a. | n.a. | n.a. | n.a. | n.a. | n.a. | n.a. | n.a. | n.a. | n.a. |
| S10_vf_120738 | n.a. | n.a. | Clustered/Singleton | VC_491_3 | A | n.a. | n.a. | n.a. | n.a. | n.a. | n.a. | n.a. | n.a. | n.a. | n.a. | n.a. |
| S10_vf_43964 | Bacteroides phage LoVEphage | MW660583 | Clustered/Singleton | VC_1525_2 | N5 | 14 | Bacteroides | Group I | Duplodnaviria | Heunggongvirae | Uroviricota | Caudoviricetes | n.a. | Unclassified | O | O |
| S10_vs_37379 | n.a. | n.a. | Clustered/Singleton | VC_1055_2 | A | n.a. | n.a. | n.a. | n.a. | n.a. | n.a. | n.a. | n.a. | n.a. | n.a. | n.a. |
| S10_vs_62776 | Bacteroides phage LoVEphage | MW660583 | Clustered/Singleton | VC_1525_0 | N2 | 32 | Bacteroides | Group I | Duplodnaviria | Heunggongvirae | Uroviricota | Caudoviricetes | n.a. | Unclassified | O | O |
| S10_vs_77004 | n.a. | n.a. | Clustered/Singleton | VC_922_0 | A | n.a. | n.a. | n.a. | n.a. | n.a. | n.a. | n.a. | n.a. | n.a. | n.a. | n.a. |
| S11_vf_104033 | Bacteroides phage SJC03 | MT074146 | Clustered/Singleton | VC_963_1 | N4 | 51 | Bacteroides | Group I | Duplodnaviria | Heunggongvirae | Uroviricota | Caudoviricetes | n.a. | Unclassified | O | O |
| S11_vf_3790 | Geobacillus virus E3 | KP144388 | Clustered/Singleton | VC_490_0 | N9 | 1 | Geobacillus | Group I | Duplodnaviria | Heunggongvirae | Uroviricota | Caudoviricetes | n.a. | Unclassified | O | O |
| S11_vf_70015 | Phage vB_RanS_PJN03 | ON245412 | Clustered/Singleton | VC_951_2 | N20 | 7 | Unspecified | Group I | Duplodnaviria | Heunggongvirae | Uroviricota | Caudoviricetes | n.a. | Unclassified | O | O |
| S11_vs_75554 | n.a. | n.a. | Clustered/Singleton | VC_1264_0 | A | n.a. | n.a. | n.a. | n.a. | n.a. | n.a. | n.a. | n.a. | n.a. | n.a. | n.a. |
| S12_vf_188819 | Clostridium phage c-st | AP008983 | Clustered/Singleton | VC_490_3 | N7 | 5 | Clostridium | Group I | Duplodnaviria | Heunggongvirae | Uroviricota | Caudoviricetes | n.a. | Unclassified | O | O |
| S12_vf_67909 | Enterococcus phage EF1 | MF001358 | Clustered/Singleton | VC_221_0 | N2 | 11 | Enterococcus | Group I | Duplodnaviria | Heunggongvirae | Uroviricota | Caudoviricetes | n.a. | Unclassified | O | O |
| S12_vs_123620 | n.a. | n.a. | Clustered/Singleton | VC_1295_0 | A | n.a. | n.a. | n.a. | n.a. | n.a. | n.a. | n.a. | n.a. | n.a. | n.a. | n.a. |
| S12_vs_172379 | n.a. | n.a. | Clustered/Singleton | VC_1477_1 | A | n.a. | n.a. | n.a. | n.a. | n.a. | n.a. | n.a. | n.a. | n.a. | n.a. | n.a. |
| S13_vf_38708 | n.a. | n.a. | Clustered/Singleton | VC_1292_0 | A | n.a. | n.a. | n.a. | n.a. | n.a. | n.a. | n.a. | n.a. | n.a. | n.a. | n.a. |
| S13_vf_4901 | Bacillus phage vB_BcoS-136 | MH884508 | Clustered/Singleton | VC_491_1 | N10 | 2 | Bacillus | Group I | Duplodnaviria | Heunggongvirae | Uroviricota | Caudoviricetes | n.a. | Unclassified | O | O |
| S13_vs_26756 | n.a. | n.a. | Clustered/Singleton | VC_1295_1 | A | n.a. | n.a. | n.a. | n.a. | n.a. | n.a. | n.a. | n.a. | n.a. | n.a. | n.a. |
| S14_vs_70985 | n.a. | n.a. | Clustered/Singleton | VC_1612_2 | A | n.a. | n.a. | n.a. | n.a. | n.a. | n.a. | n.a. | n.a. | n.a. | n.a. | n.a. |
| S14_vs_74067 | n.a. | n.a. | Clustered/Singleton | VC_1582_2 | A | n.a. | n.a. | n.a. | n.a. | n.a. | n.a. | n.a. | n.a. | n.a. | n.a. | n.a. |
| S15_vf_75924 | uncultured phage cr116_1 | MT774389 | Clustered/Singleton | VC_1079_0 | N6 | 6 | Unspecified | Group I | Duplodnaviria | Heunggongvirae | Uroviricota | Caudoviricetes | Crassvirales | Steigviridae | O | O |
| S15_vs_42043 | n.a. | n.a. | Clustered/Singleton | VC_1559_2 | A | n.a. | n.a. | n.a. | n.a. | n.a. | n.a. | n.a. | n.a. | n.a. | n.a. | n.a. |
| S16_vf_108554 | n.a. | n.a. | Clustered/Singleton | VC_1554_1 | A | n.a. | n.a. | n.a. | n.a. | n.a. | n.a. | n.a. | n.a. | n.a. | n.a. | n.a. |
| S16_vf_173099 | n.a. | n.a. | Clustered/Singleton | VC_1586_0 | A | n.a. | n.a. | n.a. | n.a. | n.a. | n.a. | n.a. | n.a. | n.a. | n.a. | n.a. |
| S16_vf_96483 | n.a. | n.a. | Clustered/Singleton | VC_1557_0 | A | n.a. | n.a. | n.a. | n.a. | n.a. | n.a. | n.a. | n.a. | n.a. | n.a. | n.a. |
| S16_vs_15093 | Butyrivibrio virus Ceridwen | MN882553 | Clustered/Singleton | VC_1434_0 | N12 | 8 | Butyrivibrio | Group I | Duplodnaviria | Heunggongvirae | Uroviricota | Caudoviricetes | n.a. | Unclassified | O | O |
| S16_vs_34751 | n.a. | n.a. | Clustered/Singleton | VC_780_0 | A | n.a. | n.a. | n.a. | n.a. | n.a. | n.a. | n.a. | n.a. | n.a. | n.a. | n.a. |
| S3_vf_108959 | n.a. | n.a. | Clustered/Singleton | VC_1254_3 | A | n.a. | n.a. | n.a. | n.a. | n.a. | n.a. | n.a. | n.a. | n.a. | n.a. | n.a. |
| S3_vf_61860 | Clostridium phage vB_CpeS-1181 | MW478292 | Clustered/Singleton | VC_923_1 | N2 | 21 | Clostridium | Group I | Duplodnaviria | Heunggongvirae | Uroviricota | Caudoviricetes | n.a. | Unclassified | O | O |
| S3_vs_16825 | Roseburia phage Jekyll | LR596902 | Clustered/Singleton | VC_1254_4 | N19 | 10 | Roseburia | Group I | Duplodnaviria | Heunggongvirae | Uroviricota | Caudoviricetes | n.a. | Unclassified | O | O |
| S3_vs_39774 | n.a. | n.a. | Clustered/Singleton | VC_1582_1 | A | n.a. | n.a. | n.a. | n.a. | n.a. | n.a. | n.a. | n.a. | n.a. | n.a. | n.a. |
| S3_vs_68031 | Faecalibacterium phage FP_oengus | MG711463 | Clustered/Singleton | VC_1242_0 | N2 | 101 | Faecalibacterium | Group I | Duplodnaviria | Heunggongvirae | Uroviricota | Caudoviricetes | n.a. | Unclassified | O | O |
| S3_vs_81793 | n.a. | n.a. | Clustered/Singleton | VC_1586_1 | A | n.a. | n.a. | n.a. | n.a. | n.a. | n.a. | n.a. | n.a. | n.a. | n.a. | n.a. |
| S4_vf_31973 | n.a. | n.a. | Clustered/Singleton | VC_490_2 | A | n.a. | n.a. | n.a. | n.a. | n.a. | n.a. | n.a. | n.a. | n.a. | n.a. | n.a. |
| S4_vs_VIRSorter_31975-circular-cat_1 | Phage vB_RanS_PJN03 | ON245412 | Clustered/Singleton | VC_948_1 | N24 | 3 | Unspecified | Group I | Duplodnaviria | Heunggongvirae | Uroviricota | Caudoviricetes | n.a. | Unclassified | O | O |
| S5_vf_145579 | uncultured phage cr85_1 | MT774390 | Clustered/Singleton | VC_1079_2 | N6 | 12 | Unspecified | Group I | Duplodnaviria | Heunggongvirae | Uroviricota | Caudoviricetes | Crassvirales | Steigviridae | O | O |
| S5_vf_67622 | IAS virus | KJ003983 | Clustered/Singleton | VC_1079_1 | N6 | 11 | Unspecified | Group I | Duplodnaviria | Heunggongvirae | Uroviricota | Caudoviricetes | Crassvirales | Steigviridae | O | O |
| S5_vs_109527 | n.a. | n.a. | Clustered/Singleton | VC_1612_3 | A | n.a. | n.a. | n.a. | n.a. | n.a. | n.a. | n.a. | n.a. | n.a. | n.a. | n.a. |
| S5_vs_122344 | Roseburia phage Jekyll | LR596902 | Clustered/Singleton | VC_1252_1 | C2 | 7 | Roseburia | Group I | Duplodnaviria | Heunggongvirae | Uroviricota | Caudoviricetes | n.a. | Unclassified | Unclassified | O |
| S5_vs_124677 | Faecalibacterium phage FP_Toutatis | MG711466 | Clustered/Singleton | VC_728_1 | C3 | 63 | Faecalibacterium | Group I | Duplodnaviria | Heunggongvirae | Uroviricota | Caudoviricetes | n.a. | Unclassified | Unclassified | O |
| S5_vs_131510 | Bacteroides phage p00 | BK010646 | Clustered/Singleton | VC_695_1 | C1 | 71 | Bacteroides | Group I | Duplodnaviria | Heunggongvirae | Uroviricota | Caudoviricetes | n.a. | Unclassified | Unclassified | O |
| S5_vs_77593 | Clostridium phage phiMMP01 | LN681541 | Clustered/Singleton | VC_151_1 | N22 | 16 | Clostridium | Group I | Duplodnaviria | Heunggongvirae | Uroviricota | Caudoviricetes | n.a. | Unclassified | O | O |
| S5_vs_97574 | Clostridium phage c-st | AP008983 | Clustered/Singleton | VC_219_1 | N8 | 13 | Clostridium | Group I | Duplodnaviria | Heunggongvirae | Uroviricota | Caudoviricetes | n.a. | Unclassified | O | O |
| S6_vs_29152 | Faecalibacterium phage FP_Brigit | MG711465 | Clustered/Singleton | VC_923_2 | N16 | 5 | Faecalibacterium | Group I | Duplodnaviria | Heunggongvirae | Uroviricota | Caudoviricetes | n.a. | Unclassified | O | O |
| S7_vf_37279 | Bacteroides phage LoVEphage | MW660583 | Clustered/Singleton | VC_1525_1 | N6 | 15 | Bacteroides | Group I | Duplodnaviria | Heunggongvirae | Uroviricota | Caudoviricetes | n.a. | Unclassified | O | O |
| S7_vs_103530 | n.a. | n.a. | Clustered/Singleton | VC_1554_2 | A | n.a. | n.a. | n.a. | n.a. | n.a. | n.a. | n.a. | n.a. | n.a. | n.a. | n.a. |
| S7_vs_135679 | n.a. | n.a. | Clustered/Singleton | VC_1594_0 | A | n.a. | n.a. | n.a. | n.a. | n.a. | n.a. | n.a. | n.a. | n.a. | n.a. | n.a. |
| S7_vs_137611 | Brevibacillus phage Sundance | KT151959 | Clustered/Singleton | VC_219_2 | N20 | 1 | Brevibacillus | Group I | Duplodnaviria | Heunggongvirae | Uroviricota | Caudoviricetes | n.a. | Unclassified | O | O |
| S7_vs_54322 | n.a. | n.a. | Clustered/Singleton | VC_1565_1 | A | n.a. | n.a. | n.a. | n.a. | n.a. | n.a. | n.a. | n.a. | n.a. | n.a. | n.a. |
| S7_vs_78507 | n.a. | n.a. | Clustered/Singleton | VC_1481_2 | A | n.a. | n.a. | n.a. | n.a. | n.a. | n.a. | n.a. | n.a. | n.a. | n.a. | n.a. |
| S8_vf_11794 | Bacillus phage vB_BcoS-136 | MH884508 | Clustered/Singleton | VC_491_2 | N17 | 3 | Bacillus | Group I | Duplodnaviria | Heunggongvirae | Uroviricota | Caudoviricetes | n.a. | Unclassified | O | O |
| S8_vf_161703 | Bacillus phage vB_BsuS_PJN02 | OM634653 | Clustered/Singleton | VC_1040_1 | N9 | 8 | Bacillus | Group I | Duplodnaviria | Heunggongvirae | Uroviricota | Caudoviricetes | n.a. | Unclassified | O | O |
| S8_vf_173225 | Geobacillus virus E3 | KP144388 | Clustered/Singleton | VC_490_1 | N10 | 3 | Geobacillus | Group I | Duplodnaviria | Heunggongvirae | Uroviricota | Caudoviricetes | n.a. | Unclassified | O | O |
| S8_vf_23955 | Faecalibacterium phage FP_oengus | MG711463 | Clustered/Singleton | VC_1085_2 | N21 | 60 | Faecalibacterium | Group I | Duplodnaviria | Heunggongvirae | Uroviricota | Caudoviricetes | n.a. | Unclassified | O | O |
| S8_vf_54514 | Butyrivibrio virus Ceridwen | MN882553 | Clustered/Singleton | VC_1554_5 | N61 | 5 | Butyrivibrio | Group I | Duplodnaviria | Heunggongvirae | Uroviricota | Caudoviricetes | n.a. | Unclassified | O | O |
| S8_vs_115958 | Streptococcus phage Javan630 | MK448997 | Clustered/Singleton | VC_649_4 | C9 | 43 | Streptococcus | Group I | Duplodnaviria | Heunggongvirae | Uroviricota | Caudoviricetes | n.a. | Unclassified | Unclassified | O |
| S8_vs_59736 | Clostridium phage c-st | AP008983 | Clustered/Singleton | VC_219_3 | N7 | 16 | Clostridium | Group I | Duplodnaviria | Heunggongvirae | Uroviricota | Caudoviricetes | n.a. | Unclassified | O | O |
| S9_vf_13507 | n.a. | n.a. | Clustered/Singleton | VC_1582_3 | A | n.a. | n.a. | n.a. | n.a. | n.a. | n.a. | n.a. | n.a. | n.a. | n.a. | n.a. |
| S9_vf_57277 | n.a. | n.a. | Clustered/Singleton | VC_1554_4 | A | n.a. | n.a. | n.a. | n.a. | n.a. | n.a. | n.a. | n.a. | n.a. | n.a. | n.a. |
| S9_vf_90847 | Enterococcus phage EF1 | MF001358 | Clustered/Singleton | VC_221_1 | N2 | 25 | Enterococcus | Group I | Duplodnaviria | Heunggongvirae | Uroviricota | Caudoviricetes | n.a. | Unclassified | O | O |
| S9_vs_10424 | n.a. | n.a. | Clustered/Singleton | VC_1421_1 | A | n.a. | n.a. | n.a. | n.a. | n.a. | n.a. | n.a. | n.a. | n.a. | n.a. | n.a. |
| S9_vs_50439 | n.a. | n.a. | Clustered/Singleton | VC_1554_3 | A | n.a. | n.a. | n.a. | n.a. | n.a. | n.a. | n.a. | n.a. | n.a. | n.a. | n.a. |
| S9_vs_90727 | Streptococcus phage Javan630 | MK448997 | Clustered/Singleton | VC_649_5 | C5 | 55 | Streptococcus | Group I | Duplodnaviria | Heunggongvirae | Uroviricota | Caudoviricetes | n.a. | Unclassified | Unclassified | O |
| S9_vs_90846 | Faecalibacterium phage FP_Lugh | MG711464 | Clustered/Singleton | VC_640_0 | N5 | 19 | Faecalibacterium | Group I | Duplodnaviria | Heunggongvirae | Uroviricota | Caudoviricetes | n.a. | Unclassified | O | O |
| S9_vs_90859 | Bacteriophage sp. | LT996078 | Clustered/Singleton | VC_278_1 | N17 | 4 | Unspecified | Unclassified | Unclassified | n.a. | n.a. | n.a. | n.a. | Unclassified | O | O |
| S10_vf_47170 | n.a. | n.a. | Outlier | n.a. | F | n.a. | n.a. | n.a. | n.a. | n.a. | n.a. | n.a. | n.a. | n.a. | n.a. | n.a. |
| S10_vf_62230 | Streptococcus phage Javan576 | MK448987 | Outlier | n.a. | N1 | 17 | Streptococcus | Group I | Duplodnaviria | Heunggongvirae | Uroviricota | Caudoviricetes | n.a. | Unclassified | O | O |
| S10_vs_106834 | Riemerella phage vB_RanS_PT03 | MZ995504 | Outlier | n.a. | N4 | 11 | Riemerella | Group I | Duplodnaviria | Heunggongvirae | Uroviricota | Caudoviricetes | n.a. | Unclassified | O | O |
| S10_vs_37694 | Cellulophaga phage Ingeline_1 | MT732435 | Outlier | n.a. | N4 | 7 | Cellulophaga | Group I | Duplodnaviria | Heunggongvirae | Uroviricota | Caudoviricetes | n.a. | Duneviridae | O | O |
| S11_vf_33433 | Tenacibaculum phage Gundel_1 | MT732474 | Outlier | n.a. | N1 | 11 | Tenacibaculum | Group I | Duplodnaviria | Heunggongvirae | Uroviricota | Caudoviricetes | n.a. | Pachyviridae | O | O |
| S11_vf_99751 | n.a. | n.a. | Outlier | n.a. | A | n.a. | n.a. | n.a. | n.a. | n.a. | n.a. | n.a. | n.a. | n.a. | n.a. | n.a. |
| S11_vs_20225 | Faecalibacterium phage FP_Lugh | MG711464 | Outlier | n.a. | N2 | 8 | Faecalibacterium | Group I | Duplodnaviria | Heunggongvirae | Uroviricota | Caudoviricetes | n.a. | Unclassified | O | O |
| S11_vs_48109 | Streptococcus phage Javan630 | MK448997 | Outlier | n.a. | N9 | 26 | Streptococcus | Group I | Duplodnaviria | Heunggongvirae | Uroviricota | Caudoviricetes | n.a. | Unclassified | O | O |
| S11_vs_53675 | n.a. | n.a. | Outlier | n.a. | A | n.a. | n.a. | n.a. | n.a. | n.a. | n.a. | n.a. | n.a. | n.a. | n.a. | n.a. |
| S11_vs_55144 | n.a. | n.a. | Outlier | n.a. | A | n.a. | n.a. | n.a. | n.a. | n.a. | n.a. | n.a. | n.a. | n.a. | n.a. | n.a. |
| S11_vs_56762 | n.a. | n.a. | Outlier | n.a. | A | n.a. | n.a. | n.a. | n.a. | n.a. | n.a. | n.a. | n.a. | n.a. | n.a. | n.a. |
| S11_vs_67573 | n.a. | n.a. | Outlier | n.a. | A | n.a. | n.a. | n.a. | n.a. | n.a. | n.a. | n.a. | n.a. | n.a. | n.a. | n.a. |
| S11_vs_68348 | Streptococcus phage IPP10 | KY065452 | Outlier | n.a. | N3 | 19 | Streptococcus | Group I | Duplodnaviria | Heunggongvirae | Uroviricota | Caudoviricetes | n.a. | Unclassified | O | O |
| S11_vs_6878 | Clostridium phage phiCT9441A | KM983329 | Outlier | n.a. | N3 | 14 | Clostridium | Group I | Duplodnaviria | Heunggongvirae | Uroviricota | Caudoviricetes | n.a. | Unclassified | O | O |
| S11_vs_79831 | n.a. | n.a. | Outlier | n.a. | A | n.a. | n.a. | n.a. | n.a. | n.a. | n.a. | n.a. | n.a. | n.a. | n.a. | n.a. |
| S11_vs_82331 | Klebsiella phage ST11-OXA245phi3.2 | MK416010 | Outlier | n.a. | N1 | 18 | Klebsiella | Group I | Duplodnaviria | Heunggongvirae | Uroviricota | Caudoviricetes | n.a. | Unclassified | O | O |
| S11_vs_89911 | n.a. | n.a. | Outlier | n.a. | A | n.a. | n.a. | n.a. | n.a. | n.a. | n.a. | n.a. | n.a. | n.a. | n.a. | n.a. |
| S12_vf_105542 | Streptococcus phage SF39 | OK173051 | Outlier | n.a. | N3 | 3 | Streptococcus | Group I | Duplodnaviria | Heunggongvirae | Uroviricota | Caudoviricetes | n.a. | Unclassified | O | O |

| | | | | | | | | | | | | | | | | |
|---|---|---|---|---|---|---|---|---|---|---|---|---|---|---|---|---|
| S12_vf_18902 | Bacillus phage PK-3 | ON881243 | Outlier | n.a. | N14 | 11 | Bacillus | Group I | Duplodnaviria | Heunggongvirae | Uroviricota | Caudoviricetes | n.a. | Unclassified | O | O |
| S12_vs_118154 | Butyrivibrio phage Idris | MN882554 | Outlier | n.a. | N20 | 4 | Butyrivibrio | Group I | Duplodnaviria | Heunggongvirae | Uroviricota | Caudoviricetes | n.a. | Unclassified | O | O |
| S12_vs_133070 | Bacillus phage BUCT082 | MZ969646 | Outlier | n.a. | N4 | 8 | Bacillus | Group I | Duplodnaviria | Heunggongvirae | Uroviricota | Caudoviricetes | n.a. | Unclassified | O | O |
| S12_vs_142625 | Elizabethkingia phage TCUEAP2 | OK632025 | Outlier | n.a. | N2 | 17 | Elizabethkingia | Group I | Duplodnaviria | Heunggongvirae | Uroviricota | Caudoviricetes | n.a. | Unclassified | O | O |
| S12_vs_14467 | n.a. | n.a. | Outlier | n.a. | A | n.a. | n.a. | n.a. | n.a. | n.a. | n.a. | n.a. | n.a. | n.a. | n.a. | n.a. |
| S12_vs_153780 | Acidithiobacillus phage AcaML1 | JX507079 | Outlier | n.a. | N1 | 25 | Acidithiobacillus | Group I | Duplodnaviria | Heunggongvirae | Uroviricota | Caudoviricetes | n.a. | Unclassified | O | O |
| S12_vs_188825 | n.a. | n.a. | Outlier | n.a. | A | n.a. | n.a. | n.a. | n.a. | n.a. | n.a. | n.a. | n.a. | n.a. | n.a. | n.a. |
| S12_vs_1980 | Elizabethkingia phage TCUEAP2 | OK632025 | Outlier | n.a. | N7 | 25 | Elizabethkingia | Group I | Duplodnaviria | Heunggongvirae | Uroviricota | Caudoviricetes | n.a. | Unclassified | O | O |
| S12_vs_22853 | Streptococcus phage SM1 | AY007505 | Outlier | n.a. | N2 | 24 | Streptococcus | Group I | Duplodnaviria | Heunggongvirae | Uroviricota | Caudoviricetes | n.a. | Unclassified | O | O |
| S12_vs_28661 | n.a. | n.a. | Outlier | n.a. | A | n.a. | n.a. | n.a. | n.a. | n.a. | n.a. | n.a. | n.a. | n.a. | n.a. | n.a. |
| S12_vs_32669 | n.a. | n.a. | Outlier | n.a. | A | n.a. | n.a. | n.a. | n.a. | n.a. | n.a. | n.a. | n.a. | n.a. | n.a. | n.a. |
| S12_vs_74280 | n.a. | n.a. | Outlier | n.a. | A | n.a. | n.a. | n.a. | n.a. | n.a. | n.a. | n.a. | n.a. | n.a. | n.a. | n.a. |
| S12_vs_75098 | Lactobacillus phage c5 | EU340421 | Outlier | n.a. | N5 | 6 | Lactobacillus | Group I | Duplodnaviria | Heunggongvirae | Uroviricota | Caudoviricetes | n.a. | Unclassified | O | O |
| S12_vs_81729 | Bacillus phage vB_BtS_BMBtp13 | KX190832 | Outlier | n.a. | N9 | 4 | Bacillus | Group I | Duplodnaviria | Heunggongvirae | Uroviricota | Caudoviricetes | n.a. | Unclassified | O | O |
| S12_vs_8204 | Bacillus phage G | JN638751 | Outlier | n.a. | N15 | 4 | Bacillus | Group I | Duplodnaviria | Heunggongvirae | Uroviricota | Caudoviricetes | n.a. | Unclassified | O | O |
| S12_vs_87088 | Faecalibacterium phage FP_Lugh | MG711464 | Outlier | n.a. | N1 | 32 | Faecalibacterium | Group I | Duplodnaviria | Heunggongvirae | Uroviricota | Caudoviricetes | n.a. | Unclassified | O | O |
| S12_vs_94042 | n.a. | n.a. | Outlier | n.a. | A | n.a. | n.a. | n.a. | n.a. | n.a. | n.a. | n.a. | n.a. | n.a. | n.a. | n.a. |
| S12_vs_99219 | n.a. | n.a. | Outlier | n.a. | A | n.a. | n.a. | n.a. | n.a. | n.a. | n.a. | n.a. | n.a. | n.a. | n.a. | n.a. |
| S13_vf_147396 | n.a. | n.a. | Outlier | n.a. | A | n.a. | n.a. | n.a. | n.a. | n.a. | n.a. | n.a. | n.a. | n.a. | n.a. | n.a. |
| S13_vf_1770 | Pectobacterium phage DU_PP_III | MF979562 | Outlier | n.a. | N2 | 13 | Pectobacterium | Group I | Duplodnaviria | Heunggongvirae | Uroviricota | Caudoviricetes | n.a. | Unclassified | O | O |
| S13_vf_54201 | Faecalibacterium phage FP_Brigit | MG711465 | Outlier | n.a. | N1 | 18 | Faecalibacterium | Group I | Duplodnaviria | Heunggongvirae | Uroviricota | Caudoviricetes | n.a. | Unclassified | O | O |
| S13_vf_66508 | Bacteroides phage ARB14 | MT074134 | Outlier | n.a. | N1 | 51 | Bacteroides | Group I | Duplodnaviria | Heunggongvirae | Uroviricota | Caudoviricetes | n.a. | Unclassified | O | O |
| S13_vs_114213 | Faecalibacterium phage FP_Mushu | MG711460 | Outlier | n.a. | N4 | 4 | Faecalibacterium | Group I | Duplodnaviria | Heunggongvirae | Uroviricota | Caudoviricetes | n.a. | Unclassified | O | O |
| S13_vs_13129 | Bacillus phage vB_BtS_BMBtp13 | KX190832 | Outlier | n.a. | N22 | 2 | Bacillus | Group I | Duplodnaviria | Heunggongvirae | Uroviricota | Caudoviricetes | n.a. | Unclassified | O | O |
| S13_vs_14794 | Aeromonas phage P5 | OM519319 | Outlier | n.a. | N1 | 2 | Aeromonas | Group I | Duplodnaviria | Heunggongvirae | Uroviricota | Caudoviricetes | n.a. | Unclassified | O | O |
| S13_vs_27462 | Streptococcus phage Dp-1 | HQ268735 | Outlier | n.a. | N1 | 35 | Streptococcus | Group I | Duplodnaviria | Heunggongvirae | Uroviricota | Caudoviricetes | n.a. | Unclassified | O | O |
| S13_vs_45488 | Curvibacter phage TJ1 | MH766655 | Outlier | n.a. | N1 | 13 | Curvibacter | Group I | Duplodnaviria | Heunggongvirae | Uroviricota | Caudoviricetes | n.a. | Unclassified | O | O |
| S13_vs_57267 | Prevotella phage Lak-C1 | MK250029 | Outlier | n.a. | N1 | 4 | Prevotella | Group I | Duplodnaviria | Heunggongvirae | Uroviricota | Caudoviricetes | n.a. | Unclassified | O | O |
| S14_vf_117147 | Bacillus phage B13 | OP066531 | Outlier | n.a. | N15 | 6 | Bacillus | Group I | Duplodnaviria | Heunggongvirae | Uroviricota | Caudoviricetes | n.a. | Unclassified | O | O |
| S14_vf_51134 | Streptococcus phage OlisA2 | OL774869 | Outlier | n.a. | N5 | 22 | Streptococcus | Group I | Duplodnaviria | Heunggongvirae | Uroviricota | Caudoviricetes | n.a. | Unclassified | O | O |
| S14_vf_57689 | Microbacterium phage Rasputia | MT639646 | Outlier | n.a. | N1 | 17 | Microbacterium | Group I | Duplodnaviria | Heunggongvirae | Uroviricota | Caudoviricetes | n.a. | Unclassified | O | O |
| S14_vs_100227 | Streptococcus phage phiARI0746 | KT337365 | Outlier | n.a. | N8 | 12 | Streptococcus | Group I | Duplodnaviria | Heunggongvirae | Uroviricota | Caudoviricetes | n.a. | Unclassified | O | O |
| S14_vs_101482 | n.a. | n.a. | Outlier | n.a. | A | n.a. | n.a. | n.a. | n.a. | n.a. | n.a. | n.a. | n.a. | n.a. | n.a. | n.a. |
| S14_vs_108376 | Clostridium phage phiCT453A | KM983327 | Outlier | n.a. | N5 | 10 | Clostridium | Group I | Duplodnaviria | Heunggongvirae | Uroviricota | Caudoviricetes | n.a. | Unclassified | O | O |
| S14_vs_112345 | n.a. | n.a. | Outlier | n.a. | A | n.a. | n.a. | n.a. | n.a. | n.a. | n.a. | n.a. | n.a. | n.a. | n.a. | n.a. |
| S14_vs_115377 | Roseburia phage Jekyll | LR596902 | Outlier | n.a. | N13 | 9 | Roseburia | Group I | Duplodnaviria | Heunggongvirae | Uroviricota | Caudoviricetes | n.a. | Unclassified | O | O |
| S14_vs_115455 | Clostridium phage phiSM101 | CP000315 | Outlier | n.a. | N2 | 16 | Clostridium | Group I | Duplodnaviria | Heunggongvirae | Uroviricota | Caudoviricetes | n.a. | Unclassified | O | O |
| S14_vs_20193 | Faecalibacterium phage FP_Lagaffe | MG711461 | Outlier | n.a. | N2 | 50 | Faecalibacterium | Group I | Duplodnaviria | Heunggongvirae | Uroviricota | Caudoviricetes | n.a. | Unclassified | O | O |
| S14_vs_26320 | Geobacillus virus E3 | KP144388 | Outlier | n.a. | N16 | 4 | Geobacillus | Group I | Duplodnaviria | Heunggongvirae | Uroviricota | Caudoviricetes | n.a. | Unclassified | O | O |
| S14_vs_30108 | n.a. | n.a. | Outlier | n.a. | A | n.a. | n.a. | n.a. | n.a. | n.a. | n.a. | n.a. | n.a. | n.a. | n.a. | n.a. |
| S14_vs_30566 | Streptococcus phage phiARI0746 | KT337365 | Outlier | n.a. | N3 | 11 | Streptococcus | Group I | Duplodnaviria | Heunggongvirae | Uroviricota | Caudoviricetes | n.a. | Unclassified | O | O |
| S14_vs_79505 | n.a. | n.a. | Outlier | n.a. | A | n.a. | n.a. | n.a. | n.a. | n.a. | n.a. | n.a. | n.a. | n.a. | n.a. | n.a. |
| S14_vs_9162 | Streptococcus phage Javan242 | MK448882 | Outlier | n.a. | N11 | 14 | Streptococcus | Group I | Duplodnaviria | Heunggongvirae | Uroviricota | Caudoviricetes | n.a. | Unclassified | O | O |
| S14_vs_93565 | Streptococcus phage Javan422 | MK448934 | Outlier | n.a. | N7 | 6 | Streptococcus | Group I | Duplodnaviria | Heunggongvirae | Uroviricota | Caudoviricetes | n.a. | Unclassified | O | O |
| S14_vs_96286 | Streptococcus phage Javan583 | MK448814 | Outlier | n.a. | N6 | 21 | Streptococcus | Group I | Duplodnaviria | Heunggongvirae | Uroviricota | Caudoviricetes | n.a. | Unclassified | O | O |
| S15_vf_58467 | Enterococcus phage 9183 | MT939241 | Outlier | n.a. | N1 | 11 | Enterococcus | Group I | Duplodnaviria | Heunggongvirae | Uroviricota | Caudoviricetes | n.a. | Unclassified | O | O |
| S16_vf_172897 | Bifidobacterium phage BigBern1 | MT006237 | Outlier | n.a. | N6 | 9 | Bifidobacterium | Group I | Duplodnaviria | Heunggongvirae | Uroviricota | Caudoviricetes | n.a. | Unclassified | O | O |
| S16_vf_32359 | n.a. | n.a. | Outlier | n.a. | A | n.a. | n.a. | n.a. | n.a. | n.a. | n.a. | n.a. | n.a. | n.a. | n.a. | n.a. |
| S16_vf_33218 | Lactococcus phage CHPC965 | MN689525 | Outlier | n.a. | N1 | 62 | Lactococcus | Group I | Duplodnaviria | Heunggongvirae | Uroviricota | Caudoviricetes | n.a. | Unclassified | O | O |
| S16_vf_9270 | n.a. | n.a. | Outlier | n.a. | A | n.a. | n.a. | n.a. | n.a. | n.a. | n.a. | n.a. | n.a. | n.a. | n.a. | n.a. |
| S16_vs_10622 | n.a. | n.a. | Outlier | n.a. | A | n.a. | n.a. | n.a. | n.a. | n.a. | n.a. | n.a. | n.a. | n.a. | n.a. | n.a. |
| S16_vs_120149 | n.a. | n.a. | Outlier | n.a. | A | n.a. | n.a. | n.a. | n.a. | n.a. | n.a. | n.a. | n.a. | n.a. | n.a. | n.a. |
| S16_vs_137222 | Faecalibacterium phage FP_Lagaffe | MG711461 | Outlier | n.a. | N3 | 48 | Faecalibacterium | Group I | Duplodnaviria | Heunggongvirae | Uroviricota | Caudoviricetes | n.a. | Unclassified | O | O |
| S16_vs_141218 | n.a. | n.a. | Outlier | n.a. | A | n.a. | n.a. | n.a. | n.a. | n.a. | n.a. | n.a. | n.a. | n.a. | n.a. | n.a. |
| S16_vs_165662 | Clostridium phage HM T | KU517658 | Outlier | n.a. | N8 | 2 | Clostridium | Group I | Duplodnaviria | Heunggongvirae | Uroviricota | Caudoviricetes | n.a. | Unclassified | O | O |
| S16_vs_18293 | Streptococcus phage Javan261 | MK448720 | Outlier | n.a. | N4 | 12 | Streptococcus | Group I | Duplodnaviria | Heunggongvirae | Uroviricota | Caudoviricetes | n.a. | Unclassified | O | O |
| S16_vs_34021 | n.a. | n.a. | Outlier | n.a. | A | n.a. | n.a. | n.a. | n.a. | n.a. | n.a. | n.a. | n.a. | n.a. | n.a. | n.a. |
| S16_vs_35031 | n.a. | n.a. | Outlier | n.a. | A | n.a. | n.a. | n.a. | n.a. | n.a. | n.a. | n.a. | n.a. | n.a. | n.a. | n.a. |
| S16_vs_38863 | Clostridium phage phiMMP01 | LN681541 | Outlier | n.a. | N9 | 19 | Clostridium | Group I | Duplodnaviria | Heunggongvirae | Uroviricota | Caudoviricetes | n.a. | Unclassified | O | O |
| S16_vs_54250 | n.a. | n.a. | Outlier | n.a. | A | n.a. | n.a. | n.a. | n.a. | n.a. | n.a. | n.a. | n.a. | n.a. | n.a. | n.a. |
| S16_vs_81805 | Faecalibacterium phage FP_Lugh | MG711464 | Outlier | n.a. | N3 | 19 | Faecalibacterium | Group I | Duplodnaviria | Heunggongvirae | Uroviricota | Caudoviricetes | n.a. | Unclassified | O | O |
| S17_vf_111931 | Clostridium phage DCp1 | OP256049 | Outlier | n.a. | N1 | 10 | Clostridium | Group I | Duplodnaviria | Heunggongvirae | Uroviricota | Caudoviricetes | n.a. | Guelinviridae | O | O |
| S17_vf_46388 | Ruminococcus phage phiRg507T2_3 | MT980837 | Outlier | n.a. | N6 | 4 | Ruminococcus | Group I | Duplodnaviria | Heunggongvirae | Uroviricota | Caudoviricetes | n.a. | Unclassified | O | O |
| S17_vs_110666 | Faecalibacterium phage FP_Brigit | MG711465 | Outlier | n.a. | N2 | 29 | Faecalibacterium | Group I | Duplodnaviria | Heunggongvirae | Uroviricota | Caudoviricetes | n.a. | Unclassified | O | O |
| S17_vs_117507 | n.a. | n.a. | Outlier | n.a. | A | n.a. | n.a. | n.a. | n.a. | n.a. | n.a. | n.a. | n.a. | n.a. | n.a. | n.a. |
| S17_vs_177202 | Faecalibacterium phage FP_Lugh | MG711464 | Outlier | n.a. | N15 | 9 | Faecalibacterium | Group I | Duplodnaviria | Heunggongvirae | Uroviricota | Caudoviricetes | n.a. | Unclassified | O | O |
| S17_vs_177203 | Acetobacter phage phiAO1 | LC644971 | Outlier | n.a. | N1 | 26 | Acetobacter | Group I | Duplodnaviria | Heunggongvirae | Uroviricota | Caudoviricetes | n.a. | Unclassified | O | O |
| S17_vs_31654 | Streptococcus phage Javan74 | MK449005 | Outlier | n.a. | N2 | 26 | Streptococcus | Group I | Duplodnaviria | Heunggongvirae | Uroviricota | Caudoviricetes | n.a. | Unclassified | O | O |
| S17_vs_52890 | n.a. | n.a. | Outlier | n.a. | A | n.a. | n.a. | n.a. | n.a. | n.a. | n.a. | n.a. | n.a. | n.a. | n.a. | n.a. |
| S1_vf_138157 | n.a. | n.a. | Outlier | n.a. | A | n.a. | n.a. | n.a. | n.a. | n.a. | n.a. | n.a. | n.a. | n.a. | n.a. | n.a. |
| S1_vf_140259 | Microviridae sp. | MG945534 | Outlier | n.a. | N1 | 6 | Unspecified | Group II | Monodnaviria | Sangervirae | Phixviricota | Malgrandaviricetes | Petitvirales | Microviridae | O | O |
| S1_vf_28398 | n.a. | n.a. | Outlier | n.a. | A | n.a. | n.a. | n.a. | n.a. | n.a. | n.a. | n.a. | n.a. | n.a. | n.a. | n.a. |
| S1_vs_133577 | Faecalibacterium phage FP_Lugh | MG711464 | Outlier | n.a. | N4 | 19 | Faecalibacterium | Group I | Duplodnaviria | Heunggongvirae | Uroviricota | Caudoviricetes | n.a. | Unclassified | O | O |
| S1_vs_23682 | Butyrivibrio virus Ceridwen | MN882553 | Outlier | n.a. | N9 | 17 | Butyrivibrio | Group I | Duplodnaviria | Heunggongvirae | Uroviricota | Caudoviricetes | n.a. | Unclassified | O | O |
| S2_vs_4368 | Butyrivibrio phage Idris | MN882554 | Outlier | n.a. | N9 | 1 | Butyrivibrio | Group I | Duplodnaviria | Heunggongvirae | Uroviricota | Caudoviricetes | n.a. | Unclassified | O | O |
| S3_vf_101591 | Gokushovirus WZ-2015a | KT264789 | Outlier | n.a. | N1 | 8 | Unspecified | Group II | Monodnaviria | Sangervirae | Phixviricota | Malgrandaviricetes | Petitvirales | Microviridae | O | O |
| S3_vf_101611 | Streptococcus phage Javan74 | MK449005 | Outlier | n.a. | N3 | 14 | Streptococcus | Group I | Duplodnaviria | Heunggongvirae | Uroviricota | Caudoviricetes | n.a. | Unclassified | O | O |
| S3_vf_111893 | Streptococcus phage Javan583 | MK448814 | Outlier | n.a. | N4 | 23 | Streptococcus | Group I | Duplodnaviria | Heunggongvirae | Uroviricota | Caudoviricetes | n.a. | Unclassified | O | O |
| S3_vf_127392 | Roseburia phage Jekyll | LR596902 | Outlier | n.a. | N28 | 7 | Roseburia | Group I | Duplodnaviria | Heunggongvirae | Uroviricota | Caudoviricetes | n.a. | Unclassified | O | O |
| S3_vf_127408 | Bifidobacterium phage BlindBasel1 | MT006235 | Outlier | n.a. | N1 | 28 | Bifidobacterium | Group I | Duplodnaviria | Heunggongvirae | Uroviricota | Caudoviricetes | n.a. | Unclassified | O | O |
| S3_vf_2186 | n.a. | n.a. | Outlier | n.a. | F | n.a. | n.a. | n.a. | n.a. | n.a. | n.a. | n.a. | n.a. | n.a. | n.a. | n.a. |
| S3_vf_23042 | Lactococcus phage P656 | MN552149 | Outlier | n.a. | N1 | 50 | Lactococcus | Group I | Duplodnaviria | Heunggongvirae | Uroviricota | Caudoviricetes | n.a. | Unclassified | O | O |
| S3_vf_5525 | n.a. | n.a. | Outlier | n.a. | A | n.a. | n.a. | n.a. | n.a. | n.a. | n.a. | n.a. | n.a. | n.a. | n.a. | n.a. |
| S3_vf_56363 | Clostridium phage phiCDKH01 | MN718463 | Outlier | n.a. | N1 | 26 | Clostridium | Group I | Duplodnaviria | Heunggongvirae | Uroviricota | Caudoviricetes | n.a. | Unclassified | O | O |
| S3_vf_99626 | Propionibacterium phage B3 | KX620749 | Outlier | n.a. | N2 | 11 | Propionibacterium | Group I | Duplodnaviria | Heunggongvirae | Uroviricota | Caudoviricetes | n.a. | Unclassified | O | O |
| S3_vs_1031 | n.a. | n.a. | Outlier | n.a. | A | n.a. | n.a. | n.a. | n.a. | n.a. | n.a. | n.a. | n.a. | n.a. | n.a. | n.a. |
| S3_vs_119771 | Faecalibacterium phage FP_Lugh | MG711464 | Outlier | n.a. | N2 | 17 | Faecalibacterium | Group I | Duplodnaviria | Heunggongvirae | Uroviricota | Caudoviricetes | n.a. | Unclassified | O | O |

| | | | | | | | | | | | | | | | | |
|---|---|---|---|---|---|---|---|---|---|---|---|---|---|---|---|---|
| S3_vs_122706 | Streptococcus phage phiARI0746 | KT337365 | Outlier | n.a. | N6 | 2 | Streptococcus | Group I | Duplodnaviria | Heunggongvirae | Uroviricota | Caudoviricetes | n.a. | Unclassified | O | O |
| S3_vs_127395 | n.a. | n.a. | Outlier | n.a. | A | n.a. | n.a. | n.a. | n.a. | n.a. | n.a. | n.a. | n.a. | n.a. | n.a. | n.a. |
| S3_vs_127397 | Streptococcus phage Javan278 | MK448896 | Outlier | n.a. | N12 | 6 | Streptococcus | Group I | Duplodnaviria | Heunggongvirae | Uroviricota | Caudoviricetes | n.a. | Unclassified | O | O |
| S3_vs_127401 | Streptococcus phage phiARI0746 | KT337365 | Outlier | n.a. | N5 | 12 | Streptococcus | Group I | Duplodnaviria | Heunggongvirae | Uroviricota | Caudoviricetes | n.a. | Unclassified | O | O |
| S3_vs_127410 | Clostridium phage phiCD38-2 | HM568888 | Outlier | n.a. | N15 | 3 | Clostridium | Group I | Duplodnaviria | Heunggongvirae | Uroviricota | Caudoviricetes | n.a. | Unclassified | O | O |
| S3_vs_15450 | Clostridium phage phiCT453A | KM983327 | Outlier | n.a. | N16 | 5 | Clostridium | Group I | Duplodnaviria | Heunggongvirae | Uroviricota | Caudoviricetes | n.a. | Unclassified | O | O |
| S3_vs_23194 | Cellulophaga phage Ingeline_1 | MT732435 | Outlier | n.a. | N2 | 7 | Cellulophaga | Group I | Duplodnaviria | Heunggongvirae | Uroviricota | Caudoviricetes | n.a. | Duneviridae | O | O |
| S3_vs_40939 | n.a. | n.a. | Outlier | n.a. | A | n.a. | n.a. | n.a. | n.a. | n.a. | n.a. | n.a. | n.a. | n.a. | n.a. | n.a. |
| S3_vs_56292 | Elizabethkingia phage TCUEAP3 | OK632026 | Outlier | n.a. | N12 | 6 | Elizabethkingia | Group I | Duplodnaviria | Heunggongvirae | Uroviricota | Caudoviricetes | n.a. | Unclassified | O | O |
| S3_vs_62410 | n.a. | n.a. | Outlier | n.a. | A | n.a. | n.a. | n.a. | n.a. | n.a. | n.a. | n.a. | n.a. | n.a. | n.a. | n.a. |
| S3_vs_6604 | Flavobacterium phage vB_FspS_laban6-1 | MN812211 | Outlier | n.a. | N4 | 12 | Flavobacterium | Group I | Duplodnaviria | Heunggongvirae | Uroviricota | Caudoviricetes | n.a. | Duneviridae | O | O |
| S3_vs_70093 | n.a. | n.a. | Outlier | n.a. | A | n.a. | n.a. | n.a. | n.a. | n.a. | n.a. | n.a. | n.a. | n.a. | n.a. | n.a. |
| S3_vs_94577 | Lactococcus phage C41431 | KX160219 | Outlier | n.a. | N2 | 26 | Lactococcus | Group I | Duplodnaviria | Heunggongvirae | Uroviricota | Caudoviricetes | n.a. | Unclassified | O | O |
| S3_vs_97827 | n.a. | n.a. | Outlier | n.a. | A | n.a. | n.a. | n.a. | n.a. | n.a. | n.a. | n.a. | n.a. | n.a. | n.a. | n.a. |
| S4_vf_13831 | n.a. | n.a. | Outlier | n.a. | A | n.a. | n.a. | n.a. | n.a. | n.a. | n.a. | n.a. | n.a. | n.a. | n.a. | n.a. |
| S4_vf_20440 | Riemerella phage RAP44 | HQ396194 | Outlier | n.a. | N11 | 5 | Riemerella | Group I | Duplodnaviria | Heunggongvirae | Uroviricota | Caudoviricetes | n.a. | Unclassified | O | O |
| S4_vf_31982 | Lactobacillus phage Ld3 | KJ564038 | Outlier | n.a. | N1 | 41 | Lactobacillus | Group I | Duplodnaviria | Heunggongvirae | Uroviricota | Caudoviricetes | n.a. | Unclassified | O | O |
| S4_vs_VIRSorter_2744-cat_3 | Bifidobacterium phage Bbif-1 | GQ141189 | Outlier | n.a. | N5 | 9 | Bifidobacterium | Group I | Duplodnaviria | Heunggongvirae | Uroviricota | Caudoviricetes | n.a. | Unclassified | O | O |
| S4_vs_VIRSorter_31972-circular-cat_2 | Faecalibacterium phage FP_Lugh | MG711464 | Outlier | n.a. | N18 | 4 | Faecalibacterium | Group I | Duplodnaviria | Heunggongvirae | Uroviricota | Caudoviricetes | n.a. | Unclassified | O | O |
| S4_vs_VIRSorter_5698-cat_2 | Bifidobacterium phage Bbif-1 | GQ141189 | Outlier | n.a. | N3 | 17 | Bifidobacterium | Group I | Duplodnaviria | Heunggongvirae | Uroviricota | Caudoviricetes | n.a. | Unclassified | O | O |
| S5_vf_103206 | n.a. | n.a. | Outlier | n.a. | F | n.a. | n.a. | n.a. | n.a. | n.a. | n.a. | n.a. | n.a. | n.a. | n.a. | n.a. |
| S5_vf_116300 | n.a. | n.a. | Outlier | n.a. | A | n.a. | n.a. | n.a. | n.a. | n.a. | n.a. | n.a. | n.a. | n.a. | n.a. | n.a. |
| S5_vf_55687 | Lactococcus phage M5938 | KX373687 | Outlier | n.a. | N1 | 58 | Lactococcus | Group I | Duplodnaviria | Heunggongvirae | Uroviricota | Caudoviricetes | n.a. | Unclassified | O | O |
| S5_vf_8808 | Lactobacillus phage c5 | EU340421 | Outlier | n.a. | N5 | 8 | Lactobacillus | Group I | Duplodnaviria | Heunggongvirae | Uroviricota | Caudoviricetes | n.a. | Unclassified | O | O |
| S5_vs_10428 | Faecalibacterium phage FP_Taranis | MG711467 | Outlier | n.a. | N3 | 17 | Faecalibacterium | Group I | Duplodnaviria | Heunggongvirae | Uroviricota | Caudoviricetes | n.a. | Unclassified | O | O |
| S5_vs_112452 | Clostridium phage phiCT453A | KM983327 | Outlier | n.a. | N7 | 16 | Clostridium | Group I | Duplodnaviria | Heunggongvirae | Uroviricota | Caudoviricetes | n.a. | Unclassified | O | O |
| S5_vs_113386 | n.a. | n.a. | Outlier | n.a. | A | n.a. | n.a. | n.a. | n.a. | n.a. | n.a. | n.a. | n.a. | n.a. | n.a. | n.a. |
| S5_vs_118168 | n.a. | n.a. | Outlier | n.a. | A | n.a. | n.a. | n.a. | n.a. | n.a. | n.a. | n.a. | n.a. | n.a. | n.a. | n.a. |
| S5_vs_120584 | Streptococcus phage Dp-1 | HQ268735 | Outlier | n.a. | N2 | 13 | Streptococcus | Group I | Duplodnaviria | Heunggongvirae | Uroviricota | Caudoviricetes | n.a. | Unclassified | O | O |
| S5_vs_121411 | n.a. | n.a. | Outlier | n.a. | A | n.a. | n.a. | n.a. | n.a. | n.a. | n.a. | n.a. | n.a. | n.a. | n.a. | n.a. |
| S5_vs_122618 | n.a. | n.a. | Outlier | n.a. | A | n.a. | n.a. | n.a. | n.a. | n.a. | n.a. | n.a. | n.a. | n.a. | n.a. | n.a. |
| S5_vs_128398 | Faecalibacterium phage FP_Epona | MG711462 | Outlier | n.a. | N10 | 30 | Faecalibacterium | Group I | Duplodnaviria | Heunggongvirae | Uroviricota | Caudoviricetes | n.a. | Unclassified | O | O |
| S5_vs_128881 | n.a. | n.a. | Outlier | n.a. | A | n.a. | n.a. | n.a. | n.a. | n.a. | n.a. | n.a. | n.a. | n.a. | n.a. | n.a. |
| S5_vs_130032 | Escherichia phage 2H10 | LR595862 | Outlier | n.a. | N1 | 65 | Escherichia | Group I | Duplodnaviria | Heunggongvirae | Uroviricota | Caudoviricetes | n.a. | Unclassified | O | O |
| S5_vs_130476 | n.a. | n.a. | Outlier | n.a. | A | n.a. | n.a. | n.a. | n.a. | n.a. | n.a. | n.a. | n.a. | n.a. | n.a. | n.a. |
| S5_vs_141621 | Clostridium phage phiCT19406B | KM983331 | Outlier | n.a. | N4 | 10 | Clostridium | Group I | Duplodnaviria | Heunggongvirae | Uroviricota | Caudoviricetes | n.a. | Unclassified | O | O |
| S5_vs_145582 | Phage WC36-2 | OL791266 | Outlier | n.a. | N4 | 23 | Unspecified | Group I | Duplodnaviria | Heunggongvirae | Uroviricota | Caudoviricetes | n.a. | Unclassified | O | O |
| S5_vs_145611 | Clostridium phage phiCT453A | KM983327 | Outlier | n.a. | N7 | 13 | Clostridium | Group I | Duplodnaviria | Heunggongvirae | Uroviricota | Caudoviricetes | n.a. | Unclassified | O | O |
| S5_vs_32885 | Faecalibacterium phage FP_Lagaffe | MG711461 | Outlier | n.a. | N3 | 13 | Faecalibacterium | Group I | Duplodnaviria | Heunggongvirae | Uroviricota | Caudoviricetes | n.a. | Unclassified | O | O |
| S5_vs_37659 | n.a. | n.a. | Outlier | n.a. | A | n.a. | n.a. | n.a. | n.a. | n.a. | n.a. | n.a. | n.a. | n.a. | n.a. | n.a. |
| S5_vs_43207 | Faecalibacterium phage FP_Lugh | MG711464 | Outlier | n.a. | N15 | 1 | Faecalibacterium | Group I | Duplodnaviria | Heunggongvirae | Uroviricota | Caudoviricetes | n.a. | Unclassified | O | O |
| S5_vs_46394 | n.a. | n.a. | Outlier | n.a. | A | n.a. | n.a. | n.a. | n.a. | n.a. | n.a. | n.a. | n.a. | n.a. | n.a. | n.a. |
| S5_vs_4737 | n.a. | n.a. | Outlier | n.a. | A | n.a. | n.a. | n.a. | n.a. | n.a. | n.a. | n.a. | n.a. | n.a. | n.a. | n.a. |
| S5_vs_51416 | Faecalibacterium phage FP_Lugh | MG711464 | Outlier | n.a. | N26 | 1 | Faecalibacterium | Group I | Duplodnaviria | Heunggongvirae | Uroviricota | Caudoviricetes | n.a. | Unclassified | O | O |
| S5_vs_59798 | Lactobacillus phage c5 | EU340421 | Outlier | n.a. | N10 | 6 | Lactobacillus | Group I | Duplodnaviria | Heunggongvirae | Uroviricota | Caudoviricetes | n.a. | Unclassified | O | O |
| S5_vs_72532 | Bacteroides phage p00 | BK010646 | Outlier | n.a. | N2 | 30 | Bacteroides | Group I | Duplodnaviria | Heunggongvirae | Uroviricota | Caudoviricetes | n.a. | Unclassified | O | O |
| S5_vs_80911 | Clostridium phage phiSM101 | CP000315 | Outlier | n.a. | N4 | 3 | Clostridium | Group I | Duplodnaviria | Heunggongvirae | Uroviricota | Caudoviricetes | n.a. | Unclassified | O | O |
| S5_vs_8125 | Clostridium phage phiSM101 | CP000315 | Outlier | n.a. | N6 | 8 | Clostridium | Group I | Duplodnaviria | Heunggongvirae | Uroviricota | Caudoviricetes | n.a. | Unclassified | O | O |
| S5_vs_850 | n.a. | n.a. | Outlier | n.a. | A | n.a. | n.a. | n.a. | n.a. | n.a. | n.a. | n.a. | n.a. | n.a. | n.a. | n.a. |
| S5_vs_89540 | Lactobacillus phage CL2 | KR905067 | Outlier | n.a. | N1 | 19 | Lactobacillus | Group I | Duplodnaviria | Heunggongvirae | Uroviricota | Caudoviricetes | n.a. | Unclassified | O | O |
| S6_vf_47238 | Pectobacterium phage DU_PP_III | MF979562 | Outlier | n.a. | N2 | 13 | Pectobacterium | Group I | Duplodnaviria | Heunggongvirae | Uroviricota | Caudoviricetes | n.a. | Unclassified | O | O |
| S6_vf_56484 | Faecalibacterium phage FP_Lugh | MG711464 | Outlier | n.a. | N9 | 9 | Faecalibacterium | Group I | Duplodnaviria | Heunggongvirae | Uroviricota | Caudoviricetes | n.a. | Unclassified | O | O |
| S6_vs_37812 | Rhodobacter phage RcOceanus | MW677520 | Outlier | n.a. | N1 | 9 | Rhodobacter | Group I | Duplodnaviria | Heunggongvirae | Uroviricota | Caudoviricetes | n.a. | Unclassified | O | O |
| S6_vs_39065 | n.a. | n.a. | Outlier | n.a. | A | n.a. | n.a. | n.a. | n.a. | n.a. | n.a. | n.a. | n.a. | n.a. | n.a. | n.a. |
| S6_vs_44070 | Bacteriophage sp. | MW202489 | Outlier | n.a. | N1 | 11 | Unspecified | Unclassified | Unclassified | n.a. | n.a. | n.a. | n.a. | Unclassified | O | O |
| S7_vf_13614 | Streptococcus phage Javan583 | MK448814 | Outlier | n.a. | N3 | 23 | Streptococcus | Group I | Duplodnaviria | Heunggongvirae | Uroviricota | Caudoviricetes | n.a. | Unclassified | O | O |
| S7_vf_138225 | Ralstonia phage RPSC1 | MF893341 | Outlier | n.a. | N3 | 4 | Ralstonia | Group I | Duplodnaviria | Heunggongvirae | Uroviricota | Caudoviricetes | n.a. | Autographiviridae | O | O |
| S7_vf_29941 | Lactococcus phage P680 | KC182551 | Outlier | n.a. | N1 | 60 | Lactococcus | Group I | Duplodnaviria | Heunggongvirae | Uroviricota | Caudoviricetes | n.a. | Unclassified | O | O |
| S7_vf_5801 | Riemerella phage RAP44 | HQ396194 | Outlier | n.a. | N1 | 20 | Riemerella | Group I | Duplodnaviria | Heunggongvirae | Uroviricota | Caudoviricetes | n.a. | Unclassified | O | O |
| S7_vf_68239 | Lactococcus phage 62601 | MF448564 | Outlier | n.a. | N1 | 46 | Lactococcus | Group I | Duplodnaviria | Heunggongvirae | Uroviricota | Caudoviricetes | n.a. | Unclassified | O | O |
| S7_vf_83624 | Clostridium phage HM2 | LT600745 | Outlier | n.a. | N28 | 4 | Clostridium | Group I | Duplodnaviria | Heunggongvirae | Uroviricota | Caudoviricetes | n.a. | Salasmaviridae | O | O |
| S7_vf_9334 | Podoviridae sp. | MW202898 | Outlier | n.a. | N1 | 9 | Unspecified | Group I | Duplodnaviria | Heunggongvirae | Uroviricota | Caudoviricetes | n.a. | Unclassified | O | O |
| S7_vs_134784 | Faecalibacterium phage FP_Lugh | MG711464 | Outlier | n.a. | N4 | 7 | Faecalibacterium | Group I | Duplodnaviria | Heunggongvirae | Uroviricota | Caudoviricetes | n.a. | Unclassified | O | O |
| S7_vs_138140 | n.a. | n.a. | Outlier | n.a. | A | n.a. | n.a. | n.a. | n.a. | n.a. | n.a. | n.a. | n.a. | n.a. | n.a. | n.a. |
| S7_vs_14275 | n.a. | n.a. | Outlier | n.a. | A | n.a. | n.a. | n.a. | n.a. | n.a. | n.a. | n.a. | n.a. | n.a. | n.a. | n.a. |
| S7_vs_16344 | Campylobacter phage PC10 | MZ047271 | Outlier | n.a. | N1 | 15 | Campylobacter | Group I | Duplodnaviria | Heunggongvirae | Uroviricota | Caudoviricetes | n.a. | Unclassified | O | O |
| S7_vs_21521 | n.a. | n.a. | Outlier | n.a. | A | n.a. | n.a. | n.a. | n.a. | n.a. | n.a. | n.a. | n.a. | n.a. | n.a. | n.a. |
| S7_vs_26235 | n.a. | n.a. | Outlier | n.a. | A | n.a. | n.a. | n.a. | n.a. | n.a. | n.a. | n.a. | n.a. | n.a. | n.a. | n.a. |
| S7_vs_29595 | Pseudomonas phage BHU-1 | OK638201 | Outlier | n.a. | N1 | 9 | Pseudomonas | Group I | Duplodnaviria | Heunggongvirae | Uroviricota | Caudoviricetes | n.a. | Unclassified | O | O |
| S7_vs_36656 | Clostridium phage phiCT453A | KM983327 | Outlier | n.a. | N1 | 16 | Clostridium | Group I | Duplodnaviria | Heunggongvirae | Uroviricota | Caudoviricetes | n.a. | Unclassified | O | O |
| S7_vs_40259 | Clostridium phage phiCDHM19 | LK985322 | Outlier | n.a. | N7 | 9 | Clostridium | Group I | Duplodnaviria | Heunggongvirae | Uroviricota | Caudoviricetes | n.a. | Unclassified | O | O |
| S7_vs_51332 | n.a. | n.a. | Outlier | n.a. | A | n.a. | n.a. | n.a. | n.a. | n.a. | n.a. | n.a. | n.a. | n.a. | n.a. | n.a. |
| S7_vs_55031 | n.a. | n.a. | Outlier | n.a. | A | n.a. | n.a. | n.a. | n.a. | n.a. | n.a. | n.a. | n.a. | n.a. | n.a. | n.a. |
| S7_vs_70265 | Clostridium phage HM2 | LT600745 | Outlier | n.a. | N32 | 4 | Clostridium | Group I | Duplodnaviria | Heunggongvirae | Uroviricota | Caudoviricetes | n.a. | Salasmaviridae | O | O |
| S7_vs_72737 | n.a. | n.a. | Outlier | n.a. | A | n.a. | n.a. | n.a. | n.a. | n.a. | n.a. | n.a. | n.a. | n.a. | n.a. | n.a. |
| S8_vf_122206 | Bacteroides phage LoVEphage | MW660583 | Outlier | n.a. | N8 | 13 | Bacteroides | Group I | Duplodnaviria | Heunggongvirae | Uroviricota | Caudoviricetes | n.a. | Unclassified | O | O |
| S8_vf_13709 | Faecalibacterium phage FP_Lugh | MG711464 | Outlier | n.a. | N8 | 6 | Faecalibacterium | Group I | Duplodnaviria | Heunggongvirae | Uroviricota | Caudoviricetes | n.a. | Unclassified | O | O |
| S8_vf_147338 | Faecalibacterium phage FP_oengus | MG711463 | Outlier | n.a. | N21 | 32 | Faecalibacterium | Group I | Duplodnaviria | Heunggongvirae | Uroviricota | Caudoviricetes | n.a. | Unclassified | O | O |
| S8_vf_192724 | Clostridium phage HM2 | LT600745 | Outlier | n.a. | N2 | 4 | Clostridium | Group I | Duplodnaviria | Heunggongvirae | Uroviricota | Caudoviricetes | n.a. | Salasmaviridae | O | O |
| S8_vf_65522 | n.a. | n.a. | Outlier | n.a. | A | n.a. | n.a. | n.a. | n.a. | n.a. | n.a. | n.a. | n.a. | n.a. | n.a. | n.a. |
| S8_vs_110692 | Streptococcus phage Javan278 | MK448896 | Outlier | n.a. | N10 | 8 | Streptococcus | Group I | Duplodnaviria | Heunggongvirae | Uroviricota | Caudoviricetes | n.a. | Unclassified | O | O |
| S8_vs_113059 | Gokushovirus WZ-2015a | KT264759 | Outlier | n.a. | N1 | 14 | Unspecified | Group II | Monodnaviria | Sangervirae | Phixviricota | Malgrandaviricetes | Petitvirales | Microviridae | O | O |
| S8_vs_126582 | Phage DP SC_6_H4_2017 | MT121964 | Outlier | n.a. | N1 | 97 | Unspecified | Group I | Duplodnaviria | Heunggongvirae | Uroviricota | Caudoviricetes | n.a. | Unclassified | O | O |
| S8_vs_132472 | n.a. | n.a. | Outlier | n.a. | A | n.a. | n.a. | n.a. | n.a. | n.a. | n.a. | n.a. | n.a. | n.a. | n.a. | n.a. |
| S8_vs_147670 | Clostridioides phage CD1801 | MW512570 | Outlier | n.a. | N1 | 9 | Clostridioides | Group I | Duplodnaviria | Heunggongvirae | Uroviricota | Caudoviricetes | n.a. | Unclassified | O | O |
| S8_vs_147697 | n.a. | n.a. | Outlier | n.a. | A | n.a. | n.a. | n.a. | n.a. | n.a. | n.a. | n.a. | n.a. | n.a. | n.a. | n.a. |
| S8_vs_185041 | Gardnerella phage vB_Gva_AB1 | MW387018 | Outlier | n.a. | N4 | 38 | Gardnerella | Group I | Duplodnaviria | Heunggongvirae | Uroviricota | Caudoviricetes | n.a. | Unclassified | O | O |

| | | | | | | | | | | | | | | | | |
|---|---|---|---|---|---|---|---|---|---|---|---|---|---|---|---|---|
| S8_vs_3224 | n.a. | n.a. | Outlier | n.a. | A | n.a. | n.a. | n.a. | n.a. | n.a. | n.a. | n.a. | n.a. | n.a. | n.a. | n.a. |
| S8_vs_63277 | Faecalibacterium phage FP_Lugh | MG711464 | Outlier | n.a. | N1 | 37 | Faecalibacterium | Group I | Duplodnaviria | Heunggongvirae | Uroviricota | Caudoviricetes | n.a. | Unclassified | O | O |
| S8_vs_72117 | n.a. | n.a. | Outlier | n.a. | A | n.a. | n.a. | n.a. | n.a. | n.a. | n.a. | n.a. | n.a. | n.a. | n.a. | n.a. |
| S9_vf_19284 | n.a. | n.a. | Outlier | n.a. | A | n.a. | n.a. | n.a. | n.a. | n.a. | n.a. | n.a. | n.a. | n.a. | n.a. | n.a. |
| S9_vf_934 | Curtobacterium phage Penoan | MZ333133 | Outlier | n.a. | N1 | 13 | Curtobacterium | Group I | Duplodnaviria | Heunggongvirae | Uroviricota | Caudoviricetes | n.a. | Unclassified | O | O |
| S9_vs_14235 | n.a. | n.a. | Outlier | n.a. | A | n.a. | n.a. | n.a. | n.a. | n.a. | n.a. | n.a. | n.a. | n.a. | n.a. | n.a. |
| S9_vs_19689 | n.a. | n.a. | Outlier | n.a. | A | n.a. | n.a. | n.a. | n.a. | n.a. | n.a. | n.a. | n.a. | n.a. | n.a. | n.a. |
| S9_vs_2799 | n.a. | n.a. | Outlier | n.a. | A | n.a. | n.a. | n.a. | n.a. | n.a. | n.a. | n.a. | n.a. | n.a. | n.a. | n.a. |
| S9_vs_41371 | Psychrobacillus phage PVJ1 | MZ983385 | Outlier | n.a. | N3 | 14 | Psychrobacillus | Group I | Duplodnaviria | Heunggongvirae | Uroviricota | Caudoviricetes | n.a. | Unclassified | O | O |
| S9_vs_44331 | Ruminococcus phage phiRg519T2 | MT980838 | Outlier | n.a. | N7 | 9 | Ruminococcus | Group I | Duplodnaviria | Heunggongvirae | Uroviricota | Caudoviricetes | n.a. | Unclassified | O | O |
| S9_vs_55738 | Streptococcus phage Javan355 | MK448738 | Outlier | n.a. | N6 | 8 | Streptococcus | Group I | Duplodnaviria | Heunggongvirae | Uroviricota | Caudoviricetes | n.a. | Unclassified | O | O |
| S9_vs_58086 | Streptococcus phage Javan290 | MK448900 | Outlier | n.a. | N15 | 5 | Streptococcus | Group I | Duplodnaviria | Heunggongvirae | Uroviricota | Caudoviricetes | n.a. | Unclassified | O | O |
| S9_vs_62803 | Faecalibacterium phage FP_Lugh | MG711464 | Outlier | n.a. | N3 | 22 | Faecalibacterium | Group I | Duplodnaviria | Heunggongvirae | Uroviricota | Caudoviricetes | n.a. | Unclassified | O | O |
| S9_vs_66877 | Faecalibacterium phage FP_Lugh | MG711464 | Outlier | n.a. | N3 | 16 | Faecalibacterium | Group I | Duplodnaviria | Heunggongvirae | Uroviricota | Caudoviricetes | n.a. | Unclassified | O | O |
| S9_vs_67792 | Phage WC36-2 | OL791266 | Outlier | n.a. | N2 | 23 | Unspecified | Group I | Duplodnaviria | Heunggongvirae | Uroviricota | Caudoviricetes | n.a. | Unclassified | O | O |
| S14_vf_117162 | Bacteroides phage EMB2 | ON721385 | Overlap (VC_1044/VC_1047) | n.a. | C7 | 57 | Bacteroides | Group I | Duplodnaviria | Heunggongvirae | Uroviricota | Caudoviricetes | n.a. | Unclassified | Unclassified | O |
| S17_vf_177200 | Bacteroides phage EMB2 | ON721385 | Overlap (VC_1044/VC_1047) | n.a. | C7 | 57 | Bacteroides | Group I | Duplodnaviria | Heunggongvirae | Uroviricota | Caudoviricetes | n.a. | Unclassified | Unclassified | O |
| S11_vf_104062 | Bacteroides phage EMB2 | ON721385 | Overlap (VC_1044/VC_1047/VC_1488) | n.a. | C4 | 143 | Bacteroides | Group I | Duplodnaviria | Heunggongvirae | Uroviricota | Caudoviricetes | n.a. | Unclassified | Unclassified | O |
| S7_vf_27309 | Bacteroides phage EMB2 | ON721385 | Overlap (VC_1044/VC_1047/VC_1488) | n.a. | C1 | 150 | Bacteroides | Group I | Duplodnaviria | Heunggongvirae | Uroviricota | Caudoviricetes | n.a. | Unclassified | Unclassified | O |
| S12_vf_145044 | n.a. | n.a. | Overlap (VC_1080/VC_1510) | n.a. | A | n.a. | n.a. | n.a. | n.a. | n.a. | n.a. | n.a. | n.a. | n.a. | n.a. | n.a. |
| S12_vf_32615 | n.a. | n.a. | Overlap (VC_1080/VC_1510) | n.a. | A | n.a. | n.a. | n.a. | n.a. | n.a. | n.a. | n.a. | n.a. | n.a. | n.a. | n.a. |
| S15_vf_33505 | uncultured phage cr53_1 | MT774396 | Overlap (VC_1080/VC_1510) | n.a. | N6 | 2 | Unspecified | Group I | Duplodnaviria | Heunggongvirae | Uroviricota | Caudoviricetes | Crassvirales | Suoliviridae | O | O |
| S5_vs_117319 | Faecalibacterium phage FP_oengus | MG711463 | Overlap (VC_1086/VC_1243) | n.a. | C1 | 149 | Faecalibacterium | Group I | Duplodnaviria | Heunggongvirae | Uroviricota | Caudoviricetes | n.a. | Unclassified | Unclassified | O |
| S6_vs_34453 | Faecalibacterium phage FP_oengus | MG711463 | Overlap (VC_1086/VC_1243) | n.a. | C1 | 143 | Faecalibacterium | Group I | Duplodnaviria | Heunggongvirae | Uroviricota | Caudoviricetes | n.a. | Unclassified | Unclassified | O |
| S7_vs_87279 | Faecalibacterium phage FP_oengus | MG711463 | Overlap (VC_1086/VC_1243) | n.a. | C1 | 178 | Faecalibacterium | Group I | Duplodnaviria | Heunggongvirae | Uroviricota | Caudoviricetes | n.a. | Unclassified | Unclassified | O |
| S3_vs_51931 | Gokushovirus WZ-2015a | KT264829 | Overlap (VC_1098/VC_1100) | n.a. | C1 | 28 | Unspecified | Group II | Monodnaviria | Sangervirae | Phixviricota | Malgrandaviricetes | Petitvirales | Microviridae | Gokushovirinae | O |
| S16_vs_101671 | Microviridae sp. | MG945704 | Overlap (VC_1160/VC_1164) | n.a. | C1 | 23 | Unspecified | Group II | Monodnaviria | Sangervirae | Phixviricota | Malgrandaviricetes | Petitvirales | Microviridae | Unclassified | O |
| S17_vs_177210 | Microviridae sp. | MG945704 | Overlap (VC_1160/VC_1164) | n.a. | C1 | 17 | Unspecified | Group II | Monodnaviria | Sangervirae | Phixviricota | Malgrandaviricetes | Petitvirales | Microviridae | Unclassified | O |
| S1_vs_3267 | Microviridae sp. | MG945704 | Overlap (VC_1160/VC_1164) | n.a. | C1 | 22 | Unspecified | Group II | Monodnaviria | Sangervirae | Phixviricota | Malgrandaviricetes | Petitvirales | Microviridae | Unclassified | O |
| S3_vs_51005 | Microviridae sp. | MG945704 | Overlap (VC_1160/VC_1164) | n.a. | C1 | 19 | Unspecified | Group II | Monodnaviria | Sangervirae | Phixviricota | Malgrandaviricetes | Petitvirales | Microviridae | Unclassified | O |
| S16_vs_118029 | Roseburia phage Jekyll | LR596902 | Overlap (VC_1254/VC_1595) | n.a. | N16 | 5 | Roseburia | Group I | Duplodnaviria | Heunggongvirae | Uroviricota | Caudoviricetes | n.a. | Unclassified | O | O |
| S7_vs_30640 | Faecalibacterium phage FP_Lugh | MG711464 | Overlap (VC_1254/VC_1595) | n.a. | N20 | 1 | Faecalibacterium | Group I | Duplodnaviria | Heunggongvirae | Uroviricota | Caudoviricetes | n.a. | Unclassified | O | O |
| S11_vs_49071 | n.a. | n.a. | Overlap (VC_1255/VC_1558) | n.a. | A | n.a. | n.a. | n.a. | n.a. | n.a. | n.a. | n.a. | n.a. | n.a. | n.a. | n.a. |
| S7_vs_101751 | n.a. | n.a. | Overlap (VC_1255/VC_1558) | n.a. | A | n.a. | n.a. | n.a. | n.a. | n.a. | n.a. | n.a. | n.a. | n.a. | n.a. | n.a. |
| S7_vs_137998 | n.a. | n.a. | Overlap (VC_1255/VC_1558) | n.a. | A | n.a. | n.a. | n.a. | n.a. | n.a. | n.a. | n.a. | n.a. | n.a. | n.a. | n.a. |
| S12_vs_26847 | n.a. | n.a. | Overlap (VC_1255/VC_1593) | n.a. | A | n.a. | n.a. | n.a. | n.a. | n.a. | n.a. | n.a. | n.a. | n.a. | n.a. | n.a. |
| S16_vf_5809 | n.a. | n.a. | Overlap (VC_1255/VC_1593) | n.a. | A | n.a. | n.a. | n.a. | n.a. | n.a. | n.a. | n.a. | n.a. | n.a. | n.a. | n.a. |
| S16_vf_84462 | n.a. | n.a. | Overlap (VC_1255/VC_1594) | n.a. | A | n.a. | n.a. | n.a. | n.a. | n.a. | n.a. | n.a. | n.a. | n.a. | n.a. | n.a. |
| S5_vf_60597 | n.a. | n.a. | Overlap (VC_1255/VC_1594) | n.a. | A | n.a. | n.a. | n.a. | n.a. | n.a. | n.a. | n.a. | n.a. | n.a. | n.a. | n.a. |
| S8_vf_201695 | n.a. | n.a. | Overlap (VC_1255/VC_1594) | n.a. | A | n.a. | n.a. | n.a. | n.a. | n.a. | n.a. | n.a. | n.a. | n.a. | n.a. | n.a. |
| S3_vs_127421 | Roseburia phage Jekyll | LR596902 | Overlap (VC_1256/VC_1564) | n.a. | N2 | 26 | Roseburia | Group I | Duplodnaviria | Heunggongvirae | Uroviricota | Caudoviricetes | n.a. | Unclassified | O | O |
| S8_vs_173961 | Roseburia phage Jekyll | LR596902 | Overlap (VC_1256/VC_1564) | n.a. | N2 | 24 | Roseburia | Group I | Duplodnaviria | Heunggongvirae | Uroviricota | Caudoviricetes | n.a. | Unclassified | O | O |
| S11_vf_58587 | n.a. | n.a. | Overlap (VC_1259/VC_1261) | n.a. | A | n.a. | n.a. | n.a. | n.a. | n.a. | n.a. | n.a. | n.a. | n.a. | n.a. | n.a. |
| S12_vf_170614 | Clostridium phage HM2 | LT600745 | Overlap (VC_1259/VC_1261) | n.a. | N34 | 4 | Clostridium | Group I | Duplodnaviria | Heunggongvirae | Uroviricota | Caudoviricetes | n.a. | Salasmaviridae | O | O |
| S16_vf_100473 | n.a. | n.a. | Overlap (VC_1259/VC_1261) | n.a. | A | n.a. | n.a. | n.a. | n.a. | n.a. | n.a. | n.a. | n.a. | n.a. | n.a. | n.a. |
| S16_vf_94866 | n.a. | n.a. | Overlap (VC_1259/VC_1261) | n.a. | A | n.a. | n.a. | n.a. | n.a. | n.a. | n.a. | n.a. | n.a. | n.a. | n.a. | n.a. |
| S5_vf_38011 | n.a. | n.a. | Overlap (VC_1259/VC_1261) | n.a. | A | n.a. | n.a. | n.a. | n.a. | n.a. | n.a. | n.a. | n.a. | n.a. | n.a. | n.a. |
| S6_vf_24553 | n.a. | n.a. | Overlap (VC_1259/VC_1261) | n.a. | A | n.a. | n.a. | n.a. | n.a. | n.a. | n.a. | n.a. | n.a. | n.a. | n.a. | n.a. |
| S7_vf_32396 | n.a. | n.a. | Overlap (VC_1259/VC_1261) | n.a. | A | n.a. | n.a. | n.a. | n.a. | n.a. | n.a. | n.a. | n.a. | n.a. | n.a. | n.a. |
| S7_vf_89668 | n.a. | n.a. | Overlap (VC_1259/VC_1261) | n.a. | A | n.a. | n.a. | n.a. | n.a. | n.a. | n.a. | n.a. | n.a. | n.a. | n.a. | n.a. |
| S8_vf_153696 | n.a. | n.a. | Overlap (VC_1259/VC_1261) | n.a. | A | n.a. | n.a. | n.a. | n.a. | n.a. | n.a. | n.a. | n.a. | n.a. | n.a. | n.a. |
| S10_vf_57705 | n.a. | n.a. | Overlap (VC_1259/VC_1563) | n.a. | A | n.a. | n.a. | n.a. | n.a. | n.a. | n.a. | n.a. | n.a. | n.a. | n.a. | n.a. |
| S12_vf_8173 | n.a. | n.a. | Overlap (VC_1259/VC_1563) | n.a. | A | n.a. | n.a. | n.a. | n.a. | n.a. | n.a. | n.a. | n.a. | n.a. | n.a. | n.a. |
| S15_vf_71616 | n.a. | n.a. | Overlap (VC_1259/VC_1563) | n.a. | A | n.a. | n.a. | n.a. | n.a. | n.a. | n.a. | n.a. | n.a. | n.a. | n.a. | n.a. |
| S7_vs_107876 | Clostridium phage HM2 | LT600745 | Overlap (VC_1260/VC_1267) | n.a. | N11 | 7 | Clostridium | Group I | Duplodnaviria | Heunggongvirae | Uroviricota | Caudoviricetes | n.a. | Salasmaviridae | O | O |
| S8_vf_3410 | Clostridium phage HM2 | LT600745 | Overlap (VC_1260/VC_1267) | n.a. | N11 | 7 | Clostridium | Group I | Duplodnaviria | Heunggongvirae | Uroviricota | Caudoviricetes | n.a. | Salasmaviridae | O | O |
| S12_vf_61299 | Clostridium phage HM2 | LT600745 | Overlap (VC_1262/VC_1263) | n.a. | N30 | 4 | Clostridium | Group I | Duplodnaviria | Heunggongvirae | Uroviricota | Caudoviricetes | n.a. | Salasmaviridae | O | O |
| S14_vf_93134 | Clostridium phage HM2 | LT600745 | Overlap (VC_1262/VC_1263) | n.a. | N30 | 4 | Clostridium | Group I | Duplodnaviria | Heunggongvirae | Uroviricota | Caudoviricetes | n.a. | Salasmaviridae | O | O |
| S17_vf_36452 | Clostridium phage HM2 | LT600745 | Overlap (VC_1262/VC_1263) | n.a. | N30 | 4 | Clostridium | Group I | Duplodnaviria | Heunggongvirae | Uroviricota | Caudoviricetes | n.a. | Salasmaviridae | O | O |
| S2_vf_17850 | Clostridium phage HM2 | LT600745 | Overlap (VC_1262/VC_1263) | n.a. | N30 | 4 | Clostridium | Group I | Duplodnaviria | Heunggongvirae | Uroviricota | Caudoviricetes | n.a. | Salasmaviridae | O | O |
| S16_vf_147326 | Clostridium phage HM2 | LT600745 | Overlap (VC_1262/VC_1265) | n.a. | N18 | 8 | Clostridium | Group I | Duplodnaviria | Heunggongvirae | Uroviricota | Caudoviricetes | n.a. | Salasmaviridae | O | O |
| S8_vf_159173 | Clostridium phage HM2 | LT600745 | Overlap (VC_1262/VC_1267) | n.a. | N30 | 4 | Clostridium | Group I | Duplodnaviria | Heunggongvirae | Uroviricota | Caudoviricetes | n.a. | Salasmaviridae | O | O |
| S13_vf_97702 | Clostridium phage HM2 | LT600745 | Overlap (VC_1264/VC_1267) | n.a. | N6 | 7 | Clostridium | Group I | Duplodnaviria | Heunggongvirae | Uroviricota | Caudoviricetes | n.a. | Salasmaviridae | O | O |
| S7_vs_13903 | Clostridium phage HM2 | LT600745 | Overlap (VC_1264/VC_1267) | n.a. | N6 | 7 | Clostridium | Group I | Duplodnaviria | Heunggongvirae | Uroviricota | Caudoviricetes | n.a. | Salasmaviridae | O | O |
| S1_vs_145792 | n.a. | n.a. | Overlap (VC_1291/VC_1578) | n.a. | A | n.a. | n.a. | n.a. | n.a. | n.a. | n.a. | n.a. | n.a. | n.a. | n.a. | n.a. |
| S3_vs_25896 | n.a. | n.a. | Overlap (VC_1291/VC_1578) | n.a. | A | n.a. | n.a. | n.a. | n.a. | n.a. | n.a. | n.a. | n.a. | n.a. | n.a. | n.a. |
| S5_vs_145591 | n.a. | n.a. | Overlap (VC_1291/VC_1578) | n.a. | A | n.a. | n.a. | n.a. | n.a. | n.a. | n.a. | n.a. | n.a. | n.a. | n.a. | n.a. |
| S5_vs_27918 | n.a. | n.a. | Overlap (VC_1291/VC_1578) | n.a. | A | n.a. | n.a. | n.a. | n.a. | n.a. | n.a. | n.a. | n.a. | n.a. | n.a. | n.a. |
| S7_vf_72493 | n.a. | n.a. | Overlap (VC_1291/VC_1578) | n.a. | A | n.a. | n.a. | n.a. | n.a. | n.a. | n.a. | n.a. | n.a. | n.a. | n.a. | n.a. |
| S9_vs_69322 | n.a. | n.a. | Overlap (VC_1291/VC_1578) | n.a. | A | n.a. | n.a. | n.a. | n.a. | n.a. | n.a. | n.a. | n.a. | n.a. | n.a. | n.a. |
| S9_vs_88286 | n.a. | n.a. | Overlap (VC_1291/VC_1578) | n.a. | A | n.a. | n.a. | n.a. | n.a. | n.a. | n.a. | n.a. | n.a. | n.a. | n.a. | n.a. |
| S13_vs_49889 | n.a. | n.a. | Overlap (VC_1292/VC_1574) | n.a. | A | n.a. | n.a. | n.a. | n.a. | n.a. | n.a. | n.a. | n.a. | n.a. | n.a. | n.a. |
| S16_vs_59697 | n.a. | n.a. | Overlap (VC_1292/VC_1574) | n.a. | A | n.a. | n.a. | n.a. | n.a. | n.a. | n.a. | n.a. | n.a. | n.a. | n.a. | n.a. |
| S8_vs_152553 | n.a. | n.a. | Overlap (VC_1292/VC_1574) | n.a. | A | n.a. | n.a. | n.a. | n.a. | n.a. | n.a. | n.a. | n.a. | n.a. | n.a. | n.a. |
| S14_vs_4887 | n.a. | n.a. | Overlap (VC_1292/VC_1575) | n.a. | A | n.a. | n.a. | n.a. | n.a. | n.a. | n.a. | n.a. | n.a. | n.a. | n.a. | n.a. |
| S7_vs_95276 | n.a. | n.a. | Overlap (VC_1292/VC_1575) | n.a. | A | n.a. | n.a. | n.a. | n.a. | n.a. | n.a. | n.a. | n.a. | n.a. | n.a. | n.a. |
| S14_vs_90934 | n.a. | n.a. | Overlap (VC_1292/VC_1610) | n.a. | A | n.a. | n.a. | n.a. | n.a. | n.a. | n.a. | n.a. | n.a. | n.a. | n.a. | n.a. |
| S5_vs_30490 | n.a. | n.a. | Overlap (VC_1292/VC_1610) | n.a. | A | n.a. | n.a. | n.a. | n.a. | n.a. | n.a. | n.a. | n.a. | n.a. | n.a. | n.a. |
| S5_vs_36605 | Faecalibacterium phage FP_Lugh | MG711464 | Overlap (VC_1294/VC_1575) | n.a. | N14 | 3 | Faecalibacterium | Group I | Duplodnaviria | Heunggongvirae | Uroviricota | Caudoviricetes | n.a. | Unclassified | O | O |
| S6_vs_22637 | n.a. | n.a. | Overlap (VC_1294/VC_1575) | n.a. | A | n.a. | n.a. | n.a. | n.a. | n.a. | n.a. | n.a. | n.a. | n.a. | n.a. | n.a. |
| S12_vf_188846 | Tortoise microvirus 19 | MK765569 | Overlap (VC_1300/VC_1301) | n.a. | N4 | 9 | Unspecified | Group II | Monodnaviria | Sangervirae | Phixviricota | Malgrandaviricetes | Petitvirales | Microviridae | O | O |
| S8_vf_208115 | Tortoise microvirus 19 | MK765569 | Overlap (VC_1300/VC_1301) | n.a. | N3 | 9 | Unspecified | Group II | Monodnaviria | Sangervirae | Phixviricota | Malgrandaviricetes | Petitvirales | Microviridae | O | O |
| S14_vf_43340 | Tortoise microvirus 14 | MK765564 | Overlap (VC_1300/VC_1302) | n.a. | N2 | 4 | Unspecified | Group II | Monodnaviria | Sangervirae | Phixviricota | Malgrandaviricetes | Petitvirales | Microviridae | O | O |
| S3_vf_59078 | Tortoise microvirus 14 | MK765564 | Overlap (VC_1300/VC_1302) | n.a. | N2 | 4 | Unspecified | Group II | Monodnaviria | Sangervirae | Phixviricota | Malgrandaviricetes | Petitvirales | Microviridae | O | O |
| S17_vf_128147 | Microvirus mar63 | MZ089809 | Overlap (VC_1335/VC_1337) | n.a. | C1 | 17 | Unspecified | Group II | Monodnaviria | Sangervirae | Phixviricota | Malgrandaviricetes | Petitvirales | Microviridae | Unclassified | O |
| S14_vf_115736 | Butyrivibrio virus Ceridwen | MN882553 | Overlap (VC_1481/VC_1482) | n.a. | C31 | 6 | Butyrivibrio | Group I | Duplodnaviria | Heunggongvirae | Uroviricota | Caudoviricetes | n.a. | Unclassified | Unclassified | O |
| S16_vs_20060 | Butyrivibrio virus Ceridwen | MN882553 | Overlap (VC_1481/VC_1482) | n.a. | C35 | 2 | Butyrivibrio | Group I | Duplodnaviria | Heunggongvirae | Uroviricota | Caudoviricetes | n.a. | Unclassified | Unclassified | O |

| | | | | | | | | | | | | | | | | |
|---|---|---|---|---|---|---|---|---|---|---|---|---|---|---|---|---|
| S1_vf_136526 | Butyrivibrio virus Ceridwen | MN882553 | Overlap (VC_1481/VC_1482) | n.a. | C35 | 3 | Butyrivibrio | Group I | Duplodnaviria | Heunggongvirae | Uroviricota | Caudoviricetes | n.a. | Unclassified | Unclassified | O |
| S1_vf_6272 | Butyrivibrio virus Ceridwen | MN882553 | Overlap (VC_1481/VC_1482) | n.a. | C36 | 6 | Butyrivibrio | Group I | Duplodnaviria | Heunggongvirae | Uroviricota | Caudoviricetes | n.a. | Unclassified | Unclassified | O |
| S1_vs_21439 | Butyrivibrio virus Ceridwen | MN882553 | Overlap (VC_1481/VC_1482) | n.a. | C32 | 2 | Butyrivibrio | Group I | Duplodnaviria | Heunggongvirae | Uroviricota | Caudoviricetes | n.a. | Unclassified | Unclassified | O |
| S3_vf_127400 | Butyrivibrio virus Ceridwen | MN882553 | Overlap (VC_1481/VC_1482) | n.a. | C36 | 2 | Butyrivibrio | Group I | Duplodnaviria | Heunggongvirae | Uroviricota | Caudoviricetes | n.a. | Unclassified | Unclassified | O |
| S8_vs_88013 | Butyrivibrio virus Ceridwen | MN882553 | Overlap (VC_1481/VC_1482) | n.a. | C30 | 7 | Butyrivibrio | Group I | Duplodnaviria | Heunggongvirae | Uroviricota | Caudoviricetes | n.a. | Unclassified | Unclassified | O |
| S14_vs_112068 | n.a. | n.a. | Overlap (VC_1482/VC_1570) | n.a. | A | n.a. | n.a. | n.a. | n.a. | n.a. | n.a. | n.a. | n.a. | n.a. | n.a. | n.a. |
| S6_vs_56492 | n.a. | n.a. | Overlap (VC_1482/VC_1570) | n.a. | A | n.a. | n.a. | n.a. | n.a. | n.a. | n.a. | n.a. | n.a. | n.a. | n.a. | n.a. |
| S7_vs_43529 | n.a. | n.a. | Overlap (VC_1482/VC_1570) | n.a. | A | n.a. | n.a. | n.a. | n.a. | n.a. | n.a. | n.a. | n.a. | n.a. | n.a. | n.a. |
| S17_vf_140653 | Bacteroides phage F4 | MT806185 | Overlap (VC_1506/VC_1507) | n.a. | N12 | 1 | Bacteroides | Group I | Duplodnaviria | Heunggongvirae | Uroviricota | Caudoviricetes | n.a. | Unclassified | O | O |
| S3_vs_9158 | n.a. | n.a. | Overlap (VC_1506/VC_1507) | n.a. | A | n.a. | n.a. | n.a. | n.a. | n.a. | n.a. | n.a. | n.a. | n.a. | n.a. | n.a. |
| S8_vf_28956 | n.a. | n.a. | Overlap (VC_1506/VC_1507) | n.a. | A | n.a. | n.a. | n.a. | n.a. | n.a. | n.a. | n.a. | n.a. | n.a. | n.a. | n.a. |
| S11_vs_45658 | Campylobacter phage PC10 | MZ047271 | Overlap (VC_1530/VC_1560) | n.a. | N8 | 6 | Campylobacter | Group I | Duplodnaviria | Heunggongvirae | Uroviricota | Caudoviricetes | n.a. | Unclassified | O | O |
| S12_vs_111286 | n.a. | n.a. | Overlap (VC_1530/VC_1560) | n.a. | A | n.a. | n.a. | n.a. | n.a. | n.a. | n.a. | n.a. | n.a. | n.a. | n.a. | n.a. |
| S5_vs_80496 | n.a. | n.a. | Overlap (VC_1530/VC_1560) | n.a. | A | n.a. | n.a. | n.a. | n.a. | n.a. | n.a. | n.a. | n.a. | n.a. | n.a. | n.a. |
| S5_vs_117417 | n.a. | n.a. | Overlap (VC_155/VC_1589) | n.a. | A | n.a. | n.a. | n.a. | n.a. | n.a. | n.a. | n.a. | n.a. | n.a. | n.a. | n.a. |
| S7_vs_28150 | n.a. | n.a. | Overlap (VC_155/VC_1589) | n.a. | A | n.a. | n.a. | n.a. | n.a. | n.a. | n.a. | n.a. | n.a. | n.a. | n.a. | n.a. |
| S11_vs_104049 | Clostridium phage phiMMP04 | JX145342 | Overlap (VC_155/VC_661) | n.a. | N7 | 9 | Clostridium | Group I | Duplodnaviria | Heunggongvirae | Uroviricota | Caudoviricetes | n.a. | Unclassified | O | O |
| S14_vs_56410 | Clostridium phage phiMMP04 | JX145342 | Overlap (VC_155/VC_661) | n.a. | N5 | 8 | Clostridium | Group I | Duplodnaviria | Heunggongvirae | Uroviricota | Caudoviricetes | n.a. | Unclassified | O | O |
| S5_vs_69980 | Clostridium phage phiCT453B | KM983328 | Overlap (VC_155/VC_661) | n.a. | N5 | 10 | Clostridium | Group I | Duplodnaviria | Heunggongvirae | Uroviricota | Caudoviricetes | n.a. | Unclassified | O | O |
| S3_vf_7137 | Bacteriophage sp. | OP549880 | Overlap (VC_1550/VC_1551) | n.a. | C3 | 24 | Unspecified | Unclassified | Unclassified | n.a. | n.a. | n.a. | n.a. | Unclassified | Unclassified | O |
| S4_vf_30103 | Bacteriophage sp. | OP549880 | Overlap (VC_1550/VC_1551) | n.a. | C3 | 25 | Unspecified | Unclassified | Unclassified | n.a. | n.a. | n.a. | n.a. | Unclassified | Unclassified | O |
| S10_vf_113489 | n.a. | n.a. | Overlap (VC_1553/VC_1555) | n.a. | A | n.a. | n.a. | n.a. | n.a. | n.a. | n.a. | n.a. | n.a. | n.a. | n.a. | n.a. |
| S16_vf_52513 | n.a. | n.a. | Overlap (VC_1555/VC_1556) | n.a. | A | n.a. | n.a. | n.a. | n.a. | n.a. | n.a. | n.a. | n.a. | n.a. | n.a. | n.a. |
| S8_vs_51368 | n.a. | n.a. | Overlap (VC_1555/VC_1556) | n.a. | A | n.a. | n.a. | n.a. | n.a. | n.a. | n.a. | n.a. | n.a. | n.a. | n.a. | n.a. |
| S16_vs_76823 | n.a. | n.a. | Overlap (VC_1555/VC_1557) | n.a. | A | n.a. | n.a. | n.a. | n.a. | n.a. | n.a. | n.a. | n.a. | n.a. | n.a. | n.a. |
| S8_vs_102608 | n.a. | n.a. | Overlap (VC_1555/VC_1557) | n.a. | A | n.a. | n.a. | n.a. | n.a. | n.a. | n.a. | n.a. | n.a. | n.a. | n.a. | n.a. |
| S11_vs_36748 | n.a. | n.a. | Overlap (VC_1566/VC_1581) | n.a. | A | n.a. | n.a. | n.a. | n.a. | n.a. | n.a. | n.a. | n.a. | n.a. | n.a. | n.a. |
| S17_vs_157209 | n.a. | n.a. | Overlap (VC_1566/VC_1581) | n.a. | A | n.a. | n.a. | n.a. | n.a. | n.a. | n.a. | n.a. | n.a. | n.a. | n.a. | n.a. |
| S3_vs_89485 | n.a. | n.a. | Overlap (VC_1566/VC_1581/VC_1585) | n.a. | A | n.a. | n.a. | n.a. | n.a. | n.a. | n.a. | n.a. | n.a. | n.a. | n.a. | n.a. |
| S8_vs_40014 | n.a. | n.a. | Overlap (VC_1566/VC_1581/VC_1585) | n.a. | A | n.a. | n.a. | n.a. | n.a. | n.a. | n.a. | n.a. | n.a. | n.a. | n.a. | n.a. |
| S5_vs_84574 | n.a. | n.a. | Overlap (VC_1571/VC_1590) | n.a. | A | n.a. | n.a. | n.a. | n.a. | n.a. | n.a. | n.a. | n.a. | n.a. | n.a. | n.a. |
| S9_vs_88847 | n.a. | n.a. | Overlap (VC_1571/VC_1590) | n.a. | A | n.a. | n.a. | n.a. | n.a. | n.a. | n.a. | n.a. | n.a. | n.a. | n.a. | n.a. |
| S13_vf_147589 | n.a. | n.a. | Overlap (VC_1601/VC_1616) | n.a. | F | n.a. | n.a. | n.a. | n.a. | n.a. | n.a. | n.a. | n.a. | n.a. | n.a. | n.a. |
| S16_vf_173133 | n.a. | n.a. | Overlap (VC_1601/VC_1616) | n.a. | F | n.a. | n.a. | n.a. | n.a. | n.a. | n.a. | n.a. | n.a. | n.a. | n.a. | n.a. |
| S5_vs_145633 | n.a. | n.a. | Overlap (VC_1601/VC_1616) | n.a. | F | n.a. | n.a. | n.a. | n.a. | n.a. | n.a. | n.a. | n.a. | n.a. | n.a. | n.a. |
| S13_vf_91189 | n.a. | n.a. | Overlap (VC_1608/VC_1609) | n.a. | F | n.a. | n.a. | n.a. | n.a. | n.a. | n.a. | n.a. | n.a. | n.a. | n.a. | n.a. |
| S4_vf_16485 | n.a. | n.a. | Overlap (VC_1608/VC_1609) | n.a. | F | n.a. | n.a. | n.a. | n.a. | n.a. | n.a. | n.a. | n.a. | n.a. | n.a. | n.a. |
| S5_vs_130294 | n.a. | n.a. | Overlap (VC_1610/VC_1611) | n.a. | A | n.a. | n.a. | n.a. | n.a. | n.a. | n.a. | n.a. | n.a. | n.a. | n.a. | n.a. |
| S7_vs_97737 | n.a. | n.a. | Overlap (VC_1610/VC_1611) | n.a. | A | n.a. | n.a. | n.a. | n.a. | n.a. | n.a. | n.a. | n.a. | n.a. | n.a. | n.a. |
| S9_vf_90856 | Streptococcus phage Javan535 | MK448799 | Overlap (VC_262/VC_427) | n.a. | C1 | 73 | Streptococcus | Group I | Duplodnaviria | Heunggongvirae | Uroviricota | Caudoviricetes | n.a. | Unclassified | Unclassified | O |
| S5_vs_60507 | Streptococcus phage Javan573 | MK448810 | Overlap (VC_265/VC_330) | n.a. | C1 | 43 | Streptococcus | Group I | Duplodnaviria | Heunggongvirae | Uroviricota | Caudoviricetes | n.a. | Unclassified | Unclassified | O |
| S5_vs_6478 | Lactobacillus phage phiadh | AJ131519 | Overlap (VC_271/VC_315) | n.a. | C1 | 86 | Lactobacillus | Group I | Duplodnaviria | Heunggongvirae | Uroviricota | Caudoviricetes | n.a. | Unclassified | Unclassified | O |
| S11_vs_13652 | Streptococcus phage Javan407 | MK448753 | Overlap (VC_326/VC_644) | n.a. | C7 | 15 | Streptococcus | Group I | Duplodnaviria | Heunggongvirae | Uroviricota | Caudoviricetes | n.a. | Unclassified | Unclassified | O |
| S11_vs_3006 | Streptococcus phage Javan407 | MK448753 | Overlap (VC_326/VC_644) | n.a. | C7 | 15 | Streptococcus | Group I | Duplodnaviria | Heunggongvirae | Uroviricota | Caudoviricetes | n.a. | Unclassified | Unclassified | O |
| S5_vs_109085 | Streptococcus phage Javan407 | MK448753 | Overlap (VC_326/VC_644) | n.a. | C7 | 11 | Streptococcus | Group I | Duplodnaviria | Heunggongvirae | Uroviricota | Caudoviricetes | n.a. | Unclassified | Unclassified | O |
| S7_vs_23770 | Streptococcus phage Javan407 | MK448753 | Overlap (VC_326/VC_644) | n.a. | C7 | 16 | Streptococcus | Group I | Duplodnaviria | Heunggongvirae | Uroviricota | Caudoviricetes | n.a. | Unclassified | Unclassified | O |
| S11_vs_38860 | n.a. | n.a. | Overlap (VC_333/VC_645) | n.a. | A | n.a. | n.a. | n.a. | n.a. | n.a. | n.a. | n.a. | n.a. | n.a. | n.a. | n.a. |
| S12_vs_98175 | Clostridium phage phiCD38-2 | HM568888 | Overlap (VC_333/VC_645) | n.a. | N24 | 2 | Clostridium | Group I | Duplodnaviria | Heunggongvirae | Uroviricota | Caudoviricetes | n.a. | Unclassified | O | O |
| S14_vs_117153 | Roseburia phage Jekyll | LR596902 | Overlap (VC_333/VC_645) | n.a. | N37 | 3 | Roseburia | Group I | Duplodnaviria | Heunggongvirae | Uroviricota | Caudoviricetes | n.a. | Unclassified | O | O |
| S16_vs_140709 | n.a. | n.a. | Overlap (VC_333/VC_645) | n.a. | A | n.a. | n.a. | n.a. | n.a. | n.a. | n.a. | n.a. | n.a. | n.a. | n.a. | n.a. |
| S16_vs_172749 | Clostridium phage phiCD38-2 | HM568888 | Overlap (VC_333/VC_645) | n.a. | N34 | 4 | Clostridium | Group I | Duplodnaviria | Heunggongvirae | Uroviricota | Caudoviricetes | n.a. | Unclassified | O | O |
| S4_vf_31682 | Bacillus phage PBC1 | JQ619704 | Overlap (VC_333/VC_645) | n.a. | N32 | 3 | Bacillus | Group I | Duplodnaviria | Heunggongvirae | Uroviricota | Caudoviricetes | n.a. | Unclassified | O | O |
| S4_vs_VIRSorter_31916-circular-cat_1 | Clostridium phage phiCD38-2 | HM568888 | Overlap (VC_333/VC_645) | n.a. | N15 | 6 | Clostridium | Group I | Duplodnaviria | Heunggongvirae | Uroviricota | Caudoviricetes | n.a. | Unclassified | O | O |
| S5_vs_145578 | Faecalibacterium phage FP_Lugh | MG711464 | Overlap (VC_333/VC_645) | n.a. | N9 | 15 | Faecalibacterium | Group I | Duplodnaviria | Heunggongvirae | Uroviricota | Caudoviricetes | n.a. | Unclassified | O | O |
| S7_vs_60775 | Clostridium phage phiCD38-2 | HM568888 | Overlap (VC_333/VC_645) | n.a. | N33 | 3 | Clostridium | Group I | Duplodnaviria | Heunggongvirae | Uroviricota | Caudoviricetes | n.a. | Unclassified | O | O |
| S10_vs_120570 | n.a. | n.a. | Overlap (VC_334/VC_1572) | n.a. | A | n.a. | n.a. | n.a. | n.a. | n.a. | n.a. | n.a. | n.a. | n.a. | n.a. | n.a. |
| S16_vs_107235 | n.a. | n.a. | Overlap (VC_334/VC_1572) | n.a. | A | n.a. | n.a. | n.a. | n.a. | n.a. | n.a. | n.a. | n.a. | n.a. | n.a. | n.a. |
| S5_vs_95307 | Solobacterium phage SMO_1P | MZ503613 | Overlap (VC_334/VC_556) | n.a. | C3 | 26 | Solobacterium | Group I | Duplodnaviria | Heunggongvirae | Uroviricota | Caudoviricetes | n.a. | Unclassified | Unclassified | O |
| S9_vs_48240 | Solobacterium phage SMO_1P | MZ503613 | Overlap (VC_334/VC_556) | n.a. | C3 | 26 | Solobacterium | Group I | Duplodnaviria | Heunggongvirae | Uroviricota | Caudoviricetes | n.a. | Unclassified | Unclassified | O |
| S16_vs_82116 | Solobacterium phage SMO_1P | MZ503613 | Overlap (VC_334/VC_556/VC_1037) | n.a. | C6 | 13 | Solobacterium | Group I | Duplodnaviria | Heunggongvirae | Uroviricota | Caudoviricetes | n.a. | Unclassified | Unclassified | O |
| S4_vf_11308 | Solobacterium phage SMO_1P | MZ503613 | Overlap (VC_334/VC_556/VC_1037) | n.a. | C8 | 15 | Solobacterium | Group I | Duplodnaviria | Heunggongvirae | Uroviricota | Caudoviricetes | n.a. | Unclassified | Unclassified | O |
| S16_vs_132979 | Faecalibacterium phage FP_Lugh | MG711464 | Overlap (VC_335/VC_558/VC_559) | n.a. | N7 | 4 | Faecalibacterium | Group I | Duplodnaviria | Heunggongvirae | Uroviricota | Caudoviricetes | n.a. | Unclassified | O | O |
| S6_vs_53966 | Faecalibacterium phage FP_Lugh | MG711464 | Overlap (VC_335/VC_558/VC_559) | n.a. | N5 | 10 | Faecalibacterium | Group I | Duplodnaviria | Heunggongvirae | Uroviricota | Caudoviricetes | n.a. | Unclassified | O | O |
| S12_vs_58148 | Geobacillus phage GBSV1 | DQ340064 | Overlap (VC_346/VC_779) | n.a. | C3 | 11 | Geobacillus | Group I | Duplodnaviria | Heunggongvirae | Uroviricota | Caudoviricetes | n.a. | Unclassified | Unclassified | O |
| S16_vs_54616 | Geobacillus phage GBSV1 | DQ340064 | Overlap (VC_346/VC_779) | n.a. | C3 | 15 | Geobacillus | Group I | Duplodnaviria | Heunggongvirae | Uroviricota | Caudoviricetes | n.a. | Unclassified | Unclassified | O |
| S11_vs_104069 | Vibrio phage vB_VspS_VS-ABTNL-3 | MF497422 | Overlap (VC_354/VC_1271) | n.a. | C1 | 11 | Vibrio | Group I | Duplodnaviria | Heunggongvirae | Uroviricota | Caudoviricetes | n.a. | Unclassified | Unclassified | O |
| S15_vs_39804 | Bacillus phage vB_BtS_BMBtp13 | KX190832 | Overlap (VC_433/VC_668) | n.a. | N2 | 18 | Bacillus | Group I | Duplodnaviria | Heunggongvirae | Uroviricota | Caudoviricetes | n.a. | Unclassified | O | O |
| S3_vs_20315 | Bacillus phage vB_BtS_BMBtp13 | KX190832 | Overlap (VC_433/VC_668) | n.a. | N2 | 21 | Bacillus | Group I | Duplodnaviria | Heunggongvirae | Uroviricota | Caudoviricetes | n.a. | Unclassified | O | O |
| S2_vf_4130 | uncultured phage cr6_1 | MT774401 | Overlap (VC_520/VC_522) | n.a. | C3 | 282 | Unspecified | Group I | Duplodnaviria | Heunggongvirae | Uroviricota | Caudoviricetes | Crassvirales | Suoliviridae | Bearivirinae | O |
| S4_vf_31825 | uncultured phage cr6_1 | MT774401 | Overlap (VC_520/VC_522) | n.a. | C5 | 216 | Unspecified | Group I | Duplodnaviria | Heunggongvirae | Uroviricota | Caudoviricetes | Crassvirales | Suoliviridae | Bearivirinae | O |
| S8_vf_49282 | uncultured phage cr6_1 | MT774401 | Overlap (VC_520/VC_522) | n.a. | C5 | 188 | Unspecified | Group I | Duplodnaviria | Heunggongvirae | Uroviricota | Caudoviricetes | Crassvirales | Suoliviridae | Bearivirinae | O |
| S10_vs_46819 | Faecalibacterium phage FP_Lugh | MG711464 | Overlap (VC_558/VC_559) | n.a. | N12 | 10 | Faecalibacterium | Group I | Duplodnaviria | Heunggongvirae | Uroviricota | Caudoviricetes | n.a. | Unclassified | O | O |
| S4_vs_VIRSorter_27316-cat_2 | Faecalibacterium phage FP_Lugh | MG711464 | Overlap (VC_558/VC_559) | n.a. | N2 | 46 | Faecalibacterium | Group I | Duplodnaviria | Heunggongvirae | Uroviricota | Caudoviricetes | n.a. | Unclassified | O | O |
| S6_vs_1146 | Streptococcus phage Javan240 | MK448881 | Overlap (VC_558/VC_559) | n.a. | N13 | 5 | Streptococcus | Group I | Duplodnaviria | Heunggongvirae | Uroviricota | Caudoviricetes | n.a. | Unclassified | O | O |
| S14_vf_85122 | Clostridium phage vB_CpeS-1181 | MW478292 | Overlap (VC_563/VC_842/VC_924) | n.a. | C4 | 25 | Clostridium | Group I | Duplodnaviria | Heunggongvirae | Uroviricota | Caudoviricetes | n.a. | Unclassified | Unclassified | O |
| S5_vf_145583 | Clostridium phage vB_CpeS-1181 | MW478292 | Overlap (VC_563/VC_842/VC_924) | n.a. | C4 | 7 | Clostridium | Group I | Duplodnaviria | Heunggongvirae | Uroviricota | Caudoviricetes | n.a. | Unclassified | Unclassified | O |
| S10_vf_46328 | Roseburia phage Jekyll | LR596902 | Overlap (VC_627/VC_1564) | n.a. | N19 | 7 | Roseburia | Group I | Duplodnaviria | Heunggongvirae | Uroviricota | Caudoviricetes | n.a. | Unclassified | O | O |
| S11_vf_34008 | Roseburia phage Jekyll | LR596902 | Overlap (VC_627/VC_1564) | n.a. | N18 | 3 | Roseburia | Group I | Duplodnaviria | Heunggongvirae | Uroviricota | Caudoviricetes | n.a. | Unclassified | O | O |
| S11_vs_82368 | Roseburia phage Jekyll | LR596902 | Overlap (VC_627/VC_1564) | n.a. | N15 | 10 | Roseburia | Group I | Duplodnaviria | Heunggongvirae | Uroviricota | Caudoviricetes | n.a. | Unclassified | O | O |
| S3_vf_11428 | Roseburia phage Jekyll | LR596902 | Overlap (VC_627/VC_1564) | n.a. | N13 | 17 | Roseburia | Group I | Duplodnaviria | Heunggongvirae | Uroviricota | Caudoviricetes | n.a. | Unclassified | O | O |
| S4_vf_13353 | Roseburia phage Jekyll | LR596902 | Overlap (VC_627/VC_1564) | n.a. | N12 | 23 | Roseburia | Group I | Duplodnaviria | Heunggongvirae | Uroviricota | Caudoviricetes | n.a. | Unclassified | O | O |
| S5_vf_105728 | Roseburia phage Jekyll | LR596902 | Overlap (VC_627/VC_1564) | n.a. | N16 | 7 | Roseburia | Group I | Duplodnaviria | Heunggongvirae | Uroviricota | Caudoviricetes | n.a. | Unclassified | O | O |
| S5_vf_145604 | Roseburia phage Jekyll | LR596902 | Overlap (VC_627/VC_1564) | n.a. | N14 | 18 | Roseburia | Group I | Duplodnaviria | Heunggongvirae | Uroviricota | Caudoviricetes | n.a. | Unclassified | O | O |
| S5_vs_68254 | Roseburia phage Jekyll | LR596902 | Overlap (VC_627/VC_1564) | n.a. | N18 | 7 | Roseburia | Group I | Duplodnaviria | Heunggongvirae | Uroviricota | Caudoviricetes | n.a. | Unclassified | O | O |
| S7_vf_49485 | Roseburia phage Jekyll | LR596902 | Overlap (VC_627/VC_1564) | n.a. | N13 | 7 | Roseburia | Group I | Duplodnaviria | Heunggongvirae | Uroviricota | Caudoviricetes | n.a. | Unclassified | O | O |
| S7_vs_65719 | Roseburia phage Jekyll | LR596902 | Overlap (VC_627/VC_1564) | n.a. | N27 | 2 | Roseburia | Group I | Duplodnaviria | Heunggongvirae | Uroviricota | Caudoviricetes | n.a. | Unclassified | O | O |
| S9_vs_90848 | Roseburia phage Jekyll | LR596902 | Overlap (VC_627/VC_1564) | n.a. | N16 | 9 | Roseburia | Group I | Duplodnaviria | Heunggongvirae | Uroviricota | Caudoviricetes | n.a. | Unclassified | O | O |

| | | | | | | | | | | | | | | | | |
|---|---|---|---|---|---|---|---|---|---|---|---|---|---|---|---|---|
| S11_vs_83666 | Faecalibacterium phage FP_Lugh | MG711464 | Overlap (VC_641/VC_1595) | n.a. | N5 | 20 | Faecalibacterium | Group I | Duplodnaviria | Heunggongvirae | Uroviricota | Caudoviricetes | n.a. | Unclassified | O | O |
| S14_vs_56251 | Clostridium phage phiCD38-2 | HM568888 | Overlap (VC_641/VC_1595) | n.a. | N13 | 4 | Clostridium | Group I | Duplodnaviria | Heunggongvirae | Uroviricota | Caudoviricetes | n.a. | Unclassified | O | O |
| S4_vs_VIRSorter_21643-cat_2 | Faecalibacterium phage FP_Lugh | MG711464 | Overlap (VC_641/VC_1595) | n.a. | N3 | 21 | Faecalibacterium | Group I | Duplodnaviria | Heunggongvirae | Uroviricota | Caudoviricetes | n.a. | Unclassified | O | O |
| S6_vs_56485 | Faecalibacterium phage FP_Lugh | MG711464 | Overlap (VC_641/VC_925) | n.a. | N18 | 3 | Faecalibacterium | Group I | Duplodnaviria | Heunggongvirae | Uroviricota | Caudoviricetes | n.a. | Unclassified | O | O |
| S9_vs_37437 | Clostridium phage phiCD111 | LN681535 | Overlap (VC_641/VC_925) | n.a. | N12 | 5 | Clostridium | Group I | Duplodnaviria | Heunggongvirae | Uroviricota | Caudoviricetes | n.a. | Unclassified | O | O |
| S14_vs_112252 | n.a. | n.a. | Overlap (VC_660/VC_734) | n.a. | A | n.a. | n.a. | n.a. | n.a. | n.a. | n.a. | n.a. | n.a. | n.a. | n.a. | n.a. |
| S5_vs_49011 | Clostridium phage phiCT19406B | KM983331 | Overlap (VC_660/VC_734) | n.a. | N9 | 5 | Clostridium | Group I | Duplodnaviria | Heunggongvirae | Uroviricota | Caudoviricetes | n.a. | Unclassified | O | O |
| S3_vf_8164 | Bacillus phage vB_BtS_BMBtp13 | KX190832 | Overlap (VC_668/VC_907) | n.a. | N12 | 7 | Bacillus | Group I | Duplodnaviria | Heunggongvirae | Uroviricota | Caudoviricetes | n.a. | Unclassified | O | O |
| S7_vf_8332 | Bacillus phage vB_BtS_BMBtp13 | KX190832 | Overlap (VC_668/VC_907) | n.a. | N9 | 8 | Bacillus | Group I | Duplodnaviria | Heunggongvirae | Uroviricota | Caudoviricetes | n.a. | Unclassified | O | O |
| S13_vf_3728 | Bacteroides phage crAss002 | MN917146 | Overlap (VC_692/VC_693) | n.a. | C1 | 233 | Bacteroides | Group I | Duplodnaviria | Heunggongvirae | Uroviricota | Caudoviricetes | Crassvirales | Intestiviridae | Churivirinae | O |
| S16_vf_173095 | Bacteroides phage crAss002 | MN917146 | Overlap (VC_692/VC_693) | n.a. | C1 | 184 | Bacteroides | Group I | Duplodnaviria | Heunggongvirae | Uroviricota | Caudoviricetes | Crassvirales | Intestiviridae | Churivirinae | O |
| S12_vf_173666 | n.a. | n.a. | Overlap (VC_725/VC_1221) | n.a. | A | n.a. | n.a. | n.a. | n.a. | n.a. | n.a. | n.a. | n.a. | n.a. | n.a. | n.a. |
| S7_vf_67910 | n.a. | n.a. | Overlap (VC_725/VC_1221) | n.a. | A | n.a. | n.a. | n.a. | n.a. | n.a. | n.a. | n.a. | n.a. | n.a. | n.a. | n.a. |
| S8_vf_195032 | Roseburia phage Shimadzu | LR596903 | Overlap (VC_725/VC_1221) | n.a. | N4 | 1 | Roseburia | Group I | Duplodnaviria | Heunggongvirae | Uroviricota | Caudoviricetes | n.a. | Unclassified | O | O |
| S11_vf_33117 | n.a. | n.a. | Overlap (VC_730/VC_1293) | n.a. | A | n.a. | n.a. | n.a. | n.a. | n.a. | n.a. | n.a. | n.a. | n.a. | n.a. | n.a. |
| S8_vs_208127 | Faecalibacterium phage FP_Lagaffe | MG711461 | Overlap (VC_730/VC_1293) | n.a. | N16 | 2 | Faecalibacterium | Group I | Duplodnaviria | Heunggongvirae | Uroviricota | Caudoviricetes | n.a. | Unclassified | O | O |
| S7_vs_135322 | Faecalibacterium phage FP_Taranis | MG711467 | Overlap (VC_730/VC_733) | n.a. | C1 | 222 | Faecalibacterium | Group I | Duplodnaviria | Heunggongvirae | Uroviricota | Caudoviricetes | n.a. | Unclassified | Unclassified | O |
| S3_vs_90331 | Faecalibacterium phage FP_Taranis | MG711467 | Overlap (VC_730/VC_733/VC_1293) | n.a. | C4 | 84 | Faecalibacterium | Group I | Duplodnaviria | Heunggongvirae | Uroviricota | Caudoviricetes | n.a. | Unclassified | Unclassified | O |
| S8_vs_179529 | Faecalibacterium phage FP_Taranis | MG711467 | Overlap (VC_730/VC_733/VC_1293) | n.a. | C6 | 73 | Faecalibacterium | Group I | Duplodnaviria | Heunggongvirae | Uroviricota | Caudoviricetes | n.a. | Unclassified | Unclassified | O |
| S3_vs_30318 | n.a. | n.a. | Overlap (VC_734/VC_1576) | n.a. | A | n.a. | n.a. | n.a. | n.a. | n.a. | n.a. | n.a. | n.a. | n.a. | n.a. | n.a. |
| S5_vf_145586 | n.a. | n.a. | Overlap (VC_734/VC_1576) | n.a. | A | n.a. | n.a. | n.a. | n.a. | n.a. | n.a. | n.a. | n.a. | n.a. | n.a. | n.a. |
| S11_vs_104042 | n.a. | n.a. | Overlap (VC_760/VC_888) | n.a. | A | n.a. | n.a. | n.a. | n.a. | n.a. | n.a. | n.a. | n.a. | n.a. | n.a. | n.a. |
| S16_vs_101072 | Stenotrophomonas phage vB_SmeS_BUCT702 | OM735687 | Overlap (VC_760/VC_888) | n.a. | N5 | 4 | Stenotrophomonas | Group I | Duplodnaviria | Heunggongvirae | Uroviricota | Caudoviricetes | n.a. | Unclassified | O | O |
| S9_vs_90850 | Stenotrophomonas phage vB_SmeS_BUCT702 | OM735687 | Overlap (VC_760/VC_888) | n.a. | N7 | 3 | Stenotrophomonas | Group I | Duplodnaviria | Heunggongvirae | Uroviricota | Caudoviricetes | n.a. | Unclassified | O | O |
| S13_vs_149860 | Bifidobacterium phage BadAztec1 | MT006233 | Overlap (VC_762/VC_763) | n.a. | C1 | 26 | Bifidobacterium | Group I | Duplodnaviria | Heunggongvirae | Uroviricota | Caudoviricetes | n.a. | Unclassified | Unclassified | O |
| S2_vf_16829 | Actinomyces phage Av-1 | DQ123818 | Overlap (VC_762/VC_763) | n.a. | C5 | 15 | Actinomyces | Group I | Duplodnaviria | Heunggongvirae | Uroviricota | Caudoviricetes | n.a. | Unclassified | Unclassified | O |
| S3_vf_33389 | Actinomyces phage Av-1 | DQ123818 | Overlap (VC_762/VC_763) | n.a. | C5 | 15 | Actinomyces | Group I | Duplodnaviria | Heunggongvirae | Uroviricota | Caudoviricetes | n.a. | Unclassified | Unclassified | O |
| S8_vf_91033 | Actinomyces phage Av-1 | DQ123818 | Overlap (VC_762/VC_763) | n.a. | C5 | 15 | Actinomyces | Group I | Duplodnaviria | Heunggongvirae | Uroviricota | Caudoviricetes | n.a. | Unclassified | Unclassified | O |
| S14_vs_30439 | Geobacillus phage GBSV1 | DQ340064 | Overlap (VC_779/VC_781) | n.a. | C18 | 6 | Geobacillus | Group I | Duplodnaviria | Heunggongvirae | Uroviricota | Caudoviricetes | n.a. | Unclassified | Unclassified | O |
| S4_vs_VIRSorter_16803-cat_2 | Geobacillus phage GBSV1 | DQ340064 | Overlap (VC_779/VC_781) | n.a. | C11 | 6 | Geobacillus | Group I | Duplodnaviria | Heunggongvirae | Uroviricota | Caudoviricetes | n.a. | Unclassified | Unclassified | O |
| S6_vs_46091 | Geobacillus phage GBSV1 | DQ340064 | Overlap (VC_779/VC_782) | n.a. | C7 | 9 | Geobacillus | Group I | Duplodnaviria | Heunggongvirae | Uroviricota | Caudoviricetes | n.a. | Unclassified | Unclassified | O |
| S9_vs_48347 | Geobacillus phage GBSV1 | DQ340064 | Overlap (VC_779/VC_782) | n.a. | C13 | 6 | Geobacillus | Group I | Duplodnaviria | Heunggongvirae | Uroviricota | Caudoviricetes | n.a. | Unclassified | Unclassified | O |
| S14_vf_117155 | Bifidobacterium phage BitterVaud1 | MT006236 | Overlap (VC_827/VC_828) | n.a. | N2 | 23 | Bifidobacterium | Group I | Duplodnaviria | Heunggongvirae | Uroviricota | Caudoviricetes | n.a. | Unclassified | O | O |
| S16_vf_16182 | Bifidobacterium phage BitterVaud1 | MT006236 | Overlap (VC_827/VC_828) | n.a. | N4 | 15 | Bifidobacterium | Group I | Duplodnaviria | Heunggongvirae | Uroviricota | Caudoviricetes | n.a. | Unclassified | O | O |
| S7_vf_37293 | Bifidobacterium phage BitterVaud1 | MT006236 | Overlap (VC_827/VC_829) | n.a. | N3 | 18 | Bifidobacterium | Group I | Duplodnaviria | Heunggongvirae | Uroviricota | Caudoviricetes | n.a. | Unclassified | O | O |
| S7_vf_49007 | Bifidobacterium phage BitterVaud1 | MT006236 | Overlap (VC_827/VC_829) | n.a. | N11 | 8 | Bifidobacterium | Group I | Duplodnaviria | Heunggongvirae | Uroviricota | Caudoviricetes | n.a. | Unclassified | O | O |
| S12_vs_167155 | Butyrivibrio virus Ceridwen | MN882553 | Overlap (VC_841/VC_1435/VC_1481) | n.a. | C22 | 17 | Butyrivibrio | Group I | Duplodnaviria | Heunggongvirae | Uroviricota | Caudoviricetes | n.a. | Unclassified | Unclassified | O |
| S4_vf_31979 | Butyrivibrio virus Ceridwen | MN882553 | Overlap (VC_841/VC_1435/VC_1481) | n.a. | C12 | 20 | Butyrivibrio | Group I | Duplodnaviria | Heunggongvirae | Uroviricota | Caudoviricetes | n.a. | Unclassified | Unclassified | O |
| S10_vs_20400 | Butyrivibrio virus Ceridwen | MN882553 | Overlap (VC_841/VC_1481) | n.a. | C23 | 18 | Butyrivibrio | Group I | Duplodnaviria | Heunggongvirae | Uroviricota | Caudoviricetes | n.a. | Unclassified | Unclassified | O |
| S11_vf_104034 | Butyrivibrio virus Ceridwen | MN882553 | Overlap (VC_841/VC_1481) | n.a. | C20 | 34 | Butyrivibrio | Group I | Duplodnaviria | Heunggongvirae | Uroviricota | Caudoviricetes | n.a. | Unclassified | Unclassified | O |
| S16_vf_12551 | Butyrivibrio virus Ceridwen | MN882553 | Overlap (VC_841/VC_1481) | n.a. | C30 | 29 | Butyrivibrio | Group I | Duplodnaviria | Heunggongvirae | Uroviricota | Caudoviricetes | n.a. | Unclassified | Unclassified | O |
| S16_vs_78429 | Butyrivibrio virus Ceridwen | MN882553 | Overlap (VC_841/VC_1481) | n.a. | C20 | 38 | Butyrivibrio | Group I | Duplodnaviria | Heunggongvirae | Uroviricota | Caudoviricetes | n.a. | Unclassified | Unclassified | O |
| S1_vs_117317 | Butyrivibrio virus Ceridwen | MN882553 | Overlap (VC_841/VC_1481) | n.a. | C25 | 34 | Butyrivibrio | Group I | Duplodnaviria | Heunggongvirae | Uroviricota | Caudoviricetes | n.a. | Unclassified | Unclassified | O |
| S1_vs_155183 | Butyrivibrio virus Ceridwen | MN882553 | Overlap (VC_841/VC_1481) | n.a. | C24 | 31 | Butyrivibrio | Group I | Duplodnaviria | Heunggongvirae | Uroviricota | Caudoviricetes | n.a. | Unclassified | Unclassified | O |
| S4_vs_VIRSorter_29332-cat_2 | Butyrivibrio virus Ceridwen | MN882553 | Overlap (VC_841/VC_1481) | n.a. | C27 | 19 | Butyrivibrio | Group I | Duplodnaviria | Heunggongvirae | Uroviricota | Caudoviricetes | n.a. | Unclassified | Unclassified | O |
| S5_vf_21869 | Butyrivibrio virus Ceridwen | MN882553 | Overlap (VC_841/VC_1481) | n.a. | C26 | 28 | Butyrivibrio | Group I | Duplodnaviria | Heunggongvirae | Uroviricota | Caudoviricetes | n.a. | Unclassified | Unclassified | O |
| S7_vs_4114 | Butyrivibrio virus Ceridwen | MN882553 | Overlap (VC_841/VC_1481) | n.a. | C28 | 14 | Butyrivibrio | Group I | Duplodnaviria | Heunggongvirae | Uroviricota | Caudoviricetes | n.a. | Unclassified | Unclassified | O |
| S7_vs_83474 | Butyrivibrio virus Ceridwen | MN882553 | Overlap (VC_841/VC_1481) | n.a. | C26 | 21 | Butyrivibrio | Group I | Duplodnaviria | Heunggongvirae | Uroviricota | Caudoviricetes | n.a. | Unclassified | Unclassified | O |
| S7_vs_94738 | Butyrivibrio virus Ceridwen | MN882553 | Overlap (VC_841/VC_1481) | n.a. | C28 | 31 | Butyrivibrio | Group I | Duplodnaviria | Heunggongvirae | Uroviricota | Caudoviricetes | n.a. | Unclassified | Unclassified | O |
| S8_vf_72329 | Butyrivibrio virus Ceridwen | MN882553 | Overlap (VC_841/VC_1481) | n.a. | C26 | 25 | Butyrivibrio | Group I | Duplodnaviria | Heunggongvirae | Uroviricota | Caudoviricetes | n.a. | Unclassified | Unclassified | O |
| S8_vs_139391 | Butyrivibrio virus Ceridwen | MN882553 | Overlap (VC_841/VC_1481) | n.a. | C25 | 32 | Butyrivibrio | Group I | Duplodnaviria | Heunggongvirae | Uroviricota | Caudoviricetes | n.a. | Unclassified | Unclassified | O |
| S8_vs_35308 | Butyrivibrio virus Ceridwen | MN882553 | Overlap (VC_841/VC_1481) | n.a. | C23 | 37 | Butyrivibrio | Group I | Duplodnaviria | Heunggongvirae | Uroviricota | Caudoviricetes | n.a. | Unclassified | Unclassified | O |
| S8_vs_54183 | Butyrivibrio virus Ceridwen | MN882553 | Overlap (VC_841/VC_1481) | n.a. | C27 | 28 | Butyrivibrio | Group I | Duplodnaviria | Heunggongvirae | Uroviricota | Caudoviricetes | n.a. | Unclassified | Unclassified | O |
| S10_vs_119298 | Croceibacter phage P2559Y | KC688701 | Overlap (VC_856/VC_857) | n.a. | N4 | 46 | Croceibacter | Group I | Duplodnaviria | Heunggongvirae | Uroviricota | Caudoviricetes | n.a. | Unclassified | O | O |
| S11_vs_104036 | Croceibacter phage P2559Y | KC688701 | Overlap (VC_856/VC_857) | n.a. | N4 | 46 | Croceibacter | Group I | Duplodnaviria | Heunggongvirae | Uroviricota | Caudoviricetes | n.a. | Unclassified | O | O |
| S16_vs_11649 | Croceibacter phage P2559Y | KC688701 | Overlap (VC_856/VC_857) | n.a. | N4 | 44 | Croceibacter | Group I | Duplodnaviria | Heunggongvirae | Uroviricota | Caudoviricetes | n.a. | Unclassified | O | O |
| S6_vs_15390 | Croceibacter phage P2559Y | KC688701 | Overlap (VC_856/VC_857) | n.a. | N4 | 41 | Croceibacter | Group I | Duplodnaviria | Heunggongvirae | Uroviricota | Caudoviricetes | n.a. | Unclassified | O | O |
| S3_vs_50557 | Bifidobacterium phage BigBern1 | MT006237 | Overlap (VC_880/VC_1487) | n.a. | N5 | 5 | Bifidobacterium | Group I | Duplodnaviria | Heunggongvirae | Uroviricota | Caudoviricetes | n.a. | Unclassified | O | O |
| S5_vf_29332 | Bifidobacterium phage BigBern1 | MT006237 | Overlap (VC_880/VC_1487) | n.a. | N4 | 8 | Bifidobacterium | Group I | Duplodnaviria | Heunggongvirae | Uroviricota | Caudoviricetes | n.a. | Unclassified | O | O |
| S7_vs_55526 | n.a. | n.a. | Overlap (VC_880/VC_1487) | n.a. | A | n.a. | n.a. | n.a. | n.a. | n.a. | n.a. | n.a. | n.a. | n.a. | n.a. | n.a. |
| S16_vs_17493 | n.a. | n.a. | Overlap (VC_921/VC_1292) | n.a. | A | n.a. | n.a. | n.a. | n.a. | n.a. | n.a. | n.a. | n.a. | n.a. | n.a. | n.a. |
| S14_vs_38905 | n.a. | n.a. | Overlap (VC_923/VC_1582) | n.a. | A | n.a. | n.a. | n.a. | n.a. | n.a. | n.a. | n.a. | n.a. | n.a. | n.a. | n.a. |
| S5_vs_13254 | n.a. | n.a. | Overlap (VC_923/VC_1582) | n.a. | A | n.a. | n.a. | n.a. | n.a. | n.a. | n.a. | n.a. | n.a. | n.a. | n.a. | n.a. |
| S14_vs_39266 | Clostridium phage vB_CpeS-1181 | MW478292 | Overlap (VC_923/VC_924) | n.a. | C13 | 2 | Clostridium | Group I | Duplodnaviria | Heunggongvirae | Uroviricota | Caudoviricetes | n.a. | Unclassified | Unclassified | O |
| S12_vf_16374 | Phage vB_RanS_PJN03 | ON245412 | Overlap (VC_952/VC_1529) | n.a. | N15 | 7 | Unspecified | Group I | Duplodnaviria | Heunggongvirae | Uroviricota | Caudoviricetes | n.a. | Unclassified | O | O |
| S15_vs_3868 | Phage vB_RanS_PJN03 | ON245412 | Overlap (VC_952/VC_1529) | n.a. | N17 | 4 | Unspecified | Group I | Duplodnaviria | Heunggongvirae | Uroviricota | Caudoviricetes | n.a. | Unclassified | O | O |
| S15_vs_43913 | Phage vB_RanS_PJN03 | ON245412 | Overlap (VC_952/VC_1529) | n.a. | N14 | 3 | Unspecified | Group I | Duplodnaviria | Heunggongvirae | Uroviricota | Caudoviricetes | n.a. | Unclassified | O | O |
| S11_vf_104046 | n.a. | n.a. | Singleton | n.a. | G | n.a. | n.a. | n.a. | n.a. | n.a. | n.a. | n.a. | n.a. | n.a. | n.a. | n.a. |
| S11_vf_89309 | n.a. | n.a. | Singleton | n.a. | G | n.a. | n.a. | n.a. | n.a. | n.a. | n.a. | n.a. | n.a. | n.a. | n.a. | n.a. |
| S11_vs_104037 | n.a. | n.a. | Singleton | n.a. | G | n.a. | n.a. | n.a. | n.a. | n.a. | n.a. | n.a. | n.a. | n.a. | n.a. | n.a. |
| S12_vf_187941 | n.a. | n.a. | Singleton | n.a. | G | n.a. | n.a. | n.a. | n.a. | n.a. | n.a. | n.a. | n.a. | n.a. | n.a. | n.a. |
| S12_vf_188827 | n.a. | n.a. | Singleton | n.a. | G | n.a. | n.a. | n.a. | n.a. | n.a. | n.a. | n.a. | n.a. | n.a. | n.a. | n.a. |
| S12_vs_164056 | n.a. | n.a. | Singleton | n.a. | G | n.a. | n.a. | n.a. | n.a. | n.a. | n.a. | n.a. | n.a. | n.a. | n.a. | n.a. |
| S12_vs_70241 | n.a. | n.a. | Singleton | n.a. | G | n.a. | n.a. | n.a. | n.a. | n.a. | n.a. | n.a. | n.a. | n.a. | n.a. | n.a. |
| S14_vs_97835 | n.a. | n.a. | Singleton | n.a. | G | n.a. | n.a. | n.a. | n.a. | n.a. | n.a. | n.a. | n.a. | n.a. | n.a. | n.a. |
| S16_vs_120605 | n.a. | n.a. | Singleton | n.a. | G | n.a. | n.a. | n.a. | n.a. | n.a. | n.a. | n.a. | n.a. | n.a. | n.a. | n.a. |
| S16_vs_140468 | n.a. | n.a. | Singleton | n.a. | G | n.a. | n.a. | n.a. | n.a. | n.a. | n.a. | n.a. | n.a. | n.a. | n.a. | n.a. |
| S16_vs_173141 | n.a. | n.a. | Singleton | n.a. | G | n.a. | n.a. | n.a. | n.a. | n.a. | n.a. | n.a. | n.a. | n.a. | n.a. | n.a. |
| S17_vf_21718 | n.a. | n.a. | Singleton | n.a. | G | n.a. | n.a. | n.a. | n.a. | n.a. | n.a. | n.a. | n.a. | n.a. | n.a. | n.a. |
| S17_vs_133231 | n.a. | n.a. | Singleton | n.a. | G | n.a. | n.a. | n.a. | n.a. | n.a. | n.a. | n.a. | n.a. | n.a. | n.a. | n.a. |
| S17_vs_8690 | n.a. | n.a. | Singleton | n.a. | G | n.a. | n.a. | n.a. | n.a. | n.a. | n.a. | n.a. | n.a. | n.a. | n.a. | n.a. |
| S3_vf_127450 | n.a. | n.a. | Singleton | n.a. | G | n.a. | n.a. | n.a. | n.a. | n.a. | n.a. | n.a. | n.a. | n.a. | n.a. | n.a. |
| S3_vf_22968 | n.a. | n.a. | Singleton | n.a. | G | n.a. | n.a. | n.a. | n.a. | n.a. | n.a. | n.a. | n.a. | n.a. | n.a. | n.a. |
| S3_vf_87896 | n.a. | n.a. | Singleton | n.a. | G | n.a. | n.a. | n.a. | n.a. | n.a. | n.a. | n.a. | n.a. | n.a. | n.a. | n.a. |
| S3_vs_127411 | n.a. | n.a. | Singleton | n.a. | G | n.a. | n.a. | n.a. | n.a. | n.a. | n.a. | n.a. | n.a. | n.a. | n.a. | n.a. |
| S3_vs_39940 | n.a. | n.a. | Singleton | n.a. | G | n.a. | n.a. | n.a. | n.a. | n.a. | n.a. | n.a. | n.a. | n.a. | n.a. | n.a. |
| S3_vs_87728 | n.a. | n.a. | Singleton | n.a. | G | n.a. | n.a. | n.a. | n.a. | n.a. | n.a. | n.a. | n.a. | n.a. | n.a. | n.a. |

| | | | | | | | | | | | | | | | | |
|---|---|---|---|---|---|---|---|---|---|---|---|---|---|---|---|---|
| S4_vf_17948 | n.a. | n.a. | Singleton | n.a. | G | n.a. | n.a. | n.a. | n.a. | n.a. | n.a. | n.a. | n.a. | n.a. | n.a. | n.a. |
| S5_vs_21778 | n.a. | n.a. | Singleton | n.a. | G | n.a. | n.a. | n.a. | n.a. | n.a. | n.a. | n.a. | n.a. | n.a. | n.a. | n.a. |
| S5_vs_41663 | n.a. | n.a. | Singleton | n.a. | G | n.a. | n.a. | n.a. | n.a. | n.a. | n.a. | n.a. | n.a. | n.a. | n.a. | n.a. |
| S5_vs_65490 | n.a. | n.a. | Singleton | n.a. | G | n.a. | n.a. | n.a. | n.a. | n.a. | n.a. | n.a. | n.a. | n.a. | n.a. | n.a. |
| S7_vf_11744 | n.a. | n.a. | Singleton | n.a. | G | n.a. | n.a. | n.a. | n.a. | n.a. | n.a. | n.a. | n.a. | n.a. | n.a. | n.a. |
| S7_vs_13370 | n.a. | n.a. | Singleton | n.a. | G | n.a. | n.a. | n.a. | n.a. | n.a. | n.a. | n.a. | n.a. | n.a. | n.a. | n.a. |
| S7_vs_138116 | n.a. | n.a. | Singleton | n.a. | G | n.a. | n.a. | n.a. | n.a. | n.a. | n.a. | n.a. | n.a. | n.a. | n.a. | n.a. |
| S7_vs_26757 | n.a. | n.a. | Singleton | n.a. | G | n.a. | n.a. | n.a. | n.a. | n.a. | n.a. | n.a. | n.a. | n.a. | n.a. | n.a. |
| S7_vs_30665 | n.a. | n.a. | Singleton | n.a. | G | n.a. | n.a. | n.a. | n.a. | n.a. | n.a. | n.a. | n.a. | n.a. | n.a. | n.a. |
| S8_vf_102508 | n.a. | n.a. | Singleton | n.a. | G | n.a. | n.a. | n.a. | n.a. | n.a. | n.a. | n.a. | n.a. | n.a. | n.a. | n.a. |
| S8_vf_208111 | n.a. | n.a. | Singleton | n.a. | G | n.a. | n.a. | n.a. | n.a. | n.a. | n.a. | n.a. | n.a. | n.a. | n.a. | n.a. |
| S8_vf_7243 | n.a. | n.a. | Singleton | n.a. | G | n.a. | n.a. | n.a. | n.a. | n.a. | n.a. | n.a. | n.a. | n.a. | n.a. | n.a. |
| S8_vs_196247 | n.a. | n.a. | Singleton | n.a. | G | n.a. | n.a. | n.a. | n.a. | n.a. | n.a. | n.a. | n.a. | n.a. | n.a. | n.a. |
| S9_vs_42609 | n.a. | n.a. | Singleton | n.a. | G | n.a. | n.a. | n.a. | n.a. | n.a. | n.a. | n.a. | n.a. | n.a. | n.a. | n.a. |

| WMS_Scaffold | Closer | Accession | Status | VC | Level | Weight | Host | BaltimoreGroup | Realm | Kingdom | Phylum | Class | Order | Family | Subfamily | Genus |
|---|---|---|---|---|---|---|---|---|---|---|---|---|---|---|---|---|
| S1-wms-combined_107073 | Burkholderia phage BcepMu | AY539836 | Clustered | VC_610_0 | N4 | 5 | Burkholderia | Group I | Duplodnaviria | Heunggongvirae | Uroviricota | Caudoviricetes | n.a. | Unclassified | O | O |
| S1-wms-combined_118167 | Clostridium phage phiMMP02 | JX145341 | Clustered | VC_825_0 | N11 | 2 | Clostridium | Group I | Duplodnaviria | Heunggongvirae | Uroviricota | Caudoviricetes | n.a. | Unclassified | O | O |
| S1-wms-combined_120533 | n.a. | n.a. | Clustered | VC_1481_0 | F | n.a. | n.a. | n.a. | n.a. | n.a. | n.a. | n.a. | n.a. | n.a. | n.a. | n.a. |
| S1-wms-combined_132934 | Faecalibacterium phage FP_Epona | MG711462 | Clustered | VC_707_0 | N10 | 46 | Faecalibacterium | Group I | Duplodnaviria | Heunggongvirae | Uroviricota | Caudoviricetes | n.a. | Unclassified | O | O |
| S1-wms-combined_151708 | n.a. | n.a. | Clustered | VC_1484_0 | A | n.a. | n.a. | n.a. | n.a. | n.a. | n.a. | n.a. | n.a. | n.a. | n.a. | n.a. |
| S1-wms-combined_156637 | n.a. | n.a. | Clustered | VC_1487_0 | F | n.a. | n.a. | n.a. | n.a. | n.a. | n.a. | n.a. | n.a. | n.a. | n.a. | n.a. |
| S1-wms-combined_159544 | Pseudomonas phage phi3 | KT887559 | Clustered | VC_5_0 | C2 | 32 | Pseudomonas | Group I | Duplodnaviria | Heunggongvirae | Uroviricota | Caudoviricetes | n.a. | Peduoviridae | Unclassified | Phitrevirus |
| S1-wms-combined_164485 | n.a. | n.a. | Clustered | VC_1484_0 | A | n.a. | n.a. | n.a. | n.a. | n.a. | n.a. | n.a. | n.a. | n.a. | n.a. | n.a. |
| S1-wms-combined_169879 | Bacillus phage BalMu-1 | KP063902 | Clustered | VC_825_0 | N9 | 12 | Bacillus | Group I | Duplodnaviria | Heunggongvirae | Uroviricota | Caudoviricetes | n.a. | Unclassified | O | O |
| S1-wms-combined_185336 | n.a. | n.a. | Clustered | VC_1491_0 | F | n.a. | n.a. | n.a. | n.a. | n.a. | n.a. | n.a. | n.a. | n.a. | n.a. | n.a. |
| S1-wms-combined_19031 | Mannheimia phage vB_MhM_3927AP2 | KP137440 | Clustered | VC_181_0 | C1 | 48 | Mannheimia | Group I | Duplodnaviria | Heunggongvirae | Uroviricota | Caudoviricetes | n.a. | Unclassified | Unclassified | Unclassified |
| S1-wms-combined_197401 | n.a. | n.a. | Clustered | VC_1408_0 | A | n.a. | n.a. | n.a. | n.a. | n.a. | n.a. | n.a. | n.a. | n.a. | n.a. | n.a. |
| S1-wms-combined_224578 | Roseburia phage Jekyll | LR596902 | Clustered | VC_418_0 | C5 | 9 | Roseburia | Group I | Duplodnaviria | Heunggongvirae | Uroviricota | Caudoviricetes | n.a. | Unclassified | Unclassified | Unclassified |
| S1-wms-combined_23248 | Faecalibacterium phage FP_Lugh | MG711464 | Clustered | VC_621_0 | C2 | 71 | Faecalibacterium | Group I | Duplodnaviria | Heunggongvirae | Uroviricota | Caudoviricetes | n.a. | Unclassified | Unclassified | Lughvirus |
| S1-wms-combined_246923 | Flavobacterium phage fF4 | MN850656 | Clustered | VC_679_0 | N12 | 13 | Flavobacterium | Group I | Duplodnaviria | Heunggongvirae | Uroviricota | Caudoviricetes | n.a. | Unclassified | O | O |
| S1-wms-combined_248622 | n.a. | n.a. | Clustered | VC_1494_0 | F | n.a. | n.a. | n.a. | n.a. | n.a. | n.a. | n.a. | n.a. | n.a. | n.a. | n.a. |
| S1-wms-combined_263562 | n.a. | n.a. | Clustered | VC_1437_0 | A | n.a. | n.a. | n.a. | n.a. | n.a. | n.a. | n.a. | n.a. | n.a. | n.a. | n.a. |
| S1-wms-combined_302166 | Faecalibacterium phage FP_Lagaffe | MG711461 | Clustered | VC_715_0 | N8 | 11 | Faecalibacterium | Group I | Duplodnaviria | Heunggongvirae | Uroviricota | Caudoviricetes | n.a. | Unclassified | O | O |
| S1-wms-combined_304377 | Faecalibacterium phage FP_oengus | MG711463 | Clustered | VC_804_0 | C1 | 134 | Faecalibacterium | Group I | Duplodnaviria | Heunggongvirae | Uroviricota | Caudoviricetes | n.a. | Unclassified | Unclassified | Oengusvirus |
| S1-wms-combined_307323 | Bifidobacterium phage PMBT6 | MH444512 | Clustered | VC_798_0 | N3 | 41 | Bifidobacterium | Group I | Duplodnaviria | Heunggongvirae | Uroviricota | Caudoviricetes | n.a. | Unclassified | O | O |
| S1-wms-combined_313851 | n.a. | n.a. | Clustered | VC_1496_0 | F | n.a. | n.a. | n.a. | n.a. | n.a. | n.a. | n.a. | n.a. | n.a. | n.a. | n.a. |
| S1-wms-combined_324053 | Myoviridae sp. | MW202455 | Clustered | VC_427_0 | C1 | 7 | Unspecified | Group I | Duplodnaviria | Heunggongvirae | Uroviricota | Caudoviricetes | n.a. | Unclassified | Unclassified | Unclassified |
| S1-wms-combined_331544 | n.a. | n.a. | Clustered | VC_1497_0 | F | n.a. | n.a. | n.a. | n.a. | n.a. | n.a. | n.a. | n.a. | n.a. | n.a. | n.a. |
| S1-wms-combined_331896 | n.a. | n.a. | Clustered | VC_1481_0 | F | n.a. | n.a. | n.a. | n.a. | n.a. | n.a. | n.a. | n.a. | n.a. | n.a. | n.a. |
| S1-wms-combined_332236 | Streptococcus phage Javan630 | MK448997 | Clustered | VC_626_0 | C5 | 55 | Streptococcus | Group I | Duplodnaviria | Heunggongvirae | Uroviricota | Caudoviricetes | n.a. | Unclassified | Unclassified | Unclassified |
| S1-wms-combined_340558 | Flavobacterium phage fF4 | MN850656 | Clustered | VC_679_0 | N18 | 4 | Flavobacterium | Group I | Duplodnaviria | Heunggongvirae | Uroviricota | Caudoviricetes | n.a. | Unclassified | O | O |
| S1-wms-combined_353757 | Clostridium phage phiCT19406B | KM983331 | Clustered | VC_650_0 | N16 | 7 | Clostridium | Group I | Duplodnaviria | Heunggongvirae | Uroviricota | Caudoviricetes | n.a. | Unclassified | O | O |
| S1-wms-combined_354899 | n.a. | n.a. | Clustered | VC_1498_0 | F | n.a. | n.a. | n.a. | n.a. | n.a. | n.a. | n.a. | n.a. | n.a. | n.a. | n.a. |
| S1-wms-combined_357373 | Tortoise microvirus 19 | MK765569 | Clustered | VC_1243_0 | C1 | 9 | Unspecified | Group II | Monodnaviria | Sangervirae | Phixviricota | Malgrandaviricetes | Petitvirales | Microviridae | Unclassified | Unclassified |
| S1-wms-combined_360285 | Exiguobacterium phage vB_EalM-137 | MH884510 | Clustered | VC_716_0 | N2 | 13 | Exiguobacterium | Group I | Duplodnaviria | Heunggongvirae | Uroviricota | Caudoviricetes | n.a. | Unclassified | O | O |
| S1-wms-combined_394574 | Parabacteroides phage PDS1 | MN929097 | Clustered | VC_831_0 | C1 | 164 | Parabacteroides | Group I | Duplodnaviria | Heunggongvirae | Uroviricota | Caudoviricetes | n.a. | Unclassified | Unclassified | Unclassified |
| S1-wms-combined_394628 | n.a. | n.a. | Clustered | VC_1500_0 | F | n.a. | n.a. | n.a. | n.a. | n.a. | n.a. | n.a. | n.a. | n.a. | n.a. | n.a. |
| S1-wms-combined_40115 | n.a. | n.a. | Clustered | VC_1491_0 | F | n.a. | n.a. | n.a. | n.a. | n.a. | n.a. | n.a. | n.a. | n.a. | n.a. | n.a. |
| S1-wms-combined_41369 | Roseburia phage Jekyll | LR596902 | Clustered | VC_1209_0 | N12 | 5 | Roseburia | Group I | Duplodnaviria | Heunggongvirae | Uroviricota | Caudoviricetes | n.a. | Unclassified | O | O |
| S1-wms-combined_52643 | Bacteroides phage p00 | BK010646 | Clustered | VC_679_0 | N6 | 14 | Bacteroides | Group I | Duplodnaviria | Heunggongvirae | Uroviricota | Caudoviricetes | n.a. | Unclassified | O | O |
| S1-wms-combined_68998 | Faecalibacterium phage FP_Taranis | MG711467 | Clustered | VC_711_0 | C1 | 223 | Faecalibacterium | Group I | Duplodnaviria | Heunggongvirae | Uroviricota | Caudoviricetes | n.a. | Unclassified | Unclassified | Taranisvirus |
| S1-wms-combined_72167 | Listeria phage LWP01 | CP011103 | Clustered | VC_420_0 | N10 | 3 | Listeria | Group I | Duplodnaviria | Heunggongvirae | Uroviricota | Caudoviricetes | n.a. | Unclassified | O | O |
| S1-wms_100010 | n.a. | n.a. | Clustered | VC_1492_0 | A | n.a. | n.a. | n.a. | n.a. | n.a. | n.a. | n.a. | n.a. | n.a. | n.a. | n.a. |
| S1-wms_130214 | Butyrivibrio virus Ceridwen | MN882553 | Clustered | VC_1209_0 | N11 | 6 | Butyrivibrio | Group I | Duplodnaviria | Heunggongvirae | Uroviricota | Caudoviricetes | n.a. | Unclassified | O | O |
| S1-wms_170115 | Clostridium phage phiCD38-2 | HM568888 | Clustered | VC_627_0 | N9 | 6 | Clostridium | Group I | Duplodnaviria | Heunggongvirae | Uroviricota | Caudoviricetes | n.a. | Unclassified | O | O |
| S1-wms_185046 | n.a. | n.a. | Clustered | VC_1239_0 | A | n.a. | n.a. | n.a. | n.a. | n.a. | n.a. | n.a. | n.a. | n.a. | n.a. | n.a. |
| S1-wms_212940 | Faecalibacterium phage FP_oengus | MG711463 | Clustered | VC_804_2 | N10 | 46 | Faecalibacterium | Group I | Duplodnaviria | Heunggongvirae | Uroviricota | Caudoviricetes | n.a. | Unclassified | O | O |
| S1-wms_225995 | n.a. | n.a. | Clustered | VC_1492_0 | A | n.a. | n.a. | n.a. | n.a. | n.a. | n.a. | n.a. | n.a. | n.a. | n.a. | n.a. |
| S1-wms_244836 | n.a. | n.a. | Clustered | VC_1473_0 | A | n.a. | n.a. | n.a. | n.a. | n.a. | n.a. | n.a. | n.a. | n.a. | n.a. | n.a. |
| S1-wms_296850 | Faecalibacterium phage FP_Toutatis | MG711466 | Clustered | VC_710_0 | C6 | 68 | Faecalibacterium | Group I | Duplodnaviria | Heunggongvirae | Uroviricota | Caudoviricetes | n.a. | Unclassified | Unclassified | Toutatisvirus |
| S1-wms_313991 | n.a. | n.a. | Clustered | VC_1496_0 | F | n.a. | n.a. | n.a. | n.a. | n.a. | n.a. | n.a. | n.a. | n.a. | n.a. | n.a. |
| S1-wms_4277 | Faecalibacterium phage FP_Mushu | MG711460 | Clustered | VC_1235_0 | C5 | 126 | Faecalibacterium | Group I | Duplodnaviria | Heunggongvirae | Uroviricota | Caudoviricetes | n.a. | Unclassified | Unclassified | Mushuvirus |
| S11-wms-combined_119215 | Clostridium phage phiCT453B | KM983328 | Clustered | VC_650_0 | N16 | 3 | Clostridium | Group I | Duplodnaviria | Heunggongvirae | Uroviricota | Caudoviricetes | n.a. | Unclassified | O | O |
| S11-wms-combined_125176 | Campylobacter phage PC10 | MZ047271 | Clustered | VC_1456_0 | N7 | 6 | Campylobacter | Group I | Duplodnaviria | Heunggongvirae | Uroviricota | Caudoviricetes | n.a. | Unclassified | O | O |
| S11-wms-combined_140009 | Faecalibacterium phage FP_Lugh | MG711464 | Clustered | VC_621_0 | C1 | 94 | Faecalibacterium | Group I | Duplodnaviria | Heunggongvirae | Uroviricota | Caudoviricetes | n.a. | Unclassified | Unclassified | Lughvirus |
| S11-wms-combined_154845 | Escherichia phage ESSI2_ev015 | LR597649 | Clustered | VC_5_0 | C1 | 105 | Escherichia | Group I | Duplodnaviria | Heunggongvirae | Uroviricota | Caudoviricetes | n.a. | Peduoviridae | Unclassified | Quadragintavirus |
| S11-wms-combined_17029 | n.a. | n.a. | Clustered | VC_1502_0 | A | n.a. | n.a. | n.a. | n.a. | n.a. | n.a. | n.a. | n.a. | n.a. | n.a. | n.a. |
| S11-wms-combined_172889 | Flavobacterium phage fF4 | MN850656 | Clustered | VC_679_0 | N17 | 7 | Flavobacterium | Group I | Duplodnaviria | Heunggongvirae | Uroviricota | Caudoviricetes | n.a. | Unclassified | O | O |
| S11-wms-combined_193148 | n.a. | n.a. | Clustered | VC_616_0 | A | n.a. | n.a. | n.a. | n.a. | n.a. | n.a. | n.a. | n.a. | n.a. | n.a. | n.a. |
| S11-wms-combined_194205 | Streptococcus phage Javan630 | MK448997 | Clustered | VC_626_0 | C3 | 66 | Streptococcus | Group I | Duplodnaviria | Heunggongvirae | Uroviricota | Caudoviricetes | n.a. | Unclassified | Unclassified | Unclassified |
| S11-wms-combined_195425 | Bacteroides phage p00 | BK010646 | Clustered | VC_677_0 | C2 | 65 | Bacteroides | Group I | Duplodnaviria | Heunggongvirae | Uroviricota | Caudoviricetes | n.a. | Unclassified | Unclassified | Unclassified |
| S11-wms-combined_198512 | Roseburia phage Shimadzu | LR596903 | Clustered | VC_715_0 | N3 | 5 | Roseburia | Group I | Duplodnaviria | Heunggongvirae | Uroviricota | Caudoviricetes | n.a. | Unclassified | O | O |
| S11-wms-combined_200084 | Bacillus phage BalMu-1 | KP063902 | Clustered | VC_825_0 | N10 | 6 | Bacillus | Group I | Duplodnaviria | Heunggongvirae | Uroviricota | Caudoviricetes | n.a. | Unclassified | O | O |
| S11-wms-combined_204522 | Burkholderia phage BcepC6B | AY605181 | Clustered | VC_616_0 | N3 | 1 | Burkholderia | Group I | Duplodnaviria | Heunggongvirae | Uroviricota | Caudoviricetes | n.a. | Unclassified | O | O |
| S11-wms-combined_217790 | n.a. | n.a. | Clustered | VC_1503_0 | A | n.a. | n.a. | n.a. | n.a. | n.a. | n.a. | n.a. | n.a. | n.a. | n.a. | n.a. |
| S11-wms-combined_221310 | n.a. | n.a. | Clustered | VC_1504_0 | A | n.a. | n.a. | n.a. | n.a. | n.a. | n.a. | n.a. | n.a. | n.a. | n.a. | n.a. |
| S11-wms-combined_268967 | Mannheimia phage vB_MhM_3927AP2 | KP137440 | Clustered | VC_181_0 | C2 | 23 | Mannheimia | Group I | Duplodnaviria | Heunggongvirae | Uroviricota | Caudoviricetes | n.a. | Unclassified | Unclassified | Unclassified |
| S11-wms-combined_283220 | Faecalibacterium phage FP_Mushu | MG711460 | Clustered | VC_1235_0 | C1 | 160 | Faecalibacterium | Group I | Duplodnaviria | Heunggongvirae | Uroviricota | Caudoviricetes | n.a. | Unclassified | Unclassified | Mushuvirus |
| S11-wms-combined_299954 | n.a. | n.a. | Clustered | VC_650_0 | A | n.a. | n.a. | n.a. | n.a. | n.a. | n.a. | n.a. | n.a. | n.a. | n.a. | n.a. |
| S11-wms-combined_304239 | Faecalibacterium phage FP_Epona | MG711462 | Clustered | VC_707_0 | N10 | 51 | Faecalibacterium | Group I | Duplodnaviria | Heunggongvirae | Uroviricota | Caudoviricetes | n.a. | Unclassified | O | O |
| S11-wms-combined_41367 | n.a. | n.a. | Clustered | VC_1505_0 | A | n.a. | n.a. | n.a. | n.a. | n.a. | n.a. | n.a. | n.a. | n.a. | n.a. | n.a. |
| S11-wms-combined_46816 | n.a. | n.a. | Clustered | VC_893_0 | A | n.a. | n.a. | n.a. | n.a. | n.a. | n.a. | n.a. | n.a. | n.a. | n.a. | n.a. |
| S11-wms-combined_48246 | Phage vB_RanS_PJN03 | ON245412 | Clustered | VC_917_0 | N10 | 5 | Unspecified | Group I | Duplodnaviria | Heunggongvirae | Uroviricota | Caudoviricetes | n.a. | Unclassified | O | O |
| S11-wms-combined_49632 | Clostridium phage phiCT453B | KM983328 | Clustered | VC_650_0 | N12 | 10 | Clostridium | Group I | Duplodnaviria | Heunggongvirae | Uroviricota | Caudoviricetes | n.a. | Unclassified | O | O |
| S11-wms-combined_58538 | Deep-sea thermophilic phage D6E | GU568037 | Clustered | VC_271_0 | N9 | 2 | Unspecified | Group I | Duplodnaviria | Heunggongvirae | Uroviricota | Caudoviricetes | n.a. | Unclassified | O | O |
| S11-wms-combined_73204 | n.a. | n.a. | Clustered | VC_1397_0 | A | n.a. | n.a. | n.a. | n.a. | n.a. | n.a. | n.a. | n.a. | n.a. | n.a. | n.a. |
| S11-wms-combined_74983 | Faecalibacterium phage FP_Toutatis | MG711466 | Clustered | VC_710_0 | C6 | 71 | Faecalibacterium | Group I | Duplodnaviria | Heunggongvirae | Uroviricota | Caudoviricetes | n.a. | Unclassified | Unclassified | Toutatisvirus |
| S11-wms-combined_83571 | Winogradskyella phage Peternella_1 | MT732475 | Clustered | VC_676_1 | N12 | 19 | Winogradskyella | Group I | Duplodnaviria | Heunggongvirae | Uroviricota | Caudoviricetes | n.a. | Winoviridae | O | O |
| S11-wms-combined_84326 | Escherichia phage vB_EcoM-613R2 | ON470591 | Clustered | VC_716_0 | N2 | 12 | Escherichia | Group I | Duplodnaviria | Heunggongvirae | Uroviricota | Caudoviricetes | n.a. | Unclassified | O | O |
| S11-wms-combined_92114 | n.a. | n.a. | Clustered | VC_1014_0 | A | n.a. | n.a. | n.a. | n.a. | n.a. | n.a. | n.a. | n.a. | n.a. | n.a. | n.a. |
| S11-wms_14390 | n.a. | n.a. | Clustered | VC_271_0 | A | n.a. | n.a. | n.a. | n.a. | n.a. | n.a. | n.a. | n.a. | n.a. | n.a. | n.a. |
| S11-wms_303398 | n.a. | n.a. | Clustered | VC_1239_0 | A | n.a. | n.a. | n.a. | n.a. | n.a. | n.a. | n.a. | n.a. | n.a. | n.a. | n.a. |
| S11-wms_3116 | Faecalibacterium phage FP_Mushu | MG711460 | Clustered | VC_1235_0 | C2 | 142 | Faecalibacterium | Group I | Duplodnaviria | Heunggongvirae | Uroviricota | Caudoviricetes | n.a. | Unclassified | Unclassified | Mushuvirus |
| S11-wms_320555 | Phage vB_RanS_PJN03 | ON245412 | Clustered | VC_918_0 | N5 | 9 | Unspecified | Group I | Duplodnaviria | Heunggongvirae | Uroviricota | Caudoviricetes | n.a. | Unclassified | O | O |
| S11-wms_330479 | Elizabethkingia phage TCUEAP1 | MN732896 | Clustered | VC_1010_0 | C1 | 8 | Elizabethkingia | Unclassified | Unclassified | n.a. | n.a. | n.a. | n.a. | Unclassified | Unclassified | Unclassified |
| S11-wms_55610 | Faecalibacterium phage FP_Brigit | MG711465 | Clustered | VC_606_0 | C2 | 149 | Faecalibacterium | Group I | Duplodnaviria | Heunggongvirae | Uroviricota | Caudoviricetes | n.a. | Unclassified | Unclassified | Brigitvirus |
| S12-wms-combined_107942 | Bifidobacterium phage BitterVaud1 | MT006236 | Clustered | VC_804_2 | N5 | 34 | Bifidobacterium | Group I | Duplodnaviria | Heunggongvirae | Uroviricota | Caudoviricetes | n.a. | Unclassified | O | O |
| S12-wms-combined_111524 | n.a. | n.a. | Clustered | VC_1503_0 | A | n.a. | n.a. | n.a. | n.a. | n.a. | n.a. | n.a. | n.a. | n.a. | n.a. | n.a. |
| S12-wms-combined_120133 | Geobacillus phage GBSV1 | DQ340064 | Clustered | VC_338_0 | N6 | 6 | Geobacillus | Group I | Duplodnaviria | Heunggongvirae | Uroviricota | Caudoviricetes | n.a. | Unclassified | O | O |
| S12-wms-combined_129554 | Faecalibacterium phage FP_Epona | MG711462 | Clustered | VC_707_1 | C3 | 50 | Faecalibacterium | Group I | Duplodnaviria | Heunggongvirae | Uroviricota | Caudoviricetes | n.a. | Unclassified | Unclassified | Eponavirus |
| S12-wms-combined_147628 | Faecalibacterium phage FP_Taranis | MG711467 | Clustered | VC_1238_0 | N1 | 24 | Faecalibacterium | Group I | Duplodnaviria | Heunggongvirae | Uroviricota | Caudoviricetes | n.a. | Unclassified | O | O |

| | | | | | | | | | | | | | | | | |
|---|---|---|---|---|---|---|---|---|---|---|---|---|---|---|---|---|
| S12-wms-combined_156234 | Faecalibacterium phage FP_Toutatis | MG711466 | Clustered | VC_710_0 | C8 | 49 | Faecalibacterium | Group I | Duplodnaviria | Heunggongvirae | Uroviricota | Caudoviricetes | n.a. | Unclassified | Unclassified | Toutatisvirus |
| S12-wms-combined_168259 | Akkermansia phage DTMo-2021a | CP084202 | Clustered | VC_736_0 | N1 | 13 | Akkermansia | Group I | Duplodnaviria | Heunggongvirae | Uroviricota | Caudoviricetes | n.a. | Unclassified | O | O |
| S12-wms-combined_17617 | Faecalibacterium phage FP_Epona | MG711462 | Clustered | VC_707_0 | N8 | 53 | Faecalibacterium | Group I | Duplodnaviria | Heunggongvirae | Uroviricota | Caudoviricetes | n.a. | Unclassified | O | O |
| S12-wms-combined_18287 | Faecalibacterium phage FP_Epona | MG711462 | Clustered | VC_707_0 | N10 | 42 | Faecalibacterium | Group I | Duplodnaviria | Heunggongvirae | Uroviricota | Caudoviricetes | n.a. | Unclassified | O | O |
| S12-wms-combined_191570 | Clostridium phage phiCT453A | KM983327 | Clustered | VC_732_0 | N4 | 9 | Clostridium | Group I | Duplodnaviria | Heunggongvirae | Uroviricota | Caudoviricetes | n.a. | Unclassified | O | O |
| S12-wms-combined_210942 | n.a. | n.a. | Clustered | VC_1487_0 | F | n.a. | n.a. | n.a. | n.a. | n.a. | n.a. | n.a. | n.a. | n.a. | n.a. | n.a. |
| S12-wms-combined_2450 | Winogradskyella phage Peternella_1 | MT732475 | Clustered | VC_676_0 | N7 | 19 | Winogradskyella | Group I | Duplodnaviria | Heunggongvirae | Uroviricota | Caudoviricetes | n.a. | Winoviridae | O | O |
| S12-wms-combined_245755 | Butyrivibrio phage Idris | MN882554 | Clustered | VC_876_0 | N6 | 38 | Butyrivibrio | Group I | Duplodnaviria | Heunggongvirae | Uroviricota | Caudoviricetes | n.a. | Unclassified | O | O |
| S12-wms-combined_250551 | Bacteroides phage p00 | BK010646 | Clustered | VC_677_0 | C1 | 130 | Bacteroides | Group I | Duplodnaviria | Heunggongvirae | Uroviricota | Caudoviricetes | n.a. | Unclassified | Unclassified | Unclassified |
| S12-wms-combined_256299 | n.a. | n.a. | Clustered | VC_1505_0 | A | n.a. | n.a. | n.a. | n.a. | n.a. | n.a. | n.a. | n.a. | n.a. | n.a. | n.a. |
| S12-wms-combined_259783 | Roseburia phage Shimadzu | LR596903 | Clustered | VC_717_0 | C1 | 228 | Roseburia | Group I | Duplodnaviria | Heunggongvirae | Uroviricota | Caudoviricetes | n.a. | Unclassified | Unclassified | Unclassified |
| S12-wms-combined_260036 | uncultured phage cr128_1 | MT774392 | Clustered | VC_1439_0 | N7 | 3 | Unspecified | Group I | Duplodnaviria | Heunggongvirae | Uroviricota | Caudoviricetes | Crassvirales | Steigviridae | O | O |
| S12-wms-combined_263377 | Clostridium phage Clo-PEP-1 | KY206887 | Clustered | VC_212_0 | N5 | 17 | Clostridium | Group I | Duplodnaviria | Heunggongvirae | Uroviricota | Caudoviricetes | n.a. | Unclassified | O | O |
| S12-wms-combined_264425 | n.a. | n.a. | Clustered | VC_1492_0 | A | n.a. | n.a. | n.a. | n.a. | n.a. | n.a. | n.a. | n.a. | n.a. | n.a. | n.a. |
| S12-wms-combined_264845 | n.a. | n.a. | Clustered | VC_1507_0 | A | n.a. | n.a. | n.a. | n.a. | n.a. | n.a. | n.a. | n.a. | n.a. | n.a. | n.a. |
| S12-wms-combined_282594 | Akkermansia phage DTMo-2021a | CP084202 | Clustered | VC_736_0 | N3 | 7 | Akkermansia | Group I | Duplodnaviria | Heunggongvirae | Uroviricota | Caudoviricetes | n.a. | Unclassified | O | O |
| S12-wms-combined_299293 | n.a. | n.a. | Clustered | VC_1492_0 | A | n.a. | n.a. | n.a. | n.a. | n.a. | n.a. | n.a. | n.a. | n.a. | n.a. | n.a. |
| S12-wms-combined_299872 | Faecalibacterium phage FP_Mushu | MG711460 | Clustered | VC_1235_0 | C3 | 71 | Faecalibacterium | Group I | Duplodnaviria | Heunggongvirae | Uroviricota | Caudoviricetes | n.a. | Unclassified | Unclassified | Mushuvirus |
| S12-wms-combined_320727 | Flavobacterium phage FPSV-S27 | MK764448 | Clustered | VC_1397_0 | N4 | 6 | Flavobacterium | Group I | Duplodnaviria | Heunggongvirae | Uroviricota | Caudoviricetes | n.a. | Unclassified | O | O |
| S12-wms-combined_321174 | Butyrivibrio phage Idris | MN882554 | Clustered | VC_915_0 | N8 | 6 | Butyrivibrio | Group I | Duplodnaviria | Heunggongvirae | Uroviricota | Caudoviricetes | n.a. | Unclassified | O | O |
| S12-wms-combined_329242 | n.a. | n.a. | Clustered | VC_1484_0 | A | n.a. | n.a. | n.a. | n.a. | n.a. | n.a. | n.a. | n.a. | n.a. | n.a. | n.a. |
| S12-wms-combined_334806 | n.a. | n.a. | Clustered | VC_1239_0 | A | n.a. | n.a. | n.a. | n.a. | n.a. | n.a. | n.a. | n.a. | n.a. | n.a. | n.a. |
| S12-wms-combined_345948 | Faecalibacterium phage FP_Lugh | MG711464 | Clustered | VC_621_0 | C5 | 43 | Faecalibacterium | Group I | Duplodnaviria | Heunggongvirae | Uroviricota | Caudoviricetes | n.a. | Unclassified | Unclassified | Lughvirus |
| S12-wms-combined_351847 | n.a. | n.a. | Clustered | VC_1436_0 | A | n.a. | n.a. | n.a. | n.a. | n.a. | n.a. | n.a. | n.a. | n.a. | n.a. | n.a. |
| S12-wms-combined_358010 | Exiguobacterium phage vB_EalM-137 | MH884510 | Clustered | VC_715_0 | N4 | 9 | Exiguobacterium | Group I | Duplodnaviria | Heunggongvirae | Uroviricota | Caudoviricetes | n.a. | Unclassified | O | O |
| S12-wms-combined_368462 | Streptococcus phage Javan630 | MK448997 | Clustered | VC_626_0 | C7 | 61 | Streptococcus | Group I | Duplodnaviria | Heunggongvirae | Uroviricota | Caudoviricetes | n.a. | Unclassified | Unclassified | Unclassified |
| S12-wms-combined_375343 | n.a. | n.a. | Clustered | VC_1502_0 | A | n.a. | n.a. | n.a. | n.a. | n.a. | n.a. | n.a. | n.a. | n.a. | n.a. | n.a. |
| S12-wms-combined_376326 | n.a. | n.a. | Clustered | VC_1076_0 | A | n.a. | n.a. | n.a. | n.a. | n.a. | n.a. | n.a. | n.a. | n.a. | n.a. | n.a. |
| S12-wms-combined_393703 | n.a. | n.a. | Clustered | VC_1481_0 | F | n.a. | n.a. | n.a. | n.a. | n.a. | n.a. | n.a. | n.a. | n.a. | n.a. | n.a. |
| S12-wms-combined_401379 | n.a. | n.a. | Clustered | VC_1500_0 | F | n.a. | n.a. | n.a. | n.a. | n.a. | n.a. | n.a. | n.a. | n.a. | n.a. | n.a. |
| S12-wms-combined_401383 | n.a. | n.a. | Clustered | VC_1510_0 | F | n.a. | n.a. | n.a. | n.a. | n.a. | n.a. | n.a. | n.a. | n.a. | n.a. | n.a. |
| S12-wms-combined_41160 | n.a. | n.a. | Clustered | VC_1238_0 | A | n.a. | n.a. | n.a. | n.a. | n.a. | n.a. | n.a. | n.a. | n.a. | n.a. | n.a. |
| S12-wms-combined_51076 | n.a. | n.a. | Clustered | VC_1479_0 | A | n.a. | n.a. | n.a. | n.a. | n.a. | n.a. | n.a. | n.a. | n.a. | n.a. | n.a. |
| S12-wms-combined_71235 | Faecalibacterium phage FP_Lugh | MG711464 | Clustered | VC_621_0 | C4 | 57 | Faecalibacterium | Group I | Duplodnaviria | Heunggongvirae | Uroviricota | Caudoviricetes | n.a. | Unclassified | Unclassified | Lughvirus |
| S12-wms-combined_7174 | Streptococcus phage Javan630 | MK448997 | Clustered | VC_626_0 | C6 | 59 | Streptococcus | Group I | Duplodnaviria | Heunggongvirae | Uroviricota | Caudoviricetes | n.a. | Unclassified | Unclassified | Unclassified |
| S12-wms-combined_91231 | n.a. | n.a. | Clustered | VC_1439_0 | A | n.a. | n.a. | n.a. | n.a. | n.a. | n.a. | n.a. | n.a. | n.a. | n.a. | n.a. |
| S12-wms-combined_99704 | Streptococcus phage Javan105 | MK448667 | Clustered | VC_626_0 | C2 | 81 | Streptococcus | Group I | Duplodnaviria | Heunggongvirae | Uroviricota | Caudoviricetes | n.a. | Unclassified | Unclassified | Unclassified |
| S12-wms_12658 | Phage vB_RanS_PJN03 | ON245412 | Clustered | VC_918_0 | N8 | 12 | Unspecified | Group I | Duplodnaviria | Heunggongvirae | Uroviricota | Caudoviricetes | n.a. | Unclassified | O | O |
| S12-wms_15700 | n.a. | n.a. | Clustered | VC_1511_0 | F | n.a. | n.a. | n.a. | n.a. | n.a. | n.a. | n.a. | n.a. | n.a. | n.a. | n.a. |
| S12-wms_179388 | n.a. | n.a. | Clustered | VC_1492_0 | A | n.a. | n.a. | n.a. | n.a. | n.a. | n.a. | n.a. | n.a. | n.a. | n.a. | n.a. |
| S12-wms_193565 | Faecalibacterium phage FP_Mushu | MG711460 | Clustered | VC_1235_0 | C1 | 150 | Faecalibacterium | Group I | Duplodnaviria | Heunggongvirae | Uroviricota | Caudoviricetes | n.a. | Unclassified | Unclassified | Mushuvirus |
| S12-wms_373775 | n.a. | n.a. | Clustered | VC_1507_0 | A | n.a. | n.a. | n.a. | n.a. | n.a. | n.a. | n.a. | n.a. | n.a. | n.a. | n.a. |
| S12-wms_383851 | n.a. | n.a. | Clustered | VC_1436_0 | A | n.a. | n.a. | n.a. | n.a. | n.a. | n.a. | n.a. | n.a. | n.a. | n.a. | n.a. |
| S12-wms_386692 | n.a. | n.a. | Clustered | VC_1456_0 | A | n.a. | n.a. | n.a. | n.a. | n.a. | n.a. | n.a. | n.a. | n.a. | n.a. | n.a. |
| S12-wms_401366 | n.a. | n.a. | Clustered | VC_1492_0 | A | n.a. | n.a. | n.a. | n.a. | n.a. | n.a. | n.a. | n.a. | n.a. | n.a. | n.a. |
| S12-wms_401372 | Phage DP SC_6_H4_2017 | MT121964 | Clustered | VC_1420_0 | C2 | 137 | Unspecified | Group I | Duplodnaviria | Heunggongvirae | Uroviricota | Caudoviricetes | n.a. | Unclassified | Unclassified | Unclassified |
| S12-wms_44817 | n.a. | n.a. | Clustered | VC_1492_0 | A | n.a. | n.a. | n.a. | n.a. | n.a. | n.a. | n.a. | n.a. | n.a. | n.a. | n.a. |
| S17-wms-combined_126600 | n.a. | n.a. | Clustered | VC_1481_0 | F | n.a. | n.a. | n.a. | n.a. | n.a. | n.a. | n.a. | n.a. | n.a. | n.a. | n.a. |
| S17-wms-combined_126774 | n.a. | n.a. | Clustered | VC_1484_0 | A | n.a. | n.a. | n.a. | n.a. | n.a. | n.a. | n.a. | n.a. | n.a. | n.a. | n.a. |
| S17-wms-combined_127688 | Faecalibacterium phage FP_Epona | MG711462 | Clustered | VC_707_1 | C2 | 106 | Faecalibacterium | Group I | Duplodnaviria | Heunggongvirae | Uroviricota | Caudoviricetes | n.a. | Unclassified | Unclassified | Eponavirus |
| S17-wms-combined_130215 | Streptococcus phage Javan261 | MK448720 | Clustered | VC_338_0 | N9 | 5 | Streptococcus | Group I | Duplodnaviria | Heunggongvirae | Uroviricota | Caudoviricetes | n.a. | Unclassified | O | O |
| S17-wms-combined_136622 | n.a. | n.a. | Clustered | VC_650_0 | A | n.a. | n.a. | n.a. | n.a. | n.a. | n.a. | n.a. | n.a. | n.a. | n.a. | n.a. |
| S17-wms-combined_146850 | n.a. | n.a. | Clustered | VC_1484_0 | A | n.a. | n.a. | n.a. | n.a. | n.a. | n.a. | n.a. | n.a. | n.a. | n.a. | n.a. |
| S17-wms-combined_157468 | n.a. | n.a. | Clustered | VC_1484_0 | A | n.a. | n.a. | n.a. | n.a. | n.a. | n.a. | n.a. | n.a. | n.a. | n.a. | n.a. |
| S17-wms-combined_158354 | n.a. | n.a. | Clustered | VC_1505_0 | A | n.a. | n.a. | n.a. | n.a. | n.a. | n.a. | n.a. | n.a. | n.a. | n.a. | n.a. |
| S17-wms-combined_162786 | n.a. | n.a. | Clustered | VC_1408_0 | A | n.a. | n.a. | n.a. | n.a. | n.a. | n.a. | n.a. | n.a. | n.a. | n.a. | n.a. |
| S17-wms-combined_16309 | Bacteroides phage F4 | MT806185 | Clustered | VC_1435_0 | N11 | 8 | Bacteroides | Group I | Duplodnaviria | Heunggongvirae | Uroviricota | Caudoviricetes | n.a. | Unclassified | O | O |
| S17-wms-combined_175924 | Faecalibacterium phage FP_Mushu | MG711460 | Clustered | VC_1235_0 | C1 | 96 | Faecalibacterium | Group I | Duplodnaviria | Heunggongvirae | Uroviricota | Caudoviricetes | n.a. | Unclassified | Unclassified | Mushuvirus |
| S17-wms-combined_179771 | n.a. | n.a. | Clustered | VC_1509_0 | A | n.a. | n.a. | n.a. | n.a. | n.a. | n.a. | n.a. | n.a. | n.a. | n.a. | n.a. |
| S17-wms-combined_193958 | Faecalibacterium phage FP_Mushu | MG711460 | Clustered | VC_1235_0 | C3 | 135 | Faecalibacterium | Group I | Duplodnaviria | Heunggongvirae | Uroviricota | Caudoviricetes | n.a. | Unclassified | Unclassified | Mushuvirus |
| S17-wms-combined_199440 | Faecalibacterium phage FP_Epona | MG711462 | Clustered | VC_707_0 | N10 | 26 | Faecalibacterium | Group I | Duplodnaviria | Heunggongvirae | Uroviricota | Caudoviricetes | n.a. | Unclassified | O | O |
| S17-wms-combined_199746 | Bacteroides phage F4 | MT806185 | Clustered | VC_1014_0 | N10 | 2 | Bacteroides | Group I | Duplodnaviria | Heunggongvirae | Uroviricota | Caudoviricetes | n.a. | Unclassified | O | O |
| S17-wms-combined_203701 | n.a. | n.a. | Clustered | VC_1508_0 | A | n.a. | n.a. | n.a. | n.a. | n.a. | n.a. | n.a. | n.a. | n.a. | n.a. | n.a. |
| S17-wms-combined_223748 | n.a. | n.a. | Clustered | VC_1512_0 | F | n.a. | n.a. | n.a. | n.a. | n.a. | n.a. | n.a. | n.a. | n.a. | n.a. | n.a. |
| S17-wms-combined_225356 | n.a. | n.a. | Clustered | VC_1513_0 | A | n.a. | n.a. | n.a. | n.a. | n.a. | n.a. | n.a. | n.a. | n.a. | n.a. | n.a. |
| S17-wms-combined_233761 | n.a. | n.a. | Clustered | VC_1495_0 | A | n.a. | n.a. | n.a. | n.a. | n.a. | n.a. | n.a. | n.a. | n.a. | n.a. | n.a. |
| S17-wms-combined_247494 | Clostridium phage phiMMP04 | JX145342 | Clustered | VC_650_0 | N15 | 6 | Clostridium | Group I | Duplodnaviria | Heunggongvirae | Uroviricota | Caudoviricetes | n.a. | Unclassified | O | O |
| S17-wms-combined_259811 | Faecalibacterium phage FP_oengus | MG711463 | Clustered | VC_804_1 | N8 | 29 | Faecalibacterium | Group I | Duplodnaviria | Heunggongvirae | Uroviricota | Caudoviricetes | n.a. | Unclassified | O | O |
| S17-wms-combined_260343 | Faecalibacterium phage FP_Toutatis | MG711466 | Clustered | VC_710_0 | C5 | 77 | Faecalibacterium | Group I | Duplodnaviria | Heunggongvirae | Uroviricota | Caudoviricetes | n.a. | Unclassified | Unclassified | Toutatisvirus |
| S17-wms-combined_30308 | n.a. | n.a. | Clustered | VC_1482_0 | F | n.a. | n.a. | n.a. | n.a. | n.a. | n.a. | n.a. | n.a. | n.a. | n.a. | n.a. |
| S17-wms-combined_47860 | Faecalibacterium phage FP_oengus | MG711463 | Clustered | VC_804_1 | N7 | 66 | Faecalibacterium | Group I | Duplodnaviria | Heunggongvirae | Uroviricota | Caudoviricetes | n.a. | Unclassified | O | O |
| S17-wms-combined_70983 | n.a. | n.a. | Clustered | VC_1494_0 | F | n.a. | n.a. | n.a. | n.a. | n.a. | n.a. | n.a. | n.a. | n.a. | n.a. | n.a. |
| S17-wms-combined_79709 | Clostridium phage phiCT453B | KM983328 | Clustered | VC_650_0 | N14 | 10 | Clostridium | Group I | Duplodnaviria | Heunggongvirae | Uroviricota | Caudoviricetes | n.a. | Unclassified | O | O |
| S17-wms-combined_90378 | Faecalibacterium phage FP_Lugh | MG711464 | Clustered | VC_621_0 | C2 | 65 | Faecalibacterium | Group I | Duplodnaviria | Heunggongvirae | Uroviricota | Caudoviricetes | n.a. | Unclassified | Unclassified | Lughvirus |
| S17-wms-combined_9864 | Streptococcus phage Javan630 | MK448997 | Clustered | VC_626_0 | C7 | 56 | Streptococcus | Group I | Duplodnaviria | Heunggongvirae | Uroviricota | Caudoviricetes | n.a. | Unclassified | Unclassified | Unclassified |
| S17-wms_124308 | Bacteroides phage F4 | MT806185 | Clustered | VC_1435_0 | N11 | 4 | Bacteroides | Group I | Duplodnaviria | Heunggongvirae | Uroviricota | Caudoviricetes | n.a. | Unclassified | O | O |
| S17-wms_254872 | Microvirus mar62 | MZ089808 | Clustered | VC_1276_0 | C1 | 18 | Unspecified | Group II | Monodnaviria | Sangervirae | Phixviricota | Malgrandaviricetes | Petitvirales | Microviridae | Unclassified | Microvirus |
| S17-wms_266511 | Enterobacter phage phiEap-2 | KT287080 | Clustered | VC_540_0 | C1 | 92 | Enterobacter | Group I | Duplodnaviria | Heunggongvirae | Uroviricota | Caudoviricetes | n.a. | Unclassified | Guernseyvirinae | Cornellvirus |
| S2-wms-combined_101826 | Paenibacillus phage PG1 | HQ332138 | Clustered | VC_734_0 | C2 | 8 | Paenibacillus | Group I | Duplodnaviria | Heunggongvirae | Uroviricota | Caudoviricetes | n.a. | Unclassified | Unclassified | Unclassified |
| S2-wms-combined_113619 | Bacteroides phage LoVEphage | MW660583 | Clustered | VC_832_0 | N12 | 3 | Bacteroides | Group I | Duplodnaviria | Heunggongvirae | Uroviricota | Caudoviricetes | n.a. | Unclassified | O | O |
| S2-wms-combined_126134 | n.a. | n.a. | Clustered | VC_650_0 | A | n.a. | n.a. | n.a. | n.a. | n.a. | n.a. | n.a. | n.a. | n.a. | n.a. | n.a. |
| S2-wms-combined_132966 | Inovirus sp. | MT135284 | Clustered | VC_1422_0 | C1 | 28 | Unspecified | Group II | Monodnaviria | Loebvirae | Hofneiviricota | Faserviricetes | Tubulavirales | Inoviridae | Unclassified | Inovirus |
| S2-wms-combined_135139 | n.a. | n.a. | Clustered | VC_738_0 | A | n.a. | n.a. | n.a. | n.a. | n.a. | n.a. | n.a. | n.a. | n.a. | n.a. | n.a. |
| S2-wms-combined_135154 | n.a. | n.a. | Clustered | VC_947_0 | A | n.a. | n.a. | n.a. | n.a. | n.a. | n.a. | n.a. | n.a. | n.a. | n.a. | n.a. |
| S2-wms-combined_165833 | Faecalibacterium phage FP_Mushu | MG711460 | Clustered | VC_1235_0 | C3 | 67 | Faecalibacterium | Group I | Duplodnaviria | Heunggongvirae | Uroviricota | Caudoviricetes | n.a. | Unclassified | Unclassified | Mushuvirus |
| S2-wms-combined_166665 | Streptococcus phage SM1 | AY007505 | Clustered | VC_324_0 | N6 | 8 | Streptococcus | Group I | Duplodnaviria | Heunggongvirae | Uroviricota | Caudoviricetes | n.a. | Unclassified | O | O |
| S2-wms-combined_174241 | Streptococcus phage Javan630 | MK448997 | Clustered | VC_626_0 | C6 | 54 | Streptococcus | Group I | Duplodnaviria | Heunggongvirae | Uroviricota | Caudoviricetes | n.a. | Unclassified | Unclassified | Unclassified |

| | | | | | | | | | | | | | | | | |
|---|---|---|---|---|---|---|---|---|---|---|---|---|---|---|---|---|
| S2-wms-combined_176376 | Clostridioides phage phiCD27 | EU719189 | Clustered | VC_825_0 | N14 | 2 | Clostridioides | Group I | Duplodnaviria | Heunggongvirae | Uroviricota | Caudoviricetes | n.a. | Unclassified | O | O |
| S2-wms-combined_180640 | Faecalibacterium phage FP_Epona | MG711462 | Clustered | VC_707_1 | C2 | 54 | Faecalibacterium | Group I | Duplodnaviria | Heunggongvirae | Uroviricota | Caudoviricetes | n.a. | Unclassified | Unclassified | Eponavirus |
| S2-wms-combined_181095 | n.a. | n.a. | Clustered | VC_1481_0 | F | n.a. | n.a. | n.a. | n.a. | n.a. | n.a. | n.a. | n.a. | n.a. | n.a. | n.a. |
| S2-wms-combined_196783 | Clostridium phage phiCT19406B | KM983331 | Clustered | VC_629_0 | N10 | 9 | Clostridium | Group I | Duplodnaviria | Heunggongvirae | Uroviricota | Caudoviricetes | n.a. | Unclassified | O | O |
| S2-wms-combined_197577 | Phage DP SC_6_H4_2017 | MT121964 | Clustered | VC_1420_0 | C1 | 138 | Unspecified | Group I | Duplodnaviria | Heunggongvirae | Uroviricota | Caudoviricetes | n.a. | Unclassified | Unclassified | Unclassified |
| S2-wms-combined_207832 | Faecalibacterium phage FP_Lagaffe | MG711461 | Clustered | VC_717_0 | C3 | 94 | Faecalibacterium | Group I | Duplodnaviria | Heunggongvirae | Uroviricota | Caudoviricetes | n.a. | Unclassified | Unclassified | Lagaffevirus |
| S2-wms-combined_210822 | n.a. | n.a. | Clustered | VC_1495_0 | A | n.a. | n.a. | n.a. | n.a. | n.a. | n.a. | n.a. | n.a. | n.a. | n.a. | n.a. |
| S2-wms-combined_218002 | Clostridioides phage CD1801 | MW512570 | Clustered | VC_148_1 | N13 | 1 | Clostridioides | Group I | Duplodnaviria | Heunggongvirae | Uroviricota | Caudoviricetes | n.a. | Unclassified | O | O |
| S2-wms-combined_227800 | Bacillus phage vB_BtS_BMBtp13 | KX190832 | Clustered | VC_420_0 | N7 | 5 | Bacillus | Group I | Duplodnaviria | Heunggongvirae | Uroviricota | Caudoviricetes | n.a. | Unclassified | O | O |
| S2-wms-combined_229529 | Phage vB_RanS_PJN03 | ON245412 | Clustered | VC_1473_0 | N6 | 2 | Unspecified | Group I | Duplodnaviria | Heunggongvirae | Uroviricota | Caudoviricetes | n.a. | Unclassified | O | O |
| S2-wms-combined_238929 | Bacillus phage Carmen17 | MG784342 | Clustered | VC_732_0 | N3 | 11 | Bacillus | Group I | Duplodnaviria | Heunggongvirae | Uroviricota | Caudoviricetes | n.a. | Unclassified | O | O |
| S2-wms-combined_244564 | Streptococcus phage Javan105 | MK448667 | Clustered | VC_626_0 | C2 | 58 | Streptococcus | Group I | Duplodnaviria | Heunggongvirae | Uroviricota | Caudoviricetes | n.a. | Unclassified | Unclassified | Unclassified |
| S2-wms-combined_246480 | Roseburia phage Jekyll | LR596902 | Clustered | VC_418_0 | C4 | 14 | Roseburia | Group I | Duplodnaviria | Heunggongvirae | Uroviricota | Caudoviricetes | n.a. | Unclassified | Unclassified | Unclassified |
| S2-wms-combined_248318 | n.a. | n.a. | Clustered | VC_1498_0 | F | n.a. | n.a. | n.a. | n.a. | n.a. | n.a. | n.a. | n.a. | n.a. | n.a. | n.a. |
| S2-wms-combined_248329 | Butyrivibrio phage Idris | MN882554 | Clustered | VC_734_0 | N11 | 3 | Butyrivibrio | Group I | Duplodnaviria | Heunggongvirae | Uroviricota | Caudoviricetes | n.a. | Unclassified | O | O |
| S2-wms-combined_31084 | Clostridium phage Clo-PEP-1 | KY206887 | Clustered | VC_212_1 | N4 | 11 | Clostridium | Group I | Duplodnaviria | Heunggongvirae | Uroviricota | Caudoviricetes | n.a. | Unclassified | O | O |
| S2-wms-combined_4685 | n.a. | n.a. | Clustered | VC_1484_0 | A | n.a. | n.a. | n.a. | n.a. | n.a. | n.a. | n.a. | n.a. | n.a. | n.a. | n.a. |
| S2-wms-combined_55495 | Geobacillus phage GBSV1 | DQ340064 | Clustered | VC_338_0 | N8 | 8 | Geobacillus | Group I | Duplodnaviria | Heunggongvirae | Uroviricota | Caudoviricetes | n.a. | Unclassified | O | O |
| S2-wms-combined_58121 | n.a. | n.a. | Clustered | VC_1436_0 | A | n.a. | n.a. | n.a. | n.a. | n.a. | n.a. | n.a. | n.a. | n.a. | n.a. | n.a. |
| S2-wms-combined_62377 | Bifidobacterium phage BitterVaud1 | MT006236 | Clustered | VC_804_2 | N7 | 33 | Bifidobacterium | Group I | Duplodnaviria | Heunggongvirae | Uroviricota | Caudoviricetes | n.a. | Unclassified | O | O |
| S2-wms-combined_62519 | Clostridioides phage CD1801 | MW512570 | Clustered | VC_148_0 | N11 | 3 | Clostridioides | Group I | Duplodnaviria | Heunggongvirae | Uroviricota | Caudoviricetes | n.a. | Unclassified | O | O |
| S2-wms-combined_6605 | n.a. | n.a. | Clustered | VC_1484_0 | A | n.a. | n.a. | n.a. | n.a. | n.a. | n.a. | n.a. | n.a. | n.a. | n.a. | n.a. |
| S2-wms-combined_85381 | Faecalibacterium phage FP_Epona | MG711462 | Clustered | VC_707_1 | C6 | 34 | Faecalibacterium | Group I | Duplodnaviria | Heunggongvirae | Uroviricota | Caudoviricetes | n.a. | Unclassified | Unclassified | Eponavirus |
| S2-wms-combined_89742 | Burkholderia phage BcepMu | AY539836 | Clustered | VC_610_0 | N4 | 4 | Burkholderia | Group I | Duplodnaviria | Heunggongvirae | Uroviricota | Caudoviricetes | n.a. | Unclassified | O | O |
| S2-wms-combined_93845 | Faecalibacterium phage FP_Mushu | MG711460 | Clustered | VC_271_0 | N8 | 8 | Faecalibacterium | Group I | Duplodnaviria | Heunggongvirae | Uroviricota | Caudoviricetes | n.a. | Unclassified | O | O |
| S2-wms_107329 | Faecalibacterium phage FP_Brigit | MG711465 | Clustered | VC_606_0 | C3 | 131 | Faecalibacterium | Group I | Duplodnaviria | Heunggongvirae | Uroviricota | Caudoviricetes | n.a. | Unclassified | Unclassified | Brigitvirus |
| S2-wms_110761 | n.a. | n.a. | Clustered | VC_1496_0 | F | n.a. | n.a. | n.a. | n.a. | n.a. | n.a. | n.a. | n.a. | n.a. | n.a. | n.a. |
| S2-wms_128831 | Phage DP SC_6_H4_2017 | MT121964 | Clustered | VC_1420_0 | C2 | 137 | Unspecified | Group I | Duplodnaviria | Heunggongvirae | Uroviricota | Caudoviricetes | n.a. | Unclassified | Unclassified | Unclassified |
| S2-wms_128962 | uncultured phage cr53_1 | MT774396 | Clustered | VC_502_0 | N2 | 228 | Unspecified | Group I | Duplodnaviria | Heunggongvirae | Uroviricota | Caudoviricetes | Crassvirales | Suoliviridae | O | O |
| S2-wms_130687 | Solobacterium phage SMO_1P | MZ503613 | Clustered | VC_417_0 | C2 | 23 | Solobacterium | Group I | Duplodnaviria | Heunggongvirae | Uroviricota | Caudoviricetes | n.a. | Unclassified | Unclassified | Unclassified |
| S2-wms_140933 | Roseburia phage Jekyll | LR596902 | Clustered | VC_418_0 | C4 | 20 | Roseburia | Group I | Duplodnaviria | Heunggongvirae | Uroviricota | Caudoviricetes | n.a. | Unclassified | Unclassified | Unclassified |
| S2-wms_156439 | Bacteroides phage F4 | MT806185 | Clustered | VC_1435_0 | N11 | 4 | Bacteroides | Group I | Duplodnaviria | Heunggongvirae | Uroviricota | Caudoviricetes | n.a. | Unclassified | O | O |
| S2-wms_23562 | n.a. | n.a. | Clustered | VC_1496_0 | F | n.a. | n.a. | n.a. | n.a. | n.a. | n.a. | n.a. | n.a. | n.a. | n.a. | n.a. |
| S2-wms_248315 | Faecalibacterium phage FP_oengus | MG711463 | Clustered | VC_804_2 | N7 | 45 | Faecalibacterium | Group I | Duplodnaviria | Heunggongvirae | Uroviricota | Caudoviricetes | n.a. | Unclassified | O | O |
| S2-wms_53881 | Bacillus phage Chedec 11 | ON042750 | Clustered | VC_821_0 | N2 | 7 | Bacillus | Group I | Duplodnaviria | Heunggongvirae | Uroviricota | Caudoviricetes | n.a. | Salasmaviridae | O | O |
| S2-wms_79304 | Actinomyces phage Av-1 | DQ123818 | Clustered | VC_744_0 | C1 | 19 | Actinomyces | Group I | Duplodnaviria | Heunggongvirae | Uroviricota | Caudoviricetes | n.a. | Unclassified | Unclassified | Dybvigvirus |
| S2-wms_79944 | Faecalibacterium phage FP_Brigit | MG711465 | Clustered | VC_825_0 | N9 | 16 | Faecalibacterium | Group I | Duplodnaviria | Heunggongvirae | Uroviricota | Caudoviricetes | n.a. | Unclassified | O | O |
| S3-wms-combined_73262 | Bacteroides phage p00 | BK010646 | Clustered | VC_677_0 | C1 | 108 | Bacteroides | Group I | Duplodnaviria | Heunggongvirae | Uroviricota | Caudoviricetes | n.a. | Unclassified | Unclassified | Unclassified |
| S3-wms_22744 | Faecalibacterium phage FP_Lagaffe | MG711461 | Clustered | VC_717_0 | C3 | 102 | Faecalibacterium | Group I | Duplodnaviria | Heunggongvirae | Uroviricota | Caudoviricetes | n.a. | Unclassified | Unclassified | Lagaffevirus |
| S3-wms_23958 | Caldibacillus phage CBP1 | MF595878 | Clustered | VC_915_0 | N18 | 5 | Caldibacillus | Group I | Duplodnaviria | Heunggongvirae | Uroviricota | Caudoviricetes | n.a. | Unclassified | O | O |
| S4-wms-combined_113721 | n.a. | n.a. | Clustered | VC_893_0 | A | n.a. | n.a. | n.a. | n.a. | n.a. | n.a. | n.a. | n.a. | n.a. | n.a. | n.a. |
| S4-wms-combined_143879 | n.a. | n.a. | Clustered | VC_1482_0 | F | n.a. | n.a. | n.a. | n.a. | n.a. | n.a. | n.a. | n.a. | n.a. | n.a. | n.a. |
| S4-wms-combined_14411 | Parabacteroides phage PDS1 | MN929097 | Clustered | VC_831_0 | C2 | 150 | Parabacteroides | Group I | Duplodnaviria | Heunggongvirae | Uroviricota | Caudoviricetes | n.a. | Unclassified | Unclassified | Unclassified |
| S4-wms-combined_17284 | Bifidobacterium phage Bbif-1 | GQ141189 | Clustered | VC_852_0 | C1 | 135 | Bifidobacterium | Group I | Duplodnaviria | Heunggongvirae | Uroviricota | Caudoviricetes | n.a. | Unclassified | Unclassified | Unclassified |
| S4-wms-combined_41490 | Burkholderia phage BcepMu | AY539836 | Clustered | VC_610_0 | N5 | 2 | Burkholderia | Group I | Duplodnaviria | Heunggongvirae | Uroviricota | Caudoviricetes | n.a. | Unclassified | O | O |
| S4-wms-combined_42647 | Faecalibacterium phage FP_Lugh | MG711464 | Clustered | VC_621_0 | C5 | 60 | Faecalibacterium | Group I | Duplodnaviria | Heunggongvirae | Uroviricota | Caudoviricetes | n.a. | Unclassified | Unclassified | Lughvirus |
| S4-wms-combined_47565 | n.a. | n.a. | Clustered | VC_1495_0 | A | n.a. | n.a. | n.a. | n.a. | n.a. | n.a. | n.a. | n.a. | n.a. | n.a. | n.a. |
| S4-wms-combined_51079 | n.a. | n.a. | Clustered | VC_1497_0 | F | n.a. | n.a. | n.a. | n.a. | n.a. | n.a. | n.a. | n.a. | n.a. | n.a. | n.a. |
| S4-wms-combined_57123 | n.a. | n.a. | Clustered | VC_1508_0 | A | n.a. | n.a. | n.a. | n.a. | n.a. | n.a. | n.a. | n.a. | n.a. | n.a. | n.a. |
| S4-wms-combined_64632 | Roseburia phage Jekyll | LR596902 | Clustered | VC_418_0 | C5 | 4 | Roseburia | Group I | Duplodnaviria | Heunggongvirae | Uroviricota | Caudoviricetes | n.a. | Unclassified | Unclassified | Unclassified |
| S4-wms_107182 | n.a. | n.a. | Clustered | VC_1513_0 | A | n.a. | n.a. | n.a. | n.a. | n.a. | n.a. | n.a. | n.a. | n.a. | n.a. | n.a. |
| S4-wms_118690 | Lactococcus phage phiQ1 | AP019527 | Clustered | VC_535_0 | N2 | 7 | Lactococcus | Group I | Duplodnaviria | Heunggongvirae | Uroviricota | Caudoviricetes | n.a. | Unclassified | O | O |
| S4-wms_141607 | Bacteroides phage F4 | MT806185 | Clustered | VC_1008_0 | C1 | 77 | Bacteroides | Group I | Duplodnaviria | Heunggongvirae | Uroviricota | Caudoviricetes | n.a. | Unclassified | Unclassified | Unclassified |
| S4-wms_161443 | Bacteriophage sp. | OP549880 | Clustered | VC_1477_0 | C2 | 25 | Unspecified | Unclassified | Unclassified | n.a. | n.a. | n.a. | n.a. | Unclassified | Unclassified | Unclassified |
| S4-wms_45128 | n.a. | n.a. | Clustered | VC_1439_0 | A | n.a. | n.a. | n.a. | n.a. | n.a. | n.a. | n.a. | n.a. | n.a. | n.a. | n.a. |
| S4-wms_62635 | Roseburia phage Jekyll | LR596902 | Clustered | VC_418_0 | C4 | 29 | Roseburia | Group I | Duplodnaviria | Heunggongvirae | Uroviricota | Caudoviricetes | n.a. | Unclassified | Unclassified | Unclassified |
| S4-wms_73990 | uncultured phage cr50_1 | MT774375 | Clustered | VC_502_1 | C1 | 130 | Unspecified | Group I | Duplodnaviria | Heunggongvirae | Uroviricota | Caudoviricetes | Crassvirales | Suoliviridae | Boorivirinae | Cohcovirus |
| S5-wms-combined_117213 | Bifidobacterium phage Bbif-1 | GQ141189 | Clustered | VC_854_0 | N3 | 19 | Bifidobacterium | Group I | Duplodnaviria | Heunggongvirae | Uroviricota | Caudoviricetes | n.a. | Unclassified | O | O |
| S5-wms-combined_134381 | Faecalibacterium phage FP_Lagaffe | MG711461 | Clustered | VC_717_0 | C3 | 84 | Faecalibacterium | Group I | Duplodnaviria | Heunggongvirae | Uroviricota | Caudoviricetes | n.a. | Unclassified | Unclassified | Lagaffevirus |
| S5-wms-combined_145067 | Clostridium phage phiCT453B | KM983328 | Clustered | VC_650_0 | N11 | 8 | Clostridium | Group I | Duplodnaviria | Heunggongvirae | Uroviricota | Caudoviricetes | n.a. | Unclassified | O | O |
| S5-wms-combined_178432 | n.a. | n.a. | Clustered | VC_1506_0 | A | n.a. | n.a. | n.a. | n.a. | n.a. | n.a. | n.a. | n.a. | n.a. | n.a. | n.a. |
| S5-wms-combined_18356 | Escherichia phage ESSI2_ev239 | LR597637 | Clustered | VC_5_0 | C1 | 58 | Escherichia | Group I | Duplodnaviria | Heunggongvirae | Uroviricota | Caudoviricetes | n.a. | Peduoviridae | Unclassified | Evevirus |
| S5-wms-combined_206050 | Streptococcus phage Javan105 | MK448667 | Clustered | VC_626_0 | C2 | 92 | Streptococcus | Group I | Duplodnaviria | Heunggongvirae | Uroviricota | Caudoviricetes | n.a. | Unclassified | Unclassified | Unclassified |
| S5-wms-combined_207291 | n.a. | n.a. | Clustered | VC_1437_0 | A | n.a. | n.a. | n.a. | n.a. | n.a. | n.a. | n.a. | n.a. | n.a. | n.a. | n.a. |
| S5-wms-combined_26971 | Butyrivibrio phage Idris | MN882554 | Clustered | VC_876_0 | N8 | 29 | Butyrivibrio | Group I | Duplodnaviria | Heunggongvirae | Uroviricota | Caudoviricetes | n.a. | Unclassified | O | O |
| S5-wms-combined_29272 | Streptococcus phage Javan630 | MK448997 | Clustered | VC_626_0 | C7 | 55 | Streptococcus | Group I | Duplodnaviria | Heunggongvirae | Uroviricota | Caudoviricetes | n.a. | Unclassified | Unclassified | Unclassified |
| S5-wms-combined_29447 | Winogradskyella phage Peternella_1 | MT732475 | Clustered | VC_676_0 | N8 | 17 | Winogradskyella | Group I | Duplodnaviria | Heunggongvirae | Uroviricota | Caudoviricetes | n.a. | Winoviridae | O | O |
| S5-wms-combined_39793 | n.a. | n.a. | Clustered | VC_1514_0 | F | n.a. | n.a. | n.a. | n.a. | n.a. | n.a. | n.a. | n.a. | n.a. | n.a. | n.a. |
| S5-wms-combined_52741 | Akkermansia phage DTMo-2021a | CP084202 | Clustered | VC_736_0 | N2 | 17 | Akkermansia | Group I | Duplodnaviria | Heunggongvirae | Uroviricota | Caudoviricetes | n.a. | Unclassified | O | O |
| S5-wms-combined_59064 | Bacteroides phage p00 | BK010646 | Clustered | VC_677_0 | C1 | 75 | Bacteroides | Group I | Duplodnaviria | Heunggongvirae | Uroviricota | Caudoviricetes | n.a. | Unclassified | Unclassified | Unclassified |
| S5-wms-combined_66017 | n.a. | n.a. | Clustered | VC_1499_0 | F | n.a. | n.a. | n.a. | n.a. | n.a. | n.a. | n.a. | n.a. | n.a. | n.a. | n.a. |
| S5-wms-combined_7292 | Caldibacillus phage CBP1 | MF595878 | Clustered | VC_734_0 | C2 | 8 | Caldibacillus | Group I | Duplodnaviria | Heunggongvirae | Uroviricota | Caudoviricetes | n.a. | Unclassified | Unclassified | Unclassified |
| S5-wms-combined_73301 | Clostridium phage phiCD38-2 | HM568888 | Clustered | VC_627_0 | N4 | 8 | Clostridium | Group I | Duplodnaviria | Heunggongvirae | Uroviricota | Caudoviricetes | n.a. | Unclassified | O | O |
| S5-wms-combined_78434 | n.a. | n.a. | Clustered | VC_1479_0 | A | n.a. | n.a. | n.a. | n.a. | n.a. | n.a. | n.a. | n.a. | n.a. | n.a. | n.a. |
| S5-wms-combined_83370 | Bacteroides phage p00 | BK010646 | Clustered | VC_680_0 | N2 | 29 | Bacteroides | Group I | Duplodnaviria | Heunggongvirae | Uroviricota | Caudoviricetes | n.a. | Unclassified | O | O |
| S5-wms-combined_83412 | Clostridium phage phiSM101 | CP000315 | Clustered | VC_19_0 | C1 | 15 | Clostridium | Group I | Duplodnaviria | Heunggongvirae | Uroviricota | Caudoviricetes | n.a. | Unclassified | Unclassified | Unclassified |
| S5-wms-combined_98202 | n.a. | n.a. | Clustered | VC_1503_0 | A | n.a. | n.a. | n.a. | n.a. | n.a. | n.a. | n.a. | n.a. | n.a. | n.a. | n.a. |
| S5-wms_210460 | n.a. | n.a. | Clustered | VC_1499_0 | F | n.a. | n.a. | n.a. | n.a. | n.a. | n.a. | n.a. | n.a. | n.a. | n.a. | n.a. |
| S5-wms_49310 | Solobacterium phage SMO_1P | MZ503613 | Clustered | VC_417_0 | C2 | 28 | Solobacterium | Group I | Duplodnaviria | Heunggongvirae | Uroviricota | Caudoviricetes | n.a. | Unclassified | Unclassified | Unclassified |
| S5-wms_52566 | n.a. | n.a. | Clustered | VC_1481_0 | F | n.a. | n.a. | n.a. | n.a. | n.a. | n.a. | n.a. | n.a. | n.a. | n.a. | n.a. |
| S5-wms_85650 | n.a. | n.a. | Clustered | VC_1505_0 | A | n.a. | n.a. | n.a. | n.a. | n.a. | n.a. | n.a. | n.a. | n.a. | n.a. | n.a. |
| S5-wms_94965 | Bifidobacterium phage BigBern1 | MT006237 | Clustered | VC_1198_0 | C2 | 16 | Bifidobacterium | Group I | Duplodnaviria | Heunggongvirae | Uroviricota | Caudoviricetes | n.a. | Unclassified | Unclassified | Unclassified |
| S6-wms-combined_113986 | Clostridium phage phiCD38-2 | HM568888 | Clustered | VC_627_0 | N3 | 9 | Clostridium | Group I | Duplodnaviria | Heunggongvirae | Uroviricota | Caudoviricetes | n.a. | Unclassified | O | O |
| S6-wms-combined_117756 | Faecalibacterium phage FP_Epona | MG711462 | Clustered | VC_707_1 | C3 | 96 | Faecalibacterium | Group I | Duplodnaviria | Heunggongvirae | Uroviricota | Caudoviricetes | n.a. | Unclassified | Unclassified | Eponavirus |
| S6-wms-combined_118484 | Bacteriophage sp. | LT985574 | Clustered | VC_338_0 | N10 | 5 | Unspecified | Unclassified | Unclassified | n.a. | n.a. | n.a. | n.a. | Unclassified | O | O |
| S6-wms-combined_119017 | Pseudomonas phage phi3 | KT887559 | Clustered | VC_5_0 | C2 | 39 | Pseudomonas | Group I | Duplodnaviria | Heunggongvirae | Uroviricota | Caudoviricetes | n.a. | Peduoviridae | Unclassified | Phitrevirus |
| S6-wms-combined_119138 | Clostridioides phage CD2301 | MW512571 | Clustered | VC_148_1 | N11 | 2 | Clostridioides | Group I | Duplodnaviria | Heunggongvirae | Uroviricota | Caudoviricetes | n.a. | Unclassified | O | O |

| | | | | | | | | | | | | | | | | |
|---|---|---|---|---|---|---|---|---|---|---|---|---|---|---|---|---|
| S6-wms-combined_126236 | Faecalibacterium phage FP_Toutatis | MG711466 | Clustered | VC_710_0 | C2 | 64 | Faecalibacterium | Group I | Duplodnaviria | Heunggongvirae | Uroviricota | Caudoviricetes | n.a. | Unclassified | Unclassified | Toutatisvirus |
| S6-wms-combined_135223 | n.a. | n.a. | Clustered | VC_1508_0 | A | n.a. | n.a. | n.a. | n.a. | n.a. | n.a. | n.a. | n.a. | n.a. | n.a. | n.a. |
| S6-wms-combined_140731 | n.a. | n.a. | Clustered | VC_825_0 | A | n.a. | n.a. | n.a. | n.a. | n.a. | n.a. | n.a. | n.a. | n.a. | n.a. | n.a. |
| S6-wms-combined_32530 | Clostridioides phage CD1801 | MW512570 | Clustered | VC_148_1 | N10 | 2 | Clostridioides | Group I | Duplodnaviria | Heunggongvirae | Uroviricota | Caudoviricetes | n.a. | Unclassified | O | O |
| S6-wms-combined_41246 | n.a. | n.a. | Clustered | VC_1484_0 | A | n.a. | n.a. | n.a. | n.a. | n.a. | n.a. | n.a. | n.a. | n.a. | n.a. | n.a. |
| S6-wms-combined_49909 | Streptococcus phage Javan35 | MK448736 | Clustered | VC_324_0 | N6 | 8 | Streptococcus | Group I | Duplodnaviria | Heunggongvirae | Uroviricota | Caudoviricetes | n.a. | Unclassified | O | O |
| S6-wms-combined_50721 | Winogradskyella phage Peternella_1 | MT732475 | Clustered | VC_676_1 | N11 | 16 | Winogradskyella | Group I | Duplodnaviria | Heunggongvirae | Uroviricota | Caudoviricetes | n.a. | Winoviridae | O | O |
| S6-wms-combined_72228 | n.a. | n.a. | Clustered | VC_1506_0 | A | n.a. | n.a. | n.a. | n.a. | n.a. | n.a. | n.a. | n.a. | n.a. | n.a. | n.a. |
| S6-wms-combined_8508 | Faecalibacterium phage FP_Epona | MG711462 | Clustered | VC_707_0 | N11 | 42 | Faecalibacterium | Group I | Duplodnaviria | Heunggongvirae | Uroviricota | Caudoviricetes | n.a. | Unclassified | O | O |
| S6-wms-combined_91039 | n.a. | n.a. | Clustered | VC_1456_0 | A | n.a. | n.a. | n.a. | n.a. | n.a. | n.a. | n.a. | n.a. | n.a. | n.a. | n.a. |
| S6-wms_132588 | Clostridium phage Clo-PEP-1 | KY206887 | Clustered | VC_212_0 | N4 | 18 | Clostridium | Group I | Duplodnaviria | Heunggongvirae | Uroviricota | Caudoviricetes | n.a. | Unclassified | O | O |
| S6-wms_137591 | n.a. | n.a. | Clustered | VC_1496_0 | F | n.a. | n.a. | n.a. | n.a. | n.a. | n.a. | n.a. | n.a. | n.a. | n.a. | n.a. |
| S6-wms_138342 | uncultured phage cr53_1 | MT774396 | Clustered | VC_502_0 | N2 | 233 | Unspecified | Group I | Duplodnaviria | Heunggongvirae | Uroviricota | Caudoviricetes | Crassvirales | Suoliviridae | O | O |
| S6-wms_54578 | Parabacteroides phage PDS1 | MN929097 | Clustered | VC_831_0 | C3 | 113 | Parabacteroides | Group I | Duplodnaviria | Heunggongvirae | Uroviricota | Caudoviricetes | n.a. | Unclassified | Unclassified | Unclassified |
| S7-wms-combined_104872 | n.a. | n.a. | Clustered | VC_1437_0 | A | n.a. | n.a. | n.a. | n.a. | n.a. | n.a. | n.a. | n.a. | n.a. | n.a. | n.a. |
| S7-wms-combined_107679_0 | Winogradskyella phage Peternella_1 | MT732475 | Clustered | VC_676_1 | N11 | 17 | Winogradskyella | Group I | Duplodnaviria | Heunggongvirae | Uroviricota | Caudoviricetes | n.a. | Winoviridae | O | O |
| S7-wms-combined_111004 | Faecalibacterium phage FP_Taranis | MG711467 | Clustered | VC_711_0 | C1 | 200 | Faecalibacterium | Group I | Duplodnaviria | Heunggongvirae | Uroviricota | Caudoviricetes | n.a. | Unclassified | Unclassified | Taranisvirus |
| S7-wms-combined_134458 | Faecalibacterium phage FP_Toutatis | MG711466 | Clustered | VC_710_0 | C4 | 73 | Faecalibacterium | Group I | Duplodnaviria | Heunggongvirae | Uroviricota | Caudoviricetes | n.a. | Unclassified | Unclassified | Toutatisvirus |
| S7-wms-combined_136642 | n.a. | n.a. | Clustered | VC_1497_0 | F | n.a. | n.a. | n.a. | n.a. | n.a. | n.a. | n.a. | n.a. | n.a. | n.a. | n.a. |
| S7-wms-combined_139683 | Butyrivibrio phage Idris | MN882554 | Clustered | VC_876_0 | N6 | 34 | Butyrivibrio | Group I | Duplodnaviria | Heunggongvirae | Uroviricota | Caudoviricetes | n.a. | Unclassified | O | O |
| S7-wms-combined_14267 | n.a. | n.a. | Clustered | VC_1456_0 | A | n.a. | n.a. | n.a. | n.a. | n.a. | n.a. | n.a. | n.a. | n.a. | n.a. | n.a. |
| S7-wms-combined_143218 | n.a. | n.a. | Clustered | VC_1514_0 | F | n.a. | n.a. | n.a. | n.a. | n.a. | n.a. | n.a. | n.a. | n.a. | n.a. | n.a. |
| S7-wms-combined_147158 | Microbacterium phage Min1 | EF579802 | Clustered | VC_798_0 | N6 | 14 | Microbacterium | Group I | Duplodnaviria | Heunggongvirae | Uroviricota | Caudoviricetes | n.a. | Unclassified | O | O |
| S7-wms-combined_163980 | n.a. | n.a. | Clustered | VC_1014_0 | A | n.a. | n.a. | n.a. | n.a. | n.a. | n.a. | n.a. | n.a. | n.a. | n.a. | n.a. |
| S7-wms-combined_185016 | Bacteroides phage F4 | MT806185 | Clustered | VC_1435_0 | N11 | 3 | Bacteroides | Group I | Duplodnaviria | Heunggongvirae | Uroviricota | Caudoviricetes | n.a. | Unclassified | O | O |
| S7-wms-combined_189250 | Bifidobacterium phage BigBern1 | MT006237 | Clustered | VC_1198_0 | C2 | 5 | Bifidobacterium | Group I | Duplodnaviria | Heunggongvirae | Uroviricota | Caudoviricetes | n.a. | Unclassified | Unclassified | Unclassified |
| S7-wms-combined_225260 | Clostridium phage HM2 | LT600745 | Clustered | VC_822_0 | C1 | 4 | Clostridium | Group I | Duplodnaviria | Heunggongvirae | Uroviricota | Caudoviricetes | n.a. | Salasmaviridae | Unclassified | Unclassified |
| S7-wms-combined_235165 | Erysipelothrix phage SE-1 | KR816341 | Clustered | VC_1109_0 | C1 | 12 | Erysipelothrix | Group I | Duplodnaviria | Heunggongvirae | Uroviricota | Caudoviricetes | n.a. | Unclassified | Unclassified | Unclassified |
| S7-wms-combined_239784 | n.a. | n.a. | Clustered | VC_1436_0 | A | n.a. | n.a. | n.a. | n.a. | n.a. | n.a. | n.a. | n.a. | n.a. | n.a. | n.a. |
| S7-wms-combined_28138 | Listeria phage LP-030-2 | JX120799 | Clustered | VC_420_0 | N8 | 6 | Listeria | Group I | Duplodnaviria | Heunggongvirae | Uroviricota | Caudoviricetes | n.a. | Unclassified | O | O |
| S7-wms-combined_31068 | n.a. | n.a. | Clustered | VC_650_0 | A | n.a. | n.a. | n.a. | n.a. | n.a. | n.a. | n.a. | n.a. | n.a. | n.a. | n.a. |
| S7-wms-combined_40779 | n.a. | n.a. | Clustered | VC_1239_0 | A | n.a. | n.a. | n.a. | n.a. | n.a. | n.a. | n.a. | n.a. | n.a. | n.a. | n.a. |
| S7-wms-combined_43950 | Bacteroides phage p00 | BK010646 | Clustered | VC_677_0 | C1 | 129 | Bacteroides | Group I | Duplodnaviria | Heunggongvirae | Uroviricota | Caudoviricetes | n.a. | Unclassified | Unclassified | Unclassified |
| S7-wms-combined_5625 | Bacteroides phage p00 | BK010646 | Clustered | VC_677_0 | C4 | 35 | Bacteroides | Group I | Duplodnaviria | Heunggongvirae | Uroviricota | Caudoviricetes | n.a. | Unclassified | Unclassified | Unclassified |
| S7-wms-combined_7205 | Clostridium phage Clo-PEP-1 | KY206887 | Clustered | VC_212_1 | N5 | 16 | Clostridium | Group I | Duplodnaviria | Heunggongvirae | Uroviricota | Caudoviricetes | n.a. | Unclassified | O | O |
| S7-wms-combined_72070 | n.a. | n.a. | Clustered | VC_1509_0 | A | n.a. | n.a. | n.a. | n.a. | n.a. | n.a. | n.a. | n.a. | n.a. | n.a. | n.a. |
| S7-wms-combined_84688 | n.a. | n.a. | Clustered | VC_1484_0 | A | n.a. | n.a. | n.a. | n.a. | n.a. | n.a. | n.a. | n.a. | n.a. | n.a. | n.a. |
| S7-wms-combined_90393 | Bacteroides phage F4 | MT806185 | Clustered | VC_1435_0 | N10 | 8 | Bacteroides | Group I | Duplodnaviria | Heunggongvirae | Uroviricota | Caudoviricetes | n.a. | Unclassified | O | O |
| S7-wms-combined_90728 | Bacteroides phage F4 | MT806185 | Clustered | VC_1435_0 | N6 | 11 | Bacteroides | Group I | Duplodnaviria | Heunggongvirae | Uroviricota | Caudoviricetes | n.a. | Unclassified | O | O |
| S7-wms_113137 | Faecalibacterium phage FP_Mushu | MG711460 | Clustered | VC_1235_0 | C1 | 170 | Faecalibacterium | Group I | Duplodnaviria | Heunggongvirae | Uroviricota | Caudoviricetes | n.a. | Unclassified | Unclassified | Mushuvirus |
| S7-wms_118607 | Faecalibacterium phage FP_Lagaffe | MG711461 | Clustered | VC_717_0 | C1 | 230 | Faecalibacterium | Group I | Duplodnaviria | Heunggongvirae | Uroviricota | Caudoviricetes | n.a. | Unclassified | Unclassified | Lagaffevirus |
| S7-wms_123954 | n.a. | n.a. | Clustered | VC_1435_0 | A | n.a. | n.a. | n.a. | n.a. | n.a. | n.a. | n.a. | n.a. | n.a. | n.a. | n.a. |
| S7-wms_127801 | Clostridium phage phiCD6356 | GU949551 | Clustered | VC_420_0 | N4 | 6 | Clostridium | Group I | Duplodnaviria | Heunggongvirae | Uroviricota | Caudoviricetes | n.a. | Unclassified | O | O |
| S7-wms_157264 | Bacteriophage sp. | OP549880 | Clustered | VC_1477_0 | C2 | 24 | Unspecified | Unclassified | Unclassified | n.a. | n.a. | n.a. | n.a. | Unclassified | Unclassified | Unclassified |
| S7-wms_159432 | Faecalibacterium phage FP_Epona | MG711462 | Clustered | VC_707_1 | C4 | 67 | Faecalibacterium | Group I | Duplodnaviria | Heunggongvirae | Uroviricota | Caudoviricetes | n.a. | Unclassified | Unclassified | Eponavirus |
| S7-wms_200936 | Faecalibacterium phage FP_Mushu | MG711460 | Clustered | VC_1235_0 | C2 | 142 | Faecalibacterium | Group I | Duplodnaviria | Heunggongvirae | Uroviricota | Caudoviricetes | n.a. | Unclassified | Unclassified | Mushuvirus |
| S7-wms_239833 | Bacillus phage vB_BsuP-Goe1 | KU831549 | Clustered | VC_821_0 | N1 | 10 | Bacillus | Group I | Duplodnaviria | Heunggongvirae | Uroviricota | Caudoviricetes | n.a. | Salasmaviridae | O | O |
| S7-wms_244425 | uncultured phage cr127_1 | MT774393 | Clustered | VC_675_0 | C1 | 70 | Unspecified | Group I | Duplodnaviria | Heunggongvirae | Uroviricota | Caudoviricetes | Crassvirales | Crevaviridae | Doltivirinae | Kahucivirus |
| S7-wms_244866 | Bacteroides phage EMB2 | ON721385 | Clustered | VC_1009_0 | C1 | 149 | Bacteroides | Group I | Duplodnaviria | Heunggongvirae | Uroviricota | Caudoviricetes | n.a. | Unclassified | Unclassified | Unclassified |
| S7-wms_53338 | Enterococcus phage Athos | LR990834 | Clustered | VC_821_0 | N1 | 9 | Enterococcus | Group I | Duplodnaviria | Heunggongvirae | Uroviricota | Caudoviricetes | n.a. | Salasmaviridae | O | O |
| S7-wms_80255 | Streptococcus phage Javan249 | MK448716 | Clustered | VC_629_0 | N2 | 10 | Streptococcus | Group I | Duplodnaviria | Heunggongvirae | Uroviricota | Caudoviricetes | n.a. | Unclassified | O | O |
| S8-wms-combined_108917 | Streptococcus phage Javan630 | MK448997 | Clustered | VC_626_0 | C7 | 54 | Streptococcus | Group I | Duplodnaviria | Heunggongvirae | Uroviricota | Caudoviricetes | n.a. | Unclassified | Unclassified | Unclassified |
| S8-wms-combined_134098 | n.a. | n.a. | Clustered | VC_1239_0 | A | n.a. | n.a. | n.a. | n.a. | n.a. | n.a. | n.a. | n.a. | n.a. | n.a. | n.a. |
| S8-wms-combined_134284 | n.a. | n.a. | Clustered | VC_1236_0 | A | n.a. | n.a. | n.a. | n.a. | n.a. | n.a. | n.a. | n.a. | n.a. | n.a. | n.a. |
| S8-wms-combined_140059 | n.a. | n.a. | Clustered | VC_420_0 | A | n.a. | n.a. | n.a. | n.a. | n.a. | n.a. | n.a. | n.a. | n.a. | n.a. | n.a. |
| S8-wms-combined_146562 | Faecalibacterium phage FP_Mushu | MG711460 | Clustered | VC_271_0 | N8 | 13 | Faecalibacterium | Group I | Duplodnaviria | Heunggongvirae | Uroviricota | Caudoviricetes | n.a. | Unclassified | O | O |
| S8-wms-combined_161181 | Clostridioides phage CD1801 | MW512570 | Clustered | VC_148_0 | N12 | 4 | Clostridioides | Group I | Duplodnaviria | Heunggongvirae | Uroviricota | Caudoviricetes | n.a. | Unclassified | O | O |
| S8-wms-combined_169835 | n.a. | n.a. | Clustered | VC_915_0 | A | n.a. | n.a. | n.a. | n.a. | n.a. | n.a. | n.a. | n.a. | n.a. | n.a. | n.a. |
| S8-wms-combined_186054 | Streptococcus phage Javan630 | MK448997 | Clustered | VC_626_0 | C5 | 72 | Streptococcus | Group I | Duplodnaviria | Heunggongvirae | Uroviricota | Caudoviricetes | n.a. | Unclassified | Unclassified | Unclassified |
| S8-wms-combined_200168 | Faecalibacterium phage FP_Epona | MG711462 | Clustered | VC_707_1 | C2 | 42 | Faecalibacterium | Group I | Duplodnaviria | Heunggongvirae | Uroviricota | Caudoviricetes | n.a. | Unclassified | Unclassified | Eponavirus |
| S8-wms-combined_211119 | n.a. | n.a. | Clustered | VC_1236_0 | A | n.a. | n.a. | n.a. | n.a. | n.a. | n.a. | n.a. | n.a. | n.a. | n.a. | n.a. |
| S8-wms-combined_215102 | n.a. | n.a. | Clustered | VC_1504_0 | A | n.a. | n.a. | n.a. | n.a. | n.a. | n.a. | n.a. | n.a. | n.a. | n.a. | n.a. |
| S8-wms-combined_217866 | Faecalibacterium phage FP_Epona | MG711462 | Clustered | VC_707_1 | C4 | 84 | Faecalibacterium | Group I | Duplodnaviria | Heunggongvirae | Uroviricota | Caudoviricetes | n.a. | Unclassified | Unclassified | Eponavirus |
| S8-wms-combined_226076 | n.a. | n.a. | Clustered | VC_1511_0 | F | n.a. | n.a. | n.a. | n.a. | n.a. | n.a. | n.a. | n.a. | n.a. | n.a. | n.a. |
| S8-wms-combined_234473 | n.a. | n.a. | Clustered | VC_1481_0 | F | n.a. | n.a. | n.a. | n.a. | n.a. | n.a. | n.a. | n.a. | n.a. | n.a. | n.a. |
| S8-wms-combined_246185 | Akkermansia phage DTMo-2021a | CP084202 | Clustered | VC_738_0 | N3 | 1 | Akkermansia | Group I | Duplodnaviria | Heunggongvirae | Uroviricota | Caudoviricetes | n.a. | Unclassified | O | O |
| S8-wms-combined_25607 | Faecalibacterium phage FP_Taranis | MG711467 | Clustered | VC_1239_0 | N20 | 2 | Faecalibacterium | Group I | Duplodnaviria | Heunggongvirae | Uroviricota | Caudoviricetes | n.a. | Unclassified | O | O |
| S8-wms-combined_26087 | Burkholderia phage BcepMu | AY539836 | Clustered | VC_610_0 | N5 | 1 | Burkholderia | Group I | Duplodnaviria | Heunggongvirae | Uroviricota | Caudoviricetes | n.a. | Unclassified | O | O |
| S8-wms-combined_264711 | n.a. | n.a. | Clustered | VC_1481_0 | F | n.a. | n.a. | n.a. | n.a. | n.a. | n.a. | n.a. | n.a. | n.a. | n.a. | n.a. |
| S8-wms-combined_270362 | Phage vB_RanS_PJN03 | ON245412 | Clustered | VC_917_0 | N10 | 6 | Unspecified | Group I | Duplodnaviria | Heunggongvirae | Uroviricota | Caudoviricetes | n.a. | Unclassified | O | O |
| S8-wms-combined_276204 | n.a. | n.a. | Clustered | VC_1501_0 | A | n.a. | n.a. | n.a. | n.a. | n.a. | n.a. | n.a. | n.a. | n.a. | n.a. | n.a. |
| S8-wms-combined_276211 | Butyrivibrio virus Ceridwen | MN882553 | Clustered | VC_1209_0 | N10 | 11 | Butyrivibrio | Group I | Duplodnaviria | Heunggongvirae | Uroviricota | Caudoviricetes | n.a. | Unclassified | O | O |
| S8-wms-combined_49369 | n.a. | n.a. | Clustered | VC_1236_0 | A | n.a. | n.a. | n.a. | n.a. | n.a. | n.a. | n.a. | n.a. | n.a. | n.a. | n.a. |
| S8-wms-combined_68780 | n.a. | n.a. | Clustered | VC_271_0 | A | n.a. | n.a. | n.a. | n.a. | n.a. | n.a. | n.a. | n.a. | n.a. | n.a. | n.a. |
| S8-wms-combined_72911 | n.a. | n.a. | Clustered | VC_1076_0 | A | n.a. | n.a. | n.a. | n.a. | n.a. | n.a. | n.a. | n.a. | n.a. | n.a. | n.a. |
| S8-wms-combined_97848 | Bacteroides phage p00 | BK010646 | Clustered | VC_677_0 | C4 | 36 | Bacteroides | Group I | Duplodnaviria | Heunggongvirae | Uroviricota | Caudoviricetes | n.a. | Unclassified | Unclassified | Unclassified |
| S8-wms_127191 | Faecalibacterium phage FP_oengus | MG711463 | Clustered | VC_804_2 | N2 | 66 | Faecalibacterium | Group I | Duplodnaviria | Heunggongvirae | Uroviricota | Caudoviricetes | n.a. | Unclassified | O | O |
| S8-wms_152831 | Faecalibacterium phage FP_Toutatis | MG711466 | Clustered | VC_710_0 | C1 | 82 | Faecalibacterium | Group I | Duplodnaviria | Heunggongvirae | Uroviricota | Caudoviricetes | n.a. | Unclassified | Unclassified | Toutatisvirus |
| S8-wms_244525 | Bacillus phage BeachBum | KY921761 | Clustered | VC_821_0 | N1 | 10 | Bacillus | Group I | Duplodnaviria | Heunggongvirae | Uroviricota | Caudoviricetes | n.a. | Salasmaviridae | O | O |
| S8-wms_30498 | Faecalibacterium phage FP_Mushu | MG711460 | Clustered | VC_1235_0 | C4 | 112 | Faecalibacterium | Group I | Duplodnaviria | Heunggongvirae | Uroviricota | Caudoviricetes | n.a. | Unclassified | Unclassified | Mushuvirus |
| S8-wms_48935 | Faecalibacterium phage FP_Toutatis | MG711466 | Clustered | VC_710_0 | C6 | 71 | Faecalibacterium | Group I | Duplodnaviria | Heunggongvirae | Uroviricota | Caudoviricetes | n.a. | Unclassified | Unclassified | Toutatisvirus |
| S8-wms_57121 | n.a. | n.a. | Clustered | VC_1501_0 | A | n.a. | n.a. | n.a. | n.a. | n.a. | n.a. | n.a. | n.a. | n.a. | n.a. | n.a. |
| S8-wms_82880 | Faecalibacterium phage FP_Brigit | MG711465 | Clustered | VC_606_0 | C3 | 104 | Faecalibacterium | Group I | Duplodnaviria | Heunggongvirae | Uroviricota | Caudoviricetes | n.a. | Unclassified | Unclassified | Brigitvirus |
| S9-wms-combined_105462 | Clostridioides phage phiCD27 | EU719189 | Clustered | VC_825_0 | N9 | 3 | Clostridioides | Group I | Duplodnaviria | Heunggongvirae | Uroviricota | Caudoviricetes | n.a. | Unclassified | O | O |
| S9-wms-combined_25007 | Streptococcus phage Javan422 | MK448934 | Clustered | VC_626_0 | C1 | 52 | Streptococcus | Group I | Duplodnaviria | Heunggongvirae | Uroviricota | Caudoviricetes | n.a. | Unclassified | Unclassified | Unclassified |
| S9-wms-combined_26410 | Faecalibacterium phage FP_oengus | MG711463 | Clustered | VC_804_2 | N8 | 48 | Faecalibacterium | Group I | Duplodnaviria | Heunggongvirae | Uroviricota | Caudoviricetes | n.a. | Unclassified | O | O |
| S9-wms-combined_28805 | Winogradskyella phage Peternella_1 | MT732475 | Clustered | VC_676_1 | N11 | 18 | Winogradskyella | Group I | Duplodnaviria | Heunggongvirae | Uroviricota | Caudoviricetes | n.a. | Winoviridae | O | O |

| | | | | | | | | | | | | | | | | |
|---|---|---|---|---|---|---|---|---|---|---|---|---|---|---|---|---|
| S9-wms-combined_31456 | Bacteroides phage p00 | BK010646 | Clustered | VC_677_0 | C4 | 33 | Bacteroides | Group I | Duplodnaviria | Heunggongvirae | Uroviricota | Caudoviricetes | n.a. | Unclassified | Unclassified | Unclassified |
| S9-wms-combined_33229 | Bacteroides phage F2 | MT806187 | Clustered | VC_1435_0 | N11 | 7 | Bacteroides | Group I | Duplodnaviria | Heunggongvirae | Uroviricota | Caudoviricetes | n.a. | Unclassified | O | O |
| S9-wms-combined_33230 | Bacteroides phage F4 | MT806185 | Clustered | VC_1435_0 | N11 | 3 | Bacteroides | Group I | Duplodnaviria | Heunggongvirae | Uroviricota | Caudoviricetes | n.a. | Unclassified | O | O |
| S9-wms-combined_36459 | Streptococcus phage Javan553 | MK448804 | Clustered | VC_338_0 | N6 | 6 | Streptococcus | Group I | Duplodnaviria | Heunggongvirae | Uroviricota | Caudoviricetes | n.a. | Unclassified | O | O |
| S9-wms-combined_37587 | Phage vB_RanS_PJN03 | ON245412 | Clustered | VC_1473_0 | N5 | 4 | Unspecified | Group I | Duplodnaviria | Heunggongvirae | Uroviricota | Caudoviricetes | n.a. | Unclassified | O | O |
| S9-wms-combined_3878 | n.a. | n.a. | Clustered | VC_1510_0 | F | n.a. | n.a. | n.a. | n.a. | n.a. | n.a. | n.a. | n.a. | n.a. | n.a. | n.a. |
| S9-wms-combined_44402 | Bacteroides phage p00 | BK010646 | Clustered | VC_680_0 | N2 | 31 | Bacteroides | Group I | Duplodnaviria | Heunggongvirae | Uroviricota | Caudoviricetes | n.a. | Unclassified | O | O |
| S9-wms-combined_53991 | Phage WC36-2 | OL791266 | Clustered | VC_947_0 | N1 | 24 | Unspecified | Group I | Duplodnaviria | Heunggongvirae | Uroviricota | Caudoviricetes | n.a. | Unclassified | O | O |
| S9-wms-combined_5526 | Bacteroides phage LoVEphage | MW660583 | Clustered | VC_832_0 | N10 | 1 | Bacteroides | Group I | Duplodnaviria | Heunggongvirae | Uroviricota | Caudoviricetes | n.a. | Unclassified | O | O |
| S9-wms-combined_56773 | n.a. | n.a. | Clustered | VC_1481_0 | F | n.a. | n.a. | n.a. | n.a. | n.a. | n.a. | n.a. | n.a. | n.a. | n.a. | n.a. |
| S9-wms-combined_64277 | Burkholderia phage phiE255 | CP000622 | Clustered | VC_250_0 | C1 | 83 | Burkholderia | Group I | Duplodnaviria | Heunggongvirae | Uroviricota | Caudoviricetes | n.a. | Unclassified | Unclassified | Bcepmuvirus |
| S9-wms-combined_71647 | Bacillus phage vB_BtS_BMBtp13 | KX190832 | Clustered | VC_420_0 | N6 | 5 | Bacillus | Group I | Duplodnaviria | Heunggongvirae | Uroviricota | Caudoviricetes | n.a. | Unclassified | O | O |
| S9-wms-combined_76274 | Bifidobacterium phage Bbif-1 | GQ141189 | Clustered | VC_854_0 | N3 | 19 | Bifidobacterium | Group I | Duplodnaviria | Heunggongvirae | Uroviricota | Caudoviricetes | n.a. | Unclassified | O | O |
| S9-wms-combined_89249 | Roseburia phage Jekyll | LR596902 | Clustered | VC_947_0 | N4 | 3 | Roseburia | Group I | Duplodnaviria | Heunggongvirae | Uroviricota | Caudoviricetes | n.a. | Unclassified | O | O |
| S9-wms_104169 | Inovirus sp. | MT135285 | Clustered | VC_1423_0 | C1 | 13 | Unspecified | Group II | Monodnaviria | Loebvirae | Hofneiviricota | Faserviricetes | Tubulavirales | Inoviridae | Unclassified | Inovirus |
| S9-wms_128232 | Tortoise microvirus 13 | MK765563 | Clustered | VC_1243_0 | C1 | 26 | Unspecified | Group II | Monodnaviria | Sangervirae | Phixviricota | Malgrandaviricetes | Petitvirales | Microviridae | Unclassified | Unclassified |
| S9-wms_19025 | Lactococcus phage phiQ1 | AP019527 | Clustered | VC_535_0 | N2 | 9 | Lactococcus | Group I | Duplodnaviria | Heunggongvirae | Uroviricota | Caudoviricetes | n.a. | Unclassified | O | O |
| S9-wms_64472 | n.a. | n.a. | Clustered | VC_1512_0 | F | n.a. | n.a. | n.a. | n.a. | n.a. | n.a. | n.a. | n.a. | n.a. | n.a. | n.a. |
| S1-wms-combined_220022 | n.a. | n.a. | Clustered/Singleton | VC_1408_1 | A | n.a. | n.a. | n.a. | n.a. | n.a. | n.a. | n.a. | n.a. | n.a. | n.a. | n.a. |
| S1-wms-combined_85225 | Propionibacterium phage Anatole | KX620748 | Clustered/Singleton | VC_798_1 | N4 | 12 | Propionibacterium | Group I | Duplodnaviria | Heunggongvirae | Uroviricota | Caudoviricetes | n.a. | Unclassified | O | O |
| S1-wms_102342 | Bacillus phage vB_BcoS-136 | MH884508 | Clustered/Singleton | VC_719_0 | N2 | 1 | Bacillus | Group I | Duplodnaviria | Heunggongvirae | Uroviricota | Caudoviricetes | n.a. | Unclassified | O | O |
| S11-wms-combined_264050 | Faecalibacterium phage FP_Epona | MG711462 | Clustered/Singleton | VC_707_3 | C1 | 64 | Faecalibacterium | Group I | Duplodnaviria | Heunggongvirae | Uroviricota | Caudoviricetes | n.a. | Unclassified | Unclassified | O |
| S11-wms-combined_267993 | Bacillus phage BalMu-1 | KP063902 | Clustered/Singleton | VC_825_1 | N11 | 4 | Bacillus | Group I | Duplodnaviria | Heunggongvirae | Uroviricota | Caudoviricetes | n.a. | Unclassified | O | O |
| S11-wms-combined_3507 | Butyrivibrio virus Ceridwen | MN882553 | Clustered/Singleton | VC_1209_2 | N8 | 17 | Butyrivibrio | Group I | Duplodnaviria | Heunggongvirae | Uroviricota | Caudoviricetes | n.a. | Unclassified | O | O |
| S12-wms-combined_129799 | Faecalibacterium phage FP_Epona | MG711462 | Clustered/Singleton | VC_707_2 | C1 | 100 | Faecalibacterium | Group I | Duplodnaviria | Heunggongvirae | Uroviricota | Caudoviricetes | n.a. | Unclassified | Unclassified | O |
| S12-wms-combined_157167 | n.a. | n.a. | Clustered/Singleton | VC_1489_0 | F | n.a. | n.a. | n.a. | n.a. | n.a. | n.a. | n.a. | n.a. | n.a. | n.a. | n.a. |
| S12-wms-combined_201125 | Phage vB_RanS_PJN03 | ON245412 | Clustered/Singleton | VC_918_2 | N12 | 10 | Unspecified | Group I | Duplodnaviria | Heunggongvirae | Uroviricota | Caudoviricetes | n.a. | Unclassified | O | O |
| S12-wms-combined_23517 | Butyrivibrio virus Ceridwen | MN882553 | Clustered/Singleton | VC_1209_1 | N11 | 7 | Butyrivibrio | Group I | Duplodnaviria | Heunggongvirae | Uroviricota | Caudoviricetes | n.a. | Unclassified | O | O |
| S12-wms-combined_323943 | Faecalibacterium phage FP_Taranis | MG711467 | Clustered/Singleton | VC_711_1 | C1 | 110 | Faecalibacterium | Group I | Duplodnaviria | Heunggongvirae | Uroviricota | Caudoviricetes | n.a. | Unclassified | Unclassified | O |
| S12-wms-combined_358797 | Elizabethkingia phage TCUEAP3 | OK632026 | Clustered/Singleton | VC_678_0 | N11 | 9 | Elizabethkingia | Group I | Duplodnaviria | Heunggongvirae | Uroviricota | Caudoviricetes | n.a. | Unclassified | O | O |
| S12-wms-combined_98046 | Streptococcus phage Javan630 | MK448997 | Clustered/Singleton | VC_626_2 | C5 | 48 | Streptococcus | Group I | Duplodnaviria | Heunggongvirae | Uroviricota | Caudoviricetes | n.a. | Unclassified | Unclassified | O |
| S17-wms-combined_186314 | Bifidobacterium phage Bbif-1 | GQ141189 | Clustered/Singleton | VC_798_2 | N5 | 15 | Bifidobacterium | Group I | Duplodnaviria | Heunggongvirae | Uroviricota | Caudoviricetes | n.a. | Unclassified | O | O |
| S17-wms-combined_2234 | Faecalibacterium phage FP_Taranis | MG711467 | Clustered/Singleton | VC_711_2 | C3 | 88 | Faecalibacterium | Group I | Duplodnaviria | Heunggongvirae | Uroviricota | Caudoviricetes | n.a. | Unclassified | Unclassified | O |
| S17-wms-combined_248321 | Faecalibacterium phage FP_Lagaffe | MG711461 | Clustered/Singleton | VC_717_1 | C4 | 90 | Faecalibacterium | Group I | Duplodnaviria | Heunggongvirae | Uroviricota | Caudoviricetes | n.a. | Unclassified | Unclassified | O |
| S17-wms-combined_95360 | Clostridium phage phiCT453A | KM983327 | Clustered/Singleton | VC_732_1 | N4 | 3 | Clostridium | Group I | Duplodnaviria | Heunggongvirae | Uroviricota | Caudoviricetes | n.a. | Unclassified | O | O |
| S2-wms-combined_116221 | Butyrivibrio phage Idris | MN882554 | Clustered/Singleton | VC_876_1 | N5 | 33 | Butyrivibrio | Group I | Duplodnaviria | Heunggongvirae | Uroviricota | Caudoviricetes | n.a. | Unclassified | O | O |
| S2-wms-combined_13246 | Faecalibacterium phage FP_Toutatis | MG711466 | Clustered/Singleton | VC_710_4 | C2 | 69 | Faecalibacterium | Group I | Duplodnaviria | Heunggongvirae | Uroviricota | Caudoviricetes | n.a. | Unclassified | Unclassified | O |
| S2-wms-combined_137119 | Faecalibacterium phage FP_Toutatis | MG711466 | Clustered/Singleton | VC_710_1 | C8 | 55 | Faecalibacterium | Group I | Duplodnaviria | Heunggongvirae | Uroviricota | Caudoviricetes | n.a. | Unclassified | Unclassified | O |
| S2-wms-combined_140996 | Faecalibacterium phage FP_Taranis | MG711467 | Clustered/Singleton | VC_711_3 | C2 | 76 | Faecalibacterium | Group I | Duplodnaviria | Heunggongvirae | Uroviricota | Caudoviricetes | n.a. | Unclassified | Unclassified | O |
| S2-wms-combined_157934 | Faecalibacterium phage FP_Mushu | MG711460 | Clustered/Singleton | VC_1235_1 | C4 | 67 | Faecalibacterium | Group I | Duplodnaviria | Heunggongvirae | Uroviricota | Caudoviricetes | n.a. | Unclassified | Unclassified | O |
| S2-wms-combined_233107 | n.a. | n.a. | Clustered/Singleton | VC_1485_0 | A | n.a. | n.a. | n.a. | n.a. | n.a. | n.a. | n.a. | n.a. | n.a. | n.a. | n.a. |
| S2-wms-combined_24549 | Clostridioides phage JD033 | MT193276 | Clustered/Singleton | VC_651_0 | N8 | 10 | Clostridioides | Group I | Duplodnaviria | Heunggongvirae | Uroviricota | Caudoviricetes | n.a. | Unclassified | O | O |
| S2-wms-combined_98888 | Faecalibacterium phage FP_oengus | MG711463 | Clustered/Singleton | VC_804_3 | C9 | 41 | Faecalibacterium | Group I | Duplodnaviria | Heunggongvirae | Uroviricota | Caudoviricetes | n.a. | Unclassified | Unclassified | O |
| S5-wms-combined_108794 | Streptococcus phage Javan422 | MK448934 | Clustered/Singleton | VC_626_3 | C5 | 55 | Streptococcus | Group I | Duplodnaviria | Heunggongvirae | Uroviricota | Caudoviricetes | n.a. | Unclassified | Unclassified | O |
| S5-wms-combined_134743 | n.a. | n.a. | Clustered/Singleton | VC_825_2 | A | n.a. | n.a. | n.a. | n.a. | n.a. | n.a. | n.a. | n.a. | n.a. | n.a. | n.a. |
| S5-wms-combined_66662 | Campylobacter phage A11a | MG065691 | Clustered/Singleton | VC_1219_1 | C1 | 8 | Campylobacter | Unclassified | Unclassified | n.a. | n.a. | n.a. | n.a. | Unclassified | Unclassified | O |
| S5-wms-combined_72410 | Faecalibacterium phage FP_oengus | MG711463 | Clustered/Singleton | VC_804_4 | C8 | 41 | Faecalibacterium | Group I | Duplodnaviria | Heunggongvirae | Uroviricota | Caudoviricetes | n.a. | Unclassified | Unclassified | O |
| S5-wms-combined_90315 | Phage vB_RanS_PJN03 | ON245412 | Clustered/Singleton | VC_917_1 | N9 | 10 | Unspecified | Group I | Duplodnaviria | Heunggongvirae | Uroviricota | Caudoviricetes | n.a. | Unclassified | O | O |
| S5-wms_67169 | Roseburia phage Shimadzu | LR596903 | Clustered/Singleton | VC_914_0 | N3 | 5 | Roseburia | Group I | Duplodnaviria | Heunggongvirae | Uroviricota | Caudoviricetes | n.a. | Unclassified | O | O |
| S6-wms-combined_118427 | Faecalibacterium phage FP_Toutatis | MG711466 | Clustered/Singleton | VC_710_3 | C8 | 60 | Faecalibacterium | Group I | Duplodnaviria | Heunggongvirae | Uroviricota | Caudoviricetes | n.a. | Unclassified | Unclassified | O |
| S6-wms-combined_82597 | n.a. | n.a. | Clustered/Singleton | VC_1435_1 | A | n.a. | n.a. | n.a. | n.a. | n.a. | n.a. | n.a. | n.a. | n.a. | n.a. | n.a. |
| S7-wms-combined_157260 | Faecalibacterium phage FP_Toutatis | MG711466 | Clustered/Singleton | VC_710_2 | C11 | 24 | Faecalibacterium | Group I | Duplodnaviria | Heunggongvirae | Uroviricota | Caudoviricetes | n.a. | Unclassified | Unclassified | O |
| S7-wms-combined_169375 | n.a. | n.a. | Clustered/Singleton | VC_1483_0 | A | n.a. | n.a. | n.a. | n.a. | n.a. | n.a. | n.a. | n.a. | n.a. | n.a. | n.a. |
| S7-wms-combined_20921 | Bacillus phage PBC2 | KT070867 | Clustered/Singleton | VC_719_1 | N2 | 5 | Bacillus | Group I | Duplodnaviria | Heunggongvirae | Uroviricota | Caudoviricetes | n.a. | Unclassified | O | O |
| S7-wms-combined_238231 | n.a. | n.a. | Clustered/Singleton | VC_271_2 | A | n.a. | n.a. | n.a. | n.a. | n.a. | n.a. | n.a. | n.a. | n.a. | n.a. | n.a. |
| S7-wms-combined_26217 | Phage vB_RanS_PJN03 | ON245412 | Clustered/Singleton | VC_918_1 | N12 | 9 | Unspecified | Group I | Duplodnaviria | Heunggongvirae | Uroviricota | Caudoviricetes | n.a. | Unclassified | O | O |
| S7-wms-combined_3697 | Campylobacter phage A11a | MG065691 | Clustered/Singleton | VC_832_1 | N13 | 2 | Campylobacter | Unclassified | Unclassified | n.a. | n.a. | n.a. | n.a. | Unclassified | O | O |
| S7-wms-combined_62119 | Bacteroides phage LoVEphage | MW660583 | Clustered/Singleton | VC_1221_1 | C1 | 248 | Bacteroides | Group I | Duplodnaviria | Heunggongvirae | Uroviricota | Caudoviricetes | n.a. | Unclassified | Unclassified | O |
| S8-wms-combined_225114 | Faecalibacterium phage FP_Taranis | MG711467 | Clustered/Singleton | VC_271_1 | N1 | 70 | Faecalibacterium | Group I | Duplodnaviria | Heunggongvirae | Uroviricota | Caudoviricetes | n.a. | Unclassified | O | O |
| S8-wms-combined_40666 | n.a. | n.a. | Clustered/Singleton | VC_1483_1 | A | n.a. | n.a. | n.a. | n.a. | n.a. | n.a. | n.a. | n.a. | n.a. | n.a. | n.a. |
| S8-wms_111056 | Bacteroides phage crAss002 | MN917146 | Clustered/Singleton | VC_674_0 | N1 | 134 | Bacteroides | Group I | Duplodnaviria | Heunggongvirae | Uroviricota | Caudoviricetes | Crassvirales | Intestiviridae | O | O |
| S9-wms-combined_20485 | Streptococcus phage P0092 | KY705252 | Clustered/Singleton | VC_255_1 | C1 | 66 | Streptococcus | Group I | Duplodnaviria | Heunggongvirae | Uroviricota | Caudoviricetes | n.a. | Unclassified | Unclassified | O |
| S9-wms-combined_29611 | Faecalibacterium phage FP_Brigit | MG711465 | Clustered/Singleton | VC_606_1 | C3 | 85 | Faecalibacterium | Group I | Duplodnaviria | Heunggongvirae | Uroviricota | Caudoviricetes | n.a. | Unclassified | Unclassified | O |
| S9-wms-combined_9931 | Erysipelothrix phage phi1605 | MF172979 | Clustered/Singleton | VC_626_1 | C6 | 58 | Erysipelothrix | Group I | Duplodnaviria | Heunggongvirae | Uroviricota | Caudoviricetes | n.a. | Unclassified | Unclassified | O |
| S1-wms-combined_102801 | n.a. | n.a. | Outlier | n.a. | A | n.a. | n.a. | n.a. | n.a. | n.a. | n.a. | n.a. | n.a. | n.a. | n.a. | n.a. |
| S1-wms-combined_116938 | Streptococcus phage Javan580 | MK448989 | Outlier | n.a. | N5 | 9 | Streptococcus | Group I | Duplodnaviria | Heunggongvirae | Uroviricota | Caudoviricetes | n.a. | Unclassified | O | O |
| S1-wms-combined_131270 | Butyrivibrio virus Ceridwen | MN882553 | Outlier | n.a. | N2 | 15 | Butyrivibrio | Group I | Duplodnaviria | Heunggongvirae | Uroviricota | Caudoviricetes | n.a. | Unclassified | O | O |
| S1-wms-combined_136097 | n.a. | n.a. | Outlier | n.a. | A | n.a. | n.a. | n.a. | n.a. | n.a. | n.a. | n.a. | n.a. | n.a. | n.a. | n.a. |
| S1-wms-combined_143459 | Pseudomonas phage vB_Pae_CF5a | MK511057 | Outlier | n.a. | N1 | 15 | Pseudomonas | Group I | Duplodnaviria | Heunggongvirae | Uroviricota | Caudoviricetes | n.a. | Unclassified | O | O |
| S1-wms-combined_164894 | Faecalibacterium phage FP_Lugh | MG711464 | Outlier | n.a. | N4 | 16 | Faecalibacterium | Group I | Duplodnaviria | Heunggongvirae | Uroviricota | Caudoviricetes | n.a. | Unclassified | O | O |
| S1-wms-combined_186878 | n.a. | n.a. | Outlier | n.a. | A | n.a. | n.a. | n.a. | n.a. | n.a. | n.a. | n.a. | n.a. | n.a. | n.a. | n.a. |
| S1-wms-combined_212892 | Butyrivibrio virus Ceridwen | MN882553 | Outlier | n.a. | N1 | 19 | Butyrivibrio | Group I | Duplodnaviria | Heunggongvirae | Uroviricota | Caudoviricetes | n.a. | Unclassified | O | O |
| S1-wms-combined_227003 | n.a. | n.a. | Outlier | n.a. | A | n.a. | n.a. | n.a. | n.a. | n.a. | n.a. | n.a. | n.a. | n.a. | n.a. | n.a. |
| S1-wms-combined_227917 | Faecalibacterium phage FP_Lugh | MG711464 | Outlier | n.a. | N2 | 15 | Faecalibacterium | Group I | Duplodnaviria | Heunggongvirae | Uroviricota | Caudoviricetes | n.a. | Unclassified | O | O |
| S1-wms-combined_252439 | Bacillus phage vB_BpsS-140 | MH884512 | Outlier | n.a. | N1 | 2 | Bacillus | Group I | Duplodnaviria | Heunggongvirae | Uroviricota | Caudoviricetes | n.a. | Unclassified | O | O |
| S1-wms-combined_279492 | Streptococcus phage Javan580 | MK448989 | Outlier | n.a. | N6 | 8 | Streptococcus | Group I | Duplodnaviria | Heunggongvirae | Uroviricota | Caudoviricetes | n.a. | Unclassified | O | O |
| S1-wms-combined_308848 | n.a. | n.a. | Outlier | n.a. | A | n.a. | n.a. | n.a. | n.a. | n.a. | n.a. | n.a. | n.a. | n.a. | n.a. | n.a. |
| S1-wms-combined_57370 | Faecalibacterium phage FP_Lagaffe | MG711461 | Outlier | n.a. | N15 | 3 | Faecalibacterium | Group I | Duplodnaviria | Heunggongvirae | Uroviricota | Caudoviricetes | n.a. | Unclassified | O | O |
| S1-wms-combined_92523 | Bacteriophage sp. | MW202489 | Outlier | n.a. | N1 | 6 | Unspecified | Unclassified | Unclassified | n.a. | n.a. | n.a. | n.a. | Unclassified | O | O |
| S1-wms-combined_94988 | n.a. | n.a. | Outlier | n.a. | A | n.a. | n.a. | n.a. | n.a. | n.a. | n.a. | n.a. | n.a. | n.a. | n.a. | n.a. |
| S1-wms_106115 | n.a. | n.a. | Outlier | n.a. | A | n.a. | n.a. | n.a. | n.a. | n.a. | n.a. | n.a. | n.a. | n.a. | n.a. | n.a. |
| S1-wms_208875 | Bacteroides phage ARB14 | MT074134 | Outlier | n.a. | N1 | 61 | Bacteroides | Group I | Duplodnaviria | Heunggongvirae | Uroviricota | Caudoviricetes | n.a. | Unclassified | O | O |
| S1-wms_236105 | Roseburia phage Jekyll | LR596902 | Outlier | n.a. | N11 | 3 | Roseburia | Group I | Duplodnaviria | Heunggongvirae | Uroviricota | Caudoviricetes | n.a. | Unclassified | O | O |
| S1-wms_29288 | Faecalibacterium phage FP_Taranis | MG711467 | Outlier | n.a. | N2 | 61 | Faecalibacterium | Group I | Duplodnaviria | Heunggongvirae | Uroviricota | Caudoviricetes | n.a. | Unclassified | O | O |
| S1-wms_394571 | Bacteroides phage SJC03 | MT074146 | Outlier | n.a. | N1 | 36 | Bacteroides | Group I | Duplodnaviria | Heunggongvirae | Uroviricota | Caudoviricetes | n.a. | Unclassified | O | O |
| S11-wms-combined_101337 | Klebsiella phage ST11-OXA245phi3.2 | MK416010 | Outlier | n.a. | N1 | 14 | Klebsiella | Group I | Duplodnaviria | Heunggongvirae | Uroviricota | Caudoviricetes | n.a. | Unclassified | O | O |
| S11-wms-combined_112147 | n.a. | n.a. | Outlier | n.a. | A | n.a. | n.a. | n.a. | n.a. | n.a. | n.a. | n.a. | n.a. | n.a. | n.a. | n.a. |

| | | | | | | | | | | | | | | | |
|---|---|---|---|---|---|---|---|---|---|---|---|---|---|---|---|
| S11-wms-combined_112733 | Alteromonadaceae phage B23 | KY939598 | Outlier | n.a. | N1 | 18 | Unspecified | Group I | Duplodnaviria | Heunggongvirae | Uroviricota | Caudoviricetes | n.a. | Unclassified | O | O |
| S11-wms-combined_116589 | n.a. | n.a. | Outlier | n.a. | A | n.a. | n.a. | n.a. | n.a. | n.a. | n.a. | n.a. | n.a. | n.a. | n.a. | n.a. |
| S11-wms-combined_14636 | Streptococcus phage Javan68 | MK449004 | Outlier | n.a. | N3 | 10 | Streptococcus | Group I | Duplodnaviria | Heunggongvirae | Uroviricota | Caudoviricetes | n.a. | Unclassified | O | O |
| S11-wms-combined_168048 | Streptococcus phage Javan1 | MK448665 | Outlier | n.a. | N2 | 5 | Streptococcus | Group I | Duplodnaviria | Heunggongvirae | Uroviricota | Caudoviricetes | n.a. | Unclassified | O | O |
| S11-wms-combined_215723 | Cellulophaga phage Ingeline_1 | MT732435 | Outlier | n.a. | N3 | 8 | Cellulophaga | Group I | Duplodnaviria | Heunggongvirae | Uroviricota | Caudoviricetes | n.a. | Duneviridae | O | O |
| S11-wms-combined_236214 | Faecalibacterium phage FP_oengus | MG711463 | Outlier | n.a. | N4 | 32 | Faecalibacterium | Group I | Duplodnaviria | Heunggongvirae | Uroviricota | Caudoviricetes | n.a. | Unclassified | O | O |
| S11-wms-combined_282227 | Streptococcus phage Javan422 | MK448934 | Outlier | n.a. | N4 | 33 | Streptococcus | Group I | Duplodnaviria | Heunggongvirae | Uroviricota | Caudoviricetes | n.a. | Unclassified | O | O |
| S11-wms-combined_329493 | n.a. | n.a. | Outlier | n.a. | A | n.a. | n.a. | n.a. | n.a. | n.a. | n.a. | n.a. | n.a. | n.a. | n.a. | n.a. |
| S11-wms-combined_63818 | Bacteroides phage LoVEphage | MW660583 | Outlier | n.a. | N11 | 8 | Bacteroides | Group I | Duplodnaviria | Heunggongvirae | Uroviricota | Caudoviricetes | n.a. | Unclassified | O | O |
| S11-wms_87293 | Yersinia phage YeP4 | MK733262 | Outlier | n.a. | N1 | 1 | Yersinia | Group I | Duplodnaviria | Heunggongvirae | Uroviricota | Caudoviricetes | n.a. | Unclassified | O | O |
| S12-wms-combined_151843 | Aeribacillus phage AP45 | KX965989 | Outlier | n.a. | N1 | 15 | Aeribacillus | Group I | Duplodnaviria | Heunggongvirae | Uroviricota | Caudoviricetes | n.a. | Unclassified | O | O |
| S12-wms-combined_164656 | n.a. | n.a. | Outlier | n.a. | A | n.a. | n.a. | n.a. | n.a. | n.a. | n.a. | n.a. | n.a. | n.a. | n.a. | n.a. |
| S12-wms-combined_190114 | Bacteroides phage p00 | BK010646 | Outlier | n.a. | N10 | 18 | Bacteroides | Group I | Duplodnaviria | Heunggongvirae | Uroviricota | Caudoviricetes | n.a. | Unclassified | O | O |
| S12-wms-combined_21504 | Acetobacter phage phiAO1 | LC644971 | Outlier | n.a. | N1 | 10 | Acetobacter | Group I | Duplodnaviria | Heunggongvirae | Uroviricota | Caudoviricetes | n.a. | Unclassified | O | O |
| S12-wms-combined_223043 | Faecalibacterium phage FP_Mushu | MG711460 | Outlier | n.a. | N6 | 47 | Faecalibacterium | Group I | Duplodnaviria | Heunggongvirae | Uroviricota | Caudoviricetes | n.a. | Unclassified | O | O |
| S12-wms-combined_264077 | Faecalibacterium phage FP_Toutatis | MG711466 | Outlier | n.a. | N1 | 57 | Faecalibacterium | Group I | Duplodnaviria | Heunggongvirae | Uroviricota | Caudoviricetes | n.a. | Unclassified | O | O |
| S12-wms-combined_293354 | Clostridium phage phiCP26F | GQ443085 | Outlier | n.a. | N4 | 2 | Clostridium | Group I | Duplodnaviria | Heunggongvirae | Uroviricota | Caudoviricetes | n.a. | Unclassified | O | O |
| S12-wms-combined_302876 | Clostridium phage phiCT453B | KM983328 | Outlier | n.a. | N1 | 22 | Clostridium | Group I | Duplodnaviria | Heunggongvirae | Uroviricota | Caudoviricetes | n.a. | Unclassified | O | O |
| S12-wms-combined_313953 | Acidithiobacillus phage AcaML1 | JX507079 | Outlier | n.a. | N1 | 25 | Acidithiobacillus | Group I | Duplodnaviria | Heunggongvirae | Uroviricota | Caudoviricetes | n.a. | Unclassified | O | O |
| S12-wms-combined_325418 | Streptococcus phage Javan422 | MK448934 | Outlier | n.a. | N9 | 13 | Streptococcus | Group I | Duplodnaviria | Heunggongvirae | Uroviricota | Caudoviricetes | n.a. | Unclassified | O | O |
| S12-wms-combined_328554 | Elizabethkingia phage TCUEAP2 | OK632025 | Outlier | n.a. | N1 | 20 | Elizabethkingia | Group I | Duplodnaviria | Heunggongvirae | Uroviricota | Caudoviricetes | n.a. | Unclassified | O | O |
| S12-wms-combined_359190 | Streptococcus phage Javan100 | MK448838 | Outlier | n.a. | N7 | 26 | Streptococcus | Group I | Duplodnaviria | Heunggongvirae | Uroviricota | Caudoviricetes | n.a. | Unclassified | O | O |
| S12-wms-combined_368562 | Bacteroides phage p00 | BK010646 | Outlier | n.a. | N7 | 10 | Bacteroides | Group I | Duplodnaviria | Heunggongvirae | Uroviricota | Caudoviricetes | n.a. | Unclassified | O | O |
| S12-wms-combined_371541 | n.a. | n.a. | Outlier | n.a. | A | n.a. | n.a. | n.a. | n.a. | n.a. | n.a. | n.a. | n.a. | n.a. | n.a. | n.a. |
| S12-wms-combined_48648 | n.a. | n.a. | Outlier | n.a. | A | n.a. | n.a. | n.a. | n.a. | n.a. | n.a. | n.a. | n.a. | n.a. | n.a. | n.a. |
| S12-wms-combined_63649 | n.a. | n.a. | Outlier | n.a. | A | n.a. | n.a. | n.a. | n.a. | n.a. | n.a. | n.a. | n.a. | n.a. | n.a. | n.a. |
| S12-wms_135291 | Streptococcus phage phi-SgaBSJ31_rum | MN270259 | Outlier | n.a. | N2 | 2 | Streptococcus | Group I | Duplodnaviria | Heunggongvirae | Uroviricota | Caudoviricetes | n.a. | Unclassified | O | O |
| S12-wms_17950 | Butyrivibrio virus Arian | MN882551 | Outlier | n.a. | N1 | 2 | Butyrivibrio | Group I | Duplodnaviria | Heunggongvirae | Uroviricota | Caudoviricetes | n.a. | Unclassified | O | O |
| S12-wms_202803 | Faecalibacterium phage FP_Lagaffe | MG711461 | Outlier | n.a. | N31 | 3 | Faecalibacterium | Group I | Duplodnaviria | Heunggongvirae | Uroviricota | Caudoviricetes | n.a. | Unclassified | O | O |
| S12-wms_235125 | n.a. | n.a. | Outlier | n.a. | A | n.a. | n.a. | n.a. | n.a. | n.a. | n.a. | n.a. | n.a. | n.a. | n.a. | n.a. |
| S17-wms-combined_106940 | Riemerella phage RAP44 | HQ396194 | Outlier | n.a. | N1 | 20 | Riemerella | Group I | Duplodnaviria | Heunggongvirae | Uroviricota | Caudoviricetes | n.a. | Unclassified | O | O |
| S17-wms-combined_140517 | Streptococcus phage Javan100 | MK448838 | Outlier | n.a. | N4 | 35 | Streptococcus | Group I | Duplodnaviria | Heunggongvirae | Uroviricota | Caudoviricetes | n.a. | Unclassified | O | O |
| S17-wms-combined_146050 | n.a. | n.a. | Outlier | n.a. | A | n.a. | n.a. | n.a. | n.a. | n.a. | n.a. | n.a. | n.a. | n.a. | n.a. | n.a. |
| S17-wms-combined_171305 | Bacteroides phage LoVEphage | MW660583 | Outlier | n.a. | N2 | 1 | Bacteroides | Group I | Duplodnaviria | Heunggongvirae | Uroviricota | Caudoviricetes | n.a. | Unclassified | O | O |
| S17-wms-combined_185239 | n.a. | n.a. | Outlier | n.a. | A | n.a. | n.a. | n.a. | n.a. | n.a. | n.a. | n.a. | n.a. | n.a. | n.a. | n.a. |
| S17-wms-combined_72837 | Bacillus phage B13 | OP066531 | Outlier | n.a. | N3 | 8 | Bacillus | Group I | Duplodnaviria | Heunggongvirae | Uroviricota | Caudoviricetes | n.a. | Unclassified | O | O |
| S17-wms_235536 | n.a. | n.a. | Outlier | n.a. | F | n.a. | n.a. | n.a. | n.a. | n.a. | n.a. | n.a. | n.a. | n.a. | n.a. | n.a. |
| S2-wms-combined_110215 | Faecalibacterium phage FP_Brigit | MG711465 | Outlier | n.a. | N7 | 23 | Faecalibacterium | Group I | Duplodnaviria | Heunggongvirae | Uroviricota | Caudoviricetes | n.a. | Unclassified | O | O |
| S2-wms-combined_126440 | Roseburia phage Jekyll | LR596902 | Outlier | n.a. | N2 | 9 | Roseburia | Group I | Duplodnaviria | Heunggongvirae | Uroviricota | Caudoviricetes | n.a. | Unclassified | O | O |
| S2-wms-combined_127080 | n.a. | n.a. | Outlier | n.a. | A | n.a. | n.a. | n.a. | n.a. | n.a. | n.a. | n.a. | n.a. | n.a. | n.a. | n.a. |
| S2-wms-combined_143737 | Cellulophaga phage Ingeline_1 | MT732435 | Outlier | n.a. | N5 | 2 | Cellulophaga | Group I | Duplodnaviria | Heunggongvirae | Uroviricota | Caudoviricetes | n.a. | Duneviridae | O | O |
| S2-wms-combined_159670 | n.a. | n.a. | Outlier | n.a. | A | n.a. | n.a. | n.a. | n.a. | n.a. | n.a. | n.a. | n.a. | n.a. | n.a. | n.a. |
| S2-wms-combined_170845 | Streptococcus phage Javan261 | MK448720 | Outlier | n.a. | N8 | 16 | Streptococcus | Group I | Duplodnaviria | Heunggongvirae | Uroviricota | Caudoviricetes | n.a. | Unclassified | O | O |
| S2-wms-combined_181391 | n.a. | n.a. | Outlier | n.a. | A | n.a. | n.a. | n.a. | n.a. | n.a. | n.a. | n.a. | n.a. | n.a. | n.a. | n.a. |
| S2-wms-combined_185494 | Delftia phage IME-DE1 | KR153873 | Outlier | n.a. | N8 | 4 | Delftia | Group I | Duplodnaviria | Heunggongvirae | Uroviricota | Caudoviricetes | n.a. | Autographiviridae | O | O |
| S2-wms-combined_190624 | n.a. | n.a. | Outlier | n.a. | A | n.a. | n.a. | n.a. | n.a. | n.a. | n.a. | n.a. | n.a. | n.a. | n.a. | n.a. |
| S2-wms-combined_191613 | n.a. | n.a. | Outlier | n.a. | A | n.a. | n.a. | n.a. | n.a. | n.a. | n.a. | n.a. | n.a. | n.a. | n.a. | n.a. |
| S2-wms-combined_199384 | Clostridioides phage phiCD08011 | MW512572 | Outlier | n.a. | N15 | 2 | Clostridioides | Group I | Duplodnaviria | Heunggongvirae | Uroviricota | Caudoviricetes | n.a. | Unclassified | O | O |
| S2-wms-combined_200390 | Clostridium phage phiCD38-2 | HM568888 | Outlier | n.a. | N4 | 4 | Clostridium | Group I | Duplodnaviria | Heunggongvirae | Uroviricota | Caudoviricetes | n.a. | Unclassified | O | O |
| S2-wms-combined_225908 | Campylobacter phage A11a | MG065691 | Outlier | n.a. | N8 | 2 | Campylobacter | Unclassified | Unclassified | n.a. | n.a. | n.a. | n.a. | Unclassified | O | O |
| S2-wms-combined_25727 | Geobacillus phage GBSV1 | DQ340064 | Outlier | n.a. | N5 | 2 | Geobacillus | Group I | Duplodnaviria | Heunggongvirae | Uroviricota | Caudoviricetes | n.a. | Unclassified | O | O |
| S2-wms-combined_51446 | n.a. | n.a. | Outlier | n.a. | F | n.a. | n.a. | n.a. | n.a. | n.a. | n.a. | n.a. | n.a. | n.a. | n.a. | n.a. |
| S2-wms-combined_52568 | n.a. | n.a. | Outlier | n.a. | A | n.a. | n.a. | n.a. | n.a. | n.a. | n.a. | n.a. | n.a. | n.a. | n.a. | n.a. |
| S2-wms-combined_5791 | Butyrivibrio phage Idris | MN882554 | Outlier | n.a. | N4 | 15 | Butyrivibrio | Group I | Duplodnaviria | Heunggongvirae | Uroviricota | Caudoviricetes | n.a. | Unclassified | O | O |
| S2-wms-combined_62839 | Bacillus phage PK-3 | ON881243 | Outlier | n.a. | N1 | 9 | Bacillus | Group I | Duplodnaviria | Heunggongvirae | Uroviricota | Caudoviricetes | n.a. | Unclassified | O | O |
| S2-wms-combined_72725 | n.a. | n.a. | Outlier | n.a. | A | n.a. | n.a. | n.a. | n.a. | n.a. | n.a. | n.a. | n.a. | n.a. | n.a. | n.a. |
| S2-wms-combined_74387 | Faecalibacterium phage FP_Lugh | MG711464 | Outlier | n.a. | N4 | 10 | Faecalibacterium | Group I | Duplodnaviria | Heunggongvirae | Uroviricota | Caudoviricetes | n.a. | Unclassified | O | O |
| S2-wms-combined_75144 | n.a. | n.a. | Outlier | n.a. | A | n.a. | n.a. | n.a. | n.a. | n.a. | n.a. | n.a. | n.a. | n.a. | n.a. | n.a. |
| S2-wms-combined_7756 | Akkermansia phage DTMo-2021a | CP084202 | Outlier | n.a. | N3 | 11 | Akkermansia | Group I | Duplodnaviria | Heunggongvirae | Uroviricota | Caudoviricetes | n.a. | Unclassified | O | O |
| S2-wms-combined_78670 | Bacillus phage vB_BtS_BMBtp15 | KX190835 | Outlier | n.a. | N3 | 6 | Bacillus | Group I | Duplodnaviria | Heunggongvirae | Uroviricota | Caudoviricetes | n.a. | Unclassified | O | O |
| S2-wms-combined_84608 | Bordetella phage BPP-1 | AY029185 | Outlier | n.a. | N1 | 30 | Bordetella | Group I | Duplodnaviria | Heunggongvirae | Uroviricota | Caudoviricetes | n.a. | Unclassified | O | O |
| S2-wms_246296 | n.a. | n.a. | Outlier | n.a. | A | n.a. | n.a. | n.a. | n.a. | n.a. | n.a. | n.a. | n.a. | n.a. | n.a. | n.a. |
| S2-wms_248324 | Clostridium phage phiCTP1 | HM159959 | Outlier | n.a. | N1 | 6 | Clostridium | Group I | Duplodnaviria | Heunggongvirae | Uroviricota | Caudoviricetes | n.a. | Unclassified | O | O |
| S2-wms_248327 | n.a. | n.a. | Outlier | n.a. | F | n.a. | n.a. | n.a. | n.a. | n.a. | n.a. | n.a. | n.a. | n.a. | n.a. | n.a. |
| S2-wms_6424 | Tenacibaculum phage Gundel_1 | MT732474 | Outlier | n.a. | N1 | 8 | Tenacibaculum | Group I | Duplodnaviria | Heunggongvirae | Uroviricota | Caudoviricetes | n.a. | Pachyviridae | O | O |
| S3-wms-combined_135428 | Propionibacterium phage Anatole | KX620748 | Outlier | n.a. | N2 | 15 | Propionibacterium | Group I | Duplodnaviria | Heunggongvirae | Uroviricota | Caudoviricetes | n.a. | Unclassified | O | O |
| S3-wms-combined_45305 | Faecalibacterium phage FP_Lugh | MG711464 | Outlier | n.a. | N2 | 19 | Faecalibacterium | Group I | Duplodnaviria | Heunggongvirae | Uroviricota | Caudoviricetes | n.a. | Unclassified | O | O |
| S3-wms_94494 | Bifidobacterium phage PMBT6 | MH444512 | Outlier | n.a. | N1 | 29 | Bifidobacterium | Group I | Duplodnaviria | Heunggongvirae | Uroviricota | Caudoviricetes | n.a. | Unclassified | O | O |
| S4-wms-combined_10758 | Cellulophaga phage phi17:2 | KC821609 | Outlier | n.a. | N1 | 7 | Cellulophaga | Group I | Duplodnaviria | Heunggongvirae | Uroviricota | Caudoviricetes | n.a. | Unclassified | O | O |
| S4-wms-combined_118531 | Podoviridae sp. | MW202722 | Outlier | n.a. | N1 | 8 | Unspecified | Group I | Duplodnaviria | Heunggongvirae | Uroviricota | Caudoviricetes | n.a. | Unclassified | O | O |
| S4-wms-combined_121978 | Gardnerella phage vB_Gva_AB1 | MW387018 | Outlier | n.a. | N5 | 36 | Gardnerella | Group I | Duplodnaviria | Heunggongvirae | Uroviricota | Caudoviricetes | n.a. | Unclassified | O | O |
| S4-wms-combined_1246 | n.a. | n.a. | Outlier | n.a. | A | n.a. | n.a. | n.a. | n.a. | n.a. | n.a. | n.a. | n.a. | n.a. | n.a. | n.a. |
| S4-wms-combined_22012 | Propionibacterium phage B3 | KX620749 | Outlier | n.a. | N3 | 16 | Propionibacterium | Group I | Duplodnaviria | Heunggongvirae | Uroviricota | Caudoviricetes | n.a. | Unclassified | O | O |
| S4-wms-combined_25201 | Streptococcus phage SM1 | AY007505 | Outlier | n.a. | N7 | 5 | Streptococcus | Group I | Duplodnaviria | Heunggongvirae | Uroviricota | Caudoviricetes | n.a. | Unclassified | O | O |
| S4-wms-combined_83492 | Bacteroides phage p00 | BK010646 | Outlier | n.a. | N9 | 18 | Bacteroides | Group I | Duplodnaviria | Heunggongvirae | Uroviricota | Caudoviricetes | n.a. | Unclassified | O | O |
| S4-wms_114120 | Pectobacterium phage DU_PP_III | MF979562 | Outlier | n.a. | N1 | 9 | Pectobacterium | Group I | Duplodnaviria | Heunggongvirae | Uroviricota | Caudoviricetes | n.a. | Unclassified | O | O |
| S4-wms_50475 | Phage vB_RanS_PJN03 | ON245412 | Outlier | n.a. | N10 | 3 | Unspecified | Group I | Duplodnaviria | Heunggongvirae | Uroviricota | Caudoviricetes | n.a. | Unclassified | O | O |
| S5-wms-combined_117095 | n.a. | n.a. | Outlier | n.a. | A | n.a. | n.a. | n.a. | n.a. | n.a. | n.a. | n.a. | n.a. | n.a. | n.a. | n.a. |
| S5-wms-combined_156329 | Geobacillus phage GBSV1 | DQ340064 | Outlier | n.a. | N7 | 4 | Geobacillus | Group I | Duplodnaviria | Heunggongvirae | Uroviricota | Caudoviricetes | n.a. | Unclassified | O | O |
| S5-wms-combined_161191 | n.a. | n.a. | Outlier | n.a. | A | n.a. | n.a. | n.a. | n.a. | n.a. | n.a. | n.a. | n.a. | n.a. | n.a. | n.a. |
| S5-wms-combined_175145 | Streptococcus phage Javan90 | MK449010 | Outlier | n.a. | N1 | 14 | Streptococcus | Group I | Duplodnaviria | Heunggongvirae | Uroviricota | Caudoviricetes | n.a. | Unclassified | O | O |
| S5-wms-combined_179241 | n.a. | n.a. | Outlier | n.a. | A | n.a. | n.a. | n.a. | n.a. | n.a. | n.a. | n.a. | n.a. | n.a. | n.a. | n.a. |
| S5-wms-combined_204782 | n.a. | n.a. | Outlier | n.a. | A | n.a. | n.a. | n.a. | n.a. | n.a. | n.a. | n.a. | n.a. | n.a. | n.a. | n.a. |
| S5-wms-combined_3201 | Yersinia phage YeP1 | MK733259 | Outlier | n.a. | N1 | 26 | Yersinia | Group I | Duplodnaviria | Heunggongvirae | Uroviricota | Caudoviricetes | n.a. | Unclassified | O | O |
| S5-wms-combined_94482 | n.a. | n.a. | Outlier | n.a. | A | n.a. | n.a. | n.a. | n.a. | n.a. | n.a. | n.a. | n.a. | n.a. | n.a. | n.a. |
| S5-wms_164032 | Lactococcus phage vB_LacS_15 | MN337887 | Outlier | n.a. | N1 | 46 | Lactococcus | Group I | Duplodnaviria | Heunggongvirae | Uroviricota | Caudoviricetes | n.a. | Unclassified | O | O |
| S6-wms-combined_56464 | Faecalibacterium phage FP_Lugh | MG711464 | Outlier | n.a. | N8 | 10 | Faecalibacterium | Group I | Duplodnaviria | Heunggongvirae | Uroviricota | Caudoviricetes | n.a. | Unclassified | O | O |

| | | | | | | | | | | | | | | | | |
|---|---|---|---|---|---|---|---|---|---|---|---|---|---|---|---|---|
| S6-wms_140743 | Bacillus phage vB_BpsS-140 | MH884512 | Outlier | n.a. | N1 | 4 | Bacillus | Group I | Duplodnaviria | Heunggongvirae | Uroviricota | Caudoviricetes | n.a. | Unclassified | O | O |
| S7-wms-combined_106253 | Listeria phage LWP01 | CP011103 | Outlier | n.a. | N5 | 5 | Listeria | Group I | Duplodnaviria | Heunggongvirae | Uroviricota | Caudoviricetes | n.a. | Unclassified | O | O |
| S7-wms-combined_107679_1 | Riemerella phage RAP44 | HQ396194 | Outlier | n.a. | N1 | 7 | Riemerella | Group I | Duplodnaviria | Heunggongvirae | Uroviricota | Caudoviricetes | n.a. | Unclassified | O | O |
| S7-wms-combined_135154 | n.a. | n.a. | Outlier | n.a. | A | n.a. | n.a. | n.a. | n.a. | n.a. | n.a. | n.a. | n.a. | n.a. | n.a. | n.a. |
| S7-wms-combined_14529 | Enterococcus phage phiFL4A | GQ478088 | Outlier | n.a. | N1 | 10 | Enterococcus | Group I | Duplodnaviria | Heunggongvirae | Uroviricota | Caudoviricetes | n.a. | Unclassified | O | O |
| S7-wms-combined_156351 | n.a. | n.a. | Outlier | n.a. | A | n.a. | n.a. | n.a. | n.a. | n.a. | n.a. | n.a. | n.a. | n.a. | n.a. | n.a. |
| S7-wms-combined_174935 | Bifidobacterium phage Bbif-1 | GQ141189 | Outlier | n.a. | N2 | 8 | Bifidobacterium | Group I | Duplodnaviria | Heunggongvirae | Uroviricota | Caudoviricetes | n.a. | Unclassified | O | O |
| S7-wms-combined_190281 | n.a. | n.a. | Outlier | n.a. | A | n.a. | n.a. | n.a. | n.a. | n.a. | n.a. | n.a. | n.a. | n.a. | n.a. | n.a. |
| S7-wms-combined_2122 | Bacteriophage sp. | LT996075 | Outlier | n.a. | N1 | 11 | Unspecified | Unclassified | Unclassified | n.a. | n.a. | n.a. | n.a. | Unclassified | O | O |
| S7-wms-combined_21693 | Streptococcus phage Javan100 | MK448838 | Outlier | n.a. | N5 | 38 | Streptococcus | Group I | Duplodnaviria | Heunggongvirae | Uroviricota | Caudoviricetes | n.a. | Unclassified | O | O |
| S7-wms-combined_217226 | Pseudomonas phage BHU-1 | OK638201 | Outlier | n.a. | N1 | 8 | Pseudomonas | Group I | Duplodnaviria | Heunggongvirae | Uroviricota | Caudoviricetes | n.a. | Unclassified | O | O |
| S7-wms-combined_218379 | Butyrivibrio virus Arian | MN882551 | Outlier | n.a. | N1 | 24 | Butyrivibrio | Group I | Duplodnaviria | Heunggongvirae | Uroviricota | Caudoviricetes | n.a. | Unclassified | O | O |
| S7-wms-combined_224523 | Clostridium phage phiCD38-2 | HM568888 | Outlier | n.a. | N8 | 2 | Clostridium | Group I | Duplodnaviria | Heunggongvirae | Uroviricota | Caudoviricetes | n.a. | Unclassified | O | O |
| S7-wms-combined_3669 | Microviridae sp. | MG945130 | Outlier | n.a. | N1 | 5 | Unspecified | Group II | Monodnaviria | Sangervirae | Phixviricota | Malgrandaviricetes | Petitvirales | Microviridae | O | O |
| S7-wms-combined_69024 | Streptococcus phage OlisA2 | OL774869 | Outlier | n.a. | N1 | 26 | Streptococcus | Group I | Duplodnaviria | Heunggongvirae | Uroviricota | Caudoviricetes | n.a. | Unclassified | O | O |
| S7-wms-combined_71592 | Streptococcus phage Javan583 | MK448814 | Outlier | n.a. | N1 | 26 | Streptococcus | Group I | Duplodnaviria | Heunggongvirae | Uroviricota | Caudoviricetes | n.a. | Unclassified | O | O |
| S7-wms-combined_74829 | Streptococcus phage Javan199 | MK448702 | Outlier | n.a. | N2 | 9 | Streptococcus | Group I | Duplodnaviria | Heunggongvirae | Uroviricota | Caudoviricetes | n.a. | Unclassified | O | O |
| S7-wms_151112 | Butyrivibrio virus Arian | MN882551 | Outlier | n.a. | N1 | 14 | Butyrivibrio | Group I | Duplodnaviria | Heunggongvirae | Uroviricota | Caudoviricetes | n.a. | Unclassified | O | O |
| S7-wms_76437 | n.a. | n.a. | Outlier | n.a. | A | n.a. | n.a. | n.a. | n.a. | n.a. | n.a. | n.a. | n.a. | n.a. | n.a. | n.a. |
| S8-wms-combined_117281 | Streptococcus phage Javan105 | MK448667 | Outlier | n.a. | N12 | 21 | Streptococcus | Group I | Duplodnaviria | Heunggongvirae | Uroviricota | Caudoviricetes | n.a. | Unclassified | O | O |
| S8-wms-combined_164289 | n.a. | n.a. | Outlier | n.a. | A | n.a. | n.a. | n.a. | n.a. | n.a. | n.a. | n.a. | n.a. | n.a. | n.a. | n.a. |
| S8-wms-combined_180957 | Bacteroides phage p00 | BK010646 | Outlier | n.a. | N9 | 15 | Bacteroides | Group I | Duplodnaviria | Heunggongvirae | Uroviricota | Caudoviricetes | n.a. | Unclassified | O | O |
| S8-wms-combined_185611 | n.a. | n.a. | Outlier | n.a. | A | n.a. | n.a. | n.a. | n.a. | n.a. | n.a. | n.a. | n.a. | n.a. | n.a. | n.a. |
| S8-wms-combined_23583 | n.a. | n.a. | Outlier | n.a. | A | n.a. | n.a. | n.a. | n.a. | n.a. | n.a. | n.a. | n.a. | n.a. | n.a. | n.a. |
| S8-wms-combined_239791 | Olleya phage Harreka_1 | MT732457 | Outlier | n.a. | N4 | 3 | Olleya | Group I | Duplodnaviria | Heunggongvirae | Uroviricota | Caudoviricetes | n.a. | Aggregaviridae | O | O |
| S8-wms-combined_48046 | n.a. | n.a. | Outlier | n.a. | A | n.a. | n.a. | n.a. | n.a. | n.a. | n.a. | n.a. | n.a. | n.a. | n.a. | n.a. |
| S8-wms-combined_74275 | Bacteroides phage F2 | MT806186 | Outlier | n.a. | N2 | 5 | Bacteroides | Group I | Duplodnaviria | Heunggongvirae | Uroviricota | Caudoviricetes | n.a. | Unclassified | O | O |
| S8-wms-combined_81597 | Bacteroides phage p00 | BK010646 | Outlier | n.a. | N9 | 20 | Bacteroides | Group I | Duplodnaviria | Heunggongvirae | Uroviricota | Caudoviricetes | n.a. | Unclassified | O | O |
| S8-wms_113693 | Streptococcus phage Javan553 | MK448804 | Outlier | n.a. | N3 | 9 | Streptococcus | Group I | Duplodnaviria | Heunggongvirae | Uroviricota | Caudoviricetes | n.a. | Unclassified | O | O |
| S8-wms_190781 | Clostridium phage phiMMP01 | LN681541 | Outlier | n.a. | N7 | 11 | Clostridium | Group I | Duplodnaviria | Heunggongvirae | Uroviricota | Caudoviricetes | n.a. | Unclassified | O | O |
| S8-wms_58515 | n.a. | n.a. | Outlier | n.a. | A | n.a. | n.a. | n.a. | n.a. | n.a. | n.a. | n.a. | n.a. | n.a. | n.a. | n.a. |
| S9-wms-combined_109322 | Streptococcus phage Javan74 | MK449005 | Outlier | n.a. | N1 | 26 | Streptococcus | Group I | Duplodnaviria | Heunggongvirae | Uroviricota | Caudoviricetes | n.a. | Unclassified | O | O |
| S9-wms-combined_122742 | n.a. | n.a. | Outlier | n.a. | A | n.a. | n.a. | n.a. | n.a. | n.a. | n.a. | n.a. | n.a. | n.a. | n.a. | n.a. |
| S9-wms-combined_124857 | n.a. | n.a. | Outlier | n.a. | A | n.a. | n.a. | n.a. | n.a. | n.a. | n.a. | n.a. | n.a. | n.a. | n.a. | n.a. |
| S9-wms-combined_2042 | n.a. | n.a. | Outlier | n.a. | A | n.a. | n.a. | n.a. | n.a. | n.a. | n.a. | n.a. | n.a. | n.a. | n.a. | n.a. |
| S9-wms-combined_3461 | n.a. | n.a. | Outlier | n.a. | A | n.a. | n.a. | n.a. | n.a. | n.a. | n.a. | n.a. | n.a. | n.a. | n.a. | n.a. |
| S9-wms-combined_93232 | Klebsiella phage ST101-KPC2phi6.3 | MK416017 | Outlier | n.a. | N1 | 72 | Klebsiella | Group I | Duplodnaviria | Heunggongvirae | Uroviricota | Caudoviricetes | n.a. | Unclassified | O | O |
| S9-wms_114971 | Clostridium phage phiMMP04 | JX145342 | Outlier | n.a. | N1 | 13 | Clostridium | Group I | Duplodnaviria | Heunggongvirae | Uroviricota | Caudoviricetes | n.a. | Unclassified | O | O |
| S12-wms-combined_148741 | n.a. | n.a. | Overlap (VC_1103/VC_1240) | n.a. | A | n.a. | n.a. | n.a. | n.a. | n.a. | n.a. | n.a. | n.a. | n.a. | n.a. | n.a. |
| S12-wms-combined_270839 | n.a. | n.a. | Overlap (VC_1103/VC_1240) | n.a. | A | n.a. | n.a. | n.a. | n.a. | n.a. | n.a. | n.a. | n.a. | n.a. | n.a. | n.a. |
| S1-wms-combined_52708 | n.a. | n.a. | Overlap (VC_1221/VC_1237) | n.a. | A | n.a. | n.a. | n.a. | n.a. | n.a. | n.a. | n.a. | n.a. | n.a. | n.a. | n.a. |
| S17-wms-combined_56061 | n.a. | n.a. | Overlap (VC_1221/VC_1237) | n.a. | A | n.a. | n.a. | n.a. | n.a. | n.a. | n.a. | n.a. | n.a. | n.a. | n.a. | n.a. |
| S2-wms-combined_181849 | n.a. | n.a. | Overlap (VC_1221/VC_1237) | n.a. | A | n.a. | n.a. | n.a. | n.a. | n.a. | n.a. | n.a. | n.a. | n.a. | n.a. | n.a. |
| S2-wms-combined_57293 | Campylobacter phage A11a | MG065691 | Overlap (VC_1221/VC_1237) | n.a. | N11 | 5 | Campylobacter | Unclassified | Unclassified | n.a. | n.a. | n.a. | n.a. | Unclassified | O | O |
| S6-wms-combined_110950 | n.a. | n.a. | Overlap (VC_1221/VC_1237) | n.a. | A | n.a. | n.a. | n.a. | n.a. | n.a. | n.a. | n.a. | n.a. | n.a. | n.a. | n.a. |
| S6-wms-combined_138710 | n.a. | n.a. | Overlap (VC_1221/VC_1237) | n.a. | A | n.a. | n.a. | n.a. | n.a. | n.a. | n.a. | n.a. | n.a. | n.a. | n.a. | n.a. |
| S7-wms-combined_137858 | n.a. | n.a. | Overlap (VC_1221/VC_1237) | n.a. | A | n.a. | n.a. | n.a. | n.a. | n.a. | n.a. | n.a. | n.a. | n.a. | n.a. | n.a. |
| S12-wms-combined_75733 | Tortoise microvirus 19 | MK765569 | Overlap (VC_1244/VC_1245) | n.a. | N2 | 9 | Unspecified | Group II | Monodnaviria | Sangervirae | Phixviricota | Malgrandaviricetes | Petitvirales | Microviridae | O | O |
| S8-wms_270 | Tortoise microvirus 19 | MK765569 | Overlap (VC_1244/VC_1245) | n.a. | N2 | 9 | Unspecified | Group II | Monodnaviria | Sangervirae | Phixviricota | Malgrandaviricetes | Petitvirales | Microviridae | O | O |
| S1-wms-combined_153670 | n.a. | n.a. | Overlap (VC_1245/VC_1487) | n.a. | A | n.a. | n.a. | n.a. | n.a. | n.a. | n.a. | n.a. | n.a. | n.a. | n.a. | n.a. |
| S1-wms_153615 | n.a. | n.a. | Overlap (VC_1245/VC_1487) | n.a. | A | n.a. | n.a. | n.a. | n.a. | n.a. | n.a. | n.a. | n.a. | n.a. | n.a. | n.a. |
| S12-wms-combined_132907 | Clostridium phage phiCT453A | KM983327 | Overlap (VC_138/VC_146) | n.a. | C5 | 31 | Clostridium | Group I | Duplodnaviria | Heunggongvirae | Uroviricota | Caudoviricetes | n.a. | Unclassified | Unclassified | O |
| S12-wms-combined_52101 | Thermus phage phi OH2 | AB823818 | Overlap (VC_138/VC_146) | n.a. | C11 | 15 | Thermus | Group I | Duplodnaviria | Heunggongvirae | Uroviricota | Caudoviricetes | n.a. | Unclassified | Unclassified | O |
| S17-wms-combined_258087 | Thermus phage phi OH2 | AB823818 | Overlap (VC_138/VC_146) | n.a. | C25 | 9 | Thermus | Group I | Duplodnaviria | Heunggongvirae | Uroviricota | Caudoviricetes | n.a. | Unclassified | Unclassified | O |
| S17-wms_106671 | Clostridium phage phiCT9441A | KM983329 | Overlap (VC_138/VC_146) | n.a. | C11 | 23 | Clostridium | Group I | Duplodnaviria | Heunggongvirae | Uroviricota | Caudoviricetes | n.a. | Unclassified | Unclassified | O |
| S2-wms-combined_135014 | Thermus phage phi OH2 | AB823818 | Overlap (VC_138/VC_146) | n.a. | C29 | 10 | Thermus | Group I | Duplodnaviria | Heunggongvirae | Uroviricota | Caudoviricetes | n.a. | Unclassified | Unclassified | O |
| S2-wms-combined_207000 | Clostridium phage phiCT9441A | KM983329 | Overlap (VC_138/VC_146) | n.a. | C11 | 22 | Clostridium | Group I | Duplodnaviria | Heunggongvirae | Uroviricota | Caudoviricetes | n.a. | Unclassified | Unclassified | O |
| S2-wms-combined_58563 | Thermus phage phi OH2 | AB823818 | Overlap (VC_138/VC_146) | n.a. | C27 | 13 | Thermus | Group I | Duplodnaviria | Heunggongvirae | Uroviricota | Caudoviricetes | n.a. | Unclassified | Unclassified | O |
| S2-wms_245232 | Thermus phage phi OH2 | AB823818 | Overlap (VC_138/VC_146) | n.a. | C17 | 19 | Thermus | Group I | Duplodnaviria | Heunggongvirae | Uroviricota | Caudoviricetes | n.a. | Unclassified | Unclassified | O |
| S8-wms-combined_219796 | Thermus phage phi OH2 | AB823818 | Overlap (VC_138/VC_146) | n.a. | C1 | 19 | Thermus | Group I | Duplodnaviria | Heunggongvirae | Uroviricota | Caudoviricetes | n.a. | Unclassified | Unclassified | O |
| S8-wms_20528 | Clostridium phage phiCT453A | KM983327 | Overlap (VC_138/VC_146) | n.a. | C15 | 28 | Clostridium | Group I | Duplodnaviria | Heunggongvirae | Uroviricota | Caudoviricetes | n.a. | Unclassified | Unclassified | O |
| S9-wms-combined_105822 | Thermus phage phi OH2 | AB823818 | Overlap (VC_138/VC_146) | n.a. | C1 | 25 | Thermus | Group I | Duplodnaviria | Heunggongvirae | Uroviricota | Caudoviricetes | n.a. | Unclassified | Unclassified | O |
| S9-wms-combined_97755 | Clostridium phage phiCT9441A | KM983329 | Overlap (VC_138/VC_146) | n.a. | C7 | 30 | Clostridium | Group I | Duplodnaviria | Heunggongvirae | Uroviricota | Caudoviricetes | n.a. | Unclassified | Unclassified | O |
| S1-wms_265285 | Thermus phage phi OH2 | AB823818 | Overlap (VC_138/VC_146/VC_147) | n.a. | C25 | 16 | Thermus | Group I | Duplodnaviria | Heunggongvirae | Uroviricota | Caudoviricetes | n.a. | Unclassified | Unclassified | O |
| S12-wms_161239 | Thermus phage phi OH2 | AB823818 | Overlap (VC_138/VC_146/VC_147) | n.a. | C21 | 4 | Thermus | Group I | Duplodnaviria | Heunggongvirae | Uroviricota | Caudoviricetes | n.a. | Unclassified | Unclassified | O |
| S17-wms_116248 | Thermus phage phi OH2 | AB823818 | Overlap (VC_138/VC_146/VC_147) | n.a. | C23 | 17 | Thermus | Group I | Duplodnaviria | Heunggongvirae | Uroviricota | Caudoviricetes | n.a. | Unclassified | Unclassified | O |
| S17-wms_29082 | Thermus phage phi OH2 | AB823818 | Overlap (VC_138/VC_146/VC_147) | n.a. | C25 | 15 | Thermus | Group I | Duplodnaviria | Heunggongvirae | Uroviricota | Caudoviricetes | n.a. | Unclassified | Unclassified | O |
| S4-wms-combined_57963 | Thermus phage phi OH2 | AB823818 | Overlap (VC_138/VC_146/VC_147) | n.a. | C25 | 11 | Thermus | Group I | Duplodnaviria | Heunggongvirae | Uroviricota | Caudoviricetes | n.a. | Unclassified | Unclassified | O |
| S17-wms-combined_66123 | Thermus phage phi OH2 | AB823818 | Overlap (VC_138/VC_146/VC_148) | n.a. | C29 | 10 | Thermus | Group I | Duplodnaviria | Heunggongvirae | Uroviricota | Caudoviricetes | n.a. | Unclassified | Unclassified | O |
| S17-wms-combined_75892 | Psychrobacillus phage PVJ1 | MZ983385 | Overlap (VC_138/VC_146/VC_148) | n.a. | C27 | 10 | Psychrobacillus | Group I | Duplodnaviria | Heunggongvirae | Uroviricota | Caudoviricetes | n.a. | Unclassified | Unclassified | O |
| S2-wms-combined_122457 | Thermus phage phi OH2 | AB823818 | Overlap (VC_138/VC_146/VC_148) | n.a. | C33 | 8 | Thermus | Group I | Duplodnaviria | Heunggongvirae | Uroviricota | Caudoviricetes | n.a. | Unclassified | Unclassified | O |
| S1-wms-combined_373308 | n.a. | n.a. | Overlap (VC_1483/VC_1490) | n.a. | F | n.a. | n.a. | n.a. | n.a. | n.a. | n.a. | n.a. | n.a. | n.a. | n.a. | n.a. |
| S5-wms-combined_163165 | n.a. | n.a. | Overlap (VC_1485/VC_1486) | n.a. | A | n.a. | n.a. | n.a. | n.a. | n.a. | n.a. | n.a. | n.a. | n.a. | n.a. | n.a. |
| S9-wms-combined_10990 | n.a. | n.a. | Overlap (VC_1489/VC_1494) | n.a. | A | n.a. | n.a. | n.a. | n.a. | n.a. | n.a. | n.a. | n.a. | n.a. | n.a. | n.a. |
| S1-wms-combined_17651 | n.a. | n.a. | Overlap (VC_1490/VC_1491) | n.a. | F | n.a. | n.a. | n.a. | n.a. | n.a. | n.a. | n.a. | n.a. | n.a. | n.a. | n.a. |
| S12-wms-combined_133608 | n.a. | n.a. | Overlap (VC_1490/VC_1491) | n.a. | F | n.a. | n.a. | n.a. | n.a. | n.a. | n.a. | n.a. | n.a. | n.a. | n.a. | n.a. |
| S12-wms-combined_117628 | Clostridium phage PhiS63 | JQ660954 | Overlap (VC_20/VC_542) | n.a. | C3 | 13 | Clostridium | Group I | Duplodnaviria | Heunggongvirae | Uroviricota | Caudoviricetes | n.a. | Unclassified | Unclassified | O |
| S2-wms_161986 | Clostridium phage PhiS63 | JQ660954 | Overlap (VC_20/VC_542) | n.a. | C5 | 14 | Clostridium | Group I | Duplodnaviria | Heunggongvirae | Uroviricota | Caudoviricetes | n.a. | Unclassified | Unclassified | O |
| S2-wms_192667 | Clostridium phage PhiS63 | JQ660954 | Overlap (VC_20/VC_542) | n.a. | C1 | 20 | Clostridium | Group I | Duplodnaviria | Heunggongvirae | Uroviricota | Caudoviricetes | n.a. | Unclassified | Unclassified | O |
| S11-wms-combined_320710 | Vibrio phage vB_VspS_VS-ABTNL-3 | MF497422 | Overlap (VC_342/VC_1215) | n.a. | C1 | 10 | Vibrio | Group I | Duplodnaviria | Heunggongvirae | Uroviricota | Caudoviricetes | n.a. | Unclassified | Unclassified | O |
| S2-wms-combined_158821 | Butyrivibrio phage Idris | MN882554 | Overlap (VC_422/VC_1413) | n.a. | C5 | 9 | Butyrivibrio | Group I | Duplodnaviria | Heunggongvirae | Uroviricota | Caudoviricetes | n.a. | Unclassified | Unclassified | O |
| S4-wms-combined_112093 | Butyrivibrio phage Idris | MN882554 | Overlap (VC_422/VC_1413) | n.a. | C5 | 7 | Butyrivibrio | Group I | Duplodnaviria | Heunggongvirae | Uroviricota | Caudoviricetes | n.a. | Unclassified | Unclassified | O |
| S17-wms-combined_56006 | Faecalibacterium phage FP_Brigit | MG711465 | Overlap (VC_607/VC_1238) | n.a. | N5 | 33 | Faecalibacterium | Group I | Duplodnaviria | Heunggongvirae | Uroviricota | Caudoviricetes | n.a. | Unclassified | O | O |
| S2-wms_124416 | Faecalibacterium phage FP_Brigit | MG711465 | Overlap (VC_607/VC_1238) | n.a. | N4 | 17 | Faecalibacterium | Group I | Duplodnaviria | Heunggongvirae | Uroviricota | Caudoviricetes | n.a. | Unclassified | O | O |
| S12-wms-combined_385270 | Lactobacillus phage c5 | EU340421 | Overlap (VC_630/VC_642) | n.a. | N5 | 6 | Lactobacillus | Group I | Duplodnaviria | Heunggongvirae | Uroviricota | Caudoviricetes | n.a. | Unclassified | O | O |
| S17-wms-combined_179363 | Lactobacillus phage c5 | EU340421 | Overlap (VC_630/VC_642) | n.a. | N4 | 9 | Lactobacillus | Group I | Duplodnaviria | Heunggongvirae | Uroviricota | Caudoviricetes | n.a. | Unclassified | O | O |
| S2-wms-combined_99446 | Clostridium phage phiCT19406B | KM983331 | Overlap (VC_630/VC_642) | n.a. | N5 | 7 | Clostridium | Group I | Duplodnaviria | Heunggongvirae | Uroviricota | Caudoviricetes | n.a. | Unclassified | O | O |

| | | | | | | | | | | | | | | | | |
|---|---|---|---|---|---|---|---|---|---|---|---|---|---|---|---|---|
| S5-wms-combined_147993 | Lactobacillus phage c5 | EU340421 | Overlap (VC_630/VC_642) | n.a. | N5 | 8 | Lactobacillus | Group I | Duplodnaviria | Heunggongvirae | Uroviricota | Caudoviricetes | n.a. | Unclassified | O | O |
| S5-wms-combined_94786 | Lactobacillus phage c5 | EU340421 | Overlap (VC_630/VC_642) | n.a. | N2 | 7 | Lactobacillus | Group I | Duplodnaviria | Heunggongvirae | Uroviricota | Caudoviricetes | n.a. | Unclassified | O | O |
| S1-wms-combined_374219 | n.a. | n.a. | Overlap (VC_651/VC_736) | n.a. | A | n.a. | n.a. | n.a. | n.a. | n.a. | n.a. | n.a. | n.a. | n.a. | n.a. | n.a. |
| S6-wms-combined_128972 | n.a. | n.a. | Overlap (VC_651/VC_736) | n.a. | A | n.a. | n.a. | n.a. | n.a. | n.a. | n.a. | n.a. | n.a. | n.a. | n.a. | n.a. |
| S1-wms-combined_107766 | n.a. | n.a. | Overlap (VC_652/VC_1481) | n.a. | A | n.a. | n.a. | n.a. | n.a. | n.a. | n.a. | n.a. | n.a. | n.a. | n.a. | n.a. |
| S12-wms-combined_25459 | n.a. | n.a. | Overlap (VC_652/VC_1481) | n.a. | A | n.a. | n.a. | n.a. | n.a. | n.a. | n.a. | n.a. | n.a. | n.a. | n.a. | n.a. |
| S12-wms-combined_320262 | n.a. | n.a. | Overlap (VC_652/VC_1481) | n.a. | A | n.a. | n.a. | n.a. | n.a. | n.a. | n.a. | n.a. | n.a. | n.a. | n.a. | n.a. |
| S2-wms-combined_208559 | n.a. | n.a. | Overlap (VC_652/VC_1481) | n.a. | A | n.a. | n.a. | n.a. | n.a. | n.a. | n.a. | n.a. | n.a. | n.a. | n.a. | n.a. |
| S5-wms-combined_183113 | n.a. | n.a. | Overlap (VC_652/VC_1481) | n.a. | A | n.a. | n.a. | n.a. | n.a. | n.a. | n.a. | n.a. | n.a. | n.a. | n.a. | n.a. |
| S12-wms-combined_184338 | Roseburia phage Jekyll | LR596902 | Overlap (VC_652/VC_894) | n.a. | N9 | 4 | Roseburia | Group I | Duplodnaviria | Heunggongvirae | Uroviricota | Caudoviricetes | n.a. | Unclassified | O | O |
| S1-wms-combined_330329 | Bacteroides phage p00 | BK010646 | Overlap (VC_678/VC_679) | n.a. | N6 | 37 | Bacteroides | Group I | Duplodnaviria | Heunggongvirae | Uroviricota | Caudoviricetes | n.a. | Unclassified | O | O |
| S11-wms-combined_226165 | Bacteroides phage p00 | BK010646 | Overlap (VC_678/VC_679) | n.a. | N10 | 22 | Bacteroides | Group I | Duplodnaviria | Heunggongvirae | Uroviricota | Caudoviricetes | n.a. | Unclassified | O | O |
| S11-wms-combined_321938 | Bacteroides phage p00 | BK010646 | Overlap (VC_678/VC_679) | n.a. | N4 | 38 | Bacteroides | Group I | Duplodnaviria | Heunggongvirae | Uroviricota | Caudoviricetes | n.a. | Unclassified | O | O |
| S11-wms-combined_64392 | Bacteroides phage p00 | BK010646 | Overlap (VC_678/VC_679) | n.a. | N2 | 56 | Bacteroides | Group I | Duplodnaviria | Heunggongvirae | Uroviricota | Caudoviricetes | n.a. | Unclassified | O | O |
| S12-wms-combined_189323 | Bacteroides phage p00 | BK010646 | Overlap (VC_678/VC_679) | n.a. | N2 | 45 | Bacteroides | Group I | Duplodnaviria | Heunggongvirae | Uroviricota | Caudoviricetes | n.a. | Unclassified | O | O |
| S2-wms-combined_147065 | Bacteroides phage p00 | BK010646 | Overlap (VC_678/VC_679) | n.a. | N7 | 30 | Bacteroides | Group I | Duplodnaviria | Heunggongvirae | Uroviricota | Caudoviricetes | n.a. | Unclassified | O | O |
| S5-wms-combined_210959 | Bacteroides phage p00 | BK010646 | Overlap (VC_678/VC_679) | n.a. | N10 | 15 | Bacteroides | Group I | Duplodnaviria | Heunggongvirae | Uroviricota | Caudoviricetes | n.a. | Unclassified | O | O |
| S5-wms-combined_67650 | Bacteroides phage p00 | BK010646 | Overlap (VC_678/VC_679) | n.a. | N11 | 14 | Bacteroides | Group I | Duplodnaviria | Heunggongvirae | Uroviricota | Caudoviricetes | n.a. | Unclassified | O | O |
| S1-wms-combined_100430 | Faecalibacterium phage FP_Taranis | MG711467 | Overlap (VC_711/VC_715) | n.a. | C8 | 25 | Faecalibacterium | Group I | Duplodnaviria | Heunggongvirae | Uroviricota | Caudoviricetes | n.a. | Unclassified | Unclassified | O |
| S1-wms-combined_285012 | Faecalibacterium phage FP_Taranis | MG711467 | Overlap (VC_711/VC_715) | n.a. | C9 | 22 | Faecalibacterium | Group I | Duplodnaviria | Heunggongvirae | Uroviricota | Caudoviricetes | n.a. | Unclassified | Unclassified | O |
| S11-wms-combined_120257 | Vibrio phage phi 2 | KJ545483 | Overlap (VC_711/VC_715) | n.a. | C9 | 8 | Vibrio | Group I | Duplodnaviria | Heunggongvirae | Uroviricota | Caudoviricetes | n.a. | Unclassified | Unclassified | O |
| S17-wms-combined_106262 | Faecalibacterium phage FP_Taranis | MG711467 | Overlap (VC_711/VC_715) | n.a. | C9 | 21 | Faecalibacterium | Group I | Duplodnaviria | Heunggongvirae | Uroviricota | Caudoviricetes | n.a. | Unclassified | Unclassified | O |
| S5-wms-combined_199135 | n.a. | n.a. | Overlap (VC_733/VC_1494) | n.a. | A | n.a. | n.a. | n.a. | n.a. | n.a. | n.a. | n.a. | n.a. | n.a. | n.a. | n.a. |
| S17-wms-combined_39687 | n.a. | n.a. | Overlap (VC_733/VC_893/VC_1494) | n.a. | A | n.a. | n.a. | n.a. | n.a. | n.a. | n.a. | n.a. | n.a. | n.a. | n.a. | n.a. |
| S12-wms_184276 | Clostridium phage HM T | KU517658 | Overlap (VC_735/VC_915) | n.a. | N11 | 4 | Clostridium | Group I | Duplodnaviria | Heunggongvirae | Uroviricota | Caudoviricetes | n.a. | Unclassified | O | O |
| S5-wms_62679 | Paenibacillus phage PG1 | HQ332138 | Overlap (VC_735/VC_915) | n.a. | N5 | 6 | Paenibacillus | Group I | Duplodnaviria | Heunggongvirae | Uroviricota | Caudoviricetes | n.a. | Unclassified | O | O |
| S2-wms-combined_201205 | Akkermansia phage DTMo-2021a | CP084202 | Overlap (VC_737/VC_738) | n.a. | C1 | 54 | Akkermansia | Group I | Duplodnaviria | Heunggongvirae | Uroviricota | Caudoviricetes | n.a. | Unclassified | Unclassified | O |
| S1-wms-combined_351889 | Butyrivibrio virus Ceridwen | MN882553 | Overlap (VC_817/VC_1210) | n.a. | C5 | 40 | Butyrivibrio | Group I | Duplodnaviria | Heunggongvirae | Uroviricota | Caudoviricetes | n.a. | Unclassified | Unclassified | O |
| S2-wms-combined_64692 | Butyrivibrio virus Ceridwen | MN882553 | Overlap (VC_817/VC_1210) | n.a. | C5 | 39 | Butyrivibrio | Group I | Duplodnaviria | Heunggongvirae | Uroviricota | Caudoviricetes | n.a. | Unclassified | Unclassified | O |
| S8-wms-combined_246389 | Butyrivibrio virus Ceridwen | MN882553 | Overlap (VC_817/VC_1210) | n.a. | C5 | 42 | Butyrivibrio | Group I | Duplodnaviria | Heunggongvirae | Uroviricota | Caudoviricetes | n.a. | Unclassified | Unclassified | O |
| S17-wms-combined_253499 | n.a. | n.a. | Overlap (VC_833/VC_1015) | n.a. | A | n.a. | n.a. | n.a. | n.a. | n.a. | n.a. | n.a. | n.a. | n.a. | n.a. | n.a. |
| S12-wms-combined_31761 | Butyrivibrio phage Idris | MN882554 | Overlap (VC_877/VC_1413) | n.a. | C1 | 67 | Butyrivibrio | Group I | Duplodnaviria | Heunggongvirae | Uroviricota | Caudoviricetes | n.a. | Unclassified | Unclassified | O |
| S5-wms-combined_26061 | Butyrivibrio phage Idris | MN882554 | Overlap (VC_877/VC_1413) | n.a. | C5 | 58 | Butyrivibrio | Group I | Duplodnaviria | Heunggongvirae | Uroviricota | Caudoviricetes | n.a. | Unclassified | Unclassified | O |
| S6-wms-combined_88414 | Butyrivibrio phage Idris | MN882554 | Overlap (VC_877/VC_1413) | n.a. | C5 | 39 | Butyrivibrio | Group I | Duplodnaviria | Heunggongvirae | Uroviricota | Caudoviricetes | n.a. | Unclassified | Unclassified | O |
| S1-wms-combined_166392 | Faecalibacterium phage FP_Lugh | MG711464 | Overlap (VC_893/VC_894) | n.a. | N9 | 2 | Faecalibacterium | Group I | Duplodnaviria | Heunggongvirae | Uroviricota | Caudoviricetes | n.a. | Unclassified | O | O |
| S1-wms_223620 | Clostridium phage phiCD38-2 | HM568888 | Overlap (VC_893/VC_894) | n.a. | N10 | 4 | Clostridium | Group I | Duplodnaviria | Heunggongvirae | Uroviricota | Caudoviricetes | n.a. | Unclassified | O | O |
| S11-wms-combined_179605 | n.a. | n.a. | Overlap (VC_893/VC_894/VC_1489/VC_1494) | n.a. | A | n.a. | n.a. | n.a. | n.a. | n.a. | n.a. | n.a. | n.a. | n.a. | n.a. | n.a. |
| S12-wms-combined_297545 | Bacteroides phage LoVEphage | MW660583 | Overlap (VC_918/VC_1456) | n.a. | N13 | 3 | Bacteroides | Group I | Duplodnaviria | Heunggongvirae | Uroviricota | Caudoviricetes | n.a. | Unclassified | O | O |
| S12-wms-combined_368190 | Phage vB_RanS_PJN03 | ON245412 | Overlap (VC_918/VC_1456) | n.a. | N12 | 7 | Unspecified | Group I | Duplodnaviria | Heunggongvirae | Uroviricota | Caudoviricetes | n.a. | Unclassified | O | O |
| S1-wms-combined_169788 | n.a. | n.a. | Singleton | n.a. | G | n.a. | n.a. | n.a. | n.a. | n.a. | n.a. | n.a. | n.a. | n.a. | n.a. | n.a. |
| S1-wms-combined_231234 | n.a. | n.a. | Singleton | n.a. | G | n.a. | n.a. | n.a. | n.a. | n.a. | n.a. | n.a. | n.a. | n.a. | n.a. | n.a. |
| S1-wms-combined_231411 | n.a. | n.a. | Singleton | n.a. | G | n.a. | n.a. | n.a. | n.a. | n.a. | n.a. | n.a. | n.a. | n.a. | n.a. | n.a. |
| S1-wms-combined_32863 | n.a. | n.a. | Singleton | n.a. | G | n.a. | n.a. | n.a. | n.a. | n.a. | n.a. | n.a. | n.a. | n.a. | n.a. | n.a. |
| S1-wms_32878 | n.a. | n.a. | Singleton | n.a. | G | n.a. | n.a. | n.a. | n.a. | n.a. | n.a. | n.a. | n.a. | n.a. | n.a. | n.a. |
| S1-wms_388292 | n.a. | n.a. | Singleton | n.a. | G | n.a. | n.a. | n.a. | n.a. | n.a. | n.a. | n.a. | n.a. | n.a. | n.a. | n.a. |
| S11-wms-combined_117756 | n.a. | n.a. | Singleton | n.a. | G | n.a. | n.a. | n.a. | n.a. | n.a. | n.a. | n.a. | n.a. | n.a. | n.a. | n.a. |
| S11-wms-combined_265453 | n.a. | n.a. | Singleton | n.a. | G | n.a. | n.a. | n.a. | n.a. | n.a. | n.a. | n.a. | n.a. | n.a. | n.a. | n.a. |
| S11-wms_32691 | n.a. | n.a. | Singleton | n.a. | G | n.a. | n.a. | n.a. | n.a. | n.a. | n.a. | n.a. | n.a. | n.a. | n.a. | n.a. |
| S11-wms_6203 | n.a. | n.a. | Singleton | n.a. | G | n.a. | n.a. | n.a. | n.a. | n.a. | n.a. | n.a. | n.a. | n.a. | n.a. | n.a. |
| S12-wms-combined_117447 | n.a. | n.a. | Singleton | n.a. | G | n.a. | n.a. | n.a. | n.a. | n.a. | n.a. | n.a. | n.a. | n.a. | n.a. | n.a. |
| S12-wms-combined_390985 | n.a. | n.a. | Singleton | n.a. | G | n.a. | n.a. | n.a. | n.a. | n.a. | n.a. | n.a. | n.a. | n.a. | n.a. | n.a. |
| S12-wms-combined_63056 | n.a. | n.a. | Singleton | n.a. | G | n.a. | n.a. | n.a. | n.a. | n.a. | n.a. | n.a. | n.a. | n.a. | n.a. | n.a. |
| S12-wms_223235 | n.a. | n.a. | Singleton | n.a. | G | n.a. | n.a. | n.a. | n.a. | n.a. | n.a. | n.a. | n.a. | n.a. | n.a. | n.a. |
| S12-wms_336780 | n.a. | n.a. | Singleton | n.a. | G | n.a. | n.a. | n.a. | n.a. | n.a. | n.a. | n.a. | n.a. | n.a. | n.a. | n.a. |
| S12-wms_352846 | n.a. | n.a. | Singleton | n.a. | G | n.a. | n.a. | n.a. | n.a. | n.a. | n.a. | n.a. | n.a. | n.a. | n.a. | n.a. |
| S2-wms-combined_115352 | n.a. | n.a. | Singleton | n.a. | G | n.a. | n.a. | n.a. | n.a. | n.a. | n.a. | n.a. | n.a. | n.a. | n.a. | n.a. |
| S2-wms-combined_204225 | n.a. | n.a. | Singleton | n.a. | G | n.a. | n.a. | n.a. | n.a. | n.a. | n.a. | n.a. | n.a. | n.a. | n.a. | n.a. |
| S2-wms-combined_222535 | n.a. | n.a. | Singleton | n.a. | G | n.a. | n.a. | n.a. | n.a. | n.a. | n.a. | n.a. | n.a. | n.a. | n.a. | n.a. |
| S2-wms-combined_230282 | n.a. | n.a. | Singleton | n.a. | G | n.a. | n.a. | n.a. | n.a. | n.a. | n.a. | n.a. | n.a. | n.a. | n.a. | n.a. |
| S2-wms-combined_30750 | n.a. | n.a. | Singleton | n.a. | G | n.a. | n.a. | n.a. | n.a. | n.a. | n.a. | n.a. | n.a. | n.a. | n.a. | n.a. |
| S2-wms-combined_48661 | n.a. | n.a. | Singleton | n.a. | G | n.a. | n.a. | n.a. | n.a. | n.a. | n.a. | n.a. | n.a. | n.a. | n.a. | n.a. |
| S2-wms-combined_52006 | n.a. | n.a. | Singleton | n.a. | G | n.a. | n.a. | n.a. | n.a. | n.a. | n.a. | n.a. | n.a. | n.a. | n.a. | n.a. |
| S2-wms-combined_73327 | n.a. | n.a. | Singleton | n.a. | G | n.a. | n.a. | n.a. | n.a. | n.a. | n.a. | n.a. | n.a. | n.a. | n.a. | n.a. |
| S2-wms-combined_95987 | n.a. | n.a. | Singleton | n.a. | G | n.a. | n.a. | n.a. | n.a. | n.a. | n.a. | n.a. | n.a. | n.a. | n.a. | n.a. |
| S4-wms-combined_24659 | n.a. | n.a. | Singleton | n.a. | G | n.a. | n.a. | n.a. | n.a. | n.a. | n.a. | n.a. | n.a. | n.a. | n.a. | n.a. |
| S5-wms-combined_106597 | n.a. | n.a. | Singleton | n.a. | G | n.a. | n.a. | n.a. | n.a. | n.a. | n.a. | n.a. | n.a. | n.a. | n.a. | n.a. |
| S5-wms-combined_197186 | n.a. | n.a. | Singleton | n.a. | G | n.a. | n.a. | n.a. | n.a. | n.a. | n.a. | n.a. | n.a. | n.a. | n.a. | n.a. |
| S5-wms-combined_205046 | n.a. | n.a. | Singleton | n.a. | G | n.a. | n.a. | n.a. | n.a. | n.a. | n.a. | n.a. | n.a. | n.a. | n.a. | n.a. |
| S5-wms_10783 | n.a. | n.a. | Singleton | n.a. | G | n.a. | n.a. | n.a. | n.a. | n.a. | n.a. | n.a. | n.a. | n.a. | n.a. | n.a. |
| S5-wms_123499 | n.a. | n.a. | Singleton | n.a. | G | n.a. | n.a. | n.a. | n.a. | n.a. | n.a. | n.a. | n.a. | n.a. | n.a. | n.a. |
| S5-wms_212596 | n.a. | n.a. | Singleton | n.a. | G | n.a. | n.a. | n.a. | n.a. | n.a. | n.a. | n.a. | n.a. | n.a. | n.a. | n.a. |
| S6-wms-combined_65225 | n.a. | n.a. | Singleton | n.a. | G | n.a. | n.a. | n.a. | n.a. | n.a. | n.a. | n.a. | n.a. | n.a. | n.a. | n.a. |
| S6-wms-combined_745 | n.a. | n.a. | Singleton | n.a. | G | n.a. | n.a. | n.a. | n.a. | n.a. | n.a. | n.a. | n.a. | n.a. | n.a. | n.a. |
| S7-wms-combined_121062 | n.a. | n.a. | Singleton | n.a. | G | n.a. | n.a. | n.a. | n.a. | n.a. | n.a. | n.a. | n.a. | n.a. | n.a. | n.a. |
| S7-wms-combined_123085 | n.a. | n.a. | Singleton | n.a. | G | n.a. | n.a. | n.a. | n.a. | n.a. | n.a. | n.a. | n.a. | n.a. | n.a. | n.a. |
| S7-wms-combined_134452 | n.a. | n.a. | Singleton | n.a. | G | n.a. | n.a. | n.a. | n.a. | n.a. | n.a. | n.a. | n.a. | n.a. | n.a. | n.a. |
| S7-wms-combined_157434 | n.a. | n.a. | Singleton | n.a. | G | n.a. | n.a. | n.a. | n.a. | n.a. | n.a. | n.a. | n.a. | n.a. | n.a. | n.a. |
| S7-wms-combined_167867 | n.a. | n.a. | Singleton | n.a. | G | n.a. | n.a. | n.a. | n.a. | n.a. | n.a. | n.a. | n.a. | n.a. | n.a. | n.a. |
| S9-wms-combined_27311 | n.a. | n.a. | Singleton | n.a. | G | n.a. | n.a. | n.a. | n.a. | n.a. | n.a. | n.a. | n.a. | n.a. | n.a. | n.a. |
| S9-wms_10480 | n.a. | n.a. | Singleton | n.a. | G | n.a. | n.a. | n.a. | n.a. | n.a. | n.a. | n.a. | n.a. | n.a. | n.a. | n.a. |

| | VLP-HQ-vOTUs | WMS-HQ-vOTUs |
|---|---|---|
| vOTUs considered as "clustered" or "clustered/singleton" | 764 | 411 |
| vOTUs considered as "singleton" | 34 | 41 |
| vOTUs considered as "outlier" | 207 | 148 |
| vOTUs considered as "overlap" between more than | 226 | 88 |
